## Supplementary material for "Estimation of Vaccine Efficacy for Variants that Emerge After the Placebo Group Is Vaccinated": GISAIDacknowledgment

We gratefully acknowledge the following Authors from the Originating laboratories responsible for obtaining the specimens, as well as the Submitting laboratories where the genome data were generated and shared via GISAID, on which this research is based.

All Submitters of data may be contacted directly via www.gisaid.org

Authors are sorted alphabetically.

| Accession ID | Originating Laboratory | Submitting Laboratory | Authors |
|---|---|---|---|
| EPI_ISL_404253 | IL Department of Public Health Chicago Laboratory | Pathogen Discovery, Respiratory Viruses Branch, Division of Viral Diseases, Centers for Dieases Control and Prevention | Ying Tao, Krista Queen, Clinton R. Paden, Jing Zhang, Yan Li, Anna Uehara, Xiaoyan Lu, Brian Lynch, Senthil Kumar K. Sakthivel, Brett L. Whitaker, Shifaq Kamili, Lijuan Wang, Janna' R. Murray, Susan I. Gerber, Stephen Lindstrom, Suxiang Tong |
| EPI_ISL_404895 | Providence Regional Medical Center | Division of Viral Diseases, Centers for Disease Control and Prevention | Queen,K., Tao,Y., Li,Y., Paden,C.R., Lu,X., Zhang,J., Gerber,S.I., Lindstrom,S., Tong,S. |
| EPI_ISL_406034, EPI_ISL_406036 | California Department of Public Health | Pathogen Discovery, Respiratory Viruses Branch, Division of Viral Diseases, Centers for Dieases Control and Prevention | Anna Uehara, Krista Queen, Ying Tao, Yan Li, Clinton R. Paden, Jing Zhang, Xiaoyan Lu, Brian Lynch, Senthil Kumar K. Sakthivel, Brett L. Whitaker, Shifaq Kamili, Lijuan Wang, Janna' R. Murray, Susan I. Gerber, Stephen Lindstrom, Suxiang Tong |
| EPI_ISL_406223 | Arizona Department of Health Services | Pathogen Discovery, Respiratory Viruses Branch, Division of Viral Diseases, Centers for Disease Control and Prevention | Ying Tao, Clinton R. Paden, Krista Queen, Anna Uehara, Yan Li, Jing Zhang, Xiaoyan Lu, Brian Lynch, Senthil Kumar K. Sakthivel, Brett L. Whitaker, Shifaq Kamili, Lijuan Wang, Janna' R. Murray, Susan I. Gerber, Stephen Lindstrom, Suxiang Tong |
| EPI_ISL_407214, EPI_ISL_407215 | Washington State Department of Health | Pathogen Discovery, Respiratory Viruses Branch, Division of Viral Diseases, Centers for Dieases Control and Prevention | Krista Queen, Azaibi Tamin, Jennifer Harcourt, Ying Tao, Clinton R. Paden, Jing Zhang, Yan Li, Anna Uehara, Xiaoyan Lu, Shifaq Kamili, Rashi Gautam, Haibin Wang, Janna' R. Murray, Susan I. Gerber, Stephen Lindstrom, Natalie Thornburg, Suxiang Tong |
| EPI_ISL_408008 | California Department of Health | Pathogen Discovery, Respiratory Viruses Branch, Division of Viral Diseases, Centers for Disease Control and Prevention | Krista Queen, Jing Zhang, Yan Li, Ying Tao, Anna Uehara, Clinton Paden, Xiaoyan Lu, Brian Lynch, Senthil Kumar K. Sakthivel, Brett L. Whitaker, Shifaq Kamili, Lijuan Wang, Janna' R. Murray, Susan I. Gerber, Stephen Lindstrom, Suxiang Tong |
| EPI_ISL_408009 | California Department of Health | Pathogen Discovery, Respiratory Viruses Branch, Division of Viral Diseases, Centers for Dieases Control and Prevention | Krista Queen, Jing Zhang, Yan Li, Ying Tao, Anna Uehara, Clinton Paden, Xiaoyan Lu, Brian Lynch, Senthil Kumar K. Sakthivel, Brett L. Whitaker, Shifaq Kamili, Lijuan Wang, Janna' R. Murray, Susan I. Gerber, Stephen Lindstrom, Suxiang Tong |
| EPI_ISL_408010 | California Department of Health | Pathogen Discovery, Respiratory Viruses Branch, Division of Viral Diseases, Centers for Dieases Control and Prevention | Ying Tao, Krista Queen, Jing Zhang, Yan Li, Anna Uehara, Clinton Paden, Xiaoyan Lu, Brian Lynch, Senthil Kumar K. Sakthivel, Brett L. Whitaker, Shifaq Kamili, Lijuan Wang, Janna' R. Murray, Susan I. Gerber, Stephen Lindstrom, Suxiang Tong |
| EPI_ISL_408670 | Wisconsin Department of Health Services | Pathogen Discovery, Respiratory Viruses Branch, Division of Viral Diseases, Centers for Dieases Control and Prevention | Jing Zhang, Anna Uehara, Krista Queen, Yan Li, Ying Tao, Clinton R. Paden, Xiaoyan Lu, Brian Lynch, Senthil Kumar K. Sakthivel, Brett L. Whitaker, Shifaq Kamili, Lijuan Wang, Janna' R. Murray, Susan I. Gerber, Stephen Lindstrom, Suxiang Tong |
| EPI_ISL_409067 | Massachusetts Department of Public Health | Pathogen Discovery, Respiratory Viruses Branch, Division of Viral Diseases, Centers for Dieases Control and Prevention | Clinton R. Paden, Jing Zhang, Krista Queen, Yan Li, Ying Tao, Anna Uehara, Xiaoyan Lu, Brian Lynch, Senthil Kumar K. Sakthivel, Brett L. Whitaker, Shifaq Kamili, Lijuan Wang, Janna' R. Murray, Susan I. Gerber, Stephen Lindstrom, Suxiang Tong |
| EPI_ISL_410044 | California Department of Public Health | Pathogen Discovery, Respiratory Viruses Branch, Division of Viral Diseases, Centers for Dieases Control and Prevention | Jing Zhang, Krista Queen, Yan Li, Ying Tao, Anna Uehara, Clinton R. Paden, Xiaoyan Lu, Brian Lynch, Senthil Kumar K. Sakthivel, Brett L. Whitaker, Shifaq Kamili, Lijuan Wang, Janna' R. Murray, Susan I. Gerber, Stephen Lindstrom, Suxiang Tong |
| EPI_ISL_410045 | IL Department of Public Health Chicago Laboratory | Pathogen Discovery, Respiratory Viruses Branch, Division of Viral Diseases, Centers for Dieases Control and Prevention | Yan Li, Jing Zhang, Krista Queen, Ying Tao, Anna Uehara, Clinton R. Paden, Xiaoyan Lu, Brian Lynch, Senthil Kumar K. Sakthivel, Brett L. Whitaker, Shifaq Kamili, Lijuan Wang, Janna' R. Murray, Susan I. Gerber, Stephen Lindstrom, Suxiang Tong |
| EPI_ISL_411954, EPI_ISL_411955 | California Department of Public Health | Pathogen Discovery, Respiratory Viruses Branch, Division of Viral Diseases, Centers for Dieases Control and Prevention | Krista Queen, Anna Uehara, Jing Zhang, Yan Li, Ying Tao, Clinton R. Paden, Haibin Wang, Shifaq Kamili, Xiaoyan Lu, Brian Lynch, Senthil Kumar K. Sakthivel, Brett L. Whitaker, Lijuan Wang, Janna' R. Murray, Susan I. Gerber, Stephen Lindstrom, Suxiang Tong |
| EPI_ISL_411956 | Texas Department of State Health Services | Pathogen Discovery, Respiratory Viruses Branch, Division of Viral Diseases, Centers for Dieases Control and Prevention | Krista Queen, Anna Uehara, Jing Zhang, Yan Li, Ying Tao, Clinton R. Paden, Haibin Wang, Shifaq Kamili, Xiaoyan Lu, Brian Lynch, Senthil Kumar K. Sakthivel, Brett L. Whitaker, Lijuan Wang, Janna' R. Murray, Susan I. Gerber, Stephen Lindstrom, Suxiang Tong |
| EPI_ISL_412862 | California Department of Public Health | Pathogen Discovery, Respiratory Viruses Branch, Division of Viral Diseases, Centers for Disease Control and Prevention | Krista Queen, Anna Uehara, Jing Zhang, Yan Li, Ying Tao, Clinton R. Paden, Haibin Wang, Shifaq Kamili, Xiaoyan Lu, Brian Lynch, Senthil Kumar K. Sakthivel, Brett L. Whitaker, Lijuan Wang, Janna' R. Murray, Jasmine Padilla, Justin Lee, Susan I. Gerber, Stephen Lindstrom, Suxiang Tong |
| EPI_ISL_412970 | Washington State Department of Health | Seattle Flu Study | Helen Chu, Michael Boeckh, Janet Englund, Michael Famulare, Barry Lutz, Deborah Nickerson, Mark Rieder, Lea Starita, Matthew Thompson, Jay Shendure, and Trevor Bedford |
| EPI_ISL_412972 | Instituto Nacional de Enfermedades Respiratorias | Instituto de Diagnostico y Referencia Epidemiologicos (INDRE) | Ramirez-Gonzalez Ernesto, Garces-Ayala Fabiola, Araiza-Rodriguez Adnan, Mendieta-Condado Edgar, Rodriguez-Maldonado Abril, Wong-Arambula Claudia, Vazquez-Perez Joel, Martinez Arturo, Boukadida Celia, Munoz-Medina Esteban, Sanchez Alejandro, Isa Pavel, Taboada Blanca, Lopez Susana, Arias Carlos, Barrera-Badillo Gisela, Hernandez-Rivas Lucia, Lopez-Martinez Irma |
| EPI_ISL_413025 | Harborview Medical Center | UW Virology Lab | Pavitra Roychoudhury, Arun Nalla, Hong Xie, Keith Jerome, Alexander Greninger |
| EPI_ISL_413455 | Washington State Public Health Lab | University of Washington Virology Lab | Pavitra Roychoudhury, Arun Nalla, Hong Xie, Keith Jerome, Alexander Greninger |
| EPI_ISL_413456 | Seattle Flu Study, University of Washington Medical Center | Seattle Flu Study, University of Washington Medical Center | Chu et al |
| EPI_ISL_413457, EPI_ISL_413458 | Washington State Public Health Lab | UW Virology Lab | Pavitra Roychoudhury, Arun Nalla, Hong Xie, Keith Jerome, Alexander Greninger |
| EPI_ISL_413486 | Valley Medical Center | University of Washington Virology Lab | Pavitra Roychoudhury, Arun Nalla, Hong Xie, Keith Jerome, Alexander Greninger |
| EPI_ISL_413487 | Harborview Medical Center | University of Washington Virology Lab | Pavitra Roychoudhury, Arun Nalla, Hong Xie, Keith Jerome, Alexander Greninger |
| EPI_ISL_413557, EPI_ISL_413558, EPI_ISL_413559 | California Department of Public Health | Chiu Laboratory, University of California, San Francisco | Xianding Deng, Scot Federman, Chao-Yang Pan, Hugo Guevara,Wei Gu, Debra A. Wadford, and Charles Y. Chiu |
| EPI_ISL_413560 | Seattle Flu Study | Seattle Flu Study | Chu et al |
| EPI_ISL_413561 | California Department of Public Health | Chiu Laboratory, University of California, San Francisco | Xianding Deng, Scot Federman, Chao-Yang Pan, Hugo Guevara,Wei Gu, Debra A. Wadford, and Charles Y. Chiu |
| EPI_ISL_413562, EPI_ISL_413563, EPI_ISL_413601 | UW Virology Lab | UW Virology Lab | Pavitra Roychoudhury, Hong Xie, Keith Jerome, Alexander Greninger |
| EPI_ISL_413606, EPI_ISL_413607, EPI_ISL_413608, EPI_ISL_413609, EPI_ISL_413610, EPI_ISL_413611 | unknown | Pathogen Discovery, Respiratory Viruses Branch, Division of Viral Diseases, Centers for Dieases Control and Prevention | Anna Uehara, Ying Tao, Clinton R. Paden, Krista Queen, Jing Zhang, Yan Li, Mary S. Keckler, Alison S Laufer Halpin, Haibin Wang, Jasmine Padilla, Justin Lee, Christopher A. Elkins, Susan I. Gerber, Suxiang Tong |
| EPI_ISL_413612, EPI_ISL_413613, EPI_ISL_413614, EPI_ISL_413615, EPI_ISL_413616, EPI_ISL_413617 | unknown | Pathogen Discovery, Respiratory Viruses Branch, Division of Viral Diseases, Centers for Dieases Control and Prevention | Ying Tao, Clinton R. Paden, Krista Queen, Anna Uehara, Jing Zhang, Yan Li, Haibin Wang, Shifaq Kamili, Xiaoyan Lu, Brian Lynch, Senthil Kumar K. Sakthivel, Brett L. Whitaker, Lijuan Wang, Janna' R. Murray, Jasmine Padilla, Justin Lee, Susan I. Gerber, Stephen Lindstrom, Suxiang Tong |
| EPI_ISL_413618, EPI_ISL_413619, EPI_ISL_413620, EPI_ISL_413621, EPI_ISL_413622, EPI_ISL_413623 | unknown | Pathogen Discovery, Respiratory Viruses Branch, Division of Viral Diseases, Centers for Dieases Control and Prevention | Clinton R. Paden, Ying Tao, Krista Queen, Anna Uehara, Jing Zhang, Yan Li, Haibin Wang, Shifaq Kamili, Xiaoyan Lu, Brian Lynch, Senthil Kumar K. Sakthivel, Brett L. Whitaker, Lijuan Wang, Janna' R. Murray, Jasmine Padilla, Justin Lee, Susan I. Gerber, Stephen Lindstrom, Suxiang Tong |
| EPI_ISL_413649, EPI_ISL_413650, EPI_ISL_413651, EPI_ISL_413652, | UW Virology Lab | UW Virology Lab | Pavitra Roychoudhury, Hong Xie, Keith Jerome, Alexander Greninger |

| | | | |
|---|---|---|---|
| EPI_ISL_413653 | | | |
| EPI_ISL_413922, EPI_ISL_413924, EPI_ISL_413925, EPI_ISL_413926, EPI_ISL_413928, EPI_ISL_413931 | California Department of Public Health | Chiu Laboratory, University of California, San Francisco | Xianding Deng, Scot Federman, Chao-Yang Pan, Hugo Guevara,Wei Gu, Debra A. Wadford, and Charles Y. Chiu |
| EPI_ISL_414363, EPI_ISL_414364, EPI_ISL_414365, EPI_ISL_414366, EPI_ISL_414367, EPI_ISL_414368, EPI_ISL_414369 | UW Virology Lab | UW Virology Lab | Pavitra Roychoudhury, Hong Xie, Keith Jerome, Alexander Greninger |
| EPI_ISL_414476 | MSHS Clinical Microbiology Laboratories | MSHS Pathogen Surveillance Program | Gopi Patel, Emilia Sordillo, Melissa Gitman, Alberto Paniz-mondolfi, Matthew Hernandez, Shelcie Fabre, Jose Polanco, Ana Sylvia Gonzalez-Reiche, Zenab Khan, Nancy Francoeur, Melissa Smith, Robert Sebra, Lisa Miorin, Wen-chun Liu, Randy Albrecht, Judith Aberg, Florian Krammer, Adolfo Garcia-Sarstre, Viviana Simon, Harm van Bakel |
| EPI_ISL_414479, EPI_ISL_414480, EPI_ISL_414481 | unknown | Pathogen Discovery, Respiratory Viruses Branch, Division of Viral Diseases, Centers for Disease Control and Prevention | Ying Tao, Krista Queen, Clinton R. Paden, Anna Uehara, Jing Zhang, Yan Li, Mary S. Keckler, Alison S. Laufer Halpin, Haibin Wang, Jasmine Padilla, Justin Lee, Christopher A. Elkins, Susan I. Gerber, Suxiang Tong |
| EPI_ISL_414482, EPI_ISL_414483, EPI_ISL_414484, EPI_ISL_414485 | unknown | Pathogen Discovery, Respiratory Viruses Branch, Division of Viral Diseases, Centers for Disease Control and Prevention | Krista Queen, Anna Uehara, Ying Tao, Clinton R. Paden, Jing Zhang, Yan Li, Haibin Wang, Shifaq Kamili, Xiaoyan Lu, Brian Lynch, Senthil Kumar K. Sakthivel, Brett L. Whitaker, Lijuan Wang, Janna' R. Murray, Jasmine Padilla, Justin Lee, Susan I. Gerber, Stephen Lindstrom, Suxiang Tong |
| EPI_ISL_414588, EPI_ISL_414589, EPI_ISL_414590 | Minnesota Department of Health, Public Health Laboratory | Minnesota Department of Health, Public Health Laboratory | Matt Plumb, Jake Garfin and Xiong Wang |
| EPI_ISL_414591, EPI_ISL_414592, EPI_ISL_414593, EPI_ISL_414594, EPI_ISL_414595, EPI_ISL_414596, EPI_ISL_414597, EPI_ISL_414616, EPI_ISL_414617, EPI_ISL_414618, EPI_ISL_414619, EPI_ISL_414620, EPI_ISL_414621, EPI_ISL_414622 | | | |
| see above | UW Virology Lab | UW Virology Lab | Pavitra Roychoudhury, Hong Xie, Keith Jerome, Alexander Greninger |
| EPI_ISL_414648 | Andersen Lab, The Scripps Research Institute | Andersen Lab, The Scripps Research Institute | Mark Zeller, Catie Anderson, Emily Spender, Sarah Topol, Raphaelle Klitting, Refugio Robles-Sikisaka, Karthik Gangavarapu, Laura Nicholson, Kristian Andersen |
| EPI_ISL_415151 | MSHS Clinical Microbiology Laboratories | MSHS Pathogen Surveillance Program | Gopi Patel, Emilia Sordillo, Melissa Gitman, Alberto Paniz-mondolfi, Matthew Hernandez, Shelcie Fabre, Jose Polanco, Ana Silvia Gonzalez-Reiche, Zenab Khan, Nancy Francoeur, Melissa Smith, Robert Sebra, Lisa Miorin, Wen-chun Liu, Randy Albrecht, Judith Aberg, Florian Krammer, Adolfo Garcia-Sarstre, Viviana Simon, Harm van Bakel |
| EPI_ISL_415539, EPI_ISL_415541, EPI_ISL_415542, EPI_ISL_415543, EPI_ISL_415544 | Utah Public Health Laboratory | Utah Public Health Laboratory | Erin Young, Kelly Oakeson |
| EPI_ISL_415591, EPI_ISL_415592, EPI_ISL_415593, EPI_ISL_415594, EPI_ISL_415595, EPI_ISL_415596, EPI_ISL_415597, EPI_ISL_415598, EPI_ISL_415599, EPI_ISL_415600, EPI_ISL_415601, EPI_ISL_415602, EPI_ISL_415603, EPI_ISL_415604, EPI_ISL_415605, EPI_ISL_415606, EPI_ISL_415607, EPI_ISL_415608, EPI_ISL_415609, EPI_ISL_415610, EPI_ISL_415611, EPI_ISL_415612, EPI_ISL_415613, EPI_ISL_415614, EPI_ISL_415615, EPI_ISL_415616, EPI_ISL_415617, EPI_ISL_415618, EPI_ISL_415619, EPI_ISL_415620, EPI_ISL_415621, EPI_ISL_415622, EPI_ISL_415623, EPI_ISL_415624, EPI_ISL_415625, EPI_ISL_415626, EPI_ISL_415627 | | | |
| see above | UW Virology Lab | UW Virology Lab | Pavitra Roychoudhury, Hong Xie, Keith Jerome, Alexander Greninger |
| EPI_ISL_416417 | Connecticut State Department of Public Health | Grubaugh Lab - Yale School of Public Health | Joseph Fauver, Chantal Vogels, Anderson Brito, Tara Alpert, Nagarjuna Cheemarla, Ellen Foxman, Anthony Muyombwe, Jafar Razeq, Richard Martinello, Albert Ko, Marie-Louise Landry, Nathan Grubaugh |
| EPI_ISL_416420, EPI_ISL_416421, EPI_ISL_416423 | Yale Clinical Virology Laboratory | Grubaugh Lab - Yale School of Public Health | Joseph Fauver, Chantal Vogels, Anderson Brito, Tara Alpert, Nagarjuna Cheemarla, Ellen Foxman, Anthony Muyombwe, Jafar Razeq, Richard Martinello, Albert Ko, Marie-Louise Landry, Nathan Grubaugh |
| EPI_ISL_416433, EPI_ISL_416434, EPI_ISL_416435, EPI_ISL_416436, EPI_ISL_416437, EPI_ISL_416438, EPI_ISL_416439, EPI_ISL_416440, EPI_ISL_416441, EPI_ISL_416442, EPI_ISL_416443, EPI_ISL_416444, EPI_ISL_416445, EPI_ISL_416446, EPI_ISL_416447, EPI_ISL_416448, EPI_ISL_416449, EPI_ISL_416450, EPI_ISL_416451, EPI_ISL_416452, EPI_ISL_416453, EPI_ISL_416454, EPI_ISL_416455, EPI_ISL_416456 | | | |
| see above | UW Virology Lab | UW Virology Lab | Pavitra Roychoudhury, Hong Xie, Keith Jerome, Alexander Greninger |
| EPI_ISL_416457 | Andersen Lab, The Scripps Research Institute | Andersen Lab, The Scripps Research Institute | Mark Zeller, Catie Anderson, Emily Spender, Sarah Topol, Raphaelle Klitting, Refugio Robles-Sikisaka, Karthik Gangavarapu, Laura Nicholson, Kristian Andersen |
| EPI_ISL_416460, EPI_ISL_416461, EPI_ISL_416462, EPI_ISL_416465, EPI_ISL_416466 | Seattle Flu Study | Seattle Flu Study | Chu et al |
| EPI_ISL_416489, EPI_ISL_416491, EPI_ISL_416492, EPI_ISL_416523 | University of Wisconsin-Madison AIDS Vaccine Research Laboratories | University of Wisconsin-Madison AIDS Vaccine Research Laboratories | Gage Moreno, Katarina Braun, et al. AIDS Vaccine Research Laboratories |
| EPI_ISL_416635, EPI_ISL_416636, EPI_ISL_416637, EPI_ISL_416638, EPI_ISL_416639, EPI_ISL_416640, EPI_ISL_416641, EPI_ISL_416642, EPI_ISL_416643, EPI_ISL_416644, EPI_ISL_416645, EPI_ISL_416646, EPI_ISL_416647, EPI_ISL_416648, EPI_ISL_416649, EPI_ISL_416650, EPI_ISL_416651, EPI_ISL_416652, EPI_ISL_416653, EPI_ISL_416654, EPI_ISL_416655, EPI_ISL_416656, EPI_ISL_416657, EPI_ISL_416658, EPI_ISL_416659, EPI_ISL_416660, EPI_ISL_416661, EPI_ISL_416662, EPI_ISL_416663, EPI_ISL_416664, EPI_ISL_416665, EPI_ISL_416666, EPI_ISL_416667, EPI_ISL_416668, EPI_ISL_416669, EPI_ISL_416670, EPI_ISL_416671, EPI_ISL_416673, EPI_ISL_416674, EPI_ISL_416675, EPI_ISL_416676, EPI_ISL_416677, EPI_ISL_416678, EPI_ISL_416679, EPI_ISL_416680, EPI_ISL_416681, EPI_ISL_416682, EPI_ISL_416683, EPI_ISL_416684, EPI_ISL_416685, EPI_ISL_416686, EPI_ISL_416687, EPI_ISL_416688, EPI_ISL_416689, EPI_ISL_416690, EPI_ISL_416691, EPI_ISL_416692, EPI_ISL_416693, EPI_ISL_416694, EPI_ISL_416695, EPI_ISL_416696, EPI_ISL_416697, EPI_ISL_416698, EPI_ISL_416699, EPI_ISL_416700, EPI_ISL_416701, EPI_ISL_416702, EPI_ISL_416703, EPI_ISL_416704, EPI_ISL_416705, EPI_ISL_416706, EPI_ISL_416707, EPI_ISL_416708, EPI_ISL_416709, EPI_ISL_416710, EPI_ISL_416711, EPI_ISL_416712, EPI_ISL_416713, EPI_ISL_416714, EPI_ISL_416715, EPI_ISL_416716, EPI_ISL_416717, EPI_ISL_416718, EPI_ISL_416719, EPI_ISL_416721, EPI_ISL_416722, EPI_ISL_416723, EPI_ISL_416724, EPI_ISL_416725, EPI_ISL_416726, EPI_ISL_416727, EPI_ISL_416728, EPI_ISL_416729 | | | |
| see above | UW Virology Lab | UW Virology Lab | Pavitra Roychoudhury, Hong Xie, Keith Jerome, Alexander Greninger |
| EPI_ISL_416830, EPI_ISL_416831, EPI_ISL_416832 | NYU Langone Health | Department of Pathology and Medicine, New York University School of Medicine | John Chen, Dacia Dimartino, Xiaojun Feng, Adriana Heguy, Megan Hogan, Emily Huang, George Jour, Christian Marier, Matt Maurano, Mark Mulligan, Peter Meyn, Marie Samanovic-Golden, Amy Rapkiewicz, Guomiao Shen, Matija Snuderl, Gael Westby, Paul Zappile |
| EPI_ISL_417026, EPI_ISL_417027, EPI_ISL_417028 | Utah Public Health Laboratory | Utah Public Health Laboratory | Erin Young, Kelly Oakeson |
| EPI_ISL_417065, EPI_ISL_417066, EPI_ISL_417067, EPI_ISL_417068, EPI_ISL_417069, EPI_ISL_417070, EPI_ISL_417071, EPI_ISL_417072, EPI_ISL_417073, EPI_ISL_417074, EPI_ISL_417075, EPI_ISL_417076, EPI_ISL_417077, EPI_ISL_417078, EPI_ISL_417079, EPI_ISL_417080, EPI_ISL_417081, EPI_ISL_417082, EPI_ISL_417084, EPI_ISL_417085, EPI_ISL_417086, EPI_ISL_417087, EPI_ISL_417088, EPI_ISL_417089, EPI_ISL_417090, EPI_ISL_417091, EPI_ISL_417092, EPI_ISL_417093, EPI_ISL_417094, EPI_ISL_417095, EPI_ISL_417096, EPI_ISL_417097, EPI_ISL_417098, EPI_ISL_417099, EPI_ISL_417100, EPI_ISL_417101, EPI_ISL_417102, EPI_ISL_417103, EPI_ISL_417104, EPI_ISL_417105, EPI_ISL_417106, EPI_ISL_417107, EPI_ISL_417108, EPI_ISL_417109, EPI_ISL_417110, EPI_ISL_417111, EPI_ISL_417112, EPI_ISL_417114, EPI_ISL_417115, EPI_ISL_417116, EPI_ISL_417117, EPI_ISL_417118, EPI_ISL_417119, EPI_ISL_417120, EPI_ISL_417121, EPI_ISL_417122, EPI_ISL_417123, EPI_ISL_417124, EPI_ISL_417125, EPI_ISL_417126, EPI_ISL_417127, EPI_ISL_417128, EPI_ISL_417129, EPI_ISL_417130, EPI_ISL_417131, EPI_ISL_417132, EPI_ISL_417133, EPI_ISL_417134, EPI_ISL_417135, EPI_ISL_417136, EPI_ISL_417137, EPI_ISL_417138, EPI_ISL_417139, EPI_ISL_417140, EPI_ISL_417141, EPI_ISL_417142, EPI_ISL_417143, EPI_ISL_417144, EPI_ISL_417145, EPI_ISL_417146, EPI_ISL_417147, EPI_ISL_417148, EPI_ISL_417149, EPI_ISL_417150, EPI_ISL_417151, EPI_ISL_417152, EPI_ISL_417153, EPI_ISL_417154, EPI_ISL_417155, EPI_ISL_417156, EPI_ISL_417157, EPI_ISL_417158, EPI_ISL_417159, EPI_ISL_417160, EPI_ISL_417161, EPI_ISL_417162 | | | |
| see above | Washington State Department of Health | Seattle Flu Study | Chu etl al |
| EPI_ISL_417163, EPI_ISL_417164, EPI_ISL_417165 | Seattle Flu Study | Seattle Flu Study | Chu etl al |
| EPI_ISL_417166, EPI_ISL_417167, EPI_ISL_417168, EPI_ISL_417169, EPI_ISL_417170, EPI_ISL_417171, EPI_ISL_417172, EPI_ISL_417174, EPI_ISL_417175 | Washington State Department of Health | Seattle Flu Study | Chu etl al |
| EPI_ISL_417189, EPI_ISL_417191, EPI_ISL_417192, EPI_ISL_417194, EPI_ISL_417196, EPI_ISL_417198 | Minnesota Department of Health, Public Health Laboratory | Minnesota Department of Health, Public Health Laboratory | Matt Plumb, Jake Garfin and Xiong Wang |
| EPI_ISL_417200, EPI_ISL_417201, EPI_ISL_417202, EPI_ISL_417203, | University of Wisconsin-Madison AIDS Vaccine Research Laboratories | University of Wisconsin-Madison AIDS Vaccine Research Laboratories | Gage Moreno, Katarina Braun, et al. AIDS Vaccine Research Laboratories |

| | | | |
|---|---|---|---|
| EPI_ISL_417204 | | | |
| EPI_ISL_417317, EPI_ISL_417318, EPI_ISL_417320 | Santa Clara County Public Health Department | Chiu Laboratory, University of California, San Francisco | Xianding Deng, Scot Federman, Wei Gu, Elsa Villarino, Brandon Bonin, Debra A. Wadford, and Charles Y. Chiu |
| EPI_ISL_417323, EPI_ISL_417324 | California Department of Public Health | Chiu Laboratory, University of California, San Francisco | Xianding Deng, Scot Federman, Chao-Yang Pan, Hugo Guevara,Wei Gu, Debra A. Wadford, and Charles Y. Chiu |
| EPI_ISL_417330, EPI_ISL_417332 | Chiu Laboratory, University of California, San Francisco | Chiu Laboratory, University of California, San Francisco | Xianding Deng, Scot Federman, Wei Gu, and Charles Y. Chiu |
| EPI_ISL_417341, EPI_ISL_417342, EPI_ISL_417343, EPI_ISL_417344, EPI_ISL_417345, EPI_ISL_417346, EPI_ISL_417347, EPI_ISL_417348, EPI_ISL_417349, EPI_ISL_417350, EPI_ISL_417351, EPI_ISL_417352, EPI_ISL_417353, EPI_ISL_417354, EPI_ISL_417355, EPI_ISL_417356, EPI_ISL_417357, EPI_ISL_417358, EPI_ISL_417359, EPI_ISL_417360, EPI_ISL_417361, EPI_ISL_417362, EPI_ISL_417363, EPI_ISL_417364, EPI_ISL_417365, EPI_ISL_417366, EPI_ISL_417367, EPI_ISL_417368, EPI_ISL_417369, EPI_ISL_417370, EPI_ISL_417371, EPI_ISL_417372, EPI_ISL_417373, EPI_ISL_417374, EPI_ISL_417375, EPI_ISL_417376, EPI_ISL_417377, EPI_ISL_417378, EPI_ISL_417379, EPI_ISL_417380, EPI_ISL_417381, EPI_ISL_417382, EPI_ISL_417448, EPI_ISL_417449, EPI_ISL_417450, EPI_ISL_417451, EPI_ISL_417452, EPI_ISL_417453, EPI_ISL_417454, EPI_ISL_417455, EPI_ISL_417456 | | | |
| see above | UW Virology Lab | UW Virology Lab | Pavitra Roychoudhury, Hong Xie, Keith Jerome, Alexander Greninger |
| EPI_ISL_417469, EPI_ISL_417470, EPI_ISL_417471, EPI_ISL_417472, EPI_ISL_417473, EPI_ISL_417474, EPI_ISL_417475, EPI_ISL_417476, EPI_ISL_417477, EPI_ISL_417478, EPI_ISL_417479, EPI_ISL_417480, EPI_ISL_417492, EPI_ISL_417493, EPI_ISL_417494, EPI_ISL_417495, EPI_ISL_417496, EPI_ISL_417497, EPI_ISL_417498, EPI_ISL_417499, EPI_ISL_417500, EPI_ISL_417501, EPI_ISL_417502, EPI_ISL_417503 | | | |
| see above | Minnesota Department of Health, Public Health Laboratory | Minnesota Department of Health, Public Health Laboratory | Matt Plumb, Jake Garfin and Xiong Wang |
| EPI_ISL_417504, EPI_ISL_417505, EPI_ISL_417506, EPI_ISL_417507, EPI_ISL_417508, EPI_ISL_417509, EPI_ISL_417510, EPI_ISL_417512, EPI_ISL_417513, EPI_ISL_417514, EPI_ISL_417515, EPI_ISL_417516, EPI_ISL_417517 | | | |
| see above | University of Wisconsin-Madison AIDS Vaccine Research Laboratories | University of Wisconsin-Madison AIDS Vaccine Research Laboratories | Gage Moreno, Katarina Braun, et al. AIDS Vaccine Research Laboratories |
| EPI_ISL_417931, EPI_ISL_417932, EPI_ISL_417933, EPI_ISL_417935, EPI_ISL_417937, EPI_ISL_417938, EPI_ISL_417939 | UCSF Clinical Microbiology Laboratory | Chan-Zuckerberg Biohub | Shaun Arevalo, Josh Batson, Olga Botvinnik, Gloria Castaneda, Angela Detweiler, David Dynerman, Samantha Hao, Jack Kamm, Amy Kistler, G. Renuka Kumar, Chaz Langelier, Lucy Li, Steve Miller, Lusajo Mwakibete, Norma Neff, Angela Pisco, Maira Phelps, Michelle Tan, Chunyu Zhao |
| EPI_ISL_417958, EPI_ISL_417959, EPI_ISL_417960, EPI_ISL_417964, EPI_ISL_417966, EPI_ISL_417970, EPI_ISL_417971, EPI_ISL_417973, EPI_ISL_417974, EPI_ISL_417976, EPI_ISL_417977 | | | |
| see above | Utah Public Health Laboratory | Utah Public Health Laboratory | Erin Young, Kelly Oakeson |
| EPI_ISL_418028, EPI_ISL_418029, EPI_ISL_418030, EPI_ISL_418032, EPI_ISL_418033, EPI_ISL_418034, EPI_ISL_418035, EPI_ISL_418036, EPI_ISL_418037, EPI_ISL_418038, EPI_ISL_418039, EPI_ISL_418040, EPI_ISL_418041, EPI_ISL_418042, EPI_ISL_418043, EPI_ISL_418044, EPI_ISL_418045, EPI_ISL_418046, EPI_ISL_418047, EPI_ISL_418048, EPI_ISL_418049, EPI_ISL_418050, EPI_ISL_418051, EPI_ISL_418053, EPI_ISL_418054, EPI_ISL_418055, EPI_ISL_418056, EPI_ISL_418057, EPI_ISL_418058, EPI_ISL_418059, EPI_ISL_418060, EPI_ISL_418061, EPI_ISL_418062, EPI_ISL_418063, EPI_ISL_418064, EPI_ISL_418065, EPI_ISL_418067, EPI_ISL_418068, EPI_ISL_418069, EPI_ISL_418070, EPI_ISL_418071, EPI_ISL_418072, EPI_ISL_418073, EPI_ISL_418074, EPI_ISL_418075, EPI_ISL_418076, EPI_ISL_418077, EPI_ISL_418078, EPI_ISL_418079, EPI_ISL_418080, EPI_ISL_418081, EPI_ISL_418082 | | | |
| see above | UW Virology Lab | UW Virology Lab | Pavitra Roychoudhury, Hong Xie, Keith Jerome, Alexander Greninger |
| EPI_ISL_418184 | Gundersen Molecular Diagnostics Laboratory | Kabara Cancer Research Institute | Craig S. Richmond & Paraic A. Kenny |
| EPI_ISL_418186 | Gundersen Molecular Diagnostic Laboratory | Kabara Cancer Research Institute | Craig S. Richmond & Paraic A. Kenny |
| EPI_ISL_418187, EPI_ISL_418188, EPI_ISL_418189 | Gundersen Molecular Diagnostics Laboratory | Kabara Cancer Research Institute | Craig S. Richmond & Paraic A. Kenny |
| EPI_ISL_418190, EPI_ISL_418192, EPI_ISL_418193, EPI_ISL_418196, EPI_ISL_418197, EPI_ISL_418198, EPI_ISL_418199, EPI_ISL_418200, EPI_ISL_418202, EPI_ISL_418203, EPI_ISL_418204 | | | |
| see above | NYU Langone Health | Department of Pathology and Medicine, New York University School of Medicine | Margaret Black, John Cadley, Paolo Cotzia, John Chen, Dacia Dimartino, Xiaojun Feng, Adriana Heguy, Megan Hogan, Emily Huang, George Jour, Christian Marier, Matthew T. Maurano, Mark J. Mulligan, Peter Meyn, Jared Pinnell, Amy Rapkiewicz, Marie Samanovic-Golden, Antonio Serrano, Guomiao Shen, Matija Snuderl, Nick Vulpescu, Gael Westby, Paul Zappile |
| EPI_ISL_418771, EPI_ISL_418772, EPI_ISL_418773, EPI_ISL_418774, EPI_ISL_418775, EPI_ISL_418776, EPI_ISL_418777 | WA State Department of Health | Pathogen Discovery, Respiratory Viruses Branch, Division of Viral Diseases, Centers for Disease Control and Prevention | Jing Zhang, Ying Tao, Clinton R. Paden, Krista Queen, Anna Uehara, Yan Li, Haibin Wang, Jesica Jacobs, Denny Russell, Brian Hiatt, Jessica Gant, Suxiang Tong |
| EPI_ISL_418778, EPI_ISL_418779, EPI_ISL_418780, EPI_ISL_418781, EPI_ISL_418782, EPI_ISL_418783, EPI_ISL_418784, EPI_ISL_418785, EPI_ISL_418786, EPI_ISL_418787, EPI_ISL_418788, EPI_ISL_418789, EPI_ISL_418790, EPI_ISL_418791 | | | |
| see above | WA State Department of Health | Pathogen Discovery, Respiratory Viruses Branch, Division of Viral Diseases, Centers for Disease Control and Prevention | Ying Tao, Jing Zhang, Clinton R. Paden, Krista Queen, Anna Uehara, Yan Li, Haibin Wang, Jesica Jacobs, Denny Russell, Brian Hiatt, Jessica Gant, Suxiang Tong |
| EPI_ISL_418864 | Virginia DCLS | Virginia DCLS | Virginia DCLS |
| EPI_ISL_418866, EPI_ISL_418867, EPI_ISL_418868, EPI_ISL_418869, EPI_ISL_418870, EPI_ISL_418871, EPI_ISL_418872, EPI_ISL_418873, EPI_ISL_418874, EPI_ISL_418875, EPI_ISL_418876, EPI_ISL_418877, EPI_ISL_418878, EPI_ISL_418879, EPI_ISL_418880, EPI_ISL_418881, EPI_ISL_418882, EPI_ISL_418883, EPI_ISL_418884, EPI_ISL_418885, EPI_ISL_418886, EPI_ISL_418887, EPI_ISL_418888, EPI_ISL_418889, EPI_ISL_418890, EPI_ISL_418891, EPI_ISL_418892, EPI_ISL_418893, EPI_ISL_418894, EPI_ISL_418895, EPI_ISL_418896, EPI_ISL_418897, EPI_ISL_418898, EPI_ISL_418899, EPI_ISL_418900, EPI_ISL_418901, EPI_ISL_418902, EPI_ISL_418903, EPI_ISL_418904, EPI_ISL_418905, EPI_ISL_418906, EPI_ISL_418907, EPI_ISL_418908, EPI_ISL_418909, EPI_ISL_418910, EPI_ISL_418911, EPI_ISL_418912, EPI_ISL_418913, EPI_ISL_418914, EPI_ISL_418915, EPI_ISL_418916, EPI_ISL_418917, EPI_ISL_418918, EPI_ISL_418919, EPI_ISL_418920, EPI_ISL_418921, EPI_ISL_418922, EPI_ISL_418923, EPI_ISL_418924, EPI_ISL_418925, EPI_ISL_418926, EPI_ISL_418927, EPI_ISL_418928, EPI_ISL_418929, EPI_ISL_418930, EPI_ISL_418931, EPI_ISL_418932, EPI_ISL_418933, EPI_ISL_418934, EPI_ISL_418935, EPI_ISL_418936, EPI_ISL_418937, EPI_ISL_418938, EPI_ISL_418939, EPI_ISL_418940, EPI_ISL_418941, EPI_ISL_418942, EPI_ISL_418943, EPI_ISL_418944, EPI_ISL_418945, EPI_ISL_418946, EPI_ISL_418947, EPI_ISL_418948, EPI_ISL_418949, EPI_ISL_418950, EPI_ISL_418951, EPI_ISL_418952, EPI_ISL_418953, EPI_ISL_418954, EPI_ISL_418955 | | | |
| see above | UW Virology Lab | UW Virology Lab | Pavitra Roychoudhury, Hong Xie, Keith Jerome, Alexander Greninger |
| EPI_ISL_418956, EPI_ISL_418957, EPI_ISL_418958 | Virginia DCLS | Virginia DCLS | Virginia DCLS |
| EPI_ISL_418962, EPI_ISL_418963, EPI_ISL_418964, EPI_ISL_418965, EPI_ISL_418967 | Utah Public Health Laboratory | Utah Public Health Laboratory | Erin Young, Kelly Oakeson |
| EPI_ISL_418971, EPI_ISL_418972, EPI_ISL_418973, EPI_ISL_418974, EPI_ISL_418975, EPI_ISL_418976, EPI_ISL_418977, EPI_ISL_418978, EPI_ISL_418979, EPI_ISL_418980 | NYU Langone Health | Department of Pathology and Medicine, New York University School of Medicine | Maria Aguero-Rosenfeld, Margaret Black, John Cadley, Paolo Cotzia, John Chen, Dacia Dimartino, Xiaojun Feng, Adriana Heguy, Megan Hogan, Emily Huang, George Jour, Christian Marier, Matthew T. Maurano, Mark J. Mulligan, Peter Meyn, Jared Pinnell, Sitharam Ramaswami, Amy Rapkiewicz, Marie Samanovic-Golden, Antonio Serrano, Guomiao Shen, Matija Snuderl, Nick Vulpescu, Gael Westby, Paul Zappile, Yutong Zhang |
| EPI_ISL_419256, EPI_ISL_419257, EPI_ISL_419258, EPI_ISL_419260, EPI_ISL_419261, EPI_ISL_419262, EPI_ISL_419263 | Virginia DCLS | Virginia DCLS | Virginia DCLS |
| EPI_ISL_419388, EPI_ISL_419390, EPI_ISL_419391, EPI_ISL_419392, EPI_ISL_419393, EPI_ISL_419394, EPI_ISL_419395, EPI_ISL_419396, EPI_ISL_419397 | Minnesota Department of Health, Public Health Laboratory | Minnesota Department of Health, Public Health Laboratory | Matt Plumb, Jake Garfin and Xiong Wang |
| EPI_ISL_419515, EPI_ISL_419516 | Yale COVID-19 Biorepository | Grubaugh Lab - Yale School of Public Health | Joseph Fauver, Tara Alpert, Anderson Brito, Anne Wyllie, Chantal Vogels, Mary Petrone, Chaney Kalinich, Isabel Ott, Arnau Casanovas, Catherine Muenker, Adam Moore, Alice Lu, Maria Tokuyama, Patrick Wong, Peiwen Lu, Saad Omer, Richard Martinello, Allison Nelson, Shelli Farhadian, Akiko Iwasaki, Charlese Dela Cruz, Albert Ko, Nathan Grubaugh |
| EPI_ISL_419517, EPI_ISL_419518, | Yale Clinical Virology Laboratory | Grubaugh Lab - Yale School of Public Health | Joseph Fauver, Anderson Brito, Tara Alpert, Chantal Vogels, Ellen Foxman, Albert Ko, Marie Landry, Nathan Grubaugh |

| | | | |
|---|---|---|---|
| EPI_ISL_419519, EPI_ISL_419520, EPI_ISL_419521, EPI_ISL_419522, EPI_ISL_419524, EPI_ISL_419525, EPI_ISL_419526, EPI_ISL_419528 | | | |
| EPI_ISL_419553 | RI State Health Laboratories | Pathogen Discovery, Respiratory Viruses Branch, Division of Viral Diseases, Centers for Disease Control and Prevention | Ying Tao, Jing Zhang, Krista Queen, Anna Uehara, Clinton R. Paden, Yan Li, Haibin Wang, Jasmine Padilla, Justin Lee, Suxiang Tong |
| EPI_ISL_419554 | California Department of Public Health | Pathogen Discovery, Respiratory Viruses Branch, Division of Viral Diseases, Centers for Disease Control and Prevention | Ying Tao, Jing Zhang, Krista Queen, Anna Uehara, Clinton R. Paden, Yan Li, Haibin Wang, Jasmine Padilla, Justin Lee, Suxiang Tong |
| EPI_ISL_419555 | WA State Department of Health | Pathogen Discovery, Respiratory Viruses Branch, Division of Viral Diseases, Centers for Disease Control and Prevention | Ying Tao, Jing Zhang, Krista Queen, Anna Uehara, Clinton R. Paden, Yan Li, Haibin Wang, Jasmine Padilla, Justin Lee, Suxiang Tong |
| EPI_ISL_419556, EPI_ISL_419557 | GA Department of Public Health Laboratory | Pathogen Discovery, Respiratory Viruses Branch, Division of Viral Diseases, Centers for Disease Control and Prevention | Ying Tao, Jing Zhang, Krista Queen, Anna Uehara, Clinton R. Paden, Yan Li, Haibin Wang, Jasmine Padilla, Justin Lee, Suxiang Tong |
| EPI_ISL_419558 | OR State PHL-Virology/Immunology Section | Pathogen Discovery, Respiratory Viruses Branch, Division of Viral Diseases, Centers for Disease Control and Prevention | Ying Tao, Jing Zhang, Krista Queen, Anna Uehara, Clinton R. Paden, Yan Li, Haibin Wang, Jasmine Padilla, Justin Lee, Suxiang Tong |
| EPI_ISL_419559, EPI_ISL_419560 | FL Bureau of Public Health Laboratories-Tampa | Pathogen Discovery, Respiratory Viruses Branch, Division of Viral Diseases, Centers for Disease Control and Prevention | Anna Uehara, Ying Tao, Jing Zhang, Krista Queen, Clinton R. Paden, Yan Li, Haibin Wang, Jasmine Padilla, Justin Lee, Suxiang Tong |
| EPI_ISL_419651, EPI_ISL_419652 | Gundersen Molecular Diagnostics Laboratory | Kabara Cancer Research Institute | Craig S. Richmond & Paraic A. Kenny |
| EPI_ISL_419696, EPI_ISL_419697, EPI_ISL_419698, EPI_ISL_419699, EPI_ISL_419700, EPI_ISL_419702, EPI_ISL_419703, EPI_ISL_419704, EPI_ISL_419705 | NYU Langone Health | Departments of Pathology and Medicine, New York University School of Medicine | Maria Aguero-Rosenfeld, Margaret Black, John Cadley, Paolo Cotzia, John Chen, Dacia Dimartino, Xiaojun Feng, Adriana Heguy, Megan Hogan, Emily Huang, George Jour, Christian Marier, Matthew T. Maurano, Mark J. Mulligan, Peter Meyn, Jared Pinnell, Sitharam Ramaswami, Amy Rapkiewicz, Marie Samanovic-Golden, Antonio Serrano, Guomiao Shen, Matija Snuderl, Nick Vulpescu, Gael Westby, Paul Zappile, Yutong Zhang |
| EPI_ISL_419706, EPI_ISL_419708, EPI_ISL_419710, EPI_ISL_419711, EPI_ISL_419712, EPI_ISL_419713, EPI_ISL_420018, EPI_ISL_420019, EPI_ISL_420020, EPI_ISL_420021, EPI_ISL_420022, EPI_ISL_420023, EPI_ISL_420024, EPI_ISL_420025, EPI_ISL_420026, EPI_ISL_420027, EPI_ISL_420028, EPI_ISL_420029 | | | |
| see above | Virginia DCLS | Virginia DCLS | Virginia DCLS |
| EPI_ISL_420086, EPI_ISL_420087, EPI_ISL_420088, EPI_ISL_420089, EPI_ISL_420090, EPI_ISL_420091, EPI_ISL_420092, EPI_ISL_420093, EPI_ISL_420094, EPI_ISL_420095, EPI_ISL_420096, EPI_ISL_420097, EPI_ISL_420098 | | | |
| see above | Yale Clinical Virology Laboratory | Grubaugh Lab - Yale School of Public Health | Joseph Fauver, Anderson Brito, Tara Alpert, Chantal Vogels, Ellen Foxman, Albert Ko, Marie Landry, Nathan Grubaugh |
| EPI_ISL_420296, EPI_ISL_420297, EPI_ISL_420298, EPI_ISL_420299, EPI_ISL_420300, EPI_ISL_420301, EPI_ISL_420302 | NYU Langone Health | Departments of Pathology and Medicine, New York University School of Medicine | Maria Aguero-Rosenfeld, Margaret Black, John Cadley, Paolo Cotzia, John Chen, Dacia Dimartino, Xiaojun Feng, Adriana Heguy, Megan Hogan, Emily Huang, George Jour, Christian Marier, Matthew T. Maurano, Mark J. Mulligan, Peter Meyn, Jared Pinnell, Sitharam Ramaswami, Amy Rapkiewicz, Marie Samanovic-Golden, Antonio Serrano, Guomiao Shen, Matija Snuderl, Nick Vulpescu, Gael Westby, Paul Zappile, Yutong Zhang |
| EPI_ISL_420303 | Alaska State Virology Laboratory | Alaska State Virology Laboratory | Chen, J |
| EPI_ISL_420304, EPI_ISL_420305, EPI_ISL_420306 | Alaska State Virology Laboratory | Alaska State Virology Laboratory | Chen, J. |
| EPI_ISL_420307, EPI_ISL_420308, EPI_ISL_420309 | NYU Langone Health | Departments of Pathology and Medicine, New York University School of Medicine | Maria Aguero-Rosenfeld, Margaret Black, John Cadley, Paolo Cotzia, John Chen, Dacia Dimartino, Xiaojun Feng, Adriana Heguy, Megan Hogan, Emily Huang, George Jour, Christian Marier, Matthew T. Maurano, Mark J. Mulligan, Peter Meyn, Jared Pinnell, Sitharam Ramaswami, Amy Rapkiewicz, Marie Samanovic-Golden, Antonio Serrano, Guomiao Shen, Matija Snuderl, Nick Vulpescu, Gael Westby, Paul Zappile, Yutong Zhang |
| EPI_ISL_420570, EPI_ISL_420571, EPI_ISL_420572, EPI_ISL_420573, EPI_ISL_420574, EPI_ISL_420575, EPI_ISL_420576, EPI_ISL_420577, EPI_ISL_420578, EPI_ISL_420579, EPI_ISL_420580, EPI_ISL_420581, EPI_ISL_420582, EPI_ISL_420584, EPI_ISL_420585, EPI_ISL_420586, EPI_ISL_420587, EPI_ISL_420588, EPI_ISL_420589, EPI_ISL_420590, EPI_ISL_420591 | | | |
| see above | NYU Langone Health | Departments of Pathology and Medicine, New York University School of Medicine | Maria Aguero-Rosenfeld, Brendan Belovarac, Margaret Black, Ludovic Boytard, John Cadley, Paolo Cotzia, John Chen, Dacia Dimartino, Xiaojun Feng, Tatyana Gindin, Adriana Heguy, Megan Hogan, Emily Huang, George Jour, Andrew Lytle, Christian Marier, Matthew T. Maurano, Mark J. Mulligan, Peter Meyn, Iman Osman, Jared Pinnell, Sitharam Ramaswami, Amy Rapkiewicz, Marie Samanovic-Golden, Antonio Serrano, Guomiao Shen, Matija Snuderl, Theodore Vougiouklakis, Nick Vulpescu, Gael Westby, Paul Zappile, Yutong Zhang |
| EPI_ISL_420628, EPI_ISL_420629, EPI_ISL_420630 | Virginia DCLS | Virginia DCLS | Virginia DCLS |
| EPI_ISL_420784 | AZ Department of Health Services | Pathogen Discovery, Respiratory Viruses Branch, Division of Viral Diseases, Centers for Disease Control and Prevention | Krista Queen, Yan Li, Ying Tao, Jing Zhang, Anne Uehara, Clinton R. Paden, Haibin Wang, Rachel Marine, Mary S. Keckler, Alison S. Laufer Halpin, Jasmine Padilla, Justin Lee, Christopher A. Elkins, Suxiang Tong |
| EPI_ISL_420785 | FL Bureau of Health Laboratories Tampa | Pathogen Discovery, Respiratory Viruses Branch, Division of Viral Diseases, Centers for Disease Control and Prevention | Krista Queen, Yan Li, Ying Tao, Jing Zhang, Anne Uehara, Clinton R. Paden, Haibin Wang, Rachel Marine, Mary S. Keckler, Alison S. Laufer Halpin, Jasmine Padilla, Justin Lee, Christopher A. Elkins, Suxiang Tong |
| EPI_ISL_420786, EPI_ISL_420787, EPI_ISL_420788 | GA Department of Public Health | Pathogen Discovery, Respiratory Viruses Branch, Division of Viral Diseases, Centers for Disease Control and Prevention | Krista Queen, Yan Li, Ying Tao, Jing Zhang, Anne Uehara, Clinton R. Paden, Haibin Wang, Rachel Marine, Mary S. Keckler, Alison S. Laufer Halpin, Jasmine Padilla, Justin Lee, Christopher A. Elkins, Suxiang Tong |
| EPI_ISL_420789, EPI_ISL_420790 | Illinois Department of Public Health Chicago Laboratory | Pathogen Discovery, Respiratory Viruses Branch, Division of Viral Diseases, Centers for Disease Control and Prevention | Krista Queen, Yan Li, Ying Tao, Jing Zhang, Anne Uehara, Clinton R. Paden, Haibin Wang, Rachel Marine, Mary S. Keckler, Alison S. Laufer Halpin, Jasmine Padilla, Justin Lee, Christopher A. Elkins, Suxiang Tong |
| EPI_ISL_420791, EPI_ISL_420792 | NH Department of Health and Human Services Public Health Labs | Pathogen Discovery, Respiratory Viruses Branch, Division of Viral Diseases, Centers for Disease Control and Prevention | Krista Queen, Yan Li, Ying Tao, Jing Zhang, Anne Uehara, Clinton R. Paden, Haibin Wang, Rachel Marine, Mary S. Keckler, Alison S. Laufer Halpin, Jasmine Padilla, Justin Lee, Christopher A. Elkins, Suxiang Tong |
| EPI_ISL_420793 | NYC Department of Health and Mental Hygiene | Pathogen Discovery, Respiratory Viruses Branch, Division of Viral Diseases, Centers for Disease Control and Prevention | Krista Queen, Yan Li, Ying Tao, Jing Zhang, Anne Uehara, Clinton R. Paden, Haibin Wang, Rachel Marine, Mary S. Keckler, Alison S. Laufer Halpin, Jasmine Padilla, Justin Lee, Christopher A. Elkins, Suxiang Tong |
| EPI_ISL_420794 | Oregon State Public Health- Virology section | Pathogen Discovery, Respiratory Viruses Branch, Division of Viral Diseases, Centers for Disease Control and Prevention | Krista Queen, Yan Li, Ying Tao, Jing Zhang, Anne Uehara, Clinton R. Paden, Haibin Wang, Rachel Marine, Mary S. Keckler, Alison S. Laufer Halpin, Jasmine Padilla, Justin Lee, Christopher A. Elkins, Suxiang Tong |
| EPI_ISL_420795 | RI State Health Laboratory | Pathogen Discovery, Respiratory Viruses Branch, Division of Viral Diseases, Centers for Disease Control and Prevention | Krista Queen, Yan Li, Ying Tao, Jing Zhang, Anne Uehara, Clinton R. Paden, Haibin Wang, Rachel Marine, Mary S. Keckler, Alison S. Laufer Halpin, Jasmine Padilla, Justin Lee, Christopher A. Elkins, Suxiang Tong |
| EPI_ISL_420796, EPI_ISL_420797, EPI_ISL_420798 | Texas DSHS Lab Services | Pathogen Discovery, Respiratory Viruses Branch, Division of Viral Diseases, Centers for Disease Control and Prevention | Krista Queen, Yan Li, Ying Tao, Jing Zhang, Anne Uehara, Clinton R. Paden, Haibin Wang, Rachel Marine, Mary S. Keckler, Alison S. Laufer Halpin, Jasmine Padilla, Justin Lee, Christopher A. Elkins, Suxiang Tong |
| EPI_ISL_420803, EPI_ISL_420804, EPI_ISL_420805, EPI_ISL_420806, EPI_ISL_420807, EPI_ISL_420808, EPI_ISL_420809, EPI_ISL_420810, EPI_ISL_420811, EPI_ISL_420812, EPI_ISL_420813, EPI_ISL_420814, EPI_ISL_420815, EPI_ISL_420816, EPI_ISL_420817, EPI_ISL_420818, EPI_ISL_420819, EPI_ISL_420820, EPI_ISL_420821, EPI_ISL_420822, EPI_ISL_420823, EPI_ISL_420824, EPI_ISL_420825 | | | |
| see above | Utah Public Health Laboratory | Utah Public Health Laboratory | Erin Young, Kelly Oakeson |
| EPI_ISL_421272 | Wyoming Public Health Laboratory | Center for Global Health, University of New Mexico Health Sciences Center | Daryl Domman, Kurt Schwalm, Rob Christensen, Wanda Manley, Cari Sloma, Noah Hull, Darrell Dinwiddie |
| EPI_ISL_421279, EPI_ISL_421281 | Clinical Diagnostics Laboratory, Diagnostic & Experimental Pathology, Lilly Research Laboratories | Clinical Diagnostics Laboratory, Diagnostic & Experimental Pathology, Lilly Research Laboratories | Tim Holzer, Mayuri Vaidya, Angie Fulford, Sam McNeely, Rachael Redmond, Phil Ebert, John Calley, Leslie O'Neill Reising, Pat Finnegan, Erin Wray, John McElwee, Jeff Fill, Joe Oakley, Andrew Schade |

| | | | |
|---|---|---|---|
| EPI_ISL_421283, EPI_ISL_421284, EPI_ISL_421285, EPI_ISL_421286, EPI_ISL_421287, EPI_ISL_421288, EPI_ISL_421289, EPI_ISL_421290, EPI_ISL_421291, EPI_ISL_421292, EPI_ISL_421293, EPI_ISL_421294, EPI_ISL_421295, EPI_ISL_421296, EPI_ISL_421297, EPI_ISL_421298, EPI_ISL_421299, EPI_ISL_421300, EPI_ISL_421301, EPI_ISL_421302, EPI_ISL_421303, EPI_ISL_421304, EPI_ISL_421305, EPI_ISL_421306, EPI_ISL_421307, EPI_ISL_421308, EPI_ISL_421309, EPI_ISL_421310, EPI_ISL_421311, EPI_ISL_421312, EPI_ISL_421313, EPI_ISL_421314, EPI_ISL_421315, EPI_ISL_421316, EPI_ISL_421317, EPI_ISL_421318, EPI_ISL_421319, EPI_ISL_421320, EPI_ISL_421321, EPI_ISL_421322, EPI_ISL_421323, EPI_ISL_421324, EPI_ISL_421325, EPI_ISL_421326, EPI_ISL_421327, EPI_ISL_421328, EPI_ISL_421329, EPI_ISL_421330, EPI_ISL_421331, EPI_ISL_421332, EPI_ISL_421333, EPI_ISL_421334, EPI_ISL_421335, EPI_ISL_421336, EPI_ISL_421338, EPI_ISL_421339, EPI_ISL_421340, EPI_ISL_421341, EPI_ISL_421342, EPI_ISL_421343 | | | |
| see above | University of Wisconsin-Madison AIDS Vaccine Research Laboratories | University of Wisconsin-Madison AIDS Vaccine Research Laboratories | Gage Moreno, Katarina Braun, et al. AIDS Vaccine Research Laboratories |
| EPI_ISL_421346, EPI_ISL_421347 | Wyoming Public Health Laboratory | Center for Global Health, University of New Mexico Health Sciences Center | Daryl Domman, Kurt Schwalm, Rob Christensen, Wanda Manley, Cari Sloma, Noah Hull, Darrell Dinwiddie |
| EPI_ISL_421348, EPI_ISL_421349, EPI_ISL_421350, EPI_ISL_421351, EPI_ISL_421352, EPI_ISL_421353, EPI_ISL_421354, EPI_ISL_421355, EPI_ISL_421356, EPI_ISL_421357, EPI_ISL_421358, EPI_ISL_421359, EPI_ISL_421360, EPI_ISL_421362, EPI_ISL_421363, EPI_ISL_421364, EPI_ISL_421365, EPI_ISL_421366, EPI_ISL_421367, EPI_ISL_421368, EPI_ISL_421369, EPI_ISL_421370, EPI_ISL_421371, EPI_ISL_421372, EPI_ISL_421373, EPI_ISL_421374, EPI_ISL_421375, EPI_ISL_421376, EPI_ISL_421377, EPI_ISL_421378, EPI_ISL_421379, EPI_ISL_421380, EPI_ISL_421381, EPI_ISL_421382, EPI_ISL_421383, EPI_ISL_421384, EPI_ISL_421385, EPI_ISL_421386, EPI_ISL_421387, EPI_ISL_421388, EPI_ISL_421389, EPI_ISL_421390, EPI_ISL_421391, EPI_ISL_421392, EPI_ISL_421393, EPI_ISL_421394, EPI_ISL_421395, EPI_ISL_421396, EPI_ISL_421397, EPI_ISL_421398, EPI_ISL_421399, EPI_ISL_421400, EPI_ISL_421401, EPI_ISL_421402, EPI_ISL_421403, EPI_ISL_421404, EPI_ISL_421405, EPI_ISL_421406, EPI_ISL_421407, EPI_ISL_421408, EPI_ISL_421409, EPI_ISL_421410, EPI_ISL_421411, EPI_ISL_421412, EPI_ISL_421413, EPI_ISL_421414, EPI_ISL_421415, EPI_ISL_421416, EPI_ISL_421417, EPI_ISL_421418, EPI_ISL_421419, EPI_ISL_421420, EPI_ISL_421421, EPI_ISL_421422, EPI_ISL_421423, EPI_ISL_421424, EPI_ISL_421425, EPI_ISL_421426, EPI_ISL_421427, EPI_ISL_421428, EPI_ISL_421429, EPI_ISL_421430, EPI_ISL_421431, EPI_ISL_421432, EPI_ISL_421433, EPI_ISL_421434, EPI_ISL_421435 | | | |
| see above | MSHS Clinical Microbiology Laboratories | MSHS Pathogen Surveillance Program | Ana S. Gonzalez-Reiche, Mitchell Sullivan, Ajay Obla, Gopi Patel, Emilia Sordillo, Melissa Gitman, Alberto Paniz-mondolfi, Matthew Hernandez, Shelcie Fabre, Jose Polanco, Zenab Khan, Bremy Albuquerque, Jayeeta Dutta, Juan Soto, Shwetha Sridhar Hara, Ying-Chih Wang, Melissa Smith, Robert Sebra, Lisa Miorin, Wen-chun Liu, Randy Albrecht, Judith Aberg, Florian Krammer, Adolfo Garcia-Sarstre, Viviana Simon, Harm van Bakel |
| EPI_ISL_421543, EPI_ISL_421544, EPI_ISL_421545, EPI_ISL_421547, EPI_ISL_421548, EPI_ISL_421549, EPI_ISL_421550, EPI_ISL_421551, EPI_ISL_421552, EPI_ISL_421553 | Wyoming Public Health Laboratory | Center for Global Health, University of New Mexico Health Sciences Center | Daryl Domman, Kurt Schwalm, Rob Christensen, Wanda Manley, Cari Sloma, Noah Hull, Darrell Dinwiddie |
| EPI_ISL_421560, EPI_ISL_421561, EPI_ISL_421562, EPI_ISL_421563 | Utah Public Health Laboratory | Utah Public Health Laboratory | Erin Young, Kelly Oakeson |
| EPI_ISL_421577, EPI_ISL_421578, EPI_ISL_421579, EPI_ISL_421580, EPI_ISL_421581, EPI_ISL_421582, EPI_ISL_421583, EPI_ISL_421584, EPI_ISL_421585, EPI_ISL_421586, EPI_ISL_421587, EPI_ISL_421588, EPI_ISL_421589, EPI_ISL_421590, EPI_ISL_421591, EPI_ISL_421592 | | | |
| see above | NYU Langone Health | Departments of Pathology and Medicine, New York University School of Medicine | Maria Aguero-Rosenfeld, Brendan Belovarac, Margaret Black, Ludovic Boytard, John Cadley, Paolo Cotzia, John Chen, Dacia Dimartino, Xiaojun Feng, Tatyana Gindin, Adriana Heguy, Megan Hogan, Emily Huang, George Jour, Andrew Lytle, Christian Marier, Matthew T. Maurano, Mark J. Mulligan, Peter Meyn, Iman Osman, Jared Pinnell, Sitharam Ramaswami, Amy Rapkiewicz, Marie Samanovic-Golden, Antonio Serrano, Guomiao Shen, Matija Snuderl, Theodore Vougiouklakis, Nick Vulpescu, Gael Westby, Paul Zappile, Yutong Zhang |
| EPI_ISL_421593, EPI_ISL_421594, EPI_ISL_421595, EPI_ISL_421596, EPI_ISL_421597, EPI_ISL_421598, EPI_ISL_421599, EPI_ISL_421600, EPI_ISL_421601, EPI_ISL_421602, EPI_ISL_421603, EPI_ISL_421604, EPI_ISL_421605, EPI_ISL_421606, EPI_ISL_421607, EPI_ISL_421608, EPI_ISL_421609, EPI_ISL_421610, EPI_ISL_421611, EPI_ISL_421612, EPI_ISL_421613, EPI_ISL_421614, EPI_ISL_421615, EPI_ISL_421616, EPI_ISL_421617, EPI_ISL_421618, EPI_ISL_421619, EPI_ISL_421620, EPI_ISL_421621, EPI_ISL_421622, EPI_ISL_421623, EPI_ISL_421624, EPI_ISL_421625, EPI_ISL_421626, EPI_ISL_421627, EPI_ISL_421628, EPI_ISL_421629, EPI_ISL_421630, EPI_ISL_421631, EPI_ISL_421632, EPI_ISL_421633, EPI_ISL_421634, EPI_ISL_421635 | | | |
| see above | MSHS Clinical Microbiology Laboratories | MSHS Pathogen Surveillance Program | Ana S. Gonzalez-Reiche, Mitchell Sullivan, Ajay Obla, Gopi Patel, Emilia Sordillo, Melissa Gitman, Alberto Paniz-mondolfi, Matthew Hernandez, Shelcie Fabre, Jose Polanco, Zenab Khan, Bremy Albuquerque, Jayeeta Dutta, Juan Soto, Shwetha Sridhar Hara, Ying-Chih Wang, Melissa Smith, Robert Sebra, Lisa Miorin, Wen-chun Liu, Randy Albrecht, Judith Aberg, Florian Krammer, Adolfo Garcia-Sarstre, Viviana Simon, Harm van Bakel |
| EPI_ISL_421660 | The Ohio State University | The Ohio State University-James Molecular Lab at Polaris | Huolin Tu, Matthew Avenarius, Preeti Panchioli, Sean Caruthers, Joan-Miquel Balada-Llasat, Jason Garee, Matt Hunt, Xiaokang Pan, Dan Jones |
| EPI_ISL_421675 | The Ohio State University Wexner Medical Center | The Ohio State University James Molecular lab | Huolin Tu, Sean Caruthers, Matthew Avenarius, Joan-Miquel Balada-Llasat, Matthew Hunt, Preeti Panchioli, Xiaokang Pen, Jason Garee, Pam Snyder, Dan Jones |
| EPI_ISL_421683, EPI_ISL_421684, EPI_ISL_421685, EPI_ISL_421686, EPI_ISL_421687, EPI_ISL_421688, EPI_ISL_421689, EPI_ISL_421690, EPI_ISL_421691, EPI_ISL_421692 | Minnesota Department of Health, Public Health Laboratory | Minnesota Department of Health, Public Health Laboratory | Matt Plumb, Jacob Garfin, Xiong Wang |
| EPI_ISL_421705, EPI_ISL_421706, EPI_ISL_421707, EPI_ISL_421708, EPI_ISL_421709, EPI_ISL_421710, EPI_ISL_421711, EPI_ISL_421712, EPI_ISL_421713, EPI_ISL_421714, EPI_ISL_421715, EPI_ISL_421716, EPI_ISL_421717, EPI_ISL_421718, EPI_ISL_421719, EPI_ISL_421720, EPI_ISL_421721, EPI_ISL_421722, EPI_ISL_421723, EPI_ISL_421724, EPI_ISL_421725, EPI_ISL_421726, EPI_ISL_421727, EPI_ISL_421728, EPI_ISL_421729, EPI_ISL_421730, EPI_ISL_421731, EPI_ISL_421732, EPI_ISL_421733 | | | |
| see above | NYU Langone Health | Departments of Pathology and Medicine, New York University School of Medicine | Maria Aguero-Rosenfeld, Brendan Belovarac, Margaret Black, Ludovic Boytard, John Cadley, Paolo Cotzia, John Chen, Dacia Dimartino, Xiaojun Feng, Tatyana Gindin, Adriana Heguy, Megan Hogan, Emily Huang, George Jour, Andrew Lytle, Christian Marier, Matthew T. Maurano, Mark J. Mulligan, Peter Meyn, Iman Osman, Jared Pinnell, Sitharam Ramaswami, Amy Rapkiewicz, Marie Samanovic-Golden, Antonio Serrano, Guomiao Shen, Matija Snuderl, Theodore Vougiouklakis, Nick Vulpescu, Gael Westby, Paul Zappile, Yutong Zhang |
| EPI_ISL_422453, EPI_ISL_422459 | Gundersen Molecular Diagnostics Laboratory | Kabara Cancer Research Institute | Craig S. Richmond & Paraic A. Kenny |
| EPI_ISL_422461, EPI_ISL_422462, EPI_ISL_422463 | Gundersen Molecular Diagnostics Laboratory | Kabara Cancer Research Institute | Craig S. Richmond, Paraic A. Kenny |
| EPI_ISL_422464 | Gundersen Molecular Diagnostics Laboratory | Kabara Cancer Research Institute | Craig S. Richmond; Paraic A. Kenny |
| EPI_ISL_422465 | Gundersen Molecular Diagnostics Laboratory | Kabara Cancer Research Institute | Craig S. Richmond, Paraic A. Kenny |
| EPI_ISL_422488, EPI_ISL_422489, EPI_ISL_422490, EPI_ISL_422491, EPI_ISL_422492, EPI_ISL_422494, EPI_ISL_422495, EPI_ISL_422496, EPI_ISL_422497, EPI_ISL_422498, EPI_ISL_422499, EPI_ISL_422500, EPI_ISL_422501, EPI_ISL_422502, EPI_ISL_422503, EPI_ISL_422504, EPI_ISL_422505, EPI_ISL_422506, EPI_ISL_422507, EPI_ISL_422508, EPI_ISL_422509, EPI_ISL_422510, EPI_ISL_422511, EPI_ISL_422512, EPI_ISL_422513, EPI_ISL_422514, EPI_ISL_422515, EPI_ISL_422516, EPI_ISL_422517, EPI_ISL_422518, EPI_ISL_422519, EPI_ISL_422520, EPI_ISL_422521, EPI_ISL_422522, EPI_ISL_422523, EPI_ISL_422524, EPI_ISL_422525, EPI_ISL_422526, EPI_ISL_422527, EPI_ISL_422528, EPI_ISL_422529, EPI_ISL_422530, EPI_ISL_422531, EPI_ISL_422532, EPI_ISL_422533, EPI_ISL_422534, EPI_ISL_422535, EPI_ISL_422536, EPI_ISL_422537, EPI_ISL_422538, EPI_ISL_422539, EPI_ISL_422540, EPI_ISL_422541, EPI_ISL_422542, EPI_ISL_422543, EPI_ISL_422544, EPI_ISL_422545, EPI_ISL_422546, EPI_ISL_422547, EPI_ISL_422548, EPI_ISL_422549, EPI_ISL_422550, EPI_ISL_422551, EPI_ISL_422552, EPI_ISL_422553, EPI_ISL_422554, EPI_ISL_422555, EPI_ISL_422556, EPI_ISL_422557, EPI_ISL_422558, EPI_ISL_422559, EPI_ISL_422560, EPI_ISL_422561, EPI_ISL_422562 | | | |
| see above | MSHS Clinical Microbiology Laboratories | MSHS Pathogen Surveillance Program | Ana S. Gonzalez-Reiche, Mitchell Sullivan, Ajay Obla, Gopi Patel, Emilia Sordillo, Melissa Gitman, Alberto Paniz-mondolfi, Matthew Hernandez, Shelcie Fabre, Jose Polanco, Zenab Khan, Bremy Albuquerque, Jayeeta Dutta, Juan Soto, Shwetha Sridhar Hara, Ying-Chih Wang, Melissa Smith, Robert Sebra, Lisa Miorin, Wen-chun Liu, Randy Albrecht, Judith Aberg, Florian Krammer, Adolfo Garcia-Sarstre, Viviana Simon, Harm van Bakel |
| EPI_ISL_422961, EPI_ISL_422962, EPI_ISL_422963, EPI_ISL_422964, EPI_ISL_422965, EPI_ISL_422966, EPI_ISL_422967, EPI_ISL_422968, EPI_ISL_422969, EPI_ISL_422970, EPI_ISL_422971, EPI_ISL_422972, EPI_ISL_422973, EPI_ISL_422974, EPI_ISL_422975, EPI_ISL_422976, EPI_ISL_422977, EPI_ISL_422978, EPI_ISL_422979, EPI_ISL_422980, EPI_ISL_422981, EPI_ISL_422982, EPI_ISL_422983, EPI_ISL_422984, EPI_ISL_422985, EPI_ISL_422986, EPI_ISL_422987, EPI_ISL_422988, EPI_ISL_422989, EPI_ISL_422990, EPI_ISL_422991, EPI_ISL_422992, EPI_ISL_422993, EPI_ISL_422994, EPI_ISL_422995, EPI_ISL_422996, EPI_ISL_422997, EPI_ISL_422998, EPI_ISL_422999, EPI_ISL_423000, EPI_ISL_423001, EPI_ISL_423002, EPI_ISL_423003, EPI_ISL_423004, EPI_ISL_423005, EPI_ISL_423006, EPI_ISL_423007, EPI_ISL_423008, EPI_ISL_423009, EPI_ISL_423010, EPI_ISL_423011, EPI_ISL_423012, EPI_ISL_423013, EPI_ISL_423014, EPI_ISL_423015, EPI_ISL_423016, EPI_ISL_423017, EPI_ISL_423018, EPI_ISL_423019, EPI_ISL_423020, EPI_ISL_423021, EPI_ISL_423022, EPI_ISL_423023, EPI_ISL_423024, EPI_ISL_423025, EPI_ISL_423026, EPI_ISL_423028, EPI_ISL_423029, EPI_ISL_423030, EPI_ISL_423031, EPI_ISL_423032, EPI_ISL_423033, EPI_ISL_424166, EPI_ISL_424167, EPI_ISL_424168, EPI_ISL_424169, EPI_ISL_424170, EPI_ISL_424171, EPI_ISL_424172, EPI_ISL_424173, EPI_ISL_424174, EPI_ISL_424175, EPI_ISL_424176, EPI_ISL_424177, EPI_ISL_424178, EPI_ISL_424179, EPI_ISL_424180, EPI_ISL_424181, EPI_ISL_424182, EPI_ISL_424183, EPI_ISL_424184, EPI_ISL_424185, EPI_ISL_424186, EPI_ISL_424187, EPI_ISL_424188, EPI_ISL_424189, EPI_ISL_424190, EPI_ISL_424191, EPI_ISL_424192, EPI_ISL_424193, EPI_ISL_424194, EPI_ISL_424195, EPI_ISL_424196, EPI_ISL_424197, EPI_ISL_424198, EPI_ISL_424199, EPI_ISL_424200, EPI_ISL_424201, EPI_ISL_424202, EPI_ISL_424203, EPI_ISL_424204, EPI_ISL_424205, EPI_ISL_424206, EPI_ISL_424207, EPI_ISL_424208, EPI_ISL_424209, EPI_ISL_424210, EPI_ISL_424211, EPI_ISL_424212, EPI_ISL_424213, EPI_ISL_424214, EPI_ISL_424215, EPI_ISL_424216, EPI_ISL_424217, EPI_ISL_424218, EPI_ISL_424219, EPI_ISL_424220, EPI_ISL_424221, EPI_ISL_424222, EPI_ISL_424223, EPI_ISL_424224, EPI_ISL_424225, EPI_ISL_424226, EPI_ISL_424227, EPI_ISL_424228, EPI_ISL_424229, EPI_ISL_424230, EPI_ISL_424231, EPI_ISL_424232, EPI_ISL_424233, EPI_ISL_424234, EPI_ISL_424235, EPI_ISL_424236, EPI_ISL_424237, EPI_ISL_424238, EPI_ISL_424239, EPI_ISL_424240, EPI_ISL_424241, EPI_ISL_424242, EPI_ISL_424243, EPI_ISL_424244, EPI_ISL_424245, EPI_ISL_424246, EPI_ISL_424247, EPI_ISL_424248, EPI_ISL_424249, EPI_ISL_424250, EPI_ISL_424251, EPI_ISL_424252, EPI_ISL_424253, EPI_ISL_424254, EPI_ISL_424255, EPI_ISL_424256, EPI_ISL_424257, EPI_ISL_424258, EPI_ISL_424259, EPI_ISL_424260, EPI_ISL_424261, EPI_ISL_424262, EPI_ISL_424263, EPI_ISL_424264, EPI_ISL_424265, EPI_ISL_424266, EPI_ISL_424267, EPI_ISL_424268, EPI_ISL_424269, EPI_ISL_424270, EPI_ISL_424271, EPI_ISL_424272, EPI_ISL_424273, EPI_ISL_424275, EPI_ISL_424276, EPI_ISL_424277, EPI_ISL_424278, EPI_ISL_424279, EPI_ISL_424280, EPI_ISL_424281, EPI_ISL_424282, EPI_ISL_424283, EPI_ISL_424284, EPI_ISL_424285, EPI_ISL_424286, EPI_ISL_424287, EPI_ISL_424288, EPI_ISL_424289, EPI_ISL_424290, EPI_ISL_424291, EPI_ISL_424292, EPI_ISL_424293, EPI_ISL_424294, EPI_ISL_424295, EPI_ISL_424296, EPI_ISL_424297, EPI_ISL_424298, EPI_ISL_424299, EPI_ISL_424300, EPI_ISL_424301, EPI_ISL_424302, EPI_ISL_424303, EPI_ISL_424304, EPI_ISL_424305, EPI_ISL_424306, EPI_ISL_424307, EPI_ISL_424308, EPI_ISL_424309, EPI_ISL_424310, EPI_ISL_424311, EPI_ISL_424312, EPI_ISL_424313, EPI_ISL_424314, EPI_ISL_424315, EPI_ISL_424316, EPI_ISL_424317, EPI_ISL_424318, EPI_ISL_424319, EPI_ISL_424320, EPI_ISL_424321, EPI_ISL_424322, EPI_ISL_424323, EPI_ISL_424324, EPI_ISL_424325, EPI_ISL_424326, EPI_ISL_424328, EPI_ISL_424329, EPI_ISL_424330, EPI_ISL_424331, EPI_ISL_424332, EPI_ISL_424333, EPI_ISL_424334, EPI_ISL_424335, EPI_ISL_424336, EPI_ISL_424337, EPI_ISL_424338, EPI_ISL_424339, EPI_ISL_424340, EPI_ISL_424341 | | | |
| see above | UW Virology Lab | UW Virology Lab | Pavitra Roychoudhury, Hong Xie, Keith Jerome, Alexander Greninger |
| EPI_ISL_424345 | Instituto Nacional de Enfermedades Respiratorias | Instituto Nacional de Enfermedades Respiratorias | Joel Armando Vázquez Pérez, Celia Boukadida, Santiago Avila Ríos, Mario Mújica Sánchez, José Arturo Martínez Orozco, Eduardo Becerril Vargas, Jorge Salas Hernández, Irma López Martínez, Lucia Hernández Rivas, Gisela Barrera Badillo, Edgar Mendieta Condado, Fabiola Garcés Ayala, Adnan Araiza Rodríguez, José Ernesto Ramírez González, Victor Hugo Borja Aburto, Concepción Grajales Muñiz, Cesar Raúl González Bonilla, Carolina González Torres, Francisco Javier Gaytán Cervantes, José Esteban Muñoz Medina, Guillermo M. Ruiz-Palacios, Pilar Ramos Cervantes, Violeta Ibarra Gonzalez, Fernando Ledesma Barrientos, Luis Alberto García Andrade, Alfredo Ponce de León Garduño, Blanca Taboada, Alejandro Sánchez, Pavel Isa, Ricardo Grande, Gloria Vázquez, Francisco Pulido, Carlos F. Arias. |
| EPI_ISL_424346, EPI_ISL_424347 | Alaska State Virology Laboratory | Alaska State Virology Laboratory | Chen, J. |
| EPI_ISL_424348 | Instituto Nacional de Enfermedades Respiratorias | Instituto Nacional de Enfermedades Respiratorias | Joel Armando Vázquez Pérez, Celia Boukadida, Santiago Avila Ríos, Mario Mújica Sánchez, José Arturo Martínez Orozco, Eduardo Becerril Vargas, Jorge Salas Hernández, Irma López Martínez, Lucia Hernández Rivas, Gisela Barrera Badillo, Edgar Mendieta Condado, Fabiola Garcés Ayala, Adnan Araiza Rodríguez, José Ernesto Ramírez González, Victor Hugo Borja Aburto, Concepción Grajales Muñiz, Cesar Raúl González Bonilla, Carolina González Torres, Francisco Javier Gaytán Cervantes, José Esteban Muñoz Medina, Guillermo M. Ruiz-Palacios, Pilar Ramos Cervantes, Violeta Ibarra Gonzalez, Fernando Ledesma Barrientos, Luis Alberto García Andrade, Alfredo Ponce de León Garduño, Blanca Taboada, Alejandro Sánchez, Pavel Isa, Ricardo Grande, Gloria Vázquez, Francisco Pulido, Carlos F. Arias. |
| EPI_ISL_424350 | Environmental and Global Health | Environmental and Global Health | Elbadry,M.A., Subramaniam,K., Waltzek,T.B., Stephenson,C.J., Gibson,J.C., Alam,M., Morris,J.G. Jr. and Lednicky,J.A. |
| EPI_ISL_424351 | Environmental and Global Health | Environmental and Global Health | Elbadry,M.A., Subramaniam,K., Waltzek,T.B., Gibson,J.C., Stephenson,C.J., Morris,J.G. Jr. and Lednicky,J.A. |
| EPI_ISL_424353 | Dirk Dittmer | Dirk Dittmer | Bailey,A.G., Caro-Vegas,C., Thompson,C., Dittmer,D., Eason,A.B., Juarez,A., Landis,J.T., McNamara,R.P., Miller,M.B., Moorad,R., Pluta,L.J., Seltzer,T.A., Villamor,F. and Vahrson,W. |
| EPI_ISL_424354 | Dirk Dittmer | Dirk Dittmer | Bailey,A.G., Caro-Vegas,C.P., Dittmer,D., Eason,A.B., Juarez,A., Landis,J.T., McNamara,R.P., Miller,M.B., Moorad,R., Pluta,L.J., Seltzer,T.A., Thompson,C., Vahrson,W. and Villamor,F. |
| EPI_ISL_424626, EPI_ISL_424627 | Instituto Nacional de Enfermedades Respiratorias | Instituto Nacional de Enfermedades Respiratorias | Joel Armando Vázquez Pérez, Celia Boukadida, Santiago Avila Ríos, Mario Mújica Sánchez, José Arturo Martínez Orozco, Eduardo Becerril Vargas, Jorge Salas Hernández, Irma López Martínez, Lucia Hernández Rivas, Gisela Barrera Badillo, Edgar Mendieta Condado, Fabiola Garcés Ayala, Adnan Araiza Rodríguez, José Ernesto Ramírez González, Victor Hugo Borja Aburto, Concepción Grajales Muñiz, Cesar Raúl González Bonilla, Carolina González Torres, Francisco Javier Gaytán Cervantes, José Esteban Muñoz Medina, Guillermo M. Ruiz-Palacios, Pilar Ramos Cervantes, Violeta Ibarra Gonzalez, Fernando Ledesma Barrientos, Luis Alberto García Andrade, Alfredo Ponce de León Garduño, Blanca Taboada, Alejandro Sánchez, Pavel Isa, Ricardo Grande, Gloria Vázquez, Francisco Pulido, Carlos F. Arias. |
| EPI_ISL_424667 | Laboratorio Estatal de Salud Publica del Estado de México | Instituto de Diagnóstico y Referencia Epidemiológicos | Irma López Martínez, José Ernesto Ramírez González, Lucia Hernández Rivas, Gisela Barrera Badillo, Edgar Mendieta Condado, Fabiola Garcés Ayala, Adnan Araiza Rodríguez, Celia Boukadida, Santiago Avila Ríos, Mario Mújica Sánchez, José Arturo Martínez Orozco,Eduardo Becerril Vargas, Joel Armando Vázquez Pérez, Victor Hugo Borja Aburto, Concepción Grajales Muñiz, Cesar Raúl González Bonilla, Carolina González Torres, Francisco Javier Gaytán Cervantes, José Esteban Muñoz Medina, Guillermo M. Ruiz-Palacios, Pilar Ramos Cervantes, Violeta Ibarra |

| | | | |
|---|---|---|---|
| | | | Gonzalez, Fernando Ledesma Barrientos, Luis Alberto García Andrade, Alfredo Ponce de León Garduño, Blanca Taboada, Alejandro Sánchez, Pavel Isa, Ricardo Grande, Gloria Vázquez, Francisco Pulido, Carlos F. Arias. |
| EPI_ISL_424668, EPI_ISL_424669 | Arizona State University Health Services | Arizona State University | Rabia Maqsood, LaRinda A. Holland, Emily A. Kaelin, Bereket Estifanos, Nicholas J. Mellor, Jason Steel, Lily I. Wu, Arvind Varsani, Rolf U. Halden, Brenda G. Hogue, Matthew Scotch, Efrem S. Lim |
| EPI_ISL_424670 | Laboratorio Estatal de Salud Publica del Estado de Queretaro | Instituto de Diagnóstico y Referencia Epidemiologicos | Gisela Barrera Badillo, Irma López Martínez, Lucia Hernández Rivas, Edgar Mendieta Condado, Fabiola Garcés Ayala, Adnan Araiza Rodríguez, Celia Boukadida, Santiago Avila Ríos, Mario Mújica Sánchez, José Arturo Martínez Orozco,Eduardo Becerril Vargas, Joel Armando Vázquez Pérez, Victor Hugo Borja Aburto, Concepción Grajales Muñiz, Cesar Raúl González Bonilla, Carolina González Torres, Francisco Javier Gaytán Cervantes, José Esteban Muñoz Medina, Guillermo M. Ruiz-Palacios, Pilar Ramos Cervantes, Violeta Ibarra Gonzalez, Fernando Ledesma Barrientos, Luis Alberto García Andrade, Alfredo Ponce de León Garduño, Blanca Taboada, Alejandro Sánchez, Pavel Isa, Ricardo Grande, Gloria Vázquez, Francisco Pulido, Carlos F. Arias,José Ernesto Ramírez González |
| EPI_ISL_424671 | Arizona State University Health Services | Arizona State University | Rabia Maqsood, LaRinda A. Holland, Emily A. Kaelin, Bereket Estifanos, Nicholas J. Mellor, Jason Steel, Lily I. Wu, Arvind Varsani, Rolf U. Halden, Brenda G. Hogue, Matthew Scotch, Efrem S. Lim |
| EPI_ISL_424672 | Laboratorio Estatal de Salud Publica del Estado de Puebla | Instituto de Diagnostico y Referencia Epidemiologicos | Fabiola Garcés Ayala, Gisela Barrera Badillo, Irma López Martínez, Lucia Hernández Rivas, Edgar Mendieta Condado, Adnan Araiza Rodríguez, Celia Boukadida, Santiago Avila Ríos, Mario Mújica Sánchez, José Arturo Martínez Orozco,Eduardo Becerril Vargas, Joel Armando Vázquez Pérez, Victor Hugo Borja Aburto, Concepción Grajales Muñiz, Cesar Raúl González Bonilla, Carolina González Torres, Francisco Javier Gaytán Cervantes, José Esteban Muñoz Medina, Guillermo M. Ruiz-Palacios, Pilar Ramos Cervantes, Violeta Ibarra Gonzalez, Fernando Ledesma Barrientos, Luis Alberto García Andrade, Alfredo Ponce de León Garduño, Blanca Taboada, Alejandro Sánchez, Pavel Isa, Ricardo Grande, Gloria Vázquez, Francisco Pulido, Carlos F. Arias,José Ernesto Ramírez González |
| EPI_ISL_424673 | Instituto de Diagnostico y Referencia Epidemiologicos | Instituto de Diagnostico y Referencia Epidemiologicos | Adnan Araiza Rodríguez, Edgar Mendieta Condado,Fabiola Garcés Ayala, Gisela Barrera Badillo, Irma López Martínez, Lucia Hernández Rivas, Celia Boukadida, Santiago Avila Ríos, Mario Mújica Sánchez, José Arturo Martínez Orozco,Eduardo Becerril Vargas, Joel Armando Vázquez Pérez, Victor Hugo Borja Aburto, Concepción Grajales Muñiz, Cesar Raúl González Bonilla, Carolina González Torres, Francisco Javier Gaytán Cervantes, José Esteban Muñoz Medina, Guillermo M. Ruiz-Palacios, Pilar Ramos Cervantes, Violeta Ibarra Gonzalez, Fernando Ledesma Barrientos, Luis Alberto García Andrade, Alfredo Ponce de León Garduño, Blanca Taboada, Alejandro Sánchez, Pavel Isa, Ricardo Grande, Gloria Vázquez, Francisco Pulido, Carlos F. Arias,José Ernesto Ramírez González |
| EPI_ISL_424841, EPI_ISL_424842 | SC Dept of Health and Env. Control-Bureau of Laboratories | Pathogen Discovery, Respiratory Viruses Branch, Division of Viral Diseases, Centers for Disease Control and Prevention | Yan Li, Krista Queen, Clinton R. Paden, Rachel Marine, Anna Uehara, Ying Tao, Jing Zhang, Haibin Wang, Mary S. Keckler, Alison S. Laufer Halpin, Christopher A. Elkins, Suxiang Tong |
| EPI_ISL_424843, EPI_ISL_424844, EPI_ISL_424845, EPI_ISL_424846, EPI_ISL_424847 | MA State Public Health Laboratory | Pathogen Discovery, Respiratory Viruses Branch, Division of Viral Diseases, Centers for Disease Control and Prevention | Yan Li, Krista Queen, Clinton R. Paden, Rachel Marine, Anna Uehara, Ying Tao, Jing Zhang, Haibin Wang, Mary S. Keckler, Alison S. Laufer Halpin, Christopher A. Elkins, Suxiang Tong |
| EPI_ISL_424848, EPI_ISL_424849 | AZ SPHL, Arizona Department of Health Services | Pathogen Discovery, Respiratory Viruses Branch, Division of Viral Diseases, Centers for Disease Control and Prevention | Yan Li, Krista Queen, Clinton R. Paden, Rachel Marine, Anna Uehara, Ying Tao, Jing Zhang, Haibin Wang, Mary S. Keckler, Alison S. Laufer Halpin, Christopher A. Elkins, Suxiang Tong |
| EPI_ISL_424850, EPI_ISL_424851 | IL Department of Public Health Chicago Laboratory | Pathogen Discovery, Respiratory Viruses Branch, Division of Viral Diseases, Centers for Disease Control and Prevention | Yan Li, Krista Queen, Clinton R. Paden, Rachel Marine, Anna Uehara, Ying Tao, Jing Zhang, Haibin Wang, Mary S. Keckler, Alison S. Laufer Halpin, Christopher A. Elkins, Suxiang Tong |
| EPI_ISL_424852 | DC Public Health Lab/ Dept. of Forensic Sciences | Pathogen Discovery, Respiratory Viruses Branch, Division of Viral Diseases, Centers for Disease Control and Prevention | Yan Li, Krista Queen, Clinton R. Paden, Rachel Marine, Anna Uehara, Ying Tao, Jing Zhang, Haibin Wang, Mary S. Keckler, Alison S. Laufer Halpin, Christopher A. Elkins, Suxiang Tong |
| EPI_ISL_424853, EPI_ISL_424854 | FL Bureau of Public Health Laboratories-Miami | Pathogen Discovery, Respiratory Viruses Branch, Division of Viral Diseases, Centers for Disease Control and Prevention | Yan Li, Krista Queen, Clinton R. Paden, Rachel Marine, Anna Uehara, Ying Tao, Jing Zhang, Haibin Wang, Mary S. Keckler, Alison S. Laufer Halpin, Christopher A. Elkins, Suxiang Tong |
| EPI_ISL_424855 | FL Bureau of Public Health Laboratories-Tampa | Pathogen Discovery, Respiratory Viruses Branch, Division of Viral Diseases, Centers for Disease Control and Prevention | Yan Li, Krista Queen, Clinton R. Paden, Rachel Marine, Anna Uehara, Ying Tao, Jing Zhang, Haibin Wang, Mary S. Keckler, Alison S. Laufer Halpin, Christopher A. Elkins, Suxiang Tong |
| EPI_ISL_424856, EPI_ISL_424857 | FL Bur. of Public Health Laboratories-Jacksonville | Pathogen Discovery, Respiratory Viruses Branch, Division of Viral Diseases, Centers for Disease Control and Prevention | Yan Li, Krista Queen, Clinton R. Paden, Rachel Marine, Anna Uehara, Ying Tao, Jing Zhang, Haibin Wang, Mary S. Keckler, Alison S. Laufer Halpin, Christopher A. Elkins, Suxiang Tong |
| EPI_ISL_424858, EPI_ISL_424859, EPI_ISL_424860, EPI_ISL_424861, EPI_ISL_424862, EPI_ISL_424863, EPI_ISL_424864 | GA Department of Public Health Laboratory | Pathogen Discovery, Respiratory Viruses Branch, Division of Viral Diseases, Centers for Disease Control and Prevention | Yan Li, Krista Queen, Clinton R. Paden, Rachel Marine, Anna Uehara, Ying Tao, Jing Zhang, Haibin Wang, Mary S. Keckler, Alison S. Laufer Halpin, Christopher A. Elkins, Suxiang Tong |
| EPI_ISL_424865 | IA State Hygienic Laboratory | Pathogen Discovery, Respiratory Viruses Branch, Division of Viral Diseases, Centers for Disease Control and Prevention | Yan Li, Krista Queen, Clinton R. Paden, Rachel Marine, Anna Uehara, Ying Tao, Jing Zhang, Haibin Wang, Mary S. Keckler, Alison S. Laufer Halpin, Christopher A. Elkins, Suxiang Tong |
| EPI_ISL_424866 | IN State Department of Health Laboratory Services | Pathogen Discovery, Respiratory Viruses Branch, Division of Viral Diseases, Centers for Disease Control and Prevention | Yan Li, Krista Queen, Clinton R. Paden, Rachel Marine, Anna Uehara, Ying Tao, Jing Zhang, Haibin Wang, Mary S. Keckler, Alison S. Laufer Halpin, Christopher A. Elkins, Suxiang Tong |
| EPI_ISL_424867 | KS Health and Environmental Laboratories | Pathogen Discovery, Respiratory Viruses Branch, Division of Viral Diseases, Centers for Disease Control and Prevention | Yan Li, Krista Queen, Clinton R. Paden, Rachel Marine, Anna Uehara, Ying Tao, Jing Zhang, Haibin Wang, Mary S. Keckler, Alison S. Laufer Halpin, Christopher A. Elkins, Suxiang Tong |
| EPI_ISL_424868 | LA Office of Public Health Laboratories | Pathogen Discovery, Respiratory Viruses Branch, Division of Viral Diseases, Centers for Disease Control and Prevention | Yan Li, Krista Queen, Clinton R. Paden, Rachel Marine, Anna Uehara, Ying Tao, Jing Zhang, Haibin Wang, Mary S. Keckler, Alison S. Laufer Halpin, Christopher A. Elkins, Suxiang Tong |
| EPI_ISL_424869 | MD DOH Laboratories Administration | Pathogen Discovery, Respiratory Viruses Branch, Division of Viral Diseases, Centers for Disease Control and Prevention | Yan Li, Krista Queen, Clinton R. Paden, Rachel Marine, Anna Uehara, Ying Tao, Jing Zhang, Haibin Wang, Mary S. Keckler, Alison S. Laufer Halpin, Christopher A. Elkins, Suxiang Tong |
| EPI_ISL_424870 | MO State Public Health Laboratory | Pathogen Discovery, Respiratory Viruses Branch, Division of Viral Diseases, Centers for Disease Control and Prevention | Yan Li, Krista Queen, Clinton R. Paden, Rachel Marine, Anna Uehara, Ying Tao, Jing Zhang, Haibin Wang, Mary S. Keckler, Alison S. Laufer Halpin, Christopher A. Elkins, Suxiang Tong |
| EPI_ISL_424871, EPI_ISL_424872, EPI_ISL_424873 | NC State Laboratory of Public Health | Pathogen Discovery, Respiratory Viruses Branch, Division of Viral Diseases, Centers for Disease Control and Prevention | Yan Li, Krista Queen, Clinton R. Paden, Rachel Marine, Anna Uehara, Ying Tao, Jing Zhang, Haibin Wang, Mary S. Keckler, Alison S. Laufer Halpin, Christopher A. Elkins, Suxiang Tong |
| EPI_ISL_424874, EPI_ISL_424875 | NE Public Health Laboratory | Pathogen Discovery, Respiratory Viruses Branch, Division of Viral Diseases, Centers for Disease Control and Prevention | Yan Li, Krista Queen, Clinton R. Paden, Rachel Marine, Anna Uehara, Ying Tao, Jing Zhang, Haibin Wang, Mary S. Keckler, Alison S. Laufer Halpin, Christopher A. Elkins, Suxiang Tong |
| EPI_ISL_424876, EPI_ISL_424877 | NH Dept. of Health and Human Services Public Health Labs | Pathogen Discovery, Respiratory Viruses Branch, Division of Viral Diseases, Centers for Disease Control and Prevention | Yan Li, Krista Queen, Clinton R. Paden, Rachel Marine, Anna Uehara, Ying Tao, Jing Zhang, Haibin Wang, Mary S. Keckler, Alison S. Laufer Halpin, Christopher A. Elkins, Suxiang Tong |
| EPI_ISL_424878 | NJ Public Health and Environmental Laboratories | Pathogen Discovery, Respiratory Viruses Branch, Division of Viral Diseases, Centers for Disease Control and Prevention | Yan Li, Krista Queen, Clinton R. Paden, Rachel Marine, Anna Uehara, Ying Tao, Jing Zhang, Haibin Wang, Mary S. Keckler, Alison S. Laufer Halpin, Christopher A. Elkins, Suxiang Tong |
| EPI_ISL_424879 | NV-Southern Nevada Public Health Laboratory | Pathogen Discovery, Respiratory Viruses Branch, Division of Viral Diseases, Centers for Disease Control and Prevention | Yan Li, Krista Queen, Clinton R. Paden, Rachel Marine, Anna Uehara, Ying Tao, Jing Zhang, Haibin Wang, Mary S. Keckler, Alison S. Laufer Halpin, Christopher A. Elkins, Suxiang Tong |
| EPI_ISL_424880 | OH Department of Health Laboratory | Pathogen Discovery, Respiratory Viruses Branch, Division of Viral Diseases, Centers for Disease Control and Prevention | Yan Li, Krista Queen, Clinton R. Paden, Rachel Marine, Anna Uehara, Ying Tao, Jing Zhang, Haibin Wang, Mary S. Keckler, Alison S. Laufer Halpin, Christopher A. Elkins, Suxiang Tong |
| EPI_ISL_424881, EPI_ISL_424882, EPI_ISL_424883, EPI_ISL_424884, EPI_ISL_424885, EPI_ISL_424886 | PA Department of Health, Bureau of Laboratories | Pathogen Discovery, Respiratory Viruses Branch, Division of Viral Diseases, Centers for Disease Control and Prevention | Yan Li, Krista Queen, Clinton R. Paden, Rachel Marine, Anna Uehara, Ying Tao, Jing Zhang, Haibin Wang, Mary S. Keckler, Alison S. Laufer Halpin, Christopher A. Elkins, Suxiang Tong |
| EPI_ISL_424887 | RI State Health Laboratories | Pathogen Discovery, Respiratory Viruses Branch, Division of Viral Diseases, Centers for Disease Control and Prevention | Yan Li, Krista Queen, Clinton R. Paden, Rachel Marine, Anna Uehara, Ying Tao, Jing Zhang, Haibin Wang, Mary S. Keckler, Alison S. Laufer Halpin, Christopher A. Elkins, Suxiang Tong |
| EPI_ISL_424888 | SC Dept of Health and Env. Control-Bureau of Laboratories | Pathogen Discovery, Respiratory Viruses Branch, Division of Viral Diseases, Centers for Disease Control and Prevention | Yan Li, Krista Queen, Clinton R. Paden, Rachel Marine, Anna Uehara, Ying Tao, Jing Zhang, Haibin Wang, Mary S. Keckler, Alison S. Laufer Halpin, Christopher A. Elkins, Suxiang Tong |
| EPI_ISL_424889, EPI_ISL_424890 | UT-Unified State Labs: Public Health Utah DOH | Pathogen Discovery, Respiratory Viruses Branch, Division of Viral Diseases, Centers for Disease Control and Prevention | Yan Li, Krista Queen, Clinton R. Paden, Rachel Marine, Anna Uehara, Ying Tao, Jing Zhang, Haibin Wang, Mary S. Keckler, Alison S. Laufer Halpin, Christopher A. Elkins, Suxiang Tong |

| | | | |
|---|---|---|---|
| EPI_ISL_424891, EPI_ISL_424892, EPI_ISL_424893, EPI_ISL_424894 | VA-Division of Consolidated Laboratory Services | Pathogen Discovery, Respiratory Viruses Branch, Division of Viral Diseases, Centers for Disease Control and Prevention | Yan Li, Krista Queen, Clinton R. Paden, Rachel Marine, Anna Uehara, Ying Tao, Jing Zhang, Haibin Wang, Mary S. Keckler, Alison S. Laufer Halpin, Christopher A. Elkins, Suxiang Tong |
| EPI_ISL_424895, EPI_ISL_424896, EPI_ISL_424897, EPI_ISL_424898, EPI_ISL_424899, EPI_ISL_424900 | IA State Hygienic Laboratory | Pathogen Discovery, Respiratory Viruses Branch, Division of Viral Diseases, Centers for Disease Control and Prevention | Ying Tao, Clinton R. Paden, Jing Zhang, Krista Queen, Anna Uehara, Yan Li, Haibin Wang, Mary S. Keckler, Alison S. Laufer Halpin, Christopher A. Elkins, Suxiang Tong |
| EPI_ISL_424901 | NJ Public Health and Environmental Laboratories | Pathogen Discovery, Respiratory Viruses Branch, Division of Viral Diseases, Centers for Disease Control and Prevention | Ying Tao, Clinton R. Paden, Jing Zhang, Krista Queen, Anna Uehara, Yan Li, Haibin Wang, Mary S. Keckler, Alison S. Laufer Halpin, Christopher A. Elkins, Suxiang Tong |
| EPI_ISL_424902, EPI_ISL_424903, EPI_ISL_424904, EPI_ISL_424905 | SC Dept of Health and Env. Control-Bureau of Laboratories | Pathogen Discovery, Respiratory Viruses Branch, Division of Viral Diseases, Centers for Disease Control and Prevention | Ying Tao, Clinton R. Paden, Jing Zhang, Krista Queen, Anna Uehara, Yan Li, Haibin Wang, Mary S. Keckler, Alison S. Laufer Halpin, Christopher A. Elkins, Suxiang Tong |
| EPI_ISL_424906 | NYC Department of Health and Mental Hygiene | Pathogen Discovery, Respiratory Viruses Branch, Division of Viral Diseases, Centers for Disease Control and Prevention | Ying Tao, Clinton R. Paden, Jing Zhang, Krista Queen, Anna Uehara, Yan Li, Haibin Wang, Mary S. Keckler, Alison S. Laufer Halpin, Christopher A. Elkins, Suxiang Tong |
| EPI_ISL_424907 | VA-Division of Consolidated Laboratory Services | Pathogen Discovery, Respiratory Viruses Branch, Division of Viral Diseases, Centers for Disease Control and Prevention | Ying Tao, Clinton R. Paden, Jing Zhang, Krista Queen, Anna Uehara, Yan Li, Haibin Wang, Mary S. Keckler, Alison S. Laufer Halpin, Christopher A. Elkins, Suxiang Tong |
| EPI_ISL_424908, EPI_ISL_424909, EPI_ISL_424910, EPI_ISL_424911, EPI_ISL_424912, EPI_ISL_424913, EPI_ISL_424914, EPI_ISL_424915, EPI_ISL_424916, EPI_ISL_424917, EPI_ISL_424918, EPI_ISL_424919, EPI_ISL_424920 | | | |
| see above | MA State Public Health Laboratory | Pathogen Discovery, Respiratory Viruses Branch, Division of Viral Diseases, Centers for Disease Control and Prevention | Ying Tao, Clinton R. Paden, Jing Zhang, Krista Queen, Anna Uehara, Yan Li, Haibin Wang, Mary S. Keckler, Alison S. Laufer Halpin, Christopher A. Elkins, Suxiang Tong |
| EPI_ISL_424929, EPI_ISL_424930, EPI_ISL_424931, EPI_ISL_424932, EPI_ISL_424933, EPI_ISL_424934, EPI_ISL_424935, EPI_ISL_424936, EPI_ISL_424937, EPI_ISL_424938, EPI_ISL_424939, EPI_ISL_424940, EPI_ISL_424941, EPI_ISL_424942, EPI_ISL_424943, EPI_ISL_424944, EPI_ISL_424945, EPI_ISL_424946, EPI_ISL_424947, EPI_ISL_424948, EPI_ISL_424949, EPI_ISL_424950, EPI_ISL_424951, EPI_ISL_424952, EPI_ISL_424953, EPI_ISL_424954, EPI_ISL_424955, EPI_ISL_424956, EPI_ISL_424957, EPI_ISL_424958, EPI_ISL_424959, EPI_ISL_424960, EPI_ISL_424961, EPI_ISL_424962, EPI_ISL_424963, EPI_ISL_424964, EPI_ISL_424965, EPI_ISL_424966, EPI_ISL_424967, EPI_ISL_424968 | | | |
| see above | NYU Langone Health | Departments of Pathology and Medicine, New York University School of Medicine | Maria Aguero-Rosenfeld, Brendan Belovarac, Margaret Black, Ludovic Boytard, John Cadley, Paolo Cotzia, John Chen, Dacia Dimartino, Xiaojun Feng, Tatyana Gindin, Adriana Heguy, Megan Hogan, Emily Huang, George Jour, Andrew Lytle, Christian Marier, Matthew T. Maurano, Mark J. Mulligan, Peter Meyn, Iman Osman, Jared Pinnell, Sitharam Ramaswami, Amy Rapkiewicz, Marie Samanovic-Golden, Antonio Serrano, Guomiao Shen, Matija Snuderl, Theodore Vougiouklakis, Nick Vulpescu, Gael Westby, Paul Zappile, Yutong Zhang |
| EPI_ISL_424981, EPI_ISL_424982 | Bureau of Laboratories, Michigan Department of Health and Human Services | Michigan Department of Health and Human Services | Blankenship HM, Riner D, Soehnlen MK |
| EPI_ISL_424983 | Dirk Dittmer | Dirk Dittmer | Bailey,A.G., Caro-Vegas,C., Dittmer,D., Eason,A.B., Juarez,A., Landis,J.T., McNamara,R.P., Miller,M.B., Moorad,R., Pluta,L.J., Seltzer,T.A., Thompson,C., Vahrson,W. and Villamor,F. |
| EPI_ISL_424984, EPI_ISL_424985, EPI_ISL_424986 | Bureau of Laboratories, Michigan Department of Health and Human Services | Michigan Department of Health and Human Services | Blankenship HM, Riner D, Soehnlen MK |
| EPI_ISL_424987 | Dirk Dittmer | Dirk Dittmer | Bailey,A.G., Caro-Vegas,C.P., Dittmer,D., Eason,A.B., Juarez,A., Landis,J.T., McNamara,R.P., Miller,M.B., Moorad,R., Pluta,L.J., Seltzer,T.A., Thompson,C., Vahrson,W. and Villamor,F. |
| EPI_ISL_424988, EPI_ISL_424989, EPI_ISL_424990, EPI_ISL_424992 | Bureau of Laboratories, Michigan Department of Health and Human Services | Michigan Department of Health and Human Services | Blankenship HM, Riner D, Soehnlen MK |
| EPI_ISL_425142, EPI_ISL_425143, EPI_ISL_425144, EPI_ISL_425145, EPI_ISL_425146, EPI_ISL_425147, EPI_ISL_425148, EPI_ISL_425149, EPI_ISL_425150, EPI_ISL_425151, EPI_ISL_425152, EPI_ISL_425153, EPI_ISL_425154, EPI_ISL_425155, EPI_ISL_425156, EPI_ISL_425157, EPI_ISL_425158, EPI_ISL_425159, EPI_ISL_425160, EPI_ISL_425161, EPI_ISL_425162, EPI_ISL_425163, EPI_ISL_425164, EPI_ISL_425165, EPI_ISL_425166, EPI_ISL_425167, EPI_ISL_425168, EPI_ISL_425169, EPI_ISL_425170, EPI_ISL_425171, EPI_ISL_425172, EPI_ISL_425173, EPI_ISL_425174, EPI_ISL_425175, EPI_ISL_425176 | | | |
| see above | University of Wisconsin-Madison AIDS Vaccine Research Laboratories | University of Wisconsin-Madison AIDS Vaccine Research Laboratories | Gage Moreno, Katarina Braun, et al. AIDS Vaccine Research Laboratories |
| EPI_ISL_425224 | Wadsworth Center, New York State Department of Health | Wadsworth Center, New York State Department of Health | Kirsten St. George, Daryl M. Lamson, Sara Griesemer, Jonathan Plitnick, Navjot Singh, Matthew D. Shudt, Erica Lasek-Nesselquist |
| EPI_ISL_426025, EPI_ISL_426026, EPI_ISL_426027, EPI_ISL_426028, EPI_ISL_426029, EPI_ISL_426030, EPI_ISL_426031, EPI_ISL_426032, EPI_ISL_426033, EPI_ISL_426034, EPI_ISL_426035, EPI_ISL_426036, EPI_ISL_426037, EPI_ISL_426038, EPI_ISL_426039, EPI_ISL_426040, EPI_ISL_426041, EPI_ISL_426042, EPI_ISL_426043, EPI_ISL_426044, EPI_ISL_426045, EPI_ISL_426046, EPI_ISL_426047, EPI_ISL_426048, EPI_ISL_426049, EPI_ISL_426050 | | | |
| see above | Wadsworth Center, New York State Department.of Health | Wadsworth Center, New York State Department.of Health | Kirsten St. George, Daryl M. Lamson, Sara Griesemer, Jonathan Plitnick, Navjot Singh, Matthew D. Shudt, Erica Lasek-Nesselquist |
| EPI_ISL_426052, EPI_ISL_426053, EPI_ISL_426054, EPI_ISL_426055, EPI_ISL_426056, EPI_ISL_426057, EPI_ISL_426058, EPI_ISL_426059, EPI_ISL_426060, EPI_ISL_426061, EPI_ISL_426062, EPI_ISL_426063, EPI_ISL_426064, EPI_ISL_426065, EPI_ISL_426066, EPI_ISL_426067, EPI_ISL_426068, EPI_ISL_426069, EPI_ISL_426070, EPI_ISL_426071, EPI_ISL_426072, EPI_ISL_426073, EPI_ISL_426074, EPI_ISL_426075, EPI_ISL_426076, EPI_ISL_426077, EPI_ISL_426078, EPI_ISL_426079, EPI_ISL_426080, EPI_ISL_426081, EPI_ISL_426082, EPI_ISL_426083, EPI_ISL_426084, EPI_ISL_426085, EPI_ISL_426086, EPI_ISL_426087, EPI_ISL_426088, EPI_ISL_426089, EPI_ISL_426090, EPI_ISL_426091, EPI_ISL_426092, EPI_ISL_426093, EPI_ISL_426094, EPI_ISL_426095, EPI_ISL_426096, EPI_ISL_426097, EPI_ISL_426098, EPI_ISL_426099, EPI_ISL_426100, EPI_ISL_426101, EPI_ISL_426102, EPI_ISL_426103, EPI_ISL_426104, EPI_ISL_426105, EPI_ISL_426106, EPI_ISL_426107, EPI_ISL_426108, EPI_ISL_426109, EPI_ISL_426110, EPI_ISL_426111, EPI_ISL_426112, EPI_ISL_426113, EPI_ISL_426114, EPI_ISL_426115, EPI_ISL_426116, EPI_ISL_426117, EPI_ISL_426118, EPI_ISL_426119, EPI_ISL_426120, EPI_ISL_426121, EPI_ISL_426122, EPI_ISL_426123, EPI_ISL_426124, EPI_ISL_426125, EPI_ISL_426126, EPI_ISL_426127, EPI_ISL_426128, EPI_ISL_426129, EPI_ISL_426130, EPI_ISL_426131, EPI_ISL_426132, EPI_ISL_426133, EPI_ISL_426135, EPI_ISL_426136, EPI_ISL_426137 | | | |
| see above | UW Virology Lab | UW Virology Lab | Pavitra Roychoudhury, Hong Xie, Keith Jerome, Alexander Greninger |
| EPI_ISL_426159, EPI_ISL_426160, EPI_ISL_426161 | Gundersen Molecular Diagnostics Laboratory | Kabara Cancer Research Institute | Craig S. Richmond, Paraic A. Kenny |
| EPI_ISL_426290 | Wadsworth Center, New York State Department of Health | Wadsworth Center, New York State Department.of Health | Kirsten St. George, Daryl M. Lamson, Sara Griesemer, Jonathan Plitnick, Navjot Singh, Matthew D. Shudt, Erica Lasek-Nesselquist |
| EPI_ISL_426291, EPI_ISL_426292, EPI_ISL_426293, EPI_ISL_426294, EPI_ISL_426295, EPI_ISL_426296, EPI_ISL_426297, EPI_ISL_426298, EPI_ISL_426299, EPI_ISL_426300, EPI_ISL_426301, EPI_ISL_426302, EPI_ISL_426303, EPI_ISL_426304, EPI_ISL_426305, EPI_ISL_426306, EPI_ISL_426307, EPI_ISL_426308, EPI_ISL_426309, EPI_ISL_426310, EPI_ISL_426311, EPI_ISL_426312, EPI_ISL_426313, EPI_ISL_426314, EPI_ISL_426315, EPI_ISL_426316, EPI_ISL_426317, EPI_ISL_426318, EPI_ISL_426319, EPI_ISL_426320, EPI_ISL_426321, EPI_ISL_426322, EPI_ISL_426323, EPI_ISL_426324, EPI_ISL_426325, EPI_ISL_426326, EPI_ISL_426327, EPI_ISL_426328 | | | |
| see above | Wadsworth Center, New York State Department.of Health | Wadsworth Center, New York State Department.of Health | Kirsten St. George, Daryl M. Lamson, Sara Griesemer, Jonathan Plitnick, Navjot Singh, Matthew D. Shudt, Erica Lasek-Nesselquist |
| EPI_ISL_426361, EPI_ISL_426362, EPI_ISL_426363 | Instituto Nacional de Ciencias Medicas y Nutricion Salvador Zubiran | Instituto Nacional de Ciencias Medicas y Nutricion Salvador Zubiran | Guillermo M. Ruiz-Palacios, Pilar Ramos Cervantes, Violeta Ibarra Gonzalez, Fernando Ledesma Barrientos, Luis Alberto García Andrade, Alfredo Ponce de León Garduño, Irma López Martínez, Lucia Hernández Rivas, Gisela Barrera Badillo, Edgar Mendieta Condado, Fabiola Garcés Ayala, Adnan Araiza Rodríguez, José Ernesto Ramírez González, Celia Boukadida, Santiago Avila Ríos, Mario Mújica Sánchez, José Arturo Martínez Orozco, Eduardo Becerril Vargas, Joel Armando Vázquez Pérez, Victor Hugo Borja Aburto, Concepción Grajales Muñiz, Cesar Raúl González Bonilla, Carolina González Torres, Francisco Javier Gaytán Cervantes, José Esteban Muñoz Medina, Blanca Taboada, Alejandro Sánchez, Pavel Isa, Ricardo Grande, Gloria Vázquez, Francisco Pulido, Carlos F. Arias |
| EPI_ISL_426364 | Instituto Nacional de Ciencias Medicas y Nutricion Salvador Zubiran | Instituto Nacional de Ciencias Medicas y Nutricion | Guillermo M. Ruiz-Palacios, Pilar Ramos Cervantes, Violeta Ibarra Gonzalez, Fernando Ledesma Barrientos, Luis Alberto García Andrade, Alfredo Ponce de León Garduño, Irma López Martínez, Lucia Hernández Rivas, Gisela Barrera Badillo, Edgar Mendieta Condado, Fabiola Garcés Ayala, Adnan Araiza Rodríguez, José Ernesto Ramírez González, Celia Boukadida, Santiago Avila Ríos, Mario Mújica Sánchez, José Arturo Martínez Orozco, Eduardo Becerril Vargas, Joel Armando Vázquez Pérez, Victor Hugo Borja Aburto, Concepción Grajales Muñiz, Cesar Raúl González Bonilla, Carolina González Torres, Francisco Javier Gaytán Cervantes, José Esteban Muñoz Medina, Blanca Taboada, Alejandro Sánchez, Pavel Isa, Ricardo Grande, Gloria Vázquez, Francisco Pulido, Carlos F. Arias |
| EPI_ISL_426365 | Instituto Nacional de Ciencias Medicas y Nutricion Salvador Zubiran | Instituto Nacional de Ciencias Medicas y Nutricion Salvador Zubiran | Guillermo M. Ruiz-Palacios, Pilar Ramos Cervantes, Violeta Ibarra Gonzalez, Fernando Ledesma Barrientos, Luis Alberto García Andrade, Alfredo Ponce de León Garduño, Irma López Martínez, Lucia Hernández Rivas, Gisela Barrera Badillo, Edgar Mendieta Condado, Fabiola Garcés Ayala, Adnan Araiza Rodríguez, José Ernesto Ramírez González, Celia Boukadida, Santiago Avila Ríos, Mario Mújica Sánchez, José Arturo Martínez Orozco, Eduardo Becerril Vargas, Joel Armando Vázquez Pérez, Victor Hugo Borja Aburto, Concepción Grajales Muñiz, Cesar Raúl González Bonilla, Carolina González Torres, Francisco Javier Gaytán Cervantes, José Esteban Muñoz Medina, Blanca Taboada, Alejandro Sánchez, Pavel Isa, Ricardo Grande, Gloria Vázquez, Francisco Pulido, Carlos F. Arias |
| EPI_ISL_426416 | CT-Dr. Katherine A. Kelley State Public Health Lab | Pathogen Discovery, Respiratory Viruses Branch, Division of Viral Diseases, Centers for Disease Control and Prevention | Anna Uehara, Yan Li, Krista Queen, Clinton R. Paden, Rachel Marine, Ying Tao, Jing Zhang, Haibin Wang, Mary S. Keckler, Alison S. Laufer Halpin, Christopher A. Elkins, Suxiang Tong |
| EPI_ISL_426417, EPI_ISL_426418, EPI_ISL_426419 | GA Department of Public Health Laboratory | Pathogen Discovery, Respiratory Viruses Branch, Division of Viral Diseases, Centers for Disease Control and Prevention | Anna Uehara, Yan Li, Krista Queen, Clinton R. Paden, Rachel Marine, Ying Tao, Jing Zhang, Haibin Wang, Mary S. Keckler, Alison S. Laufer Halpin, Christopher A. Elkins, Suxiang Tong |
| EPI_ISL_426420, EPI_ISL_426421 | HI Dept. of Health, State Laboratories Division | Pathogen Discovery, Respiratory Viruses Branch, Division of Viral Diseases, Centers for Disease Control and Prevention | Anna Uehara, Yan Li, Krista Queen, Clinton R. Paden, Rachel Marine, Ying Tao, Jing Zhang, Haibin Wang, Mary S. Keckler, Alison S. Laufer Halpin, Christopher A. Elkins, Suxiang Tong |
| EPI_ISL_426422, EPI_ISL_426423, EPI_ISL_426424 | IN State Department of Health Laboratory Services | Pathogen Discovery, Respiratory Viruses Branch, Division of Viral Diseases, Centers for Disease Control and Prevention | Krista Queen, Yan Li, Anna Uehara, Clinton R. Paden, Rachel Marine, Ying Tao, Jing Zhang, Haibin Wang, Mary S. Keckler, Alison S. Laufer Halpin, Christopher A. Elkins, Suxiang Tong |
| EPI_ISL_426425 | MD DOH Laboratories Administration | Pathogen Discovery, Respiratory Viruses Branch, Division of Viral Diseases, Centers for Disease Control and Prevention | Krista Queen, Yan Li, Anna Uehara, Clinton R. Paden, Rachel Marine, Ying Tao, Jing Zhang, Haibin Wang, Mary S. Keckler, Alison S. Laufer Halpin, Christopher A. Elkins, Suxiang Tong |
| EPI_ISL_426426, EPI_ISL_426427 | MN PHL Division, Minnesota Department of Health | Pathogen Discovery, Respiratory Viruses Branch, Division of Viral Diseases, Centers for Disease Control and Prevention | Krista Queen, Yan Li, Anna Uehara, Clinton R. Paden, Rachel Marine, Ying Tao, Jing Zhang, Haibin Wang, Mary S. Keckler, Alison S. Laufer Halpin, Christopher A. Elkins, Suxiang Tong |
| EPI_ISL_426428 | NC State Laboratory of Public Health | Pathogen Discovery, Respiratory Viruses | Krista Queen, Yan Li, Anna Uehara, Clinton R. Paden, Rachel Marine, Ying Tao, Jing Zhang, Haibin Wang, Mary S. Keckler, Alison S. Laufer Halpin, Christopher A. Elkins, Suxiang Tong |

| | | | |
|---|---|---|---|
| | | Branch, Division of Viral Diseases, Centers for Disease Control and Prevention | |
| EPI_ISL_426429 | NV State Public Health Laboratory | Pathogen Discovery, Respiratory Viruses Branch, Division of Viral Diseases, Centers for Disease Control and Prevention | Krista Queen, Yan Li, Anna Uehara, Clinton R. Paden, Rachel Marine, Ying Tao, Jing Zhang, Haibin Wang, Mary S. Keckler, Alison S. Laufer Halpin, Christopher A. Elkins, Suxiang Tong |
| EPI_ISL_426430, EPI_ISL_426431 | OH Department of Health Laboratory | Pathogen Discovery, Respiratory Viruses Branch, Division of Viral Diseases, Centers for Disease Control and Prevention | Krista Queen, Yan Li, Anna Uehara, Clinton R. Paden, Rachel Marine, Ying Tao, Jing Zhang, Haibin Wang, Mary S. Keckler, Alison S. Laufer Halpin, Christopher A. Elkins, Suxiang Tong |
| EPI_ISL_426432, EPI_ISL_426433, EPI_ISL_426434 | PA Department of Health, Bureau of Laboratories | Pathogen Discovery, Respiratory Viruses Branch, Division of Viral Diseases, Centers for Disease Control and Prevention | Krista Queen, Yan Li, Anna Uehara, Clinton R. Paden, Rachel Marine, Ying Tao, Jing Zhang, Haibin Wang, Mary S. Keckler, Alison S. Laufer Halpin, Christopher A. Elkins, Suxiang Tong |
| EPI_ISL_426435 | RI State Health Laboratories | Pathogen Discovery, Respiratory Viruses Branch, Division of Viral Diseases, Centers for Disease Control and Prevention | Krista Queen, Yan Li, Anna Uehara, Clinton R. Paden, Rachel Marine, Ying Tao, Jing Zhang, Haibin Wang, Mary S. Keckler, Alison S. Laufer Halpin, Christopher A. Elkins, Suxiang Tong |
| EPI_ISL_426436 | WA State Department of Health | Pathogen Discovery, Respiratory Viruses Branch, Division of Viral Diseases, Centers for Disease Control and Prevention | Jing Zhang, Ying Tao, Clinton R. Paden, Krista Queen, Anna Uehara, Yan Li, Haibin Wang, Jesica Jacobs, Denny Russell, Brian Hiatt, Jessica Gant, Suxiang Tong |
| EPI_ISL_426437 | WA State Department of Health | Pathogen Discovery, Respiratory Viruses Branch, Division of Viral Diseases, Centers for Disease Control and Prevention | Ying Tao, Jing Zhang, Clinton R. Paden, Krista Queen, Anna Uehara, Yan Li, Haibin Wang, Jesica Jacobs, Denny Russell, Brian Hiatt, Jessica Gant, Suxiang Tong |
| EPI_ISL_426438, EPI_ISL_426439 | WA State Department of Health | Pathogen Discovery, Respiratory Viruses Branch, Division of Viral Diseases, Centers for Disease Control and Prevention | Jing Zhang, Ying Tao, Clinton R. Paden, Krista Queen, Anna Uehara, Yan Li, Haibin Wang, Jesica Jacobs, Denny Russell, Brian Hiatt, Jessica Gant, Suxiang Tong |
| EPI_ISL_426440, EPI_ISL_426441 | WA State Department of Health | Pathogen Discovery, Respiratory Viruses Branch, Division of Viral Diseases, Centers for Disease Control and Prevention | Ying Tao, Jing Zhang, Clinton R. Paden, Krista Queen, Anna Uehara, Yan Li, Haibin Wang, Jesica Jacobs, Denny Russell, Brian Hiatt, Jessica Gant, Suxiang Tong |
| EPI_ISL_426442 | WA State Department of Health | Pathogen Discovery, Respiratory Viruses Branch, Division of Viral Diseases, Centers for Disease Control and Prevention | Jing Zhang, Ying Tao, Clinton R. Paden, Krista Queen, Anna Uehara, Yan Li, Haibin Wang, Jesica Jacobs, Denny Russell, Brian Hiatt, Jessica Gant, Suxiang Tong |
| EPI_ISL_426443, EPI_ISL_426444 | WA State Department of Health | Pathogen Discovery, Respiratory Viruses Branch, Division of Viral Diseases, Centers for Disease Control and Prevention | Ying Tao, Jing Zhang, Clinton R. Paden, Krista Queen, Anna Uehara, Yan Li, Haibin Wang, Jesica Jacobs, Denny Russell, Brian Hiatt, Jessica Gant, Suxiang Tong |
| EPI_ISL_426445 | WA State Department of Health | Pathogen Discovery, Respiratory Viruses Branch, Division of Viral Diseases, Centers for Disease Control and Prevention | Jing Zhang, Ying Tao, Clinton R. Paden, Krista Queen, Anna Uehara, Yan Li, Haibin Wang, Jesica Jacobs, Denny Russell, Brian Hiatt, Jessica Gant, Suxiang Tong |
| EPI_ISL_426446, EPI_ISL_426447, EPI_ISL_426448, EPI_ISL_426449 | WA State Department of Health | Pathogen Discovery, Respiratory Viruses Branch, Division of Viral Diseases, Centers for Disease Control and Prevention | Ying Tao, Jing Zhang, Clinton R. Paden, Krista Queen, Anna Uehara, Yan Li, Haibin Wang, Jesica Jacobs, Denny Russell, Brian Hiatt, Jessica Gant, Suxiang Tong |
| EPI_ISL_426450 | WA State Department of Health | Pathogen Discovery, Respiratory Viruses Branch, Division of Viral Diseases, Centers for Disease Control and Prevention | Jing Zhang, Ying Tao, Clinton R. Paden, Krista Queen, Anna Uehara, Yan Li, Haibin Wang, Jesica Jacobs, Denny Russell, Brian Hiatt, Jessica Gant, Suxiang Tong |
| EPI_ISL_426451, EPI_ISL_426452, EPI_ISL_426453 | WA State Department of Health | Pathogen Discovery, Respiratory Viruses Branch, Division of Viral Diseases, Centers for Disease Control and Prevention | Ying Tao, Jing Zhang, Clinton R. Paden, Krista Queen, Anna Uehara, Yan Li, Haibin Wang, Jesica Jacobs, Denny Russell, Brian Hiatt, Jessica Gant, Suxiang Tong |
| EPI_ISL_426454, EPI_ISL_426455, EPI_ISL_426456, EPI_ISL_426457, EPI_ISL_426458, EPI_ISL_426459, EPI_ISL_426460, EPI_ISL_426461, EPI_ISL_426462, EPI_ISL_426463, EPI_ISL_426464, EPI_ISL_426465, EPI_ISL_426466, EPI_ISL_426467, EPI_ISL_426468, EPI_ISL_426469, EPI_ISL_426470, EPI_ISL_426471, EPI_ISL_426472, EPI_ISL_426473, EPI_ISL_426474, EPI_ISL_426475 | | | |
| see above | Virginia DCLS | Virginia DCLS | Virginia DCLS |
| EPI_ISL_426483 | AZ SPHL, Arizona Department of Health Services | TGen North | Jolene Bowers, Megan Folkerts, Darrin Lemmer, Dave Engelthaler |
| EPI_ISL_426484 | n/a | Worobey Lab on behalf of the Arizona COVID-19 Genomics Union | Brendan B. Larsen, Megan Folkerts, Krystal Sheridan, Ashlyn Pfeiffer, Danielle Yasquez, Hayley Yaglom, Darrin Lemmer, Jolene Bowers,Evan Bolyen, Jason W. Sahl, Nicholas A. Bokulich, J. Gregory Caporaso, Crystal Hepp, Jason Ladner,David M. Engelthaler, Paul Keim, Michael Worobey |
| EPI_ISL_426485 | AZ SPHL, Arizona Department of Health Services | TGen North | Jolene Bowers, Megan Folkerts, Darrin Lemmer, Dave Engelthaler |
| EPI_ISL_426499, EPI_ISL_426500, EPI_ISL_426501, EPI_ISL_426502, EPI_ISL_426503, EPI_ISL_426504, EPI_ISL_426505, EPI_ISL_426506, EPI_ISL_426507, EPI_ISL_426508, EPI_ISL_426509, EPI_ISL_426510, EPI_ISL_426511 | | | |
| see above | TGen North | TGen North | Jolene Bowers, Megan Folkerts, Darrin Lemmer, Dave Engelthaler |
| EPI_ISL_426512, EPI_ISL_426513, EPI_ISL_426514, EPI_ISL_426515, EPI_ISL_426516, EPI_ISL_426517, EPI_ISL_426518, EPI_ISL_426519 | AZ SPHL, Arizona Department of Health Services | TGen North | Jolene Bowers, Megan Folkerts, Darrin Lemmer, Dave Engelthaler |
| EPI_ISL_426520, EPI_ISL_426521, EPI_ISL_426522, EPI_ISL_426523, EPI_ISL_426524, EPI_ISL_426525, EPI_ISL_426526 | TGen North | TGen North | Jolene Bowers, Megan Folkerts, Darrin Lemmer, Dave Engelthaler |
| EPI_ISL_426527, EPI_ISL_426528, EPI_ISL_426529, EPI_ISL_426530, EPI_ISL_426531 | AZ SPHL, Arizona Department of Health Services | TGen North | Jolene Bowers, Megan Folkerts, Darrin Lemmer, Dave Engelthaler |
| EPI_ISL_426532, EPI_ISL_426533, EPI_ISL_426534, EPI_ISL_426535, EPI_ISL_426536 | TGen North | TGen North | Jolene Bowers, Megan Folkerts, Darrin Lemmer, Dave Engelthaler |
| EPI_ISL_426537, EPI_ISL_426538, EPI_ISL_426539, EPI_ISL_426540, EPI_ISL_426541, EPI_ISL_426542, EPI_ISL_426543, EPI_ISL_426544, EPI_ISL_426545, EPI_ISL_426546, EPI_ISL_426547, EPI_ISL_426548, EPI_ISL_426549, EPI_ISL_426550, EPI_ISL_426551, EPI_ISL_426552, EPI_ISL_426553, EPI_ISL_426554, EPI_ISL_426555 | | | |
| see above | AZ SPHL, Arizona Department of Health Services | TGen North | Jolene Bowers, Megan Folkerts, Darrin Lemmer, Dave Engelthaler |
| EPI_ISL_426556, EPI_ISL_426557 | TGen North | TGen North | Jolene Bowers, Megan Folkerts, Darrin Lemmer, Dave Engelthaler |
| EPI_ISL_426558, EPI_ISL_426559, EPI_ISL_426560, EPI_ISL_426561, EPI_ISL_426562, EPI_ISL_426563, EPI_ISL_426564, EPI_ISL_426565, EPI_ISL_426566, EPI_ISL_426567, EPI_ISL_426568, EPI_ISL_426569 | | | |
| see above | AZ SPHL, Arizona Department of Health Services | TGen North | Jolene Bowers, Megan Folkerts, Darrin Lemmer, Dave Engelthaler |
| EPI_ISL_426617, EPI_ISL_426618, EPI_ISL_426619, EPI_ISL_426620, EPI_ISL_426621, EPI_ISL_426622, EPI_ISL_426623, EPI_ISL_426624, | NYU Langone Health | Departments of Pathology and Medicine, New York University School of Medicine | Maria Aguero-Rosenfeld, Brendan Belovarac, Margaret Black, Ludovic Boytard, John Cadley, Paolo Cotzia, John Chen, Dacia Dimartino, Xiaojun Feng, Tatyana Gindin, Emily Guzman, Adriana Heguy, Megan Hogan, Emily Huang, George Jour, Andrew Lytle, Christian Marier, Matthew T. Maurano, Mark J. Mulligan, Peter Meyn, Iman Osman, Jared Pinnell, Vanessa Raabe, Sitharam Ramaswami, Amy Rapkiewicz, Marie Samanovic-Golden, Antonio Serrano, Guomiao Shen, Matija Snuderl, Theodore Vougiouklakis, Nick Vulpescu, Gael Westby, Paul Zappile, Yutong Zhang |

| | | | |
|---|---|---|---|
| EPI_ISL_426625, EPI_ISL_426626 | | | |
| EPI_ISL_426627, EPI_ISL_426628 | Ochsner Health | BioInfoExperts, LLC | Amy Feehan, David Nolan, Rebecca Rose, Susanna Lamers, Sissy Cross, Julia-Garcia-Diaz, Tong Yang, Luke Caruso, David Moraga Amador, Wayra Navia, Lydia Von Borstel, Xiao Hui Zhou |
| EPI_ISL_427161, EPI_ISL_427162, EPI_ISL_427163, EPI_ISL_427164, EPI_ISL_427165, EPI_ISL_427166, EPI_ISL_427167, EPI_ISL_427168, EPI_ISL_427169, EPI_ISL_427170, EPI_ISL_427171, EPI_ISL_427172, EPI_ISL_427173, EPI_ISL_427174, EPI_ISL_427175, EPI_ISL_427176, EPI_ISL_427177, EPI_ISL_427178, EPI_ISL_427179, EPI_ISL_427180, EPI_ISL_427181, EPI_ISL_427182, EPI_ISL_427183, EPI_ISL_427184, EPI_ISL_427185, EPI_ISL_427186, EPI_ISL_427187, EPI_ISL_427188, EPI_ISL_427189, EPI_ISL_427190, EPI_ISL_427191, EPI_ISL_427192, EPI_ISL_427193, EPI_ISL_427194, EPI_ISL_427195, EPI_ISL_427196, EPI_ISL_427197, EPI_ISL_427198, EPI_ISL_427199, EPI_ISL_427200, EPI_ISL_427201, EPI_ISL_427202, EPI_ISL_427203, EPI_ISL_427204, EPI_ISL_427205, EPI_ISL_427206, EPI_ISL_427207, EPI_ISL_427208, EPI_ISL_427209, EPI_ISL_427210, EPI_ISL_427211, EPI_ISL_427212, EPI_ISL_427213, EPI_ISL_427214, EPI_ISL_427215, EPI_ISL_427216, EPI_ISL_427217, EPI_ISL_427218, EPI_ISL_427219, EPI_ISL_427220, EPI_ISL_427221, EPI_ISL_427222, EPI_ISL_427223, EPI_ISL_427224, EPI_ISL_427225, EPI_ISL_427226, EPI_ISL_427227, EPI_ISL_427228, EPI_ISL_427229, EPI_ISL_427230, EPI_ISL_427231, EPI_ISL_427232, EPI_ISL_427233, EPI_ISL_427234, EPI_ISL_427235, EPI_ISL_427236, EPI_ISL_427237, EPI_ISL_427238, EPI_ISL_427239, EPI_ISL_427240, EPI_ISL_427241, EPI_ISL_427242, EPI_ISL_427243, EPI_ISL_427244, EPI_ISL_427245, EPI_ISL_427246, EPI_ISL_427247, EPI_ISL_427248, EPI_ISL_427249, EPI_ISL_427250, EPI_ISL_427251, EPI_ISL_427252, EPI_ISL_427253, EPI_ISL_427254, EPI_ISL_427255, EPI_ISL_427256, EPI_ISL_427257, EPI_ISL_427258, EPI_ISL_427259, EPI_ISL_427260, EPI_ISL_427261, EPI_ISL_427262, EPI_ISL_427263, EPI_ISL_427264, EPI_ISL_427265, EPI_ISL_427266, EPI_ISL_427268, EPI_ISL_427269, EPI_ISL_427270 | | | |
| see above | UW Virology Lab | UW Virology Lab | Pavitra Roychoudhury, Hong Xie, Keith Jerome, Alexander Greninger |
| EPI_ISL_427271, EPI_ISL_427272 | AZ SPHL, Arizona Department of Health Services | TGen North | Jolene Bowers, Megan Folkerts, Darrin Lemmer, Dave Engelthaler |
| EPI_ISL_427273, EPI_ISL_427274, EPI_ISL_427275, EPI_ISL_427276, EPI_ISL_427277, EPI_ISL_427278, EPI_ISL_427279, EPI_ISL_427280, EPI_ISL_427281, EPI_ISL_427282, EPI_ISL_427283, EPI_ISL_427284, EPI_ISL_427285, EPI_ISL_427286, EPI_ISL_427287 | | | |
| see above | Minnesota Department of Health, Public Health Laboratory | Minnesota Department of Health, Public Health Laboratory | Matt Plumb, Jacob Garfin and Xiong Wang |
| EPI_ISL_427288 | The Ohio State University | The Ohio State University-James Molecular Lab at Polaris | Huolin Tu, Preeti Panchioli, Jason Garee, Matthew Hunt, Joan-Miquel Balada-Llasat, Erica Vincent, Weiqiang Zhao, Dan Jones |
| EPI_ISL_427290 | The Ohio State University | The Ohio State University-James Molecular Lab at Polaris | Huolin Tu, Jason Garee, Matthew Hunt, Joan-Miquel Balada-Llasat, Preeti Pancholi, Erica Vincent, Rongqin Ren, Dan Jones |
| EPI_ISL_427427, EPI_ISL_427428, EPI_ISL_427429, EPI_ISL_427430, EPI_ISL_427431, EPI_ISL_427432, EPI_ISL_427433, EPI_ISL_427434, EPI_ISL_427435, EPI_ISL_427436, EPI_ISL_427437, EPI_ISL_427438, EPI_ISL_427439, EPI_ISL_427440, EPI_ISL_427441, EPI_ISL_427442, EPI_ISL_427443, EPI_ISL_427444, EPI_ISL_427445, EPI_ISL_427446, EPI_ISL_427447, EPI_ISL_427448, EPI_ISL_427449, EPI_ISL_427450, EPI_ISL_427451, EPI_ISL_427452, EPI_ISL_427453, EPI_ISL_427454, EPI_ISL_427455, EPI_ISL_427456, EPI_ISL_427457, EPI_ISL_427458, EPI_ISL_427459, EPI_ISL_427460, EPI_ISL_427461, EPI_ISL_427462 | | | |
| see above | University of Wisconsin-Madison AIDS Vaccine Research Laboratories | University of Wisconsin-Madison AIDS Vaccine Research Laboratories | Gage Moreno, Katarina Braun, et al. AIDS Vaccine Research Laboratories |
| EPI_ISL_427469, EPI_ISL_427470, EPI_ISL_427471, EPI_ISL_427472, EPI_ISL_427473, EPI_ISL_427474, EPI_ISL_427475, EPI_ISL_427476, EPI_ISL_427477, EPI_ISL_427478, EPI_ISL_427479, EPI_ISL_427480, EPI_ISL_427481, EPI_ISL_427482, EPI_ISL_427483, EPI_ISL_427484, EPI_ISL_427485, EPI_ISL_427486, EPI_ISL_427487, EPI_ISL_427488, EPI_ISL_427489, EPI_ISL_427490, EPI_ISL_427491, EPI_ISL_427492, EPI_ISL_427493, EPI_ISL_427494, EPI_ISL_427495, EPI_ISL_427496, EPI_ISL_427497, EPI_ISL_427498, EPI_ISL_427500, EPI_ISL_427501, EPI_ISL_427502, EPI_ISL_427503, EPI_ISL_427504, EPI_ISL_427505, EPI_ISL_427506, EPI_ISL_427507, EPI_ISL_427508, EPI_ISL_427509, EPI_ISL_427510, EPI_ISL_427511, EPI_ISL_427512, EPI_ISL_427513, EPI_ISL_427514, EPI_ISL_427515, EPI_ISL_427516, EPI_ISL_427517, EPI_ISL_427518, EPI_ISL_427519, EPI_ISL_427520, EPI_ISL_427521, EPI_ISL_427524 | | | |
| see above | NYU Langone Health | Departments of Pathology and Medicine, New York University School of Medicine | Maria Aguero-Rosenfeld, Brendan Belovarac, Margaret Black, Ludovic Boytard, John Cadley, Paolo Cotzia, John Chen, Dacia Dimartino, Xiaojun Feng, Tatyana Gindin, Emily Guzman, Adriana Heguy, Megan Hogan, Emily Huang, George Jour, Andrew Lytle, Christian Marier, Matthew T. Maurano, Mark J. Mulligan, Peter Meyn, Iman Osman, Jared Pinnell, Vanessa Raabe, Sitharam Ramaswami, Amy Rapkiewicz, Marie Samanovic-Golden, Antonio Serrano, Guomiao Shen, Matija Snuderl, Theodore Vougiouklakis, Nick Vulpescu, Gael Westby, Paul Zappile, Yutong Zhang |
| EPI_ISL_427526, EPI_ISL_427527, EPI_ISL_427528, EPI_ISL_427529, EPI_ISL_427530, EPI_ISL_427531, EPI_ISL_427532, EPI_ISL_427533, EPI_ISL_427534, EPI_ISL_427535, EPI_ISL_427536, EPI_ISL_427537, EPI_ISL_427538, EPI_ISL_427539, EPI_ISL_427540, EPI_ISL_427541, EPI_ISL_427542, EPI_ISL_427543, EPI_ISL_427544, EPI_ISL_427545, EPI_ISL_427546, EPI_ISL_427547, EPI_ISL_427548, EPI_ISL_427549, EPI_ISL_427550, EPI_ISL_427551, EPI_ISL_427552, EPI_ISL_427553, EPI_ISL_427554, EPI_ISL_427555, EPI_ISL_427556, EPI_ISL_427557, EPI_ISL_427558, EPI_ISL_427559, EPI_ISL_427560, EPI_ISL_427561, EPI_ISL_427562, EPI_ISL_427563, EPI_ISL_427564, EPI_ISL_427565, EPI_ISL_427566, EPI_ISL_427567, EPI_ISL_427568, EPI_ISL_427569, EPI_ISL_427570, EPI_ISL_427571, EPI_ISL_427572, EPI_ISL_427573, EPI_ISL_427574, EPI_ISL_427575, EPI_ISL_427576, EPI_ISL_427577, EPI_ISL_427578, EPI_ISL_427579, EPI_ISL_427580, EPI_ISL_427581, EPI_ISL_427582, EPI_ISL_427583, EPI_ISL_427584, EPI_ISL_427585, EPI_ISL_427586, EPI_ISL_427587, EPI_ISL_427588, EPI_ISL_427589, EPI_ISL_427590, EPI_ISL_427591, EPI_ISL_427592, EPI_ISL_427593, EPI_ISL_427594, EPI_ISL_427595, EPI_ISL_427596, EPI_ISL_427597, EPI_ISL_427598, EPI_ISL_427599, EPI_ISL_427600, EPI_ISL_427601, EPI_ISL_427602, EPI_ISL_427603, EPI_ISL_427604, EPI_ISL_427605, EPI_ISL_427606, EPI_ISL_427607, EPI_ISL_427608, EPI_ISL_427609, EPI_ISL_427610, EPI_ISL_427611, EPI_ISL_427612, EPI_ISL_427613, EPI_ISL_427614, EPI_ISL_427615, EPI_ISL_427616, EPI_ISL_427617, EPI_ISL_427618 | | | |
| see above | NewYork-Presbyterian & Mason Lab | Mason Lab | Daniel J. Butler, Christopher Mozsary, Cem Meydan, David Danko, Jonathan Foox, Joel Rosiene, Alon Shaiber, Matthew MacKay, Ebrahim Afshinnekoo, Fritz J. Sedlazeck, Nikolay A. Ivanov, Maria Sierra, Craig D. Westover, Krista Ryon, Benjamin Young, Chandrima Bhattacharya, Phyllis Ruggiero, Justyna Gawrys, Iman Hajirasouliha, Dmitry Meleshko, Mirella Salvatore, Dong Xu, Jenny Xiang, John Sipley, Lin Cong, Arryn Craney, Priya Velu, Lars F. Westblade, Massimo Loda, Shawn Levy, Melissa Cushing, Marcin Imielinski, Hanna Rennert, Christopher E. Mason |
| EPI_ISL_427619, EPI_ISL_427620, EPI_ISL_427621, EPI_ISL_427622 | Alaska State Virology Laboratory | Alaska State Virology Laboratory | Chen, J. |
| EPI_ISL_427627, EPI_ISL_427628, EPI_ISL_427629, EPI_ISL_427630, EPI_ISL_427631, EPI_ISL_427632, EPI_ISL_427633, EPI_ISL_427634, EPI_ISL_427635, EPI_ISL_427636, EPI_ISL_427637, EPI_ISL_427638, EPI_ISL_427639, EPI_ISL_427640, EPI_ISL_427641, EPI_ISL_427642 | | | |
| see above | NYU Langone Health | Departments of Pathology and Medicine, New York University School of Medicine | Maria Aguero-Rosenfeld, Brendan Belovarac, Margaret Black, Ludovic Boytard, John Cadley, Paolo Cotzia, John Chen, Dacia Dimartino, Xiaojun Feng, Tatyana Gindin, Emily Guzman, Adriana Heguy, Megan Hogan, Emily Huang, George Jour, Andrew Lytle, Christian Marier, Matthew T. Maurano, Mark J. Mulligan, Peter Meyn, Iman Osman, Jared Pinnell, Vanessa Raabe, Sitharam Ramaswami, Amy Rapkiewicz, Marie Samanovic-Golden, Antonio Serrano, Guomiao Shen, Matija Snuderl, Theodore Vougiouklakis, Nick Vulpescu, Gael Westby, Paul Zappile, Yutong Zhang |
| EPI_ISL_428202, EPI_ISL_428203, EPI_ISL_428204, EPI_ISL_428205, EPI_ISL_428206 | Nebraska Public Health Laboratory | UNMC COVID-19 Response Team | UNMC COVID-19 Response Team |
| EPI_ISL_428252, EPI_ISL_428253, EPI_ISL_428254, EPI_ISL_428255, EPI_ISL_428256, EPI_ISL_428257, EPI_ISL_428258, EPI_ISL_428259, EPI_ISL_428260, EPI_ISL_428261, EPI_ISL_428262, EPI_ISL_428263, EPI_ISL_428264, EPI_ISL_428265, EPI_ISL_428266, EPI_ISL_428267, EPI_ISL_428268, EPI_ISL_428269, EPI_ISL_428270, EPI_ISL_428271, EPI_ISL_428272, EPI_ISL_428273, EPI_ISL_428274, EPI_ISL_428275, EPI_ISL_428276, EPI_ISL_428277, EPI_ISL_428278, EPI_ISL_428279, EPI_ISL_428280, EPI_ISL_428281, EPI_ISL_428282, EPI_ISL_428283, EPI_ISL_428284, EPI_ISL_428285, EPI_ISL_428286, EPI_ISL_428287, EPI_ISL_428288, EPI_ISL_428289, EPI_ISL_428290, EPI_ISL_428291, EPI_ISL_428292, EPI_ISL_428293, EPI_ISL_428294, EPI_ISL_428295, EPI_ISL_428296, EPI_ISL_428297, EPI_ISL_428298, EPI_ISL_428299, EPI_ISL_428300, EPI_ISL_428301, EPI_ISL_428302, EPI_ISL_428303, EPI_ISL_428304, EPI_ISL_428305, EPI_ISL_428306, EPI_ISL_428307, EPI_ISL_428308, EPI_ISL_428309, EPI_ISL_428310, EPI_ISL_428311, EPI_ISL_428312, EPI_ISL_428313, EPI_ISL_428314, EPI_ISL_428315, EPI_ISL_428316, EPI_ISL_428317, EPI_ISL_428318, EPI_ISL_428319, EPI_ISL_428320, EPI_ISL_428321, EPI_ISL_428322, EPI_ISL_428323, EPI_ISL_428324, EPI_ISL_428325, EPI_ISL_428326, EPI_ISL_428327, EPI_ISL_428328, EPI_ISL_428329, EPI_ISL_428330, EPI_ISL_428331, EPI_ISL_428332, EPI_ISL_428333, EPI_ISL_428334, EPI_ISL_428335, EPI_ISL_428336, EPI_ISL_428337, EPI_ISL_428338, EPI_ISL_428339, EPI_ISL_428340, EPI_ISL_428341, EPI_ISL_428342, EPI_ISL_428343, EPI_ISL_428344, EPI_ISL_428345 | | | |
| see above | University of Wisconsin-Madison AIDS Vaccine Research Laboratories | University of Wisconsin-Madison AIDS Vaccine Research Laboratories | Gage Moreno, Katarina Braun, et al. AIDS Vaccine Research Laboratories |
| EPI_ISL_428369, EPI_ISL_428370, EPI_ISL_428371, EPI_ISL_428372 | Yale Clinical Virology Laboratory | Grubaugh Lab - Yale School of Public Health | Joseph Fauver, Anderson Brito, Tara Alpert, Chantal Vogels, Ellen Foxman, Albert Ko, Marie Landry, Nathan Grubaugh |
| EPI_ISL_428373, EPI_ISL_428374, EPI_ISL_428375, EPI_ISL_428376, EPI_ISL_428377, EPI_ISL_428378, EPI_ISL_428379, EPI_ISL_428384, EPI_ISL_428385, EPI_ISL_428386, EPI_ISL_428387, EPI_ISL_428388, EPI_ISL_428391, EPI_ISL_428393, EPI_ISL_428394, EPI_ISL_428395, EPI_ISL_428396 | | | |
| see above | Yale COVID-19 Biorepository | Grubaugh Lab - Yale School of Public Health | Joseph Fauver, Tara Alpert, Anderson Brito, Anne Wyllie, Chantal Vogels, Mary Petrone, Chaney Kalinich, Isabel Ott, Arnau Casanovas, Catherine Muenker, Adam Moore, Alice Lu, Maria Tokuyama, Patrick Wong, Peiwen Lu, Saad Omer, Richard Martinello, Allison Nelson, Shelli Farhadian, Akiko Iwasaki, Charlese Dela Cruz, Albert Ko, Nathan Grubaugh |
| EPI_ISL_428399, EPI_ISL_428400 | Yale COVID-19 Biorepository | Grubaugh Lab - Yale School of Public Health | Joseph Fauver, Tara Alpert, Anderson Brito, Anne Wyllie, Chantal Vogels, Mary Petrone, Cole Jensen, Chaney Kalinich, Isabel Ott, Arnau Casanovas, Catherine Muenker, Adam Moore, Alice Lu, Maria Tokuyama, Patrick Wong, Peiwen Lu, Saad Omer, Richard Martinello, Allison Nelson, Shelli Farhadian, Akiko Iwasaki, Charlese Dela Cruz, Albert Ko, Nathan Grubaugh |
| EPI_ISL_428724, EPI_ISL_428725, EPI_ISL_428726, EPI_ISL_428727, EPI_ISL_428728, EPI_ISL_428729, EPI_ISL_428730, EPI_ISL_428731, EPI_ISL_428732 | University of Wisconsin-Madison AIDS Vaccine Research Laboratories | University of Wisconsin-Madison AIDS Vaccine Research Laboratories | Gage Moreno, Katarina Braun, et al. AIDS Vaccine Research Laboratories |
| EPI_ISL_428740 | Yale Clinical Virology Laboratory | Grubaugh Lab - Yale School of Public Health | Joseph Fauver, Anderson Brito, Tara Alpert, Chantal Vogels, Ellen Foxman, Albert Ko, Marie Landry, Nathan Grubaugh |
| EPI_ISL_428746, EPI_ISL_428747, EPI_ISL_428749, EPI_ISL_428750 | Yale COVID-19 Biorepository | Grubaugh Lab - Yale School of Public Health | Joseph Fauver, Tara Alpert, Anderson Brito, Anne Wyllie, Chantal Vogels, Mary Petrone, Cole Jensen, Chaney Kalinich, Isabel Ott, Arnau Casanovas, Catherine Muenker, Adam Moore, Alice Lu, Maria Tokuyama, Patrick Wong, Peiwen Lu, Saad Omer, Richard Martinello, Allison Nelson, Shelli Farhadian, Akiko Iwasaki, Charlese Dela Cruz, Albert Ko, Nathan Grubaugh |
| EPI_ISL_428757, EPI_ISL_428758, EPI_ISL_428759, EPI_ISL_428760, EPI_ISL_428761, EPI_ISL_428762, EPI_ISL_428763, EPI_ISL_428764, EPI_ISL_428765, EPI_ISL_428766, EPI_ISL_428767, EPI_ISL_428768, EPI_ISL_428769, EPI_ISL_428770, EPI_ISL_428771, EPI_ISL_428772, EPI_ISL_428773, EPI_ISL_428774, EPI_ISL_428775, EPI_ISL_428776, EPI_ISL_428777, EPI_ISL_428778, EPI_ISL_428779, EPI_ISL_428780, EPI_ISL_428781, EPI_ISL_428782, EPI_ISL_428783, EPI_ISL_428784, EPI_ISL_428785, EPI_ISL_428786, EPI_ISL_428787, EPI_ISL_428788, EPI_ISL_428789, EPI_ISL_428790, EPI_ISL_428791, EPI_ISL_428792, EPI_ISL_428793, EPI_ISL_428795, EPI_ISL_428796, EPI_ISL_428797, EPI_ISL_428798, EPI_ISL_428799, EPI_ISL_428800, EPI_ISL_428801, EPI_ISL_428802, EPI_ISL_428803, EPI_ISL_428804, EPI_ISL_428805 | | | |
| see above | NYU Langone Health | Departments of Pathology and Medicine, New York University School of Medicine | Maria Aguero-Rosenfeld, Brendan Belovarac, Margaret Black, Ludovic Boytard, John Cadley, Paolo Cotzia, John Chen, Dacia Dimartino, Xiaojun Feng, Tatyana Gindin, Emily Guzman, Adriana Heguy, Megan Hogan, Emily Huang, George Jour, Andrew Lytle, Christian Marier, Matthew T. Maurano, Mark J. Mulligan, Peter Meyn, Iman Osman, Jared Pinnell, Vanessa Raabe, Sitharam Ramaswami, Amy Rapkiewicz, Marie Samanovic-Golden, Antonio Serrano, Guomiao Shen, Matija Snuderl, Theodore Vougiouklakis, Nick Vulpescu, Gael Westby, Paul Zappile, Yutong Zhang |
| EPI_ISL_428935, EPI_ISL_428936 | University of Wisconsin-Madison AIDS Vaccine Research Laboratories | University of Wisconsin-Madison AIDS Vaccine Research Laboratories | Gage Moreno, Katarina Braun, et al. AIDS Vaccine Research Laboratories |
| EPI_ISL_428990, EPI_ISL_428991, EPI_ISL_428992, EPI_ISL_428993, EPI_ISL_428994, EPI_ISL_428995, EPI_ISL_428996, EPI_ISL_428997, EPI_ISL_428998, EPI_ISL_428999, EPI_ISL_429000, EPI_ISL_429001, EPI_ISL_429002, EPI_ISL_429003, EPI_ISL_429004, EPI_ISL_429005, EPI_ISL_429006, EPI_ISL_429007, EPI_ISL_429008, EPI_ISL_429009, EPI_ISL_429010, EPI_ISL_429011, EPI_ISL_429012, EPI_ISL_429013, EPI_ISL_429014, EPI_ISL_429015, EPI_ISL_429016, EPI_ISL_429017, EPI_ISL_429018, EPI_ISL_429019, EPI_ISL_429020, EPI_ISL_429021, EPI_ISL_429022, EPI_ISL_429023, EPI_ISL_429024, EPI_ISL_429025, EPI_ISL_429026, EPI_ISL_429027, EPI_ISL_429028, EPI_ISL_429029, EPI_ISL_429030, EPI_ISL_429031, EPI_ISL_429032, EPI_ISL_429033, EPI_ISL_429034, EPI_ISL_429035, EPI_ISL_429036, EPI_ISL_429037, EPI_ISL_429038, EPI_ISL_429039, EPI_ISL_429040, EPI_ISL_429041, EPI_ISL_429042, EPI_ISL_429043, EPI_ISL_429044, EPI_ISL_429045, EPI_ISL_429046, EPI_ISL_429047, EPI_ISL_429048, EPI_ISL_429049, EPI_ISL_429050, EPI_ISL_429051, EPI_ISL_429052, EPI_ISL_429053, EPI_ISL_429054, EPI_ISL_429055, EPI_ISL_429056, EPI_ISL_429057, EPI_ISL_429058, EPI_ISL_429059, EPI_ISL_429060, EPI_ISL_429061, EPI_ISL_429062, EPI_ISL_429063, EPI_ISL_429064, EPI_ISL_429065, EPI_ISL_429066, EPI_ISL_429067, EPI_ISL_429068, EPI_ISL_429069 | | | |
| see above | UCSF Clinical Microbiology Laboratory | Chan-Zuckerberg Biohub | CZB Cliahub Consortium |
| EPI_ISL_429597, EPI_ISL_429598, EPI_ISL_429599, EPI_ISL_429600, EPI_ISL_429601, EPI_ISL_429602, EPI_ISL_429603, EPI_ISL_429604, EPI_ISL_429605, EPI_ISL_429606, EPI_ISL_429607, EPI_ISL_429608, EPI_ISL_429609, EPI_ISL_429610, EPI_ISL_429611, EPI_ISL_429612, EPI_ISL_429613, EPI_ISL_429614, EPI_ISL_429615, EPI_ISL_429616, EPI_ISL_429617, EPI_ISL_429618, EPI_ISL_429619, EPI_ISL_429620, EPI_ISL_429621, EPI_ISL_429622, EPI_ISL_429623, EPI_ISL_429624, EPI_ISL_429625, EPI_ISL_429626, EPI_ISL_429627, EPI_ISL_429628, EPI_ISL_429629, EPI_ISL_429630, EPI_ISL_429631, EPI_ISL_429632, EPI_ISL_429633, EPI_ISL_429634, EPI_ISL_429635, EPI_ISL_429636, EPI_ISL_429637, EPI_ISL_429638, EPI_ISL_429639, EPI_ISL_429640, EPI_ISL_429641, EPI_ISL_429642, EPI_ISL_429643, EPI_ISL_429644, EPI_ISL_429645, EPI_ISL_429647, EPI_ISL_429648, EPI_ISL_429649, EPI_ISL_429650, EPI_ISL_429651, EPI_ISL_429652, EPI_ISL_429653, EPI_ISL_429654, EPI_ISL_429655, EPI_ISL_429656 | | | |
| see above | UW Virology Lab | UW Virology Lab | Pavitra Roychoudhury, Hong Xie, Keith Jerome, Alexander Greninger |
| EPI_ISL_429843 | Gundersen Molecular Diagnostics Laboratory | Kabara Cancer Research Institute | Craig S. Richmond, Paraic A. Kenny |
| EPI_ISL_429844 | Gundersen Molecular Diagnostics Laboratory | Kabara Cancer Research Institute | Craig S. Richmond; Paraic A. Kenny |

| | | | |
|---|---|---|---|
| EPI_ISL_429845, EPI_ISL_429846, EPI_ISL_429847, EPI_ISL_429848 | Gundersen Molecular Diagnostics Laboratory | Kabara Cancer Research Institute | Craig S. Richmond, Paraic A. Kenny |
| EPI_ISL_429877, EPI_ISL_429878 | Chiu Laboratory, University of California, San Francisco | Chiu Laboratory, University of California, San Francisco | Xianding Deng, Scot Federman, Wei Gu, and Charles Y. Chiu |
| EPI_ISL_429879, EPI_ISL_429880 | Santa Clara County Public Health Department | Chiu Laboratory, University of California, San Francisco | Xianding Deng, Scot Federman, Wei Gu, Elsa Villarino, Brandon Bonin, Debra A. Wadford, and Charles Y. Chiu |
| EPI_ISL_429881 | California Department of Public Health | Chiu Laboratory, University of California, San Francisco | Xianding Deng, Scot Federman, Wei Gu, and Charles Y. Chiu |
| EPI_ISL_429969, EPI_ISL_429970, EPI_ISL_429971, EPI_ISL_429972, EPI_ISL_429973, EPI_ISL_429974, EPI_ISL_429975, EPI_ISL_429976, EPI_ISL_429977, EPI_ISL_429978, EPI_ISL_429979, EPI_ISL_429980, EPI_ISL_429981, EPI_ISL_429982, EPI_ISL_429983, EPI_ISL_429984, EPI_ISL_429985, EPI_ISL_429986, EPI_ISL_429987, EPI_ISL_429988, EPI_ISL_429989 | | | |
| see above | Virginia DCLS | Virginia DCLS | Virginia DCLS |
| EPI_ISL_429990 | Rady's Childrens Hospital | Andersen lab at Scripps Research | SEARCH Alliance San Diego with Christina Clarke, Michelle Vanderpool, Teresa Mueller, Denise Malicki |
| EPI_ISL_429991 | Andersen lab at Scripps Research | Andersen lab at Scripps Research | SEARCH Alliance San Diego |
| EPI_ISL_430010 | OSU Wexner Medical Center | James Molecular Lab - OSUWMC | Huolin Tu, Preeti Pancholi, Matt Avenarius, Erica Vincent, Matt Hunt, Dan Jones |
| EPI_ISL_430016 | Andersen lab at Scripps Research | Andersen lab at Scripps Research | SEARCH Alliance San Diego |
| EPI_ISL_430019, EPI_ISL_430020, EPI_ISL_430021, EPI_ISL_430022, EPI_ISL_430023, EPI_ISL_430024, EPI_ISL_430025, EPI_ISL_430026, EPI_ISL_430027, EPI_ISL_430028, EPI_ISL_430029, EPI_ISL_430031, EPI_ISL_430032, EPI_ISL_430033, EPI_ISL_430034, EPI_ISL_430035, EPI_ISL_430036, EPI_ISL_430037, EPI_ISL_430038, EPI_ISL_430039, EPI_ISL_430041, EPI_ISL_430042, EPI_ISL_430043, EPI_ISL_430044, EPI_ISL_430045, EPI_ISL_430046, EPI_ISL_430047, EPI_ISL_430048, EPI_ISL_430050, EPI_ISL_430051, EPI_ISL_430052, EPI_ISL_430053, EPI_ISL_430054, EPI_ISL_430055, EPI_ISL_430056, EPI_ISL_430057, EPI_ISL_430058, EPI_ISL_430059, EPI_ISL_430060, EPI_ISL_430061, EPI_ISL_430062 | | | |
| see above | Utah Public Health Laboratory | Utah Public Health Laboratory | Erin Young, Kelly Oakeson |
| EPI_ISL_430113, EPI_ISL_430114, EPI_ISL_430115, EPI_ISL_430116, EPI_ISL_430117, EPI_ISL_430118, EPI_ISL_430119, EPI_ISL_430121, EPI_ISL_430122, EPI_ISL_430123, EPI_ISL_430124, EPI_ISL_430125, EPI_ISL_430126, EPI_ISL_430127, EPI_ISL_430128, EPI_ISL_430129, EPI_ISL_430130, EPI_ISL_430131, EPI_ISL_430132, EPI_ISL_430133, EPI_ISL_430134, EPI_ISL_430135, EPI_ISL_430136, EPI_ISL_430137, EPI_ISL_430139, EPI_ISL_430140, EPI_ISL_430141, EPI_ISL_430142, EPI_ISL_430143, EPI_ISL_430144, EPI_ISL_430146, EPI_ISL_430147, EPI_ISL_430148, EPI_ISL_430149, EPI_ISL_430150, EPI_ISL_430151, EPI_ISL_430152, EPI_ISL_430153, EPI_ISL_430154, EPI_ISL_430155, EPI_ISL_430156 | | | |
| see above | Seattle Flu Study | Seattle Flu Study | Chu et al |
| EPI_ISL_430160, EPI_ISL_430161, EPI_ISL_430162, EPI_ISL_430163, EPI_ISL_430164, EPI_ISL_430165, EPI_ISL_430166, EPI_ISL_430167, EPI_ISL_430168, EPI_ISL_430169, EPI_ISL_430170, EPI_ISL_430171, EPI_ISL_430172, EPI_ISL_430173, EPI_ISL_430174, EPI_ISL_430175, EPI_ISL_430176, EPI_ISL_430177, EPI_ISL_430178, EPI_ISL_430179, EPI_ISL_430180, EPI_ISL_430181, EPI_ISL_430182, EPI_ISL_430183, EPI_ISL_430184, EPI_ISL_430185, EPI_ISL_430186, EPI_ISL_430187, EPI_ISL_430188, EPI_ISL_430189, EPI_ISL_430190, EPI_ISL_430191, EPI_ISL_430192, EPI_ISL_430193, EPI_ISL_430194, EPI_ISL_430196, EPI_ISL_430197, EPI_ISL_430198, EPI_ISL_430199, EPI_ISL_430203, EPI_ISL_430204, EPI_ISL_430205, EPI_ISL_430206, EPI_ISL_430207, EPI_ISL_430208, EPI_ISL_430209, EPI_ISL_430210, EPI_ISL_430211, EPI_ISL_430212, EPI_ISL_430213, EPI_ISL_430214, EPI_ISL_430215, EPI_ISL_430216, EPI_ISL_430217, EPI_ISL_430218, EPI_ISL_430219, EPI_ISL_430220, EPI_ISL_430221, EPI_ISL_430222, EPI_ISL_430223, EPI_ISL_430224, EPI_ISL_430225, EPI_ISL_430226, EPI_ISL_430227, EPI_ISL_430228, EPI_ISL_430229, EPI_ISL_430230, EPI_ISL_430231, EPI_ISL_430232, EPI_ISL_430233, EPI_ISL_430234, EPI_ISL_430235, EPI_ISL_430236, EPI_ISL_430237, EPI_ISL_430238, EPI_ISL_430239, EPI_ISL_430240, EPI_ISL_430241, EPI_ISL_430242, EPI_ISL_430243, EPI_ISL_430244, EPI_ISL_430245, EPI_ISL_430246, EPI_ISL_430247, EPI_ISL_430248, EPI_ISL_430249, EPI_ISL_430250, EPI_ISL_430251, EPI_ISL_430252, EPI_ISL_430253, EPI_ISL_430254, EPI_ISL_430255, EPI_ISL_430256, EPI_ISL_430257, EPI_ISL_430258, EPI_ISL_430259, EPI_ISL_430261, EPI_ISL_430262, EPI_ISL_430263, EPI_ISL_430264, EPI_ISL_430265, EPI_ISL_430266, EPI_ISL_430267, EPI_ISL_430268, EPI_ISL_430269, EPI_ISL_430270, EPI_ISL_430271, EPI_ISL_430272, EPI_ISL_430273, EPI_ISL_430274, EPI_ISL_430275, EPI_ISL_430276, EPI_ISL_430277, EPI_ISL_430278, EPI_ISL_430279, EPI_ISL_430280, EPI_ISL_430281, EPI_ISL_430282, EPI_ISL_430283, EPI_ISL_430284, EPI_ISL_430285, EPI_ISL_430286, EPI_ISL_430287, EPI_ISL_430288, EPI_ISL_430289, EPI_ISL_430290, EPI_ISL_430291, EPI_ISL_430292, EPI_ISL_430293, EPI_ISL_430294, EPI_ISL_430295 | | | |
| see above | Washington State Department of Health | Seattle Flu Study | Chu et al |
| EPI_ISL_430323, EPI_ISL_430324, EPI_ISL_430325, EPI_ISL_430326, EPI_ISL_430327, EPI_ISL_430328, EPI_ISL_430329, EPI_ISL_430330, EPI_ISL_430331, EPI_ISL_430332, EPI_ISL_430333, EPI_ISL_430334, EPI_ISL_430335, EPI_ISL_430336, EPI_ISL_430337, EPI_ISL_430338, EPI_ISL_430339, EPI_ISL_430340, EPI_ISL_430341, EPI_ISL_430342, EPI_ISL_430343, EPI_ISL_430344, EPI_ISL_430345, EPI_ISL_430346, EPI_ISL_430347, EPI_ISL_430348, EPI_ISL_430349, EPI_ISL_430350, EPI_ISL_430351, EPI_ISL_430352, EPI_ISL_430353, EPI_ISL_430354, EPI_ISL_430355, EPI_ISL_430356, EPI_ISL_430357, EPI_ISL_430358, EPI_ISL_430359, EPI_ISL_430360, EPI_ISL_430361, EPI_ISL_430362, EPI_ISL_430363, EPI_ISL_430364, EPI_ISL_430365, EPI_ISL_430366, EPI_ISL_430367, EPI_ISL_430368, EPI_ISL_430369, EPI_ISL_430370, EPI_ISL_430371, EPI_ISL_430372, EPI_ISL_430373, EPI_ISL_430374, EPI_ISL_430375, EPI_ISL_430376, EPI_ISL_430377, EPI_ISL_430378, EPI_ISL_430379, EPI_ISL_430380, EPI_ISL_430381, EPI_ISL_430382, EPI_ISL_430383, EPI_ISL_430384, EPI_ISL_430385, EPI_ISL_430386, EPI_ISL_430387, EPI_ISL_430388, EPI_ISL_430389, EPI_ISL_430390, EPI_ISL_430391, EPI_ISL_430392, EPI_ISL_430393, EPI_ISL_430394, EPI_ISL_430395, EPI_ISL_430396, EPI_ISL_430397, EPI_ISL_430398, EPI_ISL_430399, EPI_ISL_430400, EPI_ISL_430401, EPI_ISL_430402, EPI_ISL_430403, EPI_ISL_430404, EPI_ISL_430405, EPI_ISL_430406, EPI_ISL_430407, EPI_ISL_430408, EPI_ISL_430409, EPI_ISL_430410, EPI_ISL_430411, EPI_ISL_430412, EPI_ISL_430413, EPI_ISL_430414, EPI_ISL_430415, EPI_ISL_430416, EPI_ISL_430417, EPI_ISL_430418, EPI_ISL_430419, EPI_ISL_430420, EPI_ISL_430421, EPI_ISL_430422, EPI_ISL_430423, EPI_ISL_430424, EPI_ISL_430425, EPI_ISL_430426, EPI_ISL_430427, EPI_ISL_430428, EPI_ISL_430429, EPI_ISL_430430, EPI_ISL_430431, EPI_ISL_430432, EPI_ISL_430433, EPI_ISL_430434 | | | |
| see above | NYU Langone Health | Departments of Pathology and Medicine, New York University School of Medicine | Maria Aguero-Rosenfeld, Brendan Belovarac, Margaret Black, Ludovic Boytard, John Cadley, Paolo Cotzia, John Chen, Dacia Dimartino, Xiaojun Feng, Tatyana Gindin, Emily Guzman, Adriana Heguy, Megan Hogan, Emily Huang, George Jour, Lawrence H. Lin, Raven Luther, Andrew Lytle, Christian Marier, Matthew T. Maurano, Mark J. Mulligan, Peter Meyn, Raquel Ordonez Ciriza, Iman Osman, Jared Pinnell, Vanessa Raabe, Sitharam Ramaswami, Amy Rapkiewicz, Andre M. Ribeiro-dos-Santos, Marie Samanovic-Golden, Antonio Serrano, Guomiao Shen, Matija Snuderl, Theodore Vougiouklakis, Nick Vulpescu, Gael Westby, Paul Zappile, Yutong Zhang |
| EPI_ISL_430791, EPI_ISL_430792 | UCSF Clinical Microbiology Laboratory | Chan-Zuckerberg Biohub | CZB Cliahub Consortium |
| EPI_ISL_430867, EPI_ISL_430868, EPI_ISL_430869, EPI_ISL_430870, EPI_ISL_430871, EPI_ISL_430872, EPI_ISL_430873, EPI_ISL_430874, EPI_ISL_430875, EPI_ISL_430876, EPI_ISL_430877, EPI_ISL_430878, EPI_ISL_430879, EPI_ISL_430880, EPI_ISL_430881, EPI_ISL_430882, EPI_ISL_430883, EPI_ISL_430884, EPI_ISL_430885, EPI_ISL_430886, EPI_ISL_430887, EPI_ISL_430888, EPI_ISL_430889, EPI_ISL_430890, EPI_ISL_430891, EPI_ISL_430892, EPI_ISL_430893, EPI_ISL_430894, EPI_ISL_430895, EPI_ISL_430897, EPI_ISL_430898, EPI_ISL_430899, EPI_ISL_430900, EPI_ISL_430901, EPI_ISL_430902, EPI_ISL_430903, EPI_ISL_430904, EPI_ISL_430905, EPI_ISL_430906, EPI_ISL_430907, EPI_ISL_430908, EPI_ISL_430909, EPI_ISL_430910, EPI_ISL_430911, EPI_ISL_430912, EPI_ISL_430913, EPI_ISL_430914, EPI_ISL_430915, EPI_ISL_430916, EPI_ISL_430917, EPI_ISL_430918, EPI_ISL_430919, EPI_ISL_430920, EPI_ISL_430921, EPI_ISL_430922, EPI_ISL_430923, EPI_ISL_430924, EPI_ISL_430925, EPI_ISL_430926, EPI_ISL_430927, EPI_ISL_430928, EPI_ISL_430929, EPI_ISL_430930, EPI_ISL_430931, EPI_ISL_430932, EPI_ISL_430933, EPI_ISL_430934, EPI_ISL_430935, EPI_ISL_430936, EPI_ISL_430937, EPI_ISL_430938, EPI_ISL_430939, EPI_ISL_430940, EPI_ISL_430941, EPI_ISL_430942, EPI_ISL_430943, EPI_ISL_430944, EPI_ISL_430945, EPI_ISL_430946, EPI_ISL_430947, EPI_ISL_430948, EPI_ISL_430949, EPI_ISL_430950, EPI_ISL_430951, EPI_ISL_430952, EPI_ISL_430953, EPI_ISL_430954, EPI_ISL_430955, EPI_ISL_430956, EPI_ISL_430957, EPI_ISL_430958, EPI_ISL_430959, EPI_ISL_430960, EPI_ISL_430961, EPI_ISL_430962, EPI_ISL_430963, EPI_ISL_430964, EPI_ISL_430965, EPI_ISL_430966, EPI_ISL_430967, EPI_ISL_430968, EPI_ISL_430969, EPI_ISL_430970, EPI_ISL_430971, EPI_ISL_430972, EPI_ISL_430973, EPI_ISL_430974, EPI_ISL_430975, EPI_ISL_430976, EPI_ISL_430977, EPI_ISL_430978, EPI_ISL_430979, EPI_ISL_430980 | | | |
| see above | UW Virology Lab | UW Virology Lab | Pavitra Roychoudhury, Hong Xie, Keith Jerome, Alexander Greninger |
| EPI_ISL_431013 | Alaska State Virology Laboratory | Alaska State Virology Laboratory | Jack Chen |
| EPI_ISL_431014, EPI_ISL_431015, EPI_ISL_431016, EPI_ISL_431017, EPI_ISL_431018, EPI_ISL_431019 | Alaska State Virology Laboratory | Alaska State Virology Laboratory | Jack Chen, Ph.D. |
| EPI_ISL_431080, EPI_ISL_431081, EPI_ISL_431082, EPI_ISL_431083, EPI_ISL_431084, EPI_ISL_431085, EPI_ISL_431086, EPI_ISL_431089, EPI_ISL_431092, EPI_ISL_431096 | Yale COVID-19 Biorepository | Grubaugh Lab - Yale School of Public Health | Joseph Fauver, Tara Alpert, Anderson Brito, Anne Wyllie, Chantal Vogels, Mary Petrone, Cole Jensen, Chaney Kalinich, Isabel Ott, Arnau Casanovas, Catherine Muenker, Adam Moore, Alice Lu, Maria Tokuyama, Patrick Wong, Peiwen Lu, Saad Omer, Richard Martinello, Allison Nelson, Shelli Farhadian, Akiko Iwasaki, Charlese Dela Cruz, Albert Ko, Nathan Grubaugh |
| EPI_ISL_434063, EPI_ISL_434064, EPI_ISL_434065, EPI_ISL_434066, EPI_ISL_434068, EPI_ISL_434069, EPI_ISL_434070, EPI_ISL_434071, EPI_ISL_434072, EPI_ISL_434073, EPI_ISL_434074, EPI_ISL_434075, EPI_ISL_434076, EPI_ISL_434077, EPI_ISL_434078, EPI_ISL_434079, EPI_ISL_434080, EPI_ISL_434081, EPI_ISL_434082, EPI_ISL_434083, EPI_ISL_434084, EPI_ISL_434085, EPI_ISL_434086, EPI_ISL_434087, EPI_ISL_434088, EPI_ISL_434089, EPI_ISL_434090, EPI_ISL_434091, EPI_ISL_434092, EPI_ISL_434093, EPI_ISL_434094, EPI_ISL_434095, EPI_ISL_434096, EPI_ISL_434097, EPI_ISL_434098, EPI_ISL_434099, EPI_ISL_434100, EPI_ISL_434101, EPI_ISL_434102, EPI_ISL_434103, EPI_ISL_434104, EPI_ISL_434105, EPI_ISL_434106, EPI_ISL_434107, EPI_ISL_434108, EPI_ISL_434109, EPI_ISL_434110, EPI_ISL_434111, EPI_ISL_434112, EPI_ISL_434113, EPI_ISL_434114, EPI_ISL_434115, EPI_ISL_434116, EPI_ISL_434117, EPI_ISL_434118, EPI_ISL_434119, EPI_ISL_434120, EPI_ISL_434121, EPI_ISL_434122, EPI_ISL_434123, EPI_ISL_434124, EPI_ISL_434128, EPI_ISL_434129, EPI_ISL_434130, EPI_ISL_434131, EPI_ISL_434132, EPI_ISL_434133, EPI_ISL_434134, EPI_ISL_434135, EPI_ISL_434136, EPI_ISL_434137, EPI_ISL_434138, EPI_ISL_434139, EPI_ISL_434140, EPI_ISL_434141, EPI_ISL_434142, EPI_ISL_434143, EPI_ISL_434144, EPI_ISL_434145, EPI_ISL_434146, EPI_ISL_434147, EPI_ISL_434148, EPI_ISL_434149, EPI_ISL_434150, EPI_ISL_434151, EPI_ISL_434152, EPI_ISL_434153, EPI_ISL_434154, EPI_ISL_434155, EPI_ISL_434156, EPI_ISL_434157, EPI_ISL_434158, EPI_ISL_434159, EPI_ISL_434160, EPI_ISL_434161, EPI_ISL_434162, EPI_ISL_434163, EPI_ISL_434164, EPI_ISL_434165, EPI_ISL_434166, EPI_ISL_434167, EPI_ISL_434168, EPI_ISL_434169, EPI_ISL_434170, EPI_ISL_434171, EPI_ISL_434172, EPI_ISL_434173, EPI_ISL_434174, EPI_ISL_434175, EPI_ISL_434176, EPI_ISL_434177, EPI_ISL_434178, EPI_ISL_434179, EPI_ISL_434180, EPI_ISL_434182, EPI_ISL_434183, EPI_ISL_434184, EPI_ISL_434185, EPI_ISL_434186, EPI_ISL_434187, EPI_ISL_434188, EPI_ISL_434189, EPI_ISL_434190, EPI_ISL_434191, EPI_ISL_434192, EPI_ISL_434193, EPI_ISL_434194, EPI_ISL_434196, EPI_ISL_434197, EPI_ISL_434198, EPI_ISL_434199, EPI_ISL_434200, EPI_ISL_434201, EPI_ISL_434202, EPI_ISL_434204, EPI_ISL_434205, EPI_ISL_434206, EPI_ISL_434207, EPI_ISL_434208, EPI_ISL_434209, EPI_ISL_434210, EPI_ISL_434211, EPI_ISL_434212, EPI_ISL_434213, EPI_ISL_434214, EPI_ISL_434215, EPI_ISL_434216, EPI_ISL_434217, EPI_ISL_434218, EPI_ISL_434219, EPI_ISL_434220, EPI_ISL_434221, EPI_ISL_434222, EPI_ISL_434223, EPI_ISL_434224, EPI_ISL_434225, EPI_ISL_434226, EPI_ISL_434227, EPI_ISL_434228, EPI_ISL_434229, EPI_ISL_434230, EPI_ISL_434231, EPI_ISL_434232, EPI_ISL_434233, EPI_ISL_434234, EPI_ISL_434235, EPI_ISL_434236, EPI_ISL_434237, EPI_ISL_434238, EPI_ISL_434239, EPI_ISL_434240, EPI_ISL_434241, EPI_ISL_434242, EPI_ISL_434243, EPI_ISL_434244, EPI_ISL_434245, EPI_ISL_434246, EPI_ISL_434247, EPI_ISL_434248, EPI_ISL_434249, EPI_ISL_434250, EPI_ISL_434251, EPI_ISL_434252, EPI_ISL_434253, EPI_ISL_434254, EPI_ISL_434255, EPI_ISL_434256, EPI_ISL_434257, EPI_ISL_434258, EPI_ISL_434259, EPI_ISL_434260, EPI_ISL_434262, EPI_ISL_434263, EPI_ISL_434264, EPI_ISL_434265, EPI_ISL_434266, EPI_ISL_434268, EPI_ISL_434269, EPI_ISL_434270, EPI_ISL_434271, EPI_ISL_434272, EPI_ISL_434273, EPI_ISL_434274, EPI_ISL_434275, EPI_ISL_434276, EPI_ISL_434277, EPI_ISL_434279, EPI_ISL_434280, EPI_ISL_434281, EPI_ISL_434282, EPI_ISL_434283, EPI_ISL_434284, EPI_ISL_434285, EPI_ISL_434286, EPI_ISL_434287, EPI_ISL_434288, EPI_ISL_434289, EPI_ISL_434290, EPI_ISL_434291, EPI_ISL_434293, EPI_ISL_434294, EPI_ISL_434296, EPI_ISL_434298, EPI_ISL_434299, EPI_ISL_434300, EPI_ISL_434301, EPI_ISL_434302, EPI_ISL_434303, EPI_ISL_434305, EPI_ISL_434306, EPI_ISL_434307, EPI_ISL_434308, EPI_ISL_434309, EPI_ISL_434310, EPI_ISL_434311, EPI_ISL_434313, EPI_ISL_434314, EPI_ISL_434315, EPI_ISL_434316, EPI_ISL_434317, EPI_ISL_434319, EPI_ISL_434320, EPI_ISL_434321, EPI_ISL_434322, EPI_ISL_434323, EPI_ISL_434324, EPI_ISL_434325, EPI_ISL_434327, EPI_ISL_434328, EPI_ISL_434329, EPI_ISL_434330, EPI_ISL_434331, EPI_ISL_434332, EPI_ISL_434333, EPI_ISL_434334, EPI_ISL_434335, EPI_ISL_434336, EPI_ISL_434337, EPI_ISL_434338, EPI_ISL_434339, EPI_ISL_434341, EPI_ISL_434342, EPI_ISL_434343, EPI_ISL_434344, EPI_ISL_434345, EPI_ISL_434346 | | | |
| see above | Washington State Department of Health | Seattle Flu Study | Chu et al |
| EPI_ISL_434517, EPI_ISL_434518, EPI_ISL_434519, EPI_ISL_434520, EPI_ISL_434521, EPI_ISL_434522, EPI_ISL_434523, EPI_ISL_434524, EPI_ISL_434525, EPI_ISL_434526, EPI_ISL_434527, EPI_ISL_434528, EPI_ISL_434529, EPI_ISL_434530, EPI_ISL_434531, EPI_ISL_434532 | | | |
| see above | Robert Garry lab | Andersen lab at Scripps Research | Allison Smither, Gilberto Sabino-Santos, Patricia Snarski, Lilia Melnik, Antoinette Bell, Kaylynn Genemaras, Arnaud Drouin, Dahlene Fusco, Robert Garry with SEARCH Alliance San Diego |
| EPI_ISL_434541, EPI_ISL_434542, EPI_ISL_434543, EPI_ISL_434545, EPI_ISL_434546, EPI_ISL_434547, EPI_ISL_434548, EPI_ISL_434549, EPI_ISL_434550, EPI_ISL_434551, EPI_ISL_434552, EPI_ISL_434553 | | | |
| see above | Puerto Rico Department of Health | Centers for Disease Control and Prevention, Dengue Branch | Gilberto A. Santiago, Glenda Gonzalez, Betzabel Flores, Keyla Charriez, Fabiola Cruz, Chaney Kalinich, Joseph Fauver, Jessica I. Falcon, Nathan Grubaugh, Jorge L. Munoz-Jordan |
| EPI_ISL_434586, EPI_ISL_434587, EPI_ISL_434588 | Johns Hopkins Hospital Department of Pathology | Johns Hopkins Hospital Department of Pathology | Peter M. Thielen, Thomas Mehoke, Shirlee Wohl, Srividya Ramakrishnan, Oluwaseun Nwulia-Falade, Amanda Ernlund, Melanie Kirsche, Paul Morris, Norah Sadowski, Nídiá Trovao, Victoria Gniazdowski, Michael Schatz, Stuart C. Ray, Winston Timp, Heba Mostafa |
| EPI_ISL_434590, EPI_ISL_434591, EPI_ISL_434592, EPI_ISL_434593, EPI_ISL_434594, EPI_ISL_434595, EPI_ISL_434596, EPI_ISL_434597, EPI_ISL_434598, EPI_ISL_434599, EPI_ISL_434600, EPI_ISL_434601, EPI_ISL_434602, EPI_ISL_434603, EPI_ISL_434604, EPI_ISL_434605, EPI_ISL_434606 | | | |
| see above | Virginia DCLS | Virginia DCLS | Virginia DCLS |
| EPI_ISL_434607, EPI_ISL_434608, EPI_ISL_434609, EPI_ISL_434610, EPI_ISL_434611, EPI_ISL_434612, EPI_ISL_434613, EPI_ISL_434614, EPI_ISL_434615 | University of Wisconsin-Madison AIDS Vaccine Research Laboratories | University of Wisconsin-Madison AIDS Vaccine Research Laboratories | Gage Moreno, Katarina Braun, et al. AIDS Vaccine Research Laboratories |
| EPI_ISL_434636 | Lednicky Laboratory, Emerging | Lednicky Laboratory at Emerging Pathogens | Elbadry,M.A., Subramaniam,K., Waltzek,T.B., Stephenson,C.J.,Gibson,J.C., Alam,M., Morris,J.G. Jr. and Lednicky,J.A. |

| | | | |
|---|---|---|---|
| | Pathogens Institute, University of Florida | Institute, University of Florida | |
| EPI_ISL_434637 | Lednicky Laboratory, Emerging Pathogens Institute, University of Florida. | Lednicky Laboratory, Emerging Pathogens Institute, University of Florida. | Elbadry,M.A.; Subramaniam,K.; Waltzek,T.B.; Gibson,J.C.; Stephenson,C.J.; Morris,J.G. Jr. and Lednicky,J.A. |
| EPI_ISL_434638, EPI_ISL_434639, EPI_ISL_434640 | Johns Hopkins Hospital Department of Pathology | Johns Hopkins Hospital Department of Pathology | Peter M. Thielen, Thomas Mehoke, Shirlee Wohl, Srividya Ramakrishnan, Oluwaseun Nwulia-Falade, Amanda Ernlund, Melanie Kirsche, Paul Morris, Norah Sadowski, Nídiá Trovao, Victoria Gniazdowski, Michael Schatz, Stuart C. Ray, Winston Timp, Heba Mostafa |
| EPI_ISL_434682, EPI_ISL_434683, EPI_ISL_434684, EPI_ISL_434685, EPI_ISL_434686, EPI_ISL_434687, EPI_ISL_434688, EPI_ISL_434689, EPI_ISL_434690, EPI_ISL_434691 | Johns Hopkins Hospital Department of Pathology | Johns Hopkins Hospital Department of Pathology | Peter M. Thielen, Thomas Mehoke, Shirlee Wohl, Srividya Ramakrishnan, Melanie Kirsche, Amanda Ernlund, Oluwaseun Falade-Nwulia, Timothy Gilpatrick, Paul Morris, Norah Sadowski, Nídiá Trovao, Victoria Gniazdowski, Michael Schatz, Stuart C. Ray, Winston Timp, Heba Mostafa |
| EPI_ISL_434712, EPI_ISL_434713, EPI_ISL_434714, EPI_ISL_434715, EPI_ISL_434716, EPI_ISL_434717, EPI_ISL_434718, EPI_ISL_434719, EPI_ISL_434720, EPI_ISL_434721, EPI_ISL_434722, EPI_ISL_434723, EPI_ISL_434724, EPI_ISL_434725, EPI_ISL_434726, EPI_ISL_434727, EPI_ISL_434728, EPI_ISL_434729, EPI_ISL_434730, EPI_ISL_434731, EPI_ISL_434732, EPI_ISL_434733, EPI_ISL_434734, EPI_ISL_434735, EPI_ISL_434736, EPI_ISL_434737, EPI_ISL_434738, EPI_ISL_434739, EPI_ISL_434740, EPI_ISL_434742, EPI_ISL_434743, EPI_ISL_434744, EPI_ISL_434745, EPI_ISL_434746, EPI_ISL_434747, EPI_ISL_434748, EPI_ISL_434749, EPI_ISL_434750, EPI_ISL_434751, EPI_ISL_434752, EPI_ISL_434753, EPI_ISL_434754, EPI_ISL_434755, EPI_ISL_434756, EPI_ISL_434757, EPI_ISL_434758, EPI_ISL_434759, EPI_ISL_434760, EPI_ISL_434761, EPI_ISL_434762, EPI_ISL_434764, EPI_ISL_434766, EPI_ISL_434768, EPI_ISL_434769, EPI_ISL_434770, EPI_ISL_434772, EPI_ISL_434775, EPI_ISL_434776, EPI_ISL_434777, EPI_ISL_434780, EPI_ISL_434781, EPI_ISL_434782, EPI_ISL_434783, EPI_ISL_434784, EPI_ISL_434785, EPI_ISL_434786, EPI_ISL_434787, EPI_ISL_434788, EPI_ISL_434789, EPI_ISL_434790, EPI_ISL_434791, EPI_ISL_434792, EPI_ISL_434793, EPI_ISL_434794, EPI_ISL_434795, EPI_ISL_434796, EPI_ISL_434797, EPI_ISL_434798, EPI_ISL_434799, EPI_ISL_434800, EPI_ISL_434801, EPI_ISL_434802, EPI_ISL_434803, EPI_ISL_434804, EPI_ISL_434805, EPI_ISL_434806, EPI_ISL_434807, EPI_ISL_434808, EPI_ISL_434809, EPI_ISL_434810, EPI_ISL_434811, EPI_ISL_434812, EPI_ISL_434813, EPI_ISL_434814, EPI_ISL_434815, EPI_ISL_434816, EPI_ISL_434817, EPI_ISL_434818, EPI_ISL_434819, EPI_ISL_434820, EPI_ISL_434821, EPI_ISL_434822, EPI_ISL_434823, EPI_ISL_434824, EPI_ISL_434825, EPI_ISL_434826, EPI_ISL_434827, EPI_ISL_434828, EPI_ISL_434829, EPI_ISL_434830, EPI_ISL_434831, EPI_ISL_434832, EPI_ISL_434833, EPI_ISL_434834, EPI_ISL_434835, EPI_ISL_434836, EPI_ISL_434837, EPI_ISL_434838, EPI_ISL_434839, EPI_ISL_434840, EPI_ISL_434841, EPI_ISL_434842, EPI_ISL_434843, EPI_ISL_434844, EPI_ISL_434845, EPI_ISL_434846, EPI_ISL_434847, EPI_ISL_434848, EPI_ISL_434849, EPI_ISL_434850, EPI_ISL_434851, EPI_ISL_434852, EPI_ISL_434853, EPI_ISL_434854, EPI_ISL_434855, EPI_ISL_434856, EPI_ISL_434857, EPI_ISL_434858, EPI_ISL_434859, EPI_ISL_434860, EPI_ISL_434861, EPI_ISL_434862, EPI_ISL_434863, EPI_ISL_434864, EPI_ISL_434865, EPI_ISL_434866, EPI_ISL_434867, EPI_ISL_434868, EPI_ISL_434869, EPI_ISL_434870, EPI_ISL_434871, EPI_ISL_434872, EPI_ISL_434873, EPI_ISL_434874, EPI_ISL_434875, EPI_ISL_434876, EPI_ISL_434877, EPI_ISL_434878, EPI_ISL_434879, EPI_ISL_434880, EPI_ISL_434881, EPI_ISL_434882, EPI_ISL_434883, EPI_ISL_434884, EPI_ISL_434885, EPI_ISL_434886, EPI_ISL_434887, EPI_ISL_434888, EPI_ISL_434889, EPI_ISL_434890, EPI_ISL_434891, EPI_ISL_434892, EPI_ISL_434893, EPI_ISL_434894, EPI_ISL_434895, EPI_ISL_434896, EPI_ISL_434897, EPI_ISL_434898, EPI_ISL_434899, EPI_ISL_434900, EPI_ISL_434901, EPI_ISL_434902, EPI_ISL_434903, EPI_ISL_434904, EPI_ISL_434905, EPI_ISL_434906, EPI_ISL_434907, EPI_ISL_434908, EPI_ISL_434909, EPI_ISL_434910, EPI_ISL_434911, EPI_ISL_434912, EPI_ISL_434913, EPI_ISL_434914, EPI_ISL_434915, EPI_ISL_434916, EPI_ISL_434917, EPI_ISL_434918, EPI_ISL_434919, EPI_ISL_434920, EPI_ISL_434921, EPI_ISL_434922, EPI_ISL_434923, EPI_ISL_434924, EPI_ISL_434925, EPI_ISL_434926, EPI_ISL_434927, EPI_ISL_434928, EPI_ISL_434929, EPI_ISL_434930, EPI_ISL_434931, EPI_ISL_434932, EPI_ISL_434933, EPI_ISL_434934, EPI_ISL_434935, EPI_ISL_434936, EPI_ISL_434937, EPI_ISL_434938, EPI_ISL_434939, EPI_ISL_434940, EPI_ISL_434941, EPI_ISL_434942, EPI_ISL_434943, EPI_ISL_434944, EPI_ISL_434945, EPI_ISL_434946, EPI_ISL_434947, EPI_ISL_434948, EPI_ISL_434949, EPI_ISL_434950, EPI_ISL_434951, EPI_ISL_434952, EPI_ISL_434953, EPI_ISL_434954, EPI_ISL_434955, EPI_ISL_434956, EPI_ISL_434957, EPI_ISL_434958, EPI_ISL_434959, EPI_ISL_434960, EPI_ISL_434961, EPI_ISL_434962, EPI_ISL_434963, EPI_ISL_434964, EPI_ISL_434965, EPI_ISL_434966, EPI_ISL_434967, EPI_ISL_434968, EPI_ISL_434969, EPI_ISL_434970, EPI_ISL_434971, EPI_ISL_434972, EPI_ISL_434973, EPI_ISL_434974, EPI_ISL_434975, EPI_ISL_434976, EPI_ISL_434977, EPI_ISL_434978, EPI_ISL_434979, EPI_ISL_434980, EPI_ISL_434981, EPI_ISL_434982, EPI_ISL_434983, EPI_ISL_434984, EPI_ISL_434985, EPI_ISL_434986, EPI_ISL_434987, EPI_ISL_434988, EPI_ISL_434989, EPI_ISL_434990, EPI_ISL_434991, EPI_ISL_434992, EPI_ISL_434993, EPI_ISL_434994, EPI_ISL_434995, EPI_ISL_434996, EPI_ISL_434997, EPI_ISL_434998, EPI_ISL_434999, EPI_ISL_435000, EPI_ISL_435001, EPI_ISL_435002, EPI_ISL_435003, EPI_ISL_435004, EPI_ISL_435005, EPI_ISL_435006, EPI_ISL_435007, EPI_ISL_435008, EPI_ISL_435009, EPI_ISL_435010, EPI_ISL_435011, EPI_ISL_435012, EPI_ISL_435013, EPI_ISL_435014, EPI_ISL_435015, EPI_ISL_435016, EPI_ISL_435017, EPI_ISL_435018, EPI_ISL_435019, EPI_ISL_435020, EPI_ISL_435021, EPI_ISL_435022, EPI_ISL_435023, EPI_ISL_435024, EPI_ISL_435025, EPI_ISL_435026, EPI_ISL_435027, EPI_ISL_435028, EPI_ISL_435029, EPI_ISL_435030, EPI_ISL_435031 | | | |
| see above | Houston Methodist Hospital | Houston Methodist Hospital | S. Wesley Long, Randall J. Olsen, Paul A. Christensen, David W. Bernard, James J. Davis, Maulik Shukla, Marcus Nguyen, Matthew Ojeda Saavedra, Concepcion C. Cantu, Prasanti Yerramilli, Layne Pruitt, Sishir Subedi, Heather Hendrickson, Ghazaleh Eskandari, Muthiah Kumaraswami, Jason S. McLellan, Hakon Jonsson, Kari Stefansson, and James M. Musser |
| EPI_ISL_435034 | LSUHS Emerging Viral Threat Laboratory | Microbial Genome Sequencing Center | Jeremy P. Kamil, John A. Vanchiere, Rona S. Scott, Camille F. Abshire, Abida Siddiqa, Byeong-Jae Lee, Chan-ki Min, Md Maksudul Alam, Monica Gestal-Cartele, Edna Ondari, Adam Greer, Malgorzata Bienkowska-Haba, Katarzyna Zwolinska, Michelle M. Arnold, Jason M. Bodily, Andrew D. Yurochko, Paul M. Weinberger, Christopher G. Kevil, Martin J. Sapp, Daniel J. Snyder, Vaughn S. Cooper |
| EPI_ISL_435035, EPI_ISL_435036, EPI_ISL_435037, EPI_ISL_435038, EPI_ISL_435039, EPI_ISL_435040, EPI_ISL_435041, EPI_ISL_435042, EPI_ISL_435043, EPI_ISL_435044 | LSUHS Emerging Viral Threat Laboratory | Microbial Genome Sequencing Center | Jeremy P. Kamil, John A. Vanchiere, Rona S. Scott, Camille F. Abshire, Abida Siddiqa, Byeong-Jae Lee, Chan-ki Min, Md Maksudul Alam, Monica Gestal-Cartele, Edna Ondari, Adam Greer, Malgorzata Bienkowska-Haba, Katarzyna Zwolinska, Jason M. Bodily, Andrew D. Yurochko, Paul M. Weinberger, Christopher G. Kevil, Martin J. Sapp, Daniel J. Snyder, Vaughn S. Cooper |
| EPI_ISL_435348, EPI_ISL_435350, EPI_ISL_435351, EPI_ISL_435352, EPI_ISL_435353, EPI_ISL_435354, EPI_ISL_435355, EPI_ISL_435356, EPI_ISL_435357, EPI_ISL_435358, EPI_ISL_435359, EPI_ISL_435360, EPI_ISL_435361, EPI_ISL_435362, EPI_ISL_435363, EPI_ISL_435364, EPI_ISL_435365, EPI_ISL_435366, EPI_ISL_435367, EPI_ISL_435368, EPI_ISL_435369, EPI_ISL_435371, EPI_ISL_435372, EPI_ISL_435373, EPI_ISL_435374, EPI_ISL_435375, EPI_ISL_435376, EPI_ISL_435377, EPI_ISL_435378, EPI_ISL_435379, EPI_ISL_435380, EPI_ISL_435381, EPI_ISL_435382, EPI_ISL_435383, EPI_ISL_435384, EPI_ISL_435385, EPI_ISL_435386, EPI_ISL_435387, EPI_ISL_435388, EPI_ISL_435389, EPI_ISL_435390, EPI_ISL_435391, EPI_ISL_435392, EPI_ISL_435393 | | | |
| see above | Utah Public Health Laboratory | Utah Public Health Laboratory | Erin Young, Kelly Oakeson |
| EPI_ISL_435394, EPI_ISL_435395, EPI_ISL_435396, EPI_ISL_435397, EPI_ISL_435398, EPI_ISL_435399, EPI_ISL_435400, EPI_ISL_435401, EPI_ISL_435402 | Gundersen Molecular Diagnostics Laboratory | Kabara Cancer Research Institute | Craig S. Richmond, Paraic A. Kenny |
| EPI_ISL_435441, EPI_ISL_435442, EPI_ISL_435443, EPI_ISL_435444 | Alaska State Virology Laboratory | Alaska State Virology Laboratory | Jack Chen, Ph.D. |
| EPI_ISL_435445, EPI_ISL_435446, EPI_ISL_435447, EPI_ISL_435448, EPI_ISL_435449, EPI_ISL_435450, EPI_ISL_435451, EPI_ISL_435452, EPI_ISL_435453, EPI_ISL_435454, EPI_ISL_435455, EPI_ISL_435456, EPI_ISL_435457, EPI_ISL_435458, EPI_ISL_435459, EPI_ISL_435460, EPI_ISL_435461, EPI_ISL_435462, EPI_ISL_435463, EPI_ISL_435464, EPI_ISL_435465, EPI_ISL_435466, EPI_ISL_435467, EPI_ISL_435468, EPI_ISL_435469, EPI_ISL_435470, EPI_ISL_435471, EPI_ISL_435472 | | | |
| see above | Robert Garry lab | Andersen lab at Scripps Research | Allison Smither, Gilberto Sabino-Santos, Patricia Snarski, Lilia Melnik, Antoinette Bell, Kaylynn Genemaras, Arnaud Drouin, Dahlene Fusco, Robert Garry with SEARCH Alliance San Diego |
| EPI_ISL_435473, EPI_ISL_435474 | Rady's Childrens Hospital | Andersen lab at Scripps Research | SEARCH Alliance San Diego |
| EPI_ISL_435475, EPI_ISL_435476, EPI_ISL_435477, EPI_ISL_435478, EPI_ISL_435479, EPI_ISL_435480, EPI_ISL_435481, EPI_ISL_435482, EPI_ISL_435483, EPI_ISL_435484, EPI_ISL_435485, EPI_ISL_435486, EPI_ISL_435487, EPI_ISL_435488, EPI_ISL_435489, EPI_ISL_435490, EPI_ISL_435491, EPI_ISL_435492, EPI_ISL_435493, EPI_ISL_435494, EPI_ISL_435495, EPI_ISL_435496, EPI_ISL_435497, EPI_ISL_435498, EPI_ISL_435499, EPI_ISL_435500, EPI_ISL_435501, EPI_ISL_435502, EPI_ISL_435503, EPI_ISL_435504, EPI_ISL_435505, EPI_ISL_435506, EPI_ISL_435507, EPI_ISL_435508, EPI_ISL_435509, EPI_ISL_435510, EPI_ISL_435511, EPI_ISL_435512, EPI_ISL_435513, EPI_ISL_435514, EPI_ISL_435515, EPI_ISL_435516, EPI_ISL_435517, EPI_ISL_435518, EPI_ISL_435519, EPI_ISL_435520, EPI_ISL_435521, EPI_ISL_435522, EPI_ISL_435523, EPI_ISL_435524, EPI_ISL_435525, EPI_ISL_435526, EPI_ISL_435527, EPI_ISL_435528, EPI_ISL_435529, EPI_ISL_435530, EPI_ISL_435531, EPI_ISL_435532, EPI_ISL_435533, EPI_ISL_435534, EPI_ISL_435535, EPI_ISL_435536, EPI_ISL_435537, EPI_ISL_435538, EPI_ISL_435539, EPI_ISL_435540, EPI_ISL_435541, EPI_ISL_435542, EPI_ISL_435543, EPI_ISL_435544, EPI_ISL_435545, EPI_ISL_435546, EPI_ISL_435547, EPI_ISL_435548, EPI_ISL_435549 | | | |
| see above | NYU Langone Health | Departments of Pathology and Medicine, New York University School of Medicine | Maria Aguero-Rosenfeld, Brendan Belovarac, Margaret Black, Ludovic Boytard, John Cadley, Paolo Cotzia, John Chen, Dacia Dimartino, Xiaojun Feng, Tatyana Gindin, Emily Guzman, Adriana Heguy, Megan Hogan, Emily Huang, George Jour, Lawrence H. Lin, Raven Luther, Andrew Lytle, Christian Marier, Matthew T. Maurano, Mark J. Mulligan, Peter Meyn, Raquel Ordonez Ciriza, Iman Osman, Jared Pinnell, Vanessa Raabe, Sitharam Ramaswami, Amy Rapkiewicz, Andre M. Ribeiro-dos-Santos, Marie Samanovic-Golden, Antonio Serrano, Guomiao Shen, Matija Snuderl, Theodore Vougiouklakis, Nick Vulpescu, Gael Westby, Paul Zappile, Yutong Zhang |
| EPI_ISL_435550, EPI_ISL_435551, EPI_ISL_435552, EPI_ISL_435553, EPI_ISL_435554 | LSUHS Emerging Viral Threat Laboratory | Microbial Genome Sequencing Center | Rona S. Scott, Jeremy P. Kamil, John A. Vanchiere, Camille F. Abshire, Abida Siddiqa, Byeong-Jae Lee, Chan-ki Min, Md Maksudul Alam, Monica Gestal-Cartele, Edna Ondari, Adam Greer, Malgorzata Bienkowska-Haba, Katarzyna Zwolinska, Jason M. Bodily, Andrew D. Yurochko, Paul M. Weinberger, Christopher G. Kevil, Martin J. Sapp, Daniel J. Snyder, Vaughn S. Cooper |
| EPI_ISL_435555, EPI_ISL_435556, EPI_ISL_435557, EPI_ISL_435558, EPI_ISL_435559, EPI_ISL_435560, EPI_ISL_435561, EPI_ISL_435562, EPI_ISL_435563, EPI_ISL_435564, EPI_ISL_435565, EPI_ISL_435566, EPI_ISL_435567, EPI_ISL_435568 | | | |
| see above | LSUHS Emerging Viral Threat Laboratory | Microbial Genome Sequencing Center | John A. Vanchiere, Jeremy P. Kamil, Rona S. Scott, Camille F. Abshire, Abida Siddiqa, Byeong-Jae Lee, Chan-ki Min, Md Maksudul Alam, Monica Gestal-Cartele, Edna Ondari, Adam Greer, Malgorzata Bienkowska-Haba, Katarzyna Zwolinska, Jason M. Bodily, Andrew D. Yurochko, Paul M. Weinberger, Christopher G. Kevil, Martin J. Sapp, Daniel J. Snyder, Vaughn S. Cooper |
| EPI_ISL_435569, EPI_ISL_435570, EPI_ISL_435571, EPI_ISL_435572, EPI_ISL_435573, EPI_ISL_435574, EPI_ISL_435575, EPI_ISL_435576, EPI_ISL_435577, EPI_ISL_435578, EPI_ISL_435579 | | | |
| see above | LSUHS Emerging Viral Threat Laboratory | Microbial Genome Sequencing Center | Jeremy P. Kamil, John A. Vanchiere, Rona S. Scott, Camille F. Abshire, Abida Siddiqa, Byeong-Jae Lee, Chan-ki Min, Md Maksudul Alam, Monica Gestal-Cartele, Edna Ondari, Adam Greer, Malgorzata Bienkowska-Haba, Katarzyna Zwolinska, Jason M. Bodily, Andrew D. Yurochko, Paul M. Weinberger, Christopher G. Kevil, Martin J. Sapp, Daniel J. Snyder, Vaughn S. Cooper |
| EPI_ISL_435580, EPI_ISL_435581, EPI_ISL_435582, EPI_ISL_435583, EPI_ISL_435584, EPI_ISL_435586, EPI_ISL_435587, EPI_ISL_435590, EPI_ISL_435591, EPI_ISL_435594, EPI_ISL_435596, EPI_ISL_435597, EPI_ISL_435598, EPI_ISL_435599, EPI_ISL_435600, EPI_ISL_435601, EPI_ISL_435608, EPI_ISL_435609, EPI_ISL_435612, EPI_ISL_435615, EPI_ISL_435618, EPI_ISL_435619, EPI_ISL_435620, EPI_ISL_435621, EPI_ISL_435627, EPI_ISL_435628, EPI_ISL_435630, EPI_ISL_435631, EPI_ISL_435633, EPI_ISL_435634, EPI_ISL_435635, EPI_ISL_435636, EPI_ISL_435637, EPI_ISL_435638, EPI_ISL_435639, EPI_ISL_435640, EPI_ISL_435641, EPI_ISL_435643, EPI_ISL_435645, EPI_ISL_435647, EPI_ISL_435649, EPI_ISL_435650, EPI_ISL_435651, EPI_ISL_435653, EPI_ISL_435654, EPI_ISL_435657, EPI_ISL_435658, EPI_ISL_435660, EPI_ISL_435661, EPI_ISL_435662, EPI_ISL_435663, EPI_ISL_435666, EPI_ISL_435667, EPI_ISL_435668, EPI_ISL_435670, EPI_ISL_435671 | | | |
| see above | Santa Clara County Public Health Department | Chiu Laboratory, University of California, San Francisco | Xianding Deng, Scot Federman, Wei Gu, Elsa Villarino, Brandon Bonin, Debra A. Wadford, and Charles Y. Chiu |
| EPI_ISL_435702, EPI_ISL_435703, EPI_ISL_435704, EPI_ISL_435705, EPI_ISL_435706, EPI_ISL_435707, EPI_ISL_435708, EPI_ISL_435709 | Yale COVID-19 Biorepository | Grubaugh Lab - Yale School of Public Health | Joseph Fauver, Tara Alpert, Anderson Brito, Anne Wyllie, Chantal Vogels, Mary Petrone, Cole Jensen, Chaney Kalinich, Isabel Ott, Arnau Casanovas, Catherine Muenker, Adam Moore, Alice Lu, Maria Tokuyama, Patrick Wong, Peiwen Lu, Saad Omer, Richard Martinello, Allison Nelson, Shelli Farhadian, Akiko Iwasaki, Charlese Dela Cruz, Albert Ko, Nathan Grubaugh |
| EPI_ISL_435710, | Connecticut State Department of Public | Grubaugh Lab - Yale School of Public Health | Joseph Fauver, Tara Alpert, Anderson Brito, Anne Wyllie, Chantal Vogels, Mary Petrone, Cole Jensen, Chaney Kalinich, Isabel Ott, Arnau Casanovas, Catherine Muenker, Adam Moore, Alice Lu, Maria Tokuyama, Patrick Wong, Peiwen Lu, Saad Omer, Richard Martinello, Allison Nelson, |

| | | | |
|---|---|---|---|
| EPI_ISL_435711, EPI_ISL_435712, EPI_ISL_435713, EPI_ISL_435714, EPI_ISL_435715, EPI_ISL_435716, EPI_ISL_435717, EPI_ISL_435718, EPI_ISL_435719 | Health | | Shelli Farhadian, Akiko Iwasaki, Charlese Dela Cruz, Albert Ko, Nathan Grubaugh |
| EPI_ISL_435720 | Yale Clinical Virology Laboratory | Grubaugh Lab - Yale School of Public Health | Joseph Fauver, Tara Alpert, Anderson Brito, Anne Wyllie, Chantal Vogels, Mary Petrone, Cole Jensen, Chaney Kalinich, Isabel Ott, Arnau Casanovas, Catherine Muenker, Adam Moore, Alice Lu, Maria Tokuyama, Patrick Wong, Peiwen Lu, Saad Omer, Richard Martinello, Allison Nelson, Shelli Farhadian, Akiko Iwasaki, Charlese Dela Cruz, Albert Ko, Nathan Grubaugh |
| EPI_ISL_435721, EPI_ISL_435722 | NYU Langone Health | Departments of Pathology and Medicine, New York University School of Medicine | Maria Aguero-Rosenfeld, Brendan Belovarac, Margaret Black, Ludovic Boytard, John Cadley, Paolo Cotzia, John Chen, Dacia Dimartino, Xiaojun Feng, Tatyana Gindin, Emily Guzman, Adriana Heguy, Megan Hogan, Emily Huang, George Jour, Lawrence H. Lin, Raven Luther, Andrew Lytle, Christian Marier, Matthew T. Maurano, Mark J. Mulligan, Peter Meyn, Raquel Ordonez Ciriza, Iman Osman, Jared Pinnell, Vanessa Raabe, Sitharam Ramaswami, Amy Rapkiewicz, Andre M. Ribeiro-dos-Santos, Marie Samanovic-Golden, Antonio Serrano, Guomiao Shen, Matija Snuderl, Theodore Vougiouklakis, Nick Vulpescu, Gael Westby, Paul Zappile, Yutong Zhang |
| EPI_ISL_436040, EPI_ISL_436041, EPI_ISL_436042, EPI_ISL_436043 | DC Public Health Lab Dept of Forensic Science | Pathogen Discovery, Respiratory Viruses Branch, Division of Viral Diseases, Centers for Disease Control and Prevention | Ying Tao, Jing Zhang, Krista Queen, Yan Li, Anna Uehara, Clinton R. Paden, Haibin Wang, Zachary Weiner, Bettina Bankamp, Suxiang Tong |
| EPI_ISL_436044 | Louisiana Office of Public Health Laboratories | Pathogen Discovery, Respiratory Viruses Branch, Division of Viral Diseases, Centers for Disease Control and Prevention | Ying Tao, Jing Zhang, Krista Queen, Yan Li, Anna Uehara, Clinton R. Paden, Haibin Wang, Zachary Weiner, Bettina Bankamp, Suxiang Tong |
| EPI_ISL_436045, EPI_ISL_436046 | US VI Department of Health | Pathogen Discovery, Respiratory Viruses Branch, Division of Viral Diseases, Centers for Disease Control and Prevention | Ying Tao, Jing Zhang, Krista Queen, Yan Li, Anna Uehara, Clinton R. Paden, Haibin Wang, Zachary Weiner, Bettina Bankamp, Suxiang Tong |
| EPI_ISL_436047, EPI_ISL_436048, EPI_ISL_436049, EPI_ISL_436050, EPI_ISL_436051, EPI_ISL_436052, EPI_ISL_436053, EPI_ISL_436054, EPI_ISL_436055, EPI_ISL_436056, EPI_ISL_436057, EPI_ISL_436058, EPI_ISL_436059, EPI_ISL_436060, EPI_ISL_436061, EPI_ISL_436062, EPI_ISL_436063, EPI_ISL_436064, EPI_ISL_436065, EPI_ISL_436066, EPI_ISL_436067, EPI_ISL_436068, EPI_ISL_436069, EPI_ISL_436070, EPI_ISL_436071, EPI_ISL_436072, EPI_ISL_436073, EPI_ISL_436074, EPI_ISL_436075, EPI_ISL_436076, EPI_ISL_436077, EPI_ISL_436078, EPI_ISL_436079, EPI_ISL_436080, EPI_ISL_436081, EPI_ISL_436082 | | | |
| see above | NYC Department of Health and Mental Hygiene | Pathogen Discovery, Respiratory Viruses Branch, Division of Viral Diseases, Centers for Disease Control and Prevention | Ying Tao, Krista Queen, Christy Harrison, Jennifer Rakeman, Clinton R. Paden, Jing Zhang, Anna Uehara, Yan Li, Haibin Wang, Jasmine Padilla, Justin Lee, Bettina Bankamp, Zachary Weiner, Suxiang Tong |
| EPI_ISL_436464 | Alaska State Virology Laboratory | Alaska State Virology Laboratory | Jack Chen |
| EPI_ISL_436466, EPI_ISL_436467, EPI_ISL_436468, EPI_ISL_436469, EPI_ISL_436470, EPI_ISL_436471, EPI_ISL_436472, EPI_ISL_436473, EPI_ISL_436474, EPI_ISL_436475, EPI_ISL_436476, EPI_ISL_436477, EPI_ISL_436478, EPI_ISL_436479, EPI_ISL_436480, EPI_ISL_436481, EPI_ISL_436482, EPI_ISL_436483, EPI_ISL_436484, EPI_ISL_436485, EPI_ISL_436486, EPI_ISL_436487, EPI_ISL_436488, EPI_ISL_436489, EPI_ISL_436490, EPI_ISL_436491, EPI_ISL_436492, EPI_ISL_436493, EPI_ISL_436494, EPI_ISL_436495, EPI_ISL_436496, EPI_ISL_436497, EPI_ISL_436498, EPI_ISL_436499, EPI_ISL_436500, EPI_ISL_436501, EPI_ISL_436502, EPI_ISL_436503, EPI_ISL_436504 | | | |
| see above | UPMC Clinical Laboratory | Microbial Genome Sequencing Center, Microbial Genomic Epidemiological Laboratory | Dan Snyder, Stephanie L Mitchell, Mustapha M Mustapha, Marissa P Griffith, Vatsala R Srinivasa, Kady D Waggle, Chinelo Ezeonwuku, Jane W. Marsh, Lee H. Harrison, Vaughn S. Cooper |
| EPI_ISL_436506, EPI_ISL_436512, EPI_ISL_436521, EPI_ISL_436522, EPI_ISL_436524, EPI_ISL_436528, EPI_ISL_436533, EPI_ISL_436537, EPI_ISL_436549, EPI_ISL_436550, EPI_ISL_436551, EPI_ISL_436552, EPI_ISL_436555, EPI_ISL_436563 | | | |
| see above | Florida Bureau of Public Health Laboratories | Florida Bureau of Public Health Laboratories | Sarah Schmedes, Jason Blanton |
| EPI_ISL_436564, EPI_ISL_436565, EPI_ISL_436566, EPI_ISL_436567, EPI_ISL_436568, EPI_ISL_436569, EPI_ISL_436570, EPI_ISL_436571, EPI_ISL_436572, EPI_ISL_436573, EPI_ISL_436574, EPI_ISL_436575, EPI_ISL_436576, EPI_ISL_436577, EPI_ISL_436578, EPI_ISL_436579, EPI_ISL_436580, EPI_ISL_436581, EPI_ISL_436582, EPI_ISL_436583, EPI_ISL_436584, EPI_ISL_436585, EPI_ISL_436586, EPI_ISL_436587, EPI_ISL_436588, EPI_ISL_436589, EPI_ISL_436590, EPI_ISL_436591, EPI_ISL_436592, EPI_ISL_436593, EPI_ISL_436594, EPI_ISL_436595, EPI_ISL_436596, EPI_ISL_436597, EPI_ISL_436598, EPI_ISL_436599, EPI_ISL_436600, EPI_ISL_436601, EPI_ISL_436602, EPI_ISL_436603, EPI_ISL_436604, EPI_ISL_436605, EPI_ISL_436606, EPI_ISL_436607, EPI_ISL_436608, EPI_ISL_436609, EPI_ISL_436610, EPI_ISL_436612, EPI_ISL_436613, EPI_ISL_436614, EPI_ISL_436615, EPI_ISL_436616, EPI_ISL_436617, EPI_ISL_436618, EPI_ISL_436619, EPI_ISL_436620, EPI_ISL_436621, EPI_ISL_436622, EPI_ISL_436623, EPI_ISL_436624, EPI_ISL_436625, EPI_ISL_436626, EPI_ISL_436627, EPI_ISL_436628, EPI_ISL_436629, EPI_ISL_436630, EPI_ISL_436631, EPI_ISL_436632, EPI_ISL_436634, EPI_ISL_436635, EPI_ISL_436636, EPI_ISL_436637, EPI_ISL_436638, EPI_ISL_436639, EPI_ISL_436640 | | | |
| see above | University of Wisconsin-Madison AIDS Vaccine Research Laboratories | University of Wisconsin-Madison AIDS Vaccine Research Laboratories | Gage Moreno, Katarina Braun, et al. AIDS Vaccine Research Laboratories |
| EPI_ISL_436641, EPI_ISL_436642, EPI_ISL_436643, EPI_ISL_436644, EPI_ISL_436645, EPI_ISL_436646, EPI_ISL_436647, EPI_ISL_436648, EPI_ISL_436650, EPI_ISL_436651, EPI_ISL_436652, EPI_ISL_436653, EPI_ISL_436654, EPI_ISL_436655, EPI_ISL_436656, EPI_ISL_436657, EPI_ISL_436658, EPI_ISL_436659, EPI_ISL_436660, EPI_ISL_436661, EPI_ISL_436662, EPI_ISL_436663, EPI_ISL_436664, EPI_ISL_436665, EPI_ISL_436666, EPI_ISL_436667, EPI_ISL_436669, EPI_ISL_436670, EPI_ISL_436671, EPI_ISL_436672, EPI_ISL_436673, EPI_ISL_436674, EPI_ISL_436675, EPI_ISL_436676, EPI_ISL_436677, EPI_ISL_436678, EPI_ISL_436679, EPI_ISL_436680, EPI_ISL_436681, EPI_ISL_436682, EPI_ISL_436683 | | | |
| see above | County of Santa Clara Public Health Department | Chan-Zuckerberg Biohub | CZB Cliahub Consortium |
| EPI_ISL_436733, EPI_ISL_436734, EPI_ISL_436735, EPI_ISL_436736, EPI_ISL_436737, EPI_ISL_436738, EPI_ISL_436739, EPI_ISL_436740, EPI_ISL_436741, EPI_ISL_436743 | NYU Langone Health | Departments of Pathology and Medicine, New York University School of Medicine | Maria Aguero-Rosenfeld, Brendan Belovarac, Margaret Black, Ludovic Boytard, John Cadley, Paolo Cotzia, John Chen, Dacia Dimartino, Xiaojun Feng, Tatyana Gindin, Emily Guzman, Adriana Heguy, Megan Hogan, Emily Huang, George Jour, Alireza Khodadadi-Jamayran, Lawrence H. Lin, Raven Luther, Andrew Lytle, Christian Marier, Matthew T. Maurano, Mark J. Mulligan, Peter Meyn, Raquel Ordonez Ciriza, Iman Osman, Jared Pinnell, Vanessa Raabe, Sitharam Ramaswami, Amy Rapkiewicz, Andre M. Ribeiro-dos-Santos, Marie Samanovic-Golden, Antonio Serrano, Guomiao Shen, Matija Snuderl, Theodore Vougiouklakis, Nick Vulpescu, Gael Westby, Paul Zappile, Yutong Zhang |
| EPI_ISL_436800, EPI_ISL_436801, EPI_ISL_436802, EPI_ISL_436803, EPI_ISL_436804, EPI_ISL_436805, EPI_ISL_436806, EPI_ISL_436807, EPI_ISL_436808, EPI_ISL_436809, EPI_ISL_436810, EPI_ISL_436811, EPI_ISL_436812, EPI_ISL_436813, EPI_ISL_436814, EPI_ISL_436815, EPI_ISL_436816, EPI_ISL_436817, EPI_ISL_436818, EPI_ISL_436819, EPI_ISL_436820, EPI_ISL_436821, EPI_ISL_436822, EPI_ISL_436823, EPI_ISL_436824, EPI_ISL_436825, EPI_ISL_436826, EPI_ISL_436827, EPI_ISL_436828, EPI_ISL_436829, EPI_ISL_436830, EPI_ISL_436831, EPI_ISL_436832, EPI_ISL_436833, EPI_ISL_436834, EPI_ISL_436835, EPI_ISL_436836, EPI_ISL_436837, EPI_ISL_436838, EPI_ISL_436839, EPI_ISL_436840, EPI_ISL_436841, EPI_ISL_436842, EPI_ISL_436843, EPI_ISL_436844, EPI_ISL_436845, EPI_ISL_436846, EPI_ISL_436847, EPI_ISL_436848, EPI_ISL_436849, EPI_ISL_436850, EPI_ISL_436851, EPI_ISL_436852, EPI_ISL_436853, EPI_ISL_436854, EPI_ISL_436855, EPI_ISL_436856, EPI_ISL_436857, EPI_ISL_436858, EPI_ISL_436859, EPI_ISL_436860, EPI_ISL_436861, EPI_ISL_436862, EPI_ISL_436863, EPI_ISL_436864, EPI_ISL_436865, EPI_ISL_436866, EPI_ISL_436867, EPI_ISL_436868, EPI_ISL_436869, EPI_ISL_436870, EPI_ISL_436871, EPI_ISL_436872, EPI_ISL_436873, EPI_ISL_436874, EPI_ISL_436875, EPI_ISL_436876, EPI_ISL_436877, EPI_ISL_436878, EPI_ISL_436879, EPI_ISL_436880, EPI_ISL_436881, EPI_ISL_436882, EPI_ISL_436883, EPI_ISL_436884, EPI_ISL_436885, EPI_ISL_436886, EPI_ISL_436887, EPI_ISL_436889, EPI_ISL_436890 | | | |
| see above | Michigan Department of Health and Human Services, Bureau of Laboratories | Michigan Department of Health and Human Services, Bureau of Laboratories | Blankenship HM, Riner D, Soehnlen MK |
| EPI_ISL_436891, EPI_ISL_436892, EPI_ISL_436893, EPI_ISL_436894, EPI_ISL_436895, EPI_ISL_436896, EPI_ISL_436897, EPI_ISL_436898, EPI_ISL_436899, EPI_ISL_436900 | Gundersen Molecular Diagnostics Laboratory | Kabara Cancer Research Institute | Craig S. Richmond, Paraic A. Kenny |
| EPI_ISL_436901, EPI_ISL_436902, EPI_ISL_436903, EPI_ISL_436904, EPI_ISL_436905, EPI_ISL_436906, EPI_ISL_436907, EPI_ISL_436909, EPI_ISL_436910, EPI_ISL_436911, EPI_ISL_436912, EPI_ISL_436914, EPI_ISL_436915, EPI_ISL_436916, EPI_ISL_436917, EPI_ISL_436918, EPI_ISL_436919, EPI_ISL_436920, EPI_ISL_436921, EPI_ISL_436922, EPI_ISL_436923, EPI_ISL_436924, EPI_ISL_436925 | | | |
| see above | Utah Public Health Laboratory | Utah Public Health Laboratory | Erin Young, Kelly Oakeson |
| EPI_ISL_436926 | $x^2$ | Utah Public Health Laboratory | Erin Young, Kelly Oakeson |
| EPI_ISL_436927, EPI_ISL_436928, EPI_ISL_436929, EPI_ISL_436930, EPI_ISL_436931, EPI_ISL_436932, EPI_ISL_436933, EPI_ISL_436934, EPI_ISL_436935, EPI_ISL_436936, EPI_ISL_436937, EPI_ISL_436938 | | | |
| see above | Utah Public Health Laboratory | Utah Public Health Laboratory | Erin Young, Kelly Oakeson |
| EPI_ISL_436939, EPI_ISL_436940, EPI_ISL_436941, EPI_ISL_436942, EPI_ISL_436943, EPI_ISL_436944, EPI_ISL_436945, EPI_ISL_436946, EPI_ISL_436947, EPI_ISL_436948, EPI_ISL_436949, EPI_ISL_436950, EPI_ISL_436952, EPI_ISL_436953, EPI_ISL_436954, EPI_ISL_436955, EPI_ISL_436956, EPI_ISL_436957, EPI_ISL_436958, EPI_ISL_436959, EPI_ISL_436960, EPI_ISL_436961 | | | |
| see above | Ochsner Health | Bioinfoexperts, LLC | Amy Feehan, David J. Nolan, Rebecca Rose, Sissy Cross, David Moraga Amador, Tong Yang, Luke Caruso, Wayra Navia, Lydia Von Borstel, Xiao Hui Zhou, Julia-Garcia-Diaz, Susanna L. Lamers |
| EPI_ISL_437043, EPI_ISL_437044, EPI_ISL_437045, EPI_ISL_437046, EPI_ISL_437047, EPI_ISL_437048, EPI_ISL_437049, EPI_ISL_437050, EPI_ISL_437051, EPI_ISL_437052, EPI_ISL_437053, EPI_ISL_437054, EPI_ISL_437055, EPI_ISL_437056, EPI_ISL_437057, EPI_ISL_437058, EPI_ISL_437059, EPI_ISL_437060, EPI_ISL_437061, EPI_ISL_437062, EPI_ISL_437063, EPI_ISL_437064, EPI_ISL_437065, EPI_ISL_437066, EPI_ISL_437067, EPI_ISL_437068, EPI_ISL_437069, EPI_ISL_437071, EPI_ISL_437072, EPI_ISL_437073, EPI_ISL_437074, EPI_ISL_437075, EPI_ISL_437076, EPI_ISL_437077, EPI_ISL_437078, EPI_ISL_437079, EPI_ISL_437080, EPI_ISL_437081, EPI_ISL_437082, EPI_ISL_437083, EPI_ISL_437084, EPI_ISL_437085, EPI_ISL_437086, EPI_ISL_437087, EPI_ISL_437088 | | | |
| see above | County of Santa Clara Public Health | Chan-Zuckerberg Biohub | CZB Cliahub Consortium |
| EPI_ISL_437097, EPI_ISL_437098, EPI_ISL_437099, EPI_ISL_437100, EPI_ISL_437101, EPI_ISL_437102, EPI_ISL_437103, EPI_ISL_437104, EPI_ISL_437105, EPI_ISL_437106, EPI_ISL_437107, EPI_ISL_437108, EPI_ISL_437109, EPI_ISL_437110, EPI_ISL_437111, EPI_ISL_437112, EPI_ISL_437113, EPI_ISL_437114, EPI_ISL_437115, EPI_ISL_437116, EPI_ISL_437117, EPI_ISL_437118, EPI_ISL_437119, EPI_ISL_437120, EPI_ISL_437121, EPI_ISL_437122, EPI_ISL_437123, EPI_ISL_437124, EPI_ISL_437125, EPI_ISL_437126, EPI_ISL_437127, EPI_ISL_437128, EPI_ISL_437129, EPI_ISL_437130, EPI_ISL_437131, EPI_ISL_437132, EPI_ISL_437133, EPI_ISL_437134, EPI_ISL_437135, EPI_ISL_437136, EPI_ISL_437137, EPI_ISL_437138, EPI_ISL_437139, EPI_ISL_437140, EPI_ISL_437141, EPI_ISL_437142, EPI_ISL_437143, EPI_ISL_437144, EPI_ISL_437145, EPI_ISL_437146, EPI_ISL_437147, EPI_ISL_437148, EPI_ISL_437149, EPI_ISL_437150, EPI_ISL_437151, EPI_ISL_437152, EPI_ISL_437153, EPI_ISL_437154, EPI_ISL_437155, EPI_ISL_437156, EPI_ISL_437157, EPI_ISL_437158, EPI_ISL_437159, EPI_ISL_437160, EPI_ISL_437161, EPI_ISL_437162, EPI_ISL_437163, EPI_ISL_437164, EPI_ISL_437165, EPI_ISL_437166, EPI_ISL_437167, EPI_ISL_437168, EPI_ISL_437169, EPI_ISL_437170, EPI_ISL_437171, EPI_ISL_437172, EPI_ISL_437173, EPI_ISL_437174, EPI_ISL_437175, EPI_ISL_437176, EPI_ISL_437177, EPI_ISL_437178, EPI_ISL_437179, EPI_ISL_437180, EPI_ISL_437181, EPI_ISL_437182, EPI_ISL_437183, EPI_ISL_437184, EPI_ISL_437185, EPI_ISL_437186 | | | |
| see above | Michigan Department of Health and Human Services, Bureau of Laboratories | Michigan Department of Health and Human Services, Bureau of Laboratories | Blankenship HM, Riner D, Soehnlen MK |
| EPI_ISL_437360, EPI_ISL_437361, EPI_ISL_437362, EPI_ISL_437363, EPI_ISL_437364, EPI_ISL_437365, EPI_ISL_437366, EPI_ISL_437367, EPI_ISL_437368, EPI_ISL_437369, EPI_ISL_437370, EPI_ISL_437371, EPI_ISL_437372, EPI_ISL_437373, EPI_ISL_437374, EPI_ISL_437375, EPI_ISL_437376, EPI_ISL_437377, EPI_ISL_437378, EPI_ISL_437379, EPI_ISL_437380, EPI_ISL_437381, EPI_ISL_437382, EPI_ISL_437383, EPI_ISL_437384, EPI_ISL_437385, EPI_ISL_437386 | | | |
| see above | Minnesota Department of Health, Public Health Laboratory | Minnesota Department of Health, Public Health Laboratory | Matt Plumb, Jacob Garfin, and Xiong Wang |
| EPI_ISL_437387, EPI_ISL_437388, EPI_ISL_437389, EPI_ISL_437390, EPI_ISL_437391, EPI_ISL_437392, EPI_ISL_437393, EPI_ISL_437394, EPI_ISL_437395, EPI_ISL_437396, EPI_ISL_437397, EPI_ISL_437398, EPI_ISL_437399, EPI_ISL_437400, EPI_ISL_437401, EPI_ISL_437402, EPI_ISL_437403, EPI_ISL_437404, EPI_ISL_437405, EPI_ISL_437406, EPI_ISL_437407, EPI_ISL_437408, EPI_ISL_437409, EPI_ISL_437410, | | | |

| | | | |
|---|---|---|---|
| EPI_ISL_437411, EPI_ISL_437412, EPI_ISL_437413, EPI_ISL_437414, EPI_ISL_437415, EPI_ISL_437416, EPI_ISL_437417, EPI_ISL_437418, EPI_ISL_437419, EPI_ISL_437420, EPI_ISL_437421, EPI_ISL_437422, EPI_ISL_437423, EPI_ISL_437424, EPI_ISL_437425, EPI_ISL_437426, EPI_ISL_437427, EPI_ISL_437428, EPI_ISL_437429, EPI_ISL_437430, EPI_ISL_437431, EPI_ISL_437432 | | | |
| see above | Virginia DCLS | Virginia DCLS | Virginia DCLS |
| EPI_ISL_437433 | Bozeman Health Deaconess Hospital | Wiedenheft lab, Montana State University | Artem Nemudryi, Anna Nemudraia, Kevin Surya, Tanner Wiegand, Murat Buyukyoruk, Royce Wilkinson, Blake Wiedenheft |
| EPI_ISL_437437 | Alaska State Virology Laboratory | Alaska State Virology Laboratory | Jack Chen, Ph.D. |
| EPI_ISL_437455, EPI_ISL_437456, EPI_ISL_437457, EPI_ISL_437458 | Clinical Diagnostics Laboratory, Diagnostic & Experimental Pathology, Lilly Research Laboratories | Clinical Diagnostics Laboratory, Diagnostic & Experimental Pathology, Lilly Research Laboratories | Tim Holzer, Mayuri Vaidya, Angie Fulford, Sam McNeely, Rachael Redmond, Phil Ebert, John Calley, Leslie ONeill Reising, Pat Finnegan, Erin Wray, John McElwee, Jeff Fill, Joe Oakley, Andrew Schade |
| EPI_ISL_437513, EPI_ISL_437514, EPI_ISL_437515, EPI_ISL_437516, EPI_ISL_437517, EPI_ISL_437518 | Alaska State Virology Laboratory | Alaska State Virology Laboratory | Jack Chen, Ph.D. |
| EPI_ISL_437520, EPI_ISL_437521, EPI_ISL_437522, EPI_ISL_437523, EPI_ISL_437524, EPI_ISL_437525, EPI_ISL_437526, EPI_ISL_437527, EPI_ISL_437528, EPI_ISL_437529, EPI_ISL_437531, EPI_ISL_437532, EPI_ISL_437533, EPI_ISL_437535 | | | |
| see above | OHSU Lab Services Molecular Microbiology Lab | Oregon SARS-CoV-2 Genome Sequencing Center | Brendan L. O'Connell, Ruth V. Nichols, Alec J. Hirsch, Guang Fan, Daniel N. Streblow, William B. Messer, Andrew C. Adey, Benjamin N. Bimber, Brian J. O'Roak |
| EPI_ISL_437540, EPI_ISL_437541, EPI_ISL_437542, EPI_ISL_437544, EPI_ISL_437545, EPI_ISL_437546, EPI_ISL_437547, EPI_ISL_437548 | Robert Garry lab | Andersen lab at Scripps Research | Allison Smither, Gilberto Sabino-Santos, Patricia Snarski, Lilia Melnik, Antoinette Bell, Kaylynn Genemaras, Arnaud Drouin, Dahlene Fusco, Robert Garry with SEARCH Alliance San Diego |
| EPI_ISL_437549, EPI_ISL_437550, EPI_ISL_437551, EPI_ISL_437552, EPI_ISL_437553, EPI_ISL_437554, EPI_ISL_437555, EPI_ISL_437556, EPI_ISL_437557, EPI_ISL_437559, EPI_ISL_437560, EPI_ISL_437561, EPI_ISL_437562, EPI_ISL_437563, EPI_ISL_437564, EPI_ISL_437565, EPI_ISL_437566, EPI_ISL_437567, EPI_ISL_437568, EPI_ISL_437569, EPI_ISL_437570, EPI_ISL_437571, EPI_ISL_437572, EPI_ISL_437573, EPI_ISL_437574, EPI_ISL_437575, EPI_ISL_437576, EPI_ISL_437577, EPI_ISL_437578, EPI_ISL_437579, EPI_ISL_437580, EPI_ISL_437581, EPI_ISL_437582, EPI_ISL_437583, EPI_ISL_437584, EPI_ISL_437585, EPI_ISL_437586, EPI_ISL_437587, EPI_ISL_437588, EPI_ISL_437589, EPI_ISL_437590, EPI_ISL_437591, EPI_ISL_437592, EPI_ISL_437593, EPI_ISL_437594, EPI_ISL_437595, EPI_ISL_437596, EPI_ISL_437598, EPI_ISL_437599, EPI_ISL_437600 | | | |
| see above | Scripps Medical Laboratory | Andersen lab at Scripps Research | SEARCH Alliance San Diego with Michael Quigley, Ellen Stefanski, Ian Mchardy |
| EPI_ISL_437763, EPI_ISL_437764, EPI_ISL_437765, EPI_ISL_437766, EPI_ISL_437767, EPI_ISL_437768, EPI_ISL_437769, EPI_ISL_437770, EPI_ISL_437771, EPI_ISL_437772, EPI_ISL_437773, EPI_ISL_437774, EPI_ISL_437775, EPI_ISL_437776, EPI_ISL_437777, EPI_ISL_437778, EPI_ISL_437779, EPI_ISL_437780, EPI_ISL_437781, EPI_ISL_437782, EPI_ISL_437783, EPI_ISL_437784, EPI_ISL_437785, EPI_ISL_437786, EPI_ISL_437787, EPI_ISL_437788, EPI_ISL_437789, EPI_ISL_437790, EPI_ISL_437791, EPI_ISL_437792, EPI_ISL_437793, EPI_ISL_437794, EPI_ISL_437795, EPI_ISL_437796, EPI_ISL_437797, EPI_ISL_437798, EPI_ISL_437799, EPI_ISL_437800, EPI_ISL_437801, EPI_ISL_437802 | | | |
| see above | Virginia DCLS | Virginia DCLS | Virginia DCLS |
| EPI_ISL_437803, EPI_ISL_437804, EPI_ISL_437805, EPI_ISL_437806, EPI_ISL_437807, EPI_ISL_437808, EPI_ISL_437809, EPI_ISL_437810, EPI_ISL_437811, EPI_ISL_437812, EPI_ISL_437813, EPI_ISL_437814, EPI_ISL_437815, EPI_ISL_437816, EPI_ISL_437817, EPI_ISL_437818, EPI_ISL_437819, EPI_ISL_437820, EPI_ISL_437821, EPI_ISL_437822, EPI_ISL_437823, EPI_ISL_437824, EPI_ISL_437825, EPI_ISL_437826, EPI_ISL_437827, EPI_ISL_437828, EPI_ISL_437829, EPI_ISL_437830, EPI_ISL_437831, EPI_ISL_437832, EPI_ISL_437833, EPI_ISL_437834, EPI_ISL_437835, EPI_ISL_437836, EPI_ISL_437837, EPI_ISL_437838, EPI_ISL_437839, EPI_ISL_437840, EPI_ISL_437841, EPI_ISL_437842, EPI_ISL_437843, EPI_ISL_437844, EPI_ISL_437845, EPI_ISL_437846, EPI_ISL_437847, EPI_ISL_437848, EPI_ISL_437849, EPI_ISL_437850, EPI_ISL_437851, EPI_ISL_437852, EPI_ISL_437853, EPI_ISL_437854, EPI_ISL_437855, EPI_ISL_437856, EPI_ISL_437857, EPI_ISL_437858, EPI_ISL_437859, EPI_ISL_437860, EPI_ISL_437861, EPI_ISL_437862, EPI_ISL_437863, EPI_ISL_437864, EPI_ISL_437865, EPI_ISL_437866, EPI_ISL_437868, EPI_ISL_437869, EPI_ISL_437870, EPI_ISL_437871, EPI_ISL_437872 | | | |
| see above | UW Virology Lab | UW Virology Lab | Pavitra Roychoudhury, Hong Xie, Keith Jerome, Alexander Greninger |
| EPI_ISL_437873 | Alaska State Virology Laboratory | Alaska State Virology Laboratory | Jack Chen, Ph.D. |
| EPI_ISL_438140, EPI_ISL_438141, EPI_ISL_438142, EPI_ISL_438143, EPI_ISL_438144, EPI_ISL_438145, EPI_ISL_438146, EPI_ISL_438147, EPI_ISL_438148, EPI_ISL_438149, EPI_ISL_438150, EPI_ISL_438151, EPI_ISL_438152, EPI_ISL_438153, EPI_ISL_438154, EPI_ISL_438155, EPI_ISL_438156, EPI_ISL_438157, EPI_ISL_438158, EPI_ISL_438159, EPI_ISL_438160, EPI_ISL_438161, EPI_ISL_438162, EPI_ISL_438163, EPI_ISL_438164, EPI_ISL_438165, EPI_ISL_438166, EPI_ISL_438167, EPI_ISL_438168, EPI_ISL_438169, EPI_ISL_438170, EPI_ISL_438171, EPI_ISL_438172, EPI_ISL_438173, EPI_ISL_438174, EPI_ISL_438175 | | | |
| see above | Seattle Flu Study | Seattle Flu Study | Chu et al |
| EPI_ISL_438176, EPI_ISL_438177, EPI_ISL_438178, EPI_ISL_438179, EPI_ISL_438180, EPI_ISL_438181, EPI_ISL_438182, EPI_ISL_438183, EPI_ISL_438184, EPI_ISL_438185, EPI_ISL_438186, EPI_ISL_438187, EPI_ISL_438188, EPI_ISL_438189, EPI_ISL_438190, EPI_ISL_438191, EPI_ISL_438192, EPI_ISL_438193, EPI_ISL_438194, EPI_ISL_438195, EPI_ISL_438196, EPI_ISL_438197, EPI_ISL_438198, EPI_ISL_438199, EPI_ISL_438200, EPI_ISL_438201, EPI_ISL_438202, EPI_ISL_438203, EPI_ISL_438204, EPI_ISL_438205, EPI_ISL_438206, EPI_ISL_438207, EPI_ISL_438208, EPI_ISL_438209, EPI_ISL_438210, EPI_ISL_438211, EPI_ISL_438212, EPI_ISL_438213, EPI_ISL_438214, EPI_ISL_438215, EPI_ISL_438216, EPI_ISL_438217, EPI_ISL_438218, EPI_ISL_438219, EPI_ISL_438220, EPI_ISL_438221 | | | |
| see above | Washington State Department of Health | Seattle Flu Study | Chu et al |
| EPI_ISL_438222, EPI_ISL_438223, EPI_ISL_438224, EPI_ISL_438225, EPI_ISL_438226, EPI_ISL_438227, EPI_ISL_438228, EPI_ISL_438229, EPI_ISL_438230, EPI_ISL_438231, EPI_ISL_438232, EPI_ISL_438233, EPI_ISL_438234 | | | |
| see above | Johns Hopkins Hospital Department of Pathology | Johns Hopkins Hospital Department of Pathology | Peter M. Thielen, Thomas Mehoke, Shirlee Wohl, Srividya Ramakrishnan, Melanie Kirsche,_Amanda Ernlund,_Oluwaseun Falade-Nwulia, Timothy Gilpatrick, Paul Morris, Norah Sadowski, N_di_ Trovao, Victoria Gniazdowski, Michael Schatz, Stuart C. Ray, Winston Timp, Heba Mostafa |
| EPI_ISL_438235, EPI_ISL_438236, EPI_ISL_438237, EPI_ISL_438238, EPI_ISL_438239, EPI_ISL_438240, EPI_ISL_438241, EPI_ISL_438242, EPI_ISL_438243, EPI_ISL_438244, EPI_ISL_438245, EPI_ISL_438246, EPI_ISL_438247 | | | |
| see above | Johns Hopkins Hospital Department of Pathology | Johns Hopkins Hospital Department of Pathology | Peter M. Thielen, Thomas Mehoke, Shirlee Wohl, Srividya Ramakrishnan, Melanie Kirsche, Amanda Ernlund, Oluwaseun Falade-Nwulia, Timothy Gilpatrick, Paul Morris, Norah Sadowski, Nidia Trovao, Victoria Gniazdowski, Michael Schatz, Stuart C. Ray, Winston Timp, Heba Mostafa |
| EPI_ISL_443183 | M Health Fairview | University of Minnesota Genomics Center | Daryl M. Gohl, John Garbe, Patrick Grady, Jerry Daniel, Ray Watson, Benjamin Auch, Andrew Nelson, Sophia Yohe, and Kenneth B. Beckman |
| EPI_ISL_443184, EPI_ISL_443185, EPI_ISL_443186 | UW Virology Lab | UW Virology Lab | Pavitra Roychoudhury, Hong Xie, Keith Jerome, Alexander Greninger |
| EPI_ISL_443253, EPI_ISL_443254, EPI_ISL_443255, EPI_ISL_443256, EPI_ISL_443257 | M Health Fairview | University of Minnesota Genomics Center | Daryl M. Gohl, John Garbe, Patrick Grady, Jerry Daniel, Ray Watson, Benjamin Auch, Andrew Nelson, Sophia Yohe, and Kenneth B. Beckman |
| EPI_ISL_444022 | Baylor College of Medicine | Baylor College of Medicine: HGSC | Vasanthi Avadhanula, Erin Nicholson, David Henke, Pedro Piedra, Harsha Doddapaneni, Donna Muzny, Qingchang Meng, Hsu Chao, Zeineen Momin, Hua Shen, George Weissenberger, Kavya Kottapalli, Yimiti Meiheerguli, Sejal Salvi, Ginger Metcalf, Vipin Menon, Sara J.J. Cregeen, Matthew C. Ross, Tulin Ayvaz, Richard Sucgang, Kristi L. Hoffman, Matthew Wong, Joseph F. Petrosino |
| EPI_ISL_444023, EPI_ISL_444024, EPI_ISL_444025, EPI_ISL_444026 | County of Santa Clara Public Health | Chan-Zuckerberg Biohub | CZB Cliahub Consortium |
| EPI_ISL_444051, EPI_ISL_444052, EPI_ISL_444053, EPI_ISL_444054, EPI_ISL_444055, EPI_ISL_444056, EPI_ISL_444057, EPI_ISL_444058, EPI_ISL_444059, EPI_ISL_444061, EPI_ISL_444062, EPI_ISL_444063, EPI_ISL_444064, EPI_ISL_444065, EPI_ISL_444066, EPI_ISL_444067, EPI_ISL_444068, EPI_ISL_444069, EPI_ISL_444070, EPI_ISL_444071, EPI_ISL_444073, EPI_ISL_444074, EPI_ISL_444075, EPI_ISL_444076, EPI_ISL_444077, EPI_ISL_444078 | | | |
| see above | UCSF Clinical Microbiology Laboratory | Chan-Zuckerberg Biohub | CZB Cliahub Consortium |
| EPI_ISL_444455 | Molecular Infectious Disease | Molecular Infectious Disease | Anderson,B.P., Rosenthal,S.H., Gerasimova,A., Kagan,R.M. and Owen,R. |
| EPI_ISL_444518 | unknown | Molecular Infectious Disease | Anderson,B.P., Rosenthal,S.H., Gerasimova,A., Kagan,R.M. and Owen,R. |
| EPI_ISL_444519 | Molecular Infectious Disease | Molecular Infectious Disease | Anderson,B.P., Rosenthal,S.H., Gerasimova,A., Kagan,R.M. and Owen,R. |
| EPI_ISL_444520, EPI_ISL_444521, EPI_ISL_444522, EPI_ISL_444523, EPI_ISL_444524, EPI_ISL_444525, EPI_ISL_444527, EPI_ISL_444528, EPI_ISL_444529, EPI_ISL_444530, EPI_ISL_444531, EPI_ISL_444532, EPI_ISL_444533, EPI_ISL_444534, EPI_ISL_444535, EPI_ISL_444536, EPI_ISL_444537, EPI_ISL_444538, EPI_ISL_444539, EPI_ISL_444540, EPI_ISL_444541, EPI_ISL_444542, EPI_ISL_444543, EPI_ISL_444544, EPI_ISL_444545, EPI_ISL_444546, EPI_ISL_444547, EPI_ISL_444548, EPI_ISL_444549, EPI_ISL_444550, EPI_ISL_444551, EPI_ISL_444552, EPI_ISL_444553, EPI_ISL_444555, EPI_ISL_444556, EPI_ISL_444558, EPI_ISL_444559, EPI_ISL_444560, EPI_ISL_444561, EPI_ISL_444562, EPI_ISL_444563, EPI_ISL_444564, EPI_ISL_444565, EPI_ISL_444566, EPI_ISL_444567, EPI_ISL_444568, EPI_ISL_444569, EPI_ISL_444570, EPI_ISL_444571, EPI_ISL_444572, EPI_ISL_444573, EPI_ISL_444574, EPI_ISL_444575, EPI_ISL_444576, EPI_ISL_444577, EPI_ISL_444578, EPI_ISL_444579, EPI_ISL_444580, EPI_ISL_444581, EPI_ISL_444582, EPI_ISL_444583, EPI_ISL_444585, EPI_ISL_444586, EPI_ISL_444587, EPI_ISL_444588, EPI_ISL_444589, EPI_ISL_444590, EPI_ISL_444591, EPI_ISL_444592, EPI_ISL_444593, EPI_ISL_444594, EPI_ISL_444595, EPI_ISL_444596, EPI_ISL_444597, EPI_ISL_444598, EPI_ISL_444599, EPI_ISL_444600, EPI_ISL_444601, EPI_ISL_444602, EPI_ISL_444603, EPI_ISL_444604, EPI_ISL_444605, EPI_ISL_444606, EPI_ISL_444607, EPI_ISL_444608, EPI_ISL_444609 | | | |
| see above | Northwestern Memorial Hospital | Ozer Lab | Ramon Lorenzo-Redondo, Hannah H. Nam, Scott C. Roberts, Lacy M. Simons, Chad J. Achenbach, Lawrence J. Jennings, Chao Qi, Alan R. Hauser, Michael G. Ison, Judd F. Hultquist, Egon A. Ozer |
| EPI_ISL_444613, EPI_ISL_444614, EPI_ISL_444615, EPI_ISL_444616, EPI_ISL_444617, EPI_ISL_444618, EPI_ISL_444619, EPI_ISL_444620, EPI_ISL_444621, EPI_ISL_444622, EPI_ISL_444623, EPI_ISL_444624, EPI_ISL_444625, EPI_ISL_444626, EPI_ISL_444627, EPI_ISL_444628, EPI_ISL_444629, EPI_ISL_444630, EPI_ISL_444631, EPI_ISL_444632, EPI_ISL_444633, EPI_ISL_444634, EPI_ISL_444635, EPI_ISL_444636, EPI_ISL_444637, EPI_ISL_444638, EPI_ISL_444639, EPI_ISL_444640, EPI_ISL_444641, EPI_ISL_444642, EPI_ISL_444643, EPI_ISL_444644, EPI_ISL_444645, EPI_ISL_444646, EPI_ISL_444647, EPI_ISL_444648, EPI_ISL_444650, EPI_ISL_444651, EPI_ISL_444652, EPI_ISL_444653, EPI_ISL_444654, EPI_ISL_444655, EPI_ISL_444656, EPI_ISL_444657, EPI_ISL_444658, EPI_ISL_444659, EPI_ISL_444660, EPI_ISL_444661, EPI_ISL_444662, EPI_ISL_444663, EPI_ISL_444664, EPI_ISL_444665, EPI_ISL_444666, EPI_ISL_444667, EPI_ISL_444668, EPI_ISL_444669, EPI_ISL_444670, EPI_ISL_444671, EPI_ISL_444673, EPI_ISL_444675, EPI_ISL_444677, EPI_ISL_444678, EPI_ISL_444679, EPI_ISL_444680, EPI_ISL_444681, EPI_ISL_444682, EPI_ISL_444683, EPI_ISL_444684, EPI_ISL_444685, EPI_ISL_444686, EPI_ISL_444687, EPI_ISL_444688, EPI_ISL_444690, EPI_ISL_444691, EPI_ISL_444692, EPI_ISL_444693, EPI_ISL_444694, EPI_ISL_444695, EPI_ISL_444696, EPI_ISL_444697, EPI_ISL_444698, EPI_ISL_444699, EPI_ISL_444700, EPI_ISL_444701, EPI_ISL_444702, EPI_ISL_444703, EPI_ISL_444704, EPI_ISL_444705, EPI_ISL_444706, EPI_ISL_444707, EPI_ISL_444708, EPI_ISL_444709, EPI_ISL_444710, EPI_ISL_444711, EPI_ISL_444712, EPI_ISL_444713, EPI_ISL_444714, EPI_ISL_444715, EPI_ISL_444716, EPI_ISL_444717, EPI_ISL_444718, EPI_ISL_444719, EPI_ISL_444720, EPI_ISL_444721, EPI_ISL_444722, EPI_ISL_444723, EPI_ISL_444724, EPI_ISL_444725, EPI_ISL_444726, EPI_ISL_444727, EPI_ISL_444728, EPI_ISL_444729, EPI_ISL_444730, EPI_ISL_444731, EPI_ISL_444732, EPI_ISL_444733, EPI_ISL_444734, EPI_ISL_444735, EPI_ISL_444736, EPI_ISL_444737, EPI_ISL_444738, EPI_ISL_444739, EPI_ISL_444740, EPI_ISL_444741, EPI_ISL_444742, EPI_ISL_444743, EPI_ISL_444744, EPI_ISL_444745, EPI_ISL_444746, EPI_ISL_444747, EPI_ISL_444748, EPI_ISL_444749, EPI_ISL_444750, EPI_ISL_444751, EPI_ISL_444752, EPI_ISL_444753, EPI_ISL_444754, EPI_ISL_444755, EPI_ISL_444756, EPI_ISL_444757, EPI_ISL_444758, EPI_ISL_444759, EPI_ISL_444760, EPI_ISL_444761, EPI_ISL_444762, EPI_ISL_444763, EPI_ISL_444764, EPI_ISL_444765, EPI_ISL_444766, EPI_ISL_444767, EPI_ISL_444768, EPI_ISL_444769, EPI_ISL_444770, EPI_ISL_444771, EPI_ISL_444772, EPI_ISL_444773, EPI_ISL_444774, EPI_ISL_444775, EPI_ISL_444776, EPI_ISL_444777, EPI_ISL_444779, EPI_ISL_444780, EPI_ISL_444781, EPI_ISL_444782, EPI_ISL_444783, EPI_ISL_444784, EPI_ISL_444786, EPI_ISL_444787, EPI_ISL_444788, EPI_ISL_444789, EPI_ISL_444790, EPI_ISL_444791 | | | |
| see above | NYU Langone Health | Departments of Pathology and Medicine, New York University School of Medicine | Maria Aguero-Rosenfeld, Brendan Belovarac, Margaret Black, Ludovic Boytard, John Cadley, Paolo Cotzia, John Chen, Dacia Dimartino, Xiaojun Feng, Tatyana Gindin, Emily Guzman, Adriana Heguy, Megan Hogan, Emily Huang, George Jour, Alireza Khodadadi-Jamayran, Lawrence H. Lin, Raven Luther, Andrew Lytle, Christian Marier, Matthew T. Maurano, Mark J. Mulligan, Peter Meyn, Raquel Ordonez Ciriza, Iman Osman, Jared Pinnell, Vanessa Raabe, Sitharam Ramaswami, Amy Rapkiewicz, Andre M. Ribeiro-dos-Santos, Marie Samanovic-Golden, Antonio Serrano, Guomiao Shen, Matija Snuderl, Theodore Vougiouklakis, Nick Vulpescu, Gael Westby, Paul Zappile, Yutong Zhang |
| EPI_ISL_444994, EPI_ISL_444995, EPI_ISL_444996, EPI_ISL_444997 | Naval Health Research Center | Naval Medical Research Center Biological Defense Research Directorate | Logan Voegtly, Regina Cer, Dessiree Pena-Gomez, Adrian Paskey,Kyle Long, Roger Pan, Melinda Balansay-Ames, Chris Myers, Ewell Hollis, Nathaniel Christy, Kimberly Bishop-Lilly |
| EPI_ISL_445006, EPI_ISL_445008, EPI_ISL_445009, EPI_ISL_445013, EPI_ISL_445015, EPI_ISL_445019, EPI_ISL_445024, EPI_ISL_445029, EPI_ISL_445030, EPI_ISL_445035, EPI_ISL_445036, EPI_ISL_445038, EPI_ISL_445041, EPI_ISL_445043, EPI_ISL_445044, EPI_ISL_445045, EPI_ISL_445046, EPI_ISL_445047, EPI_ISL_445048, EPI_ISL_445049 | | | |
| see above | Florida Bureau of Public Health | Florida Bureau of Public Health Laboratories | Sarah Schmedes, Jason Blanton |

| | | | |
|---|---|---|---|
| | Laboratories | | |
| EPI_ISL_445077 | M Health Fairview | University of Minnesota Genomics Center | Daryl M. Gohl, John Garbe, Patrick Grady, Jerry Daniel, Ray Watson, Benjamin Auch, Andrew Nelson, Sophia Yohe, and Kenneth B. Beckman |
| EPI_ISL_445078, EPI_ISL_445079, EPI_ISL_445080, EPI_ISL_445081, EPI_ISL_445082, EPI_ISL_445083, EPI_ISL_445084 | Baylor College of Medicine | Baylor College of Medicine: HGSC | Vasanthi Avadhanula, Erin Nicholson, David Henke, Pedro Piedra, Harsha Doddapaneni, Donna Muzny, Qingchang Meng, Hsu Chao, Zeineen Momin, Hua Shen, George Weissenberger, Kavya Kottapalli, Yimiti Meiheerguli, Sejal Salvi, Ginger Metcalf, Vipin Menon, Sara J.J. Cregeen, Matthew C. Ross, Tulin Ayvaz, Richard Sucgang, Kristi L. Hoffman, Matthew Wong, Joseph F. Petrosino |
| EPI_ISL_445094, EPI_ISL_445095, EPI_ISL_445096, EPI_ISL_445097, EPI_ISL_445098, EPI_ISL_445100, EPI_ISL_445101, EPI_ISL_445102, EPI_ISL_445103, EPI_ISL_445104, EPI_ISL_445105, EPI_ISL_445106, EPI_ISL_445107, EPI_ISL_445108, EPI_ISL_445109, EPI_ISL_445110, EPI_ISL_445111, EPI_ISL_445113, EPI_ISL_445114, EPI_ISL_445115, EPI_ISL_445116, EPI_ISL_445117 | | | |
| see above | UC San Diego Center for Advanced Laboratory Medicine | Andersen lab at Scripps Research | SEARCH Alliance San Diego with David Pride, Ji H Shin |
| EPI_ISL_445118 | Rady's Childrens Hospital | Andersen lab at Scripps Research | SEARCH Alliance San Diego |
| EPI_ISL_445119, EPI_ISL_445120, EPI_ISL_445121, EPI_ISL_445122, EPI_ISL_445123, EPI_ISL_445124, EPI_ISL_445125, EPI_ISL_445126, EPI_ISL_445127, EPI_ISL_445128, EPI_ISL_445129, EPI_ISL_445130, EPI_ISL_445131, EPI_ISL_445132, EPI_ISL_445133, EPI_ISL_445134, EPI_ISL_445135, EPI_ISL_445136, EPI_ISL_445137, EPI_ISL_445138, EPI_ISL_445139, EPI_ISL_445140, EPI_ISL_445141, EPI_ISL_445142, EPI_ISL_445143, EPI_ISL_445144, EPI_ISL_445145, EPI_ISL_445146, EPI_ISL_445147, EPI_ISL_445148, EPI_ISL_445149, EPI_ISL_445150, EPI_ISL_445151, EPI_ISL_445152, EPI_ISL_445153, EPI_ISL_445154, EPI_ISL_445155, EPI_ISL_445156, EPI_ISL_445157, EPI_ISL_445159, EPI_ISL_445160, EPI_ISL_445161, EPI_ISL_445162, EPI_ISL_445163 | | | |
| see above | Robert Garry lab | Andersen lab at Scripps Research | Allison Smither, Gilberto Sabino-Santos, Patricia Snarski, Lilia Melnik, Antoinette Bell, Kaylynn Genemaras, Arnaud Drouin, Dahlene Fusco, Robert Garry with SEARCH Alliance San Diego |
| EPI_ISL_445164, EPI_ISL_445166, EPI_ISL_445167, EPI_ISL_445168 | Scripps Medical Laboratory | Andersen lab at Scripps Research | SEARCH Alliance San Diego with Michael Quigley, Ellen Stefanski, Ian Mchardy |
| EPI_ISL_445169, EPI_ISL_445170, EPI_ISL_445171, EPI_ISL_445172, EPI_ISL_445173, EPI_ISL_445174, EPI_ISL_445175, EPI_ISL_445176, EPI_ISL_445177, EPI_ISL_445178, EPI_ISL_445179, EPI_ISL_445180, EPI_ISL_445181, EPI_ISL_445182 | | | |
| see above | UCSF Clinical Microbiology Laboratory | Chan-Zuckerberg Biohub | CZB Cliahub Consortium |
| EPI_ISL_447057, EPI_ISL_447058, EPI_ISL_447059, EPI_ISL_447060, EPI_ISL_447061, EPI_ISL_447062, EPI_ISL_447063, EPI_ISL_447064, EPI_ISL_447065, EPI_ISL_447066, EPI_ISL_447067, EPI_ISL_447068, EPI_ISL_447069, EPI_ISL_447070, EPI_ISL_447071, EPI_ISL_447072, EPI_ISL_447073, EPI_ISL_447074, EPI_ISL_447075, EPI_ISL_447076, EPI_ISL_447077, EPI_ISL_447078, EPI_ISL_447079, EPI_ISL_447080, EPI_ISL_447081, EPI_ISL_447082, EPI_ISL_447083, EPI_ISL_447084, EPI_ISL_447085, EPI_ISL_447086, EPI_ISL_447087, EPI_ISL_447088, EPI_ISL_447089, EPI_ISL_447090, EPI_ISL_447091, EPI_ISL_447092, EPI_ISL_447093, EPI_ISL_447094, EPI_ISL_447095, EPI_ISL_447096, EPI_ISL_447097, EPI_ISL_447098, EPI_ISL_447099, EPI_ISL_447100, EPI_ISL_447101, EPI_ISL_447102, EPI_ISL_447103, EPI_ISL_447104, EPI_ISL_447105, EPI_ISL_447106, EPI_ISL_447107, EPI_ISL_447108, EPI_ISL_447109, EPI_ISL_447110, EPI_ISL_447111, EPI_ISL_447112, EPI_ISL_447113, EPI_ISL_447114, EPI_ISL_447115, EPI_ISL_447116, EPI_ISL_447117, EPI_ISL_447118, EPI_ISL_447163, EPI_ISL_447164, EPI_ISL_447165, EPI_ISL_447166, EPI_ISL_447167, EPI_ISL_447168, EPI_ISL_447170, EPI_ISL_447171, EPI_ISL_447172, EPI_ISL_447173, EPI_ISL_447174, EPI_ISL_447175, EPI_ISL_447176, EPI_ISL_447177, EPI_ISL_447178, EPI_ISL_447179, EPI_ISL_447180, EPI_ISL_447181, EPI_ISL_447182, EPI_ISL_447183, EPI_ISL_447184, EPI_ISL_447185, EPI_ISL_447186, EPI_ISL_447187, EPI_ISL_447188, EPI_ISL_447189, EPI_ISL_447190, EPI_ISL_447191, EPI_ISL_447192, EPI_ISL_447193, EPI_ISL_447194, EPI_ISL_447195, EPI_ISL_447196, EPI_ISL_447197, EPI_ISL_447198, EPI_ISL_447199, EPI_ISL_447200, EPI_ISL_447201, EPI_ISL_447202, EPI_ISL_447203, EPI_ISL_447204, EPI_ISL_447205, EPI_ISL_447206, EPI_ISL_447207, EPI_ISL_447208, EPI_ISL_447209, EPI_ISL_447210, EPI_ISL_447211, EPI_ISL_447212, EPI_ISL_447213, EPI_ISL_447214, EPI_ISL_447215, EPI_ISL_447216, EPI_ISL_447217, EPI_ISL_447218, EPI_ISL_447219, EPI_ISL_447220, EPI_ISL_447221, EPI_ISL_447222, EPI_ISL_447223, EPI_ISL_447224, EPI_ISL_447225, EPI_ISL_447226, EPI_ISL_447227, EPI_ISL_447228, EPI_ISL_447229 | | | |
| see above | Michigan Department of Health and Human Services, Bureau of Laboratories | Michigan Department of Health and Human Services, Bureau of Laboratories | Blankenship HM, Riner D, Soehnlen MK |
| EPI_ISL_447588 | Lednicky Lab | Lednicky lab | Elbadry,M.A., Subramaniam,K., Waltzek,T.B., Gibson,J.C., Stephenson,C.J., Alam,M.M., Morris,J.G. Jr. and Lednicky,J.A. |
| EPI_ISL_447589 | University of Florida, Lednicky Lab | University of Florida, Lednicky Lab | Elbadry,M.A., Subramaniam,K., Waltzek,T.B., Gibson,J.C., Stephenson,C.J., Alam,M.M., Morris,J.G. Jr. and Lednicky,J.A. |
| EPI_ISL_447840 | DC Public Health Lab/ Dept. of Forensic Sciences | Pathogen Discovery, Respiratory Viruses Branch, Division of Viral Diseases, Centers for Disease Control and Prevention | Krista Queen, Yan Li, Anna Uehara, Jing Zhang, Ying Tao, Clinton R. Paden, Haibin Wang, Jasmine Padilla, Mary S. Keckler, Alison S. Laufer Halpin, Justin Lee, Christopher A. Elkins, Suxiang Tong |
| EPI_ISL_447841 | FL Bureau of Public Health Laboratories-Tampa | Pathogen Discovery, Respiratory Viruses Branch, Division of Viral Diseases, Centers for Disease Control and Prevention | Krista Queen, Yan Li, Anna Uehara, Jing Zhang, Ying Tao, Clinton R. Paden, Haibin Wang, Jasmine Padilla, Mary S. Keckler, Alison S. Laufer Halpin, Justin Lee, Christopher A. Elkins, Suxiang Tong |
| EPI_ISL_447842 | IA State Hygienic Laboratory | Pathogen Discovery, Respiratory Viruses Branch, Division of Viral Diseases, Centers for Disease Control and Prevention | Krista Queen, Yan Li, Anna Uehara, Jing Zhang, Ying Tao, Clinton R. Paden, Haibin Wang, Jasmine Padilla, Mary S. Keckler, Alison S. Laufer Halpin, Justin Lee, Christopher A. Elkins, Suxiang Tong |
| EPI_ISL_447843 | MD DOH Laboratories Administration | Pathogen Discovery, Respiratory Viruses Branch, Division of Viral Diseases, Centers for Disease Control and Prevention | Krista Queen, Yan Li, Anna Uehara, Jing Zhang, Ying Tao, Clinton R. Paden, Haibin Wang, Jasmine Padilla, Mary S. Keckler, Alison S. Laufer Halpin, Justin Lee, Christopher A. Elkins, Suxiang Tong |
| EPI_ISL_447844 | PA Department of Health, Bureau of Laboratories | Pathogen Discovery, Respiratory Viruses Branch, Division of Viral Diseases, Centers for Disease Control and Prevention | Krista Queen, Yan Li, Anna Uehara, Jing Zhang, Ying Tao, Clinton R. Paden, Haibin Wang, Jasmine Padilla, Mary S. Keckler, Alison S. Laufer Halpin, Justin Lee, Christopher A. Elkins, Suxiang Tong |
| EPI_ISL_447845 | PR - Biological and Chemical Emergencies Lab Office of Public Health Preparedness and Response | Pathogen Discovery, Respiratory Viruses Branch, Division of Viral Diseases, Centers for Disease Control and Prevention | Krista Queen, Yan Li, Anna Uehara, Jing Zhang, Ying Tao, Clinton R. Paden, Haibin Wang, Jasmine Padilla, Mary S. Keckler, Alison S. Laufer Halpin, Justin Lee, Christopher A. Elkins, Suxiang Tong |
| EPI_ISL_447846 | VT Dept. of Health Laboratory | Pathogen Discovery, Respiratory Viruses Branch, Division of Viral Diseases, Centers for Disease Control and Prevention | Krista Queen, Yan Li, Anna Uehara, Jing Zhang, Ying Tao, Clinton R. Paden, Haibin Wang, Jasmine Padilla, Mary S. Keckler, Alison S. Laufer Halpin, Justin Lee, Christopher A. Elkins, Suxiang Tong |
| EPI_ISL_447886 | unknown | Pathogen Discovery | Ying Tao, Yan Li, Jing Zhang, Clinton R. Paden, Krista Queen, Anna Uehara, Haibin Wang, Julu Bhatnagar, Suxiang Tong |
| EPI_ISL_447887, EPI_ISL_447888, EPI_ISL_447889, EPI_ISL_447890, EPI_ISL_447891, EPI_ISL_447892, EPI_ISL_447893, EPI_ISL_447894, EPI_ISL_447895, EPI_ISL_447896 | University of California, Davis | Chan-Zuckerberg Biohub | CZB Cliahub Consortium |
| EPI_ISL_447903 | University of Florida | University of Florida | Elbadry,M.A., Subramaniam,K., Waltzek,T.B., Lauzardo,M., Gibson,J.C., Stephenson,C.J., Alam,M.M., Morris,J.G. Jr. and Lednicky,J.A. |
| EPI_ISL_447905 | University of Florida | University of Florida | Elbadry,M.A., Subramaniam,K., Waltzek,T.B., Gibson,J.C., Stephenson,C.J., Alam,M.M., Morris,J.G. Jr. and Lednicky,J.A. |
| EPI_ISL_449801, EPI_ISL_449802, EPI_ISL_449803, EPI_ISL_449804, EPI_ISL_449805, EPI_ISL_449806, EPI_ISL_449807, EPI_ISL_449808, EPI_ISL_449809, EPI_ISL_449810, EPI_ISL_449812, EPI_ISL_449815, EPI_ISL_449816, EPI_ISL_449817, EPI_ISL_449818, EPI_ISL_449819, EPI_ISL_449820, EPI_ISL_449821, EPI_ISL_449822, EPI_ISL_449823, EPI_ISL_449824, EPI_ISL_449825, EPI_ISL_449826, EPI_ISL_449827, EPI_ISL_449828, EPI_ISL_449829, EPI_ISL_449830, EPI_ISL_449831, EPI_ISL_449832, EPI_ISL_449833, EPI_ISL_449834, EPI_ISL_449835, EPI_ISL_449836, EPI_ISL_449837, EPI_ISL_449838 | | | |
| see above | Utah Public Health Laboratory | Utah Public Health Laboratory | Erin Young, Kelly Oakeson |
| EPI_ISL_449839, EPI_ISL_449840, EPI_ISL_449841, EPI_ISL_449842, EPI_ISL_449843, EPI_ISL_449844, EPI_ISL_449845, EPI_ISL_449846, EPI_ISL_449847, EPI_ISL_449848, EPI_ISL_449849, EPI_ISL_449850, EPI_ISL_449851, EPI_ISL_449852, EPI_ISL_449853, EPI_ISL_449854, EPI_ISL_449855, EPI_ISL_449856, EPI_ISL_449857, EPI_ISL_449858, EPI_ISL_449859, EPI_ISL_449860, EPI_ISL_449861, EPI_ISL_449862, EPI_ISL_449863, EPI_ISL_449864, EPI_ISL_449865, EPI_ISL_449866, EPI_ISL_449867, EPI_ISL_449868, EPI_ISL_449869, EPI_ISL_449870, EPI_ISL_449871, EPI_ISL_449872, EPI_ISL_449873, EPI_ISL_449874, EPI_ISL_449875, EPI_ISL_449876, EPI_ISL_449877, EPI_ISL_449878, EPI_ISL_449879, EPI_ISL_449880, EPI_ISL_449881, EPI_ISL_449882, EPI_ISL_449883, EPI_ISL_449884, EPI_ISL_449885, EPI_ISL_449886, EPI_ISL_449887, EPI_ISL_449888, EPI_ISL_449889, EPI_ISL_449890, EPI_ISL_449891, EPI_ISL_449892, EPI_ISL_449893, EPI_ISL_449894, EPI_ISL_449895, EPI_ISL_449896, EPI_ISL_449897, EPI_ISL_449898, EPI_ISL_449899, EPI_ISL_449900, EPI_ISL_449901, EPI_ISL_449902, EPI_ISL_449903, EPI_ISL_449904, EPI_ISL_449905, EPI_ISL_449906, EPI_ISL_449907, EPI_ISL_449908, EPI_ISL_449909, EPI_ISL_449910, EPI_ISL_449911, EPI_ISL_449912, EPI_ISL_449913, EPI_ISL_449914, EPI_ISL_449915, EPI_ISL_449916, EPI_ISL_449917, EPI_ISL_449918, EPI_ISL_449919, EPI_ISL_449920, EPI_ISL_449921, EPI_ISL_449922, EPI_ISL_449923, EPI_ISL_449925, EPI_ISL_449926, EPI_ISL_449927, EPI_ISL_449928, EPI_ISL_449929, EPI_ISL_449930, EPI_ISL_449931, EPI_ISL_449932, EPI_ISL_449933, EPI_ISL_449934, EPI_ISL_449935, EPI_ISL_449936, EPI_ISL_449937, EPI_ISL_449938, EPI_ISL_449939, EPI_ISL_449940, EPI_ISL_449941, EPI_ISL_449943, EPI_ISL_449944, EPI_ISL_449945, EPI_ISL_449946, EPI_ISL_449947, EPI_ISL_449948, EPI_ISL_449949, EPI_ISL_449951, EPI_ISL_449952, EPI_ISL_449953, EPI_ISL_449954, EPI_ISL_449955, EPI_ISL_449956, EPI_ISL_449957, EPI_ISL_449958, EPI_ISL_449959, EPI_ISL_449960, EPI_ISL_449962, EPI_ISL_449963, EPI_ISL_449964, EPI_ISL_449965, EPI_ISL_449966, EPI_ISL_449967, EPI_ISL_449968, EPI_ISL_449969, EPI_ISL_449970, EPI_ISL_449971, EPI_ISL_449972, EPI_ISL_449973, EPI_ISL_449974, EPI_ISL_449975, EPI_ISL_449977, EPI_ISL_449978, EPI_ISL_449979, EPI_ISL_449980, EPI_ISL_449981, EPI_ISL_449982, EPI_ISL_449983, EPI_ISL_449984, EPI_ISL_449985, EPI_ISL_449986, EPI_ISL_449987, EPI_ISL_449988, EPI_ISL_449989, EPI_ISL_449990, EPI_ISL_449991, EPI_ISL_449992, EPI_ISL_449994, EPI_ISL_449995, EPI_ISL_449996, EPI_ISL_449997, EPI_ISL_449998, EPI_ISL_449999, EPI_ISL_450000, EPI_ISL_450001, EPI_ISL_450002, EPI_ISL_450003 | | | |
| see above | Washington State Department of Health | Seattle Flu Study | Chu et al |
| EPI_ISL_450004, EPI_ISL_450005, EPI_ISL_450006, EPI_ISL_450007, EPI_ISL_450008, EPI_ISL_450009, EPI_ISL_450010, EPI_ISL_450011, EPI_ISL_450012, EPI_ISL_450013, EPI_ISL_450014, EPI_ISL_450015, EPI_ISL_450016, EPI_ISL_450017, EPI_ISL_450018, EPI_ISL_450019, EPI_ISL_450020, EPI_ISL_450021, EPI_ISL_450022, EPI_ISL_450023, EPI_ISL_450024, EPI_ISL_450025, EPI_ISL_450026, EPI_ISL_450027, EPI_ISL_450028, EPI_ISL_450029, EPI_ISL_450030, EPI_ISL_450031, EPI_ISL_450032, EPI_ISL_450033, EPI_ISL_450034, EPI_ISL_450035, EPI_ISL_450036, EPI_ISL_450037, EPI_ISL_450038, EPI_ISL_450039, EPI_ISL_450040, EPI_ISL_450041, EPI_ISL_450042, EPI_ISL_450043, EPI_ISL_450044, EPI_ISL_450045, EPI_ISL_450046, EPI_ISL_450047, EPI_ISL_450048, EPI_ISL_450049, EPI_ISL_450050, EPI_ISL_450051, EPI_ISL_450052, EPI_ISL_450053, EPI_ISL_450054, EPI_ISL_450055, EPI_ISL_450056, EPI_ISL_450057, EPI_ISL_450058, EPI_ISL_450059, EPI_ISL_450060, EPI_ISL_450061, EPI_ISL_450062, EPI_ISL_450063, EPI_ISL_450064, EPI_ISL_450065, EPI_ISL_450066, EPI_ISL_450067, EPI_ISL_450068, EPI_ISL_450069, EPI_ISL_450070, EPI_ISL_450071, EPI_ISL_450072, EPI_ISL_450074, EPI_ISL_450075, EPI_ISL_450076, EPI_ISL_450077, EPI_ISL_450078, EPI_ISL_450079, EPI_ISL_450080, EPI_ISL_450081, EPI_ISL_450082, EPI_ISL_450083, EPI_ISL_450084, EPI_ISL_450085, EPI_ISL_450086, EPI_ISL_450087, EPI_ISL_450088, EPI_ISL_450089, EPI_ISL_450090, EPI_ISL_450091, EPI_ISL_450092, EPI_ISL_450093, EPI_ISL_450094, EPI_ISL_450095, EPI_ISL_450096, EPI_ISL_450097, EPI_ISL_450098, EPI_ISL_450099, EPI_ISL_450100, EPI_ISL_450101, EPI_ISL_450102, EPI_ISL_450103, EPI_ISL_450104, EPI_ISL_450105, EPI_ISL_450106, EPI_ISL_450107, EPI_ISL_450108, EPI_ISL_450109, EPI_ISL_450110, EPI_ISL_450111, EPI_ISL_450112, EPI_ISL_450113, EPI_ISL_450114, EPI_ISL_450115, EPI_ISL_450116, EPI_ISL_450117, EPI_ISL_450118, EPI_ISL_450119, EPI_ISL_450120, EPI_ISL_450121, EPI_ISL_450122, EPI_ISL_450123, EPI_ISL_450124, EPI_ISL_450125, EPI_ISL_450126, EPI_ISL_450127, EPI_ISL_450128, EPI_ISL_450129, EPI_ISL_450130, EPI_ISL_450131, EPI_ISL_450132, EPI_ISL_450133, EPI_ISL_450134, EPI_ISL_450135, EPI_ISL_450136, EPI_ISL_450137, EPI_ISL_450138, EPI_ISL_450139, EPI_ISL_450140, EPI_ISL_450141, EPI_ISL_450142, EPI_ISL_450143, EPI_ISL_450144, EPI_ISL_450145, EPI_ISL_450146, EPI_ISL_450147, EPI_ISL_450148, EPI_ISL_450149, EPI_ISL_450150, EPI_ISL_450151, EPI_ISL_450152, EPI_ISL_450153, EPI_ISL_450154, EPI_ISL_450155, EPI_ISL_450156, EPI_ISL_450157, EPI_ISL_450158, EPI_ISL_450159, EPI_ISL_450160, EPI_ISL_450161, EPI_ISL_450162, EPI_ISL_450163, EPI_ISL_450164, EPI_ISL_450165, EPI_ISL_450166, EPI_ISL_450167, EPI_ISL_450168, EPI_ISL_450169, EPI_ISL_450170 | | | |
| see above | MSHS Clinical Microbiology Laboratories | MSHS Pathogen Surveillance Program | Ana S. Gonzalez-Reiche, Mitchell Sullivan, Ajay Obla, Gopi Patel, Emilia Sordillo, Melissa Gitman, Alberto Paniz-mondolfi, Matthew Hernandez, Shelcie Fabre, Jose Polanco, Zenab Khan, Bremy Alburquerque, Jayeeta Dutta, Juan Soto, Shwetha Sridhar Hara, Ying-Chih Wang, Melissa Smith, Robert Sebra, Lisa Miorin, Wen-chun Liu, Randy Albrecht, Judith Aberg, Florian Krammer, Adolfo Garcia-Sastre, Viviana Simon, Harm van Bakel |
| EPI_ISL_450173, EPI_ISL_450174, EPI_ISL_450175, EPI_ISL_450176, EPI_ISL_450177, EPI_ISL_450178, EPI_ISL_450179, EPI_ISL_450180, EPI_ISL_450181, EPI_ISL_450182, EPI_ISL_450183, EPI_ISL_450184, EPI_ISL_450185 | | | |
| see above | Robert Garry lab | Andersen lab at Scripps Research | Allison Smither, Gilberto Sabino-Santos, Patricia Snarski, Lilia Melnik, Antoinette Bell, Kaylynn Genemaras, Arnaud Drouin, Dahlene Fusco, Robert Garry with SEARCH Alliance San Diego |
| EPI_ISL_450190 | Rady's Childrens Hospital | Andersen lab at Scripps Research | SEARCH Alliance San Diego |
| EPI_ISL_450191, EPI_ISL_450192 | Scripps Medical Laboratory | Andersen lab at Scripps Research | SEARCH Alliance San Diego with Michael Quigley, Ellen Stefanski, Ian Mchardy |
| EPI_ISL_450231 | Robert Garry lab | Andersen lab at Scripps Research | Allison Smither, Gilberto Sabino-Santos, Patricia Snarski, Lilia Melnik, Antoinette Bell, Kaylynn Genemaras, Arnaud Drouin, Dahlene Fusco, Robert Garry with SEARCH Alliance San Diego |
| EPI_ISL_450232, EPI_ISL_450233, | UCSF Clinical Microbiology Laboratory | Chiu Laboratory, University of California, San Francisco | Xianding Deng, Scot Federman, Wei Gu, and Charles Y. Chiu |

| | | | |
|---|---|---|---|
| EPI_ISL_450234, EPI_ISL_450235, EPI_ISL_450236, EPI_ISL_450237, EPI_ISL_450238, EPI_ISL_450239, EPI_ISL_450240 | | | |
| EPI_ISL_450393, EPI_ISL_450394, EPI_ISL_450395, EPI_ISL_450396, EPI_ISL_450397, EPI_ISL_450398, EPI_ISL_450399, EPI_ISL_450400, EPI_ISL_450401, EPI_ISL_450402 | NYU Langone Health | Departments of Pathology and Medicine, New York University School of Medicine | Maria Aguero-Rosenfeld, Brendan Belovarac, Margaret Black, Ludovic Boytard, John Cadley, Paolo Cotzia, John Chen, Dacia Dimartino, Xiaojun Feng, Tatyana Gindin, Emily Guzman, Adriana Heguy, Megan Hogan, Emily Huang, George Jour, Alireza Khodadadi-Jamayran, Lawrence H. Lin, Raven Luther, Andrew Lytle, Christian Marier, Matthew T. Maurano, Mark J. Mulligan, Peter Meyn, Raquel Ordonez Ciriza, Iman Osman, Jared Pinnell, Vanessa Raabe, Sitharam Ramaswami, Amy Rapkiewicz, Andre M. Ribeiro-dos-Santos, Marie Samanovic-Golden, Antonio Serrano, Guomiao Shen, Matija Snuderl, Theodore Vougiouklakis, Nick Vulpescu, Gael Westby, Paul Zappile, Yutong Zhang |
| EPI_ISL_450445, EPI_ISL_450446, EPI_ISL_450447, EPI_ISL_450448, EPI_ISL_450449, EPI_ISL_450450, EPI_ISL_450451, EPI_ISL_450452, EPI_ISL_450453, EPI_ISL_450454, EPI_ISL_450455, EPI_ISL_450456, EPI_ISL_450457, EPI_ISL_450458, EPI_ISL_450459, EPI_ISL_450460, EPI_ISL_450461, EPI_ISL_450462, EPI_ISL_450463, EPI_ISL_450464, EPI_ISL_450465, EPI_ISL_450466, EPI_ISL_450467, EPI_ISL_450468, EPI_ISL_450469, EPI_ISL_450470, EPI_ISL_450471, EPI_ISL_450472, EPI_ISL_450473, EPI_ISL_450474, EPI_ISL_450475, EPI_ISL_450476, EPI_ISL_450477, EPI_ISL_450478, EPI_ISL_450479, EPI_ISL_450480, EPI_ISL_450481 | | | |
| see above | Stanford clinical virology lab | Chan-Zuckerberg Biohub | Benjamin Pinksy, Katharine Walter, Victoria N. Parikh, John Gorzynski, Hannah N. DeJong, Matthew T. Wheeler, Jason Andrews, Manuel Rivas, Carlos Bustamante, Euan Ashley, with CZB Cliahub Consortium |
| EPI_ISL_450484, EPI_ISL_450485, EPI_ISL_450486, EPI_ISL_450487 | unknown | Data Science | Carroll,T.D., Tran,N.K., Cohen,S.H., Miller,C.J. |
| EPI_ISL_450538, EPI_ISL_450539, EPI_ISL_450540, EPI_ISL_450541, EPI_ISL_450542, EPI_ISL_450543, EPI_ISL_450544, EPI_ISL_450545, EPI_ISL_450546, EPI_ISL_450547, EPI_ISL_450548, EPI_ISL_450549, EPI_ISL_450551, EPI_ISL_450552, EPI_ISL_450553, EPI_ISL_450554, EPI_ISL_450555, EPI_ISL_450559, EPI_ISL_450560, EPI_ISL_450561, EPI_ISL_450562, EPI_ISL_450563, EPI_ISL_450564, EPI_ISL_450565, EPI_ISL_450566, EPI_ISL_450567, EPI_ISL_450568 | | | |
| see above | Utah Public Health Laboratory | Utah Public Health Laboratory | Erin Young, Kelly Oakeson |
| EPI_ISL_450570, EPI_ISL_450572, EPI_ISL_450573, EPI_ISL_450574, EPI_ISL_450575, EPI_ISL_450576, EPI_ISL_450577, EPI_ISL_450578, EPI_ISL_450579, EPI_ISL_450580, EPI_ISL_450581, EPI_ISL_450583, EPI_ISL_450584, EPI_ISL_450585, EPI_ISL_450586, EPI_ISL_450587, EPI_ISL_450588, EPI_ISL_450589, EPI_ISL_450590, EPI_ISL_450591, EPI_ISL_450592, EPI_ISL_450593, EPI_ISL_450594, EPI_ISL_450596, EPI_ISL_450597, EPI_ISL_450598, EPI_ISL_450599, EPI_ISL_450600 | | | |
| see above | Michigan Department of Health and Human Services, Bureau of Laboratories | Michigan Department of Health and Human Services, Bureau of Laboratories | Blankenship HM; Riner D; Soehnlen MK |
| EPI_ISL_450601, EPI_ISL_450602, EPI_ISL_450603, EPI_ISL_450604, EPI_ISL_450605, EPI_ISL_450606, EPI_ISL_450607, EPI_ISL_450608, EPI_ISL_450609, EPI_ISL_450610, EPI_ISL_450611, EPI_ISL_450612, EPI_ISL_450613, EPI_ISL_450614, EPI_ISL_450615, EPI_ISL_450616, EPI_ISL_450617, EPI_ISL_450618, EPI_ISL_450619, EPI_ISL_450620, EPI_ISL_450621, EPI_ISL_450622, EPI_ISL_450623, EPI_ISL_450624, EPI_ISL_450625, EPI_ISL_450626, EPI_ISL_450627, EPI_ISL_450628, EPI_ISL_450629, EPI_ISL_450630, EPI_ISL_450631, EPI_ISL_450632, EPI_ISL_450633, EPI_ISL_450634, EPI_ISL_450635, EPI_ISL_450636, EPI_ISL_450637 | | | |
| see above | Michigan Department of Health and Human Services, Bureau of Laboratories | Michigan Department of Health and Human Services, Bureau of Laboratories | Blankenship HM, Riner D, Soehnlen MK |
| EPI_ISL_450700, EPI_ISL_450701, EPI_ISL_450702, EPI_ISL_450703, EPI_ISL_450704, EPI_ISL_450705, EPI_ISL_450706, EPI_ISL_450707, EPI_ISL_450708, EPI_ISL_450709, EPI_ISL_450710, EPI_ISL_450711, EPI_ISL_450712, EPI_ISL_450713, EPI_ISL_450714, EPI_ISL_450715, EPI_ISL_450716, EPI_ISL_450717, EPI_ISL_450718, EPI_ISL_450719, EPI_ISL_450720, EPI_ISL_450721, EPI_ISL_450722 | | | |
| see above | University of Wisconsin-Madison AIDS Vaccine Research Laboratories | University of Wisconsin-Madison AIDS Vaccine Research Laboratories | Gage Moreno, Katarina Braun, et al. AIDS Vaccine Research Laboratories |
| EPI_ISL_450748, EPI_ISL_450749, EPI_ISL_450750, EPI_ISL_450751, EPI_ISL_450752, EPI_ISL_450753, EPI_ISL_450754, EPI_ISL_450755, EPI_ISL_450756, EPI_ISL_450757, EPI_ISL_450758, EPI_ISL_450759, EPI_ISL_450760, EPI_ISL_450761, EPI_ISL_450762, EPI_ISL_450763, EPI_ISL_450764, EPI_ISL_450765, EPI_ISL_450766, EPI_ISL_450767, EPI_ISL_450768, EPI_ISL_450769, EPI_ISL_450770, EPI_ISL_450771, EPI_ISL_450772, EPI_ISL_450773, EPI_ISL_450774, EPI_ISL_450775, EPI_ISL_450776, EPI_ISL_450777, EPI_ISL_450778, EPI_ISL_450779, EPI_ISL_450780 | | | |
| see above | Minnesota Department of Health, Public Health Laboratory | Minnesota Department of Health, Public Health Laboratory | Matt Plumb, Jacob Garfin, and Xiong Wang |
| EPI_ISL_450800 | AR Dept. of Health-Public Health Lab | Pathogen Discovery, Respiratory Viruses Branch, Division of Viral Diseases, Centers for Disease Control and Prevention | Yan Li, Anna Montmayeur, Ying Tao, Krista Queen, Jing Zhang, Anna Uehara, Clinton R. Paden, Rachel Marine, Haibin Wang, Zachary Weiner, Bettina Bankamp, Suxiang Tong |
| EPI_ISL_450801 | Georgia Department of Health | Pathogen Discovery, Respiratory Viruses Branch, Division of Viral Diseases, Centers for Disease Control and Prevention | Yan Li, Anna Montmayeur, Ying Tao, Krista Queen, Jing Zhang, Anna Uehara, Clinton R. Paden, Rachel Marine, Haibin Wang, Zachary Weiner, Bettina Bankamp, Suxiang Tong |
| EPI_ISL_450802, EPI_ISL_450803 | PA Department of Health, Bureau of Laboratories | Pathogen Discovery, Respiratory Viruses Branch, Division of Viral Diseases, Centers for Disease Control and Prevention | Yan Li, Anna Montmayeur, Ying Tao, Krista Queen, Jing Zhang, Anna Uehara, Clinton R. Paden, Rachel Marine, Haibin Wang, Zachary Weiner, Bettina Bankamp, Suxiang Tong |
| EPI_ISL_450804 | VI-US Virgin Islands Department of Health | Pathogen Discovery, Respiratory Viruses Branch, Division of Viral Diseases, Centers for Disease Control and Prevention | Yan Li, Anna Montmayeur, Ying Tao, Krista Queen, Jing Zhang, Anna Uehara, Clinton R. Paden, Rachel Marine, Haibin Wang, Zachary Weiner, Bettina Bankamp, Suxiang Tong |
| EPI_ISL_450805 | VI-US Virgin Islands Department of Health | Pathogen Discovery, Respiratory Viruses Branch, Division of Viral Diseases, Centers for Disease Control and Prevention | Krista Queen, Yan Li, Anna Montmayeur, Ying Tao, Jing Zhang, Anna Uehara, Clinton R. Paden, Rachel Marine, Haibin Wang, Jasmine Padilla, Justin Lee, Zachary Weiner, Bettina Bankamp, Suxiang Tong |
| EPI_ISL_450806 | VI-US Virgin Islands Department of Health | Pathogen Discovery, Respiratory Viruses Branch, Division of Viral Diseases, Centers for Disease Control and Prevention | Yan Li, Anna Montmayeur, Ying Tao, Krista Queen, Jing Zhang, Anna Uehara, Clinton R. Paden, Rachel Marine, Haibin Wang, Zachary Weiner, Bettina Bankamp, Suxiang Tong |
| EPI_ISL_450844, EPI_ISL_450846, EPI_ISL_450847, EPI_ISL_450849, EPI_ISL_450850, EPI_ISL_450851, EPI_ISL_450857, EPI_ISL_450864, EPI_ISL_450865, EPI_ISL_450869, EPI_ISL_450871 | | | |
| see above | Florida Bureau of Public Health Laboratories | Florida Bureau of Public Health Laboratories | Sarah Schmedes, Jason Blanton |
| EPI_ISL_451203, EPI_ISL_451204, EPI_ISL_451205, EPI_ISL_451206, EPI_ISL_451207, EPI_ISL_451208, EPI_ISL_451209, EPI_ISL_451210, EPI_ISL_451211, EPI_ISL_451212, EPI_ISL_451213, EPI_ISL_451214, EPI_ISL_451215, EPI_ISL_451216, EPI_ISL_451217, EPI_ISL_451218 | | | |
| see above | LSUHS Emerging Viral Threat Laboratory | Microbial Genome Sequencing Center | Jeremy P. Kamil, John A. Vanchiere, Rona S. Scott, Camille F. Abshire, Abida Siddiqa, Byeong-Jae Lee, Chan-ki Min, Md Maksudul Alam, Monica Gestal-Cartele, Edna Ondari, Adam Greer, Malgorzata Bienkowska-Haba, Katarzyna Zwolinska, Jason M. Bodily, Andrew D. Yurochko, Paul M. Weinberger, Christopher G. Kevil, Martin J. Sapp, Daniel J. Snyder, Vaughn S. Cooper |
| EPI_ISL_451219, EPI_ISL_451220, EPI_ISL_451221, EPI_ISL_451222, EPI_ISL_451223, EPI_ISL_451224, EPI_ISL_451225, EPI_ISL_451226, EPI_ISL_451227, EPI_ISL_451228, EPI_ISL_451229 | | | |
| see above | LSUHS Emerging Viral Threat Laboratory | Microbial Genome Sequencing Center | Rona S. Scott, Jeremy P. Kamil, John A. Vanchiere, Camille F. Abshire, Abida Siddiqa, Byeong-Jae Lee, Chan-ki Min, Md Maksudul Alam, Monica Gestal-Cartele, Edna Ondari, Adam Greer, Malgorzata Bienkowska-Haba, Katarzyna Zwolinska, Jason M. Bodily, Andrew D. Yurochko, Paul M. Weinberger, Christopher G. Kevil, Martin J. Sapp, Daniel J. Snyder, Vaughn S. Cooper |
| EPI_ISL_451230, EPI_ISL_451231, EPI_ISL_451232, EPI_ISL_451233, EPI_ISL_451234, EPI_ISL_451235, EPI_ISL_451236, EPI_ISL_451237, EPI_ISL_451238, EPI_ISL_451239, EPI_ISL_451240, EPI_ISL_451241, EPI_ISL_451242, EPI_ISL_451243, EPI_ISL_451244, EPI_ISL_451245, EPI_ISL_451246, EPI_ISL_451247 | | | |
| see above | LSUHS Emerging Viral Threat Laboratory | Microbial Genome Sequencing Center | John A. Vanchiere, Jeremy P. Kamil, Rona S. Scott, Camille F. Abshire, Abida Siddiqa, Byeong-Jae Lee, Chan-ki Min, Md Maksudul Alam, Monica Gestal-Cartele, Edna Ondari, Adam Greer, Malgorzata Bienkowska-Haba, Katarzyna Zwolinska, Jason M. Bodily, Andrew D. Yurochko, Paul M. Weinberger, Christopher G. Kevil, Martin J. Sapp, Daniel J. Snyder, Vaughn S. Cooper |
| EPI_ISL_451248, EPI_ISL_451249, EPI_ISL_451250, EPI_ISL_451251, EPI_ISL_451252, EPI_ISL_451253, EPI_ISL_451254, EPI_ISL_451255, EPI_ISL_451256, EPI_ISL_451257, EPI_ISL_451258, EPI_ISL_451259, EPI_ISL_451260, EPI_ISL_451261, EPI_ISL_451262, EPI_ISL_451263, EPI_ISL_451264, EPI_ISL_451265, EPI_ISL_451266, EPI_ISL_451267, EPI_ISL_451268, EPI_ISL_451269, EPI_ISL_451270, EPI_ISL_451271, EPI_ISL_451272, EPI_ISL_451273, EPI_ISL_451274, EPI_ISL_451275, EPI_ISL_451276, EPI_ISL_451277, EPI_ISL_451278, EPI_ISL_451279, EPI_ISL_451280, EPI_ISL_451281, EPI_ISL_451282, EPI_ISL_451283, EPI_ISL_451284, EPI_ISL_451285, EPI_ISL_451286, EPI_ISL_451287, EPI_ISL_451288, EPI_ISL_451289, EPI_ISL_451290, EPI_ISL_451291, EPI_ISL_451292, EPI_ISL_451293, EPI_ISL_451294, EPI_ISL_451295, EPI_ISL_451296, EPI_ISL_451297 | | | |
| see above | LSUHS Emerging Viral Threat Laboratory | Microbial Genome Sequencing Center | Jeremy P. Kamil, John A. Vanchiere, Rona S. Scott, Camille F. Abshire, Abida Siddiqa, Byeong-Jae Lee, Chan-ki Min, Md Maksudul Alam, Monica Gestal-Cartele, Edna Ondari, Adam Greer, Malgorzata Bienkowska-Haba, Katarzyna Zwolinska, Jason M. Bodily, Andrew D. Yurochko, Paul M. Weinberger, Christopher G. Kevil, Martin J. Sapp, Daniel J. Snyder, Vaughn S. Cooper |
| EPI_ISL_451401, EPI_ISL_451402, EPI_ISL_451403, EPI_ISL_451404, EPI_ISL_451405, EPI_ISL_451406, EPI_ISL_451407, EPI_ISL_451408, EPI_ISL_451409, EPI_ISL_451410, EPI_ISL_451411, EPI_ISL_451412, EPI_ISL_451413, EPI_ISL_451414, EPI_ISL_451415, EPI_ISL_451416, EPI_ISL_451417, EPI_ISL_451418, EPI_ISL_451419, EPI_ISL_451420, EPI_ISL_451421, EPI_ISL_451422, EPI_ISL_451423, EPI_ISL_451424, EPI_ISL_451426, EPI_ISL_451427, EPI_ISL_451428, EPI_ISL_451429, EPI_ISL_451430, EPI_ISL_451431, EPI_ISL_451432, EPI_ISL_451433, EPI_ISL_451434, EPI_ISL_451435, EPI_ISL_451436, EPI_ISL_451437, EPI_ISL_451438, EPI_ISL_451439, EPI_ISL_451440, EPI_ISL_451441, EPI_ISL_451442, EPI_ISL_451443, EPI_ISL_451444, EPI_ISL_451445, EPI_ISL_451446, EPI_ISL_451447, EPI_ISL_451448, EPI_ISL_451449, EPI_ISL_451450, EPI_ISL_451451, EPI_ISL_451452, EPI_ISL_451453, EPI_ISL_451454, EPI_ISL_451455, EPI_ISL_451456, EPI_ISL_451457, EPI_ISL_451458, EPI_ISL_451459, EPI_ISL_451460, EPI_ISL_451461, EPI_ISL_451462, EPI_ISL_451463, EPI_ISL_451464, EPI_ISL_451465, EPI_ISL_451466, EPI_ISL_451467, EPI_ISL_451468, EPI_ISL_451469, EPI_ISL_451470, EPI_ISL_451471, EPI_ISL_451472, EPI_ISL_451473, EPI_ISL_451474, EPI_ISL_451475, EPI_ISL_451476, EPI_ISL_451477, EPI_ISL_451478, EPI_ISL_451479, EPI_ISL_451665 | | | |
| see above | NYU Langone Health | Departments of Pathology and Medicine, New York University School of Medicine | Maria Aguero-Rosenfeld, Brendan Belovarac, Margaret Black, Ludovic Boytard, John Cadley, Paolo Cotzia, John Chen, Dacia Dimartino, Xiaojun Feng, Tatyana Gindin, Emily Guzman, Adriana Heguy, Megan Hogan, Emily Huang, George Jour, Alireza Khodadadi-Jamayran, Lawrence H. Lin, Raven Luther, Andrew Lytle, Christian Marier, Matthew T. Maurano, Mark J. Mulligan, Peter Meyn, Raquel Ordonez Ciriza, Iman Osman, Jared Pinnell, Vanessa Raabe, Sitharam Ramaswami, Amy Rapkiewicz, Andre M. Ribeiro-dos-Santos, Marie Samanovic-Golden, Antonio Serrano, Guomiao Shen, Matija Snuderl, Theodore Vougiouklakis, Nick Vulpescu, Gael Westby, Paul Zappile, Yutong Zhang |
| EPI_ISL_451960 | VI-US Virgin Islands Department of Health | Centers for Disease Control and Prevention | Queen,K., Li,Y., Montmayeur,A., Tao,Y., Zhang,J., Uehara,A., Paden,C.R., Marine,R., Wang,H., Padilla,J., Lee,J., Weiner,Z., Bankamp,B. and Tong,S. |
| EPI_ISL_452105 | Texas DSHS Lab Services | Pathogen Discovery, Respiratory Viruses Branch, Division of Viral Diseases, Centers for Disease Control and Prevention | Yan Li, Anna Montmayeur, Ying Tao, Krista Queen, Jing Zhang, Anna Uehara, Clinton R. Paden, Rachel Marine, Mary S. Keckler, Alison S. Laufer Halpin, Haibin Wang, Christopher A. Elkins, Zachary Weiner, Suxiang Tong |
| EPI_ISL_452106, EPI_ISL_452107 | LA Office of Public Health Laboratories | Pathogen Discovery, Respiratory Viruses Branch, Division of Viral Diseases, Centers for Disease Control and Prevention | Yan Li, Anna Montmayeur, Ying Tao, Krista Queen, Jing Zhang, Anna Uehara, Clinton R. Paden, Rachel Marine, Mary S. Keckler, Alison S. Laufer Halpin, Haibin Wang, Christopher A. Elkins, Zachary Weiner, Suxiang Tong |
| EPI_ISL_452108 | MN Department of Health | Pathogen Discovery, Respiratory Viruses Branch, Division of Viral Diseases, Centers | Yan Li, Anna Montmayeur, Ying Tao, Krista Queen, Jing Zhang, Anna Uehara, Clinton R. Paden, Rachel Marine, Mary S. Keckler, Alison S. Laufer Halpin, Haibin Wang, Christopher A. Elkins, Zachary Weiner, Suxiang Tong |

| | | | |
|---|---|---|---|
| | | for Disease Control and Prevention | |
| EPI_ISL_452109 | NM Department of Health | Pathogen Discovery, Respiratory Viruses Branch, Division of Viral Diseases, Centers for Disease Control and Prevention | Yan Li, Anna Montmayeur, Ying Tao, Krista Queen, Jing Zhang, Anna Uehara, Clinton R. Paden, Rachel Marine, Mary S. Keckler, Alison S. Laufer Halpin, Haibin Wang, Christopher A. Elkins, Zachary Weiner, Suxiang Tong |
| EPI_ISL_452110, EPI_ISL_452111 | FL Bureau of Public Health Laboratories | Pathogen Discovery, Respiratory Viruses Branch, Division of Viral Diseases, Centers for Disease Control and Prevention | Yan Li, Anna Montmayeur, Ying Tao, Krista Queen, Jing Zhang, Anna Uehara, Clinton R. Paden, Rachel Marine, Mary S. Keckler, Alison S. Laufer Halpin, Haibin Wang, Christopher A. Elkins, Zachary Weiner, Suxiang Tong |
| EPI_ISL_452112 | Texas DSHS Lab Services | Pathogen Discovery, Respiratory Viruses Branch, Division of Viral Diseases, Centers for Disease Control and Prevention | Yan Li, Anna Montmayeur, Ying Tao, Krista Queen, Jing Zhang, Anna Uehara, Clinton R. Paden, Rachel Marine, Mary S. Keckler, Alison S. Laufer Halpin, Haibin Wang, Christopher A. Elkins, Zachary Weiner, Suxiang Tong |
| EPI_ISL_452113 | MN Department of Health | Pathogen Discovery, Respiratory Viruses Branch, Division of Viral Diseases, Centers for Disease Control and Prevention | Yan Li, Anna Montmayeur, Ying Tao, Krista Queen, Jing Zhang, Anna Uehara, Clinton R. Paden, Rachel Marine, Mary S. Keckler, Alison S. Laufer Halpin, Haibin Wang, Christopher A. Elkins, Zachary Weiner, Suxiang Tong |
| EPI_ISL_452114 | KS Health and Environmental Laboratories | Pathogen Discovery, Respiratory Viruses Branch, Division of Viral Diseases, Centers for Disease Control and Prevention | Yan Li, Anna Montmayeur, Ying Tao, Krista Queen, Jing Zhang, Anna Uehara, Clinton R. Paden, Rachel Marine, Mary S. Keckler, Alison S. Laufer Halpin, Haibin Wang, Christopher A. Elkins, Zachary Weiner, Suxiang Tong |
| EPI_ISL_452115 | Texas DSHS Lab Services | Pathogen Discovery, Respiratory Viruses Branch, Division of Viral Diseases, Centers for Disease Control and Prevention | Yan Li, Anna Montmayeur, Ying Tao, Krista Queen, Jing Zhang, Anna Uehara, Clinton R. Paden, Rachel Marine, Mary S. Keckler, Alison S. Laufer Halpin, Haibin Wang, Christopher A. Elkins, Zachary Weiner, Suxiang Tong |
| EPI_ISL_452116, EPI_ISL_452117 | NJ Public Health and Environmental Laboratories | Pathogen Discovery, Respiratory Viruses Branch, Division of Viral Diseases, Centers for Disease Control and Prevention | Yan Li, Anna Montmayeur, Ying Tao, Krista Queen, Jing Zhang, Anna Uehara, Clinton R. Paden, Rachel Marine, Mary S. Keckler, Alison S. Laufer Halpin, Haibin Wang, Christopher A. Elkins, Zachary Weiner, Suxiang Tong |
| EPI_ISL_452118 | AZ SPHL, Arizona Department of Health Services | Pathogen Discovery, Respiratory Viruses Branch, Division of Viral Diseases, Centers for Disease Control and Prevention | Yan Li, Anna Montmayeur, Ying Tao, Krista Queen, Jing Zhang, Anna Uehara, Clinton R. Paden, Rachel Marine, Mary S. Keckler, Alison S. Laufer Halpin, Haibin Wang, Christopher A. Elkins, Zachary Weiner, Suxiang Tong |
| EPI_ISL_452119 | VI-US Virgin Islands Department of Health | Pathogen Discovery, Respiratory Viruses Branch, Division of Viral Diseases, Centers for Disease Control and Prevention | Yan Li, Anna Montmayeur, Ying Tao, Krista Queen, Jing Zhang, Anna Uehara, Clinton R. Paden, Rachel Marine, Mary S. Keckler, Alison S. Laufer Halpin, Haibin Wang, Christopher A. Elkins, Zachary Weiner, Suxiang Tong |
| EPI_ISL_452120 | Georgia Department of Health | Pathogen Discovery, Respiratory Viruses Branch, Division of Viral Diseases, Centers for Disease Control and Prevention | Jing Zhang, Anna Montmayeur, Yan Li, Ying Tao, Krista Queen, Anna Uehara, Clinton R. Paden, Rachel Marine, Mary S. Keckler, Alison S. Laufer Halpin, Haibin Wang, Christopher A. Elkins, Zachary Weiner, Suxiang Tong |
| EPI_ISL_452121, EPI_ISL_452122, EPI_ISL_452123 | VI-US Virgin Islands Department of Health | Pathogen Discovery, Respiratory Viruses Branch, Division of Viral Diseases, Centers for Disease Control and Prevention | Jing Zhang, Anna Montmayeur, Yan Li, Ying Tao, Krista Queen, Anna Uehara, Clinton R. Paden, Rachel Marine, Mary S. Keckler, Alison S. Laufer Halpin, Haibin Wang, Christopher A. Elkins, Zachary Weiner, Suxiang Tong |
| EPI_ISL_452124 | NC State Laboratory of Public Health | Pathogen Discovery, Respiratory Viruses Branch, Division of Viral Diseases, Centers for Disease Control and Prevention | Jing Zhang, Anna Montmayeur, Yan Li, Ying Tao, Krista Queen, Anna Uehara, Clinton R. Paden, Rachel Marine, Mary S. Keckler, Alison S. Laufer Halpin, Haibin Wang, Christopher A. Elkins, Zachary Weiner, Suxiang Tong |
| EPI_ISL_452125 | Georgia Department of Health | Pathogen Discovery, Respiratory Viruses Branch, Division of Viral Diseases, Centers for Disease Control and Prevention | Jing Zhang, Anna Montmayeur, Yan Li, Ying Tao, Krista Queen, Anna Uehara, Clinton R. Paden, Rachel Marine, Mary S. Keckler, Alison S. Laufer Halpin, Haibin Wang, Christopher A. Elkins, Zachary Weiner, Suxiang Tong |
| EPI_ISL_452126 | NC State Laboratory of Public Health | Pathogen Discovery, Respiratory Viruses Branch, Division of Viral Diseases, Centers for Disease Control and Prevention | Jing Zhang, Anna Montmayeur, Yan Li, Ying Tao, Krista Queen, Anna Uehara, Clinton R. Paden, Rachel Marine, Mary S. Keckler, Alison S. Laufer Halpin, Haibin Wang, Christopher A. Elkins, Zachary Weiner, Suxiang Tong |
| EPI_ISL_452127, EPI_ISL_452128, EPI_ISL_452129, EPI_ISL_452130 | CO Department of Public Health and Environment | Pathogen Discovery, Respiratory Viruses Branch, Division of Viral Diseases, Centers for Disease Control and Prevention | Jing Zhang, Anna Montmayeur, Yan Li, Ying Tao, Krista Queen, Anna Uehara, Clinton R. Paden, Rachel Marine, Mary S. Keckler, Alison S. Laufer Halpin, Haibin Wang, Christopher A. Elkins, Zachary Weiner, Suxiang Tong |
| EPI_ISL_452131 | IN State Department of Health Laboratory Services | Pathogen Discovery, Respiratory Viruses Branch, Division of Viral Diseases, Centers for Disease Control and Prevention | Jing Zhang, Anna Montmayeur, Yan Li, Ying Tao, Krista Queen, Anna Uehara, Clinton R. Paden, Rachel Marine, Mary S. Keckler, Alison S. Laufer Halpin, Haibin Wang, Christopher A. Elkins, Zachary Weiner, Suxiang Tong |
| EPI_ISL_452132 | FL Bureau of Public Health Laboratories | Pathogen Discovery, Respiratory Viruses Branch, Division of Viral Diseases, Centers for Disease Control and Prevention | Krista Queen, Yan Li, Anna Montmayeur, Ying Tao, Jing Zhang, Anna Uehara, Clinton R. Paden, Rachel Marine, Mary S. Keckler, Alison S. Laufer Halpin, Haibin Wang, Christopher A. Elkins, Zachary Weiner, Suxiang Tong |
| EPI_ISL_452133 | MN Department of Health | Pathogen Discovery, Respiratory Viruses Branch, Division of Viral Diseases, Centers for Disease Control and Prevention | Krista Queen, Yan Li, Anna Montmayeur, Ying Tao, Jing Zhang, Anna Uehara, Clinton R. Paden, Rachel Marine, Mary S. Keckler, Alison S. Laufer Halpin, Haibin Wang, Christopher A. Elkins, Zachary Weiner, Suxiang Tong |
| EPI_ISL_452134 | IL Department of Public Health Chicago Laboratory | Pathogen Discovery, Respiratory Viruses Branch, Division of Viral Diseases, Centers for Disease Control and Prevention | Krista Queen, Yan Li, Anna Montmayeur, Ying Tao, Jing Zhang, Anna Uehara, Clinton R. Paden, Rachel Marine, Mary S. Keckler, Alison S. Laufer Halpin, Haibin Wang, Christopher A. Elkins, Zachary Weiner, Suxiang Tong |
| EPI_ISL_452135, EPI_ISL_452136 | IN State Department of Health Laboratory Services | Pathogen Discovery, Respiratory Viruses Branch, Division of Viral Diseases, Centers for Disease Control and Prevention | Krista Queen, Yan Li, Anna Montmayeur, Ying Tao, Jing Zhang, Anna Uehara, Clinton R. Paden, Rachel Marine, Mary S. Keckler, Alison S. Laufer Halpin, Haibin Wang, Christopher A. Elkins, Zachary Weiner, Suxiang Tong |
| EPI_ISL_452137, EPI_ISL_452138 | VI-US Virgin Islands Department of Health | Pathogen Discovery, Respiratory Viruses Branch, Division of Viral Diseases, Centers for Disease Control and Prevention | Anna Uehara, Yan Li, Anna Montmayeur, Ying Tao, Krista Queen, Jing Zhang, Clinton R. Paden, Rachel Marine, Haibin Wang, Bettina Bankamp, Zachary Weiner, Suxiang Tong |
| EPI_ISL_452139 | Instituto de Diagnostico y Referencia Epidemiologicos (INDRE) | Instituto de diagnóstico y Referencia Epidemiologicos (INDRE) | Ramirez-Gonzalez Ernesto, Garces-Ayala Fabiola, Araiza-Rodriguez Adnan, Mendieta-Condado Edgar, Rodriguez-Maldonado Abril, Wong-Arambula Claudia, Barrera-Badillo Gisela, Hernandez-Rivas Lucia, Lopez-Martinez Irma |
| EPI_ISL_452141 | Instituto de Diagnostico y Referencia Epidemiologicos (INDRE) | Instituto de diagnóstico y Referencia Epidemiologicos (INDRE) | Ramirez-Gonzalez Ernesto, Garces-Ayala Fabiola, Araiza-Rodriguez Adnan, Mendieta-Condado Edgar, Rodriguez-Maldonado Abril, Wong-Arambula Claudia, Barrera-Badillo Gisela, Hernandez-Rivas Lucia, Lopez-Martinez Irma. |
| EPI_ISL_452144, EPI_ISL_452145 | Yale COVID-19 Biorepository | Grubaugh Lab - Yale School of Public Health | Joseph Fauver, Tara Alpert, Anderson Brito, Anne Wyllie, Chantal Vogels, Mary Petrone, Cole Jensen, Chaney Kalinich, Isabel Ott, Arnau Casanovas, Catherine Muenker, Adam Moore, Alice Lu, Maria Tokuyama, Patrick Wong, Peiwen Lu, Saad Omer, Richard Martinello, Allison Nelson, Shelli Farhadian, Akiko Iwasaki, Charlese Dela Cruz, Albert Ko, Nathan Grubaugh |
| EPI_ISL_452153, EPI_ISL_452154, EPI_ISL_452155, EPI_ISL_452156, EPI_ISL_452157, EPI_ISL_452158, EPI_ISL_452161, EPI_ISL_452163, EPI_ISL_452164, EPI_ISL_452165, EPI_ISL_452166, EPI_ISL_452167, EPI_ISL_452168, EPI_ISL_452169, EPI_ISL_452170, EPI_ISL_452171, EPI_ISL_452172, EPI_ISL_452173, EPI_ISL_452174, EPI_ISL_452175, EPI_ISL_452176, EPI_ISL_452177 | | | |
| see above | Utah Public Health Laboratory | Utah Public Health Laboratory | Erin Young, Kelly Oakeson |
| EPI_ISL_452262, EPI_ISL_452263, EPI_ISL_452264, EPI_ISL_452265, EPI_ISL_452266, EPI_ISL_452267, EPI_ISL_452268, EPI_ISL_452269, EPI_ISL_452270, EPI_ISL_452271, EPI_ISL_452272, EPI_ISL_452273, EPI_ISL_452274, EPI_ISL_452275, EPI_ISL_452276, EPI_ISL_452277, EPI_ISL_452278, EPI_ISL_452279, EPI_ISL_452280, EPI_ISL_452281, EPI_ISL_452282, EPI_ISL_452283, EPI_ISL_452284, EPI_ISL_452285, EPI_ISL_452286, EPI_ISL_452287, EPI_ISL_452288, EPI_ISL_452289, EPI_ISL_452290, EPI_ISL_452291, EPI_ISL_452292, EPI_ISL_452293, EPI_ISL_452294, EPI_ISL_452295, EPI_ISL_452296, EPI_ISL_452297, EPI_ISL_452298, EPI_ISL_452299, EPI_ISL_452300, EPI_ISL_452301, EPI_ISL_452302, EPI_ISL_452303, EPI_ISL_452304, EPI_ISL_452305, EPI_ISL_452306, EPI_ISL_452307, EPI_ISL_452308, EPI_ISL_452309, EPI_ISL_452310, EPI_ISL_452311, EPI_ISL_452312, EPI_ISL_452313, EPI_ISL_452314, EPI_ISL_452315, EPI_ISL_452316, EPI_ISL_452317, EPI_ISL_452318, EPI_ISL_452319, EPI_ISL_452320, EPI_ISL_452321, EPI_ISL_452322, EPI_ISL_452323, EPI_ISL_452324, EPI_ISL_452325, EPI_ISL_452326 | | | |
| see above | Michigan Department of Health and Human Services, Bureau of Laboratories | Michigan Department of Health and Human Services, Bureau of Laboratories | Blankenship HM, Riner D, Soehnlen MK |
| EPI_ISL_452796, EPI_ISL_452797, EPI_ISL_452798, EPI_ISL_452799, EPI_ISL_452800, EPI_ISL_452801, EPI_ISL_452802, EPI_ISL_452803, EPI_ISL_452804, EPI_ISL_452805, EPI_ISL_452806, EPI_ISL_452807, EPI_ISL_452808, EPI_ISL_452809, EPI_ISL_452810, EPI_ISL_452811, EPI_ISL_452812, EPI_ISL_452813, EPI_ISL_452814, EPI_ISL_452815, EPI_ISL_452816, EPI_ISL_452817, EPI_ISL_452818, EPI_ISL_452819, EPI_ISL_452820, EPI_ISL_452821, EPI_ISL_452822, EPI_ISL_452823, EPI_ISL_452824, EPI_ISL_452825, EPI_ISL_452826, EPI_ISL_452827, EPI_ISL_452828, EPI_ISL_452829, EPI_ISL_452830, EPI_ISL_452831, EPI_ISL_452832, EPI_ISL_452833, EPI_ISL_452834, EPI_ISL_452835, EPI_ISL_452836, EPI_ISL_452837, EPI_ISL_452838, EPI_ISL_452839, EPI_ISL_452840, EPI_ISL_452841, EPI_ISL_452842, EPI_ISL_452843, EPI_ISL_452844, EPI_ISL_452845, EPI_ISL_452846, EPI_ISL_452847, EPI_ISL_452848, EPI_ISL_452849, EPI_ISL_452850, EPI_ISL_452851, EPI_ISL_452852, EPI_ISL_452853, EPI_ISL_452854, EPI_ISL_452855, EPI_ISL_452856 | | | |
| see above | Virginia DCLS | Virginia DCLS | Virginia DCLS |
| EPI_ISL_454353 | Microbial Genome Sequencing Center; Microbial Genomic Epidemiology Laboratory | Microbial Genomic Epidemiology Laboratory, University of Pittsburgh | Mustapha M. Mustapha, Jane W. Marsh, Dan Snyder, Marissa P. Griffith, Stephanie L. Mitchell, Vatsala R. Srinivasa, Kady D. Waggle, Chinelo Ezeonwuku, Vaughn S. Cooper, Lee H. Harrison |
| EPI_ISL_454354 | UPMC Clinical Microbiology Laboratory | Microbial Genome Sequencing Center; Microbial Genomic Epidemiology Laboratory, University of Pittsburgh | Mustapha M. Mustapha, Jane W. Marsh, Dan Snyder, Marissa P. Griffith, Stephanie L. Mitchell, Vatsala R. Srinivasa, Kady D. Waggle, Chinelo Ezeonwuku, Vaughn S. Cooper, Lee H. Harrison |
| EPI_ISL_454355, EPI_ISL_454356, EPI_ISL_454357, EPI_ISL_454358, EPI_ISL_454359, EPI_ISL_454360, EPI_ISL_454361, EPI_ISL_454362, EPI_ISL_454363, EPI_ISL_454364, EPI_ISL_454365, EPI_ISL_454366, EPI_ISL_454367, EPI_ISL_454368, EPI_ISL_454369, EPI_ISL_454370, EPI_ISL_454371, EPI_ISL_454372, EPI_ISL_454373, EPI_ISL_454374, EPI_ISL_454375, EPI_ISL_454376, EPI_ISL_454377, EPI_ISL_454378, EPI_ISL_454379, EPI_ISL_454380, EPI_ISL_454381, EPI_ISL_454382, EPI_ISL_454383, EPI_ISL_454384, EPI_ISL_454385, EPI_ISL_454386, EPI_ISL_454387, EPI_ISL_454388, EPI_ISL_454389, EPI_ISL_454390, EPI_ISL_454391, EPI_ISL_454392, EPI_ISL_454393, EPI_ISL_454394, EPI_ISL_454395, EPI_ISL_454396, EPI_ISL_454397, EPI_ISL_454398, EPI_ISL_454399, EPI_ISL_454400, EPI_ISL_454401, EPI_ISL_454402, EPI_ISL_454403, EPI_ISL_454404, EPI_ISL_454405, EPI_ISL_454406, EPI_ISL_454407, EPI_ISL_454408, EPI_ISL_454409, EPI_ISL_454410, EPI_ISL_454411 | | | |
| see above | UPMC Clinical Microbiology Laboratory | Microbial Genome Sequencing Center, Microbial Genomic Epidemiological Laboratory | Mustapha M. Mustapha, Jane W. Marsh, Dan Snyder, Marissa P. Griffith, Stephanie L. Mitchell, Vatsala R. Srinivasa, Kady D. Waggle, Chinelo Ezeonwuku, Vaughn S. Cooper, Lee H. Harrison |
| EPI_ISL_454412, | Dirk Dittmer | Dirk Dittmer | Bailey,A.G., Caro-Vegas,C.P., Dittmer,D., Eason,A.B., Juarez,A., Landis,J.T., McNamara,R.P., Miller,M.B., Moorad,R., Pluta,L.J., Seltzer,T.A., Thompson,C., Vahrson,W., Villamor,F. |

| | | | |
|---|---|---|---|
| EPI_ISL_454413, EPI_ISL_454414, EPI_ISL_454415 | | | |
| EPI_ISL_454422, EPI_ISL_454423, EPI_ISL_454424, EPI_ISL_454425, EPI_ISL_454426, EPI_ISL_454427, EPI_ISL_454428, EPI_ISL_454430, EPI_ISL_454431, EPI_ISL_454432, EPI_ISL_454434, EPI_ISL_454435 | | | |
| see above | Maryland Department of Health Laboratories Administration | Maryland Department of Health Laboratories Administration | MDH Laboratories Administration |
| EPI_ISL_454607, EPI_ISL_454608, EPI_ISL_454609, EPI_ISL_454610, EPI_ISL_454611, EPI_ISL_454612, EPI_ISL_454613 | Alameda County Public Health Lab | Chan-Zuckerberg Biohub | CZB Cliahub Consortium |
| EPI_ISL_454614, EPI_ISL_454615, EPI_ISL_454616, EPI_ISL_454617, EPI_ISL_454618, EPI_ISL_454619, EPI_ISL_454620, EPI_ISL_454621, EPI_ISL_454622, EPI_ISL_454623, EPI_ISL_454624, EPI_ISL_454625, EPI_ISL_454626, EPI_ISL_454627, EPI_ISL_454628, EPI_ISL_454629, EPI_ISL_454630, EPI_ISL_454631, EPI_ISL_454632, EPI_ISL_454633, EPI_ISL_454634 | | | |
| see above | UCSF Clinical Microbiology Laboratory | Chan-Zuckerberg Biohub | CZB Cliahub Consortium |
| EPI_ISL_454635 | County Of San Luis Obispo Public Health Laboratory | Chan-Zuckerberg Biohub | CZB Cliahub Consortium |
| EPI_ISL_454636, EPI_ISL_454637, EPI_ISL_454638, EPI_ISL_454639, EPI_ISL_454640, EPI_ISL_454641 | Humboldt County Public Health Laboratory | Chan-Zuckerberg Biohub | CZB Cliahub Consortium |
| EPI_ISL_454642 | CT-Dr. Katherine A. Kelley State Public Health Lab | Pathogen Discovery, Respiratory Viruses Branch, Division of Viral Diseases, Centers for Disease Control and Prevention | Jing Zhang, Ying Tao, Clinton R. Paden, Anna Uehara, Krista Queen, Yan Li, Haibin Wang, Zachary Weiner, Bettina Bankamp, Suxiang Tong |
| EPI_ISL_454643, EPI_ISL_454644 | VI-US Virgin Islands Department of Health | Pathogen Discovery, Respiratory Viruses Branch, Division of Viral Diseases, Centers for Disease Control and Prevention | Jing Zhang, Ying Tao, Clinton R. Paden, Anna Uehara, Krista Queen, Yan Li, Haibin Wang, Zachary Weiner, Bettina Bankamp, Suxiang Tong |
| EPI_ISL_454645 | CT-Dr. Katherine A. Kelley State Public Health Lab | Pathogen Discovery, Respiratory Viruses Branch, Division of Viral Diseases, Centers for Disease Control and Prevention | Jing Zhang, Ying Tao, Clinton R. Paden, Anna Uehara, Krista Queen, Yan Li, Haibin Wang, Zachary Weiner, Bettina Bankamp, Suxiang Tong |
| EPI_ISL_454646 | City of El Paso Department of Public Health Laboratory | Pathogen Discovery, Respiratory Viruses Branch, Division of Viral Diseases, Centers for Disease Control and Prevention | Ying Tao, Clinton R. Paden, Jing Zhang, Anna Uehara, Krista Queen, Yan Li, Haibin Wang, Zachary Weiner, Michael Bowen, Suxiang Tong |
| EPI_ISL_454647, EPI_ISL_454648, EPI_ISL_454649, EPI_ISL_454650, EPI_ISL_454651, EPI_ISL_454652 | VI-US Virgin Islands Department of Health | Pathogen Discovery, Respiratory Viruses Branch, Division of Viral Diseases, Centers for Disease Control and Prevention | Ying Tao, Clinton R. Paden, Jing Zhang, Anna Uehara, Krista Queen, Yan Li, Haibin Wang, Zachary Weiner, Bettina Bankamp, Suxiang Tong |
| EPI_ISL_454653, EPI_ISL_454654, EPI_ISL_454655, EPI_ISL_454656, EPI_ISL_454657, EPI_ISL_454658, EPI_ISL_454659, EPI_ISL_454660, EPI_ISL_454661, EPI_ISL_454662, EPI_ISL_454663, EPI_ISL_454664, EPI_ISL_454665, EPI_ISL_454666, EPI_ISL_454667, EPI_ISL_454668, EPI_ISL_454670, EPI_ISL_454672, EPI_ISL_454673, EPI_ISL_454674, EPI_ISL_454675, EPI_ISL_454676, EPI_ISL_454677, EPI_ISL_454678, EPI_ISL_454679, EPI_ISL_454680, EPI_ISL_454681, EPI_ISL_454682, EPI_ISL_454683, EPI_ISL_454684, EPI_ISL_454685, EPI_ISL_454686, EPI_ISL_454687, EPI_ISL_454688, EPI_ISL_454689 | | | |
| see above | County of Santa Clara Public Health Department | Chan-Zuckerberg Biohub | CZB Cliahub Consortium |
| EPI_ISL_454690 | Emory Molecular Diagnostics Laboratory, Emory Healthcare | Piantadosi Lab, Emory Department of Pathology | Ahmed Babiker, Anne Piantadosi |
| EPI_ISL_454692 | Quest Diagnostics | Quest Diagnostics | Anderson,B.P., Rosenthal,S.H., Gerasimova,A., Kagan,R.M. and Owen, R. |
| EPI_ISL_454707, EPI_ISL_454708, EPI_ISL_454709, EPI_ISL_454710, EPI_ISL_454711, EPI_ISL_454712, EPI_ISL_454714, EPI_ISL_454715, EPI_ISL_454717, EPI_ISL_454718, EPI_ISL_454719, EPI_ISL_454720, EPI_ISL_454721, EPI_ISL_454722, EPI_ISL_454723, EPI_ISL_454725, EPI_ISL_454727, EPI_ISL_454728, EPI_ISL_454729, EPI_ISL_454730, EPI_ISL_454731 | | | |
| see above | Utah Public Health Laboratory | Utah Public Health Laboratory | Erin Young, Kelly Oakeson |
| EPI_ISL_454796, EPI_ISL_454797, EPI_ISL_454798, EPI_ISL_454799, EPI_ISL_454800, EPI_ISL_454801, EPI_ISL_454802, EPI_ISL_454803, EPI_ISL_454804, EPI_ISL_454805, EPI_ISL_454806, EPI_ISL_454807, EPI_ISL_454808, EPI_ISL_454809, EPI_ISL_454810, EPI_ISL_454811, EPI_ISL_454812, EPI_ISL_454813, EPI_ISL_454814, EPI_ISL_454815, EPI_ISL_454816, EPI_ISL_454817, EPI_ISL_454818, EPI_ISL_454819, EPI_ISL_454820, EPI_ISL_454821, EPI_ISL_454822, EPI_ISL_454823, EPI_ISL_454824, EPI_ISL_454825, EPI_ISL_454826, EPI_ISL_454827, EPI_ISL_454828, EPI_ISL_454829 | | | |
| see above | Dirk Dittmer | Dirk Dittmer | Bailey,A.G., Caro-Vegas,C.P., Dittmer,D., Eason,A.B., Juarez,A., Landis,J.T., McNamara,R.P., Miller,M.B., Moorad,R., Pluta,L.J., Seltzer,T.A., Thompson,C., Vahrson,W., Villamor,F. |
| EPI_ISL_455355, EPI_ISL_455356, EPI_ISL_455358, EPI_ISL_455359, EPI_ISL_455360, EPI_ISL_455361 | Emory Molecular Diagnostics Laboratory, Emory Healthcare | Piantadosi Lab, Emory Department of Pathology | Ahmed Babiker, Anne Piantadosi |
| EPI_ISL_455432, EPI_ISL_455434 | Instituto de Diagnostico y Referencia Epidemiologicos (INDRE) | Instituto de Diagnostico y Referencia Epidemiologicos (INDRE) | Taboada Ramírez Blanca. Ramirez-Gonzalez Ernesto, Garces-Ayala Fabiola, Araiza-Rodriguez Adnan, Mendieta-Condado Edgar, Rodriguez-Maldonado Abril, Wong-Arambula Claudia, Barrera-Badillo Gisela, Hernandez-Rivas Lucia, Lopez-Martinez Irma. |
| EPI_ISL_455435 | Instituto de Diagnostico y Referencia Epidemiologicos (INDRE) | Instituto de Diagnostico y Referencia Epidemiologicos (INDRE) | Garces-Ayala Fabiola. Taboada Ramírez Blanca. Ramirez-Gonzalez Ernesto, Araiza-Rodriguez Adnan , Mendieta-Condado Edgar, Rodriguez-Maldonado Abril, Wong-Arambula Claudia, Barrera-Badillo Gisela, Hernandez-Rivas Lucia, Lopez-Martinez Irma |
| EPI_ISL_455436 | Instituto de Diagnostico y Referencia Epidemiologicos (INDRE) | Instituto de Diagnostico y Referencia Epidemiologicos (INDRE) | Garces-Ayala Fabiola. Taboada Ramírez Blanca. Ramirez-Gonzalez Ernesto, Araiza-Rodriguez Adnan, Mendieta-Condado Edgar, Rodriguez-Maldonado Abril, Wong-Arambula Claudia, Barrera-Badillo Gisela, Hernandez-Rivas Lucia, Lopez-Martinez Irma. |
| EPI_ISL_455437, EPI_ISL_455438 | Instituto de Diagnostico y Referencia Epidemiologicos (INDRE) | Instituto de Diagnostico y Referencia Epidemiologicos (INDRE) | Araiza-Rodriguez Adnan, Garces-Ayala Fabiola. Ramirez-Gonzalez Ernesto, Mendieta-Condado Edgar, Rodriguez-Maldonado Abril, Wong-Arambula Claudia, Barrera-Badillo Gisela, Hernandez-Rivas Lucia, Lopez-Martinez Irma, Taboada Ramírez Blanca. |
| EPI_ISL_455439 | Instituto de Diagnostico y Referencia Epidemiologicos (INDRE) | Instituto de Diagnostico y Referencia Epidemiologicos (INDRE) | Mendieta-Condado Edgar, Araiza-Rodriguez Adnan, Garces-Ayala Fabiola, Rodriguez-Maldonado Abril, Wong-Arambula Claudia, Barrera-Badillo Gisela, Hernandez-Rivas Lucia, Lopez-Martinez Irma, Taboada Ramirez Blanca, Ramirez-Gonzalez Ernesto. |
| EPI_ISL_455454 | Instituto de Diagnostico y Referencia Epidemiologicos (INDRE) | Instituto de Diagnostico y Referencia Epidemiologicos (INDRE) | Mendieta-Condado Edgar, Araiza-Rodriguez Adnan, Garces-Ayala Fabiola, , Rodriguez-Maldonado Abril, Wong-Arambula Claudia, Barrera-Badillo Gisela, Taboada Ramírez Blanca, Ramirez-Gonzalez Ernesto, Hernandez-Rivas Lucia, Lopez-Martinez Irma. |
| EPI_ISL_455455 | Instituto de Diagnostico y Referencia Epidemiologicos (INDRE) | Instituto de Diagnostico y Referencia Epidemiologicos (INDRE) | Rodriguez-Maldonado Abril, Mendieta-Condado Edgar, Araiza-Rodriguez Adnan, Garces-Ayala Fabiola. Taboada Ramirez Blanca. Ramirez-Gonzalez Ernesto, Barrera-Badillo Gisela, Hernandez-Rivas Lucia, Lopez-Martinez Irma, Wong-Arambula Claudia. |
| EPI_ISL_455456 | Instituto de Diagnostico y Referencia Epidemiologicos (INDRE) | Instituto de Diagnostico y Referencia Epidemiologicos (INDRE) | Rodriguez-Maldonado Abril, Mendieta-Condado Edgar, Araiza-Rodriguez Adnan, Garces-Ayala Fabiola. Taboada Ramírez Blanca. Ramirez-Gonzalez Ernesto, , Barrera-Badillo Gisela, Hernandez-Rivas Lucia, Lopez-Martinez Irma, Wong-Arambula Claudia. |
| EPI_ISL_455568, EPI_ISL_455569, EPI_ISL_455570, EPI_ISL_455571, EPI_ISL_455572 | Gundersen Molecular Diagnostics Laboratory | Kabara Cancer Research Institute | Craig S. Richmond, Paraic A. Kenny |
| EPI_ISL_455573, EPI_ISL_455574 | Gundersen Clinical Microbiology Laboratory | Kabara Cancer Research Institute | Craig S. Richmond, Paraic A. Kenny |
| EPI_ISL_455575, EPI_ISL_455576, EPI_ISL_455577, EPI_ISL_455578, EPI_ISL_455579 | Gundersen Molecular Diagnostics Laboratory | Kabara Cancer Research Institute | Craig S. Richmond, Paraic A. Kenny |
| EPI_ISL_455580, EPI_ISL_455581, EPI_ISL_455582 | Gundersen Clinical Microbiology Laboratory | Kabara Cancer Research Institute | Craig S. Richmond, Paraic A. Kenny |
| EPI_ISL_455609, EPI_ISL_455610, EPI_ISL_455611, EPI_ISL_455612, EPI_ISL_455613, EPI_ISL_455614, EPI_ISL_455615, EPI_ISL_455616, EPI_ISL_455617, EPI_ISL_455618, EPI_ISL_455619, EPI_ISL_455620, EPI_ISL_455621, EPI_ISL_455622, EPI_ISL_455623 | | | |
| see above | Ochsner Health | Bioinfoexperts, LLC | Susanna L. Lamers, David J. Nolan, Rebecca Rose, Sissy Cross, David Moraga Amador, Tong Yang, Luke Caruso, Wayra Navia, Lydia Von Borstel, Xiao Hui Zhou, Amy Feehan, Julia-Garcia-Diaz |
| EPI_ISL_455980, EPI_ISL_455981, | LSUHS Emerging Viral Threat Laboratory | Microbial Genome Sequencing Center | Jeremy P. Kamil, John A. Vanchiere, Rona S. Scott, Camille F. Abshire, Abida Siddiqa, Byeong-Jae Lee, Chan-ki Min, Md Maksudul Alam, Monica Gestal-Cartele, Edna Ondari, Adam Greer, Malgorzata Bienkowska-Haba, Katarzyna Zwolinska, Michelle M. Arnold, Jason M. Bodily, Andrew D. Yurochko, Paul M. Weinberger, Christopher G. Kevil, Martin J. Sapp, Daniel J. Snyder, Vaughn S. Cooper |

| | | | |
|---|---|---|---|
| EPI_ISL_455982, EPI_ISL_455983, EPI_ISL_455984, EPI_ISL_455985, EPI_ISL_455986, EPI_ISL_455987, EPI_ISL_455988, EPI_ISL_455989 | | | |
| EPI_ISL_455990, EPI_ISL_455991, EPI_ISL_455992, EPI_ISL_455993, EPI_ISL_455994, EPI_ISL_455995, EPI_ISL_455996, EPI_ISL_455997, EPI_ISL_455998, EPI_ISL_455999 | LSUHS Emerging Viral Threat Laboratory | Microbial Genome Sequencing Center | John A. Vanchiere, Jeremy P. Kamil, Rona S. Scott, Camille F. Abshire, Abida Siddiqa, Byeong-Jae Lee, Chan-ki Min, Md Maksudul Alam, Monica Gestal-Cartele, Edna Ondari, Adam Greer, Malgorzata Bienkowska-Haba, Katarzyna Zwolinska, Michelle M. Arnold, Jason M. Bodily, Andrew D. Yurochko, Paul M. Weinberger, Christopher G. Kevil, Martin J. Sapp, Daniel J. Snyder, Vaughn S. Cooper |
| EPI_ISL_456000, EPI_ISL_456001, EPI_ISL_456002 | LSUHS Emerging Viral Threat Laboratory | Microbial Genome Sequencing Center | Rona S. Scott, Jeremy P. Kamil, John A. Vanchiere, Camille F. Abshire, Abida Siddiqa, Byeong-Jae Lee, Chan-ki Min, Md Maksudul Alam, Monica Gestal-Cartele, Edna Ondari, Adam Greer, Malgorzata Bienkowska-Haba, Katarzyna Zwolinska, Michelle M. Arnold, Jason M. Bodily, Andrew D. Yurochko, Paul M. Weinberger, Christopher G. Kevil, Martin J. Sapp, Daniel J. Snyder, Vaughn S. Cooper |
| EPI_ISL_456004, EPI_ISL_456006, EPI_ISL_456007, EPI_ISL_456008, EPI_ISL_456010, EPI_ISL_456011, EPI_ISL_456012, EPI_ISL_456013, EPI_ISL_456014, EPI_ISL_456015, EPI_ISL_456016, EPI_ISL_456017, EPI_ISL_456018, EPI_ISL_456019, EPI_ISL_456020, EPI_ISL_456021, EPI_ISL_456022, EPI_ISL_456023, EPI_ISL_456024, EPI_ISL_456025, EPI_ISL_456026, EPI_ISL_456027, EPI_ISL_456028, EPI_ISL_456029, EPI_ISL_456030, EPI_ISL_456031, EPI_ISL_456032, EPI_ISL_456033, EPI_ISL_456034, EPI_ISL_456035, EPI_ISL_456036, EPI_ISL_456037, EPI_ISL_456038, EPI_ISL_456039, EPI_ISL_456040, EPI_ISL_456041, EPI_ISL_456042, EPI_ISL_456043, EPI_ISL_456044, EPI_ISL_456045, EPI_ISL_456046, EPI_ISL_456047, EPI_ISL_456048, EPI_ISL_456049, EPI_ISL_456050, EPI_ISL_456051, EPI_ISL_456052, EPI_ISL_456053, EPI_ISL_456054, EPI_ISL_456055, EPI_ISL_456056, EPI_ISL_456058, EPI_ISL_456059, EPI_ISL_456060, EPI_ISL_456061, EPI_ISL_456062, EPI_ISL_456063, EPI_ISL_456064, EPI_ISL_456065, EPI_ISL_456066, EPI_ISL_456107, EPI_ISL_456108, EPI_ISL_456109, EPI_ISL_456111, EPI_ISL_456112, EPI_ISL_456113, EPI_ISL_456114 | | | |
| see above | NYU Langone Health | Departments of Pathology and Medicine, New York University School of Medicine | Maria Aguero-Rosenfeld, Brendan Belovarac, Margaret Black, Ludovic Boytard, John Cadley, Paolo Cotzia, John Chen, Dacia Dimartino, Xiaojun Feng, Tatyana Gindin, Emily Guzman, Adriana Heguy, Megan Hogan, Emily Huang, George Jour, Alireza Khodadadi-Jamayran, Lawrence H. Lin, Raven Luther, Andrew Lytle, Christian Marier, Matthew T. Maurano, Mark J. Mulligan, Peter Meyn, Raquel Ordonez Ciriza, Iman Osman, Jared Pinnell, Vanessa Raabe, Sitharam Ramaswami, Amy Rapkiewicz, Andre M. Ribeiro-dos-Santos, Marie Samanovic-Golden, Antonio Serrano, Guomiao Shen, Matija Snuderl, Theodore Vougiouklakis, Nick Vulpescu, Gael Westby, Paul Zappile, Yutong Zhang |
| EPI_ISL_456656 | American Type Culture Collection Inc. (ATCC) | American Type Culture Collection Inc. (ATCC) | Riojas, M.A., Frank,A.M., Puthuveetil, N.P., Benton,B., Peiris, J.S.M., Chu, D.K.W., King,S.P., Flores, B., Parker, M. and Rashid, S. |
| EPI_ISL_457751, EPI_ISL_457752, EPI_ISL_457753, EPI_ISL_457754, EPI_ISL_457755, EPI_ISL_457756, EPI_ISL_457757, EPI_ISL_457758, EPI_ISL_457759, EPI_ISL_457760, EPI_ISL_457761, EPI_ISL_457762, EPI_ISL_457763, EPI_ISL_457764, EPI_ISL_457765, EPI_ISL_457766, EPI_ISL_457767, EPI_ISL_457768, EPI_ISL_457769, EPI_ISL_457770, EPI_ISL_457771, EPI_ISL_457777, EPI_ISL_457780, EPI_ISL_457781, EPI_ISL_457782, EPI_ISL_457783, EPI_ISL_457784, EPI_ISL_457785, EPI_ISL_457786, EPI_ISL_457787, EPI_ISL_457788, EPI_ISL_457789, EPI_ISL_457790, EPI_ISL_457791, EPI_ISL_457792, EPI_ISL_457794, EPI_ISL_457795, EPI_ISL_457796, EPI_ISL_457797, EPI_ISL_457798, EPI_ISL_457799, EPI_ISL_457800, EPI_ISL_457801, EPI_ISL_457802, EPI_ISL_457803, EPI_ISL_457804, EPI_ISL_457805, EPI_ISL_457806, EPI_ISL_457807, EPI_ISL_457808, EPI_ISL_457809, EPI_ISL_457810, EPI_ISL_457811, EPI_ISL_457812, EPI_ISL_457813, EPI_ISL_457814, EPI_ISL_457816, EPI_ISL_457817, EPI_ISL_457818, EPI_ISL_457819, EPI_ISL_457820, EPI_ISL_457821, EPI_ISL_457822, EPI_ISL_457823 | | | |
| see above | Johns Hopkins Hospital Department of Pathology | Johns Hopkins Hospital Department of Pathology | Peter M. Thielen, Thomas Mehoke, Shirlee Wohl, Srividya Ramakrishnan, Melanie Kirsche, Amanda Ernlund, Craig Howser, Kristina Zudock, Oluwaseun Falade-Nwulia, Norah Sadowski, Paul Morris, Mark Hopkins, Yunfan Fan, Nidia Trovao, Victoria Gniazdowski, Michael C. Schatz, Stuart C. Ray, Winston Timp, Heba H. Mostafa |
| EPI_ISL_458001, EPI_ISL_458002, EPI_ISL_458003, EPI_ISL_458004, EPI_ISL_458005, EPI_ISL_458006 | Dirk Dittmer | Dirk Dittmer | Bailey,A.G., Caro-Vegas,C.P., Dittmer,D., Eason,A.B., Juarez,A., Landis,J.T., McNamara,R.P., Miller,M.B., Moorad,R., Pluta,L.J., Seltzer,T.A., Thompson,C., Vahrson,W., Villamor,F. |
| EPI_ISL_458007 | Dirk Dittmer | Dirk Dittmer | Aubrey,B.G., Caro-Vegas,C.P., Dittmer,D., Eason,A.B., Juarez,A., Landis,J.T., McNamara,R.P., Miller,M.B., Moorad,R., Pluta,L.J., Seltzer,T.A., Thompson,C., Vahrson,W., Villamor,F. |
| EPI_ISL_458017, EPI_ISL_458018, EPI_ISL_458019, EPI_ISL_458020, EPI_ISL_458021 | NYU Langone Health | Departments of Pathology and Medicine, New York University School of Medicine | Maria Aguero-Rosenfeld, Brendan Belovarac, Margaret Black, Ludovic Boytard, John Cadley, Paolo Cotzia, John Chen, Dacia Dimartino, Xiaojun Feng, Tatyana Gindin, Emily Guzman, Adriana Heguy, Megan Hogan, Emily Huang, George Jour, Alireza Khodadadi-Jamayran, Lawrence H. Lin, Raven Luther, Andrew Lytle, Christian Marier, Matthew T. Maurano, Mark J. Mulligan, Peter Meyn, Raquel Ordonez Ciriza, Iman Osman, Jared Pinnell, Vanessa Raabe, Sitharam Ramaswami, Amy Rapkiewicz, Andre M. Ribeiro-dos-Santos, Marie Samanovic-Golden, Antonio Serrano, Guomiao Shen, Matija Snuderl, Theodore Vougiouklakis, Nick Vulpescu, Gael Westby, Paul Zappile, Yutong Zhang |
| EPI_ISL_458237, EPI_ISL_458238, EPI_ISL_458239, EPI_ISL_458240, EPI_ISL_458241, EPI_ISL_458242, EPI_ISL_458243, EPI_ISL_458244, EPI_ISL_458245, EPI_ISL_458246, EPI_ISL_458247, EPI_ISL_458248, EPI_ISL_458249, EPI_ISL_458250, EPI_ISL_458251, EPI_ISL_458252, EPI_ISL_458253, EPI_ISL_458254, EPI_ISL_458255, EPI_ISL_458256, EPI_ISL_458257, EPI_ISL_458258, EPI_ISL_458259, EPI_ISL_458260, EPI_ISL_458261, EPI_ISL_458262, EPI_ISL_458263, EPI_ISL_458264, EPI_ISL_458265, EPI_ISL_458266, EPI_ISL_458267, EPI_ISL_458268, EPI_ISL_458269, EPI_ISL_458270, EPI_ISL_458271, EPI_ISL_458272, EPI_ISL_458273, EPI_ISL_458274, EPI_ISL_458275, EPI_ISL_458276, EPI_ISL_458277, EPI_ISL_458278, EPI_ISL_458279, EPI_ISL_458280, EPI_ISL_458281, EPI_ISL_458282, EPI_ISL_458283, EPI_ISL_458284 | | | |
| see above | Scripps Medical Laboratory | Andersen lab at Scripps Research | SEARCH Alliance San Diego with Michael Quigley, Ellen Stefanski, Ian Mchardy |
| EPI_ISL_458291 | Dirk Dittmer | Dirk Dittmer | Bailey,A.G., Caro-Vegas,C.P., Dittmer,D., Eason,A.B., Juarez,A., Landis,J.T., McNamara,R.P., Miller,M.B., Moorad,R., Pluta,L.J., Seltzer,T.A., Thompson,C., Vahrson,W., Villamor,F. |
| EPI_ISL_458292, EPI_ISL_458293 | Dirk Dittmer | Dirk Dittmer | Aubrey,B.G., Caro-Vegas,C.P., Dittmer,D., Eason,A.B., Juarez,A., Landis,J.T., Mcnamara,R.P., Miller,M.B., Moorad,R., Pluta,L.J., Seltzer,T.A., Thompson,C., Vahrson,W., Villamor,F. |
| EPI_ISL_458294, EPI_ISL_458295, EPI_ISL_458296, EPI_ISL_458297 | Dirk Dittmer | Dirk Dittmer | Bailey,A.G., Caro-Vegas,C.P., Dittmer,D., Eason,A.B., Juarez,A., Landis,J.T., McNamara,R.P., Miller,M.B., Moorad,R., Pluta,L.J., Seltzer,T.A., Thompson,C., Vahrson,W., Villamor,F. |
| EPI_ISL_459992, EPI_ISL_459993, EPI_ISL_459994, EPI_ISL_459995, EPI_ISL_459996, EPI_ISL_459997, EPI_ISL_459998, EPI_ISL_459999, EPI_ISL_460000, EPI_ISL_460001, EPI_ISL_460002, EPI_ISL_460003, EPI_ISL_460004, EPI_ISL_460005, EPI_ISL_460006, EPI_ISL_460008, EPI_ISL_460009, EPI_ISL_460010, EPI_ISL_460012, EPI_ISL_460013, EPI_ISL_460014, EPI_ISL_460015, EPI_ISL_460016, EPI_ISL_460017, EPI_ISL_460018, EPI_ISL_460019, EPI_ISL_460020, EPI_ISL_460021, EPI_ISL_460022, EPI_ISL_460023, EPI_ISL_460024, EPI_ISL_460025, EPI_ISL_460026, EPI_ISL_460027, EPI_ISL_460028, EPI_ISL_460029, EPI_ISL_460031, EPI_ISL_460032, EPI_ISL_460033, EPI_ISL_460034, EPI_ISL_460035, EPI_ISL_460036, EPI_ISL_460037, EPI_ISL_460038, EPI_ISL_460039, EPI_ISL_460040, EPI_ISL_460041, EPI_ISL_460042, EPI_ISL_460043 | | | |
| see above | Michigan Department of Health and Human Services, Bureau of Laboratories | Michigan Department of Health and Human Services, Bureau of Laboratories | Blankenship HM, Riner D, Soehnlen MK |
| EPI_ISL_460045, EPI_ISL_460046, EPI_ISL_460047, EPI_ISL_460048, EPI_ISL_460049, EPI_ISL_460050, EPI_ISL_460051, EPI_ISL_460052, EPI_ISL_460053, EPI_ISL_460054, EPI_ISL_460055, EPI_ISL_460056, EPI_ISL_460057, EPI_ISL_460058, EPI_ISL_460059, EPI_ISL_460060, EPI_ISL_460061, EPI_ISL_460062, EPI_ISL_460063, EPI_ISL_460064, EPI_ISL_460065, EPI_ISL_460066, EPI_ISL_460067, EPI_ISL_460068, EPI_ISL_460069, EPI_ISL_460070, EPI_ISL_460071, EPI_ISL_460072, EPI_ISL_460073, EPI_ISL_460074, EPI_ISL_460075, EPI_ISL_460076, EPI_ISL_460077, EPI_ISL_460078 | | | |
| see above | Minnesota Department of Health, Public Health Laboratory | Minnesota Department of Health, Public Health Laboratory | Matt Plumb, Jacob Garfin, and Xiong Wang |
| EPI_ISL_460097, EPI_ISL_460098, EPI_ISL_460099, EPI_ISL_460101, EPI_ISL_460102, EPI_ISL_460103, EPI_ISL_460104, EPI_ISL_460105, EPI_ISL_460106, EPI_ISL_460107, EPI_ISL_460108, EPI_ISL_460109, EPI_ISL_460110, EPI_ISL_460111, EPI_ISL_460112, EPI_ISL_460113, EPI_ISL_460114, EPI_ISL_460115, EPI_ISL_460116, EPI_ISL_460117, EPI_ISL_460118, EPI_ISL_460119, EPI_ISL_460120, EPI_ISL_460121, EPI_ISL_460122, EPI_ISL_460123, EPI_ISL_460124, EPI_ISL_460125, EPI_ISL_460126, EPI_ISL_460127, EPI_ISL_460128, EPI_ISL_460129, EPI_ISL_460130, EPI_ISL_460131, EPI_ISL_460132, EPI_ISL_460133, EPI_ISL_460134, EPI_ISL_460135, EPI_ISL_460137, EPI_ISL_460138, EPI_ISL_460139, EPI_ISL_460140, EPI_ISL_460141, EPI_ISL_460142, EPI_ISL_460143, EPI_ISL_460144, EPI_ISL_460145, EPI_ISL_460146, EPI_ISL_460147, EPI_ISL_460148, EPI_ISL_460149, EPI_ISL_460150, EPI_ISL_460151, EPI_ISL_460153, EPI_ISL_460154, EPI_ISL_460155, EPI_ISL_460156, EPI_ISL_460157, EPI_ISL_460158, EPI_ISL_460159, EPI_ISL_460160, EPI_ISL_460161, EPI_ISL_460162, EPI_ISL_460163, EPI_ISL_460164, EPI_ISL_460165, EPI_ISL_460166, EPI_ISL_460168, EPI_ISL_460169, EPI_ISL_460170, EPI_ISL_460171, EPI_ISL_460172, EPI_ISL_460173, EPI_ISL_460174, EPI_ISL_460175, EPI_ISL_460176, EPI_ISL_460177, EPI_ISL_460178, EPI_ISL_460179, EPI_ISL_460180, EPI_ISL_460181, EPI_ISL_460182, EPI_ISL_460183, EPI_ISL_460184, EPI_ISL_460185, EPI_ISL_460186, EPI_ISL_460187, EPI_ISL_460188, EPI_ISL_460189, EPI_ISL_460190, EPI_ISL_460191, EPI_ISL_460192, EPI_ISL_460193, EPI_ISL_460194, EPI_ISL_460195, EPI_ISL_460196, EPI_ISL_460197, EPI_ISL_460198, EPI_ISL_460199, EPI_ISL_460201, EPI_ISL_460202, EPI_ISL_460203, EPI_ISL_460204, EPI_ISL_460205, EPI_ISL_460206, EPI_ISL_460207, EPI_ISL_460208, EPI_ISL_460209, EPI_ISL_460210, EPI_ISL_460211, EPI_ISL_460212, EPI_ISL_460213, EPI_ISL_460214, EPI_ISL_460215, EPI_ISL_460216, EPI_ISL_460217, EPI_ISL_460218, EPI_ISL_460220, EPI_ISL_460221, EPI_ISL_460223, EPI_ISL_460224, EPI_ISL_460225, EPI_ISL_460226, EPI_ISL_460227, EPI_ISL_460229, EPI_ISL_460230, EPI_ISL_460231, EPI_ISL_460232, EPI_ISL_460233, EPI_ISL_460234, EPI_ISL_460235, EPI_ISL_460236, EPI_ISL_460237, EPI_ISL_460238, EPI_ISL_460239, EPI_ISL_460240, EPI_ISL_460241, EPI_ISL_460242, EPI_ISL_460243, EPI_ISL_460244, EPI_ISL_460245, EPI_ISL_460246, EPI_ISL_460247, EPI_ISL_460248, EPI_ISL_460249, EPI_ISL_460250, EPI_ISL_460252, EPI_ISL_460253, EPI_ISL_460254, EPI_ISL_460255, EPI_ISL_460256, EPI_ISL_460257, EPI_ISL_460258, EPI_ISL_460261, EPI_ISL_460262, EPI_ISL_460263, EPI_ISL_460264, EPI_ISL_460265, EPI_ISL_460266, EPI_ISL_460267, EPI_ISL_460268, EPI_ISL_460269, EPI_ISL_460270, EPI_ISL_460271, EPI_ISL_460272, EPI_ISL_460273, EPI_ISL_460274, EPI_ISL_460275, EPI_ISL_460276, EPI_ISL_460277, EPI_ISL_460278, EPI_ISL_460279, EPI_ISL_460280, EPI_ISL_460281, EPI_ISL_460282, EPI_ISL_460283, EPI_ISL_460284, EPI_ISL_460285, EPI_ISL_460286, EPI_ISL_460287, EPI_ISL_460288, EPI_ISL_460289, EPI_ISL_460290, EPI_ISL_460291, EPI_ISL_460292, EPI_ISL_460293, EPI_ISL_460294, EPI_ISL_460295, EPI_ISL_460296, EPI_ISL_460297, EPI_ISL_460298, EPI_ISL_460299, EPI_ISL_460300, EPI_ISL_460301, EPI_ISL_460302, EPI_ISL_460304, EPI_ISL_460305, EPI_ISL_460306, EPI_ISL_460307, EPI_ISL_460308, EPI_ISL_460309, EPI_ISL_460310, EPI_ISL_460311, EPI_ISL_460312, EPI_ISL_460313, EPI_ISL_460314, EPI_ISL_460315, EPI_ISL_460316, EPI_ISL_460317, EPI_ISL_460319, EPI_ISL_460321, EPI_ISL_460322, EPI_ISL_460323, EPI_ISL_460324, EPI_ISL_460325, EPI_ISL_460326, EPI_ISL_460327, EPI_ISL_460328, EPI_ISL_460329, EPI_ISL_460330, EPI_ISL_460331, EPI_ISL_460332, EPI_ISL_460333, EPI_ISL_460334, EPI_ISL_460335, EPI_ISL_460336, EPI_ISL_460337, EPI_ISL_460338, EPI_ISL_460339, EPI_ISL_460340, EPI_ISL_460341, EPI_ISL_460342, EPI_ISL_460343, EPI_ISL_460344, EPI_ISL_460345, EPI_ISL_460346, EPI_ISL_460347, EPI_ISL_460348, EPI_ISL_460349, EPI_ISL_460350, EPI_ISL_460351, EPI_ISL_460352, EPI_ISL_460353, EPI_ISL_460354, EPI_ISL_460355, EPI_ISL_460356, EPI_ISL_460357, EPI_ISL_460358, EPI_ISL_460359, EPI_ISL_460360, EPI_ISL_460361, EPI_ISL_460362, EPI_ISL_460363, EPI_ISL_460364, EPI_ISL_460365, EPI_ISL_460366, EPI_ISL_460367, EPI_ISL_460368, EPI_ISL_460369, EPI_ISL_460370, EPI_ISL_460371, EPI_ISL_460372, EPI_ISL_460373, EPI_ISL_460376, EPI_ISL_460377, EPI_ISL_460378, EPI_ISL_460379, EPI_ISL_460380, EPI_ISL_460381, EPI_ISL_460382, EPI_ISL_460383, EPI_ISL_460384, EPI_ISL_460385, EPI_ISL_460386, EPI_ISL_460388, EPI_ISL_460389, EPI_ISL_460390, EPI_ISL_460391, EPI_ISL_460392, EPI_ISL_460394, EPI_ISL_460395, EPI_ISL_460396, EPI_ISL_460397, EPI_ISL_460398, EPI_ISL_460399, EPI_ISL_460400, EPI_ISL_460401, EPI_ISL_460402, EPI_ISL_460403, EPI_ISL_460404, EPI_ISL_460406, EPI_ISL_460407, EPI_ISL_460408, EPI_ISL_460409, EPI_ISL_460410, EPI_ISL_460411, EPI_ISL_460412, EPI_ISL_460413, EPI_ISL_460414, EPI_ISL_460415, EPI_ISL_460417, EPI_ISL_460418, EPI_ISL_460419, EPI_ISL_460420, EPI_ISL_460421, EPI_ISL_460422, EPI_ISL_460423, EPI_ISL_460424, EPI_ISL_460425, EPI_ISL_460426, EPI_ISL_460427, EPI_ISL_460428, EPI_ISL_460429, EPI_ISL_460430, EPI_ISL_460431, EPI_ISL_460432, EPI_ISL_460433, EPI_ISL_460434, EPI_ISL_460435, EPI_ISL_460436, EPI_ISL_460437, EPI_ISL_460439, EPI_ISL_460440, EPI_ISL_460441, EPI_ISL_460442, EPI_ISL_460443, EPI_ISL_460444, EPI_ISL_460445, EPI_ISL_460446, EPI_ISL_460447, EPI_ISL_460448, EPI_ISL_460450, EPI_ISL_460451, EPI_ISL_460452, EPI_ISL_460453, EPI_ISL_460454, EPI_ISL_460455, EPI_ISL_460456, EPI_ISL_460457, EPI_ISL_460458, EPI_ISL_460459, EPI_ISL_460460, EPI_ISL_460461, EPI_ISL_460462, EPI_ISL_460463, EPI_ISL_460464, EPI_ISL_460465, EPI_ISL_460466, EPI_ISL_460467, EPI_ISL_460468, EPI_ISL_460469, EPI_ISL_460470, EPI_ISL_460471, EPI_ISL_460472 | | | |
| see above | Massachusetts General Hospital | Infectious Disease Program, Broad Institute of Harvard and MIT | Lemieux,J.E., Siddle,K.J., Shaw,B., Adams,G., Pierce,V., Turbett,S., Anahtar,M., Branda,J., Slater,D., Harris,J., Lin,A.E., Gladden-Young,A., Lagerborg,K., Rudy,M., DeRuff,K., Carter,A., Normandin,E., Bauer,M., Reilly,S., Tomkins-Tinch,C., Loreth,C., Chaluvadi,S., Neumann,A., Cusick,C., Chapman,S.B., Gnirke,A., Flowers,K., Cerrato,F., Birren,B.W., Gallagher,G., Smole,S., Park,D.J., MacInnis,B.L., Ryan,E., LaRocque,R., Rosenberg,E., Sabeti,P.C. |
| EPI_ISL_460554, EPI_ISL_460555, EPI_ISL_460556, EPI_ISL_460557, EPI_ISL_460558, EPI_ISL_460559, EPI_ISL_460560, EPI_ISL_460562, EPI_ISL_460563, EPI_ISL_460564, EPI_ISL_460565, EPI_ISL_460566, EPI_ISL_460567, EPI_ISL_460568, EPI_ISL_460569, EPI_ISL_460570, EPI_ISL_460571, EPI_ISL_460572, EPI_ISL_460573, EPI_ISL_460574, EPI_ISL_460576, EPI_ISL_460577, EPI_ISL_460578, EPI_ISL_460579, EPI_ISL_460580, EPI_ISL_460581, EPI_ISL_460582, EPI_ISL_460583, EPI_ISL_460584, EPI_ISL_460585, EPI_ISL_460586, EPI_ISL_460587, EPI_ISL_460588, EPI_ISL_460591, EPI_ISL_460592, EPI_ISL_460593, EPI_ISL_460594, EPI_ISL_460595, EPI_ISL_460596, EPI_ISL_460597, EPI_ISL_460598, EPI_ISL_460599, EPI_ISL_460600, EPI_ISL_460601, EPI_ISL_460602 | | | |
| see above | Michigan Department of Health and Human Services, Bureau of Laboratories | Michigan Department of Health and Human Services, Bureau of Laboratories | Blankenship HM, Riner D, Soehnlen MK |
| EPI_ISL_460603 | NYU Langone Health | Departments of Pathology and Medicine, New York University School of Medicine | Maria Aguero-Rosenfeld, Brendan Belovarac, Margaret Black, Ludovic Boytard, John Cadley, Paolo Cotzia, John Chen, Dacia Dimartino, Xiaojun Feng, Tatyana Gindin, Emily Guzman, Adriana Heguy, Megan Hogan, Emily Huang, George Jour, Alireza Khodadadi-Jamayran, Lawrence H. Lin, Raven Luther, Andrew Lytle, Christian Marier, Matthew T. Maurano, Mark J. Mulligan, Peter Meyn, Raquel Ordonez Ciriza, Iman Osman, Jared Pinnell, Vanessa Raabe, Sitharam Ramaswami, Amy Rapkiewicz, Andre M. Ribeiro-dos-Santos, Marie Samanovic-Golden, Antonio Serrano, Guomiao Shen, Matija Snuderl, Theodore Vougiouklakis, Nick Vulpescu, Gael Westby, Paul Zappile, Yutong Zhang |
| EPI_ISL_460621, EPI_ISL_460622, EPI_ISL_460623, EPI_ISL_460624, EPI_ISL_460625, EPI_ISL_460626, EPI_ISL_460627, EPI_ISL_460628, EPI_ISL_460629, EPI_ISL_460630, EPI_ISL_460631, EPI_ISL_460632, EPI_ISL_460633, EPI_ISL_460634, EPI_ISL_461399, EPI_ISL_461400, EPI_ISL_461401, EPI_ISL_461402, EPI_ISL_461403, EPI_ISL_461404, EPI_ISL_461405, EPI_ISL_461406, EPI_ISL_461407, EPI_ISL_461408, EPI_ISL_461409, EPI_ISL_461410, EPI_ISL_461411, EPI_ISL_461412, EPI_ISL_461413, EPI_ISL_461414, EPI_ISL_461415, EPI_ISL_461416, EPI_ISL_461417, EPI_ISL_461418, EPI_ISL_461419, EPI_ISL_461420, EPI_ISL_461421, EPI_ISL_461422, EPI_ISL_461423, EPI_ISL_461424, EPI_ISL_461425, EPI_ISL_461426, EPI_ISL_461427, EPI_ISL_461428, EPI_ISL_461429, EPI_ISL_461430, EPI_ISL_461431, EPI_ISL_461432, EPI_ISL_461433, EPI_ISL_461434, EPI_ISL_461435, EPI_ISL_461436, EPI_ISL_461437, EPI_ISL_461438, EPI_ISL_461439, EPI_ISL_461440, EPI_ISL_461441, EPI_ISL_461442, EPI_ISL_461443, EPI_ISL_461444, EPI_ISL_461445, EPI_ISL_461446, EPI_ISL_461447, EPI_ISL_461448, EPI_ISL_461449, EPI_ISL_461450, EPI_ISL_461451, EPI_ISL_461452, EPI_ISL_461453, EPI_ISL_461454, EPI_ISL_461455, EPI_ISL_461456, EPI_ISL_461457, EPI_ISL_461458, EPI_ISL_461459, EPI_ISL_461460, EPI_ISL_461461, EPI_ISL_461462, EPI_ISL_461463, EPI_ISL_461464, EPI_ISL_461465, EPI_ISL_461466, EPI_ISL_461467, EPI_ISL_461468, EPI_ISL_461469, EPI_ISL_461470, EPI_ISL_461471, EPI_ISL_461472, EPI_ISL_461473, EPI_ISL_461474, EPI_ISL_461475, EPI_ISL_461476, EPI_ISL_461477 | | | |
| see above | UW Virology Lab | UW Virology Lab | Pavitra Roychoudhury, Amin Addetia, Hong Xie, Lasata Shrestha, Truong Nguyen, Meei-Li Huang, Keith Jerome, Alexander Greninger |
| EPI_ISL_462636, EPI_ISL_462638, EPI_ISL_462639, EPI_ISL_462640, EPI_ISL_462641, EPI_ISL_462642, EPI_ISL_462643, EPI_ISL_462644, EPI_ISL_462645, EPI_ISL_462646, EPI_ISL_462647, EPI_ISL_462648, EPI_ISL_462649, EPI_ISL_462650, EPI_ISL_462651, EPI_ISL_462652, EPI_ISL_462653, EPI_ISL_462654, EPI_ISL_462655, EPI_ISL_462656, EPI_ISL_462657, EPI_ISL_462658, EPI_ISL_462659, EPI_ISL_462660, EPI_ISL_462661, EPI_ISL_462662, EPI_ISL_462663, EPI_ISL_462664, EPI_ISL_462665, EPI_ISL_462666, EPI_ISL_462667, EPI_ISL_462668, EPI_ISL_462669, EPI_ISL_462670, EPI_ISL_462671, EPI_ISL_462672, EPI_ISL_462673, EPI_ISL_462674, EPI_ISL_462675, EPI_ISL_462676, EPI_ISL_462677, EPI_ISL_462678, EPI_ISL_462679, EPI_ISL_462680, EPI_ISL_462681, EPI_ISL_462682, EPI_ISL_462683, EPI_ISL_462684, EPI_ISL_462685, EPI_ISL_462686, EPI_ISL_462687, EPI_ISL_462688, EPI_ISL_462689, EPI_ISL_462690, EPI_ISL_462691, EPI_ISL_462692, EPI_ISL_462693, EPI_ISL_462694, EPI_ISL_462695, EPI_ISL_462696, EPI_ISL_462697, EPI_ISL_462698, EPI_ISL_462699, EPI_ISL_462700, EPI_ISL_462701, EPI_ISL_462702, EPI_ISL_462703, EPI_ISL_462704, EPI_ISL_462705, EPI_ISL_462706, EPI_ISL_462707, EPI_ISL_462708, EPI_ISL_462709, EPI_ISL_462710, EPI_ISL_462711, EPI_ISL_462712, EPI_ISL_462713, EPI_ISL_462714, EPI_ISL_462715, EPI_ISL_462716, EPI_ISL_462717, EPI_ISL_462718, EPI_ISL_462719, EPI_ISL_462720, EPI_ISL_462721, EPI_ISL_462722, EPI_ISL_462723, EPI_ISL_462724, EPI_ISL_462725, EPI_ISL_462726, EPI_ISL_462727, EPI_ISL_462728, EPI_ISL_462729, EPI_ISL_462730, EPI_ISL_462731, EPI_ISL_462732, | | | |

| | | | |
|---|---|---|---|
| EPI_ISL_462733, EPI_ISL_462734, EPI_ISL_462735, EPI_ISL_462736 | | | |
| see above | Michigan Department of Health and Human Services, Bureau of Laboratories | Michigan Department of Health and Human Services, Bureau of Laboratories | Blankenship HM, Riner D, Soehnlen MK |
| EPI_ISL_462845, EPI_ISL_462846, EPI_ISL_462847, EPI_ISL_462848, EPI_ISL_462849, EPI_ISL_462850, EPI_ISL_462851, EPI_ISL_462852, EPI_ISL_462853, EPI_ISL_462854, EPI_ISL_462855, EPI_ISL_462856, EPI_ISL_462857, EPI_ISL_462858, EPI_ISL_462859, EPI_ISL_462860, EPI_ISL_462861, EPI_ISL_462862, EPI_ISL_462863, EPI_ISL_462864, EPI_ISL_462865, EPI_ISL_462866, EPI_ISL_462867, EPI_ISL_462868, EPI_ISL_462869, EPI_ISL_462870, EPI_ISL_462871, EPI_ISL_462872, EPI_ISL_462873, EPI_ISL_462874, EPI_ISL_462875, EPI_ISL_462876, EPI_ISL_462877, EPI_ISL_462878, EPI_ISL_462879, EPI_ISL_462880, EPI_ISL_462881, EPI_ISL_462882, EPI_ISL_462883, EPI_ISL_462884, EPI_ISL_462885, EPI_ISL_462886, EPI_ISL_462887, EPI_ISL_462888, EPI_ISL_462889, EPI_ISL_462890, EPI_ISL_462891, EPI_ISL_462892, EPI_ISL_462893, EPI_ISL_462894, EPI_ISL_462895, EPI_ISL_462896, EPI_ISL_462897, EPI_ISL_462898, EPI_ISL_462899, EPI_ISL_462900, EPI_ISL_462901, EPI_ISL_462902, EPI_ISL_462903, EPI_ISL_462904, EPI_ISL_462905, EPI_ISL_462906, EPI_ISL_462907, EPI_ISL_462908, EPI_ISL_462909, EPI_ISL_462910, EPI_ISL_462911 | | | |
| see above | Minnesota Department of Health, Public Health Laboratory | Minnesota Department of Health, Public Health Laboratory | Matt Plumb, Jacob Garfin, and Xiong Wang |
| EPI_ISL_462912, EPI_ISL_462913, EPI_ISL_462914, EPI_ISL_462915, EPI_ISL_462917, EPI_ISL_462918, EPI_ISL_462919, EPI_ISL_462921, EPI_ISL_462922, EPI_ISL_462924, EPI_ISL_462925, EPI_ISL_462926, EPI_ISL_462927, EPI_ISL_462929, EPI_ISL_462930, EPI_ISL_462932, EPI_ISL_462933, EPI_ISL_462935, EPI_ISL_462936, EPI_ISL_462940, EPI_ISL_462941, EPI_ISL_462943, EPI_ISL_462944, EPI_ISL_462945, EPI_ISL_462947, EPI_ISL_462948, EPI_ISL_462949, EPI_ISL_462950, EPI_ISL_462951, EPI_ISL_462952, EPI_ISL_462953, EPI_ISL_462955, EPI_ISL_462956, EPI_ISL_462957, EPI_ISL_462958, EPI_ISL_462959, EPI_ISL_462961, EPI_ISL_462963, EPI_ISL_462964, EPI_ISL_462965, EPI_ISL_462966, EPI_ISL_462967, EPI_ISL_462968, EPI_ISL_462969, EPI_ISL_462971, EPI_ISL_462973, EPI_ISL_462975, EPI_ISL_462976, EPI_ISL_462977, EPI_ISL_462979, EPI_ISL_462980, EPI_ISL_462982, EPI_ISL_462983, EPI_ISL_462987, EPI_ISL_462988 | | | |
| see above | Wyoming Public Health Laboratory | Center for Global Health, University of New Mexico Health Sciences Center | Daryl Domman, Kurt Schwalm, Rob Christensen, Wanda Manley, Cari Sloma, Noah Hull, Darrell Dinwiddie |
| EPI_ISL_462991 | Microbiology Division | Microbiology Division | Flores,H. |
| EPI_ISL_463094, EPI_ISL_463095, EPI_ISL_463096, EPI_ISL_463097, EPI_ISL_463098, EPI_ISL_463099, EPI_ISL_463100, EPI_ISL_463101, EPI_ISL_463102, EPI_ISL_463103, EPI_ISL_463104, EPI_ISL_463105, EPI_ISL_463106, EPI_ISL_463107, EPI_ISL_463108, EPI_ISL_463109, EPI_ISL_463110, EPI_ISL_463111, EPI_ISL_463112, EPI_ISL_463113, EPI_ISL_463114, EPI_ISL_463115, EPI_ISL_463116, EPI_ISL_463117, EPI_ISL_463118, EPI_ISL_463119, EPI_ISL_463120, EPI_ISL_463121, EPI_ISL_463122, EPI_ISL_463123, EPI_ISL_463124, EPI_ISL_463125, EPI_ISL_463126, EPI_ISL_463127, EPI_ISL_463128, EPI_ISL_463129, EPI_ISL_463130, EPI_ISL_463131, EPI_ISL_463132, EPI_ISL_463133, EPI_ISL_463134, EPI_ISL_463135, EPI_ISL_463136, EPI_ISL_463137 | | | |
| see above | Virginia DCLS | Virginia DCLS | Virginia DCLS |
| EPI_ISL_463138, EPI_ISL_463139, EPI_ISL_463140, EPI_ISL_463141, EPI_ISL_463143, EPI_ISL_463144, EPI_ISL_463145, EPI_ISL_463147, EPI_ISL_463148, EPI_ISL_463149, EPI_ISL_463150, EPI_ISL_463151, EPI_ISL_463152, EPI_ISL_463153, EPI_ISL_463154, EPI_ISL_463155, EPI_ISL_463156, EPI_ISL_463157, EPI_ISL_463158, EPI_ISL_463159, EPI_ISL_463160, EPI_ISL_463161, EPI_ISL_463162, EPI_ISL_463163, EPI_ISL_463164, EPI_ISL_463165, EPI_ISL_463166, EPI_ISL_463167, EPI_ISL_463168, EPI_ISL_463169, EPI_ISL_463170, EPI_ISL_463171, EPI_ISL_463172, EPI_ISL_463173 | | | |
| see above | Yale Clinical Virology Laboratory | Grubaugh Lab - Yale School of Public Health | Joseph Fauver, Tara Alpert, Anderson Brito, Anne Wyllie, Chantal Vogels, Mary Petrone, Cole Jensen, Chaney Kalinich, Isabel Ott, Arnau Casanovas, Catherine Muenker, Adam Moore, Alice Lu, Maria Tokuyama, Patrick Wong, Peiwen Lu, Saad Omer, Richard Martinello, Allison Nelson, Shelli Farhadian, Akiko Iwasaki, Charlese Dela Cruz, Albert Ko, Nathan Grubaugh |
| EPI_ISL_463277, EPI_ISL_463278, EPI_ISL_463279, EPI_ISL_463280, EPI_ISL_463281, EPI_ISL_463282, EPI_ISL_463283, EPI_ISL_463284, EPI_ISL_463285, EPI_ISL_463286, EPI_ISL_463287, EPI_ISL_463288, EPI_ISL_463289, EPI_ISL_463290, EPI_ISL_463291, EPI_ISL_463292, EPI_ISL_463293, EPI_ISL_463294, EPI_ISL_463295, EPI_ISL_463296, EPI_ISL_463297, EPI_ISL_463298, EPI_ISL_463299, EPI_ISL_463300 | | | |
| see above | Ochsner Health | Bioinfoexperts, LLC | Susanna L. Lamers, David J. Nolan, Rebecca Rose, Sissy Cross, David Moraga Amador, Tong Yang, Luke Caruso, Wayra Navia, Lydia Von Borstel, Xiao Hui Zhou, Amy Feehan, Julia-Garcia-Diaz |
| EPI_ISL_463325, EPI_ISL_463326, EPI_ISL_463327, EPI_ISL_463328, EPI_ISL_463329, EPI_ISL_463330, EPI_ISL_463331, EPI_ISL_463332, EPI_ISL_463333, EPI_ISL_463334, EPI_ISL_463335, EPI_ISL_463337, EPI_ISL_463338, EPI_ISL_463339, EPI_ISL_463340, EPI_ISL_463341, EPI_ISL_463342, EPI_ISL_463343, EPI_ISL_463344, EPI_ISL_463345, EPI_ISL_463346, EPI_ISL_463347, EPI_ISL_463348, EPI_ISL_463349, EPI_ISL_463350, EPI_ISL_463351, EPI_ISL_463352, EPI_ISL_463353, EPI_ISL_463354, EPI_ISL_463355, EPI_ISL_463356, EPI_ISL_463357, EPI_ISL_463358, EPI_ISL_463359, EPI_ISL_463360, EPI_ISL_463361, EPI_ISL_463362, EPI_ISL_463363, EPI_ISL_463364, EPI_ISL_463365, EPI_ISL_463366, EPI_ISL_463367, EPI_ISL_463368, EPI_ISL_463369, EPI_ISL_463370, EPI_ISL_463371, EPI_ISL_463378, EPI_ISL_463379, EPI_ISL_463380, EPI_ISL_463381, EPI_ISL_463382, EPI_ISL_463383, EPI_ISL_463384, EPI_ISL_463385, EPI_ISL_463386, EPI_ISL_463387, EPI_ISL_463388, EPI_ISL_463389, EPI_ISL_463390, EPI_ISL_463391, EPI_ISL_463392, EPI_ISL_463394, EPI_ISL_463395, EPI_ISL_463396, EPI_ISL_463397, EPI_ISL_463398, EPI_ISL_463399, EPI_ISL_463400, EPI_ISL_463401, EPI_ISL_463402, EPI_ISL_463403, EPI_ISL_463404, EPI_ISL_463405, EPI_ISL_463406, EPI_ISL_463407, EPI_ISL_463408, EPI_ISL_463409, EPI_ISL_463410, EPI_ISL_463411, EPI_ISL_463412, EPI_ISL_463413, EPI_ISL_463414, EPI_ISL_463415, EPI_ISL_463416, EPI_ISL_463417, EPI_ISL_463418, EPI_ISL_463419, EPI_ISL_463420, EPI_ISL_463421, EPI_ISL_463422, EPI_ISL_463423, EPI_ISL_463424, EPI_ISL_463425, EPI_ISL_463426, EPI_ISL_463427, EPI_ISL_463428, EPI_ISL_463429, EPI_ISL_463430, EPI_ISL_463431, EPI_ISL_463432, EPI_ISL_463433, EPI_ISL_463434, EPI_ISL_463435, EPI_ISL_463436, EPI_ISL_463437, EPI_ISL_463438, EPI_ISL_463439, EPI_ISL_463440, EPI_ISL_463441, EPI_ISL_463442, EPI_ISL_463443, EPI_ISL_463444, EPI_ISL_463445, EPI_ISL_463446, EPI_ISL_463449, EPI_ISL_463450, EPI_ISL_463451, EPI_ISL_463452, EPI_ISL_463453, EPI_ISL_463454, EPI_ISL_463455, EPI_ISL_463456, EPI_ISL_463457, EPI_ISL_463458, EPI_ISL_463459, EPI_ISL_463460, EPI_ISL_463461, EPI_ISL_463462, EPI_ISL_463463, EPI_ISL_463464, EPI_ISL_463465, EPI_ISL_463466, EPI_ISL_463467, EPI_ISL_463472, EPI_ISL_463478, EPI_ISL_463479, EPI_ISL_463480, EPI_ISL_463481, EPI_ISL_463482, EPI_ISL_463483, EPI_ISL_463484, EPI_ISL_463485, EPI_ISL_463486, EPI_ISL_463487, EPI_ISL_463488, EPI_ISL_463489, EPI_ISL_463490, EPI_ISL_463491, EPI_ISL_463492, EPI_ISL_463493, EPI_ISL_463494, EPI_ISL_463495, EPI_ISL_463496, EPI_ISL_463497, EPI_ISL_463498, EPI_ISL_463499, EPI_ISL_463500, EPI_ISL_463501, EPI_ISL_463502, EPI_ISL_463503, EPI_ISL_463504, EPI_ISL_463505, EPI_ISL_463506, EPI_ISL_463507, EPI_ISL_463508, EPI_ISL_463509, EPI_ISL_463510, EPI_ISL_463511, EPI_ISL_463512, EPI_ISL_463513, EPI_ISL_463514, EPI_ISL_463515, EPI_ISL_463516, EPI_ISL_463517, EPI_ISL_463518, EPI_ISL_463519, EPI_ISL_463520, EPI_ISL_463521, EPI_ISL_463522, EPI_ISL_463523, EPI_ISL_463524, EPI_ISL_463525, EPI_ISL_463526, EPI_ISL_463527, EPI_ISL_463528, EPI_ISL_463529, EPI_ISL_463530, EPI_ISL_463531, EPI_ISL_463532, EPI_ISL_463533, EPI_ISL_463534, EPI_ISL_463535, EPI_ISL_463536, EPI_ISL_463537, EPI_ISL_463538, EPI_ISL_463539, EPI_ISL_463540, EPI_ISL_463541, EPI_ISL_463542, EPI_ISL_463543, EPI_ISL_463544, EPI_ISL_463545, EPI_ISL_463546, EPI_ISL_463547, EPI_ISL_463548, EPI_ISL_463549, EPI_ISL_463550, EPI_ISL_463551, EPI_ISL_463552, EPI_ISL_463553, EPI_ISL_463554, EPI_ISL_463555, EPI_ISL_463556, EPI_ISL_463557, EPI_ISL_463558, EPI_ISL_463559, EPI_ISL_463560, EPI_ISL_463561, EPI_ISL_463562, EPI_ISL_463563, EPI_ISL_463564, EPI_ISL_463565, EPI_ISL_463566, EPI_ISL_463567, EPI_ISL_463568, EPI_ISL_463569, EPI_ISL_463570, EPI_ISL_463571, EPI_ISL_463572, EPI_ISL_463573, EPI_ISL_463574, EPI_ISL_463575, EPI_ISL_463576, EPI_ISL_463577, EPI_ISL_463578, EPI_ISL_463579, EPI_ISL_463580, EPI_ISL_463581, EPI_ISL_463582, EPI_ISL_463583, EPI_ISL_463584, EPI_ISL_463585, EPI_ISL_463586, EPI_ISL_463587, EPI_ISL_463588, EPI_ISL_463589, EPI_ISL_463590, EPI_ISL_463591, EPI_ISL_463592, EPI_ISL_463593, EPI_ISL_463594, EPI_ISL_463595, EPI_ISL_463596, EPI_ISL_463597, EPI_ISL_463598, EPI_ISL_463599, EPI_ISL_463600, EPI_ISL_463601, EPI_ISL_463602, EPI_ISL_463603, EPI_ISL_463604, EPI_ISL_463605, EPI_ISL_463606, EPI_ISL_463607, EPI_ISL_463608, EPI_ISL_463609, EPI_ISL_463610, EPI_ISL_463611, EPI_ISL_463612, EPI_ISL_463613, EPI_ISL_463614, EPI_ISL_463615, EPI_ISL_463616, EPI_ISL_463617, EPI_ISL_463618, EPI_ISL_463619, EPI_ISL_463620, EPI_ISL_463621, EPI_ISL_463622, EPI_ISL_463623, EPI_ISL_463624, EPI_ISL_463625, EPI_ISL_463626, EPI_ISL_463627, EPI_ISL_463628, EPI_ISL_463629, EPI_ISL_463630, EPI_ISL_463632, EPI_ISL_463633, EPI_ISL_463634, EPI_ISL_463635, EPI_ISL_463636, EPI_ISL_463637, EPI_ISL_463638, EPI_ISL_463639, EPI_ISL_463640, EPI_ISL_463641, EPI_ISL_463642, EPI_ISL_463643, EPI_ISL_463644, EPI_ISL_463645, EPI_ISL_463646, EPI_ISL_463647, EPI_ISL_463648, EPI_ISL_463649, EPI_ISL_463650, EPI_ISL_463651, EPI_ISL_463654, EPI_ISL_463655, EPI_ISL_463656, EPI_ISL_463657, EPI_ISL_463658, EPI_ISL_463659, EPI_ISL_463660, EPI_ISL_463661, EPI_ISL_463662, EPI_ISL_463663, EPI_ISL_463664, EPI_ISL_463665, EPI_ISL_463666, EPI_ISL_463667, EPI_ISL_463668, EPI_ISL_463669, EPI_ISL_463670, EPI_ISL_463671, EPI_ISL_463672, EPI_ISL_463673, EPI_ISL_463674, EPI_ISL_463675, EPI_ISL_463676, EPI_ISL_463677, EPI_ISL_463678, EPI_ISL_463679, EPI_ISL_463680, EPI_ISL_463681, EPI_ISL_463682, EPI_ISL_463684, EPI_ISL_463686, EPI_ISL_463687, EPI_ISL_463688, EPI_ISL_463689, EPI_ISL_463690, EPI_ISL_463691, EPI_ISL_463692, EPI_ISL_463694, EPI_ISL_463695, EPI_ISL_463696, EPI_ISL_463697, EPI_ISL_463698, EPI_ISL_463699, EPI_ISL_463700, EPI_ISL_463701, EPI_ISL_463702, EPI_ISL_463703, EPI_ISL_463704, EPI_ISL_463705 | | | |
| see above | Washington State Department of Health | Seattle Flu Study | Chu et al |
| EPI_ISL_464167 | VI-US Virgin Islands Department of Health | Pathogen Discovery, Respiratory Viruses Branch, Division of Viral Diseases, Centers for Disease Control and Prevention | Krista Queen, Ying Tao, Jing Zhang, Yan Li, Anna Uehara, Clinton R. Paden, Mary S. Keckler, Alison S. Laufer Halpin, Haibin Wang, Jasmine Padilla, Justin Lee, Christopher A. Elkins, Suxiang Tong |
| EPI_ISL_466648 | Innovative Genomics Institute, UCB | Innovative Genomics Institute, UCB | Stacia Wyman, Haridha Shivram, Liana Lareau, Shana McDevitt, Justin Choi |
| EPI_ISL_466652, EPI_ISL_466653, EPI_ISL_466654, EPI_ISL_466655, EPI_ISL_466656, EPI_ISL_466657, EPI_ISL_466658, EPI_ISL_466659, EPI_ISL_466660, EPI_ISL_466661, EPI_ISL_466662, EPI_ISL_466664, EPI_ISL_466665, EPI_ISL_466666, EPI_ISL_466667, EPI_ISL_466668, EPI_ISL_466669, EPI_ISL_466670, EPI_ISL_466671, EPI_ISL_466672, EPI_ISL_466673, EPI_ISL_466674, EPI_ISL_466675, EPI_ISL_466676, EPI_ISL_466677, EPI_ISL_466678, EPI_ISL_466679, EPI_ISL_466680, EPI_ISL_466681, EPI_ISL_466682, EPI_ISL_466683, EPI_ISL_467298 | | | |
| see above | Nebraska Public Health Laboratory | UNMC COVID-19 Response Team | UNMC COVID-19 Response Team |
| EPI_ISL_467302, EPI_ISL_467304 | Washington University in St. Louis | Washington University in St. Louis | David Wang, Carey-Ann Burnham, Scott Handley, Lindsay Droit, Stephen Tahan |
| EPI_ISL_467372, EPI_ISL_467373 | Arizona State University Health Services | Arizona State University | Peter T. Skidmore, Rabia Maqsood, LaRinda A. Holland, Emily A. Kaelin, Lily I. Wu, Arvind Varsani, Rolf U. Halden, Brenda G. Hogue, Matthew Scotch, Efrem S. Lim |
| EPI_ISL_467377, EPI_ISL_467378, EPI_ISL_467379, EPI_ISL_467380, EPI_ISL_467381, EPI_ISL_467384, EPI_ISL_467385, EPI_ISL_467387, EPI_ISL_467388, EPI_ISL_467389, EPI_ISL_467390, EPI_ISL_467391, EPI_ISL_467393, EPI_ISL_467394, EPI_ISL_467395, EPI_ISL_467396, EPI_ISL_467397, EPI_ISL_467398, EPI_ISL_467399, EPI_ISL_467400, EPI_ISL_467402, EPI_ISL_467405, EPI_ISL_467406, EPI_ISL_467407, EPI_ISL_467408, EPI_ISL_467409, EPI_ISL_467410, EPI_ISL_467411, EPI_ISL_467412, EPI_ISL_467413, EPI_ISL_467414, EPI_ISL_467416, EPI_ISL_467417, EPI_ISL_467418, EPI_ISL_467420, EPI_ISL_467421, EPI_ISL_467422 | | | |
| see above | NYU Langone Health | Departments of Pathology and Medicine, New York University School of Medicine | Maria Aguero-Rosenfeld, Brendan Belovarac, Margaret Black, Ludovic Boytard, John Cadley, Paolo Cotzia, John Chen, Dacia Dimartino, Xiaojun Feng, Tatyana Gindin, Emily Guzman, Adriana Heguy, Megan Hogan, Emily Huang, George Jour, Alireza Khodadadi-Jamayran, Lawrence H. Lin, Raven Luther, Andrew Lytle, Christian Marier, Matthew T. Maurano, Mark J. Mulligan, Peter Meyn, Raquel Ordonez Ciriza, Iman Osman, Jared Pinnell, Vanessa Raabe, Sitharam Ramaswami, Amy Rapkiewicz, Andre M. Ribeiro-dos-Santos, Marie Samanovic-Golden, Antonio Serrano, Guomiao Shen, Matija Snuderl, Theodore Vougiouklakis, Nick Vulpescu, Gael Westby, Paul Zappile, Yutong Zhang |
| EPI_ISL_467529, EPI_ISL_467532, EPI_ISL_467533, EPI_ISL_467536, EPI_ISL_467539, EPI_ISL_467540, EPI_ISL_467541, EPI_ISL_467542, EPI_ISL_467544, EPI_ISL_467545, EPI_ISL_467546, EPI_ISL_467547, EPI_ISL_467550, EPI_ISL_467551, EPI_ISL_467552, EPI_ISL_467553, EPI_ISL_467554, EPI_ISL_467555, EPI_ISL_467556, EPI_ISL_467557, EPI_ISL_467558, EPI_ISL_467559, EPI_ISL_467563, EPI_ISL_467564, EPI_ISL_467565, EPI_ISL_467566, EPI_ISL_467567, EPI_ISL_467568, EPI_ISL_467569, EPI_ISL_467570, EPI_ISL_467572, EPI_ISL_467574, EPI_ISL_467577, EPI_ISL_467582, EPI_ISL_467583, EPI_ISL_467584, EPI_ISL_467585, EPI_ISL_467586, EPI_ISL_467588, EPI_ISL_467589, EPI_ISL_467590, EPI_ISL_467591, EPI_ISL_467594, EPI_ISL_467596, EPI_ISL_467598, EPI_ISL_467599, EPI_ISL_467600, EPI_ISL_467604, EPI_ISL_467608, EPI_ISL_467609, EPI_ISL_467610, EPI_ISL_467611, EPI_ISL_467612, EPI_ISL_467613, EPI_ISL_467616, EPI_ISL_467617, EPI_ISL_467619, EPI_ISL_467620, EPI_ISL_467621, EPI_ISL_467623, EPI_ISL_467624, EPI_ISL_467625, EPI_ISL_467626, EPI_ISL_467627, EPI_ISL_467630, EPI_ISL_467632, EPI_ISL_467633, EPI_ISL_467634, EPI_ISL_467636, EPI_ISL_467638, EPI_ISL_467639, EPI_ISL_467640, EPI_ISL_467643, EPI_ISL_467644, EPI_ISL_467645, EPI_ISL_467646, EPI_ISL_467647, EPI_ISL_467648, EPI_ISL_467649, EPI_ISL_467650, EPI_ISL_467651, EPI_ISL_467652, EPI_ISL_467653, EPI_ISL_467654, EPI_ISL_467657, EPI_ISL_467659, EPI_ISL_467661, EPI_ISL_467662, EPI_ISL_467663, EPI_ISL_467664 | | | |
| see above | New Mexico Department of Health Scientific Laboratory Division | Center for Global Health, University of New Mexico Health Sciences Center | Daryl Domman, Kurt Schwalm, Twila Kunde, Joseph Hicks, Michael Edwards, Darrell Dinwiddie |
| EPI_ISL_467782, EPI_ISL_467783, EPI_ISL_467784, EPI_ISL_467785, EPI_ISL_467786, EPI_ISL_467787, EPI_ISL_467788, EPI_ISL_467789, EPI_ISL_467790, EPI_ISL_467791, EPI_ISL_467792, EPI_ISL_467793, EPI_ISL_467794, EPI_ISL_467795, EPI_ISL_467796, EPI_ISL_467797, EPI_ISL_467798, EPI_ISL_467799, EPI_ISL_467800, EPI_ISL_467801, EPI_ISL_467802, EPI_ISL_467803, EPI_ISL_467804, EPI_ISL_467805, EPI_ISL_467806, EPI_ISL_467807, EPI_ISL_467808 | | | |
| see above | Virginia DCLS | Virginia DCLS | Virginia DCLS |
| EPI_ISL_467809 | Cedars-Sinai Medical Center, Department of Pathology & Laboratory Medicine, Molecular Pathology Laboratory | Cedars-Sinai Medical Center, Molecular Pathology Laboratory of Department of Pathology & Laboratory Medicine and Genomic Core | Wenjuan Zhang, John Paul Govindavari, Brian Davis, Stephanie Chen, Jong Taek Kim, Jianbo Song, Jean Lopategui, Jasmine T Plummer, Eric Vail |
| EPI_ISL_467811, EPI_ISL_467812, EPI_ISL_467813, EPI_ISL_467814, EPI_ISL_467815, EPI_ISL_467816, EPI_ISL_467817, EPI_ISL_467818, EPI_ISL_467819, EPI_ISL_467820, EPI_ISL_467821, EPI_ISL_467822, EPI_ISL_467823, EPI_ISL_467824, EPI_ISL_467825, EPI_ISL_467826, EPI_ISL_467827, EPI_ISL_467828, EPI_ISL_467829, EPI_ISL_467830, EPI_ISL_467831, EPI_ISL_467832, EPI_ISL_467833, EPI_ISL_467834, EPI_ISL_467835, EPI_ISL_467836, EPI_ISL_467837, EPI_ISL_467838, EPI_ISL_467839, EPI_ISL_467840, EPI_ISL_467841, EPI_ISL_467842, EPI_ISL_467843, EPI_ISL_467844, EPI_ISL_467845, EPI_ISL_467846, EPI_ISL_467847, EPI_ISL_467848, EPI_ISL_467849, EPI_ISL_467850, EPI_ISL_467851, EPI_ISL_467852, EPI_ISL_467853, EPI_ISL_467854, EPI_ISL_467855, EPI_ISL_467856, EPI_ISL_467857, EPI_ISL_467858, EPI_ISL_467859, EPI_ISL_467860, EPI_ISL_467861, EPI_ISL_467862, EPI_ISL_467863, EPI_ISL_467864, EPI_ISL_467865, EPI_ISL_467866, EPI_ISL_467867, EPI_ISL_467868, EPI_ISL_467869, EPI_ISL_467870, EPI_ISL_467871, EPI_ISL_467872, EPI_ISL_467873, EPI_ISL_467874, EPI_ISL_467875, EPI_ISL_467876, EPI_ISL_467877, EPI_ISL_467878, EPI_ISL_467879, EPI_ISL_467880, EPI_ISL_467881, EPI_ISL_467882, EPI_ISL_467883, EPI_ISL_467884, EPI_ISL_467885, EPI_ISL_467886, EPI_ISL_467887, EPI_ISL_467888, EPI_ISL_467889, EPI_ISL_467890, EPI_ISL_467891, EPI_ISL_467892, EPI_ISL_467893, EPI_ISL_467894, EPI_ISL_467895, EPI_ISL_467896, EPI_ISL_467897, EPI_ISL_467898, EPI_ISL_467899, EPI_ISL_467900, EPI_ISL_467901, EPI_ISL_467902, EPI_ISL_467903, EPI_ISL_467904, EPI_ISL_467905, EPI_ISL_467906, EPI_ISL_467907, EPI_ISL_467908, EPI_ISL_467909, EPI_ISL_467910, EPI_ISL_467911, EPI_ISL_467912, EPI_ISL_467913, EPI_ISL_467914, EPI_ISL_467915, EPI_ISL_467916, EPI_ISL_467917, EPI_ISL_467918, EPI_ISL_467919, EPI_ISL_467920, EPI_ISL_467921, EPI_ISL_467922, EPI_ISL_467923, EPI_ISL_467924, EPI_ISL_467925, EPI_ISL_467926, EPI_ISL_467927 | | | |
| see above | Quest Diagnostics | Quest Diagnostics | Anderson,B.P., Rosenthal,S.H., Gerasimova,A., Kagan,R.M. and Owen, R. |
| EPI_ISL_467928, EPI_ISL_467929, EPI_ISL_467930, EPI_ISL_467931, EPI_ISL_467932, EPI_ISL_467933, EPI_ISL_467934, EPI_ISL_467935, EPI_ISL_467936, EPI_ISL_467937, EPI_ISL_467938, EPI_ISL_467939, EPI_ISL_467940, EPI_ISL_467941, EPI_ISL_467942, EPI_ISL_467943, EPI_ISL_467944 | | | |
| see above | Virginia DCLS | Virginia DCLS | Virginia DCLS |
| EPI_ISL_467945 | Montefiore Medical Center, Dept. of Pathology, Clinical Virology | Albert Einstein College of Medicine, Dept. of Microbiology & Immunology, Chandran lab | J. Maximilian Fels, Saad Khan, Ryan Forster, Karin A. Skalina, Ariel S. Wirchnianski, Denise Haslwanter, Catalina Florez, Robert H. Bortz III, M. Eugenia Dieterle, Ethan Laudermilch, Rohit K. Jangra, Amanda Mengotto, Duncan Kimmel, Shahina B. Maqbool, John M. Greally, Wendy A. Szymczak, Amy S. Fox, Michael B. Prystowsky, D. Yitzchak Goldstein, Johanna P. Daily, Libusha Kelly, Kartik Chandran |
| EPI_ISL_467946, EPI_ISL_467947, EPI_ISL_467948, EPI_ISL_467949 | Innovative Genomics Institute, UC Berkeley | Innovative Genomics Institute, UC Berkeley | Stacia Wyman, Haridha Shivram, Liana Lareau, Shana McDevitt, Justin Choi |
| EPI_ISL_467950, EPI_ISL_467951, EPI_ISL_467952, EPI_ISL_467953, EPI_ISL_467954, EPI_ISL_467955, EPI_ISL_467956, EPI_ISL_467957, EPI_ISL_467958, EPI_ISL_467959, EPI_ISL_467960, EPI_ISL_467961, EPI_ISL_467962 | | | |
| see above | San Diego County Public Health Laboratory | Andersen lab at Scripps Research | SEARCH Alliance San Diego with Tracy Basler, Jovan Shephard, Brett Austin |
| EPI_ISL_467963 | San Diego County Public Health Laboratory | Andersen lab at Scripps Research | SEARCH Alliance San Diego with Michael Quigley, Ellen Stefanski, Ian Mchardy |
| EPI_ISL_467964, EPI_ISL_467965, EPI_ISL_467966 | San Diego County Public Health Laboratory | Andersen lab at Scripps Research | SEARCH Alliance San Diego with Tracy Basler, Jovan Shephard, Brett Austin |
| EPI_ISL_467967 | Scripps Medical Laboratory | Andersen lab at Scripps Research | SEARCH Alliance San Diego with Tracy Basler, Jovan Shephard, Brett Austin |
| EPI_ISL_467968, EPI_ISL_467969, | San Diego County Public Health Laboratory | Andersen lab at Scripps Research | SEARCH Alliance San Diego with Tracy Basler, Jovan Shephard, Brett Austin |

| | | | |
|---|---|---|---|
| EPI_ISL_467970, EPI_ISL_467971 | | | |
| EPI_ISL_467972 | Scripps Medical Laboratory | Andersen lab at Scripps Research | SEARCH Alliance San Diego with Michael Quigley, Ellen Stefanski, Ian Mchardy |
| EPI_ISL_467973, EPI_ISL_467974, EPI_ISL_467975 | San Diego County Public Health Laboratory | Andersen lab at Scripps Research | SEARCH Alliance San Diego with Tracy Basler, Jovan Shephard, Brett Austin |
| EPI_ISL_467976 | Rady's Childrens Hospital | Andersen lab at Scripps Research | SEARCH Alliance San Diego |
| EPI_ISL_467977, EPI_ISL_467978, EPI_ISL_467979, EPI_ISL_467980 | San Diego County Public Health Laboratory | Andersen lab at Scripps Research | SEARCH Alliance San Diego with Tracy Basler, Jovan Shephard, Brett Austin |
| EPI_ISL_467981 | Rady's Childrens Hospital | Andersen lab at Scripps Research | SEARCH Alliance San Diego |
| EPI_ISL_467982, EPI_ISL_467983 | San Diego County Public Health Laboratory | Andersen lab at Scripps Research | SEARCH Alliance San Diego with Tracy Basler, Jovan Shephard, Brett Austin |
| EPI_ISL_467984 | Rady's Childrens Hospital | Andersen lab at Scripps Research | SEARCH Alliance San Diego |
| EPI_ISL_468063, EPI_ISL_468064, EPI_ISL_468065 | unknown | Computer Science and Engineering | Rouchka,E.C., Chariker,J.H., Chung,D., Ramirez,J., Palmer,K.E., Lasnik,A.B., Carrico,R., Arnold,F.W., Adcock,R.S., Zhang,M., Alejandro,B., Wolf,L.A., Hwang,J.Y., Park,J.W., Waigel,S., Zacharias,W. |
| EPI_ISL_468080, EPI_ISL_468081, EPI_ISL_468082, EPI_ISL_468083, EPI_ISL_468084, EPI_ISL_468086, EPI_ISL_468087, EPI_ISL_468088, EPI_ISL_468089, EPI_ISL_468091, EPI_ISL_468092, EPI_ISL_468093, EPI_ISL_468094, EPI_ISL_468095, EPI_ISL_468096, EPI_ISL_468097, EPI_ISL_468098, EPI_ISL_468099, EPI_ISL_468100, EPI_ISL_468101, EPI_ISL_468103, EPI_ISL_468104, EPI_ISL_468105, EPI_ISL_468106, EPI_ISL_468107, EPI_ISL_468108, EPI_ISL_468109, EPI_ISL_468110, EPI_ISL_468111, EPI_ISL_468112, EPI_ISL_468113, EPI_ISL_468114, EPI_ISL_468115, EPI_ISL_468116, EPI_ISL_468117, EPI_ISL_468118, EPI_ISL_468119, EPI_ISL_468120, EPI_ISL_468121, EPI_ISL_468122, EPI_ISL_468123, EPI_ISL_468124, EPI_ISL_468125, EPI_ISL_468126, EPI_ISL_468127, EPI_ISL_468128, EPI_ISL_468129 | | | |
| see above | OHSU Lab Services Molecular Microbiology Lab | Oregon SARS-CoV-2 Genome Sequencing Center | Brendan L. O'Connell, Ruth V. Nichols, Alec J. Hirsch, Guang Fan, Daniel N. Streblow, William B. Messer, Andrew C. Adey, Benjamin N. Bimber, Brian J. O'Roak |
| EPI_ISL_468345, EPI_ISL_468346, EPI_ISL_468347, EPI_ISL_468348, EPI_ISL_468349, EPI_ISL_468350, EPI_ISL_468351, EPI_ISL_468352, EPI_ISL_468353, EPI_ISL_468354, EPI_ISL_468355, EPI_ISL_468356 | | | |
| see above | County of Santa Clara Public Health Department | Chan-Zuckerberg Biohub | CZB Cliahub Consortium |
| EPI_ISL_468357, EPI_ISL_468358, EPI_ISL_468359, EPI_ISL_468360, EPI_ISL_468361, EPI_ISL_468362, EPI_ISL_468363, EPI_ISL_468364, EPI_ISL_468365, EPI_ISL_468366, EPI_ISL_468367, EPI_ISL_468368, EPI_ISL_468369, EPI_ISL_468370, EPI_ISL_468371, EPI_ISL_468372, EPI_ISL_468373, EPI_ISL_468374, EPI_ISL_468375, EPI_ISL_468376, EPI_ISL_468377, EPI_ISL_468378, EPI_ISL_468379, EPI_ISL_468380, EPI_ISL_468382, EPI_ISL_468383, EPI_ISL_468384, EPI_ISL_468385, EPI_ISL_468387 | | | |
| see above | Alameda County Public Health Lab | Chan-Zuckerberg Biohub | CZB Cliahub Consortium |
| EPI_ISL_468388, EPI_ISL_468389, EPI_ISL_468390, EPI_ISL_468391, EPI_ISL_468392, EPI_ISL_468393, EPI_ISL_468394, EPI_ISL_468395, EPI_ISL_468396, EPI_ISL_468397, EPI_ISL_468398, EPI_ISL_468399, EPI_ISL_468400, EPI_ISL_468401, EPI_ISL_468402, EPI_ISL_468403, EPI_ISL_468404, EPI_ISL_468405, EPI_ISL_468406, EPI_ISL_468407, EPI_ISL_468408, EPI_ISL_468409, EPI_ISL_468410, EPI_ISL_468411, EPI_ISL_468413, EPI_ISL_468414, EPI_ISL_468415, EPI_ISL_468416, EPI_ISL_468417, EPI_ISL_468418, EPI_ISL_468419, EPI_ISL_468420, EPI_ISL_468421, EPI_ISL_468422, EPI_ISL_468423, EPI_ISL_468424, EPI_ISL_468425, EPI_ISL_468426, EPI_ISL_468427, EPI_ISL_468428, EPI_ISL_468429, EPI_ISL_468430, EPI_ISL_468431, EPI_ISL_468432, EPI_ISL_468433, EPI_ISL_468434, EPI_ISL_468435, EPI_ISL_468436, EPI_ISL_468437 | | | |
| see above | County of San Luis Obispo Public Health Laboratory | Chan-Zuckerberg Biohub | CZB Cliahub Consortium |
| EPI_ISL_468438, EPI_ISL_468439, EPI_ISL_468440, EPI_ISL_468441, EPI_ISL_468442, EPI_ISL_468443, EPI_ISL_468444, EPI_ISL_468445, EPI_ISL_468446, EPI_ISL_468447, EPI_ISL_468448, EPI_ISL_468449, EPI_ISL_468450, EPI_ISL_468451, EPI_ISL_468452, EPI_ISL_468453, EPI_ISL_468454, EPI_ISL_468455, EPI_ISL_468456, EPI_ISL_468457, EPI_ISL_468458, EPI_ISL_468459, EPI_ISL_468460, EPI_ISL_468461 | | | |
| see above | Humboldt County Public Health Laboratory | Chan-Zuckerberg Biohub | CZB Cliahub Consortium |
| EPI_ISL_468462, EPI_ISL_468463, EPI_ISL_468464, EPI_ISL_468465, EPI_ISL_468466, EPI_ISL_468467, EPI_ISL_468468, EPI_ISL_468469, EPI_ISL_468470, EPI_ISL_468471, EPI_ISL_468472, EPI_ISL_468473, EPI_ISL_468474, EPI_ISL_468475, EPI_ISL_468476, EPI_ISL_468477, EPI_ISL_468478, EPI_ISL_468479, EPI_ISL_468480, EPI_ISL_468481, EPI_ISL_468482, EPI_ISL_468483, EPI_ISL_468484, EPI_ISL_468485, EPI_ISL_468486, EPI_ISL_468487, EPI_ISL_468488, EPI_ISL_468489, EPI_ISL_468490, EPI_ISL_468491, EPI_ISL_468492, EPI_ISL_468493, EPI_ISL_468494, EPI_ISL_468495, EPI_ISL_468496, EPI_ISL_468497, EPI_ISL_468498, EPI_ISL_468499, EPI_ISL_468500, EPI_ISL_468501, EPI_ISL_468502, EPI_ISL_468503, EPI_ISL_468504, EPI_ISL_468505 | | | |
| see above | Ventura County Public Health Lab | Chan-Zuckerberg Biohub | CZB Cliahub Consortium |
| EPI_ISL_468506, EPI_ISL_468507, EPI_ISL_468508, EPI_ISL_468509, EPI_ISL_468510, EPI_ISL_468511, EPI_ISL_468512, EPI_ISL_468513, EPI_ISL_468514, EPI_ISL_468515, EPI_ISL_468517, EPI_ISL_468518, EPI_ISL_468519, EPI_ISL_468520, EPI_ISL_468521, EPI_ISL_468522, EPI_ISL_468523, EPI_ISL_468524, EPI_ISL_468525, EPI_ISL_468526, EPI_ISL_468527, EPI_ISL_468528, EPI_ISL_468529, EPI_ISL_468530, EPI_ISL_468531, EPI_ISL_468532, EPI_ISL_468533, EPI_ISL_468534, EPI_ISL_468535, EPI_ISL_468536, EPI_ISL_468537, EPI_ISL_468538, EPI_ISL_468539, EPI_ISL_468540, EPI_ISL_468541, EPI_ISL_468542, EPI_ISL_468543, EPI_ISL_468544, EPI_ISL_468545, EPI_ISL_468546, EPI_ISL_468547, EPI_ISL_468548, EPI_ISL_468549, EPI_ISL_468550, EPI_ISL_468551, EPI_ISL_468552, EPI_ISL_468553, EPI_ISL_468554, EPI_ISL_468556, EPI_ISL_468557, EPI_ISL_468558, EPI_ISL_468559 | | | |
| see above | San Joaquin County Public Health Lab | Chan-Zuckerberg Biohub | CZB Cliahub Consortium |
| EPI_ISL_468560, EPI_ISL_468561, EPI_ISL_468562, EPI_ISL_468563, EPI_ISL_468564, EPI_ISL_468565, EPI_ISL_468566, EPI_ISL_468567, EPI_ISL_468568, EPI_ISL_468569, EPI_ISL_468570, EPI_ISL_468571, EPI_ISL_468572, EPI_ISL_468573, EPI_ISL_468574, EPI_ISL_468575, EPI_ISL_468576, EPI_ISL_468577, EPI_ISL_468578, EPI_ISL_468579, EPI_ISL_468580, EPI_ISL_468581, EPI_ISL_468582, EPI_ISL_468583, EPI_ISL_468584, EPI_ISL_468585, EPI_ISL_468586, EPI_ISL_468587, EPI_ISL_468588, EPI_ISL_468589, EPI_ISL_468590 | | | |
| see above | Quest Diagnostics | Quest Diagnostics | Anderson,B.P., Rosenthal,S.H., Gerasimova,A., Kagan,R.M. and Owen, R. |
| EPI_ISL_468592, EPI_ISL_468593, EPI_ISL_468594, EPI_ISL_468595, EPI_ISL_468596, EPI_ISL_468597, EPI_ISL_468598, EPI_ISL_468599, EPI_ISL_468600, EPI_ISL_468601, EPI_ISL_468602, EPI_ISL_468603, EPI_ISL_468604, EPI_ISL_468605, EPI_ISL_468606, EPI_ISL_468607, EPI_ISL_468608, EPI_ISL_468609, EPI_ISL_468610, EPI_ISL_468611, EPI_ISL_468612, EPI_ISL_468613, EPI_ISL_468614 | | | |
| see above | Orange County Public Health Lab | Chan-Zuckerberg Biohub | CZB Cliahub Consortium |
| EPI_ISL_468615, EPI_ISL_468616, EPI_ISL_468617, EPI_ISL_468618, EPI_ISL_468619, EPI_ISL_468620, EPI_ISL_468621, EPI_ISL_468622, EPI_ISL_468623, EPI_ISL_468624, EPI_ISL_468625, EPI_ISL_468626, EPI_ISL_468627, EPI_ISL_468628, EPI_ISL_468629, EPI_ISL_468630, EPI_ISL_468631, EPI_ISL_468632, EPI_ISL_468633, EPI_ISL_468634, EPI_ISL_468635, EPI_ISL_468636, EPI_ISL_468637, EPI_ISL_468638, EPI_ISL_468639, EPI_ISL_468640, EPI_ISL_468641, EPI_ISL_468642, EPI_ISL_468643, EPI_ISL_468645, EPI_ISL_468646, EPI_ISL_468647, EPI_ISL_468648, EPI_ISL_468649, EPI_ISL_468650, EPI_ISL_468651, EPI_ISL_468652, EPI_ISL_468654, EPI_ISL_468655 | | | |
| see above | Contra Costa Public Health Lab | Chan-Zuckerberg Biohub | CZB Cliahub Consortium |
| EPI_ISL_468701, EPI_ISL_468702, EPI_ISL_468703, EPI_ISL_468704, EPI_ISL_468705, EPI_ISL_468706, EPI_ISL_468707, EPI_ISL_468708, EPI_ISL_468709, EPI_ISL_468710, EPI_ISL_468711, EPI_ISL_468712, EPI_ISL_468713, EPI_ISL_468714, EPI_ISL_468715, EPI_ISL_468716, EPI_ISL_468717 | | | |
| see above | Ochsner Health | Bioinfoexperts, LLC | Rebecca Rose, Amy Feehan, David J. Nolan, Sissy Cross, David Moraga Amador, Tong Yang, Luke Caruso, Wayra Navia, Lydia Von Borstel, Xiao Hui Zhou, Julia-Garcia-Diaz, Susanna L. Lamers |
| EPI_ISL_468718 | Environmental and Global Health | Environmental and Global Health | Stephenson,C.J., Subramaniam,K., Waltzek,T.B., Merck,L.H., Gibson,J.C., Morris,J.G. |
| EPI_ISL_468719 | University of Florida | University of Florida | Elbadry,M.A., Subramaniam,K., Waltzek,T.B., Stephenson,C.J., Gibson,J.C., Alam,M.M., Lauzardo,M., Morris,J.G., Lednicky,J.A. |
| EPI_ISL_468720 | University of Florida | University of Florida | Stephenson,C.J., Subramaniam,K., Waltzek,T.B., Lauzardo,M., Morris,J.G., Lednicky,J.A. |
| EPI_ISL_468721 | University of Florida | University of Florida | Stephenson,C.J., Subramaniam,K., Waltzek,T.B., Lauzardo,M., Gibson,J.C., Morris,J.G., Lednicky,J.A. |
| EPI_ISL_468722 | University of Florida | University of Florida | Elbadry,M.A., Subramaniam,K., Waltzek,T.B., Lauzardo,M., Morris,J.G., Lednicky,J.A. |
| EPI_ISL_468723 | University of Florida | University of Florida | Stephenson,C.J., Subramaniam,K., Waltzek,T.B., Lauzardo,M., Morris,J.G., Lednicky,J.A. |
| EPI_ISL_469171, EPI_ISL_469172, EPI_ISL_469173, EPI_ISL_469174, EPI_ISL_469175, EPI_ISL_469176, EPI_ISL_469177, EPI_ISL_469178, EPI_ISL_469179, EPI_ISL_469180, EPI_ISL_469181, EPI_ISL_469182, EPI_ISL_469183, EPI_ISL_469184, EPI_ISL_469185, EPI_ISL_469186, EPI_ISL_469187, EPI_ISL_469188, EPI_ISL_469189, EPI_ISL_469190, EPI_ISL_469191, EPI_ISL_469192, EPI_ISL_469193, EPI_ISL_469194, EPI_ISL_469195, EPI_ISL_469196, EPI_ISL_469197, EPI_ISL_469198, EPI_ISL_469199, EPI_ISL_469200, EPI_ISL_469201, EPI_ISL_469202, EPI_ISL_469203, EPI_ISL_469204, EPI_ISL_469205, EPI_ISL_469206, EPI_ISL_469207, EPI_ISL_469208 | | | |
| see above | Yale Clinical Virology Laboratory | Grubaugh Lab - Yale School of Public Health | Joseph Fauver, Tara Alpert, Anderson Brito, Anne Wyllie, Chantal Vogels, Mary Petrone, Cole Jensen, Chaney Kalinich, Isabel Ott, Arnau Casanovas, Catherine Muenker, Adam Moore, Alice Lu, Maria Tokuyama, Patrick Wong, Peiwen Lu, Saad Omer, Richard Martinello, Allison Nelson, Shelli Farhadian, Akiko Iwasaki, Charlese Dela Cruz, Albert Ko, Nathan Grubaugh |
| EPI_ISL_470540 | Wisconsin State Laboratory of Hygiene Communicable Disease Division | Wisconsin State Laboratory of Hygiene Communicable Disease Division | Kelsey R Florek |
| EPI_ISL_470542, EPI_ISL_470543, EPI_ISL_470544, EPI_ISL_470545, EPI_ISL_470547, EPI_ISL_470548, EPI_ISL_470549, EPI_ISL_470550, EPI_ISL_470551, EPI_ISL_470552, EPI_ISL_470553, EPI_ISL_470554, EPI_ISL_470555, EPI_ISL_470558, EPI_ISL_470559, EPI_ISL_470560, EPI_ISL_470561, EPI_ISL_470563, EPI_ISL_470564, EPI_ISL_470565, EPI_ISL_470566, EPI_ISL_470567, EPI_ISL_470656, EPI_ISL_470657, EPI_ISL_470659, EPI_ISL_470660, EPI_ISL_470662, EPI_ISL_470663, EPI_ISL_470664, EPI_ISL_470665, EPI_ISL_470666, EPI_ISL_470667, EPI_ISL_470668, EPI_ISL_470669, EPI_ISL_470670, EPI_ISL_470671, EPI_ISL_470672, EPI_ISL_470673, EPI_ISL_470675, EPI_ISL_470677, EPI_ISL_470678, EPI_ISL_470679, EPI_ISL_470680, EPI_ISL_470681, EPI_ISL_470682, EPI_ISL_470683, EPI_ISL_470684, EPI_ISL_470685, EPI_ISL_470686, EPI_ISL_470687, EPI_ISL_470688, EPI_ISL_470689, EPI_ISL_470690, EPI_ISL_470691, EPI_ISL_470692, EPI_ISL_470693, EPI_ISL_470694, EPI_ISL_470696, EPI_ISL_470697, EPI_ISL_470698, EPI_ISL_470699, EPI_ISL_470700, EPI_ISL_470701, EPI_ISL_470702, EPI_ISL_470703, EPI_ISL_470704, EPI_ISL_470705, EPI_ISL_470706, EPI_ISL_470707, EPI_ISL_470708, EPI_ISL_470709, EPI_ISL_470710, EPI_ISL_470711, EPI_ISL_470712, EPI_ISL_470714, EPI_ISL_470715, EPI_ISL_470716, EPI_ISL_470717, EPI_ISL_470718 | | | |
| see above | Utah Public Health Laboratory | Utah Public Health Laboratory | Erin Young, Kelly Oakeson |
| EPI_ISL_470719, EPI_ISL_470720, EPI_ISL_470721, EPI_ISL_470722, EPI_ISL_470724, EPI_ISL_470725, EPI_ISL_470727, EPI_ISL_470728, EPI_ISL_470729, EPI_ISL_470730, EPI_ISL_470731, EPI_ISL_470732, EPI_ISL_470733, EPI_ISL_470734, EPI_ISL_470735, EPI_ISL_470736, EPI_ISL_470737, EPI_ISL_470738, EPI_ISL_470740, EPI_ISL_470741, EPI_ISL_470742, EPI_ISL_470743, EPI_ISL_470744, EPI_ISL_470745 | | | |
| see above | Utah Public Health Laboratory | Utah Public Health Laboratory | Heidi Butz, Erin Young, Kelly Oakeson |
| EPI_ISL_470747, EPI_ISL_470748, EPI_ISL_470749, EPI_ISL_470750, EPI_ISL_470751, EPI_ISL_470752, EPI_ISL_470753, EPI_ISL_470754, EPI_ISL_470755, EPI_ISL_470756, EPI_ISL_470757, EPI_ISL_470758, EPI_ISL_470759, EPI_ISL_470760, EPI_ISL_470761, EPI_ISL_470762, EPI_ISL_470763, EPI_ISL_470764, EPI_ISL_470765, EPI_ISL_470766, EPI_ISL_470767, EPI_ISL_470768, EPI_ISL_470769, EPI_ISL_470770, EPI_ISL_470771, EPI_ISL_470772, EPI_ISL_470773, EPI_ISL_470774, EPI_ISL_470775, EPI_ISL_470776, EPI_ISL_470777, EPI_ISL_470778, EPI_ISL_470779, EPI_ISL_470780, EPI_ISL_470781, EPI_ISL_470782, EPI_ISL_470783, EPI_ISL_470784, EPI_ISL_470785, EPI_ISL_470786, EPI_ISL_470787, EPI_ISL_470788, EPI_ISL_470789 | | | |
| see above | Minnesota Department of Health, Public Health Laboratory | Minnesota Department of Health, Public Health Laboratory | Matt Plumb, Jacob Garfin, and Xiong Wang |
| EPI_ISL_470790, EPI_ISL_470791, EPI_ISL_470792, EPI_ISL_470793, EPI_ISL_470794, EPI_ISL_470795, EPI_ISL_470796, EPI_ISL_470797, EPI_ISL_470798, EPI_ISL_470799, EPI_ISL_470800 | | | |
| see above | M Health Fairview | Minnesota Department of Health, Public Health Laboratory | Matt Plumb, Jacob Garfin, and Xiong Wang |
| EPI_ISL_471144, EPI_ISL_471145, EPI_ISL_471146, EPI_ISL_471147, EPI_ISL_471148, EPI_ISL_471149, EPI_ISL_471150, EPI_ISL_471151, EPI_ISL_471152, EPI_ISL_471153, EPI_ISL_471154, EPI_ISL_471155, EPI_ISL_471156 | | | |
| see above | Gundersen Molecular Diagnostics Laboratory | Kabara Cancer Research Institute | Craig S. Richmond, Paraic A. Kenny |
| EPI_ISL_471157 | Gundersen Clinical Microbiology Laboratory | Kabara Cancer Research Institute | Craig S. Richmond, Paraic A. Kenny |
| EPI_ISL_471178, EPI_ISL_471179, EPI_ISL_471180, EPI_ISL_471181, EPI_ISL_471182, EPI_ISL_471183, EPI_ISL_471184, EPI_ISL_471185, EPI_ISL_471186, EPI_ISL_471187, EPI_ISL_471188, EPI_ISL_471189, EPI_ISL_471190, EPI_ISL_471191, EPI_ISL_471192, EPI_ISL_471193, EPI_ISL_471194, EPI_ISL_471195, EPI_ISL_471196, EPI_ISL_471197, EPI_ISL_471198, EPI_ISL_471199, EPI_ISL_471200, EPI_ISL_471201, EPI_ISL_471202, EPI_ISL_471203, EPI_ISL_471204, EPI_ISL_471205, EPI_ISL_471206, EPI_ISL_471207, EPI_ISL_471208, EPI_ISL_471209, EPI_ISL_471210, EPI_ISL_471211, EPI_ISL_471212, EPI_ISL_471213, EPI_ISL_471214, EPI_ISL_471215, EPI_ISL_471216, EPI_ISL_471217, EPI_ISL_471218, EPI_ISL_471219, EPI_ISL_471220, EPI_ISL_471221, EPI_ISL_471222, EPI_ISL_471223, EPI_ISL_471224, EPI_ISL_471225, EPI_ISL_471226, EPI_ISL_471227, EPI_ISL_471228, EPI_ISL_471229, EPI_ISL_471230, EPI_ISL_471231, EPI_ISL_471232, EPI_ISL_471233, EPI_ISL_471234, EPI_ISL_471235, EPI_ISL_471236, EPI_ISL_471237, EPI_ISL_471238, EPI_ISL_471239, EPI_ISL_471240, EPI_ISL_471241, EPI_ISL_471242, EPI_ISL_471243, EPI_ISL_471244, EPI_ISL_471245, EPI_ISL_471246, EPI_ISL_471247, EPI_ISL_471248, EPI_ISL_471249, EPI_ISL_471250, EPI_ISL_471251, EPI_ISL_471252, EPI_ISL_471253, EPI_ISL_471254, EPI_ISL_471255, EPI_ISL_471256, EPI_ISL_471257, EPI_ISL_471258, EPI_ISL_471260, EPI_ISL_471261, EPI_ISL_471262, EPI_ISL_471263, EPI_ISL_471264, EPI_ISL_471265, EPI_ISL_471266 | | | |

| | | | |
|---|---|---|---|
| see above | Wisconsin State Laboratory of Hygiene Communicable Disease Division | Wisconsin State Laboratory of Hygiene Communicable Disease Division | Kelsey R. Florek, Abigail C. Shockey |
| EPI_ISL_471676, EPI_ISL_471679, EPI_ISL_471681, EPI_ISL_471683, EPI_ISL_471685, EPI_ISL_471686, EPI_ISL_471687, EPI_ISL_471688, EPI_ISL_471689, EPI_ISL_471690, EPI_ISL_471691, EPI_ISL_471692, EPI_ISL_471693, EPI_ISL_471694, EPI_ISL_471695, EPI_ISL_471696, EPI_ISL_471697, EPI_ISL_471698, EPI_ISL_471699, EPI_ISL_471700, EPI_ISL_471701, EPI_ISL_471702, EPI_ISL_471703, EPI_ISL_471704, EPI_ISL_471705, EPI_ISL_471706, EPI_ISL_471707, EPI_ISL_471708, EPI_ISL_471709, EPI_ISL_471710, EPI_ISL_471711, EPI_ISL_471712, EPI_ISL_471713, EPI_ISL_471714, EPI_ISL_471715, EPI_ISL_471716, EPI_ISL_471717, EPI_ISL_471718, EPI_ISL_471719, EPI_ISL_471720, EPI_ISL_471721, EPI_ISL_471722, EPI_ISL_471723, EPI_ISL_471724, EPI_ISL_471725, EPI_ISL_471726, EPI_ISL_471727, EPI_ISL_471728, EPI_ISL_471729, EPI_ISL_471730, EPI_ISL_471731, EPI_ISL_471732, EPI_ISL_471733, EPI_ISL_471734, EPI_ISL_471735, EPI_ISL_471736, EPI_ISL_471737, EPI_ISL_471738, EPI_ISL_471739, EPI_ISL_471740, EPI_ISL_471742, EPI_ISL_471743, EPI_ISL_471744, EPI_ISL_471745, EPI_ISL_471746, EPI_ISL_471747, EPI_ISL_471748, EPI_ISL_471749, EPI_ISL_471750, EPI_ISL_471751, EPI_ISL_471752, EPI_ISL_471753, EPI_ISL_471754, EPI_ISL_471756, EPI_ISL_471757, EPI_ISL_471758, EPI_ISL_471759, EPI_ISL_471760, EPI_ISL_471761, EPI_ISL_471762, EPI_ISL_471763, EPI_ISL_471764, EPI_ISL_471765, EPI_ISL_471766, EPI_ISL_471767, EPI_ISL_471768, EPI_ISL_471769, EPI_ISL_471770, EPI_ISL_471771, EPI_ISL_471772, EPI_ISL_471774, EPI_ISL_471775, EPI_ISL_471776, EPI_ISL_471777, EPI_ISL_471778, EPI_ISL_471779, EPI_ISL_471780, EPI_ISL_471781, EPI_ISL_471782, EPI_ISL_471783, EPI_ISL_471784, EPI_ISL_471785, EPI_ISL_471786, EPI_ISL_471787, EPI_ISL_471788, EPI_ISL_471789, EPI_ISL_471790, EPI_ISL_471791, EPI_ISL_471792, EPI_ISL_471793, EPI_ISL_471794, EPI_ISL_471795, EPI_ISL_471796, EPI_ISL_471797, EPI_ISL_471799, EPI_ISL_471800, EPI_ISL_471801, EPI_ISL_471802, EPI_ISL_471803, EPI_ISL_471804, EPI_ISL_471805, EPI_ISL_471806, EPI_ISL_471807, EPI_ISL_471808, EPI_ISL_471809, EPI_ISL_471810, EPI_ISL_471811, EPI_ISL_471812, EPI_ISL_471813, EPI_ISL_471814, EPI_ISL_471815, EPI_ISL_471816, EPI_ISL_471817, EPI_ISL_471818, EPI_ISL_471819, EPI_ISL_471820, EPI_ISL_471821, EPI_ISL_471822, EPI_ISL_471823, EPI_ISL_471824, EPI_ISL_471825, EPI_ISL_471826, EPI_ISL_471828, EPI_ISL_471829, EPI_ISL_471830, EPI_ISL_471831, EPI_ISL_471832, EPI_ISL_471833, EPI_ISL_471834, EPI_ISL_471835, EPI_ISL_471836, EPI_ISL_471837, EPI_ISL_471838, EPI_ISL_471839, EPI_ISL_471840, EPI_ISL_471841, EPI_ISL_471842, EPI_ISL_471843, EPI_ISL_471844, EPI_ISL_471845, EPI_ISL_471846, EPI_ISL_471847, EPI_ISL_471848, EPI_ISL_471849, EPI_ISL_471850, EPI_ISL_471851, EPI_ISL_471852, EPI_ISL_471853, EPI_ISL_471854, EPI_ISL_471855, EPI_ISL_471856, EPI_ISL_471857, EPI_ISL_471858, EPI_ISL_471859, EPI_ISL_471860, EPI_ISL_471861, EPI_ISL_471862, EPI_ISL_471863, EPI_ISL_471864, EPI_ISL_471865, EPI_ISL_471866, EPI_ISL_471867, EPI_ISL_471868, EPI_ISL_471869, EPI_ISL_471870, EPI_ISL_471871, EPI_ISL_471872, EPI_ISL_471873, EPI_ISL_471874, EPI_ISL_471875, EPI_ISL_471876, EPI_ISL_471877, EPI_ISL_471878, EPI_ISL_471879, EPI_ISL_471880, EPI_ISL_471881, EPI_ISL_471882, EPI_ISL_471883, EPI_ISL_471884, EPI_ISL_471885, EPI_ISL_471886, EPI_ISL_471887, EPI_ISL_471888, EPI_ISL_471889, EPI_ISL_471890, EPI_ISL_471891, EPI_ISL_471892, EPI_ISL_471893, EPI_ISL_471894, EPI_ISL_471895, EPI_ISL_471896, EPI_ISL_471897, EPI_ISL_471898, EPI_ISL_471899, EPI_ISL_471900, EPI_ISL_471901, EPI_ISL_471902, EPI_ISL_471903, EPI_ISL_471904, EPI_ISL_471905, EPI_ISL_471906, EPI_ISL_471907, EPI_ISL_471908, EPI_ISL_471909, EPI_ISL_471910 | | | |
| see above | Michigan Department of Health and Human Services, Bureau of Laboratories | Michigan Department of Health and Human Services, Bureau of Laboratories | Blankenship HM, Riner D, Soehnlen MK |
| EPI_ISL_475174, EPI_ISL_475175, EPI_ISL_475176, EPI_ISL_475177, EPI_ISL_475178, EPI_ISL_475179, EPI_ISL_475180, EPI_ISL_475181, EPI_ISL_475182, EPI_ISL_475183, EPI_ISL_475184, EPI_ISL_475185, EPI_ISL_475186, EPI_ISL_475187, EPI_ISL_475188, EPI_ISL_475189, EPI_ISL_475190, EPI_ISL_475191, EPI_ISL_475192, EPI_ISL_475193, EPI_ISL_475194, EPI_ISL_475195, EPI_ISL_475196, EPI_ISL_475197, EPI_ISL_475198, EPI_ISL_475199, EPI_ISL_475200, EPI_ISL_475201, EPI_ISL_475202, EPI_ISL_475203, EPI_ISL_475204, EPI_ISL_475205, EPI_ISL_475206, EPI_ISL_475207, EPI_ISL_475208, EPI_ISL_475209, EPI_ISL_475210, EPI_ISL_475211, EPI_ISL_475212, EPI_ISL_475213, EPI_ISL_475214, EPI_ISL_475215, EPI_ISL_475216, EPI_ISL_475217, EPI_ISL_475218, EPI_ISL_475219, EPI_ISL_475220, EPI_ISL_475221, EPI_ISL_475222, EPI_ISL_475223, EPI_ISL_475224, EPI_ISL_475225, EPI_ISL_475226, EPI_ISL_475227, EPI_ISL_475228, EPI_ISL_475229, EPI_ISL_475230, EPI_ISL_475231, EPI_ISL_475232, EPI_ISL_475233, EPI_ISL_475234, EPI_ISL_475235, EPI_ISL_475236, EPI_ISL_475237 | | | |
| see above | Nebraska Public Health Laboratory | UNMC COVID-19 Response Team | UNMC COVID-19 Response Team |
| EPI_ISL_475574, EPI_ISL_475575, EPI_ISL_475576, EPI_ISL_475577, EPI_ISL_475578, EPI_ISL_475579, EPI_ISL_475580, EPI_ISL_475581, EPI_ISL_475582, EPI_ISL_475583, EPI_ISL_475584, EPI_ISL_475585, EPI_ISL_475586, EPI_ISL_475587, EPI_ISL_475588, EPI_ISL_475589, EPI_ISL_475590, EPI_ISL_475591, EPI_ISL_475592, EPI_ISL_475593, EPI_ISL_475594, EPI_ISL_475595, EPI_ISL_475596, EPI_ISL_475597, EPI_ISL_475598, EPI_ISL_475599, EPI_ISL_475600, EPI_ISL_475601, EPI_ISL_475602, EPI_ISL_475603, EPI_ISL_475604, EPI_ISL_475605, EPI_ISL_475606, EPI_ISL_475607, EPI_ISL_475608, EPI_ISL_475609, EPI_ISL_475610, EPI_ISL_475611, EPI_ISL_475612, EPI_ISL_475613, EPI_ISL_475614, EPI_ISL_475615, EPI_ISL_475616, EPI_ISL_475617, EPI_ISL_475618, EPI_ISL_475619, EPI_ISL_475620, EPI_ISL_475621, EPI_ISL_475622, EPI_ISL_475623, EPI_ISL_475624, EPI_ISL_475625, EPI_ISL_475626, EPI_ISL_475627, EPI_ISL_475628, EPI_ISL_475629, EPI_ISL_475630, EPI_ISL_475631, EPI_ISL_475632, EPI_ISL_475633, EPI_ISL_475634, EPI_ISL_475635, EPI_ISL_475636, EPI_ISL_475637, EPI_ISL_475638, EPI_ISL_475639, EPI_ISL_475640, EPI_ISL_475641, EPI_ISL_475642, EPI_ISL_475643, EPI_ISL_475644, EPI_ISL_475645, EPI_ISL_475646, EPI_ISL_475647, EPI_ISL_475648, EPI_ISL_475649, EPI_ISL_475650, EPI_ISL_475651, EPI_ISL_475652, EPI_ISL_475653, EPI_ISL_475654, EPI_ISL_475655, EPI_ISL_475656, EPI_ISL_475657, EPI_ISL_475658, EPI_ISL_475659, EPI_ISL_475660, EPI_ISL_475661, EPI_ISL_475662, EPI_ISL_475663, EPI_ISL_475664, EPI_ISL_475665, EPI_ISL_475666, EPI_ISL_475667, EPI_ISL_475668, EPI_ISL_475669, EPI_ISL_475670, EPI_ISL_475671, EPI_ISL_475672, EPI_ISL_475673, EPI_ISL_475674, EPI_ISL_475675, EPI_ISL_475676, EPI_ISL_475677, EPI_ISL_475678, EPI_ISL_475679, EPI_ISL_475680, EPI_ISL_475681, EPI_ISL_475682, EPI_ISL_475683, EPI_ISL_475684, EPI_ISL_475685, EPI_ISL_475686, EPI_ISL_475687, EPI_ISL_475688, EPI_ISL_475689, EPI_ISL_475690, EPI_ISL_475691, EPI_ISL_475692, EPI_ISL_475693, EPI_ISL_475694, EPI_ISL_475695, EPI_ISL_475696, EPI_ISL_475697, EPI_ISL_475698, EPI_ISL_475699, EPI_ISL_475700, EPI_ISL_475701, EPI_ISL_475702, EPI_ISL_475703, EPI_ISL_475704, EPI_ISL_475705, EPI_ISL_475706, EPI_ISL_475707, EPI_ISL_475708, EPI_ISL_475709, EPI_ISL_475710, EPI_ISL_475711, EPI_ISL_475712, EPI_ISL_475713, EPI_ISL_475714, EPI_ISL_475715, EPI_ISL_475716 | | | |
| see above | Cedars-Sinai Medical Center, Department of Pathology & Laboratory Medicine, Molecular Pathology Laboratory | Cedars-Sinai Medical Center, Molecular Pathology Laboratory of Department of Pathology & Laboratory Medicine and Genomic Core | Wenjuan Zhang, John Paul Govindavari, Brian Davis, Stephanie Chen, Jong Taek Kim, Jianbo Song, Jean Lopategui, Jasmine T Plummer, Eric Vail |
| EPI_ISL_475726, EPI_ISL_475727, EPI_ISL_475728, EPI_ISL_475730, EPI_ISL_475735, EPI_ISL_475739, EPI_ISL_475743, EPI_ISL_475744 | Utah Public Health Laboratory | Utah Public Health Laboratory | Erin Young, Kelly Oakeson |
| EPI_ISL_475762 | Oklahoma State Department of Health | França Lab | Caio Martinelle B. de França, Graham Wiley, Samuel T. Dunn, and Matthew J. Miller. |
| EPI_ISL_476018, EPI_ISL_476019, EPI_ISL_476021 | Washington University in St. Louis | Washington University in St. Louis | David Wang, Carey-Ann Burnham, Scott Handley, Lindsay Droit, Stephen Tahan |
| EPI_ISL_476027, EPI_ISL_476028, EPI_ISL_476029, EPI_ISL_476030, EPI_ISL_476031, EPI_ISL_476032, EPI_ISL_476033, EPI_ISL_476034, EPI_ISL_476035, EPI_ISL_476036, EPI_ISL_476037, EPI_ISL_476038, EPI_ISL_476039, EPI_ISL_476040, EPI_ISL_476041, EPI_ISL_476042, EPI_ISL_476043, EPI_ISL_476044, EPI_ISL_476045, EPI_ISL_476046, EPI_ISL_476047, EPI_ISL_476048, EPI_ISL_476049, EPI_ISL_476050, EPI_ISL_476051, EPI_ISL_476052, EPI_ISL_476053, EPI_ISL_476054, EPI_ISL_476055, EPI_ISL_476056, EPI_ISL_476057, EPI_ISL_476058, EPI_ISL_476059, EPI_ISL_476060, EPI_ISL_476062, EPI_ISL_476063, EPI_ISL_476064, EPI_ISL_476065, EPI_ISL_476066 | | | |
| see above | Michigan Department of Health and Human Services, Bureau of Laboratories | Michigan Department of Health and Human Services, Bureau of Laboratories | Blankenship HM, Riner D, Soehnlen MK |
| EPI_ISL_476517, EPI_ISL_476518, EPI_ISL_476519, EPI_ISL_476520, EPI_ISL_476521, EPI_ISL_476522, EPI_ISL_476523, EPI_ISL_476524, EPI_ISL_476525, EPI_ISL_476526, EPI_ISL_476527, EPI_ISL_476528, EPI_ISL_476529, EPI_ISL_476530, EPI_ISL_476531, EPI_ISL_476532, EPI_ISL_476533, EPI_ISL_476534, EPI_ISL_476535, EPI_ISL_476536, EPI_ISL_476537, EPI_ISL_476538, EPI_ISL_476539, EPI_ISL_476540, EPI_ISL_476541, EPI_ISL_476542, EPI_ISL_476543, EPI_ISL_476544, EPI_ISL_476545, EPI_ISL_476546, EPI_ISL_476547, EPI_ISL_476548, EPI_ISL_476549, EPI_ISL_476550, EPI_ISL_476551, EPI_ISL_476552, EPI_ISL_476553, EPI_ISL_476554, EPI_ISL_476555, EPI_ISL_476556, EPI_ISL_476557 | | | |
| see above | Yale Clinical Virology Laboratory | Grubaugh Lab - Yale School of Public Health | Joseph Fauver, Tara Alpert, Anderson Brito, Anne Wyllie, Chantal Vogels, Mary Petrone, Cole Jensen, Chaney Kalinich, Isabel Ott, Arnau Casanovas, Catherine Muenker, Adam Moore, Alice Lu, Maria Tokuyama, Patrick Wong, Peiwen Lu, Saad Omer, Richard Martinello, Allison Nelson, Shelli Farhadian, Akiko Iwasaki, Charlese Dela Cruz, Albert Ko, Nathan Grubaugh |
| EPI_ISL_476706, EPI_ISL_476707, EPI_ISL_476708, EPI_ISL_476709, EPI_ISL_476710, EPI_ISL_476711, EPI_ISL_476712, EPI_ISL_476713, EPI_ISL_476714, EPI_ISL_476715, EPI_ISL_476716, EPI_ISL_476717, EPI_ISL_476718, EPI_ISL_476719, EPI_ISL_476720, EPI_ISL_476721, EPI_ISL_476722, EPI_ISL_476723, EPI_ISL_476724, EPI_ISL_476725, EPI_ISL_476726, EPI_ISL_476727, EPI_ISL_476728, EPI_ISL_476729, EPI_ISL_476730, EPI_ISL_476731, EPI_ISL_476732, EPI_ISL_476733, EPI_ISL_476734, EPI_ISL_476735, EPI_ISL_476736, EPI_ISL_476737, EPI_ISL_476738, EPI_ISL_476739, EPI_ISL_476740, EPI_ISL_476741, EPI_ISL_476742, EPI_ISL_476743, EPI_ISL_476744, EPI_ISL_476745, EPI_ISL_476746, EPI_ISL_476747, EPI_ISL_476748, EPI_ISL_476749, EPI_ISL_476750, EPI_ISL_476751, EPI_ISL_476752, EPI_ISL_476753, EPI_ISL_476754, EPI_ISL_476755, EPI_ISL_476756, EPI_ISL_476757, EPI_ISL_476758, EPI_ISL_476759, EPI_ISL_476760, EPI_ISL_476761, EPI_ISL_476762, EPI_ISL_476763, EPI_ISL_476764, EPI_ISL_476765, EPI_ISL_476766 | | | |
| see above | Minnesota Department of Health, Public Health Laboratory | Minnesota Department of Health, Public Health Laboratory | Matt Plumb, Jacob Garfin, and Xiong Wang |
| EPI_ISL_476768, EPI_ISL_476772, EPI_ISL_476773, EPI_ISL_476774, EPI_ISL_476775, EPI_ISL_476778, EPI_ISL_476779, EPI_ISL_476780, EPI_ISL_476781, EPI_ISL_476782, EPI_ISL_476783, EPI_ISL_476784, EPI_ISL_476785, EPI_ISL_476786, EPI_ISL_476787, EPI_ISL_476788, EPI_ISL_476789, EPI_ISL_476790, EPI_ISL_476791, EPI_ISL_476792, EPI_ISL_476793, EPI_ISL_476794 | | | |
| see above | Stanford clinical virology lab | Chan-Zuckerberg Biohub | Benjamin Pinksy, Katharine Walter, Victoria N. Parikh, John Gorzynski, Hannah N. DeJong, Matthew T. Wheeler, Jason Andrews, Manuel Rivas, Carlos Bustamante, Euan Ashley, with CZB Cliahub Consortium |
| EPI_ISL_476897 | University of South Carolina Functional Genomics Core | University of South Carolina Functional Genomics Core | Michael Shtutman |
| EPI_ISL_476898, EPI_ISL_476899 | Alaska State Virology Laboratory | Alaska State Virology Laboratory | Jack Chen, Ph.D. |
| EPI_ISL_476900, EPI_ISL_476902, EPI_ISL_476903, EPI_ISL_476904, EPI_ISL_476905, EPI_ISL_476906, EPI_ISL_476907, EPI_ISL_476908, EPI_ISL_476909, EPI_ISL_476910, EPI_ISL_476911, EPI_ISL_476912, EPI_ISL_476914, EPI_ISL_476915, EPI_ISL_476916, EPI_ISL_476917, EPI_ISL_476918, EPI_ISL_476919, EPI_ISL_476920, EPI_ISL_476921, EPI_ISL_476922, EPI_ISL_476923, EPI_ISL_476924, EPI_ISL_476925, EPI_ISL_476926, EPI_ISL_476927, EPI_ISL_476928, EPI_ISL_476929, EPI_ISL_476930, EPI_ISL_476931, EPI_ISL_476932, EPI_ISL_476933, EPI_ISL_476935, EPI_ISL_476936, EPI_ISL_476937, EPI_ISL_476938, EPI_ISL_476939, EPI_ISL_476940 | | | |
| see above | UW Virology Lab | UW Virology Lab | Pavitra Roychoudhury, Hong Xie, Lasata Shrestha, Amin Addetia, Truong Nguyen, Victoria M Rachleff, Meei-Li Huang, Keith R Jerome, Alexander Greninger |
| EPI_ISL_477163 | Laboratory of Dr. John Lednicky | University of Florida | John A. Lednicky, Maha A. Elbadry, Kuttichantran Subramaniam, Thomas B. Waltzek, John Glenn Morris, Jr. |
| EPI_ISL_477272, EPI_ISL_477273, EPI_ISL_477274, EPI_ISL_477275, EPI_ISL_477276 | Mayo Clinic & Mayo Clinic Laboratories | Minnesota Department of Health, Public Health Laboratory | Matt Plumb, Jacob Garfin, Kelly Pung, and Xiong Wang |
| EPI_ISL_477277, EPI_ISL_477278, EPI_ISL_477279, EPI_ISL_477280, EPI_ISL_477281, EPI_ISL_477282, EPI_ISL_477283, EPI_ISL_477284, EPI_ISL_477285, EPI_ISL_477286, EPI_ISL_477287, EPI_ISL_477288, EPI_ISL_477289, EPI_ISL_477290 | | | |
| see above | M Health Fairview | Minnesota Department of Health, Public Health Laboratory | Matt Plumb, Jacob Garfin, Kelly Pung, and Xiong Wang |
| EPI_ISL_477291, EPI_ISL_477292, EPI_ISL_477293, EPI_ISL_477294, EPI_ISL_477295, EPI_ISL_477296, EPI_ISL_477297, EPI_ISL_477298, EPI_ISL_477299, EPI_ISL_477300, EPI_ISL_477301, EPI_ISL_477302, EPI_ISL_477303, EPI_ISL_477304, EPI_ISL_477305, EPI_ISL_477306, EPI_ISL_477307, EPI_ISL_477308, EPI_ISL_477309, EPI_ISL_477310, EPI_ISL_477311, EPI_ISL_477312 | | | |
| see above | Mayo Clinic & Mayo Clinic Laboratories | Minnesota Department of Health, Public Health Laboratory | Matt Plumb, Jacob Garfin, Kelly Pung, and Xiong Wang |
| EPI_ISL_477631, EPI_ISL_477632, EPI_ISL_477633, EPI_ISL_477634, EPI_ISL_477635, EPI_ISL_477636, EPI_ISL_477637, EPI_ISL_477638, EPI_ISL_477639, EPI_ISL_477640, EPI_ISL_477641, EPI_ISL_477642, EPI_ISL_477643, EPI_ISL_477644, EPI_ISL_477645, EPI_ISL_477646, EPI_ISL_477647, EPI_ISL_477648, EPI_ISL_477649, EPI_ISL_477650, EPI_ISL_477651, EPI_ISL_477652, EPI_ISL_477653, EPI_ISL_477654, EPI_ISL_477655, EPI_ISL_477656, EPI_ISL_477657, EPI_ISL_477658, EPI_ISL_477659, EPI_ISL_477660, EPI_ISL_477661, EPI_ISL_477662, EPI_ISL_477663, EPI_ISL_477664, EPI_ISL_477665, EPI_ISL_477666, EPI_ISL_477667, EPI_ISL_477668, EPI_ISL_477669, EPI_ISL_477670, EPI_ISL_477671, EPI_ISL_477672 | | | |
| see above | Virginia DCLS | Virginia DCLS | Virginia DCLS |
| EPI_ISL_477673, EPI_ISL_477674, EPI_ISL_477675, EPI_ISL_477676, EPI_ISL_477677, EPI_ISL_477678, EPI_ISL_477679, EPI_ISL_477680, EPI_ISL_477681, EPI_ISL_477682, EPI_ISL_477683, EPI_ISL_477684, EPI_ISL_477685, EPI_ISL_477686, EPI_ISL_477687, EPI_ISL_477689, EPI_ISL_477690, EPI_ISL_477691, EPI_ISL_477692, EPI_ISL_477693, EPI_ISL_477694, EPI_ISL_477695, EPI_ISL_477696, EPI_ISL_477697, EPI_ISL_477698, EPI_ISL_477699, EPI_ISL_477700, EPI_ISL_477701, EPI_ISL_477702, EPI_ISL_477703, EPI_ISL_477704, EPI_ISL_477705, EPI_ISL_477706, EPI_ISL_477707, EPI_ISL_477708, EPI_ISL_477709, EPI_ISL_477710, EPI_ISL_477711, EPI_ISL_477712, EPI_ISL_477713, EPI_ISL_477714, EPI_ISL_477715, EPI_ISL_477716, EPI_ISL_477717, EPI_ISL_477718, EPI_ISL_477719, EPI_ISL_477720, EPI_ISL_477721, EPI_ISL_477723, EPI_ISL_477724 | | | |
| see above | UW Virology Lab | UW Virology Lab | Pavitra Roychoudhury, Hong Xie, Lasata Shrestha, Amin Addetia, Truong Nguyen, Victoria M Rachleff, Meei-Li Huang, Keith R Jerome, Alexander Greninger |
| EPI_ISL_479664, EPI_ISL_479665, EPI_ISL_479666, EPI_ISL_479667, EPI_ISL_479668, EPI_ISL_479671, EPI_ISL_479672, EPI_ISL_479673, EPI_ISL_479674 | Center for Genomics and System Biology, New York University | Center for Genomics and System Biology, New York University | Roder,A., Banakis,S., Johnson,K., Khalfan,M., Borenstein,E.S., Samanovic,M., Cornelius,A., Herati,R., Ulrich,R., Fleming,A., Kottkamp,A., Raabe,V., Mulligan,M.J., Gresham,D., Ghedin,E. |
| EPI_ISL_479759, EPI_ISL_479760, EPI_ISL_479762, EPI_ISL_479763, EPI_ISL_479764, EPI_ISL_479765, EPI_ISL_479766, EPI_ISL_479767, EPI_ISL_479768, EPI_ISL_479769, EPI_ISL_479770, EPI_ISL_479771, EPI_ISL_479772, EPI_ISL_479773, EPI_ISL_479774, EPI_ISL_479775 | | | |
| see above | University of Miami Immunology and Histocompatibility Laboratory | University of Miami Immunology and Histocompatibility Laboratory | Emilio Margolles-Clark, PhD and Phillip Ruiz, MD, PhD |
| EPI_ISL_480351, EPI_ISL_480352, EPI_ISL_480353, EPI_ISL_480354, EPI_ISL_480355, EPI_ISL_480356, EPI_ISL_480357, EPI_ISL_480358, EPI_ISL_480359, EPI_ISL_480360, EPI_ISL_480361, EPI_ISL_480362, EPI_ISL_480363, EPI_ISL_480364, EPI_ISL_480365, EPI_ISL_480366, EPI_ISL_480367, EPI_ISL_480368, EPI_ISL_480369, EPI_ISL_480370, EPI_ISL_480371, EPI_ISL_480372, EPI_ISL_480373, EPI_ISL_480374, | | | |

| | | | |
|---|---|---|---|
| EPI_ISL_480375, EPI_ISL_480376, EPI_ISL_480378, EPI_ISL_480379, EPI_ISL_480380, EPI_ISL_480381, EPI_ISL_480382, EPI_ISL_480383, EPI_ISL_480384, EPI_ISL_480385, EPI_ISL_480386, EPI_ISL_480387, EPI_ISL_480388, EPI_ISL_480389, EPI_ISL_480390, EPI_ISL_480391, EPI_ISL_480392, EPI_ISL_480393, EPI_ISL_480394, EPI_ISL_480395, EPI_ISL_480396, EPI_ISL_480397, EPI_ISL_480398, EPI_ISL_480399, EPI_ISL_480400, EPI_ISL_480401, EPI_ISL_480402, EPI_ISL_480403, EPI_ISL_480404, EPI_ISL_480405, EPI_ISL_480406, EPI_ISL_480407, EPI_ISL_480408, EPI_ISL_480409, EPI_ISL_480410, EPI_ISL_480411, EPI_ISL_480412, EPI_ISL_480413 | | | |
| see above | University of Wisconsin-Madison AIDS Vaccine Research Laboratories | University of Wisconsin-Madison AIDS Vaccine Research Laboratories | Gage Moreno, Katarina Braun, et al. AIDS Vaccine Research Laboratories |
| EPI_ISL_480784, EPI_ISL_480785 | NYC Department of Health and Mental Hygiene | Pathogen Discovery, Respiratory Viruses Branch, Division of Viral Diseases, Centers for Disease Control and Prevention | Krista Queen, Christine Mahl, Jennifer Rakeman, Anna Uehara, Ying Tao, Jing Zhang, Yan Li, Clinton R. Paden, Haibin Wang, Jasmine Padilla, Justin Lee, Sally Slavinski, Suxiang Tong |
| EPI_ISL_480790, EPI_ISL_480791, EPI_ISL_480792, EPI_ISL_480793, EPI_ISL_480794, EPI_ISL_480795, EPI_ISL_480796, EPI_ISL_480798, EPI_ISL_480799, EPI_ISL_480801, EPI_ISL_480802, EPI_ISL_480803, EPI_ISL_480804, EPI_ISL_480805, EPI_ISL_480806, EPI_ISL_480807, EPI_ISL_480808, EPI_ISL_480809, EPI_ISL_480810, EPI_ISL_480811, EPI_ISL_480812, EPI_ISL_480813, EPI_ISL_480814, EPI_ISL_480815, EPI_ISL_480816, EPI_ISL_480817, EPI_ISL_480818, EPI_ISL_480819, EPI_ISL_480820, EPI_ISL_480821, EPI_ISL_480822, EPI_ISL_480823, EPI_ISL_480824, EPI_ISL_480825, EPI_ISL_480826, EPI_ISL_480827, EPI_ISL_480828, EPI_ISL_480829, EPI_ISL_480830, EPI_ISL_480831, EPI_ISL_480832, EPI_ISL_480833, EPI_ISL_480834, EPI_ISL_480835, EPI_ISL_480836, EPI_ISL_480837, EPI_ISL_480838, EPI_ISL_480839, EPI_ISL_480840, EPI_ISL_480841, EPI_ISL_480842, EPI_ISL_480843, EPI_ISL_480844, EPI_ISL_480845, EPI_ISL_480846, EPI_ISL_480847, EPI_ISL_480848, EPI_ISL_480849, EPI_ISL_480850, EPI_ISL_480851, EPI_ISL_480852, EPI_ISL_480853, EPI_ISL_480854, EPI_ISL_480855, EPI_ISL_480856, EPI_ISL_480857, EPI_ISL_480858, EPI_ISL_480859, EPI_ISL_480860, EPI_ISL_480861, EPI_ISL_480862, EPI_ISL_480863, EPI_ISL_480864, EPI_ISL_480865, EPI_ISL_480866, EPI_ISL_480868, EPI_ISL_480869, EPI_ISL_480870, EPI_ISL_480871, EPI_ISL_480872, EPI_ISL_480873, EPI_ISL_480874, EPI_ISL_480875, EPI_ISL_480876, EPI_ISL_480877, EPI_ISL_480878, EPI_ISL_480879, EPI_ISL_480880, EPI_ISL_480882, EPI_ISL_480883, EPI_ISL_480884, EPI_ISL_480885, EPI_ISL_480886, EPI_ISL_480887, EPI_ISL_480888, EPI_ISL_480889, EPI_ISL_480890, EPI_ISL_480891, EPI_ISL_480892, EPI_ISL_480893, EPI_ISL_480894, EPI_ISL_480896, EPI_ISL_480897, EPI_ISL_480898, EPI_ISL_480899, EPI_ISL_480900, EPI_ISL_480901, EPI_ISL_480902, EPI_ISL_480903, EPI_ISL_480904, EPI_ISL_480905, EPI_ISL_480906, EPI_ISL_480907, EPI_ISL_480908, EPI_ISL_480909, EPI_ISL_480910, EPI_ISL_480911, EPI_ISL_480912, EPI_ISL_480913, EPI_ISL_480914, EPI_ISL_480916, EPI_ISL_480917, EPI_ISL_480918, EPI_ISL_480919, EPI_ISL_480921, EPI_ISL_480922, EPI_ISL_480923, EPI_ISL_480924, EPI_ISL_480925, EPI_ISL_480926, EPI_ISL_480927, EPI_ISL_480928, EPI_ISL_480929, EPI_ISL_480932, EPI_ISL_480933, EPI_ISL_480934, EPI_ISL_480935, EPI_ISL_480936, EPI_ISL_480938, EPI_ISL_480939, EPI_ISL_480940, EPI_ISL_480942, EPI_ISL_480943, EPI_ISL_480944, EPI_ISL_480945, EPI_ISL_480947, EPI_ISL_480948, EPI_ISL_480949, EPI_ISL_480950, EPI_ISL_480951 | | | |
| see above | Florida Bureau of Public Health Laboratories | Florida Bureau of Public Health Laboratories | Sarah Schmedes, Jason Blanton |
| EPI_ISL_481241 | M Health Fairview | Minnesota Department of Health, Public Health Laboratory | Matt Plumb, Jacob Garfin, Kelly Pung, and Xiong Wang |
| EPI_ISL_481242 | Mayo Clinic & Mayo Clinic Laboratories | Minnesota Department of Health, Public Health Laboratory | Matt Plumb, Jacob Garfin, Kelly Pung, and Xiong Wang |
| EPI_ISL_481265, EPI_ISL_481266, EPI_ISL_481267, EPI_ISL_481268, EPI_ISL_481269, EPI_ISL_481270, EPI_ISL_481271, EPI_ISL_481272, EPI_ISL_481273, EPI_ISL_481274, EPI_ISL_481275, EPI_ISL_481276, EPI_ISL_481277, EPI_ISL_481278, EPI_ISL_481279, EPI_ISL_481280, EPI_ISL_481281, EPI_ISL_481282 | | | |
| see above | Maryland Department of Health | Maryland Department of Health | Keller,E. |
| EPI_ISL_481283 | Center for Genomics and System Biology, New York University | Center for Genomics and System Biology, New York University | Roder,A., Banakis,S., Johnson,K., Khalfan,M., Borenstein,E.S., Samanovic,M., Cornelius,A., Herati,R., Ulrich,R., Fleming,A., Kottkamp,A., Raabe,V., Mulligan,M.J., Gresham,D. and Ghedin,E. |
| EPI_ISL_482292, EPI_ISL_482293, EPI_ISL_482294, EPI_ISL_482295, EPI_ISL_482296, EPI_ISL_482297, EPI_ISL_482298, EPI_ISL_482299, EPI_ISL_482300, EPI_ISL_482301, EPI_ISL_482302, EPI_ISL_482303, EPI_ISL_482304, EPI_ISL_482305, EPI_ISL_482306, EPI_ISL_482307, EPI_ISL_482308, EPI_ISL_482309, EPI_ISL_482310, EPI_ISL_482311, EPI_ISL_482312, EPI_ISL_482313, EPI_ISL_482314, EPI_ISL_482315, EPI_ISL_482316, EPI_ISL_482317, EPI_ISL_482318, EPI_ISL_482319, EPI_ISL_482320, EPI_ISL_482321, EPI_ISL_482322, EPI_ISL_482323, EPI_ISL_482324, EPI_ISL_482325, EPI_ISL_482326, EPI_ISL_482327, EPI_ISL_482328, EPI_ISL_482329, EPI_ISL_482330, EPI_ISL_482331, EPI_ISL_482332, EPI_ISL_482333, EPI_ISL_482334, EPI_ISL_482335, EPI_ISL_482336, EPI_ISL_482337, EPI_ISL_482338, EPI_ISL_482339, EPI_ISL_482340, EPI_ISL_482341, EPI_ISL_482342, EPI_ISL_482343, EPI_ISL_482344, EPI_ISL_482345, EPI_ISL_482346, EPI_ISL_482347, EPI_ISL_482348, EPI_ISL_482349, EPI_ISL_482350, EPI_ISL_482351, EPI_ISL_482352, EPI_ISL_482353, EPI_ISL_482354, EPI_ISL_482355, EPI_ISL_482356, EPI_ISL_482357, EPI_ISL_482358, EPI_ISL_482359, EPI_ISL_482360, EPI_ISL_482361, EPI_ISL_482362, EPI_ISL_482363, EPI_ISL_482364, EPI_ISL_482365, EPI_ISL_482366, EPI_ISL_482367, EPI_ISL_482368, EPI_ISL_482369, EPI_ISL_482370, EPI_ISL_482371, EPI_ISL_482372, EPI_ISL_482373, EPI_ISL_482374, EPI_ISL_482375, EPI_ISL_482376, EPI_ISL_482377, EPI_ISL_482378, EPI_ISL_482379, EPI_ISL_482380, EPI_ISL_482381, EPI_ISL_482382, EPI_ISL_482383, EPI_ISL_482384, EPI_ISL_482385, EPI_ISL_482386, EPI_ISL_482387, EPI_ISL_482388, EPI_ISL_482389, EPI_ISL_482390, EPI_ISL_482391, EPI_ISL_482392, EPI_ISL_482393, EPI_ISL_482394, EPI_ISL_482395, EPI_ISL_482396, EPI_ISL_482397, EPI_ISL_482398, EPI_ISL_482399, EPI_ISL_482400, EPI_ISL_482401, EPI_ISL_482402, EPI_ISL_482403, EPI_ISL_482404, EPI_ISL_482405, EPI_ISL_482406, EPI_ISL_482407, EPI_ISL_482408, EPI_ISL_482409, EPI_ISL_482410, EPI_ISL_482411, EPI_ISL_482412, EPI_ISL_482413, EPI_ISL_482414, EPI_ISL_482415, EPI_ISL_482416, EPI_ISL_482417, EPI_ISL_482418, EPI_ISL_482419, EPI_ISL_482420, EPI_ISL_482421, EPI_ISL_482422, EPI_ISL_482423, EPI_ISL_482424, EPI_ISL_482425, EPI_ISL_482426, EPI_ISL_482427, EPI_ISL_482428, EPI_ISL_482429, EPI_ISL_482430, EPI_ISL_482431, EPI_ISL_482432, EPI_ISL_482433, EPI_ISL_482434, EPI_ISL_482435, EPI_ISL_482436, EPI_ISL_482437, EPI_ISL_482438, EPI_ISL_482439, EPI_ISL_482440, EPI_ISL_482441, EPI_ISL_482442, EPI_ISL_482443, EPI_ISL_482444, EPI_ISL_482445, EPI_ISL_482446, EPI_ISL_482447, EPI_ISL_482448, EPI_ISL_482449, EPI_ISL_482450, EPI_ISL_482451, EPI_ISL_482452, EPI_ISL_482453, EPI_ISL_482454, EPI_ISL_482455, EPI_ISL_482456, EPI_ISL_482457, EPI_ISL_482458, EPI_ISL_482459, EPI_ISL_482460, EPI_ISL_482461, EPI_ISL_482462, EPI_ISL_482463, EPI_ISL_482464, EPI_ISL_482465, EPI_ISL_482466, EPI_ISL_482467 | | | |
| see above | Providence St. Joseph Health Molecular Genomics Laboratory | Providence St. Joseph Health Molecular Genomics Laboratory | Alexa K Dowdell, Brian D Piening, Fred L Robinson, Carlo B Bifulco, Mary Campbell |
| EPI_ISL_482946, EPI_ISL_482947, EPI_ISL_482948, EPI_ISL_482949, EPI_ISL_482950, EPI_ISL_482951, EPI_ISL_482952, EPI_ISL_482953, EPI_ISL_482954, EPI_ISL_482955, EPI_ISL_482956, EPI_ISL_482957, EPI_ISL_482958, EPI_ISL_482959, EPI_ISL_482960, EPI_ISL_482961, EPI_ISL_482962, EPI_ISL_482963, EPI_ISL_482964, EPI_ISL_482965, EPI_ISL_482966 | | | |
| see above | Minnesota Department of Health, Public Health Laboratory | Minnesota Department of Health, Public Health Laboratory | Matt Plumb, Jacob Garfin, and Xiong Wang |
| EPI_ISL_482967, EPI_ISL_482968, EPI_ISL_482969, EPI_ISL_482970, EPI_ISL_482971, EPI_ISL_482972, EPI_ISL_482973, EPI_ISL_482974, EPI_ISL_482975, EPI_ISL_482976, EPI_ISL_482977, EPI_ISL_482978, EPI_ISL_482979, EPI_ISL_482980, EPI_ISL_482981, EPI_ISL_482982, EPI_ISL_482983, EPI_ISL_482984, EPI_ISL_482985, EPI_ISL_482986, EPI_ISL_482987 | | | |
| see above | Mayo Clinic & Mayo Clinic Laboratories | Minnesota Department of Health, Public Health Laboratory | Matt Plumb, Jacob Garfin, and Xiong Wang |
| EPI_ISL_482988, EPI_ISL_482989, EPI_ISL_482990, EPI_ISL_482991, EPI_ISL_482992, EPI_ISL_482993, EPI_ISL_482994, EPI_ISL_482995, EPI_ISL_482996, EPI_ISL_482997, EPI_ISL_482998, EPI_ISL_482999, EPI_ISL_483000, EPI_ISL_483001, EPI_ISL_483002, EPI_ISL_483003, EPI_ISL_483004, EPI_ISL_483005, EPI_ISL_483006, EPI_ISL_483007, EPI_ISL_483008, EPI_ISL_483009, EPI_ISL_483010, EPI_ISL_483011, EPI_ISL_483012, EPI_ISL_483013, EPI_ISL_483014, EPI_ISL_483015, EPI_ISL_483016, EPI_ISL_483017 | | | |
| see above | Minnesota Department of Health, Public Health Laboratory | Minnesota Department of Health, Public Health Laboratory | Matt Plumb, Jacob Garfin, and Xiong Wang |
| EPI_ISL_483018, EPI_ISL_483019, EPI_ISL_483021, EPI_ISL_483022, EPI_ISL_483023, EPI_ISL_483024, EPI_ISL_483029, EPI_ISL_483031, EPI_ISL_483032 | Utah Public Health Laboratory | Utah Public Health Laboratory | Heidi Butz, Erin Young, Kelly Oakeson |
| EPI_ISL_483158, EPI_ISL_483159, EPI_ISL_483160, EPI_ISL_483161, EPI_ISL_483162, EPI_ISL_483163, EPI_ISL_483164 | San Diego County Public Health Laboratory | Andersen lab at Scripps Research | SEARCH Alliance San Diego with Tracy Basler, Jovan Shephard, Brett Austin |
| EPI_ISL_483165, EPI_ISL_483166, EPI_ISL_483167, EPI_ISL_483168, EPI_ISL_483169, EPI_ISL_483170, EPI_ISL_483171, EPI_ISL_483172, EPI_ISL_483173, EPI_ISL_483174, EPI_ISL_483175, EPI_ISL_483176, EPI_ISL_483177, EPI_ISL_483178, EPI_ISL_483179, EPI_ISL_483180, EPI_ISL_483181, EPI_ISL_483182, EPI_ISL_483183, EPI_ISL_483184, EPI_ISL_483185, EPI_ISL_483186, EPI_ISL_483187, EPI_ISL_483188, EPI_ISL_483189, EPI_ISL_483190, EPI_ISL_483191, EPI_ISL_483192, EPI_ISL_483193, EPI_ISL_483194, EPI_ISL_483195, EPI_ISL_483196, EPI_ISL_483197, EPI_ISL_483198, EPI_ISL_483199, EPI_ISL_483200, EPI_ISL_483201, EPI_ISL_483202, EPI_ISL_483203, EPI_ISL_483204, EPI_ISL_483205, EPI_ISL_483206, EPI_ISL_483207, EPI_ISL_483208, EPI_ISL_483209, EPI_ISL_483210, EPI_ISL_483211, EPI_ISL_483212, EPI_ISL_483213, EPI_ISL_483214, EPI_ISL_483215, EPI_ISL_483216, EPI_ISL_483217, EPI_ISL_483218, EPI_ISL_483219, EPI_ISL_483220, EPI_ISL_483221, EPI_ISL_483222, EPI_ISL_483223, EPI_ISL_483224, EPI_ISL_483225, EPI_ISL_483226, EPI_ISL_483227, EPI_ISL_483228, EPI_ISL_483229, EPI_ISL_483230, EPI_ISL_483231, EPI_ISL_483232, EPI_ISL_483233, EPI_ISL_483234, EPI_ISL_483235, EPI_ISL_483236, EPI_ISL_483237, EPI_ISL_483238, EPI_ISL_483239, EPI_ISL_483240, EPI_ISL_483241, EPI_ISL_483242, EPI_ISL_483243, EPI_ISL_483244, EPI_ISL_483245, EPI_ISL_483246, EPI_ISL_483247, EPI_ISL_483248, EPI_ISL_483249, EPI_ISL_483250, EPI_ISL_483251, EPI_ISL_483252, EPI_ISL_483253, EPI_ISL_483254, EPI_ISL_483255, EPI_ISL_483256, EPI_ISL_483257, EPI_ISL_483258, EPI_ISL_483259, EPI_ISL_483260, EPI_ISL_483261, EPI_ISL_483262, EPI_ISL_483263, EPI_ISL_483264, EPI_ISL_483265, EPI_ISL_483266, EPI_ISL_483267, EPI_ISL_483268, EPI_ISL_483269, EPI_ISL_483270, EPI_ISL_483271, EPI_ISL_483272, EPI_ISL_483273, EPI_ISL_483274, EPI_ISL_483275, EPI_ISL_483276, EPI_ISL_483277, EPI_ISL_483278, EPI_ISL_483279, EPI_ISL_483280, EPI_ISL_483281, EPI_ISL_483282, EPI_ISL_483283, EPI_ISL_483284, EPI_ISL_483285, EPI_ISL_483286, EPI_ISL_483287, EPI_ISL_483288, EPI_ISL_483289, EPI_ISL_483290, EPI_ISL_483291, EPI_ISL_483292, EPI_ISL_483293, EPI_ISL_483294, EPI_ISL_483295, EPI_ISL_483296, EPI_ISL_483297, EPI_ISL_483298, EPI_ISL_483299, EPI_ISL_483300, EPI_ISL_483301, EPI_ISL_483302, EPI_ISL_483303, EPI_ISL_483304, EPI_ISL_483305, EPI_ISL_483306, EPI_ISL_483307, EPI_ISL_483308, EPI_ISL_483309, EPI_ISL_483310, EPI_ISL_483311, EPI_ISL_483312, EPI_ISL_483313, EPI_ISL_483314, EPI_ISL_483315, EPI_ISL_483316, EPI_ISL_483317, EPI_ISL_483318, EPI_ISL_483319, EPI_ISL_483320, EPI_ISL_483321, EPI_ISL_483322, EPI_ISL_483323, EPI_ISL_483324, EPI_ISL_483325, EPI_ISL_483326, EPI_ISL_483327, EPI_ISL_483328, EPI_ISL_483329, EPI_ISL_483330, EPI_ISL_483331, EPI_ISL_483332, EPI_ISL_483333, EPI_ISL_483334, EPI_ISL_483335, EPI_ISL_483336, EPI_ISL_483337, EPI_ISL_483338, EPI_ISL_483339, EPI_ISL_483340, EPI_ISL_483341, EPI_ISL_483342, EPI_ISL_483343, EPI_ISL_483344, EPI_ISL_483345, EPI_ISL_483346, EPI_ISL_483347, EPI_ISL_483348, EPI_ISL_483349, EPI_ISL_483350, EPI_ISL_483351, EPI_ISL_483352, EPI_ISL_483353, EPI_ISL_483354, EPI_ISL_483355, EPI_ISL_483356, EPI_ISL_483357, EPI_ISL_483358, EPI_ISL_483359, EPI_ISL_483360, EPI_ISL_483361, EPI_ISL_483362, EPI_ISL_483363, EPI_ISL_483364, EPI_ISL_483365, EPI_ISL_483366, EPI_ISL_483367, EPI_ISL_483368, EPI_ISL_483369, EPI_ISL_483370, EPI_ISL_483371, EPI_ISL_483372, EPI_ISL_483373, EPI_ISL_483374, EPI_ISL_483375, EPI_ISL_483376, EPI_ISL_483377, EPI_ISL_483378, EPI_ISL_483380, EPI_ISL_483381, EPI_ISL_483382, EPI_ISL_483383, EPI_ISL_483384, EPI_ISL_483385, EPI_ISL_483386, EPI_ISL_483387, EPI_ISL_483388, EPI_ISL_483389, EPI_ISL_483390, EPI_ISL_483391, EPI_ISL_483392, EPI_ISL_483393, EPI_ISL_483394, EPI_ISL_483395, EPI_ISL_483396, EPI_ISL_483397 | | | |
| see above | UC San Diego Center for Advanced Laboratory Medicine | Andersen lab at Scripps Research | SEARCH Alliance San Diego with David Pride, Ji H Shin |
| EPI_ISL_483398, EPI_ISL_483399, EPI_ISL_483400, EPI_ISL_483401, EPI_ISL_483402 | UC San Diego Center for Advanced Laboratory Medicine | Andersen lab at Scripps Research | Allison Smither, Gilberto Sabino-Santos, Patricia Snarski, Lilia Melnik, Antoinette Bell, Kaylynn Genemaras, Arnaud Drouin, Dahlene Fusco, Robert Garry with SEARCH Alliance San Diego |
| EPI_ISL_483403, EPI_ISL_483404, EPI_ISL_483405, EPI_ISL_483406, EPI_ISL_483407, EPI_ISL_483408, EPI_ISL_483409, EPI_ISL_483410, EPI_ISL_483411, EPI_ISL_483412, EPI_ISL_483413, EPI_ISL_483414, EPI_ISL_483415, EPI_ISL_483416, EPI_ISL_483417, EPI_ISL_483418, EPI_ISL_483419, EPI_ISL_483420, EPI_ISL_483421, EPI_ISL_483422, EPI_ISL_483423, EPI_ISL_483425, EPI_ISL_483426, EPI_ISL_483427, EPI_ISL_483428, EPI_ISL_483429, EPI_ISL_483430, EPI_ISL_483431, EPI_ISL_483432, EPI_ISL_483433, EPI_ISL_483434, EPI_ISL_483435, EPI_ISL_483436, EPI_ISL_483437, EPI_ISL_483438, EPI_ISL_483439, EPI_ISL_483440, EPI_ISL_483441, EPI_ISL_483442, EPI_ISL_483443, EPI_ISL_483444, EPI_ISL_483445, EPI_ISL_483446, EPI_ISL_483447, EPI_ISL_483448, EPI_ISL_483449, EPI_ISL_483450, EPI_ISL_483451, EPI_ISL_483452, EPI_ISL_483453, EPI_ISL_483454, EPI_ISL_483455, EPI_ISL_483456, EPI_ISL_483457, EPI_ISL_483458, EPI_ISL_483459, EPI_ISL_483460, EPI_ISL_483461, EPI_ISL_483462, EPI_ISL_483463, EPI_ISL_483464, EPI_ISL_483465, EPI_ISL_483466, EPI_ISL_483467, EPI_ISL_483468, EPI_ISL_483469, EPI_ISL_483470, EPI_ISL_483471, EPI_ISL_483472, EPI_ISL_483473, EPI_ISL_483474, EPI_ISL_483475, EPI_ISL_483476, EPI_ISL_483477, EPI_ISL_483478, EPI_ISL_483479, EPI_ISL_483480, EPI_ISL_483481, EPI_ISL_483482, EPI_ISL_483483, EPI_ISL_483484, EPI_ISL_483485, EPI_ISL_483486, EPI_ISL_483487, EPI_ISL_483488, EPI_ISL_483489, EPI_ISL_483490, EPI_ISL_483491, EPI_ISL_483492, EPI_ISL_483493, EPI_ISL_483494, EPI_ISL_483495, EPI_ISL_483496, EPI_ISL_483497, EPI_ISL_483498, EPI_ISL_483499, EPI_ISL_483500, EPI_ISL_483501, EPI_ISL_483502, EPI_ISL_483503, EPI_ISL_483504, EPI_ISL_483505, EPI_ISL_483506, EPI_ISL_483507, EPI_ISL_483508, EPI_ISL_483509, EPI_ISL_483510, EPI_ISL_483511, EPI_ISL_483512, EPI_ISL_483513, EPI_ISL_483514, EPI_ISL_483515, EPI_ISL_483516, EPI_ISL_483517, EPI_ISL_483518, EPI_ISL_483519, EPI_ISL_483520, EPI_ISL_483521, EPI_ISL_483522, EPI_ISL_483523, EPI_ISL_483524, EPI_ISL_483525, EPI_ISL_483526, EPI_ISL_483527, EPI_ISL_483528, EPI_ISL_483529, EPI_ISL_483530 | | | |
| see above | UC San Diego Center for Advanced Laboratory Medicine | Andersen lab at Scripps Research | SEARCH Alliance San Diego with David Pride, Ji H Shin |
| EPI_ISL_483531, EPI_ISL_483532, EPI_ISL_483533, EPI_ISL_483534, EPI_ISL_483535, EPI_ISL_483536, EPI_ISL_483537, EPI_ISL_483538, EPI_ISL_483539, EPI_ISL_483540, EPI_ISL_483541 | | | |
| see above | San Diego County Public Health Laboratory | Andersen lab at Scripps Research | SEARCH Alliance San Diego with Tracy Basler, Jovan Shephard, Brett Austin |
| EPI_ISL_484692 | Minnesota Department of Health, Public Health Laboratory | Minnesota Department of Health, Public Health Laboratory | Matt Plumb, Jacob Garfin, and Xiong Wang |
| EPI_ISL_484709, EPI_ISL_484710, EPI_ISL_484711, EPI_ISL_484712, EPI_ISL_484713, EPI_ISL_484714, EPI_ISL_484715, EPI_ISL_484716, EPI_ISL_484717, EPI_ISL_484718, EPI_ISL_484719, EPI_ISL_484720, EPI_ISL_484721, EPI_ISL_484722, EPI_ISL_484723, EPI_ISL_484724, EPI_ISL_484725, EPI_ISL_484726, EPI_ISL_484727, EPI_ISL_484728, EPI_ISL_484729, EPI_ISL_484730, EPI_ISL_484731, EPI_ISL_484732, EPI_ISL_484733, EPI_ISL_484734, EPI_ISL_484735, EPI_ISL_484736, EPI_ISL_484737, EPI_ISL_484738, EPI_ISL_484739, EPI_ISL_484740, EPI_ISL_484741, EPI_ISL_484742, EPI_ISL_484743, EPI_ISL_484744, EPI_ISL_484745, EPI_ISL_484747, EPI_ISL_484748, EPI_ISL_484749, EPI_ISL_484750, EPI_ISL_484751, EPI_ISL_484752, EPI_ISL_484753, EPI_ISL_484754, EPI_ISL_484755, EPI_ISL_484756, EPI_ISL_484757, EPI_ISL_484758, EPI_ISL_484759, EPI_ISL_484760, EPI_ISL_484761, EPI_ISL_484762, EPI_ISL_484763, EPI_ISL_484765, EPI_ISL_484766, EPI_ISL_484767, EPI_ISL_484768, EPI_ISL_484769, EPI_ISL_484770, EPI_ISL_484771, EPI_ISL_484772, EPI_ISL_484773, EPI_ISL_484774, EPI_ISL_484775, EPI_ISL_484776, EPI_ISL_484777, EPI_ISL_484778, EPI_ISL_484779, EPI_ISL_484780, EPI_ISL_484781, EPI_ISL_484782, EPI_ISL_484783, EPI_ISL_484784, EPI_ISL_484785, EPI_ISL_484787, EPI_ISL_484788, EPI_ISL_484789, EPI_ISL_484790, EPI_ISL_484791, EPI_ISL_484792, EPI_ISL_484793, EPI_ISL_484794, EPI_ISL_484795, EPI_ISL_484796, EPI_ISL_484798, EPI_ISL_484799, EPI_ISL_484800, EPI_ISL_484801, EPI_ISL_484802, EPI_ISL_484803, EPI_ISL_484804, EPI_ISL_484806 | | | |
| see above | University of Michigan Clinical Microbiology Laboratory | Lauring Lab, University of Michigan, Department of Microbiology and Immunology | Valesano et al. |

| | | | |
|---|---|---|---|
| EPI_ISL_484807, EPI_ISL_484808, EPI_ISL_484809, EPI_ISL_484810, EPI_ISL_484811, EPI_ISL_484812, EPI_ISL_484813, EPI_ISL_484814, EPI_ISL_484815, EPI_ISL_484816, EPI_ISL_484817, EPI_ISL_484818, EPI_ISL_484819, EPI_ISL_484820, EPI_ISL_484821, EPI_ISL_484822, EPI_ISL_484823, EPI_ISL_484824, EPI_ISL_484825, EPI_ISL_484826, EPI_ISL_484827, EPI_ISL_484828, EPI_ISL_484829, EPI_ISL_484830, EPI_ISL_484831, EPI_ISL_484832, EPI_ISL_484833, EPI_ISL_484834, EPI_ISL_484835, EPI_ISL_484836, EPI_ISL_484837, EPI_ISL_484838, EPI_ISL_484839, EPI_ISL_484840, EPI_ISL_484841, EPI_ISL_484842, EPI_ISL_484843, EPI_ISL_484844, EPI_ISL_484845, EPI_ISL_484846, EPI_ISL_484847, EPI_ISL_484848, EPI_ISL_484849, EPI_ISL_484850, EPI_ISL_484851, EPI_ISL_484852, EPI_ISL_484853, EPI_ISL_484854, EPI_ISL_484855, EPI_ISL_484856, EPI_ISL_484857, EPI_ISL_484858, EPI_ISL_484859, EPI_ISL_484860, EPI_ISL_484861, EPI_ISL_484862, EPI_ISL_484863, EPI_ISL_484864, EPI_ISL_484865, EPI_ISL_484866, EPI_ISL_484867, EPI_ISL_484868, EPI_ISL_484869, EPI_ISL_484870, EPI_ISL_484871, EPI_ISL_484872, EPI_ISL_484873, EPI_ISL_484874, EPI_ISL_484875, EPI_ISL_484876, EPI_ISL_484877, EPI_ISL_484878, EPI_ISL_484879, EPI_ISL_484880, EPI_ISL_484881, EPI_ISL_484882, EPI_ISL_484883, EPI_ISL_484884, EPI_ISL_484885, EPI_ISL_484886, EPI_ISL_484887, EPI_ISL_484888, EPI_ISL_484889, EPI_ISL_484890, EPI_ISL_484891, EPI_ISL_484892, EPI_ISL_484893, EPI_ISL_484894, EPI_ISL_484895, EPI_ISL_484896, EPI_ISL_484897, EPI_ISL_484898, EPI_ISL_484899, EPI_ISL_484900, EPI_ISL_484901, EPI_ISL_484902, EPI_ISL_484903, EPI_ISL_484904, EPI_ISL_484905, EPI_ISL_484906, EPI_ISL_484907, EPI_ISL_484908, EPI_ISL_484909, EPI_ISL_484910, EPI_ISL_484911, EPI_ISL_484912, EPI_ISL_484913, EPI_ISL_484914, EPI_ISL_484915, EPI_ISL_484916, EPI_ISL_484917, EPI_ISL_484918, EPI_ISL_484919, EPI_ISL_484920, EPI_ISL_484921, EPI_ISL_484922, EPI_ISL_484923, EPI_ISL_484924, EPI_ISL_484925, EPI_ISL_484926, EPI_ISL_484927, EPI_ISL_484928, EPI_ISL_484929, EPI_ISL_484930, EPI_ISL_484931, EPI_ISL_484932, EPI_ISL_484933, EPI_ISL_484934, EPI_ISL_484935, EPI_ISL_484936, EPI_ISL_484937, EPI_ISL_484938, EPI_ISL_484939, EPI_ISL_484940, EPI_ISL_484941, EPI_ISL_484942, EPI_ISL_484943, EPI_ISL_484944, EPI_ISL_484945, EPI_ISL_484946, EPI_ISL_484947, EPI_ISL_484948, EPI_ISL_484949, EPI_ISL_484950, EPI_ISL_484951, EPI_ISL_484952, EPI_ISL_484953, EPI_ISL_484954, EPI_ISL_484955, EPI_ISL_484956, EPI_ISL_484957, EPI_ISL_484958, EPI_ISL_484959, EPI_ISL_484960, EPI_ISL_484961, EPI_ISL_484962, EPI_ISL_484963, EPI_ISL_484964, EPI_ISL_484965, EPI_ISL_484966, EPI_ISL_484967, EPI_ISL_484968, EPI_ISL_484969, EPI_ISL_484970, EPI_ISL_484971, EPI_ISL_484972, EPI_ISL_484973, EPI_ISL_484974, EPI_ISL_484975, EPI_ISL_484976, EPI_ISL_484977, EPI_ISL_484978, EPI_ISL_484979, EPI_ISL_484980, EPI_ISL_484981, EPI_ISL_484982, EPI_ISL_484983, EPI_ISL_484984, EPI_ISL_484985, EPI_ISL_484986, EPI_ISL_484987, EPI_ISL_484988, EPI_ISL_484989, EPI_ISL_484990, EPI_ISL_484991, EPI_ISL_484992, EPI_ISL_484993, EPI_ISL_484994, EPI_ISL_484995, EPI_ISL_484996, EPI_ISL_484997, EPI_ISL_484998, EPI_ISL_484999, EPI_ISL_485000 | | | |
| see above | University of Wisconsin-Madison AIDS Vaccine Research Laboratories | University of Wisconsin-Madison AIDS Vaccine Research Laboratories | Gage Moreno, Katarina Braun, et al. AIDS Vaccine Research Laboratories |
| EPI_ISL_485004, EPI_ISL_485005, EPI_ISL_485006, EPI_ISL_485007, EPI_ISL_485008, EPI_ISL_485009, EPI_ISL_485010, EPI_ISL_485011, EPI_ISL_485012, EPI_ISL_485013, EPI_ISL_485014, EPI_ISL_485015, EPI_ISL_485016, EPI_ISL_485017, EPI_ISL_485018, EPI_ISL_485019, EPI_ISL_485020, EPI_ISL_485021, EPI_ISL_485022, EPI_ISL_485023, EPI_ISL_485025, EPI_ISL_485026, EPI_ISL_485027, EPI_ISL_485028, EPI_ISL_485029, EPI_ISL_485030, EPI_ISL_485031, EPI_ISL_485032, EPI_ISL_485033, EPI_ISL_485034, EPI_ISL_485035, EPI_ISL_485036, EPI_ISL_485037, EPI_ISL_485038, EPI_ISL_485039, EPI_ISL_485040, EPI_ISL_485041, EPI_ISL_485042, EPI_ISL_485043, EPI_ISL_485044, EPI_ISL_485045, EPI_ISL_485046, EPI_ISL_485047, EPI_ISL_485048, EPI_ISL_485049, EPI_ISL_485051, EPI_ISL_485052, EPI_ISL_485053, EPI_ISL_485054, EPI_ISL_485055, EPI_ISL_485056, EPI_ISL_485057, EPI_ISL_485058, EPI_ISL_485059, EPI_ISL_485060, EPI_ISL_485061, EPI_ISL_485062, EPI_ISL_485063, EPI_ISL_485064, EPI_ISL_485065, EPI_ISL_485067, EPI_ISL_485068, EPI_ISL_485069, EPI_ISL_485070, EPI_ISL_485071, EPI_ISL_485072, EPI_ISL_485073, EPI_ISL_485074, EPI_ISL_485075, EPI_ISL_485076, EPI_ISL_485077, EPI_ISL_485078, EPI_ISL_485079, EPI_ISL_485080, EPI_ISL_485081, EPI_ISL_485082, EPI_ISL_485083, EPI_ISL_485084, EPI_ISL_485086, EPI_ISL_485088, EPI_ISL_485089, EPI_ISL_485090, EPI_ISL_485091, EPI_ISL_485093, EPI_ISL_485094, EPI_ISL_485095, EPI_ISL_485096, EPI_ISL_485097, EPI_ISL_485098, EPI_ISL_485099, EPI_ISL_485101, EPI_ISL_485102, EPI_ISL_485103, EPI_ISL_485104, EPI_ISL_485105, EPI_ISL_485107, EPI_ISL_485108, EPI_ISL_485109, EPI_ISL_485110, EPI_ISL_485111, EPI_ISL_485112, EPI_ISL_485114, EPI_ISL_485115, EPI_ISL_485116, EPI_ISL_485117, EPI_ISL_485118, EPI_ISL_485119, EPI_ISL_485120, EPI_ISL_485121, EPI_ISL_485122, EPI_ISL_485123, EPI_ISL_485124, EPI_ISL_485125, EPI_ISL_485126, EPI_ISL_485127, EPI_ISL_485128, EPI_ISL_485129, EPI_ISL_485130, EPI_ISL_485131, EPI_ISL_485132, EPI_ISL_485133, EPI_ISL_485134, EPI_ISL_485135, EPI_ISL_485136, EPI_ISL_485137, EPI_ISL_485138, EPI_ISL_485139, EPI_ISL_485140, EPI_ISL_485141, EPI_ISL_485142, EPI_ISL_485143, EPI_ISL_485144, EPI_ISL_485145, EPI_ISL_485146, EPI_ISL_485147, EPI_ISL_485148, EPI_ISL_485149, EPI_ISL_485150, EPI_ISL_485151, EPI_ISL_485152, EPI_ISL_485153, EPI_ISL_485155, EPI_ISL_485156, EPI_ISL_485157, EPI_ISL_485158, EPI_ISL_485159, EPI_ISL_485160, EPI_ISL_485161, EPI_ISL_485162, EPI_ISL_485163, EPI_ISL_485164, EPI_ISL_485165, EPI_ISL_485166, EPI_ISL_485167, EPI_ISL_485168, EPI_ISL_485169, EPI_ISL_485170, EPI_ISL_485171, EPI_ISL_485172, EPI_ISL_485173, EPI_ISL_485174, EPI_ISL_485175, EPI_ISL_485176, EPI_ISL_485177, EPI_ISL_485178, EPI_ISL_485179, EPI_ISL_485180, EPI_ISL_485181, EPI_ISL_485182, EPI_ISL_485183, EPI_ISL_485185, EPI_ISL_485186, EPI_ISL_485187, EPI_ISL_485189, EPI_ISL_485190, EPI_ISL_485191, EPI_ISL_485192, EPI_ISL_485193, EPI_ISL_485194, EPI_ISL_485195, EPI_ISL_485196, EPI_ISL_485197, EPI_ISL_485198, EPI_ISL_485199, EPI_ISL_485200, EPI_ISL_485201, EPI_ISL_485202, EPI_ISL_485203, EPI_ISL_485204, EPI_ISL_485205, EPI_ISL_485206, EPI_ISL_485207, EPI_ISL_485208, EPI_ISL_485209, EPI_ISL_485210, EPI_ISL_485212, EPI_ISL_485213, EPI_ISL_485214, EPI_ISL_485215, EPI_ISL_485216, EPI_ISL_485217, EPI_ISL_485218, EPI_ISL_485219, EPI_ISL_485220, EPI_ISL_485221, EPI_ISL_485222, EPI_ISL_485223, EPI_ISL_485224, EPI_ISL_485226, EPI_ISL_485227, EPI_ISL_485228, EPI_ISL_485229, EPI_ISL_485230, EPI_ISL_485231, EPI_ISL_485232, EPI_ISL_485233, EPI_ISL_485234, EPI_ISL_485235, EPI_ISL_485236, EPI_ISL_485237, EPI_ISL_485238, EPI_ISL_485240, EPI_ISL_485242, EPI_ISL_485243, EPI_ISL_485244, EPI_ISL_485245, EPI_ISL_485246, EPI_ISL_485247, EPI_ISL_485248, EPI_ISL_485249, EPI_ISL_485250, EPI_ISL_485251, EPI_ISL_485252, EPI_ISL_485253, EPI_ISL_485254, EPI_ISL_485255, EPI_ISL_485256, EPI_ISL_485257, EPI_ISL_485258, EPI_ISL_485259, EPI_ISL_485260, EPI_ISL_485261, EPI_ISL_485262, EPI_ISL_485263, EPI_ISL_485264, EPI_ISL_485265, EPI_ISL_485266, EPI_ISL_485267, EPI_ISL_485268, EPI_ISL_485269, EPI_ISL_485270, EPI_ISL_485271, EPI_ISL_485272, EPI_ISL_485273, EPI_ISL_485274, EPI_ISL_485275, EPI_ISL_485276, EPI_ISL_485277, EPI_ISL_485278, EPI_ISL_485279, EPI_ISL_485280, EPI_ISL_485281, EPI_ISL_485282, EPI_ISL_485283, EPI_ISL_485284, EPI_ISL_485285, EPI_ISL_485286, EPI_ISL_485287, EPI_ISL_485288, EPI_ISL_485289, EPI_ISL_485290, EPI_ISL_485291, EPI_ISL_485292, EPI_ISL_485293, EPI_ISL_485294, EPI_ISL_485295, EPI_ISL_485296, EPI_ISL_485297, EPI_ISL_485298, EPI_ISL_485299, EPI_ISL_485300, EPI_ISL_485301, EPI_ISL_485302, EPI_ISL_485303, EPI_ISL_485304, EPI_ISL_485305, EPI_ISL_485306, EPI_ISL_485307, EPI_ISL_485308, EPI_ISL_485309, EPI_ISL_485310, EPI_ISL_485311, EPI_ISL_485312, EPI_ISL_485313, EPI_ISL_485314, EPI_ISL_485315, EPI_ISL_485316, EPI_ISL_485317, EPI_ISL_485318, EPI_ISL_485319, EPI_ISL_485320, EPI_ISL_485321, EPI_ISL_485322, EPI_ISL_485323, EPI_ISL_485324, EPI_ISL_485325, EPI_ISL_485326, EPI_ISL_485327, EPI_ISL_485328, EPI_ISL_485329, EPI_ISL_485330, EPI_ISL_485331, EPI_ISL_485332, EPI_ISL_485333, EPI_ISL_485334, EPI_ISL_485335, EPI_ISL_485336, EPI_ISL_485337, EPI_ISL_485338, EPI_ISL_485339, EPI_ISL_485340, EPI_ISL_485341, EPI_ISL_485342, EPI_ISL_485343, EPI_ISL_485345, EPI_ISL_485346, EPI_ISL_485347, EPI_ISL_485348, EPI_ISL_485349, EPI_ISL_485351, EPI_ISL_485352, EPI_ISL_485353, EPI_ISL_485354, EPI_ISL_485355, EPI_ISL_485356, EPI_ISL_485357, EPI_ISL_485358, EPI_ISL_485359, EPI_ISL_485360, EPI_ISL_485361, EPI_ISL_485362, EPI_ISL_485363, EPI_ISL_485364, EPI_ISL_485365, EPI_ISL_485366, EPI_ISL_485367, EPI_ISL_485368, EPI_ISL_485369, EPI_ISL_485370, EPI_ISL_485371, EPI_ISL_485372, EPI_ISL_485373, EPI_ISL_485374, EPI_ISL_485375, EPI_ISL_485376, EPI_ISL_485377, EPI_ISL_485378, EPI_ISL_485379, EPI_ISL_485380, EPI_ISL_485381, EPI_ISL_485382, EPI_ISL_485383, EPI_ISL_485384, EPI_ISL_485385, EPI_ISL_485386, EPI_ISL_485387 | | | |
| see above | River Road Testing Lab | Ginkgo Bioworks Clinical Laboratory | Rebecca C. Christofferson, Stephania A. Cormier, Luan V. Dinh, E. Handly Mayton, Hollis R. O'Neil, Thaya Stoufflet, Malaika Mckenzie-Bennett, James McGann, Jim Griffin, Keith Robison, Alex Plocik, Becky Schilling, Rebecca Littlefield, Michelle Spencer, Birgitte Simen |
| EPI_ISL_485814, EPI_ISL_485815, EPI_ISL_485816, EPI_ISL_485817, EPI_ISL_485818, EPI_ISL_485819, EPI_ISL_485820, EPI_ISL_485821, EPI_ISL_485822, EPI_ISL_485823, EPI_ISL_485824, EPI_ISL_485825, EPI_ISL_485826, EPI_ISL_485827, EPI_ISL_485828, EPI_ISL_485829, EPI_ISL_485830, EPI_ISL_485831, EPI_ISL_485832, EPI_ISL_485833, EPI_ISL_485834, EPI_ISL_485835, EPI_ISL_485836, EPI_ISL_485837, EPI_ISL_485838, EPI_ISL_485839, EPI_ISL_485840, EPI_ISL_485841, EPI_ISL_485842, EPI_ISL_485843, EPI_ISL_485844, EPI_ISL_485845, EPI_ISL_485846, EPI_ISL_485847, EPI_ISL_485848, EPI_ISL_485849, EPI_ISL_485850, EPI_ISL_485851, EPI_ISL_485852, EPI_ISL_485853, EPI_ISL_485854, EPI_ISL_485855, EPI_ISL_485856, EPI_ISL_485857, EPI_ISL_485858, EPI_ISL_485859, EPI_ISL_485860, EPI_ISL_485861, EPI_ISL_485862, EPI_ISL_485863, EPI_ISL_485864, EPI_ISL_485865, EPI_ISL_485866, EPI_ISL_485867, EPI_ISL_485868, EPI_ISL_485869, EPI_ISL_485870, EPI_ISL_485871 | | | |
| see above | Virginia DCLS | Virginia DCLS | Virginia DCLS |
| EPI_ISL_485872 | New Mexico Department of Health Scientific Laboratory Division | Center for Global Health, University of New Mexico Health Sciences Center | Daryl Domman, Kurt Schwalm, Twila Kunde, Joseph Hicks, Michael Edwards, Darrell Dinwiddie |
| EPI_ISL_485873, EPI_ISL_485874, EPI_ISL_485875, EPI_ISL_485876, EPI_ISL_485878, EPI_ISL_485879, EPI_ISL_485880, EPI_ISL_485881, EPI_ISL_485882, EPI_ISL_485884, EPI_ISL_485885, EPI_ISL_485887, EPI_ISL_485888, EPI_ISL_485889, EPI_ISL_485890, EPI_ISL_485892, EPI_ISL_485895, EPI_ISL_485896, EPI_ISL_485897, EPI_ISL_485898, EPI_ISL_485901, EPI_ISL_485902, EPI_ISL_485903, EPI_ISL_485904, EPI_ISL_485906 | | | |
| see above | River Road Testing Lab | Ginkgo Bioworks Clinical Laboratory | Rebecca C. Christofferson, Stephania A. Cormier, Luan V. Dinh, E. Handly Mayton, Hollis R. O'Neil, Thaya Stoufflet, Malaika Mckenzie-Bennett, James McGann, Jim Griffin, Keith Robison, Alex Plocik, Becky Schilling, Rebecca Littlefield, Michelle Spencer, Birgitte Simen |
| EPI_ISL_485917, EPI_ISL_485918, EPI_ISL_485919, EPI_ISL_485920, EPI_ISL_485921, EPI_ISL_485922, EPI_ISL_485923, EPI_ISL_485924, EPI_ISL_485925, EPI_ISL_485926, EPI_ISL_485927, EPI_ISL_485928, EPI_ISL_485929, EPI_ISL_485930, EPI_ISL_485931, EPI_ISL_485932, EPI_ISL_485933, EPI_ISL_485934, EPI_ISL_485936, EPI_ISL_485937, EPI_ISL_485938, EPI_ISL_485939, EPI_ISL_485940, EPI_ISL_485941, EPI_ISL_485942, EPI_ISL_485944, EPI_ISL_485945, EPI_ISL_485946, EPI_ISL_485947, EPI_ISL_485948, EPI_ISL_485949, EPI_ISL_485950, EPI_ISL_485951, EPI_ISL_485952, EPI_ISL_485953, EPI_ISL_485954, EPI_ISL_485955, EPI_ISL_485956, EPI_ISL_485957, EPI_ISL_485958, EPI_ISL_485959, EPI_ISL_485960, EPI_ISL_485961, EPI_ISL_485963, EPI_ISL_485964, EPI_ISL_485967, EPI_ISL_485968, EPI_ISL_485969, EPI_ISL_485972, EPI_ISL_485973, EPI_ISL_485974, EPI_ISL_485975, EPI_ISL_485976, EPI_ISL_485977, EPI_ISL_485978, EPI_ISL_485979, EPI_ISL_485981, EPI_ISL_485982, EPI_ISL_485983, EPI_ISL_485984, EPI_ISL_485985, EPI_ISL_485986, EPI_ISL_485987, EPI_ISL_485988, EPI_ISL_485989, EPI_ISL_485990, EPI_ISL_485992, EPI_ISL_485993, EPI_ISL_485994, EPI_ISL_485995, EPI_ISL_485996, EPI_ISL_485997, EPI_ISL_485998, EPI_ISL_485999, EPI_ISL_486000, EPI_ISL_486001, EPI_ISL_486002, EPI_ISL_486003, EPI_ISL_486004, EPI_ISL_486005, EPI_ISL_486006, EPI_ISL_486007, EPI_ISL_486008, EPI_ISL_486009, EPI_ISL_486010, EPI_ISL_486011, EPI_ISL_486012, EPI_ISL_486013, EPI_ISL_486014, EPI_ISL_486015, EPI_ISL_486016, EPI_ISL_486017, EPI_ISL_486018, EPI_ISL_486019, EPI_ISL_486020, EPI_ISL_486021, EPI_ISL_486022, EPI_ISL_486023, EPI_ISL_486024, EPI_ISL_486025, EPI_ISL_486026, EPI_ISL_486027, EPI_ISL_486028, EPI_ISL_486029, EPI_ISL_486030, EPI_ISL_486032, EPI_ISL_486033, EPI_ISL_486034, EPI_ISL_486035, EPI_ISL_486036, EPI_ISL_486037, EPI_ISL_486038, EPI_ISL_486039, EPI_ISL_486040, EPI_ISL_486041, EPI_ISL_486042, EPI_ISL_486043, EPI_ISL_486044, EPI_ISL_486046, EPI_ISL_486047, EPI_ISL_486048, EPI_ISL_486049, EPI_ISL_486050, EPI_ISL_486051, EPI_ISL_486052, EPI_ISL_486053, EPI_ISL_486054, EPI_ISL_486055, EPI_ISL_486056, EPI_ISL_486057, EPI_ISL_486058, EPI_ISL_486059, EPI_ISL_486060, EPI_ISL_486061, EPI_ISL_486062, EPI_ISL_486063, EPI_ISL_486064, EPI_ISL_486065, EPI_ISL_486066, EPI_ISL_486067, EPI_ISL_486068, EPI_ISL_486069, EPI_ISL_486070, EPI_ISL_486071, EPI_ISL_486072, EPI_ISL_486074, EPI_ISL_486075, EPI_ISL_486076, EPI_ISL_486077, EPI_ISL_486078, EPI_ISL_486079, EPI_ISL_486080, EPI_ISL_486081, EPI_ISL_486082, EPI_ISL_486083, EPI_ISL_486084, EPI_ISL_486085, EPI_ISL_486086, EPI_ISL_486087, EPI_ISL_486088, EPI_ISL_486089, EPI_ISL_486093, EPI_ISL_486094, EPI_ISL_486095, EPI_ISL_486097, EPI_ISL_486098, EPI_ISL_486099, EPI_ISL_486102, EPI_ISL_486103, EPI_ISL_486105, EPI_ISL_486106, EPI_ISL_486108, EPI_ISL_486109, EPI_ISL_486110, EPI_ISL_486112, EPI_ISL_486113, EPI_ISL_486114 | | | |
| see above | UW Virology Lab | UW Virology Lab | Pavitra Roychoudhury, Hong Xie, Lasata Shrestha, Amin Addetia, Truong Nguyen, Victoria M Rachleff, Meei-Li Huang, Keith R Jerome, Alexander Greninger |
| EPI_ISL_486115, EPI_ISL_486116, EPI_ISL_486117, EPI_ISL_486118, EPI_ISL_486119 | County of Santa Clara Public Health Department | Chan-Zuckerberg Biohub | CZB Cliahub Consortium |
| EPI_ISL_486120, EPI_ISL_486121, EPI_ISL_486122, EPI_ISL_486123, EPI_ISL_486124, EPI_ISL_486125, EPI_ISL_486126, EPI_ISL_486127, EPI_ISL_486128, EPI_ISL_486129, EPI_ISL_486130, EPI_ISL_486131, EPI_ISL_486132, EPI_ISL_486133, EPI_ISL_486134, EPI_ISL_486135, EPI_ISL_486136, EPI_ISL_486137, EPI_ISL_486138, EPI_ISL_486139, EPI_ISL_486140, EPI_ISL_486141, EPI_ISL_486142, EPI_ISL_486143, EPI_ISL_486144, EPI_ISL_486145, EPI_ISL_486146, EPI_ISL_486147, EPI_ISL_486148, EPI_ISL_486149, EPI_ISL_486150, EPI_ISL_486151, EPI_ISL_486152, EPI_ISL_486153, EPI_ISL_486154, EPI_ISL_486155, EPI_ISL_486156, EPI_ISL_486157, EPI_ISL_486158, EPI_ISL_486159, EPI_ISL_486160, EPI_ISL_486161, EPI_ISL_486162, EPI_ISL_486163, EPI_ISL_486164, EPI_ISL_486165, EPI_ISL_486166, EPI_ISL_486167, EPI_ISL_486168, EPI_ISL_486169, EPI_ISL_486170, EPI_ISL_486171, EPI_ISL_486172, EPI_ISL_486173, EPI_ISL_486174, EPI_ISL_486175, EPI_ISL_486176, EPI_ISL_486177, EPI_ISL_486178, EPI_ISL_486179, EPI_ISL_486180, EPI_ISL_486181, EPI_ISL_486182, EPI_ISL_486183, EPI_ISL_486184, EPI_ISL_486185, EPI_ISL_486186, EPI_ISL_486187, EPI_ISL_486188, EPI_ISL_486189, EPI_ISL_486190, EPI_ISL_486191, EPI_ISL_486192, EPI_ISL_486193, EPI_ISL_486194, EPI_ISL_486195, EPI_ISL_486196, EPI_ISL_486197, EPI_ISL_486198, EPI_ISL_486199, EPI_ISL_486200, EPI_ISL_486201, EPI_ISL_486202, EPI_ISL_486203, EPI_ISL_486204, EPI_ISL_486205, EPI_ISL_486206, EPI_ISL_486207, EPI_ISL_486208, EPI_ISL_486209, EPI_ISL_486210, EPI_ISL_486211, EPI_ISL_486212, EPI_ISL_486213, EPI_ISL_486214, EPI_ISL_486215, EPI_ISL_486216, EPI_ISL_486217, EPI_ISL_486218, EPI_ISL_486219, EPI_ISL_486220, EPI_ISL_486221, EPI_ISL_486222, EPI_ISL_486223, EPI_ISL_486224, EPI_ISL_486225, EPI_ISL_486226, EPI_ISL_486227, EPI_ISL_486228, EPI_ISL_486229, EPI_ISL_486230, EPI_ISL_486231, EPI_ISL_486232, EPI_ISL_486233, EPI_ISL_486234, EPI_ISL_486235, EPI_ISL_486236, EPI_ISL_486237, EPI_ISL_486238, EPI_ISL_486239, EPI_ISL_486240, EPI_ISL_486241, EPI_ISL_486242, EPI_ISL_486243, EPI_ISL_486244, EPI_ISL_486245, EPI_ISL_486246, EPI_ISL_486247, EPI_ISL_486248, EPI_ISL_486249, EPI_ISL_486250, EPI_ISL_486251, EPI_ISL_486252, EPI_ISL_486253, EPI_ISL_486254, EPI_ISL_486255, EPI_ISL_486256, EPI_ISL_486257, EPI_ISL_486258, EPI_ISL_486259, EPI_ISL_486260, EPI_ISL_486261, EPI_ISL_486262, EPI_ISL_486263, EPI_ISL_486264, EPI_ISL_486265, EPI_ISL_486266, EPI_ISL_486267, EPI_ISL_486268, EPI_ISL_486269, EPI_ISL_486270, EPI_ISL_486271, EPI_ISL_486272, EPI_ISL_486273, EPI_ISL_486274, EPI_ISL_486275, EPI_ISL_486276, EPI_ISL_486277, EPI_ISL_486278, EPI_ISL_486279 | | | |
| see above | Orange County Public Health Laboratory | Chan-Zuckerberg Biohub | CZB Cliahub Consortium |
| EPI_ISL_486281, EPI_ISL_486282, EPI_ISL_486283, EPI_ISL_486284, EPI_ISL_486285, EPI_ISL_486286 | Humboldt County Public Health Laboratory | Chan-Zuckerberg Biohub | CZB Cliahub Consortium |
| EPI_ISL_486287, EPI_ISL_486288, EPI_ISL_486289, EPI_ISL_486290, EPI_ISL_486291, EPI_ISL_486292, EPI_ISL_486293, EPI_ISL_486294, EPI_ISL_486295, EPI_ISL_486296, EPI_ISL_486297, EPI_ISL_486298, EPI_ISL_486299, EPI_ISL_486300, EPI_ISL_486301, EPI_ISL_486302, EPI_ISL_486303, EPI_ISL_486304, EPI_ISL_486305, EPI_ISL_486306, EPI_ISL_486307, EPI_ISL_486308, EPI_ISL_486309, EPI_ISL_486310, EPI_ISL_486311, EPI_ISL_486312, EPI_ISL_486313, EPI_ISL_486314, EPI_ISL_486315, EPI_ISL_486316, EPI_ISL_486317, EPI_ISL_486318, EPI_ISL_486319, EPI_ISL_486320, EPI_ISL_486321, EPI_ISL_486322, EPI_ISL_486323, EPI_ISL_486324, EPI_ISL_486325, EPI_ISL_486326, EPI_ISL_486327, EPI_ISL_486328, EPI_ISL_486329, EPI_ISL_486330, EPI_ISL_486331, EPI_ISL_486332, EPI_ISL_486333, EPI_ISL_486334, EPI_ISL_486335, EPI_ISL_486336, EPI_ISL_486337, EPI_ISL_486338 | | | |
| see above | San Joaquin County Public Health Lab | Chan-Zuckerberg Biohub | CZB Cliahub Consortium |
| EPI_ISL_486339, EPI_ISL_486340, EPI_ISL_486341, EPI_ISL_486342, EPI_ISL_486343, EPI_ISL_486344, EPI_ISL_486345, EPI_ISL_486346, EPI_ISL_486347, EPI_ISL_486348, EPI_ISL_486349, EPI_ISL_486350, EPI_ISL_486351, EPI_ISL_486352, EPI_ISL_486353, EPI_ISL_486354, EPI_ISL_486355, EPI_ISL_486356, EPI_ISL_486357, EPI_ISL_486358, EPI_ISL_486359, EPI_ISL_486360, EPI_ISL_486361, EPI_ISL_486362, EPI_ISL_486363, EPI_ISL_486364, EPI_ISL_486365 | | | |
| see above | UCSF Clinical Microbiology Laboratory | Chan-Zuckerberg Biohub | CZB Cliahub Consortium |
| EPI_ISL_486830, EPI_ISL_486831 | Providence St. Joseph Health Molecular Genomics Laboratory | Providence St. Joseph Health Molecular Genomics Laboratory | Alexa K Dowdell, Brian D Piening, Fred L Robinson, Carlo B Bifulco, Mary Campbell |
| EPI_ISL_486912, EPI_ISL_486913, EPI_ISL_486914, EPI_ISL_486915, EPI_ISL_486916, EPI_ISL_486917 | Maryland Department of Health | Maryland Department of Health | Keller,E. |
| EPI_ISL_487193, EPI_ISL_487197, EPI_ISL_487198, EPI_ISL_487199, EPI_ISL_487200, EPI_ISL_487201, EPI_ISL_487202, EPI_ISL_487203, EPI_ISL_487204, EPI_ISL_487205, EPI_ISL_487206, EPI_ISL_487207, EPI_ISL_487208, EPI_ISL_487209, EPI_ISL_487210, EPI_ISL_487211, EPI_ISL_487212, EPI_ISL_487213, EPI_ISL_487214, EPI_ISL_487215, EPI_ISL_487218, EPI_ISL_487219, EPI_ISL_487220, EPI_ISL_487221, EPI_ISL_487222, EPI_ISL_487223, EPI_ISL_487224, EPI_ISL_487225, EPI_ISL_487226, EPI_ISL_487228 | | | |
| see above | Utah Public Health Laboratory | Utah Public Health Laboratory | Heidi Butz, Erin Young, Kelly Oakeson |
| EPI_ISL_487229, EPI_ISL_487230 | University of Michigan Clinical Microbiology Laboratory | Lauring Lab, University of Michigan, Department of Microbiology and Immunology | Valesano et al. |
| EPI_ISL_487246, EPI_ISL_487247, EPI_ISL_487248, EPI_ISL_487249, EPI_ISL_487250, EPI_ISL_487251, EPI_ISL_487252, EPI_ISL_487253, EPI_ISL_487254, EPI_ISL_487255, EPI_ISL_487257, EPI_ISL_487259, EPI_ISL_487260, EPI_ISL_487261, EPI_ISL_487262, EPI_ISL_487264, EPI_ISL_487266, EPI_ISL_487267, EPI_ISL_487268 | | | |
| see above | Utah Public Health Laboratory | Utah Public Health Laboratory | Heidi Butz, Erin Young, Kelly Oakeson |
| EPI_ISL_489710, EPI_ISL_489711, EPI_ISL_489712, EPI_ISL_489713, EPI_ISL_489714, EPI_ISL_489715, EPI_ISL_489716, EPI_ISL_489717, EPI_ISL_489718, EPI_ISL_489719, EPI_ISL_489720, EPI_ISL_489721, EPI_ISL_489722, EPI_ISL_489723, EPI_ISL_489724, EPI_ISL_489725, EPI_ISL_489726, EPI_ISL_489727, EPI_ISL_489728, EPI_ISL_489729, EPI_ISL_489730, EPI_ISL_489731, EPI_ISL_489732, EPI_ISL_489733, EPI_ISL_489734, EPI_ISL_489735, EPI_ISL_489736, EPI_ISL_489737, EPI_ISL_489738, EPI_ISL_489739, EPI_ISL_489740, EPI_ISL_489741, EPI_ISL_489742, EPI_ISL_489743, EPI_ISL_489745, EPI_ISL_489746, EPI_ISL_489747, EPI_ISL_489748, EPI_ISL_489751, EPI_ISL_489752, EPI_ISL_489753, EPI_ISL_489754, EPI_ISL_489755, EPI_ISL_489756, EPI_ISL_489758, EPI_ISL_489759, EPI_ISL_489760, EPI_ISL_489761, EPI_ISL_489762, EPI_ISL_489763, EPI_ISL_489764, EPI_ISL_489765, EPI_ISL_489766, EPI_ISL_489767, EPI_ISL_489768, EPI_ISL_489769, EPI_ISL_489770, EPI_ISL_489771, EPI_ISL_489772, EPI_ISL_489773, EPI_ISL_489777, EPI_ISL_489778, EPI_ISL_489780, EPI_ISL_489781, EPI_ISL_489782, EPI_ISL_489783, EPI_ISL_489784, EPI_ISL_489785, EPI_ISL_489786, EPI_ISL_489787, EPI_ISL_489788, EPI_ISL_489789, EPI_ISL_489790, EPI_ISL_489791, EPI_ISL_489792, EPI_ISL_489793, EPI_ISL_489794, EPI_ISL_489795, EPI_ISL_489796, EPI_ISL_489797, EPI_ISL_489798, EPI_ISL_489799, EPI_ISL_489800, EPI_ISL_489801, EPI_ISL_489802, EPI_ISL_489803, EPI_ISL_489805, EPI_ISL_489806, EPI_ISL_489807, EPI_ISL_489808, EPI_ISL_489810, EPI_ISL_489811, EPI_ISL_489816, EPI_ISL_489817, EPI_ISL_489818, EPI_ISL_489819, EPI_ISL_489820, EPI_ISL_489821, EPI_ISL_489822, EPI_ISL_489823, EPI_ISL_489825, EPI_ISL_489827, EPI_ISL_489828, EPI_ISL_489830 | | | |
| see above | Florida Bureau of Public Health Laboratories | Florida Bureau of Public Health Laboratories | Sarah Schmedes, Jason Blanton |
| EPI_ISL_489896, EPI_ISL_489897, EPI_ISL_489898, EPI_ISL_489899, | Gundersen Molecular Diagnostics Laboratory | Kabara Cancer Research Institute | Craig S. Richmond, Paraic A. Kenny |

| | | | |
|---|---|---|---|
| EPI_ISL_489900, EPI_ISL_489901, EPI_ISL_489902, EPI_ISL_489903 | | | |
| EPI_ISL_489904 | Gundersen Clinical Microbiology Laboratory | Kabara Cancer Research Institute | Craig S. Richmond, Paraic A. Kenny |
| EPI_ISL_489905, EPI_ISL_489906, EPI_ISL_489907, EPI_ISL_489908, EPI_ISL_489909, EPI_ISL_489910, EPI_ISL_489911, EPI_ISL_489912, EPI_ISL_489913, EPI_ISL_489914, EPI_ISL_489915, EPI_ISL_489916, EPI_ISL_489917, EPI_ISL_489918, EPI_ISL_489919, EPI_ISL_489920, EPI_ISL_489921, EPI_ISL_489922, EPI_ISL_489923, EPI_ISL_489924, EPI_ISL_489925, EPI_ISL_489926, EPI_ISL_489928, EPI_ISL_489929, EPI_ISL_489930, EPI_ISL_489931, EPI_ISL_489932, EPI_ISL_489933, EPI_ISL_489934, EPI_ISL_489935, EPI_ISL_489936, EPI_ISL_489937, EPI_ISL_489938, EPI_ISL_489939, EPI_ISL_489940, EPI_ISL_489941, EPI_ISL_489942, EPI_ISL_489943, EPI_ISL_489944, EPI_ISL_489945, EPI_ISL_489946, EPI_ISL_489947, EPI_ISL_489948, EPI_ISL_489949, EPI_ISL_489950, EPI_ISL_489951, EPI_ISL_489952, EPI_ISL_489953, EPI_ISL_489954, EPI_ISL_489955 | | | |
| see above | Gundersen Molecular Diagnostics Laboratory | Kabara Cancer Research Institute | Craig S. Richmond, Paraic A. Kenny |
| EPI_ISL_489956, EPI_ISL_489957, EPI_ISL_489958 | Gundersen Clinical Microbiology Laboratory | Kabara Cancer Research Institute | Craig S. Richmond, Paraic A. Kenny |
| EPI_ISL_490210, EPI_ISL_490211, EPI_ISL_490212, EPI_ISL_490213, EPI_ISL_490214, EPI_ISL_490215, EPI_ISL_490216, EPI_ISL_490217, EPI_ISL_490218, EPI_ISL_490219, EPI_ISL_490220, EPI_ISL_490221, EPI_ISL_490222, EPI_ISL_490223 | | | |
| see above | Quest Diagnostics | Q Squared Solutions - QRTP facility | Victor J Weigman |
| EPI_ISL_490978 | Mayo Clinic Laboratories | UW Virology Lab | Pavitra Roychoudhury, Hong Xie, Lasata Shrestha, Amin Addetia, Truong Nguyen, Victoria M Rachleff, Meei-Li Huang, Keith R Jerome, Alexander Greninger |
| EPI_ISL_490979, EPI_ISL_490980, EPI_ISL_490982, EPI_ISL_490983, EPI_ISL_490984, EPI_ISL_490985, EPI_ISL_490986, EPI_ISL_490987 | UW Virology Lab | UW Virology Lab | Pavitra Roychoudhury, Hong Xie, Lasata Shrestha, Amin Addetia, Truong Nguyen, Victoria M Rachleff, Meei-Li Huang, Keith R Jerome, Alexander Greninger |
| EPI_ISL_490988 | Mayo Clinic Laboratories | UW Virology Lab | Pavitra Roychoudhury, Hong Xie, Lasata Shrestha, Amin Addetia, Truong Nguyen, Victoria M Rachleff, Meei-Li Huang, Keith R Jerome, Alexander Greninger |
| EPI_ISL_490989, EPI_ISL_490990, EPI_ISL_490991, EPI_ISL_490992, EPI_ISL_490993, EPI_ISL_490994, EPI_ISL_490995, EPI_ISL_490996 | UW Virology Lab | UW Virology Lab | Pavitra Roychoudhury, Hong Xie, Lasata Shrestha, Amin Addetia, Truong Nguyen, Victoria M Rachleff, Meei-Li Huang, Keith R Jerome, Alexander Greninger |
| EPI_ISL_490997, EPI_ISL_490998, EPI_ISL_490999, EPI_ISL_491000 | Mayo Clinic Laboratories | UW Virology Lab | Pavitra Roychoudhury, Hong Xie, Lasata Shrestha, Amin Addetia, Truong Nguyen, Victoria M Rachleff, Meei-Li Huang, Keith R Jerome, Alexander Greninger |
| EPI_ISL_491001, EPI_ISL_491002, EPI_ISL_491003 | UW Virology Lab | UW Virology Lab | Pavitra Roychoudhury, Hong Xie, Lasata Shrestha, Amin Addetia, Truong Nguyen, Victoria M Rachleff, Meei-Li Huang, Keith R Jerome, Alexander Greninger |
| EPI_ISL_491004 | Mayo Clinic Laboratories | UW Virology Lab | Pavitra Roychoudhury, Hong Xie, Lasata Shrestha, Amin Addetia, Truong Nguyen, Victoria M Rachleff, Meei-Li Huang, Keith R Jerome, Alexander Greninger |
| EPI_ISL_491005, EPI_ISL_491006, EPI_ISL_491007, EPI_ISL_491009, EPI_ISL_491010, EPI_ISL_491011, EPI_ISL_491012, EPI_ISL_491013, EPI_ISL_491014, EPI_ISL_491016, EPI_ISL_491017, EPI_ISL_491018, EPI_ISL_491019, EPI_ISL_491020, EPI_ISL_491021, EPI_ISL_491022, EPI_ISL_491023, EPI_ISL_491024, EPI_ISL_491025, EPI_ISL_491026, EPI_ISL_491027, EPI_ISL_491028, EPI_ISL_491029, EPI_ISL_491030, EPI_ISL_491031, EPI_ISL_491032, EPI_ISL_491033, EPI_ISL_491034 | | | |
| see above | UW Virology Lab | UW Virology Lab | Pavitra Roychoudhury, Hong Xie, Lasata Shrestha, Amin Addetia, Truong Nguyen, Victoria M Rachleff, Meei-Li Huang, Keith R Jerome, Alexander Greninger |
| EPI_ISL_491097, EPI_ISL_491098, EPI_ISL_491099, EPI_ISL_491100, EPI_ISL_491101, EPI_ISL_491102, EPI_ISL_491103, EPI_ISL_491104, EPI_ISL_491105, EPI_ISL_491106, EPI_ISL_491107, EPI_ISL_491108, EPI_ISL_491109, EPI_ISL_491110 | | | |
| see above | SC Department of Health and Environmental Control | SC Department of Health and Environmental Control | Flores,H. |
| EPI_ISL_491299, EPI_ISL_491300, EPI_ISL_491301, EPI_ISL_491302, EPI_ISL_491304, EPI_ISL_491305, EPI_ISL_491306, EPI_ISL_491307, EPI_ISL_491308, EPI_ISL_491309, EPI_ISL_491310, EPI_ISL_491311, EPI_ISL_491312, EPI_ISL_491313, EPI_ISL_491314, EPI_ISL_491315, EPI_ISL_491316, EPI_ISL_491317, EPI_ISL_491318, EPI_ISL_491319, EPI_ISL_491320, EPI_ISL_491321, EPI_ISL_491322, EPI_ISL_491323, EPI_ISL_491324, EPI_ISL_491325, EPI_ISL_491326, EPI_ISL_491327, EPI_ISL_491328, EPI_ISL_491329, EPI_ISL_491330, EPI_ISL_491331, EPI_ISL_491332, EPI_ISL_491333, EPI_ISL_491334, EPI_ISL_491335, EPI_ISL_491336, EPI_ISL_491337, EPI_ISL_491338, EPI_ISL_491339, EPI_ISL_491340, EPI_ISL_491341, EPI_ISL_491342, EPI_ISL_491343, EPI_ISL_491344, EPI_ISL_491345, EPI_ISL_491346, EPI_ISL_491347, EPI_ISL_491348, EPI_ISL_491349, EPI_ISL_491350, EPI_ISL_491351, EPI_ISL_491352, EPI_ISL_491353, EPI_ISL_491354, EPI_ISL_491355, EPI_ISL_491356, EPI_ISL_491357, EPI_ISL_491358, EPI_ISL_491359, EPI_ISL_491360, EPI_ISL_491361, EPI_ISL_491362, EPI_ISL_491363, EPI_ISL_491364, EPI_ISL_491365, EPI_ISL_491366, EPI_ISL_491367, EPI_ISL_491368, EPI_ISL_491369, EPI_ISL_491370, EPI_ISL_491371, EPI_ISL_491372, EPI_ISL_491373, EPI_ISL_491374, EPI_ISL_491375, EPI_ISL_491376, EPI_ISL_491377, EPI_ISL_491378, EPI_ISL_491379, EPI_ISL_491380, EPI_ISL_491381, EPI_ISL_491382, EPI_ISL_491383, EPI_ISL_491384, EPI_ISL_491385, EPI_ISL_491386, EPI_ISL_491387, EPI_ISL_491388, EPI_ISL_491389, EPI_ISL_491390, EPI_ISL_491391, EPI_ISL_491392, EPI_ISL_491393, EPI_ISL_491394, EPI_ISL_491395, EPI_ISL_491396, EPI_ISL_491397, EPI_ISL_491398, EPI_ISL_491399, EPI_ISL_491401, EPI_ISL_491402, EPI_ISL_491403, EPI_ISL_491404, EPI_ISL_491405, EPI_ISL_491406, EPI_ISL_491407, EPI_ISL_491408, EPI_ISL_491409, EPI_ISL_491410, EPI_ISL_491411, EPI_ISL_491412, EPI_ISL_491413, EPI_ISL_491414, EPI_ISL_491415, EPI_ISL_491416, EPI_ISL_491417, EPI_ISL_491418, EPI_ISL_491419, EPI_ISL_491420, EPI_ISL_491421, EPI_ISL_491422, EPI_ISL_491424, EPI_ISL_491425 | | | |
| see above | University of Wisconsin-Madison AIDS Vaccine Research Laboratories | University of Wisconsin-Madison AIDS Vaccine Research Laboratories | Gage Moreno, Katarina Braun, et al. AIDS Vaccine Research Laboratories |
| EPI_ISL_491481 | Functional Genomics Core University of South Carolina / Prisma Health–Midlands | Functional Genomics Core, Center For Targeted Therapeutics, | Hao Ji, Diego Altomare, B.Celia Cui, Mengqian Chen, Alyssa Clay-Glimour, Michael Wyatt, Phillip Buckhaults, Helmut Albrecht, Michael Shtutman |
| EPI_ISL_491907, EPI_ISL_491908, EPI_ISL_491909, EPI_ISL_491910, EPI_ISL_491911, EPI_ISL_491912, EPI_ISL_491913, EPI_ISL_491914, EPI_ISL_491915, EPI_ISL_491916, EPI_ISL_491917, EPI_ISL_491918, EPI_ISL_491919, EPI_ISL_491920, EPI_ISL_491921, EPI_ISL_491922, EPI_ISL_491923, EPI_ISL_491924, EPI_ISL_491925, EPI_ISL_491926, EPI_ISL_491927, EPI_ISL_491928, EPI_ISL_491929, EPI_ISL_491930, EPI_ISL_491931, EPI_ISL_491942, EPI_ISL_491943 | | | |
| see above | Naval Infectious Diseases Diagnostic Laboratory | Naval Medical Research Center Biological Defense Research Directorate | Logan Voegtly, Regina Cer, Lindsay Glang, Victor Sugiharto, Francisco Malgon Bautista, Hua Wei Chen, Dessiree Pena-Gomez, Megan Schilling, Adrian Paskey, Kyle Long, Mark Simons, Kimberly Bishop-Lilly |
| EPI_ISL_492049, EPI_ISL_492050, EPI_ISL_492051, EPI_ISL_492052, EPI_ISL_492053, EPI_ISL_492055, EPI_ISL_492056, EPI_ISL_492057, EPI_ISL_492059, EPI_ISL_492060, EPI_ISL_492061, EPI_ISL_492062, EPI_ISL_492063 | | | |
| see above | Alaska State Virology Laboratory | Alaska State Virology Laboratory | Chen J et al with Pathogenomics group Dagdag R, Redlinger M, Milton E, George W, Kovalenko A, Drown DM, Bortz E |
| EPI_ISL_492074 | Functional Genomics Core University of South Carolina / Prisma Health–Midlands | Functional Genomics Core, University of South Carolina, | Hao Ji, Diego Altomare, B.Celia Cui, Mengqian Chen, Alyssa Clay-Glimour, Michael Wyatt, Phillip Buckhaults, Helmut Albrecht, Michael Shtutman |
| EPI_ISL_492087 | Alaska State Virology Laboratory | Alaska State Virology Laboratory | Chen J et al with Pathogenomics group Dagdag R, Redlinger M, Milton E, George W, Kovalenko A, Drown DM, Bortz E |
| EPI_ISL_492181, EPI_ISL_492182 | University of Arkansas for Medical Sciences (UAMS) | Department of Biomedical Informatics, University of Arkansas for Medical Sciences (UAMS) | Piroon Jenjaroenpun, David W Ussery, Thidathip Wongsurawat |
| EPI_ISL_493001 | Wyoming Public Health Laboratory | Wyoming Public Health Laboratory | Noah Hull, Rob Christensen, Jim Mildenberger, Joel Sevinsky, Cari Sloma, and Wanda Manley |
| EPI_ISL_493004, EPI_ISL_493005, EPI_ISL_493006, EPI_ISL_493007, EPI_ISL_493008, EPI_ISL_493010, EPI_ISL_493011, EPI_ISL_493012, EPI_ISL_493013, EPI_ISL_493014, EPI_ISL_493016, EPI_ISL_493017, EPI_ISL_493018, EPI_ISL_493020, EPI_ISL_493021, EPI_ISL_493023, EPI_ISL_493025, EPI_ISL_493026, EPI_ISL_493027, EPI_ISL_493030, EPI_ISL_493031, EPI_ISL_493032, EPI_ISL_493033, EPI_ISL_493035, EPI_ISL_493036, EPI_ISL_493037, EPI_ISL_493038, EPI_ISL_493039, EPI_ISL_493040, EPI_ISL_493041, EPI_ISL_493042, EPI_ISL_493043, EPI_ISL_493045, EPI_ISL_493046, EPI_ISL_493047, EPI_ISL_493048, EPI_ISL_493050, EPI_ISL_493051, EPI_ISL_493052, EPI_ISL_493056, EPI_ISL_493057, EPI_ISL_493059, EPI_ISL_493060 | | | |
| see above | Utah Public Health Laboratory | Utah Public Health Laboratory | Heidi Butz, Erin Young, Kelly Oakeson |
| EPI_ISL_493061, EPI_ISL_493062, EPI_ISL_493063, EPI_ISL_493064 | Wyoming Public Health Laboratory | Wyoming Public Health Laboratory | Noah Hull, Rob Christensen, Jim Mildenberger, Joel Sevinsky, Cari Sloma, and Wanda Manley |
| EPI_ISL_493069, EPI_ISL_493070, EPI_ISL_493071, EPI_ISL_493072, EPI_ISL_493073, EPI_ISL_493074, EPI_ISL_493075, EPI_ISL_493076, EPI_ISL_493077, EPI_ISL_493078, EPI_ISL_493079, EPI_ISL_493080, EPI_ISL_493081, EPI_ISL_493082, EPI_ISL_493083, EPI_ISL_493084, EPI_ISL_493085, EPI_ISL_493086 | | | |
| see above | Washington University in St. Louis | Washington University in St. Louis | David Wang, Carey-Ann Burnham, Scott Handley, Lindsay Droit, Stephen Tahan |
| EPI_ISL_493088, EPI_ISL_493089, EPI_ISL_493090, EPI_ISL_493091, EPI_ISL_493092, EPI_ISL_493093, EPI_ISL_493094, EPI_ISL_493096, EPI_ISL_493097, EPI_ISL_493098, EPI_ISL_493099, EPI_ISL_493100, EPI_ISL_493102, EPI_ISL_493103, EPI_ISL_493104, EPI_ISL_493105, EPI_ISL_493106, EPI_ISL_493107, EPI_ISL_493108, EPI_ISL_493109, EPI_ISL_493110, EPI_ISL_493112, EPI_ISL_493113, EPI_ISL_493114, EPI_ISL_493115, EPI_ISL_493116, EPI_ISL_493119, EPI_ISL_493120, EPI_ISL_493121, EPI_ISL_493122, EPI_ISL_493123, EPI_ISL_493124 | | | |
| see above | Utah Public Health Laboratory | Utah Public Health Laboratory | Heidi Butz, Erin Young, Kelly Oakeson |
| EPI_ISL_493125, EPI_ISL_493126, EPI_ISL_493127, EPI_ISL_493128 | Wyoming Public Health Laboratory | Wyoming Public Health Laboratory | Noah Hull, Rob Christensen, Jim Mildenberger, Joel Sevinsky, Cari Sloma, and Wanda Manley |
| EPI_ISL_493129, EPI_ISL_493130 | Functional Genomics Core University of South Carolina / Prisma Health–Midlands | Functional Genomics Core, University of South Carolina, | Hao Ji, Diego Altomare, B.Celia Cui, Mengqian Chen, Alyssa Clay-Glimour, Michael Wyatt, Phillip Buckhaults, Helmut Albrecht, Michael Shtutman |
| EPI_ISL_493209, EPI_ISL_493210, EPI_ISL_493211, EPI_ISL_493212 | Functional Genomics Core University of South Carolina / Prisma Health–Midlands | Functional Genomics Core, University of South Carolina | Hao Ji, Diego Altomare, B.Celia Cui, Mengqian Chen, Alyssa Clay-Glimour, Michael Wyatt, Phillip Buckhaults, Helmut Albrecht, Michael Shtutman |

| | | | |
|---|---|---|---|
| EPI_ISL_493334, EPI_ISL_493335, EPI_ISL_493336, EPI_ISL_493338 | Instituto de Diagnostico y Referencia Epidemiologicos (INDRE) | Instituto de Diagnostico y Referencia Epidemiologicos (INDRE) | Gisela Barrera-Badillo , Abril Rodriguez-Maldonado, Claudia Wong-Arambula , Natividad Cruz-Ortiz, Tatiana Nunez-Garcia, Dayanira Arellano-Suarez, Fabiola Garces-Ayala, Edgar Mendieta-Condado, Lucia Hernandez-Rivas, Irma Lopez-Martinez, Ernesto Ramirez-Gonzalez. |
| EPI_ISL_493339, EPI_ISL_493340, EPI_ISL_493341 | Instituto de Diagnostico y Referencia Epidemiologicos (INDRE) | Instituto de Diagnostico y Referencia Epidemiologicos (INDRE) | Gisela Barrera-Badillo , Abril Rodriguez-Maldonado, Claudia Wong-Arambula , Natividad Cruz-Ortiz, Tatiana Nunez-Garcia, Dayanira Arellano-Suarez, Adnan Araiza-Rodriguez, Edgar Mendieta-Condado, Lucia Hernandez-Rivas, Irma Lopez-Martinez, Ernesto Ramirez-Gonzalez. |
| EPI_ISL_493342, EPI_ISL_493343 | Instituto de Diagnostico y Referencia Epidemiologicos (INDRE) | Instituto de Diagnostico y Referencia Epidemiologicos (INDRE) | Ernesto Ramirez-Gonzalez, Abril Rodriguez-Maldonado, Claudia Wong-Arambula , Natividad Cruz-Ortiz, Tatiana Nunez-Garcia, Dayanira Arellano-Suarez, Adnan Araiza-Rodriguez, Edgar Mendieta-Condado, Lucia Hernandez-Rivas, Irma Lopez-Martinez, Gisela Barrera-Badillo. |
| EPI_ISL_493344, EPI_ISL_493345, EPI_ISL_493346, EPI_ISL_493347, EPI_ISL_493348, EPI_ISL_493349 | Instituto de Diagnostico y Referencia Epidemiologicos (INDRE) | Instituto de Diagnostico y Referencia Epidemiologicos (INDRE) | Ernesto Ramirez-Gonzalez, Abril Rodriguez-Maldonado, Claudia Wong-Arambula , Natividad Cruz-Ortiz, Tatiana Nunez-Garcia, Dayanira Arellano-Suarez, Adnan Araiza-Rodriguez, Fabiola Garces-Ayala, Lucia Hernandez-Rivas, Irma Lopez-Martinez, Gisela Barrera-Badillo. |
| EPI_ISL_493426 | Functional Genomics Core University of South Carolina / Prisma Health–Midlands | Functional Genomics Core, University of South Carolina, | Hao Ji, Diego Altomare, B.Celia Cui, Mengqian Chen, Alyssa Clay-Glimour, Michael Wyatt, Phillip Buckhaults, Helmut Albrecht, Michael Shtutman |
| EPI_ISL_493427 | Functional Genomics Core University of South Carolina / Prisma Health–Midlands | Functional Genomics Core, University of South Carolina | Hao Ji, Diego Altomare, B.Celia Cui, Mengqian Chen, Alyssa Clay-Glimour, Michael Wyatt, Phillip Buckhaults, Helmut Albrecht, Michael Shtutman |
| EPI_ISL_494372, EPI_ISL_494373, EPI_ISL_494374, EPI_ISL_494375, EPI_ISL_494376, EPI_ISL_494377, EPI_ISL_494378, EPI_ISL_494379, EPI_ISL_494380, EPI_ISL_494381, EPI_ISL_494382, EPI_ISL_494383, EPI_ISL_494384, EPI_ISL_494385, EPI_ISL_494386, EPI_ISL_494387, EPI_ISL_494388, EPI_ISL_494389, EPI_ISL_494390, EPI_ISL_494391, EPI_ISL_494392, EPI_ISL_494393, EPI_ISL_494394, EPI_ISL_494395, EPI_ISL_494396, EPI_ISL_494397, EPI_ISL_494398, EPI_ISL_494399, EPI_ISL_494400, EPI_ISL_494401, EPI_ISL_494402, EPI_ISL_494403, EPI_ISL_494404, EPI_ISL_494405, EPI_ISL_494406, EPI_ISL_494407, EPI_ISL_494408, EPI_ISL_494409, EPI_ISL_494410, EPI_ISL_494411, EPI_ISL_494412, EPI_ISL_494413, EPI_ISL_494414, EPI_ISL_494415, EPI_ISL_494416, EPI_ISL_494417, EPI_ISL_494418, EPI_ISL_494419, EPI_ISL_494420, EPI_ISL_494421, EPI_ISL_494422, EPI_ISL_494423, EPI_ISL_494424, EPI_ISL_494425, EPI_ISL_494426, EPI_ISL_494427, EPI_ISL_494428, EPI_ISL_494429, EPI_ISL_494430, EPI_ISL_494431, EPI_ISL_494432, EPI_ISL_494433, EPI_ISL_494434, EPI_ISL_494435, EPI_ISL_494436, EPI_ISL_494437, EPI_ISL_494438, EPI_ISL_494439, EPI_ISL_494440, EPI_ISL_494441, EPI_ISL_494442, EPI_ISL_494443, EPI_ISL_494444, EPI_ISL_494445, EPI_ISL_494446, EPI_ISL_494447, EPI_ISL_494448, EPI_ISL_494449, EPI_ISL_494450, EPI_ISL_494451, EPI_ISL_494452, EPI_ISL_494453, EPI_ISL_494454, EPI_ISL_494455, EPI_ISL_494456, EPI_ISL_494457, EPI_ISL_494458, EPI_ISL_494459, EPI_ISL_494460, EPI_ISL_494461, EPI_ISL_494462, EPI_ISL_494463, EPI_ISL_494464, EPI_ISL_494465, EPI_ISL_494466, EPI_ISL_494467, EPI_ISL_494468, EPI_ISL_494469, EPI_ISL_494470, EPI_ISL_494471, EPI_ISL_494472, EPI_ISL_494473, EPI_ISL_494474, EPI_ISL_494475, EPI_ISL_494476, EPI_ISL_494477, EPI_ISL_494478, EPI_ISL_494479, EPI_ISL_494480, EPI_ISL_494481, EPI_ISL_494482, EPI_ISL_494483, EPI_ISL_494484, EPI_ISL_494485, EPI_ISL_494486, EPI_ISL_494487, EPI_ISL_494488, EPI_ISL_494489, EPI_ISL_494490, EPI_ISL_494491, EPI_ISL_494492, EPI_ISL_494493, EPI_ISL_494494, EPI_ISL_494495, EPI_ISL_494496, EPI_ISL_494497, EPI_ISL_494498, EPI_ISL_494499, EPI_ISL_494500, EPI_ISL_494501, EPI_ISL_494502, EPI_ISL_494503 | | | |
| see above | San Diego County Public Health Laboratory | Andersen lab at Scripps Research | SEARCH Alliance San Diego with Tracy Basler, Jovan Shephard, Brett Austin |
| EPI_ISL_494504, EPI_ISL_494505, EPI_ISL_494506, EPI_ISL_494507, EPI_ISL_494508, EPI_ISL_494509, EPI_ISL_494510, EPI_ISL_494511, EPI_ISL_494512, EPI_ISL_494513, EPI_ISL_494514, EPI_ISL_494515, EPI_ISL_494516, EPI_ISL_494517, EPI_ISL_494518, EPI_ISL_494519, EPI_ISL_494520, EPI_ISL_494521, EPI_ISL_494522, EPI_ISL_494523, EPI_ISL_494524, EPI_ISL_494525, EPI_ISL_494526, EPI_ISL_494527, EPI_ISL_494528, EPI_ISL_494529, EPI_ISL_494530, EPI_ISL_494531, EPI_ISL_494532, EPI_ISL_494533, EPI_ISL_494534, EPI_ISL_494535, EPI_ISL_494536, EPI_ISL_494537, EPI_ISL_494538, EPI_ISL_494539, EPI_ISL_494540, EPI_ISL_494541, EPI_ISL_494542, EPI_ISL_494543, EPI_ISL_494544, EPI_ISL_494545, EPI_ISL_494546, EPI_ISL_494547, EPI_ISL_494548, EPI_ISL_494549, EPI_ISL_494550, EPI_ISL_494551, EPI_ISL_494552 | | | |
| see above | Quest Diagnostics | Quest Diagnostics | Anderson,B.P., Rosenthal,S.H., Gerasimova,A., Kagan,R.M. and Owen, R. |
| EPI_ISL_494553, EPI_ISL_494554, EPI_ISL_494555, EPI_ISL_494558 | Functional Genomics Core University of South Carolina / Prisma Health–Midlands | Functional Genomics Core, University of South Carolina | Hao Ji, Diego Altomare, B.Celia Cui, Mengqian Chen, Alyssa Clay-Glimour, Michael Wyatt, Phillip Buckhaults, Helmut Albrecht, Michael Shtutman |
| EPI_ISL_494568, EPI_ISL_494569 | San Diego County Public Health Laboratory | Andersen lab at Scripps Research | SEARCH Alliance San Diego with Tracy Basler, Jovan Shephard, Brett Austin |
| EPI_ISL_494570 | Rady's Childrens Hospital | Andersen lab at Scripps Research | SEARCH Alliance San Diego |
| EPI_ISL_494571, EPI_ISL_494572, EPI_ISL_494573, EPI_ISL_494574, EPI_ISL_494575, EPI_ISL_494576, EPI_ISL_494577, EPI_ISL_494578, EPI_ISL_494579, EPI_ISL_494580, EPI_ISL_494581, EPI_ISL_494582, EPI_ISL_494583, EPI_ISL_494584, EPI_ISL_494585, EPI_ISL_494586, EPI_ISL_494587, EPI_ISL_494588, EPI_ISL_494589, EPI_ISL_494590, EPI_ISL_494591, EPI_ISL_494592, EPI_ISL_494593, EPI_ISL_494594, EPI_ISL_494595 | | | |
| see above | San Diego County Public Health Laboratory | Andersen lab at Scripps Research | SEARCH Alliance San Diego with Tracy Basler, Jovan Shephard, Brett Austin |
| EPI_ISL_494596, EPI_ISL_494597, EPI_ISL_494598, EPI_ISL_494599, EPI_ISL_494600, EPI_ISL_494601, EPI_ISL_494602, EPI_ISL_494603, EPI_ISL_494604, EPI_ISL_494605, EPI_ISL_494606, EPI_ISL_494607, EPI_ISL_494608, EPI_ISL_494609, EPI_ISL_494610, EPI_ISL_494611, EPI_ISL_494612, EPI_ISL_494613, EPI_ISL_494614, EPI_ISL_494615, EPI_ISL_494616, EPI_ISL_494617, EPI_ISL_494618, EPI_ISL_494619 | | | |
| see above | Scripps Medical Laboratory | Andersen lab at Scripps Research | SEARCH Alliance San Diego with Michael Quigley, Ellen Stefanski, Ian Mchardy |
| EPI_ISL_494620, EPI_ISL_494621, EPI_ISL_494622, EPI_ISL_494623, EPI_ISL_494624, EPI_ISL_494625, EPI_ISL_494626, EPI_ISL_494627, EPI_ISL_494628, EPI_ISL_494629, EPI_ISL_494630, EPI_ISL_494631, EPI_ISL_494632, EPI_ISL_494633, EPI_ISL_494634 | | | |
| see above | San Diego County Public Health Laboratory | Andersen lab at Scripps Research | SEARCH Alliance San Diego with Tracy Basler, Jovan Shephard, Brett Austin |
| EPI_ISL_494635, EPI_ISL_494636, EPI_ISL_494637, EPI_ISL_494638, EPI_ISL_494639, EPI_ISL_494640, EPI_ISL_494641, EPI_ISL_494642, EPI_ISL_494643, EPI_ISL_494644, EPI_ISL_494645, EPI_ISL_494646, EPI_ISL_494647, EPI_ISL_494648, EPI_ISL_494649, EPI_ISL_494650, EPI_ISL_494651, EPI_ISL_494652, EPI_ISL_494653, EPI_ISL_494654, EPI_ISL_494655, EPI_ISL_494656, EPI_ISL_494657, EPI_ISL_494658, EPI_ISL_494659, EPI_ISL_494660, EPI_ISL_494661, EPI_ISL_494662, EPI_ISL_494663, EPI_ISL_494664, EPI_ISL_494665, EPI_ISL_494666, EPI_ISL_494667, EPI_ISL_494668, EPI_ISL_494669, EPI_ISL_494670, EPI_ISL_494671, EPI_ISL_494672, EPI_ISL_494673, EPI_ISL_494674, EPI_ISL_494675, EPI_ISL_494676, EPI_ISL_494677, EPI_ISL_494678, EPI_ISL_494679, EPI_ISL_494680, EPI_ISL_494681, EPI_ISL_494682, EPI_ISL_494683, EPI_ISL_494684, EPI_ISL_494685, EPI_ISL_494686, EPI_ISL_494687, EPI_ISL_494688, EPI_ISL_494689, EPI_ISL_494690, EPI_ISL_494691, EPI_ISL_494692, EPI_ISL_494693, EPI_ISL_494694, EPI_ISL_494695, EPI_ISL_494696, EPI_ISL_494697, EPI_ISL_494698, EPI_ISL_494699, EPI_ISL_494700, EPI_ISL_494701, EPI_ISL_494702, EPI_ISL_494703, EPI_ISL_494704, EPI_ISL_494705, EPI_ISL_494706, EPI_ISL_494707, EPI_ISL_494708, EPI_ISL_494709, EPI_ISL_494710, EPI_ISL_494711, EPI_ISL_494712, EPI_ISL_494713 | | | |
| see above | Scripps Medical Laboratory | Andersen lab at Scripps Research | SEARCH Alliance San Diego with Michael Quigley, Ellen Stefanski, Ian Mchardy |
| EPI_ISL_494714, EPI_ISL_494715, EPI_ISL_494716, EPI_ISL_494717, EPI_ISL_494718, EPI_ISL_494719, EPI_ISL_494720, EPI_ISL_494721, EPI_ISL_494722, EPI_ISL_494723, EPI_ISL_494724, EPI_ISL_494725, EPI_ISL_494726, EPI_ISL_494727, EPI_ISL_494728, EPI_ISL_494729, EPI_ISL_494730, EPI_ISL_494731, EPI_ISL_494732, EPI_ISL_494733, EPI_ISL_494734, EPI_ISL_494735, EPI_ISL_494736, EPI_ISL_494737, EPI_ISL_494738, EPI_ISL_494739, EPI_ISL_494740, EPI_ISL_494741, EPI_ISL_494742, EPI_ISL_494743, EPI_ISL_494744, EPI_ISL_494745, EPI_ISL_494746 | | | |
| see above | San Diego County Public Health Laboratory | Andersen lab at Scripps Research | SEARCH Alliance San Diego with Tracy Basler, Jovan Shephard, Brett Austin |
| EPI_ISL_495122, EPI_ISL_495123, EPI_ISL_495124, EPI_ISL_495125, EPI_ISL_495126, EPI_ISL_495127, EPI_ISL_495128, EPI_ISL_495129, EPI_ISL_495130, EPI_ISL_495131, EPI_ISL_495132, EPI_ISL_495133, EPI_ISL_495134, EPI_ISL_495135, EPI_ISL_495136, EPI_ISL_495137, EPI_ISL_495138, EPI_ISL_495139, EPI_ISL_495140, EPI_ISL_495141, EPI_ISL_495142, EPI_ISL_495143, EPI_ISL_495144, EPI_ISL_495145, EPI_ISL_495146, EPI_ISL_495147, EPI_ISL_495148, EPI_ISL_495149, EPI_ISL_495150, EPI_ISL_495151, EPI_ISL_495152, EPI_ISL_495153, EPI_ISL_495154, EPI_ISL_495155, EPI_ISL_495156, EPI_ISL_495157, EPI_ISL_495158, EPI_ISL_495159, EPI_ISL_495160 | | | |
| see above | Innovative Genomics Institute, UC Berkeley | Innovative Genomics Institute, UC Berkeley | Stacia Wyman, Haridha Shivram, Liana Lareau, Shana McDevitt, Justin Choi |
| EPI_ISL_495298, EPI_ISL_495299, EPI_ISL_495300, EPI_ISL_495301, EPI_ISL_495302, EPI_ISL_495303, EPI_ISL_495304, EPI_ISL_495305, EPI_ISL_495306, EPI_ISL_495307, EPI_ISL_495308, EPI_ISL_495309, EPI_ISL_495310, EPI_ISL_495311, EPI_ISL_495312, EPI_ISL_495313, EPI_ISL_495314, EPI_ISL_495315, EPI_ISL_495316, EPI_ISL_495317, EPI_ISL_495318, EPI_ISL_495319, EPI_ISL_495320, EPI_ISL_495321, EPI_ISL_495322, EPI_ISL_495323, EPI_ISL_495324, EPI_ISL_495325, EPI_ISL_495326, EPI_ISL_495327, EPI_ISL_495328, EPI_ISL_495329 | | | |
| see above | Virginia DCLS | Virginia DCLS | Virginia DCLS |
| EPI_ISL_495330, EPI_ISL_495332, EPI_ISL_495333, EPI_ISL_495334, EPI_ISL_495335, EPI_ISL_495336, EPI_ISL_495337, EPI_ISL_495339, EPI_ISL_495342, EPI_ISL_495343, EPI_ISL_495344, EPI_ISL_495345, EPI_ISL_495346, EPI_ISL_495347, EPI_ISL_495349, EPI_ISL_495350, EPI_ISL_495351, EPI_ISL_495352, EPI_ISL_495353, EPI_ISL_495355, EPI_ISL_495357, EPI_ISL_495359, EPI_ISL_495360, EPI_ISL_495361, EPI_ISL_495362, EPI_ISL_495363, EPI_ISL_495365, EPI_ISL_495369, EPI_ISL_495370, EPI_ISL_495371, EPI_ISL_495378, EPI_ISL_495379, EPI_ISL_495383, EPI_ISL_495384, EPI_ISL_495385, EPI_ISL_495386, EPI_ISL_495387, EPI_ISL_495388, EPI_ISL_495389, EPI_ISL_495390, EPI_ISL_495391, EPI_ISL_495392, EPI_ISL_495393, EPI_ISL_495394, EPI_ISL_495395, EPI_ISL_495396, EPI_ISL_495397, EPI_ISL_495398, EPI_ISL_495399, EPI_ISL_495400, EPI_ISL_495401, EPI_ISL_495402, EPI_ISL_495403, EPI_ISL_495404, EPI_ISL_495405, EPI_ISL_495406, EPI_ISL_495407, EPI_ISL_495409, EPI_ISL_495410 | | | |
| see above | Florida Bureau of Public Health Laboratories | Florida Bureau of Public Health Laboratories | Sarah Schmedes, Jason Blanton |
| EPI_ISL_495460, EPI_ISL_495461, EPI_ISL_495462, EPI_ISL_495463, EPI_ISL_495464, EPI_ISL_495465, EPI_ISL_495466, EPI_ISL_495467, EPI_ISL_495468, EPI_ISL_495469, EPI_ISL_495470, EPI_ISL_495471, EPI_ISL_495472, EPI_ISL_495473, EPI_ISL_495474, EPI_ISL_495475, EPI_ISL_495476, EPI_ISL_495477, EPI_ISL_495478, EPI_ISL_495479, EPI_ISL_495480, EPI_ISL_495481, EPI_ISL_495482, EPI_ISL_495483, EPI_ISL_495484, EPI_ISL_495485, EPI_ISL_495486, EPI_ISL_495487, EPI_ISL_495488, EPI_ISL_495489, EPI_ISL_495490, EPI_ISL_495491, EPI_ISL_495492, EPI_ISL_495493, EPI_ISL_495494, EPI_ISL_495495, EPI_ISL_495496, EPI_ISL_495497, EPI_ISL_495498, EPI_ISL_495499, EPI_ISL_495500, EPI_ISL_495501, EPI_ISL_495502, EPI_ISL_495503, EPI_ISL_495504, EPI_ISL_495505, EPI_ISL_495506, EPI_ISL_495507, EPI_ISL_495508, EPI_ISL_495509 | | | |
| see above | University of Wisconsin-Madison AIDS Vaccine Research Laboratories | University of Wisconsin-Madison AIDS Vaccine Research Laboratories | Gage Moreno, Katarina Braun, et al. AIDS Vaccine Research Laboratories |
| EPI_ISL_495563, EPI_ISL_495564, EPI_ISL_495565, EPI_ISL_495566, EPI_ISL_495567, EPI_ISL_495568, EPI_ISL_495569, EPI_ISL_495570, EPI_ISL_495571, EPI_ISL_495572, EPI_ISL_495573, EPI_ISL_495574, EPI_ISL_495575, EPI_ISL_495576, EPI_ISL_495577, EPI_ISL_495578, EPI_ISL_495579, EPI_ISL_495580, EPI_ISL_495581, EPI_ISL_495582, EPI_ISL_495583, EPI_ISL_495584, EPI_ISL_495585, EPI_ISL_495586, EPI_ISL_495587, EPI_ISL_495588, EPI_ISL_495589, EPI_ISL_495590, EPI_ISL_495591, EPI_ISL_495592, EPI_ISL_495593, EPI_ISL_495594, EPI_ISL_495595, EPI_ISL_495596 | | | |
| see above | University of Michigan Clinical Microbiology Laboratory | Lauring Lab, University of Michigan, Department of Microbiology and Immunology | Valesano et al. |
| EPI_ISL_495597, EPI_ISL_495598, EPI_ISL_495599, EPI_ISL_495600, EPI_ISL_495601, EPI_ISL_495602, EPI_ISL_495603, EPI_ISL_495604, EPI_ISL_495605, EPI_ISL_495606, EPI_ISL_495607, EPI_ISL_495608, EPI_ISL_495609 | | | |
| see above | Mayo Clinic & Mayo Clinic Laboratories | Minnesota Department of Health, Public Health Laboratory | Matt Plumb, Jacob Garfin, and Xiong Wang |
| EPI_ISL_495610, EPI_ISL_495611, EPI_ISL_495612, EPI_ISL_495613, EPI_ISL_495614, EPI_ISL_495615, EPI_ISL_495616, EPI_ISL_495617, EPI_ISL_495618, EPI_ISL_495619, EPI_ISL_495620, EPI_ISL_495621, EPI_ISL_495622, EPI_ISL_495623, EPI_ISL_495624, EPI_ISL_495625, EPI_ISL_495626, EPI_ISL_495627, EPI_ISL_495628 | | | |
| see above | Minnesota Department of Health, Public Health Laboratory | Minnesota Department of Health, Public Health Laboratory | Matt Plumb, Jacob Garfin, and Xiong Wang |
| EPI_ISL_495642, EPI_ISL_495643, EPI_ISL_495644, EPI_ISL_495645, EPI_ISL_495646, EPI_ISL_495647, EPI_ISL_495648, EPI_ISL_495649, EPI_ISL_495650, EPI_ISL_495651, EPI_ISL_495652, EPI_ISL_495653 | | | |
| see above | Washington State Department of Health | Seattle Flu Study | Deborah A. Nickerson, Chris D. Frazar, Jover Lee, Benjamin Pelle, Matthew Richardson, Amanda Adler, Elisabeth Brandstetter, Peter D. Han, Kairsten Fay, Misja Ilcisin, Kirsten Lacombe, Thomas R. Sibley, Melissa Truong, Caitlin R. Wolf, Romesh Gautom, Geoff Melly, Brian Hiatt, Philip Dykema, Scott Lindquist, Michael Boeckh, Janet A. Englund, Michael Famulare, Barry R. Lutz, Mark J. Rieder, Lea M. Starita, Matthew Thompson, Helen Y. Chu, Jay Shendure, Trevor Bedford |
| EPI_ISL_495654, EPI_ISL_495655, EPI_ISL_495656, EPI_ISL_495657, EPI_ISL_495658, EPI_ISL_495659, | Seattle Flu Study | Seattle Flu Study | Deborah A. Nickerson, Chris D. Frazar, Jover Lee, Benjamin Pelle, Matthew Richardson, Amanda Adler, Elisabeth Brandstetter, Peter D. Han, Kairsten Fay, Misja Ilcisin, Kirsten Lacombe, Thomas R. Sibley, Melissa Truong, Caitlin R. Wolf, Karen Cowgill, Stephanie Schrag, Jeff Duchin, Michael Boeckh, Janet A. Englund, Michael Famulare, Barry R. Lutz, Mark J. Rieder, Lea M. Starita, Matthew Thompson, Helen Y. Chu, Trevor Bedford, Jay Shendure |

| | | | |
|---|---|---|---|
| EPI_ISL_495660, EPI_ISL_495661, EPI_ISL_495662 | | | |
| EPI_ISL_495663 | Seattle Flu Study | Seattle Flu Study | Deborah A. Nickerson, Chris D. Frazar, Jover Lee, Benjamin Pelle, Matthew Richardson, Amanda Adler, Elisabeth Brandstetter, Peter D. Han, Kairsten Fay, Misja Ilcisin, Kirsten Lacombe, Thomas R. Sibley, Melissa Truong, Caitlin R. Wolf, Michael Boeckh, Janet A. Englund, Michael Famulare, Barry R. Lutz, Mark J. Rieder, Lea M. Starita, Matthew Thompson, Jay Shendure, Trevor Bedford, Helen Y. Chu |
| EPI_ISL_495664, EPI_ISL_495665 | Seattle Flu Study | Seattle Flu Study | Deborah A. Nickerson, Chris D. Frazar, Jover Lee, Benjamin Pelle, Matthew Richardson, Amanda Adler, Elisabeth Brandstetter, Peter D. Han, Kairsten Fay, Misja Ilcisin, Kirsten Lacombe, Thomas R. Sibley, Melissa Truong, Caitlin R. Wolf, Karen Cowgill, Stephanie Schrag, Jeff Duchin, Michael Boeckh, Janet A. Englund, Michael Famulare, Barry R. Lutz, Mark J. Rieder, Lea M. Starita, Matthew Thompson, Helen Y. Chu, Trevor Bedford, Jay Shendure |
| EPI_ISL_495666, EPI_ISL_495667, EPI_ISL_495668, EPI_ISL_495669, EPI_ISL_495670, EPI_ISL_495671, EPI_ISL_495672, EPI_ISL_495673, EPI_ISL_495674, EPI_ISL_495676, EPI_ISL_495677, EPI_ISL_495679, EPI_ISL_495680, EPI_ISL_495681, EPI_ISL_495682, EPI_ISL_495683, EPI_ISL_495684, EPI_ISL_495685, EPI_ISL_495686, EPI_ISL_495687, EPI_ISL_495688, EPI_ISL_495689, EPI_ISL_495690, EPI_ISL_495692, EPI_ISL_495693, EPI_ISL_495694, EPI_ISL_495695, EPI_ISL_495696, EPI_ISL_495697, EPI_ISL_495698, EPI_ISL_495699, EPI_ISL_495700, EPI_ISL_495701, EPI_ISL_495702, EPI_ISL_495703, EPI_ISL_495704, EPI_ISL_495705, EPI_ISL_495706, EPI_ISL_495707, EPI_ISL_495708, EPI_ISL_495709, EPI_ISL_495710, EPI_ISL_495711, EPI_ISL_495712, EPI_ISL_495713, EPI_ISL_495714, EPI_ISL_495715, EPI_ISL_495716, EPI_ISL_495717, EPI_ISL_495718, EPI_ISL_495719, EPI_ISL_495720, EPI_ISL_495721, EPI_ISL_495722, EPI_ISL_495723, EPI_ISL_495724, EPI_ISL_495725, EPI_ISL_495726, EPI_ISL_495727, EPI_ISL_495728, EPI_ISL_495729, EPI_ISL_495730, EPI_ISL_495731, EPI_ISL_495732, EPI_ISL_495733, EPI_ISL_495734, EPI_ISL_495735, EPI_ISL_495736, EPI_ISL_495737, EPI_ISL_495738, EPI_ISL_495739, EPI_ISL_495740, EPI_ISL_495741, EPI_ISL_495742, EPI_ISL_495743, EPI_ISL_495744, EPI_ISL_495745, EPI_ISL_495746, EPI_ISL_495747, EPI_ISL_495748, EPI_ISL_495749, EPI_ISL_495750, EPI_ISL_495751, EPI_ISL_495752, EPI_ISL_495753, EPI_ISL_495754, EPI_ISL_495755, EPI_ISL_495756, EPI_ISL_495757, EPI_ISL_495758, EPI_ISL_495759, EPI_ISL_495760, EPI_ISL_495761, EPI_ISL_495762, EPI_ISL_495763, EPI_ISL_495764, EPI_ISL_495765, EPI_ISL_495766, EPI_ISL_495767, EPI_ISL_495768, EPI_ISL_495769, EPI_ISL_495770, EPI_ISL_495771, EPI_ISL_495772, EPI_ISL_495773, EPI_ISL_495774, EPI_ISL_495775, EPI_ISL_495776, EPI_ISL_495777, EPI_ISL_495778, EPI_ISL_495779, EPI_ISL_495780, EPI_ISL_495781, EPI_ISL_495782, EPI_ISL_495783, EPI_ISL_495784, EPI_ISL_495785, EPI_ISL_495786, EPI_ISL_495787, EPI_ISL_495788, EPI_ISL_495789, EPI_ISL_495790, EPI_ISL_495791, EPI_ISL_495792, EPI_ISL_495793, EPI_ISL_495794, EPI_ISL_495795, EPI_ISL_495796, EPI_ISL_495797, EPI_ISL_495798, EPI_ISL_495799, EPI_ISL_495800, EPI_ISL_495801, EPI_ISL_495802, EPI_ISL_495803, EPI_ISL_495804, EPI_ISL_495805, EPI_ISL_495806, EPI_ISL_495807, EPI_ISL_495808, EPI_ISL_495809, EPI_ISL_495810, EPI_ISL_495811, EPI_ISL_495812, EPI_ISL_495813, EPI_ISL_495814, EPI_ISL_495815, EPI_ISL_495816, EPI_ISL_495817, EPI_ISL_495818, EPI_ISL_495819, EPI_ISL_495820, EPI_ISL_495821, EPI_ISL_495822, EPI_ISL_495823, EPI_ISL_495824, EPI_ISL_495825, EPI_ISL_495826, EPI_ISL_495827, EPI_ISL_495828, EPI_ISL_495829, EPI_ISL_495830, EPI_ISL_495831, EPI_ISL_495832, EPI_ISL_495833, EPI_ISL_495834, EPI_ISL_495835, EPI_ISL_495836, EPI_ISL_495837, EPI_ISL_495838, EPI_ISL_495839, EPI_ISL_495840, EPI_ISL_495841, EPI_ISL_495842, EPI_ISL_495843, EPI_ISL_495844, EPI_ISL_495845, EPI_ISL_495846, EPI_ISL_495847, EPI_ISL_495848, EPI_ISL_495849, EPI_ISL_495850, EPI_ISL_495851, EPI_ISL_495852, EPI_ISL_495853, EPI_ISL_495854, EPI_ISL_495855, EPI_ISL_495856, EPI_ISL_495857, EPI_ISL_495858, EPI_ISL_495859, EPI_ISL_495860, EPI_ISL_495861, EPI_ISL_495862, EPI_ISL_495863, EPI_ISL_495864, EPI_ISL_495865, EPI_ISL_495866, EPI_ISL_495867, EPI_ISL_495868, EPI_ISL_495869, EPI_ISL_495874, EPI_ISL_495875, EPI_ISL_495876, EPI_ISL_495877, EPI_ISL_495878, EPI_ISL_495880, EPI_ISL_495881, EPI_ISL_495882, EPI_ISL_495883, EPI_ISL_495884, EPI_ISL_495888, EPI_ISL_495889, EPI_ISL_495890, EPI_ISL_495891, EPI_ISL_495892, EPI_ISL_495893, EPI_ISL_495894, EPI_ISL_495895, EPI_ISL_495896, EPI_ISL_495897, EPI_ISL_495898, EPI_ISL_495899, EPI_ISL_495901, EPI_ISL_495902, EPI_ISL_495903, EPI_ISL_495904, EPI_ISL_495905, EPI_ISL_495906, EPI_ISL_495907, EPI_ISL_495908, EPI_ISL_495909, EPI_ISL_495910, EPI_ISL_495911, EPI_ISL_495912, EPI_ISL_495913, EPI_ISL_495914, EPI_ISL_495915, EPI_ISL_495916 | | | |
| see above | Washington State Department of Health | Seattle Flu Study | Deborah A. Nickerson, Chris D. Frazar, Jover Lee, Benjamin Pelle, Matthew Richardson, Amanda Adler, Elisabeth Brandstetter, Peter D. Han, Kairsten Fay, Misja Ilcisin, Kirsten Lacombe, Thomas R. Sibley, Melissa Truong, Caitlin R. Wolf, Romesh Gautom, Geoff |
| EPI_ISL_495918, EPI_ISL_495919, EPI_ISL_495920, EPI_ISL_495921, EPI_ISL_495922, EPI_ISL_495923, EPI_ISL_495924, EPI_ISL_495925, EPI_ISL_495926, EPI_ISL_495927, EPI_ISL_495928, EPI_ISL_495929, EPI_ISL_495930, EPI_ISL_495931, EPI_ISL_495932, EPI_ISL_495933, EPI_ISL_495934, EPI_ISL_495935, EPI_ISL_495936, EPI_ISL_495937, EPI_ISL_495938, EPI_ISL_495939, EPI_ISL_495940, EPI_ISL_495943, EPI_ISL_495944, EPI_ISL_495945, EPI_ISL_495946, EPI_ISL_495947, EPI_ISL_495948, EPI_ISL_495949, EPI_ISL_495950, EPI_ISL_495951, EPI_ISL_495952, EPI_ISL_495953, EPI_ISL_495954, EPI_ISL_495955, EPI_ISL_495956, EPI_ISL_495957, EPI_ISL_495958, EPI_ISL_495959, EPI_ISL_495961, EPI_ISL_495962, EPI_ISL_495963, EPI_ISL_495964, EPI_ISL_495965, EPI_ISL_495966, EPI_ISL_495967, EPI_ISL_495968, EPI_ISL_495969, EPI_ISL_495970, EPI_ISL_495972, EPI_ISL_495973, EPI_ISL_495974, EPI_ISL_495975, EPI_ISL_495977, EPI_ISL_495978, EPI_ISL_495979, EPI_ISL_495980, EPI_ISL_495981, EPI_ISL_495982, EPI_ISL_495983, EPI_ISL_495984, EPI_ISL_495985, EPI_ISL_495986, EPI_ISL_495987, EPI_ISL_495988, EPI_ISL_495989, EPI_ISL_495990, EPI_ISL_495991, EPI_ISL_495992, EPI_ISL_495993, EPI_ISL_495994, EPI_ISL_495995, EPI_ISL_495996, EPI_ISL_495997, EPI_ISL_495998, EPI_ISL_495999, EPI_ISL_496000, EPI_ISL_496001, EPI_ISL_496002, EPI_ISL_496003, EPI_ISL_496004, EPI_ISL_496005, EPI_ISL_496006, EPI_ISL_496007, EPI_ISL_496008, EPI_ISL_496009, EPI_ISL_496010, EPI_ISL_496011, EPI_ISL_496012, EPI_ISL_496013, EPI_ISL_496014, EPI_ISL_496015, EPI_ISL_496016, EPI_ISL_496017, EPI_ISL_496018, EPI_ISL_496019, EPI_ISL_496021, EPI_ISL_496022, EPI_ISL_496023, EPI_ISL_496024, EPI_ISL_496025, EPI_ISL_496026, EPI_ISL_496027, EPI_ISL_496028, EPI_ISL_496029, EPI_ISL_496030, EPI_ISL_496031, EPI_ISL_496032, EPI_ISL_496034, EPI_ISL_496035, EPI_ISL_496036, EPI_ISL_496037, EPI_ISL_496038, EPI_ISL_496039, EPI_ISL_496040, EPI_ISL_496041, EPI_ISL_496042, EPI_ISL_496043, EPI_ISL_496044, EPI_ISL_496045, EPI_ISL_496046, EPI_ISL_496047, EPI_ISL_496048, EPI_ISL_496049, EPI_ISL_496050, EPI_ISL_496051, EPI_ISL_496052, EPI_ISL_496053, EPI_ISL_496054, EPI_ISL_496055, EPI_ISL_496056, EPI_ISL_496057, EPI_ISL_496058, EPI_ISL_496059, EPI_ISL_496060, EPI_ISL_496061, EPI_ISL_496062, EPI_ISL_496063, EPI_ISL_496064, EPI_ISL_496065, EPI_ISL_496066, EPI_ISL_496067, EPI_ISL_496068, EPI_ISL_496069, EPI_ISL_496070, EPI_ISL_496071, EPI_ISL_496072, EPI_ISL_496073, EPI_ISL_496074, EPI_ISL_496076, EPI_ISL_496077, EPI_ISL_496078, EPI_ISL_496079, EPI_ISL_496080, EPI_ISL_496081, EPI_ISL_496082, EPI_ISL_496083, EPI_ISL_496084, EPI_ISL_496085, EPI_ISL_496086, EPI_ISL_496087, EPI_ISL_496088, EPI_ISL_496089, EPI_ISL_496090, EPI_ISL_496091, EPI_ISL_496092, EPI_ISL_496094, EPI_ISL_496095, EPI_ISL_496096, EPI_ISL_496097, EPI_ISL_496098, EPI_ISL_496099, EPI_ISL_496100, EPI_ISL_496101, EPI_ISL_496102, EPI_ISL_496103, EPI_ISL_496104, EPI_ISL_496105, EPI_ISL_496106, EPI_ISL_496107, EPI_ISL_496108, EPI_ISL_496110, EPI_ISL_496111, EPI_ISL_496112, EPI_ISL_496113, EPI_ISL_496114, EPI_ISL_496115, EPI_ISL_496116, EPI_ISL_496117, EPI_ISL_496118, EPI_ISL_496119, EPI_ISL_496120, EPI_ISL_496121, EPI_ISL_496122, EPI_ISL_496123, EPI_ISL_496124, EPI_ISL_496125, EPI_ISL_496126, EPI_ISL_496127, EPI_ISL_496128, EPI_ISL_496129, EPI_ISL_496130, EPI_ISL_496131, EPI_ISL_496132, EPI_ISL_496133, EPI_ISL_496134, EPI_ISL_496135, EPI_ISL_496136, EPI_ISL_496137, EPI_ISL_496138, EPI_ISL_496139, EPI_ISL_496140, EPI_ISL_496141, EPI_ISL_496142, EPI_ISL_496143, EPI_ISL_496144, EPI_ISL_496145, EPI_ISL_496146, EPI_ISL_496147, EPI_ISL_496148, EPI_ISL_496149, EPI_ISL_496150, EPI_ISL_496151, EPI_ISL_496152, EPI_ISL_496153, EPI_ISL_496154, EPI_ISL_496155, EPI_ISL_496156, EPI_ISL_496157, EPI_ISL_496158, EPI_ISL_496159, EPI_ISL_496160, EPI_ISL_496161, EPI_ISL_496162, EPI_ISL_496163, EPI_ISL_496164, EPI_ISL_496165, EPI_ISL_496166, EPI_ISL_496167, EPI_ISL_496168, EPI_ISL_496169, EPI_ISL_496170, EPI_ISL_496171, EPI_ISL_496173, EPI_ISL_496174, EPI_ISL_496175, EPI_ISL_496176, EPI_ISL_496177, EPI_ISL_496178, EPI_ISL_496179, EPI_ISL_496180, EPI_ISL_496181, EPI_ISL_496182, EPI_ISL_496183, EPI_ISL_496184, EPI_ISL_496185, EPI_ISL_496186, EPI_ISL_496187, EPI_ISL_496188, EPI_ISL_496189, EPI_ISL_496190, EPI_ISL_496191, EPI_ISL_496192, EPI_ISL_496193, EPI_ISL_496194, EPI_ISL_496195, EPI_ISL_496196, EPI_ISL_496197, EPI_ISL_496198, EPI_ISL_496199, EPI_ISL_496200, EPI_ISL_496201, EPI_ISL_496202, EPI_ISL_496203, EPI_ISL_496204, EPI_ISL_496205, EPI_ISL_496206, EPI_ISL_496207, EPI_ISL_496208, EPI_ISL_496209, EPI_ISL_496210, EPI_ISL_496211, EPI_ISL_496212, EPI_ISL_496213, EPI_ISL_496214, EPI_ISL_496215, EPI_ISL_496216, EPI_ISL_496217, EPI_ISL_496218, EPI_ISL_496219, EPI_ISL_496220, EPI_ISL_496221, EPI_ISL_496222, EPI_ISL_496223, EPI_ISL_496224, EPI_ISL_496225, EPI_ISL_496226, EPI_ISL_496227, EPI_ISL_496228, EPI_ISL_496229, EPI_ISL_496230, EPI_ISL_496231, EPI_ISL_496232, EPI_ISL_496233, EPI_ISL_496234, EPI_ISL_496235, EPI_ISL_496236, EPI_ISL_496237, EPI_ISL_496238, EPI_ISL_496239, EPI_ISL_496240, EPI_ISL_496241, EPI_ISL_496242, EPI_ISL_496243, EPI_ISL_496244, EPI_ISL_496245, EPI_ISL_496246, EPI_ISL_496247, EPI_ISL_496248, EPI_ISL_496249, EPI_ISL_496250, EPI_ISL_496251, EPI_ISL_496252, EPI_ISL_496253, EPI_ISL_496254, EPI_ISL_496255, EPI_ISL_496256, EPI_ISL_496257, EPI_ISL_496258, EPI_ISL_496259, EPI_ISL_496260, EPI_ISL_496261, EPI_ISL_496262, EPI_ISL_496263, EPI_ISL_496264, EPI_ISL_496265, EPI_ISL_496266, EPI_ISL_496267, EPI_ISL_496268, EPI_ISL_496269, EPI_ISL_496270, EPI_ISL_496271, EPI_ISL_496272, EPI_ISL_496273, EPI_ISL_496274, EPI_ISL_496275, EPI_ISL_496276, EPI_ISL_496277, EPI_ISL_496278, EPI_ISL_496279, EPI_ISL_496280, EPI_ISL_496281, EPI_ISL_496282, EPI_ISL_496283, EPI_ISL_496284, EPI_ISL_496285, EPI_ISL_496286, EPI_ISL_496287, EPI_ISL_496288, EPI_ISL_496289, EPI_ISL_496290, EPI_ISL_496291, EPI_ISL_496292, EPI_ISL_496294, EPI_ISL_496295, EPI_ISL_496296, EPI_ISL_496297, EPI_ISL_496298, EPI_ISL_496299, EPI_ISL_496300, EPI_ISL_496301, EPI_ISL_496302, EPI_ISL_496303, EPI_ISL_496304, EPI_ISL_496305, EPI_ISL_496306, EPI_ISL_496307, EPI_ISL_496308, EPI_ISL_496309, EPI_ISL_496310, EPI_ISL_496311, EPI_ISL_496312, EPI_ISL_496313, EPI_ISL_496314, EPI_ISL_496315, EPI_ISL_496316, EPI_ISL_496317, EPI_ISL_496318, EPI_ISL_496319, EPI_ISL_496320, EPI_ISL_496321, EPI_ISL_496322, EPI_ISL_496323, EPI_ISL_496324, EPI_ISL_496325, EPI_ISL_496326, EPI_ISL_496327, EPI_ISL_496328, EPI_ISL_496329, EPI_ISL_496330, EPI_ISL_496331, EPI_ISL_496332, EPI_ISL_496333, EPI_ISL_496334, EPI_ISL_496335, EPI_ISL_496336, EPI_ISL_496337, EPI_ISL_496338 | | | |
| see above | Washington State Department of Health | Seattle Flu Study | Deborah A. Nickerson, Chris D. Frazar, Jover Lee, Benjamin Pelle, Matthew Richardson, Amanda Adler, Elisabeth Brandstetter, Peter D. Han, Kairsten Fay, Misja Ilcisin, Kirsten Lacombe, Thomas R. Sibley, Melissa Truong, Caitlin R. Wolf, Romesh Gautom, Geoff Melly, Brian Hiatt, Philip Dykema, Scott Lindquist, Michael Boeckh, Janet A. Englund, Michael Famulare, Barry R. Lutz, Mark J. Rieder, Lea M. Starita, Matthew Thompson, Helen Y. Chu, Jay Shendure, Trevor Bedford |
| EPI_ISL_496339, EPI_ISL_496340, EPI_ISL_496341, EPI_ISL_496342, EPI_ISL_496343, EPI_ISL_496344, EPI_ISL_496345, EPI_ISL_496346, EPI_ISL_496347, EPI_ISL_496348, EPI_ISL_496349, EPI_ISL_496350, EPI_ISL_496351, EPI_ISL_496352, EPI_ISL_496353, EPI_ISL_496354, EPI_ISL_496355, EPI_ISL_496356, EPI_ISL_496357, EPI_ISL_496358, EPI_ISL_496359, EPI_ISL_496360, EPI_ISL_496361, EPI_ISL_496362, EPI_ISL_496363, EPI_ISL_496364, EPI_ISL_496365, EPI_ISL_496366, EPI_ISL_496367, EPI_ISL_496368, EPI_ISL_496369, EPI_ISL_496370, EPI_ISL_496371, EPI_ISL_496372, EPI_ISL_496373, EPI_ISL_496374, EPI_ISL_496375, EPI_ISL_496376 | | | |
| see above | Infectolab | Andersen lab at Scripps Research | SEARCH Alliance San Diego with Samuel Navarro Alvarez, Carlos A. Cota Haros, Octavio Renteria Pacheco |
| EPI_ISL_496916, EPI_ISL_496917 | Minnesota Department of Health, Public Health Laboratory | Minnesota Department of Health, Public Health Laboratory | Matt Plumb, Jacob Garfin, and Xiong Wang |
| EPI_ISL_496918 | Mayo Clinic & Mayo Clinic Laboratories | Minnesota Department of Health, Public Health Laboratory | Matt Plumb, Jacob Garfin, and Xiong Wang |
| EPI_ISL_496919, EPI_ISL_496920, EPI_ISL_496921, EPI_ISL_496922, EPI_ISL_496923, EPI_ISL_496924, EPI_ISL_496925, EPI_ISL_496926, EPI_ISL_496927, EPI_ISL_496928, EPI_ISL_496929, EPI_ISL_496930 | | | |
| see above | Minnesota Department of Health, Public Health Laboratory | Minnesota Department of Health, Public Health Laboratory | Matt Plumb, Jacob Garfin, and Xiong Wang |
| EPI_ISL_496931, EPI_ISL_496932, EPI_ISL_496933, EPI_ISL_496934, EPI_ISL_496935, EPI_ISL_496936, EPI_ISL_496937, EPI_ISL_496938, EPI_ISL_496939, EPI_ISL_496940, EPI_ISL_496941, EPI_ISL_496942, EPI_ISL_496943, EPI_ISL_496944, EPI_ISL_496945, EPI_ISL_496946, EPI_ISL_496947, EPI_ISL_496948, EPI_ISL_496949, EPI_ISL_496950, EPI_ISL_496951, EPI_ISL_496952, EPI_ISL_496953, EPI_ISL_496954, EPI_ISL_496955, EPI_ISL_496956, EPI_ISL_496957, EPI_ISL_496958, EPI_ISL_496959, EPI_ISL_496960, EPI_ISL_496961, EPI_ISL_496962, EPI_ISL_496963, EPI_ISL_496964, EPI_ISL_496965, EPI_ISL_496966, EPI_ISL_496967, EPI_ISL_496968, EPI_ISL_496969, EPI_ISL_496970, EPI_ISL_496971, EPI_ISL_496972, EPI_ISL_496974, EPI_ISL_496975, EPI_ISL_496976, EPI_ISL_496977, EPI_ISL_496978, EPI_ISL_496979, EPI_ISL_496980, EPI_ISL_496981, EPI_ISL_496982, EPI_ISL_496983, EPI_ISL_496984, EPI_ISL_496985, EPI_ISL_496986, EPI_ISL_496987, EPI_ISL_496988, EPI_ISL_496989, EPI_ISL_496990, EPI_ISL_496991, EPI_ISL_496992, EPI_ISL_496993, EPI_ISL_496994, EPI_ISL_496995, EPI_ISL_496996, EPI_ISL_496997, EPI_ISL_496998, EPI_ISL_496999, EPI_ISL_497000, EPI_ISL_497002, EPI_ISL_497003, EPI_ISL_497004, EPI_ISL_497005, EPI_ISL_497006, EPI_ISL_497007, EPI_ISL_497008, EPI_ISL_497009, EPI_ISL_497010, EPI_ISL_497011, EPI_ISL_497012, EPI_ISL_497013, EPI_ISL_497014, EPI_ISL_497015, EPI_ISL_497016, EPI_ISL_497017, EPI_ISL_497018, EPI_ISL_497019, EPI_ISL_497020, EPI_ISL_497021, EPI_ISL_497022, EPI_ISL_497023, EPI_ISL_497024, EPI_ISL_497025, EPI_ISL_497026, EPI_ISL_497027, EPI_ISL_497028, EPI_ISL_497029, EPI_ISL_497030, EPI_ISL_497031, EPI_ISL_497032, EPI_ISL_497033, EPI_ISL_497034, EPI_ISL_497035, EPI_ISL_497036, EPI_ISL_497037, EPI_ISL_497038, EPI_ISL_497039, EPI_ISL_497040, EPI_ISL_497041, EPI_ISL_497042, EPI_ISL_497043, EPI_ISL_497044, EPI_ISL_497045, EPI_ISL_497046, EPI_ISL_497047, EPI_ISL_497048, EPI_ISL_497049, EPI_ISL_497050, EPI_ISL_497051, EPI_ISL_497052, EPI_ISL_497053, EPI_ISL_497054, EPI_ISL_497055, EPI_ISL_497056, EPI_ISL_497057, EPI_ISL_497058, EPI_ISL_497059, EPI_ISL_497060, EPI_ISL_497061, EPI_ISL_497062, EPI_ISL_497063, EPI_ISL_497064, EPI_ISL_497065, EPI_ISL_497066, EPI_ISL_497067, EPI_ISL_497068, EPI_ISL_497069, EPI_ISL_497070, EPI_ISL_497071, EPI_ISL_497072, EPI_ISL_497074, EPI_ISL_497075, EPI_ISL_497076, EPI_ISL_497077, EPI_ISL_497078, EPI_ISL_497079, EPI_ISL_497080, EPI_ISL_497081, EPI_ISL_497082, EPI_ISL_497083, EPI_ISL_497084, EPI_ISL_497085, EPI_ISL_497086, EPI_ISL_497087, EPI_ISL_497088, EPI_ISL_497089, EPI_ISL_497090, EPI_ISL_497091, EPI_ISL_497092, EPI_ISL_497093, EPI_ISL_497094, EPI_ISL_497095, EPI_ISL_497096, EPI_ISL_497097, EPI_ISL_497098, EPI_ISL_497099, EPI_ISL_497100, EPI_ISL_497101, EPI_ISL_497102, EPI_ISL_497103, EPI_ISL_497104, EPI_ISL_497105, EPI_ISL_497106, EPI_ISL_497107, EPI_ISL_497109, EPI_ISL_497110, EPI_ISL_497111, EPI_ISL_497112, EPI_ISL_497113, EPI_ISL_497114, EPI_ISL_497115, EPI_ISL_497116, EPI_ISL_497117, EPI_ISL_497118, EPI_ISL_497119, EPI_ISL_497120, EPI_ISL_497121, EPI_ISL_497122, EPI_ISL_497123, EPI_ISL_497124, EPI_ISL_497125, EPI_ISL_497126, EPI_ISL_497127, EPI_ISL_497128, EPI_ISL_497129, EPI_ISL_497130, EPI_ISL_497131, EPI_ISL_497132, EPI_ISL_497133, EPI_ISL_497134, EPI_ISL_497135, EPI_ISL_497136, EPI_ISL_497137, EPI_ISL_497138, EPI_ISL_497139, EPI_ISL_497140, EPI_ISL_497141, EPI_ISL_497142, EPI_ISL_497143, EPI_ISL_497144, EPI_ISL_497145, EPI_ISL_497146, EPI_ISL_497147, EPI_ISL_497148, EPI_ISL_497149, EPI_ISL_497150, EPI_ISL_497151, EPI_ISL_497152, EPI_ISL_497153, EPI_ISL_497154, EPI_ISL_497155, EPI_ISL_497156, EPI_ISL_497157, EPI_ISL_497158, EPI_ISL_497159, EPI_ISL_497160, EPI_ISL_497161, EPI_ISL_497162, EPI_ISL_497163, EPI_ISL_497164, EPI_ISL_497165, EPI_ISL_497166, EPI_ISL_497168, EPI_ISL_497169, EPI_ISL_497170, EPI_ISL_497171, EPI_ISL_497172, EPI_ISL_497173, EPI_ISL_497174, EPI_ISL_497175, EPI_ISL_497176, EPI_ISL_497177, EPI_ISL_497178, EPI_ISL_497179, EPI_ISL_497180, EPI_ISL_497181, EPI_ISL_497182, EPI_ISL_497183, EPI_ISL_497184, EPI_ISL_497185, EPI_ISL_497186, EPI_ISL_497187, EPI_ISL_497188, EPI_ISL_497189, EPI_ISL_497190, EPI_ISL_497191, EPI_ISL_497192, EPI_ISL_497193, EPI_ISL_497194, EPI_ISL_497195, EPI_ISL_497196, EPI_ISL_497197, EPI_ISL_497198, EPI_ISL_497199, EPI_ISL_497200, EPI_ISL_497201, EPI_ISL_497202, EPI_ISL_497203, EPI_ISL_497204, EPI_ISL_497205, EPI_ISL_497206, EPI_ISL_497207, EPI_ISL_497208, EPI_ISL_497209, EPI_ISL_497210, EPI_ISL_497211, EPI_ISL_497212, EPI_ISL_497213, EPI_ISL_497214, EPI_ISL_497215, EPI_ISL_497216, EPI_ISL_497217, EPI_ISL_497218, EPI_ISL_497219, EPI_ISL_497220, EPI_ISL_497221, EPI_ISL_497222, EPI_ISL_497223, EPI_ISL_497224, EPI_ISL_497225, EPI_ISL_497226, EPI_ISL_497227, EPI_ISL_497228, EPI_ISL_497230, EPI_ISL_497231, EPI_ISL_497232, EPI_ISL_497233, EPI_ISL_497234, EPI_ISL_497235, EPI_ISL_497236, EPI_ISL_497237, EPI_ISL_497238, EPI_ISL_497239, EPI_ISL_497240, EPI_ISL_497241, EPI_ISL_497242, EPI_ISL_497243, EPI_ISL_497244, EPI_ISL_497245, EPI_ISL_497246, EPI_ISL_497247, EPI_ISL_497248, EPI_ISL_497249, EPI_ISL_497250, EPI_ISL_497251, EPI_ISL_497252, EPI_ISL_497253, EPI_ISL_497254, EPI_ISL_497255, EPI_ISL_497256, EPI_ISL_497257, EPI_ISL_497258, EPI_ISL_497259, EPI_ISL_497260, EPI_ISL_497261, EPI_ISL_497262, EPI_ISL_497263, EPI_ISL_497265, EPI_ISL_497266, EPI_ISL_497267, EPI_ISL_497268, EPI_ISL_497269, EPI_ISL_497270, EPI_ISL_497271, EPI_ISL_497272, EPI_ISL_497273, EPI_ISL_497274, EPI_ISL_497275, EPI_ISL_497276, EPI_ISL_497277, EPI_ISL_497278, EPI_ISL_497279, EPI_ISL_497280, EPI_ISL_497281, EPI_ISL_497282, EPI_ISL_497283, EPI_ISL_497284, EPI_ISL_497285, EPI_ISL_497286, EPI_ISL_497287, EPI_ISL_497288, EPI_ISL_497289, EPI_ISL_497290, EPI_ISL_497291, EPI_ISL_497292, EPI_ISL_497293, EPI_ISL_497294, EPI_ISL_497295, EPI_ISL_497296, EPI_ISL_497297, EPI_ISL_497298, EPI_ISL_497299, EPI_ISL_497300, EPI_ISL_497301, EPI_ISL_497302, EPI_ISL_497303, EPI_ISL_497304, EPI_ISL_497305, EPI_ISL_497306, EPI_ISL_497307, EPI_ISL_497308, EPI_ISL_497309, EPI_ISL_497310, EPI_ISL_497311, EPI_ISL_497312, EPI_ISL_497313, EPI_ISL_497315, EPI_ISL_497316, EPI_ISL_497317, EPI_ISL_497318, EPI_ISL_497319, EPI_ISL_497320, EPI_ISL_497321, EPI_ISL_497322, EPI_ISL_497323, EPI_ISL_497324, EPI_ISL_497325, EPI_ISL_497326, EPI_ISL_497327, EPI_ISL_497328, EPI_ISL_497329, EPI_ISL_497330, EPI_ISL_497331, EPI_ISL_497332, EPI_ISL_497333, EPI_ISL_497334, EPI_ISL_497335, EPI_ISL_497336, EPI_ISL_497337, EPI_ISL_497338, EPI_ISL_497339, EPI_ISL_497340, EPI_ISL_497341, EPI_ISL_497342, EPI_ISL_497343, EPI_ISL_497344, EPI_ISL_497345, EPI_ISL_497346, EPI_ISL_497347, EPI_ISL_497348, EPI_ISL_497349, EPI_ISL_497350, EPI_ISL_497351, EPI_ISL_497352, EPI_ISL_497353, EPI_ISL_497354, EPI_ISL_497356, EPI_ISL_497357, EPI_ISL_497358, EPI_ISL_497359, EPI_ISL_497360, EPI_ISL_497361, EPI_ISL_497362, EPI_ISL_497363, EPI_ISL_497364, EPI_ISL_497365, EPI_ISL_497366, EPI_ISL_497367, EPI_ISL_497368, EPI_ISL_497369, EPI_ISL_497370, EPI_ISL_497371, EPI_ISL_497372, EPI_ISL_497373, EPI_ISL_497374, EPI_ISL_497375, EPI_ISL_497376, EPI_ISL_497377, EPI_ISL_497378, EPI_ISL_497379, EPI_ISL_497380, EPI_ISL_497381, EPI_ISL_497382, EPI_ISL_497383, EPI_ISL_497384, EPI_ISL_497385, EPI_ISL_497386, EPI_ISL_497387, EPI_ISL_497388, EPI_ISL_497389, EPI_ISL_497390, EPI_ISL_497391, EPI_ISL_497392, EPI_ISL_497393, EPI_ISL_497394, EPI_ISL_497395, EPI_ISL_497396, EPI_ISL_497397, EPI_ISL_497398, EPI_ISL_497399, EPI_ISL_497400, EPI_ISL_497401, EPI_ISL_497402, EPI_ISL_497403, EPI_ISL_497404, EPI_ISL_497405, EPI_ISL_497406, EPI_ISL_497407, EPI_ISL_497408, EPI_ISL_497409, EPI_ISL_497410, EPI_ISL_497411, EPI_ISL_497412, EPI_ISL_497413, EPI_ISL_497414, EPI_ISL_497415, EPI_ISL_497416, EPI_ISL_497417, EPI_ISL_497418, EPI_ISL_497419, EPI_ISL_497420, EPI_ISL_497421, EPI_ISL_497422, EPI_ISL_497423, EPI_ISL_497424, EPI_ISL_497425, EPI_ISL_497426, EPI_ISL_497427, EPI_ISL_497428, EPI_ISL_497429, EPI_ISL_497430, EPI_ISL_497431, EPI_ISL_497432, EPI_ISL_497433, EPI_ISL_497434, EPI_ISL_497435, EPI_ISL_497436, EPI_ISL_497437, EPI_ISL_497438, EPI_ISL_497440, EPI_ISL_497441, EPI_ISL_497442, EPI_ISL_497443, EPI_ISL_497444, EPI_ISL_497445, EPI_ISL_497446, EPI_ISL_497447, EPI_ISL_497448, EPI_ISL_497449, EPI_ISL_497450, EPI_ISL_497451, EPI_ISL_497452, EPI_ISL_497453, EPI_ISL_497454, EPI_ISL_497455, EPI_ISL_497456, EPI_ISL_497457, EPI_ISL_497458, EPI_ISL_497459, EPI_ISL_497460, EPI_ISL_497461, EPI_ISL_497462, EPI_ISL_497463, EPI_ISL_497464, EPI_ISL_497465, EPI_ISL_497466, EPI_ISL_497467, EPI_ISL_497468, EPI_ISL_497469, EPI_ISL_497470, EPI_ISL_497471, EPI_ISL_497472, EPI_ISL_497473, EPI_ISL_497474, EPI_ISL_497475, EPI_ISL_497476, EPI_ISL_497477, EPI_ISL_497478, EPI_ISL_497479, EPI_ISL_497480, EPI_ISL_497481, EPI_ISL_497482, EPI_ISL_497483, EPI_ISL_497485, EPI_ISL_497486, EPI_ISL_497487, EPI_ISL_497488, EPI_ISL_497489, EPI_ISL_497490, EPI_ISL_497491, EPI_ISL_497492, EPI_ISL_497493, EPI_ISL_497494, EPI_ISL_497495, EPI_ISL_497496, EPI_ISL_497497, EPI_ISL_497498, EPI_ISL_497499, EPI_ISL_497500, EPI_ISL_497501, EPI_ISL_497502, EPI_ISL_497503, EPI_ISL_497504, EPI_ISL_497505, EPI_ISL_497506, EPI_ISL_497507, EPI_ISL_497508, EPI_ISL_497509, EPI_ISL_497510, EPI_ISL_497511, EPI_ISL_497512, EPI_ISL_497513, EPI_ISL_497514, EPI_ISL_497515, EPI_ISL_497516, EPI_ISL_497517, EPI_ISL_497518, EPI_ISL_497519, EPI_ISL_497520, EPI_ISL_497521, EPI_ISL_497522, EPI_ISL_497523, EPI_ISL_497524, EPI_ISL_497525, EPI_ISL_497526, EPI_ISL_497527, EPI_ISL_497528, EPI_ISL_497529, EPI_ISL_497530, EPI_ISL_497531, EPI_ISL_497532, EPI_ISL_497533, EPI_ISL_497534, EPI_ISL_497535, EPI_ISL_497536, EPI_ISL_497537, EPI_ISL_497538, EPI_ISL_497539, EPI_ISL_497540, EPI_ISL_497541, EPI_ISL_497542, EPI_ISL_497543, EPI_ISL_497544, EPI_ISL_497545, EPI_ISL_497546, EPI_ISL_497547, EPI_ISL_497548, EPI_ISL_497549, EPI_ISL_497550, EPI_ISL_497551, EPI_ISL_497552, EPI_ISL_497553, EPI_ISL_497554, EPI_ISL_497555, EPI_ISL_497556, EPI_ISL_497557, EPI_ISL_497558, EPI_ISL_497559, EPI_ISL_497560, EPI_ISL_497561, EPI_ISL_497562, EPI_ISL_497563, EPI_ISL_497564, EPI_ISL_497565, EPI_ISL_497567, EPI_ISL_497569, EPI_ISL_497570, EPI_ISL_497571, EPI_ISL_497572, EPI_ISL_497573, EPI_ISL_497574, EPI_ISL_497575, EPI_ISL_497576, EPI_ISL_497577, EPI_ISL_497578, EPI_ISL_497579, EPI_ISL_497580, EPI_ISL_497581, EPI_ISL_497582, EPI_ISL_497583, EPI_ISL_497584, EPI_ISL_497585, EPI_ISL_497586, EPI_ISL_497587, EPI_ISL_497588, EPI_ISL_497589, EPI_ISL_497590, EPI_ISL_497591, EPI_ISL_497592, EPI_ISL_497593, EPI_ISL_497594, EPI_ISL_497595, EPI_ISL_497596, EPI_ISL_497597, EPI_ISL_497598, EPI_ISL_497599, EPI_ISL_497600, EPI_ISL_497601, EPI_ISL_497602, EPI_ISL_497603, EPI_ISL_497604, EPI_ISL_497605, EPI_ISL_497607, EPI_ISL_497608, EPI_ISL_497609, EPI_ISL_497610, EPI_ISL_497611, EPI_ISL_497612, EPI_ISL_497613, EPI_ISL_497614, EPI_ISL_497615, EPI_ISL_497616, EPI_ISL_497617, EPI_ISL_497618, EPI_ISL_497619, EPI_ISL_497620, EPI_ISL_497621, EPI_ISL_497622, EPI_ISL_497623, EPI_ISL_497624, EPI_ISL_497625, EPI_ISL_497626, EPI_ISL_497627, EPI_ISL_497628, EPI_ISL_497629, EPI_ISL_497630, EPI_ISL_497631, EPI_ISL_497632, EPI_ISL_497633, EPI_ISL_497634, EPI_ISL_497635, EPI_ISL_497636, EPI_ISL_497637, EPI_ISL_497638, EPI_ISL_497639, EPI_ISL_497640, EPI_ISL_497641, EPI_ISL_497642, EPI_ISL_497643, EPI_ISL_497644, EPI_ISL_497645, EPI_ISL_497646, EPI_ISL_497647, EPI_ISL_497648, EPI_ISL_497649, EPI_ISL_497650, EPI_ISL_497651, EPI_ISL_497652, EPI_ISL_497653, EPI_ISL_497654, EPI_ISL_497655, EPI_ISL_497656, EPI_ISL_497657, EPI_ISL_497658, EPI_ISL_497659, EPI_ISL_497660, EPI_ISL_497661, EPI_ISL_497662, EPI_ISL_497663, EPI_ISL_497664, EPI_ISL_497665, EPI_ISL_497666, EPI_ISL_497667, EPI_ISL_497668, EPI_ISL_497669, EPI_ISL_497670, EPI_ISL_497671, EPI_ISL_497672, EPI_ISL_497673, EPI_ISL_497674, EPI_ISL_497675, EPI_ISL_497676, EPI_ISL_497677, EPI_ISL_497678, EPI_ISL_497679, EPI_ISL_497680, EPI_ISL_497681, EPI_ISL_497682, EPI_ISL_497683, EPI_ISL_497684, EPI_ISL_497685, EPI_ISL_497686, EPI_ISL_497687, EPI_ISL_497688, EPI_ISL_497689, EPI_ISL_497690, EPI_ISL_497691, EPI_ISL_497692, EPI_ISL_497693, EPI_ISL_497694, EPI_ISL_497695, EPI_ISL_497696, EPI_ISL_497697, EPI_ISL_497698, EPI_ISL_497699, EPI_ISL_497700, EPI_ISL_497701, EPI_ISL_497702, EPI_ISL_497703, EPI_ISL_497704, EPI_ISL_497705, EPI_ISL_497706, EPI_ISL_497707, EPI_ISL_497708, EPI_ISL_497709, EPI_ISL_497710, EPI_ISL_497711, EPI_ISL_497712, EPI_ISL_497713, EPI_ISL_497714, EPI_ISL_497715, EPI_ISL_497716, EPI_ISL_497717, EPI_ISL_497718, EPI_ISL_497719, EPI_ISL_497720, EPI_ISL_497721, EPI_ISL_497722, EPI_ISL_497723, EPI_ISL_497724, EPI_ISL_497725, EPI_ISL_497726 | | | |
| see above | Washington State Department of Health | Seattle Flu Study | Deborah A. Nickerson, Chris D. Frazar, Jover Lee, Benjamin Pelle, Matthew Richardson, Amanda Adler, Elisabeth Brandstetter, Peter D. Han, Kairsten Fay, Misja Ilcisin, Kirsten Lacombe, Thomas R. Sibley, Melissa Truong, Caitlin R. Wolf, Romesh Gautom, Geoff Melly, Brian Hiatt, Philip Dykema, Scott Lindquist, Michael Boeckh, Janet A. Englund, Michael Famulare, Barry R. Lutz, Mark J. Rieder, Lea M. Starita, Matthew Thompson, Helen Y. Chu, Jay Shendure, Trevor Bedford |
| EPI_ISL_497872 | UW Virology Lab | UW Virology Lab | Pavitra Roychoudhury, Amin Addetia, Hong Xie, Lasata Shrestha, Truong Nguyen, Meei-Li Huang, Keith Jerome, Alexander Greninger |
| EPI_ISL_498630, EPI_ISL_498631, EPI_ISL_498632, EPI_ISL_498633, EPI_ISL_498634, EPI_ISL_498635, EPI_ISL_498637, EPI_ISL_498638, EPI_ISL_498640, EPI_ISL_498641, EPI_ISL_498643, EPI_ISL_498644, EPI_ISL_498645, EPI_ISL_498646, EPI_ISL_498647, EPI_ISL_498648, EPI_ISL_498649, EPI_ISL_498650, EPI_ISL_498651, EPI_ISL_498652, EPI_ISL_498653, EPI_ISL_498654, EPI_ISL_498655, EPI_ISL_498656, EPI_ISL_498657, EPI_ISL_498658, EPI_ISL_498659, EPI_ISL_498660, EPI_ISL_498661, EPI_ISL_498662, EPI_ISL_498663, EPI_ISL_498664, EPI_ISL_498665, EPI_ISL_498666, EPI_ISL_498667, EPI_ISL_498668, EPI_ISL_498669, EPI_ISL_498670, EPI_ISL_498671, EPI_ISL_498672, EPI_ISL_498674, EPI_ISL_498677, EPI_ISL_498678, EPI_ISL_498680, EPI_ISL_498681, EPI_ISL_498682, EPI_ISL_498683, EPI_ISL_498684, EPI_ISL_498685, EPI_ISL_498686, EPI_ISL_498687, EPI_ISL_498688, EPI_ISL_498690 | | | |
| see above | Utah Public Health Laboratory | Utah Public Health Laboratory | Heidi Butz, Erin Young, Kelly Oakeson |
| EPI_ISL_498695, EPI_ISL_498696, EPI_ISL_498697, EPI_ISL_498698, EPI_ISL_498699, EPI_ISL_498700, EPI_ISL_498701, EPI_ISL_498702, EPI_ISL_498703, EPI_ISL_498704, EPI_ISL_498705, EPI_ISL_498706, EPI_ISL_498707, EPI_ISL_498708, EPI_ISL_498709, EPI_ISL_498710, EPI_ISL_498711, EPI_ISL_498712, EPI_ISL_498713, EPI_ISL_498714, EPI_ISL_498715, EPI_ISL_498716, EPI_ISL_498717, EPI_ISL_498718, EPI_ISL_498719, EPI_ISL_498720, EPI_ISL_498721, EPI_ISL_498722, EPI_ISL_498723, EPI_ISL_498724, EPI_ISL_498725, EPI_ISL_498726, EPI_ISL_498727, EPI_ISL_498728, EPI_ISL_498729, EPI_ISL_498730, EPI_ISL_498731, EPI_ISL_498732, EPI_ISL_498733, EPI_ISL_498734, EPI_ISL_498735, EPI_ISL_498736, EPI_ISL_498737, EPI_ISL_498738, EPI_ISL_498739, EPI_ISL_498740, EPI_ISL_498741, EPI_ISL_498742, EPI_ISL_498743, EPI_ISL_498744, EPI_ISL_498745, EPI_ISL_498746, EPI_ISL_498747 | | | |
| see above | Quest Diagnostics | Quest Diagnostics | Rosenthal,S.H., Gerasimova,A., Kagan,R.M. and Owen, R. |
| EPI_ISL_500459 | Quest Diagnostics | Quest Diagnostics | Rosenthal,S.H., Gerasimova,A., Kagan,R.M., Owen, R. AND Lacbawan, F. |
| EPI_ISL_500500, EPI_ISL_500501, EPI_ISL_500502 | University of Washington Virology Lab | University of Washington Virology Lab | Pavitra Roychoudhury, Hong Xie, Lasata Shrestha, Amin Addetia, Truong Nguyen, Victoria M Rachleff, Meei-Li Huang, Keith R Jerome, Alexander Greninger |
| EPI_ISL_500503, EPI_ISL_500504, EPI_ISL_500505, EPI_ISL_500506, EPI_ISL_500507, EPI_ISL_500508, EPI_ISL_500509, EPI_ISL_500510, EPI_ISL_500511, EPI_ISL_500512, EPI_ISL_500513, EPI_ISL_500514, EPI_ISL_500515, EPI_ISL_500516, EPI_ISL_500517, EPI_ISL_500518, EPI_ISL_500519, EPI_ISL_500520, EPI_ISL_500521, EPI_ISL_500522, EPI_ISL_500523, EPI_ISL_500524, EPI_ISL_500525, EPI_ISL_500526, EPI_ISL_500527, EPI_ISL_500528, EPI_ISL_500529, EPI_ISL_500530, EPI_ISL_500531, EPI_ISL_500532, EPI_ISL_500533, EPI_ISL_500534, EPI_ISL_500535, EPI_ISL_500536, EPI_ISL_500537, EPI_ISL_500538 | | | |
| see above | Mayo Clinic Laboratories | University of Washington Virology Lab | Pavitra Roychoudhury, Hong Xie, Lasata Shrestha, Amin Addetia, Truong Nguyen, Victoria M Rachleff, Meei-Li Huang, Keith R Jerome, Alexander Greninger |

| | | | |
|---|---|---|---|
| EPI_ISL_500831, EPI_ISL_500832, EPI_ISL_500833, EPI_ISL_500834, EPI_ISL_500835, EPI_ISL_500836, EPI_ISL_500837, EPI_ISL_500838, EPI_ISL_500839, EPI_ISL_500840, EPI_ISL_500841, EPI_ISL_500842, EPI_ISL_500843, EPI_ISL_500844, EPI_ISL_500845, EPI_ISL_500846, EPI_ISL_500847, EPI_ISL_500848, EPI_ISL_500849, EPI_ISL_500850, EPI_ISL_500851, EPI_ISL_500852, EPI_ISL_500853, EPI_ISL_500854, EPI_ISL_500855, EPI_ISL_500856, EPI_ISL_500857, EPI_ISL_500858, EPI_ISL_500859, EPI_ISL_500860, EPI_ISL_500861, EPI_ISL_500862, EPI_ISL_500863 | | | |
| see above | Virginia DCLS | Virginia DCLS | Virginia DCLS |
| EPI_ISL_501072, EPI_ISL_501073, EPI_ISL_501074, EPI_ISL_501075, EPI_ISL_501076, EPI_ISL_501077, EPI_ISL_501078, EPI_ISL_501079, EPI_ISL_501080, EPI_ISL_501081, EPI_ISL_501082 | | | |
| see above | Mayo Clinic Laboratories | University of Washington Virology Lab | Pavitra Roychoudhury, Hong Xie, Lasata Shrestha, Amin Addetia, Truong Nguyen, Victoria M Rachleff, Meei-Li Huang, Keith R Jerome, Alexander Greninger |
| EPI_ISL_501083, EPI_ISL_501084, EPI_ISL_501085, EPI_ISL_501086, EPI_ISL_501087, EPI_ISL_501088, EPI_ISL_501089, EPI_ISL_501090, EPI_ISL_501091, EPI_ISL_501092, EPI_ISL_501093, EPI_ISL_501094, EPI_ISL_501095, EPI_ISL_501096, EPI_ISL_501097, EPI_ISL_501098, EPI_ISL_501099, EPI_ISL_501100, EPI_ISL_501101, EPI_ISL_501102, EPI_ISL_501103, EPI_ISL_501104, EPI_ISL_501105, EPI_ISL_501106, EPI_ISL_501107, EPI_ISL_501108, EPI_ISL_501109, EPI_ISL_501110, EPI_ISL_501111, EPI_ISL_501112, EPI_ISL_501113, EPI_ISL_501114, EPI_ISL_501115, EPI_ISL_501116, EPI_ISL_501117, EPI_ISL_501118, EPI_ISL_501120, EPI_ISL_501121, EPI_ISL_501122, EPI_ISL_501123, EPI_ISL_501124, EPI_ISL_501125, EPI_ISL_501126, EPI_ISL_501127, EPI_ISL_501128, EPI_ISL_501129, EPI_ISL_501130, EPI_ISL_501131, EPI_ISL_501132, EPI_ISL_501133, EPI_ISL_501134, EPI_ISL_501135, EPI_ISL_501136, EPI_ISL_501137, EPI_ISL_501138, EPI_ISL_501139, EPI_ISL_501140, EPI_ISL_501141, EPI_ISL_501142, EPI_ISL_501143, EPI_ISL_501144, EPI_ISL_501145, EPI_ISL_501146, EPI_ISL_501147, EPI_ISL_501148, EPI_ISL_501149, EPI_ISL_501150, EPI_ISL_501151, EPI_ISL_501152, EPI_ISL_501153, EPI_ISL_501154, EPI_ISL_501155, EPI_ISL_501156, EPI_ISL_501157, EPI_ISL_501158, EPI_ISL_501159, EPI_ISL_501160, EPI_ISL_501161, EPI_ISL_501162, EPI_ISL_501163, EPI_ISL_501164 | | | |
| see above | University of Washington Virology Lab | University of Washington Virology Lab | Pavitra Roychoudhury, Hong Xie, Lasata Shrestha, Amin Addetia, Truong Nguyen, Victoria M Rachleff, Meei-Li Huang, Keith R Jerome, Alexander Greninger |
| EPI_ISL_501165 | Mayo Clinic Laboratories | University of Washington Virology Lab | Pavitra Roychoudhury, Hong Xie, Lasata Shrestha, Amin Addetia, Truong Nguyen, Victoria M Rachleff, Meei-Li Huang, Keith R Jerome, Alexander Greninger |
| EPI_ISL_501167, EPI_ISL_501168, EPI_ISL_501169, EPI_ISL_501171, EPI_ISL_501172, EPI_ISL_501173 | Baylor College of Medicine | Baylor College of Medicine: HGSC | Vasanthi Avadhanula, Erin Nicholson, David Henke, Pedro Piedra, Harsha Doddapaneni, Donna Muzny, Qingchang Meng, Hsu Chao, Zeineen Momin, Hua Shen, George Weissenberger, Kavya Kottapalli, Yimiti Meiheerguli, Sejal Salvi, Ginger Metcalf, Vipin Menon, Sara J.J. Cregeen, Matthew C. Ross, Tulin Ayvaz, Richard Sucgang, Kristi L. Hoffman, Matthew Wong, Joseph F. Petrosino |
| EPI_ISL_507419, EPI_ISL_507420, EPI_ISL_507421, EPI_ISL_507422, EPI_ISL_507423, EPI_ISL_507424, EPI_ISL_507425, EPI_ISL_507426, EPI_ISL_507427, EPI_ISL_507428, EPI_ISL_507429, EPI_ISL_507430, EPI_ISL_507431, EPI_ISL_507432, EPI_ISL_507433, EPI_ISL_507434, EPI_ISL_507435, EPI_ISL_507436, EPI_ISL_507437, EPI_ISL_507438, EPI_ISL_507439, EPI_ISL_507440, EPI_ISL_507441, EPI_ISL_507442, EPI_ISL_507443, EPI_ISL_507444, EPI_ISL_507445, EPI_ISL_507446, EPI_ISL_507449, EPI_ISL_507450, EPI_ISL_507451, EPI_ISL_507452, EPI_ISL_507453, EPI_ISL_507454, EPI_ISL_507455, EPI_ISL_507456, EPI_ISL_507457, EPI_ISL_507458, EPI_ISL_507459, EPI_ISL_507460, EPI_ISL_507461, EPI_ISL_507462, EPI_ISL_507463, EPI_ISL_507464, EPI_ISL_507465, EPI_ISL_507466, EPI_ISL_507467, EPI_ISL_507468, EPI_ISL_507469, EPI_ISL_507470, EPI_ISL_507471, EPI_ISL_507473, EPI_ISL_507475, EPI_ISL_507476, EPI_ISL_507477, EPI_ISL_507478, EPI_ISL_507479, EPI_ISL_507480, EPI_ISL_507481, EPI_ISL_507482, EPI_ISL_507483, EPI_ISL_507484, EPI_ISL_507485, EPI_ISL_507486, EPI_ISL_507487, EPI_ISL_507488, EPI_ISL_507489, EPI_ISL_507490, EPI_ISL_507491, EPI_ISL_507492, EPI_ISL_507493, EPI_ISL_507494, EPI_ISL_507495, EPI_ISL_507496, EPI_ISL_507500, EPI_ISL_507501, EPI_ISL_507502, EPI_ISL_507503, EPI_ISL_507504, EPI_ISL_507505, EPI_ISL_507506, EPI_ISL_507507, EPI_ISL_507508, EPI_ISL_507509, EPI_ISL_507510, EPI_ISL_507511, EPI_ISL_507512, EPI_ISL_507513, EPI_ISL_507514, EPI_ISL_507515, EPI_ISL_507516, EPI_ISL_507517, EPI_ISL_507518, EPI_ISL_507519, EPI_ISL_507520, EPI_ISL_507521, EPI_ISL_507522, EPI_ISL_507523, EPI_ISL_507524, EPI_ISL_507525, EPI_ISL_507526, EPI_ISL_507527, EPI_ISL_507528, EPI_ISL_507531, EPI_ISL_507532, EPI_ISL_507533, EPI_ISL_507534, EPI_ISL_507535, EPI_ISL_507536, EPI_ISL_507537, EPI_ISL_507538, EPI_ISL_507539, EPI_ISL_507540, EPI_ISL_507541, EPI_ISL_507542, EPI_ISL_507543, EPI_ISL_507544, EPI_ISL_507545, EPI_ISL_507546, EPI_ISL_507547, EPI_ISL_507548, EPI_ISL_507549, EPI_ISL_507550, EPI_ISL_507551, EPI_ISL_507552, EPI_ISL_507553, EPI_ISL_507554, EPI_ISL_507555, EPI_ISL_507556, EPI_ISL_507557, EPI_ISL_507558, EPI_ISL_507560, EPI_ISL_507562, EPI_ISL_507563, EPI_ISL_507564, EPI_ISL_507565, EPI_ISL_507566, EPI_ISL_507567, EPI_ISL_507568, EPI_ISL_507569, EPI_ISL_507570, EPI_ISL_507571, EPI_ISL_507572, EPI_ISL_507573, EPI_ISL_507574, EPI_ISL_507575, EPI_ISL_507576, EPI_ISL_507577, EPI_ISL_507578, EPI_ISL_507579, EPI_ISL_507580, EPI_ISL_507581, EPI_ISL_507582, EPI_ISL_507583, EPI_ISL_507584, EPI_ISL_507585, EPI_ISL_507586, EPI_ISL_507587, EPI_ISL_507588, EPI_ISL_507589, EPI_ISL_507590, EPI_ISL_507591, EPI_ISL_507592, EPI_ISL_507593, EPI_ISL_507594, EPI_ISL_507595, EPI_ISL_507596, EPI_ISL_507597, EPI_ISL_507598, EPI_ISL_507599, EPI_ISL_507600, EPI_ISL_507601, EPI_ISL_507602, EPI_ISL_507603, EPI_ISL_507604, EPI_ISL_507605, EPI_ISL_507606, EPI_ISL_507607, EPI_ISL_507608, EPI_ISL_507609, EPI_ISL_507610, EPI_ISL_507611, EPI_ISL_507612, EPI_ISL_507613, EPI_ISL_507614, EPI_ISL_507615, EPI_ISL_507616, EPI_ISL_507617, EPI_ISL_507618, EPI_ISL_507619, EPI_ISL_507620, EPI_ISL_507621, EPI_ISL_507622, EPI_ISL_507623, EPI_ISL_507633, EPI_ISL_507634, EPI_ISL_507635, EPI_ISL_507637, EPI_ISL_507638, EPI_ISL_507639, EPI_ISL_507643, EPI_ISL_507644, EPI_ISL_507645, EPI_ISL_507646, EPI_ISL_507647, EPI_ISL_507648, EPI_ISL_507649, EPI_ISL_507650, EPI_ISL_507651, EPI_ISL_507652, EPI_ISL_507653, EPI_ISL_507654, EPI_ISL_507655, EPI_ISL_507656, EPI_ISL_507657, EPI_ISL_507658, EPI_ISL_507659, EPI_ISL_507660, EPI_ISL_507661, EPI_ISL_507662, EPI_ISL_507663, EPI_ISL_507664, EPI_ISL_507665, EPI_ISL_507666, EPI_ISL_507667, EPI_ISL_507668, EPI_ISL_507669, EPI_ISL_507670, EPI_ISL_507671, EPI_ISL_507672, EPI_ISL_507673, EPI_ISL_507674, EPI_ISL_507675, EPI_ISL_507676, EPI_ISL_507677, EPI_ISL_507678, EPI_ISL_507679, EPI_ISL_507680, EPI_ISL_507681, EPI_ISL_507682, EPI_ISL_507683, EPI_ISL_507684, EPI_ISL_507685, EPI_ISL_507686, EPI_ISL_507687, EPI_ISL_507688, EPI_ISL_507689, EPI_ISL_507690, EPI_ISL_507691, EPI_ISL_507692, EPI_ISL_507693, EPI_ISL_507694, EPI_ISL_507695, EPI_ISL_507696, EPI_ISL_507697, EPI_ISL_507698, EPI_ISL_507699, EPI_ISL_507700, EPI_ISL_507701, EPI_ISL_507702, EPI_ISL_507703, EPI_ISL_507704, EPI_ISL_507705, EPI_ISL_507706, EPI_ISL_507707, EPI_ISL_507708, EPI_ISL_507709, EPI_ISL_507710, EPI_ISL_507711, EPI_ISL_507712, EPI_ISL_507713, EPI_ISL_507714, EPI_ISL_507715, EPI_ISL_507716, EPI_ISL_507717, EPI_ISL_507718, EPI_ISL_507719, EPI_ISL_507720, EPI_ISL_507725, EPI_ISL_507726, EPI_ISL_507727, EPI_ISL_507728, EPI_ISL_507729, EPI_ISL_507730, EPI_ISL_507731, EPI_ISL_507732, EPI_ISL_507733, EPI_ISL_507734, EPI_ISL_507735, EPI_ISL_507736, EPI_ISL_507737, EPI_ISL_507738, EPI_ISL_507739, EPI_ISL_507740, EPI_ISL_507741, EPI_ISL_507742, EPI_ISL_507743, EPI_ISL_507744, EPI_ISL_507745, EPI_ISL_507746, EPI_ISL_507747, EPI_ISL_507748, EPI_ISL_507750, EPI_ISL_507751, EPI_ISL_507752, EPI_ISL_507753, EPI_ISL_507754, EPI_ISL_507755, EPI_ISL_507756, EPI_ISL_507757, EPI_ISL_507758, EPI_ISL_507759, EPI_ISL_507760, EPI_ISL_507761, EPI_ISL_507762, EPI_ISL_507763, EPI_ISL_507764, EPI_ISL_507765, EPI_ISL_507766, EPI_ISL_507767, EPI_ISL_507768, EPI_ISL_507769, EPI_ISL_507770, EPI_ISL_507771, EPI_ISL_507772, EPI_ISL_507773, EPI_ISL_507774, EPI_ISL_507775, EPI_ISL_507776, EPI_ISL_507777, EPI_ISL_507778, EPI_ISL_507779, EPI_ISL_507780, EPI_ISL_507781, EPI_ISL_507782, EPI_ISL_507783, EPI_ISL_507784, EPI_ISL_507785, EPI_ISL_507786, EPI_ISL_507787, EPI_ISL_507788, EPI_ISL_507789, EPI_ISL_507802, EPI_ISL_507804, EPI_ISL_507808, EPI_ISL_507809, EPI_ISL_507810, EPI_ISL_507812, EPI_ISL_507813, EPI_ISL_507815, EPI_ISL_507816, EPI_ISL_507817, EPI_ISL_507818, EPI_ISL_507819, EPI_ISL_507820, EPI_ISL_507821, EPI_ISL_507822, EPI_ISL_507823, EPI_ISL_507824, EPI_ISL_507825, EPI_ISL_507826, EPI_ISL_507827, EPI_ISL_507828, EPI_ISL_507829, EPI_ISL_507831, EPI_ISL_507832, EPI_ISL_507833, EPI_ISL_507834, EPI_ISL_507835, EPI_ISL_507836, EPI_ISL_507837, EPI_ISL_507838, EPI_ISL_507839, EPI_ISL_507840, EPI_ISL_507841, EPI_ISL_507842, EPI_ISL_507843, EPI_ISL_507844, EPI_ISL_507845, EPI_ISL_507846, EPI_ISL_507847, EPI_ISL_507848, EPI_ISL_507849, EPI_ISL_507850, EPI_ISL_507851, EPI_ISL_507852, EPI_ISL_507853, EPI_ISL_507854, EPI_ISL_507855, EPI_ISL_507856, EPI_ISL_507857, EPI_ISL_507858, EPI_ISL_507859, EPI_ISL_507860, EPI_ISL_507861, EPI_ISL_507862, EPI_ISL_507863, EPI_ISL_507864, EPI_ISL_507865, EPI_ISL_507866, EPI_ISL_507867, EPI_ISL_507868, EPI_ISL_507869, EPI_ISL_507870, EPI_ISL_507871, EPI_ISL_507872, EPI_ISL_507873, EPI_ISL_507874, EPI_ISL_507875, EPI_ISL_507876, EPI_ISL_507877, EPI_ISL_507878, EPI_ISL_507879, EPI_ISL_507880, EPI_ISL_507881, EPI_ISL_507882, EPI_ISL_507883, EPI_ISL_507884, EPI_ISL_507885, EPI_ISL_507886, EPI_ISL_507887, EPI_ISL_507888, EPI_ISL_507889, EPI_ISL_507890, EPI_ISL_507891, EPI_ISL_507892, EPI_ISL_507893, EPI_ISL_507894, EPI_ISL_507895, EPI_ISL_507896, EPI_ISL_507897, EPI_ISL_507898, EPI_ISL_507899, EPI_ISL_507900, EPI_ISL_507901, EPI_ISL_507902, EPI_ISL_507903, EPI_ISL_507904, EPI_ISL_507905, EPI_ISL_507906, EPI_ISL_507907, EPI_ISL_507908, EPI_ISL_507909, EPI_ISL_507910, EPI_ISL_507911, EPI_ISL_507912, EPI_ISL_507913, EPI_ISL_507914, EPI_ISL_507915, EPI_ISL_507916, EPI_ISL_507917, EPI_ISL_507918, EPI_ISL_507919, EPI_ISL_507920, EPI_ISL_507921, EPI_ISL_507922, EPI_ISL_507923, EPI_ISL_507924, EPI_ISL_507925, EPI_ISL_507926, EPI_ISL_507927, EPI_ISL_507928, EPI_ISL_507929, EPI_ISL_507930, EPI_ISL_507931, EPI_ISL_507932, EPI_ISL_507933 | | | |
| see above | Michigan Department of Health and Human Services, Bureau of Laboratories | Michigan Department of Health and Human Services, Bureau of Laboratories | Blankenship HM, Riner D, Soehnlen MK |
| EPI_ISL_507934, EPI_ISL_507935, EPI_ISL_507936, EPI_ISL_507937, EPI_ISL_507938, EPI_ISL_507939, EPI_ISL_507940, EPI_ISL_507941, EPI_ISL_507942, EPI_ISL_507943, EPI_ISL_507944, EPI_ISL_507945, EPI_ISL_507946, EPI_ISL_507947, EPI_ISL_507948, EPI_ISL_507949, EPI_ISL_507950, EPI_ISL_507951, EPI_ISL_507952, EPI_ISL_507953, EPI_ISL_507954, EPI_ISL_507955, EPI_ISL_507956 | | | |
| see above | Minnesota Department of Health, Public Health Laboratory | Minnesota Department of Health, Public Health Laboratory | Matt Plumb, Jacob Garfin, and Xiong Wang |
| EPI_ISL_507957, EPI_ISL_507958, EPI_ISL_507959, EPI_ISL_507960, EPI_ISL_507961 | Mayo Clinic & Mayo Clinic Laboratories | Minnesota Department of Health, Public Health Laboratory | Matt Plumb, Jacob Garfin, and Xiong Wang |
| EPI_ISL_507962 | Children's Hospitals and Clinics of Minnesota | Minnesota Department of Health, Public Health Laboratory | Matt Plumb, Jacob Garfin, and Xiong Wang |
| EPI_ISL_507963, EPI_ISL_507964, EPI_ISL_507965, EPI_ISL_507966, EPI_ISL_507967, EPI_ISL_507968, EPI_ISL_507969, EPI_ISL_507970, EPI_ISL_507971, EPI_ISL_507972 | Avera Mckennan Laboratory | Minnesota Department of Health, Public Health Laboratory | Matt Plumb, Jacob Garfin, and Xiong Wang |
| EPI_ISL_507973, EPI_ISL_507974, EPI_ISL_507975, EPI_ISL_507976, EPI_ISL_507977, EPI_ISL_507978, EPI_ISL_507979, EPI_ISL_507980, EPI_ISL_507981, EPI_ISL_507982, EPI_ISL_507983 | | | |
| see above | Minnesota Department of Health, Public Health Laboratory | Minnesota Department of Health, Public Health Laboratory | Matt Plumb, Jacob Garfin, and Xiong Wang |
| EPI_ISL_507985, EPI_ISL_507986, EPI_ISL_507987, EPI_ISL_507989, EPI_ISL_507990, EPI_ISL_507991, EPI_ISL_507992, EPI_ISL_507994, EPI_ISL_507995, EPI_ISL_507999, EPI_ISL_508000, EPI_ISL_508002, EPI_ISL_508005, EPI_ISL_508008, EPI_ISL_508009, EPI_ISL_508013, EPI_ISL_508014, EPI_ISL_508018, EPI_ISL_508019, EPI_ISL_508022, EPI_ISL_508025, EPI_ISL_508026, EPI_ISL_508027, EPI_ISL_508029, EPI_ISL_508032, EPI_ISL_508035, EPI_ISL_508036, EPI_ISL_508039, EPI_ISL_508041, EPI_ISL_508047, EPI_ISL_508050, EPI_ISL_508052, EPI_ISL_508053, EPI_ISL_508055, EPI_ISL_508057, EPI_ISL_508058, EPI_ISL_508059, EPI_ISL_508060, EPI_ISL_508061, EPI_ISL_508062, EPI_ISL_508063, EPI_ISL_508066, EPI_ISL_508068, EPI_ISL_508069, EPI_ISL_508073, EPI_ISL_508075, EPI_ISL_508076, EPI_ISL_508079, EPI_ISL_508080, EPI_ISL_508081, EPI_ISL_508086, EPI_ISL_508087, EPI_ISL_508091, EPI_ISL_508092, EPI_ISL_508093, EPI_ISL_508097, EPI_ISL_508098, EPI_ISL_508099, EPI_ISL_508100, EPI_ISL_508101, EPI_ISL_508102, EPI_ISL_508106, EPI_ISL_508107, EPI_ISL_508109, EPI_ISL_508112, EPI_ISL_508121 | | | |
| see above | New Mexico Department of Health Scientific Laboratory Division | Center for Global Health, University of New Mexico Health Sciences Center | Daryl Domman, Kurt Schwalm, Twila Kunde, Joseph Hicks, Michael Edwards, Darrell Dinwiddie |
| EPI_ISL_508708, EPI_ISL_508709, EPI_ISL_508710, EPI_ISL_508711, EPI_ISL_508712, EPI_ISL_508713, EPI_ISL_508715, EPI_ISL_508716, EPI_ISL_508717, EPI_ISL_508718, EPI_ISL_508719, EPI_ISL_508720, EPI_ISL_508721, EPI_ISL_508722, EPI_ISL_508723, EPI_ISL_508724, EPI_ISL_508725, EPI_ISL_508726, EPI_ISL_508727, EPI_ISL_508728, EPI_ISL_508729, EPI_ISL_508730, EPI_ISL_508731, EPI_ISL_508732, EPI_ISL_508733, EPI_ISL_508734, EPI_ISL_508735, EPI_ISL_508736, EPI_ISL_508737, EPI_ISL_508739, EPI_ISL_508741, EPI_ISL_508742, EPI_ISL_508743, EPI_ISL_508744, EPI_ISL_508745, EPI_ISL_508746, EPI_ISL_508747, EPI_ISL_508748, EPI_ISL_508749, EPI_ISL_508750, EPI_ISL_508752, EPI_ISL_508753, EPI_ISL_508754, EPI_ISL_508755, EPI_ISL_508756, EPI_ISL_508757, EPI_ISL_508758, EPI_ISL_508759, EPI_ISL_508760, EPI_ISL_508761, EPI_ISL_508762, EPI_ISL_508763, EPI_ISL_508765, EPI_ISL_508766, EPI_ISL_508767, EPI_ISL_508769, EPI_ISL_508770, EPI_ISL_508771, EPI_ISL_508772, EPI_ISL_508773, EPI_ISL_508774, EPI_ISL_508775, EPI_ISL_508776, EPI_ISL_508777, EPI_ISL_508778, EPI_ISL_508779, EPI_ISL_508780, EPI_ISL_508781, EPI_ISL_508782, EPI_ISL_508783, EPI_ISL_508784, EPI_ISL_508785, EPI_ISL_508786, EPI_ISL_508787, EPI_ISL_508788, EPI_ISL_508789, EPI_ISL_508790, EPI_ISL_508791, EPI_ISL_508792, EPI_ISL_508793, EPI_ISL_508794, EPI_ISL_508795, EPI_ISL_508796, EPI_ISL_508797, EPI_ISL_508798, EPI_ISL_508799, EPI_ISL_508800, EPI_ISL_508801, EPI_ISL_508802, EPI_ISL_508803, EPI_ISL_508804, EPI_ISL_508805, EPI_ISL_508806, EPI_ISL_508807, EPI_ISL_508808, EPI_ISL_508813, EPI_ISL_508814, EPI_ISL_508819, EPI_ISL_508820, EPI_ISL_508821, EPI_ISL_508822, EPI_ISL_508827, EPI_ISL_508834, EPI_ISL_508835, EPI_ISL_508836, EPI_ISL_508837, EPI_ISL_508838, EPI_ISL_508839, EPI_ISL_508840, EPI_ISL_508841, EPI_ISL_508842, EPI_ISL_508845, EPI_ISL_508849, EPI_ISL_508852, EPI_ISL_508855, EPI_ISL_508860 | | | |
| see above | Florida Bureau of Public Health Laboratories | Florida Bureau of Public Health Laboratories | Sarah Schmedes, Jason Blanton |
| EPI_ISL_509059, EPI_ISL_509060, EPI_ISL_509061, EPI_ISL_509062, EPI_ISL_509063, EPI_ISL_509064, EPI_ISL_509065, EPI_ISL_509066, EPI_ISL_509067, EPI_ISL_509069, EPI_ISL_509070, EPI_ISL_509071, EPI_ISL_509072, EPI_ISL_509073, EPI_ISL_509074, EPI_ISL_509075, EPI_ISL_509076, EPI_ISL_509077, EPI_ISL_509078, EPI_ISL_509079, EPI_ISL_509080, EPI_ISL_509081, EPI_ISL_509082, EPI_ISL_509083, EPI_ISL_509084, EPI_ISL_509085, EPI_ISL_509086, EPI_ISL_509087, EPI_ISL_509088, EPI_ISL_509089, EPI_ISL_509090, EPI_ISL_509091, EPI_ISL_509092, EPI_ISL_509093, EPI_ISL_509094, EPI_ISL_509095, EPI_ISL_509096, EPI_ISL_509097, EPI_ISL_509099, EPI_ISL_509100, EPI_ISL_509101, EPI_ISL_509102, EPI_ISL_509103, EPI_ISL_509104, EPI_ISL_509105, EPI_ISL_509106, EPI_ISL_509107, EPI_ISL_509108, EPI_ISL_509109, EPI_ISL_509110, EPI_ISL_509111, EPI_ISL_509112, EPI_ISL_509113, EPI_ISL_509114, EPI_ISL_509115, EPI_ISL_509116, EPI_ISL_509117, EPI_ISL_509118, EPI_ISL_509119, EPI_ISL_509120, EPI_ISL_509121, EPI_ISL_509122, EPI_ISL_509123, EPI_ISL_509124, EPI_ISL_509125, EPI_ISL_509126, EPI_ISL_509127, EPI_ISL_509128, EPI_ISL_509129, EPI_ISL_509130, EPI_ISL_509131, EPI_ISL_509132, EPI_ISL_509133, EPI_ISL_509134, EPI_ISL_509135, EPI_ISL_509136, EPI_ISL_509137, EPI_ISL_509138, EPI_ISL_509139, EPI_ISL_509140, EPI_ISL_509141, EPI_ISL_509142, EPI_ISL_509143, EPI_ISL_509144, EPI_ISL_509145, EPI_ISL_509146, EPI_ISL_509147, EPI_ISL_509148, EPI_ISL_509149, EPI_ISL_509150, EPI_ISL_509151, EPI_ISL_509152, EPI_ISL_509153, EPI_ISL_509154, EPI_ISL_509155, EPI_ISL_509156, EPI_ISL_509157, EPI_ISL_509158, EPI_ISL_509159, EPI_ISL_509160, EPI_ISL_509161, EPI_ISL_509162, EPI_ISL_509163, EPI_ISL_509164, EPI_ISL_509165, EPI_ISL_509166, EPI_ISL_509167, EPI_ISL_509168, EPI_ISL_509169, EPI_ISL_509170, EPI_ISL_509171, EPI_ISL_509174, EPI_ISL_509175, EPI_ISL_509176, EPI_ISL_509177, EPI_ISL_509178, EPI_ISL_509179, EPI_ISL_509180, EPI_ISL_509181, EPI_ISL_509182, EPI_ISL_509183, EPI_ISL_509184, EPI_ISL_509185, EPI_ISL_509186, EPI_ISL_509187, EPI_ISL_509188, EPI_ISL_509189, EPI_ISL_509190, EPI_ISL_509191, EPI_ISL_509192, EPI_ISL_509193, EPI_ISL_509194, EPI_ISL_509195, EPI_ISL_509197, EPI_ISL_509198, EPI_ISL_509199, EPI_ISL_509200, EPI_ISL_509201, EPI_ISL_509202, EPI_ISL_509203, EPI_ISL_509204, EPI_ISL_509205, EPI_ISL_509206, EPI_ISL_509207, EPI_ISL_509208, EPI_ISL_509209, EPI_ISL_509210, EPI_ISL_509211, EPI_ISL_509212, EPI_ISL_509213, EPI_ISL_509214 | | | |
| see above | OHSU Lab Services Molecular Microbiology Lab | Oregon SARS-CoV-2 Genome Sequencing Center | Brendan L. O'Connell, Ruth V. Nichols, Sally B. Grindstaff, Alec J. Hirsch, Guang Fan, Daniel N. Streblow, William B. Messer, Andrew C. Adey, Benjamin N. Bimber, Brian J. O'Roak |
| EPI_ISL_509424, EPI_ISL_509425, EPI_ISL_509426, EPI_ISL_509427, EPI_ISL_509428, EPI_ISL_509429 | Microbiology and Immunology, University of South Alabama | Microbiology and Immunology, University of South Alabama | Wood,R.R., Roberts,R.A., Houserova,D., Borchert,G.M., Fouty,B., Rayner,J.O. |
| EPI_ISL_509444, EPI_ISL_509445, EPI_ISL_509446, EPI_ISL_509447, EPI_ISL_509448, EPI_ISL_509449, EPI_ISL_509450, EPI_ISL_509451, EPI_ISL_509452, EPI_ISL_509453, EPI_ISL_509454, EPI_ISL_509455, EPI_ISL_509456, EPI_ISL_509457, EPI_ISL_509458, EPI_ISL_509459, EPI_ISL_509460, EPI_ISL_509461, EPI_ISL_509462, EPI_ISL_509463, EPI_ISL_509464, EPI_ISL_509465, EPI_ISL_509466, EPI_ISL_509467, EPI_ISL_509468, EPI_ISL_509469, EPI_ISL_509470, EPI_ISL_509471, EPI_ISL_509472, EPI_ISL_509473, EPI_ISL_509474, EPI_ISL_509475, EPI_ISL_509476, EPI_ISL_509477, EPI_ISL_509478, EPI_ISL_509479, EPI_ISL_509480, EPI_ISL_509481, EPI_ISL_509482, EPI_ISL_509483, EPI_ISL_509484, EPI_ISL_509485, EPI_ISL_509486, EPI_ISL_509487, EPI_ISL_509488, EPI_ISL_509489, EPI_ISL_509490, EPI_ISL_509491 | | | |
| see above | Maryland Department of Health | Maryland Department of Health | Keller,E. |
| EPI_ISL_509527, EPI_ISL_509528, EPI_ISL_509529, EPI_ISL_509530, EPI_ISL_509531, EPI_ISL_509532, EPI_ISL_509533, EPI_ISL_509534, EPI_ISL_509535, EPI_ISL_509536, EPI_ISL_509537, EPI_ISL_509538, EPI_ISL_509539, EPI_ISL_509540, EPI_ISL_509541, EPI_ISL_509542, EPI_ISL_509543, EPI_ISL_509544, EPI_ISL_509545, EPI_ISL_509546, EPI_ISL_509547, EPI_ISL_509549, EPI_ISL_509550, EPI_ISL_509551, EPI_ISL_509552, EPI_ISL_509553, EPI_ISL_509554, EPI_ISL_509557, EPI_ISL_509558, EPI_ISL_509559, EPI_ISL_509560, EPI_ISL_509561, EPI_ISL_509562, EPI_ISL_509563, EPI_ISL_509567, EPI_ISL_509568, EPI_ISL_509569, EPI_ISL_509570, EPI_ISL_509571, EPI_ISL_509572, EPI_ISL_509573, EPI_ISL_509574, EPI_ISL_509575, EPI_ISL_509576, EPI_ISL_509577, EPI_ISL_509578, EPI_ISL_509579, EPI_ISL_509580, EPI_ISL_509581, EPI_ISL_509582, EPI_ISL_509583, EPI_ISL_509584, EPI_ISL_509585, EPI_ISL_509586, EPI_ISL_509587, EPI_ISL_509588, EPI_ISL_509589, EPI_ISL_509590, EPI_ISL_509591, EPI_ISL_509592, EPI_ISL_509593, EPI_ISL_509594, EPI_ISL_509595, EPI_ISL_509597, EPI_ISL_509598, EPI_ISL_509599 | | | |
| see above | Utah Public Health Laboratory | Utah Public Health Laboratory | Heidi Butz, Erin Young, Kelly Oakeson |
| EPI_ISL_509662 | Georgia Department of Health | Pathogen Discovery, Respiratory Viruses | Yan Li, Anna Montmayeur, Jing Zhang, Krista Queen, Anna Uehara, Ying Tao, Rachel Marine, Clinton R. Paden, Haibin Wang, Suxiang Tong |

| | | | |
|---|---|---|---|
| | | Branch, Division of Viral Diseases, Centers for Disease Control and Prevention | |
| EPI_ISL_509663, EPI_ISL_509664, EPI_ISL_509665, EPI_ISL_509666, EPI_ISL_509667, EPI_ISL_509668, EPI_ISL_509670, EPI_ISL_509671, EPI_ISL_509672, EPI_ISL_509673, EPI_ISL_509674, EPI_ISL_509675, EPI_ISL_509676, EPI_ISL_509677, EPI_ISL_509678, EPI_ISL_509679, EPI_ISL_509680, EPI_ISL_509681, EPI_ISL_509682 | | | |
| see above | AR Dept. of Health-Public Health Lab | Pathogen Discovery, Respiratory Viruses Branch, Division of Viral Diseases, Centers for Disease Control and Prevention | Yan Li, Anna Montmayeur, Jing Zhang, Krista Queen, Anna Uehara, Ying Tao, Rachel Marine, Clinton R. Paden, Haibin Wang, Suxiang Tong |
| EPI_ISL_509683, EPI_ISL_509684, EPI_ISL_509685 | AR Dept. of Health-Public Health Lab | Pathogen Discovery, Respiratory Viruses Branch, Division of Viral Diseases, Centers for Disease Control and Prevention | Jing Zhang, Yan Li, Anna Montmayeur, Krista Queen, Anna Uehara, Ying Tao, Rachel Marine, Clinton R. Paden, Haibin Wang, Suxiang Tong |
| EPI_ISL_509686 | M Health Fairview | Minnesota Department of Health, Public Health Laboratory | Matt Plumb, Jacob Garfin, and Xiong Wang |
| EPI_ISL_509688 | Alabama Department of Public Health Bureau of Clinical Laboratories | Pathogen Discovery, Respiratory Viruses Branch, Division of Viral Diseases, Centers for Disease Control and Prevention | Ying Tao, Jing Zhang, Krista Queen, Anna Uehara, Yan Li, Clinton Paden, Haibin Wang, Suxiang Tong |
| EPI_ISL_509689, EPI_ISL_509690, EPI_ISL_509691, EPI_ISL_509692, EPI_ISL_509693, EPI_ISL_509694 | Utah Public Health Laboratory | Pathogen Discovery, Respiratory Viruses Branch, Division of Viral Diseases, Centers for Disease Control and Prevention | Ying Tao, Jing Zhang, Krista Queen, Anna Uehara, Yan Li, Clinton Paden, Haibin Wang, Suxiang Tong |
| EPI_ISL_509704, EPI_ISL_509705, EPI_ISL_509706 | Wisconsin Department of Health Services | Pathogen Discovery, Respiratory Viruses Branch, Division of Viral Diseases, Centers for Disease Control and Prevention | Ying Tao, Jing Zhang, Krista Queen, Anna Uehara, Yan Li, Clinton Paden, Haibin Wang, Suxiang Tong |
| EPI_ISL_509707, EPI_ISL_509708, EPI_ISL_509709 | Utah Public Health Laboratory | Pathogen Discovery, Respiratory Viruses Branch, Division of Viral Diseases, Centers for Disease Control and Prevention | Jing Zhang, Ying Tao, Krista Queen, Anna Uehara, Yan Li, Clinton Paden, Haibin Wang, Suxiang Tong |
| EPI_ISL_509715 | Wisconsin Department of Health Services | Pathogen Discovery, Respiratory Viruses Branch, Division of Viral Diseases, Centers for Disease Control and Prevention | Jing Zhang, Ying Tao, Krista Queen, Anna Uehara, Yan Li, Clinton Paden, Haibin Wang, Suxiang Tong |
| EPI_ISL_509716, EPI_ISL_509717, EPI_ISL_509718, EPI_ISL_509719, EPI_ISL_509720, EPI_ISL_509721, EPI_ISL_509722, EPI_ISL_509723, EPI_ISL_509724, EPI_ISL_509725, EPI_ISL_509726, EPI_ISL_509727, EPI_ISL_509728, EPI_ISL_509729, EPI_ISL_509730, EPI_ISL_509731, EPI_ISL_509732, EPI_ISL_509733, EPI_ISL_509734, EPI_ISL_509735, EPI_ISL_509736, EPI_ISL_509737, EPI_ISL_509738, EPI_ISL_509739, EPI_ISL_509740, EPI_ISL_509741, EPI_ISL_509742, EPI_ISL_509743, EPI_ISL_509744, EPI_ISL_509745, EPI_ISL_509746, EPI_ISL_509747, EPI_ISL_509748, EPI_ISL_509749, EPI_ISL_509750, EPI_ISL_509751, EPI_ISL_509752, EPI_ISL_509753, EPI_ISL_509754, EPI_ISL_509755, EPI_ISL_509756, EPI_ISL_509757, EPI_ISL_509758, EPI_ISL_509759, EPI_ISL_509760, EPI_ISL_509761, EPI_ISL_509762, EPI_ISL_509763, EPI_ISL_509764, EPI_ISL_509765, EPI_ISL_509766, EPI_ISL_509767, EPI_ISL_509768, EPI_ISL_509769, EPI_ISL_509770, EPI_ISL_509771, EPI_ISL_509772, EPI_ISL_509773, EPI_ISL_509774, EPI_ISL_509775, EPI_ISL_509776, EPI_ISL_509777, EPI_ISL_509778, EPI_ISL_509779, EPI_ISL_509780, EPI_ISL_509781, EPI_ISL_509782, EPI_ISL_509783, EPI_ISL_509784, EPI_ISL_509785, EPI_ISL_509786, EPI_ISL_509787, EPI_ISL_509788, EPI_ISL_509789, EPI_ISL_509791, EPI_ISL_509792, EPI_ISL_509793, EPI_ISL_509794, EPI_ISL_509795, EPI_ISL_509796, EPI_ISL_509797, EPI_ISL_509798 | | | |
| see above | Florida Bureau of Public Health Laboratories | Florida Bureau of Public Health Laboratories | Sarah Schmedes, Jason Blanton |
| EPI_ISL_509799, EPI_ISL_509800, EPI_ISL_509801, EPI_ISL_509802, EPI_ISL_509803, EPI_ISL_509804, EPI_ISL_509805, EPI_ISL_509806, EPI_ISL_509807, EPI_ISL_509808, EPI_ISL_509809, EPI_ISL_509810, EPI_ISL_509811, EPI_ISL_509812, EPI_ISL_509813, EPI_ISL_509814, EPI_ISL_509815, EPI_ISL_509816, EPI_ISL_509817, EPI_ISL_509818, EPI_ISL_509819, EPI_ISL_509820, EPI_ISL_509821, EPI_ISL_509822, EPI_ISL_509823, EPI_ISL_509824, EPI_ISL_509825, EPI_ISL_509826, EPI_ISL_509827, EPI_ISL_509828, EPI_ISL_509829, EPI_ISL_509830, EPI_ISL_509831, EPI_ISL_509832, EPI_ISL_509833, EPI_ISL_509834, EPI_ISL_509835, EPI_ISL_509836, EPI_ISL_509837, EPI_ISL_509838, EPI_ISL_509839, EPI_ISL_509840, EPI_ISL_509841, EPI_ISL_509842, EPI_ISL_509843, EPI_ISL_509844, EPI_ISL_509845, EPI_ISL_509846, EPI_ISL_509847, EPI_ISL_509848, EPI_ISL_509849, EPI_ISL_509850, EPI_ISL_509851, EPI_ISL_509852, EPI_ISL_509853, EPI_ISL_509854, EPI_ISL_509855, EPI_ISL_509856, EPI_ISL_509857, EPI_ISL_509859, EPI_ISL_509860, EPI_ISL_509861, EPI_ISL_509862, EPI_ISL_509863, EPI_ISL_509864, EPI_ISL_509865, EPI_ISL_509866, EPI_ISL_509867, EPI_ISL_509868, EPI_ISL_509869, EPI_ISL_509870, EPI_ISL_509871, EPI_ISL_509872, EPI_ISL_509873, EPI_ISL_509874, EPI_ISL_509875, EPI_ISL_509876, EPI_ISL_509877, EPI_ISL_509878, EPI_ISL_509879, EPI_ISL_509880, EPI_ISL_509881, EPI_ISL_509882, EPI_ISL_509883, EPI_ISL_509884, EPI_ISL_509885, EPI_ISL_509886, EPI_ISL_509887, EPI_ISL_509888, EPI_ISL_509889, EPI_ISL_509890, EPI_ISL_509891, EPI_ISL_509892, EPI_ISL_509893, EPI_ISL_509894, EPI_ISL_509895, EPI_ISL_509896, EPI_ISL_509897, EPI_ISL_509898, EPI_ISL_509899, EPI_ISL_509900, EPI_ISL_509901, EPI_ISL_509902, EPI_ISL_509903, EPI_ISL_509904, EPI_ISL_509905, EPI_ISL_509906, EPI_ISL_509907, EPI_ISL_509908, EPI_ISL_509909, EPI_ISL_509910, EPI_ISL_509911, EPI_ISL_509912, EPI_ISL_509914, EPI_ISL_509915, EPI_ISL_509916, EPI_ISL_509917, EPI_ISL_509918, EPI_ISL_509919, EPI_ISL_509920, EPI_ISL_509921, EPI_ISL_509922, EPI_ISL_509923, EPI_ISL_509924, EPI_ISL_509925, EPI_ISL_509926, EPI_ISL_509927, EPI_ISL_509928, EPI_ISL_509929, EPI_ISL_509930, EPI_ISL_509931, EPI_ISL_509932, EPI_ISL_509933, EPI_ISL_509934, EPI_ISL_509935, EPI_ISL_509936, EPI_ISL_509937, EPI_ISL_509938, EPI_ISL_509939, EPI_ISL_509940, EPI_ISL_509941, EPI_ISL_509942, EPI_ISL_509943, EPI_ISL_509944, EPI_ISL_509945, EPI_ISL_509946, EPI_ISL_509947, EPI_ISL_509948, EPI_ISL_509949, EPI_ISL_509950, EPI_ISL_509951, EPI_ISL_509952, EPI_ISL_509953, EPI_ISL_509954, EPI_ISL_509955, EPI_ISL_509956, EPI_ISL_509957, EPI_ISL_509958, EPI_ISL_509959, EPI_ISL_509960, EPI_ISL_509961, EPI_ISL_509962, EPI_ISL_509963, EPI_ISL_509964, EPI_ISL_509965, EPI_ISL_509966, EPI_ISL_509967, EPI_ISL_509968, EPI_ISL_509969, EPI_ISL_509970, EPI_ISL_509971, EPI_ISL_509972, EPI_ISL_509973, EPI_ISL_509974, EPI_ISL_509975, EPI_ISL_509976, EPI_ISL_509977, EPI_ISL_509978, EPI_ISL_509979, EPI_ISL_509980, EPI_ISL_509981, EPI_ISL_509982, EPI_ISL_509983, EPI_ISL_509984, EPI_ISL_509985, EPI_ISL_509986, EPI_ISL_509987, EPI_ISL_509988, EPI_ISL_509989, EPI_ISL_509990, EPI_ISL_509991, EPI_ISL_509992, EPI_ISL_509993, EPI_ISL_509994, EPI_ISL_509995, EPI_ISL_509996, EPI_ISL_509997, EPI_ISL_509998, EPI_ISL_510524, EPI_ISL_510525 | | | |
| see above | University of Wisconsin-Madison AIDS Vaccine Research Laboratories | University of Wisconsin-Madison AIDS Vaccine Research Laboratories | Gage Moreno, Katarina Braun, et al. AIDS Vaccine Research Laboratories |
| EPI_ISL_510878, EPI_ISL_510879, EPI_ISL_510880, EPI_ISL_510881, EPI_ISL_510882, EPI_ISL_510883, EPI_ISL_510884, EPI_ISL_510885, EPI_ISL_510886, EPI_ISL_510887, EPI_ISL_510888 | | | |
| see above | New Mexico Department of Health Scientific Laboratory | New Mexico Department of Health Scientific Laboratory | Ellie Johnson, Anastacia Griego-Fisher |
| EPI_ISL_511805, EPI_ISL_511806, EPI_ISL_511807, EPI_ISL_511808, EPI_ISL_511809, EPI_ISL_511810, EPI_ISL_511811, EPI_ISL_511812, EPI_ISL_511813, EPI_ISL_511814, EPI_ISL_511815, EPI_ISL_511816, EPI_ISL_511817, EPI_ISL_511818, EPI_ISL_511819, EPI_ISL_511820, EPI_ISL_511821, EPI_ISL_511822, EPI_ISL_511823, EPI_ISL_511824, EPI_ISL_511825, EPI_ISL_511826, EPI_ISL_511827, EPI_ISL_511828, EPI_ISL_511829, EPI_ISL_511830, EPI_ISL_511831, EPI_ISL_511832, EPI_ISL_511833, EPI_ISL_511834, EPI_ISL_511835, EPI_ISL_511836, EPI_ISL_511837, EPI_ISL_511838, EPI_ISL_511839, EPI_ISL_511840, EPI_ISL_511841, EPI_ISL_511842, EPI_ISL_511843, EPI_ISL_511845, EPI_ISL_511846, EPI_ISL_511847, EPI_ISL_511848, EPI_ISL_511849, EPI_ISL_511850, EPI_ISL_511851 | | | |
| see above | Innovative Genomics Institute, UC Berkeley | Innovative Genomics Institute, UC Berkeley | Stacia Wyman, Haridha Shivram, Liana Lareau, Shana McDevitt, Justin Choi |
| EPI_ISL_511852, EPI_ISL_511853, EPI_ISL_511854, EPI_ISL_511855, EPI_ISL_511856, EPI_ISL_511857, EPI_ISL_511858, EPI_ISL_511859, EPI_ISL_511860, EPI_ISL_511861 | UW Virology Lab | UW Virology Lab | Pavitra Roychoudhury, Amin Addetia, Hong Xie, Lasata Shrestha, Truong Nguyen, Meei-Li Huang, Keith Jerome, Alexander Greninger |
| EPI_ISL_511862 | Innovative Genomics Institute, UC Berkeley | Innovative Genomics Institute, UC Berkeley | Stacia Wyman, Haridha Shivram, Liana Lareau, Shana McDevitt, Justin Choi |
| EPI_ISL_511863, EPI_ISL_511865, EPI_ISL_511866, EPI_ISL_511868, EPI_ISL_511869, EPI_ISL_511870, EPI_ISL_511871, EPI_ISL_511872, EPI_ISL_511873, EPI_ISL_511874, EPI_ISL_511875, EPI_ISL_511876, EPI_ISL_511877 | | | |
| see above | Johns Hopkins Hospital Department of Pathology | Johns Hopkins Hospital Department of Pathology | Peter M. Thielen, Thomas Mehoke, Shirlee Wohl, Srividya Ramakrishnan, Melanie Kirsche, Amanda Ernlund, Craig Howser, Kristina Zudock, Oluwaseun Falade-Nwulia, Norah Sadowski, Paul Morris, Mark Hopkins, Yunfan Fan, Nidia Trovao, Victoria Gniazdowski, Michael C. Schatz, Stuart C. Ray, Winston Timp, Heba H. Mostafa |
| EPI_ISL_512086 | UW Virology lab | UW Virology lab | Pavitra Roychoudhury, Amin Addetia, Hong Xie, Lasata Shrestha, Truong Nguyen, Meei-Li Huang, Keith Jerome, Alexander Greninger |
| EPI_ISL_512135, EPI_ISL_512136, EPI_ISL_512137, EPI_ISL_512138, EPI_ISL_512140, EPI_ISL_512141, EPI_ISL_512142, EPI_ISL_512143, EPI_ISL_512144, EPI_ISL_512145, EPI_ISL_512146, EPI_ISL_512147, EPI_ISL_512148, EPI_ISL_512149, EPI_ISL_512150, EPI_ISL_512151, EPI_ISL_512152, EPI_ISL_512153, EPI_ISL_512154, EPI_ISL_512155, EPI_ISL_512156, EPI_ISL_512157 | | | |
| see above | Alaska State Virology Laboratory | Alaska State Virology Laboratory | Chen J et al with Pathogenomics group Dagdag R, Redlinger M, Milton E, George W, Kovalenko A, Drown DM, Bortz E |
| EPI_ISL_512158, EPI_ISL_512159, EPI_ISL_512160, EPI_ISL_512161, EPI_ISL_512162, EPI_ISL_512163, EPI_ISL_512164, EPI_ISL_512165, EPI_ISL_512166, EPI_ISL_512167, EPI_ISL_512168, EPI_ISL_512169, EPI_ISL_512170, EPI_ISL_512171, EPI_ISL_512172, EPI_ISL_512173, EPI_ISL_512174, EPI_ISL_512175, EPI_ISL_512176, EPI_ISL_512177, EPI_ISL_512178, EPI_ISL_512179, EPI_ISL_512180, EPI_ISL_512181, EPI_ISL_512182, EPI_ISL_512183, EPI_ISL_512184, EPI_ISL_512185, EPI_ISL_512186, EPI_ISL_512187, EPI_ISL_512188, EPI_ISL_512189, EPI_ISL_512190, EPI_ISL_512191, EPI_ISL_512192, EPI_ISL_512193, EPI_ISL_512194, EPI_ISL_512195, EPI_ISL_512196, EPI_ISL_512197, EPI_ISL_512198, EPI_ISL_512199, EPI_ISL_512200, EPI_ISL_512201, EPI_ISL_512202, EPI_ISL_512203, EPI_ISL_512204, EPI_ISL_512205, EPI_ISL_512206, EPI_ISL_512207, EPI_ISL_512208, EPI_ISL_512209, EPI_ISL_512210, EPI_ISL_512211, EPI_ISL_512212, EPI_ISL_512213, EPI_ISL_512214, EPI_ISL_512215, EPI_ISL_512216, EPI_ISL_512217, EPI_ISL_512218, EPI_ISL_512219, EPI_ISL_512220, EPI_ISL_512221, EPI_ISL_512222, EPI_ISL_512223, EPI_ISL_512224, EPI_ISL_512225, EPI_ISL_512226, EPI_ISL_512227, EPI_ISL_512228, EPI_ISL_512229, EPI_ISL_512230, EPI_ISL_512231, EPI_ISL_512232, EPI_ISL_512233, EPI_ISL_512234, EPI_ISL_512235, EPI_ISL_512236, EPI_ISL_512237, EPI_ISL_512238, EPI_ISL_512239, EPI_ISL_512240, EPI_ISL_512241, EPI_ISL_512242, EPI_ISL_512243, EPI_ISL_512244, EPI_ISL_512245, EPI_ISL_512246, EPI_ISL_512247, EPI_ISL_512248, EPI_ISL_512249, EPI_ISL_512250, EPI_ISL_512251, EPI_ISL_512252, EPI_ISL_512253, EPI_ISL_512254, EPI_ISL_512255, EPI_ISL_512256, EPI_ISL_512257, EPI_ISL_512258, EPI_ISL_512259, EPI_ISL_512260, EPI_ISL_512261, EPI_ISL_512262, EPI_ISL_512263, EPI_ISL_512264, EPI_ISL_512265, EPI_ISL_512266, EPI_ISL_512267, EPI_ISL_512268, EPI_ISL_512269, EPI_ISL_512270, EPI_ISL_512271, EPI_ISL_512272, EPI_ISL_512273 | | | |
| see above | San Diego County Public Health Laboratory | Andersen lab at Scripps Research | SEARCH Alliance San Diego with Tracy Basler, Jovan Shephard, Brett Austin |
| EPI_ISL_512547, EPI_ISL_512548, EPI_ISL_512549, EPI_ISL_512550, EPI_ISL_512551, EPI_ISL_512552, EPI_ISL_512553, EPI_ISL_512554, EPI_ISL_512555, EPI_ISL_512556, EPI_ISL_512557, EPI_ISL_512558, EPI_ISL_512559, EPI_ISL_512560, EPI_ISL_512561, EPI_ISL_512562, EPI_ISL_512563, EPI_ISL_512564, EPI_ISL_512565, EPI_ISL_512566, EPI_ISL_512567, EPI_ISL_512568, EPI_ISL_512569, EPI_ISL_512570, EPI_ISL_512571, EPI_ISL_512572, EPI_ISL_512573, EPI_ISL_512574, EPI_ISL_512575, EPI_ISL_512576, EPI_ISL_512578, EPI_ISL_512579, EPI_ISL_512582, EPI_ISL_512585, EPI_ISL_512586, EPI_ISL_512593, EPI_ISL_512676, EPI_ISL_512677 | | | |
| see above | Florida Bureau of Public Health Laboratories | Florida Bureau of Public Health Laboratories | Sarah Schmedes, Jason Blanton |
| EPI_ISL_512785, EPI_ISL_512786, EPI_ISL_512787, EPI_ISL_512788, EPI_ISL_512789, EPI_ISL_512790, EPI_ISL_512791, EPI_ISL_512792, EPI_ISL_512793, EPI_ISL_512794, EPI_ISL_512795, EPI_ISL_512796, EPI_ISL_512797, EPI_ISL_512798, EPI_ISL_512799, EPI_ISL_512800, EPI_ISL_512801, EPI_ISL_512802, EPI_ISL_512803, EPI_ISL_512804, EPI_ISL_512805, EPI_ISL_512806, EPI_ISL_512807, EPI_ISL_512808, EPI_ISL_512809 | | | |
| see above | Public Health, United States Air Force School of Aerospace Medicine | Public Health, United States Air Force School of Aerospace Medicine | Fries,A.C., Purves,S.M., Meyer,J.R., Javorina,A.K., Connors,B.C., Macias,E.A., Lambert,A.W., Chapleau,R.R., Starr,C.R. |
| EPI_ISL_513265, EPI_ISL_513266, EPI_ISL_513267, EPI_ISL_513268, EPI_ISL_513269, EPI_ISL_513270, EPI_ISL_513271, EPI_ISL_513272, EPI_ISL_513273, EPI_ISL_513274, EPI_ISL_513275, EPI_ISL_513276, EPI_ISL_513277, EPI_ISL_513278, EPI_ISL_513279, EPI_ISL_513280, EPI_ISL_513281, EPI_ISL_513282, EPI_ISL_513283, EPI_ISL_513284, EPI_ISL_513285, EPI_ISL_513286, EPI_ISL_513287, EPI_ISL_513288, EPI_ISL_513289, EPI_ISL_513290, EPI_ISL_513291, EPI_ISL_513292, EPI_ISL_513293 | | | |
| see above | University of Miami Immunology and Histocompatibility Laboratory | University of Miami Immunology and Histocompatibility Laboratory | Emilio Margolles-Clark, PhD and Phillip Ruiz, MD, PhD |
| EPI_ISL_513294 | Baylor College of Medicine | Baylor College of Medicine: HGSC | Vasanthi Avadhanula, Erin Nicholson, David Henke, Pedro Piedra, Harsha Doddapaneni, Donna Muzny, Qingchang Meng, Hsu Chao, Zeineen Momin, Hua Shen, George Weissenberger, Kavya Kottapalli, Yimiti Meiheerguli, Sejal Salvi, Ginger Metcalf, Vipin Menon, Sara J.J. Cregeen, Matthew C. Ross, Tulin Ayvaz, Richard Sucgang, Kristi L. Hoffman, Matthew Wong, Joseph F. Petrosino |
| EPI_ISL_513414, EPI_ISL_513415, EPI_ISL_513416, EPI_ISL_513417, EPI_ISL_513418, EPI_ISL_513419, EPI_ISL_513420, EPI_ISL_513421, EPI_ISL_513422, EPI_ISL_513423, EPI_ISL_513424, EPI_ISL_513425, EPI_ISL_513426, EPI_ISL_513427, EPI_ISL_513428, EPI_ISL_513429, EPI_ISL_513430, EPI_ISL_513431, EPI_ISL_513432, EPI_ISL_513433, EPI_ISL_513434, EPI_ISL_513435, EPI_ISL_513436, EPI_ISL_513437, EPI_ISL_513438, EPI_ISL_513439, EPI_ISL_513440, EPI_ISL_513441, EPI_ISL_513442, EPI_ISL_513443, EPI_ISL_513444, EPI_ISL_513445, EPI_ISL_513446, EPI_ISL_513447, EPI_ISL_513448, EPI_ISL_513449, EPI_ISL_513450, EPI_ISL_513451, EPI_ISL_513452, EPI_ISL_513453, EPI_ISL_513454, EPI_ISL_513455, EPI_ISL_513456, EPI_ISL_513457, EPI_ISL_513458, EPI_ISL_513459, EPI_ISL_513460, EPI_ISL_513461, EPI_ISL_513462, EPI_ISL_513463, EPI_ISL_513464, EPI_ISL_513465, EPI_ISL_513466, EPI_ISL_513467, EPI_ISL_513468, EPI_ISL_513469, EPI_ISL_513470, EPI_ISL_513471, EPI_ISL_513472, EPI_ISL_513473, EPI_ISL_513474, EPI_ISL_513475, EPI_ISL_513476, EPI_ISL_513477, EPI_ISL_513478, EPI_ISL_513479, EPI_ISL_513480, EPI_ISL_513481, EPI_ISL_513482, EPI_ISL_513483, EPI_ISL_513484, EPI_ISL_513485, | | | |

EPI_ISL_513486, EPI_ISL_513487, EPI_ISL_513488, EPI_ISL_513489, EPI_ISL_513490, EPI_ISL_513491, EPI_ISL_513492, EPI_ISL_513493, EPI_ISL_513494, EPI_ISL_513495, EPI_ISL_513496, EPI_ISL_513497, EPI_ISL_513498, EPI_ISL_513499, EPI_ISL_513500, EPI_ISL_513501, EPI_ISL_513502, EPI_ISL_513503, EPI_ISL_513504, EPI_ISL_513505, EPI_ISL_513506, EPI_ISL_513507, EPI_ISL_513508, EPI_ISL_513509, EPI_ISL_513510

| | | | |
|---|---|---|---|
| see above | Maine HETL | Tewhey Lab, The Jackson Laboratory | Matluk,N., Dewey,H., Barter,M., Lynch,R., Munger,H. and Tewhey,R. |
| EPI_ISL_513632, EPI_ISL_513633, EPI_ISL_513634, EPI_ISL_513635, EPI_ISL_513636, EPI_ISL_513637, EPI_ISL_513638 | University of Washington Virology Lab | University of Washington Virology Lab | Pavitra Roychoudhury, Hong Xie, Lasata Shrestha, Amin Addetia, Truong Nguyen, Victoria M Rachleff, Meei-Li Huang, Keith R Jerome, Alexander Greninger |

EPI_ISL_513641, EPI_ISL_513642, EPI_ISL_513643, EPI_ISL_513644, EPI_ISL_513645, EPI_ISL_513647, EPI_ISL_513648, EPI_ISL_513649, EPI_ISL_513651, EPI_ISL_513652, EPI_ISL_513653, EPI_ISL_513654, EPI_ISL_513655, EPI_ISL_513656, EPI_ISL_513657, EPI_ISL_513658, EPI_ISL_513659, EPI_ISL_513661, EPI_ISL_513662, EPI_ISL_513663, EPI_ISL_513664, EPI_ISL_513665, EPI_ISL_513666, EPI_ISL_513667, EPI_ISL_513668, EPI_ISL_513669, EPI_ISL_513670, EPI_ISL_513671, EPI_ISL_513672, EPI_ISL_513673, EPI_ISL_513675, EPI_ISL_513676, EPI_ISL_513677, EPI_ISL_513678, EPI_ISL_513680, EPI_ISL_513681, EPI_ISL_513682, EPI_ISL_513683, EPI_ISL_513684, EPI_ISL_513685, EPI_ISL_513686, EPI_ISL_513687, EPI_ISL_513688, EPI_ISL_513689, EPI_ISL_513690, EPI_ISL_513691, EPI_ISL_513692, EPI_ISL_513693, EPI_ISL_513694, EPI_ISL_513696, EPI_ISL_513697, EPI_ISL_513698, EPI_ISL_513699, EPI_ISL_513700, EPI_ISL_513701, EPI_ISL_513702, EPI_ISL_513704, EPI_ISL_513705

| | | | |
|---|---|---|---|
| see above | Utah Public Health Laboratory | Utah Public Health Laboratory | Heidi Butz, Erin Young, Kelly Oakeson |

EPI_ISL_513707, EPI_ISL_513708, EPI_ISL_513709, EPI_ISL_513710, EPI_ISL_513711, EPI_ISL_513712, EPI_ISL_513713, EPI_ISL_513714, EPI_ISL_513715, EPI_ISL_513716, EPI_ISL_513717, EPI_ISL_513718, EPI_ISL_513719, EPI_ISL_513720, EPI_ISL_513721, EPI_ISL_513722, EPI_ISL_513723, EPI_ISL_513724, EPI_ISL_513725, EPI_ISL_513726, EPI_ISL_513727, EPI_ISL_513728, EPI_ISL_513729, EPI_ISL_513730, EPI_ISL_513731, EPI_ISL_513732, EPI_ISL_513733, EPI_ISL_513734, EPI_ISL_513735, EPI_ISL_513736, EPI_ISL_513737, EPI_ISL_513738, EPI_ISL_513739, EPI_ISL_513740, EPI_ISL_513741, EPI_ISL_513742, EPI_ISL_513743, EPI_ISL_513744, EPI_ISL_513745, EPI_ISL_513746, EPI_ISL_513747, EPI_ISL_513748, EPI_ISL_513749, EPI_ISL_513750, EPI_ISL_513751, EPI_ISL_513752, EPI_ISL_513753, EPI_ISL_513754, EPI_ISL_513755, EPI_ISL_513756, EPI_ISL_513757, EPI_ISL_513758, EPI_ISL_513760, EPI_ISL_513761, EPI_ISL_513762, EPI_ISL_513763, EPI_ISL_513764, EPI_ISL_513765, EPI_ISL_513766, EPI_ISL_513767, EPI_ISL_513768, EPI_ISL_513769, EPI_ISL_513770, EPI_ISL_513771, EPI_ISL_513772

| | | | |
|---|---|---|---|
| see above | Orange County Public Health Lab | Chan-Zuckerberg Biohub | CZB Cliahub Consortium |

EPI_ISL_513773, EPI_ISL_513774, EPI_ISL_513775, EPI_ISL_513776, EPI_ISL_513777, EPI_ISL_513778, EPI_ISL_513779, EPI_ISL_513780, EPI_ISL_513781, EPI_ISL_513782, EPI_ISL_513783, EPI_ISL_513784, EPI_ISL_513785, EPI_ISL_513786, EPI_ISL_513787, EPI_ISL_513788

| | | | |
|---|---|---|---|
| see above | County of Santa Clara Public Health Department | Chan-Zuckerberg Biohub | CZB Cliahub Consortium |

EPI_ISL_513789, EPI_ISL_513790, EPI_ISL_513791, EPI_ISL_513792, EPI_ISL_513793, EPI_ISL_513794, EPI_ISL_513795, EPI_ISL_513796, EPI_ISL_513797, EPI_ISL_513798, EPI_ISL_513799, EPI_ISL_513800, EPI_ISL_513801, EPI_ISL_513802, EPI_ISL_513803, EPI_ISL_513804, EPI_ISL_513805, EPI_ISL_513806, EPI_ISL_513807, EPI_ISL_513808, EPI_ISL_513809, EPI_ISL_513810, EPI_ISL_513811, EPI_ISL_513812, EPI_ISL_513813, EPI_ISL_513814, EPI_ISL_513815, EPI_ISL_513816, EPI_ISL_513817, EPI_ISL_513818, EPI_ISL_513819, EPI_ISL_513820, EPI_ISL_513821, EPI_ISL_513822, EPI_ISL_513823, EPI_ISL_513824, EPI_ISL_513825, EPI_ISL_513826, EPI_ISL_513827, EPI_ISL_513828, EPI_ISL_513829, EPI_ISL_513830, EPI_ISL_513831, EPI_ISL_513832, EPI_ISL_513833, EPI_ISL_513834, EPI_ISL_513835, EPI_ISL_513836, EPI_ISL_513837, EPI_ISL_513838, EPI_ISL_513839

| | | | |
|---|---|---|---|
| see above | Orange County Public Health Laboratory | Chan-Zuckerberg Biohub | CZB Cliahub Consortium |

EPI_ISL_513840, EPI_ISL_513841, EPI_ISL_513842, EPI_ISL_513843, EPI_ISL_513844, EPI_ISL_513845, EPI_ISL_513846, EPI_ISL_513847, EPI_ISL_513848, EPI_ISL_513849, EPI_ISL_513850, EPI_ISL_513851, EPI_ISL_513852, EPI_ISL_513853, EPI_ISL_513854, EPI_ISL_513855

| | | | |
|---|---|---|---|
| see above | Humboldt County Public Health Laboratory | Chan-Zuckerberg Biohub | CZB Cliahub Consortium |

EPI_ISL_513856, EPI_ISL_513857, EPI_ISL_513858, EPI_ISL_513859, EPI_ISL_513860, EPI_ISL_513861, EPI_ISL_513862, EPI_ISL_513863, EPI_ISL_513864, EPI_ISL_513865, EPI_ISL_513866, EPI_ISL_513867, EPI_ISL_513868, EPI_ISL_513869, EPI_ISL_513870, EPI_ISL_513871, EPI_ISL_513872, EPI_ISL_513873, EPI_ISL_513874, EPI_ISL_513875, EPI_ISL_513876, EPI_ISL_513877, EPI_ISL_513878, EPI_ISL_513879, EPI_ISL_513880, EPI_ISL_513881, EPI_ISL_513882

| | | | |
|---|---|---|---|
| see above | San Francisco Public Health Laboratory | Chan-Zuckerberg Biohub | CZB Cliahub Consortium |

EPI_ISL_513883, EPI_ISL_513884, EPI_ISL_513885, EPI_ISL_513886, EPI_ISL_513887, EPI_ISL_513888, EPI_ISL_513889, EPI_ISL_513891, EPI_ISL_513892, EPI_ISL_513893, EPI_ISL_513894, EPI_ISL_513895, EPI_ISL_513896, EPI_ISL_513897, EPI_ISL_513898, EPI_ISL_513899, EPI_ISL_513900, EPI_ISL_513901, EPI_ISL_513902, EPI_ISL_513903, EPI_ISL_513904, EPI_ISL_513905, EPI_ISL_513906, EPI_ISL_513907, EPI_ISL_513908, EPI_ISL_513909, EPI_ISL_513910

| | | | |
|---|---|---|---|
| see above | UCSF Clinical Microbiology Laboratory | Chan-Zuckerberg Biohub | CZB Cliahub Consortium |
| EPI_ISL_514062, EPI_ISL_514063, EPI_ISL_514064 | Navy and Marine Corps Public Health Center | Pathogen Discovery, Respiratory Viruses Branch, Division of Viral Diseases, Centers for Disease Control and Prevention | Yan Li, Anna Montmayeur, Krista Queen, Jing Zhang, Ying Tao, Anna Uehara, Rachel Marine, Clinton R. Paden, Haibin Wang, Suxiang Tong |
| EPI_ISL_514065 | Navy and Marine Corps Public Health Center | Pathogen Discovery, Respiratory Viruses Branch, Division of Viral Diseases, Centers for Disease Control and Prevention | Krista Queen, Yan Li, Anna Montmayeur, Jing Zhang, Ying Tao, Anna Uehara, Rachel Marine, Clinton R. Paden, Haibin Wang, Suxiang Tong |
| EPI_ISL_514066 | Navy and Marine Corps Public Health Center | Pathogen Discovery, Respiratory Viruses Branch, Division of Viral Diseases, Centers for Disease Control and Prevention | Yan Li, Anna Montmayeur, Krista Queen, Jing Zhang, Ying Tao, Anna Uehara, Rachel Marine, Clinton R. Paden, Haibin Wang, Suxiang Tong |
| EPI_ISL_514067, EPI_ISL_514068 | Navy and Marine Corps Public Health Center | Pathogen Discovery, Respiratory Viruses Branch, Division of Viral Diseases, Centers for Disease Control and Prevention | Krista Queen, Yan Li, Anna Montmayeur, Jing Zhang, Ying Tao, Anna Uehara, Rachel Marine, Clinton R. Paden, Haibin Wang, Suxiang Tong |
| EPI_ISL_514069, EPI_ISL_514070 | Navy and Marine Corps Public Health Center | Pathogen Discovery, Respiratory Viruses Branch, Division of Viral Diseases, Centers for Disease Control and Prevention | Yan Li, Anna Montmayeur, Krista Queen, Jing Zhang, Ying Tao, Anna Uehara, Rachel Marine, Clinton R. Paden, Haibin Wang, Suxiang Tong |
| EPI_ISL_514071, EPI_ISL_514072 | Navy and Marine Corps Public Health Center | Pathogen Discovery, Respiratory Viruses Branch, Division of Viral Diseases, Centers for Disease Control and Prevention | Krista Queen, Yan Li, Anna Montmayeur, Jing Zhang, Ying Tao, Anna Uehara, Rachel Marine, Clinton R. Paden, Haibin Wang, Suxiang Tong |
| EPI_ISL_514073, EPI_ISL_514074 | Navy and Marine Corps Public Health Center | Pathogen Discovery, Respiratory Viruses Branch, Division of Viral Diseases, Centers for Disease Control and Prevention | Yan Li, Anna Montmayeur, Krista Queen, Jing Zhang, Ying Tao, Anna Uehara, Rachel Marine, Clinton R. Paden, Haibin Wang, Suxiang Tong |
| EPI_ISL_514075, EPI_ISL_514076 | Navy and Marine Corps Public Health Center | Pathogen Discovery, Respiratory Viruses Branch, Division of Viral Diseases, Centers for Disease Control and Prevention | Krista Queen, Yan Li, Anna Montmayeur, Jing Zhang, Ying Tao, Anna Uehara, Rachel Marine, Clinton R. Paden, Haibin Wang, Suxiang Tong |
| EPI_ISL_514077, EPI_ISL_514078, EPI_ISL_514079 | Navy and Marine Corps Public Health Center | Pathogen Discovery, Respiratory Viruses Branch, Division of Viral Diseases, Centers for Disease Control and Prevention | Yan Li, Anna Montmayeur, Krista Queen, Jing Zhang, Ying Tao, Anna Uehara, Rachel Marine, Clinton R. Paden, Haibin Wang, Suxiang Tong |
| EPI_ISL_514080, EPI_ISL_514081, EPI_ISL_514082, EPI_ISL_514083 | Navy and Marine Corps Public Health Center | Pathogen Discovery, Respiratory Viruses Branch, Division of Viral Diseases, Centers for Disease Control and Prevention | Krista Queen, Yan Li, Anna Montmayeur, Jing Zhang, Ying Tao, Anna Uehara, Rachel Marine, Clinton R. Paden, Haibin Wang, Suxiang Tong |
| EPI_ISL_514084 | Navy and Marine Corps Public Health Center | Pathogen Discovery, Respiratory Viruses Branch, Division of Viral Diseases, Centers for Disease Control and Prevention | Yan Li, Anna Montmayeur, Krista Queen, Jing Zhang, Ying Tao, Anna Uehara, Rachel Marine, Clinton R. Paden, Haibin Wang, Suxiang Tong |
| EPI_ISL_514085, EPI_ISL_514086, EPI_ISL_514087 | Navy and Marine Corps Public Health Center | Pathogen Discovery, Respiratory Viruses Branch, Division of Viral Diseases, Centers for Disease Control and Prevention | Krista Queen, Yan Li, Anna Montmayeur, Jing Zhang, Ying Tao, Anna Uehara, Rachel Marine, Clinton R. Paden, Haibin Wang, Suxiang Tong |
| EPI_ISL_514088 | Navy and Marine Corps Public Health Center | Pathogen Discovery, Respiratory Viruses Branch, Division of Viral Diseases, Centers for Disease Control and Prevention | Yan Li, Anna Montmayeur, Krista Queen, Jing Zhang, Ying Tao, Anna Uehara, Rachel Marine, Clinton R. Paden, Haibin Wang, Suxiang Tong |
| EPI_ISL_514089 | Navy and Marine Corps Public Health Center | Pathogen Discovery, Respiratory Viruses Branch, Division of Viral Diseases, Centers for Disease Control and Prevention | Krista Queen, Yan Li, Anna Montmayeur, Jing Zhang, Ying Tao, Anna Uehara, Rachel Marine, Clinton R. Paden, Haibin Wang, Suxiang Tong |
| EPI_ISL_514090, EPI_ISL_514091, EPI_ISL_514092 | Navy and Marine Corps Public Health Center | Pathogen Discovery, Respiratory Viruses Branch, Division of Viral Diseases, Centers for Disease Control and Prevention | Yan Li, Anna Montmayeur, Krista Queen, Jing Zhang, Ying Tao, Anna Uehara, Rachel Marine, Clinton R. Paden, Haibin Wang, Suxiang Tong |
| EPI_ISL_514128 | Navy and Marine Corps Public Health Center | Centers for Disease Control and Prevention | Krista Queen, Yan Li, Anna Montmayeur, Jing Zhang, Ying Tao, Anna Uehara, Rachel Marine, Clinton R. Paden, Haibin Wang, Suxiang Tong |

EPI_ISL_514140, EPI_ISL_514141, EPI_ISL_514142, EPI_ISL_514143, EPI_ISL_514144, EPI_ISL_514145, EPI_ISL_514146, EPI_ISL_514147, EPI_ISL_514148, EPI_ISL_514149, EPI_ISL_514151, EPI_ISL_514152, EPI_ISL_514153, EPI_ISL_514155, EPI_ISL_514156, EPI_ISL_514157, EPI_ISL_514158, EPI_ISL_514159, EPI_ISL_514160, EPI_ISL_514161, EPI_ISL_514162, EPI_ISL_514163, EPI_ISL_514168, EPI_ISL_514169, EPI_ISL_514170, EPI_ISL_514171, EPI_ISL_514174, EPI_ISL_514175, EPI_ISL_514176, EPI_ISL_514177, EPI_ISL_514178, EPI_ISL_514179, EPI_ISL_514180, EPI_ISL_514181, EPI_ISL_514182, EPI_ISL_514183, EPI_ISL_514184, EPI_ISL_514185, EPI_ISL_514186, EPI_ISL_514187, EPI_ISL_514188, EPI_ISL_514189, EPI_ISL_514192, EPI_ISL_514194, EPI_ISL_514197, EPI_ISL_514200, EPI_ISL_514201, EPI_ISL_514202, EPI_ISL_514203, EPI_ISL_514204, EPI_ISL_514205, EPI_ISL_514207, EPI_ISL_514208, EPI_ISL_514209, EPI_ISL_514210, EPI_ISL_514211, EPI_ISL_514212, EPI_ISL_514213, EPI_ISL_514214, EPI_ISL_514215, EPI_ISL_514216, EPI_ISL_514218, EPI_ISL_514219, EPI_ISL_514220, EPI_ISL_514221, EPI_ISL_514222, EPI_ISL_514223, EPI_ISL_514224, EPI_ISL_514225

| | | | |
|---|---|---|---|
| see above | Florida Bureau of Public Health Laboratories | Florida Bureau of Public Health Laboratories | Sarah Schmedes, Jason Blanton |
| EPI_ISL_514438, EPI_ISL_514439 | Tewhey Lab, The Jackson Laboratory | The Jackson Laboratory | Matluk,N., Dewey,H., Barter,M., Lynch,R., Munger,H., Tewhey,R. |
| EPI_ISL_514616, EPI_ISL_514617, EPI_ISL_514618 | Minnesota Department of Health, Public Health Laboratory | Minnesota Department of Health, Public Health Laboratory | Matt Plumb, Jacob Garfin, and Xiong Wang |
| EPI_ISL_514619, EPI_ISL_514620, | Mayo Clinic & Mayo Clinic Laboratories | Minnesota Department of Health, Public Health Laboratory | Matt Plumb, Jacob Garfin, and Xiong Wang |

| | | | |
|---|---|---|---|
| EPI_ISL_514621, EPI_ISL_514622, EPI_ISL_514623, EPI_ISL_514624, EPI_ISL_514625, EPI_ISL_514626, EPI_ISL_514627, EPI_ISL_514628 | | | |
| EPI_ISL_514629, EPI_ISL_514630, EPI_ISL_514631, EPI_ISL_514632, EPI_ISL_514633, EPI_ISL_514634, EPI_ISL_514635 | M Health Fairview | Minnesota Department of Health, Public Health Laboratory | Matt Plumb, Jacob Garfin, and Xiong Wang |
| EPI_ISL_514636 | Mayo Clinic & Mayo Clinic Laboratories | Minnesota Department of Health, Public Health Laboratory | Matt Plumb, Jacob Garfin, and Xiong Wang |
| EPI_ISL_514637 | M Health Fairview St. Joseph's Hospital | Minnesota Department of Health, Public Health Laboratory | Matt Plumb, Jacob Garfin, and Xiong Wang |
| EPI_ISL_514638, EPI_ISL_514639, EPI_ISL_514640 | Mayo Clinic & Mayo Clinic Laboratories | Minnesota Department of Health, Public Health Laboratory | Matt Plumb, Jacob Garfin, and Xiong Wang |
| EPI_ISL_514641 | St. Luke's Hospital | Minnesota Department of Health, Public Health Laboratory | Matt Plumb, Jacob Garfin, and Xiong Wang |
| EPI_ISL_514642 | Essentia Health-St. Mary's Medical Center | Minnesota Department of Health, Public Health Laboratory | Matt Plumb, Jacob Garfin, and Xiong Wang |
| EPI_ISL_514643 | M Health Fairview St. Joseph's Hospital | Minnesota Department of Health, Public Health Laboratory | Matt Plumb, Jacob Garfin, and Xiong Wang |
| EPI_ISL_514644 | Mayo Clinic & Mayo Clinic Laboratories | Minnesota Department of Health, Public Health Laboratory | Matt Plumb, Jacob Garfin, and Xiong Wang |
| EPI_ISL_514645 | Avera Mckennan Laboratory | Minnesota Department of Health, Public Health Laboratory | Matt Plumb, Jacob Garfin, and Xiong Wang |
| EPI_ISL_514646 | St. Luke's Hospital | Minnesota Department of Health, Public Health Laboratory | Matt Plumb, Jacob Garfin, and Xiong Wang |
| EPI_ISL_514647 | Allina Health Laboratory | Minnesota Department of Health, Public Health Laboratory | Matt Plumb, Jacob Garfin, and Xiong Wang |
| EPI_ISL_514648 | Essentia Health-St. Joseph's Medical Center | Minnesota Department of Health, Public Health Laboratory | Matt Plumb, Jacob Garfin, and Xiong Wang |
| EPI_ISL_514649 | Allina Health Laboratory | Minnesota Department of Health, Public Health Laboratory | Matt Plumb, Jacob Garfin, and Xiong Wang |
| EPI_ISL_514650 | M Health Fairview St. Joseph's Hospital | Minnesota Department of Health, Public Health Laboratory | Matt Plumb, Jacob Garfin, and Xiong Wang |
| EPI_ISL_514651 | Allina Health Laboratory | Minnesota Department of Health, Public Health Laboratory | Matt Plumb, Jacob Garfin, and Xiong Wang |
| EPI_ISL_514652 | M Health Fairview St. Joseph's Hospital | Minnesota Department of Health, Public Health Laboratory | Matt Plumb, Jacob Garfin, and Xiong Wang |
| EPI_ISL_514653, EPI_ISL_514654, EPI_ISL_514655, EPI_ISL_514656 | Mayo Clinic & Mayo Clinic Laboratories | Minnesota Department of Health, Public Health Laboratory | Matt Plumb, Jacob Garfin, and Xiong Wang |
| EPI_ISL_514657 | M Health Fairview St. Joseph's Hospital | Minnesota Department of Health, Public Health Laboratory | Matt Plumb, Jacob Garfin, and Xiong Wang |
| EPI_ISL_514658, EPI_ISL_514659, EPI_ISL_514660, EPI_ISL_514661, EPI_ISL_514662, EPI_ISL_514663, EPI_ISL_514664, EPI_ISL_514665, EPI_ISL_514666, EPI_ISL_514667, EPI_ISL_514668, EPI_ISL_514669, EPI_ISL_514670, EPI_ISL_514671, EPI_ISL_514672 | | | |
| see above | Minnesota Department of Health, Public Health Laboratory | Minnesota Department of Health, Public Health Laboratory | Matt Plumb, Jacob Garfin, and Xiong Wang |
| EPI_ISL_514673 | Nevada State Public Health Laboratory | Nevada State Public Health Laboratory | Richard Tillet, Joel R. Sevinsky, Paul Hartley, Heather Kerwin, David Jackson, Subhash C. Verma, Cyprian Rosetto, Andrew Gorzalski, Chris Laverdure, Natalie Crawford, Stephanie Van Hooser, and Mark Pandori |
| EPI_ISL_515250, EPI_ISL_515251, EPI_ISL_515252, EPI_ISL_515253, EPI_ISL_515254, EPI_ISL_515255, EPI_ISL_515256, EPI_ISL_515257, EPI_ISL_515258, EPI_ISL_515259, EPI_ISL_515260, EPI_ISL_515261 | | | |
| see above | Texas Department of State Health Services | Texas Department of State Health Services | Rashmi Tuladhar, Bonnie Oh, Cara Akrout, Jenny Zhang, Maliha Rahman, Anita Pokharel, Myong Koag, Chun Wang, Rachel Lee, Grace Kubin |
| EPI_ISL_515262 | Minnesota Department of Health, Public Health Laboratory | Minnesota Department of Health, Public Health Laboratory | Matt Plumb, Jacob Garfin, and Xiong Wang |
| EPI_ISL_515263, EPI_ISL_515264, EPI_ISL_515265, EPI_ISL_515266 | Mayo Clinic & Mayo Clinic Laboratories | Minnesota Department of Health, Public Health Laboratory | Matt Plumb, Jacob Garfin, and Xiong Wang |
| EPI_ISL_515267, EPI_ISL_515268 | M Health Fairview | Minnesota Department of Health, Public Health Laboratory | Matt Plumb, Jacob Garfin, and Xiong Wang |
| EPI_ISL_515269 | Minnesota Department of Health, Public Health Laboratory | Minnesota Department of Health, Public Health Laboratory | Matt Plumb, Jacob Garfin, and Xiong Wang |
| EPI_ISL_515270, EPI_ISL_515271, EPI_ISL_515272, EPI_ISL_515273, EPI_ISL_515274, EPI_ISL_515275, EPI_ISL_515276, EPI_ISL_515277, EPI_ISL_515278, EPI_ISL_515279, EPI_ISL_515280, EPI_ISL_515281, EPI_ISL_515282, EPI_ISL_515283, EPI_ISL_515284, EPI_ISL_515285, EPI_ISL_515286 | | | |
| see above | University of Washington Virology Lab | University of Washington Virology Lab | Pavitra Roychoudhury, Hong Xie, Lasata Shrestha, Amin Addetia, Truong Nguyen, Victoria M Rachleff, Meei-Li Huang, Keith R Jerome, Alexander Greninger |
| EPI_ISL_515293, EPI_ISL_515294, EPI_ISL_515295, EPI_ISL_515296, EPI_ISL_515297, EPI_ISL_515298, EPI_ISL_515299, EPI_ISL_515300, EPI_ISL_515301, EPI_ISL_515302, EPI_ISL_515303, EPI_ISL_515304, EPI_ISL_515305, EPI_ISL_515306, EPI_ISL_515307, EPI_ISL_515308, EPI_ISL_515309, EPI_ISL_515310, EPI_ISL_515311, EPI_ISL_515312, EPI_ISL_515313, EPI_ISL_515314, EPI_ISL_515315, EPI_ISL_515316, EPI_ISL_515317, EPI_ISL_515318, EPI_ISL_515319, EPI_ISL_515320, EPI_ISL_515321, EPI_ISL_515322, EPI_ISL_515323, EPI_ISL_515324, EPI_ISL_515325, EPI_ISL_515327, EPI_ISL_515328, EPI_ISL_515329, EPI_ISL_515330, EPI_ISL_515331, EPI_ISL_515332, EPI_ISL_515333, EPI_ISL_515334, EPI_ISL_515336, EPI_ISL_515337, EPI_ISL_515338, EPI_ISL_515339, EPI_ISL_515340, EPI_ISL_515341, EPI_ISL_515342, EPI_ISL_515344, EPI_ISL_515345, EPI_ISL_515346, EPI_ISL_515347, EPI_ISL_515348, EPI_ISL_515349, EPI_ISL_515350, EPI_ISL_515351, EPI_ISL_515352, EPI_ISL_515353, EPI_ISL_515354, EPI_ISL_515355, EPI_ISL_515356, EPI_ISL_515357, EPI_ISL_515358, EPI_ISL_515359, EPI_ISL_515360, EPI_ISL_515361, EPI_ISL_515362, EPI_ISL_515363, EPI_ISL_515364, EPI_ISL_515365, EPI_ISL_515366, EPI_ISL_515367, EPI_ISL_515368, EPI_ISL_515369, EPI_ISL_515370, EPI_ISL_515371, EPI_ISL_515372, EPI_ISL_515373, EPI_ISL_515374, EPI_ISL_515375, EPI_ISL_515376, EPI_ISL_515377, EPI_ISL_515378, EPI_ISL_515379, EPI_ISL_515381, EPI_ISL_515382, EPI_ISL_515384, EPI_ISL_515385, EPI_ISL_515386, EPI_ISL_515387, EPI_ISL_515388, EPI_ISL_515389, EPI_ISL_515390, EPI_ISL_515391, EPI_ISL_515392, EPI_ISL_515393, EPI_ISL_515394, EPI_ISL_515397, EPI_ISL_515399, EPI_ISL_515400, EPI_ISL_515401, EPI_ISL_515403, EPI_ISL_515404, EPI_ISL_515405, EPI_ISL_515406, EPI_ISL_515407, EPI_ISL_515408, EPI_ISL_515409, EPI_ISL_515410, EPI_ISL_515411, EPI_ISL_515412, EPI_ISL_515413, EPI_ISL_515414, EPI_ISL_515415, EPI_ISL_515416, EPI_ISL_515417, EPI_ISL_515418, EPI_ISL_515419, EPI_ISL_515420, EPI_ISL_515421, EPI_ISL_515422, EPI_ISL_515423, EPI_ISL_515424, EPI_ISL_515425, EPI_ISL_515426, EPI_ISL_515427, EPI_ISL_515428, EPI_ISL_515429, EPI_ISL_515430, EPI_ISL_515431, EPI_ISL_515432, EPI_ISL_515433, EPI_ISL_515434, EPI_ISL_515436, EPI_ISL_515437, EPI_ISL_515438, EPI_ISL_515439, EPI_ISL_515440, EPI_ISL_515441, EPI_ISL_515442, EPI_ISL_515443, EPI_ISL_515444, EPI_ISL_515445, EPI_ISL_515446, EPI_ISL_515447, EPI_ISL_515448, EPI_ISL_515449, EPI_ISL_515450, EPI_ISL_515451, EPI_ISL_515452, EPI_ISL_515453, EPI_ISL_515454, EPI_ISL_515455, EPI_ISL_515456, EPI_ISL_515457, EPI_ISL_515458, EPI_ISL_515459, EPI_ISL_515460, EPI_ISL_515461, EPI_ISL_515462 | | | |
| see above | Nevada State Public Health Laboratory | Nevada State Public Health Laboratory | Richard Tillett, Joel R. Sevinsky, Paul Hartley, Heather Kerwin, David Jackson, Subhash C. Verma, Cyprian Rossetto, Andrew Gorzalski, Chris Laverdure, Natalie Crawford, Stephanie Van Hooser, and Mark Pandori |
| EPI_ISL_515894, EPI_ISL_515895, EPI_ISL_515896, EPI_ISL_515897, EPI_ISL_515898, EPI_ISL_515899, EPI_ISL_515900, EPI_ISL_515901, EPI_ISL_515902, EPI_ISL_515903, EPI_ISL_515905, EPI_ISL_515906, EPI_ISL_515907, EPI_ISL_515908, EPI_ISL_515909, EPI_ISL_515910, EPI_ISL_515911, EPI_ISL_515912, EPI_ISL_515913, EPI_ISL_515914, EPI_ISL_515915, EPI_ISL_515916, EPI_ISL_515917, EPI_ISL_515918, EPI_ISL_515919, EPI_ISL_515920, EPI_ISL_515921, EPI_ISL_515922, EPI_ISL_515923, EPI_ISL_515924, EPI_ISL_515925, EPI_ISL_515926, EPI_ISL_515927, EPI_ISL_515928, EPI_ISL_515929 | | | |
| see above | California Department of Public Health | California Department of Public Health | CDPH IDLB COVIDNet |
| EPI_ISL_516194, EPI_ISL_516195, EPI_ISL_516196, EPI_ISL_516197, EPI_ISL_516198, EPI_ISL_516199, EPI_ISL_516200, EPI_ISL_516201, EPI_ISL_516202, EPI_ISL_516203, EPI_ISL_516205, EPI_ISL_516206, EPI_ISL_516207, EPI_ISL_516209, EPI_ISL_516211, EPI_ISL_516212, EPI_ISL_516213, EPI_ISL_516214, EPI_ISL_516215, EPI_ISL_516216, EPI_ISL_516217, EPI_ISL_516219, EPI_ISL_516220, EPI_ISL_516221, EPI_ISL_516222, EPI_ISL_516223, EPI_ISL_516224, EPI_ISL_516225, EPI_ISL_516226, EPI_ISL_516227, EPI_ISL_516228, EPI_ISL_516229, EPI_ISL_516230, EPI_ISL_516231, EPI_ISL_516232, EPI_ISL_516233, EPI_ISL_516238, EPI_ISL_516239, EPI_ISL_516240, EPI_ISL_516241, EPI_ISL_516242, EPI_ISL_516243, EPI_ISL_516244, EPI_ISL_516245, EPI_ISL_516246, EPI_ISL_516247, EPI_ISL_516249, EPI_ISL_516250, EPI_ISL_516251, EPI_ISL_516252, EPI_ISL_516253, EPI_ISL_516254, EPI_ISL_516255, EPI_ISL_516256, EPI_ISL_516258, EPI_ISL_516259, EPI_ISL_516260, EPI_ISL_516261, EPI_ISL_516262, EPI_ISL_516263, EPI_ISL_516264, EPI_ISL_516265, EPI_ISL_516266, EPI_ISL_516267, EPI_ISL_516268, EPI_ISL_516269, EPI_ISL_516270, EPI_ISL_516271, EPI_ISL_516272, EPI_ISL_516273, EPI_ISL_516274, EPI_ISL_516275, EPI_ISL_516276, EPI_ISL_516277, EPI_ISL_516278, EPI_ISL_516279, EPI_ISL_516280, EPI_ISL_516281, EPI_ISL_516282, EPI_ISL_516283, EPI_ISL_516284, EPI_ISL_516285, EPI_ISL_516286, EPI_ISL_516287, EPI_ISL_516288, EPI_ISL_516290, EPI_ISL_516291, EPI_ISL_516292, EPI_ISL_516294, EPI_ISL_516295, EPI_ISL_516296, EPI_ISL_516297, EPI_ISL_516298, EPI_ISL_516299, EPI_ISL_516300, EPI_ISL_516301, EPI_ISL_516302, EPI_ISL_516303, EPI_ISL_516304, EPI_ISL_516305, EPI_ISL_516306, EPI_ISL_516307, EPI_ISL_516308, EPI_ISL_516309, EPI_ISL_516310, EPI_ISL_516311, EPI_ISL_516312, EPI_ISL_516314, EPI_ISL_516316, EPI_ISL_516317, EPI_ISL_516318, EPI_ISL_516320, EPI_ISL_516321, EPI_ISL_516322, EPI_ISL_516324, EPI_ISL_516325, EPI_ISL_516326, EPI_ISL_516327, EPI_ISL_516328, EPI_ISL_516329, EPI_ISL_516330, EPI_ISL_516331, EPI_ISL_516332, EPI_ISL_516333, EPI_ISL_516334, EPI_ISL_516335, EPI_ISL_516336, EPI_ISL_516337, EPI_ISL_516338, EPI_ISL_516339, EPI_ISL_516343, EPI_ISL_516344, EPI_ISL_516345, EPI_ISL_516346, EPI_ISL_516347, EPI_ISL_516348, EPI_ISL_516351, EPI_ISL_516352, EPI_ISL_516353, EPI_ISL_516354, EPI_ISL_516355, EPI_ISL_516356, EPI_ISL_516357, EPI_ISL_516358, EPI_ISL_516359, EPI_ISL_516360, EPI_ISL_516361, EPI_ISL_516362, EPI_ISL_516363, EPI_ISL_516365, EPI_ISL_516366, EPI_ISL_516367, EPI_ISL_516368, EPI_ISL_516369, EPI_ISL_516371, EPI_ISL_516372, EPI_ISL_516373, EPI_ISL_516374, EPI_ISL_516375, EPI_ISL_516376, EPI_ISL_516378, EPI_ISL_516379, EPI_ISL_516380, EPI_ISL_516384, EPI_ISL_516385, EPI_ISL_516386, EPI_ISL_516387, EPI_ISL_516388, EPI_ISL_516389, EPI_ISL_516390, EPI_ISL_516391, EPI_ISL_516392, EPI_ISL_516393, EPI_ISL_516394, EPI_ISL_516395, EPI_ISL_516396, EPI_ISL_516397, EPI_ISL_516398, EPI_ISL_516399, EPI_ISL_516400, EPI_ISL_516401, EPI_ISL_516402, EPI_ISL_516403, EPI_ISL_516404, EPI_ISL_516405, EPI_ISL_516406, EPI_ISL_516407, EPI_ISL_516408, EPI_ISL_516409, EPI_ISL_516410, EPI_ISL_516411, EPI_ISL_516412 | | | |
| see above | Michigan Department of Health and Human Services, Bureau of Laboratories | Michigan Department of Health and Human Services, Bureau of Laboratories | Blankenship HM, Riner D, Soehnlen MK |
| EPI_ISL_516432, EPI_ISL_516433, EPI_ISL_516434, EPI_ISL_516435, EPI_ISL_516436, EPI_ISL_516437, EPI_ISL_516438, EPI_ISL_516439, EPI_ISL_516440, EPI_ISL_516441, EPI_ISL_516442, EPI_ISL_516443, EPI_ISL_516444, EPI_ISL_516445, EPI_ISL_516446, EPI_ISL_516447, EPI_ISL_516448, EPI_ISL_516449, EPI_ISL_516450, EPI_ISL_516451, EPI_ISL_516452, EPI_ISL_516453, EPI_ISL_516454, EPI_ISL_516455, EPI_ISL_516456, EPI_ISL_516457, EPI_ISL_516458, EPI_ISL_516459, EPI_ISL_516460, EPI_ISL_516461, EPI_ISL_516462, EPI_ISL_516463, EPI_ISL_516464, EPI_ISL_516465, EPI_ISL_516466, EPI_ISL_516467, EPI_ISL_516468, EPI_ISL_516469, EPI_ISL_516470, EPI_ISL_516471, EPI_ISL_516472, EPI_ISL_516473, EPI_ISL_516474, EPI_ISL_516475, EPI_ISL_516476, EPI_ISL_516477, EPI_ISL_516478, EPI_ISL_516479, | | | |

| | | | |
|---|---|---|---|
| EPI_ISL_516480, EPI_ISL_516481, EPI_ISL_516482, EPI_ISL_516483, EPI_ISL_516484, EPI_ISL_516485, EPI_ISL_516486, EPI_ISL_516487, EPI_ISL_516488, EPI_ISL_516489, EPI_ISL_516490, EPI_ISL_516491, EPI_ISL_516492, EPI_ISL_516493, EPI_ISL_516494, EPI_ISL_516495, EPI_ISL_516496, EPI_ISL_516497, EPI_ISL_516498, EPI_ISL_516499, EPI_ISL_516500, EPI_ISL_516501, EPI_ISL_516502, EPI_ISL_516503, EPI_ISL_516504, EPI_ISL_516505, EPI_ISL_516506, EPI_ISL_516507, EPI_ISL_516508, EPI_ISL_516509, EPI_ISL_516510, EPI_ISL_516511, EPI_ISL_516512, EPI_ISL_516513, EPI_ISL_516514, EPI_ISL_516515, EPI_ISL_516516, EPI_ISL_516517, EPI_ISL_516518, EPI_ISL_516519, EPI_ISL_516520, EPI_ISL_516521, EPI_ISL_516522, EPI_ISL_516523, EPI_ISL_516524, EPI_ISL_516525, EPI_ISL_516526, EPI_ISL_516527, EPI_ISL_516528, EPI_ISL_516529, EPI_ISL_516530, EPI_ISL_516531, EPI_ISL_516532, EPI_ISL_516533, EPI_ISL_516534, EPI_ISL_516535, EPI_ISL_516536, EPI_ISL_516537, EPI_ISL_516538, EPI_ISL_516539, EPI_ISL_516540, EPI_ISL_516541, EPI_ISL_516542, EPI_ISL_516543, EPI_ISL_516544, EPI_ISL_516545, EPI_ISL_516546, EPI_ISL_516547, EPI_ISL_516548, EPI_ISL_516549 | | | |
| see above | University of Wisconsin-Madison AIDS Vaccine Research Laboratories | University of Wisconsin-Madison AIDS Vaccine Research Laboratories | Gage Moreno, Katarina Braun, et al. AIDS Vaccine Research Laboratories |
| EPI_ISL_516608 | Instituto de Diagnostico y Referencia Epidemiologicos (INDRE) | Instituto de Diagnostico y Referencia Epidemiologicos (INDRE) | Ernesto Ramirez-Gonzalez, Abril Rodriguez-Maldonado, Claudia Wong-Arambula , Natividad Cruz-Ortiz, Tatiana Nunez-Garcia, Dayanira Arellano-Suarez, Adnan Araiza-Rodriguez, Edgar Mendieta-Condado, Lucia Hernandez-Rivas, Irma Lopez-Martinez, Gisela Barrera-Badillo. |
| EPI_ISL_516609, EPI_ISL_516610 | Instituto de Diagnostico y Referencia Epidemiologicos (INDRE) | Instituto de Diagnostico y Referencia Epidemiologicos (INDRE) | Ernesto Ramirez-Gonzalez, Abril Rodriguez-Maldonado, Claudia Wong-Arambula , Natividad Cruz-Ortiz, Tatiana Nunez-Garcia, Dayanira Arellano-Suarez, Adnan Araiza-Rodriguez, Fabiola Garces-Ayala, Lucia Hernandez-Rivas, Irma Lopez-Martinez, Gisela Barrera-Badillo. |
| EPI_ISL_516611 | Instituto de Diagnostico y Referencia Epidemiologicos (INDRE) | Instituto de Diagnostico y Referencia Epidemiologicos (INDRE) | Gisela Barrera-Badillo , Abril Rodriguez-Maldonado, Claudia Wong-Arambula , Natividad Cruz-Ortiz, Tatiana Nunez-Garcia, Dayanira Arellano-Suarez, Fabiola Garces-Ayala, Edgar Mendieta-Condado, Lucia Hernandez-Rivas, Irma Lopez-Martinez, Ernesto Ramirez-Gonzalez. |
| EPI_ISL_516613 | Instituto de Diagnostico y Referencia Epidemiologicos (INDRE) | Instituto de Diagnostico y Referencia Epidemiologicos (INDRE) | Ernesto Ramirez-Gonzalez, Abril Rodriguez-Maldonado, Claudia Wong-Arambula , Natividad Cruz-Ortiz, Tatiana Nunez-Garcia, Dayanira Arellano-Suarez, Adnan Araiza-Rodriguez, Edgar Mendieta-Condado, Lucia Hernandez-Rivas, Irma Lopez-Martinez, Gisela Barrera-Badillo. |
| EPI_ISL_516614, EPI_ISL_516615, EPI_ISL_516616, EPI_ISL_516617 | Instituto de Diagnostico y Referencia Epidemiologicos (INDRE) | Instituto de Diagnostico y Referencia Epidemiologicos (INDRE) | Ernesto Ramirez-Gonzalez, Abril Rodriguez-Maldonado, Claudia Wong-Arambula , Natividad Cruz-Ortiz, Tatiana Nunez-Garcia, Dayanira Arellano-Suarez, Adnan Araiza-Rodriguez, Fabiola Garces-Ayala, Lucia Hernandez-Rivas, Irma Lopez-Martinez, Gisela Barrera-Badillo. |
| EPI_ISL_516618, EPI_ISL_516619 | Instituto de Diagnostico y Referencia Epidemiologicos (INDRE) | Instituto de Diagnostico y Referencia Epidemiologicos (INDRE) | Gisela Barrera-Badillo , Abril Rodriguez-Maldonado, Claudia Wong-Arambula , Natividad Cruz-Ortiz, Tatiana Nunez-Garcia, Dayanira Arellano-Suarez, Fabiola Garces-Ayala, Edgar Mendieta-Condado, Lucia Hernandez-Rivas, Irma Lopez-Martinez, Ernesto Ramirez-Gonzalez. |
| EPI_ISL_516620 | Instituto de Diagnostico y Referencia Epidemiologicos (INDRE) | Instituto de Diagnostico y Referencia Epidemiologicos (INDRE) | Gisela Barrera-Badillo , Abril Rodriguez-Maldonado, Claudia Wong-Arambula , Natividad Cruz-Ortiz, Tatiana Nunez-Garcia, Dayanira Arellano-Suarez, Adnan Araiza-Rodriguez, Edgar Mendieta-Condado, Lucia Hernandez-Rivas, Irma Lopez-Martinez, Ernesto Ramirez-Gonzalez. |
| EPI_ISL_516622 | Instituto de Diagnostico y Referencia Epidemiologicos (INDRE) | Instituto de Diagnostico y Referencia Epidemiologicos (INDRE) | Gisela Barrera-Badillo , Abril Rodriguez-Maldonado, Claudia Wong-Arambula , Natividad Cruz-Ortiz, Tatiana Nunez-Garcia, Dayanira Arellano-Suarez, Fabiola Garces-Ayala, Edgar Mendieta-Condado, Lucia Hernandez-Rivas, Irma Lopez-Martinez, Ernesto Ramirez-Gonzalez. |
| EPI_ISL_516623 | Instituto de Diagnostico y Referencia Epidemiologicos (INDRE) | Instituto de Diagnostico y Referencia Epidemiologicos (INDRE) | Gisela Barrera-Badillo , Abril Rodriguez-Maldonado, Claudia Wong-Arambula , Natividad Cruz-Ortiz, Tatiana Nunez-Garcia, Dayanira Arellano-Suarez, Adnan Araiza-Rodriguez, Edgar Mendieta-Condado, Lucia Hernandez-Rivas, Irma Lopez-Martinez, Ernesto Ramirez-Gonzalez. |
| EPI_ISL_516624, EPI_ISL_516625 | Instituto de Diagnostico y Referencia Epidemiologicos (INDRE) | Instituto de Diagnostico y Referencia Epidemiologicos (INDRE) | Gisela Barrera-Badillo , Abril Rodriguez-Maldonado, Claudia Wong-Arambula , Natividad Cruz-Ortiz, Tatiana Nunez-Garcia, Dayanira Arellano-Suarez, Fabiola Garces-Ayala, Edgar Mendieta-Condado, Lucia Hernandez-Rivas, Irma Lopez-Martinez, Ernesto Ramirez-Gonzalez. |
| EPI_ISL_516655, EPI_ISL_516656, EPI_ISL_516657, EPI_ISL_516658, EPI_ISL_516659, EPI_ISL_516660, EPI_ISL_516661, EPI_ISL_516662, EPI_ISL_516663, EPI_ISL_516664, EPI_ISL_516665, EPI_ISL_516666, EPI_ISL_516667, EPI_ISL_516668, EPI_ISL_516669, EPI_ISL_516670, EPI_ISL_516671, EPI_ISL_516672, EPI_ISL_516673, EPI_ISL_516674, EPI_ISL_516675, EPI_ISL_516676, EPI_ISL_516677, EPI_ISL_516678, EPI_ISL_516679, EPI_ISL_516680, EPI_ISL_516681, EPI_ISL_516682, EPI_ISL_516683, EPI_ISL_516684, EPI_ISL_516687, EPI_ISL_516688, EPI_ISL_516689, EPI_ISL_516690, EPI_ISL_516691, EPI_ISL_516692, EPI_ISL_516693, EPI_ISL_516694, EPI_ISL_516695, EPI_ISL_516696, EPI_ISL_516697, EPI_ISL_516698, EPI_ISL_516699, EPI_ISL_516700, EPI_ISL_516701, EPI_ISL_516702, EPI_ISL_516703, EPI_ISL_516704, EPI_ISL_516705, EPI_ISL_516706, EPI_ISL_516707, EPI_ISL_516708, EPI_ISL_516709, EPI_ISL_516710, EPI_ISL_516711, EPI_ISL_516712, EPI_ISL_516713, EPI_ISL_516714, EPI_ISL_516715, EPI_ISL_516716, EPI_ISL_516717, EPI_ISL_516718, EPI_ISL_516719, EPI_ISL_516720 | | | |
| see above | Virginia DCLS | Virginia DCLS | Virginia DCLS |
| EPI_ISL_516723, EPI_ISL_516724, EPI_ISL_516725, EPI_ISL_516726, EPI_ISL_516727, EPI_ISL_516728, EPI_ISL_516729, EPI_ISL_516730, EPI_ISL_516731, EPI_ISL_516732, EPI_ISL_516733, EPI_ISL_516734, EPI_ISL_516735, EPI_ISL_516736, EPI_ISL_516737, EPI_ISL_516738, EPI_ISL_516739, EPI_ISL_516740, EPI_ISL_516741, EPI_ISL_516742, EPI_ISL_516743, EPI_ISL_516744, EPI_ISL_516745, EPI_ISL_516746, EPI_ISL_516747 | | | |
| see above | UCSF Clinical Microbiology Laboratory | Chan-Zuckerberg Biohub | CZB Cliahub Consortium |
| EPI_ISL_516748, EPI_ISL_516749, EPI_ISL_516750, EPI_ISL_516751, EPI_ISL_516752, EPI_ISL_516753, EPI_ISL_516754, EPI_ISL_516756, EPI_ISL_516757, EPI_ISL_516758, EPI_ISL_516759, EPI_ISL_516760, EPI_ISL_516761, EPI_ISL_516762, EPI_ISL_516763, EPI_ISL_516764, EPI_ISL_516765, EPI_ISL_516766, EPI_ISL_516767, EPI_ISL_516768, EPI_ISL_516769, EPI_ISL_516770, EPI_ISL_516771, EPI_ISL_516772, EPI_ISL_516773, EPI_ISL_516774, EPI_ISL_516775, EPI_ISL_516776, EPI_ISL_516777, EPI_ISL_516778, EPI_ISL_516779, EPI_ISL_516780, EPI_ISL_516781, EPI_ISL_516782 | | | |
| see above | van Bakel Laboratory, Genetics and Genomics Sciences, Icahn School of Medicine at Mount Sinai | van Bakel Laboratory, Genetics and Genomics Sciences, Icahn School of Medicine at Mount Sinai | Andrew G. Letizia, Irene Ramos, Ajay Obla, Carl Goforth, Dawn Weir, Yongchao Ge, Marcas M. Bamman, Jayeeta Dutta, Ethan Ellis, Luis Estrella, Mary-Catherine George, Ana S. Gonzalez-Reiche, Darnell Graham, Adriana van de Guchte, Ramiro Gutierrez, Franca Jones, Aspasia Kalomoiri, Rhonda Lizewski, Stephen Lizewski, Jan Marayag, Nada Marjanovic, Eugene V. Millar, Venugopalan Nair, German Nudelman, Edgar Nunez, Brian Pike, James Regeimbal, Stas Rirak , Ernesto Santa Ana, Rachel S. Gelernter Sealfon, Robert Sebra, Mark Simons, Alessandra Soares-Schanoski, Michael Termini, Sindhu Vangeti, Carlos Williams, Harm van Bakel, Stuart C. Sealfon |
| EPI_ISL_517771, EPI_ISL_517772, EPI_ISL_517774, EPI_ISL_517776, EPI_ISL_517777, EPI_ISL_517778, EPI_ISL_517779, EPI_ISL_517780, EPI_ISL_517781, EPI_ISL_517783, EPI_ISL_517784, EPI_ISL_517786, EPI_ISL_517788, EPI_ISL_517790, EPI_ISL_517791, EPI_ISL_517792, EPI_ISL_517793, EPI_ISL_517794, EPI_ISL_517795, EPI_ISL_517796, EPI_ISL_517797, EPI_ISL_517798, EPI_ISL_517799, EPI_ISL_517800, EPI_ISL_517801, EPI_ISL_517802, EPI_ISL_517803, EPI_ISL_517804, EPI_ISL_517805, EPI_ISL_517806, EPI_ISL_517807, EPI_ISL_517808, EPI_ISL_517809, EPI_ISL_517810, EPI_ISL_517811, EPI_ISL_517812, EPI_ISL_517813, EPI_ISL_517814, EPI_ISL_517815, EPI_ISL_517816, EPI_ISL_517817, EPI_ISL_517818, EPI_ISL_517819, EPI_ISL_517821, EPI_ISL_517822, EPI_ISL_517823, EPI_ISL_517824, EPI_ISL_517825, EPI_ISL_517826, EPI_ISL_517827, EPI_ISL_517828, EPI_ISL_517829, EPI_ISL_517830, EPI_ISL_517831, EPI_ISL_517832, EPI_ISL_517833, EPI_ISL_517834, EPI_ISL_517835, EPI_ISL_517836, EPI_ISL_517839, EPI_ISL_517840, EPI_ISL_517841, EPI_ISL_517842, EPI_ISL_517843, EPI_ISL_517844, EPI_ISL_517845, EPI_ISL_517846, EPI_ISL_517847, EPI_ISL_517849, EPI_ISL_517850, EPI_ISL_517852, EPI_ISL_517853, EPI_ISL_517854, EPI_ISL_517855, EPI_ISL_517856, EPI_ISL_517857, EPI_ISL_517858, EPI_ISL_517859, EPI_ISL_517860, EPI_ISL_517861, EPI_ISL_517862, EPI_ISL_517863, EPI_ISL_517864, EPI_ISL_517865, EPI_ISL_517866, EPI_ISL_517867, EPI_ISL_517868, EPI_ISL_517869, EPI_ISL_517870, EPI_ISL_517871, EPI_ISL_517872, EPI_ISL_517873, EPI_ISL_517874, EPI_ISL_517875, EPI_ISL_517876, EPI_ISL_517877, EPI_ISL_517878, EPI_ISL_517879, EPI_ISL_517880, EPI_ISL_517881, EPI_ISL_517882, EPI_ISL_517883, EPI_ISL_517884, EPI_ISL_517885, EPI_ISL_517886, EPI_ISL_517887, EPI_ISL_517888, EPI_ISL_517889, EPI_ISL_517890, EPI_ISL_517891, EPI_ISL_517892, EPI_ISL_517893, EPI_ISL_517894, EPI_ISL_517895, EPI_ISL_517896, EPI_ISL_517897, EPI_ISL_517898, EPI_ISL_517899, EPI_ISL_517900, EPI_ISL_517901, EPI_ISL_517902, EPI_ISL_517903, EPI_ISL_517904, EPI_ISL_517905, EPI_ISL_517906, EPI_ISL_517907, EPI_ISL_517908, EPI_ISL_517909, EPI_ISL_517910, EPI_ISL_517911, EPI_ISL_517912, EPI_ISL_517913, EPI_ISL_517914, EPI_ISL_517915, EPI_ISL_517916, EPI_ISL_517917, EPI_ISL_517918, EPI_ISL_517919, EPI_ISL_517920, EPI_ISL_517921, EPI_ISL_517922, EPI_ISL_517923, EPI_ISL_517924, EPI_ISL_517925, EPI_ISL_517926, EPI_ISL_517928, EPI_ISL_517929, EPI_ISL_517930, EPI_ISL_517931, EPI_ISL_517932, EPI_ISL_517933, EPI_ISL_517934, EPI_ISL_517935, EPI_ISL_517936, EPI_ISL_517937, EPI_ISL_517938, EPI_ISL_517939, EPI_ISL_517940, EPI_ISL_517941, EPI_ISL_517942, EPI_ISL_517943, EPI_ISL_517944, EPI_ISL_517945, EPI_ISL_517946, EPI_ISL_517947, EPI_ISL_517948, EPI_ISL_517949, EPI_ISL_517950, EPI_ISL_517951, EPI_ISL_517952, EPI_ISL_517953, EPI_ISL_517954, EPI_ISL_517956, EPI_ISL_517957 | | | |
| see above | Florida Bureau of Public Health Laboratories | Florida Bureau of Public Health Laboratories | Sarah Schmedes, Jason Blanton |
| EPI_ISL_517961, EPI_ISL_517962, EPI_ISL_517963, EPI_ISL_517964, EPI_ISL_517965, EPI_ISL_517967, EPI_ISL_517968, EPI_ISL_517969, EPI_ISL_517970, EPI_ISL_517971, EPI_ISL_517974, EPI_ISL_517978, EPI_ISL_517979, EPI_ISL_517981, EPI_ISL_517982, EPI_ISL_517984, EPI_ISL_517985, EPI_ISL_517986, EPI_ISL_517987, EPI_ISL_517990, EPI_ISL_517993, EPI_ISL_517995 | | | |
| see above | Texas Department of State Health Services | Texas Department of State Health Services | Rashmi Tuladhar, Bonnie Oh, Cara Akrout, Jenny Zhang, Maliha Rahman, Anita Pokharel, Myong Koag, Chun Wang, Rachel Lee, Grace Kubin |
| EPI_ISL_518855 | Minnesota Department of Health, Public Health Laboratory | Minnesota Department of Health, Public Health Laboratory | Matt Plumb, Jacob Garfin, and Xiong Wang |
| EPI_ISL_518856, EPI_ISL_518857, EPI_ISL_518858, EPI_ISL_518859, EPI_ISL_518860, EPI_ISL_518861, EPI_ISL_518862, EPI_ISL_518863, EPI_ISL_518864, EPI_ISL_518865, EPI_ISL_518866, EPI_ISL_518867, EPI_ISL_518868, EPI_ISL_518869, EPI_ISL_518870, EPI_ISL_518871, EPI_ISL_518872, EPI_ISL_518873, EPI_ISL_518874, EPI_ISL_518875, EPI_ISL_518876, EPI_ISL_518877, EPI_ISL_518878, EPI_ISL_518879, EPI_ISL_518880, EPI_ISL_518881, EPI_ISL_518882, EPI_ISL_518883, EPI_ISL_518884, EPI_ISL_518885, EPI_ISL_518886, EPI_ISL_518887, EPI_ISL_518888, EPI_ISL_518889, EPI_ISL_518890, EPI_ISL_518891, EPI_ISL_518892, EPI_ISL_518893, EPI_ISL_518894, EPI_ISL_518895, EPI_ISL_518896, EPI_ISL_518897, EPI_ISL_518898, EPI_ISL_518899, EPI_ISL_518900, EPI_ISL_521153 | | | |
| see above | Mayo Clinic & Mayo Clinic Laboratories | Minnesota Department of Health, Public Health Laboratory | Matt Plumb, Jacob Garfin, and Xiong Wang |
| EPI_ISL_522254, EPI_ISL_522255, EPI_ISL_522257, EPI_ISL_522258, EPI_ISL_522259, EPI_ISL_522260, EPI_ISL_522261, EPI_ISL_522262, EPI_ISL_522263, EPI_ISL_522264, EPI_ISL_522265, EPI_ISL_522266, EPI_ISL_522267, EPI_ISL_522268, EPI_ISL_522269, EPI_ISL_522270, EPI_ISL_522271, EPI_ISL_522272, EPI_ISL_522274, EPI_ISL_522276, EPI_ISL_522278, EPI_ISL_522279, EPI_ISL_522281, EPI_ISL_522282, EPI_ISL_522285, EPI_ISL_522287, EPI_ISL_522288, EPI_ISL_522290, EPI_ISL_522291, EPI_ISL_522294, EPI_ISL_522295, EPI_ISL_522296, EPI_ISL_522297, EPI_ISL_522298, EPI_ISL_522299, EPI_ISL_522300, EPI_ISL_522301, EPI_ISL_522302, EPI_ISL_522303, EPI_ISL_522304, EPI_ISL_522305, EPI_ISL_522306, EPI_ISL_522307, EPI_ISL_522308, EPI_ISL_522309, EPI_ISL_522310, EPI_ISL_522311, EPI_ISL_522312, EPI_ISL_522313, EPI_ISL_522314, EPI_ISL_522315, EPI_ISL_522316, EPI_ISL_522319, EPI_ISL_522320, EPI_ISL_522321, EPI_ISL_522322, EPI_ISL_522323, EPI_ISL_522324, EPI_ISL_522326, EPI_ISL_522327, EPI_ISL_522328, EPI_ISL_522330, EPI_ISL_522331, EPI_ISL_522335, EPI_ISL_522336, EPI_ISL_522337, EPI_ISL_522338, EPI_ISL_522339, EPI_ISL_522340, EPI_ISL_522341, EPI_ISL_522343, EPI_ISL_522345, EPI_ISL_522346, EPI_ISL_522347, EPI_ISL_522348 | | | |
| see above | Utah Public Health Laboratory | Utah Public Health Laboratory | Erin Young, Kelly Oakeson |
| EPI_ISL_522352, EPI_ISL_522353, EPI_ISL_522354, EPI_ISL_522355, EPI_ISL_522356, EPI_ISL_522357, EPI_ISL_522358, EPI_ISL_522360, EPI_ISL_522362, EPI_ISL_522363, EPI_ISL_522365, EPI_ISL_522366, EPI_ISL_522367, EPI_ISL_522368, EPI_ISL_522369, EPI_ISL_522370, EPI_ISL_522371, EPI_ISL_522373, EPI_ISL_522375, EPI_ISL_522376, EPI_ISL_522378, EPI_ISL_522379, EPI_ISL_522380, EPI_ISL_522381, EPI_ISL_522382, EPI_ISL_522383, EPI_ISL_522385, EPI_ISL_522386, EPI_ISL_522387, EPI_ISL_522388, EPI_ISL_522389, EPI_ISL_522390, EPI_ISL_522391, EPI_ISL_522392, EPI_ISL_522393, EPI_ISL_522394, EPI_ISL_522395 | | | |
| see above | Texas Department of State Health Services | Texas Department of State Health Services | Rashmi Tuladhar, Bonnie Oh, Cara Akrout, Jenny Zhang, Maliha Rahman, Anita Pokharel, Myong Koag, Chun Wang, Rachel Lee, Grace Kubin |
| EPI_ISL_522396, EPI_ISL_522397, EPI_ISL_522398, EPI_ISL_522399, EPI_ISL_522400, EPI_ISL_522401, EPI_ISL_522402, EPI_ISL_522403, EPI_ISL_522404, EPI_ISL_522405 | Alaska State Virology Laboratory | Alaska State Virology Laboratory | Jack Chen, Ph.D. |
| EPI_ISL_522777, EPI_ISL_522778, EPI_ISL_522779, EPI_ISL_522780, EPI_ISL_522781, EPI_ISL_522782, EPI_ISL_522783, EPI_ISL_522784, EPI_ISL_522785, EPI_ISL_522786, EPI_ISL_522787, EPI_ISL_522788, EPI_ISL_522789, EPI_ISL_522790, EPI_ISL_522791, EPI_ISL_522792, EPI_ISL_522793, EPI_ISL_522794, EPI_ISL_522795, EPI_ISL_522796, EPI_ISL_522797, EPI_ISL_522798, EPI_ISL_522799, EPI_ISL_522800, EPI_ISL_522801, EPI_ISL_522802, EPI_ISL_522803, EPI_ISL_522804, EPI_ISL_522805, EPI_ISL_522806, EPI_ISL_522807, EPI_ISL_522808, EPI_ISL_522809, EPI_ISL_522810, EPI_ISL_522811, EPI_ISL_522812, EPI_ISL_522813, EPI_ISL_522814, EPI_ISL_522815, EPI_ISL_522816, EPI_ISL_522817, EPI_ISL_522818, EPI_ISL_522819, EPI_ISL_522820, EPI_ISL_522821, EPI_ISL_522822, EPI_ISL_522823 | | | |
| see above | Virginia DCLS | Virginia DCLS | Virginia DCLS |
| EPI_ISL_522824, EPI_ISL_522825, EPI_ISL_522826, EPI_ISL_522827, EPI_ISL_522828, EPI_ISL_522829, EPI_ISL_522830, EPI_ISL_522831, EPI_ISL_522832, EPI_ISL_522833, EPI_ISL_522834, EPI_ISL_522835, EPI_ISL_522836, EPI_ISL_522837, EPI_ISL_522838, EPI_ISL_522839, EPI_ISL_522840, EPI_ISL_522841, EPI_ISL_522842, EPI_ISL_522843, EPI_ISL_522844, EPI_ISL_522845, EPI_ISL_522846, EPI_ISL_522847, EPI_ISL_522848, EPI_ISL_522849, EPI_ISL_522850, EPI_ISL_522851, EPI_ISL_522852, EPI_ISL_522853, EPI_ISL_522854, EPI_ISL_522869, EPI_ISL_522870, EPI_ISL_522871 | | | |
| see above | Maryland Department of Health | Maryland Department of Health | Maryland Department of Health Laboratories Administration |
| EPI_ISL_522872 | Instituto Nacional de Medicina Genómica | Instituto Nacional de Medicina Genómica | Hidalgo-Miranda A, Mendoza-Vargas A, Reyes-Grajeda JP, Cisneros-Villanueva M, Cedro-Tanda A, Hurtado-Cordova E, Peñaloza-Figueroa F, Herrera-Montalvo LA |
| EPI_ISL_522873, EPI_ISL_522874, EPI_ISL_522875, EPI_ISL_522876, EPI_ISL_522877 | Instituto Nacional de Medicina Genomica | Instituto Nacional de Medicina Genomica | Hidalgo-Miranda A, Mendoza-Vargas A, Reyes-Grajeda JP, Cisneros-Villanueva M, Cedro-Tanda A, Hurtado-Cordova E, Peñaloza-Figueroa F, Herrera-Montalvo LA |
| EPI_ISL_522878 | Instituto Nacional de Medicina Genomica | Instituto Nacional de Medicina Genomica | Hidalgo-Miranda A, Mendoza-Vargas A, Reyes-Grajeda JP, Cisneros-Villanueva M, Hurtado-Cordova E, Cedro-Tanda A, Peñaloza-Figueroa F, Herrera-Montalvo LA |
| EPI_ISL_522879, | Instituto Nacional de Medicina | Instituto Nacional de Medicina Genómica | Hidalgo-Miranda A, Mendoza-Vargas A, Reyes-Grajeda JP, Cisneros-Villanueva M, Cedro-Tanda A, Hurtado-Cordova E, Peñaloza-Figueroa F, Herrera-Montalvo LA |

| | | | |
|---|---|---|---|
| EPI_ISL_522880 | Genómica | | |
| EPI_ISL_522881, EPI_ISL_522882, EPI_ISL_522883, EPI_ISL_522884, EPI_ISL_522885, EPI_ISL_522886, EPI_ISL_522887, EPI_ISL_522888, EPI_ISL_522889, EPI_ISL_522890, EPI_ISL_522891, EPI_ISL_522892, EPI_ISL_522893, EPI_ISL_522894, EPI_ISL_522895, EPI_ISL_522896, EPI_ISL_522897, EPI_ISL_522898, EPI_ISL_522899, EPI_ISL_522900, EPI_ISL_522901, EPI_ISL_522902, EPI_ISL_522903, EPI_ISL_522904, EPI_ISL_522905, EPI_ISL_522906, EPI_ISL_522907, EPI_ISL_522908, EPI_ISL_522909, EPI_ISL_522910, EPI_ISL_522911, EPI_ISL_522912, EPI_ISL_522913, EPI_ISL_522914, EPI_ISL_522915, EPI_ISL_522916, EPI_ISL_522917, EPI_ISL_522918, EPI_ISL_522919, EPI_ISL_522920, EPI_ISL_522921, EPI_ISL_522922, EPI_ISL_522923, EPI_ISL_522924, EPI_ISL_522925, EPI_ISL_522926, EPI_ISL_522927, EPI_ISL_522928, EPI_ISL_522929, EPI_ISL_522930, EPI_ISL_522931, EPI_ISL_522932, EPI_ISL_522933, EPI_ISL_522934, EPI_ISL_522935, EPI_ISL_522936, EPI_ISL_522937, EPI_ISL_522938, EPI_ISL_522939 | | | |
| see above | University of Wisconsin-Madison AIDS Vaccine Research Laboratories | University of Wisconsin-Madison AIDS Vaccine Research Laboratories | Gage Moreno, Katarina Braun, et al. AIDS Vaccine Research Laboratories |
| EPI_ISL_522940, EPI_ISL_522941, EPI_ISL_522942 | Instituto Nacional de Medicina Genómica | Instituto Nacional de Medicina Genómica | Hidalgo-Miranda A, Mendoza-Vargas A, Reyes-Grajeda JP, Cisneros-Villanueva M, Cedro-Tanda A, Hurtado-Cordova E, Peñaloza-Figueroa F, Herrera-Montalvo LA |
| EPI_ISL_522943, EPI_ISL_522944, EPI_ISL_522945, EPI_ISL_522946, EPI_ISL_522947, EPI_ISL_522948, EPI_ISL_522949, EPI_ISL_522951, EPI_ISL_522952, EPI_ISL_522953, EPI_ISL_522954, EPI_ISL_522955, EPI_ISL_522956, EPI_ISL_522957, EPI_ISL_522958, EPI_ISL_522959, EPI_ISL_522960, EPI_ISL_522961, EPI_ISL_522962, EPI_ISL_522963, EPI_ISL_522964, EPI_ISL_522965, EPI_ISL_522966, EPI_ISL_522967, EPI_ISL_522968, EPI_ISL_522969, EPI_ISL_522970, EPI_ISL_522971, EPI_ISL_522972, EPI_ISL_522973, EPI_ISL_522975, EPI_ISL_522976, EPI_ISL_522977 | | | |
| see above | Texas Department of State Health Services | Texas Department of State Health Services | Rashmi Tuladhar, Bonnie Oh, Cara Akrout, Jenny Zhang, Maliha Rahman, Anita Pokharel, Myong Koag, Chun Wang, Rachel Lee, Grace Kubin |
| EPI_ISL_522978, EPI_ISL_522979, EPI_ISL_522980, EPI_ISL_522981, EPI_ISL_522982, EPI_ISL_522983, EPI_ISL_522984, EPI_ISL_522985, EPI_ISL_522986, EPI_ISL_523500 | Instituto Nacional de Medicina Genómica | Instituto Nacional de Medicina Genómica | Hidalgo-Miranda A, Mendoza-Vargas A, Reyes-Grajeda JP, Cisneros-Villanueva M, Cedro-Tanda A, Hurtado-Cordova E, Peñaloza-Figueroa F, Herrera-Montalvo LA |
| EPI_ISL_524053, EPI_ISL_524056, EPI_ISL_524057, EPI_ISL_524059, EPI_ISL_524060, EPI_ISL_524061, EPI_ISL_524062, EPI_ISL_524063, EPI_ISL_524064, EPI_ISL_524065, EPI_ISL_524066, EPI_ISL_524067, EPI_ISL_524068, EPI_ISL_524070, EPI_ISL_524073 | | | |
| see above | Texas Department of State Health Services | Texas Department of State Health Services | Rashmi Tuladhar, Bonnie Oh, Cara Akrout, Jenny Zhang, Maliha Rahman, Anita Pokharel, Myong Koag, Chun Wang, Rachel Lee, Grace Kubin |
| EPI_ISL_524074, EPI_ISL_524075, EPI_ISL_524076, EPI_ISL_524077, EPI_ISL_524078, EPI_ISL_524079, EPI_ISL_524080, EPI_ISL_524081, EPI_ISL_524082, EPI_ISL_524083, EPI_ISL_524084, EPI_ISL_524085, EPI_ISL_524086, EPI_ISL_524087, EPI_ISL_524088, EPI_ISL_524089, EPI_ISL_524090, EPI_ISL_524092, EPI_ISL_524093, EPI_ISL_524094, EPI_ISL_524096, EPI_ISL_524097, EPI_ISL_524098, EPI_ISL_524099, EPI_ISL_524100, EPI_ISL_524101, EPI_ISL_524102, EPI_ISL_524104, EPI_ISL_524105, EPI_ISL_524106, EPI_ISL_524107, EPI_ISL_524108, EPI_ISL_524109, EPI_ISL_524110, EPI_ISL_524111, EPI_ISL_524112, EPI_ISL_524113, EPI_ISL_524114, EPI_ISL_524115, EPI_ISL_524116, EPI_ISL_524117, EPI_ISL_524118, EPI_ISL_524119, EPI_ISL_524121, EPI_ISL_524122, EPI_ISL_524123, EPI_ISL_524124, EPI_ISL_524125, EPI_ISL_524126, EPI_ISL_524127, EPI_ISL_524128, EPI_ISL_524129, EPI_ISL_524130, EPI_ISL_524131, EPI_ISL_524133, EPI_ISL_524134, EPI_ISL_524135, EPI_ISL_524136, EPI_ISL_524137, EPI_ISL_524138, EPI_ISL_524139, EPI_ISL_524141, EPI_ISL_524142, EPI_ISL_524143, EPI_ISL_524144, EPI_ISL_524145, EPI_ISL_524146, EPI_ISL_524147, EPI_ISL_524148, EPI_ISL_524149, EPI_ISL_524150, EPI_ISL_524151, EPI_ISL_524152, EPI_ISL_524153, EPI_ISL_524154, EPI_ISL_524155, EPI_ISL_524156, EPI_ISL_524157, EPI_ISL_524158, EPI_ISL_524159, EPI_ISL_524160, EPI_ISL_524161, EPI_ISL_524162, EPI_ISL_524163, EPI_ISL_524164, EPI_ISL_524165, EPI_ISL_524166, EPI_ISL_524167, EPI_ISL_524168, EPI_ISL_524169, EPI_ISL_524170, EPI_ISL_524171, EPI_ISL_524172, EPI_ISL_524173, EPI_ISL_524174, EPI_ISL_524175, EPI_ISL_524176, EPI_ISL_524177, EPI_ISL_524178, EPI_ISL_524179, EPI_ISL_524180, EPI_ISL_524181, EPI_ISL_524182, EPI_ISL_524183, EPI_ISL_524184, EPI_ISL_524185, EPI_ISL_524186, EPI_ISL_524187, EPI_ISL_524188, EPI_ISL_524189, EPI_ISL_524190, EPI_ISL_524191, EPI_ISL_524192, EPI_ISL_524193, EPI_ISL_524194, EPI_ISL_524195, EPI_ISL_524196, EPI_ISL_524197, EPI_ISL_524199, EPI_ISL_524200, EPI_ISL_524201, EPI_ISL_524202, EPI_ISL_524203, EPI_ISL_524204, EPI_ISL_524205, EPI_ISL_524206, EPI_ISL_524207, EPI_ISL_524208, EPI_ISL_524209, EPI_ISL_524210, EPI_ISL_524211, EPI_ISL_524212, EPI_ISL_524213, EPI_ISL_524214, EPI_ISL_524215, EPI_ISL_524216, EPI_ISL_524217, EPI_ISL_524218, EPI_ISL_524220, EPI_ISL_524222, EPI_ISL_524223, EPI_ISL_524224, EPI_ISL_524225, EPI_ISL_524226, EPI_ISL_524227, EPI_ISL_524228, EPI_ISL_524229, EPI_ISL_524231, EPI_ISL_524232, EPI_ISL_524233, EPI_ISL_524234, EPI_ISL_524235, EPI_ISL_524236, EPI_ISL_524237, EPI_ISL_524239, EPI_ISL_524240, EPI_ISL_524241, EPI_ISL_524242, EPI_ISL_524243, EPI_ISL_524244, EPI_ISL_524245, EPI_ISL_524246, EPI_ISL_524247, EPI_ISL_524249, EPI_ISL_524250, EPI_ISL_524251, EPI_ISL_524252, EPI_ISL_524253, EPI_ISL_524254, EPI_ISL_524255, EPI_ISL_524256, EPI_ISL_524257, EPI_ISL_524258, EPI_ISL_524260, EPI_ISL_524261, EPI_ISL_524262, EPI_ISL_524263, EPI_ISL_524264, EPI_ISL_524265, EPI_ISL_524266, EPI_ISL_524267, EPI_ISL_524268, EPI_ISL_524269, EPI_ISL_524270, EPI_ISL_524271, EPI_ISL_524272, EPI_ISL_524274, EPI_ISL_524275, EPI_ISL_524276, EPI_ISL_524277, EPI_ISL_524278, EPI_ISL_524279, EPI_ISL_524280, EPI_ISL_524281, EPI_ISL_524283, EPI_ISL_524284, EPI_ISL_524285, EPI_ISL_524287, EPI_ISL_524288, EPI_ISL_524289, EPI_ISL_524290, EPI_ISL_524291, EPI_ISL_524292, EPI_ISL_524293, EPI_ISL_524294, EPI_ISL_524295, EPI_ISL_524296, EPI_ISL_524297, EPI_ISL_524298, EPI_ISL_524299, EPI_ISL_524300, EPI_ISL_524301, EPI_ISL_524302, EPI_ISL_524303, EPI_ISL_524304, EPI_ISL_524305, EPI_ISL_524306, EPI_ISL_524307, EPI_ISL_524308, EPI_ISL_524309, EPI_ISL_524311, EPI_ISL_524313, EPI_ISL_524314, EPI_ISL_524315, EPI_ISL_524316, EPI_ISL_524318, EPI_ISL_524319, EPI_ISL_524320, EPI_ISL_524321, EPI_ISL_524322, EPI_ISL_524323, EPI_ISL_524324, EPI_ISL_524325, EPI_ISL_524327, EPI_ISL_524330, EPI_ISL_524331, EPI_ISL_524333, EPI_ISL_524334, EPI_ISL_524335, EPI_ISL_524337, EPI_ISL_524338, EPI_ISL_524339, EPI_ISL_524340, EPI_ISL_524341, EPI_ISL_524342, EPI_ISL_524343, EPI_ISL_524344, EPI_ISL_524345, EPI_ISL_524346, EPI_ISL_524347, EPI_ISL_524348, EPI_ISL_524350, EPI_ISL_524351, EPI_ISL_524352, EPI_ISL_524353, EPI_ISL_524354, EPI_ISL_524355, EPI_ISL_524356, EPI_ISL_524357, EPI_ISL_524358, EPI_ISL_524359, EPI_ISL_524360, EPI_ISL_524361, EPI_ISL_524362, EPI_ISL_524364, EPI_ISL_524365, EPI_ISL_524366, EPI_ISL_524367, EPI_ISL_524368, EPI_ISL_524369, EPI_ISL_524370, EPI_ISL_524371, EPI_ISL_524372, EPI_ISL_524373, EPI_ISL_524374, EPI_ISL_524375, EPI_ISL_524376, EPI_ISL_524377, EPI_ISL_524378, EPI_ISL_524380, EPI_ISL_524381, EPI_ISL_524382, EPI_ISL_524383, EPI_ISL_524384, EPI_ISL_524385, EPI_ISL_524386, EPI_ISL_524387, EPI_ISL_524388, EPI_ISL_524389, EPI_ISL_524391, EPI_ISL_524392, EPI_ISL_524393, EPI_ISL_524394, EPI_ISL_524395, EPI_ISL_524396, EPI_ISL_524397, EPI_ISL_524398, EPI_ISL_524400, EPI_ISL_524401, EPI_ISL_524402, EPI_ISL_524403, EPI_ISL_524404, EPI_ISL_524405, EPI_ISL_524406, EPI_ISL_524407, EPI_ISL_524408, EPI_ISL_524409, EPI_ISL_524410, EPI_ISL_524411, EPI_ISL_524412, EPI_ISL_524413, EPI_ISL_524414, EPI_ISL_524415, EPI_ISL_524416, EPI_ISL_524417, EPI_ISL_524418, EPI_ISL_524419, EPI_ISL_524420, EPI_ISL_524421, EPI_ISL_524422, EPI_ISL_524423, EPI_ISL_524424, EPI_ISL_524425 | | | |
| see above | Utah Public Health Laboratory | Utah Public Health Laboratory | Erin L. Young, Kelly Oakeson, Tara Gallagher, Michael T. Pyne, E. Susan Slechta, Melanie A. Mallory, Jeffrey B. Stevenson, Salika M. Shakir, David R. Hillyard |
| EPI_ISL_524428, EPI_ISL_524429, EPI_ISL_524430, EPI_ISL_524431, EPI_ISL_524432 | Dirk Dittmer | Dirk Dittmer | Bailey,A.G., Caro-Vegas,C.P., Dittmer,D., Eason,A.B., Juarez,A., Landis,J.T., McNamara,R.P., Miller,M.B., Moorad,R., Pluta,L.J.,Seltzer,T.A., Thompson,C., Vahrson,W. and Villamor,F. |
| EPI_ISL_524433, EPI_ISL_524434 | Environmental and Global Health, University of Florida - Gainesville | University of Florida | Elbadry,M.A., Subramaniam,K., Waltzek,T.B., Gibson,J.C., Stephenson,C.J., Alam,M.M., Morris,J.G. Jr., Lednicky,J.A. |
| EPI_ISL_524435 | Department of Immunology, The Scripps Research Institute | Department of Immunology, The Scripps Research Institute | SEARCH Alliance,S.D., Pride,D., Shin,J.H. |
| EPI_ISL_524436 | Department of Immunology, The Scripps Research Institute | Department of Immunology, The Scripps Research Institute | Smither,A., Sabino-Santos,G., Snarski,P., Melnik,L., Bell,A., Genemaras,K., Drouin,A., Fusco,D., Garry,R., SEARCH Alliance,S.D. |
| EPI_ISL_524437, EPI_ISL_524438, EPI_ISL_524439, EPI_ISL_524440, EPI_ISL_524441, EPI_ISL_524442, EPI_ISL_524443 | University of Washington Virology Lab | Laboratory Medicine, University of Washington | Roychoudhury,P., Greninger,A., Jerome,K. |
| EPI_ISL_524444 | University of Washington Virology Lab | Fred Hutchinson Cancer Research Center | Roychoudhury,P., Greninger,A., Jerome,K. |
| EPI_ISL_524450, EPI_ISL_524451, EPI_ISL_524452, EPI_ISL_524453, EPI_ISL_524454 | Microbiology & Immunology, University of North Carolina | Microbiology & Immunology, University of North Carolina | Bailey,A.G., Caro-Vegas,C.P., Dittmer,D., Eason,A.B., Juarez,A., Landis,J.T., McNamara,R.P., Miller,M.B., Moorad,R., Pluta,L.J., Seltzer,T.A., Thompson,C., Vahrson,W., Villamor,F. |
| EPI_ISL_524767, EPI_ISL_524768, EPI_ISL_524769, EPI_ISL_524770, EPI_ISL_524771, EPI_ISL_524772, EPI_ISL_524773, EPI_ISL_524774, EPI_ISL_524775, EPI_ISL_524776, EPI_ISL_524777, EPI_ISL_524778, EPI_ISL_524779, EPI_ISL_524780, EPI_ISL_524781, EPI_ISL_524802, EPI_ISL_524803, EPI_ISL_524804, EPI_ISL_524805, EPI_ISL_524806, EPI_ISL_524807, EPI_ISL_524808, EPI_ISL_524809, EPI_ISL_524810, EPI_ISL_524811, EPI_ISL_524812, EPI_ISL_524813, EPI_ISL_524814, EPI_ISL_524815, EPI_ISL_524816, EPI_ISL_524817, EPI_ISL_524818, EPI_ISL_524819, EPI_ISL_524820, EPI_ISL_524821, EPI_ISL_524822, EPI_ISL_524823, EPI_ISL_524824, EPI_ISL_524825, EPI_ISL_524826, EPI_ISL_524827, EPI_ISL_524828, EPI_ISL_524829, EPI_ISL_524830, EPI_ISL_524831, EPI_ISL_524832, EPI_ISL_524833, EPI_ISL_524834, EPI_ISL_524835, EPI_ISL_524836, EPI_ISL_524837, EPI_ISL_524838, EPI_ISL_524839, EPI_ISL_524840, EPI_ISL_524841, EPI_ISL_524842, EPI_ISL_524843, EPI_ISL_524844, EPI_ISL_524845, EPI_ISL_524846, EPI_ISL_524847, EPI_ISL_524848, EPI_ISL_524849, EPI_ISL_524850, EPI_ISL_524851, EPI_ISL_524852, EPI_ISL_524853, EPI_ISL_524854, EPI_ISL_524855, EPI_ISL_524856, EPI_ISL_524857, EPI_ISL_524858, EPI_ISL_524859, EPI_ISL_524860, EPI_ISL_524861, EPI_ISL_524862, EPI_ISL_524863, EPI_ISL_524864, EPI_ISL_524867, EPI_ISL_524868, EPI_ISL_524869, EPI_ISL_524870, EPI_ISL_524871, EPI_ISL_524872 | | | |
| see above | Utah Public Health Laboratory | Utah Public Health Laboratory | Erin L. Young, Kelly Oakeson, Tara Gallagher, Michael T. Pyne, E. Susan Slechta, Melanie A. Mallory, Jeffrey B. Stevenson, Salika M. Shakir, David R. Hillyard |
| EPI_ISL_524873, EPI_ISL_524875, EPI_ISL_524876, EPI_ISL_524877, EPI_ISL_524878, EPI_ISL_524879, EPI_ISL_524880, EPI_ISL_524881, EPI_ISL_524882, EPI_ISL_524883, EPI_ISL_524884, EPI_ISL_524885, EPI_ISL_524886, EPI_ISL_524887, EPI_ISL_524888, EPI_ISL_524889, EPI_ISL_524890, EPI_ISL_524891, EPI_ISL_524892, EPI_ISL_524893, EPI_ISL_524894, EPI_ISL_524895, EPI_ISL_524896, EPI_ISL_524897, EPI_ISL_524898, EPI_ISL_524899, EPI_ISL_524900, EPI_ISL_524901, EPI_ISL_524902, EPI_ISL_524903 | | | |
| see above | MD PHL | MD PHL | Maryland Department of Health Laboratories Administration |
| EPI_ISL_524904, EPI_ISL_524905, EPI_ISL_524906, EPI_ISL_524907, EPI_ISL_524908, EPI_ISL_524909, EPI_ISL_524910, EPI_ISL_524911, EPI_ISL_524912, EPI_ISL_524913, EPI_ISL_524915, EPI_ISL_524917, EPI_ISL_524918, EPI_ISL_524919, EPI_ISL_524920, EPI_ISL_524921, EPI_ISL_524922, EPI_ISL_524923, EPI_ISL_524924, EPI_ISL_524925, EPI_ISL_524926, EPI_ISL_524927, EPI_ISL_524928, EPI_ISL_524929, EPI_ISL_524930, EPI_ISL_524931, EPI_ISL_524932, EPI_ISL_524933, EPI_ISL_524934, EPI_ISL_524935, EPI_ISL_524936, EPI_ISL_524937, EPI_ISL_524938, EPI_ISL_524939, EPI_ISL_524940, EPI_ISL_524942, EPI_ISL_524943, EPI_ISL_524944, EPI_ISL_524945, EPI_ISL_524946, EPI_ISL_524947, EPI_ISL_524948, EPI_ISL_524949, EPI_ISL_524950, EPI_ISL_524951, EPI_ISL_524952, EPI_ISL_524954, EPI_ISL_524955, EPI_ISL_524956, EPI_ISL_524957, EPI_ISL_524958, EPI_ISL_524959, EPI_ISL_524960, EPI_ISL_524961, EPI_ISL_524962, EPI_ISL_524963, EPI_ISL_524964, EPI_ISL_524965, EPI_ISL_524966, EPI_ISL_524967, EPI_ISL_524968, EPI_ISL_524969, EPI_ISL_524970, EPI_ISL_524971, EPI_ISL_524972, EPI_ISL_524973, EPI_ISL_524974, EPI_ISL_524975, EPI_ISL_524976, EPI_ISL_524977, EPI_ISL_524979, EPI_ISL_524980, EPI_ISL_524981, EPI_ISL_524982, EPI_ISL_524983, EPI_ISL_524984, EPI_ISL_524985, EPI_ISL_524986, EPI_ISL_524987, EPI_ISL_524988, EPI_ISL_524989, EPI_ISL_524990, EPI_ISL_524991, EPI_ISL_524992, EPI_ISL_524993, EPI_ISL_524994, EPI_ISL_524995, EPI_ISL_524996, EPI_ISL_524997, EPI_ISL_524998, EPI_ISL_524999, EPI_ISL_525000, EPI_ISL_525002, EPI_ISL_525003, EPI_ISL_525004, EPI_ISL_525006, EPI_ISL_525008, EPI_ISL_525009, EPI_ISL_525010, EPI_ISL_525011, EPI_ISL_525012, EPI_ISL_525013, EPI_ISL_525014, EPI_ISL_525015, EPI_ISL_525016, EPI_ISL_525017, EPI_ISL_525018, EPI_ISL_525019, EPI_ISL_525020, EPI_ISL_525021, EPI_ISL_525022, EPI_ISL_525023, EPI_ISL_525024, EPI_ISL_525025, EPI_ISL_525026, EPI_ISL_525027, EPI_ISL_525028, EPI_ISL_525029, EPI_ISL_525030, EPI_ISL_525031, EPI_ISL_525032, EPI_ISL_525033, EPI_ISL_525034, EPI_ISL_525035, EPI_ISL_525036, EPI_ISL_525037, EPI_ISL_525038, EPI_ISL_525039, EPI_ISL_525040, EPI_ISL_525041, EPI_ISL_525042, EPI_ISL_525043, EPI_ISL_525044, EPI_ISL_525045, EPI_ISL_525046, EPI_ISL_525047, EPI_ISL_525048, EPI_ISL_525049, EPI_ISL_525050, EPI_ISL_525051, EPI_ISL_525052, EPI_ISL_525053, EPI_ISL_525054, EPI_ISL_525056, EPI_ISL_525058, EPI_ISL_525060, EPI_ISL_525061, EPI_ISL_525062, EPI_ISL_525063, EPI_ISL_525064, EPI_ISL_525065, EPI_ISL_525067, EPI_ISL_525068, EPI_ISL_525070, EPI_ISL_525071, EPI_ISL_525072, EPI_ISL_525073, EPI_ISL_525074, EPI_ISL_525075, EPI_ISL_525076, EPI_ISL_525077, EPI_ISL_525078, EPI_ISL_525079, EPI_ISL_525080, EPI_ISL_525081, EPI_ISL_525082, EPI_ISL_525083, EPI_ISL_525084, EPI_ISL_525086, EPI_ISL_525087, EPI_ISL_525088, EPI_ISL_525089, EPI_ISL_525090, EPI_ISL_525091, EPI_ISL_525092, EPI_ISL_525093, EPI_ISL_525094, EPI_ISL_525095, EPI_ISL_525096, EPI_ISL_525097, EPI_ISL_525098, EPI_ISL_525099, EPI_ISL_525100, EPI_ISL_525101, EPI_ISL_525102, EPI_ISL_525103, EPI_ISL_525104, EPI_ISL_525105, EPI_ISL_525106, EPI_ISL_525107, EPI_ISL_525108, EPI_ISL_525109, EPI_ISL_525110, EPI_ISL_525112, EPI_ISL_525113, EPI_ISL_525114, EPI_ISL_525115, EPI_ISL_525116, EPI_ISL_525117 | | | |
| see above | Utah Public Health Laboratory | Utah Public Health Laboratory | Erin L. Young, Kelly Oakeson, Tara Gallagher, Michael T. Pyne, E. Susan Slechta, Melanie A. Mallory, Jeffrey B. Stevenson, Salika M. Shakir, David R. Hillyard |
| EPI_ISL_525118 | Maryland Department of Health Laboratory | Maryland Department of Health Laboratory | Maryland Department of Health Laboratories Administration |
| EPI_ISL_525119, EPI_ISL_525120, EPI_ISL_525121, EPI_ISL_525122, EPI_ISL_525123, EPI_ISL_525124, EPI_ISL_525125, EPI_ISL_525126, EPI_ISL_525127, EPI_ISL_525128, EPI_ISL_525129, EPI_ISL_525130, EPI_ISL_525131, EPI_ISL_525132, EPI_ISL_525133, EPI_ISL_525134, EPI_ISL_525136, EPI_ISL_525137, EPI_ISL_525138, EPI_ISL_525139, EPI_ISL_525140, EPI_ISL_525141, EPI_ISL_525142, EPI_ISL_525143, EPI_ISL_525144, EPI_ISL_525145, EPI_ISL_525146, EPI_ISL_525147, EPI_ISL_525148, EPI_ISL_525149, EPI_ISL_525150, EPI_ISL_525151, EPI_ISL_525152, EPI_ISL_525153, EPI_ISL_525154, EPI_ISL_525155, EPI_ISL_525157, EPI_ISL_525158, EPI_ISL_525159, EPI_ISL_525160, EPI_ISL_525161, EPI_ISL_525162, EPI_ISL_525163, EPI_ISL_525164, EPI_ISL_525165, EPI_ISL_525166, EPI_ISL_525167, EPI_ISL_525168, EPI_ISL_525169, EPI_ISL_525170, EPI_ISL_525171, EPI_ISL_525172, EPI_ISL_525173, EPI_ISL_525174, EPI_ISL_525175, EPI_ISL_525176, EPI_ISL_525177, EPI_ISL_525179, EPI_ISL_525180, EPI_ISL_525181, EPI_ISL_525182, EPI_ISL_525183, EPI_ISL_525184, EPI_ISL_525185, EPI_ISL_525186, EPI_ISL_525187, EPI_ISL_525188 | | | |
| see above | Utah Public Health Laboratory | Utah Public Health Laboratory | Erin L. Young, Kelly Oakeson, Tara Gallagher, Michael T. Pyne, E. Susan Slechta, Melanie A. Mallory, Jeffrey B. Stevenson, Salika M. Shakir, David R. Hillyard |
| EPI_ISL_525189, EPI_ISL_525190, EPI_ISL_525191, EPI_ISL_525192, EPI_ISL_525193, EPI_ISL_525194, EPI_ISL_525195, EPI_ISL_525196, EPI_ISL_525197, EPI_ISL_525198, EPI_ISL_525199, EPI_ISL_525200, EPI_ISL_525201, EPI_ISL_525202, EPI_ISL_525203, EPI_ISL_525204, EPI_ISL_525205 | | | |
| see above | Virginia DCLS | Virginia DCLS | Virginia DCLS |
| EPI_ISL_525207, EPI_ISL_525208, EPI_ISL_525209, EPI_ISL_525211, EPI_ISL_525212, EPI_ISL_525213, EPI_ISL_525214, EPI_ISL_525215, EPI_ISL_525216, EPI_ISL_525217, EPI_ISL_525218, EPI_ISL_525219, EPI_ISL_525221, EPI_ISL_525222, EPI_ISL_525223, EPI_ISL_525224, EPI_ISL_525225, EPI_ISL_525226, EPI_ISL_525227, EPI_ISL_525228, EPI_ISL_525229, EPI_ISL_525230, EPI_ISL_525231, EPI_ISL_525232, EPI_ISL_525233, EPI_ISL_525234, EPI_ISL_525235, EPI_ISL_525236, EPI_ISL_525237, EPI_ISL_525238, EPI_ISL_525239, EPI_ISL_525241, EPI_ISL_525242, EPI_ISL_525243, EPI_ISL_525244, EPI_ISL_525246, EPI_ISL_525247, EPI_ISL_525248, EPI_ISL_525249, EPI_ISL_525250, EPI_ISL_525251, EPI_ISL_525252, EPI_ISL_525253, EPI_ISL_525254, EPI_ISL_525255, EPI_ISL_525256, EPI_ISL_525257, EPI_ISL_525258, EPI_ISL_525259, EPI_ISL_525260, EPI_ISL_525261, EPI_ISL_525262, EPI_ISL_525263, EPI_ISL_525265, EPI_ISL_525266, EPI_ISL_525267, EPI_ISL_525268, EPI_ISL_525270, EPI_ISL_525271, EPI_ISL_525272, EPI_ISL_525273, EPI_ISL_525274, EPI_ISL_525275, EPI_ISL_525277, EPI_ISL_525278, EPI_ISL_525279, EPI_ISL_525280, EPI_ISL_525281, EPI_ISL_525282, EPI_ISL_525283, EPI_ISL_525284, EPI_ISL_525285, EPI_ISL_525286, EPI_ISL_525287, EPI_ISL_525288, EPI_ISL_525289, EPI_ISL_525290, EPI_ISL_525291, EPI_ISL_525292, EPI_ISL_525293, EPI_ISL_525294, EPI_ISL_525295, EPI_ISL_525296, EPI_ISL_525297, EPI_ISL_525298, EPI_ISL_525299, EPI_ISL_525300, EPI_ISL_525301, EPI_ISL_525302, EPI_ISL_525303, EPI_ISL_525304, EPI_ISL_525305, EPI_ISL_525306, EPI_ISL_525307, EPI_ISL_525309, EPI_ISL_525310, EPI_ISL_525311, EPI_ISL_525312, EPI_ISL_525313, EPI_ISL_525314, EPI_ISL_525315, EPI_ISL_525316, EPI_ISL_525317, EPI_ISL_525318, EPI_ISL_525319, EPI_ISL_525320, EPI_ISL_525321, EPI_ISL_525322, EPI_ISL_525323, EPI_ISL_525324, EPI_ISL_525325, EPI_ISL_525326, EPI_ISL_525327, EPI_ISL_525328, EPI_ISL_525329, EPI_ISL_525330, EPI_ISL_525331, EPI_ISL_525332, EPI_ISL_525333, EPI_ISL_525334, EPI_ISL_525335 | | | |

| | | | |
|---|---|---|---|
| see above | Utah Public Health Laboratory | Utah Public Health Laboratory | Erin L. Young, Kelly Oakeson, Tara Gallagher, Michael T. Pyne, E. Susan Slechta, Melanie A. Mallory, Jeffrey B. Stevenson, Salika M. Shakir, David R. Hillyard |
| EPI_ISL_525338, EPI_ISL_525339, EPI_ISL_525340, EPI_ISL_525342, EPI_ISL_525343, EPI_ISL_525344, EPI_ISL_525345, EPI_ISL_525346, EPI_ISL_525347, EPI_ISL_525348, EPI_ISL_525350, EPI_ISL_525351 | | | |
| see above | Utah Public Health Laboratory | Utah Public Health Laboratory | Erin Young, Kelly Oakeson |
| EPI_ISL_525575, EPI_ISL_525576, EPI_ISL_525577, EPI_ISL_525578, EPI_ISL_525579, EPI_ISL_525580, EPI_ISL_525581, EPI_ISL_525582, EPI_ISL_525583, EPI_ISL_525584, EPI_ISL_525585, EPI_ISL_525586, EPI_ISL_525587, EPI_ISL_525588, EPI_ISL_525589, EPI_ISL_525590, EPI_ISL_525591, EPI_ISL_525592, EPI_ISL_525593, EPI_ISL_525594, EPI_ISL_525595, EPI_ISL_525596, EPI_ISL_525597, EPI_ISL_525598, EPI_ISL_525599, EPI_ISL_525600, EPI_ISL_525601, EPI_ISL_525602, EPI_ISL_525603, EPI_ISL_525604, EPI_ISL_525605, EPI_ISL_525606, EPI_ISL_525607, EPI_ISL_525608, EPI_ISL_525609, EPI_ISL_525610, EPI_ISL_525611, EPI_ISL_525612, EPI_ISL_525613, EPI_ISL_525614, EPI_ISL_525615, EPI_ISL_525616, EPI_ISL_525617, EPI_ISL_525618, EPI_ISL_525619, EPI_ISL_525620, EPI_ISL_525621, EPI_ISL_525622, EPI_ISL_525623, EPI_ISL_525624, EPI_ISL_525625, EPI_ISL_525626, EPI_ISL_525627, EPI_ISL_525628, EPI_ISL_525629, EPI_ISL_525630, EPI_ISL_525631, EPI_ISL_525632, EPI_ISL_525633, EPI_ISL_525634, EPI_ISL_525635, EPI_ISL_525636, EPI_ISL_525637, EPI_ISL_525638, EPI_ISL_525639, EPI_ISL_525640, EPI_ISL_525641, EPI_ISL_525642, EPI_ISL_525643, EPI_ISL_525644, EPI_ISL_525645, EPI_ISL_525646, EPI_ISL_525647, EPI_ISL_525648, EPI_ISL_525649, EPI_ISL_525650, EPI_ISL_525651, EPI_ISL_525652, EPI_ISL_525653, EPI_ISL_525654, EPI_ISL_525655, EPI_ISL_525656, EPI_ISL_525657, EPI_ISL_525658, EPI_ISL_525659, EPI_ISL_525660, EPI_ISL_525661, EPI_ISL_525662, EPI_ISL_525663, EPI_ISL_525664, EPI_ISL_525665, EPI_ISL_525666, EPI_ISL_525667, EPI_ISL_525668, EPI_ISL_525669, EPI_ISL_525670, EPI_ISL_525671, EPI_ISL_525672, EPI_ISL_525673, EPI_ISL_525674, EPI_ISL_525675, EPI_ISL_525676, EPI_ISL_525677, EPI_ISL_525678, EPI_ISL_525679, EPI_ISL_525680, EPI_ISL_525681, EPI_ISL_525682, EPI_ISL_525683, EPI_ISL_525684, EPI_ISL_525685, EPI_ISL_525686 | | | |
| see above | Wadsworth Center, New York State Department of Health | Wadsworth Center, New York State Department of Health | Kirsten St. George, Daryl M. Lamson, Sara Griesemer, Jonathan Plitnick, Navjot Singh, Matthew D. Shudt, Erica Lasek-Nesselquist |
| EPI_ISL_525699 | Seattle Flu Study | Seattle Flu Study | Deborah A. Nickerson, Chris D. Frazar, Jover Lee, Benjamin Pelle, Matthew Richardson, Amanda Adler, Elisabeth Brandstetter, Peter D. Han, Kairsten Fay, Misja Ilcisin, Kirsten Lacombe, Thomas R. Sibley, Melissa Truong, Caitlin R. Wolf, Karen Cowgill, Stephanie Schrag, Jeff Duchin, Michael Boeckh, Janet A. Englund, Michael Famulare, Barry R. Lutz, Mark J. Rieder, Lea M. Starita, Matthew Thompson, Helen Y. Chu, Trevor Bedford, Jay Shendure |
| EPI_ISL_525700, EPI_ISL_525701 | Seattle Flu Study | Seattle Flu Study | Deborah A. Nickerson, Chris D. Frazar, Jover Lee, Benjamin Pelle, Matthew Richardson, Amanda Adler, Elisabeth Brandstetter, Peter D. Han, Kairsten Fay, Misja Ilcisin, Kirsten Lacombe, Thomas R. Sibley, Melissa Truong, Caitlin R. Wolf, Michael Boeckh, Janet A. Englund, Michael Famulare, Barry R. Lutz, Mark J. Rieder, Lea M. Starita, Matthew Thompson, Jay Shendure, Trevor Bedford, Helen Y. Chu |
| EPI_ISL_525702, EPI_ISL_525703, EPI_ISL_525704, EPI_ISL_525705 | Seattle Flu Study | Seattle Flu Study | Deborah A. Nickerson, Chris D. Frazar, Jover Lee, Benjamin Pelle, Matthew Richardson, Amanda Adler, Elisabeth Brandstetter, Peter D. Han, Kairsten Fay, Misja Ilcisin, Kirsten Lacombe, Thomas R. Sibley, Melissa Truong, Caitlin R. Wolf, Karen Cowgill, Stephanie Schrag, Jeff Duchin, Michael Boeckh, Janet A. Englund, Michael Famulare, Barry R. Lutz, Mark J. Rieder, Lea M. Starita, Matthew Thompson, Helen Y. Chu, Trevor Bedford, Jay Shendure |
| EPI_ISL_525706 | Seattle Flu Study | Seattle Flu Study | Deborah A. Nickerson, Chris D. Frazar, Jover Lee, Benjamin Pelle, Matthew Richardson, Amanda Adler, Elisabeth Brandstetter, Peter D. Han, Kairsten Fay, Misja Ilcisin, Kirsten Lacombe, Thomas R. Sibley, Melissa Truong, Caitlin R. Wolf, Michael Boeckh, Janet A. Englund, Michael Famulare, Barry R. Lutz, Mark J. Rieder, Lea M. Starita, Matthew Thompson, Jay Shendure, Trevor Bedford, Helen Y. Chu |
| EPI_ISL_525707, EPI_ISL_525708, EPI_ISL_525709, EPI_ISL_525710, EPI_ISL_525711, EPI_ISL_525712, EPI_ISL_525713, EPI_ISL_525714, EPI_ISL_525715, EPI_ISL_525716, EPI_ISL_525717, EPI_ISL_525718, EPI_ISL_525719, EPI_ISL_525720, EPI_ISL_525721, EPI_ISL_525722, EPI_ISL_525723, EPI_ISL_525724, EPI_ISL_525725, EPI_ISL_525726, EPI_ISL_525727 | | | |
| see above | Seattle Flu Study | Seattle Flu Study | Deborah A. Nickerson, Chris D. Frazar, Jover Lee, Benjamin Pelle, Matthew Richardson, Amanda Adler, Elisabeth Brandstetter, Peter D. Han, Kairsten Fay, Misja Ilcisin, Kirsten Lacombe, Thomas R. Sibley, Melissa Truong, Caitlin R. Wolf, Karen Cowgill, Stephanie Schrag, Jeff Duchin, Michael Boeckh, Janet A. Englund, Michael Famulare, Barry R. Lutz, Mark J. Rieder, Lea M. Starita, Matthew Thompson, Helen Y. Chu, Trevor Bedford, Jay Shendure |
| EPI_ISL_525728 | Seattle Flu Study | Seattle Flu Study | Deborah A. Nickerson, Chris D. Frazar, Jover Lee, Benjamin Pelle, Matthew Richardson, Amanda Adler, Elisabeth Brandstetter, Peter D. Han, Kairsten Fay, Misja Ilcisin, Kirsten Lacombe, Thomas R. Sibley, Melissa Truong, Caitlin R. Wolf, Michael Boeckh, Janet A. Englund, Michael Famulare, Barry R. Lutz, Mark J. Rieder, Lea M. Starita, Matthew Thompson, Jay Shendure, Trevor Bedford, Helen Y. Chu |
| EPI_ISL_525729, EPI_ISL_525730, EPI_ISL_525731, EPI_ISL_525732, EPI_ISL_525733, EPI_ISL_525734, EPI_ISL_525735, EPI_ISL_525736, EPI_ISL_525737, EPI_ISL_525738, EPI_ISL_525739, EPI_ISL_525741 | | | |
| see above | Seattle Flu Study | Seattle Flu Study | Deborah A. Nickerson, Chris D. Frazar, Jover Lee, Benjamin Pelle, Matthew Richardson, Amanda Adler, Elisabeth Brandstetter, Peter D. Han, Kairsten Fay, Misja Ilcisin, Kirsten Lacombe, Thomas R. Sibley, Melissa Truong, Caitlin R. Wolf, Karen Cowgill, Stephanie Schrag, Jeff Duchin, Michael Boeckh, Janet A. Englund, Michael Famulare, Barry R. Lutz, Mark J. Rieder, Lea M. Starita, Matthew Thompson, Helen Y. Chu, Trevor Bedford, Jay Shendure |
| EPI_ISL_525742 | Seattle Flu Study | Seattle Flu Study | Deborah A. Nickerson, Chris D. Frazar, Jover Lee, Benjamin Pelle, Matthew Richardson, Amanda Adler, Elisabeth Brandstetter, Peter D. Han, Kairsten Fay, Misja Ilcisin, Kirsten Lacombe, Thomas R. Sibley, Melissa Truong, Caitlin R. Wolf, Michael Boeckh, Janet A. Englund, Michael Famulare, Barry R. Lutz, Mark J. Rieder, Lea M. Starita, Matthew Thompson, Jay Shendure, Trevor Bedford, Helen Y. Chu |
| EPI_ISL_525743, EPI_ISL_525744, EPI_ISL_525745, EPI_ISL_525746, EPI_ISL_525747, EPI_ISL_525748, EPI_ISL_525749, EPI_ISL_525751, EPI_ISL_525753, EPI_ISL_525754, EPI_ISL_525755, EPI_ISL_525756 | | | |
| see above | Seattle Flu Study | Seattle Flu Study | Deborah A. Nickerson, Chris D. Frazar, Jover Lee, Benjamin Pelle, Matthew Richardson, Amanda Adler, Elisabeth Brandstetter, Peter D. Han, Kairsten Fay, Misja Ilcisin, Kirsten Lacombe, Thomas R. Sibley, Melissa Truong, Caitlin R. Wolf, Karen Cowgill, Stephanie Schrag, Jeff Duchin, Michael Boeckh, Janet A. Englund, Michael Famulare, Barry R. Lutz, Mark J. Rieder, Lea M. Starita, Matthew Thompson, Helen Y. Chu, Trevor Bedford, Jay Shendure |
| EPI_ISL_525758, EPI_ISL_525759, EPI_ISL_525760 | Alaska State Virology Laboratory | Alaska State Virology Laboratory | Jack Chen, Ph.D. |
| EPI_ISL_525763, EPI_ISL_525764, EPI_ISL_525766, EPI_ISL_525767, EPI_ISL_525768, EPI_ISL_525769, EPI_ISL_525770, EPI_ISL_525771, EPI_ISL_525775, EPI_ISL_525776, EPI_ISL_525777, EPI_ISL_525779, EPI_ISL_525781, EPI_ISL_525782, EPI_ISL_525784, EPI_ISL_525785, EPI_ISL_525786, EPI_ISL_525787, EPI_ISL_525788, EPI_ISL_525789 | | | |
| see above | Texas Department of State Health Services | Texas Department of State Health Services | Jenny Zhang, Rashmi Tuladhar, Bonnie Oh, Maliha Rahman, Anita Pokharel, Myong Koag, Chun Wang, Rachel Lee, Grace Kubin |
| EPI_ISL_525802 | OHSU Lab Services Molecular Microbiology Lab | Oregon SARS-CoV-2 Genome Sequencing Center | Brendan L. O'Connell, Ruth V. Nichols, Alec J. Hirsch, Guang Fan, Daniel N. Streblow, William B. Messer, Andrew C. Adey, Benjamin N. Bimber, Brian J. O'Roak |
| EPI_ISL_525803 | OHSU Lab Services Molecular Microbiology Lab | Ginkgo Bioworks Clinical Laboratory | Brendan L. O'Connell, Ruth V. Nichols, Alec J. Hirsch, Guang Fan, Daniel N. Streblow, Malaika Mckenzie-Bennett, James McGann, Jim Griffin, Keith Robison, Alex Plocik, Becky Schilling, Rebecca Littlefield, Michelle Spencer, Birgitte Simen, William B. Messer, Andrew C. Adey, Benjamin N. Bimber, Brian J. O'Roak |
| EPI_ISL_525804, EPI_ISL_525805, EPI_ISL_525806, EPI_ISL_525807 | OHSU Lab Services Molecular Microbiology Lab | Oregon SARS-CoV-2 Genome Sequencing Center | Brendan L. O'Connell, Ruth V. Nichols, Alec J. Hirsch, Guang Fan, Daniel N. Streblow, William B. Messer, Andrew C. Adey, Benjamin N. Bimber, Brian J. O'Roak |
| EPI_ISL_525808, EPI_ISL_525809, EPI_ISL_525811, EPI_ISL_525812, EPI_ISL_525813, EPI_ISL_525814, EPI_ISL_525815, EPI_ISL_525816, EPI_ISL_525817, EPI_ISL_525818, EPI_ISL_525820, EPI_ISL_525821, EPI_ISL_525822, EPI_ISL_525823, EPI_ISL_525824 | | | |
| see above | OHSU Lab Services Molecular Microbiology Lab | Ginkgo Bioworks Clinical Laboratory | Brendan L. O'Connell, Ruth V. Nichols, Alec J. Hirsch, Guang Fan, Daniel N. Streblow, Malaika Mckenzie-Bennett, James McGann, Jim Griffin, Keith Robison, Alex Plocik, Becky Schilling, Rebecca Littlefield, Michelle Spencer, Birgitte Simen, William B. Messer, Andrew C. Adey, Benjamin N. Bimber, Brian J. O'Roak |
| EPI_ISL_525825 | OHSU Lab Services Molecular Microbiology Lab | Oregon SARS-CoV-2 Genome Sequencing Center | Brendan L. O'Connell, Ruth V. Nichols, Alec J. Hirsch, Guang Fan, Daniel N. Streblow, William B. Messer, Andrew C. Adey, Benjamin N. Bimber, Brian J. O'Roak |
| EPI_ISL_525826 | OHSU Lab Services Molecular Microbiology Lab | Ginkgo Bioworks Clinical Laboratory | Brendan L. O'Connell, Ruth V. Nichols, Alec J. Hirsch, Guang Fan, Daniel N. Streblow, Malaika Mckenzie-Bennett, James McGann, Jim Griffin, Keith Robison, Alex Plocik, Becky Schilling, Rebecca Littlefield, Michelle Spencer, Birgitte Simen, William B. Messer, Andrew C. Adey, Benjamin N. Bimber, Brian J. O'Roak |
| EPI_ISL_525827 | OHSU Lab Services Molecular Microbiology Lab | Oregon SARS-CoV-2 Genome Sequencing Center | Brendan L. O'Connell, Ruth V. Nichols, Alec J. Hirsch, Guang Fan, Daniel N. Streblow, William B. Messer, Andrew C. Adey, Benjamin N. Bimber, Brian J. O'Roak |
| EPI_ISL_525828, EPI_ISL_525829, EPI_ISL_525830, EPI_ISL_525831, EPI_ISL_525832 | OHSU Lab Services Molecular Microbiology Lab | Ginkgo Bioworks Clinical Laboratory | Brendan L. O'Connell, Ruth V. Nichols, Alec J. Hirsch, Guang Fan, Daniel N. Streblow, Malaika Mckenzie-Bennett, James McGann, Jim Griffin, Keith Robison, Alex Plocik, Becky Schilling, Rebecca Littlefield, Michelle Spencer, Birgitte Simen, William B. Messer, Andrew C. Adey, Benjamin N. Bimber, Brian J. O'Roak |
| EPI_ISL_525833 | OHSU Lab Services Molecular Microbiology Lab | Oregon SARS-CoV-2 Genome Sequencing Center | Brendan L. O'Connell, Ruth V. Nichols, Alec J. Hirsch, Guang Fan, Daniel N. Streblow, William B. Messer, Andrew C. Adey, Benjamin N. Bimber, Brian J. O'Roak |
| EPI_ISL_525835, EPI_ISL_525838, EPI_ISL_525839, EPI_ISL_525840 | OHSU Lab Services Molecular Microbiology Lab | Ginkgo Bioworks Clinical Laboratory | Brendan L. O'Connell, Ruth V. Nichols, Alec J. Hirsch, Guang Fan, Daniel N. Streblow, Malaika Mckenzie-Bennett, James McGann, Jim Griffin, Keith Robison, Alex Plocik, Becky Schilling, Rebecca Littlefield, Michelle Spencer, Birgitte Simen, William B. Messer, Andrew C. Adey, Benjamin N. Bimber, Brian J. O'Roak |
| EPI_ISL_525841 | OHSU Lab Services Molecular Microbiology Lab | Oregon SARS-CoV-2 Genome Sequencing Center | Brendan L. O'Connell, Ruth V. Nichols, Alec J. Hirsch, Guang Fan, Daniel N. Streblow, William B. Messer, Andrew C. Adey, Benjamin N. Bimber, Brian J. O'Roak |
| EPI_ISL_525842, EPI_ISL_525843, EPI_ISL_525844, EPI_ISL_525845, EPI_ISL_525846 | OHSU Lab Services Molecular Microbiology Lab | Ginkgo Bioworks Clinical Laboratory | Brendan L. O'Connell, Ruth V. Nichols, Alec J. Hirsch, Guang Fan, Daniel N. Streblow, Malaika Mckenzie-Bennett, James McGann, Jim Griffin, Keith Robison, Alex Plocik, Becky Schilling, Rebecca Littlefield, Michelle Spencer, Birgitte Simen, William B. Messer, Andrew C. Adey, Benjamin N. Bimber, Brian J. O'Roak |
| EPI_ISL_525847 | OHSU Lab Services Molecular Microbiology Lab | Oregon SARS-CoV-2 Genome Sequencing Center | Brendan L. O'Connell, Ruth V. Nichols, Alec J. Hirsch, Guang Fan, Daniel N. Streblow, William B. Messer, Andrew C. Adey, Benjamin N. Bimber, Brian J. O'Roak |
| EPI_ISL_525848 | OHSU Lab Services Molecular Microbiology Lab | Ginkgo Bioworks Clinical Laboratory | Brendan L. O'Connell, Ruth V. Nichols, Alec J. Hirsch, Guang Fan, Daniel N. Streblow, Malaika Mckenzie-Bennett, James McGann, Jim Griffin, Keith Robison, Alex Plocik, Becky Schilling, Rebecca Littlefield, Michelle Spencer, Birgitte Simen, William B. Messer, Andrew C. Adey, Benjamin N. Bimber, Brian J. O'Roak |
| EPI_ISL_525849, EPI_ISL_525851 | OHSU Lab Services Molecular Microbiology Lab | Oregon SARS-CoV-2 Genome Sequencing Center | Brendan L. O'Connell, Ruth V. Nichols, Alec J. Hirsch, Guang Fan, Daniel N. Streblow, William B. Messer, Andrew C. Adey, Benjamin N. Bimber, Brian J. O'Roak |
| EPI_ISL_525852, EPI_ISL_525853, EPI_ISL_525854, EPI_ISL_525855 | OHSU Lab Services Molecular Microbiology Lab | Ginkgo Bioworks Clinical Laboratory | Brendan L. O'Connell, Ruth V. Nichols, Alec J. Hirsch, Guang Fan, Daniel N. Streblow, Malaika Mckenzie-Bennett, James McGann, Jim Griffin, Keith Robison, Alex Plocik, Becky Schilling, Rebecca Littlefield, Michelle Spencer, Birgitte Simen, William B. Messer, Andrew C. Adey, Benjamin N. Bimber, Brian J. O'Roak |
| EPI_ISL_525856, EPI_ISL_525857, EPI_ISL_525858 | OHSU Lab Services Molecular Microbiology Lab | Oregon SARS-CoV-2 Genome Sequencing Center | Brendan L. O'Connell, Ruth V. Nichols, Alec J. Hirsch, Guang Fan, Daniel N. Streblow, William B. Messer, Andrew C. Adey, Benjamin N. Bimber, Brian J. O'Roak |
| EPI_ISL_525859 | OHSU Lab Services Molecular Microbiology Lab | Ginkgo Bioworks Clinical Laboratory | Brendan L. O'Connell, Ruth V. Nichols, Alec J. Hirsch, Guang Fan, Daniel N. Streblow, Malaika Mckenzie-Bennett, James McGann, Jim Griffin, Keith Robison, Alex Plocik, Becky Schilling, Rebecca Littlefield, Michelle Spencer, Birgitte Simen, William B. Messer, Andrew C. Adey, Benjamin N. Bimber, Brian J. O'Roak |
| EPI_ISL_525860 | OHSU Lab Services Molecular Microbiology Lab | Oregon SARS-CoV-2 Genome Sequencing Center | Brendan L. O'Connell, Ruth V. Nichols, Alec J. Hirsch, Guang Fan, Daniel N. Streblow, William B. Messer, Andrew C. Adey, Benjamin N. Bimber, Brian J. O'Roak |
| EPI_ISL_525861 | OHSU Lab Services Molecular | Ginkgo Bioworks Clinical Laboratory | Brendan L. O'Connell, Ruth V. Nichols, Alec J. Hirsch, Guang Fan, Daniel N. Streblow, Malaika Mckenzie-Bennett, James McGann, Jim Griffin, Keith Robison, Alex Plocik, Becky Schilling, Rebecca Littlefield, Michelle Spencer, Birgitte Simen, William B. Messer, Andrew C. Adey, Benjamin N. |

| | | | |
|---|---|---|---|
| | Microbiology Lab | | Bimber, Brian J. O’Roak |
| EPI_ISL_525862 | OHSU Lab Services Molecular Microbiology Lab | Oregon SARS-CoV-2 Genome Sequencing Center | Brendan L. O'Connell, Ruth V. Nichols, Alec J. Hirsch, Guang Fan, Daniel N. Streblow, William B. Messer, Andrew C. Adey, Benjamin N. Bimber, Brian J. O’Roak |
| EPI_ISL_525863, EPI_ISL_525864 | OHSU Lab Services Molecular Microbiology Lab | Ginkgo Bioworks Clinical Laboratory | Brendan L. O'Connell, Ruth V. Nichols, Alec J. Hirsch, Guang Fan, Daniel N. Streblow, Malaika Mckenzie-Bennett, James McGann, Jim Griffin, Keith Robison, Alex Plocik, Becky Schilling, Rebecca Littlefield, Michelle Spencer, Birgitte Simen, William B. Messer, Andrew C. Adey, Benjamin N. Bimber, Brian J. O’Roak |
| EPI_ISL_525865 | OHSU Lab Services Molecular Microbiology Lab | Oregon SARS-CoV-2 Genome Sequencing Center | Brendan L. O'Connell, Ruth V. Nichols, Alec J. Hirsch, Guang Fan, Daniel N. Streblow, William B. Messer, Andrew C. Adey, Benjamin N. Bimber, Brian J. O’Roak |
| EPI_ISL_525866, EPI_ISL_525867 | OHSU Lab Services Molecular Microbiology Lab | Ginkgo Bioworks Clinical Laboratory | Brendan L. O'Connell, Ruth V. Nichols, Alec J. Hirsch, Guang Fan, Daniel N. Streblow, Malaika Mckenzie-Bennett, James McGann, Jim Griffin, Keith Robison, Alex Plocik, Becky Schilling, Rebecca Littlefield, Michelle Spencer, Birgitte Simen, William B. Messer, Andrew C. Adey, Benjamin N. Bimber, Brian J. O’Roak |
| EPI_ISL_525868 | OHSU Lab Services Molecular Microbiology Lab | Oregon SARS-CoV-2 Genome Sequencing Center | Brendan L. O'Connell, Ruth V. Nichols, Alec J. Hirsch, Guang Fan, Daniel N. Streblow, William B. Messer, Andrew C. Adey, Benjamin N. Bimber, Brian J. O’Roak |
| EPI_ISL_525869 | OHSU Lab Services Molecular Microbiology Lab | Ginkgo Bioworks Clinical Laboratory | Brendan L. O'Connell, Ruth V. Nichols, Alec J. Hirsch, Guang Fan, Daniel N. Streblow, Malaika Mckenzie-Bennett, James McGann, Jim Griffin, Keith Robison, Alex Plocik, Becky Schilling, Rebecca Littlefield, Michelle Spencer, Birgitte Simen, William B. Messer, Andrew C. Adey, Benjamin N. Bimber, Brian J. O’Roak |
| EPI_ISL_525870 | OHSU Lab Services Molecular Microbiology Lab | Oregon SARS-CoV-2 Genome Sequencing Center | Brendan L. O'Connell, Ruth V. Nichols, Alec J. Hirsch, Guang Fan, Daniel N. Streblow, William B. Messer, Andrew C. Adey, Benjamin N. Bimber, Brian J. O’Roak |
| EPI_ISL_525871, EPI_ISL_525872, EPI_ISL_525873 | OHSU Lab Services Molecular Microbiology Lab | Ginkgo Bioworks Clinical Laboratory | Brendan L. O'Connell, Ruth V. Nichols, Alec J. Hirsch, Guang Fan, Daniel N. Streblow, Malaika Mckenzie-Bennett, James McGann, Jim Griffin, Keith Robison, Alex Plocik, Becky Schilling, Rebecca Littlefield, Michelle Spencer, Birgitte Simen, William B. Messer, Andrew C. Adey, Benjamin N. Bimber, Brian J. O’Roak |
| EPI_ISL_525874 | OHSU Lab Services Molecular Microbiology Lab | Oregon SARS-CoV-2 Genome Sequencing Center | Brendan L. O'Connell, Ruth V. Nichols, Alec J. Hirsch, Guang Fan, Daniel N. Streblow, William B. Messer, Andrew C. Adey, Benjamin N. Bimber, Brian J. O’Roak |
| EPI_ISL_525875 | OHSU Lab Services Molecular Microbiology Lab | Ginkgo Bioworks Clinical Laboratory | Brendan L. O'Connell, Ruth V. Nichols, Alec J. Hirsch, Guang Fan, Daniel N. Streblow, Malaika Mckenzie-Bennett, James McGann, Jim Griffin, Keith Robison, Alex Plocik, Becky Schilling, Rebecca Littlefield, Michelle Spencer, Birgitte Simen, William B. Messer, Andrew C. Adey, Benjamin N. Bimber, Brian J. O’Roak |
| EPI_ISL_525876 | OHSU Lab Services Molecular Microbiology Lab | Oregon SARS-CoV-2 Genome Sequencing Center | Brendan L. O'Connell, Ruth V. Nichols, Alec J. Hirsch, Guang Fan, Daniel N. Streblow, William B. Messer, Andrew C. Adey, Benjamin N. Bimber, Brian J. O’Roak |
| EPI_ISL_525877 | OHSU Lab Services Molecular Microbiology Lab | Ginkgo Bioworks Clinical Laboratory | Brendan L. O'Connell, Ruth V. Nichols, Alec J. Hirsch, Guang Fan, Daniel N. Streblow, Malaika Mckenzie-Bennett, James McGann, Jim Griffin, Keith Robison, Alex Plocik, Becky Schilling, Rebecca Littlefield, Michelle Spencer, Birgitte Simen, William B. Messer, Andrew C. Adey, Benjamin N. Bimber, Brian J. O’Roak |
| EPI_ISL_525878, EPI_ISL_525880, EPI_ISL_525881, EPI_ISL_525883 | OHSU Lab Services Molecular Microbiology Lab | Oregon SARS-CoV-2 Genome Sequencing Center | Brendan L. O'Connell, Ruth V. Nichols, Alec J. Hirsch, Guang Fan, Daniel N. Streblow, William B. Messer, Andrew C. Adey, Benjamin N. Bimber, Brian J. O’Roak |
| EPI_ISL_525884 | OHSU Lab Services Molecular Microbiology Lab | Ginkgo Bioworks Clinical Laboratory | Brendan L. O'Connell, Ruth V. Nichols, Alec J. Hirsch, Guang Fan, Daniel N. Streblow, Malaika Mckenzie-Bennett, James McGann, Jim Griffin, Keith Robison, Alex Plocik, Becky Schilling, Rebecca Littlefield, Michelle Spencer, Birgitte Simen, William B. Messer, Andrew C. Adey, Benjamin N. Bimber, Brian J. O’Roak |
| EPI_ISL_525885 | OHSU Lab Services Molecular Microbiology Lab | Oregon SARS-CoV-2 Genome Sequencing Center | Brendan L. O'Connell, Ruth V. Nichols, Alec J. Hirsch, Guang Fan, Daniel N. Streblow, William B. Messer, Andrew C. Adey, Benjamin N. Bimber, Brian J. O’Roak |
| EPI_ISL_525886, EPI_ISL_525887, EPI_ISL_525888, EPI_ISL_525889 | OHSU Lab Services Molecular Microbiology Lab | Ginkgo Bioworks Clinical Laboratory | Brendan L. O'Connell, Ruth V. Nichols, Alec J. Hirsch, Guang Fan, Daniel N. Streblow, Malaika Mckenzie-Bennett, James McGann, Jim Griffin, Keith Robison, Alex Plocik, Becky Schilling, Rebecca Littlefield, Michelle Spencer, Birgitte Simen, William B. Messer, Andrew C. Adey, Benjamin N. Bimber, Brian J. O’Roak |
| EPI_ISL_525891, EPI_ISL_525892, EPI_ISL_525893, EPI_ISL_525894, EPI_ISL_525895, EPI_ISL_525896, EPI_ISL_525897, EPI_ISL_525898, EPI_ISL_525899, EPI_ISL_525900, EPI_ISL_525901, EPI_ISL_525902, EPI_ISL_525903, EPI_ISL_525904, EPI_ISL_525905, EPI_ISL_525906, EPI_ISL_525907, EPI_ISL_525908, EPI_ISL_525909, EPI_ISL_525910, EPI_ISL_525911, EPI_ISL_525912, EPI_ISL_525913, EPI_ISL_525914, EPI_ISL_525915, EPI_ISL_525916, EPI_ISL_525917, EPI_ISL_525918, EPI_ISL_525919, EPI_ISL_525920, EPI_ISL_525921, EPI_ISL_525922, EPI_ISL_525923, EPI_ISL_525924, EPI_ISL_525925, EPI_ISL_525926, EPI_ISL_525927, EPI_ISL_525928, EPI_ISL_525929, EPI_ISL_525930, EPI_ISL_525931, EPI_ISL_525932, EPI_ISL_525933, EPI_ISL_525934, EPI_ISL_525935, EPI_ISL_525936, EPI_ISL_525937, EPI_ISL_525938, EPI_ISL_525939, EPI_ISL_525940, EPI_ISL_525941, EPI_ISL_525942, EPI_ISL_525943, EPI_ISL_525944, EPI_ISL_525945, EPI_ISL_525946, EPI_ISL_525947, EPI_ISL_525948, EPI_ISL_525949, EPI_ISL_525950, EPI_ISL_525951, EPI_ISL_525952, EPI_ISL_525953, EPI_ISL_525954, EPI_ISL_525955, EPI_ISL_525956, EPI_ISL_525957, EPI_ISL_525958, EPI_ISL_525959, EPI_ISL_525960, EPI_ISL_525961, EPI_ISL_525962, EPI_ISL_525963, EPI_ISL_525964, EPI_ISL_525965, EPI_ISL_525966, EPI_ISL_525967, EPI_ISL_525968, EPI_ISL_525969, EPI_ISL_525970, EPI_ISL_525971, EPI_ISL_525972, EPI_ISL_525973, EPI_ISL_525974, EPI_ISL_525975, EPI_ISL_525976, EPI_ISL_525977, EPI_ISL_525978, EPI_ISL_525979, EPI_ISL_525980, EPI_ISL_525981, EPI_ISL_525982, EPI_ISL_525983, EPI_ISL_525984, EPI_ISL_525985, EPI_ISL_525986, EPI_ISL_525987, EPI_ISL_525988, EPI_ISL_525989, EPI_ISL_525990, EPI_ISL_525991, EPI_ISL_525992, EPI_ISL_525993, EPI_ISL_525994, EPI_ISL_525995, EPI_ISL_525996, EPI_ISL_525997, EPI_ISL_525998, EPI_ISL_525999, EPI_ISL_526000, EPI_ISL_526001, EPI_ISL_526002, EPI_ISL_526003, EPI_ISL_526004, EPI_ISL_526005, EPI_ISL_526006, EPI_ISL_526007, EPI_ISL_526008, EPI_ISL_526009, EPI_ISL_526010, EPI_ISL_526011, EPI_ISL_526012, EPI_ISL_526013, EPI_ISL_526014, EPI_ISL_526015, EPI_ISL_526016, EPI_ISL_526017, EPI_ISL_526018, EPI_ISL_526019, EPI_ISL_526020, EPI_ISL_526021, EPI_ISL_526022, EPI_ISL_526023, EPI_ISL_526024, EPI_ISL_526025, EPI_ISL_526026, EPI_ISL_526027, EPI_ISL_526028, EPI_ISL_526029, EPI_ISL_526030, EPI_ISL_526031, EPI_ISL_526032, EPI_ISL_526033, EPI_ISL_526034, EPI_ISL_526035, EPI_ISL_526036, EPI_ISL_526037, EPI_ISL_526038, EPI_ISL_526039, EPI_ISL_526040, EPI_ISL_526041, EPI_ISL_526042, EPI_ISL_526043, EPI_ISL_526044, EPI_ISL_526045, EPI_ISL_526046, EPI_ISL_526047, EPI_ISL_526048, EPI_ISL_526049, EPI_ISL_526050, EPI_ISL_526051, EPI_ISL_526052, EPI_ISL_526053, EPI_ISL_526054, EPI_ISL_526055, EPI_ISL_526056, EPI_ISL_526057, EPI_ISL_526058, EPI_ISL_526059, EPI_ISL_526060, EPI_ISL_526061, EPI_ISL_526062, EPI_ISL_526063, EPI_ISL_526064, EPI_ISL_526065, EPI_ISL_526066, EPI_ISL_526067, EPI_ISL_526068, EPI_ISL_526069, EPI_ISL_526070, EPI_ISL_526071, EPI_ISL_526072, EPI_ISL_526073, EPI_ISL_526074, EPI_ISL_526075, EPI_ISL_526076, EPI_ISL_526077, EPI_ISL_526078, EPI_ISL_526079, EPI_ISL_526080, EPI_ISL_526081, EPI_ISL_526082, EPI_ISL_526083, EPI_ISL_526084, EPI_ISL_526085, EPI_ISL_526086 | | | |
| see above | OHSU Lab Services Molecular Microbiology Lab | Oregon SARS-CoV-2 Genome Sequencing Center | Brendan L. O'Connell, Ruth V. Nichols, Alec J. Hirsch, Guang Fan, Daniel N. Streblow, William B. Messer, Andrew C. Adey, Benjamin N. Bimber, Brian J. O’Roak |
| EPI_ISL_526087, EPI_ISL_526088, EPI_ISL_526089, EPI_ISL_526090 | OHSU Lab Services Molecular Microbiology Lab | Ginkgo Bioworks Clinical Laboratory | Brendan L. O'Connell, Ruth V. Nichols, Alec J. Hirsch, Guang Fan, Daniel N. Streblow, Malaika Mckenzie-Bennett, James McGann, Jim Griffin, Keith Robison, Alex Plocik, Becky Schilling, Rebecca Littlefield, Michelle Spencer, Birgitte Simen, William B. Messer, Andrew C. Adey, Benjamin N. Bimber, Brian J. O’Roak |
| EPI_ISL_526091 | OHSU Lab Services Molecular Microbiology Lab | Oregon SARS-CoV-2 Genome Sequencing Center | Brendan L. O'Connell, Ruth V. Nichols, Alec J. Hirsch, Guang Fan, Daniel N. Streblow, William B. Messer, Andrew C. Adey, Benjamin N. Bimber, Brian J. O’Roak |
| EPI_ISL_526092, EPI_ISL_526093 | OHSU Lab Services Molecular Microbiology Lab | Ginkgo Bioworks Clinical Laboratory | Brendan L. O'Connell, Ruth V. Nichols, Alec J. Hirsch, Guang Fan, Daniel N. Streblow, Malaika Mckenzie-Bennett, James McGann, Jim Griffin, Keith Robison, Alex Plocik, Becky Schilling, Rebecca Littlefield, Michelle Spencer, Birgitte Simen, William B. Messer, Andrew C. Adey, Benjamin N. Bimber, Brian J. O’Roak |
| EPI_ISL_526094, EPI_ISL_526095, EPI_ISL_526096 | OHSU Lab Services Molecular Microbiology Lab | Oregon SARS-CoV-2 Genome Sequencing Center | Brendan L. O'Connell, Ruth V. Nichols, Alec J. Hirsch, Guang Fan, Daniel N. Streblow, William B. Messer, Andrew C. Adey, Benjamin N. Bimber, Brian J. O’Roak |
| EPI_ISL_526097 | OHSU Lab Services Molecular Microbiology Lab | Ginkgo Bioworks Clinical Laboratory | Brendan L. O'Connell, Ruth V. Nichols, Alec J. Hirsch, Guang Fan, Daniel N. Streblow, Malaika Mckenzie-Bennett, James McGann, Jim Griffin, Keith Robison, Alex Plocik, Becky Schilling, Rebecca Littlefield, Michelle Spencer, Birgitte Simen, William B. Messer, Andrew C. Adey, Benjamin N. Bimber, Brian J. O’Roak |
| EPI_ISL_526098, EPI_ISL_526099, EPI_ISL_526100, EPI_ISL_526101, EPI_ISL_526102, EPI_ISL_526103, EPI_ISL_526104, EPI_ISL_526105, EPI_ISL_526106, EPI_ISL_526107, EPI_ISL_526108, EPI_ISL_526109, EPI_ISL_526110, EPI_ISL_526111, EPI_ISL_526112, EPI_ISL_526113, EPI_ISL_526114, EPI_ISL_526551, EPI_ISL_526552 | | | |
| see above | OHSU Lab Services Molecular Microbiology Lab | Oregon SARS-CoV-2 Genome Sequencing Center | Brendan L. O'Connell, Ruth V. Nichols, Alec J. Hirsch, Guang Fan, Daniel N. Streblow, William B. Messer, Andrew C. Adey, Benjamin N. Bimber, Brian J. O’Roak |
| EPI_ISL_526553, EPI_ISL_526554, EPI_ISL_526555, EPI_ISL_526556, EPI_ISL_526558, EPI_ISL_526559, EPI_ISL_526563, EPI_ISL_526564, EPI_ISL_526566, EPI_ISL_526567, EPI_ISL_526568, EPI_ISL_526569, EPI_ISL_526570, EPI_ISL_526571, EPI_ISL_526572, EPI_ISL_526573, EPI_ISL_526575, EPI_ISL_526578, EPI_ISL_526579, EPI_ISL_526580, EPI_ISL_526581, EPI_ISL_526583, EPI_ISL_526585, EPI_ISL_526586, EPI_ISL_526587, EPI_ISL_526588, EPI_ISL_526589, EPI_ISL_526590, EPI_ISL_526591, EPI_ISL_526592, EPI_ISL_526593, EPI_ISL_526594, EPI_ISL_526595, EPI_ISL_526596, EPI_ISL_526597, EPI_ISL_526598, EPI_ISL_526599, EPI_ISL_526600, EPI_ISL_526601, EPI_ISL_526602, EPI_ISL_526603, EPI_ISL_526604, EPI_ISL_526605, EPI_ISL_526606, EPI_ISL_526607, EPI_ISL_526608, EPI_ISL_526609, EPI_ISL_526610, EPI_ISL_526611, EPI_ISL_526612, EPI_ISL_526613, EPI_ISL_526614, EPI_ISL_526615, EPI_ISL_526616, EPI_ISL_526617, EPI_ISL_526618, EPI_ISL_526619, EPI_ISL_526620, EPI_ISL_526621, EPI_ISL_526622, EPI_ISL_526623, EPI_ISL_526624, EPI_ISL_526625, EPI_ISL_526626, EPI_ISL_526627, EPI_ISL_526628, EPI_ISL_526629, EPI_ISL_526630, EPI_ISL_526631, EPI_ISL_526632, EPI_ISL_526633, EPI_ISL_526634, EPI_ISL_526635, EPI_ISL_526636, EPI_ISL_526637, EPI_ISL_526638, EPI_ISL_526639, EPI_ISL_526640, EPI_ISL_526641, EPI_ISL_526642, EPI_ISL_526643, EPI_ISL_526644, EPI_ISL_526645, EPI_ISL_526646, EPI_ISL_526647, EPI_ISL_526648, EPI_ISL_526649, EPI_ISL_526650, EPI_ISL_526651, EPI_ISL_526652, EPI_ISL_526653, EPI_ISL_526655, EPI_ISL_526656, EPI_ISL_526657, EPI_ISL_526659, EPI_ISL_526660, EPI_ISL_526661, EPI_ISL_526662, EPI_ISL_526663, EPI_ISL_526664, EPI_ISL_526665, EPI_ISL_526666, EPI_ISL_526667, EPI_ISL_526668, EPI_ISL_526669, EPI_ISL_526670, EPI_ISL_526671, EPI_ISL_526672, EPI_ISL_526673, EPI_ISL_526674, EPI_ISL_526675, EPI_ISL_526676, EPI_ISL_526677, EPI_ISL_526678, EPI_ISL_526679, EPI_ISL_526680, EPI_ISL_526681, EPI_ISL_526682, EPI_ISL_526683, EPI_ISL_526684, EPI_ISL_526685 | | | |
| see above | Florida Bureau of Public Health Laboratories | Florida Bureau of Public Health Laboratories | Sarah Schmedes, Jason Blanton |
| EPI_ISL_526686, EPI_ISL_526687, EPI_ISL_526688 | Faith Laboratory, Immunology Institute, Icahn School of Medicine at Mount Sinai | van Bakel Laboratory, Genetics and Genomics Sciences, Icahn School of Medicine at Mount Sinai | Graham J. Britton, Alice Chen-Liaw, Francesca Cossarini, Alexandra Livanos, Matthew P. Spindler, Tamar Plitt, Joseph Eggers, Ilaria Mogno, Ana S. Gonzalez-Reiche, Sophia Sui, Michael Tankelevich, Lauren Tal Grinspan, Rebekah E. Dixon, Divya Jha, Gustavo Martinez-Delgado, Fatima Amanat, Daisy Hoagland, Benjamin R. tenOever, Marla C. Dubinsky, Miriam Merad, Harm Van Bakel, Florian Krammer, Gerold Bongers, Saurabh Mehandru and Jeremiah J. Faith |
| EPI_ISL_526775, EPI_ISL_526776, EPI_ISL_526777, EPI_ISL_526778, EPI_ISL_526779, EPI_ISL_526780, EPI_ISL_526781, EPI_ISL_526782, EPI_ISL_526783, EPI_ISL_526784, EPI_ISL_526785, EPI_ISL_526786, EPI_ISL_526787, EPI_ISL_526788, EPI_ISL_526789, EPI_ISL_526790, EPI_ISL_526791, EPI_ISL_526792, EPI_ISL_526793, EPI_ISL_526794, EPI_ISL_526795, EPI_ISL_526796, EPI_ISL_526797, EPI_ISL_526798, EPI_ISL_526799, EPI_ISL_526800, EPI_ISL_526801, EPI_ISL_526802, EPI_ISL_526803, EPI_ISL_526804, EPI_ISL_526805, EPI_ISL_526806, EPI_ISL_526807, EPI_ISL_526808, EPI_ISL_526809, EPI_ISL_526810, EPI_ISL_526811, EPI_ISL_526812, EPI_ISL_526813, EPI_ISL_526814, EPI_ISL_526815, EPI_ISL_526816, EPI_ISL_526817, EPI_ISL_526818, EPI_ISL_526819, EPI_ISL_526820, EPI_ISL_526821, EPI_ISL_526822, EPI_ISL_526823, EPI_ISL_526824, EPI_ISL_526825, EPI_ISL_526826, EPI_ISL_526827, EPI_ISL_526828, EPI_ISL_526829, EPI_ISL_526830, EPI_ISL_526831, EPI_ISL_526832, EPI_ISL_526833, EPI_ISL_526834, EPI_ISL_526835, EPI_ISL_526836, EPI_ISL_526837, EPI_ISL_526838, EPI_ISL_526839, EPI_ISL_526840, EPI_ISL_526841, EPI_ISL_526842, EPI_ISL_526843, EPI_ISL_526844, EPI_ISL_526845, EPI_ISL_526846, EPI_ISL_526847, EPI_ISL_526848, EPI_ISL_526849, EPI_ISL_526850, EPI_ISL_526851, EPI_ISL_526852, EPI_ISL_526853, EPI_ISL_526854, EPI_ISL_526855, EPI_ISL_526856, EPI_ISL_526857, EPI_ISL_526858, EPI_ISL_526859, EPI_ISL_526860, EPI_ISL_526861, EPI_ISL_526862, EPI_ISL_526863, EPI_ISL_526864, EPI_ISL_526865, EPI_ISL_526866, EPI_ISL_526867, EPI_ISL_526868, EPI_ISL_526869, EPI_ISL_526870, EPI_ISL_526871, EPI_ISL_526872, EPI_ISL_526873, EPI_ISL_526874, EPI_ISL_526875, EPI_ISL_526876, EPI_ISL_526877, EPI_ISL_526878, EPI_ISL_526879, EPI_ISL_526880, EPI_ISL_526881, EPI_ISL_526882, EPI_ISL_526883, EPI_ISL_526884, EPI_ISL_526885, EPI_ISL_526886, EPI_ISL_526887, EPI_ISL_526888, EPI_ISL_526889, EPI_ISL_526890, EPI_ISL_526891, EPI_ISL_526892, EPI_ISL_526893, EPI_ISL_526894, EPI_ISL_526895, EPI_ISL_526896, EPI_ISL_526897, EPI_ISL_526898, EPI_ISL_526899, EPI_ISL_526900, EPI_ISL_526901, EPI_ISL_526902, EPI_ISL_526903, EPI_ISL_526904, EPI_ISL_526905, EPI_ISL_526906, EPI_ISL_526907, EPI_ISL_526908, EPI_ISL_526909, EPI_ISL_526910, EPI_ISL_526911, EPI_ISL_526912, EPI_ISL_526913, EPI_ISL_526914, EPI_ISL_526915, EPI_ISL_526916, EPI_ISL_526917, EPI_ISL_526918, EPI_ISL_526919, EPI_ISL_526920, EPI_ISL_526921, EPI_ISL_526922, EPI_ISL_526923, EPI_ISL_526924, EPI_ISL_526925, EPI_ISL_526926, EPI_ISL_526927, EPI_ISL_526928, EPI_ISL_526929, EPI_ISL_526930, EPI_ISL_526931 | | | |
| see above | Virginia DCLS | Virginia DCLS | Virginia DCLS |
| EPI_ISL_527381, EPI_ISL_527382, EPI_ISL_527383, EPI_ISL_527384, EPI_ISL_527385, EPI_ISL_527386, EPI_ISL_527387, EPI_ISL_527388, EPI_ISL_527389, EPI_ISL_527390, EPI_ISL_527391, EPI_ISL_527392, EPI_ISL_527393, EPI_ISL_527394, EPI_ISL_527395, EPI_ISL_527396, EPI_ISL_527397, EPI_ISL_527398, EPI_ISL_527399 | | | |
| see above | University of Miami Immunology and Histocompatibility Laboratory | University of Miami Immunology and Histocompatibility Laboratory | Emilio Margolles-Clark, PhD and Phillip Ruiz, MD, PhD |
| EPI_ISL_527401, EPI_ISL_527402, EPI_ISL_527403, EPI_ISL_527404, EPI_ISL_527405, EPI_ISL_527406, EPI_ISL_527407, EPI_ISL_527408, EPI_ISL_527409, EPI_ISL_527410, EPI_ISL_527411, EPI_ISL_527412, EPI_ISL_527413, EPI_ISL_527414, EPI_ISL_527415, EPI_ISL_527416, EPI_ISL_527417, EPI_ISL_527418, EPI_ISL_527419, EPI_ISL_527420, EPI_ISL_527421, EPI_ISL_527422, EPI_ISL_527423, EPI_ISL_527424, EPI_ISL_527425, EPI_ISL_527426, EPI_ISL_527427, EPI_ISL_527428, EPI_ISL_527429, EPI_ISL_527430, EPI_ISL_527431, EPI_ISL_527432, EPI_ISL_527433, EPI_ISL_527434, EPI_ISL_527435, EPI_ISL_527436, EPI_ISL_527437, EPI_ISL_527438, EPI_ISL_527439, EPI_ISL_527440, EPI_ISL_527441, EPI_ISL_527442, EPI_ISL_527443, EPI_ISL_527444, EPI_ISL_527445, EPI_ISL_527446, EPI_ISL_527447, EPI_ISL_527448, EPI_ISL_527449, EPI_ISL_527450, EPI_ISL_527451, EPI_ISL_527452, EPI_ISL_527453, EPI_ISL_527454, EPI_ISL_527455, EPI_ISL_527456, EPI_ISL_527457, EPI_ISL_527458, EPI_ISL_527459, EPI_ISL_527460, EPI_ISL_527461, EPI_ISL_527462, EPI_ISL_527463, EPI_ISL_527464, EPI_ISL_527465, EPI_ISL_527466, EPI_ISL_527467, EPI_ISL_527468, EPI_ISL_527469, EPI_ISL_527470, EPI_ISL_527471, EPI_ISL_527472, EPI_ISL_527473, EPI_ISL_527474, EPI_ISL_527475, EPI_ISL_527476, EPI_ISL_527477, EPI_ISL_527478, EPI_ISL_527479, EPI_ISL_527480, EPI_ISL_527481, EPI_ISL_527482, EPI_ISL_527483, EPI_ISL_527484, EPI_ISL_527485, EPI_ISL_527486, EPI_ISL_527487, EPI_ISL_527488 | | | |
| see above | Colorado State University - Ebel Lab | Colorado State University - Ebel Lab | Greg Ebel et al. |
| EPI_ISL_527573, EPI_ISL_527574, EPI_ISL_527575, EPI_ISL_527576, EPI_ISL_527577, EPI_ISL_527578, EPI_ISL_527579, EPI_ISL_527580, EPI_ISL_527581, EPI_ISL_527582, EPI_ISL_527583, EPI_ISL_527584, EPI_ISL_527585, EPI_ISL_527586, EPI_ISL_527587, EPI_ISL_527588, EPI_ISL_527589, EPI_ISL_527590, EPI_ISL_527591, EPI_ISL_527592, EPI_ISL_527593, EPI_ISL_527594, EPI_ISL_527595, EPI_ISL_527596, EPI_ISL_527597, EPI_ISL_527598, EPI_ISL_527599, EPI_ISL_527600, EPI_ISL_527601, EPI_ISL_527602, EPI_ISL_527603, EPI_ISL_527604, EPI_ISL_527605, EPI_ISL_527606, EPI_ISL_527607, EPI_ISL_527608, EPI_ISL_527609, EPI_ISL_527610, EPI_ISL_527611, EPI_ISL_527612, EPI_ISL_527613, EPI_ISL_527614, EPI_ISL_527615, EPI_ISL_527616, EPI_ISL_527617, EPI_ISL_527618, EPI_ISL_527619, EPI_ISL_527620, EPI_ISL_527621, EPI_ISL_527622, EPI_ISL_527623, EPI_ISL_527624, EPI_ISL_527625, EPI_ISL_527626, EPI_ISL_527627, EPI_ISL_527628, EPI_ISL_527629 | | | |

| | | | |
|---|---|---|---|
| see above | Minnesota Department of Health, Public Health Laboratory | Minnesota Department of Health, Public Health Laboratory | Matt Plumb, Jacob Garfin, and Xiong Wang |
| EPI_ISL_527630, EPI_ISL_527631 | Mayo Clinic & Mayo Clinic Laboratories | Minnesota Department of Health, Public Health Laboratory | Matt Plumb, Jacob Garfin, and Xiong Wang |
| EPI_ISL_527632, EPI_ISL_527633, EPI_ISL_527634, EPI_ISL_527635, EPI_ISL_527636, EPI_ISL_527637, EPI_ISL_527638, EPI_ISL_527639, EPI_ISL_527640, EPI_ISL_527641, EPI_ISL_527642, EPI_ISL_527643, EPI_ISL_527644, EPI_ISL_527645, EPI_ISL_527646, EPI_ISL_527647, EPI_ISL_527648, EPI_ISL_527649, EPI_ISL_527650, EPI_ISL_527651, EPI_ISL_527652, EPI_ISL_527653, EPI_ISL_527654, EPI_ISL_527655, EPI_ISL_527656, EPI_ISL_527657 | | | |
| see above | AR Dept. of Health-Public Health Lab | Pathogen Discovery, Respiratory Viruses Branch, Division of Viral Diseases, Centers for Disease Control and Prevention | Ying Tao, Jing Zhang, Yan Li, Krista Queen, Anna Uehara, Clinton Paden, Haibin Wang, Suxiang Tong |
| EPI_ISL_527658, EPI_ISL_527659, EPI_ISL_527660, EPI_ISL_527661, EPI_ISL_527662, EPI_ISL_527663, EPI_ISL_527664, EPI_ISL_527665 | GA Department of Public Health Laboratory | Pathogen Discovery, Respiratory Viruses Branch, Division of Viral Diseases, Centers for Disease Control and Prevention | Yan Li, Anna Montmayeur, Jing Zhang, Krista Queen, Ying Tao, Anna Uehara, Rachel Marine, Clinton R. Paden, Haibin Wang, Suxiang Tong |
| EPI_ISL_527666 | IN State Department of Health Laboratory Services | Pathogen Discovery, Respiratory Viruses Branch, Division of Viral Diseases, Centers for Disease Control and Prevention | Krista Queen, Brian Lynch, Yan Li, Anna Montmayeur, Jing Zhang, Ying Tao, Anna Uehara, Rachel Marine, Clinton R. Paden, Haibin Wang, Suxiang Tong |
| EPI_ISL_527667, EPI_ISL_527668, EPI_ISL_527669, EPI_ISL_527670, EPI_ISL_527671, EPI_ISL_527672, EPI_ISL_527673, EPI_ISL_527674, EPI_ISL_527675, EPI_ISL_527676, EPI_ISL_527677, EPI_ISL_527678, EPI_ISL_527679, EPI_ISL_527680, EPI_ISL_527681, EPI_ISL_527682, EPI_ISL_527683, EPI_ISL_527684, EPI_ISL_527685, EPI_ISL_527686, EPI_ISL_527687, EPI_ISL_527688, EPI_ISL_527689, EPI_ISL_527690, EPI_ISL_527691 | | | |
| see above | MN PHL Division, Minnesota Department of Health | Pathogen Discovery, Respiratory Viruses Branch, Division of Viral Diseases, Centers for Disease Control and Prevention | Yan Li, Anna Montmayeur, Jing Zhang, Krista Queen, Ying Tao, Anna Uehara, Rachel Marine, Clinton R. Paden, Haibin Wang, Suxiang Tong |
| EPI_ISL_527692 | MN PHL Division, Minnesota Department of Health | Pathogen Discovery, Respiratory Viruses Branch, Division of Viral Diseases, Centers for Disease Control and Prevention | Krista Queen, Brian Lynch, Yan Li, Anna Montmayeur, Jing Zhang, Ying Tao, Anna Uehara, Rachel Marine, Clinton R. Paden, Haibin Wang, Suxiang Tong |
| EPI_ISL_527693, EPI_ISL_527694, EPI_ISL_527695, EPI_ISL_527696, EPI_ISL_527697, EPI_ISL_527698 | MN PHL Division, Minnesota Department of Health | Pathogen Discovery, Respiratory Viruses Branch, Division of Viral Diseases, Centers for Disease Control and Prevention | Yan Li, Anna Montmayeur, Jing Zhang, Krista Queen, Ying Tao, Anna Uehara, Rachel Marine, Clinton R. Paden, Haibin Wang, Suxiang Tong |
| EPI_ISL_527699 | MN PHL Division, Minnesota Department of Health | Pathogen Discovery, Respiratory Viruses Branch, Division of Viral Diseases, Centers for Disease Control and Prevention | Krista Queen, Brian Lynch, Yan Li, Anna Montmayeur, Jing Zhang, Ying Tao, Anna Uehara, Rachel Marine, Clinton R. Paden, Haibin Wang, Suxiang Tong |
| EPI_ISL_527700 | MN PHL Division, Minnesota Department of Health | Pathogen Discovery, Respiratory Viruses Branch, Division of Viral Diseases, Centers for Disease Control and Prevention | Yan Li, Anna Montmayeur, Jing Zhang, Krista Queen, Ying Tao, Anna Uehara, Rachel Marine, Clinton R. Paden, Haibin Wang, Suxiang Tong |
| EPI_ISL_527701 | MN PHL Division, Minnesota Department of Health | Pathogen Discovery, Respiratory Viruses Branch, Division of Viral Diseases, Centers for Disease Control and Prevention | Krista Queen, Brian Lynch, Yan Li, Anna Montmayeur, Jing Zhang, Ying Tao, Anna Uehara, Rachel Marine, Clinton R. Paden, Haibin Wang, Suxiang Tong |
| EPI_ISL_527702, EPI_ISL_527703, EPI_ISL_527704, EPI_ISL_527705, EPI_ISL_527706, EPI_ISL_527707, EPI_ISL_527708, EPI_ISL_527709, EPI_ISL_527710, EPI_ISL_527711, EPI_ISL_527712, EPI_ISL_527713 | | | |
| see above | MN PHL Division, Minnesota Department of Health | Pathogen Discovery, Respiratory Viruses Branch, Division of Viral Diseases, Centers for Disease Control and Prevention | Yan Li, Anna Montmayeur, Jing Zhang, Krista Queen, Ying Tao, Anna Uehara, Rachel Marine, Clinton R. Paden, Haibin Wang, Suxiang Tong |
| EPI_ISL_527714 | MN PHL Division, Minnesota Department of Health | Pathogen Discovery, Respiratory Viruses Branch, Division of Viral Diseases, Centers for Disease Control and Prevention | Krista Queen, Brian Lynch, Yan Li, Anna Montmayeur, Jing Zhang, Ying Tao, Anna Uehara, Rachel Marine, Clinton R. Paden, Haibin Wang, Suxiang Tong |
| EPI_ISL_527715 | MN PHL Division, Minnesota Department of Health | Pathogen Discovery, Respiratory Viruses Branch, Division of Viral Diseases, Centers for Disease Control and Prevention | Yan Li, Anna Montmayeur, Jing Zhang, Krista Queen, Ying Tao, Anna Uehara, Rachel Marine, Clinton R. Paden, Haibin Wang, Suxiang Tong |
| EPI_ISL_527716, EPI_ISL_527717 | MN PHL Division, Minnesota Department of Health | Pathogen Discovery, Respiratory Viruses Branch, Division of Viral Diseases, Centers for Disease Control and Prevention | Krista Queen, Brian Lynch, Yan Li, Anna Montmayeur, Jing Zhang, Ying Tao, Anna Uehara, Rachel Marine, Clinton R. Paden, Haibin Wang, Suxiang Tong |
| EPI_ISL_527718, EPI_ISL_527719 | MN PHL Division, Minnesota Department of Health | Pathogen Discovery, Respiratory Viruses Branch, Division of Viral Diseases, Centers for Disease Control and Prevention | Yan Li, Anna Montmayeur, Jing Zhang, Krista Queen, Ying Tao, Anna Uehara, Rachel Marine, Clinton R. Paden, Haibin Wang, Suxiang Tong |
| EPI_ISL_527720 | MN PHL Division, Minnesota Department of Health | Pathogen Discovery, Respiratory Viruses Branch, Division of Viral Diseases, Centers for Disease Control and Prevention | Krista Queen, Brian Lynch, Yan Li, Anna Montmayeur, Jing Zhang, Ying Tao, Anna Uehara, Rachel Marine, Clinton R. Paden, Haibin Wang, Suxiang Tong |
| EPI_ISL_527721, EPI_ISL_527722, EPI_ISL_527723 | MN PHL Division, Minnesota Department of Health | Pathogen Discovery, Respiratory Viruses Branch, Division of Viral Diseases, Centers for Disease Control and Prevention | Yan Li, Anna Montmayeur, Jing Zhang, Krista Queen, Ying Tao, Anna Uehara, Rachel Marine, Clinton R. Paden, Haibin Wang, Suxiang Tong |
| EPI_ISL_527724 | MN PHL Division, Minnesota Department of Health | Pathogen Discovery, Respiratory Viruses Branch, Division of Viral Diseases, Centers for Disease Control and Prevention | Krista Queen, Brian Lynch, Yan Li, Anna Montmayeur, Jing Zhang, Ying Tao, Anna Uehara, Rachel Marine, Clinton R. Paden, Haibin Wang, Suxiang Tong |
| EPI_ISL_527725, EPI_ISL_527726, EPI_ISL_527727, EPI_ISL_527728, EPI_ISL_527729, EPI_ISL_527730 | MN PHL Division, Minnesota Department of Health | Pathogen Discovery, Respiratory Viruses Branch, Division of Viral Diseases, Centers for Disease Control and Prevention | Yan Li, Anna Montmayeur, Jing Zhang, Krista Queen, Ying Tao, Anna Uehara, Rachel Marine, Clinton R. Paden, Haibin Wang, Suxiang Tong |
| EPI_ISL_527731 | MN PHL Division, Minnesota Department of Health | Pathogen Discovery, Respiratory Viruses Branch, Division of Viral Diseases, Centers for Disease Control and Prevention | Krista Queen, Brian Lynch, Yan Li, Anna Montmayeur, Jing Zhang, Ying Tao, Anna Uehara, Rachel Marine, Clinton R. Paden, Haibin Wang, Suxiang Tong |
| EPI_ISL_527732, EPI_ISL_527733 | MN PHL Division, Minnesota Department of Health | Pathogen Discovery, Respiratory Viruses Branch, Division of Viral Diseases, Centers for Disease Control and Prevention | Yan Li, Anna Montmayeur, Jing Zhang, Krista Queen, Ying Tao, Anna Uehara, Rachel Marine, Clinton R. Paden, Haibin Wang, Suxiang Tong |
| EPI_ISL_527734, EPI_ISL_527735 | MN PHL Division, Minnesota Department of Health | Pathogen Discovery, Respiratory Viruses Branch, Division of Viral Diseases, Centers for Disease Control and Prevention | Krista Queen, Brian Lynch, Yan Li, Anna Montmayeur, Jing Zhang, Ying Tao, Anna Uehara, Rachel Marine, Clinton R. Paden, Haibin Wang, Suxiang Tong |
| EPI_ISL_527736 | MN PHL Division, Minnesota Department of Health | Pathogen Discovery, Respiratory Viruses Branch, Division of Viral Diseases, Centers for Disease Control and Prevention | Yan Li, Anna Montmayeur, Jing Zhang, Krista Queen, Ying Tao, Anna Uehara, Rachel Marine, Clinton R. Paden, Haibin Wang, Suxiang Tong |
| EPI_ISL_527737 | WI State Laboratory of Hygiene | Pathogen Discovery, Respiratory Viruses Branch, Division of Viral Diseases, Centers for Disease Control and Prevention | Ying Tao, Jing Zhang, Yan Li, Krista Queen, Anna Uehara, Clinton Paden, Haibin Wang, Suxiang Tong |
| EPI_ISL_527774, EPI_ISL_527776, EPI_ISL_527777, EPI_ISL_527783, EPI_ISL_527785 | Connecticut State Department of Public Health | Grubaugh Lab - Yale School of Public Health | Joseph Fauver, Mary Petrone, Chantal Vogels, Anderson Brito, Tara Alpert, Anthony Muyombwe, Jafar Razeq, Albert Ko, Nathan Grubaugh |
| EPI_ISL_527794, EPI_ISL_527795, EPI_ISL_527796, EPI_ISL_527797, EPI_ISL_527798, EPI_ISL_527799, EPI_ISL_527800, EPI_ISL_527801, EPI_ISL_527802, EPI_ISL_527803, EPI_ISL_527804, EPI_ISL_527805, EPI_ISL_527806, EPI_ISL_527807, EPI_ISL_527808 | | | |
| see above | University of Wisconsin-Madison AIDS Vaccine Research Laboratories | University of Wisconsin-Madison AIDS Vaccine Research Laboratories | Gage Moreno, Katarina Braun, et al. AIDS Vaccine Research Laboratories |

| | | | |
|---|---|---|---|
| EPI_ISL_527820, EPI_ISL_527821, EPI_ISL_527822, EPI_ISL_527823, EPI_ISL_527824, EPI_ISL_527825, EPI_ISL_527826, EPI_ISL_527827, EPI_ISL_527828, EPI_ISL_527830, EPI_ISL_527831, EPI_ISL_527832, EPI_ISL_527833, EPI_ISL_527834, EPI_ISL_527835, EPI_ISL_527836, EPI_ISL_527838, EPI_ISL_527839, EPI_ISL_527840, EPI_ISL_527841, EPI_ISL_527842, EPI_ISL_527843, EPI_ISL_527844, EPI_ISL_527846, EPI_ISL_527847, EPI_ISL_527848, EPI_ISL_527849, EPI_ISL_527850, EPI_ISL_527851, EPI_ISL_527852, EPI_ISL_527853, EPI_ISL_527854, EPI_ISL_527855 | | | |
| see above | Texas Department of State Health Services | Texas Department of State Health Services | Bonnie Oh, Rashmi Tuladhar, Jenny Zhang, Maliha Rahman, Anita Pokharel, Myong Koag, Chun Wang, Rachel Lee, Grace Kubin |
| EPI_ISL_528485, EPI_ISL_528486, EPI_ISL_528487, EPI_ISL_528489, EPI_ISL_528490, EPI_ISL_528491, EPI_ISL_528492, EPI_ISL_528493, EPI_ISL_528494, EPI_ISL_528495, EPI_ISL_528497, EPI_ISL_528498, EPI_ISL_528499, EPI_ISL_528500, EPI_ISL_528501, EPI_ISL_528502, EPI_ISL_528503, EPI_ISL_528504, EPI_ISL_528506, EPI_ISL_528507, EPI_ISL_528508, EPI_ISL_528509, EPI_ISL_528510, EPI_ISL_528511, EPI_ISL_528513, EPI_ISL_528514, EPI_ISL_528515, EPI_ISL_528516, EPI_ISL_528521, EPI_ISL_528524, EPI_ISL_528525, EPI_ISL_528526, EPI_ISL_528527, EPI_ISL_528528, EPI_ISL_528529, EPI_ISL_528530, EPI_ISL_528531, EPI_ISL_528532, EPI_ISL_528533, EPI_ISL_528534, EPI_ISL_528535, EPI_ISL_528536, EPI_ISL_528537 | | | |
| see above | Alaska State Virology Laboratory | Alaska State Virology Laboratory | Chen J et al with Pathogenomics group Dagdag R, Redlinger M, Milton E, George W, Kovalenko A, Drown DM, Bortz E |
| EPI_ISL_528639, EPI_ISL_528640, EPI_ISL_528641, EPI_ISL_528642, EPI_ISL_528643, EPI_ISL_528644, EPI_ISL_528645, EPI_ISL_528646, EPI_ISL_528647, EPI_ISL_528648, EPI_ISL_528649, EPI_ISL_528650, EPI_ISL_528651, EPI_ISL_528652, EPI_ISL_528653, EPI_ISL_528654, EPI_ISL_528655, EPI_ISL_528656, EPI_ISL_528657, EPI_ISL_528658, EPI_ISL_528659, EPI_ISL_528660, EPI_ISL_528661, EPI_ISL_528662 | | | |
| see above | Virginia DCLS | Virginia DCLS | Virginia DCLS |
| EPI_ISL_528663, EPI_ISL_528664, EPI_ISL_528665, EPI_ISL_528666, EPI_ISL_528667, EPI_ISL_528668, EPI_ISL_528669, EPI_ISL_528670, EPI_ISL_528671, EPI_ISL_528672, EPI_ISL_528673, EPI_ISL_528674, EPI_ISL_528675, EPI_ISL_528676, EPI_ISL_528677, EPI_ISL_528678, EPI_ISL_528679, EPI_ISL_528680, EPI_ISL_528681, EPI_ISL_528682 | | | |
| see above | Virginia Division of Consolidated Laboratory Services (DCLS) | Virginia Division of Consolidated Laboratory Services (DCLS) | Virginia DCLS |
| EPI_ISL_529033, EPI_ISL_529034, EPI_ISL_529035, EPI_ISL_529036 | Wadsworth Center, New York State Department of Health | Wadsworth Center, New York State Department of Health | Kirsten St. George, Daryl M. Lamson, Sara Griesemer, Jonathan Plitnick, Navjot Singh, Matthew D. Shudt, Erica Lasek-Nesselquist |
| EPI_ISL_529080, EPI_ISL_529081, EPI_ISL_529082, EPI_ISL_529083, EPI_ISL_529084, EPI_ISL_529085, EPI_ISL_529086, EPI_ISL_529087, EPI_ISL_529088, EPI_ISL_529089, EPI_ISL_529090, EPI_ISL_529091, EPI_ISL_529092, EPI_ISL_529093, EPI_ISL_529094, EPI_ISL_529095, EPI_ISL_529096, EPI_ISL_529097, EPI_ISL_529098, EPI_ISL_529099, EPI_ISL_529100, EPI_ISL_529101, EPI_ISL_529103, EPI_ISL_529104, EPI_ISL_529105, EPI_ISL_529106, EPI_ISL_529107, EPI_ISL_529108 | | | |
| see above | Microbiology Division, SC DHEC | Microbiology Division, SC DHEC | Flores,H. |
| EPI_ISL_529151, EPI_ISL_529152, EPI_ISL_529153 | Department of Immunology, The Scripps Research Institute | Andersen lab at Scripps Research | Quigley, M., Stefanski, E., Mchardy, I. with SEARCH Alliance San Diego |
| EPI_ISL_529154 | Department of Immunology, The Scripps Research Institute | Andersen lab at Scripps Research | Basler, T., Shephard, J., Austin, B. with SEARCH Alliance San Diego |
| EPI_ISL_529155, EPI_ISL_529156, EPI_ISL_529157, EPI_ISL_529158, EPI_ISL_529159 | University of Washington, Laboratory Medicine | University of Washington, Laboratory Medicine | Roychoudhury,P., Greninger,A., Jerome,K. |
| EPI_ISL_529160, EPI_ISL_529161 | University of Washington, Laboratory Medicine | University of Washington, Laboratory Medicine | Roychoudhury, P., Greninger, A., Jerome, K. |
| EPI_ISL_529162, EPI_ISL_529163, EPI_ISL_529164, EPI_ISL_529165, EPI_ISL_529166 | Department of Immunology, The Scripps Research Institute | Andersen lab at Scripps Research | Allison Smither, Gilberto Sabino-Santos, Patricia Snarski, Lilia Melnik, Antoinette Bell, Kaylynn Genemaras, Arnaud Drouin, Dahlene Fusco, Robert Garry with SEARCH Alliance San Diego |
| EPI_ISL_529167, EPI_ISL_529168 | Maryland Department of Health Laboratory | Maryland Department of Health Laboratory | Eric Keller, Maryland Department of Health Laboratories Administration |
| EPI_ISL_529169 | Michigan Department of Health and Human Services, Bureau of Laboratories | Michigan Department of Health and Human Services, Bureau of Laboratories | Blankenship,H.M., Riner,D., Soehnlen,M. |
| EPI_ISL_529170 | University of Washington, Laboratory Medicine | University of Washington, Laboratory Medicine | Roychoudhury,P., Greninger,A., Jerome,K. |
| EPI_ISL_529171, EPI_ISL_529172, EPI_ISL_529173, EPI_ISL_529174, EPI_ISL_529175, EPI_ISL_529176, EPI_ISL_529177, EPI_ISL_529178, EPI_ISL_529180, EPI_ISL_529181, EPI_ISL_529182, EPI_ISL_529183, EPI_ISL_529184, EPI_ISL_529185, EPI_ISL_529186, EPI_ISL_529187, EPI_ISL_529188, EPI_ISL_529189, EPI_ISL_529190, EPI_ISL_529191, EPI_ISL_529192, EPI_ISL_529193, EPI_ISL_529194, EPI_ISL_529195, EPI_ISL_529196, EPI_ISL_529197, EPI_ISL_529198, EPI_ISL_529199 | | | |
| see above | South Carolina Department of Health and Environmental Control | South Carolina Department of Health and Environmental Control | Haley V. Flores |
| EPI_ISL_529200 | University of Washington, Laboratory Medicine | University of Washington, Laboratory Medicine | Roychoudhury,P., Greninger,A., Jerome,K. |
| EPI_ISL_529208, EPI_ISL_529209, EPI_ISL_529210, EPI_ISL_529211, EPI_ISL_529212 | Laboratory Medicine, University of Washington | University of Washington, Laboratory Medicine | Roychoudhury,P., Greninger,A., Jerome,K. |
| EPI_ISL_529804, EPI_ISL_529805, EPI_ISL_529806, EPI_ISL_529807, EPI_ISL_529808, EPI_ISL_529809, EPI_ISL_529810, EPI_ISL_529811, EPI_ISL_529812, EPI_ISL_529813, EPI_ISL_529814, EPI_ISL_529815, EPI_ISL_529816, EPI_ISL_529817, EPI_ISL_529818, EPI_ISL_529819, EPI_ISL_529820, EPI_ISL_529821, EPI_ISL_529823, EPI_ISL_529826, EPI_ISL_529827, EPI_ISL_529828, EPI_ISL_529829, EPI_ISL_529830, EPI_ISL_529831, EPI_ISL_529832, EPI_ISL_529833, EPI_ISL_529834, EPI_ISL_529835, EPI_ISL_529836, EPI_ISL_529837, EPI_ISL_529838, EPI_ISL_529839, EPI_ISL_529840, EPI_ISL_529841, EPI_ISL_529842, EPI_ISL_529843, EPI_ISL_529844, EPI_ISL_529845, EPI_ISL_529846, EPI_ISL_529847, EPI_ISL_529848, EPI_ISL_529849, EPI_ISL_529850, EPI_ISL_529851, EPI_ISL_529852, EPI_ISL_529853, EPI_ISL_529854, EPI_ISL_529855, EPI_ISL_529856, EPI_ISL_529857, EPI_ISL_529858, EPI_ISL_529860, EPI_ISL_529861, EPI_ISL_529862, EPI_ISL_529863, EPI_ISL_529864, EPI_ISL_529865, EPI_ISL_529866, EPI_ISL_529867, EPI_ISL_529868, EPI_ISL_529869, EPI_ISL_529870, EPI_ISL_529871, EPI_ISL_529872, EPI_ISL_529874, EPI_ISL_529876, EPI_ISL_529877, EPI_ISL_529879, EPI_ISL_529880, EPI_ISL_529881, EPI_ISL_529882, EPI_ISL_529883, EPI_ISL_529884, EPI_ISL_529885, EPI_ISL_529886, EPI_ISL_529887, EPI_ISL_529888, EPI_ISL_529889, EPI_ISL_529890, EPI_ISL_529891, EPI_ISL_529892, EPI_ISL_529893, EPI_ISL_529894, EPI_ISL_529895, EPI_ISL_529896, EPI_ISL_529897, EPI_ISL_529898, EPI_ISL_529899, EPI_ISL_529900, EPI_ISL_529902, EPI_ISL_529903, EPI_ISL_529904, EPI_ISL_529905, EPI_ISL_529906, EPI_ISL_529907 | | | |
| see above | Michigan Department of Health and Human Services, Bureau of Laboratories | Michigan Department of Health and Human Services, Bureau of Laboratories | Blankenship HM, Riner D, Soehnlen MK |
| EPI_ISL_529908, EPI_ISL_529909, EPI_ISL_529910, EPI_ISL_529911, EPI_ISL_529912, EPI_ISL_529913, EPI_ISL_529914, EPI_ISL_529915, EPI_ISL_529916, EPI_ISL_529917, EPI_ISL_529918, EPI_ISL_529919, EPI_ISL_529920, EPI_ISL_529921, EPI_ISL_529922, EPI_ISL_529923, EPI_ISL_529924, EPI_ISL_529925, EPI_ISL_529926, EPI_ISL_529927, EPI_ISL_529928, EPI_ISL_529929, EPI_ISL_529930, EPI_ISL_529931, EPI_ISL_529932, EPI_ISL_529933, EPI_ISL_529934, EPI_ISL_529935, EPI_ISL_529936 | | | |
| see above | Virginia Division of Consolidated Laboratory Services | Virginia Division of Consolidated Laboratory Services | Virginia DCLS |
| EPI_ISL_529937, EPI_ISL_529938, EPI_ISL_529939, EPI_ISL_529940, EPI_ISL_529941, EPI_ISL_529942, EPI_ISL_529943, EPI_ISL_529944, EPI_ISL_529945, EPI_ISL_529946, EPI_ISL_529947, EPI_ISL_529948, EPI_ISL_529949, EPI_ISL_529950, EPI_ISL_529951, EPI_ISL_529952, EPI_ISL_529953, EPI_ISL_529954, EPI_ISL_529955, EPI_ISL_529956, EPI_ISL_529957, EPI_ISL_529958, EPI_ISL_529959, EPI_ISL_529960 | | | |
| see above | Virginia DCLS | Virginia DCLS | Virginia DCLS |
| EPI_ISL_530124, EPI_ISL_530125 | Seattle Flu Study | Seattle Flu Study | Deborah A. Nickerson, Chris D. Frazar, Jover Lee, Benjamin Pelle, Matthew Richardson, Amanda Adler, Elisabeth Brandstetter, Peter D. Han, Kairsten Fay, Misja Ilcisin, Kirsten Lacombe, Thomas R. Sibley, Melissa Truong, Caitlin R. Wolf, Michael Boeckh, Janet A. Englund, Michael Famulare, Barry R. Lutz, Mark J. Rieder, Lea M. Starita, Matthew Thompson, Jay Shendure, Trevor Bedford, Helen Y. Chu |
| EPI_ISL_530126, EPI_ISL_530127, EPI_ISL_530128, EPI_ISL_530129, EPI_ISL_530130, EPI_ISL_530131 | Seattle Flu Study | Seattle Flu Study | Deborah A. Nickerson, Chris D. Frazar, Jover Lee, Benjamin Pelle, Matthew Richardson, Amanda Adler, Elisabeth Brandstetter, Peter D. Han, Kairsten Fay, Misja Ilcisin, Kirsten Lacombe, Thomas R. Sibley, Melissa Truong, Caitlin R. Wolf, Karen Cowgill, Stephanie Schrag, Jeff Duchin, Michael Boeckh, Janet A. Englund, Michael Famulare, Barry R. Lutz, Mark J. Rieder, Lea M. Starita, Matthew Thompson, Helen Y. Chu, Trevor Bedford, Jay Shendure |
| EPI_ISL_530132 | Seattle Flu Study | Seattle Flu Study | Deborah A. Nickerson, Chris D. Frazar, Jover Lee, Benjamin Pelle, Matthew Richardson, Amanda Adler, Elisabeth Brandstetter, Peter D. Han, Kairsten Fay, Misja Ilcisin, Kirsten Lacombe, Thomas R. Sibley, Melissa Truong, Caitlin R. Wolf, Michael Boeckh, Janet A. Englund, Michael Famulare, Barry R. Lutz, Mark J. Rieder, Lea M. Starita, Matthew Thompson, Jay Shendure, Trevor Bedford, Helen Y. Chu |
| EPI_ISL_530133, EPI_ISL_530134, EPI_ISL_530135, EPI_ISL_530136, EPI_ISL_530137, EPI_ISL_530138, EPI_ISL_530139, EPI_ISL_530140, EPI_ISL_530141, EPI_ISL_530142, EPI_ISL_530143, EPI_ISL_530144, EPI_ISL_530145, EPI_ISL_530146, EPI_ISL_530147, EPI_ISL_530148, EPI_ISL_530149, EPI_ISL_530150, EPI_ISL_530151, EPI_ISL_530152, EPI_ISL_530153, EPI_ISL_530154, EPI_ISL_530155, EPI_ISL_530156, EPI_ISL_530157, EPI_ISL_530158, EPI_ISL_530159, EPI_ISL_530161, EPI_ISL_530162 | | | |
| see above | Seattle Flu Study | Seattle Flu Study | Deborah A. Nickerson, Chris D. Frazar, Jover Lee, Benjamin Pelle, Matthew Richardson, Amanda Adler, Elisabeth Brandstetter, Peter D. Han, Kairsten Fay, Misja Ilcisin, Kirsten Lacombe, Thomas R. Sibley, Melissa Truong, Caitlin R. Wolf, Karen Cowgill, Stephanie Schrag, Jeff Duchin, Michael Boeckh, Janet A. Englund, Michael Famulare, Barry R. Lutz, Mark J. Rieder, Lea M. Starita, Matthew Thompson, Helen Y. Chu, Trevor Bedford, Jay Shendure |
| EPI_ISL_530163, EPI_ISL_530164, EPI_ISL_530165, EPI_ISL_530166, EPI_ISL_530167, EPI_ISL_530168 | M Health Fairview | Minnesota Department of Health, Public Health Laboratory | Matt Plumb, Jacob Garfin, and Xiong Wang |
| EPI_ISL_530169, EPI_ISL_530170, EPI_ISL_530171 | Hennepin County Medical Center | Minnesota Department of Health, Public Health Laboratory | Matt Plumb, Jacob Garfin, and Xiong Wang |
| EPI_ISL_530172, EPI_ISL_530173, EPI_ISL_530174, EPI_ISL_530175, EPI_ISL_530176, EPI_ISL_530177, EPI_ISL_530178, EPI_ISL_530179, EPI_ISL_530180, EPI_ISL_530181, EPI_ISL_530182, EPI_ISL_530183, EPI_ISL_530184, EPI_ISL_530185, EPI_ISL_530186, EPI_ISL_530187, EPI_ISL_530188, EPI_ISL_530189, EPI_ISL_530190, EPI_ISL_530191, EPI_ISL_530192, EPI_ISL_530193, EPI_ISL_530194, EPI_ISL_530195, EPI_ISL_530196, EPI_ISL_530197, EPI_ISL_530198, EPI_ISL_530199, EPI_ISL_530200, EPI_ISL_530201, EPI_ISL_530202, EPI_ISL_530203, EPI_ISL_530204, EPI_ISL_530205, EPI_ISL_530206, EPI_ISL_530207, EPI_ISL_530208, EPI_ISL_530209, EPI_ISL_530210, EPI_ISL_530211, EPI_ISL_530212, EPI_ISL_530213, EPI_ISL_530214, EPI_ISL_530215, EPI_ISL_530216, EPI_ISL_530217, EPI_ISL_530218, EPI_ISL_530219, EPI_ISL_530220, EPI_ISL_530221, EPI_ISL_530222, EPI_ISL_530223, EPI_ISL_530224 | | | |
| see above | Minnesota Department of Health, Public Health Laboratory | Minnesota Department of Health, Public Health Laboratory | Matt Plumb, Jacob Garfin, and Xiong Wang |

| | | | |
|---|---|---|---|
| EPI_ISL_534217 | M Health Fairview | Minnesota Department of Health, Public Health Laboratory | Matt Plumb, Jacob Garfin, and Xiong Wang |
| EPI_ISL_534261, EPI_ISL_534262, EPI_ISL_534263, EPI_ISL_534265, EPI_ISL_534266, EPI_ISL_534267, EPI_ISL_534269, EPI_ISL_534270, EPI_ISL_534271, EPI_ISL_534272, EPI_ISL_534276, EPI_ISL_534277, EPI_ISL_534279, EPI_ISL_534280, EPI_ISL_534281, EPI_ISL_534282, EPI_ISL_534283, EPI_ISL_534285, EPI_ISL_534286, EPI_ISL_534287, EPI_ISL_534288, EPI_ISL_534291, EPI_ISL_534293, EPI_ISL_534294, EPI_ISL_534295, EPI_ISL_534296, EPI_ISL_534297, EPI_ISL_534298, EPI_ISL_534299, EPI_ISL_534300, EPI_ISL_534301, EPI_ISL_534302 | | | |
| see above | Texas Department of State Health Services | Texas Department of State Health Services | Rashmi Tuladhar, Bonnie Oh, Jenny Zhang, Maliha Rahman, Anita Pokharel, Myong Koag, Chun Wang, Rachel Lee, Grace Kubin |
| EPI_ISL_534693, EPI_ISL_534694, EPI_ISL_534695, EPI_ISL_534696, EPI_ISL_534697, EPI_ISL_534698 | University of Miami Immunology and Histocompatibility Laboratory | University of Miami Immunology and Histocompatibility Laboratory | Emilio Margolles-Clark, PhD and Phillip Ruiz, MD, PhD |
| EPI_ISL_534699, EPI_ISL_534700, EPI_ISL_534701, EPI_ISL_534702, EPI_ISL_534703, EPI_ISL_534704, EPI_ISL_534705, EPI_ISL_534706, EPI_ISL_534707, EPI_ISL_534708, EPI_ISL_534709, EPI_ISL_534710, EPI_ISL_534711, EPI_ISL_534712, EPI_ISL_534713, EPI_ISL_534714, EPI_ISL_534715, EPI_ISL_534716 | | | |
| see above | MD PHL | MD PHL | Maryland Department of Health Laboratories Administration |
| EPI_ISL_534731 | Wadsworth Center, New York State Department of Health | Wadsworth Center, New York State Department of Health | Kirsten St. George, Daryl M. Lamson, Sara Griesemer, Jonathan Plitnick, Navjot Singh, Matthew D. Shudt, Erica Lasek-Nesselquist |
| EPI_ISL_535263, EPI_ISL_535264, EPI_ISL_535265, EPI_ISL_535266, EPI_ISL_535268, EPI_ISL_535269, EPI_ISL_535270, EPI_ISL_535271, EPI_ISL_535272, EPI_ISL_535273, EPI_ISL_535274, EPI_ISL_535275, EPI_ISL_535276, EPI_ISL_535277, EPI_ISL_535278, EPI_ISL_535279, EPI_ISL_535280, EPI_ISL_535281, EPI_ISL_535282, EPI_ISL_535283, EPI_ISL_535284, EPI_ISL_535285, EPI_ISL_535286, EPI_ISL_535287, EPI_ISL_535288, EPI_ISL_535291, EPI_ISL_535292, EPI_ISL_535293, EPI_ISL_535294, EPI_ISL_535295, EPI_ISL_535297, EPI_ISL_535298, EPI_ISL_535299, EPI_ISL_535300, EPI_ISL_535301, EPI_ISL_535302, EPI_ISL_535303 | | | |
| see above | New Mexico Department of Health Scientific Laboratory | New Mexico Department of Health Scientific Laboratory | Ellie Johnson, Anastacia Griego-Fisher, D'Eldra Malone |
| EPI_ISL_535305, EPI_ISL_535306, EPI_ISL_535307, EPI_ISL_535308, EPI_ISL_535309, EPI_ISL_535310, EPI_ISL_535311, EPI_ISL_535312, EPI_ISL_535313, EPI_ISL_535314, EPI_ISL_535315, EPI_ISL_535316, EPI_ISL_535317, EPI_ISL_535318, EPI_ISL_535319, EPI_ISL_535320, EPI_ISL_535321, EPI_ISL_535322, EPI_ISL_535323, EPI_ISL_535324, EPI_ISL_535325, EPI_ISL_535326, EPI_ISL_535327, EPI_ISL_535328, EPI_ISL_535329, EPI_ISL_535330, EPI_ISL_535331, EPI_ISL_535332, EPI_ISL_535333, EPI_ISL_535334, EPI_ISL_535335, EPI_ISL_535336, EPI_ISL_535337, EPI_ISL_535338, EPI_ISL_535339, EPI_ISL_535340 | | | |
| see above | LA Office of Public Health Laboratories | Pathogen Discovery, Respiratory Viruses Branch, Division of Viral Diseases, Centers for Disease Control and Prevention | Ying Tao, Jing Zhang, Yan Li, Krista Queen, Anna Uehara, Clinton Paden, Haibin Wang, Suxiang Tong |
| EPI_ISL_535341, EPI_ISL_535342 | LA Office of Public Health Laboratories | Pathogen Discovery, Respiratory Viruses Branch, Division of Viral Diseases, Centers for Disease Control and Prevention | Jing Zhang, Ying Tao, Yan Li, Krista Queen, Anna Uehara, Clinton Paden, Haibin Wang, Suxiang Tong |
| EPI_ISL_535343 | LA Office of Public Health Laboratories | Pathogen Discovery, Respiratory Viruses Branch, Division of Viral Diseases, Centers for Disease Control and Prevention | Ying Tao, Jing Zhang, Yan Li, Krista Queen, Anna Uehara, Clinton Paden, Haibin Wang, Suxiang Tong |
| EPI_ISL_535344, EPI_ISL_535345, EPI_ISL_535346 | LA Office of Public Health Laboratories | Pathogen Discovery, Respiratory Viruses Branch, Division of Viral Diseases, Centers for Disease Control and Prevention | Jing Zhang, Ying Tao, Yan Li, Krista Queen, Anna Uehara, Clinton Paden, Haibin Wang, Suxiang Tong |
| EPI_ISL_535347 | LA Office of Public Health Laboratories | Pathogen Discovery, Respiratory Viruses Branch, Division of Viral Diseases, Centers for Disease Control and Prevention | Ying Tao, Jing Zhang, Yan Li, Krista Queen, Anna Uehara, Clinton Paden, Haibin Wang, Suxiang Tong |
| EPI_ISL_535348 | LA Office of Public Health Laboratories | Pathogen Discovery, Respiratory Viruses Branch, Division of Viral Diseases, Centers for Disease Control and Prevention | Jing Zhang, Ying Tao, Yan Li, Krista Queen, Anna Uehara, Clinton Paden, Haibin Wang, Suxiang Tong |
| EPI_ISL_535349, EPI_ISL_535350 | LA Office of Public Health Laboratories | Pathogen Discovery, Respiratory Viruses Branch, Division of Viral Diseases, Centers for Disease Control and Prevention | Ying Tao, Jing Zhang, Yan Li, Krista Queen, Anna Uehara, Clinton Paden, Haibin Wang, Suxiang Tong |
| EPI_ISL_535351 | LA Office of Public Health Laboratories | Pathogen Discovery, Respiratory Viruses Branch, Division of Viral Diseases, Centers for Disease Control and Prevention | Jing Zhang, Ying Tao, Yan Li, Krista Queen, Anna Uehara, Clinton Paden, Haibin Wang, Suxiang Tong |
| EPI_ISL_535352, EPI_ISL_535353, EPI_ISL_535354, EPI_ISL_535355, EPI_ISL_535356 | LA Office of Public Health Laboratories | Pathogen Discovery, Respiratory Viruses Branch, Division of Viral Diseases, Centers for Disease Control and Prevention | Ying Tao, Jing Zhang, Yan Li, Krista Queen, Anna Uehara, Clinton Paden, Haibin Wang, Suxiang Tong |
| EPI_ISL_535357 | LA Office of Public Health Laboratories | Pathogen Discovery, Respiratory Viruses Branch, Division of Viral Diseases, Centers for Disease Control and Prevention | Jing Zhang, Ying Tao, Yan Li, Krista Queen, Anna Uehara, Clinton Paden, Haibin Wang, Suxiang Tong |
| EPI_ISL_535358 | LA Office of Public Health Laboratories | Pathogen Discovery, Respiratory Viruses Branch, Division of Viral Diseases, Centers for Disease Control and Prevention | Ying Tao, Jing Zhang, Yan Li, Krista Queen, Anna Uehara, Clinton Paden, Haibin Wang, Suxiang Tong |
| EPI_ISL_535359, EPI_ISL_535360 | RI State Health Laboratories | Pathogen Discovery, Respiratory Viruses Branch, Division of Viral Diseases, Centers for Disease Control and Prevention | Jing Zhang, Ying Tao, Yan Li, Krista Queen, Anna Uehara, Clinton Paden, Haibin Wang, Suxiang Tong |
| EPI_ISL_535361, EPI_ISL_535362, EPI_ISL_535363, EPI_ISL_535364 | Oklahoma Animal Disease Diagnostic Laboratory | Oklahoma Animal Disease Diagnostic Laboratory | Sai Narayanan, John C Ritchey, Girish Patil, Teluguakula Narasaraju, Sunil More, Jerry Malayer, Jeremiah Saliki, Anil Kaul, Akhilesh Ramachandran |
| EPI_ISL_535650 | AR Dept. of Health-Public Health Lab | Pathogen Discovery, Respiratory Viruses Branch, Division of Viral Diseases, Centers for Disease Control and Prevention | Brian Lynch, Yan Li, Jing Zhang, Ying Tao, Krista Queen, Anna Uehara, Clinton R. Paden, Rachel Marine, Haibin Wang, Suxiang Tong |
| EPI_ISL_535651 | AR Dept. of Health-Public Health Lab | Pathogen Discovery, Respiratory Viruses Branch, Division of Viral Diseases, Centers for Disease Control and Prevention | Yan Li, Jing Zhang, Ying Tao, Krista Queen, Brian Lynch, Anna Uehara, Clinton R. Paden, Rachel Marine, Haibin Wang, Suxiang Tong |
| EPI_ISL_535652 | AR Dept. of Health-Public Health Lab | Pathogen Discovery, Respiratory Viruses Branch, Division of Viral Diseases, Centers for Disease Control and Prevention | Brian Lynch, Yan Li, Jing Zhang, Ying Tao, Krista Queen, Anna Uehara, Clinton R. Paden, Rachel Marine, Haibin Wang, Suxiang Tong |
| EPI_ISL_535653 | AR Dept. of Health-Public Health Lab | Pathogen Discovery, Respiratory Viruses Branch, Division of Viral Diseases, Centers for Disease Control and Prevention | Yan Li, Jing Zhang, Ying Tao, Krista Queen, Brian Lynch, Anna Uehara, Clinton R. Paden, Rachel Marine, Haibin Wang, Suxiang Tong |
| EPI_ISL_535654 | AR Dept. of Health-Public Health Lab | Pathogen Discovery, Respiratory Viruses Branch, Division of Viral Diseases, Centers for Disease Control and Prevention | Brian Lynch, Yan Li, Jing Zhang, Ying Tao, Krista Queen, Anna Uehara, Clinton R. Paden, Rachel Marine, Haibin Wang, Suxiang Tong |
| EPI_ISL_535655, EPI_ISL_535656, EPI_ISL_535657 | AR Dept. of Health-Public Health Lab | Pathogen Discovery, Respiratory Viruses Branch, Division of Viral Diseases, Centers for Disease Control and Prevention | Yan Li, Jing Zhang, Ying Tao, Krista Queen, Brian Lynch, Anna Uehara, Clinton R. Paden, Rachel Marine, Haibin Wang, Suxiang Tong |
| EPI_ISL_535658, EPI_ISL_535659 | AR Dept. of Health-Public Health Lab | Pathogen Discovery, Respiratory Viruses Branch, Division of Viral Diseases, Centers for Disease Control and Prevention | Brian Lynch, Yan Li, Jing Zhang, Ying Tao, Krista Queen, Anna Uehara, Clinton R. Paden, Rachel Marine, Haibin Wang, Suxiang Tong |
| EPI_ISL_535660 | AR Dept. of Health-Public Health Lab | Pathogen Discovery, Respiratory Viruses Branch, Division of Viral Diseases, Centers for Disease Control and Prevention | Yan Li, Jing Zhang, Ying Tao, Krista Queen, Brian Lynch, Anna Uehara, Clinton R. Paden, Rachel Marine, Haibin Wang, Suxiang Tong |
| EPI_ISL_535661 | AR Dept. of Health-Public Health Lab | Pathogen Discovery, Respiratory Viruses Branch, Division of Viral Diseases, Centers for Disease Control and Prevention | Brian Lynch, Yan Li, Jing Zhang, Ying Tao, Krista Queen, Anna Uehara, Clinton R. Paden, Rachel Marine, Haibin Wang, Suxiang Tong |
| EPI_ISL_535662, EPI_ISL_535663, EPI_ISL_535664, EPI_ISL_535665, EPI_ISL_535666, EPI_ISL_535667, EPI_ISL_535668, EPI_ISL_535669, EPI_ISL_535670, EPI_ISL_535671, EPI_ISL_535672, EPI_ISL_535673, EPI_ISL_535674 | | | |
| see above | CDPH, Microbial Diseases Laboratory | Pathogen Discovery, Respiratory Viruses Branch, Division of Viral Diseases, Centers for Disease Control and Prevention | Yan Li, Jing Zhang, Ying Tao, Krista Queen, Brian Lynch, Anna Uehara, Clinton R. Paden, Rachel Marine, Haibin Wang, Suxiang Tong |

| | | | |
|---|---|---|---|
| EPI_ISL_535675 | CDPH, Microbial Diseases Laboratory | Pathogen Discovery, Respiratory Viruses Branch, Division of Viral Diseases, Centers for Disease Control and Prevention | Brian Lynch, Yan Li, Jing Zhang, Ying Tao, Krista Queen, Anna Uehara, Clinton R. Paden, Rachel Marine, Haibin Wang, Suxiang Tong |
| EPI_ISL_535676, EPI_ISL_535677, EPI_ISL_535678, EPI_ISL_535679, EPI_ISL_535680, EPI_ISL_535681, EPI_ISL_535682, EPI_ISL_535683, EPI_ISL_535684, EPI_ISL_535685, EPI_ISL_535686, EPI_ISL_535687, EPI_ISL_535688, EPI_ISL_535689, EPI_ISL_535690, EPI_ISL_535691, EPI_ISL_535692, EPI_ISL_535693, EPI_ISL_535694, EPI_ISL_535695, EPI_ISL_535696, EPI_ISL_535697, EPI_ISL_535698, EPI_ISL_535699, EPI_ISL_535700 | | | |
| see above | CDPH, Microbial Diseases Laboratory | Pathogen Discovery, Respiratory Viruses Branch, Division of Viral Diseases, Centers for Disease Control and Prevention | Yan Li, Jing Zhang, Ying Tao, Krista Queen, Brian Lynch, Anna Uehara, Clinton R. Paden, Rachel Marine, Haibin Wang, Suxiang Tong |
| EPI_ISL_535701 | CDPH, Microbial Diseases Laboratory | Pathogen Discovery, Respiratory Viruses Branch, Division of Viral Diseases, Centers for Disease Control and Prevention | Brian Lynch, Yan Li, Jing Zhang, Ying Tao, Krista Queen, Anna Uehara, Clinton R. Paden, Rachel Marine, Haibin Wang, Suxiang Tong |
| EPI_ISL_535702 | CDPH, Microbial Diseases Laboratory | Pathogen Discovery, Respiratory Viruses Branch, Division of Viral Diseases, Centers for Disease Control and Prevention | Yan Li, Jing Zhang, Ying Tao, Krista Queen, Brian Lynch, Anna Uehara, Clinton R. Paden, Rachel Marine, Haibin Wang, Suxiang Tong |
| EPI_ISL_535703 | CDPH, Microbial Diseases Laboratory | Pathogen Discovery, Respiratory Viruses Branch, Division of Viral Diseases, Centers for Disease Control and Prevention | Ying Tao, Jing Zhang, Yan Li, Krista Queen, Anna Uehara, Clinton R. Paden, Haibin Wang, Suxiang Tong |
| EPI_ISL_535704, EPI_ISL_535705 | CDPH, Microbial Diseases Laboratory | Pathogen Discovery, Respiratory Viruses Branch, Division of Viral Diseases, Centers for Disease Control and Prevention | Yan Li, Jing Zhang, Ying Tao, Krista Queen, Brian Lynch, Anna Uehara, Clinton R. Paden, Rachel Marine, Haibin Wang, Suxiang Tong |
| EPI_ISL_535706 | CDPH, Microbial Diseases Laboratory | Pathogen Discovery, Respiratory Viruses Branch, Division of Viral Diseases, Centers for Disease Control and Prevention | Ying Tao, Jing Zhang, Yan Li, Krista Queen, Anna Uehara, Clinton R. Paden, Haibin Wang, Suxiang Tong |
| EPI_ISL_535707 | CDPH, Microbial Diseases Laboratory | Pathogen Discovery, Respiratory Viruses Branch, Division of Viral Diseases, Centers for Disease Control and Prevention | Yan Li, Jing Zhang, Ying Tao, Krista Queen, Brian Lynch, Anna Uehara, Clinton R. Paden, Rachel Marine, Haibin Wang, Suxiang Tong |
| EPI_ISL_535708 | CDPH, Microbial Diseases Laboratory | Pathogen Discovery, Respiratory Viruses Branch, Division of Viral Diseases, Centers for Disease Control and Prevention | Brian Lynch, Yan Li, Jing Zhang, Ying Tao, Krista Queen, Anna Uehara, Clinton R. Paden, Rachel Marine, Haibin Wang, Suxiang Tong |
| EPI_ISL_535709, EPI_ISL_535710, EPI_ISL_535711, EPI_ISL_535712, EPI_ISL_535713, EPI_ISL_535714, EPI_ISL_535715 | CDPH, Microbial Diseases Laboratory | Pathogen Discovery, Respiratory Viruses Branch, Division of Viral Diseases, Centers for Disease Control and Prevention | Yan Li, Jing Zhang, Ying Tao, Krista Queen, Brian Lynch, Anna Uehara, Clinton R. Paden, Rachel Marine, Haibin Wang, Suxiang Tong |
| EPI_ISL_536572, EPI_ISL_536573, EPI_ISL_536574, EPI_ISL_536575, EPI_ISL_536576, EPI_ISL_536577, EPI_ISL_536578, EPI_ISL_536579, EPI_ISL_536580, EPI_ISL_536581, EPI_ISL_536582, EPI_ISL_536583, EPI_ISL_536584, EPI_ISL_536585, EPI_ISL_536586, EPI_ISL_536587, EPI_ISL_536588, EPI_ISL_536589, EPI_ISL_536590, EPI_ISL_536591, EPI_ISL_536592, EPI_ISL_536593, EPI_ISL_536594, EPI_ISL_536595, EPI_ISL_536596, EPI_ISL_536597, EPI_ISL_536598, EPI_ISL_536599, EPI_ISL_536600, EPI_ISL_536601, EPI_ISL_536602, EPI_ISL_536603, EPI_ISL_536604, EPI_ISL_536605, EPI_ISL_536606, EPI_ISL_536607, EPI_ISL_536608, EPI_ISL_536609, EPI_ISL_536610, EPI_ISL_536611, EPI_ISL_536612, EPI_ISL_536613, EPI_ISL_536614, EPI_ISL_536615, EPI_ISL_536616, EPI_ISL_536617, EPI_ISL_536618, EPI_ISL_536619, EPI_ISL_536620, EPI_ISL_536621, EPI_ISL_536622, EPI_ISL_536623, EPI_ISL_536624, EPI_ISL_536625, EPI_ISL_536626, EPI_ISL_536627, EPI_ISL_536628, EPI_ISL_536629, EPI_ISL_536630, EPI_ISL_536631, EPI_ISL_536632, EPI_ISL_536633, EPI_ISL_536634, EPI_ISL_536635, EPI_ISL_536636, EPI_ISL_536637, EPI_ISL_536638, EPI_ISL_536639, EPI_ISL_536640, EPI_ISL_536641, EPI_ISL_536642, EPI_ISL_536643, EPI_ISL_536644, EPI_ISL_536645, EPI_ISL_536646, EPI_ISL_536647, EPI_ISL_536648, EPI_ISL_536649, EPI_ISL_536650, EPI_ISL_536651, EPI_ISL_536652, EPI_ISL_536653, EPI_ISL_536654, EPI_ISL_536655 | | | |
| see above | University of Wisconsin-Madison AIDS Vaccine Research Laboratories | University of Wisconsin-Madison AIDS Vaccine Research Laboratories | Gage Moreno, Katarina Braun, et al. AIDS Vaccine Research Laboratories |
| EPI_ISL_536656, EPI_ISL_536657 | University of Wisconsin-Madison Campus AIDS Vaccine Research Laboratories | University of Wisconsin-Madison AIDS Vaccine Research Laboratories | Gage Moreno, Katarina Braun, et al. AIDS Vaccine Research Laboratories |
| EPI_ISL_536658, EPI_ISL_536659, EPI_ISL_536660, EPI_ISL_536661, EPI_ISL_536662, EPI_ISL_536663, EPI_ISL_536664, EPI_ISL_536665, EPI_ISL_536666, EPI_ISL_536667, EPI_ISL_536668, EPI_ISL_536669, EPI_ISL_536670, EPI_ISL_536671, EPI_ISL_536672, EPI_ISL_536673, EPI_ISL_536674, EPI_ISL_536675, EPI_ISL_536676, EPI_ISL_536677, EPI_ISL_536678, EPI_ISL_536679, EPI_ISL_536680, EPI_ISL_536681, EPI_ISL_536682, EPI_ISL_536683, EPI_ISL_536684, EPI_ISL_536685, EPI_ISL_536686, EPI_ISL_536687, EPI_ISL_536688, EPI_ISL_536689, EPI_ISL_536690, EPI_ISL_536691, EPI_ISL_536692, EPI_ISL_536693, EPI_ISL_536694, EPI_ISL_536695, EPI_ISL_536696, EPI_ISL_536697, EPI_ISL_536698, EPI_ISL_536699, EPI_ISL_536700, EPI_ISL_536701, EPI_ISL_536702, EPI_ISL_536703, EPI_ISL_536704, EPI_ISL_536705, EPI_ISL_536706, EPI_ISL_536707, EPI_ISL_536708, EPI_ISL_536709, EPI_ISL_536710, EPI_ISL_536711, EPI_ISL_536712, EPI_ISL_536713, EPI_ISL_536714, EPI_ISL_536715, EPI_ISL_536716, EPI_ISL_536717, EPI_ISL_536718, EPI_ISL_536719, EPI_ISL_536720, EPI_ISL_536721, EPI_ISL_536722, EPI_ISL_536723, EPI_ISL_536724, EPI_ISL_536725, EPI_ISL_536727, EPI_ISL_536728, EPI_ISL_536729, EPI_ISL_536730, EPI_ISL_536731, EPI_ISL_536732, EPI_ISL_536733, EPI_ISL_536734, EPI_ISL_536735, EPI_ISL_536736, EPI_ISL_536737, EPI_ISL_536738, EPI_ISL_536739, EPI_ISL_536740, EPI_ISL_536741, EPI_ISL_536742, EPI_ISL_536743, EPI_ISL_536744, EPI_ISL_536745, EPI_ISL_536746, EPI_ISL_536747, EPI_ISL_536748, EPI_ISL_536749, EPI_ISL_536750, EPI_ISL_536752, EPI_ISL_536753, EPI_ISL_536754, EPI_ISL_536755, EPI_ISL_536756, EPI_ISL_536757, EPI_ISL_536758, EPI_ISL_536759, EPI_ISL_536760, EPI_ISL_536761, EPI_ISL_536762, EPI_ISL_536763, EPI_ISL_536764, EPI_ISL_536765, EPI_ISL_536766, EPI_ISL_536767, EPI_ISL_536768, EPI_ISL_536769, EPI_ISL_536770, EPI_ISL_536771, EPI_ISL_536772, EPI_ISL_536773, EPI_ISL_536774, EPI_ISL_536775, EPI_ISL_536776, EPI_ISL_536777, EPI_ISL_536778, EPI_ISL_536779, EPI_ISL_536780, EPI_ISL_536781, EPI_ISL_536782, EPI_ISL_536783, EPI_ISL_536784 | | | |
| see above | University of Wisconsin-Madison AIDS Vaccine Research Laboratories | University of Wisconsin-Madison AIDS Vaccine Research Laboratories | Gage Moreno, Katarina Braun, et al. AIDS Vaccine Research Laboratories |
| EPI_ISL_536785 | University of Wisconsin-Madison Campus AIDS Vaccine Research Laboratories | University of Wisconsin-Madison AIDS Vaccine Research Laboratories | Gage Moreno, Katarina Braun, et al. AIDS Vaccine Research Laboratories |
| EPI_ISL_536786 | University of Wisconsin-Madison AIDS Vaccine Research Laboratories | University of Wisconsin-Madison AIDS Vaccine Research Laboratories | Gage Moreno, Katarina Braun, et al. AIDS Vaccine Research Laboratories |
| EPI_ISL_537467, EPI_ISL_537469, EPI_ISL_537470, EPI_ISL_537471, EPI_ISL_537472, EPI_ISL_537473, EPI_ISL_537474, EPI_ISL_537475, EPI_ISL_537476, EPI_ISL_537477, EPI_ISL_537478, EPI_ISL_537479, EPI_ISL_537480, EPI_ISL_537481, EPI_ISL_537482, EPI_ISL_537483, EPI_ISL_537484, EPI_ISL_537485, EPI_ISL_537486, EPI_ISL_537487, EPI_ISL_537488, EPI_ISL_537489, EPI_ISL_537490, EPI_ISL_537491, EPI_ISL_537492, EPI_ISL_537493, EPI_ISL_537494, EPI_ISL_537495, EPI_ISL_537496, EPI_ISL_537497, EPI_ISL_537498, EPI_ISL_537499, EPI_ISL_537500, EPI_ISL_537501, EPI_ISL_537502, EPI_ISL_537503, EPI_ISL_537504, EPI_ISL_537505, EPI_ISL_537506, EPI_ISL_537507, EPI_ISL_537508, EPI_ISL_537509, EPI_ISL_537510, EPI_ISL_537512, EPI_ISL_537513, EPI_ISL_537514, EPI_ISL_537515, EPI_ISL_537516, EPI_ISL_537517, EPI_ISL_537518, EPI_ISL_537519, EPI_ISL_537520, EPI_ISL_537522, EPI_ISL_537523, EPI_ISL_537524, EPI_ISL_537525, EPI_ISL_537526, EPI_ISL_537527, EPI_ISL_537528, EPI_ISL_537529, EPI_ISL_537530, EPI_ISL_537531, EPI_ISL_537532, EPI_ISL_537533, EPI_ISL_537535, EPI_ISL_537536, EPI_ISL_537537, EPI_ISL_537538, EPI_ISL_537539, EPI_ISL_537540, EPI_ISL_537541, EPI_ISL_537542, EPI_ISL_537543, EPI_ISL_537544, EPI_ISL_537545, EPI_ISL_537546, EPI_ISL_537547, EPI_ISL_537549, EPI_ISL_537550, EPI_ISL_537551, EPI_ISL_537554, EPI_ISL_537556, EPI_ISL_537557, EPI_ISL_537558, EPI_ISL_537559, EPI_ISL_537560, EPI_ISL_537561, EPI_ISL_537562, EPI_ISL_537563, EPI_ISL_537564, EPI_ISL_537565, EPI_ISL_537566, EPI_ISL_537567, EPI_ISL_537568, EPI_ISL_537569, EPI_ISL_537570, EPI_ISL_537571, EPI_ISL_537573, EPI_ISL_537575, EPI_ISL_537576, EPI_ISL_537577, EPI_ISL_537578, EPI_ISL_537579, EPI_ISL_537582, EPI_ISL_537583, EPI_ISL_537584, EPI_ISL_537585, EPI_ISL_537586, EPI_ISL_537587, EPI_ISL_537588, EPI_ISL_537589, EPI_ISL_537590, EPI_ISL_537591, EPI_ISL_537592, EPI_ISL_537593, EPI_ISL_537594, EPI_ISL_537595, EPI_ISL_537596, EPI_ISL_537597, EPI_ISL_537598, EPI_ISL_537599, EPI_ISL_537602, EPI_ISL_537603, EPI_ISL_537604, EPI_ISL_537605, EPI_ISL_537606, EPI_ISL_537607 | | | |
| see above | UCLA Pathology Clinical Microbiology Lab | Kruglyak Lab | Guo et al. |
| EPI_ISL_538233, EPI_ISL_538234, EPI_ISL_538235, EPI_ISL_538236, EPI_ISL_538239, EPI_ISL_538241, EPI_ISL_538242, EPI_ISL_538243, EPI_ISL_538245, EPI_ISL_538247, EPI_ISL_538248, EPI_ISL_538249, EPI_ISL_538264 | | | |
| see above | TriCore Reference Laboratories | Center for Global Health, University of New Mexico Health Sciences Center | Daryl Domman, Kurt Schwalm, Twila Kunde, Joseph Hicks, Michael Edwards, Darrell Dinwiddie |
| EPI_ISL_538265, EPI_ISL_538266, EPI_ISL_538267, EPI_ISL_538268, EPI_ISL_538269, EPI_ISL_538270, EPI_ISL_538271, EPI_ISL_538272, EPI_ISL_538273, EPI_ISL_538274 | Maryland Public Health Laboratory | Maryland Public Health Laboratory | Maryland Department of Health Laboratories Administration |
| EPI_ISL_538277, EPI_ISL_538279, EPI_ISL_538280, EPI_ISL_538281, EPI_ISL_538282, EPI_ISL_538283, EPI_ISL_538284, EPI_ISL_538285, EPI_ISL_538286, EPI_ISL_538288, EPI_ISL_538289, EPI_ISL_538290, EPI_ISL_538292, EPI_ISL_538293, EPI_ISL_538294, EPI_ISL_538295, EPI_ISL_538296, EPI_ISL_538297, EPI_ISL_538298, EPI_ISL_538299, EPI_ISL_538300, EPI_ISL_538301, EPI_ISL_538303, EPI_ISL_538304, EPI_ISL_538305, EPI_ISL_538306, EPI_ISL_538307, EPI_ISL_538309, EPI_ISL_538311, EPI_ISL_538313, EPI_ISL_538314, EPI_ISL_538315, EPI_ISL_538318, EPI_ISL_538319, EPI_ISL_538320 | | | |
| see above | Texas Department of State Health Services | Texas Department of State Health Services | Bonnie Oh, Rashmi Tuladhar, Jenny Zhang, Maliha Rahman, Anita Pokharel, Myong Koag, Chun Wang, Rachel Lee, Grace Kubin |
| EPI_ISL_538390, EPI_ISL_538391, EPI_ISL_538392, EPI_ISL_538393, EPI_ISL_538394, EPI_ISL_538395, EPI_ISL_538396, EPI_ISL_538397, EPI_ISL_538398, EPI_ISL_538399, EPI_ISL_538400, EPI_ISL_538401, EPI_ISL_538402, EPI_ISL_538404, EPI_ISL_538406, EPI_ISL_538407, EPI_ISL_538408, EPI_ISL_538409, EPI_ISL_538410, EPI_ISL_538411, EPI_ISL_538412, EPI_ISL_538413, EPI_ISL_538414, EPI_ISL_538415, EPI_ISL_538417, EPI_ISL_538418, EPI_ISL_538419, EPI_ISL_538420, EPI_ISL_538421, EPI_ISL_538422, EPI_ISL_538423, EPI_ISL_538424, EPI_ISL_538425, EPI_ISL_538426, EPI_ISL_538427, EPI_ISL_538428, EPI_ISL_538429, EPI_ISL_538430, EPI_ISL_538431, EPI_ISL_538432, EPI_ISL_538433, EPI_ISL_538434 | | | |
| see above | Microbiology Division, South Carolina Department of Health and Environmental Control | Microbiology Division, South Carolina Department of Health and Environmental Control | Flores,H. |
| EPI_ISL_538514, EPI_ISL_538515, EPI_ISL_538516, EPI_ISL_538517, EPI_ISL_538518, EPI_ISL_538519, EPI_ISL_538520, EPI_ISL_538521 | Infectious Diseases, North Carolina State Laboratory of Public Health COVID-19 Response Team | Infectious Diseases, North Carolina State Laboratory of Public Health COVID-19 Response Team | Chase,K. |
| EPI_ISL_538522, | Infectious Diseases, North Carolina | North Carolina State Laboratory of Public | Chase,K. |

| | | | |
|---|---|---|---|
| EPI_ISL_538523 | State Laboratory of Public Health COVID-19 Response Team | Health | |
| EPI_ISL_539495 | Centers for Disease Control and Prevention, Dengue Branch | Centers for Disease Control and Prevention, Dengue Branch | Gilberto A. Santiago, Glenda Gonzalez, Betzabel Flores, Keyla Charriez, Jorge L. Munoz-Jordan, Gabriela Paz-Bailey, Janice Perez, Vanessa Rivera-Amill, Diego Sainz de la Peña, Jorge Bertran |
| EPI_ISL_539794, EPI_ISL_539795, EPI_ISL_539796, EPI_ISL_539797, EPI_ISL_539798, EPI_ISL_539799, EPI_ISL_539800, EPI_ISL_539801, EPI_ISL_539802 | Wyoming Public Health Laboratory | Wyoming Public Health Laboratory | Noah Hull, Rob Christensen, Jim Mildenberger, Joel Sevinsky, Cari Sloma, and Wanda Manley |
| EPI_ISL_539824, EPI_ISL_539825, EPI_ISL_539826, EPI_ISL_539827, EPI_ISL_539828, EPI_ISL_539829, EPI_ISL_539830, EPI_ISL_539831, EPI_ISL_539832, EPI_ISL_539833, EPI_ISL_539834, EPI_ISL_539835, EPI_ISL_539836, EPI_ISL_539837, EPI_ISL_539838, EPI_ISL_539839, EPI_ISL_539840, EPI_ISL_539841 | | | |
| see above | Minnesota Department of Health, Public Health Laboratory | Minnesota Department of Health, Public Health Laboratory | Matt Plumb, Jacob Garfin, and Xiong Wang |
| EPI_ISL_539842, EPI_ISL_539843, EPI_ISL_539844, EPI_ISL_539845, EPI_ISL_539846, EPI_ISL_539847, EPI_ISL_539848, EPI_ISL_539849 | Mayo Clinic & Mayo Clinic Laboratories | Minnesota Department of Health, Public Health Laboratory | Matt Plumb, Jacob Garfin, and Xiong Wang |
| EPI_ISL_539879 | Microbiology Division, South Carolina Department of Health and Environmental Control | Microbiology Division, South Carolina Department of Health and Environmental Control | Flores,H. |
| EPI_ISL_540421, EPI_ISL_540422 | Wyoming Public Health Laboratory | Wyoming Public Health Laboratory | Noah Hull, Rob Christensen, Jim Mildenberger, Joel Sevinsky, Cari Sloma, and Wanda Manley |
| EPI_ISL_540431 | IN State Department of Health Laboratory Services | Pathogen Discovery, Respiratory Viruses Branch, Division of Viral Diseases, Centers for Disease Control and Prevention | Yan Li, Jing Zhang, Anna Montmayeur, Krista Queen,Ying Tao, Anna Uehara, Clinton R. Paden, Rachel Marine, Haibin Wang, Suxiang Tong |
| EPI_ISL_540432, EPI_ISL_540433 | TN Division of Laboratory Services | Pathogen Discovery, Respiratory Viruses Branch, Division of Viral Diseases, Centers for Disease Control and Prevention | Yan Li, Jing Zhang, Anna Montmayeur, Krista Queen,Ying Tao, Anna Uehara, Clinton R. Paden, Rachel Marine, Haibin Wang, Suxiang Tong |
| EPI_ISL_540434 | TN Division of Laboratory Services | Pathogen Discovery, Respiratory Viruses Branch, Division of Viral Diseases, Centers for Disease Control and Prevention | Ying Tao, Yan Li, Jing Zhang, Krista Queen, Anna Uehara, Clinton R. Paden, Haibin Wang, Suxiang Tong |
| EPI_ISL_540435 | VA-Division of Consolidated Laboratory Services | Pathogen Discovery, Respiratory Viruses Branch, Division of Viral Diseases, Centers for Disease Control and Prevention | Krista Queen, Yan Li, Jing Zhang, Anna Montmayeur, Krista Queen,Ying Tao, Anna Uehara, Clinton R. Paden, Rachel Marine, Haibin Wang, Suxiang Tong |
| EPI_ISL_540436, EPI_ISL_540437, EPI_ISL_540438 | WI State Laboratory of Hygiene | Pathogen Discovery, Respiratory Viruses Branch, Division of Viral Diseases, Centers for Disease Control and Prevention | Yan Li, Jing Zhang, Anna Montmayeur, Krista Queen,Ying Tao, Anna Uehara, Clinton R. Paden, Rachel Marine, Haibin Wang, Suxiang Tong |
| EPI_ISL_540439 | WVDHHR - Office of Laboratory Services | Pathogen Discovery, Respiratory Viruses Branch, Division of Viral Diseases, Centers for Disease Control and Prevention | Ying Tao, Yan Li, Jing Zhang, Krista Queen, Anna Uehara, Clinton R. Paden, Haibin Wang, Suxiang Tong |
| EPI_ISL_540441 | NC State Laboratory of Public Health | Pathogen Discovery, Respiratory Viruses Branch, Division of Viral Diseases, Centers for Disease Control and Prevention | Yan Li, Jing Zhang, Anna Montmayeur, Krista Queen,Ying Tao, Anna Uehara, Clinton R. Paden, Rachel Marine, Haibin Wang, Suxiang Tong |
| EPI_ISL_540921 | Wyoming Public Health Laboratory | Wyoming Public Health Laboratory | Noah Hull, Rob Christensen, Jim Mildenberger, Joel Sevinsky, Cari Sloma, and Wanda Manley |
| EPI_ISL_541140, EPI_ISL_541141, EPI_ISL_541143, EPI_ISL_541144, EPI_ISL_541145, EPI_ISL_541147, EPI_ISL_541148, EPI_ISL_541149, EPI_ISL_541150, EPI_ISL_541151, EPI_ISL_541153, EPI_ISL_541154, EPI_ISL_541155, EPI_ISL_541156, EPI_ISL_541157, EPI_ISL_541158, EPI_ISL_541159, EPI_ISL_541160, EPI_ISL_541161, EPI_ISL_541163, EPI_ISL_541164, EPI_ISL_541166, EPI_ISL_541168, EPI_ISL_541171, EPI_ISL_541172, EPI_ISL_541173, EPI_ISL_541174, EPI_ISL_541175, EPI_ISL_541176, EPI_ISL_541178, EPI_ISL_541179, EPI_ISL_541180, EPI_ISL_541181, EPI_ISL_541182, EPI_ISL_541183, EPI_ISL_541184, EPI_ISL_541185, EPI_ISL_541186, EPI_ISL_541187, EPI_ISL_541188, EPI_ISL_541189, EPI_ISL_541191, EPI_ISL_541192, EPI_ISL_541193, EPI_ISL_541195, EPI_ISL_541196, EPI_ISL_541197, EPI_ISL_541198, EPI_ISL_541199, EPI_ISL_541200, EPI_ISL_541201, EPI_ISL_541202, EPI_ISL_541203, EPI_ISL_541204, EPI_ISL_541205, EPI_ISL_541207, EPI_ISL_541208, EPI_ISL_541209, EPI_ISL_541210, EPI_ISL_541212, EPI_ISL_541213, EPI_ISL_541214, EPI_ISL_541215, EPI_ISL_541216, EPI_ISL_541217, EPI_ISL_541218, EPI_ISL_541219, EPI_ISL_541220, EPI_ISL_541221, EPI_ISL_541222, EPI_ISL_541223, EPI_ISL_541225, EPI_ISL_541226, EPI_ISL_541227, EPI_ISL_541228, EPI_ISL_541230, EPI_ISL_541231, EPI_ISL_541232, EPI_ISL_541233, EPI_ISL_541234, EPI_ISL_541235, EPI_ISL_541236, EPI_ISL_541237, EPI_ISL_541238, EPI_ISL_541239, EPI_ISL_541240, EPI_ISL_541241, EPI_ISL_541242, EPI_ISL_541244, EPI_ISL_541246, EPI_ISL_541248, EPI_ISL_541249, EPI_ISL_541250, EPI_ISL_541253, EPI_ISL_541254, EPI_ISL_541255, EPI_ISL_541256, EPI_ISL_541257, EPI_ISL_541258, EPI_ISL_541259, EPI_ISL_541260, EPI_ISL_541261, EPI_ISL_541262, EPI_ISL_541263, EPI_ISL_541264, EPI_ISL_541265, EPI_ISL_541266, EPI_ISL_541267, EPI_ISL_541268, EPI_ISL_541269, EPI_ISL_541270, EPI_ISL_541271, EPI_ISL_541272, EPI_ISL_541273, EPI_ISL_541275, EPI_ISL_541277, EPI_ISL_541278, EPI_ISL_541279, EPI_ISL_541280, EPI_ISL_541281, EPI_ISL_541282, EPI_ISL_541283, EPI_ISL_541284, EPI_ISL_541285, EPI_ISL_541286, EPI_ISL_541287, EPI_ISL_541288, EPI_ISL_541289, EPI_ISL_541290, EPI_ISL_541291, EPI_ISL_541292, EPI_ISL_541293, EPI_ISL_541294, EPI_ISL_541295, EPI_ISL_541296, EPI_ISL_541297, EPI_ISL_541299, EPI_ISL_541300, EPI_ISL_541301, EPI_ISL_541302, EPI_ISL_541303, EPI_ISL_541305, EPI_ISL_541306, EPI_ISL_541308, EPI_ISL_541309, EPI_ISL_541310, EPI_ISL_541312, EPI_ISL_541313, EPI_ISL_541314, EPI_ISL_541315, EPI_ISL_541318, EPI_ISL_541319, EPI_ISL_541320, EPI_ISL_541321, EPI_ISL_541322, EPI_ISL_541323, EPI_ISL_541324, EPI_ISL_541326, EPI_ISL_541327, EPI_ISL_541328, EPI_ISL_541329, EPI_ISL_541330, EPI_ISL_541331 | | | |
| see above | Florida Bureau of Public Health Laboratories, Florida Department of Health | Florida Bureau of Public Health Laboratories, Florida Department of Health | Schmedes,S., Blanton,J. |
| EPI_ISL_541338 | Florida Bureau of Public Health Laboratories | Florida Bureau of Public Health Laboratories | Sarah Schmedes, Jason Blanton |
| EPI_ISL_541540, EPI_ISL_541541, EPI_ISL_541542, EPI_ISL_541543, EPI_ISL_541544, EPI_ISL_541545, EPI_ISL_541546, EPI_ISL_541547, EPI_ISL_541548, EPI_ISL_541549, EPI_ISL_541550 | | | |
| see above | University of Wisconsin-Madison AIDS Vaccine Research Laboratories | University of Wisconsin-Madison AIDS Vaccine Research Laboratories | Gage Moreno, Katarina Braun, et al. AIDS Vaccine Research Laboratories |
| EPI_ISL_541551, EPI_ISL_541552, EPI_ISL_541553, EPI_ISL_541554, EPI_ISL_541555, EPI_ISL_541556, EPI_ISL_541557, EPI_ISL_541558, EPI_ISL_541559, EPI_ISL_541560, EPI_ISL_541561, EPI_ISL_541562, EPI_ISL_541563, EPI_ISL_541564, EPI_ISL_541565 | | | |
| see above | University of Wisconsin-Madison Campus AIDS Vaccine Research Laboratories | University of Wisconsin-Madison AIDS Vaccine Research Laboratories | Gage Moreno, Katarina Braun, et al. AIDS Vaccine Research Laboratories |
| EPI_ISL_541566, EPI_ISL_541567, EPI_ISL_541569, EPI_ISL_541570, EPI_ISL_541571, EPI_ISL_541572, EPI_ISL_541573, EPI_ISL_541574, EPI_ISL_541575, EPI_ISL_541576, EPI_ISL_541577, EPI_ISL_541578, EPI_ISL_541579, EPI_ISL_541580, EPI_ISL_541581, EPI_ISL_541582, EPI_ISL_541583, EPI_ISL_541584, EPI_ISL_541585, EPI_ISL_541586, EPI_ISL_541587, EPI_ISL_541588, EPI_ISL_541589, EPI_ISL_541590, EPI_ISL_541591, EPI_ISL_541592, EPI_ISL_541593, EPI_ISL_541594, EPI_ISL_541595, EPI_ISL_541596, EPI_ISL_541597, EPI_ISL_541598, EPI_ISL_541599, EPI_ISL_541600, EPI_ISL_541601, EPI_ISL_541602, EPI_ISL_541603, EPI_ISL_541604, EPI_ISL_541605, EPI_ISL_541606, EPI_ISL_541607, EPI_ISL_541608, EPI_ISL_541609, EPI_ISL_541610, EPI_ISL_541611, EPI_ISL_541612, EPI_ISL_541613, EPI_ISL_541614, EPI_ISL_541615, EPI_ISL_541616, EPI_ISL_541617, EPI_ISL_541618, EPI_ISL_541619, EPI_ISL_541620, EPI_ISL_541621, EPI_ISL_541622, EPI_ISL_541623, EPI_ISL_541624, EPI_ISL_541625, EPI_ISL_541626, EPI_ISL_541627, EPI_ISL_541628, EPI_ISL_541629, EPI_ISL_541630, EPI_ISL_541631, EPI_ISL_541632, EPI_ISL_541633, EPI_ISL_541634, EPI_ISL_541635, EPI_ISL_541636, EPI_ISL_541637, EPI_ISL_541638, EPI_ISL_541639, EPI_ISL_541640, EPI_ISL_541641, EPI_ISL_541642, EPI_ISL_541643, EPI_ISL_541644, EPI_ISL_541645, EPI_ISL_541646, EPI_ISL_541647, EPI_ISL_541648 | | | |
| see above | University of Wisconsin-Madison AIDS Vaccine Research Laboratories | University of Wisconsin-Madison AIDS Vaccine Research Laboratories | Gage Moreno, Katarina Braun, et al. AIDS Vaccine Research Laboratories |
| EPI_ISL_541663, EPI_ISL_541664, EPI_ISL_541665, EPI_ISL_541666, EPI_ISL_541667, EPI_ISL_541668, EPI_ISL_541669, EPI_ISL_541670, EPI_ISL_541671, EPI_ISL_541672, EPI_ISL_541673, EPI_ISL_541674, EPI_ISL_541675, EPI_ISL_541676, EPI_ISL_541677, EPI_ISL_541678, EPI_ISL_541679 | | | |
| see above | Microbiology Division, South Carolina Department of Health and Environmental Control | Microbiology Division, South Carolina Department of Health and Environmental Control | Flores,H. |
| EPI_ISL_541971, EPI_ISL_541974, EPI_ISL_541976, EPI_ISL_541977, EPI_ISL_541980, EPI_ISL_541981, EPI_ISL_541983, EPI_ISL_541984, EPI_ISL_541985, EPI_ISL_541987, EPI_ISL_541988, EPI_ISL_541989, EPI_ISL_541990, EPI_ISL_541991, EPI_ISL_541992, EPI_ISL_541993, EPI_ISL_541994, EPI_ISL_541995, EPI_ISL_541996, EPI_ISL_541997, EPI_ISL_541998, EPI_ISL_541999, EPI_ISL_542000, EPI_ISL_542001, EPI_ISL_542003, EPI_ISL_542004, EPI_ISL_542007, EPI_ISL_542008, EPI_ISL_542009, EPI_ISL_542010, EPI_ISL_542011 | | | |
| see above | Texas Department of State Health Services | Texas Department of State Health Services | Rashmi Tuladhar, Bonnie Oh, Jenny Zhang, Maliha Rahman, Anita Pokharel, Myong Koag, Chun Wang, Rachel Lee, Grace Kubin |
| EPI_ISL_542012, EPI_ISL_542013, EPI_ISL_542014, EPI_ISL_542015, EPI_ISL_542016, EPI_ISL_542017, EPI_ISL_542018, EPI_ISL_542019, EPI_ISL_542020, EPI_ISL_542021, EPI_ISL_542022, EPI_ISL_542023, EPI_ISL_542024, EPI_ISL_542025, EPI_ISL_542026, EPI_ISL_542027, EPI_ISL_542028, EPI_ISL_542029, EPI_ISL_542030, EPI_ISL_542031, EPI_ISL_542032, EPI_ISL_542033, EPI_ISL_542034, EPI_ISL_542035, EPI_ISL_542036, EPI_ISL_542037, EPI_ISL_542038, EPI_ISL_542039, EPI_ISL_542040, EPI_ISL_542041, EPI_ISL_542042, EPI_ISL_542043, EPI_ISL_542044, EPI_ISL_542045, EPI_ISL_542046, EPI_ISL_542047, EPI_ISL_542048, EPI_ISL_542049, EPI_ISL_542050, EPI_ISL_542051, EPI_ISL_542052, EPI_ISL_542053, EPI_ISL_542054, EPI_ISL_542055, EPI_ISL_542056, EPI_ISL_542057, EPI_ISL_542058, EPI_ISL_542059, EPI_ISL_542060, EPI_ISL_542061, EPI_ISL_542062, EPI_ISL_542063, EPI_ISL_542064, EPI_ISL_542065, EPI_ISL_542066, EPI_ISL_542067, EPI_ISL_542068, EPI_ISL_542069, EPI_ISL_542070, EPI_ISL_542071, EPI_ISL_542072, EPI_ISL_542073, EPI_ISL_542074, EPI_ISL_542076, EPI_ISL_542077, EPI_ISL_542078, EPI_ISL_542079, EPI_ISL_542080, EPI_ISL_542082, EPI_ISL_542083, EPI_ISL_542084, EPI_ISL_542085, EPI_ISL_542086, EPI_ISL_542087, EPI_ISL_542088, EPI_ISL_542089, EPI_ISL_542090, EPI_ISL_542091, EPI_ISL_542092, EPI_ISL_542093, EPI_ISL_542094, EPI_ISL_542095, EPI_ISL_542096, EPI_ISL_542097 | | | |
| see above | New Mexico Department of Health Scientific Laboratory | New Mexico Department of Health Scientific Laboratory | Ellie Johnson, Anastacia Griego-Fisher, D'Eldra Malone |
| EPI_ISL_542444, EPI_ISL_542445, EPI_ISL_542446, EPI_ISL_542447, EPI_ISL_542448, EPI_ISL_542450, EPI_ISL_542452, EPI_ISL_542453, EPI_ISL_542454, EPI_ISL_542455, EPI_ISL_542456, EPI_ISL_542457, EPI_ISL_542458, EPI_ISL_542460, EPI_ISL_542461, EPI_ISL_542463, EPI_ISL_542464, EPI_ISL_542466, EPI_ISL_542469, EPI_ISL_542470, EPI_ISL_542471, EPI_ISL_542472, EPI_ISL_542473, EPI_ISL_542474, EPI_ISL_542475, EPI_ISL_542477, EPI_ISL_542478, EPI_ISL_542479, EPI_ISL_542480, EPI_ISL_542481 | | | |
| see above | Texas Department of State Health Services | Texas Department of State Health Services | Rashmi Tuladhar, Bonnie Oh,Jenny Zhang, Maliha Rahman, Anita Pokharel, Myong Koag, Chun Wang, Rachel Lee, Grace Kubin |
| EPI_ISL_542486 | Walter Reed National Military Medical | Naval Medical Research Center Biological | Logan J. Voegtly, Regina Z. Cer, Lindsay A. Glang, Francisco J. Malagon, Kyle A. Long, Derek Larson, Sterling L. Brodniak, Kimberly A Bishop-Lilly |

| | | |
|---|---|---|
| | Center | Defense Research Directorate |

EPI_ISL_542487, EPI_ISL_542488, EPI_ISL_542489, EPI_ISL_542490, EPI_ISL_542491, EPI_ISL_542492, EPI_ISL_542493, EPI_ISL_542494, EPI_ISL_542495, EPI_ISL_542496, EPI_ISL_542497, EPI_ISL_542498, EPI_ISL_542499, EPI_ISL_542500, EPI_ISL_542501, EPI_ISL_542502, EPI_ISL_542503, EPI_ISL_542504, EPI_ISL_542505, EPI_ISL_542506, EPI_ISL_542507, EPI_ISL_542508, EPI_ISL_542509, EPI_ISL_542510,
EPI_ISL_542511, EPI_ISL_542512, EPI_ISL_542513, EPI_ISL_542514, EPI_ISL_542515, EPI_ISL_542516, EPI_ISL_542517, EPI_ISL_542518, EPI_ISL_542519, EPI_ISL_542520, EPI_ISL_542521, EPI_ISL_542522, EPI_ISL_542523, EPI_ISL_542524, EPI_ISL_542525, EPI_ISL_542526, EPI_ISL_542527, EPI_ISL_542528, EPI_ISL_542529, EPI_ISL_542530, EPI_ISL_542531, EPI_ISL_542532, EPI_ISL_542533, EPI_ISL_542534,
EPI_ISL_542535, EPI_ISL_542536, EPI_ISL_542537, EPI_ISL_542538, EPI_ISL_542539, EPI_ISL_542540, EPI_ISL_542541, EPI_ISL_542542, EPI_ISL_542543, EPI_ISL_542544, EPI_ISL_542545, EPI_ISL_542546, EPI_ISL_542547, EPI_ISL_542548, EPI_ISL_542549, EPI_ISL_542550, EPI_ISL_542551, EPI_ISL_542552, EPI_ISL_542553, EPI_ISL_542554, EPI_ISL_542555, EPI_ISL_542556, EPI_ISL_542557, EPI_ISL_542558,
EPI_ISL_542559, EPI_ISL_542560, EPI_ISL_542561, EPI_ISL_542562, EPI_ISL_542563, EPI_ISL_542564, EPI_ISL_542565, EPI_ISL_542566, EPI_ISL_542567, EPI_ISL_542568, EPI_ISL_542569, EPI_ISL_542570, EPI_ISL_542571, EPI_ISL_542572, EPI_ISL_542573, EPI_ISL_542574, EPI_ISL_542575, EPI_ISL_542576, EPI_ISL_542577, EPI_ISL_542578, EPI_ISL_542579, EPI_ISL_542580, EPI_ISL_542581, EPI_ISL_542582,
EPI_ISL_542583, EPI_ISL_542584, EPI_ISL_542585, EPI_ISL_542586, EPI_ISL_542587, EPI_ISL_542588, EPI_ISL_542589, EPI_ISL_542590, EPI_ISL_542591, EPI_ISL_542592, EPI_ISL_542593, EPI_ISL_542594, EPI_ISL_542595, EPI_ISL_542596, EPI_ISL_542597, EPI_ISL_542598, EPI_ISL_542599, EPI_ISL_542600, EPI_ISL_542601, EPI_ISL_542602, EPI_ISL_542603, EPI_ISL_542604, EPI_ISL_542605, EPI_ISL_542606,
EPI_ISL_542607, EPI_ISL_542608, EPI_ISL_542609, EPI_ISL_542610, EPI_ISL_542611, EPI_ISL_542612, EPI_ISL_542613, EPI_ISL_542614, EPI_ISL_542615, EPI_ISL_542616, EPI_ISL_542617, EPI_ISL_542618, EPI_ISL_542619, EPI_ISL_542620, EPI_ISL_542621, EPI_ISL_542622, EPI_ISL_542623, EPI_ISL_542624, EPI_ISL_542625, EPI_ISL_542626, EPI_ISL_542627, EPI_ISL_542628, EPI_ISL_542629, EPI_ISL_542630,
EPI_ISL_542631, EPI_ISL_542632, EPI_ISL_542633, EPI_ISL_542634, EPI_ISL_542635, EPI_ISL_542636, EPI_ISL_542637, EPI_ISL_542638, EPI_ISL_542639, EPI_ISL_542640, EPI_ISL_542641, EPI_ISL_542642, EPI_ISL_542643, EPI_ISL_542644, EPI_ISL_542645, EPI_ISL_542646, EPI_ISL_542647, EPI_ISL_542648, EPI_ISL_542649, EPI_ISL_542650, EPI_ISL_542651, EPI_ISL_542652, EPI_ISL_542653, EPI_ISL_542654,
EPI_ISL_542655, EPI_ISL_542656, EPI_ISL_542657, EPI_ISL_542658, EPI_ISL_542659, EPI_ISL_542660, EPI_ISL_542661, EPI_ISL_542662, EPI_ISL_542663, EPI_ISL_542664, EPI_ISL_542665, EPI_ISL_542666, EPI_ISL_542667, EPI_ISL_542668, EPI_ISL_542669, EPI_ISL_542670, EPI_ISL_542671, EPI_ISL_542672, EPI_ISL_542673, EPI_ISL_542674, EPI_ISL_542675, EPI_ISL_542676, EPI_ISL_542677, EPI_ISL_542678,
EPI_ISL_542679, EPI_ISL_542680, EPI_ISL_542681, EPI_ISL_542682, EPI_ISL_542683, EPI_ISL_542684, EPI_ISL_542685, EPI_ISL_542686, EPI_ISL_542687, EPI_ISL_542688, EPI_ISL_542689, EPI_ISL_542690, EPI_ISL_542691, EPI_ISL_542692, EPI_ISL_542693, EPI_ISL_542694, EPI_ISL_542695, EPI_ISL_542696, EPI_ISL_542697, EPI_ISL_542698, EPI_ISL_542699, EPI_ISL_542700, EPI_ISL_542701, EPI_ISL_542702,
EPI_ISL_542703, EPI_ISL_542704, EPI_ISL_542705, EPI_ISL_542706, EPI_ISL_542707, EPI_ISL_542708, EPI_ISL_542709, EPI_ISL_542710, EPI_ISL_542711, EPI_ISL_542712, EPI_ISL_542713, EPI_ISL_542714, EPI_ISL_542715, EPI_ISL_542716, EPI_ISL_542717, EPI_ISL_542718, EPI_ISL_542719, EPI_ISL_542720, EPI_ISL_542721, EPI_ISL_542722, EPI_ISL_542723, EPI_ISL_542724, EPI_ISL_542725, EPI_ISL_542726,
EPI_ISL_542727, EPI_ISL_542728, EPI_ISL_542729, EPI_ISL_542730, EPI_ISL_542731, EPI_ISL_542732, EPI_ISL_542733, EPI_ISL_542734, EPI_ISL_542735, EPI_ISL_542736, EPI_ISL_542737, EPI_ISL_542738, EPI_ISL_542739, EPI_ISL_542740, EPI_ISL_542741, EPI_ISL_542742, EPI_ISL_542743, EPI_ISL_542744, EPI_ISL_542745, EPI_ISL_542746, EPI_ISL_542747, EPI_ISL_542748, EPI_ISL_542749, EPI_ISL_542750,
EPI_ISL_542751, EPI_ISL_542752, EPI_ISL_542753, EPI_ISL_542754, EPI_ISL_542755, EPI_ISL_542756, EPI_ISL_542757, EPI_ISL_542758, EPI_ISL_542759, EPI_ISL_542760, EPI_ISL_542761, EPI_ISL_542762, EPI_ISL_542763, EPI_ISL_542764, EPI_ISL_542765, EPI_ISL_542766, EPI_ISL_542767, EPI_ISL_542768, EPI_ISL_542769, EPI_ISL_542770, EPI_ISL_542771, EPI_ISL_542772, EPI_ISL_542773, EPI_ISL_542774,
EPI_ISL_542775, EPI_ISL_542776, EPI_ISL_542777, EPI_ISL_542778, EPI_ISL_542779, EPI_ISL_542780, EPI_ISL_542781, EPI_ISL_542782, EPI_ISL_542783, EPI_ISL_542784, EPI_ISL_542785, EPI_ISL_542786, EPI_ISL_542787, EPI_ISL_542788, EPI_ISL_542789, EPI_ISL_542790, EPI_ISL_542791, EPI_ISL_542792, EPI_ISL_542793, EPI_ISL_542794, EPI_ISL_542795, EPI_ISL_542796, EPI_ISL_542797, EPI_ISL_542798,
EPI_ISL_542799, EPI_ISL_542800, EPI_ISL_542801, EPI_ISL_542802, EPI_ISL_542803, EPI_ISL_542804, EPI_ISL_542805, EPI_ISL_542806, EPI_ISL_542807, EPI_ISL_542808, EPI_ISL_542809, EPI_ISL_542810, EPI_ISL_542811, EPI_ISL_542812, EPI_ISL_542813, EPI_ISL_542814, EPI_ISL_542815, EPI_ISL_542816, EPI_ISL_542817, EPI_ISL_542818, EPI_ISL_542819, EPI_ISL_542820, EPI_ISL_542821, EPI_ISL_542822,
EPI_ISL_542823, EPI_ISL_542824, EPI_ISL_542825, EPI_ISL_542826, EPI_ISL_542827, EPI_ISL_542828, EPI_ISL_542829, EPI_ISL_542830, EPI_ISL_542831, EPI_ISL_542832, EPI_ISL_542833, EPI_ISL_542834, EPI_ISL_542835, EPI_ISL_542836, EPI_ISL_542837, EPI_ISL_542838, EPI_ISL_542839, EPI_ISL_542840, EPI_ISL_542841, EPI_ISL_542842, EPI_ISL_542843, EPI_ISL_542844, EPI_ISL_542845, EPI_ISL_542846,
EPI_ISL_542847, EPI_ISL_542848, EPI_ISL_542849, EPI_ISL_542850, EPI_ISL_542851, EPI_ISL_542852, EPI_ISL_542853, EPI_ISL_542854, EPI_ISL_542855, EPI_ISL_542856, EPI_ISL_542857, EPI_ISL_542858, EPI_ISL_542859, EPI_ISL_542860, EPI_ISL_542861, EPI_ISL_542862, EPI_ISL_542863, EPI_ISL_542864, EPI_ISL_542865, EPI_ISL_542866, EPI_ISL_542867, EPI_ISL_542868, EPI_ISL_542869, EPI_ISL_542870,
EPI_ISL_542871, EPI_ISL_542872, EPI_ISL_542873, EPI_ISL_542874, EPI_ISL_542875, EPI_ISL_542876, EPI_ISL_542877, EPI_ISL_542878, EPI_ISL_542879, EPI_ISL_542880, EPI_ISL_542881, EPI_ISL_542882, EPI_ISL_542883, EPI_ISL_542884, EPI_ISL_542885, EPI_ISL_542886, EPI_ISL_542887, EPI_ISL_542888, EPI_ISL_542889, EPI_ISL_542890, EPI_ISL_542891, EPI_ISL_542892, EPI_ISL_542893, EPI_ISL_542894,
EPI_ISL_542895, EPI_ISL_542896, EPI_ISL_542897, EPI_ISL_542898, EPI_ISL_542899, EPI_ISL_542900, EPI_ISL_542901, EPI_ISL_542902, EPI_ISL_542903, EPI_ISL_542904, EPI_ISL_542905, EPI_ISL_542906, EPI_ISL_542907, EPI_ISL_542908, EPI_ISL_542909, EPI_ISL_542910, EPI_ISL_542911, EPI_ISL_542912, EPI_ISL_542913, EPI_ISL_542914, EPI_ISL_542915, EPI_ISL_542916, EPI_ISL_542917, EPI_ISL_542918,
EPI_ISL_542919, EPI_ISL_542920, EPI_ISL_542921, EPI_ISL_542922, EPI_ISL_542923, EPI_ISL_542924, EPI_ISL_542925, EPI_ISL_542926, EPI_ISL_542927, EPI_ISL_542928, EPI_ISL_542929, EPI_ISL_542930, EPI_ISL_542931, EPI_ISL_542932, EPI_ISL_542933, EPI_ISL_542934

| | | | |
|---|---|---|---|
| see above | Houston Methodist Hospital | Houston Methodist Hospital | S. Wesley Long, Randall J. Olsen, Paul A. Christensen, David W. Bernard, James J. Davis, Maulik Shukla, Marcus Nguyen, Matthew Ojeda Saavedra, Concepcion C. Cantu, Prasanti Yerramilli, Layne Pruitt, Sishir Subedi, Hung-Che Kuo, Heather Hendrickson, Ghazaleh Eskandari, Hoang A. T. Nguyen, J. Hunter Long, Muthiah Kumaraswami, Jule Goike, Daniel Boutz, Jimmy Gollihar, Jason S. McLellan, Chia-Wei Chou, Kamyab Javanmardi, Ilya J. Finkelstein, and James M. Musser |

EPI_ISL_542935, EPI_ISL_542937, EPI_ISL_542940, EPI_ISL_542941, EPI_ISL_542942, EPI_ISL_542943, EPI_ISL_542945, EPI_ISL_542948, EPI_ISL_542949, EPI_ISL_542950, EPI_ISL_542951, EPI_ISL_542952, EPI_ISL_542955, EPI_ISL_542956, EPI_ISL_542958, EPI_ISL_542959, EPI_ISL_542960, EPI_ISL_542961, EPI_ISL_542962, EPI_ISL_542963, EPI_ISL_542964, EPI_ISL_542965, EPI_ISL_542966, EPI_ISL_542967,
EPI_ISL_542969, EPI_ISL_542970, EPI_ISL_542971, EPI_ISL_542972, EPI_ISL_542975, EPI_ISL_542977, EPI_ISL_542978, EPI_ISL_542979, EPI_ISL_542980, EPI_ISL_542981, EPI_ISL_542982, EPI_ISL_542983, EPI_ISL_542984, EPI_ISL_542985, EPI_ISL_542986, EPI_ISL_542987, EPI_ISL_542988, EPI_ISL_542989, EPI_ISL_542990, EPI_ISL_542991, EPI_ISL_542992, EPI_ISL_542993, EPI_ISL_542994, EPI_ISL_542995,
EPI_ISL_542996, EPI_ISL_542997, EPI_ISL_542998, EPI_ISL_543001, EPI_ISL_543005

| | | | |
|---|---|---|---|
| see above | TriCore Reference Laboratories | Center for Global Health, University of New Mexico Health Sciences Center | Daryl Domman, Kurt Schwalm, Twila Kunde, Joseph Hicks, Michael Edwards, Darrell Dinwiddie |

EPI_ISL_543007, EPI_ISL_543008, EPI_ISL_543009, EPI_ISL_543010, EPI_ISL_543011, EPI_ISL_543012, EPI_ISL_543013, EPI_ISL_543014, EPI_ISL_543015, EPI_ISL_543016, EPI_ISL_543017, EPI_ISL_543018, EPI_ISL_543019, EPI_ISL_543020, EPI_ISL_543021, EPI_ISL_543022, EPI_ISL_543023, EPI_ISL_543024, EPI_ISL_543025, EPI_ISL_543026, EPI_ISL_543027, EPI_ISL_543028, EPI_ISL_543029, EPI_ISL_543030,
EPI_ISL_543031, EPI_ISL_543032, EPI_ISL_543033, EPI_ISL_543034, EPI_ISL_543035, EPI_ISL_543036, EPI_ISL_543037, EPI_ISL_543038, EPI_ISL_543039, EPI_ISL_543040, EPI_ISL_543041, EPI_ISL_543042, EPI_ISL_543043, EPI_ISL_543044, EPI_ISL_543045, EPI_ISL_543046, EPI_ISL_543047, EPI_ISL_543048, EPI_ISL_543049, EPI_ISL_543050, EPI_ISL_543051, EPI_ISL_543052, EPI_ISL_543053, EPI_ISL_543054,
EPI_ISL_543055, EPI_ISL_543056, EPI_ISL_543057, EPI_ISL_543058, EPI_ISL_543059, EPI_ISL_543060, EPI_ISL_543061, EPI_ISL_543062, EPI_ISL_543063, EPI_ISL_543064, EPI_ISL_543065, EPI_ISL_543066, EPI_ISL_543067, EPI_ISL_543068, EPI_ISL_543069, EPI_ISL_543070, EPI_ISL_543071, EPI_ISL_543072, EPI_ISL_543073, EPI_ISL_543074, EPI_ISL_543075, EPI_ISL_543076, EPI_ISL_543077, EPI_ISL_543078,
EPI_ISL_543079, EPI_ISL_543080, EPI_ISL_543081, EPI_ISL_543082, EPI_ISL_543083, EPI_ISL_543084, EPI_ISL_543085, EPI_ISL_543086, EPI_ISL_543087, EPI_ISL_543088, EPI_ISL_543089, EPI_ISL_543090, EPI_ISL_543091, EPI_ISL_543092, EPI_ISL_543093, EPI_ISL_543094, EPI_ISL_543095, EPI_ISL_543096, EPI_ISL_543097, EPI_ISL_543098, EPI_ISL_543099, EPI_ISL_543100, EPI_ISL_543101, EPI_ISL_543102,
EPI_ISL_543103, EPI_ISL_543104, EPI_ISL_543105, EPI_ISL_543106, EPI_ISL_543107, EPI_ISL_543108, EPI_ISL_543109, EPI_ISL_543110, EPI_ISL_543111, EPI_ISL_543112, EPI_ISL_543113, EPI_ISL_543114, EPI_ISL_543115, EPI_ISL_543116, EPI_ISL_543117, EPI_ISL_543118, EPI_ISL_543119, EPI_ISL_543120, EPI_ISL_543121, EPI_ISL_543122, EPI_ISL_543123, EPI_ISL_543124, EPI_ISL_543125, EPI_ISL_543126,
EPI_ISL_543127, EPI_ISL_543128, EPI_ISL_543129, EPI_ISL_543130, EPI_ISL_543131, EPI_ISL_543132, EPI_ISL_543133, EPI_ISL_543134, EPI_ISL_543135, EPI_ISL_543136, EPI_ISL_543137, EPI_ISL_543138, EPI_ISL_543139, EPI_ISL_543140, EPI_ISL_543141, EPI_ISL_543142, EPI_ISL_543143, EPI_ISL_543144, EPI_ISL_543145, EPI_ISL_543146, EPI_ISL_543147, EPI_ISL_543148, EPI_ISL_543149, EPI_ISL_543150,
EPI_ISL_543151, EPI_ISL_543152, EPI_ISL_543153, EPI_ISL_543154, EPI_ISL_543155, EPI_ISL_543156, EPI_ISL_543157, EPI_ISL_543158, EPI_ISL_543159, EPI_ISL_543160, EPI_ISL_543161, EPI_ISL_543162, EPI_ISL_543163, EPI_ISL_543164, EPI_ISL_543165, EPI_ISL_543166, EPI_ISL_543167, EPI_ISL_543168, EPI_ISL_543169, EPI_ISL_543170, EPI_ISL_543171, EPI_ISL_543172, EPI_ISL_543173, EPI_ISL_543174,
EPI_ISL_543175, EPI_ISL_543176, EPI_ISL_543177, EPI_ISL_543178, EPI_ISL_543179, EPI_ISL_543180, EPI_ISL_543181, EPI_ISL_543182, EPI_ISL_543183, EPI_ISL_543184, EPI_ISL_543185, EPI_ISL_543186, EPI_ISL_543187, EPI_ISL_543188, EPI_ISL_543189, EPI_ISL_543190, EPI_ISL_543191, EPI_ISL_543192, EPI_ISL_543193, EPI_ISL_543194, EPI_ISL_543195, EPI_ISL_543196, EPI_ISL_543197, EPI_ISL_543198,
EPI_ISL_543199, EPI_ISL_543200, EPI_ISL_543201, EPI_ISL_543202, EPI_ISL_543203, EPI_ISL_543204, EPI_ISL_543205, EPI_ISL_543206, EPI_ISL_543207, EPI_ISL_543208, EPI_ISL_543209, EPI_ISL_543210, EPI_ISL_543211, EPI_ISL_543212, EPI_ISL_543213, EPI_ISL_543214, EPI_ISL_543215, EPI_ISL_543216, EPI_ISL_543217, EPI_ISL_543218, EPI_ISL_543219, EPI_ISL_543220, EPI_ISL_543221, EPI_ISL_543222,
EPI_ISL_543223, EPI_ISL_543224, EPI_ISL_543225, EPI_ISL_543226, EPI_ISL_543227, EPI_ISL_543228, EPI_ISL_543229, EPI_ISL_543230, EPI_ISL_543231, EPI_ISL_543232, EPI_ISL_543233, EPI_ISL_543234, EPI_ISL_543235, EPI_ISL_543236, EPI_ISL_543237, EPI_ISL_543238, EPI_ISL_543239, EPI_ISL_543240, EPI_ISL_543241, EPI_ISL_543242, EPI_ISL_543243, EPI_ISL_543244, EPI_ISL_543245, EPI_ISL_543246,
EPI_ISL_543247, EPI_ISL_543248, EPI_ISL_543249, EPI_ISL_543250, EPI_ISL_543251, EPI_ISL_543252, EPI_ISL_543253, EPI_ISL_543254, EPI_ISL_543255, EPI_ISL_543256, EPI_ISL_543257, EPI_ISL_543258, EPI_ISL_543259, EPI_ISL_543260, EPI_ISL_543261, EPI_ISL_543262, EPI_ISL_543263, EPI_ISL_543264, EPI_ISL_543265, EPI_ISL_543266, EPI_ISL_543267, EPI_ISL_543268, EPI_ISL_543269, EPI_ISL_543270,
EPI_ISL_543271, EPI_ISL_543272, EPI_ISL_543273, EPI_ISL_543274, EPI_ISL_543275, EPI_ISL_543276, EPI_ISL_543277, EPI_ISL_543278, EPI_ISL_543279, EPI_ISL_543280, EPI_ISL_543281, EPI_ISL_543282, EPI_ISL_543283, EPI_ISL_543284, EPI_ISL_543285, EPI_ISL_543286, EPI_ISL_543287, EPI_ISL_543288, EPI_ISL_543289, EPI_ISL_543290, EPI_ISL_543291, EPI_ISL_543292, EPI_ISL_543293, EPI_ISL_543294,
EPI_ISL_543295, EPI_ISL_543296, EPI_ISL_543297, EPI_ISL_543298, EPI_ISL_543299, EPI_ISL_543300, EPI_ISL_543301, EPI_ISL_543302, EPI_ISL_543303, EPI_ISL_543304, EPI_ISL_543305, EPI_ISL_543306, EPI_ISL_543307, EPI_ISL_543308, EPI_ISL_543309, EPI_ISL_543310, EPI_ISL_543311, EPI_ISL_543312, EPI_ISL_543313, EPI_ISL_543314, EPI_ISL_543315, EPI_ISL_543316, EPI_ISL_543317, EPI_ISL_543318,
EPI_ISL_543319, EPI_ISL_543320, EPI_ISL_543321, EPI_ISL_543322, EPI_ISL_543323, EPI_ISL_543324, EPI_ISL_543325, EPI_ISL_543326, EPI_ISL_543327, EPI_ISL_543328, EPI_ISL_543329, EPI_ISL_543330, EPI_ISL_543331, EPI_ISL_543332, EPI_ISL_543333, EPI_ISL_543334, EPI_ISL_543335, EPI_ISL_543336, EPI_ISL_543337, EPI_ISL_543338, EPI_ISL_543339, EPI_ISL_543340, EPI_ISL_543341, EPI_ISL_543342,
EPI_ISL_543343, EPI_ISL_543344, EPI_ISL_543345, EPI_ISL_543346, EPI_ISL_543347, EPI_ISL_543348, EPI_ISL_543349, EPI_ISL_543350, EPI_ISL_543351, EPI_ISL_543352, EPI_ISL_543353, EPI_ISL_543354, EPI_ISL_543355, EPI_ISL_543356, EPI_ISL_543357, EPI_ISL_543358, EPI_ISL_543359, EPI_ISL_543360, EPI_ISL_543361, EPI_ISL_543362, EPI_ISL_543363, EPI_ISL_543364, EPI_ISL_543365, EPI_ISL_543366,
EPI_ISL_543367, EPI_ISL_543368, EPI_ISL_543369, EPI_ISL_543370, EPI_ISL_543371, EPI_ISL_543372, EPI_ISL_543373, EPI_ISL_543374, EPI_ISL_543375, EPI_ISL_543376, EPI_ISL_543377, EPI_ISL_543378, EPI_ISL_543379, EPI_ISL_543380, EPI_ISL_543381, EPI_ISL_543382, EPI_ISL_543383, EPI_ISL_543384, EPI_ISL_543385, EPI_ISL_543386, EPI_ISL_543387, EPI_ISL_543388, EPI_ISL_543389, EPI_ISL_543390,
EPI_ISL_543391, EPI_ISL_543392, EPI_ISL_543393, EPI_ISL_543394, EPI_ISL_543395, EPI_ISL_543396, EPI_ISL_543397, EPI_ISL_543398, EPI_ISL_543399, EPI_ISL_543400, EPI_ISL_543401, EPI_ISL_543402, EPI_ISL_543403, EPI_ISL_543404, EPI_ISL_543405, EPI_ISL_543406, EPI_ISL_543407, EPI_ISL_543408, EPI_ISL_543409, EPI_ISL_543410, EPI_ISL_543411, EPI_ISL_543412, EPI_ISL_543413, EPI_ISL_543414,
EPI_ISL_543415, EPI_ISL_543416, EPI_ISL_543417, EPI_ISL_543418, EPI_ISL_543419, EPI_ISL_543420, EPI_ISL_543421, EPI_ISL_543422, EPI_ISL_543423, EPI_ISL_543424, EPI_ISL_543425, EPI_ISL_543426, EPI_ISL_543427, EPI_ISL_543428, EPI_ISL_543429, EPI_ISL_543430, EPI_ISL_543431, EPI_ISL_543432, EPI_ISL_543433, EPI_ISL_543434, EPI_ISL_543435, EPI_ISL_543436, EPI_ISL_543437, EPI_ISL_543438,
EPI_ISL_543439, EPI_ISL_543440, EPI_ISL_543441, EPI_ISL_543442, EPI_ISL_543443, EPI_ISL_543444, EPI_ISL_543445, EPI_ISL_543446, EPI_ISL_543447, EPI_ISL_543448, EPI_ISL_543449, EPI_ISL_543450, EPI_ISL_543451, EPI_ISL_543452, EPI_ISL_543453, EPI_ISL_543454, EPI_ISL_543455, EPI_ISL_543456, EPI_ISL_543457, EPI_ISL_543458, EPI_ISL_543459, EPI_ISL_543460, EPI_ISL_543461, EPI_ISL_543462,
EPI_ISL_543463, EPI_ISL_543464, EPI_ISL_543465, EPI_ISL_543466, EPI_ISL_543467, EPI_ISL_543468, EPI_ISL_543469, EPI_ISL_543470, EPI_ISL_543471, EPI_ISL_543472, EPI_ISL_543473, EPI_ISL_543474, EPI_ISL_543475, EPI_ISL_543476, EPI_ISL_543477, EPI_ISL_543478, EPI_ISL_543479, EPI_ISL_543480, EPI_ISL_543481, EPI_ISL_543482, EPI_ISL_543483, EPI_ISL_543484, EPI_ISL_543485, EPI_ISL_543486,
EPI_ISL_543487, EPI_ISL_543488, EPI_ISL_543489, EPI_ISL_543490, EPI_ISL_543491, EPI_ISL_543492, EPI_ISL_543493, EPI_ISL_543494, EPI_ISL_543495, EPI_ISL_543496, EPI_ISL_543497, EPI_ISL_543498, EPI_ISL_543499, EPI_ISL_543500, EPI_ISL_543501, EPI_ISL_543502, EPI_ISL_543503, EPI_ISL_543504, EPI_ISL_543505, EPI_ISL_543506, EPI_ISL_543507, EPI_ISL_543508, EPI_ISL_543509, EPI_ISL_543510,
EPI_ISL_543511, EPI_ISL_543512, EPI_ISL_543513, EPI_ISL_543514, EPI_ISL_543515, EPI_ISL_543516, EPI_ISL_543517, EPI_ISL_543518, EPI_ISL_543519, EPI_ISL_543520, EPI_ISL_543521, EPI_ISL_543522, EPI_ISL_543523, EPI_ISL_543524, EPI_ISL_543525, EPI_ISL_543526, EPI_ISL_543527, EPI_ISL_543528, EPI_ISL_543529, EPI_ISL_543530, EPI_ISL_543531, EPI_ISL_543532, EPI_ISL_543533, EPI_ISL_543534,
EPI_ISL_543535, EPI_ISL_543536, EPI_ISL_543537, EPI_ISL_543538, EPI_ISL_543539, EPI_ISL_543540, EPI_ISL_543541, EPI_ISL_543542, EPI_ISL_543543, EPI_ISL_543544, EPI_ISL_543545, EPI_ISL_543546, EPI_ISL_543547, EPI_ISL_543548, EPI_ISL_543549, EPI_ISL_543550, EPI_ISL_543551, EPI_ISL_543552, EPI_ISL_543553, EPI_ISL_543554, EPI_ISL_543555, EPI_ISL_543556, EPI_ISL_543557, EPI_ISL_543558,
EPI_ISL_543559, EPI_ISL_543560, EPI_ISL_543561, EPI_ISL_543562, EPI_ISL_543563, EPI_ISL_543564, EPI_ISL_543565, EPI_ISL_543566, EPI_ISL_543567, EPI_ISL_543568, EPI_ISL_543569, EPI_ISL_543570, EPI_ISL_543571, EPI_ISL_543572, EPI_ISL_543573, EPI_ISL_543574, EPI_ISL_543575, EPI_ISL_543576, EPI_ISL_543577, EPI_ISL_543578, EPI_ISL_543579, EPI_ISL_543580, EPI_ISL_543581, EPI_ISL_543582,
EPI_ISL_543583, EPI_ISL_543584, EPI_ISL_543585, EPI_ISL_543586, EPI_ISL_543587, EPI_ISL_543588, EPI_ISL_543589, EPI_ISL_543590, EPI_ISL_543591, EPI_ISL_543592, EPI_ISL_543593, EPI_ISL_543594, EPI_ISL_543595, EPI_ISL_543596, EPI_ISL_543597, EPI_ISL_543598, EPI_ISL_543599, EPI_ISL_543600, EPI_ISL_543601, EPI_ISL_543602, EPI_ISL_543603, EPI_ISL_543604, EPI_ISL_543605, EPI_ISL_543606,
EPI_ISL_543607, EPI_ISL_543608, EPI_ISL_543609, EPI_ISL_543610, EPI_ISL_543611, EPI_ISL_543612, EPI_ISL_543613, EPI_ISL_543614, EPI_ISL_543615, EPI_ISL_543616, EPI_ISL_543617, EPI_ISL_543618, EPI_ISL_543619, EPI_ISL_543620, EPI_ISL_543621, EPI_ISL_543622, EPI_ISL_543623, EPI_ISL_543624, EPI_ISL_543625, EPI_ISL_543626, EPI_ISL_543627, EPI_ISL_543628, EPI_ISL_543629, EPI_ISL_543630,
EPI_ISL_543631, EPI_ISL_543632, EPI_ISL_543633, EPI_ISL_543634, EPI_ISL_543635, EPI_ISL_543636, EPI_ISL_543637, EPI_ISL_543638, EPI_ISL_543639, EPI_ISL_543640, EPI_ISL_543641, EPI_ISL_543642, EPI_ISL_543643, EPI_ISL_543644, EPI_ISL_543645, EPI_ISL_543646, EPI_ISL_543647, EPI_ISL_543648, EPI_ISL_543649, EPI_ISL_543650, EPI_ISL_543651, EPI_ISL_543652, EPI_ISL_543653, EPI_ISL_543654,
EPI_ISL_543655, EPI_ISL_543656, EPI_ISL_543657, EPI_ISL_543658, EPI_ISL_543659, EPI_ISL_543660, EPI_ISL_543661, EPI_ISL_543662, EPI_ISL_543663, EPI_ISL_543664, EPI_ISL_543665, EPI_ISL_543666, EPI_ISL_543667, EPI_ISL_543668, EPI_ISL_543669, EPI_ISL_543670, EPI_ISL_543671, EPI_ISL_543672, EPI_ISL_543673, EPI_ISL_543674, EPI_ISL_543675, EPI_ISL_543676, EPI_ISL_543677, EPI_ISL_543678,
EPI_ISL_543679, EPI_ISL_543680, EPI_ISL_543681, EPI_ISL_543682, EPI_ISL_543683, EPI_ISL_543684, EPI_ISL_543685, EPI_ISL_543686, EPI_ISL_543687, EPI_ISL_543688, EPI_ISL_543689, EPI_ISL_543690, EPI_ISL_543691, EPI_ISL_543692, EPI_ISL_543693, EPI_ISL_543694, EPI_ISL_543695, EPI_ISL_543696, EPI_ISL_543697, EPI_ISL_543698, EPI_ISL_543699, EPI_ISL_543700, EPI_ISL_543701, EPI_ISL_543702,
EPI_ISL_543703, EPI_ISL_543704, EPI_ISL_543705, EPI_ISL_543706, EPI_ISL_543707, EPI_ISL_543708, EPI_ISL_543709, EPI_ISL_543710, EPI_ISL_543711, EPI_ISL_543712, EPI_ISL_543713, EPI_ISL_543714, EPI_ISL_543715, EPI_ISL_543716, EPI_ISL_543717, EPI_ISL_543718, EPI_ISL_543719, EPI_ISL_543720, EPI_ISL_543721, EPI_ISL_543722, EPI_ISL_543723, EPI_ISL_543724, EPI_ISL_543725, EPI_ISL_543726,
EPI_ISL_543727, EPI_ISL_543728, EPI_ISL_543729, EPI_ISL_543730, EPI_ISL_543731, EPI_ISL_543732, EPI_ISL_543733, EPI_ISL_543734, EPI_ISL_543735, EPI_ISL_543736, EPI_ISL_543737, EPI_ISL_543738, EPI_ISL_543739, EPI_ISL_543740, EPI_ISL_543741, EPI_ISL_543742, EPI_ISL_543743, EPI_ISL_543744, EPI_ISL_543745, EPI_ISL_543746, EPI_ISL_543747, EPI_ISL_543748, EPI_ISL_543749, EPI_ISL_543750,
EPI_ISL_543751, EPI_ISL_543752, EPI_ISL_543753, EPI_ISL_543754, EPI_ISL_543755, EPI_ISL_543756, EPI_ISL_543757, EPI_ISL_543758, EPI_ISL_543759, EPI_ISL_543760, EPI_ISL_543761, EPI_ISL_543762, EPI_ISL_543763, EPI_ISL_543764, EPI_ISL_543765, EPI_ISL_543766, EPI_ISL_543767, EPI_ISL_543768, EPI_ISL_543769, EPI_ISL_543770, EPI_ISL_543771, EPI_ISL_543772, EPI_ISL_543773, EPI_ISL_543774,
EPI_ISL_543775, EPI_ISL_543776, EPI_ISL_543777, EPI_ISL_543778, EPI_ISL_543779, EPI_ISL_543780, EPI_ISL_543781, EPI_ISL_543782, EPI_ISL_543783, EPI_ISL_543784, EPI_ISL_543785, EPI_ISL_543786, EPI_ISL_543787, EPI_ISL_543788, EPI_ISL_543789, EPI_ISL_543790, EPI_ISL_543791, EPI_ISL_543792, EPI_ISL_543793, EPI_ISL_543794, EPI_ISL_543795, EPI_ISL_543796, EPI_ISL_543797, EPI_ISL_543798,
EPI_ISL_543799, EPI_ISL_543800, EPI_ISL_543801, EPI_ISL_543802, EPI_ISL_543803, EPI_ISL_543804, EPI_ISL_543805, EPI_ISL_543806, EPI_ISL_543807, EPI_ISL_543808, EPI_ISL_543809, EPI_ISL_543810, EPI_ISL_543811, EPI_ISL_543812, EPI_ISL_543813, EPI_ISL_543814, EPI_ISL_543815, EPI_ISL_543816, EPI_ISL_543817, EPI_ISL_543818, EPI_ISL_543819, EPI_ISL_543820, EPI_ISL_543821, EPI_ISL_543822,
EPI_ISL_543823, EPI_ISL_543824, EPI_ISL_543825, EPI_ISL_543826, EPI_ISL_543827, EPI_ISL_543828, EPI_ISL_543829, EPI_ISL_543830, EPI_ISL_543831, EPI_ISL_543832, EPI_ISL_543833, EPI_ISL_543834, EPI_ISL_543835, EPI_ISL_543836, EPI_ISL_543837, EPI_ISL_543838, EPI_ISL_543839, EPI_ISL_543840, EPI_ISL_543841, EPI_ISL_543842, EPI_ISL_543843, EPI_ISL_543844, EPI_ISL_543845, EPI_ISL_543846,
EPI_ISL_543847, EPI_ISL_543848, EPI_ISL_543849, EPI_ISL_543850, EPI_ISL_543851, EPI_ISL_543852, EPI_ISL_543853, EPI_ISL_543854, EPI_ISL_543855, EPI_ISL_543856, EPI_ISL_543857, EPI_ISL_543858, EPI_ISL_543859, EPI_ISL_543860, EPI_ISL_543861, EPI_ISL_543862, EPI_ISL_543863, EPI_ISL_543864, EPI_ISL_543865, EPI_ISL_543866, EPI_ISL_543867, EPI_ISL_543868, EPI_ISL_543869, EPI_ISL_543870,
EPI_ISL_543871, EPI_ISL_543872, EPI_ISL_543873, EPI_ISL_543874, EPI_ISL_543875, EPI_ISL_543876, EPI_ISL_543877, EPI_ISL_543878, EPI_ISL_543879, EPI_ISL_543880, EPI_ISL_543881, EPI_ISL_543882, EPI_ISL_543883, EPI_ISL_543884, EPI_ISL_543885, EPI_ISL_543886, EPI_ISL_543887, EPI_ISL_543888, EPI_ISL_543889, EPI_ISL_543890, EPI_ISL_543891, EPI_ISL_543892, EPI_ISL_543893, EPI_ISL_543894,
EPI_ISL_543895, EPI_ISL_543896, EPI_ISL_543897, EPI_ISL_543898, EPI_ISL_543899, EPI_ISL_543900, EPI_ISL_543901, EPI_ISL_543902, EPI_ISL_543903, EPI_ISL_543904, EPI_ISL_543905, EPI_ISL_543906, EPI_ISL_543907, EPI_ISL_543908, EPI_ISL_543909, EPI_ISL_543910, EPI_ISL_543911, EPI_ISL_543912, EPI_ISL_543913, EPI_ISL_543914, EPI_ISL_543915, EPI_ISL_543916, EPI_ISL_543917, EPI_ISL_543918,
EPI_ISL_543919, EPI_ISL_543920, EPI_ISL_543921, EPI_ISL_543922, EPI_ISL_543923, EPI_ISL_543924, EPI_ISL_543925, EPI_ISL_543926, EPI_ISL_543927, EPI_ISL_543928, EPI_ISL_543929, EPI_ISL_543930, EPI_ISL_543931, EPI_ISL_543932, EPI_ISL_543933, EPI_ISL_543934, EPI_ISL_543935, EPI_ISL_543936, EPI_ISL_543937, EPI_ISL_543938, EPI_ISL_543939, EPI_ISL_543940, EPI_ISL_543941, EPI_ISL_543942,
EPI_ISL_543943, EPI_ISL_543944, EPI_ISL_543945, EPI_ISL_543946, EPI_ISL_543947, EPI_ISL_543948, EPI_ISL_543949, EPI_ISL_543950, EPI_ISL_543951, EPI_ISL_543952, EPI_ISL_543953, EPI_ISL_543954, EPI_ISL_543955, EPI_ISL_543956, EPI_ISL_543957, EPI_ISL_543958, EPI_ISL_543959, EPI_ISL_543960, EPI_ISL_543961, EPI_ISL_543962, EPI_ISL_543963, EPI_ISL_543964, EPI_ISL_543965, EPI_ISL_543966,
EPI_ISL_543967, EPI_ISL_543968, EPI_ISL_543969, EPI_ISL_543970, EPI_ISL_543971, EPI_ISL_543972, EPI_ISL_543973, EPI_ISL_543974, EPI_ISL_543975, EPI_ISL_543976, EPI_ISL_543977, EPI_ISL_543978, EPI_ISL_543979, EPI_ISL_543980, EPI_ISL_543981, EPI_ISL_543982, EPI_ISL_543983, EPI_ISL_543984, EPI_ISL_543985, EPI_ISL_543986, EPI_ISL_543987, EPI_ISL_543988, EPI_ISL_543989, EPI_ISL_543990,
EPI_ISL_543991, EPI_ISL_543992, EPI_ISL_543993, EPI_ISL_543994, EPI_ISL_543995, EPI_ISL_543996, EPI_ISL_543997, EPI_ISL_543998, EPI_ISL_543999, EPI_ISL_544000, EPI_ISL_544001, EPI_ISL_544002, EPI_ISL_544003, EPI_ISL_544004, EPI_ISL_544005, EPI_ISL_544006, EPI_ISL_544007, EPI_ISL_544008, EPI_ISL_544009, EPI_ISL_544010, EPI_ISL_544011, EPI_ISL_544012, EPI_ISL_544013, EPI_ISL_544014,
EPI_ISL_544015, EPI_ISL_544016, EPI_ISL_544017, EPI_ISL_544018, EPI_ISL_544019, EPI_ISL_544020, EPI_ISL_544021, EPI_ISL_544022, EPI_ISL_544023, EPI_ISL_544024, EPI_ISL_544025, EPI_ISL_544026, EPI_ISL_544027, EPI_ISL_544028, EPI_ISL_544029, EPI_ISL_544030, EPI_ISL_544031, EPI_ISL_544032, EPI_ISL_544033, EPI_ISL_544034, EPI_ISL_544035, EPI_ISL_544036, EPI_ISL_544037, EPI_ISL_544038,
EPI_ISL_544039, EPI_ISL_544040, EPI_ISL_544041, EPI_ISL_544042, EPI_ISL_544043, EPI_ISL_544044, EPI_ISL_544045, EPI_ISL_544046, EPI_ISL_544047, EPI_ISL_544048, EPI_ISL_544049, EPI_ISL_544050, EPI_ISL_544051, EPI_ISL_544052, EPI_ISL_544053, EPI_ISL_544054, EPI_ISL_544055, EPI_ISL_544056, EPI_ISL_544057, EPI_ISL_544058, EPI_ISL_544059, EPI_ISL_544060, EPI_ISL_544061, EPI_ISL_544062,
EPI_ISL_544063, EPI_ISL_544064, EPI_ISL_544065, EPI_ISL_544066, EPI_ISL_544067, EPI_ISL_544068, EPI_ISL_544069, EPI_ISL_544070, EPI_ISL_544071, EPI_ISL_544072, EPI_ISL_544073, EPI_ISL_544074, EPI_ISL_544075, EPI_ISL_544076, EPI_ISL_544077, EPI_ISL_544078, EPI_ISL_544079, EPI_ISL_544080, EPI_ISL_544081, EPI_ISL_544082, EPI_ISL_544083, EPI_ISL_544084, EPI_ISL_544085, EPI_ISL_544086,
EPI_ISL_544087, EPI_ISL_544088, EPI_ISL_544089, EPI_ISL_544090, EPI_ISL_544091, EPI_ISL_544092, EPI_ISL_544093, EPI_ISL_544094, EPI_ISL_544095, EPI_ISL_544096, EPI_ISL_544097, EPI_ISL_544098, EPI_ISL_544099, EPI_ISL_544100, EPI_ISL_544101, EPI_ISL_544102, EPI_ISL_544103, EPI_ISL_544104, EPI_ISL_544105, EPI_ISL_544106, EPI_ISL_544107, EPI_ISL_544108, EPI_ISL_544109, EPI_ISL_544110,
EPI_ISL_544111, EPI_ISL_544112, EPI_ISL_544113, EPI_ISL_544114, EPI_ISL_544115, EPI_ISL_544116, EPI_ISL_544117, EPI_ISL_544118, EPI_ISL_544119, EPI_ISL_544120, EPI_ISL_544121, EPI_ISL_544122, EPI_ISL_544123, EPI_ISL_544124, EPI_ISL_544125, EPI_ISL_544126, EPI_ISL_544127, EPI_ISL_544128, EPI_ISL_544129, EPI_ISL_544130, EPI_ISL_544131, EPI_ISL_544132, EPI_ISL_544133, EPI_ISL_544134,
EPI_ISL_544135, EPI_ISL_544136, EPI_ISL_544137, EPI_ISL_544138, EPI_ISL_544139, EPI_ISL_544140, EPI_ISL_544141, EPI_ISL_544142, EPI_ISL_544143, EPI_ISL_544144, EPI_ISL_544145, EPI_ISL_544146, EPI_ISL_544147, EPI_ISL_544148, EPI_ISL_544149, EPI_ISL_544150, EPI_ISL_544151, EPI_ISL_544152, EPI_ISL_544153, EPI_ISL_544154, EPI_ISL_544155, EPI_ISL_544156, EPI_ISL_544157, EPI_ISL_544158,
EPI_ISL_544159, EPI_ISL_544160, EPI_ISL_544161, EPI_ISL_544162, EPI_ISL_544163, EPI_ISL_544164, EPI_ISL_544165, EPI_ISL_544166, EPI_ISL_544167, EPI_ISL_544168, EPI_ISL_544169, EPI_ISL_544170, EPI_ISL_544171, EPI_ISL_544172, EPI_ISL_544173, EPI_ISL_544174, EPI_ISL_544175, EPI_ISL_544176, EPI_ISL_544177, EPI_ISL_544178, EPI_ISL_544179, EPI_ISL_544180, EPI_ISL_544181, EPI_ISL_544182,
EPI_ISL_544183, EPI_ISL_544184, EPI_ISL_544185, EPI_ISL_544186, EPI_ISL_544187, EPI_ISL_544188, EPI_ISL_544189, EPI_ISL_544190, EPI_ISL_544191, EPI_ISL_544192, EPI_ISL_544193, EPI_ISL_544194, EPI_ISL_544195, EPI_ISL_544196, EPI_ISL_544197, EPI_ISL_544198, EPI_ISL_544199, EPI_ISL_544200, EPI_ISL_544201, EPI_ISL_544202, EPI_ISL_544203, EPI_ISL_544204, EPI_ISL_544205, EPI_ISL_544206,
EPI_ISL_544207, EPI_ISL_544208, EPI_ISL_544209, EPI_ISL_544210, EPI_ISL_544211, EPI_ISL_544212, EPI_ISL_544213, EPI_ISL_544214, EPI_ISL_544215, EPI_ISL_544216, EPI_ISL_544217, EPI_ISL_544218, EPI_ISL_544219, EPI_ISL_544220, EPI_ISL_544221, EPI_ISL_544222, EPI_ISL_544223, EPI_ISL_544224, EPI_ISL_544225, EPI_ISL_544226, EPI_ISL_544227, EPI_ISL_544228, EPI_ISL_544229, EPI_ISL_544230,
EPI_ISL_544231, EPI_ISL_544232, EPI_ISL_544233, EPI_ISL_544234, EPI_ISL_544235, EPI_ISL_544236, EPI_ISL_544237, EPI_ISL_544238, EPI_ISL_544239, EPI_ISL_544240, EPI_ISL_544241, EPI_ISL_544242, EPI_ISL_544243, EPI_ISL_544244, EPI_ISL_544245, EPI_ISL_544246, EPI_ISL_544247, EPI_ISL_544248, EPI_ISL_544249, EPI_ISL_544250, EPI_ISL_544251, EPI_ISL_544252, EPI_ISL_544253, EPI_ISL_544254,
EPI_ISL_544255, EPI_ISL_544256, EPI_ISL_544257, EPI_ISL_544258, EPI_ISL_544259, EPI_ISL_544260, EPI_ISL_544261, EPI_ISL_544262, EPI_ISL_544263, EPI_ISL_544264, EPI_ISL_544265, EPI_ISL_544266, EPI_ISL_544267, EPI_ISL_544268, EPI_ISL_544269, EPI_ISL_544270, EPI_ISL_544271, EPI_ISL_544272, EPI_ISL_544273, EPI_ISL_544274, EPI_ISL_544275, EPI_ISL_544276, EPI_ISL_544277, EPI_ISL_544278,
EPI_ISL_544279, EPI_ISL_544280, EPI_ISL_544281, EPI_ISL_544282, EPI_ISL_544283, EPI_ISL_544284, EPI_ISL_544285, EPI_ISL_544286, EPI_ISL_544287, EPI_ISL_544288, EPI_ISL_544289, EPI_ISL_544290, EPI_ISL_544291, EPI_ISL_544292, EPI_ISL_544293, EPI_ISL_544294, EPI_ISL_544295, EPI_ISL_544296, EPI_ISL_544297, EPI_ISL_544298, EPI_ISL_544299, EPI_ISL_544300, EPI_ISL_544301, EPI_ISL_544302,
EPI_ISL_544303, EPI_ISL_544304, EPI_ISL_544305, EPI_ISL_544306, EPI_ISL_544307, EPI_ISL_544308, EPI_ISL_544309, EPI_ISL_544310, EPI_ISL_544311, EPI_ISL_544312, EPI_ISL_544313, EPI_ISL_544314, EPI_ISL_544315, EPI_ISL_544316, EPI_ISL_544317, EPI_ISL_544318, EPI_ISL_544319, EPI_ISL_544320, EPI_ISL_544321, EPI_ISL_544322, EPI_ISL_544323, EPI_ISL_544324, EPI_ISL_544325, EPI_ISL_544326,
EPI_ISL_544327, EPI_ISL_544328, EPI_ISL_544329, EPI_ISL_544330, EPI_ISL_544331, EPI_ISL_544332, EPI_ISL_544333, EPI_ISL_544334, EPI_ISL_544335, EPI_ISL_544336, EPI_ISL_544337, EPI_ISL_544338, EPI_ISL_544339, EPI_ISL_544340, EPI_ISL_544341, EPI_ISL_544342, EPI_ISL_544343, EPI_ISL_544344, EPI_ISL_544345, EPI_ISL_544346, EPI_ISL_544347, EPI_ISL_544348, EPI_ISL_544349, EPI_ISL_544350,
EPI_ISL_544351, EPI_ISL_544352, EPI_ISL_544353, EPI_ISL_544354, EPI_ISL_544355, EPI_ISL_544356, EPI_ISL_544357, EPI_ISL_544358, EPI_ISL_544359, EPI_ISL_544360, EPI_ISL_544361, EPI_ISL_544362, EPI_ISL_544363, EPI_ISL_544364, EPI_ISL_544365, EPI_ISL_544366, EPI_ISL_544367, EPI_ISL_544368, EPI_ISL_544369, EPI_ISL_544370, EPI_ISL_544371, EPI_ISL_544372, EPI_ISL_544373, EPI_ISL_544374,
EPI_ISL_544375, EPI_ISL_544376, EPI_ISL_544377, EPI_ISL_544378, EPI_ISL_544379, EPI_ISL_544380, EPI_ISL_544381, EPI_ISL_544382, EPI_ISL_544383, EPI_ISL_544384, EPI_ISL_544385, EPI_ISL_544386, EPI_ISL_544387, EPI_ISL_544388, EPI_ISL_544389, EPI_ISL_544390, EPI_ISL_544391, EPI_ISL_544392, EPI_ISL_544393, EPI_ISL_544394, EPI_ISL_544395, EPI_ISL_544396, EPI_ISL_544397, EPI_ISL_544398,
EPI_ISL_544399, EPI_ISL_544400, EPI_ISL_544401, EPI_ISL_544402, EPI_ISL_544403, EPI_ISL_544404, EPI_ISL_544405, EPI_ISL_544406, EPI_ISL_544407, EPI_ISL_544408, EPI_ISL_544409, EPI_ISL_544410, EPI_ISL_544411, EPI_ISL_544412, EPI_ISL_544413, EPI_ISL_544414, EPI_ISL_544415, EPI_ISL_544416, EPI_ISL_544417, EPI_ISL_544418, EPI_ISL_544419, EPI_ISL_544420, EPI_ISL_544421, EPI_ISL_544422,
EPI_ISL_544423, EPI_ISL_544424, EPI_ISL_544425, EPI_ISL_544426, EPI_ISL_544427, EPI_ISL_544428, EPI_ISL_544429, EPI_ISL_544430, EPI_ISL_544431, EPI_ISL_544432, EPI_ISL_544433, EPI_ISL_544434, EPI_ISL_544435, EPI_ISL_544436, EPI_ISL_544437, EPI_ISL_544438, EPI_ISL_544439, EPI_ISL_544440, EPI_ISL_544441, EPI_ISL_544442, EPI_ISL_544443, EPI_ISL_544444, EPI_ISL_544445, EPI_ISL_544446,
EPI_ISL_544447, EPI_ISL_544448, EPI_ISL_544449, EPI_ISL_544450, EPI_ISL_544451, EPI_ISL_544452, EPI_ISL_544453, EPI_ISL_544454, EPI_ISL_544455, EPI_ISL_544456, EPI_ISL_544457, EPI_ISL_544458, EPI_ISL_544459, EPI_ISL_544460, EPI_ISL_544461, EPI_ISL_544462, EPI_ISL_544463, EPI_ISL_544464, EPI_ISL_544465, EPI_ISL_544466, EPI_ISL_544467, EPI_ISL_544468, EPI_ISL_544469, EPI_ISL_544470,
EPI_ISL_544471, EPI_ISL_544472, EPI_ISL_544473, EPI_ISL_544474, EPI_ISL_544475, EPI_ISL_544476, EPI_ISL_544477, EPI_ISL_544478, EPI_ISL_544479, EPI_ISL_544480, EPI_ISL_544481, EPI_ISL_544482, EPI_ISL_544483, EPI_ISL_544484, EPI_ISL_544485, EPI_ISL_544486, EPI_ISL_544487, EPI_ISL_544488, EPI_ISL_544489, EPI_ISL_544490, EPI_ISL_544491, EPI_ISL_544492, EPI_ISL_544493, EPI_ISL_544494,
EPI_ISL_544495, EPI_ISL_544496, EPI_ISL_544497, EPI_ISL_544498, EPI_ISL_544499, EPI_ISL_544500, EPI_ISL_544501, EPI_ISL_544502, EPI_ISL_544503, EPI_ISL_544504, EPI_ISL_544505, EPI_ISL_544506, EPI_ISL_544507, EPI_ISL_544508, EPI_ISL_544509, EPI_ISL_544510, EPI_ISL_544511, EPI_ISL_544512, EPI_ISL_544513, EPI_ISL_544514, EPI_ISL_544515, EPI_ISL_544516, EPI_ISL_544517, EPI_ISL_544518,
EPI_ISL_544519, EPI_ISL_544520, EPI_ISL_544521, EPI_ISL_544522, EPI_ISL_544523, EPI_ISL_544524, EPI_ISL_544525, EPI_ISL_544526, EPI_ISL_544527, EPI_ISL_544528, EPI_ISL_544529, EPI_ISL_544530, EPI_ISL_544531, EPI_ISL_544532, EPI_ISL_544533, EPI_ISL_544534, EPI_ISL_544535, EPI_ISL_544536, EPI_ISL_544537, EPI_ISL_544538, EPI_ISL_544539, EPI_ISL_544540, EPI_ISL_544541, EPI_ISL_544542,
EPI_ISL_544543, EPI_ISL_544544, EPI_ISL_544545, EPI_ISL_544546, EPI_ISL_544547, EPI_ISL_544548, EPI_ISL_544549, EPI_ISL_544550, EPI_ISL_544551, EPI_ISL_544552, EPI_ISL_544553, EPI_ISL_544554, EPI_ISL_544555, EPI_ISL_544556, EPI_ISL_544557, EPI_ISL_544558, EPI_ISL_544559, EPI_ISL_544560, EPI_ISL_544561, EPI_ISL_544562, EPI_ISL_544563, EPI_ISL_544564, EPI_ISL_544565, EPI_ISL_544566,
EPI_ISL_544567, EPI_ISL_544568, EPI_ISL_544569, EPI_ISL_544570, EPI_ISL_544571, EPI_ISL_544572, EPI_ISL_544573, EPI_ISL_544574, EPI_ISL_544575, EPI_ISL_544576, EPI_ISL_544577, EPI_ISL_544578, EPI_ISL_544579, EPI_ISL_544580, EPI_ISL_544581, EPI_ISL_544582, EPI_ISL_544583, EPI_ISL_544584, EPI_ISL_544585, EPI_ISL_544586, EPI_ISL_544587, EPI_ISL_544588, EPI_ISL_544589, EPI_ISL_544590,
EPI_ISL_544591, EPI_ISL_544592, EPI_ISL_544593, EPI_ISL_544594, EPI_ISL_544595, EPI_ISL_544596, EPI_ISL_544597, EPI_ISL_544598, EPI_ISL_544599, EPI_ISL_544600, EPI_ISL_544601, EPI_ISL_544602, EPI_ISL_544603, EPI_ISL_544604, EPI_ISL_544605, EPI_ISL_544606, EPI_ISL_544607, EPI_ISL_544608, EPI_ISL_544609, EPI_ISL_544610, EPI_ISL_544611, EPI_ISL_544612, EPI_ISL_544613, EPI_ISL_544614,
EPI_ISL_544615, EPI_ISL_544616, EPI_ISL_544617, EPI_ISL_544618, EPI_ISL_544619, EPI_ISL_544620, EPI_ISL_544621, EPI_ISL_544622, EPI_ISL_544623, EPI_ISL_544624, EPI_ISL_544625, EPI_ISL_544626, EPI_ISL_544627, EPI_ISL_544628, EPI_ISL_544629, EPI_ISL_544630, EPI_ISL_544631, EPI_ISL_544632, EPI_ISL_544633, EPI_ISL_544634, EPI_ISL_544635, EPI_ISL_544636, EPI_ISL_544637, EPI_ISL_544638,
EPI_ISL_544639, EPI_ISL_544640, EPI_ISL_544641, EPI_ISL_544642, EPI_ISL_544643, EPI_ISL_544644, EPI_ISL_544645, EPI_ISL_544646, EPI_ISL_544647, EPI_ISL_544648, EPI_ISL_544649, EPI_ISL_544650, EPI_ISL_544651, EPI_ISL_544652, EPI_ISL_544653, EPI_ISL_544654, EPI_ISL_544655, EPI_ISL_544656, EPI_ISL_544657, EPI_ISL_544658, EPI_ISL_544659, EPI_ISL_544660, EPI_ISL_544661, EPI_ISL_544662,
EPI_ISL_544663, EPI_ISL_544664, EPI_ISL_544665, EPI_ISL_544666, EPI_ISL_544667, EPI_ISL_544668, EPI_ISL_544669, EPI_ISL_544670, EPI_ISL_544671, EPI_ISL_544672, EPI_ISL_544673, EPI_ISL_544674, EPI_ISL_544675, EPI_ISL_544676, EPI_ISL_544677, EPI_ISL_544678, EPI_ISL_544679, EPI_ISL_544680, EPI_ISL_544681, EPI_ISL_544682, EPI_ISL_544683, EPI_ISL_544684, EPI_ISL_544685, EPI_ISL_544686,
EPI_ISL_544687, EPI_ISL_544688, EPI_ISL_544689, EPI_ISL_544690, EPI_ISL_544691, EPI_ISL_544692, EPI_ISL_544693, EPI_ISL_544694, EPI_ISL_544695, EPI_ISL_544696, EPI_ISL_544697, EPI_ISL_544698, EPI_ISL_544699, EPI_ISL_544700, EPI_ISL_544701, EPI_ISL_544702, EPI_ISL_544703, EPI_ISL_544704, EPI_ISL_544705, EPI_ISL_544706, EPI_ISL_544707, EPI_ISL_544708, EPI_ISL_544709, EPI_ISL_544710,
EPI_ISL_544711, EPI_ISL_544712, EPI_ISL_544713, EPI_ISL_544714, EPI_ISL_544715, EPI_ISL_544716, EPI_ISL_544717, EPI_ISL_544718, EPI_ISL_544719, EPI_ISL_544720, EPI_ISL_544721, EPI_ISL_544722, EPI_ISL_544723, EPI_ISL_544724, EPI_ISL_544725, EPI_ISL_544726, EPI_ISL_544727, EPI_ISL_544728, EPI_ISL_544729, EPI_ISL_544730, EPI_ISL_544731, EPI_ISL_544732, EPI_ISL_544733, EPI_ISL_544734,
EPI_ISL_544735, EPI_ISL_544736, EPI_ISL_544737, EPI_ISL_544738, EPI_ISL_544739, EPI_ISL_544740, EPI_ISL_544741, EPI_ISL_544742, EPI_ISL_544743, EPI_ISL_544744, EPI_ISL_544745, EPI_ISL_544746, EPI_ISL_544747, EPI_ISL_544748, EPI_ISL_544749, EPI_ISL_544750, EPI_ISL_544751, EPI_ISL_544752, EPI_ISL_544753, EPI_ISL_544754, EPI_ISL_544755, EPI_ISL_544756, EPI_ISL_544757, EPI_ISL_544758,
EPI_ISL_544759, EPI_ISL_544760, EPI_ISL_544761, EPI_ISL_544762, EPI_ISL_544763, EPI_ISL_544764, EPI_ISL_544765, EPI_ISL_544766, EPI_ISL_544767, EPI_ISL_544768, EPI_ISL_544769, EPI_ISL_544770, EPI_ISL_544771, EPI_ISL_544772, EPI_ISL_544773, EPI_ISL_544774, EPI_ISL_544775, EPI_ISL_544776, EPI_ISL_544777, EPI_ISL_544778, EPI_ISL_544779, EPI_ISL_544780, EPI_ISL_544781, EPI_ISL_544782,
EPI_ISL_544783, EPI_ISL_544784, EPI_ISL_544785, EPI_ISL_544786, EPI_ISL_544787, EPI_ISL_544788, EPI_ISL_544789, EPI_ISL_544790, EPI_ISL_544791, EPI_ISL_544792, EPI_ISL_544793, EPI_ISL_544794, EPI_ISL_544795, EPI_ISL_544796, EPI_ISL_544797, EPI_ISL_544798, EPI_ISL_544799, EPI_ISL_544800, EPI_ISL_544801, EPI_ISL_544802, EPI_ISL_544803, EPI_ISL_544804, EPI_ISL_544805, EPI_ISL_544806,
EPI_ISL_544807, EPI_ISL_544808, EPI_ISL_544809, EPI_ISL_544810, EPI_ISL_544811, EPI_ISL_544812, EPI_ISL_544813, EPI_ISL_544814, EPI_ISL_544815, EPI_ISL_544816, EPI_ISL_544817, EPI_ISL_544818, EPI_ISL_544819, EPI_ISL_544820, EPI_ISL_544821, EPI_ISL_544822, EPI_ISL_544823, EPI_ISL_544824, EPI_ISL_544825, EPI_ISL_544826, EPI_ISL_544827, EPI_ISL_544828, EPI_ISL_544829, EPI_ISL_544830,
EPI_ISL_544831, EPI_ISL_544832, EPI_ISL_544833, EPI_ISL_544834, EPI_ISL_544835, EPI_ISL_544836, EPI_ISL_544837, EPI_ISL_544838, EPI_ISL_544839, EPI_ISL_544840, EPI_ISL_544841, EPI_ISL_544842, EPI_ISL_544843, EPI_ISL_544844, EPI_ISL_544845, EPI_ISL_544846, EPI_ISL_544847, EPI_ISL_544848, EPI_ISL_544849, EPI_ISL_544850, EPI_ISL_544851, EPI_ISL_544852, EPI_ISL_544853, EPI_ISL_544854,
EPI_ISL_544855, EPI_ISL_544856, EPI_ISL_544857, EPI_ISL_544858, EPI_ISL_544859, EPI_ISL_544860, EPI_ISL_544861, EPI_ISL_544862, EPI_ISL_544863, EPI_ISL_544864, EPI_ISL_544865, EPI_ISL_544866, EPI_ISL_544867, EPI_ISL_544868, EPI_ISL_544869, EPI_ISL_544870, EPI_ISL_544871, EPI_ISL_544872, EPI_ISL_544873, EPI_ISL_544874, EPI_ISL_544875, EPI_ISL_544876, EPI_ISL_544877, EPI_ISL_544878,
EPI_ISL_544879, EPI_ISL_544880, EPI_ISL_544881, EPI_ISL_544882, EPI_ISL_544883, EPI_ISL_544884, EPI_ISL_544885, EPI_ISL_544886, EPI_ISL_544887, EPI_ISL_544888, EPI_ISL_544889, EPI_ISL_544890, EPI_ISL_544891, EPI_ISL_544892, EPI_ISL_544893, EPI_ISL_544894, EPI_ISL_544895, EPI_ISL_544896, EPI_ISL_544897, EPI_ISL_544898, EPI_ISL_544899, EPI_ISL_544900, EPI_ISL_544901, EPI_ISL_544902,
EPI_ISL_544903, EPI_ISL_544904, EPI_ISL_544905, EPI_ISL_544906, EPI_ISL_544907, EPI_ISL_544908, EPI_ISL_544909, EPI_ISL_544910, EPI_ISL_544911, EPI_ISL_544912, EPI_ISL_544913, EPI_ISL_544914, EPI_ISL_544915, EPI_ISL_544916, EPI_ISL_544917, EPI_ISL_544918, EPI_ISL_544919, EPI_ISL_544920, EPI_ISL_544921, EPI_ISL_544922, EPI_ISL_544923, EPI_ISL_544924, EPI_ISL_544925, EPI_ISL_544926,

| | | | |
|---|---|---|---|
| EPI_ISL_544927, EPI_ISL_544928, EPI_ISL_544929, EPI_ISL_544930, EPI_ISL_544931, EPI_ISL_544932, EPI_ISL_544933, EPI_ISL_544934, EPI_ISL_544935, EPI_ISL_544936, EPI_ISL_544937, EPI_ISL_544938, EPI_ISL_544939, EPI_ISL_544940, EPI_ISL_544941, EPI_ISL_544942, EPI_ISL_544943, EPI_ISL_544944, EPI_ISL_544945, EPI_ISL_544946, EPI_ISL_544947, EPI_ISL_544948, EPI_ISL_544949, EPI_ISL_544950, EPI_ISL_544951, EPI_ISL_544952, EPI_ISL_544953, EPI_ISL_544954, EPI_ISL_545051, EPI_ISL_545052, EPI_ISL_545053, EPI_ISL_545054, EPI_ISL_545055, EPI_ISL_545056, EPI_ISL_545057, EPI_ISL_545058, EPI_ISL_545059, EPI_ISL_545060, EPI_ISL_545061, EPI_ISL_545062, EPI_ISL_545063, EPI_ISL_545064, EPI_ISL_545065, EPI_ISL_545066, EPI_ISL_545067, EPI_ISL_545068, EPI_ISL_545069, EPI_ISL_545070, EPI_ISL_545071, EPI_ISL_545072, EPI_ISL_545073, EPI_ISL_545074, EPI_ISL_545075, EPI_ISL_545076, EPI_ISL_545077, EPI_ISL_545078, EPI_ISL_545079, EPI_ISL_545080, EPI_ISL_545081, EPI_ISL_545082, EPI_ISL_545083, EPI_ISL_545084, EPI_ISL_545085, EPI_ISL_545086, EPI_ISL_545087, EPI_ISL_545088, EPI_ISL_545089, EPI_ISL_545090, EPI_ISL_545091, EPI_ISL_545092, EPI_ISL_545093, EPI_ISL_545094, EPI_ISL_545095, EPI_ISL_545096, EPI_ISL_545097, EPI_ISL_545098, EPI_ISL_545099, EPI_ISL_545100, EPI_ISL_545101, EPI_ISL_545102, EPI_ISL_545103, EPI_ISL_545104, EPI_ISL_545105, EPI_ISL_545106, EPI_ISL_545107, EPI_ISL_545108, EPI_ISL_545109, EPI_ISL_545110, EPI_ISL_545111, EPI_ISL_545112, EPI_ISL_545113, EPI_ISL_545114, EPI_ISL_545115, EPI_ISL_545116, EPI_ISL_545117, EPI_ISL_545118, EPI_ISL_545119, EPI_ISL_545120, EPI_ISL_545121, EPI_ISL_545122, EPI_ISL_545123, EPI_ISL_545124, EPI_ISL_545125, EPI_ISL_545126, EPI_ISL_545127, EPI_ISL_545128, EPI_ISL_545129, EPI_ISL_545130, EPI_ISL_545131, EPI_ISL_545132, EPI_ISL_545133, EPI_ISL_545134, EPI_ISL_545135, EPI_ISL_545136, EPI_ISL_545137, EPI_ISL_545138, EPI_ISL_545139, EPI_ISL_545140, EPI_ISL_545141, EPI_ISL_545142, EPI_ISL_545143, EPI_ISL_545144, EPI_ISL_545145, EPI_ISL_545146, EPI_ISL_545147, EPI_ISL_545148, EPI_ISL_545149, EPI_ISL_545150, EPI_ISL_545151, EPI_ISL_545152, EPI_ISL_545153, EPI_ISL_545154, EPI_ISL_545155, EPI_ISL_545156, EPI_ISL_545157, EPI_ISL_545158, EPI_ISL_545159, EPI_ISL_545160, EPI_ISL_545161, EPI_ISL_545162, EPI_ISL_545163, EPI_ISL_545164, EPI_ISL_545165, EPI_ISL_545166, EPI_ISL_545167, EPI_ISL_545168, EPI_ISL_545169, EPI_ISL_545170, EPI_ISL_545171, EPI_ISL_545172, EPI_ISL_545173, EPI_ISL_545174, EPI_ISL_545175, EPI_ISL_545176, EPI_ISL_545177, EPI_ISL_545178, EPI_ISL_545179, EPI_ISL_545180, EPI_ISL_545181, EPI_ISL_545182, EPI_ISL_545183, EPI_ISL_545184, EPI_ISL_545185, EPI_ISL_545186, EPI_ISL_545187, EPI_ISL_545188, EPI_ISL_545189, EPI_ISL_545190, EPI_ISL_545191, EPI_ISL_545192, EPI_ISL_545193, EPI_ISL_545194, EPI_ISL_545195, EPI_ISL_545196, EPI_ISL_545197, EPI_ISL_545198, EPI_ISL_545199, EPI_ISL_545200, EPI_ISL_545201, EPI_ISL_545202, EPI_ISL_545203, EPI_ISL_545204, EPI_ISL_545205, EPI_ISL_545206, EPI_ISL_545207, EPI_ISL_545208, EPI_ISL_545209, EPI_ISL_545210, EPI_ISL_545211, EPI_ISL_545212, EPI_ISL_545213, EPI_ISL_545214, EPI_ISL_545215, EPI_ISL_545216, EPI_ISL_545217, EPI_ISL_545218, EPI_ISL_545219, EPI_ISL_545220, EPI_ISL_545221, EPI_ISL_545222, EPI_ISL_545223, EPI_ISL_545224, EPI_ISL_545225, EPI_ISL_545226, EPI_ISL_545227, EPI_ISL_545228, EPI_ISL_545229, EPI_ISL_545230, EPI_ISL_545231, EPI_ISL_545232, EPI_ISL_545233, EPI_ISL_545234, EPI_ISL_545235, EPI_ISL_545236, EPI_ISL_545237, EPI_ISL_545238, EPI_ISL_545239, EPI_ISL_545240, EPI_ISL_545241, EPI_ISL_545242, EPI_ISL_545243, EPI_ISL_545244, EPI_ISL_545245, EPI_ISL_545246, EPI_ISL_545247, EPI_ISL_545248, EPI_ISL_545249, EPI_ISL_545250, EPI_ISL_545251, EPI_ISL_545252, EPI_ISL_545253, EPI_ISL_545254, EPI_ISL_545255, EPI_ISL_545256, EPI_ISL_545257, EPI_ISL_545258, EPI_ISL_545259, EPI_ISL_545260, EPI_ISL_545261, EPI_ISL_545262, EPI_ISL_545263, EPI_ISL_545264, EPI_ISL_545265, EPI_ISL_545266, EPI_ISL_545267, EPI_ISL_545268, EPI_ISL_545269, EPI_ISL_545270, EPI_ISL_545271, EPI_ISL_545272, EPI_ISL_545273, EPI_ISL_545274, EPI_ISL_545275, EPI_ISL_545276, EPI_ISL_545277, EPI_ISL_545278, EPI_ISL_545279, EPI_ISL_545280, EPI_ISL_545281, EPI_ISL_545282, EPI_ISL_545283, EPI_ISL_545284, EPI_ISL_545285, EPI_ISL_545286, EPI_ISL_545287, EPI_ISL_545288, EPI_ISL_545289, EPI_ISL_545290, EPI_ISL_545291, EPI_ISL_545292, EPI_ISL_545293, EPI_ISL_545294, EPI_ISL_545295, EPI_ISL_545296, EPI_ISL_545297, EPI_ISL_545298, EPI_ISL_545299, EPI_ISL_545300, EPI_ISL_545301, EPI_ISL_545302, EPI_ISL_545303, EPI_ISL_545304, EPI_ISL_545305, EPI_ISL_545306, EPI_ISL_545307, EPI_ISL_545308, EPI_ISL_545309, EPI_ISL_545310, EPI_ISL_545311, EPI_ISL_545312, EPI_ISL_545313, EPI_ISL_545314, EPI_ISL_545315, EPI_ISL_545316, EPI_ISL_545317, EPI_ISL_545318, EPI_ISL_545319, EPI_ISL_545320, EPI_ISL_545321, EPI_ISL_545322, EPI_ISL_545323, EPI_ISL_545324, EPI_ISL_545325, EPI_ISL_545326, EPI_ISL_545327, EPI_ISL_545328, EPI_ISL_545329, EPI_ISL_545330, EPI_ISL_545331, EPI_ISL_545332, EPI_ISL_545333, EPI_ISL_545334, EPI_ISL_545335, EPI_ISL_545336, EPI_ISL_545337, EPI_ISL_545338, EPI_ISL_545339, EPI_ISL_545340, EPI_ISL_545341, EPI_ISL_545342, EPI_ISL_545343, EPI_ISL_545344, EPI_ISL_545345, EPI_ISL_545346, EPI_ISL_545347, EPI_ISL_545348, EPI_ISL_545349, EPI_ISL_545350, EPI_ISL_545351, EPI_ISL_545352, EPI_ISL_545353, EPI_ISL_545354, EPI_ISL_545355, EPI_ISL_545356, EPI_ISL_545357, EPI_ISL_545358, EPI_ISL_545359, EPI_ISL_545360, EPI_ISL_545361, EPI_ISL_545362, EPI_ISL_545363, EPI_ISL_545364, EPI_ISL_545365, EPI_ISL_545366, EPI_ISL_545367, EPI_ISL_545368, EPI_ISL_545369, EPI_ISL_545370, EPI_ISL_545371, EPI_ISL_545372, EPI_ISL_545373, EPI_ISL_545374, EPI_ISL_545375, EPI_ISL_545376, EPI_ISL_545377, EPI_ISL_545378, EPI_ISL_545379, EPI_ISL_545380, EPI_ISL_545381, EPI_ISL_545382, EPI_ISL_545383, EPI_ISL_545384, EPI_ISL_545385, EPI_ISL_545386, EPI_ISL_545387, EPI_ISL_545388, EPI_ISL_545389, EPI_ISL_545390, EPI_ISL_545391, EPI_ISL_545392, EPI_ISL_545393, EPI_ISL_545394, EPI_ISL_545395, EPI_ISL_545396, EPI_ISL_545397, EPI_ISL_545398, EPI_ISL_545399, EPI_ISL_545400, EPI_ISL_545401, EPI_ISL_545402, EPI_ISL_545403, EPI_ISL_545404, EPI_ISL_545405, EPI_ISL_545406, EPI_ISL_545407, EPI_ISL_545408, EPI_ISL_545409, EPI_ISL_545410, EPI_ISL_545411, EPI_ISL_545412, EPI_ISL_545413, EPI_ISL_545414, EPI_ISL_545415, EPI_ISL_545416, EPI_ISL_545417, EPI_ISL_545418, EPI_ISL_545419, EPI_ISL_545420, EPI_ISL_545421, EPI_ISL_545422, EPI_ISL_545423, EPI_ISL_545424, EPI_ISL_545425, EPI_ISL_545426, EPI_ISL_545427, EPI_ISL_545428, EPI_ISL_545429, EPI_ISL_545430, EPI_ISL_545431, EPI_ISL_545432, EPI_ISL_545433, EPI_ISL_545434, EPI_ISL_545435, EPI_ISL_545436, EPI_ISL_545437, EPI_ISL_545438, EPI_ISL_545439, EPI_ISL_545440, EPI_ISL_545441, EPI_ISL_545442, EPI_ISL_545443, EPI_ISL_545444, EPI_ISL_545445, EPI_ISL_545446, EPI_ISL_545447, EPI_ISL_545448, EPI_ISL_545449, EPI_ISL_545450, EPI_ISL_545451, EPI_ISL_545452, EPI_ISL_545453, EPI_ISL_545454, EPI_ISL_545455, EPI_ISL_545456, EPI_ISL_545457, EPI_ISL_545458, EPI_ISL_545459, EPI_ISL_545460, EPI_ISL_545461, EPI_ISL_545462, EPI_ISL_545463, EPI_ISL_545464, EPI_ISL_545465, EPI_ISL_545466, EPI_ISL_545467, EPI_ISL_545468, EPI_ISL_545469, EPI_ISL_545470, EPI_ISL_545471, EPI_ISL_545472, EPI_ISL_545473, EPI_ISL_545474, EPI_ISL_545475, EPI_ISL_545476, EPI_ISL_545477, EPI_ISL_545478, EPI_ISL_545479, EPI_ISL_545480, EPI_ISL_545481, EPI_ISL_545482, EPI_ISL_545483, EPI_ISL_545484, EPI_ISL_545485, EPI_ISL_545486, EPI_ISL_545487, EPI_ISL_545488, EPI_ISL_545489, EPI_ISL_545490, EPI_ISL_545491, EPI_ISL_545492, EPI_ISL_545493, EPI_ISL_545494, EPI_ISL_545495, EPI_ISL_545496, EPI_ISL_545497, EPI_ISL_545498, EPI_ISL_545499, EPI_ISL_545500, EPI_ISL_545501, EPI_ISL_545502, EPI_ISL_545503, EPI_ISL_545504, EPI_ISL_545505, EPI_ISL_545506, EPI_ISL_545507, EPI_ISL_545508, EPI_ISL_545509, EPI_ISL_545510, EPI_ISL_545511, EPI_ISL_545512, EPI_ISL_545513, EPI_ISL_545514, EPI_ISL_545515, EPI_ISL_545516, EPI_ISL_545517, EPI_ISL_545518, EPI_ISL_545519, EPI_ISL_545520, EPI_ISL_545521, EPI_ISL_545522, EPI_ISL_545523, EPI_ISL_545524, EPI_ISL_545525, EPI_ISL_545526, EPI_ISL_545527, EPI_ISL_545528, EPI_ISL_545529, EPI_ISL_545530, EPI_ISL_545531, EPI_ISL_545532, EPI_ISL_545533, EPI_ISL_545534, EPI_ISL_545535, EPI_ISL_545536, EPI_ISL_545537, EPI_ISL_545538, EPI_ISL_545539, EPI_ISL_545540, EPI_ISL_545541, EPI_ISL_545542, EPI_ISL_545543, EPI_ISL_545544, EPI_ISL_545545, EPI_ISL_545546, EPI_ISL_545547, EPI_ISL_545548, EPI_ISL_545549, EPI_ISL_545550, EPI_ISL_545551, EPI_ISL_545552, EPI_ISL_545553, EPI_ISL_545554, EPI_ISL_545555, EPI_ISL_545556, EPI_ISL_545557, EPI_ISL_545558, EPI_ISL_545559, EPI_ISL_545560, EPI_ISL_545561, EPI_ISL_545562, EPI_ISL_545563, EPI_ISL_545564, EPI_ISL_545565, EPI_ISL_545566, EPI_ISL_545567, EPI_ISL_545568, EPI_ISL_545569, EPI_ISL_545570, EPI_ISL_545571, EPI_ISL_545572, EPI_ISL_545573, EPI_ISL_545574, EPI_ISL_545581, EPI_ISL_545582, EPI_ISL_545583, EPI_ISL_545584, EPI_ISL_545585, EPI_ISL_545586, EPI_ISL_545587, EPI_ISL_545588, EPI_ISL_545589, EPI_ISL_545590, EPI_ISL_545591, EPI_ISL_545592, EPI_ISL_545593, EPI_ISL_545594, EPI_ISL_545595, EPI_ISL_545596, EPI_ISL_545597, EPI_ISL_545598, EPI_ISL_545599, EPI_ISL_545600, EPI_ISL_545601, EPI_ISL_545602, EPI_ISL_545603, EPI_ISL_545604, EPI_ISL_545605, EPI_ISL_545606, EPI_ISL_545607, EPI_ISL_545608, EPI_ISL_545609, EPI_ISL_545610, EPI_ISL_545611, EPI_ISL_545612, EPI_ISL_545613, EPI_ISL_545614, EPI_ISL_545615, EPI_ISL_545616, EPI_ISL_545617, EPI_ISL_545618, EPI_ISL_545619, EPI_ISL_545620, EPI_ISL_545621, EPI_ISL_545622, EPI_ISL_545623, EPI_ISL_545624, EPI_ISL_545625, EPI_ISL_545626, EPI_ISL_545627, EPI_ISL_545628, EPI_ISL_545629, EPI_ISL_545630, EPI_ISL_545631, EPI_ISL_545632, EPI_ISL_545633, EPI_ISL_545634, EPI_ISL_545635, EPI_ISL_545636, EPI_ISL_545637, EPI_ISL_545638, EPI_ISL_545639, EPI_ISL_545640, EPI_ISL_545641, EPI_ISL_545642, EPI_ISL_545643, EPI_ISL_545644, EPI_ISL_545645, EPI_ISL_545646, EPI_ISL_545647, EPI_ISL_545648, EPI_ISL_545649, EPI_ISL_545650, EPI_ISL_545651, EPI_ISL_545652, EPI_ISL_545653, EPI_ISL_545654, EPI_ISL_545655, EPI_ISL_545656, EPI_ISL_545657, EPI_ISL_545658, EPI_ISL_545659, EPI_ISL_545660, EPI_ISL_545661, EPI_ISL_545662, EPI_ISL_545663, EPI_ISL_545664, EPI_ISL_545665, EPI_ISL_545666, EPI_ISL_545667, EPI_ISL_545668, EPI_ISL_545669, EPI_ISL_545670, EPI_ISL_545671, EPI_ISL_545672, EPI_ISL_545673, EPI_ISL_545674, EPI_ISL_545675, EPI_ISL_545676, EPI_ISL_545677, EPI_ISL_545678, EPI_ISL_545679, EPI_ISL_545680, EPI_ISL_545681, EPI_ISL_545682, EPI_ISL_545683, EPI_ISL_545684, EPI_ISL_545685, EPI_ISL_545686, EPI_ISL_545687, EPI_ISL_545688, EPI_ISL_545689, EPI_ISL_545690, EPI_ISL_545691, EPI_ISL_545692, EPI_ISL_545693, EPI_ISL_545694, EPI_ISL_545695, EPI_ISL_545696, EPI_ISL_545697, EPI_ISL_545698, EPI_ISL_545699, EPI_ISL_545700, EPI_ISL_545701, EPI_ISL_545702, EPI_ISL_545703, EPI_ISL_545704, EPI_ISL_545705, EPI_ISL_545706, EPI_ISL_545707, EPI_ISL_545708, EPI_ISL_545709, EPI_ISL_545710, EPI_ISL_545711, EPI_ISL_545712, EPI_ISL_545713, EPI_ISL_545714, EPI_ISL_545715, EPI_ISL_545716, EPI_ISL_545717, EPI_ISL_545718, EPI_ISL_545719, EPI_ISL_545720, EPI_ISL_545721, EPI_ISL_545722, EPI_ISL_545723, EPI_ISL_545724, EPI_ISL_545725, EPI_ISL_545726, EPI_ISL_545727, EPI_ISL_545728, EPI_ISL_545729, EPI_ISL_545730, EPI_ISL_545731, EPI_ISL_545732, EPI_ISL_545733, EPI_ISL_545734, EPI_ISL_545735, EPI_ISL_545736, EPI_ISL_545737, EPI_ISL_545738, EPI_ISL_545739, EPI_ISL_545740, EPI_ISL_545741, EPI_ISL_545742, EPI_ISL_545743, EPI_ISL_545744, EPI_ISL_545745, EPI_ISL_545746, EPI_ISL_545747, EPI_ISL_545748, EPI_ISL_545749, EPI_ISL_545750, EPI_ISL_545751, EPI_ISL_545752, EPI_ISL_545753, EPI_ISL_545754, EPI_ISL_545755, EPI_ISL_545756, EPI_ISL_545757, EPI_ISL_545758, EPI_ISL_545759, EPI_ISL_545760, EPI_ISL_545761, EPI_ISL_545762, EPI_ISL_545763, EPI_ISL_545764, EPI_ISL_545765, EPI_ISL_545766, EPI_ISL_545767, EPI_ISL_545768, EPI_ISL_545769, EPI_ISL_545770, EPI_ISL_545771, EPI_ISL_545772, EPI_ISL_545773, EPI_ISL_545774, EPI_ISL_545775, EPI_ISL_545776, EPI_ISL_545777, EPI_ISL_545778, EPI_ISL_545779, EPI_ISL_545780, EPI_ISL_545781, EPI_ISL_545782, EPI_ISL_545783, EPI_ISL_545784, EPI_ISL_545785, EPI_ISL_545786, EPI_ISL_545787, EPI_ISL_545788, EPI_ISL_545789, EPI_ISL_545790, EPI_ISL_545791, EPI_ISL_545792, EPI_ISL_545793, EPI_ISL_545794, EPI_ISL_545795, EPI_ISL_545796, EPI_ISL_545797, EPI_ISL_545798, EPI_ISL_545799, EPI_ISL_545800, EPI_ISL_545801, EPI_ISL_545802, EPI_ISL_545803, EPI_ISL_545804, EPI_ISL_545805, EPI_ISL_545806, EPI_ISL_545807, EPI_ISL_545808, EPI_ISL_545809, EPI_ISL_545810, EPI_ISL_545811, EPI_ISL_545812, EPI_ISL_545813, EPI_ISL_545814, EPI_ISL_545815, EPI_ISL_545816, EPI_ISL_545817, EPI_ISL_545818, EPI_ISL_545819, EPI_ISL_545820, EPI_ISL_545821, EPI_ISL_545822, EPI_ISL_545823, EPI_ISL_545824, EPI_ISL_545825, EPI_ISL_545826, EPI_ISL_545827, EPI_ISL_545828, EPI_ISL_545829, EPI_ISL_545830, EPI_ISL_545831, EPI_ISL_545832, EPI_ISL_545833, EPI_ISL_545834, EPI_ISL_545835, EPI_ISL_545836, EPI_ISL_545837, EPI_ISL_545838, EPI_ISL_545839, EPI_ISL_545840, EPI_ISL_545841, EPI_ISL_545842, EPI_ISL_545843, EPI_ISL_545844, EPI_ISL_545845, EPI_ISL_545846, EPI_ISL_545847, EPI_ISL_545848, EPI_ISL_545849, EPI_ISL_545850, EPI_ISL_545851, EPI_ISL_545852, EPI_ISL_545853, EPI_ISL_545854, EPI_ISL_545855, EPI_ISL_545856, EPI_ISL_545857, EPI_ISL_545858, EPI_ISL_545859, EPI_ISL_545860, EPI_ISL_545861, EPI_ISL_545862, EPI_ISL_545863, EPI_ISL_545864, EPI_ISL_545865, EPI_ISL_545866, EPI_ISL_545867, EPI_ISL_545868, EPI_ISL_545869, EPI_ISL_545870, EPI_ISL_545871, EPI_ISL_545872, EPI_ISL_545873, EPI_ISL_545874, EPI_ISL_545875, EPI_ISL_545876, EPI_ISL_545877, EPI_ISL_545878, EPI_ISL_545879, EPI_ISL_545880, EPI_ISL_545881, EPI_ISL_545882, EPI_ISL_545883, EPI_ISL_545884, EPI_ISL_545885, EPI_ISL_545886, EPI_ISL_545887, EPI_ISL_545888, EPI_ISL_545889, EPI_ISL_545890, EPI_ISL_545891, EPI_ISL_545892, EPI_ISL_545893, EPI_ISL_545894, EPI_ISL_545895, EPI_ISL_545896, EPI_ISL_545897, EPI_ISL_545898, EPI_ISL_545899, EPI_ISL_545900, EPI_ISL_545901, EPI_ISL_545902, EPI_ISL_545903, EPI_ISL_545904, EPI_ISL_545905, EPI_ISL_545906, EPI_ISL_545907, EPI_ISL_545908, EPI_ISL_545909, EPI_ISL_545910, EPI_ISL_545911, EPI_ISL_545912, EPI_ISL_545913, EPI_ISL_545914, EPI_ISL_545915, EPI_ISL_545916, EPI_ISL_545917, EPI_ISL_545918, EPI_ISL_545919, EPI_ISL_545920, EPI_ISL_545921, EPI_ISL_545922, EPI_ISL_545923, EPI_ISL_545924, EPI_ISL_545925, EPI_ISL_545926, EPI_ISL_545927, EPI_ISL_545928, EPI_ISL_545929, EPI_ISL_545930, EPI_ISL_545931, EPI_ISL_545932, EPI_ISL_545933, EPI_ISL_545934, EPI_ISL_545935, EPI_ISL_545936, EPI_ISL_545937, EPI_ISL_545938, EPI_ISL_545939, EPI_ISL_545940, EPI_ISL_545941, EPI_ISL_545942, EPI_ISL_545943, EPI_ISL_545944, EPI_ISL_545945, EPI_ISL_545946, EPI_ISL_545947, EPI_ISL_545948, EPI_ISL_545949, EPI_ISL_545950, EPI_ISL_545951, EPI_ISL_545952, EPI_ISL_545953, EPI_ISL_545958, EPI_ISL_545959, EPI_ISL_545960, EPI_ISL_545961, EPI_ISL_545962, EPI_ISL_545963, EPI_ISL_545964, EPI_ISL_545965, EPI_ISL_545966, EPI_ISL_545967, EPI_ISL_545968, EPI_ISL_545969, EPI_ISL_545970, EPI_ISL_545971, EPI_ISL_545972, EPI_ISL_545973, EPI_ISL_545974, EPI_ISL_545975, EPI_ISL_545976, EPI_ISL_545977, EPI_ISL_545978, EPI_ISL_545979, EPI_ISL_545980, EPI_ISL_545981, EPI_ISL_545982, EPI_ISL_545983, EPI_ISL_545984, EPI_ISL_545985, EPI_ISL_545986, EPI_ISL_545987, EPI_ISL_545988, EPI_ISL_545989, EPI_ISL_545990, EPI_ISL_545991, EPI_ISL_545992, EPI_ISL_545993, EPI_ISL_545994, EPI_ISL_545995, EPI_ISL_545996, EPI_ISL_545997, EPI_ISL_545998, EPI_ISL_545999, EPI_ISL_546000, EPI_ISL_546001, EPI_ISL_546002, EPI_ISL_546003, EPI_ISL_546004, EPI_ISL_546005, EPI_ISL_546006, EPI_ISL_546007, EPI_ISL_546008, EPI_ISL_546009, EPI_ISL_546010, EPI_ISL_546011, EPI_ISL_546012, EPI_ISL_546013, EPI_ISL_546014, EPI_ISL_546015, EPI_ISL_546016, EPI_ISL_546017, EPI_ISL_546018, EPI_ISL_546019, EPI_ISL_546020, EPI_ISL_546021, EPI_ISL_546022, EPI_ISL_546023, EPI_ISL_546024, EPI_ISL_546025, EPI_ISL_546026, EPI_ISL_546027, EPI_ISL_546028, EPI_ISL_546029, EPI_ISL_546030, EPI_ISL_546031, EPI_ISL_546032, EPI_ISL_546033, EPI_ISL_546034, EPI_ISL_546035, EPI_ISL_546036, EPI_ISL_546037, EPI_ISL_546038, EPI_ISL_546039, EPI_ISL_546040, EPI_ISL_546041, EPI_ISL_546042, EPI_ISL_546043, EPI_ISL_546044, EPI_ISL_546045, EPI_ISL_546046, EPI_ISL_546047, EPI_ISL_546048, EPI_ISL_546049, EPI_ISL_546050, EPI_ISL_546051, EPI_ISL_546052, EPI_ISL_546053, EPI_ISL_546054, EPI_ISL_546055, EPI_ISL_546056, EPI_ISL_546057, EPI_ISL_546058, EPI_ISL_546059, EPI_ISL_546060, EPI_ISL_546061, EPI_ISL_546062, EPI_ISL_546063, EPI_ISL_546064, EPI_ISL_546065, EPI_ISL_546066, EPI_ISL_546067, EPI_ISL_546068, EPI_ISL_546069, EPI_ISL_546070, EPI_ISL_546071, EPI_ISL_546072, EPI_ISL_546073, EPI_ISL_546074, EPI_ISL_546075, EPI_ISL_546076, EPI_ISL_546077, EPI_ISL_546078, EPI_ISL_546079, EPI_ISL_546080, EPI_ISL_546081, EPI_ISL_546082, EPI_ISL_546083, EPI_ISL_546084, EPI_ISL_546085, EPI_ISL_546086, EPI_ISL_546087, EPI_ISL_546088, EPI_ISL_546089, EPI_ISL_546090, EPI_ISL_546091, EPI_ISL_546092, EPI_ISL_546093, EPI_ISL_546094, EPI_ISL_546095, EPI_ISL_546096, EPI_ISL_546097, EPI_ISL_546098, EPI_ISL_546099, EPI_ISL_546100, EPI_ISL_546101, EPI_ISL_546102, EPI_ISL_546103, EPI_ISL_546104, EPI_ISL_546105, EPI_ISL_546106, EPI_ISL_546107, EPI_ISL_546108, EPI_ISL_546109, EPI_ISL_546110, EPI_ISL_546111, EPI_ISL_546112, EPI_ISL_546113, EPI_ISL_546114, EPI_ISL_546115, EPI_ISL_546116, EPI_ISL_546117, EPI_ISL_546118, EPI_ISL_546119, EPI_ISL_546120, EPI_ISL_546121, EPI_ISL_546122, EPI_ISL_546123, EPI_ISL_546124, EPI_ISL_546125, EPI_ISL_546126, EPI_ISL_546127, EPI_ISL_546128, EPI_ISL_546129, EPI_ISL_546130, EPI_ISL_546131, EPI_ISL_546132, EPI_ISL_546133, EPI_ISL_546134, EPI_ISL_546135, EPI_ISL_546136, EPI_ISL_546137, EPI_ISL_546138, EPI_ISL_546139, EPI_ISL_546140, EPI_ISL_546141, EPI_ISL_546142, EPI_ISL_546143, EPI_ISL_546144, EPI_ISL_546145, EPI_ISL_546146, EPI_ISL_546147, EPI_ISL_546148, EPI_ISL_546149, EPI_ISL_546150, EPI_ISL_546151, EPI_ISL_546152, EPI_ISL_546153, EPI_ISL_546154, EPI_ISL_546155, EPI_ISL_546156, EPI_ISL_546157, EPI_ISL_546158, EPI_ISL_546159, EPI_ISL_546160, EPI_ISL_546161, EPI_ISL_546162, EPI_ISL_546163, EPI_ISL_546164, EPI_ISL_546165, EPI_ISL_546166, EPI_ISL_546167, EPI_ISL_546168, EPI_ISL_546169, EPI_ISL_546170, EPI_ISL_546171, EPI_ISL_546172, EPI_ISL_546173, EPI_ISL_546174, EPI_ISL_546175, EPI_ISL_546176, EPI_ISL_546177, EPI_ISL_546178, EPI_ISL_546179, EPI_ISL_546180, EPI_ISL_546181, EPI_ISL_546182, EPI_ISL_546183, EPI_ISL_546184, EPI_ISL_546185, EPI_ISL_546186, EPI_ISL_546187, EPI_ISL_546188, EPI_ISL_546189, EPI_ISL_546190, EPI_ISL_546191, EPI_ISL_546192, EPI_ISL_546193, EPI_ISL_546194, EPI_ISL_546195, EPI_ISL_546196, EPI_ISL_546197, EPI_ISL_546198, EPI_ISL_546199, EPI_ISL_546200, EPI_ISL_546201, EPI_ISL_546202, EPI_ISL_546203, EPI_ISL_546204, EPI_ISL_546205, EPI_ISL_546206, EPI_ISL_546207, EPI_ISL_546208, EPI_ISL_546209, EPI_ISL_546210, EPI_ISL_546211, EPI_ISL_546212, EPI_ISL_546213, EPI_ISL_546214, EPI_ISL_546215, EPI_ISL_546216, EPI_ISL_546217, EPI_ISL_546218, EPI_ISL_546219, EPI_ISL_546220, EPI_ISL_546221, EPI_ISL_546222, EPI_ISL_546223, EPI_ISL_546224, EPI_ISL_546225, EPI_ISL_546226, EPI_ISL_546227, EPI_ISL_546228, EPI_ISL_546229, EPI_ISL_546230, EPI_ISL_546231, EPI_ISL_546232, EPI_ISL_546233, EPI_ISL_546234, EPI_ISL_546235, EPI_ISL_546236, EPI_ISL_546237, EPI_ISL_546238, EPI_ISL_546239, EPI_ISL_546240, EPI_ISL_546241, EPI_ISL_546242, EPI_ISL_546243, EPI_ISL_546244, EPI_ISL_546245, EPI_ISL_546246, EPI_ISL_546247, EPI_ISL_546248, EPI_ISL_546249, EPI_ISL_546250, EPI_ISL_546251, EPI_ISL_546252, EPI_ISL_546253, EPI_ISL_546254, EPI_ISL_546255, EPI_ISL_546256, EPI_ISL_546257, EPI_ISL_546258, EPI_ISL_546259, EPI_ISL_546260, EPI_ISL_546261, EPI_ISL_546262, EPI_ISL_546263, EPI_ISL_546264, EPI_ISL_546265, EPI_ISL_546266, EPI_ISL_546267, EPI_ISL_546268, EPI_ISL_546269, EPI_ISL_546270, EPI_ISL_546271, EPI_ISL_546272, EPI_ISL_546273, EPI_ISL_546274, EPI_ISL_546275, EPI_ISL_546276, EPI_ISL_546277, EPI_ISL_546278, EPI_ISL_546279, EPI_ISL_546280, EPI_ISL_546281, EPI_ISL_546282, EPI_ISL_546283, EPI_ISL_546284, EPI_ISL_546285, EPI_ISL_546286, EPI_ISL_546287, EPI_ISL_546288, EPI_ISL_546289, EPI_ISL_546290, EPI_ISL_546291, EPI_ISL_546292, EPI_ISL_546293, EPI_ISL_546294, EPI_ISL_546295, EPI_ISL_546296, EPI_ISL_546297, EPI_ISL_546298, EPI_ISL_546299, EPI_ISL_546300, EPI_ISL_546301, EPI_ISL_546302, EPI_ISL_546303, EPI_ISL_546304, EPI_ISL_546305, EPI_ISL_546306, EPI_ISL_546307, EPI_ISL_546308, EPI_ISL_546309, EPI_ISL_546310, EPI_ISL_546311, EPI_ISL_546312, EPI_ISL_546313, EPI_ISL_546314, EPI_ISL_546315, EPI_ISL_546316, EPI_ISL_546317, EPI_ISL_546318, EPI_ISL_546319, EPI_ISL_546320, EPI_ISL_546321, EPI_ISL_546322, EPI_ISL_546323, EPI_ISL_546324, EPI_ISL_546325, EPI_ISL_546326, EPI_ISL_546327, EPI_ISL_546328, EPI_ISL_546329, EPI_ISL_546330, EPI_ISL_546331, EPI_ISL_546332, EPI_ISL_546333, EPI_ISL_546334, EPI_ISL_546335, EPI_ISL_546336, EPI_ISL_546337, EPI_ISL_546338, EPI_ISL_546339, EPI_ISL_546340, EPI_ISL_546341, EPI_ISL_546342, EPI_ISL_546343, EPI_ISL_546344, EPI_ISL_546345, EPI_ISL_546346, EPI_ISL_546347, EPI_ISL_546348, EPI_ISL_546349, EPI_ISL_546350, EPI_ISL_546351, EPI_ISL_546352, EPI_ISL_546353, EPI_ISL_546354, EPI_ISL_546355, EPI_ISL_546356, EPI_ISL_546357, EPI_ISL_546358, EPI_ISL_546359, EPI_ISL_546360, EPI_ISL_546361, EPI_ISL_546362, EPI_ISL_546363, EPI_ISL_546364, EPI_ISL_546365, EPI_ISL_546366, EPI_ISL_546367, EPI_ISL_546368, EPI_ISL_546369, EPI_ISL_546370, EPI_ISL_546371, EPI_ISL_546372, EPI_ISL_546373, EPI_ISL_546374, EPI_ISL_546375, EPI_ISL_546376, EPI_ISL_546377, EPI_ISL_546378, EPI_ISL_546379, EPI_ISL_546380, EPI_ISL_546381, EPI_ISL_546382, EPI_ISL_546383, EPI_ISL_546384, EPI_ISL_546385, EPI_ISL_546386, EPI_ISL_546387, EPI_ISL_546388, EPI_ISL_546389, EPI_ISL_546390, EPI_ISL_546391, EPI_ISL_546392, EPI_ISL_546393, EPI_ISL_546394, EPI_ISL_546395, EPI_ISL_546396, EPI_ISL_546397, EPI_ISL_546398, EPI_ISL_546399, EPI_ISL_546400, EPI_ISL_546401, EPI_ISL_546402, EPI_ISL_546403, EPI_ISL_546404, EPI_ISL_546405, EPI_ISL_546406, EPI_ISL_546407, EPI_ISL_546408, EPI_ISL_546409, EPI_ISL_546410, EPI_ISL_546411, EPI_ISL_546412, EPI_ISL_546413, EPI_ISL_546414, EPI_ISL_546415, EPI_ISL_546416, EPI_ISL_546417, EPI_ISL_546418, EPI_ISL_546419, EPI_ISL_546420, EPI_ISL_546421, EPI_ISL_546422, EPI_ISL_546423, EPI_ISL_546424, EPI_ISL_546425, EPI_ISL_546426, EPI_ISL_546427, EPI_ISL_546428, EPI_ISL_546429, EPI_ISL_546430, EPI_ISL_546431, EPI_ISL_546432, EPI_ISL_546433, EPI_ISL_546434, EPI_ISL_546435, EPI_ISL_546437, EPI_ISL_546438, EPI_ISL_546439, EPI_ISL_546440, EPI_ISL_546441, EPI_ISL_546442, EPI_ISL_546443, EPI_ISL_546444, EPI_ISL_546445, EPI_ISL_546446, EPI_ISL_546447, EPI_ISL_546448, EPI_ISL_546449, EPI_ISL_546450, EPI_ISL_546451, EPI_ISL_546452, EPI_ISL_546453, EPI_ISL_546454, EPI_ISL_546455, EPI_ISL_546456, EPI_ISL_546457, EPI_ISL_546458, EPI_ISL_546459, EPI_ISL_546460, EPI_ISL_546461, EPI_ISL_546462, EPI_ISL_546463, EPI_ISL_546464, EPI_ISL_546465, EPI_ISL_546466, EPI_ISL_546467, EPI_ISL_546468, EPI_ISL_546469, EPI_ISL_546470, EPI_ISL_546471, EPI_ISL_546472, EPI_ISL_546473, EPI_ISL_546474, EPI_ISL_546475, EPI_ISL_546476, EPI_ISL_546477, EPI_ISL_546478, EPI_ISL_546479, EPI_ISL_546480, EPI_ISL_546481, EPI_ISL_546482, EPI_ISL_546483, EPI_ISL_546484, EPI_ISL_546485, EPI_ISL_546486, EPI_ISL_546487, EPI_ISL_546488, EPI_ISL_546489, EPI_ISL_546490, EPI_ISL_546491, EPI_ISL_546492, EPI_ISL_546493, EPI_ISL_546494, EPI_ISL_546495, EPI_ISL_546496, EPI_ISL_546497, EPI_ISL_546498, EPI_ISL_546499, EPI_ISL_546500, EPI_ISL_546501, EPI_ISL_546502, EPI_ISL_546503, EPI_ISL_546504, EPI_ISL_546505, EPI_ISL_546506, EPI_ISL_546507, EPI_ISL_546508, EPI_ISL_546509, EPI_ISL_546510, EPI_ISL_546511, EPI_ISL_546512, EPI_ISL_546513, EPI_ISL_546514, EPI_ISL_546515, EPI_ISL_546516, EPI_ISL_546517, EPI_ISL_546518, EPI_ISL_546519, EPI_ISL_546520, EPI_ISL_546521, EPI_ISL_546522, EPI_ISL_546523, EPI_ISL_546524, EPI_ISL_546525, EPI_ISL_546526, EPI_ISL_546527, EPI_ISL_546528, EPI_ISL_546529, EPI_ISL_546530, EPI_ISL_546531, EPI_ISL_546532, EPI_ISL_546533, EPI_ISL_546534, EPI_ISL_546535, EPI_ISL_546536, EPI_ISL_546537, EPI_ISL_546538, EPI_ISL_546539, EPI_ISL_546540, EPI_ISL_546541, EPI_ISL_546542, EPI_ISL_546543, EPI_ISL_546544, EPI_ISL_546545, EPI_ISL_546546, EPI_ISL_546547, EPI_ISL_546548, EPI_ISL_546549, EPI_ISL_546550, EPI_ISL_546551, EPI_ISL_546552, EPI_ISL_546553, EPI_ISL_546554, EPI_ISL_546555, EPI_ISL_546556, EPI_ISL_546557, EPI_ISL_546558, EPI_ISL_546559, EPI_ISL_546560, EPI_ISL_546561, EPI_ISL_546562, EPI_ISL_546563, EPI_ISL_546564, EPI_ISL_546565, EPI_ISL_546566, EPI_ISL_546567, EPI_ISL_546568, EPI_ISL_546569, EPI_ISL_546570, EPI_ISL_546571, EPI_ISL_546572, EPI_ISL_546573, EPI_ISL_546574, EPI_ISL_546575, EPI_ISL_546576, EPI_ISL_546577, EPI_ISL_546578, EPI_ISL_546579, EPI_ISL_546580, EPI_ISL_546581, EPI_ISL_546582, EPI_ISL_546583, EPI_ISL_546584, EPI_ISL_546585, EPI_ISL_546586, EPI_ISL_546587, EPI_ISL_546588, EPI_ISL_546589, EPI_ISL_546590, EPI_ISL_546591, EPI_ISL_546592, EPI_ISL_546593, EPI_ISL_546594, EPI_ISL_546595, EPI_ISL_546596, EPI_ISL_546597, EPI_ISL_546598, EPI_ISL_546599, EPI_ISL_546600, EPI_ISL_546601, EPI_ISL_546602, EPI_ISL_546603, EPI_ISL_546604, EPI_ISL_546605, EPI_ISL_546606, EPI_ISL_546607, EPI_ISL_546608, EPI_ISL_546609, EPI_ISL_546610, EPI_ISL_546611, EPI_ISL_546612, EPI_ISL_546613, EPI_ISL_546614, EPI_ISL_546615, EPI_ISL_546616, EPI_ISL_546617, EPI_ISL_546618, EPI_ISL_546619, EPI_ISL_546620, EPI_ISL_546621, EPI_ISL_546622, EPI_ISL_546623, EPI_ISL_546624, EPI_ISL_546625, EPI_ISL_546626, EPI_ISL_546627, EPI_ISL_546628, EPI_ISL_546629, EPI_ISL_546630, EPI_ISL_546631, EPI_ISL_546632, EPI_ISL_546633, EPI_ISL_546634, EPI_ISL_546635, EPI_ISL_546636, EPI_ISL_546637, EPI_ISL_546638, EPI_ISL_546639, EPI_ISL_546640, EPI_ISL_546641, EPI_ISL_546642, EPI_ISL_546643, EPI_ISL_546644, EPI_ISL_546645, EPI_ISL_546646, EPI_ISL_546647, EPI_ISL_546648, EPI_ISL_546649, EPI_ISL_546650, EPI_ISL_546651, EPI_ISL_546652, EPI_ISL_546653, EPI_ISL_546654, EPI_ISL_546655, EPI_ISL_546656, EPI_ISL_546657, EPI_ISL_546658, EPI_ISL_546659, EPI_ISL_546660, EPI_ISL_546661, EPI_ISL_546662, EPI_ISL_546663, EPI_ISL_546664, EPI_ISL_546665, EPI_ISL_546666, EPI_ISL_546667, EPI_ISL_546668, EPI_ISL_546669, EPI_ISL_546670, EPI_ISL_546671, EPI_ISL_546672, EPI_ISL_546673, EPI_ISL_546674, EPI_ISL_546675, EPI_ISL_546676, EPI_ISL_546677, EPI_ISL_546678, EPI_ISL_546679, EPI_ISL_546680, EPI_ISL_546681, EPI_ISL_546682, EPI_ISL_546683, EPI_ISL_546684, EPI_ISL_546685, EPI_ISL_546686, EPI_ISL_546687, EPI_ISL_546688, EPI_ISL_546689, EPI_ISL_546690, EPI_ISL_546691, EPI_ISL_546692, EPI_ISL_546693, EPI_ISL_546694, EPI_ISL_546695, EPI_ISL_546696, EPI_ISL_546697, EPI_ISL_546698, EPI_ISL_546699, EPI_ISL_546700, EPI_ISL_546701, EPI_ISL_546702, EPI_ISL_546703, EPI_ISL_546704, EPI_ISL_546705, EPI_ISL_546706, EPI_ISL_546707, EPI_ISL_546708, EPI_ISL_546709, EPI_ISL_546710, EPI_ISL_546711, EPI_ISL_546712, EPI_ISL_546713, EPI_ISL_546714, EPI_ISL_546715, EPI_ISL_546716, EPI_ISL_546717, EPI_ISL_546718, EPI_ISL_546719, EPI_ISL_546720, EPI_ISL_546721, EPI_ISL_546722, EPI_ISL_546723, EPI_ISL_546724, EPI_ISL_546725, EPI_ISL_546726, EPI_ISL_546727, EPI_ISL_546728, EPI_ISL_546729, EPI_ISL_546730, EPI_ISL_546731, EPI_ISL_546732, EPI_ISL_546733, EPI_ISL_546734, EPI_ISL_546735, EPI_ISL_546736, EPI_ISL_546737, EPI_ISL_546738, EPI_ISL_546739, EPI_ISL_546740, EPI_ISL_546741, EPI_ISL_546742, EPI_ISL_546743, EPI_ISL_546744, EPI_ISL_546745, EPI_ISL_546746, EPI_ISL_546747, EPI_ISL_546748, EPI_ISL_546749, EPI_ISL_546750, EPI_ISL_546751, EPI_ISL_546752, EPI_ISL_546753, EPI_ISL_546754, EPI_ISL_546755, EPI_ISL_546756, EPI_ISL_546757, EPI_ISL_546758, EPI_ISL_546759, EPI_ISL_546760, EPI_ISL_546761, EPI_ISL_546762, EPI_ISL_546763, EPI_ISL_546764, EPI_ISL_546765, EPI_ISL_546766, EPI_ISL_546767, EPI_ISL_546768, EPI_ISL_546769, EPI_ISL_546770, EPI_ISL_546771, EPI_ISL_546772, EPI_ISL_546773, EPI_ISL_546774, EPI_ISL_546775, EPI_ISL_546776, EPI_ISL_546777, EPI_ISL_546778, EPI_ISL_546779, EPI_ISL_546780, EPI_ISL_546781, EPI_ISL_546782, EPI_ISL_546783, EPI_ISL_546784, EPI_ISL_546785, EPI_ISL_546786, EPI_ISL_546787, EPI_ISL_546788, EPI_ISL_546789, EPI_ISL_546790, EPI_ISL_546791, EPI_ISL_546792, EPI_ISL_546793, EPI_ISL_546794, EPI_ISL_546795, EPI_ISL_546796, EPI_ISL_546797, EPI_ISL_546798, EPI_ISL_546799, EPI_ISL_546800, EPI_ISL_546801, EPI_ISL_546802, EPI_ISL_546803, EPI_ISL_546804, EPI_ISL_546805, EPI_ISL_546806, EPI_ISL_546807, EPI_ISL_546808, EPI_ISL_546809, EPI_ISL_546810, EPI_ISL_546811, EPI_ISL_546812, EPI_ISL_546813, EPI_ISL_546814, EPI_ISL_546815, EPI_ISL_546816, EPI_ISL_546817, EPI_ISL_546818, EPI_ISL_546819, EPI_ISL_546820, EPI_ISL_546821, EPI_ISL_546822, EPI_ISL_546823, EPI_ISL_546824, EPI_ISL_546825, EPI_ISL_546826, EPI_ISL_546827, EPI_ISL_546828, EPI_ISL_546829, EPI_ISL_546830, EPI_ISL_546831, EPI_ISL_546832, EPI_ISL_546833, EPI_ISL_546834, EPI_ISL_546835, EPI_ISL_546836, EPI_ISL_546837, EPI_ISL_546838, EPI_ISL_546839, EPI_ISL_546840, EPI_ISL_546841, EPI_ISL_546842, EPI_ISL_546843, EPI_ISL_546844, EPI_ISL_546845, EPI_ISL_546846, EPI_ISL_546847, EPI_ISL_546848, EPI_ISL_546849, EPI_ISL_546850, EPI_ISL_546851, EPI_ISL_546852, EPI_ISL_546853, EPI_ISL_546854, EPI_ISL_546855, EPI_ISL_546856, EPI_ISL_546857, EPI_ISL_546858, EPI_ISL_546859, EPI_ISL_546860, EPI_ISL_546861, EPI_ISL_546862, EPI_ISL_546863, EPI_ISL_546864, EPI_ISL_546865, EPI_ISL_546866, EPI_ISL_546867, EPI_ISL_546868, EPI_ISL_546869, EPI_ISL_546870, EPI_ISL_546871, EPI_ISL_546872, EPI_ISL_546873, EPI_ISL_546874, EPI_ISL_546875, EPI_ISL_546876, EPI_ISL_546877, EPI_ISL_546878, EPI_ISL_546879, EPI_ISL_546880, EPI_ISL_546881, EPI_ISL_546882, EPI_ISL_546883, EPI_ISL_546884, EPI_ISL_546885, EPI_ISL_546886, EPI_ISL_546887, EPI_ISL_546888, EPI_ISL_546889, EPI_ISL_546890, EPI_ISL_546891, EPI_ISL_546892, EPI_ISL_546893, EPI_ISL_546894, EPI_ISL_546895, EPI_ISL_546896, EPI_ISL_546897, EPI_ISL_546898, EPI_ISL_546899, EPI_ISL_546900, EPI_ISL_546901, EPI_ISL_546902, EPI_ISL_546903, EPI_ISL_546904, EPI_ISL_546905, EPI_ISL_546906, EPI_ISL_546907, EPI_ISL_546908, EPI_ISL_546909, EPI_ISL_546910, EPI_ISL_546911, EPI_ISL_546912, EPI_ISL_546913, EPI_ISL_546914, EPI_ISL_546915, EPI_ISL_546916, EPI_ISL_546917, EPI_ISL_546918, EPI_ISL_546919, EPI_ISL_546920, EPI_ISL_546921, EPI_ISL_546922, EPI_ISL_546923, EPI_ISL_546924, EPI_ISL_546925, EPI_ISL_546926, EPI_ISL_546927, EPI_ISL_546928, EPI_ISL_546929, EPI_ISL_546930, EPI_ISL_546931, EPI_ISL_546932, EPI_ISL_546933, EPI_ISL_546934, EPI_ISL_546937, EPI_ISL_546938, EPI_ISL_546939, EPI_ISL_546940, EPI_ISL_546941, EPI_ISL_546942, EPI_ISL_546943, EPI_ISL_546944, EPI_ISL_546945, EPI_ISL_546946, EPI_ISL_546947, EPI_ISL_546948, EPI_ISL_546949, EPI_ISL_546950, EPI_ISL_546951, EPI_ISL_546952, EPI_ISL_546953, EPI_ISL_546954, EPI_ISL_546955, EPI_ISL_546956, EPI_ISL_546957, EPI_ISL_546958, EPI_ISL_546959, EPI_ISL_546960, EPI_ISL_546961, EPI_ISL_546962, EPI_ISL_546963, EPI_ISL_546964, EPI_ISL_546965, EPI_ISL_546966, EPI_ISL_546967, EPI_ISL_546968, EPI_ISL_546969, EPI_ISL_546970, EPI_ISL_546971, EPI_ISL_546972, EPI_ISL_546973, EPI_ISL_546974, EPI_ISL_546975, EPI_ISL_546976, EPI_ISL_546977, EPI_ISL_546978, EPI_ISL_546979, EPI_ISL_546980, EPI_ISL_546981, EPI_ISL_546982, EPI_ISL_546983, EPI_ISL_546984, EPI_ISL_546985, EPI_ISL_546986, EPI_ISL_546987, EPI_ISL_546988, EPI_ISL_546989, EPI_ISL_546990, EPI_ISL_546991, EPI_ISL_546992, EPI_ISL_546993, EPI_ISL_546994, EPI_ISL_546995, EPI_ISL_546996, EPI_ISL_546997, EPI_ISL_546998, EPI_ISL_546999, EPI_ISL_547000, EPI_ISL_547001, EPI_ISL_547002, EPI_ISL_547003, EPI_ISL_547004, EPI_ISL_547005, EPI_ISL_547006, EPI_ISL_547007, EPI_ISL_547008, EPI_ISL_547009, EPI_ISL_547010, EPI_ISL_547011, EPI_ISL_547012, EPI_ISL_547013, EPI_ISL_547014, EPI_ISL_547015, EPI_ISL_547016, EPI_ISL_547017, EPI_ISL_547018, EPI_ISL_547019, EPI_ISL_547020, EPI_ISL_547021, EPI_ISL_547022, EPI_ISL_547023, EPI_ISL_547024, EPI_ISL_547025, EPI_ISL_547026, EPI_ISL_547027, EPI_ISL_547028, EPI_ISL_547029, EPI_ISL_547030, EPI_ISL_547031, EPI_ISL_547032, EPI_ISL_547033, EPI_ISL_547034, EPI_ISL_547035, EPI_ISL_547036, EPI_ISL_547037, EPI_ISL_547038, EPI_ISL_547039, EPI_ISL_547040, EPI_ISL_547041, EPI_ISL_547042, EPI_ISL_547043, EPI_ISL_547044, EPI_ISL_547045, EPI_ISL_547046, EPI_ISL_547047, EPI_ISL_547048, EPI_ISL_547049, EPI_ISL_547050, EPI_ISL_547051, EPI_ISL_547052, EPI_ISL_547053, EPI_ISL_547054, EPI_ISL_547055, EPI_ISL_547056, EPI_ISL_547057, EPI_ISL_547058, EPI_ISL_547059, EPI_ISL_547060, EPI_ISL_547061, EPI_ISL_547062, EPI_ISL_547063, EPI_ISL_547064, EPI_ISL_547065, EPI_ISL_547066, EPI_ISL_547067, EPI_ISL_547068, EPI_ISL_547069, EPI_ISL_547070, EPI_ISL_547071, EPI_ISL_547072, EPI_ISL_547073, EPI_ISL_547074, EPI_ISL_547075, EPI_ISL_547076, EPI_ISL_547077, EPI_ISL_547078, EPI_ISL_547079, EPI_ISL_547080, EPI_ISL_547081, EPI_ISL_547082, EPI_ISL_547083, EPI_ISL_547084, EPI_ISL_547085, EPI_ISL_547086, EPI_ISL_547087, EPI_ISL_547088, EPI_ISL_547089, EPI_ISL_547090, EPI_ISL_547091, EPI_ISL_547092, EPI_ISL_547093, EPI_ISL_547094, EPI_ISL_547095, EPI_ISL_547096, EPI_ISL_547097, EPI_ISL_547098, EPI_ISL_547099, EPI_ISL_547100, EPI_ISL_547101, EPI_ISL_547102, EPI_ISL_547103, EPI_ISL_547104, EPI_ISL_547105, EPI_ISL_547106, EPI_ISL_547107, EPI_ISL_547108, EPI_ISL_547109, EPI_ISL_547110, EPI_ISL_547111, EPI_ISL_547112, EPI_ISL_547113, EPI_ISL_547114, EPI_ISL_547115, EPI_ISL_547116, EPI_ISL_547117, EPI_ISL_547118, EPI_ISL_547119, EPI_ISL_547120, EPI_ISL_547121, EPI_ISL_547122, EPI_ISL_547123, EPI_ISL_547124, EPI_ISL_547125, EPI_ISL_547126, EPI_ISL_547127, EPI_ISL_547128, EPI_ISL_547129, EPI_ISL_547130, EPI_ISL_547131, EPI_ISL_547132, EPI_ISL_547133, EPI_ISL_547134, EPI_ISL_547135, EPI_ISL_547136, EPI_ISL_547137, EPI_ISL_547138, EPI_ISL_547139, EPI_ISL_547140, EPI_ISL_547141, EPI_ISL_547142, EPI_ISL_547143, EPI_ISL_547144, EPI_ISL_547145, EPI_ISL_547146, EPI_ISL_547147, EPI_ISL_547148, EPI_ISL_547149, EPI_ISL_547150, EPI_ISL_547151, EPI_ISL_547152, EPI_ISL_547153, EPI_ISL_547154, EPI_ISL_547155, EPI_ISL_547156, EPI_ISL_547157, EPI_ISL_547158, EPI_ISL_547159, EPI_ISL_547160, EPI_ISL_547161, EPI_ISL_547162, EPI_ISL_547163, EPI_ISL_547164, EPI_ISL_547165, EPI_ISL_547166, EPI_ISL_547167, EPI_ISL_547168, EPI_ISL_547169, EPI_ISL_547170, EPI_ISL_547171, EPI_ISL_547172, EPI_ISL_547173, EPI_ISL_547174, EPI_ISL_547175, EPI_ISL_547176, EPI_ISL_547177, EPI_ISL_547178, EPI_ISL_547179, EPI_ISL_547180, EPI_ISL_547181, EPI_ISL_547182, EPI_ISL_547183, EPI_ISL_547184, EPI_ISL_547185, EPI_ISL_547186, EPI_ISL_547187, EPI_ISL_547188, EPI_ISL_547189, EPI_ISL_547190, EPI_ISL_547191, EPI_ISL_547192, EPI_ISL_547193, EPI_ISL_547194, EPI_ISL_547195, EPI_ISL_547196, EPI_ISL_547197, EPI_ISL_547198, EPI_ISL_547199, EPI_ISL_547200, EPI_ISL_547201, EPI_ISL_547202, EPI_ISL_547203, EPI_ISL_547204, EPI_ISL_547205, EPI_ISL_547206, EPI_ISL_547207, EPI_ISL_547208, EPI_ISL_547209, EPI_ISL_547210, EPI_ISL_547211, EPI_ISL_547212, EPI_ISL_547213, EPI_ISL_547214, EPI_ISL_547215, EPI_ISL_547216, EPI_ISL_547217, EPI_ISL_547218, EPI_ISL_547219, EPI_ISL_547220, EPI_ISL_547221, EPI_ISL_547222, EPI_ISL_547223, EPI_ISL_547224, EPI_ISL_547225, EPI_ISL_547226, EPI_ISL_547227, EPI_ISL_547228, EPI_ISL_547229, EPI_ISL_547230, EPI_ISL_547231, EPI_ISL_547232, EPI_ISL_547233, EPI_ISL_547234, EPI_ISL_547235, EPI_ISL_547236, EPI_ISL_547237, EPI_ISL_547238, EPI_ISL_547239, EPI_ISL_547240, EPI_ISL_547241, EPI_ISL_547242, EPI_ISL_547243, EPI_ISL_547244, EPI_ISL_547245, EPI_ISL_547246, EPI_ISL_547247, EPI_ISL_547248, EPI_ISL_547249, EPI_ISL_547250, EPI_ISL_547251, EPI_ISL_547252, EPI_ISL_547253, EPI_ISL_547254, EPI_ISL_547255, EPI_ISL_547256, EPI_ISL_547257, EPI_ISL_547258, EPI_ISL_547259, EPI_ISL_547260, EPI_ISL_547261, EPI_ISL_547262, EPI_ISL_547263, EPI_ISL_547264, EPI_ISL_547265, EPI_ISL_547266, EPI_ISL_547267, EPI_ISL_547268, EPI_ISL_547269, EPI_ISL_547270, EPI_ISL_547271, EPI_ISL_547272, EPI_ISL_547273, EPI_ISL_547274, EPI_ISL_547275, EPI_ISL_547276, EPI_ISL_547277, EPI_ISL_547278, EPI_ISL_547279, EPI_ISL_547280, EPI_ISL_547281, EPI_ISL_547282, EPI_ISL_547283, EPI_ISL_547284, EPI_ISL_547285, EPI_ISL_547286, EPI_ISL_547287, EPI_ISL_547288, EPI_ISL_547289, EPI_ISL_547290, EPI_ISL_547291, EPI_ISL_547292, EPI_ISL_547293, EPI_ISL_547294, EPI_ISL_547295, EPI_ISL_547296, EPI_ISL_547297, EPI_ISL_547298, EPI_ISL_547299, EPI_ISL_547300, EPI_ISL_547301, EPI_ISL_547302, EPI_ISL_547303, EPI_ISL_547304, EPI_ISL_547305, EPI_ISL_547306, EPI_ISL_547307, EPI_ISL_547308, EPI_ISL_547309, EPI_ISL_547310, EPI_ISL_547311, EPI_ISL_547312, EPI_ISL_547313, EPI_ISL_547314, EPI_ISL_547315, EPI_ISL_547316, EPI_ISL_547317, EPI_ISL_547318, EPI_ISL_547319, EPI_ISL_547320, EPI_ISL_547321, EPI_ISL_547322, EPI_ISL_547323, EPI_ISL_547324, EPI_ISL_547325, EPI_ISL_547326, EPI_ISL_547327, EPI_ISL_547328, EPI_ISL_547329, EPI_ISL_547330, EPI_ISL_547331, EPI_ISL_547332, EPI_ISL_547333, EPI_ISL_547334, EPI_ISL_547335, EPI_ISL_547336, EPI_ISL_547337, EPI_ISL_547338, EPI_ISL_547339, EPI_ISL_547340, EPI_ISL_547341, EPI_ISL_547342, EPI_ISL_547343, EPI_ISL_547344, EPI_ISL_547345, EPI_ISL_547346, EPI_ISL_547347, EPI_ISL_547348, EPI_ISL_547349, EPI_ISL_547350, EPI_ISL_547351, EPI_ISL_547352, EPI_ISL_547353, EPI_ISL_547354, EPI_ISL_547355, EPI_ISL_547356, EPI_ISL_547357, EPI_ISL_547358, EPI_ISL_547359, EPI_ISL_547360, EPI_ISL_547361, EPI_ISL_547362, EPI_ISL_547363, EPI_ISL_547364, EPI_ISL_547365, EPI_ISL_547366, EPI_ISL_547367, EPI_ISL_547368, EPI_ISL_547369, EPI_ISL_547370, EPI_ISL_547371, EPI_ISL_547372, EPI_ISL_547373, EPI_ISL_547374, EPI_ISL_547375, EPI_ISL_547376, EPI_ISL_547377, EPI_ISL_547378, EPI_ISL_547379, EPI_ISL_547380, EPI_ISL_547381, EPI_ISL_547382, EPI_ISL_547383, EPI_ISL_547384, EPI_ISL_547385, EPI_ISL_547386, EPI_ISL_547387, EPI_ISL_547388, EPI_ISL_547389, EPI_ISL_547390, EPI_ISL_547391, EPI_ISL_547392, EPI_ISL_547393, EPI_ISL_547394, EPI_ISL_547395, EPI_ISL_547396, EPI_ISL_547397, EPI_ISL_547398, EPI_ISL_547399, EPI_ISL_547400, EPI_ISL_547401, EPI_ISL_547402, EPI_ISL_547403, EPI_ISL_547404, EPI_ISL_547405, EPI_ISL_547406, EPI_ISL_547407, EPI_ISL_547408, EPI_ISL_547409, EPI_ISL_547410, EPI_ISL_547411, EPI_ISL_547412, EPI_ISL_547413, EPI_ISL_547414, EPI_ISL_547415, EPI_ISL_547416, EPI_ISL_547417, EPI_ISL_547418, EPI_ISL_547419, EPI_ISL_547420, EPI_ISL_547421, EPI_ISL_547422, EPI_ISL_547423, EPI_ISL_547424, EPI_ISL_547425, EPI_ISL_547426, EPI_ISL_547427, EPI_ISL_547428, EPI_ISL_547429, EPI_ISL_547430, EPI_ISL_547431, EPI_ISL_547432 | | | |
| see above | Houston Methodist Hospital | Houston Methodist Hospital | S. Wesley Long, Randall J. Olsen, Paul A. Christensen, David W. Bernard, James J. Davis, Maulik Shukla, Marcus Nguyen, Matthew Ojeda Saavedra, Concepcion C. Cantu, Prasanti Yerramilli, Layne Pruitt, Sishir Subedi, Hung-Che Kuo, Heather Hendrickson, Ghazaleh Eskandari, Hoang A. T. Nguyen, J. Hunter Long, Muthiah Kumaraswami, Jule Goike, Daniel Boutz, Jimmy Gollihar, Jason S. McLellan, Chia-Wei Chou, Kamyab Javanmardi, Ilya J. Finkelstein, and James M. Musser |
| EPI_ISL_547581, EPI_ISL_547582, EPI_ISL_547583 | University of Wisconsin-Madison AIDS Vaccine Research Laboratories | University of Wisconsin-Madison AIDS Vaccine Research Laboratories | Gage Moreno, Katarina Braun, et al. AIDS Vaccine Research Laboratories |
| EPI_ISL_547602, EPI_ISL_547603, EPI_ISL_547604, | Gundersen Molecular Diagnostics Laboratory | Kabara Cancer Research Institute | Craig S. Richmond, Paraic A. Kenny |

| | | | |
|---|---|---|---|
| EPI_ISL_547605, EPI_ISL_547606, EPI_ISL_547607 | | | |
| EPI_ISL_547608, EPI_ISL_547609, EPI_ISL_547610 | Gundersen Clinical Microbiology Laboratory | Kabara Cancer Research Institute | Craig S. Richmond, Paraic A. Kenny |
| EPI_ISL_547611, EPI_ISL_547612, EPI_ISL_547613, EPI_ISL_547614, EPI_ISL_547615, EPI_ISL_547616, EPI_ISL_547617, EPI_ISL_547618, EPI_ISL_547619, EPI_ISL_547620, EPI_ISL_547621, EPI_ISL_547622, EPI_ISL_547623, EPI_ISL_547624, EPI_ISL_547625, EPI_ISL_547626, EPI_ISL_547627, EPI_ISL_547628, EPI_ISL_547629, EPI_ISL_547630, EPI_ISL_547631, EPI_ISL_547632, EPI_ISL_547633, EPI_ISL_547634, EPI_ISL_547635, EPI_ISL_547636, EPI_ISL_547637, EPI_ISL_547638, EPI_ISL_547639, EPI_ISL_547640, EPI_ISL_547641, EPI_ISL_547642, EPI_ISL_547643, EPI_ISL_547644, EPI_ISL_547645, EPI_ISL_547646, EPI_ISL_547647, EPI_ISL_547648, EPI_ISL_547649, EPI_ISL_547650, EPI_ISL_547651, EPI_ISL_547652, EPI_ISL_547653, EPI_ISL_547654, EPI_ISL_547655, EPI_ISL_547656, EPI_ISL_547657, EPI_ISL_547658, EPI_ISL_547659, EPI_ISL_547660, EPI_ISL_547661, EPI_ISL_547662, EPI_ISL_547663, EPI_ISL_547664, EPI_ISL_547665 | | | |
| see above | Gundersen Molecular Diagnostics Laboratory | Kabara Cancer Research Institute | Craig S. Richmond, Paraic A. Kenny |
| EPI_ISL_547666, EPI_ISL_547667, EPI_ISL_547668, EPI_ISL_547669, EPI_ISL_547670, EPI_ISL_547671, EPI_ISL_547672, EPI_ISL_547673, EPI_ISL_547674, EPI_ISL_547675, EPI_ISL_547676 | | | |
| see above | Gundersen Clinical Microbiology Laboratory | Kabara Cancer Research Institute | Craig S. Richmond, Paraic A. Kenny |
| EPI_ISL_547677, EPI_ISL_547678, EPI_ISL_547679, EPI_ISL_547680, EPI_ISL_547681, EPI_ISL_547682 | Gundersen Molecular Diagnostics Laboratory | Kabara Cancer Research Institute | Craig S. Richmond, Paraic A. Kenny |
| EPI_ISL_547683, EPI_ISL_547684 | Gundersen Clinical Microbiology Laboratory | Kabara Cancer Research Institute | Craig S. Richmond, Paraic A. Kenny |
| EPI_ISL_547685, EPI_ISL_547686, EPI_ISL_547687, EPI_ISL_547688, EPI_ISL_547689, EPI_ISL_547690, EPI_ISL_547691, EPI_ISL_547692, EPI_ISL_547693, EPI_ISL_547694, EPI_ISL_547695, EPI_ISL_547696, EPI_ISL_547697, EPI_ISL_547698, EPI_ISL_547699, EPI_ISL_547700, EPI_ISL_547701, EPI_ISL_547702, EPI_ISL_547703, EPI_ISL_547704 | | | |
| see above | Gundersen Molecular Diagnostics Laboratory | Kabara Cancer Research Institute | Craig S. Richmond, Paraic A. Kenny |
| EPI_ISL_547705 | Gundersen Clinical Microbiology Laboratory | Kabara Cancer Research Institute | Craig S. Richmond, Paraic A. Kenny |
| EPI_ISL_547706, EPI_ISL_547707, EPI_ISL_547708, EPI_ISL_547709, EPI_ISL_547710, EPI_ISL_547711, EPI_ISL_547712, EPI_ISL_547713, EPI_ISL_547714, EPI_ISL_547715, EPI_ISL_547716, EPI_ISL_547717, EPI_ISL_547718, EPI_ISL_547719, EPI_ISL_547720, EPI_ISL_547721, EPI_ISL_547722, EPI_ISL_547723, EPI_ISL_547724, EPI_ISL_547725, EPI_ISL_547726, EPI_ISL_547727, EPI_ISL_547728, EPI_ISL_547729, EPI_ISL_547730, EPI_ISL_547731, EPI_ISL_547732, EPI_ISL_547733, EPI_ISL_547734, EPI_ISL_547735, EPI_ISL_547736, EPI_ISL_547737, EPI_ISL_547738, EPI_ISL_547739, EPI_ISL_547740, EPI_ISL_547741, EPI_ISL_547742, EPI_ISL_547743, EPI_ISL_547744, EPI_ISL_547745, EPI_ISL_547746, EPI_ISL_547747, EPI_ISL_547748, EPI_ISL_547749, EPI_ISL_547750, EPI_ISL_547751, EPI_ISL_547752, EPI_ISL_547753, EPI_ISL_547754, EPI_ISL_547755, EPI_ISL_547756, EPI_ISL_547757, EPI_ISL_547758, EPI_ISL_547759, EPI_ISL_547760, EPI_ISL_547761, EPI_ISL_547762, EPI_ISL_547763, EPI_ISL_547764, EPI_ISL_547765, EPI_ISL_547766, EPI_ISL_547767, EPI_ISL_547768, EPI_ISL_547769, EPI_ISL_547770, EPI_ISL_547771, EPI_ISL_547772, EPI_ISL_547773, EPI_ISL_547774, EPI_ISL_547775, EPI_ISL_547776, EPI_ISL_547777, EPI_ISL_547778, EPI_ISL_547779, EPI_ISL_547780, EPI_ISL_547781, EPI_ISL_547782, EPI_ISL_547783, EPI_ISL_547784, EPI_ISL_547785, EPI_ISL_547786, EPI_ISL_547787, EPI_ISL_547788, EPI_ISL_547789, EPI_ISL_547790, EPI_ISL_547791, EPI_ISL_547792, EPI_ISL_547793, EPI_ISL_547794, EPI_ISL_547795, EPI_ISL_547796, EPI_ISL_547797, EPI_ISL_547798, EPI_ISL_547799, EPI_ISL_547800, EPI_ISL_547801, EPI_ISL_547802, EPI_ISL_547803, EPI_ISL_547804, EPI_ISL_547805, EPI_ISL_547806, EPI_ISL_547807, EPI_ISL_547808, EPI_ISL_547809, EPI_ISL_547810, EPI_ISL_547811, EPI_ISL_547812, EPI_ISL_547813, EPI_ISL_547814, EPI_ISL_547815, EPI_ISL_547816, EPI_ISL_547817, EPI_ISL_547818, EPI_ISL_547819, EPI_ISL_547820, EPI_ISL_547821, EPI_ISL_547822, EPI_ISL_547823, EPI_ISL_547824, EPI_ISL_547825, EPI_ISL_547826, EPI_ISL_547827, EPI_ISL_547828, EPI_ISL_547829, EPI_ISL_547830, EPI_ISL_547831, EPI_ISL_547832, EPI_ISL_547833, EPI_ISL_547834, EPI_ISL_547835, EPI_ISL_547836, EPI_ISL_547837, EPI_ISL_547838, EPI_ISL_547839, EPI_ISL_547840, EPI_ISL_547841, EPI_ISL_547842, EPI_ISL_547843, EPI_ISL_547844, EPI_ISL_547845, EPI_ISL_547846, EPI_ISL_547847, EPI_ISL_547848, EPI_ISL_547849, EPI_ISL_547850, EPI_ISL_547851, EPI_ISL_547852, EPI_ISL_547853, EPI_ISL_547854, EPI_ISL_547855, EPI_ISL_547856, EPI_ISL_547857, EPI_ISL_547858, EPI_ISL_547859, EPI_ISL_547860, EPI_ISL_547861, EPI_ISL_547862, EPI_ISL_547863, EPI_ISL_547864, EPI_ISL_547865, EPI_ISL_547866, EPI_ISL_547867, EPI_ISL_547868, EPI_ISL_547869 | | | |
| see above | Gundersen Molecular Diagnostics Laboratory | Kabara Cancer Research Institute | Craig S. Richmond, Paraic A. Kenny |
| EPI_ISL_548243, EPI_ISL_548244 | Faith Laboratory, Immunology Institute, Icahn School of Medicine at Mount Sinai | van Bakel Laboratory, Genetics and Genomics Sciences, Icahn School of Medicine at Mount Sinai | Graham J. Britton, Alice Chen-Liaw, Francesca Cossarini, Alexandra Livanos, Matthew P. Spindler, Tamar Plitt, Joseph Eggers, Ilaria Mogno, Ana S. Gonzalez-Reiche, Sophia Sui, Michael Tankelevich, Lauren Tal Grinspan, Rebekah E. Dixon, Divya Jha, Gustavo Martinez-Delgado, Fatima Amanat, Daisy Hoagland, Benjamin R. tenOever, Marla C. Dubinsky, Miriam Merad, Harm Van Bakel, Florian Krammer, Gerold Bongers, Saurabh Mehandru and Jeremiah J. Faith |
| EPI_ISL_548264 | County of Santa Clara Public Health Department | Chan-Zuckerberg Biohub | CZB Cliahub Consortium |
| EPI_ISL_548265 | County of San Luis Obispo Public Health Laboratory | Chan-Zuckerberg Biohub | CZB Cliahub Consortium |
| EPI_ISL_548266, EPI_ISL_548267 | County of Santa Clara Public Health Department | Chan-Zuckerberg Biohub | CZB Cliahub Consortium |
| EPI_ISL_548268, EPI_ISL_548269 | County of San Luis Obispo Public Health Laboratory | Chan-Zuckerberg Biohub | CZB Cliahub Consortium |
| EPI_ISL_548270, EPI_ISL_548271, EPI_ISL_548272, EPI_ISL_548273, EPI_ISL_548274, EPI_ISL_548275, EPI_ISL_548276, EPI_ISL_548278 | Orange County Public Health Laboratory | Chan-Zuckerberg Biohub | CZB Cliahub Consortium |
| EPI_ISL_548279 | County of Santa Clara Public Health Department | Chan-Zuckerberg Biohub | CZB Cliahub Consortium |
| EPI_ISL_548280, EPI_ISL_548281 | County of San Luis Obispo Public Health Laboratory | Chan-Zuckerberg Biohub | CZB Cliahub Consortium |
| EPI_ISL_548282 | County of Santa Clara Public Health Department | Chan-Zuckerberg Biohub | CZB Cliahub Consortium |
| EPI_ISL_548283 | Orange County Public Health Laboratory | Chan-Zuckerberg Biohub | CZB Cliahub Consortium |
| EPI_ISL_548284 | County of San Luis Obispo Public Health Laboratory | Chan-Zuckerberg Biohub | CZB Cliahub Consortium |
| EPI_ISL_548285 | Orange County Public Health Laboratory | Chan-Zuckerberg Biohub | CZB Cliahub Consortium |
| EPI_ISL_548286 | County of San Luis Obispo Public Health Laboratory | Chan-Zuckerberg Biohub | CZB Cliahub Consortium |
| EPI_ISL_548287 | Orange County Public Health Laboratory | Chan-Zuckerberg Biohub | CZB Cliahub Consortium |
| EPI_ISL_548288 | County of San Luis Obispo Public Health Laboratory | Chan-Zuckerberg Biohub | CZB Cliahub Consortium |
| EPI_ISL_548289 | Orange County Public Health Laboratory | Chan-Zuckerberg Biohub | CZB Cliahub Consortium |
| EPI_ISL_548290 | County of Santa Clara Public Health Department | Chan-Zuckerberg Biohub | CZB Cliahub Consortium |
| EPI_ISL_548291 | Orange County Public Health Laboratory | Chan-Zuckerberg Biohub | CZB Cliahub Consortium |
| EPI_ISL_548292 | County of San Luis Obispo Public Health Laboratory | Chan-Zuckerberg Biohub | CZB Cliahub Consortium |
| EPI_ISL_548293 | Orange County Public Health Laboratory | Chan-Zuckerberg Biohub | CZB Cliahub Consortium |
| EPI_ISL_548294, EPI_ISL_548295 | County of San Luis Obispo Public Health Laboratory | Chan-Zuckerberg Biohub | CZB Cliahub Consortium |
| EPI_ISL_548296, EPI_ISL_548297, EPI_ISL_548298 | Orange County Public Health Laboratory | Chan-Zuckerberg Biohub | CZB Cliahub Consortium |
| EPI_ISL_548299 | County of Santa Clara Public Health Department | Chan-Zuckerberg Biohub | CZB Cliahub Consortium |
| EPI_ISL_548300, EPI_ISL_548301 | Orange County Public Health Laboratory | Chan-Zuckerberg Biohub | CZB Cliahub Consortium |
| EPI_ISL_548302, EPI_ISL_548303 | County of Santa Clara Public Health Department | Chan-Zuckerberg Biohub | CZB Cliahub Consortium |
| EPI_ISL_548304 | County of San Luis Obispo Public Health Laboratory | Chan-Zuckerberg Biohub | CZB Cliahub Consortium |
| EPI_ISL_548305, EPI_ISL_548306 | Orange County Public Health Laboratory | Chan-Zuckerberg Biohub | CZB Cliahub Consortium |
| EPI_ISL_548307 | County of Santa Clara Public Health | Chan-Zuckerberg Biohub | CZB Cliahub Consortium |

| | | | |
|---|---|---|---|
| | Department | | |
| EPI_ISL_548308, EPI_ISL_548309 | Orange County Public Health Laboratory | Chan-Zuckerberg Biohub | CZB Cliahub Consortium |
| EPI_ISL_548310 | County of Santa Clara Public Health Department | Chan-Zuckerberg Biohub | CZB Cliahub Consortium |
| EPI_ISL_548311 | County of San Luis Obispo Public Health Laboratory | Chan-Zuckerberg Biohub | CZB Cliahub Consortium |
| EPI_ISL_548312, EPI_ISL_548313 | Orange County Public Health Laboratory | Chan-Zuckerberg Biohub | CZB Cliahub Consortium |
| EPI_ISL_548314 | County of Santa Clara Public Health Department | Chan-Zuckerberg Biohub | CZB Cliahub Consortium |
| EPI_ISL_548315 | Orange County Public Health Laboratory | Chan-Zuckerberg Biohub | CZB Cliahub Consortium |
| EPI_ISL_548316 | County of San Luis Obispo Public Health Laboratory | Chan-Zuckerberg Biohub | CZB Cliahub Consortium |
| EPI_ISL_548317, EPI_ISL_548318 | Orange County Public Health Laboratory | Chan-Zuckerberg Biohub | CZB Cliahub Consortium |
| EPI_ISL_548319, EPI_ISL_548320, EPI_ISL_548321 | County of Santa Clara Public Health Department | Chan-Zuckerberg Biohub | CZB Cliahub Consortium |
| EPI_ISL_548322 | County of San Luis Obispo Public Health Laboratory | Chan-Zuckerberg Biohub | CZB Cliahub Consortium |
| EPI_ISL_548323 | Orange County Public Health Laboratory | Chan-Zuckerberg Biohub | CZB Cliahub Consortium |
| EPI_ISL_548324 | County of Santa Clara Public Health Department | Chan-Zuckerberg Biohub | CZB Cliahub Consortium |
| EPI_ISL_548325 | County of San Luis Obispo Public Health Laboratory | Chan-Zuckerberg Biohub | CZB Cliahub Consortium |
| EPI_ISL_548326 | Orange County Public Health Laboratory | Chan-Zuckerberg Biohub | CZB Cliahub Consortium |
| EPI_ISL_548327 | County of Santa Clara Public Health Department | Chan-Zuckerberg Biohub | CZB Cliahub Consortium |
| EPI_ISL_548328 | Orange County Public Health Laboratory | Chan-Zuckerberg Biohub | CZB Cliahub Consortium |
| EPI_ISL_548329, EPI_ISL_548330 | County of San Luis Obispo Public Health Laboratory | Chan-Zuckerberg Biohub | CZB Cliahub Consortium |
| EPI_ISL_548331 | County of Santa Clara Public Health Department | Chan-Zuckerberg Biohub | CZB Cliahub Consortium |
| EPI_ISL_548332 | Orange County Public Health Laboratory | Chan-Zuckerberg Biohub | CZB Cliahub Consortium |
| EPI_ISL_548333 | County of Santa Clara Public Health Department | Chan-Zuckerberg Biohub | CZB Cliahub Consortium |
| EPI_ISL_548334 | Orange County Public Health Laboratory | Chan-Zuckerberg Biohub | CZB Cliahub Consortium |
| EPI_ISL_548336 | County of San Luis Obispo Public Health Laboratory | Chan-Zuckerberg Biohub | CZB Cliahub Consortium |
| EPI_ISL_548337, EPI_ISL_548338, EPI_ISL_548339, EPI_ISL_548340, EPI_ISL_548341, EPI_ISL_548342, EPI_ISL_548343, EPI_ISL_548344, EPI_ISL_548345 | County of Santa Clara Public Health Department | Chan-Zuckerberg Biohub | CZB Cliahub Consortium |
| EPI_ISL_548346 | Ventura County Public Health Lab | Chan-Zuckerberg Biohub | CZB Cliahub Consortium |
| EPI_ISL_548347 | County of San Luis Obispo Public Health Laboratory | Chan-Zuckerberg Biohub | CZB Cliahub Consortium |
| EPI_ISL_548348, EPI_ISL_548349 | Ventura County Public Health Lab | Chan-Zuckerberg Biohub | CZB Cliahub Consortium |
| EPI_ISL_548350, EPI_ISL_548351 | County of San Luis Obispo Public Health Laboratory | Chan-Zuckerberg Biohub | CZB Cliahub Consortium |
| EPI_ISL_548352, EPI_ISL_548353, EPI_ISL_548354, EPI_ISL_548355, EPI_ISL_548356 | Ventura County Public Health Lab | Chan-Zuckerberg Biohub | CZB Cliahub Consortium |
| EPI_ISL_548357 | County of San Luis Obispo Public Health Laboratory | Chan-Zuckerberg Biohub | CZB Cliahub Consortium |
| EPI_ISL_548358 | Ventura County Public Health Lab | Chan-Zuckerberg Biohub | CZB Cliahub Consortium |
| EPI_ISL_548359, EPI_ISL_548360, EPI_ISL_548361 | County of San Luis Obispo Public Health Laboratory | Chan-Zuckerberg Biohub | CZB Cliahub Consortium |
| EPI_ISL_548362 | Ventura County Public Health Lab | Chan-Zuckerberg Biohub | CZB Cliahub Consortium |
| EPI_ISL_548363 | Tuolumne County Public Health | Chan-Zuckerberg Biohub | CZB Cliahub Consortium |
| EPI_ISL_548364 | Ventura County Public Health Lab | Chan-Zuckerberg Biohub | CZB Cliahub Consortium |
| EPI_ISL_548365, EPI_ISL_548366 | County of San Luis Obispo Public Health Laboratory | Chan-Zuckerberg Biohub | CZB Cliahub Consortium |
| EPI_ISL_548367, EPI_ISL_548368 | Ventura County Public Health Lab | Chan-Zuckerberg Biohub | CZB Cliahub Consortium |
| EPI_ISL_548369, EPI_ISL_548370 | County of San Luis Obispo Public Health Laboratory | Chan-Zuckerberg Biohub | CZB Cliahub Consortium |
| EPI_ISL_548371 | County of Santa Clara Public Health Department | Chan-Zuckerberg Biohub | CZB Cliahub Consortium |
| EPI_ISL_548372 | Ventura County Public Health Lab | Chan-Zuckerberg Biohub | CZB Cliahub Consortium |
| EPI_ISL_548373 | Orange County Public Health Laboratory | Chan-Zuckerberg Biohub | CZB Cliahub Consortium |
| EPI_ISL_548374 | University of California, Davis | Chan-Zuckerberg Biohub | CZB Cliahub Consortium |
| EPI_ISL_548375 | Orange County Public Health Laboratory | Chan-Zuckerberg Biohub | CZB Cliahub Consortium |
| EPI_ISL_548376 | Ventura County Public Health Lab | Chan-Zuckerberg Biohub | CZB Cliahub Consortium |
| EPI_ISL_548377, EPI_ISL_548378 | Orange County Public Health Laboratory | Chan-Zuckerberg Biohub | CZB Cliahub Consortium |
| EPI_ISL_548379 | County of Santa Clara Public Health Department | Chan-Zuckerberg Biohub | CZB Cliahub Consortium |
| EPI_ISL_548380 | University of California, Davis | Chan-Zuckerberg Biohub | CZB Cliahub Consortium |
| EPI_ISL_548381 | Orange County Public Health Laboratory | Chan-Zuckerberg Biohub | CZB Cliahub Consortium |

| | | | |
|---|---|---|---|
| EPI_ISL_548382 | University of California, Davis | Chan-Zuckerberg Biohub | CZB Cliahub Consortium |
| EPI_ISL_548383 | Orange County Public Health Laboratory | Chan-Zuckerberg Biohub | CZB Cliahub Consortium |
| EPI_ISL_548385 | County of Santa Clara Public Health Department | Chan-Zuckerberg Biohub | CZB Cliahub Consortium |
| EPI_ISL_548386, EPI_ISL_548387 | Orange County Public Health Laboratory | Chan-Zuckerberg Biohub | CZB Cliahub Consortium |
| EPI_ISL_548388 | County of Santa Clara Public Health Department | Chan-Zuckerberg Biohub | CZB Cliahub Consortium |
| EPI_ISL_548389 | Orange County Public Health Laboratory | Chan-Zuckerberg Biohub | CZB Cliahub Consortium |
| EPI_ISL_548390 | Ventura County Public Health Lab | Chan-Zuckerberg Biohub | CZB Cliahub Consortium |
| EPI_ISL_548391 | County of San Luis Obispo Public Health Laboratory | Chan-Zuckerberg Biohub | CZB Cliahub Consortium |
| EPI_ISL_548392 | County of Santa Clara Public Health Department | Chan-Zuckerberg Biohub | CZB Cliahub Consortium |
| EPI_ISL_548393 | Orange County Public Health Laboratory | Chan-Zuckerberg Biohub | CZB Cliahub Consortium |
| EPI_ISL_548394, EPI_ISL_548395, EPI_ISL_548396 | County of Santa Clara Public Health Department | Chan-Zuckerberg Biohub | CZB Cliahub Consortium |
| EPI_ISL_548398 | Ventura County Public Health Lab | Chan-Zuckerberg Biohub | CZB Cliahub Consortium |
| EPI_ISL_548400 | Orange County Public Health Laboratory | Chan-Zuckerberg Biohub | CZB Cliahub Consortium |
| EPI_ISL_548401 | County of Santa Clara Public Health Department | Chan-Zuckerberg Biohub | CZB Cliahub Consortium |
| EPI_ISL_548402 | Ventura County Public Health Lab | Chan-Zuckerberg Biohub | CZB Cliahub Consortium |
| EPI_ISL_548403 | County of Santa Clara Public Health Department | Chan-Zuckerberg Biohub | CZB Cliahub Consortium |
| EPI_ISL_548404, EPI_ISL_548405 | Orange County Public Health Laboratory | Chan-Zuckerberg Biohub | CZB Cliahub Consortium |
| EPI_ISL_548406 | Ventura County Public Health Lab | Chan-Zuckerberg Biohub | CZB Cliahub Consortium |
| EPI_ISL_548407 | County of Santa Clara Public Health Department | Chan-Zuckerberg Biohub | CZB Cliahub Consortium |
| EPI_ISL_548408 | Orange County Public Health Laboratory | Chan-Zuckerberg Biohub | CZB Cliahub Consortium |
| EPI_ISL_548409, EPI_ISL_548410 | Ventura County Public Health Lab | Chan-Zuckerberg Biohub | CZB Cliahub Consortium |
| EPI_ISL_548411, EPI_ISL_548412, EPI_ISL_548413 | Orange County Public Health Laboratory | Chan-Zuckerberg Biohub | CZB Cliahub Consortium |
| EPI_ISL_548414 | University of California, Davis | Chan-Zuckerberg Biohub | CZB Cliahub Consortium |
| EPI_ISL_548415 | County of Santa Clara Public Health Department | Chan-Zuckerberg Biohub | CZB Cliahub Consortium |
| EPI_ISL_548416 | Orange County Public Health Laboratory | Chan-Zuckerberg Biohub | CZB Cliahub Consortium |
| EPI_ISL_548417 | County of Santa Clara Public Health Department | Chan-Zuckerberg Biohub | CZB Cliahub Consortium |
| EPI_ISL_548418 | Orange County Public Health Laboratory | Chan-Zuckerberg Biohub | CZB Cliahub Consortium |
| EPI_ISL_548419, EPI_ISL_548420, EPI_ISL_548421, EPI_ISL_548422, EPI_ISL_548423 | County of Santa Clara Public Health Department | Chan-Zuckerberg Biohub | CZB Cliahub Consortium |
| EPI_ISL_548424 | Orange County Public Health Laboratory | Chan-Zuckerberg Biohub | CZB Cliahub Consortium |
| EPI_ISL_548425 | County of San Luis Obispo Public Health Laboratory | Chan-Zuckerberg Biohub | CZB Cliahub Consortium |
| EPI_ISL_548426 | County of Santa Clara Public Health Department | Chan-Zuckerberg Biohub | CZB Cliahub Consortium |
| EPI_ISL_548427 | Ventura County Public Health Lab | Chan-Zuckerberg Biohub | CZB Cliahub Consortium |
| EPI_ISL_548428 | County of San Luis Obispo Public Health Laboratory | Chan-Zuckerberg Biohub | CZB Cliahub Consortium |
| EPI_ISL_548429 | County of Santa Clara Public Health Department | Chan-Zuckerberg Biohub | CZB Cliahub Consortium |
| EPI_ISL_548430 | Ventura County Public Health Lab | Chan-Zuckerberg Biohub | CZB Cliahub Consortium |
| EPI_ISL_548431 | Orange County Public Health Laboratory | Chan-Zuckerberg Biohub | CZB Cliahub Consortium |
| EPI_ISL_548432, EPI_ISL_548433, EPI_ISL_548434 | County of Santa Clara Public Health Department | Chan-Zuckerberg Biohub | CZB Cliahub Consortium |
| EPI_ISL_548435 | University of California, Davis | Chan-Zuckerberg Biohub | CZB Cliahub Consortium |
| EPI_ISL_548436 | County of San Luis Obispo Public Health Laboratory | Chan-Zuckerberg Biohub | CZB Cliahub Consortium |
| EPI_ISL_548437 | County of Santa Clara Public Health Department | Chan-Zuckerberg Biohub | CZB Cliahub Consortium |
| EPI_ISL_548438, EPI_ISL_548439, EPI_ISL_548440, EPI_ISL_548441 | Orange County Public Health Laboratory | Chan-Zuckerberg Biohub | CZB Cliahub Consortium |
| EPI_ISL_548442, EPI_ISL_548443 | Ventura County Public Health Lab | Chan-Zuckerberg Biohub | CZB Cliahub Consortium |
| EPI_ISL_548444, EPI_ISL_548445 | Orange County Public Health Laboratory | Chan-Zuckerberg Biohub | CZB Cliahub Consortium |
| EPI_ISL_548446 | County of Santa Clara Public Health Department | Chan-Zuckerberg Biohub | CZB Cliahub Consortium |
| EPI_ISL_548447 | University of California, Davis | Chan-Zuckerberg Biohub | CZB Cliahub Consortium |
| EPI_ISL_548448 | County of Santa Clara Public Health Department | Chan-Zuckerberg Biohub | CZB Cliahub Consortium |
| EPI_ISL_548449 | Orange County Public Health Laboratory | Chan-Zuckerberg Biohub | CZB Cliahub Consortium |
| EPI_ISL_548450 | Ventura County Public Health Lab | Chan-Zuckerberg Biohub | CZB Cliahub Consortium |
| EPI_ISL_548451 | County of Santa Clara Public Health Department | Chan-Zuckerberg Biohub | CZB Cliahub Consortium |
| EPI_ISL_548452 | University of California, Davis | Chan-Zuckerberg Biohub | CZB Cliahub Consortium |
| EPI_ISL_548453 | County of San Luis Obispo Public Health Laboratory | Chan-Zuckerberg Biohub | CZB Cliahub Consortium |

| | | | |
|---|---|---|---|
| EPI_ISL_548454 | Orange County Public Health Laboratory | Chan-Zuckerberg Biohub | CZB Cliahub Consortium |
| EPI_ISL_548455 | Ventura County Public Health Lab | Chan-Zuckerberg Biohub | CZB Cliahub Consortium |
| EPI_ISL_548456 | Orange County Public Health Laboratory | Chan-Zuckerberg Biohub | CZB Cliahub Consortium |
| EPI_ISL_548457 | County of San Luis Obispo Public Health Laboratory | Chan-Zuckerberg Biohub | CZB Cliahub Consortium |
| EPI_ISL_548458 | University of California, Davis | Chan-Zuckerberg Biohub | CZB Cliahub Consortium |
| EPI_ISL_548459 | Ventura County Public Health Lab | Chan-Zuckerberg Biohub | CZB Cliahub Consortium |
| EPI_ISL_548460 | Orange County Public Health Laboratory | Chan-Zuckerberg Biohub | CZB Cliahub Consortium |
| EPI_ISL_548461 | Ventura County Public Health Lab | Chan-Zuckerberg Biohub | CZB Cliahub Consortium |
| EPI_ISL_548462, EPI_ISL_548463, EPI_ISL_548464, EPI_ISL_548465 | Orange County Public Health Laboratory | Chan-Zuckerberg Biohub | CZB Cliahub Consortium |
| EPI_ISL_548466 | County of Santa Clara Public Health Department | Chan-Zuckerberg Biohub | CZB Cliahub Consortium |
| EPI_ISL_548467 | Orange County Public Health Laboratory | Chan-Zuckerberg Biohub | CZB Cliahub Consortium |
| EPI_ISL_548468 | County of Santa Clara Public Health Department | Chan-Zuckerberg Biohub | CZB Cliahub Consortium |
| EPI_ISL_548469 | Orange County Public Health Laboratory | Chan-Zuckerberg Biohub | CZB Cliahub Consortium |
| EPI_ISL_548470 | County of Santa Clara Public Health Department | Chan-Zuckerberg Biohub | CZB Cliahub Consortium |
| EPI_ISL_548471, EPI_ISL_548473 | Ventura County Public Health Lab | Chan-Zuckerberg Biohub | CZB Cliahub Consortium |
| EPI_ISL_548474 | University of California, Davis | Chan-Zuckerberg Biohub | CZB Cliahub Consortium |
| EPI_ISL_548475, EPI_ISL_548476 | Ventura County Public Health Lab | Chan-Zuckerberg Biohub | CZB Cliahub Consortium |
| EPI_ISL_548477 | University of California, Davis | Chan-Zuckerberg Biohub | CZB Cliahub Consortium |
| EPI_ISL_548478 | County of Santa Clara Public Health Department | Chan-Zuckerberg Biohub | CZB Cliahub Consortium |
| EPI_ISL_548479 | University of California, Davis | Chan-Zuckerberg Biohub | CZB Cliahub Consortium |
| EPI_ISL_548480, EPI_ISL_548481 | Ventura County Public Health Lab | Chan-Zuckerberg Biohub | CZB Cliahub Consortium |
| EPI_ISL_548482 | County of Santa Clara Public Health Department | Chan-Zuckerberg Biohub | CZB Cliahub Consortium |
| EPI_ISL_548483 | Ventura County Public Health Lab | Chan-Zuckerberg Biohub | CZB Cliahub Consortium |
| EPI_ISL_548484, EPI_ISL_548485 | Orange County Public Health Laboratory | Chan-Zuckerberg Biohub | CZB Cliahub Consortium |
| EPI_ISL_548486 | University of California, Davis | Chan-Zuckerberg Biohub | CZB Cliahub Consortium |
| EPI_ISL_548487 | Ventura County Public Health Lab | Chan-Zuckerberg Biohub | CZB Cliahub Consortium |
| EPI_ISL_548488 | Orange County Public Health Laboratory | Chan-Zuckerberg Biohub | CZB Cliahub Consortium |
| EPI_ISL_548489 | Ventura County Public Health Lab | Chan-Zuckerberg Biohub | CZB Cliahub Consortium |
| EPI_ISL_548490 | University of California, Davis | Chan-Zuckerberg Biohub | CZB Cliahub Consortium |
| EPI_ISL_548491, EPI_ISL_548492 | Contra Costa Public Health Lab | Chan-Zuckerberg Biohub | CZB Cliahub Consortium |
| EPI_ISL_548493 | County of San Luis Obispo Public Health Laboratory | Chan-Zuckerberg Biohub | CZB Cliahub Consortium |
| EPI_ISL_548494 | County of Santa Clara Public Health Department | Chan-Zuckerberg Biohub | CZB Cliahub Consortium |
| EPI_ISL_548495 | Contra Costa Public Health Lab | Chan-Zuckerberg Biohub | CZB Cliahub Consortium |
| EPI_ISL_548496, EPI_ISL_548497, EPI_ISL_548498 | County of Santa Clara Public Health Department | Chan-Zuckerberg Biohub | CZB Cliahub Consortium |
| EPI_ISL_548499 | Contra Costa Public Health Lab | Chan-Zuckerberg Biohub | CZB Cliahub Consortium |
| EPI_ISL_548500 | County of Santa Clara Public Health Department | Chan-Zuckerberg Biohub | CZB Cliahub Consortium |
| EPI_ISL_548501 | Contra Costa Public Health Lab | Chan-Zuckerberg Biohub | CZB Cliahub Consortium |
| EPI_ISL_548502 | County of San Luis Obispo Public Health Laboratory | Chan-Zuckerberg Biohub | CZB Cliahub Consortium |
| EPI_ISL_548503 | County of Santa Clara Public Health Department | Chan-Zuckerberg Biohub | CZB Cliahub Consortium |
| EPI_ISL_548504 | County of San Luis Obispo Public Health Laboratory | Chan-Zuckerberg Biohub | CZB Cliahub Consortium |
| EPI_ISL_548505 | UCSF Clinical Microbiology Laboratory | Chan-Zuckerberg Biohub | CZB Cliahub Consortium |
| EPI_ISL_548506, EPI_ISL_548507, EPI_ISL_548508 | Contra Costa Public Health Lab | Chan-Zuckerberg Biohub | CZB Cliahub Consortium |
| EPI_ISL_548509 | County of Santa Clara Public Health Department | Chan-Zuckerberg Biohub | CZB Cliahub Consortium |
| EPI_ISL_548510 | Contra Costa Public Health Lab | Chan-Zuckerberg Biohub | CZB Cliahub Consortium |
| EPI_ISL_548511, EPI_ISL_548512 | County of Santa Clara Public Health Department | Chan-Zuckerberg Biohub | CZB Cliahub Consortium |
| EPI_ISL_548513 | UCSF Clinical Microbiology Laboratory | Chan-Zuckerberg Biohub | CZB Cliahub Consortium |
| EPI_ISL_548514 | County of Santa Clara Public Health Department | Chan-Zuckerberg Biohub | CZB Cliahub Consortium |
| EPI_ISL_548515, EPI_ISL_548516 | Contra Costa Public Health Lab | Chan-Zuckerberg Biohub | CZB Cliahub Consortium |
| EPI_ISL_548517 | County of San Luis Obispo Public Health Laboratory | Chan-Zuckerberg Biohub | CZB Cliahub Consortium |
| EPI_ISL_548518 | County of Santa Clara Public Health Department | Chan-Zuckerberg Biohub | CZB Cliahub Consortium |
| EPI_ISL_548519 | County of San Luis Obispo Public Health Laboratory | Chan-Zuckerberg Biohub | CZB Cliahub Consortium |
| EPI_ISL_548520 | Contra Costa Public Health Lab | Chan-Zuckerberg Biohub | CZB Cliahub Consortium |
| EPI_ISL_548521 | County of San Luis Obispo Public Health Laboratory | Chan-Zuckerberg Biohub | CZB Cliahub Consortium |
| EPI_ISL_548522 | UCSF Clinical Microbiology Laboratory | Chan-Zuckerberg Biohub | CZB Cliahub Consortium |

| | | | |
|---|---|---|---|
| EPI_ISL_548523 | County of Santa Clara Public Health Department | Chan-Zuckerberg Biohub | CZB Cliahub Consortium |
| EPI_ISL_548524 | Contra Costa Public Health Lab | Chan-Zuckerberg Biohub | CZB Cliahub Consortium |
| EPI_ISL_548525, EPI_ISL_548526 | County of Santa Clara Public Health Department | Chan-Zuckerberg Biohub | CZB Cliahub Consortium |
| EPI_ISL_548527, EPI_ISL_548528 | Contra Costa Public Health Lab | Chan-Zuckerberg Biohub | CZB Cliahub Consortium |
| EPI_ISL_548529, EPI_ISL_548530 | County of Santa Clara Public Health Department | Chan-Zuckerberg Biohub | CZB Cliahub Consortium |
| EPI_ISL_548531 | Contra Costa Public Health Lab | Chan-Zuckerberg Biohub | CZB Cliahub Consortium |
| EPI_ISL_548532 | County of San Luis Obispo Public Health Laboratory | Chan-Zuckerberg Biohub | CZB Cliahub Consortium |
| EPI_ISL_548533 | County of Santa Clara Public Health Department | Chan-Zuckerberg Biohub | CZB Cliahub Consortium |
| EPI_ISL_548534, EPI_ISL_548535 | Contra Costa Public Health Lab | Chan-Zuckerberg Biohub | CZB Cliahub Consortium |
| EPI_ISL_548536 | County of Santa Clara Public Health Department | Chan-Zuckerberg Biohub | CZB Cliahub Consortium |
| EPI_ISL_548537, EPI_ISL_548538, EPI_ISL_548539, EPI_ISL_548540 | Contra Costa Public Health Lab | Chan-Zuckerberg Biohub | CZB Cliahub Consortium |
| EPI_ISL_548541 | County of Santa Clara Public Health Department | Chan-Zuckerberg Biohub | CZB Cliahub Consortium |
| EPI_ISL_548542 | County of San Luis Obispo Public Health Laboratory | Chan-Zuckerberg Biohub | CZB Cliahub Consortium |
| EPI_ISL_548543 | County of Santa Clara Public Health Department | Chan-Zuckerberg Biohub | CZB Cliahub Consortium |
| EPI_ISL_548544 | County of San Luis Obispo Public Health Laboratory | Chan-Zuckerberg Biohub | CZB Cliahub Consortium |
| EPI_ISL_548545 | Contra Costa Public Health Lab | Chan-Zuckerberg Biohub | CZB Cliahub Consortium |
| EPI_ISL_548546, EPI_ISL_548547 | County of Santa Clara Public Health Department | Chan-Zuckerberg Biohub | CZB Cliahub Consortium |
| EPI_ISL_548548, EPI_ISL_548549 | Contra Costa Public Health Lab | Chan-Zuckerberg Biohub | CZB Cliahub Consortium |
| EPI_ISL_548550, EPI_ISL_548551 | County of Santa Clara Public Health Department | Chan-Zuckerberg Biohub | CZB Cliahub Consortium |
| EPI_ISL_548552 | Contra Costa Public Health Lab | Chan-Zuckerberg Biohub | CZB Cliahub Consortium |
| EPI_ISL_548553 | County of Santa Clara Public Health Department | Chan-Zuckerberg Biohub | CZB Cliahub Consortium |
| EPI_ISL_548554 | Contra Costa Public Health Lab | Chan-Zuckerberg Biohub | CZB Cliahub Consortium |
| EPI_ISL_548555 | UCSF Clinical Microbiology Laboratory | Chan-Zuckerberg Biohub | CZB Cliahub Consortium |
| EPI_ISL_548556, EPI_ISL_548557, EPI_ISL_548558 | Contra Costa Public Health Lab | Chan-Zuckerberg Biohub | CZB Cliahub Consortium |
| EPI_ISL_548559 | County of Santa Clara Public Health Department | Chan-Zuckerberg Biohub | CZB Cliahub Consortium |
| EPI_ISL_548560 | County of San Luis Obispo Public Health Laboratory | Chan-Zuckerberg Biohub | CZB Cliahub Consortium |
| EPI_ISL_548561 | Contra Costa Public Health Lab | Chan-Zuckerberg Biohub | CZB Cliahub Consortium |
| EPI_ISL_548562, EPI_ISL_548563 | County of Santa Clara Public Health Department | Chan-Zuckerberg Biohub | CZB Cliahub Consortium |
| EPI_ISL_548564, EPI_ISL_548565, EPI_ISL_548566, EPI_ISL_548567 | Contra Costa Public Health Lab | Chan-Zuckerberg Biohub | CZB Cliahub Consortium |
| EPI_ISL_548568 | County of Santa Clara Public Health Department | Chan-Zuckerberg Biohub | CZB Cliahub Consortium |
| EPI_ISL_548569 | Contra Costa Public Health Lab | Chan-Zuckerberg Biohub | CZB Cliahub Consortium |
| EPI_ISL_548570 | County of Santa Clara Public Health Department | Chan-Zuckerberg Biohub | CZB Cliahub Consortium |
| EPI_ISL_548571 | UCSF Clinical Microbiology Laboratory | Chan-Zuckerberg Biohub | CZB Cliahub Consortium |
| EPI_ISL_548572, EPI_ISL_548573, EPI_ISL_548574, EPI_ISL_548575 | Contra Costa Public Health Lab | Chan-Zuckerberg Biohub | CZB Cliahub Consortium |
| EPI_ISL_548576 | County of Santa Clara Public Health Department | Chan-Zuckerberg Biohub | CZB Cliahub Consortium |
| EPI_ISL_548577 | Contra Costa Public Health Lab | Chan-Zuckerberg Biohub | CZB Cliahub Consortium |
| EPI_ISL_548578 | County of Santa Clara Public Health Department | Chan-Zuckerberg Biohub | CZB Cliahub Consortium |
| EPI_ISL_548579 | Contra Costa Public Health Lab | Chan-Zuckerberg Biohub | CZB Cliahub Consortium |
| EPI_ISL_548580, EPI_ISL_548581 | Orange County Public Health Laboratory | Chan-Zuckerberg Biohub | CZB Cliahub Consortium |
| EPI_ISL_548582, EPI_ISL_548583, EPI_ISL_548584 | County of Santa Clara Public Health Department | Chan-Zuckerberg Biohub | CZB Cliahub Consortium |
| EPI_ISL_548585, EPI_ISL_548586, EPI_ISL_548587 | Orange County Public Health Laboratory | Chan-Zuckerberg Biohub | CZB Cliahub Consortium |
| EPI_ISL_548588 | County of Santa Clara Public Health Department | Chan-Zuckerberg Biohub | CZB Cliahub Consortium |
| EPI_ISL_548589, EPI_ISL_548590, EPI_ISL_548591, EPI_ISL_548592 | Orange County Public Health Laboratory | Chan-Zuckerberg Biohub | CZB Cliahub Consortium |
| EPI_ISL_548593 | County of Santa Clara Public Health Department | Chan-Zuckerberg Biohub | CZB Cliahub Consortium |
| EPI_ISL_548594 | Orange County Public Health Laboratory | Chan-Zuckerberg Biohub | CZB Cliahub Consortium |
| EPI_ISL_548595, | County of Santa Clara Public Health | Chan-Zuckerberg Biohub | CZB Cliahub Consortium |

| | | | |
|---|---|---|---|
| EPI_ISL_548596, EPI_ISL_548597, EPI_ISL_548598 | Department | | |
| EPI_ISL_548599 | Tuolumne County Public Health | Chan-Zuckerberg Biohub | CZB Cliahub Consortium |
| EPI_ISL_548600 | County of Santa Clara Public Health Department | Chan-Zuckerberg Biohub | CZB Cliahub Consortium |
| EPI_ISL_548601 | Orange County Public Health Laboratory | Chan-Zuckerberg Biohub | CZB Cliahub Consortium |
| EPI_ISL_548602 | County of Santa Clara Public Health Department | Chan-Zuckerberg Biohub | CZB Cliahub Consortium |
| EPI_ISL_548603 | County of San Luis Obispo Public Health Laboratory | Chan-Zuckerberg Biohub | CZB Cliahub Consortium |
| EPI_ISL_548604, EPI_ISL_548605, EPI_ISL_548606, EPI_ISL_548607 | County of Santa Clara Public Health Department | Chan-Zuckerberg Biohub | CZB Cliahub Consortium |
| EPI_ISL_548608, EPI_ISL_548609 | Orange County Public Health Laboratory | Chan-Zuckerberg Biohub | CZB Cliahub Consortium |
| EPI_ISL_548610 | County of Santa Clara Public Health Department | Chan-Zuckerberg Biohub | CZB Cliahub Consortium |
| EPI_ISL_548611 | Orange County Public Health Laboratory | Chan-Zuckerberg Biohub | CZB Cliahub Consortium |
| EPI_ISL_548612 | County of Santa Clara Public Health Department | Chan-Zuckerberg Biohub | CZB Cliahub Consortium |
| EPI_ISL_548613, EPI_ISL_548614, EPI_ISL_548615 | Orange County Public Health Laboratory | Chan-Zuckerberg Biohub | CZB Cliahub Consortium |
| EPI_ISL_548616, EPI_ISL_548617, EPI_ISL_548618, EPI_ISL_548619 | County of Santa Clara Public Health Department | Chan-Zuckerberg Biohub | CZB Cliahub Consortium |
| EPI_ISL_548620 | Orange County Public Health Laboratory | Chan-Zuckerberg Biohub | CZB Cliahub Consortium |
| EPI_ISL_548621, EPI_ISL_548622 | County of Santa Clara Public Health Department | Chan-Zuckerberg Biohub | CZB Cliahub Consortium |
| EPI_ISL_548623 | Orange County Public Health Laboratory | Chan-Zuckerberg Biohub | CZB Cliahub Consortium |
| EPI_ISL_548624 | County of Santa Clara Public Health Department | Chan-Zuckerberg Biohub | CZB Cliahub Consortium |
| EPI_ISL_548625 | Orange County Public Health Laboratory | Chan-Zuckerberg Biohub | CZB Cliahub Consortium |
| EPI_ISL_548626 | Tuolumne County Public Health | Chan-Zuckerberg Biohub | CZB Cliahub Consortium |
| EPI_ISL_548627, EPI_ISL_548628 | Orange County Public Health Laboratory | Chan-Zuckerberg Biohub | CZB Cliahub Consortium |
| EPI_ISL_548629, EPI_ISL_548630 | County of Santa Clara Public Health Department | Chan-Zuckerberg Biohub | CZB Cliahub Consortium |
| EPI_ISL_548631 | County of San Luis Obispo Public Health Laboratory | Chan-Zuckerberg Biohub | CZB Cliahub Consortium |
| EPI_ISL_548632, EPI_ISL_548633 | County of Santa Clara Public Health Department | Chan-Zuckerberg Biohub | CZB Cliahub Consortium |
| EPI_ISL_548634, EPI_ISL_548635 | Orange County Public Health Laboratory | Chan-Zuckerberg Biohub | CZB Cliahub Consortium |
| EPI_ISL_548636 | County of Santa Clara Public Health Department | Chan-Zuckerberg Biohub | CZB Cliahub Consortium |
| EPI_ISL_548637, EPI_ISL_548638 | Orange County Public Health Laboratory | Chan-Zuckerberg Biohub | CZB Cliahub Consortium |
| EPI_ISL_548639 | County of Santa Clara Public Health Department | Chan-Zuckerberg Biohub | CZB Cliahub Consortium |
| EPI_ISL_548640, EPI_ISL_548641 | Orange County Public Health Laboratory | Chan-Zuckerberg Biohub | CZB Cliahub Consortium |
| EPI_ISL_548642, EPI_ISL_548643 | County of Santa Clara Public Health Department | Chan-Zuckerberg Biohub | CZB Cliahub Consortium |
| EPI_ISL_548644 | County of San Luis Obispo Public Health Laboratory | Chan-Zuckerberg Biohub | CZB Cliahub Consortium |
| EPI_ISL_548645 | County of Santa Clara Public Health Department | Chan-Zuckerberg Biohub | CZB Cliahub Consortium |
| EPI_ISL_548646 | Orange County Public Health Laboratory | Chan-Zuckerberg Biohub | CZB Cliahub Consortium |
| EPI_ISL_548647 | County of Santa Clara Public Health Department | Chan-Zuckerberg Biohub | CZB Cliahub Consortium |
| EPI_ISL_548648 | Orange County Public Health Laboratory | Chan-Zuckerberg Biohub | CZB Cliahub Consortium |
| EPI_ISL_548649, EPI_ISL_548650 | County of Santa Clara Public Health Department | Chan-Zuckerberg Biohub | CZB Cliahub Consortium |
| EPI_ISL_548651 | Orange County Public Health Laboratory | Chan-Zuckerberg Biohub | CZB Cliahub Consortium |
| EPI_ISL_548652 | County of Santa Clara Public Health Department | Chan-Zuckerberg Biohub | CZB Cliahub Consortium |
| EPI_ISL_548653, EPI_ISL_548654 | Orange County Public Health Laboratory | Chan-Zuckerberg Biohub | CZB Cliahub Consortium |
| EPI_ISL_548655, EPI_ISL_548656, EPI_ISL_548657, EPI_ISL_548658, EPI_ISL_548659 | County of Santa Clara Public Health Department | Chan-Zuckerberg Biohub | CZB Cliahub Consortium |
| EPI_ISL_548660 | Orange County Public Health Laboratory | Chan-Zuckerberg Biohub | CZB Cliahub Consortium |
| EPI_ISL_548661 | County of Santa Clara Public Health Department | Chan-Zuckerberg Biohub | CZB Cliahub Consortium |
| EPI_ISL_548662 | Orange County Public Health Laboratory | Chan-Zuckerberg Biohub | CZB Cliahub Consortium |
| EPI_ISL_548663 | County of San Luis Obispo Public Health Laboratory | Chan-Zuckerberg Biohub | CZB Cliahub Consortium |
| EPI_ISL_548664 | County of Santa Clara Public Health Department | Chan-Zuckerberg Biohub | CZB Cliahub Consortium |
| EPI_ISL_548665, EPI_ISL_548666, EPI_ISL_548667, EPI_ISL_548668 | Orange County Public Health Laboratory | Chan-Zuckerberg Biohub | CZB Cliahub Consortium |
| EPI_ISL_548669, | County of Santa Clara Public Health | Chan-Zuckerberg Biohub | CZB Cliahub Consortium |

| | | | |
|---|---|---|---|
| EPI_ISL_548670 | Department | | |
| EPI_ISL_548671 | County of San Luis Obispo Public Health Laboratory | Chan-Zuckerberg Biohub | CZB Cliahub Consortium |
| EPI_ISL_548672, EPI_ISL_548673 | County of Santa Clara Public Health Department | Chan-Zuckerberg Biohub | CZB Cliahub Consortium |
| EPI_ISL_548674, EPI_ISL_548675 | Orange County Public Health Laboratory | Chan-Zuckerberg Biohub | CZB Cliahub Consortium |
| EPI_ISL_548676 | County of Santa Clara Public Health Department | Chan-Zuckerberg Biohub | CZB Cliahub Consortium |
| EPI_ISL_548677 | Orange County Public Health Laboratory | Chan-Zuckerberg Biohub | CZB Cliahub Consortium |
| EPI_ISL_548678, EPI_ISL_548679 | County of Santa Clara Public Health Department | Chan-Zuckerberg Biohub | CZB Cliahub Consortium |
| EPI_ISL_549178, EPI_ISL_549181, EPI_ISL_549182, EPI_ISL_549183, EPI_ISL_549184, EPI_ISL_549186, EPI_ISL_549187, EPI_ISL_549189, EPI_ISL_549190, EPI_ISL_549191, EPI_ISL_549192, EPI_ISL_549193, EPI_ISL_549194, EPI_ISL_549195, EPI_ISL_549196, EPI_ISL_549197, EPI_ISL_549199, EPI_ISL_549200, EPI_ISL_549201, EPI_ISL_549202, EPI_ISL_549203, EPI_ISL_549204, EPI_ISL_549205, EPI_ISL_549206, EPI_ISL_549207, EPI_ISL_549208, EPI_ISL_549210, EPI_ISL_549211, EPI_ISL_549212, EPI_ISL_549213, EPI_ISL_549214, EPI_ISL_549215, EPI_ISL_549216, EPI_ISL_549217, EPI_ISL_549218, EPI_ISL_549219, EPI_ISL_549220, EPI_ISL_549221, EPI_ISL_549222, EPI_ISL_549223, EPI_ISL_549224, EPI_ISL_549225, EPI_ISL_549226, EPI_ISL_549227, EPI_ISL_549228, EPI_ISL_549230, EPI_ISL_549231, EPI_ISL_549232, EPI_ISL_549234, EPI_ISL_549235, EPI_ISL_549236, EPI_ISL_549237, EPI_ISL_549238, EPI_ISL_549239, EPI_ISL_549240, EPI_ISL_549241, EPI_ISL_549242, EPI_ISL_549243, EPI_ISL_549244, EPI_ISL_549245, EPI_ISL_549246, EPI_ISL_549247, EPI_ISL_549248, EPI_ISL_549249, EPI_ISL_549250, EPI_ISL_549251, EPI_ISL_549252, EPI_ISL_549253, EPI_ISL_549254, EPI_ISL_549255, EPI_ISL_549257, EPI_ISL_549258, EPI_ISL_549259, EPI_ISL_549261, EPI_ISL_549262, EPI_ISL_549263, EPI_ISL_549264, EPI_ISL_549265, EPI_ISL_549266, EPI_ISL_549267, EPI_ISL_549268, EPI_ISL_549269, EPI_ISL_549270 | | | |
| see above | Florida Bureau of Public Health Laboratories | Florida Bureau of Public Health Laboratories | Sarah Schmedes, Jason Blanton |
| EPI_ISL_560307 | Delaware Public Health Lab | Delaware Public Health Lab | Gregory Hovan |
| EPI_ISL_560325, EPI_ISL_560326, EPI_ISL_560327, EPI_ISL_560329, EPI_ISL_560330, EPI_ISL_560331, EPI_ISL_560332, EPI_ISL_560333, EPI_ISL_560334, EPI_ISL_560336, EPI_ISL_560337, EPI_ISL_560338, EPI_ISL_560339, EPI_ISL_560340, EPI_ISL_560342, EPI_ISL_560343, EPI_ISL_560344, EPI_ISL_560345, EPI_ISL_560346, EPI_ISL_560347, EPI_ISL_560348, EPI_ISL_560349, EPI_ISL_560350, EPI_ISL_560351, EPI_ISL_560352, EPI_ISL_560353, EPI_ISL_560354, EPI_ISL_560356, EPI_ISL_560357 | | | |
| see above | TriCore Reference Laboratories | Center for Global Health, University of New Mexico Health Sciences Center | Daryl Domman, Kurt Schwalm, Twila Kunde, Joseph Hicks, Michael Edwards, Darrell Dinwiddie |
| EPI_ISL_560406 | Delaware Public Health Lab | Delaware Public Health Lab | Gregory Hovan |
| EPI_ISL_560554, EPI_ISL_560555, EPI_ISL_560556, EPI_ISL_560557, EPI_ISL_560558, EPI_ISL_560559, EPI_ISL_560560, EPI_ISL_560561, EPI_ISL_560562, EPI_ISL_560563, EPI_ISL_560564, EPI_ISL_560565, EPI_ISL_560566 | | | |
| see above | Alaska State Virology Laboratory | Alaska State Virology Laboratory | Jack Chen, Ph.D. |
| EPI_ISL_560567, EPI_ISL_560647, EPI_ISL_560648, EPI_ISL_560741, EPI_ISL_560742 | Delaware Public Health Lab | Delaware Public Health Lab | Gregory Hovan |
| EPI_ISL_560743, EPI_ISL_560744, EPI_ISL_560745, EPI_ISL_560746, EPI_ISL_560747, EPI_ISL_560748, EPI_ISL_560749, EPI_ISL_560750, EPI_ISL_560751, EPI_ISL_560752, EPI_ISL_560753, EPI_ISL_560754, EPI_ISL_560755, EPI_ISL_560756, EPI_ISL_560757, EPI_ISL_560758, EPI_ISL_560759, EPI_ISL_560760, EPI_ISL_560761, EPI_ISL_560762, EPI_ISL_560763, EPI_ISL_560764, EPI_ISL_560765, EPI_ISL_560766, EPI_ISL_560767, EPI_ISL_560768, EPI_ISL_560769, EPI_ISL_560770, EPI_ISL_560771, EPI_ISL_560772, EPI_ISL_560773, EPI_ISL_560774, EPI_ISL_560775, EPI_ISL_560776, EPI_ISL_560777, EPI_ISL_560778, EPI_ISL_560779, EPI_ISL_560780, EPI_ISL_560781, EPI_ISL_560782, EPI_ISL_560783, EPI_ISL_560784, EPI_ISL_560785, EPI_ISL_560786, EPI_ISL_560787, EPI_ISL_560788, EPI_ISL_560789, EPI_ISL_560790, EPI_ISL_560791 | | | |
| see above | Minnesota Department of Health, Public Health Laboratory | Minnesota Department of Health, Public Health Laboratory | Matt Plumb, Jacob Garfin, and Xiong Wang |
| EPI_ISL_560792, EPI_ISL_560793, EPI_ISL_560794, EPI_ISL_560795, EPI_ISL_560796, EPI_ISL_560797 | Mayo Clinic & Mayo Clinic Laboratories | Minnesota Department of Health, Public Health Laboratory | Matt Plumb, Jacob Garfin, and Xiong Wang |
| EPI_ISL_560798, EPI_ISL_560799, EPI_ISL_560800, EPI_ISL_560801, EPI_ISL_560802, EPI_ISL_560803, EPI_ISL_560804, EPI_ISL_560805 | M Health Fairview | Minnesota Department of Health, Public Health Laboratory | Matt Plumb, Jacob Garfin, and Xiong Wang |
| EPI_ISL_560806, EPI_ISL_560807, EPI_ISL_560808, EPI_ISL_560809, EPI_ISL_560810, EPI_ISL_560811, EPI_ISL_560812, EPI_ISL_560813, EPI_ISL_560814, EPI_ISL_560815, EPI_ISL_560816, EPI_ISL_560817, EPI_ISL_560818, EPI_ISL_560819, EPI_ISL_560820, EPI_ISL_560821, EPI_ISL_560822, EPI_ISL_560823, EPI_ISL_560824, EPI_ISL_560825, EPI_ISL_560826, EPI_ISL_560827, EPI_ISL_560828, EPI_ISL_560829, EPI_ISL_560830, EPI_ISL_560831, EPI_ISL_560832 | | | |
| see above | Maryland Public Health Laboratory | Maryland Public Health Laboratory | Maryland Department of Health Laboratories Administration |
| EPI_ISL_560833, EPI_ISL_560834, EPI_ISL_560837, EPI_ISL_560838, EPI_ISL_560840, EPI_ISL_560841, EPI_ISL_560842, EPI_ISL_560843, EPI_ISL_560844, EPI_ISL_560845, EPI_ISL_560846, EPI_ISL_560847, EPI_ISL_560849, EPI_ISL_560850, EPI_ISL_560851, EPI_ISL_560852, EPI_ISL_560853, EPI_ISL_560856, EPI_ISL_560857, EPI_ISL_560858, EPI_ISL_560859, EPI_ISL_560860, EPI_ISL_560861, EPI_ISL_560862, EPI_ISL_560863, EPI_ISL_560864, EPI_ISL_560865, EPI_ISL_560868, EPI_ISL_560869, EPI_ISL_560870, EPI_ISL_560871, EPI_ISL_560872, EPI_ISL_560873, EPI_ISL_560874, EPI_ISL_560875, EPI_ISL_560876, EPI_ISL_560877, EPI_ISL_560878, EPI_ISL_560879, EPI_ISL_560880, EPI_ISL_560882, EPI_ISL_560884, EPI_ISL_560886, EPI_ISL_560887, EPI_ISL_560888, EPI_ISL_560889, EPI_ISL_560890, EPI_ISL_560891, EPI_ISL_560892, EPI_ISL_560893, EPI_ISL_560894, EPI_ISL_560897, EPI_ISL_560899, EPI_ISL_560903, EPI_ISL_560904, EPI_ISL_560905, EPI_ISL_560907, EPI_ISL_560908, EPI_ISL_560911, EPI_ISL_560912, EPI_ISL_560913, EPI_ISL_560914, EPI_ISL_560916, EPI_ISL_560917, EPI_ISL_560918, EPI_ISL_560919, EPI_ISL_560920, EPI_ISL_560921, EPI_ISL_560922, EPI_ISL_560923, EPI_ISL_560924, EPI_ISL_560925 | | | |
| see above | Utah Public Health Laboratory | Utah Public Health Laboratory | Erin Young, Kelly Oakeson |
| EPI_ISL_560926, EPI_ISL_560927, EPI_ISL_560928, EPI_ISL_560929, EPI_ISL_560930, EPI_ISL_560931, EPI_ISL_560932, EPI_ISL_560933, EPI_ISL_560934, EPI_ISL_560935, EPI_ISL_560936, EPI_ISL_560937, EPI_ISL_560938, EPI_ISL_560939, EPI_ISL_560940, EPI_ISL_560941, EPI_ISL_560942, EPI_ISL_560943, EPI_ISL_560944, EPI_ISL_560945, EPI_ISL_560946, EPI_ISL_560947, EPI_ISL_560948, EPI_ISL_560949, EPI_ISL_560950, EPI_ISL_560951, EPI_ISL_560952, EPI_ISL_560953, EPI_ISL_560954, EPI_ISL_560955, EPI_ISL_560956, EPI_ISL_560957, EPI_ISL_560958, EPI_ISL_560959, EPI_ISL_560960, EPI_ISL_560961, EPI_ISL_560962, EPI_ISL_560963, EPI_ISL_560964, EPI_ISL_560965, EPI_ISL_560966, EPI_ISL_560967, EPI_ISL_560968, EPI_ISL_560969, EPI_ISL_560970 | | | |
| see above | Texas Department of State Health Services | Texas Department of State Health Services | Rashmi Tuladhar, Bonnie Oh, Jenny Zhang, Maliha Rahman, Anita Pokharel, Myong Koag, Chun Wang, Rachel Lee, Grace Kubin |
| EPI_ISL_561335, EPI_ISL_561336, EPI_ISL_561345, EPI_ISL_561346, EPI_ISL_561351, EPI_ISL_561352, EPI_ISL_561353, EPI_ISL_561356, EPI_ISL_561358, EPI_ISL_561360, EPI_ISL_565830, EPI_ISL_565831, EPI_ISL_565832 | | | |
| see above | Delaware Public Health Lab | Delaware Public Health Lab | Gregory Hovan |
| EPI_ISL_565833, EPI_ISL_565834, EPI_ISL_565835, EPI_ISL_565836, EPI_ISL_565837, EPI_ISL_565838, EPI_ISL_565839, EPI_ISL_565841, EPI_ISL_565842, EPI_ISL_565843, EPI_ISL_565845, EPI_ISL_565846, EPI_ISL_565847, EPI_ISL_565848, EPI_ISL_565849, EPI_ISL_565850, EPI_ISL_565851, EPI_ISL_565852, EPI_ISL_565853, EPI_ISL_565855, EPI_ISL_565856, EPI_ISL_565857, EPI_ISL_565858, EPI_ISL_565859, EPI_ISL_565861, EPI_ISL_565863, EPI_ISL_565864, EPI_ISL_565865, EPI_ISL_565866, EPI_ISL_565869, EPI_ISL_565870, EPI_ISL_565872, EPI_ISL_565873, EPI_ISL_565874, EPI_ISL_565875, EPI_ISL_565876, EPI_ISL_565878, EPI_ISL_565879, EPI_ISL_565880, EPI_ISL_565881, EPI_ISL_565882, EPI_ISL_565883, EPI_ISL_565884, EPI_ISL_565886, EPI_ISL_565888, EPI_ISL_565890, EPI_ISL_565891, EPI_ISL_565892, EPI_ISL_565893, EPI_ISL_565894, EPI_ISL_565895, EPI_ISL_565898, EPI_ISL_565902, EPI_ISL_565903, EPI_ISL_565906, EPI_ISL_565907, EPI_ISL_565929, EPI_ISL_565932, EPI_ISL_565933, EPI_ISL_565934, EPI_ISL_565935, EPI_ISL_565936, EPI_ISL_565937, EPI_ISL_565938, EPI_ISL_565939, EPI_ISL_565940, EPI_ISL_565941, EPI_ISL_565942, EPI_ISL_565943, EPI_ISL_565944, EPI_ISL_565945, EPI_ISL_565946, EPI_ISL_565947, EPI_ISL_565948, EPI_ISL_565949, EPI_ISL_565950, EPI_ISL_565951, EPI_ISL_565952, EPI_ISL_565953, EPI_ISL_565954, EPI_ISL_565955, EPI_ISL_565956, EPI_ISL_565957, EPI_ISL_565958, EPI_ISL_565959, EPI_ISL_565960, EPI_ISL_565961, EPI_ISL_565962, EPI_ISL_565963, EPI_ISL_565964, EPI_ISL_565965, EPI_ISL_565966, EPI_ISL_565967, EPI_ISL_565968, EPI_ISL_565969, EPI_ISL_565970, EPI_ISL_565971, EPI_ISL_565972, EPI_ISL_565973, EPI_ISL_565974, EPI_ISL_565975, EPI_ISL_565976, EPI_ISL_565977, EPI_ISL_565978, EPI_ISL_565979, EPI_ISL_565980, EPI_ISL_565981, EPI_ISL_565982, EPI_ISL_565983, EPI_ISL_565984, EPI_ISL_565985, EPI_ISL_565986, EPI_ISL_565987, EPI_ISL_565988, EPI_ISL_565989, EPI_ISL_565999, EPI_ISL_566000, EPI_ISL_566001, EPI_ISL_566002, EPI_ISL_566003, EPI_ISL_566004, EPI_ISL_566007, EPI_ISL_566008, EPI_ISL_566009, EPI_ISL_566010, EPI_ISL_566011, EPI_ISL_566012, EPI_ISL_566013, EPI_ISL_566014, EPI_ISL_566015, EPI_ISL_566016, EPI_ISL_566017, EPI_ISL_566018, EPI_ISL_566019, EPI_ISL_566020, EPI_ISL_566021, EPI_ISL_566022, EPI_ISL_566023, EPI_ISL_566024, EPI_ISL_566025, EPI_ISL_566026, EPI_ISL_566027, EPI_ISL_566028, EPI_ISL_566029, EPI_ISL_566030, EPI_ISL_566031, EPI_ISL_566032, EPI_ISL_566033, EPI_ISL_566034, EPI_ISL_566035 | | | |
| see above | Michigan Department of Health and Human Services, Bureau of Laboratories | Michigan Department of Health and Human Services, Bureau of Laboratories | Blankenship HM, Riner D, Soehnlen MK |
| EPI_ISL_566084 | Delaware Public Health Lab | Delaware Public Health Lab | Gregory Hovan |
| EPI_ISL_566085, EPI_ISL_566086, EPI_ISL_566087, EPI_ISL_566088, EPI_ISL_566089, EPI_ISL_566090, EPI_ISL_566091, EPI_ISL_566092, EPI_ISL_566093, EPI_ISL_566094, EPI_ISL_566095, EPI_ISL_566096, EPI_ISL_566097, EPI_ISL_566098, EPI_ISL_566099, EPI_ISL_566100, EPI_ISL_566101, EPI_ISL_566102, EPI_ISL_566103, EPI_ISL_566104, EPI_ISL_566105, EPI_ISL_566106, EPI_ISL_566107 | | | |
| see above | GA Department of Public Health Laboratory | Pathogen Discovery, Respiratory Viruses Branch, Division of Viral Diseases, Centers for Disease Control and Prevention | Jing Zhang, Brian Lynch, Yan Li, Anna Montmayeur, Krista Queen, Ying Tao, Anna Uehara, Clinton R. Paden, Rachel Marine, Haibin Wang, Suxiang Tong |
| EPI_ISL_566108 | Delaware Public Health Lab | Delaware Public Health Lab | Gregory Hovan |
| EPI_ISL_568587, EPI_ISL_568589, EPI_ISL_568590, EPI_ISL_568591, EPI_ISL_568592, EPI_ISL_568593, EPI_ISL_568594, EPI_ISL_568595, EPI_ISL_568596, EPI_ISL_568597, EPI_ISL_568598, EPI_ISL_568599, EPI_ISL_568600, EPI_ISL_568601, EPI_ISL_568602, EPI_ISL_568603, EPI_ISL_568604, EPI_ISL_568606, EPI_ISL_568607, EPI_ISL_568608, EPI_ISL_568609, EPI_ISL_568610, EPI_ISL_568612, EPI_ISL_568614, EPI_ISL_568616, EPI_ISL_568617, EPI_ISL_568618, EPI_ISL_568619, EPI_ISL_568620, EPI_ISL_568621, EPI_ISL_568622, EPI_ISL_568623, EPI_ISL_568624, EPI_ISL_568625, EPI_ISL_568626, EPI_ISL_568627, EPI_ISL_568628, EPI_ISL_568630, EPI_ISL_568631, EPI_ISL_568632, EPI_ISL_568633, EPI_ISL_568635, EPI_ISL_568637, EPI_ISL_568638, EPI_ISL_568639, EPI_ISL_568640, EPI_ISL_568644, EPI_ISL_568645, EPI_ISL_568648, EPI_ISL_568649, EPI_ISL_568650, EPI_ISL_568651, EPI_ISL_568652, EPI_ISL_568654, EPI_ISL_568656, EPI_ISL_568657, EPI_ISL_568658, EPI_ISL_568659, EPI_ISL_568660, EPI_ISL_568661, EPI_ISL_568662, EPI_ISL_568663, EPI_ISL_568664, EPI_ISL_568665, EPI_ISL_568666, EPI_ISL_568667, EPI_ISL_568668, EPI_ISL_568669, EPI_ISL_568671, EPI_ISL_568672, EPI_ISL_568673, EPI_ISL_568674, EPI_ISL_568675, EPI_ISL_568676, EPI_ISL_568677, EPI_ISL_568678, EPI_ISL_568679, EPI_ISL_568681, EPI_ISL_568683, EPI_ISL_568684, EPI_ISL_568876 | | | |
| see above | Florida Bureau of Public Health Laboratories | Florida Bureau of Public Health Laboratories | Sarah Schmedes, Jason Blanton |
| EPI_ISL_569609 | Bennet County Hospital | South Dakota Public Health Laboratory | Matt Plumb, Jacob Garfin, Xiong Wang, and Chris Carlson |
| EPI_ISL_569610 | Avera McKennan Hospital | South Dakota Public Health Laboratory | Matt Plumb, Jacob Garfin, Xiong Wang, and Chris Carlson |
| EPI_ISL_569611 | Sioux Falls Urgent Care | South Dakota Public Health Laboratory | Matt Plumb, Jacob Garfin, Xiong Wang, and Chris Carlson |
| EPI_ISL_569612, EPI_ISL_569613 | Cheyenne River Health Center | South Dakota Public Health Laboratory | Matt Plumb, Jacob Garfin, Xiong Wang, and Chris Carlson |
| EPI_ISL_569614 | IHS Fort Yates Hospital | South Dakota Public Health Laboratory | Matt Plumb, Jacob Garfin, Xiong Wang, and Chris Carlson |
| EPI_ISL_569615 | Quick Care Huron | South Dakota Public Health Laboratory | Matt Plumb, Jacob Garfin, Xiong Wang, and Chris Carlson |

| | | | |
|---|---|---|---|
| EPI_ISL_569616 | Aurora County Clinic | South Dakota Public Health Laboratory | Matt Plumb, Jacob Garfin, Xiong Wang, and Chris Carlson |
| EPI_ISL_569617 | SD Urban Indian Health Pierre | South Dakota Public Health Laboratory | Matt Plumb, Jacob Garfin, Xiong Wang, and Chris Carlson |
| EPI_ISL_569618, EPI_ISL_569619 | Bethel Lutheran Home | South Dakota Public Health Laboratory | Matt Plumb, Jacob Garfin, Xiong Wang, and Chris Carlson |
| EPI_ISL_569620 | McCrossan Boys Ranch | South Dakota Public Health Laboratory | Matt Plumb, Jacob Garfin, Xiong Wang, and Chris Carlson |
| EPI_ISL_569621 | Quick Care Watertown | South Dakota Public Health Laboratory | Matt Plumb, Jacob Garfin, Xiong Wang, and Chris Carlson |
| EPI_ISL_569622 | Woodrow Wilson Keeble Memorial Healthcare | South Dakota Public Health Laboratory | Matt Plumb, Jacob Garfin, Xiong Wang, and Chris Carlson |
| EPI_ISL_569623 | Quick Care Watertown | South Dakota Public Health Laboratory | Matt Plumb, Jacob Garfin, Xiong Wang, and Chris Carlson |
| EPI_ISL_569624 | Avera McKennan Hospital | South Dakota Public Health Laboratory | Matt Plumb, Jacob Garfin, Xiong Wang, and Chris Carlson |
| EPI_ISL_569625, EPI_ISL_569626, EPI_ISL_569627, EPI_ISL_569628, EPI_ISL_569631, EPI_ISL_569632, EPI_ISL_569633, EPI_ISL_569634, EPI_ISL_569635, EPI_ISL_569636, EPI_ISL_569637, EPI_ISL_569638, EPI_ISL_569639, EPI_ISL_569640, EPI_ISL_569641, EPI_ISL_569642, EPI_ISL_569643, EPI_ISL_569644, EPI_ISL_569645, EPI_ISL_569646, EPI_ISL_569647, EPI_ISL_569648, EPI_ISL_569649, EPI_ISL_569650, EPI_ISL_569651, EPI_ISL_569652, EPI_ISL_569653, EPI_ISL_569654, EPI_ISL_569655, EPI_ISL_569656, EPI_ISL_569657, EPI_ISL_569658, EPI_ISL_569659, EPI_ISL_569660, EPI_ISL_569661, EPI_ISL_569662, EPI_ISL_569663 | | | |
| see above | New Mexico Department of Health Scientific Laboratory | New Mexico Department of Health Scientific Laboratory | Ellie Johnson, Anastacia Griego-Fisher, D'Eldra Malone |
| EPI_ISL_569689, EPI_ISL_569692, EPI_ISL_569693, EPI_ISL_569694, EPI_ISL_569695, EPI_ISL_569697, EPI_ISL_569699, EPI_ISL_569700, EPI_ISL_569701, EPI_ISL_569702, EPI_ISL_569703, EPI_ISL_569704, EPI_ISL_569705, EPI_ISL_569706, EPI_ISL_569708, EPI_ISL_569709, EPI_ISL_569710, EPI_ISL_569711, EPI_ISL_569712, EPI_ISL_569713, EPI_ISL_569714, EPI_ISL_569715, EPI_ISL_569716, EPI_ISL_569719, EPI_ISL_569720, EPI_ISL_569721, EPI_ISL_569722, EPI_ISL_569724, EPI_ISL_569725, EPI_ISL_569726, EPI_ISL_569727, EPI_ISL_569728, EPI_ISL_569729, EPI_ISL_569731, EPI_ISL_569732, EPI_ISL_569733, EPI_ISL_569734 | | | |
| see above | Texas Department of State Health Services | Texas Department of State Health Services | Rashmi Tuladhar, Bonnie Oh,Jenny Zhang, Maliha Rahman, Anita Pokharel, Myong Koag, Chun Wang, Rachel Lee, Grace Kubin |
| EPI_ISL_569887, EPI_ISL_569888, EPI_ISL_569889, EPI_ISL_569890, EPI_ISL_569891, EPI_ISL_569892, EPI_ISL_569893, EPI_ISL_569894, EPI_ISL_569895, EPI_ISL_569896, EPI_ISL_569897, EPI_ISL_569898, EPI_ISL_569899, EPI_ISL_569900, EPI_ISL_569901, EPI_ISL_569902, EPI_ISL_569903, EPI_ISL_569904, EPI_ISL_569905, EPI_ISL_569906, EPI_ISL_569907, EPI_ISL_569908, EPI_ISL_569909, EPI_ISL_569910, EPI_ISL_569911, EPI_ISL_569912, EPI_ISL_569913, EPI_ISL_569914, EPI_ISL_569915, EPI_ISL_569916, EPI_ISL_569917, EPI_ISL_569918, EPI_ISL_569919, EPI_ISL_569920, EPI_ISL_569921, EPI_ISL_569922, EPI_ISL_569923, EPI_ISL_569924, EPI_ISL_569925, EPI_ISL_569926, EPI_ISL_569927, EPI_ISL_569929, EPI_ISL_569930, EPI_ISL_569931, EPI_ISL_569932, EPI_ISL_569933, EPI_ISL_569934, EPI_ISL_569935, EPI_ISL_569936, EPI_ISL_569937, EPI_ISL_569938, EPI_ISL_569939, EPI_ISL_569940, EPI_ISL_569941, EPI_ISL_569942, EPI_ISL_569943, EPI_ISL_569944, EPI_ISL_569945, EPI_ISL_569946, EPI_ISL_569947, EPI_ISL_569948 | | | |
| see above | Innovative Genomics Institute, UC Berkeley | Innovative Genomics Institute, UC Berkeley | Stacia Wyman, Haridha Shivram, Phil Frankino, Liana Lareau, Shana McDevitt, Justin Choi |
| EPI_ISL_570027, EPI_ISL_570028, EPI_ISL_570029, EPI_ISL_570030, EPI_ISL_570031, EPI_ISL_570032, EPI_ISL_570033, EPI_ISL_570034, EPI_ISL_570035, EPI_ISL_570036, EPI_ISL_570037, EPI_ISL_570038, EPI_ISL_570039, EPI_ISL_570040, EPI_ISL_570041, EPI_ISL_570042, EPI_ISL_570043, EPI_ISL_570044, EPI_ISL_570045, EPI_ISL_570046, EPI_ISL_570047, EPI_ISL_570048, EPI_ISL_570049, EPI_ISL_570050, EPI_ISL_570051, EPI_ISL_570052, EPI_ISL_570053, EPI_ISL_570054, EPI_ISL_570055, EPI_ISL_570056, EPI_ISL_570057, EPI_ISL_570058, EPI_ISL_570059, EPI_ISL_570060, EPI_ISL_570061, EPI_ISL_570062, EPI_ISL_570063, EPI_ISL_570064, EPI_ISL_570065, EPI_ISL_570066, EPI_ISL_570067, EPI_ISL_570068, EPI_ISL_570069, EPI_ISL_570070, EPI_ISL_570071, EPI_ISL_570072, EPI_ISL_570073, EPI_ISL_570074, EPI_ISL_570075, EPI_ISL_570076, EPI_ISL_570077, EPI_ISL_570078, EPI_ISL_570079, EPI_ISL_570080, EPI_ISL_570081, EPI_ISL_570082, EPI_ISL_570083, EPI_ISL_570084, EPI_ISL_570085, EPI_ISL_570086, EPI_ISL_570087, EPI_ISL_570088, EPI_ISL_570089, EPI_ISL_570090, EPI_ISL_570091, EPI_ISL_570092, EPI_ISL_570093, EPI_ISL_570094, EPI_ISL_570095, EPI_ISL_570096, EPI_ISL_570097, EPI_ISL_570098, EPI_ISL_570099, EPI_ISL_570100, EPI_ISL_570101, EPI_ISL_570102, EPI_ISL_570103, EPI_ISL_570104, EPI_ISL_570105, EPI_ISL_570106, EPI_ISL_570107, EPI_ISL_570108, EPI_ISL_570109, EPI_ISL_570110, EPI_ISL_570111, EPI_ISL_570112, EPI_ISL_570113, EPI_ISL_570114, EPI_ISL_570115, EPI_ISL_570116, EPI_ISL_570118, EPI_ISL_570119, EPI_ISL_570120, EPI_ISL_570121, EPI_ISL_570122, EPI_ISL_570123, EPI_ISL_570124, EPI_ISL_570125, EPI_ISL_570126, EPI_ISL_570127, EPI_ISL_570128, EPI_ISL_570129, EPI_ISL_570130, EPI_ISL_570131, EPI_ISL_570132, EPI_ISL_570133, EPI_ISL_570134, EPI_ISL_570135, EPI_ISL_570136, EPI_ISL_570137, EPI_ISL_570138, EPI_ISL_570139, EPI_ISL_570140, EPI_ISL_570141, EPI_ISL_570142, EPI_ISL_570143, EPI_ISL_570144, EPI_ISL_570145, EPI_ISL_570146, EPI_ISL_570147, EPI_ISL_570148, EPI_ISL_570149, EPI_ISL_570150, EPI_ISL_570151, EPI_ISL_570152, EPI_ISL_570153, EPI_ISL_570154, EPI_ISL_570155, EPI_ISL_570156, EPI_ISL_570157, EPI_ISL_570158, EPI_ISL_570159, EPI_ISL_570160, EPI_ISL_570161, EPI_ISL_570162, EPI_ISL_570163, EPI_ISL_570164, EPI_ISL_570165, EPI_ISL_570166, EPI_ISL_570167, EPI_ISL_570168, EPI_ISL_570169, EPI_ISL_570170, EPI_ISL_570171, EPI_ISL_570172, EPI_ISL_570173, EPI_ISL_570174, EPI_ISL_570175, EPI_ISL_570176, EPI_ISL_570177, EPI_ISL_570178, EPI_ISL_570179, EPI_ISL_570180, EPI_ISL_570181, EPI_ISL_570182, EPI_ISL_570184, EPI_ISL_570185, EPI_ISL_570186, EPI_ISL_570187, EPI_ISL_570188, EPI_ISL_570189, EPI_ISL_570190, EPI_ISL_570191, EPI_ISL_570192, EPI_ISL_570193, EPI_ISL_570194, EPI_ISL_570195, EPI_ISL_570196, EPI_ISL_570197, EPI_ISL_570198, EPI_ISL_570199, EPI_ISL_570200, EPI_ISL_570201, EPI_ISL_570202, EPI_ISL_570203, EPI_ISL_570204, EPI_ISL_570205, EPI_ISL_570206, EPI_ISL_570207, EPI_ISL_570208, EPI_ISL_570209, EPI_ISL_570210, EPI_ISL_570211, EPI_ISL_570212, EPI_ISL_570213, EPI_ISL_570214, EPI_ISL_570215, EPI_ISL_570216, EPI_ISL_570217, EPI_ISL_570218, EPI_ISL_570219, EPI_ISL_570220, EPI_ISL_570221, EPI_ISL_570222, EPI_ISL_570223, EPI_ISL_570224, EPI_ISL_570225, EPI_ISL_570226, EPI_ISL_570227, EPI_ISL_570228, EPI_ISL_570229, EPI_ISL_570230, EPI_ISL_570231, EPI_ISL_570232, EPI_ISL_570233, EPI_ISL_570234, EPI_ISL_570235, EPI_ISL_570236, EPI_ISL_570237, EPI_ISL_570238, EPI_ISL_570239, EPI_ISL_570240, EPI_ISL_570241, EPI_ISL_570242, EPI_ISL_570243, EPI_ISL_570244, EPI_ISL_570245, EPI_ISL_570246, EPI_ISL_570247, EPI_ISL_570248, EPI_ISL_570249, EPI_ISL_570250, EPI_ISL_570251, EPI_ISL_570252, EPI_ISL_570253, EPI_ISL_570254, EPI_ISL_570255, EPI_ISL_570256, EPI_ISL_570257, EPI_ISL_570258, EPI_ISL_570259, EPI_ISL_570260, EPI_ISL_570261, EPI_ISL_570262, EPI_ISL_570263, EPI_ISL_570264, EPI_ISL_570265, EPI_ISL_570266, EPI_ISL_570267, EPI_ISL_570268, EPI_ISL_570269, EPI_ISL_570270, EPI_ISL_570271, EPI_ISL_570272, EPI_ISL_570273, EPI_ISL_570274, EPI_ISL_570275, EPI_ISL_570276, EPI_ISL_570277, EPI_ISL_570278, EPI_ISL_570279, EPI_ISL_570280, EPI_ISL_570281, EPI_ISL_570282, EPI_ISL_570283, EPI_ISL_570284, EPI_ISL_570285, EPI_ISL_570286, EPI_ISL_570287, EPI_ISL_570288, EPI_ISL_570289, EPI_ISL_570290, EPI_ISL_570291, EPI_ISL_570292, EPI_ISL_570293, EPI_ISL_570294, EPI_ISL_570295, EPI_ISL_570296, EPI_ISL_570297, EPI_ISL_570298, EPI_ISL_570299, EPI_ISL_570300, EPI_ISL_570301, EPI_ISL_570302, EPI_ISL_570303, EPI_ISL_570304, EPI_ISL_570305, EPI_ISL_570306, EPI_ISL_570307, EPI_ISL_570308, EPI_ISL_570309, EPI_ISL_570310, EPI_ISL_570311, EPI_ISL_570312, EPI_ISL_570313, EPI_ISL_570314, EPI_ISL_570315, EPI_ISL_570316, EPI_ISL_570317, EPI_ISL_570318, EPI_ISL_570319, EPI_ISL_570320, EPI_ISL_570321, EPI_ISL_570322, EPI_ISL_570323, EPI_ISL_570324, EPI_ISL_570325, EPI_ISL_570326, EPI_ISL_570327, EPI_ISL_570328, EPI_ISL_570329, EPI_ISL_570330, EPI_ISL_570331, EPI_ISL_570332, EPI_ISL_570333, EPI_ISL_570334, EPI_ISL_570335, EPI_ISL_570336, EPI_ISL_570337, EPI_ISL_570338, EPI_ISL_570339, EPI_ISL_570340, EPI_ISL_570341, EPI_ISL_570342, EPI_ISL_570343, EPI_ISL_570344, EPI_ISL_570345, EPI_ISL_570346, EPI_ISL_570347, EPI_ISL_570348, EPI_ISL_570349, EPI_ISL_570350, EPI_ISL_570351, EPI_ISL_570352, EPI_ISL_570353, EPI_ISL_570354, EPI_ISL_570355, EPI_ISL_570356, EPI_ISL_570357, EPI_ISL_570358, EPI_ISL_570359, EPI_ISL_570360, EPI_ISL_570361, EPI_ISL_570362, EPI_ISL_570363, EPI_ISL_570364, EPI_ISL_570365, EPI_ISL_570366, EPI_ISL_570367, EPI_ISL_570368, EPI_ISL_570369, EPI_ISL_570370, EPI_ISL_570371, EPI_ISL_570372, EPI_ISL_570373, EPI_ISL_570374, EPI_ISL_570375, EPI_ISL_570376, EPI_ISL_570377, EPI_ISL_570378, EPI_ISL_570379, EPI_ISL_570380, EPI_ISL_570381, EPI_ISL_570382, EPI_ISL_570383, EPI_ISL_570384, EPI_ISL_570385, EPI_ISL_570386, EPI_ISL_570387, EPI_ISL_570388, EPI_ISL_570389, EPI_ISL_570390, EPI_ISL_570391, EPI_ISL_570392, EPI_ISL_570393, EPI_ISL_570394, EPI_ISL_570395, EPI_ISL_570396, EPI_ISL_570397, EPI_ISL_570398, EPI_ISL_570399, EPI_ISL_570400, EPI_ISL_570401, EPI_ISL_570402, EPI_ISL_570403, EPI_ISL_570404, EPI_ISL_570405, EPI_ISL_570406, EPI_ISL_570407, EPI_ISL_570408, EPI_ISL_570409, EPI_ISL_570410, EPI_ISL_570411, EPI_ISL_570412, EPI_ISL_570413, EPI_ISL_570414, EPI_ISL_570415, EPI_ISL_570416, EPI_ISL_570417, EPI_ISL_570418, EPI_ISL_570419, EPI_ISL_570420, EPI_ISL_570421, EPI_ISL_570422, EPI_ISL_570423, EPI_ISL_570424, EPI_ISL_570425, EPI_ISL_570426, EPI_ISL_570427, EPI_ISL_570428, EPI_ISL_570429, EPI_ISL_570430, EPI_ISL_570431, EPI_ISL_570432, EPI_ISL_570433, EPI_ISL_570434, EPI_ISL_570435, EPI_ISL_570436, EPI_ISL_570437, EPI_ISL_570438, EPI_ISL_570439, EPI_ISL_570440, EPI_ISL_570441, EPI_ISL_570442, EPI_ISL_570443, EPI_ISL_570444, EPI_ISL_570445, EPI_ISL_570446, EPI_ISL_570447, EPI_ISL_570448, EPI_ISL_570449, EPI_ISL_570450, EPI_ISL_570451, EPI_ISL_570452, EPI_ISL_570453, EPI_ISL_570454, EPI_ISL_570455, EPI_ISL_570456, EPI_ISL_570457, EPI_ISL_570458, EPI_ISL_570459, EPI_ISL_570460, EPI_ISL_570461, EPI_ISL_570462, EPI_ISL_570463, EPI_ISL_570464, EPI_ISL_570465, EPI_ISL_570466, EPI_ISL_570467, EPI_ISL_570468, EPI_ISL_570469, EPI_ISL_570470, EPI_ISL_570471, EPI_ISL_570472, EPI_ISL_570473, EPI_ISL_570474, EPI_ISL_570475, EPI_ISL_570476, EPI_ISL_570477, EPI_ISL_570478, EPI_ISL_570479, EPI_ISL_570480, EPI_ISL_570481, EPI_ISL_570482, EPI_ISL_570483, EPI_ISL_570484, EPI_ISL_570485, EPI_ISL_570486, EPI_ISL_570487, EPI_ISL_570488, EPI_ISL_570489, EPI_ISL_570490, EPI_ISL_570491, EPI_ISL_570492, EPI_ISL_570493, EPI_ISL_570494, EPI_ISL_570495, EPI_ISL_570496, EPI_ISL_570497, EPI_ISL_570498, EPI_ISL_570499, EPI_ISL_570500, EPI_ISL_570501, EPI_ISL_570502, EPI_ISL_570503, EPI_ISL_570504, EPI_ISL_570505, EPI_ISL_570506, EPI_ISL_570507, EPI_ISL_570508, EPI_ISL_570509, EPI_ISL_570510, EPI_ISL_570511, EPI_ISL_570512, EPI_ISL_570513, EPI_ISL_570514, EPI_ISL_570515, EPI_ISL_570516, EPI_ISL_570517, EPI_ISL_570518, EPI_ISL_570519, EPI_ISL_570520, EPI_ISL_570521, EPI_ISL_570522, EPI_ISL_570523, EPI_ISL_570524, EPI_ISL_570525, EPI_ISL_570526, EPI_ISL_570527, EPI_ISL_570528, EPI_ISL_570529, EPI_ISL_570530, EPI_ISL_570531, EPI_ISL_570532, EPI_ISL_570533, EPI_ISL_570534, EPI_ISL_570535, EPI_ISL_570536, EPI_ISL_570537, EPI_ISL_570538, EPI_ISL_570539, EPI_ISL_570540, EPI_ISL_570541, EPI_ISL_570542, EPI_ISL_570543, EPI_ISL_570544, EPI_ISL_570545, EPI_ISL_570546, EPI_ISL_570547, EPI_ISL_570548, EPI_ISL_570549, EPI_ISL_570550, EPI_ISL_570551, EPI_ISL_570552, EPI_ISL_570553, EPI_ISL_570554, EPI_ISL_570555, EPI_ISL_570556, EPI_ISL_570557, EPI_ISL_570558, EPI_ISL_570559, EPI_ISL_570560, EPI_ISL_570561, EPI_ISL_570562, EPI_ISL_570563, EPI_ISL_570564, EPI_ISL_570565, EPI_ISL_570566, EPI_ISL_570567, EPI_ISL_570568, EPI_ISL_570569, EPI_ISL_570570, EPI_ISL_570571, EPI_ISL_570572, EPI_ISL_570573, EPI_ISL_570574, EPI_ISL_570575, EPI_ISL_570576, EPI_ISL_570577, EPI_ISL_570578, EPI_ISL_570579, EPI_ISL_570580, EPI_ISL_570581, EPI_ISL_570582, EPI_ISL_570583, EPI_ISL_570584, EPI_ISL_570585, EPI_ISL_570586, EPI_ISL_570587, EPI_ISL_570588, EPI_ISL_570589, EPI_ISL_570590, EPI_ISL_570591, EPI_ISL_570592, EPI_ISL_570593, EPI_ISL_570594, EPI_ISL_570595, EPI_ISL_570596, EPI_ISL_570597, EPI_ISL_570598, EPI_ISL_570599, EPI_ISL_570600, EPI_ISL_570601, EPI_ISL_570602, EPI_ISL_570603, EPI_ISL_570604, EPI_ISL_570605, EPI_ISL_570606, EPI_ISL_570607, EPI_ISL_570608, EPI_ISL_570609, EPI_ISL_570610, EPI_ISL_570611, EPI_ISL_570612, EPI_ISL_570613, EPI_ISL_570614, EPI_ISL_570615, EPI_ISL_570616, EPI_ISL_570617, EPI_ISL_570618, EPI_ISL_570619, EPI_ISL_570620, EPI_ISL_570621, EPI_ISL_570622, EPI_ISL_570623, EPI_ISL_570624, EPI_ISL_570625, EPI_ISL_570626, EPI_ISL_570627, EPI_ISL_570628, EPI_ISL_570629, EPI_ISL_570630, EPI_ISL_570631, EPI_ISL_570632, EPI_ISL_570633, EPI_ISL_570634, EPI_ISL_570635, EPI_ISL_570636, EPI_ISL_570637, EPI_ISL_570638, EPI_ISL_570639, EPI_ISL_570640, EPI_ISL_570641, EPI_ISL_570642, EPI_ISL_570643, EPI_ISL_570644, EPI_ISL_570645, EPI_ISL_570646, EPI_ISL_570647, EPI_ISL_570648, EPI_ISL_570649, EPI_ISL_570650, EPI_ISL_570651, EPI_ISL_570652, EPI_ISL_570653, EPI_ISL_570654, EPI_ISL_570655, EPI_ISL_570656, EPI_ISL_570657, EPI_ISL_570658, EPI_ISL_570659, EPI_ISL_570660, EPI_ISL_570661, EPI_ISL_570662, EPI_ISL_570663, EPI_ISL_570664, EPI_ISL_570665, EPI_ISL_570666, EPI_ISL_570667, EPI_ISL_570668, EPI_ISL_570669, EPI_ISL_570670, EPI_ISL_570671, EPI_ISL_570672, EPI_ISL_570673, EPI_ISL_570674, EPI_ISL_570675, EPI_ISL_570676, EPI_ISL_570677, EPI_ISL_570678, EPI_ISL_570679, EPI_ISL_570680, EPI_ISL_570681, EPI_ISL_570682, EPI_ISL_570683, EPI_ISL_570684, EPI_ISL_570685, EPI_ISL_570686, EPI_ISL_570687, EPI_ISL_570688, EPI_ISL_570689, EPI_ISL_570690, EPI_ISL_570691, EPI_ISL_570692, EPI_ISL_570693, EPI_ISL_570694, EPI_ISL_570695, EPI_ISL_570696, EPI_ISL_570697, EPI_ISL_570698, EPI_ISL_570699, EPI_ISL_570700, EPI_ISL_570701, EPI_ISL_570702, EPI_ISL_570703, EPI_ISL_570704, EPI_ISL_570705, EPI_ISL_570706, EPI_ISL_570707, EPI_ISL_570708, EPI_ISL_570709, EPI_ISL_570710, EPI_ISL_570711, EPI_ISL_570712, EPI_ISL_570713, EPI_ISL_570714, EPI_ISL_570715, EPI_ISL_570716, EPI_ISL_570717, EPI_ISL_570718, EPI_ISL_570719, EPI_ISL_570720, EPI_ISL_570721, EPI_ISL_570722, EPI_ISL_570723, EPI_ISL_570724, EPI_ISL_570725, EPI_ISL_570726, EPI_ISL_570727, EPI_ISL_570728, EPI_ISL_570729, EPI_ISL_570730, EPI_ISL_570731, EPI_ISL_570732, EPI_ISL_570733, EPI_ISL_570734, EPI_ISL_570735, EPI_ISL_570736, EPI_ISL_570737, EPI_ISL_570738, EPI_ISL_570739, EPI_ISL_570740, EPI_ISL_570741, EPI_ISL_570742, EPI_ISL_570743, EPI_ISL_570744, EPI_ISL_570745, EPI_ISL_570746, EPI_ISL_570747, EPI_ISL_570748, EPI_ISL_570749, EPI_ISL_570750, EPI_ISL_570751, EPI_ISL_570752, EPI_ISL_570753, EPI_ISL_570754, EPI_ISL_570755, EPI_ISL_570756, EPI_ISL_570757, EPI_ISL_570758, EPI_ISL_570759, EPI_ISL_570760, EPI_ISL_570761, EPI_ISL_570762, EPI_ISL_570763, EPI_ISL_570764, EPI_ISL_570765, EPI_ISL_570766, EPI_ISL_570767, EPI_ISL_570768, EPI_ISL_570769, EPI_ISL_570770, EPI_ISL_570771, EPI_ISL_570772, EPI_ISL_570773, EPI_ISL_570774, EPI_ISL_570775, EPI_ISL_570776, EPI_ISL_570777, EPI_ISL_570778, EPI_ISL_570779, EPI_ISL_570780, EPI_ISL_570781, EPI_ISL_570782, EPI_ISL_570783, EPI_ISL_570784, EPI_ISL_570785, EPI_ISL_570786, EPI_ISL_570787, EPI_ISL_570788, EPI_ISL_570789, EPI_ISL_570790, EPI_ISL_570791, EPI_ISL_570792, EPI_ISL_570793, EPI_ISL_570794, EPI_ISL_570795, EPI_ISL_570796, EPI_ISL_570797, EPI_ISL_570798, EPI_ISL_570799, EPI_ISL_570800, EPI_ISL_570801, EPI_ISL_570802, EPI_ISL_570803, EPI_ISL_570804, EPI_ISL_570805, EPI_ISL_570806, EPI_ISL_570807, EPI_ISL_570808, EPI_ISL_570809, EPI_ISL_570810, EPI_ISL_570811, EPI_ISL_570812, EPI_ISL_570813, EPI_ISL_570814, EPI_ISL_570815, EPI_ISL_570816, EPI_ISL_570817, EPI_ISL_570818, EPI_ISL_570819, EPI_ISL_570820, EPI_ISL_570821, EPI_ISL_570822, EPI_ISL_570823, EPI_ISL_570824, EPI_ISL_570825, EPI_ISL_570826, EPI_ISL_570827, EPI_ISL_570828, EPI_ISL_570829, EPI_ISL_570830, EPI_ISL_570831, EPI_ISL_570832, EPI_ISL_570833, EPI_ISL_570834, EPI_ISL_570835, EPI_ISL_570836, EPI_ISL_570837, EPI_ISL_570838, EPI_ISL_570839, EPI_ISL_570840, EPI_ISL_570841, EPI_ISL_570842, EPI_ISL_570843, EPI_ISL_570844, EPI_ISL_570846, EPI_ISL_570847, EPI_ISL_570848, EPI_ISL_570849, EPI_ISL_570850, EPI_ISL_570851, EPI_ISL_570852, EPI_ISL_570853, EPI_ISL_570854, EPI_ISL_570855, EPI_ISL_570856, EPI_ISL_570857, EPI_ISL_570858, EPI_ISL_570859, EPI_ISL_570860, EPI_ISL_570861, EPI_ISL_570862, EPI_ISL_570863, EPI_ISL_570864, EPI_ISL_570865, EPI_ISL_570866, EPI_ISL_570867, EPI_ISL_570868, EPI_ISL_570869, EPI_ISL_570870, EPI_ISL_570871, EPI_ISL_570872, EPI_ISL_570873, EPI_ISL_570874, EPI_ISL_570875, EPI_ISL_570876, EPI_ISL_570877, EPI_ISL_570878, EPI_ISL_570879, EPI_ISL_570880, EPI_ISL_570881, EPI_ISL_570882, EPI_ISL_570883, EPI_ISL_570884, EPI_ISL_570885, EPI_ISL_570886, EPI_ISL_570887, EPI_ISL_570888, EPI_ISL_570889, EPI_ISL_570890, EPI_ISL_570891, EPI_ISL_570892, EPI_ISL_570893, EPI_ISL_570894, EPI_ISL_570895, EPI_ISL_570896, EPI_ISL_570897, EPI_ISL_570898, EPI_ISL_570899, EPI_ISL_570900, EPI_ISL_570901, EPI_ISL_570902, EPI_ISL_570903, EPI_ISL_570904, EPI_ISL_570905, EPI_ISL_570906, EPI_ISL_570907, EPI_ISL_570908, EPI_ISL_570909, EPI_ISL_570910, EPI_ISL_570911, EPI_ISL_570912, EPI_ISL_570913, EPI_ISL_570914, EPI_ISL_570915, EPI_ISL_570916, EPI_ISL_570917, EPI_ISL_570918, EPI_ISL_570919, EPI_ISL_570920, EPI_ISL_570921, EPI_ISL_570922, EPI_ISL_570923, EPI_ISL_570924, EPI_ISL_570925, EPI_ISL_570926, EPI_ISL_570927, EPI_ISL_570928, EPI_ISL_570929, EPI_ISL_570930, EPI_ISL_570931, EPI_ISL_570932, EPI_ISL_570933, EPI_ISL_570934, EPI_ISL_570935, EPI_ISL_570936, EPI_ISL_570937, EPI_ISL_570938, EPI_ISL_570939, EPI_ISL_570940, EPI_ISL_570941, EPI_ISL_570942, EPI_ISL_570943, EPI_ISL_570944, EPI_ISL_570945, EPI_ISL_570946, EPI_ISL_570947, EPI_ISL_570948, EPI_ISL_570949, EPI_ISL_570950, EPI_ISL_570951, EPI_ISL_570952, EPI_ISL_570953, EPI_ISL_570954, EPI_ISL_570955, EPI_ISL_570956, EPI_ISL_570957, EPI_ISL_570958, EPI_ISL_570959, EPI_ISL_570960, EPI_ISL_570961, EPI_ISL_570962, EPI_ISL_570963, EPI_ISL_570964, EPI_ISL_570965, EPI_ISL_570966, EPI_ISL_570967, EPI_ISL_570968, EPI_ISL_570969, EPI_ISL_570970, EPI_ISL_570971, EPI_ISL_570972, EPI_ISL_570973, EPI_ISL_570974, EPI_ISL_570975, EPI_ISL_570976, EPI_ISL_570977, EPI_ISL_570978, EPI_ISL_570979, EPI_ISL_570980, EPI_ISL_570981, EPI_ISL_570982, EPI_ISL_570983, EPI_ISL_570984, EPI_ISL_570985, EPI_ISL_570986, EPI_ISL_570987, EPI_ISL_570988, EPI_ISL_570989, EPI_ISL_570990, EPI_ISL_570991, EPI_ISL_570992, EPI_ISL_570993, EPI_ISL_570994, EPI_ISL_570995, EPI_ISL_570996, EPI_ISL_570997, EPI_ISL_570998, EPI_ISL_570999, EPI_ISL_571000, EPI_ISL_571001, EPI_ISL_571002, EPI_ISL_571003, EPI_ISL_571004, EPI_ISL_571005, EPI_ISL_571006, EPI_ISL_571007, EPI_ISL_571008, EPI_ISL_571009, EPI_ISL_571010, EPI_ISL_571011, EPI_ISL_571012, EPI_ISL_571013, EPI_ISL_571014, EPI_ISL_571015 | | | |
| see above | UW Virology Lab | UW Virology Lab | Pavitra Roychoudhury, Hong Xie, Lasata Shrestha, Amin Addetia, Victoria M Rachleff, Meei-Li Huang, Keith R Jerome, Alexander Greninger |
| EPI_ISL_571016, EPI_ISL_571017, EPI_ISL_571018, EPI_ISL_571019, EPI_ISL_571020, EPI_ISL_571021, EPI_ISL_571022, EPI_ISL_571023, EPI_ISL_571024, EPI_ISL_571025, EPI_ISL_571026, EPI_ISL_571027, EPI_ISL_571028, EPI_ISL_571029, EPI_ISL_571030, EPI_ISL_571031, EPI_ISL_571032, EPI_ISL_571033, EPI_ISL_571034, EPI_ISL_571035, EPI_ISL_571036, EPI_ISL_571037, EPI_ISL_571038, EPI_ISL_571039, EPI_ISL_571040, EPI_ISL_571041, EPI_ISL_571042, EPI_ISL_571043, EPI_ISL_571044, EPI_ISL_571045, EPI_ISL_571046, EPI_ISL_571047, EPI_ISL_571048, EPI_ISL_571049, EPI_ISL_571050, EPI_ISL_571051, EPI_ISL_571052, EPI_ISL_571053, EPI_ISL_571054, EPI_ISL_571055, EPI_ISL_571056, EPI_ISL_571057, EPI_ISL_571058, EPI_ISL_571059, EPI_ISL_571060, EPI_ISL_571061, EPI_ISL_571062, EPI_ISL_571063, EPI_ISL_571064, EPI_ISL_571065, EPI_ISL_571066, EPI_ISL_571067, EPI_ISL_571068, EPI_ISL_571069, EPI_ISL_571070, EPI_ISL_571071, EPI_ISL_571072, EPI_ISL_571073, EPI_ISL_571074, EPI_ISL_571075, EPI_ISL_571076, EPI_ISL_571077, EPI_ISL_571078, EPI_ISL_571079, EPI_ISL_571080, EPI_ISL_571081, EPI_ISL_571082, EPI_ISL_571083, EPI_ISL_571084, EPI_ISL_571085, EPI_ISL_571086, EPI_ISL_571087, EPI_ISL_571088, EPI_ISL_571089, EPI_ISL_571090, EPI_ISL_571091, EPI_ISL_571092, EPI_ISL_571093, EPI_ISL_571094, EPI_ISL_571095, EPI_ISL_571096, EPI_ISL_571097, EPI_ISL_571098, EPI_ISL_571099, EPI_ISL_571100, EPI_ISL_571101, EPI_ISL_571102, EPI_ISL_571103, EPI_ISL_571104, EPI_ISL_571105, EPI_ISL_571106, EPI_ISL_571107, EPI_ISL_571108, EPI_ISL_571109, EPI_ISL_571110, EPI_ISL_571111, EPI_ISL_571112, EPI_ISL_571113, EPI_ISL_571114, EPI_ISL_571115, EPI_ISL_571116, EPI_ISL_571117, EPI_ISL_571118, EPI_ISL_571119, EPI_ISL_571120, EPI_ISL_571121, EPI_ISL_571122, EPI_ISL_571123, EPI_ISL_571124, EPI_ISL_571125, EPI_ISL_571126, EPI_ISL_571127, EPI_ISL_571128, EPI_ISL_571129, EPI_ISL_571130, EPI_ISL_571131, EPI_ISL_571132, EPI_ISL_571133, EPI_ISL_571134, EPI_ISL_571135, EPI_ISL_571136, EPI_ISL_571137, EPI_ISL_571138, EPI_ISL_571139, EPI_ISL_571140, EPI_ISL_571141, EPI_ISL_571142, EPI_ISL_571143, EPI_ISL_571144, EPI_ISL_571145, EPI_ISL_571146, EPI_ISL_571147, EPI_ISL_571148, EPI_ISL_571149, EPI_ISL_571150, EPI_ISL_571151, EPI_ISL_571152, EPI_ISL_571153, EPI_ISL_571154, EPI_ISL_571155, EPI_ISL_571156, EPI_ISL_571157, EPI_ISL_571158, EPI_ISL_571159, EPI_ISL_571160, EPI_ISL_571161, EPI_ISL_571162, EPI_ISL_571163, EPI_ISL_571164, EPI_ISL_571165, EPI_ISL_571166, EPI_ISL_571167, EPI_ISL_571168, EPI_ISL_571169, EPI_ISL_571170, EPI_ISL_571171, EPI_ISL_571172, EPI_ISL_571173, EPI_ISL_571174, EPI_ISL_571175, EPI_ISL_571176, EPI_ISL_571177, EPI_ISL_571178, EPI_ISL_571179, EPI_ISL_571180, EPI_ISL_571181, EPI_ISL_571182, EPI_ISL_571183, EPI_ISL_571184, EPI_ISL_571185, EPI_ISL_571186, EPI_ISL_571187, EPI_ISL_571188, EPI_ISL_571189, EPI_ISL_571190, EPI_ISL_571191, EPI_ISL_571192, EPI_ISL_571193, EPI_ISL_571194, EPI_ISL_571195, EPI_ISL_571196, EPI_ISL_571197, EPI_ISL_571198, EPI_ISL_571199, EPI_ISL_571200, EPI_ISL_571201, EPI_ISL_571202, EPI_ISL_571203, EPI_ISL_571204, EPI_ISL_571205, EPI_ISL_571206, EPI_ISL_571207, EPI_ISL_571208, EPI_ISL_571209, EPI_ISL_571210, EPI_ISL_571211, EPI_ISL_571212, EPI_ISL_571213, EPI_ISL_571214, EPI_ISL_571215, EPI_ISL_571216, EPI_ISL_571217, EPI_ISL_571218, EPI_ISL_571219, EPI_ISL_571220, EPI_ISL_571221, EPI_ISL_571222, EPI_ISL_571223, EPI_ISL_571224, EPI_ISL_571225, EPI_ISL_571226, EPI_ISL_571227, EPI_ISL_571228, EPI_ISL_571229, EPI_ISL_571230, EPI_ISL_571231, EPI_ISL_571232, EPI_ISL_571233, EPI_ISL_571234, EPI_ISL_571235, EPI_ISL_571236, EPI_ISL_571237, EPI_ISL_571238, EPI_ISL_571239, EPI_ISL_571240, EPI_ISL_571241, EPI_ISL_571242, EPI_ISL_571243, EPI_ISL_571244, EPI_ISL_571245, EPI_ISL_571246, EPI_ISL_571247, EPI_ISL_571248, EPI_ISL_571249, EPI_ISL_571250, EPI_ISL_571251, EPI_ISL_571252, EPI_ISL_571253, EPI_ISL_571254, EPI_ISL_571255, EPI_ISL_571256, EPI_ISL_571257, EPI_ISL_571258, EPI_ISL_571259, EPI_ISL_571260, EPI_ISL_571261, EPI_ISL_571262, EPI_ISL_571263, EPI_ISL_571264, EPI_ISL_571265, EPI_ISL_571266, EPI_ISL_571267, EPI_ISL_571268, EPI_ISL_571269, EPI_ISL_571270, EPI_ISL_571271, EPI_ISL_571272, EPI_ISL_571273, EPI_ISL_571274, EPI_ISL_571275, EPI_ISL_571276, EPI_ISL_571277, EPI_ISL_571278, EPI_ISL_571279, EPI_ISL_571280, EPI_ISL_571281, EPI_ISL_571282, EPI_ISL_571283, EPI_ISL_571284, EPI_ISL_571285, EPI_ISL_571286, EPI_ISL_571287, EPI_ISL_571288, EPI_ISL_571289, EPI_ISL_571290, EPI_ISL_571291, EPI_ISL_571292, EPI_ISL_571293, EPI_ISL_571294, EPI_ISL_571295, EPI_ISL_571296, EPI_ISL_571297, EPI_ISL_571298, EPI_ISL_571299, EPI_ISL_571300, EPI_ISL_571301, EPI_ISL_571302, EPI_ISL_571303, EPI_ISL_571304, EPI_ISL_571305, EPI_ISL_571306, EPI_ISL_571307, EPI_ISL_571308, EPI_ISL_571309, EPI_ISL_571310, EPI_ISL_571311, EPI_ISL_571312, EPI_ISL_571313, EPI_ISL_571314, EPI_ISL_571315, EPI_ISL_571316, EPI_ISL_571317, EPI_ISL_571318, EPI_ISL_571319, EPI_ISL_571320, EPI_ISL_571321, EPI_ISL_571322, EPI_ISL_571323, EPI_ISL_571324, EPI_ISL_571325, EPI_ISL_571326, EPI_ISL_571327, EPI_ISL_571328, EPI_ISL_571329, EPI_ISL_571330, EPI_ISL_571331, EPI_ISL_571332, EPI_ISL_571333, EPI_ISL_571334, EPI_ISL_571335, EPI_ISL_571336, EPI_ISL_571337, EPI_ISL_571338, EPI_ISL_571339, EPI_ISL_571340, EPI_ISL_571341, EPI_ISL_571342, EPI_ISL_571343, EPI_ISL_571344, EPI_ISL_571345, EPI_ISL_571346, EPI_ISL_571347, EPI_ISL_571348, EPI_ISL_571349, EPI_ISL_571350, EPI_ISL_571351, EPI_ISL_571352, EPI_ISL_571353, EPI_ISL_571354, EPI_ISL_571355, EPI_ISL_571356, EPI_ISL_571357, EPI_ISL_571358, EPI_ISL_571359, EPI_ISL_571360, EPI_ISL_571361, EPI_ISL_571362, EPI_ISL_571363, EPI_ISL_571364, EPI_ISL_571365, EPI_ISL_571366, EPI_ISL_571367, EPI_ISL_571368, EPI_ISL_571369, EPI_ISL_571370, EPI_ISL_571371, EPI_ISL_571372, EPI_ISL_571373, EPI_ISL_571374, EPI_ISL_571375, EPI_ISL_571376, EPI_ISL_571377, EPI_ISL_571378, EPI_ISL_571379, EPI_ISL_571380, EPI_ISL_571381, EPI_ISL_571382, EPI_ISL_571383, EPI_ISL_571384, EPI_ISL_571385, EPI_ISL_571386, EPI_ISL_571387, EPI_ISL_571388, EPI_ISL_571389, EPI_ISL_571390, EPI_ISL_571391, EPI_ISL_571392, EPI_ISL_571393, EPI_ISL_571394, EPI_ISL_571395, EPI_ISL_571396, EPI_ISL_571397, EPI_ISL_571398, EPI_ISL_571399, EPI_ISL_571400, EPI_ISL_571401, EPI_ISL_571402, EPI_ISL_571403, EPI_ISL_571404, EPI_ISL_571405, EPI_ISL_571406, EPI_ISL_571407, EPI_ISL_571408, EPI_ISL_571409, EPI_ISL_571410, EPI_ISL_571411, EPI_ISL_571412, EPI_ISL_571413, EPI_ISL_571414, EPI_ISL_571415, EPI_ISL_571416, EPI_ISL_571417, EPI_ISL_571418, EPI_ISL_571419, EPI_ISL_571420, EPI_ISL_571421, EPI_ISL_571422, EPI_ISL_571423, EPI_ISL_571424, EPI_ISL_571425, EPI_ISL_571426, EPI_ISL_571427, EPI_ISL_571428, EPI_ISL_571429, EPI_ISL_571430, EPI_ISL_571431, EPI_ISL_571432, EPI_ISL_571433, EPI_ISL_571434, EPI_ISL_571435, EPI_ISL_571436, EPI_ISL_571437, EPI_ISL_571438, EPI_ISL_571439, EPI_ISL_571440, EPI_ISL_571441, EPI_ISL_571442, EPI_ISL_571443, EPI_ISL_571444, EPI_ISL_571445, EPI_ISL_571446, EPI_ISL_571447, EPI_ISL_571448, EPI_ISL_571449, EPI_ISL_571450, EPI_ISL_571451, EPI_ISL_571452, EPI_ISL_571453, EPI_ISL_571454, EPI_ISL_571455, EPI_ISL_571456, EPI_ISL_571457, EPI_ISL_571458, EPI_ISL_571459, EPI_ISL_571460, EPI_ISL_571461, EPI_ISL_571462, EPI_ISL_571463, EPI_ISL_571464, EPI_ISL_571465, EPI_ISL_571466, EPI_ISL_571467, EPI_ISL_571468, EPI_ISL_571469, EPI_ISL_571470, EPI_ISL_571471, EPI_ISL_571472, EPI_ISL_571473, EPI_ISL_571474, EPI_ISL_571475, EPI_ISL_571476, EPI_ISL_571477, EPI_ISL_571478, EPI_ISL_571479, EPI_ISL_571480, EPI_ISL_571481, EPI_ISL_571482, EPI_ISL_571483, EPI_ISL_571484, EPI_ISL_571485, EPI_ISL_571486, EPI_ISL_571487, EPI_ISL_571488, EPI_ISL_571489, EPI_ISL_571490, EPI_ISL_571491, EPI_ISL_571492, EPI_ISL_571493, EPI_ISL_571494, EPI_ISL_571495, EPI_ISL_571496, EPI_ISL_571497, EPI_ISL_571498, EPI_ISL_571499, EPI_ISL_571500, EPI_ISL_571501, EPI_ISL_571502, EPI_ISL_571503, EPI_ISL_571504, EPI_ISL_571505, EPI_ISL_571506, EPI_ISL_571507, EPI_ISL_571508, EPI_ISL_571509, EPI_ISL_571510, EPI_ISL_571511, EPI_ISL_571512, EPI_ISL_571513, EPI_ISL_571514, EPI_ISL_571515, EPI_ISL_571516, EPI_ISL_571517, EPI_ISL_571518, EPI_ISL_571519, EPI_ISL_571520, EPI_ISL_571521, EPI_ISL_571522, EPI_ISL_571523, EPI_ISL_571524, EPI_ISL_571525, EPI_ISL_571526, EPI_ISL_571527, EPI_ISL_571528, EPI_ISL_571529, EPI_ISL_571530, EPI_ISL_571531, EPI_ISL_571532, EPI_ISL_571533, EPI_ISL_571534, EPI_ISL_571535, EPI_ISL_571536, EPI_ISL_571537, EPI_ISL_571538, EPI_ISL_571539, EPI_ISL_571540, EPI_ISL_571541, EPI_ISL_571542, EPI_ISL_571543, EPI_ISL_571544, EPI_ISL_571545, EPI_ISL_571546, EPI_ISL_571547, EPI_ISL_571548, EPI_ISL_571549, EPI_ISL_571550, EPI_ISL_571551, EPI_ISL_571552, EPI_ISL_571553, EPI_ISL_571554, EPI_ISL_571555, EPI_ISL_571556, EPI_ISL_571557, EPI_ISL_571558, EPI_ISL_571559, EPI_ISL_571560, EPI_ISL_571561, EPI_ISL_571562, EPI_ISL_571563, EPI_ISL_571564, EPI_ISL_571565, EPI_ISL_571566, EPI_ISL_571567, EPI_ISL_571568, EPI_ISL_571569, EPI_ISL_571570, EPI_ISL_571571, EPI_ISL_571572, EPI_ISL_571573, EPI_ISL_571574, EPI_ISL_571575, EPI_ISL_571576, EPI_ISL_571577, EPI_ISL_571578, EPI_ISL_571579, EPI_ISL_571580, EPI_ISL_571581, EPI_ISL_571582, EPI_ISL_571583, EPI_ISL_571584, EPI_ISL_571585, EPI_ISL_571586, EPI_ISL_571587, EPI_ISL_571588, EPI_ISL_571589, EPI_ISL_571590, EPI_ISL_571591, EPI_ISL_571592, EPI_ISL_571593, EPI_ISL_571594, EPI_ISL_571595, EPI_ISL_571596, EPI_ISL_571597, EPI_ISL_571598, EPI_ISL_571599, EPI_ISL_571600, EPI_ISL_571601, EPI_ISL_571602, EPI_ISL_571603, EPI_ISL_571604, EPI_ISL_571605, EPI_ISL_571606, EPI_ISL_571607, EPI_ISL_571608, EPI_ISL_571609, EPI_ISL_571610, EPI_ISL_571611, EPI_ISL_571612, EPI_ISL_571613, EPI_ISL_571614, EPI_ISL_571615, EPI_ISL_571616, EPI_ISL_571617, EPI_ISL_571618, EPI_ISL_571619, EPI_ISL_571620, EPI_ISL_571621, EPI_ISL_571622, EPI_ISL_571623, EPI_ISL_571624, EPI_ISL_571625, EPI_ISL_571626, EPI_ISL_571627, EPI_ISL_571628, EPI_ISL_571629, EPI_ISL_571630, EPI_ISL_571631, EPI_ISL_571632, EPI_ISL_571633, EPI_ISL_571634, EPI_ISL_571635, EPI_ISL_571636, EPI_ISL_571637, EPI_ISL_571638, EPI_ISL_571639, EPI_ISL_571640, EPI_ISL_571641, EPI_ISL_571642, EPI_ISL_571643, EPI_ISL_571644, EPI_ISL_571645, EPI_ISL_571646, EPI_ISL_571647, EPI_ISL_571648, EPI_ISL_571649, EPI_ISL_571650, EPI_ISL_571651, EPI_ISL_571652, EPI_ISL_571653, EPI_ISL_571654, EPI_ISL_571655, EPI_ISL_571656, EPI_ISL_571657, EPI_ISL_571658, EPI_ISL_571659, EPI_ISL_571660, EPI_ISL_571661, EPI_ISL_571662, EPI_ISL_571663, EPI_ISL_571664, EPI_ISL_571665, EPI_ISL_571666, EPI_ISL_571667, EPI_ISL_571668, EPI_ISL_571669, EPI_ISL_571670, EPI_ISL_571671, EPI_ISL_571672, EPI_ISL_571673, EPI_ISL_571674, EPI_ISL_571675, EPI_ISL_571676, EPI_ISL_571677, EPI_ISL_571678, EPI_ISL_571679, EPI_ISL_571680, EPI_ISL_571681, EPI_ISL_571682, EPI_ISL_571683, EPI_ISL_571684, EPI_ISL_571685, EPI_ISL_571686, EPI_ISL_571687, EPI_ISL_571688, EPI_ISL_571689, EPI_ISL_571690, EPI_ISL_571691, EPI_ISL_571692, EPI_ISL_571693, EPI_ISL_571694, EPI_ISL_571695, EPI_ISL_571696, EPI_ISL_571697, EPI_ISL_571698, EPI_ISL_571699, EPI_ISL_571700, EPI_ISL_571701, EPI_ISL_571702, EPI_ISL_571703, EPI_ISL_571704, EPI_ISL_571705, EPI_ISL_571706, EPI_ISL_571707, EPI_ISL_571708, EPI_ISL_571709, EPI_ISL_571710, EPI_ISL_571711, EPI_ISL_571712, EPI_ISL_571713, EPI_ISL_571714, EPI_ISL_571715, EPI_ISL_571716, EPI_ISL_571717, EPI_ISL_571718, EPI_ISL_571719, EPI_ISL_571720, EPI_ISL_571721, EPI_ISL_571722, EPI_ISL_571723, EPI_ISL_571724, EPI_ISL_571725, EPI_ISL_571726, EPI_ISL_571727, EPI_ISL_571728, EPI_ISL_571729, EPI_ISL_571730, EPI_ISL_571731, EPI_ISL_571732, EPI_ISL_571733, EPI_ISL_571734, EPI_ISL_571735, EPI_ISL_571736, EPI_ISL_571737, EPI_ISL_571738, EPI_ISL_571739, EPI_ISL_571740, EPI_ISL_571741, EPI_ISL_571742, EPI_ISL_571743, EPI_ISL_571744, EPI_ISL_571745, EPI_ISL_571746, EPI_ISL_571747, EPI_ISL_571748, EPI_ISL_571749, EPI_ISL_571750, EPI_ISL_571751, EPI_ISL_571752, EPI_ISL_571753, EPI_ISL_571754, EPI_ISL_571755, EPI_ISL_571756, EPI_ISL_571757, EPI_ISL_571758, EPI_ISL_571759, EPI_ISL_571760, EPI_ISL_571761, EPI_ISL_571762, EPI_ISL_571763, EPI_ISL_571764, EPI_ISL_571765, EPI_ISL_571766, EPI_ISL_571767, EPI_ISL_571768, EPI_ISL_571769, EPI_ISL_571770, EPI_ISL_571771, EPI_ISL_571772, EPI_ISL_571773, EPI_ISL_571774, EPI_ISL_571775, EPI_ISL_571776, EPI_ISL_571777, EPI_ISL_571778, EPI_ISL_571779, EPI_ISL_571780, EPI_ISL_571781, EPI_ISL_571782, EPI_ISL_571783, EPI_ISL_571784, EPI_ISL_571785, EPI_ISL_571786, EPI_ISL_571787, EPI_ISL_571788, EPI_ISL_571789, EPI_ISL_571790, EPI_ISL_571791, EPI_ISL_571792, EPI_ISL_571793, EPI_ISL_571794, EPI_ISL_571795, EPI_ISL_571796, EPI_ISL_571797, EPI_ISL_571798, EPI_ISL_571799, EPI_ISL_571800, EPI_ISL_571801, EPI_ISL_571802, EPI_ISL_571803, EPI_ISL_571804, EPI_ISL_571805, EPI_ISL_571806, EPI_ISL_571807, EPI_ISL_571808, EPI_ISL_571809, EPI_ISL_571810, EPI_ISL_571811, EPI_ISL_571812, EPI_ISL_571813, EPI_ISL_571814, EPI_ISL_571815, EPI_ISL_571816, EPI_ISL_571817, EPI_ISL_571818, EPI_ISL_571819, EPI_ISL_571820, EPI_ISL_571821, EPI_ISL_571822, EPI_ISL_571823, EPI_ISL_571824, EPI_ISL_571825, EPI_ISL_571826, EPI_ISL_571827, EPI_ISL_571828, EPI_ISL_571829, EPI_ISL_571830, EPI_ISL_571831, EPI_ISL_571832, EPI_ISL_571833, EPI_ISL_571834, EPI_ISL_571835, EPI_ISL_571836, EPI_ISL_571837, EPI_ISL_571838, EPI_ISL_571839, EPI_ISL_571840, EPI_ISL_571841, EPI_ISL_571842, EPI_ISL_571843, EPI_ISL_571844, EPI_ISL_571845, EPI_ISL_571846, EPI_ISL_571847, EPI_ISL_571848, EPI_ISL_571849, EPI_ISL_571850, EPI_ISL_571851, EPI_ISL_571852, EPI_ISL_571853, EPI_ISL_571854, EPI_ISL_571855, EPI_ISL_571856, EPI_ISL_571857, EPI_ISL_571858, EPI_ISL_571859, EPI_ISL_571860, EPI_ISL_571861, EPI_ISL_571862, EPI_ISL_571863, EPI_ISL_571864, EPI_ISL_571865, EPI_ISL_571866, EPI_ISL_571867, EPI_ISL_571868, EPI_ISL_571869, EPI_ISL_571870, EPI_ISL_571871, EPI_ISL_571872, EPI_ISL_571873, EPI_ISL_571874, EPI_ISL_571875, EPI_ISL_571876, EPI_ISL_571877, EPI_ISL_571878, EPI_ISL_571879, EPI_ISL_571880, EPI_ISL_571881, EPI_ISL_571882, EPI_ISL_571883, EPI_ISL_571884, EPI_ISL_571885, EPI_ISL_571886, EPI_ISL_571887, EPI_ISL_571888, EPI_ISL_571889, EPI_ISL_571890, EPI_ISL_571891, EPI_ISL_571892, EPI_ISL_571893, EPI_ISL_571894, EPI_ISL_571895, EPI_ISL_571896, EPI_ISL_571897, EPI_ISL_571898, EPI_ISL_571899, EPI_ISL_571900, EPI_ISL_571901, EPI_ISL_571902, EPI_ISL_571903, | | | |

| | | | |
|---|---|---|---|
| EPI_ISL_571904, EPI_ISL_571905, EPI_ISL_571906, EPI_ISL_571907, EPI_ISL_571908, EPI_ISL_571909, EPI_ISL_571910, EPI_ISL_571911, EPI_ISL_571912, EPI_ISL_571913, EPI_ISL_571914, EPI_ISL_571915, EPI_ISL_571916, EPI_ISL_571917, EPI_ISL_571918, EPI_ISL_571919, EPI_ISL_571920, EPI_ISL_571921, EPI_ISL_571922, EPI_ISL_571923, EPI_ISL_571924, EPI_ISL_571925, EPI_ISL_571926, EPI_ISL_571927, EPI_ISL_571928, EPI_ISL_571929, EPI_ISL_571930, EPI_ISL_571931, EPI_ISL_571932, EPI_ISL_571933, EPI_ISL_571934, EPI_ISL_571935, EPI_ISL_571936, EPI_ISL_571937, EPI_ISL_571938, EPI_ISL_571939, EPI_ISL_571940, EPI_ISL_571941, EPI_ISL_571942, EPI_ISL_571943, EPI_ISL_571944, EPI_ISL_571945, EPI_ISL_571946, EPI_ISL_571947, EPI_ISL_571948, EPI_ISL_571949, EPI_ISL_571950, EPI_ISL_571951, EPI_ISL_571952, EPI_ISL_571953, EPI_ISL_571954, EPI_ISL_571955, EPI_ISL_571956, EPI_ISL_571957, EPI_ISL_571958, EPI_ISL_571959, EPI_ISL_571960, EPI_ISL_571961, EPI_ISL_571962, EPI_ISL_571963, EPI_ISL_571964, EPI_ISL_571965, EPI_ISL_571966, EPI_ISL_571967, EPI_ISL_571968, EPI_ISL_571969, EPI_ISL_571970, EPI_ISL_571971, EPI_ISL_571972, EPI_ISL_571973, EPI_ISL_571974, EPI_ISL_571975, EPI_ISL_571976, EPI_ISL_571977, EPI_ISL_571978, EPI_ISL_571979, EPI_ISL_571980, EPI_ISL_571981, EPI_ISL_571982, EPI_ISL_571983, EPI_ISL_571984, EPI_ISL_571985, EPI_ISL_571986, EPI_ISL_571987, EPI_ISL_571988, EPI_ISL_571989, EPI_ISL_571990, EPI_ISL_571991, EPI_ISL_571992, EPI_ISL_571993, EPI_ISL_571994, EPI_ISL_571995, EPI_ISL_571996, EPI_ISL_571997, EPI_ISL_571998, EPI_ISL_571999, EPI_ISL_572000, EPI_ISL_572001, EPI_ISL_572002, EPI_ISL_572003, EPI_ISL_572004, EPI_ISL_572005, EPI_ISL_572006, EPI_ISL_572007, EPI_ISL_572008, EPI_ISL_572009, EPI_ISL_572010, EPI_ISL_572011, EPI_ISL_572012, EPI_ISL_572013, EPI_ISL_572014, EPI_ISL_572015, EPI_ISL_572016, EPI_ISL_572017, EPI_ISL_572018, EPI_ISL_572019, EPI_ISL_572020, EPI_ISL_572021, EPI_ISL_572022, EPI_ISL_572023, EPI_ISL_572024, EPI_ISL_572025, EPI_ISL_572026, EPI_ISL_572027, EPI_ISL_572028, EPI_ISL_572029, EPI_ISL_572030, EPI_ISL_572031, EPI_ISL_572032, EPI_ISL_572033, EPI_ISL_572034, EPI_ISL_572035, EPI_ISL_572036, EPI_ISL_572037, EPI_ISL_572038, EPI_ISL_572039, EPI_ISL_572040, EPI_ISL_572041, EPI_ISL_572042, EPI_ISL_572043, EPI_ISL_572044, EPI_ISL_572045, EPI_ISL_572046, EPI_ISL_572047, EPI_ISL_572048, EPI_ISL_572049, EPI_ISL_572050, EPI_ISL_572051, EPI_ISL_572052, EPI_ISL_572053, EPI_ISL_572054, EPI_ISL_572055, EPI_ISL_572056, EPI_ISL_572057, EPI_ISL_572058, EPI_ISL_572059, EPI_ISL_572060, EPI_ISL_572061, EPI_ISL_572062, EPI_ISL_572063, EPI_ISL_572064, EPI_ISL_572065, EPI_ISL_572066, EPI_ISL_572067, EPI_ISL_572068, EPI_ISL_572069, EPI_ISL_572070, EPI_ISL_572071, EPI_ISL_572072, EPI_ISL_572073, EPI_ISL_572074, EPI_ISL_572075, EPI_ISL_572076, EPI_ISL_572077, EPI_ISL_572078, EPI_ISL_572079, EPI_ISL_572080, EPI_ISL_572081, EPI_ISL_572082, EPI_ISL_572083, EPI_ISL_572084, EPI_ISL_572085, EPI_ISL_572086, EPI_ISL_572087, EPI_ISL_572088, EPI_ISL_572089, EPI_ISL_572090, EPI_ISL_572091, EPI_ISL_572092, EPI_ISL_572093, EPI_ISL_572094, EPI_ISL_572095, EPI_ISL_572096, EPI_ISL_572097, EPI_ISL_572098, EPI_ISL_572099, EPI_ISL_572100, EPI_ISL_572101, EPI_ISL_572102, EPI_ISL_572103, EPI_ISL_572104, EPI_ISL_572105, EPI_ISL_572106, EPI_ISL_572107, EPI_ISL_572108, EPI_ISL_572109, EPI_ISL_572110, EPI_ISL_572111, EPI_ISL_572112, EPI_ISL_572113, EPI_ISL_572114, EPI_ISL_572115, EPI_ISL_572116, EPI_ISL_572117, EPI_ISL_572118, EPI_ISL_572119, EPI_ISL_572120, EPI_ISL_572121, EPI_ISL_572122, EPI_ISL_572123, EPI_ISL_572124, EPI_ISL_572125, EPI_ISL_572126, EPI_ISL_572127, EPI_ISL_572128, EPI_ISL_572129, EPI_ISL_572130, EPI_ISL_572131, EPI_ISL_572132, EPI_ISL_572133, EPI_ISL_572134, EPI_ISL_572135, EPI_ISL_572136, EPI_ISL_572137, EPI_ISL_572138, EPI_ISL_572139, EPI_ISL_572140, EPI_ISL_572141, EPI_ISL_572142, EPI_ISL_572143, EPI_ISL_572144, EPI_ISL_572145, EPI_ISL_572146, EPI_ISL_572147, EPI_ISL_572148, EPI_ISL_572149, EPI_ISL_572150, EPI_ISL_572151, EPI_ISL_572152, EPI_ISL_572153, EPI_ISL_572154, EPI_ISL_572155, EPI_ISL_572156, EPI_ISL_572157, EPI_ISL_572158, EPI_ISL_572159, EPI_ISL_572160, EPI_ISL_572161, EPI_ISL_572162, EPI_ISL_572163, EPI_ISL_572164, EPI_ISL_572165, EPI_ISL_572166, EPI_ISL_572167, EPI_ISL_572168, EPI_ISL_572169, EPI_ISL_572170, EPI_ISL_572171, EPI_ISL_572172, EPI_ISL_572173, EPI_ISL_572174, EPI_ISL_572175, EPI_ISL_572176, EPI_ISL_572177, EPI_ISL_572178, EPI_ISL_572179, EPI_ISL_572180, EPI_ISL_572181, EPI_ISL_572182, EPI_ISL_572183, EPI_ISL_572184, EPI_ISL_572185 | | | |
| see above | Quest Diagnostics | Quest Diagnostics | Rosenthal,S.H., Gerasimova,A., Kagan,R.M., Anderson, B., Grover, D., Livingston, K.E., Hua, M., Liu Y., Shalhout, D.F., Owen, R., Lacbawan, F. |
| EPI_ISL_572195, EPI_ISL_572196, EPI_ISL_572197, EPI_ISL_572198, EPI_ISL_572199, EPI_ISL_572200, EPI_ISL_572201, EPI_ISL_572202, EPI_ISL_572203, EPI_ISL_572204, EPI_ISL_572205, EPI_ISL_572206, EPI_ISL_572207, EPI_ISL_572208, EPI_ISL_572209, EPI_ISL_572210, EPI_ISL_572211, EPI_ISL_572212, EPI_ISL_572213, EPI_ISL_572214, EPI_ISL_572215, EPI_ISL_572216, EPI_ISL_572217, EPI_ISL_572218, EPI_ISL_572219, EPI_ISL_572220, EPI_ISL_572221, EPI_ISL_572222, EPI_ISL_572223, EPI_ISL_572224, EPI_ISL_572225, EPI_ISL_572226, EPI_ISL_572227, EPI_ISL_572228, EPI_ISL_572229, EPI_ISL_572230, EPI_ISL_572231, EPI_ISL_572232, EPI_ISL_572233, EPI_ISL_572234, EPI_ISL_572235, EPI_ISL_572236, EPI_ISL_572237, EPI_ISL_572238, EPI_ISL_572239, EPI_ISL_572240, EPI_ISL_572241, EPI_ISL_572242, EPI_ISL_572243, EPI_ISL_572244, EPI_ISL_572245, EPI_ISL_572246, EPI_ISL_572247, EPI_ISL_572248, EPI_ISL_572249, EPI_ISL_572250, EPI_ISL_572251, EPI_ISL_572252, EPI_ISL_572253, EPI_ISL_572254, EPI_ISL_572255, EPI_ISL_572256, EPI_ISL_572257, EPI_ISL_572258, EPI_ISL_572259, EPI_ISL_572260, EPI_ISL_572261, EPI_ISL_572262, EPI_ISL_572263, EPI_ISL_572264, EPI_ISL_572265, EPI_ISL_572266, EPI_ISL_572267, EPI_ISL_572268, EPI_ISL_572269, EPI_ISL_572270, EPI_ISL_572271, EPI_ISL_572272, EPI_ISL_572273, EPI_ISL_572274, EPI_ISL_572275, EPI_ISL_572276, EPI_ISL_572277, EPI_ISL_572278, EPI_ISL_572279, EPI_ISL_572280, EPI_ISL_572281, EPI_ISL_572282, EPI_ISL_572283, EPI_ISL_572284, EPI_ISL_572285, EPI_ISL_572286, EPI_ISL_572287, EPI_ISL_572288, EPI_ISL_572289, EPI_ISL_572290, EPI_ISL_572291, EPI_ISL_572292, EPI_ISL_572293, EPI_ISL_572294, EPI_ISL_572295, EPI_ISL_572296, EPI_ISL_572297, EPI_ISL_572298, EPI_ISL_572299, EPI_ISL_572300, EPI_ISL_572301, EPI_ISL_572302, EPI_ISL_572303, EPI_ISL_572304, EPI_ISL_572305, EPI_ISL_572306, EPI_ISL_572307, EPI_ISL_572308, EPI_ISL_572309, EPI_ISL_572310, EPI_ISL_572311, EPI_ISL_572312, EPI_ISL_572313, EPI_ISL_572314, EPI_ISL_572315, EPI_ISL_572316, EPI_ISL_572317, EPI_ISL_572318 | | | |
| see above | Virginia DCLS | Virginia DCLS | Virginia DCLS |
| EPI_ISL_574260, EPI_ISL_574261, EPI_ISL_574262, EPI_ISL_574263, EPI_ISL_574264, EPI_ISL_574265, EPI_ISL_574266, EPI_ISL_574267, EPI_ISL_574268, EPI_ISL_574269, EPI_ISL_574270, EPI_ISL_574271, EPI_ISL_574272, EPI_ISL_574273, EPI_ISL_574274, EPI_ISL_574275, EPI_ISL_574276, EPI_ISL_574277, EPI_ISL_574278, EPI_ISL_574279, EPI_ISL_574280, EPI_ISL_574281, EPI_ISL_574282, EPI_ISL_574283, EPI_ISL_574284, EPI_ISL_574285, EPI_ISL_574286, EPI_ISL_574287, EPI_ISL_574288, EPI_ISL_574289, EPI_ISL_574290, EPI_ISL_574291 | | | |
| see above | New Mexico Department of Health Scientific Laboratory | New Mexico Department of Health Scientific Laboratory | Ellie Johnson, Anastacia Griego-Fisher, D'Eldra Malone |
| EPI_ISL_574295, EPI_ISL_574296, EPI_ISL_574297, EPI_ISL_574298, EPI_ISL_574299, EPI_ISL_574300 | LSUHS Emerging Viral Threat Laboratory | Microbial Genome Sequencing Center | Jeremy P. Kamil, Rona S. Scott, Maarten Van Diest, Malgorzata Bienkowska-Haba, Katarzyna Zwolinska, Andrew D. Yurochko, Christopher G. Kevil, Martin J. Sapp, Daniel J. Snyder, Vaughn S. Cooper, John A. Vanchiere |
| EPI_ISL_574301, EPI_ISL_574302, EPI_ISL_574303, EPI_ISL_574304, EPI_ISL_574305, EPI_ISL_574306, EPI_ISL_574307, EPI_ISL_574308, EPI_ISL_574309, EPI_ISL_574310, EPI_ISL_574311, EPI_ISL_574312, EPI_ISL_574313, EPI_ISL_574314, EPI_ISL_574315, EPI_ISL_574316, EPI_ISL_574317, EPI_ISL_574318, EPI_ISL_574319 | | | |
| see above | LSUHS Emerging Viral Threat Laboratory | Microbial Genome Sequencing Center | Rona S. Scott, Jeremy P. Kamil, Maarten Van Diest, Malgorzata Bienkowska-Haba, Katarzyna Zwolinska, Andrew D. Yurochko, Christopher G. Kevil, Martin J. Sapp, Daniel J. Snyder, Vaughn S. Cooper, John A. Vanchiere |
| EPI_ISL_574320, EPI_ISL_574321, EPI_ISL_574322, EPI_ISL_574323, EPI_ISL_574324, EPI_ISL_574325, EPI_ISL_574326, EPI_ISL_574327, EPI_ISL_574328, EPI_ISL_574329, EPI_ISL_574330, EPI_ISL_574331, EPI_ISL_574332, EPI_ISL_574333, EPI_ISL_574334, EPI_ISL_574335, EPI_ISL_574336, EPI_ISL_574337, EPI_ISL_574338, EPI_ISL_574339, EPI_ISL_574340, EPI_ISL_574341, EPI_ISL_574342, EPI_ISL_574343, EPI_ISL_574344, EPI_ISL_574345, EPI_ISL_574346, EPI_ISL_574347, EPI_ISL_574348, EPI_ISL_574349, EPI_ISL_574350, EPI_ISL_574351, EPI_ISL_574352, EPI_ISL_574353, EPI_ISL_574354, EPI_ISL_574355, EPI_ISL_574356, EPI_ISL_574357, EPI_ISL_574358, EPI_ISL_574359, EPI_ISL_574360, EPI_ISL_574361, EPI_ISL_574362, EPI_ISL_574363, EPI_ISL_574364, EPI_ISL_574365, EPI_ISL_574366, EPI_ISL_574367, EPI_ISL_574368, EPI_ISL_574369, EPI_ISL_574370, EPI_ISL_574371, EPI_ISL_574372, EPI_ISL_574373, EPI_ISL_574374, EPI_ISL_574375, EPI_ISL_574376, EPI_ISL_574377, EPI_ISL_574378, EPI_ISL_574379, EPI_ISL_574380, EPI_ISL_574381, EPI_ISL_574382, EPI_ISL_574383, EPI_ISL_574384, EPI_ISL_574385, EPI_ISL_574386, EPI_ISL_574387, EPI_ISL_574388, EPI_ISL_574389, EPI_ISL_574390, EPI_ISL_574391, EPI_ISL_574392, EPI_ISL_574393, EPI_ISL_574394, EPI_ISL_574395, EPI_ISL_574396, EPI_ISL_574397, EPI_ISL_574398, EPI_ISL_574399, EPI_ISL_574400, EPI_ISL_574401, EPI_ISL_574402, EPI_ISL_574403, EPI_ISL_574404, EPI_ISL_574405, EPI_ISL_574406, EPI_ISL_574407, EPI_ISL_574408, EPI_ISL_574409, EPI_ISL_574410, EPI_ISL_574411, EPI_ISL_574412, EPI_ISL_574413, EPI_ISL_574414, EPI_ISL_574415, EPI_ISL_574416, EPI_ISL_574417, EPI_ISL_574418, EPI_ISL_574419, EPI_ISL_574420, EPI_ISL_574421, EPI_ISL_574422, EPI_ISL_574423, EPI_ISL_574424, EPI_ISL_574425, EPI_ISL_574426, EPI_ISL_574427, EPI_ISL_574428, EPI_ISL_574429, EPI_ISL_574430 | | | |
| see above | LSUHS Emerging Viral Threat Laboratory | Microbial Genome Sequencing Center | Jeremy P. Kamil, Rona S. Scott, Maarten Van Diest, Malgorzata Bienkowska-Haba, Katarzyna Zwolinska, Andrew D. Yurochko, Christopher G. Kevil, Martin J. Sapp, Daniel J. Snyder, Vaughn S. Cooper, John A. Vanchiere |
| EPI_ISL_574434, EPI_ISL_574435, EPI_ISL_574436, EPI_ISL_574437, EPI_ISL_574438, EPI_ISL_574439, EPI_ISL_574440, EPI_ISL_574441, EPI_ISL_574442, EPI_ISL_574443, EPI_ISL_574445, EPI_ISL_574446, EPI_ISL_574447, EPI_ISL_574448, EPI_ISL_574449, EPI_ISL_574450, EPI_ISL_574451, EPI_ISL_574452, EPI_ISL_574453, EPI_ISL_574454, EPI_ISL_574455, EPI_ISL_574456, EPI_ISL_574457, EPI_ISL_574458, EPI_ISL_574459, EPI_ISL_574460, EPI_ISL_574461, EPI_ISL_574462, EPI_ISL_574463, EPI_ISL_574464, EPI_ISL_574465, EPI_ISL_574466, EPI_ISL_574467, EPI_ISL_574468, EPI_ISL_574469, EPI_ISL_574470, EPI_ISL_574471, EPI_ISL_574472, EPI_ISL_574473, EPI_ISL_574474, EPI_ISL_574475, EPI_ISL_574476, EPI_ISL_574477, EPI_ISL_574478 | | | |
| see above | Texas Department of State Health Services | Texas Department of State Health Services | Rashmi Tuladhar, Bonnie Oh, Jenny Zhang, Maliha Rahman, Anita Pokharel, Myong Koag, Chun Wang, Rachel Lee, Grace Kubin |
| EPI_ISL_574543, EPI_ISL_574544, EPI_ISL_574545, EPI_ISL_574546, EPI_ISL_574547, EPI_ISL_574548, EPI_ISL_574549, EPI_ISL_574550, EPI_ISL_574551, EPI_ISL_574552, EPI_ISL_574553, EPI_ISL_574554, EPI_ISL_574555, EPI_ISL_574556, EPI_ISL_574557, EPI_ISL_574558, EPI_ISL_574559, EPI_ISL_574560, EPI_ISL_574561, EPI_ISL_574562, EPI_ISL_574563, EPI_ISL_574564, EPI_ISL_574565, EPI_ISL_574566, EPI_ISL_574567, EPI_ISL_574568, EPI_ISL_574569, EPI_ISL_574570, EPI_ISL_574571, EPI_ISL_574572, EPI_ISL_574573, EPI_ISL_574574, EPI_ISL_574575 | | | |
| see above | Microbiology Division, South Carolina Department of Health and Environmental Control | Microbiology Division, South Carolina Department of Health and Environmental Control | Flores,H. |
| EPI_ISL_574581, EPI_ISL_574584, EPI_ISL_574585, EPI_ISL_574586, EPI_ISL_574599 | Delaware Public Health Lab | Delaware Public Health Lab | Gregory Hovan |
| EPI_ISL_574648 | Seattle Flu Study | Seattle Flu Study | Deborah A. Nickerson, Chris D. Frazar, Jover Lee, Benjamin Pelle, Matthew Richardson, Amanda Adler, Elisabeth Brandstetter, Peter D. Han, Kairsten Fay, Misja Ilcisin, Kirsten Lacombe, Thomas R. Sibley, Melissa Truong, Caitlin R. Wolf, Michael Boeckh, Janet A. Englund, Michael Famulare, Barry R. Lutz, Mark J. Rieder, Lea M. Starita, Matthew Thompson, Jay Shendure, Trevor Bedford, Helen Y. Chu |
| EPI_ISL_574649, EPI_ISL_574650, EPI_ISL_574651, EPI_ISL_574652, EPI_ISL_574653, EPI_ISL_574654, EPI_ISL_574655, EPI_ISL_574656, EPI_ISL_574657, EPI_ISL_574658, EPI_ISL_574659, EPI_ISL_574660, EPI_ISL_574661, EPI_ISL_574662 | | | |
| see above | Seattle Flu Study | Seattle Flu Study | Deborah A. Nickerson, Chris D. Frazar, Jover Lee, Benjamin Pelle, Matthew Richardson, Amanda Adler, Elisabeth Brandstetter, Peter D. Han, Kairsten Fay, Misja Ilcisin, Kirsten Lacombe, Thomas R. Sibley, Melissa Truong, Caitlin R. Wolf, Karen Cowgill, Stephanie Schrag, Jeff Duchin, Michael Boeckh, Janet A. Englund, Michael Famulare, Barry R. Lutz, Mark J. Rieder, Lea M. Starita, Matthew Thompson, Helen Y. Chu, Trevor Bedford, Jay Shendure |
| EPI_ISL_574663, EPI_ISL_574664, EPI_ISL_574665, EPI_ISL_574666, EPI_ISL_574667 | Seattle Flu Study | Seattle Flu Study | Deborah A. Nickerson, Chris D. Frazar, Jover Lee, Benjamin Pelle, Matthew Richardson, Amanda Adler, Elisabeth Brandstetter, Peter D. Han, Kairsten Fay, Misja Ilcisin, Kirsten Lacombe, Thomas R. Sibley, Melissa Truong, Caitlin R. Wolf, Michael Boeckh, Janet A. Englund, Michael Famulare, Barry R. Lutz, Mark J. Rieder, Lea M. Starita, Matthew Thompson, Jay Shendure, Trevor Bedford, Helen Y. Chu |
| EPI_ISL_574668, EPI_ISL_574669, EPI_ISL_574670, EPI_ISL_574671, EPI_ISL_574672, EPI_ISL_574673, EPI_ISL_574674, EPI_ISL_574675, EPI_ISL_574676, EPI_ISL_574677, EPI_ISL_574678 | | | |
| see above | Seattle Flu Study | Seattle Flu Study | Deborah A. Nickerson, Chris D. Frazar, Jover Lee, Benjamin Pelle, Matthew Richardson, Amanda Adler, Elisabeth Brandstetter, Peter D. Han, Kairsten Fay, Misja Ilcisin, Kirsten Lacombe, Thomas R. Sibley, Melissa Truong, Caitlin R. Wolf, Karen Cowgill, Stephanie Schrag, Jeff Duchin, Michael Boeckh, Janet A. Englund, Michael Famulare, Barry R. Lutz, Mark J. Rieder, Lea M. Starita, Matthew Thompson, Helen Y. Chu, Trevor Bedford, Jay Shendure |
| EPI_ISL_574679 | Seattle Flu Study | Seattle Flu Study | Deborah A. Nickerson, Chris D. Frazar, Jover Lee, Benjamin Pelle, Matthew Richardson, Amanda Adler, Elisabeth Brandstetter, Peter D. Han, Kairsten Fay, Misja Ilcisin, Kirsten Lacombe, Thomas R. Sibley, Melissa Truong, Caitlin R. Wolf, Michael Boeckh, Janet A. Englund, Michael Famulare, Barry R. Lutz, Mark J. Rieder, Lea M. Starita, Matthew Thompson, Jay Shendure, Trevor Bedford, Helen Y. Chu |
| EPI_ISL_574680, EPI_ISL_574681 | Seattle Flu Study | Seattle Flu Study | Deborah A. Nickerson, Chris D. Frazar, Jover Lee, Benjamin Pelle, Matthew Richardson, Amanda Adler, Elisabeth Brandstetter, Peter D. Han, Kairsten Fay, Misja Ilcisin, Kirsten Lacombe, Thomas R. Sibley, Melissa Truong, Caitlin R. Wolf, Karen Cowgill, Stephanie Schrag, Jeff Duchin, Michael Boeckh, Janet A. Englund, Michael Famulare, Barry R. Lutz, Mark J. Rieder, Lea M. Starita, Matthew Thompson, Helen Y. Chu, Trevor Bedford, Jay Shendure |
| EPI_ISL_574682 | Seattle Flu Study | Seattle Flu Study | Deborah A. Nickerson, Chris D. Frazar, Jover Lee, Benjamin Pelle, Matthew Richardson, Amanda Adler, Elisabeth Brandstetter, Peter D. Han, Kairsten Fay, Misja Ilcisin, Kirsten Lacombe, Thomas R. Sibley, Melissa Truong, Caitlin R. Wolf, Michael Boeckh, Janet A. Englund, Michael Famulare, Barry R. Lutz, Mark J. Rieder, Lea M. Starita, Matthew Thompson, Jay Shendure, Trevor Bedford, Helen Y. Chu |
| EPI_ISL_574683 | Seattle Flu Study | Seattle Flu Study | Deborah A. Nickerson, Chris D. Frazar, Jover Lee, Benjamin Pelle, Matthew Richardson, Amanda Adler, Elisabeth Brandstetter, Peter D. Han, Kairsten Fay, Misja Ilcisin, Kirsten Lacombe, Thomas R. Sibley, Melissa Truong, Caitlin R. Wolf, Karen Cowgill, Stephanie Schrag, Jeff Duchin, Michael Boeckh, Janet A. Englund, Michael Famulare, Barry R. Lutz, Mark J. Rieder, Lea M. Starita, Matthew Thompson, Helen Y. Chu, Trevor Bedford, Jay Shendure |
| EPI_ISL_574684 | Seattle Flu Study | Seattle Flu Study | Deborah A. Nickerson, Chris D. Frazar, Jover Lee, Benjamin Pelle, Matthew Richardson, Amanda Adler, Elisabeth Brandstetter, Peter D. Han, Kairsten Fay, Misja Ilcisin, Kirsten Lacombe, Thomas R. Sibley, Melissa Truong, Caitlin R. Wolf, Michael Boeckh, Janet A. Englund, Michael Famulare, Barry R. Lutz, Mark J. Rieder, Lea M. Starita, Matthew Thompson, Jay Shendure, Trevor Bedford, Helen Y. Chu |
| EPI_ISL_574685, EPI_ISL_574686, EPI_ISL_574687, EPI_ISL_574688, EPI_ISL_574689, EPI_ISL_574690, EPI_ISL_574691, EPI_ISL_575028, EPI_ISL_575029, EPI_ISL_575030, EPI_ISL_575031, EPI_ISL_575032 | | | |
| see above | Seattle Flu Study | Seattle Flu Study | Deborah A. Nickerson, Chris D. Frazar, Jover Lee, Benjamin Pelle, Matthew Richardson, Amanda Adler, Elisabeth Brandstetter, Peter D. Han, Kairsten Fay, Misja Ilcisin, Kirsten Lacombe, Thomas R. Sibley, Melissa Truong, Caitlin R. Wolf, Karen Cowgill, Stephanie Schrag, Jeff Duchin, Michael Boeckh, Janet A. Englund, Michael Famulare, Barry R. Lutz, Mark J. Rieder, Lea M. Starita, Matthew Thompson, Helen Y. Chu, Trevor Bedford, Jay Shendure |
| EPI_ISL_575033 | Seattle Flu Study | Seattle Flu Study | Deborah A. Nickerson, Chris D. Frazar, Jover Lee, Benjamin Pelle, Matthew Richardson, Amanda Adler, Elisabeth Brandstetter, Peter D. Han, Kairsten Fay, Misja Ilcisin, Kirsten Lacombe, Thomas R. Sibley, Melissa Truong, Caitlin R. Wolf, Romesh Gautom, Geoff Melly, Brian Hiatt, Philip Dykema, Scott Lindquist, Michael Boeckh, Janet A. Englund, Michael Famulare, Barry R. Lutz, Mark J. Rieder, Lea M. Starita, Matthew Thompson, Helen Y. Chu, Jay Shendure, Trevor Bedford |
| EPI_ISL_575034, EPI_ISL_575035, EPI_ISL_575036, EPI_ISL_575037, EPI_ISL_575038, EPI_ISL_575039, EPI_ISL_575040, EPI_ISL_575041, EPI_ISL_575043, EPI_ISL_575044, EPI_ISL_575045, EPI_ISL_575046, EPI_ISL_575047, EPI_ISL_575048, EPI_ISL_575049, EPI_ISL_575050, EPI_ISL_575051, EPI_ISL_575052, EPI_ISL_575053, EPI_ISL_575054, EPI_ISL_575055, EPI_ISL_575057, EPI_ISL_575058, EPI_ISL_575059, EPI_ISL_575060, EPI_ISL_575061, EPI_ISL_575062, EPI_ISL_575063, EPI_ISL_575065, EPI_ISL_575067, EPI_ISL_575068, EPI_ISL_575069, EPI_ISL_575070, EPI_ISL_575071, EPI_ISL_575074, EPI_ISL_575075, EPI_ISL_575076, EPI_ISL_575077, EPI_ISL_575078, EPI_ISL_575079, EPI_ISL_575080, EPI_ISL_575081, EPI_ISL_575083, EPI_ISL_575084, EPI_ISL_575085, EPI_ISL_575086, EPI_ISL_575087, EPI_ISL_575088, EPI_ISL_575089, EPI_ISL_575090, EPI_ISL_575091, EPI_ISL_575092, EPI_ISL_575093, EPI_ISL_575099, EPI_ISL_575100, EPI_ISL_575101, EPI_ISL_575102, EPI_ISL_575103, EPI_ISL_575104, EPI_ISL_575105, EPI_ISL_575106, EPI_ISL_575107, EPI_ISL_575108, EPI_ISL_575109, EPI_ISL_575110, EPI_ISL_575111, EPI_ISL_575112, EPI_ISL_575114, EPI_ISL_575115, EPI_ISL_575119, EPI_ISL_575120, EPI_ISL_575122, EPI_ISL_575123, EPI_ISL_575125, EPI_ISL_575126, EPI_ISL_575127, EPI_ISL_575129, EPI_ISL_575130, EPI_ISL_575131, EPI_ISL_575132, EPI_ISL_575133, EPI_ISL_575134, EPI_ISL_575136, EPI_ISL_575137, EPI_ISL_575139, EPI_ISL_575140, EPI_ISL_575141, EPI_ISL_575142, EPI_ISL_575143, EPI_ISL_575144, EPI_ISL_575145, EPI_ISL_575146, EPI_ISL_575147, EPI_ISL_575148, EPI_ISL_575149, EPI_ISL_575151, EPI_ISL_575154, EPI_ISL_575156, EPI_ISL_575157, EPI_ISL_575158, EPI_ISL_575160, EPI_ISL_575161, EPI_ISL_575162, EPI_ISL_575163, EPI_ISL_575164, EPI_ISL_575165, EPI_ISL_575166, EPI_ISL_575167, EPI_ISL_575168, EPI_ISL_575172, EPI_ISL_575173, EPI_ISL_575174, EPI_ISL_575175, EPI_ISL_575177, EPI_ISL_575178, EPI_ISL_575179, EPI_ISL_575180, EPI_ISL_575181, EPI_ISL_575182, EPI_ISL_575183, EPI_ISL_575185, EPI_ISL_575186, EPI_ISL_575187, EPI_ISL_575188, EPI_ISL_575189, EPI_ISL_575190, EPI_ISL_575191, EPI_ISL_575192, EPI_ISL_575193, EPI_ISL_575194, EPI_ISL_575196, EPI_ISL_575197, EPI_ISL_575198, EPI_ISL_575199, EPI_ISL_575200, EPI_ISL_575202, EPI_ISL_575203, EPI_ISL_575204, EPI_ISL_575206, EPI_ISL_575207, EPI_ISL_575208, EPI_ISL_575209, EPI_ISL_575210, EPI_ISL_575211, EPI_ISL_575213, EPI_ISL_575214, EPI_ISL_575215, EPI_ISL_575217, EPI_ISL_575221, EPI_ISL_575222, EPI_ISL_575223, EPI_ISL_575225, EPI_ISL_575227, EPI_ISL_575228, EPI_ISL_575229, EPI_ISL_575230, EPI_ISL_575231, EPI_ISL_575232, EPI_ISL_575233, EPI_ISL_575234, EPI_ISL_575236, EPI_ISL_575237, EPI_ISL_575239, EPI_ISL_575242, EPI_ISL_575244, EPI_ISL_575245, EPI_ISL_575246, EPI_ISL_575249, EPI_ISL_575251, EPI_ISL_575252, EPI_ISL_575254, EPI_ISL_575255, EPI_ISL_575256, EPI_ISL_575257, EPI_ISL_575258, EPI_ISL_575259, EPI_ISL_575260, EPI_ISL_575263, EPI_ISL_575265, EPI_ISL_575266, EPI_ISL_575267, EPI_ISL_575268, EPI_ISL_575269, EPI_ISL_575271, EPI_ISL_575274, EPI_ISL_575276, EPI_ISL_575277, EPI_ISL_575279, EPI_ISL_575280, EPI_ISL_575281, EPI_ISL_575282, EPI_ISL_575286, EPI_ISL_575287 | | | |
| see above | Utah Public Health Laboratory | Utah Public Health Laboratory | Erin Young, Kelly Oakeson |
| EPI_ISL_575335 | Delaware Public Health Lab | Delaware Public Health Lab | Gregory Hovan |
| EPI_ISL_576111, EPI_ISL_576112 | Alaska State Virology Laboratory | Alaska State Virology Laboratory | Jack Chen, Ph.D. |

| | | | |
|---|---|---|---|
| EPI_ISL_576150, EPI_ISL_576151, EPI_ISL_576152, EPI_ISL_576153, EPI_ISL_576154, EPI_ISL_576155, EPI_ISL_576156, EPI_ISL_576157, EPI_ISL_576158, EPI_ISL_576159, EPI_ISL_576160, EPI_ISL_576161, EPI_ISL_576162, EPI_ISL_576163, EPI_ISL_576164, EPI_ISL_576165, EPI_ISL_576166, EPI_ISL_576167, EPI_ISL_576168, EPI_ISL_576169 | | | |
| see above | Delaware Public Health Lab | Delaware Public Health Lab | Gregory Hovan |
| EPI_ISL_576170, EPI_ISL_576171 | PA Department of Health, Bureau of Laboratories | Pathogen Discovery, Respiratory Viruses Branch, Division of Viral Diseases, Centers for Disease Control and Prevention | Yan Li, Anna Montmayeur, Brian Lynch, Jing Zhang, Krista Queen, Ying Tao, Anna Uehara, Rachel Marine, Clinton R. Paden, Peter Cook, Haibin Wang, Suxiang Tong |
| EPI_ISL_576172 | TN Division of Laboratory Services | Pathogen Discovery, Respiratory Viruses Branch, Division of Viral Diseases, Centers for Disease Control and Prevention | Yan Li, Anna Montmayeur, Brian Lynch, Jing Zhang, Krista Queen, Ying Tao, Anna Uehara, Rachel Marine, Clinton R. Paden, Peter Cook, Haibin Wang, Suxiang Tong |
| EPI_ISL_576173 | FL Department of Health-Bureau of Epidemiology | Pathogen Discovery, Respiratory Viruses Branch, Division of Viral Diseases, Centers for Disease Control and Prevention | Ying Tao, Jing Zhang, Brian Lynch, Yan Li, Krista Queen, Anna Uehara, Clinton R. Paden, Peter Cook, Haibin Wang, Suxiang Tong |
| EPI_ISL_576174 | VA-Division of Consolidated Laboratory Services | Pathogen Discovery, Respiratory Viruses Branch, Division of Viral Diseases, Centers for Disease Control and Prevention | Ying Tao, Jing Zhang, Brian Lynch, Yan Li, Krista Queen, Anna Uehara, Clinton R. Paden, Peter Cook, Haibin Wang, Suxiang Tong |
| EPI_ISL_576175, EPI_ISL_576176 | TN Division of Laboratory Services | Pathogen Discovery, Respiratory Viruses Branch, Division of Viral Diseases, Centers for Disease Control and Prevention | Yan Li, Anna Montmayeur, Brian Lynch, Jing Zhang, Krista Queen, Ying Tao, Anna Uehara, Rachel Marine, Clinton R. Paden, Peter Cook, Haibin Wang, Suxiang Tong |
| EPI_ISL_576177 | CA, CDPH, Viral and Rickettsial Disease Laboratory | Pathogen Discovery, Respiratory Viruses Branch, Division of Viral Diseases, Centers for Disease Control and Prevention | Ying Tao, Jing Zhang, Brian Lynch, Yan Li, Krista Queen, Anna Uehara, Clinton R. Paden, Peter Cook, Haibin Wang, Suxiang Tong |
| EPI_ISL_576178 | DC Public Health Lab/ Dept. of Forensic Sciences | Pathogen Discovery, Respiratory Viruses Branch, Division of Viral Diseases, Centers for Disease Control and Prevention | Ying Tao, Jing Zhang, Brian Lynch, Yan Li, Krista Queen, Anna Uehara, Clinton R. Paden, Peter Cook, Haibin Wang, Suxiang Tong |
| EPI_ISL_576179, EPI_ISL_576180, EPI_ISL_576181, EPI_ISL_576182, EPI_ISL_576183, EPI_ISL_576184, EPI_ISL_576185, EPI_ISL_576186, EPI_ISL_576187, EPI_ISL_576188 | WI State Laboratory of Hygiene | Pathogen Discovery, Respiratory Viruses Branch, Division of Viral Diseases, Centers for Disease Control and Prevention | Ying Tao, Jing Zhang, Brian Lynch, Yan Li, Krista Queen, Anna Uehara, Clinton R. Paden, Peter Cook, Haibin Wang, Suxiang Tong |
| EPI_ISL_576189, EPI_ISL_576190, EPI_ISL_576191, EPI_ISL_576192, EPI_ISL_576193, EPI_ISL_576194, EPI_ISL_576195 | GA Department of Public Health Laboratory | Pathogen Discovery, Respiratory Viruses Branch, Division of Viral Diseases, Centers for Disease Control and Prevention | Ying Tao, Jing Zhang, Brian Lynch, Yan Li, Krista Queen, Anna Uehara, Clinton R. Paden, Peter Cook, Haibin Wang, Suxiang Tong |
| EPI_ISL_576196, EPI_ISL_576197 | AR Dept. of Health-Public Health Lab | Pathogen Discovery, Respiratory Viruses Branch, Division of Viral Diseases, Centers for Disease Control and Prevention | Ying Tao, Jing Zhang, Brian Lynch, Yan Li, Krista Queen, Anna Uehara, Clinton R. Paden, Peter Cook, Haibin Wang, Suxiang Tong |
| EPI_ISL_576198, EPI_ISL_576199, EPI_ISL_576200, EPI_ISL_576201, EPI_ISL_576202, EPI_ISL_576203, EPI_ISL_576204, EPI_ISL_576205, EPI_ISL_576206, EPI_ISL_576207, EPI_ISL_576208, EPI_ISL_576209, EPI_ISL_576210, EPI_ISL_576211, EPI_ISL_576212, EPI_ISL_576213, EPI_ISL_576214, EPI_ISL_576215, EPI_ISL_576216, EPI_ISL_576217, EPI_ISL_576218, EPI_ISL_576219, EPI_ISL_576220, EPI_ISL_576221 | | | |
| see above | Michigan Department of Health and Human Services, Bureau of Laboratories | Michigan Department of Health and Human Services, Bureau of Laboratories | Blankenship HM, Riner D, Soehnlen MK |
| EPI_ISL_576223, EPI_ISL_576224, EPI_ISL_576225, EPI_ISL_576226, EPI_ISL_576227 | Delaware Public Health Lab | Delaware Public Health Lab | Gregory Hovan |
| EPI_ISL_576228, EPI_ISL_576229, EPI_ISL_576230, EPI_ISL_576231, EPI_ISL_576232, EPI_ISL_576233, EPI_ISL_576234, EPI_ISL_576235, EPI_ISL_576236, EPI_ISL_576237, EPI_ISL_576238, EPI_ISL_576239, EPI_ISL_576240, EPI_ISL_576241, EPI_ISL_576242, EPI_ISL_576243, EPI_ISL_576244, EPI_ISL_576245, EPI_ISL_576246, EPI_ISL_576247, EPI_ISL_576248, EPI_ISL_576249, EPI_ISL_576250, EPI_ISL_576251, EPI_ISL_576252, EPI_ISL_576253, EPI_ISL_576254, EPI_ISL_576255, EPI_ISL_576256 | | | |
| see above | Minnesota Department of Health, Public Health Laboratory | Minnesota Department of Health, Public Health Laboratory | Matt Plumb, Jacob Garfin, Alexandra Lorentz, and Xiong Wang |
| EPI_ISL_576257, EPI_ISL_576258, EPI_ISL_576259, EPI_ISL_576260 | Instituto de Diagnostico y Referencia Epidemiologicos (INDRE) | Instituto de Diagnostico y Referencia Epidemiologicos (INDRE) | Gisela Barrera-Badillo , Abril Rodriguez-Maldonado, Claudia Wong-Arambula , Natividad Cruz-Ortiz, Tatiana Nunez-Garcia, Dayanira Arellano-Suarez, Fabiola Garces-Ayala, Edgar Mendieta-Condado, Lucia Hernandez-Rivas, Irma Lopez-Martinez, Ernesto Ramirez-Gonzalez. |
| EPI_ISL_576261, EPI_ISL_576262, EPI_ISL_576263 | Instituto de Diagnostico y Referencia Epidemiologicos (INDRE) | Instituto de Diagnostico y Referencia Epidemiologicos (INDRE) | Gisela Barrera-Badillo , Abril Rodriguez-Maldonado, Claudia Wong-Arambula , Natividad Cruz-Ortiz, Tatiana Nunez-Garcia, Dayanira Arellano-Suarez, Adnan Araiza-Rodriguez, Edgar Mendieta-Condado, Lucia Hernandez-Rivas, Irma Lopez-Martinez, Ernesto Ramirez-Gonzalez. |
| EPI_ISL_576264, EPI_ISL_576265 | Instituto de Diagnostico y Referencia Epidemiologicos (INDRE) | Instituto de Diagnostico y Referencia Epidemiologicos (INDRE) | Ernesto Ramirez-Gonzalez, Abril Rodriguez-Maldonado, Claudia Wong-Arambula , Natividad Cruz-Ortiz, Tatiana Nunez-Garcia, Dayanira Arellano-Suarez, Adnan Araiza-Rodriguez, Edgar Mendieta-Condado, Lucia Hernandez-Rivas, Irma Lopez-Martinez, Gisela Barrera-Badillo. |
| EPI_ISL_576266, EPI_ISL_576267, EPI_ISL_576268, EPI_ISL_576269, EPI_ISL_576270, EPI_ISL_576271 | Instituto de Diagnostico y Referencia Epidemiologicos (INDRE) | Instituto de Diagnostico y Referencia Epidemiologicos (INDRE) | Ernesto Ramirez-Gonzalez, Abril Rodriguez-Maldonado, Claudia Wong-Arambula , Natividad Cruz-Ortiz, Tatiana Nunez-Garcia, Dayanira Arellano-Suarez, Adnan Araiza-Rodriguez, Fabiola Garces-Ayala, Lucia Hernandez-Rivas, Irma Lopez-Martinez, Gisela Barrera-Badillo. |
| EPI_ISL_576272 | Instituto de Diagnostico y Referencia Epidemiologicos (INDRE) | Instituto de Diagnostico y Referencia Epidemiologicos (INDRE) | Gisela Barrera-Badillo , Abril Rodriguez-Maldonado, Claudia Wong-Arambula , Natividad Cruz-Ortiz, Tatiana Nunez-Garcia, Dayanira Arellano-Suarez, Fabiola Garces-Ayala, Edgar Mendieta-Condado, Lucia Hernandez-Rivas, Irma Lopez-Martinez, Ernesto Ramirez-Gonzalez. |
| EPI_ISL_576273 | Instituto de Diagnostico y Referencia Epidemiologicos (INDRE) | Instituto de Diagnostico y Referencia Epidemiologicos (INDRE) | Ernesto Ramirez-Gonzalez, Abril Rodriguez-Maldonado, Claudia Wong-Arambula , Natividad Cruz-Ortiz, Tatiana Nunez-Garcia, Dayanira Arellano-Suarez, Adnan Araiza-Rodriguez, Fabiola Garces-Ayala, Lucia Hernandez-Rivas, Irma Lopez-Martinez, Gisela Barrera-Badillo. |
| EPI_ISL_576274, EPI_ISL_576275, EPI_ISL_576276 | Instituto de Diagnostico y Referencia Epidemiologicos (INDRE) | Instituto de Diagnostico y Referencia Epidemiologicos (INDRE) | Gisela Barrera-Badillo , Abril Rodriguez-Maldonado, Claudia Wong-Arambula , Natividad Cruz-Ortiz, Tatiana Nunez-Garcia, Dayanira Arellano-Suarez, Adnan Araiza-Rodriguez, Edgar Mendieta-Condado, Lucia Hernandez-Rivas, Irma Lopez-Martinez, Ernesto Ramirez-Gonzalez. |
| EPI_ISL_576277, EPI_ISL_576278 | Instituto de Diagnostico y Referencia Epidemiologicos (INDRE) | Instituto de Diagnostico y Referencia Epidemiologicos (INDRE) | Ernesto Ramirez-Gonzalez, Abril Rodriguez-Maldonado, Claudia Wong-Arambula , Natividad Cruz-Ortiz, Tatiana Nunez-Garcia, Dayanira Arellano-Suarez, Adnan Araiza-Rodriguez, Edgar Mendieta-Condado, Lucia Hernandez-Rivas, Irma Lopez-Martinez, Gisela Barrera-Badillo. |
| EPI_ISL_576279 | Instituto de Diagnostico y Referencia Epidemiologicos (INDRE) | Instituto de Diagnostico y Referencia Epidemiologicos (INDRE) | Ernesto Ramirez-Gonzalez, Abril Rodriguez-Maldonado, Claudia Wong-Arambula , Natividad Cruz-Ortiz, Tatiana Nunez-Garcia, Dayanira Arellano-Suarez, Adnan Araiza-Rodriguez, Fabiola Garces-Ayala, Lucia Hernandez-Rivas, Irma Lopez-Martinez, Gisela Barrera-Badillo. |
| EPI_ISL_576283, EPI_ISL_576285, EPI_ISL_576286, EPI_ISL_576287, EPI_ISL_576288, EPI_ISL_576289, EPI_ISL_576290, EPI_ISL_576292, EPI_ISL_576293, EPI_ISL_576297, EPI_ISL_576298, EPI_ISL_576299, EPI_ISL_576300, EPI_ISL_576301, EPI_ISL_576302, EPI_ISL_576304, EPI_ISL_576305, EPI_ISL_576306, EPI_ISL_576307, EPI_ISL_576308, EPI_ISL_576309, EPI_ISL_576310, EPI_ISL_576311, EPI_ISL_576312, EPI_ISL_576313, EPI_ISL_576315, EPI_ISL_576316, EPI_ISL_576317, EPI_ISL_576318, EPI_ISL_576319, EPI_ISL_576320, EPI_ISL_576321, EPI_ISL_576324, EPI_ISL_576325 | | | |
| see above | Texas Department of State Health Services | Texas Department of State Health Services | Rashmi Tuladhar, Bonnie Oh, Jenny Zhang, Maliha Rahman, Anita Pokharel, Myong Koag, Chun Wang, Rachel Lee, Grace Kubin |
| EPI_ISL_576331, EPI_ISL_576332, EPI_ISL_576333, EPI_ISL_576334, EPI_ISL_576347, EPI_ISL_576348, EPI_ISL_576349, EPI_ISL_576351, EPI_ISL_576353, EPI_ISL_576354, EPI_ISL_576355, EPI_ISL_576356, EPI_ISL_576358, EPI_ISL_576359, EPI_ISL_576360, EPI_ISL_576361 | | | |
| see above | Texas Department of State Health Services | Texas Department of State Health Services | Rashmi Tuladhar, Bonnie Oh, Mayela Pedrueza, Jenny Zhang, Maliha Rahman, Anita Pokharel, Myong Koag, Chun Wang, Rachel Lee, Grace Kubin |
| EPI_ISL_576399, EPI_ISL_576400, EPI_ISL_576401, EPI_ISL_576402, EPI_ISL_576403, EPI_ISL_576404, EPI_ISL_576405, EPI_ISL_576406, EPI_ISL_576407, EPI_ISL_576408, EPI_ISL_576409, EPI_ISL_576410, EPI_ISL_576411, EPI_ISL_576412, EPI_ISL_576414, EPI_ISL_576415, EPI_ISL_576416, EPI_ISL_576417, EPI_ISL_576418, EPI_ISL_576419, EPI_ISL_576420, EPI_ISL_576422, EPI_ISL_576423, EPI_ISL_576424, EPI_ISL_576425, EPI_ISL_576426, EPI_ISL_576427, EPI_ISL_576428, EPI_ISL_576429, EPI_ISL_576430, EPI_ISL_576431, EPI_ISL_576432, EPI_ISL_576433, EPI_ISL_576434, EPI_ISL_576435, EPI_ISL_576436, EPI_ISL_576437, EPI_ISL_576438, EPI_ISL_576439, EPI_ISL_576440, EPI_ISL_576441, EPI_ISL_576442, EPI_ISL_576443, EPI_ISL_576444, EPI_ISL_576445, EPI_ISL_576446, EPI_ISL_576447, EPI_ISL_576448, EPI_ISL_576449, EPI_ISL_576450, EPI_ISL_576451, EPI_ISL_576452, EPI_ISL_576453, EPI_ISL_576454, EPI_ISL_576455, EPI_ISL_576456, EPI_ISL_576457, EPI_ISL_576458, EPI_ISL_576459, EPI_ISL_576460, EPI_ISL_576461, EPI_ISL_576462, EPI_ISL_576463, EPI_ISL_576464, EPI_ISL_576465, EPI_ISL_576466, EPI_ISL_576467, EPI_ISL_576468, EPI_ISL_576469, EPI_ISL_576470, EPI_ISL_576471, EPI_ISL_576472, EPI_ISL_576473, EPI_ISL_576474, EPI_ISL_576475, EPI_ISL_576476, EPI_ISL_576477, EPI_ISL_576478, EPI_ISL_576479, EPI_ISL_576480, EPI_ISL_576481, EPI_ISL_576482, EPI_ISL_576483, EPI_ISL_576484, EPI_ISL_576485, EPI_ISL_576486, EPI_ISL_576487, EPI_ISL_576488, EPI_ISL_576489, EPI_ISL_576490, EPI_ISL_576491, EPI_ISL_576492, EPI_ISL_576493, EPI_ISL_576494, EPI_ISL_576495, EPI_ISL_576496, EPI_ISL_576497, EPI_ISL_576498, EPI_ISL_576499, EPI_ISL_576500, EPI_ISL_576501, EPI_ISL_576502, EPI_ISL_576503, EPI_ISL_576504, EPI_ISL_576505, EPI_ISL_576506, EPI_ISL_576507, EPI_ISL_576508, EPI_ISL_576509, EPI_ISL_576510, EPI_ISL_576511, EPI_ISL_576512, EPI_ISL_576513, EPI_ISL_576514, EPI_ISL_576515, EPI_ISL_576516, EPI_ISL_576517 | | | |
| see above | UW Virology Lab | UW Virology Lab | Pavitra Roychoudhury, Hong Xie, Lasata Shrestha, Amin Addetia, Victoria M Rachleff, Meei-Li Huang, Keith R Jerome, Alexander Greninger |
| EPI_ISL_576518, EPI_ISL_576519, EPI_ISL_576520, EPI_ISL_576521, EPI_ISL_576522, EPI_ISL_576523, EPI_ISL_576524, EPI_ISL_576525, EPI_ISL_576526, EPI_ISL_576527, EPI_ISL_576528, EPI_ISL_576529, EPI_ISL_576530, EPI_ISL_576531, EPI_ISL_576532, EPI_ISL_576533, EPI_ISL_576534, EPI_ISL_576535, EPI_ISL_576536, EPI_ISL_576537, EPI_ISL_576538, EPI_ISL_576539, EPI_ISL_576540, EPI_ISL_576541, EPI_ISL_576542, EPI_ISL_576543, EPI_ISL_576544, EPI_ISL_576545, EPI_ISL_576546, EPI_ISL_576547, EPI_ISL_576548, EPI_ISL_576549, EPI_ISL_576550, EPI_ISL_576551, EPI_ISL_576552, EPI_ISL_576553, EPI_ISL_576554, EPI_ISL_576555, EPI_ISL_576556 | | | |

| | | | |
|---|---|---|---|
| see above | Innovative Genomics Institute, UC Berkeley | Innovative Genomics Institute, UC Berkeley | Stacia Wyman, Haridha Shivram, Phil Frankino, Liana Lareau, Shana McDevitt, Justin Choi |
| EPI_ISL_576557, EPI_ISL_576558, EPI_ISL_576559, EPI_ISL_576560, EPI_ISL_576561, EPI_ISL_576562, EPI_ISL_576563, EPI_ISL_576564 | UW Virology Lab | UW Virology Lab | Pavitra Roychoudhury, Hong Xie, Lasata Shrestha, Amin Addetia, Victoria M Rachleff, Meei-Li Huang, Keith R Jerome, Alexander Greninger |
| EPI_ISL_577547, EPI_ISL_577548, EPI_ISL_577549, EPI_ISL_577550, EPI_ISL_577551, EPI_ISL_577552, EPI_ISL_577553, EPI_ISL_577554, EPI_ISL_577555, EPI_ISL_577556, EPI_ISL_577557, EPI_ISL_577558, EPI_ISL_577559, EPI_ISL_577560, EPI_ISL_577561, EPI_ISL_577562, EPI_ISL_577563, EPI_ISL_577565, EPI_ISL_577566, EPI_ISL_577567, EPI_ISL_577568, EPI_ISL_577569, EPI_ISL_577570, EPI_ISL_577571, EPI_ISL_577572, EPI_ISL_577573, EPI_ISL_577574, EPI_ISL_577575, EPI_ISL_577576, EPI_ISL_577577, EPI_ISL_577578, EPI_ISL_577579, EPI_ISL_577580, EPI_ISL_577581, EPI_ISL_577582, EPI_ISL_577583, EPI_ISL_577584, EPI_ISL_577585, EPI_ISL_577586, EPI_ISL_577587, EPI_ISL_577588, EPI_ISL_577589, EPI_ISL_577590, EPI_ISL_577591, EPI_ISL_577592, EPI_ISL_577593, EPI_ISL_577594, EPI_ISL_577595, EPI_ISL_577596 | | | |
| see above | Michigan Department of Health and Human Services, Bureau of Laboratories | Michigan Department of Health and Human Services, Bureau of Laboratories | Blankenship HM, Riner D, Soehnlen MK |
| EPI_ISL_578082, EPI_ISL_578083, EPI_ISL_578084, EPI_ISL_578085, EPI_ISL_578086, EPI_ISL_578087, EPI_ISL_578088, EPI_ISL_578089, EPI_ISL_578090, EPI_ISL_578091, EPI_ISL_578092, EPI_ISL_578093, EPI_ISL_578094, EPI_ISL_578095, EPI_ISL_578096, EPI_ISL_578097, EPI_ISL_578098, EPI_ISL_578099, EPI_ISL_578100, EPI_ISL_578101, EPI_ISL_578102, EPI_ISL_578103, EPI_ISL_578104, EPI_ISL_578105, EPI_ISL_578106, EPI_ISL_578107, EPI_ISL_578108, EPI_ISL_578109, EPI_ISL_578110, EPI_ISL_578111, EPI_ISL_578112, EPI_ISL_578113, EPI_ISL_578114, EPI_ISL_578115, EPI_ISL_578116, EPI_ISL_578117, EPI_ISL_578118, EPI_ISL_578119, EPI_ISL_578120, EPI_ISL_578121, EPI_ISL_578122, EPI_ISL_578123, EPI_ISL_578124, EPI_ISL_578125, EPI_ISL_578126, EPI_ISL_578127, EPI_ISL_578128, EPI_ISL_578129, EPI_ISL_578130, EPI_ISL_578131, EPI_ISL_578132, EPI_ISL_578133, EPI_ISL_578134, EPI_ISL_578135, EPI_ISL_578136, EPI_ISL_578137, EPI_ISL_578138, EPI_ISL_578139, EPI_ISL_578140, EPI_ISL_578141, EPI_ISL_578142, EPI_ISL_578143, EPI_ISL_578144, EPI_ISL_578145, EPI_ISL_578146, EPI_ISL_578147, EPI_ISL_578148, EPI_ISL_578149, EPI_ISL_578150, EPI_ISL_578151, EPI_ISL_578152, EPI_ISL_578153, EPI_ISL_578154, EPI_ISL_578155, EPI_ISL_578156, EPI_ISL_578157, EPI_ISL_578158, EPI_ISL_578159, EPI_ISL_578160, EPI_ISL_578161, EPI_ISL_578162, EPI_ISL_578163, EPI_ISL_578164, EPI_ISL_578165, EPI_ISL_578166, EPI_ISL_578167, EPI_ISL_578169, EPI_ISL_578170, EPI_ISL_578171, EPI_ISL_578172, EPI_ISL_578173, EPI_ISL_578174, EPI_ISL_578185 | | | |
| see above | University of Michigan Clinical Microbiology Laboratory | Lauring Lab, University of Michigan, Department of Microbiology and Immunology | Valesano |
| EPI_ISL_578335, EPI_ISL_578336, EPI_ISL_578337, EPI_ISL_578338, EPI_ISL_578339, EPI_ISL_578340, EPI_ISL_578341, EPI_ISL_578342, EPI_ISL_578343, EPI_ISL_578344, EPI_ISL_578345, EPI_ISL_578346, EPI_ISL_578347, EPI_ISL_578348, EPI_ISL_578349, EPI_ISL_578350, EPI_ISL_578351, EPI_ISL_578352, EPI_ISL_578353, EPI_ISL_578354, EPI_ISL_578355, EPI_ISL_578356, EPI_ISL_578357, EPI_ISL_578358, EPI_ISL_578359, EPI_ISL_578360, EPI_ISL_578361, EPI_ISL_578362, EPI_ISL_578363, EPI_ISL_578364, EPI_ISL_578365, EPI_ISL_578366, EPI_ISL_578367, EPI_ISL_578368, EPI_ISL_578369, EPI_ISL_578370, EPI_ISL_578371, EPI_ISL_578372, EPI_ISL_578373, EPI_ISL_578374, EPI_ISL_578375, EPI_ISL_578376, EPI_ISL_578377, EPI_ISL_578378, EPI_ISL_578379, EPI_ISL_578380, EPI_ISL_578381, EPI_ISL_578382, EPI_ISL_578383, EPI_ISL_578384, EPI_ISL_578385, EPI_ISL_578386, EPI_ISL_578387, EPI_ISL_578388, EPI_ISL_578389, EPI_ISL_578390, EPI_ISL_578391, EPI_ISL_578392, EPI_ISL_578393, EPI_ISL_578394, EPI_ISL_578395, EPI_ISL_578396, EPI_ISL_578397, EPI_ISL_578398, EPI_ISL_578399, EPI_ISL_578400, EPI_ISL_578401, EPI_ISL_578402, EPI_ISL_578403, EPI_ISL_578404, EPI_ISL_578405, EPI_ISL_578406, EPI_ISL_578407, EPI_ISL_578408, EPI_ISL_578409, EPI_ISL_578410, EPI_ISL_578411, EPI_ISL_578412, EPI_ISL_578413, EPI_ISL_578414, EPI_ISL_578415, EPI_ISL_578416, EPI_ISL_578417, EPI_ISL_578418, EPI_ISL_578419, EPI_ISL_578420, EPI_ISL_578421, EPI_ISL_578422, EPI_ISL_578423, EPI_ISL_578424, EPI_ISL_578425, EPI_ISL_578426, EPI_ISL_578427, EPI_ISL_578428, EPI_ISL_578429, EPI_ISL_578430, EPI_ISL_578431, EPI_ISL_578432, EPI_ISL_578433, EPI_ISL_578434, EPI_ISL_578435, EPI_ISL_578436, EPI_ISL_578437, EPI_ISL_578438, EPI_ISL_578439, EPI_ISL_578440, EPI_ISL_578441, EPI_ISL_578442, EPI_ISL_578443, EPI_ISL_578444, EPI_ISL_578445, EPI_ISL_578446, EPI_ISL_578447, EPI_ISL_578448, EPI_ISL_578449, EPI_ISL_578450, EPI_ISL_578451, EPI_ISL_578452, EPI_ISL_578453, EPI_ISL_578454, EPI_ISL_578455, EPI_ISL_578456, EPI_ISL_578457, EPI_ISL_578458, EPI_ISL_578459, EPI_ISL_578460, EPI_ISL_578461, EPI_ISL_578462, EPI_ISL_578463, EPI_ISL_578464, EPI_ISL_578465, EPI_ISL_578466, EPI_ISL_578467, EPI_ISL_578468, EPI_ISL_578469, EPI_ISL_578470, EPI_ISL_578471, EPI_ISL_578472, EPI_ISL_578473, EPI_ISL_578474, EPI_ISL_578475, EPI_ISL_578476, EPI_ISL_578477, EPI_ISL_578478, EPI_ISL_578479, EPI_ISL_578480, EPI_ISL_578481, EPI_ISL_578482, EPI_ISL_578483, EPI_ISL_578484, EPI_ISL_578485, EPI_ISL_578486, EPI_ISL_578487, EPI_ISL_578488, EPI_ISL_578489, EPI_ISL_578490, EPI_ISL_578491, EPI_ISL_578492, EPI_ISL_578493, EPI_ISL_578494, EPI_ISL_578495, EPI_ISL_578496, EPI_ISL_578497, EPI_ISL_578498, EPI_ISL_578499, EPI_ISL_578500, EPI_ISL_578501, EPI_ISL_578502, EPI_ISL_578503, EPI_ISL_578504, EPI_ISL_578505, EPI_ISL_578506, EPI_ISL_578507, EPI_ISL_578508, EPI_ISL_578509, EPI_ISL_578510, EPI_ISL_578511, EPI_ISL_578512, EPI_ISL_578513, EPI_ISL_578514, EPI_ISL_578515, EPI_ISL_578516, EPI_ISL_578517, EPI_ISL_578518, EPI_ISL_578519, EPI_ISL_578520, EPI_ISL_578521, EPI_ISL_578522, EPI_ISL_578523, EPI_ISL_578524, EPI_ISL_578525, EPI_ISL_578526, EPI_ISL_578527, EPI_ISL_578528, EPI_ISL_578529, EPI_ISL_578530, EPI_ISL_578531, EPI_ISL_578532, EPI_ISL_578533, EPI_ISL_578534, EPI_ISL_578535, EPI_ISL_578536, EPI_ISL_578537, EPI_ISL_578538, EPI_ISL_578539, EPI_ISL_578540, EPI_ISL_578541, EPI_ISL_578542, EPI_ISL_578543, EPI_ISL_578544, EPI_ISL_578545, EPI_ISL_578546, EPI_ISL_578547, EPI_ISL_578548, EPI_ISL_578549, EPI_ISL_578550, EPI_ISL_578551, EPI_ISL_578552, EPI_ISL_578553, EPI_ISL_578554, EPI_ISL_578555, EPI_ISL_578556, EPI_ISL_578557, EPI_ISL_578558, EPI_ISL_578559, EPI_ISL_578560, EPI_ISL_578561, EPI_ISL_578562, EPI_ISL_578563, EPI_ISL_578564, EPI_ISL_578565, EPI_ISL_578566, EPI_ISL_578567, EPI_ISL_578568, EPI_ISL_578569, EPI_ISL_578570, EPI_ISL_578571, EPI_ISL_578572, EPI_ISL_578573, EPI_ISL_578574, EPI_ISL_578575, EPI_ISL_578576, EPI_ISL_578577, EPI_ISL_578578, EPI_ISL_578579, EPI_ISL_578580, EPI_ISL_578581, EPI_ISL_578582, EPI_ISL_578583, EPI_ISL_578584, EPI_ISL_578585, EPI_ISL_578586, EPI_ISL_578587, EPI_ISL_578588, EPI_ISL_578589, EPI_ISL_578590, EPI_ISL_578591, EPI_ISL_578592, EPI_ISL_578593, EPI_ISL_578594, EPI_ISL_578595, EPI_ISL_578596, EPI_ISL_578597, EPI_ISL_578598, EPI_ISL_578599, EPI_ISL_578600, EPI_ISL_578601, EPI_ISL_578602, EPI_ISL_578603, EPI_ISL_578604, EPI_ISL_578605, EPI_ISL_578606, EPI_ISL_578607, EPI_ISL_578608, EPI_ISL_578609, EPI_ISL_578610, EPI_ISL_578611, EPI_ISL_578612, EPI_ISL_578613, EPI_ISL_578614, EPI_ISL_578615, EPI_ISL_578616, EPI_ISL_578617, EPI_ISL_578618, EPI_ISL_578619, EPI_ISL_578620, EPI_ISL_578621, EPI_ISL_578622, EPI_ISL_578623, EPI_ISL_578624, EPI_ISL_578625, EPI_ISL_578626, EPI_ISL_578627, EPI_ISL_578628, EPI_ISL_578629, EPI_ISL_578630, EPI_ISL_578631, EPI_ISL_578632, EPI_ISL_578633, EPI_ISL_578634, EPI_ISL_578635, EPI_ISL_578636, EPI_ISL_578637, EPI_ISL_578638, EPI_ISL_578639, EPI_ISL_578640, EPI_ISL_578641, EPI_ISL_578642, EPI_ISL_578643, EPI_ISL_578644, EPI_ISL_578645, EPI_ISL_578646, EPI_ISL_578647, EPI_ISL_578648, EPI_ISL_578649, EPI_ISL_578650, EPI_ISL_578651, EPI_ISL_578652, EPI_ISL_578653, EPI_ISL_578654, EPI_ISL_578655, EPI_ISL_578656, EPI_ISL_578657, EPI_ISL_578658, EPI_ISL_578659, EPI_ISL_578660, EPI_ISL_578661, EPI_ISL_578662, EPI_ISL_578663, EPI_ISL_578664, EPI_ISL_578665, EPI_ISL_578666, EPI_ISL_578667, EPI_ISL_578668, EPI_ISL_578669, EPI_ISL_578670, EPI_ISL_578671, EPI_ISL_578672, EPI_ISL_578673, EPI_ISL_578674, EPI_ISL_578675, EPI_ISL_578676, EPI_ISL_578677, EPI_ISL_578678, EPI_ISL_578679, EPI_ISL_578680, EPI_ISL_578681, EPI_ISL_578682, EPI_ISL_578683, EPI_ISL_578684, EPI_ISL_578685, EPI_ISL_578686, EPI_ISL_578687, EPI_ISL_578688, EPI_ISL_578689, EPI_ISL_578690, EPI_ISL_578691, EPI_ISL_578692, EPI_ISL_578693, EPI_ISL_578694, EPI_ISL_578695 | | | |
| see above | Wisconsin State Laboratory of Hygiene Communicable Disease Division | Wisconsin State Laboratory of Hygiene Communicable Disease Division | Kelsey R. Florek, Abigail C. Shockey |
| EPI_ISL_578698 | LSUHS Emerging Viral Threat Laboratory | Microbial Genome Sequencing Center | Maarten Van Diest, Jeremy P. Kamil, Rona S. Scott, Malgorzata Bienkowska-Haba, Katarzyna Zwolinska, Andrew D. Yurochko, Christopher G. Kevil, Martin J. Sapp, Daniel J. Snyder, Vaughn S. Cooper, John A. Vanchiere |
| EPI_ISL_578699, EPI_ISL_578700 | LSUHS Emerging Viral Threat Laboratory | Microbial Genome Sequencing Center | Jeremy P. Kamil, Rona S. Scott, Maarten Van Diest, Malgorzata Bienkowska-Haba, Katarzyna Zwolinska, Andrew D. Yurochko, Christopher G. Kevil, Martin J. Sapp, Daniel J. Snyder, Vaughn S. Cooper, John A. Vanchiere |
| EPI_ISL_578704, EPI_ISL_578705, EPI_ISL_578706, EPI_ISL_578707, EPI_ISL_578708, EPI_ISL_578709, EPI_ISL_578710, EPI_ISL_578711, EPI_ISL_578712, EPI_ISL_578713, EPI_ISL_578714, EPI_ISL_578715, EPI_ISL_578716, EPI_ISL_578717, EPI_ISL_578718, EPI_ISL_578719, EPI_ISL_578720, EPI_ISL_578721, EPI_ISL_578722, EPI_ISL_578723, EPI_ISL_578724, EPI_ISL_578725, EPI_ISL_578726, EPI_ISL_578727 | | | |
| see above | LSUHS Emerging Viral Threat Laboratory | Microbial Genome Sequencing Center | Maarten Van Diest, Jeremy P. Kamil, Rona S. Scott, Malgorzata Bienkowska-Haba, Katarzyna Zwolinska, Andrew D. Yurochko, Christopher G. Kevil, Martin J. Sapp, Daniel J. Snyder, Vaughn S. Cooper, John A. Vanchiere |
| EPI_ISL_578728, EPI_ISL_578729, EPI_ISL_578730, EPI_ISL_578731, EPI_ISL_578732, EPI_ISL_578733, EPI_ISL_578734, EPI_ISL_578735, EPI_ISL_578736, EPI_ISL_578737, EPI_ISL_578738, EPI_ISL_578739, EPI_ISL_578740, EPI_ISL_578741, EPI_ISL_578742, EPI_ISL_578743, EPI_ISL_578744, EPI_ISL_578745, EPI_ISL_578746, EPI_ISL_578747, EPI_ISL_578748, EPI_ISL_578749, EPI_ISL_578750, EPI_ISL_578751, EPI_ISL_578752 | | | |
| see above | LSUHS Emerging Viral Threat Laboratory | Microbial Genome Sequencing Center | Rona S. Scott, Jeremy P. Kamil, Maarten Van Diest, Malgorzata Bienkowska-Haba, Katarzyna Zwolinska, Andrew D. Yurochko, Christopher G. Kevil, Martin J. Sapp, Daniel J. Snyder, Vaughn S. Cooper, John A. Vanchiere |
| EPI_ISL_578753, EPI_ISL_578754, EPI_ISL_578755, EPI_ISL_578756, EPI_ISL_578757, EPI_ISL_578758, EPI_ISL_578759, EPI_ISL_578760, EPI_ISL_578761, EPI_ISL_578762, EPI_ISL_578763, EPI_ISL_578764, EPI_ISL_578765, EPI_ISL_578766, EPI_ISL_578767, EPI_ISL_578768, EPI_ISL_578769, EPI_ISL_578770, EPI_ISL_578771, EPI_ISL_578772, EPI_ISL_578773, EPI_ISL_578774, EPI_ISL_578775, EPI_ISL_578776, EPI_ISL_578777, EPI_ISL_578778, EPI_ISL_578779, EPI_ISL_578780, EPI_ISL_578781, EPI_ISL_578782, EPI_ISL_578783, EPI_ISL_578784, EPI_ISL_578785, EPI_ISL_578786, EPI_ISL_578787, EPI_ISL_578788, EPI_ISL_578789, EPI_ISL_578790, EPI_ISL_578791, EPI_ISL_578792, EPI_ISL_578793, EPI_ISL_578794, EPI_ISL_578795, EPI_ISL_578796, EPI_ISL_578797, EPI_ISL_578798, EPI_ISL_578799, EPI_ISL_578800, EPI_ISL_578801, EPI_ISL_578802, EPI_ISL_578803, EPI_ISL_578804, EPI_ISL_578805, EPI_ISL_578806, EPI_ISL_578807, EPI_ISL_578808, EPI_ISL_578809, EPI_ISL_578810, EPI_ISL_578811, EPI_ISL_578812, EPI_ISL_578813, EPI_ISL_578814, EPI_ISL_578815, EPI_ISL_578816, EPI_ISL_578817, EPI_ISL_578818, EPI_ISL_578819, EPI_ISL_578820, EPI_ISL_578821, EPI_ISL_578822, EPI_ISL_578823, EPI_ISL_578824, EPI_ISL_578825, EPI_ISL_578826, EPI_ISL_578827, EPI_ISL_578828, EPI_ISL_578829, EPI_ISL_578830, EPI_ISL_578831, EPI_ISL_578832, EPI_ISL_578833, EPI_ISL_578834, EPI_ISL_578835, EPI_ISL_578836, EPI_ISL_578837, EPI_ISL_578838, EPI_ISL_578839, EPI_ISL_578840, EPI_ISL_578841, EPI_ISL_578842, EPI_ISL_578843, EPI_ISL_578844, EPI_ISL_578845, EPI_ISL_578846, EPI_ISL_578847, EPI_ISL_578848, EPI_ISL_578849, EPI_ISL_578850, EPI_ISL_578851, EPI_ISL_578852, EPI_ISL_578853, EPI_ISL_578854, EPI_ISL_578855, EPI_ISL_578856, EPI_ISL_578857, EPI_ISL_578858, EPI_ISL_578859, EPI_ISL_578860, EPI_ISL_578861, EPI_ISL_578862, EPI_ISL_578863, EPI_ISL_578864, EPI_ISL_578865, EPI_ISL_578866, EPI_ISL_578867, EPI_ISL_578868, EPI_ISL_578869, EPI_ISL_578870, EPI_ISL_578871, EPI_ISL_578872, EPI_ISL_578873, EPI_ISL_578874, EPI_ISL_578875, EPI_ISL_578876, EPI_ISL_578877, EPI_ISL_578878, EPI_ISL_578879, EPI_ISL_578880, EPI_ISL_578881, EPI_ISL_578882, EPI_ISL_578883, EPI_ISL_578884, EPI_ISL_578885, EPI_ISL_578886, EPI_ISL_578887, EPI_ISL_578888, EPI_ISL_578889, EPI_ISL_578890, EPI_ISL_578891, EPI_ISL_578892, EPI_ISL_578893, EPI_ISL_578894, EPI_ISL_578895, EPI_ISL_578896, EPI_ISL_578897, EPI_ISL_578898, EPI_ISL_578899, EPI_ISL_578900, EPI_ISL_578901, EPI_ISL_578902, EPI_ISL_578903, EPI_ISL_578904, EPI_ISL_578905, EPI_ISL_578906, EPI_ISL_578907, EPI_ISL_578908, EPI_ISL_578909, EPI_ISL_578910, EPI_ISL_578911, EPI_ISL_578912, EPI_ISL_578913, EPI_ISL_578914, EPI_ISL_578915, EPI_ISL_578916, EPI_ISL_578917, EPI_ISL_578918, EPI_ISL_578919, EPI_ISL_578920, EPI_ISL_578921, EPI_ISL_578922, EPI_ISL_578923, EPI_ISL_578924, EPI_ISL_578925, EPI_ISL_578926, EPI_ISL_578927, EPI_ISL_578928, EPI_ISL_578929, EPI_ISL_578930, EPI_ISL_578931, EPI_ISL_578932, EPI_ISL_578933, EPI_ISL_578934, EPI_ISL_578935, EPI_ISL_578936, EPI_ISL_578937, EPI_ISL_578938, EPI_ISL_578939, EPI_ISL_578940, EPI_ISL_578941, EPI_ISL_578942, EPI_ISL_578943, EPI_ISL_578944, EPI_ISL_578945, EPI_ISL_578946, EPI_ISL_578947, EPI_ISL_578948, EPI_ISL_578949, EPI_ISL_578950, EPI_ISL_578951, EPI_ISL_578952, EPI_ISL_578953, EPI_ISL_578954, EPI_ISL_578955, EPI_ISL_578956, EPI_ISL_578957, EPI_ISL_578958, EPI_ISL_578959, EPI_ISL_578960, EPI_ISL_578961, EPI_ISL_578962, EPI_ISL_578963, EPI_ISL_578964, EPI_ISL_578965, EPI_ISL_578966, EPI_ISL_578967, EPI_ISL_578968, EPI_ISL_578969, EPI_ISL_578970, EPI_ISL_578971, EPI_ISL_578972, EPI_ISL_578973, EPI_ISL_578974, EPI_ISL_578975, EPI_ISL_578976, EPI_ISL_578977, EPI_ISL_578978, EPI_ISL_578979, EPI_ISL_578980, EPI_ISL_578981, EPI_ISL_578982, EPI_ISL_578983, EPI_ISL_578984, EPI_ISL_578985, EPI_ISL_578986, EPI_ISL_578987, EPI_ISL_578988, EPI_ISL_578989, EPI_ISL_578990, EPI_ISL_578991, EPI_ISL_578992, EPI_ISL_578993, EPI_ISL_578994, EPI_ISL_578995, EPI_ISL_578996, EPI_ISL_578997, EPI_ISL_578998, EPI_ISL_578999, EPI_ISL_579000, EPI_ISL_579001, EPI_ISL_579002, EPI_ISL_579003, EPI_ISL_579004, EPI_ISL_579005, EPI_ISL_579006, EPI_ISL_579007, EPI_ISL_579008, EPI_ISL_579009, EPI_ISL_579010, EPI_ISL_579011, EPI_ISL_579012, EPI_ISL_579013, EPI_ISL_579014, EPI_ISL_579015, EPI_ISL_579016, EPI_ISL_579017, EPI_ISL_579018, EPI_ISL_579019, EPI_ISL_579020, EPI_ISL_579021, EPI_ISL_579022, EPI_ISL_579023, EPI_ISL_579024, EPI_ISL_579025, EPI_ISL_579026, EPI_ISL_579027, EPI_ISL_579028, EPI_ISL_579029, EPI_ISL_579030, EPI_ISL_579031, EPI_ISL_579032, EPI_ISL_579033, EPI_ISL_579034, EPI_ISL_579035, EPI_ISL_579036, EPI_ISL_579037, EPI_ISL_579038, EPI_ISL_579039, EPI_ISL_579040, EPI_ISL_579041, EPI_ISL_579042, EPI_ISL_579043, EPI_ISL_579044, EPI_ISL_579045, EPI_ISL_579046, EPI_ISL_579047, EPI_ISL_579048, EPI_ISL_579049, EPI_ISL_579050, EPI_ISL_579051, EPI_ISL_579052, EPI_ISL_579053, EPI_ISL_579054, EPI_ISL_579055, EPI_ISL_579056, EPI_ISL_579057 | | | |
| see above | LSUHS Emerging Viral Threat Laboratory | Microbial Genome Sequencing Center | Jeremy P. Kamil, Rona S. Scott, Maarten Van Diest, Malgorzata Bienkowska-Haba, Katarzyna Zwolinska, Andrew D. Yurochko, Christopher G. Kevil, Martin J. Sapp, Daniel J. Snyder, Vaughn S. Cooper, John A. Vanchiere |
| EPI_ISL_581508, EPI_ISL_581509, EPI_ISL_581510, EPI_ISL_581511, EPI_ISL_581512, EPI_ISL_581513, EPI_ISL_581514, EPI_ISL_581515, EPI_ISL_581516, EPI_ISL_581517, EPI_ISL_581518, EPI_ISL_581520, EPI_ISL_581522, EPI_ISL_581523, EPI_ISL_581524, EPI_ISL_581525, EPI_ISL_581526, EPI_ISL_581527, EPI_ISL_581528, EPI_ISL_581529, EPI_ISL_581530, EPI_ISL_581531, EPI_ISL_581532, EPI_ISL_581533, EPI_ISL_581534, EPI_ISL_581535, EPI_ISL_581536, EPI_ISL_581537, EPI_ISL_581538, EPI_ISL_581539, EPI_ISL_581540, EPI_ISL_581541, EPI_ISL_581542, EPI_ISL_581543, EPI_ISL_581544, EPI_ISL_581545, EPI_ISL_581546, EPI_ISL_581547, EPI_ISL_581548, EPI_ISL_581549, EPI_ISL_581550, EPI_ISL_581551, EPI_ISL_581552, EPI_ISL_581553, EPI_ISL_581554, EPI_ISL_581555, EPI_ISL_581556, EPI_ISL_581557, EPI_ISL_581558, EPI_ISL_581559, EPI_ISL_581560, EPI_ISL_581561, EPI_ISL_581562, EPI_ISL_581563, EPI_ISL_581564, EPI_ISL_581565, EPI_ISL_581566, EPI_ISL_581567, EPI_ISL_581568, EPI_ISL_581569, EPI_ISL_581570, EPI_ISL_581571, EPI_ISL_581572, EPI_ISL_581573, EPI_ISL_581574 | | | |
| see above | Virginia DCLS | Virginia DCLS | Virginia DCLS |
| EPI_ISL_582133 | Delaware Public Health Lab | Delaware Public Health Lab | Gregory Hovan |
| EPI_ISL_582148, EPI_ISL_582149, EPI_ISL_582150, EPI_ISL_582151, EPI_ISL_582152, EPI_ISL_582153, EPI_ISL_582155, EPI_ISL_582156, EPI_ISL_582157, EPI_ISL_582158, EPI_ISL_582161, EPI_ISL_582163, EPI_ISL_582166, EPI_ISL_582167, EPI_ISL_582168, EPI_ISL_582170, EPI_ISL_582171, EPI_ISL_582178, EPI_ISL_582183, EPI_ISL_582187 | | | |
| see above | TriCore Reference Laboratories | Center for Global Health, University of New Mexico Health Sciences Center | Daryl Domman, Kurt Schwalm, Twila Kunde, Joseph Hicks, Michael Edwards, Darrell Dinwiddie |
| EPI_ISL_582222, EPI_ISL_582223, EPI_ISL_582224, EPI_ISL_582225, EPI_ISL_582226, EPI_ISL_582228, EPI_ISL_582229, EPI_ISL_582230, EPI_ISL_582233, EPI_ISL_582235, EPI_ISL_582236, EPI_ISL_582237, EPI_ISL_582238 | | | |
| see above | Wyoming Public Health Laboratory | Center for Global Health, University of New Mexico Health Sciences Center | Daryl Domman, Kurt Schwalm, Rob Christensen, Wanda Manley, Cari Sloma, Noah Hull, Darrell Dinwiddie |
| EPI_ISL_582851, EPI_ISL_582852, EPI_ISL_582853, EPI_ISL_582854, EPI_ISL_582855, EPI_ISL_582856, EPI_ISL_582857, EPI_ISL_582858, EPI_ISL_582859, EPI_ISL_582860, EPI_ISL_582861, EPI_ISL_582862, EPI_ISL_582863, EPI_ISL_582864, EPI_ISL_582865, EPI_ISL_582866, EPI_ISL_582867, EPI_ISL_582868, EPI_ISL_582869, EPI_ISL_582870, EPI_ISL_582871, EPI_ISL_582872, EPI_ISL_582873, EPI_ISL_582874, EPI_ISL_582875, EPI_ISL_582876, EPI_ISL_582877, EPI_ISL_582878, EPI_ISL_582879, EPI_ISL_582880, EPI_ISL_582881, EPI_ISL_582882, EPI_ISL_582883, EPI_ISL_582884, EPI_ISL_582885, EPI_ISL_582886, EPI_ISL_582887, EPI_ISL_582888, EPI_ISL_582889, EPI_ISL_582890, EPI_ISL_582891, EPI_ISL_582892, EPI_ISL_582893, EPI_ISL_582894, EPI_ISL_582895, EPI_ISL_582896, EPI_ISL_582897, EPI_ISL_582898, EPI_ISL_582899, EPI_ISL_582900, EPI_ISL_582901, EPI_ISL_582902, EPI_ISL_582903, EPI_ISL_582904, EPI_ISL_582905, EPI_ISL_582906, EPI_ISL_582907, EPI_ISL_582908, EPI_ISL_582909, EPI_ISL_582910, EPI_ISL_582911, EPI_ISL_582912, EPI_ISL_582913, EPI_ISL_582914, EPI_ISL_582915, EPI_ISL_582916, EPI_ISL_582917, EPI_ISL_582918, EPI_ISL_582919, EPI_ISL_582920, EPI_ISL_582921, EPI_ISL_582922, EPI_ISL_582923, EPI_ISL_582924, EPI_ISL_582925, EPI_ISL_582926, EPI_ISL_582927, EPI_ISL_582928, EPI_ISL_582929, EPI_ISL_582930, EPI_ISL_582931, EPI_ISL_582932, EPI_ISL_582933, EPI_ISL_582934, EPI_ISL_582935, EPI_ISL_582936, EPI_ISL_582937, EPI_ISL_582938, EPI_ISL_582939, EPI_ISL_582940, EPI_ISL_582941, EPI_ISL_582942, EPI_ISL_582943, EPI_ISL_582944, EPI_ISL_582945, EPI_ISL_582946, EPI_ISL_582947, EPI_ISL_582948, EPI_ISL_582949, EPI_ISL_582950, EPI_ISL_582951, EPI_ISL_582952, EPI_ISL_582953, EPI_ISL_582954, EPI_ISL_582955, EPI_ISL_582956, EPI_ISL_582957, EPI_ISL_582958 | | | |
| see above | County of Santa Clara Public Health Department | Chan-Zuckerberg Biohub | CZB Cliahub Consortium |
| EPI_ISL_582959, EPI_ISL_582960, EPI_ISL_582961, EPI_ISL_582962, EPI_ISL_582963, EPI_ISL_582964, EPI_ISL_582965, EPI_ISL_582966, EPI_ISL_582967, EPI_ISL_582968, EPI_ISL_582969, EPI_ISL_582970, EPI_ISL_582971, EPI_ISL_582972, EPI_ISL_582973, EPI_ISL_582974 | | | |
| see above | San Luis Obispo Public Health Department | Chan-Zuckerberg Biohub | CZB Cliahub Consortium |
| EPI_ISL_582975, EPI_ISL_582976, EPI_ISL_582977, EPI_ISL_582978, EPI_ISL_582979, EPI_ISL_582980, EPI_ISL_582981, EPI_ISL_582982, EPI_ISL_582983, EPI_ISL_582984, EPI_ISL_582985, EPI_ISL_582986, EPI_ISL_582987, EPI_ISL_582988, EPI_ISL_582989, EPI_ISL_582990, EPI_ISL_582991, EPI_ISL_582992, EPI_ISL_582993, EPI_ISL_582994, EPI_ISL_582995, EPI_ISL_582996, EPI_ISL_582997, EPI_ISL_582998, EPI_ISL_582999, EPI_ISL_583000, EPI_ISL_583001, EPI_ISL_583002, EPI_ISL_583003, EPI_ISL_583004, EPI_ISL_583005, EPI_ISL_583006, EPI_ISL_583007, EPI_ISL_583008, EPI_ISL_583009, EPI_ISL_583010, EPI_ISL_583011, EPI_ISL_583012, EPI_ISL_583013, EPI_ISL_583014, EPI_ISL_583015, EPI_ISL_583016, EPI_ISL_583017, EPI_ISL_583018, EPI_ISL_583019, EPI_ISL_583020, EPI_ISL_583021, EPI_ISL_583022, EPI_ISL_583023, EPI_ISL_583024, EPI_ISL_583025, EPI_ISL_583026, EPI_ISL_583027, EPI_ISL_583028, EPI_ISL_583029, EPI_ISL_583030, EPI_ISL_583031, EPI_ISL_583032, EPI_ISL_583033, EPI_ISL_583034, EPI_ISL_583035, EPI_ISL_583036, EPI_ISL_583037, EPI_ISL_583038, EPI_ISL_583039, EPI_ISL_583040, EPI_ISL_583041, EPI_ISL_583042, EPI_ISL_583043, EPI_ISL_583044, EPI_ISL_583045 | | | |
| see above | Orange County Public Health Lab | Chan-Zuckerberg Biohub | CZB Cliahub Consortium |
| EPI_ISL_583046, EPI_ISL_583047, EPI_ISL_583048, EPI_ISL_583049, EPI_ISL_583050, EPI_ISL_583051, EPI_ISL_583052, EPI_ISL_583053, EPI_ISL_583054, EPI_ISL_583055, EPI_ISL_583056, EPI_ISL_583057, EPI_ISL_583058, EPI_ISL_583059, EPI_ISL_583060, EPI_ISL_583061, EPI_ISL_583062, EPI_ISL_583063, EPI_ISL_583064, EPI_ISL_583065, EPI_ISL_583066, EPI_ISL_583067, EPI_ISL_583068, EPI_ISL_583069, EPI_ISL_583070, EPI_ISL_583071, EPI_ISL_583072, EPI_ISL_583073, EPI_ISL_583074, EPI_ISL_583075, EPI_ISL_583076, EPI_ISL_583077, EPI_ISL_583078, EPI_ISL_583079, EPI_ISL_583080, EPI_ISL_583081, EPI_ISL_583082, EPI_ISL_583083, EPI_ISL_583084, EPI_ISL_583085, EPI_ISL_583086, EPI_ISL_583087, EPI_ISL_583088, EPI_ISL_583089, EPI_ISL_583090, EPI_ISL_583091, EPI_ISL_583092, EPI_ISL_583093, EPI_ISL_583094, EPI_ISL_583095, EPI_ISL_583096, EPI_ISL_583097, EPI_ISL_583098, EPI_ISL_583099, EPI_ISL_583100, EPI_ISL_583101, EPI_ISL_583102, EPI_ISL_583103, EPI_ISL_583105, EPI_ISL_583106, EPI_ISL_583107, EPI_ISL_583108, EPI_ISL_583109, EPI_ISL_583110, EPI_ISL_583111, EPI_ISL_583112, EPI_ISL_583113, EPI_ISL_583114, EPI_ISL_583115, EPI_ISL_583116, EPI_ISL_583117, EPI_ISL_583118, EPI_ISL_583119, EPI_ISL_583120, EPI_ISL_583121, EPI_ISL_583122, EPI_ISL_583123, EPI_ISL_583124, EPI_ISL_583125, EPI_ISL_583126, EPI_ISL_583127, EPI_ISL_583128, EPI_ISL_583129, EPI_ISL_583130, EPI_ISL_583131, EPI_ISL_583132, EPI_ISL_583133, EPI_ISL_583134, EPI_ISL_583135, EPI_ISL_583136, EPI_ISL_583137, EPI_ISL_583138, EPI_ISL_583139, EPI_ISL_583140, EPI_ISL_583141, EPI_ISL_583142, EPI_ISL_583143, EPI_ISL_583144, EPI_ISL_583145, EPI_ISL_583146, EPI_ISL_583147, EPI_ISL_583148, EPI_ISL_583149, EPI_ISL_583150, EPI_ISL_583151, EPI_ISL_583152, EPI_ISL_583153, EPI_ISL_583154, EPI_ISL_583155, EPI_ISL_583156, EPI_ISL_583157, EPI_ISL_583158, EPI_ISL_583159, EPI_ISL_583160, EPI_ISL_583161, EPI_ISL_583162, EPI_ISL_583163, EPI_ISL_583164, EPI_ISL_583165, EPI_ISL_583166, EPI_ISL_583167, EPI_ISL_583168, EPI_ISL_583169, EPI_ISL_583170 | | | |
| see above | Humboldt County Public Health | Chan-Zuckerberg Biohub | CZB Cliahub Consortium |

| | | | |
|---|---|---|---|
| | Laboratory | | |
| EPI_ISL_583171, EPI_ISL_583172 | Tulare County Public Health Lab | Chan-Zuckerberg Biohub | CZB Cliahub Consortium |
| EPI_ISL_583173, EPI_ISL_583174, EPI_ISL_583175, EPI_ISL_583176, EPI_ISL_583177, EPI_ISL_583178, EPI_ISL_583179, EPI_ISL_583180, EPI_ISL_583181, EPI_ISL_583182, EPI_ISL_583183, EPI_ISL_583184, EPI_ISL_583185, EPI_ISL_583186, EPI_ISL_583187, EPI_ISL_583188, EPI_ISL_583189 | | | |
| see above | San Bernardino County Public Health Lab | Chan-Zuckerberg Biohub | CZB Cliahub Consortium |
| EPI_ISL_583190, EPI_ISL_583191, EPI_ISL_583192, EPI_ISL_583193, EPI_ISL_583194, EPI_ISL_583195, EPI_ISL_583196, EPI_ISL_583197 | San Francisco Public Health Laboratory | Chan-Zuckerberg Biohub | CZB Cliahub Consortium |
| EPI_ISL_583198 | Marin County Public Health Department | Chan-Zuckerberg Biohub | CZB Cliahub Consortium |
| EPI_ISL_583199, EPI_ISL_583200, EPI_ISL_583201, EPI_ISL_583202, EPI_ISL_583203, EPI_ISL_583204, EPI_ISL_583205, EPI_ISL_583206, EPI_ISL_583207, EPI_ISL_583208, EPI_ISL_583209, EPI_ISL_583210, EPI_ISL_583211, EPI_ISL_583212, EPI_ISL_583213, EPI_ISL_583214, EPI_ISL_583215, EPI_ISL_583216, EPI_ISL_583217, EPI_ISL_583218, EPI_ISL_583219, EPI_ISL_583220, EPI_ISL_583221, EPI_ISL_583222, EPI_ISL_583223, EPI_ISL_583224, EPI_ISL_583225, EPI_ISL_583226, EPI_ISL_583227, EPI_ISL_583228, EPI_ISL_583229, EPI_ISL_583230, EPI_ISL_583231 | | | |
| see above | UCSF Clinical Microbiology Laboratory | Chan-Zuckerberg Biohub | CZB Cliahub Consortium |
| EPI_ISL_583246, EPI_ISL_583247, EPI_ISL_583248 | Delaware Public Health Lab | Delaware Public Health Lab | Gregory Hovan |
| EPI_ISL_583249, EPI_ISL_583250, EPI_ISL_583251, EPI_ISL_583252, EPI_ISL_583253, EPI_ISL_583254, EPI_ISL_583255, EPI_ISL_583256, EPI_ISL_583257, EPI_ISL_583258, EPI_ISL_583259, EPI_ISL_583260, EPI_ISL_583261, EPI_ISL_583262, EPI_ISL_583263, EPI_ISL_583264, EPI_ISL_583265, EPI_ISL_583266, EPI_ISL_583267, EPI_ISL_583268, EPI_ISL_583269, EPI_ISL_583270, EPI_ISL_583271, EPI_ISL_583272, EPI_ISL_583273, EPI_ISL_583274, EPI_ISL_583275, EPI_ISL_583276, EPI_ISL_583277, EPI_ISL_583278, EPI_ISL_583279, EPI_ISL_583280, EPI_ISL_583281, EPI_ISL_583282, EPI_ISL_583283, EPI_ISL_583284, EPI_ISL_583285, EPI_ISL_583286, EPI_ISL_583287, EPI_ISL_583288, EPI_ISL_583289, EPI_ISL_583290, EPI_ISL_583291, EPI_ISL_583292, EPI_ISL_583293, EPI_ISL_583294, EPI_ISL_583295, EPI_ISL_583296, EPI_ISL_583297, EPI_ISL_583298, EPI_ISL_583299, EPI_ISL_583300, EPI_ISL_583301, EPI_ISL_583302, EPI_ISL_583303, EPI_ISL_583304, EPI_ISL_583305, EPI_ISL_583306, EPI_ISL_583307, EPI_ISL_583308, EPI_ISL_583309, EPI_ISL_583310, EPI_ISL_583311, EPI_ISL_583312, EPI_ISL_583313, EPI_ISL_583314, EPI_ISL_583315, EPI_ISL_583316, EPI_ISL_583317, EPI_ISL_583318, EPI_ISL_583319, EPI_ISL_583320, EPI_ISL_583321, EPI_ISL_583322, EPI_ISL_583323, EPI_ISL_583324, EPI_ISL_583325, EPI_ISL_583326, EPI_ISL_583327, EPI_ISL_583328, EPI_ISL_583329, EPI_ISL_583330, EPI_ISL_583331, EPI_ISL_583332, EPI_ISL_583333, EPI_ISL_583334, EPI_ISL_583335, EPI_ISL_583336, EPI_ISL_583337, EPI_ISL_583338, EPI_ISL_583339, EPI_ISL_583340, EPI_ISL_583341, EPI_ISL_583342, EPI_ISL_583343, EPI_ISL_583344, EPI_ISL_583345, EPI_ISL_583346, EPI_ISL_583347, EPI_ISL_583348, EPI_ISL_583349, EPI_ISL_583350, EPI_ISL_583351, EPI_ISL_583352, EPI_ISL_583353, EPI_ISL_583354, EPI_ISL_583355, EPI_ISL_583356, EPI_ISL_583357, EPI_ISL_583358, EPI_ISL_583359, EPI_ISL_583360, EPI_ISL_583361, EPI_ISL_583362, EPI_ISL_583363, EPI_ISL_583364, EPI_ISL_583365, EPI_ISL_583366, EPI_ISL_583367, EPI_ISL_583368, EPI_ISL_583369, EPI_ISL_583370, EPI_ISL_583371, EPI_ISL_583372, EPI_ISL_583373, EPI_ISL_583374, EPI_ISL_583375, EPI_ISL_583376, EPI_ISL_583377, EPI_ISL_583378, EPI_ISL_583379, EPI_ISL_583380, EPI_ISL_583381, EPI_ISL_583382, EPI_ISL_583383, EPI_ISL_583384, EPI_ISL_583385, EPI_ISL_583386, EPI_ISL_583387, EPI_ISL_583388, EPI_ISL_583389, EPI_ISL_583390, EPI_ISL_583391, EPI_ISL_583392, EPI_ISL_583393, EPI_ISL_583394, EPI_ISL_583395, EPI_ISL_583396, EPI_ISL_583397, EPI_ISL_583398, EPI_ISL_583399, EPI_ISL_583400, EPI_ISL_583401, EPI_ISL_583402, EPI_ISL_583403, EPI_ISL_583404, EPI_ISL_583405, EPI_ISL_583406, EPI_ISL_583407, EPI_ISL_583408, EPI_ISL_583409, EPI_ISL_583410, EPI_ISL_583411, EPI_ISL_583412, EPI_ISL_583413, EPI_ISL_583414, EPI_ISL_583415, EPI_ISL_583416, EPI_ISL_583417, EPI_ISL_583418, EPI_ISL_583419, EPI_ISL_583420, EPI_ISL_583421, EPI_ISL_583422, EPI_ISL_583423, EPI_ISL_583424, EPI_ISL_583425 | | | |
| see above | University of Michigan Clinical Microbiology Laboratory | Lauring Lab, University of Michigan, Department of Microbiology and Immunology | Valesano |
| EPI_ISL_583426 | Garcia-Sastre Laboratory, Department of Microbiology, Icahn School of Medicine at Mount Sinai | van Bakel Laboratory, Genetics and Genomics Sciences, Icahn School of Medicine at Mount Sinai | Teresa Aydillo, Ana S. Gonzalez-Reiche, Sadaf Aslam, Adriana van de Guchte, Zenab Khan, Ajay Obla, Jayeeta Dutta, Harm van Bakel, Judith Aberg, Adolfo García-Sastre, Gunjan Shah, Tobias Hohl, Genovefa Papanicolaou, Miguel-Angel Perales, Kent Sepkowitz, Ngolela Esther Babady, and Mini Kamboj |
| EPI_ISL_583427, EPI_ISL_583428 | Memorial Sloan Kettering Cancer Center | van Bakel Laboratory, Genetics and Genomics Sciences, Icahn School of Medicine at Mount Sinai | Teresa Aydillo, Ana S. Gonzalez-Reiche, Sadaf Aslam, Adriana van de Guchte, Zenab Khan, Ajay Obla, Jayeeta Dutta, Harm van Bakel, Judith Aberg, Adolfo García-Sastre, Gunjan Shah, Tobias Hohl, Genovefa Papanicolaou, Miguel-Angel Perales, Kent Sepkowitz, Ngolela Esther Babady, and Mini Kamboj |
| EPI_ISL_583429 | Garcia-Sastre Laboratory, Department of Microbiology, Icahn School of Medicine at Mount Sinai | van Bakel Laboratory, Genetics and Genomics Sciences, Icahn School of Medicine at Mount Sinai | Teresa Aydillo, Ana S. Gonzalez-Reiche, Sadaf Aslam, Adriana van de Guchte, Zenab Khan, Ajay Obla, Jayeeta Dutta, Harm van Bakel, Judith Aberg, Adolfo García-Sastre, Gunjan Shah, Tobias Hohl, Genovefa Papanicolaou, Miguel-Angel Perales, Kent Sepkowitz, Ngolela Esther Babady, and Mini Kamboj |
| EPI_ISL_583430, EPI_ISL_583431, EPI_ISL_583432, EPI_ISL_583433 | Memorial Sloan Kettering Cancer Center | van Bakel Laboratory, Genetics and Genomics Sciences, Icahn School of Medicine at Mount Sinai | Teresa Aydillo, Ana S. Gonzalez-Reiche, Sadaf Aslam, Adriana van de Guchte, Zenab Khan, Ajay Obla, Jayeeta Dutta, Harm van Bakel, Judith Aberg, Adolfo García-Sastre, Gunjan Shah, Tobias Hohl, Genovefa Papanicolaou, Miguel-Angel Perales, Kent Sepkowitz, Ngolela Esther Babady, and Mini Kamboj |
| EPI_ISL_583434, EPI_ISL_583435, EPI_ISL_583436 | Garcia-Sastre Laboratory, Department of Microbiology, Icahn School of Medicine at Mount Sinai | van Bakel Laboratory, Genetics and Genomics Sciences, Icahn School of Medicine at Mount Sinai | Teresa Aydillo, Ana S. Gonzalez-Reiche, Sadaf Aslam, Adriana van de Guchte, Zenab Khan, Ajay Obla, Jayeeta Dutta, Harm van Bakel, Judith Aberg, Adolfo García-Sastre, Gunjan Shah, Tobias Hohl, Genovefa Papanicolaou, Miguel-Angel Perales, Kent Sepkowitz, Ngolela Esther Babady, and Mini Kamboj |
| EPI_ISL_583437, EPI_ISL_583438, EPI_ISL_583439, EPI_ISL_583440 | Memorial Sloan Kettering Cancer Center | van Bakel Laboratory, Genetics and Genomics Sciences, Icahn School of Medicine at Mount Sinai | Teresa Aydillo, Ana S. Gonzalez-Reiche, Sadaf Aslam, Adriana van de Guchte, Zenab Khan, Ajay Obla, Jayeeta Dutta, Harm van Bakel, Judith Aberg, Adolfo García-Sastre, Gunjan Shah, Tobias Hohl, Genovefa Papanicolaou, Miguel-Angel Perales, Kent Sepkowitz, Ngolela Esther Babady, and Mini Kamboj |
| EPI_ISL_583441, EPI_ISL_583442 | Garcia-Sastre Laboratory, Department of Microbiology, Icahn School of Medicine at Mount Sinai | van Bakel Laboratory, Genetics and Genomics Sciences, Icahn School of Medicine at Mount Sinai | Teresa Aydillo, Ana S. Gonzalez-Reiche, Sadaf Aslam, Adriana van de Guchte, Zenab Khan, Ajay Obla, Jayeeta Dutta, Harm van Bakel, Judith Aberg, Adolfo García-Sastre, Gunjan Shah, Tobias Hohl, Genovefa Papanicolaou, Miguel-Angel Perales, Kent Sepkowitz, Ngolela Esther Babady, and Mini Kamboj |
| EPI_ISL_583443, EPI_ISL_583444, EPI_ISL_583445, EPI_ISL_583446, EPI_ISL_583447, EPI_ISL_583448, EPI_ISL_583449, EPI_ISL_583450, EPI_ISL_583451 | Memorial Sloan Kettering Cancer Center | van Bakel Laboratory, Genetics and Genomics Sciences, Icahn School of Medicine at Mount Sinai | Teresa Aydillo, Ana S. Gonzalez-Reiche, Sadaf Aslam, Adriana van de Guchte, Zenab Khan, Ajay Obla, Jayeeta Dutta, Harm van Bakel, Judith Aberg, Adolfo García-Sastre, Gunjan Shah, Tobias Hohl, Genovefa Papanicolaou, Miguel-Angel Perales, Kent Sepkowitz, Ngolela Esther Babady, and Mini Kamboj |
| EPI_ISL_583452, EPI_ISL_583453, EPI_ISL_583454 | Garcia-Sastre Laboratory, Department of Microbiology, Icahn School of Medicine at Mount Sinai | van Bakel Laboratory, Genetics and Genomics Sciences, Icahn School of Medicine at Mount Sinai | Teresa Aydillo, Ana S. Gonzalez-Reiche, Sadaf Aslam, Adriana van de Guchte, Zenab Khan, Ajay Obla, Jayeeta Dutta, Harm van Bakel, Judith Aberg, Adolfo García-Sastre, Gunjan Shah, Tobias Hohl, Genovefa Papanicolaou, Miguel-Angel Perales, Kent Sepkowitz, Ngolela Esther Babady, and Mini Kamboj |
| EPI_ISL_583455, EPI_ISL_583456, EPI_ISL_583457 | Memorial Sloan Kettering Cancer Center | van Bakel Laboratory, Genetics and Genomics Sciences, Icahn School of Medicine at Mount Sinai | Teresa Aydillo, Ana S. Gonzalez-Reiche, Sadaf Aslam, Adriana van de Guchte, Zenab Khan, Ajay Obla, Jayeeta Dutta, Harm van Bakel, Judith Aberg, Adolfo García-Sastre, Gunjan Shah, Tobias Hohl, Genovefa Papanicolaou, Miguel-Angel Perales, Kent Sepkowitz, Ngolela Esther Babady, and Mini Kamboj |
| EPI_ISL_583458, EPI_ISL_583459 | Garcia-Sastre Laboratory, Department of Microbiology, Icahn School of Medicine at Mount Sinai | van Bakel Laboratory, Genetics and Genomics Sciences, Icahn School of Medicine at Mount Sinai | Teresa Aydillo, Ana S. Gonzalez-Reiche, Sadaf Aslam, Adriana van de Guchte, Zenab Khan, Ajay Obla, Jayeeta Dutta, Harm van Bakel, Judith Aberg, Adolfo García-Sastre, Gunjan Shah, Tobias Hohl, Genovefa Papanicolaou, Miguel-Angel Perales, Kent Sepkowitz, Ngolela Esther Babady, and Mini Kamboj |
| EPI_ISL_583460, EPI_ISL_583461, EPI_ISL_583462 | Memorial Sloan Kettering Cancer Center | van Bakel Laboratory, Genetics and Genomics Sciences, Icahn School of Medicine at Mount Sinai | Teresa Aydillo, Ana S. Gonzalez-Reiche, Sadaf Aslam, Adriana van de Guchte, Zenab Khan, Ajay Obla, Jayeeta Dutta, Harm van Bakel, Judith Aberg, Adolfo García-Sastre, Gunjan Shah, Tobias Hohl, Genovefa Papanicolaou, Miguel-Angel Perales, Kent Sepkowitz, Ngolela Esther Babady, and Mini Kamboj |
| EPI_ISL_583463, EPI_ISL_583464 | Garcia-Sastre Laboratory, Department of Microbiology, Icahn School of Medicine at Mount Sinai | van Bakel Laboratory, Genetics and Genomics Sciences, Icahn School of Medicine at Mount Sinai | Teresa Aydillo, Ana S. Gonzalez-Reiche, Sadaf Aslam, Adriana van de Guchte, Zenab Khan, Ajay Obla, Jayeeta Dutta, Harm van Bakel, Judith Aberg, Adolfo García-Sastre, Gunjan Shah, Tobias Hohl, Genovefa Papanicolaou, Miguel-Angel Perales, Kent Sepkowitz, Ngolela Esther Babady, and Mini Kamboj |
| EPI_ISL_583465, EPI_ISL_583466 | Memorial Sloan Kettering Cancer Center | van Bakel Laboratory, Genetics and Genomics Sciences, Icahn School of Medicine at Mount Sinai | Teresa Aydillo, Ana S. Gonzalez-Reiche, Sadaf Aslam, Adriana van de Guchte, Zenab Khan, Ajay Obla, Jayeeta Dutta, Harm van Bakel, Judith Aberg, Adolfo García-Sastre, Gunjan Shah, Tobias Hohl, Genovefa Papanicolaou, Miguel-Angel Perales, Kent Sepkowitz, Ngolela Esther Babady, and Mini Kamboj |
| EPI_ISL_583467 | Garcia-Sastre Laboratory, Department of Microbiology, Icahn School of Medicine at Mount Sinai | van Bakel Laboratory, Genetics and Genomics Sciences, Icahn School of Medicine at Mount Sinai | Teresa Aydillo, Ana S. Gonzalez-Reiche, Sadaf Aslam, Adriana van de Guchte, Zenab Khan, Ajay Obla, Jayeeta Dutta, Harm van Bakel, Judith Aberg, Adolfo García-Sastre, Gunjan Shah, Tobias Hohl, Genovefa Papanicolaou, Miguel-Angel Perales, Kent Sepkowitz, Ngolela Esther Babady, and Mini Kamboj |
| EPI_ISL_583468, EPI_ISL_583469, EPI_ISL_583470 | Memorial Sloan Kettering Cancer Center | van Bakel Laboratory, Genetics and Genomics Sciences, Icahn School of Medicine at Mount Sinai | Teresa Aydillo, Ana S. Gonzalez-Reiche, Sadaf Aslam, Adriana van de Guchte, Zenab Khan, Ajay Obla, Jayeeta Dutta, Harm van Bakel, Judith Aberg, Adolfo García-Sastre, Gunjan Shah, Tobias Hohl, Genovefa Papanicolaou, Miguel-Angel Perales, Kent Sepkowitz, Ngolela Esther Babady, and Mini Kamboj |
| EPI_ISL_583471 | Garcia-Sastre Laboratory, Department of Microbiology, Icahn School of Medicine at Mount Sinai | van Bakel Laboratory, Genetics and Genomics Sciences, Icahn School of Medicine at Mount Sinai | Teresa Aydillo, Ana S. Gonzalez-Reiche, Sadaf Aslam, Adriana van de Guchte, Zenab Khan, Ajay Obla, Jayeeta Dutta, Harm van Bakel, Judith Aberg, Adolfo García-Sastre, Gunjan Shah, Tobias Hohl, Genovefa Papanicolaou, Miguel-Angel Perales, Kent Sepkowitz, Ngolela Esther Babady, and Mini Kamboj |
| EPI_ISL_583472 | Memorial Sloan Kettering Cancer Center | van Bakel Laboratory, Genetics and Genomics Sciences, Icahn School of Medicine at Mount Sinai | Teresa Aydillo, Ana S. Gonzalez-Reiche, Sadaf Aslam, Adriana van de Guchte, Zenab Khan, Ajay Obla, Jayeeta Dutta, Harm van Bakel, Judith Aberg, Adolfo García-Sastre, Gunjan Shah, Tobias Hohl, Genovefa Papanicolaou, Miguel-Angel Perales, Kent Sepkowitz, Ngolela Esther Babady, and Mini Kamboj |
| EPI_ISL_583473 | Garcia-Sastre Laboratory, Department of Microbiology, Icahn School of Medicine at Mount Sinai | van Bakel Laboratory, Genetics and Genomics Sciences, Icahn School of Medicine at Mount Sinai | Teresa Aydillo, Ana S. Gonzalez-Reiche, Sadaf Aslam, Adriana van de Guchte, Zenab Khan, Ajay Obla, Jayeeta Dutta, Harm van Bakel, Judith Aberg, Adolfo García-Sastre, Gunjan Shah, Tobias Hohl, Genovefa Papanicolaou, Miguel-Angel Perales, Kent Sepkowitz, Ngolela Esther Babady, and Mini Kamboj |
| EPI_ISL_583474, | Memorial Sloan Kettering Cancer Center | van Bakel Laboratory, Genetics and | Teresa Aydillo, Ana S. Gonzalez-Reiche, Sadaf Aslam, Adriana van de Guchte, Zenab Khan, Ajay Obla, Jayeeta Dutta, Harm van Bakel, Judith Aberg, Adolfo García-Sastre, Gunjan Shah, Tobias Hohl, Genovefa Papanicolaou, Miguel-Angel Perales, Kent Sepkowitz, Ngolela Esther Babady, |

| | | | |
|---|---|---|---|
| EPI_ISL_583475, EPI_ISL_583476 | | Genomics Sciences, Icahn School of Medicine at Mount Sinai | and Mini Kamboj |
| EPI_ISL_583477 | Garcia-Sastre Laboratory, Department of Microbiology, Icahn School of Medicine at Mount Sinai | van Bakel Laboratory, Genetics and Genomics Sciences, Icahn School of Medicine at Mount Sinai | Teresa Aydillo, Ana S. Gonzalez-Reiche, Sadaf Aslam, Adriana van de Guchte, Zenab Khan, Ajay Obla, Jayeeta Dutta, Harm van Bakel, Judith Aberg, Adolfo García-Sastre, Gunjan Shah, Tobias Hohl, Genovefa Papanicolaou, Miguel-Angel Perales, Kent Sepkowitz, Ngolela Esther Babady, and Mini Kamboj |
| EPI_ISL_583478, EPI_ISL_583479, EPI_ISL_583480 | Memorial Sloan Kettering Cancer Center | van Bakel Laboratory, Genetics and Genomics Sciences, Icahn School of Medicine at Mount Sinai | Teresa Aydillo, Ana S. Gonzalez-Reiche, Sadaf Aslam, Adriana van de Guchte, Zenab Khan, Ajay Obla, Jayeeta Dutta, Harm van Bakel, Judith Aberg, Adolfo García-Sastre, Gunjan Shah, Tobias Hohl, Genovefa Papanicolaou, Miguel-Angel Perales, Kent Sepkowitz, Ngolela Esther Babady, and Mini Kamboj |
| EPI_ISL_583506, EPI_ISL_583507, EPI_ISL_583508, EPI_ISL_583509, EPI_ISL_583510, EPI_ISL_583511, EPI_ISL_583512, EPI_ISL_583513, EPI_ISL_583514, EPI_ISL_583515, EPI_ISL_583516, EPI_ISL_583517, EPI_ISL_583518, EPI_ISL_583519, EPI_ISL_583520, EPI_ISL_583521, EPI_ISL_583522, EPI_ISL_583523, EPI_ISL_583524, EPI_ISL_583525, EPI_ISL_583526, EPI_ISL_583527, EPI_ISL_583528, EPI_ISL_583529, EPI_ISL_583530, EPI_ISL_583531, EPI_ISL_583532, EPI_ISL_583533, EPI_ISL_583534 | | | |
| see above | Michigan Department of Health and Human Services, Bureau of Laboratories | Michigan Department of Health and Human Services, Bureau of Laboratories | Blankenship HM, Riner D, Soehnlen MK |
| EPI_ISL_583994, EPI_ISL_583995, EPI_ISL_584013, EPI_ISL_584014 | Delaware Public Health Lab | Delaware Public Health Lab | Gregory Hovan |
| EPI_ISL_584095, EPI_ISL_584096, EPI_ISL_584097 | University of Michigan Clinical Microbiology Laboratory | Lauring Lab, University of Michigan, Department of Microbiology and Immunology | Valesano |
| EPI_ISL_584123, EPI_ISL_584127, EPI_ISL_584128, EPI_ISL_584129, EPI_ISL_584131, EPI_ISL_584132, EPI_ISL_584133, EPI_ISL_584136, EPI_ISL_584137, EPI_ISL_584138, EPI_ISL_584139, EPI_ISL_584140, EPI_ISL_584141, EPI_ISL_584142, EPI_ISL_584143, EPI_ISL_584145, EPI_ISL_584146, EPI_ISL_584147, EPI_ISL_584148, EPI_ISL_584149, EPI_ISL_584150, EPI_ISL_584151, EPI_ISL_584152, EPI_ISL_584154, EPI_ISL_584155, EPI_ISL_584156, EPI_ISL_584157, EPI_ISL_584158, EPI_ISL_584160, EPI_ISL_584162, EPI_ISL_584163, EPI_ISL_584164 | | | |
| see above | Texas Department of State Health Services | Texas Department of State Health Services | Rashmi Tuladhar, Bonnie Oh, Jenny Zhang, Maliha Rahman, Anita Pokharel, Mayela Pedrueza, Myong Koag, Chun Wang, Rachel Lee, Grace Kubin |
| EPI_ISL_586243, EPI_ISL_586244, EPI_ISL_586245, EPI_ISL_586246, EPI_ISL_586247, EPI_ISL_586248, EPI_ISL_586249, EPI_ISL_586250, EPI_ISL_586251, EPI_ISL_586252, EPI_ISL_586253, EPI_ISL_586254, EPI_ISL_586255, EPI_ISL_586256, EPI_ISL_586257, EPI_ISL_586258, EPI_ISL_586259, EPI_ISL_586260, EPI_ISL_586261, EPI_ISL_586262, EPI_ISL_586263, EPI_ISL_586264, EPI_ISL_586265, EPI_ISL_586266 | | | |
| see above | Alaska State Virology Laboratory | Alaska State Virology Laboratory | Jack Chen, Ph.D. |
| EPI_ISL_590699, EPI_ISL_590700, EPI_ISL_590701, EPI_ISL_590702, EPI_ISL_590703, EPI_ISL_590704, EPI_ISL_590705, EPI_ISL_590706, EPI_ISL_590707, EPI_ISL_590708, EPI_ISL_590709, EPI_ISL_590710, EPI_ISL_590711, EPI_ISL_590712, EPI_ISL_590713, EPI_ISL_590714, EPI_ISL_590715, EPI_ISL_590716, EPI_ISL_590717, EPI_ISL_590718, EPI_ISL_590719, EPI_ISL_590720, EPI_ISL_590721, EPI_ISL_590722, EPI_ISL_590723, EPI_ISL_590724, EPI_ISL_590725, EPI_ISL_590726, EPI_ISL_590727, EPI_ISL_590728, EPI_ISL_590729, EPI_ISL_590730, EPI_ISL_590731, EPI_ISL_590732, EPI_ISL_590733, EPI_ISL_590734, EPI_ISL_590735, EPI_ISL_590736, EPI_ISL_590737, EPI_ISL_590738, EPI_ISL_590739, EPI_ISL_590740, EPI_ISL_590741, EPI_ISL_590742, EPI_ISL_590743, EPI_ISL_590744, EPI_ISL_590745, EPI_ISL_590746, EPI_ISL_590747, EPI_ISL_590748, EPI_ISL_590749, EPI_ISL_590750, EPI_ISL_590751, EPI_ISL_590752 | | | |
| see above | University of Michigan Clinical Microbiology Laboratory | Lauring Lab, University of Michigan, Department of Microbiology and Immunology | Valesano |
| EPI_ISL_590753, EPI_ISL_590754, EPI_ISL_590755, EPI_ISL_590756, EPI_ISL_590757, EPI_ISL_590758, EPI_ISL_590759, EPI_ISL_590760, EPI_ISL_590761, EPI_ISL_590762, EPI_ISL_590763, EPI_ISL_590764, EPI_ISL_590765, EPI_ISL_590766, EPI_ISL_590767, EPI_ISL_590768, EPI_ISL_590769, EPI_ISL_590770, EPI_ISL_590771, EPI_ISL_590772, EPI_ISL_590773, EPI_ISL_590774, EPI_ISL_590775, EPI_ISL_590776, EPI_ISL_590777, EPI_ISL_590778, EPI_ISL_590779, EPI_ISL_590780, EPI_ISL_590781, EPI_ISL_590782, EPI_ISL_590783, EPI_ISL_590784, EPI_ISL_590785, EPI_ISL_590786, EPI_ISL_590787, EPI_ISL_590788, EPI_ISL_590789, EPI_ISL_590790, EPI_ISL_590791, EPI_ISL_590792, EPI_ISL_590793, EPI_ISL_590794, EPI_ISL_590795, EPI_ISL_590796, EPI_ISL_590797, EPI_ISL_590798, EPI_ISL_590799, EPI_ISL_590800, EPI_ISL_590801, EPI_ISL_590802, EPI_ISL_590803, EPI_ISL_590804, EPI_ISL_590805, EPI_ISL_590806, EPI_ISL_590807, EPI_ISL_590808, EPI_ISL_590809, EPI_ISL_590810, EPI_ISL_590811, EPI_ISL_590812, EPI_ISL_590813, EPI_ISL_590814, EPI_ISL_590815, EPI_ISL_590816, EPI_ISL_590817, EPI_ISL_590818, EPI_ISL_590819, EPI_ISL_590820, EPI_ISL_590821, EPI_ISL_590822, EPI_ISL_590837 | | | |
| see above | Minnesota Department of Health, Public Health Laboratory | Minnesota Department of Health, Public Health Laboratory | Matt Plumb, Jacob Garfin, Alexandra Lorentz, and Xiong Wang |
| EPI_ISL_590839, EPI_ISL_590840, EPI_ISL_590843, EPI_ISL_590851, EPI_ISL_590852, EPI_ISL_590853, EPI_ISL_590854, EPI_ISL_590860, EPI_ISL_590865, EPI_ISL_590867, EPI_ISL_590869, EPI_ISL_590871, EPI_ISL_590872, EPI_ISL_590874 | | | |
| see above | Texas Department of State Health Services | Texas Department of State Health Services | Rashmi Tuladhar, Bonnie Oh, Jenny Zhang, Maliha Rahman, Anita Pokharel, Myong Koag, Chung Wang, Rachel Lee, Grace Kubin, Mayela Pedrueza |
| EPI_ISL_591022, EPI_ISL_591023, EPI_ISL_591024, EPI_ISL_591025, EPI_ISL_591026, EPI_ISL_591027, EPI_ISL_591028, EPI_ISL_591029, EPI_ISL_591030, EPI_ISL_591031, EPI_ISL_591032, EPI_ISL_591033, EPI_ISL_591034, EPI_ISL_591035, EPI_ISL_591036 | | | |
| see above | MD PHL | MD PHL | Maryland Department of Health Laboratories Administration |
| EPI_ISL_591054, EPI_ISL_591055, EPI_ISL_591056 | Virus Ecology, Rocky Mountain Laboratories, National Institutes of Health | Virus Ecology, Rocky Mountain Laboratories, National Institutes of Health | Van Doremalen,N., Holbrook,M.G., Barbian,K.D., Bushmaker,C., Bushmaker,T., Martens,C.A., Munster,V.J. |
| EPI_ISL_591281, EPI_ISL_591282, EPI_ISL_591284, EPI_ISL_591285, EPI_ISL_591287, EPI_ISL_591288, EPI_ISL_591289, EPI_ISL_591290, EPI_ISL_591291, EPI_ISL_591292, EPI_ISL_591293, EPI_ISL_591294, EPI_ISL_591295, EPI_ISL_591297, EPI_ISL_591298, EPI_ISL_591300, EPI_ISL_591301, EPI_ISL_591302, EPI_ISL_591303, EPI_ISL_591304, EPI_ISL_591305, EPI_ISL_591306, EPI_ISL_591307, EPI_ISL_591308, EPI_ISL_591309, EPI_ISL_591310, EPI_ISL_591312, EPI_ISL_591315 | | | |
| see above | Utah Public Health Laboratory | Utah Public Health Laboratory | Erin Young, Kelly Oakeson |
| EPI_ISL_593478, EPI_ISL_593479, EPI_ISL_593480, EPI_ISL_593553, EPI_ISL_593554, EPI_ISL_593555, EPI_ISL_593556, EPI_ISL_593557, EPI_ISL_593558 | Brigham and Women's Hospital | Jonathan Li Laboratory | Manish C. Choudhary, James Regan, Jonathan Z. Li |
| EPI_ISL_593563, EPI_ISL_593564, EPI_ISL_593565, EPI_ISL_593566, EPI_ISL_593568, EPI_ISL_593569, EPI_ISL_593571, EPI_ISL_593572, EPI_ISL_593574, EPI_ISL_593575, EPI_ISL_593576, EPI_ISL_593577, EPI_ISL_593578, EPI_ISL_593579, EPI_ISL_593581, EPI_ISL_593582, EPI_ISL_593584, EPI_ISL_593587, EPI_ISL_593588, EPI_ISL_593589, EPI_ISL_593591, EPI_ISL_593592, EPI_ISL_593593, EPI_ISL_593594, EPI_ISL_593595 | | | |
| see above | Texas Department of State Health Services | Texas Department of State Health Services | Rashmi Tuladhar, Bonnie Oh, Jenny Zhang, Maliha Rahman, Anita Pokharel, Myong Koag, Chung Wang, Rachel Lee, Grace Kubin, Mayela Pedrueza |
| EPI_ISL_593596 | Swedish Medical Center, Seattle, WA, USA | Institute for Systems Biology | Jason D. Goldman, Kai Wang, Katharina Röltgen, Sandra C. A. Nielsen, Jared C. Roach, Samia N. Naccache, Fan Yang, Oliver F. Wirz, Kathryn E. Yost, Ji-Yeun Lee, Kelly Chun, Terri Wrin,, Christos J. Petropoulos, Inyoul Lee, Shannon Fallen, Paula M. Manner, Julie A. Wallick, Heather A. Algren, RN, Kim M. Murray, Yapeng Su, Jennifer Hadlock, Joshua Jeharajah, William R. Berrington, George P. Pappas, Sonam T. Nyatsatsang, Alexander L. Greninger, Ansuman T. Satpathy, John S. Pauk, Scott D. Boyd, James R. Heath |
| EPI_ISL_593938, EPI_ISL_593939, EPI_ISL_593940, EPI_ISL_593941, EPI_ISL_593942, EPI_ISL_593943, EPI_ISL_593944, EPI_ISL_593945, EPI_ISL_593947, EPI_ISL_593948, EPI_ISL_593949, EPI_ISL_593950, EPI_ISL_593951, EPI_ISL_593952, EPI_ISL_593953, EPI_ISL_593954, EPI_ISL_593955, EPI_ISL_593956, EPI_ISL_593957, EPI_ISL_593958, EPI_ISL_593960, EPI_ISL_593961, EPI_ISL_593962, EPI_ISL_593963, EPI_ISL_593964, EPI_ISL_593965, EPI_ISL_593966, EPI_ISL_593967, EPI_ISL_593968, EPI_ISL_593969, EPI_ISL_593970, EPI_ISL_593971, EPI_ISL_593972, EPI_ISL_593973, EPI_ISL_593974, EPI_ISL_593975, EPI_ISL_593976, EPI_ISL_593977, EPI_ISL_593978, EPI_ISL_593979, EPI_ISL_593980, EPI_ISL_593981 | | | |
| see above | Delaware Public Health Lab | Delaware Public Health Lab | Gregory Hovan |
| EPI_ISL_593999, EPI_ISL_594002, EPI_ISL_594005, EPI_ISL_594007, EPI_ISL_594008, EPI_ISL_594009, EPI_ISL_594010, EPI_ISL_594011, EPI_ISL_594012, EPI_ISL_594013, EPI_ISL_594014, EPI_ISL_594015, EPI_ISL_594016, EPI_ISL_594017, EPI_ISL_594018, EPI_ISL_594019, EPI_ISL_594020, EPI_ISL_594021, EPI_ISL_594023, EPI_ISL_594026, EPI_ISL_594027, EPI_ISL_594028, EPI_ISL_594030, EPI_ISL_594031, EPI_ISL_594032, EPI_ISL_594034, EPI_ISL_594035, EPI_ISL_594038, EPI_ISL_594039, EPI_ISL_594040, EPI_ISL_594041, EPI_ISL_594042, EPI_ISL_594043, EPI_ISL_594044, EPI_ISL_594045, EPI_ISL_594046, EPI_ISL_594047, EPI_ISL_594048, EPI_ISL_594049, EPI_ISL_594050, EPI_ISL_594051, EPI_ISL_594052, EPI_ISL_594053, EPI_ISL_594054, EPI_ISL_594055, EPI_ISL_594056, EPI_ISL_594057, EPI_ISL_594058, EPI_ISL_594059, EPI_ISL_594060, EPI_ISL_594061, EPI_ISL_594062, EPI_ISL_594063, EPI_ISL_594064, EPI_ISL_594065, EPI_ISL_594066, EPI_ISL_594067, EPI_ISL_594068, EPI_ISL_594069, EPI_ISL_594070, EPI_ISL_594071, EPI_ISL_594072, EPI_ISL_594073, EPI_ISL_594074, EPI_ISL_594075, EPI_ISL_594076, EPI_ISL_594079, EPI_ISL_594080, EPI_ISL_594081, EPI_ISL_594083, EPI_ISL_594084, EPI_ISL_594086, EPI_ISL_594087, EPI_ISL_594088, EPI_ISL_594089, EPI_ISL_594090, EPI_ISL_594091, EPI_ISL_594093, EPI_ISL_594094, EPI_ISL_594095, EPI_ISL_594097, EPI_ISL_594098, EPI_ISL_594099, EPI_ISL_594101, EPI_ISL_594102, EPI_ISL_594103, EPI_ISL_594104, EPI_ISL_594105, EPI_ISL_594106, EPI_ISL_594107, EPI_ISL_594108, EPI_ISL_594110, EPI_ISL_594111, EPI_ISL_594112, EPI_ISL_594113, EPI_ISL_594114, EPI_ISL_594115, EPI_ISL_594116 | | | |
| see above | Utah Public Health Laboratory | Utah Public Health Laboratory | Erin Young, Kelly Oakeson |
| EPI_ISL_594121, EPI_ISL_594124, EPI_ISL_594127, EPI_ISL_594128, EPI_ISL_594131, EPI_ISL_594133 | Yale COVID-19 Biorepository | Grubaugh Lab - Yale School of Public Health | Joseph Fauver, Tara Alpert, Anderson Brito, Anne Wyllie, Chantal Vogels, Mary Petrone, Chaney Kalinich, Isabel Ott, Arnau Casanovas, Catherine Muenker, Adam Moore, Alice Lu, Maria Tokuyama, Patrick Wong, Peiwen Lu, Saad Omer, Richard Martinello, Allison Nelson, Shelli Farhadian, Akiko Iwasaki, Charlese Dela Cruz, Albert Ko, Nathan Grubaugh |
| EPI_ISL_594248, EPI_ISL_594249, EPI_ISL_594251, EPI_ISL_594252, EPI_ISL_594253, EPI_ISL_594255, EPI_ISL_594256, EPI_ISL_594258, EPI_ISL_594259, EPI_ISL_594260, EPI_ISL_594262, EPI_ISL_594263, EPI_ISL_594265, EPI_ISL_594266, EPI_ISL_594267, EPI_ISL_594268, EPI_ISL_594269, EPI_ISL_594270, EPI_ISL_594271, EPI_ISL_594272, EPI_ISL_594273, EPI_ISL_594274, EPI_ISL_594275 | | | |
| see above | Utah Public Health Laboratory | Utah Public Health Laboratory | Erin Young, Kelly Oakeson |
| EPI_ISL_594279, EPI_ISL_594280, EPI_ISL_594281, EPI_ISL_594282, EPI_ISL_594283, EPI_ISL_594284, EPI_ISL_594286, EPI_ISL_594287, EPI_ISL_594288, EPI_ISL_594289, EPI_ISL_594290, EPI_ISL_594291, EPI_ISL_594292, EPI_ISL_594293, EPI_ISL_594294, EPI_ISL_594295, EPI_ISL_594296, EPI_ISL_594297, EPI_ISL_594298, EPI_ISL_594299, EPI_ISL_594300, EPI_ISL_594301, EPI_ISL_594302, EPI_ISL_594303, EPI_ISL_594304, EPI_ISL_594305, EPI_ISL_594306, EPI_ISL_594307, EPI_ISL_594308, EPI_ISL_594309, EPI_ISL_594310, EPI_ISL_594311, EPI_ISL_594312, EPI_ISL_594313, EPI_ISL_594314, EPI_ISL_594315, EPI_ISL_594316, EPI_ISL_594317, EPI_ISL_594318, EPI_ISL_594319, EPI_ISL_594320, EPI_ISL_594321, EPI_ISL_594322, EPI_ISL_594324, EPI_ISL_594325, EPI_ISL_594326, EPI_ISL_594327, EPI_ISL_594328, EPI_ISL_594329, EPI_ISL_594330, EPI_ISL_594331, EPI_ISL_594332, EPI_ISL_594333, EPI_ISL_594334, EPI_ISL_594335, EPI_ISL_594336, EPI_ISL_594337, EPI_ISL_594338, EPI_ISL_594339, EPI_ISL_594340, EPI_ISL_594341, EPI_ISL_594342, EPI_ISL_594343, EPI_ISL_594344, EPI_ISL_594345, EPI_ISL_594346, EPI_ISL_594347, EPI_ISL_594348, EPI_ISL_594349, EPI_ISL_594350, EPI_ISL_594351, EPI_ISL_594352, EPI_ISL_594353, EPI_ISL_594354, EPI_ISL_594355, EPI_ISL_594356, EPI_ISL_594357, EPI_ISL_594358, EPI_ISL_594359, EPI_ISL_594360, EPI_ISL_594361, EPI_ISL_594362, EPI_ISL_594363, EPI_ISL_594364, EPI_ISL_594365, EPI_ISL_594366, EPI_ISL_594367, EPI_ISL_594368, EPI_ISL_594369, EPI_ISL_594370, EPI_ISL_594371, EPI_ISL_594373, EPI_ISL_594374, EPI_ISL_594375, EPI_ISL_594376, EPI_ISL_594377, EPI_ISL_594378, EPI_ISL_594379, EPI_ISL_594380, EPI_ISL_594381, EPI_ISL_594382, EPI_ISL_594383, EPI_ISL_594384, EPI_ISL_594385, EPI_ISL_594386, EPI_ISL_594388, EPI_ISL_594389, EPI_ISL_594391, EPI_ISL_594393 | | | |
| see above | Florida Bureau of Public Health Laboratories | Florida Bureau of Public Health Laboratories | Sarah Schmedes, Jason Blanton |
| EPI_ISL_594394, EPI_ISL_594395, EPI_ISL_594396, EPI_ISL_594397, EPI_ISL_594398, EPI_ISL_594399, EPI_ISL_594400, EPI_ISL_594401, EPI_ISL_594402, EPI_ISL_594403, EPI_ISL_594404, EPI_ISL_594406, EPI_ISL_594407, EPI_ISL_594409, EPI_ISL_594410, EPI_ISL_594411, EPI_ISL_594412, EPI_ISL_594413, EPI_ISL_594414, EPI_ISL_594415, EPI_ISL_594416, EPI_ISL_594417, EPI_ISL_594418, EPI_ISL_594419, EPI_ISL_594420, EPI_ISL_594421, EPI_ISL_594422, EPI_ISL_594423, EPI_ISL_594425, EPI_ISL_594427, EPI_ISL_594428, EPI_ISL_594429, EPI_ISL_594430, EPI_ISL_594431, EPI_ISL_594432, EPI_ISL_594433, EPI_ISL_594438, EPI_ISL_594439, EPI_ISL_594441, EPI_ISL_594442, EPI_ISL_594443, EPI_ISL_594444 | | | |
| see above | Utah Public Health Laboratory | Utah Public Health Laboratory | Erin Young, Kelly Oakeson |
| EPI_ISL_594447, EPI_ISL_594448, EPI_ISL_594449, EPI_ISL_594450, EPI_ISL_594451, | Washington State Public Health Laboratories | Pathogen Discovery, Respiratory Viruses Branch, Division of Viral Diseases, Centers for Disease Control and Prevention | Ying Tao, Yan Li, Clinton Paden, Jing Zhang, Krista Queen, Anna Uehara, Haibin Wang, Julu Bhatnagar, Suxiang Tong |

| | | | |
|---|---|---|---|
| EPI_ISL_594452, EPI_ISL_594453, EPI_ISL_594454 | | | |
| EPI_ISL_594455 | Texas Department of State Health Services Public Health Laboratory | Pathogen Discovery, Respiratory Viruses Branch, Division of Viral Diseases, Centers for Disease Control and Prevention | Ying Tao, Yan Li, Clinton Paden, Jing Zhang, Krista Queen, Anna Uehara, Haibin Wang, Julu Bhatnagar, Suxiang Tong |
| EPI_ISL_594456 | Georgia Public Health Laboratory | Pathogen Discovery, Respiratory Viruses Branch, Division of Viral Diseases, Centers for Disease Control and Prevention | Ying Tao, Yan Li, Clinton Paden, Jing Zhang, Krista Queen, Anna Uehara, Haibin Wang, Julu Bhatnagar, Suxiang Tong |
| EPI_ISL_594457 | NYS Department of Health | Pathogen Discovery, Respiratory Viruses Branch, Division of Viral Diseases, Centers for Disease Control and Prevention | Ying Tao, Yan Li, Clinton Paden, Jing Zhang, Krista Queen, Anna Uehara, Haibin Wang, Julu Bhatnagar, Suxiang Tong |
| EPI_ISL_594458 | California Department of Public Health | Pathogen Discovery, Respiratory Viruses Branch, Division of Viral Diseases, Centers for Disease Control and Prevention | Ying Tao, Yan Li, Clinton Paden, Jing Zhang, Krista Queen, Anna Uehara, Haibin Wang, Julu Bhatnagar, Suxiang Tong |
| EPI_ISL_594459 | Washington State Public Health Laboratories | Pathogen Discovery, Respiratory Viruses Branch, Division of Viral Diseases, Centers for Disease Control and Prevention | Ying Tao, Yan Li, Clinton Paden, Jing Zhang, Krista Queen, Anna Uehara, Haibin Wang, Julu Bhatnagar, Suxiang Tong |
| EPI_ISL_594460 | DC Department of Forensic Sciences | Pathogen Discovery, Respiratory Viruses Branch, Division of Viral Diseases, Centers for Disease Control and Prevention | Ying Tao, Yan Li, Clinton Paden, Jing Zhang, Krista Queen, Anna Uehara, Haibin Wang, Julu Bhatnagar, Suxiang Tong |
| EPI_ISL_594461 | FL Bureau of Public Health Laboratories | Pathogen Discovery, Respiratory Viruses Branch, Division of Viral Diseases, Centers for Disease Control and Prevention | Ying Tao, Yan Li, Clinton Paden, Jing Zhang, Krista Queen, Anna Uehara, Haibin Wang, Julu Bhatnagar, Suxiang Tong |
| EPI_ISL_594462 | NM Department of Health | Pathogen Discovery, Respiratory Viruses Branch, Division of Viral Diseases, Centers for Disease Control and Prevention | Ying Tao, Yan Li, Clinton Paden, Jing Zhang, Krista Queen, Anna Uehara, Haibin Wang, Julu Bhatnagar, Suxiang Tong |
| EPI_ISL_594463 | MN Department of Health | Pathogen Discovery, Respiratory Viruses Branch, Division of Viral Diseases, Centers for Disease Control and Prevention | Ying Tao, Yan Li, Clinton Paden, Jing Zhang, Krista Queen, Anna Uehara, Haibin Wang, Julu Bhatnagar, Suxiang Tong |
| EPI_ISL_594464 | Texas Department of State Health Services Public Health Laboratory | Pathogen Discovery, Respiratory Viruses Branch, Division of Viral Diseases, Centers for Disease Control and Prevention | Ying Tao, Yan Li, Clinton Paden, Jing Zhang, Krista Queen, Anna Uehara, Haibin Wang, Julu Bhatnagar, Suxiang Tong |
| EPI_ISL_594465 | FL Bureau of Public Health Laboratories | Pathogen Discovery, Respiratory Viruses Branch, Division of Viral Diseases, Centers for Disease Control and Prevention | Ying Tao, Yan Li, Clinton Paden, Jing Zhang, Krista Queen, Anna Uehara, Haibin Wang, Julu Bhatnagar, Suxiang Tong |
| EPI_ISL_596386 | National Institute for Allergy and Infectious Diseases Integrated Research Facility - Frederick (NIAID IRF- Frederick), National Institutes of Health (NIH) | National Institute for Allergy and Infectious Diseases Integrated Research Facility - Frederick (NIAID IRF- Frederick), National Institutes of Health (NIH) | Kocher,G., Kugelman,J.R., Beitzel,B. and Palacios,G. |
| EPI_ISL_596388, EPI_ISL_596389, EPI_ISL_596390, EPI_ISL_596391 | Seattle Flu Study | Seattle Flu Study | Deborah A. Nickerson, Chris D. Frazar, Jover Lee, Benjamin Pelle, Matthew Richardson, Amanda Adler, Elisabeth Brandstetter, Peter D. Han, Kairsten Fay, Misja Ilcisin, Kirsten Lacombe, Thomas R. Sibley, Melissa Truong, Caitlin R. Wolf, Karen Cowgill, Stephanie Schrag, Jeff Duchin, Michael Boeckh, Janet A. Englund, Michael Famulare, Barry R. Lutz, Mark J. Rieder, Lea M. Starita, Matthew Thompson, Helen Y. Chu, Trevor Bedford, Jay Shendure |
| EPI_ISL_596393 | Seattle Flu Study | Seattle Flu Study | Deborah A. Nickerson, Chris D. Frazar, Jover Lee, Benjamin Pelle, Matthew Richardson, Amanda Adler, Elisabeth Brandstetter, Peter D. Han, Kairsten Fay, Misja Ilcisin, Kirsten Lacombe, Thomas R. Sibley, Melissa Truong, Caitlin R. Wolf, Michael Boeckh, Janet A. Englund, Michael Famulare, Barry R. Lutz, Mark J. Rieder, Lea M. Starita, Matthew Thompson, Jay Shendure, Trevor Bedford, Helen Y. Chu |
| EPI_ISL_596394, EPI_ISL_596395, EPI_ISL_596396, EPI_ISL_596397, EPI_ISL_596398, EPI_ISL_596400, EPI_ISL_596401, EPI_ISL_596402, EPI_ISL_596403, EPI_ISL_596405, EPI_ISL_596406, EPI_ISL_596407, EPI_ISL_596408, EPI_ISL_596409 | | | |
| see above | Seattle Flu Study | Seattle Flu Study | Deborah A. Nickerson, Chris D. Frazar, Jover Lee, Benjamin Pelle, Matthew Richardson, Amanda Adler, Elisabeth Brandstetter, Peter D. Han, Kairsten Fay, Misja Ilcisin, Kirsten Lacombe, Thomas R. Sibley, Melissa Truong, Caitlin R. Wolf, Karen Cowgill, Stephanie Schrag, Jeff Duchin, Michael Boeckh, Janet A. Englund, Michael Famulare, Barry R. Lutz, Mark J. Rieder, Lea M. Starita, Matthew Thompson, Helen Y. Chu, Trevor Bedford, Jay Shendure |
| EPI_ISL_596410 | Seattle Flu Study | Seattle Flu Study | Deborah A. Nickerson, Chris D. Frazar, Jover Lee, Benjamin Pelle, Matthew Richardson, Amanda Adler, Elisabeth Brandstetter, Peter D. Han, Kairsten Fay, Misja Ilcisin, Kirsten Lacombe, Thomas R. Sibley, Melissa Truong, Caitlin R. Wolf, Michael Boeckh, Janet A. Englund, Michael Famulare, Barry R. Lutz, Mark J. Rieder, Lea M. Starita, Matthew Thompson, Jay Shendure, Trevor Bedford, Helen Y. Chu |
| EPI_ISL_596415, EPI_ISL_596416 | Seattle Flu Study | Seattle Flu Study | Deborah A. Nickerson, Chris D. Frazar, Jover Lee, Benjamin Pelle, Matthew Richardson, Amanda Adler, Elisabeth Brandstetter, Peter D. Han, Kairsten Fay, Misja Ilcisin, Kirsten Lacombe, Thomas R. Sibley, Melissa Truong, Caitlin R. Wolf, Karen Cowgill, Stephanie Schrag, Jeff Duchin, Michael Boeckh, Janet A. Englund, Michael Famulare, Barry R. Lutz, Mark J. Rieder, Lea M. Starita, Matthew Thompson, Helen Y. Chu, Trevor Bedford, Jay Shendure |
| EPI_ISL_596417, EPI_ISL_596418, EPI_ISL_596419, EPI_ISL_596420 | Seattle Flu Study | Seattle Flu Study | Deborah A. Nickerson, Chris D. Frazar, Jover Lee, Benjamin Pelle, Matthew Richardson, Amanda Adler, Elisabeth Brandstetter, Peter D. Han, Kairsten Fay, Misja Ilcisin, Kirsten Lacombe, Thomas R. Sibley, Melissa Truong, Caitlin R. Wolf, Michael Boeckh, Janet A. Englund, Michael Famulare, Barry R. Lutz, Mark J. Rieder, Lea M. Starita, Matthew Thompson, Jay Shendure, Trevor Bedford, Helen Y. Chu |
| EPI_ISL_596421, EPI_ISL_596422, EPI_ISL_596423, EPI_ISL_596424 | Seattle Flu Study | Seattle Flu Study | Deborah A. Nickerson, Chris D. Frazar, Jover Lee, Benjamin Pelle, Matthew Richardson, Amanda Adler, Elisabeth Brandstetter, Peter D. Han, Kairsten Fay, Misja Ilcisin, Kirsten Lacombe, Thomas R. Sibley, Melissa Truong, Caitlin R. Wolf, Karen Cowgill, Stephanie Schrag, Jeff Duchin, Michael Boeckh, Janet A. Englund, Michael Famulare, Barry R. Lutz, Mark J. Rieder, Lea M. Starita, Matthew Thompson, Helen Y. Chu, Trevor Bedford, Jay Shendure |
| EPI_ISL_596426, EPI_ISL_596427, EPI_ISL_596428, EPI_ISL_596429, EPI_ISL_596430, EPI_ISL_596431, EPI_ISL_596432, EPI_ISL_596434, EPI_ISL_596435, EPI_ISL_596436 | Seattle Flu Study | Seattle Flu Study | Deborah A. Nickerson, Chris D. Frazar, Jover Lee, Benjamin Pelle, Matthew Richardson, Amanda Adler, Elisabeth Brandstetter, Peter D. Han, Kairsten Fay, Misja Ilcisin, Kirsten Lacombe, Thomas R. Sibley, Melissa Truong, Caitlin R. Wolf, Michael Boeckh, Janet A. Englund, Michael Famulare, Barry R. Lutz, Mark J. Rieder, Lea M. Starita, Matthew Thompson, Jay Shendure, Trevor Bedford, Helen Y. Chu |
| EPI_ISL_596437, EPI_ISL_596439 | Seattle Flu Study | Seattle Flu Study | Deborah A. Nickerson, Chris D. Frazar, Jover Lee, Benjamin Pelle, Matthew Richardson, Amanda Adler, Elisabeth Brandstetter, Peter D. Han, Kairsten Fay, Misja Ilcisin, Kirsten Lacombe, Thomas R. Sibley, Melissa Truong, Caitlin R. Wolf, Karen Cowgill, Stephanie Schrag, Jeff Duchin, Michael Boeckh, Janet A. Englund, Michael Famulare, Barry R. Lutz, Mark J. Rieder, Lea M. Starita, Matthew Thompson, Helen Y. Chu, Trevor Bedford, Jay Shendure |
| EPI_ISL_596440, EPI_ISL_596441, EPI_ISL_596442, EPI_ISL_596443, EPI_ISL_596444, EPI_ISL_596445 | Seattle Flu Study | Seattle Flu Study | Deborah A. Nickerson, Chris D. Frazar, Jover Lee, Benjamin Pelle, Matthew Richardson, Amanda Adler, Elisabeth Brandstetter, Peter D. Han, Kairsten Fay, Misja Ilcisin, Kirsten Lacombe, Thomas R. Sibley, Melissa Truong, Caitlin R. Wolf, Michael Boeckh, Janet A. Englund, Michael Famulare, Barry R. Lutz, Mark J. Rieder, Lea M. Starita, Matthew Thompson, Jay Shendure, Trevor Bedford, Helen Y. Chu |
| EPI_ISL_596446 | Seattle Flu Study | Seattle Flu Study | Deborah A. Nickerson, Chris D. Frazar, Jover Lee, Benjamin Pelle, Matthew Richardson, Amanda Adler, Elisabeth Brandstetter, Peter D. Han, Kairsten Fay, Misja Ilcisin, Kirsten Lacombe, Thomas R. Sibley, Melissa Truong, Caitlin R. Wolf, Karen Cowgill, Stephanie Schrag, Jeff Duchin, Michael Boeckh, Janet A. Englund, Michael Famulare, Barry R. Lutz, Mark J. Rieder, Lea M. Starita, Matthew Thompson, Helen Y. Chu, Trevor Bedford, Jay Shendure |
| EPI_ISL_596447, EPI_ISL_596448 | New Mexico Department of Health Scientific Laboratory Division | Los Alamos National Laboratory Bioscience Division | Chien-Chi Lo, Migun Shakya, Cheryl Gleasner, Kim McMurry, Alina Deshpande, Twila Kunde, Joseph Hicks, Michael Edwards, Patrick Chain |
| EPI_ISL_596569, EPI_ISL_596570, EPI_ISL_596571, EPI_ISL_596572, EPI_ISL_596573, EPI_ISL_596574, EPI_ISL_596575, EPI_ISL_596576, EPI_ISL_596577, EPI_ISL_596578, EPI_ISL_596579, EPI_ISL_596580, EPI_ISL_596581, EPI_ISL_596582, EPI_ISL_596583, EPI_ISL_596584, EPI_ISL_596585, EPI_ISL_596586, EPI_ISL_596587, EPI_ISL_596588, EPI_ISL_596589, EPI_ISL_596590, EPI_ISL_596591, EPI_ISL_596592, EPI_ISL_596593, EPI_ISL_596594, EPI_ISL_596595, EPI_ISL_596596, EPI_ISL_596597, EPI_ISL_596598, EPI_ISL_596599, EPI_ISL_596600, EPI_ISL_596601, EPI_ISL_596602, EPI_ISL_596603, EPI_ISL_596604, EPI_ISL_596605, EPI_ISL_596606, EPI_ISL_596607, EPI_ISL_596608, EPI_ISL_596609, EPI_ISL_596610, EPI_ISL_596611, EPI_ISL_596612, EPI_ISL_596613, EPI_ISL_596614, EPI_ISL_596615, EPI_ISL_596616, EPI_ISL_596617, EPI_ISL_596618, EPI_ISL_596619, EPI_ISL_596620, EPI_ISL_596621, EPI_ISL_596622, EPI_ISL_596623, EPI_ISL_596624, EPI_ISL_596625, EPI_ISL_596626 | | | |
| see above | University of Michigan Clinical Microbiology Laboratory | Lauring Lab, University of Michigan, Department of Microbiology and Immunology | Valesano |
| EPI_ISL_602162, EPI_ISL_602163, EPI_ISL_602164, EPI_ISL_602165, EPI_ISL_602166, EPI_ISL_602167, EPI_ISL_602168, EPI_ISL_602169, EPI_ISL_602170, EPI_ISL_602171, EPI_ISL_602172, EPI_ISL_602173, EPI_ISL_602174, EPI_ISL_602175, EPI_ISL_602176, EPI_ISL_602177, EPI_ISL_602178, EPI_ISL_602179, EPI_ISL_602180, EPI_ISL_602181, EPI_ISL_602182, EPI_ISL_602183, EPI_ISL_602184, EPI_ISL_602185, EPI_ISL_602186, EPI_ISL_602187, EPI_ISL_602188, EPI_ISL_602189, EPI_ISL_602190, EPI_ISL_602191, EPI_ISL_602192, EPI_ISL_602193, EPI_ISL_602194, EPI_ISL_602195, EPI_ISL_602196, EPI_ISL_602197, EPI_ISL_602198, EPI_ISL_602199, EPI_ISL_602200, EPI_ISL_602201, EPI_ISL_602202, EPI_ISL_602203, EPI_ISL_602204, EPI_ISL_602205, EPI_ISL_602206, EPI_ISL_602207 | | | |
| see above | New Mexico Department of Health Scientific Laboratory | New Mexico Department of Health Scientific Laboratory | Ellie Johnson, Anastacia Griego-Fisher, D'Eldra Malone |
| EPI_ISL_602208, EPI_ISL_602209 | New Mexico Department of Health Scientific Laboratory Division | Los Alamos National Laboratory Bioscience Division | Chien-Chi Lo, Migun Shakya, Cheryl Gleasner, Kim McMurry, Alina Deshpande, Twila Kunde, Joseph Hicks, Michael Edwards, Patrick Chain |
| EPI_ISL_602210, EPI_ISL_602211, EPI_ISL_602212, EPI_ISL_602213, EPI_ISL_602216, EPI_ISL_602217, EPI_ISL_602218, EPI_ISL_602219, EPI_ISL_602220, EPI_ISL_602223, EPI_ISL_602224, EPI_ISL_602225, EPI_ISL_602226, EPI_ISL_602228, EPI_ISL_602231, EPI_ISL_602233, EPI_ISL_602234, EPI_ISL_602235, EPI_ISL_602237, EPI_ISL_602238, EPI_ISL_602239, EPI_ISL_602240, EPI_ISL_602241, EPI_ISL_602242, | | | |

| | | | |
|---|---|---|---|
| EPI_ISL_602243, EPI_ISL_602244, EPI_ISL_602245, EPI_ISL_602246, EPI_ISL_602249, EPI_ISL_602253, EPI_ISL_602255 | | | |
| see above | Texas Department of State Health Services | Texas Department of State Health Services | Rashmi Tuladhar, Bonnie Oh, Jenny Zhang, Maliha Rahman, Anita Pokharel, Myong Koag, Chung Wang, Rachel Lee, Grace Kubin, Mayela Pedrueza |
| EPI_ISL_602258, EPI_ISL_602259, EPI_ISL_602260, EPI_ISL_602261, EPI_ISL_602262, EPI_ISL_602263, EPI_ISL_602264, EPI_ISL_602265, EPI_ISL_602266, EPI_ISL_602267, EPI_ISL_602268, EPI_ISL_602269, EPI_ISL_602270, EPI_ISL_602271, EPI_ISL_602272, EPI_ISL_602273, EPI_ISL_602274, EPI_ISL_602275, EPI_ISL_602276, EPI_ISL_602277 | | | |
| see above | MD PHL | MD PHL | Maryland Department of Health Laboratories Administration |
| EPI_ISL_602278, EPI_ISL_602279, EPI_ISL_602280, EPI_ISL_602281 | New Mexico Department of Health Scientific Laboratory Division | Los Alamos National Laboratory Bioscience Division | Chien-Chi Lo, Migun Shakya, Cheryl Gleasner, Kim McMurry, Alina Deshpande, Twila Kunde, Joseph Hicks, Michael Edwards, Patrick Chain |
| EPI_ISL_602306, EPI_ISL_602307, EPI_ISL_602308, EPI_ISL_602309, EPI_ISL_602310, EPI_ISL_602311, EPI_ISL_602312, EPI_ISL_602313, EPI_ISL_602314, EPI_ISL_602315, EPI_ISL_602316, EPI_ISL_602317, EPI_ISL_602318, EPI_ISL_602319, EPI_ISL_602320, EPI_ISL_602321, EPI_ISL_602322, EPI_ISL_602323, EPI_ISL_602324, EPI_ISL_602325, EPI_ISL_602326, EPI_ISL_602327, EPI_ISL_602328 | | | |
| see above | University of Miami Immunology and Histocompatibility Laboratory | University of Miami Immunology and Histocompatibility Laboratory | Emilio Margolles-Clark, PhD and Phillip Ruiz, MD, PhD |
| EPI_ISL_602559, EPI_ISL_602560, EPI_ISL_602561 | Alaska State Virology Laboratory | Alaska State Virology Laboratory | DeRonde,S., Deuling,H., Chen,J. |
| EPI_ISL_602931, EPI_ISL_602934, EPI_ISL_602935, EPI_ISL_602936, EPI_ISL_602937, EPI_ISL_602942, EPI_ISL_602944, EPI_ISL_602946, EPI_ISL_602947, EPI_ISL_602948, EPI_ISL_602949, EPI_ISL_602950, EPI_ISL_602951, EPI_ISL_602952, EPI_ISL_602954, EPI_ISL_602955 | | | |
| see above | Utah Public Health Laboratory | Utah Public Health Laboratory | Erin Young, Kelly Oakeson |
| EPI_ISL_602956, EPI_ISL_602957, EPI_ISL_602958, EPI_ISL_602959, EPI_ISL_602960, EPI_ISL_602961, EPI_ISL_602962, EPI_ISL_602963, EPI_ISL_602964, EPI_ISL_602965, EPI_ISL_602966, EPI_ISL_602967, EPI_ISL_602968, EPI_ISL_602969, EPI_ISL_602970, EPI_ISL_602971, EPI_ISL_602972, EPI_ISL_602973, EPI_ISL_602974, EPI_ISL_602975, EPI_ISL_602976, EPI_ISL_602977, EPI_ISL_602978, EPI_ISL_602979, EPI_ISL_602980, EPI_ISL_602981, EPI_ISL_602982, EPI_ISL_602983, EPI_ISL_602984, EPI_ISL_602985, EPI_ISL_602986, EPI_ISL_602987, EPI_ISL_602988, EPI_ISL_602989, EPI_ISL_602990, EPI_ISL_602991, EPI_ISL_602992, EPI_ISL_602993, EPI_ISL_602994, EPI_ISL_602995, EPI_ISL_602996, EPI_ISL_602997, EPI_ISL_602998, EPI_ISL_602999 | | | |
| see above | Minnesota Department of Health, Public Health Laboratory | Minnesota Department of Health, Public Health Laboratory | Matt Plumb, Jacob Garfin, Alexandra Lorentz, and Xiong Wang |
| EPI_ISL_603000, EPI_ISL_603001, EPI_ISL_603002, EPI_ISL_603003 | Sanford South University Medical Center | Minnesota Department of Health, Public Health Laboratory | Matt Plumb, Jacob Garfin, Alexandra Lorentz, and Xiong Wang |
| EPI_ISL_603004, EPI_ISL_603005 | Essentia Health-St. Mary's Medical Center | Minnesota Department of Health, Public Health Laboratory | Matt Plumb, Jacob Garfin, Alexandra Lorentz, and Xiong Wang |
| EPI_ISL_603006, EPI_ISL_603007, EPI_ISL_603008, EPI_ISL_603009 | Minnesota Department of Health, Public Health Laboratory | Minnesota Department of Health, Public Health Laboratory | Matt Plumb, Jacob Garfin, Alexandra Lorentz, and Xiong Wang |
| EPI_ISL_603010, EPI_ISL_603011 | Mayo Clinic & Mayo Clinic Laboratories | Minnesota Department of Health, Public Health Laboratory | Matt Plumb, Jacob Garfin, Alexandra Lorentz, and Xiong Wang |
| EPI_ISL_603012, EPI_ISL_603013, EPI_ISL_603014 | Essentia Health-St. Mary's Medical Center | Minnesota Department of Health, Public Health Laboratory | Matt Plumb, Jacob Garfin, Alexandra Lorentz, and Xiong Wang |
| EPI_ISL_603015 | Minnesota Department of Health, Public Health Laboratory | Minnesota Department of Health, Public Health Laboratory | Matt Plumb, Jacob Garfin, Alexandra Lorentz, and Xiong Wang |
| EPI_ISL_603016, EPI_ISL_603017 | Essentia Health-St. Mary's Medical Center | Minnesota Department of Health, Public Health Laboratory | Matt Plumb, Jacob Garfin, Alexandra Lorentz, and Xiong Wang |
| EPI_ISL_603018, EPI_ISL_603019, EPI_ISL_603020 | Minnesota Department of Health, Public Health Laboratory | Minnesota Department of Health, Public Health Laboratory | Matt Plumb, Jacob Garfin, Alexandra Lorentz, and Xiong Wang |
| EPI_ISL_603049, EPI_ISL_603050, EPI_ISL_603051, EPI_ISL_603052 | Utah Public Health Laboratory, Utah Public Health Laboratory Infectious Disease submission group | Utah Public Health Laboratory, Utah Public Health Laboratory Infectious Disease submission group | Young,E.L., Oakeson,K. |
| EPI_ISL_603248 | Brotman Baty Institute for Precision Medicine | Brotman Baty Institute for Precision Medicine | Trevor Bedford, Jennifer K. Logue, Peter D. Han, Caitlin R. Wolf, Chris D. Frazar, Benjamin Pelle, Mark J. Rieder, Deborah A. Nickerson, Christina M. Lockwood, Lea M. Starita, Helen Y. Chu, Jay Shendure |
| EPI_ISL_603251, EPI_ISL_603252, EPI_ISL_603253, EPI_ISL_603254, EPI_ISL_603255 | UW Virology Lab | UW Virology Lab | Pavitra Roychoudhury, Hong Xie, Lasata Shrestha, Meei-Li Huang, Keith R Jerome, Alexander Greninger |
| EPI_ISL_603759, EPI_ISL_603760, EPI_ISL_603761, EPI_ISL_603762, EPI_ISL_603763, EPI_ISL_603764, EPI_ISL_603765, EPI_ISL_603766, EPI_ISL_603767, EPI_ISL_603768, EPI_ISL_603769, EPI_ISL_603770, EPI_ISL_603771, EPI_ISL_603772, EPI_ISL_603773, EPI_ISL_603774, EPI_ISL_603775, EPI_ISL_603776, EPI_ISL_603777, EPI_ISL_603778, EPI_ISL_603779, EPI_ISL_603780, EPI_ISL_603781, EPI_ISL_603782, EPI_ISL_603783, EPI_ISL_603784, EPI_ISL_603785, EPI_ISL_603786, EPI_ISL_603787, EPI_ISL_603788, EPI_ISL_603789, EPI_ISL_603790, EPI_ISL_603791, EPI_ISL_603792, EPI_ISL_603793, EPI_ISL_603794, EPI_ISL_603795, EPI_ISL_603796, EPI_ISL_603797, EPI_ISL_603798, EPI_ISL_603799, EPI_ISL_603800, EPI_ISL_603801, EPI_ISL_603802, EPI_ISL_603803, EPI_ISL_603804, EPI_ISL_603805, EPI_ISL_603806, EPI_ISL_603807, EPI_ISL_603808, EPI_ISL_603809, EPI_ISL_603810, EPI_ISL_603811, EPI_ISL_603812, EPI_ISL_603813, EPI_ISL_603814, EPI_ISL_603815, EPI_ISL_603816, EPI_ISL_603817, EPI_ISL_603818, EPI_ISL_603819, EPI_ISL_603820, EPI_ISL_603821, EPI_ISL_603822, EPI_ISL_603823, EPI_ISL_603824, EPI_ISL_603825, EPI_ISL_603826, EPI_ISL_603827, EPI_ISL_603828, EPI_ISL_603829, EPI_ISL_603830, EPI_ISL_603831, EPI_ISL_603832, EPI_ISL_603833, EPI_ISL_603834, EPI_ISL_603835, EPI_ISL_603836, EPI_ISL_603837, EPI_ISL_603838, EPI_ISL_603839, EPI_ISL_603840, EPI_ISL_603841, EPI_ISL_603842, EPI_ISL_603843, EPI_ISL_603844, EPI_ISL_603845, EPI_ISL_603846, EPI_ISL_603847, EPI_ISL_603848, EPI_ISL_603849, EPI_ISL_603850, EPI_ISL_603851, EPI_ISL_603852, EPI_ISL_603853, EPI_ISL_603854, EPI_ISL_603855, EPI_ISL_603856, EPI_ISL_603857, EPI_ISL_603858, EPI_ISL_603859, EPI_ISL_603860, EPI_ISL_603861, EPI_ISL_603862, EPI_ISL_603863, EPI_ISL_603864, EPI_ISL_603865, EPI_ISL_603866, EPI_ISL_603867, EPI_ISL_603868, EPI_ISL_603869, EPI_ISL_603870, EPI_ISL_603871, EPI_ISL_603872, EPI_ISL_603873, EPI_ISL_603874, EPI_ISL_603875, EPI_ISL_603876, EPI_ISL_603877, EPI_ISL_603878, EPI_ISL_603879, EPI_ISL_603880, EPI_ISL_603881, EPI_ISL_603882, EPI_ISL_603883, EPI_ISL_603884, EPI_ISL_603885, EPI_ISL_603886, EPI_ISL_603887, EPI_ISL_603888, EPI_ISL_603889, EPI_ISL_603890, EPI_ISL_603891, EPI_ISL_603892, EPI_ISL_603893, EPI_ISL_603894, EPI_ISL_603895, EPI_ISL_603896, EPI_ISL_603897, EPI_ISL_603898, EPI_ISL_603899, EPI_ISL_603900, EPI_ISL_603901, EPI_ISL_603902, EPI_ISL_603903, EPI_ISL_603904, EPI_ISL_603905, EPI_ISL_603906, EPI_ISL_603907, EPI_ISL_603908, EPI_ISL_603909, EPI_ISL_603910, EPI_ISL_603911, EPI_ISL_603912, EPI_ISL_603913, EPI_ISL_603914, EPI_ISL_603915, EPI_ISL_603916, EPI_ISL_603917, EPI_ISL_603918, EPI_ISL_603919, EPI_ISL_603920, EPI_ISL_603921, EPI_ISL_603922, EPI_ISL_603923, EPI_ISL_603924, EPI_ISL_603925, EPI_ISL_603926, EPI_ISL_603927, EPI_ISL_603928, EPI_ISL_603929, EPI_ISL_603930, EPI_ISL_603931, EPI_ISL_603932, EPI_ISL_603933, EPI_ISL_603934, EPI_ISL_603935, EPI_ISL_603936, EPI_ISL_603937, EPI_ISL_603938, EPI_ISL_603939, EPI_ISL_603940, EPI_ISL_603941, EPI_ISL_603942, EPI_ISL_603943, EPI_ISL_603944, EPI_ISL_603945, EPI_ISL_603946, EPI_ISL_603947, EPI_ISL_603948, EPI_ISL_603949, EPI_ISL_603950, EPI_ISL_603951, EPI_ISL_603952, EPI_ISL_603953, EPI_ISL_603954, EPI_ISL_603955, EPI_ISL_603956, EPI_ISL_603957, EPI_ISL_603958, EPI_ISL_603959, EPI_ISL_603960, EPI_ISL_603961, EPI_ISL_603962, EPI_ISL_603963, EPI_ISL_603964, EPI_ISL_603965, EPI_ISL_603966, EPI_ISL_603967, EPI_ISL_603968, EPI_ISL_603969, EPI_ISL_603970, EPI_ISL_603971, EPI_ISL_603972, EPI_ISL_603973, EPI_ISL_603974, EPI_ISL_603975, EPI_ISL_603976, EPI_ISL_603977, EPI_ISL_603978, EPI_ISL_603979, EPI_ISL_603980, EPI_ISL_603981, EPI_ISL_603982, EPI_ISL_603983, EPI_ISL_603984, EPI_ISL_603985, EPI_ISL_603986, EPI_ISL_603987, EPI_ISL_603988, EPI_ISL_603989, EPI_ISL_603990, EPI_ISL_603991, EPI_ISL_603992, EPI_ISL_603993, EPI_ISL_603994, EPI_ISL_603995, EPI_ISL_603996, EPI_ISL_603997, EPI_ISL_603998, EPI_ISL_603999, EPI_ISL_604000, EPI_ISL_604001, EPI_ISL_604002, EPI_ISL_604003, EPI_ISL_604004, EPI_ISL_604005, EPI_ISL_604006, EPI_ISL_604007, EPI_ISL_604008, EPI_ISL_604009, EPI_ISL_604010, EPI_ISL_604011, EPI_ISL_604012, EPI_ISL_604013, EPI_ISL_604014, EPI_ISL_604015, EPI_ISL_604016, EPI_ISL_604017, EPI_ISL_604018, EPI_ISL_604019, EPI_ISL_604020, EPI_ISL_604021, EPI_ISL_604022, EPI_ISL_604023, EPI_ISL_604024, EPI_ISL_604025, EPI_ISL_604026, EPI_ISL_604027, EPI_ISL_604028, EPI_ISL_604029, EPI_ISL_604030, EPI_ISL_604031, EPI_ISL_604032, EPI_ISL_604033, EPI_ISL_604034, EPI_ISL_604035, EPI_ISL_604036, EPI_ISL_604037, EPI_ISL_604038, EPI_ISL_604039, EPI_ISL_604040, EPI_ISL_604041, EPI_ISL_604042, EPI_ISL_604043, EPI_ISL_604044, EPI_ISL_604045, EPI_ISL_604046, EPI_ISL_604047, EPI_ISL_604048, EPI_ISL_604049, EPI_ISL_604050, EPI_ISL_604051, EPI_ISL_604052, EPI_ISL_604053, EPI_ISL_604054, EPI_ISL_604055, EPI_ISL_604056, EPI_ISL_604057, EPI_ISL_604058, EPI_ISL_604059, EPI_ISL_604060, EPI_ISL_604061, EPI_ISL_604062, EPI_ISL_604063, EPI_ISL_604064, EPI_ISL_604065, EPI_ISL_604066, EPI_ISL_604067, EPI_ISL_604068, EPI_ISL_604069, EPI_ISL_604070, EPI_ISL_604071, EPI_ISL_604072, EPI_ISL_604073, EPI_ISL_604074, EPI_ISL_604075, EPI_ISL_604076, EPI_ISL_604077, EPI_ISL_604078, EPI_ISL_604079, EPI_ISL_604080, EPI_ISL_604081, EPI_ISL_604082, EPI_ISL_604083, EPI_ISL_604084, EPI_ISL_604085, EPI_ISL_604086, EPI_ISL_604087, EPI_ISL_604088, EPI_ISL_604089, EPI_ISL_604090, EPI_ISL_604091, EPI_ISL_604092, EPI_ISL_604093, EPI_ISL_604094, EPI_ISL_604095, EPI_ISL_604096, EPI_ISL_604097, EPI_ISL_604098, EPI_ISL_604099, EPI_ISL_604100, EPI_ISL_604101, EPI_ISL_604102, EPI_ISL_604103, EPI_ISL_604104, EPI_ISL_604105, EPI_ISL_604106, EPI_ISL_604107, EPI_ISL_604108, EPI_ISL_604109, EPI_ISL_604110, EPI_ISL_604111, EPI_ISL_604112, EPI_ISL_604113, EPI_ISL_604114, EPI_ISL_604115, EPI_ISL_604116, EPI_ISL_604117, EPI_ISL_604118, EPI_ISL_604119, EPI_ISL_604120, EPI_ISL_604121, EPI_ISL_604122, EPI_ISL_604123, EPI_ISL_604124, EPI_ISL_604125, EPI_ISL_604126, EPI_ISL_604127, EPI_ISL_604128, EPI_ISL_604129, EPI_ISL_604130, EPI_ISL_604131, EPI_ISL_604132, EPI_ISL_604133, EPI_ISL_604134, EPI_ISL_604135, EPI_ISL_604136, EPI_ISL_604137, EPI_ISL_604138, EPI_ISL_604139, EPI_ISL_604140, EPI_ISL_604141, EPI_ISL_604142, EPI_ISL_604143, EPI_ISL_604144, EPI_ISL_604145, EPI_ISL_604146, EPI_ISL_604147, EPI_ISL_604148, EPI_ISL_604149, EPI_ISL_604150, EPI_ISL_604151, EPI_ISL_604152, EPI_ISL_604153, EPI_ISL_604154, EPI_ISL_604155, EPI_ISL_604156, EPI_ISL_604157, EPI_ISL_604158, EPI_ISL_604159, EPI_ISL_604160, EPI_ISL_604161, EPI_ISL_604162, EPI_ISL_604163, EPI_ISL_604164, EPI_ISL_604165, EPI_ISL_604166, EPI_ISL_604167, EPI_ISL_604168, EPI_ISL_604169, EPI_ISL_604170, EPI_ISL_604171, EPI_ISL_604172, EPI_ISL_604173, EPI_ISL_604174, EPI_ISL_604175, EPI_ISL_604176, EPI_ISL_604177, EPI_ISL_604178, EPI_ISL_604179, EPI_ISL_604180, EPI_ISL_604181, EPI_ISL_604182, EPI_ISL_604183, EPI_ISL_604184, EPI_ISL_604185, EPI_ISL_604186, EPI_ISL_604187, EPI_ISL_604188, EPI_ISL_604189, EPI_ISL_604190, EPI_ISL_604191, EPI_ISL_604192, EPI_ISL_604193, EPI_ISL_604194, EPI_ISL_604195, EPI_ISL_604196, EPI_ISL_604197, EPI_ISL_604198, EPI_ISL_604199, EPI_ISL_604200, EPI_ISL_604201, EPI_ISL_604202, EPI_ISL_604203, EPI_ISL_604204, EPI_ISL_604205, EPI_ISL_604206, EPI_ISL_604207, EPI_ISL_604208, EPI_ISL_604209, EPI_ISL_604210, EPI_ISL_604211, EPI_ISL_604212, EPI_ISL_604213, EPI_ISL_604214, EPI_ISL_604215, EPI_ISL_604216, EPI_ISL_604217, EPI_ISL_604218, EPI_ISL_604219, EPI_ISL_604220, EPI_ISL_604221, EPI_ISL_604222, EPI_ISL_604223, EPI_ISL_604224, EPI_ISL_604225, EPI_ISL_604226, EPI_ISL_604227, EPI_ISL_604228, EPI_ISL_604229, EPI_ISL_604230, EPI_ISL_604231, EPI_ISL_604232, EPI_ISL_604233, EPI_ISL_604234, EPI_ISL_604235, EPI_ISL_604236, EPI_ISL_604237, EPI_ISL_604238, EPI_ISL_604239, EPI_ISL_604240, EPI_ISL_604241, EPI_ISL_604242, EPI_ISL_604243, EPI_ISL_604244, EPI_ISL_604245, EPI_ISL_604246, EPI_ISL_604247, EPI_ISL_604248, EPI_ISL_604249, EPI_ISL_604250, EPI_ISL_604251, EPI_ISL_604252, EPI_ISL_604253, EPI_ISL_604254, EPI_ISL_604255, EPI_ISL_604256, EPI_ISL_604257, EPI_ISL_604258, EPI_ISL_604259, EPI_ISL_604260, EPI_ISL_604261, EPI_ISL_604262, EPI_ISL_604263, EPI_ISL_604264, EPI_ISL_604265, EPI_ISL_604266, EPI_ISL_604267, EPI_ISL_604268, EPI_ISL_604269, EPI_ISL_604270, EPI_ISL_604271, EPI_ISL_604272, EPI_ISL_604273, EPI_ISL_604274, EPI_ISL_604275, EPI_ISL_604276, EPI_ISL_604277, EPI_ISL_604278, EPI_ISL_604279, EPI_ISL_604280, EPI_ISL_604281, EPI_ISL_604282, EPI_ISL_604283, EPI_ISL_604284, EPI_ISL_604285, EPI_ISL_604286, EPI_ISL_604287, EPI_ISL_604288, EPI_ISL_604289, EPI_ISL_604290, EPI_ISL_604291, EPI_ISL_604292, EPI_ISL_604293, EPI_ISL_604294, EPI_ISL_604295, EPI_ISL_604296, EPI_ISL_604297, EPI_ISL_604298, EPI_ISL_604299, EPI_ISL_604300, EPI_ISL_604301, EPI_ISL_604302, EPI_ISL_604303, EPI_ISL_604304, EPI_ISL_604305, EPI_ISL_604306, EPI_ISL_604307, EPI_ISL_604308, EPI_ISL_604309, EPI_ISL_604310, EPI_ISL_604311, EPI_ISL_604312, EPI_ISL_604313, EPI_ISL_604314, EPI_ISL_604315, EPI_ISL_604316, EPI_ISL_604317, EPI_ISL_604318, EPI_ISL_604319, EPI_ISL_604320, EPI_ISL_604321, EPI_ISL_604322, EPI_ISL_604323, EPI_ISL_604324, EPI_ISL_604325, EPI_ISL_604326, EPI_ISL_604327, EPI_ISL_604328, EPI_ISL_604329, EPI_ISL_604330, EPI_ISL_604331, EPI_ISL_604332, EPI_ISL_604333, EPI_ISL_604334, EPI_ISL_604335, EPI_ISL_604336, EPI_ISL_604337, EPI_ISL_604338, EPI_ISL_604339, EPI_ISL_604340, EPI_ISL_604341, EPI_ISL_604342, EPI_ISL_604343, EPI_ISL_604344, EPI_ISL_604345, EPI_ISL_604346, EPI_ISL_604347, EPI_ISL_604348, EPI_ISL_604349, EPI_ISL_604350, EPI_ISL_604351, EPI_ISL_604352, EPI_ISL_604353, EPI_ISL_604354, EPI_ISL_604355, EPI_ISL_604356, EPI_ISL_604357, EPI_ISL_604358, EPI_ISL_604359, EPI_ISL_604360, EPI_ISL_604361, EPI_ISL_604362, EPI_ISL_604363, EPI_ISL_604364, EPI_ISL_604365, EPI_ISL_604366, EPI_ISL_604367, EPI_ISL_604368, EPI_ISL_604369, EPI_ISL_604370, EPI_ISL_604371, EPI_ISL_604372, EPI_ISL_604373, EPI_ISL_604374, EPI_ISL_604375, EPI_ISL_604376, EPI_ISL_604377, EPI_ISL_604378, EPI_ISL_604379, EPI_ISL_604380, EPI_ISL_604381, EPI_ISL_604382, EPI_ISL_604383, EPI_ISL_604384, EPI_ISL_604385, EPI_ISL_604386, EPI_ISL_604387, EPI_ISL_604388, EPI_ISL_604389, EPI_ISL_604390, EPI_ISL_604391, EPI_ISL_604392, EPI_ISL_604393, EPI_ISL_604394, EPI_ISL_604395, EPI_ISL_604396, EPI_ISL_604397, EPI_ISL_604398, EPI_ISL_604399, EPI_ISL_604400, EPI_ISL_604401, EPI_ISL_604402, EPI_ISL_604403, EPI_ISL_604404, EPI_ISL_604405, EPI_ISL_604406, EPI_ISL_604407, EPI_ISL_604408, EPI_ISL_604409, EPI_ISL_604410, EPI_ISL_604411, EPI_ISL_604412, EPI_ISL_604413, EPI_ISL_604414, EPI_ISL_604415, EPI_ISL_604416, EPI_ISL_604417, EPI_ISL_604418, EPI_ISL_604419, EPI_ISL_604420, EPI_ISL_604421, EPI_ISL_604422, EPI_ISL_604423, EPI_ISL_604424, EPI_ISL_604425, EPI_ISL_604426, EPI_ISL_604427, EPI_ISL_604428, EPI_ISL_604429, EPI_ISL_604430, EPI_ISL_604431, EPI_ISL_604432, EPI_ISL_604433, EPI_ISL_604434, EPI_ISL_604435, EPI_ISL_604436, EPI_ISL_604437, EPI_ISL_604438, EPI_ISL_604439, EPI_ISL_604440, EPI_ISL_604441, EPI_ISL_604442, EPI_ISL_604443, EPI_ISL_604444, EPI_ISL_604445, EPI_ISL_604446, EPI_ISL_604447, EPI_ISL_604448, EPI_ISL_604449, EPI_ISL_604450, EPI_ISL_604451, EPI_ISL_604452, EPI_ISL_604453, EPI_ISL_604454, EPI_ISL_604455, EPI_ISL_604456, EPI_ISL_604457, EPI_ISL_604458, EPI_ISL_604459, EPI_ISL_604460, EPI_ISL_604461, EPI_ISL_604462, EPI_ISL_604463, EPI_ISL_604464, EPI_ISL_604465, EPI_ISL_604466, EPI_ISL_604467, EPI_ISL_604468, EPI_ISL_604469, EPI_ISL_604470, EPI_ISL_604471, EPI_ISL_604472, EPI_ISL_604473, EPI_ISL_604474, EPI_ISL_604475, EPI_ISL_604476, EPI_ISL_604477, EPI_ISL_604478, EPI_ISL_604479, EPI_ISL_604480, EPI_ISL_604481, EPI_ISL_604482, EPI_ISL_604483, EPI_ISL_604484, EPI_ISL_604485, EPI_ISL_604486, EPI_ISL_604487, EPI_ISL_604488, EPI_ISL_604489, EPI_ISL_604490, EPI_ISL_604491, EPI_ISL_604492, EPI_ISL_604493, EPI_ISL_604494, EPI_ISL_604495, EPI_ISL_604496, EPI_ISL_604497, EPI_ISL_604498, EPI_ISL_604499, EPI_ISL_604500, EPI_ISL_604501, EPI_ISL_604502, EPI_ISL_604503, EPI_ISL_604504, EPI_ISL_604505, EPI_ISL_604506, EPI_ISL_604507, EPI_ISL_604508, EPI_ISL_604509, EPI_ISL_604510, EPI_ISL_604511, EPI_ISL_604512, EPI_ISL_604513, EPI_ISL_604514, EPI_ISL_604515, EPI_ISL_604516, EPI_ISL_604517, EPI_ISL_604518, EPI_ISL_604519, EPI_ISL_604520, EPI_ISL_604521, EPI_ISL_604522, EPI_ISL_604523, EPI_ISL_604524, EPI_ISL_604525, EPI_ISL_604526, EPI_ISL_604527, EPI_ISL_604528, EPI_ISL_604529, EPI_ISL_604530, EPI_ISL_604531, EPI_ISL_604532, EPI_ISL_604533, EPI_ISL_604534, EPI_ISL_604535, EPI_ISL_604536, EPI_ISL_604537, EPI_ISL_604538, EPI_ISL_604539, EPI_ISL_604540, EPI_ISL_604541, EPI_ISL_604542, EPI_ISL_604543, EPI_ISL_604544, EPI_ISL_604545, EPI_ISL_604546, EPI_ISL_604547, EPI_ISL_604548, EPI_ISL_604549, EPI_ISL_604550, EPI_ISL_604551, EPI_ISL_604552, EPI_ISL_604553, EPI_ISL_604554, EPI_ISL_604555, EPI_ISL_604556, EPI_ISL_604557, EPI_ISL_604558, EPI_ISL_604559, EPI_ISL_604560, EPI_ISL_604561, EPI_ISL_604562, EPI_ISL_604563, EPI_ISL_604564, EPI_ISL_604565, EPI_ISL_604566, EPI_ISL_604567, EPI_ISL_604568, EPI_ISL_604569, EPI_ISL_604570, EPI_ISL_604571, EPI_ISL_604572, EPI_ISL_604573, EPI_ISL_604574, EPI_ISL_604575, EPI_ISL_604576, EPI_ISL_604577, EPI_ISL_604578, EPI_ISL_604579, EPI_ISL_604580, EPI_ISL_604581, EPI_ISL_604582, EPI_ISL_604583, EPI_ISL_604584, EPI_ISL_604585, EPI_ISL_604586, EPI_ISL_604587, EPI_ISL_604588, EPI_ISL_604589, EPI_ISL_604590, EPI_ISL_604591, EPI_ISL_604592, EPI_ISL_604593, EPI_ISL_604594, EPI_ISL_604595, EPI_ISL_604596, EPI_ISL_604597, EPI_ISL_604598, EPI_ISL_604599, EPI_ISL_604600, EPI_ISL_604601, EPI_ISL_604602, EPI_ISL_604603, EPI_ISL_604604, EPI_ISL_604605, EPI_ISL_604606, EPI_ISL_604607, EPI_ISL_604608, EPI_ISL_604609, EPI_ISL_604610, EPI_ISL_604611, EPI_ISL_604612, EPI_ISL_604613, EPI_ISL_604614, EPI_ISL_604615, EPI_ISL_604616, EPI_ISL_604617, EPI_ISL_604618, EPI_ISL_604619, EPI_ISL_604620, EPI_ISL_604621, EPI_ISL_604622, EPI_ISL_604623, EPI_ISL_604624, EPI_ISL_604625, EPI_ISL_604626, EPI_ISL_604627, EPI_ISL_604628, EPI_ISL_604629, EPI_ISL_604630, EPI_ISL_604631, EPI_ISL_604632, EPI_ISL_604633, EPI_ISL_604634, EPI_ISL_604635, EPI_ISL_604636, EPI_ISL_604637, EPI_ISL_604638, EPI_ISL_604639, EPI_ISL_604640, EPI_ISL_604641, EPI_ISL_604642, EPI_ISL_604643, EPI_ISL_604644, EPI_ISL_604645, EPI_ISL_604646, EPI_ISL_604647, EPI_ISL_604648, EPI_ISL_604649, EPI_ISL_604650, EPI_ISL_604651, EPI_ISL_604652, EPI_ISL_604653, EPI_ISL_604654, EPI_ISL_604655, EPI_ISL_604656, EPI_ISL_604657, EPI_ISL_604658, EPI_ISL_604659, EPI_ISL_604660, EPI_ISL_604661, EPI_ISL_604662, EPI_ISL_604663, EPI_ISL_604664, EPI_ISL_604665, EPI_ISL_604666, EPI_ISL_604667, EPI_ISL_604668, EPI_ISL_604669, EPI_ISL_604670, EPI_ISL_604671, EPI_ISL_604672, EPI_ISL_604673, EPI_ISL_604674, EPI_ISL_604675, EPI_ISL_604676, EPI_ISL_604677, EPI_ISL_604678, EPI_ISL_604679, EPI_ISL_604680, EPI_ISL_604681, EPI_ISL_604682, EPI_ISL_604683, EPI_ISL_604684, EPI_ISL_604685, EPI_ISL_604686, EPI_ISL_604687, EPI_ISL_604688, EPI_ISL_604689, EPI_ISL_604690, EPI_ISL_604691, EPI_ISL_604692, EPI_ISL_604693, EPI_ISL_604694, EPI_ISL_604695, EPI_ISL_604696, EPI_ISL_604697, EPI_ISL_604698, EPI_ISL_604699, EPI_ISL_604700, EPI_ISL_604701, EPI_ISL_604702, EPI_ISL_604703, EPI_ISL_604704, EPI_ISL_604705, EPI_ISL_604706, EPI_ISL_604707, EPI_ISL_604708, EPI_ISL_604709, EPI_ISL_604710, EPI_ISL_604711, EPI_ISL_604712, EPI_ISL_604713, EPI_ISL_604714, EPI_ISL_604715, EPI_ISL_604716, EPI_ISL_604717, EPI_ISL_604718, EPI_ISL_604719, EPI_ISL_604720, EPI_ISL_604721, EPI_ISL_604722, EPI_ISL_604723, EPI_ISL_604724, EPI_ISL_604725, EPI_ISL_604726, EPI_ISL_604727, EPI_ISL_604728, EPI_ISL_604729, EPI_ISL_604730, EPI_ISL_604731, EPI_ISL_604732, EPI_ISL_604733, EPI_ISL_604734, EPI_ISL_604735, EPI_ISL_604736, EPI_ISL_604737, EPI_ISL_604738, EPI_ISL_604739, EPI_ISL_604740, EPI_ISL_604741, EPI_ISL_604742, EPI_ISL_604743, EPI_ISL_604744, EPI_ISL_604745, EPI_ISL_604746, EPI_ISL_604747, EPI_ISL_604748, EPI_ISL_604749, EPI_ISL_604750, EPI_ISL_604751, EPI_ISL_604752, EPI_ISL_604753, EPI_ISL_604754, EPI_ISL_604755, EPI_ISL_604756, EPI_ISL_604757, EPI_ISL_604758, EPI_ISL_604759, EPI_ISL_604760, EPI_ISL_604761, EPI_ISL_604762, EPI_ISL_604763, EPI_ISL_604764, EPI_ISL_604765, EPI_ISL_604766, EPI_ISL_604767, EPI_ISL_604768, EPI_ISL_604769, EPI_ISL_604770, EPI_ISL_604771, EPI_ISL_604772, EPI_ISL_604773, EPI_ISL_604774, EPI_ISL_604775, EPI_ISL_604776, EPI_ISL_604777, EPI_ISL_604778, EPI_ISL_604779, EPI_ISL_604780, EPI_ISL_604781, EPI_ISL_604782, EPI_ISL_604783, EPI_ISL_604784, EPI_ISL_604785, EPI_ISL_604786, EPI_ISL_604787, EPI_ISL_604788, EPI_ISL_604789, EPI_ISL_604790, EPI_ISL_604791, EPI_ISL_604792, EPI_ISL_604793, EPI_ISL_604794, EPI_ISL_604795, EPI_ISL_604796, EPI_ISL_604797, EPI_ISL_604798, EPI_ISL_604799, EPI_ISL_604800, EPI_ISL_604801, EPI_ISL_604802, EPI_ISL_604803, EPI_ISL_604804, EPI_ISL_604805, EPI_ISL_604806, EPI_ISL_604807, EPI_ISL_604808, EPI_ISL_604809, EPI_ISL_604810, EPI_ISL_604811, EPI_ISL_604812, EPI_ISL_604813, EPI_ISL_604814, EPI_ISL_604815, EPI_ISL_604816, EPI_ISL_604817, EPI_ISL_604818, EPI_ISL_604819, EPI_ISL_604820, EPI_ISL_604821, EPI_ISL_604822, EPI_ISL_604823, EPI_ISL_604824, EPI_ISL_604825, EPI_ISL_604826, EPI_ISL_604827, EPI_ISL_604828, EPI_ISL_604829, EPI_ISL_604830, EPI_ISL_604831, EPI_ISL_604832, EPI_ISL_604833, EPI_ISL_604834, EPI_ISL_604835, EPI_ISL_604836, EPI_ISL_604837, EPI_ISL_604838, | | | |

| | | | |
|---|---|---|---|
| EPI_ISL_604839, EPI_ISL_604840, EPI_ISL_604841, EPI_ISL_604842, EPI_ISL_604843, EPI_ISL_604844, EPI_ISL_604845, EPI_ISL_604846, EPI_ISL_604847, EPI_ISL_604848, EPI_ISL_604849, EPI_ISL_604850, EPI_ISL_604851, EPI_ISL_604852, EPI_ISL_604853, EPI_ISL_604854, EPI_ISL_604855, EPI_ISL_604856, EPI_ISL_604857, EPI_ISL_604858, EPI_ISL_604859, EPI_ISL_604860, EPI_ISL_604861, EPI_ISL_604862, EPI_ISL_604863, EPI_ISL_604864, EPI_ISL_604865, EPI_ISL_604866, EPI_ISL_604867, EPI_ISL_604868, EPI_ISL_604869, EPI_ISL_604870, EPI_ISL_604871, EPI_ISL_604872, EPI_ISL_604873, EPI_ISL_604874, EPI_ISL_604875, EPI_ISL_604876, EPI_ISL_604877, EPI_ISL_604878, EPI_ISL_604879, EPI_ISL_604880, EPI_ISL_604881, EPI_ISL_604882, EPI_ISL_604883, EPI_ISL_604884, EPI_ISL_604885, EPI_ISL_604886, EPI_ISL_604887, EPI_ISL_604888, EPI_ISL_604889, EPI_ISL_604890, EPI_ISL_604891, EPI_ISL_604892, EPI_ISL_604893, EPI_ISL_604894, EPI_ISL_604895, EPI_ISL_604896, EPI_ISL_604897, EPI_ISL_604898, EPI_ISL_604899, EPI_ISL_604900, EPI_ISL_604901, EPI_ISL_604902, EPI_ISL_604903, EPI_ISL_604904, EPI_ISL_604905, EPI_ISL_604906, EPI_ISL_604907, EPI_ISL_604908, EPI_ISL_604909, EPI_ISL_604910, EPI_ISL_604911, EPI_ISL_604912, EPI_ISL_604913, EPI_ISL_604914, EPI_ISL_604915, EPI_ISL_604916, EPI_ISL_604917, EPI_ISL_604918, EPI_ISL_604919, EPI_ISL_604920, EPI_ISL_604921, EPI_ISL_604922, EPI_ISL_604923, EPI_ISL_604924, EPI_ISL_604925, EPI_ISL_604926, EPI_ISL_604927, EPI_ISL_604928, EPI_ISL_604929, EPI_ISL_604930, EPI_ISL_604931, EPI_ISL_604932, EPI_ISL_604933, EPI_ISL_604934, EPI_ISL_604935, EPI_ISL_604936, EPI_ISL_604937, EPI_ISL_604938, EPI_ISL_604939, EPI_ISL_604940, EPI_ISL_604941, EPI_ISL_604942, EPI_ISL_604943, EPI_ISL_604944, EPI_ISL_604945, EPI_ISL_604946, EPI_ISL_604947, EPI_ISL_604948, EPI_ISL_604949, EPI_ISL_604950, EPI_ISL_604951, EPI_ISL_604952, EPI_ISL_604953, EPI_ISL_604954, EPI_ISL_604955, EPI_ISL_604956, EPI_ISL_604957, EPI_ISL_604958, EPI_ISL_604959, EPI_ISL_604960, EPI_ISL_604961, EPI_ISL_604962, EPI_ISL_604963, EPI_ISL_604964, EPI_ISL_604965, EPI_ISL_604966, EPI_ISL_604967, EPI_ISL_604968, EPI_ISL_604969, EPI_ISL_604970, EPI_ISL_604971, EPI_ISL_604972, EPI_ISL_604973, EPI_ISL_604974, EPI_ISL_604975, EPI_ISL_604976, EPI_ISL_604977, EPI_ISL_604978, EPI_ISL_604979, EPI_ISL_604980, EPI_ISL_604981, EPI_ISL_604982, EPI_ISL_604983, EPI_ISL_604984, EPI_ISL_604985, EPI_ISL_604986, EPI_ISL_604987, EPI_ISL_604988, EPI_ISL_604989, EPI_ISL_604990, EPI_ISL_604991, EPI_ISL_604992, EPI_ISL_604993, EPI_ISL_604994, EPI_ISL_604995, EPI_ISL_604996, EPI_ISL_604997, EPI_ISL_604998, EPI_ISL_604999, EPI_ISL_605000, EPI_ISL_605001, EPI_ISL_605002, EPI_ISL_605003, EPI_ISL_605004, EPI_ISL_605005, EPI_ISL_605006, EPI_ISL_605007, EPI_ISL_605008, EPI_ISL_605009, EPI_ISL_605010, EPI_ISL_605011, EPI_ISL_605012, EPI_ISL_605013, EPI_ISL_605014, EPI_ISL_605015, EPI_ISL_605016, EPI_ISL_605017, EPI_ISL_605018, EPI_ISL_605019, EPI_ISL_605020, EPI_ISL_605021, EPI_ISL_605022, EPI_ISL_605023, EPI_ISL_605024, EPI_ISL_605025, EPI_ISL_605026, EPI_ISL_605027, EPI_ISL_605028, EPI_ISL_605029, EPI_ISL_605030, EPI_ISL_605031, EPI_ISL_605032, EPI_ISL_605033, EPI_ISL_605034, EPI_ISL_605035, EPI_ISL_605036, EPI_ISL_605037, EPI_ISL_605038, EPI_ISL_605039, EPI_ISL_605040, EPI_ISL_605041, EPI_ISL_605042, EPI_ISL_605043, EPI_ISL_605044, EPI_ISL_605045, EPI_ISL_605046, EPI_ISL_605047, EPI_ISL_605048, EPI_ISL_605049, EPI_ISL_605050, EPI_ISL_605051, EPI_ISL_605052, EPI_ISL_605053, EPI_ISL_605054, EPI_ISL_605055, EPI_ISL_605056, EPI_ISL_605057 | | | |
| see above | Quest Diagnostics | Quest Diagnostics | Rosenthal,S.H., Gerasimova,A., Kagan,R.M., Anderson, B., Grover, D., Livingston, K.E., Hua, M., Liu Y., Shalhout, D.F., Owen, R., Lacbawan, F. |
| EPI_ISL_605147 | University of Miami Immunology and Histocompatibility Laboratory | University of Miami Immunology and Histocompatibility Laboratory | Emilio Margolles-Clark, PhD and Phillip Ruiz, MD, PhD |
| EPI_ISL_605148, EPI_ISL_605150, EPI_ISL_605151, EPI_ISL_605153, EPI_ISL_605155, EPI_ISL_605156, EPI_ISL_605157, EPI_ISL_605158, EPI_ISL_605159, EPI_ISL_605160, EPI_ISL_605161, EPI_ISL_605162, EPI_ISL_605164, EPI_ISL_605165, EPI_ISL_605166, EPI_ISL_605170, EPI_ISL_605171, EPI_ISL_605172, EPI_ISL_605173, EPI_ISL_605176, EPI_ISL_605177, EPI_ISL_605180, EPI_ISL_605181, EPI_ISL_605182, EPI_ISL_605183, EPI_ISL_605184, EPI_ISL_605186, EPI_ISL_605187, EPI_ISL_605188, EPI_ISL_605192, EPI_ISL_605193, EPI_ISL_605194, EPI_ISL_605197, EPI_ISL_605198, EPI_ISL_605199, EPI_ISL_605200, EPI_ISL_605201, EPI_ISL_605202, EPI_ISL_605204, EPI_ISL_605205, EPI_ISL_605206, EPI_ISL_605207, EPI_ISL_605208, EPI_ISL_605209, EPI_ISL_605210, EPI_ISL_605211, EPI_ISL_605212, EPI_ISL_605213, EPI_ISL_605215, EPI_ISL_605216, EPI_ISL_605217, EPI_ISL_605218, EPI_ISL_605220, EPI_ISL_605221, EPI_ISL_605223, EPI_ISL_605225, EPI_ISL_605226, EPI_ISL_605227, EPI_ISL_605229, EPI_ISL_605230, EPI_ISL_605231, EPI_ISL_605232, EPI_ISL_605234, EPI_ISL_605235, EPI_ISL_605236, EPI_ISL_605237, EPI_ISL_605238, EPI_ISL_605239, EPI_ISL_605240, EPI_ISL_605241, EPI_ISL_605242, EPI_ISL_605243, EPI_ISL_605244, EPI_ISL_605245, EPI_ISL_605247, EPI_ISL_605248, EPI_ISL_605249, EPI_ISL_605250, EPI_ISL_605251, EPI_ISL_605252, EPI_ISL_605253, EPI_ISL_605254, EPI_ISL_605255, EPI_ISL_605256, EPI_ISL_605257, EPI_ISL_605258, EPI_ISL_605259, EPI_ISL_605260, EPI_ISL_605261, EPI_ISL_605262, EPI_ISL_605263, EPI_ISL_605264, EPI_ISL_605265, EPI_ISL_605266, EPI_ISL_605267, EPI_ISL_605268, EPI_ISL_605269, EPI_ISL_605270, EPI_ISL_605271, EPI_ISL_605273, EPI_ISL_605274, EPI_ISL_605275, EPI_ISL_605276, EPI_ISL_605277, EPI_ISL_605278, EPI_ISL_605279, EPI_ISL_605280, EPI_ISL_605281, EPI_ISL_605282, EPI_ISL_605283, EPI_ISL_605284, EPI_ISL_605285, EPI_ISL_605286, EPI_ISL_605287, EPI_ISL_605288, EPI_ISL_605289, EPI_ISL_605290, EPI_ISL_605291, EPI_ISL_605292, EPI_ISL_605293, EPI_ISL_605294, EPI_ISL_605295, EPI_ISL_605296, EPI_ISL_605297, EPI_ISL_605298, EPI_ISL_605300, EPI_ISL_605301, EPI_ISL_605302, EPI_ISL_605303, EPI_ISL_605304, EPI_ISL_605305, EPI_ISL_605306, EPI_ISL_605307, EPI_ISL_605308, EPI_ISL_605309, EPI_ISL_605310, EPI_ISL_605311, EPI_ISL_605312, EPI_ISL_605313, EPI_ISL_605314, EPI_ISL_605315, EPI_ISL_605316, EPI_ISL_605317, EPI_ISL_605318, EPI_ISL_605319, EPI_ISL_605320, EPI_ISL_605321, EPI_ISL_605322, EPI_ISL_605323, EPI_ISL_605324, EPI_ISL_605325, EPI_ISL_605326, EPI_ISL_605327, EPI_ISL_605328, EPI_ISL_605331, EPI_ISL_605332, EPI_ISL_605335, EPI_ISL_605336, EPI_ISL_605337, EPI_ISL_605338, EPI_ISL_605339, EPI_ISL_605341, EPI_ISL_605343, EPI_ISL_605344, EPI_ISL_605345, EPI_ISL_605347, EPI_ISL_605348, EPI_ISL_605349, EPI_ISL_605350, EPI_ISL_605351, EPI_ISL_605352, EPI_ISL_605354, EPI_ISL_605355, EPI_ISL_605356, EPI_ISL_605357, EPI_ISL_605358, EPI_ISL_605359, EPI_ISL_605360, EPI_ISL_605361, EPI_ISL_605362, EPI_ISL_605363, EPI_ISL_605364, EPI_ISL_605365, EPI_ISL_605366, EPI_ISL_605367, EPI_ISL_605368, EPI_ISL_605369, EPI_ISL_605370, EPI_ISL_605372, EPI_ISL_605373, EPI_ISL_605374, EPI_ISL_605375, EPI_ISL_605376, EPI_ISL_605377, EPI_ISL_605378, EPI_ISL_605379, EPI_ISL_605380, EPI_ISL_605382, EPI_ISL_605383, EPI_ISL_605384, EPI_ISL_605385, EPI_ISL_605386, EPI_ISL_605387, EPI_ISL_605388, EPI_ISL_605389, EPI_ISL_605390, EPI_ISL_605391, EPI_ISL_605392, EPI_ISL_605393, EPI_ISL_605394, EPI_ISL_605395, EPI_ISL_605396, EPI_ISL_605397, EPI_ISL_605398, EPI_ISL_605399, EPI_ISL_605400, EPI_ISL_605401, EPI_ISL_605402, EPI_ISL_605403 | | | |
| see above | Utah Public Health Laboratory | Utah Public Health Laboratory | Erin L. Young, Kelly Oakeson, Tara Gallagher, Michael T. Pyne, E. Susan Slechta, Melanie A. Mallory, Jeffrey B. Stevenson, Salika M. Shakir, David R. Hillyard |
| EPI_ISL_605405, EPI_ISL_605406, EPI_ISL_605407, EPI_ISL_605408, EPI_ISL_605409, EPI_ISL_605410, EPI_ISL_605411, EPI_ISL_605412, EPI_ISL_605413, EPI_ISL_605414, EPI_ISL_605415, EPI_ISL_605416, EPI_ISL_605417, EPI_ISL_605418, EPI_ISL_605419, EPI_ISL_605420, EPI_ISL_605421, EPI_ISL_605422, EPI_ISL_605423, EPI_ISL_605424, EPI_ISL_605425, EPI_ISL_605426, EPI_ISL_605427, EPI_ISL_605428, EPI_ISL_605429, EPI_ISL_605430, EPI_ISL_605431, EPI_ISL_605432, EPI_ISL_605433, EPI_ISL_605434, EPI_ISL_605435, EPI_ISL_605436, EPI_ISL_605437, EPI_ISL_605438, EPI_ISL_605439, EPI_ISL_605440, EPI_ISL_605441, EPI_ISL_605442, EPI_ISL_605443, EPI_ISL_605444, EPI_ISL_605445, EPI_ISL_605446, EPI_ISL_605447, EPI_ISL_605448, EPI_ISL_605449, EPI_ISL_605450, EPI_ISL_605451, EPI_ISL_605452, EPI_ISL_605454, EPI_ISL_605455, EPI_ISL_605456, EPI_ISL_605457, EPI_ISL_605458, EPI_ISL_605459, EPI_ISL_605460, EPI_ISL_605461, EPI_ISL_605462, EPI_ISL_605463, EPI_ISL_605464, EPI_ISL_605465, EPI_ISL_605466, EPI_ISL_605467, EPI_ISL_605468, EPI_ISL_605469, EPI_ISL_605470, EPI_ISL_605471, EPI_ISL_605472, EPI_ISL_605473, EPI_ISL_605474, EPI_ISL_605475, EPI_ISL_605476, EPI_ISL_605477, EPI_ISL_605478, EPI_ISL_605479, EPI_ISL_605480, EPI_ISL_605481, EPI_ISL_605482, EPI_ISL_605483, EPI_ISL_605484, EPI_ISL_605485, EPI_ISL_605486, EPI_ISL_605487, EPI_ISL_605488, EPI_ISL_605489, EPI_ISL_605490, EPI_ISL_605491, EPI_ISL_605492, EPI_ISL_605493, EPI_ISL_605494, EPI_ISL_605495, EPI_ISL_605496, EPI_ISL_605497, EPI_ISL_605498, EPI_ISL_605499, EPI_ISL_605500, EPI_ISL_605501, EPI_ISL_605502, EPI_ISL_605503, EPI_ISL_605505, EPI_ISL_605506, EPI_ISL_605507, EPI_ISL_605508, EPI_ISL_605509, EPI_ISL_605510, EPI_ISL_605511, EPI_ISL_605512, EPI_ISL_605513, EPI_ISL_605514, EPI_ISL_605515, EPI_ISL_605516, EPI_ISL_605517, EPI_ISL_605518, EPI_ISL_605519, EPI_ISL_605520, EPI_ISL_605521, EPI_ISL_605522, EPI_ISL_605523, EPI_ISL_605524, EPI_ISL_605525, EPI_ISL_605526, EPI_ISL_605527, EPI_ISL_605528, EPI_ISL_605529, EPI_ISL_605530, EPI_ISL_605531, EPI_ISL_605532, EPI_ISL_605533, EPI_ISL_605534, EPI_ISL_605535, EPI_ISL_605536, EPI_ISL_605537, EPI_ISL_605538, EPI_ISL_605539, EPI_ISL_605540, EPI_ISL_605541, EPI_ISL_605542, EPI_ISL_605543, EPI_ISL_605544, EPI_ISL_605545, EPI_ISL_605546, EPI_ISL_605547, EPI_ISL_605548, EPI_ISL_605549, EPI_ISL_605550, EPI_ISL_605551, EPI_ISL_605552, EPI_ISL_605554, EPI_ISL_605555, EPI_ISL_605556, EPI_ISL_605557, EPI_ISL_605558, EPI_ISL_605559, EPI_ISL_605560, EPI_ISL_605561, EPI_ISL_605562, EPI_ISL_605563, EPI_ISL_605564, EPI_ISL_605565, EPI_ISL_605566, EPI_ISL_605567, EPI_ISL_605568, EPI_ISL_605569, EPI_ISL_605570, EPI_ISL_605571, EPI_ISL_605572, EPI_ISL_605573, EPI_ISL_605574, EPI_ISL_605575, EPI_ISL_605576, EPI_ISL_605577, EPI_ISL_605578, EPI_ISL_605579, EPI_ISL_605580, EPI_ISL_605581, EPI_ISL_605582, EPI_ISL_605583, EPI_ISL_605584, EPI_ISL_605585, EPI_ISL_605586, EPI_ISL_605587, EPI_ISL_605588, EPI_ISL_605589, EPI_ISL_605590, EPI_ISL_605591, EPI_ISL_605592, EPI_ISL_605593, EPI_ISL_605594, EPI_ISL_605595, EPI_ISL_605596, EPI_ISL_605598, EPI_ISL_605599, EPI_ISL_605600, EPI_ISL_605601, EPI_ISL_605602, EPI_ISL_605603, EPI_ISL_605604, EPI_ISL_605605, EPI_ISL_605606, EPI_ISL_605607, EPI_ISL_605608, EPI_ISL_605609, EPI_ISL_605610, EPI_ISL_605611, EPI_ISL_605612, EPI_ISL_605613, EPI_ISL_605614, EPI_ISL_605615, EPI_ISL_605616, EPI_ISL_605617, EPI_ISL_605618, EPI_ISL_605619, EPI_ISL_605620, EPI_ISL_605621, EPI_ISL_605622, EPI_ISL_605623, EPI_ISL_605624, EPI_ISL_605625, EPI_ISL_605626, EPI_ISL_605627, EPI_ISL_605628, EPI_ISL_605629, EPI_ISL_605630, EPI_ISL_605631, EPI_ISL_605632, EPI_ISL_605633, EPI_ISL_605634, EPI_ISL_605635, EPI_ISL_605636, EPI_ISL_605637, EPI_ISL_605638, EPI_ISL_605639, EPI_ISL_605640, EPI_ISL_605641, EPI_ISL_605642, EPI_ISL_605643, EPI_ISL_605644, EPI_ISL_605645, EPI_ISL_605646, EPI_ISL_605647, EPI_ISL_605648, EPI_ISL_605649, EPI_ISL_605650, EPI_ISL_605651, EPI_ISL_605652, EPI_ISL_605653, EPI_ISL_605654, EPI_ISL_605655, EPI_ISL_605656, EPI_ISL_605657, EPI_ISL_605658, EPI_ISL_605659, EPI_ISL_605660, EPI_ISL_605661, EPI_ISL_605662, EPI_ISL_605663, EPI_ISL_605664, EPI_ISL_605665, EPI_ISL_605666, EPI_ISL_605667, EPI_ISL_605668, EPI_ISL_605669, EPI_ISL_605670, EPI_ISL_605671, EPI_ISL_605672, EPI_ISL_605673, EPI_ISL_605674, EPI_ISL_605675, EPI_ISL_605676, EPI_ISL_605677, EPI_ISL_605678, EPI_ISL_605679, EPI_ISL_605680, EPI_ISL_605681, EPI_ISL_605682, EPI_ISL_605683, EPI_ISL_605684, EPI_ISL_605685, EPI_ISL_605686, EPI_ISL_605687, EPI_ISL_605688, EPI_ISL_605689, EPI_ISL_605690, EPI_ISL_605691, EPI_ISL_605693, EPI_ISL_605694, EPI_ISL_605695, EPI_ISL_605696, EPI_ISL_605697, EPI_ISL_605698, EPI_ISL_605699, EPI_ISL_605700, EPI_ISL_605701, EPI_ISL_605702, EPI_ISL_605703, EPI_ISL_605704, EPI_ISL_605705, EPI_ISL_605706, EPI_ISL_605707, EPI_ISL_605708, EPI_ISL_605709, EPI_ISL_605710, EPI_ISL_605711, EPI_ISL_605712, EPI_ISL_605713, EPI_ISL_605714, EPI_ISL_605715, EPI_ISL_605716, EPI_ISL_605717, EPI_ISL_605718, EPI_ISL_605719, EPI_ISL_605720, EPI_ISL_605721, EPI_ISL_605722, EPI_ISL_605723, EPI_ISL_605724, EPI_ISL_605725, EPI_ISL_605726, EPI_ISL_605727, EPI_ISL_605728, EPI_ISL_605729, EPI_ISL_605730, EPI_ISL_605731, EPI_ISL_605732, EPI_ISL_605733, EPI_ISL_605734, EPI_ISL_605735, EPI_ISL_605736, EPI_ISL_605737, EPI_ISL_605738, EPI_ISL_605739, EPI_ISL_605740, EPI_ISL_605741, EPI_ISL_605742, EPI_ISL_605743, EPI_ISL_605745, EPI_ISL_605746, EPI_ISL_605747, EPI_ISL_605748, EPI_ISL_605749, EPI_ISL_605750, EPI_ISL_605751, EPI_ISL_605752, EPI_ISL_605753, EPI_ISL_605754, EPI_ISL_605755, EPI_ISL_605756, EPI_ISL_605757, EPI_ISL_605758, EPI_ISL_605759, EPI_ISL_605760, EPI_ISL_605761, EPI_ISL_605762, EPI_ISL_605763, EPI_ISL_605764, EPI_ISL_605765, EPI_ISL_605766, EPI_ISL_605767, EPI_ISL_605768, EPI_ISL_605769, EPI_ISL_605770, EPI_ISL_605771, EPI_ISL_605772, EPI_ISL_605773, EPI_ISL_605774, EPI_ISL_605775, EPI_ISL_605776, EPI_ISL_605777, EPI_ISL_605778, EPI_ISL_605779 | | | |
| see above | University of Wisconsin-Madison AIDS Vaccine Research Laboratories | University of Wisconsin-Madison AIDS Vaccine Research Laboratories | Gage Moreno, Katarina Braun, et al. AIDS Vaccine Research Laboratories |
| EPI_ISL_605791 | Brotman Baty Institute for Precision Medicine | Brotman Baty Institute for Precision Medicine | Trevor Bedford, Jennifer K. Logue, Peter D. Han, Caitlin R. Wolf, Chris D. Frazar, Benjamin Pelle, Erica Ryke, Jover Lee, Mark J. Rieder, Deborah A. Nickerson, Christina M. Lockwood, Lea M. Starita, Helen Y. Chu, Jay Shendure |
| EPI_ISL_609971, EPI_ISL_609972, EPI_ISL_609973, EPI_ISL_609974, EPI_ISL_609975, EPI_ISL_609976, EPI_ISL_609977, EPI_ISL_609978, EPI_ISL_609979, EPI_ISL_609980, EPI_ISL_609981, EPI_ISL_609982, EPI_ISL_609983, EPI_ISL_609984, EPI_ISL_609985, EPI_ISL_609986, EPI_ISL_609987, EPI_ISL_609988 | | | |
| see above | Virginia DCLS | Virginia DCLS | Virginia DCLS |
| EPI_ISL_610000, EPI_ISL_610002, EPI_ISL_610003, EPI_ISL_610005, EPI_ISL_610008, EPI_ISL_610009, EPI_ISL_610010, EPI_ISL_610011, EPI_ISL_610012, EPI_ISL_610013, EPI_ISL_610015, EPI_ISL_610016, EPI_ISL_610018, EPI_ISL_610019, EPI_ISL_610022, EPI_ISL_610023, EPI_ISL_610025, EPI_ISL_610026, EPI_ISL_610028, EPI_ISL_610029, EPI_ISL_610030, EPI_ISL_610031, EPI_ISL_610032, EPI_ISL_610033, EPI_ISL_610034, EPI_ISL_610035, EPI_ISL_610037, EPI_ISL_610038, EPI_ISL_610040, EPI_ISL_610043, EPI_ISL_610044, EPI_ISL_610045, EPI_ISL_610047 | | | |
| see above | Texas Department of State Health Services | Texas Department of State Health Services | Rashmi Tuladhar, Bonnie Oh, Jenny Zhang, Maliha Rahman, Anita Pokharel, Myong Koag, Chung Wang, Rachel Lee, Grace Kubin, Mayela Pedrueza |
| EPI_ISL_610048, EPI_ISL_610049, EPI_ISL_610050, EPI_ISL_610051, EPI_ISL_610052, EPI_ISL_610053, EPI_ISL_610054, EPI_ISL_610055, EPI_ISL_610056, EPI_ISL_610057, EPI_ISL_610058, EPI_ISL_610059, EPI_ISL_610060, EPI_ISL_610061, EPI_ISL_610062, EPI_ISL_610063, EPI_ISL_610064, EPI_ISL_610065, EPI_ISL_610066, EPI_ISL_610067, EPI_ISL_610068, EPI_ISL_610069, EPI_ISL_610070, EPI_ISL_610071, EPI_ISL_610072, EPI_ISL_610073, EPI_ISL_610074, EPI_ISL_610075, EPI_ISL_610076, EPI_ISL_610077, EPI_ISL_610078, EPI_ISL_610079, EPI_ISL_610080, EPI_ISL_610081, EPI_ISL_610082, EPI_ISL_610083, EPI_ISL_610084, EPI_ISL_610085, EPI_ISL_610086, EPI_ISL_610087, EPI_ISL_610088, EPI_ISL_610089, EPI_ISL_610090, EPI_ISL_610091, EPI_ISL_610092, EPI_ISL_610093, EPI_ISL_610094, EPI_ISL_610095, EPI_ISL_610096, EPI_ISL_610097, EPI_ISL_610098, EPI_ISL_610099, EPI_ISL_610100, EPI_ISL_610101, EPI_ISL_610102, EPI_ISL_610103, EPI_ISL_610104, EPI_ISL_610105, EPI_ISL_610106, EPI_ISL_610107, EPI_ISL_610108, EPI_ISL_610109, EPI_ISL_610110, EPI_ISL_610111, EPI_ISL_610112, EPI_ISL_610113, EPI_ISL_610114, EPI_ISL_610115, EPI_ISL_610116, EPI_ISL_610117 | | | |
| see above | University of Michigan Clinical Microbiology Laboratory | Lauring Lab, University of Michigan, Department of Microbiology and Immunology | Valesano |
| EPI_ISL_610118, EPI_ISL_610119, EPI_ISL_610120, EPI_ISL_610121, EPI_ISL_610122, EPI_ISL_610123, EPI_ISL_610124, EPI_ISL_610125, EPI_ISL_610126, EPI_ISL_610127, EPI_ISL_610128, EPI_ISL_610129, EPI_ISL_610130, EPI_ISL_610131, EPI_ISL_610132, EPI_ISL_610133, EPI_ISL_610134, EPI_ISL_610135, EPI_ISL_610136, EPI_ISL_610137, EPI_ISL_610138, EPI_ISL_610139, EPI_ISL_610140, EPI_ISL_610141, EPI_ISL_610142, EPI_ISL_610143, EPI_ISL_610144, EPI_ISL_610145, EPI_ISL_610146, EPI_ISL_610147, EPI_ISL_610148, EPI_ISL_610149, EPI_ISL_610150, EPI_ISL_610151 | | | |
| see above | Virginia DCLS | Virginia DCLS | Virginia DCLS |
| EPI_ISL_610157, EPI_ISL_610159, EPI_ISL_610160 | Washington University in St. Louis | Washington University in St. Louis | David Wang, Carey-Ann Burnham, Bijal Parikh, Scott Handley, Lindsay Droit, Stephen Tahan |
| EPI_ISL_610246, EPI_ISL_610247, EPI_ISL_610248 | University of Michigan Clinical Microbiology Laboratory | Lauring Lab, University of Michigan, Department of Microbiology and Immunology | Valesano |
| EPI_ISL_610250 | New Mexico Department of Health Scientific Laboratory Division | Center for Global Health, University of New Mexico Health Sciences Center | Daryl Domman, Kurt Schwalm, Twila Kunde, Joseph Hicks, Michael Edwards, Darrell Dinwiddie |
| EPI_ISL_610251, EPI_ISL_610252 | TriCore Reference Laboratories | Center for Global Health, University of New Mexico Health Sciences Center | Daryl Domman, Kurt Schwalm, Twila Kunde, Joseph Hicks, Michael Edwards, Darrell Dinwiddie |
| EPI_ISL_610253 | New Mexico Department of Health Scientific Laboratory Division | Center for Global Health, University of New Mexico Health Sciences Center | Daryl Domman, Kurt Schwalm, Twila Kunde, Joseph Hicks, Michael Edwards, Darrell Dinwiddie |
| EPI_ISL_610254, EPI_ISL_610255, EPI_ISL_610256, EPI_ISL_610258, EPI_ISL_610259, EPI_ISL_610260, EPI_ISL_610261, EPI_ISL_610262, EPI_ISL_610263, EPI_ISL_610264, EPI_ISL_610265, EPI_ISL_610266, EPI_ISL_610267 | | | |
| see above | TriCore Reference Laboratories | Center for Global Health, University of New Mexico Health Sciences Center | Daryl Domman, Kurt Schwalm, Twila Kunde, Joseph Hicks, Michael Edwards, Darrell Dinwiddie |
| EPI_ISL_610268, EPI_ISL_610269, EPI_ISL_610270, EPI_ISL_610271, EPI_ISL_610272, EPI_ISL_610273, EPI_ISL_610274, EPI_ISL_610275, EPI_ISL_610276, EPI_ISL_610277, EPI_ISL_610278, EPI_ISL_610279, EPI_ISL_610280, EPI_ISL_610281 | | | |
| see above | New Mexico Department of Health Scientific Laboratory Division | Center for Global Health, University of New Mexico Health Sciences Center | Daryl Domman, Kurt Schwalm, Twila Kunde, Joseph Hicks, Michael Edwards, Darrell Dinwiddie |
| EPI_ISL_610282 | TriCore Reference Laboratories | Center for Global Health, University of New Mexico Health Sciences Center | Daryl Domman, Kurt Schwalm, Twila Kunde, Joseph Hicks, Michael Edwards, Darrell Dinwiddie |
| EPI_ISL_610283, EPI_ISL_610284, EPI_ISL_610285 | UW Virology Lab | UW Virology Lab | Pavitra Roychoudhury, Hong Xie, Lasata Shrestha, Meei-Li Huang, Keith R Jerome, Alexander Greninger |
| EPI_ISL_611491, EPI_ISL_611493, EPI_ISL_611494, EPI_ISL_611497, EPI_ISL_611499, EPI_ISL_611500, EPI_ISL_611501, EPI_ISL_611502, EPI_ISL_611503, EPI_ISL_611504, EPI_ISL_611505, EPI_ISL_611506, EPI_ISL_611507, EPI_ISL_611513 | | | |
| see above | Utah Public Health Laboratory | Utah Public Health Laboratory | Erin Young, Kelly Oakeson |
| EPI_ISL_613463, EPI_ISL_613464, EPI_ISL_613465, EPI_ISL_613466, EPI_ISL_613467, EPI_ISL_613468, EPI_ISL_613469, EPI_ISL_613470, EPI_ISL_613471, EPI_ISL_613472, EPI_ISL_613473, EPI_ISL_613474, EPI_ISL_613475, EPI_ISL_613476, EPI_ISL_613477, EPI_ISL_613478, EPI_ISL_613479, EPI_ISL_613480, EPI_ISL_613481, EPI_ISL_613482, EPI_ISL_613483, EPI_ISL_613484, EPI_ISL_613485, EPI_ISL_613486, EPI_ISL_613487, EPI_ISL_613488, EPI_ISL_613489, EPI_ISL_613490, EPI_ISL_613491, EPI_ISL_613492, EPI_ISL_613493, EPI_ISL_613494, EPI_ISL_613495, EPI_ISL_613496, EPI_ISL_613497, EPI_ISL_613498, EPI_ISL_613499, EPI_ISL_613500, EPI_ISL_613501, EPI_ISL_613502, EPI_ISL_613503, EPI_ISL_613504, EPI_ISL_613505, EPI_ISL_613506, EPI_ISL_613507, EPI_ISL_613508, EPI_ISL_613509, EPI_ISL_613510, EPI_ISL_613511, EPI_ISL_613512, EPI_ISL_613513, EPI_ISL_613514, EPI_ISL_613515, EPI_ISL_613516, EPI_ISL_613517, EPI_ISL_613518, EPI_ISL_613519, EPI_ISL_613520, EPI_ISL_613521, EPI_ISL_613522, EPI_ISL_613523, EPI_ISL_613524, EPI_ISL_613525, EPI_ISL_613526, EPI_ISL_613527, EPI_ISL_613528, EPI_ISL_613529, EPI_ISL_613530, EPI_ISL_613531, EPI_ISL_613532, EPI_ISL_613533, EPI_ISL_613534, EPI_ISL_613535, EPI_ISL_613536, EPI_ISL_613537, EPI_ISL_613538, EPI_ISL_613539, EPI_ISL_613540 | | | |
| see above | Public Health Laboratory - Infectious Disease Lab, Minnesota Department of Health Infectious Disease Laboratory Submission Group | Minnesota Department of Health, Public Health Laboratory | Plumb,M., Garfin,J., Lorentz,A., Wang,X. |
| EPI_ISL_613712, EPI_ISL_613713, EPI_ISL_613714, EPI_ISL_613715, EPI_ISL_613716, EPI_ISL_613717, EPI_ISL_613718, EPI_ISL_613719, EPI_ISL_613720, EPI_ISL_613721, EPI_ISL_613722, EPI_ISL_613723, EPI_ISL_613724, EPI_ISL_613725, EPI_ISL_613726, EPI_ISL_613727, EPI_ISL_613728, EPI_ISL_613729, EPI_ISL_613730, EPI_ISL_613731, EPI_ISL_613732, EPI_ISL_613733, EPI_ISL_613734, EPI_ISL_613735, EPI_ISL_613736, EPI_ISL_613737, EPI_ISL_613738, EPI_ISL_613739, EPI_ISL_613740, EPI_ISL_613741, EPI_ISL_613742, EPI_ISL_613743, EPI_ISL_613744, EPI_ISL_613745, EPI_ISL_613746, EPI_ISL_613747, EPI_ISL_613748, EPI_ISL_613749, EPI_ISL_613750, EPI_ISL_613751, EPI_ISL_613752, EPI_ISL_613753, EPI_ISL_613754, EPI_ISL_613755, EPI_ISL_613756, EPI_ISL_613757, EPI_ISL_613758, EPI_ISL_613759, EPI_ISL_613760, EPI_ISL_613761, EPI_ISL_613762, EPI_ISL_613763, EPI_ISL_613764, EPI_ISL_613765, EPI_ISL_613766, EPI_ISL_613767, EPI_ISL_613768, EPI_ISL_613769, EPI_ISL_613770, EPI_ISL_613771, EPI_ISL_613829, EPI_ISL_613830, EPI_ISL_613831, EPI_ISL_613832, EPI_ISL_613833, EPI_ISL_613834, EPI_ISL_613835, EPI_ISL_613836, EPI_ISL_613837, EPI_ISL_613838, EPI_ISL_613839, EPI_ISL_613840, EPI_ISL_613841, EPI_ISL_613842, EPI_ISL_613843, EPI_ISL_613844, EPI_ISL_613845, EPI_ISL_613846, EPI_ISL_613847, EPI_ISL_613848, EPI_ISL_613849, EPI_ISL_613850, EPI_ISL_613851, EPI_ISL_613852, EPI_ISL_613853, EPI_ISL_613854, EPI_ISL_613855, EPI_ISL_613856, EPI_ISL_613857, EPI_ISL_613858, EPI_ISL_613859, EPI_ISL_613860, EPI_ISL_613861, EPI_ISL_613862, EPI_ISL_613863, EPI_ISL_613864, EPI_ISL_613865, EPI_ISL_613866, EPI_ISL_613867, EPI_ISL_613868, EPI_ISL_613869, EPI_ISL_613870, EPI_ISL_613871, EPI_ISL_613872, EPI_ISL_613873, EPI_ISL_613874, EPI_ISL_613875, EPI_ISL_613876, EPI_ISL_613877, EPI_ISL_613878, EPI_ISL_613879, EPI_ISL_613880, EPI_ISL_613881, EPI_ISL_613882, EPI_ISL_613883, EPI_ISL_613884, EPI_ISL_613885, EPI_ISL_613886, EPI_ISL_613887, EPI_ISL_613888, | | | |

| | | | |
|---|---|---|---|
| EPI_ISL_613889, EPI_ISL_613890, EPI_ISL_613891, EPI_ISL_613892, EPI_ISL_613893, EPI_ISL_613894, EPI_ISL_613895, EPI_ISL_613896, EPI_ISL_613897, EPI_ISL_613898, EPI_ISL_613899, EPI_ISL_613900, EPI_ISL_613901, EPI_ISL_613902, EPI_ISL_613903, EPI_ISL_613904, EPI_ISL_613905, EPI_ISL_613906, EPI_ISL_613907, EPI_ISL_613908, EPI_ISL_613909, EPI_ISL_613910, EPI_ISL_613911, EPI_ISL_613912, EPI_ISL_613913, EPI_ISL_613914, EPI_ISL_613915, EPI_ISL_613916, EPI_ISL_613917, EPI_ISL_613918, EPI_ISL_613919, EPI_ISL_613920, EPI_ISL_613921, EPI_ISL_613922, EPI_ISL_613923, EPI_ISL_613924, EPI_ISL_613925, EPI_ISL_613926, EPI_ISL_613927, EPI_ISL_613928, EPI_ISL_613929, EPI_ISL_613930, EPI_ISL_613931, EPI_ISL_613932, EPI_ISL_613933, EPI_ISL_613934, EPI_ISL_613935, EPI_ISL_613936, EPI_ISL_613937, EPI_ISL_613938, EPI_ISL_613939, EPI_ISL_613940, EPI_ISL_613941, EPI_ISL_613942, EPI_ISL_613943, EPI_ISL_613944, EPI_ISL_613945, EPI_ISL_613946, EPI_ISL_613947, EPI_ISL_613948, EPI_ISL_613949, EPI_ISL_613950 | | | |
| see above | Florida Bureau of Public Health Laboratories | Florida Bureau of Public Health Laboratories | Sarah Schmedes, Jason Blanton |
| EPI_ISL_613976, EPI_ISL_613977, EPI_ISL_613978, EPI_ISL_613979, EPI_ISL_613980, EPI_ISL_613981, EPI_ISL_613982, EPI_ISL_613983, EPI_ISL_613984, EPI_ISL_613985, EPI_ISL_613986, EPI_ISL_613987, EPI_ISL_613988, EPI_ISL_613989, EPI_ISL_613990, EPI_ISL_613991, EPI_ISL_613992, EPI_ISL_613993, EPI_ISL_613994, EPI_ISL_613995, EPI_ISL_613996, EPI_ISL_613997, EPI_ISL_613998, EPI_ISL_613999, EPI_ISL_614000, EPI_ISL_614001, EPI_ISL_614002, EPI_ISL_614003, EPI_ISL_614004, EPI_ISL_614005, EPI_ISL_614006, EPI_ISL_614007, EPI_ISL_614008, EPI_ISL_614009, EPI_ISL_614010 | | | |
| see above | Texas Department of State Health Services | Texas Department of State Health Services | Rashmi Tuladhar, Bonnie Oh, Jenny Zhang, Maliha Rahman, Anita Pokharel, Myong Koag, Chung Wang, Rachel Lee, Grace Kubin, Mayela Pedrueza |
| EPI_ISL_614012, EPI_ISL_614013, EPI_ISL_614014, EPI_ISL_614015, EPI_ISL_614016, EPI_ISL_614017, EPI_ISL_614018, EPI_ISL_614019, EPI_ISL_614020, EPI_ISL_614021, EPI_ISL_614022, EPI_ISL_614023, EPI_ISL_614024, EPI_ISL_614025, EPI_ISL_614026, EPI_ISL_614027, EPI_ISL_614028, EPI_ISL_614029, EPI_ISL_614030, EPI_ISL_614031, EPI_ISL_614032, EPI_ISL_614033, EPI_ISL_614034, EPI_ISL_614035, EPI_ISL_614036, EPI_ISL_614037, EPI_ISL_614038, EPI_ISL_614039, EPI_ISL_614040, EPI_ISL_614041, EPI_ISL_614042, EPI_ISL_614043, EPI_ISL_614044, EPI_ISL_614045, EPI_ISL_614046, EPI_ISL_614047, EPI_ISL_614048, EPI_ISL_614049, EPI_ISL_614050, EPI_ISL_614051, EPI_ISL_614052, EPI_ISL_614053, EPI_ISL_614054, EPI_ISL_614055, EPI_ISL_614056, EPI_ISL_614057, EPI_ISL_614058, EPI_ISL_614059, EPI_ISL_614060, EPI_ISL_614061, EPI_ISL_614062, EPI_ISL_614063, EPI_ISL_614064, EPI_ISL_614065, EPI_ISL_614066, EPI_ISL_614067, EPI_ISL_614068, EPI_ISL_614069, EPI_ISL_614070, EPI_ISL_614071, EPI_ISL_614072, EPI_ISL_614073, EPI_ISL_614074, EPI_ISL_614075, EPI_ISL_614076, EPI_ISL_614077, EPI_ISL_614078, EPI_ISL_614079, EPI_ISL_614080, EPI_ISL_614081, EPI_ISL_614082, EPI_ISL_614083, EPI_ISL_614084, EPI_ISL_614085, EPI_ISL_614086, EPI_ISL_614087, EPI_ISL_614088, EPI_ISL_614089, EPI_ISL_614090, EPI_ISL_614091, EPI_ISL_614092, EPI_ISL_614093, EPI_ISL_614094, EPI_ISL_614096, EPI_ISL_614097, EPI_ISL_614098, EPI_ISL_614099, EPI_ISL_614100, EPI_ISL_614101, EPI_ISL_614102, EPI_ISL_614103, EPI_ISL_614104, EPI_ISL_614105, EPI_ISL_614106, EPI_ISL_614107, EPI_ISL_614108, EPI_ISL_614109, EPI_ISL_614110, EPI_ISL_614111, EPI_ISL_614112, EPI_ISL_614113, EPI_ISL_614114, EPI_ISL_614115, EPI_ISL_614116, EPI_ISL_614117, EPI_ISL_614118, EPI_ISL_614119, EPI_ISL_614120, EPI_ISL_614121, EPI_ISL_614122, EPI_ISL_614123, EPI_ISL_614124, EPI_ISL_614125, EPI_ISL_614126, EPI_ISL_614127, EPI_ISL_614128, EPI_ISL_614129, EPI_ISL_614130, EPI_ISL_614131, EPI_ISL_614132, EPI_ISL_614133, EPI_ISL_614134, EPI_ISL_614135, EPI_ISL_614136, EPI_ISL_614137, EPI_ISL_614138, EPI_ISL_614139, EPI_ISL_614140, EPI_ISL_614141, EPI_ISL_614142, EPI_ISL_614143, EPI_ISL_614144, EPI_ISL_614145, EPI_ISL_614146, EPI_ISL_614147, EPI_ISL_614148, EPI_ISL_614149, EPI_ISL_614150, EPI_ISL_614151, EPI_ISL_614152, EPI_ISL_614153, EPI_ISL_614154 | | | |
| see above | Virginia DCLS | Virginia DCLS | Virginia DCLS |
| EPI_ISL_614160, EPI_ISL_614161, EPI_ISL_614162, EPI_ISL_614163, EPI_ISL_614164, EPI_ISL_614165, EPI_ISL_614166, EPI_ISL_614167, EPI_ISL_614168, EPI_ISL_614169, EPI_ISL_614170, EPI_ISL_614171, EPI_ISL_614172, EPI_ISL_614173, EPI_ISL_614174, EPI_ISL_614175, EPI_ISL_614176, EPI_ISL_614177, EPI_ISL_614178, EPI_ISL_614179, EPI_ISL_614180, EPI_ISL_614183, EPI_ISL_614184, EPI_ISL_614185, EPI_ISL_614186, EPI_ISL_614187, EPI_ISL_614188, EPI_ISL_614189, EPI_ISL_614190, EPI_ISL_614191, EPI_ISL_614192, EPI_ISL_614193, EPI_ISL_614194, EPI_ISL_614195, EPI_ISL_614196, EPI_ISL_614197, EPI_ISL_614198, EPI_ISL_614199, EPI_ISL_614200, EPI_ISL_614201, EPI_ISL_614202, EPI_ISL_614203, EPI_ISL_614204, EPI_ISL_614205, EPI_ISL_614206, EPI_ISL_614207, EPI_ISL_614208, EPI_ISL_614209, EPI_ISL_614210, EPI_ISL_614211, EPI_ISL_614212, EPI_ISL_614213, EPI_ISL_614214, EPI_ISL_614215, EPI_ISL_614216, EPI_ISL_614217, EPI_ISL_614218, EPI_ISL_614219, EPI_ISL_614220, EPI_ISL_614221, EPI_ISL_614222, EPI_ISL_614223, EPI_ISL_614224, EPI_ISL_614225, EPI_ISL_614226, EPI_ISL_614227, EPI_ISL_614228, EPI_ISL_614229, EPI_ISL_614230, EPI_ISL_614231, EPI_ISL_614232, EPI_ISL_614233, EPI_ISL_614234, EPI_ISL_614235, EPI_ISL_614236, EPI_ISL_614237, EPI_ISL_614238, EPI_ISL_614239, EPI_ISL_614240, EPI_ISL_614241, EPI_ISL_614242, EPI_ISL_614243, EPI_ISL_614244, EPI_ISL_614245, EPI_ISL_614246 | | | |
| see above | Michigan Department of Health and Human Services, Bureau of Laboratories | Michigan Department of Health and Human Services, Bureau of Laboratories | Blankenship HM, Riner D, Soehnlen MK |
| EPI_ISL_614248, EPI_ISL_614249, EPI_ISL_614250, EPI_ISL_614251, EPI_ISL_614252, EPI_ISL_614253, EPI_ISL_614254, EPI_ISL_614255, EPI_ISL_614256, EPI_ISL_614257, EPI_ISL_614258, EPI_ISL_614259, EPI_ISL_614260, EPI_ISL_614261, EPI_ISL_614262, EPI_ISL_614263, EPI_ISL_614264 | | | |
| see above | Wyoming Public Health Laboratory | Center for Global Health, University of New Mexico Health Sciences Center | Daryl Domman, Kurt Schwalm, Rob Christensen, Wanda Manley, Cari Sloma, Noah Hull, Darrell Dinwiddie |
| EPI_ISL_614294 | Maimonides Medical Center | New York City Public Health Laboratory | Jade Wang, et al. |
| EPI_ISL_615122, EPI_ISL_615123, EPI_ISL_615124, EPI_ISL_615125, EPI_ISL_615126, EPI_ISL_615127, EPI_ISL_615128, EPI_ISL_615129, EPI_ISL_615130, EPI_ISL_615131, EPI_ISL_615132, EPI_ISL_615133, EPI_ISL_615134, EPI_ISL_615135, EPI_ISL_615137, EPI_ISL_615139, EPI_ISL_615140, EPI_ISL_615141, EPI_ISL_615142, EPI_ISL_615143, EPI_ISL_615144, EPI_ISL_615146, EPI_ISL_615147, EPI_ISL_615148, EPI_ISL_615149, EPI_ISL_615150, EPI_ISL_615153, EPI_ISL_615154, EPI_ISL_615155, EPI_ISL_615156, EPI_ISL_615157, EPI_ISL_615158, EPI_ISL_615159, EPI_ISL_615160, EPI_ISL_615161, EPI_ISL_615162, EPI_ISL_615165 | | | |
| see above | Texas Department of State Health Services | Texas Department of State Health Services | Rashmi Tuladhar, Bonnie Oh, Jenny Zhang, Maliha Rahman, Anita Pokharel, Myong Koag, Chung Wang, Rachel Lee, Grace Kubin, Mayela Pedrueza |
| EPI_ISL_623171, EPI_ISL_623172, EPI_ISL_623174, EPI_ISL_623175, EPI_ISL_623176, EPI_ISL_623177, EPI_ISL_623178, EPI_ISL_623179, EPI_ISL_623180, EPI_ISL_623181, EPI_ISL_623182, EPI_ISL_623183, EPI_ISL_623185, EPI_ISL_623186, EPI_ISL_623187, EPI_ISL_623188, EPI_ISL_623189, EPI_ISL_623190, EPI_ISL_623191, EPI_ISL_623192, EPI_ISL_623193, EPI_ISL_623194, EPI_ISL_623195, EPI_ISL_623196, EPI_ISL_623197, EPI_ISL_623198, EPI_ISL_623199, EPI_ISL_623200, EPI_ISL_623202, EPI_ISL_623203, EPI_ISL_623204, EPI_ISL_623205, EPI_ISL_623206, EPI_ISL_623207, EPI_ISL_623208, EPI_ISL_623209, EPI_ISL_623210 | | | |
| see above | Utah Public Health Laboratory | Utah Public Health Laboratory | Erin Young, Kelly Oakeson |
| EPI_ISL_625478, EPI_ISL_625479, EPI_ISL_625480, EPI_ISL_625481, EPI_ISL_625482, EPI_ISL_625483, EPI_ISL_625484, EPI_ISL_625485, EPI_ISL_625486, EPI_ISL_625487, EPI_ISL_625488, EPI_ISL_625489, EPI_ISL_625490, EPI_ISL_625491, EPI_ISL_625492, EPI_ISL_625493, EPI_ISL_625494, EPI_ISL_625495, EPI_ISL_625496, EPI_ISL_625497, EPI_ISL_625498, EPI_ISL_625499 | | | |
| see above | Santa Clara County Public Health Laboratory | Chan-Zuckerberg Biohub | CZB Cliahub Consortium |
| EPI_ISL_625500, EPI_ISL_625501, EPI_ISL_625502, EPI_ISL_625503, EPI_ISL_625504, EPI_ISL_625505, EPI_ISL_625506, EPI_ISL_625507, EPI_ISL_625508, EPI_ISL_625509, EPI_ISL_625510, EPI_ISL_625511, EPI_ISL_625512, EPI_ISL_625513, EPI_ISL_625514, EPI_ISL_625515, EPI_ISL_625516, EPI_ISL_625517, EPI_ISL_625518, EPI_ISL_625519, EPI_ISL_625520, EPI_ISL_625521, EPI_ISL_625522, EPI_ISL_625523, EPI_ISL_625524, EPI_ISL_625525, EPI_ISL_625526, EPI_ISL_625527, EPI_ISL_625528, EPI_ISL_625529, EPI_ISL_625530, EPI_ISL_625531, EPI_ISL_625532, EPI_ISL_625533, EPI_ISL_625534, EPI_ISL_625535, EPI_ISL_625536, EPI_ISL_625537, EPI_ISL_625538, EPI_ISL_625539, EPI_ISL_625540, EPI_ISL_625541, EPI_ISL_625542, EPI_ISL_625543, EPI_ISL_625544, EPI_ISL_625545, EPI_ISL_625546 | | | |
| see above | Alameda County Public Health Lab | Chan-Zuckerberg Biohub | CZB Cliahub Consortium |
| EPI_ISL_625547, EPI_ISL_625548, EPI_ISL_625549 | County of San Luis Obispo Public Health Laboratory | Chan-Zuckerberg Biohub | CZB Cliahub Consortium |
| EPI_ISL_625550, EPI_ISL_625551, EPI_ISL_625552, EPI_ISL_625553, EPI_ISL_625554, EPI_ISL_625555, EPI_ISL_625556, EPI_ISL_625557, EPI_ISL_625558 | UCSF Clinical Microbiology Laboratory | Chan-Zuckerberg Biohub | CZB Cliahub Consortium |
| EPI_ISL_625559, EPI_ISL_625560, EPI_ISL_625561, EPI_ISL_625562, EPI_ISL_625563, EPI_ISL_625564, EPI_ISL_625565, EPI_ISL_625566, EPI_ISL_625567, EPI_ISL_625568, EPI_ISL_625569, EPI_ISL_625570, EPI_ISL_625571, EPI_ISL_625572, EPI_ISL_625573, EPI_ISL_625574, EPI_ISL_625575, EPI_ISL_625576, EPI_ISL_625577, EPI_ISL_625578, EPI_ISL_625579, EPI_ISL_625580, EPI_ISL_625581, EPI_ISL_625582, EPI_ISL_625583, EPI_ISL_625584, EPI_ISL_625585, EPI_ISL_625586, EPI_ISL_625587, EPI_ISL_625588, EPI_ISL_625589, EPI_ISL_625590, EPI_ISL_625591, EPI_ISL_625592, EPI_ISL_625593 | | | |
| see above | NaN | Chan-Zuckerberg Biohub | CZB Cliahub Consortium |
| EPI_ISL_625594, EPI_ISL_625595, EPI_ISL_625596, EPI_ISL_625597, EPI_ISL_625598, EPI_ISL_625599, EPI_ISL_625600, EPI_ISL_625601, EPI_ISL_625602, EPI_ISL_625603, EPI_ISL_625604, EPI_ISL_625605, EPI_ISL_625606, EPI_ISL_625607, EPI_ISL_625608, EPI_ISL_625609, EPI_ISL_625610 | | | |
| see above | Santa Clara County Public Health Laboratory | Chan-Zuckerberg Biohub | CZB Cliahub Consortium |
| EPI_ISL_625611, EPI_ISL_625612, EPI_ISL_625613, EPI_ISL_625614, EPI_ISL_625615, EPI_ISL_625616, EPI_ISL_625617, EPI_ISL_625618, EPI_ISL_625619, EPI_ISL_625620, EPI_ISL_625621, EPI_ISL_625622 | | | |
| see above | Alameda County Public Health Lab | Chan-Zuckerberg Biohub | CZB Cliahub Consortium |
| EPI_ISL_625623, EPI_ISL_625624, EPI_ISL_625625, EPI_ISL_625626 | County of San Luis Obispo Public Health Laboratory | Chan-Zuckerberg Biohub | CZB Cliahub Consortium |
| EPI_ISL_625627, EPI_ISL_625628, EPI_ISL_625629, EPI_ISL_625630, EPI_ISL_625632, EPI_ISL_625633, EPI_ISL_625634, EPI_ISL_625635, EPI_ISL_625636, EPI_ISL_625637, EPI_ISL_625638, EPI_ISL_625639, EPI_ISL_625640, EPI_ISL_625641, EPI_ISL_625642, EPI_ISL_625643, EPI_ISL_625644, EPI_ISL_625645, EPI_ISL_625646, EPI_ISL_625647, EPI_ISL_625648, EPI_ISL_625649, EPI_ISL_625650, EPI_ISL_625651, EPI_ISL_625652, EPI_ISL_625653, EPI_ISL_625654, EPI_ISL_625655, EPI_ISL_625656, EPI_ISL_625657, EPI_ISL_625658, EPI_ISL_625659 | | | |
| see above | Orange County Public Health Lab | Chan-Zuckerberg Biohub | CZB Cliahub Consortium |
| EPI_ISL_625660 | San Joaquin County Public Health Lab | Chan-Zuckerberg Biohub | CZB Cliahub Consortium |
| EPI_ISL_625662, EPI_ISL_625663, EPI_ISL_625664, EPI_ISL_625665, EPI_ISL_625666, EPI_ISL_625667, EPI_ISL_625668, EPI_ISL_625669, EPI_ISL_625670 | UCSF Clinical Microbiology Laboratory | Chan-Zuckerberg Biohub | CZB Cliahub Consortium |
| EPI_ISL_626357, EPI_ISL_626358, EPI_ISL_626359, EPI_ISL_626360, EPI_ISL_626361, EPI_ISL_626362, EPI_ISL_626363, EPI_ISL_626364, EPI_ISL_626365, EPI_ISL_626366, EPI_ISL_626367, EPI_ISL_626368, EPI_ISL_626369, EPI_ISL_626370, EPI_ISL_626371, EPI_ISL_626372, EPI_ISL_626373, EPI_ISL_626374, EPI_ISL_626375, EPI_ISL_626376, EPI_ISL_626377, EPI_ISL_626378, EPI_ISL_626379, EPI_ISL_626381, EPI_ISL_626382, EPI_ISL_626383, EPI_ISL_626384, EPI_ISL_626385, EPI_ISL_626386, EPI_ISL_626387, EPI_ISL_626388, EPI_ISL_626389, EPI_ISL_626390, EPI_ISL_626391, EPI_ISL_626392, EPI_ISL_626393, EPI_ISL_626394, EPI_ISL_626395, EPI_ISL_626396, EPI_ISL_626397, EPI_ISL_626398, EPI_ISL_626399, EPI_ISL_626400, EPI_ISL_626401, EPI_ISL_626402, EPI_ISL_626403, EPI_ISL_626404, EPI_ISL_626405, EPI_ISL_626406, EPI_ISL_626407, EPI_ISL_626408, EPI_ISL_626409, EPI_ISL_626410, EPI_ISL_626411, EPI_ISL_626413, EPI_ISL_626414, EPI_ISL_626415, EPI_ISL_626416, EPI_ISL_626417, EPI_ISL_626418, EPI_ISL_626419, EPI_ISL_626420, EPI_ISL_626421, EPI_ISL_626422, EPI_ISL_626423, EPI_ISL_626424, EPI_ISL_626425, EPI_ISL_626427, EPI_ISL_626428, EPI_ISL_626430, EPI_ISL_626431, EPI_ISL_626432, EPI_ISL_626433, EPI_ISL_626434, EPI_ISL_626435, EPI_ISL_626436, EPI_ISL_626437, EPI_ISL_626438, EPI_ISL_626439, EPI_ISL_626440, EPI_ISL_626441, EPI_ISL_626442, EPI_ISL_626443, EPI_ISL_626444, EPI_ISL_626445, EPI_ISL_626446, EPI_ISL_626447, EPI_ISL_626448, EPI_ISL_626449, EPI_ISL_626450, EPI_ISL_626451, EPI_ISL_626452, EPI_ISL_626453, EPI_ISL_626454, EPI_ISL_626455, EPI_ISL_626457, EPI_ISL_626458, EPI_ISL_626459, EPI_ISL_626460, EPI_ISL_626461, EPI_ISL_626462, EPI_ISL_626463, EPI_ISL_626464, EPI_ISL_626465, EPI_ISL_626466, EPI_ISL_626467, EPI_ISL_626468, EPI_ISL_626469, EPI_ISL_626470, EPI_ISL_626471, EPI_ISL_626472, EPI_ISL_626473, EPI_ISL_626474, EPI_ISL_626475, EPI_ISL_626476, EPI_ISL_626477, EPI_ISL_626478, EPI_ISL_626479, EPI_ISL_626480, EPI_ISL_626481, EPI_ISL_626482, EPI_ISL_626483, EPI_ISL_626484, EPI_ISL_626485, EPI_ISL_626486, EPI_ISL_626487, EPI_ISL_626488, EPI_ISL_626489, EPI_ISL_626491, EPI_ISL_626492, EPI_ISL_626493, EPI_ISL_626494, EPI_ISL_626495, EPI_ISL_626497, EPI_ISL_626498, EPI_ISL_626500, EPI_ISL_626501, EPI_ISL_626502, EPI_ISL_626503, EPI_ISL_626505, EPI_ISL_626506, EPI_ISL_626507, EPI_ISL_626508, EPI_ISL_626509, EPI_ISL_626510, EPI_ISL_626511, EPI_ISL_626512, EPI_ISL_626514, EPI_ISL_626515, EPI_ISL_626516, EPI_ISL_626517, EPI_ISL_626518, EPI_ISL_626520, EPI_ISL_626521, EPI_ISL_626522, EPI_ISL_626523, EPI_ISL_626524, EPI_ISL_626525, EPI_ISL_626526, EPI_ISL_626527, EPI_ISL_626528, EPI_ISL_626529, EPI_ISL_626530, EPI_ISL_626531, EPI_ISL_626532, EPI_ISL_626533, EPI_ISL_626534, EPI_ISL_626536, EPI_ISL_626537, EPI_ISL_626538, EPI_ISL_626539, EPI_ISL_626540, EPI_ISL_626541, EPI_ISL_626542, EPI_ISL_626543, EPI_ISL_626544, EPI_ISL_626545, EPI_ISL_626546, EPI_ISL_626547, EPI_ISL_626548 | | | |
| see above | Northwestern Memorial Hospital | Ozer Lab | Ramon Lorenzo-Redondo, Hannah H. Nam, Scott C. Roberts, Lacy M. Simons, Chad J. Achenbach, Lawrence J. Jennings, Chao Qi, Alan R. Hauser, Michael G. Ison, Judd F. Hultquist, Egon A. Ozer |
| EPI_ISL_628701, EPI_ISL_628702, EPI_ISL_628703, EPI_ISL_628704, EPI_ISL_628705, EPI_ISL_628706, EPI_ISL_628707, EPI_ISL_628708, EPI_ISL_628709, EPI_ISL_628710, EPI_ISL_628711, EPI_ISL_628712, EPI_ISL_628713, EPI_ISL_628714, EPI_ISL_628715, EPI_ISL_628716, EPI_ISL_628717, EPI_ISL_628718, EPI_ISL_628719, EPI_ISL_628720, EPI_ISL_628721, EPI_ISL_628722, EPI_ISL_628723, EPI_ISL_628724, EPI_ISL_628725, EPI_ISL_628726, EPI_ISL_628727, EPI_ISL_628728, EPI_ISL_628729 | | | |
| see above | Texas Department of State Health Services | Texas Department of State Health Services | Rashmi Tuladhar, Bonnie Oh, Jenny Zhang, Maliha Rahman, Anita Pokharel, Myong Koag, Chung Wang, Rachel Lee, Grace Kubin, Mayela Pedrueza |
| EPI_ISL_628804, EPI_ISL_628805, EPI_ISL_628806, EPI_ISL_628807, EPI_ISL_628808, EPI_ISL_628809, EPI_ISL_628810, EPI_ISL_628811, EPI_ISL_628812, EPI_ISL_628813, EPI_ISL_628814, EPI_ISL_628815, EPI_ISL_628816, EPI_ISL_628817, EPI_ISL_628818, EPI_ISL_628819, EPI_ISL_628820, EPI_ISL_628821, EPI_ISL_628822, EPI_ISL_628823, EPI_ISL_628824, EPI_ISL_628825, EPI_ISL_628826, EPI_ISL_628827, EPI_ISL_628828, EPI_ISL_628829, EPI_ISL_628830, EPI_ISL_628831, EPI_ISL_628832, EPI_ISL_628833, EPI_ISL_628834, EPI_ISL_628835, EPI_ISL_628836, EPI_ISL_628837, EPI_ISL_628838, EPI_ISL_628839, EPI_ISL_628840, EPI_ISL_628841, EPI_ISL_628842, EPI_ISL_628843, EPI_ISL_628844, EPI_ISL_628845, EPI_ISL_628846, EPI_ISL_628847, EPI_ISL_628848, EPI_ISL_628849, EPI_ISL_628850, EPI_ISL_628851, EPI_ISL_628852, EPI_ISL_628853, EPI_ISL_628854, EPI_ISL_628855, EPI_ISL_628856, EPI_ISL_628857, EPI_ISL_628858, EPI_ISL_628859, EPI_ISL_628860, EPI_ISL_628861, EPI_ISL_628862, EPI_ISL_628863, EPI_ISL_628864, EPI_ISL_628865, EPI_ISL_628866, EPI_ISL_628867, EPI_ISL_628868, EPI_ISL_628869, EPI_ISL_628870, EPI_ISL_628871, EPI_ISL_628872, EPI_ISL_628873, EPI_ISL_628874, EPI_ISL_628875, EPI_ISL_628876, EPI_ISL_628877, EPI_ISL_628878, EPI_ISL_628879, EPI_ISL_628880, EPI_ISL_628881, EPI_ISL_628882, EPI_ISL_628883, EPI_ISL_628884, EPI_ISL_628885, EPI_ISL_628886 | | | |
| see above | University of Michigan Clinical | Lauring Lab, University of Michigan, | Valesano |

| | | | |
|---|---|---|---|
| | Microbiology Laboratory | Department of Microbiology and Immunology | |
| EPI_ISL_628932, EPI_ISL_628933, EPI_ISL_628934, EPI_ISL_628935, EPI_ISL_628936, EPI_ISL_628937, EPI_ISL_628938, EPI_ISL_628939, EPI_ISL_628940, EPI_ISL_628941, EPI_ISL_628943, EPI_ISL_628944, EPI_ISL_628945, EPI_ISL_628946, EPI_ISL_628947, EPI_ISL_628948, EPI_ISL_628949, EPI_ISL_628950, EPI_ISL_628951, EPI_ISL_628952, EPI_ISL_628953, EPI_ISL_628954, EPI_ISL_628955, EPI_ISL_628958, EPI_ISL_628959, EPI_ISL_628960, EPI_ISL_628961, EPI_ISL_628962, EPI_ISL_628963, EPI_ISL_628964, EPI_ISL_628966, EPI_ISL_628967, EPI_ISL_628968, EPI_ISL_628969, EPI_ISL_628971, EPI_ISL_628972, EPI_ISL_628973, EPI_ISL_628975, EPI_ISL_628976, EPI_ISL_628977, EPI_ISL_628978, EPI_ISL_628979, EPI_ISL_628981, EPI_ISL_628982, EPI_ISL_628983, EPI_ISL_628984, EPI_ISL_628985, EPI_ISL_628986, EPI_ISL_628987, EPI_ISL_628988, EPI_ISL_628989, EPI_ISL_628990, EPI_ISL_628991, EPI_ISL_628992, EPI_ISL_628993, EPI_ISL_628994, EPI_ISL_628996, EPI_ISL_628997, EPI_ISL_628998, EPI_ISL_628999 | | | |
| see above | Utah Public Health Laboratory | Utah Public Health Laboratory | Erin Young, Kelly Oakeson |
| EPI_ISL_629036, EPI_ISL_629037, EPI_ISL_629038, EPI_ISL_629039, EPI_ISL_629040, EPI_ISL_629041, EPI_ISL_629042, EPI_ISL_629043, EPI_ISL_629044, EPI_ISL_629045, EPI_ISL_629046, EPI_ISL_629047, EPI_ISL_629048, EPI_ISL_629049, EPI_ISL_629050, EPI_ISL_629051, EPI_ISL_629052, EPI_ISL_629053, EPI_ISL_629054, EPI_ISL_629055, EPI_ISL_629056, EPI_ISL_629057, EPI_ISL_629058, EPI_ISL_629059, EPI_ISL_629060, EPI_ISL_629061, EPI_ISL_629062, EPI_ISL_629063, EPI_ISL_629064, EPI_ISL_629065, EPI_ISL_629066, EPI_ISL_629067, EPI_ISL_629068, EPI_ISL_629069, EPI_ISL_629070, EPI_ISL_629071, EPI_ISL_629072, EPI_ISL_629073, EPI_ISL_629074, EPI_ISL_629075, EPI_ISL_629076, EPI_ISL_629077, EPI_ISL_629078, EPI_ISL_629079, EPI_ISL_629080 | | | |
| see above | Maryland Public Health Laboratory | Maryland Public Health Laboratory | Maryland Department of Health Laboratories Administration |
| EPI_ISL_631389, EPI_ISL_631390, EPI_ISL_631391, EPI_ISL_631392, EPI_ISL_631393, EPI_ISL_631394, EPI_ISL_631395, EPI_ISL_631396, EPI_ISL_631397, EPI_ISL_631398, EPI_ISL_631399, EPI_ISL_631400, EPI_ISL_631401, EPI_ISL_631402, EPI_ISL_631403, EPI_ISL_631404, EPI_ISL_631405, EPI_ISL_631406, EPI_ISL_631407, EPI_ISL_631408, EPI_ISL_631409, EPI_ISL_631410, EPI_ISL_631411, EPI_ISL_631412, EPI_ISL_631413, EPI_ISL_631414, EPI_ISL_631415, EPI_ISL_631416, EPI_ISL_631417, EPI_ISL_631418, EPI_ISL_631419, EPI_ISL_631420, EPI_ISL_631421, EPI_ISL_631422, EPI_ISL_631423, EPI_ISL_631424, EPI_ISL_631425, EPI_ISL_631426, EPI_ISL_631427, EPI_ISL_631428, EPI_ISL_631429, EPI_ISL_631430, EPI_ISL_631431, EPI_ISL_631432, EPI_ISL_631433, EPI_ISL_631434, EPI_ISL_631435, EPI_ISL_631436, EPI_ISL_631437, EPI_ISL_631438, EPI_ISL_631439, EPI_ISL_631440, EPI_ISL_631441, EPI_ISL_631442, EPI_ISL_631443, EPI_ISL_631444, EPI_ISL_631445, EPI_ISL_631446, EPI_ISL_631447, EPI_ISL_631448, EPI_ISL_631449, EPI_ISL_631450, EPI_ISL_631451, EPI_ISL_631452, EPI_ISL_631453, EPI_ISL_631454, EPI_ISL_631455, EPI_ISL_631456, EPI_ISL_631457, EPI_ISL_631458, EPI_ISL_631459, EPI_ISL_631460, EPI_ISL_631461, EPI_ISL_631462, EPI_ISL_631463, EPI_ISL_631464, EPI_ISL_631465, EPI_ISL_631466, EPI_ISL_631467, EPI_ISL_631468, EPI_ISL_631469, EPI_ISL_631470, EPI_ISL_631471, EPI_ISL_631472, EPI_ISL_631473, EPI_ISL_631474, EPI_ISL_631475, EPI_ISL_631476, EPI_ISL_631477, EPI_ISL_631478, EPI_ISL_631479, EPI_ISL_631480, EPI_ISL_631481, EPI_ISL_631482, EPI_ISL_631483, EPI_ISL_631484, EPI_ISL_631485, EPI_ISL_631486, EPI_ISL_631487, EPI_ISL_631488, EPI_ISL_631489, EPI_ISL_631490, EPI_ISL_631491, EPI_ISL_631492, EPI_ISL_631493, EPI_ISL_631494, EPI_ISL_631495, EPI_ISL_631496, EPI_ISL_631497, EPI_ISL_631498, EPI_ISL_631499, EPI_ISL_631500, EPI_ISL_631501 | | | |
| see above | Wisconsin State Laboratory of Hygiene Communicable Disease Division | Wisconsin State Laboratory of Hygiene Communicable Disease Division | Kelsey R. Florek, Abigail C. Shockey |
| EPI_ISL_631502, EPI_ISL_631503 | New York-Presbyterian-Columbia University Medical Center | New York City Public Health Laboratory | Jade Wang, et al. |
| EPI_ISL_631504 | Wyckoff Heights Medical Center | New York City Public Health Laboratory | Jade Wang, et al. |
| EPI_ISL_631505 | Northwell Health-GoHealth Urgent Care 10025 | New York City Public Health Laboratory | Jade Wang, et al. |
| EPI_ISL_631506 | NYC Department Of Health And Mental Hygiene | New York City Public Health Laboratory | Jade Wang, et al. |
| EPI_ISL_631507 | Mount Sinai West | New York City Public Health Laboratory | Jade Wang, et al. |
| EPI_ISL_631508 | Brooklyn Hospital Center | New York City Public Health Laboratory | Jade Wang, et al. |
| EPI_ISL_631509 | New York Presbyterian Lower Manhattan | New York City Public Health Laboratory | Jade Wang, et al. |
| EPI_ISL_631510, EPI_ISL_631511 | New York Presbyterian/ Weill Cornell Medical Center | New York City Public Health Laboratory | Jade Wang, et al. |
| EPI_ISL_631512 | Mount Sinai Hospital | New York City Public Health Laboratory | Jade Wang, et al. |
| EPI_ISL_631513 | Montefiore Medical Center | New York City Public Health Laboratory | Jade Wang, et al. |
| EPI_ISL_631514 | New York Community Hospital | New York City Public Health Laboratory | Jade Wang, et al. |
| EPI_ISL_631515 | NYU Langone Health | New York City Public Health Laboratory | Jade Wang, et al. |
| EPI_ISL_631516 | Jamaica Hospital Medical Center | New York City Public Health Laboratory | Jade Wang, et al. |
| EPI_ISL_631517, EPI_ISL_631518, EPI_ISL_631519 | New York Presbyterian-Brooklyn Methodist Hospital | New York City Public Health Laboratory | Jade Wang, et al. |
| EPI_ISL_631520 | NYC HH Lincoln Medical And Mental Health Center | New York City Public Health Laboratory | Jade Wang, et al. |
| EPI_ISL_631521 | Mount Sinai Hospital | New York City Public Health Laboratory | Jade Wang, et al. |
| EPI_ISL_631522 | Robert Kutnick | New York City Public Health Laboratory | Jade Wang, et al. |
| EPI_ISL_631523 | NYU Langone Health | New York City Public Health Laboratory | Jade Wang, et al. |
| EPI_ISL_631524 | Jamaica Hospital Medical Center | New York City Public Health Laboratory | Jade Wang, et al. |
| EPI_ISL_631525 | Mount Sinai Hospital | New York City Public Health Laboratory | Jade Wang, et al. |
| EPI_ISL_631526, EPI_ISL_631527 | NYU Langone Health | New York City Public Health Laboratory | Jade Wang, et al. |
| EPI_ISL_631528 | Maimonides Medical Center | New York City Public Health Laboratory | Jade Wang, et al. |
| EPI_ISL_631529 | St. Johns Episcopal Hospital | New York City Public Health Laboratory | Jade Wang, et al. |
| EPI_ISL_631530 | New York Presbyterian Queens | New York City Public Health Laboratory | Jade Wang, et al. |
| EPI_ISL_631531 | NYU Langone Health | New York City Public Health Laboratory | Jade Wang, et al. |
| EPI_ISL_631532 | Mount Sinai Hospital | New York City Public Health Laboratory | Jade Wang, et al. |
| EPI_ISL_631533 | New York Presbyterian Lower Manhattan | New York City Public Health Laboratory | Jade Wang, et al. |
| EPI_ISL_631534 | Mount Sinai Hospital | New York City Public Health Laboratory | Jade Wang, et al. |
| EPI_ISL_631535 | NYC Department Of Health And Mental Hygiene | New York City Public Health Laboratory | Jade Wang, et al. |
| EPI_ISL_631536 | New York Presbyterian Lower Manhattan | New York City Public Health Laboratory | Jade Wang, et al. |
| EPI_ISL_631537 | Mount Sinai Hospital | New York City Public Health Laboratory | Jade Wang, et al. |
| EPI_ISL_631538 | Maimonides Medical Center | New York City Public Health Laboratory | Jade Wang, et al. |
| EPI_ISL_631539, EPI_ISL_631540 | New York Presbyterian Lower Manhattan | New York City Public Health Laboratory | Jade Wang, et al. |
| EPI_ISL_631541 | NYC Department Of Health And Mental Hygiene | New York City Public Health Laboratory | Jade Wang, et al. |
| EPI_ISL_631542 | New York Presbyterian Lower Manhattan | New York City Public Health Laboratory | Jade Wang, et al. |
| EPI_ISL_631543 | Robert Kutnick | New York City Public Health Laboratory | Jade Wang, et al. |
| EPI_ISL_631544, EPI_ISL_631545 | New York Presbyterian-Brooklyn Methodist Hospital | New York City Public Health Laboratory | Jade Wang, et al. |
| EPI_ISL_631546 | Maimonides Medical Center | New York City Public Health Laboratory | Jade Wang, et al. |
| EPI_ISL_631547 | New York Presbyterian Queens | New York City Public Health Laboratory | Jade Wang, et al. |
| EPI_ISL_631548 | New York Presbyterian-Brooklyn Methodist Hospital | New York City Public Health Laboratory | Jade Wang, et al. |
| EPI_ISL_631549 | Mount Sinai Hospital | New York City Public Health Laboratory | Jade Wang, et al. |
| EPI_ISL_631550, EPI_ISL_631551 | Jamaica Hospital Medical Center | New York City Public Health Laboratory | Jade Wang, et al. |
| EPI_ISL_631552, EPI_ISL_631553 | Brookdale University Hospital Medical Center | New York City Public Health Laboratory | Jade Wang, et al. |
| EPI_ISL_631554 | Jamaica Hospital Medical Center | New York City Public Health Laboratory | Jade Wang, et al. |
| EPI_ISL_631555 | Richmond University Medical Center | New York City Public Health Laboratory | Jade Wang, et al. |
| EPI_ISL_631556 | NYC HH Jacobi Medical Center | New York City Public Health Laboratory | Jade Wang, et al. |
| EPI_ISL_631557 | NYC HH Lincoln Medical And Mental Health Center | New York City Public Health Laboratory | Jade Wang, et al. |
| EPI_ISL_631558, EPI_ISL_631559 | NYU Langone Health | New York City Public Health Laboratory | Jade Wang, et al. |
| EPI_ISL_631560, | Montefiore Medical Center | New York City Public Health Laboratory | Jade Wang, et al. |

| | | | |
|---|---|---|---|
| EPI_ISL_631561 | | | |
| EPI_ISL_631562 | Brookdale University Hospital Medical Center | New York City Public Health Laboratory | Jade Wang, et al. |
| EPI_ISL_631563 | Flushing Hospital Medical Center | New York City Public Health Laboratory | Jade Wang, et al. |
| EPI_ISL_631564 | NYC HH Elmhurst Hospital Medical Center | New York City Public Health Laboratory | Jade Wang, et al. |
| EPI_ISL_631565 | Brookdale University Hospital Medical Center | New York City Public Health Laboratory | Jade Wang, et al. |
| EPI_ISL_631566, EPI_ISL_631567 | New York Presbyterian-Brooklyn Methodist Hospital | New York City Public Health Laboratory | Jade Wang, et al. |
| EPI_ISL_631568, EPI_ISL_631569 | New York Presbyterian Queens | New York City Public Health Laboratory | Jade Wang, et al. |
| EPI_ISL_631570 | OCME Office Of Chief Medical Examiner | New York City Public Health Laboratory | Jade Wang, et al. |
| EPI_ISL_631571 | Jamaica Hospital Medical Center | New York City Public Health Laboratory | Jade Wang, et al. |
| EPI_ISL_631572 | NYC Department Of Health And Mental Hygiene | New York City Public Health Laboratory | Jade Wang, et al. |
| EPI_ISL_631573 | Brookdale University Hospital Medical Center | New York City Public Health Laboratory | Jade Wang, et al. |
| EPI_ISL_631574, EPI_ISL_631575 | Jamaica Hospital Medical Center | New York City Public Health Laboratory | Jade Wang, et al. |
| EPI_ISL_631576 | Richmond University Medical Center | New York City Public Health Laboratory | Jade Wang, et al. |
| EPI_ISL_631577 | NYC HH Lincoln Medical And Mental Health Center | New York City Public Health Laboratory | Jade Wang, et al. |
| EPI_ISL_631578 | Jamaica Hospital Medical Center | New York City Public Health Laboratory | Jade Wang, et al. |
| EPI_ISL_631579 | Flushing Hospital Medical Center | New York City Public Health Laboratory | Jade Wang, et al. |
| EPI_ISL_631580 | Bellevue Hospital Center | New York City Public Health Laboratory | Jade Wang, et al. |
| EPI_ISL_631581 | SUNY Downstate Medical Center | New York City Public Health Laboratory | Jade Wang, et al. |
| EPI_ISL_631582 | Jamaica Hospital Medical Center | New York City Public Health Laboratory | Jade Wang, et al. |
| EPI_ISL_631583 | Richmond University Medical Center | New York City Public Health Laboratory | Jade Wang, et al. |
| EPI_ISL_631584 | NYC HH Lincoln Medical And Mental Health Center | New York City Public Health Laboratory | Jade Wang, et al. |
| EPI_ISL_631585 | Jamaica Hospital Medical Center | New York City Public Health Laboratory | Jade Wang, et al. |
| EPI_ISL_631586 | Flushing Hospital Medical Center | New York City Public Health Laboratory | Jade Wang, et al. |
| EPI_ISL_631587 | NYC HH Lincoln Medical And Mental Health Center | New York City Public Health Laboratory | Jade Wang, et al. |
| EPI_ISL_631588 | SUNY Downstate Medical Center | New York City Public Health Laboratory | Jade Wang, et al. |
| EPI_ISL_631589 | Flushing Hospital Medical Center | New York City Public Health Laboratory | Jade Wang, et al. |
| EPI_ISL_631590 | Jamaica Hospital Medical Center | New York City Public Health Laboratory | Jade Wang, et al. |
| EPI_ISL_631591 | OCME Office Of Chief Medical Examiner | New York City Public Health Laboratory | Jade Wang, et al. |
| EPI_ISL_631592, EPI_ISL_631593 | Jamaica Hospital Medical Center | New York City Public Health Laboratory | Jade Wang, et al. |
| EPI_ISL_631594 | OCME Office Of Chief Medical Examiner | New York City Public Health Laboratory | Jade Wang, et al. |
| EPI_ISL_631595 | Jamaica Hospital Medical Center | New York City Public Health Laboratory | Jade Wang, et al. |
| EPI_ISL_631596 | James J. Peters VA Medical Center | New York City Public Health Laboratory | Jade Wang, et al. |
| EPI_ISL_631597 | Jamaica Hospital Medical Center | New York City Public Health Laboratory | Jade Wang, et al. |
| EPI_ISL_631598 | OCME Office Of Chief Medical Examiner | New York City Public Health Laboratory | Jade Wang, et al. |
| EPI_ISL_631599 | Jamaica Hospital Medical Center | New York City Public Health Laboratory | Jade Wang, et al. |
| EPI_ISL_631600 | James J. Peters VA Medical Center | New York City Public Health Laboratory | Jade Wang, et al. |
| EPI_ISL_631601 | Jamaica Hospital Medical Center | New York City Public Health Laboratory | Jade Wang, et al. |
| EPI_ISL_631602 | Flushing Hospital Medical Center | New York City Public Health Laboratory | Jade Wang, et al. |
| EPI_ISL_631603 | NYC HH Lincoln Medical And Mental Health Center | New York City Public Health Laboratory | Jade Wang, et al. |
| EPI_ISL_631604 | Jamaica Hospital Medical Center | New York City Public Health Laboratory | Jade Wang, et al. |
| EPI_ISL_631605 | OCME Office Of Chief Medical Examiner | New York City Public Health Laboratory | Jade Wang, et al. |
| EPI_ISL_631606 | Flushing Hospital Medical Center | New York City Public Health Laboratory | Jade Wang, et al. |
| EPI_ISL_631607 | NYC HH Lincoln Medical And Mental Health Center | New York City Public Health Laboratory | Jade Wang, et al. |
| EPI_ISL_631608, EPI_ISL_631609, EPI_ISL_631610 | Jamaica Hospital Medical Center | New York City Public Health Laboratory | Jade Wang, et al. |
| EPI_ISL_631611 | OCME Office Of Chief Medical Examiner | New York City Public Health Laboratory | Jade Wang, et al. |
| EPI_ISL_631612, EPI_ISL_631613 | Jamaica Hospital Medical Center | New York City Public Health Laboratory | Jade Wang, et al. |
| EPI_ISL_631614 | Flushing Hospital Medical Center | New York City Public Health Laboratory | Jade Wang, et al. |
| EPI_ISL_631615 | Jamaica Hospital Medical Center | New York City Public Health Laboratory | Jade Wang, et al. |
| EPI_ISL_631616 | Flushing Hospital Medical Center | New York City Public Health Laboratory | Jade Wang, et al. |
| EPI_ISL_631617, EPI_ISL_631618, EPI_ISL_631619 | OCME Office Of Chief Medical Examiner | New York City Public Health Laboratory | Jade Wang, et al. |
| EPI_ISL_631620 | Flushing Hospital Medical Center | New York City Public Health Laboratory | Jade Wang, et al. |
| EPI_ISL_631621 | Jamaica Hospital Medical Center | New York City Public Health Laboratory | Jade Wang, et al. |
| EPI_ISL_631622, EPI_ISL_631623, EPI_ISL_631624, EPI_ISL_631625, EPI_ISL_631626, EPI_ISL_631627, EPI_ISL_631628, EPI_ISL_631629, EPI_ISL_631630, EPI_ISL_631631, EPI_ISL_631632, EPI_ISL_631633 | | | |
| see above | Richmond University Medical Center | New York City Public Health Laboratory | Jade Wang, et al. |
| EPI_ISL_631634 | Jamaica Hospital Medical Center | New York City Public Health Laboratory | Jade Wang, et al. |
| EPI_ISL_631635 | OCME Office Of Chief Medical Examiner | New York City Public Health Laboratory | Jade Wang, et al. |
| EPI_ISL_631636, EPI_ISL_631637 | Jamaica Hospital Medical Center | New York City Public Health Laboratory | Jade Wang, et al. |
| EPI_ISL_631638 | Flushing Hospital Medical Center | New York City Public Health Laboratory | Jade Wang, et al. |
| EPI_ISL_631639 | OCME Office Of Chief Medical Examiner | New York City Public Health Laboratory | Jade Wang, et al. |
| EPI_ISL_631640, EPI_ISL_631641, EPI_ISL_631642, EPI_ISL_631645, EPI_ISL_631646, EPI_ISL_631647, EPI_ISL_631648, EPI_ISL_631649, EPI_ISL_631651, EPI_ISL_631652, EPI_ISL_631653, EPI_ISL_631655, EPI_ISL_631656, EPI_ISL_631657, EPI_ISL_631659, EPI_ISL_631660, EPI_ISL_631661, EPI_ISL_631662, EPI_ISL_631663, EPI_ISL_631665, EPI_ISL_631666, EPI_ISL_631667, EPI_ISL_631669, EPI_ISL_631670, EPI_ISL_631674, EPI_ISL_631677, EPI_ISL_631678, EPI_ISL_631679, EPI_ISL_631682, EPI_ISL_631683, EPI_ISL_631684 | | | |
| see above | Texas Department of State Health Services | Texas Department of State Health Services | Rashmi Tuladhar, Bonnie Oh, Jenny Zhang, Maliha Rahman, Anita Pokharel, Myong Koag, Chung Wang, Rachel Lee, Grace Kubin, Mayela Pedrueza |

| | | | |
|---|---|---|---|
| EPI_ISL_631685 | OCME Office Of Chief Medical Examiner | New York City Public Health Laboratory | Jade Wang, et al. |
| EPI_ISL_631686 | NYC HH Lincoln Medical And Mental Health Center | New York City Public Health Laboratory | Jade Wang, et al. |
| EPI_ISL_631687 | OCME Office Of Chief Medical Examiner | New York City Public Health Laboratory | Jade Wang, et al. |
| EPI_ISL_631688 | Wildlife Conservation Society - Bronx Zoo | New York City Public Health Laboratory | Jade Wang, et al. |
| EPI_ISL_631689, EPI_ISL_631690 | Lincoln Medical Center | New York City Public Health Laboratory | Jade Wang, et al. |
| EPI_ISL_631691 | NYC HH Lincoln Medical And Mental Health Center | New York City Public Health Laboratory | Jade Wang, et al. |
| EPI_ISL_631692 | Flushing Hospital Medical Center | New York City Public Health Laboratory | Jade Wang, et al. |
| EPI_ISL_631693 | NYC HH Lincoln Medical And Mental Health Center | New York City Public Health Laboratory | Jade Wang, et al. |
| EPI_ISL_631694, EPI_ISL_631695 | OCME Office Of Chief Medical Examiner | New York City Public Health Laboratory | Jade Wang, et al. |
| EPI_ISL_631696 | Flushing Hospital Medical Center | New York City Public Health Laboratory | Jade Wang, et al. |
| EPI_ISL_631697, EPI_ISL_631698 | Jamaica Hospital Medical Center | New York City Public Health Laboratory | Jade Wang, et al. |
| EPI_ISL_631699, EPI_ISL_631700, EPI_ISL_631701 | Flushing Hospital Medical Center | New York City Public Health Laboratory | Jade Wang, et al. |
| EPI_ISL_631702, EPI_ISL_631703, EPI_ISL_631704, EPI_ISL_631705, EPI_ISL_631706 | Jamaica Hospital Medical Center | New York City Public Health Laboratory | Jade Wang, et al. |
| EPI_ISL_631707, EPI_ISL_631708 | Flushing Hospital Medical Center | New York City Public Health Laboratory | Jade Wang, et al. |
| EPI_ISL_631709 | Jamaica Hospital Medical Center | New York City Public Health Laboratory | Jade Wang, et al. |
| EPI_ISL_631710 | Flushing Hospital Medical Center | New York City Public Health Laboratory | Jade Wang, et al. |
| EPI_ISL_631711, EPI_ISL_631712, EPI_ISL_631713 | Jamaica Hospital Medical Center | New York City Public Health Laboratory | Jade Wang, et al. |
| EPI_ISL_631714, EPI_ISL_631715 | Flushing Hospital Medical Center | New York City Public Health Laboratory | Jade Wang, et al. |
| EPI_ISL_631716, EPI_ISL_631717, EPI_ISL_631718, EPI_ISL_631719, EPI_ISL_631720, EPI_ISL_631721, EPI_ISL_631722, EPI_ISL_631723 | Jamaica Hospital Medical Center | New York City Public Health Laboratory | Jade Wang, et al. |
| EPI_ISL_631724, EPI_ISL_631725 | Flushing Hospital Medical Center | New York City Public Health Laboratory | Jade Wang, et al. |
| EPI_ISL_631726 | Jamaica Hospital Medical Center | New York City Public Health Laboratory | Jade Wang, et al. |
| EPI_ISL_631727 | Flushing Hospital Medical Center | New York City Public Health Laboratory | Jade Wang, et al. |
| EPI_ISL_631728, EPI_ISL_631729, EPI_ISL_631730, EPI_ISL_631731 | Jamaica Hospital Medical Center | New York City Public Health Laboratory | Jade Wang, et al. |
| EPI_ISL_631732 | Flushing Hospital Medical Center | New York City Public Health Laboratory | Jade Wang, et al. |
| EPI_ISL_631733, EPI_ISL_631734, EPI_ISL_631735, EPI_ISL_631736, EPI_ISL_631737, EPI_ISL_631738, EPI_ISL_631739, EPI_ISL_631740, EPI_ISL_631741, EPI_ISL_631742, EPI_ISL_631743, EPI_ISL_631744, EPI_ISL_631745, EPI_ISL_631746, EPI_ISL_631747 | | | |
| see above | Jamaica Hospital Medical Center | New York City Public Health Laboratory | Jade Wang, et al. |
| EPI_ISL_631748, EPI_ISL_631749, EPI_ISL_631750, EPI_ISL_631751 | St Barnabas Hospital | New York City Public Health Laboratory | Jade Wang, et al. |
| EPI_ISL_631752, EPI_ISL_631753, EPI_ISL_631754 | NYC HH Lincoln Medical And Mental Health Center | New York City Public Health Laboratory | Jade Wang, et al. |
| EPI_ISL_631755 | Richmond University Medical Center | New York City Public Health Laboratory | Jade Wang, et al. |
| EPI_ISL_631756 | Flushing Hospital Medical Center | New York City Public Health Laboratory | Jade Wang, et al. |
| EPI_ISL_631757 | Richmond University Medical Center | New York City Public Health Laboratory | Jade Wang, et al. |
| EPI_ISL_631758, EPI_ISL_631759, EPI_ISL_631760, EPI_ISL_631761, EPI_ISL_631762, EPI_ISL_631763 | OCME Office Of Chief Medical Examiner | New York City Public Health Laboratory | Jade Wang, et al. |
| EPI_ISL_631764 | Mount Sinai Hospital | New York City Public Health Laboratory | Jade Wang, et al. |
| EPI_ISL_631765 | Bellevue Hospital Center | New York City Public Health Laboratory | Jade Wang, et al. |
| EPI_ISL_631766 | Flushing Hospital Medical Center | New York City Public Health Laboratory | Jade Wang, et al. |
| EPI_ISL_631767, EPI_ISL_631768 | Jamaica Hospital Medical Center | New York City Public Health Laboratory | Jade Wang, et al. |
| EPI_ISL_631769, EPI_ISL_631770, EPI_ISL_631771 | Flushing Hospital Medical Center | New York City Public Health Laboratory | Jade Wang, et al. |
| EPI_ISL_631772 | Jamaica Hospital Medical Center | New York City Public Health Laboratory | Jade Wang, et al. |
| EPI_ISL_631773, EPI_ISL_631774 | Flushing Hospital Medical Center | New York City Public Health Laboratory | Jade Wang, et al. |
| EPI_ISL_631775, EPI_ISL_631776, EPI_ISL_631777 | Jamaica Hospital Medical Center | New York City Public Health Laboratory | Jade Wang, et al. |
| EPI_ISL_631778, EPI_ISL_631779, EPI_ISL_631780, EPI_ISL_631781, EPI_ISL_631782, EPI_ISL_631783, EPI_ISL_631784 | St Barnabas Hospital | New York City Public Health Laboratory | Jade Wang, et al. |

| | | | |
|---|---|---|---|
| EPI_ISL_631785, EPI_ISL_631786 | Mount Sinai Hospital | New York City Public Health Laboratory | Jade Wang, et al. |
| EPI_ISL_631787 | Flushing Hospital Medical Center | New York City Public Health Laboratory | Jade Wang, et al. |
| EPI_ISL_631788, EPI_ISL_631789, EPI_ISL_631790, EPI_ISL_631791 | Jamaica Hospital Medical Center | New York City Public Health Laboratory | Jade Wang, et al. |
| EPI_ISL_631792 | Richmond University Medical Center | New York City Public Health Laboratory | Jade Wang, et al. |
| EPI_ISL_631793, EPI_ISL_631794 | SUNY Downstate Medical Center | New York City Public Health Laboratory | Jade Wang, et al. |
| EPI_ISL_631795 | Jamaica Hospital Medical Center | New York City Public Health Laboratory | Jade Wang, et al. |
| EPI_ISL_631796 | Maimonides Medical Center | New York City Public Health Laboratory | Jade Wang, et al. |
| EPI_ISL_631797 | OCME Office Of Chief Medical Examiner | New York City Public Health Laboratory | Jade Wang, et al. |
| EPI_ISL_631798 | Jamaica Hospital Medical Center | New York City Public Health Laboratory | Jade Wang, et al. |
| EPI_ISL_631799 | NYC HH Lincoln Medical And Mental Health Center | New York City Public Health Laboratory | Jade Wang, et al. |
| EPI_ISL_631800 | Brookdale University Hospital Medical Center | New York City Public Health Laboratory | Jade Wang, et al. |
| EPI_ISL_631801 | Richmond University Medical Center | New York City Public Health Laboratory | Jade Wang, et al. |
| EPI_ISL_631802 | SUNY Downstate Medical Center | New York City Public Health Laboratory | Jade Wang, et al. |
| EPI_ISL_631803 | Brookdale University Hospital Medical Center | New York City Public Health Laboratory | Jade Wang, et al. |
| EPI_ISL_631804 | Bellevue Hospital Center | New York City Public Health Laboratory | Jade Wang, et al. |
| EPI_ISL_631805, EPI_ISL_631806, EPI_ISL_631807 | Brookdale University Hospital Medical Center | New York City Public Health Laboratory | Jade Wang, et al. |
| EPI_ISL_631808 | Wyckoff Heights Medical Center | New York City Public Health Laboratory | Jade Wang, et al. |
| EPI_ISL_631809, EPI_ISL_631810 | Jamaica Hospital Medical Center | New York City Public Health Laboratory | Jade Wang, et al. |
| EPI_ISL_631811 | Wyckoff Heights Medical Center | New York City Public Health Laboratory | Jade Wang, et al. |
| EPI_ISL_631812, EPI_ISL_631813 | Jamaica Hospital Medical Center | New York City Public Health Laboratory | Jade Wang, et al. |
| EPI_ISL_631814, EPI_ISL_631815, EPI_ISL_631816, EPI_ISL_631817, EPI_ISL_631818 | Richmond University Medical Center | New York City Public Health Laboratory | Jade Wang, et al. |
| EPI_ISL_631819 | Brookdale University Hospital Medical Center | New York City Public Health Laboratory | Jade Wang, et al. |
| EPI_ISL_631820 | Jamaica Hospital Medical Center | New York City Public Health Laboratory | Jade Wang, et al. |
| EPI_ISL_631821 | Richmond University Medical Center | New York City Public Health Laboratory | Jade Wang, et al. |
| EPI_ISL_631822, EPI_ISL_631823, EPI_ISL_631825 | Jamaica Hospital Medical Center | New York City Public Health Laboratory | Jade Wang, et al. |
| EPI_ISL_631826 | Flushing Hospital Medical Center | New York City Public Health Laboratory | Jade Wang, et al. |
| EPI_ISL_631827 | NYC HH Lincoln Medical And Mental Health Center | New York City Public Health Laboratory | Jade Wang, et al. |
| EPI_ISL_631828 | Richmond University Medical Center | New York City Public Health Laboratory | Jade Wang, et al. |
| EPI_ISL_631829 | Brookdale University Hospital Medical Center | New York City Public Health Laboratory | Jade Wang, et al. |
| EPI_ISL_631830 | Mount Sinai Hospital | New York City Public Health Laboratory | Jade Wang, et al. |
| EPI_ISL_631831 | VA NY Harbor Healthcare System | New York City Public Health Laboratory | Jade Wang, et al. |
| EPI_ISL_631832 | Metro Urgicare | New York City Public Health Laboratory | Jade Wang, et al. |
| EPI_ISL_631833 | Jamaica Hospital Medical Center | New York City Public Health Laboratory | Jade Wang, et al. |
| EPI_ISL_631834 | Mount Sinai Hospital | New York City Public Health Laboratory | Jade Wang, et al. |
| EPI_ISL_631835, EPI_ISL_631836 | Jamaica Hospital Medical Center | New York City Public Health Laboratory | Jade Wang, et al. |
| EPI_ISL_631837 | Richmond University Medical Center | New York City Public Health Laboratory | Jade Wang, et al. |
| EPI_ISL_631838 | Bellevue Hospital Center | New York City Public Health Laboratory | Jade Wang, et al. |
| EPI_ISL_631839 | Mount Sinai Hospital | New York City Public Health Laboratory | Jade Wang, et al. |
| EPI_ISL_631840, EPI_ISL_631841, EPI_ISL_631842, EPI_ISL_631843, EPI_ISL_631844, EPI_ISL_631845, EPI_ISL_631846 | OCME Office Of Chief Medical Examiner | New York City Public Health Laboratory | Jade Wang, et al. |
| EPI_ISL_631847 | Jamaica Hospital Medical Center | New York City Public Health Laboratory | Jade Wang, et al. |
| EPI_ISL_631848 | Flushing Hospital Medical Center | New York City Public Health Laboratory | Jade Wang, et al. |
| EPI_ISL_631849, EPI_ISL_631850 | Jamaica Hospital Medical Center | New York City Public Health Laboratory | Jade Wang, et al. |
| EPI_ISL_631851 | Flushing Hospital Medical Center | New York City Public Health Laboratory | Jade Wang, et al. |
| EPI_ISL_631852, EPI_ISL_631853, EPI_ISL_631854 | Jamaica Hospital Medical Center | New York City Public Health Laboratory | Jade Wang, et al. |
| EPI_ISL_631855 | Flushing Hospital Medical Center | New York City Public Health Laboratory | Jade Wang, et al. |
| EPI_ISL_631856, EPI_ISL_631857, EPI_ISL_631858, EPI_ISL_631859 | St Barnabas Hospital | New York City Public Health Laboratory | Jade Wang, et al. |
| EPI_ISL_631860 | New York Presbyterian/ Weill Cornell Medical Center | New York City Public Health Laboratory | Jade Wang, et al. |
| EPI_ISL_631861 | New York Presbyterian Queens | New York City Public Health Laboratory | Jade Wang, et al. |
| EPI_ISL_631862 | NYC Department Of Health And Mental Hygiene | New York City Public Health Laboratory | Jade Wang, et al. |
| EPI_ISL_631863 | Maimonides Medical Center | New York City Public Health Laboratory | Jade Wang, et al. |
| EPI_ISL_631864 | Brookdale University Hospital Medical Center | New York City Public Health Laboratory | Jade Wang, et al. |

| | | | |
|---|---|---|---|
| EPI_ISL_631865 | NYC HH Elmhurst Hospital Medical Center | New York City Public Health Laboratory | Jade Wang, et al. |
| EPI_ISL_631866, EPI_ISL_631867 | Jamaica Hospital Medical Center | New York City Public Health Laboratory | Jade Wang, et al. |
| EPI_ISL_631868, EPI_ISL_631869 | Richmond University Medical Center | New York City Public Health Laboratory | Jade Wang, et al. |
| EPI_ISL_631870, EPI_ISL_631871, EPI_ISL_631872 | Jamaica Hospital Medical Center | New York City Public Health Laboratory | Jade Wang, et al. |
| EPI_ISL_631873 | Richmond University Medical Center | New York City Public Health Laboratory | Jade Wang, et al. |
| EPI_ISL_631874 | James J. Peters VA Medical Center | New York City Public Health Laboratory | Jade Wang, et al. |
| EPI_ISL_631875 | Richmond University Medical Center | New York City Public Health Laboratory | Jade Wang, et al. |
| EPI_ISL_631876, EPI_ISL_631877, EPI_ISL_631878, EPI_ISL_631879 | Jamaica Hospital Medical Center | New York City Public Health Laboratory | Jade Wang, et al. |
| EPI_ISL_631880 | Mount Sinai West | New York City Public Health Laboratory | Jade Wang, et al. |
| EPI_ISL_631881 | Maimonides Medical Center | New York City Public Health Laboratory | Jade Wang, et al. |
| EPI_ISL_631882, EPI_ISL_631883 | Jamaica Hospital Medical Center | New York City Public Health Laboratory | Jade Wang, et al. |
| EPI_ISL_631884 | Brookdale University Hospital Medical Center | New York City Public Health Laboratory | Jade Wang, et al. |
| EPI_ISL_631885 | Flushing Hospital Medical Center | New York City Public Health Laboratory | Jade Wang, et al. |
| EPI_ISL_631886 | NYC HH Lincoln Medical And Mental Health Center | New York City Public Health Laboratory | Jade Wang, et al. |
| EPI_ISL_631887 | Jamaica Hospital Medical Center | New York City Public Health Laboratory | Jade Wang, et al. |
| EPI_ISL_631888 | St Barnabas Hospital | New York City Public Health Laboratory | Jade Wang, et al. |
| EPI_ISL_631889, EPI_ISL_631890 | Jamaica Hospital Medical Center | New York City Public Health Laboratory | Jade Wang, et al. |
| EPI_ISL_631891 | Flushing Hospital Medical Center | New York City Public Health Laboratory | Jade Wang, et al. |
| EPI_ISL_631892 | Richmond University Medical Center | New York City Public Health Laboratory | Jade Wang, et al. |
| EPI_ISL_631893, EPI_ISL_631894 | Flushing Hospital Medical Center | New York City Public Health Laboratory | Jade Wang, et al. |
| EPI_ISL_631895, EPI_ISL_631896 | Jamaica Hospital Medical Center | New York City Public Health Laboratory | Jade Wang, et al. |
| EPI_ISL_631897 | Richmond University Medical Center | New York City Public Health Laboratory | Jade Wang, et al. |
| EPI_ISL_631898 | OCME Office Of Chief Medical Examiner | New York City Public Health Laboratory | Jade Wang, et al. |
| EPI_ISL_631899 | Richmond University Medical Center | New York City Public Health Laboratory | Jade Wang, et al. |
| EPI_ISL_631900 | St Barnabas Hospital | New York City Public Health Laboratory | Jade Wang, et al. |
| EPI_ISL_631901, EPI_ISL_631902 | Jamaica Hospital Medical Center | New York City Public Health Laboratory | Jade Wang, et al. |
| EPI_ISL_631903 | Richmond University Medical Center | New York City Public Health Laboratory | Jade Wang, et al. |
| EPI_ISL_631904 | Jamaica Hospital Medical Center | New York City Public Health Laboratory | Jade Wang, et al. |
| EPI_ISL_631905 | Richmond University Medical Center | New York City Public Health Laboratory | Jade Wang, et al. |
| EPI_ISL_631906, EPI_ISL_631907, EPI_ISL_631908 | Flushing Hospital Medical Center | New York City Public Health Laboratory | Jade Wang, et al. |
| EPI_ISL_631909, EPI_ISL_631910 | Jamaica Hospital Medical Center | New York City Public Health Laboratory | Jade Wang, et al. |
| EPI_ISL_631911, EPI_ISL_631912 | St Barnabas Hospital | New York City Public Health Laboratory | Jade Wang, et al. |
| EPI_ISL_631913, EPI_ISL_631914 | Mount Sinai Hospital | New York City Public Health Laboratory | Jade Wang, et al. |
| EPI_ISL_631915 | Maimonides Medical Center | New York City Public Health Laboratory | Jade Wang, et al. |
| EPI_ISL_631916 | Flushing Hospital Medical Center | New York City Public Health Laboratory | Jade Wang, et al. |
| EPI_ISL_631917 | St Barnabas Hospital | New York City Public Health Laboratory | Jade Wang, et al. |
| EPI_ISL_631918 | Flushing Hospital Medical Center | New York City Public Health Laboratory | Jade Wang, et al. |
| EPI_ISL_631919, EPI_ISL_631920 | Richmond University Medical Center | New York City Public Health Laboratory | Jade Wang, et al. |
| EPI_ISL_631921, EPI_ISL_631922 | Jamaica Hospital Medical Center | New York City Public Health Laboratory | Jade Wang, et al. |
| EPI_ISL_631923 | Flushing Hospital Medical Center | New York City Public Health Laboratory | Jade Wang, et al. |
| EPI_ISL_631924 | St Barnabas Hospital | New York City Public Health Laboratory | Jade Wang, et al. |
| EPI_ISL_631925, EPI_ISL_631926, EPI_ISL_631927, EPI_ISL_631928 | Jamaica Hospital Medical Center | New York City Public Health Laboratory | Jade Wang, et al. |
| EPI_ISL_631929 | Flushing Hospital Medical Center | New York City Public Health Laboratory | Jade Wang, et al. |
| EPI_ISL_631930 | Jamaica Hospital Medical Center | New York City Public Health Laboratory | Jade Wang, et al. |
| EPI_ISL_631931 | Flushing Hospital Medical Center | New York City Public Health Laboratory | Jade Wang, et al. |
| EPI_ISL_631932 | OCME Office Of Chief Medical Examiner | New York City Public Health Laboratory | Jade Wang, et al. |
| EPI_ISL_631933 | St Barnabas Hospital | New York City Public Health Laboratory | Jade Wang, et al. |
| EPI_ISL_631934 | OCME Office Of Chief Medical Examiner | New York City Public Health Laboratory | Jade Wang, et al. |
| EPI_ISL_631935 | St Barnabas Hospital | New York City Public Health Laboratory | Jade Wang, et al. |
| EPI_ISL_631936, EPI_ISL_631937, EPI_ISL_631938 | OCME Office Of Chief Medical Examiner | New York City Public Health Laboratory | Jade Wang, et al. |
| EPI_ISL_631939, EPI_ISL_631940, EPI_ISL_631941, EPI_ISL_631942, EPI_ISL_631943, EPI_ISL_631944, EPI_ISL_631945, EPI_ISL_631946, EPI_ISL_631947, EPI_ISL_631948, EPI_ISL_631949 | | | |
| see above | St Barnabas Hospital | New York City Public Health Laboratory | Jade Wang, et al. |
| EPI_ISL_631950, EPI_ISL_631951 | Jamaica Hospital Medical Center | New York City Public Health Laboratory | Jade Wang, et al. |
| EPI_ISL_631952 | St Barnabas Hospital | New York City Public Health Laboratory | Jade Wang, et al. |
| EPI_ISL_631953 | NYC HH Lincoln Medical And Mental Health Center | New York City Public Health Laboratory | Jade Wang, et al. |
| EPI_ISL_631954 | Richmond University Medical Center | New York City Public Health Laboratory | Jade Wang, et al. |

| | | | |
|---|---|---|---|
| EPI_ISL_631955 | Flushing Hospital Medical Center | New York City Public Health Laboratory | Jade Wang, et al. |
| EPI_ISL_631956 | OCME Office Of Chief Medical Examiner | New York City Public Health Laboratory | Jade Wang, et al. |
| EPI_ISL_631957, EPI_ISL_631958 | St Barnabas Hospital | New York City Public Health Laboratory | Jade Wang, et al. |
| EPI_ISL_631959, EPI_ISL_631960, EPI_ISL_631961 | Flushing Hospital Medical Center | New York City Public Health Laboratory | Jade Wang, et al. |
| EPI_ISL_631962 | Richmond University Medical Center | New York City Public Health Laboratory | Jade Wang, et al. |
| EPI_ISL_631963 | Jamaica Hospital Medical Center | New York City Public Health Laboratory | Jade Wang, et al. |
| EPI_ISL_631964 | Flushing Hospital Medical Center | New York City Public Health Laboratory | Jade Wang, et al. |
| EPI_ISL_631965 | Richmond University Medical Center | New York City Public Health Laboratory | Jade Wang, et al. |
| EPI_ISL_631966 | Jamaica Hospital Medical Center | New York City Public Health Laboratory | Jade Wang, et al. |
| EPI_ISL_631967 | Richmond University Medical Center | New York City Public Health Laboratory | Jade Wang, et al. |
| EPI_ISL_631968 | Flushing Hospital Medical Center | New York City Public Health Laboratory | Jade Wang, et al. |
| EPI_ISL_631969, EPI_ISL_631970 | Jamaica Hospital Medical Center | New York City Public Health Laboratory | Jade Wang, et al. |
| EPI_ISL_631971 | St Barnabas Hospital | New York City Public Health Laboratory | Jade Wang, et al. |
| EPI_ISL_631972 | Richmond University Medical Center | New York City Public Health Laboratory | Jade Wang, et al. |
| EPI_ISL_631973 | Jamaica Hospital Medical Center | New York City Public Health Laboratory | Jade Wang, et al. |
| EPI_ISL_631974 | NYC HH Coney Island Hospital | New York City Public Health Laboratory | Jade Wang, et al. |
| EPI_ISL_631975 | Richmond University Medical Center | New York City Public Health Laboratory | Jade Wang, et al. |
| EPI_ISL_631976 | Brookdale University Hospital Medical Center | New York City Public Health Laboratory | Jade Wang, et al. |
| EPI_ISL_631977 | SUNY Downstate Medical Center | New York City Public Health Laboratory | Jade Wang, et al. |
| EPI_ISL_631978 | Brookdale University Hospital Medical Center | New York City Public Health Laboratory | Jade Wang, et al. |
| EPI_ISL_631979 | Jamaica Hospital Medical Center | New York City Public Health Laboratory | Jade Wang, et al. |
| EPI_ISL_631980 | St Barnabas Hospital | New York City Public Health Laboratory | Jade Wang, et al. |
| EPI_ISL_631981 | Brookdale University Hospital Medical Center | New York City Public Health Laboratory | Jade Wang, et al. |
| EPI_ISL_631982 | Montefiore Medical Center | New York City Public Health Laboratory | Jade Wang, et al. |
| EPI_ISL_631983 | Mount Sinai Hospital | New York City Public Health Laboratory | Jade Wang, et al. |
| EPI_ISL_631984 | Brookdale University Hospital Medical Center | New York City Public Health Laboratory | Jade Wang, et al. |
| EPI_ISL_631985 | St Barnabas Hospital | New York City Public Health Laboratory | Jade Wang, et al. |
| EPI_ISL_631986 | Flushing Hospital Medical Center | New York City Public Health Laboratory | Jade Wang, et al. |
| EPI_ISL_631987, EPI_ISL_631988 | St Barnabas Hospital | New York City Public Health Laboratory | Jade Wang, et al. |
| EPI_ISL_631989 | Flushing Hospital Medical Center | New York City Public Health Laboratory | Jade Wang, et al. |
| EPI_ISL_631990 | St Barnabas Hospital | New York City Public Health Laboratory | Jade Wang, et al. |
| EPI_ISL_631991 | Flushing Hospital Medical Center | New York City Public Health Laboratory | Jade Wang, et al. |
| EPI_ISL_631992 | Richmond University Medical Center | New York City Public Health Laboratory | Jade Wang, et al. |
| EPI_ISL_631993 | Maimonides Medical Center | New York City Public Health Laboratory | Jade Wang, et al. |
| EPI_ISL_631994 | Jamaica Hospital Medical Center | New York City Public Health Laboratory | Jade Wang, et al. |
| EPI_ISL_631995 | Wyckoff Heights Medical Center | New York City Public Health Laboratory | Jade Wang, et al. |
| EPI_ISL_631996, EPI_ISL_631997 | Jamaica Hospital Medical Center | New York City Public Health Laboratory | Jade Wang, et al. |
| EPI_ISL_631998, EPI_ISL_631999, EPI_ISL_632000, EPI_ISL_632001, EPI_ISL_632002 | Richmond University Medical Center | New York City Public Health Laboratory | Jade Wang, et al. |
| EPI_ISL_632003, EPI_ISL_632004 | Flushing Hospital Medical Center | New York City Public Health Laboratory | Jade Wang, et al. |
| EPI_ISL_632005 | Jamaica Hospital Medical Center | New York City Public Health Laboratory | Jade Wang, et al. |
| EPI_ISL_632006 | OCME Office Of Chief Medical Examiner | New York City Public Health Laboratory | Jade Wang, et al. |
| EPI_ISL_632007, EPI_ISL_632008, EPI_ISL_632009 | Jamaica Hospital Medical Center | New York City Public Health Laboratory | Jade Wang, et al. |
| EPI_ISL_632010 | Flushing Hospital Medical Center | New York City Public Health Laboratory | Jade Wang, et al. |
| EPI_ISL_632011, EPI_ISL_632012 | Jamaica Hospital Medical Center | New York City Public Health Laboratory | Jade Wang, et al. |
| EPI_ISL_632013, EPI_ISL_632014, EPI_ISL_632015, EPI_ISL_632016 | Richmond University Medical Center | New York City Public Health Laboratory | Jade Wang, et al. |
| EPI_ISL_632017 | Jamaica Hospital Medical Center | New York City Public Health Laboratory | Jade Wang, et al. |
| EPI_ISL_632018, EPI_ISL_632019, EPI_ISL_632020 | Flushing Hospital Medical Center | New York City Public Health Laboratory | Jade Wang, et al. |
| EPI_ISL_632021 | Jamaica Hospital Medical Center | New York City Public Health Laboratory | Jade Wang, et al. |
| EPI_ISL_632022 | Flushing Hospital Medical Center | New York City Public Health Laboratory | Jade Wang, et al. |
| EPI_ISL_632023 | Jamaica Hospital Medical Center | New York City Public Health Laboratory | Jade Wang, et al. |
| EPI_ISL_632024 | Flushing Hospital Medical Center | New York City Public Health Laboratory | Jade Wang, et al. |
| EPI_ISL_632025 | Jamaica Hospital Medical Center | New York City Public Health Laboratory | Jade Wang, et al. |
| EPI_ISL_632026 | NYC HH Lincoln Medical And Mental Health Center | New York City Public Health Laboratory | Jade Wang, et al. |
| EPI_ISL_632027 | Flushing Hospital Medical Center | New York City Public Health Laboratory | Jade Wang, et al. |
| EPI_ISL_632028, EPI_ISL_632029 | Jamaica Hospital Medical Center | New York City Public Health Laboratory | Jade Wang, et al. |
| EPI_ISL_632030, EPI_ISL_632031, EPI_ISL_632032 | Richmond University Medical Center | New York City Public Health Laboratory | Jade Wang, et al. |
| EPI_ISL_632034 | Flushing Hospital Medical Center | New York City Public Health Laboratory | Jade Wang, et al. |

| | | | |
|---|---|---|---|
| EPI_ISL_632035 | Jamaica Hospital Medical Center | New York City Public Health Laboratory | Jade Wang, et al. |
| EPI_ISL_632036 | Richmond University Medical Center | New York City Public Health Laboratory | Jade Wang, et al. |
| EPI_ISL_632037 | Jamaica Hospital Medical Center | New York City Public Health Laboratory | Jade Wang, et al. |
| EPI_ISL_632038 | New York Presbyterian Lower Manhattan | New York City Public Health Laboratory | Jade Wang, et al. |
| EPI_ISL_632039 | NYC Department Of Health And Mental Hygiene | New York City Public Health Laboratory | Jade Wang, et al. |
| EPI_ISL_632040 | New York Presbyterian Queens | New York City Public Health Laboratory | Jade Wang, et al. |
| EPI_ISL_632041, EPI_ISL_632042 | New York Presbyterian Lower Manhattan | New York City Public Health Laboratory | Jade Wang, et al. |
| EPI_ISL_632043, EPI_ISL_632044 | NYU Langone Health | New York City Public Health Laboratory | Jade Wang, et al. |
| EPI_ISL_632045 | New York Presbyterian Lower Manhattan | New York City Public Health Laboratory | Jade Wang, et al. |
| EPI_ISL_632046 | New York Presbyterian-Brooklyn Methodist Hospital | New York City Public Health Laboratory | Jade Wang, et al. |
| EPI_ISL_632047 | New York Presbyterian Lower Manhattan | New York City Public Health Laboratory | Jade Wang, et al. |
| EPI_ISL_632048 | New York Presbyterian-Brooklyn Methodist Hospital | New York City Public Health Laboratory | Jade Wang, et al. |
| EPI_ISL_632049 | New York Presbyterian Queens | New York City Public Health Laboratory | Jade Wang, et al. |
| EPI_ISL_632050 | Long Island Jewish Medical Center | New York City Public Health Laboratory | Jade Wang, et al. |
| EPI_ISL_632051 | Jamaica Hospital Medical Center | New York City Public Health Laboratory | Jade Wang, et al. |
| EPI_ISL_632052, EPI_ISL_632053 | New York Presbyterian Queens | New York City Public Health Laboratory | Jade Wang, et al. |
| EPI_ISL_632054 | New York Presbyterian-Brooklyn Methodist Hospital | New York City Public Health Laboratory | Jade Wang, et al. |
| EPI_ISL_632055, EPI_ISL_632056 | New York Community Hospital | New York City Public Health Laboratory | Jade Wang, et al. |
| EPI_ISL_632057 | New York-Presbyterian-Columbia University Medical Center | New York City Public Health Laboratory | Jade Wang, et al. |
| EPI_ISL_632058, EPI_ISL_632059 | Proximedical Urgent Care | New York City Public Health Laboratory | Jade Wang, et al. |
| EPI_ISL_632060 | NYC HH Queens Hospital Center | New York City Public Health Laboratory | Jade Wang, et al. |
| EPI_ISL_632061 | Jamaica Hospital Medical Center | New York City Public Health Laboratory | Jade Wang, et al. |
| EPI_ISL_632062 | NYC Department Of Health And Mental Hygiene | New York City Public Health Laboratory | Jade Wang, et al. |
| EPI_ISL_632063, EPI_ISL_632064 | New York Presbyterian Queens | New York City Public Health Laboratory | Jade Wang, et al. |
| EPI_ISL_632065, EPI_ISL_632066 | Jamaica Hospital Medical Center | New York City Public Health Laboratory | Jade Wang, et al. |
| EPI_ISL_632067 | NYU Langone Health | New York City Public Health Laboratory | Jade Wang, et al. |
| EPI_ISL_632068, EPI_ISL_632069 | Bellevue Hospital Center | New York City Public Health Laboratory | Jade Wang, et al. |
| EPI_ISL_632070, EPI_ISL_632071 | New York Presbyterian Queens | New York City Public Health Laboratory | Jade Wang, et al. |
| EPI_ISL_632072 | NYC HH Lincoln Medical And Mental Health Center | New York City Public Health Laboratory | Jade Wang, et al. |
| EPI_ISL_632073 | OCME Office Of Chief Medical Examiner | New York City Public Health Laboratory | Jade Wang, et al. |
| EPI_ISL_632074 | NYC Department Of Health And Mental Hygiene | New York City Public Health Laboratory | Jade Wang, et al. |
| EPI_ISL_632075 | Jamaica Hospital Medical Center | New York City Public Health Laboratory | Jade Wang, et al. |
| EPI_ISL_632076 | NYU Langone Health | New York City Public Health Laboratory | Jade Wang, et al. |
| EPI_ISL_632077, EPI_ISL_632078 | Brookdale University Hospital Medical Center | New York City Public Health Laboratory | Jade Wang, et al. |
| EPI_ISL_632079 | NYU Langone Health | New York City Public Health Laboratory | Jade Wang, et al. |
| EPI_ISL_632080 | SUNY Downstate Medical Center | New York City Public Health Laboratory | Jade Wang, et al. |
| EPI_ISL_632081 | New York Presbyterian-Brooklyn Methodist Hospital | New York City Public Health Laboratory | Jade Wang, et al. |
| EPI_ISL_632082 | New York Presbyterian/ Weill Cornell Medical Center | New York City Public Health Laboratory | Jade Wang, et al. |
| EPI_ISL_632083, EPI_ISL_632084 | NYC Department Of Health And Mental Hygiene | New York City Public Health Laboratory | Jade Wang, et al. |
| EPI_ISL_632085 | Richmond University Medical Center | New York City Public Health Laboratory | Jade Wang, et al. |
| EPI_ISL_632086 | Jamaica Hospital Medical Center | New York City Public Health Laboratory | Jade Wang, et al. |
| EPI_ISL_632087 | Flushing Hospital Medical Center | New York City Public Health Laboratory | Jade Wang, et al. |
| EPI_ISL_632088 | Jamaica Hospital Medical Center | New York City Public Health Laboratory | Jade Wang, et al. |
| EPI_ISL_632089 | Brookdale University Hospital Medical Center | New York City Public Health Laboratory | Jade Wang, et al. |
| EPI_ISL_632090 | Jamaica Hospital Medical Center | New York City Public Health Laboratory | Jade Wang, et al. |
| EPI_ISL_632091, EPI_ISL_632092 | SUNY Downstate Medical Center | New York City Public Health Laboratory | Jade Wang, et al. |
| EPI_ISL_632093, EPI_ISL_632094, EPI_ISL_632095, EPI_ISL_632096 | Jamaica Hospital Medical Center | New York City Public Health Laboratory | Jade Wang, et al. |
| EPI_ISL_632097 | Flushing Hospital Medical Center | New York City Public Health Laboratory | Jade Wang, et al. |
| EPI_ISL_632098 | Bellevue Hospital Center | New York City Public Health Laboratory | Jade Wang, et al. |
| EPI_ISL_632099 | VA NY Harbor Healthcare System | New York City Public Health Laboratory | Jade Wang, et al. |
| EPI_ISL_632100 | Brookdale University Hospital Medical Center | New York City Public Health Laboratory | Jade Wang, et al. |
| EPI_ISL_632101, EPI_ISL_632102 | Mount Sinai Queens | New York City Public Health Laboratory | Jade Wang, et al. |
| EPI_ISL_632103, EPI_ISL_632104, EPI_ISL_632105 | Flushing Hospital Medical Center | New York City Public Health Laboratory | Jade Wang, et al. |
| EPI_ISL_632106, | Jamaica Hospital Medical Center | New York City Public Health Laboratory | Jade Wang, et al. |

| | | | |
|---|---|---|---|
| EPI_ISL_632107, EPI_ISL_632108, EPI_ISL_632109, EPI_ISL_632110 | | | |
| EPI_ISL_632111 | Montefiore Medical Center | New York City Public Health Laboratory | Jade Wang, et al. |
| EPI_ISL_632112, EPI_ISL_632113 | Richmond University Medical Center | New York City Public Health Laboratory | Jade Wang, et al. |
| EPI_ISL_632114 | Brookdale University Hospital Medical Center | New York City Public Health Laboratory | Jade Wang, et al. |
| EPI_ISL_632115 | Richmond University Medical Center | New York City Public Health Laboratory | Jade Wang, et al. |
| EPI_ISL_632116 | NYC HH Lincoln Medical And Mental Health Center | New York City Public Health Laboratory | Jade Wang, et al. |
| EPI_ISL_632117, EPI_ISL_632118 | Richmond University Medical Center | New York City Public Health Laboratory | Jade Wang, et al. |
| EPI_ISL_632119, EPI_ISL_632120 | Flushing Hospital Medical Center | New York City Public Health Laboratory | Jade Wang, et al. |
| EPI_ISL_632121 | Bellevue Hospital Center | New York City Public Health Laboratory | Jade Wang, et al. |
| EPI_ISL_632122 | Flushing Hospital Medical Center | New York City Public Health Laboratory | Jade Wang, et al. |
| EPI_ISL_632123 | Brookdale University Hospital Medical Center | New York City Public Health Laboratory | Jade Wang, et al. |
| EPI_ISL_632124, EPI_ISL_632125, EPI_ISL_632126, EPI_ISL_632127 | Flushing Hospital Medical Center | New York City Public Health Laboratory | Jade Wang, et al. |
| EPI_ISL_632128 | Jamaica Hospital Medical Center | New York City Public Health Laboratory | Jade Wang, et al. |
| EPI_ISL_632129 | Wyckoff Heights Medical Center | New York City Public Health Laboratory | Jade Wang, et al. |
| EPI_ISL_632130 | Jamaica Hospital Medical Center | New York City Public Health Laboratory | Jade Wang, et al. |
| EPI_ISL_632131 | NYC HH Lincoln Medical And Mental Health Center | New York City Public Health Laboratory | Jade Wang, et al. |
| EPI_ISL_632132, EPI_ISL_632133, EPI_ISL_632134 | Flushing Hospital Medical Center | New York City Public Health Laboratory | Jade Wang, et al. |
| EPI_ISL_632135 | SUNY Downstate Medical Center | New York City Public Health Laboratory | Jade Wang, et al. |
| EPI_ISL_632136 | Bellevue Hospital Center | New York City Public Health Laboratory | Jade Wang, et al. |
| EPI_ISL_632137, EPI_ISL_632138, EPI_ISL_632139, EPI_ISL_632140 | Flushing Hospital Medical Center | New York City Public Health Laboratory | Jade Wang, et al. |
| EPI_ISL_632141 | Wyckoff Heights Medical Center | New York City Public Health Laboratory | Jade Wang, et al. |
| EPI_ISL_632142 | Brookdale University Hospital Medical Center | New York City Public Health Laboratory | Jade Wang, et al. |
| EPI_ISL_632143 | Jamaica Hospital Medical Center | New York City Public Health Laboratory | Jade Wang, et al. |
| EPI_ISL_632144 | Flushing Hospital Medical Center | New York City Public Health Laboratory | Jade Wang, et al. |
| EPI_ISL_632145 | Jamaica Hospital Medical Center | New York City Public Health Laboratory | Jade Wang, et al. |
| EPI_ISL_632146, EPI_ISL_632147 | NYC HH Lincoln Medical And Mental Health Center | New York City Public Health Laboratory | Jade Wang, et al. |
| EPI_ISL_632148 | SUNY Downstate Medical Center | New York City Public Health Laboratory | Jade Wang, et al. |
| EPI_ISL_632149, EPI_ISL_632150, EPI_ISL_632151, EPI_ISL_632152 | Jamaica Hospital Medical Center | New York City Public Health Laboratory | Jade Wang, et al. |
| EPI_ISL_632153 | SUNY Downstate Medical Center | New York City Public Health Laboratory | Jade Wang, et al. |
| EPI_ISL_632154 | Richmond University Medical Center | New York City Public Health Laboratory | Jade Wang, et al. |
| EPI_ISL_632155, EPI_ISL_632156 | Brookdale University Hospital Medical Center | New York City Public Health Laboratory | Jade Wang, et al. |
| EPI_ISL_632157 | Wyckoff Heights Medical Center | New York City Public Health Laboratory | Jade Wang, et al. |
| EPI_ISL_632158 | SUNY Downstate Medical Center | New York City Public Health Laboratory | Jade Wang, et al. |
| EPI_ISL_632159 | Wyckoff Heights Medical Center | New York City Public Health Laboratory | Jade Wang, et al. |
| EPI_ISL_632160, EPI_ISL_632161 | SUNY Downstate Medical Center | New York City Public Health Laboratory | Jade Wang, et al. |
| EPI_ISL_632162 | Jamaica Hospital Medical Center | New York City Public Health Laboratory | Jade Wang, et al. |
| EPI_ISL_632163, EPI_ISL_632164, EPI_ISL_632165 | Richmond University Medical Center | New York City Public Health Laboratory | Jade Wang, et al. |
| EPI_ISL_632166, EPI_ISL_632167, EPI_ISL_632168, EPI_ISL_632169, EPI_ISL_632170, EPI_ISL_632171, EPI_ISL_632172 | Jamaica Hospital Medical Center | New York City Public Health Laboratory | Jade Wang, et al. |
| EPI_ISL_632173 | Richmond University Medical Center | New York City Public Health Laboratory | Jade Wang, et al. |
| EPI_ISL_632174 | Flushing Hospital Medical Center | New York City Public Health Laboratory | Jade Wang, et al. |
| EPI_ISL_632175, EPI_ISL_632176 | OCME Office Of Chief Medical Examiner | New York City Public Health Laboratory | Jade Wang, et al. |
| EPI_ISL_632177 | NYC HH Lincoln Medical And Mental Health Center | New York City Public Health Laboratory | Jade Wang, et al. |
| EPI_ISL_632178 | OCME Office Of Chief Medical Examiner | New York City Public Health Laboratory | Jade Wang, et al. |
| EPI_ISL_632179 | James J. Peters VA Medical Center | New York City Public Health Laboratory | Jade Wang, et al. |
| EPI_ISL_632180 | Jamaica Hospital Medical Center | New York City Public Health Laboratory | Jade Wang, et al. |
| EPI_ISL_632181 | Flushing Hospital Medical Center | New York City Public Health Laboratory | Jade Wang, et al. |
| EPI_ISL_632182 | Jamaica Hospital Medical Center | New York City Public Health Laboratory | Jade Wang, et al. |
| EPI_ISL_632183 | OCME Office Of Chief Medical Examiner | New York City Public Health Laboratory | Jade Wang, et al. |
| EPI_ISL_632184 | Jamaica Hospital Medical Center | New York City Public Health Laboratory | Jade Wang, et al. |
| EPI_ISL_632185 | Flushing Hospital Medical Center | New York City Public Health Laboratory | Jade Wang, et al. |
| EPI_ISL_632186, EPI_ISL_632187 | NYC HH Lincoln Medical And Mental Health Center | New York City Public Health Laboratory | Jade Wang, et al. |
| EPI_ISL_632188, | Jamaica Hospital Medical Center | New York City Public Health Laboratory | Jade Wang, et al. |

| | | | |
|---|---|---|---|
| EPI_ISL_632189 | | | |
| EPI_ISL_632190 | Flushing Hospital Medical Center | New York City Public Health Laboratory | Jade Wang, et al. |
| EPI_ISL_632191 | Jamaica Hospital Medical Center | New York City Public Health Laboratory | Jade Wang, et al. |
| EPI_ISL_632192, EPI_ISL_632193 | NYC HH Lincoln Medical And Mental Health Center | New York City Public Health Laboratory | Jade Wang, et al. |
| EPI_ISL_632194 | Flushing Hospital Medical Center | New York City Public Health Laboratory | Jade Wang, et al. |
| EPI_ISL_632195 | Jamaica Hospital Medical Center | New York City Public Health Laboratory | Jade Wang, et al. |
| EPI_ISL_632196, EPI_ISL_632197 | Flushing Hospital Medical Center | New York City Public Health Laboratory | Jade Wang, et al. |
| EPI_ISL_632198 | Jamaica Hospital Medical Center | New York City Public Health Laboratory | Jade Wang, et al. |
| EPI_ISL_632199 | Richmond University Medical Center | New York City Public Health Laboratory | Jade Wang, et al. |
| EPI_ISL_632200 | James J. Peters VA Medical Center | New York City Public Health Laboratory | Jade Wang, et al. |
| EPI_ISL_632201 | Richmond University Medical Center | New York City Public Health Laboratory | Jade Wang, et al. |
| EPI_ISL_632202 | OCME Office Of Chief Medical Examiner | New York City Public Health Laboratory | Jade Wang, et al. |
| EPI_ISL_632203 | Richmond University Medical Center | New York City Public Health Laboratory | Jade Wang, et al. |
| EPI_ISL_632204 | Jamaica Hospital Medical Center | New York City Public Health Laboratory | Jade Wang, et al. |
| EPI_ISL_632205, EPI_ISL_632206, EPI_ISL_632207, EPI_ISL_632208 | Richmond University Medical Center | New York City Public Health Laboratory | Jade Wang, et al. |
| EPI_ISL_632209, EPI_ISL_632210 | Jamaica Hospital Medical Center | New York City Public Health Laboratory | Jade Wang, et al. |
| EPI_ISL_632211, EPI_ISL_632212, EPI_ISL_632213 | Flushing Hospital Medical Center | New York City Public Health Laboratory | Jade Wang, et al. |
| EPI_ISL_632214 | Jamaica Hospital Medical Center | New York City Public Health Laboratory | Jade Wang, et al. |
| EPI_ISL_632215, EPI_ISL_632216, EPI_ISL_632217, EPI_ISL_632218 | Richmond University Medical Center | New York City Public Health Laboratory | Jade Wang, et al. |
| EPI_ISL_632219 | Jamaica Hospital Medical Center | New York City Public Health Laboratory | Jade Wang, et al. |
| EPI_ISL_632220 | Flushing Hospital Medical Center | New York City Public Health Laboratory | Jade Wang, et al. |
| EPI_ISL_632221 | Jamaica Hospital Medical Center | New York City Public Health Laboratory | Jade Wang, et al. |
| EPI_ISL_632222, EPI_ISL_632223, EPI_ISL_632224, EPI_ISL_632225, EPI_ISL_632226 | Flushing Hospital Medical Center | New York City Public Health Laboratory | Jade Wang, et al. |
| EPI_ISL_632227, EPI_ISL_632228, EPI_ISL_632229 | Jamaica Hospital Medical Center | New York City Public Health Laboratory | Jade Wang, et al. |
| EPI_ISL_632230, EPI_ISL_632231, EPI_ISL_632232, EPI_ISL_632233, EPI_ISL_632234 | Richmond University Medical Center | New York City Public Health Laboratory | Jade Wang, et al. |
| EPI_ISL_632235 | NYC HH Lincoln Medical And Mental Health Center | New York City Public Health Laboratory | Jade Wang, et al. |
| EPI_ISL_632236, EPI_ISL_632237 | Flushing Hospital Medical Center | New York City Public Health Laboratory | Jade Wang, et al. |
| EPI_ISL_632238 | OCME Office Of Chief Medical Examiner | New York City Public Health Laboratory | Jade Wang, et al. |
| EPI_ISL_632239, EPI_ISL_632240, EPI_ISL_632241 | Jamaica Hospital Medical Center | New York City Public Health Laboratory | Jade Wang, et al. |
| EPI_ISL_632286 | Laboratory Services Section, Texas Department of State Health Services | Laboratory Services Section, Texas Department of State Health Services | Tuladhar,R., Oh,B., Zhang,J., Rahman,M., Pokharel,A., Pedrueza,M.,Bonser,J., Koag,M., Wang,C., Lee,R. and Kubin,G. |
| EPI_ISL_632287 | Environmental and Global Health, University of Florida | Environmental and Global Health, University of Florida | Alam,M.M., Lauzardo,M., Elbadry,M.A., Stephenson,C.J., Gibson,J.C., Morris,J.G. and Lednicky,J.A. |
| EPI_ISL_632288, EPI_ISL_632289, EPI_ISL_632290, EPI_ISL_632291, EPI_ISL_632292, EPI_ISL_632293, EPI_ISL_632294, EPI_ISL_632295, EPI_ISL_632296, EPI_ISL_632297, EPI_ISL_632298, EPI_ISL_632299, EPI_ISL_632300, EPI_ISL_632301, EPI_ISL_632302, EPI_ISL_632303, EPI_ISL_632304, EPI_ISL_632305, EPI_ISL_632306, EPI_ISL_632307, EPI_ISL_632308, EPI_ISL_632309 | | | |
| see above | Brigham and Women's Hospital | Jonathan Li laboratory | Choudhary MC, Esmaeilzadeh E, Etemad B, Mohammadi A, Regan J, Li JZ |
| EPI_ISL_632823, EPI_ISL_632824, EPI_ISL_632825, EPI_ISL_632826, EPI_ISL_632827, EPI_ISL_632828, EPI_ISL_632829, EPI_ISL_632830, EPI_ISL_632831, EPI_ISL_632832, EPI_ISL_632833, EPI_ISL_632834, EPI_ISL_632835, EPI_ISL_632836, EPI_ISL_632837, EPI_ISL_632838, EPI_ISL_632839, EPI_ISL_632840, EPI_ISL_632841, EPI_ISL_632842, EPI_ISL_632843, EPI_ISL_632844, EPI_ISL_632845 | | | |
| see above | New Mexico Department of Health Scientific Laboratory | New Mexico Department of Health Scientific Laboratory | Ellie Johnson, Anastacia Griego-Fisher, D'Eldra Malone |
| EPI_ISL_632847, EPI_ISL_632848, EPI_ISL_632849, EPI_ISL_632850, EPI_ISL_632851, EPI_ISL_632852, EPI_ISL_632853, EPI_ISL_632854, EPI_ISL_632855, EPI_ISL_632856, EPI_ISL_632857, EPI_ISL_632858, EPI_ISL_632859, EPI_ISL_632860, EPI_ISL_632861, EPI_ISL_632862, EPI_ISL_632863, EPI_ISL_632864, EPI_ISL_632865, EPI_ISL_632866, EPI_ISL_632867, EPI_ISL_632868, EPI_ISL_632869, EPI_ISL_632870, EPI_ISL_632871, EPI_ISL_632872, EPI_ISL_632873, EPI_ISL_632874, EPI_ISL_632875, EPI_ISL_632876, EPI_ISL_632877, EPI_ISL_632878, EPI_ISL_632879, EPI_ISL_632880, EPI_ISL_632881, EPI_ISL_632882, EPI_ISL_632883, EPI_ISL_632884, EPI_ISL_632885, EPI_ISL_632886, EPI_ISL_632887, EPI_ISL_632888, EPI_ISL_632889 | | | |
| see above | Idaho Bureau of Laboratories | Center for Global Health, University of New Mexico Health Sciences Center | Daryl Domman, Kurt Schwalm, Matthew Burns, Robert Voermans, Christopher Ball, Darrell Dinwiddie |
| EPI_ISL_632890, EPI_ISL_632891, EPI_ISL_632892, EPI_ISL_632893, EPI_ISL_632894, EPI_ISL_632895, EPI_ISL_632896, EPI_ISL_632897, EPI_ISL_632898 | University of Wisconsin-Madison AIDS Vaccine Research Laboratories | University of Wisconsin-Madison AIDS Vaccine Research Laboratories | Gage Moreno, Katarina Braun, et al. AIDS Vaccine Research Laboratories |
| EPI_ISL_632939 | Jamaica Hospital Medical Center | New York City Public Health Laboratory | Jade Wang, et al. |
| EPI_ISL_632940 | Richmond University Medical Center | New York City Public Health Laboratory | Jade Wang, et al. |
| EPI_ISL_632941 | Jamaica Hospital Medical Center | New York City Public Health Laboratory | Jade Wang, et al. |
| EPI_ISL_632942, EPI_ISL_632943 | Richmond University Medical Center | New York City Public Health Laboratory | Jade Wang, et al. |
| EPI_ISL_632944 | Jamaica Hospital Medical Center | New York City Public Health Laboratory | Jade Wang, et al. |
| EPI_ISL_632945, EPI_ISL_632946 | OCME Office Of Chief Medical Examiner | New York City Public Health Laboratory | Jade Wang, et al. |
| EPI_ISL_632947 | Flushing Hospital Medical Center | New York City Public Health Laboratory | Jade Wang, et al. |
| EPI_ISL_632948 | Richmond University Medical Center | New York City Public Health Laboratory | Jade Wang, et al. |
| EPI_ISL_632949, EPI_ISL_632950 | Jamaica Hospital Medical Center | New York City Public Health Laboratory | Jade Wang, et al. |
| EPI_ISL_632951, | Flushing Hospital Medical Center | New York City Public Health Laboratory | Jade Wang, et al. |

| | | | |
|---|---|---|---|
| EPI_ISL_632952 | | | |
| EPI_ISL_632953 | Jamaica Hospital Medical Center | New York City Public Health Laboratory | Jade Wang, et al. |
| EPI_ISL_632954, EPI_ISL_632955, EPI_ISL_632956 | Flushing Hospital Medical Center | New York City Public Health Laboratory | Jade Wang, et al. |
| EPI_ISL_632957, EPI_ISL_632958 | Jamaica Hospital Medical Center | New York City Public Health Laboratory | Jade Wang, et al. |
| EPI_ISL_632959 | Richmond University Medical Center | New York City Public Health Laboratory | Jade Wang, et al. |
| EPI_ISL_632960 | Flushing Hospital Medical Center | New York City Public Health Laboratory | Jade Wang, et al. |
| EPI_ISL_632961 | Jamaica Hospital Medical Center | New York City Public Health Laboratory | Jade Wang, et al. |
| EPI_ISL_632962 | Richmond University Medical Center | New York City Public Health Laboratory | Jade Wang, et al. |
| EPI_ISL_632963 | NYC Department Of Health And Mental Hygiene | New York City Public Health Laboratory | Jade Wang, et al. |
| EPI_ISL_632964 | Department of Homeless Services | New York City Public Health Laboratory | Jade Wang, et al. |
| EPI_ISL_632965 | Richmond University Medical Center | New York City Public Health Laboratory | Jade Wang, et al. |
| EPI_ISL_632966, EPI_ISL_632967, EPI_ISL_632968 | Flushing Hospital Medical Center | New York City Public Health Laboratory | Jade Wang, et al. |
| EPI_ISL_632969, EPI_ISL_632970, EPI_ISL_632971, EPI_ISL_632972 | Jamaica Hospital Medical Center | New York City Public Health Laboratory | Jade Wang, et al. |
| EPI_ISL_632973 | Flushing Hospital Medical Center | New York City Public Health Laboratory | Jade Wang, et al. |
| EPI_ISL_632974 | Jamaica Hospital Medical Center | New York City Public Health Laboratory | Jade Wang, et al. |
| EPI_ISL_632975 | Richmond University Medical Center | New York City Public Health Laboratory | Jade Wang, et al. |
| EPI_ISL_632976, EPI_ISL_632977, EPI_ISL_632978, EPI_ISL_632979 | Empire City Laboratories | New York City Public Health Laboratory | Jade Wang, et al. |
| EPI_ISL_632980, EPI_ISL_632981, EPI_ISL_632982, EPI_ISL_632983, EPI_ISL_632984, EPI_ISL_632985, EPI_ISL_632986, EPI_ISL_632987, EPI_ISL_632988, EPI_ISL_632989, EPI_ISL_632990, EPI_ISL_632991, EPI_ISL_632992, EPI_ISL_632993 | | | |
| see above | DOHMH Corona | New York City Public Health Laboratory | Jade Wang, et al. |
| EPI_ISL_632994, EPI_ISL_632995, EPI_ISL_632996, EPI_ISL_632997, EPI_ISL_632998 | DOHMH Crown Heights | New York City Public Health Laboratory | Jade Wang, et al. |
| EPI_ISL_632999, EPI_ISL_633000, EPI_ISL_633001, EPI_ISL_633002, EPI_ISL_633003, EPI_ISL_633004, EPI_ISL_633005, EPI_ISL_633006, EPI_ISL_633007, EPI_ISL_633008 | DOHMH Jamaica | New York City Public Health Laboratory | Jade Wang, et al. |
| EPI_ISL_633009, EPI_ISL_633010 | DOHMH Central Harlem | New York City Public Health Laboratory | Jade Wang, et al. |
| EPI_ISL_633011 | DOHMH PHL | New York City Public Health Laboratory | Jade Wang, et al. |
| EPI_ISL_633012 | DOHMH Riverside | New York City Public Health Laboratory | Jade Wang, et al. |
| EPI_ISL_633013 | DOHMH PHL | New York City Public Health Laboratory | Jade Wang, et al. |
| EPI_ISL_633014 | DOHMH Chelsea | New York City Public Health Laboratory | Jade Wang, et al. |
| EPI_ISL_633015 | DOHMH Morrisania | New York City Public Health Laboratory | Jade Wang, et al. |
| EPI_ISL_633016, EPI_ISL_633017 | DOHMH Chelsea | New York City Public Health Laboratory | Jade Wang, et al. |
| EPI_ISL_633018 | DOHMH Central Harlem | New York City Public Health Laboratory | Jade Wang, et al. |
| EPI_ISL_633019, EPI_ISL_633020, EPI_ISL_633021, EPI_ISL_633022 | DOHMH Morrisania | New York City Public Health Laboratory | Jade Wang, et al. |
| EPI_ISL_633023 | DOHMH Chelsea | New York City Public Health Laboratory | Jade Wang, et al. |
| EPI_ISL_633024 | DOHMH Morrisania | New York City Public Health Laboratory | Jade Wang, et al. |
| EPI_ISL_633025 | DOHMH Chelsea | New York City Public Health Laboratory | Jade Wang, et al. |
| EPI_ISL_633026 | DOHMH PHL | New York City Public Health Laboratory | Jade Wang, et al. |
| EPI_ISL_633027, EPI_ISL_633028, EPI_ISL_633029, EPI_ISL_633030 | DOHMH Morrisania | New York City Public Health Laboratory | Jade Wang, et al. |
| EPI_ISL_633031 | DOHMH Chelsea | New York City Public Health Laboratory | Jade Wang, et al. |
| EPI_ISL_633032 | DOHMH Morrisania | New York City Public Health Laboratory | Jade Wang, et al. |
| EPI_ISL_633033 | DOHMH Chelsea | New York City Public Health Laboratory | Jade Wang, et al. |
| EPI_ISL_633034 | DOHMH Riverside | New York City Public Health Laboratory | Jade Wang, et al. |
| EPI_ISL_633035 | DOHMH PHL | New York City Public Health Laboratory | Jade Wang, et al. |
| EPI_ISL_633036 | DOHMH Morrisania | New York City Public Health Laboratory | Jade Wang, et al. |
| EPI_ISL_633037 | DOHMH Central Harlem | New York City Public Health Laboratory | Jade Wang, et al. |
| EPI_ISL_633038 | DOHMH PHL | New York City Public Health Laboratory | Jade Wang, et al. |
| EPI_ISL_633039, EPI_ISL_633040 | DOHMH Chelsea | New York City Public Health Laboratory | Jade Wang, et al. |
| EPI_ISL_633041, EPI_ISL_633042, EPI_ISL_633043 | DOHMH Riverside | New York City Public Health Laboratory | Jade Wang, et al. |
| EPI_ISL_633044 | DOHMH Morrisania | New York City Public Health Laboratory | Jade Wang, et al. |
| EPI_ISL_633045 | DOHMH PHL | New York City Public Health Laboratory | Jade Wang, et al. |
| EPI_ISL_633046, EPI_ISL_633047 | DOHMH Chelsea | New York City Public Health Laboratory | Jade Wang, et al. |
| EPI_ISL_633048, EPI_ISL_633049 | DOHMH Morrisania | New York City Public Health Laboratory | Jade Wang, et al. |
| EPI_ISL_633050, | DOHMH PHL | New York City Public Health Laboratory | Jade Wang, et al. |

| | | | |
|---|---|---|---|
| EPI_ISL_633051 | | | |
| EPI_ISL_633052 | DOHMH Chelsea | New York City Public Health Laboratory | Jade Wang, et al. |
| EPI_ISL_633053 | DOHMH Morrisania | New York City Public Health Laboratory | Jade Wang, et al. |
| EPI_ISL_633054, EPI_ISL_633055 | DOHMH PHL | New York City Public Health Laboratory | Jade Wang, et al. |
| EPI_ISL_633056 | DOHMH Central Harlem | New York City Public Health Laboratory | Jade Wang, et al. |
| EPI_ISL_633057, EPI_ISL_633058 | DOHMH Morrisania | New York City Public Health Laboratory | Jade Wang, et al. |
| EPI_ISL_633059 | DOHMH Corona | New York City Public Health Laboratory | Jade Wang, et al. |
| EPI_ISL_633060, EPI_ISL_633061, EPI_ISL_633062 | DOHMH Jamaica | New York City Public Health Laboratory | Jade Wang, et al. |
| EPI_ISL_633063 | DOHMH Crown Heights | New York City Public Health Laboratory | Jade Wang, et al. |
| EPI_ISL_633064 | DOHMH Jamaica | New York City Public Health Laboratory | Jade Wang, et al. |
| EPI_ISL_633065 | DOHMH Morrisania | New York City Public Health Laboratory | Jade Wang, et al. |
| EPI_ISL_633066 | DOHMH Chelsea | New York City Public Health Laboratory | Jade Wang, et al. |
| EPI_ISL_633067, EPI_ISL_633068 | DOHMH Morrisania | New York City Public Health Laboratory | Jade Wang, et al. |
| EPI_ISL_633069 | DOHMH Chelsea | New York City Public Health Laboratory | Jade Wang, et al. |
| EPI_ISL_633070 | Flushing Hospital Medical Center | New York City Public Health Laboratory | Jade Wang, et al. |
| EPI_ISL_633071 | DOHMH PHL | New York City Public Health Laboratory | Jade Wang, et al. |
| EPI_ISL_633072 | DOHMH Riverside | New York City Public Health Laboratory | Jade Wang, et al. |
| EPI_ISL_633073 | DOHMH Fort Greene | New York City Public Health Laboratory | Jade Wang, et al. |
| EPI_ISL_633074 | DOHMH Corona | New York City Public Health Laboratory | Jade Wang, et al. |
| EPI_ISL_633075 | DOHMH Morrisania | New York City Public Health Laboratory | Jade Wang, et al. |
| EPI_ISL_633076, EPI_ISL_633077, EPI_ISL_633078 | DOHMH Chelsea | New York City Public Health Laboratory | Jade Wang, et al. |
| EPI_ISL_633079 | DOHMH PHL | New York City Public Health Laboratory | Jade Wang, et al. |
| EPI_ISL_633080, EPI_ISL_633081, EPI_ISL_633082 | DOHMH Morrisania | New York City Public Health Laboratory | Jade Wang, et al. |
| EPI_ISL_633083, EPI_ISL_633084, EPI_ISL_633085, EPI_ISL_633086, EPI_ISL_633087, EPI_ISL_633088, EPI_ISL_633089, EPI_ISL_633090, EPI_ISL_633091, EPI_ISL_633092, EPI_ISL_633093, EPI_ISL_633094, EPI_ISL_633095, EPI_ISL_633096, EPI_ISL_633097, EPI_ISL_633098, EPI_ISL_633099 | | | |
| see above | MD PHL | MD PHL | Maryland Department of Health Laboratories Administration |
| EPI_ISL_634838, EPI_ISL_634839, EPI_ISL_634840, EPI_ISL_634841, EPI_ISL_634842, EPI_ISL_634843, EPI_ISL_634844, EPI_ISL_634845, EPI_ISL_634846, EPI_ISL_634847, EPI_ISL_634848, EPI_ISL_634849, EPI_ISL_634850, EPI_ISL_634851, EPI_ISL_634852, EPI_ISL_634853, EPI_ISL_634854, EPI_ISL_634855, EPI_ISL_634856 | | | |
| see above | Minnesota Department of Health, Public Health Laboratory | Minnesota Department of Health, Public Health Laboratory | Matt Plumb, Jacob Garfin, Alexandra Lorentz, and Xiong Wang |
| EPI_ISL_634857 | Minneapolis VA Health Care System | Minnesota Department of Health, Public Health Laboratory | Matt Plumb, Jacob Garfin, Alexandra Lorentz, and Xiong Wang |
| EPI_ISL_634858, EPI_ISL_634859, EPI_ISL_634860, EPI_ISL_634861, EPI_ISL_634862, EPI_ISL_634863 | Minnesota Department of Health, Public Health Laboratory | Minnesota Department of Health, Public Health Laboratory | Matt Plumb, Jacob Garfin, Alexandra Lorentz, and Xiong Wang |
| EPI_ISL_634864, EPI_ISL_634865 | HealthPartners Central Lab | Minnesota Department of Health, Public Health Laboratory | Matt Plumb, Jacob Garfin, Alexandra Lorentz, and Xiong Wang |
| EPI_ISL_634866, EPI_ISL_634867, EPI_ISL_634868, EPI_ISL_634869, EPI_ISL_634870, EPI_ISL_634871, EPI_ISL_634872, EPI_ISL_634873, EPI_ISL_634874, EPI_ISL_634875 | Minnesota Department of Health, Public Health Laboratory | Minnesota Department of Health, Public Health Laboratory | Matt Plumb, Jacob Garfin, Alexandra Lorentz, and Xiong Wang |
| EPI_ISL_634896, EPI_ISL_634897, EPI_ISL_634898, EPI_ISL_634899, EPI_ISL_634900, EPI_ISL_634901, EPI_ISL_634902, EPI_ISL_634903, EPI_ISL_634904, EPI_ISL_634905, EPI_ISL_634906, EPI_ISL_634907, EPI_ISL_634908, EPI_ISL_634909, EPI_ISL_634911, EPI_ISL_634912, EPI_ISL_634913, EPI_ISL_634914, EPI_ISL_634915, EPI_ISL_634916, EPI_ISL_634918, EPI_ISL_634919, EPI_ISL_634920, EPI_ISL_634921, EPI_ISL_634922, EPI_ISL_634923, EPI_ISL_634924, EPI_ISL_634925, EPI_ISL_634926, EPI_ISL_634927, EPI_ISL_634928, EPI_ISL_634929, EPI_ISL_634931, EPI_ISL_634932, EPI_ISL_634933 | | | |
| see above | Utah Public Health Laboratory | Utah Public Health Laboratory | Erin L. Young, Kelly F. Oakeson |
| EPI_ISL_634934, EPI_ISL_634935, EPI_ISL_634937, EPI_ISL_634938, EPI_ISL_634941, EPI_ISL_634942, EPI_ISL_634943, EPI_ISL_634944, EPI_ISL_634946, EPI_ISL_634947, EPI_ISL_634948, EPI_ISL_634949, EPI_ISL_634950, EPI_ISL_634951, EPI_ISL_634952, EPI_ISL_634953, EPI_ISL_634954, EPI_ISL_634955, EPI_ISL_634956, EPI_ISL_634958, EPI_ISL_634959, EPI_ISL_634961, EPI_ISL_634962, EPI_ISL_634964, EPI_ISL_634966, EPI_ISL_634967, EPI_ISL_634968, EPI_ISL_634969, EPI_ISL_634970, EPI_ISL_634971, EPI_ISL_634972, EPI_ISL_634973, EPI_ISL_634975 | | | |
| see above | Texas Department of State Health Services | Texas Department of State Health Services | Rashmi Tuladhar, Bonnie Oh, Jenny Zhang, Maliha Rahman, Anita Pokharel, Myong Koag, Chung Wang, Rachel Lee, Grace Kubin, Mayela Pedrueza |
| EPI_ISL_635300, EPI_ISL_635301, EPI_ISL_635302, EPI_ISL_635303, EPI_ISL_635304, EPI_ISL_635305, EPI_ISL_635306, EPI_ISL_635307, EPI_ISL_635308, EPI_ISL_635309, EPI_ISL_635310, EPI_ISL_635311, EPI_ISL_635312, EPI_ISL_635313, EPI_ISL_635314, EPI_ISL_635315, EPI_ISL_635316, EPI_ISL_635317, EPI_ISL_635318, EPI_ISL_635319, EPI_ISL_635320, EPI_ISL_635321, EPI_ISL_635322, EPI_ISL_635323, EPI_ISL_635324, EPI_ISL_635325, EPI_ISL_635326, EPI_ISL_635327, EPI_ISL_635328, EPI_ISL_635329, EPI_ISL_635330, EPI_ISL_635331, EPI_ISL_635332, EPI_ISL_635333, EPI_ISL_635334, EPI_ISL_635335, EPI_ISL_635336, EPI_ISL_635337, EPI_ISL_635338, EPI_ISL_635339, EPI_ISL_635340, EPI_ISL_635341, EPI_ISL_635342, EPI_ISL_635343, EPI_ISL_635344, EPI_ISL_635345, EPI_ISL_635346, EPI_ISL_635347, EPI_ISL_635348, EPI_ISL_635349, EPI_ISL_635350, EPI_ISL_635351, EPI_ISL_635352, EPI_ISL_635353, EPI_ISL_635354, EPI_ISL_635355, EPI_ISL_635356, EPI_ISL_635357, EPI_ISL_635358, EPI_ISL_635359, EPI_ISL_635360, EPI_ISL_635361, EPI_ISL_635362, EPI_ISL_635363, EPI_ISL_635364, EPI_ISL_635365, EPI_ISL_635366, EPI_ISL_635367, EPI_ISL_635368, EPI_ISL_635369, EPI_ISL_635370, EPI_ISL_635371, EPI_ISL_635372, EPI_ISL_635373, EPI_ISL_635374, EPI_ISL_635375, EPI_ISL_635376, EPI_ISL_635377, EPI_ISL_635378, EPI_ISL_635379, EPI_ISL_635380, EPI_ISL_635381, EPI_ISL_635382, EPI_ISL_635383, EPI_ISL_635384, EPI_ISL_635385, EPI_ISL_635386, EPI_ISL_635387, EPI_ISL_635388, EPI_ISL_635389, EPI_ISL_635390, EPI_ISL_635391, EPI_ISL_635392, EPI_ISL_635393, EPI_ISL_635394, EPI_ISL_635395, EPI_ISL_635396, EPI_ISL_635397, EPI_ISL_635398, EPI_ISL_635399, EPI_ISL_635400, EPI_ISL_635401, EPI_ISL_635402, EPI_ISL_635403, EPI_ISL_635404, EPI_ISL_635405, EPI_ISL_635406, EPI_ISL_635407, EPI_ISL_635408, EPI_ISL_635409, EPI_ISL_635410, EPI_ISL_635411, EPI_ISL_635412, EPI_ISL_635413, EPI_ISL_635414, EPI_ISL_635415, EPI_ISL_635416, EPI_ISL_635417, EPI_ISL_635418, EPI_ISL_635419, EPI_ISL_635420, EPI_ISL_635421, EPI_ISL_635422, EPI_ISL_635423, EPI_ISL_635424, EPI_ISL_635425, EPI_ISL_635426, EPI_ISL_635427, EPI_ISL_635428, EPI_ISL_635429, EPI_ISL_635430, EPI_ISL_635431, EPI_ISL_635432, EPI_ISL_635433, EPI_ISL_635434, EPI_ISL_635435, EPI_ISL_635436, EPI_ISL_635437, EPI_ISL_635438, EPI_ISL_635439, EPI_ISL_635440, EPI_ISL_635441, EPI_ISL_635442, EPI_ISL_635443, EPI_ISL_635444, EPI_ISL_635445, EPI_ISL_635446, EPI_ISL_635447, EPI_ISL_635448, EPI_ISL_635449, EPI_ISL_635450, EPI_ISL_635451, EPI_ISL_635452, EPI_ISL_635453, EPI_ISL_635454, EPI_ISL_635455, EPI_ISL_635456, EPI_ISL_635457, EPI_ISL_635458, EPI_ISL_635459, EPI_ISL_635460, EPI_ISL_635461, EPI_ISL_635462, EPI_ISL_635463, EPI_ISL_635464, EPI_ISL_635465, EPI_ISL_635466, EPI_ISL_635467, EPI_ISL_635468, EPI_ISL_635469, EPI_ISL_635470, EPI_ISL_635471, EPI_ISL_635472, EPI_ISL_635473, EPI_ISL_635474, EPI_ISL_635475, EPI_ISL_635476, EPI_ISL_635477, EPI_ISL_635478 | | | |
| see above | San Diego County Public Health Laboratory | Andersen lab at Scripps Research | SEARCH Alliance San Diego with Tracy Basler, Jovan Shephard, Brett Austin |
| EPI_ISL_635479, EPI_ISL_635480, EPI_ISL_635481, EPI_ISL_635482, EPI_ISL_635483, EPI_ISL_635484, EPI_ISL_635485, EPI_ISL_635486, EPI_ISL_635487, EPI_ISL_635488, EPI_ISL_635489, EPI_ISL_635490, EPI_ISL_635491, EPI_ISL_635492, EPI_ISL_635493, EPI_ISL_635494, EPI_ISL_635495, EPI_ISL_635496, EPI_ISL_635497, EPI_ISL_635498, EPI_ISL_635499, EPI_ISL_635500, EPI_ISL_635501, EPI_ISL_635502, EPI_ISL_635503, EPI_ISL_635504, EPI_ISL_635505, EPI_ISL_635506, EPI_ISL_635507, EPI_ISL_635508, EPI_ISL_635509, EPI_ISL_635510, EPI_ISL_635511, EPI_ISL_635512, EPI_ISL_635513, EPI_ISL_635514, EPI_ISL_635515, EPI_ISL_635516, EPI_ISL_635517, EPI_ISL_635518, EPI_ISL_635519, EPI_ISL_635520, EPI_ISL_635521, EPI_ISL_635522, EPI_ISL_635523, EPI_ISL_635524, EPI_ISL_635525, EPI_ISL_635526, EPI_ISL_635527, EPI_ISL_635528, EPI_ISL_635529, EPI_ISL_635530, EPI_ISL_635531, EPI_ISL_635532, EPI_ISL_635533, EPI_ISL_635534, EPI_ISL_635535, EPI_ISL_635536, EPI_ISL_635537, EPI_ISL_635538, EPI_ISL_635539, EPI_ISL_635540, EPI_ISL_635541, EPI_ISL_635542, EPI_ISL_635543, EPI_ISL_635544, EPI_ISL_635545, EPI_ISL_635546, EPI_ISL_635547, EPI_ISL_635548, EPI_ISL_635549, EPI_ISL_635550, EPI_ISL_635551, EPI_ISL_635552, EPI_ISL_635553, EPI_ISL_635554, EPI_ISL_635555, EPI_ISL_635556, EPI_ISL_635557, EPI_ISL_635558, EPI_ISL_635559, EPI_ISL_635560, EPI_ISL_635561, EPI_ISL_635562, EPI_ISL_635563, EPI_ISL_635564, EPI_ISL_635565, EPI_ISL_635566, EPI_ISL_635567, EPI_ISL_635568, EPI_ISL_635569, EPI_ISL_635570, EPI_ISL_635571, EPI_ISL_635572, EPI_ISL_635573, EPI_ISL_635574, EPI_ISL_635575, EPI_ISL_635576 | | | |
| see above | Centro de Diagn√≥stico COVID-19 UABC Tijuana | Andersen lab at Scripps Research | SEARCH Alliance San Diego with Idanya Rubí Serafín Higuera, Manuel Sánchez Alavez, Jorge Luis Jiménez Niebla, Germán Ibarra, Jonathan Vincent Baena, Oscar Efrén Zazueta Fierro |
| EPI_ISL_635577, EPI_ISL_635578, EPI_ISL_635579, EPI_ISL_635580, EPI_ISL_635581, EPI_ISL_635582, EPI_ISL_635583, EPI_ISL_635584, EPI_ISL_635585, EPI_ISL_635586, EPI_ISL_635587, EPI_ISL_635588, EPI_ISL_635589, EPI_ISL_635590, EPI_ISL_635591, EPI_ISL_635592, EPI_ISL_635593, EPI_ISL_635594, EPI_ISL_635595, EPI_ISL_635596, EPI_ISL_635597, EPI_ISL_635598, EPI_ISL_635599, EPI_ISL_635600, EPI_ISL_635601, EPI_ISL_635602, EPI_ISL_635603, EPI_ISL_635604, EPI_ISL_635605, EPI_ISL_635606, EPI_ISL_635607, EPI_ISL_635608, EPI_ISL_635609, EPI_ISL_635610, EPI_ISL_635611, EPI_ISL_635612, EPI_ISL_635613, EPI_ISL_635614, EPI_ISL_635615, EPI_ISL_635616, EPI_ISL_635617, EPI_ISL_635618, EPI_ISL_635619, EPI_ISL_635620, EPI_ISL_635621, EPI_ISL_635622, EPI_ISL_635623, EPI_ISL_635624, EPI_ISL_635625, EPI_ISL_635626, EPI_ISL_635627, EPI_ISL_635628, EPI_ISL_635629, EPI_ISL_635630, EPI_ISL_635631, EPI_ISL_635632, EPI_ISL_635633, EPI_ISL_635634, EPI_ISL_635635, EPI_ISL_635636, EPI_ISL_635637, EPI_ISL_635638, EPI_ISL_635639, EPI_ISL_635640, EPI_ISL_635641, EPI_ISL_635642, EPI_ISL_635643, EPI_ISL_635644, EPI_ISL_635645, EPI_ISL_635646, EPI_ISL_635647, EPI_ISL_635648, EPI_ISL_635649, EPI_ISL_635650, EPI_ISL_635651, EPI_ISL_635652, EPI_ISL_635653, EPI_ISL_635654, EPI_ISL_635655, EPI_ISL_635656, EPI_ISL_635657, EPI_ISL_635658, EPI_ISL_635659, EPI_ISL_635660, EPI_ISL_635661, EPI_ISL_635662, EPI_ISL_635663, EPI_ISL_635664, EPI_ISL_635665, EPI_ISL_635666, EPI_ISL_635667, EPI_ISL_635668, EPI_ISL_635669, EPI_ISL_635670, EPI_ISL_635671, EPI_ISL_635672, EPI_ISL_635673, EPI_ISL_635674, EPI_ISL_635675, EPI_ISL_635676, EPI_ISL_635677, EPI_ISL_635678, EPI_ISL_635679, EPI_ISL_635680, EPI_ISL_635681, EPI_ISL_635682, EPI_ISL_635683, EPI_ISL_635684, EPI_ISL_635685, EPI_ISL_635686, EPI_ISL_635687, EPI_ISL_635688, EPI_ISL_635689, EPI_ISL_635690, EPI_ISL_635691, EPI_ISL_635692, EPI_ISL_635693, EPI_ISL_635694, EPI_ISL_635695, EPI_ISL_635696, EPI_ISL_635697, EPI_ISL_635698, EPI_ISL_635699, EPI_ISL_635700, EPI_ISL_635701, EPI_ISL_635702, EPI_ISL_635703, EPI_ISL_635704, EPI_ISL_635705, EPI_ISL_635706, EPI_ISL_635707, EPI_ISL_635708, EPI_ISL_635709, EPI_ISL_635710, EPI_ISL_635711, EPI_ISL_635712, EPI_ISL_635713, EPI_ISL_635714, EPI_ISL_635715, EPI_ISL_635716, EPI_ISL_635717, EPI_ISL_635718, EPI_ISL_635719, EPI_ISL_635720, EPI_ISL_635721, EPI_ISL_635722, EPI_ISL_635723, EPI_ISL_635724, EPI_ISL_635725, EPI_ISL_635726, EPI_ISL_635727, EPI_ISL_635728, EPI_ISL_635729, EPI_ISL_635730, EPI_ISL_635731, EPI_ISL_635732, EPI_ISL_635733, EPI_ISL_635734, EPI_ISL_635735, EPI_ISL_635736, EPI_ISL_635737, EPI_ISL_635738, EPI_ISL_635739, EPI_ISL_635740, EPI_ISL_635741, EPI_ISL_635742, EPI_ISL_635743, EPI_ISL_635744, EPI_ISL_635745, EPI_ISL_635746, EPI_ISL_635747, EPI_ISL_635748, EPI_ISL_635749, EPI_ISL_635750, EPI_ISL_635751, EPI_ISL_635752, EPI_ISL_635753, EPI_ISL_635754, EPI_ISL_635755, EPI_ISL_635756, EPI_ISL_635757, EPI_ISL_635758, EPI_ISL_635759, EPI_ISL_635760, EPI_ISL_635761, EPI_ISL_635762, EPI_ISL_635763, EPI_ISL_635764, EPI_ISL_635765, EPI_ISL_635766, EPI_ISL_635767, EPI_ISL_635768, EPI_ISL_635769, EPI_ISL_635770, EPI_ISL_635771, EPI_ISL_635772, EPI_ISL_635773, EPI_ISL_635774, EPI_ISL_635775, EPI_ISL_635776, EPI_ISL_635783, EPI_ISL_635784, EPI_ISL_635785, EPI_ISL_635786, EPI_ISL_635787, EPI_ISL_635788, EPI_ISL_635789, EPI_ISL_635790, EPI_ISL_635791, EPI_ISL_635792, EPI_ISL_635793, EPI_ISL_635794, EPI_ISL_635795, EPI_ISL_635796, EPI_ISL_635797, EPI_ISL_635798, EPI_ISL_635799, EPI_ISL_635800, EPI_ISL_635801, EPI_ISL_635802, EPI_ISL_635803, EPI_ISL_635804, EPI_ISL_635805, EPI_ISL_635806, EPI_ISL_635807, EPI_ISL_635808, EPI_ISL_635809, EPI_ISL_635810, EPI_ISL_635811, EPI_ISL_635812, EPI_ISL_635813, EPI_ISL_635814, EPI_ISL_635815, EPI_ISL_635816, EPI_ISL_635817, EPI_ISL_635818, EPI_ISL_635819, EPI_ISL_635820, EPI_ISL_635821, EPI_ISL_635822, EPI_ISL_635823, EPI_ISL_635824, EPI_ISL_635825, EPI_ISL_635826, EPI_ISL_635827, EPI_ISL_635828, EPI_ISL_635829, EPI_ISL_635830, EPI_ISL_635831, EPI_ISL_635832, EPI_ISL_635833, EPI_ISL_635834, EPI_ISL_635835, EPI_ISL_635836, EPI_ISL_635837, EPI_ISL_635838, EPI_ISL_635839, EPI_ISL_635840, EPI_ISL_635841, EPI_ISL_635842, EPI_ISL_635843, EPI_ISL_635844, EPI_ISL_635845, EPI_ISL_635846, EPI_ISL_635847, EPI_ISL_635848, EPI_ISL_635849, EPI_ISL_635850, EPI_ISL_635851, EPI_ISL_635852, EPI_ISL_635853, EPI_ISL_635854, EPI_ISL_635855, EPI_ISL_635856, EPI_ISL_635857, EPI_ISL_635858, EPI_ISL_635859, EPI_ISL_635860, EPI_ISL_635861, EPI_ISL_635862, EPI_ISL_635863, EPI_ISL_635864, EPI_ISL_635865, EPI_ISL_635866, EPI_ISL_635867, EPI_ISL_635868, EPI_ISL_635869, EPI_ISL_635870, | | | |

| | | | |
|---|---|---|---|
| EPI_ISL_635871, EPI_ISL_635872, EPI_ISL_635873, EPI_ISL_635874, EPI_ISL_635875, EPI_ISL_635876, EPI_ISL_635877, EPI_ISL_635878, EPI_ISL_635879, EPI_ISL_635880, EPI_ISL_635881, EPI_ISL_635882, EPI_ISL_635883, EPI_ISL_635884, EPI_ISL_635885, EPI_ISL_635886, EPI_ISL_635887, EPI_ISL_635888, EPI_ISL_635889, EPI_ISL_635890, EPI_ISL_635891, EPI_ISL_635892, EPI_ISL_635893, EPI_ISL_635894, EPI_ISL_635895, EPI_ISL_635896, EPI_ISL_635897, EPI_ISL_635898, EPI_ISL_635899, EPI_ISL_635900, EPI_ISL_635901, EPI_ISL_635902, EPI_ISL_635903, EPI_ISL_635904, EPI_ISL_635905, EPI_ISL_635906, EPI_ISL_635907, EPI_ISL_635908, EPI_ISL_635909, EPI_ISL_635910, EPI_ISL_635911, EPI_ISL_635912, EPI_ISL_635913, EPI_ISL_635914, EPI_ISL_635915, EPI_ISL_635916, EPI_ISL_635917, EPI_ISL_635918, EPI_ISL_635919, EPI_ISL_635920, EPI_ISL_635921, EPI_ISL_635922, EPI_ISL_635923, EPI_ISL_635924, EPI_ISL_635925, EPI_ISL_635926, EPI_ISL_635927, EPI_ISL_635928, EPI_ISL_635929, EPI_ISL_635930, EPI_ISL_635931, EPI_ISL_635932, EPI_ISL_635933, EPI_ISL_635934, EPI_ISL_635935, EPI_ISL_635936, EPI_ISL_635937, EPI_ISL_635938, EPI_ISL_635939, EPI_ISL_635940, EPI_ISL_635941, EPI_ISL_635942, EPI_ISL_635943, EPI_ISL_635944, EPI_ISL_635945, EPI_ISL_635946, EPI_ISL_635947, EPI_ISL_635948, EPI_ISL_635949, EPI_ISL_635950, EPI_ISL_635951, EPI_ISL_635952, EPI_ISL_635953, EPI_ISL_635954, EPI_ISL_635955, EPI_ISL_635956, EPI_ISL_635957, EPI_ISL_635958, EPI_ISL_635959, EPI_ISL_635960, EPI_ISL_635961, EPI_ISL_635962, EPI_ISL_635963, EPI_ISL_635964, EPI_ISL_635965, EPI_ISL_635966, EPI_ISL_635967, EPI_ISL_635968, EPI_ISL_635969, EPI_ISL_635970, EPI_ISL_635971, EPI_ISL_635972, EPI_ISL_635973, EPI_ISL_635974, EPI_ISL_635975, EPI_ISL_635976, EPI_ISL_635977, EPI_ISL_635978, EPI_ISL_635979, EPI_ISL_635980, EPI_ISL_635981, EPI_ISL_635982, EPI_ISL_635983, EPI_ISL_635984, EPI_ISL_635985, EPI_ISL_635986, EPI_ISL_635987, EPI_ISL_635988, EPI_ISL_635989, EPI_ISL_635990, EPI_ISL_635991, EPI_ISL_635992, EPI_ISL_635993, EPI_ISL_635994, EPI_ISL_635995, EPI_ISL_635996, EPI_ISL_635997, EPI_ISL_635998, EPI_ISL_635999, EPI_ISL_636000, EPI_ISL_636001, EPI_ISL_636002, EPI_ISL_636003, EPI_ISL_636004, EPI_ISL_636005, EPI_ISL_636006, EPI_ISL_636007, EPI_ISL_636008, EPI_ISL_636009, EPI_ISL_636010, EPI_ISL_636011, EPI_ISL_636012, EPI_ISL_636013, EPI_ISL_636014, EPI_ISL_636015, EPI_ISL_636016, EPI_ISL_636017, EPI_ISL_636018, EPI_ISL_636019, EPI_ISL_636020, EPI_ISL_636021, EPI_ISL_636022, EPI_ISL_636023, EPI_ISL_636024, EPI_ISL_636025, EPI_ISL_636026, EPI_ISL_636027, EPI_ISL_636028, EPI_ISL_636029, EPI_ISL_636030, EPI_ISL_636031, EPI_ISL_636032, EPI_ISL_636033, EPI_ISL_636034, EPI_ISL_636035, EPI_ISL_636036, EPI_ISL_636037, EPI_ISL_636038, EPI_ISL_636039, EPI_ISL_636040, EPI_ISL_636041, EPI_ISL_636042, EPI_ISL_636043, EPI_ISL_636044, EPI_ISL_636045, EPI_ISL_636046, EPI_ISL_636047, EPI_ISL_636048, EPI_ISL_636049, EPI_ISL_636050, EPI_ISL_636051, EPI_ISL_636052, EPI_ISL_636053, EPI_ISL_636054, EPI_ISL_636055, EPI_ISL_636056, EPI_ISL_636057, EPI_ISL_636058, EPI_ISL_636059, EPI_ISL_636060, EPI_ISL_636061, EPI_ISL_636062, EPI_ISL_636063, EPI_ISL_636064, EPI_ISL_636065, EPI_ISL_636066, EPI_ISL_636067, EPI_ISL_636068, EPI_ISL_636069, EPI_ISL_636070, EPI_ISL_636071, EPI_ISL_636072, EPI_ISL_636073, EPI_ISL_636074, EPI_ISL_636075, EPI_ISL_636076, EPI_ISL_636077, EPI_ISL_636078, EPI_ISL_636079, EPI_ISL_636080, EPI_ISL_636081, EPI_ISL_636082, EPI_ISL_636083, EPI_ISL_636084, EPI_ISL_636085, EPI_ISL_636086, EPI_ISL_636087, EPI_ISL_636088, EPI_ISL_636089, EPI_ISL_636090, EPI_ISL_636091, EPI_ISL_636092, EPI_ISL_636093, EPI_ISL_636094, EPI_ISL_636095, EPI_ISL_636096, EPI_ISL_636097, EPI_ISL_636098, EPI_ISL_636099, EPI_ISL_636100, EPI_ISL_636101, EPI_ISL_636102, EPI_ISL_636103, EPI_ISL_636104, EPI_ISL_636105, EPI_ISL_636106, EPI_ISL_636107, EPI_ISL_636108, EPI_ISL_636109, EPI_ISL_636110, EPI_ISL_636111, EPI_ISL_636112 | | | |
| see above | San Diego County Public Health Laboratory | Andersen lab at Scripps Research | SEARCH Alliance San Diego with Tracy Basler, Jovan Shephard, Brett Austin |
| EPI_ISL_636113, EPI_ISL_636114, EPI_ISL_636115, EPI_ISL_636116, EPI_ISL_636117, EPI_ISL_636118, EPI_ISL_636119, EPI_ISL_636120 | Sharp HealthCare Laboratory | Andersen lab at Scripps Research | SEARCH Alliance San Diego with Aaron Harding, Jacquelyn Berumen, Cathy Woerle, Liam McGinnis |
| EPI_ISL_636121, EPI_ISL_636122, EPI_ISL_636123, EPI_ISL_636124, EPI_ISL_636125, EPI_ISL_636126, EPI_ISL_636127, EPI_ISL_636128, EPI_ISL_636129, EPI_ISL_636130, EPI_ISL_636131, EPI_ISL_636132, EPI_ISL_636133, EPI_ISL_636134, EPI_ISL_636135, EPI_ISL_636136, EPI_ISL_636137, EPI_ISL_636138, EPI_ISL_636139, EPI_ISL_636140, EPI_ISL_636141, EPI_ISL_636142, EPI_ISL_636143, EPI_ISL_636144, EPI_ISL_636145, EPI_ISL_636146, EPI_ISL_636147, EPI_ISL_636148, EPI_ISL_636149, EPI_ISL_636150, EPI_ISL_636151, EPI_ISL_636152, EPI_ISL_636153, EPI_ISL_636154, EPI_ISL_636155, EPI_ISL_636156, EPI_ISL_636157, EPI_ISL_636158, EPI_ISL_636159, EPI_ISL_636160, EPI_ISL_636161, EPI_ISL_636162, EPI_ISL_636163, EPI_ISL_636164, EPI_ISL_636165, EPI_ISL_636166, EPI_ISL_636167, EPI_ISL_636168, EPI_ISL_636169, EPI_ISL_636170, EPI_ISL_636171, EPI_ISL_636172, EPI_ISL_636173, EPI_ISL_636174, EPI_ISL_636175, EPI_ISL_636176, EPI_ISL_636177, EPI_ISL_636178, EPI_ISL_636179, EPI_ISL_636180, EPI_ISL_636181, EPI_ISL_636182, EPI_ISL_636183, EPI_ISL_636184, EPI_ISL_636185, EPI_ISL_636186, EPI_ISL_636187, EPI_ISL_636188, EPI_ISL_636189, EPI_ISL_636190, EPI_ISL_636191, EPI_ISL_636192, EPI_ISL_636193, EPI_ISL_636194, EPI_ISL_636195, EPI_ISL_636196, EPI_ISL_636197, EPI_ISL_636198, EPI_ISL_636199, EPI_ISL_636200, EPI_ISL_636201, EPI_ISL_636202, EPI_ISL_636203, EPI_ISL_636204, EPI_ISL_636205, EPI_ISL_636206, EPI_ISL_636207, EPI_ISL_636208, EPI_ISL_636209, EPI_ISL_636210, EPI_ISL_636211, EPI_ISL_636212, EPI_ISL_636213, EPI_ISL_636214, EPI_ISL_636215, EPI_ISL_636216, EPI_ISL_636217, EPI_ISL_636218, EPI_ISL_636219, EPI_ISL_636220, EPI_ISL_636221, EPI_ISL_636222, EPI_ISL_636223, EPI_ISL_636224, EPI_ISL_636225, EPI_ISL_636226, EPI_ISL_636227, EPI_ISL_636228, EPI_ISL_636229, EPI_ISL_636230, EPI_ISL_636231, EPI_ISL_636232, EPI_ISL_636233, EPI_ISL_636234, EPI_ISL_636235, EPI_ISL_636236, EPI_ISL_636237, EPI_ISL_636238, EPI_ISL_636239, EPI_ISL_636240, EPI_ISL_636241, EPI_ISL_636242, EPI_ISL_636243, EPI_ISL_636244, EPI_ISL_636245, EPI_ISL_636246, EPI_ISL_636247, EPI_ISL_636248, EPI_ISL_636249, EPI_ISL_636250, EPI_ISL_636251, EPI_ISL_636252, EPI_ISL_636253, EPI_ISL_636254, EPI_ISL_636255, EPI_ISL_636256, EPI_ISL_636257, EPI_ISL_636258, EPI_ISL_636259, EPI_ISL_636260, EPI_ISL_636261, EPI_ISL_636262, EPI_ISL_636263, EPI_ISL_636264, EPI_ISL_636265, EPI_ISL_636266 | | | |
| see above | San Diego County Public Health Laboratory | Andersen lab at Scripps Research | SEARCH Alliance San Diego with Tracy Basler, Jovan Shephard, Brett Austin |
| EPI_ISL_640142, EPI_ISL_640143, EPI_ISL_640144, EPI_ISL_640145, EPI_ISL_640146, EPI_ISL_640147, EPI_ISL_640148, EPI_ISL_640149, EPI_ISL_640150, EPI_ISL_640151, EPI_ISL_640152, EPI_ISL_640153, EPI_ISL_640154, EPI_ISL_640155, EPI_ISL_640156, EPI_ISL_640157, EPI_ISL_640158, EPI_ISL_640159, EPI_ISL_640160, EPI_ISL_640161, EPI_ISL_640162, EPI_ISL_640163, EPI_ISL_640164, EPI_ISL_640165, EPI_ISL_640166, EPI_ISL_640167, EPI_ISL_640168, EPI_ISL_640169, EPI_ISL_640170, EPI_ISL_640171, EPI_ISL_640172, EPI_ISL_640173, EPI_ISL_640174, EPI_ISL_640175, EPI_ISL_640176, EPI_ISL_640177, EPI_ISL_640178, EPI_ISL_640179, EPI_ISL_640180, EPI_ISL_640181, EPI_ISL_640182, EPI_ISL_640183, EPI_ISL_640184, EPI_ISL_640185, EPI_ISL_640186, EPI_ISL_640187, EPI_ISL_640188, EPI_ISL_640189, EPI_ISL_640190, EPI_ISL_640191, EPI_ISL_640192, EPI_ISL_640193, EPI_ISL_640194, EPI_ISL_640195, EPI_ISL_640196, EPI_ISL_640197, EPI_ISL_640198, EPI_ISL_640199, EPI_ISL_640200, EPI_ISL_640201, EPI_ISL_640202, EPI_ISL_640203, EPI_ISL_640204, EPI_ISL_640205, EPI_ISL_640206, EPI_ISL_640207, EPI_ISL_640208, EPI_ISL_640209, EPI_ISL_640210, EPI_ISL_640211, EPI_ISL_640212, EPI_ISL_640213, EPI_ISL_640214, EPI_ISL_640215, EPI_ISL_640216, EPI_ISL_640217, EPI_ISL_640218, EPI_ISL_640273, EPI_ISL_640274, EPI_ISL_640275 | | | |
| see above | University of Michigan Clinical Microbiology Laboratory | Lauring Lab, University of Michigan, Department of Microbiology and Immunology | Valesano |
| EPI_ISL_640279, EPI_ISL_640280, EPI_ISL_640281, EPI_ISL_640288, EPI_ISL_640289, EPI_ISL_640290, EPI_ISL_640298, EPI_ISL_640299, EPI_ISL_640300 | Utah Public Health Laboratory | Utah Public Health Laboratory | Erin Young, Kelly Oakeson |
| EPI_ISL_641311 | Respiratory virus Laboratory, Chinese Academy of Medical Science | Respiratory virus Laboratory, Chinese Academy of Medical Science | Li,J., Zhen,H., Chen,Y. and Liu,L. |
| EPI_ISL_641312, EPI_ISL_641313, EPI_ISL_641314, EPI_ISL_641315, EPI_ISL_641316, EPI_ISL_641317 | Rocky Mountain Laboratories, RTS Genomics Unit, National Institute of Allergy and Infectious Diseases, National Institutes of Health | Rocky Mountain Laboratories, RTS Genomics Unit, National Institute of Allergy and Infectious Diseases, National Institutes of Health | Avanzato,V.A., Matson,M.J., Seifert,S.N., Pryce,R., Williamson,B.N., Anzick,S.L., Barbian,K., Judson,S.D., Fischer,E.R., Martens,C., Bowden,T.A., de Wit,E., Riedo,F.X., Munster,V.J., Siefert,S.N., WIlliamson,B.N., Anzick,S., Martens,C.A. |
| EPI_ISL_641318 | Environmental and Global Health, University of Florida | Environmental and Global Health, University of Florida | Nannu-Shankar,S., Witanachchi,C.T., Alam,M.M., Loeb,J.C., Stephenson,C.J., Fan,Z.H., Wu,C.-Y., Lednicky,J.A. |
| EPI_ISL_641319 | Environmental and Global Health, University of Florida | Environmental and Global Health, University of Florida | Elbadry,M.A., Subramaniam,K., Waltzek,T.B., Loeb,J.C., Stephenson,C.J., Lauzardo,M., Morris,J.G., Lednicky,J.A. |
| EPI_ISL_644163, EPI_ISL_644164, EPI_ISL_644165, EPI_ISL_644166, EPI_ISL_644167, EPI_ISL_644168, EPI_ISL_644169, EPI_ISL_644170, EPI_ISL_644171, EPI_ISL_644172, EPI_ISL_644173, EPI_ISL_644174, EPI_ISL_644176, EPI_ISL_644177, EPI_ISL_644178, EPI_ISL_644179, EPI_ISL_644180, EPI_ISL_644181, EPI_ISL_644184, EPI_ISL_644185, EPI_ISL_644186, EPI_ISL_644188 | | | |
| see above | Texas Department of State Health Services | Texas Department of State Health Services | Rashmi Tuladhar, Bonnie Oh, Jenny Zhang, Maliha Rahman, Anita Pokharel, Myong Koag, Chung Wang, Rachel Lee, Grace Kubin, Mayela Pedrueza |
| EPI_ISL_644189, EPI_ISL_644190, EPI_ISL_644192, EPI_ISL_644193, EPI_ISL_644194, EPI_ISL_644196, EPI_ISL_644199, EPI_ISL_644200, EPI_ISL_644201, EPI_ISL_644203, EPI_ISL_644204, EPI_ISL_644207, EPI_ISL_644208 | | | |
| see above | Texas Department of State Health Services | Texas Department of State Health Services | Rashmi Tuladhar, Bonnie Oh, Jenny Zhang, Maliha Rahman, Anita Pokharel, Myong Koag, Chung Wang, Rachel Lee, Grace Kubin, Mayela Pedrueza, James Daniel Bonser |
| EPI_ISL_644346, EPI_ISL_644347, EPI_ISL_644348, EPI_ISL_644349, EPI_ISL_644350, EPI_ISL_644351, EPI_ISL_644352, EPI_ISL_644353, EPI_ISL_644354, EPI_ISL_644355, EPI_ISL_644356, EPI_ISL_644357, EPI_ISL_644358, EPI_ISL_644359, EPI_ISL_644360, EPI_ISL_644361, EPI_ISL_644362, EPI_ISL_644363, EPI_ISL_644364, EPI_ISL_644365, EPI_ISL_644366, EPI_ISL_644367, EPI_ISL_644368, EPI_ISL_644369, EPI_ISL_644370, EPI_ISL_644371, EPI_ISL_644372, EPI_ISL_644373, EPI_ISL_644374 | | | |
| see above | Michigan Department of Health and Human Services, Bureau of Laboratories | Michigan Department of Health and Human Services, Bureau of Laboratories | Blankenship HM, Riner D, Soehnlen MK |
| EPI_ISL_644584, EPI_ISL_644585, EPI_ISL_644586, EPI_ISL_644587, EPI_ISL_644588, EPI_ISL_644589, EPI_ISL_644590, EPI_ISL_644591, EPI_ISL_644592, EPI_ISL_644593, EPI_ISL_644594, EPI_ISL_644595, EPI_ISL_644596, EPI_ISL_644597, EPI_ISL_644598, EPI_ISL_644599, EPI_ISL_644600, EPI_ISL_644601, EPI_ISL_644602, EPI_ISL_644603, EPI_ISL_644604, EPI_ISL_644605, EPI_ISL_644606, EPI_ISL_644607, EPI_ISL_644608, EPI_ISL_644609 | | | |
| see above | New Mexico Department of Health Scientific Laboratory | New Mexico Department of Health Scientific Laboratory | Ellie Johnson, Anastacia Griego-Fisher, D'Eldra Malone |
| EPI_ISL_644823 | Madigan Army Medical Center | U.S. Air Force School of Aerospace Medicine | Emily Parsons, Matthew Timlin, Clarise Starr, Anthony Fries, Ronald Wells, Mattew Studer, Rebecca Sainato |
| EPI_ISL_644825, EPI_ISL_644826, EPI_ISL_644827, EPI_ISL_644828, EPI_ISL_644829, EPI_ISL_644830, EPI_ISL_644831, EPI_ISL_644832, EPI_ISL_644833, EPI_ISL_644834, EPI_ISL_644835, EPI_ISL_644836, EPI_ISL_644837, EPI_ISL_644838, EPI_ISL_644839, EPI_ISL_644840, EPI_ISL_644841, EPI_ISL_644842, EPI_ISL_644843, EPI_ISL_644844, EPI_ISL_644845, EPI_ISL_644846, EPI_ISL_644847, EPI_ISL_644848, EPI_ISL_644849, EPI_ISL_644850, EPI_ISL_644851, EPI_ISL_644852, EPI_ISL_644853, EPI_ISL_644854, EPI_ISL_644855, EPI_ISL_644856, EPI_ISL_644857, EPI_ISL_644858, EPI_ISL_644859, EPI_ISL_644860, EPI_ISL_644861, EPI_ISL_644862, EPI_ISL_644863, EPI_ISL_644864, EPI_ISL_644865, EPI_ISL_644866, EPI_ISL_644867, EPI_ISL_644868, EPI_ISL_644869, EPI_ISL_644870, EPI_ISL_644871, EPI_ISL_644872, EPI_ISL_644873, EPI_ISL_644874, EPI_ISL_644875, EPI_ISL_644876, EPI_ISL_644877, EPI_ISL_644878, EPI_ISL_644879, EPI_ISL_644880, EPI_ISL_644881, EPI_ISL_644882, EPI_ISL_644883, EPI_ISL_644884, EPI_ISL_644885, EPI_ISL_644886, EPI_ISL_644887, EPI_ISL_644888, EPI_ISL_644889, EPI_ISL_644890, EPI_ISL_644891, EPI_ISL_644892, EPI_ISL_644893, EPI_ISL_644894, EPI_ISL_644895, EPI_ISL_644896, EPI_ISL_644897, EPI_ISL_644898, EPI_ISL_644899, EPI_ISL_644900, EPI_ISL_644901, EPI_ISL_644902, EPI_ISL_644903, EPI_ISL_644904, EPI_ISL_644905, EPI_ISL_644906, EPI_ISL_644907, EPI_ISL_644908, EPI_ISL_644909, EPI_ISL_644910, EPI_ISL_644911, EPI_ISL_644912, EPI_ISL_644913, EPI_ISL_644914, EPI_ISL_644915, EPI_ISL_644916, EPI_ISL_644917, EPI_ISL_644918, EPI_ISL_644919, EPI_ISL_644920, EPI_ISL_644921, EPI_ISL_644922, EPI_ISL_644923, EPI_ISL_644924, EPI_ISL_644925, EPI_ISL_644926, EPI_ISL_644927, EPI_ISL_644928, EPI_ISL_644929, EPI_ISL_644930, EPI_ISL_644931, EPI_ISL_644932 | | | |
| see above | Virginia DCLS | Virginia DCLS | Virginia DCLS |
| EPI_ISL_644933, EPI_ISL_644934, EPI_ISL_644935 | Essentia Health-St. Mary's Medical Center | Minnesota Department of Health, Public Health Laboratory | Matt Plumb, Jacob Garfin, Alexandra Lorentz, and Xiong Wang |
| EPI_ISL_644936, EPI_ISL_644937, EPI_ISL_644938, EPI_ISL_644939, EPI_ISL_644940 | Mayo Clinic & Mayo Clinic Laboratories | Minnesota Department of Health, Public Health Laboratory | Matt Plumb, Jacob Garfin, Alexandra Lorentz, and Xiong Wang |
| EPI_ISL_644941 | Allina Health Laboratory | Minnesota Department of Health, Public Health Laboratory | Matt Plumb, Jacob Garfin, Alexandra Lorentz, and Xiong Wang |
| EPI_ISL_644942 | Hennepin County Medical Center | Minnesota Department of Health, Public Health Laboratory | Matt Plumb, Jacob Garfin, Alexandra Lorentz, and Xiong Wang |
| EPI_ISL_644943 | Essentia Health-St. Mary's Medical Center | Minnesota Department of Health, Public Health Laboratory | Matt Plumb, Jacob Garfin, Alexandra Lorentz, and Xiong Wang |
| EPI_ISL_644944 | M Health Fairview | Minnesota Department of Health, Public Health Laboratory | Matt Plumb, Jacob Garfin, Alexandra Lorentz, and Xiong Wang |
| EPI_ISL_647985 | RI State Health Laboratories | Pathogen Discovery, Respiratory Viruses Branch, Division of Viral Diseases, Centers for Disease Control and Prevention | Yan Li, Jing Zhang, Ying Tao, Brian Lynch, Krista Queen, Anna Montmayeur, Anna Uehara, Clinton R. Paden, Rachel Marine, Haibin Wang, Suxiang Tong |

| | | | |
|---|---|---|---|
| EPI_ISL_647986 | Indiana State Department of Health | Pathogen Discovery, Respiratory Viruses Branch, Division of Viral Diseases, Centers for Disease Control and Prevention | Yan Li, Jing Zhang, Ying Tao, Brian Lynch, Krista Queen, Anna Montmayeur, Anna Uehara, Clinton R. Paden, Rachel Marine, Haibin Wang, Suxiang Tong |
| EPI_ISL_647987, EPI_ISL_647988, EPI_ISL_647989 | GA Department of Public Health Laboratory | Pathogen Discovery, Respiratory Viruses Branch, Division of Viral Diseases, Centers for Disease Control and Prevention | Yan Li, Jing Zhang, Ying Tao, Brian Lynch, Krista Queen, Anna Montmayeur, Anna Uehara, Clinton R. Paden, Rachel Marine, Haibin Wang, Suxiang Tong |
| EPI_ISL_647990, EPI_ISL_647991, EPI_ISL_647992, EPI_ISL_647993, EPI_ISL_647994, EPI_ISL_647995, EPI_ISL_647996, EPI_ISL_647997, EPI_ISL_647998, EPI_ISL_647999, EPI_ISL_648000, EPI_ISL_648001 | | | |
| see above | IA State Hygienic Laboratory | Pathogen Discovery, Respiratory Viruses Branch, Division of Viral Diseases, Centers for Disease Control and Prevention | Ying Tao, Yan Li, Jing Zhang, Brian Lynch, Krista Queen, Anna Uehara, Clinton R. Paden, Haibin Wang, Suxiang Tong |
| EPI_ISL_648002, EPI_ISL_648003 | GA Department of Public Health Laboratory | Pathogen Discovery, Respiratory Viruses Branch, Division of Viral Diseases, Centers for Disease Control and Prevention | Yan Li, Jing Zhang, Ying Tao, Brian Lynch, Krista Queen, Anna Montmayeur, Anna Uehara, Clinton R. Paden, Rachel Marine, Haibin Wang, Suxiang Tong |
| EPI_ISL_648004, EPI_ISL_648005, EPI_ISL_648006, EPI_ISL_648007, EPI_ISL_648008, EPI_ISL_648009, EPI_ISL_648010, EPI_ISL_648011, EPI_ISL_648012, EPI_ISL_648013, EPI_ISL_648014, EPI_ISL_648015, EPI_ISL_648016, EPI_ISL_648017, EPI_ISL_648018, EPI_ISL_648019, EPI_ISL_648020, EPI_ISL_648021, EPI_ISL_648022, EPI_ISL_648023, EPI_ISL_648024, EPI_ISL_648025, EPI_ISL_648026, EPI_ISL_648027, EPI_ISL_648028, EPI_ISL_648029, EPI_ISL_648030, EPI_ISL_648031, EPI_ISL_648032, EPI_ISL_648033, EPI_ISL_648034 | | | |
| see above | MS Public Health Laboratory | Pathogen Discovery, Respiratory Viruses Branch, Division of Viral Diseases, Centers for Disease Control and Prevention | Yan Li, Jing Zhang, Ying Tao, Brian Lynch, Krista Queen, Anna Montmayeur, Anna Uehara, Clinton R. Paden, Rachel Marine, Haibin Wang, Suxiang Tong |
| EPI_ISL_648035, EPI_ISL_648036 | WI State Laboratory of Hygiene | Pathogen Discovery, Respiratory Viruses Branch, Division of Viral Diseases, Centers for Disease Control and Prevention | Yan Li, Jing Zhang, Ying Tao, Brian Lynch, Krista Queen, Anna Montmayeur, Anna Uehara, Clinton R. Paden, Rachel Marine, Haibin Wang, Suxiang Tong |
| EPI_ISL_648037, EPI_ISL_648038, EPI_ISL_648039, EPI_ISL_648040, EPI_ISL_648041, EPI_ISL_648042 | MS Public Health Laboratory | Pathogen Discovery, Respiratory Viruses Branch, Division of Viral Diseases, Centers for Disease Control and Prevention | Yan Li, Jing Zhang, Ying Tao, Brian Lynch, Krista Queen, Anna Montmayeur, Anna Uehara, Clinton R. Paden, Rachel Marine, Haibin Wang, Suxiang Tong |
| EPI_ISL_648122 | VA-Division of Consolidated Laboratory Services | Pathogen Discovery, Respiratory Viruses Branch, Division of Viral Diseases, Centers for Disease Control and Prevention | Yan Li, Jing Zhang, Ying Tao, Brian Lynch, Krista Queen, Anna Montmayeur, Anna Uehara, Clinton R. Paden, Rachel Marine, Haibin Wang, Suxiang Tong |
| EPI_ISL_648123 | GA Department of Public Health Laboratory | Pathogen Discovery, Respiratory Viruses Branch, Division of Viral Diseases, Centers for Disease Control and Prevention | Yan Li, Jing Zhang, Ying Tao, Brian Lynch, Krista Queen, Anna Montmayeur, Anna Uehara, Clinton R. Paden, Rachel Marine, Haibin Wang, Suxiang Tong |
| EPI_ISL_648219, EPI_ISL_648220, EPI_ISL_648221, EPI_ISL_648222, EPI_ISL_648223, EPI_ISL_648224, EPI_ISL_648225, EPI_ISL_648226, EPI_ISL_648227, EPI_ISL_648228, EPI_ISL_648229, EPI_ISL_648230, EPI_ISL_648231, EPI_ISL_648232, EPI_ISL_648233, EPI_ISL_648234, EPI_ISL_648235, EPI_ISL_648236, EPI_ISL_648237, EPI_ISL_648238, EPI_ISL_648239, EPI_ISL_648240, EPI_ISL_648241, EPI_ISL_648242, EPI_ISL_648243, EPI_ISL_648244, EPI_ISL_648245, EPI_ISL_648256 | | | |
| see above | Texas Department of State Health Services | Texas Department of State Health Services | Rashmi Tuladhar, Bonnie Oh, Jenny Zhang, Maliha Rahman, Anita Pokharel, Myong Koag, Chung Wang, Rachel Lee, Grace Kubin, Mayela Pedrueza, James Daniel Bonser |
| EPI_ISL_648265, EPI_ISL_648267, EPI_ISL_648270 | Brigham and Women's Hospital | Jonathan Li laboratory | Jonathan Z. Li, Manish C. Choudhary, Upasana D. Adhikari, George Eng, Douglas S Kwon |
| EPI_ISL_648271, EPI_ISL_648272, EPI_ISL_648273, EPI_ISL_648274, EPI_ISL_648275, EPI_ISL_648276, EPI_ISL_648277, EPI_ISL_648278, EPI_ISL_648279, EPI_ISL_648280, EPI_ISL_648281, EPI_ISL_648282, EPI_ISL_648283, EPI_ISL_648284, EPI_ISL_648285, EPI_ISL_648286, EPI_ISL_648287, EPI_ISL_648288, EPI_ISL_648289, EPI_ISL_648290, EPI_ISL_648291, EPI_ISL_648292, EPI_ISL_648293, EPI_ISL_648294, EPI_ISL_648295, EPI_ISL_648296, EPI_ISL_648297, EPI_ISL_648298, EPI_ISL_648299, EPI_ISL_648300, EPI_ISL_648301, EPI_ISL_648302 | | | |
| see above | MD PHL | MD PHL | Maryland Department of Health Laboratories Administration |
| EPI_ISL_648381, EPI_ISL_648382, EPI_ISL_648383, EPI_ISL_648384, EPI_ISL_648385, EPI_ISL_648386, EPI_ISL_648387, EPI_ISL_648388, EPI_ISL_648389, EPI_ISL_648390, EPI_ISL_648391, EPI_ISL_648392, EPI_ISL_648393, EPI_ISL_648394, EPI_ISL_648395, EPI_ISL_648396, EPI_ISL_648397, EPI_ISL_648399, EPI_ISL_648400, EPI_ISL_648401, EPI_ISL_648402, EPI_ISL_648403, EPI_ISL_648404, EPI_ISL_648405, EPI_ISL_648406, EPI_ISL_648407, EPI_ISL_648408, EPI_ISL_648409, EPI_ISL_648410, EPI_ISL_648411, EPI_ISL_648412, EPI_ISL_648413, EPI_ISL_648414, EPI_ISL_648415, EPI_ISL_648416, EPI_ISL_648417, EPI_ISL_648418, EPI_ISL_648419, EPI_ISL_648420, EPI_ISL_648421, EPI_ISL_648422, EPI_ISL_648423, EPI_ISL_648424, EPI_ISL_648425, EPI_ISL_648426, EPI_ISL_648427, EPI_ISL_648428, EPI_ISL_648429, EPI_ISL_648430, EPI_ISL_648431, EPI_ISL_648432, EPI_ISL_648433, EPI_ISL_648434, EPI_ISL_648435, EPI_ISL_648436, EPI_ISL_648437, EPI_ISL_648438, EPI_ISL_648439, EPI_ISL_648440, EPI_ISL_648441, EPI_ISL_648442, EPI_ISL_648443, EPI_ISL_648444, EPI_ISL_648445, EPI_ISL_648446, EPI_ISL_648447, EPI_ISL_648448, EPI_ISL_648449, EPI_ISL_648450, EPI_ISL_648451, EPI_ISL_648452, EPI_ISL_648453, EPI_ISL_648454, EPI_ISL_648455, EPI_ISL_648456, EPI_ISL_648457, EPI_ISL_648458, EPI_ISL_648459, EPI_ISL_648460, EPI_ISL_648461, EPI_ISL_648462, EPI_ISL_648463, EPI_ISL_648464, EPI_ISL_648465, EPI_ISL_648466, EPI_ISL_648467, EPI_ISL_648468, EPI_ISL_648469, EPI_ISL_648470, EPI_ISL_648471, EPI_ISL_648472, EPI_ISL_648473, EPI_ISL_648474, EPI_ISL_648475, EPI_ISL_648476, EPI_ISL_648477, EPI_ISL_648479, EPI_ISL_648480, EPI_ISL_648481, EPI_ISL_648482, EPI_ISL_648483, EPI_ISL_648484, EPI_ISL_648485, EPI_ISL_648486, EPI_ISL_648487, EPI_ISL_648488, EPI_ISL_648489, EPI_ISL_648490, EPI_ISL_648492, EPI_ISL_648493, EPI_ISL_648494, EPI_ISL_648495 | | | |
| see above | Santa Clara County Public Health Laboratory | Chan-Zuckerberg Biohub | CZB Cliahub Consortium |
| EPI_ISL_648496, EPI_ISL_648497, EPI_ISL_648498, EPI_ISL_648499, EPI_ISL_648500, EPI_ISL_648501, EPI_ISL_648502, EPI_ISL_648503, EPI_ISL_648504, EPI_ISL_648505, EPI_ISL_648506, EPI_ISL_648508, EPI_ISL_648509, EPI_ISL_648510, EPI_ISL_648511, EPI_ISL_648512, EPI_ISL_648513, EPI_ISL_648514, EPI_ISL_648515, EPI_ISL_648516, EPI_ISL_648517, EPI_ISL_648518, EPI_ISL_648519, EPI_ISL_648520, EPI_ISL_648521, EPI_ISL_648522, EPI_ISL_648523, EPI_ISL_648524, EPI_ISL_648525, EPI_ISL_648526, EPI_ISL_648527, EPI_ISL_648528, EPI_ISL_648529, EPI_ISL_648530 | | | |
| see above | Orange County Public Health Lab | Chan-Zuckerberg Biohub | CZB Cliahub Consortium |
| EPI_ISL_648531, EPI_ISL_648532, EPI_ISL_648533, EPI_ISL_648534, EPI_ISL_648535, EPI_ISL_648536, EPI_ISL_648537, EPI_ISL_648538, EPI_ISL_648539, EPI_ISL_648540, EPI_ISL_648541 | | | |
| see above | Tulare County Public Health Lab | Chan-Zuckerberg Biohub | CZB Cliahub Consortium |
| EPI_ISL_648542 | Santa Clara County Public Health Laboratory | Chan-Zuckerberg Biohub | CZB Cliahub Consortium |
| EPI_ISL_648543, EPI_ISL_648544 | UCSF Clinical Microbiology Laboratory | Chan-Zuckerberg Biohub | CZB Cliahub Consortium |
| EPI_ISL_648545, EPI_ISL_648546, EPI_ISL_648547, EPI_ISL_648548, EPI_ISL_648549, EPI_ISL_648550, EPI_ISL_648551, EPI_ISL_648552, EPI_ISL_648553, EPI_ISL_648554 | Madera County Department of Public Health | Chan-Zuckerberg Biohub | CZB Cliahub Consortium |
| EPI_ISL_648555, EPI_ISL_648556, EPI_ISL_648558, EPI_ISL_648560, EPI_ISL_648561, EPI_ISL_648562, EPI_ISL_648563, EPI_ISL_648564, EPI_ISL_648565, EPI_ISL_648568, EPI_ISL_648569, EPI_ISL_648570, EPI_ISL_648571, EPI_ISL_648572, EPI_ISL_648573, EPI_ISL_648574, EPI_ISL_648575, EPI_ISL_648576, EPI_ISL_648577, EPI_ISL_648578, EPI_ISL_648579, EPI_ISL_648580, EPI_ISL_648581, EPI_ISL_648582, EPI_ISL_648583, EPI_ISL_648585, EPI_ISL_648586, EPI_ISL_648588, EPI_ISL_648590, EPI_ISL_648591, EPI_ISL_648592, EPI_ISL_648593, EPI_ISL_648594 | | | |
| see above | Utah Public Health Laboratory | Utah Public Health Laboratory | Erin Young, Kelly Oakeson |
| EPI_ISL_648595, EPI_ISL_648602, EPI_ISL_648603 | Laboratorio de Infectología Servicio de Infectología Hospital Universitario Dr. José Eleuterio González - Universidad Autónoma de Nuevo León | Laboratorio de Infectología Molecular Departamento de Bioquímica y Medicina Molecular Facultad de Medicina - Universidad Autónoma de Nuevo León | Kame A. Galán-Huerta, María F. Herrera-Saldivar, Natalia Martínez-Acuña, Sonia A. Lozano-Sepúlveda, Daniel Arellanos-Soto, Ana M. Rivas-Estilla, Paola Bocanegra-Ibarias, Samantha M. Flores-Treviño, Elvira Garza-González, Eduardo Perez-Alba, Laura Nuzzolo-Shihadeh, Adrian Camacho-Ortiz, Roberto Montes-de-Oca, Consuelo Treviño-Garza, Manuel E. de-la-O-Cavazos |
| EPI_ISL_648607 | Laboratorio de Infectología, Servicio de Infectología, Hospital Universitario Dr. José Eleuterio González - Universidad Autónoma de Nuevo León | Laboratorio de Infectología Molecular, Departamento de Bioquímica y Medicina Molecular,Facultad de Medicina - Universidad Autónoma de Nuevo León | Kame A. Galán-Huerta, María F. Herrera-Saldivar, Natalia Martínez-Acuña, Sonia A. Lozano-Sepúlveda, Daniel Arellanos-Soto, Ana M. Rivas-Estilla, Paola Bocanegra-Ibarias, Samantha M. Flores-Treviño, Elvira Garza-González, Eduardo Perez-Alba, Laura Nuzzolo-Shihadeh, Adrian Camacho-Ortiz, Roberto Montes-de-Oca, Consuelo Treviño-Garza, Manuel E. de-la-O-Cavazos |
| EPI_ISL_648822 | Virus Ecology Section, RML | Virus Ecology Section, RML | van Doremalen,N., Holbrook,M.G., Barbian,K.D., Bushmaker,C., Bushmaker,T., Martens,C.A. and Munster,V.J |
| EPI_ISL_648823, EPI_ISL_648824, EPI_ISL_648825, EPI_ISL_648826, EPI_ISL_648827, EPI_ISL_648828, EPI_ISL_648829, EPI_ISL_648830, EPI_ISL_648831, EPI_ISL_648832, EPI_ISL_648833, EPI_ISL_648834, EPI_ISL_648835, EPI_ISL_648836, EPI_ISL_648837, EPI_ISL_648838, EPI_ISL_648839, EPI_ISL_648840, EPI_ISL_648841, EPI_ISL_648842, EPI_ISL_648843, EPI_ISL_648844, EPI_ISL_648845, EPI_ISL_648846, EPI_ISL_648847, EPI_ISL_648848, EPI_ISL_648849, EPI_ISL_648850, EPI_ISL_648851, EPI_ISL_648852, EPI_ISL_648853, EPI_ISL_648854, EPI_ISL_648855, EPI_ISL_648856, EPI_ISL_648857, EPI_ISL_648858, EPI_ISL_648859, EPI_ISL_648860, EPI_ISL_648861, EPI_ISL_648862, EPI_ISL_648863, EPI_ISL_648864, EPI_ISL_648865, EPI_ISL_648866, EPI_ISL_648867, EPI_ISL_648868, EPI_ISL_648869, EPI_ISL_648870, EPI_ISL_648871, EPI_ISL_648872, EPI_ISL_648873, EPI_ISL_648874, EPI_ISL_648875, EPI_ISL_648876, EPI_ISL_648877, EPI_ISL_648878, EPI_ISL_648879, EPI_ISL_648880, EPI_ISL_648881, EPI_ISL_648882, EPI_ISL_648883, EPI_ISL_648884, EPI_ISL_648885, EPI_ISL_648886, EPI_ISL_648887, EPI_ISL_648888, EPI_ISL_648889, EPI_ISL_648890, EPI_ISL_648891, EPI_ISL_648892, EPI_ISL_648893, EPI_ISL_648894, EPI_ISL_648895, EPI_ISL_648896, EPI_ISL_648897, EPI_ISL_648898, EPI_ISL_648899, EPI_ISL_648900, EPI_ISL_648901, EPI_ISL_648902, EPI_ISL_648903, EPI_ISL_648904, EPI_ISL_648905, EPI_ISL_648906, EPI_ISL_648907, EPI_ISL_648908, EPI_ISL_648909, EPI_ISL_648910, EPI_ISL_648911, EPI_ISL_648912, EPI_ISL_648913, EPI_ISL_648914, EPI_ISL_648915, EPI_ISL_648916, EPI_ISL_648917, EPI_ISL_648918, EPI_ISL_648919, EPI_ISL_648920, EPI_ISL_648921, EPI_ISL_648922, EPI_ISL_648923, EPI_ISL_648924, EPI_ISL_648925, EPI_ISL_648926, EPI_ISL_648927, EPI_ISL_648928, EPI_ISL_648929, EPI_ISL_648930, EPI_ISL_648931, EPI_ISL_648932, EPI_ISL_648933, EPI_ISL_648934, EPI_ISL_648935, EPI_ISL_648936, EPI_ISL_648937, EPI_ISL_648938, EPI_ISL_648939, EPI_ISL_648940, EPI_ISL_648941, EPI_ISL_648942, EPI_ISL_648943, EPI_ISL_648944, EPI_ISL_648945, EPI_ISL_648946, EPI_ISL_648947, EPI_ISL_648948, EPI_ISL_648949, EPI_ISL_648950, EPI_ISL_648951, EPI_ISL_648952, EPI_ISL_648953, EPI_ISL_648954, EPI_ISL_648955, EPI_ISL_648956, EPI_ISL_648957, EPI_ISL_648958, EPI_ISL_648959, EPI_ISL_648960, EPI_ISL_648961, EPI_ISL_648962, EPI_ISL_648963, EPI_ISL_648964, EPI_ISL_648965, EPI_ISL_648966, EPI_ISL_648967, EPI_ISL_648968, EPI_ISL_648969, EPI_ISL_648970, EPI_ISL_648971, EPI_ISL_648972, EPI_ISL_648973, EPI_ISL_648974, EPI_ISL_648975, EPI_ISL_648976, EPI_ISL_648977, EPI_ISL_648978, EPI_ISL_648979, EPI_ISL_648980, EPI_ISL_648981, EPI_ISL_648982, EPI_ISL_648983, EPI_ISL_648984, EPI_ISL_648985, EPI_ISL_648986, EPI_ISL_648987, EPI_ISL_648988, EPI_ISL_648989, EPI_ISL_648990, EPI_ISL_648991, EPI_ISL_648992, EPI_ISL_648993, EPI_ISL_648994, EPI_ISL_648995, EPI_ISL_648996, EPI_ISL_648997, EPI_ISL_648998, EPI_ISL_648999, EPI_ISL_649000, EPI_ISL_649001, EPI_ISL_649002, EPI_ISL_649003, EPI_ISL_649004, EPI_ISL_649005, EPI_ISL_649006, EPI_ISL_649007, EPI_ISL_649008, EPI_ISL_649009, EPI_ISL_649010, EPI_ISL_649011, EPI_ISL_649012, EPI_ISL_649013, EPI_ISL_649014, EPI_ISL_649015, EPI_ISL_649016, EPI_ISL_649017, EPI_ISL_649018, EPI_ISL_649019, EPI_ISL_649020, EPI_ISL_649021, EPI_ISL_649022, EPI_ISL_649023, EPI_ISL_649024, EPI_ISL_649025, EPI_ISL_649026, EPI_ISL_649027, EPI_ISL_649028, EPI_ISL_649029, EPI_ISL_649030, EPI_ISL_649031, EPI_ISL_649032, EPI_ISL_649033, EPI_ISL_649034, EPI_ISL_649035, EPI_ISL_649036, EPI_ISL_649037, EPI_ISL_649038, EPI_ISL_649039, EPI_ISL_649040, EPI_ISL_649041, EPI_ISL_649042, EPI_ISL_649043, EPI_ISL_649044, EPI_ISL_649045, EPI_ISL_649046, EPI_ISL_649047, EPI_ISL_649048, EPI_ISL_649049, EPI_ISL_649050, EPI_ISL_649051, EPI_ISL_649052, EPI_ISL_649053, EPI_ISL_649054, EPI_ISL_649055, EPI_ISL_649056 | | | |
| see above | San Diego County Public Health Laboratory | Andersen lab at Scripps Research | SEARCH Alliance San Diego with Tracy Basler, Jovan Shephard, Brett Austin |
| EPI_ISL_649057, EPI_ISL_649058, EPI_ISL_649059 | University of Michigan Clinical Microbiology Laboratory | Lauring Lab, University of Michigan, Department of Microbiology and Immunology | Valesano |

| | | | |
|---|---|---|---|
| EPI_ISL_649152 | Infectious diseases, Chan Zuckerberg Biohub | Infectious diseases, Chan Zuckerberg Biohub | Bhatt,K.D. |
| EPI_ISL_649972, EPI_ISL_649973, EPI_ISL_649974, EPI_ISL_649975, EPI_ISL_649976, EPI_ISL_649977, EPI_ISL_649978, EPI_ISL_649979, EPI_ISL_649980, EPI_ISL_649981, EPI_ISL_649982, EPI_ISL_649983, EPI_ISL_649984, EPI_ISL_649985, EPI_ISL_649986, EPI_ISL_649987, EPI_ISL_649988, EPI_ISL_649989, EPI_ISL_649990, EPI_ISL_649991, EPI_ISL_649992, EPI_ISL_649993, EPI_ISL_649994, EPI_ISL_649995, EPI_ISL_649996, EPI_ISL_649997, EPI_ISL_649998, EPI_ISL_649999, EPI_ISL_650000, EPI_ISL_650001, EPI_ISL_650002, EPI_ISL_650003, EPI_ISL_650004, EPI_ISL_650005, EPI_ISL_650006, EPI_ISL_650007, EPI_ISL_650008, EPI_ISL_650009, EPI_ISL_650010, EPI_ISL_650011, EPI_ISL_650012, EPI_ISL_650013, EPI_ISL_650014, EPI_ISL_650015, EPI_ISL_650016, EPI_ISL_650017, EPI_ISL_650018, EPI_ISL_650019, EPI_ISL_650020, EPI_ISL_650021, EPI_ISL_650022, EPI_ISL_650023, EPI_ISL_650024, EPI_ISL_650025, EPI_ISL_650026, EPI_ISL_650027, EPI_ISL_650028, EPI_ISL_650029, EPI_ISL_650030, EPI_ISL_650031, EPI_ISL_650032, EPI_ISL_650033, EPI_ISL_650034, EPI_ISL_650035, EPI_ISL_650036, EPI_ISL_650037, EPI_ISL_650038, EPI_ISL_650039, EPI_ISL_650040, EPI_ISL_650041, EPI_ISL_650042, EPI_ISL_650043, EPI_ISL_650044, EPI_ISL_650045, EPI_ISL_650046, EPI_ISL_650047, EPI_ISL_650048, EPI_ISL_650049, EPI_ISL_650050, EPI_ISL_650051, EPI_ISL_650052, EPI_ISL_650053, EPI_ISL_650054, EPI_ISL_650055, EPI_ISL_650056, EPI_ISL_650057, EPI_ISL_650058, EPI_ISL_650059, EPI_ISL_650060, EPI_ISL_650061, EPI_ISL_650062, EPI_ISL_650063, EPI_ISL_650064, EPI_ISL_650065, EPI_ISL_650066, EPI_ISL_650067, EPI_ISL_650068, EPI_ISL_650069, EPI_ISL_650070, EPI_ISL_650071, EPI_ISL_650072, EPI_ISL_650073, EPI_ISL_650074, EPI_ISL_650075, EPI_ISL_650076, EPI_ISL_650077, EPI_ISL_650078, EPI_ISL_650079, EPI_ISL_650080, EPI_ISL_650081, EPI_ISL_650082, EPI_ISL_650083, EPI_ISL_650084, EPI_ISL_650085, EPI_ISL_650086, EPI_ISL_650087, EPI_ISL_650088, EPI_ISL_650089, EPI_ISL_650090, EPI_ISL_650091, EPI_ISL_650092, EPI_ISL_650093, EPI_ISL_650094, EPI_ISL_650095, EPI_ISL_650096, EPI_ISL_650097, EPI_ISL_650098, EPI_ISL_650099, EPI_ISL_650100, EPI_ISL_650101, EPI_ISL_650102, EPI_ISL_650103, EPI_ISL_650104, EPI_ISL_650105, EPI_ISL_650106, EPI_ISL_650107, EPI_ISL_650108, EPI_ISL_650109, EPI_ISL_650110, EPI_ISL_650111 | | | |
| see above | University of Michigan Clinical Microbiology Laboratory | Lauring Lab, University of Michigan, Department of Microbiology and Immunology | Valesano |
| EPI_ISL_653098, EPI_ISL_653100, EPI_ISL_653101, EPI_ISL_653102, EPI_ISL_653103, EPI_ISL_653104, EPI_ISL_653105, EPI_ISL_653106, EPI_ISL_653107, EPI_ISL_653108, EPI_ISL_653109, EPI_ISL_653110, EPI_ISL_653111, EPI_ISL_653112, EPI_ISL_653113, EPI_ISL_653114, EPI_ISL_653115, EPI_ISL_653117, EPI_ISL_653118, EPI_ISL_653119, EPI_ISL_653120, EPI_ISL_653121, EPI_ISL_653122, EPI_ISL_653123, EPI_ISL_653124, EPI_ISL_653125, EPI_ISL_653126, EPI_ISL_653127, EPI_ISL_653128, EPI_ISL_653129, EPI_ISL_653130, EPI_ISL_653131, EPI_ISL_653132, EPI_ISL_653133, EPI_ISL_653134, EPI_ISL_653137, EPI_ISL_653138, EPI_ISL_653139, EPI_ISL_653140, EPI_ISL_653141, EPI_ISL_653143, EPI_ISL_653145, EPI_ISL_653146, EPI_ISL_653147, EPI_ISL_653148, EPI_ISL_653149, EPI_ISL_653150, EPI_ISL_653151, EPI_ISL_653153, EPI_ISL_653154, EPI_ISL_653156, EPI_ISL_653157, EPI_ISL_653158, EPI_ISL_653161, EPI_ISL_653162, EPI_ISL_653163, EPI_ISL_653164, EPI_ISL_653165, EPI_ISL_653166, EPI_ISL_653169, EPI_ISL_653170, EPI_ISL_653174, EPI_ISL_653175, EPI_ISL_653178, EPI_ISL_653179, EPI_ISL_653180, EPI_ISL_653181, EPI_ISL_653182, EPI_ISL_653183, EPI_ISL_653185, EPI_ISL_653190, EPI_ISL_653191, EPI_ISL_653193, EPI_ISL_653194, EPI_ISL_653195, EPI_ISL_653200, EPI_ISL_653201, EPI_ISL_653202, EPI_ISL_653203, EPI_ISL_653204, EPI_ISL_653205, EPI_ISL_653206, EPI_ISL_653207, EPI_ISL_653208, EPI_ISL_653209, EPI_ISL_653211, EPI_ISL_653213, EPI_ISL_653214, EPI_ISL_653215, EPI_ISL_653216, EPI_ISL_653217, EPI_ISL_653218, EPI_ISL_653219, EPI_ISL_653220, EPI_ISL_653221, EPI_ISL_653222, EPI_ISL_653224, EPI_ISL_653226, EPI_ISL_653227, EPI_ISL_653228, EPI_ISL_653230, EPI_ISL_653231, EPI_ISL_653232, EPI_ISL_653233, EPI_ISL_653236, EPI_ISL_653241, EPI_ISL_653244, EPI_ISL_653245, EPI_ISL_653246, EPI_ISL_653247, EPI_ISL_653248, EPI_ISL_653249, EPI_ISL_653251, EPI_ISL_653253, EPI_ISL_653254, EPI_ISL_653255, EPI_ISL_653256, EPI_ISL_653257, EPI_ISL_653258, EPI_ISL_653259, EPI_ISL_653260, EPI_ISL_653261, EPI_ISL_653262, EPI_ISL_653263, EPI_ISL_653264, EPI_ISL_653265, EPI_ISL_653266, EPI_ISL_653267, EPI_ISL_653268, EPI_ISL_653269, EPI_ISL_653271, EPI_ISL_653272, EPI_ISL_653273, EPI_ISL_653274, EPI_ISL_653275, EPI_ISL_653276, EPI_ISL_653277, EPI_ISL_653278, EPI_ISL_653280, EPI_ISL_653281, EPI_ISL_653284, EPI_ISL_653288, EPI_ISL_653294, EPI_ISL_653295, EPI_ISL_653296, EPI_ISL_653297, EPI_ISL_653298, EPI_ISL_653300, EPI_ISL_653302, EPI_ISL_653303, EPI_ISL_653304, EPI_ISL_653306, EPI_ISL_653307, EPI_ISL_653310, EPI_ISL_653311, EPI_ISL_653313, EPI_ISL_653314, EPI_ISL_653317, EPI_ISL_653320, EPI_ISL_653321, EPI_ISL_653322, EPI_ISL_653323, EPI_ISL_653324, EPI_ISL_653325, EPI_ISL_653326, EPI_ISL_653329, EPI_ISL_653331, EPI_ISL_653333, EPI_ISL_653334, EPI_ISL_653336, EPI_ISL_653337, EPI_ISL_653339, EPI_ISL_653340, EPI_ISL_653341, EPI_ISL_653344 | | | |
| see above | Florida Bureau of Public Health Laboratories | Florida Bureau of Public Health Laboratories | Sarah Schmedes, Jason Blanton |
| EPI_ISL_653345, EPI_ISL_653346, EPI_ISL_653347, EPI_ISL_653348, EPI_ISL_653349, EPI_ISL_653350, EPI_ISL_653351, EPI_ISL_653352, EPI_ISL_653353, EPI_ISL_653354, EPI_ISL_653355, EPI_ISL_653356, EPI_ISL_653357, EPI_ISL_653358, EPI_ISL_653359, EPI_ISL_653360, EPI_ISL_653361, EPI_ISL_653362, EPI_ISL_653363, EPI_ISL_653364, EPI_ISL_653365, EPI_ISL_653366, EPI_ISL_653367, EPI_ISL_653368, EPI_ISL_653369 | | | |
| see above | LSUHS Emerging Viral Threat Laboratory | Microbial Genome Sequencing Center | Maarten Van Diest, Jeremy P. Kamil, Rona S. Scott, Malgorzata Bienkowska-Haba, Katarzyna Zwolinska, Andrew D. Yurochko, Christopher G. Kevil, Martin J. Sapp, Daniel J. Snyder, Vaughn S. Cooper, John A. Vanchiere |
| EPI_ISL_653370, EPI_ISL_653371, EPI_ISL_653372, EPI_ISL_653373, EPI_ISL_653374, EPI_ISL_653375, EPI_ISL_653376, EPI_ISL_653377, EPI_ISL_653378, EPI_ISL_653379, EPI_ISL_653380, EPI_ISL_653381, EPI_ISL_653382, EPI_ISL_653383, EPI_ISL_653384, EPI_ISL_653385, EPI_ISL_653386, EPI_ISL_653387, EPI_ISL_653388, EPI_ISL_653389, EPI_ISL_653390, EPI_ISL_653391, EPI_ISL_653392, EPI_ISL_653393, EPI_ISL_653394 | | | |
| see above | LSUHS Emerging Viral Threat Laboratory | Microbial Genome Sequencing Center | Rona S. Scott, Jeremy P. Kamil, Maarten Van Diest, Malgorzata Bienkowska-Haba, Katarzyna Zwolinska, Andrew D. Yurochko, Christopher G. Kevil, Martin J. Sapp, Daniel J. Snyder, Vaughn S. Cooper, John A. Vanchiere |
| EPI_ISL_653395, EPI_ISL_653396, EPI_ISL_653397, EPI_ISL_653398, EPI_ISL_653399, EPI_ISL_653400, EPI_ISL_653401, EPI_ISL_653402, EPI_ISL_653403, EPI_ISL_653404, EPI_ISL_653405, EPI_ISL_653406, EPI_ISL_653407, EPI_ISL_653408, EPI_ISL_653409, EPI_ISL_653410, EPI_ISL_653411, EPI_ISL_653412, EPI_ISL_653413, EPI_ISL_653414, EPI_ISL_653415, EPI_ISL_653416, EPI_ISL_653417, EPI_ISL_653418, EPI_ISL_653419, EPI_ISL_653420, EPI_ISL_653421, EPI_ISL_653422, EPI_ISL_653423, EPI_ISL_653424, EPI_ISL_653425, EPI_ISL_653426, EPI_ISL_653427, EPI_ISL_653428, EPI_ISL_653429, EPI_ISL_653430, EPI_ISL_653431, EPI_ISL_653432, EPI_ISL_653433, EPI_ISL_653434, EPI_ISL_653435, EPI_ISL_653436, EPI_ISL_653437, EPI_ISL_653438, EPI_ISL_653439, EPI_ISL_653440, EPI_ISL_653441, EPI_ISL_653442, EPI_ISL_653443, EPI_ISL_653444, EPI_ISL_653445, EPI_ISL_653446, EPI_ISL_653447, EPI_ISL_653448, EPI_ISL_653449, EPI_ISL_653450, EPI_ISL_653451, EPI_ISL_653452, EPI_ISL_653453, EPI_ISL_653454, EPI_ISL_653455, EPI_ISL_653456, EPI_ISL_653457, EPI_ISL_653458, EPI_ISL_653459, EPI_ISL_653460, EPI_ISL_653461, EPI_ISL_653462, EPI_ISL_653463, EPI_ISL_653464, EPI_ISL_653465, EPI_ISL_653466, EPI_ISL_653467, EPI_ISL_653468, EPI_ISL_653469, EPI_ISL_653470, EPI_ISL_653471, EPI_ISL_653472, EPI_ISL_653473, EPI_ISL_653474, EPI_ISL_653475, EPI_ISL_653476, EPI_ISL_653477, EPI_ISL_653478, EPI_ISL_653479, EPI_ISL_653480, EPI_ISL_653481, EPI_ISL_653482, EPI_ISL_653483, EPI_ISL_653484, EPI_ISL_653485, EPI_ISL_653486, EPI_ISL_653487, EPI_ISL_653488, EPI_ISL_653489, EPI_ISL_653490, EPI_ISL_653491, EPI_ISL_653492, EPI_ISL_653493, EPI_ISL_653494, EPI_ISL_653495, EPI_ISL_653496, EPI_ISL_653497, EPI_ISL_653498, EPI_ISL_653499, EPI_ISL_653500, EPI_ISL_653501, EPI_ISL_653502, EPI_ISL_653503, EPI_ISL_653504, EPI_ISL_653505, EPI_ISL_653506, EPI_ISL_653507, EPI_ISL_653508, EPI_ISL_653509, EPI_ISL_653510, EPI_ISL_653511, EPI_ISL_653512, EPI_ISL_653513, EPI_ISL_653514, EPI_ISL_653515, EPI_ISL_653516, EPI_ISL_653517, EPI_ISL_653518, EPI_ISL_653519, EPI_ISL_653520, EPI_ISL_653521, EPI_ISL_653522, EPI_ISL_653523, EPI_ISL_653524, EPI_ISL_653525, EPI_ISL_653526, EPI_ISL_653527, EPI_ISL_653528, EPI_ISL_653529, EPI_ISL_653530, EPI_ISL_653531, EPI_ISL_653532, EPI_ISL_653533, EPI_ISL_653534, EPI_ISL_653535, EPI_ISL_653536, EPI_ISL_653537, EPI_ISL_653538, EPI_ISL_653539, EPI_ISL_653540, EPI_ISL_653541, EPI_ISL_653542, EPI_ISL_653543, EPI_ISL_653544, EPI_ISL_653545, EPI_ISL_653546, EPI_ISL_653547, EPI_ISL_653548, EPI_ISL_653549, EPI_ISL_653550, EPI_ISL_653551, EPI_ISL_653552, EPI_ISL_653553, EPI_ISL_653554, EPI_ISL_653555, EPI_ISL_653556, EPI_ISL_653557, EPI_ISL_653558, EPI_ISL_653559, EPI_ISL_653560, EPI_ISL_653561, EPI_ISL_653562, EPI_ISL_653563, EPI_ISL_653564, EPI_ISL_653565, EPI_ISL_653566, EPI_ISL_653567, EPI_ISL_653568, EPI_ISL_653569, EPI_ISL_653570, EPI_ISL_653571, EPI_ISL_653572, EPI_ISL_653573, EPI_ISL_653574, EPI_ISL_653575, EPI_ISL_653576, EPI_ISL_653577, EPI_ISL_653578, EPI_ISL_653579, EPI_ISL_653580, EPI_ISL_653581, EPI_ISL_653582, EPI_ISL_653583, EPI_ISL_653584, EPI_ISL_653585, EPI_ISL_653586, EPI_ISL_653587, EPI_ISL_653588, EPI_ISL_653589, EPI_ISL_653590, EPI_ISL_653591, EPI_ISL_653592, EPI_ISL_653593, EPI_ISL_653594, EPI_ISL_653595, EPI_ISL_653596, EPI_ISL_653597, EPI_ISL_653598, EPI_ISL_653599, EPI_ISL_653600, EPI_ISL_653601, EPI_ISL_653602, EPI_ISL_653603, EPI_ISL_653604, EPI_ISL_653605, EPI_ISL_653606, EPI_ISL_653607, EPI_ISL_653608, EPI_ISL_653609, EPI_ISL_653610, EPI_ISL_653611, EPI_ISL_653612, EPI_ISL_653613, EPI_ISL_653614, EPI_ISL_653615, EPI_ISL_653616, EPI_ISL_653617, EPI_ISL_653618, EPI_ISL_653619, EPI_ISL_653620, EPI_ISL_653621, EPI_ISL_653622, EPI_ISL_653623, EPI_ISL_653624, EPI_ISL_653625, EPI_ISL_653626, EPI_ISL_653627, EPI_ISL_653628, EPI_ISL_653629, EPI_ISL_653630, EPI_ISL_653631, EPI_ISL_653632, EPI_ISL_653633, EPI_ISL_653634, EPI_ISL_653635, EPI_ISL_653636, EPI_ISL_653637, EPI_ISL_653638, EPI_ISL_653639, EPI_ISL_653640, EPI_ISL_653641, EPI_ISL_653642, EPI_ISL_653643, EPI_ISL_653644, EPI_ISL_653645, EPI_ISL_653646, EPI_ISL_653647, EPI_ISL_653648, EPI_ISL_653649, EPI_ISL_653650, EPI_ISL_653651, EPI_ISL_653652, EPI_ISL_653653, EPI_ISL_653654, EPI_ISL_653655, EPI_ISL_653656, EPI_ISL_653657, EPI_ISL_653658, EPI_ISL_653659, EPI_ISL_653660, EPI_ISL_653661, EPI_ISL_653662, EPI_ISL_653663, EPI_ISL_653664, EPI_ISL_653665, EPI_ISL_653666, EPI_ISL_653667, EPI_ISL_653668, EPI_ISL_653669, EPI_ISL_653670, EPI_ISL_653671, EPI_ISL_653672, EPI_ISL_653673, EPI_ISL_653674, EPI_ISL_653675, EPI_ISL_653676, EPI_ISL_653677, EPI_ISL_653678, EPI_ISL_653679, EPI_ISL_653680, EPI_ISL_653681, EPI_ISL_653682, EPI_ISL_653683, EPI_ISL_653684, EPI_ISL_653685, EPI_ISL_653686, EPI_ISL_653687, EPI_ISL_653688, EPI_ISL_653689, EPI_ISL_653690, EPI_ISL_653691, EPI_ISL_653692, EPI_ISL_653693, EPI_ISL_653694, EPI_ISL_653695, EPI_ISL_653696, EPI_ISL_653697, EPI_ISL_653698, EPI_ISL_653699, EPI_ISL_653700, EPI_ISL_653701, EPI_ISL_653702, EPI_ISL_653703, EPI_ISL_653704, EPI_ISL_653705, EPI_ISL_653706, EPI_ISL_653707, EPI_ISL_653708, EPI_ISL_653709, EPI_ISL_653710, EPI_ISL_653711, EPI_ISL_653712, EPI_ISL_653713, EPI_ISL_653714, EPI_ISL_653715, EPI_ISL_653716, EPI_ISL_653717, EPI_ISL_653718, EPI_ISL_653719, EPI_ISL_653720, EPI_ISL_653721, EPI_ISL_653722, EPI_ISL_653723, EPI_ISL_653724, EPI_ISL_653725, EPI_ISL_653726, EPI_ISL_653727, EPI_ISL_653728, EPI_ISL_653729, EPI_ISL_653730, EPI_ISL_653731, EPI_ISL_653732, EPI_ISL_653733, EPI_ISL_653734, EPI_ISL_653735, EPI_ISL_653736, EPI_ISL_653737, EPI_ISL_653738, EPI_ISL_653739, EPI_ISL_653740, EPI_ISL_653741, EPI_ISL_653742, EPI_ISL_653743, EPI_ISL_653744 | | | |
| see above | LSUHS Emerging Viral Threat Laboratory | Microbial Genome Sequencing Center | Jeremy P. Kamil, Rona S. Scott, Maarten Van Diest, Malgorzata Bienkowska-Haba, Katarzyna Zwolinska, Andrew D. Yurochko, Christopher G. Kevil, Martin J. Sapp, Daniel J. Snyder, Vaughn S. Cooper, John A. Vanchiere |
| EPI_ISL_654695 | Minnesota Department of Health, Public Health Laboratory | Minnesota Department of Health, Public Health Laboratory | Matt Plumb, Jacob Garfin, Alexandra Lorentz, and Xiong Wang |
| EPI_ISL_654696, EPI_ISL_654697 | Essentia Health-St. Mary's Medical Center | Minnesota Department of Health, Public Health Laboratory | Matt Plumb, Jacob Garfin, Alexandra Lorentz, and Xiong Wang |
| EPI_ISL_654698 | Mayo Clinic & Mayo Clinic Laboratories | Minnesota Department of Health, Public Health Laboratory | Matt Plumb, Jacob Garfin, Alexandra Lorentz, and Xiong Wang |
| EPI_ISL_654699 | Essentia Health-St. Mary's Medical Center | Minnesota Department of Health, Public Health Laboratory | Matt Plumb, Jacob Garfin, Alexandra Lorentz, and Xiong Wang |
| EPI_ISL_654700, EPI_ISL_654701, EPI_ISL_654702, EPI_ISL_654703, EPI_ISL_654704, EPI_ISL_654705, EPI_ISL_654706, EPI_ISL_654707, EPI_ISL_654708, EPI_ISL_654709, EPI_ISL_654710, EPI_ISL_654711, EPI_ISL_654712, EPI_ISL_654713, EPI_ISL_654714, EPI_ISL_654715, EPI_ISL_654716, EPI_ISL_654717, EPI_ISL_654718, EPI_ISL_654719, EPI_ISL_654720, EPI_ISL_654721, EPI_ISL_654722, EPI_ISL_654723, EPI_ISL_654724, EPI_ISL_654725, EPI_ISL_654726, EPI_ISL_654727, EPI_ISL_654728, EPI_ISL_654729 | | | |
| see above | Minnesota Department of Health, Public Health Laboratory | Minnesota Department of Health, Public Health Laboratory | Matt Plumb, Jacob Garfin, Alexandra Lorentz, and Xiong Wang |
| EPI_ISL_654731, EPI_ISL_654732, EPI_ISL_654733, EPI_ISL_654734, EPI_ISL_654735, EPI_ISL_654736, EPI_ISL_654738, EPI_ISL_654739, EPI_ISL_654740, EPI_ISL_654741, EPI_ISL_654742, EPI_ISL_654743, EPI_ISL_654744, EPI_ISL_654745, EPI_ISL_654746, EPI_ISL_654747, EPI_ISL_654748, EPI_ISL_654749, EPI_ISL_654750, EPI_ISL_654751, EPI_ISL_654752, EPI_ISL_654753, EPI_ISL_654754, EPI_ISL_654755, EPI_ISL_654756, EPI_ISL_654757, EPI_ISL_654758, EPI_ISL_654759, EPI_ISL_654769, EPI_ISL_654770, EPI_ISL_654771, EPI_ISL_654772, EPI_ISL_654773, EPI_ISL_654774, EPI_ISL_654900, EPI_ISL_654901, EPI_ISL_654904, EPI_ISL_654905, EPI_ISL_654906, EPI_ISL_654908, EPI_ISL_654909, EPI_ISL_654911, EPI_ISL_654912, EPI_ISL_654913, EPI_ISL_654914, EPI_ISL_654915, EPI_ISL_654917, EPI_ISL_654918, EPI_ISL_654922, EPI_ISL_654923, EPI_ISL_654925, EPI_ISL_654926, EPI_ISL_654927, EPI_ISL_654929, EPI_ISL_654930, EPI_ISL_654931, EPI_ISL_654932, EPI_ISL_654933, EPI_ISL_654934, EPI_ISL_654937, EPI_ISL_654938, EPI_ISL_654939, EPI_ISL_654940 | | | |
| see above | Texas Department of State Health Services | Texas Department of State Health Services | Rashmi Tuladhar, Bonnie Oh, Jenny Zhang, Maliha Rahman, Anita Pokharel, Myong Koag, Chung Wang, Rachel Lee, Grace Kubin, Mayela Pedrueza, James Daniel Bonser |
| EPI_ISL_658863, EPI_ISL_658865, EPI_ISL_658868, EPI_ISL_658870 | Instituto de Diagnostico y Referencia Epidemiologicos (INDRE) | Instituto de Diagnostico y Referencia Epidemiologicos (INDRE) | Gisela Barrera-Badillo , Abril Rodriguez-Maldonado, Claudia Wong-Arambula , Natividad Cruz-Ortiz, Tatiana Nunez-Garcia, Dayanira Arellano-Suarez, Fabiola Garces-Ayala, Lucia Hernandez-Rivas, Irma Lopez-Martinez, Ernesto Ramirez-Gonzalez. |
| EPI_ISL_658873, EPI_ISL_658875 | Instituto de Diagnostico y Referencia Epidemiologicos (INDRE) | Instituto de Diagnostico y Referencia Epidemiologicos (INDRE) | Gisela Barrera-Badillo , Abril Rodriguez-Maldonado, Claudia Wong-Arambula , Natividad Cruz-Ortiz, Tatiana Nunez-Garcia, Dayanira Arellano-Suarez, Fabiola Garces-Ayala, Lucia Hernandez-Rivas, Irma Lopez-Martinez, Ernesto Ramirez-Gonzalez. |
| EPI_ISL_658878, EPI_ISL_658880, EPI_ISL_658883, EPI_ISL_658886, EPI_ISL_658888, EPI_ISL_658891, EPI_ISL_658893, EPI_ISL_658896 | Instituto de Diagnostico y Referencia Epidemiologicos (INDRE) | Instituto de Diagnostico y Referencia Epidemiologicos (INDRE) | Ernesto Ramirez-Gonzalez, Abril Rodriguez-Maldonado, Claudia Wong-Arambula , Natividad Cruz-Ortiz, Tatiana Nunez-Garcia, Dayanira Arellano-Suarez, Fabiola Garces-Ayala, Lucia Hernandez-Rivas, Irma Lopez-Martinez, Gisela Barrera-Badillo. |
| EPI_ISL_658899 | Instituto de Diagnostico y Referencia Epidemiologicos (INDRE) | Instituto de Diagnostico y Referencia Epidemiologicos (INDRE) | Gisela Barrera-Badillo , Abril Rodriguez-Maldonado, Claudia Wong-Arambula , Natividad Cruz-Ortiz, Tatiana Nunez-Garcia, Dayanira Arellano-Suarez, Fabiola Garces-Ayala, Lucia Hernandez-Rivas, Irma Lopez-Martinez, Ernesto Ramirez-Gonzalez. |
| EPI_ISL_658901 | Instituto de Diagnostico y Referencia Epidemiologicos (INDRE) | Instituto de Diagnostico y Referencia Epidemiologicos (INDRE) | Ernesto Ramirez-Gonzalez, Abril Rodriguez-Maldonado, Claudia Wong-Arambula , Natividad Cruz-Ortiz, Tatiana Nunez-Garcia, Dayanira Arellano-Suarez, Fabiola Garces-Ayala, Lucia Hernandez-Rivas, Irma Lopez-Martinez, Gisela Barrera-Badillo. |
| EPI_ISL_658904 | Instituto de Diagnostico y Referencia Epidemiologicos (INDRE) | Instituto de Diagnostico y Referencia Epidemiologicos (INDRE) | Ernesto Ramirez-Gonzalez, Abril Rodriguez-Maldonado, Claudia Wong-Arambula , Natividad Cruz-Ortiz, Tatiana Nunez-Garcia, Dayanira Arellano-Suarez, Fabiola Garces-Ayala, Lucia Hernandez-Rivas, Irma Lopez-Martinez, Gisela Barrera-Badillo, Marisol Galindo-Galindo, Linda Andarade-Sanchez, Octavio Ruiz-Muñiz, Pamela Ramirez-Medina, Roman Canul-Aguilar, Gaudalupe Herrera-Ramirez, Jazmin Galicia-Hernandez. |
| EPI_ISL_660068 | Instituto de Diagnostico y Referencia Epidemiologicos (INDRE) | Instituto de diagnóstico y Referencia Epidemiologicos (INDRE) Departamento de Virología | Gisela Barrera-Badillo , Abril Rodriguez-Maldonado, Claudia Wong-Arambula , Natividad Cruz-Ortiz, Tatiana Nunez-Garcia, Dayanira Arellano-Suarez, Fabiola Garces-Ayala, Lucia Hernandez-Rivas, Irma Lopez-Martinez, Ernesto Ramirez-Gonzalez. |
| EPI_ISL_660071, EPI_ISL_660072, EPI_ISL_660073, EPI_ISL_660074, EPI_ISL_660075, EPI_ISL_660076, EPI_ISL_660077, EPI_ISL_660078, EPI_ISL_660079, EPI_ISL_660080, EPI_ISL_660081, EPI_ISL_660082, EPI_ISL_660083, EPI_ISL_660084, EPI_ISL_660085, EPI_ISL_660086, EPI_ISL_660087, EPI_ISL_660088, EPI_ISL_660089, EPI_ISL_660090, EPI_ISL_660091, EPI_ISL_660092, EPI_ISL_660093, EPI_ISL_660094, EPI_ISL_660095, EPI_ISL_660096, EPI_ISL_660097, EPI_ISL_660098, EPI_ISL_660099, EPI_ISL_660100, EPI_ISL_660101, EPI_ISL_660102, EPI_ISL_660103, EPI_ISL_660104, EPI_ISL_660105 | | | |
| see above | MD PHL | MD PHL | Maryland Department of Health Laboratories Administration |
| EPI_ISL_660795, EPI_ISL_660796, EPI_ISL_660797, EPI_ISL_660798, EPI_ISL_660799, EPI_ISL_660800, EPI_ISL_660801 | Gundersen Molecular Diagnostics Laboratory | Kabara Cancer Research Institute | Craig S. Richmond, Paraic A. Kenny |
| EPI_ISL_660802, EPI_ISL_660803, | Gundersen Clinical Microbiology Laboratory | Kabara Cancer Research Institute | Craig S. Richmond, Paraic A. Kenny |

| | | | |
|---|---|---|---|
| EPI_ISL_660804, EPI_ISL_660805 | | | |
| EPI_ISL_660806, EPI_ISL_660807, EPI_ISL_660808, EPI_ISL_660809, EPI_ISL_660810, EPI_ISL_660811, EPI_ISL_660812, EPI_ISL_660813, EPI_ISL_660814, EPI_ISL_660815, EPI_ISL_660816, EPI_ISL_660817, EPI_ISL_660818, EPI_ISL_660819, EPI_ISL_660820, EPI_ISL_660821, EPI_ISL_660822, EPI_ISL_660823, EPI_ISL_660824, EPI_ISL_660825, EPI_ISL_660826, EPI_ISL_660827, EPI_ISL_660828, EPI_ISL_660829, EPI_ISL_660830, EPI_ISL_660831, EPI_ISL_660832, EPI_ISL_660833, EPI_ISL_660834, EPI_ISL_660835, EPI_ISL_660836, EPI_ISL_660837, EPI_ISL_660838, EPI_ISL_660839, EPI_ISL_660840, EPI_ISL_660841, EPI_ISL_660842, EPI_ISL_660843, EPI_ISL_660844, EPI_ISL_660845, EPI_ISL_660846, EPI_ISL_660847, EPI_ISL_660848, EPI_ISL_660849, EPI_ISL_660850, EPI_ISL_660851, EPI_ISL_660852, EPI_ISL_660853, EPI_ISL_660854, EPI_ISL_660855, EPI_ISL_660856, EPI_ISL_660857, EPI_ISL_660858, EPI_ISL_660859, EPI_ISL_660860, EPI_ISL_660861, EPI_ISL_660862, EPI_ISL_660863, EPI_ISL_660864, EPI_ISL_660865, EPI_ISL_660866, EPI_ISL_660867, EPI_ISL_660868 | | | |
| see above | Gundersen Molecular Diagnostics Laboratory | Kabara Cancer Research Institute | Craig S. Richmond, Paraic A. Kenny |
| EPI_ISL_660869, EPI_ISL_660870, EPI_ISL_660871, EPI_ISL_660872, EPI_ISL_660873, EPI_ISL_660874, EPI_ISL_660875, EPI_ISL_660876, EPI_ISL_660877, EPI_ISL_660878, EPI_ISL_660879, EPI_ISL_660880, EPI_ISL_660881 | | | |
| see above | Gundersen Clinical Microbiology Laboratory | Kabara Cancer Research Institute | Craig S. Richmond, Paraic A. Kenny |
| EPI_ISL_660882, EPI_ISL_660883, EPI_ISL_660884, EPI_ISL_660885, EPI_ISL_660886, EPI_ISL_660887, EPI_ISL_660888, EPI_ISL_660889, EPI_ISL_660890, EPI_ISL_660891 | Gundersen Molecular Diagnostics Laboratory | Kabara Cancer Research Institute | Craig S. Richmond, Paraic A. Kenny |
| EPI_ISL_660892, EPI_ISL_660893 | Gundersen Clinical Microbiology Laboratory | Kabara Cancer Research Institute | Craig S. Richmond, Paraic A. Kenny |
| EPI_ISL_660894, EPI_ISL_660895, EPI_ISL_660896, EPI_ISL_660897, EPI_ISL_660898, EPI_ISL_660899, EPI_ISL_660900, EPI_ISL_660901, EPI_ISL_660902, EPI_ISL_660903, EPI_ISL_660904, EPI_ISL_660905, EPI_ISL_660906, EPI_ISL_660907, EPI_ISL_660908, EPI_ISL_660909, EPI_ISL_660910, EPI_ISL_660911, EPI_ISL_660912, EPI_ISL_660913, EPI_ISL_660914, EPI_ISL_660915, EPI_ISL_660916, EPI_ISL_660917, EPI_ISL_660918, EPI_ISL_660919, EPI_ISL_660920, EPI_ISL_660921, EPI_ISL_660922, EPI_ISL_660923, EPI_ISL_660924, EPI_ISL_660925, EPI_ISL_660926, EPI_ISL_660927, EPI_ISL_660928, EPI_ISL_660929, EPI_ISL_660930, EPI_ISL_660931, EPI_ISL_660932, EPI_ISL_660933, EPI_ISL_660934, EPI_ISL_660935, EPI_ISL_660936, EPI_ISL_660937, EPI_ISL_660938, EPI_ISL_660939, EPI_ISL_660940, EPI_ISL_660941, EPI_ISL_660942, EPI_ISL_660943, EPI_ISL_660944, EPI_ISL_660945, EPI_ISL_660946, EPI_ISL_660947, EPI_ISL_660948, EPI_ISL_660949, EPI_ISL_660950, EPI_ISL_660951, EPI_ISL_660952, EPI_ISL_660953, EPI_ISL_660954, EPI_ISL_660955, EPI_ISL_660956, EPI_ISL_660957, EPI_ISL_660958, EPI_ISL_660959, EPI_ISL_660960, EPI_ISL_660961, EPI_ISL_660962, EPI_ISL_660963, EPI_ISL_660964, EPI_ISL_660965, EPI_ISL_660966, EPI_ISL_660967, EPI_ISL_660968, EPI_ISL_660969, EPI_ISL_660970, EPI_ISL_660971, EPI_ISL_660972, EPI_ISL_660973, EPI_ISL_660974, EPI_ISL_660975, EPI_ISL_660976, EPI_ISL_660977, EPI_ISL_660978, EPI_ISL_660979, EPI_ISL_660980, EPI_ISL_660981, EPI_ISL_660982, EPI_ISL_660983, EPI_ISL_660984, EPI_ISL_660985, EPI_ISL_660986, EPI_ISL_660987, EPI_ISL_660988, EPI_ISL_660989, EPI_ISL_660990, EPI_ISL_660991, EPI_ISL_660992, EPI_ISL_660993, EPI_ISL_660994, EPI_ISL_660995, EPI_ISL_660996, EPI_ISL_660997, EPI_ISL_660998, EPI_ISL_660999, EPI_ISL_661000, EPI_ISL_661001, EPI_ISL_661002, EPI_ISL_661003, EPI_ISL_661004, EPI_ISL_661005, EPI_ISL_661006, EPI_ISL_661007, EPI_ISL_661008, EPI_ISL_661009, EPI_ISL_661010, EPI_ISL_661011, EPI_ISL_661012, EPI_ISL_661013, EPI_ISL_661014, EPI_ISL_661015, EPI_ISL_661016, EPI_ISL_661017, EPI_ISL_661018, EPI_ISL_661019, EPI_ISL_661020, EPI_ISL_661021, EPI_ISL_661022, EPI_ISL_661023, EPI_ISL_661024, EPI_ISL_661025, EPI_ISL_661026, EPI_ISL_661027, EPI_ISL_661028, EPI_ISL_661029, EPI_ISL_661030, EPI_ISL_661031, EPI_ISL_661032, EPI_ISL_661033, EPI_ISL_661034, EPI_ISL_661035, EPI_ISL_661036, EPI_ISL_661037, EPI_ISL_661038, EPI_ISL_661039, EPI_ISL_661040, EPI_ISL_661041, EPI_ISL_661042, EPI_ISL_661043, EPI_ISL_661044, EPI_ISL_661045, EPI_ISL_661046, EPI_ISL_661047, EPI_ISL_661048, EPI_ISL_661049, EPI_ISL_661050, EPI_ISL_661051, EPI_ISL_661052, EPI_ISL_661053, EPI_ISL_661054, EPI_ISL_661055, EPI_ISL_661056, EPI_ISL_661057, EPI_ISL_661058, EPI_ISL_661059, EPI_ISL_661060, EPI_ISL_661061, EPI_ISL_661062, EPI_ISL_661063, EPI_ISL_661064, EPI_ISL_661065, EPI_ISL_661066, EPI_ISL_661067, EPI_ISL_661068, EPI_ISL_661069, EPI_ISL_661070, EPI_ISL_661071, EPI_ISL_661072, EPI_ISL_661073, EPI_ISL_661074, EPI_ISL_661075, EPI_ISL_661076, EPI_ISL_661077, EPI_ISL_661078, EPI_ISL_661079, EPI_ISL_661080, EPI_ISL_661081, EPI_ISL_661082, EPI_ISL_661083, EPI_ISL_661084, EPI_ISL_661085, EPI_ISL_661086, EPI_ISL_661087, EPI_ISL_661088, EPI_ISL_661089, EPI_ISL_661090, EPI_ISL_661091, EPI_ISL_661092, EPI_ISL_661093, EPI_ISL_661094, EPI_ISL_661095, EPI_ISL_661096, EPI_ISL_661097, EPI_ISL_661098, EPI_ISL_661099, EPI_ISL_661100, EPI_ISL_661101, EPI_ISL_661102, EPI_ISL_661103, EPI_ISL_661104, EPI_ISL_661105, EPI_ISL_661106, EPI_ISL_661107, EPI_ISL_661108, EPI_ISL_661109, EPI_ISL_661110, EPI_ISL_661111, EPI_ISL_661112, EPI_ISL_661113, EPI_ISL_661114, EPI_ISL_661115, EPI_ISL_661116, EPI_ISL_661117, EPI_ISL_661118, EPI_ISL_661119, EPI_ISL_661120, EPI_ISL_661121, EPI_ISL_661122, EPI_ISL_661123, EPI_ISL_661124, EPI_ISL_661125, EPI_ISL_661126, EPI_ISL_661127, EPI_ISL_661128, EPI_ISL_661129, EPI_ISL_661130, EPI_ISL_661131, EPI_ISL_661132, EPI_ISL_661133, EPI_ISL_661134, EPI_ISL_661135, EPI_ISL_661136, EPI_ISL_661137, EPI_ISL_661138, EPI_ISL_661139, EPI_ISL_661140, EPI_ISL_661141, EPI_ISL_661142, EPI_ISL_661143, EPI_ISL_661144, EPI_ISL_661145, EPI_ISL_661146, EPI_ISL_661147, EPI_ISL_661148, EPI_ISL_661149, EPI_ISL_661150, EPI_ISL_661151, EPI_ISL_661152, EPI_ISL_661153, EPI_ISL_661154, EPI_ISL_661155, EPI_ISL_661156, EPI_ISL_661157, EPI_ISL_661158, EPI_ISL_661159, EPI_ISL_661160, EPI_ISL_661161, EPI_ISL_661162, EPI_ISL_661163, EPI_ISL_661164, EPI_ISL_661165, EPI_ISL_661166, EPI_ISL_661167, EPI_ISL_661168, EPI_ISL_661169, EPI_ISL_661170, EPI_ISL_661171, EPI_ISL_661172, EPI_ISL_661173 | | | |
| see above | Gundersen Molecular Diagnostics Laboratory | Kabara Cancer Research Institute | Craig S. Richmond, Paraic A. Kenny |
| EPI_ISL_661252, EPI_ISL_661253, EPI_ISL_661254 | Quest Diagnostics | Quest Diagnostics | Rosenthal,S.H., Gerasimova,A., Kagan,R.M., Anderson, B., Owen, R., Lacbawan, F. |
| EPI_ISL_666609, EPI_ISL_666613 | LSUHS Emerging Viral Threat Laboratory | Microbial Genome Sequencing Center | Jeremy P. Kamil, Rona S. Scott, Maarten Van Diest, Malgorzata Bienkowska-Haba, Katarzyna Zwolinska, Andrew D. Yurochko, Christopher G. Kevil, Martin J. Sapp, Daniel J. Snyder, Vaughn S. Cooper, John A. Vanchiere |
| EPI_ISL_666617, EPI_ISL_666618, EPI_ISL_666619, EPI_ISL_666620, EPI_ISL_666621 | Environmental and Global Health, University of Florida | Environmental and Global Health, University of Florida | Loeb,J.C., Silva,L.O., Elbadry,M.A., Stephenson,C.J., Morris,J.G., Lednicky,J.A. |
| EPI_ISL_666622 | Environmental and Global Health, University of Florida | Environmental and Global Health, University of Florida | Stephenson,C.J., Elbadry,M.A., Loeb,J.C., Silva,L.O., Morris,J.G., Lednicky,J.A. |
| EPI_ISL_666771, EPI_ISL_666772, EPI_ISL_666773, EPI_ISL_666774, EPI_ISL_666775, EPI_ISL_666776, EPI_ISL_666777, EPI_ISL_666778, EPI_ISL_666779, EPI_ISL_666780, EPI_ISL_666781, EPI_ISL_666782, EPI_ISL_666783, EPI_ISL_666784, EPI_ISL_666785, EPI_ISL_666786, EPI_ISL_666787, EPI_ISL_666788, EPI_ISL_666789, EPI_ISL_666790, EPI_ISL_666791, EPI_ISL_666792, EPI_ISL_666793, EPI_ISL_666794, EPI_ISL_666795, EPI_ISL_666796, EPI_ISL_666797, EPI_ISL_666798, EPI_ISL_666799, EPI_ISL_666800, EPI_ISL_666801, EPI_ISL_666802, EPI_ISL_666803, EPI_ISL_666804, EPI_ISL_666805, EPI_ISL_666806, EPI_ISL_666807, EPI_ISL_666808, EPI_ISL_666809, EPI_ISL_666810, EPI_ISL_666811, EPI_ISL_666812, EPI_ISL_666813 | | | |
| see above | Maryland Public Health Laboratory | Maryland Public Health Laboratory | Maryland Department of Health Laboratories Administration |
| EPI_ISL_666815, EPI_ISL_666818, EPI_ISL_666819, EPI_ISL_666821, EPI_ISL_666825, EPI_ISL_666826, EPI_ISL_666828, EPI_ISL_666829, EPI_ISL_666830, EPI_ISL_666831, EPI_ISL_666833, EPI_ISL_666834, EPI_ISL_666836, EPI_ISL_666837, EPI_ISL_666840, EPI_ISL_666842, EPI_ISL_666844, EPI_ISL_666846, EPI_ISL_666847, EPI_ISL_666848, EPI_ISL_666849, EPI_ISL_666850, EPI_ISL_666851, EPI_ISL_666852, EPI_ISL_666853, EPI_ISL_666854, EPI_ISL_666859, EPI_ISL_666867 | | | |
| see above | Florida Bureau of Public Health Laboratories | Florida Bureau of Public Health Laboratories | Sarah Schmedes, Jason Blanton |
| EPI_ISL_666897, EPI_ISL_666898, EPI_ISL_666899, EPI_ISL_666900, EPI_ISL_666901, EPI_ISL_666902, EPI_ISL_666903, EPI_ISL_666904, EPI_ISL_666905, EPI_ISL_666906, EPI_ISL_666907, EPI_ISL_666908, EPI_ISL_666909, EPI_ISL_666910, EPI_ISL_666911, EPI_ISL_666912, EPI_ISL_666913, EPI_ISL_666914, EPI_ISL_666915, EPI_ISL_666916, EPI_ISL_666917, EPI_ISL_666918, EPI_ISL_666919, EPI_ISL_666920, EPI_ISL_666921, EPI_ISL_666922, EPI_ISL_666923, EPI_ISL_666924, EPI_ISL_666925, EPI_ISL_666926, EPI_ISL_666927, EPI_ISL_666928, EPI_ISL_666929, EPI_ISL_666930, EPI_ISL_666931, EPI_ISL_666932, EPI_ISL_666933, EPI_ISL_666934, EPI_ISL_666935, EPI_ISL_666936, EPI_ISL_666937, EPI_ISL_666938, EPI_ISL_666939, EPI_ISL_666940, EPI_ISL_666941, EPI_ISL_666942, EPI_ISL_666943, EPI_ISL_666944, EPI_ISL_666945, EPI_ISL_666946, EPI_ISL_666947, EPI_ISL_666948, EPI_ISL_666949, EPI_ISL_666950, EPI_ISL_666951, EPI_ISL_666952, EPI_ISL_666953, EPI_ISL_666954, EPI_ISL_666955, EPI_ISL_666956, EPI_ISL_666957, EPI_ISL_666958, EPI_ISL_666959, EPI_ISL_666960, EPI_ISL_666961, EPI_ISL_666962, EPI_ISL_666963 | | | |
| see above | Michigan Department of Health and Human Services, Bureau of Laboratories | Michigan Department of Health and Human Services, Bureau of Laboratories | Blankenship HM, Riner D, Soehnlen MK |
| EPI_ISL_666964 | Quest Diagnostics | Quest Diagnostics | Rosenthal,S.H., Gerasimova,A., Kagan,R.M., Anderson, B., Owen, R., Lacbawan, F. |
| EPI_ISL_666965, EPI_ISL_666966, EPI_ISL_666967, EPI_ISL_666968, EPI_ISL_666969, EPI_ISL_666970, EPI_ISL_666971, EPI_ISL_666972, EPI_ISL_666973, EPI_ISL_666974, EPI_ISL_666975, EPI_ISL_666976, EPI_ISL_666977, EPI_ISL_666978, EPI_ISL_666979, EPI_ISL_666980, EPI_ISL_666981, EPI_ISL_666982, EPI_ISL_666983, EPI_ISL_666984, EPI_ISL_666985, EPI_ISL_666986, EPI_ISL_666987, EPI_ISL_666988, EPI_ISL_666989, EPI_ISL_666990, EPI_ISL_666991, EPI_ISL_666992, EPI_ISL_666993, EPI_ISL_666994, EPI_ISL_666995, EPI_ISL_666996, EPI_ISL_666997, EPI_ISL_666998, EPI_ISL_666999, EPI_ISL_667000, EPI_ISL_667001, EPI_ISL_667002, EPI_ISL_667003, EPI_ISL_667004, EPI_ISL_667005, EPI_ISL_667006, EPI_ISL_667007, EPI_ISL_667008, EPI_ISL_667009, EPI_ISL_667010, EPI_ISL_667011, EPI_ISL_667012, EPI_ISL_667013, EPI_ISL_667014, EPI_ISL_667015, EPI_ISL_667016, EPI_ISL_667017, EPI_ISL_667018, EPI_ISL_667019, EPI_ISL_667020, EPI_ISL_667021, EPI_ISL_667022, EPI_ISL_667023, EPI_ISL_667024, EPI_ISL_667025, EPI_ISL_667026, EPI_ISL_667027, EPI_ISL_667028, EPI_ISL_667029, EPI_ISL_667030, EPI_ISL_667031, EPI_ISL_667032, EPI_ISL_667033, EPI_ISL_667034, EPI_ISL_667035, EPI_ISL_667036, EPI_ISL_667037, EPI_ISL_667038, EPI_ISL_667039, EPI_ISL_667040, EPI_ISL_667041, EPI_ISL_667042, EPI_ISL_667043, EPI_ISL_667044, EPI_ISL_667045, EPI_ISL_667046, EPI_ISL_667047, EPI_ISL_667048, EPI_ISL_667049, EPI_ISL_667050, EPI_ISL_667051, EPI_ISL_667052, EPI_ISL_667053, EPI_ISL_667054, EPI_ISL_667055, EPI_ISL_667056, EPI_ISL_667057, EPI_ISL_667058, EPI_ISL_667059, EPI_ISL_667060, EPI_ISL_667061, EPI_ISL_667062, EPI_ISL_667063, EPI_ISL_667064, EPI_ISL_667065, EPI_ISL_667066, EPI_ISL_667067, EPI_ISL_667068, EPI_ISL_667069, EPI_ISL_667070, EPI_ISL_667071, EPI_ISL_667072, EPI_ISL_667073, EPI_ISL_667074, EPI_ISL_667075, EPI_ISL_667076, EPI_ISL_667077, EPI_ISL_667078 | | | |
| see above | San Diego County Public Health Laboratory | Andersen lab at Scripps Research | SEARCH Alliance San Diego with Tracy Basler, Jovan Shephard, Brett Austin |
| EPI_ISL_667079, EPI_ISL_667082, EPI_ISL_667083, EPI_ISL_667085, EPI_ISL_667086, EPI_ISL_667087, EPI_ISL_667088, EPI_ISL_667089, EPI_ISL_667090, EPI_ISL_667091, EPI_ISL_667092, EPI_ISL_667093, EPI_ISL_667094, EPI_ISL_667095, EPI_ISL_667096, EPI_ISL_667097, EPI_ISL_667098, EPI_ISL_667099, EPI_ISL_667100, EPI_ISL_667101, EPI_ISL_667102, EPI_ISL_667104, EPI_ISL_667105, EPI_ISL_667106, EPI_ISL_667107, EPI_ISL_667108, EPI_ISL_667109, EPI_ISL_667110, EPI_ISL_667112, EPI_ISL_667113, EPI_ISL_667114, EPI_ISL_667115, EPI_ISL_667116, EPI_ISL_667117, EPI_ISL_667118, EPI_ISL_667119, EPI_ISL_667120, EPI_ISL_667121, EPI_ISL_667122, EPI_ISL_667123, EPI_ISL_667124, EPI_ISL_667125, EPI_ISL_667126, EPI_ISL_667127, EPI_ISL_667128, EPI_ISL_667129, EPI_ISL_667130, EPI_ISL_667131, EPI_ISL_667132, EPI_ISL_667133, EPI_ISL_667134, EPI_ISL_667135, EPI_ISL_667136, EPI_ISL_667137, EPI_ISL_667138, EPI_ISL_667139, EPI_ISL_667140, EPI_ISL_667142, EPI_ISL_667143, EPI_ISL_667144, EPI_ISL_667145, EPI_ISL_667146, EPI_ISL_667147, EPI_ISL_667149, EPI_ISL_667150, EPI_ISL_667151, EPI_ISL_667152, EPI_ISL_667153, EPI_ISL_667154, EPI_ISL_667155, EPI_ISL_667156, EPI_ISL_667157, EPI_ISL_667158, EPI_ISL_667159, EPI_ISL_667160, EPI_ISL_667161, EPI_ISL_667162, EPI_ISL_667163, EPI_ISL_667164, EPI_ISL_667165, EPI_ISL_667166, EPI_ISL_667167, EPI_ISL_667168, EPI_ISL_667169, EPI_ISL_667170, EPI_ISL_667171, EPI_ISL_667172, EPI_ISL_667173, EPI_ISL_667174, EPI_ISL_667175, EPI_ISL_667176, EPI_ISL_667177, EPI_ISL_667178, EPI_ISL_667179, EPI_ISL_667180, EPI_ISL_667181, EPI_ISL_667182, EPI_ISL_667183, EPI_ISL_667184, EPI_ISL_667185, EPI_ISL_667187, EPI_ISL_667188, EPI_ISL_667189, EPI_ISL_667190, EPI_ISL_667191, EPI_ISL_667192, EPI_ISL_667193, EPI_ISL_667194, EPI_ISL_667195, EPI_ISL_667196, EPI_ISL_667197, EPI_ISL_667198, EPI_ISL_667199, EPI_ISL_667200, EPI_ISL_667201, EPI_ISL_667202, EPI_ISL_667203, EPI_ISL_667205, EPI_ISL_667206, EPI_ISL_667207, EPI_ISL_667208, EPI_ISL_667209, EPI_ISL_667210, EPI_ISL_667212, EPI_ISL_667213, EPI_ISL_667214, EPI_ISL_667215, EPI_ISL_667216, EPI_ISL_667217, EPI_ISL_667218, EPI_ISL_667219, EPI_ISL_667220, EPI_ISL_667222, EPI_ISL_667223, EPI_ISL_667224, EPI_ISL_667225, EPI_ISL_667226, EPI_ISL_667227, EPI_ISL_667228, EPI_ISL_667229, EPI_ISL_667230, EPI_ISL_667231, EPI_ISL_667232, EPI_ISL_667233, EPI_ISL_667234, EPI_ISL_667235, EPI_ISL_667236, EPI_ISL_667237, EPI_ISL_667238, EPI_ISL_667239, EPI_ISL_667240, EPI_ISL_667241, EPI_ISL_667242, EPI_ISL_667243, EPI_ISL_667244, EPI_ISL_667245, EPI_ISL_667246, EPI_ISL_667247, EPI_ISL_667248, EPI_ISL_667251, EPI_ISL_667252, EPI_ISL_667253, EPI_ISL_667254, EPI_ISL_667255, EPI_ISL_667256, EPI_ISL_667257, EPI_ISL_667258, EPI_ISL_667259, EPI_ISL_667260, EPI_ISL_667261, EPI_ISL_667262, EPI_ISL_667263, EPI_ISL_667264, EPI_ISL_667265, EPI_ISL_667266, EPI_ISL_667267, EPI_ISL_667268, EPI_ISL_667269, EPI_ISL_667270, EPI_ISL_667271, EPI_ISL_667272, EPI_ISL_667273, EPI_ISL_667274, EPI_ISL_667275, EPI_ISL_667276, EPI_ISL_667277, EPI_ISL_667278, EPI_ISL_667279, EPI_ISL_667280, EPI_ISL_667281, EPI_ISL_667282, EPI_ISL_667283, EPI_ISL_667285, EPI_ISL_667286, EPI_ISL_667289, EPI_ISL_667290, EPI_ISL_667291, EPI_ISL_667292, EPI_ISL_667293, EPI_ISL_667294, EPI_ISL_667295, EPI_ISL_667296, EPI_ISL_667297, EPI_ISL_667298, EPI_ISL_667299, EPI_ISL_667301, EPI_ISL_667302, EPI_ISL_667303, EPI_ISL_667304, EPI_ISL_667305, EPI_ISL_667306, EPI_ISL_667307, EPI_ISL_667308, EPI_ISL_667309, EPI_ISL_667310, EPI_ISL_667311, EPI_ISL_667312, EPI_ISL_667313, EPI_ISL_667314, EPI_ISL_667315, EPI_ISL_667316, EPI_ISL_667317, EPI_ISL_667318, EPI_ISL_667319, EPI_ISL_667320, EPI_ISL_667321, EPI_ISL_667322, EPI_ISL_667323, EPI_ISL_667324, EPI_ISL_667325, EPI_ISL_667326, EPI_ISL_667327, EPI_ISL_667328, EPI_ISL_667329, EPI_ISL_667330, EPI_ISL_667331, EPI_ISL_667332, EPI_ISL_667333, EPI_ISL_667334, EPI_ISL_667335, EPI_ISL_667336, EPI_ISL_667337, EPI_ISL_667338, EPI_ISL_667339, EPI_ISL_667340, EPI_ISL_667341, EPI_ISL_667342, EPI_ISL_667343, EPI_ISL_667344, EPI_ISL_667345, EPI_ISL_667347, EPI_ISL_667348, EPI_ISL_667350, EPI_ISL_667351, EPI_ISL_667352, EPI_ISL_667354, EPI_ISL_667355, EPI_ISL_667356, EPI_ISL_667357, EPI_ISL_667358, EPI_ISL_667359, EPI_ISL_667360, EPI_ISL_667361, EPI_ISL_667363, EPI_ISL_667364, EPI_ISL_667365, EPI_ISL_667366, EPI_ISL_667367, EPI_ISL_667368, EPI_ISL_667369, EPI_ISL_667370, EPI_ISL_667371, EPI_ISL_667372, EPI_ISL_667373, EPI_ISL_667374, EPI_ISL_667375, EPI_ISL_667376, EPI_ISL_667377, EPI_ISL_667378, EPI_ISL_667380, EPI_ISL_667381, EPI_ISL_667383, EPI_ISL_667384, EPI_ISL_667385, EPI_ISL_667386, EPI_ISL_667387, EPI_ISL_667388, EPI_ISL_667389, EPI_ISL_667390, EPI_ISL_667391, EPI_ISL_667392, EPI_ISL_667393, EPI_ISL_667394, EPI_ISL_667395, EPI_ISL_667396, EPI_ISL_667397, EPI_ISL_667398, EPI_ISL_667400, EPI_ISL_667401, EPI_ISL_667402, EPI_ISL_667403, EPI_ISL_667405, EPI_ISL_667406, EPI_ISL_667407, EPI_ISL_667408, EPI_ISL_667409, EPI_ISL_667410, EPI_ISL_667411, EPI_ISL_667413, EPI_ISL_667414, EPI_ISL_667415, EPI_ISL_667417, EPI_ISL_667418, EPI_ISL_667419, EPI_ISL_667420, EPI_ISL_667421, EPI_ISL_667422, EPI_ISL_667423, EPI_ISL_667424, EPI_ISL_667425, EPI_ISL_667426, EPI_ISL_667427, EPI_ISL_667429, EPI_ISL_667430, EPI_ISL_667431, EPI_ISL_667432, EPI_ISL_667433, EPI_ISL_667434, EPI_ISL_667435, EPI_ISL_667436, EPI_ISL_667437, EPI_ISL_667438, EPI_ISL_667439, EPI_ISL_667440, EPI_ISL_667441, EPI_ISL_667442, EPI_ISL_667444, EPI_ISL_667445, EPI_ISL_667446, EPI_ISL_667447, EPI_ISL_667448, EPI_ISL_667449, EPI_ISL_667450, EPI_ISL_667451, EPI_ISL_667452, EPI_ISL_667453, EPI_ISL_667454, EPI_ISL_667455, EPI_ISL_667456, EPI_ISL_667457, EPI_ISL_667458, EPI_ISL_667459, EPI_ISL_667460, EPI_ISL_667461, EPI_ISL_667462, EPI_ISL_667463, EPI_ISL_667465, EPI_ISL_667466, EPI_ISL_667467, EPI_ISL_667468, EPI_ISL_667470, EPI_ISL_667471, EPI_ISL_667472, EPI_ISL_667473, EPI_ISL_667474, EPI_ISL_667475, EPI_ISL_667476, EPI_ISL_667477, EPI_ISL_667478, EPI_ISL_667479, EPI_ISL_667480, EPI_ISL_667481, EPI_ISL_667482, EPI_ISL_667483, EPI_ISL_667484, EPI_ISL_667485, EPI_ISL_667486, EPI_ISL_667487, EPI_ISL_667488, EPI_ISL_667489, EPI_ISL_667490, EPI_ISL_667491, EPI_ISL_667492, EPI_ISL_667493, EPI_ISL_667495, EPI_ISL_667496, EPI_ISL_667497, EPI_ISL_667498, EPI_ISL_667500, EPI_ISL_667501, EPI_ISL_667502, EPI_ISL_667503, EPI_ISL_667505, EPI_ISL_667506, EPI_ISL_667507, EPI_ISL_667508, EPI_ISL_667509, EPI_ISL_667510, EPI_ISL_667511, EPI_ISL_667512, EPI_ISL_667513, EPI_ISL_667514, EPI_ISL_667515, EPI_ISL_667516, EPI_ISL_667518, EPI_ISL_667519, EPI_ISL_667520, EPI_ISL_667521, EPI_ISL_667522, EPI_ISL_667523, EPI_ISL_667524, EPI_ISL_667525, EPI_ISL_667526, EPI_ISL_667527, EPI_ISL_667528, EPI_ISL_667529, EPI_ISL_667530, EPI_ISL_667531, EPI_ISL_667532, EPI_ISL_667533, EPI_ISL_667534, EPI_ISL_667535, EPI_ISL_667536, EPI_ISL_667537, EPI_ISL_667538, EPI_ISL_667539, EPI_ISL_667540, EPI_ISL_667541, EPI_ISL_667542, EPI_ISL_667543, EPI_ISL_667544, EPI_ISL_667545, EPI_ISL_667546, EPI_ISL_667547, EPI_ISL_667548, EPI_ISL_667551, EPI_ISL_667552, EPI_ISL_667553, EPI_ISL_667554, EPI_ISL_667555, EPI_ISL_667556, EPI_ISL_667557 | | | |
| see above | OHSU Lab Services Molecular Microbiology Lab | Oregon SARS-CoV-2 Genome Sequencing Center | Brendan L. O'Connell, Ruth V. Nichols, Sally Grindstaff, Alec J. Hirsch, Donna Hansel, Guang Fan, Daniel N. Streblow, William B. Messer, Andrew C. Adey, Benjamin N. Bimber, Brian J. O’Roak |
| EPI_ISL_667776 | Texas Department of State Health Services | Texas Department of State Health Services | Pedrueza,M., Oh,B., Zhang,J., Rahman,M., Pokharel,A., Tuladhar,R., Koag,M., Wang,C., Lee,R., Kubin,G. and Bonser,J. |
| EPI_ISL_671607, EPI_ISL_671608, EPI_ISL_671609, EPI_ISL_671610, EPI_ISL_671611, EPI_ISL_671612, EPI_ISL_671613, EPI_ISL_671614, EPI_ISL_671615, EPI_ISL_671616, EPI_ISL_671617, EPI_ISL_671618, EPI_ISL_671619, EPI_ISL_671620, EPI_ISL_671621, EPI_ISL_671622, EPI_ISL_671623, EPI_ISL_671624, EPI_ISL_671625, EPI_ISL_671626, EPI_ISL_671627, EPI_ISL_671628, EPI_ISL_671629, EPI_ISL_671630, EPI_ISL_671631, EPI_ISL_671632, EPI_ISL_671633, EPI_ISL_671634, EPI_ISL_671635, EPI_ISL_671636, EPI_ISL_671637, EPI_ISL_671638, EPI_ISL_671639, EPI_ISL_671640, EPI_ISL_671641, EPI_ISL_671650, EPI_ISL_671651 | | | |
| see above | Texas Department of State Health Services | Texas Department of State Health Services | Rashmi Tuladhar, Bonnie Oh, Jenny Zhang, Maliha Rahman, Anita Pokharel, Myong Koag, Chung Wang, Rachel Lee, Grace Kubin, Mayela Pedrueza, James Daniel Bonser |
| EPI_ISL_671674 | DOHMH Corona | New York City Public Health Laboratory | Jade Wang, et al. |
| EPI_ISL_671675 | DOHMH Jamaica | New York City Public Health Laboratory | Jade Wang, et al. |

| | | | |
|---|---|---|---|
| EPI_ISL_671676 | DOHMH PHL | New York City Public Health Laboratory | Jade Wang, et al. |
| EPI_ISL_671677 | DOHMH Corona | New York City Public Health Laboratory | Jade Wang, et al. |
| EPI_ISL_671678 | DOHMH Fort Greene | New York City Public Health Laboratory | Jade Wang, et al. |
| EPI_ISL_671679, EPI_ISL_671680 | DOHMH Morrisania | New York City Public Health Laboratory | Jade Wang, et al. |
| EPI_ISL_671681, EPI_ISL_671682, EPI_ISL_671683 | DOHMH Corona | New York City Public Health Laboratory | Jade Wang, et al. |
| EPI_ISL_671684 | DOHMH Chelsea | New York City Public Health Laboratory | Jade Wang, et al. |
| EPI_ISL_671685 | DOHMH Corona | New York City Public Health Laboratory | Jade Wang, et al. |
| EPI_ISL_671686, EPI_ISL_671687, EPI_ISL_671688 | DOHMH Morrisania | New York City Public Health Laboratory | Jade Wang, et al. |
| EPI_ISL_671689 | DOHMH Central Harlem | New York City Public Health Laboratory | Jade Wang, et al. |
| EPI_ISL_671690 | DOHMH Crown Heights | New York City Public Health Laboratory | Jade Wang, et al. |
| EPI_ISL_671691 | DOHMH Corona | New York City Public Health Laboratory | Jade Wang, et al. |
| EPI_ISL_671692 | DOHMH Jamaica | New York City Public Health Laboratory | Jade Wang, et al. |
| EPI_ISL_671693 | DOHMH Chelsea | New York City Public Health Laboratory | Jade Wang, et al. |
| EPI_ISL_671694 | DOHMH Riverside | New York City Public Health Laboratory | Jade Wang, et al. |
| EPI_ISL_671695 | DOHMH Corona | New York City Public Health Laboratory | Jade Wang, et al. |
| EPI_ISL_671696 | DOHMH PHL | New York City Public Health Laboratory | Jade Wang, et al. |
| EPI_ISL_671697 | DOHMH Corona | New York City Public Health Laboratory | Jade Wang, et al. |
| EPI_ISL_671698 | DOHMH Jamaica | New York City Public Health Laboratory | Jade Wang, et al. |
| EPI_ISL_671699, EPI_ISL_671700 | DOHMH Corona | New York City Public Health Laboratory | Jade Wang, et al. |
| EPI_ISL_671701 | DOHMH Riverside | New York City Public Health Laboratory | Jade Wang, et al. |
| EPI_ISL_671702 | DOHMH Chelsea | New York City Public Health Laboratory | Jade Wang, et al. |
| EPI_ISL_671703 | DOHMH Corona | New York City Public Health Laboratory | Jade Wang, et al. |
| EPI_ISL_671704 | DOHMH Central Harlem | New York City Public Health Laboratory | Jade Wang, et al. |
| EPI_ISL_671705 | DOHMH Corona | New York City Public Health Laboratory | Jade Wang, et al. |
| EPI_ISL_671706, EPI_ISL_671707 | DOHMH Jamaica | New York City Public Health Laboratory | Jade Wang, et al. |
| EPI_ISL_671708 | DOHMH Morrisania | New York City Public Health Laboratory | Jade Wang, et al. |
| EPI_ISL_671709 | DOHMH Riverside | New York City Public Health Laboratory | Jade Wang, et al. |
| EPI_ISL_671710, EPI_ISL_671711 | DOHMH Corona | New York City Public Health Laboratory | Jade Wang, et al. |
| EPI_ISL_671712 | DOHMH Jamaica | New York City Public Health Laboratory | Jade Wang, et al. |
| EPI_ISL_671713 | DOHMH Chelsea | New York City Public Health Laboratory | Jade Wang, et al. |
| EPI_ISL_671714 | DOHMH Jamaica | New York City Public Health Laboratory | Jade Wang, et al. |
| EPI_ISL_671715, EPI_ISL_671716 | DOHMH PHL | New York City Public Health Laboratory | Jade Wang, et al. |
| EPI_ISL_671717, EPI_ISL_671718, EPI_ISL_671719, EPI_ISL_671720, EPI_ISL_671721 | DOHMH Corona | New York City Public Health Laboratory | Jade Wang, et al. |
| EPI_ISL_671722 | DOHMH Chelsea | New York City Public Health Laboratory | Jade Wang, et al. |
| EPI_ISL_671723, EPI_ISL_671724 | DOHMH Jamaica | New York City Public Health Laboratory | Jade Wang, et al. |
| EPI_ISL_671725 | DOHMH Riverside | New York City Public Health Laboratory | Jade Wang, et al. |
| EPI_ISL_671726, EPI_ISL_671727, EPI_ISL_671728, EPI_ISL_671729 | DOHMH Morrisania | New York City Public Health Laboratory | Jade Wang, et al. |
| EPI_ISL_671730 | DOHMH Jamaica | New York City Public Health Laboratory | Jade Wang, et al. |
| EPI_ISL_671731, EPI_ISL_671732 | DOHMH Corona | New York City Public Health Laboratory | Jade Wang, et al. |
| EPI_ISL_671733 | DOHMH Riverside | New York City Public Health Laboratory | Jade Wang, et al. |
| EPI_ISL_671734 | DOHMH PHL | New York City Public Health Laboratory | Jade Wang, et al. |
| EPI_ISL_671735 | DOHMH Jamaica | New York City Public Health Laboratory | Jade Wang, et al. |
| EPI_ISL_671736 | DOHMH Morrisania | New York City Public Health Laboratory | Jade Wang, et al. |
| EPI_ISL_671737 | DOHMH PHL | New York City Public Health Laboratory | Jade Wang, et al. |
| EPI_ISL_671738 | DOHMH Fort Greene | New York City Public Health Laboratory | Jade Wang, et al. |
| EPI_ISL_671739 | DOHMH Jamaica | New York City Public Health Laboratory | Jade Wang, et al. |
| EPI_ISL_671740, EPI_ISL_671741, EPI_ISL_671742 | DOHMH Morrisania | New York City Public Health Laboratory | Jade Wang, et al. |
| EPI_ISL_671743 | DOHMH Jamaica | New York City Public Health Laboratory | Jade Wang, et al. |
| EPI_ISL_671744 | DOHMH PHL | New York City Public Health Laboratory | Jade Wang, et al. |
| EPI_ISL_671745 | DOHMH Riverside | New York City Public Health Laboratory | Jade Wang, et al. |
| EPI_ISL_671746, EPI_ISL_671747 | DOHMH Corona | New York City Public Health Laboratory | Jade Wang, et al. |
| EPI_ISL_671748, EPI_ISL_671749, EPI_ISL_671750 | DOHMH Riverside | New York City Public Health Laboratory | Jade Wang, et al. |
| EPI_ISL_671751, EPI_ISL_671752, EPI_ISL_671753, EPI_ISL_671754, EPI_ISL_671755, EPI_ISL_671756, EPI_ISL_671757, EPI_ISL_671758 | DOHMH Morrisania | New York City Public Health Laboratory | Jade Wang, et al. |
| EPI_ISL_671759 | DOHMH Fort Greene | New York City Public Health Laboratory | Jade Wang, et al. |
| EPI_ISL_671760 | DOHMH Crown Heights | New York City Public Health Laboratory | Jade Wang, et al. |
| EPI_ISL_671761 | DOHMH Corona | New York City Public Health Laboratory | Jade Wang, et al. |

| | | | |
|---|---|---|---|
| EPI_ISL_671762 | DOHMH Riverside | New York City Public Health Laboratory | Jade Wang, et al. |
| EPI_ISL_671763 | DOHMH Chelsea | New York City Public Health Laboratory | Jade Wang, et al. |
| EPI_ISL_671765, EPI_ISL_671767, EPI_ISL_671768 | Santa Clara County Public Health Department | Chiu Laboratory, University of California, San Francisco | Xianding Deng, Scot Federman, Wei Gu, Elsa Villarino, Brandon Bonin, Debra A. Wadford, and Charles Y. Chiu |
| EPI_ISL_672013 | Santa Clara County Public Health Laboratory | Chan-Zuckerberg Biohub | CZB Cliahub Consortium |
| EPI_ISL_672014 | Alameda County Public Health Lab | Chan-Zuckerberg Biohub | CZB Cliahub Consortium |
| EPI_ISL_672015, EPI_ISL_672016 | UCSF Clinical Microbiology Laboratory | Chan-Zuckerberg Biohub | CZB Cliahub Consortium |
| EPI_ISL_672017 | The Ashley Laboratory, Stanford University | Chan-Zuckerberg Biohub | CZB Cliahub Consortium |
| EPI_ISL_672018 | UCSF Clinical Microbiology Laboratory | Chan-Zuckerberg Biohub | CZB Cliahub Consortium |
| EPI_ISL_672019 | Madera County Department of Public Health | Chan-Zuckerberg Biohub | CZB Cliahub Consortium |
| EPI_ISL_672020, EPI_ISL_672021 | UCSF Clinical Microbiology Laboratory | Chan-Zuckerberg Biohub | CZB Cliahub Consortium |
| EPI_ISL_672022 | Madera County Department of Public Health | Chan-Zuckerberg Biohub | CZB Cliahub Consortium |
| EPI_ISL_672023, EPI_ISL_672024 | Alameda County Public Health Lab | Chan-Zuckerberg Biohub | CZB Cliahub Consortium |
| EPI_ISL_672025 | UCSF Clinical Microbiology Laboratory | Chan-Zuckerberg Biohub | CZB Cliahub Consortium |
| EPI_ISL_672026 | Madera County Department of Public Health | Chan-Zuckerberg Biohub | CZB Cliahub Consortium |
| EPI_ISL_672027 | The Ashley Laboratory, Stanford University | Chan-Zuckerberg Biohub | CZB Cliahub Consortium |
| EPI_ISL_672028 | County of San Luis Obispo Public Health Laboratory | Chan-Zuckerberg Biohub | CZB Cliahub Consortium |
| EPI_ISL_672029 | Alameda County Public Health Lab | Chan-Zuckerberg Biohub | CZB Cliahub Consortium |
| EPI_ISL_672030 | The Ashley Laboratory, Stanford University | Chan-Zuckerberg Biohub | CZB Cliahub Consortium |
| EPI_ISL_672031 | Santa Clara County Public Health Laboratory | Chan-Zuckerberg Biohub | CZB Cliahub Consortium |
| EPI_ISL_672032 | Alameda County Public Health Lab | Chan-Zuckerberg Biohub | CZB Cliahub Consortium |
| EPI_ISL_672033, EPI_ISL_672035 | Santa Clara County Public Health Laboratory | Chan-Zuckerberg Biohub | CZB Cliahub Consortium |
| EPI_ISL_672036 | Alameda County Public Health Lab | Chan-Zuckerberg Biohub | CZB Cliahub Consortium |
| EPI_ISL_672037 | Madera County Department of Public Health | Chan-Zuckerberg Biohub | CZB Cliahub Consortium |
| EPI_ISL_672038, EPI_ISL_672039, EPI_ISL_672040 | Alameda County Public Health Lab | Chan-Zuckerberg Biohub | CZB Cliahub Consortium |
| EPI_ISL_672041 | Madera County Department of Public Health | Chan-Zuckerberg Biohub | CZB Cliahub Consortium |
| EPI_ISL_672042 | The Ashley Laboratory, Stanford University | Chan-Zuckerberg Biohub | CZB Cliahub Consortium |
| EPI_ISL_672043, EPI_ISL_672044 | Santa Clara County Public Health Laboratory | Chan-Zuckerberg Biohub | CZB Cliahub Consortium |
| EPI_ISL_672045 | Alameda County Public Health Lab | Chan-Zuckerberg Biohub | CZB Cliahub Consortium |
| EPI_ISL_672046 | Madera County Department of Public Health | Chan-Zuckerberg Biohub | CZB Cliahub Consortium |
| EPI_ISL_672047 | The Ashley Laboratory, Stanford University | Chan-Zuckerberg Biohub | CZB Cliahub Consortium |
| EPI_ISL_672048 | Madera County Department of Public Health | Chan-Zuckerberg Biohub | CZB Cliahub Consortium |
| EPI_ISL_672049 | The Ashley Laboratory, Stanford University | Chan-Zuckerberg Biohub | CZB Cliahub Consortium |
| EPI_ISL_672050 | Alameda County Public Health Lab | Chan-Zuckerberg Biohub | CZB Cliahub Consortium |
| EPI_ISL_672051 | The Ashley Laboratory, Stanford University | Chan-Zuckerberg Biohub | CZB Cliahub Consortium |
| EPI_ISL_672052 | Santa Clara County Public Health Laboratory | Chan-Zuckerberg Biohub | CZB Cliahub Consortium |
| EPI_ISL_672053, EPI_ISL_672054 | Alameda County Public Health Lab | Chan-Zuckerberg Biohub | CZB Cliahub Consortium |
| EPI_ISL_672055, EPI_ISL_672056 | The Ashley Laboratory, Stanford University | Chan-Zuckerberg Biohub | CZB Cliahub Consortium |
| EPI_ISL_672057 | Santa Clara County Public Health Laboratory | Chan-Zuckerberg Biohub | CZB Cliahub Consortium |
| EPI_ISL_672058 | The Ashley Laboratory, Stanford University | Chan-Zuckerberg Biohub | CZB Cliahub Consortium |
| EPI_ISL_672059 | Madera County Department of Public Health | Chan-Zuckerberg Biohub | CZB Cliahub Consortium |
| EPI_ISL_672060 | Alameda County Public Health Lab | Chan-Zuckerberg Biohub | CZB Cliahub Consortium |
| EPI_ISL_672061 | The Ashley Laboratory, Stanford University | Chan-Zuckerberg Biohub | CZB Cliahub Consortium |
| EPI_ISL_672062 | Alameda County Public Health Lab | Chan-Zuckerberg Biohub | CZB Cliahub Consortium |
| EPI_ISL_672063 | The Ashley Laboratory, Stanford University | Chan-Zuckerberg Biohub | CZB Cliahub Consortium |
| EPI_ISL_672064 | Alameda County Public Health Lab | Chan-Zuckerberg Biohub | CZB Cliahub Consortium |
| EPI_ISL_672065 | Santa Clara County Public Health Laboratory | Chan-Zuckerberg Biohub | CZB Cliahub Consortium |
| EPI_ISL_672066 | Alameda County Public Health Lab | Chan-Zuckerberg Biohub | CZB Cliahub Consortium |
| EPI_ISL_672067 | Madera County Department of Public Health | Chan-Zuckerberg Biohub | CZB Cliahub Consortium |
| EPI_ISL_672068 | County of San Luis Obispo Public Health Laboratory | Chan-Zuckerberg Biohub | CZB Cliahub Consortium |
| EPI_ISL_672069, EPI_ISL_672070 | Santa Clara County Public Health Laboratory | Chan-Zuckerberg Biohub | CZB Cliahub Consortium |

| | | | |
|---|---|---|---|
| EPI_ISL_672071 | The Ashley Laboratory, Stanford University | Chan-Zuckerberg Biohub | CZB Cliahub Consortium |
| EPI_ISL_672072, EPI_ISL_672073 | Santa Clara County Public Health Laboratory | Chan-Zuckerberg Biohub | CZB Cliahub Consortium |
| EPI_ISL_672074 | The Ashley Laboratory, Stanford University | Chan-Zuckerberg Biohub | CZB Cliahub Consortium |
| EPI_ISL_672075 | Orange County Public Health Lab | Chan-Zuckerberg Biohub | CZB Cliahub Consortium |
| EPI_ISL_672076 | Alameda County Public Health Lab | Chan-Zuckerberg Biohub | CZB Cliahub Consortium |
| EPI_ISL_672077 | San Francisco Public Health Laboratory | Chan-Zuckerberg Biohub | CZB Cliahub Consortium |
| EPI_ISL_672078 | Alameda County Public Health Lab | Chan-Zuckerberg Biohub | CZB Cliahub Consortium |
| EPI_ISL_672079 | The Ashley Laboratory, Stanford University | Chan-Zuckerberg Biohub | CZB Cliahub Consortium |
| EPI_ISL_672080 | Santa Clara County Public Health Laboratory | Chan-Zuckerberg Biohub | CZB Cliahub Consortium |
| EPI_ISL_672081 | Madera County Department of Public Health | Chan-Zuckerberg Biohub | CZB Cliahub Consortium |
| EPI_ISL_672082 | Alameda County Public Health Lab | Chan-Zuckerberg Biohub | CZB Cliahub Consortium |
| EPI_ISL_672083 | The Ashley Laboratory, Stanford University | Chan-Zuckerberg Biohub | CZB Cliahub Consortium |
| EPI_ISL_672084 | Orange County Public Health Lab | Chan-Zuckerberg Biohub | CZB Cliahub Consortium |
| EPI_ISL_672085 | Santa Clara County Public Health Laboratory | Chan-Zuckerberg Biohub | CZB Cliahub Consortium |
| EPI_ISL_672086 | Alameda County Public Health Lab | Chan-Zuckerberg Biohub | CZB Cliahub Consortium |
| EPI_ISL_672087 | County of San Luis Obispo Public Health Laboratory | Chan-Zuckerberg Biohub | CZB Cliahub Consortium |
| EPI_ISL_672088, EPI_ISL_672089 | Santa Clara County Public Health Laboratory | Chan-Zuckerberg Biohub | CZB Cliahub Consortium |
| EPI_ISL_672098, EPI_ISL_672099, EPI_ISL_672100, EPI_ISL_672101, EPI_ISL_672102 | Madera County Department of Public Health | Chan-Zuckerberg Biohub | CZB Cliahub Consortium |
| EPI_ISL_672103 | Orange County Public Health Lab | Chan-Zuckerberg Biohub | CZB Cliahub Consortium |
| EPI_ISL_672104 | Santa Clara County Public Health Laboratory | Chan-Zuckerberg Biohub | CZB Cliahub Consortium |
| EPI_ISL_672105 | The Ashley Laboratory, Stanford University | Chan-Zuckerberg Biohub | CZB Cliahub Consortium |
| EPI_ISL_672106, EPI_ISL_672107, EPI_ISL_672108 | Santa Clara County Public Health Laboratory | Chan-Zuckerberg Biohub | CZB Cliahub Consortium |
| EPI_ISL_672109, EPI_ISL_672110 | Orange County Public Health Lab | Chan-Zuckerberg Biohub | CZB Cliahub Consortium |
| EPI_ISL_672111 | Santa Clara County Public Health Laboratory | Chan-Zuckerberg Biohub | CZB Cliahub Consortium |
| EPI_ISL_672112 | The Ashley Laboratory, Stanford University | Chan-Zuckerberg Biohub | CZB Cliahub Consortium |
| EPI_ISL_672113 | Alameda County Public Health Lab | Chan-Zuckerberg Biohub | CZB Cliahub Consortium |
| EPI_ISL_672114, EPI_ISL_672115, EPI_ISL_672116, EPI_ISL_672117, EPI_ISL_672118, EPI_ISL_672119, EPI_ISL_672120, EPI_ISL_672121, EPI_ISL_672122, EPI_ISL_672123, EPI_ISL_672124, EPI_ISL_672125, EPI_ISL_672126, EPI_ISL_672127, EPI_ISL_672128, EPI_ISL_672129, EPI_ISL_672130, EPI_ISL_672131, EPI_ISL_672132, EPI_ISL_672133, EPI_ISL_672134, EPI_ISL_672135, EPI_ISL_672136, EPI_ISL_672137, EPI_ISL_672138, EPI_ISL_672139, EPI_ISL_672140, EPI_ISL_672141, EPI_ISL_672142, EPI_ISL_672143, EPI_ISL_672144, EPI_ISL_672145, EPI_ISL_672146, EPI_ISL_672147, EPI_ISL_672148, EPI_ISL_672149, EPI_ISL_672150, EPI_ISL_672151, EPI_ISL_672152, EPI_ISL_672153, EPI_ISL_672154, EPI_ISL_672155, EPI_ISL_672156, EPI_ISL_672157, EPI_ISL_672158, EPI_ISL_672159, EPI_ISL_672160, EPI_ISL_672161, EPI_ISL_672162, EPI_ISL_672163, EPI_ISL_672164, EPI_ISL_672165, EPI_ISL_672166, EPI_ISL_672167, EPI_ISL_672168, EPI_ISL_672169, EPI_ISL_672170, EPI_ISL_672171, EPI_ISL_672172, EPI_ISL_672173, EPI_ISL_672174, EPI_ISL_672175, EPI_ISL_672176, EPI_ISL_672177, EPI_ISL_672178, EPI_ISL_672179, EPI_ISL_672180, EPI_ISL_672181, EPI_ISL_672182, EPI_ISL_672183, EPI_ISL_672184, EPI_ISL_672185, EPI_ISL_672186, EPI_ISL_672187, EPI_ISL_672188, EPI_ISL_672189, EPI_ISL_672190, EPI_ISL_672191, EPI_ISL_672192, EPI_ISL_672193, EPI_ISL_672194, EPI_ISL_672195, EPI_ISL_672196, EPI_ISL_672197, EPI_ISL_672198, EPI_ISL_672199, EPI_ISL_672200, EPI_ISL_672201, EPI_ISL_672202, EPI_ISL_672203, EPI_ISL_672204, EPI_ISL_672205, EPI_ISL_672206, EPI_ISL_672207, EPI_ISL_672208, EPI_ISL_672209, EPI_ISL_672210, EPI_ISL_672211, EPI_ISL_672212, EPI_ISL_672213, EPI_ISL_672214, EPI_ISL_672215, EPI_ISL_672216, EPI_ISL_672217, EPI_ISL_672218, EPI_ISL_672219, EPI_ISL_672220, EPI_ISL_672221, EPI_ISL_672222, EPI_ISL_672223, EPI_ISL_672224, EPI_ISL_672225, EPI_ISL_672226, EPI_ISL_672227, EPI_ISL_672228, EPI_ISL_672229, EPI_ISL_672230, EPI_ISL_672231, EPI_ISL_672232, EPI_ISL_672233, EPI_ISL_672234, EPI_ISL_672235, EPI_ISL_672236, EPI_ISL_672237, EPI_ISL_672238, EPI_ISL_672239, EPI_ISL_672240, EPI_ISL_672241, EPI_ISL_672242, EPI_ISL_672243, EPI_ISL_672244, EPI_ISL_672245, EPI_ISL_672246, EPI_ISL_672247, EPI_ISL_672248, EPI_ISL_672249, EPI_ISL_672250, EPI_ISL_672251, EPI_ISL_672252, EPI_ISL_672253, EPI_ISL_672254, EPI_ISL_672255, EPI_ISL_672256, EPI_ISL_672257, EPI_ISL_672258, EPI_ISL_672259, EPI_ISL_672260, EPI_ISL_672261, EPI_ISL_672262, EPI_ISL_672263, EPI_ISL_672264, EPI_ISL_672265, EPI_ISL_672266, EPI_ISL_672267, EPI_ISL_672268, EPI_ISL_672269, EPI_ISL_672270, EPI_ISL_672271, EPI_ISL_672272, EPI_ISL_672273, EPI_ISL_672274 | | | |
| see above | The Ashley Laboratory, Stanford University | Chan-Zuckerberg Biohub | CZB Cliahub Consortium |
| EPI_ISL_672275, EPI_ISL_672276, EPI_ISL_672277, EPI_ISL_672278, EPI_ISL_672279, EPI_ISL_672280, EPI_ISL_672281, EPI_ISL_672282, EPI_ISL_672283, EPI_ISL_672284, EPI_ISL_672285, EPI_ISL_672286, EPI_ISL_672287, EPI_ISL_672288, EPI_ISL_672289, EPI_ISL_672290, EPI_ISL_672291, EPI_ISL_672292, EPI_ISL_672293, EPI_ISL_672294, EPI_ISL_672295, EPI_ISL_672296, EPI_ISL_672297, EPI_ISL_672298, EPI_ISL_672299, EPI_ISL_672300, EPI_ISL_672301, EPI_ISL_672302, EPI_ISL_672303, EPI_ISL_672304 | | | |
| see above | Santa Clara County Public Health Laboratory | Chan-Zuckerberg Biohub | CZB Cliahub Consortium |
| EPI_ISL_672305, EPI_ISL_672306, EPI_ISL_672307, EPI_ISL_672308, EPI_ISL_672309, EPI_ISL_672310, EPI_ISL_672311, EPI_ISL_672312, EPI_ISL_672313, EPI_ISL_672314, EPI_ISL_672315, EPI_ISL_672316, EPI_ISL_672317, EPI_ISL_672318, EPI_ISL_672319, EPI_ISL_672320, EPI_ISL_672321, EPI_ISL_672322, EPI_ISL_672323, EPI_ISL_672324, EPI_ISL_672325, EPI_ISL_672326, EPI_ISL_672327, EPI_ISL_672328, EPI_ISL_672329, EPI_ISL_672330, EPI_ISL_672331, EPI_ISL_672332, EPI_ISL_672333, EPI_ISL_672334, EPI_ISL_672335, EPI_ISL_672336, EPI_ISL_672337, EPI_ISL_672338, EPI_ISL_672339, EPI_ISL_672340, EPI_ISL_672341, EPI_ISL_672342, EPI_ISL_672343, EPI_ISL_672344, EPI_ISL_672345, EPI_ISL_672346, EPI_ISL_672347, EPI_ISL_672348, EPI_ISL_672349, EPI_ISL_672350, EPI_ISL_672351, EPI_ISL_672352, EPI_ISL_672353 | | | |
| see above | Alameda County Public Health Lab | Chan-Zuckerberg Biohub | CZB Cliahub Consortium |
| EPI_ISL_672354, EPI_ISL_672355, EPI_ISL_672356, EPI_ISL_672357, EPI_ISL_672358, EPI_ISL_672359 | County of San Luis Obispo Public Health Laboratory | Chan-Zuckerberg Biohub | CZB Cliahub Consortium |
| EPI_ISL_672360, EPI_ISL_672361, EPI_ISL_672362, EPI_ISL_672363, EPI_ISL_672364, EPI_ISL_672365, EPI_ISL_672366, EPI_ISL_672367, EPI_ISL_672368, EPI_ISL_672369, EPI_ISL_672370, EPI_ISL_672371, EPI_ISL_672372, EPI_ISL_672373, EPI_ISL_672374 | | | |
| see above | Orange County Public Health Lab | Chan-Zuckerberg Biohub | CZB Cliahub Consortium |
| EPI_ISL_672375, EPI_ISL_672376, EPI_ISL_672377, EPI_ISL_672378, EPI_ISL_672379, EPI_ISL_672380, EPI_ISL_672381, EPI_ISL_672382, EPI_ISL_672383, EPI_ISL_672384, EPI_ISL_672385 | | | |
| see above | San Francisco Public Health Laboratory | Chan-Zuckerberg Biohub | CZB Cliahub Consortium |
| EPI_ISL_672386, EPI_ISL_672387, EPI_ISL_672388, EPI_ISL_672389, EPI_ISL_672390, EPI_ISL_672391, EPI_ISL_672392, EPI_ISL_672393 | UCSF Clinical Microbiology Laboratory | Chan-Zuckerberg Biohub | CZB Cliahub Consortium |
| EPI_ISL_672394, EPI_ISL_672395, EPI_ISL_672396, EPI_ISL_672397, EPI_ISL_672398, EPI_ISL_672399, EPI_ISL_672400, EPI_ISL_672401, EPI_ISL_672402, EPI_ISL_672403, EPI_ISL_672404, EPI_ISL_672405, EPI_ISL_672406, EPI_ISL_672407, EPI_ISL_672408, EPI_ISL_672409, EPI_ISL_672410, EPI_ISL_672411, EPI_ISL_672412, EPI_ISL_672413, EPI_ISL_672414, EPI_ISL_672415, EPI_ISL_672416, EPI_ISL_672417, EPI_ISL_672418, EPI_ISL_672419, EPI_ISL_672420, EPI_ISL_672421, EPI_ISL_672422 | | | |
| see above | Madera County Department of Public Health | Chan-Zuckerberg Biohub | CZB Cliahub Consortium |
| EPI_ISL_672423, EPI_ISL_672424, EPI_ISL_672425 | Fresno County Public Health Laboratory | Chan-Zuckerberg Biohub | CZB Cliahub Consortium |
| EPI_ISL_672426, EPI_ISL_672427, EPI_ISL_672428 | UCSF Clinical Microbiology Laboratory | Chan-Zuckerberg Biohub | CZB Cliahub Consortium |

| | | | |
|---|---|---|---|
| EPI_ISL_672429, EPI_ISL_672430, EPI_ISL_672431, EPI_ISL_672432, EPI_ISL_672433, EPI_ISL_672434, EPI_ISL_672435, EPI_ISL_672436, EPI_ISL_672437, EPI_ISL_672438 | Santa Clara County Public Health Laboratory | Chan-Zuckerberg Biohub | CZB Cliahub Consortium |
| EPI_ISL_672439, EPI_ISL_672440, EPI_ISL_672441, EPI_ISL_672442, EPI_ISL_672443, EPI_ISL_672444, EPI_ISL_672445, EPI_ISL_672446, EPI_ISL_672447, EPI_ISL_672448, EPI_ISL_672449, EPI_ISL_672450, EPI_ISL_672451, EPI_ISL_672452, EPI_ISL_672453, EPI_ISL_672454, EPI_ISL_672455, EPI_ISL_672456, EPI_ISL_672457, EPI_ISL_672458, EPI_ISL_672459, EPI_ISL_672460, EPI_ISL_672461, EPI_ISL_672462, EPI_ISL_672463, EPI_ISL_672464, EPI_ISL_672465, EPI_ISL_672466, EPI_ISL_672467, EPI_ISL_672468, EPI_ISL_672469, EPI_ISL_672470, EPI_ISL_672471, EPI_ISL_672472, EPI_ISL_672473, EPI_ISL_672474, EPI_ISL_672475, EPI_ISL_672476 | | | |
| see above | Alameda County Public Health Lab | Chan-Zuckerberg Biohub | CZB Cliahub Consortium |
| EPI_ISL_672477, EPI_ISL_672478, EPI_ISL_672479, EPI_ISL_672480, EPI_ISL_672481, EPI_ISL_672482, EPI_ISL_672483, EPI_ISL_672484, EPI_ISL_672485, EPI_ISL_672486, EPI_ISL_672487 | | | |
| see above | Orange County Public Health Lab | Chan-Zuckerberg Biohub | CZB Cliahub Consortium |
| EPI_ISL_672488, EPI_ISL_672489 | Contra Costa County Public Health Lab | Chan-Zuckerberg Biohub | CZB Cliahub Consortium |
| EPI_ISL_672490, EPI_ISL_672491, EPI_ISL_672492, EPI_ISL_672493, EPI_ISL_672494, EPI_ISL_672495, EPI_ISL_672496, EPI_ISL_672497, EPI_ISL_672498, EPI_ISL_672499, EPI_ISL_672500, EPI_ISL_672501 | | | |
| see above | UCSF Clinical Microbiology Laboratory | Chan-Zuckerberg Biohub | CZB Cliahub Consortium |
| EPI_ISL_672502, EPI_ISL_672503, EPI_ISL_672504, EPI_ISL_672505, EPI_ISL_672506, EPI_ISL_672507, EPI_ISL_672508, EPI_ISL_672509, EPI_ISL_672510, EPI_ISL_672511, EPI_ISL_672512, EPI_ISL_672513, EPI_ISL_672514, EPI_ISL_672515, EPI_ISL_672516, EPI_ISL_672517, EPI_ISL_672518, EPI_ISL_672519, EPI_ISL_672520, EPI_ISL_672521, EPI_ISL_672522, EPI_ISL_672523, EPI_ISL_672524, EPI_ISL_672525, EPI_ISL_672526, EPI_ISL_672527, EPI_ISL_672528, EPI_ISL_672529, EPI_ISL_672530 | | | |
| see above | Madera County Department of Public Health | Chan-Zuckerberg Biohub | CZB Cliahub Consortium |
| EPI_ISL_672531, EPI_ISL_672532, EPI_ISL_672533, EPI_ISL_672534, EPI_ISL_672535, EPI_ISL_672536, EPI_ISL_672537, EPI_ISL_672538, EPI_ISL_672539, EPI_ISL_672540, EPI_ISL_672541, EPI_ISL_672542, EPI_ISL_672548, EPI_ISL_672549, EPI_ISL_672550, EPI_ISL_672551, EPI_ISL_672552, EPI_ISL_672553, EPI_ISL_672554, EPI_ISL_672555, EPI_ISL_672556, EPI_ISL_672557, EPI_ISL_672558, EPI_ISL_672559, EPI_ISL_672560, EPI_ISL_672561, EPI_ISL_672562, EPI_ISL_672563, EPI_ISL_672564, EPI_ISL_672565, EPI_ISL_672566, EPI_ISL_672567, EPI_ISL_672568, EPI_ISL_672569 | | | |
| see above | Utah Public Health Laboratory | Utah Public Health Laboratory | Erin Young, Kelly Oakeson |
| EPI_ISL_676604, EPI_ISL_676605, EPI_ISL_676606, EPI_ISL_676607, EPI_ISL_676608, EPI_ISL_676609, EPI_ISL_676610, EPI_ISL_676611, EPI_ISL_676612, EPI_ISL_676613, EPI_ISL_676614, EPI_ISL_676615, EPI_ISL_676616, EPI_ISL_676617, EPI_ISL_676618, EPI_ISL_676619, EPI_ISL_676620, EPI_ISL_676621, EPI_ISL_676622, EPI_ISL_676623, EPI_ISL_676624, EPI_ISL_676625, EPI_ISL_676626, EPI_ISL_676627, EPI_ISL_676628, EPI_ISL_676629, EPI_ISL_676630, EPI_ISL_676631, EPI_ISL_676632, EPI_ISL_676633, EPI_ISL_676634, EPI_ISL_676635, EPI_ISL_676636, EPI_ISL_676637, EPI_ISL_676638, EPI_ISL_676642, EPI_ISL_676643, EPI_ISL_676644, EPI_ISL_676645, EPI_ISL_676646, EPI_ISL_676647 | | | |
| see above | Texas Department of State Health Services | Texas Department of State Health Services | Rashmi Tuladhar, Bonnie Oh, Jenny Zhang, Maliha Rahman, Anita Pokharel, Myong Koag, Chung Wang, Rachel Lee, Grace Kubin, Mayela Pedrueza, James Daniel Bonser |
| EPI_ISL_676648, EPI_ISL_676649, EPI_ISL_676650 | Wadsworth Center, New York State Department of Health | Wadsworth Center, New York State Department of Health | Kirsten St. George, Daryl M. Lamson, Sara Griesemer, Jonathan Plitnick, Navjot Singh, Matthew D. Shudt, Erica Lasek-Nesselquist |
| EPI_ISL_676651 | Wadsworth Center, New York State Department.of Health | Wadsworth Center, New York State Department.of Health | Kirsten St. George, Daryl M. Lamson, Alexis Russel, Jonathan Plitnick, Navjot Singh, John Kelly, Sara Griesemer, Erasmus Schneider, Erica Lasek-Nesselquist |
| EPI_ISL_676652 | Masonic Medical Research Institute | Wadsworth Center, New York State Department of Health | Nathan Tucker, Kirsten St. George, Daryl M. Lamson, Alexis Russel, Jonathan Plitnick, Navjot Singh, John Kelly, Sara Griesemer, Erasmus Schneider, Erica Lasek-Nesselquist |
| EPI_ISL_676653, EPI_ISL_676654, EPI_ISL_676655, EPI_ISL_676656, EPI_ISL_676657, EPI_ISL_676658 | Wadsworth Center, New York State Department.of Health | Wadsworth Center, New York State Department.of Health | Kirsten St. George, Daryl M. Lamson, Alexis Russel, Jonathan Plitnick, Navjot Singh, John Kelly, Sara Griesemer, Erasmus Schneider, Erica Lasek-Nesselquist |
| EPI_ISL_676659 | Masonic Medical Research Institute | Wadsworth Center, New York State Department.of Health | Nathan Tucker, Kirsten St. George, Daryl M. Lamson, Alexis Russel, Jonathan Plitnick, Navjot Singh, John Kelly, Sara Griesemer, Erasmus Schneider, Erica Lasek-Nesselquist |
| EPI_ISL_676660 | Wadsworth Center, New York State Department.of Health | Wadsworth Center, New York State Department.of Health | Kirsten St. George, Daryl M. Lamson, Alexis Russel, Jonathan Plitnick, Navjot Singh, John Kelly, Sara Griesemer, Erasmus Schneider, Erica Lasek-Nesselquist |
| EPI_ISL_676661 | Masonic Medical Research Institute | Wadsworth Center, New York State Department.of Health | Nathan Tucker, Kirsten St. George, Daryl M. Lamson, Alexis Russel, Jonathan Plitnick, Navjot Singh, John Kelly, Sara Griesemer, Erasmus Schneider, Erica Lasek-Nesselquist |
| EPI_ISL_676662 | Wadsworth Center, New York State Department.of Health | Wadsworth Center, New York State Department.of Health | Kirsten St. George, Daryl M. Lamson, Alexis Russel, Jonathan Plitnick, Navjot Singh, John Kelly, Sara Griesemer, Erasmus Schneider, Erica Lasek-Nesselquist |
| EPI_ISL_676663 | Masonic Medical Research Institute | Wadsworth Center, New York State Department.of Health | Nathan Tucker, Kirsten St. George, Daryl M. Lamson, Alexis Russel, Jonathan Plitnick, Navjot Singh, John Kelly, Sara Griesemer, Erasmus Schneider, Erica Lasek-Nesselquist |
| EPI_ISL_676664 | Wadsworth Center, New York State Department.of Health | Wadsworth Center, New York State Department.of Health | Kirsten St. George, Daryl M. Lamson, Alexis Russel, Jonathan Plitnick, Navjot Singh, John Kelly, Sara Griesemer, Erasmus Schneider, Erica Lasek-Nesselquist |
| EPI_ISL_676665 | Masonic Medical Research Institute | Wadsworth Center, New York State Department.of Health | Nathan Tucker, Kirsten St. George, Daryl M. Lamson, Alexis Russel, Jonathan Plitnick, Navjot Singh, John Kelly, Sara Griesemer, Erasmus Schneider, Erica Lasek-Nesselquist |
| EPI_ISL_676666, EPI_ISL_676667 | Wadsworth Center, New York State Department.of Health | Wadsworth Center, New York State Department.of Health | Kirsten St. George, Daryl M. Lamson, Alexis Russel, Jonathan Plitnick, Navjot Singh, John Kelly, Sara Griesemer, Erasmus Schneider, Erica Lasek-Nesselquist |
| EPI_ISL_676668 | Masonic Medical Research Institute | Wadsworth Center, New York State Department.of Health | Nathan Tucker, Kirsten St. George, Daryl M. Lamson, Alexis Russel, Jonathan Plitnick, Navjot Singh, John Kelly, Sara Griesemer, Erasmus Schneider, Erica Lasek-Nesselquist |
| EPI_ISL_676669, EPI_ISL_676670, EPI_ISL_676671, EPI_ISL_676672 | Wadsworth Center, New York State Department.of Health | Wadsworth Center, New York State Department.of Health | Kirsten St. George, Daryl M. Lamson, Alexis Russel, Jonathan Plitnick, Navjot Singh, John Kelly, Sara Griesemer, Erasmus Schneider, Erica Lasek-Nesselquist |
| EPI_ISL_676673 | Masonic Medical Research Institute | Wadsworth Center, New York State Department.of Health | Nathan Tucker, Kirsten St. George, Daryl M. Lamson, Alexis Russel, Jonathan Plitnick, Navjot Singh, John Kelly, Sara Griesemer, Erasmus Schneider, Erica Lasek-Nesselquist |
| EPI_ISL_676674, EPI_ISL_676675, EPI_ISL_676676, EPI_ISL_676677, EPI_ISL_676678, EPI_ISL_676679 | Wadsworth Center, New York State Department.of Health | Wadsworth Center, New York State Department.of Health | Kirsten St. George, Daryl M. Lamson, Alexis Russel, Jonathan Plitnick, Navjot Singh, John Kelly, Sara Griesemer, Erasmus Schneider, Erica Lasek-Nesselquist |
| EPI_ISL_676680 | Masonic Medical Research Institute | Wadsworth Center, New York State Department.of Health | Nathan Tucker, Kirsten St. George, Daryl M. Lamson, Alexis Russel, Jonathan Plitnick, Navjot Singh, John Kelly, Sara Griesemer, Erasmus Schneider, Erica Lasek-Nesselquist |
| EPI_ISL_676681, EPI_ISL_676682, EPI_ISL_676683, EPI_ISL_676684, EPI_ISL_676685, EPI_ISL_676686, EPI_ISL_676687, EPI_ISL_676688, EPI_ISL_676689, EPI_ISL_676690, EPI_ISL_676691, EPI_ISL_676692, EPI_ISL_676693, EPI_ISL_676694, EPI_ISL_676695, EPI_ISL_676696, EPI_ISL_676697, EPI_ISL_676698, EPI_ISL_676699, EPI_ISL_676700, EPI_ISL_676701, EPI_ISL_676702, EPI_ISL_676703, EPI_ISL_676704, EPI_ISL_676705, EPI_ISL_676706, EPI_ISL_676707, EPI_ISL_676708 | | | |
| see above | Wadsworth Center, New York State Department.of Health | Wadsworth Center, New York State Department.of Health | Kirsten St. George, Daryl M. Lamson, Alexis Russel, Jonathan Plitnick, Navjot Singh, John Kelly, Sara Griesemer, Erasmus Schneider, Erica Lasek-Nesselquist |
| EPI_ISL_676709 | Masonic Medical Research Institute | Wadsworth Center, New York State Department.of Health | Nathan Tucker, Kirsten St. George, Daryl M. Lamson, Alexis Russel, Jonathan Plitnick, Navjot Singh, John Kelly, Sara Griesemer, Erasmus Schneider, Erica Lasek-Nesselquist |
| EPI_ISL_676710, EPI_ISL_676711, EPI_ISL_676712 | Wadsworth Center, New York State Department.of Health | Wadsworth Center, New York State Department.of Health | Kirsten St. George, Daryl M. Lamson, Alexis Russel, Jonathan Plitnick, Navjot Singh, John Kelly, Sara Griesemer, Erasmus Schneider, Erica Lasek-Nesselquist |
| EPI_ISL_676713 | Masonic Medical Research Institute | Wadsworth Center, New York State Department.of Health | Nathan Tucker, Kirsten St. George, Daryl M. Lamson, Alexis Russel, Jonathan Plitnick, Navjot Singh, John Kelly, Sara Griesemer, Erasmus Schneider, Erica Lasek-Nesselquist |
| EPI_ISL_676714, EPI_ISL_676715, EPI_ISL_676716, EPI_ISL_676717, EPI_ISL_676718, EPI_ISL_676719, EPI_ISL_676720, EPI_ISL_676721, EPI_ISL_676722, EPI_ISL_676723, EPI_ISL_676724, EPI_ISL_676725 | | | |
| see above | Wadsworth Center, New York State Department.of Health | Wadsworth Center, New York State Department.of Health | Kirsten St. George, Daryl M. Lamson, Alexis Russel, Jonathan Plitnick, Navjot Singh, John Kelly, Sara Griesemer, Erasmus Schneider, Erica Lasek-Nesselquist |
| EPI_ISL_676726 | Masonic Medical Research Institute | Wadsworth Center, New York State Department.of Health | Nathan Tucker, Kirsten St. George, Daryl M. Lamson, Alexis Russel, Jonathan Plitnick, Navjot Singh, John Kelly, Sara Griesemer, Erasmus Schneider, Erica Lasek-Nesselquist |
| EPI_ISL_676727, EPI_ISL_676728, EPI_ISL_676729, EPI_ISL_676730, EPI_ISL_676731, EPI_ISL_676732, EPI_ISL_676733, EPI_ISL_676734, EPI_ISL_676735, EPI_ISL_676736, EPI_ISL_676737, EPI_ISL_676738, EPI_ISL_676739, EPI_ISL_676740, EPI_ISL_676741, EPI_ISL_676742, EPI_ISL_676743, EPI_ISL_676744, EPI_ISL_676745, EPI_ISL_676746, EPI_ISL_676747, EPI_ISL_676748, EPI_ISL_676749, EPI_ISL_676750, | | | |

EPI_ISL_676751, EPI_ISL_676752, EPI_ISL_676753, EPI_ISL_676754, EPI_ISL_676755, EPI_ISL_676756, EPI_ISL_676757, EPI_ISL_676758, EPI_ISL_676759, EPI_ISL_676760, EPI_ISL_676761, EPI_ISL_676762, EPI_ISL_676763, EPI_ISL_676764, EPI_ISL_676765, EPI_ISL_676766, EPI_ISL_676767, EPI_ISL_676768, EPI_ISL_676769, EPI_ISL_676770, EPI_ISL_676771, EPI_ISL_676772, EPI_ISL_676773, EPI_ISL_676774, EPI_ISL_676775, EPI_ISL_676776, EPI_ISL_676777, EPI_ISL_676778, EPI_ISL_676779, EPI_ISL_676780, EPI_ISL_676781, EPI_ISL_676782, EPI_ISL_676783, EPI_ISL_676784, EPI_ISL_676785, EPI_ISL_676786, EPI_ISL_676787, EPI_ISL_676788, EPI_ISL_676789, EPI_ISL_676790, EPI_ISL_676791, EPI_ISL_676792, EPI_ISL_676793, EPI_ISL_676794, EPI_ISL_676795, EPI_ISL_676796, EPI_ISL_676797, EPI_ISL_676798, EPI_ISL_676799, EPI_ISL_676800, EPI_ISL_676801, EPI_ISL_676802, EPI_ISL_676803, EPI_ISL_676804, EPI_ISL_676805, EPI_ISL_676806, EPI_ISL_676807, EPI_ISL_676808, EPI_ISL_676809, EPI_ISL_676810, EPI_ISL_676811, EPI_ISL_676812, EPI_ISL_676813, EPI_ISL_676814, EPI_ISL_676815, EPI_ISL_676816, EPI_ISL_676817, EPI_ISL_676818, EPI_ISL_676819, EPI_ISL_676820, EPI_ISL_676821, EPI_ISL_676822, EPI_ISL_676823, EPI_ISL_676824, EPI_ISL_676825, EPI_ISL_676826, EPI_ISL_676827, EPI_ISL_676828, EPI_ISL_676829, EPI_ISL_676830, EPI_ISL_676831, EPI_ISL_676832, EPI_ISL_676833, EPI_ISL_676834, EPI_ISL_676835, EPI_ISL_676836, EPI_ISL_676837, EPI_ISL_676838, EPI_ISL_676839, EPI_ISL_676840, EPI_ISL_676841, EPI_ISL_676842, EPI_ISL_676843, EPI_ISL_676844, EPI_ISL_676845, EPI_ISL_676846, EPI_ISL_676847, EPI_ISL_676848, EPI_ISL_676849, EPI_ISL_676850, EPI_ISL_676851, EPI_ISL_676852, EPI_ISL_676853, EPI_ISL_676854, EPI_ISL_676855, EPI_ISL_676856, EPI_ISL_676857, EPI_ISL_676858, EPI_ISL_676859, EPI_ISL_676860, EPI_ISL_676861, EPI_ISL_676862, EPI_ISL_676863, EPI_ISL_676864, EPI_ISL_676865, EPI_ISL_676866, EPI_ISL_676867, EPI_ISL_676868, EPI_ISL_676869, EPI_ISL_676870, EPI_ISL_676871, EPI_ISL_676872, EPI_ISL_676873, EPI_ISL_676874, EPI_ISL_676875, EPI_ISL_676876, EPI_ISL_676877, EPI_ISL_676878, EPI_ISL_676879, EPI_ISL_676880, EPI_ISL_676881, EPI_ISL_676882, EPI_ISL_676883, EPI_ISL_676884, EPI_ISL_676885, EPI_ISL_676886, EPI_ISL_676887, EPI_ISL_676888, EPI_ISL_676889, EPI_ISL_676890, EPI_ISL_676891, EPI_ISL_676892, EPI_ISL_676893, EPI_ISL_676894, EPI_ISL_676895, EPI_ISL_676896, EPI_ISL_676897, EPI_ISL_676898, EPI_ISL_676899, EPI_ISL_676900, EPI_ISL_676901, EPI_ISL_676902, EPI_ISL_676903, EPI_ISL_676904, EPI_ISL_676905, EPI_ISL_676906, EPI_ISL_676907, EPI_ISL_676908, EPI_ISL_676909, EPI_ISL_676910, EPI_ISL_676911, EPI_ISL_676912, EPI_ISL_676913, EPI_ISL_676914, EPI_ISL_676915, EPI_ISL_676916, EPI_ISL_676917, EPI_ISL_676918, EPI_ISL_676919, EPI_ISL_676920, EPI_ISL_676921, EPI_ISL_676922, EPI_ISL_676923, EPI_ISL_676924, EPI_ISL_676925, EPI_ISL_676926, EPI_ISL_676927, EPI_ISL_676928, EPI_ISL_676929, EPI_ISL_676930, EPI_ISL_676931, EPI_ISL_676932, EPI_ISL_676933, EPI_ISL_676934, EPI_ISL_676935, EPI_ISL_676936, EPI_ISL_676937, EPI_ISL_676938, EPI_ISL_676939, EPI_ISL_676940, EPI_ISL_676941, EPI_ISL_676942, EPI_ISL_676943, EPI_ISL_676944, EPI_ISL_676945, EPI_ISL_676946, EPI_ISL_676947, EPI_ISL_676948, EPI_ISL_676949, EPI_ISL_676950, EPI_ISL_676951, EPI_ISL_676952, EPI_ISL_676953, EPI_ISL_676954, EPI_ISL_676955, EPI_ISL_676956, EPI_ISL_676957, EPI_ISL_676958, EPI_ISL_676959, EPI_ISL_676960, EPI_ISL_676961, EPI_ISL_676962, EPI_ISL_676963, EPI_ISL_676964, EPI_ISL_676965, EPI_ISL_676966, EPI_ISL_676967, EPI_ISL_676968, EPI_ISL_676969, EPI_ISL_676970, EPI_ISL_676971, EPI_ISL_676972, EPI_ISL_676973, EPI_ISL_676974, EPI_ISL_676975, EPI_ISL_676976, EPI_ISL_676977, EPI_ISL_676978, EPI_ISL_676979, EPI_ISL_676980, EPI_ISL_676981, EPI_ISL_676982, EPI_ISL_676983, EPI_ISL_676984, EPI_ISL_676985, EPI_ISL_676986, EPI_ISL_676987, EPI_ISL_676988, EPI_ISL_676989, EPI_ISL_676990, EPI_ISL_676991, EPI_ISL_676992, EPI_ISL_676993, EPI_ISL_676994

see above | Wadsworth Center, New York State Department.of Health | Wadsworth Center, New York State Department.of Health | Kirsten St. George, Daryl M. Lamson, Alexis Russel, Jonathan Plitnick, Navjot Singh, John Kelly, Sara Griesemer, Erasmus Schneider, Erica Lasek-Nesselquist

EPI_ISL_676995, EPI_ISL_676996, EPI_ISL_676997, EPI_ISL_676998, EPI_ISL_676999, EPI_ISL_677000, EPI_ISL_677001, EPI_ISL_677002, EPI_ISL_677003, EPI_ISL_677004, EPI_ISL_677005, EPI_ISL_677006, EPI_ISL_677007, EPI_ISL_677008, EPI_ISL_677009, EPI_ISL_677010, EPI_ISL_677011, EPI_ISL_677012, EPI_ISL_677013, EPI_ISL_677014, EPI_ISL_677015, EPI_ISL_677016, EPI_ISL_677017, EPI_ISL_677018, EPI_ISL_677019, EPI_ISL_677020, EPI_ISL_677021, EPI_ISL_677022, EPI_ISL_677023, EPI_ISL_677024, EPI_ISL_677025

see above | Masonic Medical Research Institute | Wadsworth Center, New York State Department.of Health | Nathan Tucker, Kirsten St. George, Daryl M. Lamson, Alexis Russel, Jonathan Plitnick, Navjot Singh, John Kelly, Sara Griesemer, Erasmus Schneider, Erica Lasek-Nesselquist

EPI_ISL_677026, EPI_ISL_677027, EPI_ISL_677028, EPI_ISL_677029, EPI_ISL_677030, EPI_ISL_677031, EPI_ISL_677032, EPI_ISL_677033, EPI_ISL_677034, EPI_ISL_677035, EPI_ISL_677036, EPI_ISL_677037, EPI_ISL_677038, EPI_ISL_677039, EPI_ISL_677040, EPI_ISL_677041, EPI_ISL_677042, EPI_ISL_677043, EPI_ISL_677044, EPI_ISL_677045, EPI_ISL_677046, EPI_ISL_677047, EPI_ISL_677048

see above | Wadsworth Center, New York State Department.of Health | Wadsworth Center, New York State Department.of Health | Kirsten St. George, Daryl M. Lamson, Alexis Russel, Jonathan Plitnick, Navjot Singh, John Kelly, Sara Griesemer, Erasmus Schneider, Erica Lasek-Nesselquist

EPI_ISL_677049, EPI_ISL_677050, EPI_ISL_677051, EPI_ISL_677052, EPI_ISL_677053, EPI_ISL_677054, EPI_ISL_677055, EPI_ISL_677056, EPI_ISL_677057, EPI_ISL_677058, EPI_ISL_677059, EPI_ISL_677060, EPI_ISL_677061, EPI_ISL_677062, EPI_ISL_677063, EPI_ISL_677064, EPI_ISL_677065, EPI_ISL_677066, EPI_ISL_677067, EPI_ISL_677068, EPI_ISL_677069, EPI_ISL_677070, EPI_ISL_677071, EPI_ISL_677072, EPI_ISL_677073, EPI_ISL_677074, EPI_ISL_677075, EPI_ISL_677076

see above | Masonic Medical Research Institute | Wadsworth Center, New York State Department.of Health | Nathan Tucker, Kirsten St. George, Daryl M. Lamson, Alexis Russel, Jonathan Plitnick, Navjot Singh, John Kelly, Sara Griesemer, Erasmus Schneider, Erica Lasek-Nesselquist

EPI_ISL_677077, EPI_ISL_677078, EPI_ISL_677079, EPI_ISL_677080, EPI_ISL_677081, EPI_ISL_677082, EPI_ISL_677083, EPI_ISL_677084, EPI_ISL_677085, EPI_ISL_677086, EPI_ISL_677087, EPI_ISL_677088, EPI_ISL_677089, EPI_ISL_677090, EPI_ISL_677091, EPI_ISL_677092, EPI_ISL_677093, EPI_ISL_677094, EPI_ISL_677095, EPI_ISL_677096, EPI_ISL_677097, EPI_ISL_677098, EPI_ISL_677099, EPI_ISL_677100, EPI_ISL_677101, EPI_ISL_677102, EPI_ISL_677103, EPI_ISL_677104, EPI_ISL_677105, EPI_ISL_677106, EPI_ISL_677107, EPI_ISL_677108, EPI_ISL_677109, EPI_ISL_677110

see above | Wadsworth Center, New York State Department.of Health | Wadsworth Center, New York State Department.of Health | Kirsten St. George, Daryl M. Lamson, Alexis Russel, Jonathan Plitnick, Navjot Singh, John Kelly, Sara Griesemer, Erasmus Schneider, Erica Lasek-Nesselquist

EPI_ISL_677111, EPI_ISL_677112, EPI_ISL_677113, EPI_ISL_677114, EPI_ISL_677115, EPI_ISL_677116, EPI_ISL_677117, EPI_ISL_677118, EPI_ISL_677119, EPI_ISL_677120, EPI_ISL_677121, EPI_ISL_677122, EPI_ISL_677123, EPI_ISL_677124, EPI_ISL_677125, EPI_ISL_677126, EPI_ISL_677127, EPI_ISL_677128, EPI_ISL_677129

see above | Masonic Medical Research Institute | Wadsworth Center, New York State Department.of Health | Nathan Tucker, Kirsten St. George, Daryl M. Lamson, Alexis Russel, Jonathan Plitnick, Navjot Singh, John Kelly, Sara Griesemer, Erasmus Schneider, Erica Lasek-Nesselquist

EPI_ISL_677130, EPI_ISL_677131, EPI_ISL_677132, EPI_ISL_677133 | Wadsworth Center, New York State Department.of Health | Wadsworth Center, New York State Department.of Health | Kirsten St. George, Daryl M. Lamson, Alexis Russel, Jonathan Plitnick, Navjot Singh, John Kelly, Sara Griesemer, Erasmus Schneider, Erica Lasek-Nesselquist

EPI_ISL_677134, EPI_ISL_677135, EPI_ISL_677136, EPI_ISL_677137, EPI_ISL_677138, EPI_ISL_677139, EPI_ISL_677140, EPI_ISL_677141, EPI_ISL_677142, EPI_ISL_677143, EPI_ISL_677144, EPI_ISL_677145, EPI_ISL_677146, EPI_ISL_677147, EPI_ISL_677148, EPI_ISL_677149, EPI_ISL_677150, EPI_ISL_677151, EPI_ISL_677152, EPI_ISL_677153, EPI_ISL_677154, EPI_ISL_677155, EPI_ISL_677156, EPI_ISL_677157, EPI_ISL_677158, EPI_ISL_677159, EPI_ISL_677160, EPI_ISL_677161, EPI_ISL_677162, EPI_ISL_677163, EPI_ISL_677164, EPI_ISL_677165, EPI_ISL_677166, EPI_ISL_677167, EPI_ISL_677168, EPI_ISL_677169, EPI_ISL_677170, EPI_ISL_677171, EPI_ISL_677172, EPI_ISL_677173, EPI_ISL_677174, EPI_ISL_677175, EPI_ISL_677176, EPI_ISL_677177, EPI_ISL_677178, EPI_ISL_677179, EPI_ISL_677180, EPI_ISL_677181, EPI_ISL_677182, EPI_ISL_677183, EPI_ISL_677184, EPI_ISL_677185, EPI_ISL_677186, EPI_ISL_677187, EPI_ISL_677188, EPI_ISL_677189, EPI_ISL_677190, EPI_ISL_677191, EPI_ISL_677192, EPI_ISL_677193, EPI_ISL_677194, EPI_ISL_677195, EPI_ISL_677196, EPI_ISL_677197, EPI_ISL_677198, EPI_ISL_677199, EPI_ISL_677200, EPI_ISL_677201, EPI_ISL_677202, EPI_ISL_677203, EPI_ISL_677204, EPI_ISL_677205, EPI_ISL_677206, EPI_ISL_677207, EPI_ISL_677208, EPI_ISL_677209, EPI_ISL_677210, EPI_ISL_677211, EPI_ISL_677212, EPI_ISL_677213, EPI_ISL_677214, EPI_ISL_677215, EPI_ISL_677216, EPI_ISL_677217, EPI_ISL_677218, EPI_ISL_677219, EPI_ISL_677220, EPI_ISL_677221, EPI_ISL_677222, EPI_ISL_677223, EPI_ISL_677224, EPI_ISL_677225, EPI_ISL_677226, EPI_ISL_677227, EPI_ISL_677228, EPI_ISL_677229, EPI_ISL_677230, EPI_ISL_677231, EPI_ISL_677232, EPI_ISL_677233, EPI_ISL_677234, EPI_ISL_677235, EPI_ISL_677236, EPI_ISL_677237, EPI_ISL_677238, EPI_ISL_677239, EPI_ISL_677240

see above | Virginia Division of Consolidated Laboratory Services | Virginia Division of Consolidated Laboratory Services | Virginia DCLS

EPI_ISL_677242, EPI_ISL_677243, EPI_ISL_677244, EPI_ISL_677245, EPI_ISL_677246, EPI_ISL_677247, EPI_ISL_677248, EPI_ISL_677249, EPI_ISL_677250, EPI_ISL_677251, EPI_ISL_677252, EPI_ISL_677253, EPI_ISL_677254, EPI_ISL_677255, EPI_ISL_677256, EPI_ISL_677257, EPI_ISL_677258, EPI_ISL_677259, EPI_ISL_677260, EPI_ISL_677261, EPI_ISL_677262, EPI_ISL_677263, EPI_ISL_677264, EPI_ISL_677265, EPI_ISL_677285, EPI_ISL_677286, EPI_ISL_677287, EPI_ISL_677288, EPI_ISL_677289, EPI_ISL_677290, EPI_ISL_677291, EPI_ISL_677292, EPI_ISL_677293, EPI_ISL_677294, EPI_ISL_677295, EPI_ISL_677296, EPI_ISL_677297, EPI_ISL_677298, EPI_ISL_677299, EPI_ISL_677300, EPI_ISL_677301, EPI_ISL_677302, EPI_ISL_677303, EPI_ISL_677304, EPI_ISL_677305, EPI_ISL_677306, EPI_ISL_677307, EPI_ISL_677308, EPI_ISL_677309, EPI_ISL_677310, EPI_ISL_677311, EPI_ISL_677312, EPI_ISL_677313, EPI_ISL_677314, EPI_ISL_677315, EPI_ISL_677316, EPI_ISL_677317, EPI_ISL_677318, EPI_ISL_677319

see above | Colorado Department of Public Health and Environment | Colorado Department of Puplic Health and Environment | Laura Bankers, Molly Hetherington-Rauth, Shannon Ely, Shannon R. Matzinger, Sarah Elizabeth Totten, Emily A. Travanty

EPI_ISL_677320, EPI_ISL_677321, EPI_ISL_677322, EPI_ISL_677323, EPI_ISL_677324, EPI_ISL_677325, EPI_ISL_677326, EPI_ISL_677327, EPI_ISL_677328, EPI_ISL_677329, EPI_ISL_677330, EPI_ISL_677331, EPI_ISL_677332, EPI_ISL_677333, EPI_ISL_677334, EPI_ISL_677335, EPI_ISL_677336, EPI_ISL_677338, EPI_ISL_677339, EPI_ISL_677340, EPI_ISL_677341, EPI_ISL_677342, EPI_ISL_677343, EPI_ISL_677344, EPI_ISL_677345, EPI_ISL_677346, EPI_ISL_677347, EPI_ISL_677348, EPI_ISL_677349, EPI_ISL_677350, EPI_ISL_677351, EPI_ISL_677352, EPI_ISL_677353, EPI_ISL_677354, EPI_ISL_677355, EPI_ISL_677356, EPI_ISL_677357, EPI_ISL_677358, EPI_ISL_677359, EPI_ISL_677360, EPI_ISL_677361, EPI_ISL_677362, EPI_ISL_677363, EPI_ISL_677364, EPI_ISL_677365, EPI_ISL_677366, EPI_ISL_677367, EPI_ISL_677368, EPI_ISL_677369, EPI_ISL_677370, EPI_ISL_677371, EPI_ISL_677372, EPI_ISL_677373, EPI_ISL_677374, EPI_ISL_677375, EPI_ISL_677376, EPI_ISL_677377, EPI_ISL_677378, EPI_ISL_677379, EPI_ISL_677380, EPI_ISL_677381, EPI_ISL_677382, EPI_ISL_677383, EPI_ISL_677384, EPI_ISL_677385, EPI_ISL_677387, EPI_ISL_677388, EPI_ISL_677389, EPI_ISL_677390, EPI_ISL_677391, EPI_ISL_677392, EPI_ISL_677393, EPI_ISL_677394, EPI_ISL_677395, EPI_ISL_677396, EPI_ISL_677397, EPI_ISL_677398, EPI_ISL_677399, EPI_ISL_677400, EPI_ISL_677401, EPI_ISL_677402, EPI_ISL_677403, EPI_ISL_677404, EPI_ISL_677405, EPI_ISL_677406, EPI_ISL_677407, EPI_ISL_677408, EPI_ISL_677409, EPI_ISL_677410, EPI_ISL_677411, EPI_ISL_677412, EPI_ISL_677413, EPI_ISL_677414, EPI_ISL_677415, EPI_ISL_677416, EPI_ISL_677417, EPI_ISL_677418, EPI_ISL_677419, EPI_ISL_677420, EPI_ISL_677421, EPI_ISL_677422, EPI_ISL_677423, EPI_ISL_677424, EPI_ISL_677425, EPI_ISL_677426, EPI_ISL_677427, EPI_ISL_677428, EPI_ISL_677429, EPI_ISL_677430, EPI_ISL_677431, EPI_ISL_677432, EPI_ISL_677433, EPI_ISL_677434, EPI_ISL_677435, EPI_ISL_677436, EPI_ISL_677437, EPI_ISL_677438, EPI_ISL_677439, EPI_ISL_677440, EPI_ISL_677441, EPI_ISL_677442, EPI_ISL_677443, EPI_ISL_677444, EPI_ISL_677445, EPI_ISL_677446, EPI_ISL_677447, EPI_ISL_677448, EPI_ISL_677449, EPI_ISL_677450, EPI_ISL_677451, EPI_ISL_677452, EPI_ISL_677453, EPI_ISL_677454, EPI_ISL_677455, EPI_ISL_677456, EPI_ISL_677457, EPI_ISL_677458, EPI_ISL_677459, EPI_ISL_677460, EPI_ISL_677461, EPI_ISL_677462, EPI_ISL_677463, EPI_ISL_677464, EPI_ISL_677465, EPI_ISL_677466, EPI_ISL_677467, EPI_ISL_677468, EPI_ISL_677469, EPI_ISL_677470, EPI_ISL_677471, EPI_ISL_677472, EPI_ISL_677473, EPI_ISL_677474, EPI_ISL_677475, EPI_ISL_677476, EPI_ISL_677477, EPI_ISL_677478, EPI_ISL_677479, EPI_ISL_677480, EPI_ISL_677481, EPI_ISL_677482, EPI_ISL_677483, EPI_ISL_677484, EPI_ISL_677485, EPI_ISL_677486, EPI_ISL_677487, EPI_ISL_677488, EPI_ISL_677489, EPI_ISL_677490, EPI_ISL_677491, EPI_ISL_677492, EPI_ISL_677493, EPI_ISL_677494, EPI_ISL_677495, EPI_ISL_677496, EPI_ISL_677497, EPI_ISL_677498, EPI_ISL_677499, EPI_ISL_677500, EPI_ISL_677501, EPI_ISL_677502, EPI_ISL_677503, EPI_ISL_677504, EPI_ISL_677505, EPI_ISL_677506, EPI_ISL_677507, EPI_ISL_677508, EPI_ISL_677509, EPI_ISL_677510, EPI_ISL_677511, EPI_ISL_677512, EPI_ISL_677513, EPI_ISL_677514, EPI_ISL_677515, EPI_ISL_677516, EPI_ISL_677517, EPI_ISL_677518, EPI_ISL_677519, EPI_ISL_677520, EPI_ISL_677521, EPI_ISL_677522, EPI_ISL_677523, EPI_ISL_677524, EPI_ISL_677525, EPI_ISL_677526, EPI_ISL_677527, EPI_ISL_677528, EPI_ISL_677529, EPI_ISL_677530, EPI_ISL_677531, EPI_ISL_677532, EPI_ISL_677533, EPI_ISL_677534, EPI_ISL_677535, EPI_ISL_677536, EPI_ISL_677537, EPI_ISL_677538, EPI_ISL_677539, EPI_ISL_677540, EPI_ISL_677542, EPI_ISL_677543, EPI_ISL_677544, EPI_ISL_677545, EPI_ISL_677546, EPI_ISL_677547, EPI_ISL_677548, EPI_ISL_677549, EPI_ISL_677550, EPI_ISL_677551, EPI_ISL_677552, EPI_ISL_677554, EPI_ISL_677555, EPI_ISL_677556, EPI_ISL_677557, EPI_ISL_677558, EPI_ISL_677559, EPI_ISL_677560, EPI_ISL_677561, EPI_ISL_677562, EPI_ISL_677564, EPI_ISL_677565, EPI_ISL_677566, EPI_ISL_677567, EPI_ISL_677568, EPI_ISL_677570, EPI_ISL_677571, EPI_ISL_677572, EPI_ISL_677573, EPI_ISL_677574, EPI_ISL_677576, EPI_ISL_677577, EPI_ISL_677578, EPI_ISL_677579, EPI_ISL_677580, EPI_ISL_677581, EPI_ISL_677582, EPI_ISL_677583, EPI_ISL_677585, EPI_ISL_677586, EPI_ISL_677587, EPI_ISL_677588, EPI_ISL_677589, EPI_ISL_677591, EPI_ISL_677592, EPI_ISL_677593, EPI_ISL_677594, EPI_ISL_677595, EPI_ISL_677596, EPI_ISL_677597, EPI_ISL_677598, EPI_ISL_677599, EPI_ISL_677600, EPI_ISL_677601, EPI_ISL_677602, EPI_ISL_677603, EPI_ISL_677604, EPI_ISL_677605, EPI_ISL_677606, EPI_ISL_677607, EPI_ISL_677608, EPI_ISL_677609, EPI_ISL_677610, EPI_ISL_677611, EPI_ISL_677612, EPI_ISL_677613, EPI_ISL_677614, EPI_ISL_677615, EPI_ISL_677616, EPI_ISL_677617, EPI_ISL_677618, EPI_ISL_677619, EPI_ISL_677620, EPI_ISL_677621, EPI_ISL_677622, EPI_ISL_677623, EPI_ISL_677624, EPI_ISL_677625, EPI_ISL_677626, EPI_ISL_677627, EPI_ISL_677628, EPI_ISL_677629, EPI_ISL_677630, EPI_ISL_677631, EPI_ISL_677632, EPI_ISL_677633

see above | University of Wisconsin-Madison AIDS Vaccine Research Laboratories | University of Wisconsin-Madison AIDS Vaccine Research Laboratories | Gage Moreno, Katarina Braun, et al. AIDS Vaccine Research Laboratories

EPI_ISL_677638, EPI_ISL_677639, EPI_ISL_677640, EPI_ISL_677641, EPI_ISL_677642, EPI_ISL_677643, EPI_ISL_677644, EPI_ISL_677645, EPI_ISL_677646, EPI_ISL_677647, EPI_ISL_677648, EPI_ISL_677649, EPI_ISL_677650, EPI_ISL_677652, EPI_ISL_677653, EPI_ISL_677654, EPI_ISL_677655, EPI_ISL_677656, EPI_ISL_677657

see above | Colorado Department of Public Health and Environment | Colorado Department of Puplic Health and Environment | Laura Bankers, Molly C. Hetherington-Rauth, Shannon Ely, Shannon R. Matzinger, Sarah Elizabeth Totten, Emily A. Travanty

EPI_ISL_677658, EPI_ISL_677659, EPI_ISL_677660, EPI_ISL_677661, EPI_ISL_677662, EPI_ISL_677663 | University of Wisconsin-Madison AIDS Vaccine Research Laboratories | University of Wisconsin-Madison AIDS Vaccine Research Laboratories | Gage Moreno, Katarina Braun, et al. AIDS Vaccine Research Laboratories

EPI_ISL_677664, EPI_ISL_677665, EPI_ISL_677666, EPI_ISL_677667, EPI_ISL_677668 | Wadsworth Center, New York State Department.of Health | Wadsworth Center, New York State Department.of Health | Kirsten St. George, Daryl M. Lamson, Alexis Russel, Jonathan Plitnick, Navjot Singh, John Kelly, Sara Griesemer, Erasmus Schneider, Erica Lasek-Nesselquist

EPI_ISL_677669 | Masonic Medical Research Institute | Wadsworth Center, New York State Department.of Health | Nathan Tucker, Kirsten St. George, Daryl M. Lamson, Alexis Russel, Jonathan Plitnick, Navjot Singh, John Kelly, Sara Griesemer, Erasmus Schneider, Erica Lasek-Nesselquist

EPI_ISL_677670, EPI_ISL_677671, EPI_ISL_677673 | Wadsworth Center, New York State Department.of Health | Wadsworth Center, New York State Department.of Health | Kirsten St. George, Daryl M. Lamson, Alexis Russel, Jonathan Plitnick, Navjot Singh, John Kelly, Sara Griesemer, Erasmus Schneider, Erica Lasek-Nesselquist

EPI_ISL_677678, EPI_ISL_677679, EPI_ISL_677680, EPI_ISL_677681, EPI_ISL_677682, EPI_ISL_677683, EPI_ISL_677684, EPI_ISL_677685, EPI_ISL_677686, EPI_ISL_677687, EPI_ISL_677688, EPI_ISL_677689, EPI_ISL_677692, EPI_ISL_677695, EPI_ISL_677696, EPI_ISL_677699

see above | Vanda Pharmaceuticals Clinical Site | Vanda Pharmaceuticals | Vanda Pharmaceuticals

EPI_ISL_677821, EPI_ISL_677823, EPI_ISL_677824, EPI_ISL_677825, EPI_ISL_677826, EPI_ISL_677827, EPI_ISL_677828, EPI_ISL_677829, EPI_ISL_677830, EPI_ISL_677831, EPI_ISL_677832, EPI_ISL_677833, EPI_ISL_677834, EPI_ISL_677835, EPI_ISL_677836, EPI_ISL_677837, EPI_ISL_677838, EPI_ISL_677839, EPI_ISL_677840, EPI_ISL_677841, EPI_ISL_677842, EPI_ISL_677843, EPI_ISL_677844, EPI_ISL_677845, EPI_ISL_677846, EPI_ISL_677847, EPI_ISL_677848, EPI_ISL_677849, EPI_ISL_677850, EPI_ISL_677851, EPI_ISL_677852, EPI_ISL_677853, EPI_ISL_677854, EPI_ISL_677855, EPI_ISL_677856, EPI_ISL_677857, EPI_ISL_677858, EPI_ISL_677859, EPI_ISL_677860, EPI_ISL_677861, EPI_ISL_677862, EPI_ISL_677863, EPI_ISL_677864, EPI_ISL_677865, EPI_ISL_677866, EPI_ISL_677867, EPI_ISL_677868, EPI_ISL_677869, EPI_ISL_677870, EPI_ISL_677871, EPI_ISL_677872, EPI_ISL_677873, EPI_ISL_677874, EPI_ISL_677875, EPI_ISL_677876, EPI_ISL_677877, EPI_ISL_677878, EPI_ISL_677879, EPI_ISL_677880, EPI_ISL_677881, EPI_ISL_677882, EPI_ISL_677883, EPI_ISL_677884, EPI_ISL_677885, EPI_ISL_677886, EPI_ISL_677887, EPI_ISL_677889, EPI_ISL_677890, EPI_ISL_677891, EPI_ISL_677892, EPI_ISL_677893, EPI_ISL_677894, EPI_ISL_677896, EPI_ISL_677897, EPI_ISL_677898, EPI_ISL_677899, EPI_ISL_677900, EPI_ISL_677901, EPI_ISL_677904, EPI_ISL_677905, EPI_ISL_677906, EPI_ISL_677907

see above | Innovative Genomics Institute, UC Berkeley | Innovative Genomics Institute, UC Berkeley | Stacia Wyman, Haridha Shivram, Phil Frankino, Liana Lareau, Shana McDevitt, Justin Choi

EPI_ISL_678509, EPI_ISL_678511, EPI_ISL_678515, EPI_ISL_678516, EPI_ISL_678517, EPI_ISL_678519, EPI_ISL_678520, EPI_ISL_678521, EPI_ISL_678522, EPI_ISL_678523, EPI_ISL_678524, EPI_ISL_678525, EPI_ISL_678526, EPI_ISL_678527, EPI_ISL_678529, EPI_ISL_678530, EPI_ISL_678531, EPI_ISL_678532, EPI_ISL_678533

see above | Utah Public Health Laboratory | Utah Public Health Laboratory | Erin Young, Kelly Oakeson

EPI_ISL_681320, EPI_ISL_681321 | Pathology and Laboratory Medicine, UW-Madison | Pathology and Laboratory Medicine, UW-Madison | Moreno,G., Braun,K., Baczenas,J.J., Baker,D.

EPI_ISL_681322 | Environmental and Global Health, University of Florida | Environmental and Global Health, University of Florida | Loeb,J.C., Stephenson,C.J., Merck,L., Morris,J.G. and Lednicky,J.A.

EPI_ISL_681776, EPI_ISL_681777, EPI_ISL_681778, EPI_ISL_681779, EPI_ISL_681780, EPI_ISL_681781, EPI_ISL_681782, EPI_ISL_681783, EPI_ISL_681784, EPI_ISL_681785, EPI_ISL_681786, EPI_ISL_681787, EPI_ISL_681788, EPI_ISL_681789, EPI_ISL_681790, EPI_ISL_681791, EPI_ISL_681792, EPI_ISL_681793, EPI_ISL_681794, EPI_ISL_681795, EPI_ISL_681796, EPI_ISL_681797, EPI_ISL_681798, EPI_ISL_681799,

| | | | |
|---|---|---|---|
| EPI_ISL_681800, EPI_ISL_681801, EPI_ISL_681802, EPI_ISL_681803, EPI_ISL_681804, EPI_ISL_681805, EPI_ISL_681806, EPI_ISL_681807, EPI_ISL_681808, EPI_ISL_681809, EPI_ISL_681810, EPI_ISL_681811, EPI_ISL_681812, EPI_ISL_681813, EPI_ISL_681814, EPI_ISL_681815, EPI_ISL_681816, EPI_ISL_681817, EPI_ISL_681818, EPI_ISL_681819, EPI_ISL_681820, EPI_ISL_681821, EPI_ISL_681822, EPI_ISL_681823, EPI_ISL_681824, EPI_ISL_681825, EPI_ISL_681826, EPI_ISL_681827, EPI_ISL_681828 | | | |
| see above | GA Department of Public Health Laboratory | Pathogen Discovery, Respiratory Viruses Branch, Division of Viral Diseases, Centers for Disease Control and Prevention | Yan Li, Ying Tao, Jing Zhang, Krista Queen, Anna Uehara, Peter Cook, Clinton R. Paden, Haibin Wang, Suxiang Tong |
| EPI_ISL_681843, EPI_ISL_681844, EPI_ISL_681845, EPI_ISL_681846, EPI_ISL_681847, EPI_ISL_681848, EPI_ISL_681849, EPI_ISL_681850, EPI_ISL_681851, EPI_ISL_681852, EPI_ISL_681853, EPI_ISL_681854, EPI_ISL_681855, EPI_ISL_681856, EPI_ISL_681857, EPI_ISL_681858, EPI_ISL_681859, EPI_ISL_681860, EPI_ISL_681861, EPI_ISL_681862, EPI_ISL_681863, EPI_ISL_681864, EPI_ISL_681865, EPI_ISL_681866, EPI_ISL_681867, EPI_ISL_681876, EPI_ISL_681877, EPI_ISL_681878, EPI_ISL_681879, EPI_ISL_681880, EPI_ISL_681881, EPI_ISL_681882, EPI_ISL_681883 | | | |
| see above | Texas Department of State Health Services | Texas Department of State Health Services | Rashmi Tuladhar, Bonnie Oh, Jenny Zhang, Maliha Rahman, Anita Pokharel, Myong Koag, Chung Wang, Rachel Lee, Grace Kubin, Mayela Pedrueza, James Daniel Bonser |
| EPI_ISL_681929, EPI_ISL_681930, EPI_ISL_681931, EPI_ISL_681932, EPI_ISL_681933, EPI_ISL_681934, EPI_ISL_681935, EPI_ISL_681936, EPI_ISL_681937, EPI_ISL_681938, EPI_ISL_681939, EPI_ISL_681940, EPI_ISL_681941, EPI_ISL_681942, EPI_ISL_681943, EPI_ISL_681944, EPI_ISL_681945, EPI_ISL_681946, EPI_ISL_681947, EPI_ISL_681948, EPI_ISL_681949, EPI_ISL_681950, EPI_ISL_681951, EPI_ISL_681952, EPI_ISL_681953, EPI_ISL_681954, EPI_ISL_681955, EPI_ISL_681956, EPI_ISL_681957, EPI_ISL_681958, EPI_ISL_681959, EPI_ISL_681960, EPI_ISL_681961, EPI_ISL_681962, EPI_ISL_681963, EPI_ISL_681964, EPI_ISL_681965, EPI_ISL_681966, EPI_ISL_681967, EPI_ISL_681968, EPI_ISL_681969, EPI_ISL_681970, EPI_ISL_681971, EPI_ISL_681972, EPI_ISL_681973, EPI_ISL_681974, EPI_ISL_681975, EPI_ISL_681976, EPI_ISL_681977, EPI_ISL_681978, EPI_ISL_681979, EPI_ISL_681980, EPI_ISL_681981, EPI_ISL_681982, EPI_ISL_681983, EPI_ISL_681984, EPI_ISL_681985, EPI_ISL_681986, EPI_ISL_681987, EPI_ISL_681988, EPI_ISL_681989, EPI_ISL_681990, EPI_ISL_681991, EPI_ISL_681992, EPI_ISL_681993, EPI_ISL_681994, EPI_ISL_681995, EPI_ISL_681996, EPI_ISL_681997, EPI_ISL_681998, EPI_ISL_681999, EPI_ISL_682000, EPI_ISL_682001, EPI_ISL_682002, EPI_ISL_682003, EPI_ISL_682004, EPI_ISL_682005, EPI_ISL_682006, EPI_ISL_682007, EPI_ISL_682008, EPI_ISL_682009, EPI_ISL_682010, EPI_ISL_682011, EPI_ISL_682012, EPI_ISL_682013, EPI_ISL_682014, EPI_ISL_682015, EPI_ISL_682016, EPI_ISL_682017, EPI_ISL_682018, EPI_ISL_682019, EPI_ISL_682020, EPI_ISL_682021, EPI_ISL_682022, EPI_ISL_682023, EPI_ISL_682024, EPI_ISL_682025, EPI_ISL_682026, EPI_ISL_682027, EPI_ISL_682028, EPI_ISL_682029, EPI_ISL_682030, EPI_ISL_682031, EPI_ISL_682032, EPI_ISL_682033, EPI_ISL_682034, EPI_ISL_682035, EPI_ISL_682036, EPI_ISL_682037, EPI_ISL_682038, EPI_ISL_682039, EPI_ISL_682040, EPI_ISL_682041, EPI_ISL_682042, EPI_ISL_682043, EPI_ISL_682044, EPI_ISL_682045, EPI_ISL_682046, EPI_ISL_682047, EPI_ISL_682048, EPI_ISL_682049, EPI_ISL_682050, EPI_ISL_682051, EPI_ISL_682052, EPI_ISL_682053, EPI_ISL_682054, EPI_ISL_682055, EPI_ISL_682056 | | | |
| see above | UPMC Clinical Microbiology Laboratory | Microbial Genomic Epidemiology Laboratory, University of Pittsburgh | Mustapha M. Mustapha, Jane W. Marsh, Dan Snyder, Marissa P. Griffith, Stephanie L. Mitchell, Vatsala R. Srinivasa, Kady D. Waggle, Chinelo Ezeonwuku, Vaughn S. Cooper, Lee H. Harrison |
| EPI_ISL_682060, EPI_ISL_682061, EPI_ISL_682062, EPI_ISL_682063, EPI_ISL_682064, EPI_ISL_682065, EPI_ISL_682066, EPI_ISL_682067, EPI_ISL_682068, EPI_ISL_682069, EPI_ISL_682070, EPI_ISL_682071, EPI_ISL_682072, EPI_ISL_682073, EPI_ISL_682074, EPI_ISL_682075, EPI_ISL_682076, EPI_ISL_682077, EPI_ISL_682078 | | | |
| see above | Maryland Public Health Laboratory | Maryland Public Health Laboratory | Maryland Department of Health Laboratories Administration |
| EPI_ISL_682079, EPI_ISL_682080, EPI_ISL_682081, EPI_ISL_682082, EPI_ISL_682083, EPI_ISL_682084, EPI_ISL_682085, EPI_ISL_682086, EPI_ISL_682087, EPI_ISL_682088, EPI_ISL_682089, EPI_ISL_682090, EPI_ISL_682091, EPI_ISL_682092, EPI_ISL_682093, EPI_ISL_682094, EPI_ISL_682095, EPI_ISL_682096, EPI_ISL_682097, EPI_ISL_682098, EPI_ISL_682099, EPI_ISL_682100, EPI_ISL_682101, EPI_ISL_682102, EPI_ISL_682103, EPI_ISL_682104, EPI_ISL_682105, EPI_ISL_682106, EPI_ISL_682107, EPI_ISL_682108, EPI_ISL_682109, EPI_ISL_682110, EPI_ISL_682111, EPI_ISL_682112, EPI_ISL_682113, EPI_ISL_682114, EPI_ISL_682115, EPI_ISL_682116, EPI_ISL_682117, EPI_ISL_682118, EPI_ISL_682119, EPI_ISL_682120, EPI_ISL_682121, EPI_ISL_682122, EPI_ISL_682123, EPI_ISL_682124, EPI_ISL_682125, EPI_ISL_682126, EPI_ISL_682127, EPI_ISL_682128, EPI_ISL_682129, EPI_ISL_682130, EPI_ISL_682131, EPI_ISL_682132, EPI_ISL_682133, EPI_ISL_682134, EPI_ISL_682135, EPI_ISL_682136, EPI_ISL_682137, EPI_ISL_682138, EPI_ISL_682139, EPI_ISL_682140, EPI_ISL_682141, EPI_ISL_682142, EPI_ISL_682143, EPI_ISL_682144, EPI_ISL_682145, EPI_ISL_682146, EPI_ISL_682147, EPI_ISL_682148, EPI_ISL_682149, EPI_ISL_682150, EPI_ISL_682151, EPI_ISL_682152, EPI_ISL_682153, EPI_ISL_682154, EPI_ISL_682155, EPI_ISL_682156, EPI_ISL_682157, EPI_ISL_682158, EPI_ISL_682159, EPI_ISL_682160, EPI_ISL_682161, EPI_ISL_682162, EPI_ISL_682163, EPI_ISL_682164, EPI_ISL_682165, EPI_ISL_682166, EPI_ISL_682167, EPI_ISL_682168, EPI_ISL_682169, EPI_ISL_682170, EPI_ISL_682171, EPI_ISL_682172, EPI_ISL_682173, EPI_ISL_682174, EPI_ISL_682175, EPI_ISL_682176, EPI_ISL_682177, EPI_ISL_682178, EPI_ISL_682179, EPI_ISL_682180, EPI_ISL_682181, EPI_ISL_682182, EPI_ISL_682183, EPI_ISL_682184, EPI_ISL_682185, EPI_ISL_682186, EPI_ISL_682187, EPI_ISL_682188, EPI_ISL_682189, EPI_ISL_682190, EPI_ISL_682191, EPI_ISL_682192, EPI_ISL_682193, EPI_ISL_682194, EPI_ISL_682195, EPI_ISL_682196, EPI_ISL_682197, EPI_ISL_682198, EPI_ISL_682199, EPI_ISL_682200, EPI_ISL_682201, EPI_ISL_682202, EPI_ISL_682203, EPI_ISL_682204, EPI_ISL_682205, EPI_ISL_682206, EPI_ISL_682207, EPI_ISL_682208, EPI_ISL_682209, EPI_ISL_682210, EPI_ISL_682211, EPI_ISL_682212, EPI_ISL_682213, EPI_ISL_682214, EPI_ISL_682215, EPI_ISL_682216, EPI_ISL_682217, EPI_ISL_682218, EPI_ISL_682219, EPI_ISL_682220, EPI_ISL_682221, EPI_ISL_682222, EPI_ISL_682223, EPI_ISL_682224, EPI_ISL_682225, EPI_ISL_682226, EPI_ISL_682227, EPI_ISL_682228, EPI_ISL_682229, EPI_ISL_682230, EPI_ISL_682231, EPI_ISL_682232, EPI_ISL_682233, EPI_ISL_682234 | | | |
| see above | University of Michigan Clinical Microbiology Laboratory | Lauring Lab, University of Michigan, Department of Microbiology and Immunology | Valesano |
| EPI_ISL_683404, EPI_ISL_683407, EPI_ISL_683408, EPI_ISL_683409, EPI_ISL_683410, EPI_ISL_683411, EPI_ISL_683412, EPI_ISL_683413, EPI_ISL_683415, EPI_ISL_683417, EPI_ISL_683418, EPI_ISL_683419, EPI_ISL_683420, EPI_ISL_683421, EPI_ISL_683422, EPI_ISL_683423, EPI_ISL_683424, EPI_ISL_683425, EPI_ISL_683426, EPI_ISL_683427, EPI_ISL_683430, EPI_ISL_683433, EPI_ISL_683434, EPI_ISL_683435, EPI_ISL_683436, EPI_ISL_683439, EPI_ISL_683440, EPI_ISL_683441, EPI_ISL_683442, EPI_ISL_683443, EPI_ISL_683444, EPI_ISL_683445, EPI_ISL_683447 | | | |
| see above | Texas Department of State Health Services | Texas Department of State Health Services | Rashmi Tuladhar, Bonnie Oh, Jenny Zhang, Maliha Rahman, Anita Pokharel, Myong Koag, Chung Wang, Rachel Lee, Grace Kubin, Mayela Pedrueza, James Daniel Bonser |
| EPI_ISL_683678, EPI_ISL_683679, EPI_ISL_683680, EPI_ISL_683681, EPI_ISL_683682, EPI_ISL_683683, EPI_ISL_683684, EPI_ISL_683685, EPI_ISL_683686, EPI_ISL_683687, EPI_ISL_683688 | | | |
| see above | M Health Fairview | Minnesota Department of Health, Public Health Laboratory | Alexandra Lorentz, Jacob Garfin, Matt Plumb, and Xiong Wang |
| EPI_ISL_683689 | Hennepin County Medical Center | Minnesota Department of Health, Public Health Laboratory | Alexandra Lorentz, Jacob Garfin, Matt Plumb, and Xiong Wang |
| EPI_ISL_683690, EPI_ISL_683691, EPI_ISL_683692, EPI_ISL_683693, EPI_ISL_683694 | Essentia Health-St. Mary's Medical Center | Minnesota Department of Health, Public Health Laboratory | Alexandra Lorentz, Jacob Garfin, Matt Plumb, and Xiong Wang |
| EPI_ISL_683695 | Mayo Clinic & Mayo Clinic Laboratories | Minnesota Department of Health, Public Health Laboratory | Alexandra Lorentz, Jacob Garfin, Matt Plumb, and Xiong Wang |
| EPI_ISL_683696, EPI_ISL_683697, EPI_ISL_683698, EPI_ISL_683699, EPI_ISL_683700, EPI_ISL_683701, EPI_ISL_683702, EPI_ISL_683703, EPI_ISL_683704, EPI_ISL_683705, EPI_ISL_683706, EPI_ISL_683707, EPI_ISL_683708, EPI_ISL_683709, EPI_ISL_683710, EPI_ISL_683711, EPI_ISL_683712, EPI_ISL_683713 | | | |
| see above | Essentia Health-St. Mary's Medical Center | Minnesota Department of Health, Public Health Laboratory | Alexandra Lorentz, Jacob Garfin, Matt Plumb, and Xiong Wang |
| EPI_ISL_683714, EPI_ISL_683715, EPI_ISL_683716, EPI_ISL_683717, EPI_ISL_683718, EPI_ISL_683719, EPI_ISL_683720, EPI_ISL_683721, EPI_ISL_683722, EPI_ISL_683723, EPI_ISL_683724, EPI_ISL_683725, EPI_ISL_683726, EPI_ISL_683727, EPI_ISL_683728 | | | |
| see above | Mayo Clinic & Mayo Clinic Laboratories | Minnesota Department of Health, Public Health Laboratory | Alexandra Lorentz, Jacob Garfin, Matt Plumb, and Xiong Wang |
| EPI_ISL_683729, EPI_ISL_683730, EPI_ISL_683731, EPI_ISL_683732, EPI_ISL_683733, EPI_ISL_683734, EPI_ISL_683735, EPI_ISL_683736, EPI_ISL_683737, EPI_ISL_683738, EPI_ISL_683739, EPI_ISL_683740, EPI_ISL_683741, EPI_ISL_683742, EPI_ISL_683743, EPI_ISL_683744, EPI_ISL_683745, EPI_ISL_683746, EPI_ISL_683747, EPI_ISL_683748, EPI_ISL_683749, EPI_ISL_683750 | | | |
| see above | Minnesota Department of Health, Public Health Laboratory | Minnesota Department of Health, Public Health Laboratory | Alexandra Lorentz, Jacob Garfin, Matt Plumb, and Xiong Wang |
| EPI_ISL_683753 | DOHMH Corona | New York City Public Health Laboratory | Jade Wang, et al. |
| EPI_ISL_683754 | DOHMH Morrisania | New York City Public Health Laboratory | Jade Wang, et al. |
| EPI_ISL_683755 | DOHMH PHL | New York City Public Health Laboratory | Jade Wang, et al. |
| EPI_ISL_683756 | DOHMH Central Harlem | New York City Public Health Laboratory | Jade Wang, et al. |
| EPI_ISL_683757, EPI_ISL_683758 | DOHMH Corona | New York City Public Health Laboratory | Jade Wang, et al. |
| EPI_ISL_683759 | DOHMH PHL | New York City Public Health Laboratory | Jade Wang, et al. |
| EPI_ISL_683760 | DOHMH Morrisania | New York City Public Health Laboratory | Jade Wang, et al. |
| EPI_ISL_683761 | DOHMH Corona | New York City Public Health Laboratory | Jade Wang, et al. |
| EPI_ISL_683762, EPI_ISL_683763 | DOHMH Morrisania | New York City Public Health Laboratory | Jade Wang, et al. |
| EPI_ISL_683764 | DOHMH Corona | New York City Public Health Laboratory | Jade Wang, et al. |
| EPI_ISL_683765 | DOHMH Chelsea | New York City Public Health Laboratory | Jade Wang, et al. |
| EPI_ISL_683766 | DOHMH Corona | New York City Public Health Laboratory | Jade Wang, et al. |
| EPI_ISL_683767, EPI_ISL_683768, EPI_ISL_683769 | DOHMH Riverside | New York City Public Health Laboratory | Jade Wang, et al. |
| EPI_ISL_683770 | DOHMH Corona | New York City Public Health Laboratory | Jade Wang, et al. |
| EPI_ISL_683771 | DOHMH Fort Greene | New York City Public Health Laboratory | Jade Wang, et al. |
| EPI_ISL_683772 | DOHMH Morrisania | New York City Public Health Laboratory | Jade Wang, et al. |
| EPI_ISL_683773 | DOHMH Corona | New York City Public Health Laboratory | Jade Wang, et al. |
| EPI_ISL_683774 | DOHMH PHL | New York City Public Health Laboratory | Jade Wang, et al. |
| EPI_ISL_683775, EPI_ISL_683776 | DOHMH Jamaica | New York City Public Health Laboratory | Jade Wang, et al. |
| EPI_ISL_683777 | DOHMH Corona | New York City Public Health Laboratory | Jade Wang, et al. |
| EPI_ISL_683778 | DOHMH Morrisania | New York City Public Health Laboratory | Jade Wang, et al. |
| EPI_ISL_683779 | DOHMH Corona | New York City Public Health Laboratory | Jade Wang, et al. |
| EPI_ISL_683780 | DOHMH Chelsea | New York City Public Health Laboratory | Jade Wang, et al. |
| EPI_ISL_683781, EPI_ISL_683782 | DOHMH Jamaica | New York City Public Health Laboratory | Jade Wang, et al. |
| EPI_ISL_683783 | DOHMH PHL | New York City Public Health Laboratory | Jade Wang, et al. |
| EPI_ISL_683784 | DOHMH Riverside | New York City Public Health Laboratory | Jade Wang, et al. |
| EPI_ISL_683785 | DOHMH Corona | New York City Public Health Laboratory | Jade Wang, et al. |
| EPI_ISL_683786 | DOHMH Jamaica | New York City Public Health Laboratory | Jade Wang, et al. |
| EPI_ISL_683787 | DOHMH PHL | New York City Public Health Laboratory | Jade Wang, et al. |

| | | | |
|---|---|---|---|
| EPI_ISL_683788 | DOHMH Chelsea | New York City Public Health Laboratory | Jade Wang, et al. |
| EPI_ISL_683789 | DOHMH PHL | New York City Public Health Laboratory | Jade Wang, et al. |
| EPI_ISL_683790 | DOHMH Morrisania | New York City Public Health Laboratory | Jade Wang, et al. |
| EPI_ISL_683791 | DOHMH Jamaica | New York City Public Health Laboratory | Jade Wang, et al. |
| EPI_ISL_683792 | DOHMH Chelsea | New York City Public Health Laboratory | Jade Wang, et al. |
| EPI_ISL_683793, EPI_ISL_683794 | DOHMH Corona | New York City Public Health Laboratory | Jade Wang, et al. |
| EPI_ISL_683795 | DOHMH Central Harlem | New York City Public Health Laboratory | Jade Wang, et al. |
| EPI_ISL_683796 | DOHMH Corona | New York City Public Health Laboratory | Jade Wang, et al. |
| EPI_ISL_683797 | DOHMH PHL | New York City Public Health Laboratory | Jade Wang, et al. |
| EPI_ISL_683798, EPI_ISL_683799, EPI_ISL_683800 | DOHMH Corona | New York City Public Health Laboratory | Jade Wang, et al. |
| EPI_ISL_683801 | DOHMH Morrisania | New York City Public Health Laboratory | Jade Wang, et al. |
| EPI_ISL_683802 | DOHMH Corona | New York City Public Health Laboratory | Jade Wang, et al. |
| EPI_ISL_683803, EPI_ISL_683804, EPI_ISL_683805, EPI_ISL_683806 | DOHMH Morrisania | New York City Public Health Laboratory | Jade Wang, et al. |
| EPI_ISL_683807, EPI_ISL_683808 | DOHMH Chelsea | New York City Public Health Laboratory | Jade Wang, et al. |
| EPI_ISL_683809 | DOHMH Corona | New York City Public Health Laboratory | Jade Wang, et al. |
| EPI_ISL_683810 | DOHMH PHL | New York City Public Health Laboratory | Jade Wang, et al. |
| EPI_ISL_683811, EPI_ISL_683812 | DOHMH Corona | New York City Public Health Laboratory | Jade Wang, et al. |
| EPI_ISL_683813 | DOHMH Central Harlem | New York City Public Health Laboratory | Jade Wang, et al. |
| EPI_ISL_683814 | DOHMH Morrisania | New York City Public Health Laboratory | Jade Wang, et al. |
| EPI_ISL_683815 | DOHMH Corona | New York City Public Health Laboratory | Jade Wang, et al. |
| EPI_ISL_683816 | DOHMH Fort Greene | New York City Public Health Laboratory | Jade Wang, et al. |
| EPI_ISL_683817, EPI_ISL_683818 | DOHMH Corona | New York City Public Health Laboratory | Jade Wang, et al. |
| EPI_ISL_683819, EPI_ISL_683820 | DOHMH Riverside | New York City Public Health Laboratory | Jade Wang, et al. |
| EPI_ISL_683821 | DOHMH Crown Heights | New York City Public Health Laboratory | Jade Wang, et al. |
| EPI_ISL_683822 | DOHMH Jamaica | New York City Public Health Laboratory | Jade Wang, et al. |
| EPI_ISL_683823 | DOHMH Morrisania | New York City Public Health Laboratory | Jade Wang, et al. |
| EPI_ISL_683824 | DOHMH Corona | New York City Public Health Laboratory | Jade Wang, et al. |
| EPI_ISL_683825, EPI_ISL_683826 | DOHMH Morrisania | New York City Public Health Laboratory | Jade Wang, et al. |
| EPI_ISL_683827 | DOHMH Crown Heights | New York City Public Health Laboratory | Jade Wang, et al. |
| EPI_ISL_683828 | DOHMH Chelsea | New York City Public Health Laboratory | Jade Wang, et al. |
| EPI_ISL_683829 | DOHMH Central Harlem | New York City Public Health Laboratory | Jade Wang, et al. |
| EPI_ISL_683830 | DOHMH Chelsea | New York City Public Health Laboratory | Jade Wang, et al. |
| EPI_ISL_683831 | DOHMH Corona | New York City Public Health Laboratory | Jade Wang, et al. |
| EPI_ISL_683832 | DOHMH Fort Greene | New York City Public Health Laboratory | Jade Wang, et al. |
| EPI_ISL_683833 | DOHMH Corona | New York City Public Health Laboratory | Jade Wang, et al. |
| EPI_ISL_683834 | DOHMH Central Harlem | New York City Public Health Laboratory | Jade Wang, et al. |
| EPI_ISL_683836 | DOHMH PHL | New York City Public Health Laboratory | Jade Wang, et al. |
| EPI_ISL_683837, EPI_ISL_683838 | DOHMH Jamaica | New York City Public Health Laboratory | Jade Wang, et al. |
| EPI_ISL_683839, EPI_ISL_683840 | DOHMH Morrisania | New York City Public Health Laboratory | Jade Wang, et al. |
| EPI_ISL_683841 | DOHMH Corona | New York City Public Health Laboratory | Jade Wang, et al. |
| EPI_ISL_683842 | DOHMH Central Harlem | New York City Public Health Laboratory | Jade Wang, et al. |
| EPI_ISL_683843, EPI_ISL_683844, EPI_ISL_683845 | DOHMH Morrisania | New York City Public Health Laboratory | Jade Wang, et al. |
| EPI_ISL_683846 | DOHMH PHL | New York City Public Health Laboratory | Jade Wang, et al. |
| EPI_ISL_683847 | DOHMH Corona | New York City Public Health Laboratory | Jade Wang, et al. |
| EPI_ISL_683848 | DOHMH Central Harlem | New York City Public Health Laboratory | Jade Wang, et al. |
| EPI_ISL_683849 | DOHMH Corona | New York City Public Health Laboratory | Jade Wang, et al. |
| EPI_ISL_683850 | DOHMH Jamaica | New York City Public Health Laboratory | Jade Wang, et al. |
| EPI_ISL_683851 | DOHMH Corona | New York City Public Health Laboratory | Jade Wang, et al. |
| EPI_ISL_683852 | DOHMH PHL | New York City Public Health Laboratory | Jade Wang, et al. |
| EPI_ISL_683853, EPI_ISL_683854 | DOHMH Morrisania | New York City Public Health Laboratory | Jade Wang, et al. |
| EPI_ISL_683855 | DOHMH Corona | New York City Public Health Laboratory | Jade Wang, et al. |
| EPI_ISL_683856 | DOHMH Crown Heights | New York City Public Health Laboratory | Jade Wang, et al. |
| EPI_ISL_683857 | DOHMH Corona | New York City Public Health Laboratory | Jade Wang, et al. |
| EPI_ISL_683858 | DOHMH Jamaica | New York City Public Health Laboratory | Jade Wang, et al. |
| EPI_ISL_683859 | DOHMH Central Harlem | New York City Public Health Laboratory | Jade Wang, et al. |
| EPI_ISL_683860 | DOHMH Corona | New York City Public Health Laboratory | Jade Wang, et al. |
| EPI_ISL_683861, EPI_ISL_683862 | DOHMH Jamaica | New York City Public Health Laboratory | Jade Wang, et al. |
| EPI_ISL_683863 | DOHMH Morrisania | New York City Public Health Laboratory | Jade Wang, et al. |
| EPI_ISL_683864 | DOHMH Jamaica | New York City Public Health Laboratory | Jade Wang, et al. |
| EPI_ISL_683865 | DOHMH Chelsea | New York City Public Health Laboratory | Jade Wang, et al. |
| EPI_ISL_683866 | DOHMH Riverside | New York City Public Health Laboratory | Jade Wang, et al. |
| EPI_ISL_683867 | DOHMH Corona | New York City Public Health Laboratory | Jade Wang, et al. |
| EPI_ISL_683868 | DOHMH Fort Greene | New York City Public Health Laboratory | Jade Wang, et al. |
| EPI_ISL_683869 | DOHMH Morrisania | New York City Public Health Laboratory | Jade Wang, et al. |
| EPI_ISL_683870 | DOHMH Corona | New York City Public Health Laboratory | Jade Wang, et al. |

| | | | |
|---|---|---|---|
| EPI_ISL_683871 | DOHMH PHL | New York City Public Health Laboratory | Jade Wang, et al. |
| EPI_ISL_683872,<br>EPI_ISL_683873 | DOHMH Corona | New York City Public Health Laboratory | Jade Wang, et al. |
| EPI_ISL_683874 | DOHMH Jamaica | New York City Public Health Laboratory | Jade Wang, et al. |
| EPI_ISL_683875 | DOHMH PHL | New York City Public Health Laboratory | Jade Wang, et al. |
| EPI_ISL_683876 | DOHMH Corona | New York City Public Health Laboratory | Jade Wang, et al. |
| EPI_ISL_683878 | DOHMH Jamaica | New York City Public Health Laboratory | Jade Wang, et al. |
| EPI_ISL_683879,<br>EPI_ISL_683880 | DOHMH Morrisania | New York City Public Health Laboratory | Jade Wang, et al. |
| EPI_ISL_683881 | DOHMH Corona | New York City Public Health Laboratory | Jade Wang, et al. |
| EPI_ISL_683882,<br>EPI_ISL_683883 | DOHMH Central Harlem | New York City Public Health Laboratory | Jade Wang, et al. |
| EPI_ISL_683884 | DOHMH Morrisania | New York City Public Health Laboratory | Jade Wang, et al. |
| EPI_ISL_683885 | DOHMH Jamaica | New York City Public Health Laboratory | Jade Wang, et al. |
| EPI_ISL_683886 | DOHMH Morrisania | New York City Public Health Laboratory | Jade Wang, et al. |
| EPI_ISL_683887,<br>EPI_ISL_683888 | DOHMH Fort Greene | New York City Public Health Laboratory | Jade Wang, et al. |
| EPI_ISL_683889 | DOHMH Morrisania | New York City Public Health Laboratory | Jade Wang, et al. |
| EPI_ISL_683890 | DOHMH PHL | New York City Public Health Laboratory | Jade Wang, et al. |
| EPI_ISL_683891 | DOHMH Crown Heights | New York City Public Health Laboratory | Jade Wang, et al. |
| EPI_ISL_683892 | DOHMH Jamaica | New York City Public Health Laboratory | Jade Wang, et al. |
| EPI_ISL_683893 | DOHMH Corona | New York City Public Health Laboratory | Jade Wang, et al. |
| EPI_ISL_683894,<br>EPI_ISL_683895 | DOHMH Riverside | New York City Public Health Laboratory | Jade Wang, et al. |
| EPI_ISL_683896,<br>EPI_ISL_683897,<br>EPI_ISL_683898,<br>EPI_ISL_683899,<br>EPI_ISL_683900 | DOHMH Corona | New York City Public Health Laboratory | Jade Wang, et al. |
| EPI_ISL_683901 | DOHMH Jamaica | New York City Public Health Laboratory | Jade Wang, et al. |
| EPI_ISL_683902 | DOHMH Riverside | New York City Public Health Laboratory | Jade Wang, et al. |
| EPI_ISL_683903 | DOHMH Jamaica | New York City Public Health Laboratory | Jade Wang, et al. |
| EPI_ISL_683904 | DOHMH Crown Heights | New York City Public Health Laboratory | Jade Wang, et al. |
| EPI_ISL_683905 | DOHMH Morrisania | New York City Public Health Laboratory | Jade Wang, et al. |
| EPI_ISL_683906 | DOHMH Fort Greene | New York City Public Health Laboratory | Jade Wang, et al. |
| EPI_ISL_683907 | DOHMH Riverside | New York City Public Health Laboratory | Jade Wang, et al. |
| EPI_ISL_683908,<br>EPI_ISL_683909 | DOHMH Corona | New York City Public Health Laboratory | Jade Wang, et al. |
| EPI_ISL_683910 | DOHMH Crown Heights | New York City Public Health Laboratory | Jade Wang, et al. |
| EPI_ISL_683911,<br>EPI_ISL_683912 | DOHMH Corona | New York City Public Health Laboratory | Jade Wang, et al. |
| EPI_ISL_683913 | DOHMH Fort Greene | New York City Public Health Laboratory | Jade Wang, et al. |
| EPI_ISL_683914,<br>EPI_ISL_683915,<br>EPI_ISL_683916 | DOHMH Jamaica | New York City Public Health Laboratory | Jade Wang, et al. |
| EPI_ISL_683917,<br>EPI_ISL_683918,<br>EPI_ISL_683919 | DOHMH Morrisania | New York City Public Health Laboratory | Jade Wang, et al. |
| EPI_ISL_683920,<br>EPI_ISL_683921 | DOHMH Jamaica | New York City Public Health Laboratory | Jade Wang, et al. |
| EPI_ISL_683922 | DOHMH Fort Greene | New York City Public Health Laboratory | Jade Wang, et al. |
| EPI_ISL_683923 | DOHMH Chelsea | New York City Public Health Laboratory | Jade Wang, et al. |
| EPI_ISL_683924 | DOHMH Central Harlem | New York City Public Health Laboratory | Jade Wang, et al. |
| EPI_ISL_683925 | DOHMH Jamaica | New York City Public Health Laboratory | Jade Wang, et al. |
| EPI_ISL_683926 | DOHMH Corona | New York City Public Health Laboratory | Jade Wang, et al. |
| EPI_ISL_683927,<br>EPI_ISL_683928 | DOHMH Central Harlem | New York City Public Health Laboratory | Jade Wang, et al. |
| EPI_ISL_683929 | DOHMH Corona | New York City Public Health Laboratory | Jade Wang, et al. |
| EPI_ISL_683930 | DOHMH Jamaica | New York City Public Health Laboratory | Jade Wang, et al. |
| EPI_ISL_683931 | DOHMH Chelsea | New York City Public Health Laboratory | Jade Wang, et al. |
| EPI_ISL_683932 | DOHMH Corona | New York City Public Health Laboratory | Jade Wang, et al. |
| EPI_ISL_683933 | DOHMH Central Harlem | New York City Public Health Laboratory | Jade Wang, et al. |
| EPI_ISL_683934 | DOHMH Corona | New York City Public Health Laboratory | Jade Wang, et al. |
| EPI_ISL_683935 | DOHMH Crown Heights | New York City Public Health Laboratory | Jade Wang, et al. |
| EPI_ISL_683936 | DOHMH Chelsea | New York City Public Health Laboratory | Jade Wang, et al. |
| EPI_ISL_683937 | DOHMH Central Harlem | New York City Public Health Laboratory | Jade Wang, et al. |
| EPI_ISL_683938 | DOHMH Morrisania | New York City Public Health Laboratory | Jade Wang, et al. |
| EPI_ISL_683939 | DOHMH Jamaica | New York City Public Health Laboratory | Jade Wang, et al. |
| EPI_ISL_683940 | DOHMH Morrisania | New York City Public Health Laboratory | Jade Wang, et al. |
| EPI_ISL_683941 | DOHMH Chelsea | New York City Public Health Laboratory | Jade Wang, et al. |
| EPI_ISL_683942 | DOHMH Riverside | New York City Public Health Laboratory | Jade Wang, et al. |
| EPI_ISL_683943 | DOHMH PHL | New York City Public Health Laboratory | Jade Wang, et al. |
| EPI_ISL_683944,<br>EPI_ISL_683945 | DOHMH Jamaica | New York City Public Health Laboratory | Jade Wang, et al. |
| EPI_ISL_683946,<br>EPI_ISL_683947 | DOHMH Morrisania | New York City Public Health Laboratory | Jade Wang, et al. |
| EPI_ISL_683948 | DOHMH Jamaica | New York City Public Health Laboratory | Jade Wang, et al. |
| EPI_ISL_683949,<br>EPI_ISL_683950 | DOHMH Riverside | New York City Public Health Laboratory | Jade Wang, et al. |
| EPI_ISL_683951 | DOHMH Jamaica | New York City Public Health Laboratory | Jade Wang, et al. |
| EPI_ISL_683952,<br>EPI_ISL_683953 | DOHMH Corona | New York City Public Health Laboratory | Jade Wang, et al. |

| | | | |
|---|---|---|---|
| EPI_ISL_683954 | DOHMH PHL | New York City Public Health Laboratory | Jade Wang, et al. |
| EPI_ISL_683955 | DOHMH Riverside | New York City Public Health Laboratory | Jade Wang, et al. |
| EPI_ISL_683956 | DOHMH Jamaica | New York City Public Health Laboratory | Jade Wang, et al. |
| EPI_ISL_683957 | DOHMH Chelsea | New York City Public Health Laboratory | Jade Wang, et al. |
| EPI_ISL_683958 | DOHMH Jamaica | New York City Public Health Laboratory | Jade Wang, et al. |
| EPI_ISL_683959 | DOHMH Morrisania | New York City Public Health Laboratory | Jade Wang, et al. |
| EPI_ISL_683960 | DOHMH Corona | New York City Public Health Laboratory | Jade Wang, et al. |
| EPI_ISL_683961 | DOHMH Chelsea | New York City Public Health Laboratory | Jade Wang, et al. |
| EPI_ISL_683962 | DOHMH Central Harlem | New York City Public Health Laboratory | Jade Wang, et al. |
| EPI_ISL_683963 | DOHMH Fort Greene | New York City Public Health Laboratory | Jade Wang, et al. |
| EPI_ISL_683964 | DOHMH Central Harlem | New York City Public Health Laboratory | Jade Wang, et al. |
| EPI_ISL_683965, EPI_ISL_683966 | DOHMH Morrisania | New York City Public Health Laboratory | Jade Wang, et al. |
| EPI_ISL_683967 | DOHMH Chelsea | New York City Public Health Laboratory | Jade Wang, et al. |
| EPI_ISL_683968 | DOHMH Central Harlem | New York City Public Health Laboratory | Jade Wang, et al. |
| EPI_ISL_683969 | DOHMH Morrisania | New York City Public Health Laboratory | Jade Wang, et al. |
| EPI_ISL_683970 | DOHMH Crown Heights | New York City Public Health Laboratory | Jade Wang, et al. |
| EPI_ISL_683971, EPI_ISL_683972 | DOHMH Chelsea | New York City Public Health Laboratory | Jade Wang, et al. |
| EPI_ISL_683973 | DOHMH Fort Greene | New York City Public Health Laboratory | Jade Wang, et al. |
| EPI_ISL_683974 | DOHMH Central Harlem | New York City Public Health Laboratory | Jade Wang, et al. |
| EPI_ISL_683975 | DOHMH Jamaica | New York City Public Health Laboratory | Jade Wang, et al. |
| EPI_ISL_683976, EPI_ISL_683977 | DOHMH Central Harlem | New York City Public Health Laboratory | Jade Wang, et al. |
| EPI_ISL_683978, EPI_ISL_683979 | DOHMH Corona | New York City Public Health Laboratory | Jade Wang, et al. |
| EPI_ISL_683980, EPI_ISL_683981 | DOHMH Chelsea | New York City Public Health Laboratory | Jade Wang, et al. |
| EPI_ISL_683982, EPI_ISL_683983, EPI_ISL_683984, EPI_ISL_683985 | DOHMH Central Harlem | New York City Public Health Laboratory | Jade Wang, et al. |
| EPI_ISL_683986 | DOHMH Jamaica | New York City Public Health Laboratory | Jade Wang, et al. |
| EPI_ISL_683987, EPI_ISL_683988 | DOHMH PHL | New York City Public Health Laboratory | Jade Wang, et al. |
| EPI_ISL_683989 | DOHMH Chelsea | New York City Public Health Laboratory | Jade Wang, et al. |
| EPI_ISL_683990 | DOHMH Jamaica | New York City Public Health Laboratory | Jade Wang, et al. |
| EPI_ISL_683991 | DOHMH Central Harlem | New York City Public Health Laboratory | Jade Wang, et al. |
| EPI_ISL_683992 | DOHMH Morrisania | New York City Public Health Laboratory | Jade Wang, et al. |
| EPI_ISL_683993 | DOHMH PHL | New York City Public Health Laboratory | Jade Wang, et al. |
| EPI_ISL_683994 | DOHMH Riverside | New York City Public Health Laboratory | Jade Wang, et al. |
| EPI_ISL_683995 | DOHMH PHL | New York City Public Health Laboratory | Jade Wang, et al. |
| EPI_ISL_683996 | DOHMH Fort Greene | New York City Public Health Laboratory | Jade Wang, et al. |
| EPI_ISL_684000, EPI_ISL_684002, EPI_ISL_684003, EPI_ISL_684004, EPI_ISL_684005, EPI_ISL_684006, EPI_ISL_684007, EPI_ISL_684008, EPI_ISL_684010, EPI_ISL_684011, EPI_ISL_684012, EPI_ISL_684013, EPI_ISL_684015, EPI_ISL_684016, EPI_ISL_684017, EPI_ISL_684018, EPI_ISL_684020, EPI_ISL_684021, EPI_ISL_684023, EPI_ISL_684024, EPI_ISL_684026, EPI_ISL_684027 | | | |
| see above | Utah Public Health Laboratory | Utah Public Health Laboratory | Erin Young, Kelly Oakeson |
| EPI_ISL_692769, EPI_ISL_692770, EPI_ISL_692771, EPI_ISL_692772, EPI_ISL_692773, EPI_ISL_692774, EPI_ISL_692776, EPI_ISL_692778, EPI_ISL_692780, EPI_ISL_692782, EPI_ISL_692783, EPI_ISL_692784, EPI_ISL_692785, EPI_ISL_692786, EPI_ISL_692787, EPI_ISL_692788, EPI_ISL_692789, EPI_ISL_692790, EPI_ISL_692791, EPI_ISL_692792, EPI_ISL_692794, EPI_ISL_692795, EPI_ISL_692796, EPI_ISL_692798, EPI_ISL_692799, EPI_ISL_692801, EPI_ISL_692803, EPI_ISL_692804, EPI_ISL_692809, EPI_ISL_692810, EPI_ISL_692811, EPI_ISL_692812, EPI_ISL_692813, EPI_ISL_692814, EPI_ISL_692815, EPI_ISL_692816, EPI_ISL_692817, EPI_ISL_692818, EPI_ISL_692819, EPI_ISL_692820, EPI_ISL_692822, EPI_ISL_692831, EPI_ISL_692832, EPI_ISL_692834, EPI_ISL_692835, EPI_ISL_692836, EPI_ISL_692837, EPI_ISL_692838, EPI_ISL_692839, EPI_ISL_692840, EPI_ISL_692841, EPI_ISL_692842, EPI_ISL_692843, EPI_ISL_692844, EPI_ISL_692845, EPI_ISL_692846, EPI_ISL_692847, EPI_ISL_692848, EPI_ISL_692849, EPI_ISL_692850, EPI_ISL_692851, EPI_ISL_692852, EPI_ISL_692853, EPI_ISL_692854, EPI_ISL_692855, EPI_ISL_692858, EPI_ISL_692859, EPI_ISL_692860, EPI_ISL_692863, EPI_ISL_692865, EPI_ISL_692866, EPI_ISL_692867, EPI_ISL_692868, EPI_ISL_692869, EPI_ISL_692870, EPI_ISL_692871, EPI_ISL_692873, EPI_ISL_692874, EPI_ISL_692875, EPI_ISL_692876, EPI_ISL_692878, EPI_ISL_692880, EPI_ISL_692881, EPI_ISL_692884, EPI_ISL_692885, EPI_ISL_692886, EPI_ISL_692887, EPI_ISL_692888, EPI_ISL_692889, EPI_ISL_692890, EPI_ISL_692891, EPI_ISL_692892, EPI_ISL_692893, EPI_ISL_692894, EPI_ISL_692895, EPI_ISL_692896, EPI_ISL_692897, EPI_ISL_692898, EPI_ISL_692899, EPI_ISL_692900, EPI_ISL_692901, EPI_ISL_692902, EPI_ISL_692903, EPI_ISL_692904, EPI_ISL_692905, EPI_ISL_692908, EPI_ISL_692909, EPI_ISL_692910, EPI_ISL_692911, EPI_ISL_692912, EPI_ISL_692913, EPI_ISL_692915, EPI_ISL_692916, EPI_ISL_692917, EPI_ISL_692918, EPI_ISL_692919, EPI_ISL_692920, EPI_ISL_692921, EPI_ISL_692922, EPI_ISL_692923, EPI_ISL_692924, EPI_ISL_692925, EPI_ISL_692926, EPI_ISL_692927, EPI_ISL_692928, EPI_ISL_692929, EPI_ISL_692930, EPI_ISL_692931, EPI_ISL_692932, EPI_ISL_692933, EPI_ISL_692934, EPI_ISL_692935, EPI_ISL_692936, EPI_ISL_692937, EPI_ISL_692938, EPI_ISL_692939, EPI_ISL_692940, EPI_ISL_692941, EPI_ISL_692942, EPI_ISL_692943, EPI_ISL_692944, EPI_ISL_692945, EPI_ISL_692946, EPI_ISL_692947, EPI_ISL_692949, EPI_ISL_692950, EPI_ISL_692952, EPI_ISL_692953, EPI_ISL_692954, EPI_ISL_692955, EPI_ISL_692956, EPI_ISL_692957, EPI_ISL_692958, EPI_ISL_692961, EPI_ISL_692962, EPI_ISL_692963, EPI_ISL_692964, EPI_ISL_692965, EPI_ISL_692966, EPI_ISL_692968, EPI_ISL_692970, EPI_ISL_692971, EPI_ISL_692972, EPI_ISL_692974, EPI_ISL_692975, EPI_ISL_692976, EPI_ISL_692977, EPI_ISL_692978, EPI_ISL_692979, EPI_ISL_692980, EPI_ISL_692981, EPI_ISL_692982, EPI_ISL_692983, EPI_ISL_692984, EPI_ISL_692985, EPI_ISL_692986, EPI_ISL_692987, EPI_ISL_692988, EPI_ISL_692989, EPI_ISL_692991, EPI_ISL_692992, EPI_ISL_692993, EPI_ISL_692994, EPI_ISL_692996, EPI_ISL_692998, EPI_ISL_692999, EPI_ISL_693000, EPI_ISL_693001, EPI_ISL_693002, EPI_ISL_693003, EPI_ISL_693004, EPI_ISL_693005, EPI_ISL_693006, EPI_ISL_693007, EPI_ISL_693008, EPI_ISL_693009, EPI_ISL_693010, EPI_ISL_693011, EPI_ISL_693012, EPI_ISL_693013, EPI_ISL_693015, EPI_ISL_693016, EPI_ISL_693018, EPI_ISL_693019, EPI_ISL_693020, EPI_ISL_693021, EPI_ISL_693023, EPI_ISL_693027, EPI_ISL_693028, EPI_ISL_693030, EPI_ISL_693031, EPI_ISL_693032, EPI_ISL_693035, EPI_ISL_693036, EPI_ISL_693037, EPI_ISL_693038, EPI_ISL_693039, EPI_ISL_693040, EPI_ISL_693042, EPI_ISL_693043, EPI_ISL_693044, EPI_ISL_693045, EPI_ISL_693046, EPI_ISL_693047, EPI_ISL_693048, EPI_ISL_693049, EPI_ISL_693050, EPI_ISL_693051, EPI_ISL_693052, EPI_ISL_693054, EPI_ISL_693055, EPI_ISL_693057, EPI_ISL_693058, EPI_ISL_693059, EPI_ISL_693060, EPI_ISL_693061, EPI_ISL_693062, EPI_ISL_693063, EPI_ISL_693064, EPI_ISL_693065, EPI_ISL_693066, EPI_ISL_693067, EPI_ISL_693069, EPI_ISL_693070, EPI_ISL_693071, EPI_ISL_693072, EPI_ISL_693073, EPI_ISL_693074, EPI_ISL_693075, EPI_ISL_693076, EPI_ISL_693077, EPI_ISL_693078, EPI_ISL_693080, EPI_ISL_693082, EPI_ISL_693083, EPI_ISL_693084, EPI_ISL_693086, EPI_ISL_693089, EPI_ISL_693090, EPI_ISL_693091, EPI_ISL_693097, EPI_ISL_693098, EPI_ISL_693099, EPI_ISL_693101, EPI_ISL_693102, EPI_ISL_693103, EPI_ISL_693104, EPI_ISL_693105, EPI_ISL_693106, EPI_ISL_693107, EPI_ISL_693108, EPI_ISL_693109, EPI_ISL_693110, EPI_ISL_693113, EPI_ISL_693114, EPI_ISL_693115, EPI_ISL_693117, EPI_ISL_693119, EPI_ISL_693120, EPI_ISL_693121, EPI_ISL_693122, EPI_ISL_693124, EPI_ISL_693125, EPI_ISL_693126, EPI_ISL_693127, EPI_ISL_693128, EPI_ISL_693129, EPI_ISL_693130, EPI_ISL_693131, EPI_ISL_693133, EPI_ISL_693134, EPI_ISL_693135, EPI_ISL_693136, EPI_ISL_693137, EPI_ISL_693139, EPI_ISL_693142, EPI_ISL_693143, EPI_ISL_693144, EPI_ISL_693145, EPI_ISL_693146, EPI_ISL_693147, EPI_ISL_693148, EPI_ISL_693149, EPI_ISL_693150, EPI_ISL_693151, EPI_ISL_693152, EPI_ISL_693153, EPI_ISL_693155, EPI_ISL_693156, EPI_ISL_693157, EPI_ISL_693158, EPI_ISL_693159, EPI_ISL_693162, EPI_ISL_693164, EPI_ISL_693165, EPI_ISL_693166, EPI_ISL_693167, EPI_ISL_693170, EPI_ISL_693171, EPI_ISL_693172, EPI_ISL_693173, EPI_ISL_693174, EPI_ISL_693176, EPI_ISL_693177, EPI_ISL_693178, EPI_ISL_693179, EPI_ISL_693180, EPI_ISL_693181, EPI_ISL_693182, EPI_ISL_693183, EPI_ISL_693185, EPI_ISL_693186, EPI_ISL_693187, EPI_ISL_693188, EPI_ISL_693189, EPI_ISL_693190, EPI_ISL_693191 | | | |
| see above | Massachusetts State Public Health Laboratory | Massachusetts State Public Health Laboratory | Andrew Lang, Timelia Fink, Glen Gallagher, Sandra Smole |
| EPI_ISL_693658, EPI_ISL_693659 | Washington University in St. Louis | Washington University in St. Louis | David Wang, Carey-Ann Burnham, Bijal Parikh, Scott Handley, Lindsay Droit, Stephen Tahan |
| EPI_ISL_693688, EPI_ISL_693689, EPI_ISL_693690, EPI_ISL_693691, EPI_ISL_693692, EPI_ISL_693693, EPI_ISL_693694, EPI_ISL_693695, EPI_ISL_693696, EPI_ISL_693697, EPI_ISL_693698, EPI_ISL_693699, EPI_ISL_693700, EPI_ISL_693701, EPI_ISL_693702, EPI_ISL_693703, EPI_ISL_693704, EPI_ISL_693705, EPI_ISL_693706, EPI_ISL_693707, EPI_ISL_693708, EPI_ISL_693709, EPI_ISL_693710, EPI_ISL_693711, EPI_ISL_693712, EPI_ISL_693713, EPI_ISL_693714, EPI_ISL_693715, EPI_ISL_693727, EPI_ISL_693728, EPI_ISL_693729, EPI_ISL_693730, EPI_ISL_693731, EPI_ISL_693732, EPI_ISL_693733, EPI_ISL_693734, EPI_ISL_693735, EPI_ISL_693736, EPI_ISL_693737, EPI_ISL_693738, EPI_ISL_693739, EPI_ISL_693740, EPI_ISL_693741, EPI_ISL_693742, EPI_ISL_693743, EPI_ISL_693744, EPI_ISL_693745, EPI_ISL_693746, EPI_ISL_693747, EPI_ISL_693748, EPI_ISL_693749, EPI_ISL_693750, EPI_ISL_693751, EPI_ISL_693752, EPI_ISL_693753, EPI_ISL_693754, EPI_ISL_693755, EPI_ISL_693756, EPI_ISL_693757 | | | |
| see above | Delaware Public Health Laboratory | Delaware Public Health Laboratory | Gregory Hovan |
| EPI_ISL_694009, EPI_ISL_694010, EPI_ISL_694011, EPI_ISL_694013, EPI_ISL_694014, EPI_ISL_694016, EPI_ISL_694017, EPI_ISL_694019, EPI_ISL_694020, EPI_ISL_694021, EPI_ISL_694026, EPI_ISL_694027, EPI_ISL_694030, EPI_ISL_694031, EPI_ISL_694032, EPI_ISL_694033, EPI_ISL_694034, EPI_ISL_694035, EPI_ISL_694037, EPI_ISL_694039 | | | |
| see above | TGen North | TGen North | Hayley D Yaglom, Marette Gebhardt, Ashlyn Pfeiffer, Mary Ellen Ormsby, Daniel Jasso-Selles, Darrin Lemmer, Megan Folkerts, Chris French, Matthew Maurer, Jolene R Bowers, David M Engelthaler |
| EPI_ISL_694046 | Los Angeles County Public Health Laboratories | Los Angeles County Public Health Laboratories | Peera Hemarajata et al. |
| EPI_ISL_694047, EPI_ISL_694048 | TGen North | TGen North | Jolene Bowers, Megan Folkerts, Chris French, Hayley Yaglom, Ashlyn Pfeiffer, Darrin Lemmer, Dave Engelthaler, The Arizona COVID Genomics Union (ACGU) |
| EPI_ISL_694049 | AZ SPHL, Arizona Department of Health Services | TGen North | Jolene Bowers, Megan Folkerts, Chris French, Hayley Yaglom, Ashlyn Pfeiffer, Darrin Lemmer, Dave Engelthaler, The Arizona COVID Genomics Union (ACGU) |
| EPI_ISL_694050, EPI_ISL_694051, EPI_ISL_694052 | TGen North | TGen North | Jolene Bowers, Megan Folkerts, Chris French, Hayley Yaglom, Ashlyn Pfeiffer, Darrin Lemmer, Dave Engelthaler, The Arizona COVID Genomics Union (ACGU) |
| EPI_ISL_694053, EPI_ISL_694054, EPI_ISL_694055, EPI_ISL_694056, EPI_ISL_694057, EPI_ISL_694058, EPI_ISL_694059, EPI_ISL_694060, EPI_ISL_694061, EPI_ISL_694062, EPI_ISL_694063, EPI_ISL_694064, EPI_ISL_694065, EPI_ISL_694066, EPI_ISL_694067, EPI_ISL_694068, EPI_ISL_694069, EPI_ISL_694070, EPI_ISL_694071, EPI_ISL_694072, EPI_ISL_694073, EPI_ISL_694074, EPI_ISL_694075, EPI_ISL_694076, EPI_ISL_694077, EPI_ISL_694078, EPI_ISL_694079, EPI_ISL_694080, EPI_ISL_694081, EPI_ISL_694082, EPI_ISL_694083, EPI_ISL_694084, EPI_ISL_694085, EPI_ISL_694086, EPI_ISL_694087, EPI_ISL_694088, EPI_ISL_694089, EPI_ISL_694090, EPI_ISL_694092, EPI_ISL_694093, EPI_ISL_694094, EPI_ISL_694095, EPI_ISL_694096, EPI_ISL_694097, EPI_ISL_694098, EPI_ISL_694099, EPI_ISL_694100, EPI_ISL_694101, EPI_ISL_694102, EPI_ISL_694103, EPI_ISL_694104, EPI_ISL_694105, EPI_ISL_694106, EPI_ISL_694107, EPI_ISL_694110, EPI_ISL_694111, EPI_ISL_694112, EPI_ISL_694113, EPI_ISL_694115, EPI_ISL_694116, EPI_ISL_694117, EPI_ISL_694119, EPI_ISL_694120, EPI_ISL_694123, EPI_ISL_694124, EPI_ISL_694125, EPI_ISL_694126, EPI_ISL_694127, EPI_ISL_694128, EPI_ISL_694130, EPI_ISL_694131, EPI_ISL_694132, EPI_ISL_694133, EPI_ISL_694134, EPI_ISL_694135, EPI_ISL_694136, EPI_ISL_694137, EPI_ISL_694138 | | | |
| see above | AZ SPHL, Arizona Department of Health Services | TGen North | Jolene Bowers, Megan Folkerts, Chris French, Hayley Yaglom, Ashlyn Pfeiffer, Darrin Lemmer, Dave Engelthaler, The Arizona COVID Genomics Union (ACGU) |
| EPI_ISL_694139, EPI_ISL_694140, EPI_ISL_694141, EPI_ISL_694142, | TGen North | TGen North | Jolene Bowers, Megan Folkerts, Chris French, Hayley Yaglom, Ashlyn Pfeiffer, Darrin Lemmer, Dave Engelthaler, The Arizona COVID Genomics Union (ACGU) |

EPI_ISL_694143, EPI_ISL_694144, EPI_ISL_694145, EPI_ISL_694146, EPI_ISL_694147

EPI_ISL_694148, EPI_ISL_694149, EPI_ISL_694151, EPI_ISL_694152, EPI_ISL_694155, EPI_ISL_694156, EPI_ISL_694157, EPI_ISL_694158, EPI_ISL_694159, EPI_ISL_694160, EPI_ISL_694161, EPI_ISL_694162, EPI_ISL_694163, EPI_ISL_694164, EPI_ISL_694165, EPI_ISL_694166, EPI_ISL_694167, EPI_ISL_694168, EPI_ISL_694170, EPI_ISL_694171, EPI_ISL_694173, EPI_ISL_694174, EPI_ISL_694175, EPI_ISL_694176, EPI_ISL_694177, EPI_ISL_694178, EPI_ISL_694179, EPI_ISL_694180, EPI_ISL_694181, EPI_ISL_694182, EPI_ISL_694183, EPI_ISL_694184, EPI_ISL_694185, EPI_ISL_694186, EPI_ISL_694187, EPI_ISL_694188, EPI_ISL_694189, EPI_ISL_694191, EPI_ISL_694193, EPI_ISL_694194, EPI_ISL_694195, EPI_ISL_694196, EPI_ISL_694198, EPI_ISL_694199, EPI_ISL_694200, EPI_ISL_694201, EPI_ISL_694202, EPI_ISL_694203, EPI_ISL_694204, EPI_ISL_694205, EPI_ISL_694207, EPI_ISL_694208, EPI_ISL_694209, EPI_ISL_694210, EPI_ISL_694211, EPI_ISL_694212, EPI_ISL_694213, EPI_ISL_694214, EPI_ISL_694215, EPI_ISL_694216, EPI_ISL_694217, EPI_ISL_694218

| see above | AZ SPHL, Arizona Department of Health Services | TGen North | Jolene Bowers, Megan Folkerts, Chris French, Hayley Yaglom, Ashlyn Pfeiffer, Darrin Lemmer, Dave Engelthaler, The Arizona COVID Genomics Union (ACGU) |
|---|---|---|---|

EPI_ISL_694220, EPI_ISL_694221, EPI_ISL_694222, EPI_ISL_694223, EPI_ISL_694224, EPI_ISL_694226, EPI_ISL_694228, EPI_ISL_694230, EPI_ISL_694231, EPI_ISL_694232, EPI_ISL_694233, EPI_ISL_694235, EPI_ISL_694236, EPI_ISL_694237, EPI_ISL_694238, EPI_ISL_694239, EPI_ISL_694241, EPI_ISL_694242, EPI_ISL_694243, EPI_ISL_694244, EPI_ISL_694245, EPI_ISL_694246, EPI_ISL_694247

| see above | TGen North | TGen North | Jolene Bowers, Megan Folkerts, Chris French, Hayley Yaglom, Ashlyn Pfeiffer, Darrin Lemmer, Dave Engelthaler, The Arizona COVID Genomics Union (ACGU) |
|---|---|---|---|

EPI_ISL_694248, EPI_ISL_694249, EPI_ISL_694250, EPI_ISL_694251, EPI_ISL_694252, EPI_ISL_694253, EPI_ISL_694254, EPI_ISL_694255, EPI_ISL_694256, EPI_ISL_694257, EPI_ISL_694258, EPI_ISL_694259, EPI_ISL_694261, EPI_ISL_694264, EPI_ISL_694265, EPI_ISL_694266, EPI_ISL_694267, EPI_ISL_694268, EPI_ISL_694269, EPI_ISL_694270, EPI_ISL_694271, EPI_ISL_694272, EPI_ISL_694273, EPI_ISL_694274, EPI_ISL_694275, EPI_ISL_694276, EPI_ISL_694277, EPI_ISL_694278, EPI_ISL_694279, EPI_ISL_694280, EPI_ISL_694281, EPI_ISL_694282, EPI_ISL_694283, EPI_ISL_694284, EPI_ISL_694285, EPI_ISL_694286, EPI_ISL_694287, EPI_ISL_694288, EPI_ISL_694289, EPI_ISL_694290, EPI_ISL_694291, EPI_ISL_694292, EPI_ISL_694293, EPI_ISL_694294, EPI_ISL_694295, EPI_ISL_694296, EPI_ISL_694297, EPI_ISL_694298, EPI_ISL_694299, EPI_ISL_694301, EPI_ISL_694302, EPI_ISL_694303, EPI_ISL_694304, EPI_ISL_694305, EPI_ISL_694306, EPI_ISL_694307, EPI_ISL_694308, EPI_ISL_694309, EPI_ISL_694311, EPI_ISL_694312, EPI_ISL_694313, EPI_ISL_694314, EPI_ISL_694315

| see above | AZ SPHL, Arizona Department of Health Services | TGen North | Jolene Bowers, Megan Folkerts, Chris French, Hayley Yaglom, Ashlyn Pfeiffer, Darrin Lemmer, Dave Engelthaler, The Arizona COVID Genomics Union (ACGU) |
|---|---|---|---|

EPI_ISL_694316, EPI_ISL_694317, EPI_ISL_694318, EPI_ISL_694319, EPI_ISL_694320, EPI_ISL_694321, EPI_ISL_694322, EPI_ISL_694323, EPI_ISL_694324, EPI_ISL_694325, EPI_ISL_694326, EPI_ISL_694327, EPI_ISL_694328, EPI_ISL_694329, EPI_ISL_694330, EPI_ISL_694331, EPI_ISL_694332, EPI_ISL_694333, EPI_ISL_694335, EPI_ISL_694336, EPI_ISL_694337, EPI_ISL_694338, EPI_ISL_694339, EPI_ISL_694340, EPI_ISL_694341, EPI_ISL_694343, EPI_ISL_694344, EPI_ISL_694345, EPI_ISL_694346, EPI_ISL_694347, EPI_ISL_694348, EPI_ISL_694349, EPI_ISL_694350, EPI_ISL_694351, EPI_ISL_694352, EPI_ISL_694353, EPI_ISL_694354, EPI_ISL_694356, EPI_ISL_694357, EPI_ISL_694359, EPI_ISL_694360, EPI_ISL_694361, EPI_ISL_694362, EPI_ISL_694363, EPI_ISL_694364, EPI_ISL_694366, EPI_ISL_694367, EPI_ISL_694368, EPI_ISL_694369, EPI_ISL_694370, EPI_ISL_694371, EPI_ISL_694372, EPI_ISL_694373, EPI_ISL_694374, EPI_ISL_694375, EPI_ISL_694376, EPI_ISL_694377, EPI_ISL_694378, EPI_ISL_694379, EPI_ISL_694380, EPI_ISL_694381, EPI_ISL_694382, EPI_ISL_694383, EPI_ISL_694384, EPI_ISL_694385, EPI_ISL_694386, EPI_ISL_694387, EPI_ISL_694388, EPI_ISL_694389, EPI_ISL_694390, EPI_ISL_694391, EPI_ISL_694392, EPI_ISL_694393, EPI_ISL_694394, EPI_ISL_694396, EPI_ISL_694397, EPI_ISL_694398, EPI_ISL_694399, EPI_ISL_694400, EPI_ISL_694401, EPI_ISL_694403, EPI_ISL_694404

| see above | TGen North | TGen North | Jolene Bowers, Megan Folkerts, Chris French, Hayley Yaglom, Ashlyn Pfeiffer, Darrin Lemmer, Dave Engelthaler, The Arizona COVID Genomics Union (ACGU) |
|---|---|---|---|

EPI_ISL_694405, EPI_ISL_694406, EPI_ISL_694408, EPI_ISL_694411, EPI_ISL_694412, EPI_ISL_694413, EPI_ISL_694414, EPI_ISL_694415, EPI_ISL_694416, EPI_ISL_694420, EPI_ISL_694421, EPI_ISL_694422, EPI_ISL_694423, EPI_ISL_694424, EPI_ISL_694425, EPI_ISL_694427

| see above | AZ SPHL, Arizona Department of Health Services | TGen North | Jolene Bowers, Megan Folkerts, Chris French, Hayley Yaglom, Ashlyn Pfeiffer, Darrin Lemmer, Dave Engelthaler, The Arizona COVID Genomics Union (ACGU) |
|---|---|---|---|

EPI_ISL_694428, EPI_ISL_694429, EPI_ISL_694430, EPI_ISL_694431, EPI_ISL_694432, EPI_ISL_694433, EPI_ISL_694434, EPI_ISL_694435, EPI_ISL_694436, EPI_ISL_694437, EPI_ISL_694438, EPI_ISL_694439, EPI_ISL_694440, EPI_ISL_694441, EPI_ISL_694442, EPI_ISL_694443, EPI_ISL_694445, EPI_ISL_694446, EPI_ISL_694448, EPI_ISL_694449, EPI_ISL_694450, EPI_ISL_694451, EPI_ISL_694452, EPI_ISL_694453, EPI_ISL_694454, EPI_ISL_694455, EPI_ISL_694456, EPI_ISL_694457

| see above | TGen North | TGen North | Jolene Bowers, Megan Folkerts, Chris French, Hayley Yaglom, Ashlyn Pfeiffer, Darrin Lemmer, Dave Engelthaler, The Arizona COVID Genomics Union (ACGU) |
|---|---|---|---|

EPI_ISL_694458, EPI_ISL_694459, EPI_ISL_694460, EPI_ISL_694461, EPI_ISL_694462, EPI_ISL_694464, EPI_ISL_694465, EPI_ISL_694467, EPI_ISL_694468, EPI_ISL_694469, EPI_ISL_694470, EPI_ISL_694471, EPI_ISL_694472, EPI_ISL_694473, EPI_ISL_694474, EPI_ISL_694475, EPI_ISL_694476, EPI_ISL_694477, EPI_ISL_694479, EPI_ISL_694480, EPI_ISL_694481, EPI_ISL_694483, EPI_ISL_694484, EPI_ISL_694485, EPI_ISL_694486, EPI_ISL_694487, EPI_ISL_694488, EPI_ISL_694489, EPI_ISL_694490, EPI_ISL_694491, EPI_ISL_694492, EPI_ISL_694493, EPI_ISL_694494, EPI_ISL_694495, EPI_ISL_694496, EPI_ISL_694497, EPI_ISL_694498, EPI_ISL_694499, EPI_ISL_694500, EPI_ISL_694501, EPI_ISL_694502, EPI_ISL_694503, EPI_ISL_694504, EPI_ISL_694505, EPI_ISL_694506, EPI_ISL_694508, EPI_ISL_694509, EPI_ISL_694510, EPI_ISL_694511, EPI_ISL_694512, EPI_ISL_694513, EPI_ISL_694514, EPI_ISL_694515, EPI_ISL_694516, EPI_ISL_694517, EPI_ISL_694518, EPI_ISL_694519, EPI_ISL_694520, EPI_ISL_694521, EPI_ISL_694522, EPI_ISL_694523, EPI_ISL_694524, EPI_ISL_694525, EPI_ISL_694526, EPI_ISL_694527, EPI_ISL_694529, EPI_ISL_694530, EPI_ISL_694531, EPI_ISL_694532, EPI_ISL_694533, EPI_ISL_694534, EPI_ISL_694535, EPI_ISL_694538, EPI_ISL_694539, EPI_ISL_694541, EPI_ISL_694542, EPI_ISL_694543, EPI_ISL_694544, EPI_ISL_694545, EPI_ISL_694546, EPI_ISL_694547, EPI_ISL_694548, EPI_ISL_694550, EPI_ISL_694551, EPI_ISL_694552, EPI_ISL_694553, EPI_ISL_694554, EPI_ISL_694555, EPI_ISL_694556, EPI_ISL_694558, EPI_ISL_694560, EPI_ISL_694561, EPI_ISL_694562, EPI_ISL_694564, EPI_ISL_694565, EPI_ISL_694566, EPI_ISL_694567, EPI_ISL_694568, EPI_ISL_694569, EPI_ISL_694570, EPI_ISL_694571, EPI_ISL_694572, EPI_ISL_694574, EPI_ISL_694575, EPI_ISL_694576, EPI_ISL_694577, EPI_ISL_694578, EPI_ISL_694579, EPI_ISL_694580, EPI_ISL_694581, EPI_ISL_694582, EPI_ISL_694583, EPI_ISL_694584, EPI_ISL_694585, EPI_ISL_694586, EPI_ISL_694587, EPI_ISL_694588, EPI_ISL_694589, EPI_ISL_694591, EPI_ISL_694592, EPI_ISL_694593, EPI_ISL_694594, EPI_ISL_694595, EPI_ISL_694596, EPI_ISL_694597, EPI_ISL_694598, EPI_ISL_694599, EPI_ISL_694600

| see above | AZ SPHL, Arizona Department of Health Services | TGen North | Jolene Bowers, Megan Folkerts, Chris French, Hayley Yaglom, Ashlyn Pfeiffer, Darrin Lemmer, Dave Engelthaler, The Arizona COVID Genomics Union (ACGU) |
|---|---|---|---|

EPI_ISL_694601, EPI_ISL_694602, EPI_ISL_694603, EPI_ISL_694604, EPI_ISL_694605, EPI_ISL_694606, EPI_ISL_694607, EPI_ISL_694608, EPI_ISL_694609, EPI_ISL_694610, EPI_ISL_694611, EPI_ISL_694612, EPI_ISL_694614, EPI_ISL_694615, EPI_ISL_694616, EPI_ISL_694617, EPI_ISL_694618, EPI_ISL_694619, EPI_ISL_694620, EPI_ISL_694621, EPI_ISL_694622, EPI_ISL_694623, EPI_ISL_694624, EPI_ISL_694625, EPI_ISL_694626, EPI_ISL_694627, EPI_ISL_694628, EPI_ISL_694629, EPI_ISL_694630, EPI_ISL_694631, EPI_ISL_694632, EPI_ISL_694635, EPI_ISL_694636, EPI_ISL_694637, EPI_ISL_694638, EPI_ISL_694639, EPI_ISL_694640, EPI_ISL_694641, EPI_ISL_694642, EPI_ISL_694643, EPI_ISL_694645, EPI_ISL_694646, EPI_ISL_694647, EPI_ISL_694648, EPI_ISL_694649, EPI_ISL_694650, EPI_ISL_694651, EPI_ISL_694652, EPI_ISL_694653, EPI_ISL_694654, EPI_ISL_694655, EPI_ISL_694656, EPI_ISL_694657, EPI_ISL_694658, EPI_ISL_694659, EPI_ISL_694660, EPI_ISL_694661, EPI_ISL_694662, EPI_ISL_694663, EPI_ISL_694664, EPI_ISL_694665, EPI_ISL_694666, EPI_ISL_694667, EPI_ISL_694668, EPI_ISL_694669, EPI_ISL_694670, EPI_ISL_694671, EPI_ISL_694672, EPI_ISL_694673, EPI_ISL_694674, EPI_ISL_694675, EPI_ISL_694676, EPI_ISL_694677, EPI_ISL_694678, EPI_ISL_694679, EPI_ISL_694680, EPI_ISL_694681, EPI_ISL_694682, EPI_ISL_694683, EPI_ISL_694684, EPI_ISL_694685, EPI_ISL_694686, EPI_ISL_694687, EPI_ISL_694688, EPI_ISL_694689, EPI_ISL_694690, EPI_ISL_694691, EPI_ISL_694692, EPI_ISL_694693, EPI_ISL_694695, EPI_ISL_694696, EPI_ISL_694697, EPI_ISL_694698, EPI_ISL_694699, EPI_ISL_694701, EPI_ISL_694702, EPI_ISL_694703, EPI_ISL_694704, EPI_ISL_694706, EPI_ISL_694707, EPI_ISL_694708, EPI_ISL_694709, EPI_ISL_694710, EPI_ISL_694711, EPI_ISL_694712, EPI_ISL_694713, EPI_ISL_694714, EPI_ISL_694716, EPI_ISL_694717, EPI_ISL_694718, EPI_ISL_694719, EPI_ISL_694720, EPI_ISL_694721, EPI_ISL_694722, EPI_ISL_694723

| see above | TGen North | TGen North | Jolene Bowers, Megan Folkerts, Chris French, Hayley Yaglom, Ashlyn Pfeiffer, Darrin Lemmer, Dave Engelthaler, The Arizona COVID Genomics Union (ACGU) |
|---|---|---|---|

EPI_ISL_694724, EPI_ISL_694726, EPI_ISL_694727, EPI_ISL_694728, EPI_ISL_694729, EPI_ISL_694730, EPI_ISL_694731, EPI_ISL_694732, EPI_ISL_694733, EPI_ISL_694734, EPI_ISL_694736, EPI_ISL_694737, EPI_ISL_694738, EPI_ISL_694739, EPI_ISL_694740, EPI_ISL_694741, EPI_ISL_694742, EPI_ISL_694743, EPI_ISL_694744, EPI_ISL_694745, EPI_ISL_694746, EPI_ISL_694747, EPI_ISL_694748, EPI_ISL_694750, EPI_ISL_694751, EPI_ISL_694752, EPI_ISL_694753, EPI_ISL_694754, EPI_ISL_694755, EPI_ISL_694756, EPI_ISL_694757, EPI_ISL_694758, EPI_ISL_694759, EPI_ISL_694761, EPI_ISL_694762, EPI_ISL_694763, EPI_ISL_694764, EPI_ISL_694765, EPI_ISL_694766, EPI_ISL_694767, EPI_ISL_694768, EPI_ISL_694769, EPI_ISL_694771, EPI_ISL_694772, EPI_ISL_694773, EPI_ISL_694774, EPI_ISL_694775, EPI_ISL_694776, EPI_ISL_694779, EPI_ISL_694780, EPI_ISL_694781, EPI_ISL_694782, EPI_ISL_694783, EPI_ISL_694784, EPI_ISL_694785, EPI_ISL_694786, EPI_ISL_694787, EPI_ISL_694788, EPI_ISL_694789, EPI_ISL_694790, EPI_ISL_694791, EPI_ISL_694792, EPI_ISL_694793, EPI_ISL_694794, EPI_ISL_694796

| see above | AZ SPHL, Arizona Department of Health Services | TGen North | Jolene Bowers, Megan Folkerts, Chris French, Hayley Yaglom, Ashlyn Pfeiffer, Darrin Lemmer, Dave Engelthaler, The Arizona COVID Genomics Union (ACGU) |
|---|---|---|---|
| EPI_ISL_694797, EPI_ISL_694798 | TGen North | TGen North | Jolene Bowers, Megan Folkerts, Chris French, Hayley Yaglom, Ashlyn Pfeiffer, Darrin Lemmer, Dave Engelthaler, The Arizona COVID Genomics Union (ACGU) |

EPI_ISL_694799, EPI_ISL_694800, EPI_ISL_694801, EPI_ISL_694802, EPI_ISL_694805, EPI_ISL_694806, EPI_ISL_694808, EPI_ISL_694809, EPI_ISL_694810, EPI_ISL_694811, EPI_ISL_694812, EPI_ISL_694813, EPI_ISL_694814, EPI_ISL_694816, EPI_ISL_694818, EPI_ISL_694819, EPI_ISL_694821, EPI_ISL_694822, EPI_ISL_694823, EPI_ISL_694824, EPI_ISL_694825, EPI_ISL_694826, EPI_ISL_694827, EPI_ISL_694828, EPI_ISL_694829, EPI_ISL_694830, EPI_ISL_694831, EPI_ISL_694832, EPI_ISL_694833, EPI_ISL_694834, EPI_ISL_694835, EPI_ISL_694836, EPI_ISL_694837, EPI_ISL_694838, EPI_ISL_694839, EPI_ISL_694840, EPI_ISL_694841, EPI_ISL_694842, EPI_ISL_694844

| see above | AZ SPHL, Arizona Department of Health Services | TGen North | Jolene Bowers, Megan Folkerts, Chris French, Hayley Yaglom, Ashlyn Pfeiffer, Darrin Lemmer, Dave Engelthaler, The Arizona COVID Genomics Union (ACGU) |
|---|---|---|---|

EPI_ISL_694845, EPI_ISL_694846, EPI_ISL_694847, EPI_ISL_694848, EPI_ISL_694849, EPI_ISL_694850, EPI_ISL_694851, EPI_ISL_694853, EPI_ISL_694855, EPI_ISL_694856, EPI_ISL_694857, EPI_ISL_694858, EPI_ISL_694859, EPI_ISL_694860, EPI_ISL_694861, EPI_ISL_694862, EPI_ISL_694864, EPI_ISL_694865, EPI_ISL_694866, EPI_ISL_694867, EPI_ISL_694868, EPI_ISL_694869, EPI_ISL_694870, EPI_ISL_694871, EPI_ISL_694872, EPI_ISL_694873, EPI_ISL_694876, EPI_ISL_694879, EPI_ISL_694880, EPI_ISL_694881, EPI_ISL_694882, EPI_ISL_694883, EPI_ISL_694884

| see above | TGen North | TGen North | Jolene Bowers, Megan Folkerts, Chris French, Hayley Yaglom, Ashlyn Pfeiffer, Darrin Lemmer, Dave Engelthaler, The Arizona COVID Genomics Union (ACGU) |
|---|---|---|---|

EPI_ISL_694885, EPI_ISL_694886, EPI_ISL_694887, EPI_ISL_694888, EPI_ISL_694889, EPI_ISL_694890, EPI_ISL_694891, EPI_ISL_694892, EPI_ISL_694893, EPI_ISL_694894, EPI_ISL_694895, EPI_ISL_694896, EPI_ISL_694897, EPI_ISL_694898, EPI_ISL_694899, EPI_ISL_694900, EPI_ISL_694901, EPI_ISL_694902, EPI_ISL_694903, EPI_ISL_694905, EPI_ISL_694906, EPI_ISL_694907, EPI_ISL_694908, EPI_ISL_694909, EPI_ISL_694910, EPI_ISL_694911, EPI_ISL_694912, EPI_ISL_694913, EPI_ISL_694914, EPI_ISL_694916, EPI_ISL_694917, EPI_ISL_694918, EPI_ISL_694919, EPI_ISL_694920, EPI_ISL_694921, EPI_ISL_694922, EPI_ISL_694923, EPI_ISL_694924, EPI_ISL_694925, EPI_ISL_694926, EPI_ISL_694927, EPI_ISL_694928, EPI_ISL_694929, EPI_ISL_694930, EPI_ISL_694931, EPI_ISL_694932, EPI_ISL_694933, EPI_ISL_694934, EPI_ISL_694935, EPI_ISL_694936, EPI_ISL_694937, EPI_ISL_694938, EPI_ISL_694939, EPI_ISL_694940, EPI_ISL_694941, EPI_ISL_694942, EPI_ISL_694943, EPI_ISL_694945, EPI_ISL_694946, EPI_ISL_694947, EPI_ISL_694948, EPI_ISL_694949, EPI_ISL_694950

| see above | AZ SPHL, Arizona Department of Health Services | TGen North | Jolene Bowers, Megan Folkerts, Chris French, Hayley Yaglom, Ashlyn Pfeiffer, Darrin Lemmer, Dave Engelthaler, The Arizona COVID Genomics Union (ACGU) |
|---|---|---|---|

EPI_ISL_694951, EPI_ISL_694953, EPI_ISL_694956, EPI_ISL_694957, EPI_ISL_694959, EPI_ISL_694960, EPI_ISL_694961, EPI_ISL_694962, EPI_ISL_694963, EPI_ISL_694964, EPI_ISL_694965, EPI_ISL_694966, EPI_ISL_694969, EPI_ISL_694970, EPI_ISL_694971, EPI_ISL_694972, EPI_ISL_694973, EPI_ISL_694974, EPI_ISL_694975, EPI_ISL_694976, EPI_ISL_694977, EPI_ISL_694978, EPI_ISL_694980, EPI_ISL_694981, EPI_ISL_694982, EPI_ISL_694983, EPI_ISL_694984, EPI_ISL_694987, EPI_ISL_694988, EPI_ISL_694989, EPI_ISL_694991, EPI_ISL_694994, EPI_ISL_694995, EPI_ISL_694996, EPI_ISL_694997, EPI_ISL_694998, EPI_ISL_694999, EPI_ISL_695000, EPI_ISL_695001, EPI_ISL_695002, EPI_ISL_695003, EPI_ISL_695004, EPI_ISL_695005, EPI_ISL_695006, EPI_ISL_695007, EPI_ISL_695009, EPI_ISL_695011, EPI_ISL_695012, EPI_ISL_695013, EPI_ISL_695014, EPI_ISL_695015, EPI_ISL_695017, EPI_ISL_695018, EPI_ISL_695019, EPI_ISL_695020, EPI_ISL_695021, EPI_ISL_695022, EPI_ISL_695024, EPI_ISL_695025, EPI_ISL_695026, EPI_ISL_695027, EPI_ISL_695028, EPI_ISL_695029, EPI_ISL_695030, EPI_ISL_695031, EPI_ISL_695032, EPI_ISL_695033, EPI_ISL_695034, EPI_ISL_695035, EPI_ISL_695036, EPI_ISL_695037, EPI_ISL_695039, EPI_ISL_695041, EPI_ISL_695042, EPI_ISL_695043, EPI_ISL_695045, EPI_ISL_695047, EPI_ISL_695049, EPI_ISL_695050, EPI_ISL_695051, EPI_ISL_695053, EPI_ISL_695054, EPI_ISL_695055, EPI_ISL_695056, EPI_ISL_695057, EPI_ISL_695061, EPI_ISL_695062, EPI_ISL_695063, EPI_ISL_695064, EPI_ISL_695065, EPI_ISL_695066, EPI_ISL_695068, EPI_ISL_695069, EPI_ISL_695071, EPI_ISL_695072, EPI_ISL_695073, EPI_ISL_695074, EPI_ISL_695075, EPI_ISL_695076, EPI_ISL_695077, EPI_ISL_695078, EPI_ISL_695080, EPI_ISL_695081, EPI_ISL_695082, EPI_ISL_695083, EPI_ISL_695084, EPI_ISL_695085, EPI_ISL_695086, EPI_ISL_695087, EPI_ISL_695088, EPI_ISL_695089, EPI_ISL_695091, EPI_ISL_695092, EPI_ISL_695093, EPI_ISL_695095, EPI_ISL_695096, EPI_ISL_695097, EPI_ISL_695099, EPI_ISL_695100, EPI_ISL_695102

| see above | TGen North | TGen North | Jolene Bowers, Megan Folkerts, Chris French, Hayley Yaglom, Ashlyn Pfeiffer, Darrin Lemmer, Dave Engelthaler, The Arizona COVID Genomics Union (ACGU) |
|---|---|---|---|

EPI_ISL_695103, EPI_ISL_695104, EPI_ISL_695105, EPI_ISL_695106, EPI_ISL_695107, EPI_ISL_695108, EPI_ISL_695109, EPI_ISL_695110, EPI_ISL_695111, EPI_ISL_695112, EPI_ISL_695113, EPI_ISL_695114, EPI_ISL_695115, EPI_ISL_695116, EPI_ISL_695117, EPI_ISL_695118, EPI_ISL_695119, EPI_ISL_695120, EPI_ISL_695121, EPI_ISL_695122, EPI_ISL_695124, EPI_ISL_695126, EPI_ISL_695127, EPI_ISL_695128, EPI_ISL_695129, EPI_ISL_695130, EPI_ISL_695131, EPI_ISL_695132, EPI_ISL_695133, EPI_ISL_695134, EPI_ISL_695135, EPI_ISL_695136, EPI_ISL_695137, EPI_ISL_695138, EPI_ISL_695139, EPI_ISL_695140, EPI_ISL_695143, EPI_ISL_695144, EPI_ISL_695145, EPI_ISL_695146, EPI_ISL_695147, EPI_ISL_695148, EPI_ISL_695149, EPI_ISL_695150, EPI_ISL_695151, EPI_ISL_695152, EPI_ISL_695153, EPI_ISL_695154, EPI_ISL_695155, EPI_ISL_695156, EPI_ISL_695157, EPI_ISL_695158, EPI_ISL_695159, EPI_ISL_695160, EPI_ISL_695161, EPI_ISL_695162, EPI_ISL_695163, EPI_ISL_695164, EPI_ISL_695165, EPI_ISL_695166, EPI_ISL_695167, EPI_ISL_695168, EPI_ISL_695169, EPI_ISL_695170, EPI_ISL_695171, EPI_ISL_695172, EPI_ISL_695173, EPI_ISL_695176, EPI_ISL_695177, EPI_ISL_695178, EPI_ISL_695181, EPI_ISL_695184, EPI_ISL_695185, EPI_ISL_695186, EPI_ISL_695189, EPI_ISL_695190, EPI_ISL_695191, EPI_ISL_695192, EPI_ISL_695193, EPI_ISL_695194, EPI_ISL_695198, EPI_ISL_695202, EPI_ISL_695203, EPI_ISL_695205, EPI_ISL_695206, EPI_ISL_695207, EPI_ISL_695209, EPI_ISL_695211, EPI_ISL_695212, EPI_ISL_695214, EPI_ISL_695215, EPI_ISL_695220, EPI_ISL_695221, EPI_ISL_695222, EPI_ISL_695223, EPI_ISL_695224, EPI_ISL_695225, EPI_ISL_695226, EPI_ISL_695228

| see above | AZ SPHL, Arizona Department of Health Services | TGen North | Jolene Bowers, Megan Folkerts, Chris French, Hayley Yaglom, Ashlyn Pfeiffer, Darrin Lemmer, Dave Engelthaler, The Arizona COVID Genomics Union (ACGU) |
|---|---|---|---|

EPI_ISL_695230, EPI_ISL_695231, EPI_ISL_695232, EPI_ISL_695233, EPI_ISL_695235, EPI_ISL_695236, EPI_ISL_695237, EPI_ISL_695238, EPI_ISL_695239, EPI_ISL_695240, EPI_ISL_695241, EPI_ISL_695242, EPI_ISL_695243, EPI_ISL_695244, EPI_ISL_695245, EPI_ISL_695246, EPI_ISL_695247, EPI_ISL_695248, EPI_ISL_695249, EPI_ISL_695250, EPI_ISL_695251, EPI_ISL_695252, EPI_ISL_695253, EPI_ISL_695254, EPI_ISL_695255, EPI_ISL_695256, EPI_ISL_695257, EPI_ISL_695258, EPI_ISL_695259, EPI_ISL_695260, EPI_ISL_695261, EPI_ISL_695263, EPI_ISL_695264, EPI_ISL_695265, EPI_ISL_695267, EPI_ISL_695268, EPI_ISL_695269, EPI_ISL_695270, EPI_ISL_695271, EPI_ISL_695272, EPI_ISL_695273, EPI_ISL_695274, EPI_ISL_695275, EPI_ISL_695276, EPI_ISL_695277, EPI_ISL_695278, EPI_ISL_695279, EPI_ISL_695280, EPI_ISL_695281, EPI_ISL_695282, EPI_ISL_695283, EPI_ISL_695284, EPI_ISL_695285, EPI_ISL_695286, EPI_ISL_695287, EPI_ISL_695288, EPI_ISL_695289, EPI_ISL_695290, EPI_ISL_695291, EPI_ISL_695293, EPI_ISL_695294, EPI_ISL_695295, EPI_ISL_695296, EPI_ISL_695297, EPI_ISL_695298, EPI_ISL_695299, EPI_ISL_695300, EPI_ISL_695301, EPI_ISL_695302, EPI_ISL_695303, EPI_ISL_695304, EPI_ISL_695305, EPI_ISL_695306, EPI_ISL_695307, EPI_ISL_695308, EPI_ISL_695309, EPI_ISL_695310, EPI_ISL_695311, EPI_ISL_695312, EPI_ISL_695313, EPI_ISL_695315

| see above | TGen North | TGen North | Jolene Bowers, Megan Folkerts, Chris French, Hayley Yaglom, Ashlyn Pfeiffer, Darrin Lemmer, Dave Engelthaler, The Arizona COVID Genomics Union (ACGU) |
|---|---|---|---|

EPI_ISL_695318, EPI_ISL_695319, EPI_ISL_695322, EPI_ISL_695323, EPI_ISL_695325, EPI_ISL_695326, EPI_ISL_695328, EPI_ISL_695330, EPI_ISL_695333, EPI_ISL_695334, EPI_ISL_695335, EPI_ISL_695336, EPI_ISL_695338, EPI_ISL_695339, EPI_ISL_695340, EPI_ISL_695342, EPI_ISL_695343, EPI_ISL_695345, EPI_ISL_695346, EPI_ISL_695350, EPI_ISL_695351, EPI_ISL_695352, EPI_ISL_695354, EPI_ISL_695355, EPI_ISL_695356, EPI_ISL_695359, EPI_ISL_695360, EPI_ISL_695361, EPI_ISL_695362, EPI_ISL_695364, EPI_ISL_695365, EPI_ISL_695367, EPI_ISL_695368, EPI_ISL_695369, EPI_ISL_695370

| see above | AZ SPHL, Arizona Department of Health Services | TGen North | Jolene Bowers, Megan Folkerts, Chris French, Hayley Yaglom, Ashlyn Pfeiffer, Darrin Lemmer, Dave Engelthaler, The Arizona COVID Genomics Union (ACGU) |
|---|---|---|---|

EPI_ISL_695372, EPI_ISL_695373, EPI_ISL_695374, EPI_ISL_695376, EPI_ISL_695377, EPI_ISL_695378, EPI_ISL_695379, EPI_ISL_695380, EPI_ISL_695381, EPI_ISL_695383, EPI_ISL_695384, EPI_ISL_695386, EPI_ISL_695387, EPI_ISL_695389, EPI_ISL_695392

| see above | TGen North | TGen North | Jolene Bowers, Megan Folkerts, Chris French, Hayley Yaglom, Ashlyn Pfeiffer, Darrin Lemmer, Dave Engelthaler, The Arizona COVID Genomics Union (ACGU) |
|---|---|---|---|

EPI_ISL_695394, EPI_ISL_695395, EPI_ISL_695396, EPI_ISL_695397, EPI_ISL_695398, EPI_ISL_695399, EPI_ISL_695401, EPI_ISL_695402, EPI_ISL_695403, EPI_ISL_695406, EPI_ISL_695408, EPI_ISL_695410, EPI_ISL_695411, EPI_ISL_695412, EPI_ISL_695415, EPI_ISL_695416, EPI_ISL_695417, EPI_ISL_695419, EPI_ISL_695422, EPI_ISL_695423, EPI_ISL_695425, EPI_ISL_695426, EPI_ISL_695427, EPI_ISL_695428, EPI_ISL_695429, EPI_ISL_695430, EPI_ISL_695431, EPI_ISL_695432, EPI_ISL_695433, EPI_ISL_695434, EPI_ISL_695435, EPI_ISL_695436, EPI_ISL_695437, EPI_ISL_695438, EPI_ISL_695440, EPI_ISL_695442, EPI_ISL_695445, EPI_ISL_695446, EPI_ISL_695447, EPI_ISL_695448, EPI_ISL_695449, EPI_ISL_695450, EPI_ISL_695451, EPI_ISL_695452, EPI_ISL_695453, EPI_ISL_695454, EPI_ISL_695455, EPI_ISL_695456, EPI_ISL_695458, EPI_ISL_695459, EPI_ISL_695460, EPI_ISL_695461, EPI_ISL_695462, EPI_ISL_695463, EPI_ISL_695465, EPI_ISL_695466, EPI_ISL_695467, EPI_ISL_695468, EPI_ISL_695469, EPI_ISL_695470, EPI_ISL_695472, EPI_ISL_695473, EPI_ISL_695474, EPI_ISL_695475, EPI_ISL_695476, EPI_ISL_695477, EPI_ISL_695478, EPI_ISL_695479, EPI_ISL_695480, EPI_ISL_695481, EPI_ISL_695482, EPI_ISL_695483, EPI_ISL_695484, EPI_ISL_695485, EPI_ISL_695486, EPI_ISL_695487, EPI_ISL_695488, EPI_ISL_695489, EPI_ISL_695490, EPI_ISL_695491, EPI_ISL_695492, EPI_ISL_695493, EPI_ISL_695494, EPI_ISL_695495, EPI_ISL_695496, EPI_ISL_695497, EPI_ISL_695498, EPI_ISL_695500, EPI_ISL_695501, EPI_ISL_695502, EPI_ISL_695503, EPI_ISL_695506, EPI_ISL_695508, EPI_ISL_695509, EPI_ISL_695510, EPI_ISL_695512, EPI_ISL_695513, EPI_ISL_695514, EPI_ISL_695515, EPI_ISL_695516, EPI_ISL_695517, EPI_ISL_695519

| see above | AZ SPHL, Arizona Department of Health Services | TGen North | Jolene Bowers, Megan Folkerts, Chris French, Hayley Yaglom, Ashlyn Pfeiffer, Darrin Lemmer, Dave Engelthaler, The Arizona COVID Genomics Union (ACGU) |
|---|---|---|---|

EPI_ISL_695521, EPI_ISL_695522, EPI_ISL_695523, EPI_ISL_695524, EPI_ISL_695525, EPI_ISL_695526, EPI_ISL_695527, EPI_ISL_695528, EPI_ISL_695529, EPI_ISL_695531, EPI_ISL_695532, EPI_ISL_695533, EPI_ISL_695535, EPI_ISL_695536, EPI_ISL_695537, EPI_ISL_695538, EPI_ISL_695539, EPI_ISL_695541, EPI_ISL_695542, EPI_ISL_695543, EPI_ISL_695544, EPI_ISL_695545, EPI_ISL_695546, EPI_ISL_695547,

EPI_ISL_695548, EPI_ISL_695549, EPI_ISL_695550, EPI_ISL_695552, EPI_ISL_695553, EPI_ISL_695555, EPI_ISL_695556, EPI_ISL_695557, EPI_ISL_695558, EPI_ISL_695559, EPI_ISL_695560, EPI_ISL_695561, EPI_ISL_695562

| | | | |
|---|---|---|---|
| see above | TGen North | TGen North | Jolene Bowers, Megan Folkerts, Chris French, Hayley Yaglom, Ashlyn Pfeiffer, Darrin Lemmer, Dave Engelthaler, The Arizona COVID Genomics Union (ACGU) |

EPI_ISL_695563, EPI_ISL_695564, EPI_ISL_695565, EPI_ISL_695566, EPI_ISL_695567, EPI_ISL_695568, EPI_ISL_695570, EPI_ISL_695573, EPI_ISL_695574, EPI_ISL_695575, EPI_ISL_695576, EPI_ISL_695577, EPI_ISL_695578, EPI_ISL_695579, EPI_ISL_695581, EPI_ISL_695582, EPI_ISL_695583, EPI_ISL_695584, EPI_ISL_695585, EPI_ISL_695586, EPI_ISL_695587, EPI_ISL_695588, EPI_ISL_695589, EPI_ISL_695590, EPI_ISL_695591, EPI_ISL_695592, EPI_ISL_695593, EPI_ISL_695594, EPI_ISL_695595, EPI_ISL_695596, EPI_ISL_695598, EPI_ISL_695599, EPI_ISL_695600, EPI_ISL_695601, EPI_ISL_695602, EPI_ISL_695603, EPI_ISL_695605, EPI_ISL_695606, EPI_ISL_695607, EPI_ISL_695608, EPI_ISL_695610, EPI_ISL_695611, EPI_ISL_695612, EPI_ISL_695613

| | | | |
|---|---|---|---|
| see above | AZ SPHL, Arizona Department of Health Services | TGen North | Jolene Bowers, Megan Folkerts, Chris French, Hayley Yaglom, Ashlyn Pfeiffer, Darrin Lemmer, Dave Engelthaler, The Arizona COVID Genomics Union (ACGU) |

EPI_ISL_695616, EPI_ISL_695618, EPI_ISL_695621, EPI_ISL_695622, EPI_ISL_695623, EPI_ISL_695624, EPI_ISL_695626, EPI_ISL_695627, EPI_ISL_695628, EPI_ISL_695629, EPI_ISL_695632, EPI_ISL_695633, EPI_ISL_695634, EPI_ISL_695635, EPI_ISL_695637, EPI_ISL_695638, EPI_ISL_695639

| | | | |
|---|---|---|---|
| see above | TGen North | TGen North | Jolene Bowers, Megan Folkerts, Chris French, Hayley Yaglom, Ashlyn Pfeiffer, Darrin Lemmer, Dave Engelthaler, The Arizona COVID Genomics Union (ACGU) |
| EPI_ISL_695642, EPI_ISL_695644, EPI_ISL_695646, EPI_ISL_695647, EPI_ISL_695649, EPI_ISL_695650, EPI_ISL_695653, EPI_ISL_695654, EPI_ISL_695655 | AZ SPHL, Arizona Department of Health Services | TGen North | Jolene Bowers, Megan Folkerts, Chris French, Hayley Yaglom, Ashlyn Pfeiffer, Darrin Lemmer, Dave Engelthaler, The Arizona COVID Genomics Union (ACGU) |
| EPI_ISL_695656, EPI_ISL_695657, EPI_ISL_695659, EPI_ISL_695660, EPI_ISL_695662, EPI_ISL_695663, EPI_ISL_695664 | TGen North | TGen North | Jolene Bowers, Megan Folkerts, Chris French, Hayley Yaglom, Ashlyn Pfeiffer, Darrin Lemmer, Dave Engelthaler, The Arizona COVID Genomics Union (ACGU) |

EPI_ISL_695665, EPI_ISL_695666, EPI_ISL_695667, EPI_ISL_695668, EPI_ISL_695669, EPI_ISL_695671, EPI_ISL_695672, EPI_ISL_695673, EPI_ISL_695675, EPI_ISL_695676, EPI_ISL_695679, EPI_ISL_695680, EPI_ISL_695683, EPI_ISL_695684, EPI_ISL_695685, EPI_ISL_695686, EPI_ISL_695687, EPI_ISL_695688, EPI_ISL_695689, EPI_ISL_695690, EPI_ISL_695691, EPI_ISL_695692, EPI_ISL_695693, EPI_ISL_695694, EPI_ISL_695696, EPI_ISL_695697, EPI_ISL_695700, EPI_ISL_695701

| | | | |
|---|---|---|---|
| see above | AZ SPHL, Arizona Department of Health Services | TGen North | Jolene Bowers, Megan Folkerts, Chris French, Hayley Yaglom, Ashlyn Pfeiffer, Darrin Lemmer, Dave Engelthaler, The Arizona COVID Genomics Union (ACGU) |
| EPI_ISL_695702, EPI_ISL_695705, EPI_ISL_695706, EPI_ISL_695707, EPI_ISL_695708, EPI_ISL_695710 | TGen North | TGen North | Jolene Bowers, Megan Folkerts, Chris French, Hayley Yaglom, Ashlyn Pfeiffer, Darrin Lemmer, Dave Engelthaler, The Arizona COVID Genomics Union (ACGU) |

EPI_ISL_695712, EPI_ISL_695713, EPI_ISL_695714, EPI_ISL_695715, EPI_ISL_695716, EPI_ISL_695717, EPI_ISL_695718, EPI_ISL_695719, EPI_ISL_695720, EPI_ISL_695721, EPI_ISL_695722, EPI_ISL_695723, EPI_ISL_695724, EPI_ISL_695725, EPI_ISL_695726, EPI_ISL_695727, EPI_ISL_695728, EPI_ISL_695730, EPI_ISL_695731, EPI_ISL_695735, EPI_ISL_695736, EPI_ISL_695737, EPI_ISL_695738, EPI_ISL_695739, EPI_ISL_695740, EPI_ISL_695741, EPI_ISL_695742, EPI_ISL_695743, EPI_ISL_695744, EPI_ISL_695745, EPI_ISL_695746, EPI_ISL_695747, EPI_ISL_695749, EPI_ISL_695752

| | | | |
|---|---|---|---|
| see above | AZ SPHL, Arizona Department of Health Services | TGen North | Jolene Bowers, Megan Folkerts, Chris French, Hayley Yaglom, Ashlyn Pfeiffer, Darrin Lemmer, Dave Engelthaler, The Arizona COVID Genomics Union (ACGU) |
| EPI_ISL_695753, EPI_ISL_695754 | TGen North | TGen North | Jolene Bowers, Megan Folkerts, Chris French, Hayley Yaglom, Ashlyn Pfeiffer, Darrin Lemmer, Dave Engelthaler, The Arizona COVID Genomics Union (ACGU) |

EPI_ISL_695755, EPI_ISL_695756, EPI_ISL_695757, EPI_ISL_695759, EPI_ISL_695760, EPI_ISL_695761, EPI_ISL_695762, EPI_ISL_695763, EPI_ISL_695764, EPI_ISL_695765, EPI_ISL_695766, EPI_ISL_695767, EPI_ISL_695768, EPI_ISL_695769, EPI_ISL_695770, EPI_ISL_695772, EPI_ISL_695773, EPI_ISL_695774, EPI_ISL_695775, EPI_ISL_695776, EPI_ISL_695777, EPI_ISL_695778, EPI_ISL_695779, EPI_ISL_695780, EPI_ISL_695781, EPI_ISL_695782, EPI_ISL_695783, EPI_ISL_695784, EPI_ISL_695785, EPI_ISL_695786, EPI_ISL_695787, EPI_ISL_695788, EPI_ISL_695789, EPI_ISL_695790, EPI_ISL_695791, EPI_ISL_695792, EPI_ISL_695793, EPI_ISL_695794, EPI_ISL_695795, EPI_ISL_695796, EPI_ISL_695797, EPI_ISL_695798, EPI_ISL_695799, EPI_ISL_695800, EPI_ISL_695801, EPI_ISL_695802, EPI_ISL_695803, EPI_ISL_695804, EPI_ISL_695806, EPI_ISL_695807, EPI_ISL_695808, EPI_ISL_695809, EPI_ISL_695810, EPI_ISL_695811, EPI_ISL_695812, EPI_ISL_695813, EPI_ISL_695814, EPI_ISL_695815, EPI_ISL_695816, EPI_ISL_695817, EPI_ISL_695818, EPI_ISL_695819, EPI_ISL_695820, EPI_ISL_695821, EPI_ISL_695822, EPI_ISL_695823, EPI_ISL_695824, EPI_ISL_695825, EPI_ISL_695826, EPI_ISL_695827, EPI_ISL_695828, EPI_ISL_695829

| | | | |
|---|---|---|---|
| see above | Sonora Quest Laboratories, Laboratory Sciences of Arizona | TGen North | Jolene Bowers, Megan Folkerts, Chris French, Hayley Yaglom, Ashlyn Pfeiffer, Darrin Lemmer, Dave Engelthaler, The Arizona COVID Genomics Union (ACGU) |
| EPI_ISL_695830, EPI_ISL_695831, EPI_ISL_695832, EPI_ISL_695833, EPI_ISL_695834, EPI_ISL_695835, EPI_ISL_695836, EPI_ISL_695837, EPI_ISL_695838 | AZ SPHL, Arizona Department of Health Services | TGen North | Jolene Bowers, Megan Folkerts, Chris French, Hayley Yaglom, Ashlyn Pfeiffer, Darrin Lemmer, Dave Engelthaler, The Arizona COVID Genomics Union (ACGU) |

EPI_ISL_695840, EPI_ISL_695841, EPI_ISL_695842, EPI_ISL_695843, EPI_ISL_695844, EPI_ISL_695845, EPI_ISL_695846, EPI_ISL_695847, EPI_ISL_695848, EPI_ISL_695849, EPI_ISL_695850, EPI_ISL_695851, EPI_ISL_695852, EPI_ISL_695853, EPI_ISL_695854, EPI_ISL_695855, EPI_ISL_695856, EPI_ISL_695857, EPI_ISL_695858, EPI_ISL_695859, EPI_ISL_695861, EPI_ISL_695862, EPI_ISL_695863, EPI_ISL_695865, EPI_ISL_695866, EPI_ISL_695867, EPI_ISL_695868, EPI_ISL_695869, EPI_ISL_695870, EPI_ISL_695871, EPI_ISL_695872, EPI_ISL_695873, EPI_ISL_695874, EPI_ISL_695875, EPI_ISL_695876, EPI_ISL_695877, EPI_ISL_695879, EPI_ISL_695880, EPI_ISL_695881, EPI_ISL_695882, EPI_ISL_695883, EPI_ISL_695885, EPI_ISL_695886, EPI_ISL_695887, EPI_ISL_695888, EPI_ISL_695889, EPI_ISL_695890, EPI_ISL_695891, EPI_ISL_695892, EPI_ISL_695893, EPI_ISL_695894, EPI_ISL_695895, EPI_ISL_695896, EPI_ISL_695897, EPI_ISL_695898, EPI_ISL_695900, EPI_ISL_695901, EPI_ISL_695902, EPI_ISL_695903, EPI_ISL_695904, EPI_ISL_695907, EPI_ISL_695908, EPI_ISL_695909, EPI_ISL_695910, EPI_ISL_695911, EPI_ISL_695912, EPI_ISL_695913, EPI_ISL_695915, EPI_ISL_695916, EPI_ISL_695917, EPI_ISL_695918, EPI_ISL_695920, EPI_ISL_695921, EPI_ISL_695922, EPI_ISL_695923, EPI_ISL_695924, EPI_ISL_695926, EPI_ISL_695927, EPI_ISL_695928, EPI_ISL_695929, EPI_ISL_695930, EPI_ISL_695933, EPI_ISL_695934, EPI_ISL_695936, EPI_ISL_695937, EPI_ISL_695938, EPI_ISL_695939, EPI_ISL_695940, EPI_ISL_695941, EPI_ISL_695943, EPI_ISL_695944, EPI_ISL_695945, EPI_ISL_695946, EPI_ISL_695947, EPI_ISL_695948, EPI_ISL_695949, EPI_ISL_695950, EPI_ISL_695951, EPI_ISL_695952, EPI_ISL_695953, EPI_ISL_695954, EPI_ISL_695956, EPI_ISL_695957, EPI_ISL_695958, EPI_ISL_695959, EPI_ISL_695960, EPI_ISL_695961, EPI_ISL_695962, EPI_ISL_695963, EPI_ISL_695964, EPI_ISL_695965, EPI_ISL_695966, EPI_ISL_695967, EPI_ISL_695968, EPI_ISL_695969, EPI_ISL_695970, EPI_ISL_695972, EPI_ISL_695973, EPI_ISL_695975, EPI_ISL_695976, EPI_ISL_695977, EPI_ISL_695978, EPI_ISL_695979, EPI_ISL_695980, EPI_ISL_695981, EPI_ISL_695982, EPI_ISL_695983, EPI_ISL_695984, EPI_ISL_695985, EPI_ISL_695986, EPI_ISL_695987, EPI_ISL_695988, EPI_ISL_695989, EPI_ISL_695990, EPI_ISL_695991, EPI_ISL_695992, EPI_ISL_695993, EPI_ISL_695994, EPI_ISL_695996, EPI_ISL_695998, EPI_ISL_695999, EPI_ISL_696000, EPI_ISL_696001, EPI_ISL_696002, EPI_ISL_696003, EPI_ISL_696004, EPI_ISL_696005, EPI_ISL_696006, EPI_ISL_696008, EPI_ISL_696009, EPI_ISL_696010, EPI_ISL_696011, EPI_ISL_696013, EPI_ISL_696014, EPI_ISL_696015, EPI_ISL_696017, EPI_ISL_696018, EPI_ISL_696019, EPI_ISL_696020, EPI_ISL_696021, EPI_ISL_696022, EPI_ISL_696023, EPI_ISL_696025, EPI_ISL_696026, EPI_ISL_696027, EPI_ISL_696028, EPI_ISL_696029, EPI_ISL_696031, EPI_ISL_696033, EPI_ISL_696034, EPI_ISL_696035, EPI_ISL_696036, EPI_ISL_696037, EPI_ISL_696038, EPI_ISL_696039, EPI_ISL_696040, EPI_ISL_696041, EPI_ISL_696042, EPI_ISL_696043, EPI_ISL_696044, EPI_ISL_696045, EPI_ISL_696046, EPI_ISL_696047, EPI_ISL_696048, EPI_ISL_696049, EPI_ISL_696051, EPI_ISL_696052, EPI_ISL_696053, EPI_ISL_696055, EPI_ISL_696056, EPI_ISL_696057, EPI_ISL_696058, EPI_ISL_696059, EPI_ISL_696060, EPI_ISL_696061, EPI_ISL_696062, EPI_ISL_696064, EPI_ISL_696065, EPI_ISL_696066, EPI_ISL_696067, EPI_ISL_696068, EPI_ISL_696069, EPI_ISL_696070, EPI_ISL_696071, EPI_ISL_696072, EPI_ISL_696073, EPI_ISL_696074, EPI_ISL_696075, EPI_ISL_696076, EPI_ISL_696077, EPI_ISL_696078, EPI_ISL_696079, EPI_ISL_696080, EPI_ISL_696081, EPI_ISL_696082, EPI_ISL_696085, EPI_ISL_696086, EPI_ISL_696087, EPI_ISL_696088, EPI_ISL_696090, EPI_ISL_696091, EPI_ISL_696092, EPI_ISL_696093, EPI_ISL_696094, EPI_ISL_696095, EPI_ISL_696097, EPI_ISL_696098, EPI_ISL_696099, EPI_ISL_696100, EPI_ISL_696103, EPI_ISL_696104, EPI_ISL_696106, EPI_ISL_696107, EPI_ISL_696109, EPI_ISL_696110, EPI_ISL_696111, EPI_ISL_696113, EPI_ISL_696114, EPI_ISL_696115, EPI_ISL_696116, EPI_ISL_696117, EPI_ISL_696119, EPI_ISL_696120, EPI_ISL_696121, EPI_ISL_696122, EPI_ISL_696123, EPI_ISL_696125, EPI_ISL_696126, EPI_ISL_696127, EPI_ISL_696128, EPI_ISL_696130, EPI_ISL_696131, EPI_ISL_696132, EPI_ISL_696133, EPI_ISL_696134, EPI_ISL_696135, EPI_ISL_696136, EPI_ISL_696137, EPI_ISL_696138, EPI_ISL_696139, EPI_ISL_696140, EPI_ISL_696141, EPI_ISL_696142, EPI_ISL_696143, EPI_ISL_696144, EPI_ISL_696145, EPI_ISL_696146, EPI_ISL_696147, EPI_ISL_696149, EPI_ISL_696150, EPI_ISL_696151, EPI_ISL_696152, EPI_ISL_696153, EPI_ISL_696156, EPI_ISL_696157, EPI_ISL_696158, EPI_ISL_696159, EPI_ISL_696160, EPI_ISL_696162, EPI_ISL_696163, EPI_ISL_696164, EPI_ISL_696165, EPI_ISL_696166, EPI_ISL_696167, EPI_ISL_696168, EPI_ISL_696169, EPI_ISL_696170, EPI_ISL_696171, EPI_ISL_696172, EPI_ISL_696173, EPI_ISL_696174, EPI_ISL_696175, EPI_ISL_696176, EPI_ISL_696178, EPI_ISL_696179, EPI_ISL_696181, EPI_ISL_696182, EPI_ISL_696183, EPI_ISL_696184, EPI_ISL_696185, EPI_ISL_696186, EPI_ISL_696187, EPI_ISL_696189, EPI_ISL_696190, EPI_ISL_696192, EPI_ISL_696193, EPI_ISL_696194, EPI_ISL_696195, EPI_ISL_696196, EPI_ISL_696197, EPI_ISL_696199, EPI_ISL_696201, EPI_ISL_696202, EPI_ISL_696203, EPI_ISL_696204, EPI_ISL_696205, EPI_ISL_696206, EPI_ISL_696207, EPI_ISL_696208, EPI_ISL_696209, EPI_ISL_696210, EPI_ISL_696212, EPI_ISL_696213, EPI_ISL_696214, EPI_ISL_696215, EPI_ISL_696216, EPI_ISL_696217, EPI_ISL_696218, EPI_ISL_696219, EPI_ISL_696220, EPI_ISL_696221, EPI_ISL_696222, EPI_ISL_696223, EPI_ISL_696224, EPI_ISL_696225, EPI_ISL_696226, EPI_ISL_696227, EPI_ISL_696228, EPI_ISL_696229, EPI_ISL_696230, EPI_ISL_696231, EPI_ISL_696232, EPI_ISL_696233, EPI_ISL_696234, EPI_ISL_696235, EPI_ISL_696236, EPI_ISL_696237, EPI_ISL_696238, EPI_ISL_696239, EPI_ISL_696240, EPI_ISL_696241, EPI_ISL_696242, EPI_ISL_696243, EPI_ISL_696244, EPI_ISL_696245, EPI_ISL_696246, EPI_ISL_696248, EPI_ISL_696249, EPI_ISL_696250, EPI_ISL_696251, EPI_ISL_696252, EPI_ISL_696253, EPI_ISL_696254, EPI_ISL_696255, EPI_ISL_696256, EPI_ISL_696257, EPI_ISL_696259, EPI_ISL_696260, EPI_ISL_696261, EPI_ISL_696262, EPI_ISL_696263, EPI_ISL_696265, EPI_ISL_696266, EPI_ISL_696268, EPI_ISL_696269, EPI_ISL_696270, EPI_ISL_696271, EPI_ISL_696272, EPI_ISL_696274, EPI_ISL_696275, EPI_ISL_696276, EPI_ISL_696277, EPI_ISL_696278, EPI_ISL_696279, EPI_ISL_696282, EPI_ISL_696283, EPI_ISL_696284, EPI_ISL_696285, EPI_ISL_696287, EPI_ISL_696288, EPI_ISL_696290, EPI_ISL_696293, EPI_ISL_696294, EPI_ISL_696296, EPI_ISL_696297, EPI_ISL_696298, EPI_ISL_696300, EPI_ISL_696301, EPI_ISL_696302, EPI_ISL_696303, EPI_ISL_696304, EPI_ISL_696305, EPI_ISL_696308, EPI_ISL_696310, EPI_ISL_696311, EPI_ISL_696313, EPI_ISL_696314, EPI_ISL_696315, EPI_ISL_696316, EPI_ISL_696317, EPI_ISL_696318, EPI_ISL_696320, EPI_ISL_696321, EPI_ISL_696322, EPI_ISL_696323, EPI_ISL_696324, EPI_ISL_696325, EPI_ISL_696326, EPI_ISL_696327, EPI_ISL_696328, EPI_ISL_696329, EPI_ISL_696330, EPI_ISL_696331, EPI_ISL_696332, EPI_ISL_696333, EPI_ISL_696334, EPI_ISL_696335, EPI_ISL_696336, EPI_ISL_696338, EPI_ISL_696339, EPI_ISL_696340, EPI_ISL_696341, EPI_ISL_696342, EPI_ISL_696343, EPI_ISL_696344, EPI_ISL_696345, EPI_ISL_696348, EPI_ISL_696350, EPI_ISL_696351, EPI_ISL_696352, EPI_ISL_696353, EPI_ISL_696354, EPI_ISL_696356, EPI_ISL_696357, EPI_ISL_696358, EPI_ISL_696359, EPI_ISL_696362, EPI_ISL_696363, EPI_ISL_696364, EPI_ISL_696365, EPI_ISL_696366, EPI_ISL_696367, EPI_ISL_696369, EPI_ISL_696370, EPI_ISL_696371, EPI_ISL_696372, EPI_ISL_696373, EPI_ISL_696374, EPI_ISL_696375, EPI_ISL_696376, EPI_ISL_696377, EPI_ISL_696378, EPI_ISL_696379, EPI_ISL_696380, EPI_ISL_696382, EPI_ISL_696383, EPI_ISL_696384, EPI_ISL_696385, EPI_ISL_696386, EPI_ISL_696387, EPI_ISL_696388, EPI_ISL_696389, EPI_ISL_696390, EPI_ISL_696391, EPI_ISL_696392, EPI_ISL_696393, EPI_ISL_696394, EPI_ISL_696395, EPI_ISL_696396, EPI_ISL_696397, EPI_ISL_696398, EPI_ISL_696399, EPI_ISL_696400, EPI_ISL_696401, EPI_ISL_696404, EPI_ISL_696405, EPI_ISL_696406, EPI_ISL_696407, EPI_ISL_696408, EPI_ISL_696409, EPI_ISL_696410, EPI_ISL_696411, EPI_ISL_696412, EPI_ISL_696413, EPI_ISL_696414, EPI_ISL_696415, EPI_ISL_696416, EPI_ISL_696418, EPI_ISL_696419, EPI_ISL_696420, EPI_ISL_696421, EPI_ISL_696422, EPI_ISL_696423, EPI_ISL_696424, EPI_ISL_696425, EPI_ISL_696427, EPI_ISL_696429, EPI_ISL_696430, EPI_ISL_696432, EPI_ISL_696433, EPI_ISL_696434, EPI_ISL_696435, EPI_ISL_696436, EPI_ISL_696439, EPI_ISL_696440, EPI_ISL_696441, EPI_ISL_696442, EPI_ISL_696443, EPI_ISL_696446, EPI_ISL_696447, EPI_ISL_696448, EPI_ISL_696449, EPI_ISL_696450

| | | | |
|---|---|---|---|
| see above | Sonora Quest Laboratories, Laboratory Sciences of Arizona | TGen North | Jolene Bowers, Megan Folkerts, Chris French, Hayley Yaglom, Ashlyn Pfeiffer, Darrin Lemmer, Dave Engelthaler, The Arizona COVID Genomics Union (ACGU) |

EPI_ISL_700699, EPI_ISL_700700, EPI_ISL_700701, EPI_ISL_700702, EPI_ISL_700703, EPI_ISL_700704, EPI_ISL_700705, EPI_ISL_700706, EPI_ISL_700707, EPI_ISL_700708, EPI_ISL_700709, EPI_ISL_700710, EPI_ISL_700711, EPI_ISL_700712, EPI_ISL_700713, EPI_ISL_700714, EPI_ISL_700715, EPI_ISL_700716, EPI_ISL_700717, EPI_ISL_700718, EPI_ISL_700719, EPI_ISL_700720, EPI_ISL_700721, EPI_ISL_700722, EPI_ISL_700723, EPI_ISL_700724, EPI_ISL_700725, EPI_ISL_700726, EPI_ISL_700727, EPI_ISL_700728, EPI_ISL_700729, EPI_ISL_700730, EPI_ISL_700739, EPI_ISL_700740, EPI_ISL_700741, EPI_ISL_700742

| | | | |
|---|---|---|---|
| see above | Texas Department of State Health Services | Texas Department of State Health Services | Rashmi Tuladhar, Bonnie Oh, Jenny Zhang, Maliha Rahman, Anita Pokharel, Myong Koag, Chung Wang, Rachel Lee, Grace Kubin, Mayela Pedrueza, James Daniel Bonser |
| EPI_ISL_707693, EPI_ISL_707694, EPI_ISL_707695 | Los Angeles County Public Health Laboratories | Los Angeles County Public Health Laboratories | P. Hemarajata et al. |

EPI_ISL_707808, EPI_ISL_707809, EPI_ISL_707810, EPI_ISL_707811, EPI_ISL_707812, EPI_ISL_707813, EPI_ISL_707814, EPI_ISL_707815, EPI_ISL_707816, EPI_ISL_707817, EPI_ISL_707818, EPI_ISL_707819, EPI_ISL_707820, EPI_ISL_707821, EPI_ISL_707822, EPI_ISL_707823, EPI_ISL_707824, EPI_ISL_707825, EPI_ISL_707826, EPI_ISL_707827, EPI_ISL_707828, EPI_ISL_707829, EPI_ISL_707830, EPI_ISL_707831, EPI_ISL_707832, EPI_ISL_707833, EPI_ISL_707834, EPI_ISL_707835, EPI_ISL_707836, EPI_ISL_707837, EPI_ISL_707838, EPI_ISL_707839, EPI_ISL_707840, EPI_ISL_707841, EPI_ISL_707842, EPI_ISL_707843, EPI_ISL_707844, EPI_ISL_707845, EPI_ISL_707846, EPI_ISL_707847, EPI_ISL_707848, EPI_ISL_707849, EPI_ISL_707850, EPI_ISL_707851, EPI_ISL_707852, EPI_ISL_707853, EPI_ISL_707854, EPI_ISL_707855, EPI_ISL_707856, EPI_ISL_707857, EPI_ISL_707858, EPI_ISL_707859, EPI_ISL_707860, EPI_ISL_707861, EPI_ISL_707862, EPI_ISL_707863, EPI_ISL_707864, EPI_ISL_707865, EPI_ISL_707866, EPI_ISL_707867, EPI_ISL_707868, EPI_ISL_707869, EPI_ISL_707870, EPI_ISL_707871, EPI_ISL_707872, EPI_ISL_707873, EPI_ISL_707874, EPI_ISL_707875, EPI_ISL_707876, EPI_ISL_707877, EPI_ISL_707878, EPI_ISL_707879, EPI_ISL_707880, EPI_ISL_707881, EPI_ISL_707882

| | | | |
|---|---|---|---|
| see above | University of Michigan Clinical Microbiology Laboratory | Lauring Lab, University of Michigan, Department of Microbiology and Immunology | Valesano |

EPI_ISL_707883, EPI_ISL_707885, EPI_ISL_707886, EPI_ISL_707887, EPI_ISL_707888, EPI_ISL_707889, EPI_ISL_707903, EPI_ISL_707904, EPI_ISL_707905, EPI_ISL_707906, EPI_ISL_707907, EPI_ISL_707908, EPI_ISL_707909, EPI_ISL_707910, EPI_ISL_707911, EPI_ISL_707912, EPI_ISL_707913, EPI_ISL_707914, EPI_ISL_707915, EPI_ISL_707916, EPI_ISL_707917, EPI_ISL_707918, EPI_ISL_707919, EPI_ISL_707920, EPI_ISL_707921, EPI_ISL_707922, EPI_ISL_707923, EPI_ISL_707924, EPI_ISL_707925, EPI_ISL_707926, EPI_ISL_707927, EPI_ISL_707929, EPI_ISL_707930, EPI_ISL_707931, EPI_ISL_707932, EPI_ISL_708182, EPI_ISL_708183

| | | | |
|---|---|---|---|
| see above | Los Angeles County Public Health Laboratories | Los Angeles County Public Health Laboratories | P. Hemarajata et al. |

EPI_ISL_708200, EPI_ISL_708201, EPI_ISL_708202, EPI_ISL_708203, EPI_ISL_708204, EPI_ISL_708205, EPI_ISL_708206, EPI_ISL_708207, EPI_ISL_708208, EPI_ISL_708209, EPI_ISL_708210, EPI_ISL_708211, EPI_ISL_708212, EPI_ISL_708213, EPI_ISL_708214, EPI_ISL_708215, EPI_ISL_708216, EPI_ISL_708217, EPI_ISL_708218, EPI_ISL_708219, EPI_ISL_708220, EPI_ISL_708221, EPI_ISL_708222, EPI_ISL_708223, EPI_ISL_708224, EPI_ISL_708225, EPI_ISL_708226, EPI_ISL_708227, EPI_ISL_708228, EPI_ISL_708229, EPI_ISL_708230, EPI_ISL_708231, EPI_ISL_708232, EPI_ISL_708233, EPI_ISL_708234, EPI_ISL_708235, EPI_ISL_708236, EPI_ISL_708237, EPI_ISL_708238, EPI_ISL_708239, EPI_ISL_708240, EPI_ISL_708241, EPI_ISL_708242, EPI_ISL_708243, EPI_ISL_708244, EPI_ISL_708245, EPI_ISL_708246, EPI_ISL_708247, EPI_ISL_708248, EPI_ISL_708249, EPI_ISL_708250, EPI_ISL_708251, EPI_ISL_708252, EPI_ISL_708253, EPI_ISL_708254, EPI_ISL_708255, EPI_ISL_708256, EPI_ISL_708257, EPI_ISL_708258, EPI_ISL_708259, EPI_ISL_708260, EPI_ISL_708261, EPI_ISL_708262, EPI_ISL_708263, EPI_ISL_708264, EPI_ISL_708265, EPI_ISL_708266, EPI_ISL_708267, EPI_ISL_708268, EPI_ISL_708269, EPI_ISL_708270, EPI_ISL_708271,

| | | | |
|---|---|---|---|
| EPI_ISL_708272, EPI_ISL_708273, EPI_ISL_708277, EPI_ISL_708278, EPI_ISL_708279, EPI_ISL_708280, EPI_ISL_708281, EPI_ISL_708282, EPI_ISL_708283, EPI_ISL_708284, EPI_ISL_708285, EPI_ISL_708286, EPI_ISL_708287, EPI_ISL_708288, EPI_ISL_708289, EPI_ISL_708290, EPI_ISL_708291, EPI_ISL_708292, EPI_ISL_708293, EPI_ISL_708294, EPI_ISL_708295, EPI_ISL_708296, EPI_ISL_708297, EPI_ISL_708298, EPI_ISL_708299, EPI_ISL_708300, EPI_ISL_708301, EPI_ISL_708302, EPI_ISL_708303, EPI_ISL_708304, EPI_ISL_708305, EPI_ISL_708317, EPI_ISL_708318, EPI_ISL_708319, EPI_ISL_708320, EPI_ISL_708321, EPI_ISL_708322, EPI_ISL_708323, EPI_ISL_708324, EPI_ISL_708325, EPI_ISL_708326, EPI_ISL_708327, EPI_ISL_708328, EPI_ISL_708329, EPI_ISL_708330, EPI_ISL_708331, EPI_ISL_708332, EPI_ISL_708333, EPI_ISL_708334, EPI_ISL_708335, EPI_ISL_708336, EPI_ISL_708337, EPI_ISL_708338, EPI_ISL_708339, EPI_ISL_708340, EPI_ISL_708341, EPI_ISL_708342, EPI_ISL_708343, EPI_ISL_708344, EPI_ISL_708345, EPI_ISL_708346, EPI_ISL_708347, EPI_ISL_708348, EPI_ISL_708349, EPI_ISL_708350, EPI_ISL_708351, EPI_ISL_708352, EPI_ISL_708353, EPI_ISL_708354, EPI_ISL_708355, EPI_ISL_708356, EPI_ISL_708357, EPI_ISL_708358, EPI_ISL_708359, EPI_ISL_708360, EPI_ISL_708361, EPI_ISL_708362, EPI_ISL_708363, EPI_ISL_708364, EPI_ISL_708365, EPI_ISL_708366, EPI_ISL_708367, EPI_ISL_708368, EPI_ISL_708369, EPI_ISL_708370, EPI_ISL_708371, EPI_ISL_708372, EPI_ISL_708373, EPI_ISL_708374, EPI_ISL_708375, EPI_ISL_708376, EPI_ISL_708377, EPI_ISL_708378 | | | |
| see above | Michigan Department of Health and Human Services, Bureau of Laboratories | Michigan Department of Health and Human Services, Bureau of Laboratories | Blankenship HM, Riner D, Soehnlen MK |
| EPI_ISL_708382, EPI_ISL_708383, EPI_ISL_708384, EPI_ISL_708385, EPI_ISL_708386, EPI_ISL_708387, EPI_ISL_708388, EPI_ISL_708389, EPI_ISL_708390, EPI_ISL_708391, EPI_ISL_708392, EPI_ISL_708393, EPI_ISL_708394, EPI_ISL_708395, EPI_ISL_708396, EPI_ISL_708397, EPI_ISL_708398, EPI_ISL_708399, EPI_ISL_708400, EPI_ISL_708401, EPI_ISL_708402, EPI_ISL_708403, EPI_ISL_708404, EPI_ISL_708405, EPI_ISL_708406, EPI_ISL_708407, EPI_ISL_708408, EPI_ISL_708409, EPI_ISL_708410, EPI_ISL_708411, EPI_ISL_708433, EPI_ISL_708434, EPI_ISL_708435, EPI_ISL_708436, EPI_ISL_708437, EPI_ISL_708438, EPI_ISL_708439, EPI_ISL_708440, EPI_ISL_708441, EPI_ISL_708442, EPI_ISL_708443, EPI_ISL_708444, EPI_ISL_708445, EPI_ISL_708446, EPI_ISL_708447, EPI_ISL_708448, EPI_ISL_708449, EPI_ISL_708450, EPI_ISL_708451, EPI_ISL_708452 | | | |
| see above | Delaware Public Health Lab | Delaware Public Health Lab | Gregory Hovan |
| EPI_ISL_708460, EPI_ISL_708461, EPI_ISL_708462, EPI_ISL_708463, EPI_ISL_708464, EPI_ISL_708465, EPI_ISL_708466, EPI_ISL_708467, EPI_ISL_708468, EPI_ISL_708469, EPI_ISL_708470, EPI_ISL_708471, EPI_ISL_708472, EPI_ISL_708473, EPI_ISL_708474, EPI_ISL_708475, EPI_ISL_708476, EPI_ISL_708477, EPI_ISL_708478, EPI_ISL_708479, EPI_ISL_708480, EPI_ISL_708481, EPI_ISL_708482 | | | |
| see above | Minnesota Department of Health, Public Health Laboratory | Minnesota Department of Health, Public Health Laboratory | Alexandra Lorentz, Jacob Garfin, Matt Plumb, and Xiong Wang |
| EPI_ISL_708483 | Mayo Clinic & Mayo Clinic Laboratories | Minnesota Department of Health, Public Health Laboratory | Alexandra Lorentz, Jacob Garfin, Matt Plumb, and Xiong Wang |
| EPI_ISL_708484 | Essentia Health-St. Mary's Medical Center | Minnesota Department of Health, Public Health Laboratory | Alexandra Lorentz, Jacob Garfin, Matt Plumb, and Xiong Wang |
| EPI_ISL_708485, EPI_ISL_708486, EPI_ISL_708487, EPI_ISL_708488, EPI_ISL_708489, EPI_ISL_708490, EPI_ISL_708491, EPI_ISL_708492, EPI_ISL_708493, EPI_ISL_708494, EPI_ISL_708495, EPI_ISL_708496, EPI_ISL_708497, EPI_ISL_708498, EPI_ISL_708499, EPI_ISL_708500, EPI_ISL_708501, EPI_ISL_708502, EPI_ISL_708503 | | | |
| see above | Minnesota Department of Health, Public Health Laboratory | Minnesota Department of Health, Public Health Laboratory | Alexandra Lorentz, Jacob Garfin, Matt Plumb, and Xiong Wang |
| EPI_ISL_708504 | M Health Fairview | Minnesota Department of Health, Public Health Laboratory | Alexandra Lorentz, Jacob Garfin, Matt Plumb, and Xiong Wang |
| EPI_ISL_708505, EPI_ISL_708506, EPI_ISL_708507 | Los Angeles County Public Health Laboratories | Los Angeles County Public Health Laboratories | P. Hemarajata et al. |
| EPI_ISL_708531, EPI_ISL_708532, EPI_ISL_708533, EPI_ISL_708534, EPI_ISL_708535, EPI_ISL_708536, EPI_ISL_708537, EPI_ISL_708538, EPI_ISL_708539, EPI_ISL_708540, EPI_ISL_708541, EPI_ISL_708542, EPI_ISL_708543, EPI_ISL_708544, EPI_ISL_708545, EPI_ISL_708546, EPI_ISL_708547, EPI_ISL_708548, EPI_ISL_708549, EPI_ISL_708550, EPI_ISL_708551, EPI_ISL_708552, EPI_ISL_708553, EPI_ISL_708554, EPI_ISL_708555, EPI_ISL_708556, EPI_ISL_708557, EPI_ISL_708558, EPI_ISL_708559, EPI_ISL_708560, EPI_ISL_708561, EPI_ISL_708562, EPI_ISL_708563, EPI_ISL_708568, EPI_ISL_708569, EPI_ISL_708570, EPI_ISL_708571, EPI_ISL_708572, EPI_ISL_708573, EPI_ISL_708574, EPI_ISL_708575, EPI_ISL_708576, EPI_ISL_708577, EPI_ISL_708578, EPI_ISL_708579, EPI_ISL_708580, EPI_ISL_708581, EPI_ISL_708582, EPI_ISL_708583, EPI_ISL_708584, EPI_ISL_708585, EPI_ISL_708586, EPI_ISL_708587, EPI_ISL_708588, EPI_ISL_708589, EPI_ISL_708590, EPI_ISL_708591, EPI_ISL_708592, EPI_ISL_708593, EPI_ISL_708594, EPI_ISL_708595, EPI_ISL_708596, EPI_ISL_708597, EPI_ISL_708598, EPI_ISL_708599, EPI_ISL_708600, EPI_ISL_708601, EPI_ISL_708602, EPI_ISL_708603, EPI_ISL_708604, EPI_ISL_708605, EPI_ISL_708606 | | | |
| see above | Michigan Department of Health and Human Services, Bureau of Laboratories | Michigan Department of Health and Human Services, Bureau of Laboratories | Blankenship HM, Riner D, Soehnlen MK |
| EPI_ISL_708607, EPI_ISL_708608, EPI_ISL_708609, EPI_ISL_708610, EPI_ISL_708611, EPI_ISL_708612, EPI_ISL_708613, EPI_ISL_708614, EPI_ISL_708615, EPI_ISL_708616, EPI_ISL_708617, EPI_ISL_708618, EPI_ISL_708619, EPI_ISL_708620, EPI_ISL_708621, EPI_ISL_708622, EPI_ISL_708623, EPI_ISL_708624, EPI_ISL_708625, EPI_ISL_708626, EPI_ISL_708627, EPI_ISL_708628, EPI_ISL_708629, EPI_ISL_708630, EPI_ISL_708631, EPI_ISL_708632, EPI_ISL_708633, EPI_ISL_708634, EPI_ISL_708635, EPI_ISL_708636, EPI_ISL_708637, EPI_ISL_708646, EPI_ISL_708647, EPI_ISL_708648, EPI_ISL_708649, EPI_ISL_708650, EPI_ISL_708651 | | | |
| see above | Texas Department of State Health Services | Texas Department of State Health Services | Rashmi Tuladhar, Bonnie Oh, Jenny Zhang, Maliha Rahman, Anita Pokharel, Myong Koag, Chung Wang, Rachel Lee, Grace Kubin, Mayela Pedrueza, James Daniel Bonser |
| EPI_ISL_708724, EPI_ISL_708725, EPI_ISL_708726, EPI_ISL_708727, EPI_ISL_708728, EPI_ISL_708729, EPI_ISL_708730, EPI_ISL_708731, EPI_ISL_708732, EPI_ISL_708733, EPI_ISL_708734 | | | |
| see above | Los Angeles County Public Health Laboratories | Los Angeles County Public Health Laboratories | P. Hemarajata et al. |
| EPI_ISL_710035, EPI_ISL_710036, EPI_ISL_710037, EPI_ISL_710038, EPI_ISL_710039 | University of Michigan Clinical Microbiology Laboratory | Lauring Lab, University of Michigan, Department of Microbiology and Immunology | Valesano |
| EPI_ISL_710043, EPI_ISL_710044, EPI_ISL_710045, EPI_ISL_710046, EPI_ISL_710047, EPI_ISL_710048, EPI_ISL_710049, EPI_ISL_710050, EPI_ISL_710051, EPI_ISL_710052, EPI_ISL_710053, EPI_ISL_710054, EPI_ISL_710055, EPI_ISL_710056, EPI_ISL_710057, EPI_ISL_710058, EPI_ISL_710059, EPI_ISL_710060, EPI_ISL_710061, EPI_ISL_710062, EPI_ISL_710063, EPI_ISL_710064, EPI_ISL_710065, EPI_ISL_710066, EPI_ISL_710067, EPI_ISL_710068, EPI_ISL_710069, EPI_ISL_710070, EPI_ISL_710071, EPI_ISL_710072, EPI_ISL_710073, EPI_ISL_710074, EPI_ISL_710075, EPI_ISL_710076, EPI_ISL_710077, EPI_ISL_710078, EPI_ISL_710079, EPI_ISL_710080, EPI_ISL_710081, EPI_ISL_710087, EPI_ISL_710088, EPI_ISL_710089 | | | |
| see above | Texas Department of State Health Services | Texas Department of State Health Services | Anita Pokharel, Bonnie Oh, James Daniel Bonser, Rashmi Tuladhar, Mayela Pedrueza, Jenny Zhang, Maliha Rahman, Myong Koag, Chung Wang, Rachel Lee, Grace Kubin |
| EPI_ISL_710090, EPI_ISL_710091, EPI_ISL_710092, EPI_ISL_710093, EPI_ISL_710094, EPI_ISL_710095, EPI_ISL_710096, EPI_ISL_710097, EPI_ISL_710098 | Maryland Public Health Laboratory | Maryland Public Health Laboratory | Maryland Department of Health Laboratories Administration |
| EPI_ISL_710099, EPI_ISL_710100, EPI_ISL_710101, EPI_ISL_710102, EPI_ISL_710103, EPI_ISL_710104, EPI_ISL_710105, EPI_ISL_710106, EPI_ISL_710107, EPI_ISL_710108, EPI_ISL_710109, EPI_ISL_710110, EPI_ISL_710111, EPI_ISL_710112, EPI_ISL_710113, EPI_ISL_710114, EPI_ISL_710115, EPI_ISL_710116, EPI_ISL_710117, EPI_ISL_710118 | | | |
| see above | Los Angeles County Public Health Laboratories | Los Angeles County Public Health Laboratories | P. Hemarajata et al. |
| EPI_ISL_710130, EPI_ISL_710131, EPI_ISL_710132, EPI_ISL_710133, EPI_ISL_710134, EPI_ISL_710135, EPI_ISL_710136, EPI_ISL_710137, EPI_ISL_710138, EPI_ISL_710139, EPI_ISL_710140, EPI_ISL_710141, EPI_ISL_710142, EPI_ISL_710143, EPI_ISL_710144, EPI_ISL_710145, EPI_ISL_710146, EPI_ISL_710147, EPI_ISL_710148, EPI_ISL_710149, EPI_ISL_710150, EPI_ISL_710151, EPI_ISL_710152, EPI_ISL_710153, EPI_ISL_710154, EPI_ISL_710155, EPI_ISL_710156, EPI_ISL_710157, EPI_ISL_710180, EPI_ISL_710181, EPI_ISL_710182, EPI_ISL_710183, EPI_ISL_710184, EPI_ISL_710185, EPI_ISL_710186, EPI_ISL_710187, EPI_ISL_710188, EPI_ISL_710189, EPI_ISL_710190, EPI_ISL_710191, EPI_ISL_710192, EPI_ISL_710193, EPI_ISL_710194, EPI_ISL_710195, EPI_ISL_710196, EPI_ISL_710197, EPI_ISL_710198, EPI_ISL_710199, EPI_ISL_710200, EPI_ISL_710201, EPI_ISL_710202, EPI_ISL_710203, EPI_ISL_710204, EPI_ISL_710205, EPI_ISL_710206, EPI_ISL_710207, EPI_ISL_710208, EPI_ISL_710209, EPI_ISL_710210, EPI_ISL_710211, EPI_ISL_710212, EPI_ISL_710213, EPI_ISL_710214, EPI_ISL_710215, EPI_ISL_710216, EPI_ISL_710217, EPI_ISL_710218, EPI_ISL_710219, EPI_ISL_710220, EPI_ISL_710221, EPI_ISL_710222, EPI_ISL_710223, EPI_ISL_710224, EPI_ISL_710225, EPI_ISL_710226, EPI_ISL_710227, EPI_ISL_710228, EPI_ISL_710229, EPI_ISL_710230, EPI_ISL_710231, EPI_ISL_710232, EPI_ISL_710233, EPI_ISL_710234, EPI_ISL_710235, EPI_ISL_710236, EPI_ISL_710237, EPI_ISL_710238, EPI_ISL_710239, EPI_ISL_710240, EPI_ISL_710241, EPI_ISL_710242, EPI_ISL_710243, EPI_ISL_710244, EPI_ISL_710245, EPI_ISL_710246, EPI_ISL_710247, EPI_ISL_710248, EPI_ISL_710249, EPI_ISL_710250, EPI_ISL_710251, EPI_ISL_710252, EPI_ISL_710253, EPI_ISL_710254, EPI_ISL_710255, EPI_ISL_710256, EPI_ISL_710257, EPI_ISL_710258, EPI_ISL_710259, EPI_ISL_710260, EPI_ISL_710261, EPI_ISL_710262, EPI_ISL_710263, EPI_ISL_710264, EPI_ISL_710265, EPI_ISL_710266, EPI_ISL_710267, EPI_ISL_710268, EPI_ISL_710269, EPI_ISL_710270, EPI_ISL_710301, EPI_ISL_710302, EPI_ISL_710303, EPI_ISL_710304, EPI_ISL_710305, EPI_ISL_710306, EPI_ISL_710307, EPI_ISL_710308, EPI_ISL_710309, EPI_ISL_710310, EPI_ISL_710311, EPI_ISL_710312, EPI_ISL_710313, EPI_ISL_710314, EPI_ISL_710315, EPI_ISL_710316, EPI_ISL_710317, EPI_ISL_710318, EPI_ISL_710319, EPI_ISL_710320, EPI_ISL_710321, EPI_ISL_710322, EPI_ISL_710323, EPI_ISL_710324, EPI_ISL_710325, EPI_ISL_710326, EPI_ISL_710327, EPI_ISL_710328, EPI_ISL_710329, EPI_ISL_710330, EPI_ISL_710331, EPI_ISL_710332, EPI_ISL_710333, EPI_ISL_710334, EPI_ISL_710335, EPI_ISL_710336, EPI_ISL_710337, EPI_ISL_710338, EPI_ISL_710339, EPI_ISL_710340, EPI_ISL_710341, EPI_ISL_710342, EPI_ISL_710343, EPI_ISL_710344, EPI_ISL_710345, EPI_ISL_710346, EPI_ISL_710347, EPI_ISL_710348, EPI_ISL_710349, EPI_ISL_710350, EPI_ISL_710351, EPI_ISL_710352, EPI_ISL_710353, EPI_ISL_710354, EPI_ISL_710355, EPI_ISL_710356, EPI_ISL_710357, EPI_ISL_710358, EPI_ISL_710359, EPI_ISL_710360, EPI_ISL_710361, EPI_ISL_710362, EPI_ISL_710363, EPI_ISL_710364, EPI_ISL_710365, EPI_ISL_710366, EPI_ISL_710367, EPI_ISL_710368, EPI_ISL_710369, EPI_ISL_710370, EPI_ISL_710371, EPI_ISL_710372, EPI_ISL_710373 | | | |
| see above | Colorado Department of Public Health and Environment | Colorado Department of Puplic Health and Environment | Laura Bankers, Molly C. Hetherington-Rauth, Shannon Ely, Shannon R. Matzinger, Sarah Elizabeth Totten, Emily A. Travanty |
| EPI_ISL_710374, EPI_ISL_710375, EPI_ISL_710376, EPI_ISL_710377, EPI_ISL_710378, EPI_ISL_710379, EPI_ISL_710380, EPI_ISL_710381, EPI_ISL_710382, EPI_ISL_710383, EPI_ISL_710384, EPI_ISL_710385, EPI_ISL_710387, EPI_ISL_710388, EPI_ISL_710389, EPI_ISL_710390, EPI_ISL_710391, EPI_ISL_710392, EPI_ISL_710393, EPI_ISL_710394, EPI_ISL_710395, EPI_ISL_710396, EPI_ISL_710397, EPI_ISL_710398, EPI_ISL_710399, EPI_ISL_710400, EPI_ISL_710401, EPI_ISL_710402, EPI_ISL_710403, EPI_ISL_710404, EPI_ISL_710405, EPI_ISL_710406, EPI_ISL_710407, EPI_ISL_710408, EPI_ISL_710409, EPI_ISL_710410, EPI_ISL_710411, EPI_ISL_710412, EPI_ISL_710413, EPI_ISL_710414, EPI_ISL_710415 | | | |
| see above | Texas Department of State Health Services | Texas Department of State Health Services | Rashmi Tuladhar, Bonnie Oh, Jenny Zhang, Maliha Rahman, Anita Pokharel, Myong Koag, Chung Wang, Rachel Lee, Grace Kubin, Mayela Pedrueza, James Daniel Bonser |
| EPI_ISL_710416, EPI_ISL_710417, EPI_ISL_710418, EPI_ISL_710419, EPI_ISL_710420, EPI_ISL_710421, EPI_ISL_710422, EPI_ISL_710423, EPI_ISL_710430, EPI_ISL_710431, EPI_ISL_710432 | | | |
| see above | Los Angeles County Public Health Laboratories | Los Angeles County Public Health Laboratories | P. Hemarajata et al. |
| EPI_ISL_717766, EPI_ISL_717767, EPI_ISL_717768, EPI_ISL_717769, EPI_ISL_717770, EPI_ISL_717771, EPI_ISL_717772, EPI_ISL_717773, EPI_ISL_717774, EPI_ISL_717775, EPI_ISL_717776, EPI_ISL_717777, EPI_ISL_717778, EPI_ISL_717779, EPI_ISL_717780, EPI_ISL_717781, EPI_ISL_717782, EPI_ISL_717783, EPI_ISL_717784 | | | |
| see above | UW Virology Lab | UW Virology Lab | Pavitra Roychoudhury, Hong Xie, Lasata Shrestha, Michelle Lin, Meei-Li Huang, Keith R Jerome, Alexander Greninger |
| EPI_ISL_723118, EPI_ISL_723119, EPI_ISL_723120, EPI_ISL_723121, EPI_ISL_723122, EPI_ISL_723123, EPI_ISL_723124, EPI_ISL_723125, EPI_ISL_723126, EPI_ISL_723127, EPI_ISL_723128, EPI_ISL_723129, EPI_ISL_723130, EPI_ISL_723131, EPI_ISL_723132, EPI_ISL_723133, EPI_ISL_723134, EPI_ISL_723135, EPI_ISL_723136, EPI_ISL_723137, EPI_ISL_723138, EPI_ISL_723139, EPI_ISL_723140 | | | |
| see above | Minnesota Department of Health, Public Health Laboratory | Minnesota Department of Health, Public Health Laboratory | Alexandra Lorentz, Jacob Garfin, Matt Plumb, and Xiong Wang |
| EPI_ISL_723141, EPI_ISL_723142, EPI_ISL_723143, EPI_ISL_723144, EPI_ISL_723145 | Mayo Clinic & Mayo Clinic Laboratories | Minnesota Department of Health, Public Health Laboratory | Alexandra Lorentz, Jacob Garfin, Matt Plumb, and Xiong Wang |
| EPI_ISL_723146 | Minnesota Department of Health, Public Health Laboratory | Minnesota Department of Health, Public Health Laboratory | Alexandra Lorentz, Jacob Garfin, Matt Plumb, and Xiong Wang |
| EPI_ISL_723147, EPI_ISL_723148, EPI_ISL_723149, EPI_ISL_723150, EPI_ISL_723151, EPI_ISL_723152, EPI_ISL_723153, | Vault Health | Minnesota Department of Health, Public Health Laboratory | Alexandra Lorentz, Jacob Garfin, Matt Plumb, and Xiong Wang |

| | | | |
|---|---|---|---|
| EPI_ISL_723154 | | | |
| EPI_ISL_723470, EPI_ISL_723471, EPI_ISL_723472, EPI_ISL_723473, EPI_ISL_723474, EPI_ISL_723475, EPI_ISL_723476, EPI_ISL_723477, EPI_ISL_723478, EPI_ISL_723479, EPI_ISL_723480, EPI_ISL_723481, EPI_ISL_723482, EPI_ISL_723483, EPI_ISL_723484, EPI_ISL_723485, EPI_ISL_723486, EPI_ISL_723487, EPI_ISL_723488, EPI_ISL_723489, EPI_ISL_723490, EPI_ISL_723491 | | | |
| see above | Virginia DCLS | Virginia DCLS | Virginia DCLS |
| EPI_ISL_723492, EPI_ISL_723493, EPI_ISL_723494, EPI_ISL_723495, EPI_ISL_723496, EPI_ISL_723497, EPI_ISL_723498, EPI_ISL_723499, EPI_ISL_723500, EPI_ISL_723501, EPI_ISL_723502, EPI_ISL_723503, EPI_ISL_723504, EPI_ISL_723505, EPI_ISL_723506, EPI_ISL_723507, EPI_ISL_723508, EPI_ISL_723509, EPI_ISL_723510, EPI_ISL_723511, EPI_ISL_723512, EPI_ISL_723513, EPI_ISL_723514, EPI_ISL_723515, EPI_ISL_723516, EPI_ISL_723517, EPI_ISL_723518, EPI_ISL_723519, EPI_ISL_723520 | | | |
| see above | Virginia Division of Consolidated Laboratory Services (DCLS) | Virginia Division of Consolidated Laboratory Services (DCLS) | Virginia DCLS |
| EPI_ISL_728001, EPI_ISL_728002, EPI_ISL_728003, EPI_ISL_728004, EPI_ISL_728005, EPI_ISL_728006, EPI_ISL_728007, EPI_ISL_728008, EPI_ISL_728009, EPI_ISL_728010, EPI_ISL_728011, EPI_ISL_728012, EPI_ISL_728013, EPI_ISL_728014, EPI_ISL_728015, EPI_ISL_728016, EPI_ISL_728017, EPI_ISL_728018, EPI_ISL_728019, EPI_ISL_728020, EPI_ISL_728021, EPI_ISL_728022, EPI_ISL_728023, EPI_ISL_728024, EPI_ISL_728025, EPI_ISL_728026, EPI_ISL_728027, EPI_ISL_728028, EPI_ISL_728029, EPI_ISL_728030, EPI_ISL_728031, EPI_ISL_728032, EPI_ISL_728033, EPI_ISL_728034, EPI_ISL_728035, EPI_ISL_728036, EPI_ISL_728037, EPI_ISL_728038, EPI_ISL_728039, EPI_ISL_728040, EPI_ISL_728041, EPI_ISL_728042, EPI_ISL_728043, EPI_ISL_728044, EPI_ISL_728045, EPI_ISL_728046, EPI_ISL_728047, EPI_ISL_728048, EPI_ISL_728049, EPI_ISL_728050, EPI_ISL_728051, EPI_ISL_728052, EPI_ISL_728053, EPI_ISL_728054, EPI_ISL_728055, EPI_ISL_728056, EPI_ISL_728057, EPI_ISL_728058, EPI_ISL_728059, EPI_ISL_728060, EPI_ISL_728061, EPI_ISL_728062, EPI_ISL_728063, EPI_ISL_728064, EPI_ISL_728065, EPI_ISL_728066, EPI_ISL_728067, EPI_ISL_728068, EPI_ISL_728069, EPI_ISL_728070, EPI_ISL_728071, EPI_ISL_728072, EPI_ISL_728073, EPI_ISL_728074, EPI_ISL_728075, EPI_ISL_728076, EPI_ISL_728077, EPI_ISL_728078, EPI_ISL_728079, EPI_ISL_728080, EPI_ISL_728081, EPI_ISL_728082, EPI_ISL_728083, EPI_ISL_728084, EPI_ISL_728085, EPI_ISL_728086, EPI_ISL_728087, EPI_ISL_728088, EPI_ISL_728089, EPI_ISL_728090, EPI_ISL_728091, EPI_ISL_728092, EPI_ISL_728093, EPI_ISL_728094, EPI_ISL_728095, EPI_ISL_728096, EPI_ISL_728097, EPI_ISL_728098, EPI_ISL_728099, EPI_ISL_728100, EPI_ISL_728101, EPI_ISL_728102, EPI_ISL_728103, EPI_ISL_728104, EPI_ISL_728105, EPI_ISL_728106, EPI_ISL_728107, EPI_ISL_728108, EPI_ISL_728109, EPI_ISL_728110, EPI_ISL_728112, EPI_ISL_728113, EPI_ISL_728114, EPI_ISL_728115, EPI_ISL_728116, EPI_ISL_728117, EPI_ISL_728118, EPI_ISL_728119, EPI_ISL_728120, EPI_ISL_728122, EPI_ISL_728124, EPI_ISL_728125, EPI_ISL_728126, EPI_ISL_728127, EPI_ISL_728128, EPI_ISL_728129, EPI_ISL_728130, EPI_ISL_728131, EPI_ISL_728132, EPI_ISL_728133, EPI_ISL_728134, EPI_ISL_728135, EPI_ISL_728136, EPI_ISL_728137, EPI_ISL_728138, EPI_ISL_728139, EPI_ISL_728140, EPI_ISL_728141, EPI_ISL_728142, EPI_ISL_728143, EPI_ISL_728144, EPI_ISL_728145, EPI_ISL_728146, EPI_ISL_728147, EPI_ISL_728148, EPI_ISL_728149, EPI_ISL_728150, EPI_ISL_728151, EPI_ISL_728152 | | | |
| see above | University of Wisconsin-Madison AIDS Vaccine Research Laboratories | University of Wisconsin-Madison AIDS Vaccine Research Laboratories | Gage Moreno, Katarina Braun, et al. AIDS Vaccine Research Laboratories |
| EPI_ISL_728368, EPI_ISL_728369, EPI_ISL_728370, EPI_ISL_728371, EPI_ISL_728372, EPI_ISL_728373, EPI_ISL_728374, EPI_ISL_728375, EPI_ISL_728376, EPI_ISL_728377, EPI_ISL_728378, EPI_ISL_728379, EPI_ISL_728380, EPI_ISL_728381, EPI_ISL_728382, EPI_ISL_728383, EPI_ISL_728384, EPI_ISL_728385, EPI_ISL_728386, EPI_ISL_728387, EPI_ISL_728388, EPI_ISL_728389, EPI_ISL_728390, EPI_ISL_728391, EPI_ISL_728392, EPI_ISL_728393, EPI_ISL_728394, EPI_ISL_728395, EPI_ISL_728396, EPI_ISL_728397, EPI_ISL_728398, EPI_ISL_728399, EPI_ISL_728400, EPI_ISL_728401, EPI_ISL_728402, EPI_ISL_728403, EPI_ISL_728404, EPI_ISL_728405, EPI_ISL_728406, EPI_ISL_728407, EPI_ISL_728408, EPI_ISL_728409, EPI_ISL_728410, EPI_ISL_728411, EPI_ISL_728412, EPI_ISL_728413, EPI_ISL_728414, EPI_ISL_728415, EPI_ISL_728416, EPI_ISL_728417, EPI_ISL_728418, EPI_ISL_728419, EPI_ISL_728420, EPI_ISL_728421, EPI_ISL_728422, EPI_ISL_728423, EPI_ISL_728424, EPI_ISL_728425, EPI_ISL_728426, EPI_ISL_728427, EPI_ISL_728428, EPI_ISL_728429, EPI_ISL_728430, EPI_ISL_728431, EPI_ISL_728432, EPI_ISL_728433, EPI_ISL_728434, EPI_ISL_728435, EPI_ISL_728436, EPI_ISL_728437, EPI_ISL_728438, EPI_ISL_728439, EPI_ISL_728440, EPI_ISL_728441, EPI_ISL_728442, EPI_ISL_728443, EPI_ISL_728444, EPI_ISL_728445, EPI_ISL_728446, EPI_ISL_728447, EPI_ISL_728448, EPI_ISL_728449, EPI_ISL_728450, EPI_ISL_728451, EPI_ISL_728452, EPI_ISL_728453, EPI_ISL_728454, EPI_ISL_728455, EPI_ISL_728456, EPI_ISL_728457, EPI_ISL_728458, EPI_ISL_728459, EPI_ISL_728460, EPI_ISL_728461, EPI_ISL_728462, EPI_ISL_728463, EPI_ISL_728464, EPI_ISL_728465, EPI_ISL_728466, EPI_ISL_728467, EPI_ISL_728468, EPI_ISL_728469, EPI_ISL_728470, EPI_ISL_728471, EPI_ISL_728472, EPI_ISL_728473, EPI_ISL_728474, EPI_ISL_728475, EPI_ISL_728476, EPI_ISL_728477, EPI_ISL_728478, EPI_ISL_728479, EPI_ISL_728480, EPI_ISL_728481, EPI_ISL_728482, EPI_ISL_728483, EPI_ISL_728484, EPI_ISL_728485, EPI_ISL_728486, EPI_ISL_728487, EPI_ISL_728488, EPI_ISL_728489, EPI_ISL_728490, EPI_ISL_728491, EPI_ISL_728492, EPI_ISL_728493, EPI_ISL_728494, EPI_ISL_728495, EPI_ISL_728496, EPI_ISL_728497, EPI_ISL_728498, EPI_ISL_728499, EPI_ISL_728500, EPI_ISL_728501, EPI_ISL_728502, EPI_ISL_728503, EPI_ISL_728504, EPI_ISL_728505, EPI_ISL_728506, EPI_ISL_728507, EPI_ISL_728508, EPI_ISL_728509, EPI_ISL_728510, EPI_ISL_728511, EPI_ISL_728512, EPI_ISL_728513, EPI_ISL_728514, EPI_ISL_728515 | | | |
| see above | University of Michigan Clinical Microbiology Laboratory | Lauring Lab, University of Michigan, Department of Microbiology and Immunology | Valesano |
| EPI_ISL_728552, EPI_ISL_728553, EPI_ISL_728554, EPI_ISL_728555 | University of Wisconsin-Madison AIDS Vaccine Research Laboratories | University of Wisconsin-Madison AIDS Vaccine Research Laboratories | Gage Moreno, Katarina Braun, et al. AIDS Vaccine Research Laboratories |
| EPI_ISL_729735, EPI_ISL_729736 | Connecticut Department of Health | Grubaugh Lab - Yale School of Public Health | Joseph Fauver, Tara Alpert, Anderson Brito, Annie Watkins, Anne Wyllie, Chantal Vogels, Mary Petrone, Chaney Kalinich, Isabel Ott, Arnau Casanovas, Catherine Muenker, Adam Moore, Alice Lu, Maria Tokuyama, Patrick Wong, Peiwen Lu, Saad Omer, Richard Martinello, Allison Nelson, Shelli Farhadian, Akiko Iwasaki, Charlese Dela Cruz, Albert Ko, Nathan Grubaugh |
| EPI_ISL_729737 | Grubaugh Lab - Yale School of Public Health | Grubaugh Lab - Yale School of Public Health | Joseph Fauver, Tara Alpert, Anderson Brito, Annie Watkins, Anne Wyllie, Chantal Vogels, Mary Petrone, Chaney Kalinich, Isabel Ott, Arnau Casanovas, Catherine Muenker, Adam Moore, Alice Lu, Maria Tokuyama, Patrick Wong, Peiwen Lu, Saad Omer, Richard Martinello, Allison Nelson, Shelli Farhadian, Akiko Iwasaki, Charlese Dela Cruz, Albert Ko, Nathan Grubaugh |
| EPI_ISL_729738 | Yale Pathology Lab | Grubaugh Lab - Yale School of Public Health | Joseph Fauver, Tara Alpert, Anderson Brito, Annie Watkins, Anne Wyllie, Chantal Vogels, Mary Petrone, Chaney Kalinich, Isabel Ott, Arnau Casanovas, Catherine Muenker, Adam Moore, Alice Lu, Maria Tokuyama, Patrick Wong, Peiwen Lu, Saad Omer, Richard Martinello, Allison Nelson, Shelli Farhadian, Akiko Iwasaki, Charlese Dela Cruz, Albert Ko, Nathan Grubaugh |
| EPI_ISL_729739, EPI_ISL_729740 | Grubaugh Lab - Yale School of Public Health | Grubaugh Lab - Yale School of Public Health | Joseph Fauver, Tara Alpert, Anderson Brito, Annie Watkins, Anne Wyllie, Chantal Vogels, Mary Petrone, Chaney Kalinich, Isabel Ott, Arnau Casanovas, Catherine Muenker, Adam Moore, Alice Lu, Maria Tokuyama, Patrick Wong, Peiwen Lu, Saad Omer, Richard Martinello, Allison Nelson, Shelli Farhadian, Akiko Iwasaki, Charlese Dela Cruz, Albert Ko, Nathan Grubaugh |
| EPI_ISL_729741 | Connecticut Department of Health | Grubaugh Lab - Yale School of Public Health | Joseph Fauver, Tara Alpert, Anderson Brito, Annie Watkins, Anne Wyllie, Chantal Vogels, Mary Petrone, Chaney Kalinich, Isabel Ott, Arnau Casanovas, Catherine Muenker, Adam Moore, Alice Lu, Maria Tokuyama, Patrick Wong, Peiwen Lu, Saad Omer, Richard Martinello, Allison Nelson, Shelli Farhadian, Akiko Iwasaki, Charlese Dela Cruz, Albert Ko, Nathan Grubaugh |
| EPI_ISL_729742 | Yale Pathology Lab | Grubaugh Lab - Yale School of Public Health | Joseph Fauver, Tara Alpert, Anderson Brito, Annie Watkins, Anne Wyllie, Chantal Vogels, Mary Petrone, Chaney Kalinich, Isabel Ott, Arnau Casanovas, Catherine Muenker, Adam Moore, Alice Lu, Maria Tokuyama, Patrick Wong, Peiwen Lu, Saad Omer, Richard Martinello, Allison Nelson, Shelli Farhadian, Akiko Iwasaki, Charlese Dela Cruz, Albert Ko, Nathan Grubaugh |
| EPI_ISL_729743, EPI_ISL_729744, EPI_ISL_729745, EPI_ISL_729746, EPI_ISL_729747, EPI_ISL_729748, EPI_ISL_729749, EPI_ISL_729750 | Connecticut Department of Health | Grubaugh Lab - Yale School of Public Health | Joseph Fauver, Tara Alpert, Anderson Brito, Annie Watkins, Anne Wyllie, Chantal Vogels, Mary Petrone, Chaney Kalinich, Isabel Ott, Arnau Casanovas, Catherine Muenker, Adam Moore, Alice Lu, Maria Tokuyama, Patrick Wong, Peiwen Lu, Saad Omer, Richard Martinello, Allison Nelson, Shelli Farhadian, Akiko Iwasaki, Charlese Dela Cruz, Albert Ko, Nathan Grubaugh |
| EPI_ISL_729751 | Yale Pathology Lab | Grubaugh Lab - Yale School of Public Health | Joseph Fauver, Tara Alpert, Anderson Brito, Annie Watkins, Anne Wyllie, Chantal Vogels, Mary Petrone, Chaney Kalinich, Isabel Ott, Arnau Casanovas, Catherine Muenker, Adam Moore, Alice Lu, Maria Tokuyama, Patrick Wong, Peiwen Lu, Saad Omer, Richard Martinello, Allison Nelson, Shelli Farhadian, Akiko Iwasaki, Charlese Dela Cruz, Albert Ko, Nathan Grubaugh |
| EPI_ISL_729752, EPI_ISL_729753, EPI_ISL_729754, EPI_ISL_729755, EPI_ISL_729756 | Connecticut Department of Health | Grubaugh Lab - Yale School of Public Health | Joseph Fauver, Tara Alpert, Anderson Brito, Annie Watkins, Anne Wyllie, Chantal Vogels, Mary Petrone, Chaney Kalinich, Isabel Ott, Arnau Casanovas, Catherine Muenker, Adam Moore, Alice Lu, Maria Tokuyama, Patrick Wong, Peiwen Lu, Saad Omer, Richard Martinello, Allison Nelson, Shelli Farhadian, Akiko Iwasaki, Charlese Dela Cruz, Albert Ko, Nathan Grubaugh |
| EPI_ISL_729757 | Yale Pathology Lab | Grubaugh Lab - Yale School of Public Health | Joseph Fauver, Tara Alpert, Anderson Brito, Annie Watkins, Anne Wyllie, Chantal Vogels, Mary Petrone, Chaney Kalinich, Isabel Ott, Arnau Casanovas, Catherine Muenker, Adam Moore, Alice Lu, Maria Tokuyama, Patrick Wong, Peiwen Lu, Saad Omer, Richard Martinello, Allison Nelson, Shelli Farhadian, Akiko Iwasaki, Charlese Dela Cruz, Albert Ko, Nathan Grubaugh |
| EPI_ISL_729758, EPI_ISL_729759, EPI_ISL_729760 | Connecticut Department of Health | Grubaugh Lab - Yale School of Public Health | Joseph Fauver, Tara Alpert, Anderson Brito, Annie Watkins, Anne Wyllie, Chantal Vogels, Mary Petrone, Chaney Kalinich, Isabel Ott, Arnau Casanovas, Catherine Muenker, Adam Moore, Alice Lu, Maria Tokuyama, Patrick Wong, Peiwen Lu, Saad Omer, Richard Martinello, Allison Nelson, Shelli Farhadian, Akiko Iwasaki, Charlese Dela Cruz, Albert Ko, Nathan Grubaugh |
| EPI_ISL_729761, EPI_ISL_729762, EPI_ISL_729763, EPI_ISL_729764 | Yale Pathology Lab | Grubaugh Lab - Yale School of Public Health | Joseph Fauver, Tara Alpert, Anderson Brito, Annie Watkins, Anne Wyllie, Chantal Vogels, Mary Petrone, Chaney Kalinich, Isabel Ott, Arnau Casanovas, Catherine Muenker, Adam Moore, Alice Lu, Maria Tokuyama, Patrick Wong, Peiwen Lu, Saad Omer, Richard Martinello, Allison Nelson, Shelli Farhadian, Akiko Iwasaki, Charlese Dela Cruz, Albert Ko, Nathan Grubaugh |
| EPI_ISL_729765 | Connecticut Department of Health | Grubaugh Lab - Yale School of Public Health | Joseph Fauver, Tara Alpert, Anderson Brito, Annie Watkins, Anne Wyllie, Chantal Vogels, Mary Petrone, Chaney Kalinich, Isabel Ott, Arnau Casanovas, Catherine Muenker, Adam Moore, Alice Lu, Maria Tokuyama, Patrick Wong, Peiwen Lu, Saad Omer, Richard Martinello, Allison Nelson, Shelli Farhadian, Akiko Iwasaki, Charlese Dela Cruz, Albert Ko, Nathan Grubaugh |
| EPI_ISL_729766, EPI_ISL_729767, EPI_ISL_729768, EPI_ISL_729769 | Yale Pathology Lab | Grubaugh Lab - Yale School of Public Health | Joseph Fauver, Tara Alpert, Anderson Brito, Annie Watkins, Anne Wyllie, Chantal Vogels, Mary Petrone, Chaney Kalinich, Isabel Ott, Arnau Casanovas, Catherine Muenker, Adam Moore, Alice Lu, Maria Tokuyama, Patrick Wong, Peiwen Lu, Saad Omer, Richard Martinello, Allison Nelson, Shelli Farhadian, Akiko Iwasaki, Charlese Dela Cruz, Albert Ko, Nathan Grubaugh |
| EPI_ISL_730062, EPI_ISL_730064, EPI_ISL_730065, EPI_ISL_730067, EPI_ISL_730070, EPI_ISL_730074, EPI_ISL_730075, EPI_ISL_730076, EPI_ISL_730080, EPI_ISL_730082 | Yale Clinical Virology Laboratory | Grubaugh Lab - Yale School of Public Health | Joseph Fauver, Tara Alpert, Anderson Brito, Annie Watkins, Anne Wyllie, Chantal Vogels, Mary Petrone, Chaney Kalinich, Isabel Ott, Arnau Casanovas, Catherine Muenker, Adam Moore, Alice Lu, Maria Tokuyama, Patrick Wong, Peiwen Lu, Saad Omer, Richard Martinello, Allison Nelson, Shelli Farhadian, Akiko Iwasaki, Charlese Dela Cruz, Albert Ko, Nathan Grubaugh |
| EPI_ISL_730085, EPI_ISL_730086, EPI_ISL_730087, EPI_ISL_730088, EPI_ISL_730089, EPI_ISL_730090, EPI_ISL_730091, EPI_ISL_730092, EPI_ISL_730093, EPI_ISL_730094, EPI_ISL_730095, EPI_ISL_730096, EPI_ISL_730097, EPI_ISL_730098, EPI_ISL_730099, EPI_ISL_730100, EPI_ISL_730101, EPI_ISL_730102, EPI_ISL_730103, EPI_ISL_730104, EPI_ISL_730105, EPI_ISL_730106, EPI_ISL_730107, EPI_ISL_730108, EPI_ISL_730109, EPI_ISL_730110, EPI_ISL_730111, EPI_ISL_730112, EPI_ISL_730113, EPI_ISL_730114, EPI_ISL_730115, EPI_ISL_730116, EPI_ISL_730117, EPI_ISL_730118, EPI_ISL_730119, EPI_ISL_730120, EPI_ISL_730121, EPI_ISL_730122, EPI_ISL_730123, EPI_ISL_730124, EPI_ISL_730125, EPI_ISL_730126 | | | |
| see above | San Diego County Public Health Laboratory | Andersen lab at Scripps Research | SEARCH Alliance San Diego with Tracy Basler, Jovan Shephard, Brett Austin |
| EPI_ISL_730127, EPI_ISL_730128, EPI_ISL_730129, EPI_ISL_730130, EPI_ISL_730131, EPI_ISL_730132, EPI_ISL_730133, EPI_ISL_730134, EPI_ISL_730135, EPI_ISL_730136 | Sharp HealthCare Laboratory | Andersen lab at Scripps Research | SEARCH Alliance San Diego with Aaron Harding, Jacquelyn Berumen, Cathy Woerle, Liam McGinnis, Art Mendoza, Omid Bakhtar |
| EPI_ISL_730137, EPI_ISL_730138, EPI_ISL_730139, EPI_ISL_730140, EPI_ISL_730141, EPI_ISL_730142, EPI_ISL_730143, EPI_ISL_730144, EPI_ISL_730145, EPI_ISL_730146, EPI_ISL_730147, EPI_ISL_730148, EPI_ISL_730149, EPI_ISL_730150, EPI_ISL_730151, EPI_ISL_730152, EPI_ISL_730153, EPI_ISL_730154, EPI_ISL_730155, EPI_ISL_730156, EPI_ISL_730157, EPI_ISL_730158, EPI_ISL_730159, EPI_ISL_730160 | | | |
| see above | Scripps Medical Laboratory | Andersen lab at Scripps Research | SEARCH Alliance San Diego with Michael Quigley, Ellen Stefanski, Ian Mchardy |

| | | | |
|---|---|---|---|
| EPI_ISL_730161, EPI_ISL_730162, EPI_ISL_730163, EPI_ISL_730164, EPI_ISL_730165, EPI_ISL_730166, EPI_ISL_730167, EPI_ISL_730168, EPI_ISL_730169, EPI_ISL_730170, EPI_ISL_730171, EPI_ISL_730172, EPI_ISL_730173, EPI_ISL_730174, EPI_ISL_730175, EPI_ISL_730176, EPI_ISL_730177, EPI_ISL_730178, EPI_ISL_730179, EPI_ISL_730180, EPI_ISL_730181, EPI_ISL_730182, EPI_ISL_730183, EPI_ISL_730184, EPI_ISL_730185, EPI_ISL_730186, EPI_ISL_730187, EPI_ISL_730188, EPI_ISL_730189, EPI_ISL_730190, EPI_ISL_730191, EPI_ISL_730192, EPI_ISL_730193, EPI_ISL_730194, EPI_ISL_730195, EPI_ISL_730196 | | | |
| see above | San Diego County Public Health Laboratory | Andersen lab at Scripps Research | SEARCH Alliance San Diego with Tracy Basler, Jovan Shephard, Brett Austin |
| EPI_ISL_730197, EPI_ISL_730198, EPI_ISL_730199, EPI_ISL_730200, EPI_ISL_730201, EPI_ISL_730202, EPI_ISL_730203, EPI_ISL_730204, EPI_ISL_730205, EPI_ISL_730206, EPI_ISL_730207, EPI_ISL_730208, EPI_ISL_730209, EPI_ISL_730210, EPI_ISL_730211, EPI_ISL_730212, EPI_ISL_730213, EPI_ISL_730214, EPI_ISL_730215, EPI_ISL_730216, EPI_ISL_730217, EPI_ISL_730218, EPI_ISL_730219, EPI_ISL_730220, EPI_ISL_730221, EPI_ISL_730222, EPI_ISL_730223, EPI_ISL_730224, EPI_ISL_730225, EPI_ISL_730226, EPI_ISL_730227, EPI_ISL_730228 | | | |
| see above | Genomica Lab Molecular, M√©xico | Andersen lab at Scripps Research | SEARCH Alliance San Diego with Jonathan Gonzalez Garcia, Jose Roman Chavez Mendez, Jose Horacio Reyna Verdugo, Martin Gonzalez Ibarra, Luis Alberto Rangel Gonzalez |
| EPI_ISL_730291, EPI_ISL_730292, EPI_ISL_730293, EPI_ISL_730294, EPI_ISL_730295, EPI_ISL_730296, EPI_ISL_730297, EPI_ISL_730298, EPI_ISL_730299, EPI_ISL_730300, EPI_ISL_730301, EPI_ISL_730302, EPI_ISL_730303, EPI_ISL_730304, EPI_ISL_730305, EPI_ISL_730306, EPI_ISL_730307, EPI_ISL_730308, EPI_ISL_730309, EPI_ISL_730310, EPI_ISL_730311, EPI_ISL_730312, EPI_ISL_730313, EPI_ISL_730314, EPI_ISL_730315, EPI_ISL_730316, EPI_ISL_730317, EPI_ISL_730318, EPI_ISL_730319, EPI_ISL_730320, EPI_ISL_730321, EPI_ISL_730322, EPI_ISL_730323, EPI_ISL_730324, EPI_ISL_730325, EPI_ISL_730326, EPI_ISL_730327, EPI_ISL_730328, EPI_ISL_730329, EPI_ISL_730330, EPI_ISL_730331, EPI_ISL_730332, EPI_ISL_730333, EPI_ISL_730334, EPI_ISL_730335, EPI_ISL_730336, EPI_ISL_730337, EPI_ISL_730338, EPI_ISL_730339, EPI_ISL_730340, EPI_ISL_730341, EPI_ISL_730342, EPI_ISL_730343, EPI_ISL_730344, EPI_ISL_730345, EPI_ISL_730346, EPI_ISL_730347, EPI_ISL_730348, EPI_ISL_730349, EPI_ISL_730350, EPI_ISL_730351, EPI_ISL_730352, EPI_ISL_730353, EPI_ISL_730354, EPI_ISL_730355, EPI_ISL_730356, EPI_ISL_730357, EPI_ISL_730358, EPI_ISL_730359, EPI_ISL_730360, EPI_ISL_730361, EPI_ISL_730362, EPI_ISL_730363, EPI_ISL_730364, EPI_ISL_730365, EPI_ISL_730366, EPI_ISL_730367, EPI_ISL_730368, EPI_ISL_730369, EPI_ISL_730370, EPI_ISL_730371 | | | |
| see above | San Diego County Public Health Laboratory | Andersen lab at Scripps Research | SEARCH Alliance San Diego with Tracy Basler, Jovan Shephard, Brett Austin |
| EPI_ISL_730564, EPI_ISL_730565, EPI_ISL_730566 | University of Michigan Clinical Microbiology Laboratory | Lauring Lab, University of Michigan, Department of Microbiology and Immunology | Valesano |
| EPI_ISL_732704, EPI_ISL_732705, EPI_ISL_732706, EPI_ISL_732707, EPI_ISL_732708, EPI_ISL_732709, EPI_ISL_732710, EPI_ISL_732711, EPI_ISL_732712, EPI_ISL_732713, EPI_ISL_732714, EPI_ISL_732715, EPI_ISL_732716, EPI_ISL_732717, EPI_ISL_732718, EPI_ISL_732719, EPI_ISL_732720, EPI_ISL_732721, EPI_ISL_732722, EPI_ISL_732723, EPI_ISL_732724, EPI_ISL_732725, EPI_ISL_732726, EPI_ISL_732727, EPI_ISL_732728, EPI_ISL_732729, EPI_ISL_732730, EPI_ISL_732731, EPI_ISL_732732, EPI_ISL_732733, EPI_ISL_732734, EPI_ISL_732735, EPI_ISL_732736, EPI_ISL_732737, EPI_ISL_732738, EPI_ISL_732739, EPI_ISL_732740, EPI_ISL_732741, EPI_ISL_732742, EPI_ISL_732743, EPI_ISL_732744, EPI_ISL_732746, EPI_ISL_732747 | | | |
| see above | New Mexico Department of Health Scientific Laboratory | New Mexico Department of Health Scientific Laboratory | Ellie Johnson, Anastacia Griego-Fisher, D'Eldra Malone |
| EPI_ISL_732820, EPI_ISL_732821, EPI_ISL_732822, EPI_ISL_732823, EPI_ISL_732824, EPI_ISL_732825, EPI_ISL_732826 | Nebraska Public Health Laboratory | UNMC COVID-19 Response Team | UNMC COVID-19 Response Team |
| EPI_ISL_734170, EPI_ISL_734171, EPI_ISL_734172, EPI_ISL_734173, EPI_ISL_734174, EPI_ISL_734175, EPI_ISL_734176, EPI_ISL_734177, EPI_ISL_734178, EPI_ISL_734179, EPI_ISL_734180, EPI_ISL_734181, EPI_ISL_734182, EPI_ISL_734183, EPI_ISL_734184, EPI_ISL_734185, EPI_ISL_734186, EPI_ISL_734187, EPI_ISL_734188, EPI_ISL_734189, EPI_ISL_734190, EPI_ISL_734191, EPI_ISL_734192, EPI_ISL_734193, EPI_ISL_734194, EPI_ISL_734195, EPI_ISL_734196, EPI_ISL_734197, EPI_ISL_734198, EPI_ISL_734199, EPI_ISL_734200, EPI_ISL_734201, EPI_ISL_734202, EPI_ISL_734203, EPI_ISL_734204, EPI_ISL_734205, EPI_ISL_734206, EPI_ISL_734207, EPI_ISL_734208, EPI_ISL_734209, EPI_ISL_734210, EPI_ISL_734211, EPI_ISL_734212, EPI_ISL_734213, EPI_ISL_734214, EPI_ISL_734215, EPI_ISL_734216, EPI_ISL_734217, EPI_ISL_734218, EPI_ISL_734219, EPI_ISL_734220, EPI_ISL_734221, EPI_ISL_734222, EPI_ISL_734223, EPI_ISL_734224, EPI_ISL_734225, EPI_ISL_734226, EPI_ISL_734227, EPI_ISL_734228, EPI_ISL_734229, EPI_ISL_734230, EPI_ISL_734231, EPI_ISL_734232, EPI_ISL_734233, EPI_ISL_734234, EPI_ISL_734235, EPI_ISL_734236, EPI_ISL_734237, EPI_ISL_734238, EPI_ISL_734239, EPI_ISL_734240, EPI_ISL_734241, EPI_ISL_734242, EPI_ISL_734243, EPI_ISL_734244, EPI_ISL_734245, EPI_ISL_734246, EPI_ISL_734247, EPI_ISL_734248, EPI_ISL_734249, EPI_ISL_734250, EPI_ISL_734251, EPI_ISL_734252, EPI_ISL_734253, EPI_ISL_734254, EPI_ISL_734255, EPI_ISL_734256, EPI_ISL_734257, EPI_ISL_734258, EPI_ISL_734259, EPI_ISL_734260, EPI_ISL_734261, EPI_ISL_734262, EPI_ISL_734263, EPI_ISL_734264, EPI_ISL_734265, EPI_ISL_734266, EPI_ISL_734267, EPI_ISL_734268, EPI_ISL_734269, EPI_ISL_734270, EPI_ISL_734271, EPI_ISL_734272, EPI_ISL_734273, EPI_ISL_734274, EPI_ISL_734275, EPI_ISL_734276, EPI_ISL_734277, EPI_ISL_734278, EPI_ISL_734279, EPI_ISL_734280, EPI_ISL_734281, EPI_ISL_734282, EPI_ISL_734283, EPI_ISL_734284, EPI_ISL_734285, EPI_ISL_734286 | | | |
| see above | Virginia Division of Consolidated Laboratory Services (DCLS) | Virginia Division of Consolidated Laboratory Services (DCLS) | Virginia DCLS |
| EPI_ISL_734287, EPI_ISL_734288, EPI_ISL_734289, EPI_ISL_734290, EPI_ISL_734291, EPI_ISL_734292, EPI_ISL_734293, EPI_ISL_734294, EPI_ISL_734295, EPI_ISL_734296, EPI_ISL_734297, EPI_ISL_734298, EPI_ISL_734299, EPI_ISL_734300, EPI_ISL_734301, EPI_ISL_734302, EPI_ISL_734303, EPI_ISL_734304, EPI_ISL_734305, EPI_ISL_734306, EPI_ISL_734307, EPI_ISL_734308, EPI_ISL_734309, EPI_ISL_734310, EPI_ISL_734311, EPI_ISL_734312, EPI_ISL_734313, EPI_ISL_734314, EPI_ISL_734315, EPI_ISL_734316, EPI_ISL_734317, EPI_ISL_734318, EPI_ISL_734319, EPI_ISL_734320, EPI_ISL_734321, EPI_ISL_734322, EPI_ISL_734323, EPI_ISL_734324, EPI_ISL_734325, EPI_ISL_734326, EPI_ISL_734327, EPI_ISL_734328, EPI_ISL_734329, EPI_ISL_734330, EPI_ISL_734331, EPI_ISL_734332, EPI_ISL_734333, EPI_ISL_734334, EPI_ISL_734335, EPI_ISL_734336, EPI_ISL_734337, EPI_ISL_734338, EPI_ISL_734339, EPI_ISL_734340, EPI_ISL_734341, EPI_ISL_734342, EPI_ISL_734343, EPI_ISL_734344, EPI_ISL_734345, EPI_ISL_734346, EPI_ISL_734347, EPI_ISL_734348, EPI_ISL_734349, EPI_ISL_734350, EPI_ISL_734351, EPI_ISL_734352, EPI_ISL_734353, EPI_ISL_734354, EPI_ISL_734355, EPI_ISL_734356, EPI_ISL_734357, EPI_ISL_734358, EPI_ISL_734359, EPI_ISL_734360, EPI_ISL_734361, EPI_ISL_734362, EPI_ISL_734363, EPI_ISL_734364, EPI_ISL_734365, EPI_ISL_734366, EPI_ISL_734367, EPI_ISL_734368, EPI_ISL_734369, EPI_ISL_734370, EPI_ISL_734371, EPI_ISL_734372, EPI_ISL_734373, EPI_ISL_734374, EPI_ISL_734375, EPI_ISL_734376, EPI_ISL_734377, EPI_ISL_734378, EPI_ISL_734379, EPI_ISL_734380, EPI_ISL_734381, EPI_ISL_734382, EPI_ISL_734383, EPI_ISL_734384, EPI_ISL_734385, EPI_ISL_734386, EPI_ISL_734387, EPI_ISL_734388, EPI_ISL_734389, EPI_ISL_734390, EPI_ISL_734391, EPI_ISL_734392, EPI_ISL_734393, EPI_ISL_734394, EPI_ISL_734395, EPI_ISL_734396, EPI_ISL_734397, EPI_ISL_734398, EPI_ISL_734399, EPI_ISL_734400, EPI_ISL_734401, EPI_ISL_734402, EPI_ISL_734403, EPI_ISL_734404, EPI_ISL_734405, EPI_ISL_734406, EPI_ISL_734407, EPI_ISL_734408, EPI_ISL_734409, EPI_ISL_734410, EPI_ISL_734411, EPI_ISL_734412, EPI_ISL_734413, EPI_ISL_734414, EPI_ISL_734415, EPI_ISL_734416, EPI_ISL_734417, EPI_ISL_734418, EPI_ISL_734419, EPI_ISL_734420, EPI_ISL_734421, EPI_ISL_734422, EPI_ISL_734423, EPI_ISL_734424, EPI_ISL_734425, EPI_ISL_734426, EPI_ISL_734427, EPI_ISL_734428, EPI_ISL_734429, EPI_ISL_734430, EPI_ISL_734431, EPI_ISL_734432, EPI_ISL_734433, EPI_ISL_734434, EPI_ISL_734435, EPI_ISL_734436, EPI_ISL_734437, EPI_ISL_734438, EPI_ISL_734439, EPI_ISL_734440, EPI_ISL_734441, EPI_ISL_734442, EPI_ISL_734443, EPI_ISL_734444, EPI_ISL_734445, EPI_ISL_734446 | | | |
| see above | Wadsworth Center, New York State Department.of Health | Wadsworth Center, New York State Department.of Health | Kirsten St. George, Daryl M. Lamson, Alexis Russel, Jonathan Plitnick, Navjot Singh, John Kelly, Sara Griesemer, Erasmus Schneider, Erica Lasek-Nesselquist |
| EPI_ISL_734447, EPI_ISL_734448, EPI_ISL_734449, EPI_ISL_734450, EPI_ISL_734451, EPI_ISL_734452, EPI_ISL_734453, EPI_ISL_734454, EPI_ISL_734455, EPI_ISL_734456, EPI_ISL_734457, EPI_ISL_734458, EPI_ISL_734459, EPI_ISL_734460, EPI_ISL_734461, EPI_ISL_734462, EPI_ISL_734463, EPI_ISL_734464, EPI_ISL_734465, EPI_ISL_734466, EPI_ISL_734467, EPI_ISL_734468, EPI_ISL_734469, EPI_ISL_734470, EPI_ISL_734471, EPI_ISL_734472, EPI_ISL_734473, EPI_ISL_734474, EPI_ISL_734475, EPI_ISL_734476, EPI_ISL_734477, EPI_ISL_734478, EPI_ISL_734479, EPI_ISL_734480, EPI_ISL_734481 | | | |
| see above | Masonic Medical Research Institute | Wadsworth Center, New York State Department.of Health | Kirsten St. George, Nathan Tucker, Ryan D. Pfeiffer, Daryl M. Lamson, Alexis Russel, Jonathan Plitnick, Navjot Singh, John Kelly, Sara Griesemer, Erasmus Schneider, Erica Lasek-Nesselquist |
| EPI_ISL_735374, EPI_ISL_735375, EPI_ISL_735376, EPI_ISL_735377, EPI_ISL_735378, EPI_ISL_735379, EPI_ISL_735380, EPI_ISL_735381, EPI_ISL_735382, EPI_ISL_735383 | Los Angeles County Public Health Laboratories | Los Angeles County Public Health Laboratories | P. Hemarajata et al. |
| EPI_ISL_735449, EPI_ISL_735450, EPI_ISL_735451, EPI_ISL_735452, EPI_ISL_735453, EPI_ISL_735454, EPI_ISL_735457, EPI_ISL_735458, EPI_ISL_735459, EPI_ISL_735461, EPI_ISL_735462, EPI_ISL_735463, EPI_ISL_735464, EPI_ISL_735465, EPI_ISL_735466, EPI_ISL_735467, EPI_ISL_735468, EPI_ISL_735469, EPI_ISL_735470, EPI_ISL_735471, EPI_ISL_735472, EPI_ISL_735473, EPI_ISL_735474, EPI_ISL_735475, EPI_ISL_735476, EPI_ISL_735477, EPI_ISL_735478, EPI_ISL_735479, EPI_ISL_735482, EPI_ISL_735484, EPI_ISL_735485, EPI_ISL_735486, EPI_ISL_735487, EPI_ISL_735488, EPI_ISL_735489, EPI_ISL_736775, EPI_ISL_736776, EPI_ISL_737081, EPI_ISL_737082, EPI_ISL_737083, EPI_ISL_737084, EPI_ISL_737085, EPI_ISL_737086, EPI_ISL_737087, EPI_ISL_737088, EPI_ISL_737089, EPI_ISL_737090, EPI_ISL_737091, EPI_ISL_737092, EPI_ISL_737093, EPI_ISL_737094, EPI_ISL_737095, EPI_ISL_737096, EPI_ISL_737097, EPI_ISL_737098, EPI_ISL_737099 | | | |
| see above | UW Virology Lab | UW Virology Lab | Pavitra Roychoudhury, Hong Xie, Lasata Shrestha, Meei-Li Huang, Keith R Jerome, Alexander Greninger |
| EPI_ISL_737100, EPI_ISL_737101, EPI_ISL_737102, EPI_ISL_737103, EPI_ISL_737104, EPI_ISL_737105, EPI_ISL_737106, EPI_ISL_737107, EPI_ISL_737108, EPI_ISL_737109, EPI_ISL_737110, EPI_ISL_737111, EPI_ISL_737112, EPI_ISL_737113, EPI_ISL_737114, EPI_ISL_737115, EPI_ISL_737116, EPI_ISL_737117, EPI_ISL_737118, EPI_ISL_737119, EPI_ISL_737120, EPI_ISL_737121, EPI_ISL_737122, EPI_ISL_737123, EPI_ISL_737124, EPI_ISL_737125, EPI_ISL_737126, EPI_ISL_737127, EPI_ISL_737128, EPI_ISL_737129, EPI_ISL_737130, EPI_ISL_737131, EPI_ISL_737132, EPI_ISL_737133, EPI_ISL_737134, EPI_ISL_737135, EPI_ISL_737136, EPI_ISL_737137, EPI_ISL_737138, EPI_ISL_737139, EPI_ISL_737140, EPI_ISL_737141, EPI_ISL_737142, EPI_ISL_737143, EPI_ISL_737144, EPI_ISL_737145, EPI_ISL_737146, EPI_ISL_737147, EPI_ISL_737148, EPI_ISL_737153, EPI_ISL_737154, EPI_ISL_737155, EPI_ISL_737156, EPI_ISL_737157, EPI_ISL_737158, EPI_ISL_737159, EPI_ISL_737160, EPI_ISL_737161, EPI_ISL_737162, EPI_ISL_737163, EPI_ISL_737164, EPI_ISL_737165, EPI_ISL_737166, EPI_ISL_737167, EPI_ISL_737168, EPI_ISL_737169, EPI_ISL_737170, EPI_ISL_737171, EPI_ISL_737172, EPI_ISL_737173, EPI_ISL_737174, EPI_ISL_737175, EPI_ISL_737176, EPI_ISL_737177, EPI_ISL_737178, EPI_ISL_737179, EPI_ISL_737180, EPI_ISL_737181, EPI_ISL_737182, EPI_ISL_737183, EPI_ISL_737184, EPI_ISL_737185, EPI_ISL_737186, EPI_ISL_737187, EPI_ISL_737188, EPI_ISL_737189, EPI_ISL_737190, EPI_ISL_737191, EPI_ISL_737192, EPI_ISL_737193, EPI_ISL_737194, EPI_ISL_737195, EPI_ISL_737196, EPI_ISL_737197, EPI_ISL_737198 | | | |
| see above | Michigan Department of Health and Human Services, Bureau of Laboratories | Michigan Department of Health and Human Services, Bureau of Laboratories | Blankenship HM, Riner D, Soehnlen MK |
| EPI_ISL_737236, EPI_ISL_737237, EPI_ISL_737238, EPI_ISL_737239, EPI_ISL_737240, EPI_ISL_737241, EPI_ISL_737242, EPI_ISL_737243, EPI_ISL_737244, EPI_ISL_737245, EPI_ISL_737246, EPI_ISL_737247, EPI_ISL_737248, EPI_ISL_737249, EPI_ISL_737250, EPI_ISL_737251, EPI_ISL_737252, EPI_ISL_737253, EPI_ISL_737254, EPI_ISL_737255, EPI_ISL_737256, EPI_ISL_737257, EPI_ISL_737258, EPI_ISL_737259, EPI_ISL_737260, EPI_ISL_737261 | | | |
| see above | Los Angeles County Public Health Laboratories | Los Angeles County Public Health Laboratories | P. Hemarajata et al. |
| EPI_ISL_737268, EPI_ISL_737269, EPI_ISL_737270, EPI_ISL_737271, EPI_ISL_737272, EPI_ISL_737273, EPI_ISL_737274, EPI_ISL_737275, EPI_ISL_737276, EPI_ISL_737277, EPI_ISL_737278, EPI_ISL_737279, EPI_ISL_737280, EPI_ISL_737282, EPI_ISL_737283, EPI_ISL_737284, EPI_ISL_737285, EPI_ISL_737286, EPI_ISL_737287, EPI_ISL_737288, EPI_ISL_737289, EPI_ISL_737290, EPI_ISL_737291, EPI_ISL_737292, EPI_ISL_737293, EPI_ISL_737294, EPI_ISL_737295, EPI_ISL_737296, EPI_ISL_737297, EPI_ISL_737298, EPI_ISL_737299, EPI_ISL_737300, EPI_ISL_737301, EPI_ISL_737302, EPI_ISL_737303, EPI_ISL_737304, EPI_ISL_737305, EPI_ISL_737306, EPI_ISL_737307, EPI_ISL_737308 | | | |
| see above | Michigan Department of Health and Human Services, Bureau of Laboratories | Michigan Department of Health and Human Services, Bureau of Laboratories | Blankenship HM, Riner D, Soehnlen MK |
| EPI_ISL_737309 | UW Virology Lab | UW Virology Lab | Pavitra Roychoudhury, Hong Xie, Lasata Shrestha, Meei-Li Huang, Keith R Jerome, Alexander Greninger |
| EPI_ISL_738080 | Laboratory Services Section, Texas Department of State Health Services | Laboratory Services Section, Texas Department of State Health Services | Pokharel,A., Oh,B., Bonser,J.D., Tuladhar,R., Pedrueza,M., Zhang,J., Rahman,M., Koag,M., Wang,C., Lee,R. and Kubin,G. |
| EPI_ISL_738148, EPI_ISL_738149, EPI_ISL_738150, EPI_ISL_738151, EPI_ISL_738152, EPI_ISL_738153, EPI_ISL_738154, EPI_ISL_738155, EPI_ISL_738156, EPI_ISL_738157, EPI_ISL_738158, EPI_ISL_738159, EPI_ISL_738160, EPI_ISL_738161, EPI_ISL_738162, EPI_ISL_738163, EPI_ISL_738164, EPI_ISL_738165, EPI_ISL_738166, EPI_ISL_738167, EPI_ISL_738168, EPI_ISL_738169, EPI_ISL_738170, EPI_ISL_738171, EPI_ISL_738172, EPI_ISL_738173, EPI_ISL_738174, EPI_ISL_738175, EPI_ISL_738176, EPI_ISL_738177, EPI_ISL_738178, EPI_ISL_738179, EPI_ISL_738180, EPI_ISL_738181, EPI_ISL_738182, EPI_ISL_738183, EPI_ISL_738184, EPI_ISL_738185, EPI_ISL_738186, EPI_ISL_738187, EPI_ISL_738188, EPI_ISL_738189, EPI_ISL_738190, EPI_ISL_738191, EPI_ISL_738192, EPI_ISL_738193 | | | |
| see above | Texas Department of State Health Services | Texas Department of State Health Services | Anita Pokharel, Bonnie Oh, James Daniel Bonser, Rashmi Tuladhar, Mayela Pedrueza, Jenny Zhang, Maliha Rahman, Myong Koag, Chung Wang, Rachel Lee, Grace Kubin |
| EPI_ISL_738244, EPI_ISL_738246, EPI_ISL_738247, EPI_ISL_738248, EPI_ISL_738249, EPI_ISL_738250, EPI_ISL_738252, EPI_ISL_738253, EPI_ISL_738254, EPI_ISL_738255, EPI_ISL_738256, EPI_ISL_738257, EPI_ISL_738258, EPI_ISL_738259, EPI_ISL_738260, EPI_ISL_738261, EPI_ISL_738262, EPI_ISL_738263, EPI_ISL_738264, EPI_ISL_738265, EPI_ISL_738267, EPI_ISL_738268, EPI_ISL_738283, EPI_ISL_738287, EPI_ISL_738288 | | | |
| see above | Connecticut Veterans' Affairs Hospital | Grubaugh Lab - Yale School of Public Health | Joseph Fauver, Tara Alpert, Chantal Vogels, Mary Petrone, Isabel Ott, Ellen Foxman, Shaili Gupta, Danielle Plank, Nathan Grubaugh |
| EPI_ISL_738290, EPI_ISL_738291, EPI_ISL_738292, EPI_ISL_738293, EPI_ISL_738294, EPI_ISL_738295, EPI_ISL_738296, EPI_ISL_738297 | Yale Pathology Lab | Grubaugh Lab - Yale School of Public Health | Joseph Fauver, Tara Alpert, Chantal Vogels, Mary Petrone, Isabel Ott, Ellen Foxman, Shaili Gupta, Danielle Plank, Nathan Grubaugh |
| EPI_ISL_738298, EPI_ISL_738300, EPI_ISL_738302, | Connecticut Veterans' Affairs Hospital | Grubaugh Lab - Yale School of Public Health | Joseph Fauver, Tara Alpert, Chantal Vogels, Mary Petrone, Isabel Ott, Ellen Foxman, Shaili Gupta, Danielle Plank, Nathan Grubaugh |

| | | | |
|---|---|---|---|
| EPI_ISL_738303, EPI_ISL_738305, EPI_ISL_738307, EPI_ISL_738309, EPI_ISL_738310, EPI_ISL_738311, EPI_ISL_738312 | | | |
| EPI_ISL_738316, EPI_ISL_738350, EPI_ISL_738351, EPI_ISL_738352, EPI_ISL_738353, EPI_ISL_738354, EPI_ISL_738355, EPI_ISL_738356, EPI_ISL_738357, EPI_ISL_738358, EPI_ISL_738359 | | | |
| see above | Utah Public Health Laboratory | Utah Public Health Laboratory | Erin Young, Kelly Oakeson, Tara Gallagher |
| EPI_ISL_738500, EPI_ISL_738501, EPI_ISL_738502 | Santa Clara County Public Health Laboratory | Chan-Zuckerberg Biohub | CZB Cliahub Consortium |
| EPI_ISL_738503 | Orange County Public Health Lab | Chan-Zuckerberg Biohub | CZB Cliahub Consortium |
| EPI_ISL_738504, EPI_ISL_738505 | Humboldt County Public Health Laboratory | Chan-Zuckerberg Biohub | CZB Cliahub Consortium |
| EPI_ISL_738506 | Monterey County Public Health Lab | Chan-Zuckerberg Biohub | CZB Cliahub Consortium |
| EPI_ISL_738507 | Orange County Public Health Lab | Chan-Zuckerberg Biohub | CZB Cliahub Consortium |
| EPI_ISL_738508 | Santa Clara County Public Health Laboratory | Chan-Zuckerberg Biohub | CZB Cliahub Consortium |
| EPI_ISL_738509, EPI_ISL_738510 | Alameda County Public Health Lab | Chan-Zuckerberg Biohub | CZB Cliahub Consortium |
| EPI_ISL_738511 | Santa Clara County Public Health Laboratory | Chan-Zuckerberg Biohub | CZB Cliahub Consortium |
| EPI_ISL_738512 | Napa-Solano-Yolo- Marin County (NSYM) Public Health Laboratories | Chan-Zuckerberg Biohub | CZB Cliahub Consortium |
| EPI_ISL_738513, EPI_ISL_738514 | Alameda County Public Health Lab | Chan-Zuckerberg Biohub | CZB Cliahub Consortium |
| EPI_ISL_738515 | Orange County Public Health Lab | Chan-Zuckerberg Biohub | CZB Cliahub Consortium |
| EPI_ISL_738516, EPI_ISL_738517 | Santa Clara County Public Health Laboratory | Chan-Zuckerberg Biohub | CZB Cliahub Consortium |
| EPI_ISL_738518 | Orange County Public Health Lab | Chan-Zuckerberg Biohub | CZB Cliahub Consortium |
| EPI_ISL_738519 | Alameda County Public Health Lab | Chan-Zuckerberg Biohub | CZB Cliahub Consortium |
| EPI_ISL_738520, EPI_ISL_738521 | Santa Clara County Public Health Laboratory | Chan-Zuckerberg Biohub | CZB Cliahub Consortium |
| EPI_ISL_738522, EPI_ISL_738523, EPI_ISL_738524 | Alameda County Public Health Lab | Chan-Zuckerberg Biohub | CZB Cliahub Consortium |
| EPI_ISL_738525, EPI_ISL_738526 | Santa Clara County Public Health Laboratory | Chan-Zuckerberg Biohub | CZB Cliahub Consortium |
| EPI_ISL_738527, EPI_ISL_738528 | Alameda County Public Health Lab | Chan-Zuckerberg Biohub | CZB Cliahub Consortium |
| EPI_ISL_738529 | Orange County Public Health Lab | Chan-Zuckerberg Biohub | CZB Cliahub Consortium |
| EPI_ISL_738530 | Humboldt County Public Health Laboratory | Chan-Zuckerberg Biohub | CZB Cliahub Consortium |
| EPI_ISL_738531 | Alameda County Public Health Lab | Chan-Zuckerberg Biohub | CZB Cliahub Consortium |
| EPI_ISL_738532, EPI_ISL_738533, EPI_ISL_738534 | Santa Clara County Public Health Laboratory | Chan-Zuckerberg Biohub | CZB Cliahub Consortium |
| EPI_ISL_738535 | Alameda County Public Health Lab | Chan-Zuckerberg Biohub | CZB Cliahub Consortium |
| EPI_ISL_738536 | Santa Clara County Public Health Laboratory | Chan-Zuckerberg Biohub | CZB Cliahub Consortium |
| EPI_ISL_738537, EPI_ISL_738538 | Alameda County Public Health Lab | Chan-Zuckerberg Biohub | CZB Cliahub Consortium |
| EPI_ISL_738539 | Humboldt County Public Health Laboratory | Chan-Zuckerberg Biohub | CZB Cliahub Consortium |
| EPI_ISL_738540 | Alameda County Public Health Lab | Chan-Zuckerberg Biohub | CZB Cliahub Consortium |
| EPI_ISL_738541 | Napa-Solano-Yolo- Marin County (NSYM) Public Health Laboratories | Chan-Zuckerberg Biohub | CZB Cliahub Consortium |
| EPI_ISL_738542, EPI_ISL_738543 | Santa Clara County Public Health Laboratory | Chan-Zuckerberg Biohub | CZB Cliahub Consortium |
| EPI_ISL_738544 | UCSF Clinical Microbiology Laboratory | Chan-Zuckerberg Biohub | CZB Cliahub Consortium |
| EPI_ISL_738545 | Alameda County Public Health Lab | Chan-Zuckerberg Biohub | CZB Cliahub Consortium |
| EPI_ISL_738546 | Santa Clara County Public Health Laboratory | Chan-Zuckerberg Biohub | CZB Cliahub Consortium |
| EPI_ISL_738547 | Alameda County Public Health Lab | Chan-Zuckerberg Biohub | CZB Cliahub Consortium |
| EPI_ISL_738548 | Orange County Public Health Lab | Chan-Zuckerberg Biohub | CZB Cliahub Consortium |
| EPI_ISL_738549, EPI_ISL_738550, EPI_ISL_738551, EPI_ISL_738552 | Santa Clara County Public Health Laboratory | Chan-Zuckerberg Biohub | CZB Cliahub Consortium |
| EPI_ISL_738553, EPI_ISL_738554 | Alameda County Public Health Lab | Chan-Zuckerberg Biohub | CZB Cliahub Consortium |
| EPI_ISL_738555, EPI_ISL_738556, EPI_ISL_738557, EPI_ISL_738558 | Santa Clara County Public Health Laboratory | Chan-Zuckerberg Biohub | CZB Cliahub Consortium |
| EPI_ISL_738559 | Orange County Public Health Lab | Chan-Zuckerberg Biohub | CZB Cliahub Consortium |
| EPI_ISL_738560 | Santa Clara County Public Health Laboratory | Chan-Zuckerberg Biohub | CZB Cliahub Consortium |
| EPI_ISL_738561, EPI_ISL_738562 | Alameda County Public Health Lab | Chan-Zuckerberg Biohub | CZB Cliahub Consortium |
| EPI_ISL_738563 | Humboldt County Public Health Laboratory | Chan-Zuckerberg Biohub | CZB Cliahub Consortium |
| EPI_ISL_738564, EPI_ISL_738565 | Alameda County Public Health Lab | Chan-Zuckerberg Biohub | CZB Cliahub Consortium |
| EPI_ISL_738566 | Santa Clara County Public Health Laboratory | Chan-Zuckerberg Biohub | CZB Cliahub Consortium |
| EPI_ISL_738567 | Renegade | Chan-Zuckerberg Biohub | CZB Cliahub Consortium |
| EPI_ISL_738568 | Santa Clara County Public Health | Chan-Zuckerberg Biohub | CZB Cliahub Consortium |

| | | | |
|---|---|---|---|
| | Laboratory | | |
| EPI_ISL_738569, EPI_ISL_738570, EPI_ISL_738571 | Orange County Public Health Lab | Chan-Zuckerberg Biohub | CZB Cliahub Consortium |
| EPI_ISL_738572, EPI_ISL_738573 | Alameda County Public Health Lab | Chan-Zuckerberg Biohub | CZB Cliahub Consortium |
| EPI_ISL_738574, EPI_ISL_738575, EPI_ISL_738576 | Renegade | Chan-Zuckerberg Biohub | CZB Cliahub Consortium |
| EPI_ISL_738577 | Alameda County Public Health Lab | Chan-Zuckerberg Biohub | CZB Cliahub Consortium |
| EPI_ISL_738578, EPI_ISL_738579 | Santa Clara County Public Health Laboratory | Chan-Zuckerberg Biohub | CZB Cliahub Consortium |
| EPI_ISL_738580 | Alameda County Public Health Lab | Chan-Zuckerberg Biohub | CZB Cliahub Consortium |
| EPI_ISL_738581, EPI_ISL_738582, EPI_ISL_738583 | Santa Clara County Public Health Laboratory | Chan-Zuckerberg Biohub | CZB Cliahub Consortium |
| EPI_ISL_738584, EPI_ISL_738585 | Alameda County Public Health Lab | Chan-Zuckerberg Biohub | CZB Cliahub Consortium |
| EPI_ISL_738586 | Santa Clara County Public Health Laboratory | Chan-Zuckerberg Biohub | CZB Cliahub Consortium |
| EPI_ISL_738587 | Alameda County Public Health Lab | Chan-Zuckerberg Biohub | CZB Cliahub Consortium |
| EPI_ISL_738588 | Orange County Public Health Lab | Chan-Zuckerberg Biohub | CZB Cliahub Consortium |
| EPI_ISL_738589, EPI_ISL_738590 | Santa Clara County Public Health Laboratory | Chan-Zuckerberg Biohub | CZB Cliahub Consortium |
| EPI_ISL_738591 | Renegade | Chan-Zuckerberg Biohub | CZB Cliahub Consortium |
| EPI_ISL_738592 | Alameda County Public Health Lab | Chan-Zuckerberg Biohub | CZB Cliahub Consortium |
| EPI_ISL_738593 | Santa Clara County Public Health Laboratory | Chan-Zuckerberg Biohub | CZB Cliahub Consortium |
| EPI_ISL_738594 | Orange County Public Health Lab | Chan-Zuckerberg Biohub | CZB Cliahub Consortium |
| EPI_ISL_738595, EPI_ISL_738596, EPI_ISL_738597, EPI_ISL_738598 | Santa Clara County Public Health Laboratory | Chan-Zuckerberg Biohub | CZB Cliahub Consortium |
| EPI_ISL_738599 | Orange County Public Health Lab | Chan-Zuckerberg Biohub | CZB Cliahub Consortium |
| EPI_ISL_738600 | Humboldt County Public Health Laboratory | Chan-Zuckerberg Biohub | CZB Cliahub Consortium |
| EPI_ISL_738601 | Santa Clara County Public Health Laboratory | Chan-Zuckerberg Biohub | CZB Cliahub Consortium |
| EPI_ISL_738602, EPI_ISL_738603 | Orange County Public Health Lab | Chan-Zuckerberg Biohub | CZB Cliahub Consortium |
| EPI_ISL_738604, EPI_ISL_738605, EPI_ISL_738606, EPI_ISL_738607 | Alameda County Public Health Lab | Chan-Zuckerberg Biohub | CZB Cliahub Consortium |
| EPI_ISL_738608 | Santa Clara County Public Health Laboratory | Chan-Zuckerberg Biohub | CZB Cliahub Consortium |
| EPI_ISL_738609 | Alameda County Public Health Lab | Chan-Zuckerberg Biohub | CZB Cliahub Consortium |
| EPI_ISL_738610, EPI_ISL_738611, EPI_ISL_738612 | Santa Clara County Public Health Laboratory | Chan-Zuckerberg Biohub | CZB Cliahub Consortium |
| EPI_ISL_738613, EPI_ISL_738614, EPI_ISL_738615, EPI_ISL_738616 | Alameda County Public Health Lab | Chan-Zuckerberg Biohub | CZB Cliahub Consortium |
| EPI_ISL_738617, EPI_ISL_738618 | Santa Clara County Public Health Laboratory | Chan-Zuckerberg Biohub | CZB Cliahub Consortium |
| EPI_ISL_738619 | Alameda County Public Health Lab | Chan-Zuckerberg Biohub | CZB Cliahub Consortium |
| EPI_ISL_738620, EPI_ISL_738621 | Orange County Public Health Lab | Chan-Zuckerberg Biohub | CZB Cliahub Consortium |
| EPI_ISL_738622, EPI_ISL_738623 | Alameda County Public Health Lab | Chan-Zuckerberg Biohub | CZB Cliahub Consortium |
| EPI_ISL_738624 | Orange County Public Health Lab | Chan-Zuckerberg Biohub | CZB Cliahub Consortium |
| EPI_ISL_738625, EPI_ISL_738626, EPI_ISL_738627 | Alameda County Public Health Lab | Chan-Zuckerberg Biohub | CZB Cliahub Consortium |
| EPI_ISL_738628 | Santa Clara County Public Health Laboratory | Chan-Zuckerberg Biohub | CZB Cliahub Consortium |
| EPI_ISL_738629 | Alameda County Public Health Lab | Chan-Zuckerberg Biohub | CZB Cliahub Consortium |
| EPI_ISL_738630 | Orange County Public Health Lab | Chan-Zuckerberg Biohub | CZB Cliahub Consortium |
| EPI_ISL_738631 | Santa Clara County Public Health Laboratory | Chan-Zuckerberg Biohub | CZB Cliahub Consortium |
| EPI_ISL_738632 | Orange County Public Health Lab | Chan-Zuckerberg Biohub | CZB Cliahub Consortium |
| EPI_ISL_738633 | Alameda County Public Health Lab | Chan-Zuckerberg Biohub | CZB Cliahub Consortium |
| EPI_ISL_738634, EPI_ISL_738635, EPI_ISL_738636, EPI_ISL_738637, EPI_ISL_738638, EPI_ISL_738639, EPI_ISL_738640, EPI_ISL_738641 | Orange County Public Health Lab | Chan-Zuckerberg Biohub | CZB Cliahub Consortium |
| EPI_ISL_738642 | Alameda County Public Health Lab | Chan-Zuckerberg Biohub | CZB Cliahub Consortium |
| EPI_ISL_738643, EPI_ISL_738644, EPI_ISL_738645 | Santa Clara County Public Health Laboratory | Chan-Zuckerberg Biohub | CZB Cliahub Consortium |
| EPI_ISL_738646 | Orange County Public Health Lab | Chan-Zuckerberg Biohub | CZB Cliahub Consortium |
| EPI_ISL_738647 | Humboldt County Public Health Laboratory | Chan-Zuckerberg Biohub | CZB Cliahub Consortium |
| EPI_ISL_738648 | Alameda County Public Health Lab | Chan-Zuckerberg Biohub | CZB Cliahub Consortium |
| EPI_ISL_738649 | Humboldt County Public Health | Chan-Zuckerberg Biohub | CZB Cliahub Consortium |

| | | | |
|---|---|---|---|
| | Laboratory | | |
| EPI_ISL_738650 | Santa Clara County Public Health Laboratory | Chan-Zuckerberg Biohub | CZB Cliahub Consortium |
| EPI_ISL_738651 | Humboldt County Public Health Laboratory | Chan-Zuckerberg Biohub | CZB Cliahub Consortium |
| EPI_ISL_738652, EPI_ISL_738653 | Santa Clara County Public Health Laboratory | Chan-Zuckerberg Biohub | CZB Cliahub Consortium |
| EPI_ISL_738654 | Alameda County Public Health Lab | Chan-Zuckerberg Biohub | CZB Cliahub Consortium |
| EPI_ISL_738655 | Orange County Public Health Lab | Chan-Zuckerberg Biohub | CZB Cliahub Consortium |
| EPI_ISL_738656 | Alameda County Public Health Lab | Chan-Zuckerberg Biohub | CZB Cliahub Consortium |
| EPI_ISL_738657 | Santa Clara County Public Health Laboratory | Chan-Zuckerberg Biohub | CZB Cliahub Consortium |
| EPI_ISL_738658 | Monterey County Public Health Lab | Chan-Zuckerberg Biohub | CZB Cliahub Consortium |
| EPI_ISL_738659 | Alameda County Public Health Lab | Chan-Zuckerberg Biohub | CZB Cliahub Consortium |
| EPI_ISL_738660 | Renegade | Chan-Zuckerberg Biohub | CZB Cliahub Consortium |
| EPI_ISL_738661 | Santa Clara County Public Health Laboratory | Chan-Zuckerberg Biohub | CZB Cliahub Consortium |
| EPI_ISL_738662 | Renegade | Chan-Zuckerberg Biohub | CZB Cliahub Consortium |
| EPI_ISL_738663 | Santa Clara County Public Health Laboratory | Chan-Zuckerberg Biohub | CZB Cliahub Consortium |
| EPI_ISL_738664 | Orange County Public Health Lab | Chan-Zuckerberg Biohub | CZB Cliahub Consortium |
| EPI_ISL_738665 | Alameda County Public Health Lab | Chan-Zuckerberg Biohub | CZB Cliahub Consortium |
| EPI_ISL_738666 | Madera County Department of Public Health | Chan-Zuckerberg Biohub | CZB Cliahub Consortium |
| EPI_ISL_738667 | Santa Clara County Public Health Laboratory | Chan-Zuckerberg Biohub | CZB Cliahub Consortium |
| EPI_ISL_738668 | Alameda County Public Health Lab | Chan-Zuckerberg Biohub | CZB Cliahub Consortium |
| EPI_ISL_738669 | Orange County Public Health Lab | Chan-Zuckerberg Biohub | CZB Cliahub Consortium |
| EPI_ISL_738670 | Santa Clara County Public Health Laboratory | Chan-Zuckerberg Biohub | CZB Cliahub Consortium |
| EPI_ISL_738671 | Alameda County Public Health Lab | Chan-Zuckerberg Biohub | CZB Cliahub Consortium |
| EPI_ISL_738672, EPI_ISL_738673 | Santa Clara County Public Health Laboratory | Chan-Zuckerberg Biohub | CZB Cliahub Consortium |
| EPI_ISL_738674, EPI_ISL_738675, EPI_ISL_738676, EPI_ISL_738677, EPI_ISL_738678 | Alameda County Public Health Lab | Chan-Zuckerberg Biohub | CZB Cliahub Consortium |
| EPI_ISL_738707, EPI_ISL_738708 | Madera County Department of Public Health | Chan-Zuckerberg Biohub | CZB Cliahub Consortium |
| EPI_ISL_738709 | Orange County Public Health Lab | Chan-Zuckerberg Biohub | CZB Cliahub Consortium |
| EPI_ISL_738710 | Santa Clara County Public Health Laboratory | Chan-Zuckerberg Biohub | CZB Cliahub Consortium |
| EPI_ISL_738711 | Alameda County Public Health Lab | Chan-Zuckerberg Biohub | CZB Cliahub Consortium |
| EPI_ISL_738712, EPI_ISL_738713, EPI_ISL_738714 | Renegade | Chan-Zuckerberg Biohub | CZB Cliahub Consortium |
| EPI_ISL_738715 | Alameda County Public Health Lab | Chan-Zuckerberg Biohub | CZB Cliahub Consortium |
| EPI_ISL_738716, EPI_ISL_738717, EPI_ISL_738718, EPI_ISL_738719, EPI_ISL_738720 | Santa Clara County Public Health Laboratory | Chan-Zuckerberg Biohub | CZB Cliahub Consortium |
| EPI_ISL_738721 | Alameda County Public Health Lab | Chan-Zuckerberg Biohub | CZB Cliahub Consortium |
| EPI_ISL_738722 | Orange County Public Health Lab | Chan-Zuckerberg Biohub | CZB Cliahub Consortium |
| EPI_ISL_738723 | Renegade | Chan-Zuckerberg Biohub | CZB Cliahub Consortium |
| EPI_ISL_738724 | Alameda County Public Health Lab | Chan-Zuckerberg Biohub | CZB Cliahub Consortium |
| EPI_ISL_738725 | Renegade | Chan-Zuckerberg Biohub | CZB Cliahub Consortium |
| EPI_ISL_738726 | Santa Clara County Public Health Laboratory | Chan-Zuckerberg Biohub | CZB Cliahub Consortium |
| EPI_ISL_738727 | Renegade | Chan-Zuckerberg Biohub | CZB Cliahub Consortium |
| EPI_ISL_738728 | Santa Clara County Public Health Laboratory | Chan-Zuckerberg Biohub | CZB Cliahub Consortium |
| EPI_ISL_738729 | Orange County Public Health Lab | Chan-Zuckerberg Biohub | CZB Cliahub Consortium |
| EPI_ISL_738730 | Santa Clara County Public Health Laboratory | Chan-Zuckerberg Biohub | CZB Cliahub Consortium |
| EPI_ISL_738731, EPI_ISL_738732, EPI_ISL_738733, EPI_ISL_738734, EPI_ISL_738735, EPI_ISL_738736, EPI_ISL_738737, EPI_ISL_738738, EPI_ISL_738739, EPI_ISL_738740, EPI_ISL_738741 | | | |
| see above | Orange County Public Health Lab | Chan-Zuckerberg Biohub | CZB Cliahub Consortium |
| EPI_ISL_738742 | Humboldt County Public Health Laboratory | Chan-Zuckerberg Biohub | CZB Cliahub Consortium |
| EPI_ISL_738743, EPI_ISL_738744, EPI_ISL_738745 | Alameda County Public Health Lab | Chan-Zuckerberg Biohub | CZB Cliahub Consortium |
| EPI_ISL_738746, EPI_ISL_738747 | Santa Clara County Public Health Laboratory | Chan-Zuckerberg Biohub | CZB Cliahub Consortium |
| EPI_ISL_738748, EPI_ISL_738749 | Alameda County Public Health Lab | Chan-Zuckerberg Biohub | CZB Cliahub Consortium |
| EPI_ISL_738750 | Santa Clara County Public Health Laboratory | Chan-Zuckerberg Biohub | CZB Cliahub Consortium |
| EPI_ISL_738751 | Alameda County Public Health Lab | Chan-Zuckerberg Biohub | CZB Cliahub Consortium |
| EPI_ISL_738752 | Orange County Public Health Lab | Chan-Zuckerberg Biohub | CZB Cliahub Consortium |
| EPI_ISL_738753 | Alameda County Public Health Lab | Chan-Zuckerberg Biohub | CZB Cliahub Consortium |
| EPI_ISL_738754 | Napa-Solano-Yolo- Marin County (NSYM) Public Health Laboratories | Chan-Zuckerberg Biohub | CZB Cliahub Consortium |
| EPI_ISL_738755 | Orange County Public Health Lab | Chan-Zuckerberg Biohub | CZB Cliahub Consortium |
| EPI_ISL_738756 | Humboldt County Public Health Laboratory | Chan-Zuckerberg Biohub | CZB Cliahub Consortium |

| | | | |
|---|---|---|---|
| EPI_ISL_738757, EPI_ISL_738758 | Orange County Public Health Lab | Chan-Zuckerberg Biohub | CZB Cliahub Consortium |
| EPI_ISL_738759 | Humboldt County Public Health Laboratory | Chan-Zuckerberg Biohub | CZB Cliahub Consortium |
| EPI_ISL_738760 | Alameda County Public Health Lab | Chan-Zuckerberg Biohub | CZB Cliahub Consortium |
| EPI_ISL_738761, EPI_ISL_738762 | Orange County Public Health Lab | Chan-Zuckerberg Biohub | CZB Cliahub Consortium |
| EPI_ISL_738763, EPI_ISL_738764, EPI_ISL_738765 | Santa Clara County Public Health Laboratory | Chan-Zuckerberg Biohub | CZB Cliahub Consortium |
| EPI_ISL_738766, EPI_ISL_738767, EPI_ISL_738768 | Alameda County Public Health Lab | Chan-Zuckerberg Biohub | CZB Cliahub Consortium |
| EPI_ISL_738769 | Santa Clara County Public Health Laboratory | Chan-Zuckerberg Biohub | CZB Cliahub Consortium |
| EPI_ISL_738770 | Renegade | Chan-Zuckerberg Biohub | CZB Cliahub Consortium |
| EPI_ISL_738771, EPI_ISL_738772 | Santa Clara County Public Health Laboratory | Chan-Zuckerberg Biohub | CZB Cliahub Consortium |
| EPI_ISL_738773, EPI_ISL_738774, EPI_ISL_738775, EPI_ISL_738776 | Alameda County Public Health Lab | Chan-Zuckerberg Biohub | CZB Cliahub Consortium |
| EPI_ISL_738777 | Napa-Solano-Yolo- Marin County (NSYM) Public Health Laboratories | Chan-Zuckerberg Biohub | CZB Cliahub Consortium |
| EPI_ISL_738778, EPI_ISL_738779 | Humboldt County Public Health Laboratory | Chan-Zuckerberg Biohub | CZB Cliahub Consortium |
| EPI_ISL_738780 | Santa Clara County Public Health Laboratory | Chan-Zuckerberg Biohub | CZB Cliahub Consortium |
| EPI_ISL_738781 | Alameda County Public Health Lab | Chan-Zuckerberg Biohub | CZB Cliahub Consortium |
| EPI_ISL_738782 | Orange County Public Health Lab | Chan-Zuckerberg Biohub | CZB Cliahub Consortium |
| EPI_ISL_738783 | Alameda County Public Health Lab | Chan-Zuckerberg Biohub | CZB Cliahub Consortium |
| EPI_ISL_738784 | Orange County Public Health Lab | Chan-Zuckerberg Biohub | CZB Cliahub Consortium |
| EPI_ISL_738785, EPI_ISL_738786 | Humboldt County Public Health Laboratory | Chan-Zuckerberg Biohub | CZB Cliahub Consortium |
| EPI_ISL_738787 | Orange County Public Health Lab | Chan-Zuckerberg Biohub | CZB Cliahub Consortium |
| EPI_ISL_738788 | Alameda County Public Health Lab | Chan-Zuckerberg Biohub | CZB Cliahub Consortium |
| EPI_ISL_738789 | Santa Clara County Public Health Laboratory | Chan-Zuckerberg Biohub | CZB Cliahub Consortium |
| EPI_ISL_738790 | Orange County Public Health Lab | Chan-Zuckerberg Biohub | CZB Cliahub Consortium |
| EPI_ISL_738791 | Santa Clara County Public Health Laboratory | Chan-Zuckerberg Biohub | CZB Cliahub Consortium |
| EPI_ISL_738792, EPI_ISL_738793 | Alameda County Public Health Lab | Chan-Zuckerberg Biohub | CZB Cliahub Consortium |
| EPI_ISL_738794 | Santa Clara County Public Health Laboratory | Chan-Zuckerberg Biohub | CZB Cliahub Consortium |
| EPI_ISL_738795 | Humboldt County Public Health Laboratory | Chan-Zuckerberg Biohub | CZB Cliahub Consortium |
| EPI_ISL_738796, EPI_ISL_738797 | Alameda County Public Health Lab | Chan-Zuckerberg Biohub | CZB Cliahub Consortium |
| EPI_ISL_738798 | Santa Clara County Public Health Laboratory | Chan-Zuckerberg Biohub | CZB Cliahub Consortium |
| EPI_ISL_738799 | Orange County Public Health Lab | Chan-Zuckerberg Biohub | CZB Cliahub Consortium |
| EPI_ISL_738800 | Alameda County Public Health Lab | Chan-Zuckerberg Biohub | CZB Cliahub Consortium |
| EPI_ISL_738801 | Humboldt County Public Health Laboratory | Chan-Zuckerberg Biohub | CZB Cliahub Consortium |
| EPI_ISL_738802 | Alameda County Public Health Lab | Chan-Zuckerberg Biohub | CZB Cliahub Consortium |
| EPI_ISL_738803 | Santa Clara County Public Health Laboratory | Chan-Zuckerberg Biohub | CZB Cliahub Consortium |
| EPI_ISL_738804 | Alameda County Public Health Lab | Chan-Zuckerberg Biohub | CZB Cliahub Consortium |
| EPI_ISL_738805 | Santa Clara County Public Health Laboratory | Chan-Zuckerberg Biohub | CZB Cliahub Consortium |
| EPI_ISL_738806, EPI_ISL_738807 | Humboldt County Public Health Laboratory | Chan-Zuckerberg Biohub | CZB Cliahub Consortium |
| EPI_ISL_738808 | Orange County Public Health Lab | Chan-Zuckerberg Biohub | CZB Cliahub Consortium |
| EPI_ISL_738809, EPI_ISL_738810 | Humboldt County Public Health Laboratory | Chan-Zuckerberg Biohub | CZB Cliahub Consortium |
| EPI_ISL_738811 | Orange County Public Health Lab | Chan-Zuckerberg Biohub | CZB Cliahub Consortium |
| EPI_ISL_738812 | Santa Clara County Public Health Laboratory | Chan-Zuckerberg Biohub | CZB Cliahub Consortium |
| EPI_ISL_738813, EPI_ISL_738814 | Alameda County Public Health Lab | Chan-Zuckerberg Biohub | CZB Cliahub Consortium |
| EPI_ISL_738815, EPI_ISL_738816 | Orange County Public Health Lab | Chan-Zuckerberg Biohub | CZB Cliahub Consortium |
| EPI_ISL_738817 | Alameda County Public Health Lab | Chan-Zuckerberg Biohub | CZB Cliahub Consortium |
| EPI_ISL_738818 | Orange County Public Health Lab | Chan-Zuckerberg Biohub | CZB Cliahub Consortium |
| EPI_ISL_738819 | Alameda County Public Health Lab | Chan-Zuckerberg Biohub | CZB Cliahub Consortium |
| EPI_ISL_738820 | Napa-Solano-Yolo- Marin County (NSYM) Public Health Laboratories | Chan-Zuckerberg Biohub | CZB Cliahub Consortium |
| EPI_ISL_738821, EPI_ISL_738822 | Alameda County Public Health Lab | Chan-Zuckerberg Biohub | CZB Cliahub Consortium |
| EPI_ISL_738823, EPI_ISL_738824 | Orange County Public Health Lab | Chan-Zuckerberg Biohub | CZB Cliahub Consortium |
| EPI_ISL_738825 | Alameda County Public Health Lab | Chan-Zuckerberg Biohub | CZB Cliahub Consortium |
| EPI_ISL_738826 | Orange County Public Health Lab | Chan-Zuckerberg Biohub | CZB Cliahub Consortium |
| EPI_ISL_738827 | Humboldt County Public Health Laboratory | Chan-Zuckerberg Biohub | CZB Cliahub Consortium |
| EPI_ISL_738828 | Alameda County Public Health Lab | Chan-Zuckerberg Biohub | CZB Cliahub Consortium |

| | | | |
|---|---|---|---|
| EPI_ISL_738829, EPI_ISL_738830 | Orange County Public Health Lab | Chan-Zuckerberg Biohub | CZB Cliahub Consortium |
| EPI_ISL_738831 | Alameda County Public Health Lab | Chan-Zuckerberg Biohub | CZB Cliahub Consortium |
| EPI_ISL_738832 | Humboldt County Public Health Laboratory | Chan-Zuckerberg Biohub | CZB Cliahub Consortium |
| EPI_ISL_738833 | Alameda County Public Health Lab | Chan-Zuckerberg Biohub | CZB Cliahub Consortium |
| EPI_ISL_738834 | Santa Clara County Public Health Laboratory | Chan-Zuckerberg Biohub | CZB Cliahub Consortium |
| EPI_ISL_738835 | Alameda County Public Health Lab | Chan-Zuckerberg Biohub | CZB Cliahub Consortium |
| EPI_ISL_738836 | Napa-Solano-Yolo- Marin County (NSYM) Public Health Laboratories | Chan-Zuckerberg Biohub | CZB Cliahub Consortium |
| EPI_ISL_738837, EPI_ISL_738838 | Alameda County Public Health Lab | Chan-Zuckerberg Biohub | CZB Cliahub Consortium |
| EPI_ISL_738839 | Santa Clara County Public Health Laboratory | Chan-Zuckerberg Biohub | CZB Cliahub Consortium |
| EPI_ISL_738840, EPI_ISL_738841 | Orange County Public Health Lab | Chan-Zuckerberg Biohub | CZB Cliahub Consortium |
| EPI_ISL_738842 | Santa Clara County Public Health Laboratory | Chan-Zuckerberg Biohub | CZB Cliahub Consortium |
| EPI_ISL_738843 | Alameda County Public Health Lab | Chan-Zuckerberg Biohub | CZB Cliahub Consortium |
| EPI_ISL_738844 | Humboldt County Public Health Laboratory | Chan-Zuckerberg Biohub | CZB Cliahub Consortium |
| EPI_ISL_738845 | Orange County Public Health Lab | Chan-Zuckerberg Biohub | CZB Cliahub Consortium |
| EPI_ISL_738846, EPI_ISL_738847, EPI_ISL_738848 | Alameda County Public Health Lab | Chan-Zuckerberg Biohub | CZB Cliahub Consortium |
| EPI_ISL_738849 | Humboldt County Public Health Laboratory | Chan-Zuckerberg Biohub | CZB Cliahub Consortium |
| EPI_ISL_738850 | Orange County Public Health Lab | Chan-Zuckerberg Biohub | CZB Cliahub Consortium |
| EPI_ISL_738851 | Renegade | Chan-Zuckerberg Biohub | CZB Cliahub Consortium |
| EPI_ISL_738852 | Humboldt County Public Health Laboratory | Chan-Zuckerberg Biohub | CZB Cliahub Consortium |
| EPI_ISL_738853 | Santa Clara County Public Health Laboratory | Chan-Zuckerberg Biohub | CZB Cliahub Consortium |
| EPI_ISL_738854 | Humboldt County Public Health Laboratory | Chan-Zuckerberg Biohub | CZB Cliahub Consortium |
| EPI_ISL_738855 | Santa Clara County Public Health Laboratory | Chan-Zuckerberg Biohub | CZB Cliahub Consortium |
| EPI_ISL_738856, EPI_ISL_738857 | Orange County Public Health Lab | Chan-Zuckerberg Biohub | CZB Cliahub Consortium |
| EPI_ISL_738858 | Alameda County Public Health Lab | Chan-Zuckerberg Biohub | CZB Cliahub Consortium |
| EPI_ISL_738859 | Santa Clara County Public Health Laboratory | Chan-Zuckerberg Biohub | CZB Cliahub Consortium |
| EPI_ISL_738860 | Orange County Public Health Lab | Chan-Zuckerberg Biohub | CZB Cliahub Consortium |
| EPI_ISL_738861 | Santa Clara County Public Health Laboratory | Chan-Zuckerberg Biohub | CZB Cliahub Consortium |
| EPI_ISL_738862, EPI_ISL_738863 | Orange County Public Health Lab | Chan-Zuckerberg Biohub | CZB Cliahub Consortium |
| EPI_ISL_738864 | Santa Clara County Public Health Laboratory | Chan-Zuckerberg Biohub | CZB Cliahub Consortium |
| EPI_ISL_738865 | Humboldt County Public Health Laboratory | Chan-Zuckerberg Biohub | CZB Cliahub Consortium |
| EPI_ISL_738866 | Orange County Public Health Lab | Chan-Zuckerberg Biohub | CZB Cliahub Consortium |
| EPI_ISL_738867 | Santa Clara County Public Health Laboratory | Chan-Zuckerberg Biohub | CZB Cliahub Consortium |
| EPI_ISL_738868 | Alameda County Public Health Lab | Chan-Zuckerberg Biohub | CZB Cliahub Consortium |
| EPI_ISL_738869 | Orange County Public Health Lab | Chan-Zuckerberg Biohub | CZB Cliahub Consortium |
| EPI_ISL_738870 | Napa-Solano-Yolo- Marin County (NSYM) Public Health Laboratories | Chan-Zuckerberg Biohub | CZB Cliahub Consortium |
| EPI_ISL_738871, EPI_ISL_738872, EPI_ISL_738873, EPI_ISL_738874 | Orange County Public Health Lab | Chan-Zuckerberg Biohub | CZB Cliahub Consortium |
| EPI_ISL_738875 | Humboldt County Public Health Laboratory | Chan-Zuckerberg Biohub | CZB Cliahub Consortium |
| EPI_ISL_738876 | Alameda County Public Health Lab | Chan-Zuckerberg Biohub | CZB Cliahub Consortium |
| EPI_ISL_738877 | Renegade | Chan-Zuckerberg Biohub | CZB Cliahub Consortium |
| EPI_ISL_738878 | Orange County Public Health Lab | Chan-Zuckerberg Biohub | CZB Cliahub Consortium |
| EPI_ISL_738879 | Humboldt County Public Health Laboratory | Chan-Zuckerberg Biohub | CZB Cliahub Consortium |
| EPI_ISL_738880 | Orange County Public Health Lab | Chan-Zuckerberg Biohub | CZB Cliahub Consortium |
| EPI_ISL_738881 | Alameda County Public Health Lab | Chan-Zuckerberg Biohub | CZB Cliahub Consortium |
| EPI_ISL_738882 | Orange County Public Health Lab | Chan-Zuckerberg Biohub | CZB Cliahub Consortium |
| EPI_ISL_738883 | Santa Clara County Public Health Laboratory | Chan-Zuckerberg Biohub | CZB Cliahub Consortium |
| EPI_ISL_738884, EPI_ISL_738885 | Orange County Public Health Lab | Chan-Zuckerberg Biohub | CZB Cliahub Consortium |
| EPI_ISL_738886 | Alameda County Public Health Lab | Chan-Zuckerberg Biohub | CZB Cliahub Consortium |
| EPI_ISL_738887 | Santa Clara County Public Health Laboratory | Chan-Zuckerberg Biohub | CZB Cliahub Consortium |
| EPI_ISL_738888 | Orange County Public Health Lab | Chan-Zuckerberg Biohub | CZB Cliahub Consortium |
| EPI_ISL_738889 | Santa Clara County Public Health Laboratory | Chan-Zuckerberg Biohub | CZB Cliahub Consortium |
| EPI_ISL_738890 | County of San Luis Obispo Public Health Laboratory | Chan-Zuckerberg Biohub | CZB Cliahub Consortium |
| EPI_ISL_738891, EPI_ISL_738892 | Alameda County Public Health Lab | Chan-Zuckerberg Biohub | CZB Cliahub Consortium |

| | | | |
|---|---|---|---|
| EPI_ISL_738893 | Orange County Public Health Lab | Chan-Zuckerberg Biohub | CZB Cliahub Consortium |
| EPI_ISL_738894 | Santa Clara County Public Health Laboratory | Chan-Zuckerberg Biohub | CZB Cliahub Consortium |
| EPI_ISL_738895, EPI_ISL_738896 | Alameda County Public Health Lab | Chan-Zuckerberg Biohub | CZB Cliahub Consortium |
| EPI_ISL_738897 | Napa-Solano-Yolo- Marin County (NSYM) Public Health Laboratories | Chan-Zuckerberg Biohub | CZB Cliahub Consortium |
| EPI_ISL_738898 | Santa Clara County Public Health Laboratory | Chan-Zuckerberg Biohub | CZB Cliahub Consortium |
| EPI_ISL_738899, EPI_ISL_738900 | Orange County Public Health Lab | Chan-Zuckerberg Biohub | CZB Cliahub Consortium |
| EPI_ISL_738901 | Renegade | Chan-Zuckerberg Biohub | CZB Cliahub Consortium |
| EPI_ISL_738902 | Humboldt County Public Health Laboratory | Chan-Zuckerberg Biohub | CZB Cliahub Consortium |
| EPI_ISL_738903 | Santa Clara County Public Health Laboratory | Chan-Zuckerberg Biohub | CZB Cliahub Consortium |
| EPI_ISL_738904 | Alameda County Public Health Lab | Chan-Zuckerberg Biohub | CZB Cliahub Consortium |
| EPI_ISL_738905 | Orange County Public Health Lab | Chan-Zuckerberg Biohub | CZB Cliahub Consortium |
| EPI_ISL_738906 | Santa Clara County Public Health Laboratory | Chan-Zuckerberg Biohub | CZB Cliahub Consortium |
| EPI_ISL_738907, EPI_ISL_738908 | Orange County Public Health Lab | Chan-Zuckerberg Biohub | CZB Cliahub Consortium |
| EPI_ISL_738909, EPI_ISL_738910 | Santa Clara County Public Health Laboratory | Chan-Zuckerberg Biohub | CZB Cliahub Consortium |
| EPI_ISL_738911 | Napa-Solano-Yolo- Marin County (NSYM) Public Health Laboratories | Chan-Zuckerberg Biohub | CZB Cliahub Consortium |
| EPI_ISL_738912, EPI_ISL_738913 | Humboldt County Public Health Laboratory | Chan-Zuckerberg Biohub | CZB Cliahub Consortium |
| EPI_ISL_738914 | Alameda County Public Health Lab | Chan-Zuckerberg Biohub | CZB Cliahub Consortium |
| EPI_ISL_738915 | Santa Clara County Public Health Laboratory | Chan-Zuckerberg Biohub | CZB Cliahub Consortium |
| EPI_ISL_738916 | Madera County Department of Public Health | Chan-Zuckerberg Biohub | CZB Cliahub Consortium |
| EPI_ISL_738917, EPI_ISL_738918 | Humboldt County Public Health Laboratory | Chan-Zuckerberg Biohub | CZB Cliahub Consortium |
| EPI_ISL_738919 | Alameda County Public Health Lab | Chan-Zuckerberg Biohub | CZB Cliahub Consortium |
| EPI_ISL_738920 | Santa Clara County Public Health Laboratory | Chan-Zuckerberg Biohub | CZB Cliahub Consortium |
| EPI_ISL_738921 | Orange County Public Health Lab | Chan-Zuckerberg Biohub | CZB Cliahub Consortium |
| EPI_ISL_738922 | Santa Clara County Public Health Laboratory | Chan-Zuckerberg Biohub | CZB Cliahub Consortium |
| EPI_ISL_738923 | Humboldt County Public Health Laboratory | Chan-Zuckerberg Biohub | CZB Cliahub Consortium |
| EPI_ISL_738924 | Santa Clara County Public Health Laboratory | Chan-Zuckerberg Biohub | CZB Cliahub Consortium |
| EPI_ISL_738925 | Alameda County Public Health Lab | Chan-Zuckerberg Biohub | CZB Cliahub Consortium |
| EPI_ISL_738926, EPI_ISL_738927 | Santa Clara County Public Health Laboratory | Chan-Zuckerberg Biohub | CZB Cliahub Consortium |
| EPI_ISL_738928 | Orange County Public Health Lab | Chan-Zuckerberg Biohub | CZB Cliahub Consortium |
| EPI_ISL_738929 | Humboldt County Public Health Laboratory | Chan-Zuckerberg Biohub | CZB Cliahub Consortium |
| EPI_ISL_738930 | Orange County Public Health Lab | Chan-Zuckerberg Biohub | CZB Cliahub Consortium |
| EPI_ISL_738931 | Santa Clara County Public Health Laboratory | Chan-Zuckerberg Biohub | CZB Cliahub Consortium |
| EPI_ISL_738932 | Alameda County Public Health Lab | Chan-Zuckerberg Biohub | CZB Cliahub Consortium |
| EPI_ISL_738933 | Santa Clara County Public Health Laboratory | Chan-Zuckerberg Biohub | CZB Cliahub Consortium |
| EPI_ISL_738934 | Alameda County Public Health Lab | Chan-Zuckerberg Biohub | CZB Cliahub Consortium |
| EPI_ISL_738935 | Orange County Public Health Lab | Chan-Zuckerberg Biohub | CZB Cliahub Consortium |
| EPI_ISL_738936 | Santa Clara County Public Health Laboratory | Chan-Zuckerberg Biohub | CZB Cliahub Consortium |
| EPI_ISL_738937 | Orange County Public Health Lab | Chan-Zuckerberg Biohub | CZB Cliahub Consortium |
| EPI_ISL_738938 | Humboldt County Public Health Laboratory | Chan-Zuckerberg Biohub | CZB Cliahub Consortium |
| EPI_ISL_738939 | Orange County Public Health Lab | Chan-Zuckerberg Biohub | CZB Cliahub Consortium |
| EPI_ISL_738940 | Alameda County Public Health Lab | Chan-Zuckerberg Biohub | CZB Cliahub Consortium |
| EPI_ISL_738941, EPI_ISL_738942, EPI_ISL_738943 | Santa Clara County Public Health Laboratory | Chan-Zuckerberg Biohub | CZB Cliahub Consortium |
| EPI_ISL_738944 | Orange County Public Health Lab | Chan-Zuckerberg Biohub | CZB Cliahub Consortium |
| EPI_ISL_738945 | Alameda County Public Health Lab | Chan-Zuckerberg Biohub | CZB Cliahub Consortium |
| EPI_ISL_738946 | Orange County Public Health Lab | Chan-Zuckerberg Biohub | CZB Cliahub Consortium |
| EPI_ISL_738947 | Humboldt County Public Health Laboratory | Chan-Zuckerberg Biohub | CZB Cliahub Consortium |
| EPI_ISL_738948 | Santa Clara County Public Health Laboratory | Chan-Zuckerberg Biohub | CZB Cliahub Consortium |
| EPI_ISL_738949 | Orange County Public Health Lab | Chan-Zuckerberg Biohub | CZB Cliahub Consortium |
| EPI_ISL_738950 | Alameda County Public Health Lab | Chan-Zuckerberg Biohub | CZB Cliahub Consortium |
| EPI_ISL_738951 | Humboldt County Public Health Laboratory | Chan-Zuckerberg Biohub | CZB Cliahub Consortium |
| EPI_ISL_738952 | Napa-Solano-Yolo- Marin County (NSYM) Public Health Laboratories | Chan-Zuckerberg Biohub | CZB Cliahub Consortium |
| EPI_ISL_738953 | Humboldt County Public Health Laboratory | Chan-Zuckerberg Biohub | CZB Cliahub Consortium |
| EPI_ISL_738954 | Orange County Public Health Lab | Chan-Zuckerberg Biohub | CZB Cliahub Consortium |
| EPI_ISL_738955, | Santa Clara County Public Health | Chan-Zuckerberg Biohub | CZB Cliahub Consortium |

| | | | |
|---|---|---|---|
| EPI_ISL_738956 | Laboratory | | |
| EPI_ISL_738957 | Alameda County Public Health Lab | Chan-Zuckerberg Biohub | CZB Cliahub Consortium |
| EPI_ISL_738958 | Humboldt County Public Health Laboratory | Chan-Zuckerberg Biohub | CZB Cliahub Consortium |
| EPI_ISL_738959 | Napa-Solano-Yolo- Marin County (NSYM) Public Health Laboratories | Chan-Zuckerberg Biohub | CZB Cliahub Consortium |
| EPI_ISL_738960 | Santa Clara County Public Health Laboratory | Chan-Zuckerberg Biohub | CZB Cliahub Consortium |
| EPI_ISL_738961 | Humboldt County Public Health Laboratory | Chan-Zuckerberg Biohub | CZB Cliahub Consortium |
| EPI_ISL_738962 | Santa Clara County Public Health Laboratory | Chan-Zuckerberg Biohub | CZB Cliahub Consortium |
| EPI_ISL_738963 | Alameda County Public Health Lab | Chan-Zuckerberg Biohub | CZB Cliahub Consortium |
| EPI_ISL_738964 | Madera County Department of Public Health | Chan-Zuckerberg Biohub | CZB Cliahub Consortium |
| EPI_ISL_738965 | Santa Clara County Public Health Laboratory | Chan-Zuckerberg Biohub | CZB Cliahub Consortium |
| EPI_ISL_738966 | Orange County Public Health Lab | Chan-Zuckerberg Biohub | CZB Cliahub Consortium |
| EPI_ISL_738967 | Santa Clara County Public Health Laboratory | Chan-Zuckerberg Biohub | CZB Cliahub Consortium |
| EPI_ISL_738968 | Orange County Public Health Lab | Chan-Zuckerberg Biohub | CZB Cliahub Consortium |
| EPI_ISL_738969 | Humboldt County Public Health Laboratory | Chan-Zuckerberg Biohub | CZB Cliahub Consortium |
| EPI_ISL_738970 | Orange County Public Health Lab | Chan-Zuckerberg Biohub | CZB Cliahub Consortium |
| EPI_ISL_738971, EPI_ISL_738972 | Santa Clara County Public Health Laboratory | Chan-Zuckerberg Biohub | CZB Cliahub Consortium |
| EPI_ISL_738973 | Orange County Public Health Lab | Chan-Zuckerberg Biohub | CZB Cliahub Consortium |
| EPI_ISL_738974 | Alameda County Public Health Lab | Chan-Zuckerberg Biohub | CZB Cliahub Consortium |
| EPI_ISL_738975 | Orange County Public Health Lab | Chan-Zuckerberg Biohub | CZB Cliahub Consortium |
| EPI_ISL_738976 | Alameda County Public Health Lab | Chan-Zuckerberg Biohub | CZB Cliahub Consortium |
| EPI_ISL_738977 | Renegade | Chan-Zuckerberg Biohub | CZB Cliahub Consortium |
| EPI_ISL_738978 | Orange County Public Health Lab | Chan-Zuckerberg Biohub | CZB Cliahub Consortium |
| EPI_ISL_738979 | Santa Clara County Public Health Laboratory | Chan-Zuckerberg Biohub | CZB Cliahub Consortium |
| EPI_ISL_738980 | Humboldt County Public Health Laboratory | Chan-Zuckerberg Biohub | CZB Cliahub Consortium |
| EPI_ISL_738981 | Orange County Public Health Lab | Chan-Zuckerberg Biohub | CZB Cliahub Consortium |
| EPI_ISL_738982, EPI_ISL_738983 | Alameda County Public Health Lab | Chan-Zuckerberg Biohub | CZB Cliahub Consortium |
| EPI_ISL_738984 | Santa Clara County Public Health Laboratory | Chan-Zuckerberg Biohub | CZB Cliahub Consortium |
| EPI_ISL_738985 | Humboldt County Public Health Laboratory | Chan-Zuckerberg Biohub | CZB Cliahub Consortium |
| EPI_ISL_738986 | Alameda County Public Health Lab | Chan-Zuckerberg Biohub | CZB Cliahub Consortium |
| EPI_ISL_738987 | Santa Clara County Public Health Laboratory | Chan-Zuckerberg Biohub | CZB Cliahub Consortium |
| EPI_ISL_738988 | Humboldt County Public Health Laboratory | Chan-Zuckerberg Biohub | CZB Cliahub Consortium |
| EPI_ISL_738989 | Orange County Public Health Lab | Chan-Zuckerberg Biohub | CZB Cliahub Consortium |
| EPI_ISL_738990 | County of San Luis Obispo Public Health Laboratory | Chan-Zuckerberg Biohub | CZB Cliahub Consortium |
| EPI_ISL_738991 | Orange County Public Health Lab | Chan-Zuckerberg Biohub | CZB Cliahub Consortium |
| EPI_ISL_738992 | Alameda County Public Health Lab | Chan-Zuckerberg Biohub | CZB Cliahub Consortium |
| EPI_ISL_738993 | Orange County Public Health Lab | Chan-Zuckerberg Biohub | CZB Cliahub Consortium |
| EPI_ISL_738994 | Santa Clara County Public Health Laboratory | Chan-Zuckerberg Biohub | CZB Cliahub Consortium |
| EPI_ISL_738995 | Orange County Public Health Lab | Chan-Zuckerberg Biohub | CZB Cliahub Consortium |
| EPI_ISL_738996, EPI_ISL_738997, EPI_ISL_738998 | Santa Clara County Public Health Laboratory | Chan-Zuckerberg Biohub | CZB Cliahub Consortium |
| EPI_ISL_738999, EPI_ISL_739000, EPI_ISL_739001 | Humboldt County Public Health Laboratory | Chan-Zuckerberg Biohub | CZB Cliahub Consortium |
| EPI_ISL_739002 | Orange County Public Health Lab | Chan-Zuckerberg Biohub | CZB Cliahub Consortium |
| EPI_ISL_739003 | Alameda County Public Health Lab | Chan-Zuckerberg Biohub | CZB Cliahub Consortium |
| EPI_ISL_739004 | UCSF Clinical Microbiology Laboratory | Chan-Zuckerberg Biohub | CZB Cliahub Consortium |
| EPI_ISL_739005, EPI_ISL_739006 | Santa Clara County Public Health Laboratory | Chan-Zuckerberg Biohub | CZB Cliahub Consortium |
| EPI_ISL_739007 | Humboldt County Public Health Laboratory | Chan-Zuckerberg Biohub | CZB Cliahub Consortium |
| EPI_ISL_739008, EPI_ISL_739009, EPI_ISL_739010 | Santa Clara County Public Health Laboratory | Chan-Zuckerberg Biohub | CZB Cliahub Consortium |
| EPI_ISL_739011 | Humboldt County Public Health Laboratory | Chan-Zuckerberg Biohub | CZB Cliahub Consortium |
| EPI_ISL_739012 | Santa Clara County Public Health Laboratory | Chan-Zuckerberg Biohub | CZB Cliahub Consortium |
| EPI_ISL_739013 | Orange County Public Health Lab | Chan-Zuckerberg Biohub | CZB Cliahub Consortium |
| EPI_ISL_739014 | Renegade | Chan-Zuckerberg Biohub | CZB Cliahub Consortium |
| EPI_ISL_739015 | Santa Clara County Public Health Laboratory | Chan-Zuckerberg Biohub | CZB Cliahub Consortium |
| EPI_ISL_739016 | Orange County Public Health Lab | Chan-Zuckerberg Biohub | CZB Cliahub Consortium |
| EPI_ISL_739017 | County of San Luis Obispo Public Health Laboratory | Chan-Zuckerberg Biohub | CZB Cliahub Consortium |
| EPI_ISL_739018 | Alameda County Public Health Lab | Chan-Zuckerberg Biohub | CZB Cliahub Consortium |
| EPI_ISL_739019, | Santa Clara County Public Health | Chan-Zuckerberg Biohub | CZB Cliahub Consortium |

| | | | |
|---|---|---|---|
| EPI_ISL_739020, EPI_ISL_739021 | Laboratory | | |
| EPI_ISL_739022 | Renegade | Chan-Zuckerberg Biohub | CZB Cliahub Consortium |
| EPI_ISL_739023 | Alameda County Public Health Lab | Chan-Zuckerberg Biohub | CZB Cliahub Consortium |
| EPI_ISL_739024, EPI_ISL_739025, EPI_ISL_739026, EPI_ISL_739027 | Santa Clara County Public Health Laboratory | Chan-Zuckerberg Biohub | CZB Cliahub Consortium |
| EPI_ISL_739028 | Orange County Public Health Lab | Chan-Zuckerberg Biohub | CZB Cliahub Consortium |
| EPI_ISL_739029 | Alameda County Public Health Lab | Chan-Zuckerberg Biohub | CZB Cliahub Consortium |
| EPI_ISL_739030, EPI_ISL_739031 | Santa Clara County Public Health Laboratory | Chan-Zuckerberg Biohub | CZB Cliahub Consortium |
| EPI_ISL_739032 | Humboldt County Public Health Laboratory | Chan-Zuckerberg Biohub | CZB Cliahub Consortium |
| EPI_ISL_739033 | Alameda County Public Health Lab | Chan-Zuckerberg Biohub | CZB Cliahub Consortium |
| EPI_ISL_739034 | Orange County Public Health Lab | Chan-Zuckerberg Biohub | CZB Cliahub Consortium |
| EPI_ISL_739035 | Alameda County Public Health Lab | Chan-Zuckerberg Biohub | CZB Cliahub Consortium |
| EPI_ISL_739036, EPI_ISL_739037 | Santa Clara County Public Health Laboratory | Chan-Zuckerberg Biohub | CZB Cliahub Consortium |
| EPI_ISL_739038 | County of San Luis Obispo Public Health Laboratory | Chan-Zuckerberg Biohub | CZB Cliahub Consortium |
| EPI_ISL_739039, EPI_ISL_739040, EPI_ISL_739041 | Santa Clara County Public Health Laboratory | Chan-Zuckerberg Biohub | CZB Cliahub Consortium |
| EPI_ISL_739042 | Orange County Public Health Lab | Chan-Zuckerberg Biohub | CZB Cliahub Consortium |
| EPI_ISL_739043 | Humboldt County Public Health Laboratory | Chan-Zuckerberg Biohub | CZB Cliahub Consortium |
| EPI_ISL_739044 | Orange County Public Health Lab | Chan-Zuckerberg Biohub | CZB Cliahub Consortium |
| EPI_ISL_739045 | Santa Clara County Public Health Laboratory | Chan-Zuckerberg Biohub | CZB Cliahub Consortium |
| EPI_ISL_739046, EPI_ISL_739047 | Alameda County Public Health Lab | Chan-Zuckerberg Biohub | CZB Cliahub Consortium |
| EPI_ISL_739048 | Orange County Public Health Lab | Chan-Zuckerberg Biohub | CZB Cliahub Consortium |
| EPI_ISL_739049 | Alameda County Public Health Lab | Chan-Zuckerberg Biohub | CZB Cliahub Consortium |
| EPI_ISL_739050 | Santa Clara County Public Health Laboratory | Chan-Zuckerberg Biohub | CZB Cliahub Consortium |
| EPI_ISL_739051, EPI_ISL_739052 | Humboldt County Public Health Laboratory | Chan-Zuckerberg Biohub | CZB Cliahub Consortium |
| EPI_ISL_739053 | Santa Clara County Public Health Laboratory | Chan-Zuckerberg Biohub | CZB Cliahub Consortium |
| EPI_ISL_739054 | Alameda County Public Health Lab | Chan-Zuckerberg Biohub | CZB Cliahub Consortium |
| EPI_ISL_739055 | Orange County Public Health Lab | Chan-Zuckerberg Biohub | CZB Cliahub Consortium |
| EPI_ISL_739056, EPI_ISL_739057 | Alameda County Public Health Lab | Chan-Zuckerberg Biohub | CZB Cliahub Consortium |
| EPI_ISL_739058 | Santa Clara County Public Health Laboratory | Chan-Zuckerberg Biohub | CZB Cliahub Consortium |
| EPI_ISL_739059 | Alameda County Public Health Lab | Chan-Zuckerberg Biohub | CZB Cliahub Consortium |
| EPI_ISL_739060 | Santa Clara County Public Health Laboratory | Chan-Zuckerberg Biohub | CZB Cliahub Consortium |
| EPI_ISL_739061 | Madera County Department of Public Health | Chan-Zuckerberg Biohub | CZB Cliahub Consortium |
| EPI_ISL_739062 | Alameda County Public Health Lab | Chan-Zuckerberg Biohub | CZB Cliahub Consortium |
| EPI_ISL_739063 | Orange County Public Health Lab | Chan-Zuckerberg Biohub | CZB Cliahub Consortium |
| EPI_ISL_739064 | Alameda County Public Health Lab | Chan-Zuckerberg Biohub | CZB Cliahub Consortium |
| EPI_ISL_739065, EPI_ISL_739066, EPI_ISL_739067, EPI_ISL_739068 | Santa Clara County Public Health Laboratory | Chan-Zuckerberg Biohub | CZB Cliahub Consortium |
| EPI_ISL_739069 | Orange County Public Health Lab | Chan-Zuckerberg Biohub | CZB Cliahub Consortium |
| EPI_ISL_739070 | Santa Clara County Public Health Laboratory | Chan-Zuckerberg Biohub | CZB Cliahub Consortium |
| EPI_ISL_739071, EPI_ISL_739072 | Orange County Public Health Lab | Chan-Zuckerberg Biohub | CZB Cliahub Consortium |
| EPI_ISL_739073 | Alameda County Public Health Lab | Chan-Zuckerberg Biohub | CZB Cliahub Consortium |
| EPI_ISL_739074 | Santa Clara County Public Health Laboratory | Chan-Zuckerberg Biohub | CZB Cliahub Consortium |
| EPI_ISL_739075 | Alameda County Public Health Lab | Chan-Zuckerberg Biohub | CZB Cliahub Consortium |
| EPI_ISL_739076 | Santa Clara County Public Health Laboratory | Chan-Zuckerberg Biohub | CZB Cliahub Consortium |
| EPI_ISL_739077 | Alameda County Public Health Lab | Chan-Zuckerberg Biohub | CZB Cliahub Consortium |
| EPI_ISL_739078 | Orange County Public Health Lab | Chan-Zuckerberg Biohub | CZB Cliahub Consortium |
| EPI_ISL_739079 | Humboldt County Public Health Laboratory | Chan-Zuckerberg Biohub | CZB Cliahub Consortium |
| EPI_ISL_739080 | Santa Clara County Public Health Laboratory | Chan-Zuckerberg Biohub | CZB Cliahub Consortium |
| EPI_ISL_739081 | Orange County Public Health Lab | Chan-Zuckerberg Biohub | CZB Cliahub Consortium |
| EPI_ISL_739082, EPI_ISL_739083, EPI_ISL_739084 | Alameda County Public Health Lab | Chan-Zuckerberg Biohub | CZB Cliahub Consortium |
| EPI_ISL_739085 | Santa Clara County Public Health Laboratory | Chan-Zuckerberg Biohub | CZB Cliahub Consortium |
| EPI_ISL_739086 | Humboldt County Public Health Laboratory | Chan-Zuckerberg Biohub | CZB Cliahub Consortium |
| EPI_ISL_739087 | Santa Clara County Public Health Laboratory | Chan-Zuckerberg Biohub | CZB Cliahub Consortium |
| EPI_ISL_739088 | Alameda County Public Health Lab | Chan-Zuckerberg Biohub | CZB Cliahub Consortium |
| EPI_ISL_739089 | Orange County Public Health Lab | Chan-Zuckerberg Biohub | CZB Cliahub Consortium |

| | | | |
|---|---|---|---|
| EPI_ISL_739090 | Alameda County Public Health Lab | Chan-Zuckerberg Biohub | CZB Cliahub Consortium |
| EPI_ISL_739091 | Santa Clara County Public Health Laboratory | Chan-Zuckerberg Biohub | CZB Cliahub Consortium |
| EPI_ISL_739092 | Orange County Public Health Lab | Chan-Zuckerberg Biohub | CZB Cliahub Consortium |
| EPI_ISL_739093 | Alameda County Public Health Lab | Chan-Zuckerberg Biohub | CZB Cliahub Consortium |
| EPI_ISL_739094 | Orange County Public Health Lab | Chan-Zuckerberg Biohub | CZB Cliahub Consortium |
| EPI_ISL_739095 | County of San Luis Obispo Public Health Laboratory | Chan-Zuckerberg Biohub | CZB Cliahub Consortium |
| EPI_ISL_739096 | Alameda County Public Health Lab | Chan-Zuckerberg Biohub | CZB Cliahub Consortium |
| EPI_ISL_739097 | Orange County Public Health Lab | Chan-Zuckerberg Biohub | CZB Cliahub Consortium |
| EPI_ISL_739098 | Santa Clara County Public Health Laboratory | Chan-Zuckerberg Biohub | CZB Cliahub Consortium |
| EPI_ISL_739099 | Orange County Public Health Lab | Chan-Zuckerberg Biohub | CZB Cliahub Consortium |
| EPI_ISL_739100 | Alameda County Public Health Lab | Chan-Zuckerberg Biohub | CZB Cliahub Consortium |
| EPI_ISL_739101 | Monterey County Public Health Lab | Chan-Zuckerberg Biohub | CZB Cliahub Consortium |
| EPI_ISL_739102 | County of San Luis Obispo Public Health Laboratory | Chan-Zuckerberg Biohub | CZB Cliahub Consortium |
| EPI_ISL_739103, EPI_ISL_739104 | Santa Clara County Public Health Laboratory | Chan-Zuckerberg Biohub | CZB Cliahub Consortium |
| EPI_ISL_739105 | Orange County Public Health Lab | Chan-Zuckerberg Biohub | CZB Cliahub Consortium |
| EPI_ISL_739106 | Humboldt County Public Health Laboratory | Chan-Zuckerberg Biohub | CZB Cliahub Consortium |
| EPI_ISL_739107 | Santa Clara County Public Health Laboratory | Chan-Zuckerberg Biohub | CZB Cliahub Consortium |
| EPI_ISL_739108 | Orange County Public Health Lab | Chan-Zuckerberg Biohub | CZB Cliahub Consortium |
| EPI_ISL_739109, EPI_ISL_739110, EPI_ISL_739111 | Alameda County Public Health Lab | Chan-Zuckerberg Biohub | CZB Cliahub Consortium |
| EPI_ISL_739112 | Humboldt County Public Health Laboratory | Chan-Zuckerberg Biohub | CZB Cliahub Consortium |
| EPI_ISL_739113 | Alameda County Public Health Lab | Chan-Zuckerberg Biohub | CZB Cliahub Consortium |
| EPI_ISL_739114 | Humboldt County Public Health Laboratory | Chan-Zuckerberg Biohub | CZB Cliahub Consortium |
| EPI_ISL_739115 | Alameda County Public Health Lab | Chan-Zuckerberg Biohub | CZB Cliahub Consortium |
| EPI_ISL_739116 | Orange County Public Health Lab | Chan-Zuckerberg Biohub | CZB Cliahub Consortium |
| EPI_ISL_739117 | Santa Clara County Public Health Laboratory | Chan-Zuckerberg Biohub | CZB Cliahub Consortium |
| EPI_ISL_739118 | Alameda County Public Health Lab | Chan-Zuckerberg Biohub | CZB Cliahub Consortium |
| EPI_ISL_739119 | Tulare County Public Health Lab | Chan-Zuckerberg Biohub | CZB Cliahub Consortium |
| EPI_ISL_739120 | Alameda County Public Health Lab | Chan-Zuckerberg Biohub | CZB Cliahub Consortium |
| EPI_ISL_739121, EPI_ISL_739122 | Santa Clara County Public Health Laboratory | Chan-Zuckerberg Biohub | CZB Cliahub Consortium |
| EPI_ISL_739123 | Humboldt County Public Health Laboratory | Chan-Zuckerberg Biohub | CZB Cliahub Consortium |
| EPI_ISL_739124 | Alameda County Public Health Lab | Chan-Zuckerberg Biohub | CZB Cliahub Consortium |
| EPI_ISL_739125 | Orange County Public Health Lab | Chan-Zuckerberg Biohub | CZB Cliahub Consortium |
| EPI_ISL_739126 | Humboldt County Public Health Laboratory | Chan-Zuckerberg Biohub | CZB Cliahub Consortium |
| EPI_ISL_739127 | Alameda County Public Health Lab | Chan-Zuckerberg Biohub | CZB Cliahub Consortium |
| EPI_ISL_739128, EPI_ISL_739129, EPI_ISL_739130 | Santa Clara County Public Health Laboratory | Chan-Zuckerberg Biohub | CZB Cliahub Consortium |
| EPI_ISL_739131 | Humboldt County Public Health Laboratory | Chan-Zuckerberg Biohub | CZB Cliahub Consortium |
| EPI_ISL_739132 | Orange County Public Health Lab | Chan-Zuckerberg Biohub | CZB Cliahub Consortium |
| EPI_ISL_739133 | Madera County Department of Public Health | Chan-Zuckerberg Biohub | CZB Cliahub Consortium |
| EPI_ISL_739134, EPI_ISL_739135, EPI_ISL_739136, EPI_ISL_739137 | Alameda County Public Health Lab | Chan-Zuckerberg Biohub | CZB Cliahub Consortium |
| EPI_ISL_739138 | Santa Clara County Public Health Laboratory | Chan-Zuckerberg Biohub | CZB Cliahub Consortium |
| EPI_ISL_739139 | Orange County Public Health Lab | Chan-Zuckerberg Biohub | CZB Cliahub Consortium |
| EPI_ISL_739140 | Alameda County Public Health Lab | Chan-Zuckerberg Biohub | CZB Cliahub Consortium |
| EPI_ISL_739141 | Santa Clara County Public Health Laboratory | Chan-Zuckerberg Biohub | CZB Cliahub Consortium |
| EPI_ISL_739142, EPI_ISL_739143 | Orange County Public Health Lab | Chan-Zuckerberg Biohub | CZB Cliahub Consortium |
| EPI_ISL_739144 | Alameda County Public Health Lab | Chan-Zuckerberg Biohub | CZB Cliahub Consortium |
| EPI_ISL_739145, EPI_ISL_739146, EPI_ISL_739147 | Santa Clara County Public Health Laboratory | Chan-Zuckerberg Biohub | CZB Cliahub Consortium |
| EPI_ISL_739148 | Humboldt County Public Health Laboratory | Chan-Zuckerberg Biohub | CZB Cliahub Consortium |
| EPI_ISL_739149, EPI_ISL_739150 | Alameda County Public Health Lab | Chan-Zuckerberg Biohub | CZB Cliahub Consortium |
| EPI_ISL_739151 | Orange County Public Health Lab | Chan-Zuckerberg Biohub | CZB Cliahub Consortium |
| EPI_ISL_739152, EPI_ISL_739153 | Santa Clara County Public Health Laboratory | Chan-Zuckerberg Biohub | CZB Cliahub Consortium |
| EPI_ISL_739154 | Orange County Public Health Lab | Chan-Zuckerberg Biohub | CZB Cliahub Consortium |
| EPI_ISL_739155 | Humboldt County Public Health Laboratory | Chan-Zuckerberg Biohub | CZB Cliahub Consortium |
| EPI_ISL_739156 | Orange County Public Health Lab | Chan-Zuckerberg Biohub | CZB Cliahub Consortium |
| EPI_ISL_739157 | Santa Clara County Public Health Laboratory | Chan-Zuckerberg Biohub | CZB Cliahub Consortium |

| | | | |
|---|---|---|---|
| EPI_ISL_739158 | Alameda County Public Health Lab | Chan-Zuckerberg Biohub | CZB Cliahub Consortium |
| EPI_ISL_739159 | Humboldt County Public Health Laboratory | Chan-Zuckerberg Biohub | CZB Cliahub Consortium |
| EPI_ISL_739160, EPI_ISL_739161 | Alameda County Public Health Lab | Chan-Zuckerberg Biohub | CZB Cliahub Consortium |
| EPI_ISL_739162 | Orange County Public Health Lab | Chan-Zuckerberg Biohub | CZB Cliahub Consortium |
| EPI_ISL_739163, EPI_ISL_739164 | Alameda County Public Health Lab | Chan-Zuckerberg Biohub | CZB Cliahub Consortium |
| EPI_ISL_739165 | Humboldt County Public Health Laboratory | Chan-Zuckerberg Biohub | CZB Cliahub Consortium |
| EPI_ISL_739166, EPI_ISL_739167, EPI_ISL_739168 | Santa Clara County Public Health Laboratory | Chan-Zuckerberg Biohub | CZB Cliahub Consortium |
| EPI_ISL_739169 | Napa-Solano-Yolo- Marin County (NSYM) Public Health Laboratories | Chan-Zuckerberg Biohub | CZB Cliahub Consortium |
| EPI_ISL_739170 | Humboldt County Public Health Laboratory | Chan-Zuckerberg Biohub | CZB Cliahub Consortium |
| EPI_ISL_739171 | Alameda County Public Health Lab | Chan-Zuckerberg Biohub | CZB Cliahub Consortium |
| EPI_ISL_739172 | Madera County Department of Public Health | Chan-Zuckerberg Biohub | CZB Cliahub Consortium |
| EPI_ISL_739173, EPI_ISL_739174 | Alameda County Public Health Lab | Chan-Zuckerberg Biohub | CZB Cliahub Consortium |
| EPI_ISL_739175, EPI_ISL_739176 | Orange County Public Health Lab | Chan-Zuckerberg Biohub | CZB Cliahub Consortium |
| EPI_ISL_739177 | Humboldt County Public Health Laboratory | Chan-Zuckerberg Biohub | CZB Cliahub Consortium |
| EPI_ISL_739178, EPI_ISL_739179 | Alameda County Public Health Lab | Chan-Zuckerberg Biohub | CZB Cliahub Consortium |
| EPI_ISL_739180 | Santa Clara County Public Health Laboratory | Chan-Zuckerberg Biohub | CZB Cliahub Consortium |
| EPI_ISL_739181 | Napa-Solano-Yolo- Marin County (NSYM) Public Health Laboratories | Chan-Zuckerberg Biohub | CZB Cliahub Consortium |
| EPI_ISL_739182 | Orange County Public Health Lab | Chan-Zuckerberg Biohub | CZB Cliahub Consortium |
| EPI_ISL_739183 | Humboldt County Public Health Laboratory | Chan-Zuckerberg Biohub | CZB Cliahub Consortium |
| EPI_ISL_739184 | Santa Clara County Public Health Laboratory | Chan-Zuckerberg Biohub | CZB Cliahub Consortium |
| EPI_ISL_739185 | Orange County Public Health Lab | Chan-Zuckerberg Biohub | CZB Cliahub Consortium |
| EPI_ISL_739186, EPI_ISL_739187, EPI_ISL_739188 | Santa Clara County Public Health Laboratory | Chan-Zuckerberg Biohub | CZB Cliahub Consortium |
| EPI_ISL_739189, EPI_ISL_739190 | Alameda County Public Health Lab | Chan-Zuckerberg Biohub | CZB Cliahub Consortium |
| EPI_ISL_739191 | Orange County Public Health Lab | Chan-Zuckerberg Biohub | CZB Cliahub Consortium |
| EPI_ISL_739192 | Alameda County Public Health Lab | Chan-Zuckerberg Biohub | CZB Cliahub Consortium |
| EPI_ISL_739193 | Tulare County Public Health Lab | Chan-Zuckerberg Biohub | CZB Cliahub Consortium |
| EPI_ISL_739194 | Humboldt County Public Health Laboratory | Chan-Zuckerberg Biohub | CZB Cliahub Consortium |
| EPI_ISL_739195 | Santa Clara County Public Health Laboratory | Chan-Zuckerberg Biohub | CZB Cliahub Consortium |
| EPI_ISL_739196 | Alameda County Public Health Lab | Chan-Zuckerberg Biohub | CZB Cliahub Consortium |
| EPI_ISL_739197 | Madera County Department of Public Health | Chan-Zuckerberg Biohub | CZB Cliahub Consortium |
| EPI_ISL_739198 | Humboldt County Public Health Laboratory | Chan-Zuckerberg Biohub | CZB Cliahub Consortium |
| EPI_ISL_739199 | Orange County Public Health Lab | Chan-Zuckerberg Biohub | CZB Cliahub Consortium |
| EPI_ISL_739200, EPI_ISL_739201, EPI_ISL_739202 | Alameda County Public Health Lab | Chan-Zuckerberg Biohub | CZB Cliahub Consortium |
| EPI_ISL_739203 | UCSF Clinical Microbiology Laboratory | Chan-Zuckerberg Biohub | CZB Cliahub Consortium |
| EPI_ISL_739204 | Santa Clara County Public Health Laboratory | Chan-Zuckerberg Biohub | CZB Cliahub Consortium |
| EPI_ISL_739205 | Orange County Public Health Lab | Chan-Zuckerberg Biohub | CZB Cliahub Consortium |
| EPI_ISL_739206, EPI_ISL_739207, EPI_ISL_739208, EPI_ISL_739209 | Alameda County Public Health Lab | Chan-Zuckerberg Biohub | CZB Cliahub Consortium |
| EPI_ISL_739210 | Humboldt County Public Health Laboratory | Chan-Zuckerberg Biohub | CZB Cliahub Consortium |
| EPI_ISL_739211 | Orange County Public Health Lab | Chan-Zuckerberg Biohub | CZB Cliahub Consortium |
| EPI_ISL_739212, EPI_ISL_739213 | Alameda County Public Health Lab | Chan-Zuckerberg Biohub | CZB Cliahub Consortium |
| EPI_ISL_739214 | Santa Clara County Public Health Laboratory | Chan-Zuckerberg Biohub | CZB Cliahub Consortium |
| EPI_ISL_739215 | Alameda County Public Health Lab | Chan-Zuckerberg Biohub | CZB Cliahub Consortium |
| EPI_ISL_739216 | Orange County Public Health Lab | Chan-Zuckerberg Biohub | CZB Cliahub Consortium |
| EPI_ISL_739217, EPI_ISL_739218 | Alameda County Public Health Lab | Chan-Zuckerberg Biohub | CZB Cliahub Consortium |
| EPI_ISL_739219 | Santa Clara County Public Health Laboratory | Chan-Zuckerberg Biohub | CZB Cliahub Consortium |
| EPI_ISL_739220 | Alameda County Public Health Lab | Chan-Zuckerberg Biohub | CZB Cliahub Consortium |
| EPI_ISL_739221 | Santa Clara County Public Health Laboratory | Chan-Zuckerberg Biohub | CZB Cliahub Consortium |
| EPI_ISL_739222 | Orange County Public Health Lab | Chan-Zuckerberg Biohub | CZB Cliahub Consortium |
| EPI_ISL_739223 | Napa-Solano-Yolo- Marin County (NSYM) Public Health Laboratories | Chan-Zuckerberg Biohub | CZB Cliahub Consortium |
| EPI_ISL_739224 | Santa Clara County Public Health Laboratory | Chan-Zuckerberg Biohub | CZB Cliahub Consortium |

| | | | |
|---|---|---|---|
| EPI_ISL_739225 | Humboldt County Public Health Laboratory | Chan-Zuckerberg Biohub | CZB Cliahub Consortium |
| EPI_ISL_739226 | Alameda County Public Health Lab | Chan-Zuckerberg Biohub | CZB Cliahub Consortium |
| EPI_ISL_739227 | Orange County Public Health Lab | Chan-Zuckerberg Biohub | CZB Cliahub Consortium |
| EPI_ISL_739228 | Humboldt County Public Health Laboratory | Chan-Zuckerberg Biohub | CZB Cliahub Consortium |
| EPI_ISL_739229 | Alameda County Public Health Lab | Chan-Zuckerberg Biohub | CZB Cliahub Consortium |
| EPI_ISL_739230 | Orange County Public Health Lab | Chan-Zuckerberg Biohub | CZB Cliahub Consortium |
| EPI_ISL_739231 | Alameda County Public Health Lab | Chan-Zuckerberg Biohub | CZB Cliahub Consortium |
| EPI_ISL_739232 | Santa Clara County Public Health Laboratory | Chan-Zuckerberg Biohub | CZB Cliahub Consortium |
| EPI_ISL_739233 | Alameda County Public Health Lab | Chan-Zuckerberg Biohub | CZB Cliahub Consortium |
| EPI_ISL_739234 | Santa Clara County Public Health Laboratory | Chan-Zuckerberg Biohub | CZB Cliahub Consortium |
| EPI_ISL_739235 | Humboldt County Public Health Laboratory | Chan-Zuckerberg Biohub | CZB Cliahub Consortium |
| EPI_ISL_739236 | Alameda County Public Health Lab | Chan-Zuckerberg Biohub | CZB Cliahub Consortium |
| EPI_ISL_739237 | Santa Clara County Public Health Laboratory | Chan-Zuckerberg Biohub | CZB Cliahub Consortium |
| EPI_ISL_739238, EPI_ISL_739239 | Orange County Public Health Lab | Chan-Zuckerberg Biohub | CZB Cliahub Consortium |
| EPI_ISL_739240 | Humboldt County Public Health Laboratory | Chan-Zuckerberg Biohub | CZB Cliahub Consortium |
| EPI_ISL_739241 | Alameda County Public Health Lab | Chan-Zuckerberg Biohub | CZB Cliahub Consortium |
| EPI_ISL_739242 | Humboldt County Public Health Laboratory | Chan-Zuckerberg Biohub | CZB Cliahub Consortium |
| EPI_ISL_739243, EPI_ISL_739244 | Alameda County Public Health Lab | Chan-Zuckerberg Biohub | CZB Cliahub Consortium |
| EPI_ISL_739245 | Orange County Public Health Lab | Chan-Zuckerberg Biohub | CZB Cliahub Consortium |
| EPI_ISL_739246, EPI_ISL_739247, EPI_ISL_739248, EPI_ISL_739249 | Santa Clara County Public Health Laboratory | Chan-Zuckerberg Biohub | CZB Cliahub Consortium |
| EPI_ISL_739250 | Humboldt County Public Health Laboratory | Chan-Zuckerberg Biohub | CZB Cliahub Consortium |
| EPI_ISL_739251 | Napa-Solano-Yolo- Marin County (NSYM) Public Health Laboratories | Chan-Zuckerberg Biohub | CZB Cliahub Consortium |
| EPI_ISL_739252, EPI_ISL_739253, EPI_ISL_739254 | Santa Clara County Public Health Laboratory | Chan-Zuckerberg Biohub | CZB Cliahub Consortium |
| EPI_ISL_739255 | Orange County Public Health Lab | Chan-Zuckerberg Biohub | CZB Cliahub Consortium |
| EPI_ISL_739256 | Santa Clara County Public Health Laboratory | Chan-Zuckerberg Biohub | CZB Cliahub Consortium |
| EPI_ISL_739257 | Alameda County Public Health Lab | Chan-Zuckerberg Biohub | CZB Cliahub Consortium |
| EPI_ISL_739258, EPI_ISL_739259 | Orange County Public Health Lab | Chan-Zuckerberg Biohub | CZB Cliahub Consortium |
| EPI_ISL_739260 | Santa Clara County Public Health Laboratory | Chan-Zuckerberg Biohub | CZB Cliahub Consortium |
| EPI_ISL_739261, EPI_ISL_739262 | Alameda County Public Health Lab | Chan-Zuckerberg Biohub | CZB Cliahub Consortium |
| EPI_ISL_739263 | Santa Clara County Public Health Laboratory | Chan-Zuckerberg Biohub | CZB Cliahub Consortium |
| EPI_ISL_739264 | Humboldt County Public Health Laboratory | Chan-Zuckerberg Biohub | CZB Cliahub Consortium |
| EPI_ISL_739265, EPI_ISL_739266, EPI_ISL_739267 | Orange County Public Health Lab | Chan-Zuckerberg Biohub | CZB Cliahub Consortium |
| EPI_ISL_739268 | Alameda County Public Health Lab | Chan-Zuckerberg Biohub | CZB Cliahub Consortium |
| EPI_ISL_739269 | Humboldt County Public Health Laboratory | Chan-Zuckerberg Biohub | CZB Cliahub Consortium |
| EPI_ISL_739270 | Alameda County Public Health Lab | Chan-Zuckerberg Biohub | CZB Cliahub Consortium |
| EPI_ISL_739271 | Orange County Public Health Lab | Chan-Zuckerberg Biohub | CZB Cliahub Consortium |
| EPI_ISL_739272 | Humboldt County Public Health Laboratory | Chan-Zuckerberg Biohub | CZB Cliahub Consortium |
| EPI_ISL_739273 | Santa Clara County Public Health Laboratory | Chan-Zuckerberg Biohub | CZB Cliahub Consortium |
| EPI_ISL_739274 | Humboldt County Public Health Laboratory | Chan-Zuckerberg Biohub | CZB Cliahub Consortium |
| EPI_ISL_739275 | Santa Clara County Public Health Laboratory | Chan-Zuckerberg Biohub | CZB Cliahub Consortium |
| EPI_ISL_739276 | Orange County Public Health Lab | Chan-Zuckerberg Biohub | CZB Cliahub Consortium |
| EPI_ISL_739277, EPI_ISL_739278 | Santa Clara County Public Health Laboratory | Chan-Zuckerberg Biohub | CZB Cliahub Consortium |
| EPI_ISL_739279 | Orange County Public Health Lab | Chan-Zuckerberg Biohub | CZB Cliahub Consortium |
| EPI_ISL_739280 | Alameda County Public Health Lab | Chan-Zuckerberg Biohub | CZB Cliahub Consortium |
| EPI_ISL_739281, EPI_ISL_739282 | Santa Clara County Public Health Laboratory | Chan-Zuckerberg Biohub | CZB Cliahub Consortium |
| EPI_ISL_739283 | Alameda County Public Health Lab | Chan-Zuckerberg Biohub | CZB Cliahub Consortium |
| EPI_ISL_739284 | Santa Clara County Public Health Laboratory | Chan-Zuckerberg Biohub | CZB Cliahub Consortium |
| EPI_ISL_739285, EPI_ISL_739286 | Alameda County Public Health Lab | Chan-Zuckerberg Biohub | CZB Cliahub Consortium |
| EPI_ISL_739287 | Humboldt County Public Health Laboratory | Chan-Zuckerberg Biohub | CZB Cliahub Consortium |
| EPI_ISL_739288 | Alameda County Public Health Lab | Chan-Zuckerberg Biohub | CZB Cliahub Consortium |
| EPI_ISL_739289 | Orange County Public Health Lab | Chan-Zuckerberg Biohub | CZB Cliahub Consortium |
| EPI_ISL_739290 | Alameda County Public Health Lab | Chan-Zuckerberg Biohub | CZB Cliahub Consortium |

| | | | |
|---|---|---|---|
| EPI_ISL_739291 | Orange County Public Health Lab | Chan-Zuckerberg Biohub | CZB Cliahub Consortium |
| EPI_ISL_739292 | Humboldt County Public Health Laboratory | Chan-Zuckerberg Biohub | CZB Cliahub Consortium |
| EPI_ISL_739293 | Santa Clara County Public Health Laboratory | Chan-Zuckerberg Biohub | CZB Cliahub Consortium |
| EPI_ISL_739294 | Alameda County Public Health Lab | Chan-Zuckerberg Biohub | CZB Cliahub Consortium |
| EPI_ISL_739295 | UCSF Clinical Microbiology Laboratory | Chan-Zuckerberg Biohub | CZB Cliahub Consortium |
| EPI_ISL_739296 | Orange County Public Health Lab | Chan-Zuckerberg Biohub | CZB Cliahub Consortium |
| EPI_ISL_739297 | Humboldt County Public Health Laboratory | Chan-Zuckerberg Biohub | CZB Cliahub Consortium |
| EPI_ISL_739298 | Santa Clara County Public Health Laboratory | Chan-Zuckerberg Biohub | CZB Cliahub Consortium |
| EPI_ISL_739299 | Humboldt County Public Health Laboratory | Chan-Zuckerberg Biohub | CZB Cliahub Consortium |
| EPI_ISL_739300 | Alameda County Public Health Lab | Chan-Zuckerberg Biohub | CZB Cliahub Consortium |
| EPI_ISL_739301 | Napa-Solano-Yolo- Marin County (NSYM) Public Health Laboratories | Chan-Zuckerberg Biohub | CZB Cliahub Consortium |
| EPI_ISL_739302, EPI_ISL_739303 | Alameda County Public Health Lab | Chan-Zuckerberg Biohub | CZB Cliahub Consortium |
| EPI_ISL_739304 | Orange County Public Health Lab | Chan-Zuckerberg Biohub | CZB Cliahub Consortium |
| EPI_ISL_739305 | Santa Clara County Public Health Laboratory | Chan-Zuckerberg Biohub | CZB Cliahub Consortium |
| EPI_ISL_739306, EPI_ISL_739307, EPI_ISL_739308, EPI_ISL_739309 | Alameda County Public Health Lab | Chan-Zuckerberg Biohub | CZB Cliahub Consortium |
| EPI_ISL_739310 | Santa Clara County Public Health Laboratory | Chan-Zuckerberg Biohub | CZB Cliahub Consortium |
| EPI_ISL_739311 | Humboldt County Public Health Laboratory | Chan-Zuckerberg Biohub | CZB Cliahub Consortium |
| EPI_ISL_739312 | Alameda County Public Health Lab | Chan-Zuckerberg Biohub | CZB Cliahub Consortium |
| EPI_ISL_739313 | Humboldt County Public Health Laboratory | Chan-Zuckerberg Biohub | CZB Cliahub Consortium |
| EPI_ISL_739314 | Orange County Public Health Lab | Chan-Zuckerberg Biohub | CZB Cliahub Consortium |
| EPI_ISL_739315 | Napa-Solano-Yolo- Marin County (NSYM) Public Health Laboratories | Chan-Zuckerberg Biohub | CZB Cliahub Consortium |
| EPI_ISL_739316 | Renegade | Chan-Zuckerberg Biohub | CZB Cliahub Consortium |
| EPI_ISL_739317 | Santa Clara County Public Health Laboratory | Chan-Zuckerberg Biohub | CZB Cliahub Consortium |
| EPI_ISL_739318 | Humboldt County Public Health Laboratory | Chan-Zuckerberg Biohub | CZB Cliahub Consortium |
| EPI_ISL_739319, EPI_ISL_739320, EPI_ISL_739321 | Santa Clara County Public Health Laboratory | Chan-Zuckerberg Biohub | CZB Cliahub Consortium |
| EPI_ISL_739322 | Orange County Public Health Lab | Chan-Zuckerberg Biohub | CZB Cliahub Consortium |
| EPI_ISL_739323, EPI_ISL_739324 | Alameda County Public Health Lab | Chan-Zuckerberg Biohub | CZB Cliahub Consortium |
| EPI_ISL_739325 | Napa-Solano-Yolo- Marin County (NSYM) Public Health Laboratories | Chan-Zuckerberg Biohub | CZB Cliahub Consortium |
| EPI_ISL_739326 | Alameda County Public Health Lab | Chan-Zuckerberg Biohub | CZB Cliahub Consortium |
| EPI_ISL_739327 | Santa Clara County Public Health Laboratory | Chan-Zuckerberg Biohub | CZB Cliahub Consortium |
| EPI_ISL_739328 | Alameda County Public Health Lab | Chan-Zuckerberg Biohub | CZB Cliahub Consortium |
| EPI_ISL_739329 | Monterey County Public Health Lab | Chan-Zuckerberg Biohub | CZB Cliahub Consortium |
| EPI_ISL_739330 | Humboldt County Public Health Laboratory | Chan-Zuckerberg Biohub | CZB Cliahub Consortium |
| EPI_ISL_739331 | Orange County Public Health Lab | Chan-Zuckerberg Biohub | CZB Cliahub Consortium |
| EPI_ISL_739332 | Santa Clara County Public Health Laboratory | Chan-Zuckerberg Biohub | CZB Cliahub Consortium |
| EPI_ISL_739333 | Alameda County Public Health Lab | Chan-Zuckerberg Biohub | CZB Cliahub Consortium |
| EPI_ISL_739334 | Orange County Public Health Lab | Chan-Zuckerberg Biohub | CZB Cliahub Consortium |
| EPI_ISL_739335 | Santa Clara County Public Health Laboratory | Chan-Zuckerberg Biohub | CZB Cliahub Consortium |
| EPI_ISL_739336 | Monterey County Public Health Lab | Chan-Zuckerberg Biohub | CZB Cliahub Consortium |
| EPI_ISL_739337, EPI_ISL_739338 | Alameda County Public Health Lab | Chan-Zuckerberg Biohub | CZB Cliahub Consortium |
| EPI_ISL_739339 | Napa-Solano-Yolo- Marin County (NSYM) Public Health Laboratories | Chan-Zuckerberg Biohub | CZB Cliahub Consortium |
| EPI_ISL_739340 | Alameda County Public Health Lab | Chan-Zuckerberg Biohub | CZB Cliahub Consortium |
| EPI_ISL_739341, EPI_ISL_739342 | Orange County Public Health Lab | Chan-Zuckerberg Biohub | CZB Cliahub Consortium |
| EPI_ISL_739343, EPI_ISL_739344 | Santa Clara County Public Health Laboratory | Chan-Zuckerberg Biohub | CZB Cliahub Consortium |
| EPI_ISL_739345 | Orange County Public Health Lab | Chan-Zuckerberg Biohub | CZB Cliahub Consortium |
| EPI_ISL_739346 | Santa Clara County Public Health Laboratory | Chan-Zuckerberg Biohub | CZB Cliahub Consortium |
| EPI_ISL_739347 | Orange County Public Health Lab | Chan-Zuckerberg Biohub | CZB Cliahub Consortium |
| EPI_ISL_739348 | Renegade | Chan-Zuckerberg Biohub | CZB Cliahub Consortium |
| EPI_ISL_739349 | Alameda County Public Health Lab | Chan-Zuckerberg Biohub | CZB Cliahub Consortium |
| EPI_ISL_739350 | Orange County Public Health Lab | Chan-Zuckerberg Biohub | CZB Cliahub Consortium |
| EPI_ISL_739351 | Alameda County Public Health Lab | Chan-Zuckerberg Biohub | CZB Cliahub Consortium |
| EPI_ISL_739352 | Santa Clara County Public Health Laboratory | Chan-Zuckerberg Biohub | CZB Cliahub Consortium |
| EPI_ISL_739353 | Orange County Public Health Lab | Chan-Zuckerberg Biohub | CZB Cliahub Consortium |
| EPI_ISL_739354 | Santa Clara County Public Health Laboratory | Chan-Zuckerberg Biohub | CZB Cliahub Consortium |

| | | | |
|---|---|---|---|
| EPI_ISL_739355 | Renegade | Chan-Zuckerberg Biohub | CZB Cliahub Consortium |
| EPI_ISL_739356 | Alameda County Public Health Lab | Chan-Zuckerberg Biohub | CZB Cliahub Consortium |
| EPI_ISL_739357 | Santa Clara County Public Health Laboratory | Chan-Zuckerberg Biohub | CZB Cliahub Consortium |
| EPI_ISL_739358 | Alameda County Public Health Lab | Chan-Zuckerberg Biohub | CZB Cliahub Consortium |
| EPI_ISL_739359 | Orange County Public Health Lab | Chan-Zuckerberg Biohub | CZB Cliahub Consortium |
| EPI_ISL_739360 | Santa Clara County Public Health Laboratory | Chan-Zuckerberg Biohub | CZB Cliahub Consortium |
| EPI_ISL_739361 | Alameda County Public Health Lab | Chan-Zuckerberg Biohub | CZB Cliahub Consortium |
| EPI_ISL_739362 | Orange County Public Health Lab | Chan-Zuckerberg Biohub | CZB Cliahub Consortium |
| EPI_ISL_739363 | County of San Luis Obispo Public Health Laboratory | Chan-Zuckerberg Biohub | CZB Cliahub Consortium |
| EPI_ISL_739364 | Monterey County Public Health Lab | Chan-Zuckerberg Biohub | CZB Cliahub Consortium |
| EPI_ISL_739365 | Santa Clara County Public Health Laboratory | Chan-Zuckerberg Biohub | CZB Cliahub Consortium |
| EPI_ISL_739366, EPI_ISL_739367 | Humboldt County Public Health Laboratory | Chan-Zuckerberg Biohub | CZB Cliahub Consortium |
| EPI_ISL_739368 | Alameda County Public Health Lab | Chan-Zuckerberg Biohub | CZB Cliahub Consortium |
| EPI_ISL_739369 | Orange County Public Health Lab | Chan-Zuckerberg Biohub | CZB Cliahub Consortium |
| EPI_ISL_739370, EPI_ISL_739371 | Santa Clara County Public Health Laboratory | Chan-Zuckerberg Biohub | CZB Cliahub Consortium |
| EPI_ISL_739372 | Monterey County Public Health Lab | Chan-Zuckerberg Biohub | CZB Cliahub Consortium |
| EPI_ISL_739373 | Orange County Public Health Lab | Chan-Zuckerberg Biohub | CZB Cliahub Consortium |
| EPI_ISL_739374 | Santa Clara County Public Health Laboratory | Chan-Zuckerberg Biohub | CZB Cliahub Consortium |
| EPI_ISL_739375 | Orange County Public Health Lab | Chan-Zuckerberg Biohub | CZB Cliahub Consortium |
| EPI_ISL_739376, EPI_ISL_739377 | Santa Clara County Public Health Laboratory | Chan-Zuckerberg Biohub | CZB Cliahub Consortium |
| EPI_ISL_739378 | Alameda County Public Health Lab | Chan-Zuckerberg Biohub | CZB Cliahub Consortium |
| EPI_ISL_739379 | Orange County Public Health Lab | Chan-Zuckerberg Biohub | CZB Cliahub Consortium |
| EPI_ISL_739380, EPI_ISL_739381, EPI_ISL_739382 | Santa Clara County Public Health Laboratory | Chan-Zuckerberg Biohub | CZB Cliahub Consortium |
| EPI_ISL_739383 | Alameda County Public Health Lab | Chan-Zuckerberg Biohub | CZB Cliahub Consortium |
| EPI_ISL_739384 | Orange County Public Health Lab | Chan-Zuckerberg Biohub | CZB Cliahub Consortium |
| EPI_ISL_739385 | Alameda County Public Health Lab | Chan-Zuckerberg Biohub | CZB Cliahub Consortium |
| EPI_ISL_739386 | Orange County Public Health Lab | Chan-Zuckerberg Biohub | CZB Cliahub Consortium |
| EPI_ISL_739387 | Alameda County Public Health Lab | Chan-Zuckerberg Biohub | CZB Cliahub Consortium |
| EPI_ISL_739388 | Santa Clara County Public Health Laboratory | Chan-Zuckerberg Biohub | CZB Cliahub Consortium |
| EPI_ISL_739389 | Humboldt County Public Health Laboratory | Chan-Zuckerberg Biohub | CZB Cliahub Consortium |
| EPI_ISL_739390 | Madera County Department of Public Health | Chan-Zuckerberg Biohub | CZB Cliahub Consortium |
| EPI_ISL_739391 | Alameda County Public Health Lab | Chan-Zuckerberg Biohub | CZB Cliahub Consortium |
| EPI_ISL_739392 | Santa Clara County Public Health Laboratory | Chan-Zuckerberg Biohub | CZB Cliahub Consortium |
| EPI_ISL_739393 | Madera County Department of Public Health | Chan-Zuckerberg Biohub | CZB Cliahub Consortium |
| EPI_ISL_739394 | Renegade | Chan-Zuckerberg Biohub | CZB Cliahub Consortium |
| EPI_ISL_739395 | Alameda County Public Health Lab | Chan-Zuckerberg Biohub | CZB Cliahub Consortium |
| EPI_ISL_739396, EPI_ISL_739397 | Orange County Public Health Lab | Chan-Zuckerberg Biohub | CZB Cliahub Consortium |
| EPI_ISL_739398 | Santa Clara County Public Health Laboratory | Chan-Zuckerberg Biohub | CZB Cliahub Consortium |
| EPI_ISL_739399 | County of San Luis Obispo Public Health Laboratory | Chan-Zuckerberg Biohub | CZB Cliahub Consortium |
| EPI_ISL_739400 | Santa Clara County Public Health Laboratory | Chan-Zuckerberg Biohub | CZB Cliahub Consortium |
| EPI_ISL_739401 | Orange County Public Health Lab | Chan-Zuckerberg Biohub | CZB Cliahub Consortium |
| EPI_ISL_739402 | Santa Clara County Public Health Laboratory | Chan-Zuckerberg Biohub | CZB Cliahub Consortium |
| EPI_ISL_739403 | Humboldt County Public Health Laboratory | Chan-Zuckerberg Biohub | CZB Cliahub Consortium |
| EPI_ISL_739404 | Santa Clara County Public Health Laboratory | Chan-Zuckerberg Biohub | CZB Cliahub Consortium |
| EPI_ISL_739405 | Humboldt County Public Health Laboratory | Chan-Zuckerberg Biohub | CZB Cliahub Consortium |
| EPI_ISL_739406, EPI_ISL_739407, EPI_ISL_739408 | Alameda County Public Health Lab | Chan-Zuckerberg Biohub | CZB Cliahub Consortium |
| EPI_ISL_739409 | Santa Clara County Public Health Laboratory | Chan-Zuckerberg Biohub | CZB Cliahub Consortium |
| EPI_ISL_739410 | Orange County Public Health Lab | Chan-Zuckerberg Biohub | CZB Cliahub Consortium |
| EPI_ISL_739411 | Humboldt County Public Health Laboratory | Chan-Zuckerberg Biohub | CZB Cliahub Consortium |
| EPI_ISL_739412 | Orange County Public Health Lab | Chan-Zuckerberg Biohub | CZB Cliahub Consortium |
| EPI_ISL_739413, EPI_ISL_739414, EPI_ISL_739415, EPI_ISL_739416 | Santa Clara County Public Health Laboratory | Chan-Zuckerberg Biohub | CZB Cliahub Consortium |
| EPI_ISL_739417, EPI_ISL_739418 | Humboldt County Public Health Laboratory | Chan-Zuckerberg Biohub | CZB Cliahub Consortium |
| EPI_ISL_739419 | Santa Clara County Public Health Laboratory | Chan-Zuckerberg Biohub | CZB Cliahub Consortium |
| EPI_ISL_739420, | Orange County Public Health Lab | Chan-Zuckerberg Biohub | CZB Cliahub Consortium |

| | | | |
|---|---|---|---|
| EPI_ISL_739421 | | | |
| EPI_ISL_739422 | Alameda County Public Health Lab | Chan-Zuckerberg Biohub | CZB Cliahub Consortium |
| EPI_ISL_739423 | Orange County Public Health Lab | Chan-Zuckerberg Biohub | CZB Cliahub Consortium |
| EPI_ISL_739424 | Alameda County Public Health Lab | Chan-Zuckerberg Biohub | CZB Cliahub Consortium |
| EPI_ISL_739425 | Humboldt County Public Health Laboratory | Chan-Zuckerberg Biohub | CZB Cliahub Consortium |
| EPI_ISL_739426, EPI_ISL_739427 | Alameda County Public Health Lab | Chan-Zuckerberg Biohub | CZB Cliahub Consortium |
| EPI_ISL_739428 | Orange County Public Health Lab | Chan-Zuckerberg Biohub | CZB Cliahub Consortium |
| EPI_ISL_739429 | Madera County Department of Public Health | Chan-Zuckerberg Biohub | CZB Cliahub Consortium |
| EPI_ISL_739430 | Humboldt County Public Health Laboratory | Chan-Zuckerberg Biohub | CZB Cliahub Consortium |
| EPI_ISL_739431 | Santa Clara County Public Health Laboratory | Chan-Zuckerberg Biohub | CZB Cliahub Consortium |
| EPI_ISL_739432 | Orange County Public Health Lab | Chan-Zuckerberg Biohub | CZB Cliahub Consortium |
| EPI_ISL_739433 | Humboldt County Public Health Laboratory | Chan-Zuckerberg Biohub | CZB Cliahub Consortium |
| EPI_ISL_739434, EPI_ISL_739435 | Alameda County Public Health Lab | Chan-Zuckerberg Biohub | CZB Cliahub Consortium |
| EPI_ISL_739436 | Humboldt County Public Health Laboratory | Chan-Zuckerberg Biohub | CZB Cliahub Consortium |
| EPI_ISL_739437, EPI_ISL_739438 | Orange County Public Health Lab | Chan-Zuckerberg Biohub | CZB Cliahub Consortium |
| EPI_ISL_739439 | Alameda County Public Health Lab | Chan-Zuckerberg Biohub | CZB Cliahub Consortium |
| EPI_ISL_739440 | Santa Clara County Public Health Laboratory | Chan-Zuckerberg Biohub | CZB Cliahub Consortium |
| EPI_ISL_739441 | Orange County Public Health Lab | Chan-Zuckerberg Biohub | CZB Cliahub Consortium |
| EPI_ISL_739442 | Humboldt County Public Health Laboratory | Chan-Zuckerberg Biohub | CZB Cliahub Consortium |
| EPI_ISL_739443 | Santa Clara County Public Health Laboratory | Chan-Zuckerberg Biohub | CZB Cliahub Consortium |
| EPI_ISL_739444 | Alameda County Public Health Lab | Chan-Zuckerberg Biohub | CZB Cliahub Consortium |
| EPI_ISL_739445 | Orange County Public Health Lab | Chan-Zuckerberg Biohub | CZB Cliahub Consortium |
| EPI_ISL_739446 | Alameda County Public Health Lab | Chan-Zuckerberg Biohub | CZB Cliahub Consortium |
| EPI_ISL_739447 | Humboldt County Public Health Laboratory | Chan-Zuckerberg Biohub | CZB Cliahub Consortium |
| EPI_ISL_739448, EPI_ISL_739449 | Alameda County Public Health Lab | Chan-Zuckerberg Biohub | CZB Cliahub Consortium |
| EPI_ISL_739450 | Orange County Public Health Lab | Chan-Zuckerberg Biohub | CZB Cliahub Consortium |
| EPI_ISL_739451 | Santa Clara County Public Health Laboratory | Chan-Zuckerberg Biohub | CZB Cliahub Consortium |
| EPI_ISL_739452 | Humboldt County Public Health Laboratory | Chan-Zuckerberg Biohub | CZB Cliahub Consortium |
| EPI_ISL_739453 | Santa Clara County Public Health Laboratory | Chan-Zuckerberg Biohub | CZB Cliahub Consortium |
| EPI_ISL_739454 | Alameda County Public Health Lab | Chan-Zuckerberg Biohub | CZB Cliahub Consortium |
| EPI_ISL_739455 | Orange County Public Health Lab | Chan-Zuckerberg Biohub | CZB Cliahub Consortium |
| EPI_ISL_739456, EPI_ISL_739457, EPI_ISL_739458 | Santa Clara County Public Health Laboratory | Chan-Zuckerberg Biohub | CZB Cliahub Consortium |
| EPI_ISL_739459 | Alameda County Public Health Lab | Chan-Zuckerberg Biohub | CZB Cliahub Consortium |
| EPI_ISL_739460 | Orange County Public Health Lab | Chan-Zuckerberg Biohub | CZB Cliahub Consortium |
| EPI_ISL_739461 | Humboldt County Public Health Laboratory | Chan-Zuckerberg Biohub | CZB Cliahub Consortium |
| EPI_ISL_739462 | Orange County Public Health Lab | Chan-Zuckerberg Biohub | CZB Cliahub Consortium |
| EPI_ISL_739463 | Alameda County Public Health Lab | Chan-Zuckerberg Biohub | CZB Cliahub Consortium |
| EPI_ISL_739464, EPI_ISL_739465 | Orange County Public Health Lab | Chan-Zuckerberg Biohub | CZB Cliahub Consortium |
| EPI_ISL_739466, EPI_ISL_739467 | Renegade | Chan-Zuckerberg Biohub | CZB Cliahub Consortium |
| EPI_ISL_739468 | Alameda County Public Health Lab | Chan-Zuckerberg Biohub | CZB Cliahub Consortium |
| EPI_ISL_739469 | Napa-Solano-Yolo- Marin County (NSYM) Public Health Laboratories | Chan-Zuckerberg Biohub | CZB Cliahub Consortium |
| EPI_ISL_739470 | Alameda County Public Health Lab | Chan-Zuckerberg Biohub | CZB Cliahub Consortium |
| EPI_ISL_739471 | Napa-Solano-Yolo- Marin County (NSYM) Public Health Laboratories | Chan-Zuckerberg Biohub | CZB Cliahub Consortium |
| EPI_ISL_739472, EPI_ISL_739473 | Alameda County Public Health Lab | Chan-Zuckerberg Biohub | CZB Cliahub Consortium |
| EPI_ISL_739474 | Napa-Solano-Yolo- Marin County (NSYM) Public Health Laboratories | Chan-Zuckerberg Biohub | CZB Cliahub Consortium |
| EPI_ISL_739475 | Humboldt County Public Health Laboratory | Chan-Zuckerberg Biohub | CZB Cliahub Consortium |
| EPI_ISL_739476 | Santa Clara County Public Health Laboratory | Chan-Zuckerberg Biohub | CZB Cliahub Consortium |
| EPI_ISL_739477 | Orange County Public Health Lab | Chan-Zuckerberg Biohub | CZB Cliahub Consortium |
| EPI_ISL_739478 | Renegade | Chan-Zuckerberg Biohub | CZB Cliahub Consortium |
| EPI_ISL_739479 | Santa Clara County Public Health Laboratory | Chan-Zuckerberg Biohub | CZB Cliahub Consortium |
| EPI_ISL_739480 | County of San Luis Obispo Public Health Laboratory | Chan-Zuckerberg Biohub | CZB Cliahub Consortium |
| EPI_ISL_739481 | Santa Clara County Public Health Laboratory | Chan-Zuckerberg Biohub | CZB Cliahub Consortium |
| EPI_ISL_739482 | Orange County Public Health Lab | Chan-Zuckerberg Biohub | CZB Cliahub Consortium |
| EPI_ISL_739483, EPI_ISL_739484 | Alameda County Public Health Lab | Chan-Zuckerberg Biohub | CZB Cliahub Consortium |

| | | | |
|---|---|---|---|
| EPI_ISL_739485 | Santa Clara County Public Health Laboratory | Chan-Zuckerberg Biohub | CZB Cliahub Consortium |
| EPI_ISL_739486 | Orange County Public Health Lab | Chan-Zuckerberg Biohub | CZB Cliahub Consortium |
| EPI_ISL_739487 | Santa Clara County Public Health Laboratory | Chan-Zuckerberg Biohub | CZB Cliahub Consortium |
| EPI_ISL_739488 | Orange County Public Health Lab | Chan-Zuckerberg Biohub | CZB Cliahub Consortium |
| EPI_ISL_739489 | Humboldt County Public Health Laboratory | Chan-Zuckerberg Biohub | CZB Cliahub Consortium |
| EPI_ISL_739490 | Orange County Public Health Lab | Chan-Zuckerberg Biohub | CZB Cliahub Consortium |
| EPI_ISL_739491 | Santa Clara County Public Health Laboratory | Chan-Zuckerberg Biohub | CZB Cliahub Consortium |
| EPI_ISL_739492 | County of San Luis Obispo Public Health Laboratory | Chan-Zuckerberg Biohub | CZB Cliahub Consortium |
| EPI_ISL_739493, EPI_ISL_739494 | Orange County Public Health Lab | Chan-Zuckerberg Biohub | CZB Cliahub Consortium |
| EPI_ISL_739495 | Alameda County Public Health Lab | Chan-Zuckerberg Biohub | CZB Cliahub Consortium |
| EPI_ISL_739496, EPI_ISL_739497 | Orange County Public Health Lab | Chan-Zuckerberg Biohub | CZB Cliahub Consortium |
| EPI_ISL_739498 | Santa Clara County Public Health Laboratory | Chan-Zuckerberg Biohub | CZB Cliahub Consortium |
| EPI_ISL_739499 | Humboldt County Public Health Laboratory | Chan-Zuckerberg Biohub | CZB Cliahub Consortium |
| EPI_ISL_739500 | Orange County Public Health Lab | Chan-Zuckerberg Biohub | CZB Cliahub Consortium |
| EPI_ISL_739501, EPI_ISL_739502 | Santa Clara County Public Health Laboratory | Chan-Zuckerberg Biohub | CZB Cliahub Consortium |
| EPI_ISL_739503 | Alameda County Public Health Lab | Chan-Zuckerberg Biohub | CZB Cliahub Consortium |
| EPI_ISL_739504, EPI_ISL_739505 | Santa Clara County Public Health Laboratory | Chan-Zuckerberg Biohub | CZB Cliahub Consortium |
| EPI_ISL_739506, EPI_ISL_739507 | Alameda County Public Health Lab | Chan-Zuckerberg Biohub | CZB Cliahub Consortium |
| EPI_ISL_739508, EPI_ISL_739509, EPI_ISL_739510 | Santa Clara County Public Health Laboratory | Chan-Zuckerberg Biohub | CZB Cliahub Consortium |
| EPI_ISL_739511 | Alameda County Public Health Lab | Chan-Zuckerberg Biohub | CZB Cliahub Consortium |
| EPI_ISL_739512 | Santa Clara County Public Health Laboratory | Chan-Zuckerberg Biohub | CZB Cliahub Consortium |
| EPI_ISL_739513 | Orange County Public Health Lab | Chan-Zuckerberg Biohub | CZB Cliahub Consortium |
| EPI_ISL_739514 | Napa-Solano-Yolo- Marin County (NSYM) Public Health Laboratories | Chan-Zuckerberg Biohub | CZB Cliahub Consortium |
| EPI_ISL_739515, EPI_ISL_739516 | Alameda County Public Health Lab | Chan-Zuckerberg Biohub | CZB Cliahub Consortium |
| EPI_ISL_739517 | Orange County Public Health Lab | Chan-Zuckerberg Biohub | CZB Cliahub Consortium |
| EPI_ISL_739518, EPI_ISL_739519 | Alameda County Public Health Lab | Chan-Zuckerberg Biohub | CZB Cliahub Consortium |
| EPI_ISL_739520, EPI_ISL_739521 | Santa Clara County Public Health Laboratory | Chan-Zuckerberg Biohub | CZB Cliahub Consortium |
| EPI_ISL_739522, EPI_ISL_739523 | Alameda County Public Health Lab | Chan-Zuckerberg Biohub | CZB Cliahub Consortium |
| EPI_ISL_739524 | Orange County Public Health Lab | Chan-Zuckerberg Biohub | CZB Cliahub Consortium |
| EPI_ISL_739525 | Humboldt County Public Health Laboratory | Chan-Zuckerberg Biohub | CZB Cliahub Consortium |
| EPI_ISL_739526 | Alameda County Public Health Lab | Chan-Zuckerberg Biohub | CZB Cliahub Consortium |
| EPI_ISL_739527 | Humboldt County Public Health Laboratory | Chan-Zuckerberg Biohub | CZB Cliahub Consortium |
| EPI_ISL_739528 | Alameda County Public Health Lab | Chan-Zuckerberg Biohub | CZB Cliahub Consortium |
| EPI_ISL_739529 | Orange County Public Health Lab | Chan-Zuckerberg Biohub | CZB Cliahub Consortium |
| EPI_ISL_739530, EPI_ISL_739531 | Humboldt County Public Health Laboratory | Chan-Zuckerberg Biohub | CZB Cliahub Consortium |
| EPI_ISL_739532 | Orange County Public Health Lab | Chan-Zuckerberg Biohub | CZB Cliahub Consortium |
| EPI_ISL_739533 | Humboldt County Public Health Laboratory | Chan-Zuckerberg Biohub | CZB Cliahub Consortium |
| EPI_ISL_739534 | Orange County Public Health Lab | Chan-Zuckerberg Biohub | CZB Cliahub Consortium |
| EPI_ISL_739535 | Alameda County Public Health Lab | Chan-Zuckerberg Biohub | CZB Cliahub Consortium |
| EPI_ISL_739536, EPI_ISL_739537 | Santa Clara County Public Health Laboratory | Chan-Zuckerberg Biohub | CZB Cliahub Consortium |
| EPI_ISL_739538 | Alameda County Public Health Lab | Chan-Zuckerberg Biohub | CZB Cliahub Consortium |
| EPI_ISL_739539 | Madera County Department of Public Health | Chan-Zuckerberg Biohub | CZB Cliahub Consortium |
| EPI_ISL_739540 | Orange County Public Health Lab | Chan-Zuckerberg Biohub | CZB Cliahub Consortium |
| EPI_ISL_739541 | Alameda County Public Health Lab | Chan-Zuckerberg Biohub | CZB Cliahub Consortium |
| EPI_ISL_739542 | Humboldt County Public Health Laboratory | Chan-Zuckerberg Biohub | CZB Cliahub Consortium |
| EPI_ISL_739543 | Santa Clara County Public Health Laboratory | Chan-Zuckerberg Biohub | CZB Cliahub Consortium |
| EPI_ISL_739544 | Alameda County Public Health Lab | Chan-Zuckerberg Biohub | CZB Cliahub Consortium |
| EPI_ISL_739545 | Orange County Public Health Lab | Chan-Zuckerberg Biohub | CZB Cliahub Consortium |
| EPI_ISL_739546 | Monterey County Public Health Lab | Chan-Zuckerberg Biohub | CZB Cliahub Consortium |
| EPI_ISL_739547 | Orange County Public Health Lab | Chan-Zuckerberg Biohub | CZB Cliahub Consortium |
| EPI_ISL_739548, EPI_ISL_739549, EPI_ISL_739550 | Santa Clara County Public Health Laboratory | Chan-Zuckerberg Biohub | CZB Cliahub Consortium |
| EPI_ISL_739551, EPI_ISL_739552 | Alameda County Public Health Lab | Chan-Zuckerberg Biohub | CZB Cliahub Consortium |
| EPI_ISL_739553 | Humboldt County Public Health Laboratory | Chan-Zuckerberg Biohub | CZB Cliahub Consortium |
| EPI_ISL_739554 | Santa Clara County Public Health Laboratory | Chan-Zuckerberg Biohub | CZB Cliahub Consortium |

| | | | |
|---|---|---|---|
| EPI_ISL_739555 | Alameda County Public Health Lab | Chan-Zuckerberg Biohub | CZB Cliahub Consortium |
| EPI_ISL_739556 | Orange County Public Health Lab | Chan-Zuckerberg Biohub | CZB Cliahub Consortium |
| EPI_ISL_739557 | Napa-Solano-Yolo- Marin County (NSYM) Public Health Laboratories | Chan-Zuckerberg Biohub | CZB Cliahub Consortium |
| EPI_ISL_739558 | Humboldt County Public Health Laboratory | Chan-Zuckerberg Biohub | CZB Cliahub Consortium |
| EPI_ISL_739559 | Orange County Public Health Lab | Chan-Zuckerberg Biohub | CZB Cliahub Consortium |
| EPI_ISL_739560, EPI_ISL_739561, EPI_ISL_739562 | Santa Clara County Public Health Laboratory | Chan-Zuckerberg Biohub | CZB Cliahub Consortium |
| EPI_ISL_739563 | Alameda County Public Health Lab | Chan-Zuckerberg Biohub | CZB Cliahub Consortium |
| EPI_ISL_739564 | Renegade | Chan-Zuckerberg Biohub | CZB Cliahub Consortium |
| EPI_ISL_739565 | Alameda County Public Health Lab | Chan-Zuckerberg Biohub | CZB Cliahub Consortium |
| EPI_ISL_739566 | Orange County Public Health Lab | Chan-Zuckerberg Biohub | CZB Cliahub Consortium |
| EPI_ISL_739567 | Madera County Department of Public Health | Chan-Zuckerberg Biohub | CZB Cliahub Consortium |
| EPI_ISL_739568 | Orange County Public Health Lab | Chan-Zuckerberg Biohub | CZB Cliahub Consortium |
| EPI_ISL_739569 | Alameda County Public Health Lab | Chan-Zuckerberg Biohub | CZB Cliahub Consortium |
| EPI_ISL_739570 | Orange County Public Health Lab | Chan-Zuckerberg Biohub | CZB Cliahub Consortium |
| EPI_ISL_739571 | Humboldt County Public Health Laboratory | Chan-Zuckerberg Biohub | CZB Cliahub Consortium |
| EPI_ISL_739572 | Alameda County Public Health Lab | Chan-Zuckerberg Biohub | CZB Cliahub Consortium |
| EPI_ISL_739573 | Orange County Public Health Lab | Chan-Zuckerberg Biohub | CZB Cliahub Consortium |
| EPI_ISL_739574 | Santa Clara County Public Health Laboratory | Chan-Zuckerberg Biohub | CZB Cliahub Consortium |
| EPI_ISL_739575, EPI_ISL_739576 | Alameda County Public Health Lab | Chan-Zuckerberg Biohub | CZB Cliahub Consortium |
| EPI_ISL_739577 | Orange County Public Health Lab | Chan-Zuckerberg Biohub | CZB Cliahub Consortium |
| EPI_ISL_739578 | Alameda County Public Health Lab | Chan-Zuckerberg Biohub | CZB Cliahub Consortium |
| EPI_ISL_739579, EPI_ISL_739580 | Santa Clara County Public Health Laboratory | Chan-Zuckerberg Biohub | CZB Cliahub Consortium |
| EPI_ISL_739581, EPI_ISL_739582 | Orange County Public Health Lab | Chan-Zuckerberg Biohub | CZB Cliahub Consortium |
| EPI_ISL_739583 | Alameda County Public Health Lab | Chan-Zuckerberg Biohub | CZB Cliahub Consortium |
| EPI_ISL_739584 | Santa Clara County Public Health Laboratory | Chan-Zuckerberg Biohub | CZB Cliahub Consortium |
| EPI_ISL_739585 | Humboldt County Public Health Laboratory | Chan-Zuckerberg Biohub | CZB Cliahub Consortium |
| EPI_ISL_739586 | Orange County Public Health Lab | Chan-Zuckerberg Biohub | CZB Cliahub Consortium |
| EPI_ISL_739587 | Alameda County Public Health Lab | Chan-Zuckerberg Biohub | CZB Cliahub Consortium |
| EPI_ISL_739588 | County of San Luis Obispo Public Health Laboratory | Chan-Zuckerberg Biohub | CZB Cliahub Consortium |
| EPI_ISL_739589 | Santa Clara County Public Health Laboratory | Chan-Zuckerberg Biohub | CZB Cliahub Consortium |
| EPI_ISL_739590 | Alameda County Public Health Lab | Chan-Zuckerberg Biohub | CZB Cliahub Consortium |
| EPI_ISL_739591, EPI_ISL_739592 | Orange County Public Health Lab | Chan-Zuckerberg Biohub | CZB Cliahub Consortium |
| EPI_ISL_739593, EPI_ISL_739594 | Santa Clara County Public Health Laboratory | Chan-Zuckerberg Biohub | CZB Cliahub Consortium |
| EPI_ISL_739595 | Orange County Public Health Lab | Chan-Zuckerberg Biohub | CZB Cliahub Consortium |
| EPI_ISL_739596, EPI_ISL_739597 | Santa Clara County Public Health Laboratory | Chan-Zuckerberg Biohub | CZB Cliahub Consortium |
| EPI_ISL_739598 | Alameda County Public Health Lab | Chan-Zuckerberg Biohub | CZB Cliahub Consortium |
| EPI_ISL_739599 | Orange County Public Health Lab | Chan-Zuckerberg Biohub | CZB Cliahub Consortium |
| EPI_ISL_739600, EPI_ISL_739601 | Alameda County Public Health Lab | Chan-Zuckerberg Biohub | CZB Cliahub Consortium |
| EPI_ISL_739602 | Santa Clara County Public Health Laboratory | Chan-Zuckerberg Biohub | CZB Cliahub Consortium |
| EPI_ISL_739603 | San Francisco Public Health Laboratory | Chan-Zuckerberg Biohub | CZB Cliahub Consortium |
| EPI_ISL_739604 | Alameda County Public Health Lab | Chan-Zuckerberg Biohub | CZB Cliahub Consortium |
| EPI_ISL_739605 | Santa Clara County Public Health Laboratory | Chan-Zuckerberg Biohub | CZB Cliahub Consortium |
| EPI_ISL_739606 | Alameda County Public Health Lab | Chan-Zuckerberg Biohub | CZB Cliahub Consortium |
| EPI_ISL_739607 | Orange County Public Health Lab | Chan-Zuckerberg Biohub | CZB Cliahub Consortium |
| EPI_ISL_739608 | Humboldt County Public Health Laboratory | Chan-Zuckerberg Biohub | CZB Cliahub Consortium |
| EPI_ISL_739609 | Orange County Public Health Lab | Chan-Zuckerberg Biohub | CZB Cliahub Consortium |
| EPI_ISL_739610 | Alameda County Public Health Lab | Chan-Zuckerberg Biohub | CZB Cliahub Consortium |
| EPI_ISL_739611 | Humboldt County Public Health Laboratory | Chan-Zuckerberg Biohub | CZB Cliahub Consortium |
| EPI_ISL_739612 | Santa Clara County Public Health Laboratory | Chan-Zuckerberg Biohub | CZB Cliahub Consortium |
| EPI_ISL_739613 | UCSF Clinical Microbiology Laboratory | Chan-Zuckerberg Biohub | CZB Cliahub Consortium |
| EPI_ISL_739614 | Orange County Public Health Lab | Chan-Zuckerberg Biohub | CZB Cliahub Consortium |
| EPI_ISL_739615 | Santa Clara County Public Health Laboratory | Chan-Zuckerberg Biohub | CZB Cliahub Consortium |
| EPI_ISL_739616 | Alameda County Public Health Lab | Chan-Zuckerberg Biohub | CZB Cliahub Consortium |
| EPI_ISL_739617, EPI_ISL_739618 | Santa Clara County Public Health Laboratory | Chan-Zuckerberg Biohub | CZB Cliahub Consortium |
| EPI_ISL_739619 | Alameda County Public Health Lab | Chan-Zuckerberg Biohub | CZB Cliahub Consortium |
| EPI_ISL_739620 | Orange County Public Health Lab | Chan-Zuckerberg Biohub | CZB Cliahub Consortium |
| EPI_ISL_739621 | Humboldt County Public Health Laboratory | Chan-Zuckerberg Biohub | CZB Cliahub Consortium |
| EPI_ISL_739622, | Alameda County Public Health Lab | Chan-Zuckerberg Biohub | CZB Cliahub Consortium |

| | | | |
|---|---|---|---|
| EPI_ISL_739623 | | | |
| EPI_ISL_739624 | Santa Clara County Public Health Laboratory | Chan-Zuckerberg Biohub | CZB Cliahub Consortium |
| EPI_ISL_739625 | Orange County Public Health Lab | Chan-Zuckerberg Biohub | CZB Cliahub Consortium |
| EPI_ISL_739626 | Santa Clara County Public Health Laboratory | Chan-Zuckerberg Biohub | CZB Cliahub Consortium |
| EPI_ISL_739627 | Orange County Public Health Lab | Chan-Zuckerberg Biohub | CZB Cliahub Consortium |
| EPI_ISL_739628 | Napa-Solano-Yolo- Marin County (NSYM) Public Health Laboratories | Chan-Zuckerberg Biohub | CZB Cliahub Consortium |
| EPI_ISL_739629 | Santa Clara County Public Health Laboratory | Chan-Zuckerberg Biohub | CZB Cliahub Consortium |
| EPI_ISL_739630 | Renegade | Chan-Zuckerberg Biohub | CZB Cliahub Consortium |
| EPI_ISL_739631 | Alameda County Public Health Lab | Chan-Zuckerberg Biohub | CZB Cliahub Consortium |
| EPI_ISL_739632, EPI_ISL_739633 | UCSF Clinical Microbiology Laboratory | Chan-Zuckerberg Biohub | CZB Cliahub Consortium |
| EPI_ISL_739634 | Alameda County Public Health Lab | Chan-Zuckerberg Biohub | CZB Cliahub Consortium |
| EPI_ISL_739635 | Napa-Solano-Yolo- Marin County (NSYM) Public Health Laboratories | Chan-Zuckerberg Biohub | CZB Cliahub Consortium |
| EPI_ISL_739636, EPI_ISL_739637 | Orange County Public Health Lab | Chan-Zuckerberg Biohub | CZB Cliahub Consortium |
| EPI_ISL_739638 | Alameda County Public Health Lab | Chan-Zuckerberg Biohub | CZB Cliahub Consortium |
| EPI_ISL_739639 | Tulare County Public Health Lab | Chan-Zuckerberg Biohub | CZB Cliahub Consortium |
| EPI_ISL_739640 | Alameda County Public Health Lab | Chan-Zuckerberg Biohub | CZB Cliahub Consortium |
| EPI_ISL_739641 | Orange County Public Health Lab | Chan-Zuckerberg Biohub | CZB Cliahub Consortium |
| EPI_ISL_739642 | Santa Clara County Public Health Laboratory | Chan-Zuckerberg Biohub | CZB Cliahub Consortium |
| EPI_ISL_739643, EPI_ISL_739644 | Alameda County Public Health Lab | Chan-Zuckerberg Biohub | CZB Cliahub Consortium |
| EPI_ISL_739645 | Napa-Solano-Yolo- Marin County (NSYM) Public Health Laboratories | Chan-Zuckerberg Biohub | CZB Cliahub Consortium |
| EPI_ISL_739646 | Orange County Public Health Lab | Chan-Zuckerberg Biohub | CZB Cliahub Consortium |
| EPI_ISL_739647 | Santa Clara County Public Health Laboratory | Chan-Zuckerberg Biohub | CZB Cliahub Consortium |
| EPI_ISL_739648 | Alameda County Public Health Lab | Chan-Zuckerberg Biohub | CZB Cliahub Consortium |
| EPI_ISL_739649, EPI_ISL_739650 | Santa Clara County Public Health Laboratory | Chan-Zuckerberg Biohub | CZB Cliahub Consortium |
| EPI_ISL_739651 | Humboldt County Public Health Laboratory | Chan-Zuckerberg Biohub | CZB Cliahub Consortium |
| EPI_ISL_739652 | Alameda County Public Health Lab | Chan-Zuckerberg Biohub | CZB Cliahub Consortium |
| EPI_ISL_739653 | Santa Clara County Public Health Laboratory | Chan-Zuckerberg Biohub | CZB Cliahub Consortium |
| EPI_ISL_739654 | Orange County Public Health Lab | Chan-Zuckerberg Biohub | CZB Cliahub Consortium |
| EPI_ISL_739655 | Renegade | Chan-Zuckerberg Biohub | CZB Cliahub Consortium |
| EPI_ISL_739656 | Alameda County Public Health Lab | Chan-Zuckerberg Biohub | CZB Cliahub Consortium |
| EPI_ISL_739657 | Santa Clara County Public Health Laboratory | Chan-Zuckerberg Biohub | CZB Cliahub Consortium |
| EPI_ISL_739658 | Orange County Public Health Lab | Chan-Zuckerberg Biohub | CZB Cliahub Consortium |
| EPI_ISL_745261, EPI_ISL_745262, EPI_ISL_745263, EPI_ISL_745264, EPI_ISL_745265, EPI_ISL_745266, EPI_ISL_745267, EPI_ISL_745268, EPI_ISL_745269, EPI_ISL_745270, EPI_ISL_745271, EPI_ISL_745272, EPI_ISL_745273, EPI_ISL_745274, EPI_ISL_745276, EPI_ISL_745277, EPI_ISL_745278, EPI_ISL_745279, EPI_ISL_745280, EPI_ISL_745281, EPI_ISL_745282, EPI_ISL_745283, EPI_ISL_745284, EPI_ISL_745285, EPI_ISL_745286, EPI_ISL_745287, EPI_ISL_745288, EPI_ISL_745289, EPI_ISL_745290, EPI_ISL_745291, EPI_ISL_745292, EPI_ISL_745293, EPI_ISL_745294, EPI_ISL_745295, EPI_ISL_745296, EPI_ISL_745297, EPI_ISL_745298, EPI_ISL_745299, EPI_ISL_745300, EPI_ISL_745301, EPI_ISL_745302, EPI_ISL_745303, EPI_ISL_745304, EPI_ISL_745305, EPI_ISL_745306 | | | |
| see above | Texas Department of State Health Services | Texas Department of State Health Services | Rashmi Tuladhar, Bonnie Oh, Jenny Zhang, Maliha Rahman, Anita Pokharel, Myong Koag, Chung Wang, Rachel Lee, Grace Kubin, Mayela Pedrueza, James Daniel Bonser |
| EPI_ISL_745398 | DOHMH Jamaica | New York City Public Health Laboratory | Jade Wang, et al. |
| EPI_ISL_745399 | DOHMH Morrisania | New York City Public Health Laboratory | Jade Wang, et al. |
| EPI_ISL_745400 | DOHMH Central Harlem | New York City Public Health Laboratory | Jade Wang, et al. |
| EPI_ISL_745401 | DOHMH Corona | New York City Public Health Laboratory | Jade Wang, et al. |
| EPI_ISL_745402 | DOHMH Jamaica | New York City Public Health Laboratory | Jade Wang, et al. |
| EPI_ISL_745403 | DOHMH Corona | New York City Public Health Laboratory | Jade Wang, et al. |
| EPI_ISL_745404 | DOHMH Central Harlem | New York City Public Health Laboratory | Jade Wang, et al. |
| EPI_ISL_745405 | DOHMH Jamaica | New York City Public Health Laboratory | Jade Wang, et al. |
| EPI_ISL_745406 | DOHMH Morrisania | New York City Public Health Laboratory | Jade Wang, et al. |
| EPI_ISL_745407 | DOHMH Jamaica | New York City Public Health Laboratory | Jade Wang, et al. |
| EPI_ISL_745408 | DOHMH Riverside | New York City Public Health Laboratory | Jade Wang, et al. |
| EPI_ISL_745409 | DOHMH Corona | New York City Public Health Laboratory | Jade Wang, et al. |
| EPI_ISL_745410 | DOHMH Chelsea | New York City Public Health Laboratory | Jade Wang, et al. |
| EPI_ISL_745411 | DOHMH Corona | New York City Public Health Laboratory | Jade Wang, et al. |
| EPI_ISL_745412 | DOHMH Morrisania | New York City Public Health Laboratory | Jade Wang, et al. |
| EPI_ISL_745413 | DOHMH Central Harlem | New York City Public Health Laboratory | Jade Wang, et al. |
| EPI_ISL_745414, EPI_ISL_745415, EPI_ISL_745416, EPI_ISL_745417, EPI_ISL_745418 | DOHMH Jamaica | New York City Public Health Laboratory | Jade Wang, et al. |
| EPI_ISL_745419, EPI_ISL_745420 | DOHMH Crown Heights | New York City Public Health Laboratory | Jade Wang, et al. |
| EPI_ISL_745421, EPI_ISL_745422 | DOHMH Chelsea | New York City Public Health Laboratory | Jade Wang, et al. |
| EPI_ISL_745423, EPI_ISL_745424 | DOHMH Corona | New York City Public Health Laboratory | Jade Wang, et al. |
| EPI_ISL_745425 | DOHMH Central Harlem | New York City Public Health Laboratory | Jade Wang, et al. |
| EPI_ISL_745426, EPI_ISL_745427 | DOHMH Corona | New York City Public Health Laboratory | Jade Wang, et al. |
| EPI_ISL_745428, EPI_ISL_745429 | DOHMH Riverside | New York City Public Health Laboratory | Jade Wang, et al. |
| EPI_ISL_745430, EPI_ISL_745431, | DOHMH Morrisania | New York City Public Health Laboratory | Jade Wang, et al. |

| | | | |
|---|---|---|---|
| EPI_ISL_745432, EPI_ISL_745433 | | | |
| EPI_ISL_745434, EPI_ISL_745435 | DOHMH Corona | New York City Public Health Laboratory | Jade Wang, et al. |
| EPI_ISL_745436 | DOHMH Chelsea | New York City Public Health Laboratory | Jade Wang, et al. |
| EPI_ISL_745437, EPI_ISL_745438 | DOHMH PHL | New York City Public Health Laboratory | Jade Wang, et al. |
| EPI_ISL_745439 | DOHMH Riverside | New York City Public Health Laboratory | Jade Wang, et al. |
| EPI_ISL_745440 | DOHMH Jamaica | New York City Public Health Laboratory | Jade Wang, et al. |
| EPI_ISL_745441, EPI_ISL_745442, EPI_ISL_745443 | DOHMH Morrisania | New York City Public Health Laboratory | Jade Wang, et al. |
| EPI_ISL_745444 | DOHMH Jamaica | New York City Public Health Laboratory | Jade Wang, et al. |
| EPI_ISL_745445 | DOHMH Crown Heights | New York City Public Health Laboratory | Jade Wang, et al. |
| EPI_ISL_745446, EPI_ISL_745447 | DOHMH Corona | New York City Public Health Laboratory | Jade Wang, et al. |
| EPI_ISL_745448 | DOHMH Crown Heights | New York City Public Health Laboratory | Jade Wang, et al. |
| EPI_ISL_745449, EPI_ISL_745450 | DOHMH Morrisania | New York City Public Health Laboratory | Jade Wang, et al. |
| EPI_ISL_745451 | DOHMH Central Harlem | New York City Public Health Laboratory | Jade Wang, et al. |
| EPI_ISL_745452 | DOHMH Crown Heights | New York City Public Health Laboratory | Jade Wang, et al. |
| EPI_ISL_745453 | DOHMH Chelsea | New York City Public Health Laboratory | Jade Wang, et al. |
| EPI_ISL_745454 | DOHMH Crown Heights | New York City Public Health Laboratory | Jade Wang, et al. |
| EPI_ISL_745455 | DOHMH Corona | New York City Public Health Laboratory | Jade Wang, et al. |
| EPI_ISL_745456 | DOHMH PHL | New York City Public Health Laboratory | Jade Wang, et al. |
| EPI_ISL_745457 | DOHMH Chelsea | New York City Public Health Laboratory | Jade Wang, et al. |
| EPI_ISL_745458, EPI_ISL_745459, EPI_ISL_745460, EPI_ISL_745461, EPI_ISL_745462 | DOHMH Corona | New York City Public Health Laboratory | Jade Wang, et al. |
| EPI_ISL_745463 | DOHMH Fort Greene | New York City Public Health Laboratory | Jade Wang, et al. |
| EPI_ISL_745464 | DOHMH Corona | New York City Public Health Laboratory | Jade Wang, et al. |
| EPI_ISL_745465 | DOHMH PHL | New York City Public Health Laboratory | Jade Wang, et al. |
| EPI_ISL_745466, EPI_ISL_745467 | DOHMH Corona | New York City Public Health Laboratory | Jade Wang, et al. |
| EPI_ISL_745468 | DOHMH Riverside | New York City Public Health Laboratory | Jade Wang, et al. |
| EPI_ISL_745469 | DOHMH Central Harlem | New York City Public Health Laboratory | Jade Wang, et al. |
| EPI_ISL_745470 | DOHMH Morrisania | New York City Public Health Laboratory | Jade Wang, et al. |
| EPI_ISL_745471, EPI_ISL_745472, EPI_ISL_745473, EPI_ISL_745474, EPI_ISL_745475, EPI_ISL_745476, EPI_ISL_745477, EPI_ISL_745478, EPI_ISL_745479, EPI_ISL_745480, EPI_ISL_745481, EPI_ISL_745482, EPI_ISL_745483, EPI_ISL_745484, EPI_ISL_745485, EPI_ISL_745486, EPI_ISL_745487, EPI_ISL_745488, EPI_ISL_745489, EPI_ISL_745490, EPI_ISL_745491, EPI_ISL_745492, EPI_ISL_745493, EPI_ISL_745494, EPI_ISL_745495, EPI_ISL_745496, EPI_ISL_745497, EPI_ISL_745498, EPI_ISL_745499, EPI_ISL_745500, EPI_ISL_745501, EPI_ISL_745502, EPI_ISL_745503, EPI_ISL_745504, EPI_ISL_745505, EPI_ISL_745506, EPI_ISL_745507, EPI_ISL_745508, EPI_ISL_745509, EPI_ISL_745510, EPI_ISL_745511, EPI_ISL_745512, EPI_ISL_745513, EPI_ISL_745514, EPI_ISL_745515, EPI_ISL_745516, EPI_ISL_745517, EPI_ISL_745518, EPI_ISL_745519, EPI_ISL_745520, EPI_ISL_745521, EPI_ISL_745522, EPI_ISL_745523, EPI_ISL_745524, EPI_ISL_745525, EPI_ISL_745526, EPI_ISL_745527, EPI_ISL_745528, EPI_ISL_745529, EPI_ISL_745530, EPI_ISL_745531, EPI_ISL_745532, EPI_ISL_745533, EPI_ISL_745534, EPI_ISL_745535, EPI_ISL_745536, EPI_ISL_745537, EPI_ISL_745538, EPI_ISL_745539, EPI_ISL_745540, EPI_ISL_745541, EPI_ISL_745542, EPI_ISL_745543, EPI_ISL_745544, EPI_ISL_745545, EPI_ISL_745546, EPI_ISL_745547, EPI_ISL_745548, EPI_ISL_745549, EPI_ISL_745550, EPI_ISL_745551, EPI_ISL_745552, EPI_ISL_745553, EPI_ISL_745554, EPI_ISL_745555, EPI_ISL_745556, EPI_ISL_745557, EPI_ISL_745558, EPI_ISL_745559, EPI_ISL_745560, EPI_ISL_745561, EPI_ISL_745562, EPI_ISL_745563, EPI_ISL_745564, EPI_ISL_745565, EPI_ISL_745566, EPI_ISL_745567, EPI_ISL_745568, EPI_ISL_745569, EPI_ISL_745570, EPI_ISL_745571, EPI_ISL_745572, EPI_ISL_745573, EPI_ISL_745574, EPI_ISL_745575, EPI_ISL_745576, EPI_ISL_745577, EPI_ISL_745578, EPI_ISL_745579, EPI_ISL_745580, EPI_ISL_745581, EPI_ISL_745582, EPI_ISL_745583, EPI_ISL_745584, EPI_ISL_745585, EPI_ISL_745586, EPI_ISL_745587, EPI_ISL_745588, EPI_ISL_745589, EPI_ISL_745590, EPI_ISL_745591, EPI_ISL_745592, EPI_ISL_745593, EPI_ISL_745594, EPI_ISL_745595, EPI_ISL_745596, EPI_ISL_745597, EPI_ISL_745598, EPI_ISL_745659, EPI_ISL_745660, EPI_ISL_745661, EPI_ISL_745662, EPI_ISL_745663, EPI_ISL_745664, EPI_ISL_745665, EPI_ISL_745666, EPI_ISL_745667, EPI_ISL_745668, EPI_ISL_745669, EPI_ISL_745670, EPI_ISL_745671, EPI_ISL_745672, EPI_ISL_745673, EPI_ISL_745674, EPI_ISL_745675, EPI_ISL_745676, EPI_ISL_745677, EPI_ISL_745678, EPI_ISL_745679, EPI_ISL_745680, EPI_ISL_745681, EPI_ISL_745682, EPI_ISL_745683, EPI_ISL_745684, EPI_ISL_745685, EPI_ISL_745686, EPI_ISL_745687, EPI_ISL_745688, EPI_ISL_745689, EPI_ISL_745690, EPI_ISL_745691, EPI_ISL_745692, EPI_ISL_745693, EPI_ISL_745694, EPI_ISL_745695, EPI_ISL_745696, EPI_ISL_745697, EPI_ISL_745698, EPI_ISL_745699, EPI_ISL_745700, EPI_ISL_745701, EPI_ISL_745702, EPI_ISL_745703, EPI_ISL_745704, EPI_ISL_745705, EPI_ISL_745706, EPI_ISL_745707, EPI_ISL_745708, EPI_ISL_745709, EPI_ISL_745710, EPI_ISL_745711, EPI_ISL_745712, EPI_ISL_745713, EPI_ISL_745714, EPI_ISL_745715, EPI_ISL_745716, EPI_ISL_745717, EPI_ISL_745718, EPI_ISL_745719, EPI_ISL_745720, EPI_ISL_745721, EPI_ISL_745722, EPI_ISL_745723, EPI_ISL_745724, EPI_ISL_745725, EPI_ISL_745726, EPI_ISL_745727, EPI_ISL_745728, EPI_ISL_745729, EPI_ISL_745730, EPI_ISL_745731, EPI_ISL_745732, EPI_ISL_745733, EPI_ISL_745734, EPI_ISL_745735, EPI_ISL_745736, EPI_ISL_745737, EPI_ISL_745738, EPI_ISL_745739, EPI_ISL_745740, EPI_ISL_745741, EPI_ISL_745742, EPI_ISL_745743, EPI_ISL_745744, EPI_ISL_745745, EPI_ISL_745746, EPI_ISL_745747, EPI_ISL_745748, EPI_ISL_745749, EPI_ISL_745750, EPI_ISL_745751, EPI_ISL_745752, EPI_ISL_745753, EPI_ISL_745754, EPI_ISL_745755, EPI_ISL_745756, EPI_ISL_745757, EPI_ISL_745758, EPI_ISL_745759, EPI_ISL_745760, EPI_ISL_745761, EPI_ISL_745762, EPI_ISL_745763, EPI_ISL_745764, EPI_ISL_745765, EPI_ISL_745766, EPI_ISL_745767, EPI_ISL_745768, EPI_ISL_745769, EPI_ISL_745770, EPI_ISL_745771, EPI_ISL_745772, EPI_ISL_745773, EPI_ISL_745774, EPI_ISL_745775, EPI_ISL_745776, EPI_ISL_745777, EPI_ISL_745778, EPI_ISL_745779, EPI_ISL_745780, EPI_ISL_745781, EPI_ISL_745782, EPI_ISL_745783, EPI_ISL_745784, EPI_ISL_745785, EPI_ISL_745786, EPI_ISL_745787, EPI_ISL_745788, EPI_ISL_745789, EPI_ISL_745790, EPI_ISL_745791, EPI_ISL_745792, EPI_ISL_745793, EPI_ISL_745794, EPI_ISL_745795, EPI_ISL_745796, EPI_ISL_745797, EPI_ISL_745798, EPI_ISL_745799, EPI_ISL_745800, EPI_ISL_745801, EPI_ISL_745802, EPI_ISL_745803, EPI_ISL_745804, EPI_ISL_745805, EPI_ISL_745806, EPI_ISL_745807, EPI_ISL_745808, EPI_ISL_745809, EPI_ISL_745810, EPI_ISL_745811, EPI_ISL_745812, EPI_ISL_745813, EPI_ISL_745814, EPI_ISL_745815, EPI_ISL_745816, EPI_ISL_745817, EPI_ISL_745818, EPI_ISL_745819, EPI_ISL_745820, EPI_ISL_745821, EPI_ISL_745822, EPI_ISL_745823, EPI_ISL_745824, EPI_ISL_745825, EPI_ISL_745826, EPI_ISL_745827, EPI_ISL_745828, EPI_ISL_745829, EPI_ISL_745830, EPI_ISL_745831, EPI_ISL_745832, EPI_ISL_745833, EPI_ISL_745834, EPI_ISL_745835, EPI_ISL_745836, EPI_ISL_745837, EPI_ISL_745838, EPI_ISL_745839, EPI_ISL_745840, EPI_ISL_745841, EPI_ISL_745842, EPI_ISL_745843, EPI_ISL_745844, EPI_ISL_745845, EPI_ISL_745846, EPI_ISL_745847, EPI_ISL_745848, EPI_ISL_745849, EPI_ISL_745850, EPI_ISL_745851, EPI_ISL_745852, EPI_ISL_745853, EPI_ISL_745854, EPI_ISL_745855, EPI_ISL_745856, EPI_ISL_745857, EPI_ISL_745858, EPI_ISL_745859, EPI_ISL_745860, EPI_ISL_745861, EPI_ISL_745862, EPI_ISL_745863, EPI_ISL_745864, EPI_ISL_745865, EPI_ISL_745866, EPI_ISL_745867, EPI_ISL_745868, EPI_ISL_745869, EPI_ISL_745870, EPI_ISL_745871, EPI_ISL_745872, EPI_ISL_745873, EPI_ISL_745874, EPI_ISL_745875, EPI_ISL_745876, EPI_ISL_745877, EPI_ISL_745878, EPI_ISL_745879, EPI_ISL_745880, EPI_ISL_745881, EPI_ISL_745882, EPI_ISL_745883, EPI_ISL_745884, EPI_ISL_745885, EPI_ISL_745886, EPI_ISL_745887, EPI_ISL_745888, EPI_ISL_745889, EPI_ISL_745890, EPI_ISL_745891, EPI_ISL_745892, EPI_ISL_745893, EPI_ISL_745894, EPI_ISL_745895, EPI_ISL_745896, EPI_ISL_745897, EPI_ISL_745898, EPI_ISL_745899, EPI_ISL_745900, EPI_ISL_745901, EPI_ISL_745902, EPI_ISL_745903, EPI_ISL_745904, EPI_ISL_745905, EPI_ISL_745906, EPI_ISL_745907, EPI_ISL_745908, EPI_ISL_745909, EPI_ISL_745910, EPI_ISL_745911, EPI_ISL_745912, EPI_ISL_745913, EPI_ISL_745914, EPI_ISL_745915, EPI_ISL_745916, EPI_ISL_745917, EPI_ISL_745918, EPI_ISL_745919, EPI_ISL_745920, EPI_ISL_745921, EPI_ISL_745922, EPI_ISL_745923, EPI_ISL_745924, EPI_ISL_745925, EPI_ISL_745926, EPI_ISL_745927, EPI_ISL_745928, EPI_ISL_745929, EPI_ISL_745930, EPI_ISL_745931, EPI_ISL_745932, EPI_ISL_745933, EPI_ISL_745934, EPI_ISL_745935, EPI_ISL_745936, EPI_ISL_745937, EPI_ISL_745938, EPI_ISL_745939, EPI_ISL_745940, EPI_ISL_745941, EPI_ISL_745942, EPI_ISL_745943, EPI_ISL_745944, EPI_ISL_745945, EPI_ISL_745946, EPI_ISL_745947, EPI_ISL_745948, EPI_ISL_745949, EPI_ISL_745950, EPI_ISL_745951, EPI_ISL_745952, EPI_ISL_745953, EPI_ISL_745954, EPI_ISL_745955, EPI_ISL_745956, EPI_ISL_745957, EPI_ISL_745958, EPI_ISL_745959, EPI_ISL_745960, EPI_ISL_745961, EPI_ISL_745962, EPI_ISL_745963, EPI_ISL_745964, EPI_ISL_745965, EPI_ISL_745966, EPI_ISL_745967, EPI_ISL_745968, EPI_ISL_745969, EPI_ISL_745970, EPI_ISL_745971, EPI_ISL_745972, EPI_ISL_745973, EPI_ISL_745974, EPI_ISL_745975, EPI_ISL_745976, EPI_ISL_745977, EPI_ISL_745978, EPI_ISL_745979, EPI_ISL_745980, EPI_ISL_745981, EPI_ISL_745982, EPI_ISL_745983, EPI_ISL_745984, EPI_ISL_745985, EPI_ISL_745986, EPI_ISL_745987, EPI_ISL_745988, EPI_ISL_745989, EPI_ISL_745990, EPI_ISL_745991, EPI_ISL_745992, EPI_ISL_745993, EPI_ISL_745994, EPI_ISL_745995, EPI_ISL_745996, EPI_ISL_745997, EPI_ISL_745998, EPI_ISL_745999, EPI_ISL_746000, EPI_ISL_746001, EPI_ISL_746002, EPI_ISL_746003, EPI_ISL_746004, EPI_ISL_746005, EPI_ISL_746006, EPI_ISL_746007, EPI_ISL_746008, EPI_ISL_746009, EPI_ISL_746010, EPI_ISL_746011, EPI_ISL_746012, EPI_ISL_746013, EPI_ISL_746014, EPI_ISL_746015, EPI_ISL_746016, EPI_ISL_746017, EPI_ISL_746018, EPI_ISL_746019, EPI_ISL_746020, EPI_ISL_746021, EPI_ISL_746022, EPI_ISL_746023, EPI_ISL_746024, EPI_ISL_746025, EPI_ISL_746026, EPI_ISL_746027, EPI_ISL_746028, EPI_ISL_746029, EPI_ISL_746030, EPI_ISL_746031, EPI_ISL_746032, EPI_ISL_746033, EPI_ISL_746034, EPI_ISL_746035, EPI_ISL_746036, EPI_ISL_746037, EPI_ISL_746038, EPI_ISL_746039, EPI_ISL_746040, EPI_ISL_746041, EPI_ISL_746042, EPI_ISL_746043, EPI_ISL_746044, EPI_ISL_746045, EPI_ISL_746046, EPI_ISL_746047, EPI_ISL_746048, EPI_ISL_746049, EPI_ISL_746050, EPI_ISL_746051, EPI_ISL_746052, EPI_ISL_746053, EPI_ISL_746054, EPI_ISL_746055, EPI_ISL_746056, EPI_ISL_746057, EPI_ISL_746058, EPI_ISL_746059, EPI_ISL_746060, EPI_ISL_746061, EPI_ISL_746062, EPI_ISL_746063, EPI_ISL_746064, EPI_ISL_746065, EPI_ISL_746066, EPI_ISL_746067, EPI_ISL_746068, EPI_ISL_746069, EPI_ISL_746070, EPI_ISL_746071, EPI_ISL_746072, EPI_ISL_746073, EPI_ISL_746074, EPI_ISL_746075, EPI_ISL_746076, EPI_ISL_746077, EPI_ISL_746078, EPI_ISL_746079, EPI_ISL_746080, EPI_ISL_746081, EPI_ISL_746082, EPI_ISL_746083, EPI_ISL_746084, EPI_ISL_746085, EPI_ISL_746086, EPI_ISL_746087, EPI_ISL_746088, EPI_ISL_746089, EPI_ISL_746090, EPI_ISL_746091, EPI_ISL_746092, EPI_ISL_746093, EPI_ISL_746094, EPI_ISL_746095, EPI_ISL_746096, EPI_ISL_746097, EPI_ISL_746098, EPI_ISL_746099, EPI_ISL_746100, EPI_ISL_746101, EPI_ISL_746102, EPI_ISL_746103, EPI_ISL_746104, EPI_ISL_746105, EPI_ISL_746106, EPI_ISL_746107, EPI_ISL_746108, EPI_ISL_746109, EPI_ISL_746110, EPI_ISL_746111, EPI_ISL_746112, EPI_ISL_746113, EPI_ISL_746114, EPI_ISL_746115, EPI_ISL_746116, EPI_ISL_746117, EPI_ISL_746118, EPI_ISL_746119, EPI_ISL_746120, EPI_ISL_746121, EPI_ISL_746122, EPI_ISL_746123, EPI_ISL_746124, EPI_ISL_746125, EPI_ISL_746126, EPI_ISL_746127, EPI_ISL_746128, EPI_ISL_746129, EPI_ISL_746130, EPI_ISL_746131, EPI_ISL_746132, EPI_ISL_746133, EPI_ISL_746134, EPI_ISL_746135, EPI_ISL_746136, EPI_ISL_746137, EPI_ISL_746138, EPI_ISL_746139, EPI_ISL_746140, EPI_ISL_746141, EPI_ISL_746142, EPI_ISL_746143, EPI_ISL_746144, EPI_ISL_746145, EPI_ISL_746146, EPI_ISL_746147, EPI_ISL_746148, EPI_ISL_746149, EPI_ISL_746150, EPI_ISL_746151, EPI_ISL_746152, EPI_ISL_746153, EPI_ISL_746154, EPI_ISL_746155, EPI_ISL_746156, EPI_ISL_746157, EPI_ISL_746158, EPI_ISL_746159, EPI_ISL_746160, EPI_ISL_746161, EPI_ISL_746162, EPI_ISL_746163, EPI_ISL_746164, EPI_ISL_746165, EPI_ISL_746166, EPI_ISL_746167, EPI_ISL_746168, EPI_ISL_746169, EPI_ISL_746170, EPI_ISL_746171, EPI_ISL_746172, EPI_ISL_746173, EPI_ISL_746174, EPI_ISL_746175, EPI_ISL_746176, EPI_ISL_746177, EPI_ISL_746178, EPI_ISL_746179, EPI_ISL_746180, EPI_ISL_746181, EPI_ISL_746182, EPI_ISL_746183, EPI_ISL_746184, EPI_ISL_746185, EPI_ISL_746186, EPI_ISL_746187, EPI_ISL_746188, EPI_ISL_746189, EPI_ISL_746190, EPI_ISL_746191, EPI_ISL_746192, EPI_ISL_746193, EPI_ISL_746194, EPI_ISL_746195, EPI_ISL_746196, EPI_ISL_746197, EPI_ISL_746198, EPI_ISL_746199, EPI_ISL_746200, EPI_ISL_746201, EPI_ISL_746202, EPI_ISL_746203, EPI_ISL_746204, EPI_ISL_746205, EPI_ISL_746206, EPI_ISL_746207, EPI_ISL_746208, EPI_ISL_746209, EPI_ISL_746210, EPI_ISL_746211, EPI_ISL_746212, EPI_ISL_746213, EPI_ISL_746214, EPI_ISL_746215, EPI_ISL_746216, EPI_ISL_746217, EPI_ISL_746218, EPI_ISL_746219, EPI_ISL_746220, EPI_ISL_746221, EPI_ISL_746222, EPI_ISL_746223, EPI_ISL_746224, EPI_ISL_746225, EPI_ISL_746226, EPI_ISL_746227, EPI_ISL_746228, EPI_ISL_746229, EPI_ISL_746230, EPI_ISL_746231, EPI_ISL_746232, EPI_ISL_746233, EPI_ISL_746234, EPI_ISL_746235, EPI_ISL_746236, EPI_ISL_746237, EPI_ISL_746238, EPI_ISL_746239, EPI_ISL_746240, EPI_ISL_746241, EPI_ISL_746242, EPI_ISL_746243, EPI_ISL_746244, EPI_ISL_746245, EPI_ISL_746246, EPI_ISL_746247, EPI_ISL_746248, EPI_ISL_746249, EPI_ISL_746250, EPI_ISL_746251, EPI_ISL_746252, EPI_ISL_746253, EPI_ISL_746254, EPI_ISL_746255, EPI_ISL_746256, EPI_ISL_746257, EPI_ISL_746258, EPI_ISL_746259, EPI_ISL_746260, EPI_ISL_746261, EPI_ISL_746262, EPI_ISL_746263, EPI_ISL_746264, EPI_ISL_746265, EPI_ISL_746266, EPI_ISL_746267, EPI_ISL_746268, EPI_ISL_746269, EPI_ISL_746270, EPI_ISL_746271, EPI_ISL_746272, EPI_ISL_746273, EPI_ISL_746274, EPI_ISL_746275, EPI_ISL_746276, EPI_ISL_746277, EPI_ISL_746278, EPI_ISL_746279, EPI_ISL_746280, EPI_ISL_746281, EPI_ISL_746282, EPI_ISL_746283, EPI_ISL_746284, EPI_ISL_746285, EPI_ISL_746286, EPI_ISL_746287, EPI_ISL_746288, EPI_ISL_746289, EPI_ISL_746290, EPI_ISL_746291, EPI_ISL_746292, EPI_ISL_746293, EPI_ISL_746294, EPI_ISL_746295, EPI_ISL_746296, EPI_ISL_746297, EPI_ISL_746298, EPI_ISL_746299, EPI_ISL_746300, EPI_ISL_746301, EPI_ISL_746302, EPI_ISL_746303, EPI_ISL_746304, EPI_ISL_746305, EPI_ISL_746306, EPI_ISL_746307, EPI_ISL_746308, EPI_ISL_746309, EPI_ISL_746310, EPI_ISL_746311, EPI_ISL_746312, EPI_ISL_746313, EPI_ISL_746314, EPI_ISL_746315, EPI_ISL_746316, EPI_ISL_746317 | | | |
| see above | Ginkgo Bioworks Clinical Laboratory | Utah Public Health Laboratory | Erin L. Young, Kelly Oakeson, Tara Gallagher, Michael T. Pyne, E. Susan Slechta, Melanie A. Mallory, Jeffrey B. Stevenson, Salika M. Shakir, David R. Hillyard, Malaika McKenzie-Bennett, James McGann, Jim Griffin, Keith Robison, Alex Plocik, Becky Schilling, Martha Pierson, Rebecca Littlefield, Michelle Spencer, Birgitte Simen |
| EPI_ISL_746325, EPI_ISL_746326, EPI_ISL_746327, EPI_ISL_746328, EPI_ISL_746329, EPI_ISL_746330, EPI_ISL_746331, EPI_ISL_746332, EPI_ISL_746333, EPI_ISL_746334, EPI_ISL_746335, EPI_ISL_746336, EPI_ISL_746337, EPI_ISL_746338, EPI_ISL_746339, EPI_ISL_746340, EPI_ISL_746341, EPI_ISL_746360, EPI_ISL_746361, EPI_ISL_746362, EPI_ISL_746363, EPI_ISL_746364, EPI_ISL_746365, EPI_ISL_746366, EPI_ISL_746367, EPI_ISL_746368, EPI_ISL_746369, EPI_ISL_746370, EPI_ISL_746371, EPI_ISL_746372, EPI_ISL_746373, EPI_ISL_746374, EPI_ISL_746375, EPI_ISL_746376, EPI_ISL_746377, EPI_ISL_746378, EPI_ISL_746379, EPI_ISL_746380, EPI_ISL_746381, EPI_ISL_746382, EPI_ISL_746383, EPI_ISL_746384, EPI_ISL_746385, EPI_ISL_746386, EPI_ISL_746387, EPI_ISL_746388, EPI_ISL_746389, EPI_ISL_746390, EPI_ISL_746391, EPI_ISL_746392, EPI_ISL_746393, EPI_ISL_746394, EPI_ISL_746395, EPI_ISL_746396, EPI_ISL_746397, EPI_ISL_746398, EPI_ISL_746399, EPI_ISL_746400, EPI_ISL_746401, EPI_ISL_746402, EPI_ISL_746403, EPI_ISL_746404, EPI_ISL_746405, EPI_ISL_746406, EPI_ISL_746424, EPI_ISL_746425, EPI_ISL_746426, EPI_ISL_746427, EPI_ISL_746428, EPI_ISL_746429, EPI_ISL_746430, EPI_ISL_746431, EPI_ISL_746432, EPI_ISL_746433, EPI_ISL_746434, EPI_ISL_746435, EPI_ISL_746436, EPI_ISL_746437, EPI_ISL_746438, EPI_ISL_746439, EPI_ISL_746440, EPI_ISL_746441, EPI_ISL_746442, EPI_ISL_746443, EPI_ISL_746444, EPI_ISL_746445, EPI_ISL_746446, EPI_ISL_746447, EPI_ISL_746448, EPI_ISL_746449, EPI_ISL_746450, EPI_ISL_746451, EPI_ISL_746452, EPI_ISL_746453, EPI_ISL_746454, EPI_ISL_746455, EPI_ISL_746456, EPI_ISL_746457, EPI_ISL_746458, EPI_ISL_746459, EPI_ISL_746460, EPI_ISL_746461, EPI_ISL_746462, EPI_ISL_746463, EPI_ISL_746464, EPI_ISL_746465, EPI_ISL_746466, EPI_ISL_746467, EPI_ISL_746468, EPI_ISL_746469, EPI_ISL_746470, EPI_ISL_746471, EPI_ISL_746472, EPI_ISL_746473, EPI_ISL_746474, EPI_ISL_746475, EPI_ISL_746476, EPI_ISL_746477, EPI_ISL_746836, EPI_ISL_746837, EPI_ISL_746838, EPI_ISL_746839, EPI_ISL_746840, EPI_ISL_746841, EPI_ISL_746842, EPI_ISL_746843, EPI_ISL_746844, EPI_ISL_746845, EPI_ISL_746846, EPI_ISL_746847, EPI_ISL_746848, EPI_ISL_746849, EPI_ISL_746850, EPI_ISL_746851, EPI_ISL_746852, EPI_ISL_746853, EPI_ISL_746854, EPI_ISL_746855, EPI_ISL_746856, EPI_ISL_746857, EPI_ISL_746858, EPI_ISL_746859, EPI_ISL_746860, EPI_ISL_746861, EPI_ISL_746862, EPI_ISL_746863, EPI_ISL_746864, EPI_ISL_746865, EPI_ISL_746866, EPI_ISL_746867, EPI_ISL_746868, EPI_ISL_746869, EPI_ISL_746870, EPI_ISL_746871, EPI_ISL_746872, EPI_ISL_746873, EPI_ISL_746874, EPI_ISL_746875, EPI_ISL_746876, EPI_ISL_746877, EPI_ISL_746878, EPI_ISL_746879, EPI_ISL_746880, EPI_ISL_746881, EPI_ISL_746882, EPI_ISL_746883, EPI_ISL_746884, EPI_ISL_746885, EPI_ISL_746886, EPI_ISL_746887, EPI_ISL_746888, EPI_ISL_746889, EPI_ISL_746890, EPI_ISL_746948, EPI_ISL_746949, EPI_ISL_746950, EPI_ISL_746951, EPI_ISL_746952, EPI_ISL_746953, EPI_ISL_746954, EPI_ISL_746955, EPI_ISL_746956, EPI_ISL_746957, EPI_ISL_746958, EPI_ISL_746959, EPI_ISL_746960, EPI_ISL_746961, EPI_ISL_746962, EPI_ISL_746963, EPI_ISL_746964, EPI_ISL_746965, EPI_ISL_746966, EPI_ISL_746967, EPI_ISL_746968, EPI_ISL_746969, EPI_ISL_746970, EPI_ISL_746971, EPI_ISL_746972, EPI_ISL_746973, EPI_ISL_746974, EPI_ISL_746975, EPI_ISL_746976, EPI_ISL_746977, EPI_ISL_746978, EPI_ISL_746979, EPI_ISL_746980, EPI_ISL_746981, EPI_ISL_746982, EPI_ISL_746983, EPI_ISL_746984, EPI_ISL_746985, EPI_ISL_746986, EPI_ISL_746987, EPI_ISL_746988, EPI_ISL_746989, EPI_ISL_746990, EPI_ISL_746991, EPI_ISL_746992, EPI_ISL_746993, EPI_ISL_746994, EPI_ISL_746995, EPI_ISL_746996, EPI_ISL_746997, EPI_ISL_746998, EPI_ISL_746999, EPI_ISL_747000, EPI_ISL_747001, EPI_ISL_747002, EPI_ISL_747003, EPI_ISL_747004, EPI_ISL_747005, EPI_ISL_747006, EPI_ISL_747007, EPI_ISL_747008, EPI_ISL_747009, EPI_ISL_747010, EPI_ISL_747011, EPI_ISL_747012, EPI_ISL_747013, EPI_ISL_747014, EPI_ISL_747015, EPI_ISL_747016, EPI_ISL_747017, EPI_ISL_747018, EPI_ISL_747019, EPI_ISL_747020, EPI_ISL_747021, EPI_ISL_747022, EPI_ISL_747023, EPI_ISL_747024, EPI_ISL_747025, EPI_ISL_747026, EPI_ISL_747027, EPI_ISL_747028, EPI_ISL_747029, EPI_ISL_747030, EPI_ISL_747031 | | | |
| see above | Utah Public Health Laboratory | Utah Public Health Laboratory | Erin Young, Kelly Oakeson, Tara Gallagher |
| EPI_ISL_747032, EPI_ISL_747033, EPI_ISL_747034, EPI_ISL_747035, EPI_ISL_747036, EPI_ISL_747038, EPI_ISL_747039, EPI_ISL_747040, EPI_ISL_747041, EPI_ISL_747042, EPI_ISL_747043, EPI_ISL_747044, EPI_ISL_747045, EPI_ISL_747046, EPI_ISL_747047, EPI_ISL_747048, EPI_ISL_747049, EPI_ISL_747050, EPI_ISL_747051, EPI_ISL_747052, EPI_ISL_747053, EPI_ISL_747054, EPI_ISL_747055, EPI_ISL_747056, EPI_ISL_747057, EPI_ISL_747058, EPI_ISL_747059, EPI_ISL_747060, EPI_ISL_747061, EPI_ISL_747062, EPI_ISL_747063, EPI_ISL_747064, EPI_ISL_747065, EPI_ISL_747066, EPI_ISL_747067, EPI_ISL_747068, EPI_ISL_747069, EPI_ISL_747070, EPI_ISL_747071, EPI_ISL_747072, EPI_ISL_747073, EPI_ISL_747074, EPI_ISL_747075, EPI_ISL_747076, EPI_ISL_747077, EPI_ISL_747078, EPI_ISL_747079, EPI_ISL_747080, EPI_ISL_747081, EPI_ISL_747082, EPI_ISL_747083, EPI_ISL_747084, EPI_ISL_747085, EPI_ISL_747086, EPI_ISL_747087, EPI_ISL_747088, EPI_ISL_747089, EPI_ISL_747090, EPI_ISL_747091, EPI_ISL_747092, EPI_ISL_747093, EPI_ISL_747094, EPI_ISL_747095, EPI_ISL_747096, EPI_ISL_747097, EPI_ISL_747098, EPI_ISL_747099, EPI_ISL_747100, EPI_ISL_747101, EPI_ISL_747102, EPI_ISL_747103, EPI_ISL_747104, EPI_ISL_747105, EPI_ISL_747106, EPI_ISL_747107, EPI_ISL_747108, EPI_ISL_747110, EPI_ISL_747111, EPI_ISL_747112, EPI_ISL_747113, EPI_ISL_747114, EPI_ISL_747115, EPI_ISL_747116, EPI_ISL_747117, EPI_ISL_747118, EPI_ISL_747119, EPI_ISL_747120, EPI_ISL_747121, EPI_ISL_747122, EPI_ISL_747123, EPI_ISL_747124, EPI_ISL_747125, EPI_ISL_747126, EPI_ISL_747127, EPI_ISL_747128, EPI_ISL_747129, EPI_ISL_747130, EPI_ISL_747131, EPI_ISL_747132, EPI_ISL_747133, EPI_ISL_747134, EPI_ISL_747135, EPI_ISL_747136, EPI_ISL_747137, EPI_ISL_747138, EPI_ISL_747139, EPI_ISL_747140, EPI_ISL_747141, EPI_ISL_747142, EPI_ISL_747143, EPI_ISL_747144, EPI_ISL_747145, EPI_ISL_747146, EPI_ISL_747147, EPI_ISL_747149, EPI_ISL_747150, EPI_ISL_747151, EPI_ISL_747152, EPI_ISL_747153, EPI_ISL_747154, EPI_ISL_747155, EPI_ISL_747156, EPI_ISL_747157, EPI_ISL_747158, EPI_ISL_747159, EPI_ISL_747160, EPI_ISL_747161, EPI_ISL_747162, EPI_ISL_747163, EPI_ISL_747164, EPI_ISL_747165, EPI_ISL_747166, EPI_ISL_747167, EPI_ISL_747168, EPI_ISL_747169, EPI_ISL_747170, EPI_ISL_747171, EPI_ISL_747172, EPI_ISL_747173, EPI_ISL_747174, EPI_ISL_747175, EPI_ISL_747176, EPI_ISL_747177, EPI_ISL_747178, EPI_ISL_747179, EPI_ISL_747180, EPI_ISL_747181, EPI_ISL_747182, EPI_ISL_747183, EPI_ISL_747184, EPI_ISL_747185, EPI_ISL_747186, EPI_ISL_747187, EPI_ISL_747188, EPI_ISL_747189, EPI_ISL_747190, EPI_ISL_747191, EPI_ISL_747192, EPI_ISL_747193 | | | |
| see above | Respiratory Viruses Branch, Centers for Disease Control and Prevention | Respiratory Viruses Branch, Centers for Disease Control and Prevention | Queen,K., Li,Y., Tao,Y., Uehara,A., Montmayeur,A., Paden,C.R., Cook,P.W., Marine,R., Sheth,M., Wang,H., Lee,J., Tong,S. |

| | | | |
|---|---|---|---|
| EPI_ISL_747195, EPI_ISL_747196, EPI_ISL_747197, EPI_ISL_747198, EPI_ISL_747199, EPI_ISL_747200, EPI_ISL_747201, EPI_ISL_747202, EPI_ISL_747203, EPI_ISL_747204, EPI_ISL_747205, EPI_ISL_747206, EPI_ISL_747207, EPI_ISL_747208, EPI_ISL_747209, EPI_ISL_747210, EPI_ISL_747211, EPI_ISL_747212, EPI_ISL_747213, EPI_ISL_747214, EPI_ISL_747215, EPI_ISL_747216, EPI_ISL_747217, EPI_ISL_747218, EPI_ISL_747219, EPI_ISL_747220, EPI_ISL_747221, EPI_ISL_747222, EPI_ISL_747223, EPI_ISL_747224, EPI_ISL_747225, EPI_ISL_747226, EPI_ISL_747227, EPI_ISL_747228, EPI_ISL_747229, EPI_ISL_747230, EPI_ISL_747231, EPI_ISL_747232 | | | |
| see above | UW Virology Lab | UW Virology Lab | Pavitra Roychoudhury, Hong Xie, Lasata Shrestha, Michelle Lin, Meei-Li Huang, Keith R Jerome, Alexander Greninger |
| EPI_ISL_747242 | Pathogen Lab (BSL3), Biomedical Innovation Department, Applied and Experimental Biology Division, Scientific Research Center and High Education from Ensenada (CICESE) | Pathogen Laboratory (BSL3), Biomedical Innovation Department, Experimental and Applied Biology Division, Scientific Research Center and High Education from Ensenada (CICESE) | Cervantes-Luevano K, Martinez M, Saavedra A, Galindo C and Licea-Navarro A |
| EPI_ISL_747243 | General Hospital Laboratory | Pathogen Laboratory (BSL3), Biomedical Innovation Department, Experimental and Applied Biology Division, Scientific Research Center and High Education from Ensenada (CICESE) | Cervantes-Luevano K., Martinez M.A., Saavedra-Flores A., Galindo C. and Licea-Navarro A. |
| EPI_ISL_751504, EPI_ISL_751505, EPI_ISL_751506, EPI_ISL_751507, EPI_ISL_751508, EPI_ISL_751509, EPI_ISL_751510, EPI_ISL_751511, EPI_ISL_751512, EPI_ISL_751513, EPI_ISL_751514, EPI_ISL_751515, EPI_ISL_751516, EPI_ISL_751517, EPI_ISL_751518, EPI_ISL_751519 | | | |
| see above | Texas Department of State Health Services | Texas Department of State Health Services | Rashmi Tuladhar, Bonnie Oh, Jenny Zhang, Maliha Rahman, Anita Pokharel, Myong Koag, Chung Wang, Rachel Lee, Grace Kubin, Mayela Pedrueza, James Daniel Bonser |
| EPI_ISL_751542 | SC Dept of Health and Env. Control-Bureau of Laboratories | Genomics and Discovery, Respiratory Viruses Branch, Division of Viral Diseases, Centers for Disease Control and Prevention | Krista Queen, Yan Li, Ying Tao, Jing Zhang, Anna Uehara, Anna Montmayeur, Clinton R. Paden, Peter W. Cook,Rachel Marine, Mili Sheth, Haibin Wang, Justin Lee, Suxiang Tong |
| EPI_ISL_751543 | DE Public Health Laboratory | Genomics and Discovery, Respiratory Viruses Branch, Division of Viral Diseases, Centers for Disease Control and Prevention | Krista Queen, Yan Li, Ying Tao, Jing Zhang, Anna Uehara, Anna Montmayeur, Clinton R. Paden, Peter W. Cook,Rachel Marine, Mili Sheth, Haibin Wang, Justin Lee, Suxiang Tong |
| EPI_ISL_751544 | NH Dept. of Health and Human Services Public Health Labs | Genomics and Discovery, Respiratory Viruses Branch, Division of Viral Diseases, Centers for Disease Control and Prevention | Krista Queen, Yan Li, Ying Tao, Jing Zhang, Anna Uehara, Anna Montmayeur, Clinton R. Paden, Peter W. Cook,Rachel Marine, Mili Sheth, Haibin Wang, Justin Lee, Suxiang Tong |
| EPI_ISL_751545 | NYSDOH Wadsworth Center, Virology Lab | Genomics and Discovery, Respiratory Viruses Branch, Division of Viral Diseases, Centers for Disease Control and Prevention | Krista Queen, Yan Li, Ying Tao, Jing Zhang, Anna Uehara, Anna Montmayeur, Clinton R. Paden, Peter W. Cook,Rachel Marine, Mili Sheth, Haibin Wang, Justin Lee, Suxiang Tong |
| EPI_ISL_751546 | VA-Division of Consolidated Laboratory Services | Genomics and Discovery, Respiratory Viruses Branch, Division of Viral Diseases, Centers for Disease Control and Prevention | Krista Queen, Yan Li, Ying Tao, Jing Zhang, Anna Uehara, Anna Montmayeur, Clinton R. Paden, Peter W. Cook,Rachel Marine, Mili Sheth, Haibin Wang, Justin Lee, Suxiang Tong |
| EPI_ISL_751547 | DE Public Health Laboratory | Genomics and Discovery, Respiratory Viruses Branch, Division of Viral Diseases, Centers for Disease Control and Prevention | Krista Queen, Yan Li, Ying Tao, Jing Zhang, Anna Uehara, Anna Montmayeur, Clinton R. Paden, Peter W. Cook,Rachel Marine, Mili Sheth, Haibin Wang, Justin Lee, Suxiang Tong |
| EPI_ISL_751548 | NYSDOH Wadsworth Center, Virology Lab | Genomics and Discovery, Respiratory Viruses Branch, Division of Viral Diseases, Centers for Disease Control and Prevention | Krista Queen, Yan Li, Ying Tao, Jing Zhang, Anna Uehara, Anna Montmayeur, Clinton R. Paden, Peter W. Cook,Rachel Marine, Mili Sheth, Haibin Wang, Justin Lee, Suxiang Tong |
| EPI_ISL_751554 | CT-Dr. Katherine A. Kelley State Public Health Lab | Genomics and Discovery, Respiratory Viruses Branch, Division of Viral Diseases, Centers for Disease Control and Prevention | Krista Queen, Yan Li, Ying Tao, Jing Zhang, Anna Uehara, Anna Montmayeur, Clinton R. Paden, Peter W. Cook,Rachel Marine, Mili Sheth, Haibin Wang, Justin Lee, Suxiang Tong |
| EPI_ISL_751555 | WY Public Health Laboratory | Genomics and Discovery, Respiratory Viruses Branch, Division of Viral Diseases, Centers for Disease Control and Prevention | Krista Queen, Yan Li, Ying Tao, Jing Zhang, Anna Uehara, Anna Montmayeur, Clinton R. Paden, Peter W. Cook,Rachel Marine, Mili Sheth, Haibin Wang, Justin Lee, Suxiang Tong |
| EPI_ISL_751556, EPI_ISL_751557 | NV State Public Health Laboratory | Genomics and Discovery, Respiratory Viruses Branch, Division of Viral Diseases, Centers for Disease Control and Prevention | Krista Queen, Yan Li, Ying Tao, Jing Zhang, Anna Uehara, Anna Montmayeur, Clinton R. Paden, Peter W. Cook,Rachel Marine, Mili Sheth, Haibin Wang, Justin Lee, Suxiang Tong |
| EPI_ISL_751558 | NH Dept. of Health and Human Services Public Health Labs | Genomics and Discovery, Respiratory Viruses Branch, Division of Viral Diseases, Centers for Disease Control and Prevention | Krista Queen, Yan Li, Ying Tao, Jing Zhang, Anna Uehara, Anna Montmayeur, Clinton R. Paden, Peter W. Cook,Rachel Marine, Mili Sheth, Haibin Wang, Justin Lee, Suxiang Tong |
| EPI_ISL_751559 | MD DOH Laboratories Administration | Genomics and Discovery, Respiratory Viruses Branch, Division of Viral Diseases, Centers for Disease Control and Prevention | Krista Queen, Yan Li, Ying Tao, Jing Zhang, Anna Uehara, Anna Montmayeur, Clinton R. Paden, Peter W. Cook,Rachel Marine, Mili Sheth, Haibin Wang, Justin Lee, Suxiang Tong |
| EPI_ISL_751560 | NYSDOH Wadsworth Center, Virology Lab | Genomics and Discovery, Respiratory Viruses Branch, Division of Viral Diseases, Centers for Disease Control and Prevention | Krista Queen, Yan Li, Ying Tao, Jing Zhang, Anna Uehara, Anna Montmayeur, Clinton R. Paden, Peter W. Cook,Rachel Marine, Mili Sheth, Haibin Wang, Justin Lee, Suxiang Tong |
| EPI_ISL_751561 | MT Public Health Laboratory | Genomics and Discovery, Respiratory Viruses Branch, Division of Viral Diseases, Centers for Disease Control and Prevention | Krista Queen, Yan Li, Ying Tao, Jing Zhang, Anna Uehara, Anna Montmayeur, Clinton R. Paden, Peter W. Cook,Rachel Marine, Mili Sheth, Haibin Wang, Justin Lee, Suxiang Tong |
| EPI_ISL_751562 | NYSDOH Wadsworth Center, Virology Lab | Genomics and Discovery, Respiratory Viruses Branch, Division of Viral Diseases, Centers for Disease Control and Prevention | Krista Queen, Yan Li, Ying Tao, Jing Zhang, Anna Uehara, Anna Montmayeur, Clinton R. Paden, Peter W. Cook,Rachel Marine, Mili Sheth, Haibin Wang, Justin Lee, Suxiang Tong |
| EPI_ISL_751563 | NE Public Health Laboratory | Genomics and Discovery, Respiratory Viruses Branch, Division of Viral Diseases, Centers for Disease Control and Prevention | Krista Queen, Yan Li, Ying Tao, Jing Zhang, Anna Uehara, Anna Montmayeur, Clinton R. Paden, Peter W. Cook,Rachel Marine, Mili Sheth, Haibin Wang, Justin Lee, Suxiang Tong |
| EPI_ISL_751564 | VA-Division of Consolidated Laboratory Services | Genomics and Discovery, Respiratory Viruses Branch, Division of Viral Diseases, Centers for Disease Control and Prevention | Krista Queen, Yan Li, Ying Tao, Jing Zhang, Anna Uehara, Anna Montmayeur, Clinton R. Paden, Peter W. Cook,Rachel Marine, Mili Sheth, Haibin Wang, Justin Lee, Suxiang Tong |
| EPI_ISL_751565 | IA State Hygienic Laboratory | Genomics and Discovery, Respiratory Viruses Branch, Division of Viral Diseases, Centers for Disease Control and Prevention | Krista Queen, Yan Li, Ying Tao, Jing Zhang, Anna Uehara, Anna Montmayeur, Clinton R. Paden, Peter W. Cook,Rachel Marine, Mili Sheth, Haibin Wang, Justin Lee, Suxiang Tong |
| EPI_ISL_751566 | WVDHHR - Office of Laboratory Services | Genomics and Discovery, Respiratory Viruses Branch, Division of Viral Diseases, Centers for Disease Control and Prevention | Krista Queen, Yan Li, Ying Tao, Jing Zhang, Anna Uehara, Anna Montmayeur, Clinton R. Paden, Peter W. Cook,Rachel Marine, Mili Sheth, Haibin Wang, Justin Lee, Suxiang Tong |
| EPI_ISL_751567 | ID Bureau of Laboratories | Genomics and Discovery, Respiratory Viruses Branch, Division of Viral Diseases, Centers for Disease Control and Prevention | Krista Queen, Yan Li, Ying Tao, Jing Zhang, Anna Uehara, Anna Montmayeur, Clinton R. Paden, Peter W. Cook,Rachel Marine, Mili Sheth, Haibin Wang, Justin Lee, Suxiang Tong |
| EPI_ISL_751568 | MS Public Health Laboratory | Genomics and Discovery, Respiratory Viruses Branch, Division of Viral Diseases, Centers for Disease Control and Prevention | Krista Queen, Yan Li, Ying Tao, Jing Zhang, Anna Uehara, Anna Montmayeur, Clinton R. Paden, Peter W. Cook,Rachel Marine, Mili Sheth, Haibin Wang, Justin Lee, Suxiang Tong |
| EPI_ISL_751569 | MI - Michigan Department of Health and Human Services - Bureau of Laboratories | Genomics and Discovery, Respiratory Viruses Branch, Division of Viral Diseases, Centers for Disease Control and Prevention | Krista Queen, Yan Li, Ying Tao, Jing Zhang, Anna Uehara, Anna Montmayeur, Clinton R. Paden, Peter W. Cook,Rachel Marine, Mili Sheth, Haibin Wang, Justin Lee, Suxiang Tong |
| EPI_ISL_751570 | KY State Public Health Lab | Genomics and Discovery, Respiratory Viruses Branch, Division of Viral Diseases, Centers for Disease Control and Prevention | Krista Queen, Yan Li, Ying Tao, Jing Zhang, Anna Uehara, Anna Montmayeur, Clinton R. Paden, Peter W. Cook,Rachel Marine, Mili Sheth, Haibin Wang, Justin Lee, Suxiang Tong |
| EPI_ISL_751571 | TX DSHS, Lab Services Section MC 1947 | Genomics and Discovery, Respiratory Viruses Branch, Division of Viral Diseases, Centers for Disease Control and Prevention | Krista Queen, Yan Li, Ying Tao, Jing Zhang, Anna Uehara, Anna Montmayeur, Clinton R. Paden, Peter W. Cook,Rachel Marine, Mili Sheth, Haibin Wang, Justin Lee, Suxiang Tong |
| EPI_ISL_751572 | KS Health and Environmental Laboratories | Genomics and Discovery, Respiratory Viruses Branch, Division of Viral Diseases, Centers for Disease Control and Prevention | Krista Queen, Yan Li, Ying Tao, Jing Zhang, Anna Uehara, Anna Montmayeur, Clinton R. Paden, Peter W. Cook,Rachel Marine, Mili Sheth, Haibin Wang, Justin Lee, Suxiang Tong |
| EPI_ISL_751573 | MN PHL Division, Minnesota Department of Health | Genomics and Discovery, Respiratory Viruses Branch, Division of Viral Diseases, Centers for Disease Control and Prevention | Krista Queen, Yan Li, Ying Tao, Jing Zhang, Anna Uehara, Anna Montmayeur, Clinton R. Paden, Peter W. Cook,Rachel Marine, Mili Sheth, Haibin Wang, Justin Lee, Suxiang Tong |
| EPI_ISL_751574 | AR Dept. of Health-Public Health Lab | Genomics and Discovery, Respiratory | Krista Queen, Yan Li, Ying Tao, Jing Zhang, Anna Uehara, Anna Montmayeur, Clinton R. Paden, Peter W. Cook,Rachel Marine, Mili Sheth, Haibin Wang, Justin Lee, Suxiang Tong |

| | | | |
|---|---|---|---|
| | | Viruses Branch, Division of Viral Diseases, Centers for Disease Control and Prevention | |
| EPI_ISL_751575 | CO Dept. of Public Health and Environment, Lab Services Division | Genomics and Discovery, Respiratory Viruses Branch, Division of Viral Diseases, Centers for Disease Control and Prevention | Krista Queen, Yan Li, Ying Tao, Jing Zhang, Anna Uehara, Anna Montmayeur, Clinton R. Paden, Peter W. Cook,Rachel Marine, Mili Sheth, Haibin Wang, Justin Lee, Suxiang Tong |
| EPI_ISL_751576 | MN PHL Division, Minnesota Department of Health | Genomics and Discovery, Respiratory Viruses Branch, Division of Viral Diseases, Centers for Disease Control and Prevention | Krista Queen, Yan Li, Ying Tao, Jing Zhang, Anna Uehara, Anna Montmayeur, Clinton R. Paden, Peter W. Cook,Rachel Marine, Mili Sheth, Haibin Wang, Justin Lee, Suxiang Tong |
| EPI_ISL_751577 | MD DOH Laboratories Administration | Genomics and Discovery, Respiratory Viruses Branch, Division of Viral Diseases, Centers for Disease Control and Prevention | Krista Queen, Yan Li, Ying Tao, Jing Zhang, Anna Uehara, Anna Montmayeur, Clinton R. Paden, Peter W. Cook,Rachel Marine, Mili Sheth, Haibin Wang, Justin Lee, Suxiang Tong |
| EPI_ISL_751578 | HI Dept. of Health, State Laboratories Division | Genomics and Discovery, Respiratory Viruses Branch, Division of Viral Diseases, Centers for Disease Control and Prevention | Krista Queen, Yan Li, Ying Tao, Jing Zhang, Anna Uehara, Anna Montmayeur, Clinton R. Paden, Peter W. Cook,Rachel Marine, Mili Sheth, Haibin Wang, Justin Lee, Suxiang Tong |
| EPI_ISL_751579 | IN State Department of Health Laboratory Services | Genomics and Discovery, Respiratory Viruses Branch, Division of Viral Diseases, Centers for Disease Control and Prevention | Krista Queen, Yan Li, Ying Tao, Jing Zhang, Anna Uehara, Anna Montmayeur, Clinton R. Paden, Peter W. Cook,Rachel Marine, Mili Sheth, Haibin Wang, Justin Lee, Suxiang Tong |
| EPI_ISL_751580, EPI_ISL_751581 | NYSDOH Wadsworth Center, Virology Lab | Genomics and Discovery, Respiratory Viruses Branch, Division of Viral Diseases, Centers for Disease Control and Prevention | Krista Queen, Yan Li, Ying Tao, Jing Zhang, Anna Uehara, Anna Montmayeur, Clinton R. Paden, Peter W. Cook,Rachel Marine, Mili Sheth, Haibin Wang, Justin Lee, Suxiang Tong |
| EPI_ISL_751582 | NH Dept. of Health and Human Services Public Health Labs | Genomics and Discovery, Respiratory Viruses Branch, Division of Viral Diseases, Centers for Disease Control and Prevention | Krista Queen, Yan Li, Ying Tao, Jing Zhang, Anna Uehara, Anna Montmayeur, Clinton R. Paden, Peter W. Cook,Rachel Marine, Mili Sheth, Haibin Wang, Justin Lee, Suxiang Tong |
| EPI_ISL_751583 | NE Public Health Laboratory | Genomics and Discovery, Respiratory Viruses Branch, Division of Viral Diseases, Centers for Disease Control and Prevention | Krista Queen, Yan Li, Ying Tao, Jing Zhang, Anna Uehara, Anna Montmayeur, Clinton R. Paden, Peter W. Cook,Rachel Marine, Mili Sheth, Haibin Wang, Justin Lee, Suxiang Tong |
| EPI_ISL_751584 | NH Dept. of Health and Human Services Public Health Labs | Genomics and Discovery, Respiratory Viruses Branch, Division of Viral Diseases, Centers for Disease Control and Prevention | Krista Queen, Yan Li, Ying Tao, Jing Zhang, Anna Uehara, Anna Montmayeur, Clinton R. Paden, Peter W. Cook,Rachel Marine, Mili Sheth, Haibin Wang, Justin Lee, Suxiang Tong |
| EPI_ISL_751585 | NE Public Health Laboratory | Genomics and Discovery, Respiratory Viruses Branch, Division of Viral Diseases, Centers for Disease Control and Prevention | Krista Queen, Yan Li, Ying Tao, Jing Zhang, Anna Uehara, Anna Montmayeur, Clinton R. Paden, Peter W. Cook,Rachel Marine, Mili Sheth, Haibin Wang, Justin Lee, Suxiang Tong |
| EPI_ISL_751586 | OH Department of Health Laboratory | Genomics and Discovery, Respiratory Viruses Branch, Division of Viral Diseases, Centers for Disease Control and Prevention | Krista Queen, Yan Li, Ying Tao, Jing Zhang, Anna Uehara, Anna Montmayeur, Clinton R. Paden, Peter W. Cook,Rachel Marine, Mili Sheth, Haibin Wang, Justin Lee, Suxiang Tong |
| EPI_ISL_751587 | NH Dept. of Health and Human Services Public Health Labs | Genomics and Discovery, Respiratory Viruses Branch, Division of Viral Diseases, Centers for Disease Control and Prevention | Krista Queen, Yan Li, Ying Tao, Jing Zhang, Anna Uehara, Anna Montmayeur, Clinton R. Paden, Peter W. Cook,Rachel Marine, Mili Sheth, Haibin Wang, Justin Lee, Suxiang Tong |
| EPI_ISL_751588 | VA-Division of Consolidated Laboratory Services | Genomics and Discovery, Respiratory Viruses Branch, Division of Viral Diseases, Centers for Disease Control and Prevention | Krista Queen, Yan Li, Ying Tao, Jing Zhang, Anna Uehara, Anna Montmayeur, Clinton R. Paden, Peter W. Cook,Rachel Marine, Mili Sheth, Haibin Wang, Justin Lee, Suxiang Tong |
| EPI_ISL_751589 | WVDHHR - Office of Laboratory Services | Genomics and Discovery, Respiratory Viruses Branch, Division of Viral Diseases, Centers for Disease Control and Prevention | Krista Queen, Yan Li, Ying Tao, Jing Zhang, Anna Uehara, Anna Montmayeur, Clinton R. Paden, Peter W. Cook,Rachel Marine, Mili Sheth, Haibin Wang, Justin Lee, Suxiang Tong |
| EPI_ISL_751590 | ID Bureau of Laboratories | Genomics and Discovery, Respiratory Viruses Branch, Division of Viral Diseases, Centers for Disease Control and Prevention | Krista Queen, Yan Li, Ying Tao, Jing Zhang, Anna Uehara, Anna Montmayeur, Clinton R. Paden, Peter W. Cook,Rachel Marine, Mili Sheth, Haibin Wang, Justin Lee, Suxiang Tong |
| EPI_ISL_751591 | WY Public Health Laboratory | Genomics and Discovery, Respiratory Viruses Branch, Division of Viral Diseases, Centers for Disease Control and Prevention | Krista Queen, Yan Li, Ying Tao, Jing Zhang, Anna Uehara, Anna Montmayeur, Clinton R. Paden, Peter W. Cook,Rachel Marine, Mili Sheth, Haibin Wang, Justin Lee, Suxiang Tong |
| EPI_ISL_751592, EPI_ISL_751593 | AK State Public Health Lab, State Health Department | Genomics and Discovery, Respiratory Viruses Branch, Division of Viral Diseases, Centers for Disease Control and Prevention | Krista Queen, Yan Li, Ying Tao, Jing Zhang, Anna Uehara, Anna Montmayeur, Clinton R. Paden, Peter W. Cook,Rachel Marine, Mili Sheth, Haibin Wang, Justin Lee, Suxiang Tong |
| EPI_ISL_751594 | KY State Public Health Lab | Genomics and Discovery, Respiratory Viruses Branch, Division of Viral Diseases, Centers for Disease Control and Prevention | Krista Queen, Yan Li, Ying Tao, Jing Zhang, Anna Uehara, Anna Montmayeur, Clinton R. Paden, Peter W. Cook,Rachel Marine, Mili Sheth, Haibin Wang, Justin Lee, Suxiang Tong |
| EPI_ISL_751595 | RI State Health Laboratories | Genomics and Discovery, Respiratory Viruses Branch, Division of Viral Diseases, Centers for Disease Control and Prevention | Krista Queen, Yan Li, Ying Tao, Jing Zhang, Anna Uehara, Anna Montmayeur, Clinton R. Paden, Peter W. Cook,Rachel Marine, Mili Sheth, Haibin Wang, Justin Lee, Suxiang Tong |
| EPI_ISL_751596 | KY State Public Health Lab | Genomics and Discovery, Respiratory Viruses Branch, Division of Viral Diseases, Centers for Disease Control and Prevention | Krista Queen, Yan Li, Ying Tao, Jing Zhang, Anna Uehara, Anna Montmayeur, Clinton R. Paden, Peter W. Cook,Rachel Marine, Mili Sheth, Haibin Wang, Justin Lee, Suxiang Tong |
| EPI_ISL_751597, EPI_ISL_751598 | AK State Public Health Lab, State Health Department | Genomics and Discovery, Respiratory Viruses Branch, Division of Viral Diseases, Centers for Disease Control and Prevention | Krista Queen, Yan Li, Ying Tao, Jing Zhang, Anna Uehara, Anna Montmayeur, Clinton R. Paden, Peter W. Cook,Rachel Marine, Mili Sheth, Haibin Wang, Justin Lee, Suxiang Tong |
| EPI_ISL_751599, EPI_ISL_751600 | IN State Department of Health Laboratory Services | Genomics and Discovery, Respiratory Viruses Branch, Division of Viral Diseases, Centers for Disease Control and Prevention | Krista Queen, Yan Li, Ying Tao, Jing Zhang, Anna Uehara, Anna Montmayeur, Clinton R. Paden, Peter W. Cook,Rachel Marine, Mili Sheth, Haibin Wang, Justin Lee, Suxiang Tong |
| EPI_ISL_751601 | MI - Michigan Department of Health and Human Services - Bureau of Laboratories | Genomics and Discovery, Respiratory Viruses Branch, Division of Viral Diseases, Centers for Disease Control and Prevention | Krista Queen, Yan Li, Ying Tao, Jing Zhang, Anna Uehara, Anna Montmayeur, Clinton R. Paden, Peter W. Cook,Rachel Marine, Mili Sheth, Haibin Wang, Justin Lee, Suxiang Tong |
| EPI_ISL_751602 | DE Public Health Laboratory | Genomics and Discovery, Respiratory Viruses Branch, Division of Viral Diseases, Centers for Disease Control and Prevention | Krista Queen, Yan Li, Ying Tao, Jing Zhang, Anna Uehara, Anna Montmayeur, Clinton R. Paden, Peter W. Cook,Rachel Marine, Mili Sheth, Haibin Wang, Justin Lee, Suxiang Tong |
| EPI_ISL_751603 | HI Dept. of Health, State Laboratories Division | Genomics and Discovery, Respiratory Viruses Branch, Division of Viral Diseases, Centers for Disease Control and Prevention | Krista Queen, Yan Li, Ying Tao, Jing Zhang, Anna Uehara, Anna Montmayeur, Clinton R. Paden, Peter W. Cook,Rachel Marine, Mili Sheth, Haibin Wang, Justin Lee, Suxiang Tong |
| EPI_ISL_751604 | MD DOH Laboratories Administration | Genomics and Discovery, Respiratory Viruses Branch, Division of Viral Diseases, Centers for Disease Control and Prevention | Krista Queen, Yan Li, Ying Tao, Jing Zhang, Anna Uehara, Anna Montmayeur, Clinton R. Paden, Peter W. Cook,Rachel Marine, Mili Sheth, Haibin Wang, Justin Lee, Suxiang Tong |
| EPI_ISL_751605 | DE Public Health Laboratory | Genomics and Discovery, Respiratory Viruses Branch, Division of Viral Diseases, Centers for Disease Control and Prevention | Krista Queen, Yan Li, Ying Tao, Jing Zhang, Anna Uehara, Anna Montmayeur, Clinton R. Paden, Peter W. Cook,Rachel Marine, Mili Sheth, Haibin Wang, Justin Lee, Suxiang Tong |
| EPI_ISL_751606, EPI_ISL_751607 | MD DOH Laboratories Administration | Genomics and Discovery, Respiratory Viruses Branch, Division of Viral Diseases, Centers for Disease Control and Prevention | Krista Queen, Yan Li, Ying Tao, Jing Zhang, Anna Uehara, Anna Montmayeur, Clinton R. Paden, Peter W. Cook,Rachel Marine, Mili Sheth, Haibin Wang, Justin Lee, Suxiang Tong |
| EPI_ISL_751608 | NE Public Health Laboratory | Genomics and Discovery, Respiratory Viruses Branch, Division of Viral Diseases, Centers for Disease Control and Prevention | Krista Queen, Yan Li, Ying Tao, Jing Zhang, Anna Uehara, Anna Montmayeur, Clinton R. Paden, Peter W. Cook,Rachel Marine, Mili Sheth, Haibin Wang, Justin Lee, Suxiang Tong |
| EPI_ISL_751609, EPI_ISL_751610 | OH Department of Health Laboratory | Genomics and Discovery, Respiratory Viruses Branch, Division of Viral Diseases, Centers for Disease Control and Prevention | Krista Queen, Yan Li, Ying Tao, Jing Zhang, Anna Uehara, Anna Montmayeur, Clinton R. Paden, Peter W. Cook,Rachel Marine, Mili Sheth, Haibin Wang, Justin Lee, Suxiang Tong |
| EPI_ISL_751611 | SC Dept of Health and Env. Control-Bureau of Laboratories | Genomics and Discovery, Respiratory Viruses Branch, Division of Viral Diseases, Centers for Disease Control and Prevention | Krista Queen, Yan Li, Ying Tao, Jing Zhang, Anna Uehara, Anna Montmayeur, Clinton R. Paden, Peter W. Cook,Rachel Marine, Mili Sheth, Haibin Wang, Justin Lee, Suxiang Tong |

| | | | |
|---|---|---|---|
| EPI_ISL_751612 | ID Bureau of Laboratories | Genomics and Discovery, Respiratory Viruses Branch, Division of Viral Diseases, Centers for Disease Control and Prevention | Krista Queen, Yan Li, Ying Tao, Jing Zhang, Anna Uehara, Anna Montmayeur, Clinton R. Paden, Peter W. Cook,Rachel Marine, Mili Sheth, Haibin Wang, Justin Lee, Suxiang Tong |
| EPI_ISL_751613 | OH Department of Health Laboratory | Genomics and Discovery, Respiratory Viruses Branch, Division of Viral Diseases, Centers for Disease Control and Prevention | Krista Queen, Yan Li, Ying Tao, Jing Zhang, Anna Uehara, Anna Montmayeur, Clinton R. Paden, Peter W. Cook,Rachel Marine, Mili Sheth, Haibin Wang, Justin Lee, Suxiang Tong |
| EPI_ISL_751614 | ID Bureau of Laboratories | Genomics and Discovery, Respiratory Viruses Branch, Division of Viral Diseases, Centers for Disease Control and Prevention | Krista Queen, Yan Li, Ying Tao, Jing Zhang, Anna Uehara, Anna Montmayeur, Clinton R. Paden, Peter W. Cook,Rachel Marine, Mili Sheth, Haibin Wang, Justin Lee, Suxiang Tong |
| EPI_ISL_751615 | OR State PHL-Virology/Immunology Section | Genomics and Discovery, Respiratory Viruses Branch, Division of Viral Diseases, Centers for Disease Control and Prevention | Krista Queen, Yan Li, Ying Tao, Jing Zhang, Anna Uehara, Anna Montmayeur, Clinton R. Paden, Peter W. Cook,Rachel Marine, Mili Sheth, Haibin Wang, Justin Lee, Suxiang Tong |
| EPI_ISL_751616 | OH Department of Health Laboratory | Genomics and Discovery, Respiratory Viruses Branch, Division of Viral Diseases, Centers for Disease Control and Prevention | Krista Queen, Yan Li, Ying Tao, Jing Zhang, Anna Uehara, Anna Montmayeur, Clinton R. Paden, Peter W. Cook,Rachel Marine, Mili Sheth, Haibin Wang, Justin Lee, Suxiang Tong |
| EPI_ISL_751617 | TX DSHS, Lab Services Section MC 1947 | Genomics and Discovery, Respiratory Viruses Branch, Division of Viral Diseases, Centers for Disease Control and Prevention | Krista Queen, Yan Li, Ying Tao, Jing Zhang, Anna Uehara, Anna Montmayeur, Clinton R. Paden, Peter W. Cook,Rachel Marine, Mili Sheth, Haibin Wang, Justin Lee, Suxiang Tong |
| EPI_ISL_751618 | ID Bureau of Laboratories | Genomics and Discovery, Respiratory Viruses Branch, Division of Viral Diseases, Centers for Disease Control and Prevention | Krista Queen, Yan Li, Ying Tao, Jing Zhang, Anna Uehara, Anna Montmayeur, Clinton R. Paden, Peter W. Cook,Rachel Marine, Mili Sheth, Haibin Wang, Justin Lee, Suxiang Tong |
| EPI_ISL_751619 | WY Public Health Laboratory | Genomics and Discovery, Respiratory Viruses Branch, Division of Viral Diseases, Centers for Disease Control and Prevention | Krista Queen, Yan Li, Ying Tao, Jing Zhang, Anna Uehara, Anna Montmayeur, Clinton R. Paden, Peter W. Cook,Rachel Marine, Mili Sheth, Haibin Wang, Justin Lee, Suxiang Tong |
| EPI_ISL_751620 | LA Office of Public Health Laboratories | Genomics and Discovery, Respiratory Viruses Branch, Division of Viral Diseases, Centers for Disease Control and Prevention | Krista Queen, Yan Li, Ying Tao, Jing Zhang, Anna Uehara, Anna Montmayeur, Clinton R. Paden, Peter W. Cook,Rachel Marine, Mili Sheth, Haibin Wang, Justin Lee, Suxiang Tong |
| EPI_ISL_751621 | DE Public Health Laboratory | Genomics and Discovery, Respiratory Viruses Branch, Division of Viral Diseases, Centers for Disease Control and Prevention | Krista Queen, Yan Li, Ying Tao, Jing Zhang, Anna Uehara, Anna Montmayeur, Clinton R. Paden, Peter W. Cook,Rachel Marine, Mili Sheth, Haibin Wang, Justin Lee, Suxiang Tong |
| EPI_ISL_751622 | MI - Michigan Department of Health and Human Services - Bureau of Laboratories | Genomics and Discovery, Respiratory Viruses Branch, Division of Viral Diseases, Centers for Disease Control and Prevention | Krista Queen, Yan Li, Ying Tao, Jing Zhang, Anna Uehara, Anna Montmayeur, Clinton R. Paden, Peter W. Cook,Rachel Marine, Mili Sheth, Haibin Wang, Justin Lee, Suxiang Tong |
| EPI_ISL_751623 | DE Public Health Laboratory | Genomics and Discovery, Respiratory Viruses Branch, Division of Viral Diseases, Centers for Disease Control and Prevention | Krista Queen, Yan Li, Ying Tao, Jing Zhang, Anna Uehara, Anna Montmayeur, Clinton R. Paden, Peter W. Cook,Rachel Marine, Mili Sheth, Haibin Wang, Justin Lee, Suxiang Tong |
| EPI_ISL_751624 | CT-Dr. Katherine A. Kelley State Public Health Lab | Genomics and Discovery, Respiratory Viruses Branch, Division of Viral Diseases, Centers for Disease Control and Prevention | Krista Queen, Yan Li, Ying Tao, Jing Zhang, Anna Uehara, Anna Montmayeur, Clinton R. Paden, Peter W. Cook,Rachel Marine, Mili Sheth, Haibin Wang, Justin Lee, Suxiang Tong |
| EPI_ISL_751625 | MD DOH Laboratories Administration | Genomics and Discovery, Respiratory Viruses Branch, Division of Viral Diseases, Centers for Disease Control and Prevention | Krista Queen, Yan Li, Ying Tao, Jing Zhang, Anna Uehara, Anna Montmayeur, Clinton R. Paden, Peter W. Cook,Rachel Marine, Mili Sheth, Haibin Wang, Justin Lee, Suxiang Tong |
| EPI_ISL_751626 | IN State Department of Health Laboratory Services | Genomics and Discovery, Respiratory Viruses Branch, Division of Viral Diseases, Centers for Disease Control and Prevention | Krista Queen, Yan Li, Ying Tao, Jing Zhang, Anna Uehara, Anna Montmayeur, Clinton R. Paden, Peter W. Cook,Rachel Marine, Mili Sheth, Haibin Wang, Justin Lee, Suxiang Tong |
| EPI_ISL_751627 | DE Public Health Laboratory | Genomics and Discovery, Respiratory Viruses Branch, Division of Viral Diseases, Centers for Disease Control and Prevention | Krista Queen, Yan Li, Ying Tao, Jing Zhang, Anna Uehara, Anna Montmayeur, Clinton R. Paden, Peter W. Cook,Rachel Marine, Mili Sheth, Haibin Wang, Justin Lee, Suxiang Tong |
| EPI_ISL_751628, EPI_ISL_751629 | MD DOH Laboratories Administration | Genomics and Discovery, Respiratory Viruses Branch, Division of Viral Diseases, Centers for Disease Control and Prevention | Krista Queen, Yan Li, Ying Tao, Jing Zhang, Anna Uehara, Anna Montmayeur, Clinton R. Paden, Peter W. Cook,Rachel Marine, Mili Sheth, Haibin Wang, Justin Lee, Suxiang Tong |
| EPI_ISL_751630 | CO Dept. of Public Health and Environment, Lab Services Division | Genomics and Discovery, Respiratory Viruses Branch, Division of Viral Diseases, Centers for Disease Control and Prevention | Krista Queen, Yan Li, Ying Tao, Jing Zhang, Anna Uehara, Anna Montmayeur, Clinton R. Paden, Peter W. Cook,Rachel Marine, Mili Sheth, Haibin Wang, Justin Lee, Suxiang Tong |
| EPI_ISL_751631 | DE Public Health Laboratory | Genomics and Discovery, Respiratory Viruses Branch, Division of Viral Diseases, Centers for Disease Control and Prevention | Krista Queen, Yan Li, Ying Tao, Jing Zhang, Anna Uehara, Anna Montmayeur, Clinton R. Paden, Peter W. Cook,Rachel Marine, Mili Sheth, Haibin Wang, Justin Lee, Suxiang Tong |
| EPI_ISL_751632 | IN State Department of Health Laboratory Services | Genomics and Discovery, Respiratory Viruses Branch, Division of Viral Diseases, Centers for Disease Control and Prevention | Krista Queen, Yan Li, Ying Tao, Jing Zhang, Anna Uehara, Anna Montmayeur, Clinton R. Paden, Peter W. Cook,Rachel Marine, Mili Sheth, Haibin Wang, Justin Lee, Suxiang Tong |
| EPI_ISL_751633 | MT Public Health Laboratory | Genomics and Discovery, Respiratory Viruses Branch, Division of Viral Diseases, Centers for Disease Control and Prevention | Krista Queen, Yan Li, Ying Tao, Jing Zhang, Anna Uehara, Anna Montmayeur, Clinton R. Paden, Peter W. Cook,Rachel Marine, Mili Sheth, Haibin Wang, Justin Lee, Suxiang Tong |
| EPI_ISL_751634, EPI_ISL_751635 | MO State Public Health Laboratory | Genomics and Discovery, Respiratory Viruses Branch, Division of Viral Diseases, Centers for Disease Control and Prevention | Krista Queen, Yan Li, Ying Tao, Jing Zhang, Anna Uehara, Anna Montmayeur, Clinton R. Paden, Peter W. Cook,Rachel Marine, Mili Sheth, Haibin Wang, Justin Lee, Suxiang Tong |
| EPI_ISL_751636, EPI_ISL_751637 | NV State Public Health Laboratory | Genomics and Discovery, Respiratory Viruses Branch, Division of Viral Diseases, Centers for Disease Control and Prevention | Krista Queen, Yan Li, Ying Tao, Jing Zhang, Anna Uehara, Anna Montmayeur, Clinton R. Paden, Peter W. Cook,Rachel Marine, Mili Sheth, Haibin Wang, Justin Lee, Suxiang Tong |
| EPI_ISL_751638 | NE Public Health Laboratory | Genomics and Discovery, Respiratory Viruses Branch, Division of Viral Diseases, Centers for Disease Control and Prevention | Krista Queen, Yan Li, Ying Tao, Jing Zhang, Anna Uehara, Anna Montmayeur, Clinton R. Paden, Peter W. Cook,Rachel Marine, Mili Sheth, Haibin Wang, Justin Lee, Suxiang Tong |
| EPI_ISL_751639 | NV State Public Health Laboratory | Genomics and Discovery, Respiratory Viruses Branch, Division of Viral Diseases, Centers for Disease Control and Prevention | Krista Queen, Yan Li, Ying Tao, Jing Zhang, Anna Uehara, Anna Montmayeur, Clinton R. Paden, Peter W. Cook,Rachel Marine, Mili Sheth, Haibin Wang, Justin Lee, Suxiang Tong |
| EPI_ISL_751640 | MT Public Health Laboratory | Genomics and Discovery, Respiratory Viruses Branch, Division of Viral Diseases, Centers for Disease Control and Prevention | Krista Queen, Yan Li, Ying Tao, Jing Zhang, Anna Uehara, Anna Montmayeur, Clinton R. Paden, Peter W. Cook,Rachel Marine, Mili Sheth, Haibin Wang, Justin Lee, Suxiang Tong |
| EPI_ISL_751641 | MN PHL Division, Minnesota Department of Health | Genomics and Discovery, Respiratory Viruses Branch, Division of Viral Diseases, Centers for Disease Control and Prevention | Krista Queen, Yan Li, Ying Tao, Jing Zhang, Anna Uehara, Anna Montmayeur, Clinton R. Paden, Peter W. Cook,Rachel Marine, Mili Sheth, Haibin Wang, Justin Lee, Suxiang Tong |
| EPI_ISL_751642 | NH Dept. of Health and Human Services Public Health Labs | Genomics and Discovery, Respiratory Viruses Branch, Division of Viral Diseases, Centers for Disease Control and Prevention | Krista Queen, Yan Li, Ying Tao, Jing Zhang, Anna Uehara, Anna Montmayeur, Clinton R. Paden, Peter W. Cook,Rachel Marine, Mili Sheth, Haibin Wang, Justin Lee, Suxiang Tong |
| EPI_ISL_751643 | WY Public Health Laboratory | Genomics and Discovery, Respiratory Viruses Branch, Division of Viral Diseases, Centers for Disease Control and Prevention | Krista Queen, Yan Li, Ying Tao, Jing Zhang, Anna Uehara, Anna Montmayeur, Clinton R. Paden, Peter W. Cook,Rachel Marine, Mili Sheth, Haibin Wang, Justin Lee, Suxiang Tong |
| EPI_ISL_751644, EPI_ISL_751645, EPI_ISL_751646 | TX DSHS, Lab Services Section MC 1947 | Genomics and Discovery, Respiratory Viruses Branch, Division of Viral Diseases, Centers for Disease Control and Prevention | Krista Queen, Yan Li, Ying Tao, Jing Zhang, Anna Uehara, Anna Montmayeur, Clinton R. Paden, Peter W. Cook,Rachel Marine, Mili Sheth, Haibin Wang, Justin Lee, Suxiang Tong |
| EPI_ISL_751647 | AL Dept. of Public Health Bureau of Clinical Laboratories | Genomics and Discovery, Respiratory Viruses Branch, Division of Viral Diseases, Centers for Disease Control and Prevention | Krista Queen, Yan Li, Ying Tao, Jing Zhang, Anna Uehara, Anna Montmayeur, Clinton R. Paden, Peter W. Cook,Rachel Marine, Mili Sheth, Haibin Wang, Justin Lee, Suxiang Tong |
| EPI_ISL_751648 | KY State Public Health Lab | Genomics and Discovery, Respiratory Viruses Branch, Division of Viral Diseases, Centers for Disease Control and Prevention | Krista Queen, Yan Li, Ying Tao, Jing Zhang, Anna Uehara, Anna Montmayeur, Clinton R. Paden, Peter W. Cook,Rachel Marine, Mili Sheth, Haibin Wang, Justin Lee, Suxiang Tong |

| | | | |
|---|---|---|---|
| EPI_ISL_751649, EPI_ISL_751650 | CT-Dr. Katherine A. Kelley State Public Health Lab | Genomics and Discovery, Respiratory Viruses Branch, Division of Viral Diseases, Centers for Disease Control and Prevention | Krista Queen, Yan Li, Ying Tao, Jing Zhang, Anna Uehara, Anna Montmayeur, Clinton R. Paden, Peter W. Cook,Rachel Marine, Mili Sheth, Haibin Wang, Justin Lee, Suxiang Tong |
| EPI_ISL_751651 | HI Dept. of Health, State Laboratories Division | Genomics and Discovery, Respiratory Viruses Branch, Division of Viral Diseases, Centers for Disease Control and Prevention | Krista Queen, Yan Li, Ying Tao, Jing Zhang, Anna Uehara, Anna Montmayeur, Clinton R. Paden, Peter W. Cook,Rachel Marine, Mili Sheth, Haibin Wang, Justin Lee, Suxiang Tong |
| EPI_ISL_751652 | AR Dept. of Health-Public Health Lab | Genomics and Discovery, Respiratory Viruses Branch, Division of Viral Diseases, Centers for Disease Control and Prevention | Krista Queen, Yan Li, Ying Tao, Jing Zhang, Anna Uehara, Anna Montmayeur, Clinton R. Paden, Peter W. Cook,Rachel Marine, Mili Sheth, Haibin Wang, Justin Lee, Suxiang Tong |
| EPI_ISL_751653, EPI_ISL_751654, EPI_ISL_751655 | MN PHL Division, Minnesota Department of Health | Genomics and Discovery, Respiratory Viruses Branch, Division of Viral Diseases, Centers for Disease Control and Prevention | Krista Queen, Yan Li, Ying Tao, Jing Zhang, Anna Uehara, Anna Montmayeur, Clinton R. Paden, Peter W. Cook,Rachel Marine, Mili Sheth, Haibin Wang, Justin Lee, Suxiang Tong |
| EPI_ISL_751656, EPI_ISL_751657 | MO State Public Health Laboratory | Genomics and Discovery, Respiratory Viruses Branch, Division of Viral Diseases, Centers for Disease Control and Prevention | Krista Queen, Yan Li, Ying Tao, Jing Zhang, Anna Uehara, Anna Montmayeur, Clinton R. Paden, Peter W. Cook,Rachel Marine, Mili Sheth, Haibin Wang, Justin Lee, Suxiang Tong |
| EPI_ISL_751658 | NYSDOH Wadsworth Center, Virology Lab | Genomics and Discovery, Respiratory Viruses Branch, Division of Viral Diseases, Centers for Disease Control and Prevention | Krista Queen, Yan Li, Ying Tao, Jing Zhang, Anna Uehara, Anna Montmayeur, Clinton R. Paden, Peter W. Cook,Rachel Marine, Mili Sheth, Haibin Wang, Justin Lee, Suxiang Tong |
| EPI_ISL_751659 | NJ Public Health and Environmental Laboratories | Genomics and Discovery, Respiratory Viruses Branch, Division of Viral Diseases, Centers for Disease Control and Prevention | Krista Queen, Yan Li, Ying Tao, Jing Zhang, Anna Uehara, Anna Montmayeur, Clinton R. Paden, Peter W. Cook,Rachel Marine, Mili Sheth, Haibin Wang, Justin Lee, Suxiang Tong |
| EPI_ISL_751660 | MN PHL Division, Minnesota Department of Health | Genomics and Discovery, Respiratory Viruses Branch, Division of Viral Diseases, Centers for Disease Control and Prevention | Krista Queen, Yan Li, Ying Tao, Jing Zhang, Anna Uehara, Anna Montmayeur, Clinton R. Paden, Peter W. Cook,Rachel Marine, Mili Sheth, Haibin Wang, Justin Lee, Suxiang Tong |
| EPI_ISL_751661 | MS Public Health Laboratory | Genomics and Discovery, Respiratory Viruses Branch, Division of Viral Diseases, Centers for Disease Control and Prevention | Krista Queen, Yan Li, Ying Tao, Jing Zhang, Anna Uehara, Anna Montmayeur, Clinton R. Paden, Peter W. Cook,Rachel Marine, Mili Sheth, Haibin Wang, Justin Lee, Suxiang Tong |
| EPI_ISL_751662 | NJ Public Health and Environmental Laboratories | Genomics and Discovery, Respiratory Viruses Branch, Division of Viral Diseases, Centers for Disease Control and Prevention | Krista Queen, Yan Li, Ying Tao, Jing Zhang, Anna Uehara, Anna Montmayeur, Clinton R. Paden, Peter W. Cook,Rachel Marine, Mili Sheth, Haibin Wang, Justin Lee, Suxiang Tong |
| EPI_ISL_751663 | WY Public Health Laboratory | Genomics and Discovery, Respiratory Viruses Branch, Division of Viral Diseases, Centers for Disease Control and Prevention | Krista Queen, Yan Li, Ying Tao, Jing Zhang, Anna Uehara, Anna Montmayeur, Clinton R. Paden, Peter W. Cook,Rachel Marine, Mili Sheth, Haibin Wang, Justin Lee, Suxiang Tong |
| EPI_ISL_751664, EPI_ISL_751665 | SC Dept of Health and Env. Control-Bureau of Laboratories | Genomics and Discovery, Respiratory Viruses Branch, Division of Viral Diseases, Centers for Disease Control and Prevention | Krista Queen, Yan Li, Ying Tao, Jing Zhang, Anna Uehara, Anna Montmayeur, Clinton R. Paden, Peter W. Cook,Rachel Marine, Mili Sheth, Haibin Wang, Justin Lee, Suxiang Tong |
| EPI_ISL_751666 | IA State Hygienic Laboratory | Genomics and Discovery, Respiratory Viruses Branch, Division of Viral Diseases, Centers for Disease Control and Prevention | Krista Queen, Yan Li, Ying Tao, Jing Zhang, Anna Uehara, Anna Montmayeur, Clinton R. Paden, Peter W. Cook,Rachel Marine, Mili Sheth, Haibin Wang, Justin Lee, Suxiang Tong |
| EPI_ISL_751667 | RI State Health Laboratories | Genomics and Discovery, Respiratory Viruses Branch, Division of Viral Diseases, Centers for Disease Control and Prevention | Krista Queen, Yan Li, Ying Tao, Jing Zhang, Anna Uehara, Anna Montmayeur, Clinton R. Paden, Peter W. Cook,Rachel Marine, Mili Sheth, Haibin Wang, Justin Lee, Suxiang Tong |
| EPI_ISL_751668 | AL Dept. of Public Health Bureau of Clinical Laboratories | Genomics and Discovery, Respiratory Viruses Branch, Division of Viral Diseases, Centers for Disease Control and Prevention | Krista Queen, Yan Li, Ying Tao, Jing Zhang, Anna Uehara, Anna Montmayeur, Clinton R. Paden, Peter W. Cook,Rachel Marine, Mili Sheth, Haibin Wang, Justin Lee, Suxiang Tong |
| EPI_ISL_751669 | LA Office of Public Health Laboratories | Genomics and Discovery, Respiratory Viruses Branch, Division of Viral Diseases, Centers for Disease Control and Prevention | Krista Queen, Yan Li, Ying Tao, Jing Zhang, Anna Uehara, Anna Montmayeur, Clinton R. Paden, Peter W. Cook,Rachel Marine, Mili Sheth, Haibin Wang, Justin Lee, Suxiang Tong |
| EPI_ISL_751670 | KY State Public Health Lab | Genomics and Discovery, Respiratory Viruses Branch, Division of Viral Diseases, Centers for Disease Control and Prevention | Krista Queen, Yan Li, Ying Tao, Jing Zhang, Anna Uehara, Anna Montmayeur, Clinton R. Paden, Peter W. Cook,Rachel Marine, Mili Sheth, Haibin Wang, Justin Lee, Suxiang Tong |
| EPI_ISL_751671 | RI State Health Laboratories | Genomics and Discovery, Respiratory Viruses Branch, Division of Viral Diseases, Centers for Disease Control and Prevention | Krista Queen, Yan Li, Ying Tao, Jing Zhang, Anna Uehara, Anna Montmayeur, Clinton R. Paden, Peter W. Cook,Rachel Marine, Mili Sheth, Haibin Wang, Justin Lee, Suxiang Tong |
| EPI_ISL_751672 | IN State Department of Health Laboratory Services | Genomics and Discovery, Respiratory Viruses Branch, Division of Viral Diseases, Centers for Disease Control and Prevention | Krista Queen, Yan Li, Ying Tao, Jing Zhang, Anna Uehara, Anna Montmayeur, Clinton R. Paden, Peter W. Cook,Rachel Marine, Mili Sheth, Haibin Wang, Justin Lee, Suxiang Tong |
| EPI_ISL_751673, EPI_ISL_751674, EPI_ISL_751675 | HI Dept. of Health, State Laboratories Division | Genomics and Discovery, Respiratory Viruses Branch, Division of Viral Diseases, Centers for Disease Control and Prevention | Krista Queen, Yan Li, Ying Tao, Jing Zhang, Anna Uehara, Anna Montmayeur, Clinton R. Paden, Peter W. Cook,Rachel Marine, Mili Sheth, Haibin Wang, Justin Lee, Suxiang Tong |
| EPI_ISL_751676 | IN State Department of Health Laboratory Services | Genomics and Discovery, Respiratory Viruses Branch, Division of Viral Diseases, Centers for Disease Control and Prevention | Krista Queen, Yan Li, Ying Tao, Jing Zhang, Anna Uehara, Anna Montmayeur, Clinton R. Paden, Peter W. Cook,Rachel Marine, Mili Sheth, Haibin Wang, Justin Lee, Suxiang Tong |
| EPI_ISL_751677 | KS Health and Environmental Laboratories | Genomics and Discovery, Respiratory Viruses Branch, Division of Viral Diseases, Centers for Disease Control and Prevention | Krista Queen, Yan Li, Ying Tao, Jing Zhang, Anna Uehara, Anna Montmayeur, Clinton R. Paden, Peter W. Cook,Rachel Marine, Mili Sheth, Haibin Wang, Justin Lee, Suxiang Tong |
| EPI_ISL_751678 | NH Dept. of Health and Human Services Public Health Labs | Genomics and Discovery, Respiratory Viruses Branch, Division of Viral Diseases, Centers for Disease Control and Prevention | Krista Queen, Yan Li, Ying Tao, Jing Zhang, Anna Uehara, Anna Montmayeur, Clinton R. Paden, Peter W. Cook,Rachel Marine, Mili Sheth, Haibin Wang, Justin Lee, Suxiang Tong |
| EPI_ISL_751679 | MS Public Health Laboratory | Genomics and Discovery, Respiratory Viruses Branch, Division of Viral Diseases, Centers for Disease Control and Prevention | Krista Queen, Yan Li, Ying Tao, Jing Zhang, Anna Uehara, Anna Montmayeur, Clinton R. Paden, Peter W. Cook,Rachel Marine, Mili Sheth, Haibin Wang, Justin Lee, Suxiang Tong |
| EPI_ISL_751680 | NJ Public Health and Environmental Laboratories | Genomics and Discovery, Respiratory Viruses Branch, Division of Viral Diseases, Centers for Disease Control and Prevention | Krista Queen, Yan Li, Ying Tao, Jing Zhang, Anna Uehara, Anna Montmayeur, Clinton R. Paden, Peter W. Cook,Rachel Marine, Mili Sheth, Haibin Wang, Justin Lee, Suxiang Tong |
| EPI_ISL_751681 | NE Public Health Laboratory | Genomics and Discovery, Respiratory Viruses Branch, Division of Viral Diseases, Centers for Disease Control and Prevention | Krista Queen, Yan Li, Ying Tao, Jing Zhang, Anna Uehara, Anna Montmayeur, Clinton R. Paden, Peter W. Cook,Rachel Marine, Mili Sheth, Haibin Wang, Justin Lee, Suxiang Tong |
| EPI_ISL_751682 | NYSDOH Wadsworth Center, Virology Lab | Genomics and Discovery, Respiratory Viruses Branch, Division of Viral Diseases, Centers for Disease Control and Prevention | Krista Queen, Yan Li, Ying Tao, Jing Zhang, Anna Uehara, Anna Montmayeur, Clinton R. Paden, Peter W. Cook,Rachel Marine, Mili Sheth, Haibin Wang, Justin Lee, Suxiang Tong |
| EPI_ISL_751683 | MS Public Health Laboratory | Genomics and Discovery, Respiratory Viruses Branch, Division of Viral Diseases, Centers for Disease Control and Prevention | Krista Queen, Yan Li, Ying Tao, Jing Zhang, Anna Uehara, Anna Montmayeur, Clinton R. Paden, Peter W. Cook,Rachel Marine, Mili Sheth, Haibin Wang, Justin Lee, Suxiang Tong |
| EPI_ISL_751684 | TX DSHS, Lab Services Section MC 1947 | Genomics and Discovery, Respiratory Viruses Branch, Division of Viral Diseases, Centers for Disease Control and Prevention | Krista Queen, Yan Li, Ying Tao, Jing Zhang, Anna Uehara, Anna Montmayeur, Clinton R. Paden, Peter W. Cook,Rachel Marine, Mili Sheth, Haibin Wang, Justin Lee, Suxiang Tong |
| EPI_ISL_751685, EPI_ISL_751686 | VA-Division of Consolidated Laboratory Services | Genomics and Discovery, Respiratory Viruses Branch, Division of Viral Diseases, Centers for Disease Control and Prevention | Krista Queen, Yan Li, Ying Tao, Jing Zhang, Anna Uehara, Anna Montmayeur, Clinton R. Paden, Peter W. Cook,Rachel Marine, Mili Sheth, Haibin Wang, Justin Lee, Suxiang Tong |
| EPI_ISL_751687 | IA State Hygienic Laboratory | Genomics and Discovery, Respiratory Viruses Branch, Division of Viral Diseases, Centers for Disease Control and Prevention | Krista Queen, Yan Li, Ying Tao, Jing Zhang, Anna Uehara, Anna Montmayeur, Clinton R. Paden, Peter W. Cook,Rachel Marine, Mili Sheth, Haibin Wang, Justin Lee, Suxiang Tong |
| EPI_ISL_751688 | WY Public Health Laboratory | Genomics and Discovery, Respiratory Viruses Branch, Division of Viral Diseases, Centers for Disease Control and Prevention | Krista Queen, Yan Li, Ying Tao, Jing Zhang, Anna Uehara, Anna Montmayeur, Clinton R. Paden, Peter W. Cook,Rachel Marine, Mili Sheth, Haibin Wang, Justin Lee, Suxiang Tong |

| | | | |
|---|---|---|---|
| EPI_ISL_751689 | OR State PHL-Virology/Immunology Section | Genomics and Discovery, Respiratory Viruses Branch, Division of Viral Diseases, Centers for Disease Control and Prevention | Krista Queen, Yan Li, Ying Tao, Jing Zhang, Anna Uehara, Anna Montmayeur, Clinton R. Paden, Peter W. Cook,Rachel Marine, Mili Sheth, Haibin Wang, Justin Lee, Suxiang Tong |
| EPI_ISL_751690 | VA-Division of Consolidated Laboratory Services | Genomics and Discovery, Respiratory Viruses Branch, Division of Viral Diseases, Centers for Disease Control and Prevention | Krista Queen, Yan Li, Ying Tao, Jing Zhang, Anna Uehara, Anna Montmayeur, Clinton R. Paden, Peter W. Cook,Rachel Marine, Mili Sheth, Haibin Wang, Justin Lee, Suxiang Tong |
| EPI_ISL_751691 | WY Public Health Laboratory | Genomics and Discovery, Respiratory Viruses Branch, Division of Viral Diseases, Centers for Disease Control and Prevention | Krista Queen, Yan Li, Ying Tao, Jing Zhang, Anna Uehara, Anna Montmayeur, Clinton R. Paden, Peter W. Cook,Rachel Marine, Mili Sheth, Haibin Wang, Justin Lee, Suxiang Tong |
| EPI_ISL_751692 | IA State Hygienic Laboratory | Genomics and Discovery, Respiratory Viruses Branch, Division of Viral Diseases, Centers for Disease Control and Prevention | Krista Queen, Yan Li, Ying Tao, Jing Zhang, Anna Uehara, Anna Montmayeur, Clinton R. Paden, Peter W. Cook,Rachel Marine, Mili Sheth, Haibin Wang, Justin Lee, Suxiang Tong |
| EPI_ISL_751693 | OR State PHL-Virology/Immunology Section | Genomics and Discovery, Respiratory Viruses Branch, Division of Viral Diseases, Centers for Disease Control and Prevention | Krista Queen, Yan Li, Ying Tao, Jing Zhang, Anna Uehara, Anna Montmayeur, Clinton R. Paden, Peter W. Cook,Rachel Marine, Mili Sheth, Haibin Wang, Justin Lee, Suxiang Tong |
| EPI_ISL_751694 | AK State Public Health Lab, State Health Department | Genomics and Discovery, Respiratory Viruses Branch, Division of Viral Diseases, Centers for Disease Control and Prevention | Krista Queen, Yan Li, Ying Tao, Jing Zhang, Anna Uehara, Anna Montmayeur, Clinton R. Paden, Peter W. Cook,Rachel Marine, Mili Sheth, Haibin Wang, Justin Lee, Suxiang Tong |
| EPI_ISL_751695 | RI State Health Laboratories | Genomics and Discovery, Respiratory Viruses Branch, Division of Viral Diseases, Centers for Disease Control and Prevention | Krista Queen, Yan Li, Ying Tao, Jing Zhang, Anna Uehara, Anna Montmayeur, Clinton R. Paden, Peter W. Cook,Rachel Marine, Mili Sheth, Haibin Wang, Justin Lee, Suxiang Tong |
| EPI_ISL_751696 | LA Office of Public Health Laboratories | Genomics and Discovery, Respiratory Viruses Branch, Division of Viral Diseases, Centers for Disease Control and Prevention | Krista Queen, Yan Li, Ying Tao, Jing Zhang, Anna Uehara, Anna Montmayeur, Clinton R. Paden, Peter W. Cook,Rachel Marine, Mili Sheth, Haibin Wang, Justin Lee, Suxiang Tong |
| EPI_ISL_751697, EPI_ISL_751698, EPI_ISL_751699 | AR Dept. of Health-Public Health Lab | Genomics and Discovery, Respiratory Viruses Branch, Division of Viral Diseases, Centers for Disease Control and Prevention | Krista Queen, Yan Li, Ying Tao, Jing Zhang, Anna Uehara, Anna Montmayeur, Clinton R. Paden, Peter W. Cook,Rachel Marine, Mili Sheth, Haibin Wang, Justin Lee, Suxiang Tong |
| EPI_ISL_751700 | MD DOH Laboratories Administration | Genomics and Discovery, Respiratory Viruses Branch, Division of Viral Diseases, Centers for Disease Control and Prevention | Krista Queen, Yan Li, Ying Tao, Jing Zhang, Anna Uehara, Anna Montmayeur, Clinton R. Paden, Peter W. Cook,Rachel Marine, Mili Sheth, Haibin Wang, Justin Lee, Suxiang Tong |
| EPI_ISL_751701 | CO Dept. of Public Health and Environment, Lab Services Division | Genomics and Discovery, Respiratory Viruses Branch, Division of Viral Diseases, Centers for Disease Control and Prevention | Krista Queen, Yan Li, Ying Tao, Jing Zhang, Anna Uehara, Anna Montmayeur, Clinton R. Paden, Peter W. Cook,Rachel Marine, Mili Sheth, Haibin Wang, Justin Lee, Suxiang Tong |
| EPI_ISL_751702, EPI_ISL_751703 | MD DOH Laboratories Administration | Genomics and Discovery, Respiratory Viruses Branch, Division of Viral Diseases, Centers for Disease Control and Prevention | Krista Queen, Yan Li, Ying Tao, Jing Zhang, Anna Uehara, Anna Montmayeur, Clinton R. Paden, Peter W. Cook,Rachel Marine, Mili Sheth, Haibin Wang, Justin Lee, Suxiang Tong |
| EPI_ISL_751704, EPI_ISL_751705 | CO Dept. of Public Health and Environment, Lab Services Division | Genomics and Discovery, Respiratory Viruses Branch, Division of Viral Diseases, Centers for Disease Control and Prevention | Krista Queen, Yan Li, Ying Tao, Jing Zhang, Anna Uehara, Anna Montmayeur, Clinton R. Paden, Peter W. Cook,Rachel Marine, Mili Sheth, Haibin Wang, Justin Lee, Suxiang Tong |
| EPI_ISL_751706 | CT-Dr. Katherine A. Kelley State Public Health Lab | Genomics and Discovery, Respiratory Viruses Branch, Division of Viral Diseases, Centers for Disease Control and Prevention | Krista Queen, Yan Li, Ying Tao, Jing Zhang, Anna Uehara, Anna Montmayeur, Clinton R. Paden, Peter W. Cook,Rachel Marine, Mili Sheth, Haibin Wang, Justin Lee, Suxiang Tong |
| EPI_ISL_751707 | KS Health and Environmental Laboratories | Genomics and Discovery, Respiratory Viruses Branch, Division of Viral Diseases, Centers for Disease Control and Prevention | Krista Queen, Yan Li, Ying Tao, Jing Zhang, Anna Uehara, Anna Montmayeur, Clinton R. Paden, Peter W. Cook,Rachel Marine, Mili Sheth, Haibin Wang, Justin Lee, Suxiang Tong |
| EPI_ISL_751708 | NV State Public Health Laboratory | Genomics and Discovery, Respiratory Viruses Branch, Division of Viral Diseases, Centers for Disease Control and Prevention | Krista Queen, Yan Li, Ying Tao, Jing Zhang, Anna Uehara, Anna Montmayeur, Clinton R. Paden, Peter W. Cook,Rachel Marine, Mili Sheth, Haibin Wang, Justin Lee, Suxiang Tong |
| EPI_ISL_751709 | NJ Public Health and Environmental Laboratories | Genomics and Discovery, Respiratory Viruses Branch, Division of Viral Diseases, Centers for Disease Control and Prevention | Krista Queen, Yan Li, Ying Tao, Jing Zhang, Anna Uehara, Anna Montmayeur, Clinton R. Paden, Peter W. Cook,Rachel Marine, Mili Sheth, Haibin Wang, Justin Lee, Suxiang Tong |
| EPI_ISL_751710, EPI_ISL_751711 | MT Public Health Laboratory | Genomics and Discovery, Respiratory Viruses Branch, Division of Viral Diseases, Centers for Disease Control and Prevention | Krista Queen, Yan Li, Ying Tao, Jing Zhang, Anna Uehara, Anna Montmayeur, Clinton R. Paden, Peter W. Cook,Rachel Marine, Mili Sheth, Haibin Wang, Justin Lee, Suxiang Tong |
| EPI_ISL_751712 | NH Dept. of Health and Human Services Public Health Labs | Genomics and Discovery, Respiratory Viruses Branch, Division of Viral Diseases, Centers for Disease Control and Prevention | Krista Queen, Yan Li, Ying Tao, Jing Zhang, Anna Uehara, Anna Montmayeur, Clinton R. Paden, Peter W. Cook,Rachel Marine, Mili Sheth, Haibin Wang, Justin Lee, Suxiang Tong |
| EPI_ISL_751713 | MT Public Health Laboratory | Genomics and Discovery, Respiratory Viruses Branch, Division of Viral Diseases, Centers for Disease Control and Prevention | Krista Queen, Yan Li, Ying Tao, Jing Zhang, Anna Uehara, Anna Montmayeur, Clinton R. Paden, Peter W. Cook,Rachel Marine, Mili Sheth, Haibin Wang, Justin Lee, Suxiang Tong |
| EPI_ISL_751714 | NV State Public Health Laboratory | Genomics and Discovery, Respiratory Viruses Branch, Division of Viral Diseases, Centers for Disease Control and Prevention | Krista Queen, Yan Li, Ying Tao, Jing Zhang, Anna Uehara, Anna Montmayeur, Clinton R. Paden, Peter W. Cook,Rachel Marine, Mili Sheth, Haibin Wang, Justin Lee, Suxiang Tong |
| EPI_ISL_751715 | MO State Public Health Laboratory | Genomics and Discovery, Respiratory Viruses Branch, Division of Viral Diseases, Centers for Disease Control and Prevention | Krista Queen, Yan Li, Ying Tao, Jing Zhang, Anna Uehara, Anna Montmayeur, Clinton R. Paden, Peter W. Cook,Rachel Marine, Mili Sheth, Haibin Wang, Justin Lee, Suxiang Tong |
| EPI_ISL_751716 | MN PHL Division, Minnesota Department of Health | Genomics and Discovery, Respiratory Viruses Branch, Division of Viral Diseases, Centers for Disease Control and Prevention | Krista Queen, Yan Li, Ying Tao, Jing Zhang, Anna Uehara, Anna Montmayeur, Clinton R. Paden, Peter W. Cook,Rachel Marine, Mili Sheth, Haibin Wang, Justin Lee, Suxiang Tong |
| EPI_ISL_751717 | NYSDOH Wadsworth Center, Virology Lab | Genomics and Discovery, Respiratory Viruses Branch, Division of Viral Diseases, Centers for Disease Control and Prevention | Krista Queen, Yan Li, Ying Tao, Jing Zhang, Anna Uehara, Anna Montmayeur, Clinton R. Paden, Peter W. Cook,Rachel Marine, Mili Sheth, Haibin Wang, Justin Lee, Suxiang Tong |
| EPI_ISL_751718 | SC Dept of Health and Env. Control-Bureau of Laboratories | Genomics and Discovery, Respiratory Viruses Branch, Division of Viral Diseases, Centers for Disease Control and Prevention | Krista Queen, Yan Li, Ying Tao, Jing Zhang, Anna Uehara, Anna Montmayeur, Clinton R. Paden, Peter W. Cook,Rachel Marine, Mili Sheth, Haibin Wang, Justin Lee, Suxiang Tong |
| EPI_ISL_751719 | ID Bureau of Laboratories | Genomics and Discovery, Respiratory Viruses Branch, Division of Viral Diseases, Centers for Disease Control and Prevention | Krista Queen, Yan Li, Ying Tao, Jing Zhang, Anna Uehara, Anna Montmayeur, Clinton R. Paden, Peter W. Cook,Rachel Marine, Mili Sheth, Haibin Wang, Justin Lee, Suxiang Tong |
| EPI_ISL_751720 | TX DSHS, Lab Services Section MC 1947 | Genomics and Discovery, Respiratory Viruses Branch, Division of Viral Diseases, Centers for Disease Control and Prevention | Krista Queen, Yan Li, Ying Tao, Jing Zhang, Anna Uehara, Anna Montmayeur, Clinton R. Paden, Peter W. Cook,Rachel Marine, Mili Sheth, Haibin Wang, Justin Lee, Suxiang Tong |
| EPI_ISL_751721 | IA State Hygienic Laboratory | Genomics and Discovery, Respiratory Viruses Branch, Division of Viral Diseases, Centers for Disease Control and Prevention | Krista Queen, Yan Li, Ying Tao, Jing Zhang, Anna Uehara, Anna Montmayeur, Clinton R. Paden, Peter W. Cook,Rachel Marine, Mili Sheth, Haibin Wang, Justin Lee, Suxiang Tong |
| EPI_ISL_751722 | LA Office of Public Health Laboratories | Genomics and Discovery, Respiratory Viruses Branch, Division of Viral Diseases, Centers for Disease Control and Prevention | Krista Queen, Yan Li, Ying Tao, Jing Zhang, Anna Uehara, Anna Montmayeur, Clinton R. Paden, Peter W. Cook,Rachel Marine, Mili Sheth, Haibin Wang, Justin Lee, Suxiang Tong |
| EPI_ISL_751723 | AR Dept. of Health-Public Health Lab | Genomics and Discovery, Respiratory Viruses Branch, Division of Viral Diseases, Centers for Disease Control and Prevention | Krista Queen, Yan Li, Ying Tao, Jing Zhang, Anna Uehara, Anna Montmayeur, Clinton R. Paden, Peter W. Cook,Rachel Marine, Mili Sheth, Haibin Wang, Justin Lee, Suxiang Tong |
| EPI_ISL_751724 | KS Health and Environmental Laboratories | Genomics and Discovery, Respiratory Viruses Branch, Division of Viral Diseases, Centers for Disease Control and Prevention | Krista Queen, Yan Li, Ying Tao, Jing Zhang, Anna Uehara, Anna Montmayeur, Clinton R. Paden, Peter W. Cook,Rachel Marine, Mili Sheth, Haibin Wang, Justin Lee, Suxiang Tong |
| EPI_ISL_751725 | MD DOH Laboratories Administration | Genomics and Discovery, Respiratory Viruses Branch, Division of Viral Diseases, Centers for Disease Control and Prevention | Krista Queen, Yan Li, Ying Tao, Jing Zhang, Anna Uehara, Anna Montmayeur, Clinton R. Paden, Peter W. Cook,Rachel Marine, Mili Sheth, Haibin Wang, Justin Lee, Suxiang Tong |

| | | | |
|---|---|---|---|
| EPI_ISL_751726 | DE Public Health Laboratory | Genomics and Discovery, Respiratory Viruses Branch, Division of Viral Diseases, Centers for Disease Control and Prevention | Krista Queen, Yan Li, Ying Tao, Jing Zhang, Anna Uehara, Anna Montmayeur, Clinton R. Paden, Peter W. Cook,Rachel Marine, Mili Sheth, Haibin Wang, Justin Lee, Suxiang Tong |
| EPI_ISL_751727 | IN State Department of Health Laboratory Services | Genomics and Discovery, Respiratory Viruses Branch, Division of Viral Diseases, Centers for Disease Control and Prevention | Krista Queen, Yan Li, Ying Tao, Jing Zhang, Anna Uehara, Anna Montmayeur, Clinton R. Paden, Peter W. Cook,Rachel Marine, Mili Sheth, Haibin Wang, Justin Lee, Suxiang Tong |
| EPI_ISL_751728 | CO Dept. of Public Health and Environment, Lab Services Division | Genomics and Discovery, Respiratory Viruses Branch, Division of Viral Diseases, Centers for Disease Control and Prevention | Krista Queen, Yan Li, Ying Tao, Jing Zhang, Anna Uehara, Anna Montmayeur, Clinton R. Paden, Peter W. Cook,Rachel Marine, Mili Sheth, Haibin Wang, Justin Lee, Suxiang Tong |
| EPI_ISL_751729 | CT-Dr. Katherine A. Kelley State Public Health Lab | Genomics and Discovery, Respiratory Viruses Branch, Division of Viral Diseases, Centers for Disease Control and Prevention | Krista Queen, Yan Li, Ying Tao, Jing Zhang, Anna Uehara, Anna Montmayeur, Clinton R. Paden, Peter W. Cook,Rachel Marine, Mili Sheth, Haibin Wang, Justin Lee, Suxiang Tong |
| EPI_ISL_751730 | KS Health and Environmental Laboratories | Genomics and Discovery, Respiratory Viruses Branch, Division of Viral Diseases, Centers for Disease Control and Prevention | Krista Queen, Yan Li, Ying Tao, Jing Zhang, Anna Uehara, Anna Montmayeur, Clinton R. Paden, Peter W. Cook,Rachel Marine, Mili Sheth, Haibin Wang, Justin Lee, Suxiang Tong |
| EPI_ISL_751731 | AR Dept. of Health-Public Health Lab | Genomics and Discovery, Respiratory Viruses Branch, Division of Viral Diseases, Centers for Disease Control and Prevention | Krista Queen, Yan Li, Ying Tao, Jing Zhang, Anna Uehara, Anna Montmayeur, Clinton R. Paden, Peter W. Cook,Rachel Marine, Mili Sheth, Haibin Wang, Justin Lee, Suxiang Tong |
| EPI_ISL_751732 | NE Public Health Laboratory | Genomics and Discovery, Respiratory Viruses Branch, Division of Viral Diseases, Centers for Disease Control and Prevention | Krista Queen, Yan Li, Ying Tao, Jing Zhang, Anna Uehara, Anna Montmayeur, Clinton R. Paden, Peter W. Cook,Rachel Marine, Mili Sheth, Haibin Wang, Justin Lee, Suxiang Tong |
| EPI_ISL_751733 | MN PHL Division, Minnesota Department of Health | Genomics and Discovery, Respiratory Viruses Branch, Division of Viral Diseases, Centers for Disease Control and Prevention | Krista Queen, Yan Li, Ying Tao, Jing Zhang, Anna Uehara, Anna Montmayeur, Clinton R. Paden, Peter W. Cook,Rachel Marine, Mili Sheth, Haibin Wang, Justin Lee, Suxiang Tong |
| EPI_ISL_751734 | NE Public Health Laboratory | Genomics and Discovery, Respiratory Viruses Branch, Division of Viral Diseases, Centers for Disease Control and Prevention | Krista Queen, Yan Li, Ying Tao, Jing Zhang, Anna Uehara, Anna Montmayeur, Clinton R. Paden, Peter W. Cook,Rachel Marine, Mili Sheth, Haibin Wang, Justin Lee, Suxiang Tong |
| EPI_ISL_751735 | NH Dept. of Health and Human Services Public Health Labs | Genomics and Discovery, Respiratory Viruses Branch, Division of Viral Diseases, Centers for Disease Control and Prevention | Krista Queen, Yan Li, Ying Tao, Jing Zhang, Anna Uehara, Anna Montmayeur, Clinton R. Paden, Peter W. Cook,Rachel Marine, Mili Sheth, Haibin Wang, Justin Lee, Suxiang Tong |
| EPI_ISL_751736 | NV State Public Health Laboratory | Genomics and Discovery, Respiratory Viruses Branch, Division of Viral Diseases, Centers for Disease Control and Prevention | Krista Queen, Yan Li, Ying Tao, Jing Zhang, Anna Uehara, Anna Montmayeur, Clinton R. Paden, Peter W. Cook,Rachel Marine, Mili Sheth, Haibin Wang, Justin Lee, Suxiang Tong |
| EPI_ISL_751737 | MO State Public Health Laboratory | Genomics and Discovery, Respiratory Viruses Branch, Division of Viral Diseases, Centers for Disease Control and Prevention | Krista Queen, Yan Li, Ying Tao, Jing Zhang, Anna Uehara, Anna Montmayeur, Clinton R. Paden, Peter W. Cook,Rachel Marine, Mili Sheth, Haibin Wang, Justin Lee, Suxiang Tong |
| EPI_ISL_751738 | MT Public Health Laboratory | Genomics and Discovery, Respiratory Viruses Branch, Division of Viral Diseases, Centers for Disease Control and Prevention | Krista Queen, Yan Li, Ying Tao, Jing Zhang, Anna Uehara, Anna Montmayeur, Clinton R. Paden, Peter W. Cook,Rachel Marine, Mili Sheth, Haibin Wang, Justin Lee, Suxiang Tong |
| EPI_ISL_751739 | MN PHL Division, Minnesota Department of Health | Genomics and Discovery, Respiratory Viruses Branch, Division of Viral Diseases, Centers for Disease Control and Prevention | Krista Queen, Yan Li, Ying Tao, Jing Zhang, Anna Uehara, Anna Montmayeur, Clinton R. Paden, Peter W. Cook,Rachel Marine, Mili Sheth, Haibin Wang, Justin Lee, Suxiang Tong |
| EPI_ISL_751740 | MO State Public Health Laboratory | Genomics and Discovery, Respiratory Viruses Branch, Division of Viral Diseases, Centers for Disease Control and Prevention | Krista Queen, Yan Li, Ying Tao, Jing Zhang, Anna Uehara, Anna Montmayeur, Clinton R. Paden, Peter W. Cook,Rachel Marine, Mili Sheth, Haibin Wang, Justin Lee, Suxiang Tong |
| EPI_ISL_751741 | OH Department of Health Laboratory | Genomics and Discovery, Respiratory Viruses Branch, Division of Viral Diseases, Centers for Disease Control and Prevention | Krista Queen, Yan Li, Ying Tao, Jing Zhang, Anna Uehara, Anna Montmayeur, Clinton R. Paden, Peter W. Cook,Rachel Marine, Mili Sheth, Haibin Wang, Justin Lee, Suxiang Tong |
| EPI_ISL_751742 | IA State Hygienic Laboratory | Genomics and Discovery, Respiratory Viruses Branch, Division of Viral Diseases, Centers for Disease Control and Prevention | Krista Queen, Yan Li, Ying Tao, Jing Zhang, Anna Uehara, Anna Montmayeur, Clinton R. Paden, Peter W. Cook,Rachel Marine, Mili Sheth, Haibin Wang, Justin Lee, Suxiang Tong |
| EPI_ISL_751743 | WY Public Health Laboratory | Genomics and Discovery, Respiratory Viruses Branch, Division of Viral Diseases, Centers for Disease Control and Prevention | Krista Queen, Yan Li, Ying Tao, Jing Zhang, Anna Uehara, Anna Montmayeur, Clinton R. Paden, Peter W. Cook,Rachel Marine, Mili Sheth, Haibin Wang, Justin Lee, Suxiang Tong |
| EPI_ISL_751744 | SC Dept of Health and Env. Control-Bureau of Laboratories | Genomics and Discovery, Respiratory Viruses Branch, Division of Viral Diseases, Centers for Disease Control and Prevention | Krista Queen, Yan Li, Ying Tao, Jing Zhang, Anna Uehara, Anna Montmayeur, Clinton R. Paden, Peter W. Cook,Rachel Marine, Mili Sheth, Haibin Wang, Justin Lee, Suxiang Tong |
| EPI_ISL_751745 | VA-Division of Consolidated Laboratory Services | Genomics and Discovery, Respiratory Viruses Branch, Division of Viral Diseases, Centers for Disease Control and Prevention | Krista Queen, Yan Li, Ying Tao, Jing Zhang, Anna Uehara, Anna Montmayeur, Clinton R. Paden, Peter W. Cook,Rachel Marine, Mili Sheth, Haibin Wang, Justin Lee, Suxiang Tong |
| EPI_ISL_751746 | WVDHHR - Office of Laboratory Services | Genomics and Discovery, Respiratory Viruses Branch, Division of Viral Diseases, Centers for Disease Control and Prevention | Krista Queen, Yan Li, Ying Tao, Jing Zhang, Anna Uehara, Anna Montmayeur, Clinton R. Paden, Peter W. Cook,Rachel Marine, Mili Sheth, Haibin Wang, Justin Lee, Suxiang Tong |
| EPI_ISL_751747 | VA-Division of Consolidated Laboratory Services | Genomics and Discovery, Respiratory Viruses Branch, Division of Viral Diseases, Centers for Disease Control and Prevention | Krista Queen, Yan Li, Ying Tao, Jing Zhang, Anna Uehara, Anna Montmayeur, Clinton R. Paden, Peter W. Cook,Rachel Marine, Mili Sheth, Haibin Wang, Justin Lee, Suxiang Tong |
| EPI_ISL_751748 | ID Bureau of Laboratories | Genomics and Discovery, Respiratory Viruses Branch, Division of Viral Diseases, Centers for Disease Control and Prevention | Krista Queen, Yan Li, Ying Tao, Jing Zhang, Anna Uehara, Anna Montmayeur, Clinton R. Paden, Peter W. Cook,Rachel Marine, Mili Sheth, Haibin Wang, Justin Lee, Suxiang Tong |
| EPI_ISL_751749 | TX DSHS, Lab Services Section MC 1947 | Genomics and Discovery, Respiratory Viruses Branch, Division of Viral Diseases, Centers for Disease Control and Prevention | Krista Queen, Yan Li, Ying Tao, Jing Zhang, Anna Uehara, Anna Montmayeur, Clinton R. Paden, Peter W. Cook,Rachel Marine, Mili Sheth, Haibin Wang, Justin Lee, Suxiang Tong |
| EPI_ISL_751750 | ID Bureau of Laboratories | Genomics and Discovery, Respiratory Viruses Branch, Division of Viral Diseases, Centers for Disease Control and Prevention | Krista Queen, Yan Li, Ying Tao, Jing Zhang, Anna Uehara, Anna Montmayeur, Clinton R. Paden, Peter W. Cook,Rachel Marine, Mili Sheth, Haibin Wang, Justin Lee, Suxiang Tong |
| EPI_ISL_751751 | RI State Health Laboratories | Genomics and Discovery, Respiratory Viruses Branch, Division of Viral Diseases, Centers for Disease Control and Prevention | Krista Queen, Yan Li, Ying Tao, Jing Zhang, Anna Uehara, Anna Montmayeur, Clinton R. Paden, Peter W. Cook,Rachel Marine, Mili Sheth, Haibin Wang, Justin Lee, Suxiang Tong |
| EPI_ISL_751752 | LA Office of Public Health Laboratories | Genomics and Discovery, Respiratory Viruses Branch, Division of Viral Diseases, Centers for Disease Control and Prevention | Krista Queen, Yan Li, Ying Tao, Jing Zhang, Anna Uehara, Anna Montmayeur, Clinton R. Paden, Peter W. Cook,Rachel Marine, Mili Sheth, Haibin Wang, Justin Lee, Suxiang Tong |
| EPI_ISL_751753 | AL Dept. of Public Health Bureau of Clinical Laboratories | Genomics and Discovery, Respiratory Viruses Branch, Division of Viral Diseases, Centers for Disease Control and Prevention | Krista Queen, Yan Li, Ying Tao, Jing Zhang, Anna Uehara, Anna Montmayeur, Clinton R. Paden, Peter W. Cook,Rachel Marine, Mili Sheth, Haibin Wang, Justin Lee, Suxiang Tong |
| EPI_ISL_751754 | KY State Public Health Lab | Genomics and Discovery, Respiratory Viruses Branch, Division of Viral Diseases, Centers for Disease Control and Prevention | Krista Queen, Yan Li, Ying Tao, Jing Zhang, Anna Uehara, Anna Montmayeur, Clinton R. Paden, Peter W. Cook,Rachel Marine, Mili Sheth, Haibin Wang, Justin Lee, Suxiang Tong |
| EPI_ISL_751755, EPI_ISL_751756 | AL Dept. of Public Health Bureau of Clinical Laboratories | Genomics and Discovery, Respiratory Viruses Branch, Division of Viral Diseases, Centers for Disease Control and Prevention | Krista Queen, Yan Li, Ying Tao, Jing Zhang, Anna Uehara, Anna Montmayeur, Clinton R. Paden, Peter W. Cook,Rachel Marine, Mili Sheth, Haibin Wang, Justin Lee, Suxiang Tong |
| EPI_ISL_751757 | RI State Health Laboratories | Genomics and Discovery, Respiratory Viruses Branch, Division of Viral Diseases, Centers for Disease Control and Prevention | Krista Queen, Yan Li, Ying Tao, Jing Zhang, Anna Uehara, Anna Montmayeur, Clinton R. Paden, Peter W. Cook,Rachel Marine, Mili Sheth, Haibin Wang, Justin Lee, Suxiang Tong |
| EPI_ISL_751758 | HI Dept. of Health, State Laboratories Division | Genomics and Discovery, Respiratory Viruses Branch, Division of Viral Diseases, Centers for Disease Control and Prevention | Krista Queen, Yan Li, Ying Tao, Jing Zhang, Anna Uehara, Anna Montmayeur, Clinton R. Paden, Peter W. Cook,Rachel Marine, Mili Sheth, Haibin Wang, Justin Lee, Suxiang Tong |

| | | | |
|---|---|---|---|
| EPI_ISL_751759 | CT-Dr. Katherine A. Kelley State Public Health Lab | Genomics and Discovery, Respiratory Viruses Branch, Division of Viral Diseases, Centers for Disease Control and Prevention | Krista Queen, Yan Li, Ying Tao, Jing Zhang, Anna Uehara, Anna Montmayeur, Clinton R. Paden, Peter W. Cook,Rachel Marine, Mili Sheth, Haibin Wang, Justin Lee, Suxiang Tong |
| EPI_ISL_751760 | AR Dept. of Health-Public Health Lab | Genomics and Discovery, Respiratory Viruses Branch, Division of Viral Diseases, Centers for Disease Control and Prevention | Krista Queen, Yan Li, Ying Tao, Jing Zhang, Anna Uehara, Anna Montmayeur, Clinton R. Paden, Peter W. Cook,Rachel Marine, Mili Sheth, Haibin Wang, Justin Lee, Suxiang Tong |
| EPI_ISL_751761 | HI Dept. of Health, State Laboratories Division | Genomics and Discovery, Respiratory Viruses Branch, Division of Viral Diseases, Centers for Disease Control and Prevention | Krista Queen, Yan Li, Ying Tao, Jing Zhang, Anna Uehara, Anna Montmayeur, Clinton R. Paden, Peter W. Cook,Rachel Marine, Mili Sheth, Haibin Wang, Justin Lee, Suxiang Tong |
| EPI_ISL_751762 | CT-Dr. Katherine A. Kelley State Public Health Lab | Genomics and Discovery, Respiratory Viruses Branch, Division of Viral Diseases, Centers for Disease Control and Prevention | Krista Queen, Yan Li, Ying Tao, Jing Zhang, Anna Uehara, Anna Montmayeur, Clinton R. Paden, Peter W. Cook,Rachel Marine, Mili Sheth, Haibin Wang, Justin Lee, Suxiang Tong |
| EPI_ISL_751763 | CO Dept. of Public Health and Environment, Lab Services Division | Genomics and Discovery, Respiratory Viruses Branch, Division of Viral Diseases, Centers for Disease Control and Prevention | Krista Queen, Yan Li, Ying Tao, Jing Zhang, Anna Uehara, Anna Montmayeur, Clinton R. Paden, Peter W. Cook,Rachel Marine, Mili Sheth, Haibin Wang, Justin Lee, Suxiang Tong |
| EPI_ISL_751764, EPI_ISL_751765 | AR Dept. of Health-Public Health Lab | Genomics and Discovery, Respiratory Viruses Branch, Division of Viral Diseases, Centers for Disease Control and Prevention | Krista Queen, Yan Li, Ying Tao, Jing Zhang, Anna Uehara, Anna Montmayeur, Clinton R. Paden, Peter W. Cook,Rachel Marine, Mili Sheth, Haibin Wang, Justin Lee, Suxiang Tong |
| EPI_ISL_751766 | CO Dept. of Public Health and Environment, Lab Services Division | Genomics and Discovery, Respiratory Viruses Branch, Division of Viral Diseases, Centers for Disease Control and Prevention | Krista Queen, Yan Li, Ying Tao, Jing Zhang, Anna Uehara, Anna Montmayeur, Clinton R. Paden, Peter W. Cook,Rachel Marine, Mili Sheth, Haibin Wang, Justin Lee, Suxiang Tong |
| EPI_ISL_751767 | HI Dept. of Health, State Laboratories Division | Genomics and Discovery, Respiratory Viruses Branch, Division of Viral Diseases, Centers for Disease Control and Prevention | Krista Queen, Yan Li, Ying Tao, Jing Zhang, Anna Uehara, Anna Montmayeur, Clinton R. Paden, Peter W. Cook,Rachel Marine, Mili Sheth, Haibin Wang, Justin Lee, Suxiang Tong |
| EPI_ISL_751768 | IN State Department of Health Laboratory Services | Genomics and Discovery, Respiratory Viruses Branch, Division of Viral Diseases, Centers for Disease Control and Prevention | Krista Queen, Yan Li, Ying Tao, Jing Zhang, Anna Uehara, Anna Montmayeur, Clinton R. Paden, Peter W. Cook,Rachel Marine, Mili Sheth, Haibin Wang, Justin Lee, Suxiang Tong |
| EPI_ISL_751769 | MI - Michigan Department of Health and Human Services - Bureau of Laboratories | Genomics and Discovery, Respiratory Viruses Branch, Division of Viral Diseases, Centers for Disease Control and Prevention | Krista Queen, Yan Li, Ying Tao, Jing Zhang, Anna Uehara, Anna Montmayeur, Clinton R. Paden, Peter W. Cook,Rachel Marine, Mili Sheth, Haibin Wang, Justin Lee, Suxiang Tong |
| EPI_ISL_751770 | IN State Department of Health Laboratory Services | Genomics and Discovery, Respiratory Viruses Branch, Division of Viral Diseases, Centers for Disease Control and Prevention | Krista Queen, Yan Li, Ying Tao, Jing Zhang, Anna Uehara, Anna Montmayeur, Clinton R. Paden, Peter W. Cook,Rachel Marine, Mili Sheth, Haibin Wang, Justin Lee, Suxiang Tong |
| EPI_ISL_751771 | MD DOH Laboratories Administration | Genomics and Discovery, Respiratory Viruses Branch, Division of Viral Diseases, Centers for Disease Control and Prevention | Krista Queen, Yan Li, Ying Tao, Jing Zhang, Anna Uehara, Anna Montmayeur, Clinton R. Paden, Peter W. Cook,Rachel Marine, Mili Sheth, Haibin Wang, Justin Lee, Suxiang Tong |
| EPI_ISL_751772 | HI Dept. of Health, State Laboratories Division | Genomics and Discovery, Respiratory Viruses Branch, Division of Viral Diseases, Centers for Disease Control and Prevention | Krista Queen, Yan Li, Ying Tao, Jing Zhang, Anna Uehara, Anna Montmayeur, Clinton R. Paden, Peter W. Cook,Rachel Marine, Mili Sheth, Haibin Wang, Justin Lee, Suxiang Tong |
| EPI_ISL_751773 | NJ Public Health and Environmental Laboratories | Genomics and Discovery, Respiratory Viruses Branch, Division of Viral Diseases, Centers for Disease Control and Prevention | Krista Queen, Yan Li, Ying Tao, Jing Zhang, Anna Uehara, Anna Montmayeur, Clinton R. Paden, Peter W. Cook,Rachel Marine, Mili Sheth, Haibin Wang, Justin Lee, Suxiang Tong |
| EPI_ISL_751774 | MO State Public Health Laboratory | Genomics and Discovery, Respiratory Viruses Branch, Division of Viral Diseases, Centers for Disease Control and Prevention | Krista Queen, Yan Li, Ying Tao, Jing Zhang, Anna Uehara, Anna Montmayeur, Clinton R. Paden, Peter W. Cook,Rachel Marine, Mili Sheth, Haibin Wang, Justin Lee, Suxiang Tong |
| EPI_ISL_751775 | MT Public Health Laboratory | Genomics and Discovery, Respiratory Viruses Branch, Division of Viral Diseases, Centers for Disease Control and Prevention | Krista Queen, Yan Li, Ying Tao, Jing Zhang, Anna Uehara, Anna Montmayeur, Clinton R. Paden, Peter W. Cook,Rachel Marine, Mili Sheth, Haibin Wang, Justin Lee, Suxiang Tong |
| EPI_ISL_751776 | MS Public Health Laboratory | Genomics and Discovery, Respiratory Viruses Branch, Division of Viral Diseases, Centers for Disease Control and Prevention | Krista Queen, Yan Li, Ying Tao, Jing Zhang, Anna Uehara, Anna Montmayeur, Clinton R. Paden, Peter W. Cook,Rachel Marine, Mili Sheth, Haibin Wang, Justin Lee, Suxiang Tong |
| EPI_ISL_751777 | OH Department of Health Laboratory | Genomics and Discovery, Respiratory Viruses Branch, Division of Viral Diseases, Centers for Disease Control and Prevention | Krista Queen, Yan Li, Ying Tao, Jing Zhang, Anna Uehara, Anna Montmayeur, Clinton R. Paden, Peter W. Cook,Rachel Marine, Mili Sheth, Haibin Wang, Justin Lee, Suxiang Tong |
| EPI_ISL_751778 | NV State Public Health Laboratory | Genomics and Discovery, Respiratory Viruses Branch, Division of Viral Diseases, Centers for Disease Control and Prevention | Krista Queen, Yan Li, Ying Tao, Jing Zhang, Anna Uehara, Anna Montmayeur, Clinton R. Paden, Peter W. Cook,Rachel Marine, Mili Sheth, Haibin Wang, Justin Lee, Suxiang Tong |
| EPI_ISL_751779 | IA State Hygienic Laboratory | Genomics and Discovery, Respiratory Viruses Branch, Division of Viral Diseases, Centers for Disease Control and Prevention | Krista Queen, Yan Li, Ying Tao, Jing Zhang, Anna Uehara, Anna Montmayeur, Clinton R. Paden, Peter W. Cook,Rachel Marine, Mili Sheth, Haibin Wang, Justin Lee, Suxiang Tong |
| EPI_ISL_751780 | ID Bureau of Laboratories | Genomics and Discovery, Respiratory Viruses Branch, Division of Viral Diseases, Centers for Disease Control and Prevention | Krista Queen, Yan Li, Ying Tao, Jing Zhang, Anna Uehara, Anna Montmayeur, Clinton R. Paden, Peter W. Cook,Rachel Marine, Mili Sheth, Haibin Wang, Justin Lee, Suxiang Tong |
| EPI_ISL_751781 | WY Public Health Laboratory | Genomics and Discovery, Respiratory Viruses Branch, Division of Viral Diseases, Centers for Disease Control and Prevention | Krista Queen, Yan Li, Ying Tao, Jing Zhang, Anna Uehara, Anna Montmayeur, Clinton R. Paden, Peter W. Cook,Rachel Marine, Mili Sheth, Haibin Wang, Justin Lee, Suxiang Tong |
| EPI_ISL_751782 | ID Bureau of Laboratories | Genomics and Discovery, Respiratory Viruses Branch, Division of Viral Diseases, Centers for Disease Control and Prevention | Krista Queen, Yan Li, Ying Tao, Jing Zhang, Anna Uehara, Anna Montmayeur, Clinton R. Paden, Peter W. Cook,Rachel Marine, Mili Sheth, Haibin Wang, Justin Lee, Suxiang Tong |
| EPI_ISL_751783 | IA State Hygienic Laboratory | Genomics and Discovery, Respiratory Viruses Branch, Division of Viral Diseases, Centers for Disease Control and Prevention | Krista Queen, Yan Li, Ying Tao, Jing Zhang, Anna Uehara, Anna Montmayeur, Clinton R. Paden, Peter W. Cook,Rachel Marine, Mili Sheth, Haibin Wang, Justin Lee, Suxiang Tong |
| EPI_ISL_751784 | VA-Division of Consolidated Laboratory Services | Genomics and Discovery, Respiratory Viruses Branch, Division of Viral Diseases, Centers for Disease Control and Prevention | Krista Queen, Yan Li, Ying Tao, Jing Zhang, Anna Uehara, Anna Montmayeur, Clinton R. Paden, Peter W. Cook,Rachel Marine, Mili Sheth, Haibin Wang, Justin Lee, Suxiang Tong |
| EPI_ISL_751785, EPI_ISL_751786 | WVDHHR - Office of Laboratory Services | Genomics and Discovery, Respiratory Viruses Branch, Division of Viral Diseases, Centers for Disease Control and Prevention | Krista Queen, Yan Li, Ying Tao, Jing Zhang, Anna Uehara, Anna Montmayeur, Clinton R. Paden, Peter W. Cook,Rachel Marine, Mili Sheth, Haibin Wang, Justin Lee, Suxiang Tong |
| EPI_ISL_751787 | TX DSHS, Lab Services Section MC 1947 | Genomics and Discovery, Respiratory Viruses Branch, Division of Viral Diseases, Centers for Disease Control and Prevention | Krista Queen, Yan Li, Ying Tao, Jing Zhang, Anna Uehara, Anna Montmayeur, Clinton R. Paden, Peter W. Cook,Rachel Marine, Mili Sheth, Haibin Wang, Justin Lee, Suxiang Tong |
| EPI_ISL_751788 | OH Department of Health Laboratory | Genomics and Discovery, Respiratory Viruses Branch, Division of Viral Diseases, Centers for Disease Control and Prevention | Krista Queen, Yan Li, Ying Tao, Jing Zhang, Anna Uehara, Anna Montmayeur, Clinton R. Paden, Peter W. Cook,Rachel Marine, Mili Sheth, Haibin Wang, Justin Lee, Suxiang Tong |
| EPI_ISL_751789 | ID Bureau of Laboratories | Genomics and Discovery, Respiratory Viruses Branch, Division of Viral Diseases, Centers for Disease Control and Prevention | Krista Queen, Yan Li, Ying Tao, Jing Zhang, Anna Uehara, Anna Montmayeur, Clinton R. Paden, Peter W. Cook,Rachel Marine, Mili Sheth, Haibin Wang, Justin Lee, Suxiang Tong |
| EPI_ISL_751790 | RI State Health Laboratories | Genomics and Discovery, Respiratory Viruses Branch, Division of Viral Diseases, Centers for Disease Control and Prevention | Krista Queen, Yan Li, Ying Tao, Jing Zhang, Anna Uehara, Anna Montmayeur, Clinton R. Paden, Peter W. Cook,Rachel Marine, Mili Sheth, Haibin Wang, Justin Lee, Suxiang Tong |
| EPI_ISL_751791, EPI_ISL_751792 | AL Dept. of Public Health Bureau of Clinical Laboratories | Genomics and Discovery, Respiratory Viruses Branch, Division of Viral Diseases, Centers for Disease Control and Prevention | Krista Queen, Yan Li, Ying Tao, Jing Zhang, Anna Uehara, Anna Montmayeur, Clinton R. Paden, Peter W. Cook,Rachel Marine, Mili Sheth, Haibin Wang, Justin Lee, Suxiang Tong |
| EPI_ISL_751793 | AK State Public Health Lab, State Health Department | Genomics and Discovery, Respiratory Viruses Branch, Division of Viral Diseases, | Krista Queen, Yan Li, Ying Tao, Jing Zhang, Anna Uehara, Anna Montmayeur, Clinton R. Paden, Peter W. Cook,Rachel Marine, Mili Sheth, Haibin Wang, Justin Lee, Suxiang Tong |

| | | | |
|---|---|---|---|
| | | Centers for Disease Control and Prevention | |
| EPI_ISL_751794 | KY State Public Health Lab | Genomics and Discovery, Respiratory Viruses Branch, Division of Viral Diseases, Centers for Disease Control and Prevention | Krista Queen, Yan Li, Ying Tao, Jing Zhang, Anna Uehara, Anna Montmayeur, Clinton R. Paden, Peter W. Cook,Rachel Marine, Mili Sheth, Haibin Wang, Justin Lee, Suxiang Tong |
| EPI_ISL_751795 | RI State Health Laboratories | Genomics and Discovery, Respiratory Viruses Branch, Division of Viral Diseases, Centers for Disease Control and Prevention | Krista Queen, Yan Li, Ying Tao, Jing Zhang, Anna Uehara, Anna Montmayeur, Clinton R. Paden, Peter W. Cook,Rachel Marine, Mili Sheth, Haibin Wang, Justin Lee, Suxiang Tong |
| EPI_ISL_751800 | Colorado Department of Public Health and Environment | Colorado Department of Public Health & Environment | Laura Bankers, Molly C. Hetherington-Rauth, Diana Ir, Shannon Ely, Shannon R. Matzinger, Sarah Elizabeth Totten, Emily A. Travanty |
| EPI_ISL_751801 | UCSD EXCITE | Andersen lab at Scripps Research | SEARCH Alliance San Diego |
| EPI_ISL_751802, EPI_ISL_751803, EPI_ISL_751804, EPI_ISL_751805, EPI_ISL_751806, EPI_ISL_751807, EPI_ISL_751808, EPI_ISL_751809, EPI_ISL_751810, EPI_ISL_751811, EPI_ISL_751812, EPI_ISL_751813, EPI_ISL_751814, EPI_ISL_751815, EPI_ISL_751816, EPI_ISL_751817, EPI_ISL_751818, EPI_ISL_751819, EPI_ISL_751820, EPI_ISL_751821, EPI_ISL_751822, EPI_ISL_751823, EPI_ISL_751824, EPI_ISL_751825, EPI_ISL_751826, EPI_ISL_751827, EPI_ISL_751828, EPI_ISL_751829, EPI_ISL_751830, EPI_ISL_751831, EPI_ISL_751832, EPI_ISL_751833, EPI_ISL_751834, EPI_ISL_751835, EPI_ISL_751836, EPI_ISL_751837, EPI_ISL_751838, EPI_ISL_751839, EPI_ISL_751840, EPI_ISL_751841, EPI_ISL_751842, EPI_ISL_751843, EPI_ISL_751844, EPI_ISL_751845, EPI_ISL_751846 | | | |
| see above | New Mexico Department of Health Scientific Laboratory | New Mexico Department of Health Scientific Laboratory | D'eldra Malone, Ellie Johnson, Anastacia Griego-Fisher |
| EPI_ISL_752606 | Yale Pathology Lab | Grubaugh Lab - Yale School of Public Health | Tara Alpert, Joseph Fauver, Anderson Brito, Chen Liu, Pei Hui, Jianhui Wang, Kien Pham, Nathan Grubaugh |
| EPI_ISL_752607, EPI_ISL_752608, EPI_ISL_752609, EPI_ISL_752610, EPI_ISL_752611, EPI_ISL_752612, EPI_ISL_752613, EPI_ISL_752614, EPI_ISL_752615, EPI_ISL_752616, EPI_ISL_752617, EPI_ISL_752618, EPI_ISL_752619, EPI_ISL_752620, EPI_ISL_752621, EPI_ISL_752622, EPI_ISL_752623, EPI_ISL_752624, EPI_ISL_752625, EPI_ISL_752626, EPI_ISL_752627, EPI_ISL_752628, EPI_ISL_752629, EPI_ISL_752630, EPI_ISL_752631, EPI_ISL_752632, EPI_ISL_752633, EPI_ISL_752634, EPI_ISL_752635, EPI_ISL_752636, EPI_ISL_752637, EPI_ISL_752638, EPI_ISL_752639, EPI_ISL_752640, EPI_ISL_752641, EPI_ISL_752642, EPI_ISL_752643, EPI_ISL_752644, EPI_ISL_752645, EPI_ISL_752646, EPI_ISL_752647, EPI_ISL_752648, EPI_ISL_752649, EPI_ISL_752650, EPI_ISL_752651, EPI_ISL_752652, EPI_ISL_752653, EPI_ISL_752654, EPI_ISL_752655, EPI_ISL_752656, EPI_ISL_752657, EPI_ISL_752658, EPI_ISL_752659, EPI_ISL_752660, EPI_ISL_752661, EPI_ISL_752662, EPI_ISL_752663, EPI_ISL_752664, EPI_ISL_752665, EPI_ISL_752666, EPI_ISL_752667, EPI_ISL_752668, EPI_ISL_752669, EPI_ISL_752670, EPI_ISL_752671, EPI_ISL_752672, EPI_ISL_752673, EPI_ISL_752674, EPI_ISL_752675, EPI_ISL_752676, EPI_ISL_752677, EPI_ISL_752678, EPI_ISL_752679, EPI_ISL_752680, EPI_ISL_752681, EPI_ISL_752682, EPI_ISL_752683, EPI_ISL_752684, EPI_ISL_752685, EPI_ISL_752686, EPI_ISL_752687, EPI_ISL_752688, EPI_ISL_752689, EPI_ISL_752690, EPI_ISL_752691, EPI_ISL_752692, EPI_ISL_752693, EPI_ISL_752694, EPI_ISL_752695, EPI_ISL_752696, EPI_ISL_752697, EPI_ISL_752698, EPI_ISL_752699, EPI_ISL_752700, EPI_ISL_752701, EPI_ISL_752702, EPI_ISL_752703, EPI_ISL_752704, EPI_ISL_752705, EPI_ISL_752706, EPI_ISL_752707, EPI_ISL_752708, EPI_ISL_752709, EPI_ISL_752710, EPI_ISL_752711, EPI_ISL_752712, EPI_ISL_752713, EPI_ISL_752714, EPI_ISL_752715, EPI_ISL_752716, EPI_ISL_752717, EPI_ISL_752718, EPI_ISL_752719, EPI_ISL_752720, EPI_ISL_752721, EPI_ISL_752722, EPI_ISL_752723, EPI_ISL_752724, EPI_ISL_752725, EPI_ISL_752726, EPI_ISL_752727, EPI_ISL_752728, EPI_ISL_752729, EPI_ISL_752730, EPI_ISL_752731, EPI_ISL_752732, EPI_ISL_752733, EPI_ISL_752734, EPI_ISL_752735, EPI_ISL_752736, EPI_ISL_752737, EPI_ISL_752738, EPI_ISL_752739, EPI_ISL_752740, EPI_ISL_752741, EPI_ISL_752742, EPI_ISL_752743, EPI_ISL_752744, EPI_ISL_752745, EPI_ISL_752746, EPI_ISL_752747, EPI_ISL_752748, EPI_ISL_752749, EPI_ISL_752750, EPI_ISL_752751, EPI_ISL_752752, EPI_ISL_752753, EPI_ISL_752754, EPI_ISL_752755, EPI_ISL_752756, EPI_ISL_752757, EPI_ISL_752758, EPI_ISL_752759, EPI_ISL_752760, EPI_ISL_752761, EPI_ISL_752762, EPI_ISL_752763, EPI_ISL_752764, EPI_ISL_752765, EPI_ISL_752766, EPI_ISL_752767, EPI_ISL_752768, EPI_ISL_752769, EPI_ISL_752770, EPI_ISL_752771, EPI_ISL_752772, EPI_ISL_752773, EPI_ISL_752774, EPI_ISL_752775, EPI_ISL_752776, EPI_ISL_752777, EPI_ISL_752778, EPI_ISL_752779, EPI_ISL_752780, EPI_ISL_752781, EPI_ISL_752782, EPI_ISL_752783, EPI_ISL_752784, EPI_ISL_752785, EPI_ISL_752786, EPI_ISL_752787, EPI_ISL_752788, EPI_ISL_752789, EPI_ISL_752790, EPI_ISL_752791, EPI_ISL_752792, EPI_ISL_752793, EPI_ISL_752794, EPI_ISL_752795, EPI_ISL_752796, EPI_ISL_752797, EPI_ISL_752798, EPI_ISL_752799, EPI_ISL_752800, EPI_ISL_752801, EPI_ISL_752802, EPI_ISL_752803, EPI_ISL_752804, EPI_ISL_752805, EPI_ISL_752806, EPI_ISL_752807, EPI_ISL_752808, EPI_ISL_752809, EPI_ISL_752810, EPI_ISL_752811, EPI_ISL_752812, EPI_ISL_752813, EPI_ISL_752814, EPI_ISL_752815, EPI_ISL_752816, EPI_ISL_752817, EPI_ISL_752818, EPI_ISL_752819, EPI_ISL_752820, EPI_ISL_752821, EPI_ISL_752822, EPI_ISL_752823, EPI_ISL_752824, EPI_ISL_752825, EPI_ISL_752826, EPI_ISL_752827, EPI_ISL_752828, EPI_ISL_752829, EPI_ISL_752830, EPI_ISL_752831, EPI_ISL_752832, EPI_ISL_752833, EPI_ISL_752834, EPI_ISL_752835, EPI_ISL_752836, EPI_ISL_752837, EPI_ISL_752838, EPI_ISL_752839, EPI_ISL_752840, EPI_ISL_752841, EPI_ISL_752842, EPI_ISL_752843, EPI_ISL_752844, EPI_ISL_752845, EPI_ISL_752846, EPI_ISL_752847, EPI_ISL_752848, EPI_ISL_752849, EPI_ISL_752850, EPI_ISL_752851, EPI_ISL_752852, EPI_ISL_752853, EPI_ISL_752854, EPI_ISL_752855, EPI_ISL_752856, EPI_ISL_752857, EPI_ISL_752858, EPI_ISL_752859, EPI_ISL_752860, EPI_ISL_752861, EPI_ISL_752862, EPI_ISL_752863, EPI_ISL_752864, EPI_ISL_752865, EPI_ISL_752866, EPI_ISL_752867, EPI_ISL_752868, EPI_ISL_752869, EPI_ISL_752870, EPI_ISL_752871, EPI_ISL_752872, EPI_ISL_752873, EPI_ISL_752874, EPI_ISL_752875, EPI_ISL_752876, EPI_ISL_752877, EPI_ISL_752878, EPI_ISL_752879, EPI_ISL_752880, EPI_ISL_752881, EPI_ISL_752882, EPI_ISL_752883, EPI_ISL_752884, EPI_ISL_752885, EPI_ISL_752886, EPI_ISL_752887, EPI_ISL_752888, EPI_ISL_752889, EPI_ISL_752890, EPI_ISL_752891, EPI_ISL_752892, EPI_ISL_752893, EPI_ISL_752894, EPI_ISL_752895, EPI_ISL_752896, EPI_ISL_752897, EPI_ISL_752898, EPI_ISL_752899, EPI_ISL_752900, EPI_ISL_752901, EPI_ISL_752902, EPI_ISL_752903, EPI_ISL_752904, EPI_ISL_752905, EPI_ISL_752906, EPI_ISL_752907, EPI_ISL_752908, EPI_ISL_752909, EPI_ISL_752910, EPI_ISL_752911, EPI_ISL_752912, EPI_ISL_752913, EPI_ISL_752914, EPI_ISL_752915, EPI_ISL_752916, EPI_ISL_752917, EPI_ISL_752918, EPI_ISL_752919, EPI_ISL_752920, EPI_ISL_752921, EPI_ISL_752922, EPI_ISL_752923, EPI_ISL_752924, EPI_ISL_752925, EPI_ISL_752926, EPI_ISL_752927, EPI_ISL_752928, EPI_ISL_752929, EPI_ISL_752930, EPI_ISL_752931, EPI_ISL_752932, EPI_ISL_752933, EPI_ISL_752934, EPI_ISL_752935, EPI_ISL_752936, EPI_ISL_752937, EPI_ISL_752938, EPI_ISL_752939, EPI_ISL_752940, EPI_ISL_752941, EPI_ISL_752942, EPI_ISL_752944, EPI_ISL_752945, EPI_ISL_752946, EPI_ISL_752947, EPI_ISL_752948, EPI_ISL_752949, EPI_ISL_752950, EPI_ISL_752951, EPI_ISL_752952, EPI_ISL_752953, EPI_ISL_752954, EPI_ISL_752955, EPI_ISL_752956, EPI_ISL_752957, EPI_ISL_752958, EPI_ISL_752959, EPI_ISL_752960, EPI_ISL_752961, EPI_ISL_752962, EPI_ISL_752963, EPI_ISL_752964, EPI_ISL_752965, EPI_ISL_752966, EPI_ISL_752967, EPI_ISL_752968, EPI_ISL_752969, EPI_ISL_752970, EPI_ISL_752971, EPI_ISL_752972, EPI_ISL_752973, EPI_ISL_752974, EPI_ISL_752975, EPI_ISL_752976, EPI_ISL_752977, EPI_ISL_752978, EPI_ISL_752979, EPI_ISL_752980, EPI_ISL_752981, EPI_ISL_752982, EPI_ISL_752983, EPI_ISL_752984, EPI_ISL_752985, EPI_ISL_752986, EPI_ISL_752987, EPI_ISL_752988, EPI_ISL_752989, EPI_ISL_752990, EPI_ISL_752991, EPI_ISL_752992, EPI_ISL_752993, EPI_ISL_752994, EPI_ISL_752995, EPI_ISL_752996, EPI_ISL_752997, EPI_ISL_752998, EPI_ISL_752999, EPI_ISL_753000, EPI_ISL_753001, EPI_ISL_753002, EPI_ISL_753003, EPI_ISL_753004, EPI_ISL_753005, EPI_ISL_753006, EPI_ISL_753007, EPI_ISL_753008, EPI_ISL_753009, EPI_ISL_753010, EPI_ISL_753011, EPI_ISL_753012, EPI_ISL_753013, EPI_ISL_753014, EPI_ISL_753015, EPI_ISL_753016, EPI_ISL_753017, EPI_ISL_753018, EPI_ISL_753019, EPI_ISL_753020, EPI_ISL_753021, EPI_ISL_753022, EPI_ISL_753023, EPI_ISL_753024, EPI_ISL_753026, EPI_ISL_753027, EPI_ISL_753028, EPI_ISL_753029, EPI_ISL_753030, EPI_ISL_753031, EPI_ISL_753032, EPI_ISL_753033, EPI_ISL_753034, EPI_ISL_753035, EPI_ISL_753036, EPI_ISL_753037, EPI_ISL_753038, EPI_ISL_753039, EPI_ISL_753040, EPI_ISL_753041, EPI_ISL_753042, EPI_ISL_753043, EPI_ISL_753044, EPI_ISL_753045, EPI_ISL_753046, EPI_ISL_753047, EPI_ISL_753048, EPI_ISL_753049, EPI_ISL_753050, EPI_ISL_753051, EPI_ISL_753052, EPI_ISL_753053, EPI_ISL_753054, EPI_ISL_753055, EPI_ISL_753056, EPI_ISL_753057, EPI_ISL_753058, EPI_ISL_753059, EPI_ISL_753060, EPI_ISL_753061, EPI_ISL_753062, EPI_ISL_753063, EPI_ISL_753064, EPI_ISL_753065, EPI_ISL_753066, EPI_ISL_753067, EPI_ISL_753068, EPI_ISL_753069, EPI_ISL_753070, EPI_ISL_753071, EPI_ISL_753072, EPI_ISL_753073, EPI_ISL_753074, EPI_ISL_753075, EPI_ISL_753076, EPI_ISL_753077, EPI_ISL_753078, EPI_ISL_753079, EPI_ISL_753080, EPI_ISL_753081, EPI_ISL_753082, EPI_ISL_753083, EPI_ISL_753084, EPI_ISL_753085, EPI_ISL_753086, EPI_ISL_753087, EPI_ISL_753088, EPI_ISL_753089, EPI_ISL_753090, EPI_ISL_753091, EPI_ISL_753092, EPI_ISL_753093, EPI_ISL_753094, EPI_ISL_753095, EPI_ISL_753096, EPI_ISL_753097, EPI_ISL_753098, EPI_ISL_753099, EPI_ISL_753100, EPI_ISL_753101, EPI_ISL_753102, EPI_ISL_753103, EPI_ISL_753104, EPI_ISL_753105, EPI_ISL_753106, EPI_ISL_753107, EPI_ISL_753108, EPI_ISL_753109, EPI_ISL_753110, EPI_ISL_753111, EPI_ISL_753112, EPI_ISL_753113, EPI_ISL_753114, EPI_ISL_753115, EPI_ISL_753116, EPI_ISL_753117, EPI_ISL_753118, EPI_ISL_753119, EPI_ISL_753120, EPI_ISL_753121, EPI_ISL_753122, EPI_ISL_753123, EPI_ISL_753124, EPI_ISL_753125, EPI_ISL_753126, EPI_ISL_753127, EPI_ISL_753128, EPI_ISL_753129, EPI_ISL_753130, EPI_ISL_753131, EPI_ISL_753132, EPI_ISL_753133, EPI_ISL_753134, EPI_ISL_753135, EPI_ISL_753136, EPI_ISL_753137, EPI_ISL_753138, EPI_ISL_753139, EPI_ISL_753140, EPI_ISL_753141, EPI_ISL_753142, EPI_ISL_753143, EPI_ISL_753144, EPI_ISL_753145, EPI_ISL_753146, EPI_ISL_753147, EPI_ISL_753148, EPI_ISL_753149, EPI_ISL_753150, EPI_ISL_753151, EPI_ISL_753152, EPI_ISL_753153, EPI_ISL_753154, EPI_ISL_753155, EPI_ISL_753156, EPI_ISL_753157, EPI_ISL_753158, EPI_ISL_753159, EPI_ISL_753160, EPI_ISL_753161, EPI_ISL_753162, EPI_ISL_753163, EPI_ISL_753164, EPI_ISL_753165, EPI_ISL_753166, EPI_ISL_753167, EPI_ISL_753168, EPI_ISL_753169, EPI_ISL_753170, EPI_ISL_753171, EPI_ISL_753172, EPI_ISL_753173, EPI_ISL_753174, EPI_ISL_753175, EPI_ISL_753176, EPI_ISL_753177, EPI_ISL_753178, EPI_ISL_753179, EPI_ISL_753180, EPI_ISL_753181, EPI_ISL_753182, EPI_ISL_753183, EPI_ISL_753184, EPI_ISL_753185, EPI_ISL_753186, EPI_ISL_753187, EPI_ISL_753188, EPI_ISL_753189, EPI_ISL_753190, EPI_ISL_753191, EPI_ISL_753192, EPI_ISL_753193, EPI_ISL_753194, EPI_ISL_753195, EPI_ISL_753196, EPI_ISL_753197, EPI_ISL_753198, EPI_ISL_753199, EPI_ISL_753200, EPI_ISL_753201, EPI_ISL_753202, EPI_ISL_753203, EPI_ISL_753204, EPI_ISL_753205, EPI_ISL_753206, EPI_ISL_753207, EPI_ISL_753208, EPI_ISL_753209, EPI_ISL_753210, EPI_ISL_753211, EPI_ISL_753212, EPI_ISL_753213, EPI_ISL_753214, EPI_ISL_753215, EPI_ISL_753216, EPI_ISL_753217, EPI_ISL_753218, EPI_ISL_753219, EPI_ISL_753220, EPI_ISL_753221, EPI_ISL_753222, EPI_ISL_753223, EPI_ISL_753224, EPI_ISL_753225, EPI_ISL_753226, EPI_ISL_753227, EPI_ISL_753228, EPI_ISL_753229, EPI_ISL_753230, EPI_ISL_753231, EPI_ISL_753232, EPI_ISL_753233, EPI_ISL_753234, EPI_ISL_753235, EPI_ISL_753236, EPI_ISL_753237, EPI_ISL_753238, EPI_ISL_753239, EPI_ISL_753240, EPI_ISL_753241, EPI_ISL_753242, EPI_ISL_753243, EPI_ISL_753244, EPI_ISL_753245, EPI_ISL_753246, EPI_ISL_753247, EPI_ISL_753248, EPI_ISL_753249, EPI_ISL_753250, EPI_ISL_753251, EPI_ISL_753252, EPI_ISL_753253, EPI_ISL_753254, EPI_ISL_753255, EPI_ISL_753256, EPI_ISL_753257, EPI_ISL_753258, EPI_ISL_753259, EPI_ISL_753260, EPI_ISL_753261 | | | |
| see above | State Laboratories Division, Hawaii State Department of Health | State Laboratories Division, Hawaii State Department of Health | Pamela O'Brien, Sabrina Diemert, Drew Kuwazaki, Razvan Sultana, Edward Desmond |
| EPI_ISL_753262, EPI_ISL_753263, EPI_ISL_753264, EPI_ISL_753265, EPI_ISL_753266, EPI_ISL_753267, EPI_ISL_753268, EPI_ISL_753269, EPI_ISL_753270, EPI_ISL_753271, EPI_ISL_753272, EPI_ISL_753273, EPI_ISL_753274, EPI_ISL_753275, EPI_ISL_753276, EPI_ISL_753277, EPI_ISL_753278, EPI_ISL_753279, EPI_ISL_753280, EPI_ISL_753281, EPI_ISL_753282, EPI_ISL_753283, EPI_ISL_753284, EPI_ISL_753285, EPI_ISL_753286, EPI_ISL_753287, EPI_ISL_753288, EPI_ISL_753289, EPI_ISL_753290, EPI_ISL_753291, EPI_ISL_753292, EPI_ISL_753293, EPI_ISL_753294, EPI_ISL_753295, EPI_ISL_753296, EPI_ISL_753297, EPI_ISL_753298, EPI_ISL_753299, EPI_ISL_753300, EPI_ISL_753301, EPI_ISL_753302, EPI_ISL_753303, EPI_ISL_753304, EPI_ISL_753305, EPI_ISL_753306, EPI_ISL_753307, EPI_ISL_753308, EPI_ISL_753309, EPI_ISL_753310, EPI_ISL_753311, EPI_ISL_753312, EPI_ISL_753313, EPI_ISL_753314, EPI_ISL_753315, EPI_ISL_753316, EPI_ISL_753317, EPI_ISL_753318, EPI_ISL_753319, EPI_ISL_753320, EPI_ISL_753321, EPI_ISL_753322, EPI_ISL_753323, EPI_ISL_753324, EPI_ISL_753325, EPI_ISL_753326, EPI_ISL_753327, EPI_ISL_753328, EPI_ISL_753329, EPI_ISL_753330, EPI_ISL_753331, EPI_ISL_753332, EPI_ISL_753333, EPI_ISL_753334, EPI_ISL_753335, EPI_ISL_753336, EPI_ISL_753337, EPI_ISL_753338, EPI_ISL_753339, EPI_ISL_753340, EPI_ISL_753341, EPI_ISL_753342, EPI_ISL_753343, EPI_ISL_753344, EPI_ISL_753345, EPI_ISL_753346, EPI_ISL_753347, EPI_ISL_753348, EPI_ISL_753349, EPI_ISL_753350, EPI_ISL_753351, EPI_ISL_753352, EPI_ISL_753353, EPI_ISL_753354, EPI_ISL_753355, EPI_ISL_753356, EPI_ISL_753357, EPI_ISL_753358, EPI_ISL_753359, EPI_ISL_753360, EPI_ISL_753361, EPI_ISL_753362, EPI_ISL_753363, EPI_ISL_753364, EPI_ISL_753365, EPI_ISL_753366, EPI_ISL_753367, EPI_ISL_753368, EPI_ISL_753369, EPI_ISL_753370, EPI_ISL_753371, EPI_ISL_753372, EPI_ISL_753373, EPI_ISL_753374, EPI_ISL_753375, EPI_ISL_753376, EPI_ISL_753377, EPI_ISL_753378, EPI_ISL_753379, EPI_ISL_753380, EPI_ISL_753381, EPI_ISL_753382, EPI_ISL_753383, EPI_ISL_753384, EPI_ISL_753385, EPI_ISL_753386, EPI_ISL_753387, EPI_ISL_753388, EPI_ISL_753389, EPI_ISL_753390, EPI_ISL_753391, EPI_ISL_753392, EPI_ISL_753393, EPI_ISL_753394, EPI_ISL_753395, EPI_ISL_753396, EPI_ISL_753397, EPI_ISL_753398, EPI_ISL_753399, EPI_ISL_753400, EPI_ISL_753401, EPI_ISL_753402, EPI_ISL_753403, EPI_ISL_753404, EPI_ISL_753405, EPI_ISL_753406, EPI_ISL_753407, EPI_ISL_753408, EPI_ISL_753409, EPI_ISL_753410, EPI_ISL_753411, EPI_ISL_753412, EPI_ISL_753413, EPI_ISL_753414, EPI_ISL_753415, EPI_ISL_753416, EPI_ISL_753417, EPI_ISL_753418, EPI_ISL_753419, EPI_ISL_753420, EPI_ISL_753421, EPI_ISL_753422, EPI_ISL_753423, EPI_ISL_753424, EPI_ISL_753425, EPI_ISL_753426, EPI_ISL_753427, EPI_ISL_753428, EPI_ISL_753429, EPI_ISL_753430, EPI_ISL_753431, EPI_ISL_753432, EPI_ISL_753433, EPI_ISL_753434, EPI_ISL_753435, EPI_ISL_753436, EPI_ISL_753437, EPI_ISL_753438, EPI_ISL_753439, EPI_ISL_753440, EPI_ISL_753441, EPI_ISL_753442, EPI_ISL_753443, EPI_ISL_753444, EPI_ISL_753445, EPI_ISL_753446, EPI_ISL_753447, EPI_ISL_753448, EPI_ISL_753449, EPI_ISL_753450, EPI_ISL_753451, EPI_ISL_753452, EPI_ISL_753453, EPI_ISL_753454, EPI_ISL_753455, EPI_ISL_753456, EPI_ISL_753457, EPI_ISL_753458, EPI_ISL_753459, EPI_ISL_753460, EPI_ISL_753461, EPI_ISL_753462, EPI_ISL_753463, EPI_ISL_753464, EPI_ISL_753465, EPI_ISL_753466, EPI_ISL_753467, EPI_ISL_753468, EPI_ISL_753469, EPI_ISL_753470, EPI_ISL_753471, EPI_ISL_753472, EPI_ISL_753473, EPI_ISL_753474, EPI_ISL_753475, EPI_ISL_753476, EPI_ISL_753477, EPI_ISL_753478, EPI_ISL_753479, EPI_ISL_753480, EPI_ISL_753481, EPI_ISL_753482, EPI_ISL_753483, EPI_ISL_753484, EPI_ISL_753485, EPI_ISL_753486, EPI_ISL_753487, EPI_ISL_753488, EPI_ISL_753489, EPI_ISL_753490, EPI_ISL_753491, EPI_ISL_753492, EPI_ISL_753493, EPI_ISL_753494, EPI_ISL_753495, EPI_ISL_753496, EPI_ISL_753497, EPI_ISL_753498, EPI_ISL_753499, EPI_ISL_753500, EPI_ISL_753501, EPI_ISL_753502, EPI_ISL_753503, EPI_ISL_753504, EPI_ISL_753505, EPI_ISL_753506, EPI_ISL_753507, EPI_ISL_753508, EPI_ISL_753509, EPI_ISL_753510, EPI_ISL_753511, EPI_ISL_753512, EPI_ISL_753513, EPI_ISL_753514, EPI_ISL_753515, EPI_ISL_753516, EPI_ISL_753517, EPI_ISL_753518, EPI_ISL_753519, EPI_ISL_753520, EPI_ISL_753521, EPI_ISL_753522, EPI_ISL_753523, EPI_ISL_753524, EPI_ISL_753525, EPI_ISL_753526, EPI_ISL_753527, EPI_ISL_753528, EPI_ISL_753529, EPI_ISL_753530, EPI_ISL_753531, EPI_ISL_753532, EPI_ISL_753533, EPI_ISL_753534, EPI_ISL_753535, EPI_ISL_753536, EPI_ISL_753537, EPI_ISL_753538, EPI_ISL_753539, EPI_ISL_753540, EPI_ISL_753541, EPI_ISL_753542, EPI_ISL_753543, EPI_ISL_753544, EPI_ISL_753545, EPI_ISL_753546, EPI_ISL_753547, EPI_ISL_753548, EPI_ISL_753549, EPI_ISL_753550, EPI_ISL_753551, EPI_ISL_753552, EPI_ISL_753553, EPI_ISL_753554, EPI_ISL_753555, EPI_ISL_753556, EPI_ISL_753557, EPI_ISL_753558, EPI_ISL_753559, EPI_ISL_753560, EPI_ISL_753561, EPI_ISL_753562, EPI_ISL_753563, EPI_ISL_753564, EPI_ISL_753565, EPI_ISL_753566, EPI_ISL_753567, EPI_ISL_753568, EPI_ISL_753569, EPI_ISL_753570, EPI_ISL_753571, EPI_ISL_753572, EPI_ISL_753573, EPI_ISL_753574, EPI_ISL_753575, EPI_ISL_753576, EPI_ISL_753577, EPI_ISL_753578, EPI_ISL_753579, EPI_ISL_753580, EPI_ISL_753581, EPI_ISL_753582, EPI_ISL_753583, EPI_ISL_753584, EPI_ISL_753585, EPI_ISL_753586, EPI_ISL_753587, EPI_ISL_753588, EPI_ISL_753589, EPI_ISL_753590, EPI_ISL_753591, EPI_ISL_753592, EPI_ISL_753593, EPI_ISL_753594, EPI_ISL_753595, EPI_ISL_753596, EPI_ISL_753597, EPI_ISL_753598, EPI_ISL_753599, EPI_ISL_753600, EPI_ISL_753601, EPI_ISL_753602, EPI_ISL_753603, EPI_ISL_753604, EPI_ISL_753605, EPI_ISL_753606, EPI_ISL_753607, EPI_ISL_753608, EPI_ISL_753609, EPI_ISL_753610, EPI_ISL_753611, EPI_ISL_753612, EPI_ISL_753613, EPI_ISL_753614, EPI_ISL_753615, EPI_ISL_753616, EPI_ISL_753617, EPI_ISL_753618, EPI_ISL_753619, EPI_ISL_753620, EPI_ISL_753621, EPI_ISL_753622, EPI_ISL_753623, EPI_ISL_753624, EPI_ISL_753625, EPI_ISL_753626, EPI_ISL_753627, EPI_ISL_753628, EPI_ISL_753629, EPI_ISL_753630, EPI_ISL_753631, EPI_ISL_753632, EPI_ISL_753633, EPI_ISL_753634, EPI_ISL_753635, EPI_ISL_753636, EPI_ISL_753637, EPI_ISL_753638, EPI_ISL_753639, EPI_ISL_753640, EPI_ISL_753641, EPI_ISL_753642, EPI_ISL_753643, EPI_ISL_753644, EPI_ISL_753645, EPI_ISL_753646, EPI_ISL_753647, EPI_ISL_753648, EPI_ISL_753649, EPI_ISL_753650, EPI_ISL_753651, EPI_ISL_753652, EPI_ISL_753653, EPI_ISL_753654, EPI_ISL_753655, EPI_ISL_753656, EPI_ISL_753657, EPI_ISL_753658, EPI_ISL_753659, EPI_ISL_753660, EPI_ISL_753661, EPI_ISL_753662, EPI_ISL_753663, EPI_ISL_753664, EPI_ISL_753665, EPI_ISL_753666, EPI_ISL_753667, EPI_ISL_753668, EPI_ISL_753669, EPI_ISL_753670, EPI_ISL_753671, EPI_ISL_753672, EPI_ISL_753673, EPI_ISL_753674, EPI_ISL_753675, EPI_ISL_753676, EPI_ISL_753677, EPI_ISL_753678, EPI_ISL_753679, EPI_ISL_753680, EPI_ISL_753681, EPI_ISL_753682, EPI_ISL_753683, EPI_ISL_753684, EPI_ISL_753685, EPI_ISL_753686, EPI_ISL_753687, EPI_ISL_753688, EPI_ISL_753689, EPI_ISL_753690, EPI_ISL_753691, EPI_ISL_753692 | | | |
| see above | Clinical virology Laboratory, Children's Hospital Los Angeles | Center for Personalized Medicine, Children's Hospital Los Angeles | Gai et al |
| EPI_ISL_754110, EPI_ISL_754111, EPI_ISL_754112, EPI_ISL_754113, EPI_ISL_754114, EPI_ISL_754115, EPI_ISL_754116, EPI_ISL_754117, EPI_ISL_754118, EPI_ISL_754119, EPI_ISL_754120, EPI_ISL_754121, EPI_ISL_754122, EPI_ISL_754123 | | | |
| see above | UCLA Clinical Micro Lab | Los Angeles County Public Health Laboratories | P. Hemarajata et al. |
| EPI_ISL_754124, EPI_ISL_754125, EPI_ISL_754126, EPI_ISL_754127, EPI_ISL_754128, EPI_ISL_754129 | USC Clinical Lab | Los Angeles County Public Health Laboratories | P. Hemarajata et al. |
| EPI_ISL_754130, EPI_ISL_754131, EPI_ISL_754132 | Los Angeles County Public Health Laboratories | Los Angeles County Public Health Laboratories | P. Hemarajata et al. |
| EPI_ISL_754133, EPI_ISL_754134, EPI_ISL_754135 | USC Clinical Lab | Los Angeles County Public Health Laboratories | P. Hemarajata et al. |
| EPI_ISL_754182 | Wyoming Public Health Laboratory | Wyoming Public Health Laboratory | Noah Hull, Taylor Fearing, Channing Weber, Ashley Norberg, Bailey Bowcutt, and Wanda Manley |
| EPI_ISL_754183 | Wyoming Public Health Laboratory | Wyoming Public Health Laborator | Noah Hull, Taylor Fearing, Channing Weber, Ashley Norberg, Bailey Bowcutt, and Wanda Manley |
| EPI_ISL_754215, EPI_ISL_754216, EPI_ISL_754217, EPI_ISL_754218, EPI_ISL_754219, EPI_ISL_754220, EPI_ISL_754221, EPI_ISL_754222, EPI_ISL_754223, EPI_ISL_754224, EPI_ISL_754225, EPI_ISL_754226, EPI_ISL_754227, EPI_ISL_754228, EPI_ISL_754388, EPI_ISL_754389 | | | |
| see above | Wyoming Public Health Laboratory | Wyoming Public Health Laboratory | Noah Hull, Taylor Fearing, Channing Weber, Ashley Norberg, Bailey Bowcutt, and Wanda Manley |
| EPI_ISL_754390 | USC Clinical Lab | Los Angeles County Public Health Laboratories | P. Hemarajata et al. |
| EPI_ISL_754394, EPI_ISL_754395, EPI_ISL_754396, EPI_ISL_754397, EPI_ISL_754398, EPI_ISL_754399, EPI_ISL_754400, EPI_ISL_754401, EPI_ISL_754402, EPI_ISL_754403, EPI_ISL_754404, EPI_ISL_754405, EPI_ISL_754406, EPI_ISL_754407, EPI_ISL_754408, EPI_ISL_754409, EPI_ISL_754410, EPI_ISL_754411, EPI_ISL_754412, EPI_ISL_754413, EPI_ISL_754414, EPI_ISL_754415, EPI_ISL_754416, EPI_ISL_754417, EPI_ISL_754418, EPI_ISL_754419, EPI_ISL_754420, EPI_ISL_754421, EPI_ISL_754422, EPI_ISL_754423, EPI_ISL_754424, EPI_ISL_754425, EPI_ISL_754426, EPI_ISL_754427, EPI_ISL_754428, EPI_ISL_754429, EPI_ISL_754430, EPI_ISL_754431, EPI_ISL_754432, EPI_ISL_754433, EPI_ISL_754434, EPI_ISL_754435, EPI_ISL_754436, EPI_ISL_754437, EPI_ISL_754438, EPI_ISL_754439, EPI_ISL_754440, EPI_ISL_754441, EPI_ISL_754442, EPI_ISL_754443, EPI_ISL_754444, EPI_ISL_754445, EPI_ISL_754446, EPI_ISL_754447, EPI_ISL_754448, EPI_ISL_754449, EPI_ISL_754450, EPI_ISL_754451, EPI_ISL_754452, EPI_ISL_754453, EPI_ISL_754454, EPI_ISL_754455, EPI_ISL_754456, EPI_ISL_754457, EPI_ISL_754458, EPI_ISL_754459, EPI_ISL_754460, EPI_ISL_754461, EPI_ISL_754462, EPI_ISL_754463, EPI_ISL_754464, EPI_ISL_754465, EPI_ISL_754466, EPI_ISL_754467, EPI_ISL_754468, EPI_ISL_754469, EPI_ISL_754470, EPI_ISL_754471, EPI_ISL_754472, EPI_ISL_754473, EPI_ISL_754474, EPI_ISL_754475, EPI_ISL_754476, EPI_ISL_754477, EPI_ISL_754478, EPI_ISL_754479, EPI_ISL_754480, EPI_ISL_754481, EPI_ISL_754482, EPI_ISL_754483, EPI_ISL_754484, EPI_ISL_754485, EPI_ISL_754486, EPI_ISL_754487, EPI_ISL_754488, EPI_ISL_754489, EPI_ISL_754490, EPI_ISL_754491, EPI_ISL_754492, EPI_ISL_754493, EPI_ISL_754494, EPI_ISL_754495, EPI_ISL_754496, EPI_ISL_754497, EPI_ISL_754498, EPI_ISL_754499, EPI_ISL_754500, EPI_ISL_754501, EPI_ISL_754502, EPI_ISL_754503, EPI_ISL_754504, EPI_ISL_754505, EPI_ISL_754506, EPI_ISL_754507, EPI_ISL_754508, EPI_ISL_754509, EPI_ISL_754510, EPI_ISL_754511, EPI_ISL_754512, EPI_ISL_754513, EPI_ISL_754514, EPI_ISL_754515, EPI_ISL_754516, EPI_ISL_754517, EPI_ISL_754518, EPI_ISL_754519, EPI_ISL_754520, EPI_ISL_754521, EPI_ISL_754522, EPI_ISL_754523, EPI_ISL_754524, EPI_ISL_754525, EPI_ISL_754526, EPI_ISL_754527, EPI_ISL_754528, EPI_ISL_754529, EPI_ISL_754530, EPI_ISL_754531, EPI_ISL_754532, EPI_ISL_754533, EPI_ISL_754534, EPI_ISL_754535, EPI_ISL_754536, EPI_ISL_754537, EPI_ISL_754538, EPI_ISL_754539, EPI_ISL_754540, EPI_ISL_754541, EPI_ISL_754542, EPI_ISL_754543, EPI_ISL_754544, EPI_ISL_754545, EPI_ISL_754546, EPI_ISL_754547, EPI_ISL_754548, EPI_ISL_754549, EPI_ISL_754550, EPI_ISL_754551, EPI_ISL_754552, EPI_ISL_754553, EPI_ISL_754554, EPI_ISL_754555, EPI_ISL_754556, EPI_ISL_754557, EPI_ISL_754558, EPI_ISL_754559, EPI_ISL_754560, EPI_ISL_754561, EPI_ISL_754562, EPI_ISL_754563, EPI_ISL_754564, EPI_ISL_754565, EPI_ISL_754566, EPI_ISL_754567, EPI_ISL_754568, EPI_ISL_754569, EPI_ISL_754570, EPI_ISL_754571, EPI_ISL_754572, EPI_ISL_754573, EPI_ISL_754574, EPI_ISL_754575, EPI_ISL_754576, EPI_ISL_754577, EPI_ISL_754578, EPI_ISL_754579, EPI_ISL_754580, EPI_ISL_754581, EPI_ISL_754582, EPI_ISL_754583, EPI_ISL_754584, EPI_ISL_754585, EPI_ISL_754586, EPI_ISL_754587, EPI_ISL_754588, EPI_ISL_754589, EPI_ISL_754590, EPI_ISL_754591, EPI_ISL_754592, EPI_ISL_754593, EPI_ISL_754594, EPI_ISL_754595, EPI_ISL_754596, EPI_ISL_754597, EPI_ISL_754598, EPI_ISL_754599, EPI_ISL_754600, EPI_ISL_754601, EPI_ISL_754602, EPI_ISL_754603, EPI_ISL_754604, EPI_ISL_754605, EPI_ISL_754606, EPI_ISL_754607, EPI_ISL_754608, EPI_ISL_754609, EPI_ISL_754610, EPI_ISL_754611, EPI_ISL_754612, EPI_ISL_754613, EPI_ISL_754614, EPI_ISL_754615, EPI_ISL_754616, EPI_ISL_754617, EPI_ISL_754618 | | | |
| see above | Wadsworth Center, New York State Department.of Health | Wadsworth Center, New York State Department.of Health | Kirsten St. George, Daryl M. Lamson, Alexis Russel, Matthew Shudt, Melissa A Leisner, Jonathan Plitnick, Navjot Singh, John Kelly, Sara Griesemer, Erasmus Schneider, Erica Lasek-Nesselquist |
| EPI_ISL_754619, EPI_ISL_754620, EPI_ISL_754621, EPI_ISL_754622, EPI_ISL_754623, EPI_ISL_754625, EPI_ISL_754626, EPI_ISL_754627, EPI_ISL_754628, EPI_ISL_754629, EPI_ISL_754630, EPI_ISL_754631, EPI_ISL_754632, EPI_ISL_754633, EPI_ISL_754634, EPI_ISL_754635, EPI_ISL_754636, EPI_ISL_754637, EPI_ISL_754638, EPI_ISL_754639, EPI_ISL_754640, EPI_ISL_754641, EPI_ISL_754642, EPI_ISL_754643, EPI_ISL_754644, EPI_ISL_754645, EPI_ISL_754646, EPI_ISL_754647, EPI_ISL_754648, EPI_ISL_754649, EPI_ISL_754650, EPI_ISL_754651, EPI_ISL_754652, EPI_ISL_754653, EPI_ISL_754654, EPI_ISL_754655, EPI_ISL_754656, EPI_ISL_754657, EPI_ISL_754658, EPI_ISL_754659, EPI_ISL_754660, EPI_ISL_754661, EPI_ISL_754662, EPI_ISL_754663, EPI_ISL_754664, EPI_ISL_754665, EPI_ISL_754666, EPI_ISL_754667, | | | |

| | | | |
|---|---|---|---|
| EPI_ISL_754668, EPI_ISL_754669, EPI_ISL_754670, EPI_ISL_754671, EPI_ISL_754672, EPI_ISL_754673, EPI_ISL_754674, EPI_ISL_754675, EPI_ISL_754676 | | | |
| see above | University of Wisconsin-Madison AIDS Vaccine Research Laboratories | University of Wisconsin-Madison AIDS Vaccine Research Laboratories | Gage Moreno, Katarina Braun, et al. AIDS Vaccine Research Laboratories |
| EPI_ISL_754677, EPI_ISL_754678, EPI_ISL_754679, EPI_ISL_754680, EPI_ISL_754681, EPI_ISL_754682, EPI_ISL_754683, EPI_ISL_754684, EPI_ISL_754685, EPI_ISL_754686, EPI_ISL_754687, EPI_ISL_754688, EPI_ISL_754689, EPI_ISL_754690, EPI_ISL_754691, EPI_ISL_754692, EPI_ISL_754693, EPI_ISL_754694, EPI_ISL_754695, EPI_ISL_754696, EPI_ISL_754697, EPI_ISL_754698, EPI_ISL_754699, EPI_ISL_754700, EPI_ISL_754701, EPI_ISL_754702, EPI_ISL_754703, EPI_ISL_754704, EPI_ISL_754705, EPI_ISL_754706, EPI_ISL_754707, EPI_ISL_754708, EPI_ISL_754709, EPI_ISL_754710, EPI_ISL_754711, EPI_ISL_754712, EPI_ISL_754713, EPI_ISL_754714, EPI_ISL_754715, EPI_ISL_754716, EPI_ISL_754717, EPI_ISL_754718, EPI_ISL_754719, EPI_ISL_754720, EPI_ISL_754721, EPI_ISL_754722, EPI_ISL_754723, EPI_ISL_754724, EPI_ISL_754725, EPI_ISL_754726, EPI_ISL_754727, EPI_ISL_754728, EPI_ISL_754729, EPI_ISL_754730, EPI_ISL_754731, EPI_ISL_754732, EPI_ISL_754733, EPI_ISL_754734, EPI_ISL_754735, EPI_ISL_754736, EPI_ISL_754737, EPI_ISL_754738, EPI_ISL_754739, EPI_ISL_754740, EPI_ISL_754741, EPI_ISL_754742, EPI_ISL_754743, EPI_ISL_754744, EPI_ISL_754745 | | | |
| see above | Wadsworth Center, New York State Department.of Health | Wadsworth Center, New York State Department.of Health | Kirsten St. George, Daryl M. Lamson, Alexis Russel, Matthew Shudt, Melissa A Leisner, Jonathan Plitnick, Navjot Singh, John Kelly, Sara Griesemer, Erasmus Schneider, Erica Lasek-Nesselquist |
| EPI_ISL_754746, EPI_ISL_754747, EPI_ISL_754748, EPI_ISL_754749, EPI_ISL_754750, EPI_ISL_754751, EPI_ISL_754752, EPI_ISL_754753, EPI_ISL_754754, EPI_ISL_754755, EPI_ISL_754756, EPI_ISL_754757, EPI_ISL_754758, EPI_ISL_754759, EPI_ISL_754760, EPI_ISL_754761, EPI_ISL_754762, EPI_ISL_754763, EPI_ISL_754764, EPI_ISL_754765, EPI_ISL_754766, EPI_ISL_754767, EPI_ISL_754768, EPI_ISL_754769, EPI_ISL_754770, EPI_ISL_754771, EPI_ISL_754772, EPI_ISL_754773, EPI_ISL_754774, EPI_ISL_754775, EPI_ISL_754776, EPI_ISL_754777, EPI_ISL_754778 | | | |
| see above | Emory Molecular Diagnostics Laboratory, Emory Healthcare | Piantadosi Lab, Emory Department of Pathology | Ahmed Babiker, Anne Piantadosi |
| EPI_ISL_754779, EPI_ISL_754780, EPI_ISL_754781, EPI_ISL_754782, EPI_ISL_754783, EPI_ISL_754784, EPI_ISL_754785, EPI_ISL_754786, EPI_ISL_754787, EPI_ISL_754788, EPI_ISL_754789, EPI_ISL_754790, EPI_ISL_754791, EPI_ISL_754792, EPI_ISL_754793, EPI_ISL_754794, EPI_ISL_754795, EPI_ISL_754796, EPI_ISL_754797, EPI_ISL_754798, EPI_ISL_754799, EPI_ISL_754800, EPI_ISL_754801, EPI_ISL_754802, EPI_ISL_754803, EPI_ISL_754804, EPI_ISL_754805, EPI_ISL_754806, EPI_ISL_754807, EPI_ISL_754808, EPI_ISL_754809, EPI_ISL_754810, EPI_ISL_754811, EPI_ISL_754812, EPI_ISL_754813, EPI_ISL_754814, EPI_ISL_754815, EPI_ISL_754816, EPI_ISL_754817, EPI_ISL_754818, EPI_ISL_754819, EPI_ISL_754820, EPI_ISL_754821, EPI_ISL_754822, EPI_ISL_754823, EPI_ISL_754824, EPI_ISL_754825, EPI_ISL_754826, EPI_ISL_754827, EPI_ISL_754828, EPI_ISL_754829, EPI_ISL_754830, EPI_ISL_754831, EPI_ISL_754832, EPI_ISL_754833, EPI_ISL_754834, EPI_ISL_754835, EPI_ISL_754836, EPI_ISL_754837, EPI_ISL_754838, EPI_ISL_754839, EPI_ISL_754840, EPI_ISL_754841 | | | |
| see above | Wadsworth Center, New York State Department.of Health | Wadsworth Center, New York State Department.of Health | Kirsten St. George, Daryl M. Lamson, Alexis Russel, Matthew Shudt, Melissa A Leisner, Jonathan Plitnick, Navjot Singh, John Kelly, Sara Griesemer, Erasmus Schneider, Erica Lasek-Nesselquist |
| EPI_ISL_754857, EPI_ISL_754858, EPI_ISL_754859, EPI_ISL_754860, EPI_ISL_754864, EPI_ISL_754865, EPI_ISL_754866, EPI_ISL_754867, EPI_ISL_754868, EPI_ISL_754869, EPI_ISL_754870, EPI_ISL_754871, EPI_ISL_754872, EPI_ISL_754873, EPI_ISL_754874, EPI_ISL_754875, EPI_ISL_754876, EPI_ISL_754877, EPI_ISL_754878, EPI_ISL_754879, EPI_ISL_754880, EPI_ISL_754881, EPI_ISL_754882, EPI_ISL_754883, EPI_ISL_754884, EPI_ISL_754885, EPI_ISL_754886, EPI_ISL_754887, EPI_ISL_754888, EPI_ISL_754889, EPI_ISL_754890, EPI_ISL_754891, EPI_ISL_754892, EPI_ISL_754893, EPI_ISL_754894, EPI_ISL_754895, EPI_ISL_754896, EPI_ISL_754897, EPI_ISL_754898, EPI_ISL_754899, EPI_ISL_754900, EPI_ISL_754901 | | | |
| see above | Innovative Genomics Institute, UC Berkeley | Innovative Genomics Institute, UC Berkeley | Stacia Wyman, Haridha Shivram, Phil Frankino, Liana Lareau, Shana McDevitt, Justin Choi |
| EPI_ISL_754914, EPI_ISL_754915, EPI_ISL_754916, EPI_ISL_754917, EPI_ISL_754918, EPI_ISL_754919, EPI_ISL_754920, EPI_ISL_754921, EPI_ISL_754922, EPI_ISL_754923, EPI_ISL_754924, EPI_ISL_754925, EPI_ISL_754926, EPI_ISL_754927, EPI_ISL_754928, EPI_ISL_754929, EPI_ISL_754930, EPI_ISL_754931, EPI_ISL_754932, EPI_ISL_754933, EPI_ISL_754934, EPI_ISL_754935, EPI_ISL_754936, EPI_ISL_754937, EPI_ISL_754938, EPI_ISL_754939, EPI_ISL_754940, EPI_ISL_754941, EPI_ISL_754942, EPI_ISL_754943, EPI_ISL_754944, EPI_ISL_754945, EPI_ISL_754946, EPI_ISL_754947, EPI_ISL_754948, EPI_ISL_754949, EPI_ISL_754950, EPI_ISL_754951, EPI_ISL_754952, EPI_ISL_754953, EPI_ISL_754954, EPI_ISL_754955, EPI_ISL_754956, EPI_ISL_754957, EPI_ISL_754958, EPI_ISL_754959, EPI_ISL_754960, EPI_ISL_754961, EPI_ISL_754962, EPI_ISL_754963, EPI_ISL_754964, EPI_ISL_754965, EPI_ISL_754966, EPI_ISL_754967, EPI_ISL_754968, EPI_ISL_754969, EPI_ISL_754970, EPI_ISL_754971, EPI_ISL_754972, EPI_ISL_754973, EPI_ISL_754974, EPI_ISL_754975, EPI_ISL_754976, EPI_ISL_754977, EPI_ISL_754978, EPI_ISL_754979, EPI_ISL_754980, EPI_ISL_754981, EPI_ISL_754982, EPI_ISL_754983, EPI_ISL_754984, EPI_ISL_754985, EPI_ISL_754986, EPI_ISL_754987, EPI_ISL_754988, EPI_ISL_754989, EPI_ISL_754990, EPI_ISL_754991, EPI_ISL_754992, EPI_ISL_754993, EPI_ISL_754994, EPI_ISL_754995, EPI_ISL_754996, EPI_ISL_754997, EPI_ISL_754998, EPI_ISL_754999, EPI_ISL_755000, EPI_ISL_755001, EPI_ISL_755002, EPI_ISL_755003, EPI_ISL_755004, EPI_ISL_755005, EPI_ISL_755006, EPI_ISL_755007, EPI_ISL_755008, EPI_ISL_755009, EPI_ISL_755010, EPI_ISL_755011, EPI_ISL_755012, EPI_ISL_755013, EPI_ISL_755014, EPI_ISL_755015, EPI_ISL_755030, EPI_ISL_755031, EPI_ISL_755032, EPI_ISL_755033, EPI_ISL_755034, EPI_ISL_755035, EPI_ISL_755036, EPI_ISL_755037, EPI_ISL_755038, EPI_ISL_755039, EPI_ISL_755040, EPI_ISL_755041, EPI_ISL_755042, EPI_ISL_755043, EPI_ISL_755044, EPI_ISL_755045, EPI_ISL_755046, EPI_ISL_755047, EPI_ISL_755048, EPI_ISL_755049, EPI_ISL_755050, EPI_ISL_755051, EPI_ISL_755052, EPI_ISL_755053, EPI_ISL_755054, EPI_ISL_755055, EPI_ISL_755056, EPI_ISL_755057, EPI_ISL_755058, EPI_ISL_755059, EPI_ISL_755060, EPI_ISL_755061, EPI_ISL_755062, EPI_ISL_755063, EPI_ISL_755064, EPI_ISL_755065, EPI_ISL_755066, EPI_ISL_755067, EPI_ISL_755068, EPI_ISL_755069, EPI_ISL_755070, EPI_ISL_755071, EPI_ISL_755072, EPI_ISL_755073, EPI_ISL_755074, EPI_ISL_755075, EPI_ISL_755076, EPI_ISL_755077, EPI_ISL_755078, EPI_ISL_755079, EPI_ISL_755080, EPI_ISL_755081, EPI_ISL_755082, EPI_ISL_755083, EPI_ISL_755084, EPI_ISL_755085, EPI_ISL_755086, EPI_ISL_755087, EPI_ISL_755088, EPI_ISL_755089, EPI_ISL_755090, EPI_ISL_755091, EPI_ISL_755092, EPI_ISL_755093, EPI_ISL_755094, EPI_ISL_755095, EPI_ISL_755096, EPI_ISL_755097, EPI_ISL_755098, EPI_ISL_755099, EPI_ISL_755100, EPI_ISL_755101, EPI_ISL_755102, EPI_ISL_755103, EPI_ISL_755104, EPI_ISL_755105, EPI_ISL_755106, EPI_ISL_755107, EPI_ISL_755108, EPI_ISL_755109, EPI_ISL_755110, EPI_ISL_755111, EPI_ISL_755112, EPI_ISL_755113, EPI_ISL_755114 | | | |
| see above | California Department of Public Health | California Department of Public Health | CDPH IDLB COVIDNet |
| EPI_ISL_755115, EPI_ISL_755116, EPI_ISL_755117, EPI_ISL_755119, EPI_ISL_755130, EPI_ISL_755132 | San Diego County Public Health Laboratory | Andersen lab at Scripps Research | SEARCH Alliance San Diego with Tracy Basler, Jovan Shephard, Brett Austin |
| EPI_ISL_755133 | UCSD EXCITE lab | Andersen lab at Scripps Research | SEARCH Alliance San Diego |
| EPI_ISL_755134, EPI_ISL_755135, EPI_ISL_755136 | San Diego County Public Health Laboratory | Andersen lab at Scripps Research | SEARCH Alliance San Diego with Tracy Basler, Jovan Shephard, Brett Austin |
| EPI_ISL_755137 | UCSD EXCITE lab | Andersen lab at Scripps Research | SEARCH Alliance San Diego |
| EPI_ISL_755138, EPI_ISL_755139 | San Diego County Public Health Laboratory | Andersen lab at Scripps Research | SEARCH Alliance San Diego with Tracy Basler, Jovan Shephard, Brett Austin |
| EPI_ISL_755140, EPI_ISL_755146, EPI_ISL_755147, EPI_ISL_755148, EPI_ISL_755149, EPI_ISL_755150, EPI_ISL_755151, EPI_ISL_755152, EPI_ISL_755153, EPI_ISL_755154, EPI_ISL_755155, EPI_ISL_755156, EPI_ISL_755157, EPI_ISL_755158, EPI_ISL_755159, EPI_ISL_755160, EPI_ISL_755161, EPI_ISL_755162, EPI_ISL_755163, EPI_ISL_755164, EPI_ISL_755165, EPI_ISL_755166, EPI_ISL_755167, EPI_ISL_755168, EPI_ISL_755169, EPI_ISL_755170, EPI_ISL_755171, EPI_ISL_755172, EPI_ISL_755173, EPI_ISL_755174, EPI_ISL_755175, EPI_ISL_755176, EPI_ISL_755177, EPI_ISL_755178, EPI_ISL_755179, EPI_ISL_755180, EPI_ISL_755181, EPI_ISL_755182, EPI_ISL_755183, EPI_ISL_755184, EPI_ISL_755185, EPI_ISL_755186, EPI_ISL_755187, EPI_ISL_755188, EPI_ISL_755189, EPI_ISL_755190, EPI_ISL_755191, EPI_ISL_755192, EPI_ISL_755193, EPI_ISL_755194, EPI_ISL_755195, EPI_ISL_755196, EPI_ISL_755197, EPI_ISL_755198, EPI_ISL_755199, EPI_ISL_755200, EPI_ISL_755201, EPI_ISL_755202, EPI_ISL_755203, EPI_ISL_755204, EPI_ISL_755205, EPI_ISL_755206, EPI_ISL_755207, EPI_ISL_755208, EPI_ISL_755209, EPI_ISL_755215, EPI_ISL_755216, EPI_ISL_755217, EPI_ISL_755218, EPI_ISL_755219, EPI_ISL_755220, EPI_ISL_755221, EPI_ISL_755222, EPI_ISL_755223, EPI_ISL_755224, EPI_ISL_755225, EPI_ISL_755226, EPI_ISL_755227, EPI_ISL_755228, EPI_ISL_755229, EPI_ISL_755230, EPI_ISL_755231 | | | |
| see above | UCSD EXCITE lab | Andersen lab at Scripps Research | SEARCH Alliance San Diego |
| EPI_ISL_755269, EPI_ISL_755270, EPI_ISL_755271, EPI_ISL_755272, EPI_ISL_755273, EPI_ISL_755274, EPI_ISL_755275, EPI_ISL_755276, EPI_ISL_755277, EPI_ISL_755278, EPI_ISL_755279, EPI_ISL_755280, EPI_ISL_755281, EPI_ISL_755282, EPI_ISL_755283, EPI_ISL_755284, EPI_ISL_755285, EPI_ISL_755286, EPI_ISL_755287, EPI_ISL_755288, EPI_ISL_755289, EPI_ISL_755290, EPI_ISL_755291, EPI_ISL_755292, EPI_ISL_755293, EPI_ISL_755294, EPI_ISL_755295, EPI_ISL_755296, EPI_ISL_755297, EPI_ISL_755298, EPI_ISL_755299, EPI_ISL_755300 | | | |
| see above | San Diego County Public Health Laboratory | Andersen lab at Scripps Research | SEARCH Alliance San Diego with Tracy Basler, Jovan Shephard, Brett Austin |
| EPI_ISL_755306, EPI_ISL_755307, EPI_ISL_755308, EPI_ISL_755309, EPI_ISL_755311, EPI_ISL_755312, EPI_ISL_755313, EPI_ISL_755314, EPI_ISL_755315, EPI_ISL_755316, EPI_ISL_755317, EPI_ISL_755318, EPI_ISL_755319, EPI_ISL_755320, EPI_ISL_755321, EPI_ISL_755322, EPI_ISL_755323, EPI_ISL_755324, EPI_ISL_755325, EPI_ISL_755326, EPI_ISL_755327, EPI_ISL_755328, EPI_ISL_755329, EPI_ISL_755330, EPI_ISL_755331, EPI_ISL_755332, EPI_ISL_755333, EPI_ISL_755334, EPI_ISL_755335, EPI_ISL_755336, EPI_ISL_755337, EPI_ISL_755338, EPI_ISL_755339, EPI_ISL_755340, EPI_ISL_755341, EPI_ISL_755342, EPI_ISL_755343, EPI_ISL_755344, EPI_ISL_755345, EPI_ISL_755346, EPI_ISL_755347, EPI_ISL_755348, EPI_ISL_755349, EPI_ISL_755350, EPI_ISL_755351, EPI_ISL_755352, EPI_ISL_755353, EPI_ISL_755354, EPI_ISL_755355, EPI_ISL_755356, EPI_ISL_755357, EPI_ISL_755358, EPI_ISL_755359, EPI_ISL_755360, EPI_ISL_755361, EPI_ISL_755363, EPI_ISL_755364, EPI_ISL_755365, EPI_ISL_755366, EPI_ISL_755367, EPI_ISL_755368, EPI_ISL_755369, EPI_ISL_755370, EPI_ISL_755371, EPI_ISL_755372, EPI_ISL_755373, EPI_ISL_755374, EPI_ISL_755375, EPI_ISL_755376, EPI_ISL_755377, EPI_ISL_755378, EPI_ISL_755379, EPI_ISL_755380, EPI_ISL_755381, EPI_ISL_755382, EPI_ISL_755383, EPI_ISL_755384, EPI_ISL_755385, EPI_ISL_755386, EPI_ISL_755387, EPI_ISL_755388, EPI_ISL_755389, EPI_ISL_755390, EPI_ISL_755391, EPI_ISL_755392, EPI_ISL_755393, EPI_ISL_755394, EPI_ISL_755395, EPI_ISL_755396, EPI_ISL_755397, EPI_ISL_755398, EPI_ISL_755399, EPI_ISL_755400, EPI_ISL_755401, EPI_ISL_755402, EPI_ISL_755403, EPI_ISL_755404, EPI_ISL_755405, EPI_ISL_755406, EPI_ISL_755407, EPI_ISL_755408, EPI_ISL_755409, EPI_ISL_755410, EPI_ISL_755411, EPI_ISL_755412, EPI_ISL_755413, EPI_ISL_755414, EPI_ISL_755415, EPI_ISL_755416, EPI_ISL_755417, EPI_ISL_755418, EPI_ISL_755419, EPI_ISL_755420, EPI_ISL_755421, EPI_ISL_755422, EPI_ISL_755423, EPI_ISL_755424, EPI_ISL_755425, EPI_ISL_755427, EPI_ISL_755428, EPI_ISL_755429, EPI_ISL_755430, EPI_ISL_755431, EPI_ISL_755432, EPI_ISL_755433, EPI_ISL_755434, EPI_ISL_755435, EPI_ISL_755436, EPI_ISL_755437, EPI_ISL_755438, EPI_ISL_755439, EPI_ISL_755440, EPI_ISL_755441, EPI_ISL_755442, EPI_ISL_755443, EPI_ISL_755444, EPI_ISL_755445, EPI_ISL_755446, EPI_ISL_755447, EPI_ISL_755448, EPI_ISL_755449, EPI_ISL_755450, EPI_ISL_755451, EPI_ISL_755452, EPI_ISL_755453, EPI_ISL_755454, EPI_ISL_755455, EPI_ISL_755457, EPI_ISL_755458, EPI_ISL_755460, EPI_ISL_755461, EPI_ISL_755462, EPI_ISL_755463, EPI_ISL_755464, EPI_ISL_755465, EPI_ISL_755466, EPI_ISL_755467, EPI_ISL_755468, EPI_ISL_755469, EPI_ISL_755470, EPI_ISL_755471, EPI_ISL_755472, EPI_ISL_755473, EPI_ISL_755474, EPI_ISL_755475, EPI_ISL_755476, EPI_ISL_755477, EPI_ISL_755478, EPI_ISL_755479, EPI_ISL_755480, EPI_ISL_755481, EPI_ISL_755482, EPI_ISL_755483, EPI_ISL_755484, EPI_ISL_755485, EPI_ISL_755486, EPI_ISL_755487, EPI_ISL_755488, EPI_ISL_755489, EPI_ISL_755490, EPI_ISL_755491, EPI_ISL_755492, EPI_ISL_755493, EPI_ISL_755494, EPI_ISL_755495, EPI_ISL_755496, EPI_ISL_755497, EPI_ISL_755498, EPI_ISL_755499, EPI_ISL_755500, EPI_ISL_755501, EPI_ISL_755502, EPI_ISL_755503, EPI_ISL_755504, EPI_ISL_755505, EPI_ISL_755506, EPI_ISL_755507, EPI_ISL_755508, EPI_ISL_755509, EPI_ISL_755510, EPI_ISL_755511, EPI_ISL_755512, EPI_ISL_755513, EPI_ISL_755514, EPI_ISL_755517, EPI_ISL_755518, EPI_ISL_755519, EPI_ISL_755520, EPI_ISL_755522, EPI_ISL_755523, EPI_ISL_755524, EPI_ISL_755525, EPI_ISL_755526, EPI_ISL_755527, EPI_ISL_755528, EPI_ISL_755529, EPI_ISL_755530, EPI_ISL_755531, EPI_ISL_755532, EPI_ISL_755533, EPI_ISL_755534, EPI_ISL_755535, EPI_ISL_755536, EPI_ISL_755537, EPI_ISL_755538, EPI_ISL_755539, EPI_ISL_755540, EPI_ISL_755541, EPI_ISL_755542, EPI_ISL_755543, EPI_ISL_755544, EPI_ISL_755545, EPI_ISL_755546, EPI_ISL_755547, EPI_ISL_755548, EPI_ISL_755549, EPI_ISL_755550, EPI_ISL_755551, EPI_ISL_755552, EPI_ISL_755553, EPI_ISL_755554, EPI_ISL_755555, EPI_ISL_755556, EPI_ISL_755557, EPI_ISL_755558, EPI_ISL_755559, EPI_ISL_755560, EPI_ISL_755561, EPI_ISL_755562, EPI_ISL_755563, EPI_ISL_755564, EPI_ISL_755565, EPI_ISL_755566, EPI_ISL_755567 | | | |
| see above | Maine Health and Environmental Testing Laboratory | Tewhey Lab, The Jackson Laboratory | Matluk,N., Dewey,H., Iosue,F., Barter,M., Lynch,R., Munger,H. and Tewhey,R. |
| EPI_ISL_755590, EPI_ISL_755591, EPI_ISL_755592, EPI_ISL_755593, EPI_ISL_755594, EPI_ISL_755595, EPI_ISL_755596, EPI_ISL_755597, EPI_ISL_755598, EPI_ISL_755599, EPI_ISL_755600, EPI_ISL_755601, EPI_ISL_755602, EPI_ISL_755603, EPI_ISL_755604, EPI_ISL_755605, EPI_ISL_755606, EPI_ISL_755607, EPI_ISL_755608, EPI_ISL_755609, EPI_ISL_755610, EPI_ISL_755611, EPI_ISL_755612, EPI_ISL_755613, EPI_ISL_755616, EPI_ISL_755617, EPI_ISL_755618 | | | |
| see above | Helix/Illumina | Genomics and Discovery, Respiratory Viruses Branch, Division of Viral Diseases, Centers for Disease Control and Prevention | Peter W. Cook, Dhwani Batra, Eileen de Feo, Jan Antico, Christine Tran, Matthew Tolentino, Shannon Wickline, Kim Gietzen, Brad Sickler, Jingtao Liu, Eric Allen, Phil Febbo, Summer Galloway, Nicole L. Washington, Simon White, Geraint Levan, Kelly Schiabor Barrett, Elizabeth Cirulli, Alexandre Bolze, Ary Ascencio, Charlotte Rivera-Garcia, Ryan Cho, Jason Nguyen, Sherry Wang, Jimmy Ramirez, Tyler Cassens, Efren Sandoval, Magnus Isaksson, William Lee, David Becker, Marc Laurent, James Lu, Clinton R. Paden, Suxiang Tong, Duncan MacCannell |
| EPI_ISL_755940, EPI_ISL_755942 | California Department of Public Health | Chiu Laboratory, University of California, San Francisco | Charles Chiu, Xianding (Wayne) Deng, Candace Wang, Jill Hacker, Debra Wadford |
| EPI_ISL_756126, EPI_ISL_756128, EPI_ISL_756129, EPI_ISL_756130, EPI_ISL_756131, EPI_ISL_756132, EPI_ISL_756133, EPI_ISL_756134, EPI_ISL_756164, EPI_ISL_756184, EPI_ISL_756185, EPI_ISL_756186, EPI_ISL_756187, EPI_ISL_756188, EPI_ISL_756189, EPI_ISL_756190 | | | |
| see above | Wyoming Public Health Laboratory | Wyoming Public Health Laboratory | Noah Hull, Taylor Fearing, Channing Weber, Ashley Norberg, Bailey Bowcutt, and Wanda Manley |
| EPI_ISL_756191, EPI_ISL_756192, EPI_ISL_756193, EPI_ISL_756194, EPI_ISL_756195, EPI_ISL_756196, EPI_ISL_756197, EPI_ISL_756198, EPI_ISL_756199, EPI_ISL_756200, EPI_ISL_756201, EPI_ISL_756202, EPI_ISL_756203, EPI_ISL_756204, EPI_ISL_756205, EPI_ISL_756206, EPI_ISL_756207, EPI_ISL_756208, EPI_ISL_756209, EPI_ISL_756210, EPI_ISL_756211, EPI_ISL_756212, EPI_ISL_756213, EPI_ISL_756214, EPI_ISL_756215, EPI_ISL_756216, EPI_ISL_756217, EPI_ISL_756218, EPI_ISL_756219, EPI_ISL_756220, EPI_ISL_756221, EPI_ISL_756223, EPI_ISL_756224, EPI_ISL_756225, EPI_ISL_756226, EPI_ISL_756227, EPI_ISL_756228, EPI_ISL_756229, EPI_ISL_756230, EPI_ISL_756231, EPI_ISL_756232, EPI_ISL_756233, EPI_ISL_756234, EPI_ISL_756235, EPI_ISL_756236, EPI_ISL_756237, EPI_ISL_756238, EPI_ISL_756239, EPI_ISL_756240 | | | |
| see above | UW Virology Lab | UW Virology Lab | Pavitra Roychoudhury, Hong Xie, Lasata Shrestha, Meei-Li Huang, Keith R Jerome, Alexander Greninger |
| EPI_ISL_756241, EPI_ISL_756242, EPI_ISL_756243, EPI_ISL_756244, EPI_ISL_756245, EPI_ISL_756246, EPI_ISL_756253, EPI_ISL_756254, EPI_ISL_756255, EPI_ISL_756256, EPI_ISL_756257, EPI_ISL_756258, EPI_ISL_756259, EPI_ISL_756260, EPI_ISL_756261, EPI_ISL_756262, EPI_ISL_756267, EPI_ISL_756268, EPI_ISL_756269, EPI_ISL_756270 | | | |
| see above | Wyoming Public Health Laboratory | Wyoming Public Health Laboratory | Noah Hull, Taylor Fearing, Channing Weber, Ashley Norberg, Bailey Bowcutt, and Wanda Manley |
| EPI_ISL_756275, EPI_ISL_756276, EPI_ISL_756277, EPI_ISL_756278, EPI_ISL_756279, EPI_ISL_756280, EPI_ISL_756281 | State Laboratories Division, Hawaii State Department of Health | State Laboratories Division, Hawaii State Department of Health | Pamela O'Brien, Sabrina Diemert, Drew Kuwazaki, Razvan Sultana, Edward Desmond |
| EPI_ISL_756295, EPI_ISL_756296, EPI_ISL_756297, EPI_ISL_756298, EPI_ISL_756299, EPI_ISL_756300, EPI_ISL_756301, EPI_ISL_756302, EPI_ISL_756303, EPI_ISL_756304, EPI_ISL_756305 | | | |
| see above | UW Virology Lab | UW Virology Lab | Pavitra Roychoudhury, Hong Xie, Lasata Shrestha, Meei-Li Huang, Keith R Jerome, Alexander Greninger |
| EPI_ISL_756312, EPI_ISL_756313, EPI_ISL_756314, EPI_ISL_756315, EPI_ISL_756316, EPI_ISL_756317, EPI_ISL_756318, EPI_ISL_756319, EPI_ISL_756320, EPI_ISL_756321, EPI_ISL_756322, EPI_ISL_756323, EPI_ISL_756324, EPI_ISL_756325, EPI_ISL_756326, EPI_ISL_756327, EPI_ISL_756330, EPI_ISL_756332, EPI_ISL_756333, EPI_ISL_756334, EPI_ISL_756336, EPI_ISL_756337, EPI_ISL_756338, EPI_ISL_756339, EPI_ISL_756340, EPI_ISL_756341, EPI_ISL_756342, EPI_ISL_756343, EPI_ISL_756344, EPI_ISL_756346, EPI_ISL_756347, EPI_ISL_756348, EPI_ISL_756349, EPI_ISL_756350, EPI_ISL_756351, EPI_ISL_756352, EPI_ISL_756353, EPI_ISL_756354, EPI_ISL_756355, EPI_ISL_756356 | | | |
| see above | Innovative Genomics Institute, UC Berkeley | Innovative Genomics Institute, UC Berkeley | Stacia Wyman, Haridha Shivram, Phil Frankino, Liana Lareau, Shana McDevitt, Justin Choi |
| EPI_ISL_759706, EPI_ISL_759724, EPI_ISL_759725, EPI_ISL_759726, EPI_ISL_759727, EPI_ISL_759728, EPI_ISL_759729, EPI_ISL_759730, EPI_ISL_759731, EPI_ISL_759733, EPI_ISL_759734, EPI_ISL_759735, EPI_ISL_759736, EPI_ISL_759737, EPI_ISL_759739, EPI_ISL_759740, EPI_ISL_759741, EPI_ISL_759742, EPI_ISL_759743, EPI_ISL_759744, EPI_ISL_759745, EPI_ISL_759746, EPI_ISL_759747, EPI_ISL_759748, | | | |

| | | | |
|---|---|---|---|
| EPI_ISL_759749, EPI_ISL_759750, EPI_ISL_759751, EPI_ISL_759752, EPI_ISL_759753, EPI_ISL_759754, EPI_ISL_759755, EPI_ISL_759756, EPI_ISL_759758, EPI_ISL_759759 | | | |
| see above | University of Wisconsin-Madison AIDS Vaccine Research Laboratories | University of Wisconsin-Madison AIDS Vaccine Research Laboratories | Gage Moreno, Katarina Braun, et al. AIDS Vaccine Research Laboratories |
| EPI_ISL_759860, EPI_ISL_759861, EPI_ISL_759862, EPI_ISL_759863, EPI_ISL_759864, EPI_ISL_759865 | Molecular, ProgenaBiome | Molecular, ProgenaBiome | Papoutsis,A.J. |
| EPI_ISL_763076 | Ohio Department Of Health | Ohio Department of Health | Jennifer Holmes, Eric Brandt, Keoni Omura, Glen McGillivary, Caitlin McDonnell, Kirtana Ramadugu, Kelsey Florek, Heather Blankenship, Quanta Brown, and Tammy Bannerman |
| EPI_ISL_763319 | Ohio Department of Health Laboratory | Ohio Department of Health laboratory | Jennifer Holmes, Eric Brandt, Keoni Omura, Glen McGillivary, Caitlin McDonnell, Kirtana Ramadugu, Kelsey Florek, Heather Blankenship, Quanta Brown, and Tammy Bannerman |
| EPI_ISL_763321 | Ohio Department of Health Laboratory | Ohio Depart of Health Laboratory | Jennifer Holmes, Eric Brandt, Keoni Omura, Glen McGillivary, Caitlin McDonnell, Kirtana Ramadugu, Kelsey Florek, Heather Blankenship, Quanta Brown, and Tammy Bannerman |
| EPI_ISL_763333, EPI_ISL_763339, EPI_ISL_763342, EPI_ISL_763343 | Florida Bureau of Public Health Laboratories | Florida Bureau of Public Health Laboratories | Sarah Schmedes, Jason Blanton |
| EPI_ISL_765209 | Ohio Department of Health Laboratory | Ohio Department of Health Laboratory | Holmes, Jennifer; Eric Brandt, Glen McGillivary, Keoni Omura, Caitlin McDonnell, Heather Blankenship, Kelsey Florek, Kirtana Ramadugu, Erica Leasure, Quanta Brown and Tammy Bannerman |
| EPI_ISL_765222 | Ohio Department of Health Laboratory | Ohio Department of Health Laboratory | Holmes, Jennifer; Eric Brandt, Keoni Omura, Glen McGillivary, Caitiln McDonnell, Kirtana Ramadugu, Erica Leasure, Kelsey Florek, Heather Blankenship, Quanta Brown, and Tammy Bannerman |
| EPI_ISL_765480 | Wadsworth Center, New York State Department.of Health | Wadsworth Center, New York State Department.of Health | Kirsten St. George, Daryl M. Lamson, Alexis Russel, Matthew Shudt, Melissa A Leisner, Jonathan Plitnick, Navjot Singh, John Kelly, Sara Griesemer, Erasmus Schneider, Erica Lasek-Nesselquist |
| EPI_ISL_765481 | SARATOGA HOSPITAL LABORATORY | Wadsworth Center, New York State Department.of Health | Kirsten St. George, Daryl M. Lamson, Alexis Russel, Matthew Shudt, Melissa A Leisner, Jonathan Plitnick, Navjot Singh, John Kelly, Sara Griesemer, Erasmus Schneider, Erica Lasek-Nesselquist |
| EPI_ISL_765482 | Wadsworth Center, New York State Department.of Health | Wadsworth Center, New York State Department.of Health | Kirsten St. George, Daryl M. Lamson, Alexis Russel, Matthew Shudt, Melissa A Leisner, Jonathan Plitnick, Navjot Singh, John Kelly, Sara Griesemer, Erasmus Schneider, Erica Lasek-Nesselquist |
| EPI_ISL_765483 | SARATOGA HOSPITAL LABORATORY | Wadsworth Center, New York State Department.of Health | Kirsten St. George, Daryl M. Lamson, Alexis Russel, Matthew Shudt, Melissa A Leisner, Jonathan Plitnick, Navjot Singh, John Kelly, Sara Griesemer, Erasmus Schneider, Erica Lasek-Nesselquist |
| EPI_ISL_765484, EPI_ISL_765485, EPI_ISL_765486, EPI_ISL_765487, EPI_ISL_765488 | Wadsworth Center, New York State Department.of Health | Wadsworth Center, New York State Department.of Health | Kirsten St. George, Daryl M. Lamson, Alexis Russel, Matthew Shudt, Melissa A Leisner, Jonathan Plitnick, Navjot Singh, John Kelly, Sara Griesemer, Erasmus Schneider, Erica Lasek-Nesselquist |
| EPI_ISL_765489, EPI_ISL_765490, EPI_ISL_765491, EPI_ISL_765492, EPI_ISL_765493, EPI_ISL_765494, EPI_ISL_765495 | MONTEFIORE MEDICAL CENTER LABORATORIES | Wadsworth Center, New York State Department.of Health | Kirsten St. George, Daryl M. Lamson, Alexis Russel, Matthew Shudt, Melissa A Leisner, Jonathan Plitnick, Navjot Singh, John Kelly, Sara Griesemer, Erasmus Schneider, Erica Lasek-Nesselquist |
| EPI_ISL_765496, EPI_ISL_765497, EPI_ISL_765498, EPI_ISL_765499, EPI_ISL_765500, EPI_ISL_765501, EPI_ISL_765502, EPI_ISL_765503, EPI_ISL_765504, EPI_ISL_765505, EPI_ISL_765506, EPI_ISL_765507, EPI_ISL_765508, EPI_ISL_765509, EPI_ISL_765510, EPI_ISL_765511, EPI_ISL_765512, EPI_ISL_765513, EPI_ISL_765514, EPI_ISL_765515, EPI_ISL_765516 | | | |
| see above | Wadsworth Center, New York State Department.of Health | Wadsworth Center, New York State Department.of Health | Kirsten St. George, Daryl M. Lamson, Alexis Russel, Matthew Shudt, Melissa A Leisner, Jonathan Plitnick, Navjot Singh, John Kelly, Sara Griesemer, Erasmus Schneider, Erica Lasek-Nesselquist |
| EPI_ISL_765517, EPI_ISL_765518, EPI_ISL_765519, EPI_ISL_765520, EPI_ISL_765521, EPI_ISL_765522, EPI_ISL_765523, EPI_ISL_765524, EPI_ISL_765525, EPI_ISL_765526, EPI_ISL_765527, EPI_ISL_765528, EPI_ISL_765529 | | | |
| see above | SARATOGA HOSPITAL LABORATORY | Wadsworth Center, New York State Department.of Health | Kirsten St. George, Daryl M. Lamson, Alexis Russel, Matthew Shudt, Melissa A Leisner, Jonathan Plitnick, Navjot Singh, John Kelly, Sara Griesemer, Erasmus Schneider, Erica Lasek-Nesselquist |
| EPI_ISL_765530, EPI_ISL_765531, EPI_ISL_765532, EPI_ISL_765533, EPI_ISL_765534, EPI_ISL_765535, EPI_ISL_765536, EPI_ISL_765537, EPI_ISL_765538, EPI_ISL_765539, EPI_ISL_765540, EPI_ISL_765541, EPI_ISL_765542, EPI_ISL_765543, EPI_ISL_765544, EPI_ISL_765545, EPI_ISL_765546, EPI_ISL_765547, EPI_ISL_765548, EPI_ISL_765549, EPI_ISL_765550, EPI_ISL_765551, EPI_ISL_765552, EPI_ISL_765553, EPI_ISL_765554, EPI_ISL_765555, EPI_ISL_765556, EPI_ISL_765557, EPI_ISL_765558, EPI_ISL_765559, EPI_ISL_765560, EPI_ISL_765561, EPI_ISL_765562, EPI_ISL_765563, EPI_ISL_765564, EPI_ISL_765565, EPI_ISL_765566 | | | |
| see above | MONTEFIORE MEDICAL CENTER LABORATORIES | Wadsworth Center, New York State Department.of Health | Kirsten St. George, Daryl M. Lamson, Alexis Russel, Matthew Shudt, Melissa A Leisner, Jonathan Plitnick, Navjot Singh, John Kelly, Sara Griesemer, Erasmus Schneider, Erica Lasek-Nesselquist |
| EPI_ISL_765574, EPI_ISL_765575, EPI_ISL_765576, EPI_ISL_765577, EPI_ISL_765578, EPI_ISL_765579, EPI_ISL_765580, EPI_ISL_765581, EPI_ISL_765582 | Colorado Mesa University | Infectious Disease Program, Broad Institute of Harvard and MIT | Lemieux,J.E., Siddle,K.J., Shaw,B., Adams,G., Pierce,V., Turbett,S., Anahtar,M., Branda,J., Slater,D., Harris,J., Lin,A.E., Gladden-Young,A., Lagerborg,K., Rudy,M., DeRuff,K., Carter,A., Normandin,E., Bauer,M., Reilly,S., Tomkins-Tinch,C., Loreth,C., Chaluvadi,S., Neumann,A., Cusick,C., Chapman,S.B., Gnirke,A., Flowers,K., Cerrato,F., Birren,B.W., Gallagher,G., Smole,S., Park,D.J., MacInnis,B.L., Ryan,E., LaRocque,R., Rosenberg,E. and Sabeti,P.C. |
| EPI_ISL_765583, EPI_ISL_765584, EPI_ISL_765585, EPI_ISL_765586, EPI_ISL_765587, EPI_ISL_765588, EPI_ISL_765589, EPI_ISL_765590, EPI_ISL_765591, EPI_ISL_765592, EPI_ISL_765593, EPI_ISL_765594, EPI_ISL_765595, EPI_ISL_765596, EPI_ISL_765597, EPI_ISL_765598, EPI_ISL_765599, EPI_ISL_765600, EPI_ISL_765601, EPI_ISL_765602, EPI_ISL_765603, EPI_ISL_765604, EPI_ISL_765605, EPI_ISL_765606, EPI_ISL_765607, EPI_ISL_765608, EPI_ISL_765609, EPI_ISL_765610, EPI_ISL_765611, EPI_ISL_765612 | | | |
| see above | Brigham and Womens Hospital | Infectious Disease Program, Broad Institute of Harvard and MIT | Lemieux,J.E., Siddle,K.J., Shaw,B., Adams,G., Pierce,V., Turbett,S., Anahtar,M., Branda,J., Slater,D., Harris,J., Lin,A.E., Gladden-Young,A., Lagerborg,K., Rudy,M., DeRuff,K., Carter,A., Normandin,E., Bauer,M., Reilly,S., Tomkins-Tinch,C., Loreth,C., Chaluvadi,S., Neumann,A., Cusick,C., Chapman,S.B., Gnirke,A., Flowers,K., Cerrato,F., Birren,B.W., Gallagher,G., Smole,S., Park,D.J., MacInnis,B.L., Ryan,E., LaRocque,R., Rosenberg,E. and Sabeti,P.C. |
| EPI_ISL_765613, EPI_ISL_765614, EPI_ISL_765615, EPI_ISL_765616, EPI_ISL_765617, EPI_ISL_765618, EPI_ISL_765619, EPI_ISL_765620, EPI_ISL_765621, EPI_ISL_765622, EPI_ISL_765623, EPI_ISL_765624, EPI_ISL_765625, EPI_ISL_765626, EPI_ISL_765627, EPI_ISL_765628, EPI_ISL_765629, EPI_ISL_765630, EPI_ISL_765631, EPI_ISL_765632, EPI_ISL_765633, EPI_ISL_765634, EPI_ISL_765635, EPI_ISL_765636, EPI_ISL_765637, EPI_ISL_765638, EPI_ISL_765639, EPI_ISL_765640, EPI_ISL_765641, EPI_ISL_765642, EPI_ISL_765643, EPI_ISL_765644, EPI_ISL_765645, EPI_ISL_765646, EPI_ISL_765647, EPI_ISL_765648, EPI_ISL_765649, EPI_ISL_765650, EPI_ISL_765651, EPI_ISL_765652, EPI_ISL_765653, EPI_ISL_765654, EPI_ISL_765655, EPI_ISL_765656, EPI_ISL_765657, EPI_ISL_765658, EPI_ISL_765659, EPI_ISL_765660, EPI_ISL_765661, EPI_ISL_765662, EPI_ISL_765663, EPI_ISL_765664, EPI_ISL_765665, EPI_ISL_765666, EPI_ISL_765667, EPI_ISL_765668, EPI_ISL_765669, EPI_ISL_765670, EPI_ISL_765671, EPI_ISL_765672, EPI_ISL_765673, EPI_ISL_765674, EPI_ISL_765675, EPI_ISL_765676, EPI_ISL_765677, EPI_ISL_765678, EPI_ISL_765679, EPI_ISL_765680, EPI_ISL_765681, EPI_ISL_765682, EPI_ISL_765683, EPI_ISL_765684, EPI_ISL_765685, EPI_ISL_765686, EPI_ISL_765687, EPI_ISL_765688, EPI_ISL_765689, EPI_ISL_765690, EPI_ISL_765691, EPI_ISL_765692, EPI_ISL_765693, EPI_ISL_765694, EPI_ISL_765695, EPI_ISL_765696, EPI_ISL_765697, EPI_ISL_765698, EPI_ISL_765699, EPI_ISL_765700, EPI_ISL_765701, EPI_ISL_765702, EPI_ISL_765703, EPI_ISL_765704, EPI_ISL_765705, EPI_ISL_765706, EPI_ISL_765707, EPI_ISL_765708, EPI_ISL_765709, EPI_ISL_765710, EPI_ISL_765711, EPI_ISL_765712, EPI_ISL_765713, EPI_ISL_765714, EPI_ISL_765715, EPI_ISL_765716, EPI_ISL_765717, EPI_ISL_765718, EPI_ISL_765719, EPI_ISL_765720, EPI_ISL_765721, EPI_ISL_765722, EPI_ISL_765723, EPI_ISL_765724, EPI_ISL_765725, EPI_ISL_765726, EPI_ISL_765727, EPI_ISL_765728, EPI_ISL_765729, EPI_ISL_765730, EPI_ISL_765731, EPI_ISL_765732, EPI_ISL_765733, EPI_ISL_765734, EPI_ISL_765735, EPI_ISL_765736, EPI_ISL_765737, EPI_ISL_765738, EPI_ISL_765739, EPI_ISL_765740, EPI_ISL_765741, EPI_ISL_765742, EPI_ISL_765743, EPI_ISL_765744, EPI_ISL_765745, EPI_ISL_765746, EPI_ISL_765747, EPI_ISL_765748, EPI_ISL_765749, EPI_ISL_765750, EPI_ISL_765751, EPI_ISL_765752, EPI_ISL_765753, EPI_ISL_765754, EPI_ISL_765755, EPI_ISL_765756, EPI_ISL_765757, EPI_ISL_765758, EPI_ISL_765759, EPI_ISL_765760, EPI_ISL_765761, EPI_ISL_765762, EPI_ISL_765763, EPI_ISL_765764, EPI_ISL_765765, EPI_ISL_765766, EPI_ISL_765767, EPI_ISL_765768, EPI_ISL_765769, EPI_ISL_765770, EPI_ISL_765771, EPI_ISL_765772, EPI_ISL_765773, EPI_ISL_765774, EPI_ISL_765775, EPI_ISL_765776, EPI_ISL_765777, EPI_ISL_765778, EPI_ISL_765779, EPI_ISL_765780, EPI_ISL_765781, EPI_ISL_765782, EPI_ISL_765783, EPI_ISL_765784, EPI_ISL_765785, EPI_ISL_765786, EPI_ISL_765787, EPI_ISL_765788, EPI_ISL_765789, EPI_ISL_765790, EPI_ISL_765791, EPI_ISL_765792, EPI_ISL_765793, EPI_ISL_765794, EPI_ISL_765795, EPI_ISL_765796, EPI_ISL_765797, EPI_ISL_765798, EPI_ISL_765799, EPI_ISL_765800, EPI_ISL_765801, EPI_ISL_765802, EPI_ISL_765803, EPI_ISL_765804, EPI_ISL_765805, EPI_ISL_765806, EPI_ISL_765807, EPI_ISL_765808, EPI_ISL_765809, EPI_ISL_765810, EPI_ISL_765811, EPI_ISL_765812, EPI_ISL_765813, EPI_ISL_765814, EPI_ISL_765815, EPI_ISL_765816, EPI_ISL_765817 | | | |
| see above | Massachusetts General Hospital | Infectious Disease Program, Broad Institute of Harvard and MIT | Lemieux,J.E., Siddle,K.J., Shaw,B., Adams,G., Pierce,V., Turbett,S., Anahtar,M., Branda,J., Slater,D., Harris,J., Lin,A.E., Gladden-Young,A., Lagerborg,K., Rudy,M., DeRuff,K., Carter,A., Normandin,E., Bauer,M., Reilly,S., Tomkins-Tinch,C., Loreth,C., Chaluvadi,S., Neumann,A., Cusick,C., Chapman,S.B., Gnirke,A., Flowers,K., Cerrato,F., Birren,B.W., Gallagher,G., Smole,S., Park,D.J., MacInnis,B.L., Ryan,E., LaRocque,R., Rosenberg,E. and Sabeti,P.C. |
| EPI_ISL_765818, EPI_ISL_765819, EPI_ISL_765820, EPI_ISL_765821, EPI_ISL_765822, EPI_ISL_765823 | North Shore Medical Center | Infectious Disease Program, Broad Institute of Harvard and MIT | Lemieux,J.E., Siddle,K.J., Shaw,B., Adams,G., Pierce,V., Turbett,S., Anahtar,M., Branda,J., Slater,D., Harris,J., Lin,A.E., Gladden-Young,A., Lagerborg,K., Rudy,M., DeRuff,K., Carter,A., Normandin,E., Bauer,M., Reilly,S., Tomkins-Tinch,C., Loreth,C., Chaluvadi,S., Neumann,A., Cusick,C., Chapman,S.B., Gnirke,A., Flowers,K., Cerrato,F., Birren,B.W., Gallagher,G., Smole,S., Park,D.J., MacInnis,B.L., Ryan,E., LaRocque,R., Rosenberg,E. and Sabeti,P.C. |
| EPI_ISL_765824, EPI_ISL_765825, EPI_ISL_765826, EPI_ISL_765827, EPI_ISL_765828, EPI_ISL_765829, EPI_ISL_765830, EPI_ISL_765831, EPI_ISL_765832, EPI_ISL_765833, EPI_ISL_765834, EPI_ISL_765835, EPI_ISL_765836, EPI_ISL_765837, EPI_ISL_765838, EPI_ISL_765839, EPI_ISL_765840, EPI_ISL_765841, EPI_ISL_765842, EPI_ISL_765843, EPI_ISL_765844, EPI_ISL_765845, EPI_ISL_765846, EPI_ISL_765847, EPI_ISL_765848, EPI_ISL_765849, EPI_ISL_765850, EPI_ISL_765851, EPI_ISL_765852, EPI_ISL_765853, EPI_ISL_765854, EPI_ISL_765855, EPI_ISL_765856, EPI_ISL_765857, EPI_ISL_765858, EPI_ISL_765859, EPI_ISL_765860, EPI_ISL_765861, EPI_ISL_765862, EPI_ISL_765863, EPI_ISL_765864, EPI_ISL_765865, EPI_ISL_765866, EPI_ISL_765867, EPI_ISL_765868, EPI_ISL_765869, EPI_ISL_765870, EPI_ISL_765871, EPI_ISL_765872, EPI_ISL_765873, EPI_ISL_765874, EPI_ISL_765875, EPI_ISL_765876, EPI_ISL_765877, EPI_ISL_765878, EPI_ISL_765879, EPI_ISL_765880, EPI_ISL_765881, EPI_ISL_765882, EPI_ISL_765883, EPI_ISL_765884, EPI_ISL_765885, EPI_ISL_765886, EPI_ISL_765887, EPI_ISL_765888, EPI_ISL_765889, EPI_ISL_765890, EPI_ISL_765891, EPI_ISL_765892 | | | |
| see above | Massachusetts General Hospital | Infectious Disease Program, Broad Institute of Harvard and MIT | Lemieux,J.E., Siddle,K.J., Shaw,B., Adams,G., Pierce,V., Turbett,S., Anahtar,M., Branda,J., Slater,D., Harris,J., Lin,A.E., Gladden-Young,A., Lagerborg,K., Rudy,M., DeRuff,K., Carter,A., Normandin,E., Bauer,M., Reilly,S., Tomkins-Tinch,C., Loreth,C., Chaluvadi,S., Neumann,A., Cusick,C., Chapman,S.B., Gnirke,A., Flowers,K., Cerrato,F., Birren,B.W., Gallagher,G., Smole,S., Park,D.J., MacInnis,B.L., Ryan,E., LaRocque,R., Rosenberg,E. and Sabeti,P.C. |
| EPI_ISL_765896, EPI_ISL_765897, EPI_ISL_765898, EPI_ISL_765899, EPI_ISL_765900, EPI_ISL_765901, EPI_ISL_765902, EPI_ISL_765903, EPI_ISL_765904, EPI_ISL_765905, EPI_ISL_765906, EPI_ISL_765907, EPI_ISL_765908, EPI_ISL_765909, EPI_ISL_765910, EPI_ISL_765911, EPI_ISL_765912, EPI_ISL_765913, EPI_ISL_765914, EPI_ISL_765915, EPI_ISL_765916, EPI_ISL_765917, EPI_ISL_765918, EPI_ISL_765919, EPI_ISL_765920, EPI_ISL_765921, EPI_ISL_765922, EPI_ISL_765923, EPI_ISL_765924, EPI_ISL_765925, EPI_ISL_765926, EPI_ISL_765934, EPI_ISL_765935, EPI_ISL_765936, EPI_ISL_765937, EPI_ISL_765938, EPI_ISL_765939, EPI_ISL_765940, EPI_ISL_765941 | | | |
| see above | TXDSHS | TXDSHS | Rashmi Tuladhar, Bonnie Oh, Jenny Zhang, Maliha Rahman, Anita Pokharel, Myong Koag, Chung Wang, Rachel Lee, Grace Kubin, Mayela Pedrueza, James Daniel Bonser |
| EPI_ISL_765942, EPI_ISL_765944, EPI_ISL_765945, EPI_ISL_765946, EPI_ISL_765947, EPI_ISL_765948, EPI_ISL_765949, EPI_ISL_765950, EPI_ISL_765951, EPI_ISL_765952, EPI_ISL_765953, EPI_ISL_765954, EPI_ISL_765955, EPI_ISL_765956, EPI_ISL_765957, EPI_ISL_765959 | | | |
| see above | Worobey Lab, Department of Ecology and Evolutionary Biology, University of Arizona | Worobey Lab, Department of Ecology and Evolutionary Biology, University of Arizona | Brendan Larsen, Grace Quirk, Thomas Watts, David Baltrus, Michael Worobey |
| EPI_ISL_765960, EPI_ISL_765961, EPI_ISL_765962, EPI_ISL_765963, EPI_ISL_765964, EPI_ISL_765965, EPI_ISL_765966, EPI_ISL_765967, EPI_ISL_765968, EPI_ISL_765969, EPI_ISL_765970, EPI_ISL_765971, EPI_ISL_765972, EPI_ISL_765973, EPI_ISL_765974, EPI_ISL_765975, EPI_ISL_765976, EPI_ISL_765977, EPI_ISL_765978, EPI_ISL_765979, EPI_ISL_765980, EPI_ISL_765981, EPI_ISL_765982, EPI_ISL_765983, EPI_ISL_765984, EPI_ISL_765985 | | | |
| see above | Wyoming Public Health Laboratory | Wyoming Public Health Laboratory | Noah Hull, Taylor Fearing, Channing Weber, Ashley Norberg, Bailey Bowcutt, and Wanda Manley |
| EPI_ISL_765986 | Massachusetts General Hospital | Infectious Disease Program, Broad Institute | Lemieux,J.E., Siddle,K.J., Shaw,B., Adams,G., Pierce,V., Turbett,S., Anahtar,M., Branda,J., Slater,D., Harris,J., Lin,A.E., Gladden-Young,A., Lagerborg,K., Rudy,M., DeRuff,K., Carter,A., Normandin,E., Bauer,M., Reilly,S., Tomkins-Tinch,C., Loreth,C., Chaluvadi,S., Neumann,A., Cusick,C., |

| | | | |
|---|---|---|---|
| | | of Harvard and MIT | Chapman,S.B., Gnirke,A., Flowers,K., Cerrato,F., Birren,B.W., Gallagher,G., Smole,S., Park,D.J., MacInnis,B.L., Ryan,E., LaRocque,R., Rosenberg,E. and Sabeti,P.C. |
| EPI_ISL_765987, EPI_ISL_765988, EPI_ISL_765989, EPI_ISL_765990, EPI_ISL_765991, EPI_ISL_765992, EPI_ISL_765993 | UCLA Clinical Micro Lab | Los Angeles County Public Health Laboratories | P. Hemarajata et al. |
| EPI_ISL_765994, EPI_ISL_765995, EPI_ISL_765996, EPI_ISL_765997 | USC Clinical Lab | Los Angeles County Public Health Laboratories | P. Hemarajata et al. |
| EPI_ISL_765998, EPI_ISL_765999, EPI_ISL_766000, EPI_ISL_766001, EPI_ISL_766002, EPI_ISL_766003, EPI_ISL_766004, EPI_ISL_766005, EPI_ISL_766006, EPI_ISL_766007, EPI_ISL_766008, EPI_ISL_766009, EPI_ISL_766010, EPI_ISL_766011, EPI_ISL_766012, EPI_ISL_766013, EPI_ISL_766014 | | | |
| see above | Los Angeles County Public Health Laboratories | Los Angeles County Public Health Laboratories | P. Hemarajata et al. |
| EPI_ISL_766015 | USC Clinical Lab | Los Angeles County Public Health Laboratories | P. Hemarajata el al. |
| EPI_ISL_766646, EPI_ISL_766647, EPI_ISL_766648, EPI_ISL_766649, EPI_ISL_766650, EPI_ISL_766651, EPI_ISL_766652, EPI_ISL_766653, EPI_ISL_766654, EPI_ISL_766655, EPI_ISL_766656, EPI_ISL_766657, EPI_ISL_766658, EPI_ISL_766659, EPI_ISL_766660, EPI_ISL_766661, EPI_ISL_766662, EPI_ISL_766663, EPI_ISL_766664, EPI_ISL_766665, EPI_ISL_766666, EPI_ISL_766667, EPI_ISL_766668, EPI_ISL_766669, EPI_ISL_766670, EPI_ISL_766671, EPI_ISL_766686, EPI_ISL_766687, EPI_ISL_766688, EPI_ISL_766689, EPI_ISL_766690, EPI_ISL_766691, EPI_ISL_766731, EPI_ISL_766732, EPI_ISL_766733, EPI_ISL_766734, EPI_ISL_766735, EPI_ISL_766736, EPI_ISL_766737, EPI_ISL_766738, EPI_ISL_766739, EPI_ISL_766740, EPI_ISL_766741, EPI_ISL_766742, EPI_ISL_766743, EPI_ISL_766744, EPI_ISL_766745, EPI_ISL_766746, EPI_ISL_766747, EPI_ISL_766748, EPI_ISL_766749, EPI_ISL_766750, EPI_ISL_766751, EPI_ISL_766752, EPI_ISL_766753, EPI_ISL_766754, EPI_ISL_766762, EPI_ISL_766763, EPI_ISL_766764, EPI_ISL_766765, EPI_ISL_766766, EPI_ISL_766767, EPI_ISL_766768, EPI_ISL_766769, EPI_ISL_766770, EPI_ISL_766771, EPI_ISL_766772, EPI_ISL_766773 | | | |
| see above | Texas Department of State Health Services | Texas Department of State Health Services | Rashmi Tuladhar, Bonnie Oh, Jenny Zhang, Maliha Rahman, Anita Pokharel, Myong Koag, Chung Wang, Rachel Lee, Grace Kubin, Mayela Pedrueza, James Daniel Bonser |
| EPI_ISL_766775, EPI_ISL_766776, EPI_ISL_766777, EPI_ISL_766778, EPI_ISL_766779, EPI_ISL_766780, EPI_ISL_766781, EPI_ISL_766782, EPI_ISL_766783, EPI_ISL_766784, EPI_ISL_766785, EPI_ISL_766786, EPI_ISL_766787, EPI_ISL_766788, EPI_ISL_766790 | | | |
| see above | Wyoming Public Health Laboratory | Wyoming Public Health Laboratory | Noah Hull, Taylor Fearing, Channing Weber, Ashley Norberg, Bailey Bowcutt, and Wanda Manley |
| EPI_ISL_766877, EPI_ISL_766878, EPI_ISL_766879, EPI_ISL_766880, EPI_ISL_766881, EPI_ISL_766882 | Wyoming Public Health Laboratory | Wyoming Public Health Laboratory | Noah Hull, Taylor Fearing, Lynette Gumbleton, Channing Weber, Ashley Norberg, Bailey Bowcutt, and Wanda Manley |
| EPI_ISL_766883, EPI_ISL_766884, EPI_ISL_766885, EPI_ISL_766886, EPI_ISL_766887, EPI_ISL_766888, EPI_ISL_766889, EPI_ISL_766890, EPI_ISL_766891, EPI_ISL_766892, EPI_ISL_766893, EPI_ISL_766894, EPI_ISL_766895, EPI_ISL_766896, EPI_ISL_766898, EPI_ISL_766899, EPI_ISL_766900, EPI_ISL_766901, EPI_ISL_766902, EPI_ISL_766903, EPI_ISL_766904, EPI_ISL_766905, EPI_ISL_766906, EPI_ISL_766907, EPI_ISL_766908, EPI_ISL_766909, EPI_ISL_766910, EPI_ISL_766911, EPI_ISL_766912, EPI_ISL_766913, EPI_ISL_766914, EPI_ISL_766915, EPI_ISL_766916, EPI_ISL_766917, EPI_ISL_766918, EPI_ISL_766919, EPI_ISL_766920, EPI_ISL_766921, EPI_ISL_766922, EPI_ISL_766923 | | | |
| see above | New Mexico Department of Health Scientific Laboratory | New Mexico Department of Health Scientific Laboratory | D'eldra Malone, Ellie Johnson, Anastacia Griego-Fisher |
| EPI_ISL_766938 | Nevada State Health Lab | Nevada State Health Lab | Andrew Gorzalski |
| EPI_ISL_766971, EPI_ISL_766972, EPI_ISL_766973, EPI_ISL_766974, EPI_ISL_766975, EPI_ISL_766976, EPI_ISL_766977, EPI_ISL_766978, EPI_ISL_766979, EPI_ISL_766980, EPI_ISL_766981, EPI_ISL_766982, EPI_ISL_766983, EPI_ISL_766984, EPI_ISL_766985, EPI_ISL_766986, EPI_ISL_766987, EPI_ISL_766989, EPI_ISL_766990, EPI_ISL_766991, EPI_ISL_766992, EPI_ISL_766993, EPI_ISL_766994, EPI_ISL_766995, EPI_ISL_766996, EPI_ISL_766997, EPI_ISL_766998, EPI_ISL_766999, EPI_ISL_767000, EPI_ISL_767001, EPI_ISL_767002, EPI_ISL_767003, EPI_ISL_767004, EPI_ISL_767005, EPI_ISL_767006, EPI_ISL_767007, EPI_ISL_767008, EPI_ISL_767009, EPI_ISL_767010, EPI_ISL_767011, EPI_ISL_767012 | | | |
| see above | Delaware Public Health Laboratory | Delaware Public Health Laboratory | Gregory Hovan |
| EPI_ISL_767044, EPI_ISL_767045, EPI_ISL_767046, EPI_ISL_767047, EPI_ISL_767048, EPI_ISL_767049, EPI_ISL_767050, EPI_ISL_767051, EPI_ISL_767052, EPI_ISL_767053, EPI_ISL_767054, EPI_ISL_767055, EPI_ISL_767056, EPI_ISL_767057, EPI_ISL_767058, EPI_ISL_767059, EPI_ISL_767060, EPI_ISL_767061, EPI_ISL_767062, EPI_ISL_767063, EPI_ISL_767064, EPI_ISL_767065, EPI_ISL_767066, EPI_ISL_767067, EPI_ISL_767068, EPI_ISL_767069, EPI_ISL_767070, EPI_ISL_767071, EPI_ISL_767072, EPI_ISL_767073, EPI_ISL_767074, EPI_ISL_767075, EPI_ISL_767076, EPI_ISL_767077, EPI_ISL_767078, EPI_ISL_767079, EPI_ISL_767080, EPI_ISL_767081, EPI_ISL_767082, EPI_ISL_767083, EPI_ISL_767084, EPI_ISL_767085, EPI_ISL_767086, EPI_ISL_767087, EPI_ISL_767088 | | | |
| see above | New Mexico Department of Health Scientific Laboratory | New Mexico Department of Health Scientific Laboratory | D'eldra Malone, Ellie Johnson, Anastacia Griego-Fisher |
| EPI_ISL_767347, EPI_ISL_767348, EPI_ISL_767349, EPI_ISL_767350, EPI_ISL_767351, EPI_ISL_767352, EPI_ISL_767353, EPI_ISL_767354, EPI_ISL_767355, EPI_ISL_767356, EPI_ISL_767357, EPI_ISL_767358, EPI_ISL_767359, EPI_ISL_767360, EPI_ISL_767361 | | | |
| see above | Michigan Department of Health and Human Services, Bureau of Laboratories | Michigan Department of Health and Human Services, Bureau of Laboratories | Blankenship HM, Riner D, Soehnlen MK |
| EPI_ISL_767404, EPI_ISL_767405, EPI_ISL_767407, EPI_ISL_767408 | Ochsner Health | BioInfo Experts, LLC | Rebecca Rose, Amy Feehan, David J. Nolan, Sissy Cross, David Moraga Amador, Tong Yang, Luke Caruso, Wayra Navia, Lydia Von Borstel, Xiao Hui Zhou, Julia-Garcia-Diaz, Susanna L. Lamers |
| EPI_ISL_767416 | Florida Bureau of Public Health Laboratories | Florida Bureau of Public Health Laboratories | Sarah Schmedes, Jason Blanton |
| EPI_ISL_767417 | URMC LABS | Wadsworth Center, New York State Department.of Health | Kirsten St. George, Daryl M. Lamson, Alexis Russel, Matthew Shudt, Melissa A Leisner, Jonathan Plitnick, Navjot Singh, John Kelly, Sara Griesemer, Erasmus Schneider, Erica Lasek-Nesselquist |
| EPI_ISL_767418 | Wadsworth Center, New York State Department.of Health | Wadsworth Center, New York State Department.of Health | Kirsten St. George, Daryl M. Lamson, Alexis Russel, Matthew Shudt, Melissa A Leisner, Jonathan Plitnick, Navjot Singh, John Kelly, Sara Griesemer, Erasmus Schneider, Erica Lasek-Nesselquist |
| EPI_ISL_767419 | URMC LABS | Wadsworth Center, New York State Department.of Health | Kirsten St. George, Daryl M. Lamson, Alexis Russel, Matthew Shudt, Melissa A Leisner, Jonathan Plitnick, Navjot Singh, John Kelly, Sara Griesemer, Erasmus Schneider, Erica Lasek-Nesselquist |
| EPI_ISL_767420, EPI_ISL_767421, EPI_ISL_767422 | Wadsworth Center, New York State Department.of Health | Wadsworth Center, New York State Department.of Health | Kirsten St. George, Daryl M. Lamson, Alexis Russel, Matthew Shudt, Melissa A Leisner, Jonathan Plitnick, Navjot Singh, John Kelly, Sara Griesemer, Erasmus Schneider, Erica Lasek-Nesselquist |
| EPI_ISL_767423, EPI_ISL_767424 | URMC LABS | Wadsworth Center, New York State Department.of Health | Kirsten St. George, Daryl M. Lamson, Alexis Russel, Matthew Shudt, Melissa A Leisner, Jonathan Plitnick, Navjot Singh, John Kelly, Sara Griesemer, Erasmus Schneider, Erica Lasek-Nesselquist |
| EPI_ISL_767425, EPI_ISL_767426, EPI_ISL_767427, EPI_ISL_767428, EPI_ISL_767429, EPI_ISL_767430, EPI_ISL_767431, EPI_ISL_767432, EPI_ISL_767433, EPI_ISL_767434, EPI_ISL_767435, EPI_ISL_767436, EPI_ISL_767437, EPI_ISL_767438, EPI_ISL_767439, EPI_ISL_767440, EPI_ISL_767441, EPI_ISL_767442, EPI_ISL_767443, EPI_ISL_767444, EPI_ISL_767445, EPI_ISL_767446, EPI_ISL_767447, EPI_ISL_767448, EPI_ISL_767449, EPI_ISL_767450, EPI_ISL_767451, EPI_ISL_767452, EPI_ISL_767453, EPI_ISL_767454, EPI_ISL_767455, EPI_ISL_767456, EPI_ISL_767457, EPI_ISL_767458, EPI_ISL_767459, EPI_ISL_767460, EPI_ISL_767461, EPI_ISL_767462, EPI_ISL_767463, EPI_ISL_767464, EPI_ISL_767465, EPI_ISL_767466, EPI_ISL_767467, EPI_ISL_767468, EPI_ISL_767469, EPI_ISL_767470, EPI_ISL_767471, EPI_ISL_767472, EPI_ISL_767473, EPI_ISL_767474, EPI_ISL_767475, EPI_ISL_767476, EPI_ISL_767477, EPI_ISL_767478, EPI_ISL_767479, EPI_ISL_767480, EPI_ISL_767481, EPI_ISL_767482, EPI_ISL_767483, EPI_ISL_767484, EPI_ISL_767485, EPI_ISL_767486, EPI_ISL_767487, EPI_ISL_767488, EPI_ISL_767489, EPI_ISL_767490, EPI_ISL_767491, EPI_ISL_767492, EPI_ISL_767493, EPI_ISL_767494, EPI_ISL_767495, EPI_ISL_767496, EPI_ISL_767497, EPI_ISL_767498, EPI_ISL_767499, EPI_ISL_767500, EPI_ISL_767501, EPI_ISL_767502, EPI_ISL_767503, EPI_ISL_767504, EPI_ISL_767505, EPI_ISL_767506, EPI_ISL_767507, EPI_ISL_767508, EPI_ISL_767509, EPI_ISL_767510, EPI_ISL_767511, EPI_ISL_767512, EPI_ISL_767513, EPI_ISL_767514, EPI_ISL_767515, EPI_ISL_767516, EPI_ISL_767517, EPI_ISL_767518, EPI_ISL_767519, EPI_ISL_767520, EPI_ISL_767521, EPI_ISL_767522, EPI_ISL_767523, EPI_ISL_767524, EPI_ISL_767525, EPI_ISL_767526, EPI_ISL_767527, EPI_ISL_767528, EPI_ISL_767529, EPI_ISL_767530, EPI_ISL_767531, EPI_ISL_767532, EPI_ISL_767533, EPI_ISL_767534, EPI_ISL_767535, EPI_ISL_767536, EPI_ISL_767537 | | | |
| see above | Wadsworth Center, New York State Department.of Health | Wadsworth Center, New York State Department.of Health | Kirsten St. George, Daryl M. Lamson, Alexis Russel, Matthew Shudt, Melissa A Leisner, Jonathan Plitnick, Navjot Singh, John Kelly, Sara Griesemer, Erasmus Schneider, Erica Lasek-Nesselquist |
| EPI_ISL_767538, EPI_ISL_767539, EPI_ISL_767540, EPI_ISL_767541, EPI_ISL_767542, EPI_ISL_767543, EPI_ISL_767544, EPI_ISL_767545, EPI_ISL_767546, EPI_ISL_767547, EPI_ISL_767548, EPI_ISL_767549, EPI_ISL_767550, EPI_ISL_767551, EPI_ISL_767552, EPI_ISL_767553, EPI_ISL_767554, EPI_ISL_767555, EPI_ISL_767556, EPI_ISL_767557, EPI_ISL_767558, EPI_ISL_767559, EPI_ISL_767560, EPI_ISL_767561, EPI_ISL_767562, EPI_ISL_767563, EPI_ISL_767564, EPI_ISL_767565, EPI_ISL_767566, EPI_ISL_767567, EPI_ISL_767568, EPI_ISL_767569, EPI_ISL_767570, EPI_ISL_767571, EPI_ISL_767572, EPI_ISL_767573, EPI_ISL_767574, EPI_ISL_767575, EPI_ISL_767576, EPI_ISL_767577 | | | |
| see above | URMC LABS | Wadsworth Center, New York State Department.of Health | Kirsten St. George, Daryl M. Lamson, Alexis Russel, Matthew Shudt, Melissa A Leisner, Jonathan Plitnick, Navjot Singh, John Kelly, Sara Griesemer, Erasmus Schneider, Erica Lasek-Nesselquist |
| EPI_ISL_767578 | Wadsworth Center, New York State Department.of Health | Wadsworth Center, New York State Department.of Health | Kirsten St. George, Daryl M. Lamson, Alexis Russel, Matthew Shudt, Melissa A Leisner, Jonathan Plitnick, Navjot Singh, John Kelly, Sara Griesemer, Erasmus Schneider, Erica Lasek-Nesselquist |
| EPI_ISL_767579 | URMC LABS | Wadsworth Center, New York State Department.of Health | Kirsten St. George, Daryl M. Lamson, Alexis Russel, Matthew Shudt, Melissa A Leisner, Jonathan Plitnick, Navjot Singh, John Kelly, Sara Griesemer, Erasmus Schneider, Erica Lasek-Nesselquist |
| EPI_ISL_767580, EPI_ISL_767581, EPI_ISL_767582 | WHITE PLAINS HOSPITAL CENTER LABORATORY | Wadsworth Center, New York State Department.of Health | Kirsten St. George, Daryl M. Lamson, Alexis Russel, Matthew Shudt, Melissa A Leisner, Jonathan Plitnick, Navjot Singh, John Kelly, Sara Griesemer, Erasmus Schneider, Erica Lasek-Nesselquist |
| EPI_ISL_767583 | MEMORIAL SLOAN KETTERING CANCER CENTER | Wadsworth Center, New York State Department.of Health | Kirsten St. George, Daryl M. Lamson, Alexis Russel, Matthew Shudt, Melissa A Leisner, Jonathan Plitnick, Navjot Singh, John Kelly, Sara Griesemer, Erasmus Schneider, Erica Lasek-Nesselquist |
| EPI_ISL_767584, EPI_ISL_767585, EPI_ISL_767586, EPI_ISL_767587 | WHITE PLAINS HOSPITAL CENTER LABORATORY | Wadsworth Center, New York State Department.of Health | Kirsten St. George, Daryl M. Lamson, Alexis Russel, Matthew Shudt, Melissa A Leisner, Jonathan Plitnick, Navjot Singh, John Kelly, Sara Griesemer, Erasmus Schneider, Erica Lasek-Nesselquist |
| EPI_ISL_767588 | MEMORIAL SLOAN KETTERING CANCER CENTER | Wadsworth Center, New York State Department.of Health | Kirsten St. George, Daryl M. Lamson, Alexis Russel, Matthew Shudt, Melissa A Leisner, Jonathan Plitnick, Navjot Singh, John Kelly, Sara Griesemer, Erasmus Schneider, Erica Lasek-Nesselquist |
| EPI_ISL_767589 | SARATOGA HOSPITAL LABORATORY | Wadsworth Center, New York State Department.of Health | Kirsten St. George, Daryl M. Lamson, Alexis Russel, Matthew Shudt, Melissa A Leisner, Jonathan Plitnick, Navjot Singh, John Kelly, Sara Griesemer, Erasmus Schneider, Erica Lasek-Nesselquist |
| EPI_ISL_767590 | WHITE PLAINS HOSPITAL CENTER LABORATORY | Wadsworth Center, New York State Department.of Health | Kirsten St. George, Daryl M. Lamson, Alexis Russel, Matthew Shudt, Melissa A Leisner, Jonathan Plitnick, Navjot Singh, John Kelly, Sara Griesemer, Erasmus Schneider, Erica Lasek-Nesselquist |
| EPI_ISL_767591, | MEMORIAL SLOAN KETTERING CANCER | Wadsworth Center, New York State | Kirsten St. George, Daryl M. Lamson, Alexis Russel, Matthew Shudt, Melissa A Leisner, Jonathan Plitnick, Navjot Singh, John Kelly, Sara Griesemer, Erasmus Schneider, Erica Lasek-Nesselquist |

| | | | |
|---|---|---|---|
| EPI_ISL_767592, EPI_ISL_767593 | CENTER | Department.of Health | |
| EPI_ISL_767594 | SARATOGA HOSPITAL LABORATORY | Wadsworth Center, New York State Department.of Health | Kirsten St. George, Daryl M. Lamson, Alexis Russel, Matthew Shudt, Melissa A Leisner, Jonathan Plitnick, Navjot Singh, John Kelly, Sara Griesemer, Erasmus Schneider, Erica Lasek-Nesselquist |
| EPI_ISL_767595 | WHITE PLAINS HOSPITAL CENTER LABORATORY | Wadsworth Center, New York State Department.of Health | Kirsten St. George, Daryl M. Lamson, Alexis Russel, Matthew Shudt, Melissa A Leisner, Jonathan Plitnick, Navjot Singh, John Kelly, Sara Griesemer, Erasmus Schneider, Erica Lasek-Nesselquist |
| EPI_ISL_767596, EPI_ISL_767597 | MEMORIAL SLOAN KETTERING CANCER CENTER | Wadsworth Center, New York State Department.of Health | Kirsten St. George, Daryl M. Lamson, Alexis Russel, Matthew Shudt, Melissa A Leisner, Jonathan Plitnick, Navjot Singh, John Kelly, Sara Griesemer, Erasmus Schneider, Erica Lasek-Nesselquist |
| EPI_ISL_767598 | WHITE PLAINS HOSPITAL CENTER LABORATORY | Wadsworth Center, New York State Department.of Health | Kirsten St. George, Daryl M. Lamson, Alexis Russel, Matthew Shudt, Melissa A Leisner, Jonathan Plitnick, Navjot Singh, John Kelly, Sara Griesemer, Erasmus Schneider, Erica Lasek-Nesselquist |
| EPI_ISL_767599, EPI_ISL_767600, EPI_ISL_767601, EPI_ISL_767602, EPI_ISL_767603, EPI_ISL_767604, EPI_ISL_767605, EPI_ISL_767606, EPI_ISL_767607, EPI_ISL_767608, EPI_ISL_767609, EPI_ISL_767610, EPI_ISL_767611, EPI_ISL_767612, EPI_ISL_767613, EPI_ISL_767614 | | | |
| see above | SARATOGA HOSPITAL LABORATORY | Wadsworth Center, New York State Department.of Health | Kirsten St. George, Daryl M. Lamson, Alexis Russel, Matthew Shudt, Melissa A Leisner, Jonathan Plitnick, Navjot Singh, John Kelly, Sara Griesemer, Erasmus Schneider, Erica Lasek-Nesselquist |
| EPI_ISL_767615, EPI_ISL_767616, EPI_ISL_767617, EPI_ISL_767618, EPI_ISL_767619, EPI_ISL_767620, EPI_ISL_767621, EPI_ISL_767622, EPI_ISL_767623, EPI_ISL_767624, EPI_ISL_767625, EPI_ISL_767626, EPI_ISL_767627, EPI_ISL_767628 | | | |
| see above | WHITE PLAINS HOSPITAL CENTER LABORATORY | Wadsworth Center, New York State Department.of Health | Kirsten St. George, Daryl M. Lamson, Alexis Russel, Matthew Shudt, Melissa A Leisner, Jonathan Plitnick, Navjot Singh, John Kelly, Sara Griesemer, Erasmus Schneider, Erica Lasek-Nesselquist |
| EPI_ISL_767629, EPI_ISL_767630, EPI_ISL_767631, EPI_ISL_767632, EPI_ISL_767633, EPI_ISL_767634, EPI_ISL_767635, EPI_ISL_767636, EPI_ISL_767637, EPI_ISL_767638, EPI_ISL_767639, EPI_ISL_767640, EPI_ISL_767641, EPI_ISL_767642, EPI_ISL_767643, EPI_ISL_767644, EPI_ISL_767645, EPI_ISL_767646, EPI_ISL_767647 | | | |
| see above | MEMORIAL SLOAN KETTERING CANCER CENTER | Wadsworth Center, New York State Department.of Health | Kirsten St. George, Daryl M. Lamson, Alexis Russel, Matthew Shudt, Melissa A Leisner, Jonathan Plitnick, Navjot Singh, John Kelly, Sara Griesemer, Erasmus Schneider, Erica Lasek-Nesselquist |
| EPI_ISL_767648 | BIO-REFERENCE LABORATORIES | Wadsworth Center, New York State Department.of Health | Kirsten St. George, Daryl M. Lamson, Alexis Russel, Matthew Shudt, Melissa A Leisner, Jonathan Plitnick, Navjot Singh, John Kelly, Sara Griesemer, Erasmus Schneider, Erica Lasek-Nesselquist |
| EPI_ISL_767649 | MEMORIAL SLOAN KETTERING CANCER CENTER | Wadsworth Center, New York State Department.of Health | Kirsten St. George, Daryl M. Lamson, Alexis Russel, Matthew Shudt, Melissa A Leisner, Jonathan Plitnick, Navjot Singh, John Kelly, Sara Griesemer, Erasmus Schneider, Erica Lasek-Nesselquist |
| EPI_ISL_767650, EPI_ISL_767651, EPI_ISL_767652, EPI_ISL_767653 | BIO-REFERENCE LABORATORIES | Wadsworth Center, New York State Department.of Health | Kirsten St. George, Daryl M. Lamson, Alexis Russel, Matthew Shudt, Melissa A Leisner, Jonathan Plitnick, Navjot Singh, John Kelly, Sara Griesemer, Erasmus Schneider, Erica Lasek-Nesselquist |
| EPI_ISL_767655, EPI_ISL_767656, EPI_ISL_767657, EPI_ISL_767658, EPI_ISL_767659, EPI_ISL_767660, EPI_ISL_767661, EPI_ISL_767662, EPI_ISL_767663, EPI_ISL_767664 | MEMORIAL SLOAN KETTERING CANCER CENTER | Wadsworth Center, New York State Department.of Health | Kirsten St. George, Daryl M. Lamson, Alexis Russel, Matthew Shudt, Melissa A Leisner, Jonathan Plitnick, Navjot Singh, John Kelly, Sara Griesemer, Erasmus Schneider, Erica Lasek-Nesselquist |
| EPI_ISL_767665, EPI_ISL_767666, EPI_ISL_767667, EPI_ISL_767668, EPI_ISL_767669, EPI_ISL_767670, EPI_ISL_767671, EPI_ISL_767672, EPI_ISL_767673, EPI_ISL_767674, EPI_ISL_767675, EPI_ISL_767676, EPI_ISL_767677, EPI_ISL_767678, EPI_ISL_767679, EPI_ISL_767680 | | | |
| see above | BIO-REFERENCE LABORATORIES | Wadsworth Center, New York State Department.of Health | Kirsten St. George, Daryl M. Lamson, Alexis Russel, Matthew Shudt, Melissa A Leisner, Jonathan Plitnick, Navjot Singh, John Kelly, Sara Griesemer, Erasmus Schneider, Erica Lasek-Nesselquist |
| EPI_ISL_767681, EPI_ISL_767682, EPI_ISL_767683, EPI_ISL_767684, EPI_ISL_767685, EPI_ISL_767686, EPI_ISL_767687, EPI_ISL_767688, EPI_ISL_767689, EPI_ISL_767690, EPI_ISL_767691, EPI_ISL_767692, EPI_ISL_767693, EPI_ISL_767694, EPI_ISL_767695, EPI_ISL_767696, EPI_ISL_767697, EPI_ISL_767698, EPI_ISL_767699, EPI_ISL_767700, EPI_ISL_767701, EPI_ISL_767702, EPI_ISL_767703, EPI_ISL_767704, EPI_ISL_767705, EPI_ISL_767706, EPI_ISL_767707, EPI_ISL_767708, EPI_ISL_767709, EPI_ISL_767710, EPI_ISL_767711, EPI_ISL_767712, EPI_ISL_767713, EPI_ISL_767714, EPI_ISL_767715 | | | |
| see above | MEMORIAL SLOAN KETTERING CANCER CENTER | Wadsworth Center, New York State Department.of Health | Kirsten St. George, Daryl M. Lamson, Alexis Russel, Matthew Shudt, Melissa A Leisner, Jonathan Plitnick, Navjot Singh, John Kelly, Sara Griesemer, Erasmus Schneider, Erica Lasek-Nesselquist |
| EPI_ISL_768325, EPI_ISL_768326, EPI_ISL_768327, EPI_ISL_768328, EPI_ISL_768329, EPI_ISL_768330, EPI_ISL_768331, EPI_ISL_768332, EPI_ISL_768333, EPI_ISL_768334, EPI_ISL_768335, EPI_ISL_768336, EPI_ISL_768337, EPI_ISL_768338, EPI_ISL_768339, EPI_ISL_768340, EPI_ISL_768341, EPI_ISL_768342, EPI_ISL_768343, EPI_ISL_768344, EPI_ISL_768345, EPI_ISL_768346, EPI_ISL_768347, EPI_ISL_768348, EPI_ISL_768349, EPI_ISL_768350, EPI_ISL_768351, EPI_ISL_768352, EPI_ISL_768353, EPI_ISL_768354, EPI_ISL_768355, EPI_ISL_768356, EPI_ISL_768357, EPI_ISL_768358, EPI_ISL_768359, EPI_ISL_768360, EPI_ISL_768361, EPI_ISL_768362, EPI_ISL_768363, EPI_ISL_768364, EPI_ISL_768365, EPI_ISL_768366, EPI_ISL_768367, EPI_ISL_768368, EPI_ISL_768369, EPI_ISL_768370, EPI_ISL_768371, EPI_ISL_768372, EPI_ISL_768373, EPI_ISL_768374, EPI_ISL_768375, EPI_ISL_768376, EPI_ISL_768377, EPI_ISL_768378, EPI_ISL_768379, EPI_ISL_768380, EPI_ISL_768381, EPI_ISL_768382, EPI_ISL_768383, EPI_ISL_768384, EPI_ISL_768385, EPI_ISL_768386, EPI_ISL_768387, EPI_ISL_768388, EPI_ISL_768389, EPI_ISL_768390, EPI_ISL_768391, EPI_ISL_768392, EPI_ISL_768393, EPI_ISL_768394, EPI_ISL_768395, EPI_ISL_768396, EPI_ISL_768397, EPI_ISL_768398, EPI_ISL_768399, EPI_ISL_768400, EPI_ISL_768401, EPI_ISL_768402, EPI_ISL_768403, EPI_ISL_768404, EPI_ISL_768405, EPI_ISL_768406, EPI_ISL_768407, EPI_ISL_768408, EPI_ISL_768409, EPI_ISL_768410, EPI_ISL_768411, EPI_ISL_768412, EPI_ISL_768413, EPI_ISL_768414, EPI_ISL_768415, EPI_ISL_768416, EPI_ISL_768417, EPI_ISL_768418, EPI_ISL_768419, EPI_ISL_768420, EPI_ISL_768421, EPI_ISL_768422, EPI_ISL_768423, EPI_ISL_768424, EPI_ISL_768425, EPI_ISL_768426, EPI_ISL_768427, EPI_ISL_768428, EPI_ISL_768429, EPI_ISL_768430, EPI_ISL_768431, EPI_ISL_768432, EPI_ISL_768433, EPI_ISL_768434, EPI_ISL_768435, EPI_ISL_768436, EPI_ISL_768437, EPI_ISL_768438, EPI_ISL_768439, EPI_ISL_768440, EPI_ISL_768441, EPI_ISL_768442, EPI_ISL_768443, EPI_ISL_768444, EPI_ISL_768445, EPI_ISL_768446, EPI_ISL_768447, EPI_ISL_768448, EPI_ISL_768449, EPI_ISL_768450, EPI_ISL_768451, EPI_ISL_768452, EPI_ISL_768453, EPI_ISL_768454, EPI_ISL_768455, EPI_ISL_768456, EPI_ISL_768457, EPI_ISL_768458, EPI_ISL_768459, EPI_ISL_768460, EPI_ISL_768461, EPI_ISL_768462, EPI_ISL_768463, EPI_ISL_768464, EPI_ISL_768465, EPI_ISL_768466, EPI_ISL_768467, EPI_ISL_768468, EPI_ISL_768469, EPI_ISL_768470, EPI_ISL_768471, EPI_ISL_768472, EPI_ISL_768473, EPI_ISL_768474, EPI_ISL_768475, EPI_ISL_768476, EPI_ISL_768477, EPI_ISL_768478, EPI_ISL_768479, EPI_ISL_768480, EPI_ISL_768481, EPI_ISL_768482, EPI_ISL_768483, EPI_ISL_768484, EPI_ISL_768485, EPI_ISL_768486, EPI_ISL_768487, EPI_ISL_768488, EPI_ISL_768489, EPI_ISL_768490, EPI_ISL_768491, EPI_ISL_768492, EPI_ISL_768493, EPI_ISL_768494, EPI_ISL_768495, EPI_ISL_768496 | | | |
| see above | LSUHS Emerging Viral Threat Laboratory | Microbial Genome Sequencing Center | Jeremy P. Kamil, Jennifer L. Carroll, Camille F. Abshire, Maarten Van Diest, Andrew D. Yurochko, Martin J. Sapp, Rona S. Scott, Christopher G. Kevil, Daniel J. Snyder, Vaughn S. Cooper, John A. Vanchiere |
| EPI_ISL_768497, EPI_ISL_768498, EPI_ISL_768499, EPI_ISL_768500, EPI_ISL_768501, EPI_ISL_768502, EPI_ISL_768503, EPI_ISL_768504, EPI_ISL_768505, EPI_ISL_768506, EPI_ISL_768507, EPI_ISL_768508 | | | |
| see above | LSUHS Emerging Viral Threat Laboratory | Microbial Genome Sequencing Center | Jennifer L. Carroll, Jeremy P. Kamil, Camille F. Abshire, Maarten Van Diest, Andrew D. Yurochko, Martin J. Sapp, Rona S. Scott, Christopher G. Kevil, Daniel J. Snyder, Vaughn S. Cooper, John A. Vanchiere |
| EPI_ISL_768509, EPI_ISL_768510, EPI_ISL_768511, EPI_ISL_768512, EPI_ISL_768513, EPI_ISL_768514, EPI_ISL_768515, EPI_ISL_768516, EPI_ISL_768517, EPI_ISL_768518, EPI_ISL_768519, EPI_ISL_768520 | | | |
| see above | LSUHS Emerging Viral Threat Laboratory | Microbial Genome Sequencing Center | Maarten Van Diest, Jeremy P. Kamil, Jennifer L. Carroll, Camille F. Abshire, Andrew D. Yurochko, Martin J. Sapp, Rona S. Scott, Christopher G. Kevil, Daniel J. Snyder, Vaughn S. Cooper, John A. Vanchiere |
| EPI_ISL_768525 | LSUHS Emerging Viral Threat Laboratory | Microbial Genome Sequencing Center | Jeremy P. Kamil, Jennifer L. Carroll, Camille F. Abshire, Maarten Van Diest, Andrew D. Yurochko, Martin J. Sapp, Rona S. Scott, Christopher G. Kevil, Daniel J. Snyder, Vaughn S. Cooper, John A. Vanchiere |
| EPI_ISL_769879, EPI_ISL_769880, EPI_ISL_769881, EPI_ISL_769882, EPI_ISL_769883, EPI_ISL_769884, EPI_ISL_769885, EPI_ISL_769886, EPI_ISL_769887, EPI_ISL_769888, EPI_ISL_769889, EPI_ISL_769890, EPI_ISL_769891, EPI_ISL_769892, EPI_ISL_769893, EPI_ISL_769894, EPI_ISL_769895, EPI_ISL_769896, EPI_ISL_769897, EPI_ISL_769898, EPI_ISL_769899, EPI_ISL_769900, EPI_ISL_769901, EPI_ISL_769902, EPI_ISL_769903, EPI_ISL_769904, EPI_ISL_769905, EPI_ISL_769906, EPI_ISL_769907, EPI_ISL_769908, EPI_ISL_769909, EPI_ISL_769910, EPI_ISL_769911, EPI_ISL_769912, EPI_ISL_769913, EPI_ISL_769914, EPI_ISL_769915 | | | |
| see above | Wyoming Public Health Laboratory | Wyoming Public Health Laboratory | Noah Hull, Taylor Fearing, Lynette Gumbleton, Channing Weber, Ashley Norberg, Bailey Bowcutt, and Wanda Manley |
| EPI_ISL_769916, EPI_ISL_769917, EPI_ISL_769918, EPI_ISL_769919, EPI_ISL_769920, EPI_ISL_769921, EPI_ISL_769922, EPI_ISL_769923, EPI_ISL_769924, EPI_ISL_769925, EPI_ISL_769926, EPI_ISL_769927, EPI_ISL_769928, EPI_ISL_769929, EPI_ISL_769930, EPI_ISL_769931, EPI_ISL_769932, EPI_ISL_769933, EPI_ISL_769934, EPI_ISL_769935, EPI_ISL_769936, EPI_ISL_769937, EPI_ISL_769938, EPI_ISL_769939, EPI_ISL_769940, EPI_ISL_769941, EPI_ISL_769942, EPI_ISL_769943, EPI_ISL_769944, EPI_ISL_769945, EPI_ISL_769946, EPI_ISL_769947, EPI_ISL_769948, EPI_ISL_769949, EPI_ISL_769950, EPI_ISL_769951, EPI_ISL_769952, EPI_ISL_769953, EPI_ISL_769954, EPI_ISL_769955, EPI_ISL_769956, EPI_ISL_769957, EPI_ISL_769958, EPI_ISL_769959, EPI_ISL_769960, EPI_ISL_769961, EPI_ISL_769962, EPI_ISL_769963, EPI_ISL_769964, EPI_ISL_769965, EPI_ISL_769966, EPI_ISL_769967, EPI_ISL_769968, EPI_ISL_769969, EPI_ISL_769970, EPI_ISL_769971, EPI_ISL_769972, EPI_ISL_769973, EPI_ISL_769974, EPI_ISL_769975, EPI_ISL_769976, EPI_ISL_769977, EPI_ISL_769978, EPI_ISL_769979, EPI_ISL_769980, EPI_ISL_769981, EPI_ISL_769982, EPI_ISL_769983, EPI_ISL_769984, EPI_ISL_769985 | | | |
| see above | Albany Medical Center Hospital Clinical Laboratories | Wadsworth Center, New York State Department.of Health | Kirsten St. George, Daryl M. Lamson, Alexis Russel, Matthew Shudt, Melissa A Leisner, Jonathan Plitnick, Navjot Singh, John Kelly, Sara Griesemer, Erasmus Schneider, Erica Lasek-Nesselquist |
| EPI_ISL_770063, EPI_ISL_770064, EPI_ISL_770065, EPI_ISL_770066, EPI_ISL_770067, EPI_ISL_770069, EPI_ISL_770070, EPI_ISL_770071, EPI_ISL_770072, EPI_ISL_770073, EPI_ISL_770074, EPI_ISL_770075, EPI_ISL_770076, EPI_ISL_770077, EPI_ISL_770078, EPI_ISL_770079, EPI_ISL_770080, EPI_ISL_770081, EPI_ISL_770082, EPI_ISL_770083, EPI_ISL_770084, EPI_ISL_770085, EPI_ISL_770086, EPI_ISL_770087, EPI_ISL_770088, EPI_ISL_770089, EPI_ISL_770090, EPI_ISL_770091, EPI_ISL_770092, EPI_ISL_770093, EPI_ISL_770094, EPI_ISL_770095, EPI_ISL_770096, EPI_ISL_770097, EPI_ISL_770098, EPI_ISL_770099, EPI_ISL_770100, EPI_ISL_770101, EPI_ISL_770102, EPI_ISL_770103, EPI_ISL_770104, EPI_ISL_770105, EPI_ISL_770106, EPI_ISL_770107, EPI_ISL_770108, EPI_ISL_770109, EPI_ISL_770110, EPI_ISL_770111, EPI_ISL_770112, EPI_ISL_770113, EPI_ISL_770114, EPI_ISL_770115, EPI_ISL_770116, EPI_ISL_770117, EPI_ISL_770118, EPI_ISL_770119, EPI_ISL_770120, EPI_ISL_770121, EPI_ISL_770124, EPI_ISL_770127, EPI_ISL_770128, EPI_ISL_770444, EPI_ISL_770445, EPI_ISL_770447, EPI_ISL_770448, EPI_ISL_770449, EPI_ISL_770451, EPI_ISL_770452, EPI_ISL_770453, EPI_ISL_770454, EPI_ISL_770455, EPI_ISL_770456, EPI_ISL_770457, EPI_ISL_770458, EPI_ISL_770459, EPI_ISL_770460, EPI_ISL_770461, EPI_ISL_770476, EPI_ISL_770477, EPI_ISL_770478, EPI_ISL_770479, EPI_ISL_770480, EPI_ISL_770481, EPI_ISL_770482, EPI_ISL_770483, EPI_ISL_770484, EPI_ISL_770485, EPI_ISL_770486, EPI_ISL_770487, EPI_ISL_770488, EPI_ISL_770489, EPI_ISL_770490, EPI_ISL_770491, EPI_ISL_770492, EPI_ISL_770493, EPI_ISL_770494, EPI_ISL_770495, EPI_ISL_770496, EPI_ISL_770497 | | | |
| see above | Wyoming Public Health Laboratory | Wyoming Public Health Laboratory | Noah Hull, Taylor Fearing, Lynette Gumbleton, Channing Weber, Ashley Norberg, Bailey Bowcutt, and Wanda Manley |
| EPI_ISL_770632, EPI_ISL_770775, EPI_ISL_770776, EPI_ISL_770777 | Ohio Department of Health Laboratory | Ohio Department of Health Laboratory | Holmes, Jennifer; Eric Brandt, Keoni Omura, Glen McGillivary, Caitiln McDonnell, Kirtana Ramadugu, Erica Leasure, Kelsey Florek, Heather Blankenship, Quanta Brown, and Tammy Bannerman |
| EPI_ISL_770778, EPI_ISL_770779, EPI_ISL_770780, EPI_ISL_770781, EPI_ISL_770782, EPI_ISL_770783, EPI_ISL_770784, EPI_ISL_770785, EPI_ISL_770786, EPI_ISL_770787, EPI_ISL_770788, EPI_ISL_770789, EPI_ISL_770790, EPI_ISL_770791, EPI_ISL_770792 | | | |
| see above | Minnesota Department of Health, Public Health Laboratory | Minnesota Department of Health, Public Health Laboratory | Alexandra Lorentz, Jacob Garfin, Matt Plumb, and Xiong Wang |
| EPI_ISL_770793, EPI_ISL_770794 | Hennepin County Medical Center | Minnesota Department of Health, Public Health Laboratory | Alexandra Lorentz, Jacob Garfin, Matt Plumb, and Xiong Wang |
| EPI_ISL_770795, EPI_ISL_770796 | Minnesota Department of Health, Public Health Laboratory | Minnesota Department of Health, Public Health Laboratory | Alexandra Lorentz, Jacob Garfin, Matt Plumb, and Xiong Wang |
| EPI_ISL_770797 | Hennepin County Medical Center | Minnesota Department of Health, Public Health Laboratory | Alexandra Lorentz, Jacob Garfin, Matt Plumb, and Xiong Wang |
| EPI_ISL_770798, EPI_ISL_770799 | Minnesota Department of Health, Public Health Laboratory | Minnesota Department of Health, Public Health Laboratory | Alexandra Lorentz, Jacob Garfin, Matt Plumb, and Xiong Wang |
| EPI_ISL_770800, EPI_ISL_770801, | Vault Health | Minnesota Department of Health, Public Health Laboratory | Alexandra Lorentz, Jacob Garfin, Matt Plumb, and Xiong Wang |

| | | | |
|---|---|---|---|
| EPI_ISL_770802, EPI_ISL_770803, EPI_ISL_770804, EPI_ISL_770805, EPI_ISL_770806, EPI_ISL_770807 | | | |
| EPI_ISL_770808, EPI_ISL_770809, EPI_ISL_770810, EPI_ISL_770811, EPI_ISL_770812, EPI_ISL_770813 | Essentia Health-St. Mary's Medical Center | Minnesota Department of Health, Public Health Laboratory | Alexandra Lorentz, Jacob Garfin, Matt Plumb, and Xiong Wang |
| EPI_ISL_770814, EPI_ISL_770815 | Vault Health | Minnesota Department of Health, Public Health Laboratory | Alexandra Lorentz, Jacob Garfin, Matt Plumb, and Xiong Wang |
| EPI_ISL_770816, EPI_ISL_770817 | Minnesota Department of Health, Public Health Laboratory | Minnesota Department of Health, Public Health Laboratory | Alexandra Lorentz, Jacob Garfin, Matt Plumb, and Xiong Wang |
| EPI_ISL_770818 | North Memorial | Minnesota Department of Health, Public Health Laboratory | Alexandra Lorentz, Jacob Garfin, Matt Plumb, and Xiong Wang |
| EPI_ISL_770819 | Minnesota Department of Health, Public Health Laboratory | Minnesota Department of Health, Public Health Laboratory | Alexandra Lorentz, Jacob Garfin, Matt Plumb, and Xiong Wang |
| EPI_ISL_771152, EPI_ISL_771153, EPI_ISL_771154, EPI_ISL_771155, EPI_ISL_771156, EPI_ISL_771157, EPI_ISL_771158, EPI_ISL_771159, EPI_ISL_771160, EPI_ISL_771161, EPI_ISL_771162, EPI_ISL_771163, EPI_ISL_771164, EPI_ISL_771165, EPI_ISL_771166, EPI_ISL_771167, EPI_ISL_771168, EPI_ISL_771169, EPI_ISL_771170, EPI_ISL_771171, EPI_ISL_771172, EPI_ISL_771173, EPI_ISL_771174, EPI_ISL_771175, EPI_ISL_771176, EPI_ISL_771177, EPI_ISL_771178, EPI_ISL_771179, EPI_ISL_771180, EPI_ISL_771181, EPI_ISL_771182, EPI_ISL_771183, EPI_ISL_771184, EPI_ISL_771185, EPI_ISL_771186, EPI_ISL_771187, EPI_ISL_771189, EPI_ISL_771190, EPI_ISL_771191, EPI_ISL_771222, EPI_ISL_771223, EPI_ISL_771225, EPI_ISL_771226, EPI_ISL_771227, EPI_ISL_771228, EPI_ISL_771229, EPI_ISL_771230, EPI_ISL_771231, EPI_ISL_771232, EPI_ISL_771233, EPI_ISL_771245, EPI_ISL_771246, EPI_ISL_771247, EPI_ISL_771248, EPI_ISL_771249, EPI_ISL_771250, EPI_ISL_771251, EPI_ISL_771252, EPI_ISL_771253, EPI_ISL_771254, EPI_ISL_771255, EPI_ISL_771256, EPI_ISL_771257, EPI_ISL_771258, EPI_ISL_771259, EPI_ISL_771260, EPI_ISL_771261, EPI_ISL_771262, EPI_ISL_771263, EPI_ISL_771264, EPI_ISL_771265, EPI_ISL_771266, EPI_ISL_771267, EPI_ISL_771268, EPI_ISL_771269, EPI_ISL_771270, EPI_ISL_771271, EPI_ISL_771272, EPI_ISL_771273, EPI_ISL_771274, EPI_ISL_771275, EPI_ISL_771276, EPI_ISL_771277, EPI_ISL_771278, EPI_ISL_771279, EPI_ISL_771280, EPI_ISL_771281, EPI_ISL_771282, EPI_ISL_771283, EPI_ISL_771284, EPI_ISL_771285, EPI_ISL_771286, EPI_ISL_771287, EPI_ISL_771288, EPI_ISL_771289, EPI_ISL_771290, EPI_ISL_771291, EPI_ISL_771292, EPI_ISL_771293, EPI_ISL_771294, EPI_ISL_771295, EPI_ISL_771296, EPI_ISL_771297, EPI_ISL_771298, EPI_ISL_771299, EPI_ISL_771300, EPI_ISL_771301, EPI_ISL_771302, EPI_ISL_771303, EPI_ISL_771304, EPI_ISL_771305, EPI_ISL_771306, EPI_ISL_771307, EPI_ISL_771308, EPI_ISL_771309, EPI_ISL_771310, EPI_ISL_771311, EPI_ISL_771312, EPI_ISL_771313, EPI_ISL_771314, EPI_ISL_771315, EPI_ISL_771316, EPI_ISL_771317, EPI_ISL_771318, EPI_ISL_771319, EPI_ISL_771320, EPI_ISL_771321, EPI_ISL_771322, EPI_ISL_771323, EPI_ISL_771324, EPI_ISL_771325, EPI_ISL_771326, EPI_ISL_771327, EPI_ISL_771328, EPI_ISL_771329, EPI_ISL_771330, EPI_ISL_771331, EPI_ISL_771332, EPI_ISL_771333, EPI_ISL_771334, EPI_ISL_771335, EPI_ISL_771336, EPI_ISL_771337, EPI_ISL_771338, EPI_ISL_771339, EPI_ISL_771340, EPI_ISL_771341, EPI_ISL_771342, EPI_ISL_771343, EPI_ISL_771344, EPI_ISL_771345, EPI_ISL_771346, EPI_ISL_771347, EPI_ISL_771348, EPI_ISL_771349, EPI_ISL_771350, EPI_ISL_771351, EPI_ISL_771352, EPI_ISL_771353, EPI_ISL_771354, EPI_ISL_771355, EPI_ISL_771356, EPI_ISL_771357, EPI_ISL_771358, EPI_ISL_771359, EPI_ISL_771360 | | | |
| see above | Colorado Department of Public Health and Environment | Colorado Department of Puplic Health and Environment | Laura Bankers, Molly C. Hetherington-Rauth, Diana Ir, Shannon Ely, Shannon R. Matzinger, Sarah Elizabeth Totten, Emily A. Travanty |
| EPI_ISL_771361, EPI_ISL_771362, EPI_ISL_771364, EPI_ISL_771365 | Washington State Department of Health | Seattle Flu Study | Deborah A. Nickerson, Chris D. Frazar, Jover Lee, Benjamin Pelle, Matthew Richardson, Amanda Adler, Elisabeth Brandstetter, Peter D. Han, Kairsten Fay, Misja Ilcisin, Kirsten Lacombe, Thomas R. Sibley, Melissa Truong, Caitlin R. Wolf, Romesh Gautom, Geoff Melly, Brian Hiatt, Philip Dykema, Scott Lindquist, Michael Boeckh, Janet A. Englund, Michael Famulare, Barry R. Lutz, Mark J. Rieder, Lea M. Starita, Matthew Thompson, Helen Y. Chu, Jay Shendure, Trevor Bedford |
| EPI_ISL_775600, EPI_ISL_775601, EPI_ISL_775602 | Ohio Department of Health Laboratory | Ohio Department of Health Laboratory | Holmes, Jennifer; Eric Brandt, Keoni Omura, Glen McGillivary, Caitiln McDonnell, Kirtana Ramadugu, Erica Leasure, Kelsey Florek, Heather Blankenship, Quanta Brown, and Tammy Bannerman |
| EPI_ISL_775603, EPI_ISL_775605, EPI_ISL_775606, EPI_ISL_776567, EPI_ISL_776568 | Ohio Department of Health Laboratory | Ohio Department of Health Laboratory | Holmes, Jennifer; Eric Brandt, Keoni Omura, Glen McGillivary, Caitlin McDonnell, Kirtana Ramadugu, Erica Leasure, Kelsey Florek, Heather Blankenship, Quanta Brown, and Tammy Bannerman |
| EPI_ISL_776657, EPI_ISL_776658, EPI_ISL_776659, EPI_ISL_776660, EPI_ISL_776661, EPI_ISL_776662, EPI_ISL_776663, EPI_ISL_776667, EPI_ISL_776668, EPI_ISL_776669, EPI_ISL_776670, EPI_ISL_776671, EPI_ISL_776672, EPI_ISL_776673, EPI_ISL_776674, EPI_ISL_776675, EPI_ISL_776676, EPI_ISL_776677, EPI_ISL_776678, EPI_ISL_776679, EPI_ISL_776680, EPI_ISL_776681, EPI_ISL_776682, EPI_ISL_776683, EPI_ISL_776684, EPI_ISL_776685, EPI_ISL_776686, EPI_ISL_776687, EPI_ISL_776688, EPI_ISL_776689, EPI_ISL_776690, EPI_ISL_776691, EPI_ISL_776692, EPI_ISL_776693, EPI_ISL_776694, EPI_ISL_776695, EPI_ISL_776696, EPI_ISL_776697, EPI_ISL_776698, EPI_ISL_776699, EPI_ISL_776700, EPI_ISL_776701, EPI_ISL_776702, EPI_ISL_776703, EPI_ISL_776704, EPI_ISL_776705, EPI_ISL_776706, EPI_ISL_776707, EPI_ISL_776708, EPI_ISL_776709, EPI_ISL_776710, EPI_ISL_776712, EPI_ISL_776713, EPI_ISL_776714, EPI_ISL_776715, EPI_ISL_776716, EPI_ISL_776717, EPI_ISL_776718, EPI_ISL_776719, EPI_ISL_776720, EPI_ISL_776721, EPI_ISL_776722, EPI_ISL_776723, EPI_ISL_776724, EPI_ISL_776725, EPI_ISL_776726, EPI_ISL_776727, EPI_ISL_776728, EPI_ISL_776729, EPI_ISL_776730, EPI_ISL_776731, EPI_ISL_776732, EPI_ISL_776733, EPI_ISL_776734, EPI_ISL_776735, EPI_ISL_776736, EPI_ISL_776737, EPI_ISL_776738, EPI_ISL_776739, EPI_ISL_776740, EPI_ISL_776741, EPI_ISL_776742, EPI_ISL_776743, EPI_ISL_776744, EPI_ISL_776745, EPI_ISL_776746, EPI_ISL_776747, EPI_ISL_776748, EPI_ISL_776749 | | | |
| see above | UW Virology Lab | UW Virology Lab | Pavitra Roychoudhury, Hong Xie, Lasata Shrestha, Meei-Li Huang, Keith R Jerome, Alexander Greninger |
| EPI_ISL_776773, EPI_ISL_776774, EPI_ISL_776869, EPI_ISL_776872, EPI_ISL_776874, EPI_ISL_776876, EPI_ISL_776994 | Ohio Department of Health Laboratory | Ohio Department of Health Laboratory | Holmes, Jennifer; Eric Brandt, Keoni Omura, Glen McGillivary, Caitlin McDonnell, Kirtana Ramadugu, Erica Leasure, Kelsey Florek, Heather Blankenship, Quanta Brown, and Tammy Bannerman |
| EPI_ISL_778842 | Clinical Pathology Labs | Centers for Disease Control and Prevention Division of Viral Diseases, Pathogen Discovery | Ying Tao Yan Li Jing Zhang Krista Queen Anna Uehara Peter Cook Clinton R. Paden Haibin Wang Suxiang Tong |
| EPI_ISL_778856, EPI_ISL_778857, EPI_ISL_778858, EPI_ISL_778859, EPI_ISL_778861, EPI_ISL_778862, EPI_ISL_778864 | Ohio Department of Health Laboratory | Ohio Department of Health Laboratory | Holmes, Jennifer; Eric Brandt, Keoni Omura, Glen McGillivary, Caitlin McDonnell, Kirtana Ramadugu, Erica Leasure, Kelsey Florek, Heather Blankenship, Quanta Brown, and Tammy Bannerman |
| EPI_ISL_778865, EPI_ISL_778866 | Clinical Pathology Labs | Centers for Disease Control and Prevention Division of Viral Diseases, Pathogen Discovery | Ying Tao Yan Li Jing Zhang Krista Queen Anna Uehara Peter Cook Clinton R. Paden Haibin Wang Suxiang Tong |
| EPI_ISL_778867 | Synergy Laboratories | Synergy Laboratories | Megan Cornwell |
| EPI_ISL_778871, EPI_ISL_778872, EPI_ISL_778873, EPI_ISL_778874, EPI_ISL_778875, EPI_ISL_778876, EPI_ISL_778877, EPI_ISL_778878, EPI_ISL_778879, EPI_ISL_778880, EPI_ISL_778881, EPI_ISL_778882, EPI_ISL_778883, EPI_ISL_778884, EPI_ISL_778885, EPI_ISL_778886, EPI_ISL_778887, EPI_ISL_778888, EPI_ISL_778889, EPI_ISL_778890, EPI_ISL_778891, EPI_ISL_778892, EPI_ISL_778893, EPI_ISL_778894, EPI_ISL_778895, EPI_ISL_778896, EPI_ISL_778897, EPI_ISL_778898, EPI_ISL_778899, EPI_ISL_778900, EPI_ISL_778901, EPI_ISL_778902, EPI_ISL_778903, EPI_ISL_778904, EPI_ISL_778905, EPI_ISL_778906, EPI_ISL_778907, EPI_ISL_778908, EPI_ISL_778909, EPI_ISL_778910, EPI_ISL_778911, EPI_ISL_778912, EPI_ISL_778913, EPI_ISL_778914, EPI_ISL_778915, EPI_ISL_778916, EPI_ISL_778917 | | | |
| see above | Maryland Public Health Laboratory | Maryland Public Health Laboratory | Maryland Department of Health Laboratories Administration |
| EPI_ISL_778918, EPI_ISL_778919, EPI_ISL_778920, EPI_ISL_778921, EPI_ISL_778922, EPI_ISL_778923, EPI_ISL_778924, EPI_ISL_778925, EPI_ISL_778926, EPI_ISL_778927, EPI_ISL_778928, EPI_ISL_778929, EPI_ISL_778930, EPI_ISL_778931, EPI_ISL_778932, EPI_ISL_778933, EPI_ISL_778934, EPI_ISL_778935, EPI_ISL_778936, EPI_ISL_778937, EPI_ISL_778938, EPI_ISL_778939, EPI_ISL_778940, EPI_ISL_778941, EPI_ISL_778942, EPI_ISL_778943, EPI_ISL_778944, EPI_ISL_778945, EPI_ISL_778946, EPI_ISL_778947, EPI_ISL_778948, EPI_ISL_778949, EPI_ISL_778950, EPI_ISL_778951, EPI_ISL_778952, EPI_ISL_778953, EPI_ISL_778954, EPI_ISL_778955, EPI_ISL_778956, EPI_ISL_778957, EPI_ISL_778958, EPI_ISL_778959, EPI_ISL_778960, EPI_ISL_778961, EPI_ISL_778962, EPI_ISL_778963, EPI_ISL_778964, EPI_ISL_778965, EPI_ISL_778966, EPI_ISL_778967, EPI_ISL_778968, EPI_ISL_778969, EPI_ISL_778970, EPI_ISL_778971, EPI_ISL_778972, EPI_ISL_778973, EPI_ISL_778974, EPI_ISL_778975, EPI_ISL_778976, EPI_ISL_778977, EPI_ISL_778978, EPI_ISL_778979, EPI_ISL_778980, EPI_ISL_778981, EPI_ISL_778982, EPI_ISL_778983, EPI_ISL_778984, EPI_ISL_778985, EPI_ISL_778986, EPI_ISL_778987, EPI_ISL_778988, EPI_ISL_778989, EPI_ISL_778990, EPI_ISL_778991, EPI_ISL_778992, EPI_ISL_778993, EPI_ISL_778994, EPI_ISL_778995, EPI_ISL_778996, EPI_ISL_778997, EPI_ISL_778998, EPI_ISL_778999, EPI_ISL_779000, EPI_ISL_779001, EPI_ISL_779002, EPI_ISL_779003, EPI_ISL_779004, EPI_ISL_779005, EPI_ISL_779006, EPI_ISL_779007, EPI_ISL_779008, EPI_ISL_779009, EPI_ISL_779010, EPI_ISL_779011, EPI_ISL_779012, EPI_ISL_779013, EPI_ISL_779014, EPI_ISL_779015, EPI_ISL_779016, EPI_ISL_779017, EPI_ISL_779018, EPI_ISL_779019, EPI_ISL_779020, EPI_ISL_779021, EPI_ISL_779022, EPI_ISL_779023, EPI_ISL_779024, EPI_ISL_779025, EPI_ISL_779026, EPI_ISL_779027, EPI_ISL_779028, EPI_ISL_779029, EPI_ISL_779030, EPI_ISL_779031, EPI_ISL_779032, EPI_ISL_779033, EPI_ISL_779034, EPI_ISL_779035, EPI_ISL_779036, EPI_ISL_779037, EPI_ISL_779038, EPI_ISL_779039, EPI_ISL_779040, EPI_ISL_779041, EPI_ISL_779042, EPI_ISL_779043, EPI_ISL_779044, EPI_ISL_779045, EPI_ISL_779046, EPI_ISL_779047, EPI_ISL_779048, EPI_ISL_779049, EPI_ISL_779050, EPI_ISL_779051, EPI_ISL_779052, EPI_ISL_779053, EPI_ISL_779054, EPI_ISL_779055, EPI_ISL_779056, EPI_ISL_779057, EPI_ISL_779058, EPI_ISL_779059, EPI_ISL_779060, EPI_ISL_779061, EPI_ISL_779062, EPI_ISL_779063, EPI_ISL_779064, EPI_ISL_779065, EPI_ISL_779066, EPI_ISL_779067, EPI_ISL_779068, EPI_ISL_779069, EPI_ISL_779070, EPI_ISL_779071, EPI_ISL_779072, EPI_ISL_779073, EPI_ISL_779074, EPI_ISL_779075, EPI_ISL_779076, EPI_ISL_779077, EPI_ISL_779078, EPI_ISL_779079, EPI_ISL_779080, EPI_ISL_779081, EPI_ISL_779082, EPI_ISL_779083, EPI_ISL_779084, EPI_ISL_779085, EPI_ISL_779086, EPI_ISL_779087, EPI_ISL_779088, EPI_ISL_779089, EPI_ISL_779090, EPI_ISL_779091, EPI_ISL_779092, EPI_ISL_779093, EPI_ISL_779094, EPI_ISL_779095, EPI_ISL_779096, EPI_ISL_779097, EPI_ISL_779098, EPI_ISL_779099, EPI_ISL_779100, EPI_ISL_779101, EPI_ISL_779102, EPI_ISL_779103, EPI_ISL_779104 | | | |
| see above | LSUHS Emerging Viral Threat Laboratory | Microbial Genome Sequencing Center | Jeremy P. Kamil, Jennifer L. Carroll, Camille F. Abshire, Maarten Van Diest, Andrew D. Yurochko, Martin J. Sapp, Rona S. Scott, Christopher G. Kevil, Daniel J. Snyder, Vaughn S. Cooper, John A. Vanchiere |
| EPI_ISL_779105, EPI_ISL_779106, EPI_ISL_779107, EPI_ISL_779108, EPI_ISL_779109, EPI_ISL_779110, EPI_ISL_779111, EPI_ISL_779112, EPI_ISL_779113, EPI_ISL_779114, EPI_ISL_779115, EPI_ISL_779116 | | | |
| see above | LSUHS Emerging Viral Threat Laboratory | Microbial Genome Sequencing Center | Jennifer L. Carroll, Jeremy P. Kamil, Camille F. Abshire, Maarten Van Diest, Andrew D. Yurochko, Martin J. Sapp, Rona S. Scott, Christopher G. Kevil, Daniel J. Snyder, Vaughn S. Cooper, John A. Vanchiere |
| EPI_ISL_779117, EPI_ISL_779118, EPI_ISL_779119, EPI_ISL_779120, EPI_ISL_779121, EPI_ISL_779122, EPI_ISL_779123, EPI_ISL_779124, EPI_ISL_779125, EPI_ISL_779126, EPI_ISL_779127, EPI_ISL_779128 | | | |
| see above | LSUHS Emerging Viral Threat Laboratory | Microbial Genome Sequencing Center | Maarten Van Diest, Jeremy P. Kamil, Jennifer L. Carroll, Camille F. Abshire, Andrew D. Yurochko, Martin J. Sapp, Rona S. Scott, Christopher G. Kevil, Daniel J. Snyder, Vaughn S. Cooper, John A. Vanchiere |
| EPI_ISL_779135, EPI_ISL_779136 | Wyoming Public Health Laboratory | Wyoming Public Health Laboratory | Noah Hull, Taylor Fearing, Lynette Gumbleton, Channing Weber, Ashley Norberg, Bailey Bowcutt, and Wanda Manley |
| EPI_ISL_779169, EPI_ISL_779170, EPI_ISL_779171, EPI_ISL_779172, EPI_ISL_779173, EPI_ISL_779174, EPI_ISL_779175, EPI_ISL_779176, EPI_ISL_779177, EPI_ISL_779178, EPI_ISL_779179, EPI_ISL_779180, EPI_ISL_779181, EPI_ISL_779182, EPI_ISL_779183, EPI_ISL_779184 | | | |
| see above | Laboratorio de Infectología, Servicio de Infectología, Hospital Universitario Dr. José Eleuterio González - Universidad Autónoma de Nuevo León | Laboratorio de Infectología Molecular, Departamento de Bioquímica y Medicina Molecular,Facultad de Medicina - Universidad Autónoma de Nuevo León | Kame A. Galán-Huerta, María F. Herrera-Saldivar, Natalia Martínez-Acuña, Sonia A. Lozano-Sepúlveda, Daniel Arellanos-Soto, Ana M. Rivas-Estilla, Paola Bocanegra-Ibarias, Samantha M. Flores-Treviño, Elvira Garza-González, Eduardo Perez-Alba, Laura Nuzzolo-Shihadeh, Adrian Camacho-Ortiz |
| EPI_ISL_779185, EPI_ISL_779186, EPI_ISL_779187, EPI_ISL_779188, EPI_ISL_779189, EPI_ISL_779190, EPI_ISL_779191, EPI_ISL_779192, EPI_ISL_779193, EPI_ISL_779194, EPI_ISL_779195, EPI_ISL_779196, EPI_ISL_779197, EPI_ISL_779198 | | | |
| see above | Laboratorio Estatal de Salud Pública de Nuevo León | Laboratorio de Infectología Molecular, Departamento de Bioquímica y Medicina Molecular,Facultad de Medicina - Universidad | Kame A. Galán-Huerta, María F. Herrera-Saldivar, Natalia Martínez-Acuña, Sonia A. Lozano-Sepúlveda, Daniel Arellanos-Soto, Ana M. Rivas-Estilla, Samuel Buentello-Wong, Else del Carmen García-García, Gloria A. Jasso-de-la-Peña, Roberto Montes-de-Oca, Consuelo Treviño-Garza, Manuel E. de-la-O-Cavazos |

Autónoma de Nuevo León

EPI_ISL_779319, EPI_ISL_779320, EPI_ISL_779321, EPI_ISL_779322, EPI_ISL_779323, EPI_ISL_779324, EPI_ISL_779325, EPI_ISL_779326, EPI_ISL_779327, EPI_ISL_779328, EPI_ISL_779329, EPI_ISL_779330, EPI_ISL_779331, EPI_ISL_779332, EPI_ISL_779333, EPI_ISL_779334, EPI_ISL_779335, EPI_ISL_779336, EPI_ISL_779337, EPI_ISL_779338, EPI_ISL_779339, EPI_ISL_779340, EPI_ISL_779341, EPI_ISL_779342, EPI_ISL_779343, EPI_ISL_779344, EPI_ISL_779345, EPI_ISL_779346, EPI_ISL_779347, EPI_ISL_779348, EPI_ISL_779349, EPI_ISL_779350, EPI_ISL_779351, EPI_ISL_779352, EPI_ISL_779353, EPI_ISL_779354, EPI_ISL_779355, EPI_ISL_779356, EPI_ISL_779357, EPI_ISL_779358, EPI_ISL_779359, EPI_ISL_779360, EPI_ISL_779361, EPI_ISL_779362, EPI_ISL_779363, EPI_ISL_779364, EPI_ISL_779365, EPI_ISL_779366, EPI_ISL_779367, EPI_ISL_779368, EPI_ISL_779369, EPI_ISL_779370, EPI_ISL_779371, EPI_ISL_779372, EPI_ISL_779373, EPI_ISL_779374, EPI_ISL_779375, EPI_ISL_779376, EPI_ISL_779377, EPI_ISL_779378, EPI_ISL_779379, EPI_ISL_779380, EPI_ISL_779381, EPI_ISL_779382

see above | University of Wisconsin-Madison AIDS Vaccine Research Laboratories | University of Wisconsin-Madison AIDS Vaccine Research Laboratories | Gage Moreno, Katarina Braun, et al. AIDS Vaccine Research Laboratories

EPI_ISL_780097, EPI_ISL_780098, EPI_ISL_780099, EPI_ISL_780100, EPI_ISL_780101, EPI_ISL_780102, EPI_ISL_780103, EPI_ISL_780104, EPI_ISL_780105, EPI_ISL_780106, EPI_ISL_780107, EPI_ISL_780108, EPI_ISL_780109, EPI_ISL_780110, EPI_ISL_780111, EPI_ISL_780112, EPI_ISL_780113, EPI_ISL_780114, EPI_ISL_780115, EPI_ISL_780116, EPI_ISL_780117, EPI_ISL_780118, EPI_ISL_780119, EPI_ISL_780120, EPI_ISL_780121, EPI_ISL_780122, EPI_ISL_780123, EPI_ISL_780124, EPI_ISL_780125, EPI_ISL_780126, EPI_ISL_780127, EPI_ISL_780128, EPI_ISL_780129, EPI_ISL_780130, EPI_ISL_780131, EPI_ISL_780132, EPI_ISL_780133, EPI_ISL_780134, EPI_ISL_780135, EPI_ISL_780136, EPI_ISL_780137, EPI_ISL_780138, EPI_ISL_780139, EPI_ISL_780140, EPI_ISL_780141, EPI_ISL_780142, EPI_ISL_780143, EPI_ISL_780144, EPI_ISL_780145, EPI_ISL_780146, EPI_ISL_780147, EPI_ISL_780148, EPI_ISL_780149, EPI_ISL_780150, EPI_ISL_780151, EPI_ISL_780152, EPI_ISL_780153, EPI_ISL_780154, EPI_ISL_780155, EPI_ISL_780156, EPI_ISL_780157, EPI_ISL_780158, EPI_ISL_780159, EPI_ISL_780160, EPI_ISL_780161, EPI_ISL_780162, EPI_ISL_780163, EPI_ISL_780164, EPI_ISL_780165, EPI_ISL_780166, EPI_ISL_780167, EPI_ISL_780168, EPI_ISL_780169, EPI_ISL_780170, EPI_ISL_780171, EPI_ISL_780172, EPI_ISL_780173, EPI_ISL_780174, EPI_ISL_780175, EPI_ISL_780176, EPI_ISL_780177, EPI_ISL_780178, EPI_ISL_780179, EPI_ISL_780180, EPI_ISL_780181, EPI_ISL_780182, EPI_ISL_780183, EPI_ISL_780184, EPI_ISL_780185, EPI_ISL_780186, EPI_ISL_780187, EPI_ISL_780188, EPI_ISL_780189, EPI_ISL_780190, EPI_ISL_780191, EPI_ISL_780192, EPI_ISL_780193, EPI_ISL_780194, EPI_ISL_780195, EPI_ISL_780196, EPI_ISL_780197, EPI_ISL_780198, EPI_ISL_780199, EPI_ISL_780200, EPI_ISL_780201, EPI_ISL_780202, EPI_ISL_780203, EPI_ISL_780204, EPI_ISL_780205, EPI_ISL_780206, EPI_ISL_780207, EPI_ISL_780208, EPI_ISL_780209, EPI_ISL_780210, EPI_ISL_780211, EPI_ISL_780212, EPI_ISL_780213, EPI_ISL_780214, EPI_ISL_780215, EPI_ISL_780216, EPI_ISL_780217, EPI_ISL_780218, EPI_ISL_780219, EPI_ISL_780220, EPI_ISL_780221, EPI_ISL_780222, EPI_ISL_780223, EPI_ISL_780224, EPI_ISL_780225, EPI_ISL_780226, EPI_ISL_780227, EPI_ISL_780228, EPI_ISL_780229, EPI_ISL_780230, EPI_ISL_780231, EPI_ISL_780232, EPI_ISL_780233, EPI_ISL_780234, EPI_ISL_780235, EPI_ISL_780236, EPI_ISL_780237, EPI_ISL_780238, EPI_ISL_780239, EPI_ISL_780240, EPI_ISL_780241, EPI_ISL_780242, EPI_ISL_780243, EPI_ISL_780244, EPI_ISL_780245, EPI_ISL_780246, EPI_ISL_780247, EPI_ISL_780248, EPI_ISL_780249, EPI_ISL_780250, EPI_ISL_780251, EPI_ISL_780252, EPI_ISL_780253, EPI_ISL_780254, EPI_ISL_780255, EPI_ISL_780256, EPI_ISL_780257, EPI_ISL_780258, EPI_ISL_780259, EPI_ISL_780260, EPI_ISL_780261, EPI_ISL_780262, EPI_ISL_780263, EPI_ISL_780264, EPI_ISL_780265, EPI_ISL_780266, EPI_ISL_780267, EPI_ISL_780268, EPI_ISL_780269, EPI_ISL_780270, EPI_ISL_780271, EPI_ISL_780272, EPI_ISL_780273, EPI_ISL_780274, EPI_ISL_780275, EPI_ISL_780276, EPI_ISL_780277, EPI_ISL_780278, EPI_ISL_780279, EPI_ISL_780280, EPI_ISL_780281, EPI_ISL_780282, EPI_ISL_780283, EPI_ISL_780284, EPI_ISL_780285, EPI_ISL_780286, EPI_ISL_780287, EPI_ISL_780288, EPI_ISL_780289, EPI_ISL_780290, EPI_ISL_780291, EPI_ISL_780292, EPI_ISL_780293, EPI_ISL_780294, EPI_ISL_780295, EPI_ISL_780296, EPI_ISL_780297, EPI_ISL_780298, EPI_ISL_780299, EPI_ISL_780300, EPI_ISL_780301, EPI_ISL_780302, EPI_ISL_780303, EPI_ISL_780304, EPI_ISL_780305, EPI_ISL_780306, EPI_ISL_780307, EPI_ISL_780308, EPI_ISL_780309, EPI_ISL_780310, EPI_ISL_780311, EPI_ISL_780312, EPI_ISL_780313, EPI_ISL_780314, EPI_ISL_780315, EPI_ISL_780316, EPI_ISL_780317, EPI_ISL_780318, EPI_ISL_780319, EPI_ISL_780320, EPI_ISL_780321, EPI_ISL_780322, EPI_ISL_780323, EPI_ISL_780324, EPI_ISL_780325, EPI_ISL_780326, EPI_ISL_780327, EPI_ISL_780328, EPI_ISL_780329, EPI_ISL_780330, EPI_ISL_780331, EPI_ISL_780332, EPI_ISL_780333, EPI_ISL_780334, EPI_ISL_780335, EPI_ISL_780336, EPI_ISL_780337, EPI_ISL_780338, EPI_ISL_780339, EPI_ISL_780340, EPI_ISL_780341, EPI_ISL_780342, EPI_ISL_780343, EPI_ISL_780344, EPI_ISL_780345, EPI_ISL_780346, EPI_ISL_780347, EPI_ISL_780348, EPI_ISL_780349, EPI_ISL_780350, EPI_ISL_780351, EPI_ISL_780352, EPI_ISL_780353, EPI_ISL_780354, EPI_ISL_780355, EPI_ISL_780356, EPI_ISL_780357, EPI_ISL_780358, EPI_ISL_780359, EPI_ISL_780360, EPI_ISL_780361, EPI_ISL_780362, EPI_ISL_780363, EPI_ISL_780364, EPI_ISL_780365, EPI_ISL_780366, EPI_ISL_780367, EPI_ISL_780368, EPI_ISL_780369, EPI_ISL_780370, EPI_ISL_780371, EPI_ISL_780372, EPI_ISL_780373, EPI_ISL_780374

see above | Houston Methodist Hospital | Houston Methodist Hospital | S. Wesley Long, Randall J. Olsen, Paul A. Christensen, David W. Bernard, James J. Davis, Maulik Shukla, Marcus Nguyen, Matthew Ojeda Saavedra, Prasanti Yerramilli, Layne Pruitt, Sishir Subedi, Heather Hendrickson, and James M. Musser

EPI_ISL_780377, EPI_ISL_780379, EPI_ISL_780380, EPI_ISL_780417, EPI_ISL_780419, EPI_ISL_780421, EPI_ISL_780422, EPI_ISL_780423, EPI_ISL_780424, EPI_ISL_780425, EPI_ISL_780426

see above | Ohio Department of Health Laboratory | Ohio Department of Health Laboratory | Holmes, Jennifer; Eric Brandt, Keoni Omura, Glen McGillivary, Caitlin McDonnell, Kirtana Ramadugu, Erica Leasure, Kelsey Florek, Heather Blankenship, Quanta Brown, and Tammy Bannerman

EPI_ISL_780427, EPI_ISL_780428, EPI_ISL_780429, EPI_ISL_780430, EPI_ISL_780431, EPI_ISL_780432, EPI_ISL_780433, EPI_ISL_780434, EPI_ISL_780435, EPI_ISL_780436, EPI_ISL_780437, EPI_ISL_780438, EPI_ISL_780439, EPI_ISL_780440, EPI_ISL_780441, EPI_ISL_780442, EPI_ISL_780443, EPI_ISL_780444, EPI_ISL_780445, EPI_ISL_780446, EPI_ISL_780447, EPI_ISL_780448, EPI_ISL_780449, EPI_ISL_780450, EPI_ISL_780451, EPI_ISL_780452, EPI_ISL_780453, EPI_ISL_780454, EPI_ISL_780455, EPI_ISL_780456, EPI_ISL_780457, EPI_ISL_780458, EPI_ISL_780459, EPI_ISL_780460, EPI_ISL_780461, EPI_ISL_780462, EPI_ISL_780463, EPI_ISL_780464, EPI_ISL_780465, EPI_ISL_780466, EPI_ISL_780467, EPI_ISL_780468, EPI_ISL_780469, EPI_ISL_780470, EPI_ISL_780471, EPI_ISL_780472, EPI_ISL_780473, EPI_ISL_780474, EPI_ISL_780475, EPI_ISL_780476, EPI_ISL_780477, EPI_ISL_780478, EPI_ISL_780479, EPI_ISL_780480, EPI_ISL_780481, EPI_ISL_780482, EPI_ISL_780483, EPI_ISL_780484, EPI_ISL_780485, EPI_ISL_780486, EPI_ISL_780487, EPI_ISL_780488, EPI_ISL_780489, EPI_ISL_780490, EPI_ISL_780491, EPI_ISL_780492, EPI_ISL_780493, EPI_ISL_780494, EPI_ISL_780495, EPI_ISL_780496, EPI_ISL_780497, EPI_ISL_780498, EPI_ISL_780499, EPI_ISL_780500, EPI_ISL_780501, EPI_ISL_780502, EPI_ISL_780503, EPI_ISL_780504, EPI_ISL_780505, EPI_ISL_780506, EPI_ISL_780507, EPI_ISL_780508, EPI_ISL_780509, EPI_ISL_780510, EPI_ISL_780511, EPI_ISL_780512, EPI_ISL_780513, EPI_ISL_780514, EPI_ISL_780515, EPI_ISL_780516, EPI_ISL_780517, EPI_ISL_780518, EPI_ISL_780519, EPI_ISL_780520, EPI_ISL_780521, EPI_ISL_780522, EPI_ISL_780523, EPI_ISL_780524, EPI_ISL_780525, EPI_ISL_780526, EPI_ISL_780527, EPI_ISL_780528, EPI_ISL_780529, EPI_ISL_780530, EPI_ISL_780531, EPI_ISL_780532, EPI_ISL_780533, EPI_ISL_780534, EPI_ISL_780535, EPI_ISL_780536, EPI_ISL_780537, EPI_ISL_780538, EPI_ISL_780539, EPI_ISL_780540, EPI_ISL_780541, EPI_ISL_780542, EPI_ISL_780543, EPI_ISL_780544, EPI_ISL_780545, EPI_ISL_780546, EPI_ISL_780547, EPI_ISL_780548, EPI_ISL_780549, EPI_ISL_780550, EPI_ISL_780551, EPI_ISL_780552, EPI_ISL_780553, EPI_ISL_780554, EPI_ISL_780555, EPI_ISL_780556, EPI_ISL_780557, EPI_ISL_780558, EPI_ISL_780559, EPI_ISL_780560, EPI_ISL_780561, EPI_ISL_780562, EPI_ISL_780563, EPI_ISL_780564, EPI_ISL_780565, EPI_ISL_780566, EPI_ISL_780567, EPI_ISL_780568, EPI_ISL_780569, EPI_ISL_780570, EPI_ISL_780571, EPI_ISL_780572, EPI_ISL_780573, EPI_ISL_780574, EPI_ISL_780575, EPI_ISL_780576, EPI_ISL_780577, EPI_ISL_780578, EPI_ISL_780579, EPI_ISL_780580, EPI_ISL_780581, EPI_ISL_780582, EPI_ISL_780583, EPI_ISL_780584, EPI_ISL_780585, EPI_ISL_780586, EPI_ISL_780587, EPI_ISL_780588, EPI_ISL_780589, EPI_ISL_780590, EPI_ISL_780591, EPI_ISL_780592, EPI_ISL_780593, EPI_ISL_780594, EPI_ISL_780595, EPI_ISL_780596, EPI_ISL_780597, EPI_ISL_780598, EPI_ISL_780599, EPI_ISL_780600, EPI_ISL_780601, EPI_ISL_780602, EPI_ISL_780603, EPI_ISL_780604, EPI_ISL_780605, EPI_ISL_780606, EPI_ISL_780607, EPI_ISL_780608, EPI_ISL_780609, EPI_ISL_780610, EPI_ISL_780611, EPI_ISL_780612, EPI_ISL_780613, EPI_ISL_780614, EPI_ISL_780615, EPI_ISL_780616, EPI_ISL_780617, EPI_ISL_780618, EPI_ISL_780619, EPI_ISL_780620, EPI_ISL_780621, EPI_ISL_780622, EPI_ISL_780623, EPI_ISL_780624, EPI_ISL_780625, EPI_ISL_780626, EPI_ISL_780627, EPI_ISL_780628, EPI_ISL_780629, EPI_ISL_780630, EPI_ISL_780631, EPI_ISL_780632, EPI_ISL_780633, EPI_ISL_780634, EPI_ISL_780635, EPI_ISL_780636, EPI_ISL_780637, EPI_ISL_780638, EPI_ISL_780639, EPI_ISL_780640, EPI_ISL_780641, EPI_ISL_780642, EPI_ISL_780643, EPI_ISL_780644, EPI_ISL_780645, EPI_ISL_780646, EPI_ISL_780647, EPI_ISL_780648, EPI_ISL_780649, EPI_ISL_780650, EPI_ISL_780651, EPI_ISL_780652, EPI_ISL_780653, EPI_ISL_780654, EPI_ISL_780655, EPI_ISL_780656, EPI_ISL_780657, EPI_ISL_780658, EPI_ISL_780659, EPI_ISL_780660, EPI_ISL_780661, EPI_ISL_780662, EPI_ISL_780663, EPI_ISL_780664, EPI_ISL_780665, EPI_ISL_780666, EPI_ISL_780667, EPI_ISL_780668, EPI_ISL_780669

see above | Houston Methodist Hospital | Houston Methodist Hospital | S. Wesley Long, Randall J. Olsen, Paul A. Christensen, David W. Bernard, James J. Davis, Maulik Shukla, Marcus Nguyen, Matthew Ojeda Saavedra, Prasanti Yerramilli, Layne Pruitt, Sishir Subedi, Heather Hendrickson, and James M. Musser

EPI_ISL_780670 | Ohio Department of Health Laboratory | Ohio Department of Health Laboratory | Holmes, Jennifer; Eric Brandt, Keoni Omura, Glen McGillivary, Caitlin McDonnell, Kirtana Ramadugu, Erica Leasure, Kelsey Florek, Heather Blankenship, Quanta Brown, and Tammy Bannerman

EPI_ISL_780671, EPI_ISL_780672, EPI_ISL_780673, EPI_ISL_780674, EPI_ISL_780675, EPI_ISL_780676, EPI_ISL_780677, EPI_ISL_780678, EPI_ISL_780679, EPI_ISL_780680, EPI_ISL_780681, EPI_ISL_780682, EPI_ISL_780683, EPI_ISL_780684, EPI_ISL_780685, EPI_ISL_780686, EPI_ISL_780687, EPI_ISL_780688, EPI_ISL_780689, EPI_ISL_780690, EPI_ISL_780691, EPI_ISL_780692, EPI_ISL_780693, EPI_ISL_780694, EPI_ISL_780695, EPI_ISL_780696, EPI_ISL_780697, EPI_ISL_780698, EPI_ISL_780699, EPI_ISL_780700, EPI_ISL_780701, EPI_ISL_780702, EPI_ISL_780703, EPI_ISL_780704, EPI_ISL_780705, EPI_ISL_780706, EPI_ISL_780707, EPI_ISL_780708, EPI_ISL_780709, EPI_ISL_780710, EPI_ISL_780711, EPI_ISL_780712, EPI_ISL_780713, EPI_ISL_780714, EPI_ISL_780715, EPI_ISL_780716, EPI_ISL_780717, EPI_ISL_780718, EPI_ISL_780719, EPI_ISL_780720, EPI_ISL_780721, EPI_ISL_780722, EPI_ISL_780723, EPI_ISL_780724, EPI_ISL_780725, EPI_ISL_780726, EPI_ISL_780727, EPI_ISL_780728, EPI_ISL_780729, EPI_ISL_780730, EPI_ISL_780731, EPI_ISL_780732, EPI_ISL_780733, EPI_ISL_780734, EPI_ISL_780735, EPI_ISL_780736, EPI_ISL_780737, EPI_ISL_780738, EPI_ISL_780739, EPI_ISL_780740, EPI_ISL_780741, EPI_ISL_780742, EPI_ISL_780743, EPI_ISL_780744, EPI_ISL_780745, EPI_ISL_780746, EPI_ISL_780747, EPI_ISL_780748, EPI_ISL_780749, EPI_ISL_780750, EPI_ISL_780751, EPI_ISL_780752, EPI_ISL_780753, EPI_ISL_780754, EPI_ISL_780755, EPI_ISL_780756, EPI_ISL_780757, EPI_ISL_780758, EPI_ISL_780759, EPI_ISL_780760, EPI_ISL_780761, EPI_ISL_780762, EPI_ISL_780763, EPI_ISL_780764, EPI_ISL_780765, EPI_ISL_780766, EPI_ISL_780767, EPI_ISL_780768, EPI_ISL_780769, EPI_ISL_780770, EPI_ISL_780771, EPI_ISL_780772, EPI_ISL_780773, EPI_ISL_780774, EPI_ISL_780775, EPI_ISL_780776, EPI_ISL_780777, EPI_ISL_780778, EPI_ISL_780779, EPI_ISL_780780, EPI_ISL_780781, EPI_ISL_780782, EPI_ISL_780783, EPI_ISL_780784, EPI_ISL_780785, EPI_ISL_780786, EPI_ISL_780787, EPI_ISL_780788, EPI_ISL_780789, EPI_ISL_780790, EPI_ISL_780791, EPI_ISL_780792, EPI_ISL_780793, EPI_ISL_780794, EPI_ISL_780795, EPI_ISL_780796, EPI_ISL_780797, EPI_ISL_780798, EPI_ISL_780799, EPI_ISL_780800, EPI_ISL_780801, EPI_ISL_780802, EPI_ISL_780803, EPI_ISL_780804, EPI_ISL_780805, EPI_ISL_780806, EPI_ISL_780807, EPI_ISL_780808, EPI_ISL_780809, EPI_ISL_780810, EPI_ISL_780811, EPI_ISL_780812, EPI_ISL_780813, EPI_ISL_780814, EPI_ISL_780815, EPI_ISL_780816, EPI_ISL_780817, EPI_ISL_780818, EPI_ISL_780819, EPI_ISL_780820, EPI_ISL_780821, EPI_ISL_780822, EPI_ISL_780823, EPI_ISL_780824, EPI_ISL_780825, EPI_ISL_780826, EPI_ISL_780827, EPI_ISL_780828, EPI_ISL_780829, EPI_ISL_780830, EPI_ISL_780831, EPI_ISL_780832, EPI_ISL_780833, EPI_ISL_780834, EPI_ISL_780835, EPI_ISL_780836, EPI_ISL_780837, EPI_ISL_780838, EPI_ISL_780839, EPI_ISL_780840, EPI_ISL_780841, EPI_ISL_780842, EPI_ISL_780843, EPI_ISL_780844, EPI_ISL_780845, EPI_ISL_780846, EPI_ISL_780847, EPI_ISL_780848, EPI_ISL_780849, EPI_ISL_780850, EPI_ISL_780851, EPI_ISL_780852, EPI_ISL_780853, EPI_ISL_780854, EPI_ISL_780855, EPI_ISL_780856, EPI_ISL_780857, EPI_ISL_780858, EPI_ISL_780859, EPI_ISL_780860, EPI_ISL_780861, EPI_ISL_780862, EPI_ISL_780863, EPI_ISL_780864, EPI_ISL_780865, EPI_ISL_780866, EPI_ISL_780867, EPI_ISL_780868, EPI_ISL_780869, EPI_ISL_780870, EPI_ISL_780871, EPI_ISL_780872, EPI_ISL_780873, EPI_ISL_780874, EPI_ISL_780875, EPI_ISL_780876, EPI_ISL_780877, EPI_ISL_780878, EPI_ISL_780879, EPI_ISL_780880, EPI_ISL_780881, EPI_ISL_780882, EPI_ISL_780883, EPI_ISL_780884, EPI_ISL_780885, EPI_ISL_780886, EPI_ISL_780887, EPI_ISL_780888, EPI_ISL_780889, EPI_ISL_780890, EPI_ISL_780891, EPI_ISL_780892, EPI_ISL_780893, EPI_ISL_780894, EPI_ISL_780895, EPI_ISL_780896, EPI_ISL_780897, EPI_ISL_780898, EPI_ISL_780899, EPI_ISL_780900, EPI_ISL_780901, EPI_ISL_780902, EPI_ISL_780903, EPI_ISL_780904, EPI_ISL_780905, EPI_ISL_780906, EPI_ISL_780907, EPI_ISL_780908, EPI_ISL_780909, EPI_ISL_780910, EPI_ISL_780911, EPI_ISL_780912, EPI_ISL_780913, EPI_ISL_780914, EPI_ISL_780915, EPI_ISL_780916, EPI_ISL_780917, EPI_ISL_780918, EPI_ISL_780919, EPI_ISL_780920, EPI_ISL_780921, EPI_ISL_780922, EPI_ISL_780923, EPI_ISL_780924, EPI_ISL_780925, EPI_ISL_780926, EPI_ISL_780927, EPI_ISL_780928, EPI_ISL_780929, EPI_ISL_780930, EPI_ISL_780931, EPI_ISL_780932, EPI_ISL_780933, EPI_ISL_780934, EPI_ISL_780935, EPI_ISL_780936, EPI_ISL_780937, EPI_ISL_780938, EPI_ISL_780939, EPI_ISL_780940, EPI_ISL_780941, EPI_ISL_780942, EPI_ISL_780943, EPI_ISL_780944, EPI_ISL_780945, EPI_ISL_780946, EPI_ISL_780947, EPI_ISL_780948, EPI_ISL_780949, EPI_ISL_780950, EPI_ISL_780951, EPI_ISL_780952, EPI_ISL_780953, EPI_ISL_780954, EPI_ISL_780955, EPI_ISL_780956, EPI_ISL_780957, EPI_ISL_780958, EPI_ISL_780959, EPI_ISL_780960, EPI_ISL_780961, EPI_ISL_780962, EPI_ISL_780963, EPI_ISL_780964, EPI_ISL_780965, EPI_ISL_780966, EPI_ISL_780967, EPI_ISL_780968, EPI_ISL_780969, EPI_ISL_780970, EPI_ISL_780971, EPI_ISL_780972, EPI_ISL_780973, EPI_ISL_780974, EPI_ISL_780975, EPI_ISL_780976, EPI_ISL_780977, EPI_ISL_780978, EPI_ISL_780979, EPI_ISL_780980, EPI_ISL_780981, EPI_ISL_780982, EPI_ISL_780983, EPI_ISL_780984, EPI_ISL_780985, EPI_ISL_780986, EPI_ISL_780987, EPI_ISL_780988, EPI_ISL_780989, EPI_ISL_780990, EPI_ISL_780991, EPI_ISL_780992, EPI_ISL_780993, EPI_ISL_780994, EPI_ISL_780995, EPI_ISL_780996, EPI_ISL_780997, EPI_ISL_780998, EPI_ISL_780999, EPI_ISL_781000, EPI_ISL_781001, EPI_ISL_781002, EPI_ISL_781003, EPI_ISL_781004, EPI_ISL_781005, EPI_ISL_781006, EPI_ISL_781007, EPI_ISL_781008, EPI_ISL_781009, EPI_ISL_781010, EPI_ISL_781011, EPI_ISL_781012, EPI_ISL_781013, EPI_ISL_781014, EPI_ISL_781015, EPI_ISL_781016, EPI_ISL_781017, EPI_ISL_781018, EPI_ISL_781019, EPI_ISL_781020, EPI_ISL_781021, EPI_ISL_781022, EPI_ISL_781023, EPI_ISL_781024, EPI_ISL_781025, EPI_ISL_781026, EPI_ISL_781027, EPI_ISL_781028, EPI_ISL_781029, EPI_ISL_781030, EPI_ISL_781031, EPI_ISL_781032, EPI_ISL_781033, EPI_ISL_781034, EPI_ISL_781035, EPI_ISL_781036, EPI_ISL_781037, EPI_ISL_781038, EPI_ISL_781039, EPI_ISL_781040, EPI_ISL_781041, EPI_ISL_781042, EPI_ISL_781043, EPI_ISL_781044, EPI_ISL_781045, EPI_ISL_781046, EPI_ISL_781047, EPI_ISL_781048, EPI_ISL_781049, EPI_ISL_781050, EPI_ISL_781051, EPI_ISL_781052, EPI_ISL_781053, EPI_ISL_781054, EPI_ISL_781055, EPI_ISL_781056, EPI_ISL_781057, EPI_ISL_781058, EPI_ISL_781059, EPI_ISL_781060, EPI_ISL_781061, EPI_ISL_781062, EPI_ISL_781063, EPI_ISL_781064, EPI_ISL_781065, EPI_ISL_781066, EPI_ISL_781067, EPI_ISL_781068, EPI_ISL_781069, EPI_ISL_781070, EPI_ISL_781071, EPI_ISL_781072, EPI_ISL_781073, EPI_ISL_781074, EPI_ISL_781075, EPI_ISL_781076, EPI_ISL_781077, EPI_ISL_781078, EPI_ISL_781079, EPI_ISL_781080, EPI_ISL_781081, EPI_ISL_781082, EPI_ISL_781083, EPI_ISL_781084, EPI_ISL_781085, EPI_ISL_781086, EPI_ISL_781087, EPI_ISL_781088, EPI_ISL_781089, EPI_ISL_781090, EPI_ISL_781091, EPI_ISL_781092, EPI_ISL_781093, EPI_ISL_781094, EPI_ISL_781095, EPI_ISL_781096, EPI_ISL_781097, EPI_ISL_781098, EPI_ISL_781099, EPI_ISL_781100, EPI_ISL_781101, EPI_ISL_781102, EPI_ISL_781103, EPI_ISL_781104, EPI_ISL_781105, EPI_ISL_781106, EPI_ISL_781107, EPI_ISL_781108, EPI_ISL_781109, EPI_ISL_781110, EPI_ISL_781111, EPI_ISL_781112

see above | Houston Methodist Hospital | Houston Methodist Hospital | S. Wesley Long, Randall J. Olsen, Paul A. Christensen, David W. Bernard, James J. Davis, Maulik Shukla, Marcus Nguyen, Matthew Ojeda Saavedra, Prasanti Yerramilli, Layne Pruitt, Sishir Subedi, Heather Hendrickson, and James M. Musser

EPI_ISL_781113 | Ohio Department of Health Laboratory | Ohio Department of Health Laboratory | Holmes, Jennifer; Eric Brandt, Keoni Omura, Glen McGillivary, Caitlin McDonnell, Kirtana Ramadugu, Erica Leasure, Kelsey Florek, Heather Blankenship, Quanta Brown, and Tammy Bannerman

EPI_ISL_781114, EPI_ISL_781115, EPI_ISL_781116, EPI_ISL_781117, EPI_ISL_781118, EPI_ISL_781119, EPI_ISL_781120, EPI_ISL_781121, EPI_ISL_781122, EPI_ISL_781123, EPI_ISL_781124, EPI_ISL_781125, EPI_ISL_781126, EPI_ISL_781127, EPI_ISL_781128, EPI_ISL_781129, EPI_ISL_781130, EPI_ISL_781131, EPI_ISL_781132, EPI_ISL_781133, EPI_ISL_781134, EPI_ISL_781135, EPI_ISL_781136, EPI_ISL_781137, EPI_ISL_781138, EPI_ISL_781139, EPI_ISL_781140, EPI_ISL_781141, EPI_ISL_781142, EPI_ISL_781143, EPI_ISL_781144, EPI_ISL_781145, EPI_ISL_781146, EPI_ISL_781147, EPI_ISL_781148, EPI_ISL_781149, EPI_ISL_781150, EPI_ISL_781151, EPI_ISL_781152, EPI_ISL_781153, EPI_ISL_781154, EPI_ISL_781155, EPI_ISL_781156, EPI_ISL_781157, EPI_ISL_781158, EPI_ISL_781159, EPI_ISL_781160, EPI_ISL_781161, EPI_ISL_781162, EPI_ISL_781163, EPI_ISL_781164, EPI_ISL_781165, EPI_ISL_781166, EPI_ISL_781167, EPI_ISL_781168, EPI_ISL_781169, EPI_ISL_781170, EPI_ISL_781171, EPI_ISL_781172, EPI_ISL_781173, EPI_ISL_781174, EPI_ISL_781175, EPI_ISL_781176, EPI_ISL_781177, EPI_ISL_783536, EPI_ISL_783537, EPI_ISL_783538, EPI_ISL_783539, EPI_ISL_783540, EPI_ISL_783541, EPI_ISL_783542, EPI_ISL_783543, EPI_ISL_783544, EPI_ISL_783545, EPI_ISL_783546, EPI_ISL_783547, EPI_ISL_783548, EPI_ISL_783549, EPI_ISL_783550, EPI_ISL_783551, EPI_ISL_783552, EPI_ISL_783553, EPI_ISL_783554, EPI_ISL_783555, EPI_ISL_783556, EPI_ISL_783557, EPI_ISL_783558, EPI_ISL_783559, EPI_ISL_783560, EPI_ISL_783561, EPI_ISL_783562, EPI_ISL_783563, EPI_ISL_783564, EPI_ISL_783565, EPI_ISL_783566, EPI_ISL_783567, EPI_ISL_783568, EPI_ISL_783569, EPI_ISL_783570, EPI_ISL_783571, EPI_ISL_783572, EPI_ISL_783573, EPI_ISL_783574, EPI_ISL_783575, EPI_ISL_783576, EPI_ISL_783577, EPI_ISL_783578, EPI_ISL_783579, EPI_ISL_783580, EPI_ISL_783581, EPI_ISL_783582, EPI_ISL_783583, EPI_ISL_783584, EPI_ISL_783585, EPI_ISL_783586, EPI_ISL_783587, EPI_ISL_783588, EPI_ISL_783589, EPI_ISL_783590, EPI_ISL_783591, EPI_ISL_783592, EPI_ISL_783593, EPI_ISL_783594, EPI_ISL_783595, EPI_ISL_783596, EPI_ISL_783597, EPI_ISL_783598, EPI_ISL_783599, EPI_ISL_783600, EPI_ISL_783601, EPI_ISL_783602, EPI_ISL_783603, EPI_ISL_783604, EPI_ISL_783605, EPI_ISL_783606, EPI_ISL_783607, EPI_ISL_783608, EPI_ISL_783609, EPI_ISL_783610, EPI_ISL_783611, EPI_ISL_783612, EPI_ISL_783613, EPI_ISL_783614, EPI_ISL_783615, EPI_ISL_783616, EPI_ISL_783617, EPI_ISL_783618, EPI_ISL_783619, EPI_ISL_783620, EPI_ISL_783621, EPI_ISL_783622, EPI_ISL_783623, EPI_ISL_783624, EPI_ISL_783625, EPI_ISL_783626, EPI_ISL_783627, EPI_ISL_783628, EPI_ISL_783629, EPI_ISL_783630, EPI_ISL_783631, EPI_ISL_783632, EPI_ISL_783633, EPI_ISL_783634, EPI_ISL_783635, EPI_ISL_783636, EPI_ISL_783637, EPI_ISL_783638, EPI_ISL_783639, EPI_ISL_783640, EPI_ISL_783641, EPI_ISL_783642, EPI_ISL_783643, EPI_ISL_783644, EPI_ISL_783645, EPI_ISL_783646, EPI_ISL_783647, EPI_ISL_783648, EPI_ISL_783649, EPI_ISL_783650, EPI_ISL_783651, EPI_ISL_783652, EPI_ISL_783653, EPI_ISL_783654, EPI_ISL_783655, EPI_ISL_783656, EPI_ISL_783657, EPI_ISL_783658, EPI_ISL_783659, EPI_ISL_783660, EPI_ISL_783661, EPI_ISL_783662, EPI_ISL_783663, EPI_ISL_783664, EPI_ISL_783665, EPI_ISL_783666, EPI_ISL_783667, EPI_ISL_783668, EPI_ISL_783669, EPI_ISL_783670, EPI_ISL_783671, EPI_ISL_783672, EPI_ISL_783673, EPI_ISL_783674, EPI_ISL_783675, EPI_ISL_783676, EPI_ISL_783677, EPI_ISL_783678, EPI_ISL_783679, EPI_ISL_783680, EPI_ISL_783681, EPI_ISL_783682, EPI_ISL_783683, EPI_ISL_783684, EPI_ISL_783685, EPI_ISL_783686, EPI_ISL_783687, EPI_ISL_783688, EPI_ISL_783689, EPI_ISL_783690, EPI_ISL_783691, EPI_ISL_783692, EPI_ISL_783693, EPI_ISL_783694, EPI_ISL_783695, EPI_ISL_783696, EPI_ISL_783697, EPI_ISL_783698, EPI_ISL_783699, EPI_ISL_783700, EPI_ISL_783701, EPI_ISL_783702, EPI_ISL_783703, EPI_ISL_783704, EPI_ISL_783705, EPI_ISL_783706, EPI_ISL_783707, EPI_ISL_783708, EPI_ISL_783709, EPI_ISL_783710, EPI_ISL_783711, EPI_ISL_783712, EPI_ISL_783713, EPI_ISL_783714, EPI_ISL_783715, EPI_ISL_783716, EPI_ISL_783717, EPI_ISL_783718, EPI_ISL_783719, EPI_ISL_783720, EPI_ISL_783721, EPI_ISL_783722, EPI_ISL_783723, EPI_ISL_783724, EPI_ISL_783725, EPI_ISL_783726, EPI_ISL_783727, EPI_ISL_783728, EPI_ISL_783729, EPI_ISL_783730, EPI_ISL_783731, EPI_ISL_783732, EPI_ISL_783733, EPI_ISL_783734, EPI_ISL_783735, EPI_ISL_783736, EPI_ISL_783737, EPI_ISL_783738, EPI_ISL_783739, EPI_ISL_783740, EPI_ISL_783741, EPI_ISL_783742, EPI_ISL_783743, EPI_ISL_783744, EPI_ISL_783745, EPI_ISL_783746, EPI_ISL_783747, EPI_ISL_783748, EPI_ISL_783749, EPI_ISL_783750, EPI_ISL_783751, EPI_ISL_783752, EPI_ISL_783753, EPI_ISL_783754, EPI_ISL_783755, EPI_ISL_783756, EPI_ISL_783757, EPI_ISL_783758, EPI_ISL_783759, EPI_ISL_783760, EPI_ISL_783761, EPI_ISL_783762, EPI_ISL_783763, EPI_ISL_783764, EPI_ISL_783765, EPI_ISL_783766, EPI_ISL_783767, EPI_ISL_783768, EPI_ISL_783769, EPI_ISL_783770, EPI_ISL_783771, EPI_ISL_783772, EPI_ISL_783773, EPI_ISL_783774, EPI_ISL_783775, EPI_ISL_783776, EPI_ISL_783777, EPI_ISL_783778, EPI_ISL_783779, EPI_ISL_783780, EPI_ISL_783781, EPI_ISL_783782, EPI_ISL_783783, EPI_ISL_783784, EPI_ISL_783785, EPI_ISL_783786, EPI_ISL_783787, EPI_ISL_783788, EPI_ISL_783789, EPI_ISL_783790, EPI_ISL_783791, EPI_ISL_783792, EPI_ISL_783793, EPI_ISL_783794, EPI_ISL_783795, EPI_ISL_783796, EPI_ISL_783797, EPI_ISL_783798, EPI_ISL_783799, EPI_ISL_783800, EPI_ISL_783801, EPI_ISL_783802, EPI_ISL_783803, EPI_ISL_783804, EPI_ISL_783805, EPI_ISL_783806, EPI_ISL_783807, EPI_ISL_783808, EPI_ISL_783809, EPI_ISL_783810, EPI_ISL_783811, EPI_ISL_783812, EPI_ISL_783813, EPI_ISL_783814, EPI_ISL_783815, EPI_ISL_783816, EPI_ISL_783817, EPI_ISL_783818, EPI_ISL_783819, EPI_ISL_783820, EPI_ISL_783821, EPI_ISL_783822, EPI_ISL_783823, EPI_ISL_783824, EPI_ISL_783825, EPI_ISL_783826, EPI_ISL_783827, EPI_ISL_783828, EPI_ISL_783829, EPI_ISL_783830, EPI_ISL_783831, EPI_ISL_783832, EPI_ISL_783833, EPI_ISL_783834, EPI_ISL_783835, EPI_ISL_783836, EPI_ISL_783837, EPI_ISL_783838, EPI_ISL_783839, EPI_ISL_783840, EPI_ISL_783841, EPI_ISL_783842, EPI_ISL_783843, EPI_ISL_783844, EPI_ISL_783845, EPI_ISL_783846, EPI_ISL_783847, EPI_ISL_783848, EPI_ISL_783849, EPI_ISL_783851, EPI_ISL_783852, EPI_ISL_783853, EPI_ISL_783854, EPI_ISL_783855, EPI_ISL_783856, EPI_ISL_783857, EPI_ISL_783858, EPI_ISL_783859, EPI_ISL_783860, EPI_ISL_783861, EPI_ISL_783862, EPI_ISL_783863, EPI_ISL_783864, EPI_ISL_783865, EPI_ISL_783866, EPI_ISL_783867, EPI_ISL_783868, EPI_ISL_783869, EPI_ISL_783870, EPI_ISL_783871, EPI_ISL_783872, EPI_ISL_783873, EPI_ISL_783874, EPI_ISL_783875, EPI_ISL_783876, EPI_ISL_783877, EPI_ISL_783878, EPI_ISL_783879, EPI_ISL_783880, EPI_ISL_783881, EPI_ISL_783882, EPI_ISL_783883, EPI_ISL_783884, EPI_ISL_783885, EPI_ISL_783886, EPI_ISL_783887, EPI_ISL_783888, EPI_ISL_783889, EPI_ISL_783890, EPI_ISL_783891, EPI_ISL_783892, EPI_ISL_783893, EPI_ISL_783894, EPI_ISL_783895, EPI_ISL_783896, EPI_ISL_783897, EPI_ISL_783898, EPI_ISL_783899, EPI_ISL_783900, EPI_ISL_783901, EPI_ISL_783902, EPI_ISL_783903, EPI_ISL_783904, EPI_ISL_783905, EPI_ISL_783906, EPI_ISL_783907, EPI_ISL_783908, EPI_ISL_783909, EPI_ISL_783910, EPI_ISL_783911, EPI_ISL_783912, EPI_ISL_783913, EPI_ISL_783914, EPI_ISL_783915, EPI_ISL_783916, EPI_ISL_783917, EPI_ISL_783918, EPI_ISL_783919, EPI_ISL_783920, EPI_ISL_783921, EPI_ISL_783922, EPI_ISL_783923, EPI_ISL_783924, EPI_ISL_783925, EPI_ISL_783926, EPI_ISL_783927, EPI_ISL_783928, EPI_ISL_783929, EPI_ISL_783930, EPI_ISL_783931, EPI_ISL_783932, EPI_ISL_783933, EPI_ISL_783934, EPI_ISL_783935, EPI_ISL_783936, EPI_ISL_783937, EPI_ISL_783938, EPI_ISL_783939, EPI_ISL_783940, EPI_ISL_783941, EPI_ISL_783942, EPI_ISL_783943, EPI_ISL_783944, EPI_ISL_783945, EPI_ISL_783946, EPI_ISL_783947, EPI_ISL_783948, EPI_ISL_783949, EPI_ISL_783950, EPI_ISL_783951, EPI_ISL_783952, EPI_ISL_783953, EPI_ISL_783954, EPI_ISL_783955, EPI_ISL_783956, EPI_ISL_783957, EPI_ISL_783958, EPI_ISL_783959, EPI_ISL_783960, EPI_ISL_783961, EPI_ISL_783962, EPI_ISL_783963, EPI_ISL_783964, EPI_ISL_783965, EPI_ISL_783966, EPI_ISL_783967, EPI_ISL_783968, EPI_ISL_783969, EPI_ISL_783970, EPI_ISL_783971, EPI_ISL_783972, EPI_ISL_783973, EPI_ISL_783974, EPI_ISL_783975, EPI_ISL_783976, EPI_ISL_783977, EPI_ISL_783978, EPI_ISL_783979, EPI_ISL_783980, EPI_ISL_783981, EPI_ISL_783982, EPI_ISL_783983, EPI_ISL_783984, EPI_ISL_783985, EPI_ISL_783986, EPI_ISL_783987, EPI_ISL_783988, EPI_ISL_783989, EPI_ISL_783990, EPI_ISL_783991, EPI_ISL_783992, EPI_ISL_783993, EPI_ISL_783994, EPI_ISL_783995, EPI_ISL_783996, EPI_ISL_783997, EPI_ISL_783998, EPI_ISL_783999, EPI_ISL_784000, EPI_ISL_784001, EPI_ISL_784002, EPI_ISL_784003, EPI_ISL_784004, EPI_ISL_784005, EPI_ISL_784006, EPI_ISL_784007, EPI_ISL_784008, EPI_ISL_784009, EPI_ISL_784010, EPI_ISL_784011, EPI_ISL_784012, EPI_ISL_784013, EPI_ISL_784014, EPI_ISL_784015, EPI_ISL_784016, EPI_ISL_784017, EPI_ISL_784018, EPI_ISL_784019, EPI_ISL_784020, EPI_ISL_784021, EPI_ISL_784022, EPI_ISL_784023, EPI_ISL_784024, EPI_ISL_784025, EPI_ISL_784026, EPI_ISL_784027, EPI_ISL_784028, EPI_ISL_784029, EPI_ISL_784030, EPI_ISL_784031, EPI_ISL_784032, EPI_ISL_784033, EPI_ISL_784034, EPI_ISL_784035, EPI_ISL_784036, EPI_ISL_784037, EPI_ISL_784038, EPI_ISL_784039, EPI_ISL_784040, EPI_ISL_784041, EPI_ISL_784042, EPI_ISL_784043, EPI_ISL_784044, EPI_ISL_784045, EPI_ISL_784046, EPI_ISL_784047, EPI_ISL_784048, EPI_ISL_784049, EPI_ISL_784050, EPI_ISL_784051, EPI_ISL_784052, EPI_ISL_784053, EPI_ISL_784054, EPI_ISL_784055, EPI_ISL_784056, EPI_ISL_784057, EPI_ISL_784058, EPI_ISL_784059, EPI_ISL_784060, EPI_ISL_784061, EPI_ISL_784062, EPI_ISL_784063, EPI_ISL_784064, EPI_ISL_784065, EPI_ISL_784066, EPI_ISL_784067, EPI_ISL_784068, EPI_ISL_784069, EPI_ISL_784070, EPI_ISL_784071, EPI_ISL_784072, EPI_ISL_784073, EPI_ISL_784074, EPI_ISL_784075, EPI_ISL_784076, EPI_ISL_784077, EPI_ISL_784078, EPI_ISL_784079, EPI_ISL_784080, EPI_ISL_784081, EPI_ISL_784082, EPI_ISL_784083, EPI_ISL_784084, EPI_ISL_784085, EPI_ISL_784086, EPI_ISL_784087, EPI_ISL_784088, EPI_ISL_784089, EPI_ISL_784090, EPI_ISL_784091, EPI_ISL_784092, EPI_ISL_784093, EPI_ISL_784094, EPI_ISL_784095, EPI_ISL_784096, EPI_ISL_784097, EPI_ISL_784098, EPI_ISL_784099, EPI_ISL_784100, EPI_ISL_784101, EPI_ISL_784102, EPI_ISL_784103, EPI_ISL_784104, EPI_ISL_784105, EPI_ISL_784106, EPI_ISL_784107, EPI_ISL_784108, EPI_ISL_784109, EPI_ISL_784110, EPI_ISL_784111, EPI_ISL_784112, EPI_ISL_784113, EPI_ISL_784114, EPI_ISL_784115, EPI_ISL_784116, EPI_ISL_784117, EPI_ISL_784118, EPI_ISL_784119, EPI_ISL_784120, EPI_ISL_784121, EPI_ISL_784122, EPI_ISL_784123, EPI_ISL_784124, EPI_ISL_784125, EPI_ISL_784126, EPI_ISL_784127, EPI_ISL_784128, EPI_ISL_784129, EPI_ISL_784130, EPI_ISL_784131, EPI_ISL_784132, EPI_ISL_784133, EPI_ISL_784134, EPI_ISL_784135, EPI_ISL_784136, EPI_ISL_784137, EPI_ISL_784138, EPI_ISL_784139, EPI_ISL_784140, EPI_ISL_784141, EPI_ISL_784142, EPI_ISL_784143, EPI_ISL_784144, EPI_ISL_784145, EPI_ISL_784146, EPI_ISL_784147, EPI_ISL_784148, EPI_ISL_784149, EPI_ISL_784150, EPI_ISL_784151, EPI_ISL_784152, EPI_ISL_784153, EPI_ISL_784154, EPI_ISL_784155, EPI_ISL_784156, EPI_ISL_784157, EPI_ISL_784158, EPI_ISL_784159, EPI_ISL_784160, EPI_ISL_784161, EPI_ISL_784162, EPI_ISL_784163, EPI_ISL_784164, EPI_ISL_784165, EPI_ISL_784166, EPI_ISL_784167, EPI_ISL_784168, EPI_ISL_784169, EPI_ISL_784170, EPI_ISL_784171, EPI_ISL_784172, EPI_ISL_784173, EPI_ISL_784174, EPI_ISL_784175, EPI_ISL_784176, EPI_ISL_784177, EPI_ISL_784178, EPI_ISL_784179, EPI_ISL_784180, EPI_ISL_784181, EPI_ISL_784182, EPI_ISL_784183, EPI_ISL_784184, EPI_ISL_784185, EPI_ISL_784186, EPI_ISL_784187, EPI_ISL_784188, EPI_ISL_784189, EPI_ISL_784190, EPI_ISL_784191, EPI_ISL_784192, EPI_ISL_784193, EPI_ISL_784194, EPI_ISL_784195, EPI_ISL_784196, EPI_ISL_784197, EPI_ISL_784198, EPI_ISL_784199, EPI_ISL_784200, EPI_ISL_784201, EPI_ISL_784202, EPI_ISL_784203, EPI_ISL_784204, EPI_ISL_784205, EPI_ISL_784206, EPI_ISL_784207, EPI_ISL_784208, EPI_ISL_784209, EPI_ISL_784210, EPI_ISL_784211, EPI_ISL_784212, EPI_ISL_784213, EPI_ISL_784214, EPI_ISL_784215, EPI_ISL_784216, EPI_ISL_784217, EPI_ISL_784218, EPI_ISL_784219, EPI_ISL_784220, EPI_ISL_784221, EPI_ISL_784222, EPI_ISL_784223, EPI_ISL_784224, EPI_ISL_784225, EPI_ISL_784226, EPI_ISL_784227, EPI_ISL_784228, EPI_ISL_784229, EPI_ISL_784230, EPI_ISL_784231, EPI_ISL_784232, EPI_ISL_784233, EPI_ISL_784234, EPI_ISL_784235, EPI_ISL_784236, EPI_ISL_784237, EPI_ISL_784238, EPI_ISL_784239, EPI_ISL_784240, EPI_ISL_784241, EPI_ISL_784242, EPI_ISL_784243, EPI_ISL_784244, EPI_ISL_784245, EPI_ISL_784246, EPI_ISL_784247, EPI_ISL_784248, EPI_ISL_784249, EPI_ISL_784250, EPI_ISL_784251, EPI_ISL_784252, EPI_ISL_784253, EPI_ISL_784254, EPI_ISL_784255, EPI_ISL_784256, EPI_ISL_784257, EPI_ISL_784258, EPI_ISL_784259, EPI_ISL_784260, EPI_ISL_784261, EPI_ISL_784262, EPI_ISL_784263, EPI_ISL_784264, EPI_ISL_784265, EPI_ISL_784266, EPI_ISL_784267, EPI_ISL_784268, EPI_ISL_784269, EPI_ISL_784270, EPI_ISL_784271, EPI_ISL_784272, EPI_ISL_784273, EPI_ISL_784274, EPI_ISL_784275, EPI_ISL_784276, EPI_ISL_784277, EPI_ISL_784278, EPI_ISL_784279, EPI_ISL_784280, EPI_ISL_784281, EPI_ISL_784282, EPI_ISL_784283, EPI_ISL_784284, EPI_ISL_784285, EPI_ISL_784286, EPI_ISL_784287, EPI_ISL_784288, EPI_ISL_784289, EPI_ISL_784290, EPI_ISL_784291, EPI_ISL_784292, EPI_ISL_784293, EPI_ISL_784294, EPI_ISL_784295, EPI_ISL_784296, EPI_ISL_784297, EPI_ISL_784298, EPI_ISL_784299, EPI_ISL_784300, EPI_ISL_784301, EPI_ISL_784302, EPI_ISL_784303, EPI_ISL_784304, EPI_ISL_784305, EPI_ISL_784306, EPI_ISL_784307, EPI_ISL_784308, EPI_ISL_784309, EPI_ISL_784310, EPI_ISL_784311, EPI_ISL_784312, EPI_ISL_784313, EPI_ISL_784314, EPI_ISL_784315, EPI_ISL_784316, EPI_ISL_784317, EPI_ISL_784318, EPI_ISL_784319, EPI_ISL_784320, EPI_ISL_784321, EPI_ISL_784322, EPI_ISL_784323, EPI_ISL_784324, EPI_ISL_784325, EPI_ISL_784326, EPI_ISL_784327, EPI_ISL_784328, EPI_ISL_784329, EPI_ISL_784330, EPI_ISL_784331, EPI_ISL_784332, EPI_ISL_784333, EPI_ISL_784334, EPI_ISL_784335, EPI_ISL_784336, EPI_ISL_784337, EPI_ISL_784338, EPI_ISL_784339, EPI_ISL_784340, EPI_ISL_784341, EPI_ISL_784342, EPI_ISL_784343, EPI_ISL_784344, EPI_ISL_784345, EPI_ISL_784346, EPI_ISL_784347, EPI_ISL_784348, EPI_ISL_784349, EPI_ISL_784350, EPI_ISL_784351, EPI_ISL_784352, EPI_ISL_784353, EPI_ISL_784354, EPI_ISL_784355, EPI_ISL_784356, EPI_ISL_784357, EPI_ISL_784358, EPI_ISL_784359, EPI_ISL_784360, EPI_ISL_784361, EPI_ISL_784362, EPI_ISL_784363, EPI_ISL_784364, EPI_ISL_784365, EPI_ISL_784366, EPI_ISL_784367, EPI_ISL_784368, EPI_ISL_784369, EPI_ISL_784370, EPI_ISL_784371, EPI_ISL_784372, EPI_ISL_784373, EPI_ISL_784374, EPI_ISL_784375, EPI_ISL_784376, EPI_ISL_784377, EPI_ISL_784378, EPI_ISL_784379, EPI_ISL_784380, EPI_ISL_784381, EPI_ISL_784382, EPI_ISL_784383, EPI_ISL_784384, EPI_ISL_784385, EPI_ISL_784386, EPI_ISL_784387, EPI_ISL_784388, EPI_ISL_784389, EPI_ISL_784390, EPI_ISL_784391, EPI_ISL_784392, EPI_ISL_784393, EPI_ISL_784394, EPI_ISL_784395, EPI_ISL_784396, EPI_ISL_784397, EPI_ISL_784398, EPI_ISL_784399, EPI_ISL_784400, EPI_ISL_784401, EPI_ISL_784402, EPI_ISL_784403, EPI_ISL_784404, EPI_ISL_784405, EPI_ISL_784406, EPI_ISL_784407, EPI_ISL_784408, EPI_ISL_784409, EPI_ISL_784410, EPI_ISL_784411, EPI_ISL_784412, EPI_ISL_784413, EPI_ISL_784414, EPI_ISL_784415, EPI_ISL_784416, EPI_ISL_784417, EPI_ISL_784418, EPI_ISL_784419, EPI_ISL_784420, EPI_ISL_784421, EPI_ISL_784422, EPI_ISL_784423, EPI_ISL_784424, EPI_ISL_784425, EPI_ISL_784426, EPI_ISL_784427, EPI_ISL_784428, EPI_ISL_784429, EPI_ISL_784430, EPI_ISL_784431, EPI_ISL_784432, EPI_ISL_784433, EPI_ISL_784434, EPI_ISL_784435, EPI_ISL_784436, EPI_ISL_784437, EPI_ISL_784438, EPI_ISL_784439, EPI_ISL_784440, EPI_ISL_784441, EPI_ISL_784442, EPI_ISL_784443, EPI_ISL_784444, EPI_ISL_784445, EPI_ISL_784446, EPI_ISL_784447, EPI_ISL_784448, EPI_ISL_784449, EPI_ISL_784450, EPI_ISL_784451, EPI_ISL_784452, EPI_ISL_784453, EPI_ISL_784454, EPI_ISL_784455, EPI_ISL_784456, EPI_ISL_784457, EPI_ISL_784458, EPI_ISL_784459, EPI_ISL_784460, EPI_ISL_784461, EPI_ISL_784462, EPI_ISL_784463, EPI_ISL_784464, EPI_ISL_784465, EPI_ISL_784466, EPI_ISL_784467, EPI_ISL_784468, EPI_ISL_784469, EPI_ISL_784470, EPI_ISL_784471, EPI_ISL_784472, EPI_ISL_784473, EPI_ISL_784474, EPI_ISL_784475, EPI_ISL_784476, EPI_ISL_784477, EPI_ISL_784478, EPI_ISL_784479, EPI_ISL_784480, EPI_ISL_784481, EPI_ISL_784482, EPI_ISL_784483, EPI_ISL_784484, EPI_ISL_784485, EPI_ISL_784486, EPI_ISL_784487, EPI_ISL_784488, EPI_ISL_784489, EPI_ISL_784490, EPI_ISL_784491, EPI_ISL_784492, EPI_ISL_784493, EPI_ISL_784494, EPI_ISL_784495, EPI_ISL_784496, EPI_ISL_784497, EPI_ISL_784498, EPI_ISL_784499, EPI_ISL_784500, EPI_ISL_784501, EPI_ISL_784502, EPI_ISL_784503, EPI_ISL_784504, EPI_ISL_784505, EPI_ISL_784506, EPI_ISL_784507, EPI_ISL_784508, EPI_ISL_784509, EPI_ISL_784510, EPI_ISL_784511, EPI_ISL_784512, EPI_ISL_784513, EPI_ISL_784514, EPI_ISL_784515, EPI_ISL_784516, EPI_ISL_784517, EPI_ISL_784518, EPI_ISL_784519, EPI_ISL_784520, EPI_ISL_784521, EPI_ISL_784522, EPI_ISL_784523, EPI_ISL_784524, EPI_ISL_784525, EPI_ISL_784526, EPI_ISL_784527, EPI_ISL_784528, EPI_ISL_784529, EPI_ISL_784530, EPI_ISL_784531, EPI_ISL_784532, EPI_ISL_784533, EPI_ISL_784534, EPI_ISL_784535, EPI_ISL_784536, EPI_ISL_784537, EPI_ISL_784538, EPI_ISL_784539, EPI_ISL_784540, EPI_ISL_784541, EPI_ISL_784542, EPI_ISL_784543, EPI_ISL_784544, EPI_ISL_784545, EPI_ISL_784546, EPI_ISL_784547, EPI_ISL_784548, EPI_ISL_784549, EPI_ISL_784550, EPI_ISL_784551, EPI_ISL_784552, EPI_ISL_784553, EPI_ISL_784554, EPI_ISL_784555, EPI_ISL_784556, EPI_ISL_784557, EPI_ISL_784558, EPI_ISL_784559, EPI_ISL_784560, EPI_ISL_784561, EPI_ISL_784562, EPI_ISL_784563, EPI_ISL_784564, EPI_ISL_784565, EPI_ISL_784566, EPI_ISL_784567, EPI_ISL_784568, EPI_ISL_784569, EPI_ISL_784570, EPI_ISL_784571, EPI_ISL_784572, EPI_ISL_784573, EPI_ISL_784574, EPI_ISL_784575, EPI_ISL_784576, EPI_ISL_784577, EPI_ISL_784578, EPI_ISL_784579, EPI_ISL_784580, EPI_ISL_784581, EPI_ISL_784582, EPI_ISL_784583, EPI_ISL_784584, EPI_ISL_784585, EPI_ISL_784586, EPI_ISL_784587, EPI_ISL_784588, EPI_ISL_784589, EPI_ISL_784590, EPI_ISL_784591, EPI_ISL_784592, EPI_ISL_784593, EPI_ISL_784594, EPI_ISL_784595, EPI_ISL_784596, EPI_ISL_784597, EPI_ISL_784598, EPI_ISL_784599, EPI_ISL_784600, EPI_ISL_784601, EPI_ISL_784602, EPI_ISL_784603, EPI_ISL_784604, EPI_ISL_784605, EPI_ISL_784606, EPI_ISL_784607, EPI_ISL_784608, EPI_ISL_784609, EPI_ISL_784610, EPI_ISL_784611, EPI_ISL_784612, EPI_ISL_784613, EPI_ISL_784614, EPI_ISL_784615, EPI_ISL_784616, EPI_ISL_784617, EPI_ISL_784618, EPI_ISL_784619, EPI_ISL_784620, EPI_ISL_784621, EPI_ISL_784622, EPI_ISL_784623, EPI_ISL_784624, EPI_ISL_784625, EPI_ISL_784626, EPI_ISL_784627, EPI_ISL_784628, EPI_ISL_784629, EPI_ISL_784630, EPI_ISL_784631, EPI_ISL_784632, EPI_ISL_784633, EPI_ISL_784634, EPI_ISL_784635, EPI_ISL_784636, EPI_ISL_784637, EPI_ISL_784638, EPI_ISL_784639, EPI_ISL_784640, EPI_ISL_784641, EPI_ISL_784642, EPI_ISL_784643, EPI_ISL_784644, EPI_ISL_784645, EPI_ISL_784646, EPI_ISL_784647, EPI_ISL_784648, EPI_ISL_784649, EPI_ISL_784650, EPI_ISL_784651, EPI_ISL_784652, EPI_ISL_784653, EPI_ISL_784654, EPI_ISL_784655, EPI_ISL_784656, EPI_ISL_784657, EPI_ISL_784658, EPI_ISL_784659, EPI_ISL_784660, EPI_ISL_784661, EPI_ISL_784662, EPI_ISL_784663, EPI_ISL_784664, EPI_ISL_784665, EPI_ISL_784666, EPI_ISL_784667, EPI_ISL_784668, EPI_ISL_784669, EPI_ISL_784670, EPI_ISL_784671, EPI_ISL_784672,

EPI_ISL_784673, EPI_ISL_784674, EPI_ISL_784675, EPI_ISL_784676, EPI_ISL_784677, EPI_ISL_784678, EPI_ISL_784679, EPI_ISL_784680, EPI_ISL_784681, EPI_ISL_784682, EPI_ISL_784683, EPI_ISL_784684, EPI_ISL_784685, EPI_ISL_784686, EPI_ISL_784687, EPI_ISL_784688, EPI_ISL_784689, EPI_ISL_784690, EPI_ISL_784691, EPI_ISL_784692, EPI_ISL_784693, EPI_ISL_784694, EPI_ISL_784695, EPI_ISL_784696,
EPI_ISL_784697, EPI_ISL_784698, EPI_ISL_784699, EPI_ISL_784700, EPI_ISL_784701, EPI_ISL_784702, EPI_ISL_784703, EPI_ISL_784704, EPI_ISL_784705, EPI_ISL_784706, EPI_ISL_784707, EPI_ISL_784708, EPI_ISL_784709, EPI_ISL_784710, EPI_ISL_784711, EPI_ISL_784712, EPI_ISL_784713, EPI_ISL_784714, EPI_ISL_784715, EPI_ISL_784716, EPI_ISL_784717, EPI_ISL_784718, EPI_ISL_784719, EPI_ISL_784720,
EPI_ISL_784721, EPI_ISL_784722, EPI_ISL_784723, EPI_ISL_784724, EPI_ISL_784725, EPI_ISL_784726, EPI_ISL_784727, EPI_ISL_784728, EPI_ISL_784729, EPI_ISL_784730, EPI_ISL_784731, EPI_ISL_784732, EPI_ISL_784733, EPI_ISL_784734, EPI_ISL_784735, EPI_ISL_784736, EPI_ISL_784737, EPI_ISL_784738, EPI_ISL_784739, EPI_ISL_784740, EPI_ISL_784741, EPI_ISL_784742, EPI_ISL_784743, EPI_ISL_784744,
EPI_ISL_784745, EPI_ISL_784746, EPI_ISL_784747, EPI_ISL_784748, EPI_ISL_784749, EPI_ISL_784750, EPI_ISL_784751, EPI_ISL_784752, EPI_ISL_784753, EPI_ISL_784754, EPI_ISL_784755, EPI_ISL_784756, EPI_ISL_784757, EPI_ISL_784758, EPI_ISL_784759, EPI_ISL_784760, EPI_ISL_784761, EPI_ISL_784762, EPI_ISL_784763, EPI_ISL_784764, EPI_ISL_784765, EPI_ISL_784766, EPI_ISL_784767, EPI_ISL_784768,
EPI_ISL_784769, EPI_ISL_784770, EPI_ISL_784771, EPI_ISL_784772, EPI_ISL_784773, EPI_ISL_784774, EPI_ISL_784775, EPI_ISL_784776, EPI_ISL_784777, EPI_ISL_784778, EPI_ISL_784779, EPI_ISL_784780, EPI_ISL_784781, EPI_ISL_784782, EPI_ISL_784783, EPI_ISL_784784, EPI_ISL_784785, EPI_ISL_784786, EPI_ISL_784787, EPI_ISL_784788, EPI_ISL_784789, EPI_ISL_784790, EPI_ISL_784791, EPI_ISL_784792,
EPI_ISL_784793, EPI_ISL_784794, EPI_ISL_784795, EPI_ISL_784796, EPI_ISL_784797, EPI_ISL_784798, EPI_ISL_784799, EPI_ISL_784800, EPI_ISL_784801, EPI_ISL_784802, EPI_ISL_784803, EPI_ISL_784804, EPI_ISL_784805, EPI_ISL_784806, EPI_ISL_784807, EPI_ISL_784808, EPI_ISL_784809, EPI_ISL_784810, EPI_ISL_784811, EPI_ISL_784812, EPI_ISL_784813, EPI_ISL_784814, EPI_ISL_784815, EPI_ISL_784816,
EPI_ISL_784817, EPI_ISL_784818, EPI_ISL_784819, EPI_ISL_784820, EPI_ISL_784821, EPI_ISL_784822, EPI_ISL_784823, EPI_ISL_784824, EPI_ISL_784825, EPI_ISL_784826, EPI_ISL_784827, EPI_ISL_784828, EPI_ISL_784829, EPI_ISL_784830, EPI_ISL_784831, EPI_ISL_784832, EPI_ISL_784833, EPI_ISL_784834, EPI_ISL_784835, EPI_ISL_784836, EPI_ISL_784837, EPI_ISL_784838, EPI_ISL_784839, EPI_ISL_784840,
EPI_ISL_784841, EPI_ISL_784842, EPI_ISL_784843, EPI_ISL_784844, EPI_ISL_784845, EPI_ISL_784846, EPI_ISL_784847, EPI_ISL_784848, EPI_ISL_784849, EPI_ISL_784850, EPI_ISL_784851, EPI_ISL_784852, EPI_ISL_784853, EPI_ISL_784854, EPI_ISL_784855, EPI_ISL_784856, EPI_ISL_784857, EPI_ISL_784858, EPI_ISL_784859, EPI_ISL_784860, EPI_ISL_784861, EPI_ISL_784862, EPI_ISL_784863, EPI_ISL_784864,
EPI_ISL_784865, EPI_ISL_784866, EPI_ISL_784867, EPI_ISL_784868, EPI_ISL_784869, EPI_ISL_784870, EPI_ISL_784871, EPI_ISL_784872, EPI_ISL_784873, EPI_ISL_784874, EPI_ISL_784875, EPI_ISL_784876, EPI_ISL_784877, EPI_ISL_784878, EPI_ISL_784879, EPI_ISL_784880, EPI_ISL_784881, EPI_ISL_784882, EPI_ISL_784883, EPI_ISL_784884, EPI_ISL_784885, EPI_ISL_784886, EPI_ISL_784887, EPI_ISL_784888,
EPI_ISL_784889, EPI_ISL_784890, EPI_ISL_784891, EPI_ISL_784892, EPI_ISL_784893, EPI_ISL_784894, EPI_ISL_784895, EPI_ISL_784896, EPI_ISL_784897, EPI_ISL_784898, EPI_ISL_784899, EPI_ISL_784900, EPI_ISL_784901, EPI_ISL_784902, EPI_ISL_784903, EPI_ISL_784904, EPI_ISL_784905, EPI_ISL_784906, EPI_ISL_784907, EPI_ISL_784908, EPI_ISL_784909, EPI_ISL_784910, EPI_ISL_784911, EPI_ISL_784912,
EPI_ISL_784913, EPI_ISL_784914, EPI_ISL_784915, EPI_ISL_784916, EPI_ISL_784917, EPI_ISL_784918, EPI_ISL_784919, EPI_ISL_784920, EPI_ISL_784921, EPI_ISL_784922, EPI_ISL_784923, EPI_ISL_784924, EPI_ISL_784925, EPI_ISL_784926, EPI_ISL_784927, EPI_ISL_784928, EPI_ISL_784929, EPI_ISL_784930, EPI_ISL_784931, EPI_ISL_784932, EPI_ISL_784933, EPI_ISL_784934, EPI_ISL_784935, EPI_ISL_784936,
EPI_ISL_784937, EPI_ISL_784938, EPI_ISL_784939, EPI_ISL_784940, EPI_ISL_784941, EPI_ISL_784942, EPI_ISL_784943, EPI_ISL_784944, EPI_ISL_784945, EPI_ISL_784946, EPI_ISL_784947, EPI_ISL_784948, EPI_ISL_784949, EPI_ISL_784950, EPI_ISL_784951, EPI_ISL_784952, EPI_ISL_784953, EPI_ISL_784954, EPI_ISL_784955, EPI_ISL_784956, EPI_ISL_784957, EPI_ISL_784958, EPI_ISL_784959, EPI_ISL_784960,
EPI_ISL_784961, EPI_ISL_784962, EPI_ISL_784963, EPI_ISL_784964, EPI_ISL_784965, EPI_ISL_784966, EPI_ISL_784967, EPI_ISL_784968, EPI_ISL_784969, EPI_ISL_784970, EPI_ISL_784971, EPI_ISL_784972, EPI_ISL_784973, EPI_ISL_784974, EPI_ISL_784975, EPI_ISL_784976, EPI_ISL_784977, EPI_ISL_784978, EPI_ISL_784979, EPI_ISL_784980, EPI_ISL_784981, EPI_ISL_784982, EPI_ISL_784983, EPI_ISL_784984,
EPI_ISL_784985, EPI_ISL_784986, EPI_ISL_784987, EPI_ISL_784988, EPI_ISL_784989, EPI_ISL_784990, EPI_ISL_784991, EPI_ISL_784992, EPI_ISL_784993, EPI_ISL_784994, EPI_ISL_784995, EPI_ISL_784996, EPI_ISL_784997, EPI_ISL_784998, EPI_ISL_784999, EPI_ISL_785000, EPI_ISL_785001, EPI_ISL_785002, EPI_ISL_785003, EPI_ISL_785004, EPI_ISL_785005, EPI_ISL_785006, EPI_ISL_785007, EPI_ISL_785008,
EPI_ISL_785009, EPI_ISL_785010, EPI_ISL_785011, EPI_ISL_785012, EPI_ISL_785013, EPI_ISL_785014, EPI_ISL_785015, EPI_ISL_785016, EPI_ISL_785017, EPI_ISL_785018, EPI_ISL_785019, EPI_ISL_785020, EPI_ISL_785021, EPI_ISL_785022, EPI_ISL_785023, EPI_ISL_785024, EPI_ISL_785025, EPI_ISL_785026, EPI_ISL_785027, EPI_ISL_785028, EPI_ISL_785029, EPI_ISL_785030, EPI_ISL_785031, EPI_ISL_785032,
EPI_ISL_785033, EPI_ISL_785034, EPI_ISL_785035, EPI_ISL_785036, EPI_ISL_785037, EPI_ISL_785038, EPI_ISL_785039, EPI_ISL_785040, EPI_ISL_785041, EPI_ISL_785042, EPI_ISL_785043, EPI_ISL_785044, EPI_ISL_785045, EPI_ISL_785046, EPI_ISL_785047, EPI_ISL_785048, EPI_ISL_785049, EPI_ISL_785050, EPI_ISL_785051, EPI_ISL_785052, EPI_ISL_785053, EPI_ISL_785054, EPI_ISL_785055, EPI_ISL_785056,
EPI_ISL_785057, EPI_ISL_785058, EPI_ISL_785059, EPI_ISL_785060, EPI_ISL_785061, EPI_ISL_785062, EPI_ISL_785063, EPI_ISL_785064, EPI_ISL_785065, EPI_ISL_785066, EPI_ISL_785067, EPI_ISL_785068, EPI_ISL_785069, EPI_ISL_785070, EPI_ISL_785071, EPI_ISL_785072, EPI_ISL_785073, EPI_ISL_785074, EPI_ISL_785075, EPI_ISL_785076, EPI_ISL_785077, EPI_ISL_785078, EPI_ISL_785079, EPI_ISL_785080,
EPI_ISL_785081, EPI_ISL_785082, EPI_ISL_785083, EPI_ISL_785084, EPI_ISL_785085, EPI_ISL_785086, EPI_ISL_785087, EPI_ISL_785088, EPI_ISL_785089, EPI_ISL_785090, EPI_ISL_785091, EPI_ISL_785092, EPI_ISL_785093, EPI_ISL_785094, EPI_ISL_785095, EPI_ISL_785096, EPI_ISL_785097, EPI_ISL_785098, EPI_ISL_785099, EPI_ISL_785100, EPI_ISL_785101, EPI_ISL_785102, EPI_ISL_785103, EPI_ISL_785104,
EPI_ISL_785105, EPI_ISL_785106, EPI_ISL_785107, EPI_ISL_785108, EPI_ISL_785109, EPI_ISL_785110, EPI_ISL_785111, EPI_ISL_785112, EPI_ISL_785113, EPI_ISL_785114, EPI_ISL_785115, EPI_ISL_785116, EPI_ISL_785117, EPI_ISL_785118, EPI_ISL_785119, EPI_ISL_785120, EPI_ISL_785121, EPI_ISL_785122, EPI_ISL_785123, EPI_ISL_785124, EPI_ISL_785125, EPI_ISL_785126, EPI_ISL_785127, EPI_ISL_785128,
EPI_ISL_785129, EPI_ISL_785130, EPI_ISL_785131, EPI_ISL_785132, EPI_ISL_785133, EPI_ISL_785134, EPI_ISL_785135, EPI_ISL_785136, EPI_ISL_785137, EPI_ISL_785138, EPI_ISL_785139, EPI_ISL_785140, EPI_ISL_785141, EPI_ISL_785142, EPI_ISL_785143, EPI_ISL_785144, EPI_ISL_785145, EPI_ISL_785146, EPI_ISL_785147, EPI_ISL_785148, EPI_ISL_785149, EPI_ISL_785150, EPI_ISL_785151, EPI_ISL_785152,
EPI_ISL_785153, EPI_ISL_785154, EPI_ISL_785155, EPI_ISL_785156, EPI_ISL_785157, EPI_ISL_785158, EPI_ISL_785159, EPI_ISL_785160, EPI_ISL_785161, EPI_ISL_785162, EPI_ISL_785163, EPI_ISL_785164, EPI_ISL_785165, EPI_ISL_785166, EPI_ISL_785167, EPI_ISL_785168, EPI_ISL_785169, EPI_ISL_785170, EPI_ISL_785171, EPI_ISL_785172, EPI_ISL_785173, EPI_ISL_785174, EPI_ISL_785175, EPI_ISL_785176,
EPI_ISL_785177, EPI_ISL_785178, EPI_ISL_785179, EPI_ISL_785180, EPI_ISL_785181, EPI_ISL_785182, EPI_ISL_785183, EPI_ISL_785184, EPI_ISL_785185, EPI_ISL_785186, EPI_ISL_785187, EPI_ISL_785188, EPI_ISL_785189, EPI_ISL_785190, EPI_ISL_785191, EPI_ISL_785192, EPI_ISL_785193, EPI_ISL_785194, EPI_ISL_785195, EPI_ISL_785196, EPI_ISL_785197, EPI_ISL_785198, EPI_ISL_785199, EPI_ISL_785200,
EPI_ISL_785201, EPI_ISL_785202, EPI_ISL_785203, EPI_ISL_785204, EPI_ISL_785205, EPI_ISL_785206, EPI_ISL_785207, EPI_ISL_785208, EPI_ISL_785209, EPI_ISL_785210, EPI_ISL_785211, EPI_ISL_785212, EPI_ISL_785213, EPI_ISL_785214, EPI_ISL_785215, EPI_ISL_785216, EPI_ISL_785217, EPI_ISL_785218, EPI_ISL_785219, EPI_ISL_785220, EPI_ISL_785221, EPI_ISL_785222, EPI_ISL_785223, EPI_ISL_785224,
EPI_ISL_785225, EPI_ISL_785226, EPI_ISL_785227, EPI_ISL_785228, EPI_ISL_785229, EPI_ISL_785230, EPI_ISL_785231, EPI_ISL_785232, EPI_ISL_785233, EPI_ISL_785234, EPI_ISL_785235, EPI_ISL_785236, EPI_ISL_785237, EPI_ISL_785238, EPI_ISL_785239, EPI_ISL_785240, EPI_ISL_785241, EPI_ISL_785242, EPI_ISL_785243, EPI_ISL_785244, EPI_ISL_785245, EPI_ISL_785246, EPI_ISL_785247, EPI_ISL_785248,
EPI_ISL_785249, EPI_ISL_785250, EPI_ISL_785251, EPI_ISL_785252, EPI_ISL_785253, EPI_ISL_785254, EPI_ISL_785255, EPI_ISL_785256, EPI_ISL_785257, EPI_ISL_785258, EPI_ISL_785259, EPI_ISL_785260, EPI_ISL_785261, EPI_ISL_785262, EPI_ISL_785263, EPI_ISL_785264, EPI_ISL_785265, EPI_ISL_785266, EPI_ISL_785267, EPI_ISL_785268, EPI_ISL_785269, EPI_ISL_785270, EPI_ISL_785271, EPI_ISL_785272,
EPI_ISL_785273, EPI_ISL_785274, EPI_ISL_785275, EPI_ISL_785276, EPI_ISL_785277, EPI_ISL_785278, EPI_ISL_785279, EPI_ISL_785280, EPI_ISL_785281, EPI_ISL_785282, EPI_ISL_785283, EPI_ISL_785284, EPI_ISL_785285, EPI_ISL_785286, EPI_ISL_785287, EPI_ISL_785288, EPI_ISL_785289, EPI_ISL_785290, EPI_ISL_785291, EPI_ISL_785292, EPI_ISL_785293, EPI_ISL_785294, EPI_ISL_785295, EPI_ISL_785296,
EPI_ISL_785297, EPI_ISL_785298, EPI_ISL_785299, EPI_ISL_785300, EPI_ISL_785301, EPI_ISL_785302, EPI_ISL_785303, EPI_ISL_785304, EPI_ISL_785305, EPI_ISL_785306, EPI_ISL_785307, EPI_ISL_785308, EPI_ISL_785309, EPI_ISL_785310, EPI_ISL_785311, EPI_ISL_785312, EPI_ISL_785313, EPI_ISL_785314, EPI_ISL_785315, EPI_ISL_785316, EPI_ISL_785317, EPI_ISL_785318, EPI_ISL_785319, EPI_ISL_785320,
EPI_ISL_785321, EPI_ISL_785322, EPI_ISL_785323, EPI_ISL_785324, EPI_ISL_785325, EPI_ISL_785326, EPI_ISL_785327, EPI_ISL_785328, EPI_ISL_785329, EPI_ISL_785330, EPI_ISL_785331, EPI_ISL_785332, EPI_ISL_785333, EPI_ISL_785334, EPI_ISL_785335, EPI_ISL_785336, EPI_ISL_785337, EPI_ISL_785338, EPI_ISL_785339, EPI_ISL_785340, EPI_ISL_785341, EPI_ISL_785342, EPI_ISL_785343, EPI_ISL_785344,
EPI_ISL_785345, EPI_ISL_785346, EPI_ISL_785347, EPI_ISL_785348, EPI_ISL_785349, EPI_ISL_785350, EPI_ISL_785351, EPI_ISL_785352, EPI_ISL_785353, EPI_ISL_785354, EPI_ISL_785355, EPI_ISL_785356, EPI_ISL_785357, EPI_ISL_785358, EPI_ISL_785359, EPI_ISL_785360, EPI_ISL_785361, EPI_ISL_785362, EPI_ISL_785363, EPI_ISL_785364, EPI_ISL_785365, EPI_ISL_785366, EPI_ISL_785367, EPI_ISL_785368,
EPI_ISL_785369, EPI_ISL_785370, EPI_ISL_785371, EPI_ISL_785372, EPI_ISL_785373, EPI_ISL_785374, EPI_ISL_785375, EPI_ISL_785376, EPI_ISL_785377, EPI_ISL_785378, EPI_ISL_785379, EPI_ISL_785380, EPI_ISL_785381, EPI_ISL_785382, EPI_ISL_785383, EPI_ISL_785384, EPI_ISL_785385, EPI_ISL_785386, EPI_ISL_785387, EPI_ISL_785388, EPI_ISL_785389, EPI_ISL_785390, EPI_ISL_785391, EPI_ISL_785392,
EPI_ISL_785393, EPI_ISL_785394, EPI_ISL_785395, EPI_ISL_785396, EPI_ISL_785397, EPI_ISL_785398, EPI_ISL_785399, EPI_ISL_785400, EPI_ISL_785401, EPI_ISL_785402, EPI_ISL_785403, EPI_ISL_785404, EPI_ISL_785405, EPI_ISL_785406, EPI_ISL_785407, EPI_ISL_785408, EPI_ISL_785409, EPI_ISL_785410, EPI_ISL_785411, EPI_ISL_785412, EPI_ISL_785413, EPI_ISL_785414, EPI_ISL_785415, EPI_ISL_785416,
EPI_ISL_785417, EPI_ISL_785418, EPI_ISL_785419, EPI_ISL_785420, EPI_ISL_785421, EPI_ISL_785422, EPI_ISL_785423, EPI_ISL_785424, EPI_ISL_785425, EPI_ISL_785426, EPI_ISL_785427, EPI_ISL_785428, EPI_ISL_785429, EPI_ISL_785430, EPI_ISL_785431, EPI_ISL_785432, EPI_ISL_785433, EPI_ISL_785434, EPI_ISL_785435, EPI_ISL_785436, EPI_ISL_785437, EPI_ISL_785438, EPI_ISL_785439, EPI_ISL_785440,
EPI_ISL_785441, EPI_ISL_785442, EPI_ISL_785443, EPI_ISL_785444, EPI_ISL_785445, EPI_ISL_785446, EPI_ISL_785447, EPI_ISL_785448, EPI_ISL_785449, EPI_ISL_785450, EPI_ISL_785451, EPI_ISL_785452, EPI_ISL_785453, EPI_ISL_785454, EPI_ISL_785455, EPI_ISL_785456, EPI_ISL_785457, EPI_ISL_785458, EPI_ISL_785459, EPI_ISL_785460, EPI_ISL_785461, EPI_ISL_785462, EPI_ISL_785463, EPI_ISL_785464,
EPI_ISL_785465, EPI_ISL_785466, EPI_ISL_785467, EPI_ISL_785468, EPI_ISL_785469, EPI_ISL_785470, EPI_ISL_785471, EPI_ISL_785472, EPI_ISL_785473, EPI_ISL_785474, EPI_ISL_785475, EPI_ISL_785476, EPI_ISL_785477, EPI_ISL_785478, EPI_ISL_785479, EPI_ISL_785480, EPI_ISL_785481, EPI_ISL_785482, EPI_ISL_785483, EPI_ISL_785484, EPI_ISL_785485, EPI_ISL_785486, EPI_ISL_785487, EPI_ISL_785488,
EPI_ISL_785489, EPI_ISL_785490, EPI_ISL_785491, EPI_ISL_785492, EPI_ISL_785493, EPI_ISL_785494, EPI_ISL_785495, EPI_ISL_785496, EPI_ISL_785497, EPI_ISL_785498, EPI_ISL_785499, EPI_ISL_785500, EPI_ISL_785501, EPI_ISL_785502, EPI_ISL_785503, EPI_ISL_785504, EPI_ISL_785505, EPI_ISL_785506, EPI_ISL_785507, EPI_ISL_785508, EPI_ISL_785509, EPI_ISL_785510, EPI_ISL_785511, EPI_ISL_785512,
EPI_ISL_785513, EPI_ISL_785514, EPI_ISL_785515, EPI_ISL_785516, EPI_ISL_785517, EPI_ISL_785518, EPI_ISL_785519, EPI_ISL_785520, EPI_ISL_785521, EPI_ISL_785522, EPI_ISL_785523, EPI_ISL_785524, EPI_ISL_785525, EPI_ISL_785526, EPI_ISL_785527, EPI_ISL_785528, EPI_ISL_785529, EPI_ISL_785530, EPI_ISL_785531, EPI_ISL_785532, EPI_ISL_785533, EPI_ISL_785534, EPI_ISL_785535, EPI_ISL_785536,
EPI_ISL_785537, EPI_ISL_785538, EPI_ISL_785539, EPI_ISL_785540, EPI_ISL_785541, EPI_ISL_785542, EPI_ISL_785543, EPI_ISL_785544, EPI_ISL_785545, EPI_ISL_785546, EPI_ISL_785547, EPI_ISL_785548, EPI_ISL_785549, EPI_ISL_785550, EPI_ISL_785551, EPI_ISL_785552, EPI_ISL_785553, EPI_ISL_785554, EPI_ISL_785555, EPI_ISL_785556, EPI_ISL_785557, EPI_ISL_785558, EPI_ISL_785559, EPI_ISL_785560,
EPI_ISL_785561, EPI_ISL_785562, EPI_ISL_785563, EPI_ISL_785564, EPI_ISL_785565, EPI_ISL_785566, EPI_ISL_785567, EPI_ISL_785568, EPI_ISL_785569, EPI_ISL_785570, EPI_ISL_785571, EPI_ISL_785572, EPI_ISL_785573, EPI_ISL_785574, EPI_ISL_785575, EPI_ISL_785576, EPI_ISL_785577, EPI_ISL_785578, EPI_ISL_785579, EPI_ISL_785580, EPI_ISL_785581, EPI_ISL_785582, EPI_ISL_785583, EPI_ISL_785584,
EPI_ISL_785585, EPI_ISL_785586, EPI_ISL_785587, EPI_ISL_785588, EPI_ISL_785589, EPI_ISL_785590, EPI_ISL_785591, EPI_ISL_785592, EPI_ISL_785593, EPI_ISL_785594, EPI_ISL_785595, EPI_ISL_785596, EPI_ISL_785597, EPI_ISL_785598, EPI_ISL_785599, EPI_ISL_785600, EPI_ISL_785601, EPI_ISL_785602, EPI_ISL_785603, EPI_ISL_785604, EPI_ISL_785605, EPI_ISL_785606, EPI_ISL_785607, EPI_ISL_785608,
EPI_ISL_785609, EPI_ISL_785610, EPI_ISL_785611, EPI_ISL_785612, EPI_ISL_785613, EPI_ISL_785614, EPI_ISL_785615, EPI_ISL_785616, EPI_ISL_785617, EPI_ISL_785618, EPI_ISL_785619, EPI_ISL_785620, EPI_ISL_785621, EPI_ISL_785622, EPI_ISL_785623, EPI_ISL_785624, EPI_ISL_785625, EPI_ISL_785626, EPI_ISL_785627, EPI_ISL_785628, EPI_ISL_785629, EPI_ISL_785630, EPI_ISL_785631, EPI_ISL_785632,
EPI_ISL_785633, EPI_ISL_785634, EPI_ISL_785635, EPI_ISL_785636, EPI_ISL_785637, EPI_ISL_785638, EPI_ISL_785639, EPI_ISL_785640, EPI_ISL_785641, EPI_ISL_785642, EPI_ISL_785643, EPI_ISL_785644, EPI_ISL_785645, EPI_ISL_785646, EPI_ISL_785647, EPI_ISL_785648, EPI_ISL_785649, EPI_ISL_785650, EPI_ISL_785651, EPI_ISL_785652, EPI_ISL_785653, EPI_ISL_785654, EPI_ISL_785655, EPI_ISL_785656,
EPI_ISL_785657, EPI_ISL_785658, EPI_ISL_785659, EPI_ISL_785660, EPI_ISL_785661, EPI_ISL_785662, EPI_ISL_785663, EPI_ISL_785664, EPI_ISL_785665, EPI_ISL_785666, EPI_ISL_785667, EPI_ISL_785668, EPI_ISL_785669, EPI_ISL_785670, EPI_ISL_785671, EPI_ISL_785672, EPI_ISL_785673, EPI_ISL_785674, EPI_ISL_785675, EPI_ISL_785676, EPI_ISL_785677, EPI_ISL_785678, EPI_ISL_785679, EPI_ISL_785680,
EPI_ISL_785681, EPI_ISL_785682, EPI_ISL_785683, EPI_ISL_785684, EPI_ISL_785685, EPI_ISL_785686, EPI_ISL_785687, EPI_ISL_785688, EPI_ISL_785689, EPI_ISL_785690, EPI_ISL_785691, EPI_ISL_785692, EPI_ISL_785693, EPI_ISL_785694, EPI_ISL_785695, EPI_ISL_785696, EPI_ISL_785697, EPI_ISL_785698, EPI_ISL_785699, EPI_ISL_785700, EPI_ISL_785701, EPI_ISL_785702, EPI_ISL_785703, EPI_ISL_785704,
EPI_ISL_785705, EPI_ISL_785706, EPI_ISL_785707, EPI_ISL_785708, EPI_ISL_785709, EPI_ISL_785710, EPI_ISL_785711, EPI_ISL_785712, EPI_ISL_785713, EPI_ISL_785714, EPI_ISL_785715, EPI_ISL_785716, EPI_ISL_785717, EPI_ISL_785718, EPI_ISL_785719, EPI_ISL_785720, EPI_ISL_785721, EPI_ISL_785722, EPI_ISL_785723, EPI_ISL_785724, EPI_ISL_785725, EPI_ISL_785726, EPI_ISL_785727, EPI_ISL_785728,
EPI_ISL_785729, EPI_ISL_785730, EPI_ISL_785731, EPI_ISL_785732, EPI_ISL_785733, EPI_ISL_785734, EPI_ISL_785735, EPI_ISL_785736, EPI_ISL_785737, EPI_ISL_785738, EPI_ISL_785739, EPI_ISL_785740, EPI_ISL_785741, EPI_ISL_785742, EPI_ISL_785743, EPI_ISL_785744, EPI_ISL_785745, EPI_ISL_785746, EPI_ISL_785747, EPI_ISL_785748, EPI_ISL_785749, EPI_ISL_785750, EPI_ISL_785751, EPI_ISL_785752,
EPI_ISL_785753, EPI_ISL_785754, EPI_ISL_785755, EPI_ISL_785756, EPI_ISL_785757, EPI_ISL_785758, EPI_ISL_785759, EPI_ISL_785760, EPI_ISL_785761, EPI_ISL_785762, EPI_ISL_785763, EPI_ISL_785764, EPI_ISL_785765, EPI_ISL_785766, EPI_ISL_785767, EPI_ISL_785768, EPI_ISL_785769, EPI_ISL_785770, EPI_ISL_785771, EPI_ISL_785772, EPI_ISL_785773, EPI_ISL_785774, EPI_ISL_785775, EPI_ISL_785776,
EPI_ISL_785777, EPI_ISL_785778, EPI_ISL_785779, EPI_ISL_785780, EPI_ISL_785781, EPI_ISL_785782, EPI_ISL_785783, EPI_ISL_785784, EPI_ISL_785785, EPI_ISL_785786, EPI_ISL_785787, EPI_ISL_785788, EPI_ISL_785789, EPI_ISL_785790, EPI_ISL_785791, EPI_ISL_785792, EPI_ISL_785793, EPI_ISL_785794, EPI_ISL_785795, EPI_ISL_785796, EPI_ISL_785797, EPI_ISL_785798, EPI_ISL_785799, EPI_ISL_785800,
EPI_ISL_785801, EPI_ISL_785802, EPI_ISL_785803, EPI_ISL_785804, EPI_ISL_785805, EPI_ISL_785806, EPI_ISL_785807, EPI_ISL_785808, EPI_ISL_785809, EPI_ISL_785810, EPI_ISL_785811, EPI_ISL_785812, EPI_ISL_785813, EPI_ISL_785814, EPI_ISL_785815, EPI_ISL_785816, EPI_ISL_785817, EPI_ISL_785818, EPI_ISL_785819, EPI_ISL_785820, EPI_ISL_785821, EPI_ISL_785822, EPI_ISL_785823, EPI_ISL_785824,
EPI_ISL_785825, EPI_ISL_785826, EPI_ISL_785827, EPI_ISL_785828, EPI_ISL_785829, EPI_ISL_785830, EPI_ISL_785831, EPI_ISL_785832, EPI_ISL_785833, EPI_ISL_785834, EPI_ISL_785835, EPI_ISL_785836, EPI_ISL_785837, EPI_ISL_785838, EPI_ISL_785839, EPI_ISL_785840, EPI_ISL_785841, EPI_ISL_785842, EPI_ISL_785843, EPI_ISL_785844, EPI_ISL_785845, EPI_ISL_785846, EPI_ISL_785847, EPI_ISL_785848,
EPI_ISL_785849, EPI_ISL_785850, EPI_ISL_785851, EPI_ISL_785852, EPI_ISL_785853, EPI_ISL_785854, EPI_ISL_785855, EPI_ISL_785856, EPI_ISL_785857, EPI_ISL_785858, EPI_ISL_785859, EPI_ISL_785860, EPI_ISL_785861, EPI_ISL_785862, EPI_ISL_785863, EPI_ISL_785864, EPI_ISL_785865, EPI_ISL_785866, EPI_ISL_785867, EPI_ISL_785868, EPI_ISL_785869, EPI_ISL_785870, EPI_ISL_785871, EPI_ISL_785872,
EPI_ISL_785873, EPI_ISL_785874, EPI_ISL_785875, EPI_ISL_785876, EPI_ISL_785877, EPI_ISL_785878, EPI_ISL_785879, EPI_ISL_785880, EPI_ISL_785881, EPI_ISL_785882, EPI_ISL_785883, EPI_ISL_785884, EPI_ISL_785885, EPI_ISL_785886, EPI_ISL_785887, EPI_ISL_785888, EPI_ISL_785889, EPI_ISL_785890, EPI_ISL_785891, EPI_ISL_785892, EPI_ISL_785893, EPI_ISL_785894, EPI_ISL_785895, EPI_ISL_785896,
EPI_ISL_785897, EPI_ISL_785898, EPI_ISL_785899, EPI_ISL_785900, EPI_ISL_785901, EPI_ISL_785902, EPI_ISL_785903, EPI_ISL_785904, EPI_ISL_785905, EPI_ISL_785906, EPI_ISL_785907, EPI_ISL_785908, EPI_ISL_785909, EPI_ISL_785910, EPI_ISL_785911, EPI_ISL_785912, EPI_ISL_785913, EPI_ISL_785914, EPI_ISL_785915, EPI_ISL_785916, EPI_ISL_785917, EPI_ISL_785918, EPI_ISL_785919, EPI_ISL_785920,
EPI_ISL_785921, EPI_ISL_785922, EPI_ISL_785923, EPI_ISL_785924, EPI_ISL_785925, EPI_ISL_785926, EPI_ISL_785927, EPI_ISL_785928, EPI_ISL_785929, EPI_ISL_785930, EPI_ISL_785931, EPI_ISL_785932, EPI_ISL_785933, EPI_ISL_785934, EPI_ISL_785935, EPI_ISL_785936, EPI_ISL_785937, EPI_ISL_785938, EPI_ISL_785939, EPI_ISL_785940, EPI_ISL_785941, EPI_ISL_785942, EPI_ISL_785943, EPI_ISL_785944,
EPI_ISL_785945, EPI_ISL_785946, EPI_ISL_785947, EPI_ISL_785948, EPI_ISL_785949, EPI_ISL_785950, EPI_ISL_785951, EPI_ISL_785952, EPI_ISL_785953, EPI_ISL_785954, EPI_ISL_785955, EPI_ISL_785956, EPI_ISL_785957, EPI_ISL_785958, EPI_ISL_785959, EPI_ISL_785960, EPI_ISL_785961, EPI_ISL_785962, EPI_ISL_785963, EPI_ISL_785964, EPI_ISL_785965, EPI_ISL_785966, EPI_ISL_785967, EPI_ISL_785968,
EPI_ISL_785969, EPI_ISL_785970, EPI_ISL_785971, EPI_ISL_785972, EPI_ISL_785973, EPI_ISL_785974, EPI_ISL_785975, EPI_ISL_785976, EPI_ISL_785977, EPI_ISL_785978, EPI_ISL_785979, EPI_ISL_785980, EPI_ISL_785981, EPI_ISL_785982, EPI_ISL_785983, EPI_ISL_785984, EPI_ISL_785985, EPI_ISL_785986, EPI_ISL_785987, EPI_ISL_785988, EPI_ISL_785989, EPI_ISL_785990, EPI_ISL_785991, EPI_ISL_785992,
EPI_ISL_785993, EPI_ISL_785994, EPI_ISL_785995, EPI_ISL_785996, EPI_ISL_785997, EPI_ISL_785998, EPI_ISL_785999, EPI_ISL_786000, EPI_ISL_786001, EPI_ISL_786002, EPI_ISL_786003, EPI_ISL_786004, EPI_ISL_786005, EPI_ISL_786006, EPI_ISL_786007, EPI_ISL_786008, EPI_ISL_786009, EPI_ISL_786010, EPI_ISL_786011, EPI_ISL_786012, EPI_ISL_786013, EPI_ISL_786014, EPI_ISL_786015, EPI_ISL_786016,
EPI_ISL_786017, EPI_ISL_786018, EPI_ISL_786019, EPI_ISL_786020, EPI_ISL_786021, EPI_ISL_786022, EPI_ISL_786023, EPI_ISL_786024, EPI_ISL_786025, EPI_ISL_786026, EPI_ISL_786027, EPI_ISL_786028, EPI_ISL_786029, EPI_ISL_786030, EPI_ISL_786031, EPI_ISL_786032, EPI_ISL_786033, EPI_ISL_786034, EPI_ISL_786035, EPI_ISL_786036, EPI_ISL_786037, EPI_ISL_786038, EPI_ISL_786039, EPI_ISL_786040,
EPI_ISL_786041, EPI_ISL_786042, EPI_ISL_786043, EPI_ISL_786044, EPI_ISL_786045, EPI_ISL_786046, EPI_ISL_786047, EPI_ISL_786048, EPI_ISL_786049, EPI_ISL_786050, EPI_ISL_786051, EPI_ISL_786052, EPI_ISL_786053, EPI_ISL_786054, EPI_ISL_786055, EPI_ISL_786056, EPI_ISL_786057, EPI_ISL_786058, EPI_ISL_786059, EPI_ISL_786060, EPI_ISL_786061, EPI_ISL_786062, EPI_ISL_786063, EPI_ISL_786064,
EPI_ISL_786065, EPI_ISL_786066, EPI_ISL_786067, EPI_ISL_786068, EPI_ISL_786069, EPI_ISL_786070, EPI_ISL_786071, EPI_ISL_786072, EPI_ISL_786073, EPI_ISL_786074, EPI_ISL_786075, EPI_ISL_786076, EPI_ISL_786077, EPI_ISL_786078, EPI_ISL_786079, EPI_ISL_786080, EPI_ISL_786081, EPI_ISL_786082, EPI_ISL_786083, EPI_ISL_786084, EPI_ISL_786085, EPI_ISL_786086, EPI_ISL_786087, EPI_ISL_786088,
EPI_ISL_786089, EPI_ISL_786090, EPI_ISL_786091, EPI_ISL_786092, EPI_ISL_786093, EPI_ISL_786094, EPI_ISL_786095, EPI_ISL_786096, EPI_ISL_786097, EPI_ISL_786098, EPI_ISL_786099, EPI_ISL_786100, EPI_ISL_786101, EPI_ISL_786102, EPI_ISL_786103, EPI_ISL_786104, EPI_ISL_786105, EPI_ISL_786106, EPI_ISL_786107, EPI_ISL_786108, EPI_ISL_786109, EPI_ISL_786110, EPI_ISL_786111, EPI_ISL_786112,
EPI_ISL_786113, EPI_ISL_786114, EPI_ISL_786115, EPI_ISL_786116, EPI_ISL_786117, EPI_ISL_786118, EPI_ISL_786119, EPI_ISL_786120, EPI_ISL_786121, EPI_ISL_786122, EPI_ISL_786123, EPI_ISL_786124, EPI_ISL_786125, EPI_ISL_786126, EPI_ISL_786127, EPI_ISL_786128, EPI_ISL_786129, EPI_ISL_786130, EPI_ISL_786131, EPI_ISL_786132, EPI_ISL_786133, EPI_ISL_786134, EPI_ISL_786135, EPI_ISL_786136,
EPI_ISL_786137, EPI_ISL_786138, EPI_ISL_786139, EPI_ISL_786140, EPI_ISL_786141, EPI_ISL_786142, EPI_ISL_786143, EPI_ISL_786144, EPI_ISL_786145, EPI_ISL_786146, EPI_ISL_786147, EPI_ISL_786148, EPI_ISL_786149, EPI_ISL_786150, EPI_ISL_786151, EPI_ISL_786152, EPI_ISL_786153, EPI_ISL_786154, EPI_ISL_786155, EPI_ISL_786156, EPI_ISL_786157, EPI_ISL_786158, EPI_ISL_786159, EPI_ISL_786160,
EPI_ISL_786161, EPI_ISL_786162, EPI_ISL_786163, EPI_ISL_786164, EPI_ISL_786165, EPI_ISL_786166, EPI_ISL_786167, EPI_ISL_786168, EPI_ISL_786169, EPI_ISL_786170, EPI_ISL_786171, EPI_ISL_786172, EPI_ISL_786173, EPI_ISL_786174, EPI_ISL_786175, EPI_ISL_786176, EPI_ISL_786177, EPI_ISL_786178, EPI_ISL_786179, EPI_ISL_786180, EPI_ISL_786181, EPI_ISL_786182, EPI_ISL_786183, EPI_ISL_786184,
EPI_ISL_786185, EPI_ISL_786186, EPI_ISL_786187, EPI_ISL_786188, EPI_ISL_786189, EPI_ISL_786190, EPI_ISL_786191, EPI_ISL_786192, EPI_ISL_786193, EPI_ISL_786194, EPI_ISL_786195, EPI_ISL_786196, EPI_ISL_786197, EPI_ISL_786198, EPI_ISL_786199, EPI_ISL_786200, EPI_ISL_786201, EPI_ISL_786202, EPI_ISL_786203, EPI_ISL_786204, EPI_ISL_786205, EPI_ISL_786206, EPI_ISL_786207, EPI_ISL_786208,
EPI_ISL_786209, EPI_ISL_786210, EPI_ISL_786211, EPI_ISL_786212, EPI_ISL_786213, EPI_ISL_786214, EPI_ISL_786215, EPI_ISL_786216, EPI_ISL_786217, EPI_ISL_786218, EPI_ISL_786219, EPI_ISL_786220, EPI_ISL_786221, EPI_ISL_786222, EPI_ISL_786223, EPI_ISL_786224, EPI_ISL_786225, EPI_ISL_786226, EPI_ISL_786227, EPI_ISL_786228, EPI_ISL_786229, EPI_ISL_786230, EPI_ISL_786231, EPI_ISL_786232,
EPI_ISL_786233, EPI_ISL_786234, EPI_ISL_786235, EPI_ISL_786236, EPI_ISL_786237, EPI_ISL_786238, EPI_ISL_786239, EPI_ISL_786240, EPI_ISL_786241, EPI_ISL_786242, EPI_ISL_786243, EPI_ISL_786244, EPI_ISL_786245, EPI_ISL_786246, EPI_ISL_786247, EPI_ISL_786248, EPI_ISL_786249, EPI_ISL_786250, EPI_ISL_786251, EPI_ISL_786252, EPI_ISL_786253, EPI_ISL_786254, EPI_ISL_786255, EPI_ISL_786256,
EPI_ISL_786257, EPI_ISL_786258, EPI_ISL_786259, EPI_ISL_786260, EPI_ISL_786261, EPI_ISL_786262, EPI_ISL_786263, EPI_ISL_786264, EPI_ISL_786265, EPI_ISL_786266, EPI_ISL_786267, EPI_ISL_786268, EPI_ISL_786269, EPI_ISL_786270, EPI_ISL_786271, EPI_ISL_786272, EPI_ISL_786273, EPI_ISL_786274, EPI_ISL_786275, EPI_ISL_786276, EPI_ISL_786277, EPI_ISL_786278, EPI_ISL_786279, EPI_ISL_786280,
EPI_ISL_786281, EPI_ISL_786282, EPI_ISL_786283, EPI_ISL_786284, EPI_ISL_786285, EPI_ISL_786286, EPI_ISL_786287, EPI_ISL_786288, EPI_ISL_786289, EPI_ISL_786290, EPI_ISL_786291, EPI_ISL_786292, EPI_ISL_786293, EPI_ISL_786294, EPI_ISL_786295, EPI_ISL_786296, EPI_ISL_786297, EPI_ISL_786298, EPI_ISL_786299, EPI_ISL_786300, EPI_ISL_786301, EPI_ISL_786302, EPI_ISL_786303, EPI_ISL_786304,
EPI_ISL_786305, EPI_ISL_786306, EPI_ISL_786307, EPI_ISL_786308, EPI_ISL_786309, EPI_ISL_786310, EPI_ISL_786311, EPI_ISL_786312, EPI_ISL_786313, EPI_ISL_786314, EPI_ISL_786315, EPI_ISL_786316, EPI_ISL_786317, EPI_ISL_786318, EPI_ISL_786319, EPI_ISL_786320, EPI_ISL_786321, EPI_ISL_786322, EPI_ISL_786323, EPI_ISL_786324, EPI_ISL_786325, EPI_ISL_786326, EPI_ISL_786327, EPI_ISL_786328,
EPI_ISL_786329, EPI_ISL_786330, EPI_ISL_786331, EPI_ISL_786332, EPI_ISL_786333, EPI_ISL_786334, EPI_ISL_786335, EPI_ISL_786336, EPI_ISL_786337, EPI_ISL_786338, EPI_ISL_786339, EPI_ISL_786340, EPI_ISL_786341, EPI_ISL_786342, EPI_ISL_786343, EPI_ISL_786344, EPI_ISL_786345, EPI_ISL_786346, EPI_ISL_786347, EPI_ISL_786348, EPI_ISL_786349, EPI_ISL_786350, EPI_ISL_786351, EPI_ISL_786352,
EPI_ISL_786353, EPI_ISL_786354, EPI_ISL_786355, EPI_ISL_786356, EPI_ISL_786357, EPI_ISL_786358, EPI_ISL_786359, EPI_ISL_786360, EPI_ISL_786361, EPI_ISL_786362, EPI_ISL_786363, EPI_ISL_786364, EPI_ISL_786365, EPI_ISL_786366, EPI_ISL_786367, EPI_ISL_786368, EPI_ISL_786369, EPI_ISL_786370, EPI_ISL_786371, EPI_ISL_786372, EPI_ISL_786373, EPI_ISL_786374, EPI_ISL_786375, EPI_ISL_786376,
EPI_ISL_786377, EPI_ISL_786378, EPI_ISL_786379, EPI_ISL_786380, EPI_ISL_786381, EPI_ISL_786382, EPI_ISL_786383, EPI_ISL_786384, EPI_ISL_786385, EPI_ISL_786386, EPI_ISL_786387, EPI_ISL_786388, EPI_ISL_786389, EPI_ISL_786390, EPI_ISL_786391, EPI_ISL_786392, EPI_ISL_786393, EPI_ISL_786394, EPI_ISL_786395, EPI_ISL_786396, EPI_ISL_786397, EPI_ISL_786398, EPI_ISL_786399, EPI_ISL_786400,
EPI_ISL_786401, EPI_ISL_786402, EPI_ISL_786403, EPI_ISL_786404, EPI_ISL_786405, EPI_ISL_786406, EPI_ISL_786407, EPI_ISL_786408, EPI_ISL_786409, EPI_ISL_786410, EPI_ISL_786411, EPI_ISL_786412, EPI_ISL_786413, EPI_ISL_786414, EPI_ISL_786415, EPI_ISL_786416, EPI_ISL_786417, EPI_ISL_786418, EPI_ISL_786419, EPI_ISL_786420, EPI_ISL_786421, EPI_ISL_786422, EPI_ISL_786423, EPI_ISL_786424,
EPI_ISL_786425, EPI_ISL_786426, EPI_ISL_786427, EPI_ISL_786428, EPI_ISL_786429, EPI_ISL_786430, EPI_ISL_786431, EPI_ISL_786432, EPI_ISL_786433, EPI_ISL_786434, EPI_ISL_786435, EPI_ISL_786436, EPI_ISL_786437, EPI_ISL_786438, EPI_ISL_786439, EPI_ISL_786440, EPI_ISL_786441, EPI_ISL_786442, EPI_ISL_786443, EPI_ISL_786444, EPI_ISL_786445, EPI_ISL_786446, EPI_ISL_786447, EPI_ISL_786448,
EPI_ISL_786449, EPI_ISL_786450, EPI_ISL_786451, EPI_ISL_786452, EPI_ISL_786453, EPI_ISL_786454, EPI_ISL_786455, EPI_ISL_786456, EPI_ISL_786457, EPI_ISL_786458, EPI_ISL_786459, EPI_ISL_786460, EPI_ISL_786461, EPI_ISL_786462, EPI_ISL_786463, EPI_ISL_786464, EPI_ISL_786465, EPI_ISL_786466, EPI_ISL_786467, EPI_ISL_786468, EPI_ISL_786469, EPI_ISL_786470, EPI_ISL_786471, EPI_ISL_786472,
EPI_ISL_786473, EPI_ISL_786474, EPI_ISL_786475, EPI_ISL_786476, EPI_ISL_786477, EPI_ISL_786478, EPI_ISL_786479, EPI_ISL_786480, EPI_ISL_786481, EPI_ISL_786482, EPI_ISL_786483, EPI_ISL_786484, EPI_ISL_786485, EPI_ISL_786486, EPI_ISL_786487, EPI_ISL_786488, EPI_ISL_786489, EPI_ISL_786490, EPI_ISL_786491, EPI_ISL_786492, EPI_ISL_786493, EPI_ISL_786494, EPI_ISL_786495, EPI_ISL_786496,
EPI_ISL_786497, EPI_ISL_786498, EPI_ISL_786499, EPI_ISL_786500, EPI_ISL_786501, EPI_ISL_786502, EPI_ISL_786503, EPI_ISL_786504, EPI_ISL_786505, EPI_ISL_786506, EPI_ISL_786507, EPI_ISL_786508, EPI_ISL_786509, EPI_ISL_786510, EPI_ISL_786511, EPI_ISL_786512, EPI_ISL_786513, EPI_ISL_786514, EPI_ISL_786515, EPI_ISL_786516, EPI_ISL_786517, EPI_ISL_786518, EPI_ISL_786519, EPI_ISL_786520,
EPI_ISL_786521, EPI_ISL_786522, EPI_ISL_786523, EPI_ISL_786524, EPI_ISL_786525, EPI_ISL_786526, EPI_ISL_786527, EPI_ISL_786528, EPI_ISL_786529, EPI_ISL_786530, EPI_ISL_786531, EPI_ISL_786532, EPI_ISL_786533, EPI_ISL_786534, EPI_ISL_786535, EPI_ISL_786536, EPI_ISL_786537, EPI_ISL_786538, EPI_ISL_786539, EPI_ISL_786540, EPI_ISL_786541, EPI_ISL_786542, EPI_ISL_786543, EPI_ISL_786544,
EPI_ISL_786545, EPI_ISL_786546, EPI_ISL_786547, EPI_ISL_786548, EPI_ISL_786549, EPI_ISL_786550, EPI_ISL_786551, EPI_ISL_786552, EPI_ISL_786553, EPI_ISL_786554, EPI_ISL_786555, EPI_ISL_786556, EPI_ISL_786557, EPI_ISL_786558, EPI_ISL_786559, EPI_ISL_786560, EPI_ISL_786561, EPI_ISL_786562, EPI_ISL_786563, EPI_ISL_786564, EPI_ISL_786565, EPI_ISL_786566, EPI_ISL_786567, EPI_ISL_786568,
EPI_ISL_786569, EPI_ISL_786570, EPI_ISL_786571, EPI_ISL_786572, EPI_ISL_786573, EPI_ISL_786574, EPI_ISL_786575, EPI_ISL_786576, EPI_ISL_786577, EPI_ISL_786578, EPI_ISL_786579, EPI_ISL_786580, EPI_ISL_786581, EPI_ISL_786582, EPI_ISL_786583, EPI_ISL_786584, EPI_ISL_786585, EPI_ISL_786586, EPI_ISL_786587, EPI_ISL_786588, EPI_ISL_786589, EPI_ISL_786590, EPI_ISL_786591, EPI_ISL_786592,
EPI_ISL_786593, EPI_ISL_786594, EPI_ISL_786595, EPI_ISL_786596, EPI_ISL_786597, EPI_ISL_786598, EPI_ISL_786599, EPI_ISL_786600, EPI_ISL_786601, EPI_ISL_786602, EPI_ISL_786603, EPI_ISL_786604, EPI_ISL_786605, EPI_ISL_786606, EPI_ISL_786607, EPI_ISL_786608, EPI_ISL_786609, EPI_ISL_786610, EPI_ISL_786611, EPI_ISL_786612, EPI_ISL_786613, EPI_ISL_786614, EPI_ISL_786615, EPI_ISL_786616,
EPI_ISL_786617, EPI_ISL_786618, EPI_ISL_786619, EPI_ISL_786620, EPI_ISL_786621, EPI_ISL_786622, EPI_ISL_786623, EPI_ISL_786624, EPI_ISL_786625, EPI_ISL_786626, EPI_ISL_786627, EPI_ISL_786628, EPI_ISL_786629, EPI_ISL_786630, EPI_ISL_786631, EPI_ISL_786632, EPI_ISL_786633, EPI_ISL_786634, EPI_ISL_786635, EPI_ISL_786636, EPI_ISL_786637, EPI_ISL_786638, EPI_ISL_786639, EPI_ISL_786640,
EPI_ISL_786641, EPI_ISL_786642, EPI_ISL_786643, EPI_ISL_786644, EPI_ISL_786645, EPI_ISL_786646, EPI_ISL_786647, EPI_ISL_786648, EPI_ISL_786649, EPI_ISL_786650, EPI_ISL_786651, EPI_ISL_786652, EPI_ISL_786653, EPI_ISL_786654, EPI_ISL_786655, EPI_ISL_786656, EPI_ISL_786657, EPI_ISL_786658, EPI_ISL_786659, EPI_ISL_786660, EPI_ISL_786661, EPI_ISL_786662, EPI_ISL_786663, EPI_ISL_786664,
EPI_ISL_786665, EPI_ISL_786666, EPI_ISL_786667, EPI_ISL_786668, EPI_ISL_786669, EPI_ISL_786670, EPI_ISL_786671, EPI_ISL_786672, EPI_ISL_786673, EPI_ISL_786674, EPI_ISL_786675, EPI_ISL_786676, EPI_ISL_786677, EPI_ISL_786678, EPI_ISL_786679, EPI_ISL_786680, EPI_ISL_786681, EPI_ISL_786682, EPI_ISL_786683, EPI_ISL_786684, EPI_ISL_786685, EPI_ISL_786686, EPI_ISL_786687, EPI_ISL_786688,
EPI_ISL_786689, EPI_ISL_786690, EPI_ISL_786691, EPI_ISL_786692, EPI_ISL_786693, EPI_ISL_786694, EPI_ISL_786695, EPI_ISL_786696, EPI_ISL_786697, EPI_ISL_786698, EPI_ISL_786699, EPI_ISL_786700, EPI_ISL_786701, EPI_ISL_786702, EPI_ISL_786703, EPI_ISL_786704, EPI_ISL_786705, EPI_ISL_786706, EPI_ISL_786707, EPI_ISL_786708, EPI_ISL_786709, EPI_ISL_786710, EPI_ISL_786711, EPI_ISL_786712,
EPI_ISL_786713, EPI_ISL_786714, EPI_ISL_786715, EPI_ISL_786716, EPI_ISL_786717, EPI_ISL_786718, EPI_ISL_786719, EPI_ISL_786720, EPI_ISL_786721, EPI_ISL_786722, EPI_ISL_786723, EPI_ISL_786724, EPI_ISL_786725, EPI_ISL_786726, EPI_ISL_786727, EPI_ISL_786728, EPI_ISL_786729, EPI_ISL_786730, EPI_ISL_786731, EPI_ISL_786732, EPI_ISL_786733, EPI_ISL_786734, EPI_ISL_786735, EPI_ISL_786736,
EPI_ISL_786737, EPI_ISL_786738, EPI_ISL_786739, EPI_ISL_786740, EPI_ISL_786741, EPI_ISL_786742, EPI_ISL_786743, EPI_ISL_786744, EPI_ISL_786745, EPI_ISL_786746, EPI_ISL_786747, EPI_ISL_786748, EPI_ISL_786749, EPI_ISL_786750, EPI_ISL_786751, EPI_ISL_786752, EPI_ISL_786753, EPI_ISL_786754, EPI_ISL_786755, EPI_ISL_786756, EPI_ISL_786757, EPI_ISL_786758, EPI_ISL_786759, EPI_ISL_786760,
EPI_ISL_786761, EPI_ISL_786762, EPI_ISL_786763, EPI_ISL_786764, EPI_ISL_786765, EPI_ISL_786766, EPI_ISL_786767, EPI_ISL_786768, EPI_ISL_786769, EPI_ISL_786770, EPI_ISL_786771, EPI_ISL_786772, EPI_ISL_786773, EPI_ISL_786774, EPI_ISL_786775, EPI_ISL_786776, EPI_ISL_786777, EPI_ISL_786778, EPI_ISL_786779, EPI_ISL_786780, EPI_ISL_786781, EPI_ISL_786782, EPI_ISL_786783, EPI_ISL_786784,
EPI_ISL_786785, EPI_ISL_786786, EPI_ISL_786787, EPI_ISL_786788, EPI_ISL_786789, EPI_ISL_786790, EPI_ISL_786791, EPI_ISL_786792, EPI_ISL_786793, EPI_ISL_786794, EPI_ISL_786795, EPI_ISL_786796, EPI_ISL_786797, EPI_ISL_786798, EPI_ISL_786799, EPI_ISL_786800, EPI_ISL_786801, EPI_ISL_786802, EPI_ISL_786803, EPI_ISL_786804, EPI_ISL_786805, EPI_ISL_786806, EPI_ISL_786807, EPI_ISL_786808,
EPI_ISL_786809, EPI_ISL_786810, EPI_ISL_786811, EPI_ISL_786812, EPI_ISL_786813, EPI_ISL_786814, EPI_ISL_786815, EPI_ISL_786816, EPI_ISL_786817, EPI_ISL_786818, EPI_ISL_786819, EPI_ISL_786820, EPI_ISL_786821, EPI_ISL_786822, EPI_ISL_786823, EPI_ISL_786824, EPI_ISL_786825, EPI_ISL_786826, EPI_ISL_786827, EPI_ISL_786828, EPI_ISL_786829, EPI_ISL_786830, EPI_ISL_786831, EPI_ISL_786832,
EPI_ISL_786833, EPI_ISL_786834, EPI_ISL_786835, EPI_ISL_786836, EPI_ISL_786837, EPI_ISL_786838, EPI_ISL_786839, EPI_ISL_786840, EPI_ISL_786841, EPI_ISL_786842, EPI_ISL_786843, EPI_ISL_786844, EPI_ISL_786845, EPI_ISL_786846, EPI_ISL_786847, EPI_ISL_786848, EPI_ISL_786849, EPI_ISL_786850, EPI_ISL_786851, EPI_ISL_786852, EPI_ISL_786853, EPI_ISL_786854, EPI_ISL_786855, EPI_ISL_786856,
EPI_ISL_786857, EPI_ISL_786858, EPI_ISL_786859, EPI_ISL_786860, EPI_ISL_786861, EPI_ISL_786862, EPI_ISL_786863, EPI_ISL_786864, EPI_ISL_786865, EPI_ISL_786866, EPI_ISL_786867, EPI_ISL_786868, EPI_ISL_786869, EPI_ISL_786870, EPI_ISL_786871, EPI_ISL_786872, EPI_ISL_786873, EPI_ISL_786874, EPI_ISL_786875, EPI_ISL_786876, EPI_ISL_786877, EPI_ISL_786878, EPI_ISL_786879, EPI_ISL_786880,
EPI_ISL_786881, EPI_ISL_786882, EPI_ISL_786883, EPI_ISL_786884, EPI_ISL_786885, EPI_ISL_786886, EPI_ISL_786887, EPI_ISL_786888, EPI_ISL_786889, EPI_ISL_786890, EPI_ISL_786891, EPI_ISL_786892, EPI_ISL_786893, EPI_ISL_786894, EPI_ISL_786895, EPI_ISL_786896, EPI_ISL_786897, EPI_ISL_786898, EPI_ISL_786899, EPI_ISL_786900, EPI_ISL_786901, EPI_ISL_786902, EPI_ISL_786903, EPI_ISL_786904,
EPI_ISL_786905, EPI_ISL_786906, EPI_ISL_786907, EPI_ISL_786908, EPI_ISL_786909, EPI_ISL_786910, EPI_ISL_786911, EPI_ISL_786912, EPI_ISL_786913, EPI_ISL_786914, EPI_ISL_786915, EPI_ISL_786916, EPI_ISL_786917, EPI_ISL_786918, EPI_ISL_786919, EPI_ISL_786920, EPI_ISL_786921, EPI_ISL_786922, EPI_ISL_786923, EPI_ISL_786924, EPI_ISL_786925, EPI_ISL_786926, EPI_ISL_786927, EPI_ISL_786928,
EPI_ISL_786929, EPI_ISL_786930, EPI_ISL_786931, EPI_ISL_786932, EPI_ISL_786933, EPI_ISL_786934, EPI_ISL_786935, EPI_ISL_786936, EPI_ISL_786937, EPI_ISL_786938, EPI_ISL_786939, EPI_ISL_786940, EPI_ISL_786941, EPI_ISL_786942, EPI_ISL_786943, EPI_ISL_786944, EPI_ISL_786945, EPI_ISL_786946, EPI_ISL_786947, EPI_ISL_786948, EPI_ISL_786949, EPI_ISL_786950, EPI_ISL_786951, EPI_ISL_786952,
EPI_ISL_786953, EPI_ISL_786954, EPI_ISL_786955, EPI_ISL_786956, EPI_ISL_786957, EPI_ISL_786958, EPI_ISL_786959, EPI_ISL_786960, EPI_ISL_786961, EPI_ISL_786962, EPI_ISL_786963, EPI_ISL_786964, EPI_ISL_786965, EPI_ISL_786966, EPI_ISL_786967, EPI_ISL_786968, EPI_ISL_786969, EPI_ISL_786970, EPI_ISL_786971, EPI_ISL_786972, EPI_ISL_786973, EPI_ISL_786974, EPI_ISL_786975, EPI_ISL_786976,
EPI_ISL_786977, EPI_ISL_786978, EPI_ISL_786979, EPI_ISL_786980, EPI_ISL_786981, EPI_ISL_786982, EPI_ISL_786983, EPI_ISL_786984, EPI_ISL_786985, EPI_ISL_786986, EPI_ISL_786987, EPI_ISL_786988, EPI_ISL_786989, EPI_ISL_786990, EPI_ISL_786991, EPI_ISL_786992, EPI_ISL_786993, EPI_ISL_786994, EPI_ISL_786995, EPI_ISL_786996, EPI_ISL_786997, EPI_ISL_786998, EPI_ISL_786999, EPI_ISL_787000,
EPI_ISL_787001, EPI_ISL_787002, EPI_ISL_787003, EPI_ISL_787004, EPI_ISL_787005, EPI_ISL_787006, EPI_ISL_787007, EPI_ISL_787008, EPI_ISL_787009, EPI_ISL_787010, EPI_ISL_787011, EPI_ISL_787012, EPI_ISL_787013, EPI_ISL_787014, EPI_ISL_787015, EPI_ISL_787016, EPI_ISL_787017, EPI_ISL_787018, EPI_ISL_787019, EPI_ISL_787020, EPI_ISL_787021, EPI_ISL_787022, EPI_ISL_787023, EPI_ISL_787024,
EPI_ISL_787025, EPI_ISL_787026, EPI_ISL_787027, EPI_ISL_787028, EPI_ISL_787029, EPI_ISL_787030, EPI_ISL_787031, EPI_ISL_787032, EPI_ISL_787033, EPI_ISL_787034, EPI_ISL_787035, EPI_ISL_787036, EPI_ISL_787037, EPI_ISL_787038, EPI_ISL_787039, EPI_ISL_787040, EPI_ISL_787041, EPI_ISL_787042, EPI_ISL_787043, EPI_ISL_787044, EPI_ISL_787045, EPI_ISL_787046, EPI_ISL_787047, EPI_ISL_787048,
EPI_ISL_787049, EPI_ISL_787050, EPI_ISL_787051, EPI_ISL_787052, EPI_ISL_787053, EPI_ISL_787054, EPI_ISL_787055, EPI_ISL_787056, EPI_ISL_787057, EPI_ISL_787058, EPI_ISL_787059, EPI_ISL_787060, EPI_ISL_787061, EPI_ISL_787062, EPI_ISL_787063, EPI_ISL_787064, EPI_ISL_787065, EPI_ISL_787066, EPI_ISL_787067, EPI_ISL_787068, EPI_ISL_787069, EPI_ISL_787070, EPI_ISL_787071, EPI_ISL_787072,
EPI_ISL_787073, EPI_ISL_787074, EPI_ISL_787075, EPI_ISL_787076, EPI_ISL_787077, EPI_ISL_787078, EPI_ISL_787079, EPI_ISL_787080, EPI_ISL_787081, EPI_ISL_787082, EPI_ISL_787083, EPI_ISL_787084, EPI_ISL_787085, EPI_ISL_787086, EPI_ISL_787087, EPI_ISL_787088, EPI_ISL_787089, EPI_ISL_787090, EPI_ISL_787091, EPI_ISL_787092, EPI_ISL_787093, EPI_ISL_787094, EPI_ISL_787095, EPI_ISL_787096,
EPI_ISL_787097, EPI_ISL_787098, EPI_ISL_787099, EPI_ISL_787100, EPI_ISL_787101, EPI_ISL_787102, EPI_ISL_787103, EPI_ISL_787104, EPI_ISL_787105, EPI_ISL_787106, EPI_ISL_787107, EPI_ISL_787108, EPI_ISL_787109, EPI_ISL_787110, EPI_ISL_787111, EPI_ISL_787112, EPI_ISL_787113, EPI_ISL_787114, EPI_ISL_787115, EPI_ISL_787116, EPI_ISL_787117, EPI_ISL_787118, EPI_ISL_787119, EPI_ISL_787120,
EPI_ISL_787121, EPI_ISL_787122, EPI_ISL_787123, EPI_ISL_787124, EPI_ISL_787125, EPI_ISL_787126, EPI_ISL_787127, EPI_ISL_787128, EPI_ISL_787129, EPI_ISL_787130, EPI_ISL_787131, EPI_ISL_787132, EPI_ISL_787133, EPI_ISL_787134, EPI_ISL_787135, EPI_ISL_787136, EPI_ISL_787137, EPI_ISL_787138, EPI_ISL_787139, EPI_ISL_787140, EPI_ISL_787141, EPI_ISL_787142, EPI_ISL_787143, EPI_ISL_787144,
EPI_ISL_787145, EPI_ISL_787146, EPI_ISL_787147, EPI_ISL_787148, EPI_ISL_787149, EPI_ISL_787150, EPI_ISL_787151, EPI_ISL_787152, EPI_ISL_787153, EPI_ISL_787154, EPI_ISL_787155, EPI_ISL_787156, EPI_ISL_787157, EPI_ISL_787158, EPI_ISL_787159, EPI_ISL_787160, EPI_ISL_787161, EPI_ISL_787162, EPI_ISL_787163, EPI_ISL_787164, EPI_ISL_787165, EPI_ISL_787166, EPI_ISL_787167, EPI_ISL_787168,
EPI_ISL_787169, EPI_ISL_787170, EPI_ISL_787171, EPI_ISL_787172, EPI_ISL_787173, EPI_ISL_787174, EPI_ISL_787175, EPI_ISL_787176, EPI_ISL_787177, EPI_ISL_787178, EPI_ISL_787179, EPI_ISL_787180, EPI_ISL_787181, EPI_ISL_787182, EPI_ISL_787183, EPI_ISL_787184, EPI_ISL_787185, EPI_ISL_787186, EPI_ISL_787187, EPI_ISL_787188, EPI_ISL_787189, EPI_ISL_787190, EPI_ISL_787191, EPI_ISL_787192,
EPI_ISL_787193, EPI_ISL_787194, EPI_ISL_787195, EPI_ISL_787196, EPI_ISL_787197, EPI_ISL_787198, EPI_ISL_787199, EPI_ISL_787200, EPI_ISL_787201, EPI_ISL_787202, EPI_ISL_787203, EPI_ISL_787204, EPI_ISL_787205, EPI_ISL_787206, EPI_ISL_787207, EPI_ISL_787208, EPI_ISL_787209, EPI_ISL_787210, EPI_ISL_787211, EPI_ISL_787212, EPI_ISL_787213, EPI_ISL_787214, EPI_ISL_787215, EPI_ISL_787216,
EPI_ISL_787217, EPI_ISL_787218, EPI_ISL_787219, EPI_ISL_787220, EPI_ISL_787221, EPI_ISL_787222, EPI_ISL_787223, EPI_ISL_787224, EPI_ISL_787225, EPI_ISL_787226, EPI_ISL_787227, EPI_ISL_787228, EPI_ISL_787229, EPI_ISL_787230, EPI_ISL_787231, EPI_ISL_787232, EPI_ISL_787233, EPI_ISL_787234, EPI_ISL_787235, EPI_ISL_787236, EPI_ISL_787237, EPI_ISL_787238, EPI_ISL_787239, EPI_ISL_787240,
EPI_ISL_787241, EPI_ISL_787242, EPI_ISL_787243, EPI_ISL_787244, EPI_ISL_787245, EPI_ISL_787246, EPI_ISL_787247, EPI_ISL_787248, EPI_ISL_787249, EPI_ISL_787250, EPI_ISL_787251, EPI_ISL_787252, EPI_ISL_787253, EPI_ISL_787254, EPI_ISL_787255, EPI_ISL_787256, EPI_ISL_787257, EPI_ISL_787258, EPI_ISL_787259, EPI_ISL_787260, EPI_ISL_787261, EPI_ISL_787262, EPI_ISL_787263, EPI_ISL_787264,
EPI_ISL_787265, EPI_ISL_787266, EPI_ISL_787267, EPI_ISL_787268, EPI_ISL_787269, EPI_ISL_787270, EPI_ISL_787271, EPI_ISL_787272, EPI_ISL_787273, EPI_ISL_787274, EPI_ISL_787275, EPI_ISL_787276, EPI_ISL_787277, EPI_ISL_787278, EPI_ISL_787279, EPI_ISL_787280, EPI_ISL_787281, EPI_ISL_787282, EPI_ISL_787283, EPI_ISL_787284, EPI_ISL_787285, EPI_ISL_787286, EPI_ISL_787287, EPI_ISL_787288,
EPI_ISL_787289, EPI_ISL_787290, EPI_ISL_787291, EPI_ISL_787292, EPI_ISL_787293, EPI_ISL_787294, EPI_ISL_787295, EPI_ISL_787296, EPI_ISL_787297, EPI_ISL_787298, EPI_ISL_787299, EPI_ISL_787300, EPI_ISL_787301, EPI_ISL_787302, EPI_ISL_787303, EPI_ISL_787304, EPI_ISL_787305, EPI_ISL_787306, EPI_ISL_787307, EPI_ISL_787308, EPI_ISL_787309, EPI_ISL_787310, EPI_ISL_787311, EPI_ISL_787312,

EPI_ISL_787313, EPI_ISL_787314, EPI_ISL_787315, EPI_ISL_787316, EPI_ISL_787317, EPI_ISL_787318, EPI_ISL_787319, EPI_ISL_787320, EPI_ISL_787321, EPI_ISL_787322, EPI_ISL_787323, EPI_ISL_787324, EPI_ISL_787325, EPI_ISL_787326, EPI_ISL_787327, EPI_ISL_787328, EPI_ISL_787329, EPI_ISL_787330, EPI_ISL_787331, EPI_ISL_787332, EPI_ISL_787333, EPI_ISL_787334, EPI_ISL_787335, EPI_ISL_787336,
EPI_ISL_787337, EPI_ISL_787338, EPI_ISL_787339, EPI_ISL_787340, EPI_ISL_787341, EPI_ISL_787342, EPI_ISL_787343, EPI_ISL_787344, EPI_ISL_787345, EPI_ISL_787346, EPI_ISL_787347, EPI_ISL_787348, EPI_ISL_787349, EPI_ISL_787350, EPI_ISL_787351, EPI_ISL_787352, EPI_ISL_787353, EPI_ISL_787354, EPI_ISL_787355, EPI_ISL_787356, EPI_ISL_787357, EPI_ISL_787358, EPI_ISL_787359, EPI_ISL_787360,
EPI_ISL_787361, EPI_ISL_787362, EPI_ISL_787363, EPI_ISL_787364, EPI_ISL_787365, EPI_ISL_787366, EPI_ISL_787367, EPI_ISL_787368, EPI_ISL_787369, EPI_ISL_787370, EPI_ISL_787371, EPI_ISL_787372, EPI_ISL_787373, EPI_ISL_787374, EPI_ISL_787375, EPI_ISL_787376, EPI_ISL_787377, EPI_ISL_787378, EPI_ISL_787379, EPI_ISL_787380, EPI_ISL_787381, EPI_ISL_787382, EPI_ISL_787383, EPI_ISL_787384,
EPI_ISL_787385, EPI_ISL_787386, EPI_ISL_787387, EPI_ISL_787388, EPI_ISL_787389, EPI_ISL_787390, EPI_ISL_787391, EPI_ISL_787392, EPI_ISL_787393, EPI_ISL_787394, EPI_ISL_787395, EPI_ISL_787396, EPI_ISL_787397, EPI_ISL_787398, EPI_ISL_787399, EPI_ISL_787400, EPI_ISL_787401, EPI_ISL_787402, EPI_ISL_787403, EPI_ISL_787404, EPI_ISL_787405, EPI_ISL_787406, EPI_ISL_787407, EPI_ISL_787408,
EPI_ISL_787409, EPI_ISL_787410, EPI_ISL_787411, EPI_ISL_787412, EPI_ISL_787413, EPI_ISL_787414, EPI_ISL_787415, EPI_ISL_787416, EPI_ISL_787417, EPI_ISL_787418, EPI_ISL_787419, EPI_ISL_787420, EPI_ISL_787421, EPI_ISL_787422, EPI_ISL_787423, EPI_ISL_787424, EPI_ISL_787425, EPI_ISL_787426, EPI_ISL_787427, EPI_ISL_787428, EPI_ISL_787429, EPI_ISL_787430, EPI_ISL_787431, EPI_ISL_787432,
EPI_ISL_787433, EPI_ISL_787434, EPI_ISL_787435, EPI_ISL_787436, EPI_ISL_787437, EPI_ISL_787438, EPI_ISL_787439, EPI_ISL_787440, EPI_ISL_787441, EPI_ISL_787442, EPI_ISL_787443, EPI_ISL_787444, EPI_ISL_787445, EPI_ISL_787446, EPI_ISL_787447, EPI_ISL_787448, EPI_ISL_787449, EPI_ISL_787450, EPI_ISL_787451, EPI_ISL_787452, EPI_ISL_787453, EPI_ISL_787454, EPI_ISL_787455, EPI_ISL_787456,
EPI_ISL_787457, EPI_ISL_787458, EPI_ISL_787459, EPI_ISL_787460, EPI_ISL_787461, EPI_ISL_787462, EPI_ISL_787463, EPI_ISL_787464, EPI_ISL_787465, EPI_ISL_787466, EPI_ISL_787467, EPI_ISL_787468, EPI_ISL_787469, EPI_ISL_787470, EPI_ISL_787471, EPI_ISL_787472, EPI_ISL_787473, EPI_ISL_787474, EPI_ISL_787475, EPI_ISL_787476, EPI_ISL_787477, EPI_ISL_787478, EPI_ISL_787479, EPI_ISL_787480,
EPI_ISL_787481, EPI_ISL_787482, EPI_ISL_787483, EPI_ISL_787484, EPI_ISL_787485, EPI_ISL_787486, EPI_ISL_787487, EPI_ISL_787488, EPI_ISL_787489, EPI_ISL_787490, EPI_ISL_787491, EPI_ISL_787492, EPI_ISL_787493, EPI_ISL_787494, EPI_ISL_787495, EPI_ISL_787496, EPI_ISL_787497, EPI_ISL_787498, EPI_ISL_787499, EPI_ISL_787500, EPI_ISL_787501, EPI_ISL_787502, EPI_ISL_787503, EPI_ISL_787504,
EPI_ISL_787505, EPI_ISL_787506, EPI_ISL_787507, EPI_ISL_787508, EPI_ISL_787509, EPI_ISL_787510, EPI_ISL_787511, EPI_ISL_787512, EPI_ISL_787513, EPI_ISL_787514, EPI_ISL_787515, EPI_ISL_787516, EPI_ISL_787517, EPI_ISL_787518, EPI_ISL_787519, EPI_ISL_787520, EPI_ISL_787521, EPI_ISL_787522, EPI_ISL_787523, EPI_ISL_787524, EPI_ISL_787525, EPI_ISL_787526, EPI_ISL_787527, EPI_ISL_787528,
EPI_ISL_787529, EPI_ISL_787530, EPI_ISL_787531, EPI_ISL_787532, EPI_ISL_787533, EPI_ISL_787534, EPI_ISL_787535, EPI_ISL_787536, EPI_ISL_787537, EPI_ISL_787538, EPI_ISL_787539, EPI_ISL_787540, EPI_ISL_787541, EPI_ISL_787542, EPI_ISL_787543, EPI_ISL_787544, EPI_ISL_787545, EPI_ISL_787546, EPI_ISL_787547, EPI_ISL_787548, EPI_ISL_787549, EPI_ISL_787550, EPI_ISL_787551, EPI_ISL_787552,
EPI_ISL_787553, EPI_ISL_787554, EPI_ISL_787555, EPI_ISL_787556, EPI_ISL_787557, EPI_ISL_787558, EPI_ISL_787559, EPI_ISL_787560, EPI_ISL_787561, EPI_ISL_787562, EPI_ISL_787563, EPI_ISL_787564, EPI_ISL_787565, EPI_ISL_787566, EPI_ISL_787567, EPI_ISL_787568, EPI_ISL_787569, EPI_ISL_787570, EPI_ISL_787571, EPI_ISL_787572, EPI_ISL_787573, EPI_ISL_787574, EPI_ISL_787575, EPI_ISL_787576,
EPI_ISL_787577, EPI_ISL_787578, EPI_ISL_787579, EPI_ISL_787580, EPI_ISL_787581, EPI_ISL_787582, EPI_ISL_787583, EPI_ISL_787584, EPI_ISL_787585, EPI_ISL_787586, EPI_ISL_787587, EPI_ISL_787588, EPI_ISL_787589, EPI_ISL_787590, EPI_ISL_787591, EPI_ISL_787592, EPI_ISL_787593, EPI_ISL_787594, EPI_ISL_787595, EPI_ISL_787596, EPI_ISL_787597, EPI_ISL_787598, EPI_ISL_787599, EPI_ISL_787600,
EPI_ISL_787601, EPI_ISL_787602, EPI_ISL_787603, EPI_ISL_787604, EPI_ISL_787605, EPI_ISL_787606, EPI_ISL_787607, EPI_ISL_787608, EPI_ISL_787609, EPI_ISL_787610, EPI_ISL_787611, EPI_ISL_787612, EPI_ISL_787613, EPI_ISL_787614, EPI_ISL_787615, EPI_ISL_787616, EPI_ISL_787617, EPI_ISL_787618, EPI_ISL_787619, EPI_ISL_787620, EPI_ISL_787621, EPI_ISL_787622, EPI_ISL_787623, EPI_ISL_787624,
EPI_ISL_787625, EPI_ISL_787626, EPI_ISL_787627, EPI_ISL_787628, EPI_ISL_787629, EPI_ISL_787630, EPI_ISL_787631, EPI_ISL_787632, EPI_ISL_787633, EPI_ISL_787634, EPI_ISL_787635, EPI_ISL_787636, EPI_ISL_787637, EPI_ISL_787638, EPI_ISL_787639, EPI_ISL_787640, EPI_ISL_787641, EPI_ISL_787642, EPI_ISL_787643, EPI_ISL_787644, EPI_ISL_787645, EPI_ISL_787646, EPI_ISL_787647, EPI_ISL_787648,
EPI_ISL_787649, EPI_ISL_787650, EPI_ISL_787651, EPI_ISL_787652, EPI_ISL_787653, EPI_ISL_787654, EPI_ISL_787655, EPI_ISL_787656, EPI_ISL_787657, EPI_ISL_787658, EPI_ISL_787659, EPI_ISL_787660, EPI_ISL_787661, EPI_ISL_787662, EPI_ISL_787663, EPI_ISL_787664, EPI_ISL_787665, EPI_ISL_787666, EPI_ISL_787667, EPI_ISL_787668, EPI_ISL_787669, EPI_ISL_787670, EPI_ISL_787671, EPI_ISL_787672,
EPI_ISL_787673, EPI_ISL_787674, EPI_ISL_787675, EPI_ISL_787676, EPI_ISL_787677, EPI_ISL_787678, EPI_ISL_787679, EPI_ISL_787680, EPI_ISL_787681, EPI_ISL_787682, EPI_ISL_787683, EPI_ISL_787684, EPI_ISL_787685, EPI_ISL_787686, EPI_ISL_787687, EPI_ISL_787688, EPI_ISL_787689, EPI_ISL_787690, EPI_ISL_787691, EPI_ISL_787692, EPI_ISL_787693, EPI_ISL_787694, EPI_ISL_787695, EPI_ISL_787696,
EPI_ISL_787697, EPI_ISL_787698, EPI_ISL_787699, EPI_ISL_787700, EPI_ISL_787701, EPI_ISL_787702, EPI_ISL_787703, EPI_ISL_787704, EPI_ISL_787705, EPI_ISL_787706, EPI_ISL_787707, EPI_ISL_787708, EPI_ISL_787709, EPI_ISL_787710, EPI_ISL_787711, EPI_ISL_787712, EPI_ISL_787713, EPI_ISL_787714, EPI_ISL_787715, EPI_ISL_787716, EPI_ISL_787717, EPI_ISL_787718, EPI_ISL_787719, EPI_ISL_787720,
EPI_ISL_787721, EPI_ISL_787722, EPI_ISL_787723, EPI_ISL_787724, EPI_ISL_787725, EPI_ISL_787726, EPI_ISL_787727, EPI_ISL_787728, EPI_ISL_787729, EPI_ISL_787730, EPI_ISL_787731, EPI_ISL_787732, EPI_ISL_787733, EPI_ISL_787734, EPI_ISL_787735, EPI_ISL_787736, EPI_ISL_787737, EPI_ISL_787738, EPI_ISL_787739, EPI_ISL_787740, EPI_ISL_787741, EPI_ISL_787742, EPI_ISL_787743, EPI_ISL_787744,
EPI_ISL_787745, EPI_ISL_787746, EPI_ISL_787747, EPI_ISL_787748, EPI_ISL_787749, EPI_ISL_787750, EPI_ISL_787751, EPI_ISL_787752, EPI_ISL_787753, EPI_ISL_787754, EPI_ISL_787755, EPI_ISL_787756, EPI_ISL_787757, EPI_ISL_787758, EPI_ISL_787759, EPI_ISL_787760, EPI_ISL_787761, EPI_ISL_787762, EPI_ISL_787763, EPI_ISL_787764, EPI_ISL_787765, EPI_ISL_787766, EPI_ISL_787767, EPI_ISL_787768,
EPI_ISL_787769, EPI_ISL_787770, EPI_ISL_787771, EPI_ISL_787772, EPI_ISL_787773, EPI_ISL_787774, EPI_ISL_787775, EPI_ISL_787776, EPI_ISL_787777, EPI_ISL_787778, EPI_ISL_787779, EPI_ISL_787780, EPI_ISL_787781, EPI_ISL_787782, EPI_ISL_787783, EPI_ISL_787784, EPI_ISL_787785, EPI_ISL_787786, EPI_ISL_787787, EPI_ISL_787788, EPI_ISL_787789, EPI_ISL_787790, EPI_ISL_787791, EPI_ISL_787792,
EPI_ISL_787793, EPI_ISL_787794, EPI_ISL_787795, EPI_ISL_787796, EPI_ISL_787797, EPI_ISL_787798, EPI_ISL_787799, EPI_ISL_787800, EPI_ISL_787801, EPI_ISL_787802, EPI_ISL_787803, EPI_ISL_787804, EPI_ISL_787805, EPI_ISL_787806, EPI_ISL_787807, EPI_ISL_787808, EPI_ISL_787809, EPI_ISL_787810, EPI_ISL_787811, EPI_ISL_787812, EPI_ISL_787813, EPI_ISL_787814, EPI_ISL_787815, EPI_ISL_787816,
EPI_ISL_787817, EPI_ISL_787818, EPI_ISL_787819, EPI_ISL_787820, EPI_ISL_787821, EPI_ISL_787822, EPI_ISL_787823, EPI_ISL_787824, EPI_ISL_787825, EPI_ISL_787826, EPI_ISL_787827, EPI_ISL_787828, EPI_ISL_787829, EPI_ISL_787830, EPI_ISL_787831, EPI_ISL_787832, EPI_ISL_787833, EPI_ISL_787834, EPI_ISL_787835, EPI_ISL_787836, EPI_ISL_787837, EPI_ISL_787838, EPI_ISL_787839, EPI_ISL_787840,
EPI_ISL_787841, EPI_ISL_787842, EPI_ISL_787843, EPI_ISL_787844, EPI_ISL_787845, EPI_ISL_787846, EPI_ISL_787847, EPI_ISL_787848, EPI_ISL_787849, EPI_ISL_787850, EPI_ISL_787851, EPI_ISL_787852, EPI_ISL_787853, EPI_ISL_787854, EPI_ISL_787855, EPI_ISL_787856, EPI_ISL_787857, EPI_ISL_787858, EPI_ISL_787859, EPI_ISL_787860, EPI_ISL_787861, EPI_ISL_787862, EPI_ISL_787863, EPI_ISL_787864,
EPI_ISL_787865, EPI_ISL_787866, EPI_ISL_787867, EPI_ISL_787868, EPI_ISL_787869, EPI_ISL_787870, EPI_ISL_787871, EPI_ISL_787872, EPI_ISL_787873, EPI_ISL_787874, EPI_ISL_787875, EPI_ISL_787876, EPI_ISL_787877, EPI_ISL_787878, EPI_ISL_787879, EPI_ISL_787880, EPI_ISL_787881, EPI_ISL_787882, EPI_ISL_787883, EPI_ISL_787884, EPI_ISL_787885, EPI_ISL_787886, EPI_ISL_787887, EPI_ISL_787888,
EPI_ISL_787889, EPI_ISL_787890, EPI_ISL_787891, EPI_ISL_787892, EPI_ISL_787893, EPI_ISL_787894, EPI_ISL_787895, EPI_ISL_787896, EPI_ISL_787897, EPI_ISL_787898, EPI_ISL_787899, EPI_ISL_787900, EPI_ISL_787901, EPI_ISL_787902, EPI_ISL_787903, EPI_ISL_787904, EPI_ISL_787905, EPI_ISL_787906, EPI_ISL_787907, EPI_ISL_787908, EPI_ISL_787909, EPI_ISL_787910, EPI_ISL_787911, EPI_ISL_787912,
EPI_ISL_787913, EPI_ISL_787914, EPI_ISL_787915, EPI_ISL_787916, EPI_ISL_787917, EPI_ISL_787918, EPI_ISL_787919, EPI_ISL_787920, EPI_ISL_787921, EPI_ISL_787922, EPI_ISL_787923, EPI_ISL_787924, EPI_ISL_787925, EPI_ISL_787926, EPI_ISL_787927, EPI_ISL_787928, EPI_ISL_787929, EPI_ISL_787930, EPI_ISL_787931, EPI_ISL_787932, EPI_ISL_787933, EPI_ISL_787934, EPI_ISL_787935, EPI_ISL_787936,
EPI_ISL_787937, EPI_ISL_787938, EPI_ISL_787939, EPI_ISL_787940, EPI_ISL_787941, EPI_ISL_787942, EPI_ISL_787943, EPI_ISL_787944, EPI_ISL_787945, EPI_ISL_787946, EPI_ISL_787947, EPI_ISL_787948, EPI_ISL_787949, EPI_ISL_787950, EPI_ISL_787951, EPI_ISL_787952, EPI_ISL_787953, EPI_ISL_787954, EPI_ISL_787955, EPI_ISL_787956, EPI_ISL_787957, EPI_ISL_787958, EPI_ISL_787959, EPI_ISL_787960,
EPI_ISL_787961, EPI_ISL_787962, EPI_ISL_787963, EPI_ISL_787964, EPI_ISL_787965, EPI_ISL_787966, EPI_ISL_787967, EPI_ISL_787968, EPI_ISL_787969, EPI_ISL_787970, EPI_ISL_787971, EPI_ISL_787972, EPI_ISL_787973, EPI_ISL_787974, EPI_ISL_787975, EPI_ISL_787976, EPI_ISL_787977, EPI_ISL_787978, EPI_ISL_787979, EPI_ISL_787980, EPI_ISL_787981, EPI_ISL_787982, EPI_ISL_787983, EPI_ISL_787984,
EPI_ISL_787985, EPI_ISL_787986, EPI_ISL_787987, EPI_ISL_787988, EPI_ISL_787989, EPI_ISL_787990, EPI_ISL_787991, EPI_ISL_787992, EPI_ISL_787993, EPI_ISL_787994, EPI_ISL_787995, EPI_ISL_787996, EPI_ISL_787997, EPI_ISL_787998, EPI_ISL_787999, EPI_ISL_788000, EPI_ISL_788001, EPI_ISL_788002, EPI_ISL_788003, EPI_ISL_788004, EPI_ISL_788005, EPI_ISL_788006, EPI_ISL_788007, EPI_ISL_788008,
EPI_ISL_788009, EPI_ISL_788010, EPI_ISL_788011, EPI_ISL_788012, EPI_ISL_788013, EPI_ISL_788014, EPI_ISL_788015, EPI_ISL_788016, EPI_ISL_788017, EPI_ISL_788018, EPI_ISL_788019, EPI_ISL_788020, EPI_ISL_788021, EPI_ISL_788022, EPI_ISL_788023, EPI_ISL_788024, EPI_ISL_788025, EPI_ISL_788026, EPI_ISL_788027, EPI_ISL_788028, EPI_ISL_788029, EPI_ISL_788030, EPI_ISL_788031, EPI_ISL_788032,
EPI_ISL_788033, EPI_ISL_788034, EPI_ISL_788035, EPI_ISL_788036, EPI_ISL_788037, EPI_ISL_788038, EPI_ISL_788039, EPI_ISL_788040, EPI_ISL_788041, EPI_ISL_788042, EPI_ISL_788043, EPI_ISL_788044, EPI_ISL_788045, EPI_ISL_788046, EPI_ISL_788047, EPI_ISL_788048, EPI_ISL_788049, EPI_ISL_788050, EPI_ISL_788051, EPI_ISL_788052, EPI_ISL_788053, EPI_ISL_788054, EPI_ISL_788055, EPI_ISL_788056,
EPI_ISL_788057, EPI_ISL_788058, EPI_ISL_788059, EPI_ISL_788060, EPI_ISL_788061, EPI_ISL_788062, EPI_ISL_788063, EPI_ISL_788064, EPI_ISL_788065, EPI_ISL_788066, EPI_ISL_788067, EPI_ISL_788068, EPI_ISL_788069, EPI_ISL_788070, EPI_ISL_788071, EPI_ISL_788072, EPI_ISL_788073, EPI_ISL_788074, EPI_ISL_788075, EPI_ISL_788076, EPI_ISL_788077, EPI_ISL_788078, EPI_ISL_788079, EPI_ISL_788080,
EPI_ISL_788081, EPI_ISL_788082, EPI_ISL_788083, EPI_ISL_788084, EPI_ISL_788085, EPI_ISL_788086, EPI_ISL_788087, EPI_ISL_788088, EPI_ISL_788089, EPI_ISL_788090, EPI_ISL_788091, EPI_ISL_788092, EPI_ISL_788093, EPI_ISL_788094, EPI_ISL_788095, EPI_ISL_788096, EPI_ISL_788097, EPI_ISL_788098, EPI_ISL_788099, EPI_ISL_788100, EPI_ISL_788101, EPI_ISL_788102, EPI_ISL_788103, EPI_ISL_788104,
EPI_ISL_788105, EPI_ISL_788106, EPI_ISL_788107, EPI_ISL_788108, EPI_ISL_788109, EPI_ISL_788110, EPI_ISL_788111, EPI_ISL_788112, EPI_ISL_788113, EPI_ISL_788114, EPI_ISL_788115, EPI_ISL_788116, EPI_ISL_788117, EPI_ISL_788118, EPI_ISL_788119, EPI_ISL_788120, EPI_ISL_788121, EPI_ISL_788122, EPI_ISL_788123, EPI_ISL_788124, EPI_ISL_788125, EPI_ISL_788126, EPI_ISL_788127, EPI_ISL_788128,
EPI_ISL_788129, EPI_ISL_788130, EPI_ISL_788131, EPI_ISL_788132, EPI_ISL_788133, EPI_ISL_788134, EPI_ISL_788135, EPI_ISL_788136, EPI_ISL_788137, EPI_ISL_788138, EPI_ISL_788139, EPI_ISL_788140, EPI_ISL_788141, EPI_ISL_788142, EPI_ISL_788143, EPI_ISL_788144, EPI_ISL_788145, EPI_ISL_788146, EPI_ISL_788147, EPI_ISL_788148, EPI_ISL_788149, EPI_ISL_788150, EPI_ISL_788151, EPI_ISL_788152,
EPI_ISL_788153, EPI_ISL_788154, EPI_ISL_788155, EPI_ISL_788156, EPI_ISL_788157, EPI_ISL_788158, EPI_ISL_788159, EPI_ISL_788160, EPI_ISL_788161, EPI_ISL_788162, EPI_ISL_788163, EPI_ISL_788164, EPI_ISL_788165, EPI_ISL_788166, EPI_ISL_788167, EPI_ISL_788168, EPI_ISL_788169, EPI_ISL_788170, EPI_ISL_788171, EPI_ISL_788172, EPI_ISL_788173, EPI_ISL_788174, EPI_ISL_788175, EPI_ISL_788176,
EPI_ISL_788177, EPI_ISL_788178, EPI_ISL_788179, EPI_ISL_788180, EPI_ISL_788181, EPI_ISL_788182, EPI_ISL_788183, EPI_ISL_788184, EPI_ISL_788185, EPI_ISL_788186, EPI_ISL_788187, EPI_ISL_788188, EPI_ISL_788189, EPI_ISL_788190, EPI_ISL_788191, EPI_ISL_788192, EPI_ISL_788193, EPI_ISL_788194, EPI_ISL_788195, EPI_ISL_788196, EPI_ISL_788197, EPI_ISL_788198, EPI_ISL_788199, EPI_ISL_788200,
EPI_ISL_788201, EPI_ISL_788202, EPI_ISL_788203, EPI_ISL_788204, EPI_ISL_788205, EPI_ISL_788206, EPI_ISL_788207, EPI_ISL_788208, EPI_ISL_788209, EPI_ISL_788210, EPI_ISL_788211, EPI_ISL_788212, EPI_ISL_788213, EPI_ISL_788214, EPI_ISL_788215, EPI_ISL_788216, EPI_ISL_788217, EPI_ISL_788218, EPI_ISL_788219, EPI_ISL_788220, EPI_ISL_788221, EPI_ISL_788222, EPI_ISL_788223, EPI_ISL_788224,
EPI_ISL_788225, EPI_ISL_788226, EPI_ISL_788227, EPI_ISL_788228, EPI_ISL_788229, EPI_ISL_788230, EPI_ISL_788231, EPI_ISL_788232, EPI_ISL_788233, EPI_ISL_788234, EPI_ISL_788235, EPI_ISL_788236, EPI_ISL_788237, EPI_ISL_788238, EPI_ISL_788239, EPI_ISL_788240, EPI_ISL_788241, EPI_ISL_788242, EPI_ISL_788243, EPI_ISL_788244, EPI_ISL_788245, EPI_ISL_788246, EPI_ISL_788247, EPI_ISL_788248,
EPI_ISL_788249, EPI_ISL_788250, EPI_ISL_788251, EPI_ISL_788252, EPI_ISL_788253, EPI_ISL_788254, EPI_ISL_788255, EPI_ISL_788256, EPI_ISL_788257, EPI_ISL_788258, EPI_ISL_788259, EPI_ISL_788260, EPI_ISL_788261, EPI_ISL_788262, EPI_ISL_788263, EPI_ISL_788264, EPI_ISL_788265, EPI_ISL_788266, EPI_ISL_788267, EPI_ISL_788268, EPI_ISL_788269, EPI_ISL_788270, EPI_ISL_788271, EPI_ISL_788272,
EPI_ISL_788273, EPI_ISL_788274, EPI_ISL_788275, EPI_ISL_788276, EPI_ISL_788277, EPI_ISL_788278, EPI_ISL_788279, EPI_ISL_788280, EPI_ISL_788281, EPI_ISL_788282, EPI_ISL_788283, EPI_ISL_788284, EPI_ISL_788285, EPI_ISL_788286, EPI_ISL_788287, EPI_ISL_788288, EPI_ISL_788289, EPI_ISL_788290, EPI_ISL_788291, EPI_ISL_788292, EPI_ISL_788293, EPI_ISL_788294, EPI_ISL_788295, EPI_ISL_788296,
EPI_ISL_788297, EPI_ISL_788298, EPI_ISL_788299, EPI_ISL_788300, EPI_ISL_788301, EPI_ISL_788302, EPI_ISL_788303, EPI_ISL_788304, EPI_ISL_788305, EPI_ISL_788306, EPI_ISL_788307, EPI_ISL_788308, EPI_ISL_788309, EPI_ISL_788310, EPI_ISL_788311, EPI_ISL_788312, EPI_ISL_788313, EPI_ISL_788314, EPI_ISL_788315, EPI_ISL_788316, EPI_ISL_788317, EPI_ISL_788318, EPI_ISL_788319, EPI_ISL_788320,
EPI_ISL_788321, EPI_ISL_788322, EPI_ISL_788323, EPI_ISL_788324, EPI_ISL_788325, EPI_ISL_788326, EPI_ISL_788327, EPI_ISL_788328, EPI_ISL_788329, EPI_ISL_788330, EPI_ISL_788331, EPI_ISL_788332, EPI_ISL_788333, EPI_ISL_788334, EPI_ISL_788335, EPI_ISL_788336, EPI_ISL_788337, EPI_ISL_788338, EPI_ISL_788339, EPI_ISL_788340, EPI_ISL_788341, EPI_ISL_788342, EPI_ISL_788343, EPI_ISL_788344,
EPI_ISL_788345, EPI_ISL_788346, EPI_ISL_788347, EPI_ISL_788348, EPI_ISL_788349, EPI_ISL_788350, EPI_ISL_788351, EPI_ISL_788352, EPI_ISL_788353, EPI_ISL_788354, EPI_ISL_788355, EPI_ISL_788356, EPI_ISL_788357, EPI_ISL_788358, EPI_ISL_788359, EPI_ISL_788360, EPI_ISL_788361, EPI_ISL_788362, EPI_ISL_788363, EPI_ISL_788364, EPI_ISL_788365, EPI_ISL_788366, EPI_ISL_788367, EPI_ISL_788368,
EPI_ISL_788369, EPI_ISL_788370, EPI_ISL_788371, EPI_ISL_788372, EPI_ISL_788373, EPI_ISL_788374, EPI_ISL_788375, EPI_ISL_788376, EPI_ISL_788377, EPI_ISL_788378, EPI_ISL_788379, EPI_ISL_788380, EPI_ISL_788381, EPI_ISL_788382, EPI_ISL_788383, EPI_ISL_788384, EPI_ISL_788385, EPI_ISL_788386, EPI_ISL_788387, EPI_ISL_788388, EPI_ISL_788389, EPI_ISL_788390, EPI_ISL_788391, EPI_ISL_788392,
EPI_ISL_788393, EPI_ISL_788394, EPI_ISL_788395, EPI_ISL_788396, EPI_ISL_788397, EPI_ISL_788398, EPI_ISL_788399, EPI_ISL_788400, EPI_ISL_788401, EPI_ISL_788402, EPI_ISL_788403, EPI_ISL_788404, EPI_ISL_788405, EPI_ISL_788406, EPI_ISL_788407, EPI_ISL_788408, EPI_ISL_788409, EPI_ISL_788410, EPI_ISL_788411, EPI_ISL_788412, EPI_ISL_788413, EPI_ISL_788414, EPI_ISL_788415, EPI_ISL_788416,
EPI_ISL_788417, EPI_ISL_788418, EPI_ISL_788419, EPI_ISL_788420, EPI_ISL_788421, EPI_ISL_788422, EPI_ISL_788423, EPI_ISL_788424, EPI_ISL_788425, EPI_ISL_788426, EPI_ISL_788427, EPI_ISL_788428, EPI_ISL_788429, EPI_ISL_788430, EPI_ISL_788431, EPI_ISL_788432, EPI_ISL_788433, EPI_ISL_788434, EPI_ISL_788435, EPI_ISL_788436, EPI_ISL_788437, EPI_ISL_788438, EPI_ISL_788439, EPI_ISL_788440,
EPI_ISL_788441, EPI_ISL_788442, EPI_ISL_788443, EPI_ISL_788444, EPI_ISL_788445, EPI_ISL_788446, EPI_ISL_788447, EPI_ISL_788448, EPI_ISL_788449, EPI_ISL_788450, EPI_ISL_788451, EPI_ISL_788452, EPI_ISL_788453, EPI_ISL_788454, EPI_ISL_788455, EPI_ISL_788456, EPI_ISL_788457, EPI_ISL_788458, EPI_ISL_788459, EPI_ISL_788460, EPI_ISL_788461, EPI_ISL_788462, EPI_ISL_788463, EPI_ISL_788464,
EPI_ISL_788465, EPI_ISL_788466, EPI_ISL_788467, EPI_ISL_788468, EPI_ISL_788469, EPI_ISL_788470, EPI_ISL_788471, EPI_ISL_788472, EPI_ISL_788473, EPI_ISL_788474, EPI_ISL_788475, EPI_ISL_788476, EPI_ISL_788477, EPI_ISL_788478, EPI_ISL_788479, EPI_ISL_788480, EPI_ISL_788481, EPI_ISL_788482, EPI_ISL_788483, EPI_ISL_788484, EPI_ISL_788485, EPI_ISL_788486, EPI_ISL_788487, EPI_ISL_788488,
EPI_ISL_788489, EPI_ISL_788490, EPI_ISL_788491, EPI_ISL_788492, EPI_ISL_788493, EPI_ISL_788494, EPI_ISL_788495, EPI_ISL_788496, EPI_ISL_788497, EPI_ISL_788498, EPI_ISL_788499, EPI_ISL_788500, EPI_ISL_788501, EPI_ISL_788502, EPI_ISL_788503, EPI_ISL_788504, EPI_ISL_788505, EPI_ISL_788506, EPI_ISL_788507, EPI_ISL_788508, EPI_ISL_788509, EPI_ISL_788510, EPI_ISL_788511, EPI_ISL_788512,
EPI_ISL_788513, EPI_ISL_788514, EPI_ISL_788515, EPI_ISL_788516, EPI_ISL_788517, EPI_ISL_788518, EPI_ISL_788519, EPI_ISL_788520, EPI_ISL_788521, EPI_ISL_788522, EPI_ISL_788523, EPI_ISL_788524, EPI_ISL_788525, EPI_ISL_788526, EPI_ISL_788527, EPI_ISL_788528, EPI_ISL_788529, EPI_ISL_788530, EPI_ISL_788531, EPI_ISL_788532, EPI_ISL_788533, EPI_ISL_788534, EPI_ISL_788535, EPI_ISL_788536,
EPI_ISL_788537, EPI_ISL_788538, EPI_ISL_788539, EPI_ISL_788540, EPI_ISL_788541, EPI_ISL_788542, EPI_ISL_788543, EPI_ISL_788544, EPI_ISL_788545, EPI_ISL_788546, EPI_ISL_788547, EPI_ISL_788548, EPI_ISL_788549, EPI_ISL_788550, EPI_ISL_788551, EPI_ISL_788552, EPI_ISL_788553, EPI_ISL_788554, EPI_ISL_788555, EPI_ISL_788556, EPI_ISL_788557, EPI_ISL_788558, EPI_ISL_788559, EPI_ISL_788560,
EPI_ISL_788561, EPI_ISL_788562, EPI_ISL_788563, EPI_ISL_788564, EPI_ISL_788565, EPI_ISL_788566, EPI_ISL_788567, EPI_ISL_788568, EPI_ISL_788569, EPI_ISL_788570, EPI_ISL_788571, EPI_ISL_788572, EPI_ISL_788573, EPI_ISL_788574, EPI_ISL_788575, EPI_ISL_788576, EPI_ISL_788577, EPI_ISL_788578, EPI_ISL_788579, EPI_ISL_788580, EPI_ISL_788581, EPI_ISL_788582, EPI_ISL_788583, EPI_ISL_788584,
EPI_ISL_788585, EPI_ISL_788586, EPI_ISL_788587, EPI_ISL_788588, EPI_ISL_788589, EPI_ISL_788590, EPI_ISL_788591, EPI_ISL_788592, EPI_ISL_788593, EPI_ISL_788594, EPI_ISL_788595, EPI_ISL_788596, EPI_ISL_788597, EPI_ISL_788598, EPI_ISL_788599, EPI_ISL_788600, EPI_ISL_788601, EPI_ISL_788602, EPI_ISL_788603, EPI_ISL_788604, EPI_ISL_788605, EPI_ISL_788606, EPI_ISL_788607, EPI_ISL_788608,
EPI_ISL_788609, EPI_ISL_788610, EPI_ISL_788611, EPI_ISL_788612, EPI_ISL_788613, EPI_ISL_788614, EPI_ISL_788615, EPI_ISL_788616, EPI_ISL_788617, EPI_ISL_788618, EPI_ISL_788619, EPI_ISL_788620, EPI_ISL_788621, EPI_ISL_788622, EPI_ISL_788623, EPI_ISL_788624, EPI_ISL_788625, EPI_ISL_788626, EPI_ISL_788627, EPI_ISL_788628, EPI_ISL_788629, EPI_ISL_788630, EPI_ISL_788631, EPI_ISL_788632,
EPI_ISL_788633, EPI_ISL_788634, EPI_ISL_788635, EPI_ISL_788636, EPI_ISL_788637, EPI_ISL_788638, EPI_ISL_788639, EPI_ISL_788640, EPI_ISL_788641, EPI_ISL_788642, EPI_ISL_788643, EPI_ISL_788644, EPI_ISL_788645, EPI_ISL_788646, EPI_ISL_788647, EPI_ISL_788648, EPI_ISL_788649, EPI_ISL_788650, EPI_ISL_788651, EPI_ISL_788652, EPI_ISL_788653, EPI_ISL_788654, EPI_ISL_788655, EPI_ISL_788656,
EPI_ISL_788657, EPI_ISL_788658, EPI_ISL_788659, EPI_ISL_788660, EPI_ISL_788661, EPI_ISL_788662, EPI_ISL_788663, EPI_ISL_788664, EPI_ISL_788665, EPI_ISL_788666, EPI_ISL_788667, EPI_ISL_788668, EPI_ISL_788669, EPI_ISL_788670, EPI_ISL_788671, EPI_ISL_788672, EPI_ISL_788673, EPI_ISL_788674, EPI_ISL_788675, EPI_ISL_788676, EPI_ISL_788677, EPI_ISL_788678, EPI_ISL_788679, EPI_ISL_788680,
EPI_ISL_788681, EPI_ISL_788682, EPI_ISL_788683, EPI_ISL_788684, EPI_ISL_788685, EPI_ISL_788686, EPI_ISL_788687, EPI_ISL_788688, EPI_ISL_788689, EPI_ISL_788690, EPI_ISL_788691, EPI_ISL_788692, EPI_ISL_788693, EPI_ISL_788694, EPI_ISL_788695, EPI_ISL_788696, EPI_ISL_788697, EPI_ISL_788698, EPI_ISL_788699, EPI_ISL_788700, EPI_ISL_788701, EPI_ISL_788702, EPI_ISL_788703, EPI_ISL_788704,
EPI_ISL_788705, EPI_ISL_788706, EPI_ISL_788707, EPI_ISL_788708, EPI_ISL_788709, EPI_ISL_788710, EPI_ISL_788711, EPI_ISL_788712, EPI_ISL_788713, EPI_ISL_788714, EPI_ISL_788715, EPI_ISL_788716, EPI_ISL_788717, EPI_ISL_788718, EPI_ISL_788719, EPI_ISL_788720, EPI_ISL_788721, EPI_ISL_788722, EPI_ISL_788723, EPI_ISL_788724, EPI_ISL_788725, EPI_ISL_788726, EPI_ISL_788727, EPI_ISL_788728,
EPI_ISL_788729, EPI_ISL_788730, EPI_ISL_788731, EPI_ISL_788732, EPI_ISL_788733, EPI_ISL_788734, EPI_ISL_788735, EPI_ISL_788736, EPI_ISL_788737, EPI_ISL_788738, EPI_ISL_788739, EPI_ISL_788740, EPI_ISL_788741, EPI_ISL_788742, EPI_ISL_788743, EPI_ISL_788744, EPI_ISL_788745, EPI_ISL_788746, EPI_ISL_788747, EPI_ISL_788748, EPI_ISL_788749, EPI_ISL_788750, EPI_ISL_788751, EPI_ISL_788752,
EPI_ISL_788753, EPI_ISL_788754, EPI_ISL_788755, EPI_ISL_788756, EPI_ISL_788757, EPI_ISL_788758, EPI_ISL_788759, EPI_ISL_788760, EPI_ISL_788761, EPI_ISL_788762, EPI_ISL_788763, EPI_ISL_788764, EPI_ISL_788765, EPI_ISL_788766, EPI_ISL_788767, EPI_ISL_788768, EPI_ISL_788769, EPI_ISL_788770, EPI_ISL_788771, EPI_ISL_788772, EPI_ISL_788773, EPI_ISL_788774, EPI_ISL_788775, EPI_ISL_788776,
EPI_ISL_788777, EPI_ISL_788778, EPI_ISL_788779, EPI_ISL_788780, EPI_ISL_788781, EPI_ISL_788782, EPI_ISL_788783, EPI_ISL_788784, EPI_ISL_788785, EPI_ISL_788786, EPI_ISL_788787, EPI_ISL_788788, EPI_ISL_788789, EPI_ISL_788790, EPI_ISL_788791, EPI_ISL_788792, EPI_ISL_788793, EPI_ISL_788794, EPI_ISL_788795, EPI_ISL_788796, EPI_ISL_788797, EPI_ISL_788798, EPI_ISL_788799, EPI_ISL_788800,
EPI_ISL_788801, EPI_ISL_788802, EPI_ISL_788803, EPI_ISL_788804, EPI_ISL_788805, EPI_ISL_788806, EPI_ISL_788807, EPI_ISL_788808, EPI_ISL_788809, EPI_ISL_788810, EPI_ISL_788811, EPI_ISL_788812, EPI_ISL_788813, EPI_ISL_788814, EPI_ISL_788815, EPI_ISL_788816, EPI_ISL_788817, EPI_ISL_788818, EPI_ISL_788819, EPI_ISL_788820, EPI_ISL_788821, EPI_ISL_788822, EPI_ISL_788823, EPI_ISL_788824,
EPI_ISL_788825, EPI_ISL_788826, EPI_ISL_788827, EPI_ISL_788828, EPI_ISL_788829, EPI_ISL_788830, EPI_ISL_788831, EPI_ISL_788832, EPI_ISL_788833, EPI_ISL_788834, EPI_ISL_788835, EPI_ISL_788836, EPI_ISL_788837, EPI_ISL_788838, EPI_ISL_788839, EPI_ISL_788840, EPI_ISL_788841, EPI_ISL_788842, EPI_ISL_788843, EPI_ISL_788844, EPI_ISL_788845, EPI_ISL_788846, EPI_ISL_788847, EPI_ISL_788848,
EPI_ISL_788849, EPI_ISL_788850, EPI_ISL_788851, EPI_ISL_788852, EPI_ISL_788853, EPI_ISL_788854, EPI_ISL_788855, EPI_ISL_788856, EPI_ISL_788857, EPI_ISL_788858, EPI_ISL_788859, EPI_ISL_788860, EPI_ISL_788861, EPI_ISL_788862, EPI_ISL_788863, EPI_ISL_788864, EPI_ISL_788865, EPI_ISL_788866, EPI_ISL_788867, EPI_ISL_788868, EPI_ISL_788869, EPI_ISL_788870, EPI_ISL_788871, EPI_ISL_788872,
EPI_ISL_788873, EPI_ISL_788874, EPI_ISL_788875, EPI_ISL_788876, EPI_ISL_788877, EPI_ISL_788878, EPI_ISL_788879, EPI_ISL_788880, EPI_ISL_788881, EPI_ISL_788882, EPI_ISL_788883, EPI_ISL_788884, EPI_ISL_788885, EPI_ISL_788886, EPI_ISL_788887, EPI_ISL_788888, EPI_ISL_788889, EPI_ISL_788890, EPI_ISL_788891, EPI_ISL_788892, EPI_ISL_788893, EPI_ISL_788894, EPI_ISL_788895, EPI_ISL_788896,
EPI_ISL_788897

| | | | |
|---|---|---|---|
| see above | Houston Methodist Hospital | Houston Methodist Hospital | S. Wesley Long, Randall J. Olsen, Paul A. Christensen, David W. Bernard, James J. Davis, Maulik Shukla, Marcus Nguyen, Matthew Ojeda Saavedra, Prasanti Yerramilli, Layne Pruitt, Sishir Subedi, Heather Hendrickson, and James M. Musser |

EPI_ISL_788898, EPI_ISL_788899, EPI_ISL_788900, EPI_ISL_788901, EPI_ISL_788902, EPI_ISL_788903, EPI_ISL_788904, EPI_ISL_788905, EPI_ISL_788906, EPI_ISL_788907, EPI_ISL_788908, EPI_ISL_788909, EPI_ISL_788910, EPI_ISL_788911, EPI_ISL_788912, EPI_ISL_788913, EPI_ISL_788914, EPI_ISL_788915, EPI_ISL_788916, EPI_ISL_788917, EPI_ISL_788918, EPI_ISL_788919, EPI_ISL_788939, EPI_ISL_788940,
EPI_ISL_788941, EPI_ISL_788942

| | | | |
|---|---|---|---|
| see above | University of Wisconsin-Madison AIDS Vaccine Research Laboratories | University of Wisconsin-Madison AIDS Vaccine Research Laboratories | Gage Moreno, Katarina Braun, et al. AIDS Vaccine Research Laboratories |
| EPI_ISL_789102, EPI_ISL_789103, EPI_ISL_789104 | Ohio Department of Health Laboratory | Ohio Department of Health Laboratory | Holmes, Jennifer; Eric Brandt, Keoni Omura, Glen McGillivary, Caitlin McDonnell, Kirtana Ramadugu, Erica Leasure, Kelsey Florek, Heather Blankenship, Quanta Brown, and Tammy Bannerman |

EPI_ISL_789105, EPI_ISL_789106, EPI_ISL_789107, EPI_ISL_789108, EPI_ISL_789109, EPI_ISL_789110, EPI_ISL_789111, EPI_ISL_789112, EPI_ISL_789113, EPI_ISL_789114, EPI_ISL_789115, EPI_ISL_789116, EPI_ISL_789117, EPI_ISL_789118, EPI_ISL_789119, EPI_ISL_789120, EPI_ISL_789121, EPI_ISL_789122, EPI_ISL_789123, EPI_ISL_789124, EPI_ISL_789125, EPI_ISL_789126, EPI_ISL_789127, EPI_ISL_789128,
EPI_ISL_789129, EPI_ISL_789130, EPI_ISL_789131, EPI_ISL_789132, EPI_ISL_789133, EPI_ISL_789134, EPI_ISL_789135, EPI_ISL_789136, EPI_ISL_789137, EPI_ISL_789138, EPI_ISL_789139, EPI_ISL_789140, EPI_ISL_789141, EPI_ISL_789142, EPI_ISL_789143, EPI_ISL_789144, EPI_ISL_789145, EPI_ISL_789146, EPI_ISL_789147, EPI_ISL_789148, EPI_ISL_789149, EPI_ISL_789150, EPI_ISL_789151, EPI_ISL_789152,
EPI_ISL_789153, EPI_ISL_789154, EPI_ISL_789155, EPI_ISL_789156, EPI_ISL_789157, EPI_ISL_789158, EPI_ISL_789159, EPI_ISL_789160, EPI_ISL_789161, EPI_ISL_789162, EPI_ISL_789163, EPI_ISL_789164, EPI_ISL_789165, EPI_ISL_789166, EPI_ISL_789167, EPI_ISL_789168, EPI_ISL_789169, EPI_ISL_789170, EPI_ISL_789171, EPI_ISL_789172, EPI_ISL_789173, EPI_ISL_789174, EPI_ISL_789175, EPI_ISL_789176,
EPI_ISL_789177, EPI_ISL_789178, EPI_ISL_789179, EPI_ISL_789180, EPI_ISL_789181, EPI_ISL_789182, EPI_ISL_789183, EPI_ISL_789184, EPI_ISL_789185, EPI_ISL_789186, EPI_ISL_789187, EPI_ISL_789188, EPI_ISL_789189, EPI_ISL_789190, EPI_ISL_789191, EPI_ISL_789192, EPI_ISL_789193, EPI_ISL_789194, EPI_ISL_789195, EPI_ISL_789196, EPI_ISL_789197, EPI_ISL_789198, EPI_ISL_789199, EPI_ISL_789200,
EPI_ISL_789201, EPI_ISL_789202, EPI_ISL_789203, EPI_ISL_789204, EPI_ISL_789205, EPI_ISL_789206, EPI_ISL_789207, EPI_ISL_789208, EPI_ISL_789209, EPI_ISL_789210, EPI_ISL_789211, EPI_ISL_789212, EPI_ISL_789213, EPI_ISL_789214, EPI_ISL_789215, EPI_ISL_789216, EPI_ISL_789217, EPI_ISL_789218, EPI_ISL_789219, EPI_ISL_789220, EPI_ISL_789221, EPI_ISL_789222, EPI_ISL_789223, EPI_ISL_789224,
EPI_ISL_789225, EPI_ISL_789226, EPI_ISL_789227, EPI_ISL_789228, EPI_ISL_789229, EPI_ISL_789230, EPI_ISL_789231, EPI_ISL_789232, EPI_ISL_789233, EPI_ISL_789234, EPI_ISL_789235, EPI_ISL_789236, EPI_ISL_789237, EPI_ISL_789238, EPI_ISL_789239, EPI_ISL_789240, EPI_ISL_789241, EPI_ISL_789242, EPI_ISL_789243, EPI_ISL_789244, EPI_ISL_789245, EPI_ISL_789246, EPI_ISL_789247, EPI_ISL_789248,
EPI_ISL_789249, EPI_ISL_789250, EPI_ISL_789251, EPI_ISL_789252, EPI_ISL_789253, EPI_ISL_789254, EPI_ISL_789255, EPI_ISL_789256, EPI_ISL_789257, EPI_ISL_789258, EPI_ISL_789259, EPI_ISL_789260, EPI_ISL_789261, EPI_ISL_789262, EPI_ISL_789263, EPI_ISL_789264, EPI_ISL_789265, EPI_ISL_789266, EPI_ISL_789267, EPI_ISL_789268, EPI_ISL_789269, EPI_ISL_789270, EPI_ISL_789271, EPI_ISL_789272,
EPI_ISL_789273, EPI_ISL_789274, EPI_ISL_789275, EPI_ISL_789276, EPI_ISL_789277, EPI_ISL_789278, EPI_ISL_789279, EPI_ISL_789280, EPI_ISL_789281, EPI_ISL_789282, EPI_ISL_789283, EPI_ISL_789284, EPI_ISL_789285, EPI_ISL_789286, EPI_ISL_789287, EPI_ISL_789288, EPI_ISL_789289, EPI_ISL_789290, EPI_ISL_789291, EPI_ISL_789292, EPI_ISL_789293, EPI_ISL_789294, EPI_ISL_789295, EPI_ISL_789296,
EPI_ISL_789297, EPI_ISL_789298, EPI_ISL_789299, EPI_ISL_789300, EPI_ISL_789301, EPI_ISL_789302, EPI_ISL_789303, EPI_ISL_789304, EPI_ISL_789305, EPI_ISL_789306, EPI_ISL_789307, EPI_ISL_789308, EPI_ISL_789309, EPI_ISL_789310, EPI_ISL_789311, EPI_ISL_789312, EPI_ISL_789313, EPI_ISL_789314, EPI_ISL_789315, EPI_ISL_789316, EPI_ISL_789317, EPI_ISL_789318, EPI_ISL_789319, EPI_ISL_789320,
EPI_ISL_789321, EPI_ISL_789322, EPI_ISL_789323, EPI_ISL_789324, EPI_ISL_789325, EPI_ISL_789326, EPI_ISL_789327, EPI_ISL_789328, EPI_ISL_789329, EPI_ISL_789330, EPI_ISL_789331, EPI_ISL_789332, EPI_ISL_789333, EPI_ISL_789334, EPI_ISL_789335, EPI_ISL_789336, EPI_ISL_789337, EPI_ISL_789338, EPI_ISL_789339, EPI_ISL_789340, EPI_ISL_789341, EPI_ISL_789342, EPI_ISL_789343, EPI_ISL_789344,
EPI_ISL_789345, EPI_ISL_789346, EPI_ISL_789347, EPI_ISL_789348, EPI_ISL_789349, EPI_ISL_789350, EPI_ISL_789351, EPI_ISL_789352, EPI_ISL_789353, EPI_ISL_789354, EPI_ISL_789355, EPI_ISL_789356, EPI_ISL_789357, EPI_ISL_789358, EPI_ISL_789359, EPI_ISL_789360, EPI_ISL_789361, EPI_ISL_789362, EPI_ISL_789363, EPI_ISL_789364, EPI_ISL_789365, EPI_ISL_789366, EPI_ISL_789367, EPI_ISL_789368,
EPI_ISL_789369, EPI_ISL_789370, EPI_ISL_789371, EPI_ISL_789372, EPI_ISL_789373, EPI_ISL_789374, EPI_ISL_789375, EPI_ISL_789376, EPI_ISL_789377, EPI_ISL_789378, EPI_ISL_789379, EPI_ISL_789380, EPI_ISL_789381, EPI_ISL_789382, EPI_ISL_789383, EPI_ISL_789384, EPI_ISL_789385, EPI_ISL_789386, EPI_ISL_789387, EPI_ISL_789388, EPI_ISL_789389, EPI_ISL_789390, EPI_ISL_789391, EPI_ISL_789392,
EPI_ISL_789393, EPI_ISL_789394, EPI_ISL_789395, EPI_ISL_789396, EPI_ISL_789397, EPI_ISL_789398, EPI_ISL_789399, EPI_ISL_789400, EPI_ISL_789401, EPI_ISL_789402, EPI_ISL_789403, EPI_ISL_789404, EPI_ISL_789405, EPI_ISL_789406, EPI_ISL_789407, EPI_ISL_789408, EPI_ISL_789409, EPI_ISL_789410, EPI_ISL_789411, EPI_ISL_789412, EPI_ISL_789413, EPI_ISL_789414, EPI_ISL_789415, EPI_ISL_789416,
EPI_ISL_789417, EPI_ISL_789418, EPI_ISL_789419, EPI_ISL_789420, EPI_ISL_789421, EPI_ISL_789422, EPI_ISL_789423, EPI_ISL_789424, EPI_ISL_789425, EPI_ISL_789426, EPI_ISL_789427, EPI_ISL_789428, EPI_ISL_789429, EPI_ISL_789430, EPI_ISL_789431, EPI_ISL_789432, EPI_ISL_789433, EPI_ISL_789434, EPI_ISL_789435, EPI_ISL_789436, EPI_ISL_789437, EPI_ISL_789438, EPI_ISL_789439, EPI_ISL_789440,
EPI_ISL_789441, EPI_ISL_789442, EPI_ISL_789443, EPI_ISL_789444, EPI_ISL_789445, EPI_ISL_789446, EPI_ISL_789447, EPI_ISL_789448, EPI_ISL_789449, EPI_ISL_789450, EPI_ISL_789451, EPI_ISL_789452, EPI_ISL_789453, EPI_ISL_789454, EPI_ISL_789455, EPI_ISL_789456, EPI_ISL_789457, EPI_ISL_789458, EPI_ISL_789459, EPI_ISL_789460, EPI_ISL_789461, EPI_ISL_789462, EPI_ISL_789463, EPI_ISL_789464,
EPI_ISL_789465, EPI_ISL_789466, EPI_ISL_789467, EPI_ISL_789468, EPI_ISL_789469, EPI_ISL_789470, EPI_ISL_789471, EPI_ISL_789472, EPI_ISL_789473, EPI_ISL_789474, EPI_ISL_789475, EPI_ISL_789476, EPI_ISL_789477, EPI_ISL_789478, EPI_ISL_789479, EPI_ISL_789480, EPI_ISL_789481, EPI_ISL_789482, EPI_ISL_789483, EPI_ISL_789484, EPI_ISL_789485, EPI_ISL_789486, EPI_ISL_789487, EPI_ISL_789488,
EPI_ISL_789489, EPI_ISL_789490, EPI_ISL_789491, EPI_ISL_789492, EPI_ISL_789493, EPI_ISL_789494, EPI_ISL_789495, EPI_ISL_789496, EPI_ISL_789497, EPI_ISL_789498, EPI_ISL_789499, EPI_ISL_789500, EPI_ISL_789501, EPI_ISL_789502, EPI_ISL_789503, EPI_ISL_789504, EPI_ISL_789505, EPI_ISL_789506, EPI_ISL_789507, EPI_ISL_789508, EPI_ISL_789509, EPI_ISL_789510, EPI_ISL_789511, EPI_ISL_789512,
EPI_ISL_789513, EPI_ISL_789514, EPI_ISL_789515, EPI_ISL_789516, EPI_ISL_789517, EPI_ISL_789518, EPI_ISL_789519, EPI_ISL_789520, EPI_ISL_789521, EPI_ISL_789522, EPI_ISL_789523, EPI_ISL_789524, EPI_ISL_789525, EPI_ISL_789526, EPI_ISL_789527, EPI_ISL_789528, EPI_ISL_789529, EPI_ISL_789530, EPI_ISL_789531, EPI_ISL_789532, EPI_ISL_789533, EPI_ISL_789534, EPI_ISL_789535, EPI_ISL_789536,
EPI_ISL_789537, EPI_ISL_789538, EPI_ISL_789539, EPI_ISL_789540, EPI_ISL_789541, EPI_ISL_789542, EPI_ISL_789543, EPI_ISL_789544, EPI_ISL_789545, EPI_ISL_789546, EPI_ISL_789547, EPI_ISL_789548, EPI_ISL_789549, EPI_ISL_789550, EPI_ISL_789551, EPI_ISL_789552, EPI_ISL_789553, EPI_ISL_789554, EPI_ISL_789555, EPI_ISL_789556, EPI_ISL_789557, EPI_ISL_789558, EPI_ISL_789559, EPI_ISL_789560,
EPI_ISL_789561, EPI_ISL_789562, EPI_ISL_789563, EPI_ISL_789564, EPI_ISL_789565, EPI_ISL_789566, EPI_ISL_789567, EPI_ISL_789568, EPI_ISL_789569, EPI_ISL_789570, EPI_ISL_789571, EPI_ISL_789572, EPI_ISL_789573, EPI_ISL_789574, EPI_ISL_789575, EPI_ISL_789576, EPI_ISL_789577, EPI_ISL_789578, EPI_ISL_789579, EPI_ISL_789580, EPI_ISL_789581, EPI_ISL_789582, EPI_ISL_789583, EPI_ISL_789584,
EPI_ISL_789585, EPI_ISL_789586, EPI_ISL_789587, EPI_ISL_789588, EPI_ISL_789589, EPI_ISL_789590, EPI_ISL_789591, EPI_ISL_789592, EPI_ISL_789593, EPI_ISL_789594, EPI_ISL_789595, EPI_ISL_789596, EPI_ISL_789597, EPI_ISL_789598, EPI_ISL_789599, EPI_ISL_789600, EPI_ISL_789601, EPI_ISL_789602, EPI_ISL_789603, EPI_ISL_789604, EPI_ISL_789605, EPI_ISL_789606, EPI_ISL_789607, EPI_ISL_789608,
EPI_ISL_789609, EPI_ISL_789610, EPI_ISL_789611, EPI_ISL_789612, EPI_ISL_789613, EPI_ISL_789614, EPI_ISL_789615, EPI_ISL_789616, EPI_ISL_789617, EPI_ISL_789618, EPI_ISL_789619, EPI_ISL_789620, EPI_ISL_789621, EPI_ISL_789622, EPI_ISL_789623, EPI_ISL_789624, EPI_ISL_789625, EPI_ISL_789626, EPI_ISL_789627, EPI_ISL_789628, EPI_ISL_789629, EPI_ISL_789630, EPI_ISL_789631, EPI_ISL_789632,
EPI_ISL_789633, EPI_ISL_789634, EPI_ISL_789635, EPI_ISL_789636, EPI_ISL_789637, EPI_ISL_789638, EPI_ISL_789639, EPI_ISL_789640, EPI_ISL_789641, EPI_ISL_789642, EPI_ISL_789643, EPI_ISL_789644, EPI_ISL_789645, EPI_ISL_789646, EPI_ISL_789647, EPI_ISL_789648, EPI_ISL_789649, EPI_ISL_789650, EPI_ISL_789651, EPI_ISL_789652, EPI_ISL_789653, EPI_ISL_789654, EPI_ISL_789655, EPI_ISL_789656,
EPI_ISL_789657, EPI_ISL_789658, EPI_ISL_789659, EPI_ISL_789660, EPI_ISL_789661, EPI_ISL_789662, EPI_ISL_789663, EPI_ISL_789664, EPI_ISL_789665, EPI_ISL_789666, EPI_ISL_789667, EPI_ISL_789668, EPI_ISL_789669, EPI_ISL_789670, EPI_ISL_789671, EPI_ISL_789672, EPI_ISL_789673, EPI_ISL_789674, EPI_ISL_789675, EPI_ISL_789676, EPI_ISL_789677, EPI_ISL_789678, EPI_ISL_789679, EPI_ISL_789680,
EPI_ISL_789681, EPI_ISL_789682, EPI_ISL_789683, EPI_ISL_789684, EPI_ISL_789685, EPI_ISL_789686, EPI_ISL_789687, EPI_ISL_789688, EPI_ISL_789689, EPI_ISL_789690, EPI_ISL_789691, EPI_ISL_789692, EPI_ISL_789693, EPI_ISL_789694, EPI_ISL_789695, EPI_ISL_789696, EPI_ISL_789697, EPI_ISL_789698, EPI_ISL_789699, EPI_ISL_789700, EPI_ISL_789701, EPI_ISL_789702, EPI_ISL_789703, EPI_ISL_789704,
EPI_ISL_789705, EPI_ISL_789706, EPI_ISL_789707, EPI_ISL_789708, EPI_ISL_789709, EPI_ISL_789710, EPI_ISL_789711, EPI_ISL_789712, EPI_ISL_789713, EPI_ISL_789714, EPI_ISL_789715, EPI_ISL_789716, EPI_ISL_789717, EPI_ISL_789718, EPI_ISL_789719, EPI_ISL_789720, EPI_ISL_789721, EPI_ISL_789722, EPI_ISL_789723, EPI_ISL_789724, EPI_ISL_789725, EPI_ISL_789726, EPI_ISL_789727, EPI_ISL_789728,
EPI_ISL_789729, EPI_ISL_789730, EPI_ISL_789731, EPI_ISL_789732, EPI_ISL_789733, EPI_ISL_789734, EPI_ISL_789735, EPI_ISL_789736, EPI_ISL_789737, EPI_ISL_789738, EPI_ISL_789739, EPI_ISL_789740, EPI_ISL_789741, EPI_ISL_789742, EPI_ISL_789743, EPI_ISL_789744, EPI_ISL_789745, EPI_ISL_789746, EPI_ISL_789747, EPI_ISL_789748, EPI_ISL_789749, EPI_ISL_789750, EPI_ISL_789751, EPI_ISL_789752,
EPI_ISL_789753, EPI_ISL_789754, EPI_ISL_789755, EPI_ISL_789756, EPI_ISL_789757, EPI_ISL_789758, EPI_ISL_789759, EPI_ISL_789760, EPI_ISL_789761, EPI_ISL_789762, EPI_ISL_789763, EPI_ISL_789764, EPI_ISL_789765, EPI_ISL_789766, EPI_ISL_789767, EPI_ISL_789768, EPI_ISL_789769, EPI_ISL_789770, EPI_ISL_789771, EPI_ISL_789772, EPI_ISL_789773, EPI_ISL_789774, EPI_ISL_789775, EPI_ISL_789776,
EPI_ISL_789777, EPI_ISL_789778, EPI_ISL_789779, EPI_ISL_789780, EPI_ISL_789781, EPI_ISL_789782, EPI_ISL_789783, EPI_ISL_789784, EPI_ISL_789785, EPI_ISL_789786, EPI_ISL_789787, EPI_ISL_789788, EPI_ISL_789789, EPI_ISL_789790, EPI_ISL_789791, EPI_ISL_789792, EPI_ISL_789793, EPI_ISL_789794, EPI_ISL_789795, EPI_ISL_789796, EPI_ISL_789797, EPI_ISL_789798, EPI_ISL_789799, EPI_ISL_789800,
EPI_ISL_789801, EPI_ISL_789802, EPI_ISL_789803, EPI_ISL_789804, EPI_ISL_789805, EPI_ISL_789806, EPI_ISL_789807, EPI_ISL_789808, EPI_ISL_789809, EPI_ISL_789810, EPI_ISL_789811, EPI_ISL_789812, EPI_ISL_789813, EPI_ISL_789814, EPI_ISL_789815, EPI_ISL_789816, EPI_ISL_789817, EPI_ISL_789818, EPI_ISL_789819, EPI_ISL_789820, EPI_ISL_789821, EPI_ISL_789822, EPI_ISL_789823, EPI_ISL_789824,
EPI_ISL_789825, EPI_ISL_789826, EPI_ISL_789827, EPI_ISL_789828, EPI_ISL_789829, EPI_ISL_789830, EPI_ISL_789831, EPI_ISL_789832, EPI_ISL_789833, EPI_ISL_789834, EPI_ISL_789835, EPI_ISL_789836, EPI_ISL_789837, EPI_ISL_789838, EPI_ISL_789839, EPI_ISL_789840, EPI_ISL_789841, EPI_ISL_789842, EPI_ISL_789843, EPI_ISL_789844, EPI_ISL_789845, EPI_ISL_789846, EPI_ISL_789847, EPI_ISL_789848,
EPI_ISL_789849, EPI_ISL_789850, EPI_ISL_789851, EPI_ISL_789852, EPI_ISL_789853, EPI_ISL_789854, EPI_ISL_789855, EPI_ISL_789856, EPI_ISL_789857, EPI_ISL_789858, EPI_ISL_789859, EPI_ISL_789860, EPI_ISL_789861, EPI_ISL_789862, EPI_ISL_789863, EPI_ISL_789864, EPI_ISL_789865, EPI_ISL_789866, EPI_ISL_789867, EPI_ISL_789868, EPI_ISL_789869, EPI_ISL_789870, EPI_ISL_789871, EPI_ISL_789872,
EPI_ISL_789873, EPI_ISL_789874, EPI_ISL_789875, EPI_ISL_789876, EPI_ISL_789877, EPI_ISL_789878, EPI_ISL_789879, EPI_ISL_789880, EPI_ISL_789881, EPI_ISL_789882, EPI_ISL_789883, EPI_ISL_789884, EPI_ISL_789885, EPI_ISL_789886, EPI_ISL_789887, EPI_ISL_789888, EPI_ISL_789889, EPI_ISL_789890, EPI_ISL_789891, EPI_ISL_789892, EPI_ISL_789893, EPI_ISL_789894, EPI_ISL_789895, EPI_ISL_789896,

| | | | |
|---|---|---|---|
| EPI_ISL_789897, EPI_ISL_789898, EPI_ISL_789899, EPI_ISL_789900, EPI_ISL_789901, EPI_ISL_789902, EPI_ISL_789903, EPI_ISL_789904, EPI_ISL_789905, EPI_ISL_789906, EPI_ISL_789907, EPI_ISL_789908, EPI_ISL_789909, EPI_ISL_789910, EPI_ISL_789911, EPI_ISL_789912, EPI_ISL_789913, EPI_ISL_789914, EPI_ISL_789915, EPI_ISL_789916, EPI_ISL_789917, EPI_ISL_789918, EPI_ISL_789919, EPI_ISL_789920, EPI_ISL_789921, EPI_ISL_789922, EPI_ISL_789923, EPI_ISL_789924, EPI_ISL_789925, EPI_ISL_789926, EPI_ISL_789927, EPI_ISL_789928, EPI_ISL_789929, EPI_ISL_789930, EPI_ISL_789931, EPI_ISL_789932, EPI_ISL_789933, EPI_ISL_789934, EPI_ISL_789935, EPI_ISL_789936, EPI_ISL_789937, EPI_ISL_789938, EPI_ISL_789939, EPI_ISL_789940, EPI_ISL_789941, EPI_ISL_789942, EPI_ISL_789943, EPI_ISL_789944, EPI_ISL_789945, EPI_ISL_789946, EPI_ISL_789947, EPI_ISL_789948, EPI_ISL_789949, EPI_ISL_789950, EPI_ISL_789951, EPI_ISL_789952, EPI_ISL_789953, EPI_ISL_789954, EPI_ISL_789955, EPI_ISL_789956, EPI_ISL_789957, EPI_ISL_789958, EPI_ISL_789959, EPI_ISL_789960, EPI_ISL_789961, EPI_ISL_789962, EPI_ISL_789963, EPI_ISL_789964, EPI_ISL_789965, EPI_ISL_789966, EPI_ISL_789967, EPI_ISL_789968, EPI_ISL_789969, EPI_ISL_789970, EPI_ISL_789971, EPI_ISL_789972, EPI_ISL_789973, EPI_ISL_789974, EPI_ISL_789975, EPI_ISL_789976, EPI_ISL_789977, EPI_ISL_789978, EPI_ISL_789979, EPI_ISL_789980, EPI_ISL_789981, EPI_ISL_789982, EPI_ISL_789983, EPI_ISL_789984, EPI_ISL_789985, EPI_ISL_789986, EPI_ISL_789987, EPI_ISL_789988, EPI_ISL_789989, EPI_ISL_789990, EPI_ISL_789991, EPI_ISL_789992, EPI_ISL_789993, EPI_ISL_789994, EPI_ISL_789995, EPI_ISL_789996, EPI_ISL_789997, EPI_ISL_789998, EPI_ISL_789999, EPI_ISL_790000, EPI_ISL_790001, EPI_ISL_790002, EPI_ISL_790003, EPI_ISL_790004, EPI_ISL_790005, EPI_ISL_790006, EPI_ISL_790007, EPI_ISL_790008, EPI_ISL_790009, EPI_ISL_790010, EPI_ISL_790011, EPI_ISL_790012, EPI_ISL_790013, EPI_ISL_790014, EPI_ISL_790015, EPI_ISL_790016, EPI_ISL_790017, EPI_ISL_790018, EPI_ISL_790019, EPI_ISL_790020, EPI_ISL_790021, EPI_ISL_790022, EPI_ISL_790023, EPI_ISL_790024, EPI_ISL_790025, EPI_ISL_790026, EPI_ISL_790027, EPI_ISL_790028, EPI_ISL_790029, EPI_ISL_790030, EPI_ISL_790031, EPI_ISL_790032, EPI_ISL_790033, EPI_ISL_790034, EPI_ISL_790035, EPI_ISL_790036, EPI_ISL_790037, EPI_ISL_790038, EPI_ISL_790039, EPI_ISL_790040, EPI_ISL_790041, EPI_ISL_790042, EPI_ISL_790043, EPI_ISL_790044, EPI_ISL_790045, EPI_ISL_790046, EPI_ISL_790047, EPI_ISL_790048, EPI_ISL_790049, EPI_ISL_790050, EPI_ISL_790051, EPI_ISL_790052, EPI_ISL_790053, EPI_ISL_790054, EPI_ISL_790055, EPI_ISL_790056, EPI_ISL_790057, EPI_ISL_790058, EPI_ISL_790059, EPI_ISL_790060, EPI_ISL_790061, EPI_ISL_790062, EPI_ISL_790063, EPI_ISL_790064, EPI_ISL_790065, EPI_ISL_790066, EPI_ISL_790067, EPI_ISL_790068, EPI_ISL_790069, EPI_ISL_790070, EPI_ISL_790071, EPI_ISL_790072, EPI_ISL_790073, EPI_ISL_790074, EPI_ISL_790075, EPI_ISL_790076, EPI_ISL_790077, EPI_ISL_790078, EPI_ISL_790079, EPI_ISL_790080, EPI_ISL_790081, EPI_ISL_790082, EPI_ISL_790083, EPI_ISL_790084, EPI_ISL_790085, EPI_ISL_790086, EPI_ISL_790087, EPI_ISL_790088, EPI_ISL_790089, EPI_ISL_790090, EPI_ISL_790091, EPI_ISL_790092, EPI_ISL_790093, EPI_ISL_790094, EPI_ISL_790095, EPI_ISL_790096, EPI_ISL_790097, EPI_ISL_790098, EPI_ISL_790099, EPI_ISL_790100, EPI_ISL_790101, EPI_ISL_790102, EPI_ISL_790103, EPI_ISL_790104, EPI_ISL_790105, EPI_ISL_790106, EPI_ISL_790107, EPI_ISL_790108, EPI_ISL_790109, EPI_ISL_790110, EPI_ISL_790111, EPI_ISL_790112, EPI_ISL_790113, EPI_ISL_790114, EPI_ISL_790115, EPI_ISL_790116, EPI_ISL_790117, EPI_ISL_790118, EPI_ISL_790119, EPI_ISL_790120, EPI_ISL_790121, EPI_ISL_790122, EPI_ISL_790123, EPI_ISL_790124, EPI_ISL_790125, EPI_ISL_790126, EPI_ISL_790127, EPI_ISL_790128, EPI_ISL_790129, EPI_ISL_790130, EPI_ISL_790131, EPI_ISL_790132, EPI_ISL_790133, EPI_ISL_790134, EPI_ISL_790135, EPI_ISL_790136, EPI_ISL_790137, EPI_ISL_790138, EPI_ISL_790139, EPI_ISL_790140, EPI_ISL_790141, EPI_ISL_790142, EPI_ISL_790143, EPI_ISL_790144, EPI_ISL_790145, EPI_ISL_790146, EPI_ISL_790147, EPI_ISL_790148, EPI_ISL_790149, EPI_ISL_790150, EPI_ISL_790151, EPI_ISL_790152, EPI_ISL_790153, EPI_ISL_790154, EPI_ISL_790155, EPI_ISL_790156, EPI_ISL_790157, EPI_ISL_790158, EPI_ISL_790159, EPI_ISL_790160, EPI_ISL_790161, EPI_ISL_790162, EPI_ISL_790163, EPI_ISL_790164, EPI_ISL_790165, EPI_ISL_790166, EPI_ISL_790167, EPI_ISL_790168, EPI_ISL_790169, EPI_ISL_790170, EPI_ISL_790171, EPI_ISL_790172, EPI_ISL_790173, EPI_ISL_790174, EPI_ISL_790175, EPI_ISL_790176, EPI_ISL_790177, EPI_ISL_790178, EPI_ISL_790179, EPI_ISL_790180, EPI_ISL_790181, EPI_ISL_790182, EPI_ISL_790183, EPI_ISL_790184, EPI_ISL_790185, EPI_ISL_790186, EPI_ISL_790187, EPI_ISL_790188, EPI_ISL_790189, EPI_ISL_790190, EPI_ISL_790191, EPI_ISL_790192, EPI_ISL_790193, EPI_ISL_790194, EPI_ISL_790195, EPI_ISL_790196, EPI_ISL_790197, EPI_ISL_790198, EPI_ISL_790199, EPI_ISL_790200, EPI_ISL_790201, EPI_ISL_790202, EPI_ISL_790203, EPI_ISL_790204, EPI_ISL_790205, EPI_ISL_790206, EPI_ISL_790207, EPI_ISL_790208, EPI_ISL_790209, EPI_ISL_790210, EPI_ISL_790211, EPI_ISL_790212, EPI_ISL_790213, EPI_ISL_790214, EPI_ISL_790215, EPI_ISL_790216, EPI_ISL_790217, EPI_ISL_790218, EPI_ISL_790219, EPI_ISL_790220, EPI_ISL_790221, EPI_ISL_790222, EPI_ISL_790223, EPI_ISL_790224, EPI_ISL_790225, EPI_ISL_790226, EPI_ISL_790227, EPI_ISL_790228, EPI_ISL_790229, EPI_ISL_790230, EPI_ISL_790231, EPI_ISL_790232, EPI_ISL_790233, EPI_ISL_790234, EPI_ISL_790235, EPI_ISL_790236, EPI_ISL_790237, EPI_ISL_790238, EPI_ISL_790239, EPI_ISL_790240, EPI_ISL_790241, EPI_ISL_790242, EPI_ISL_790243, EPI_ISL_790244, EPI_ISL_790245, EPI_ISL_790246, EPI_ISL_790247, EPI_ISL_790248, EPI_ISL_790249, EPI_ISL_790250, EPI_ISL_790251, EPI_ISL_790252, EPI_ISL_790253, EPI_ISL_790254, EPI_ISL_790255, EPI_ISL_790256, EPI_ISL_790257, EPI_ISL_790258, EPI_ISL_790259, EPI_ISL_790260, EPI_ISL_790261, EPI_ISL_790262, EPI_ISL_790263, EPI_ISL_790264, EPI_ISL_790265, EPI_ISL_790266, EPI_ISL_790267, EPI_ISL_790268, EPI_ISL_790269, EPI_ISL_790270, EPI_ISL_790271, EPI_ISL_790272, EPI_ISL_790273, EPI_ISL_790274, EPI_ISL_790275, EPI_ISL_790276, EPI_ISL_790277, EPI_ISL_790278, EPI_ISL_790279, EPI_ISL_790280, EPI_ISL_790281, EPI_ISL_790282, EPI_ISL_790283, EPI_ISL_790284, EPI_ISL_790285, EPI_ISL_790286, EPI_ISL_790287, EPI_ISL_790288, EPI_ISL_790289, EPI_ISL_790290, EPI_ISL_790291, EPI_ISL_790292, EPI_ISL_790293, EPI_ISL_790294, EPI_ISL_790295, EPI_ISL_790296, EPI_ISL_790297, EPI_ISL_790298, EPI_ISL_790299, EPI_ISL_790300, EPI_ISL_790301, EPI_ISL_790302, EPI_ISL_790303, EPI_ISL_790304, EPI_ISL_790305, EPI_ISL_790306, EPI_ISL_790307, EPI_ISL_790308, EPI_ISL_790309, EPI_ISL_790310, EPI_ISL_790311, EPI_ISL_790312, EPI_ISL_790313, EPI_ISL_790314, EPI_ISL_790315, EPI_ISL_790316, EPI_ISL_790317, EPI_ISL_790318, EPI_ISL_790319, EPI_ISL_790320, EPI_ISL_790321, EPI_ISL_790322, EPI_ISL_790323, EPI_ISL_790324, EPI_ISL_790325, EPI_ISL_790326, EPI_ISL_790327, EPI_ISL_790328, EPI_ISL_790329, EPI_ISL_790330, EPI_ISL_790331, EPI_ISL_790332, EPI_ISL_790333, EPI_ISL_790334, EPI_ISL_790335, EPI_ISL_790336, EPI_ISL_790337, EPI_ISL_790338, EPI_ISL_790339, EPI_ISL_790340, EPI_ISL_790341, EPI_ISL_790342, EPI_ISL_790343, EPI_ISL_790344, EPI_ISL_790345, EPI_ISL_790346, EPI_ISL_790347, EPI_ISL_790348, EPI_ISL_790349, EPI_ISL_790350, EPI_ISL_790351, EPI_ISL_790352, EPI_ISL_790353, EPI_ISL_790354, EPI_ISL_790355, EPI_ISL_790356, EPI_ISL_790357, EPI_ISL_790358, EPI_ISL_790359, EPI_ISL_790360, EPI_ISL_790361, EPI_ISL_790362, EPI_ISL_790363, EPI_ISL_790364, EPI_ISL_790365, EPI_ISL_790366, EPI_ISL_790367, EPI_ISL_790368, EPI_ISL_790369, EPI_ISL_790370, EPI_ISL_790371, EPI_ISL_790372, EPI_ISL_790373, EPI_ISL_790374, EPI_ISL_790375, EPI_ISL_790376, EPI_ISL_790377, EPI_ISL_790378, EPI_ISL_790379, EPI_ISL_790380, EPI_ISL_790381, EPI_ISL_790382, EPI_ISL_790383, EPI_ISL_790384, EPI_ISL_790385, EPI_ISL_790386, EPI_ISL_790387, EPI_ISL_790388, EPI_ISL_790389, EPI_ISL_790390, EPI_ISL_790391, EPI_ISL_790392, EPI_ISL_790393, EPI_ISL_790394, EPI_ISL_790395, EPI_ISL_790396, EPI_ISL_790397, EPI_ISL_790398, EPI_ISL_790399, EPI_ISL_790400, EPI_ISL_790401, EPI_ISL_790402, EPI_ISL_790403, EPI_ISL_790404, EPI_ISL_790405, EPI_ISL_790406, EPI_ISL_790407, EPI_ISL_790408, EPI_ISL_790409, EPI_ISL_790410, EPI_ISL_790411, EPI_ISL_790412, EPI_ISL_790413, EPI_ISL_790414, EPI_ISL_790415, EPI_ISL_790416, EPI_ISL_790417, EPI_ISL_790418, EPI_ISL_790419, EPI_ISL_790420, EPI_ISL_790421, EPI_ISL_790422, EPI_ISL_790423, EPI_ISL_790424, EPI_ISL_790425, EPI_ISL_790426, EPI_ISL_790427, EPI_ISL_790428, EPI_ISL_790429, EPI_ISL_790430, EPI_ISL_790431, EPI_ISL_790432, EPI_ISL_790433, EPI_ISL_790434, EPI_ISL_790435, EPI_ISL_790436, EPI_ISL_790437, EPI_ISL_790438, EPI_ISL_790439, EPI_ISL_790440, EPI_ISL_790441, EPI_ISL_790442, EPI_ISL_790443, EPI_ISL_790444, EPI_ISL_790445, EPI_ISL_790446, EPI_ISL_790447, EPI_ISL_790448, EPI_ISL_790449, EPI_ISL_790450, EPI_ISL_790451, EPI_ISL_790452, EPI_ISL_790453, EPI_ISL_790454, EPI_ISL_790455, EPI_ISL_790456, EPI_ISL_790457, EPI_ISL_790458, EPI_ISL_790459, EPI_ISL_790460, EPI_ISL_790461, EPI_ISL_790462, EPI_ISL_790463, EPI_ISL_790464, EPI_ISL_790465, EPI_ISL_790466, EPI_ISL_790467, EPI_ISL_790468, EPI_ISL_790469, EPI_ISL_790470, EPI_ISL_790471, EPI_ISL_790472, EPI_ISL_790473, EPI_ISL_790474, EPI_ISL_790475, EPI_ISL_790476, EPI_ISL_790477, EPI_ISL_790478, EPI_ISL_790479, EPI_ISL_790480, EPI_ISL_790481, EPI_ISL_790482, EPI_ISL_790483, EPI_ISL_790484, EPI_ISL_790485, EPI_ISL_790486, EPI_ISL_790487, EPI_ISL_790488, EPI_ISL_790489, EPI_ISL_790490, EPI_ISL_790491, EPI_ISL_790492, EPI_ISL_790493, EPI_ISL_790494, EPI_ISL_790495, EPI_ISL_790496, EPI_ISL_790497, EPI_ISL_790498, EPI_ISL_790499, EPI_ISL_790500, EPI_ISL_790501, EPI_ISL_790502, EPI_ISL_790503, EPI_ISL_790504, EPI_ISL_790505, EPI_ISL_790506, EPI_ISL_790507, EPI_ISL_790508, EPI_ISL_790509, EPI_ISL_790510, EPI_ISL_790511, EPI_ISL_790512, EPI_ISL_790513, EPI_ISL_790514, EPI_ISL_790515, EPI_ISL_790516, EPI_ISL_790517, EPI_ISL_790518, EPI_ISL_790519, EPI_ISL_790520, EPI_ISL_790521, EPI_ISL_790522, EPI_ISL_790523, EPI_ISL_790524, EPI_ISL_790525, EPI_ISL_790526, EPI_ISL_790527, EPI_ISL_790528, EPI_ISL_790529, EPI_ISL_790530, EPI_ISL_790531, EPI_ISL_790532, EPI_ISL_790533, EPI_ISL_790534, EPI_ISL_790535, EPI_ISL_790536, EPI_ISL_790537, EPI_ISL_790538, EPI_ISL_790539, EPI_ISL_790540, EPI_ISL_790541, EPI_ISL_790542, EPI_ISL_790543, EPI_ISL_790544, EPI_ISL_790545, EPI_ISL_790546, EPI_ISL_790547, EPI_ISL_790548, EPI_ISL_790549, EPI_ISL_790550, EPI_ISL_790551, EPI_ISL_790552, EPI_ISL_790553, EPI_ISL_790554, EPI_ISL_790555, EPI_ISL_790556, EPI_ISL_790557, EPI_ISL_790558, EPI_ISL_790559, EPI_ISL_790560, EPI_ISL_790561, EPI_ISL_790562, EPI_ISL_790563, EPI_ISL_790564, EPI_ISL_790565, EPI_ISL_790566 | | | |
| see above | Houston Methodist Hospital | Houston Methodist Hospital | S. Wesley Long, Randall J. Olsen, Paul A. Christensen, David W. Bernard, James J. Davis, Maulik Shukla, Marcus Nguyen, Matthew Ojeda Saavedra, Prasanti Yerramilli, Layne Pruitt, Sishir Subedi, Heather Hendrickson, and James M. Musser |
| EPI_ISL_790567, EPI_ISL_790568, EPI_ISL_790569 | Florida Bureau of Public Health Laboratories | Florida Bureau of Public Health Laboratories | Sarah Schmedes, Jason Blanton |
| EPI_ISL_791097, EPI_ISL_791098, EPI_ISL_791099, EPI_ISL_791100, EPI_ISL_791101, EPI_ISL_791102, EPI_ISL_791103, EPI_ISL_791104, EPI_ISL_791105, EPI_ISL_791106, EPI_ISL_791107, EPI_ISL_791108, EPI_ISL_791109, EPI_ISL_791110, EPI_ISL_791111, EPI_ISL_791112, EPI_ISL_791113, EPI_ISL_791114, EPI_ISL_791115, EPI_ISL_791116, EPI_ISL_791117, EPI_ISL_791118, EPI_ISL_791119, EPI_ISL_791120, EPI_ISL_791121, EPI_ISL_791122, EPI_ISL_791123, EPI_ISL_791124, EPI_ISL_791125, EPI_ISL_791126, EPI_ISL_791127, EPI_ISL_791128, EPI_ISL_791129, EPI_ISL_791130, EPI_ISL_791131, EPI_ISL_791132, EPI_ISL_791133, EPI_ISL_791134, EPI_ISL_791135, EPI_ISL_791136, EPI_ISL_791137, EPI_ISL_791138, EPI_ISL_791139, EPI_ISL_791140, EPI_ISL_791141, EPI_ISL_791143, EPI_ISL_791144, EPI_ISL_791145, EPI_ISL_791146, EPI_ISL_791147, EPI_ISL_791148, EPI_ISL_791149, EPI_ISL_791150, EPI_ISL_791151, EPI_ISL_791152, EPI_ISL_791153, EPI_ISL_791154, EPI_ISL_791155, EPI_ISL_791156, EPI_ISL_791157, EPI_ISL_791158, EPI_ISL_791159, EPI_ISL_791160, EPI_ISL_791161, EPI_ISL_791162, EPI_ISL_791163, EPI_ISL_791164, EPI_ISL_791165, EPI_ISL_791166, EPI_ISL_791167, EPI_ISL_791168, EPI_ISL_791169, EPI_ISL_791170, EPI_ISL_791171, EPI_ISL_791172, EPI_ISL_791173, EPI_ISL_791174, EPI_ISL_791175, EPI_ISL_791177, EPI_ISL_791178, EPI_ISL_791179, EPI_ISL_791180, EPI_ISL_791181, EPI_ISL_791182, EPI_ISL_791327, EPI_ISL_791328, EPI_ISL_791329, EPI_ISL_791331 | | | |
| see above | University of Wisconsin-Madison AIDS Vaccine Research Laboratories | University of Wisconsin-Madison AIDS Vaccine Research Laboratories | Gage Moreno, Katarina Braun, et al. AIDS Vaccine Research Laboratories |
| EPI_ISL_791334 | Johns Hopkins Hospital Department of Pathology | Johns Hopkins Hospital Department of Pathology | C. Paul Morris, Chun Huai Luo, Heba H. Mostafa |
| EPI_ISL_791335 | Johns Hopkins Hospital Department of Pathology | Johns Hopkins Hospital Department of Pathology | C. Paul Morris, Chun Huai Luo, Adannaya Amadi, Nicholas Gallagher, Heba H. Mostafa |
| EPI_ISL_791336 | Johns Hopkins Hospital Department of Pathology | Johns Hopkins Hospital Department of Pathology | C. Paul Morris, Chun Huai Luo, Heba H. Mostafa |
| EPI_ISL_791337 | Johns Hopkins Hospital Department of Pathology | Johns Hopkins Hospital Department of Pathology | C. Paul Morris, Chun Huai Luo, Adannaya Amadi, Nicholas Gallagher, Heba H. Mostafa |
| EPI_ISL_791338 | Johns Hopkins Hospital Department of Pathology | Johns Hopkins Hospital Department of Pathology | C. Paul Morris, Chun Huai Luo, Heba H. Mostafa |
| EPI_ISL_791339, EPI_ISL_791340 | Johns Hopkins Hospital Department of Pathology | Johns Hopkins Hospital Department of Pathology | C. Paul Morris, Chun Huai Luo, Adannaya Amadi, Nicholas Gallagher, Heba H. Mostafa |
| EPI_ISL_791341, EPI_ISL_791342, EPI_ISL_791343 | Johns Hopkins Hospital Department of Pathology | Johns Hopkins Hospital Department of Pathology | C. Paul Morris, Chun Huai Luo, Heba H. Mostafa |
| EPI_ISL_791344 | Johns Hopkins Hospital Department of Pathology | Johns Hopkins Hospital Department of Pathology | C. Paul Morris, Chun Huai Luo, Adannaya Amadi, Nicholas Gallagher, Heba H. Mostafa |
| EPI_ISL_791345 | Johns Hopkins Hospital Department of Pathology | Johns Hopkins Hospital Department of Pathology | C. Paul Morris, Chun Huai Luo, Heba H. Mostafa |
| EPI_ISL_791346 | Johns Hopkins Hospital Department of Pathology | Johns Hopkins Hospital Department of Pathology | C. Paul Morris, Chun Huai Luo, Adannaya Amadi, Nicholas Gallagher, Heba H. Mostafa |
| EPI_ISL_791347, EPI_ISL_791348, EPI_ISL_791349, EPI_ISL_791350, EPI_ISL_791351, EPI_ISL_791352, EPI_ISL_791353, EPI_ISL_791354, EPI_ISL_791355, EPI_ISL_791356, EPI_ISL_791357, EPI_ISL_791358, EPI_ISL_791359, EPI_ISL_791360, EPI_ISL_791361, EPI_ISL_791362, EPI_ISL_791363, EPI_ISL_791364, EPI_ISL_791365, EPI_ISL_791366, EPI_ISL_791367, EPI_ISL_791368, EPI_ISL_791369, EPI_ISL_791370, EPI_ISL_791371, EPI_ISL_791372, EPI_ISL_791373, EPI_ISL_791374, EPI_ISL_791375, EPI_ISL_791376, EPI_ISL_791377, EPI_ISL_791378, EPI_ISL_791379, EPI_ISL_791380, EPI_ISL_791381, EPI_ISL_791382, EPI_ISL_791383, EPI_ISL_791384, EPI_ISL_791385, EPI_ISL_791386, EPI_ISL_791387, EPI_ISL_791388, EPI_ISL_791389, EPI_ISL_791390, EPI_ISL_791391, EPI_ISL_791392, EPI_ISL_791393, EPI_ISL_791394, EPI_ISL_791395, EPI_ISL_791396, EPI_ISL_791397, EPI_ISL_791398, EPI_ISL_791399, EPI_ISL_791400, EPI_ISL_791401, EPI_ISL_791402, EPI_ISL_791403, EPI_ISL_791404, EPI_ISL_791405, EPI_ISL_791406, EPI_ISL_791407, EPI_ISL_791408, EPI_ISL_791409, EPI_ISL_791410, EPI_ISL_791411, EPI_ISL_791412, EPI_ISL_791413, EPI_ISL_791414 | | | |
| see above | Johns Hopkins Hospital Department of Pathology | Johns Hopkins Hospital Department of Pathology | C. Paul Morris, Chun Huai Luo, Heba H. Mostafa |
| EPI_ISL_791415, EPI_ISL_791416, EPI_ISL_791417, EPI_ISL_791418, EPI_ISL_791419, EPI_ISL_791420, EPI_ISL_791421, EPI_ISL_791422, EPI_ISL_791423, EPI_ISL_791424, EPI_ISL_791425, EPI_ISL_791426, EPI_ISL_791427, EPI_ISL_791428, EPI_ISL_791429, EPI_ISL_791430, EPI_ISL_791431, EPI_ISL_791432, EPI_ISL_791433, EPI_ISL_791434, EPI_ISL_791435, EPI_ISL_791436, EPI_ISL_791437, EPI_ISL_791438, EPI_ISL_791439, EPI_ISL_791440, EPI_ISL_791441, EPI_ISL_791442, EPI_ISL_791443, EPI_ISL_791444, EPI_ISL_791445, EPI_ISL_791446, EPI_ISL_791447, EPI_ISL_791448, EPI_ISL_791449, EPI_ISL_791450, EPI_ISL_791451, EPI_ISL_791452, EPI_ISL_791453, EPI_ISL_791454, EPI_ISL_791455, EPI_ISL_791456, EPI_ISL_791457, EPI_ISL_791458, EPI_ISL_791459, EPI_ISL_791460, EPI_ISL_791461, EPI_ISL_791462, EPI_ISL_791463, EPI_ISL_791464, EPI_ISL_791465, EPI_ISL_791466, EPI_ISL_791467, EPI_ISL_791468, EPI_ISL_791469, EPI_ISL_791470, EPI_ISL_791471, EPI_ISL_791472, EPI_ISL_791473, EPI_ISL_791474, EPI_ISL_791475, EPI_ISL_791476, EPI_ISL_791477, EPI_ISL_791478, EPI_ISL_791479, EPI_ISL_791480, EPI_ISL_791481, EPI_ISL_791482, EPI_ISL_791483, EPI_ISL_791484, EPI_ISL_791485, EPI_ISL_791486, EPI_ISL_791487 | | | |
| see above | Johns Hopkins Hospital Department of Pathology | Johns Hopkins Hospital Department of Pathology | C. Paul Morris, Chun Huai Luo, Adannaya Amadi, Nicholas Gallagher, Heba H. Mostafa |
| EPI_ISL_791514, EPI_ISL_791515, EPI_ISL_791518, EPI_ISL_791519, EPI_ISL_791520, EPI_ISL_791521, EPI_ISL_791522, EPI_ISL_791523, EPI_ISL_791524, EPI_ISL_791525, EPI_ISL_791526, EPI_ISL_791527, EPI_ISL_791529, EPI_ISL_791530, EPI_ISL_791531, EPI_ISL_791532, EPI_ISL_791533, EPI_ISL_791534, EPI_ISL_791535, EPI_ISL_791536, EPI_ISL_791537, EPI_ISL_791538, EPI_ISL_791539, EPI_ISL_791540, EPI_ISL_791541, EPI_ISL_791542, EPI_ISL_791543, EPI_ISL_791544, EPI_ISL_791545, EPI_ISL_791546, EPI_ISL_791548, EPI_ISL_791549, EPI_ISL_791550, EPI_ISL_791551, EPI_ISL_791552, EPI_ISL_791553, EPI_ISL_791554, EPI_ISL_791556, EPI_ISL_791557, EPI_ISL_791558, EPI_ISL_791559, EPI_ISL_791560, EPI_ISL_791561, EPI_ISL_791562, EPI_ISL_791563, EPI_ISL_791564, EPI_ISL_791565, EPI_ISL_791566, EPI_ISL_791569, EPI_ISL_791570, EPI_ISL_791571, EPI_ISL_791572, EPI_ISL_791573, EPI_ISL_791574, EPI_ISL_791575, EPI_ISL_791576, EPI_ISL_791577, EPI_ISL_791578, EPI_ISL_791579, EPI_ISL_791580, EPI_ISL_791581, EPI_ISL_791582, EPI_ISL_791584, EPI_ISL_791585, EPI_ISL_791586, EPI_ISL_791587, EPI_ISL_791588, EPI_ISL_791589, EPI_ISL_791590, EPI_ISL_791591, EPI_ISL_791592, EPI_ISL_791593, EPI_ISL_791594, EPI_ISL_791595, EPI_ISL_791596, EPI_ISL_791597, EPI_ISL_791598, EPI_ISL_791600, EPI_ISL_791601, EPI_ISL_791602, EPI_ISL_791603, EPI_ISL_791604, EPI_ISL_791605, EPI_ISL_791606, EPI_ISL_791607, EPI_ISL_791608, EPI_ISL_791609, EPI_ISL_791610, EPI_ISL_791611, EPI_ISL_791612, EPI_ISL_791613, EPI_ISL_791614, EPI_ISL_791615, EPI_ISL_791616, EPI_ISL_791617, EPI_ISL_791618, EPI_ISL_791619, EPI_ISL_791620, EPI_ISL_791621, EPI_ISL_791622, EPI_ISL_791624, EPI_ISL_791625, EPI_ISL_791626, EPI_ISL_791627, EPI_ISL_791628, EPI_ISL_791629, EPI_ISL_791630, EPI_ISL_791631, EPI_ISL_791632, EPI_ISL_791633, EPI_ISL_791634, EPI_ISL_791635, EPI_ISL_791636, EPI_ISL_791637, EPI_ISL_791638, EPI_ISL_791639, EPI_ISL_791640, EPI_ISL_791641, EPI_ISL_791642, EPI_ISL_791643, EPI_ISL_791644, EPI_ISL_791645, EPI_ISL_791646, EPI_ISL_791647, EPI_ISL_791648, EPI_ISL_791649, EPI_ISL_791650, EPI_ISL_791651, EPI_ISL_791652, EPI_ISL_791653, EPI_ISL_791654, EPI_ISL_791655, EPI_ISL_791656, EPI_ISL_791657, EPI_ISL_791658, EPI_ISL_791659, EPI_ISL_791660, EPI_ISL_791661, EPI_ISL_791662, EPI_ISL_791663, EPI_ISL_791664, EPI_ISL_791665, EPI_ISL_791666, EPI_ISL_791667, EPI_ISL_791669, EPI_ISL_791670, EPI_ISL_791671, EPI_ISL_791672, EPI_ISL_791673, EPI_ISL_791674, EPI_ISL_791675, EPI_ISL_791676, EPI_ISL_791677, EPI_ISL_791678, EPI_ISL_791679, EPI_ISL_791680, EPI_ISL_791681, EPI_ISL_791682, EPI_ISL_791683, EPI_ISL_791684, EPI_ISL_791685, EPI_ISL_791686, EPI_ISL_791687, EPI_ISL_791688, EPI_ISL_791689, EPI_ISL_791690, EPI_ISL_791691, EPI_ISL_791692, EPI_ISL_791693, EPI_ISL_791694, EPI_ISL_791695, EPI_ISL_791696, EPI_ISL_791697, EPI_ISL_791698, EPI_ISL_791699, EPI_ISL_791700, EPI_ISL_791701, EPI_ISL_791702, EPI_ISL_791703, EPI_ISL_791704, EPI_ISL_791705, EPI_ISL_791706, EPI_ISL_791707, EPI_ISL_791708, EPI_ISL_791709, EPI_ISL_791711, EPI_ISL_791712, EPI_ISL_791713, EPI_ISL_791714, EPI_ISL_791715, EPI_ISL_791716, EPI_ISL_791717, EPI_ISL_791719, EPI_ISL_791720, EPI_ISL_791722, EPI_ISL_791723, EPI_ISL_791724, EPI_ISL_791725, EPI_ISL_791726 | | | |
| see above | Massachusetts State Public Health Laboratory | Infectious Disease Program, Broad Institute of Harvard and MIT | Lemieux,J.E., Siddle,K.J., Shaw,B., Adams,G., Pierce,V., Turbett,S., Anahtar,M., Branda,J., Slater,D., Harris,J., Lin,A.E., Gladden-Young,A., Lagerborg,K., Rudy,M., DeRuff,K., Carter,A., Normandin,E., Bauer,M., Reilly,S., Tomkins-Tinch,C., Loreth,C., Chaluvadi,S., Neumann,A., Cusick,C., Chapman,S.B., Gnirke,A., Flowers,K., Cerrato,F., Birren,B.W., Gallagher,G., Smole,S., Park,D.J., MacInnis,B.L., Ryan,E., LaRocque,R., Rosenberg,E. and Sabeti,P.C. |
| EPI_ISL_791729, EPI_ISL_791730, EPI_ISL_791731, EPI_ISL_791732, EPI_ISL_791734, EPI_ISL_791735, EPI_ISL_791736, EPI_ISL_791737, EPI_ISL_791738, EPI_ISL_791739, EPI_ISL_791740, EPI_ISL_791741, EPI_ISL_791742, EPI_ISL_791743, EPI_ISL_791744, EPI_ISL_791745, EPI_ISL_791746, EPI_ISL_791747, EPI_ISL_791748, EPI_ISL_791749, EPI_ISL_791750, EPI_ISL_791751, EPI_ISL_791752, EPI_ISL_791753, EPI_ISL_791754, EPI_ISL_791755, EPI_ISL_791756, EPI_ISL_791757, EPI_ISL_791758, EPI_ISL_791759, EPI_ISL_791760, EPI_ISL_791761, EPI_ISL_791762, EPI_ISL_791763, EPI_ISL_791766, EPI_ISL_791767, EPI_ISL_791768, EPI_ISL_791769, EPI_ISL_791770, EPI_ISL_791771, EPI_ISL_791772, EPI_ISL_791773, EPI_ISL_791774, EPI_ISL_791775, EPI_ISL_791776, EPI_ISL_791777, EPI_ISL_791778, EPI_ISL_791779, EPI_ISL_791780, EPI_ISL_791781, EPI_ISL_791782, EPI_ISL_791783, EPI_ISL_791784, EPI_ISL_791785, EPI_ISL_791786, EPI_ISL_791787, EPI_ISL_791788, EPI_ISL_791789, EPI_ISL_791790, EPI_ISL_791792, EPI_ISL_791793, EPI_ISL_791794, EPI_ISL_791795, EPI_ISL_791796, EPI_ISL_791797, EPI_ISL_791798, EPI_ISL_791799, EPI_ISL_791801, EPI_ISL_791802, EPI_ISL_791803, EPI_ISL_791805, EPI_ISL_791807, EPI_ISL_791808, EPI_ISL_791809, EPI_ISL_791810, EPI_ISL_791811, EPI_ISL_791812, EPI_ISL_791813, EPI_ISL_791814, EPI_ISL_791815, EPI_ISL_791816, EPI_ISL_791817, EPI_ISL_791818, EPI_ISL_791819, EPI_ISL_791821, EPI_ISL_791822, EPI_ISL_791823, EPI_ISL_791824, EPI_ISL_791825, EPI_ISL_791826, EPI_ISL_791828, EPI_ISL_791829, EPI_ISL_791830, EPI_ISL_791831, EPI_ISL_791832, EPI_ISL_791833, EPI_ISL_791834, EPI_ISL_791835, EPI_ISL_791836, EPI_ISL_791837, EPI_ISL_791838, EPI_ISL_791839, EPI_ISL_791841, EPI_ISL_791842, EPI_ISL_791843, EPI_ISL_791844, EPI_ISL_791845, EPI_ISL_791847, EPI_ISL_791848, EPI_ISL_791849, EPI_ISL_791850, EPI_ISL_791851, EPI_ISL_791852, EPI_ISL_791853, EPI_ISL_791854, EPI_ISL_791855, EPI_ISL_791856, EPI_ISL_791857, EPI_ISL_791858, EPI_ISL_791859, EPI_ISL_791860, EPI_ISL_791861, EPI_ISL_791863, EPI_ISL_791864, EPI_ISL_791865, EPI_ISL_791866, EPI_ISL_791867, EPI_ISL_791868, EPI_ISL_791869, EPI_ISL_791871, EPI_ISL_791872, EPI_ISL_791873, EPI_ISL_791874, EPI_ISL_791875, EPI_ISL_791876, EPI_ISL_791877, EPI_ISL_791878, EPI_ISL_791879, EPI_ISL_791881, EPI_ISL_791882, EPI_ISL_791883, EPI_ISL_791884, EPI_ISL_791885, EPI_ISL_791886, EPI_ISL_791887, EPI_ISL_791888, EPI_ISL_791889, EPI_ISL_791891, EPI_ISL_791892, EPI_ISL_791893, EPI_ISL_791894, EPI_ISL_791895, EPI_ISL_791896, EPI_ISL_791897, EPI_ISL_791898, EPI_ISL_791899, EPI_ISL_791900, EPI_ISL_791901, EPI_ISL_791902, EPI_ISL_791903, EPI_ISL_791904, EPI_ISL_791905, EPI_ISL_791906, EPI_ISL_791907, EPI_ISL_791908, EPI_ISL_791909, EPI_ISL_791910, EPI_ISL_791911, EPI_ISL_791912, EPI_ISL_791914, EPI_ISL_791915, EPI_ISL_791916, EPI_ISL_791918, EPI_ISL_791919, EPI_ISL_791920, EPI_ISL_791921, EPI_ISL_791922, EPI_ISL_791923, EPI_ISL_791924, EPI_ISL_791925, EPI_ISL_791926, EPI_ISL_791927, EPI_ISL_791928, EPI_ISL_791929, EPI_ISL_791930, EPI_ISL_791931, EPI_ISL_791932, EPI_ISL_791933, EPI_ISL_791934, EPI_ISL_791935, EPI_ISL_791936, EPI_ISL_791937, EPI_ISL_791938, EPI_ISL_791939, EPI_ISL_791940, EPI_ISL_791942, EPI_ISL_791943, EPI_ISL_791944, EPI_ISL_791945, EPI_ISL_791946, EPI_ISL_791947, EPI_ISL_791948, EPI_ISL_791949, EPI_ISL_791950, EPI_ISL_791951, EPI_ISL_791952, EPI_ISL_791953, EPI_ISL_791954, EPI_ISL_791955, EPI_ISL_791956, EPI_ISL_791957, EPI_ISL_791958, EPI_ISL_791959, EPI_ISL_791960, EPI_ISL_791961, EPI_ISL_791962, EPI_ISL_791963, EPI_ISL_791964, EPI_ISL_791966, EPI_ISL_791968, EPI_ISL_791969, EPI_ISL_791970, EPI_ISL_791971, EPI_ISL_791973, EPI_ISL_791975, EPI_ISL_791976 | | | |
| see above | Massachusetts General Hospital | Infectious Disease Program, Broad Institute of Harvard and MIT | Lemieux,J.E., Siddle,K.J., Shaw,B., Adams,G., Pierce,V., Turbett,S., Anahtar,M., Branda,J., Slater,D., Harris,J., Lin,A.E., Gladden-Young,A., Lagerborg,K., Rudy,M., DeRuff,K., Carter,A., Normandin,E., Bauer,M., Reilly,S., Tomkins-Tinch,C., Loreth,C., Chaluvadi,S., Neumann,A., Cusick,C., Chapman,S.B., Gnirke,A., Flowers,K., Cerrato,F., Birren,B.W., Gallagher,G., Smole,S., Park,D.J., MacInnis,B.L., Ryan,E., LaRocque,R., Rosenberg,E. and Sabeti,P.C. |
| EPI_ISL_792092, EPI_ISL_792093, EPI_ISL_792094, EPI_ISL_792095, EPI_ISL_792096, EPI_ISL_792097, | UW Virology Lab | UW Virology Lab | Pavitra Roychoudhury, Hong Xie, Lasata Shrestha, Meei-Li Huang, Keith R Jerome, Alexander Greninger |

| | | | |
|---|---|---|---|
| EPI_ISL_792098, EPI_ISL_792099, EPI_ISL_792100 | | | |
| EPI_ISL_792655, EPI_ISL_792656, EPI_ISL_792657, EPI_ISL_792658, EPI_ISL_792659, EPI_ISL_792660, EPI_ISL_792661, EPI_ISL_792662, EPI_ISL_792663, EPI_ISL_792664, EPI_ISL_792665, EPI_ISL_792666, EPI_ISL_792667, EPI_ISL_792668, EPI_ISL_792669, EPI_ISL_792670, EPI_ISL_792671, EPI_ISL_792672, EPI_ISL_792673, EPI_ISL_792674, EPI_ISL_792675, EPI_ISL_792676, EPI_ISL_792677, EPI_ISL_792678 | | | |
| see above | Los Angeles County Public Health Laboratories | Los Angeles County Public Health Laboratories | P. Hemarajata et al. |
| EPI_ISL_792679 | Olive View Medical Center | Los Angeles County Public Health Laboratories | P. Hemarajata et al. |
| EPI_ISL_794014, EPI_ISL_794015, EPI_ISL_794016, EPI_ISL_794017, EPI_ISL_794018, EPI_ISL_794019, EPI_ISL_794020, EPI_ISL_794021, EPI_ISL_794022, EPI_ISL_794023, EPI_ISL_794024, EPI_ISL_794025, EPI_ISL_794026, EPI_ISL_794027, EPI_ISL_794028, EPI_ISL_794029, EPI_ISL_794030, EPI_ISL_794031, EPI_ISL_794032, EPI_ISL_794033, EPI_ISL_794034, EPI_ISL_794035, EPI_ISL_794036, EPI_ISL_794037, EPI_ISL_794038, EPI_ISL_794039, EPI_ISL_794040, EPI_ISL_794041, EPI_ISL_794042, EPI_ISL_794043, EPI_ISL_794044, EPI_ISL_794045, EPI_ISL_794046, EPI_ISL_794047, EPI_ISL_794048, EPI_ISL_794049, EPI_ISL_794050, EPI_ISL_794051, EPI_ISL_794052, EPI_ISL_794053, EPI_ISL_794054, EPI_ISL_794055, EPI_ISL_794056, EPI_ISL_794057, EPI_ISL_794058, EPI_ISL_794059, EPI_ISL_794060, EPI_ISL_794061, EPI_ISL_794062, EPI_ISL_794063, EPI_ISL_794064, EPI_ISL_794065, EPI_ISL_794066, EPI_ISL_794067, EPI_ISL_794068, EPI_ISL_794069, EPI_ISL_794070, EPI_ISL_794071, EPI_ISL_794072, EPI_ISL_794073, EPI_ISL_794074, EPI_ISL_794075, EPI_ISL_794076, EPI_ISL_794077, EPI_ISL_794078, EPI_ISL_794079, EPI_ISL_794080, EPI_ISL_794081, EPI_ISL_794082 | | | |
| see above | Wadsworth Center, New York State Department.of Health | Wadsworth Center, New York State Department.of Health | Kirsten St. George, Daryl M. Lamson, Alexis Russel, Matthew Shudt, Melissa A Leisner, Jonathan Plitnick, Navjot Singh, John Kelly, Sara Griesemer, Erasmus Schneider, Erica Lasek-Nesselquist |
| EPI_ISL_794083, EPI_ISL_794084, EPI_ISL_794085, EPI_ISL_794086, EPI_ISL_794087, EPI_ISL_794088, EPI_ISL_794089, EPI_ISL_794090, EPI_ISL_794091, EPI_ISL_794092, EPI_ISL_794093, EPI_ISL_794094, EPI_ISL_794095, EPI_ISL_794096, EPI_ISL_794097, EPI_ISL_794098, EPI_ISL_794099, EPI_ISL_794100, EPI_ISL_794101, EPI_ISL_794102, EPI_ISL_794103, EPI_ISL_794104, EPI_ISL_794105, EPI_ISL_794106 | | | |
| see above | URMC LABS | Wadsworth Center, New York State Department.of Health | Kirsten St. George, Daryl M. Lamson, Alexis Russel, Matthew Shudt, Melissa A Leisner, Jonathan Plitnick, Navjot Singh, John Kelly, Sara Griesemer, Erasmus Schneider, Erica Lasek-Nesselquist |
| EPI_ISL_794107, EPI_ISL_794108, EPI_ISL_794109, EPI_ISL_794110, EPI_ISL_794111, EPI_ISL_794112, EPI_ISL_794113, EPI_ISL_794114, EPI_ISL_794115, EPI_ISL_794116, EPI_ISL_794117, EPI_ISL_794118 | | | |
| see above | GLENS FALLS HOSPITAL LABORATORY | Wadsworth Center, New York State Department.of Health | Kirsten St. George, Daryl M. Lamson, Alexis Russel, Matthew Shudt, Melissa A Leisner, Jonathan Plitnick, Navjot Singh, John Kelly, Sara Griesemer, Erasmus Schneider, Erica Lasek-Nesselquist |
| EPI_ISL_794119, EPI_ISL_794120, EPI_ISL_794121, EPI_ISL_794122, EPI_ISL_794123, EPI_ISL_794124, EPI_ISL_794125, EPI_ISL_794126, EPI_ISL_794127, EPI_ISL_794128, EPI_ISL_794129, EPI_ISL_794130, EPI_ISL_794131, EPI_ISL_794132, EPI_ISL_794133, EPI_ISL_794134, EPI_ISL_794135, EPI_ISL_794136, EPI_ISL_794137, EPI_ISL_794138, EPI_ISL_794139, EPI_ISL_794140, EPI_ISL_794141, EPI_ISL_794142, EPI_ISL_794143, EPI_ISL_794144, EPI_ISL_794145, EPI_ISL_794146, EPI_ISL_794147, EPI_ISL_794148, EPI_ISL_794149, EPI_ISL_794150, EPI_ISL_794151, EPI_ISL_794152, EPI_ISL_794153, EPI_ISL_794154, EPI_ISL_794155, EPI_ISL_794156, EPI_ISL_794157, EPI_ISL_794158 | | | |
| see above | NORTHWELL HEALTH LABORATORIES | Wadsworth Center, New York State Department.of Health | Kirsten St. George, Daryl M. Lamson, Alexis Russel, Matthew Shudt, Melissa A Leisner, Jonathan Plitnick, Navjot Singh, John Kelly, Sara Griesemer, Erasmus Schneider, Erica Lasek-Nesselquist |
| EPI_ISL_794159, EPI_ISL_794160, EPI_ISL_794161, EPI_ISL_794162, EPI_ISL_794163, EPI_ISL_794164, EPI_ISL_794165, EPI_ISL_794166, EPI_ISL_794167, EPI_ISL_794168, EPI_ISL_794169, EPI_ISL_794170, EPI_ISL_794171, EPI_ISL_794172, EPI_ISL_794173, EPI_ISL_794174, EPI_ISL_794175, EPI_ISL_794176, EPI_ISL_794177, EPI_ISL_794178, EPI_ISL_794179, EPI_ISL_794180, EPI_ISL_794181 | | | |
| see above | Wadsworth Center, New York State Department.of Health | Wadsworth Center, New York State Department.of Health | Kirsten St. George, Daryl M. Lamson, Alexis Russel, Matthew Shudt, Melissa A Leisner, Jonathan Plitnick, Navjot Singh, John Kelly, Sara Griesemer, Erasmus Schneider, Erica Lasek-Nesselquist |
| EPI_ISL_794182, EPI_ISL_794183, EPI_ISL_794184, EPI_ISL_794185, EPI_ISL_794186, EPI_ISL_794187, EPI_ISL_794188, EPI_ISL_794189, EPI_ISL_794190, EPI_ISL_794191, EPI_ISL_794192, EPI_ISL_794193, EPI_ISL_794194, EPI_ISL_794195, EPI_ISL_794196, EPI_ISL_794197, EPI_ISL_794198, EPI_ISL_794199, EPI_ISL_794200, EPI_ISL_794201, EPI_ISL_794202, EPI_ISL_794203, EPI_ISL_794204, EPI_ISL_794205 | | | |
| see above | URMC LABS | Wadsworth Center, New York State Department.of Health | Kirsten St. George, Daryl M. Lamson, Alexis Russel, Matthew Shudt, Melissa A Leisner, Jonathan Plitnick, Navjot Singh, John Kelly, Sara Griesemer, Erasmus Schneider, Erica Lasek-Nesselquist |
| EPI_ISL_794206, EPI_ISL_794207 | Wadsworth Center, New York State Department.of Health | Wadsworth Center, New York State Department.of Health | Kirsten St. George, Daryl M. Lamson, Alexis Russel, Matthew Shudt, Melissa A Leisner, Jonathan Plitnick, Navjot Singh, John Kelly, Sara Griesemer, Erasmus Schneider, Erica Lasek-Nesselquist |
| EPI_ISL_794208, EPI_ISL_794209, EPI_ISL_794210, EPI_ISL_794211, EPI_ISL_794212, EPI_ISL_794213, EPI_ISL_794214, EPI_ISL_794215, EPI_ISL_794216, EPI_ISL_794217, EPI_ISL_794218, EPI_ISL_794219, EPI_ISL_794220, EPI_ISL_794221, EPI_ISL_794222, EPI_ISL_794223, EPI_ISL_794224, EPI_ISL_794225, EPI_ISL_794226, EPI_ISL_794227, EPI_ISL_794228, EPI_ISL_794229, EPI_ISL_794230, EPI_ISL_794231, EPI_ISL_794232, EPI_ISL_794233, EPI_ISL_794234, EPI_ISL_794235, EPI_ISL_794236, EPI_ISL_794237, EPI_ISL_794238, EPI_ISL_794239, EPI_ISL_794240, EPI_ISL_794241, EPI_ISL_794242, EPI_ISL_794243, EPI_ISL_794244, EPI_ISL_794245, EPI_ISL_794246, EPI_ISL_794247, EPI_ISL_794248, EPI_ISL_794249, EPI_ISL_794250, EPI_ISL_794251, EPI_ISL_794252, EPI_ISL_794253, EPI_ISL_794254, EPI_ISL_794255, EPI_ISL_794256, EPI_ISL_794257, EPI_ISL_794258, EPI_ISL_794259, EPI_ISL_794260, EPI_ISL_794261, EPI_ISL_794262, EPI_ISL_794263, EPI_ISL_794264, EPI_ISL_794265, EPI_ISL_794266, EPI_ISL_794267, EPI_ISL_794268, EPI_ISL_794269, EPI_ISL_794270, EPI_ISL_794271, EPI_ISL_794272, EPI_ISL_794273, EPI_ISL_794274, EPI_ISL_794275, EPI_ISL_794276, EPI_ISL_794277, EPI_ISL_794278, EPI_ISL_794279, EPI_ISL_794280, EPI_ISL_794281, EPI_ISL_794282 | | | |
| see above | WESTCHESTER MEDICAL CENTER | Wadsworth Center, New York State Department.of Health | Kirsten St. George, Daryl M. Lamson, Alexis Russel, Matthew Shudt, Melissa A Leisner, Jonathan Plitnick, Navjot Singh, John Kelly, Sara Griesemer, Erasmus Schneider, Erica Lasek-Nesselquist |
| EPI_ISL_794283, EPI_ISL_794284, EPI_ISL_794285, EPI_ISL_794286, EPI_ISL_794287, EPI_ISL_794288, EPI_ISL_794289, EPI_ISL_794290, EPI_ISL_794291, EPI_ISL_794292, EPI_ISL_794293, EPI_ISL_794294, EPI_ISL_794295, EPI_ISL_794296, EPI_ISL_794297, EPI_ISL_794298, EPI_ISL_794299, EPI_ISL_794300, EPI_ISL_794301, EPI_ISL_794302, EPI_ISL_794303, EPI_ISL_794304, EPI_ISL_794305, EPI_ISL_794306, EPI_ISL_794307 | | | |
| see above | Columbia University Irving Medical Center | Wadsworth Center, New York State Department.of Health | Kirsten St. George, Daryl M. Lamson, Alexis Russel, Matthew Shudt, Melissa A Leisner, Jonathan Plitnick, Navjot Singh, John Kelly, Sara Griesemer, Erasmus Schneider, Erica Lasek-Nesselquist |
| EPI_ISL_794308, EPI_ISL_794309, EPI_ISL_794310, EPI_ISL_794311, EPI_ISL_794312, EPI_ISL_794313, EPI_ISL_794314, EPI_ISL_794315, EPI_ISL_794316, EPI_ISL_794317, EPI_ISL_794318, EPI_ISL_794319, EPI_ISL_794320 | | | |
| see above | Wadsworth Center, New York State Department.of Health | Wadsworth Center, New York State Department.of Health | Kirsten St. George, Daryl M. Lamson, Alexis Russel, Matthew Shudt, Melissa A Leisner, Jonathan Plitnick, Navjot Singh, John Kelly, Sara Griesemer, Erasmus Schneider, Erica Lasek-Nesselquist |
| EPI_ISL_794592 | Laboratorio Estatal de Salud Pública de Tamaulipas | Instituto de diagnóstico y Referencia Epidemiologicos (INDRE) | Abril Rodriguez-Maldonado, Claudia Wong-Arambula, Fabiola Garces-Ayala, Gisela Barrera-Badillo, Ana Maria Cortez-Calderon, Bernardita Reyes-Berrones, Hilda del Carmen-Selvera, Gloria Molina-Gamboa, Lucia Hernandez-Rivas, Irma Lopez-Martinez, Celia Alpuche-Aranda, Jose Luis Alomia-Zegarra, Hugo Lopez Gatell-Ramirez, Ernesto Ramirez-Gonzalez. |
| EPI_ISL_794635, EPI_ISL_794636, EPI_ISL_794638, EPI_ISL_794639, EPI_ISL_794640 | USC Clinical Lab | Los Angeles County Public Health Laboratories | P. Hemarajata et al. |
| EPI_ISL_794641, EPI_ISL_794642, EPI_ISL_794643, EPI_ISL_794644, EPI_ISL_794645, EPI_ISL_794646, EPI_ISL_794647, EPI_ISL_794648 | UCLA Clinical Micro Lab | Los Angeles County Public Health Laboratories | P. Hemarajata et al. |
| EPI_ISL_801522, EPI_ISL_801523, EPI_ISL_801524, EPI_ISL_801525, EPI_ISL_801526, EPI_ISL_801527, EPI_ISL_801528, EPI_ISL_801529, EPI_ISL_801530, EPI_ISL_801531, EPI_ISL_801532, EPI_ISL_801533, EPI_ISL_801534, EPI_ISL_801535, EPI_ISL_801536 | | | |
| see above | Sonic Reference Laboratory | Pathogen Discovery, Respiratory Viruses Branch, Division of Viral Diseases, Centers for Disease Control and Prevention | Ying Tao, Yan Li, Jing Zhang, Krista Queen, Anna Uehara, Peter Cook, Clinton R. Paden, Haibin Wang, Suxiang Tong |
| EPI_ISL_801537 | Sunrise Medical Labs | Pathogen Discovery, Respiratory Viruses Branch, Division of Viral Diseases, Centers for Disease Control and Prevention | Ying Tao, Yan Li, Jing Zhang, Krista Queen, Anna Uehara, Peter Cook, Clinton R. Paden, Haibin Wang, Suxiang Tong |
| EPI_ISL_801861, EPI_ISL_801862, EPI_ISL_801863, EPI_ISL_801864, EPI_ISL_801865, EPI_ISL_801866, EPI_ISL_801867, EPI_ISL_801868, EPI_ISL_801869, EPI_ISL_801870, EPI_ISL_801871, EPI_ISL_801872, EPI_ISL_801873, EPI_ISL_801874, EPI_ISL_801875, EPI_ISL_801876, EPI_ISL_801877, EPI_ISL_801878, EPI_ISL_801879, EPI_ISL_801880, EPI_ISL_801881, EPI_ISL_801882, EPI_ISL_801883, EPI_ISL_801884, EPI_ISL_801885, EPI_ISL_801886, EPI_ISL_801887, EPI_ISL_801888, EPI_ISL_801889, EPI_ISL_801890, EPI_ISL_801891, EPI_ISL_801892, EPI_ISL_801893, EPI_ISL_801894, EPI_ISL_801895, EPI_ISL_801896, EPI_ISL_801897, EPI_ISL_801898, EPI_ISL_801899, EPI_ISL_801900, EPI_ISL_801901, EPI_ISL_801902, EPI_ISL_801903, EPI_ISL_801904, EPI_ISL_801905, EPI_ISL_801906, EPI_ISL_801907, EPI_ISL_801908, EPI_ISL_801909, EPI_ISL_801910, EPI_ISL_801911, EPI_ISL_801912, EPI_ISL_801913, EPI_ISL_801914, EPI_ISL_801915, EPI_ISL_801916, EPI_ISL_801917, EPI_ISL_801918, EPI_ISL_801919, EPI_ISL_801920, EPI_ISL_801921, EPI_ISL_801922, EPI_ISL_801923, EPI_ISL_801924, EPI_ISL_801925, EPI_ISL_801926, EPI_ISL_801927, EPI_ISL_801928, EPI_ISL_801929, EPI_ISL_801930, EPI_ISL_801931, EPI_ISL_801932, EPI_ISL_801933, EPI_ISL_801934, EPI_ISL_801935, EPI_ISL_801936, EPI_ISL_801937, EPI_ISL_801938, EPI_ISL_801939, EPI_ISL_801940, EPI_ISL_801941, EPI_ISL_801942, EPI_ISL_801943, EPI_ISL_801944, EPI_ISL_801947, EPI_ISL_801948, EPI_ISL_801949, EPI_ISL_801950, EPI_ISL_801951, EPI_ISL_801952, EPI_ISL_801953, EPI_ISL_801954, EPI_ISL_801955, EPI_ISL_801956, EPI_ISL_801957, EPI_ISL_801958, EPI_ISL_801959, EPI_ISL_801960, EPI_ISL_801961, EPI_ISL_801962, EPI_ISL_801963, EPI_ISL_801964, EPI_ISL_801965, EPI_ISL_801966, EPI_ISL_801967, EPI_ISL_801968, EPI_ISL_801969, EPI_ISL_801970, EPI_ISL_801971, EPI_ISL_801972, EPI_ISL_801973, EPI_ISL_801974, EPI_ISL_801975, EPI_ISL_801976, EPI_ISL_801977, EPI_ISL_801978, EPI_ISL_801979, EPI_ISL_801980, EPI_ISL_801981, EPI_ISL_801982, EPI_ISL_801983, EPI_ISL_801984, EPI_ISL_801985, EPI_ISL_801986, EPI_ISL_801987, EPI_ISL_801988, EPI_ISL_801989, EPI_ISL_801990, EPI_ISL_801991, EPI_ISL_801992, EPI_ISL_801993, EPI_ISL_801994, EPI_ISL_801995, EPI_ISL_801996, EPI_ISL_801997, EPI_ISL_801998, EPI_ISL_801999, EPI_ISL_802000, EPI_ISL_802001, EPI_ISL_802002, EPI_ISL_802003, EPI_ISL_802004, EPI_ISL_802005, EPI_ISL_802006, EPI_ISL_802007, EPI_ISL_802008, EPI_ISL_802009, EPI_ISL_802010, EPI_ISL_802011, EPI_ISL_802012, EPI_ISL_802013, EPI_ISL_802014, EPI_ISL_802015, EPI_ISL_802016, EPI_ISL_802017, EPI_ISL_802018, EPI_ISL_802019, EPI_ISL_802020, EPI_ISL_802021, EPI_ISL_802022, EPI_ISL_802023, EPI_ISL_802024, EPI_ISL_802025, EPI_ISL_802026, EPI_ISL_802027, EPI_ISL_802028, EPI_ISL_802029, EPI_ISL_802030, EPI_ISL_802031, EPI_ISL_802032, EPI_ISL_802033, EPI_ISL_802034, EPI_ISL_802035, EPI_ISL_802036, EPI_ISL_802037, EPI_ISL_802038, EPI_ISL_802039, EPI_ISL_802040, EPI_ISL_802041, EPI_ISL_802042, EPI_ISL_802043, EPI_ISL_802044, EPI_ISL_802045, EPI_ISL_802046, EPI_ISL_802047, EPI_ISL_802048, EPI_ISL_802049, EPI_ISL_802050, EPI_ISL_802051, EPI_ISL_802052, EPI_ISL_802053, EPI_ISL_802054, EPI_ISL_802055, EPI_ISL_802056, EPI_ISL_802057, EPI_ISL_802058, EPI_ISL_802059, EPI_ISL_802060, EPI_ISL_802061, EPI_ISL_802062, EPI_ISL_802063, EPI_ISL_802064, EPI_ISL_802065, EPI_ISL_802066, EPI_ISL_802067, EPI_ISL_802068, EPI_ISL_802069, EPI_ISL_802070, EPI_ISL_802071, EPI_ISL_802072, EPI_ISL_802073, EPI_ISL_802074, EPI_ISL_802075, EPI_ISL_802076, EPI_ISL_802077, EPI_ISL_802078, EPI_ISL_802079, EPI_ISL_802080, EPI_ISL_802081, EPI_ISL_802082, EPI_ISL_802083, EPI_ISL_802084, EPI_ISL_802085, EPI_ISL_802086, EPI_ISL_802087, EPI_ISL_802088, EPI_ISL_802089, EPI_ISL_802090, EPI_ISL_802091, EPI_ISL_802092, EPI_ISL_802093, EPI_ISL_802094, EPI_ISL_802095, EPI_ISL_802096, EPI_ISL_802097, EPI_ISL_802098, EPI_ISL_802099, EPI_ISL_802100, EPI_ISL_802101, EPI_ISL_802102, EPI_ISL_802103, EPI_ISL_802104, EPI_ISL_802105, EPI_ISL_802106, EPI_ISL_802107, EPI_ISL_802108, EPI_ISL_802109, EPI_ISL_802110, EPI_ISL_802111, EPI_ISL_802112, EPI_ISL_802113, EPI_ISL_802114, EPI_ISL_802115, EPI_ISL_802116, EPI_ISL_802117, EPI_ISL_802118, EPI_ISL_802119, EPI_ISL_802120, EPI_ISL_802121, EPI_ISL_802122, EPI_ISL_802123, EPI_ISL_802124, EPI_ISL_802125, EPI_ISL_802126, EPI_ISL_802127, EPI_ISL_802128, EPI_ISL_802129, EPI_ISL_802130, EPI_ISL_802131, EPI_ISL_802132, EPI_ISL_802133, EPI_ISL_802134, EPI_ISL_802135, EPI_ISL_802136, EPI_ISL_802137, EPI_ISL_802138, EPI_ISL_802139, EPI_ISL_802140, EPI_ISL_802141, EPI_ISL_802142, EPI_ISL_802143, EPI_ISL_802144, EPI_ISL_802145, EPI_ISL_802146, EPI_ISL_802147, EPI_ISL_802148, EPI_ISL_802149, EPI_ISL_802150, EPI_ISL_802151, EPI_ISL_802152, EPI_ISL_802153, EPI_ISL_802154, EPI_ISL_802155, EPI_ISL_802156, EPI_ISL_802157, EPI_ISL_802158, EPI_ISL_802159, EPI_ISL_802160, EPI_ISL_802161, EPI_ISL_802162, EPI_ISL_802163, EPI_ISL_802164, EPI_ISL_802165, EPI_ISL_802166, EPI_ISL_802167, EPI_ISL_802168, EPI_ISL_802169, EPI_ISL_802170, EPI_ISL_802171, EPI_ISL_802172, EPI_ISL_802173, EPI_ISL_802174, EPI_ISL_802175, EPI_ISL_802176, EPI_ISL_802177, EPI_ISL_802178, EPI_ISL_802179, EPI_ISL_802180, EPI_ISL_802181, EPI_ISL_802182, EPI_ISL_802183, EPI_ISL_802184, EPI_ISL_802185, EPI_ISL_802186, EPI_ISL_802187, EPI_ISL_802188, EPI_ISL_802189, EPI_ISL_802190, EPI_ISL_802191, EPI_ISL_802192, EPI_ISL_802193, EPI_ISL_802194, EPI_ISL_802195, EPI_ISL_802196, EPI_ISL_802197, EPI_ISL_802198, EPI_ISL_802199, EPI_ISL_802200, EPI_ISL_802201, EPI_ISL_802202, EPI_ISL_802203, EPI_ISL_802204, EPI_ISL_802205, EPI_ISL_802206, EPI_ISL_802207, EPI_ISL_802208, EPI_ISL_802209, EPI_ISL_802210, EPI_ISL_802211, EPI_ISL_802212, EPI_ISL_802213, EPI_ISL_802214, EPI_ISL_802215, EPI_ISL_802216, EPI_ISL_802217, EPI_ISL_802218, EPI_ISL_802219, EPI_ISL_802220, EPI_ISL_802221, EPI_ISL_802222, EPI_ISL_802223, EPI_ISL_802224, EPI_ISL_802225, EPI_ISL_802226, EPI_ISL_802227, EPI_ISL_802228, EPI_ISL_802229, EPI_ISL_802230, EPI_ISL_802231, EPI_ISL_802232, EPI_ISL_802233, EPI_ISL_802234, EPI_ISL_802235, EPI_ISL_802236, EPI_ISL_802237, EPI_ISL_802238, EPI_ISL_802239, EPI_ISL_802240, EPI_ISL_802241, EPI_ISL_802242, EPI_ISL_802243, EPI_ISL_802244, EPI_ISL_802245, EPI_ISL_802246, EPI_ISL_802247, EPI_ISL_802248, EPI_ISL_802249, EPI_ISL_802250, EPI_ISL_802251, EPI_ISL_802252, EPI_ISL_802253, EPI_ISL_802254, EPI_ISL_802255, EPI_ISL_802256, EPI_ISL_802257, EPI_ISL_802258, EPI_ISL_802259, EPI_ISL_802260, EPI_ISL_802261, EPI_ISL_802262, EPI_ISL_802263, EPI_ISL_802264, EPI_ISL_802265, EPI_ISL_802266, EPI_ISL_802267, EPI_ISL_802268, EPI_ISL_802269, EPI_ISL_802270, EPI_ISL_802271, EPI_ISL_802272, EPI_ISL_802273, EPI_ISL_802274, EPI_ISL_802275, EPI_ISL_802276, EPI_ISL_802277, EPI_ISL_802278, EPI_ISL_802279, EPI_ISL_802280, EPI_ISL_802281, EPI_ISL_802282, EPI_ISL_802283, EPI_ISL_802284, EPI_ISL_802285, EPI_ISL_802286, EPI_ISL_802287, EPI_ISL_802288, EPI_ISL_802289, EPI_ISL_802290, EPI_ISL_802291, EPI_ISL_802292, EPI_ISL_802293, EPI_ISL_802294, EPI_ISL_802295, EPI_ISL_802296, EPI_ISL_802297, EPI_ISL_802298, EPI_ISL_802299, EPI_ISL_802300, EPI_ISL_802301, EPI_ISL_802302, EPI_ISL_802303, EPI_ISL_802304, EPI_ISL_802305, EPI_ISL_802306, EPI_ISL_802307, EPI_ISL_802308, EPI_ISL_802309, EPI_ISL_802310, EPI_ISL_802311, EPI_ISL_802312, EPI_ISL_802313, EPI_ISL_802314, EPI_ISL_802315, EPI_ISL_802316, EPI_ISL_802317, EPI_ISL_802318, EPI_ISL_802319, EPI_ISL_802320, EPI_ISL_802321, EPI_ISL_802322, EPI_ISL_802323, EPI_ISL_802324, EPI_ISL_802325, EPI_ISL_802326, EPI_ISL_802327, EPI_ISL_802328, EPI_ISL_802329, EPI_ISL_802330, EPI_ISL_802331, EPI_ISL_802332, EPI_ISL_802333, EPI_ISL_802334, EPI_ISL_802335, EPI_ISL_802336, EPI_ISL_802337, EPI_ISL_802338, EPI_ISL_802339, EPI_ISL_802340, EPI_ISL_802341, EPI_ISL_802342, EPI_ISL_802343, EPI_ISL_802344, EPI_ISL_802345, EPI_ISL_802346, EPI_ISL_802347, EPI_ISL_802348, EPI_ISL_802349, EPI_ISL_802350, EPI_ISL_802351, EPI_ISL_802352, EPI_ISL_802353, EPI_ISL_802354, EPI_ISL_802355, EPI_ISL_802356, EPI_ISL_802357, EPI_ISL_802358, EPI_ISL_802359, EPI_ISL_802360, EPI_ISL_802361, EPI_ISL_802362, EPI_ISL_802363, EPI_ISL_802364, EPI_ISL_802365, EPI_ISL_802366, EPI_ISL_802367, EPI_ISL_802368, EPI_ISL_802369, EPI_ISL_802370, EPI_ISL_802371, EPI_ISL_802372, EPI_ISL_802373, EPI_ISL_802374, EPI_ISL_802375, EPI_ISL_802376, EPI_ISL_802377, EPI_ISL_802378, EPI_ISL_802379, EPI_ISL_802380, EPI_ISL_802381, EPI_ISL_802382, EPI_ISL_802383, EPI_ISL_802384, EPI_ISL_802385, EPI_ISL_802386, EPI_ISL_802387, EPI_ISL_802388, EPI_ISL_802389, EPI_ISL_802390, EPI_ISL_802391, EPI_ISL_802392, EPI_ISL_802393, EPI_ISL_802394, EPI_ISL_802395, EPI_ISL_802396, EPI_ISL_802397, EPI_ISL_802398, EPI_ISL_802399, EPI_ISL_802400, EPI_ISL_802401, EPI_ISL_802402, EPI_ISL_802403, EPI_ISL_802404, EPI_ISL_802405, EPI_ISL_802406, EPI_ISL_802407, EPI_ISL_802408, EPI_ISL_802409, EPI_ISL_802410, EPI_ISL_802411, EPI_ISL_802412, EPI_ISL_802413, EPI_ISL_802414, EPI_ISL_802415, EPI_ISL_802416, EPI_ISL_802417, EPI_ISL_802418, EPI_ISL_802419, EPI_ISL_802420, EPI_ISL_802421, EPI_ISL_802422, EPI_ISL_802423, EPI_ISL_802424, EPI_ISL_802425, EPI_ISL_802426, EPI_ISL_802427 | | | |
| see above | MSHS Clinical Microbiology Laboratories | MSHS Pathogen Surveillance Program | Ana S. Gonzalez-Reiche, Hala Alshammary, Mitchell J. Sullivan, Brianne Ciferri, Ajay Obla, Angela Amoako, Mahmoud Awawda, Elena Hirsch, Ashley S. Salimbangon, Levy Sominsky, Katherine Beach, Kayla Russo, Charles Gleason, Shelcie Fabre, Giulio Kleiner, Zenab Khan, Bremy Alburquerque, Adriana van de Guchte, Komal Srivastava, Matthew M. Hernandez, Jayeeta Dutta, Denise Jurczyszak, Emily Ferreri, Rachel Chernet, Nancy Francoeur, Betsaida Salom Melo, Irina Oussenko, Gintaras Deikus, Juan Soto, Shwetha Hara Sridhar, Ying-Chih Wang, Kathryn Twyman, Andrew Kasarskis, Deena R. Altman, Robert Sebra, Adolfo García-Sastre, Marta Luksza, Gopi Patel, Sarah Schaefer, Melissa Gitman, Michael D. Nowak, Alberto Paniz-Mondolfi, Emilia Mia Sordillo, Viviana Simon, Harm van Bakel |
| EPI_ISL_802428 | SARATOGA HOSPITAL LABORATORY | Wadsworth Center, New York State Department.of Health | Kirsten St. George, Daryl M. Lamson, Alexis Russel, Matthew Shudt, Melissa A Leisner, Jonathan Plitnick, Navjot Singh, John Kelly, Sara Griesemer, Erasmus Schneider, Erica Lasek-Nesselquist |

| | | | |
|---|---|---|---|
| EPI_ISL_802429 | Wadsworth Center, New York State Department.of Health | Wadsworth Center, New York State Department.of Health | Kirsten St. George, Daryl M. Lamson, Alexis Russel, Matthew Shudt, Melissa A Leisner, Jonathan Plitnick, Navjot Singh, John Kelly, Sara Griesemer, Erasmus Schneider, Erica Lasek-Nesselquist |
| EPI_ISL_802430 | BIO-REFERENCE LABORATORIES | Wadsworth Center, New York State Department.of Health | Kirsten St. George, Daryl M. Lamson, Alexis Russel, Matthew Shudt, Melissa A Leisner, Jonathan Plitnick, Navjot Singh, John Kelly, Sara Griesemer, Erasmus Schneider, Erica Lasek-Nesselquist |
| EPI_ISL_802431 | Wadsworth Center, New York State Department.of Health | Wadsworth Center, New York State Department.of Health | Kirsten St. George, Daryl M. Lamson, Alexis Russel, Matthew Shudt, Melissa A Leisner, Jonathan Plitnick, Navjot Singh, John Kelly, Sara Griesemer, Erasmus Schneider, Erica Lasek-Nesselquist |
| EPI_ISL_802432 | SARATOGA HOSPITAL LABORATORY | Wadsworth Center, New York State Department.of Health | Kirsten St. George, Daryl M. Lamson, Alexis Russel, Matthew Shudt, Melissa A Leisner, Jonathan Plitnick, Navjot Singh, John Kelly, Sara Griesemer, Erasmus Schneider, Erica Lasek-Nesselquist |
| EPI_ISL_802433 | Wadsworth Center, New York State Department.of Health | Wadsworth Center, New York State Department.of Health | Kirsten St. George, Daryl M. Lamson, Alexis Russel, Matthew Shudt, Melissa A Leisner, Jonathan Plitnick, Navjot Singh, John Kelly, Sara Griesemer, Erasmus Schneider, Erica Lasek-Nesselquist |
| EPI_ISL_802434, EPI_ISL_802435 | WESTCHESTER MEDICAL CENTER | Wadsworth Center, New York State Department.of Health | Kirsten St. George, Daryl M. Lamson, Alexis Russel, Matthew Shudt, Melissa A Leisner, Jonathan Plitnick, Navjot Singh, John Kelly, Sara Griesemer, Erasmus Schneider, Erica Lasek-Nesselquist |
| EPI_ISL_802436 | TEMPUS LABS INC | Wadsworth Center, New York State Department.of Health | Kirsten St. George, Daryl M. Lamson, Alexis Russel, Matthew Shudt, Melissa A Leisner, Jonathan Plitnick, Navjot Singh, John Kelly, Sara Griesemer, Erasmus Schneider, Erica Lasek-Nesselquist |
| EPI_ISL_802437 | SARATOGA HOSPITAL LABORATORY | Wadsworth Center, New York State Department.of Health | Kirsten St. George, Daryl M. Lamson, Alexis Russel, Matthew Shudt, Melissa A Leisner, Jonathan Plitnick, Navjot Singh, John Kelly, Sara Griesemer, Erasmus Schneider, Erica Lasek-Nesselquist |
| EPI_ISL_802438 | Wadsworth Center, New York State Department.of Health | Wadsworth Center, New York State Department.of Health | Kirsten St. George, Daryl M. Lamson, Alexis Russel, Matthew Shudt, Melissa A Leisner, Jonathan Plitnick, Navjot Singh, John Kelly, Sara Griesemer, Erasmus Schneider, Erica Lasek-Nesselquist |
| EPI_ISL_802439 | SARATOGA HOSPITAL LABORATORY | Wadsworth Center, New York State Department.of Health | Kirsten St. George, Daryl M. Lamson, Alexis Russel, Matthew Shudt, Melissa A Leisner, Jonathan Plitnick, Navjot Singh, John Kelly, Sara Griesemer, Erasmus Schneider, Erica Lasek-Nesselquist |
| EPI_ISL_802440, EPI_ISL_802441 | WESTCHESTER MEDICAL CENTER | Wadsworth Center, New York State Department.of Health | Kirsten St. George, Daryl M. Lamson, Alexis Russel, Matthew Shudt, Melissa A Leisner, Jonathan Plitnick, Navjot Singh, John Kelly, Sara Griesemer, Erasmus Schneider, Erica Lasek-Nesselquist |
| EPI_ISL_802442 | Wadsworth Center, New York State Department.of Health | Wadsworth Center, New York State Department.of Health | Kirsten St. George, Daryl M. Lamson, Alexis Russel, Matthew Shudt, Melissa A Leisner, Jonathan Plitnick, Navjot Singh, John Kelly, Sara Griesemer, Erasmus Schneider, Erica Lasek-Nesselquist |
| EPI_ISL_802443 | WESTCHESTER MEDICAL CENTER | Wadsworth Center, New York State Department.of Health | Kirsten St. George, Daryl M. Lamson, Alexis Russel, Matthew Shudt, Melissa A Leisner, Jonathan Plitnick, Navjot Singh, John Kelly, Sara Griesemer, Erasmus Schneider, Erica Lasek-Nesselquist |
| EPI_ISL_802444 | SUNRISE MEDICAL LABORATORIES | Wadsworth Center, New York State Department.of Health | Kirsten St. George, Daryl M. Lamson, Alexis Russel, Matthew Shudt, Melissa A Leisner, Jonathan Plitnick, Navjot Singh, John Kelly, Sara Griesemer, Erasmus Schneider, Erica Lasek-Nesselquist |
| EPI_ISL_802445, EPI_ISL_802446, EPI_ISL_802447 | BIO-REFERENCE LABORATORIES | Wadsworth Center, New York State Department.of Health | Kirsten St. George, Daryl M. Lamson, Alexis Russel, Matthew Shudt, Melissa A Leisner, Jonathan Plitnick, Navjot Singh, John Kelly, Sara Griesemer, Erasmus Schneider, Erica Lasek-Nesselquist |
| EPI_ISL_802448 | WESTCHESTER MEDICAL CENTER | Wadsworth Center, New York State Department.of Health | Kirsten St. George, Daryl M. Lamson, Alexis Russel, Matthew Shudt, Melissa A Leisner, Jonathan Plitnick, Navjot Singh, John Kelly, Sara Griesemer, Erasmus Schneider, Erica Lasek-Nesselquist |
| EPI_ISL_802449, EPI_ISL_802450, EPI_ISL_802451, EPI_ISL_802452, EPI_ISL_802453, EPI_ISL_802454, EPI_ISL_802455, EPI_ISL_802456, EPI_ISL_802457, EPI_ISL_802458, EPI_ISL_802459, EPI_ISL_802460, EPI_ISL_802461, EPI_ISL_802462, EPI_ISL_802463, EPI_ISL_802464, EPI_ISL_802465, EPI_ISL_802466 | | | |
| see above | Wadsworth Center, New York State Department.of Health | Wadsworth Center, New York State Department.of Health | Kirsten St. George, Daryl M. Lamson, Alexis Russel, Matthew Shudt, Melissa A Leisner, Jonathan Plitnick, Navjot Singh, John Kelly, Sara Griesemer, Erasmus Schneider, Erica Lasek-Nesselquist |
| EPI_ISL_802467, EPI_ISL_802468 | SARATOGA HOSPITAL LABORATORY | Wadsworth Center, New York State Department.of Health | Kirsten St. George, Daryl M. Lamson, Alexis Russel, Matthew Shudt, Melissa A Leisner, Jonathan Plitnick, Navjot Singh, John Kelly, Sara Griesemer, Erasmus Schneider, Erica Lasek-Nesselquist |
| EPI_ISL_802469, EPI_ISL_802470, EPI_ISL_802471, EPI_ISL_802472, EPI_ISL_802473, EPI_ISL_802474, EPI_ISL_802475, EPI_ISL_802476, EPI_ISL_802477, EPI_ISL_802478, EPI_ISL_802479, EPI_ISL_802480, EPI_ISL_802481, EPI_ISL_802482, EPI_ISL_802483, EPI_ISL_802484, EPI_ISL_802485, EPI_ISL_802486, EPI_ISL_802487, EPI_ISL_802488, EPI_ISL_802489, EPI_ISL_802490 | | | |
| see above | WESTCHESTER MEDICAL CENTER | Wadsworth Center, New York State Department.of Health | Kirsten St. George, Daryl M. Lamson, Alexis Russel, Matthew Shudt, Melissa A Leisner, Jonathan Plitnick, Navjot Singh, John Kelly, Sara Griesemer, Erasmus Schneider, Erica Lasek-Nesselquist |
| EPI_ISL_802491, EPI_ISL_802492, EPI_ISL_802493, EPI_ISL_802494, EPI_ISL_802495, EPI_ISL_802496, EPI_ISL_802497 | SARATOGA HOSPITAL LABORATORY | Wadsworth Center, New York State Department.of Health | Kirsten St. George, Daryl M. Lamson, Alexis Russel, Matthew Shudt, Melissa A Leisner, Jonathan Plitnick, Navjot Singh, John Kelly, Sara Griesemer, Erasmus Schneider, Erica Lasek-Nesselquist |
| EPI_ISL_802498, EPI_ISL_802499, EPI_ISL_802500 | Wadsworth Center, New York State Department.of Health | Wadsworth Center, New York State Department.of Health | Kirsten St. George, Daryl M. Lamson, Alexis Russel, Matthew Shudt, Melissa A Leisner, Jonathan Plitnick, Navjot Singh, John Kelly, Sara Griesemer, Erasmus Schneider, Erica Lasek-Nesselquist |
| EPI_ISL_802501 | Ohio Department of Health Laboratory | Ohio Department of Health Laboratory | Holmes, Jennifer; Eric Brandt, Keoni Omura, Glen McGillivary, Caitlin McDonnell, Kirtana Ramadugu, Erica Leasure, Kelsey Florek, Heather Blankenship, Quanta Brown, and Tammy Bannerman |
| EPI_ISL_802502, EPI_ISL_802503 | Connecticut Department of Health | Grubaugh Lab - Yale School of Public Health | Tara Alpert, Joseph Fauver, Anderson Brito, Mallery Breban, Anne Wyllie, Chantal Vogels, Mary Petrone, Chaney Kalinich, Isabel Ott, Nathan Grubaugh |
| EPI_ISL_802567, EPI_ISL_802568, EPI_ISL_802569, EPI_ISL_802570, EPI_ISL_802571 | Essentia Health-St. Mary's Medical Center | Minnesota Department of Health, Public Health Laboratory | Alexandra Lorentz, Jacob Garfin, Matt Plumb, and Xiong Wang |
| EPI_ISL_802572 | UMN Advanced Research and Diagnostics | Minnesota Department of Health, Public Health Laboratory | Alexandra Lorentz, Jacob Garfin, Matt Plumb, and Xiong Wang |
| EPI_ISL_802573 | M Health Fairview | Minnesota Department of Health, Public Health Laboratory | Alexandra Lorentz, Jacob Garfin, Matt Plumb, and Xiong Wang |
| EPI_ISL_802574, EPI_ISL_802575, EPI_ISL_802576, EPI_ISL_802577, EPI_ISL_802578, EPI_ISL_802579, EPI_ISL_802580, EPI_ISL_802581, EPI_ISL_802582, EPI_ISL_802583, EPI_ISL_802584, EPI_ISL_802585, EPI_ISL_802586, EPI_ISL_802587, EPI_ISL_802588, EPI_ISL_802589, EPI_ISL_802590, EPI_ISL_802591, EPI_ISL_802592, EPI_ISL_802593, EPI_ISL_802594, EPI_ISL_802595, EPI_ISL_802596, EPI_ISL_802597, EPI_ISL_802598 | | | |
| see above | Vault Health | Minnesota Department of Health, Public Health Laboratory | Alexandra Lorentz, Jacob Garfin, Matt Plumb, and Xiong Wang |
| EPI_ISL_802599, EPI_ISL_802600, EPI_ISL_802601, EPI_ISL_802602, EPI_ISL_802603, EPI_ISL_802604, EPI_ISL_802605, EPI_ISL_802606, EPI_ISL_802607, EPI_ISL_802608, EPI_ISL_802609, EPI_ISL_802610, EPI_ISL_802611, EPI_ISL_802612, EPI_ISL_802613, EPI_ISL_802614, EPI_ISL_802615, EPI_ISL_802616, EPI_ISL_802617, EPI_ISL_802618, EPI_ISL_802619, EPI_ISL_802620, EPI_ISL_802621, EPI_ISL_802622, EPI_ISL_802623, EPI_ISL_802624, EPI_ISL_802625, EPI_ISL_802626, EPI_ISL_802627, EPI_ISL_802628, EPI_ISL_802629, EPI_ISL_802630, EPI_ISL_802631, EPI_ISL_802632, EPI_ISL_802633, EPI_ISL_802634, EPI_ISL_802635, EPI_ISL_802636, EPI_ISL_802637, EPI_ISL_802638, EPI_ISL_802639, EPI_ISL_802640, EPI_ISL_802641, EPI_ISL_802642, EPI_ISL_802643, EPI_ISL_802644, EPI_ISL_802645, EPI_ISL_802646, EPI_ISL_802647, EPI_ISL_802648, EPI_ISL_802649, EPI_ISL_802650, EPI_ISL_802651, EPI_ISL_802654, EPI_ISL_802655, EPI_ISL_802656, EPI_ISL_802657, EPI_ISL_802658, EPI_ISL_802659, EPI_ISL_802660, EPI_ISL_802661, EPI_ISL_802662, EPI_ISL_802663, EPI_ISL_802664, EPI_ISL_802665, EPI_ISL_802666, EPI_ISL_802667, EPI_ISL_802668, EPI_ISL_802669, EPI_ISL_802670, EPI_ISL_802671, EPI_ISL_802672, EPI_ISL_802673, EPI_ISL_802674, EPI_ISL_802675, EPI_ISL_802676, EPI_ISL_802677, EPI_ISL_802678, EPI_ISL_802679, EPI_ISL_802680, EPI_ISL_802681, EPI_ISL_802682, EPI_ISL_802683, EPI_ISL_802684, EPI_ISL_802685, EPI_ISL_802686, EPI_ISL_802687, EPI_ISL_802688, EPI_ISL_802689, EPI_ISL_802690, EPI_ISL_802691, EPI_ISL_802692, EPI_ISL_802693, EPI_ISL_802694, EPI_ISL_802695, EPI_ISL_802696, EPI_ISL_802697, EPI_ISL_802698, EPI_ISL_802699, EPI_ISL_802700, EPI_ISL_802701, EPI_ISL_802702, EPI_ISL_802703, EPI_ISL_802704, EPI_ISL_802705, EPI_ISL_802706, EPI_ISL_802707, EPI_ISL_802708, EPI_ISL_802709, EPI_ISL_802710, EPI_ISL_802711, EPI_ISL_802712, EPI_ISL_802713, EPI_ISL_802714, EPI_ISL_802715, EPI_ISL_802716, EPI_ISL_802717, EPI_ISL_802718, EPI_ISL_802719, EPI_ISL_802720, EPI_ISL_802721, EPI_ISL_802722, EPI_ISL_802723 | | | |
| see above | Helix/Illumina | Genomics and Discovery, Respiratory Viruses Branch, Division of Viral Diseases, Centers for Disease Control and Prevention | Peter W. Cook, Dhwani Batra, Ben L. Rambo-Martin Eileen de Feo, Jan Antico, Christine Tran, Matthew Tolentino, Shannon Wickline, Kim Gietzen, Brad Sickler, Jingtao Liu, Eric Allen, Phil Febbo, Summer Galloway, Nicole L. Washington, Simon White, Geraint Levan, Kelly Schiabor Barrett, Elizabeth Cirulli, Alexandre Bolze, Ary Ascencio, Charlotte Rivera-Garcia, Ryan Cho, Jason Nguyen, Sherry Wang, Jimmy Ramirez, Tyler Cassens, Efren Sandoval, Magnus Isaksson, William Lee, David Becker, Marc Laurent, James Lu, Clinton R. Paden, Suxiang Tong, Duncan MacCannell |
| EPI_ISL_802725 | Florida Bureau of Public Health Laboratories | Florida Bureau of Public Health Laboratories | Sarah Schmedes, Jason Blanton |
| EPI_ISL_802777, EPI_ISL_802778, EPI_ISL_802779, EPI_ISL_802780, EPI_ISL_802781, EPI_ISL_802782, EPI_ISL_802783, EPI_ISL_802784, EPI_ISL_802785, EPI_ISL_802786, EPI_ISL_802787, EPI_ISL_802788, EPI_ISL_802789 | | | |
| see above | Columbia University Irving Medical Center | Wadsworth Center, New York State Department.of Health | Kirsten St. George, Daryl M. Lamson, Alexis Russel, Matthew Shudt, Melissa A Leisner, Jonathan Plitnick, Navjot Singh, John Kelly, Sara Griesemer, Erasmus Schneider, Erica Lasek-Nesselquist |
| EPI_ISL_802790, EPI_ISL_802791, EPI_ISL_802792, EPI_ISL_802793 | BIO-REFERENCE LABORATORIES | Wadsworth Center, New York State Department.of Health | Kirsten St. George, Daryl M. Lamson, Alexis Russel, Matthew Shudt, Melissa A Leisner, Jonathan Plitnick, Navjot Singh, John Kelly, Sara Griesemer, Erasmus Schneider, Erica Lasek-Nesselquist |
| EPI_ISL_802864, EPI_ISL_802865, EPI_ISL_802866, EPI_ISL_802867, EPI_ISL_802868, EPI_ISL_802869, EPI_ISL_802870, EPI_ISL_802871, EPI_ISL_802872, EPI_ISL_802873, EPI_ISL_802874, EPI_ISL_802875, EPI_ISL_802876, EPI_ISL_802877, EPI_ISL_802878, EPI_ISL_802879, EPI_ISL_802880, EPI_ISL_802881, EPI_ISL_802882, EPI_ISL_802883, EPI_ISL_802884, EPI_ISL_802885, EPI_ISL_802886, EPI_ISL_802887, EPI_ISL_802888, EPI_ISL_802889, EPI_ISL_802890, EPI_ISL_802891, EPI_ISL_802892, EPI_ISL_802893, EPI_ISL_802894, EPI_ISL_802895, EPI_ISL_802896, EPI_ISL_802918, EPI_ISL_802919, EPI_ISL_802920, EPI_ISL_802921, EPI_ISL_802922, EPI_ISL_802923, EPI_ISL_802924, EPI_ISL_802925, EPI_ISL_802926, EPI_ISL_802927, EPI_ISL_802928, EPI_ISL_802929, EPI_ISL_802930, EPI_ISL_802931, EPI_ISL_802932, EPI_ISL_802933, EPI_ISL_802934, EPI_ISL_802935, EPI_ISL_802936, EPI_ISL_802937, EPI_ISL_802938, EPI_ISL_802939, EPI_ISL_802940, EPI_ISL_802941, EPI_ISL_802942, EPI_ISL_802943, EPI_ISL_802944, EPI_ISL_802945, EPI_ISL_802946, EPI_ISL_802947, EPI_ISL_802948, EPI_ISL_802949, EPI_ISL_802950, EPI_ISL_802951, EPI_ISL_802952, EPI_ISL_802953, EPI_ISL_802954, EPI_ISL_802955, EPI_ISL_802956, EPI_ISL_802957, EPI_ISL_802958, EPI_ISL_802959, EPI_ISL_802981, EPI_ISL_802982, EPI_ISL_802983, EPI_ISL_802984, EPI_ISL_802985, EPI_ISL_802986, EPI_ISL_802987, EPI_ISL_802988, EPI_ISL_802989, EPI_ISL_802990, EPI_ISL_802991, EPI_ISL_802992, EPI_ISL_802993 | | | |
| see above | Utah Public Health Laboratory | Utah Public Health Laboratory | Erin Young, Kelly Oakeson, Tara Gallagher |
| EPI_ISL_802996, EPI_ISL_802997, EPI_ISL_802998, EPI_ISL_802999, EPI_ISL_803000, EPI_ISL_803001, EPI_ISL_803002, EPI_ISL_803003, EPI_ISL_803004, EPI_ISL_803005, EPI_ISL_803006, EPI_ISL_803007, EPI_ISL_803008, EPI_ISL_803009, EPI_ISL_803010, EPI_ISL_803011, EPI_ISL_803012, EPI_ISL_803013, EPI_ISL_803014, EPI_ISL_803015, EPI_ISL_803016, EPI_ISL_803017, EPI_ISL_803018, EPI_ISL_803019, EPI_ISL_803020, EPI_ISL_803021, EPI_ISL_803022, EPI_ISL_803023, EPI_ISL_803024, EPI_ISL_803025, EPI_ISL_803026, EPI_ISL_803027, EPI_ISL_803028, EPI_ISL_803029, EPI_ISL_803030, EPI_ISL_803031, EPI_ISL_803032, EPI_ISL_803033, EPI_ISL_803034, EPI_ISL_803035, EPI_ISL_803036, EPI_ISL_803037, EPI_ISL_803038, EPI_ISL_803039, EPI_ISL_803040, EPI_ISL_803041, EPI_ISL_803042, EPI_ISL_803043, EPI_ISL_803044, EPI_ISL_803045, EPI_ISL_803046, EPI_ISL_803047, EPI_ISL_803048, EPI_ISL_803049, EPI_ISL_803050, EPI_ISL_803051, EPI_ISL_803052, EPI_ISL_803053, EPI_ISL_803054, EPI_ISL_803055, EPI_ISL_803056, EPI_ISL_803057, EPI_ISL_803058, EPI_ISL_803059, EPI_ISL_803060, EPI_ISL_803061, EPI_ISL_803062, EPI_ISL_803063, EPI_ISL_803064, EPI_ISL_803065, EPI_ISL_803066, EPI_ISL_803067, | | | |

| | | | |
|---|---|---|---|
| EPI_ISL_803068, EPI_ISL_803069, EPI_ISL_803070, EPI_ISL_803071, EPI_ISL_803072, EPI_ISL_803073, EPI_ISL_803074, EPI_ISL_803075, EPI_ISL_803076, EPI_ISL_803077, EPI_ISL_803078, EPI_ISL_803079, EPI_ISL_803080, EPI_ISL_803081, EPI_ISL_803082, EPI_ISL_803083, EPI_ISL_803084, EPI_ISL_803085, EPI_ISL_803086, EPI_ISL_803087, EPI_ISL_803088, EPI_ISL_803089, EPI_ISL_803090, EPI_ISL_803091, EPI_ISL_803092, EPI_ISL_803093, EPI_ISL_803094, EPI_ISL_803095, EPI_ISL_803096, EPI_ISL_803097 | | | |
| see above | Quest Diagnostics | Quest Diagnostics | Rosenthal,S.H., Gerasimova,A., Kagan,R.M., Anderson, B., Livingston, K.E., Hua, M., Liu Y., Shalhout, D.F., Owen, R., Lacbawan, F. |
| EPI_ISL_803099, EPI_ISL_803100, EPI_ISL_803101, EPI_ISL_803102, EPI_ISL_803103, EPI_ISL_803104, EPI_ISL_803105, EPI_ISL_803106 | Respiratory Viruses Branch, Centers for Disease Control and Prevention | Respiratory Viruses Branch, Centers for Disease Control and Prevention | Queen,K., Li,Y., Tao,Y., Uehara,A., Montmayeur,A., Paden,C.R., Cook,P.W., Marine,R., Sheth,M., Wang,H., Lee,J., Tong,S. |
| EPI_ISL_803122, EPI_ISL_803123, EPI_ISL_803124, EPI_ISL_803125, EPI_ISL_803126, EPI_ISL_803127, EPI_ISL_803128, EPI_ISL_803129, EPI_ISL_803130, EPI_ISL_803131, EPI_ISL_803132, EPI_ISL_803133, EPI_ISL_803134, EPI_ISL_803135, EPI_ISL_803136, EPI_ISL_803137, EPI_ISL_803138, EPI_ISL_803139, EPI_ISL_803140, EPI_ISL_803141, EPI_ISL_803142, EPI_ISL_803143, EPI_ISL_803144, EPI_ISL_803145, EPI_ISL_803146, EPI_ISL_803147, EPI_ISL_803148, EPI_ISL_803149, EPI_ISL_803150, EPI_ISL_803151, EPI_ISL_803152, EPI_ISL_803153, EPI_ISL_803154, EPI_ISL_803155, EPI_ISL_803156, EPI_ISL_803157, EPI_ISL_803158, EPI_ISL_803159, EPI_ISL_803160, EPI_ISL_803161, EPI_ISL_803162, EPI_ISL_803163, EPI_ISL_803164, EPI_ISL_803165, EPI_ISL_803166, EPI_ISL_803167, EPI_ISL_803168, EPI_ISL_803169, EPI_ISL_803170, EPI_ISL_803171, EPI_ISL_803172, EPI_ISL_803173, EPI_ISL_803174, EPI_ISL_803175, EPI_ISL_803176, EPI_ISL_803177, EPI_ISL_803178, EPI_ISL_803179, EPI_ISL_803180, EPI_ISL_803181, EPI_ISL_803182, EPI_ISL_803183, EPI_ISL_803184, EPI_ISL_803185, EPI_ISL_803186, EPI_ISL_803187, EPI_ISL_803188, EPI_ISL_803189, EPI_ISL_803190, EPI_ISL_803191, EPI_ISL_803192, EPI_ISL_803193, EPI_ISL_803194, EPI_ISL_803195, EPI_ISL_803196, EPI_ISL_803197, EPI_ISL_803198, EPI_ISL_803199, EPI_ISL_803200, EPI_ISL_803201, EPI_ISL_803202, EPI_ISL_803203, EPI_ISL_803204, EPI_ISL_803205, EPI_ISL_803206, EPI_ISL_803207, EPI_ISL_803208, EPI_ISL_803209, EPI_ISL_803210, EPI_ISL_803211, EPI_ISL_803212, EPI_ISL_803213, EPI_ISL_803214, EPI_ISL_803215, EPI_ISL_803216, EPI_ISL_803217, EPI_ISL_803218, EPI_ISL_803219, EPI_ISL_803220, EPI_ISL_803221, EPI_ISL_803222, EPI_ISL_803223, EPI_ISL_803224, EPI_ISL_803225, EPI_ISL_803226, EPI_ISL_803227, EPI_ISL_803228, EPI_ISL_803229, EPI_ISL_803230, EPI_ISL_803231, EPI_ISL_803232, EPI_ISL_803233, EPI_ISL_803234, EPI_ISL_803235, EPI_ISL_803236, EPI_ISL_803237, EPI_ISL_803238, EPI_ISL_803239, EPI_ISL_803240, EPI_ISL_803241, EPI_ISL_803242, EPI_ISL_803243, EPI_ISL_803244, EPI_ISL_803245, EPI_ISL_803246, EPI_ISL_803247, EPI_ISL_803248, EPI_ISL_803249, EPI_ISL_803250, EPI_ISL_803251, EPI_ISL_803252, EPI_ISL_803253, EPI_ISL_803254, EPI_ISL_803255, EPI_ISL_803256, EPI_ISL_803257, EPI_ISL_803258, EPI_ISL_803259, EPI_ISL_803260, EPI_ISL_803261, EPI_ISL_803262, EPI_ISL_803263, EPI_ISL_803264, EPI_ISL_803265, EPI_ISL_803266, EPI_ISL_803267, EPI_ISL_803268, EPI_ISL_803269, EPI_ISL_803270, EPI_ISL_803271, EPI_ISL_803272, EPI_ISL_803273, EPI_ISL_803274, EPI_ISL_803275, EPI_ISL_803276, EPI_ISL_803277, EPI_ISL_803278, EPI_ISL_803279, EPI_ISL_803280, EPI_ISL_803281, EPI_ISL_803282, EPI_ISL_803283, EPI_ISL_803284, EPI_ISL_803285, EPI_ISL_803286, EPI_ISL_803287, EPI_ISL_803288, EPI_ISL_803289, EPI_ISL_803290, EPI_ISL_803291, EPI_ISL_803292, EPI_ISL_803293, EPI_ISL_803294, EPI_ISL_803295, EPI_ISL_803296, EPI_ISL_803297, EPI_ISL_803298, EPI_ISL_803299, EPI_ISL_803300, EPI_ISL_803301, EPI_ISL_803302, EPI_ISL_803303, EPI_ISL_803304, EPI_ISL_803305, EPI_ISL_803306, EPI_ISL_803307, EPI_ISL_803308, EPI_ISL_803309, EPI_ISL_803310, EPI_ISL_803311, EPI_ISL_803312, EPI_ISL_803313, EPI_ISL_803314, EPI_ISL_803315, EPI_ISL_803316, EPI_ISL_803317, EPI_ISL_803318, EPI_ISL_803319, EPI_ISL_803320, EPI_ISL_803321, EPI_ISL_803322, EPI_ISL_803323, EPI_ISL_803324, EPI_ISL_803325, EPI_ISL_803326, EPI_ISL_803327, EPI_ISL_803328, EPI_ISL_803329, EPI_ISL_803330, EPI_ISL_803331, EPI_ISL_803332, EPI_ISL_803333, EPI_ISL_803334, EPI_ISL_803335, EPI_ISL_803336, EPI_ISL_803337, EPI_ISL_803338, EPI_ISL_803339, EPI_ISL_803340, EPI_ISL_803341, EPI_ISL_803342, EPI_ISL_803343, EPI_ISL_803344, EPI_ISL_803345, EPI_ISL_803346, EPI_ISL_803347, EPI_ISL_803348, EPI_ISL_803349, EPI_ISL_803350, EPI_ISL_803351, EPI_ISL_803352, EPI_ISL_803353, EPI_ISL_803354, EPI_ISL_803355, EPI_ISL_803356, EPI_ISL_803357, EPI_ISL_803358, EPI_ISL_803359, EPI_ISL_803360, EPI_ISL_803361, EPI_ISL_803362, EPI_ISL_803363, EPI_ISL_803364, EPI_ISL_803365, EPI_ISL_803366, EPI_ISL_803367, EPI_ISL_803368, EPI_ISL_803369, EPI_ISL_803370, EPI_ISL_803371, EPI_ISL_803372, EPI_ISL_803373, EPI_ISL_803374, EPI_ISL_803375, EPI_ISL_803376, EPI_ISL_803377, EPI_ISL_803378, EPI_ISL_803379, EPI_ISL_803380, EPI_ISL_803381, EPI_ISL_803382, EPI_ISL_803383, EPI_ISL_803384, EPI_ISL_803385, EPI_ISL_803386, EPI_ISL_803387, EPI_ISL_803388, EPI_ISL_803389, EPI_ISL_803390, EPI_ISL_803391, EPI_ISL_803392, EPI_ISL_803393, EPI_ISL_803394, EPI_ISL_803395, EPI_ISL_803396, EPI_ISL_803397, EPI_ISL_803398, EPI_ISL_803399, EPI_ISL_803400, EPI_ISL_803401, EPI_ISL_803402, EPI_ISL_803403, EPI_ISL_803404, EPI_ISL_803405, EPI_ISL_803406, EPI_ISL_803407, EPI_ISL_803408, EPI_ISL_803409, EPI_ISL_803410, EPI_ISL_803411, EPI_ISL_803412, EPI_ISL_803413, EPI_ISL_803414, EPI_ISL_803415, EPI_ISL_803416, EPI_ISL_803417, EPI_ISL_803418, EPI_ISL_803419, EPI_ISL_803420, EPI_ISL_803421, EPI_ISL_803422, EPI_ISL_803423, EPI_ISL_803424, EPI_ISL_803425, EPI_ISL_803426, EPI_ISL_803427, EPI_ISL_803428, EPI_ISL_803429, EPI_ISL_803431, EPI_ISL_803432, EPI_ISL_803433, EPI_ISL_803434, EPI_ISL_803435, EPI_ISL_803436, EPI_ISL_803437, EPI_ISL_803438, EPI_ISL_803439, EPI_ISL_803440, EPI_ISL_803441, EPI_ISL_803442, EPI_ISL_803443, EPI_ISL_803444, EPI_ISL_803445, EPI_ISL_803446, EPI_ISL_803447, EPI_ISL_803448, EPI_ISL_803449, EPI_ISL_803450, EPI_ISL_803451, EPI_ISL_803452, EPI_ISL_803453, EPI_ISL_803454, EPI_ISL_803455, EPI_ISL_803456, EPI_ISL_803457, EPI_ISL_803458, EPI_ISL_803459, EPI_ISL_803460, EPI_ISL_803461, EPI_ISL_803462, EPI_ISL_803463, EPI_ISL_803464, EPI_ISL_803465, EPI_ISL_803466, EPI_ISL_803467, EPI_ISL_803468, EPI_ISL_803469, EPI_ISL_803470, EPI_ISL_803471, EPI_ISL_803472, EPI_ISL_803473, EPI_ISL_803474, EPI_ISL_803475, EPI_ISL_803476, EPI_ISL_803477, EPI_ISL_803478, EPI_ISL_803479, EPI_ISL_803480, EPI_ISL_803481, EPI_ISL_803482, EPI_ISL_803483, EPI_ISL_803484, EPI_ISL_803485, EPI_ISL_803486, EPI_ISL_803487, EPI_ISL_803488, EPI_ISL_803489, EPI_ISL_803490, EPI_ISL_803491, EPI_ISL_803492, EPI_ISL_803493, EPI_ISL_803494, EPI_ISL_803495, EPI_ISL_803496, EPI_ISL_803497, EPI_ISL_803498, EPI_ISL_803499, EPI_ISL_803500, EPI_ISL_803501, EPI_ISL_803502, EPI_ISL_803503, EPI_ISL_803504, EPI_ISL_803505, EPI_ISL_803506, EPI_ISL_803507, EPI_ISL_803508, EPI_ISL_803509, EPI_ISL_803510, EPI_ISL_803511, EPI_ISL_803512, EPI_ISL_803513, EPI_ISL_803514, EPI_ISL_803515, EPI_ISL_803516, EPI_ISL_803517, EPI_ISL_803518, EPI_ISL_803519, EPI_ISL_803520, EPI_ISL_803521, EPI_ISL_803522, EPI_ISL_803523, EPI_ISL_803524, EPI_ISL_803525, EPI_ISL_803526, EPI_ISL_803527, EPI_ISL_803528, EPI_ISL_803529, EPI_ISL_803530, EPI_ISL_803531, EPI_ISL_803532, EPI_ISL_803533, EPI_ISL_803534, EPI_ISL_803535, EPI_ISL_803536, EPI_ISL_803537, EPI_ISL_803538, EPI_ISL_803539, EPI_ISL_803540, EPI_ISL_803541, EPI_ISL_803542, EPI_ISL_803543, EPI_ISL_803544, EPI_ISL_803545, EPI_ISL_803546, EPI_ISL_803547, EPI_ISL_803548, EPI_ISL_803549, EPI_ISL_803550, EPI_ISL_803551, EPI_ISL_803552, EPI_ISL_803553, EPI_ISL_803554, EPI_ISL_803555, EPI_ISL_803556, EPI_ISL_803557, EPI_ISL_803558, EPI_ISL_803559, EPI_ISL_803560, EPI_ISL_803561, EPI_ISL_803562, EPI_ISL_803563, EPI_ISL_803564, EPI_ISL_803565, EPI_ISL_803566, EPI_ISL_803567, EPI_ISL_803568, EPI_ISL_803569, EPI_ISL_803570, EPI_ISL_803571, EPI_ISL_803572, EPI_ISL_803573, EPI_ISL_803574, EPI_ISL_803575, EPI_ISL_803576, EPI_ISL_803577, EPI_ISL_803578, EPI_ISL_803579, EPI_ISL_803580, EPI_ISL_803581, EPI_ISL_803582, EPI_ISL_803583, EPI_ISL_803584, EPI_ISL_803585, EPI_ISL_803586, EPI_ISL_803587, EPI_ISL_803588, EPI_ISL_803589, EPI_ISL_803590, EPI_ISL_803591, EPI_ISL_803592, EPI_ISL_803593, EPI_ISL_803594, EPI_ISL_803595, EPI_ISL_803596, EPI_ISL_803597, EPI_ISL_803598, EPI_ISL_803599, EPI_ISL_803600, EPI_ISL_803601, EPI_ISL_803602, EPI_ISL_803603, EPI_ISL_803604, EPI_ISL_803605, EPI_ISL_803606, EPI_ISL_803607, EPI_ISL_803608, EPI_ISL_803609, EPI_ISL_803610, EPI_ISL_803611, EPI_ISL_803612, EPI_ISL_803613, EPI_ISL_803614, EPI_ISL_803615, EPI_ISL_803616, EPI_ISL_803617, EPI_ISL_803618, EPI_ISL_803619, EPI_ISL_803620, EPI_ISL_803621, EPI_ISL_803622, EPI_ISL_803623, EPI_ISL_803624, EPI_ISL_803625, EPI_ISL_803626, EPI_ISL_803627, EPI_ISL_803628, EPI_ISL_803629, EPI_ISL_803630, EPI_ISL_803631, EPI_ISL_803632, EPI_ISL_803633, EPI_ISL_803634, EPI_ISL_803635, EPI_ISL_803636, EPI_ISL_803637, EPI_ISL_803638, EPI_ISL_803639, EPI_ISL_803640, EPI_ISL_803641, EPI_ISL_803642, EPI_ISL_803643, EPI_ISL_803644, EPI_ISL_803645, EPI_ISL_803646, EPI_ISL_803647, EPI_ISL_803648, EPI_ISL_803649, EPI_ISL_803650, EPI_ISL_803651, EPI_ISL_803652, EPI_ISL_803653, EPI_ISL_803654, EPI_ISL_803655, EPI_ISL_803656, EPI_ISL_803657, EPI_ISL_803658, EPI_ISL_803659, EPI_ISL_803660, EPI_ISL_803661, EPI_ISL_803662, EPI_ISL_803663, EPI_ISL_803664, EPI_ISL_803665, EPI_ISL_803666, EPI_ISL_803667, EPI_ISL_803668, EPI_ISL_803669, EPI_ISL_803670, EPI_ISL_803671, EPI_ISL_803672, EPI_ISL_803673, EPI_ISL_803674, EPI_ISL_803675, EPI_ISL_803676, EPI_ISL_803677, EPI_ISL_803678, EPI_ISL_803679, EPI_ISL_803680, EPI_ISL_803681, EPI_ISL_803682, EPI_ISL_803683, EPI_ISL_803684, EPI_ISL_803685, EPI_ISL_803686, EPI_ISL_803687, EPI_ISL_803688, EPI_ISL_803689, EPI_ISL_803690, EPI_ISL_803691, EPI_ISL_803692, EPI_ISL_803693, EPI_ISL_803694, EPI_ISL_803695, EPI_ISL_803696, EPI_ISL_803697, EPI_ISL_803698, EPI_ISL_803699, EPI_ISL_803700, EPI_ISL_803701, EPI_ISL_803702, EPI_ISL_803703, EPI_ISL_803704, EPI_ISL_803705, EPI_ISL_803706, EPI_ISL_803707, EPI_ISL_803708, EPI_ISL_803709, EPI_ISL_803710, EPI_ISL_803711, EPI_ISL_803712, EPI_ISL_803713, EPI_ISL_803714, EPI_ISL_803715, EPI_ISL_803716, EPI_ISL_803717, EPI_ISL_803718, EPI_ISL_803719, EPI_ISL_803720, EPI_ISL_803721, EPI_ISL_803722, EPI_ISL_803723, EPI_ISL_803724, EPI_ISL_803725, EPI_ISL_803726, EPI_ISL_803727, EPI_ISL_803728, EPI_ISL_803729, EPI_ISL_803730, EPI_ISL_803731, EPI_ISL_803732, EPI_ISL_803733, EPI_ISL_803734, EPI_ISL_803735, EPI_ISL_803736, EPI_ISL_803737, EPI_ISL_803738, EPI_ISL_803739, EPI_ISL_803740, EPI_ISL_803741, EPI_ISL_803742, EPI_ISL_803743, EPI_ISL_803744, EPI_ISL_803745, EPI_ISL_803746, EPI_ISL_803747, EPI_ISL_803748, EPI_ISL_803749, EPI_ISL_803750, EPI_ISL_803751, EPI_ISL_803752, EPI_ISL_803753, EPI_ISL_803754, EPI_ISL_803755, EPI_ISL_803756, EPI_ISL_803757, EPI_ISL_803758, EPI_ISL_803759, EPI_ISL_803760, EPI_ISL_803761, EPI_ISL_803762, EPI_ISL_803763, EPI_ISL_803764, EPI_ISL_803765, EPI_ISL_803766, EPI_ISL_803767, EPI_ISL_803768, EPI_ISL_803769, EPI_ISL_803770, EPI_ISL_803771, EPI_ISL_803772, EPI_ISL_803773, EPI_ISL_803774, EPI_ISL_803775, EPI_ISL_803776, EPI_ISL_803777, EPI_ISL_803778, EPI_ISL_803779, EPI_ISL_803780, EPI_ISL_803781, EPI_ISL_803782, EPI_ISL_803783, EPI_ISL_803784, EPI_ISL_803785, EPI_ISL_803786, EPI_ISL_803787, EPI_ISL_803788, EPI_ISL_803789, EPI_ISL_803790, EPI_ISL_803791, EPI_ISL_803792, EPI_ISL_803793, EPI_ISL_803794, EPI_ISL_803795, EPI_ISL_803796, EPI_ISL_803797, EPI_ISL_803798, EPI_ISL_803799, EPI_ISL_803800, EPI_ISL_803801, EPI_ISL_803802, EPI_ISL_803803, EPI_ISL_803804, EPI_ISL_803805, EPI_ISL_803806, EPI_ISL_803807, EPI_ISL_803808, EPI_ISL_803809, EPI_ISL_803810, EPI_ISL_803811, EPI_ISL_803812, EPI_ISL_803813, EPI_ISL_803814, EPI_ISL_803815, EPI_ISL_803816, EPI_ISL_803818, EPI_ISL_803819, EPI_ISL_803820, EPI_ISL_803821, EPI_ISL_803822, EPI_ISL_803823, EPI_ISL_803824, EPI_ISL_803825, EPI_ISL_803826, EPI_ISL_803827, EPI_ISL_803828, EPI_ISL_803829, EPI_ISL_803830, EPI_ISL_803831, EPI_ISL_803832, EPI_ISL_803833, EPI_ISL_803834, EPI_ISL_803835, EPI_ISL_803836, EPI_ISL_803837, EPI_ISL_803838, EPI_ISL_803839, EPI_ISL_803840, EPI_ISL_803841, EPI_ISL_803842, EPI_ISL_803843, EPI_ISL_803844, EPI_ISL_803845, EPI_ISL_803846, EPI_ISL_803847, EPI_ISL_803848, EPI_ISL_803849 | | | |
| see above | Wisconsin State Laboratory of Hygiene Communicable Disease Division | Wisconsin State Laboratory of Hygiene Communicable Disease Division | Kelsey R. Florek, Abigail C. Shockey |
| EPI_ISL_804019, EPI_ISL_804020, EPI_ISL_804021, EPI_ISL_804022, EPI_ISL_804023, EPI_ISL_804024, EPI_ISL_804025, EPI_ISL_804026, EPI_ISL_804027, EPI_ISL_804028, EPI_ISL_804029, EPI_ISL_804030, EPI_ISL_804031, EPI_ISL_804032, EPI_ISL_804033, EPI_ISL_804034, EPI_ISL_804035, EPI_ISL_804036, EPI_ISL_804463, EPI_ISL_804464, EPI_ISL_804465, EPI_ISL_804592, EPI_ISL_804593, EPI_ISL_804594, EPI_ISL_804595, EPI_ISL_804596, EPI_ISL_804597, EPI_ISL_804598 | | | |
| see above | Maryland Public Health Laboratory | Maryland Public Health Laboratory | Maryland Department of Health Laboratories Administration |
| EPI_ISL_804599, EPI_ISL_804600, EPI_ISL_804601, EPI_ISL_804602, EPI_ISL_804603, EPI_ISL_804604, EPI_ISL_804605, EPI_ISL_804606, EPI_ISL_804607, EPI_ISL_804608, EPI_ISL_804609, EPI_ISL_804610, EPI_ISL_804611, EPI_ISL_804612, EPI_ISL_804613, EPI_ISL_804614, EPI_ISL_804615, EPI_ISL_804616, EPI_ISL_804617, EPI_ISL_804618, EPI_ISL_804619, EPI_ISL_804620, EPI_ISL_804621, EPI_ISL_804622, EPI_ISL_804623, EPI_ISL_804624, EPI_ISL_804625, EPI_ISL_804626, EPI_ISL_804627, EPI_ISL_804628, EPI_ISL_804629, EPI_ISL_804630, EPI_ISL_804631, EPI_ISL_804632, EPI_ISL_804633, EPI_ISL_804634, EPI_ISL_804635, EPI_ISL_804636, EPI_ISL_804637, EPI_ISL_804638, EPI_ISL_804639, EPI_ISL_804640, EPI_ISL_804641, EPI_ISL_804642, EPI_ISL_804643, EPI_ISL_804644, EPI_ISL_804645, EPI_ISL_804646, EPI_ISL_804647, EPI_ISL_804648, EPI_ISL_804649, EPI_ISL_804650, EPI_ISL_804651, EPI_ISL_804652, EPI_ISL_804653, EPI_ISL_804654, EPI_ISL_804655, EPI_ISL_804656, EPI_ISL_804657, EPI_ISL_804658, EPI_ISL_804659, EPI_ISL_804660, EPI_ISL_804661, EPI_ISL_804662, EPI_ISL_804663, EPI_ISL_804664, EPI_ISL_804665, EPI_ISL_804666, EPI_ISL_804667, EPI_ISL_804668, EPI_ISL_804669, EPI_ISL_804670, EPI_ISL_804671, EPI_ISL_804672, EPI_ISL_804673, EPI_ISL_804674, EPI_ISL_804675, EPI_ISL_804676, EPI_ISL_804677, EPI_ISL_804678, EPI_ISL_804679, EPI_ISL_804680, EPI_ISL_804681, EPI_ISL_804682, EPI_ISL_804683, EPI_ISL_804684, EPI_ISL_804685, EPI_ISL_804686, EPI_ISL_804687, EPI_ISL_804688, EPI_ISL_804689, EPI_ISL_804690, EPI_ISL_804691, EPI_ISL_804692, EPI_ISL_804693, EPI_ISL_804694, EPI_ISL_804695, EPI_ISL_804696, EPI_ISL_804706, EPI_ISL_804707, EPI_ISL_804708, EPI_ISL_804709, EPI_ISL_804710, EPI_ISL_804711, EPI_ISL_804712, EPI_ISL_804713, EPI_ISL_804714, EPI_ISL_804715, EPI_ISL_804716, EPI_ISL_804717, EPI_ISL_804718, EPI_ISL_804719, EPI_ISL_804720, EPI_ISL_804721, EPI_ISL_804722, EPI_ISL_804723, EPI_ISL_804724, EPI_ISL_804725, EPI_ISL_804726, EPI_ISL_804727, EPI_ISL_804728, EPI_ISL_804729, EPI_ISL_804730, EPI_ISL_804731, EPI_ISL_804732, EPI_ISL_804733, EPI_ISL_804734, EPI_ISL_804735, EPI_ISL_804736, EPI_ISL_804737, EPI_ISL_804738, EPI_ISL_804739, EPI_ISL_804740, EPI_ISL_804741, EPI_ISL_804742, EPI_ISL_804743, EPI_ISL_804744, EPI_ISL_804745, EPI_ISL_804746, EPI_ISL_804747, EPI_ISL_804748, EPI_ISL_804749, EPI_ISL_804750, EPI_ISL_804751, EPI_ISL_804752, EPI_ISL_804753, EPI_ISL_804754, EPI_ISL_804755, EPI_ISL_804756, EPI_ISL_804757, EPI_ISL_804758, EPI_ISL_804759, EPI_ISL_804760, EPI_ISL_804761, EPI_ISL_804762, EPI_ISL_804763, EPI_ISL_804764, EPI_ISL_804765, EPI_ISL_804766, EPI_ISL_804767, EPI_ISL_804768, EPI_ISL_804769, EPI_ISL_804770, EPI_ISL_804771, EPI_ISL_804772, EPI_ISL_804773, EPI_ISL_804774, EPI_ISL_804775, EPI_ISL_804776, EPI_ISL_804777, EPI_ISL_804778, EPI_ISL_804779, EPI_ISL_804780, EPI_ISL_804781, EPI_ISL_804782, EPI_ISL_804783, EPI_ISL_804784, EPI_ISL_804785, EPI_ISL_804786, EPI_ISL_804787, EPI_ISL_804788, EPI_ISL_804789, EPI_ISL_804790, EPI_ISL_804791, EPI_ISL_804792, EPI_ISL_804793, EPI_ISL_804794, EPI_ISL_804795, EPI_ISL_804796, EPI_ISL_804797, EPI_ISL_804798, EPI_ISL_804799, EPI_ISL_804800, EPI_ISL_804801, EPI_ISL_804802, EPI_ISL_804803, EPI_ISL_804804, EPI_ISL_804805, EPI_ISL_804806, EPI_ISL_804807, EPI_ISL_804808, EPI_ISL_804809, EPI_ISL_804810, EPI_ISL_804811 | | | |
| see above | Michigan Department of Health and Human Services, Bureau of Laboratories | Michigan Department of Health and Human Services, Bureau of Laboratories | Blankenship HM, Riner D, Soehnlen MK |
| EPI_ISL_804845, EPI_ISL_804846, EPI_ISL_804847, EPI_ISL_804848, EPI_ISL_804849, EPI_ISL_804850, EPI_ISL_804851, EPI_ISL_804852, EPI_ISL_804853, EPI_ISL_804854, EPI_ISL_804855, EPI_ISL_804856, EPI_ISL_804857, EPI_ISL_804858, EPI_ISL_804859, EPI_ISL_804860, EPI_ISL_804861, EPI_ISL_804862, EPI_ISL_804863, EPI_ISL_804864, EPI_ISL_804865, EPI_ISL_804866, EPI_ISL_804867, EPI_ISL_804868, EPI_ISL_804869, EPI_ISL_804870, EPI_ISL_804871, EPI_ISL_804872, EPI_ISL_804873, EPI_ISL_804874, EPI_ISL_804875, EPI_ISL_804876, EPI_ISL_804877, EPI_ISL_804878, EPI_ISL_804879, EPI_ISL_804880, EPI_ISL_804894, EPI_ISL_804895, EPI_ISL_804896, EPI_ISL_804897, EPI_ISL_804898, EPI_ISL_804899, EPI_ISL_804900, EPI_ISL_804901, EPI_ISL_804902, EPI_ISL_804903, EPI_ISL_804904, EPI_ISL_804905, EPI_ISL_804906, EPI_ISL_804907, EPI_ISL_804908, EPI_ISL_804909, EPI_ISL_804910, EPI_ISL_804911, EPI_ISL_804912, EPI_ISL_804913, EPI_ISL_804914, EPI_ISL_804915, EPI_ISL_804916, EPI_ISL_804917, EPI_ISL_804918, EPI_ISL_804919, EPI_ISL_804920, EPI_ISL_804921, EPI_ISL_804922, EPI_ISL_804923, EPI_ISL_804924, EPI_ISL_804925, EPI_ISL_804926, EPI_ISL_804927, EPI_ISL_804928, EPI_ISL_804929, EPI_ISL_804930, EPI_ISL_804931, EPI_ISL_804932, EPI_ISL_804933, EPI_ISL_804934, EPI_ISL_804935, EPI_ISL_804936, EPI_ISL_804937, EPI_ISL_804938, EPI_ISL_804939, EPI_ISL_804940, EPI_ISL_804941, EPI_ISL_804942, EPI_ISL_804943, EPI_ISL_804944 | | | |
| see above | DC Public Health Lab/ Dept. of Forensic Sciences | DC Public Health Lab/ Dept. of Forensic Sciences | Scott Nguyen, Elizabeth Zelaya, Connie Maza, Monica Mann, Brittany Hamilton, David Payne, Jocelyn Hauser |
| EPI_ISL_804945 | Wadsworth Center, New York State Department.of Health | Wadsworth Center, New York State Department.of Health | Kirsten St. George, Daryl M. Lamson, Alexis Russel, Matthew Shudt, Melissa A Leisner, Jonathan Plitnick, Navjot Singh, John Kelly, Sara Griesemer, Erasmus Schneider, Erica Lasek-Nesselquist |
| EPI_ISL_804946 | BIO-REFERENCE LABORATORIES | Wadsworth Center, New York State Department.of Health | Kirsten St. George, Daryl M. Lamson, Alexis Russel, Matthew Shudt, Melissa A Leisner, Jonathan Plitnick, Navjot Singh, John Kelly, Sara Griesemer, Erasmus Schneider, Erica Lasek-Nesselquist |
| EPI_ISL_804947, EPI_ISL_804948 | Wadsworth Center, New York State Department.of Health | Wadsworth Center, New York State Department.of Health | Kirsten St. George, Daryl M. Lamson, Alexis Russel, Matthew Shudt, Melissa A Leisner, Jonathan Plitnick, Navjot Singh, John Kelly, Sara Griesemer, Erasmus Schneider, Erica Lasek-Nesselquist |
| EPI_ISL_804980 | Columbia University Irving Medical Center | Wadsworth Center, New York State Department of Health | Kirsten St. George, Daryl M. Lamson, Alexis Russel, Matthew Shudt, Melissa A Leisner, Jonathan Plitnick, Navjot Singh, John Kelly, Erasmus Schneider, Erica Lasek-Nesselquist |
| EPI_ISL_804981, EPI_ISL_804982, EPI_ISL_804983, EPI_ISL_804984 | BIO-REFERENCE LABORATORIES | Wadsworth Center, New York State Department of Health | Kirsten St. George, Daryl M. Lamson, Alexis Russel, Matthew Shudt, Melissa A Leisner, Jonathan Plitnick, Navjot Singh, John Kelly, Erasmus Schneider, Erica Lasek-Nesselquist |
| EPI_ISL_806543 | Santa Clara County Public Health Laboratory | Santa Clara County Public Health Laboratory | Santa Clara Co Public Health Department |
| EPI_ISL_806545 | Santa Clara County Public Health Laboratory | Santa Clara County Public Health Laboratory | Santa CLara County Public Health Department |
| EPI_ISL_806807, EPI_ISL_806808 | Gundersen Molecular Diagnostics Laboratory | Kabara Cancer Research Institute | Craig S. Richmond, Paraic A. Kenny |
| EPI_ISL_806809 | Seattle Flu Study | Seattle Flu Study | Deborah A. Nickerson, Chris D. Frazar, Jover Lee, Benjamin Pelle, Matthew Richardson, Amanda Adler, Elisabeth Brandstetter, Peter D. Han, Kairsten Fay, Misja Ilcisin, Kirsten Lacombe, Thomas R. Sibley, Melissa Truong, Caitlin R. Wolf, Michael Boeckh, Janet A. Englund, Michael Famulare, Barry R. Lutz, Mark J. Rieder, Lea M. Starita, Matthew Thompson, Jay Shendure, Trevor Bedford, Helen Y. Chu |
| EPI_ISL_806811, EPI_ISL_806814, EPI_ISL_806821, EPI_ISL_806822, EPI_ISL_806823, EPI_ISL_806824, EPI_ISL_806828, EPI_ISL_806830, EPI_ISL_806832, EPI_ISL_806834, EPI_ISL_806836, EPI_ISL_806837, EPI_ISL_806838, EPI_ISL_806839, EPI_ISL_806843, EPI_ISL_806845, EPI_ISL_806850, EPI_ISL_806852 | | | |
| see above | Alaska State Virology Laboratory | Alaska State Virology Laboratory | Stephanie DeRonde, Lisa Smith, Ph.D., Devin M. Drown, Ph.D., Jack Chen, Ph.D. |
| EPI_ISL_806853, EPI_ISL_806854, EPI_ISL_806855, EPI_ISL_806856, EPI_ISL_806857, EPI_ISL_806858, EPI_ISL_806859, EPI_ISL_806860, EPI_ISL_806861, EPI_ISL_806862, EPI_ISL_806863, EPI_ISL_806864, EPI_ISL_806865, EPI_ISL_806866, EPI_ISL_806867, EPI_ISL_806868, EPI_ISL_806869, EPI_ISL_806870, EPI_ISL_806871, EPI_ISL_806872, EPI_ISL_806873, EPI_ISL_806874, EPI_ISL_806875, EPI_ISL_806876, EPI_ISL_806877, EPI_ISL_806878, EPI_ISL_806879, EPI_ISL_806880, EPI_ISL_806881, EPI_ISL_806882, EPI_ISL_806883, EPI_ISL_806884, EPI_ISL_806885, EPI_ISL_806886, EPI_ISL_806887, EPI_ISL_806888, EPI_ISL_806889, EPI_ISL_806890, EPI_ISL_806891, EPI_ISL_806892, EPI_ISL_806893, EPI_ISL_806894, EPI_ISL_806895, EPI_ISL_806896, EPI_ISL_806897, EPI_ISL_806898, EPI_ISL_806899, EPI_ISL_806900, EPI_ISL_806901, EPI_ISL_806902, EPI_ISL_806903, EPI_ISL_806904, EPI_ISL_806905, EPI_ISL_806906, EPI_ISL_806907, EPI_ISL_806908, EPI_ISL_806909, EPI_ISL_806910, EPI_ISL_806911, EPI_ISL_806912, EPI_ISL_806913, EPI_ISL_806914, EPI_ISL_806915, EPI_ISL_806916, EPI_ISL_806917, EPI_ISL_806918, EPI_ISL_806919, EPI_ISL_806920, EPI_ISL_806921, EPI_ISL_806922, EPI_ISL_806923, EPI_ISL_806924, EPI_ISL_806925, EPI_ISL_806926, EPI_ISL_806927, EPI_ISL_806928, EPI_ISL_806929, EPI_ISL_806930, EPI_ISL_806931, EPI_ISL_806932, EPI_ISL_806933, EPI_ISL_806934, EPI_ISL_806935, EPI_ISL_806936, EPI_ISL_806937, EPI_ISL_806938, EPI_ISL_806939, EPI_ISL_806940, EPI_ISL_806941, EPI_ISL_806942, EPI_ISL_806943, EPI_ISL_806944, EPI_ISL_806945, EPI_ISL_806946, EPI_ISL_806947, EPI_ISL_806948, EPI_ISL_806949, EPI_ISL_806950, EPI_ISL_806951, EPI_ISL_806952, EPI_ISL_806953, EPI_ISL_806954, EPI_ISL_806955, EPI_ISL_806956, EPI_ISL_806957, EPI_ISL_806958, EPI_ISL_806959, EPI_ISL_806960, EPI_ISL_806961, EPI_ISL_806962, EPI_ISL_806963, EPI_ISL_806964, EPI_ISL_806965, EPI_ISL_806966, EPI_ISL_806967, EPI_ISL_806968, EPI_ISL_806969, EPI_ISL_806970, EPI_ISL_806971, EPI_ISL_806972, EPI_ISL_806973, EPI_ISL_806974, EPI_ISL_806975, EPI_ISL_806976, EPI_ISL_806977, EPI_ISL_806978, EPI_ISL_806980, EPI_ISL_806981, EPI_ISL_806982, EPI_ISL_806983, EPI_ISL_806984, EPI_ISL_806985, EPI_ISL_806986, EPI_ISL_806987, EPI_ISL_806988, EPI_ISL_806989, EPI_ISL_806990, EPI_ISL_806991, EPI_ISL_806992, EPI_ISL_806993, EPI_ISL_806994, EPI_ISL_806995, EPI_ISL_806996, EPI_ISL_806997, EPI_ISL_806999, EPI_ISL_807000, EPI_ISL_807001, EPI_ISL_807002, EPI_ISL_807003, EPI_ISL_807004, EPI_ISL_807005, EPI_ISL_807006, EPI_ISL_807007, EPI_ISL_807008, EPI_ISL_807010, EPI_ISL_807011, EPI_ISL_807012, EPI_ISL_807014, EPI_ISL_807015, EPI_ISL_807016, EPI_ISL_807018, EPI_ISL_807019, EPI_ISL_807020, EPI_ISL_807021, EPI_ISL_807022, EPI_ISL_807023, EPI_ISL_807024, EPI_ISL_807025, EPI_ISL_807026, EPI_ISL_807027, EPI_ISL_807028, EPI_ISL_807029, EPI_ISL_807030, EPI_ISL_807031, EPI_ISL_807032, EPI_ISL_807033, EPI_ISL_807034, EPI_ISL_807035, EPI_ISL_807036, EPI_ISL_807037, EPI_ISL_807038, EPI_ISL_807039, EPI_ISL_807040, EPI_ISL_807041, EPI_ISL_807042, EPI_ISL_807043, EPI_ISL_807044, EPI_ISL_807045, EPI_ISL_807046, EPI_ISL_807047, EPI_ISL_807049, EPI_ISL_807050, EPI_ISL_807051, EPI_ISL_807052, EPI_ISL_807054, EPI_ISL_807055, EPI_ISL_807056, EPI_ISL_807057, EPI_ISL_807058, EPI_ISL_807059, EPI_ISL_807060, EPI_ISL_807061, EPI_ISL_807063, EPI_ISL_807064, EPI_ISL_807065, EPI_ISL_807066, EPI_ISL_807067, EPI_ISL_807068, EPI_ISL_807069, EPI_ISL_807071, EPI_ISL_807072, EPI_ISL_807074, EPI_ISL_807075, EPI_ISL_807077, EPI_ISL_807078, EPI_ISL_807079, EPI_ISL_807080, EPI_ISL_807081, EPI_ISL_807082, EPI_ISL_807083, EPI_ISL_807084, EPI_ISL_807085, EPI_ISL_807086, EPI_ISL_807087, EPI_ISL_807089, EPI_ISL_807090, EPI_ISL_807091, EPI_ISL_807092, EPI_ISL_807093, EPI_ISL_807094, EPI_ISL_807095, EPI_ISL_807096, EPI_ISL_807097, EPI_ISL_807098, EPI_ISL_807099, EPI_ISL_807100, EPI_ISL_807101, EPI_ISL_807102, EPI_ISL_807103, EPI_ISL_807104, EPI_ISL_807105, EPI_ISL_807106, EPI_ISL_807107, EPI_ISL_807108, EPI_ISL_807109, EPI_ISL_807110, EPI_ISL_807111, EPI_ISL_807112, EPI_ISL_807113, EPI_ISL_807114, EPI_ISL_807115, EPI_ISL_807116, EPI_ISL_807117, EPI_ISL_807118, EPI_ISL_807119, EPI_ISL_807120, EPI_ISL_807121, EPI_ISL_807122, EPI_ISL_807123, EPI_ISL_807124, EPI_ISL_807125, EPI_ISL_807126, EPI_ISL_807127, EPI_ISL_807128, EPI_ISL_807129, EPI_ISL_807130, EPI_ISL_807131, EPI_ISL_807132, EPI_ISL_807133, EPI_ISL_807134, EPI_ISL_807135, EPI_ISL_807137, EPI_ISL_807138, EPI_ISL_807139, EPI_ISL_807140, EPI_ISL_807141, EPI_ISL_807144, EPI_ISL_807145, EPI_ISL_807146, EPI_ISL_807147 | | | |
| see above | Washington State Department of Health | Seattle Flu Study | Deborah A. Nickerson, Chris D. Frazar, Jover Lee, Benjamin Pelle, Matthew Richardson, Amanda Adler, Elisabeth Brandstetter, Peter D. Han, Kairsten Fay, Misja Ilcisin, Kirsten Lacombe, Thomas R. Sibley, Melissa Truong, Caitlin R. Wolf, Romesh Gautom, Geoff Melly, Brian Hiatt, Philip Dykema, Scott Lindquist, Michael Boeckh, Janet A. Englund, Michael Famulare, Barry R. Lutz, Mark J. Rieder, Lea M. Starita, Matthew Thompson, Helen Y. Chu, Jay Shendure, Trevor Bedford |

| | | | |
|---|---|---|---|
| EPI_ISL_812114 | Arizona State University | Arizona State University | LaRinda A. Holland, Peter T. Skidmore, Emily A. Kaelin, Nicholas J. Mellor, Kristina Buss, Joy M. Blain, Valerie Harris, Joshua LaBaer, Vel Murugan, Efrem S. Lim |
| EPI_ISL_812115, EPI_ISL_812116, EPI_ISL_812119, EPI_ISL_812120, EPI_ISL_812121 | Santa Clara County Public Health Laboratory | Santa Clara County Public Health Laboratory | Santa Clara County Public Health Department |
| EPI_ISL_812131 | GA Department of Public Health Laboratory | Pathogen Discovery, Respiratory Viruses Branch, Division of Viral Diseases, Centers for Disease Control and Prevention | Yan Li, Ying Tao, Anna Montmayeur, Jing Zhang, Brian Lynch, Krista Queen, Anna Uehara, Rachel Marine, Peter Cook, Clinton R. Paden, Haibin Wang, Suxiang Tong |
| EPI_ISL_812132 | TN Division of Laboratory Services | Pathogen Discovery, Respiratory Viruses Branch, Division of Viral Diseases, Centers for Disease Control and Prevention | Yan Li, Ying Tao, Anna Montmayeur, Jing Zhang, Brian Lynch, Krista Queen, Anna Uehara, Rachel Marine, Peter Cook, Clinton R. Paden, Haibin Wang, Suxiang Tong |
| EPI_ISL_812138 | FL Bureau of Public Health Laboratories-Tampa | Pathogen Discovery, Respiratory Viruses Branch, Division of Viral Diseases, Centers for Disease Control and Prevention | Yan Li, Ying Tao, Anna Montmayeur, Jing Zhang, Brian Lynch, Krista Queen, Anna Uehara, Rachel Marine, Peter Cook, Clinton R. Paden, Haibin Wang, Suxiang Tong |
| EPI_ISL_812139 | TN Division of Laboratory Services | Pathogen Discovery, Respiratory Viruses Branch, Division of Viral Diseases, Centers for Disease Control and Prevention | Yan Li, Ying Tao, Anna Montmayeur, Jing Zhang, Brian Lynch, Krista Queen, Anna Uehara, Rachel Marine, Peter Cook, Clinton R. Paden, Haibin Wang, Suxiang Tong |
| EPI_ISL_812140 | GA Department of Public Health Laboratory | Pathogen Discovery, Respiratory Viruses Branch, Division of Viral Diseases, Centers for Disease Control and Prevention | Yan Li, Ying Tao, Anna Montmayeur, Jing Zhang, Brian Lynch, Krista Queen, Anna Uehara, Rachel Marine, Peter Cook, Clinton R. Paden, Haibin Wang, Suxiang Tong |
| EPI_ISL_812141 | CT-Dr. Katherine A. Kelley State Public Health Lab | Pathogen Discovery, Respiratory Viruses Branch, Division of Viral Diseases, Centers for Disease Control and Prevention | Yan Li, Ying Tao, Anna Montmayeur, Jing Zhang, Brian Lynch, Krista Queen, Anna Uehara, Rachel Marine, Peter Cook, Clinton R. Paden, Haibin Wang, Suxiang Tong |
| EPI_ISL_812142 | FL Bureau of Public Health Laboratories-Tampa | Pathogen Discovery, Respiratory Viruses Branch, Division of Viral Diseases, Centers for Disease Control and Prevention | Yan Li, Ying Tao, Anna Montmayeur, Jing Zhang, Brian Lynch, Krista Queen, Anna Uehara, Rachel Marine, Peter Cook, Clinton R. Paden, Haibin Wang, Suxiang Tong |
| EPI_ISL_812143, EPI_ISL_812144, EPI_ISL_812145, EPI_ISL_812146, EPI_ISL_812147, EPI_ISL_812148, EPI_ISL_812149, EPI_ISL_812150, EPI_ISL_812151, EPI_ISL_812152, EPI_ISL_812153, EPI_ISL_812154, EPI_ISL_812155, EPI_ISL_812156, EPI_ISL_812157, EPI_ISL_812158, EPI_ISL_812159, EPI_ISL_812160, EPI_ISL_812161, EPI_ISL_812162, EPI_ISL_812163, EPI_ISL_812164, EPI_ISL_812165, EPI_ISL_812166, EPI_ISL_812167, EPI_ISL_812168, EPI_ISL_812169, EPI_ISL_812170, EPI_ISL_812171, EPI_ISL_812172, EPI_ISL_812173, EPI_ISL_812174, EPI_ISL_812175, EPI_ISL_812176, EPI_ISL_812177, EPI_ISL_812178, EPI_ISL_812179, EPI_ISL_812180, EPI_ISL_812181, EPI_ISL_812182, EPI_ISL_812183, EPI_ISL_812184, EPI_ISL_812185, EPI_ISL_812186, EPI_ISL_812187, EPI_ISL_812188, EPI_ISL_812189, EPI_ISL_812190, EPI_ISL_812191, EPI_ISL_812192, EPI_ISL_812193, EPI_ISL_812194, EPI_ISL_812195, EPI_ISL_812196, EPI_ISL_812197 | | | |
| see above | GA Department of Public Health Laboratory | Pathogen Discovery, Respiratory Viruses Branch, Division of Viral Diseases, Centers for Disease Control and Prevention | Yan Li, Ying Tao, Anna Montmayeur, Jing Zhang, Brian Lynch, Krista Queen, Anna Uehara, Rachel Marine, Peter Cook, Clinton R. Paden, Haibin Wang, Suxiang Tong |
| EPI_ISL_812198 | MN PHL Division, Minnesota Department of Health | Pathogen Discovery, Respiratory Viruses Branch, Division of Viral Diseases, Centers for Disease Control and Prevention | Yan Li, Ying Tao, Anna Montmayeur, Jing Zhang, Brian Lynch, Krista Queen, Anna Uehara, Rachel Marine, Peter Cook, Clinton R. Paden, Haibin Wang, Suxiang Tong |
| EPI_ISL_812199, EPI_ISL_812200 | NM Dept. Health, Scientific Laboratory Division | Pathogen Discovery, Respiratory Viruses Branch, Division of Viral Diseases, Centers for Disease Control and Prevention | Yan Li, Ying Tao, Anna Montmayeur, Jing Zhang, Brian Lynch, Krista Queen, Anna Uehara, Rachel Marine, Peter Cook, Clinton R. Paden, Haibin Wang, Suxiang Tong |
| EPI_ISL_812201 | NYC Department of Health and Mental Hygiene | Pathogen Discovery, Respiratory Viruses Branch, Division of Viral Diseases, Centers for Disease Control and Prevention | Yan Li, Ying Tao, Anna Montmayeur, Jing Zhang, Brian Lynch, Krista Queen, Anna Uehara, Rachel Marine, Peter Cook, Clinton R. Paden, Haibin Wang, Suxiang Tong |
| EPI_ISL_812202, EPI_ISL_812203 | RI State Health Laboratories | Pathogen Discovery, Respiratory Viruses Branch, Division of Viral Diseases, Centers for Disease Control and Prevention | Yan Li, Ying Tao, Anna Montmayeur, Jing Zhang, Brian Lynch, Krista Queen, Anna Uehara, Rachel Marine, Peter Cook, Clinton R. Paden, Haibin Wang, Suxiang Tong |
| EPI_ISL_812204, EPI_ISL_812205, EPI_ISL_812206, EPI_ISL_812207, EPI_ISL_812208, EPI_ISL_812209, EPI_ISL_812210, EPI_ISL_812211, EPI_ISL_812212, EPI_ISL_812213, EPI_ISL_812214, EPI_ISL_812215, EPI_ISL_812216, EPI_ISL_812217, EPI_ISL_812218, EPI_ISL_812219, EPI_ISL_812220, EPI_ISL_812221, EPI_ISL_812222, EPI_ISL_812223, EPI_ISL_812224, EPI_ISL_812225, EPI_ISL_812226, EPI_ISL_812227, EPI_ISL_812228, EPI_ISL_812229, EPI_ISL_812230, EPI_ISL_812231 | | | |
| see above | TN Division of Laboratory Services | Pathogen Discovery, Respiratory Viruses Branch, Division of Viral Diseases, Centers for Disease Control and Prevention | Yan Li, Ying Tao, Anna Montmayeur, Jing Zhang, Brian Lynch, Krista Queen, Anna Uehara, Rachel Marine, Peter Cook, Clinton R. Paden, Haibin Wang, Suxiang Tong |
| EPI_ISL_812232 | UT-Unified State Labs: Public Health Utah DOH | Pathogen Discovery, Respiratory Viruses Branch, Division of Viral Diseases, Centers for Disease Control and Prevention | Yan Li, Ying Tao, Anna Montmayeur, Jing Zhang, Brian Lynch, Krista Queen, Anna Uehara, Rachel Marine, Peter Cook, Clinton R. Paden, Haibin Wang, Suxiang Tong |
| EPI_ISL_812233, EPI_ISL_812234, EPI_ISL_812235, EPI_ISL_812236, EPI_ISL_812237, EPI_ISL_812238, EPI_ISL_812239, EPI_ISL_812240, EPI_ISL_812241, EPI_ISL_812242, EPI_ISL_812243, EPI_ISL_812244, EPI_ISL_812245, EPI_ISL_812246, EPI_ISL_812247, EPI_ISL_812248 | | | |
| see above | North Dakota Department of Health, Public Health Laboratory | North Dakota Department of Health, Public Health Laboratory | Lisa Wingerter |
| EPI_ISL_812286, EPI_ISL_812291, EPI_ISL_812292, EPI_ISL_812293, EPI_ISL_812294, EPI_ISL_812295, EPI_ISL_812296, EPI_ISL_812297, EPI_ISL_812298, EPI_ISL_812299, EPI_ISL_812300, EPI_ISL_812301, EPI_ISL_812303, EPI_ISL_812304, EPI_ISL_812306, EPI_ISL_812307 | | | |
| see above | Delaware Public Health Lab | Delaware Public Health Lab | Gregory Hovan |
| EPI_ISL_812308, EPI_ISL_812309, EPI_ISL_812310, EPI_ISL_812311, EPI_ISL_812312, EPI_ISL_812313, EPI_ISL_812314, EPI_ISL_812315, EPI_ISL_812316, EPI_ISL_812317, EPI_ISL_812318, EPI_ISL_812319, EPI_ISL_812320, EPI_ISL_812321, EPI_ISL_812322, EPI_ISL_812323, EPI_ISL_812324, EPI_ISL_812325, EPI_ISL_812326, EPI_ISL_812327, EPI_ISL_812328, EPI_ISL_812329, EPI_ISL_812330, EPI_ISL_812331, EPI_ISL_812332, EPI_ISL_812333, EPI_ISL_812334, EPI_ISL_812335, EPI_ISL_812336, EPI_ISL_812337, EPI_ISL_812338, EPI_ISL_812339, EPI_ISL_812340, EPI_ISL_812341, EPI_ISL_812342, EPI_ISL_812343, EPI_ISL_812344, EPI_ISL_812345 | | | |
| see above | United States Air Force School of Aerospace Medicine | United States Air Force School of Aerospace Medicine | Anthony Fries, Jennifer Meyer, Amanda Javorina, Sarah Purves, William Gruner, Clarise Starr, Elizabeth Macias |
| EPI_ISL_812346 | Oklahoma Animal Disease Diagnostic Laboratory | Oklahoma Animal Disease Diagnostic Laboratory | Sai Narayanan, Girish Patil, Sunil More, Jeremiah Saliki, Akhilesh Ramachandran |
| EPI_ISL_812348 | University of Wisconsin-Madison AIDS Vaccine Research Laboratories | University of Wisconsin-Madison AIDS Vaccine Research Laboratories | Gage Moreno, Katarina Braun, et al. AIDS Vaccine Research Laboratories |
| EPI_ISL_812350 | Santa Clara County Public Health Laboratory | Santa Clara County Public Health Laboratory | Santa Clara County Public Health Department |
| EPI_ISL_812361, EPI_ISL_812362, EPI_ISL_812363, EPI_ISL_812364, EPI_ISL_812366, EPI_ISL_812367, EPI_ISL_812368, EPI_ISL_812369, EPI_ISL_812372, EPI_ISL_812373, EPI_ISL_812374, EPI_ISL_812375, EPI_ISL_812376, EPI_ISL_812377, EPI_ISL_812378, EPI_ISL_812379, EPI_ISL_812382, EPI_ISL_812383, EPI_ISL_812384, EPI_ISL_812385, EPI_ISL_812386, EPI_ISL_812387, EPI_ISL_812388, EPI_ISL_812390, EPI_ISL_812391, EPI_ISL_812393, EPI_ISL_812394, EPI_ISL_812395, EPI_ISL_812396, EPI_ISL_812397, EPI_ISL_812398, EPI_ISL_812400, EPI_ISL_812401, EPI_ISL_812402, EPI_ISL_812405, EPI_ISL_812406, EPI_ISL_812409, EPI_ISL_812410, EPI_ISL_812411, EPI_ISL_812414, EPI_ISL_812415, EPI_ISL_812416, EPI_ISL_812418, EPI_ISL_812419, EPI_ISL_812420 | | | |
| see above | Utah Public Health Laboratory | Utah Public Health Laboratory | Erin L. Young, Kelly F. Oakeson, Tara Gallagher |
| EPI_ISL_812421, EPI_ISL_812422 | University of Wisconsin-Madison AIDS Vaccine Research Laboratories | University of Wisconsin-Madison AIDS Vaccine Research Laboratories | Gage Moreno, Katarina Braun, et al. AIDS Vaccine Research Laboratories |
| EPI_ISL_812526, EPI_ISL_812527, EPI_ISL_812528, EPI_ISL_812529, EPI_ISL_812530, EPI_ISL_812531, EPI_ISL_812532, EPI_ISL_812533, EPI_ISL_812534, EPI_ISL_812535, EPI_ISL_812536, EPI_ISL_812537, EPI_ISL_812538, EPI_ISL_812539, EPI_ISL_812540, EPI_ISL_812541, EPI_ISL_812542, EPI_ISL_812543, EPI_ISL_812544, EPI_ISL_812545, EPI_ISL_812546, EPI_ISL_812547, EPI_ISL_812548, EPI_ISL_812549, EPI_ISL_812550, EPI_ISL_812551, EPI_ISL_812552, EPI_ISL_812553, EPI_ISL_812554, EPI_ISL_812555, EPI_ISL_812556, EPI_ISL_812557, EPI_ISL_812558, EPI_ISL_812559, EPI_ISL_812560, EPI_ISL_812561, EPI_ISL_812562, EPI_ISL_812563, EPI_ISL_812564, EPI_ISL_812565, EPI_ISL_812566, EPI_ISL_812567, EPI_ISL_812568, EPI_ISL_812569, EPI_ISL_812570, EPI_ISL_812571, EPI_ISL_812572, EPI_ISL_812573, EPI_ISL_812574, EPI_ISL_812575, EPI_ISL_812576, EPI_ISL_812577, EPI_ISL_812578, EPI_ISL_812579, EPI_ISL_812580, EPI_ISL_812581, EPI_ISL_812582, EPI_ISL_812583, EPI_ISL_812584, EPI_ISL_812585, EPI_ISL_812586, EPI_ISL_812587, EPI_ISL_812588, EPI_ISL_812589, EPI_ISL_812590, EPI_ISL_812591, EPI_ISL_812592, EPI_ISL_812593, EPI_ISL_812594, EPI_ISL_812595, EPI_ISL_812596, EPI_ISL_812597, EPI_ISL_812598, EPI_ISL_812599, EPI_ISL_812600, EPI_ISL_812601, EPI_ISL_812602, EPI_ISL_812603, EPI_ISL_812604, EPI_ISL_812605, EPI_ISL_812606, EPI_ISL_812607, EPI_ISL_812608, EPI_ISL_812609, EPI_ISL_812610, EPI_ISL_812611, EPI_ISL_812612, EPI_ISL_812613, EPI_ISL_812614, EPI_ISL_812615, EPI_ISL_812616, EPI_ISL_812617, EPI_ISL_812618, EPI_ISL_812619, EPI_ISL_812621, EPI_ISL_812622, EPI_ISL_812623, EPI_ISL_812624, EPI_ISL_812625, EPI_ISL_812626, EPI_ISL_812627, EPI_ISL_812628, EPI_ISL_812629, EPI_ISL_812630, EPI_ISL_812631, EPI_ISL_812633, EPI_ISL_812634, EPI_ISL_812635, EPI_ISL_812636, EPI_ISL_812637, EPI_ISL_812638, EPI_ISL_812639, EPI_ISL_812640, EPI_ISL_812641, EPI_ISL_812642, EPI_ISL_812643, EPI_ISL_812644, EPI_ISL_812645, EPI_ISL_812646, EPI_ISL_812647, EPI_ISL_812648, EPI_ISL_812649, EPI_ISL_812650, EPI_ISL_812651, EPI_ISL_812653, EPI_ISL_812654, EPI_ISL_812655, EPI_ISL_812656, EPI_ISL_812657, EPI_ISL_812658, EPI_ISL_812659, EPI_ISL_812660, EPI_ISL_812661, EPI_ISL_812662, EPI_ISL_812663, EPI_ISL_812664, EPI_ISL_812665, EPI_ISL_812666, EPI_ISL_812667, EPI_ISL_812668, EPI_ISL_812669, EPI_ISL_812670, EPI_ISL_812671, EPI_ISL_812672, EPI_ISL_812673, EPI_ISL_812674, EPI_ISL_812675, EPI_ISL_812676, EPI_ISL_812677, EPI_ISL_812678, EPI_ISL_812680, EPI_ISL_812681, EPI_ISL_812682, EPI_ISL_812683, EPI_ISL_812684, EPI_ISL_812685, EPI_ISL_812686, EPI_ISL_812687, EPI_ISL_812688, EPI_ISL_812689, EPI_ISL_812690, EPI_ISL_812691, EPI_ISL_812692, EPI_ISL_812694, EPI_ISL_812695, EPI_ISL_812696, EPI_ISL_812697, EPI_ISL_812698, EPI_ISL_812699, EPI_ISL_812700, EPI_ISL_812701, EPI_ISL_812702, EPI_ISL_812703, EPI_ISL_812704, EPI_ISL_812705, EPI_ISL_812706, EPI_ISL_812707, EPI_ISL_812708, EPI_ISL_812709, EPI_ISL_812710, EPI_ISL_812711 | | | |
| see above | United States Air Force School of Aerospace Medicine | United States Air Force School of Aerospace Medicine | Anthony Fries, Jennifer Meyer, Amanda Javorina, Sarah Purves, William Gruner, Clarise Starr, Elizabeth Macias |
| EPI_ISL_812722, EPI_ISL_812723, EPI_ISL_812724 | DOHMH PHL | New York City Public Health Laboratory | Jade Wang, et al. |
| EPI_ISL_812725 | DOHMH Crown Heights | New York City Public Health Laboratory | Jade Wang, et al. |
| EPI_ISL_812726 | DOHMH Corona | New York City Public Health Laboratory | Jade Wang, et al. |
| EPI_ISL_812727 | DOHMH Jamaica | New York City Public Health Laboratory | Jade Wang, et al. |
| EPI_ISL_812728 | DOHMH Riverside | New York City Public Health Laboratory | Jade Wang, et al. |
| EPI_ISL_812729 | DOHMH Central Harlem | New York City Public Health Laboratory | Jade Wang, et al. |
| EPI_ISL_812730 | DOHMH Crown Heights | New York City Public Health Laboratory | Jade Wang, et al. |
| EPI_ISL_812731 | DOHMH Jamaica | New York City Public Health Laboratory | Jade Wang, et al. |

| | | | |
|---|---|---|---|
| EPI_ISL_812732, EPI_ISL_812733 | DOHMH Corona | New York City Public Health Laboratory | Jade Wang, et al. |
| EPI_ISL_812734 | DOHMH Crown Heights | New York City Public Health Laboratory | Jade Wang, et al. |
| EPI_ISL_812735, EPI_ISL_812736, EPI_ISL_812737 | DOHMH Morrisania | New York City Public Health Laboratory | Jade Wang, et al. |
| EPI_ISL_812738 | DOHMH Crown Heights | New York City Public Health Laboratory | Jade Wang, et al. |
| EPI_ISL_812739 | DOHMH Central Harlem | New York City Public Health Laboratory | Jade Wang, et al. |
| EPI_ISL_812740 | DOHMH Corona | New York City Public Health Laboratory | Jade Wang, et al. |
| EPI_ISL_812741 | DOHMH Central Harlem | New York City Public Health Laboratory | Jade Wang, et al. |
| EPI_ISL_812742, EPI_ISL_812743, EPI_ISL_812744 | DOHMH Corona | New York City Public Health Laboratory | Jade Wang, et al. |
| EPI_ISL_812745 | DOHMH Riverside | New York City Public Health Laboratory | Jade Wang, et al. |
| EPI_ISL_812746, EPI_ISL_812747, EPI_ISL_812748 | DOHMH Corona | New York City Public Health Laboratory | Jade Wang, et al. |
| EPI_ISL_812749 | DOHMH Central Harlem | New York City Public Health Laboratory | Jade Wang, et al. |
| EPI_ISL_812750, EPI_ISL_812751 | DOHMH Corona | New York City Public Health Laboratory | Jade Wang, et al. |
| EPI_ISL_812752 | DOHMH Crown Heights | New York City Public Health Laboratory | Jade Wang, et al. |
| EPI_ISL_812753, EPI_ISL_812754, EPI_ISL_812755, EPI_ISL_812756, EPI_ISL_812757 | DOHMH Morrisania | New York City Public Health Laboratory | Jade Wang, et al. |
| EPI_ISL_812758 | DOHMH Crown Heights | New York City Public Health Laboratory | Jade Wang, et al. |
| EPI_ISL_812759 | DOHMH Riverside | New York City Public Health Laboratory | Jade Wang, et al. |
| EPI_ISL_812760 | DOHMH Corona | New York City Public Health Laboratory | Jade Wang, et al. |
| EPI_ISL_816135, EPI_ISL_816136, EPI_ISL_816432 | Ohio Department of Health Laboratory | Ohio Department of Health Laboratory | Holmes, Jennifer; Eric Brandt, Keoni Omura, Glen McGillivary, Caitlin McDonnell, Kirtana Ramadugu, Erica Leasure, Kelsey Florek, Heather Blankenship, Quanta Brown, and Tammy Bannerman |
| EPI_ISL_819125 | Wyoming Public Health Laboratory | Wyoming Public Health Laboratory | Noah Hull, Taylor Fearing, Lynette Gumbleton, Channing Weber, Ashley Norberg, Bailey Bowcutt, and Wanda Manley |
| EPI_ISL_819127 | Ohio Department of Health Laboratory | Ohio Department of Health Laboratory | Holmes, Jennifer; Eric Brandt, Keoni Omura, Glen McGillivary, Caitlin McDonnell, Kirtana Ramadugu, Erica Leasure, Kelsey Florek, Heather Blankenship, Quanta Brown, and Tammy Bannerman |
| EPI_ISL_819204, EPI_ISL_819205, EPI_ISL_819206, EPI_ISL_819207, EPI_ISL_819208, EPI_ISL_819209, EPI_ISL_819210, EPI_ISL_819211, EPI_ISL_819216, EPI_ISL_819217, EPI_ISL_819218, EPI_ISL_819219, EPI_ISL_819220, EPI_ISL_819221, EPI_ISL_819222, EPI_ISL_819223, EPI_ISL_819224, EPI_ISL_819225, EPI_ISL_819226, EPI_ISL_819227, EPI_ISL_819228, EPI_ISL_819229, EPI_ISL_819230, EPI_ISL_819231, EPI_ISL_819232, EPI_ISL_819233, EPI_ISL_819234, EPI_ISL_819235, EPI_ISL_819236, EPI_ISL_819237, EPI_ISL_819238, EPI_ISL_819239, EPI_ISL_819240, EPI_ISL_819242, EPI_ISL_819243, EPI_ISL_819244, EPI_ISL_819245, EPI_ISL_819246, EPI_ISL_819247, EPI_ISL_819248, EPI_ISL_819249, EPI_ISL_819250, EPI_ISL_819251, EPI_ISL_819252, EPI_ISL_819253, EPI_ISL_819254, EPI_ISL_819255, EPI_ISL_819256, EPI_ISL_819257, EPI_ISL_819258, EPI_ISL_819259, EPI_ISL_819260, EPI_ISL_819261, EPI_ISL_819262, EPI_ISL_819263, EPI_ISL_819264, EPI_ISL_819265, EPI_ISL_819266, EPI_ISL_819267, EPI_ISL_819268, EPI_ISL_819269, EPI_ISL_819270, EPI_ISL_819271, EPI_ISL_819272, EPI_ISL_819273, EPI_ISL_819274, EPI_ISL_819275, EPI_ISL_819276, EPI_ISL_819277, EPI_ISL_819278, EPI_ISL_819279, EPI_ISL_819280, EPI_ISL_819281, EPI_ISL_819282, EPI_ISL_819283, EPI_ISL_819284, EPI_ISL_819285, EPI_ISL_819286, EPI_ISL_819287, EPI_ISL_819288, EPI_ISL_819289, EPI_ISL_819290, EPI_ISL_819291, EPI_ISL_819292 | | | |
| see above | Wyoming Public Health Laboratory | Wyoming Public Health Laboratory | Noah Hull, Taylor Fearing, Lynette Gumbleton, Channing Weber, Ashley Norberg, Bailey Bowcutt, and Wanda Manley |
| EPI_ISL_823813 | DOHMH Jamaica | New York City Public Health Laboratory | Jade Wang, et al. |
| EPI_ISL_823814 | DOHMH Riverside | New York City Public Health Laboratory | Jade Wang, et al. |
| EPI_ISL_823815, EPI_ISL_823816 | DOHMH Chelsea | New York City Public Health Laboratory | Jade Wang, et al. |
| EPI_ISL_823817, EPI_ISL_823818, EPI_ISL_823819 | DOHMH Corona | New York City Public Health Laboratory | Jade Wang, et al. |
| EPI_ISL_823820 | DOHMH PHL | New York City Public Health Laboratory | Jade Wang, et al. |
| EPI_ISL_823821, EPI_ISL_823822 | DOHMH Jamaica | New York City Public Health Laboratory | Jade Wang, et al. |
| EPI_ISL_823823, EPI_ISL_823824, EPI_ISL_823825 | DOHMH Riverside | New York City Public Health Laboratory | Jade Wang, et al. |
| EPI_ISL_823826, EPI_ISL_823827, EPI_ISL_823828 | DOHMH Corona | New York City Public Health Laboratory | Jade Wang, et al. |
| EPI_ISL_823829, EPI_ISL_823830, EPI_ISL_823831 | DOHMH Morrisania | New York City Public Health Laboratory | Jade Wang, et al. |
| EPI_ISL_823832, EPI_ISL_823833, EPI_ISL_823834 | DOHMH Central Harlem | New York City Public Health Laboratory | Jade Wang, et al. |
| EPI_ISL_823835, EPI_ISL_823836, EPI_ISL_823837 | DOHMH Corona | New York City Public Health Laboratory | Jade Wang, et al. |
| EPI_ISL_823838, EPI_ISL_823839, EPI_ISL_823840 | DOHMH Central Harlem | New York City Public Health Laboratory | Jade Wang, et al. |
| EPI_ISL_823841 | DOHMH Riverside | New York City Public Health Laboratory | Jade Wang, et al. |
| EPI_ISL_823842 | DOHMH PHL | New York City Public Health Laboratory | Jade Wang, et al. |
| EPI_ISL_823843, EPI_ISL_823844 | DOHMH Central Harlem | New York City Public Health Laboratory | Jade Wang, et al. |
| EPI_ISL_823845 | DOHMH Morrisania | New York City Public Health Laboratory | Jade Wang, et al. |
| EPI_ISL_823846, EPI_ISL_823847, EPI_ISL_823848 | DOHMH Jamaica | New York City Public Health Laboratory | Jade Wang, et al. |
| EPI_ISL_823849 | DOHMH Fort Greene | New York City Public Health Laboratory | Jade Wang, et al. |
| EPI_ISL_823850 | DOHMH Corona | New York City Public Health Laboratory | Jade Wang, et al. |
| EPI_ISL_823851 | DOHMH Fort Greene | New York City Public Health Laboratory | Jade Wang, et al. |
| EPI_ISL_823852, EPI_ISL_823853 | DOHMH Crown Heights | New York City Public Health Laboratory | Jade Wang, et al. |
| EPI_ISL_823854, EPI_ISL_823855, EPI_ISL_823856, EPI_ISL_823857, EPI_ISL_823858, EPI_ISL_823859, EPI_ISL_823860 | DOHMH Morrisania | New York City Public Health Laboratory | Jade Wang, et al. |
| EPI_ISL_823861, EPI_ISL_823862, EPI_ISL_823863 | DOHMH Corona | New York City Public Health Laboratory | Jade Wang, et al. |

| | | | |
|---|---|---|---|
| EPI_ISL_823864 | DOHMH Chelsea | New York City Public Health Laboratory | Jade Wang, et al. |
| EPI_ISL_823865 | DOHMH Central Harlem | New York City Public Health Laboratory | Jade Wang, et al. |
| EPI_ISL_823866, EPI_ISL_823867 | DOHMH Fort Greene | New York City Public Health Laboratory | Jade Wang, et al. |
| EPI_ISL_823868 | DOHMH Corona | New York City Public Health Laboratory | Jade Wang, et al. |
| EPI_ISL_823869 | DOHMH Central Harlem | New York City Public Health Laboratory | Jade Wang, et al. |
| EPI_ISL_823870, EPI_ISL_823871 | DOHMH Jamaica | New York City Public Health Laboratory | Jade Wang, et al. |
| EPI_ISL_823872, EPI_ISL_823873, EPI_ISL_823874 | DOHMH Morrisania | New York City Public Health Laboratory | Jade Wang, et al. |
| EPI_ISL_823875 | DOHMH PHL | New York City Public Health Laboratory | Jade Wang, et al. |
| EPI_ISL_823876, EPI_ISL_823877 | DOHMH Jamaica | New York City Public Health Laboratory | Jade Wang, et al. |
| EPI_ISL_823878, EPI_ISL_823879, EPI_ISL_823880, EPI_ISL_823881, EPI_ISL_823882 | DOHMH Corona | New York City Public Health Laboratory | Jade Wang, et al. |
| EPI_ISL_823883 | DOHMH Riverside | New York City Public Health Laboratory | Jade Wang, et al. |
| EPI_ISL_823884, EPI_ISL_823885, EPI_ISL_823886, EPI_ISL_823887 | DOHMH Central Harlem | New York City Public Health Laboratory | Jade Wang, et al. |
| EPI_ISL_823888 | DOHMH Riverside | New York City Public Health Laboratory | Jade Wang, et al. |
| EPI_ISL_823889, EPI_ISL_823890, EPI_ISL_823891 | DOHMH PHL | New York City Public Health Laboratory | Jade Wang, et al. |
| EPI_ISL_823892, EPI_ISL_823893 | DOHMH Morrisania | New York City Public Health Laboratory | Jade Wang, et al. |
| EPI_ISL_823894, EPI_ISL_823895 | DOHMH Jamaica | New York City Public Health Laboratory | Jade Wang, et al. |
| EPI_ISL_823896, EPI_ISL_823897 | DOHMH PHL | New York City Public Health Laboratory | Jade Wang, et al. |
| EPI_ISL_823898, EPI_ISL_823899 | DOHMH Corona | New York City Public Health Laboratory | Jade Wang, et al. |
| EPI_ISL_823900 | DOHMH Riverside | New York City Public Health Laboratory | Jade Wang, et al. |
| EPI_ISL_823901, EPI_ISL_823902 | DOHMH Central Harlem | New York City Public Health Laboratory | Jade Wang, et al. |
| EPI_ISL_823903 | DOHMH Morrisania | New York City Public Health Laboratory | Jade Wang, et al. |
| EPI_ISL_823904 | DOHMH Fort Greene | New York City Public Health Laboratory | Jade Wang, et al. |
| EPI_ISL_823905, EPI_ISL_823906 | DOHMH Corona | New York City Public Health Laboratory | Jade Wang, et al. |
| EPI_ISL_823907, EPI_ISL_823908 | DOHMH Jamaica | New York City Public Health Laboratory | Jade Wang, et al. |
| EPI_ISL_823909 | DOHMH PHL | New York City Public Health Laboratory | Jade Wang, et al. |
| EPI_ISL_823910, EPI_ISL_823911 | DOHMH Fort Greene | New York City Public Health Laboratory | Jade Wang, et al. |
| EPI_ISL_823912 | DOHMH Central Harlem | New York City Public Health Laboratory | Jade Wang, et al. |
| EPI_ISL_823913, EPI_ISL_823914 | DOHMH PHL | New York City Public Health Laboratory | Jade Wang, et al. |
| EPI_ISL_823915, EPI_ISL_823916, EPI_ISL_823917 | DOHMH Jamaica | New York City Public Health Laboratory | Jade Wang, et al. |
| EPI_ISL_823918, EPI_ISL_823919, EPI_ISL_823920, EPI_ISL_823921, EPI_ISL_823922 | DOHMH Corona | New York City Public Health Laboratory | Jade Wang, et al. |
| EPI_ISL_823923 | DOHMH Fort Greene | New York City Public Health Laboratory | Jade Wang, et al. |
| EPI_ISL_823924 | DOHMH Chelsea | New York City Public Health Laboratory | Jade Wang, et al. |
| EPI_ISL_823925 | DOHMH Crown Heights | New York City Public Health Laboratory | Jade Wang, et al. |
| EPI_ISL_823926 | DOHMH Jamaica | New York City Public Health Laboratory | Jade Wang, et al. |
| EPI_ISL_823927 | DOHMH Central Harlem | New York City Public Health Laboratory | Jade Wang, et al. |
| EPI_ISL_823928 | DOHMH Jamaica | New York City Public Health Laboratory | Jade Wang, et al. |
| EPI_ISL_823929 | DOHMH Morrisania | New York City Public Health Laboratory | Jade Wang, et al. |
| EPI_ISL_823930 | DOHMH Chelsea | New York City Public Health Laboratory | Jade Wang, et al. |
| EPI_ISL_823931 | DOHMH Corona | New York City Public Health Laboratory | Jade Wang, et al. |
| EPI_ISL_823932 | DOHMH Morrisania | New York City Public Health Laboratory | Jade Wang, et al. |
| EPI_ISL_823933 | DOHMH Corona | New York City Public Health Laboratory | Jade Wang, et al. |
| EPI_ISL_823934 | DOHMH Chelsea | New York City Public Health Laboratory | Jade Wang, et al. |
| EPI_ISL_823935, EPI_ISL_823936 | DOHMH Crown Heights | New York City Public Health Laboratory | Jade Wang, et al. |
| EPI_ISL_823937 | DOHMH PHL | New York City Public Health Laboratory | Jade Wang, et al. |
| EPI_ISL_823938 | DOHMH Fort Greene | New York City Public Health Laboratory | Jade Wang, et al. |
| EPI_ISL_823939 | DOHMH Riverside | New York City Public Health Laboratory | Jade Wang, et al. |
| EPI_ISL_823940, EPI_ISL_823941 | DOHMH Jamaica | New York City Public Health Laboratory | Jade Wang, et al. |
| EPI_ISL_823942 | DOHMH Crown Heights | New York City Public Health Laboratory | Jade Wang, et al. |
| EPI_ISL_823943, EPI_ISL_823944, EPI_ISL_823945 | DOHMH Corona | New York City Public Health Laboratory | Jade Wang, et al. |
| EPI_ISL_823946 | DOHMH Jamaica | New York City Public Health Laboratory | Jade Wang, et al. |
| EPI_ISL_823947 | DOHMH Morrisania | New York City Public Health Laboratory | Jade Wang, et al. |
| EPI_ISL_823948 | DOHMH Chelsea | New York City Public Health Laboratory | Jade Wang, et al. |
| EPI_ISL_823949 | DOHMH Jamaica | New York City Public Health Laboratory | Jade Wang, et al. |

| | | | |
|---|---|---|---|
| EPI_ISL_823950 | DOHMH Central Harlem | New York City Public Health Laboratory | Jade Wang, et al. |
| EPI_ISL_823951, EPI_ISL_823952 | DOHMH Riverside | New York City Public Health Laboratory | Jade Wang, et al. |
| EPI_ISL_823953 | DOHMH Corona | New York City Public Health Laboratory | Jade Wang, et al. |
| EPI_ISL_823954 | DOHMH Chelsea | New York City Public Health Laboratory | Jade Wang, et al. |
| EPI_ISL_823955, EPI_ISL_823956 | DOHMH Jamaica | New York City Public Health Laboratory | Jade Wang, et al. |
| EPI_ISL_823957 | DOHMH Corona | New York City Public Health Laboratory | Jade Wang, et al. |
| EPI_ISL_823958 | DOHMH PHL | New York City Public Health Laboratory | Jade Wang, et al. |
| EPI_ISL_823959, EPI_ISL_823960 | DOHMH Morrisania | New York City Public Health Laboratory | Jade Wang, et al. |
| EPI_ISL_823961 | DOHMH Jamaica | New York City Public Health Laboratory | Jade Wang, et al. |
| EPI_ISL_823962 | DOHMH Chelsea | New York City Public Health Laboratory | Jade Wang, et al. |
| EPI_ISL_823963, EPI_ISL_823964, EPI_ISL_823965, EPI_ISL_823966, EPI_ISL_823967 | DOHMH Corona | New York City Public Health Laboratory | Jade Wang, et al. |
| EPI_ISL_823968, EPI_ISL_823969 | DOHMH Jamaica | New York City Public Health Laboratory | Jade Wang, et al. |
| EPI_ISL_823970 | Department of Homeless Services | New York City Public Health Laboratory | Jade Wang, et al. |
| EPI_ISL_823971, EPI_ISL_823972 | OCME Office Of Chief Medical Examiner | New York City Public Health Laboratory | Jade Wang, et al. |
| EPI_ISL_823973 | Department of Homeless Services | New York City Public Health Laboratory | Jade Wang, et al. |
| EPI_ISL_823974 | OCME Office Of Chief Medical Examiner | New York City Public Health Laboratory | Jade Wang, et al. |
| EPI_ISL_823975 | Department of Homeless Services | New York City Public Health Laboratory | Jade Wang, et al. |
| EPI_ISL_824295 | DOHMH Morrisania | New York City Public Health Laboratory | Jade Wang, et al. |
| EPI_ISL_824296, EPI_ISL_824297 | DOHMH Jamaica | New York City Public Health Laboratory | Jade Wang, et al. |
| EPI_ISL_824298 | DOHMH Central Harlem | New York City Public Health Laboratory | Jade Wang, et al. |
| EPI_ISL_824299 | DOHMH Morrisania | New York City Public Health Laboratory | Jade Wang, et al. |
| EPI_ISL_824300 | DOHMH Chelsea | New York City Public Health Laboratory | Jade Wang, et al. |
| EPI_ISL_824301 | DOHMH Crown Heights | New York City Public Health Laboratory | Jade Wang, et al. |
| EPI_ISL_824302 | DOHMH Jamaica | New York City Public Health Laboratory | Jade Wang, et al. |
| EPI_ISL_824303 | DOHMH Morrisania | New York City Public Health Laboratory | Jade Wang, et al. |
| EPI_ISL_824304, EPI_ISL_824305 | DOHMH Jamaica | New York City Public Health Laboratory | Jade Wang, et al. |
| EPI_ISL_824306, EPI_ISL_824307 | DOHMH PHL | New York City Public Health Laboratory | Jade Wang, et al. |
| EPI_ISL_824308, EPI_ISL_824309 | DOHMH Jamaica | New York City Public Health Laboratory | Jade Wang, et al. |
| EPI_ISL_824310 | DOHMH Crown Heights | New York City Public Health Laboratory | Jade Wang, et al. |
| EPI_ISL_824311, EPI_ISL_824312 | DOHMH Fort Greene | New York City Public Health Laboratory | Jade Wang, et al. |
| EPI_ISL_824313 | DOHMH Jamaica | New York City Public Health Laboratory | Jade Wang, et al. |
| EPI_ISL_824314, EPI_ISL_824315 | DOHMH Corona | New York City Public Health Laboratory | Jade Wang, et al. |
| EPI_ISL_824316 | DOHMH Crown Heights | New York City Public Health Laboratory | Jade Wang, et al. |
| EPI_ISL_824317, EPI_ISL_824318 | DOHMH Central Harlem | New York City Public Health Laboratory | Jade Wang, et al. |
| EPI_ISL_824319 | DOHMH Chelsea | New York City Public Health Laboratory | Jade Wang, et al. |
| EPI_ISL_824320 | DOHMH Corona | New York City Public Health Laboratory | Jade Wang, et al. |
| EPI_ISL_824321, EPI_ISL_824322 | DOHMH Fort Greene | New York City Public Health Laboratory | Jade Wang, et al. |
| EPI_ISL_824323, EPI_ISL_824324, EPI_ISL_824325, EPI_ISL_824326, EPI_ISL_824327 | DOHMH Morrisania | New York City Public Health Laboratory | Jade Wang, et al. |
| EPI_ISL_824328 | DOHMH Corona | New York City Public Health Laboratory | Jade Wang, et al. |
| EPI_ISL_824329, EPI_ISL_824330 | DOHMH Jamaica | New York City Public Health Laboratory | Jade Wang, et al. |
| EPI_ISL_824331 | DOHMH Corona | New York City Public Health Laboratory | Jade Wang, et al. |
| EPI_ISL_824332, EPI_ISL_824333 | Ohio Department of Health Laboratory | Ohio Department of Health Laboratory | Holmes, Jennifer; Eric Brandt, Keoni Omura, Glen McGillivary, Caitlin McDonnell, Kirtana Ramadugu, Erica Leasure, Kelsey Florek, Heather Blankenship, Quanta Brown, and Tammy Bannerman |
| EPI_ISL_824340, EPI_ISL_824342, EPI_ISL_824345, EPI_ISL_824346, EPI_ISL_824347, EPI_ISL_824348, EPI_ISL_824349, EPI_ISL_824350, EPI_ISL_824351, EPI_ISL_824352, EPI_ISL_824353, EPI_ISL_824354, EPI_ISL_824355, EPI_ISL_824356, EPI_ISL_824357, EPI_ISL_824358, EPI_ISL_824359, EPI_ISL_824360, EPI_ISL_824361, EPI_ISL_824362, EPI_ISL_824363, EPI_ISL_824364, EPI_ISL_824365, EPI_ISL_824366, EPI_ISL_824367, EPI_ISL_824368, EPI_ISL_824369, EPI_ISL_824370, EPI_ISL_824371, EPI_ISL_824372, EPI_ISL_824373, EPI_ISL_824374, EPI_ISL_824375, EPI_ISL_824376, EPI_ISL_824377, EPI_ISL_824378, EPI_ISL_824379, EPI_ISL_824380, EPI_ISL_824381, EPI_ISL_824382, EPI_ISL_824383 | | | |
| see above | Michigan Department of Health and Human Services, Bureau of Laboratories | Michigan Department of Health and Human Services, Bureau of Laboratories | Blankenship HM, Riner D, Soehnlen MK |
| EPI_ISL_824384, EPI_ISL_824385, EPI_ISL_824386, EPI_ISL_824387, EPI_ISL_824389 | Ohio Department of Health Laboratory | Ohio Department of Health Laboratory | Holmes, Jennifer; Eric Brandt, Keoni Omura, Glen McGillivary, Caitlin McDonnell, Kirtana Ramadugu, Erica Leasure, Kelsey Florek, Heather Blankenship, Quanta Brown, and Tammy Bannerman |
| EPI_ISL_824390, EPI_ISL_824392, EPI_ISL_824393, EPI_ISL_824394, EPI_ISL_824395, EPI_ISL_824396, EPI_ISL_824397, EPI_ISL_824398, EPI_ISL_824399, EPI_ISL_824400, EPI_ISL_824401, EPI_ISL_824402, EPI_ISL_824403, EPI_ISL_824404, EPI_ISL_824405, EPI_ISL_824406 | | | |
| see above | California Department of Public Health | California Department of Public Health | CDPH IDLB COVIDNet |
| EPI_ISL_824513, EPI_ISL_824514, EPI_ISL_824515, EPI_ISL_824516, EPI_ISL_824517, EPI_ISL_824518, EPI_ISL_824519, EPI_ISL_824520, EPI_ISL_824521, EPI_ISL_824522, EPI_ISL_824523, EPI_ISL_824524, EPI_ISL_824525, EPI_ISL_824526, EPI_ISL_824527, EPI_ISL_824528, EPI_ISL_824529, EPI_ISL_824530, EPI_ISL_824531, EPI_ISL_824532, EPI_ISL_824533, EPI_ISL_824534, EPI_ISL_824536, EPI_ISL_824537, EPI_ISL_824538, EPI_ISL_824539, EPI_ISL_824540, EPI_ISL_824541, EPI_ISL_824542, EPI_ISL_824543, EPI_ISL_824544, EPI_ISL_824545, EPI_ISL_824546, EPI_ISL_824547, EPI_ISL_824548, EPI_ISL_824549, EPI_ISL_824550, EPI_ISL_824551, EPI_ISL_824552, EPI_ISL_824553, EPI_ISL_824554 | | | |
| see above | New Mexico Department of Health Scientific Laboratory | New Mexico Department of Health Scientific Laboratory | Ellie Johnson, Anastacia Griego-Fisher, D'eldra Malone |
| EPI_ISL_824555, EPI_ISL_824556, EPI_ISL_824557, EPI_ISL_824558, EPI_ISL_824559, EPI_ISL_824560, EPI_ISL_824561, EPI_ISL_824562, EPI_ISL_824563, EPI_ISL_824564, EPI_ISL_824565, EPI_ISL_824566, EPI_ISL_824567, EPI_ISL_824568, EPI_ISL_824569, EPI_ISL_824570, EPI_ISL_824571, EPI_ISL_824572, EPI_ISL_824573, EPI_ISL_824574, EPI_ISL_824575, EPI_ISL_824576, EPI_ISL_824577, EPI_ISL_824578, EPI_ISL_824579, EPI_ISL_824580, EPI_ISL_824581, EPI_ISL_824582, EPI_ISL_824583, EPI_ISL_824584, EPI_ISL_824585, EPI_ISL_824586, EPI_ISL_824587, EPI_ISL_824588, EPI_ISL_824589, EPI_ISL_824590, EPI_ISL_824591, EPI_ISL_824592, EPI_ISL_824593, EPI_ISL_824594, EPI_ISL_824595, EPI_ISL_824596, EPI_ISL_824597, EPI_ISL_824598, EPI_ISL_824599, EPI_ISL_824600, EPI_ISL_824601, EPI_ISL_824602, EPI_ISL_824603, EPI_ISL_824604, EPI_ISL_824605, EPI_ISL_824606, EPI_ISL_824607, EPI_ISL_824608, EPI_ISL_824609, EPI_ISL_824610, EPI_ISL_824611, EPI_ISL_824612, EPI_ISL_824613, EPI_ISL_824614, EPI_ISL_824615, EPI_ISL_824616, EPI_ISL_824618, EPI_ISL_824619, EPI_ISL_824620, EPI_ISL_824621, EPI_ISL_824622, EPI_ISL_824623, EPI_ISL_824624, EPI_ISL_824625, EPI_ISL_824626, EPI_ISL_824627, EPI_ISL_824628, EPI_ISL_824629, EPI_ISL_824630, EPI_ISL_824631, EPI_ISL_824632, EPI_ISL_824633, EPI_ISL_824634, EPI_ISL_824635, EPI_ISL_824636, EPI_ISL_824637, EPI_ISL_824638, EPI_ISL_824639, EPI_ISL_824640, EPI_ISL_824641, EPI_ISL_824642, EPI_ISL_824643, EPI_ISL_824644, EPI_ISL_824645, EPI_ISL_824646, EPI_ISL_824647, EPI_ISL_824648, EPI_ISL_824649, EPI_ISL_824650, EPI_ISL_824651, EPI_ISL_824652, EPI_ISL_824653, EPI_ISL_824654, EPI_ISL_824655, EPI_ISL_824656, EPI_ISL_824658, EPI_ISL_824659, EPI_ISL_824660, EPI_ISL_824661, EPI_ISL_824662, EPI_ISL_824663, EPI_ISL_824664, EPI_ISL_824665, EPI_ISL_824666, EPI_ISL_824667, EPI_ISL_824668, EPI_ISL_824669, EPI_ISL_824670, EPI_ISL_824671, EPI_ISL_824672, EPI_ISL_824673, EPI_ISL_824674, EPI_ISL_824675, EPI_ISL_824676, EPI_ISL_824677, EPI_ISL_824678, EPI_ISL_824679, EPI_ISL_824680, EPI_ISL_824681, EPI_ISL_824682, EPI_ISL_824683, EPI_ISL_824684, EPI_ISL_824685, EPI_ISL_824686, EPI_ISL_824687, EPI_ISL_824688, EPI_ISL_824689, EPI_ISL_824690, EPI_ISL_824691, EPI_ISL_824692, EPI_ISL_824693, EPI_ISL_824694, EPI_ISL_824695, EPI_ISL_824696, EPI_ISL_824697, EPI_ISL_824698, EPI_ISL_824699, EPI_ISL_824700, EPI_ISL_824701, EPI_ISL_824702, EPI_ISL_824703, EPI_ISL_824704, EPI_ISL_824705, EPI_ISL_824706, EPI_ISL_824707, EPI_ISL_824708, EPI_ISL_824709, EPI_ISL_824710, EPI_ISL_824711, EPI_ISL_824712, EPI_ISL_824713, EPI_ISL_824714, EPI_ISL_824715, EPI_ISL_824716, EPI_ISL_824717, EPI_ISL_824718, EPI_ISL_824719, EPI_ISL_824720, EPI_ISL_824721, EPI_ISL_824722, EPI_ISL_824723, EPI_ISL_824724, EPI_ISL_824725, EPI_ISL_824726, EPI_ISL_824727, EPI_ISL_824728, EPI_ISL_824729, EPI_ISL_824730, EPI_ISL_824731, EPI_ISL_824732, EPI_ISL_824733, EPI_ISL_824734, EPI_ISL_824735, EPI_ISL_824736, EPI_ISL_824737, EPI_ISL_824738, EPI_ISL_824739, EPI_ISL_824740, EPI_ISL_824741 | | | |

| | | | |
|---|---|---|---|
| see above | Cedars-Sinai Medical Center, Department of Pathology & Laboratory Medicine, Molecular Pathology Laboratory | Cedars-Sinai Medical Center, Molecular Pathology Laboratory of Department of Pathology & Laboratory Medicine and Genomic Core | Wenjuan Zhang, Brian Davis, Stephanie Chen, Jorge Mario Sincuir Martinez, Jasmine T Plummer, Eric Vail |
| EPI_ISL_824787, EPI_ISL_824788, EPI_ISL_824789 | Arizona State Public Health Laboratory | Arizona State Public Health Laboratory | Trung Huynh, Jessica Escobar, Katherine Fullerton, Nobuko Fukushima, Stacy White, Linda Getsinger, Victor Waddell |
| EPI_ISL_824865, EPI_ISL_824866, EPI_ISL_824867, EPI_ISL_824868, EPI_ISL_824869, EPI_ISL_824870, EPI_ISL_824871, EPI_ISL_824872, EPI_ISL_824873, EPI_ISL_824874, EPI_ISL_824875, EPI_ISL_824876, EPI_ISL_824877, EPI_ISL_824878, EPI_ISL_824879, EPI_ISL_824880, EPI_ISL_824881, EPI_ISL_824882, EPI_ISL_824883, EPI_ISL_824884, EPI_ISL_824885, EPI_ISL_824886, EPI_ISL_824887, EPI_ISL_824888, EPI_ISL_824889, EPI_ISL_824890, EPI_ISL_824891, EPI_ISL_824892, EPI_ISL_824893, EPI_ISL_824894, EPI_ISL_824895, EPI_ISL_824896, EPI_ISL_824897, EPI_ISL_824898, EPI_ISL_824899, EPI_ISL_824900, EPI_ISL_824901, EPI_ISL_824902, EPI_ISL_824903, EPI_ISL_824904, EPI_ISL_824905, EPI_ISL_824906, EPI_ISL_824907, EPI_ISL_824908, EPI_ISL_824909, EPI_ISL_824910, EPI_ISL_824911, EPI_ISL_824912, EPI_ISL_824913, EPI_ISL_824914, EPI_ISL_824915, EPI_ISL_824916, EPI_ISL_824917, EPI_ISL_824918, EPI_ISL_824919 | | | |
| see above | UW Virology Lab | UW Virology Lab | Pavitra Roychoudhury, Hong Xie, Lasata Shrestha, Meei-Li Huang, Keith R Jerome, Alexander Greninger |
| EPI_ISL_824922, EPI_ISL_824923, EPI_ISL_824924, EPI_ISL_824925, EPI_ISL_824926, EPI_ISL_824927, EPI_ISL_824928, EPI_ISL_824929, EPI_ISL_824930, EPI_ISL_824931, EPI_ISL_824932, EPI_ISL_824933, EPI_ISL_824934, EPI_ISL_824935, EPI_ISL_824937, EPI_ISL_824938, EPI_ISL_824939, EPI_ISL_824940, EPI_ISL_824941 | | | |
| see above | Arizona State Public Health Laboratory | Arizona State Public Health Laboratory | Trung Huynh, Jessica Escobar, Katherine Fullerton, Nobuko Fukushima, Stacy White, Linda Getsinger, Victor Waddell |
| EPI_ISL_824945, EPI_ISL_824946, EPI_ISL_824947, EPI_ISL_824948, EPI_ISL_824949, EPI_ISL_824950, EPI_ISL_824951, EPI_ISL_824952, EPI_ISL_824953, EPI_ISL_824954, EPI_ISL_824955, EPI_ISL_824956, EPI_ISL_824957, EPI_ISL_824958, EPI_ISL_824959, EPI_ISL_824960, EPI_ISL_824961, EPI_ISL_824962, EPI_ISL_824963, EPI_ISL_824964, EPI_ISL_824965, EPI_ISL_824966, EPI_ISL_824967, EPI_ISL_824968, EPI_ISL_824969, EPI_ISL_824970, EPI_ISL_824971, EPI_ISL_824972, EPI_ISL_824973, EPI_ISL_824974, EPI_ISL_824975, EPI_ISL_824976, EPI_ISL_824977, EPI_ISL_824978, EPI_ISL_824979, EPI_ISL_824980, EPI_ISL_824981, EPI_ISL_824982, EPI_ISL_824983, EPI_ISL_824984 | | | |
| see above | Maryland Public Health Laboratory | Maryland Public Health Laboratory | Maryland Department of Health Laboratories Administration |
| EPI_ISL_824987, EPI_ISL_824988, EPI_ISL_824989, EPI_ISL_824990, EPI_ISL_824991, EPI_ISL_824992, EPI_ISL_824993, EPI_ISL_824994, EPI_ISL_824995, EPI_ISL_824996, EPI_ISL_824997, EPI_ISL_824998, EPI_ISL_824999, EPI_ISL_825000, EPI_ISL_825001, EPI_ISL_825002, EPI_ISL_825003, EPI_ISL_825004, EPI_ISL_825005, EPI_ISL_825006, EPI_ISL_825007, EPI_ISL_825008, EPI_ISL_825009, EPI_ISL_825010, EPI_ISL_825011, EPI_ISL_825012 | | | |
| see above | Arizona State Public Health Laboratory | Arizona State Public Health Laboratory | Trung Huynh, Jessica Escobar, Katherine Fullerton, Nobuko Fukushima, Stacy White, Linda Getsinger, Victor Waddell |
| EPI_ISL_825013 | Johns Hopkins Pathology | Johns Hopkins Hospital Department of Pathology | C. Paul Morris, Chun Huai Luo, Adannaya Amadi, Nicholas Gallagher, Heba H. Mostafa |
| EPI_ISL_825014 | Northwestern Memorial Hospital | Ozer Lab | Ramon Lorenzo-Redondo, Lacy M. Simons, Lawrence J. Jennings, Michael G. Ison, Judd F. Hultquist, Egon A. Ozer |
| EPI_ISL_825015 | UW Virology Lab | UW Virology Lab | Pavitra Roychoudhury, Hong Xie, Lasata Shrestha, Meei-Li Huang, Keith R Jerome, Alexander Greninger |
| EPI_ISL_825028, EPI_ISL_825029, EPI_ISL_825030, EPI_ISL_825031, EPI_ISL_825032 | Infectious Diseases, NC SLPH COVID-19 Response Team | Infectious Diseases, NC SLPH COVID-19 Response Team | Miller,M.C., Chase,K. |
| EPI_ISL_826286, EPI_ISL_826287, EPI_ISL_826288, EPI_ISL_826289, EPI_ISL_826290, EPI_ISL_826291, EPI_ISL_826292, EPI_ISL_826293, EPI_ISL_826294, EPI_ISL_826295 | The Jackson Laboratory | The Jackson Laboratory | Lloyd M, Maurya R, Renzette N, Omerza G, Kelly K, Li L, Wei C L, Adams M |
| EPI_ISL_826297, EPI_ISL_826298, EPI_ISL_826299, EPI_ISL_826300, EPI_ISL_826301, EPI_ISL_826302, EPI_ISL_826303, EPI_ISL_826304, EPI_ISL_826306, EPI_ISL_826307, EPI_ISL_826308, EPI_ISL_826309, EPI_ISL_826310, EPI_ISL_826311, EPI_ISL_826312, EPI_ISL_826313, EPI_ISL_826314, EPI_ISL_826315, EPI_ISL_826316, EPI_ISL_826317, EPI_ISL_826318 | | | |
| see above | Wyoming Public Health Laboratory | Wyoming Public Health Laboratory | Noah Hull, Taylor Fearing, Lynette Gumbleton, Channing Weber, Ashley Norberg, Bailey Bowcutt, and Wanda Manley |
| EPI_ISL_826319, EPI_ISL_826320, EPI_ISL_826321, EPI_ISL_826322, EPI_ISL_826323, EPI_ISL_826324, EPI_ISL_826325, EPI_ISL_826326, EPI_ISL_826327, EPI_ISL_826328, EPI_ISL_826329, EPI_ISL_826330, EPI_ISL_826331, EPI_ISL_826332, EPI_ISL_826333, EPI_ISL_826334, EPI_ISL_826335, EPI_ISL_826336, EPI_ISL_826337, EPI_ISL_826338, EPI_ISL_826339, EPI_ISL_826340, EPI_ISL_826341, EPI_ISL_826342, EPI_ISL_826343, EPI_ISL_826344, EPI_ISL_826345, EPI_ISL_826346, EPI_ISL_826347, EPI_ISL_826348, EPI_ISL_826349, EPI_ISL_826350, EPI_ISL_826351, EPI_ISL_826352, EPI_ISL_826353, EPI_ISL_826354, EPI_ISL_826355, EPI_ISL_826356, EPI_ISL_826357, EPI_ISL_826358, EPI_ISL_826359, EPI_ISL_826360, EPI_ISL_826361, EPI_ISL_826362, EPI_ISL_826363, EPI_ISL_826364, EPI_ISL_826365, EPI_ISL_826366, EPI_ISL_826367, EPI_ISL_826368, EPI_ISL_826369, EPI_ISL_826370, EPI_ISL_826371, EPI_ISL_826372, EPI_ISL_826373, EPI_ISL_826374, EPI_ISL_826375, EPI_ISL_826376, EPI_ISL_826377, EPI_ISL_826378, EPI_ISL_826379, EPI_ISL_826380, EPI_ISL_826381, EPI_ISL_826382, EPI_ISL_826383, EPI_ISL_826384, EPI_ISL_826385, EPI_ISL_826386, EPI_ISL_826387, EPI_ISL_826388, EPI_ISL_826389, EPI_ISL_826390, EPI_ISL_826391, EPI_ISL_826392, EPI_ISL_826393, EPI_ISL_826394, EPI_ISL_826395, EPI_ISL_826396, EPI_ISL_826397, EPI_ISL_826398, EPI_ISL_826399, EPI_ISL_826400, EPI_ISL_826401, EPI_ISL_826402, EPI_ISL_826403, EPI_ISL_826404, EPI_ISL_826405, EPI_ISL_826406, EPI_ISL_826407, EPI_ISL_826408, EPI_ISL_826409, EPI_ISL_826410, EPI_ISL_826411, EPI_ISL_826412, EPI_ISL_826413, EPI_ISL_826414, EPI_ISL_826415, EPI_ISL_826416, EPI_ISL_826417, EPI_ISL_826418, EPI_ISL_826419, EPI_ISL_826420, EPI_ISL_826421, EPI_ISL_826422, EPI_ISL_826423, EPI_ISL_826424, EPI_ISL_826425, EPI_ISL_826426, EPI_ISL_826427, EPI_ISL_826428, EPI_ISL_826429, EPI_ISL_826430, EPI_ISL_826431, EPI_ISL_826432, EPI_ISL_826433, EPI_ISL_826434, EPI_ISL_826435, EPI_ISL_826436, EPI_ISL_826437, EPI_ISL_826438, EPI_ISL_826439, EPI_ISL_826440, EPI_ISL_826441, EPI_ISL_826442, EPI_ISL_826443, EPI_ISL_826444, EPI_ISL_826445, EPI_ISL_826446, EPI_ISL_826447, EPI_ISL_826448, EPI_ISL_826449, EPI_ISL_826450, EPI_ISL_826451, EPI_ISL_826452, EPI_ISL_826453, EPI_ISL_826454, EPI_ISL_826455, EPI_ISL_826456 | | | |
| see above | University of Michigan Clinical Microbiology Laboratory | Lauring Lab, University of Michigan, Department of Microbiology and Immunology | Valesano |
| EPI_ISL_826461 | Montana Public Health Laboratory | Wyoming Public Health Laboratory | Noah Hull, Joy Ritter, Taylor Fearing, Lynette Gumbleton, Channing Weber, Ashley Norberg, Bailey Bowcutt, Wanda Manley, Deborah Gibson |
| EPI_ISL_826463 | The Ohio State University | OSU Polaris Molecular Laboratory | Huolin Tu, Matthew R Avenarius, Laura Kubatko, Matthew Hunt, Xiaokang Pan, Peng Ru, Jason Garee, Keelie Thomas, Peter Mohler, Preeti Pancholi, Dan Jones |
| EPI_ISL_826521 | The Ohio State University | James Molecular Laboratory | Huolin Tu, Matthew R Avenarius, Laura Kubatko, Matthew Hunt, Xiaokang Pan, Peng Ru, Jason Garee, Keelie Thomas, Peter Mohler, Preeti Pancholi, Dan Jones |
| EPI_ISL_826522 | The Jackson Laboratory | The Jackson Laboratory | Lloyd M, Maurya R, Renzette N, Omerza G, Kelly K, Li L, Wei C L, Adams M |
| EPI_ISL_826537, EPI_ISL_826538, EPI_ISL_826539, EPI_ISL_826540, EPI_ISL_826541, EPI_ISL_826542, EPI_ISL_826543, EPI_ISL_826544, EPI_ISL_826545, EPI_ISL_826546, EPI_ISL_826547, EPI_ISL_826548, EPI_ISL_826549, EPI_ISL_826550, EPI_ISL_826551, EPI_ISL_826552, EPI_ISL_826553, EPI_ISL_826554, EPI_ISL_826555, EPI_ISL_826556, EPI_ISL_826557, EPI_ISL_826558, EPI_ISL_826559, EPI_ISL_826560, EPI_ISL_826561, EPI_ISL_826562, EPI_ISL_826563, EPI_ISL_826564, EPI_ISL_826565, EPI_ISL_826566, EPI_ISL_826567, EPI_ISL_826568, EPI_ISL_826577 | | | |
| see above | Texas Department of State Health Services | Texas Department of State Health Services | Rashmi Tuladhar, Bonnie Oh, Jenny Zhang, Maliha Rahman, Anita Pokharel, Myong Koag, Chung Wang, Rachel Lee, Grace Kubin, Mayela Pedrueza, James Daniel Bonser |
| EPI_ISL_826578, EPI_ISL_826579, EPI_ISL_826580, EPI_ISL_826581, EPI_ISL_826582, EPI_ISL_826583, EPI_ISL_826584, EPI_ISL_826585, EPI_ISL_826586, EPI_ISL_826587, EPI_ISL_826588, EPI_ISL_826589, EPI_ISL_826590, EPI_ISL_826591, EPI_ISL_826592, EPI_ISL_826593, EPI_ISL_826594, EPI_ISL_826595, EPI_ISL_826596, EPI_ISL_826597, EPI_ISL_826598, EPI_ISL_826599, EPI_ISL_826600, EPI_ISL_826601, EPI_ISL_826602, EPI_ISL_826603, EPI_ISL_826604, EPI_ISL_826605, EPI_ISL_826606, EPI_ISL_826607, EPI_ISL_826608, EPI_ISL_826609, EPI_ISL_826610, EPI_ISL_826611, EPI_ISL_826612, EPI_ISL_826613, EPI_ISL_826614, EPI_ISL_826615, EPI_ISL_826616, EPI_ISL_826617, EPI_ISL_826618, EPI_ISL_826619, EPI_ISL_826620, EPI_ISL_826621, EPI_ISL_826622, EPI_ISL_826623, EPI_ISL_826624, EPI_ISL_826625, EPI_ISL_826626, EPI_ISL_826627, EPI_ISL_826628, EPI_ISL_826629, EPI_ISL_826630, EPI_ISL_826631, EPI_ISL_826632, EPI_ISL_826633, EPI_ISL_826634, EPI_ISL_826635, EPI_ISL_826636, EPI_ISL_826637, EPI_ISL_826638, EPI_ISL_826639, EPI_ISL_826640, EPI_ISL_826641, EPI_ISL_826642, EPI_ISL_826643, EPI_ISL_826644, EPI_ISL_826645, EPI_ISL_826646, EPI_ISL_826647, EPI_ISL_826648, EPI_ISL_826649, EPI_ISL_826650, EPI_ISL_826651, EPI_ISL_826652, EPI_ISL_826653, EPI_ISL_826654, EPI_ISL_826655, EPI_ISL_826656, EPI_ISL_826657, EPI_ISL_826658, EPI_ISL_826659, EPI_ISL_826660, EPI_ISL_826661, EPI_ISL_826662, EPI_ISL_826663, EPI_ISL_826664, EPI_ISL_826665, EPI_ISL_826666, EPI_ISL_826667, EPI_ISL_826668, EPI_ISL_826669, EPI_ISL_826670 | | | |
| see above | Montefiore Medical Center | Albert Einstein College of Medicine, Dept. of Microbiology & Immunology, Chandran lab | J. Maximilian Fels, Saad Khan, Ryan Forster, Karin A. Skalina, Surksha Sirichand, Amy S. Fox, Aviv Bergman, William B. Mitchell, Lucia R. Wolgast, Wendy Szymczak, Robert H. Bortz III, M. Eugenia Dieterle, Catalina Florez, Denise Haslwanter, Rohit K. Jangra, Ethan Laudermilch, Ariel S. Wirchnianski, Jason Barnhill, David L. Goldman, Hnin Khine, D. Yitzchak Goldstein, Johanna P. Daily, Kartik Chandran, Libusha Kelly |
| EPI_ISL_827438, EPI_ISL_827439, EPI_ISL_827441, EPI_ISL_827443, EPI_ISL_827445, EPI_ISL_827446 | University of Michigan Clinical Microbiology Laboratory | Lauring Lab, University of Michigan, Department of Microbiology and Immunology | Valesano |
| EPI_ISL_830208, EPI_ISL_830210, EPI_ISL_830212 | KALEIDA CENTER FOR LABORATORY MEDICINE | Wadsworth Center, New York State Department of Health | Kirsten St. George, Daryl M. Lamson, Alexis Russel, Matthew Shudt, Melissa A Leisner, Jonathan Plitnick, Navjot Singh, John Kelly, Erasmus Schneider, Erica Lasek-Nesselquist |
| EPI_ISL_830214 | BOSTON HEART DIAGNOSTICS CORP | Wadsworth Center, New York State Department of Health | Kirsten St. George, Daryl M. Lamson, Alexis Russel, Matthew Shudt, Melissa A Leisner, Jonathan Plitnick, Navjot Singh, John Kelly, Erasmus Schneider, Erica Lasek-Nesselquist |
| EPI_ISL_830216, EPI_ISL_830218, EPI_ISL_830220, EPI_ISL_830221 | Wadsworth Center, New York State Department of Health | Wadsworth Center, New York State Department of Health | Kirsten St. George, Daryl M. Lamson, Alexis Russel, Matthew Shudt, Melissa A Leisner, Jonathan Plitnick, Navjot Singh, John Kelly, Erasmus Schneider, Erica Lasek-Nesselquist |
| EPI_ISL_830223, EPI_ISL_830225, EPI_ISL_830227, EPI_ISL_830229, EPI_ISL_830230, EPI_ISL_830232, EPI_ISL_830234, EPI_ISL_830236, EPI_ISL_830238, EPI_ISL_830239, EPI_ISL_830241, EPI_ISL_830243 | | | |
| see above | KALEIDA CENTER FOR LABORATORY MEDICINE | Wadsworth Center, New York State Department of Health | Kirsten St. George, Daryl M. Lamson, Alexis Russel, Matthew Shudt, Melissa A Leisner, Jonathan Plitnick, Navjot Singh, John Kelly, Erasmus Schneider, Erica Lasek-Nesselquist |
| EPI_ISL_830244, EPI_ISL_830246, EPI_ISL_830248, EPI_ISL_830250, EPI_ISL_830252, EPI_ISL_830253, EPI_ISL_830255, EPI_ISL_830257, EPI_ISL_830259, EPI_ISL_830261, EPI_ISL_830263, EPI_ISL_830265, EPI_ISL_830267, EPI_ISL_830269, EPI_ISL_830270, EPI_ISL_830272, EPI_ISL_830274, EPI_ISL_830276, EPI_ISL_830278, EPI_ISL_830280, EPI_ISL_830282, EPI_ISL_830284, EPI_ISL_830286, EPI_ISL_830288, EPI_ISL_830289, EPI_ISL_830291, EPI_ISL_830293, EPI_ISL_830294, EPI_ISL_830296, EPI_ISL_830298, EPI_ISL_830300, EPI_ISL_830302, EPI_ISL_830304, EPI_ISL_830306, EPI_ISL_830308, EPI_ISL_830310, EPI_ISL_830312, EPI_ISL_830313, EPI_ISL_830315, EPI_ISL_830317, EPI_ISL_830319, EPI_ISL_830321, EPI_ISL_830323, EPI_ISL_830325, EPI_ISL_830326, EPI_ISL_830329, EPI_ISL_830331, EPI_ISL_830333, EPI_ISL_830334, EPI_ISL_830335, EPI_ISL_830337, EPI_ISL_830338, EPI_ISL_830340, EPI_ISL_830342, EPI_ISL_830344, EPI_ISL_830346, EPI_ISL_830348, EPI_ISL_830349 | | | |
| see above | Wadsworth Center, New York State Department of Health | Wadsworth Center, New York State Department of Health | Kirsten St. George, Daryl M. Lamson, Alexis Russel, Matthew Shudt, Melissa A Leisner, Jonathan Plitnick, Navjot Singh, John Kelly, Erasmus Schneider, Erica Lasek-Nesselquist |
| EPI_ISL_830571 | KALEIDA CENTER FOR LABORATORY MEDICINE | Wadsworth Center, New York State Department of Health | Kirsten St. George, Daryl M. Lamson, Alexis Russel, Matthew Shudt, Melissa A Leisner, Jonathan Plitnick, Navjot Singh, John Kelly, Erasmus Schneider, Erica Lasek-Nesselquist |
| EPI_ISL_830572 | NORTHWELL HEALTH LABORATORIES | Wadsworth Center, New York State Department of Health | Kirsten St. George, Daryl M. Lamson, Alexis Russel, Matthew Shudt, Melissa A Leisner, Jonathan Plitnick, Navjot Singh, John Kelly, Erasmus Schneider, Erica Lasek-Nesselquist |
| EPI_ISL_830573 | KALEIDA CENTER FOR LABORATORY MEDICINE | Wadsworth Center, New York State Department of Health | Kirsten St. George, Daryl M. Lamson, Alexis Russel, Matthew Shudt, Melissa A Leisner, Jonathan Plitnick, Navjot Singh, John Kelly, Erasmus Schneider, Erica Lasek-Nesselquist |
| EPI_ISL_830574 | NORTHWELL HEALTH LABORATORIES | Wadsworth Center, New York State Department of Health | Kirsten St. George, Daryl M. Lamson, Alexis Russel, Matthew Shudt, Melissa A Leisner, Jonathan Plitnick, Navjot Singh, John Kelly, Erasmus Schneider, Erica Lasek-Nesselquist |

| | | | |
|---|---|---|---|
| EPI_ISL_830575 | KALEIDA CENTER FOR LABORATORY MEDICINE | Wadsworth Center, New York State Department of Health | Kirsten St. George, Daryl M. Lamson, Alexis Russel, Matthew Shudt, Melissa A Leisner, Jonathan Plitnick, Navjot Singh, John Kelly, Erasmus Schneider, Erica Lasek-Nesselquist |
| EPI_ISL_830576, EPI_ISL_830577, EPI_ISL_830578 | NORTHWELL HEALTH LABORATORIES | Wadsworth Center, New York State Department of Health | Kirsten St. George, Daryl M. Lamson, Alexis Russel, Matthew Shudt, Melissa A Leisner, Jonathan Plitnick, Navjot Singh, John Kelly, Erasmus Schneider, Erica Lasek-Nesselquist |
| EPI_ISL_830579, EPI_ISL_830580, EPI_ISL_830581, EPI_ISL_830582, EPI_ISL_830583, EPI_ISL_830584, EPI_ISL_830585, EPI_ISL_830586, EPI_ISL_830587, EPI_ISL_830588, EPI_ISL_830589, EPI_ISL_830590, EPI_ISL_830591, EPI_ISL_830592, EPI_ISL_830593, EPI_ISL_830594, EPI_ISL_830595, EPI_ISL_830596, EPI_ISL_830597, EPI_ISL_830598, EPI_ISL_830599, EPI_ISL_830600, EPI_ISL_830601, EPI_ISL_830602, EPI_ISL_830603, EPI_ISL_830604, EPI_ISL_830605, EPI_ISL_830606, EPI_ISL_830607, EPI_ISL_830608, EPI_ISL_830609, EPI_ISL_830610, EPI_ISL_830611, EPI_ISL_830612, EPI_ISL_830613, EPI_ISL_830614, EPI_ISL_830615, EPI_ISL_830616, EPI_ISL_830617, EPI_ISL_830618, EPI_ISL_830619, EPI_ISL_830620, EPI_ISL_830621, EPI_ISL_830622, EPI_ISL_830623, EPI_ISL_830624, EPI_ISL_830625, EPI_ISL_830626, EPI_ISL_830627 | | | |
| see above | KALEIDA CENTER FOR LABORATORY MEDICINE | Wadsworth Center, New York State Department of Health | Kirsten St. George, Daryl M. Lamson, Alexis Russel, Matthew Shudt, Melissa A Leisner, Jonathan Plitnick, Navjot Singh, John Kelly, Erasmus Schneider, Erica Lasek-Nesselquist |
| EPI_ISL_830628, EPI_ISL_830629, EPI_ISL_830630, EPI_ISL_830631, EPI_ISL_830632, EPI_ISL_830633 | NORTHWELL HEALTH LABORATORIES | Wadsworth Center, New York State Department of Health | Kirsten St. George, Daryl M. Lamson, Alexis Russel, Matthew Shudt, Melissa A Leisner, Jonathan Plitnick, Navjot Singh, John Kelly, Erasmus Schneider, Erica Lasek-Nesselquist |
| EPI_ISL_830634 | MARY IMOGENE BASSETT HOSPITAL | Wadsworth Center, New York State Department of Health | Kirsten St. George, Daryl M. Lamson, Alexis Russel, Matthew Shudt, Melissa A Leisner, Jonathan Plitnick, Navjot Singh, John Kelly, Erasmus Schneider, Erica Lasek-Nesselquist |
| EPI_ISL_830635, EPI_ISL_830636, EPI_ISL_830637, EPI_ISL_830638, EPI_ISL_830639, EPI_ISL_830640, EPI_ISL_830641, EPI_ISL_830642 | NORTHWELL HEALTH LABORATORIES | Wadsworth Center, New York State Department of Health | Kirsten St. George, Daryl M. Lamson, Alexis Russel, Matthew Shudt, Melissa A Leisner, Jonathan Plitnick, Navjot Singh, John Kelly, Erasmus Schneider, Erica Lasek-Nesselquist |
| EPI_ISL_830643, EPI_ISL_830644, EPI_ISL_830645, EPI_ISL_830646, EPI_ISL_830647 | Wadsworth Center, New York State Department of Health | Wadsworth Center, New York State Department of Health | Kirsten St. George, Daryl M. Lamson, Alexis Russel, Matthew Shudt, Melissa A Leisner, Jonathan Plitnick, Navjot Singh, John Kelly, Erasmus Schneider, Erica Lasek-Nesselquist |
| EPI_ISL_830649 | Gundersen Molecular Diagnostics Laboratory | Kabara Cancer Research Institute | Craig S. Richmond, Paraic A. Kenny |
| EPI_ISL_830650, EPI_ISL_830651, EPI_ISL_830652, EPI_ISL_830653, EPI_ISL_830654, EPI_ISL_830655, EPI_ISL_830656, EPI_ISL_830657, EPI_ISL_830658, EPI_ISL_830659, EPI_ISL_830660, EPI_ISL_830661, EPI_ISL_830662, EPI_ISL_830663, EPI_ISL_830664, EPI_ISL_830665, EPI_ISL_830666, EPI_ISL_830667, EPI_ISL_830668, EPI_ISL_830669, EPI_ISL_830670, EPI_ISL_830671, EPI_ISL_830672, EPI_ISL_830673, EPI_ISL_830674, EPI_ISL_830675, EPI_ISL_830676, EPI_ISL_830677, EPI_ISL_830678, EPI_ISL_830679, EPI_ISL_830680, EPI_ISL_830681, EPI_ISL_830682 | | | |
| see above | SUNY UPSTATE MEDICAL UNIVERSITY | Wadsworth Center, New York State Department of Health | Kirsten St. George, Daryl M. Lamson, Alexis Russel, Matthew Shudt, Melissa A Leisner, Jonathan Plitnick, Navjot Singh, John Kelly, Erasmus Schneider, Erica Lasek-Nesselquist |
| EPI_ISL_830683, EPI_ISL_830684, EPI_ISL_830685, EPI_ISL_830686, EPI_ISL_830687, EPI_ISL_830688, EPI_ISL_830689, EPI_ISL_830690, EPI_ISL_830691, EPI_ISL_830692, EPI_ISL_830693, EPI_ISL_830694, EPI_ISL_830695, EPI_ISL_830696, EPI_ISL_830697, EPI_ISL_830698, EPI_ISL_830700, EPI_ISL_830701, EPI_ISL_830702, EPI_ISL_830703, EPI_ISL_830704, EPI_ISL_830705, EPI_ISL_830706, EPI_ISL_830707, EPI_ISL_830708, EPI_ISL_830709, EPI_ISL_830710, EPI_ISL_830711, EPI_ISL_830712, EPI_ISL_830713, EPI_ISL_830714, EPI_ISL_830715, EPI_ISL_830716, EPI_ISL_830717, EPI_ISL_830718, EPI_ISL_830719 | | | |
| see above | ALBANY MEDICAL CENTER HOSPITAL CLINICAL LABORATORIES | Wadsworth Center, New York State Department of Health | Kirsten St. George, Daryl M. Lamson, Alexis Russel, Matthew Shudt, Melissa A Leisner, Jonathan Plitnick, Navjot Singh, John Kelly, Erasmus Schneider, Erica Lasek-Nesselquist |
| EPI_ISL_830720, EPI_ISL_830721, EPI_ISL_830722, EPI_ISL_830723 | BIO-REFERENCE LABORATORIES | Wadsworth Center, New York State Department of Health | Kirsten St. George, Daryl M. Lamson, Alexis Russel, Matthew Shudt, Melissa A Leisner, Jonathan Plitnick, Navjot Singh, John Kelly, Erasmus Schneider, Erica Lasek-Nesselquist |
| EPI_ISL_830724, EPI_ISL_830725 | ACUTIS DIAGNOSTICS | Wadsworth Center, New York State Department of Health | Kirsten St. George, Daryl M. Lamson, Alexis Russel, Matthew Shudt, Melissa A Leisner, Jonathan Plitnick, Navjot Singh, John Kelly, Erasmus Schneider, Erica Lasek-Nesselquist |
| EPI_ISL_830726 | Wadsworth Center, New York State Department of Health | Wadsworth Center, New York State Department of Health | Kirsten St. George, Daryl M. Lamson, Alexis Russel, Matthew Shudt, Melissa A Leisner, Jonathan Plitnick, Navjot Singh, John Kelly, Erasmus Schneider, Erica Lasek-Nesselquist |
| EPI_ISL_831289, EPI_ISL_831290, EPI_ISL_831291, EPI_ISL_831292, EPI_ISL_831293, EPI_ISL_831294, EPI_ISL_831295, EPI_ISL_831296, EPI_ISL_831297, EPI_ISL_831298, EPI_ISL_831299, EPI_ISL_831300, EPI_ISL_831301, EPI_ISL_831302, EPI_ISL_831303, EPI_ISL_831304, EPI_ISL_831305, EPI_ISL_831306, EPI_ISL_831307, EPI_ISL_831308, EPI_ISL_831309, EPI_ISL_831310, EPI_ISL_831311, EPI_ISL_831312, EPI_ISL_831313, EPI_ISL_831314, EPI_ISL_831315, EPI_ISL_831316, EPI_ISL_831317, EPI_ISL_831318, EPI_ISL_831319, EPI_ISL_831320, EPI_ISL_831321, EPI_ISL_831331, EPI_ISL_831332 | | | |
| see above | Texas Department of State Health Services | Texas Department of State Health Services | Anita Pokharel, Bonnie Oh, James Daniel Bonser, Rashmi Tuladhar, Mayela Pedrueza, Jenny Zhang, Maliha Rahman, Myong Koag, Chung Wang, Rachel Lee, Grace Kubin |
| EPI_ISL_831334, EPI_ISL_831335, EPI_ISL_831336, EPI_ISL_831337, EPI_ISL_831338, EPI_ISL_831339, EPI_ISL_831340, EPI_ISL_831341, EPI_ISL_831342, EPI_ISL_831343, EPI_ISL_831344, EPI_ISL_831345, EPI_ISL_831346, EPI_ISL_831347, EPI_ISL_831348, EPI_ISL_831349, EPI_ISL_831350, EPI_ISL_831351, EPI_ISL_831352, EPI_ISL_831353, EPI_ISL_831354, EPI_ISL_831355, EPI_ISL_831356, EPI_ISL_831357, EPI_ISL_831358, EPI_ISL_831359, EPI_ISL_831360 | | | |
| see above | Clinical Molecular Microbiology Laboratory, UNC Hospitals | Jeremy Wang | Jeremy Wang, Alexander Rubinsteyn, Cecilia Thompson, Clark Cunningham, Kaylee Gentry, Jonathan Juliano, Benjamin Vincent, Melissa Miller, Robert Hagan, Corbin Jones |
| EPI_ISL_831361, EPI_ISL_831362, EPI_ISL_831363, EPI_ISL_831364, EPI_ISL_831365, EPI_ISL_831366, EPI_ISL_831367, EPI_ISL_831368, EPI_ISL_831369 | Clinical Molecular Microbiology Laboratory, UNC Hospitals | Jeremy Wang | Jeremy Wang, Alexander Rubinsteyn, Colleen Rice, Jason Smedberg, Melissa Miller, Corbin Jones, Robert Hagan |
| EPI_ISL_831475, EPI_ISL_831476, EPI_ISL_831477, EPI_ISL_831478, EPI_ISL_831479, EPI_ISL_831480, EPI_ISL_831481, EPI_ISL_831482, EPI_ISL_831483, EPI_ISL_831484, EPI_ISL_831485, EPI_ISL_831488, EPI_ISL_831489, EPI_ISL_831490, EPI_ISL_831491, EPI_ISL_831492, EPI_ISL_831493, EPI_ISL_831494, EPI_ISL_831495, EPI_ISL_831496, EPI_ISL_831497, EPI_ISL_831498, EPI_ISL_831499, EPI_ISL_831500, EPI_ISL_831501, EPI_ISL_831502, EPI_ISL_831503, EPI_ISL_831504, EPI_ISL_831505, EPI_ISL_831506, EPI_ISL_831507, EPI_ISL_831508, EPI_ISL_831509, EPI_ISL_831510, EPI_ISL_831511, EPI_ISL_831512, EPI_ISL_831513, EPI_ISL_831514, EPI_ISL_831515, EPI_ISL_831516, EPI_ISL_831517, EPI_ISL_831518, EPI_ISL_831519, EPI_ISL_831520, EPI_ISL_831521, EPI_ISL_831522, EPI_ISL_831523, EPI_ISL_831524, EPI_ISL_831525, EPI_ISL_831526, EPI_ISL_831527, EPI_ISL_831528, EPI_ISL_831529, EPI_ISL_831530, EPI_ISL_831531, EPI_ISL_831532, EPI_ISL_831533, EPI_ISL_831534, EPI_ISL_831535, EPI_ISL_831536, EPI_ISL_831537, EPI_ISL_831538, EPI_ISL_831539, EPI_ISL_831540, EPI_ISL_831541, EPI_ISL_831542, EPI_ISL_831543, EPI_ISL_831544, EPI_ISL_831545, EPI_ISL_831546, EPI_ISL_831547, EPI_ISL_831548, EPI_ISL_831549, EPI_ISL_831550, EPI_ISL_831551, EPI_ISL_831552, EPI_ISL_831553, EPI_ISL_831554, EPI_ISL_831555, EPI_ISL_831556, EPI_ISL_831557, EPI_ISL_831558, EPI_ISL_831559, EPI_ISL_831560, EPI_ISL_831561, EPI_ISL_831562, EPI_ISL_831563, EPI_ISL_831564, EPI_ISL_831565, EPI_ISL_831566, EPI_ISL_831567, EPI_ISL_831568, EPI_ISL_831569, EPI_ISL_831570, EPI_ISL_831571, EPI_ISL_831572, EPI_ISL_831573, EPI_ISL_831574, EPI_ISL_831575, EPI_ISL_831576, EPI_ISL_831577, EPI_ISL_831578, EPI_ISL_831579, EPI_ISL_831580, EPI_ISL_831581, EPI_ISL_831582, EPI_ISL_831583, EPI_ISL_831584, EPI_ISL_831585, EPI_ISL_831586, EPI_ISL_831587, EPI_ISL_831588, EPI_ISL_831589, EPI_ISL_831590, EPI_ISL_831591, EPI_ISL_831592, EPI_ISL_831593, EPI_ISL_831594, EPI_ISL_831595, EPI_ISL_831596, EPI_ISL_831597, EPI_ISL_831598, EPI_ISL_831599, EPI_ISL_831600, EPI_ISL_831601, EPI_ISL_831602, EPI_ISL_831603, EPI_ISL_831604, EPI_ISL_831605, EPI_ISL_831606, EPI_ISL_831607, EPI_ISL_831608, EPI_ISL_831609, EPI_ISL_831610, EPI_ISL_831611, EPI_ISL_831612, EPI_ISL_831613, EPI_ISL_831614, EPI_ISL_831615, EPI_ISL_831616, EPI_ISL_831617, EPI_ISL_831618, EPI_ISL_831619, EPI_ISL_831620, EPI_ISL_831622, EPI_ISL_831623, EPI_ISL_831624, EPI_ISL_831625, EPI_ISL_831626, EPI_ISL_831627, EPI_ISL_831628, EPI_ISL_831629, EPI_ISL_831630, EPI_ISL_831631, EPI_ISL_831632, EPI_ISL_831633, EPI_ISL_831634, EPI_ISL_831635, EPI_ISL_831636, EPI_ISL_831637, EPI_ISL_831638, EPI_ISL_831639, EPI_ISL_831640, EPI_ISL_831641, EPI_ISL_831642, EPI_ISL_831643 | | | |
| see above | University of Wisconsin-Madison AIDS Vaccine Research Laboratories | University of Wisconsin-Madison AIDS Vaccine Research Laboratories | Gage Moreno, Katarina Braun, et al. AIDS Vaccine Research Laboratories |
| EPI_ISL_831648, EPI_ISL_831662, EPI_ISL_831687 | Santa Clara County Public Health Laboratory | Santa Clara County Public Health Laboratory | Santa Clara County Public Health Department |
| EPI_ISL_831690, EPI_ISL_831691, EPI_ISL_831692, EPI_ISL_831693, EPI_ISL_831694, EPI_ISL_831695, EPI_ISL_831696, EPI_ISL_831697, EPI_ISL_831698, EPI_ISL_831699, EPI_ISL_831700, EPI_ISL_831701, EPI_ISL_831702, EPI_ISL_831703, EPI_ISL_831704, EPI_ISL_831705, EPI_ISL_831706, EPI_ISL_831707, EPI_ISL_831708, EPI_ISL_831709, EPI_ISL_831710, EPI_ISL_831711, EPI_ISL_831712, EPI_ISL_831713, EPI_ISL_831714, EPI_ISL_831715, EPI_ISL_831716, EPI_ISL_831717, EPI_ISL_831718, EPI_ISL_831719, EPI_ISL_831720, EPI_ISL_831721, EPI_ISL_831722, EPI_ISL_831723, EPI_ISL_831724, EPI_ISL_831725, EPI_ISL_831726, EPI_ISL_831727, EPI_ISL_831728, EPI_ISL_831729, EPI_ISL_831748, EPI_ISL_831749, EPI_ISL_831750, EPI_ISL_831751, EPI_ISL_831752, EPI_ISL_831753, EPI_ISL_831754, EPI_ISL_831755, EPI_ISL_831756, EPI_ISL_831757, EPI_ISL_831758, EPI_ISL_831759, EPI_ISL_831760, EPI_ISL_831761, EPI_ISL_831762, EPI_ISL_831763, EPI_ISL_831764, EPI_ISL_831765, EPI_ISL_831766, EPI_ISL_831767, EPI_ISL_831768, EPI_ISL_831769, EPI_ISL_831770, EPI_ISL_831771, EPI_ISL_831772, EPI_ISL_831773, EPI_ISL_831774, EPI_ISL_831775, EPI_ISL_831776, EPI_ISL_831777, EPI_ISL_831778, EPI_ISL_831779, EPI_ISL_831780, EPI_ISL_831781, EPI_ISL_831782, EPI_ISL_831783, EPI_ISL_831784, EPI_ISL_831785, EPI_ISL_831786, EPI_ISL_831787, EPI_ISL_831788, EPI_ISL_831789, EPI_ISL_831790, EPI_ISL_831791, EPI_ISL_831792, EPI_ISL_831793, EPI_ISL_831794, EPI_ISL_831795, EPI_ISL_831796, EPI_ISL_831797, EPI_ISL_831798, EPI_ISL_831799, EPI_ISL_831800, EPI_ISL_831801, EPI_ISL_831802, EPI_ISL_831803, EPI_ISL_831804, EPI_ISL_831805, EPI_ISL_831806, EPI_ISL_831807, EPI_ISL_831808, EPI_ISL_831809, EPI_ISL_831810, EPI_ISL_831811, EPI_ISL_831812, EPI_ISL_831813, EPI_ISL_831814, EPI_ISL_831815, EPI_ISL_831816, EPI_ISL_831817, EPI_ISL_831818, EPI_ISL_831819, EPI_ISL_831820, EPI_ISL_831821, EPI_ISL_831822, EPI_ISL_831823, EPI_ISL_831824, EPI_ISL_831825, EPI_ISL_831826, EPI_ISL_831827, EPI_ISL_831828, EPI_ISL_831829, EPI_ISL_831830, EPI_ISL_831831, EPI_ISL_831832, EPI_ISL_831833, EPI_ISL_831834, EPI_ISL_831835, EPI_ISL_831836, EPI_ISL_831837, EPI_ISL_831838, EPI_ISL_831839, EPI_ISL_831840, EPI_ISL_831841, EPI_ISL_831842, EPI_ISL_831843, EPI_ISL_831844, EPI_ISL_831845, EPI_ISL_831846, EPI_ISL_831847, EPI_ISL_831848, EPI_ISL_831849, EPI_ISL_831850, EPI_ISL_831851, EPI_ISL_831852, EPI_ISL_831853, EPI_ISL_831854, EPI_ISL_831855, EPI_ISL_831856, EPI_ISL_831857, EPI_ISL_831858, EPI_ISL_831859, EPI_ISL_831860, EPI_ISL_831861, EPI_ISL_831862, EPI_ISL_831863, EPI_ISL_831864, EPI_ISL_831865, EPI_ISL_831866, EPI_ISL_831867, EPI_ISL_831868, EPI_ISL_831869, EPI_ISL_831870, EPI_ISL_831871, EPI_ISL_831872, EPI_ISL_831873, EPI_ISL_831874, EPI_ISL_831875, EPI_ISL_831876, EPI_ISL_831877, EPI_ISL_831878, EPI_ISL_831879, EPI_ISL_831880, EPI_ISL_831881, EPI_ISL_831882, EPI_ISL_831883, EPI_ISL_831884, EPI_ISL_831885, EPI_ISL_831886, EPI_ISL_831887, EPI_ISL_831888, EPI_ISL_831889, EPI_ISL_831890, EPI_ISL_831891 | | | |
| see above | United States Air Force School of Aerospace Medicine | United States Air Force School of Aerospace Medicine | Anthony Fries, Jennifer Meyer, William Gruner, Amanda Javorina, Sarah Purves, Clarise Starr, Elizabeth Macias |
| EPI_ISL_831893, EPI_ISL_831894, EPI_ISL_831895, EPI_ISL_831896, EPI_ISL_831897, EPI_ISL_831899 | Santa Clara County Public Health Laboratory | Santa Clara County Public Health Laboratory | Santa Clara County Public Health Department |
| EPI_ISL_831900, EPI_ISL_831901, EPI_ISL_831902, EPI_ISL_831903, EPI_ISL_831904, EPI_ISL_831905, EPI_ISL_831906, EPI_ISL_831907, EPI_ISL_831908, EPI_ISL_831909, EPI_ISL_831910, EPI_ISL_831911, EPI_ISL_831912, EPI_ISL_831914, EPI_ISL_831915, EPI_ISL_831916, EPI_ISL_831917, EPI_ISL_831918, EPI_ISL_831919, EPI_ISL_831920, EPI_ISL_831921, EPI_ISL_831922, EPI_ISL_831923, EPI_ISL_831924, EPI_ISL_831925, EPI_ISL_831926, EPI_ISL_831927, EPI_ISL_831928, EPI_ISL_831929, EPI_ISL_831930, EPI_ISL_831931, EPI_ISL_831932, EPI_ISL_831933, EPI_ISL_831934, EPI_ISL_831935, EPI_ISL_831936, EPI_ISL_831937 | | | |
| see above | New Mexico Department of Health | New Mexico Department of Health Scientific | Ellie Johnson, Anastacia Griego-Fisher, D'eldra Malone |

| | | | |
|---|---|---|---|
| | Scientific Laboratory | Laboratory | |
| EPI_ISL_832014 | Utah Public Health Laboratory | Utah Public Health Laboratory | Erin L. Young, Kelly F. Oakeson, Tara Gallagher |
| EPI_ISL_832015, EPI_ISL_832016, EPI_ISL_832017, EPI_ISL_832018 | University of Wisconsin-Madison AIDS Vaccine Research Laboratories | University of Wisconsin-Madison AIDS Vaccine Research Laboratories | Gage Moreno, Katarina Braun, et al. AIDS Vaccine Research Laboratories |
| EPI_ISL_832019 | Southern Nevada Public Health Laboratory | Southern Nevada Public Health Laboratory | Michael Picker, Holly Hansen, Erin Buttery, Joseph Benson |
| EPI_ISL_832023, EPI_ISL_832024, EPI_ISL_832025, EPI_ISL_832026, EPI_ISL_832027, EPI_ISL_832028, EPI_ISL_832029, EPI_ISL_832030, EPI_ISL_832031, EPI_ISL_832032, EPI_ISL_832034, EPI_ISL_832035, EPI_ISL_832036, EPI_ISL_832037, EPI_ISL_832038, EPI_ISL_832039, EPI_ISL_832040, EPI_ISL_832041, EPI_ISL_832042, EPI_ISL_832043, EPI_ISL_832044, EPI_ISL_832045, EPI_ISL_832046, EPI_ISL_832047, EPI_ISL_832048, EPI_ISL_832049, EPI_ISL_832050, EPI_ISL_832051, EPI_ISL_832052, EPI_ISL_832053, EPI_ISL_832054, EPI_ISL_832055, EPI_ISL_832056, EPI_ISL_832057, EPI_ISL_832058, EPI_ISL_832059, EPI_ISL_832060, EPI_ISL_832061, EPI_ISL_832062, EPI_ISL_832063, EPI_ISL_832064 | | | |
| see above | Wyoming Public Health Laboratory | Wyoming Public Health Laboratory | Noah Hull, Taylor Fearing, Lynette Gumbleton, Channing Weber, Ashley Norberg, Bailey Bowcutt, and Wanda Manley |
| EPI_ISL_832066, EPI_ISL_832068, EPI_ISL_832075, EPI_ISL_832076, EPI_ISL_832079, EPI_ISL_832080, EPI_ISL_832081 | Santa Clara County Public Health Laboratory | Santa Clara County Public Health Laboratory | Santa Clara County Public Health Department |
| EPI_ISL_832082, EPI_ISL_832083, EPI_ISL_832084, EPI_ISL_832085, EPI_ISL_832086, EPI_ISL_832088, EPI_ISL_832089 | Clinical Molecular Microbiology Laboratory, UNC Hospitals | Jeremy Wang | Jeremy Wang, Alexander Rubinsteyn, Cecilia Thompson, Clark Cunningham, Kaylee Gentry, Jonathan Juliano, Benjamin Vincent, Melissa Miller, Robert Hagan, Corbin Jones |
| EPI_ISL_832090 | Santa Clara County Public Health Laboratory | Santa Clara County Public Health Laboratory | Santa Clara County Public Health Department |
| EPI_ISL_832091, EPI_ISL_832092, EPI_ISL_832093, EPI_ISL_832094 | Clinical Molecular Microbiology Laboratory, UNC Hospitals | Jeremy Wang | Jeremy Wang, Alexander Rubinsteyn, Cecilia Thompson, Clark Cunningham, Kaylee Gentry, Jonathan Juliano, Benjamin Vincent, Melissa Miller, Robert Hagan, Corbin Jones |
| EPI_ISL_832095, EPI_ISL_832097 | Santa Clara County Public Health Laboratory | Santa Clara County Public Health Laboratory | Santa Clara County Public Health Department |
| EPI_ISL_832105, EPI_ISL_832106, EPI_ISL_832107, EPI_ISL_832108, EPI_ISL_832109, EPI_ISL_832110, EPI_ISL_832111, EPI_ISL_832112, EPI_ISL_832113, EPI_ISL_832114, EPI_ISL_832115, EPI_ISL_832116, EPI_ISL_832117, EPI_ISL_832118, EPI_ISL_832119, EPI_ISL_832120 | | | |
| see above | MD Laboratories | Los Angeles County Public Health Laboratories | P. Hemarajata et al. |
| EPI_ISL_832121, EPI_ISL_832122, EPI_ISL_832123, EPI_ISL_832124, EPI_ISL_832125, EPI_ISL_832126 | Clinical Molecular Microbiology Laboratory, UNC Hospitals | Jeremy Wang | Jeremy Wang, Alexander Rubinsteyn, Colleen Rice, Jason Smedberg, Melissa Miller, Corbin Jones, Robert Hagan |
| EPI_ISL_832127 | Michigan Department of Health and Human Services, Bureau of Laboratories | Michigan Department of Health and Human Services, Bureau of Laboratories | Blankenship HM, Riner D, Soehnlen MK |
| EPI_ISL_832203, EPI_ISL_832204 | Northwest Pathology | UW Virology Lab | Pavitra Roychoudhury, Hong Xie, Lasata Shrestha, Michelle Lin, Meei-Li Huang, Keith R Jerome, Alexander Greninger |
| EPI_ISL_832243 | DOHMH Corona | New York City Public Health Laboratory | Jade Wang, et al. |
| EPI_ISL_832244 | DOHMH Crown Heights | New York City Public Health Laboratory | Jade Wang, et al. |
| EPI_ISL_832245 | DOHMH Corona | New York City Public Health Laboratory | Jade Wang, et al. |
| EPI_ISL_832246 | DOHMH Jamaica | New York City Public Health Laboratory | Jade Wang, et al. |
| EPI_ISL_832247, EPI_ISL_832248 | DOHMH Morrisania | New York City Public Health Laboratory | Jade Wang, et al. |
| EPI_ISL_832249 | DOHMH PHL | New York City Public Health Laboratory | Jade Wang, et al. |
| EPI_ISL_832250 | DOHMH Jamaica | New York City Public Health Laboratory | Jade Wang, et al. |
| EPI_ISL_832251, EPI_ISL_832252, EPI_ISL_832253, EPI_ISL_832254 | DOHMH Corona | New York City Public Health Laboratory | Jade Wang, et al. |
| EPI_ISL_832255 | DOHMH Morrisania | New York City Public Health Laboratory | Jade Wang, et al. |
| EPI_ISL_832256 | DOHMH Central Harlem | New York City Public Health Laboratory | Jade Wang, et al. |
| EPI_ISL_832257 | DOHMH Morrisania | New York City Public Health Laboratory | Jade Wang, et al. |
| EPI_ISL_832258, EPI_ISL_832259 | DOHMH PHL | New York City Public Health Laboratory | Jade Wang, et al. |
| EPI_ISL_832260, EPI_ISL_832261, EPI_ISL_832262 | DOHMH Corona | New York City Public Health Laboratory | Jade Wang, et al. |
| EPI_ISL_832263 | DOHMH Jamaica | New York City Public Health Laboratory | Jade Wang, et al. |
| EPI_ISL_832264 | DOHMH Morrisania | New York City Public Health Laboratory | Jade Wang, et al. |
| EPI_ISL_832265 | DOHMH Jamaica | New York City Public Health Laboratory | Jade Wang, et al. |
| EPI_ISL_832266, EPI_ISL_832267 | DOHMH Central Harlem | New York City Public Health Laboratory | Jade Wang, et al. |
| EPI_ISL_832268, EPI_ISL_832269 | DOHMH Fort Greene | New York City Public Health Laboratory | Jade Wang, et al. |
| EPI_ISL_832270, EPI_ISL_832271 | DOHMH Riverside | New York City Public Health Laboratory | Jade Wang, et al. |
| EPI_ISL_832272, EPI_ISL_832273, EPI_ISL_832274 | DOHMH Jamaica | New York City Public Health Laboratory | Jade Wang, et al. |
| EPI_ISL_832275 | DOHMH Crown Heights | New York City Public Health Laboratory | Jade Wang, et al. |
| EPI_ISL_832276 | DOHMH Jamaica | New York City Public Health Laboratory | Jade Wang, et al. |
| EPI_ISL_832277, EPI_ISL_832278, EPI_ISL_832279 | DOHMH Corona | New York City Public Health Laboratory | Jade Wang, et al. |
| EPI_ISL_832280, EPI_ISL_832281 | DOHMH Morrisania | New York City Public Health Laboratory | Jade Wang, et al. |
| EPI_ISL_832282 | DOHMH Chelsea | New York City Public Health Laboratory | Jade Wang, et al. |
| EPI_ISL_832283 | DOHMH Jamaica | New York City Public Health Laboratory | Jade Wang, et al. |
| EPI_ISL_832284 | DOHMH Central Harlem | New York City Public Health Laboratory | Jade Wang, et al. |
| EPI_ISL_832285 | DOHMH Morrisania | New York City Public Health Laboratory | Jade Wang, et al. |
| EPI_ISL_832286 | DOHMH Fort Greene | New York City Public Health Laboratory | Jade Wang, et al. |

| | | | |
|---|---|---|---|
| EPI_ISL_832287, EPI_ISL_832288 | DOHMH Crown Heights | New York City Public Health Laboratory | Jade Wang, et al. |
| EPI_ISL_832289 | DOHMH Morrisania | New York City Public Health Laboratory | Jade Wang, et al. |
| EPI_ISL_832290, EPI_ISL_832291 | DOHMH Corona | New York City Public Health Laboratory | Jade Wang, et al. |
| EPI_ISL_832292, EPI_ISL_832293 | DOHMH Crown Heights | New York City Public Health Laboratory | Jade Wang, et al. |
| EPI_ISL_832294 | DOHMH Jamaica | New York City Public Health Laboratory | Jade Wang, et al. |
| EPI_ISL_832295 | DOHMH Crown Heights | New York City Public Health Laboratory | Jade Wang, et al. |
| EPI_ISL_832296 | DOHMH Riverside | New York City Public Health Laboratory | Jade Wang, et al. |
| EPI_ISL_832297, EPI_ISL_832298 | DOHMH Jamaica | New York City Public Health Laboratory | Jade Wang, et al. |
| EPI_ISL_832299 | DOHMH Morrisania | New York City Public Health Laboratory | Jade Wang, et al. |
| EPI_ISL_832300 | DOHMH Central Harlem | New York City Public Health Laboratory | Jade Wang, et al. |
| EPI_ISL_832301, EPI_ISL_832302 | DOHMH Chelsea | New York City Public Health Laboratory | Jade Wang, et al. |
| EPI_ISL_832303, EPI_ISL_832304 | DOHMH Corona | New York City Public Health Laboratory | Jade Wang, et al. |
| EPI_ISL_832305 | DOHMH Riverside | New York City Public Health Laboratory | Jade Wang, et al. |
| EPI_ISL_832306, EPI_ISL_832307, EPI_ISL_832308, EPI_ISL_832309, EPI_ISL_832310, EPI_ISL_832311 | DOHMH Jamaica | New York City Public Health Laboratory | Jade Wang, et al. |
| EPI_ISL_832312 | DOHMH Central Harlem | New York City Public Health Laboratory | Jade Wang, et al. |
| EPI_ISL_832313, EPI_ISL_832314 | DOHMH Chelsea | New York City Public Health Laboratory | Jade Wang, et al. |
| EPI_ISL_832315, EPI_ISL_832316 | DOHMH Morrisania | New York City Public Health Laboratory | Jade Wang, et al. |
| EPI_ISL_832317, EPI_ISL_832318, EPI_ISL_832319 | DOHMH Jamaica | New York City Public Health Laboratory | Jade Wang, et al. |
| EPI_ISL_832320, EPI_ISL_832321, EPI_ISL_832322 | DOHMH Corona | New York City Public Health Laboratory | Jade Wang, et al. |
| EPI_ISL_832323, EPI_ISL_832324 | DOHMH Jamaica | New York City Public Health Laboratory | Jade Wang, et al. |
| EPI_ISL_832325 | DOHMH Corona | New York City Public Health Laboratory | Jade Wang, et al. |
| EPI_ISL_832326 | DOHMH Riverside | New York City Public Health Laboratory | Jade Wang, et al. |
| EPI_ISL_832327 | DOHMH Morrisania | New York City Public Health Laboratory | Jade Wang, et al. |
| EPI_ISL_832328, EPI_ISL_832329, EPI_ISL_832330 | DOHMH Jamaica | New York City Public Health Laboratory | Jade Wang, et al. |
| EPI_ISL_832331, EPI_ISL_832332, EPI_ISL_832333, EPI_ISL_832334, EPI_ISL_832335 | DOHMH Corona | New York City Public Health Laboratory | Jade Wang, et al. |
| EPI_ISL_832336, EPI_ISL_832337 | DOHMH Central Harlem | New York City Public Health Laboratory | Jade Wang, et al. |
| EPI_ISL_832338 | DOHMH Crown Heights | New York City Public Health Laboratory | Jade Wang, et al. |
| EPI_ISL_832339 | DOHMH Riverside | New York City Public Health Laboratory | Jade Wang, et al. |
| EPI_ISL_832340 | DOHMH Crown Heights | New York City Public Health Laboratory | Jade Wang, et al. |
| EPI_ISL_832341 | DOHMH Chelsea | New York City Public Health Laboratory | Jade Wang, et al. |
| EPI_ISL_832342, EPI_ISL_832343, EPI_ISL_832344, EPI_ISL_832345 | DOHMH Morrisania | New York City Public Health Laboratory | Jade Wang, et al. |
| EPI_ISL_832346, EPI_ISL_832347, EPI_ISL_832348, EPI_ISL_832349, EPI_ISL_832350 | DOHMH Central Harlem | New York City Public Health Laboratory | Jade Wang, et al. |
| EPI_ISL_832351 | DOHMH Corona | New York City Public Health Laboratory | Jade Wang, et al. |
| EPI_ISL_832352, EPI_ISL_832353 | DOHMH PHL | New York City Public Health Laboratory | Jade Wang, et al. |
| EPI_ISL_832354 | DOHMH Jamaica | New York City Public Health Laboratory | Jade Wang, et al. |
| EPI_ISL_832355, EPI_ISL_832356 | DOHMH Corona | New York City Public Health Laboratory | Jade Wang, et al. |
| EPI_ISL_832357, EPI_ISL_832358 | DOHMH Jamaica | New York City Public Health Laboratory | Jade Wang, et al. |
| EPI_ISL_832359, EPI_ISL_832360 | DOHMH Corona | New York City Public Health Laboratory | Jade Wang, et al. |
| EPI_ISL_832361 | DOHMH Jamaica | New York City Public Health Laboratory | Jade Wang, et al. |
| EPI_ISL_832362, EPI_ISL_832363 | DOHMH Chelsea | New York City Public Health Laboratory | Jade Wang, et al. |
| EPI_ISL_832375, EPI_ISL_832376, EPI_ISL_832377 | Santa Clara County Public Health Laboratory | Santa Clara County Public Health Laboratory | Santa Clara County Public Health Department |
| EPI_ISL_832378 | Ohio State | James Molecular Lab | Huolin Tu, Matthew R Avenarius, Laura Kubatko, Matthew Hunt, Xiaokang Pan, Peng Ru, Jason Garee, Keelie Thomas, Peter Mohler, Preeti Pancholi, Dan Jones |
| EPI_ISL_832379, EPI_ISL_832380, EPI_ISL_832381, EPI_ISL_832382, EPI_ISL_832383, EPI_ISL_832388, EPI_ISL_832391, EPI_ISL_832392, EPI_ISL_832394, EPI_ISL_832395, EPI_ISL_832396, EPI_ISL_832398, EPI_ISL_832399, EPI_ISL_832400, EPI_ISL_832401, EPI_ISL_832402, EPI_ISL_832403, EPI_ISL_832404 | | | |
| see above | Santa Clara County Public Health Laboratory | Santa Clara County Public Health Laboratory | Santa Clara County Public Health Department |
| EPI_ISL_832405, EPI_ISL_832407, EPI_ISL_832408, EPI_ISL_832409, EPI_ISL_832410, EPI_ISL_832411, EPI_ISL_832412, EPI_ISL_832413, EPI_ISL_832414, EPI_ISL_832415, EPI_ISL_832416, EPI_ISL_832417, EPI_ISL_832418, EPI_ISL_832419, EPI_ISL_832420, EPI_ISL_832421, EPI_ISL_832422, EPI_ISL_832423, EPI_ISL_832424, EPI_ISL_832425, EPI_ISL_832426, EPI_ISL_832427, EPI_ISL_832428, EPI_ISL_832429, EPI_ISL_832430, EPI_ISL_832431, EPI_ISL_832432, EPI_ISL_832433, EPI_ISL_832434, EPI_ISL_832435, EPI_ISL_832436, EPI_ISL_832437, EPI_ISL_832438, EPI_ISL_832439, EPI_ISL_832440, EPI_ISL_832441, EPI_ISL_832442, EPI_ISL_832443, EPI_ISL_832444, EPI_ISL_832445, EPI_ISL_832446, EPI_ISL_832447, EPI_ISL_832448, EPI_ISL_832449, EPI_ISL_832450, EPI_ISL_832452, EPI_ISL_832453, EPI_ISL_832454, EPI_ISL_832455, EPI_ISL_832456, EPI_ISL_832457, EPI_ISL_832458, EPI_ISL_832459, EPI_ISL_832460, EPI_ISL_832461, EPI_ISL_832462, EPI_ISL_832463, EPI_ISL_832464, EPI_ISL_832465, EPI_ISL_832466, EPI_ISL_832467, EPI_ISL_832468, EPI_ISL_832469, EPI_ISL_832470, EPI_ISL_832471, EPI_ISL_832472, EPI_ISL_832473, EPI_ISL_832474, EPI_ISL_832475, EPI_ISL_832476, EPI_ISL_832477, EPI_ISL_832478, EPI_ISL_832479, EPI_ISL_832480, EPI_ISL_832481, EPI_ISL_832482, EPI_ISL_832483, EPI_ISL_832486, EPI_ISL_832487, EPI_ISL_832488, EPI_ISL_832489, EPI_ISL_832490, EPI_ISL_832491, EPI_ISL_832492, EPI_ISL_832493, EPI_ISL_832494, EPI_ISL_832495, EPI_ISL_832496, EPI_ISL_832497, EPI_ISL_832498, EPI_ISL_832499, EPI_ISL_832500, EPI_ISL_832501, EPI_ISL_832502, EPI_ISL_832503, EPI_ISL_832504, EPI_ISL_832505, EPI_ISL_832506, EPI_ISL_832507, EPI_ISL_832508, EPI_ISL_832509, EPI_ISL_832510, EPI_ISL_832512, EPI_ISL_832513, EPI_ISL_832514, EPI_ISL_832515, EPI_ISL_832516, EPI_ISL_832517, EPI_ISL_832518, EPI_ISL_832519, EPI_ISL_832520, EPI_ISL_832521, EPI_ISL_832522, EPI_ISL_832523, EPI_ISL_832524, EPI_ISL_832525, EPI_ISL_832526, EPI_ISL_832527, EPI_ISL_832528, EPI_ISL_832529, EPI_ISL_832530, EPI_ISL_832531, EPI_ISL_832532, EPI_ISL_832534, EPI_ISL_832535, EPI_ISL_832536, EPI_ISL_832537, EPI_ISL_832538, EPI_ISL_832539, EPI_ISL_832540, EPI_ISL_832541, EPI_ISL_832542, EPI_ISL_832543, EPI_ISL_832544, EPI_ISL_832545, EPI_ISL_832546, EPI_ISL_832547, EPI_ISL_832548, EPI_ISL_832549, EPI_ISL_832550, EPI_ISL_832551, EPI_ISL_832552, EPI_ISL_832553, EPI_ISL_832554, EPI_ISL_832555, EPI_ISL_832556, EPI_ISL_832557, EPI_ISL_832558, EPI_ISL_832559, EPI_ISL_832560, EPI_ISL_832561, EPI_ISL_832562, EPI_ISL_832563, EPI_ISL_832564, EPI_ISL_832565, EPI_ISL_832566, EPI_ISL_832567, EPI_ISL_832568, EPI_ISL_832569, EPI_ISL_832570, EPI_ISL_832571, EPI_ISL_832572, EPI_ISL_832573, EPI_ISL_832574, EPI_ISL_832575, EPI_ISL_832576, EPI_ISL_832577, EPI_ISL_832578, | | | |

| | | | |
|---|---|---|---|
| EPI_ISL_832579, EPI_ISL_832580, EPI_ISL_832581, EPI_ISL_832582, EPI_ISL_832583, EPI_ISL_832584, EPI_ISL_832585, EPI_ISL_832586, EPI_ISL_832587, EPI_ISL_832588, EPI_ISL_832589, EPI_ISL_832590, EPI_ISL_832591, EPI_ISL_832592, EPI_ISL_832593, EPI_ISL_832594, EPI_ISL_832595, EPI_ISL_832596, EPI_ISL_832597, EPI_ISL_832598, EPI_ISL_832599, EPI_ISL_832600, EPI_ISL_832601, EPI_ISL_832602, EPI_ISL_832603, EPI_ISL_832604, EPI_ISL_832605, EPI_ISL_832606, EPI_ISL_832607, EPI_ISL_832608, EPI_ISL_832609, EPI_ISL_832610, EPI_ISL_832612, EPI_ISL_832613, EPI_ISL_832614, EPI_ISL_832615, EPI_ISL_832616, EPI_ISL_832618, EPI_ISL_832619, EPI_ISL_832621, EPI_ISL_832622, EPI_ISL_832623, EPI_ISL_832624, EPI_ISL_832625, EPI_ISL_832626, EPI_ISL_832627, EPI_ISL_832628, EPI_ISL_832629, EPI_ISL_832630, EPI_ISL_832631, EPI_ISL_832632, EPI_ISL_832633, EPI_ISL_832634, EPI_ISL_832635, EPI_ISL_832636, EPI_ISL_832637, EPI_ISL_832638, EPI_ISL_832639, EPI_ISL_832640, EPI_ISL_832641, EPI_ISL_832642, EPI_ISL_832643, EPI_ISL_832644, EPI_ISL_832645, EPI_ISL_832646, EPI_ISL_832647, EPI_ISL_832648, EPI_ISL_832649, EPI_ISL_832650, EPI_ISL_832651, EPI_ISL_832652, EPI_ISL_832653, EPI_ISL_832654, EPI_ISL_832655, EPI_ISL_832656, EPI_ISL_832657, EPI_ISL_832658, EPI_ISL_832659, EPI_ISL_832660, EPI_ISL_832661, EPI_ISL_832662, EPI_ISL_832663, EPI_ISL_832664, EPI_ISL_832665, EPI_ISL_832666, EPI_ISL_832667, EPI_ISL_832668, EPI_ISL_832670, EPI_ISL_832671, EPI_ISL_832672, EPI_ISL_832673, EPI_ISL_832674, EPI_ISL_832676, EPI_ISL_832677, EPI_ISL_832678, EPI_ISL_832679, EPI_ISL_832680, EPI_ISL_832681, EPI_ISL_832682, EPI_ISL_832683, EPI_ISL_832684, EPI_ISL_832685, EPI_ISL_832687, EPI_ISL_832688, EPI_ISL_832689, EPI_ISL_832690, EPI_ISL_832691, EPI_ISL_832692, EPI_ISL_832693, EPI_ISL_832694, EPI_ISL_832695, EPI_ISL_832696, EPI_ISL_832697, EPI_ISL_832698, EPI_ISL_832699, EPI_ISL_832700, EPI_ISL_832701, EPI_ISL_832702, EPI_ISL_832703, EPI_ISL_832704, EPI_ISL_832706, EPI_ISL_832707, EPI_ISL_832708, EPI_ISL_832709, EPI_ISL_832710, EPI_ISL_832711, EPI_ISL_832712, EPI_ISL_832713, EPI_ISL_832714, EPI_ISL_832716, EPI_ISL_832717, EPI_ISL_832718, EPI_ISL_832719, EPI_ISL_832720, EPI_ISL_832721, EPI_ISL_832722, EPI_ISL_832723, EPI_ISL_832724, EPI_ISL_832726, EPI_ISL_832727, EPI_ISL_832728, EPI_ISL_832729, EPI_ISL_832730, EPI_ISL_832731, EPI_ISL_832732, EPI_ISL_832733, EPI_ISL_832734, EPI_ISL_832735, EPI_ISL_832736, EPI_ISL_832737, EPI_ISL_832738, EPI_ISL_832739, EPI_ISL_832740, EPI_ISL_832741, EPI_ISL_832742, EPI_ISL_832743, EPI_ISL_832744, EPI_ISL_832745, EPI_ISL_832746, EPI_ISL_832747, EPI_ISL_832748, EPI_ISL_832749, EPI_ISL_832752, EPI_ISL_832753, EPI_ISL_832755, EPI_ISL_832756, EPI_ISL_832757, EPI_ISL_832758, EPI_ISL_832759, EPI_ISL_832760, EPI_ISL_832761, EPI_ISL_832762, EPI_ISL_832763, EPI_ISL_832764, EPI_ISL_832765, EPI_ISL_832766, EPI_ISL_832767, EPI_ISL_832768, EPI_ISL_832769, EPI_ISL_832770, EPI_ISL_832771, EPI_ISL_832772, EPI_ISL_832773, EPI_ISL_832774, EPI_ISL_832775, EPI_ISL_832776, EPI_ISL_832777, EPI_ISL_832778, EPI_ISL_832779, EPI_ISL_832780, EPI_ISL_832781, EPI_ISL_832782, EPI_ISL_832783, EPI_ISL_832784, EPI_ISL_832786, EPI_ISL_832787, EPI_ISL_832788, EPI_ISL_832789, EPI_ISL_832790, EPI_ISL_832791, EPI_ISL_832792, EPI_ISL_832793, EPI_ISL_832794, EPI_ISL_832795, EPI_ISL_832796, EPI_ISL_832797, EPI_ISL_832798, EPI_ISL_832800, EPI_ISL_832801, EPI_ISL_832803, EPI_ISL_832805, EPI_ISL_832806, EPI_ISL_832807, EPI_ISL_832809, EPI_ISL_832811, EPI_ISL_832812, EPI_ISL_832813, EPI_ISL_832814 | | | |
| see above | OHSU Lab Services Molecular Microbiology Lab | Oregon SARS-CoV-2 Genome Sequencing Center | Brendan L. O'Connell, Ruth V. Nichols, Sally Grindstaff, Alec J. Hirsch, Donna Hansel, Guang Fan, Daniel N. Streblow, William B. Messer, Andrew C. Adey, Benjamin N. Bimber, Brian J. O’Roak |
| EPI_ISL_832823, EPI_ISL_832824, EPI_ISL_832825, EPI_ISL_832826, EPI_ISL_832827, EPI_ISL_832828, EPI_ISL_832829, EPI_ISL_832830, EPI_ISL_832831, EPI_ISL_832832, EPI_ISL_832833, EPI_ISL_832834, EPI_ISL_832835, EPI_ISL_832836, EPI_ISL_832837, EPI_ISL_832838, EPI_ISL_832839, EPI_ISL_832840, EPI_ISL_832841, EPI_ISL_832842, EPI_ISL_832843, EPI_ISL_832844, EPI_ISL_832845, EPI_ISL_832846, EPI_ISL_832848, EPI_ISL_832849, EPI_ISL_832850, EPI_ISL_832851, EPI_ISL_832852, EPI_ISL_832853, EPI_ISL_832854, EPI_ISL_832855, EPI_ISL_832856, EPI_ISL_832857, EPI_ISL_832858, EPI_ISL_832859, EPI_ISL_832860, EPI_ISL_832861, EPI_ISL_832862, EPI_ISL_832863, EPI_ISL_832865, EPI_ISL_832866, EPI_ISL_832867, EPI_ISL_832868, EPI_ISL_832869, EPI_ISL_832871, EPI_ISL_832872, EPI_ISL_832873, EPI_ISL_832874, EPI_ISL_832875, EPI_ISL_832876, EPI_ISL_832878, EPI_ISL_832879, EPI_ISL_832880, EPI_ISL_832881, EPI_ISL_832882, EPI_ISL_832883, EPI_ISL_832884, EPI_ISL_832885, EPI_ISL_832886, EPI_ISL_832887, EPI_ISL_832888, EPI_ISL_832889, EPI_ISL_832890, EPI_ISL_832891, EPI_ISL_832892, EPI_ISL_832893, EPI_ISL_832894, EPI_ISL_832895, EPI_ISL_832896, EPI_ISL_832897, EPI_ISL_832898, EPI_ISL_832899, EPI_ISL_832900, EPI_ISL_832901, EPI_ISL_832902, EPI_ISL_832903, EPI_ISL_832904, EPI_ISL_832905, EPI_ISL_832906, EPI_ISL_832907, EPI_ISL_832908, EPI_ISL_832909, EPI_ISL_832910, EPI_ISL_832911, EPI_ISL_832912, EPI_ISL_832913, EPI_ISL_832914, EPI_ISL_832915, EPI_ISL_832916, EPI_ISL_832917, EPI_ISL_832918, EPI_ISL_832919, EPI_ISL_832920, EPI_ISL_832921, EPI_ISL_832922, EPI_ISL_832923, EPI_ISL_832924, EPI_ISL_832925, EPI_ISL_832926, EPI_ISL_832927, EPI_ISL_832928, EPI_ISL_832929, EPI_ISL_832930, EPI_ISL_832931, EPI_ISL_832932, EPI_ISL_832933, EPI_ISL_832934, EPI_ISL_832935, EPI_ISL_832936, EPI_ISL_832937, EPI_ISL_832938, EPI_ISL_832939, EPI_ISL_832940, EPI_ISL_832941, EPI_ISL_832942, EPI_ISL_832943, EPI_ISL_832944, EPI_ISL_832945, EPI_ISL_832946, EPI_ISL_832947, EPI_ISL_832948, EPI_ISL_832949, EPI_ISL_832950, EPI_ISL_832951, EPI_ISL_832952, EPI_ISL_832953, EPI_ISL_832954, EPI_ISL_832955, EPI_ISL_832956, EPI_ISL_832957, EPI_ISL_832958, EPI_ISL_832959, EPI_ISL_832960, EPI_ISL_832961, EPI_ISL_832962, EPI_ISL_832963, EPI_ISL_832964, EPI_ISL_832965, EPI_ISL_832966, EPI_ISL_832967, EPI_ISL_832968, EPI_ISL_832969, EPI_ISL_832970, EPI_ISL_832971, EPI_ISL_832972, EPI_ISL_832973, EPI_ISL_832974, EPI_ISL_832975, EPI_ISL_832976, EPI_ISL_832977, EPI_ISL_832978, EPI_ISL_832979, EPI_ISL_832980, EPI_ISL_832981, EPI_ISL_832982, EPI_ISL_832983, EPI_ISL_832984, EPI_ISL_832985, EPI_ISL_832986, EPI_ISL_832987, EPI_ISL_832988, EPI_ISL_832989, EPI_ISL_832990, EPI_ISL_832991, EPI_ISL_832992, EPI_ISL_832993, EPI_ISL_832994, EPI_ISL_832995, EPI_ISL_832996, EPI_ISL_832997, EPI_ISL_832998, EPI_ISL_832999, EPI_ISL_833000, EPI_ISL_833001, EPI_ISL_833002, EPI_ISL_833003, EPI_ISL_833004, EPI_ISL_833005, EPI_ISL_833006, EPI_ISL_833008, EPI_ISL_833009, EPI_ISL_833010, EPI_ISL_833011, EPI_ISL_833012, EPI_ISL_833013, EPI_ISL_833014, EPI_ISL_833015, EPI_ISL_833016, EPI_ISL_833017, EPI_ISL_833018, EPI_ISL_833019, EPI_ISL_833020, EPI_ISL_833021, EPI_ISL_833022, EPI_ISL_833023, EPI_ISL_833024, EPI_ISL_833025, EPI_ISL_833026, EPI_ISL_833027, EPI_ISL_833028, EPI_ISL_833029, EPI_ISL_833030, EPI_ISL_833031, EPI_ISL_833032, EPI_ISL_833033, EPI_ISL_833034, EPI_ISL_833035, EPI_ISL_833036, EPI_ISL_833037 | | | |
| see above | Maine HETL | Tewhey Lab, The Jackson Laboratory | Matluk,N., Dewey,H., Iosue,F., Barter,M., Lynch,R., Munger,H. and Tewhey,R. |
| EPI_ISL_833345, EPI_ISL_833346, EPI_ISL_833347, EPI_ISL_833348, EPI_ISL_833349, EPI_ISL_833350, EPI_ISL_833351, EPI_ISL_833352, EPI_ISL_833353, EPI_ISL_833354, EPI_ISL_833355, EPI_ISL_833356, EPI_ISL_833357, EPI_ISL_833358, EPI_ISL_833359, EPI_ISL_833360, EPI_ISL_833361, EPI_ISL_833362, EPI_ISL_833363 | | | |
| see above | Microbiology Division, South Carolina Department of Health and Environmental Control (SC DHEC) | Microbiology Division, South Carolina Department of Health and Environmental Control (SC DHEC) | Flores,H., Freeman,J. |
| EPI_ISL_833394, EPI_ISL_833395, EPI_ISL_833396, EPI_ISL_833397, EPI_ISL_833398, EPI_ISL_833399, EPI_ISL_833400, EPI_ISL_833401, EPI_ISL_833402, EPI_ISL_833403, EPI_ISL_833404 | | | |
| see above | Saint Francis Medical Center | Los Angeles County Public Health Laboratories | P. Hemarajata et al. |
| EPI_ISL_833405, EPI_ISL_833406, EPI_ISL_833407, EPI_ISL_833408, EPI_ISL_833409, EPI_ISL_833410, EPI_ISL_833411, EPI_ISL_833412 | Beverly Hospital | Los Angeles County Public Health Laboratories | P. Hemarajata et al. |
| EPI_ISL_833413 | City of Hope Comprehensive Cancer Center | Los Angeles County Public Health Laboratories | P. Hemarajata et al. |
| EPI_ISL_833414, EPI_ISL_833415, EPI_ISL_833416 | USC Clinical Lab | Los Angeles County Public Health Laboratories | P. Hemarajata et al. |
| EPI_ISL_833417, EPI_ISL_833419, EPI_ISL_833420, EPI_ISL_833421, EPI_ISL_833422, EPI_ISL_833423, EPI_ISL_833424, EPI_ISL_833425, EPI_ISL_833426, EPI_ISL_833427 | MD Laboratories | Los Angeles County Public Health Laboratories | P. Hemarajata et al. |
| EPI_ISL_833428, EPI_ISL_833429, EPI_ISL_833430, EPI_ISL_833431 | UCLA Clinical Micro Lab | Los Angeles County Public Health Laboratories | P. Hemarajata et al. |
| EPI_ISL_833432 | Torrance Memorial Medical Center | Los Angeles County Public Health Laboratories | P. Hemarajata et al. |
| EPI_ISL_833489 | Saint Francis Medical Center | Los Angeles County Public Health Laboratories | P. Hemarajata et al. |
| EPI_ISL_833490, EPI_ISL_833491 | MD Laboratories | Los Angeles County Public Health Laboratories | P. Hemarajata et al. |
| EPI_ISL_837505, EPI_ISL_837506, EPI_ISL_837507, EPI_ISL_837508, EPI_ISL_837509, EPI_ISL_837510, EPI_ISL_837511, EPI_ISL_837512, EPI_ISL_837513, EPI_ISL_837514, EPI_ISL_837515, EPI_ISL_837517, EPI_ISL_837518, EPI_ISL_837519, EPI_ISL_837520, EPI_ISL_837521, EPI_ISL_837522, EPI_ISL_837523, EPI_ISL_837524, EPI_ISL_837525, EPI_ISL_837526, EPI_ISL_837527, EPI_ISL_837528, EPI_ISL_837529, EPI_ISL_837530, EPI_ISL_837531, EPI_ISL_837532, EPI_ISL_837533, EPI_ISL_837534, EPI_ISL_837535, EPI_ISL_837536, EPI_ISL_837537, EPI_ISL_837538, EPI_ISL_837539, EPI_ISL_837540, EPI_ISL_837541, EPI_ISL_837542, EPI_ISL_837543, EPI_ISL_837544, EPI_ISL_837545, EPI_ISL_837546, EPI_ISL_837547, EPI_ISL_837548, EPI_ISL_837549 | | | |
| see above | UW Virology Lab | UW Virology Lab | Pavitra Roychoudhury, Hong Xie, Lasata Shrestha, Meei-Li Huang, Keith R Jerome, Alexander Greninger |
| EPI_ISL_837600, EPI_ISL_837601, EPI_ISL_837602, EPI_ISL_837603, EPI_ISL_837604, EPI_ISL_837605, EPI_ISL_837606, EPI_ISL_837607, EPI_ISL_837608, EPI_ISL_837609, EPI_ISL_837610, EPI_ISL_837611, EPI_ISL_837612, EPI_ISL_837613, EPI_ISL_837614, EPI_ISL_837615, EPI_ISL_837616, EPI_ISL_837617, EPI_ISL_837618, EPI_ISL_837619, EPI_ISL_837620, EPI_ISL_837621, EPI_ISL_837622, EPI_ISL_837623, EPI_ISL_837624, EPI_ISL_837625, EPI_ISL_837626, EPI_ISL_837627, EPI_ISL_837628, EPI_ISL_837629, EPI_ISL_837630, EPI_ISL_837631, EPI_ISL_837632, EPI_ISL_837633, EPI_ISL_837634, EPI_ISL_837635, EPI_ISL_837636, EPI_ISL_837637, EPI_ISL_837638, EPI_ISL_837639, EPI_ISL_837640, EPI_ISL_837641, EPI_ISL_837642, EPI_ISL_837643, EPI_ISL_837644, EPI_ISL_837645, EPI_ISL_837646, EPI_ISL_837647, EPI_ISL_837648, EPI_ISL_837649, EPI_ISL_837650, EPI_ISL_837651, EPI_ISL_837652, EPI_ISL_837653, EPI_ISL_837654, EPI_ISL_837655, EPI_ISL_837656, EPI_ISL_837657, EPI_ISL_837658, EPI_ISL_837659, EPI_ISL_837660, EPI_ISL_837661, EPI_ISL_837662, EPI_ISL_837663, EPI_ISL_837664, EPI_ISL_837665, EPI_ISL_837666, EPI_ISL_837667, EPI_ISL_837668, EPI_ISL_837669, EPI_ISL_837670, EPI_ISL_837671, EPI_ISL_837672, EPI_ISL_837673, EPI_ISL_837674, EPI_ISL_837675, EPI_ISL_837676, EPI_ISL_837677, EPI_ISL_837678, EPI_ISL_837679, EPI_ISL_837680, EPI_ISL_837681, EPI_ISL_837682, EPI_ISL_837683, EPI_ISL_837684, EPI_ISL_837685, EPI_ISL_837686, EPI_ISL_837687, EPI_ISL_837688, EPI_ISL_837689, EPI_ISL_837690, EPI_ISL_837691, EPI_ISL_837692, EPI_ISL_837693, EPI_ISL_837694, EPI_ISL_837695, EPI_ISL_837696, EPI_ISL_837697, EPI_ISL_837698, EPI_ISL_837699, EPI_ISL_837700, EPI_ISL_837701, EPI_ISL_837702, EPI_ISL_837703, EPI_ISL_837704, EPI_ISL_837705, EPI_ISL_837706, EPI_ISL_837707, EPI_ISL_837708, EPI_ISL_837709, EPI_ISL_837710, EPI_ISL_837711, EPI_ISL_837712, EPI_ISL_837713, EPI_ISL_837714, EPI_ISL_837715, EPI_ISL_837716, EPI_ISL_837717, EPI_ISL_837718, EPI_ISL_837719, EPI_ISL_837720, EPI_ISL_837721, EPI_ISL_837722, EPI_ISL_837723, EPI_ISL_837724, EPI_ISL_837725, EPI_ISL_837726, EPI_ISL_837727, EPI_ISL_837728, EPI_ISL_837729, EPI_ISL_837730, EPI_ISL_837731, EPI_ISL_837732, EPI_ISL_837733, EPI_ISL_837734, EPI_ISL_837735, EPI_ISL_837736, EPI_ISL_837737, EPI_ISL_837738, EPI_ISL_837739, EPI_ISL_837740, EPI_ISL_837741, EPI_ISL_837742, EPI_ISL_837743, EPI_ISL_837744, EPI_ISL_837745, EPI_ISL_837746, EPI_ISL_837747, EPI_ISL_837748, EPI_ISL_837749, EPI_ISL_837750, EPI_ISL_837751, EPI_ISL_837752, EPI_ISL_837753, EPI_ISL_837754, EPI_ISL_837755, EPI_ISL_837756, EPI_ISL_837757, EPI_ISL_837758, EPI_ISL_837759, EPI_ISL_837760, EPI_ISL_837761, EPI_ISL_837762, EPI_ISL_837763, EPI_ISL_837764, EPI_ISL_837765, EPI_ISL_837766, EPI_ISL_837767, EPI_ISL_837768, EPI_ISL_837769, EPI_ISL_837770, EPI_ISL_837771, EPI_ISL_837772, EPI_ISL_837773, EPI_ISL_837774, EPI_ISL_837775, EPI_ISL_837776, EPI_ISL_837777, EPI_ISL_837778, EPI_ISL_837779, EPI_ISL_837780, EPI_ISL_837781, EPI_ISL_837782, EPI_ISL_837783, EPI_ISL_837784, EPI_ISL_837785, EPI_ISL_837786, EPI_ISL_837787, EPI_ISL_837788, EPI_ISL_837789, EPI_ISL_837790, EPI_ISL_837791, EPI_ISL_837792, EPI_ISL_837793, EPI_ISL_837794, EPI_ISL_837795, EPI_ISL_837796, EPI_ISL_837797, EPI_ISL_837798, EPI_ISL_837799, EPI_ISL_837800, EPI_ISL_837801, EPI_ISL_837802, EPI_ISL_837803, EPI_ISL_837804, EPI_ISL_837805, EPI_ISL_837806, EPI_ISL_837807, EPI_ISL_837808, EPI_ISL_837809, EPI_ISL_837810, EPI_ISL_837811, EPI_ISL_837812, EPI_ISL_837813, EPI_ISL_837814 | | | |
| see above | Instituto Nacional de Enfermedades Respiratorias (INER) | Instituto Nacional de Enfermedades Respiratorias (INER) | Celia Boukadida, Margarita Matías-Florentino, Alma Rincón-Rubio, Hector Esteban Paz-Juárez, Olivia Briceño, Edgar Sevilla-Reyes, Fidencio Mejía-Nepomuceno, Mario Mújica-Sánchez, Eduardo Becerril-Vargas, José Arturo Martínez-Orozco, Alejandra Hernández-Terán, Jorge Salas-Hernández, Santiago Ávila-Ríos, Joel Armando Vázquez-Pérez |
| EPI_ISL_837815, EPI_ISL_842609, EPI_ISL_842610, EPI_ISL_842611, EPI_ISL_842612 | Wyoming Public Health Laboratory | Wyoming Public Health Laboratory | Noah Hull, Taylor Fearing, Lynette Gumbleton, Channing Weber, Ashley Norberg, Bailey Bowcutt, and Wanda Manley |
| EPI_ISL_843170, EPI_ISL_843171, EPI_ISL_843172, EPI_ISL_843173, EPI_ISL_843174, EPI_ISL_843175, EPI_ISL_843176, EPI_ISL_843177, EPI_ISL_843178, EPI_ISL_843179, EPI_ISL_843180, EPI_ISL_843181, EPI_ISL_843182, EPI_ISL_843183, EPI_ISL_843184, EPI_ISL_843185, EPI_ISL_843186, EPI_ISL_843187, EPI_ISL_843188, EPI_ISL_843189, EPI_ISL_843190 | | | |
| see above | Maryland Public Health Laboratory | Maryland Public Health Laboratory | Maryland Department of Health Laboratories Administration |
| EPI_ISL_843191, EPI_ISL_843192 | Instituto Nacional de Enfermedades Respiratorias (INER) | Instituto Nacional de Enfermedades Respiratorias (INER) | Celia Boukadida, Margarita Matías-Florentino, Alma Rincón-Rubio, Hector Esteban Paz-Juárez, Olivia Briceño, Edgar Sevilla-Reyes, Fidencio Mejía-Nepomuceno, Mario Mújica-Sánchez, Eduardo Becerril-Vargas, José Arturo Martínez-Orozco, Alejandra Hernández-Terán, Jorge Salas-Hernández, Santiago Ávila-Ríos, Joel Armando Vázquez-Pérez |
| EPI_ISL_845544 | Santa Clara County Public Health Laboratory | Santa Clara County Public Health Laboratory | Santa Clara County Public Health Department |
| EPI_ISL_845657, EPI_ISL_845658, EPI_ISL_845659, EPI_ISL_845660, EPI_ISL_845661, EPI_ISL_845662, EPI_ISL_845663, EPI_ISL_845664, EPI_ISL_845665, EPI_ISL_845666, EPI_ISL_845667, EPI_ISL_845668, EPI_ISL_845669, EPI_ISL_845670, EPI_ISL_845671, EPI_ISL_845672, EPI_ISL_845673, EPI_ISL_845674, EPI_ISL_845675, EPI_ISL_845676, EPI_ISL_845677, EPI_ISL_845678, EPI_ISL_845679 | | | |
| see above | Quest Diagnostics | Quest Diagnostics | Rosenthal,S.H., Gerasimova,A., Kagan,R.M., Anderson, B., Bernstein, L.E., Livingston, K.E., Hua, M., Liu Y., Shalhout, D.F., Shlyakhter, I.A., Owen, R., Lacbawan, F. |
| EPI_ISL_845754, EPI_ISL_845755, EPI_ISL_845756, EPI_ISL_845757, EPI_ISL_845758, EPI_ISL_845759, EPI_ISL_845760, EPI_ISL_845761, EPI_ISL_845762, EPI_ISL_845763, EPI_ISL_845764, EPI_ISL_845765, EPI_ISL_845766, EPI_ISL_845767, EPI_ISL_845768, EPI_ISL_845769, EPI_ISL_845770, EPI_ISL_845771, EPI_ISL_845772, EPI_ISL_845773, EPI_ISL_845774, EPI_ISL_845775, EPI_ISL_845776, EPI_ISL_845777, EPI_ISL_845778, EPI_ISL_845779, EPI_ISL_845780, EPI_ISL_845781, EPI_ISL_845782 | | | |
| see above | Emory Molecular Diagnostics Laboratory, Emory Healthcare | Piantadosi Lab, Emory Department of Pathology | Ahmed Babiker, Anne Piantadosi |
| EPI_ISL_845783 | Quest Diagnostics | Quest Diagnostics | Rosenthal,S.H., Gerasimova,A., Kagan,R.M., Anderson, B., Bernstein, L.E., Livingston, K.E., Hua, M., Liu Y., Shalhout, D.F., Shlyakhter, I.A., Owen, R., Lacbawan, F. |
| EPI_ISL_845811, EPI_ISL_845812, EPI_ISL_845813, EPI_ISL_845814, EPI_ISL_845815, EPI_ISL_845816, EPI_ISL_845818, EPI_ISL_845819, EPI_ISL_845821, EPI_ISL_845822, EPI_ISL_845823, EPI_ISL_845826, EPI_ISL_845827, EPI_ISL_845828, EPI_ISL_845829, EPI_ISL_845831, EPI_ISL_845832, EPI_ISL_845834, EPI_ISL_845835, EPI_ISL_845836, EPI_ISL_845837, EPI_ISL_845838, EPI_ISL_845839, EPI_ISL_845840, EPI_ISL_845841, EPI_ISL_845842, EPI_ISL_845843, EPI_ISL_845844, EPI_ISL_845845, EPI_ISL_845846, EPI_ISL_845847, EPI_ISL_845848, EPI_ISL_845849, EPI_ISL_845850, EPI_ISL_845851, EPI_ISL_845852, EPI_ISL_845853, EPI_ISL_845854, EPI_ISL_845855, EPI_ISL_845856, EPI_ISL_845857, EPI_ISL_845858, EPI_ISL_845859, EPI_ISL_845860, EPI_ISL_845861, EPI_ISL_845862, EPI_ISL_845863, EPI_ISL_845866, EPI_ISL_845867, EPI_ISL_845868, EPI_ISL_845869, EPI_ISL_845870, EPI_ISL_845871, EPI_ISL_845872 | | | |

| | | | |
|---|---|---|---|
| see above | TGen North | TGen North | Jolene Bowers, Megan Folkerts, Chris French, Hayley Yaglom, Ashlyn Pfeiffer, Darrin Lemmer, Dave Engelthaler, The Arizona COVID Genomics Union (ACGU) |
| EPI_ISL_845891, EPI_ISL_845892 | Benaroya Research Institute | UW Virology Lab | Pavitra Roychoudhury, Hong Xie, Lasata Shrestha, Michelle Lin, Meei-Li Huang, Keith R Jerome, Alexander Greninger |
| EPI_ISL_846599, EPI_ISL_846600, EPI_ISL_846628, EPI_ISL_846629, EPI_ISL_846630, EPI_ISL_846651, EPI_ISL_846655 | Ohio Department of Health Laboratory | Ohio Department of Health Laboratory | Holmes, Jennifer; Eric Brandt, Keoni Omura, Glen McGillivary, Caitlin McDonnell, Kirtana Ramadugu, Erica Leasure, Kelsey Florek, Heather Blankenship, Quanta Brown, and Tammy Bannerman |
| EPI_ISL_846656, EPI_ISL_846657, EPI_ISL_846658, EPI_ISL_846659, EPI_ISL_846660, EPI_ISL_846661, EPI_ISL_846662, EPI_ISL_846663, EPI_ISL_846664, EPI_ISL_846665, EPI_ISL_846666, EPI_ISL_846667, EPI_ISL_846668, EPI_ISL_846669, EPI_ISL_846670, EPI_ISL_846671, EPI_ISL_846672, EPI_ISL_846673, EPI_ISL_846674, EPI_ISL_846675, EPI_ISL_846676, EPI_ISL_846677, EPI_ISL_846678, EPI_ISL_846679, EPI_ISL_846680, EPI_ISL_846681, EPI_ISL_846682, EPI_ISL_846683, EPI_ISL_846684, EPI_ISL_846685, EPI_ISL_846686, EPI_ISL_846687, EPI_ISL_846688, EPI_ISL_846689, EPI_ISL_846690, EPI_ISL_846691, EPI_ISL_846692, EPI_ISL_846693, EPI_ISL_846694, EPI_ISL_846695, EPI_ISL_846696, EPI_ISL_846697, EPI_ISL_846698, EPI_ISL_846699, EPI_ISL_846700, EPI_ISL_846701, EPI_ISL_846702, EPI_ISL_846703, EPI_ISL_846704, EPI_ISL_846705, EPI_ISL_846706, EPI_ISL_846707, EPI_ISL_846708, EPI_ISL_846709, EPI_ISL_846710, EPI_ISL_846711, EPI_ISL_846712, EPI_ISL_846713, EPI_ISL_846714, EPI_ISL_846715, EPI_ISL_846716, EPI_ISL_846717, EPI_ISL_846718, EPI_ISL_846719, EPI_ISL_846720, EPI_ISL_846721, EPI_ISL_846722, EPI_ISL_846723, EPI_ISL_846724, EPI_ISL_846725, EPI_ISL_846726, EPI_ISL_846727, EPI_ISL_846728, EPI_ISL_846729, EPI_ISL_846730, EPI_ISL_846731, EPI_ISL_846732, EPI_ISL_846733, EPI_ISL_846734, EPI_ISL_846735, EPI_ISL_846736, EPI_ISL_846737, EPI_ISL_846738, EPI_ISL_846739, EPI_ISL_846740, EPI_ISL_846741, EPI_ISL_846742, EPI_ISL_846743, EPI_ISL_846744, EPI_ISL_846745, EPI_ISL_846747, EPI_ISL_846748, EPI_ISL_846749, EPI_ISL_846750, EPI_ISL_846751, EPI_ISL_846752, EPI_ISL_846753, EPI_ISL_846754, EPI_ISL_846755, EPI_ISL_846756, EPI_ISL_846757, EPI_ISL_846758, EPI_ISL_846759, EPI_ISL_846760, EPI_ISL_846761, EPI_ISL_846762, EPI_ISL_846763, EPI_ISL_846764, EPI_ISL_846765, EPI_ISL_846766, EPI_ISL_846767, EPI_ISL_846768, EPI_ISL_846769, EPI_ISL_846770, EPI_ISL_846771, EPI_ISL_846772, EPI_ISL_846773 | | | |
| see above | University of Michigan Clinical Microbiology Laboratory | Lauring Lab, University of Michigan, Department of Microbiology and Immunology | Valesano |
| EPI_ISL_847515 | Chiu Laboratory, University of California, San Francisco | Chiu Laboratory, University of California, San Francisco | Charles Chiu, Xianding (Wayne) Deng, Candace Wang, Brian Bushnell, Scot Federman, Jill Hacker, Debra Wadford |
| EPI_ISL_847516 | California Department of Public Health | Chiu Laboratory, University of California, San Francisco | Charles Chiu, Xianding (Wayne) Deng, Candace Wang, Brian Bushnell, Scot Federman, Jill Hacker, Debra Wadford |
| EPI_ISL_847517 | Chiu Laboratory, University of California, San Francisco | Chiu Laboratory, University of California, San Francisco | Charles Chiu, Xianding (Wayne) Deng, Candace Wang, Brian Bushnell, Scot Federman, Jill Hacker, Debra Wadford |
| EPI_ISL_847518, EPI_ISL_847519, EPI_ISL_847520, EPI_ISL_847521, EPI_ISL_847522, EPI_ISL_847523, EPI_ISL_847524, EPI_ISL_847525, EPI_ISL_847526, EPI_ISL_847527, EPI_ISL_847528, EPI_ISL_847529, EPI_ISL_847530, EPI_ISL_847531, EPI_ISL_847532, EPI_ISL_847533, EPI_ISL_847534, EPI_ISL_847535 | | | |
| see above | California Department of Public Health | Chiu Laboratory, University of California, San Francisco | Charles Chiu, Xianding (Wayne) Deng, Candace Wang, Brian Bushnell, Scot Federman, Jill Hacker, Debra Wadford |
| EPI_ISL_847536 | Chiu Laboratory, University of California, San Francisco | Chiu Laboratory, University of California, San Francisco | Charles Chiu, Xianding (Wayne) Deng, Candace Wang, Brian Bushnell, Scot Federman, Jill Hacker, Debra Wadford |
| EPI_ISL_847537, EPI_ISL_847538, EPI_ISL_847539 | California Department of Public Health | Chiu Laboratory, University of California, San Francisco | Charles Chiu, Xianding (Wayne) Deng, Candace Wang, Brian Bushnell, Scot Federman, Jill Hacker, Debra Wadford |
| EPI_ISL_847540 | Chiu Laboratory, University of California, San Francisco | Chiu Laboratory, University of California, San Francisco | Charles Chiu, Xianding (Wayne) Deng, Candace Wang, Brian Bushnell, Scot Federman, Jill Hacker, Debra Wadford |
| EPI_ISL_847541, EPI_ISL_847542, EPI_ISL_847543, EPI_ISL_847544 | California Department of Public Health | Chiu Laboratory, University of California, San Francisco | Charles Chiu, Xianding (Wayne) Deng, Candace Wang, Brian Bushnell, Scot Federman, Jill Hacker, Debra Wadford |
| EPI_ISL_847545, EPI_ISL_847546 | Chiu Laboratory, University of California, San Francisco | Chiu Laboratory, University of California, San Francisco | Charles Chiu, Xianding (Wayne) Deng, Candace Wang, Brian Bushnell, Scot Federman, Jill Hacker, Debra Wadford |
| EPI_ISL_847547, EPI_ISL_847548, EPI_ISL_847549, EPI_ISL_847550, EPI_ISL_847551, EPI_ISL_847552 | California Department of Public Health | Chiu Laboratory, University of California, San Francisco | Charles Chiu, Xianding (Wayne) Deng, Candace Wang, Brian Bushnell, Scot Federman, Jill Hacker, Debra Wadford |
| EPI_ISL_847553 | Chiu Laboratory, University of California, San Francisco | Chiu Laboratory, University of California, San Francisco | Charles Chiu, Xianding (Wayne) Deng, Candace Wang, Brian Bushnell, Scot Federman, Jill Hacker, Debra Wadford |
| EPI_ISL_847554, EPI_ISL_847555, EPI_ISL_847556, EPI_ISL_847557, EPI_ISL_847558, EPI_ISL_847559, EPI_ISL_847560, EPI_ISL_847561, EPI_ISL_847562 | California Department of Public Health | Chiu Laboratory, University of California, San Francisco | Charles Chiu, Xianding (Wayne) Deng, Candace Wang, Brian Bushnell, Scot Federman, Jill Hacker, Debra Wadford |
| EPI_ISL_847563 | Chiu Laboratory, University of California, San Francisco | Chiu Laboratory, University of California, San Francisco | Charles Chiu, Xianding (Wayne) Deng, Candace Wang, Brian Bushnell, Scot Federman, Jill Hacker, Debra Wadford |
| EPI_ISL_847564, EPI_ISL_847565, EPI_ISL_847566, EPI_ISL_847567, EPI_ISL_847568, EPI_ISL_847569, EPI_ISL_847570, EPI_ISL_847571, EPI_ISL_847572, EPI_ISL_847573, EPI_ISL_847574, EPI_ISL_847575, EPI_ISL_847576, EPI_ISL_847577, EPI_ISL_847578 | | | |
| see above | California Department of Public Health | Chiu Laboratory, University of California, San Francisco | Charles Chiu, Xianding (Wayne) Deng, Candace Wang, Brian Bushnell, Scot Federman, Jill Hacker, Debra Wadford |
| EPI_ISL_847579 | Chiu Laboratory, University of California, San Francisco | Chiu Laboratory, University of California, San Francisco | Charles Chiu, Xianding (Wayne) Deng, Candace Wang, Brian Bushnell, Scot Federman, Jill Hacker, Debra Wadford |
| EPI_ISL_847580, EPI_ISL_847581, EPI_ISL_847582, EPI_ISL_847583, EPI_ISL_847585, EPI_ISL_847586, EPI_ISL_847587, EPI_ISL_847588, EPI_ISL_847589, EPI_ISL_847590, EPI_ISL_847591, EPI_ISL_847592, EPI_ISL_847593, EPI_ISL_847594, EPI_ISL_847595, EPI_ISL_847596, EPI_ISL_847597, EPI_ISL_847598, EPI_ISL_847599, EPI_ISL_847600, EPI_ISL_847601, EPI_ISL_847602, EPI_ISL_847603, EPI_ISL_847604, EPI_ISL_847605, EPI_ISL_847606, EPI_ISL_847607, EPI_ISL_847608, EPI_ISL_847609, EPI_ISL_847610, EPI_ISL_847611, EPI_ISL_847612, EPI_ISL_847613, EPI_ISL_847614, EPI_ISL_847615, EPI_ISL_847616, EPI_ISL_847617, EPI_ISL_847618 | | | |
| see above | California Department of Public Health | Chiu Laboratory, University of California, San Francisco | Charles Chiu, Xianding (Wayne) Deng, Candace Wang, Brian Bushnell, Scot Federman, Jill Hacker, Debra Wadford |
| EPI_ISL_847619, EPI_ISL_847620, EPI_ISL_847621, EPI_ISL_847622 | Chiu Laboratory, University of California, San Francisco | Chiu Laboratory, University of California, San Francisco | Charles Chiu, Xianding (Wayne) Deng, Candace Wang, Brian Bushnell, Scot Federman, Jill Hacker, Debra Wadford |
| EPI_ISL_847623, EPI_ISL_847624, EPI_ISL_847625, EPI_ISL_847626, EPI_ISL_847627, EPI_ISL_847628, EPI_ISL_847629, EPI_ISL_847630, EPI_ISL_847631, EPI_ISL_847632, EPI_ISL_847633, EPI_ISL_847634, EPI_ISL_847635, EPI_ISL_847636, EPI_ISL_847637, EPI_ISL_847638, EPI_ISL_847639 | | | |
| see above | California Department of Public Health | Chiu Laboratory, University of California, San Francisco | Charles Chiu, Xianding (Wayne) Deng, Candace Wang, Brian Bushnell, Scot Federman, Jill Hacker, Debra Wadford |
| EPI_ISL_847640, EPI_ISL_847641, EPI_ISL_847642, EPI_ISL_847643, EPI_ISL_847644 | Chiu Laboratory, University of California, San Francisco | Chiu Laboratory, University of California, San Francisco | Charles Chiu, Xianding (Wayne) Deng, Candace Wang, Brian Bushnell, Scot Federman, Jill Hacker, Debra Wadford |
| EPI_ISL_847645, EPI_ISL_847646, EPI_ISL_847647, EPI_ISL_847648, EPI_ISL_847649, EPI_ISL_847650, EPI_ISL_847651, EPI_ISL_847652, EPI_ISL_847653, EPI_ISL_847654, EPI_ISL_847655, EPI_ISL_847656, EPI_ISL_847657, EPI_ISL_847658, EPI_ISL_847659, EPI_ISL_847660, EPI_ISL_847661, EPI_ISL_847662, EPI_ISL_847663, EPI_ISL_847664, EPI_ISL_847665, EPI_ISL_847666, EPI_ISL_847667, EPI_ISL_847668, EPI_ISL_847669, EPI_ISL_847670, EPI_ISL_847671, EPI_ISL_847672, EPI_ISL_847673, EPI_ISL_847674, EPI_ISL_847675, EPI_ISL_847676, EPI_ISL_847677, EPI_ISL_847678, EPI_ISL_847679, EPI_ISL_847680, EPI_ISL_847681, EPI_ISL_847682, EPI_ISL_847683, EPI_ISL_847684, EPI_ISL_847685, EPI_ISL_847686, EPI_ISL_847687, EPI_ISL_847688, EPI_ISL_847689, EPI_ISL_847690, EPI_ISL_847691, EPI_ISL_847692, EPI_ISL_847693, EPI_ISL_847694, EPI_ISL_847695, EPI_ISL_847696, EPI_ISL_847697, EPI_ISL_847698, EPI_ISL_847699, EPI_ISL_847700, EPI_ISL_847701, EPI_ISL_847702, EPI_ISL_847703, EPI_ISL_847704, EPI_ISL_847705, EPI_ISL_847706, EPI_ISL_847707, EPI_ISL_847708, EPI_ISL_847709, EPI_ISL_847754, EPI_ISL_847755, EPI_ISL_847756, EPI_ISL_847757, EPI_ISL_847758, EPI_ISL_847759, EPI_ISL_847760, EPI_ISL_847761, EPI_ISL_847762, EPI_ISL_847763, EPI_ISL_847764, EPI_ISL_847765, EPI_ISL_847766, EPI_ISL_847767, EPI_ISL_847768, EPI_ISL_847769, EPI_ISL_847770, EPI_ISL_847771, EPI_ISL_847772, EPI_ISL_847773, EPI_ISL_847774, EPI_ISL_847775, EPI_ISL_847776, EPI_ISL_847777, EPI_ISL_847778, EPI_ISL_847779, EPI_ISL_847780, EPI_ISL_847781 | | | |
| see above | California Department of Public Health | Chiu Laboratory, University of California, San Francisco | Charles Chiu, Xianding (Wayne) Deng, Candace Wang, Brian Bushnell, Scot Federman, Jill Hacker, Debra Wadford |
| EPI_ISL_847782 | Chiu Laboratory, University of California, San Francisco | Chiu Laboratory, University of California, San Francisco | Charles Chiu, Xianding (Wayne) Deng, Candace Wang, Brian Bushnell, Scot Federman, Jill Hacker, Debra Wadford |
| EPI_ISL_847783, EPI_ISL_847784, EPI_ISL_847785, EPI_ISL_847786, EPI_ISL_847787, EPI_ISL_847788, EPI_ISL_847789, EPI_ISL_847790, EPI_ISL_847791, EPI_ISL_847792, EPI_ISL_847793, EPI_ISL_847794, EPI_ISL_847795, EPI_ISL_847796, EPI_ISL_847797, EPI_ISL_847798, EPI_ISL_847799, EPI_ISL_847800, EPI_ISL_847801, EPI_ISL_847802, EPI_ISL_847803, EPI_ISL_847804, EPI_ISL_847805, EPI_ISL_847806, EPI_ISL_847807, EPI_ISL_847808, EPI_ISL_847809, EPI_ISL_847810, EPI_ISL_847811, EPI_ISL_847812, EPI_ISL_847813, EPI_ISL_847814, EPI_ISL_847815, EPI_ISL_847816, EPI_ISL_847817, EPI_ISL_847818 | | | |
| see above | California Department of Public Health | Chiu Laboratory, University of California, San Francisco | Charles Chiu, Xianding (Wayne) Deng, Candace Wang, Brian Bushnell, Scot Federman, Jill Hacker, Debra Wadford |
| EPI_ISL_847819 | Ohio Department of Health Laboratory | Ohio Department of Health Laboratory | Holmes, Jennifer; Eric Brandt, Keoni Omura, Glen McGillivary, Caitlin McDonnell, Kirtana Ramadugu, Erica Leasure, Kelsey Florek, Heather Blankenship, Quanta Brown, and Tammy Bannerman |
| EPI_ISL_847821, EPI_ISL_847822 | University of Michigan Clinical Microbiology Laboratory | Lauring Lab, University of Michigan, Department of Microbiology and Immunology | Valesano |

| | | | |
|---|---|---|---|
| EPI_ISL_847823, EPI_ISL_847824, EPI_ISL_847828 | Ohio Department of Health Laboratory | Ohio Department of Health Laboratory | Holmes, Jennifer; Eric Brandt, Keoni Omura, Glen McGillivary, Caitlin McDonnell, Kirtana Ramadugu, Erica Leasure, Kelsey Florek, Heather Blankenship, Quanta Brown, and Tammy Bannerman |
| EPI_ISL_847829, EPI_ISL_847830 | Tempus | Grubaugh Lab - Yale School of Public Health | Tara Alpert, Joseph Fauver, Anderson Brito, Mallery Breban, Anne Wyllie, Chantal Vogels, Mary Petrone, Chaney Kalinich, Isabel Ott, Nathan Grubaugh |
| EPI_ISL_847831, EPI_ISL_847832 | Yale New Haven Hospital | Grubaugh Lab - Yale School of Public Health | Tara Alpert, Joseph Fauver, Anderson Brito, Mallery Breban, Anne Wyllie, Chantal Vogels, Mary Petrone, Chaney Kalinich, Isabel Ott, Nathan Grubaugh |
| EPI_ISL_847842 | Ohio Department of Health Laboratory | Ohio Department of Health Laboratory | Holmes, Jennifer; Eric Brandt, Keoni Omura, Glen McGillivary, Caitlin McDonnell, Kirtana Ramadugu, Erica Leasure, Kelsey Florek, Heather Blankenship, Quanta Brown, and Tammy Bannerman |

EPI_ISL_847978, EPI_ISL_847979, EPI_ISL_847980, EPI_ISL_847981, EPI_ISL_847982, EPI_ISL_847983, EPI_ISL_847984, EPI_ISL_847985, EPI_ISL_847986, EPI_ISL_847987, EPI_ISL_847988, EPI_ISL_847989, EPI_ISL_847990, EPI_ISL_847991, EPI_ISL_847992, EPI_ISL_847993, EPI_ISL_847994, EPI_ISL_847995, EPI_ISL_847996, EPI_ISL_847997, EPI_ISL_847998, EPI_ISL_847999, EPI_ISL_848000, EPI_ISL_848001, EPI_ISL_848002, EPI_ISL_848003, EPI_ISL_848004, EPI_ISL_848005, EPI_ISL_848006, EPI_ISL_848007, EPI_ISL_848008, EPI_ISL_848009, EPI_ISL_848010, EPI_ISL_848011, EPI_ISL_848012, EPI_ISL_848013, EPI_ISL_848014, EPI_ISL_848015, EPI_ISL_848016, EPI_ISL_848017, EPI_ISL_848018, EPI_ISL_848019, EPI_ISL_848020, EPI_ISL_848021, EPI_ISL_848022, EPI_ISL_848023, EPI_ISL_848024, EPI_ISL_848025, EPI_ISL_848026, EPI_ISL_848027, EPI_ISL_848028, EPI_ISL_848029, EPI_ISL_848030, EPI_ISL_848031, EPI_ISL_848032, EPI_ISL_848033, EPI_ISL_848034, EPI_ISL_848035, EPI_ISL_848036, EPI_ISL_848037, EPI_ISL_848038, EPI_ISL_848039, EPI_ISL_848040, EPI_ISL_848041, EPI_ISL_848042, EPI_ISL_848043, EPI_ISL_848055, EPI_ISL_848056, EPI_ISL_848057, EPI_ISL_848058, EPI_ISL_848059, EPI_ISL_848060, EPI_ISL_848061, EPI_ISL_848062

| | | | |
|---|---|---|---|
| see above | Michigan Department of Health and Human Services, Bureau of Laboratories | Michigan Department of Health and Human Services, Bureau of Laboratories | Blankenship HM, Riner D, Soehnlen MK |
| EPI_ISL_848069 | Tulane Medical Center | Centers for Disease Control and Prevention Division of Viral Diseases, Pathogen Discovery | Ying Tao, Yan Li, Jing Zhang, Krista Queen, Anna Uehara, Peter Cook, Clinton R. Paden, Haibin Wang, Suxiang Tong |

EPI_ISL_848172, EPI_ISL_848173, EPI_ISL_848174, EPI_ISL_848175, EPI_ISL_848176, EPI_ISL_848177, EPI_ISL_848178, EPI_ISL_848179, EPI_ISL_848180, EPI_ISL_848181, EPI_ISL_848182, EPI_ISL_848183, EPI_ISL_848184, EPI_ISL_848185, EPI_ISL_848186, EPI_ISL_848187

| | | | |
|---|---|---|---|
| see above | North Dakota Department of Health, Public Health Laboratory | North Dakota Department of Health, Public Health Laboratory | Lisa Wingerter |

EPI_ISL_848203, EPI_ISL_848204, EPI_ISL_848205, EPI_ISL_848206, EPI_ISL_848207, EPI_ISL_848208, EPI_ISL_848209, EPI_ISL_848210, EPI_ISL_848211, EPI_ISL_848212, EPI_ISL_848213, EPI_ISL_848214, EPI_ISL_848215, EPI_ISL_848216, EPI_ISL_848217, EPI_ISL_848218, EPI_ISL_848219, EPI_ISL_848220, EPI_ISL_848221, EPI_ISL_848222, EPI_ISL_848223, EPI_ISL_848224, EPI_ISL_848225, EPI_ISL_848226, EPI_ISL_848227, EPI_ISL_848228, EPI_ISL_848229, EPI_ISL_848230, EPI_ISL_848231, EPI_ISL_848232, EPI_ISL_848233, EPI_ISL_848234, EPI_ISL_848235, EPI_ISL_848236, EPI_ISL_848237, EPI_ISL_848238, EPI_ISL_848239, EPI_ISL_848240, EPI_ISL_848241, EPI_ISL_848242, EPI_ISL_848243, EPI_ISL_848244, EPI_ISL_848245, EPI_ISL_848246, EPI_ISL_848247, EPI_ISL_848248, EPI_ISL_848249, EPI_ISL_848250, EPI_ISL_848251, EPI_ISL_848252, EPI_ISL_848253, EPI_ISL_848254, EPI_ISL_848255, EPI_ISL_848256, EPI_ISL_848257, EPI_ISL_848258, EPI_ISL_848259, EPI_ISL_848260, EPI_ISL_848261, EPI_ISL_848262, EPI_ISL_848263, EPI_ISL_848264, EPI_ISL_848265, EPI_ISL_848266, EPI_ISL_848267, EPI_ISL_848268, EPI_ISL_848269, EPI_ISL_848270, EPI_ISL_848271, EPI_ISL_848272, EPI_ISL_848273, EPI_ISL_848274, EPI_ISL_848275, EPI_ISL_848276, EPI_ISL_848277, EPI_ISL_848278, EPI_ISL_848279, EPI_ISL_848280, EPI_ISL_848281, EPI_ISL_848282, EPI_ISL_848283, EPI_ISL_848284, EPI_ISL_848285, EPI_ISL_848286, EPI_ISL_848287, EPI_ISL_848288, EPI_ISL_848289, EPI_ISL_848290, EPI_ISL_848291, EPI_ISL_848292, EPI_ISL_848293, EPI_ISL_848294, EPI_ISL_848295, EPI_ISL_848296, EPI_ISL_848297, EPI_ISL_848298, EPI_ISL_848299, EPI_ISL_848300, EPI_ISL_848301, EPI_ISL_848302, EPI_ISL_848303, EPI_ISL_848304, EPI_ISL_848305, EPI_ISL_848306, EPI_ISL_848307, EPI_ISL_848308, EPI_ISL_848309, EPI_ISL_848310, EPI_ISL_848311, EPI_ISL_848312, EPI_ISL_848313, EPI_ISL_848314, EPI_ISL_848315, EPI_ISL_848316, EPI_ISL_848317, EPI_ISL_848318, EPI_ISL_848319, EPI_ISL_848320, EPI_ISL_848321, EPI_ISL_848322, EPI_ISL_848323, EPI_ISL_848324, EPI_ISL_848325, EPI_ISL_848326, EPI_ISL_848327, EPI_ISL_848328, EPI_ISL_848329, EPI_ISL_848330, EPI_ISL_848331, EPI_ISL_848332, EPI_ISL_848333, EPI_ISL_848334, EPI_ISL_848335, EPI_ISL_848336, EPI_ISL_848337, EPI_ISL_848338, EPI_ISL_848339, EPI_ISL_848340, EPI_ISL_848341, EPI_ISL_848342, EPI_ISL_848343, EPI_ISL_848344, EPI_ISL_848345, EPI_ISL_848346, EPI_ISL_848347, EPI_ISL_848348, EPI_ISL_848349, EPI_ISL_848350, EPI_ISL_848351, EPI_ISL_848352, EPI_ISL_848353, EPI_ISL_848354, EPI_ISL_848355, EPI_ISL_848356, EPI_ISL_848357, EPI_ISL_848358, EPI_ISL_848359, EPI_ISL_848360, EPI_ISL_848361, EPI_ISL_848362, EPI_ISL_848363, EPI_ISL_848364, EPI_ISL_848365, EPI_ISL_848366, EPI_ISL_848367, EPI_ISL_848368, EPI_ISL_848369, EPI_ISL_848370, EPI_ISL_848371, EPI_ISL_848372, EPI_ISL_848373, EPI_ISL_848374, EPI_ISL_848375, EPI_ISL_848376, EPI_ISL_848377, EPI_ISL_848378, EPI_ISL_848379, EPI_ISL_848380, EPI_ISL_848381, EPI_ISL_848382, EPI_ISL_848383, EPI_ISL_848384, EPI_ISL_848385, EPI_ISL_848386, EPI_ISL_848387, EPI_ISL_848388, EPI_ISL_848389, EPI_ISL_848390, EPI_ISL_848391, EPI_ISL_848392, EPI_ISL_848393, EPI_ISL_848394, EPI_ISL_848395, EPI_ISL_848396, EPI_ISL_848397, EPI_ISL_848398, EPI_ISL_848399, EPI_ISL_848400, EPI_ISL_848401, EPI_ISL_848402, EPI_ISL_848403, EPI_ISL_848404, EPI_ISL_848405, EPI_ISL_848406, EPI_ISL_848407, EPI_ISL_848408, EPI_ISL_848409, EPI_ISL_848410, EPI_ISL_848411, EPI_ISL_848412, EPI_ISL_848413, EPI_ISL_848414, EPI_ISL_848415, EPI_ISL_848416, EPI_ISL_848417, EPI_ISL_848418, EPI_ISL_848419, EPI_ISL_848420, EPI_ISL_848421, EPI_ISL_848422, EPI_ISL_848423, EPI_ISL_848424, EPI_ISL_848425, EPI_ISL_848426, EPI_ISL_848427, EPI_ISL_848428, EPI_ISL_848429, EPI_ISL_848430, EPI_ISL_848431, EPI_ISL_848432, EPI_ISL_848433, EPI_ISL_848434, EPI_ISL_848435, EPI_ISL_848436, EPI_ISL_848437, EPI_ISL_848438, EPI_ISL_848439, EPI_ISL_848453, EPI_ISL_848454, EPI_ISL_848455, EPI_ISL_848456, EPI_ISL_848457, EPI_ISL_848458, EPI_ISL_848459, EPI_ISL_848460, EPI_ISL_848461, EPI_ISL_848462, EPI_ISL_848463, EPI_ISL_848464, EPI_ISL_848465, EPI_ISL_848466, EPI_ISL_848467, EPI_ISL_848468, EPI_ISL_848469, EPI_ISL_848470, EPI_ISL_848471, EPI_ISL_848472, EPI_ISL_848473, EPI_ISL_848474, EPI_ISL_848475, EPI_ISL_848476, EPI_ISL_848477, EPI_ISL_848478, EPI_ISL_848479, EPI_ISL_848480, EPI_ISL_848481, EPI_ISL_848482, EPI_ISL_848483, EPI_ISL_848484, EPI_ISL_848485, EPI_ISL_848486, EPI_ISL_848487, EPI_ISL_848488, EPI_ISL_848489, EPI_ISL_848490, EPI_ISL_848491, EPI_ISL_848492, EPI_ISL_848493, EPI_ISL_848494, EPI_ISL_848495, EPI_ISL_848496, EPI_ISL_848497, EPI_ISL_848498, EPI_ISL_848499, EPI_ISL_848500, EPI_ISL_848501, EPI_ISL_848502, EPI_ISL_848503, EPI_ISL_848504, EPI_ISL_848505, EPI_ISL_848506, EPI_ISL_848507, EPI_ISL_848508, EPI_ISL_848509, EPI_ISL_848510, EPI_ISL_848511, EPI_ISL_848512, EPI_ISL_848513, EPI_ISL_848514, EPI_ISL_848515, EPI_ISL_848516, EPI_ISL_848517, EPI_ISL_848518, EPI_ISL_848519, EPI_ISL_848520, EPI_ISL_848521, EPI_ISL_848522, EPI_ISL_848523, EPI_ISL_848524, EPI_ISL_848525, EPI_ISL_848526, EPI_ISL_848527, EPI_ISL_848528, EPI_ISL_848529, EPI_ISL_848530, EPI_ISL_848531, EPI_ISL_848532, EPI_ISL_848533, EPI_ISL_848534, EPI_ISL_848535, EPI_ISL_848536, EPI_ISL_848537, EPI_ISL_848538, EPI_ISL_848539, EPI_ISL_848540, EPI_ISL_848541, EPI_ISL_848542, EPI_ISL_848543, EPI_ISL_848544, EPI_ISL_848545

| | | | |
|---|---|---|---|
| see above | Illinois Department of Public Health | Gagnon Lab, Southern Illinois University | Keith Gagnon |
| EPI_ISL_848546, EPI_ISL_848547, EPI_ISL_848548, EPI_ISL_848549, EPI_ISL_848550, EPI_ISL_848551, EPI_ISL_848552, EPI_ISL_848553, EPI_ISL_848554 | North Dakota Department of Health, Public Health Laboratory | North Dakota Department of Health, Public Health Laboratory | Lisa Wingerter |
| EPI_ISL_848630, EPI_ISL_848631, EPI_ISL_848632, EPI_ISL_848633 | Montana Public Health Laboratory | Wyoming Public Health Laboratory | Noah Hull, Joy Ritter, Taylor Fearing, Lynette Gumbleton, Channing Weber, Ashley Norberg, Bailey Bowcutt, Wanda Manley, Deborah Gibson |
| EPI_ISL_848634, EPI_ISL_848635, EPI_ISL_848636, EPI_ISL_848637, EPI_ISL_848638, EPI_ISL_848639, EPI_ISL_848640, EPI_ISL_848641 | North Dakota Department of Health, Public Health Laboratory | North Dakota Department of Health, Public Health Laboratory | Lisa Wingerter |

EPI_ISL_848643, EPI_ISL_848644, EPI_ISL_848645, EPI_ISL_848646, EPI_ISL_848648, EPI_ISL_848651, EPI_ISL_848652, EPI_ISL_848653, EPI_ISL_848654, EPI_ISL_848655, EPI_ISL_848656, EPI_ISL_848657, EPI_ISL_848658, EPI_ISL_848659, EPI_ISL_848660, EPI_ISL_848661, EPI_ISL_848662, EPI_ISL_848663, EPI_ISL_848664, EPI_ISL_848665, EPI_ISL_848671, EPI_ISL_848672, EPI_ISL_848673, EPI_ISL_848674, EPI_ISL_848675, EPI_ISL_848676, EPI_ISL_848677, EPI_ISL_848679, EPI_ISL_848680, EPI_ISL_848681, EPI_ISL_848682, EPI_ISL_848683, EPI_ISL_848684, EPI_ISL_848685, EPI_ISL_848686, EPI_ISL_848687, EPI_ISL_848688, EPI_ISL_848689, EPI_ISL_848690, EPI_ISL_848691, EPI_ISL_848692, EPI_ISL_848693, EPI_ISL_848694, EPI_ISL_848697, EPI_ISL_848698, EPI_ISL_848699, EPI_ISL_848700, EPI_ISL_848701, EPI_ISL_848702, EPI_ISL_848703, EPI_ISL_848704, EPI_ISL_848705, EPI_ISL_848706, EPI_ISL_848707, EPI_ISL_848708, EPI_ISL_848709, EPI_ISL_848714, EPI_ISL_848715, EPI_ISL_848716, EPI_ISL_848717, EPI_ISL_848718, EPI_ISL_848719, EPI_ISL_848720, EPI_ISL_848721, EPI_ISL_848722, EPI_ISL_848723, EPI_ISL_848724, EPI_ISL_848726, EPI_ISL_848727, EPI_ISL_848728, EPI_ISL_848729, EPI_ISL_848730, EPI_ISL_848731, EPI_ISL_848732, EPI_ISL_848734, EPI_ISL_848735, EPI_ISL_848736, EPI_ISL_848737, EPI_ISL_848738, EPI_ISL_848739, EPI_ISL_848740, EPI_ISL_848741, EPI_ISL_848742, EPI_ISL_848743, EPI_ISL_848744, EPI_ISL_848745, EPI_ISL_848746, EPI_ISL_848747, EPI_ISL_848748, EPI_ISL_848749, EPI_ISL_848750, EPI_ISL_848751, EPI_ISL_848752, EPI_ISL_848753, EPI_ISL_848754, EPI_ISL_848755, EPI_ISL_848756, EPI_ISL_848757, EPI_ISL_848758, EPI_ISL_848759, EPI_ISL_848760, EPI_ISL_848761, EPI_ISL_848762, EPI_ISL_848763, EPI_ISL_848764, EPI_ISL_848765, EPI_ISL_848766, EPI_ISL_848767, EPI_ISL_848768, EPI_ISL_848769, EPI_ISL_848770, EPI_ISL_848772, EPI_ISL_848773, EPI_ISL_848774, EPI_ISL_848775, EPI_ISL_848776, EPI_ISL_848777, EPI_ISL_848778, EPI_ISL_848779, EPI_ISL_848781, EPI_ISL_848782, EPI_ISL_848783, EPI_ISL_848784, EPI_ISL_848785, EPI_ISL_848786, EPI_ISL_848787, EPI_ISL_848788, EPI_ISL_848789, EPI_ISL_848790, EPI_ISL_848791, EPI_ISL_848792, EPI_ISL_848793, EPI_ISL_848794, EPI_ISL_848795, EPI_ISL_848796, EPI_ISL_848797, EPI_ISL_848798, EPI_ISL_848799, EPI_ISL_848800, EPI_ISL_848801, EPI_ISL_848802, EPI_ISL_848803, EPI_ISL_848805, EPI_ISL_848806, EPI_ISL_848807, EPI_ISL_848808, EPI_ISL_848809, EPI_ISL_848810, EPI_ISL_848811, EPI_ISL_848812, EPI_ISL_848813, EPI_ISL_848814, EPI_ISL_848815, EPI_ISL_848816, EPI_ISL_848817, EPI_ISL_848818, EPI_ISL_848819, EPI_ISL_848820, EPI_ISL_848821, EPI_ISL_848822, EPI_ISL_848823, EPI_ISL_848824, EPI_ISL_848825, EPI_ISL_848826, EPI_ISL_848827, EPI_ISL_848828, EPI_ISL_848829, EPI_ISL_848830, EPI_ISL_848831, EPI_ISL_848832, EPI_ISL_848833, EPI_ISL_848835, EPI_ISL_848836, EPI_ISL_848837, EPI_ISL_848838, EPI_ISL_848839, EPI_ISL_848840, EPI_ISL_848841, EPI_ISL_848842, EPI_ISL_848843, EPI_ISL_848844, EPI_ISL_848845, EPI_ISL_848846, EPI_ISL_848847, EPI_ISL_848848, EPI_ISL_848849, EPI_ISL_848850, EPI_ISL_848851, EPI_ISL_848852, EPI_ISL_848853, EPI_ISL_848854, EPI_ISL_848855, EPI_ISL_848856, EPI_ISL_848857, EPI_ISL_848858, EPI_ISL_848859, EPI_ISL_848860, EPI_ISL_848861, EPI_ISL_848863, EPI_ISL_848864, EPI_ISL_848865, EPI_ISL_848866, EPI_ISL_848867, EPI_ISL_848868, EPI_ISL_848869, EPI_ISL_848870, EPI_ISL_848871, EPI_ISL_848872, EPI_ISL_848873, EPI_ISL_848874, EPI_ISL_848875, EPI_ISL_848876, EPI_ISL_848877, EPI_ISL_848878, EPI_ISL_848879, EPI_ISL_848880, EPI_ISL_848881, EPI_ISL_848882, EPI_ISL_848883, EPI_ISL_848884, EPI_ISL_848885, EPI_ISL_848886, EPI_ISL_848887, EPI_ISL_848888, EPI_ISL_848889, EPI_ISL_848890, EPI_ISL_848891, EPI_ISL_848892, EPI_ISL_848893, EPI_ISL_848894, EPI_ISL_848895, EPI_ISL_848896, EPI_ISL_848897, EPI_ISL_848898, EPI_ISL_848899, EPI_ISL_848900, EPI_ISL_848901, EPI_ISL_848902, EPI_ISL_848903, EPI_ISL_848904, EPI_ISL_848905, EPI_ISL_848906, EPI_ISL_848907, EPI_ISL_848908, EPI_ISL_848909, EPI_ISL_848910, EPI_ISL_848911, EPI_ISL_848912, EPI_ISL_848913, EPI_ISL_848914, EPI_ISL_848915, EPI_ISL_848916, EPI_ISL_848917, EPI_ISL_848918, EPI_ISL_848919, EPI_ISL_848920, EPI_ISL_848921, EPI_ISL_848922, EPI_ISL_848928, EPI_ISL_848929, EPI_ISL_848930, EPI_ISL_848931, EPI_ISL_848932, EPI_ISL_848933, EPI_ISL_848934, EPI_ISL_848935, EPI_ISL_848936, EPI_ISL_848937, EPI_ISL_848938, EPI_ISL_848939, EPI_ISL_848940, EPI_ISL_848941, EPI_ISL_848943, EPI_ISL_848944, EPI_ISL_848945, EPI_ISL_848946, EPI_ISL_848947, EPI_ISL_848948, EPI_ISL_848950, EPI_ISL_848951, EPI_ISL_848953, EPI_ISL_848954, EPI_ISL_848955, EPI_ISL_848956, EPI_ISL_848957, EPI_ISL_848958, EPI_ISL_848959, EPI_ISL_848960, EPI_ISL_848961, EPI_ISL_848962, EPI_ISL_848963, EPI_ISL_848964, EPI_ISL_848965, EPI_ISL_848966, EPI_ISL_848967, EPI_ISL_848968, EPI_ISL_848969, EPI_ISL_848970, EPI_ISL_848971, EPI_ISL_848972, EPI_ISL_848973, EPI_ISL_848974, EPI_ISL_848975, EPI_ISL_848976, EPI_ISL_848977, EPI_ISL_848978, EPI_ISL_848979, EPI_ISL_848980, EPI_ISL_848981, EPI_ISL_848982, EPI_ISL_848983, EPI_ISL_848984, EPI_ISL_848985, EPI_ISL_848986, EPI_ISL_848987, EPI_ISL_848988, EPI_ISL_848989, EPI_ISL_848990, EPI_ISL_848991, EPI_ISL_848992, EPI_ISL_848993, EPI_ISL_848994, EPI_ISL_848995, EPI_ISL_848996, EPI_ISL_848997, EPI_ISL_848998, EPI_ISL_848999, EPI_ISL_849000, EPI_ISL_849001, EPI_ISL_849002, EPI_ISL_849003, EPI_ISL_849004, EPI_ISL_849005, EPI_ISL_849006, EPI_ISL_849007, EPI_ISL_849008, EPI_ISL_849009, EPI_ISL_849010, EPI_ISL_849011, EPI_ISL_849012, EPI_ISL_849013, EPI_ISL_849014, EPI_ISL_849015, EPI_ISL_849016, EPI_ISL_849017, EPI_ISL_849018, EPI_ISL_849019, EPI_ISL_849020, EPI_ISL_849021, EPI_ISL_849022, EPI_ISL_849023, EPI_ISL_849024, EPI_ISL_849025, EPI_ISL_849026, EPI_ISL_849027, EPI_ISL_849028, EPI_ISL_849029, EPI_ISL_849030, EPI_ISL_849032, EPI_ISL_849033, EPI_ISL_849034, EPI_ISL_849035, EPI_ISL_849036, EPI_ISL_849037, EPI_ISL_849041, EPI_ISL_849042, EPI_ISL_849043, EPI_ISL_849044, EPI_ISL_849045, EPI_ISL_849046, EPI_ISL_849047, EPI_ISL_849048, EPI_ISL_849049, EPI_ISL_849050, EPI_ISL_849051, EPI_ISL_849052, EPI_ISL_849053, EPI_ISL_849054, EPI_ISL_849055, EPI_ISL_849056, EPI_ISL_849057, EPI_ISL_849065, EPI_ISL_849066, EPI_ISL_849067, EPI_ISL_849068, EPI_ISL_849069, EPI_ISL_849070, EPI_ISL_849071, EPI_ISL_849072, EPI_ISL_849073, EPI_ISL_849074, EPI_ISL_849075, EPI_ISL_849076, EPI_ISL_849077, EPI_ISL_849078, EPI_ISL_849079, EPI_ISL_849080, EPI_ISL_849081, EPI_ISL_849082, EPI_ISL_849083, EPI_ISL_849084, EPI_ISL_849085, EPI_ISL_849086, EPI_ISL_849087, EPI_ISL_849088, EPI_ISL_849089, EPI_ISL_849090, EPI_ISL_849091, EPI_ISL_849095, EPI_ISL_849096, EPI_ISL_849097, EPI_ISL_849098, EPI_ISL_849099, EPI_ISL_849100, EPI_ISL_849101, EPI_ISL_849102, EPI_ISL_849103, EPI_ISL_849104, EPI_ISL_849105, EPI_ISL_849106, EPI_ISL_849107, EPI_ISL_849109, EPI_ISL_849110, EPI_ISL_849111, EPI_ISL_849112, EPI_ISL_849113, EPI_ISL_849114, EPI_ISL_849120, EPI_ISL_849121, EPI_ISL_849122, EPI_ISL_849123, EPI_ISL_849124, EPI_ISL_849125, EPI_ISL_849126, EPI_ISL_849127, EPI_ISL_849128, EPI_ISL_849129, EPI_ISL_849130, EPI_ISL_849131, EPI_ISL_849132, EPI_ISL_849133, EPI_ISL_849134, EPI_ISL_849135, EPI_ISL_849136, EPI_ISL_849137, EPI_ISL_849138, EPI_ISL_849141, EPI_ISL_849142, EPI_ISL_849143, EPI_ISL_849144, EPI_ISL_849145, EPI_ISL_849146, EPI_ISL_849147

| | | | |
|---|---|---|---|
| see above | Florida Bureau of Public Health Laboratories | Florida Bureau of Public Health Laboratories | Sarah Schmedes, Jason Blanton |

EPI_ISL_849205, EPI_ISL_849206, EPI_ISL_849207, EPI_ISL_849208, EPI_ISL_849209, EPI_ISL_849210, EPI_ISL_849211, EPI_ISL_849212, EPI_ISL_849213, EPI_ISL_849214, EPI_ISL_849215, EPI_ISL_849216, EPI_ISL_849217, EPI_ISL_849218, EPI_ISL_849219, EPI_ISL_849220, EPI_ISL_849221, EPI_ISL_849222, EPI_ISL_849223, EPI_ISL_849224, EPI_ISL_849225, EPI_ISL_849226, EPI_ISL_849227, EPI_ISL_849228, EPI_ISL_849229, EPI_ISL_849230, EPI_ISL_849231, EPI_ISL_849232, EPI_ISL_849233, EPI_ISL_849234, EPI_ISL_849235, EPI_ISL_849236, EPI_ISL_849237, EPI_ISL_849238, EPI_ISL_849239, EPI_ISL_849240, EPI_ISL_849241, EPI_ISL_849242, EPI_ISL_849243, EPI_ISL_849244, EPI_ISL_849245, EPI_ISL_849246, EPI_ISL_849247, EPI_ISL_849248, EPI_ISL_849249, EPI_ISL_849250, EPI_ISL_849251, EPI_ISL_849252, EPI_ISL_849275, EPI_ISL_849276, EPI_ISL_849277, EPI_ISL_849278

| | | | |
|---|---|---|---|
| see above | Utah Public Health Laboratory | Utah Public Health Laboratory | Erin L. Young, Kelly F. Oakeson, Tara Gallagher |

EPI_ISL_849322, EPI_ISL_849323, EPI_ISL_849324, EPI_ISL_849325, EPI_ISL_849327, EPI_ISL_849328, EPI_ISL_849329, EPI_ISL_849330, EPI_ISL_849331, EPI_ISL_849332, EPI_ISL_849333, EPI_ISL_849335, EPI_ISL_849336, EPI_ISL_849337, EPI_ISL_849338, EPI_ISL_849339, EPI_ISL_849340, EPI_ISL_849341, EPI_ISL_849342, EPI_ISL_849343, EPI_ISL_849344, EPI_ISL_849345, EPI_ISL_849346, EPI_ISL_849348, EPI_ISL_849349

| | | | |
|---|---|---|---|
| see above | Delaware Public Health Lab | Delaware Public Health Lab | Gregory Hovan |

EPI_ISL_849383, EPI_ISL_849385, EPI_ISL_849386, EPI_ISL_849387, EPI_ISL_849388, EPI_ISL_849389, EPI_ISL_849390, EPI_ISL_849392, EPI_ISL_849393, EPI_ISL_849394, EPI_ISL_849395, EPI_ISL_849396, EPI_ISL_849397, EPI_ISL_849398, EPI_ISL_849399, EPI_ISL_849400, EPI_ISL_849401, EPI_ISL_849402, EPI_ISL_849403, EPI_ISL_849405, EPI_ISL_849406, EPI_ISL_849407

| | | | |
|---|---|---|---|
| see above | Washington State Department of Health | Seattle Flu Study | Deborah A. Nickerson, Chris D. Frazar, Jover Lee, Benjamin Pelle, Matthew Richardson, Amanda Adler, Elisabeth Brandstetter, Peter D. Han, Kairsten Fay, Misja Ilcisin, Kirsten Lacombe, Thomas R. Sibley, Melissa Truong, Caitlin R. Wolf, Romesh Gautom, Geoff Melly, Brian Hiatt, Philip Dykema, Scott Lindquist, Michael Boeckh, Janet A. Englund, Michael Famulare, Barry R. Lutz, Mark J. Rieder, Lea M. Starita, Matthew Thompson, Helen Y. Chu, Jay Shendure, Trevor Bedford |
| EPI_ISL_849408 | Seattle Flu Study | Seattle Flu Study | Deborah A. Nickerson, Chris D. Frazar, Jover Lee, Benjamin Pelle, Matthew Richardson, Amanda Adler, Elisabeth Brandstetter, Peter D. Han, Kairsten Fay, Misja Ilcisin, Kirsten Lacombe, Thomas R. Sibley, Melissa Truong, Caitlin R. Wolf, Michael Boeckh, Janet A. Englund, Michael Famulare, Barry R. Lutz, Mark J. Rieder, Lea M. Starita, Matthew Thompson, Jay Shendure, Trevor Bedford, Helen Y. Chu |

EPI_ISL_849409, EPI_ISL_849410, EPI_ISL_849411, EPI_ISL_849412, EPI_ISL_849413, EPI_ISL_849414, EPI_ISL_849415, EPI_ISL_849416, EPI_ISL_849417, EPI_ISL_849418, EPI_ISL_849419, EPI_ISL_849420, EPI_ISL_849421, EPI_ISL_849422, EPI_ISL_849423

| | | | |
|---|---|---|---|
| see above | Washington State Department of Health | Seattle Flu Study | Deborah A. Nickerson, Chris D. Frazar, Jover Lee, Benjamin Pelle, Matthew Richardson, Amanda Adler, Elisabeth Brandstetter, Peter D. Han, Kairsten Fay, Misja Ilcisin, Kirsten Lacombe, Thomas R. Sibley, Melissa Truong, Caitlin R. Wolf, Romesh Gautom, Geoff Melly, Brian Hiatt, Philip Dykema, Scott Lindquist, Michael Boeckh, Janet A. Englund, Michael Famulare, Barry R. Lutz, Mark J. Rieder, Lea M. Starita, Matthew Thompson, Helen Y. Chu, Jay Shendure, Trevor Bedford |
| EPI_ISL_849424 | Seattle Flu Study | Seattle Flu Study | Deborah A. Nickerson, Chris D. Frazar, Jover Lee, Benjamin Pelle, Matthew Richardson, Amanda Adler, Elisabeth Brandstetter, Peter D. Han, Kairsten Fay, Misja Ilcisin, Kirsten Lacombe, Thomas R. Sibley, Melissa Truong, Caitlin R. Wolf, Michael Boeckh, Janet A. Englund, Michael Famulare, Barry R. Lutz, Mark J. Rieder, Lea M. Starita, Matthew Thompson, Jay Shendure, Trevor Bedford, Helen Y. Chu |
| EPI_ISL_849425, EPI_ISL_849426, EPI_ISL_849427, EPI_ISL_849428, EPI_ISL_849429 | Washington State Department of Health | Seattle Flu Study | Deborah A. Nickerson, Chris D. Frazar, Jover Lee, Benjamin Pelle, Matthew Richardson, Amanda Adler, Elisabeth Brandstetter, Peter D. Han, Kairsten Fay, Misja Ilcisin, Kirsten Lacombe, Thomas R. Sibley, Melissa Truong, Caitlin R. Wolf, Romesh Gautom, Geoff Melly, Brian Hiatt, Philip Dykema, Scott Lindquist, Michael Boeckh, Janet A. Englund, Michael Famulare, Barry R. Lutz, Mark J. Rieder, Lea M. Starita, Matthew Thompson, Helen Y. Chu, Jay Shendure, Trevor Bedford |

| | | | |
|---|---|---|---|
| EPI_ISL_849430 | Seattle Flu Study | Seattle Flu Study | Deborah A. Nickerson, Chris D. Frazar, Jover Lee, Benjamin Pelle, Matthew Richardson, Amanda Adler, Elisabeth Brandstetter, Peter D. Han, Kairsten Fay, Misja Ilcisin, Kirsten Lacombe, Thomas R. Sibley, Melissa Truong, Caitlin R. Wolf, Michael Boeckh, Janet A. Englund, Michael Famulare, Barry R. Lutz, Mark J. Rieder, Lea M. Starita, Matthew Thompson, Jay Shendure, Trevor Bedford, Helen Y. Chu |
| EPI_ISL_849431, EPI_ISL_849432, EPI_ISL_849433, EPI_ISL_849434, EPI_ISL_849435, EPI_ISL_849436, EPI_ISL_849437, EPI_ISL_849438, EPI_ISL_849439 | Washington State Department of Health | Seattle Flu Study | Deborah A. Nickerson, Chris D. Frazar, Jover Lee, Benjamin Pelle, Matthew Richardson, Amanda Adler, Elisabeth Brandstetter, Peter D. Han, Kairsten Fay, Misja Ilcisin, Kirsten Lacombe, Thomas R. Sibley, Melissa Truong, Caitlin R. Wolf, Romesh Gautom, Geoff Melly, Brian Hiatt, Philip Dykema, Scott Lindquist, Michael Boeckh, Janet A. Englund, Michael Famulare, Barry R. Lutz, Mark J. Rieder, Lea M. Starita, Matthew Thompson, Helen Y. Chu, Jay Shendure, Trevor Bedford |
| EPI_ISL_849440, EPI_ISL_849441, EPI_ISL_849442 | Seattle Flu Study | Seattle Flu Study | Deborah A. Nickerson, Chris D. Frazar, Jover Lee, Benjamin Pelle, Matthew Richardson, Amanda Adler, Elisabeth Brandstetter, Peter D. Han, Kairsten Fay, Misja Ilcisin, Kirsten Lacombe, Thomas R. Sibley, Melissa Truong, Caitlin R. Wolf, Michael Boeckh, Janet A. Englund, Michael Famulare, Barry R. Lutz, Mark J. Rieder, Lea M. Starita, Matthew Thompson, Jay Shendure, Trevor Bedford, Helen Y. Chu |
| EPI_ISL_849443 | Washington State Department of Health | Seattle Flu Study | Deborah A. Nickerson, Chris D. Frazar, Jover Lee, Benjamin Pelle, Matthew Richardson, Amanda Adler, Elisabeth Brandstetter, Peter D. Han, Kairsten Fay, Misja Ilcisin, Kirsten Lacombe, Thomas R. Sibley, Melissa Truong, Caitlin R. Wolf, Romesh Gautom, Geoff Melly, Brian Hiatt, Philip Dykema, Scott Lindquist, Michael Boeckh, Janet A. Englund, Michael Famulare, Barry R. Lutz, Mark J. Rieder, Lea M. Starita, Matthew Thompson, Helen Y. Chu, Jay Shendure, Trevor Bedford |
| EPI_ISL_849444 | Seattle Flu Study | Seattle Flu Study | Deborah A. Nickerson, Chris D. Frazar, Jover Lee, Benjamin Pelle, Matthew Richardson, Amanda Adler, Elisabeth Brandstetter, Peter D. Han, Kairsten Fay, Misja Ilcisin, Kirsten Lacombe, Thomas R. Sibley, Melissa Truong, Caitlin R. Wolf, Michael Boeckh, Janet A. Englund, Michael Famulare, Barry R. Lutz, Mark J. Rieder, Lea M. Starita, Matthew Thompson, Jay Shendure, Trevor Bedford, Helen Y. Chu |
| EPI_ISL_849445, EPI_ISL_849446, EPI_ISL_849447, EPI_ISL_849448, EPI_ISL_849449, EPI_ISL_849450, EPI_ISL_849451, EPI_ISL_849452, EPI_ISL_849453, EPI_ISL_849454, EPI_ISL_849455, EPI_ISL_849456, EPI_ISL_849457, EPI_ISL_849458, EPI_ISL_849459, EPI_ISL_849460, EPI_ISL_849461 | | | |
| see above | Washington State Department of Health | Seattle Flu Study | Deborah A. Nickerson, Chris D. Frazar, Jover Lee, Benjamin Pelle, Matthew Richardson, Amanda Adler, Elisabeth Brandstetter, Peter D. Han, Kairsten Fay, Misja Ilcisin, Kirsten Lacombe, Thomas R. Sibley, Melissa Truong, Caitlin R. Wolf, Romesh Gautom, Geoff Melly, Brian Hiatt, Philip Dykema, Scott Lindquist, Michael Boeckh, Janet A. Englund, Michael Famulare, Barry R. Lutz, Mark J. Rieder, Lea M. Starita, Matthew Thompson, Helen Y. Chu, Jay Shendure, Trevor Bedford |
| EPI_ISL_849462 | Seattle Flu Study | Seattle Flu Study | Deborah A. Nickerson, Chris D. Frazar, Jover Lee, Benjamin Pelle, Matthew Richardson, Amanda Adler, Elisabeth Brandstetter, Peter D. Han, Kairsten Fay, Misja Ilcisin, Kirsten Lacombe, Thomas R. Sibley, Melissa Truong, Caitlin R. Wolf, Michael Boeckh, Janet A. Englund, Michael Famulare, Barry R. Lutz, Mark J. Rieder, Lea M. Starita, Matthew Thompson, Jay Shendure, Trevor Bedford, Helen Y. Chu |
| EPI_ISL_849463, EPI_ISL_849464, EPI_ISL_849465, EPI_ISL_849466 | Washington State Department of Health | Seattle Flu Study | Deborah A. Nickerson, Chris D. Frazar, Jover Lee, Benjamin Pelle, Matthew Richardson, Amanda Adler, Elisabeth Brandstetter, Peter D. Han, Kairsten Fay, Misja Ilcisin, Kirsten Lacombe, Thomas R. Sibley, Melissa Truong, Caitlin R. Wolf, Romesh Gautom, Geoff Melly, Brian Hiatt, Philip Dykema, Scott Lindquist, Michael Boeckh, Janet A. Englund, Michael Famulare, Barry R. Lutz, Mark J. Rieder, Lea M. Starita, Matthew Thompson, Helen Y. Chu, Jay Shendure, Trevor Bedford |
| EPI_ISL_849479, EPI_ISL_849480 | Seattle Flu Study | Seattle Flu Study | Deborah A. Nickerson, Chris D. Frazar, Jover Lee, Benjamin Pelle, Matthew Richardson, Amanda Adler, Elisabeth Brandstetter, Peter D. Han, Kairsten Fay, Misja Ilcisin, Kirsten Lacombe, Thomas R. Sibley, Melissa Truong, Caitlin R. Wolf, Karen Cowgill, Stephanie Schrag, Jeff Duchin, Michael Boeckh, Janet A. Englund, Michael Famulare, Barry R. Lutz, Mark J. Rieder, Lea M. Starita, Matthew Thompson, Helen Y. Chu, Trevor Bedford, Jay Shendure |
| EPI_ISL_849481, EPI_ISL_849482, EPI_ISL_849483, EPI_ISL_849484, EPI_ISL_849485 | Seattle Flu Study | Seattle Flu Study | Deborah A. Nickerson, Chris D. Frazar, Jover Lee, Benjamin Pelle, Matthew Richardson, Amanda Adler, Elisabeth Brandstetter, Peter D. Han, Kairsten Fay, Misja Ilcisin, Kirsten Lacombe, Thomas R. Sibley, Melissa Truong, Caitlin R. Wolf, Michael Boeckh, Janet A. Englund, Michael Famulare, Barry R. Lutz, Mark J. Rieder, Lea M. Starita, Matthew Thompson, Jay Shendure, Trevor Bedford, Helen Y. Chu |
| EPI_ISL_849486, EPI_ISL_849487, EPI_ISL_849488 | Seattle Flu Study | Seattle Flu Study | Deborah A. Nickerson, Chris D. Frazar, Jover Lee, Benjamin Pelle, Matthew Richardson, Amanda Adler, Elisabeth Brandstetter, Peter D. Han, Kairsten Fay, Misja Ilcisin, Kirsten Lacombe, Thomas R. Sibley, Melissa Truong, Caitlin R. Wolf, Karen Cowgill, Stephanie Schrag, Jeff Duchin, Michael Boeckh, Janet A. Englund, Michael Famulare, Barry R. Lutz, Mark J. Rieder, Lea M. Starita, Matthew Thompson, Helen Y. Chu, Trevor Bedford, Jay Shendure |
| EPI_ISL_849489, EPI_ISL_849490, EPI_ISL_849491, EPI_ISL_849492, EPI_ISL_849493, EPI_ISL_849494, EPI_ISL_849495, EPI_ISL_849496, EPI_ISL_849497, EPI_ISL_849498, EPI_ISL_849499, EPI_ISL_849500, EPI_ISL_849501, EPI_ISL_849502, EPI_ISL_849503, EPI_ISL_849504, EPI_ISL_849505, EPI_ISL_849506, EPI_ISL_849507, EPI_ISL_849508, EPI_ISL_849509, EPI_ISL_849510, EPI_ISL_849511, EPI_ISL_849512, EPI_ISL_849513, EPI_ISL_849514, EPI_ISL_849515, EPI_ISL_849516, EPI_ISL_849517 | | | |
| see above | Seattle Flu Study | Seattle Flu Study | Deborah A. Nickerson, Chris D. Frazar, Jover Lee, Benjamin Pelle, Matthew Richardson, Amanda Adler, Elisabeth Brandstetter, Peter D. Han, Kairsten Fay, Misja Ilcisin, Kirsten Lacombe, Thomas R. Sibley, Melissa Truong, Caitlin R. Wolf, Michael Boeckh, Janet A. Englund, Michael Famulare, Barry R. Lutz, Mark J. Rieder, Lea M. Starita, Matthew Thompson, Jay Shendure, Trevor Bedford, Helen Y. Chu |
| EPI_ISL_849518, EPI_ISL_849519, EPI_ISL_849520, EPI_ISL_849521, EPI_ISL_849522, EPI_ISL_849523, EPI_ISL_849524, EPI_ISL_849525, EPI_ISL_849526, EPI_ISL_849527, EPI_ISL_849528, EPI_ISL_849529, EPI_ISL_849530, EPI_ISL_849531, EPI_ISL_849532, EPI_ISL_849533, EPI_ISL_849534, EPI_ISL_849535, EPI_ISL_849536, EPI_ISL_849537, EPI_ISL_849538, EPI_ISL_849539, EPI_ISL_849540, EPI_ISL_849541, EPI_ISL_849542, EPI_ISL_849543, EPI_ISL_849544, EPI_ISL_849545, EPI_ISL_849546, EPI_ISL_849547, EPI_ISL_849548, EPI_ISL_849549, EPI_ISL_849550, EPI_ISL_849551, EPI_ISL_849554, EPI_ISL_849556, EPI_ISL_849557, EPI_ISL_849558, EPI_ISL_849559, EPI_ISL_849560 | | | |
| see above | Washington State Department of Health | Seattle Flu Study | Deborah A. Nickerson, Chris D. Frazar, Jover Lee, Benjamin Pelle, Matthew Richardson, Amanda Adler, Elisabeth Brandstetter, Peter D. Han, Kairsten Fay, Misja Ilcisin, Kirsten Lacombe, Thomas R. Sibley, Melissa Truong, Caitlin R. Wolf, Romesh Gautom, Geoff Melly, Brian Hiatt, Philip Dykema, Scott Lindquist, Michael Boeckh, Janet A. Englund, Michael Famulare, Barry R. Lutz, Mark J. Rieder, Lea M. Starita, Matthew Thompson, Helen Y. Chu, Jay Shendure, Trevor Bedford |
| EPI_ISL_849593 | Seattle Flu Study | Seattle Flu Study | Deborah A. Nickerson, Chris D. Frazar, Jover Lee, Benjamin Pelle, Matthew Richardson, Amanda Adler, Elisabeth Brandstetter, Peter D. Han, Kairsten Fay, Misja Ilcisin, Kirsten Lacombe, Thomas R. Sibley, Melissa Truong, Caitlin R. Wolf, Karen Cowgill, Stephanie Schrag, Jeff Duchin, Michael Boeckh, Janet A. Englund, Michael Famulare, Barry R. Lutz, Mark J. Rieder, Lea M. Starita, Matthew Thompson, Helen Y. Chu, Trevor Bedford, Jay Shendure |
| EPI_ISL_849594, EPI_ISL_849595 | Seattle Flu Study | Seattle Flu Study | Deborah A. Nickerson, Chris D. Frazar, Jover Lee, Benjamin Pelle, Matthew Richardson, Amanda Adler, Elisabeth Brandstetter, Peter D. Han, Kairsten Fay, Misja Ilcisin, Kirsten Lacombe, Thomas R. Sibley, Melissa Truong, Caitlin R. Wolf, Michael Boeckh, Janet A. Englund, Michael Famulare, Barry R. Lutz, Mark J. Rieder, Lea M. Starita, Matthew Thompson, Jay Shendure, Trevor Bedford, Helen Y. Chu |
| EPI_ISL_849596 | Seattle Flu Study | Seattle Flu Study | Deborah A. Nickerson, Chris D. Frazar, Jover Lee, Benjamin Pelle, Matthew Richardson, Amanda Adler, Elisabeth Brandstetter, Peter D. Han, Kairsten Fay, Misja Ilcisin, Kirsten Lacombe, Thomas R. Sibley, Melissa Truong, Caitlin R. Wolf, Karen Cowgill, Stephanie Schrag, Jeff Duchin, Michael Boeckh, Janet A. Englund, Michael Famulare, Barry R. Lutz, Mark J. Rieder, Lea M. Starita, Matthew Thompson, Helen Y. Chu, Trevor Bedford, Jay Shendure |
| EPI_ISL_849597, EPI_ISL_849598, EPI_ISL_849599, EPI_ISL_849600, EPI_ISL_849601, EPI_ISL_849602, EPI_ISL_849603, EPI_ISL_849604, EPI_ISL_849605, EPI_ISL_849606, EPI_ISL_849607, EPI_ISL_849608, EPI_ISL_849609, EPI_ISL_849610, EPI_ISL_849611, EPI_ISL_849612, EPI_ISL_849613, EPI_ISL_849614, EPI_ISL_849615 | | | |
| see above | Seattle Flu Study | Seattle Flu Study | Deborah A. Nickerson, Chris D. Frazar, Jover Lee, Benjamin Pelle, Matthew Richardson, Amanda Adler, Elisabeth Brandstetter, Peter D. Han, Kairsten Fay, Misja Ilcisin, Kirsten Lacombe, Thomas R. Sibley, Melissa Truong, Caitlin R. Wolf, Michael Boeckh, Janet A. Englund, Michael Famulare, Barry R. Lutz, Mark J. Rieder, Lea M. Starita, Matthew Thompson, Jay Shendure, Trevor Bedford, Helen Y. Chu |
| EPI_ISL_849616, EPI_ISL_849617, EPI_ISL_849618, EPI_ISL_849619, EPI_ISL_849620, EPI_ISL_849621, EPI_ISL_849622, EPI_ISL_849623, EPI_ISL_849624, EPI_ISL_849625, EPI_ISL_849626, EPI_ISL_849627, EPI_ISL_849628, EPI_ISL_849629, EPI_ISL_849630, EPI_ISL_849631, EPI_ISL_849632, EPI_ISL_849633 | | | |
| see above | Washington State Department of Health | Seattle Flu Study | Deborah A. Nickerson, Chris D. Frazar, Jover Lee, Benjamin Pelle, Matthew Richardson, Amanda Adler, Elisabeth Brandstetter, Peter D. Han, Kairsten Fay, Misja Ilcisin, Kirsten Lacombe, Thomas R. Sibley, Melissa Truong, Caitlin R. Wolf, Romesh Gautom, Geoff Melly, Brian Hiatt, Philip Dykema, Scott Lindquist, Michael Boeckh, Janet A. Englund, Michael Famulare, Barry R. Lutz, Mark J. Rieder, Lea M. Starita, Matthew Thompson, Helen Y. Chu, Jay Shendure, Trevor Bedford |
| EPI_ISL_849764, EPI_ISL_849766, EPI_ISL_849767, EPI_ISL_849768, EPI_ISL_849769, EPI_ISL_849771, EPI_ISL_849772, EPI_ISL_849774, EPI_ISL_849775, EPI_ISL_849776, EPI_ISL_849777, EPI_ISL_849778, EPI_ISL_849779, EPI_ISL_849780, EPI_ISL_849781, EPI_ISL_849782, EPI_ISL_849783, EPI_ISL_849784, EPI_ISL_849785, EPI_ISL_849786, EPI_ISL_849787, EPI_ISL_849789, EPI_ISL_849790, EPI_ISL_849792, EPI_ISL_849793, EPI_ISL_849867, EPI_ISL_849868, EPI_ISL_849869, EPI_ISL_849870, EPI_ISL_849871, EPI_ISL_849872, EPI_ISL_849874, EPI_ISL_849875, EPI_ISL_849876, EPI_ISL_849877, EPI_ISL_849879, EPI_ISL_849880, EPI_ISL_849881, EPI_ISL_849883, EPI_ISL_849884, EPI_ISL_849885, EPI_ISL_849889, EPI_ISL_849890, EPI_ISL_849891, EPI_ISL_849892, EPI_ISL_849893, EPI_ISL_849894, EPI_ISL_849895, EPI_ISL_849896, EPI_ISL_849897, EPI_ISL_849898, EPI_ISL_849899, EPI_ISL_849900, EPI_ISL_849901, EPI_ISL_849902, EPI_ISL_849903, EPI_ISL_849905, EPI_ISL_849906, EPI_ISL_849907, EPI_ISL_849908, EPI_ISL_849909, EPI_ISL_849910, EPI_ISL_849911, EPI_ISL_849912, EPI_ISL_849913, EPI_ISL_849914, EPI_ISL_849915, EPI_ISL_849916, EPI_ISL_849917, EPI_ISL_849918, EPI_ISL_849920 | | | |
| see above | Utah Public Health Laboratory | Utah Public Health Laboratory | Erin L. Young, Kelly F. Oakeson, Tara Gallagher |
| EPI_ISL_849922 | Orange County Public Health Lab | Chan-Zuckerberg Biohub | CZB Cliahub Consortium |
| EPI_ISL_849923, EPI_ISL_849924, EPI_ISL_849925, EPI_ISL_849926, EPI_ISL_849927 | UCSF Clinical Microbiology Laboratory | Chan-Zuckerberg Biohub | CZB Cliahub Consortium |
| EPI_ISL_849928, EPI_ISL_849929, EPI_ISL_849930, EPI_ISL_849931, EPI_ISL_849932, EPI_ISL_849933, EPI_ISL_849934, EPI_ISL_849935, EPI_ISL_849936, EPI_ISL_849937, EPI_ISL_849938, EPI_ISL_849939, EPI_ISL_849940, EPI_ISL_849941, EPI_ISL_849942, EPI_ISL_849943, EPI_ISL_849944, EPI_ISL_849945, EPI_ISL_849946, EPI_ISL_849947, EPI_ISL_849948, EPI_ISL_849949, EPI_ISL_849950, EPI_ISL_849951, EPI_ISL_849952 | | | |
| see above | UC Davis- Department of Pathology and Laboratory Medicine | Chan-Zuckerberg Biohub | CZB Cliahub Consortium |
| EPI_ISL_849953, EPI_ISL_849954, EPI_ISL_849955, EPI_ISL_849957, EPI_ISL_849958, EPI_ISL_849959, EPI_ISL_849960, EPI_ISL_849961, EPI_ISL_849962, EPI_ISL_849963, EPI_ISL_849964, EPI_ISL_849965, EPI_ISL_849966, EPI_ISL_849967, EPI_ISL_849968, EPI_ISL_849969, EPI_ISL_849970, EPI_ISL_849971, EPI_ISL_849972, EPI_ISL_849973, EPI_ISL_849974, EPI_ISL_849975, EPI_ISL_849976, EPI_ISL_849977, EPI_ISL_849978, EPI_ISL_849979, EPI_ISL_849980, EPI_ISL_849981, EPI_ISL_849982, EPI_ISL_849983, EPI_ISL_849984, EPI_ISL_849985, EPI_ISL_849986, EPI_ISL_849987, EPI_ISL_849988, EPI_ISL_849989, EPI_ISL_849990, EPI_ISL_849991, EPI_ISL_849992, EPI_ISL_849993, EPI_ISL_849994, EPI_ISL_849995, EPI_ISL_849996, EPI_ISL_849997, EPI_ISL_849998, EPI_ISL_849999, EPI_ISL_850000, EPI_ISL_850001, EPI_ISL_850002, EPI_ISL_850003, EPI_ISL_850004, EPI_ISL_850005, EPI_ISL_850006, EPI_ISL_850007, EPI_ISL_850008, EPI_ISL_850009, EPI_ISL_850010, EPI_ISL_850011, EPI_ISL_850012, EPI_ISL_850013, EPI_ISL_850014, EPI_ISL_850015, EPI_ISL_850016, EPI_ISL_850017, EPI_ISL_850018 | | | |
| see above | Santa Clara County Public Health Laboratory | Chan-Zuckerberg Biohub | CZB Cliahub Consortium |
| EPI_ISL_850019, EPI_ISL_850020, EPI_ISL_850021, EPI_ISL_850022, EPI_ISL_850023, EPI_ISL_850024, EPI_ISL_850025, EPI_ISL_850026, EPI_ISL_850027, EPI_ISL_850029, EPI_ISL_850030, EPI_ISL_850031, EPI_ISL_850032, EPI_ISL_850033, EPI_ISL_850034, EPI_ISL_850035, EPI_ISL_850036, EPI_ISL_850037, EPI_ISL_850038, EPI_ISL_850039, EPI_ISL_850040, EPI_ISL_850041, EPI_ISL_850042, EPI_ISL_850043, EPI_ISL_850044, EPI_ISL_850045, EPI_ISL_850046, EPI_ISL_850047, EPI_ISL_850048, EPI_ISL_850049, EPI_ISL_850050, EPI_ISL_850051, EPI_ISL_850052, EPI_ISL_850053, EPI_ISL_850054, EPI_ISL_850055, EPI_ISL_850056, EPI_ISL_850057, EPI_ISL_850058, EPI_ISL_850059, EPI_ISL_850060, EPI_ISL_850061, EPI_ISL_850062, EPI_ISL_850063, EPI_ISL_850064, EPI_ISL_850065, EPI_ISL_850066, EPI_ISL_850067, EPI_ISL_850068, EPI_ISL_850069, EPI_ISL_850070, EPI_ISL_850071, EPI_ISL_850072, EPI_ISL_850073, EPI_ISL_850074, EPI_ISL_850075, EPI_ISL_850076, EPI_ISL_850077, EPI_ISL_850078, EPI_ISL_850079, EPI_ISL_850080, EPI_ISL_850081, EPI_ISL_850082, EPI_ISL_850083, EPI_ISL_850084, EPI_ISL_850085, EPI_ISL_850086, EPI_ISL_850087, EPI_ISL_850088, EPI_ISL_850089, EPI_ISL_850090, EPI_ISL_850091, EPI_ISL_850092, EPI_ISL_850093, EPI_ISL_850094, EPI_ISL_850095, EPI_ISL_850096, EPI_ISL_850097, EPI_ISL_850098, EPI_ISL_850099, EPI_ISL_850100, EPI_ISL_850101, EPI_ISL_850102, EPI_ISL_850103, EPI_ISL_850104, EPI_ISL_850105, EPI_ISL_850106, EPI_ISL_850107, EPI_ISL_850108, EPI_ISL_850109, EPI_ISL_850110, EPI_ISL_850111, EPI_ISL_850112, EPI_ISL_850113, EPI_ISL_850114, EPI_ISL_850115, EPI_ISL_850116, EPI_ISL_850117, EPI_ISL_850118, EPI_ISL_850119, EPI_ISL_850120, EPI_ISL_850121, EPI_ISL_850122, EPI_ISL_850123, EPI_ISL_850124, EPI_ISL_850125, EPI_ISL_850126, EPI_ISL_850127, EPI_ISL_850128, EPI_ISL_850129, EPI_ISL_850130, EPI_ISL_850131, EPI_ISL_850132, EPI_ISL_850133, EPI_ISL_850134, EPI_ISL_850135, EPI_ISL_850136, EPI_ISL_850137, EPI_ISL_850138, EPI_ISL_850139, EPI_ISL_850140, EPI_ISL_850141, EPI_ISL_850142, EPI_ISL_850143, EPI_ISL_850144, EPI_ISL_850145, EPI_ISL_850146, EPI_ISL_850147, EPI_ISL_850148, EPI_ISL_850149, EPI_ISL_850150, EPI_ISL_850151, EPI_ISL_850152, EPI_ISL_850153, EPI_ISL_850154, EPI_ISL_850155, EPI_ISL_850156, EPI_ISL_850157, EPI_ISL_850158, EPI_ISL_850159, EPI_ISL_850160, EPI_ISL_850161, EPI_ISL_850162, EPI_ISL_850163, EPI_ISL_850164, EPI_ISL_850165, EPI_ISL_850166, EPI_ISL_850167, EPI_ISL_850168, EPI_ISL_850169, EPI_ISL_850170, EPI_ISL_850171, EPI_ISL_850172, EPI_ISL_850173, EPI_ISL_850174, EPI_ISL_850175, EPI_ISL_850176, EPI_ISL_850177, EPI_ISL_850178, EPI_ISL_850179 | | | |
| see above | Renegade | Chan-Zuckerberg Biohub | CZB Cliahub Consortium |
| EPI_ISL_850517, EPI_ISL_850518, EPI_ISL_850519, EPI_ISL_850520, EPI_ISL_850521, EPI_ISL_850522, EPI_ISL_850523, EPI_ISL_850524, EPI_ISL_850525, EPI_ISL_850526, EPI_ISL_850527, EPI_ISL_850528, EPI_ISL_850529, EPI_ISL_850530, EPI_ISL_850531, EPI_ISL_850532, EPI_ISL_850533, EPI_ISL_850534, EPI_ISL_850535, EPI_ISL_850536, EPI_ISL_850537, EPI_ISL_850538, EPI_ISL_850539, EPI_ISL_850540, EPI_ISL_850541, EPI_ISL_850543, EPI_ISL_850544, EPI_ISL_850545, EPI_ISL_850546, EPI_ISL_850547, EPI_ISL_850548, EPI_ISL_850549, EPI_ISL_850550, EPI_ISL_850552, EPI_ISL_850553, EPI_ISL_850554, EPI_ISL_850555, EPI_ISL_850557, EPI_ISL_850558, EPI_ISL_850559, EPI_ISL_850560, EPI_ISL_850561, EPI_ISL_850562, EPI_ISL_850563, EPI_ISL_850564, EPI_ISL_850565, EPI_ISL_850566, EPI_ISL_850567, EPI_ISL_850568, EPI_ISL_850569, EPI_ISL_850570, EPI_ISL_850571, EPI_ISL_850572, EPI_ISL_850573, EPI_ISL_850574, EPI_ISL_850575, EPI_ISL_850576, EPI_ISL_850577, EPI_ISL_850578, EPI_ISL_850579, EPI_ISL_850580, EPI_ISL_850581, EPI_ISL_850582, EPI_ISL_850583, EPI_ISL_850584, EPI_ISL_850585, EPI_ISL_850586, EPI_ISL_850587, EPI_ISL_850588, EPI_ISL_850589, EPI_ISL_850590, EPI_ISL_850591, EPI_ISL_850592, EPI_ISL_850593, EPI_ISL_850594, EPI_ISL_850595, EPI_ISL_850596, EPI_ISL_850597, EPI_ISL_850598, EPI_ISL_850599, EPI_ISL_850600, EPI_ISL_850601, EPI_ISL_850602, EPI_ISL_850603, EPI_ISL_850604, EPI_ISL_850605, EPI_ISL_850606, EPI_ISL_850607, EPI_ISL_850608, EPI_ISL_850609, EPI_ISL_850610, EPI_ISL_850611, EPI_ISL_850612, EPI_ISL_850614, EPI_ISL_850615, EPI_ISL_850616, EPI_ISL_850617, EPI_ISL_850618, EPI_ISL_850619, EPI_ISL_850620, EPI_ISL_850621, EPI_ISL_850622, EPI_ISL_850623, EPI_ISL_850624, EPI_ISL_850625, EPI_ISL_850626, EPI_ISL_850627, EPI_ISL_850628, EPI_ISL_850629, EPI_ISL_850630, EPI_ISL_850631, EPI_ISL_850632, EPI_ISL_850633, EPI_ISL_850634, EPI_ISL_850635, EPI_ISL_850636, EPI_ISL_850637, EPI_ISL_850638, EPI_ISL_850639, EPI_ISL_850640, EPI_ISL_850641, EPI_ISL_850642, EPI_ISL_850643, EPI_ISL_850644, EPI_ISL_850645, EPI_ISL_850646, EPI_ISL_850647, EPI_ISL_850648, EPI_ISL_850649, EPI_ISL_850650, EPI_ISL_850651 | | | |
| see above | Helix/Illumina | Genomics and Discovery, Respiratory Viruses Branch, Division of Viral Diseases, Centers for Disease Control and Prevention | Peter W. Cook, Dhwani Batra, Ben L. Rambo-Martin Eileen de Feo, Jan Antico, Christine Tran, Matthew Tolentino, Shannon Wickline, Kim Gietzen, Brad Sickler, Jingtao Liu, Eric Allen, Phil Febbo, Summer Galloway, Nicole L. Washington, Simon White, Geraint Levan, Kelly Schiabor Barrett, Elizabeth Cirulli, Alexandre Bolze, Ary Ascencio, Charlotte Rivera-Garcia, Ryan Cho, Jason Nguyen, Sherry Wang, Jimmy Ramirez, Tyler Cassens, Efren Sandoval, Magnus Isaksson, William Lee, David Becker, Marc Laurent, James Lu, Clinton R. Paden, Suxiang Tong, Duncan MacCannell |
| EPI_ISL_850689, EPI_ISL_850690, EPI_ISL_850691, EPI_ISL_850692, EPI_ISL_850693, EPI_ISL_850694, EPI_ISL_850695, EPI_ISL_850696, EPI_ISL_850697, EPI_ISL_850698, EPI_ISL_850699, EPI_ISL_850700, EPI_ISL_850701, EPI_ISL_850702, EPI_ISL_850703, EPI_ISL_850704, EPI_ISL_850705, EPI_ISL_850706, EPI_ISL_850707, EPI_ISL_850709, EPI_ISL_850710, EPI_ISL_850711, EPI_ISL_850712, EPI_ISL_850713, | | | |

| | | | |
|---|---|---|---|
| EPI_ISL_850714, EPI_ISL_850715, EPI_ISL_850716, EPI_ISL_850717, EPI_ISL_850718, EPI_ISL_850719, EPI_ISL_850720, EPI_ISL_850722, EPI_ISL_850723, EPI_ISL_850724, EPI_ISL_850725, EPI_ISL_850726, EPI_ISL_850727, EPI_ISL_850728, EPI_ISL_850729, EPI_ISL_850730, EPI_ISL_850731, EPI_ISL_850732, EPI_ISL_850733, EPI_ISL_850734, EPI_ISL_850735, EPI_ISL_850736, EPI_ISL_850738, EPI_ISL_850739, EPI_ISL_850740, EPI_ISL_850741, EPI_ISL_850742, EPI_ISL_850743, EPI_ISL_850744, EPI_ISL_850745, EPI_ISL_850746, EPI_ISL_850747, EPI_ISL_850748, EPI_ISL_850749, EPI_ISL_850750, EPI_ISL_850751, EPI_ISL_850752, EPI_ISL_850753, EPI_ISL_850754, EPI_ISL_850755, EPI_ISL_850756, EPI_ISL_850757, EPI_ISL_850758, EPI_ISL_850759, EPI_ISL_850760, EPI_ISL_850761, EPI_ISL_850762, EPI_ISL_850763, EPI_ISL_850764, EPI_ISL_850765, EPI_ISL_850766, EPI_ISL_850767, EPI_ISL_850768, EPI_ISL_850769, EPI_ISL_850770, EPI_ISL_850771, EPI_ISL_850772, EPI_ISL_850773, EPI_ISL_850774, EPI_ISL_850775, EPI_ISL_850776, EPI_ISL_850777, EPI_ISL_850778, EPI_ISL_850779, EPI_ISL_850780, EPI_ISL_850781, EPI_ISL_850782, EPI_ISL_850783, EPI_ISL_850784, EPI_ISL_850785, EPI_ISL_850786, EPI_ISL_850787, EPI_ISL_850788, EPI_ISL_850789, EPI_ISL_850790, EPI_ISL_850791, EPI_ISL_850792, EPI_ISL_850793, EPI_ISL_850794, EPI_ISL_850795, EPI_ISL_850796, EPI_ISL_850797, EPI_ISL_850798, EPI_ISL_850799, EPI_ISL_850800, EPI_ISL_850801, EPI_ISL_850802, EPI_ISL_850803, EPI_ISL_850804, EPI_ISL_850805, EPI_ISL_850806, EPI_ISL_850807, EPI_ISL_850808, EPI_ISL_850809, EPI_ISL_850810, EPI_ISL_850811, EPI_ISL_850812, EPI_ISL_850813, EPI_ISL_850814, EPI_ISL_850815, EPI_ISL_850816, EPI_ISL_850817, EPI_ISL_850818, EPI_ISL_850819, EPI_ISL_850820, EPI_ISL_850821, EPI_ISL_850822, EPI_ISL_850823, EPI_ISL_850824, EPI_ISL_850825, EPI_ISL_850826, EPI_ISL_850827, EPI_ISL_850828, EPI_ISL_850829, EPI_ISL_850830, EPI_ISL_850831, EPI_ISL_850832, EPI_ISL_850833, EPI_ISL_850834, EPI_ISL_850835, EPI_ISL_850836, EPI_ISL_850837, EPI_ISL_850838, EPI_ISL_850839, EPI_ISL_850840, EPI_ISL_850841, EPI_ISL_850842, EPI_ISL_850843, EPI_ISL_850844, EPI_ISL_850845, EPI_ISL_850846, EPI_ISL_850847, EPI_ISL_850848, EPI_ISL_850849, EPI_ISL_850850, EPI_ISL_850851, EPI_ISL_850852, EPI_ISL_850853, EPI_ISL_850854, EPI_ISL_850855, EPI_ISL_850856, EPI_ISL_850857, EPI_ISL_850858, EPI_ISL_850859, EPI_ISL_850860, EPI_ISL_850861, EPI_ISL_850862, EPI_ISL_850863, EPI_ISL_850864, EPI_ISL_850865, EPI_ISL_850866, EPI_ISL_850867, EPI_ISL_850868, EPI_ISL_850869, EPI_ISL_850870, EPI_ISL_850871, EPI_ISL_850873, EPI_ISL_850874, EPI_ISL_850875, EPI_ISL_850876, EPI_ISL_850877, EPI_ISL_850878, EPI_ISL_850879, EPI_ISL_850880, EPI_ISL_850881, EPI_ISL_850882, EPI_ISL_850883, EPI_ISL_850884, EPI_ISL_850885, EPI_ISL_850886, EPI_ISL_850888, EPI_ISL_850889, EPI_ISL_850890, EPI_ISL_850891, EPI_ISL_850892, EPI_ISL_850893, EPI_ISL_850894, EPI_ISL_850895, EPI_ISL_850896, EPI_ISL_850897, EPI_ISL_850898, EPI_ISL_850899, EPI_ISL_850900, EPI_ISL_850901, EPI_ISL_850902, EPI_ISL_850903, EPI_ISL_850904, EPI_ISL_850905, EPI_ISL_850906, EPI_ISL_850907, EPI_ISL_850908, EPI_ISL_850909, EPI_ISL_850910, EPI_ISL_850911, EPI_ISL_850912, EPI_ISL_850913, EPI_ISL_850914, EPI_ISL_850915, EPI_ISL_850916, EPI_ISL_850917, EPI_ISL_850918, EPI_ISL_850919, EPI_ISL_850920, EPI_ISL_850921, EPI_ISL_850922, EPI_ISL_850923, EPI_ISL_850924, EPI_ISL_850925, EPI_ISL_850926, EPI_ISL_850927, EPI_ISL_850928, EPI_ISL_850929, EPI_ISL_850930, EPI_ISL_850931, EPI_ISL_850932, EPI_ISL_850933, EPI_ISL_850934, EPI_ISL_850935, EPI_ISL_850936, EPI_ISL_850937, EPI_ISL_850938, EPI_ISL_850939, EPI_ISL_850940, EPI_ISL_850941, EPI_ISL_850942, EPI_ISL_850943, EPI_ISL_850944 | | | |
| see above | Helix / Illumina | Genomics and Discovery, Respiratory Viruses Branch, Division of Viral Diseases, Centers for Disease Control and Prevention | Peter W. Cook, Dhwani Batra, Ben L. Rambo-Martin Eileen de Feo, Jan Antico, Christine Tran, Matthew Tolentino, Shannon Wickline, Kim Gietzen, Brad Sickler, Jingtao Liu, Eric Allen, Phil Febbo, Summer Galloway, Nicole L. Washington, Simon White, Geraint Levan, Kelly Schiabor Barrett, Elizabeth Cirulli, Alexandre Bolze, Ary Ascencio, Charlotte Rivera-Garcia, Ryan Cho, Jason Nguyen, Sherry Wang, Jimmy Ramirez, Tyler Cassens, Efren Sandoval, Magnus Isaksson, William Lee, David Becker, Marc Laurent, James Lu, Clinton R. Paden, Suxiang Tong, Duncan MacCannell |
| EPI_ISL_850952, EPI_ISL_850953, EPI_ISL_850954, EPI_ISL_850955, EPI_ISL_850956, EPI_ISL_850957, EPI_ISL_850958, EPI_ISL_850959, EPI_ISL_850960, EPI_ISL_850961, EPI_ISL_850962, EPI_ISL_850963, EPI_ISL_850964, EPI_ISL_850965, EPI_ISL_850966, EPI_ISL_850967, EPI_ISL_850968, EPI_ISL_850969, EPI_ISL_850970, EPI_ISL_850971, EPI_ISL_850972, EPI_ISL_850973, EPI_ISL_850974, EPI_ISL_850975, EPI_ISL_850976, EPI_ISL_850977, EPI_ISL_850978, EPI_ISL_850979, EPI_ISL_850980, EPI_ISL_850981, EPI_ISL_850982, EPI_ISL_850983, EPI_ISL_850984, EPI_ISL_850985, EPI_ISL_850986, EPI_ISL_850987, EPI_ISL_850988, EPI_ISL_850989, EPI_ISL_850990, EPI_ISL_850991, EPI_ISL_850992, EPI_ISL_850993, EPI_ISL_850994, EPI_ISL_850995, EPI_ISL_850996, EPI_ISL_850997, EPI_ISL_850998, EPI_ISL_850999, EPI_ISL_851000, EPI_ISL_851001, EPI_ISL_851002, EPI_ISL_851003, EPI_ISL_851004, EPI_ISL_851005, EPI_ISL_851006, EPI_ISL_851007, EPI_ISL_851008, EPI_ISL_851009, EPI_ISL_851010, EPI_ISL_851011, EPI_ISL_851012, EPI_ISL_851013, EPI_ISL_851014, EPI_ISL_851015, EPI_ISL_851016, EPI_ISL_851017, EPI_ISL_851018, EPI_ISL_851019, EPI_ISL_851020, EPI_ISL_851021, EPI_ISL_851022, EPI_ISL_851023, EPI_ISL_851024, EPI_ISL_851025, EPI_ISL_851026, EPI_ISL_851027, EPI_ISL_851028, EPI_ISL_851029, EPI_ISL_851030, EPI_ISL_851031, EPI_ISL_851032, EPI_ISL_851033, EPI_ISL_851034, EPI_ISL_851035, EPI_ISL_851036, EPI_ISL_851037, EPI_ISL_851038, EPI_ISL_851039, EPI_ISL_851040 | | | |
| see above | Helix/Illumina | Genomics and Discovery, Respiratory Viruses Branch, Division of Viral Diseases, Centers for Disease Control and Prevention | Peter W. Cook, Dhwani Batra, Ben L. Rambo-Martin Eileen de Feo, Jan Antico, Christine Tran, Matthew Tolentino, Shannon Wickline, Kim Gietzen, Brad Sickler, Jingtao Liu, Eric Allen, Phil Febbo, Summer Galloway, Nicole L. Washington, Simon White, Geraint Levan, Kelly Schiabor Barrett, Elizabeth Cirulli, Alexandre Bolze, Ary Ascencio, Charlotte Rivera-Garcia, Ryan Cho, Jason Nguyen, Sherry Wang, Jimmy Ramirez, Tyler Cassens, Efren Sandoval, Magnus Isaksson, William Lee, David Becker, Marc Laurent, James Lu, Clinton R. Paden, Suxiang Tong, Duncan MacCannell |
| EPI_ISL_852617, EPI_ISL_852618, EPI_ISL_852620, EPI_ISL_852621, EPI_ISL_852623, EPI_ISL_852625, EPI_ISL_852626 | Ohio Department of Health Laboratory | Ohio Department of Health Laboratory | Holmes, Jennifer; Eric Brandt, Keoni Omura, Glen McGillivary, Caitlin McDonnell, Kirtana Ramadugu, Erica Leasure, Kelsey Florek, Heather Blankenship, Quanta Brown, and Tammy Bannerman |
| EPI_ISL_852858, EPI_ISL_852859, EPI_ISL_852860, EPI_ISL_852861 | Florida Bureau of Public Health Laboratories | Florida Bureau of Public Health Laboratories | Sarah Schmedes, Jason Blanton |
| EPI_ISL_853275, EPI_ISL_853276, EPI_ISL_853277, EPI_ISL_853278, EPI_ISL_853279, EPI_ISL_853280, EPI_ISL_853281, EPI_ISL_853282, EPI_ISL_853283, EPI_ISL_853284, EPI_ISL_853285, EPI_ISL_853286, EPI_ISL_853287, EPI_ISL_853288, EPI_ISL_853289, EPI_ISL_853290, EPI_ISL_853291, EPI_ISL_853292, EPI_ISL_853293, EPI_ISL_853294, EPI_ISL_853295, EPI_ISL_853296, EPI_ISL_853297, EPI_ISL_853298, EPI_ISL_853299, EPI_ISL_853300, EPI_ISL_853301, EPI_ISL_853302, EPI_ISL_853303, EPI_ISL_853304, EPI_ISL_853305, EPI_ISL_853306, EPI_ISL_853307, EPI_ISL_853308, EPI_ISL_853309, EPI_ISL_853310, EPI_ISL_853311, EPI_ISL_853312, EPI_ISL_853313, EPI_ISL_853314, EPI_ISL_853316, EPI_ISL_853317, EPI_ISL_853318, EPI_ISL_853319, EPI_ISL_853320, EPI_ISL_853321, EPI_ISL_853322, EPI_ISL_853323, EPI_ISL_853324, EPI_ISL_853325, EPI_ISL_853326, EPI_ISL_853327, EPI_ISL_853328, EPI_ISL_853329, EPI_ISL_853330, EPI_ISL_853331, EPI_ISL_853332, EPI_ISL_853333, EPI_ISL_853334, EPI_ISL_853335, EPI_ISL_853336, EPI_ISL_853337, EPI_ISL_853338, EPI_ISL_853339, EPI_ISL_853340, EPI_ISL_853341, EPI_ISL_853342, EPI_ISL_853343, EPI_ISL_853344, EPI_ISL_853345, EPI_ISL_853346, EPI_ISL_853347, EPI_ISL_853348, EPI_ISL_853349, EPI_ISL_853350, EPI_ISL_853351, EPI_ISL_853352, EPI_ISL_853353, EPI_ISL_853354, EPI_ISL_853355, EPI_ISL_853356, EPI_ISL_853357, EPI_ISL_853358, EPI_ISL_853359, EPI_ISL_853360, EPI_ISL_853361, EPI_ISL_853362, EPI_ISL_853363, EPI_ISL_853364, EPI_ISL_853365, EPI_ISL_853366, EPI_ISL_853367, EPI_ISL_853368, EPI_ISL_853369, EPI_ISL_853370, EPI_ISL_853371, EPI_ISL_853372, EPI_ISL_853373, EPI_ISL_853374, EPI_ISL_853375, EPI_ISL_853376, EPI_ISL_853377, EPI_ISL_853378, EPI_ISL_853379, EPI_ISL_853380, EPI_ISL_853381, EPI_ISL_853382, EPI_ISL_853383, EPI_ISL_853384, EPI_ISL_853385, EPI_ISL_853386, EPI_ISL_853387, EPI_ISL_853388, EPI_ISL_853389, EPI_ISL_853390, EPI_ISL_853391, EPI_ISL_853392, EPI_ISL_853393 | | | |
| see above | UPMC Clinical Microbiology Laboratory | Microbial Genome Sequencing Center; Microbial Genomic Epidemiology Laboratory | Mustapha M. Mustapha, Jane W. Marsh, Dan Snyder, Marissa P. Griffith, Stephanie L. Mitchell, Vatsala R. Srinivasa, Kady D. Waggle, Chinelo Ezeonwuku, Vaughn S. Cooper, Lee H. Harrison |
| EPI_ISL_853416, EPI_ISL_853417, EPI_ISL_853418, EPI_ISL_853419, EPI_ISL_853420, EPI_ISL_853421, EPI_ISL_853422, EPI_ISL_853423, EPI_ISL_853424, EPI_ISL_853425, EPI_ISL_853426, EPI_ISL_853427, EPI_ISL_853428, EPI_ISL_853429, EPI_ISL_853430, EPI_ISL_853431, EPI_ISL_853432, EPI_ISL_853433, EPI_ISL_853434, EPI_ISL_853435, EPI_ISL_853436, EPI_ISL_853437, EPI_ISL_853438, EPI_ISL_853439, EPI_ISL_853440, EPI_ISL_853441, EPI_ISL_853442, EPI_ISL_853443, EPI_ISL_853444, EPI_ISL_853445, EPI_ISL_853446, EPI_ISL_853447, EPI_ISL_853448, EPI_ISL_853449, EPI_ISL_853450, EPI_ISL_853451, EPI_ISL_853452, EPI_ISL_853453, EPI_ISL_853454, EPI_ISL_853455, EPI_ISL_853456, EPI_ISL_853457, EPI_ISL_853458, EPI_ISL_853459, EPI_ISL_853460, EPI_ISL_853461, EPI_ISL_853462, EPI_ISL_853463, EPI_ISL_853464, EPI_ISL_853465, EPI_ISL_853466, EPI_ISL_853467, EPI_ISL_853468, EPI_ISL_853469, EPI_ISL_853470, EPI_ISL_853471, EPI_ISL_853472, EPI_ISL_853473, EPI_ISL_853474, EPI_ISL_853475, EPI_ISL_853476, EPI_ISL_853477, EPI_ISL_853478, EPI_ISL_853479, EPI_ISL_853480, EPI_ISL_853481, EPI_ISL_853482, EPI_ISL_853483, EPI_ISL_853484, EPI_ISL_853485, EPI_ISL_853486, EPI_ISL_853487, EPI_ISL_853488, EPI_ISL_853489, EPI_ISL_853490, EPI_ISL_853491, EPI_ISL_853492, EPI_ISL_853493, EPI_ISL_853494, EPI_ISL_853495, EPI_ISL_853496, EPI_ISL_853497, EPI_ISL_853498, EPI_ISL_853499, EPI_ISL_853500, EPI_ISL_853501, EPI_ISL_853502, EPI_ISL_853503, EPI_ISL_853504, EPI_ISL_853505, EPI_ISL_853506, EPI_ISL_853507, EPI_ISL_853508, EPI_ISL_853509, EPI_ISL_853510, EPI_ISL_853511, EPI_ISL_853512, EPI_ISL_853513, EPI_ISL_853514, EPI_ISL_853515, EPI_ISL_853516, EPI_ISL_853517, EPI_ISL_853518, EPI_ISL_853519, EPI_ISL_853520, EPI_ISL_853521, EPI_ISL_853522, EPI_ISL_853523, EPI_ISL_853524, EPI_ISL_853525, EPI_ISL_853526, EPI_ISL_853527, EPI_ISL_853528, EPI_ISL_853529, EPI_ISL_853530, EPI_ISL_853531, EPI_ISL_853532, EPI_ISL_853533, EPI_ISL_853534, EPI_ISL_853535, EPI_ISL_853536, EPI_ISL_853537, EPI_ISL_853538, EPI_ISL_853539, EPI_ISL_853540, EPI_ISL_853541, EPI_ISL_853542, EPI_ISL_853543, EPI_ISL_853544, EPI_ISL_853545, EPI_ISL_853546, EPI_ISL_853547, EPI_ISL_853548, EPI_ISL_853549, EPI_ISL_853550, EPI_ISL_853551, EPI_ISL_853552, EPI_ISL_853553, EPI_ISL_853554, EPI_ISL_853555, EPI_ISL_853556, EPI_ISL_853557, EPI_ISL_853558, EPI_ISL_853559, EPI_ISL_853560, EPI_ISL_853561, EPI_ISL_853562, EPI_ISL_853563, EPI_ISL_853564, EPI_ISL_853565 | | | |
| see above | Santa Clara County Public Health Laboratory | Chan-Zuckerberg Biohub | CZB Cliahub Consortium |
| EPI_ISL_853648, EPI_ISL_853649 | THE MARY IMOGENE BASSETT HOSPITAL | Wadsworth Center, New York State Department of Health | Kirsten St. George, Daryl M. Lamson, Alexis Russel, Matthew Shudt, Melissa A Leisner, Jonathan Plitnick, Navjot Singh, John Kelly, Erasmus Schneider, Erica Lasek-Nesselquist |
| EPI_ISL_853650, EPI_ISL_853651 | NORTHWELL HEALTH LABORATORIES | Wadsworth Center, New York State Department of Health | Kirsten St. George, Daryl M. Lamson, Alexis Russel, Matthew Shudt, Melissa A Leisner, Jonathan Plitnick, Navjot Singh, John Kelly, Erasmus Schneider, Erica Lasek-Nesselquist |
| EPI_ISL_853652 | THE MARY IMOGENE BASSETT HOSPITAL | Wadsworth Center, New York State Department of Health | Kirsten St. George, Daryl M. Lamson, Alexis Russel, Matthew Shudt, Melissa A Leisner, Jonathan Plitnick, Navjot Singh, John Kelly, Erasmus Schneider, Erica Lasek-Nesselquist |
| EPI_ISL_853653, EPI_ISL_853654, EPI_ISL_853655, EPI_ISL_853656, EPI_ISL_853657, EPI_ISL_853658, EPI_ISL_853659, EPI_ISL_853660, EPI_ISL_853661, EPI_ISL_853662, EPI_ISL_853663, EPI_ISL_853664, EPI_ISL_853665, EPI_ISL_853666 | | | |
| see above | NORTHWELL HEALTH LABORATORIES | Wadsworth Center, New York State Department of Health | Kirsten St. George, Daryl M. Lamson, Alexis Russel, Matthew Shudt, Melissa A Leisner, Jonathan Plitnick, Navjot Singh, John Kelly, Erasmus Schneider, Erica Lasek-Nesselquist |
| EPI_ISL_853667, EPI_ISL_853668, EPI_ISL_853669 | THE MARY IMOGENE BASSETT HOSPITAL | Wadsworth Center, New York State Department of Health | Kirsten St. George, Daryl M. Lamson, Alexis Russel, Matthew Shudt, Melissa A Leisner, Jonathan Plitnick, Navjot Singh, John Kelly, Erasmus Schneider, Erica Lasek-Nesselquist |
| EPI_ISL_853670, EPI_ISL_853671, EPI_ISL_853672, EPI_ISL_853673, EPI_ISL_853674, EPI_ISL_853675 | NORTHWELL HEALTH LABORATORIES | Wadsworth Center, New York State Department of Health | Kirsten St. George, Daryl M. Lamson, Alexis Russel, Matthew Shudt, Melissa A Leisner, Jonathan Plitnick, Navjot Singh, John Kelly, Erasmus Schneider, Erica Lasek-Nesselquist |
| EPI_ISL_853676, EPI_ISL_853677, EPI_ISL_853678, EPI_ISL_853679, EPI_ISL_853680, EPI_ISL_853681, EPI_ISL_853682 | ACUTIS DIAGNOSTICS | Wadsworth Center, New York State Department of Health | Kirsten St. George, Daryl M. Lamson, Alexis Russel, Matthew Shudt, Melissa A Leisner, Jonathan Plitnick, Navjot Singh, John Kelly, Erasmus Schneider, Erica Lasek-Nesselquist |
| EPI_ISL_853683 | NORTHWELL HEALTH LABORATORIES | Wadsworth Center, New York State Department of Health | Kirsten St. George, Daryl M. Lamson, Alexis Russel, Matthew Shudt, Melissa A Leisner, Jonathan Plitnick, Navjot Singh, John Kelly, Erasmus Schneider, Erica Lasek-Nesselquist |
| EPI_ISL_853684, EPI_ISL_853685, EPI_ISL_853686, EPI_ISL_853687 | ALBANY MEDICAL CENTER HOSPITAL CLINICAL LABORATORIES | Wadsworth Center, New York State Department of Health | Kirsten St. George, Daryl M. Lamson, Alexis Russel, Matthew Shudt, Melissa A Leisner, Jonathan Plitnick, Navjot Singh, John Kelly, Erasmus Schneider, Erica Lasek-Nesselquist |
| EPI_ISL_853688 | THE MARY IMOGENE BASSETT HOSPITAL | Wadsworth Center, New York State Department of Health | Kirsten St. George, Daryl M. Lamson, Alexis Russel, Matthew Shudt, Melissa A Leisner, Jonathan Plitnick, Navjot Singh, John Kelly, Erasmus Schneider, Erica Lasek-Nesselquist |
| EPI_ISL_853689 | SUNY UPSTATE MEDICAL UNIVERSITY | Wadsworth Center, New York State Department of Health | Kirsten St. George, Daryl M. Lamson, Alexis Russel, Matthew Shudt, Melissa A Leisner, Jonathan Plitnick, Navjot Singh, John Kelly, Erasmus Schneider, Erica Lasek-Nesselquist |
| EPI_ISL_853690, EPI_ISL_853691, EPI_ISL_853692, EPI_ISL_853693, EPI_ISL_853694, EPI_ISL_853695, EPI_ISL_853696, EPI_ISL_853697, EPI_ISL_853698, EPI_ISL_853699, EPI_ISL_853700, EPI_ISL_853701, EPI_ISL_853702, EPI_ISL_853703, EPI_ISL_853704 | | | |
| see above | ALBANY MEDICAL CENTER HOSPITAL CLINICAL LABORATORIES | Wadsworth Center, New York State Department of Health | Kirsten St. George, Daryl M. Lamson, Alexis Russel, Matthew Shudt, Melissa A Leisner, Jonathan Plitnick, Navjot Singh, John Kelly, Erasmus Schneider, Erica Lasek-Nesselquist |
| EPI_ISL_853705 | NORTHWELL HEALTH LABORATORIES | Wadsworth Center, New York State Department of Health | Kirsten St. George, Daryl M. Lamson, Alexis Russel, Matthew Shudt, Melissa A Leisner, Jonathan Plitnick, Navjot Singh, John Kelly, Erasmus Schneider, Erica Lasek-Nesselquist |
| EPI_ISL_853706, EPI_ISL_853707, EPI_ISL_853708 | SUNY UPSTATE MEDICAL UNIVERSITY | Wadsworth Center, New York State Department of Health | Kirsten St. George, Daryl M. Lamson, Alexis Russel, Matthew Shudt, Melissa A Leisner, Jonathan Plitnick, Navjot Singh, John Kelly, Erasmus Schneider, Erica Lasek-Nesselquist |
| EPI_ISL_853709, EPI_ISL_853710, EPI_ISL_853711 | ALBANY MEDICAL CENTER HOSPITAL CLINICAL LABORATORIES | Wadsworth Center, New York State Department of Health | Kirsten St. George, Daryl M. Lamson, Alexis Russel, Matthew Shudt, Melissa A Leisner, Jonathan Plitnick, Navjot Singh, John Kelly, Erasmus Schneider, Erica Lasek-Nesselquist |
| EPI_ISL_853712, EPI_ISL_853713, EPI_ISL_853714 | NORTHWELL HEALTH LABORATORIES | Wadsworth Center, New York State Department of Health | Kirsten St. George, Daryl M. Lamson, Alexis Russel, Matthew Shudt, Melissa A Leisner, Jonathan Plitnick, Navjot Singh, John Kelly, Erasmus Schneider, Erica Lasek-Nesselquist |
| EPI_ISL_853715 | SUNY UPSTATE MEDICAL UNIVERSITY | Wadsworth Center, New York State Department of Health | Kirsten St. George, Daryl M. Lamson, Alexis Russel, Matthew Shudt, Melissa A Leisner, Jonathan Plitnick, Navjot Singh, John Kelly, Erasmus Schneider, Erica Lasek-Nesselquist |

| | | | |
|---|---|---|---|
| EPI_ISL_853716, EPI_ISL_853717 | NORTHWELL HEALTH LABORATORIES | Wadsworth Center, New York State Department of Health | Kirsten St. George, Daryl M. Lamson, Alexis Russel, Matthew Shudt, Melissa A Leisner, Jonathan Plitnick, Navjot Singh, John Kelly, Erasmus Schneider, Erica Lasek-Nesselquist |
| EPI_ISL_854305 | ESOTERIX GENETIC LABORATORIES LLC | Wadsworth Center, New York State Department of Health | Kirsten St. George, Daryl M. Lamson, Alexis Russel, Matthew Shudt, Melissa A Leisner, Jonathan Plitnick, Navjot Singh, John Kelly, Erasmus Schneider, Erica Lasek-Nesselquist |
| EPI_ISL_854306, EPI_ISL_854307, EPI_ISL_854308, EPI_ISL_854309, EPI_ISL_854310, EPI_ISL_854311, EPI_ISL_854312, EPI_ISL_854313, EPI_ISL_854314, EPI_ISL_854315, EPI_ISL_854316, EPI_ISL_854317, EPI_ISL_854318, EPI_ISL_854319, EPI_ISL_854320, EPI_ISL_854321, EPI_ISL_854322, EPI_ISL_854323, EPI_ISL_854324, EPI_ISL_854325, EPI_ISL_854326, EPI_ISL_854327, EPI_ISL_854328, EPI_ISL_854329, EPI_ISL_854330, EPI_ISL_854331, EPI_ISL_854332, EPI_ISL_854333, EPI_ISL_854334, EPI_ISL_854335, EPI_ISL_854336, EPI_ISL_854337, EPI_ISL_854338, EPI_ISL_854339, EPI_ISL_854340, EPI_ISL_854341, EPI_ISL_854342, EPI_ISL_854343, EPI_ISL_854344, EPI_ISL_854345, EPI_ISL_854346, EPI_ISL_854347, EPI_ISL_854348, EPI_ISL_854349 | | | |
| see above | URMC LABS | Wadsworth Center, New York State Department of Health | Kirsten St. George, Daryl M. Lamson, Alexis Russel, Matthew Shudt, Melissa A Leisner, Jonathan Plitnick, Navjot Singh, John Kelly, Erasmus Schneider, Erica Lasek-Nesselquist |
| EPI_ISL_854350, EPI_ISL_854351, EPI_ISL_854352, EPI_ISL_854353, EPI_ISL_854354, EPI_ISL_854355, EPI_ISL_854356, EPI_ISL_854357, EPI_ISL_854358, EPI_ISL_854359, EPI_ISL_854360, EPI_ISL_854361, EPI_ISL_854362, EPI_ISL_854363, EPI_ISL_854364, EPI_ISL_854365, EPI_ISL_854366, EPI_ISL_854367 | | | |
| see above | BIO-REFERENCE LABORATORIES | Wadsworth Center, New York State Department of Health | Kirsten St. George, Daryl M. Lamson, Alexis Russel, Matthew Shudt, Melissa A Leisner, Jonathan Plitnick, Navjot Singh, John Kelly, Erasmus Schneider, Erica Lasek-Nesselquist |
| EPI_ISL_854368 | Wadsworth Center, New York State Department of Health | Wadsworth Center, New York State Department of Health | Kirsten St. George, Daryl M. Lamson, Alexis Russel, Matthew Shudt, Melissa A Leisner, Jonathan Plitnick, Navjot Singh, John Kelly, Erasmus Schneider, Erica Lasek-Nesselquist |
| EPI_ISL_854369 | BIO-REFERENCE LABORATORIES | Wadsworth Center, New York State Department of Health | Kirsten St. George, Daryl M. Lamson, Alexis Russel, Matthew Shudt, Melissa A Leisner, Jonathan Plitnick, Navjot Singh, John Kelly, Erasmus Schneider, Erica Lasek-Nesselquist |
| EPI_ISL_854370, EPI_ISL_854371 | URMC LABS | Wadsworth Center, New York State Department of Health | Kirsten St. George, Daryl M. Lamson, Alexis Russel, Matthew Shudt, Melissa A Leisner, Jonathan Plitnick, Navjot Singh, John Kelly, Erasmus Schneider, Erica Lasek-Nesselquist |
| EPI_ISL_854372 | BIO-REFERENCE LABORATORIES | Wadsworth Center, New York State Department of Health | Kirsten St. George, Daryl M. Lamson, Alexis Russel, Matthew Shudt, Melissa A Leisner, Jonathan Plitnick, Navjot Singh, John Kelly, Erasmus Schneider, Erica Lasek-Nesselquist |
| EPI_ISL_854373 | MONTEFIORE MEDICAL CENTER LABORATORIES | Wadsworth Center, New York State Department of Health | Kirsten St. George, Daryl M. Lamson, Alexis Russel, Matthew Shudt, Melissa A Leisner, Jonathan Plitnick, Navjot Singh, John Kelly, Erasmus Schneider, Erica Lasek-Nesselquist |
| EPI_ISL_854374, EPI_ISL_854375, EPI_ISL_854376, EPI_ISL_854377, EPI_ISL_854378, EPI_ISL_854379, EPI_ISL_854380, EPI_ISL_854381, EPI_ISL_854382, EPI_ISL_854383, EPI_ISL_854384, EPI_ISL_854385, EPI_ISL_854386, EPI_ISL_854387, EPI_ISL_854388, EPI_ISL_854389, EPI_ISL_854390, EPI_ISL_854391, EPI_ISL_854392, EPI_ISL_854393, EPI_ISL_854394, EPI_ISL_854395, EPI_ISL_854396, EPI_ISL_854397, EPI_ISL_854398 | | | |
| see above | WESTCHESTER MEDICAL CENTER | Wadsworth Center, New York State Department of Health | Kirsten St. George, Daryl M. Lamson, Alexis Russel, Matthew Shudt, Melissa A Leisner, Jonathan Plitnick, Navjot Singh, John Kelly, Erasmus Schneider, Erica Lasek-Nesselquist |
| EPI_ISL_854399 | URMC LABS | Wadsworth Center, New York State Department of Health | Kirsten St. George, Daryl M. Lamson, Alexis Russel, Matthew Shudt, Melissa A Leisner, Jonathan Plitnick, Navjot Singh, John Kelly, Erasmus Schneider, Erica Lasek-Nesselquist |
| EPI_ISL_854400, EPI_ISL_854401, EPI_ISL_854402, EPI_ISL_854403 | WESTCHESTER MEDICAL CENTER | Wadsworth Center, New York State Department of Health | Kirsten St. George, Daryl M. Lamson, Alexis Russel, Matthew Shudt, Melissa A Leisner, Jonathan Plitnick, Navjot Singh, John Kelly, Erasmus Schneider, Erica Lasek-Nesselquist |
| EPI_ISL_854404 | URMC LABS | Wadsworth Center, New York State Department of Health | Kirsten St. George, Daryl M. Lamson, Alexis Russel, Matthew Shudt, Melissa A Leisner, Jonathan Plitnick, Navjot Singh, John Kelly, Erasmus Schneider, Erica Lasek-Nesselquist |
| EPI_ISL_854405, EPI_ISL_854406, EPI_ISL_854407, EPI_ISL_854408, EPI_ISL_854409, EPI_ISL_854410, EPI_ISL_854411, EPI_ISL_854412, EPI_ISL_854413, EPI_ISL_854414, EPI_ISL_854415, EPI_ISL_854416, EPI_ISL_854417, EPI_ISL_854418, EPI_ISL_854419, EPI_ISL_854420 | | | |
| see above | MONTEFIORE MEDICAL CENTER LABORATORIES | Wadsworth Center, New York State Department of Health | Kirsten St. George, Daryl M. Lamson, Alexis Russel, Matthew Shudt, Melissa A Leisner, Jonathan Plitnick, Navjot Singh, John Kelly, Erasmus Schneider, Erica Lasek-Nesselquist |
| EPI_ISL_854421 | SARATOGA HOSPITAL LABORATORY | Wadsworth Center, New York State Department of Health | Kirsten St. George, Daryl M. Lamson, Alexis Russel, Matthew Shudt, Melissa A Leisner, Jonathan Plitnick, Navjot Singh, John Kelly, Erasmus Schneider, Erica Lasek-Nesselquist |
| EPI_ISL_854422, EPI_ISL_854423, EPI_ISL_854424, EPI_ISL_854425, EPI_ISL_854426, EPI_ISL_854427, EPI_ISL_854428, EPI_ISL_854429, EPI_ISL_854430, EPI_ISL_854431, EPI_ISL_854432, EPI_ISL_854433, EPI_ISL_854434, EPI_ISL_854435 | | | |
| see above | MONTEFIORE MEDICAL CENTER LABORATORIES | Wadsworth Center, New York State Department of Health | Kirsten St. George, Daryl M. Lamson, Alexis Russel, Matthew Shudt, Melissa A Leisner, Jonathan Plitnick, Navjot Singh, John Kelly, Erasmus Schneider, Erica Lasek-Nesselquist |
| EPI_ISL_854436, EPI_ISL_854437, EPI_ISL_854438, EPI_ISL_854439, EPI_ISL_854440, EPI_ISL_854441, EPI_ISL_854442, EPI_ISL_854443, EPI_ISL_854444, EPI_ISL_854445 | SARATOGA HOSPITAL LABORATORY | Wadsworth Center, New York State Department of Health | Kirsten St. George, Daryl M. Lamson, Alexis Russel, Matthew Shudt, Melissa A Leisner, Jonathan Plitnick, Navjot Singh, John Kelly, Erasmus Schneider, Erica Lasek-Nesselquist |
| EPI_ISL_854446 | BOSTON HEART DIAGNOSTICS CORP | Wadsworth Center, New York State Department of Health | Kirsten St. George, Daryl M. Lamson, Alexis Russel, Matthew Shudt, Melissa A Leisner, Jonathan Plitnick, Navjot Singh, John Kelly, Erasmus Schneider, Erica Lasek-Nesselquist |
| EPI_ISL_854447, EPI_ISL_854448 | TEMPUS LABS INC | Wadsworth Center, New York State Department of Health | Kirsten St. George, Daryl M. Lamson, Alexis Russel, Matthew Shudt, Melissa A Leisner, Jonathan Plitnick, Navjot Singh, John Kelly, Erasmus Schneider, Erica Lasek-Nesselquist |
| EPI_ISL_854449 | WESTCHESTER MEDICAL CENTER | Wadsworth Center, New York State Department of Health | Kirsten St. George, Daryl M. Lamson, Alexis Russel, Matthew Shudt, Melissa A Leisner, Jonathan Plitnick, Navjot Singh, John Kelly, Erasmus Schneider, Erica Lasek-Nesselquist |
| EPI_ISL_854450 | MONTEFIORE MEDICAL CENTER LABORATORIES | Wadsworth Center, New York State Department of Health | Kirsten St. George, Daryl M. Lamson, Alexis Russel, Matthew Shudt, Melissa A Leisner, Jonathan Plitnick, Navjot Singh, John Kelly, Erasmus Schneider, Erica Lasek-Nesselquist |
| EPI_ISL_854451 | WESTCHESTER MEDICAL CENTER | Wadsworth Center, New York State Department of Health | Kirsten St. George, Daryl M. Lamson, Alexis Russel, Matthew Shudt, Melissa A Leisner, Jonathan Plitnick, Navjot Singh, John Kelly, Erasmus Schneider, Erica Lasek-Nesselquist |
| EPI_ISL_854452, EPI_ISL_854453, EPI_ISL_854454, EPI_ISL_854455, EPI_ISL_854456, EPI_ISL_854457, EPI_ISL_854458, EPI_ISL_854459 | MONTEFIORE MEDICAL CENTER LABORATORIES | Wadsworth Center, New York State Department of Health | Kirsten St. George, Daryl M. Lamson, Alexis Russel, Matthew Shudt, Melissa A Leisner, Jonathan Plitnick, Navjot Singh, John Kelly, Erasmus Schneider, Erica Lasek-Nesselquist |
| EPI_ISL_854460 | Northwestern Memorial Hospital | Ozer Lab | Ramon Lorenzo-Redondo, Lacy M. Simons, Lawrence J. Jennings, Michael G. Ison, Judd F. Hultquist, Egon A. Ozer |
| EPI_ISL_854810, EPI_ISL_854811, EPI_ISL_854812, EPI_ISL_854813, EPI_ISL_854814, EPI_ISL_854815, EPI_ISL_854816, EPI_ISL_854817, EPI_ISL_854818, EPI_ISL_854819, EPI_ISL_854820, EPI_ISL_854821, EPI_ISL_854822, EPI_ISL_854823, EPI_ISL_854824, EPI_ISL_854825, EPI_ISL_854826, EPI_ISL_854827, EPI_ISL_854828, EPI_ISL_854829, EPI_ISL_854830, EPI_ISL_854831, EPI_ISL_854832, EPI_ISL_854833, EPI_ISL_854834, EPI_ISL_854835, EPI_ISL_854836, EPI_ISL_854837, EPI_ISL_854838, EPI_ISL_854839, EPI_ISL_854840, EPI_ISL_854841, EPI_ISL_854842, EPI_ISL_854843, EPI_ISL_854844, EPI_ISL_854845, EPI_ISL_854846, EPI_ISL_854847, EPI_ISL_854848, EPI_ISL_854849, EPI_ISL_854850, EPI_ISL_854851, EPI_ISL_854852, EPI_ISL_854853, EPI_ISL_854854, EPI_ISL_854855, EPI_ISL_854856, EPI_ISL_854857, EPI_ISL_854858, EPI_ISL_854859, EPI_ISL_854860, EPI_ISL_854861, EPI_ISL_854862, EPI_ISL_854863, EPI_ISL_854864, EPI_ISL_854865, EPI_ISL_854866, EPI_ISL_854867, EPI_ISL_854868, EPI_ISL_854869, EPI_ISL_854870, EPI_ISL_854871, EPI_ISL_854872, EPI_ISL_854873, EPI_ISL_854874, EPI_ISL_854875, EPI_ISL_854876, EPI_ISL_854877, EPI_ISL_854878, EPI_ISL_854879, EPI_ISL_854880, EPI_ISL_854881, EPI_ISL_854882, EPI_ISL_854883, EPI_ISL_854884, EPI_ISL_854885, EPI_ISL_854886, EPI_ISL_854887, EPI_ISL_854888, EPI_ISL_854889, EPI_ISL_854890, EPI_ISL_854891, EPI_ISL_854892, EPI_ISL_854893, EPI_ISL_854894, EPI_ISL_854895, EPI_ISL_854896, EPI_ISL_854897, EPI_ISL_854898, EPI_ISL_854899, EPI_ISL_854900, EPI_ISL_854901, EPI_ISL_854902, EPI_ISL_854903, EPI_ISL_854904, EPI_ISL_854905, EPI_ISL_854906, EPI_ISL_854907, EPI_ISL_854908, EPI_ISL_854909, EPI_ISL_854910, EPI_ISL_854911, EPI_ISL_854912, EPI_ISL_854913, EPI_ISL_854914, EPI_ISL_854915, EPI_ISL_854916, EPI_ISL_854917, EPI_ISL_854918, EPI_ISL_854919, EPI_ISL_854920, EPI_ISL_854921, EPI_ISL_854922, EPI_ISL_854923, EPI_ISL_854924, EPI_ISL_854925, EPI_ISL_854926, EPI_ISL_854927, EPI_ISL_854928, EPI_ISL_854929, EPI_ISL_854930, EPI_ISL_854931, EPI_ISL_854932, EPI_ISL_854933, EPI_ISL_854934, EPI_ISL_854935, EPI_ISL_854936, EPI_ISL_854937, EPI_ISL_854938, EPI_ISL_854939, EPI_ISL_854940, EPI_ISL_854941, EPI_ISL_854942, EPI_ISL_854943, EPI_ISL_854944, EPI_ISL_854945, EPI_ISL_854946, EPI_ISL_854947, EPI_ISL_854948, EPI_ISL_854949, EPI_ISL_854950, EPI_ISL_854951, EPI_ISL_854952, EPI_ISL_854953, EPI_ISL_854954, EPI_ISL_854955, EPI_ISL_854956, EPI_ISL_854957, EPI_ISL_854958, EPI_ISL_854959, EPI_ISL_854960, EPI_ISL_854961, EPI_ISL_854962, EPI_ISL_854963, EPI_ISL_854964, EPI_ISL_854965, EPI_ISL_854966, EPI_ISL_854967, EPI_ISL_854968, EPI_ISL_854969, EPI_ISL_854970, EPI_ISL_854971, EPI_ISL_854972, EPI_ISL_854973, EPI_ISL_854974, EPI_ISL_854975, EPI_ISL_854976, EPI_ISL_855014, EPI_ISL_855015, EPI_ISL_855016, EPI_ISL_855017, EPI_ISL_855018, EPI_ISL_855019, EPI_ISL_855020, EPI_ISL_855021, EPI_ISL_855022, EPI_ISL_855023, EPI_ISL_855024, EPI_ISL_855025, EPI_ISL_855026, EPI_ISL_855027, EPI_ISL_855028, EPI_ISL_855029, EPI_ISL_855030, EPI_ISL_855031, EPI_ISL_855032, EPI_ISL_855033, EPI_ISL_855034, EPI_ISL_855035, EPI_ISL_855036, EPI_ISL_855037, EPI_ISL_855038, EPI_ISL_855039, EPI_ISL_855040, EPI_ISL_855041, EPI_ISL_855042, EPI_ISL_855043, EPI_ISL_855044, EPI_ISL_855045, EPI_ISL_855046, EPI_ISL_855047, EPI_ISL_855048, EPI_ISL_855049, EPI_ISL_855050, EPI_ISL_855051, EPI_ISL_855052, EPI_ISL_855053, EPI_ISL_855054, EPI_ISL_855055, EPI_ISL_855056, EPI_ISL_855057, EPI_ISL_855058, EPI_ISL_855059, EPI_ISL_855060, EPI_ISL_855061, EPI_ISL_855062, EPI_ISL_855063, EPI_ISL_855064, EPI_ISL_855065, EPI_ISL_855066, EPI_ISL_855067, EPI_ISL_855068, EPI_ISL_855069, EPI_ISL_855070, EPI_ISL_855071, EPI_ISL_855072, EPI_ISL_855073, EPI_ISL_855074, EPI_ISL_855075, EPI_ISL_855076, EPI_ISL_855077, EPI_ISL_855078, EPI_ISL_855079, EPI_ISL_855080, EPI_ISL_855081, EPI_ISL_855082, EPI_ISL_855083, EPI_ISL_855084, EPI_ISL_855085, EPI_ISL_855086, EPI_ISL_855087, EPI_ISL_855088, EPI_ISL_855089, EPI_ISL_855090, EPI_ISL_855091, EPI_ISL_855092, EPI_ISL_855093, EPI_ISL_855094, EPI_ISL_855095, EPI_ISL_855096, EPI_ISL_855097, EPI_ISL_855098, EPI_ISL_855099, EPI_ISL_855100, EPI_ISL_855101, EPI_ISL_855102, EPI_ISL_855103, EPI_ISL_855104, EPI_ISL_855105, EPI_ISL_855106, EPI_ISL_855107, EPI_ISL_855108, EPI_ISL_855109, EPI_ISL_855110, EPI_ISL_855111, EPI_ISL_855112, EPI_ISL_855113, EPI_ISL_855114, EPI_ISL_855115, EPI_ISL_855116, EPI_ISL_855117, EPI_ISL_855118, EPI_ISL_855119, EPI_ISL_855120, EPI_ISL_855121, EPI_ISL_855122, EPI_ISL_855123, EPI_ISL_855124, EPI_ISL_855125, EPI_ISL_855126, EPI_ISL_855127, EPI_ISL_855128, EPI_ISL_855129, EPI_ISL_855130, EPI_ISL_855131, EPI_ISL_855132, EPI_ISL_855133, EPI_ISL_855134, EPI_ISL_855135, EPI_ISL_855136, EPI_ISL_855137, EPI_ISL_855138, EPI_ISL_855139, EPI_ISL_855140, EPI_ISL_855141, EPI_ISL_855142, EPI_ISL_855143, EPI_ISL_855144, EPI_ISL_855145, EPI_ISL_855146, EPI_ISL_855147, EPI_ISL_855148, EPI_ISL_855149, EPI_ISL_855150, EPI_ISL_855151, EPI_ISL_855152, EPI_ISL_855153, EPI_ISL_855154, EPI_ISL_855155, EPI_ISL_855156, EPI_ISL_855157, EPI_ISL_855158, EPI_ISL_855159, EPI_ISL_855160, EPI_ISL_855161, EPI_ISL_855162, EPI_ISL_855163, EPI_ISL_855164, EPI_ISL_855165, EPI_ISL_855166, EPI_ISL_855167, EPI_ISL_855168, EPI_ISL_855169, EPI_ISL_855170, EPI_ISL_855171, EPI_ISL_855172, EPI_ISL_855173, EPI_ISL_855174, EPI_ISL_855175, EPI_ISL_855176, EPI_ISL_855177, EPI_ISL_855178, EPI_ISL_855179, EPI_ISL_855180, EPI_ISL_855181, EPI_ISL_855182, EPI_ISL_855183, EPI_ISL_855184, EPI_ISL_855185 | | | |
| see above | Quest Diagnostics | Quest Diagnostics | Rosenthal,S.H., Gerasimova,A., Kagan,R.M., Anderson, B., Hua, M., Liu Y., Bernstein, L.E., Livingston, K.E., Perez, A., Shalhout, D.F., Shlyakhter, I.A., Owen, R., Tanpaiboon, P., Lacbawan, F. |
| EPI_ISL_855186, EPI_ISL_855187, EPI_ISL_855188, EPI_ISL_855189, EPI_ISL_855190, EPI_ISL_855191, EPI_ISL_855192, EPI_ISL_855193, EPI_ISL_855194, EPI_ISL_855195, EPI_ISL_855196, EPI_ISL_855197, EPI_ISL_855198, EPI_ISL_855199, EPI_ISL_855200, EPI_ISL_855201, EPI_ISL_855202, EPI_ISL_855203, EPI_ISL_855204, EPI_ISL_855205, EPI_ISL_855206, EPI_ISL_855207, EPI_ISL_855208, EPI_ISL_855209, EPI_ISL_855210, EPI_ISL_855211, EPI_ISL_855212, EPI_ISL_855213, EPI_ISL_855214, EPI_ISL_855215, EPI_ISL_855216, EPI_ISL_855217, EPI_ISL_855218, EPI_ISL_855219, EPI_ISL_855220, EPI_ISL_855221, EPI_ISL_855222, EPI_ISL_855223, EPI_ISL_855224, EPI_ISL_855225, EPI_ISL_855226, EPI_ISL_855227, EPI_ISL_855228, EPI_ISL_855229, EPI_ISL_855230, EPI_ISL_855231, EPI_ISL_855232, EPI_ISL_855233, EPI_ISL_855234, EPI_ISL_855235, EPI_ISL_855236, EPI_ISL_855237, EPI_ISL_855238, EPI_ISL_855239, EPI_ISL_855240, EPI_ISL_855241, EPI_ISL_855242, EPI_ISL_855243, EPI_ISL_855244, EPI_ISL_855245, EPI_ISL_855246, EPI_ISL_855247, EPI_ISL_855248, EPI_ISL_855249, EPI_ISL_855250, EPI_ISL_855251, EPI_ISL_855252, EPI_ISL_855253, EPI_ISL_855254, EPI_ISL_855255, EPI_ISL_855256, EPI_ISL_855257, EPI_ISL_855258, EPI_ISL_855259, EPI_ISL_855260, EPI_ISL_855261, EPI_ISL_855262, EPI_ISL_855263, EPI_ISL_855264, EPI_ISL_855265, EPI_ISL_855266, EPI_ISL_855267, EPI_ISL_855268, EPI_ISL_855269, EPI_ISL_855270, EPI_ISL_855271, EPI_ISL_855272, EPI_ISL_855273, EPI_ISL_855274, EPI_ISL_855275, EPI_ISL_855276, EPI_ISL_855277, EPI_ISL_855278, EPI_ISL_855279, EPI_ISL_855280, EPI_ISL_855281, EPI_ISL_855282, EPI_ISL_855283, EPI_ISL_855284, EPI_ISL_855285, EPI_ISL_855286, EPI_ISL_855287, EPI_ISL_855288, EPI_ISL_855289, EPI_ISL_855290, EPI_ISL_855291, EPI_ISL_855292, EPI_ISL_855293, EPI_ISL_855294, EPI_ISL_855295, EPI_ISL_855296, EPI_ISL_855297, EPI_ISL_855298, EPI_ISL_855299, EPI_ISL_855300, EPI_ISL_855301, EPI_ISL_855302, EPI_ISL_855303, EPI_ISL_855304, EPI_ISL_855305, EPI_ISL_855306, EPI_ISL_855307, EPI_ISL_855308, EPI_ISL_855309, EPI_ISL_855310, EPI_ISL_855311, EPI_ISL_855312, EPI_ISL_855313, EPI_ISL_855314, EPI_ISL_855315, EPI_ISL_855316, EPI_ISL_855317, EPI_ISL_855318, EPI_ISL_855319, EPI_ISL_855320, EPI_ISL_855321, EPI_ISL_855322, EPI_ISL_855323, EPI_ISL_855324, EPI_ISL_855325, EPI_ISL_855326, EPI_ISL_855327, EPI_ISL_855328, EPI_ISL_855329, EPI_ISL_855330, EPI_ISL_855331, EPI_ISL_855332, EPI_ISL_855333, EPI_ISL_855334, EPI_ISL_855335, EPI_ISL_855336, EPI_ISL_855337, EPI_ISL_855338, EPI_ISL_855339 | | | |
| see above | Institute for Computational Biomedicine, Weill Cornell Medicine | Institute for Computational Biomedicine, Weill Cornell Medicine | Butler,D.J., Mozsary,C., Meydan,C., Danko,D., Foox,J., Rosiene,J., Shaiber,A., Afshinnekoo,E., MacKay,M., Sedlazeck,F.J., Ivanov,N.A., Sierra,M., Pohle,D., Zietz,M., Gisladottir,U., Ramlall,V., Westover,C.D., Ryon,K., Young,B., Bhattacharya,C., Ruggiero,P., Langhorst,B.W., Tanner,N., Gawrys,J., Meleshko,D., Xu,D., Steel,P.A.D., Shemesh,A.J., Xiang,J., Thierry-Mieg,J., Thierry-Mieg,D., Schwartz,R.E., Iftner,A., Bezdan,D., Sipley,J., Cong,L., Craney,A., Velu,P., Melnick,A.M., Hajirasouliha,I., Horner,S.M., Iftner,T., Salvatore,M., Loda,M., Westblade,L.F., Cushing,M., Levy,S., Wu,S., Tatonetti,N., Imielinski,M., Rennert,H., Mason,C.E. |

| | | | |
|---|---|---|---|
| EPI_ISL_855391, EPI_ISL_855393, EPI_ISL_855394, EPI_ISL_855397, EPI_ISL_855398 | California Department of Public Health | Chiu Laboratory, University of California, San Francisco | Charles Chiu, Xianding (Wayne) Deng, Candace Wang, Brian Bushnell, Scot Federman, Jill Hacker, Debra Wadford |
| EPI_ISL_855400 | Chiu Laboratory, University of California, San Francisco | Chiu Laboratory, University of California, San Francisco | Charles Chiu, Xianding (Wayne) Deng, Candace Wang, Brian Bushnell, Scot Federman, Jill Hacker, Debra Wadford |
| EPI_ISL_856696, EPI_ISL_856697, EPI_ISL_856699, EPI_ISL_856701, EPI_ISL_856702, EPI_ISL_856755 | Ohio Department of Health Laboratory | Ohio Department of Health Laboratory | Holmes, Jennifer; Eric Brandt, Keoni Omura, Glen McGillivary, Caitlin McDonnell, Kirtana Ramadugu, Erica Leasure, Kelsey Florek, Heather Blankenship, Quanta Brown, and Tammy Bannerman |
| EPI_ISL_856802, EPI_ISL_856803, EPI_ISL_856804, EPI_ISL_856805, EPI_ISL_856806, EPI_ISL_856807, EPI_ISL_856808, EPI_ISL_856809, EPI_ISL_856810, EPI_ISL_856811, EPI_ISL_856812, EPI_ISL_856813, EPI_ISL_856814, EPI_ISL_856815, EPI_ISL_856816, EPI_ISL_856817, EPI_ISL_856818, EPI_ISL_856819, EPI_ISL_856820, EPI_ISL_856821, EPI_ISL_856822, EPI_ISL_856823, EPI_ISL_856824, EPI_ISL_856825, EPI_ISL_856826, EPI_ISL_856827, EPI_ISL_856828, EPI_ISL_856829, EPI_ISL_856830, EPI_ISL_856831, EPI_ISL_856832, EPI_ISL_856833, EPI_ISL_856834, EPI_ISL_856835, EPI_ISL_856836, EPI_ISL_856837, EPI_ISL_856838, EPI_ISL_856858, EPI_ISL_856859, EPI_ISL_856860, EPI_ISL_856861, EPI_ISL_856862, EPI_ISL_856863, EPI_ISL_856864, EPI_ISL_856865, EPI_ISL_856866, EPI_ISL_856867, EPI_ISL_856868 | | | |
| see above | Utah Public Health Laboratory | Utah Public Health Laboratory | Erin L. Young, Kelly F. Oakeson, Tara Gallagher |
| EPI_ISL_856912, EPI_ISL_856913, EPI_ISL_856914, EPI_ISL_856915, EPI_ISL_856916, EPI_ISL_856917, EPI_ISL_856918, EPI_ISL_856919, EPI_ISL_856920, EPI_ISL_856921, EPI_ISL_856922, EPI_ISL_856923, EPI_ISL_856924, EPI_ISL_856925, EPI_ISL_856926, EPI_ISL_856927, EPI_ISL_856928, EPI_ISL_856929, EPI_ISL_856930, EPI_ISL_856931, EPI_ISL_856932, EPI_ISL_856933, EPI_ISL_856934, EPI_ISL_856935, EPI_ISL_856936, EPI_ISL_856937, EPI_ISL_856939, EPI_ISL_856940, EPI_ISL_856941, EPI_ISL_856942, EPI_ISL_856943, EPI_ISL_856944, EPI_ISL_856945, EPI_ISL_856946, EPI_ISL_856947, EPI_ISL_856948, EPI_ISL_856949, EPI_ISL_856950, EPI_ISL_856951, EPI_ISL_856952, EPI_ISL_856953, EPI_ISL_856954, EPI_ISL_856955, EPI_ISL_856956, EPI_ISL_856957, EPI_ISL_856958, EPI_ISL_856959, EPI_ISL_856960, EPI_ISL_856961, EPI_ISL_856962, EPI_ISL_856963, EPI_ISL_856964, EPI_ISL_856965, EPI_ISL_856966, EPI_ISL_856967, EPI_ISL_856968, EPI_ISL_856969, EPI_ISL_856970 | | | |
| see above | Wyoming Public Health Laboratory | Wyoming Public Health Laboratory | Noah Hull, Taylor Fearing, Lynette Gumbleton, Channing Weber, Ashley Norberg, Bailey Bowcutt, and Wanda Manley |
| EPI_ISL_856974, EPI_ISL_856975, EPI_ISL_856976, EPI_ISL_856977, EPI_ISL_856978, EPI_ISL_856979, EPI_ISL_856980, EPI_ISL_856981, EPI_ISL_856982, EPI_ISL_856983, EPI_ISL_856984, EPI_ISL_856985, EPI_ISL_856986, EPI_ISL_856987, EPI_ISL_856988, EPI_ISL_856989, EPI_ISL_856990, EPI_ISL_856991, EPI_ISL_856992, EPI_ISL_856993, EPI_ISL_856994, EPI_ISL_856995, EPI_ISL_856996, EPI_ISL_856997, EPI_ISL_856998, EPI_ISL_856999, EPI_ISL_857000, EPI_ISL_857001, EPI_ISL_857002, EPI_ISL_857003, EPI_ISL_857004, EPI_ISL_857005, EPI_ISL_857006, EPI_ISL_857007, EPI_ISL_857008, EPI_ISL_857009, EPI_ISL_857010, EPI_ISL_857011, EPI_ISL_857012, EPI_ISL_857013, EPI_ISL_857014, EPI_ISL_857015, EPI_ISL_857016, EPI_ISL_857017, EPI_ISL_857018, EPI_ISL_857019, EPI_ISL_857020, EPI_ISL_857021, EPI_ISL_857022, EPI_ISL_857023, EPI_ISL_857024, EPI_ISL_857025, EPI_ISL_857026, EPI_ISL_857027, EPI_ISL_857028, EPI_ISL_857029, EPI_ISL_857030, EPI_ISL_857031, EPI_ISL_857032, EPI_ISL_857033, EPI_ISL_857034, EPI_ISL_857035, EPI_ISL_857036, EPI_ISL_857037, EPI_ISL_857038, EPI_ISL_857039 | | | |
| see above | The Ashley Laboratory, Stanford University | Chan-Zuckerberg Biohub | CZB Cliahub Consortium |
| EPI_ISL_857043, EPI_ISL_857044, EPI_ISL_857045, EPI_ISL_857046, EPI_ISL_857048, EPI_ISL_857049 | Colorado Department of Public Health and Environment | Colorado Department of Puplic Health and Environment | Laura Bankers, Molly C. Hetherington-Rauth, Diana Ir, Shannon Ely, Shannon R. Matzinger, Sarah Elizabeth Totten, Emily A. Travanty |
| EPI_ISL_857056, EPI_ISL_857057, EPI_ISL_857058, EPI_ISL_857059, EPI_ISL_857060, EPI_ISL_857061, EPI_ISL_857062 | OCME Office Of Chief Medical Examiner | New York City Public Health Laboratory | Jade Wang, et al. |
| EPI_ISL_857063, EPI_ISL_857064, EPI_ISL_857065 | DOHMH PHL | New York City Public Health Laboratory | Jade Wang, et al. |
| EPI_ISL_857066 | OCME Office Of Chief Medical Examiner | New York City Public Health Laboratory | Jade Wang, et al. |
| EPI_ISL_857067 | DOHMH Corona | New York City Public Health Laboratory | Jade Wang, et al. |
| EPI_ISL_857068 | DOHMH PHL | New York City Public Health Laboratory | Jade Wang, et al. |
| EPI_ISL_857069 | DOHMH Jamaica | New York City Public Health Laboratory | Jade Wang, et al. |
| EPI_ISL_857070 | DOHMH PHL | New York City Public Health Laboratory | Jade Wang, et al. |
| EPI_ISL_857071 | DOHMH Morrisania | New York City Public Health Laboratory | Jade Wang, et al. |
| EPI_ISL_857072 | DOHMH PHL | New York City Public Health Laboratory | Jade Wang, et al. |
| EPI_ISL_857073 | DOHMH Corona | New York City Public Health Laboratory | Jade Wang, et al. |
| EPI_ISL_857074 | DOHMH Morrisania | New York City Public Health Laboratory | Jade Wang, et al. |
| EPI_ISL_857075 | DOHMH Jamaica | New York City Public Health Laboratory | Jade Wang, et al. |
| EPI_ISL_857076 | DOHMH Chelsea | New York City Public Health Laboratory | Jade Wang, et al. |
| EPI_ISL_857077, EPI_ISL_857078, EPI_ISL_857079, EPI_ISL_857080 | DOHMH Central Harlem | New York City Public Health Laboratory | Jade Wang, et al. |
| EPI_ISL_857081 | DOHMH Morrisania | New York City Public Health Laboratory | Jade Wang, et al. |
| EPI_ISL_857082, EPI_ISL_857083, EPI_ISL_857084 | DOHMH Corona | New York City Public Health Laboratory | Jade Wang, et al. |
| EPI_ISL_857085, EPI_ISL_857086 | DOHMH Central Harlem | New York City Public Health Laboratory | Jade Wang, et al. |
| EPI_ISL_857087, EPI_ISL_857088 | DOHMH PHL | New York City Public Health Laboratory | Jade Wang, et al. |
| EPI_ISL_857089 | DOHMH Crown Heights | New York City Public Health Laboratory | Jade Wang, et al. |
| EPI_ISL_857090 | DOHMH Riverside | New York City Public Health Laboratory | Jade Wang, et al. |
| EPI_ISL_857091 | DOHMH Jamaica | New York City Public Health Laboratory | Jade Wang, et al. |
| EPI_ISL_857092 | DOHMH Riverside | New York City Public Health Laboratory | Jade Wang, et al. |
| EPI_ISL_857093 | DOHMH Morrisania | New York City Public Health Laboratory | Jade Wang, et al. |
| EPI_ISL_857094 | DOHMH Central Harlem | New York City Public Health Laboratory | Jade Wang, et al. |
| EPI_ISL_857095 | DOHMH Fort Greene | New York City Public Health Laboratory | Jade Wang, et al. |
| EPI_ISL_857096 | DOHMH Jamaica | New York City Public Health Laboratory | Jade Wang, et al. |
| EPI_ISL_857097 | DOHMH Crown Heights | New York City Public Health Laboratory | Jade Wang, et al. |
| EPI_ISL_857098, EPI_ISL_857099 | DOHMH Morrisania | New York City Public Health Laboratory | Jade Wang, et al. |
| EPI_ISL_857100 | DOHMH Central Harlem | New York City Public Health Laboratory | Jade Wang, et al. |
| EPI_ISL_857101, EPI_ISL_857102, EPI_ISL_857103, EPI_ISL_857104, EPI_ISL_857105 | DOHMH Corona | New York City Public Health Laboratory | Jade Wang, et al. |
| EPI_ISL_857106, EPI_ISL_857107, EPI_ISL_857108, EPI_ISL_857109, EPI_ISL_857110 | DOHMH Chelsea | New York City Public Health Laboratory | Jade Wang, et al. |
| EPI_ISL_857111 | DOHMH Riverside | New York City Public Health Laboratory | Jade Wang, et al. |
| EPI_ISL_857112, EPI_ISL_857113, EPI_ISL_857114, EPI_ISL_857115 | DOHMH Fort Greene | New York City Public Health Laboratory | Jade Wang, et al. |

| | | | |
|---|---|---|---|
| EPI_ISL_857116 | DOHMH Central Harlem | New York City Public Health Laboratory | Jade Wang, et al. |
| EPI_ISL_857117 | DOHMH Chelsea | New York City Public Health Laboratory | Jade Wang, et al. |
| EPI_ISL_857118 | DOHMH Central Harlem | New York City Public Health Laboratory | Jade Wang, et al. |
| EPI_ISL_857119 | DOHMH Morrisania | New York City Public Health Laboratory | Jade Wang, et al. |
| EPI_ISL_857120 | DOHMH Central Harlem | New York City Public Health Laboratory | Jade Wang, et al. |
| EPI_ISL_857121 | DOHMH PHL | New York City Public Health Laboratory | Jade Wang, et al. |
| EPI_ISL_857122, EPI_ISL_857123, EPI_ISL_857124, EPI_ISL_857125 | DOHMH Morrisania | New York City Public Health Laboratory | Jade Wang, et al. |
| EPI_ISL_857126 | DOHMH Central Harlem | New York City Public Health Laboratory | Jade Wang, et al. |
| EPI_ISL_857127, EPI_ISL_857128, EPI_ISL_857129 | DOHMH PHL | New York City Public Health Laboratory | Jade Wang, et al. |
| EPI_ISL_857130, EPI_ISL_857131 | DOHMH Chelsea | New York City Public Health Laboratory | Jade Wang, et al. |
| EPI_ISL_857132, EPI_ISL_857133 | DOHMH Central Harlem | New York City Public Health Laboratory | Jade Wang, et al. |
| EPI_ISL_857134 | DOHMH PHL | New York City Public Health Laboratory | Jade Wang, et al. |
| EPI_ISL_857135 | DOHMH Corona | New York City Public Health Laboratory | Jade Wang, et al. |
| EPI_ISL_857136, EPI_ISL_857137, EPI_ISL_857138, EPI_ISL_857139 | DOHMH Chelsea | New York City Public Health Laboratory | Jade Wang, et al. |
| EPI_ISL_857140, EPI_ISL_857141 | DOHMH Morrisania | New York City Public Health Laboratory | Jade Wang, et al. |
| EPI_ISL_857142 | DOHMH Central Harlem | New York City Public Health Laboratory | Jade Wang, et al. |
| EPI_ISL_857143, EPI_ISL_857144 | DOHMH Riverside | New York City Public Health Laboratory | Jade Wang, et al. |
| EPI_ISL_857145, EPI_ISL_857146, EPI_ISL_857147 | DOHMH Corona | New York City Public Health Laboratory | Jade Wang, et al. |
| EPI_ISL_857148 | DOHMH Fort Greene | New York City Public Health Laboratory | Jade Wang, et al. |
| EPI_ISL_857149, EPI_ISL_857150, EPI_ISL_857151, EPI_ISL_857152 | DOHMH Jamaica | New York City Public Health Laboratory | Jade Wang, et al. |
| EPI_ISL_857153 | DOHMH Corona | New York City Public Health Laboratory | Jade Wang, et al. |
| EPI_ISL_857154, EPI_ISL_857155 | DOHMH Jamaica | New York City Public Health Laboratory | Jade Wang, et al. |
| EPI_ISL_857156 | DOHMH Corona | New York City Public Health Laboratory | Jade Wang, et al. |
| EPI_ISL_857157 | DOHMH PHL | New York City Public Health Laboratory | Jade Wang, et al. |
| EPI_ISL_857158 | DOHMH Corona | New York City Public Health Laboratory | Jade Wang, et al. |
| EPI_ISL_857159, EPI_ISL_857160 | DOHMH Morrisania | New York City Public Health Laboratory | Jade Wang, et al. |
| EPI_ISL_857161, EPI_ISL_857162 | DOHMH PHL | New York City Public Health Laboratory | Jade Wang, et al. |
| EPI_ISL_857163 | DOHMH Central Harlem | New York City Public Health Laboratory | Jade Wang, et al. |
| EPI_ISL_857164 | DOHMH Morrisania | New York City Public Health Laboratory | Jade Wang, et al. |
| EPI_ISL_857165 | DOHMH Corona | New York City Public Health Laboratory | Jade Wang, et al. |
| EPI_ISL_857166, EPI_ISL_857167 | DOHMH Fort Greene | New York City Public Health Laboratory | Jade Wang, et al. |
| EPI_ISL_857168 | DOHMH Corona | New York City Public Health Laboratory | Jade Wang, et al. |
| EPI_ISL_857169, EPI_ISL_857170, EPI_ISL_857171 | DOHMH Morrisania | New York City Public Health Laboratory | Jade Wang, et al. |
| EPI_ISL_857172, EPI_ISL_857173 | DOHMH Crown Heights | New York City Public Health Laboratory | Jade Wang, et al. |
| EPI_ISL_857174, EPI_ISL_857175, EPI_ISL_857176, EPI_ISL_857177 | DOHMH Jamaica | New York City Public Health Laboratory | Jade Wang, et al. |
| EPI_ISL_857178 | DOHMH Riverside | New York City Public Health Laboratory | Jade Wang, et al. |
| EPI_ISL_857179 | DOHMH Jamaica | New York City Public Health Laboratory | Jade Wang, et al. |
| EPI_ISL_857180, EPI_ISL_857181, EPI_ISL_857182, EPI_ISL_857183 | DOHMH Central Harlem | New York City Public Health Laboratory | Jade Wang, et al. |
| EPI_ISL_857184 | DOHMH Riverside | New York City Public Health Laboratory | Jade Wang, et al. |
| EPI_ISL_857185 | DOHMH PHL | New York City Public Health Laboratory | Jade Wang, et al. |
| EPI_ISL_857186 | OCME Office Of Chief Medical Examiner | New York City Public Health Laboratory | Jade Wang, et al. |
| EPI_ISL_857187 | DOHMH PHL | New York City Public Health Laboratory | Jade Wang, et al. |
| EPI_ISL_857188 | Department of Homeless Services | New York City Public Health Laboratory | Jade Wang, et al. |
| EPI_ISL_857189 | DOHMH Corona | New York City Public Health Laboratory | Jade Wang, et al. |
| EPI_ISL_857190 | DOHMH PHL | New York City Public Health Laboratory | Jade Wang, et al. |
| EPI_ISL_857191 | DOHMH Morrisania | New York City Public Health Laboratory | Jade Wang, et al. |
| EPI_ISL_857192 | DOHMH PHL | New York City Public Health Laboratory | Jade Wang, et al. |
| EPI_ISL_857193 | OCME Office Of Chief Medical Examiner | New York City Public Health Laboratory | Jade Wang, et al. |
| EPI_ISL_857194 | DOHMH Central Harlem | New York City Public Health Laboratory | Jade Wang, et al. |
| EPI_ISL_857195 | DOHMH Riverside | New York City Public Health Laboratory | Jade Wang, et al. |
| EPI_ISL_857196 | DOHMH Crown Heights | New York City Public Health Laboratory | Jade Wang, et al. |
| EPI_ISL_857197 | DOHMH Jamaica | New York City Public Health Laboratory | Jade Wang, et al. |
| EPI_ISL_857198 | Department of Homeless Services | New York City Public Health Laboratory | Jade Wang, et al. |
| EPI_ISL_857199 | OCME Office Of Chief Medical Examiner | New York City Public Health Laboratory | Jade Wang, et al. |
| EPI_ISL_857200 | DOHMH Central Harlem | New York City Public Health Laboratory | Jade Wang, et al. |
| EPI_ISL_857201 | DOHMH Jamaica | New York City Public Health Laboratory | Jade Wang, et al. |

| | | | |
|---|---|---|---|
| EPI_ISL_857202 | DOHMH PHL | New York City Public Health Laboratory | Jade Wang, et al. |
| EPI_ISL_857203 | DOHMH Riverside | New York City Public Health Laboratory | Jade Wang, et al. |
| EPI_ISL_857204, EPI_ISL_857205 | DOHMH Central Harlem | New York City Public Health Laboratory | Jade Wang, et al. |
| EPI_ISL_857206 | DOHMH PHL | New York City Public Health Laboratory | Jade Wang, et al. |
| EPI_ISL_857207, EPI_ISL_857208 | DOHMH Morrisania | New York City Public Health Laboratory | Jade Wang, et al. |
| EPI_ISL_857209 | DOHMH Central Harlem | New York City Public Health Laboratory | Jade Wang, et al. |
| EPI_ISL_857210, EPI_ISL_857211, EPI_ISL_857212, EPI_ISL_857213, EPI_ISL_857214, EPI_ISL_857215, EPI_ISL_857216, EPI_ISL_857217 | DOHMH Jamaica | New York City Public Health Laboratory | Jade Wang, et al. |
| EPI_ISL_857218, EPI_ISL_857219, EPI_ISL_857220 | DOHMH Corona | New York City Public Health Laboratory | Jade Wang, et al. |
| EPI_ISL_857221 | DOHMH Fort Greene | New York City Public Health Laboratory | Jade Wang, et al. |
| EPI_ISL_857222, EPI_ISL_857223, EPI_ISL_857224 | DOHMH Jamaica | New York City Public Health Laboratory | Jade Wang, et al. |
| EPI_ISL_857225 | DOHMH Corona | New York City Public Health Laboratory | Jade Wang, et al. |
| EPI_ISL_857226 | DOHMH Jamaica | New York City Public Health Laboratory | Jade Wang, et al. |
| EPI_ISL_857227, EPI_ISL_857228 | DOHMH Riverside | New York City Public Health Laboratory | Jade Wang, et al. |
| EPI_ISL_857229 | DOHMH Morrisania | New York City Public Health Laboratory | Jade Wang, et al. |
| EPI_ISL_857230 | DOHMH Chelsea | New York City Public Health Laboratory | Jade Wang, et al. |
| EPI_ISL_857231, EPI_ISL_857232, EPI_ISL_857233 | DOHMH Central Harlem | New York City Public Health Laboratory | Jade Wang, et al. |
| EPI_ISL_857234, EPI_ISL_857235, EPI_ISL_857236, EPI_ISL_857237 | DOHMH Jamaica | New York City Public Health Laboratory | Jade Wang, et al. |
| EPI_ISL_857238, EPI_ISL_857239, EPI_ISL_857240 | DOHMH Corona | New York City Public Health Laboratory | Jade Wang, et al. |
| EPI_ISL_857241, EPI_ISL_857242, EPI_ISL_857243, EPI_ISL_857244, EPI_ISL_857245, EPI_ISL_857246, EPI_ISL_857247, EPI_ISL_857248, EPI_ISL_857249, EPI_ISL_857250 | DOHMH Jamaica | New York City Public Health Laboratory | Jade Wang, et al. |
| EPI_ISL_857251, EPI_ISL_857252 | DOHMH Morrisania | New York City Public Health Laboratory | Jade Wang, et al. |
| EPI_ISL_857253, EPI_ISL_857254 | DOHMH Chelsea | New York City Public Health Laboratory | Jade Wang, et al. |
| EPI_ISL_857255 | DOHMH Central Harlem | New York City Public Health Laboratory | Jade Wang, et al. |
| EPI_ISL_857256 | DOHMH Fort Greene | New York City Public Health Laboratory | Jade Wang, et al. |
| EPI_ISL_857257 | DOHMH PHL | New York City Public Health Laboratory | Jade Wang, et al. |
| EPI_ISL_857258, EPI_ISL_857259 | DOHMH Central Harlem | New York City Public Health Laboratory | Jade Wang, et al. |
| EPI_ISL_857260, EPI_ISL_857261, EPI_ISL_857262 | DOHMH Chelsea | New York City Public Health Laboratory | Jade Wang, et al. |
| EPI_ISL_857263, EPI_ISL_857264 | DOHMH Corona | New York City Public Health Laboratory | Jade Wang, et al. |
| EPI_ISL_857265 | DOHMH Fort Greene | New York City Public Health Laboratory | Jade Wang, et al. |
| EPI_ISL_857266 | DOHMH PHL | New York City Public Health Laboratory | Jade Wang, et al. |
| EPI_ISL_857267, EPI_ISL_857268, EPI_ISL_857269, EPI_ISL_857270 | DOHMH Jamaica | New York City Public Health Laboratory | Jade Wang, et al. |
| EPI_ISL_857271, EPI_ISL_857272, EPI_ISL_857273 | DOHMH Morrisania | New York City Public Health Laboratory | Jade Wang, et al. |
| EPI_ISL_857274, EPI_ISL_857275, EPI_ISL_857276 | DOHMH Jamaica | New York City Public Health Laboratory | Jade Wang, et al. |
| EPI_ISL_857277 | DOHMH Corona | New York City Public Health Laboratory | Jade Wang, et al. |
| EPI_ISL_857278 | DOHMH PHL | New York City Public Health Laboratory | Jade Wang, et al. |
| EPI_ISL_857279, EPI_ISL_857280 | DOHMH Fort Greene | New York City Public Health Laboratory | Jade Wang, et al. |
| EPI_ISL_857281 | DOHMH Riverside | New York City Public Health Laboratory | Jade Wang, et al. |
| EPI_ISL_857282, EPI_ISL_857283 | DOHMH Morrisania | New York City Public Health Laboratory | Jade Wang, et al. |
| EPI_ISL_857284 | DOHMH PHL | New York City Public Health Laboratory | Jade Wang, et al. |
| EPI_ISL_857285, EPI_ISL_857286 | DOHMH Central Harlem | New York City Public Health Laboratory | Jade Wang, et al. |
| EPI_ISL_857287, EPI_ISL_857288 | Department of Homeless Services | New York City Public Health Laboratory | Jade Wang, et al. |
| EPI_ISL_857289 | OCME Office Of Chief Medical Examiner | New York City Public Health Laboratory | Jade Wang, et al. |
| EPI_ISL_857290 | Department of Homeless Services | New York City Public Health Laboratory | Jade Wang, et al. |
| EPI_ISL_857291, EPI_ISL_857292 | OCME Office Of Chief Medical Examiner | New York City Public Health Laboratory | Jade Wang, et al. |

| | | | |
|---|---|---|---|
| EPI_ISL_857293 | Department of Homeless Services | New York City Public Health Laboratory | Jade Wang, et al. |
| EPI_ISL_857294, EPI_ISL_857295, EPI_ISL_857296, EPI_ISL_857297 | OCME Office Of Chief Medical Examiner | New York City Public Health Laboratory | Jade Wang, et al. |
| EPI_ISL_857298 | Department of Homeless Services | New York City Public Health Laboratory | Jade Wang, et al. |
| EPI_ISL_857299, EPI_ISL_857300 | OCME Office Of Chief Medical Examiner | New York City Public Health Laboratory | Jade Wang, et al. |
| EPI_ISL_857301 | Department of Homeless Services | New York City Public Health Laboratory | Jade Wang, et al. |
| EPI_ISL_857302, EPI_ISL_857303, EPI_ISL_857304, EPI_ISL_857305 | OCME Office Of Chief Medical Examiner | New York City Public Health Laboratory | Jade Wang, et al. |
| EPI_ISL_857306 | Department of Homeless Services | New York City Public Health Laboratory | Jade Wang, et al. |
| EPI_ISL_857307 | OCME Office Of Chief Medical Examiner | New York City Public Health Laboratory | Jade Wang, et al. |
| EPI_ISL_857349, EPI_ISL_857351, EPI_ISL_857352, EPI_ISL_857355, EPI_ISL_857357, EPI_ISL_857358, EPI_ISL_857360, EPI_ISL_857361, EPI_ISL_857362, EPI_ISL_857363, EPI_ISL_857364, EPI_ISL_857365, EPI_ISL_857366, EPI_ISL_857367, EPI_ISL_857368, EPI_ISL_857369, EPI_ISL_857370, EPI_ISL_857372, EPI_ISL_857374, EPI_ISL_857375, EPI_ISL_857376, EPI_ISL_857377, EPI_ISL_857378, EPI_ISL_857379, EPI_ISL_857381, EPI_ISL_857382, EPI_ISL_857384, EPI_ISL_857385, EPI_ISL_857386, EPI_ISL_857387, EPI_ISL_857388, EPI_ISL_857389, EPI_ISL_857391, EPI_ISL_857392, EPI_ISL_857393, EPI_ISL_857394 | | | |
| see above | Texas Department of State Health Services (TXDSHS) | Texas Department of State Health Services (TXDSHS) | Anita Pokharel, Bonnie Oh, James Daniel Bonser, Rashmi Tuladhar, Mayela Pedrueza, Jenny Zhang, Maliha Rahman, Myong Koag, Chung Wang, Rachel Lee, Grace Kubin |
| EPI_ISL_857396, EPI_ISL_857397, EPI_ISL_857398, EPI_ISL_857399, EPI_ISL_857400, EPI_ISL_857401, EPI_ISL_857402, EPI_ISL_857403, EPI_ISL_857404, EPI_ISL_857405, EPI_ISL_857406, EPI_ISL_857407, EPI_ISL_857408, EPI_ISL_857409, EPI_ISL_857410, EPI_ISL_857411, EPI_ISL_857412, EPI_ISL_857413, EPI_ISL_857414, EPI_ISL_857415, EPI_ISL_857416, EPI_ISL_857417, EPI_ISL_857418, EPI_ISL_857419, EPI_ISL_857420, EPI_ISL_857421, EPI_ISL_857422, EPI_ISL_857423, EPI_ISL_857424, EPI_ISL_857425, EPI_ISL_857426, EPI_ISL_857427, EPI_ISL_857428, EPI_ISL_857429, EPI_ISL_857431, EPI_ISL_857432, EPI_ISL_857433, EPI_ISL_857434, EPI_ISL_857435, EPI_ISL_857436, EPI_ISL_857437, EPI_ISL_857438, EPI_ISL_857439, EPI_ISL_857440, EPI_ISL_857441, EPI_ISL_857442, EPI_ISL_857443, EPI_ISL_857444, EPI_ISL_857445, EPI_ISL_857446, EPI_ISL_857447, EPI_ISL_857448, EPI_ISL_857449, EPI_ISL_857450, EPI_ISL_857451, EPI_ISL_857452, EPI_ISL_857453, EPI_ISL_857454, EPI_ISL_857455, EPI_ISL_857456, EPI_ISL_857457, EPI_ISL_857458, EPI_ISL_857459, EPI_ISL_857460, EPI_ISL_857461, EPI_ISL_857462, EPI_ISL_857463, EPI_ISL_857464 | | | |
| see above | Maine HETL | Tewhey Lab, The Jackson Laboratory | Matluk,N., Dewey,H., Iosue,F., Barter,M., Lynch,R., Munger,H. and Tewhey,R. |
| EPI_ISL_860256, EPI_ISL_860258, EPI_ISL_860296, EPI_ISL_860549, EPI_ISL_860552, EPI_ISL_860710, EPI_ISL_860711, EPI_ISL_860714, EPI_ISL_860717, EPI_ISL_860718 | Ohio Department of Health Laboratory | Ohio Department of Health Laboratory | Holmes, Jennifer; Eric Brandt, Keoni Omura, Glen McGillivary, Caitlin McDonnell, Kirtana Ramadugu, Erica Leasure, Kelsey Florek, Heather Blankenship, Quanta Brown, and Tammy Bannerman |
| EPI_ISL_860727, EPI_ISL_860728 | University of Michigan Clinical Microbiology Laboratory | Lauring Lab, University of Michigan, Department of Microbiology and Immunology | Valesano |
| EPI_ISL_860729, EPI_ISL_860791, EPI_ISL_860792, EPI_ISL_860793, EPI_ISL_860795, EPI_ISL_860796, EPI_ISL_860819 | Ohio Department of Health Laboratory | Ohio Department of Health Laboratory | Holmes, Jennifer; Eric Brandt, Keoni Omura, Glen McGillivary, Caitlin McDonnell, Kirtana Ramadugu, Erica Leasure, Kelsey Florek, Heather Blankenship, Quanta Brown, and Tammy Bannerman |
| EPI_ISL_860932, EPI_ISL_860933, EPI_ISL_860934, EPI_ISL_860935, EPI_ISL_860936, EPI_ISL_860937, EPI_ISL_860938, EPI_ISL_860939, EPI_ISL_860940, EPI_ISL_860941, EPI_ISL_860942, EPI_ISL_860943, EPI_ISL_860944, EPI_ISL_860945, EPI_ISL_860946, EPI_ISL_860947, EPI_ISL_860948, EPI_ISL_860949, EPI_ISL_860950, EPI_ISL_860951, EPI_ISL_860952, EPI_ISL_860953, EPI_ISL_860954, EPI_ISL_860955, EPI_ISL_860956, EPI_ISL_860957, EPI_ISL_860958, EPI_ISL_860959, EPI_ISL_860960, EPI_ISL_860961, EPI_ISL_860962, EPI_ISL_860963, EPI_ISL_860964, EPI_ISL_860965, EPI_ISL_860966, EPI_ISL_860967, EPI_ISL_860968, EPI_ISL_860969, EPI_ISL_860970, EPI_ISL_860971, EPI_ISL_860972, EPI_ISL_860973, EPI_ISL_860974, EPI_ISL_860975, EPI_ISL_860976, EPI_ISL_860977, EPI_ISL_860978, EPI_ISL_860979, EPI_ISL_860980, EPI_ISL_860981, EPI_ISL_860982, EPI_ISL_860983, EPI_ISL_860984, EPI_ISL_860985, EPI_ISL_860986, EPI_ISL_860987, EPI_ISL_860988, EPI_ISL_860989, EPI_ISL_860990, EPI_ISL_860991, EPI_ISL_860992, EPI_ISL_860993, EPI_ISL_860994, EPI_ISL_860995, EPI_ISL_860996, EPI_ISL_860997, EPI_ISL_860998, EPI_ISL_860999, EPI_ISL_861000, EPI_ISL_861001, EPI_ISL_861002, EPI_ISL_861003, EPI_ISL_861004, EPI_ISL_861005, EPI_ISL_861006, EPI_ISL_861007, EPI_ISL_861008, EPI_ISL_861009, EPI_ISL_861010, EPI_ISL_861011, EPI_ISL_861012, EPI_ISL_861013, EPI_ISL_861014, EPI_ISL_861015, EPI_ISL_861016, EPI_ISL_861017, EPI_ISL_861018, EPI_ISL_861019, EPI_ISL_861020, EPI_ISL_861021, EPI_ISL_861022, EPI_ISL_861023, EPI_ISL_861024, EPI_ISL_861025, EPI_ISL_861026, EPI_ISL_861027, EPI_ISL_861028, EPI_ISL_861029, EPI_ISL_861030, EPI_ISL_861031, EPI_ISL_861032, EPI_ISL_861033, EPI_ISL_861034, EPI_ISL_861035, EPI_ISL_861036, EPI_ISL_861037, EPI_ISL_861038, EPI_ISL_861039, EPI_ISL_861040, EPI_ISL_861041, EPI_ISL_861042, EPI_ISL_861043, EPI_ISL_861044, EPI_ISL_861045, EPI_ISL_861046, EPI_ISL_861047, EPI_ISL_861048, EPI_ISL_861049, EPI_ISL_861050, EPI_ISL_861051, EPI_ISL_861052, EPI_ISL_861053, EPI_ISL_861054, EPI_ISL_861055, EPI_ISL_861056, EPI_ISL_861057, EPI_ISL_861058, EPI_ISL_861059, EPI_ISL_861060, EPI_ISL_861061, EPI_ISL_861062, EPI_ISL_861063, EPI_ISL_861064, EPI_ISL_861065, EPI_ISL_861066, EPI_ISL_861067, EPI_ISL_861068, EPI_ISL_861069, EPI_ISL_861070, EPI_ISL_861071, EPI_ISL_861072, EPI_ISL_861073, EPI_ISL_861074, EPI_ISL_861075, EPI_ISL_861076, EPI_ISL_861077, EPI_ISL_861078, EPI_ISL_861079, EPI_ISL_861080, EPI_ISL_861081, EPI_ISL_861082, EPI_ISL_861083, EPI_ISL_861084, EPI_ISL_861085, EPI_ISL_861086, EPI_ISL_861087, EPI_ISL_861088, EPI_ISL_861089, EPI_ISL_861090, EPI_ISL_861091, EPI_ISL_861092, EPI_ISL_861093, EPI_ISL_861094, EPI_ISL_861095, EPI_ISL_861096, EPI_ISL_861097, EPI_ISL_861098, EPI_ISL_861099, EPI_ISL_861100, EPI_ISL_861101, EPI_ISL_861102, EPI_ISL_861103, EPI_ISL_861104 | | | |
| see above | Johns Hopkins Hospital Department of Pathology | Johns Hopkins Hospital Department of Pathology | C. Paul Morris, Chun Huai Luo, Adannaya Amadi, Nicholas Gallagher, Heba H. Mostafa |
| EPI_ISL_861105 | Ohio Department of Health Laboratory | Ohio Department of Health Laboratory | Holmes, Jennifer; Eric Brandt, Keoni Omura, Glen McGillivary, Caitlin McDonnell, Kirtana Ramadugu, Erica Leasure, Kelsey Florek, Heather Blankenship, Quanta Brown, and Tammy Bannerman |
| EPI_ISL_861106, EPI_ISL_861107, EPI_ISL_861108 | KALEIDA CENTER FOR LABORATORY MEDICINE | Wadsworth Center, New York State Department of Health | Kirsten St. George, Daryl M. Lamson, Alexis Russel, Matthew Shudt, Melissa A Leisner, Jonathan Plitnick, Navjot Singh, John Kelly, Erasmus Schneider, Erica Lasek-Nesselquist |
| EPI_ISL_861109, EPI_ISL_861110, EPI_ISL_861111, EPI_ISL_861112, EPI_ISL_861113, EPI_ISL_861114 | New York Presbyterian Hospital | Wadsworth Center, New York State Department of Health | Kirsten St. George, Daryl M. Lamson, Alexis Russel, Matthew Shudt, Melissa A Leisner, Jonathan Plitnick, Navjot Singh, John Kelly, Erasmus Schneider, Erica Lasek-Nesselquist |
| EPI_ISL_861115, EPI_ISL_861116, EPI_ISL_861117 | Wadsworth Center, New York State Department of Health | Wadsworth Center, New York State Department of Health | Kirsten St. George, Daryl M. Lamson, Alexis Russel, Matthew Shudt, Melissa A Leisner, Jonathan Plitnick, Navjot Singh, John Kelly, Erasmus Schneider, Erica Lasek-Nesselquist |
| EPI_ISL_861118, EPI_ISL_861119, EPI_ISL_861120, EPI_ISL_861121, EPI_ISL_861122, EPI_ISL_861123, EPI_ISL_861124, EPI_ISL_861125, EPI_ISL_861126, EPI_ISL_861127, EPI_ISL_861128, EPI_ISL_861129, EPI_ISL_861130, EPI_ISL_861131, EPI_ISL_861132, EPI_ISL_861133 | | | |
| see above | New York Presbyterian Hospital | Wadsworth Center, New York State Department of Health | Kirsten St. George, Daryl M. Lamson, Alexis Russel, Matthew Shudt, Melissa A Leisner, Jonathan Plitnick, Navjot Singh, John Kelly, Erasmus Schneider, Erica Lasek-Nesselquist |
| EPI_ISL_861134 | KALEIDA CENTER FOR LABORATORY MEDICINE | Wadsworth Center, New York State Department of Health | Kirsten St. George, Daryl M. Lamson, Alexis Russel, Matthew Shudt, Melissa A Leisner, Jonathan Plitnick, Navjot Singh, John Kelly, Erasmus Schneider, Erica Lasek-Nesselquist |
| EPI_ISL_861135, EPI_ISL_861136, EPI_ISL_861137, EPI_ISL_861138, EPI_ISL_861139, EPI_ISL_861140, EPI_ISL_861141, EPI_ISL_861142, EPI_ISL_861143 | ADIRONDACK MEDICAL CENTER | Wadsworth Center, New York State Department of Health | Kirsten St. George, Daryl M. Lamson, Alexis Russel, Matthew Shudt, Melissa A Leisner, Jonathan Plitnick, Navjot Singh, John Kelly, Erasmus Schneider, Erica Lasek-Nesselquist |
| EPI_ISL_861144, EPI_ISL_861145, EPI_ISL_861146, EPI_ISL_861147, EPI_ISL_861148, EPI_ISL_861149, EPI_ISL_861150, EPI_ISL_861151, EPI_ISL_861152, EPI_ISL_861153, EPI_ISL_861154, EPI_ISL_861155, EPI_ISL_861156, EPI_ISL_861157, EPI_ISL_861158, EPI_ISL_861159, EPI_ISL_861160, EPI_ISL_861161, EPI_ISL_861162, EPI_ISL_861163, EPI_ISL_861164, EPI_ISL_861165, EPI_ISL_861166, EPI_ISL_861167, EPI_ISL_861168, EPI_ISL_861169, EPI_ISL_861170, EPI_ISL_861171, EPI_ISL_861172, EPI_ISL_861173, EPI_ISL_861174 | | | |
| see above | KALEIDA CENTER FOR LABORATORY MEDICINE | Wadsworth Center, New York State Department of Health | Kirsten St. George, Daryl M. Lamson, Alexis Russel, Matthew Shudt, Melissa A Leisner, Jonathan Plitnick, Navjot Singh, John Kelly, Erasmus Schneider, Erica Lasek-Nesselquist |
| EPI_ISL_861175, EPI_ISL_861176, EPI_ISL_861177, EPI_ISL_861178, EPI_ISL_861179, EPI_ISL_861180, EPI_ISL_861181, EPI_ISL_861182 | BIO-REFERENCE LABORATORIES | Wadsworth Center, New York State Department of Health | Kirsten St. George, Daryl M. Lamson, Alexis Russel, Matthew Shudt, Melissa A Leisner, Jonathan Plitnick, Navjot Singh, John Kelly, Erasmus Schneider, Erica Lasek-Nesselquist |
| EPI_ISL_861183, EPI_ISL_861184 | URMC LABS | Wadsworth Center, New York State Department of Health | Kirsten St. George, Daryl M. Lamson, Alexis Russel, Matthew Shudt, Melissa A Leisner, Jonathan Plitnick, Navjot Singh, John Kelly, Erasmus Schneider, Erica Lasek-Nesselquist |
| EPI_ISL_861185 | BIO-REFERENCE LABORATORIES | Wadsworth Center, New York State Department of Health | Kirsten St. George, Daryl M. Lamson, Alexis Russel, Matthew Shudt, Melissa A Leisner, Jonathan Plitnick, Navjot Singh, John Kelly, Erasmus Schneider, Erica Lasek-Nesselquist |
| EPI_ISL_861186 | NORTH SHORE UNIVERSITY HOSPITAL | Wadsworth Center, New York State Department of Health | Kirsten St. George, Daryl M. Lamson, Alexis Russel, Matthew Shudt, Melissa A Leisner, Jonathan Plitnick, Navjot Singh, John Kelly, Erasmus Schneider, Erica Lasek-Nesselquist |

| | | | |
|---|---|---|---|
| EPI_ISL_861187 | BIO-REFERENCE LABORATORIES | Wadsworth Center, New York State Department of Health | Kirsten St. George, Daryl M. Lamson, Alexis Russel, Matthew Shudt, Melissa A Leisner, Jonathan Plitnick, Navjot Singh, John Kelly, Erasmus Schneider, Erica Lasek-Nesselquist |
| EPI_ISL_861188, EPI_ISL_861189, EPI_ISL_861190, EPI_ISL_861191, EPI_ISL_861192, EPI_ISL_861193, EPI_ISL_861194, EPI_ISL_861195, EPI_ISL_861196, EPI_ISL_861197, EPI_ISL_861198 | | | |
| see above | NORTH SHORE UNIVERSITY HOSPITAL | Wadsworth Center, New York State Department of Health | Kirsten St. George, Daryl M. Lamson, Alexis Russel, Matthew Shudt, Melissa A Leisner, Jonathan Plitnick, Navjot Singh, John Kelly, Erasmus Schneider, Erica Lasek-Nesselquist |
| EPI_ISL_861199, EPI_ISL_861200, EPI_ISL_861201, EPI_ISL_861202, EPI_ISL_861203, EPI_ISL_861204, EPI_ISL_861205, EPI_ISL_861206, EPI_ISL_861207, EPI_ISL_861208, EPI_ISL_861209 | | | |
| see above | URMC LABS | Wadsworth Center, New York State Department of Health | Kirsten St. George, Daryl M. Lamson, Alexis Russel, Matthew Shudt, Melissa A Leisner, Jonathan Plitnick, Navjot Singh, John Kelly, Erasmus Schneider, Erica Lasek-Nesselquist |
| EPI_ISL_861210, EPI_ISL_861211, EPI_ISL_861212, EPI_ISL_861213, EPI_ISL_861214, EPI_ISL_861215, EPI_ISL_861216, EPI_ISL_861217, EPI_ISL_861218, EPI_ISL_861219, EPI_ISL_861220, EPI_ISL_861221, EPI_ISL_861222, EPI_ISL_861223, EPI_ISL_861224, EPI_ISL_861225, EPI_ISL_861226, EPI_ISL_861227, EPI_ISL_861228, EPI_ISL_861229, EPI_ISL_861230, EPI_ISL_861231, EPI_ISL_861232 | | | |
| see above | BIO-REFERENCE LABORATORIES | Wadsworth Center, New York State Department of Health | Kirsten St. George, Daryl M. Lamson, Alexis Russel, Matthew Shudt, Melissa A Leisner, Jonathan Plitnick, Navjot Singh, John Kelly, Erasmus Schneider, Erica Lasek-Nesselquist |
| EPI_ISL_861233, EPI_ISL_861240, EPI_ISL_861241, EPI_ISL_861243 | Ohio Department of Health Laboratory | Ohio Department of Health Laboratory | Holmes, Jennifer; Eric Brandt, Keoni Omura, Glen McGillivary, Caitlin McDonnell, Kirtana Ramadugu, Erica Leasure, Kelsey Florek, Heather Blankenship, Quanta Brown, and Tammy Bannerman |
| EPI_ISL_861244, EPI_ISL_861245, EPI_ISL_861246, EPI_ISL_861247, EPI_ISL_861248, EPI_ISL_861249, EPI_ISL_861250, EPI_ISL_861251, EPI_ISL_861252, EPI_ISL_861253, EPI_ISL_861254, EPI_ISL_861255, EPI_ISL_861256, EPI_ISL_861257, EPI_ISL_861258, EPI_ISL_861259, EPI_ISL_861260, EPI_ISL_861261, EPI_ISL_861262, EPI_ISL_861263, EPI_ISL_861264, EPI_ISL_861265, EPI_ISL_861266, EPI_ISL_861267, EPI_ISL_861268, EPI_ISL_861269, EPI_ISL_861270, EPI_ISL_861271, EPI_ISL_861272, EPI_ISL_861273, EPI_ISL_861274, EPI_ISL_861275, EPI_ISL_861276, EPI_ISL_861277, EPI_ISL_861278, EPI_ISL_861279, EPI_ISL_861280, EPI_ISL_861281, EPI_ISL_861282, EPI_ISL_861283, EPI_ISL_861284, EPI_ISL_861285, EPI_ISL_861286, EPI_ISL_861287, EPI_ISL_861288, EPI_ISL_861289, EPI_ISL_861290, EPI_ISL_861291, EPI_ISL_861292, EPI_ISL_861293, EPI_ISL_861294, EPI_ISL_861295, EPI_ISL_861296, EPI_ISL_861297, EPI_ISL_861298, EPI_ISL_861299, EPI_ISL_861300, EPI_ISL_861301, EPI_ISL_861302, EPI_ISL_861303, EPI_ISL_861304, EPI_ISL_861305, EPI_ISL_861306, EPI_ISL_861307, EPI_ISL_861308, EPI_ISL_861309, EPI_ISL_861310, EPI_ISL_861311, EPI_ISL_861312, EPI_ISL_861313, EPI_ISL_861314, EPI_ISL_861315, EPI_ISL_861316, EPI_ISL_861317, EPI_ISL_861318, EPI_ISL_861319, EPI_ISL_861320, EPI_ISL_861321, EPI_ISL_861322, EPI_ISL_861323, EPI_ISL_861324, EPI_ISL_861325, EPI_ISL_861326, EPI_ISL_861327, EPI_ISL_861328, EPI_ISL_861329, EPI_ISL_861330, EPI_ISL_861331 | | | |
| see above | MONTEFIORE MEDICAL CENTER LABORATORIES | Wadsworth Center, New York State Department of Health | Kirsten St. George, Daryl M. Lamson, Alexis Russel, Matthew Shudt, Melissa A Leisner, Jonathan Plitnick, Navjot Singh, John Kelly, Erasmus Schneider, Erica Lasek-Nesselquist |
| EPI_ISL_861332 | DOHMH Central Harlem | New York City Public Health Laboratory | Jade Wang, et al. |
| EPI_ISL_861333, EPI_ISL_861334, EPI_ISL_861335, EPI_ISL_861336, EPI_ISL_861337, EPI_ISL_861338, EPI_ISL_861339, EPI_ISL_861340, EPI_ISL_861341, EPI_ISL_861342, EPI_ISL_861343, EPI_ISL_861344, EPI_ISL_861345, EPI_ISL_861346, EPI_ISL_861347, EPI_ISL_861348, EPI_ISL_861349, EPI_ISL_861350, EPI_ISL_861351, EPI_ISL_861352, EPI_ISL_861353, EPI_ISL_861354, EPI_ISL_861355, EPI_ISL_861356, EPI_ISL_861357, EPI_ISL_861358, EPI_ISL_861359, EPI_ISL_861360, EPI_ISL_861361, EPI_ISL_861362, EPI_ISL_861363, EPI_ISL_861364, EPI_ISL_861365, EPI_ISL_861366, EPI_ISL_861367, EPI_ISL_861368, EPI_ISL_861369, EPI_ISL_861370, EPI_ISL_861371, EPI_ISL_861372, EPI_ISL_861373, EPI_ISL_861374, EPI_ISL_861375, EPI_ISL_861376, EPI_ISL_861377, EPI_ISL_861378, EPI_ISL_861379, EPI_ISL_861380, EPI_ISL_861381, EPI_ISL_861382, EPI_ISL_861383, EPI_ISL_861384, EPI_ISL_861385, EPI_ISL_861386, EPI_ISL_861387, EPI_ISL_861388, EPI_ISL_861389, EPI_ISL_861390, EPI_ISL_861391, EPI_ISL_861392, EPI_ISL_861393, EPI_ISL_861394, EPI_ISL_861395, EPI_ISL_861396, EPI_ISL_861397, EPI_ISL_861398, EPI_ISL_861399, EPI_ISL_861400, EPI_ISL_861401, EPI_ISL_861402, EPI_ISL_861403, EPI_ISL_861404, EPI_ISL_861405, EPI_ISL_861406, EPI_ISL_861407, EPI_ISL_861408, EPI_ISL_861409, EPI_ISL_861410, EPI_ISL_861411, EPI_ISL_861412 | | | |
| see above | WESTCHESTER MEDICAL CENTER | Wadsworth Center, New York State Department of Health | Kirsten St. George, Daryl M. Lamson, Alexis Russel, Matthew Shudt, Melissa A Leisner, Jonathan Plitnick, Navjot Singh, John Kelly, Erasmus Schneider, Erica Lasek-Nesselquist |
| EPI_ISL_861413, EPI_ISL_861414, EPI_ISL_861415, EPI_ISL_861416, EPI_ISL_861417, EPI_ISL_861418 | NORTH SHORE UNIVERSITY HOSPITAL | Wadsworth Center, New York State Department of Health | Kirsten St. George, Daryl M. Lamson, Alexis Russel, Matthew Shudt, Melissa A Leisner, Jonathan Plitnick, Navjot Singh, John Kelly, Erasmus Schneider, Erica Lasek-Nesselquist |
| EPI_ISL_861419 | Instituto de Diagnostico y Referencia Epidemiologicos (INDRE) | Instituto de Diagnostico y Referencia Epidemiologicos (INDRE) | Fabiola Garces-Ayala, Gisela Barrera-Badillo , Abril Rodriguez-Maldonado, Claudia Wong-Arambula , Natividad Cruz-Ortiz, Tatiana Nunez-Garcia, Dayanira Arellano-Suarez, Adnan-Araiza Rodríguez, Edgar Mendieta-Condado, Lucia Hernandez-Rivas, Irma Lopez-Martinez, Ernesto Ramirez-Gonzalez. |
| EPI_ISL_861420 | Instituto de Diagnostico y Referencia Epidemiologicos (INDRE) | Instituto de Diagnostico y Referencia Epidemiologicos (INDRE) | Abril Rodriguez-Maldonado, Gisela Barrera-Badillo ,Claudia Wong-Arambula , Natividad Cruz-Ortiz, Tatiana Nunez-Garcia, Dayanira Arellano-Suarez, Fabiola Garces-Ayala, Adnan-Araiza Rodríguez, Edgar Mendieta-Condado, Lucia Hernandez-Rivas, Irma Lopez-Martinez, Ernesto Ramirez-Gonzalez. |
| EPI_ISL_861421 | Instituto de Diagnostico y Referencia Epidemiologicos (INDRE) | Instituto de Diagnostico y Referencia Epidemiologicos (INDRE) | Claudia Wong-Arambula, Gisela Barrera-Badillo , Abril Rodriguez-Maldonado, Natividad Cruz-Ortiz, Tatiana Nunez-Garcia, Dayanira Arellano-Suarez, Fabiola Garces-Ayala, Adnan-Araiza Rodríguez, Edgar Mendieta-Condado, Lucia Hernandez-Rivas, Irma Lopez-Martinez, Ernesto Ramirez-Gonzalez. |
| EPI_ISL_861422, EPI_ISL_861423, EPI_ISL_861424, EPI_ISL_861425, EPI_ISL_861426 | Oklahoma Animal Disease Diagnostic Laboratory | Oklahoma Animal Disease Diagnostic Laboratory | Sai Narayanan, Girish Patil, Sunil More, Jeremiah Saliki, Akhilesh Ramachandran |
| EPI_ISL_861427, EPI_ISL_861428, EPI_ISL_861429, EPI_ISL_861431, EPI_ISL_861432 | Wyoming Public Health Laboratory | Wyoming Public Health Laboratory | Noah Hull, Taylor Fearing, Lynette Gumbleton, Channing Weber, Ashley Norberg, Bailey Bowcutt, and Wanda Manley |
| EPI_ISL_861433, EPI_ISL_861434, EPI_ISL_861435 | Ohio State University | James Molecular Laboratory | Huolin Tu, Matthew R Avenarius, Laura Kubatko, Matthew Hunt, Xiaokang Pan, Peng Ru, Jason Garee, Keelie Thomas, Peter Mohler, Preeti Pancholi, Dan Jones |
| EPI_ISL_861439, EPI_ISL_861440, EPI_ISL_861441, EPI_ISL_861442 | The Ohio State University | James Molecular Laboratory | Huolin Tu, Matthew R Avenarius, Laura Kubatko, Matthew Hunt, Xiaokang Pan, Peng Ru, Jason Garee, Keelie Thomas, Peter Mohler, Preeti Pancholi, Dan Jones |
| EPI_ISL_861443, EPI_ISL_861444, EPI_ISL_861445, EPI_ISL_861446, EPI_ISL_861447, EPI_ISL_861448, EPI_ISL_861449, EPI_ISL_861450, EPI_ISL_861451, EPI_ISL_861452, EPI_ISL_861453, EPI_ISL_861454, EPI_ISL_861455 | | | |
| see above | Clinical Molecular Microbiology Laboratory, UNC Hospitals | Jeremy Wang | Jeremy Wang, Alexander Rubinsteyn, Colleen Rice, Jason Smedberg, Melissa Miller, Corbin Jones, Robert Hagan |
| EPI_ISL_861472, EPI_ISL_861473, EPI_ISL_861474, EPI_ISL_861475 | Gundersen Molecular Diagnostics Laboratory | Kabara Cancer Research Institute | Craig S. Richmond, Paraic A. Kenny |
| EPI_ISL_861476, EPI_ISL_861477, EPI_ISL_861478, EPI_ISL_861480, EPI_ISL_861485, EPI_ISL_861486 | Wyoming Public Health Laboratory | Wyoming Public Health Laboratory | Noah Hull, Taylor Fearing, Lynette Gumbleton, Channing Weber, Ashley Norberg, Bailey Bowcutt, and Wanda Manley |
| EPI_ISL_861686, EPI_ISL_861687, EPI_ISL_861688, EPI_ISL_861689, EPI_ISL_861690, EPI_ISL_861691, EPI_ISL_861692, EPI_ISL_861693, EPI_ISL_861694, EPI_ISL_861695, EPI_ISL_861696, EPI_ISL_861697, EPI_ISL_861698, EPI_ISL_861699 | | | |
| see above | Los Angeles County Public Health Laboratories | Los Angeles County Public Health Laboratories | P. Hemarajata et al. |
| EPI_ISL_861700, EPI_ISL_861701, EPI_ISL_861702, EPI_ISL_861703 | MD Laboratories | Los Angeles County Public Health Laboratories | P. Hemarajata et al. |
| EPI_ISL_861704, EPI_ISL_861705 | UCLA Clinical Micro Lab | Los Angeles County Public Health Laboratories | P. Hemarajata et al. |
| EPI_ISL_861730, EPI_ISL_861731 | UW Virology Lab | UW Virology Lab | Pavitra Roychoudhury, Hong Xie, Lasata Shrestha, Michelle Lin, Meei-Li Huang, Keith R Jerome, Alexander Greninger |
| EPI_ISL_861732, EPI_ISL_861733 | Tempus | Grubaugh Lab - Yale School of Public Health | Tara Alpert, Joseph Fauver, Anderson Brito, Mallery Breban, Anne Wyllie, Chantal Vogels, Mary Petrone, Annie Watkins, Chaney Kalinich, Isabel Ott, Nathan Grubaugh |
| EPI_ISL_861734 | Yale Pathology Lab | Grubaugh Lab - Yale School of Public Health | Tara Alpert, Joseph Fauver, Chen Liu, Pei Hui, Jianhui Wang, Susan Bell and Han Zhou, Anderson Brito, Mallery Breban, Anne Wyllie, Chantal Vogels, Mary Petrone, Chaney Kalinich, Isabel Ott, Arnau Casanovas, Catherine Muenker, Adam Moore, Alice Lu, Maria Tokuyama, Patrick Wong, Peiwen Lu, Saad Omer, Richard Martinello, Allison Nelson, Shelli Farhadian, Akiko Iwasaki, Charlese Dela Cruz, Albert Ko, Nathan Grubaugh |
| EPI_ISL_861735 | Tempus | Grubaugh Lab - Yale School of Public Health | Tara Alpert, Joseph Fauver, Anderson Brito, Mallery Breban, Anne Wyllie, Chantal Vogels, Mary Petrone, Annie Watkins, Chaney Kalinich, Isabel Ott, Nathan Grubaugh |
| EPI_ISL_861736 | Yale Pathology Lab | Grubaugh Lab - Yale School of Public Health | Tara Alpert, Joseph Fauver, Chen Liu, Pei Hui, Jianhui Wang, Susan Bell and Han Zhou, Anderson Brito, Mallery Breban, Anne Wyllie, Chantal Vogels, Mary Petrone, Chaney Kalinich, Isabel Ott, Arnau Casanovas, Catherine Muenker, Adam Moore, Alice Lu, Maria Tokuyama, Patrick Wong, Peiwen Lu, Saad Omer, Richard Martinello, Allison Nelson, Shelli Farhadian, Akiko Iwasaki, Charlese Dela Cruz, Albert Ko, Nathan Grubaugh |
| EPI_ISL_861737, EPI_ISL_861738, EPI_ISL_861739, EPI_ISL_861740, EPI_ISL_861741, | Tempus | Grubaugh Lab - Yale School of Public Health | Tara Alpert, Joseph Fauver, Anderson Brito, Mallery Breban, Anne Wyllie, Chantal Vogels, Mary Petrone, Annie Watkins, Chaney Kalinich, Isabel Ott, Nathan Grubaugh |

| | | | |
|---|---|---|---|
| EPI_ISL_861742, EPI_ISL_861743, EPI_ISL_861744 | | | |
| EPI_ISL_861745 | Yale Pathology Lab | Grubaugh Lab - Yale School of Public Health | Tara Alpert, Joseph Fauver, Chen Liu, Pei Hui, Jianhui Wang, Susan Bell and Han Zhou, Anderson Brito, Mallery Breban, Anne Wyllie, Chantal Vogels, Mary Petrone, Chaney Kalinich, Isabel Ott, Arnau Casanovas, Catherine Muenker, Adam Moore, Alice Lu, Maria Tokuyama, Patrick Wong, Peiwen Lu, Saad Omer, Richard Martinello, Allison Nelson, Shelli Farhadian, Akiko Iwasaki, Charlese Dela Cruz, Albert Ko, Nathan Grubaugh |
| EPI_ISL_861746 | Tempus | Grubaugh Lab - Yale School of Public Health | Tara Alpert, Joseph Fauver, Anderson Brito, Mallery Breban, Anne Wyllie, Chantal Vogels, Mary Petrone, Annie Watkins, Chaney Kalinich, Isabel Ott, Nathan Grubaugh |
| EPI_ISL_861916, EPI_ISL_861917, EPI_ISL_861918, EPI_ISL_861919, EPI_ISL_861921, EPI_ISL_861922, EPI_ISL_861923, EPI_ISL_861924, EPI_ISL_861925, EPI_ISL_861926, EPI_ISL_861927, EPI_ISL_861928, EPI_ISL_861929, EPI_ISL_861930, EPI_ISL_861931, EPI_ISL_861933, EPI_ISL_861934, EPI_ISL_861935, EPI_ISL_861936, EPI_ISL_861937, EPI_ISL_861938, EPI_ISL_861939, EPI_ISL_861940, EPI_ISL_861942, EPI_ISL_861943, EPI_ISL_861944, EPI_ISL_861945, EPI_ISL_861946, EPI_ISL_861947, EPI_ISL_861948, EPI_ISL_861949, EPI_ISL_861950, EPI_ISL_861951, EPI_ISL_861952, EPI_ISL_861953, EPI_ISL_861954, EPI_ISL_861955, EPI_ISL_861956, EPI_ISL_861957, EPI_ISL_861958, EPI_ISL_861959, EPI_ISL_861960, EPI_ISL_861961, EPI_ISL_861962, EPI_ISL_861963, EPI_ISL_861964, EPI_ISL_861965, EPI_ISL_861966, EPI_ISL_861967, EPI_ISL_861968, EPI_ISL_861969, EPI_ISL_861970, EPI_ISL_861971, EPI_ISL_861972, EPI_ISL_861973, EPI_ISL_861974, EPI_ISL_861975, EPI_ISL_861976, EPI_ISL_861977, EPI_ISL_861978, EPI_ISL_861979, EPI_ISL_861980, EPI_ISL_861981, EPI_ISL_861982, EPI_ISL_861983, EPI_ISL_861984, EPI_ISL_861985, EPI_ISL_861986, EPI_ISL_861987, EPI_ISL_861988, EPI_ISL_861989, EPI_ISL_861990, EPI_ISL_861991, EPI_ISL_861993, EPI_ISL_861994, EPI_ISL_861995, EPI_ISL_861996, EPI_ISL_861998, EPI_ISL_862000, EPI_ISL_862001, EPI_ISL_862002, EPI_ISL_862003, EPI_ISL_862004, EPI_ISL_862005, EPI_ISL_862006, EPI_ISL_862007, EPI_ISL_862008, EPI_ISL_862009, EPI_ISL_862010, EPI_ISL_862011, EPI_ISL_862012, EPI_ISL_862013, EPI_ISL_862014, EPI_ISL_862015, EPI_ISL_862016, EPI_ISL_862017, EPI_ISL_862018, EPI_ISL_862019, EPI_ISL_862020, EPI_ISL_862021, EPI_ISL_862022, EPI_ISL_862023, EPI_ISL_862024, EPI_ISL_862027, EPI_ISL_862028, EPI_ISL_862029, EPI_ISL_862030, EPI_ISL_862031, EPI_ISL_862032, EPI_ISL_862034, EPI_ISL_862035 | | | |
| see above | OHSU Lab Services Molecular Microbiology Lab | Oregon SARS-CoV-2 Genome Sequencing Center | Brendan L. O'Connell, Sally Grindstaff, Kayla Carter, Ruth V. Nichols, Alec J. Hirsch, Donna Hansel, Guang Fan, Xuan Qin, Daniel N. Streblow, William B. Messer, Andrew C. Adey, Benjamin N. Bimber, Brian J. O'Roak |
| EPI_ISL_862036, EPI_ISL_862037, EPI_ISL_862038 | Johns Hopkins Hospital Department of Pathology | Johns Hopkins Hospital Department of Pathology | C. Paul Morris, Chun Huai Luo, Adannaya Amadi, Matthew Schwartz, Nicholas Gallagher, Heba H. Mostafa |
| EPI_ISL_862121 | MD Laboratories | Los Angeles County Public Health Laboratories | P. Hemarajata et al. |
| EPI_ISL_862122, EPI_ISL_862123 | UCLA Clinical Micro Lab | Los Angeles County Public Health Laboratories | P. Hemarajata et al. |
| EPI_ISL_862665, EPI_ISL_862666, EPI_ISL_862667, EPI_ISL_862668 | University of Michigan Clinical Microbiology Laboratory | Lauring Lab, University of Michigan, Department of Microbiology and Immunology | Valesano |
| EPI_ISL_862669, EPI_ISL_862670, EPI_ISL_862671, EPI_ISL_862672, EPI_ISL_862674, EPI_ISL_862675, EPI_ISL_862676, EPI_ISL_862677, EPI_ISL_862678, EPI_ISL_862679, EPI_ISL_862680, EPI_ISL_862681, EPI_ISL_862682, EPI_ISL_862683, EPI_ISL_862684, EPI_ISL_862685, EPI_ISL_862686, EPI_ISL_862687, EPI_ISL_862688, EPI_ISL_862689, EPI_ISL_862690, EPI_ISL_862691, EPI_ISL_862692, EPI_ISL_862693, EPI_ISL_862694, EPI_ISL_862695, EPI_ISL_862696, EPI_ISL_862697, EPI_ISL_862698, EPI_ISL_862699, EPI_ISL_862700, EPI_ISL_862701, EPI_ISL_862702, EPI_ISL_862703, EPI_ISL_862704, EPI_ISL_862705, EPI_ISL_862706, EPI_ISL_862707, EPI_ISL_862708, EPI_ISL_862709, EPI_ISL_862710, EPI_ISL_862712, EPI_ISL_862713 | | | |
| see above | New Mexico Department of Health Scientific Laboratory | New Mexico Department of Health Scientific Laboratory | D'eldra Malone, Ellie Johnson, Anastacia Griego-Fisher |
| EPI_ISL_862714, EPI_ISL_862715 | University of Michigan Clinical Microbiology Laboratory | Lauring Lab, University of Michigan, Department of Microbiology and Immunology | Valesano |
| EPI_ISL_862723 | San Diego County Public Health Laboratory | Andersen lab at Scripps Research | SEARCH Alliance San Diego with Tracy Basler, Jovan Shephard, Brett Austin |
| EPI_ISL_862741, EPI_ISL_862742, EPI_ISL_862744, EPI_ISL_862745, EPI_ISL_862746, EPI_ISL_862747, EPI_ISL_862748, EPI_ISL_862749, EPI_ISL_862750, EPI_ISL_862752, EPI_ISL_862754, EPI_ISL_862756, EPI_ISL_862757, EPI_ISL_862758, EPI_ISL_862759, EPI_ISL_862762, EPI_ISL_862763, EPI_ISL_862764, EPI_ISL_862766, EPI_ISL_862767, EPI_ISL_862769, EPI_ISL_862770, EPI_ISL_862771, EPI_ISL_862772, EPI_ISL_862773, EPI_ISL_862774, EPI_ISL_862775, EPI_ISL_862776, EPI_ISL_862777, EPI_ISL_862778 | | | |
| see above | Utah Public Health Laboratory, Utah Public Health Laboratory Infectious Disease submission group | Utah Public Health Laboratory, Utah Public Health Laboratory Infectious Disease submission group | Young,E.L., Oakeson,K.F., Gallagher,T. |
| EPI_ISL_869079, EPI_ISL_869080, EPI_ISL_869081 | Ohio Department of Health Laboratory | Ohio Department of Health Laboratory | Holmes, Jennifer; Eric Brandt, Keoni Omura, Glen McGillivary, Caitlin McDonnell, Kirtana Ramadugu, Erica Leasure, Kelsey Florek, Heather Blankenship, Quanta Brown, and Tammy Bannerman |
| EPI_ISL_869167, EPI_ISL_869168, EPI_ISL_869169, EPI_ISL_869170, EPI_ISL_869171, EPI_ISL_869172, EPI_ISL_869173, EPI_ISL_869174, EPI_ISL_869175, EPI_ISL_869176, EPI_ISL_869177, EPI_ISL_869178, EPI_ISL_869179, EPI_ISL_869180, EPI_ISL_869181, EPI_ISL_869182, EPI_ISL_869183, EPI_ISL_869184, EPI_ISL_869185, EPI_ISL_869186, EPI_ISL_869187, EPI_ISL_869188, EPI_ISL_869189, EPI_ISL_869190, EPI_ISL_869191, EPI_ISL_869192, EPI_ISL_869193, EPI_ISL_869194, EPI_ISL_869195, EPI_ISL_869196, EPI_ISL_869197, EPI_ISL_869198, EPI_ISL_869199, EPI_ISL_869200, EPI_ISL_869201, EPI_ISL_869202, EPI_ISL_869203, EPI_ISL_869204, EPI_ISL_869205, EPI_ISL_869206, EPI_ISL_869207, EPI_ISL_869208, EPI_ISL_869209, EPI_ISL_869210, EPI_ISL_869211, EPI_ISL_869212, EPI_ISL_869213, EPI_ISL_869214, EPI_ISL_869215, EPI_ISL_869216, EPI_ISL_869217, EPI_ISL_869218, EPI_ISL_869219, EPI_ISL_869220, EPI_ISL_869221, EPI_ISL_869222, EPI_ISL_869223, EPI_ISL_869224, EPI_ISL_869225, EPI_ISL_869226, EPI_ISL_869227, EPI_ISL_869228, EPI_ISL_869229, EPI_ISL_869230, EPI_ISL_869231, EPI_ISL_869232, EPI_ISL_869233, EPI_ISL_869234, EPI_ISL_869235, EPI_ISL_869236, EPI_ISL_869237, EPI_ISL_869238, EPI_ISL_869239 | | | |
| see above | New Mexico Department of Health Scientific Laboratory | Center for Global Health, University of New Mexico Health Sciences Center | Daryl Domman, Kurt Schwalm, Twila Kunde, Joseph Hicks, Anastacia Griego, Michael Edwards, Darrell Dinwiddie |
| EPI_ISL_870065 | Ohio Department of Health Laboratory | Ohio Department of Health Laboratory | Holmes, Jennifer; Eric Brandt, Keoni Omura, Glen McGillivary, Caitlin McDonnell, Kirtana Ramadugu, Erica Leasure, Kelsey Florek, Heather Blankenship, Quanta Brown, and Tammy Bannerman |
| EPI_ISL_870317, EPI_ISL_870319, EPI_ISL_870321, EPI_ISL_870322 | University of Michigan Clinical Microbiology Laboratory | Lauring Lab, University of Michigan, Department of Microbiology and Immunology | Valesano |
| EPI_ISL_871325, EPI_ISL_871788, EPI_ISL_871791, EPI_ISL_871816 | Ohio Department of Health Laboratory | Ohio Department of Health Laboratory | Holmes, Jennifer; Eric Brandt, Keoni Omura, Glen McGillivary, Caitlin McDonnell, Kirtana Ramadugu, Erica Leasure, Kelsey Florek, Heather Blankenship, Quanta Brown, and Tammy Bannerman |
| EPI_ISL_871817 | Eurofins Diatherix | Hudsonalpha Genome Sequencing Center | Jane Grimwood, Melissa Williams, Lori H. Handley, Joshua Stough, Leslie Malone, Stefan Brzezinski, Ada Stewart, Teresa Jones, Jenell Webber, John T. Lovell, Jennifer Cart, and Jeremy Schmutz |
| EPI_ISL_871818, EPI_ISL_871819 | Ohio Department of Health Laboratory | Ohio Department of Health Laboratory | Holmes, Jennifer; Eric Brandt, Keoni Omura, Glen McGillivary, Caitlin McDonnell, Kirtana Ramadugu, Erica Leasure, Kelsey Florek, Heather Blankenship, Quanta Brown, and Tammy Bannerman |
| EPI_ISL_871840, EPI_ISL_871841, EPI_ISL_871842, EPI_ISL_871843, EPI_ISL_871850, EPI_ISL_871851, EPI_ISL_871852, EPI_ISL_871853, EPI_ISL_871854, EPI_ISL_871855, EPI_ISL_871856, EPI_ISL_871857, EPI_ISL_871858, EPI_ISL_871859, EPI_ISL_871860, EPI_ISL_871861, EPI_ISL_871862, EPI_ISL_871863, EPI_ISL_871864, EPI_ISL_871865, EPI_ISL_871866, EPI_ISL_871867, EPI_ISL_871868, EPI_ISL_871869, EPI_ISL_871870, EPI_ISL_871871, EPI_ISL_871873, EPI_ISL_871877, EPI_ISL_871878 | | | |
| see above | Wyoming Public Health Laboratory | Wyoming Public Health Laboratory | Noah Hull, Taylor Fearing, Lynette Gumbleton, Channing Weber, Ashley Norberg, Bailey Bowcutt, and Wanda Manley |
| EPI_ISL_871965, EPI_ISL_871966, EPI_ISL_871967, EPI_ISL_871968, EPI_ISL_871969 | Ohio Department of Health Laboratory | Ohio Department of Health Laboratory | Holmes, Jennifer; Eric Brandt, Keoni Omura, Glen McGillivary, Caitlin McDonnell, Kirtana Ramadugu, Erica Leasure, Kelsey Florek, Heather Blankenship, Quanta Brown, and Tammy Bannerman |
| EPI_ISL_871970, EPI_ISL_871971 | Wyoming Public Health Laboratory | Wyoming Public Health Laboratory | Noah Hull, Taylor Fearing, Lynette Gumbleton, Channing Weber, Ashley Norberg, Bailey Bowcutt, and Wanda Manley |
| EPI_ISL_872002 | Ohio Department of Health Laboratory | Ohio Department of Health Laboratory | Holmes, Jennifer; Eric Brandt, Keoni Omura, Glen McGillivary, Caitlin McDonnell, Kirtana Ramadugu, Erica Leasure, Kelsey Florek, Heather Blankenship, Quanta Brown, and Tammy Bannerman |
| EPI_ISL_872088 | Instituto de Diagnostico y Referencia Epidemiologicos (INDRE) | Instituto de Diagnostico y Referencia Epidemiologicos (INDRE) | Fabiola Garces-Ayala, Gisela Barrera-Badillo , Abril Rodriguez-Maldonado, Claudia Wong-Arambula , Natividad Cruz-Ortiz, Tatiana Nunez-Garcia, Dayanira Arellano-Suarez, Adnan-Araiza Rodríguez, Edgar Mendieta-Condado, Lucia Hernandez-Rivas, Irma Lopez-Martinez, Ernesto Ramirez-Gonzalez. |
| EPI_ISL_872089 | Instituto de Diagnostico y Referencia Epidemiologicos (INDRE) | Instituto de Diagnostico y Referencia Epidemiologicos (INDRE) | Abril Rodriguez-Maldonado, Gisela Barrera-Badillo ,Claudia Wong-Arambula , Natividad Cruz-Ortiz, Tatiana Nunez-Garcia, Dayanira Arellano-Suarez, Fabiola Garces-Ayala, Adnan-Araiza Rodríguez, Edgar Mendieta-Condado, Lucia Hernandez-Rivas, Irma Lopez-Martinez, Ernesto Ramirez-Gonzalez. |
| EPI_ISL_872090 | Instituto de Diagnostico y Referencia Epidemiologicos (INDRE) | Instituto de Diagnostico y Referencia Epidemiologicos (INDRE) | Claudia Wong-Arambula, Gisela Barrera-Badillo , Abril Rodriguez-Maldonado, Natividad Cruz-Ortiz, Tatiana Nunez-Garcia, Dayanira Arellano-Suarez, Fabiola Garces-Ayala, Adnan-Araiza Rodríguez, Edgar Mendieta-Condado, Lucia Hernandez-Rivas, Irma Lopez-Martinez, Ernesto Ramirez-Gonzalez. |
| EPI_ISL_872091 | Instituto de Diagnostico y Referencia Epidemiologicos (INDRE) | Instituto de Diagnostico y Referencia Epidemiologicos (INDRE) | Adnan-Araiza Rodríguez,Claudia Wong-Arambula, Gisela Barrera-Badillo , Abril Rodriguez-Maldonado, Natividad Cruz-Ortiz, Tatiana Nunez-Garcia, Dayanira Arellano-Suarez, Fabiola Garces-Ayala, Edgar Mendieta-Condado, Lucia Hernandez-Rivas, Irma Lopez-Martinez, Ernesto Ramirez-Gonzalez. |
| EPI_ISL_872092 | Instituto de Diagnostico y Referencia Epidemiologicos (INDRE) | Instituto de Diagnostico y Referencia Epidemiologicos (INDRE) | Ernesto Ramirez-Gonzalez, Adnan-Araiza Rodríguez,Claudia Wong-Arambula, Gisela Barrera-Badillo , Abril Rodriguez-Maldonado, Natividad Cruz-Ortiz, Tatiana Nunez-Garcia, Dayanira Arellano-Suarez, Fabiola Garces-Ayala, Edgar Mendieta-Condado, Lucia Hernandez-Rivas, Irma Lopez-Martinez. |
| EPI_ISL_872093 | Instituto de Diagnostico y Referencia Epidemiologicos (INDRE) | Instituto de Diagnostico y Referencia Epidemiologicos (INDRE) | Edgar Mendieta-Condado, Ernesto Ramirez-Gonzalez, Adnan-Araiza Rodríguez,Claudia Wong-Arambula, Gisela Barrera-Badillo , Abril Rodriguez-Maldonado, Natividad Cruz-Ortiz, Tatiana Nunez-Garcia, Dayanira Arellano-Suarez, Fabiola Garces-Ayala, Lucia Hernandez-Rivas, Irma Lopez-Martinez. |
| EPI_ISL_872097 | Instituto de Diagnostico y Referencia Epidemiologicos (INDRE) | Instituto de Diagnostico y Referencia Epidemiologicos (INDRE) | Abril Rodriguez-Maldonado, Gisela Barrera-Badillo ,Claudia Wong-Arambula , Natividad Cruz-Ortiz, Tatiana Nunez-Garcia, Dayanira Arellano-Suarez, Fabiola Garces-Ayala, Adnan-Araiza Rodríguez, Edgar Mendieta-Condado, Lucia Hernandez-Rivas, Irma Lopez-Martinez, Ernesto Ramirez-Gonzalez. |
| EPI_ISL_872098 | Instituto de Diagnostico y Referencia Epidemiologicos (INDRE) | Instituto de Diagnostico y Referencia Epidemiologicos (INDRE) | Claudia Wong-Arambula, Gisela Barrera-Badillo , Abril Rodriguez-Maldonado, Natividad Cruz-Ortiz, Tatiana Nunez-Garcia, Dayanira Arellano-Suarez, Fabiola Garces-Ayala, Adnan-Araiza Rodríguez, Edgar Mendieta-Condado, Lucia Hernandez-Rivas, Irma Lopez-Martinez, Ernesto Ramirez-Gonzalez. |
| EPI_ISL_872099 | Instituto de Diagnostico y Referencia Epidemiologicos (INDRE) | Instituto de Diagnostico y Referencia Epidemiologicos (INDRE) | Fabiola Garces-Ayala, Gisela Barrera-Badillo , Abril Rodriguez-Maldonado, Claudia Wong-Arambula , Natividad Cruz-Ortiz, Tatiana Nunez-Garcia, Dayanira Arellano-Suarez, Adnan-Araiza Rodríguez, Edgar Mendieta-Condado, Lucia Hernandez-Rivas, Irma Lopez-Martinez, Ernesto Ramirez-Gonzalez. |
| EPI_ISL_872349 | Alaska State Virology Lab | Alaska State Virology Lab | Stephanie DeRonde, Lisa Smith, Ph.D., Jack Chen, Ph.D. |
| EPI_ISL_872350, EPI_ISL_872351, EPI_ISL_872352, EPI_ISL_872355, EPI_ISL_872356, EPI_ISL_872357, EPI_ISL_872358, EPI_ISL_872359, EPI_ISL_872360, EPI_ISL_872361, EPI_ISL_872363, EPI_ISL_872365, EPI_ISL_872366, EPI_ISL_872367, EPI_ISL_872369, EPI_ISL_872370, EPI_ISL_872371, EPI_ISL_872372, EPI_ISL_872373, EPI_ISL_872374, EPI_ISL_872375, EPI_ISL_872376, EPI_ISL_872377, EPI_ISL_872378, EPI_ISL_872379, EPI_ISL_872380, EPI_ISL_872381, EPI_ISL_872382, EPI_ISL_872383, EPI_ISL_872387, EPI_ISL_872388, EPI_ISL_872389, EPI_ISL_872390, EPI_ISL_872393 | | | |
| see above | Texas Department of State Health Services (TXDSHS) | Texas Department of State Health Services (TXDSHS) | Bonnie Oh, Anita Pokharel, James Daniel Bonser, Myong Koag, Chung Wang, Rachel Lee, Grace Kubin, Rashmi Tuladhar, Mayela Pedrueza, Maliha Rahman, Jenny Zhang |
| EPI_ISL_872395, EPI_ISL_872396, EPI_ISL_872397, EPI_ISL_872398, EPI_ISL_872399, EPI_ISL_872400, EPI_ISL_872401, EPI_ISL_872402, EPI_ISL_872403, EPI_ISL_872404, EPI_ISL_872405, EPI_ISL_872406, EPI_ISL_872407, EPI_ISL_872408, EPI_ISL_872409, EPI_ISL_872410, EPI_ISL_872411, EPI_ISL_872412, EPI_ISL_872413, EPI_ISL_872414, EPI_ISL_872415, EPI_ISL_872416, EPI_ISL_872417, EPI_ISL_872418, | | | |

| | | | |
|---|---|---|---|
| EPI_ISL_872419, EPI_ISL_872420, EPI_ISL_872421, EPI_ISL_872422, EPI_ISL_872423, EPI_ISL_872424, EPI_ISL_872425, EPI_ISL_872426, EPI_ISL_872427, EPI_ISL_872428, EPI_ISL_872429, EPI_ISL_872430, EPI_ISL_872431, EPI_ISL_872432, EPI_ISL_872433, EPI_ISL_872434, EPI_ISL_872435, EPI_ISL_872436, EPI_ISL_872437, EPI_ISL_872438, EPI_ISL_872439, EPI_ISL_872440, EPI_ISL_872441, EPI_ISL_872442, EPI_ISL_872443, EPI_ISL_872444, EPI_ISL_872445, EPI_ISL_872446, EPI_ISL_872447, EPI_ISL_872448, EPI_ISL_872449, EPI_ISL_872450, EPI_ISL_872451, EPI_ISL_872452, EPI_ISL_872453, EPI_ISL_872454, EPI_ISL_872455, EPI_ISL_872456, EPI_ISL_872457, EPI_ISL_872458, EPI_ISL_872459, EPI_ISL_872460, EPI_ISL_872461, EPI_ISL_872462, EPI_ISL_872463, EPI_ISL_872464, EPI_ISL_872465, EPI_ISL_872466, EPI_ISL_872467, EPI_ISL_872468, EPI_ISL_872469, EPI_ISL_872470, EPI_ISL_872471, EPI_ISL_872472, EPI_ISL_872473, EPI_ISL_872474, EPI_ISL_872475, EPI_ISL_872476, EPI_ISL_872477 | | | |
| see above | Eurofins Diatherix | Hudsonalpha Genome Sequencing Center | Jane Grimwood, Melissa Williams, Lori H. Handley, Joshua Stough, Leslie Malone, Stefan Brzezinski, Ada Stewart, Teresa Jones, Jenell Webber, John Lovell, Jennifer Cart, and Jeremy Schmutz |
| EPI_ISL_872479, EPI_ISL_872480, EPI_ISL_872481, EPI_ISL_872482, EPI_ISL_872483, EPI_ISL_872484, EPI_ISL_872485, EPI_ISL_872486, EPI_ISL_872487, EPI_ISL_872488, EPI_ISL_872489, EPI_ISL_872490, EPI_ISL_872491, EPI_ISL_872492, EPI_ISL_872493, EPI_ISL_872495, EPI_ISL_872496, EPI_ISL_872497, EPI_ISL_872498, EPI_ISL_872499, EPI_ISL_872500, EPI_ISL_872501, EPI_ISL_872502, EPI_ISL_872503, EPI_ISL_872504, EPI_ISL_872505, EPI_ISL_872506, EPI_ISL_872507, EPI_ISL_872508, EPI_ISL_872509, EPI_ISL_872510, EPI_ISL_872511, EPI_ISL_872512, EPI_ISL_872513, EPI_ISL_872515, EPI_ISL_872517, EPI_ISL_872518, EPI_ISL_872519, EPI_ISL_872520, EPI_ISL_872521, EPI_ISL_872524, EPI_ISL_872525, EPI_ISL_872526, EPI_ISL_872527, EPI_ISL_872528, EPI_ISL_872529, EPI_ISL_872530, EPI_ISL_872531, EPI_ISL_872532, EPI_ISL_872533, EPI_ISL_872534, EPI_ISL_872535, EPI_ISL_872536, EPI_ISL_872537, EPI_ISL_872539, EPI_ISL_872540, EPI_ISL_872541, EPI_ISL_872542, EPI_ISL_872543, EPI_ISL_872544, EPI_ISL_872545, EPI_ISL_872546, EPI_ISL_872547, EPI_ISL_872548, EPI_ISL_872549, EPI_ISL_872550, EPI_ISL_872551, EPI_ISL_872552, EPI_ISL_872553, EPI_ISL_872554, EPI_ISL_872555, EPI_ISL_872557, EPI_ISL_872558, EPI_ISL_872559, EPI_ISL_872560, EPI_ISL_872561, EPI_ISL_872562, EPI_ISL_872564, EPI_ISL_872565, EPI_ISL_872566 | | | |
| see above | New Mexico Department of Health Scientific Laboratory | Center for Global Health, University of New Mexico Health Sciences Center | Daryl Domman, Kurt Schwalm, Twila Kunde, Joseph Hicks, Anastacia Griego, Michael Edwards, Darrell Dinwiddie |
| EPI_ISL_872644, EPI_ISL_872645, EPI_ISL_872647, EPI_ISL_872648, EPI_ISL_872652, EPI_ISL_872654, EPI_ISL_872656, EPI_ISL_872658, EPI_ISL_872662, EPI_ISL_872663, EPI_ISL_872665, EPI_ISL_872667, EPI_ISL_872668, EPI_ISL_872669, EPI_ISL_872671, EPI_ISL_872676, EPI_ISL_872677 | | | |
| see above | University of Massachusetts Medical School | Infectious Disease Program, Broad Institute of Harvard and MIT | Lemieux,J.E., Siddle,K.J., Ward,D., Ellison,R., Adams,G., Gladden-Young,A., Lagerborg,K., Rudy,M., DeRuff,K., Carter,A., Normandin,E., Bauer,M., Reilly,S., Tomkins-Tinch,C., Loreth,C., Chaluvadi,S., Birren,B.W., Gallagher,G., Smole,S., Park,D.J., MacInnis,B.L., and Sabeti,P.C. |
| EPI_ISL_872680, EPI_ISL_872681, EPI_ISL_872682, EPI_ISL_872684, EPI_ISL_872689, EPI_ISL_872690, EPI_ISL_872691, EPI_ISL_872692, EPI_ISL_872694, EPI_ISL_872697, EPI_ISL_872698, EPI_ISL_872700, EPI_ISL_872702, EPI_ISL_872704, EPI_ISL_872708, EPI_ISL_872709, EPI_ISL_872712, EPI_ISL_872713, EPI_ISL_872715, EPI_ISL_872716, EPI_ISL_872717, EPI_ISL_872718, EPI_ISL_872719, EPI_ISL_872721, EPI_ISL_872723, EPI_ISL_872725, EPI_ISL_872726, EPI_ISL_872727, EPI_ISL_872728, EPI_ISL_872729, EPI_ISL_872730, EPI_ISL_872731, EPI_ISL_872732, EPI_ISL_872733, EPI_ISL_872734, EPI_ISL_872736, EPI_ISL_872737, EPI_ISL_872738, EPI_ISL_872739, EPI_ISL_872741, EPI_ISL_872743, EPI_ISL_872744 | | | |
| see above | Rhode Island Department of Health | Infectious Disease Program, Broad Institute of Harvard and MIT | Lemieux,J.E., Siddle,K.J., Huard,R., King,E., Azevedo,K., Miller,A., Adams,G., Gladden-Young,A., Lagerborg,K., Rudy,M., DeRuff,K., Carter,A., Normandin,E., Bauer,M., Reilly,S., Tomkins-Tinch,C., Loreth,C., Chaluvadi,S., Birren,B.W., Gallagher,G., Smole,S., Park,D.J., MacInnis,B.L., and Sabeti,P.C. |
| EPI_ISL_872745, EPI_ISL_872746, EPI_ISL_872747, EPI_ISL_872748, EPI_ISL_872749, EPI_ISL_872751, EPI_ISL_872755, EPI_ISL_872759, EPI_ISL_872760, EPI_ISL_872761, EPI_ISL_872762, EPI_ISL_872763, EPI_ISL_872766, EPI_ISL_872767, EPI_ISL_872769, EPI_ISL_872770, EPI_ISL_872771, EPI_ISL_872772, EPI_ISL_872773, EPI_ISL_872774, EPI_ISL_872775, EPI_ISL_872776, EPI_ISL_872777 | | | |
| see above | Colorado Mesa University | Infectious Disease Program, Broad Institute of Harvard and MIT | Lemieux,J.E., Siddle,K.J., Marshall,J., ONeill,M., Bronson,A., Adams,G., Gladden-Young,A., Lagerborg,K., Rudy,M., DeRuff,K., Carter,A., Normandin,E., Bauer,M., Reilly,S., Tomkins-Tinch,C., Loreth,C., Chaluvadi,S., Birren,B.W., Gallagher,G., Smole,S., Park,D.J., MacInnis,B.L., and Sabeti,P.C. |
| EPI_ISL_872778, EPI_ISL_872779, EPI_ISL_872780, EPI_ISL_872781, EPI_ISL_872782, EPI_ISL_872783, EPI_ISL_872784, EPI_ISL_872785, EPI_ISL_872786, EPI_ISL_872787, EPI_ISL_872788, EPI_ISL_872789, EPI_ISL_872790, EPI_ISL_872791, EPI_ISL_872792, EPI_ISL_872793, EPI_ISL_872794, EPI_ISL_872795, EPI_ISL_872796, EPI_ISL_872797, EPI_ISL_872798, EPI_ISL_872799, EPI_ISL_872800, EPI_ISL_872801, EPI_ISL_872802, EPI_ISL_872803, EPI_ISL_872804, EPI_ISL_872805, EPI_ISL_872806, EPI_ISL_872807, EPI_ISL_872808, EPI_ISL_872809, EPI_ISL_872810, EPI_ISL_872811, EPI_ISL_872812, EPI_ISL_872813, EPI_ISL_872814, EPI_ISL_872815, EPI_ISL_872816, EPI_ISL_872817, EPI_ISL_872818, EPI_ISL_872819, EPI_ISL_872820, EPI_ISL_872821, EPI_ISL_872822, EPI_ISL_872823, EPI_ISL_872824, EPI_ISL_872825, EPI_ISL_872826, EPI_ISL_872827, EPI_ISL_872828, EPI_ISL_872829, EPI_ISL_872830, EPI_ISL_872831, EPI_ISL_872832, EPI_ISL_872833, EPI_ISL_872834, EPI_ISL_872835, EPI_ISL_872836, EPI_ISL_872837, EPI_ISL_872838, EPI_ISL_872839, EPI_ISL_872840, EPI_ISL_872841, EPI_ISL_872842, EPI_ISL_872843, EPI_ISL_872844, EPI_ISL_872845, EPI_ISL_872846, EPI_ISL_872847, EPI_ISL_872848, EPI_ISL_872849, EPI_ISL_872850, EPI_ISL_872851, EPI_ISL_872852, EPI_ISL_872853, EPI_ISL_872854, EPI_ISL_872855, EPI_ISL_872856, EPI_ISL_872857, EPI_ISL_872858, EPI_ISL_872859, EPI_ISL_872860, EPI_ISL_872861, EPI_ISL_872862, EPI_ISL_872863, EPI_ISL_872864, EPI_ISL_872865, EPI_ISL_872866, EPI_ISL_872867, EPI_ISL_872868, EPI_ISL_872869, EPI_ISL_872870, EPI_ISL_872871, EPI_ISL_872872, EPI_ISL_872873, EPI_ISL_872874, EPI_ISL_872875, EPI_ISL_872876, EPI_ISL_872877, EPI_ISL_872878, EPI_ISL_872879, EPI_ISL_872880, EPI_ISL_872881, EPI_ISL_872882, EPI_ISL_872883, EPI_ISL_872884, EPI_ISL_872885, EPI_ISL_872886, EPI_ISL_872887, EPI_ISL_872888, EPI_ISL_872889, EPI_ISL_872890, EPI_ISL_872891, EPI_ISL_872892, EPI_ISL_872893, EPI_ISL_872894, EPI_ISL_872895, EPI_ISL_872896 | | | |
| see above | University of Wisconsin-Madison AIDS Vaccine Research Laboratories | University of Wisconsin-Madison AIDS Vaccine Research Laboratories | Gage Moreno, Katarina Braun, et al. AIDS Vaccine Research Laboratories |
| EPI_ISL_873041, EPI_ISL_873042, EPI_ISL_873043, EPI_ISL_873044, EPI_ISL_873045, EPI_ISL_873046, EPI_ISL_873047, EPI_ISL_873048, EPI_ISL_873049, EPI_ISL_873050, EPI_ISL_873051, EPI_ISL_873052, EPI_ISL_873053, EPI_ISL_873054, EPI_ISL_873055, EPI_ISL_873056, EPI_ISL_873057, EPI_ISL_873058, EPI_ISL_873059, EPI_ISL_873060, EPI_ISL_873061, EPI_ISL_873062, EPI_ISL_873063, EPI_ISL_873064, EPI_ISL_873065, EPI_ISL_873066, EPI_ISL_873067, EPI_ISL_873068, EPI_ISL_873069, EPI_ISL_873070, EPI_ISL_873071, EPI_ISL_873072, EPI_ISL_873073, EPI_ISL_873074, EPI_ISL_873075, EPI_ISL_873076, EPI_ISL_873077, EPI_ISL_873078, EPI_ISL_873079, EPI_ISL_873080, EPI_ISL_873081, EPI_ISL_873082, EPI_ISL_873083, EPI_ISL_873084, EPI_ISL_873085, EPI_ISL_873086, EPI_ISL_873087, EPI_ISL_873088, EPI_ISL_873089, EPI_ISL_873090, EPI_ISL_873091, EPI_ISL_873092, EPI_ISL_873093, EPI_ISL_873094, EPI_ISL_873095, EPI_ISL_873096, EPI_ISL_873097, EPI_ISL_873098, EPI_ISL_873099, EPI_ISL_873100, EPI_ISL_873101, EPI_ISL_873102, EPI_ISL_873103, EPI_ISL_873104, EPI_ISL_873105, EPI_ISL_873106, EPI_ISL_873107, EPI_ISL_873108, EPI_ISL_873109, EPI_ISL_873110, EPI_ISL_873111, EPI_ISL_873112, EPI_ISL_873113, EPI_ISL_873114, EPI_ISL_873115, EPI_ISL_873116, EPI_ISL_873117, EPI_ISL_873118, EPI_ISL_873119, EPI_ISL_873120, EPI_ISL_873121, EPI_ISL_873122, EPI_ISL_873123, EPI_ISL_873124, EPI_ISL_873125, EPI_ISL_873126, EPI_ISL_873127, EPI_ISL_873128, EPI_ISL_873129, EPI_ISL_873130, EPI_ISL_873131, EPI_ISL_873132, EPI_ISL_873133, EPI_ISL_873134, EPI_ISL_873135, EPI_ISL_873136, EPI_ISL_873137, EPI_ISL_873138, EPI_ISL_873139, EPI_ISL_873140, EPI_ISL_873141, EPI_ISL_873142, EPI_ISL_873143, EPI_ISL_873144, EPI_ISL_873145, EPI_ISL_873146, EPI_ISL_873147, EPI_ISL_873148, EPI_ISL_873149, EPI_ISL_873150, EPI_ISL_873151, EPI_ISL_873152, EPI_ISL_873153, EPI_ISL_873154, EPI_ISL_873155, EPI_ISL_873156, EPI_ISL_873157, EPI_ISL_873158, EPI_ISL_873159, EPI_ISL_873160, EPI_ISL_873161, EPI_ISL_873162, EPI_ISL_873163 | | | |
| see above | University of Michigan Clinical Microbiology Laboratory | Lauring Lab, University of Michigan, Department of Microbiology and Immunology | Valesano |
| EPI_ISL_873166, EPI_ISL_873167, EPI_ISL_873168, EPI_ISL_873170, EPI_ISL_873171, EPI_ISL_873172, EPI_ISL_873173, EPI_ISL_873175, EPI_ISL_873176, EPI_ISL_873177, EPI_ISL_873178, EPI_ISL_873179, EPI_ISL_873180, EPI_ISL_873182, EPI_ISL_873183, EPI_ISL_873184, EPI_ISL_873185, EPI_ISL_873186, EPI_ISL_873187, EPI_ISL_873188, EPI_ISL_873189, EPI_ISL_873190, EPI_ISL_873191, EPI_ISL_873192, EPI_ISL_873193 | | | |
| see above | Microbiology Division, South Carolina Department of Health and Environmental Control (SC DHEC) | Microbiology Division, South Carolina Department of Health and Environmental Control (SC DHEC) | Flores,H., Freeman,J. |
| EPI_ISL_873194, EPI_ISL_873195, EPI_ISL_873196, EPI_ISL_873197, EPI_ISL_873198, EPI_ISL_873199, EPI_ISL_873200, EPI_ISL_873201, EPI_ISL_873202, EPI_ISL_873203, EPI_ISL_873204, EPI_ISL_873205, EPI_ISL_873206, EPI_ISL_873207, EPI_ISL_873208 | | | |
| see above | North Dakota Department of Health, Public Health Laboratory | North Dakota Department of Health, Public Health Laboratory | Lisa Wingerter |
| EPI_ISL_873211, EPI_ISL_873212, EPI_ISL_873213, EPI_ISL_873214, EPI_ISL_873215, EPI_ISL_873216, EPI_ISL_873217, EPI_ISL_873218, EPI_ISL_873219, EPI_ISL_873220, EPI_ISL_873221, EPI_ISL_873222, EPI_ISL_873223, EPI_ISL_873224, EPI_ISL_873225, EPI_ISL_873226, EPI_ISL_873227, EPI_ISL_873228, EPI_ISL_873229, EPI_ISL_873230, EPI_ISL_873231, EPI_ISL_873232, EPI_ISL_873233, EPI_ISL_873234, EPI_ISL_873235, EPI_ISL_873236, EPI_ISL_873237, EPI_ISL_873238, EPI_ISL_873239, EPI_ISL_873240, EPI_ISL_873241, EPI_ISL_873242, EPI_ISL_873243, EPI_ISL_873244, EPI_ISL_873245, EPI_ISL_873246, EPI_ISL_873247, EPI_ISL_873248, EPI_ISL_873249, EPI_ISL_873250, EPI_ISL_873251, EPI_ISL_873252, EPI_ISL_873253, EPI_ISL_873254, EPI_ISL_873255, EPI_ISL_873256, EPI_ISL_873257, EPI_ISL_873258, EPI_ISL_873259, EPI_ISL_873260, EPI_ISL_873261, EPI_ISL_873262, EPI_ISL_873263, EPI_ISL_873264, EPI_ISL_873265, EPI_ISL_873266, EPI_ISL_873267, EPI_ISL_873268, EPI_ISL_873269 | | | |
| see above | M Health Fairview | Minnesota Department of Health, Public Health Laboratory | Alexandra Lorentz, Jacob Garfin, Matt Plumb, and Xiong Wang |
| EPI_ISL_873270, EPI_ISL_873271 | Minnesota Department of Health, Public Health Laboratory | Minnesota Department of Health, Public Health Laboratory | Alexandra Lorentz, Jacob Garfin, Matt Plumb, and Xiong Wang |
| EPI_ISL_873272, EPI_ISL_873273, EPI_ISL_873274, EPI_ISL_873275 | Vault Health | Minnesota Department of Health, Public Health Laboratory | Alexandra Lorentz, Jacob Garfin, Matt Plumb, and Xiong Wang |
| EPI_ISL_873276 | Hennepin County Medical Center | Minnesota Department of Health, Public Health Laboratory | Alexandra Lorentz, Jacob Garfin, Matt Plumb, and Xiong Wang |
| EPI_ISL_873277 | Environmental and Global Health, University of Florida | Environmental and Global Health, University of Florida | Iovine,N.M., Waltzek,T.B., Subramaniam,K., Loeb,J.C.,Stephenson,C.J., Lessard,K., Reeves,M., Wilkerson,G., Morris,J.G. and Lednicky,J.A. |
| EPI_ISL_875347, EPI_ISL_875348, EPI_ISL_875561, EPI_ISL_875562, EPI_ISL_875565 | Ohio Department of Health Laboratory | Ohio Department of Health Laboratory | Holmes, Jennifer; Eric Brandt, Keoni Omura, Glen McGillivary, Caitlin McDonnell, Kirtana Ramadugu, Erica Leasure, Kelsey Florek, Heather Blankenship, Quanta Brown, and Tammy Bannerman |
| EPI_ISL_875570, EPI_ISL_875571, EPI_ISL_875572, EPI_ISL_875573, EPI_ISL_875574, EPI_ISL_875577, EPI_ISL_875578, EPI_ISL_875579, EPI_ISL_875580, EPI_ISL_875581, EPI_ISL_875582, EPI_ISL_875583, EPI_ISL_875584, EPI_ISL_875585, EPI_ISL_875586, EPI_ISL_875587, EPI_ISL_875588, EPI_ISL_875589, EPI_ISL_875590, EPI_ISL_875591, EPI_ISL_875592, EPI_ISL_875593, EPI_ISL_875594, EPI_ISL_875595, EPI_ISL_875596, EPI_ISL_875597, EPI_ISL_875598, EPI_ISL_875599, EPI_ISL_875600, EPI_ISL_875601, EPI_ISL_875602, EPI_ISL_875603, EPI_ISL_875604, EPI_ISL_875605, EPI_ISL_875606, EPI_ISL_875607, EPI_ISL_875608, EPI_ISL_875609, EPI_ISL_875610, EPI_ISL_875611, EPI_ISL_875612, EPI_ISL_875613, EPI_ISL_875614, EPI_ISL_875615, EPI_ISL_875616, EPI_ISL_875617, EPI_ISL_875618, EPI_ISL_875619, EPI_ISL_875620, EPI_ISL_875621, EPI_ISL_875622, EPI_ISL_875623, EPI_ISL_875624, EPI_ISL_875625, EPI_ISL_875626, EPI_ISL_875627, EPI_ISL_875628, EPI_ISL_875629, EPI_ISL_875630, EPI_ISL_875631, EPI_ISL_875632, EPI_ISL_875633, EPI_ISL_875634, EPI_ISL_875635, EPI_ISL_875636, EPI_ISL_875637, EPI_ISL_875638, EPI_ISL_875639, EPI_ISL_875640, EPI_ISL_875641, EPI_ISL_875642, EPI_ISL_875643, EPI_ISL_875644, EPI_ISL_875645, EPI_ISL_875646, EPI_ISL_875647, EPI_ISL_875648, EPI_ISL_875649, EPI_ISL_875650, EPI_ISL_875651, EPI_ISL_875652, EPI_ISL_875653, EPI_ISL_875654, EPI_ISL_875655, EPI_ISL_875656, EPI_ISL_875657, EPI_ISL_875658, EPI_ISL_875659 | | | |
| see above | Eurofins Diatherix | Hudsonalpha Genome Sequencing Center | Jane Grimwood, Melissa Williams, Lori H. Handley, Joshua Stough, Leslie Malone, Stefan Brzezinski, Ada Stewart, Teresa Jones, Jenell Webber, John Lovell, Jennifer Cart, and Jeremy Schmutz |
| EPI_ISL_875660, EPI_ISL_875661, EPI_ISL_875662, EPI_ISL_875663, EPI_ISL_875664, EPI_ISL_875665 | University of Michigan Clinical Microbiology Laboratory | Lauring Lab, University of Michigan, Department of Microbiology and Immunology | Valesano |
| EPI_ISL_875675, EPI_ISL_875676, EPI_ISL_875677, EPI_ISL_875678, EPI_ISL_875679, EPI_ISL_875680, EPI_ISL_875681, EPI_ISL_875682, EPI_ISL_875683, EPI_ISL_875684 | Kansas Health and Environmental Lab | Kansas Health and Environmental Lab | Mike Grose, Carissa Robertson, Ben Olsen, and Phil Adam |
| EPI_ISL_875871, EPI_ISL_875872 | Ohio Department of Health Laboratory | Ohio Department of Health Laboratory | Holmes, Jennifer; Eric Brandt, Keoni Omura, Glen McGillivary, Caitlin McDonnell, Kirtana Ramadugu, Erica Leasure, Kelsey Florek, Heather Blankenship, Quanta Brown, and Tammy Bannerman |
| EPI_ISL_875874, EPI_ISL_875877, EPI_ISL_875878, EPI_ISL_875879, EPI_ISL_875880, EPI_ISL_875881, EPI_ISL_875882, EPI_ISL_875883, EPI_ISL_875884, EPI_ISL_875885, EPI_ISL_875886, EPI_ISL_875887, EPI_ISL_875888, EPI_ISL_875889, EPI_ISL_875890, EPI_ISL_875891, EPI_ISL_875892, EPI_ISL_875893, EPI_ISL_875894, EPI_ISL_875895, EPI_ISL_875896, EPI_ISL_875897, EPI_ISL_875898, EPI_ISL_875899, EPI_ISL_875900, EPI_ISL_875901, EPI_ISL_875902, EPI_ISL_875903, EPI_ISL_875904, EPI_ISL_875905, EPI_ISL_875906, EPI_ISL_875907, EPI_ISL_875908, EPI_ISL_875909, EPI_ISL_875910, EPI_ISL_875911, EPI_ISL_875912, EPI_ISL_875913, EPI_ISL_875914, EPI_ISL_875915, EPI_ISL_875916, EPI_ISL_875917, EPI_ISL_875918, EPI_ISL_875919, EPI_ISL_875920, EPI_ISL_875921, EPI_ISL_875922, EPI_ISL_875923, EPI_ISL_875924, EPI_ISL_875925, EPI_ISL_875926, EPI_ISL_875927, EPI_ISL_875928, EPI_ISL_875929, EPI_ISL_875930, EPI_ISL_875931, EPI_ISL_875932, EPI_ISL_875933, EPI_ISL_875934, EPI_ISL_875935, EPI_ISL_875936, EPI_ISL_875937, EPI_ISL_875938, EPI_ISL_875939, EPI_ISL_875940, EPI_ISL_875941, EPI_ISL_875942, EPI_ISL_875943, EPI_ISL_875944, EPI_ISL_875945, EPI_ISL_875946, EPI_ISL_875947, EPI_ISL_875948, EPI_ISL_875949, EPI_ISL_875950, EPI_ISL_875951, EPI_ISL_875952, EPI_ISL_875953, EPI_ISL_875954, EPI_ISL_875955, EPI_ISL_875956, EPI_ISL_875957, EPI_ISL_875958 | | | |
| see above | Eurofins Diatherix | Hudsonalpha Genome Sequencing Center | Jane Grimwood, Melissa Williams, Lori H. Handley, Joshua Stough, Leslie Malone, Stefan Brzezinski, Ada Stewart, Teresa Jones, Jenell Webber, John Lovell, Jennifer Cart, and Jeremy Schmutz |
| EPI_ISL_876032, EPI_ISL_876033, EPI_ISL_876034, EPI_ISL_876035, EPI_ISL_876036, | Montefiore Medical Center | Albert Einstein College of Medicine, Dept. of Microbiology & Immunology, Chandran lab | J. Maximilian Fels, Saad Khan, Ryan Forster, Karin A. Skalina, Surksha Sirichand, Amy S. Fox, Aviv Bergman, William B. Mitchell, Lucia R. Wolgast, Wendy Szymczak, Robert H. Bortz III, M. Eugenia Dieterle, Catalina Florez, Denise Haslwanter, Rohit K. Jangra, Ethan Laudermilch, Ariel S. Wirchnianski, Jason Barnhill, David L. Goldman, Hnin Khine, D. Yitzchak Goldstein, Johanna P. Daily, Kartik Chandran, Libusha Kelly |

| | | | |
|---|---|---|---|
| EPI_ISL_876037, EPI_ISL_876038, EPI_ISL_876039, EPI_ISL_876040, EPI_ISL_876041 | | | |
| EPI_ISL_876044 | M Health Fairview | Minnesota Department of Health, Public Health Laboratory | Alexandra Lorentz, Jacob Garfin, Matt Plumb, and Xiong Wang |
| EPI_ISL_876045, EPI_ISL_876046, EPI_ISL_876047, EPI_ISL_876048, EPI_ISL_876049, EPI_ISL_876050, EPI_ISL_876051, EPI_ISL_876052, EPI_ISL_876053, EPI_ISL_876054, EPI_ISL_876055, EPI_ISL_876056, EPI_ISL_876057, EPI_ISL_876058, EPI_ISL_876059, EPI_ISL_876060, EPI_ISL_876061, EPI_ISL_876062, EPI_ISL_876063, EPI_ISL_876064, EPI_ISL_876065, EPI_ISL_876066, EPI_ISL_876067, EPI_ISL_876068, EPI_ISL_876069, EPI_ISL_876070, EPI_ISL_876071, EPI_ISL_876072, EPI_ISL_876073, EPI_ISL_876074, EPI_ISL_876075, EPI_ISL_876076, EPI_ISL_876077, EPI_ISL_876078, EPI_ISL_876079, EPI_ISL_876080, EPI_ISL_876081, EPI_ISL_876082, EPI_ISL_876083, EPI_ISL_876084, EPI_ISL_876085, EPI_ISL_876086, EPI_ISL_876087, EPI_ISL_876088, EPI_ISL_876089, EPI_ISL_876090, EPI_ISL_876091, EPI_ISL_876092, EPI_ISL_876093, EPI_ISL_876094, EPI_ISL_876095, EPI_ISL_876096, EPI_ISL_876097, EPI_ISL_876098, EPI_ISL_876099, EPI_ISL_876100, EPI_ISL_876101, EPI_ISL_876102, EPI_ISL_876103, EPI_ISL_876104, EPI_ISL_876105, EPI_ISL_876106, EPI_ISL_876107, EPI_ISL_876108, EPI_ISL_876109, EPI_ISL_876110, EPI_ISL_876111, EPI_ISL_876112, EPI_ISL_876113, EPI_ISL_876114, EPI_ISL_876115, EPI_ISL_876116, EPI_ISL_876117, EPI_ISL_876118, EPI_ISL_876119, EPI_ISL_876120, EPI_ISL_876121, EPI_ISL_876122, EPI_ISL_876123, EPI_ISL_876124, EPI_ISL_876125, EPI_ISL_876126, EPI_ISL_876127, EPI_ISL_876128, EPI_ISL_876129, EPI_ISL_876130, EPI_ISL_876131, EPI_ISL_876132, EPI_ISL_876133, EPI_ISL_876134, EPI_ISL_876135, EPI_ISL_876136, EPI_ISL_876137, EPI_ISL_876138, EPI_ISL_876139, EPI_ISL_876140, EPI_ISL_876141, EPI_ISL_876142, EPI_ISL_876143, EPI_ISL_876144, EPI_ISL_876145, EPI_ISL_876146, EPI_ISL_876147, EPI_ISL_876148, EPI_ISL_876149, EPI_ISL_876150, EPI_ISL_876151, EPI_ISL_876152, EPI_ISL_876153, EPI_ISL_876154, EPI_ISL_876155, EPI_ISL_876156, EPI_ISL_876157, EPI_ISL_876158, EPI_ISL_876159, EPI_ISL_876160, EPI_ISL_876161, EPI_ISL_876162, EPI_ISL_876163, EPI_ISL_876202, EPI_ISL_876203, EPI_ISL_876204, EPI_ISL_876205, EPI_ISL_876206, EPI_ISL_876207, EPI_ISL_876208, EPI_ISL_876209, EPI_ISL_876210, EPI_ISL_876211, EPI_ISL_876212, EPI_ISL_876213, EPI_ISL_876214, EPI_ISL_876215, EPI_ISL_876216, EPI_ISL_876217, EPI_ISL_876218, EPI_ISL_876219, EPI_ISL_876220, EPI_ISL_876221, EPI_ISL_876222, EPI_ISL_876223, EPI_ISL_876224, EPI_ISL_876225, EPI_ISL_876226, EPI_ISL_876227, EPI_ISL_876228, EPI_ISL_876229, EPI_ISL_876230, EPI_ISL_876231, EPI_ISL_876232, EPI_ISL_876233, EPI_ISL_876234, EPI_ISL_876235, EPI_ISL_876236, EPI_ISL_876237, EPI_ISL_876238, EPI_ISL_876239, EPI_ISL_876240, EPI_ISL_876241, EPI_ISL_876242, EPI_ISL_876243, EPI_ISL_876244, EPI_ISL_876245, EPI_ISL_876246, EPI_ISL_876247, EPI_ISL_876248, EPI_ISL_876249, EPI_ISL_876250, EPI_ISL_876251, EPI_ISL_876252, EPI_ISL_876253, EPI_ISL_876254, EPI_ISL_876255, EPI_ISL_876256, EPI_ISL_876257, EPI_ISL_876258, EPI_ISL_876259, EPI_ISL_876260, EPI_ISL_876261, EPI_ISL_876262, EPI_ISL_876263, EPI_ISL_876264, EPI_ISL_876265, EPI_ISL_876266, EPI_ISL_876267, EPI_ISL_876268, EPI_ISL_876269, EPI_ISL_876270, EPI_ISL_876271, EPI_ISL_876272, EPI_ISL_876273, EPI_ISL_876274, EPI_ISL_876275, EPI_ISL_876276, EPI_ISL_876277, EPI_ISL_876278, EPI_ISL_876279, EPI_ISL_876280, EPI_ISL_876281, EPI_ISL_876282, EPI_ISL_876283, EPI_ISL_876284, EPI_ISL_876285, EPI_ISL_876286, EPI_ISL_876287, EPI_ISL_876288, EPI_ISL_876289, EPI_ISL_876290, EPI_ISL_876291, EPI_ISL_876292, EPI_ISL_876293, EPI_ISL_876294, EPI_ISL_876295, EPI_ISL_876296, EPI_ISL_876297, EPI_ISL_876298, EPI_ISL_876299, EPI_ISL_876300, EPI_ISL_876301, EPI_ISL_876302, EPI_ISL_876303, EPI_ISL_876304, EPI_ISL_876305, EPI_ISL_876306, EPI_ISL_876307, EPI_ISL_876308, EPI_ISL_876309, EPI_ISL_876310, EPI_ISL_876311, EPI_ISL_876312, EPI_ISL_876313, EPI_ISL_876314, EPI_ISL_876315, EPI_ISL_876316, EPI_ISL_876317, EPI_ISL_876318, EPI_ISL_876319, EPI_ISL_876320, EPI_ISL_876321, EPI_ISL_876322, EPI_ISL_876323, EPI_ISL_876324, EPI_ISL_876325, EPI_ISL_876326, EPI_ISL_876327, EPI_ISL_876328, EPI_ISL_876329, EPI_ISL_876330 | | | |
| see above | Massachusetts State Public Health Laboratory | Massachusetts State Public Health Laboratory | Andrew Lang, Timelia Fink, Glen Gallagher, Sandra Smole |
| EPI_ISL_876331, EPI_ISL_876332, EPI_ISL_876333, EPI_ISL_876334, EPI_ISL_876335, EPI_ISL_876336, EPI_ISL_876337, EPI_ISL_876338, EPI_ISL_876339, EPI_ISL_876340, EPI_ISL_876341, EPI_ISL_876342, EPI_ISL_876343, EPI_ISL_876344, EPI_ISL_876345, EPI_ISL_876346, EPI_ISL_876347, EPI_ISL_876348, EPI_ISL_876349, EPI_ISL_876350, EPI_ISL_876351, EPI_ISL_876352, EPI_ISL_876353, EPI_ISL_876354, EPI_ISL_876355, EPI_ISL_876356, EPI_ISL_876357, EPI_ISL_876358, EPI_ISL_876359, EPI_ISL_876360, EPI_ISL_876361, EPI_ISL_876362, EPI_ISL_876363, EPI_ISL_876364, EPI_ISL_876365, EPI_ISL_876366, EPI_ISL_876367, EPI_ISL_876368, EPI_ISL_876369, EPI_ISL_876370, EPI_ISL_876371, EPI_ISL_876372, EPI_ISL_876373, EPI_ISL_876374, EPI_ISL_876375, EPI_ISL_876376, EPI_ISL_876377, EPI_ISL_876378, EPI_ISL_876379, EPI_ISL_876380, EPI_ISL_876381, EPI_ISL_876382, EPI_ISL_876383, EPI_ISL_876384, EPI_ISL_876385, EPI_ISL_876386, EPI_ISL_876387, EPI_ISL_876388, EPI_ISL_876389, EPI_ISL_876390, EPI_ISL_876391, EPI_ISL_876392, EPI_ISL_876393, EPI_ISL_876394, EPI_ISL_876395, EPI_ISL_876396, EPI_ISL_876397, EPI_ISL_876398, EPI_ISL_876399, EPI_ISL_876400, EPI_ISL_876401, EPI_ISL_876402, EPI_ISL_876403, EPI_ISL_876404, EPI_ISL_876405, EPI_ISL_876406, EPI_ISL_876407, EPI_ISL_876408, EPI_ISL_876409, EPI_ISL_876410, EPI_ISL_876411, EPI_ISL_876412, EPI_ISL_876413, EPI_ISL_876414, EPI_ISL_876415, EPI_ISL_876416, EPI_ISL_876417, EPI_ISL_876418, EPI_ISL_876419, EPI_ISL_876420, EPI_ISL_876421, EPI_ISL_876422, EPI_ISL_876423, EPI_ISL_876424, EPI_ISL_876425 | | | |
| see above | Eurofins Diatherix | Hudsonalpha Genome Sequencing Center | Jane Grimwood, Melissa Williams, Lori H. Handley, Joshua Stough, Leslie Malone, Stefan Brzezinski, Ada Stewart, Teresa Jones, Jenell Webber, John Lovell, Jennifer Cart, and Jeremy Schmutz |
| EPI_ISL_876427 | Ohio Department of Health Laboratory | Ohio Department of Health Laboratory | Holmes, Jennifer; Eric Brandt, Keoni Omura, Glen McGillivary, Caitlin McDonnell, Kirtana Ramadugu, Erica Leasure, Kelsey Florek, Heather Blankenship, Quanta Brown, and Tammy Bannerman |
| EPI_ISL_876428, EPI_ISL_876429, EPI_ISL_876431, EPI_ISL_876432, EPI_ISL_876433, EPI_ISL_876434, EPI_ISL_876435, EPI_ISL_876436, EPI_ISL_876437, EPI_ISL_876438, EPI_ISL_876439, EPI_ISL_876440, EPI_ISL_876441, EPI_ISL_876442, EPI_ISL_876443, EPI_ISL_876444, EPI_ISL_876445, EPI_ISL_876446, EPI_ISL_876447, EPI_ISL_876448, EPI_ISL_876449, EPI_ISL_876450, EPI_ISL_876451, EPI_ISL_876452, EPI_ISL_876453, EPI_ISL_876454, EPI_ISL_876455, EPI_ISL_876456, EPI_ISL_876457, EPI_ISL_876458, EPI_ISL_876459, EPI_ISL_876460, EPI_ISL_876461, EPI_ISL_876462, EPI_ISL_876463, EPI_ISL_876464, EPI_ISL_876465, EPI_ISL_876466, EPI_ISL_876467, EPI_ISL_876468, EPI_ISL_876469, EPI_ISL_876470, EPI_ISL_876471, EPI_ISL_876472, EPI_ISL_876473, EPI_ISL_876474, EPI_ISL_876475, EPI_ISL_876476, EPI_ISL_876477, EPI_ISL_876478, EPI_ISL_876479, EPI_ISL_876480, EPI_ISL_876481, EPI_ISL_876482, EPI_ISL_876483, EPI_ISL_876484, EPI_ISL_876485, EPI_ISL_876486, EPI_ISL_876487, EPI_ISL_876488, EPI_ISL_876489, EPI_ISL_876490, EPI_ISL_876491, EPI_ISL_876492, EPI_ISL_876493, EPI_ISL_876494, EPI_ISL_876495, EPI_ISL_876496, EPI_ISL_876497, EPI_ISL_876498, EPI_ISL_876499, EPI_ISL_876500, EPI_ISL_876501, EPI_ISL_876502, EPI_ISL_876503, EPI_ISL_876504, EPI_ISL_876505, EPI_ISL_876506, EPI_ISL_876507, EPI_ISL_876508, EPI_ISL_876509, EPI_ISL_876510, EPI_ISL_876511, EPI_ISL_876512, EPI_ISL_876513, EPI_ISL_876514, EPI_ISL_876515, EPI_ISL_876516, EPI_ISL_876517 | | | |
| see above | Eurofins Diatherix | Hudsonalpha Genome Sequencing Center | Jane Grimwood, Melissa Williams, Lori H. Handley, Joshua Stough, Leslie Malone, Stefan Brzezinski, Ada Stewart, Teresa Jones, Jenell Webber, John Lovell, Jennifer Cart, and Jeremy Schmutz |
| EPI_ISL_876518 | Ohio Department of Health Laboratory | Ohio Department of Health Laboratory | Holmes, Jennifer; Eric Brandt, Keoni Omura, Glen McGillivary, Caitlin McDonnell, Kirtana Ramadugu, Erica Leasure, Kelsey Florek, Heather Blankenship, Quanta Brown, and Tammy Bannerman |
| EPI_ISL_876519 | DOHMH Jamaica | New York City Public Health Laboratory | Jade Wang, et al. |
| EPI_ISL_876520, EPI_ISL_876521, EPI_ISL_876522, EPI_ISL_876523, EPI_ISL_876524, EPI_ISL_876525, EPI_ISL_876526, EPI_ISL_876527, EPI_ISL_876528, EPI_ISL_876529, EPI_ISL_876532, EPI_ISL_876533, EPI_ISL_876534, EPI_ISL_876535, EPI_ISL_876536, EPI_ISL_876537, EPI_ISL_876538, EPI_ISL_876539, EPI_ISL_876540, EPI_ISL_876541, EPI_ISL_876542, EPI_ISL_876543, EPI_ISL_876544, EPI_ISL_876545, EPI_ISL_876546, EPI_ISL_876547, EPI_ISL_876548, EPI_ISL_876549, EPI_ISL_876550, EPI_ISL_876551, EPI_ISL_876552, EPI_ISL_876553, EPI_ISL_876554, EPI_ISL_876555, EPI_ISL_876556, EPI_ISL_876557, EPI_ISL_876558, EPI_ISL_876559, EPI_ISL_876560, EPI_ISL_876561, EPI_ISL_876562, EPI_ISL_876563, EPI_ISL_876564, EPI_ISL_876565, EPI_ISL_876566, EPI_ISL_876567, EPI_ISL_876568, EPI_ISL_876569, EPI_ISL_876570, EPI_ISL_876571, EPI_ISL_876572, EPI_ISL_876573, EPI_ISL_876574, EPI_ISL_876575, EPI_ISL_876576, EPI_ISL_876577, EPI_ISL_876578, EPI_ISL_876579, EPI_ISL_876580, EPI_ISL_876581, EPI_ISL_876582, EPI_ISL_876583, EPI_ISL_876584, EPI_ISL_876585, EPI_ISL_876586, EPI_ISL_876587 | | | |
| see above | Florida Bureau of Public Health Laboratories | Florida Bureau of Public Health Laboratories | Sarah Schmedes, Jason Blanton |
| EPI_ISL_876588, EPI_ISL_876589 | Blackhawk Genomics | Pathogen Discovery, Respiratory Viruses Branch, Division of Viral Diseases, Centers for Disease Control and Prevention | Ying Tao, Yan Li, Jing Zhang, Krista Queen, Anna Uehara, Peter Cook, Clinton R. Paden, Haibin Wang, Suxiang Tong |
| EPI_ISL_876590, EPI_ISL_876591, EPI_ISL_876592, EPI_ISL_876593, EPI_ISL_876594, EPI_ISL_876595, EPI_ISL_876596 | TN Division of Laboratory Services | Pathogen Discovery, Respiratory Viruses Branch, Division of Viral Diseases, Centers for Disease Control and Prevention | Ying Tao, Yan Li, Jing Zhang, Krista Queen, Anna Uehara, Peter Cook, Clinton R. Paden, Haibin Wang, Suxiang Tong |
| EPI_ISL_876625, EPI_ISL_876626, EPI_ISL_876627, EPI_ISL_876628, EPI_ISL_876629, EPI_ISL_876630, EPI_ISL_876631, EPI_ISL_876632, EPI_ISL_876633, EPI_ISL_876634, EPI_ISL_876635, EPI_ISL_876636, EPI_ISL_876637, EPI_ISL_876638, EPI_ISL_876639, EPI_ISL_876644, EPI_ISL_876645, EPI_ISL_876646, EPI_ISL_876647, EPI_ISL_876648, EPI_ISL_876649, EPI_ISL_876650, EPI_ISL_876651, EPI_ISL_876652, EPI_ISL_876653, EPI_ISL_876654, EPI_ISL_876655, EPI_ISL_876656, EPI_ISL_876657, EPI_ISL_876658, EPI_ISL_876659, EPI_ISL_876660, EPI_ISL_876661, EPI_ISL_876662, EPI_ISL_876663, EPI_ISL_876664, EPI_ISL_876665, EPI_ISL_876666, EPI_ISL_876667, EPI_ISL_876668, EPI_ISL_876669, EPI_ISL_876670, EPI_ISL_876671, EPI_ISL_876672, EPI_ISL_876673, EPI_ISL_876674, EPI_ISL_876675, EPI_ISL_876676, EPI_ISL_876677, EPI_ISL_876678, EPI_ISL_876679, EPI_ISL_876680, EPI_ISL_876681, EPI_ISL_876682, EPI_ISL_876683, EPI_ISL_876684, EPI_ISL_876685, EPI_ISL_876686, EPI_ISL_876687, EPI_ISL_876688, EPI_ISL_876689, EPI_ISL_876690, EPI_ISL_876691, EPI_ISL_876692, EPI_ISL_876693, EPI_ISL_876694, EPI_ISL_876695, EPI_ISL_876696, EPI_ISL_876697, EPI_ISL_876698, EPI_ISL_876699, EPI_ISL_876700, EPI_ISL_876701, EPI_ISL_876702, EPI_ISL_876703, EPI_ISL_876704, EPI_ISL_876705, EPI_ISL_876706, EPI_ISL_876707, EPI_ISL_876708, EPI_ISL_876709, EPI_ISL_876710, EPI_ISL_876711, EPI_ISL_876712, EPI_ISL_876713, EPI_ISL_876714, EPI_ISL_876715, EPI_ISL_876716, EPI_ISL_876717, EPI_ISL_876718, EPI_ISL_876719, EPI_ISL_876720, EPI_ISL_876721, EPI_ISL_876722, EPI_ISL_876723, EPI_ISL_876724, EPI_ISL_876725, EPI_ISL_876726, EPI_ISL_876727, EPI_ISL_876728, EPI_ISL_876729, EPI_ISL_876730, EPI_ISL_876731, EPI_ISL_876732, EPI_ISL_876733, EPI_ISL_876734, EPI_ISL_876735, EPI_ISL_876736 | | | |
| see above | Helix/Illumina | Genomics and Discovery, Respiratory Viruses Branch, Division of Viral Diseases, Centers for Disease Control and Prevention | Peter W. Cook,Dhwani Batra,Ben L. Rambo-Martin,Eileen de Feo,Jan Antico,Christine Tran,Matthew Tolentino,Shannon Wickline,Kim Gietzen,Brad Sickler,Jingtao Liu,Eric Allen,Phil Febbo,Summer Galloway,Nicole L. Washington,Simon White,Geraint Levan,Kelly Schiabor Barrett,Elizabeth Cirulli,Alexandre Bolze,Ary Ascencio,Charlotte Rivera-Garcia,Ryan Cho,Jason Nguyen,Sherry Wang,Jimmy Ramirez,Tyler Cassens,Efren Sandoval,Magnus Isaksson,William Lee,David Becker,Marc Laurent,James Lu,Clinton R. Paden,Suxiang Tong,Duncan MacCannell, |
| EPI_ISL_876737 | Florida Bureau of Public Health Laboratories | Florida Bureau of Public Health Laboratories | Sarah Schmedes, Jason Blanton |
| EPI_ISL_876814, EPI_ISL_876815, EPI_ISL_876816, EPI_ISL_876817, EPI_ISL_876818, EPI_ISL_876819, EPI_ISL_876820, EPI_ISL_876821, EPI_ISL_876822, EPI_ISL_876823, EPI_ISL_876824, EPI_ISL_876825, EPI_ISL_876826, EPI_ISL_876827, EPI_ISL_876828, EPI_ISL_876829, EPI_ISL_876830, EPI_ISL_876831, EPI_ISL_876832, EPI_ISL_876833, EPI_ISL_876834, EPI_ISL_876835, EPI_ISL_876836, EPI_ISL_876837, EPI_ISL_876838, EPI_ISL_876839, EPI_ISL_876840, EPI_ISL_876841, EPI_ISL_876842, EPI_ISL_876843, EPI_ISL_876844, EPI_ISL_876845, EPI_ISL_876846, EPI_ISL_876847, EPI_ISL_876848, EPI_ISL_876849, EPI_ISL_876850, EPI_ISL_876851, EPI_ISL_876852, EPI_ISL_876853, EPI_ISL_876854, EPI_ISL_876855, EPI_ISL_876856, EPI_ISL_876857, EPI_ISL_876858, EPI_ISL_876859, EPI_ISL_876860, EPI_ISL_876861, EPI_ISL_876862, EPI_ISL_876863, EPI_ISL_876864, EPI_ISL_876865, EPI_ISL_876866, EPI_ISL_876867, EPI_ISL_876868, EPI_ISL_876869, EPI_ISL_876870, EPI_ISL_876871, EPI_ISL_876872, EPI_ISL_876873, EPI_ISL_876874, EPI_ISL_876875, EPI_ISL_876876, EPI_ISL_876877, EPI_ISL_876878, EPI_ISL_876879, EPI_ISL_876880, EPI_ISL_876881, EPI_ISL_876882, EPI_ISL_876883, EPI_ISL_876884, EPI_ISL_876885, EPI_ISL_876886, EPI_ISL_876887, EPI_ISL_876888, EPI_ISL_876889, EPI_ISL_876890, EPI_ISL_876891, EPI_ISL_876892, EPI_ISL_876893, EPI_ISL_876894, EPI_ISL_876895, EPI_ISL_876896, EPI_ISL_876897, EPI_ISL_876898, EPI_ISL_876899, EPI_ISL_876900, EPI_ISL_876901, EPI_ISL_876902, EPI_ISL_876903, EPI_ISL_876904, EPI_ISL_876905, EPI_ISL_876906, EPI_ISL_876907, EPI_ISL_876908, EPI_ISL_876909, EPI_ISL_876910, EPI_ISL_876911, EPI_ISL_876912, EPI_ISL_876913, EPI_ISL_876914, EPI_ISL_876915, EPI_ISL_876916, EPI_ISL_876917, EPI_ISL_876918, EPI_ISL_876919, EPI_ISL_876920, EPI_ISL_876921, EPI_ISL_876922, EPI_ISL_876923, EPI_ISL_876924, EPI_ISL_876925, EPI_ISL_876926, EPI_ISL_876927, EPI_ISL_876928, EPI_ISL_876929, EPI_ISL_876930, EPI_ISL_876931, EPI_ISL_876932, EPI_ISL_876933, EPI_ISL_876934, EPI_ISL_876935, EPI_ISL_876936, EPI_ISL_876937, EPI_ISL_876938, EPI_ISL_876939, EPI_ISL_876940, EPI_ISL_876941, EPI_ISL_876942, EPI_ISL_876943, EPI_ISL_876944, EPI_ISL_876945, EPI_ISL_876946, EPI_ISL_876947, EPI_ISL_876948, EPI_ISL_876949, EPI_ISL_876950, EPI_ISL_876951, EPI_ISL_876952, EPI_ISL_876953, EPI_ISL_876954, EPI_ISL_876955, EPI_ISL_876956, EPI_ISL_876957, EPI_ISL_876958, EPI_ISL_876959, EPI_ISL_876960, EPI_ISL_876961, EPI_ISL_876962, EPI_ISL_876963, EPI_ISL_876964, EPI_ISL_876965, EPI_ISL_876966, EPI_ISL_876967, EPI_ISL_876968, EPI_ISL_877037, EPI_ISL_877038, EPI_ISL_877039, EPI_ISL_877040, EPI_ISL_877041, EPI_ISL_877042, EPI_ISL_877043, EPI_ISL_877045, EPI_ISL_877046, EPI_ISL_877047, EPI_ISL_877048, EPI_ISL_877049, EPI_ISL_877050, EPI_ISL_877051, EPI_ISL_877052, EPI_ISL_877053, EPI_ISL_877054, EPI_ISL_877055, EPI_ISL_877056, EPI_ISL_877057, EPI_ISL_877058, EPI_ISL_877059, EPI_ISL_877060, EPI_ISL_877061, EPI_ISL_877062, EPI_ISL_877063, EPI_ISL_877064, EPI_ISL_877065, EPI_ISL_877066, EPI_ISL_877067, EPI_ISL_877068, EPI_ISL_877069, EPI_ISL_877070, EPI_ISL_877071, EPI_ISL_877072, EPI_ISL_877073, EPI_ISL_877074, EPI_ISL_877075, EPI_ISL_877076, EPI_ISL_877077, EPI_ISL_877078, EPI_ISL_877079, EPI_ISL_877080, EPI_ISL_877081, EPI_ISL_877082, EPI_ISL_877083, EPI_ISL_877084, EPI_ISL_877085, EPI_ISL_877086, EPI_ISL_877087, EPI_ISL_877088, EPI_ISL_877089, EPI_ISL_877090, EPI_ISL_877091, EPI_ISL_877092, EPI_ISL_877093, EPI_ISL_877094, EPI_ISL_877095, EPI_ISL_877096, EPI_ISL_877097, EPI_ISL_877098, EPI_ISL_877099, EPI_ISL_877100, EPI_ISL_877101, EPI_ISL_877102, EPI_ISL_877103, EPI_ISL_877104, EPI_ISL_877105, EPI_ISL_877106, EPI_ISL_877107, EPI_ISL_877108, EPI_ISL_877109, EPI_ISL_877110, EPI_ISL_877111, EPI_ISL_877112, EPI_ISL_877113, EPI_ISL_877114, EPI_ISL_877115, EPI_ISL_877116, EPI_ISL_877117, EPI_ISL_877118, EPI_ISL_877119, EPI_ISL_877120, EPI_ISL_877121, EPI_ISL_877122, EPI_ISL_877123, EPI_ISL_877124, EPI_ISL_877125 | | | |
| see above | Quest Diagnostics | Quest Diagnostics | Rosenthal,S.H., Gerasimova,A., Kagan,R.M., Anderson, B., Hua, M., Liu Y., Bernstein, L.E., Livingston, K.E., Perez, A., Shalhout, D.F., Shlyakhter, I.A., Owen, R., Tanpaiboon, P., Lacbawan, F. |
| EPI_ISL_877132 | The Ohio State University Wexner Medical Center | James Molecular Lab | Huolin Tu, Matthew R Avenarius, Laura Kubatko, Matthew Hunt, Xiaokang Pan, Peng Ru, Jason Garee, Keelie Thomas, Peter Mohler, Preeti Pancholi, Dan Jones |
| EPI_ISL_877133, EPI_ISL_877134, EPI_ISL_877135, EPI_ISL_877136, EPI_ISL_877137, EPI_ISL_877138, EPI_ISL_877139, EPI_ISL_877140, EPI_ISL_877141, EPI_ISL_877142, EPI_ISL_877143, EPI_ISL_877144, EPI_ISL_877145, EPI_ISL_877146, EPI_ISL_877147, EPI_ISL_877148, EPI_ISL_877149, EPI_ISL_877150, EPI_ISL_877151, EPI_ISL_877152, EPI_ISL_877153, EPI_ISL_877154, EPI_ISL_877155, EPI_ISL_877156, EPI_ISL_877157, EPI_ISL_877158, EPI_ISL_877159, EPI_ISL_877160, EPI_ISL_877161, EPI_ISL_877162, EPI_ISL_877163, EPI_ISL_877164, EPI_ISL_877165, EPI_ISL_877166, EPI_ISL_877167, EPI_ISL_877168, EPI_ISL_877169, EPI_ISL_877170, EPI_ISL_877171, EPI_ISL_877172, EPI_ISL_877173, EPI_ISL_877174, EPI_ISL_877175, EPI_ISL_877176, EPI_ISL_877177, EPI_ISL_877178, EPI_ISL_877179, EPI_ISL_877180, EPI_ISL_877181, EPI_ISL_877182, EPI_ISL_877184 | | | |
| see above | Univeristy of New Mexico Hospital | Center for Global Health, University of New Mexico Health Sciences Center | Daryl Domman, Kurt Schwalm, Justin Bacca, Jon Femling, Darrell Dinwiddie |
| EPI_ISL_877186, EPI_ISL_877187, EPI_ISL_877188, EPI_ISL_877189, EPI_ISL_877190, EPI_ISL_877191, EPI_ISL_877192, EPI_ISL_877193, EPI_ISL_877194, EPI_ISL_877195, EPI_ISL_877196, EPI_ISL_877197, EPI_ISL_877198, EPI_ISL_877199, EPI_ISL_877200, EPI_ISL_877201, EPI_ISL_877202, EPI_ISL_877203, EPI_ISL_877204 | | | |
| see above | Quest Diagnostics | Quest Diagnostics | Rosenthal,S.H., Gerasimova,A., Kagan,R.M., Anderson, B., Hua, M., Liu Y., Bernstein, L.E., Livingston, K.E., Perez, A., Shalhout, D.F., Shlyakhter, I.A., Owen, R., Tanpaiboon, P., Lacbawan, F. |
| EPI_ISL_877237, EPI_ISL_877238, EPI_ISL_877239, EPI_ISL_877240, EPI_ISL_877241, EPI_ISL_877242, EPI_ISL_877243, EPI_ISL_877244, EPI_ISL_877245, EPI_ISL_877246, EPI_ISL_877247, EPI_ISL_877248, EPI_ISL_877249, EPI_ISL_877250, EPI_ISL_877251, EPI_ISL_877252, EPI_ISL_877253, EPI_ISL_877254, EPI_ISL_877255, EPI_ISL_877256, EPI_ISL_877257, EPI_ISL_877258, EPI_ISL_877259, EPI_ISL_877260, EPI_ISL_877261, EPI_ISL_877262, EPI_ISL_877263, EPI_ISL_877264, EPI_ISL_877265, EPI_ISL_877266, EPI_ISL_877267, EPI_ISL_877268, EPI_ISL_877269, EPI_ISL_877270, EPI_ISL_877271, EPI_ISL_877272, EPI_ISL_877273, EPI_ISL_877274, EPI_ISL_877275, EPI_ISL_877276, EPI_ISL_877277, EPI_ISL_877278, EPI_ISL_877279, EPI_ISL_877280, EPI_ISL_877281, EPI_ISL_877282, EPI_ISL_877283, EPI_ISL_877284, EPI_ISL_877285, EPI_ISL_877286, EPI_ISL_877287, EPI_ISL_877288, EPI_ISL_877289, EPI_ISL_877290, EPI_ISL_877291, EPI_ISL_877292, EPI_ISL_877293, EPI_ISL_877294, EPI_ISL_877295, EPI_ISL_877296, EPI_ISL_877297, EPI_ISL_877298, EPI_ISL_877299, EPI_ISL_877300, EPI_ISL_877301, EPI_ISL_877302, EPI_ISL_877303, EPI_ISL_877304, EPI_ISL_877305, EPI_ISL_877306, EPI_ISL_877307, EPI_ISL_877308, EPI_ISL_877309, EPI_ISL_877310, EPI_ISL_877311, EPI_ISL_877312, EPI_ISL_877313, EPI_ISL_877314, EPI_ISL_877315, EPI_ISL_877316, EPI_ISL_877317, EPI_ISL_877318, EPI_ISL_877319, EPI_ISL_877320, EPI_ISL_877321, EPI_ISL_877322, EPI_ISL_877323, EPI_ISL_877324, EPI_ISL_877325, EPI_ISL_877326, EPI_ISL_877327, EPI_ISL_877328, EPI_ISL_877331, EPI_ISL_877332, EPI_ISL_877333, EPI_ISL_877334, EPI_ISL_877335, EPI_ISL_877336, EPI_ISL_877337, EPI_ISL_877338, EPI_ISL_877339, EPI_ISL_877340, EPI_ISL_877341, EPI_ISL_877342, EPI_ISL_877343, EPI_ISL_877344, EPI_ISL_877345, EPI_ISL_877346, EPI_ISL_877347, EPI_ISL_877348, EPI_ISL_877349, EPI_ISL_877350, EPI_ISL_877351, EPI_ISL_877352, EPI_ISL_877353, EPI_ISL_877354, EPI_ISL_877355, EPI_ISL_877356, EPI_ISL_877357, EPI_ISL_877358, EPI_ISL_877359, EPI_ISL_877360, EPI_ISL_877361, EPI_ISL_877362, EPI_ISL_877363, EPI_ISL_877364, EPI_ISL_877365, EPI_ISL_877366, EPI_ISL_877367, EPI_ISL_877368, EPI_ISL_877369, EPI_ISL_877370, EPI_ISL_877371, EPI_ISL_877372, EPI_ISL_877373, EPI_ISL_877374, EPI_ISL_877375, EPI_ISL_877376, EPI_ISL_877377, EPI_ISL_877378, EPI_ISL_877379, EPI_ISL_877380, EPI_ISL_877381, EPI_ISL_877382, EPI_ISL_877383, EPI_ISL_877384, EPI_ISL_877385, EPI_ISL_877386, EPI_ISL_877387, EPI_ISL_877388, EPI_ISL_877389, EPI_ISL_877390, EPI_ISL_877391, EPI_ISL_877392, EPI_ISL_877393, EPI_ISL_877394, EPI_ISL_877395, EPI_ISL_877396, EPI_ISL_877397, EPI_ISL_877398, EPI_ISL_877399, EPI_ISL_877400, EPI_ISL_877401, EPI_ISL_877402, EPI_ISL_877403, EPI_ISL_877404, EPI_ISL_877405, EPI_ISL_877406, EPI_ISL_877407, EPI_ISL_877408, EPI_ISL_877409, EPI_ISL_877410, EPI_ISL_877411, EPI_ISL_877413, EPI_ISL_877414, EPI_ISL_877415, EPI_ISL_877416, EPI_ISL_877417, EPI_ISL_877418, EPI_ISL_877461, EPI_ISL_877462, EPI_ISL_877463, EPI_ISL_877464, EPI_ISL_877465, EPI_ISL_877466, EPI_ISL_877467, EPI_ISL_877468, EPI_ISL_877469, EPI_ISL_877470, EPI_ISL_877471, EPI_ISL_877472, EPI_ISL_877473, EPI_ISL_877474, EPI_ISL_877475, EPI_ISL_877476, EPI_ISL_877477, EPI_ISL_877478, EPI_ISL_877479, EPI_ISL_877480, EPI_ISL_877481, EPI_ISL_877482, EPI_ISL_877483, EPI_ISL_877484, EPI_ISL_877485, EPI_ISL_877486, EPI_ISL_877487, EPI_ISL_877488, EPI_ISL_877489, EPI_ISL_877490, EPI_ISL_877491, EPI_ISL_877492, EPI_ISL_877493, EPI_ISL_877494, EPI_ISL_877495, EPI_ISL_877496, EPI_ISL_877497, EPI_ISL_877498, EPI_ISL_877499, EPI_ISL_877500, EPI_ISL_877501, EPI_ISL_877502, EPI_ISL_877503, EPI_ISL_877504, EPI_ISL_877505, EPI_ISL_877506, EPI_ISL_877507, EPI_ISL_877508, EPI_ISL_877509, EPI_ISL_877510, EPI_ISL_877511, EPI_ISL_877512, EPI_ISL_877513, EPI_ISL_877514, EPI_ISL_877515, EPI_ISL_877516, EPI_ISL_877517, EPI_ISL_877518, EPI_ISL_877519, EPI_ISL_877520, EPI_ISL_877521, EPI_ISL_877522, EPI_ISL_877523, EPI_ISL_877524, EPI_ISL_877525, EPI_ISL_877526, EPI_ISL_877527, EPI_ISL_877528, EPI_ISL_877529, EPI_ISL_877530, EPI_ISL_877531, EPI_ISL_877532, EPI_ISL_877533, EPI_ISL_877534, EPI_ISL_877535, EPI_ISL_877536, EPI_ISL_877537, EPI_ISL_877539, EPI_ISL_877540, EPI_ISL_877541, EPI_ISL_877542, EPI_ISL_877543, EPI_ISL_877544, EPI_ISL_877545, EPI_ISL_877546, EPI_ISL_877547, EPI_ISL_877548, EPI_ISL_877549, EPI_ISL_877550 | | | |
| see above | Eurofins Diatherix | Hudsonalpha Genome Sequencing Center | Jane Grimwood, Melissa Williams, Lori H. Handley, Joshua Stough, Leslie Malone, Stefan Brzezinski, Ada Stewart, Teresa Jones, Jenell Webber, John Lovell, Jennifer Cart, and Jeremy Schmutz |
| EPI_ISL_877585, EPI_ISL_877586, EPI_ISL_877587, EPI_ISL_877588, EPI_ISL_877589, EPI_ISL_877590, EPI_ISL_877591, EPI_ISL_877592, EPI_ISL_877593, EPI_ISL_877594, EPI_ISL_877595, EPI_ISL_877596, EPI_ISL_877597, EPI_ISL_877598, EPI_ISL_877599, EPI_ISL_877600, EPI_ISL_877601, EPI_ISL_877602, EPI_ISL_877603, EPI_ISL_877604, EPI_ISL_877605, EPI_ISL_877606, EPI_ISL_877607, EPI_ISL_877608, EPI_ISL_877609, EPI_ISL_877610, EPI_ISL_877611, EPI_ISL_877612, EPI_ISL_877613, EPI_ISL_877614, EPI_ISL_877615, EPI_ISL_877616, EPI_ISL_877617, EPI_ISL_877618, EPI_ISL_877619, EPI_ISL_877620, EPI_ISL_877621, EPI_ISL_877622, EPI_ISL_877623, EPI_ISL_877624, EPI_ISL_877625, EPI_ISL_877626, EPI_ISL_877627, EPI_ISL_877628, EPI_ISL_877629, EPI_ISL_877630, EPI_ISL_877631, EPI_ISL_877632, EPI_ISL_877633, EPI_ISL_877634, EPI_ISL_877635, EPI_ISL_877636, EPI_ISL_877637, EPI_ISL_877638, EPI_ISL_877639, EPI_ISL_877640, EPI_ISL_877641, EPI_ISL_877642, EPI_ISL_877643, EPI_ISL_877644, EPI_ISL_877645, EPI_ISL_877646, EPI_ISL_877647, EPI_ISL_877648, EPI_ISL_877649, EPI_ISL_877650, EPI_ISL_877651, EPI_ISL_877652, EPI_ISL_877653, EPI_ISL_877654, EPI_ISL_877655, EPI_ISL_877656, EPI_ISL_877657, EPI_ISL_877658, EPI_ISL_877659, EPI_ISL_877660, EPI_ISL_877661, EPI_ISL_877662, EPI_ISL_877663, EPI_ISL_877664, EPI_ISL_877665, EPI_ISL_877666, EPI_ISL_877667, EPI_ISL_877668, EPI_ISL_877669, EPI_ISL_877670, EPI_ISL_877671, EPI_ISL_877672, EPI_ISL_877673, EPI_ISL_877674, EPI_ISL_877675, EPI_ISL_877676, EPI_ISL_877677, EPI_ISL_877678, EPI_ISL_877679, EPI_ISL_877680, EPI_ISL_877681, EPI_ISL_877682, EPI_ISL_877683, EPI_ISL_877684, EPI_ISL_877685, EPI_ISL_877686, EPI_ISL_877687, EPI_ISL_877688, EPI_ISL_877689, EPI_ISL_877690, EPI_ISL_877691, EPI_ISL_877692, EPI_ISL_877693, EPI_ISL_877694, EPI_ISL_877695, EPI_ISL_877696, EPI_ISL_877697, EPI_ISL_877698, EPI_ISL_877699, EPI_ISL_877700, EPI_ISL_877701, EPI_ISL_877702, EPI_ISL_877703, EPI_ISL_877704, EPI_ISL_877705, EPI_ISL_877706, EPI_ISL_877707, EPI_ISL_877708, EPI_ISL_877709, EPI_ISL_877710, EPI_ISL_877711, EPI_ISL_877712, EPI_ISL_877713, EPI_ISL_877714, EPI_ISL_877715, EPI_ISL_877716, EPI_ISL_877717, EPI_ISL_877718, EPI_ISL_877719, EPI_ISL_877720, EPI_ISL_877721, EPI_ISL_877722, EPI_ISL_877723, EPI_ISL_877724, EPI_ISL_877725, EPI_ISL_877726, EPI_ISL_877727, EPI_ISL_877728, | | | |

| | | | |
|---|---|---|---|
| EPI_ISL_877729, EPI_ISL_877730, EPI_ISL_877731, EPI_ISL_877732, EPI_ISL_877733, EPI_ISL_877734, EPI_ISL_877735, EPI_ISL_877736, EPI_ISL_877737, EPI_ISL_877738, EPI_ISL_877739, EPI_ISL_877740, EPI_ISL_877741, EPI_ISL_877742, EPI_ISL_877743, EPI_ISL_877744, EPI_ISL_877745, EPI_ISL_877746, EPI_ISL_877747, EPI_ISL_877748, EPI_ISL_877749, EPI_ISL_877750, EPI_ISL_877751, EPI_ISL_877752, EPI_ISL_877753, EPI_ISL_877754, EPI_ISL_877755, EPI_ISL_877756, EPI_ISL_877757, EPI_ISL_877758, EPI_ISL_877759, EPI_ISL_877760, EPI_ISL_877761, EPI_ISL_877762, EPI_ISL_877763 | | | |
| see above | Clinical Molecular Microbiology Laboratory, UNC Hospital | Dirk Dittmer | Razia Moorad , Justin T. Landis , Brent A. Eason, Melissa B. Miller, Linda Pluta, Dirk Dittmer, Angelica Juarez, Cecilia Thompson , Cameroon Grant, Evelyn Hoffman, Patricio Cano, Jason Wong, Carolina Caro-Vegas, Blossom Damania. |
| EPI_ISL_878152, EPI_ISL_878155, EPI_ISL_878157, EPI_ISL_878159, EPI_ISL_878162, EPI_ISL_878165, EPI_ISL_878167, EPI_ISL_878170, EPI_ISL_878175, EPI_ISL_878178, EPI_ISL_878180, EPI_ISL_878183, EPI_ISL_878186, EPI_ISL_878189, EPI_ISL_878191, EPI_ISL_878194, EPI_ISL_878197, EPI_ISL_878199, EPI_ISL_878202, EPI_ISL_878205, EPI_ISL_878207, EPI_ISL_878209, EPI_ISL_878212, EPI_ISL_878215, EPI_ISL_878217, EPI_ISL_878221, EPI_ISL_878223, EPI_ISL_878226, EPI_ISL_878229, EPI_ISL_878232, EPI_ISL_878234, EPI_ISL_878237, EPI_ISL_878239, EPI_ISL_878242, EPI_ISL_878245, EPI_ISL_878248, EPI_ISL_878250, EPI_ISL_878253, EPI_ISL_878256, EPI_ISL_878258, EPI_ISL_878260, EPI_ISL_878263, EPI_ISL_878266, EPI_ISL_878268, EPI_ISL_878271, EPI_ISL_878274, EPI_ISL_878276, EPI_ISL_878279, EPI_ISL_878281, EPI_ISL_878284, EPI_ISL_878287, EPI_ISL_878290, EPI_ISL_878292, EPI_ISL_878295, EPI_ISL_878298, EPI_ISL_878300, EPI_ISL_878303, EPI_ISL_878305, EPI_ISL_878308, EPI_ISL_878311, EPI_ISL_878314, EPI_ISL_878316, EPI_ISL_878319, EPI_ISL_878322, EPI_ISL_878325, EPI_ISL_878327, EPI_ISL_878330, EPI_ISL_878333, EPI_ISL_878336, EPI_ISL_878338, EPI_ISL_878341, EPI_ISL_878342, EPI_ISL_878343, EPI_ISL_878344, EPI_ISL_878345, EPI_ISL_878346, EPI_ISL_878347, EPI_ISL_878348, EPI_ISL_878349, EPI_ISL_878350, EPI_ISL_878351, EPI_ISL_878352, EPI_ISL_878353, EPI_ISL_878354, EPI_ISL_878355, EPI_ISL_878356, EPI_ISL_878357, EPI_ISL_878358, EPI_ISL_878359, EPI_ISL_878360, EPI_ISL_878361, EPI_ISL_878362, EPI_ISL_878363, EPI_ISL_878364, EPI_ISL_878365, EPI_ISL_878366, EPI_ISL_878367, EPI_ISL_878368, EPI_ISL_878369, EPI_ISL_878370, EPI_ISL_878371, EPI_ISL_878372, EPI_ISL_878373, EPI_ISL_878374, EPI_ISL_878375, EPI_ISL_878376, EPI_ISL_878377, EPI_ISL_878378, EPI_ISL_878379, EPI_ISL_878380, EPI_ISL_878381, EPI_ISL_878382, EPI_ISL_878383, EPI_ISL_878384, EPI_ISL_878385, EPI_ISL_878386, EPI_ISL_878387, EPI_ISL_878388, EPI_ISL_878389, EPI_ISL_878390, EPI_ISL_878391, EPI_ISL_878392, EPI_ISL_878393, EPI_ISL_878394, EPI_ISL_878395, EPI_ISL_878396, EPI_ISL_878397, EPI_ISL_878398, EPI_ISL_878399, EPI_ISL_878400, EPI_ISL_878401, EPI_ISL_878402, EPI_ISL_878403, EPI_ISL_878404, EPI_ISL_878405, EPI_ISL_878406, EPI_ISL_878407, EPI_ISL_878408, EPI_ISL_878411, EPI_ISL_878413, EPI_ISL_878416, EPI_ISL_878418, EPI_ISL_878421, EPI_ISL_878423, EPI_ISL_878426, EPI_ISL_878429, EPI_ISL_878431, EPI_ISL_878434, EPI_ISL_878437 | | | |
| see above | San Diego County Public Health Laboratory | Andersen lab at Scripps Research | SEARCH Alliance San Diego with Tracy Basler, Jovan Shephard, Brett Austin |
| EPI_ISL_878529, EPI_ISL_878531, EPI_ISL_878534, EPI_ISL_878537, EPI_ISL_878539, EPI_ISL_878541, EPI_ISL_878544, EPI_ISL_878547, EPI_ISL_878549, EPI_ISL_878552, EPI_ISL_878554, EPI_ISL_878557, EPI_ISL_878560, EPI_ISL_878563, EPI_ISL_878565, EPI_ISL_878568 | | | |
| see above | Robert Garry lab | Andersen lab at Scripps Research | Allison Smither, Gilberto Sabino-Santos, Patricia Snarski, Lilia Melnik, Antoinette Bell, Kaylynn Genemaras, Arnaud Drouin, Dahlene Fusco, Robert Garry with SEARCH Alliance San Diego |
| EPI_ISL_878584, EPI_ISL_878587, EPI_ISL_878589, EPI_ISL_878592, EPI_ISL_878595, EPI_ISL_878597, EPI_ISL_878600, EPI_ISL_878603, EPI_ISL_878605, EPI_ISL_878608, EPI_ISL_878611, EPI_ISL_878613, EPI_ISL_878616, EPI_ISL_878619, EPI_ISL_878622, EPI_ISL_878624, EPI_ISL_878627, EPI_ISL_878629, EPI_ISL_878632, EPI_ISL_878635 | | | |
| see above | San Diego County Public Health Laboratory | Andersen lab at Scripps Research | SEARCH Alliance San Diego with Tracy Basler, Jovan Shephard, Brett Austin |
| EPI_ISL_878637, EPI_ISL_878640, EPI_ISL_878643, EPI_ISL_878645, EPI_ISL_878648, EPI_ISL_878651, EPI_ISL_878653, EPI_ISL_878656, EPI_ISL_878658, EPI_ISL_878661, EPI_ISL_878664, EPI_ISL_878666, EPI_ISL_878669, EPI_ISL_878671, EPI_ISL_878674, EPI_ISL_878677, EPI_ISL_878679, EPI_ISL_878682, EPI_ISL_878684, EPI_ISL_878687, EPI_ISL_878690, EPI_ISL_878693, EPI_ISL_878695, EPI_ISL_878697, EPI_ISL_878700, EPI_ISL_878702, EPI_ISL_878705, EPI_ISL_878707, EPI_ISL_878710, EPI_ISL_878713, EPI_ISL_878716, EPI_ISL_878719, EPI_ISL_878721, EPI_ISL_878724, EPI_ISL_878726, EPI_ISL_878729, EPI_ISL_878732, EPI_ISL_878734, EPI_ISL_878736, EPI_ISL_878739, EPI_ISL_878742, EPI_ISL_878744, EPI_ISL_878747, EPI_ISL_878750, EPI_ISL_878752, EPI_ISL_878755, EPI_ISL_878757, EPI_ISL_878760, EPI_ISL_878763, EPI_ISL_878765, EPI_ISL_878768, EPI_ISL_878771, EPI_ISL_878773, EPI_ISL_878776, EPI_ISL_878779, EPI_ISL_878781, EPI_ISL_878784, EPI_ISL_878786, EPI_ISL_878789, EPI_ISL_878791, EPI_ISL_878794, EPI_ISL_878797 | | | |
| see above | Rady's Childrens Hospital | Andersen lab at Scripps Research | SEARCH Alliance San Diego with Nanda Radamchar, David Dimmock, Linda Luo, Christina Clarke, Kathryn Bouic, Teresa Mueller, Denise Malicki |
| EPI_ISL_879924, EPI_ISL_879926, EPI_ISL_879929, EPI_ISL_879932, EPI_ISL_879934, EPI_ISL_879937, EPI_ISL_879939, EPI_ISL_879942, EPI_ISL_879945, EPI_ISL_879947, EPI_ISL_879950, EPI_ISL_879953, EPI_ISL_879956, EPI_ISL_879959, EPI_ISL_879961, EPI_ISL_879964, EPI_ISL_879967, EPI_ISL_879969, EPI_ISL_879972, EPI_ISL_879975, EPI_ISL_879977, EPI_ISL_879980, EPI_ISL_879982, EPI_ISL_879985, EPI_ISL_879987, EPI_ISL_879990, EPI_ISL_879993, EPI_ISL_879995, EPI_ISL_879998, EPI_ISL_880000, EPI_ISL_880003, EPI_ISL_880006, EPI_ISL_880009, EPI_ISL_880011, EPI_ISL_880014, EPI_ISL_880017, EPI_ISL_880019, EPI_ISL_880022, EPI_ISL_880024, EPI_ISL_880027, EPI_ISL_880029, EPI_ISL_880032, EPI_ISL_880034 | | | |
| see above | San Diego County Public Health Laboratory | Andersen lab at Scripps Research | SEARCH Alliance San Diego with Tracy Basler, Jovan Shephard, Brett Austin |
| EPI_ISL_880037, EPI_ISL_880040, EPI_ISL_880042, EPI_ISL_880045, EPI_ISL_880048, EPI_ISL_880050, EPI_ISL_880053, EPI_ISL_880056, EPI_ISL_880058, EPI_ISL_880061, EPI_ISL_880063, EPI_ISL_880066, EPI_ISL_880069, EPI_ISL_880071, EPI_ISL_880074, EPI_ISL_880077, EPI_ISL_880079, EPI_ISL_880082, EPI_ISL_880085, EPI_ISL_880087, EPI_ISL_880090, EPI_ISL_880092, EPI_ISL_880095, EPI_ISL_880098, EPI_ISL_880101, EPI_ISL_880103, EPI_ISL_880106, EPI_ISL_880109, EPI_ISL_880111, EPI_ISL_880114, EPI_ISL_880117, EPI_ISL_880119, EPI_ISL_880122, EPI_ISL_880125, EPI_ISL_880128, EPI_ISL_880130, EPI_ISL_880133, EPI_ISL_880135, EPI_ISL_880138, EPI_ISL_880140, EPI_ISL_880141, EPI_ISL_880142, EPI_ISL_880143, EPI_ISL_880144, EPI_ISL_880145, EPI_ISL_880146, EPI_ISL_880147, EPI_ISL_880148, EPI_ISL_880149, EPI_ISL_880150, EPI_ISL_880151, EPI_ISL_880152, EPI_ISL_880153, EPI_ISL_880154, EPI_ISL_880155, EPI_ISL_880156, EPI_ISL_880157, EPI_ISL_880158, EPI_ISL_880159, EPI_ISL_880160, EPI_ISL_880161, EPI_ISL_880162, EPI_ISL_880163, EPI_ISL_880164, EPI_ISL_880165, EPI_ISL_880166, EPI_ISL_880167, EPI_ISL_880168, EPI_ISL_880169, EPI_ISL_880170 | | | |
| see above | Sharp HealthCare Laboratory | Andersen lab at Scripps Research | SEARCH Alliance San Diego with Aaron Harding, Jacquelyn Berumen, Cathy Woerle, Liam McGinnis, Art Mendoza, Omid Bakhtar |
| EPI_ISL_880171, EPI_ISL_880172, EPI_ISL_880173, EPI_ISL_880174, EPI_ISL_880175, EPI_ISL_880176, EPI_ISL_880177, EPI_ISL_880178, EPI_ISL_880179, EPI_ISL_880180, EPI_ISL_880181, EPI_ISL_880182, EPI_ISL_880183, EPI_ISL_880184, EPI_ISL_880185, EPI_ISL_880186, EPI_ISL_880187, EPI_ISL_880188, EPI_ISL_880189, EPI_ISL_880190, EPI_ISL_880191, EPI_ISL_880192, EPI_ISL_880193, EPI_ISL_880194, EPI_ISL_880195, EPI_ISL_880196, EPI_ISL_880197, EPI_ISL_880198, EPI_ISL_880199, EPI_ISL_880200 | | | |
| see above | Rady's Childrens Hospital | Andersen lab at Scripps Research | SEARCH Alliance San Diego with Nanda Radamchar, David Dimmock, Linda Luo, Christina Clarke, Kathryn Bouic, Teresa Mueller, Denise Malicki |
| EPI_ISL_880201, EPI_ISL_880202, EPI_ISL_880203, EPI_ISL_880204, EPI_ISL_880205, EPI_ISL_880206, EPI_ISL_880207, EPI_ISL_880208, EPI_ISL_880209, EPI_ISL_880212, EPI_ISL_880215, EPI_ISL_880218, EPI_ISL_880220, EPI_ISL_880223, EPI_ISL_880225, EPI_ISL_880228, EPI_ISL_880231, EPI_ISL_880233, EPI_ISL_880236, EPI_ISL_880239, EPI_ISL_880241, EPI_ISL_880244, EPI_ISL_880246, EPI_ISL_880249, EPI_ISL_880251, EPI_ISL_880253, EPI_ISL_880256, EPI_ISL_880259, EPI_ISL_880261, EPI_ISL_880263, EPI_ISL_880266, EPI_ISL_880268, EPI_ISL_880271, EPI_ISL_880274, EPI_ISL_880277, EPI_ISL_880279, EPI_ISL_880282, EPI_ISL_880284, EPI_ISL_880287, EPI_ISL_880290, EPI_ISL_880292, EPI_ISL_880294, EPI_ISL_880297, EPI_ISL_880300, EPI_ISL_880303, EPI_ISL_880305, EPI_ISL_880308, EPI_ISL_880310, EPI_ISL_880313, EPI_ISL_880316, EPI_ISL_880319, EPI_ISL_880321, EPI_ISL_880324, EPI_ISL_880326, EPI_ISL_880329 | | | |
| see above | Scripps Medical Laboratory | Andersen lab at Scripps Research | SEARCH Alliance San Diego with Michael Quigley, Ellen Stefanski, Ian Mchardy |
| EPI_ISL_882647, EPI_ISL_882650 | Ohio Department of Health Laboratory | Ohio Department of Health Laboratory | Holmes, Jennifer; Eric Brandt, Keoni Omura, Glen McGillivary, Caitlin McDonnell, Kirtana Ramadugu, Erica Leasure, Kelsey Florek, Heather Blankenship, Quanta Brown, and Tammy Bannerman |
| EPI_ISL_882805 | MSHS Clinical Microbiology Laboratories | MSHS Pathogen Surveillance Program | Ana S. Gonzalez-Reiche, Hala Alshammary, Mitchell J. Sullivan, Brianne Ciferri, Ajay Obla, Angela Amoako, Mahmoud Awawda, Elena Hirsch, Ashley S. Salimbangon, Levy Sominsky, Katherine Beach, Kayla Russo, Charles Gleason, Shelcie Fabre, Giulio Kleiner, Zenab Khan, Bremy Alburquerque, Adriana van de Guchte, Komal Srivastava, Matthew M. Hernandez, Jayeeta Dutta, Denise Jurczyszak, Emily Ferreri, Rachel Chernet, Nancy Francoeur, Betsaida Salom Melo, Irina Oussenko, Gintaras Deikus, Juan Soto, Shwetha Hara Sridhar, Ying-Chih Wang, Kathryn Twyman, Andrew Kasarskis, Deena R. Altman, Robert Sebra, Adolfo García-Sastre, Marta Luksza, Gopi Patel, Sarah Schaefer, Melissa Gitman, Michael D. Nowak, Alberto Paniz-Mondolfi, Emilia Mia Sordillo, Viviana Simon, Harm van Bakel |
| EPI_ISL_882963, EPI_ISL_882964, EPI_ISL_882965, EPI_ISL_882966 | MD Laboratories | Los Angeles County Public Health Laboratories | P. Hemarajata et al. |
| EPI_ISL_882967, EPI_ISL_882968, EPI_ISL_882969, EPI_ISL_882970, EPI_ISL_882971, EPI_ISL_882972, EPI_ISL_882973, EPI_ISL_882974, EPI_ISL_882975, EPI_ISL_882976, EPI_ISL_882977, EPI_ISL_882978, EPI_ISL_882979, EPI_ISL_882980, EPI_ISL_882981, EPI_ISL_882982, EPI_ISL_882983, EPI_ISL_882984 | | | |
| see above | UCLA Clinical Micro Lab | Los Angeles County Public Health Laboratories | P. Hemarajata et al. |
| EPI_ISL_882985, EPI_ISL_882986, EPI_ISL_882987, EPI_ISL_882988, EPI_ISL_882989, EPI_ISL_882990, EPI_ISL_882991, EPI_ISL_882992, EPI_ISL_882993, EPI_ISL_882994, EPI_ISL_882995, EPI_ISL_882996 | | | |
| see above | MD Laboratories | Los Angeles County Public Health Laboratories | P. Hemarajata et al. |
| EPI_ISL_882997, EPI_ISL_882998 | Motion Picture Television Fund Hospital Lab | Los Angeles County Public Health Laboratories | P. Hemarajata et al. |
| EPI_ISL_882999, EPI_ISL_883000, EPI_ISL_883001, EPI_ISL_883002, EPI_ISL_883003, EPI_ISL_883004, EPI_ISL_883005, EPI_ISL_883006, EPI_ISL_883007, EPI_ISL_883008, EPI_ISL_883009, EPI_ISL_883010, EPI_ISL_883011, EPI_ISL_883012, EPI_ISL_883013, EPI_ISL_883014, EPI_ISL_883015, EPI_ISL_883016, EPI_ISL_883017, EPI_ISL_883018, EPI_ISL_883019, EPI_ISL_883020, EPI_ISL_883021, EPI_ISL_883022, EPI_ISL_883023, EPI_ISL_883024, EPI_ISL_883025, EPI_ISL_883026, EPI_ISL_883027, EPI_ISL_883028, EPI_ISL_883029, EPI_ISL_883030, EPI_ISL_883031, EPI_ISL_883032, EPI_ISL_883033, EPI_ISL_883034, EPI_ISL_883035 | | | |
| see above | Maryland Public Health Laboratory | Maryland Public Health Laboratory | Maryland Department of Health Laboratories Administration |
| EPI_ISL_883037, EPI_ISL_883038, EPI_ISL_883039, EPI_ISL_883040, EPI_ISL_883041, EPI_ISL_883042, EPI_ISL_883043, EPI_ISL_883044, EPI_ISL_883045, EPI_ISL_883046, EPI_ISL_883047, EPI_ISL_883048, EPI_ISL_883049, EPI_ISL_883050, EPI_ISL_883051, EPI_ISL_883052, EPI_ISL_883053, EPI_ISL_883054, EPI_ISL_883055, EPI_ISL_883056, EPI_ISL_883057, EPI_ISL_883058, EPI_ISL_883059, EPI_ISL_883060, EPI_ISL_883061, EPI_ISL_883062, EPI_ISL_883063, EPI_ISL_883064, EPI_ISL_883065 | | | |
| see above | Kansas Health and Environmental Lab | Kansas Health and Environmental Lab | Mike Grose, Carissa Robertson, Ben Olsen, and Phil Adam |
| EPI_ISL_883066, EPI_ISL_883067, EPI_ISL_883068, EPI_ISL_883069, EPI_ISL_883070, EPI_ISL_883071, EPI_ISL_883072, EPI_ISL_883073, EPI_ISL_883074, EPI_ISL_883075, EPI_ISL_883076, EPI_ISL_883077, EPI_ISL_883078, EPI_ISL_883079, EPI_ISL_883080, EPI_ISL_883081, EPI_ISL_883082, EPI_ISL_883083, EPI_ISL_883084, EPI_ISL_883085, EPI_ISL_883086, EPI_ISL_883087, EPI_ISL_883088, EPI_ISL_883089, EPI_ISL_883090, EPI_ISL_883091, EPI_ISL_883092, EPI_ISL_883093, EPI_ISL_883094, EPI_ISL_883095, EPI_ISL_883096, EPI_ISL_883097, EPI_ISL_883098, EPI_ISL_883099, EPI_ISL_883100, EPI_ISL_883101, EPI_ISL_883102, EPI_ISL_883103, EPI_ISL_883104, EPI_ISL_883105, EPI_ISL_883106, EPI_ISL_883107, EPI_ISL_883108, EPI_ISL_883109, EPI_ISL_883110, EPI_ISL_883111, EPI_ISL_883112, EPI_ISL_883113, EPI_ISL_883114, EPI_ISL_883115, EPI_ISL_883116, EPI_ISL_883117, EPI_ISL_883118, EPI_ISL_883119, EPI_ISL_883120, EPI_ISL_883121, EPI_ISL_883122, EPI_ISL_883123, EPI_ISL_883124, EPI_ISL_883125, EPI_ISL_883126, EPI_ISL_883127, EPI_ISL_883128, EPI_ISL_883129, EPI_ISL_883130, EPI_ISL_883131, EPI_ISL_883132, EPI_ISL_883133, EPI_ISL_883134, EPI_ISL_883135, EPI_ISL_883136, EPI_ISL_883137, EPI_ISL_883138, EPI_ISL_883139, EPI_ISL_883140, EPI_ISL_883141, EPI_ISL_883142, EPI_ISL_883143, EPI_ISL_883144, EPI_ISL_883145, EPI_ISL_883146, EPI_ISL_883147, EPI_ISL_883148, EPI_ISL_883149, EPI_ISL_883150, EPI_ISL_883151, EPI_ISL_883152, EPI_ISL_883153, EPI_ISL_883154, EPI_ISL_883195, EPI_ISL_883197, EPI_ISL_883198, EPI_ISL_883199, EPI_ISL_883200, EPI_ISL_883201, EPI_ISL_883202, EPI_ISL_883203, EPI_ISL_883204, EPI_ISL_883205, EPI_ISL_883206, EPI_ISL_883207, EPI_ISL_883208, EPI_ISL_883209, EPI_ISL_883210, EPI_ISL_883211, EPI_ISL_883212, EPI_ISL_883213, EPI_ISL_883214, EPI_ISL_883215, EPI_ISL_883216, EPI_ISL_883217, EPI_ISL_883218, EPI_ISL_883219, EPI_ISL_883220, EPI_ISL_883221, EPI_ISL_883222, EPI_ISL_883223, EPI_ISL_883224, EPI_ISL_883225, EPI_ISL_883226, EPI_ISL_883227, EPI_ISL_883228, EPI_ISL_883229, EPI_ISL_883230, EPI_ISL_883231, EPI_ISL_883232, EPI_ISL_883233, EPI_ISL_883234, EPI_ISL_883235, EPI_ISL_883236, EPI_ISL_883237, EPI_ISL_883238, EPI_ISL_883239, EPI_ISL_883240, EPI_ISL_883241, EPI_ISL_883242, EPI_ISL_883243, EPI_ISL_883244, EPI_ISL_883245, EPI_ISL_883246, EPI_ISL_883247, EPI_ISL_883248, EPI_ISL_883249, EPI_ISL_883250, EPI_ISL_883251, EPI_ISL_883252, EPI_ISL_883253, EPI_ISL_883254, EPI_ISL_883255, EPI_ISL_883256, EPI_ISL_883257, EPI_ISL_883258, EPI_ISL_883259, EPI_ISL_883260, EPI_ISL_883261, EPI_ISL_883262, EPI_ISL_883263, EPI_ISL_883264, EPI_ISL_883265, EPI_ISL_883266, EPI_ISL_883267, EPI_ISL_883268, EPI_ISL_883269, EPI_ISL_883270, EPI_ISL_883271, EPI_ISL_883272, EPI_ISL_883273, EPI_ISL_883274, EPI_ISL_883275, EPI_ISL_883276, EPI_ISL_883277, EPI_ISL_883278, EPI_ISL_883279, EPI_ISL_883280, EPI_ISL_883281, EPI_ISL_883282, EPI_ISL_883283, EPI_ISL_883284 | | | |
| see above | Eurofins Diatherix | Hudsonalpha Genome Sequencing Center | Jane Grimwood, Melissa Williams, Lori H. Handley, Joshua Stough, Leslie Malone, Stefan Brzezinski, Ada Stewart, Teresa Jones, Jenell Webber, John Lovell, Jennifer Cart, and Jeremy Schmutz |
| EPI_ISL_883307, EPI_ISL_883308 | Pandemic Response Laboratory | New York City Public Health Laboratory | Jade Wang, et al. |
| EPI_ISL_883309 | DOHMH Central Harlem | New York City Public Health Laboratory | Jade Wang, et al. |
| EPI_ISL_883310 | DOHMH Jamaica | New York City Public Health Laboratory | Jade Wang, et al. |
| EPI_ISL_883311 | Pandemic Response Laboratory | New York City Public Health Laboratory | Jade Wang, et al. |
| EPI_ISL_883312, EPI_ISL_883313 | DOHMH Corona | New York City Public Health Laboratory | Jade Wang, et al. |
| EPI_ISL_883314 | DOHMH Jamaica | New York City Public Health Laboratory | Jade Wang, et al. |
| EPI_ISL_883315, EPI_ISL_883316 | OCME Office Of Chief Medical Examiner | New York City Public Health Laboratory | Jade Wang, et al. |
| EPI_ISL_883317 | Pandemic Response Laboratory | New York City Public Health Laboratory | Jade Wang, et al. |
| EPI_ISL_883318, EPI_ISL_883319, EPI_ISL_883320, EPI_ISL_883321, EPI_ISL_883322, EPI_ISL_883323 | DOHMH Jamaica | New York City Public Health Laboratory | Jade Wang, et al. |
| EPI_ISL_883324 | DOHMH Central Harlem | New York City Public Health Laboratory | Jade Wang, et al. |
| EPI_ISL_883325 | OCME Office Of Chief Medical Examiner | New York City Public Health Laboratory | Jade Wang, et al. |
| EPI_ISL_883326 | Department of Homeless Services | New York City Public Health Laboratory | Jade Wang, et al. |
| EPI_ISL_883327 | DOHMH Central Harlem | New York City Public Health Laboratory | Jade Wang, et al. |

| | | | |
|---|---|---|---|
| EPI_ISL_883328 | OCME Office Of Chief Medical Examiner | New York City Public Health Laboratory | Jade Wang, et al. |
| EPI_ISL_883329 | DOHMH Central Harlem | New York City Public Health Laboratory | Jade Wang, et al. |
| EPI_ISL_883330 | OCME Office Of Chief Medical Examiner | New York City Public Health Laboratory | Jade Wang, et al. |
| EPI_ISL_883331 | DOHMH Morrisania | New York City Public Health Laboratory | Jade Wang, et al. |
| EPI_ISL_883332 | Department of Homeless Services | New York City Public Health Laboratory | Jade Wang, et al. |
| EPI_ISL_883333, EPI_ISL_883334 | DOHMH Riverside | New York City Public Health Laboratory | Jade Wang, et al. |
| EPI_ISL_883335 | DOHMH Chelsea | New York City Public Health Laboratory | Jade Wang, et al. |
| EPI_ISL_883336 | OCME Office Of Chief Medical Examiner | New York City Public Health Laboratory | Jade Wang, et al. |
| EPI_ISL_883337 | DOHMH Crown Heights | New York City Public Health Laboratory | Jade Wang, et al. |
| EPI_ISL_883338 | DOHMH Morrisania | New York City Public Health Laboratory | Jade Wang, et al. |
| EPI_ISL_883339 | DOHMH Chelsea | New York City Public Health Laboratory | Jade Wang, et al. |
| EPI_ISL_883340, EPI_ISL_883341, EPI_ISL_883342, EPI_ISL_883343 | DOHMH PHL | New York City Public Health Laboratory | Jade Wang, et al. |
| EPI_ISL_883344, EPI_ISL_883345 | OCME Office Of Chief Medical Examiner | New York City Public Health Laboratory | Jade Wang, et al. |
| EPI_ISL_883346 | DOHMH PHL | New York City Public Health Laboratory | Jade Wang, et al. |
| EPI_ISL_883347 | DOHMH Morrisania | New York City Public Health Laboratory | Jade Wang, et al. |
| EPI_ISL_883348 | DOHMH Corona | New York City Public Health Laboratory | Jade Wang, et al. |
| EPI_ISL_883349 | DOHMH Jamaica | New York City Public Health Laboratory | Jade Wang, et al. |
| EPI_ISL_883350, EPI_ISL_883351, EPI_ISL_883352, EPI_ISL_883353 | OCME Office Of Chief Medical Examiner | New York City Public Health Laboratory | Jade Wang, et al. |
| EPI_ISL_883354, EPI_ISL_883355, EPI_ISL_883356 | DOHMH Corona | New York City Public Health Laboratory | Jade Wang, et al. |
| EPI_ISL_883357, EPI_ISL_883358, EPI_ISL_883359, EPI_ISL_883360 | DOHMH Crown Heights | New York City Public Health Laboratory | Jade Wang, et al. |
| EPI_ISL_883361, EPI_ISL_883362, EPI_ISL_883363 | DOHMH PHL | New York City Public Health Laboratory | Jade Wang, et al. |
| EPI_ISL_883364 | DOHMH Crown Heights | New York City Public Health Laboratory | Jade Wang, et al. |
| EPI_ISL_883365, EPI_ISL_883366, EPI_ISL_883367 | DOHMH Morrisania | New York City Public Health Laboratory | Jade Wang, et al. |
| EPI_ISL_883368, EPI_ISL_883369 | DOHMH Chelsea | New York City Public Health Laboratory | Jade Wang, et al. |
| EPI_ISL_883370 | DOHMH Crown Heights | New York City Public Health Laboratory | Jade Wang, et al. |
| EPI_ISL_883371, EPI_ISL_883372 | DOHMH Central Harlem | New York City Public Health Laboratory | Jade Wang, et al. |
| EPI_ISL_883373, EPI_ISL_883374, EPI_ISL_883375 | DOHMH Jamaica | New York City Public Health Laboratory | Jade Wang, et al. |
| EPI_ISL_883376, EPI_ISL_883377, EPI_ISL_883378, EPI_ISL_883379 | DOHMH Central Harlem | New York City Public Health Laboratory | Jade Wang, et al. |
| EPI_ISL_883380, EPI_ISL_883381, EPI_ISL_883382 | DOHMH PHL | New York City Public Health Laboratory | Jade Wang, et al. |
| EPI_ISL_883383, EPI_ISL_883384, EPI_ISL_883385, EPI_ISL_883386, EPI_ISL_883387, EPI_ISL_883388, EPI_ISL_883389 | Pandemic Response Laboratory | New York City Public Health Laboratory | Jade Wang, et al. |
| EPI_ISL_883390, EPI_ISL_883391, EPI_ISL_883392, EPI_ISL_883393, EPI_ISL_883394 | OCME Office Of Chief Medical Examiner | New York City Public Health Laboratory | Jade Wang, et al. |
| EPI_ISL_883395, EPI_ISL_883396 | Department of Homeless Services | New York City Public Health Laboratory | Jade Wang, et al. |
| EPI_ISL_883397, EPI_ISL_883398, EPI_ISL_883399, EPI_ISL_883400, EPI_ISL_883401, EPI_ISL_883402 | OCME Office Of Chief Medical Examiner | New York City Public Health Laboratory | Jade Wang, et al. |
| EPI_ISL_883403 | Department of Homeless Services | New York City Public Health Laboratory | Jade Wang, et al. |
| EPI_ISL_883404, EPI_ISL_883405, EPI_ISL_883406, EPI_ISL_883407, EPI_ISL_883408 | OCME Office Of Chief Medical Examiner | New York City Public Health Laboratory | Jade Wang, et al. |
| EPI_ISL_883409 | DOHMH Central Harlem | New York City Public Health Laboratory | Jade Wang, et al. |
| EPI_ISL_883410, EPI_ISL_883411 | DOHMH Corona | New York City Public Health Laboratory | Jade Wang, et al. |
| EPI_ISL_883412 | DOHMH Crown Heights | New York City Public Health Laboratory | Jade Wang, et al. |
| EPI_ISL_883413, EPI_ISL_883414, EPI_ISL_883415 | DOHMH Central Harlem | New York City Public Health Laboratory | Jade Wang, et al. |
| EPI_ISL_883416, EPI_ISL_883417 | DOHMH Fort Greene | New York City Public Health Laboratory | Jade Wang, et al. |
| EPI_ISL_883418, EPI_ISL_883419, | DOHMH Jamaica | New York City Public Health Laboratory | Jade Wang, et al. |

| | | | |
|---|---|---|---|
| EPI_ISL_883420, EPI_ISL_883421, EPI_ISL_883422, EPI_ISL_883423 | | | |
| EPI_ISL_883424 | DOHMH PHL | New York City Public Health Laboratory | Jade Wang, et al. |
| EPI_ISL_883425 | BOSTON HEART DIAGNOSTICS CORP | Wadsworth Center, New York State Department of Health | Kirsten St. George, Daryl M. Lamson, Alexis Russel, Matthew Shudt, Melissa A Leisner, Jonathan Plitnick, Navjot Singh, John Kelly, Erasmus Schneider, Erica Lasek-Nesselquist |
| EPI_ISL_883426, EPI_ISL_883427, EPI_ISL_883428 | NORTHWELL HEALTH LABORATORIES | Wadsworth Center, New York State Department of Health | Kirsten St. George, Daryl M. Lamson, Alexis Russel, Matthew Shudt, Melissa A Leisner, Jonathan Plitnick, Navjot Singh, John Kelly, Erasmus Schneider, Erica Lasek-Nesselquist |
| EPI_ISL_883429, EPI_ISL_883430 | BOSTON HEART DIAGNOSTICS CORP | Wadsworth Center, New York State Department of Health | Kirsten St. George, Daryl M. Lamson, Alexis Russel, Matthew Shudt, Melissa A Leisner, Jonathan Plitnick, Navjot Singh, John Kelly, Erasmus Schneider, Erica Lasek-Nesselquist |
| EPI_ISL_883431, EPI_ISL_883432, EPI_ISL_883433, EPI_ISL_883434, EPI_ISL_883435 | NORTHWELL HEALTH LABORATORIES | Wadsworth Center, New York State Department of Health | Kirsten St. George, Daryl M. Lamson, Alexis Russel, Matthew Shudt, Melissa A Leisner, Jonathan Plitnick, Navjot Singh, John Kelly, Erasmus Schneider, Erica Lasek-Nesselquist |
| EPI_ISL_883436, EPI_ISL_883437, EPI_ISL_883438, EPI_ISL_883439, EPI_ISL_883440, EPI_ISL_883441, EPI_ISL_883442, EPI_ISL_883443, EPI_ISL_883444 | ADIRONDACK MEDICAL CENTER | Wadsworth Center, New York State Department of Health | Kirsten St. George, Daryl M. Lamson, Alexis Russel, Matthew Shudt, Melissa A Leisner, Jonathan Plitnick, Navjot Singh, John Kelly, Erasmus Schneider, Erica Lasek-Nesselquist |
| EPI_ISL_883445, EPI_ISL_883446, EPI_ISL_883447, EPI_ISL_883448, EPI_ISL_883449, EPI_ISL_883450, EPI_ISL_883451, EPI_ISL_883452, EPI_ISL_883453, EPI_ISL_883454, EPI_ISL_883455, EPI_ISL_883456, EPI_ISL_883457, EPI_ISL_883458, EPI_ISL_883459, EPI_ISL_883460, EPI_ISL_883461, EPI_ISL_883462, EPI_ISL_883463, EPI_ISL_883464, EPI_ISL_883465, EPI_ISL_883466, EPI_ISL_883467, EPI_ISL_883468, EPI_ISL_883469, EPI_ISL_883470, EPI_ISL_883471, EPI_ISL_883472 | | | |
| see above | NORTHWELL HEALTH LABORATORIES | Wadsworth Center, New York State Department of Health | Kirsten St. George, Daryl M. Lamson, Alexis Russel, Matthew Shudt, Melissa A Leisner, Jonathan Plitnick, Navjot Singh, John Kelly, Erasmus Schneider, Erica Lasek-Nesselquist |
| EPI_ISL_883473 | ADIRONDACK MEDICAL CENTER | Wadsworth Center, New York State Department of Health | Kirsten St. George, Daryl M. Lamson, Alexis Russel, Matthew Shudt, Melissa A Leisner, Jonathan Plitnick, Navjot Singh, John Kelly, Erasmus Schneider, Erica Lasek-Nesselquist |
| EPI_ISL_883474, EPI_ISL_883475, EPI_ISL_883476, EPI_ISL_883477, EPI_ISL_883478, EPI_ISL_883479, EPI_ISL_883480, EPI_ISL_883481, EPI_ISL_883482, EPI_ISL_883483, EPI_ISL_883484, EPI_ISL_883485, EPI_ISL_883486, EPI_ISL_883487, EPI_ISL_883488, EPI_ISL_883489, EPI_ISL_883490, EPI_ISL_883491, EPI_ISL_883492, EPI_ISL_883493, EPI_ISL_883494, EPI_ISL_883495, EPI_ISL_883496, EPI_ISL_883497, EPI_ISL_883498 | | | |
| see above | NORTHWELL HEALTH LABORATORIES | Wadsworth Center, New York State Department of Health | Kirsten St. George, Daryl M. Lamson, Alexis Russel, Matthew Shudt, Melissa A Leisner, Jonathan Plitnick, Navjot Singh, John Kelly, Erasmus Schneider, Erica Lasek-Nesselquist |
| EPI_ISL_883501, EPI_ISL_883502, EPI_ISL_883503, EPI_ISL_883504, EPI_ISL_883505, EPI_ISL_883506, EPI_ISL_883507, EPI_ISL_883508, EPI_ISL_883509, EPI_ISL_883510, EPI_ISL_883511, EPI_ISL_883512, EPI_ISL_883513, EPI_ISL_883514, EPI_ISL_883515, EPI_ISL_883516, EPI_ISL_883517, EPI_ISL_883518, EPI_ISL_883519, EPI_ISL_883520, EPI_ISL_883521, EPI_ISL_883522, EPI_ISL_883523, EPI_ISL_883524, EPI_ISL_883525, EPI_ISL_883526, EPI_ISL_883527, EPI_ISL_883528, EPI_ISL_883529, EPI_ISL_883530, EPI_ISL_883531, EPI_ISL_883532, EPI_ISL_883533, EPI_ISL_883534, EPI_ISL_883535, EPI_ISL_883536, EPI_ISL_883537, EPI_ISL_883538, EPI_ISL_883539, EPI_ISL_883540, EPI_ISL_883541, EPI_ISL_883542, EPI_ISL_883543, EPI_ISL_883544, EPI_ISL_883545, EPI_ISL_883546, EPI_ISL_883547, EPI_ISL_883548, EPI_ISL_883549, EPI_ISL_883550, EPI_ISL_883551, EPI_ISL_883552, EPI_ISL_883553, EPI_ISL_883554, EPI_ISL_883555, EPI_ISL_883556, EPI_ISL_883557, EPI_ISL_883558, EPI_ISL_883559, EPI_ISL_883560, EPI_ISL_883561, EPI_ISL_883562, EPI_ISL_883563, EPI_ISL_883564, EPI_ISL_883565, EPI_ISL_883566, EPI_ISL_883567, EPI_ISL_883568, EPI_ISL_883569, EPI_ISL_883570, EPI_ISL_883571, EPI_ISL_883572, EPI_ISL_883573, EPI_ISL_883574, EPI_ISL_883575, EPI_ISL_883576, EPI_ISL_883577, EPI_ISL_883578, EPI_ISL_883579, EPI_ISL_883580, EPI_ISL_883581, EPI_ISL_883582, EPI_ISL_883583, EPI_ISL_883584, EPI_ISL_883585, EPI_ISL_883586, EPI_ISL_883587, EPI_ISL_883588, EPI_ISL_883589, EPI_ISL_883590, EPI_ISL_883591, EPI_ISL_883593, EPI_ISL_883594, EPI_ISL_883595, EPI_ISL_883596, EPI_ISL_883597, EPI_ISL_883598, EPI_ISL_883599, EPI_ISL_883600, EPI_ISL_883601, EPI_ISL_883602, EPI_ISL_883603, EPI_ISL_883604, EPI_ISL_883605, EPI_ISL_883606, EPI_ISL_883607, EPI_ISL_883608, EPI_ISL_883609, EPI_ISL_883610, EPI_ISL_883611, EPI_ISL_883612, EPI_ISL_883613, EPI_ISL_883614, EPI_ISL_883615, EPI_ISL_883616, EPI_ISL_883617, EPI_ISL_883618, EPI_ISL_883619, EPI_ISL_883620, EPI_ISL_883621, EPI_ISL_883622, EPI_ISL_883623, EPI_ISL_883624, EPI_ISL_883625, EPI_ISL_883626, EPI_ISL_883627, EPI_ISL_883628, EPI_ISL_883629, EPI_ISL_883630, EPI_ISL_883631, EPI_ISL_883632, EPI_ISL_883633, EPI_ISL_883634, EPI_ISL_883635, EPI_ISL_883636, EPI_ISL_883637, EPI_ISL_883638, EPI_ISL_883639, EPI_ISL_883640, EPI_ISL_883641, EPI_ISL_883642, EPI_ISL_883643, EPI_ISL_883644, EPI_ISL_883645, EPI_ISL_883646, EPI_ISL_883647, EPI_ISL_883648, EPI_ISL_883649, EPI_ISL_883650, EPI_ISL_883651, EPI_ISL_883652, EPI_ISL_883653, EPI_ISL_883654, EPI_ISL_883655, EPI_ISL_883656, EPI_ISL_883657, EPI_ISL_883658, EPI_ISL_883659, EPI_ISL_883660, EPI_ISL_883661, EPI_ISL_883662, EPI_ISL_883663, EPI_ISL_883664, EPI_ISL_883665, EPI_ISL_883666, EPI_ISL_883667, EPI_ISL_883668, EPI_ISL_883669, EPI_ISL_883670, EPI_ISL_883671, EPI_ISL_883672, EPI_ISL_883673, EPI_ISL_883674, EPI_ISL_883675, EPI_ISL_883676, EPI_ISL_883677, EPI_ISL_883678, EPI_ISL_883679, EPI_ISL_883680, EPI_ISL_883681, EPI_ISL_883682, EPI_ISL_883683, EPI_ISL_883684, EPI_ISL_883685, EPI_ISL_883686, EPI_ISL_883687, EPI_ISL_883688, EPI_ISL_883689, EPI_ISL_883690, EPI_ISL_883691, EPI_ISL_883692, EPI_ISL_883693, EPI_ISL_883694, EPI_ISL_883695, EPI_ISL_883696, EPI_ISL_883697, EPI_ISL_883698, EPI_ISL_883699, EPI_ISL_883700, EPI_ISL_883701, EPI_ISL_883702, EPI_ISL_883703, EPI_ISL_883704, EPI_ISL_883705, EPI_ISL_883706, EPI_ISL_883707, EPI_ISL_883708, EPI_ISL_883709, EPI_ISL_883710, EPI_ISL_883711, EPI_ISL_883712, EPI_ISL_883713, EPI_ISL_883714, EPI_ISL_883715, EPI_ISL_883716, EPI_ISL_883717, EPI_ISL_883718, EPI_ISL_883719, EPI_ISL_883720, EPI_ISL_883721, EPI_ISL_883722, EPI_ISL_883723, EPI_ISL_883724, EPI_ISL_883725, EPI_ISL_883726, EPI_ISL_883727, EPI_ISL_883728, EPI_ISL_883729, EPI_ISL_883730, EPI_ISL_883731, EPI_ISL_883732, EPI_ISL_883733, EPI_ISL_883734, EPI_ISL_883735, EPI_ISL_883736, EPI_ISL_883737, EPI_ISL_883738, EPI_ISL_883739, EPI_ISL_883740, EPI_ISL_883741, EPI_ISL_883742, EPI_ISL_883743, EPI_ISL_883744, EPI_ISL_883745, EPI_ISL_883746, EPI_ISL_883747, EPI_ISL_883748, EPI_ISL_883749, EPI_ISL_883750, EPI_ISL_883751, EPI_ISL_883752, EPI_ISL_883753, EPI_ISL_883754, EPI_ISL_883755, EPI_ISL_883756, EPI_ISL_883757, EPI_ISL_883758, EPI_ISL_883759, EPI_ISL_883760, EPI_ISL_883761, EPI_ISL_883762, EPI_ISL_883763, EPI_ISL_883764, EPI_ISL_883765, EPI_ISL_883766, EPI_ISL_883767, EPI_ISL_883768, EPI_ISL_883769, EPI_ISL_883770, EPI_ISL_883771, EPI_ISL_883772, EPI_ISL_883773, EPI_ISL_883774, EPI_ISL_883777, EPI_ISL_883778, EPI_ISL_883779, EPI_ISL_883780, EPI_ISL_883781, EPI_ISL_883782, EPI_ISL_883783, EPI_ISL_883784, EPI_ISL_883785, EPI_ISL_883786, EPI_ISL_883787, EPI_ISL_883788, EPI_ISL_883789, EPI_ISL_883790, EPI_ISL_883791, EPI_ISL_883792, EPI_ISL_883793, EPI_ISL_883794, EPI_ISL_883795, EPI_ISL_883796, EPI_ISL_883797, EPI_ISL_883798, EPI_ISL_883799, EPI_ISL_883800, EPI_ISL_883801, EPI_ISL_883802, EPI_ISL_883803, EPI_ISL_883804, EPI_ISL_883805, EPI_ISL_883806, EPI_ISL_883807, EPI_ISL_883808, EPI_ISL_883809, EPI_ISL_883810, EPI_ISL_883811, EPI_ISL_883812, EPI_ISL_883813, EPI_ISL_883814, EPI_ISL_883815, EPI_ISL_883816, EPI_ISL_883817, EPI_ISL_883818, EPI_ISL_883819, EPI_ISL_883820, EPI_ISL_883821, EPI_ISL_883822, EPI_ISL_883823, EPI_ISL_883824, EPI_ISL_883825, EPI_ISL_883826, EPI_ISL_883827, EPI_ISL_883828, EPI_ISL_883829, EPI_ISL_883830, EPI_ISL_883831, EPI_ISL_883832, EPI_ISL_883833, EPI_ISL_883834, EPI_ISL_883835, EPI_ISL_883836, EPI_ISL_883837, EPI_ISL_883838, EPI_ISL_883839, EPI_ISL_883840, EPI_ISL_883841, EPI_ISL_883842, EPI_ISL_883843, EPI_ISL_883844, EPI_ISL_883845, EPI_ISL_883846, EPI_ISL_883847, EPI_ISL_883848, EPI_ISL_883849, EPI_ISL_883850, EPI_ISL_883851, EPI_ISL_883852, EPI_ISL_883853, EPI_ISL_883854, EPI_ISL_883855, EPI_ISL_883856, EPI_ISL_883857, EPI_ISL_883858, EPI_ISL_883859, EPI_ISL_883860, EPI_ISL_883861, EPI_ISL_883867, EPI_ISL_883868, EPI_ISL_883869, EPI_ISL_883870, EPI_ISL_883871, EPI_ISL_883872, EPI_ISL_883873, EPI_ISL_883874, EPI_ISL_883875, EPI_ISL_883876, EPI_ISL_883877, EPI_ISL_883878, EPI_ISL_883879, EPI_ISL_883880, EPI_ISL_883881, EPI_ISL_883882, EPI_ISL_883883, EPI_ISL_883884, EPI_ISL_883885, EPI_ISL_883886, EPI_ISL_883887, EPI_ISL_883888, EPI_ISL_883889, EPI_ISL_883890, EPI_ISL_883891, EPI_ISL_883892, EPI_ISL_883893, EPI_ISL_883894, EPI_ISL_883895, EPI_ISL_883896, EPI_ISL_883897, EPI_ISL_883898, EPI_ISL_883899, EPI_ISL_883900, EPI_ISL_883901, EPI_ISL_883902, EPI_ISL_883903, EPI_ISL_883904, EPI_ISL_883905, EPI_ISL_883906, EPI_ISL_883907, EPI_ISL_883908, EPI_ISL_883909, EPI_ISL_883910, EPI_ISL_883911, EPI_ISL_883912, EPI_ISL_883913, EPI_ISL_883914, EPI_ISL_883915, EPI_ISL_883916, EPI_ISL_883917, EPI_ISL_883918, EPI_ISL_883919, EPI_ISL_883920, EPI_ISL_883921, EPI_ISL_883922, EPI_ISL_883923, EPI_ISL_883924, EPI_ISL_883925, EPI_ISL_883926, EPI_ISL_883927, EPI_ISL_883928, EPI_ISL_883929, EPI_ISL_883930, EPI_ISL_883931, EPI_ISL_883932, EPI_ISL_883933, EPI_ISL_883934, EPI_ISL_883935, EPI_ISL_883936, EPI_ISL_883937, EPI_ISL_883938, EPI_ISL_883939, EPI_ISL_883940, EPI_ISL_883941, EPI_ISL_883942, EPI_ISL_883943, EPI_ISL_883944, EPI_ISL_883945, EPI_ISL_883946, EPI_ISL_883947, EPI_ISL_883948, EPI_ISL_883949, EPI_ISL_883950, EPI_ISL_883951, EPI_ISL_883952, EPI_ISL_883953, EPI_ISL_883954, EPI_ISL_883955, EPI_ISL_883956 | | | |
| see above | Eurofins Diatherix | Hudsonalpha Genome Sequencing Center | Jane Grimwood, Melissa Williams, Lori H. Handley, Joshua Stough, Leslie Malone, Stefan Brzezinski, Ada Stewart, Teresa Jones, Jenell Webber, John Lovell, Jennifer Cart, and Jeremy Schmutz |
| EPI_ISL_884008, EPI_ISL_884009, EPI_ISL_884010, EPI_ISL_884011, EPI_ISL_884012 | ALBANY MEDICAL CENTER HOSPITAL CLINICAL LABORATORIES | Wadsworth Center, New York State Department of Health | Kirsten St. George, Daryl M. Lamson, Alexis Russel, Matthew Shudt, Melissa A Leisner, Jonathan Plitnick, Navjot Singh, John Kelly, Erasmus Schneider, Erica Lasek-Nesselquist |
| EPI_ISL_884013, EPI_ISL_884014, EPI_ISL_884015, EPI_ISL_884016 | Wadsworth Center, New York State Department of Health | Wadsworth Center, New York State Department of Health | Kirsten St. George, Daryl M. Lamson, Alexis Russel, Matthew Shudt, Melissa A Leisner, Jonathan Plitnick, Navjot Singh, John Kelly, Erasmus Schneider, Erica Lasek-Nesselquist |
| EPI_ISL_884017, EPI_ISL_884018, EPI_ISL_884019, EPI_ISL_884020, EPI_ISL_884021, EPI_ISL_884023, EPI_ISL_884024, EPI_ISL_884025, EPI_ISL_884026, EPI_ISL_884027, EPI_ISL_884028, EPI_ISL_884029, EPI_ISL_884030, EPI_ISL_884031 | | | |
| see above | ALBANY MEDICAL CENTER HOSPITAL CLINICAL LABORATORIES | Wadsworth Center, New York State Department of Health | Kirsten St. George, Daryl M. Lamson, Alexis Russel, Matthew Shudt, Melissa A Leisner, Jonathan Plitnick, Navjot Singh, John Kelly, Erasmus Schneider, Erica Lasek-Nesselquist |
| EPI_ISL_884032, EPI_ISL_884033, EPI_ISL_884034, EPI_ISL_884035, EPI_ISL_884036, EPI_ISL_884037, EPI_ISL_884038, EPI_ISL_884039, EPI_ISL_884040, EPI_ISL_884041 | Wadsworth Center, New York State Department of Health | Wadsworth Center, New York State Department of Health | Kirsten St. George, Daryl M. Lamson, Alexis Russel, Matthew Shudt, Melissa A Leisner, Jonathan Plitnick, Navjot Singh, John Kelly, Erasmus Schneider, Erica Lasek-Nesselquist |
| EPI_ISL_884042, EPI_ISL_884043, EPI_ISL_884044, EPI_ISL_884045, EPI_ISL_884046 | ALBANY MEDICAL CENTER HOSPITAL CLINICAL LABORATORIES | Wadsworth Center, New York State Department of Health | Kirsten St. George, Daryl M. Lamson, Alexis Russel, Matthew Shudt, Melissa A Leisner, Jonathan Plitnick, Navjot Singh, John Kelly, Erasmus Schneider, Erica Lasek-Nesselquist |
| EPI_ISL_884047 | Wadsworth Center, New York State Department of Health | Wadsworth Center, New York State Department of Health | Kirsten St. George, Daryl M. Lamson, Alexis Russel, Matthew Shudt, Melissa A Leisner, Jonathan Plitnick, Navjot Singh, John Kelly, Erasmus Schneider, Erica Lasek-Nesselquist |
| EPI_ISL_884048, EPI_ISL_884049, EPI_ISL_884050, EPI_ISL_884051, EPI_ISL_884052, EPI_ISL_884053 | ALBANY MEDICAL CENTER HOSPITAL CLINICAL LABORATORIES | Wadsworth Center, New York State Department of Health | Kirsten St. George, Daryl M. Lamson, Alexis Russel, Matthew Shudt, Melissa A Leisner, Jonathan Plitnick, Navjot Singh, John Kelly, Erasmus Schneider, Erica Lasek-Nesselquist |
| EPI_ISL_884054 | TEMPUS LABS INC | Wadsworth Center, New York State Department of Health | Kirsten St. George, Daryl M. Lamson, Alexis Russel, Matthew Shudt, Melissa A Leisner, Jonathan Plitnick, Navjot Singh, John Kelly, Erasmus Schneider, Erica Lasek-Nesselquist |
| EPI_ISL_884055, EPI_ISL_884056, EPI_ISL_884057, EPI_ISL_884058 | Wadsworth Center, New York State Department of Health | Wadsworth Center, New York State Department of Health | Kirsten St. George, Daryl M. Lamson, Alexis Russel, Matthew Shudt, Melissa A Leisner, Jonathan Plitnick, Navjot Singh, John Kelly, Erasmus Schneider, Erica Lasek-Nesselquist |
| EPI_ISL_884059, EPI_ISL_884060, EPI_ISL_884061, EPI_ISL_884062, EPI_ISL_884063, EPI_ISL_884064, EPI_ISL_884065, EPI_ISL_884066, EPI_ISL_884067, EPI_ISL_884068, EPI_ISL_884069, EPI_ISL_884070, EPI_ISL_884071, EPI_ISL_884072, EPI_ISL_884073, EPI_ISL_884074, EPI_ISL_884075, EPI_ISL_884076, EPI_ISL_884077, EPI_ISL_884078, EPI_ISL_884079 | | | |
| see above | ALBANY MEDICAL CENTER HOSPITAL | Wadsworth Center, New York State | Kirsten St. George, Daryl M. Lamson, Alexis Russel, Matthew Shudt, Melissa A Leisner, Jonathan Plitnick, Navjot Singh, John Kelly, Erasmus Schneider, Erica Lasek-Nesselquist |

| | | | |
|---|---|---|---|
| | CLINICAL LABORATORIES | Department of Health | |
| EPI_ISL_884080 | Wadsworth Center, New York State Department of Health | Wadsworth Center, New York State Department of Health | Kirsten St. George, Daryl M. Lamson, Alexis Russel, Matthew Shudt, Melissa A Leisner, Jonathan Plitnick, Navjot Singh, John Kelly, Erasmus Schneider, Erica Lasek-Nesselquist |
| EPI_ISL_884081, EPI_ISL_884082, EPI_ISL_884083, EPI_ISL_884084, EPI_ISL_884085 | ALBANY MEDICAL CENTER HOSPITAL CLINICAL LABORATORIES | Wadsworth Center, New York State Department of Health | Kirsten St. George, Daryl M. Lamson, Alexis Russel, Matthew Shudt, Melissa A Leisner, Jonathan Plitnick, Navjot Singh, John Kelly, Erasmus Schneider, Erica Lasek-Nesselquist |
| EPI_ISL_884086 | Wadsworth Center, New York State Department of Health | Wadsworth Center, New York State Department of Health | Kirsten St. George, Daryl M. Lamson, Alexis Russel, Matthew Shudt, Melissa A Leisner, Jonathan Plitnick, Navjot Singh, John Kelly, Erasmus Schneider, Erica Lasek-Nesselquist |
| EPI_ISL_884089, EPI_ISL_884090, EPI_ISL_884091, EPI_ISL_884092, EPI_ISL_884093, EPI_ISL_884094, EPI_ISL_884095, EPI_ISL_884096, EPI_ISL_884097, EPI_ISL_884098, EPI_ISL_884099, EPI_ISL_884100, EPI_ISL_884101, EPI_ISL_884102, EPI_ISL_884103, EPI_ISL_884104, EPI_ISL_884105, EPI_ISL_884106, EPI_ISL_884107, EPI_ISL_884108, EPI_ISL_884109, EPI_ISL_884110, EPI_ISL_884111, EPI_ISL_884112, EPI_ISL_884113, EPI_ISL_884114, EPI_ISL_884115, EPI_ISL_884116, EPI_ISL_884117, EPI_ISL_884118, EPI_ISL_884119, EPI_ISL_884120, EPI_ISL_884121, EPI_ISL_884122, EPI_ISL_884123, EPI_ISL_884124, EPI_ISL_884125, EPI_ISL_884126, EPI_ISL_884127, EPI_ISL_884128, EPI_ISL_884129, EPI_ISL_884130, EPI_ISL_884131, EPI_ISL_884132, EPI_ISL_884133, EPI_ISL_884134, EPI_ISL_884135, EPI_ISL_884136, EPI_ISL_884137, EPI_ISL_884138, EPI_ISL_884139, EPI_ISL_884140, EPI_ISL_884141, EPI_ISL_884142, EPI_ISL_884143, EPI_ISL_884144, EPI_ISL_884145, EPI_ISL_884146, EPI_ISL_884147, EPI_ISL_884148, EPI_ISL_884149, EPI_ISL_884150, EPI_ISL_884151, EPI_ISL_884152, EPI_ISL_884153, EPI_ISL_884154, EPI_ISL_884155, EPI_ISL_884156, EPI_ISL_884157, EPI_ISL_884158, EPI_ISL_884159, EPI_ISL_884160, EPI_ISL_884161, EPI_ISL_884162, EPI_ISL_884163, EPI_ISL_884164, EPI_ISL_884165, EPI_ISL_884166, EPI_ISL_884167, EPI_ISL_884168, EPI_ISL_884169, EPI_ISL_884170, EPI_ISL_884171, EPI_ISL_884172, EPI_ISL_884173, EPI_ISL_884174, EPI_ISL_884175, EPI_ISL_884176, EPI_ISL_884177, EPI_ISL_884178, EPI_ISL_884179, EPI_ISL_884180, EPI_ISL_884181 | | | |
| see above | Eurofins Diatherix | Hudsonalpha Genome Sequencing Center | Jane Grimwood, Melissa Williams, Lori H. Handley, Joshua Stough, Leslie Malone, Stefan Brzezinski, Ada Stewart, Teresa Jones, Jenell Webber, John Lovell, Jennifer Cart, and Jeremy Schmutz |
| EPI_ISL_884182, EPI_ISL_884183, EPI_ISL_884184, EPI_ISL_884192, EPI_ISL_884193, EPI_ISL_884194, EPI_ISL_884195 | Wyoming Public Health Laboratory | Wyoming Public Health Laboratory | Noah Hull, Taylor Fearing, Lynette Gumbleton, Channing Weber, Ashley Norberg, Bailey Bowcutt, and Wanda Manley |
| EPI_ISL_884214, EPI_ISL_884215, EPI_ISL_884216, EPI_ISL_884217, EPI_ISL_884218, EPI_ISL_884221 | Alaska State Virology Laboratory | Alaska State Virology Laboratory | Stephanie DeRonde, Lisa Smith, PhD. Jack Chen, PhD. |
| EPI_ISL_884223, EPI_ISL_884224, EPI_ISL_884225, EPI_ISL_884226, EPI_ISL_884227, EPI_ISL_884228, EPI_ISL_884229, EPI_ISL_884230, EPI_ISL_884231, EPI_ISL_884232, EPI_ISL_884233, EPI_ISL_884234, EPI_ISL_884235, EPI_ISL_884236, EPI_ISL_884237, EPI_ISL_884238, EPI_ISL_884239, EPI_ISL_884240, EPI_ISL_884241, EPI_ISL_884242, EPI_ISL_884243, EPI_ISL_884244, EPI_ISL_884245, EPI_ISL_884246, EPI_ISL_884247, EPI_ISL_884248 | | | |
| see above | Kansas Health and Environmental Lab | Kansas Health and Environmental Lab | Mike Grose, Paige Drury, Carissa Robertson, Ben Olsen, and Phil Adam |
| EPI_ISL_884251, EPI_ISL_884252, EPI_ISL_884253, EPI_ISL_884254, EPI_ISL_884255, EPI_ISL_884256, EPI_ISL_884257, EPI_ISL_884258, EPI_ISL_884259 | Maryland Public Health Laboratory | Maryland Public Health Laboratory | Maryland Department of Health Laboratories Administration |
| EPI_ISL_884291, EPI_ISL_884292, EPI_ISL_884293 | Clinical Molecular Microbiology Laboratory, UNC Hospital | Dirk Dittmer | Razia Moorad , Justin T. Landis , Brent A. Eason, Melissa B. Miller, Linda Pluta, Dirk Dittmer, Angelica Juarez, Cecilia Thompson , Cameroon Grant, Evelyn Hoffman, Patricio Cano, Jason Wong, Carolina Caro-Vegas, Blossom Damania. |
| EPI_ISL_884295 | Microbiology Division, South Carolina Department of Health and Environmental Control Public Health Laboratory (SC DHEC PHL) | Microbiology Division, South Carolina Department of Health and Environmental Control Public Health Laboratory (SC DHEC PHL) | Haley Flores, Jessica Freeman |
| EPI_ISL_884297, EPI_ISL_884298, EPI_ISL_884299, EPI_ISL_884300, EPI_ISL_884301, EPI_ISL_884302 | Santa Clara County Public Health Laboratory | Santa Clara County Public Health Laboratory | Santa Clara County Public Health Department |
| EPI_ISL_884303, EPI_ISL_884304, EPI_ISL_884305, EPI_ISL_884306, EPI_ISL_884307, EPI_ISL_884308, EPI_ISL_884309, EPI_ISL_884310, EPI_ISL_884311, EPI_ISL_884312, EPI_ISL_884313, EPI_ISL_884314, EPI_ISL_884315, EPI_ISL_884316, EPI_ISL_884317, EPI_ISL_884318, EPI_ISL_884319, EPI_ISL_884320, EPI_ISL_884321, EPI_ISL_884322, EPI_ISL_884323, EPI_ISL_884324, EPI_ISL_884325, EPI_ISL_884326, EPI_ISL_884327, EPI_ISL_884328, EPI_ISL_884329, EPI_ISL_884330, EPI_ISL_884331, EPI_ISL_884332, EPI_ISL_884333, EPI_ISL_884334, EPI_ISL_884335, EPI_ISL_884336, EPI_ISL_884337, EPI_ISL_884338, EPI_ISL_884339, EPI_ISL_884340, EPI_ISL_884341, EPI_ISL_884342, EPI_ISL_884343, EPI_ISL_884344, EPI_ISL_884345, EPI_ISL_884346, EPI_ISL_884347, EPI_ISL_884348, EPI_ISL_884349, EPI_ISL_884350, EPI_ISL_884351, EPI_ISL_884352, EPI_ISL_884353, EPI_ISL_884354, EPI_ISL_884355, EPI_ISL_884356, EPI_ISL_884357, EPI_ISL_884358, EPI_ISL_884359, EPI_ISL_884360, EPI_ISL_884361, EPI_ISL_884362, EPI_ISL_884363, EPI_ISL_884364, EPI_ISL_884365, EPI_ISL_884366, EPI_ISL_884367, EPI_ISL_884368, EPI_ISL_884369, EPI_ISL_884370, EPI_ISL_884371, EPI_ISL_884372, EPI_ISL_884373, EPI_ISL_884374, EPI_ISL_884375, EPI_ISL_884376, EPI_ISL_884377, EPI_ISL_884378, EPI_ISL_884379, EPI_ISL_884380, EPI_ISL_884381, EPI_ISL_884382, EPI_ISL_884383, EPI_ISL_884384, EPI_ISL_884385, EPI_ISL_884386, EPI_ISL_884387, EPI_ISL_884388, EPI_ISL_884389, EPI_ISL_884390, EPI_ISL_884391, EPI_ISL_884392, EPI_ISL_884393, EPI_ISL_884394, EPI_ISL_884395, EPI_ISL_884396, EPI_ISL_884397, EPI_ISL_884398, EPI_ISL_884399, EPI_ISL_884400, EPI_ISL_884401, EPI_ISL_884402, EPI_ISL_884403, EPI_ISL_884404, EPI_ISL_884405, EPI_ISL_884406, EPI_ISL_884407, EPI_ISL_884408, EPI_ISL_884409, EPI_ISL_884410, EPI_ISL_884411, EPI_ISL_884412, EPI_ISL_884413, EPI_ISL_884414, EPI_ISL_884415, EPI_ISL_884416, EPI_ISL_884417, EPI_ISL_884418, EPI_ISL_884419, EPI_ISL_884420, EPI_ISL_884421, EPI_ISL_884422, EPI_ISL_884423, EPI_ISL_884424, EPI_ISL_884425, EPI_ISL_884426, EPI_ISL_884427, EPI_ISL_884428, EPI_ISL_884429, EPI_ISL_884430, EPI_ISL_884431, EPI_ISL_884432, EPI_ISL_884433, EPI_ISL_884434, EPI_ISL_884435, EPI_ISL_884436, EPI_ISL_884437, EPI_ISL_884438, EPI_ISL_884439, EPI_ISL_884440, EPI_ISL_884441, EPI_ISL_884442, EPI_ISL_884443, EPI_ISL_884444 | | | |
| see above | Infectious Diseases, Quest Diagnostics | Infectious Diseases, Quest Diagnostics | Rosenthal,S.H., Gerasimova,A., Kagan,R.M., Anderson,B., Bernstein,L.E., Livingston,K.E., Hua,M., Liu,Y., Shalhout,D.F., Owen,R., Lacbawan,F. |
| EPI_ISL_884445, EPI_ISL_884446, EPI_ISL_884447, EPI_ISL_884448, EPI_ISL_884449, EPI_ISL_884450, EPI_ISL_884451, EPI_ISL_884452, EPI_ISL_884453, EPI_ISL_884454, EPI_ISL_884455, EPI_ISL_884456, EPI_ISL_884457, EPI_ISL_884458, EPI_ISL_884459, EPI_ISL_884460, EPI_ISL_884461, EPI_ISL_884462, EPI_ISL_884463, EPI_ISL_884464, EPI_ISL_884466, EPI_ISL_884468, EPI_ISL_884469, EPI_ISL_884470, EPI_ISL_884471, EPI_ISL_884472, EPI_ISL_884473, EPI_ISL_884474, EPI_ISL_884475, EPI_ISL_884476, EPI_ISL_884477, EPI_ISL_884478, EPI_ISL_884479, EPI_ISL_884480, EPI_ISL_884481, EPI_ISL_884482, EPI_ISL_884483, EPI_ISL_884484, EPI_ISL_884485, EPI_ISL_884486, EPI_ISL_884487, EPI_ISL_884488, EPI_ISL_884489, EPI_ISL_884490, EPI_ISL_884491, EPI_ISL_884492, EPI_ISL_884493, EPI_ISL_884494, EPI_ISL_884495, EPI_ISL_884496, EPI_ISL_884497, EPI_ISL_884498, EPI_ISL_884499, EPI_ISL_884500, EPI_ISL_884501, EPI_ISL_884502, EPI_ISL_884503, EPI_ISL_884504, EPI_ISL_884505, EPI_ISL_884506, EPI_ISL_884507, EPI_ISL_884508, EPI_ISL_884509, EPI_ISL_884510, EPI_ISL_884511, EPI_ISL_884512, EPI_ISL_884513, EPI_ISL_884514, EPI_ISL_884515, EPI_ISL_884516, EPI_ISL_884517, EPI_ISL_884518, EPI_ISL_884519, EPI_ISL_884520, EPI_ISL_884521, EPI_ISL_884522, EPI_ISL_884523, EPI_ISL_884524, EPI_ISL_884525, EPI_ISL_884526, EPI_ISL_884527, EPI_ISL_884528, EPI_ISL_884529, EPI_ISL_884530, EPI_ISL_884532, EPI_ISL_884533, EPI_ISL_884534, EPI_ISL_884535, EPI_ISL_884536, EPI_ISL_884537, EPI_ISL_884538, EPI_ISL_884539, EPI_ISL_884540, EPI_ISL_884541, EPI_ISL_884542, EPI_ISL_884543, EPI_ISL_884544, EPI_ISL_884545, EPI_ISL_884546, EPI_ISL_884547, EPI_ISL_884548, EPI_ISL_884549, EPI_ISL_884550, EPI_ISL_884551, EPI_ISL_884552, EPI_ISL_884553, EPI_ISL_884554, EPI_ISL_884555, EPI_ISL_884556, EPI_ISL_884558, EPI_ISL_884560, EPI_ISL_884561, EPI_ISL_884563, EPI_ISL_884564, EPI_ISL_884565, EPI_ISL_884567, EPI_ISL_884568, EPI_ISL_884569, EPI_ISL_884570, EPI_ISL_884571, EPI_ISL_884572, EPI_ISL_884573, EPI_ISL_884576, EPI_ISL_884577, EPI_ISL_884579 | | | |
| see above | Molecular Microbiology & Immunology, University of Missouri | Molecular Microbiology & Immunology, University of Missouri | Tang,C.Y., Li,T., Hang,J., Lidl,G.M., Wan,X.-F. |
| EPI_ISL_884580, EPI_ISL_884581, EPI_ISL_884582, EPI_ISL_884583, EPI_ISL_884584, EPI_ISL_884585, EPI_ISL_884586, EPI_ISL_884587, EPI_ISL_884588, EPI_ISL_884589, EPI_ISL_884590, EPI_ISL_884591, EPI_ISL_884592, EPI_ISL_884593, EPI_ISL_884594, EPI_ISL_884595, EPI_ISL_884596, EPI_ISL_884597, EPI_ISL_884598, EPI_ISL_884599, EPI_ISL_884600, EPI_ISL_884601, EPI_ISL_884602, EPI_ISL_884603, EPI_ISL_884604, EPI_ISL_884605, EPI_ISL_884606, EPI_ISL_884607, EPI_ISL_884609, EPI_ISL_884610, EPI_ISL_884611, EPI_ISL_884612, EPI_ISL_884613, EPI_ISL_884614, EPI_ISL_884615, EPI_ISL_884616, EPI_ISL_884617, EPI_ISL_884618, EPI_ISL_884619, EPI_ISL_884620, EPI_ISL_884621, EPI_ISL_884622, EPI_ISL_884623, EPI_ISL_884624, EPI_ISL_884625, EPI_ISL_884626, EPI_ISL_884627, EPI_ISL_884628, EPI_ISL_884629, EPI_ISL_884630, EPI_ISL_884631, EPI_ISL_884632, EPI_ISL_884633, EPI_ISL_884634, EPI_ISL_884635, EPI_ISL_884636, EPI_ISL_884637, EPI_ISL_884638, EPI_ISL_884639, EPI_ISL_884640, EPI_ISL_884641, EPI_ISL_884642, EPI_ISL_884643, EPI_ISL_884644, EPI_ISL_884645, EPI_ISL_884646, EPI_ISL_884647, EPI_ISL_884648, EPI_ISL_884649, EPI_ISL_884650, EPI_ISL_884651, EPI_ISL_884652, EPI_ISL_884653, EPI_ISL_884654, EPI_ISL_884655, EPI_ISL_884656, EPI_ISL_884657, EPI_ISL_884658, EPI_ISL_884659, EPI_ISL_884660, EPI_ISL_884661, EPI_ISL_884662, EPI_ISL_884663, EPI_ISL_884664, EPI_ISL_884665, EPI_ISL_884666, EPI_ISL_884667, EPI_ISL_884668, EPI_ISL_884669, EPI_ISL_884670, EPI_ISL_884671, EPI_ISL_884672, EPI_ISL_884673, EPI_ISL_884675, EPI_ISL_884676, EPI_ISL_884677, EPI_ISL_884678, EPI_ISL_884679, EPI_ISL_884680, EPI_ISL_884681, EPI_ISL_884682, EPI_ISL_884683, EPI_ISL_884684, EPI_ISL_884685, EPI_ISL_884686, EPI_ISL_884687, EPI_ISL_884688, EPI_ISL_884689, EPI_ISL_884690, EPI_ISL_884691, EPI_ISL_884692, EPI_ISL_884693, EPI_ISL_884694, EPI_ISL_884695, EPI_ISL_884696, EPI_ISL_884697, EPI_ISL_884698, EPI_ISL_884699, EPI_ISL_884700, EPI_ISL_884701, EPI_ISL_884702, EPI_ISL_884703, EPI_ISL_884704, EPI_ISL_884705, EPI_ISL_884706, EPI_ISL_884707, EPI_ISL_884710, EPI_ISL_884711, EPI_ISL_884712, EPI_ISL_884713, EPI_ISL_884714, EPI_ISL_884715, EPI_ISL_884716, EPI_ISL_884717, EPI_ISL_884718, EPI_ISL_884719, EPI_ISL_884720, EPI_ISL_884721, EPI_ISL_884722, EPI_ISL_884723, EPI_ISL_884724, EPI_ISL_884725, EPI_ISL_884726, EPI_ISL_884727, EPI_ISL_884728, EPI_ISL_884729, EPI_ISL_884730, EPI_ISL_884731, EPI_ISL_884732, EPI_ISL_884733, EPI_ISL_884734, EPI_ISL_884735, EPI_ISL_884736, EPI_ISL_884737, EPI_ISL_884738, EPI_ISL_884740, EPI_ISL_884741, EPI_ISL_884742, EPI_ISL_884743, EPI_ISL_884744, EPI_ISL_884745, EPI_ISL_884746, EPI_ISL_884747, EPI_ISL_884748, EPI_ISL_884749, EPI_ISL_884750, EPI_ISL_884751, EPI_ISL_884752, EPI_ISL_884753, EPI_ISL_884754, EPI_ISL_884756, EPI_ISL_884757, EPI_ISL_884758, EPI_ISL_884759, EPI_ISL_884760, EPI_ISL_884761, EPI_ISL_884762, EPI_ISL_884763, EPI_ISL_884764, EPI_ISL_884765, EPI_ISL_884766, EPI_ISL_884767, EPI_ISL_884768, EPI_ISL_884769, EPI_ISL_884770, EPI_ISL_884771, EPI_ISL_884772, EPI_ISL_884773, EPI_ISL_884774, EPI_ISL_884775, EPI_ISL_884776, EPI_ISL_884777, EPI_ISL_884778, EPI_ISL_884779, EPI_ISL_884780, EPI_ISL_884781, EPI_ISL_884782, EPI_ISL_884783, EPI_ISL_884784, EPI_ISL_884785, EPI_ISL_884786, EPI_ISL_884787, EPI_ISL_884788, EPI_ISL_884789, EPI_ISL_884790, EPI_ISL_884791, EPI_ISL_884792, EPI_ISL_884793, EPI_ISL_884794, EPI_ISL_884795, EPI_ISL_884796, EPI_ISL_884798, EPI_ISL_884799, EPI_ISL_884800, EPI_ISL_884801, EPI_ISL_884802, EPI_ISL_884803, EPI_ISL_884804, EPI_ISL_884805, EPI_ISL_884806, EPI_ISL_884807, EPI_ISL_884808, EPI_ISL_884809, EPI_ISL_884810, EPI_ISL_884811, EPI_ISL_884812, EPI_ISL_884813, EPI_ISL_884814, EPI_ISL_884815, EPI_ISL_884816, EPI_ISL_884817, EPI_ISL_884818, EPI_ISL_884819, EPI_ISL_884820, EPI_ISL_884821, EPI_ISL_884822, EPI_ISL_884823, EPI_ISL_884824, EPI_ISL_884825 | | | |
| see above | Respiratory Viruses Branch, Centers for Disease Control and Prevention | Respiratory Viruses Branch, Centers for Disease Control and Prevention | Cook,P.W., Batra,D., Rambo-Martin,B.L., de Feo,E., Antico,J., Tran,C., Tolentino,M., Wickline,S., Gietzen,K., Sickler,B., Liu,J., Allen,E., Febbo,P., Galloway,S., Washington,N.L., White,S., Levan,G., Barret,K.S., Cirulli,E., Bolze,A., Ascencio,A., Rivera-Garcia,C., Cho,R., Nguyen,J., Wang,S., Ramirez,J., Cassens,T., Sandoval,E., Isaksson,M., Lee,W., Becker,D., Laurent,M., Lu,J., Paden,C.R., Tong,S., MacCannell,D. |
| EPI_ISL_884857, EPI_ISL_884858, EPI_ISL_884859, EPI_ISL_884860, EPI_ISL_884861, EPI_ISL_884862, EPI_ISL_884863 | Bacteriology, Georgia Public Health Laboratory (GPHL) | Bacteriology, Georgia Public Health Laboratory (GPHL) | Reeves,S., Edwards,J., Dixey,C., Parrott,T. |
| EPI_ISL_884866, EPI_ISL_884867, EPI_ISL_884868, EPI_ISL_884869, EPI_ISL_884870 | CA DPH Viral and Rickettsial Disease Laboratory | Chan-Zuckerberg Biohub | CZB Cliahub Consortium |
| EPI_ISL_884871, EPI_ISL_884872, EPI_ISL_884873, EPI_ISL_884874, EPI_ISL_884875, EPI_ISL_884876, EPI_ISL_884877, EPI_ISL_884878, EPI_ISL_884879, EPI_ISL_884880, EPI_ISL_884881, EPI_ISL_884882, EPI_ISL_884883, EPI_ISL_884884, EPI_ISL_884885, EPI_ISL_884886, EPI_ISL_884887, EPI_ISL_884888, EPI_ISL_884889, EPI_ISL_884890, EPI_ISL_884891, EPI_ISL_884892, EPI_ISL_884893, EPI_ISL_884894, EPI_ISL_884895, EPI_ISL_884896, EPI_ISL_884897, EPI_ISL_884898, EPI_ISL_884899, EPI_ISL_884900, EPI_ISL_884901, EPI_ISL_884902, EPI_ISL_884903, EPI_ISL_884904, EPI_ISL_884905, EPI_ISL_884906, EPI_ISL_884907, EPI_ISL_884908, EPI_ISL_884910, EPI_ISL_884911, EPI_ISL_884912, EPI_ISL_884913, EPI_ISL_884914, EPI_ISL_884915, EPI_ISL_884917, EPI_ISL_884918, EPI_ISL_884919, EPI_ISL_884920, EPI_ISL_884921, EPI_ISL_884922, EPI_ISL_884923, EPI_ISL_884924, EPI_ISL_884925, EPI_ISL_884926, EPI_ISL_884927, EPI_ISL_884928, EPI_ISL_884929, EPI_ISL_884930, EPI_ISL_884931, EPI_ISL_884932, EPI_ISL_884933, EPI_ISL_884934, EPI_ISL_884935, EPI_ISL_884936, EPI_ISL_884937, EPI_ISL_884938, EPI_ISL_884939, EPI_ISL_884940, EPI_ISL_884941, EPI_ISL_884942, EPI_ISL_884943, EPI_ISL_884944, EPI_ISL_884945, EPI_ISL_884946, EPI_ISL_884947, EPI_ISL_884948, EPI_ISL_884949, EPI_ISL_884950, EPI_ISL_884951, EPI_ISL_884952, EPI_ISL_884953, EPI_ISL_884954, EPI_ISL_884955, EPI_ISL_884956, EPI_ISL_884957, EPI_ISL_884958, EPI_ISL_884959, EPI_ISL_884960, EPI_ISL_884961, EPI_ISL_884962, EPI_ISL_884963, EPI_ISL_884964, EPI_ISL_884965, EPI_ISL_884966, EPI_ISL_884967, EPI_ISL_884968, EPI_ISL_884969, EPI_ISL_884970, EPI_ISL_884971, EPI_ISL_884972, EPI_ISL_884973, EPI_ISL_884974, EPI_ISL_884975, EPI_ISL_884976, EPI_ISL_884977, EPI_ISL_884978, EPI_ISL_884979, EPI_ISL_884980, EPI_ISL_884981, EPI_ISL_884982, EPI_ISL_884983, EPI_ISL_884984, EPI_ISL_884985, EPI_ISL_884986, EPI_ISL_884987, EPI_ISL_884988, EPI_ISL_884989, EPI_ISL_884990, EPI_ISL_884991, EPI_ISL_884992, EPI_ISL_884993, EPI_ISL_884994, EPI_ISL_884995, EPI_ISL_884996, EPI_ISL_884997, EPI_ISL_884998, EPI_ISL_884999, EPI_ISL_885000, EPI_ISL_885001, EPI_ISL_885002, EPI_ISL_885003, EPI_ISL_885004, EPI_ISL_885005, EPI_ISL_885006, EPI_ISL_885007, EPI_ISL_885008, EPI_ISL_885009, EPI_ISL_885010, EPI_ISL_885011, EPI_ISL_885012, EPI_ISL_885013, EPI_ISL_885014, EPI_ISL_885015, EPI_ISL_885016, EPI_ISL_885017, EPI_ISL_885018, EPI_ISL_885019, EPI_ISL_885020, EPI_ISL_885021, EPI_ISL_885022, EPI_ISL_885023, EPI_ISL_885024, EPI_ISL_885025, EPI_ISL_885026, EPI_ISL_885027, EPI_ISL_885028, EPI_ISL_885029, EPI_ISL_885030, EPI_ISL_885031, EPI_ISL_885032, EPI_ISL_885033, EPI_ISL_885034, EPI_ISL_885035, EPI_ISL_885036, EPI_ISL_885037, EPI_ISL_885038, EPI_ISL_885039, EPI_ISL_885040, EPI_ISL_885041, EPI_ISL_885042, EPI_ISL_885043, EPI_ISL_885044, EPI_ISL_885045, EPI_ISL_885046, EPI_ISL_885047, EPI_ISL_885048, EPI_ISL_885049, EPI_ISL_885050, EPI_ISL_885051 | | | |

see above | Santa Clara County Public Health Laboratory | Chan-Zuckerberg Biohub | CZB Cliahub Consortium

EPI_ISL_885052, EPI_ISL_885053, EPI_ISL_885054, EPI_ISL_885055, EPI_ISL_885056, EPI_ISL_885057, EPI_ISL_885058, EPI_ISL_885059, EPI_ISL_885060, EPI_ISL_885061, EPI_ISL_885062, EPI_ISL_885063, EPI_ISL_885064, EPI_ISL_885065, EPI_ISL_885066, EPI_ISL_885067, EPI_ISL_885068, EPI_ISL_885069, EPI_ISL_885070, EPI_ISL_885071, EPI_ISL_885072, EPI_ISL_885073, EPI_ISL_885074, EPI_ISL_885075, EPI_ISL_885076, EPI_ISL_885077, EPI_ISL_885078, EPI_ISL_885079, EPI_ISL_885080, EPI_ISL_885081, EPI_ISL_885082, EPI_ISL_885083, EPI_ISL_885084, EPI_ISL_885085, EPI_ISL_885086, EPI_ISL_885087, EPI_ISL_885088, EPI_ISL_885089, EPI_ISL_885090, EPI_ISL_885091, EPI_ISL_885092, EPI_ISL_885093, EPI_ISL_885094, EPI_ISL_885096, EPI_ISL_885097, EPI_ISL_885098, EPI_ISL_885099, EPI_ISL_885100, EPI_ISL_885101, EPI_ISL_885102, EPI_ISL_885103, EPI_ISL_885104, EPI_ISL_885105, EPI_ISL_885106, EPI_ISL_885107, EPI_ISL_885108, EPI_ISL_885109, EPI_ISL_885110, EPI_ISL_885111, EPI_ISL_885112, EPI_ISL_885113, EPI_ISL_885114, EPI_ISL_885115, EPI_ISL_885116, EPI_ISL_885118, EPI_ISL_885119, EPI_ISL_885120, EPI_ISL_885121, EPI_ISL_885122, EPI_ISL_885123, EPI_ISL_885124, EPI_ISL_885125, EPI_ISL_885126, EPI_ISL_885127

see above | Orange County Public Health Lab | Chan-Zuckerberg Biohub | CZB Cliahub Consortium

EPI_ISL_885128, EPI_ISL_885129, EPI_ISL_885130, EPI_ISL_885131, EPI_ISL_885132, EPI_ISL_885133, EPI_ISL_885134, EPI_ISL_885135, EPI_ISL_885136, EPI_ISL_885137, EPI_ISL_885138

see above | California Institute of Technology | Chan-Zuckerberg Biohub | CZB Cliahub Consortium

EPI_ISL_886160, EPI_ISL_886161, EPI_ISL_886162, EPI_ISL_886163, EPI_ISL_886164, EPI_ISL_886165, EPI_ISL_886166, EPI_ISL_886167, EPI_ISL_886168, EPI_ISL_886173, EPI_ISL_886179, EPI_ISL_886180, EPI_ISL_886181, EPI_ISL_886182, EPI_ISL_886183, EPI_ISL_886186, EPI_ISL_886187, EPI_ISL_886188, EPI_ISL_886190, EPI_ISL_886191, EPI_ISL_886192, EPI_ISL_886193, EPI_ISL_886194, EPI_ISL_886196, EPI_ISL_886197, EPI_ISL_886198, EPI_ISL_886199, EPI_ISL_886200, EPI_ISL_886201, EPI_ISL_886202, EPI_ISL_886203, EPI_ISL_886204, EPI_ISL_886205, EPI_ISL_886206, EPI_ISL_886207, EPI_ISL_886208, EPI_ISL_886209, EPI_ISL_886210, EPI_ISL_886211, EPI_ISL_886213, EPI_ISL_886214, EPI_ISL_886215, EPI_ISL_886216, EPI_ISL_886217, EPI_ISL_886218, EPI_ISL_886220, EPI_ISL_886222, EPI_ISL_886223, EPI_ISL_886224, EPI_ISL_886225, EPI_ISL_886226, EPI_ISL_886230, EPI_ISL_886232, EPI_ISL_886234, EPI_ISL_886235, EPI_ISL_886236, EPI_ISL_886237, EPI_ISL_886239, EPI_ISL_886240, EPI_ISL_886244, EPI_ISL_886246, EPI_ISL_886248, EPI_ISL_886249, EPI_ISL_886251, EPI_ISL_886252, EPI_ISL_886253, EPI_ISL_886254, EPI_ISL_886256, EPI_ISL_886257, EPI_ISL_886258, EPI_ISL_886259, EPI_ISL_886261, EPI_ISL_886262, EPI_ISL_886263, EPI_ISL_886266, EPI_ISL_886267, EPI_ISL_886269, EPI_ISL_886271, EPI_ISL_886273, EPI_ISL_886274, EPI_ISL_886275, EPI_ISL_886276, EPI_ISL_886277, EPI_ISL_886278, EPI_ISL_886279, EPI_ISL_886280, EPI_ISL_886282, EPI_ISL_886283, EPI_ISL_886284, EPI_ISL_886287, EPI_ISL_886288, EPI_ISL_886290, EPI_ISL_886291, EPI_ISL_886292, EPI_ISL_886293, EPI_ISL_886294, EPI_ISL_886295, EPI_ISL_886296, EPI_ISL_886298, EPI_ISL_886299, EPI_ISL_886300, EPI_ISL_886302, EPI_ISL_886303, EPI_ISL_886305, EPI_ISL_886306, EPI_ISL_886307, EPI_ISL_886308, EPI_ISL_886309, EPI_ISL_886310, EPI_ISL_886312, EPI_ISL_886313, EPI_ISL_886314, EPI_ISL_886316, EPI_ISL_886318, EPI_ISL_886319, EPI_ISL_886320, EPI_ISL_886321, EPI_ISL_886322, EPI_ISL_886323, EPI_ISL_886325, EPI_ISL_886326, EPI_ISL_886327, EPI_ISL_886330, EPI_ISL_886331, EPI_ISL_886332, EPI_ISL_886333, EPI_ISL_886334, EPI_ISL_886335, EPI_ISL_886337, EPI_ISL_886339, EPI_ISL_886340, EPI_ISL_886341, EPI_ISL_886342, EPI_ISL_886343, EPI_ISL_886344, EPI_ISL_886345, EPI_ISL_886346, EPI_ISL_886347, EPI_ISL_886348, EPI_ISL_886350, EPI_ISL_886351, EPI_ISL_886358, EPI_ISL_886359, EPI_ISL_886360, EPI_ISL_886361, EPI_ISL_886362, EPI_ISL_886363, EPI_ISL_886364, EPI_ISL_886366, EPI_ISL_886368, EPI_ISL_886371, EPI_ISL_886372, EPI_ISL_886373, EPI_ISL_886374, EPI_ISL_886375, EPI_ISL_886376, EPI_ISL_886377, EPI_ISL_886378, EPI_ISL_886379, EPI_ISL_886386, EPI_ISL_886388, EPI_ISL_886389, EPI_ISL_886390, EPI_ISL_886392, EPI_ISL_886394, EPI_ISL_886396, EPI_ISL_886397, EPI_ISL_886398, EPI_ISL_886399, EPI_ISL_886400, EPI_ISL_886402, EPI_ISL_886403, EPI_ISL_886404, EPI_ISL_886405, EPI_ISL_886406, EPI_ISL_886407, EPI_ISL_886408, EPI_ISL_886409, EPI_ISL_886410, EPI_ISL_886411, EPI_ISL_886413, EPI_ISL_886414, EPI_ISL_886415, EPI_ISL_886418, EPI_ISL_886421, EPI_ISL_886422, EPI_ISL_886423, EPI_ISL_886424, EPI_ISL_886425, EPI_ISL_886426, EPI_ISL_886427, EPI_ISL_886428, EPI_ISL_886429, EPI_ISL_886430, EPI_ISL_886431, EPI_ISL_886433, EPI_ISL_886434, EPI_ISL_886436, EPI_ISL_886437, EPI_ISL_886440, EPI_ISL_886442, EPI_ISL_886443, EPI_ISL_886444, EPI_ISL_886446, EPI_ISL_886447, EPI_ISL_886448, EPI_ISL_886449, EPI_ISL_886450, EPI_ISL_886452, EPI_ISL_886453, EPI_ISL_886456, EPI_ISL_886457, EPI_ISL_886458, EPI_ISL_886459, EPI_ISL_886460, EPI_ISL_886461, EPI_ISL_886462, EPI_ISL_886463, EPI_ISL_886464, EPI_ISL_886465, EPI_ISL_886466, EPI_ISL_886467, EPI_ISL_886470, EPI_ISL_886472, EPI_ISL_886473, EPI_ISL_886475, EPI_ISL_886476, EPI_ISL_886477, EPI_ISL_886479, EPI_ISL_886480, EPI_ISL_886481, EPI_ISL_886485, EPI_ISL_886487, EPI_ISL_886488, EPI_ISL_886489, EPI_ISL_886490, EPI_ISL_886491, EPI_ISL_886492, EPI_ISL_886493, EPI_ISL_886494, EPI_ISL_886495, EPI_ISL_886498, EPI_ISL_886499, EPI_ISL_886501, EPI_ISL_886503, EPI_ISL_886504, EPI_ISL_886505, EPI_ISL_886506, EPI_ISL_886507, EPI_ISL_886508, EPI_ISL_886509, EPI_ISL_886510, EPI_ISL_886511, EPI_ISL_886513, EPI_ISL_886514, EPI_ISL_886515, EPI_ISL_886516, EPI_ISL_886517, EPI_ISL_886519, EPI_ISL_886520, EPI_ISL_886521, EPI_ISL_886522, EPI_ISL_886523, EPI_ISL_886524, EPI_ISL_886526, EPI_ISL_886527, EPI_ISL_886528, EPI_ISL_886529, EPI_ISL_886530, EPI_ISL_886531, EPI_ISL_886533, EPI_ISL_886534, EPI_ISL_886538, EPI_ISL_886539, EPI_ISL_886540, EPI_ISL_886543, EPI_ISL_886545, EPI_ISL_886546, EPI_ISL_886547, EPI_ISL_886548, EPI_ISL_886549, EPI_ISL_886550, EPI_ISL_886552, EPI_ISL_886553, EPI_ISL_886554, EPI_ISL_886555, EPI_ISL_886556, EPI_ISL_886559, EPI_ISL_886560, EPI_ISL_886561, EPI_ISL_886562, EPI_ISL_886563, EPI_ISL_886564, EPI_ISL_886566, EPI_ISL_886568, EPI_ISL_886569, EPI_ISL_886571, EPI_ISL_886572, EPI_ISL_886573, EPI_ISL_886575, EPI_ISL_886576, EPI_ISL_886579, EPI_ISL_886580, EPI_ISL_886581, EPI_ISL_886582, EPI_ISL_886585, EPI_ISL_886586, EPI_ISL_886588, EPI_ISL_886590, EPI_ISL_886591, EPI_ISL_886592, EPI_ISL_886593, EPI_ISL_886594, EPI_ISL_886595, EPI_ISL_886596, EPI_ISL_886597, EPI_ISL_886598, EPI_ISL_886599, EPI_ISL_886600, EPI_ISL_886601, EPI_ISL_886602, EPI_ISL_886604, EPI_ISL_886605, EPI_ISL_886606, EPI_ISL_886609, EPI_ISL_886610, EPI_ISL_886611, EPI_ISL_886612, EPI_ISL_886615, EPI_ISL_886618, EPI_ISL_886620, EPI_ISL_886621, EPI_ISL_886622, EPI_ISL_886624, EPI_ISL_886625, EPI_ISL_886626, EPI_ISL_886627, EPI_ISL_886628, EPI_ISL_886630, EPI_ISL_886631, EPI_ISL_886634, EPI_ISL_886635, EPI_ISL_886638, EPI_ISL_886639, EPI_ISL_886640, EPI_ISL_886641, EPI_ISL_886642, EPI_ISL_886643, EPI_ISL_886644, EPI_ISL_886646, EPI_ISL_886647, EPI_ISL_886648, EPI_ISL_886650, EPI_ISL_886652, EPI_ISL_886653, EPI_ISL_886654, EPI_ISL_886655, EPI_ISL_886656, EPI_ISL_886657, EPI_ISL_886659, EPI_ISL_886660, EPI_ISL_886661, EPI_ISL_886663, EPI_ISL_886664, EPI_ISL_886665, EPI_ISL_886666, EPI_ISL_886667, EPI_ISL_886668, EPI_ISL_886669, EPI_ISL_886670, EPI_ISL_886671, EPI_ISL_886673, EPI_ISL_886674, EPI_ISL_886675, EPI_ISL_886676, EPI_ISL_886678, EPI_ISL_886680, EPI_ISL_886681, EPI_ISL_886685, EPI_ISL_886686, EPI_ISL_886688, EPI_ISL_886689, EPI_ISL_886690, EPI_ISL_886692, EPI_ISL_886693, EPI_ISL_886694, EPI_ISL_886698, EPI_ISL_886699, EPI_ISL_886701, EPI_ISL_886702, EPI_ISL_886703, EPI_ISL_886704, EPI_ISL_886706, EPI_ISL_886707, EPI_ISL_886708, EPI_ISL_886710, EPI_ISL_886711, EPI_ISL_886712, EPI_ISL_886713, EPI_ISL_886714, EPI_ISL_886715, EPI_ISL_886716, EPI_ISL_886717, EPI_ISL_886718, EPI_ISL_886719, EPI_ISL_886720, EPI_ISL_886721, EPI_ISL_886722, EPI_ISL_886725, EPI_ISL_886726, EPI_ISL_886727, EPI_ISL_886729, EPI_ISL_886731, EPI_ISL_886732, EPI_ISL_886733, EPI_ISL_886734, EPI_ISL_886736, EPI_ISL_886737, EPI_ISL_886738, EPI_ISL_886739, EPI_ISL_886740, EPI_ISL_886741, EPI_ISL_886742, EPI_ISL_886745, EPI_ISL_886746, EPI_ISL_886749, EPI_ISL_886750, EPI_ISL_886751, EPI_ISL_886752, EPI_ISL_886753, EPI_ISL_886754, EPI_ISL_886755, EPI_ISL_886756, EPI_ISL_886757, EPI_ISL_886758, EPI_ISL_886759, EPI_ISL_886760, EPI_ISL_886761, EPI_ISL_886762, EPI_ISL_886763, EPI_ISL_886765, EPI_ISL_886766, EPI_ISL_886767, EPI_ISL_886768, EPI_ISL_886770, EPI_ISL_886771, EPI_ISL_886773, EPI_ISL_886774, EPI_ISL_886775, EPI_ISL_886776, EPI_ISL_886777, EPI_ISL_886778, EPI_ISL_886781, EPI_ISL_886782, EPI_ISL_886783, EPI_ISL_886784, EPI_ISL_886785, EPI_ISL_886786, EPI_ISL_886787, EPI_ISL_886788, EPI_ISL_886790, EPI_ISL_886791, EPI_ISL_886792, EPI_ISL_886793, EPI_ISL_886794, EPI_ISL_886795, EPI_ISL_886796, EPI_ISL_886797, EPI_ISL_886798, EPI_ISL_886799, EPI_ISL_886800, EPI_ISL_886801, EPI_ISL_886802, EPI_ISL_886803, EPI_ISL_886807, EPI_ISL_886808, EPI_ISL_886809, EPI_ISL_886810, EPI_ISL_886812, EPI_ISL_886814, EPI_ISL_886820, EPI_ISL_886822, EPI_ISL_886823, EPI_ISL_886825, EPI_ISL_886826, EPI_ISL_886827, EPI_ISL_886829, EPI_ISL_886830, EPI_ISL_886832, EPI_ISL_886834, EPI_ISL_886835, EPI_ISL_886837, EPI_ISL_886838, EPI_ISL_886840, EPI_ISL_886841, EPI_ISL_886842, EPI_ISL_886843, EPI_ISL_886844, EPI_ISL_886845, EPI_ISL_886846, EPI_ISL_886847, EPI_ISL_886849, EPI_ISL_886851, EPI_ISL_886852, EPI_ISL_886853, EPI_ISL_886854, EPI_ISL_886855, EPI_ISL_886859, EPI_ISL_886860, EPI_ISL_886861, EPI_ISL_886862, EPI_ISL_886864, EPI_ISL_886865, EPI_ISL_886866, EPI_ISL_886867, EPI_ISL_886868, EPI_ISL_886869, EPI_ISL_886873, EPI_ISL_886875, EPI_ISL_886876, EPI_ISL_886877, EPI_ISL_886878, EPI_ISL_886879, EPI_ISL_886880, EPI_ISL_886881, EPI_ISL_886882, EPI_ISL_886883, EPI_ISL_886884, EPI_ISL_886885, EPI_ISL_886886, EPI_ISL_886887, EPI_ISL_886888, EPI_ISL_886889, EPI_ISL_886890, EPI_ISL_886891, EPI_ISL_886892, EPI_ISL_886894, EPI_ISL_886898, EPI_ISL_886899, EPI_ISL_886900, EPI_ISL_886901, EPI_ISL_886902, EPI_ISL_886903, EPI_ISL_886904, EPI_ISL_886905, EPI_ISL_886906, EPI_ISL_886907, EPI_ISL_886908, EPI_ISL_886911, EPI_ISL_886912, EPI_ISL_886913, EPI_ISL_886914, EPI_ISL_886915, EPI_ISL_886917, EPI_ISL_886918, EPI_ISL_886919, EPI_ISL_886920, EPI_ISL_886921, EPI_ISL_886926, EPI_ISL_886927, EPI_ISL_886928, EPI_ISL_886929, EPI_ISL_886930, EPI_ISL_886931, EPI_ISL_886932, EPI_ISL_886933, EPI_ISL_886934, EPI_ISL_886935, EPI_ISL_886936, EPI_ISL_886937, EPI_ISL_886938, EPI_ISL_886940, EPI_ISL_886941, EPI_ISL_886942, EPI_ISL_886943, EPI_ISL_886944, EPI_ISL_886947, EPI_ISL_886948, EPI_ISL_886949, EPI_ISL_886950, EPI_ISL_886951, EPI_ISL_886952, EPI_ISL_886953, EPI_ISL_886954, EPI_ISL_886955, EPI_ISL_886956, EPI_ISL_886957, EPI_ISL_886961, EPI_ISL_886962, EPI_ISL_886963, EPI_ISL_886964, EPI_ISL_886965, EPI_ISL_886967, EPI_ISL_886968, EPI_ISL_886969, EPI_ISL_886971, EPI_ISL_886972, EPI_ISL_886973, EPI_ISL_886974, EPI_ISL_886975, EPI_ISL_886976, EPI_ISL_886979, EPI_ISL_886980, EPI_ISL_886982, EPI_ISL_886983, EPI_ISL_886984, EPI_ISL_886985, EPI_ISL_886986, EPI_ISL_886987, EPI_ISL_886989, EPI_ISL_886990, EPI_ISL_886991, EPI_ISL_886992, EPI_ISL_886995, EPI_ISL_886997, EPI_ISL_886998, EPI_ISL_886999, EPI_ISL_887000, EPI_ISL_887002, EPI_ISL_887003, EPI_ISL_887004, EPI_ISL_887005, EPI_ISL_887006, EPI_ISL_887008, EPI_ISL_887009, EPI_ISL_887011, EPI_ISL_887012, EPI_ISL_887013, EPI_ISL_887014, EPI_ISL_887015, EPI_ISL_887016, EPI_ISL_887017, EPI_ISL_887018, EPI_ISL_887019, EPI_ISL_887020, EPI_ISL_887021, EPI_ISL_887022, EPI_ISL_887023, EPI_ISL_887024, EPI_ISL_887028, EPI_ISL_887030, EPI_ISL_887031, EPI_ISL_887032, EPI_ISL_887033, EPI_ISL_887034, EPI_ISL_887035, EPI_ISL_887037, EPI_ISL_887038, EPI_ISL_887039, EPI_ISL_887040, EPI_ISL_887041, EPI_ISL_887042, EPI_ISL_887049, EPI_ISL_887050, EPI_ISL_887051, EPI_ISL_887052, EPI_ISL_887078, EPI_ISL_887079, EPI_ISL_887081, EPI_ISL_887082, EPI_ISL_887083, EPI_ISL_887084, EPI_ISL_887085, EPI_ISL_887086, EPI_ISL_887087, EPI_ISL_887089, EPI_ISL_887090, EPI_ISL_887091, EPI_ISL_887095, EPI_ISL_887097, EPI_ISL_887098, EPI_ISL_887099, EPI_ISL_887101, EPI_ISL_887102, EPI_ISL_887103

see above | Labcorp | Genomics and Discovery, Respiratory Viruses Branch, Division of Viral Diseases, Centers for Disease Control and Prevention | Peter W. Cook,Dhwani Batra,Ben L. Rambo-Martin,Summer Galloway,Brian Krueger,Minoo Agarwal,Eyad Almasri,Debbie Boles,Ayla Burns,Nuthawin Charoensri,Oren Cohen,Susan Countryman,Mary Ann Cristobal,Bobbi Croy,Suzanne Dale,Hrushikesh Deshmukh,Amanda Douglas,Vincent Drouillon,Marcia Eisenberg,Howard Engler,Rama Ghatti,Prashant Gupta,Susan Hicks,Jake Humphrey,Lax Iyer,Manoj Jain,Mohan Kolli,Tim Kuphal,Stanley Letovsky,Michael Levandoski,Craig Lukasik,Jonathan Meltzer,Brian Norvell,Mindy Nye,Scott Parker,Christos Petropoulos,John Pruitt,Steven Ragan,Scott Ryan,Mike Sapeta,Jana Schroth,Suresh Babu Selvaraju,Goran Stevovic,Amanda Suchanek,Andrea Throop,Lyndon Tilson,Thomas Urban,Joe Voshell,Kimberly Wagner,Jonathan Williams,Mary Williamson,Qian Zeng,Tricia Zwiefelhofer,Clinton R. Paden,Suxiang Tong,Duncan MacCannell,

EPI_ISL_887145, EPI_ISL_887146, EPI_ISL_887147, EPI_ISL_887149, EPI_ISL_887150, EPI_ISL_887151, EPI_ISL_887152, EPI_ISL_887153, EPI_ISL_887154, EPI_ISL_887155, EPI_ISL_887156, EPI_ISL_887157, EPI_ISL_887158, EPI_ISL_887159, EPI_ISL_887160, EPI_ISL_887161, EPI_ISL_887162, EPI_ISL_887163, EPI_ISL_887164, EPI_ISL_887165

see above | Massachusetts General Hospital | Infectious Disease Program, Broad Institute of Harvard and MIT | Lemieux,J.E., Siddle,K.J., Shaw,B., Adams,G., Pierce,V., Turbett,S., Anahtar,M., Branda,J., Slater,D., Harris,J., Lin,A.E., Gladden-Young,A., Lagerborg,K., Rudy,M., DeRuff,K., Carter,A., Normandin,E., Bauer,M., Reilly,S., Tomkins-Tinch,C., Loreth,C., Chaluvadi,S., Neumann,A., Cusick,C., Chapman,S.B., Gnirke,A., Flowers,K., Cerrato,F., Birren,B.W., Gallagher,G., Smole,S., Park,D.J., MacInnis,B.L., Ryan,E., LaRocque,R., Rosenberg,E. and Sabeti,P.C.

EPI_ISL_887508, EPI_ISL_887509, EPI_ISL_887510, EPI_ISL_887511, EPI_ISL_887512, EPI_ISL_887513, EPI_ISL_887514, EPI_ISL_887515, EPI_ISL_887516, EPI_ISL_887517, EPI_ISL_887518, EPI_ISL_887519, EPI_ISL_887520, EPI_ISL_887521, EPI_ISL_887522, EPI_ISL_887523, EPI_ISL_887524, EPI_ISL_887525, EPI_ISL_887526, EPI_ISL_887527, EPI_ISL_887528, EPI_ISL_887529, EPI_ISL_887532, EPI_ISL_887533, EPI_ISL_887534, EPI_ISL_887535, EPI_ISL_887536, EPI_ISL_887537, EPI_ISL_887538, EPI_ISL_887539, EPI_ISL_887540, EPI_ISL_887541, EPI_ISL_887542, EPI_ISL_887543, EPI_ISL_887544, EPI_ISL_887545, EPI_ISL_887546, EPI_ISL_887547, EPI_ISL_887548, EPI_ISL_887549, EPI_ISL_887550, EPI_ISL_887551, EPI_ISL_887552, EPI_ISL_887553, EPI_ISL_887554, EPI_ISL_887555, EPI_ISL_887556, EPI_ISL_887557, EPI_ISL_887558, EPI_ISL_887559, EPI_ISL_887560, EPI_ISL_887561, EPI_ISL_887562, EPI_ISL_887563, EPI_ISL_887564, EPI_ISL_887565, EPI_ISL_887566, EPI_ISL_887567, EPI_ISL_887568, EPI_ISL_887569, EPI_ISL_887570, EPI_ISL_887571, EPI_ISL_887572, EPI_ISL_887573, EPI_ISL_887574, EPI_ISL_887575, EPI_ISL_887576, EPI_ISL_887577, EPI_ISL_887578, EPI_ISL_887579, EPI_ISL_887580, EPI_ISL_887581, EPI_ISL_887582, EPI_ISL_887583, EPI_ISL_887584, EPI_ISL_887585, EPI_ISL_887586

see above | Johns Hopkins Hospital Department of Pathology | Johns Hopkins Hospital Department of Pathology | C. Paul Morris, Chun Huai Luo, Adannaya Amadi, Matthew Schwartz, Nicholas Gallagher, Heba H. Mostafa

EPI_ISL_887589 | SC Dept of Health and Env. Control-Bureau of Laboratories | Centers for Disease Control and Prevention Division of Viral Diseases, Pathogen Discovery | Yan Li, Anna Montmayeur, Ying Tao, Jing Zhang, Krista Queen, Anna Uehara, Rachel Marine, Peter Cook, Clinton R. Paden, Haibin Wang, Suxiang Tong

EPI_ISL_887591, EPI_ISL_887592, EPI_ISL_887593, EPI_ISL_887594, EPI_ISL_887595, EPI_ISL_887596, EPI_ISL_887597, EPI_ISL_887598, EPI_ISL_887599, EPI_ISL_887600, EPI_ISL_887601, EPI_ISL_887602, EPI_ISL_887603, EPI_ISL_887604, EPI_ISL_887605, EPI_ISL_887606, EPI_ISL_887607, EPI_ISL_887608, EPI_ISL_887609, EPI_ISL_887610, EPI_ISL_887611, EPI_ISL_887612, EPI_ISL_887613, EPI_ISL_887614, EPI_ISL_887615, EPI_ISL_887616, EPI_ISL_887617, EPI_ISL_887618, EPI_ISL_887619, EPI_ISL_887620, EPI_ISL_887621, EPI_ISL_887622, EPI_ISL_887623, EPI_ISL_887624, EPI_ISL_887625, EPI_ISL_887626, EPI_ISL_887627, EPI_ISL_887628, EPI_ISL_887629, EPI_ISL_887630, EPI_ISL_887632, EPI_ISL_887633, EPI_ISL_887634, EPI_ISL_887635, EPI_ISL_887636, EPI_ISL_887637, EPI_ISL_887638, EPI_ISL_887639, EPI_ISL_887640, EPI_ISL_887641, EPI_ISL_887642, EPI_ISL_887643, EPI_ISL_887644, EPI_ISL_887645, EPI_ISL_887646, EPI_ISL_887647, EPI_ISL_887648, EPI_ISL_887649, EPI_ISL_887650, EPI_ISL_887651, EPI_ISL_887652, EPI_ISL_887653, EPI_ISL_887654, EPI_ISL_887655, EPI_ISL_887656, EPI_ISL_887657, EPI_ISL_887658, EPI_ISL_887659, EPI_ISL_887660, EPI_ISL_887661, EPI_ISL_887662, EPI_ISL_887663, EPI_ISL_887664, EPI_ISL_887665, EPI_ISL_887666, EPI_ISL_887667, EPI_ISL_887668, EPI_ISL_887669, EPI_ISL_887670, EPI_ISL_887671, EPI_ISL_887672, EPI_ISL_887673, EPI_ISL_887674, EPI_ISL_887675, EPI_ISL_887676, EPI_ISL_887677, EPI_ISL_887678, EPI_ISL_887679, EPI_ISL_887680, EPI_ISL_887681, EPI_ISL_887682, EPI_ISL_887683, EPI_ISL_887684, EPI_ISL_887685, EPI_ISL_887686, EPI_ISL_887687, EPI_ISL_887688, EPI_ISL_887689, EPI_ISL_887690, EPI_ISL_887691, EPI_ISL_887692, EPI_ISL_887693, EPI_ISL_887694, EPI_ISL_887695, EPI_ISL_887696, EPI_ISL_887697, EPI_ISL_887698, EPI_ISL_887699, EPI_ISL_887700, EPI_ISL_887701, EPI_ISL_887702, EPI_ISL_887703, EPI_ISL_887704, EPI_ISL_887705, EPI_ISL_887706, EPI_ISL_887707, EPI_ISL_887708, EPI_ISL_887709, EPI_ISL_887710, EPI_ISL_887711, EPI_ISL_887712, EPI_ISL_887713, EPI_ISL_887714, EPI_ISL_887715, EPI_ISL_887716, EPI_ISL_887717, EPI_ISL_887718, EPI_ISL_887719, EPI_ISL_887720, EPI_ISL_887721, EPI_ISL_887722, EPI_ISL_887723, EPI_ISL_887724, EPI_ISL_887725, EPI_ISL_887726, EPI_ISL_887727, EPI_ISL_887728, EPI_ISL_887729, EPI_ISL_887730, EPI_ISL_887731, EPI_ISL_887732, EPI_ISL_887733, EPI_ISL_887734, EPI_ISL_887735, EPI_ISL_887736, EPI_ISL_887737, EPI_ISL_887738, EPI_ISL_887739, EPI_ISL_887740, EPI_ISL_887741, EPI_ISL_887879, EPI_ISL_887880, EPI_ISL_887881, EPI_ISL_887883, EPI_ISL_887884, EPI_ISL_887885, EPI_ISL_887886, EPI_ISL_887887, EPI_ISL_887888, EPI_ISL_887889, EPI_ISL_887890, EPI_ISL_887891, EPI_ISL_887892, EPI_ISL_887893, EPI_ISL_887894, EPI_ISL_887895, EPI_ISL_887896, EPI_ISL_887897, EPI_ISL_887898, EPI_ISL_887899, EPI_ISL_887900, EPI_ISL_887901, EPI_ISL_887902, EPI_ISL_887903, EPI_ISL_887904, EPI_ISL_887945, EPI_ISL_887946, EPI_ISL_887947, EPI_ISL_887948, EPI_ISL_887949, EPI_ISL_887950, EPI_ISL_887951, EPI_ISL_887952, EPI_ISL_887953, EPI_ISL_887954, EPI_ISL_887955, EPI_ISL_887956, EPI_ISL_887957, EPI_ISL_887958, EPI_ISL_887959, EPI_ISL_887960, EPI_ISL_887961, EPI_ISL_887962, EPI_ISL_887963, EPI_ISL_887964, EPI_ISL_887965, EPI_ISL_887966, EPI_ISL_887967, EPI_ISL_887968, EPI_ISL_887969, EPI_ISL_887970, EPI_ISL_887971, EPI_ISL_887972, EPI_ISL_887973, EPI_ISL_887974, EPI_ISL_887975, EPI_ISL_887976, EPI_ISL_887977, EPI_ISL_887978, EPI_ISL_887979, EPI_ISL_887980, EPI_ISL_887981, EPI_ISL_887982, EPI_ISL_887983, EPI_ISL_887984, EPI_ISL_887985, EPI_ISL_887986, EPI_ISL_887987, EPI_ISL_887988, EPI_ISL_887989, EPI_ISL_887990, EPI_ISL_887991, EPI_ISL_887992, EPI_ISL_887993, EPI_ISL_887994, EPI_ISL_887995, EPI_ISL_887996, EPI_ISL_887997, EPI_ISL_887998, EPI_ISL_887999, EPI_ISL_888000, EPI_ISL_888001, EPI_ISL_888002, EPI_ISL_888003, EPI_ISL_888004, EPI_ISL_888005, EPI_ISL_888006, EPI_ISL_888007, EPI_ISL_888008, EPI_ISL_888009, EPI_ISL_888010, EPI_ISL_888011, EPI_ISL_888012, EPI_ISL_888013, EPI_ISL_888014, EPI_ISL_888015, EPI_ISL_888016, EPI_ISL_888017, EPI_ISL_888018, EPI_ISL_888019, EPI_ISL_888020, EPI_ISL_888021, EPI_ISL_888022, EPI_ISL_888023, EPI_ISL_888024, EPI_ISL_888025, EPI_ISL_888026, EPI_ISL_888027, EPI_ISL_888028, EPI_ISL_888029, EPI_ISL_888030, EPI_ISL_888031, EPI_ISL_888032, EPI_ISL_888033, EPI_ISL_888034, EPI_ISL_888035, EPI_ISL_888036, EPI_ISL_888037, EPI_ISL_888038, EPI_ISL_888039, EPI_ISL_888040, EPI_ISL_888041, EPI_ISL_888043, EPI_ISL_888044, EPI_ISL_888045, EPI_ISL_888046, EPI_ISL_888047, EPI_ISL_888048, EPI_ISL_888049, EPI_ISL_888050, EPI_ISL_888051, EPI_ISL_888052, EPI_ISL_888053, EPI_ISL_888054, EPI_ISL_888055, EPI_ISL_888056, EPI_ISL_888057, EPI_ISL_888058, EPI_ISL_888059, EPI_ISL_888060, EPI_ISL_888061, EPI_ISL_888062, EPI_ISL_888063, EPI_ISL_888064, EPI_ISL_888065, EPI_ISL_888066, EPI_ISL_888067, EPI_ISL_888068, EPI_ISL_888069, EPI_ISL_888070, EPI_ISL_888071, EPI_ISL_888072, EPI_ISL_888073, EPI_ISL_888074, EPI_ISL_888075, EPI_ISL_888076, EPI_ISL_888077, EPI_ISL_888078, EPI_ISL_888079, EPI_ISL_888080, EPI_ISL_888081, EPI_ISL_888082, EPI_ISL_888083, EPI_ISL_888084, EPI_ISL_888085, EPI_ISL_888086, EPI_ISL_888087, EPI_ISL_888088, EPI_ISL_888089, EPI_ISL_888090, EPI_ISL_888091, EPI_ISL_888092, EPI_ISL_888093, EPI_ISL_888094, EPI_ISL_888095, EPI_ISL_888096, EPI_ISL_888097, EPI_ISL_888098, EPI_ISL_888099, EPI_ISL_888100, EPI_ISL_888101, EPI_ISL_888102, EPI_ISL_888103, EPI_ISL_888104, EPI_ISL_888105, EPI_ISL_888106, EPI_ISL_888107, EPI_ISL_888108, EPI_ISL_888109, EPI_ISL_888110, EPI_ISL_888111, EPI_ISL_888112, EPI_ISL_888113, EPI_ISL_888114, EPI_ISL_888115, EPI_ISL_888116, EPI_ISL_888117, EPI_ISL_888118, EPI_ISL_888119

see above | Labcorp | Genomics and Discovery, Respiratory Viruses Branch, Division of Viral Diseases, Centers for Disease Control and Prevention | Peter W. Cook,Dhwani Batra,Ben L. Rambo-Martin,Summer Galloway,Brian Krueger,Minoo Agarwal,Eyad Almasri,Debbie Boles,Ayla Burns,Nuthawin Charoensri,Oren Cohen,Susan Countryman,Mary Ann Cristobal,Bobbi Croy,Suzanne Dale,Hrushikesh Deshmukh,Amanda Douglas,Vincent Drouillon,Marcia Eisenberg,Howard Engler,Rama Ghatti,Prashant Gupta,Susan Hicks,Jake Humphrey,Lax Iyer,Manoj Jain,Mohan Kolli,Tim Kuphal,Stanley Letovsky,Michael Levandoski,Craig Lukasik,Jonathan Meltzer,Brian Norvell,Mindy Nye,Scott Parker,Christos Petropoulos,John Pruitt,Steven Ragan,Scott Ryan,Mike Sapeta,Jana Schroth,Suresh Babu Selvaraju,Goran Stevovic,Amanda Suchanek,Andrea Throop,Lyndon Tilson,Thomas Urban,Joe Voshell,Kimberly Wagner,Jonathan Williams,Mary Williamson,Qian Zeng,Tricia Zwiefelhofer,Clinton R. Paden,Suxiang Tong,Duncan MacCannell,

EPI_ISL_888120 | SC Dept of Health and Env. Control-Bureau of Laboratories | Centers for Disease Control and Prevention Division of Viral Diseases, Pathogen Discovery | Yan Li, Anna Montmayeur, Ying Tao, Jing Zhang, Krista Queen, Anna Uehara, Rachel Marine, Peter Cook, Clinton R. Paden, Haibin Wang, Suxiang Tong

EPI_ISL_888121, EPI_ISL_888122, EPI_ISL_888123, EPI_ISL_888124, EPI_ISL_888125, EPI_ISL_888126, EPI_ISL_888127, EPI_ISL_888128, EPI_ISL_888129, EPI_ISL_888130, EPI_ISL_888131, EPI_ISL_888132, EPI_ISL_888133, EPI_ISL_888134, EPI_ISL_888135, EPI_ISL_888136, EPI_ISL_888137, EPI_ISL_888138, EPI_ISL_888140, EPI_ISL_888141, EPI_ISL_888142, EPI_ISL_888143, EPI_ISL_888144, EPI_ISL_888145, EPI_ISL_888146, EPI_ISL_888147, EPI_ISL_888148, EPI_ISL_888149, EPI_ISL_888150, EPI_ISL_888151, EPI_ISL_888152, EPI_ISL_888153, EPI_ISL_888154, EPI_ISL_888155, EPI_ISL_888156, EPI_ISL_888157, EPI_ISL_888158, EPI_ISL_888159, EPI_ISL_888160, EPI_ISL_888161, EPI_ISL_888162, EPI_ISL_888163, EPI_ISL_888164, EPI_ISL_888165, EPI_ISL_888166, EPI_ISL_888167, EPI_ISL_888168, EPI_ISL_888169, EPI_ISL_888170, EPI_ISL_888171, EPI_ISL_888172, EPI_ISL_888173, EPI_ISL_888174, EPI_ISL_888175, EPI_ISL_888176, EPI_ISL_888177, EPI_ISL_888178, EPI_ISL_888179, EPI_ISL_888180, EPI_ISL_888181, EPI_ISL_888182, EPI_ISL_888183, EPI_ISL_888184, EPI_ISL_888185, EPI_ISL_888186, EPI_ISL_888187, EPI_ISL_888188, EPI_ISL_888189, EPI_ISL_888190, EPI_ISL_888191, EPI_ISL_888192, EPI_ISL_888193, EPI_ISL_888194, EPI_ISL_888195, EPI_ISL_888196, EPI_ISL_888197, EPI_ISL_888198, EPI_ISL_888199, EPI_ISL_888200, EPI_ISL_888201, EPI_ISL_888202, EPI_ISL_888203, EPI_ISL_888204, EPI_ISL_888205, EPI_ISL_888206, EPI_ISL_888207, EPI_ISL_888208, EPI_ISL_888209, EPI_ISL_888210, EPI_ISL_888211, EPI_ISL_888212, EPI_ISL_888213, EPI_ISL_888214, EPI_ISL_888215, EPI_ISL_888216, EPI_ISL_888217, EPI_ISL_888218, EPI_ISL_888219, EPI_ISL_888220, EPI_ISL_888221, EPI_ISL_888222, EPI_ISL_888223, EPI_ISL_888224, EPI_ISL_888225, EPI_ISL_888226, EPI_ISL_888227, EPI_ISL_888228, EPI_ISL_888229, EPI_ISL_888230, EPI_ISL_888231, EPI_ISL_888232, EPI_ISL_888233, EPI_ISL_888234, EPI_ISL_888235, EPI_ISL_888236, EPI_ISL_888237, EPI_ISL_888238, EPI_ISL_888239, EPI_ISL_888240, EPI_ISL_888241, EPI_ISL_888242, EPI_ISL_888243, EPI_ISL_888244, EPI_ISL_888245, EPI_ISL_888246, EPI_ISL_888247, EPI_ISL_888248, EPI_ISL_888249, EPI_ISL_888250, EPI_ISL_888251, EPI_ISL_888252, EPI_ISL_888253, EPI_ISL_888254, EPI_ISL_888255, EPI_ISL_888256, EPI_ISL_888257, EPI_ISL_888258, EPI_ISL_888259, EPI_ISL_888260, EPI_ISL_888261, EPI_ISL_888262, EPI_ISL_888263, EPI_ISL_888264, EPI_ISL_888265, EPI_ISL_888266, EPI_ISL_888267, EPI_ISL_888268, EPI_ISL_888269, EPI_ISL_888270, EPI_ISL_888271, EPI_ISL_888272, EPI_ISL_888273, EPI_ISL_888274, EPI_ISL_888275, EPI_ISL_888276, EPI_ISL_888312, EPI_ISL_888313, EPI_ISL_888314, EPI_ISL_888315, EPI_ISL_888316, EPI_ISL_888317, EPI_ISL_888318, EPI_ISL_888319, EPI_ISL_888320, EPI_ISL_888321, EPI_ISL_888322, EPI_ISL_888323, EPI_ISL_888324, EPI_ISL_888325, EPI_ISL_888326, EPI_ISL_888327, EPI_ISL_888328, EPI_ISL_888329, EPI_ISL_888330, EPI_ISL_888331, EPI_ISL_888332, EPI_ISL_888333, EPI_ISL_888334, EPI_ISL_888335, EPI_ISL_888336, EPI_ISL_888337, EPI_ISL_888338, EPI_ISL_888339, EPI_ISL_888340, EPI_ISL_888341, EPI_ISL_888342, EPI_ISL_888343, EPI_ISL_888344, EPI_ISL_888345, EPI_ISL_888346, EPI_ISL_888347, EPI_ISL_888348, EPI_ISL_888349, EPI_ISL_888350, EPI_ISL_888351, EPI_ISL_888352, EPI_ISL_888353, EPI_ISL_888354, EPI_ISL_888355, EPI_ISL_888356, EPI_ISL_888357, EPI_ISL_888358, EPI_ISL_888359, EPI_ISL_888360, EPI_ISL_888361, EPI_ISL_888362, EPI_ISL_888363, EPI_ISL_888364, EPI_ISL_888365, EPI_ISL_888366, EPI_ISL_888367, EPI_ISL_888368, EPI_ISL_888369, EPI_ISL_888370, EPI_ISL_888371, EPI_ISL_888372, EPI_ISL_888373, EPI_ISL_888374, EPI_ISL_888375, EPI_ISL_888376, EPI_ISL_888377, EPI_ISL_888378, EPI_ISL_888379, EPI_ISL_888380, EPI_ISL_888381, EPI_ISL_888382, EPI_ISL_888383, EPI_ISL_888384, EPI_ISL_888385, EPI_ISL_888386, EPI_ISL_888387, EPI_ISL_888388, EPI_ISL_888389, EPI_ISL_888390, EPI_ISL_888391, EPI_ISL_888392, EPI_ISL_888393, EPI_ISL_888394, EPI_ISL_888395, EPI_ISL_888396, EPI_ISL_888397, EPI_ISL_888398, EPI_ISL_888399, EPI_ISL_888400, EPI_ISL_888401, EPI_ISL_888402, EPI_ISL_888403, EPI_ISL_888404, EPI_ISL_888405, EPI_ISL_888406, EPI_ISL_888407, EPI_ISL_888408, EPI_ISL_888409, EPI_ISL_888410, EPI_ISL_888411, EPI_ISL_888412, EPI_ISL_888413, EPI_ISL_888414, EPI_ISL_888415, EPI_ISL_888416, EPI_ISL_888417, EPI_ISL_888418, EPI_ISL_888419, EPI_ISL_888420, EPI_ISL_888421, EPI_ISL_888422, EPI_ISL_888423, EPI_ISL_888424, EPI_ISL_888425, EPI_ISL_888426, EPI_ISL_888427, EPI_ISL_888428, EPI_ISL_888429, EPI_ISL_888430, EPI_ISL_888431, EPI_ISL_888432, EPI_ISL_888433, EPI_ISL_888434, EPI_ISL_888448, EPI_ISL_888449, EPI_ISL_888450, EPI_ISL_888451, EPI_ISL_888452, EPI_ISL_888453, EPI_ISL_888454, EPI_ISL_888455, EPI_ISL_888456, EPI_ISL_888457, EPI_ISL_888458, EPI_ISL_888459, EPI_ISL_888460, EPI_ISL_888461, EPI_ISL_888462, EPI_ISL_888463, EPI_ISL_888464, EPI_ISL_888465, EPI_ISL_888466, EPI_ISL_888467, EPI_ISL_888468, EPI_ISL_888469, EPI_ISL_888470, EPI_ISL_888471, EPI_ISL_888472, EPI_ISL_888473, EPI_ISL_888474, EPI_ISL_888475, EPI_ISL_888476, EPI_ISL_888477, EPI_ISL_888478, EPI_ISL_888479, EPI_ISL_888480, EPI_ISL_888481, EPI_ISL_888482, EPI_ISL_888483, EPI_ISL_888485, EPI_ISL_888486, EPI_ISL_888487, EPI_ISL_888488, EPI_ISL_888489, EPI_ISL_888490, EPI_ISL_888491, EPI_ISL_888492, EPI_ISL_888493, EPI_ISL_888494, EPI_ISL_888495, EPI_ISL_888496, EPI_ISL_888497, EPI_ISL_888498, EPI_ISL_888499, EPI_ISL_888502, EPI_ISL_888503, EPI_ISL_888504, EPI_ISL_888505, EPI_ISL_888506, EPI_ISL_888507, EPI_ISL_888508, EPI_ISL_888509, EPI_ISL_888510, EPI_ISL_888511, EPI_ISL_888512, EPI_ISL_888513, EPI_ISL_888514, EPI_ISL_888515, EPI_ISL_888516, EPI_ISL_888517, EPI_ISL_888518, EPI_ISL_888519, EPI_ISL_888520, EPI_ISL_888521, EPI_ISL_888522, EPI_ISL_888523, EPI_ISL_888524, EPI_ISL_888525, EPI_ISL_888526, EPI_ISL_888527, EPI_ISL_888528, EPI_ISL_888529, EPI_ISL_888530, EPI_ISL_888531, EPI_ISL_888532, EPI_ISL_888533, EPI_ISL_888534, EPI_ISL_888535, EPI_ISL_888536, EPI_ISL_888537, EPI_ISL_888538, EPI_ISL_888539, EPI_ISL_888540, EPI_ISL_888541, EPI_ISL_888542, EPI_ISL_888543, EPI_ISL_888544, EPI_ISL_888545, EPI_ISL_888546, EPI_ISL_888547, EPI_ISL_888548, EPI_ISL_888549, EPI_ISL_888550, EPI_ISL_888551, EPI_ISL_888552, EPI_ISL_888553, EPI_ISL_888554, EPI_ISL_888555, EPI_ISL_888556, EPI_ISL_888557, EPI_ISL_888558, EPI_ISL_888559, EPI_ISL_888560, EPI_ISL_888561, EPI_ISL_888562, EPI_ISL_888563, EPI_ISL_888564, EPI_ISL_888565, EPI_ISL_888566, EPI_ISL_888567, EPI_ISL_888568, EPI_ISL_888569, EPI_ISL_888570, EPI_ISL_888571, EPI_ISL_888572, EPI_ISL_888573, EPI_ISL_888574, EPI_ISL_888575, EPI_ISL_888576, EPI_ISL_888577, EPI_ISL_888578, EPI_ISL_888579, EPI_ISL_888580, EPI_ISL_888581, EPI_ISL_888582, EPI_ISL_888583, EPI_ISL_888584, EPI_ISL_888585, EPI_ISL_888586, EPI_ISL_888587, EPI_ISL_888588

see above | Labcorp | Genomics and Discovery, Respiratory Viruses Branch, Division of Viral Diseases, Centers for Disease Control and Prevention | Peter W. Cook,Dhwani Batra,Ben L. Rambo-Martin,Summer Galloway,Brian Krueger,Minoo Agarwal,Eyad Almasri,Debbie Boles,Ayla Burns,Nuthawin Charoensri,Oren Cohen,Susan Countryman,Mary Ann Cristobal,Bobbi Croy,Suzanne Dale,Hrushikesh Deshmukh,Amanda Douglas,Vincent Drouillon,Marcia Eisenberg,Howard Engler,Rama Ghatti,Prashant Gupta,Susan Hicks,Jake Humphrey,Lax Iyer,Manoj Jain,Mohan Kolli,Tim Kuphal,Stanley Letovsky,Michael Levandoski,Craig Lukasik,Jonathan Meltzer,Brian Norvell,Mindy Nye,Scott Parker,Christos Petropoulos,John Pruitt,Steven Ragan,Scott Ryan,Mike Sapeta,Jana Schroth,Suresh Babu Selvaraju,Goran Stevovic,Amanda Suchanek,Andrea Throop,Lyndon Tilson,Thomas Urban,Joe Voshell,Kimberly Wagner,Jonathan Williams,Mary Williamson,Qian Zeng,Tricia Zwiefelhofer,Clinton R. Paden,Suxiang Tong,Duncan MacCannell,

EPI_ISL_888597, EPI_ISL_888598, EPI_ISL_888599, EPI_ISL_888600, EPI_ISL_888602, EPI_ISL_888605, EPI_ISL_888606, EPI_ISL_888607, EPI_ISL_888608, EPI_ISL_888609, EPI_ISL_888611, EPI_ISL_888614, EPI_ISL_888615, EPI_ISL_888616, EPI_ISL_888617, EPI_ISL_888618, EPI_ISL_888619, EPI_ISL_888620, EPI_ISL_888621, EPI_ISL_888622, EPI_ISL_888624, EPI_ISL_888625

see above | Wyoming Public Health Laboratory | Wyoming Public Health Laboratory | Noah Hull, Taylor Fearing, Lynette Gumbleton, Channing Weber, Ashley Norberg, Bailey Bowcutt, and Wanda Manley

EPI_ISL_888628, EPI_ISL_888629, EPI_ISL_888630, EPI_ISL_888631, EPI_ISL_888632, EPI_ISL_888633, EPI_ISL_888634, EPI_ISL_888635, EPI_ISL_888636, EPI_ISL_888637, EPI_ISL_888638, EPI_ISL_888639, EPI_ISL_888640, EPI_ISL_888641, EPI_ISL_888642, EPI_ISL_888643, EPI_ISL_888644, EPI_ISL_888645, EPI_ISL_888646, EPI_ISL_888647, EPI_ISL_888648, EPI_ISL_888649, EPI_ISL_888650, EPI_ISL_888651

| | | | |
|---|---|---|---|
| see above | Univeristy of New Mexico Hospital | Center for Global Health, University of New Mexico Health Sciences Center | Daryl Domman, Kurt Schwalm, Justin Bacca, Jon Femling, Darrell Dinwiddie |
| EPI_ISL_888653, EPI_ISL_888654, EPI_ISL_888655, EPI_ISL_888656, EPI_ISL_888657, EPI_ISL_888658, EPI_ISL_888659, EPI_ISL_888660, EPI_ISL_888661, EPI_ISL_888662, EPI_ISL_888663, EPI_ISL_888664, EPI_ISL_888665, EPI_ISL_888666, EPI_ISL_888667, EPI_ISL_888668, EPI_ISL_888669, EPI_ISL_888670 | | | |
| see above | University of Michigan Clinical Microbiology Laboratory | Lauring Lab, University of Michigan, Department of Microbiology and Immunology | Valesano |
| EPI_ISL_888673 | Montana Public Health Laboratory | Wyoming Public Health Laboratory | Noah Hull, Joy Ritter, Taylor Fearing, Lynette Gumbleton, Channing Weber, Ashley Norberg, Bailey Bowcutt, Wanda Manley, Deborah Gibson |
| EPI_ISL_888791, EPI_ISL_888792, EPI_ISL_888793, EPI_ISL_888794, EPI_ISL_888795, EPI_ISL_888796, EPI_ISL_888797, EPI_ISL_888798, EPI_ISL_888799, EPI_ISL_888800, EPI_ISL_888801, EPI_ISL_888802, EPI_ISL_888803, EPI_ISL_888804, EPI_ISL_888805, EPI_ISL_888806, EPI_ISL_888807, EPI_ISL_888808, EPI_ISL_888809, EPI_ISL_888810, EPI_ISL_888811, EPI_ISL_888812, EPI_ISL_888813, EPI_ISL_888814, EPI_ISL_888815, EPI_ISL_888816, EPI_ISL_888817 | | | |
| see above | Univeristy of New Mexico Hospital | Center for Global Health, University of New Mexico Health Sciences Center | Daryl Domman, Kurt Schwalm, Justin Bacca, Jon Femling, Darrell Dinwiddie |
| EPI_ISL_888835, EPI_ISL_888836, EPI_ISL_888837, EPI_ISL_888838, EPI_ISL_888839, EPI_ISL_888841, EPI_ISL_888842, EPI_ISL_888843, EPI_ISL_888844, EPI_ISL_888846, EPI_ISL_888847, EPI_ISL_888848, EPI_ISL_888849, EPI_ISL_888850, EPI_ISL_888851, EPI_ISL_888852, EPI_ISL_888853, EPI_ISL_888854, EPI_ISL_888855, EPI_ISL_888856, EPI_ISL_888858, EPI_ISL_888859, EPI_ISL_888860, EPI_ISL_888861, EPI_ISL_888862, EPI_ISL_888864, EPI_ISL_888865, EPI_ISL_888866, EPI_ISL_888869, EPI_ISL_888870, EPI_ISL_888871, EPI_ISL_888872, EPI_ISL_888875, EPI_ISL_888876, EPI_ISL_888877, EPI_ISL_888878, EPI_ISL_888879, EPI_ISL_888881, EPI_ISL_888882, EPI_ISL_888883, EPI_ISL_888886, EPI_ISL_888887, EPI_ISL_888888, EPI_ISL_888889, EPI_ISL_888890, EPI_ISL_888891, EPI_ISL_888893, EPI_ISL_888894, EPI_ISL_888895, EPI_ISL_888897, EPI_ISL_888899, EPI_ISL_888901, EPI_ISL_888902, EPI_ISL_888904, EPI_ISL_888906 | | | |
| see above | Michigan Department of Health and Human Services, Bureau of Laboratories | Michigan Department of Health and Human Services, Bureau of Laboratories | Blankenship HM, Riner D, Soehnlen MK |
| EPI_ISL_888907, EPI_ISL_888908, EPI_ISL_888916, EPI_ISL_888917, EPI_ISL_888918, EPI_ISL_888919, EPI_ISL_888920, EPI_ISL_888921, EPI_ISL_888922, EPI_ISL_888923, EPI_ISL_888924, EPI_ISL_888925, EPI_ISL_888926, EPI_ISL_888927, EPI_ISL_888928, EPI_ISL_888929, EPI_ISL_888930, EPI_ISL_888931, EPI_ISL_888932, EPI_ISL_888933, EPI_ISL_888934, EPI_ISL_888935, EPI_ISL_888936, EPI_ISL_888937, EPI_ISL_888938, EPI_ISL_888940, EPI_ISL_888941, EPI_ISL_888942, EPI_ISL_888943, EPI_ISL_888944, EPI_ISL_888945, EPI_ISL_888946, EPI_ISL_888947, EPI_ISL_888948, EPI_ISL_888949, EPI_ISL_888950, EPI_ISL_888951, EPI_ISL_888952, EPI_ISL_888953, EPI_ISL_888954, EPI_ISL_888955, EPI_ISL_888956, EPI_ISL_888957, EPI_ISL_888958, EPI_ISL_888959, EPI_ISL_888960, EPI_ISL_888963, EPI_ISL_888964, EPI_ISL_888965, EPI_ISL_888966, EPI_ISL_888967, EPI_ISL_888968, EPI_ISL_888970, EPI_ISL_888971, EPI_ISL_888972, EPI_ISL_888973, EPI_ISL_888974, EPI_ISL_888975, EPI_ISL_888976, EPI_ISL_888977, EPI_ISL_888978, EPI_ISL_889440 | | | |
| see above | Wyoming Public Health Laboratory | Wyoming Public Health Laboratory | Noah Hull, Taylor Fearing, Lynette Gumbleton, Channing Weber, Ashley Norberg, Bailey Bowcutt, and Wanda Manley |
| EPI_ISL_889443 | Ohio Department of Health Laboratory | Ohio Department of Health Laboratory | Holmes, Jennifer; Eric Brandt, Keoni Omura, Glen McGillivary, Caitlin McDonnell, Jade Mowery, Stephanie Mccracken, Tyler Payne, Kirtana Ramadugu, Erica Leasure, Brent Lee, Kelsey Florek, Heather Blankenship, Quanta Brown, and Tammy Bannerman |
| EPI_ISL_889444, EPI_ISL_889445, EPI_ISL_889446, EPI_ISL_889447, EPI_ISL_889448, EPI_ISL_889449, EPI_ISL_889450, EPI_ISL_889451, EPI_ISL_889452, EPI_ISL_889453, EPI_ISL_889454, EPI_ISL_889455, EPI_ISL_889456, EPI_ISL_889457, EPI_ISL_889458, EPI_ISL_889459, EPI_ISL_889460, EPI_ISL_889461, EPI_ISL_889462, EPI_ISL_889463, EPI_ISL_889464, EPI_ISL_889465, EPI_ISL_889466, EPI_ISL_889467, EPI_ISL_889468, EPI_ISL_889469, EPI_ISL_889470, EPI_ISL_889471, EPI_ISL_889472, EPI_ISL_889473, EPI_ISL_889474, EPI_ISL_889475, EPI_ISL_889476, EPI_ISL_889477, EPI_ISL_889478, EPI_ISL_889479, EPI_ISL_889480, EPI_ISL_889481, EPI_ISL_889482, EPI_ISL_889483, EPI_ISL_889484, EPI_ISL_889485, EPI_ISL_889486, EPI_ISL_889487, EPI_ISL_889488, EPI_ISL_889489, EPI_ISL_889490, EPI_ISL_889491, EPI_ISL_889492, EPI_ISL_889493, EPI_ISL_889494, EPI_ISL_889495, EPI_ISL_889496, EPI_ISL_889497, EPI_ISL_889498, EPI_ISL_889499, EPI_ISL_889500, EPI_ISL_889501, EPI_ISL_889502, EPI_ISL_889503, EPI_ISL_889504, EPI_ISL_889505, EPI_ISL_889506, EPI_ISL_889507, EPI_ISL_889508, EPI_ISL_889509, EPI_ISL_889510, EPI_ISL_889511, EPI_ISL_889512, EPI_ISL_889513, EPI_ISL_889514, EPI_ISL_889515, EPI_ISL_889516, EPI_ISL_889517, EPI_ISL_889518, EPI_ISL_889519, EPI_ISL_889520, EPI_ISL_889521, EPI_ISL_889522, EPI_ISL_889523, EPI_ISL_889524, EPI_ISL_889525, EPI_ISL_889526, EPI_ISL_889527, EPI_ISL_889528, EPI_ISL_889529, EPI_ISL_889530, EPI_ISL_889531, EPI_ISL_889532, EPI_ISL_889533, EPI_ISL_889534, EPI_ISL_889535, EPI_ISL_889536, EPI_ISL_889537, EPI_ISL_889538, EPI_ISL_889539, EPI_ISL_889540, EPI_ISL_889541 | | | |
| see above | LSUHS Emerging Viral Threat Laboratory | Microbial Genome Sequencing Center | Jeremy P. Kamil, Jennifer L. Carroll, Camille F. Abshire, Maarten Van Diest, Mohammed N.A. Siddiquey, Andrew D. Yurochko, Martin J. Sapp, Rona S. Scott, Christopher G. Kevil, Daniel J. Snyder, Vaughn S. Cooper, John A. Vanchiere |
| EPI_ISL_889542 | Ohio Department of Health Laboratory | Ohio Department of Health Laboratory | Holmes, Jennifer; Eric Brandt, Keoni Omura, Glen McGillivary, Caitlin McDonnell, Jade Mowery, Stephanie Mccracken, Tyler Payne, Kirtana Ramadugu, Erica Leasure, Brent Lee, Kelsey Florek, Heather Blankenship, Quanta Brown, and Tammy Bannerman |
| EPI_ISL_889543, EPI_ISL_889544, EPI_ISL_889545, EPI_ISL_889546, EPI_ISL_889547, EPI_ISL_889548, EPI_ISL_889549, EPI_ISL_889550, EPI_ISL_889551, EPI_ISL_889552, EPI_ISL_889553, EPI_ISL_889554, EPI_ISL_889555, EPI_ISL_889556, EPI_ISL_889557, EPI_ISL_889558, EPI_ISL_889559, EPI_ISL_889560, EPI_ISL_889561, EPI_ISL_889562, EPI_ISL_889563, EPI_ISL_889564, EPI_ISL_889565, EPI_ISL_889566, EPI_ISL_889567, EPI_ISL_889568, EPI_ISL_889569, EPI_ISL_889570, EPI_ISL_889571, EPI_ISL_889572, EPI_ISL_889573, EPI_ISL_889574, EPI_ISL_889575, EPI_ISL_889576, EPI_ISL_889577, EPI_ISL_889578, EPI_ISL_889579, EPI_ISL_889580, EPI_ISL_889581, EPI_ISL_889582, EPI_ISL_889583, EPI_ISL_889584, EPI_ISL_889585, EPI_ISL_889586, EPI_ISL_889587, EPI_ISL_889588, EPI_ISL_889589, EPI_ISL_889590, EPI_ISL_889591, EPI_ISL_889592, EPI_ISL_889593, EPI_ISL_889594, EPI_ISL_889595, EPI_ISL_889596, EPI_ISL_889597, EPI_ISL_889598, EPI_ISL_889599, EPI_ISL_889600, EPI_ISL_889601, EPI_ISL_889602, EPI_ISL_889603, EPI_ISL_889604, EPI_ISL_889605, EPI_ISL_889606, EPI_ISL_889607, EPI_ISL_889608, EPI_ISL_889609, EPI_ISL_889610, EPI_ISL_889611, EPI_ISL_889612, EPI_ISL_889613, EPI_ISL_889614, EPI_ISL_889615, EPI_ISL_889616, EPI_ISL_889617, EPI_ISL_889618, EPI_ISL_889619, EPI_ISL_889620, EPI_ISL_889621, EPI_ISL_889622, EPI_ISL_889623, EPI_ISL_889624, EPI_ISL_889625, EPI_ISL_889626, EPI_ISL_889627, EPI_ISL_889628, EPI_ISL_889629, EPI_ISL_889630, EPI_ISL_889631, EPI_ISL_889632, EPI_ISL_889633, EPI_ISL_889634, EPI_ISL_889635, EPI_ISL_889636, EPI_ISL_889637, EPI_ISL_889638, EPI_ISL_889639, EPI_ISL_889640, EPI_ISL_889641, EPI_ISL_889642, EPI_ISL_889643, EPI_ISL_889644, EPI_ISL_889645, EPI_ISL_889646, EPI_ISL_889647, EPI_ISL_889648, EPI_ISL_889649, EPI_ISL_889650, EPI_ISL_889651, EPI_ISL_889652, EPI_ISL_889653, EPI_ISL_889654, EPI_ISL_889655, EPI_ISL_889656, EPI_ISL_889657, EPI_ISL_889658, EPI_ISL_889659, EPI_ISL_889660, EPI_ISL_889661, EPI_ISL_889662, EPI_ISL_889663, EPI_ISL_889664, EPI_ISL_889665, EPI_ISL_889666, EPI_ISL_889667, EPI_ISL_889668, EPI_ISL_889669, EPI_ISL_889670, EPI_ISL_889671, EPI_ISL_889672, EPI_ISL_889673, EPI_ISL_889674, EPI_ISL_889675, EPI_ISL_889676, EPI_ISL_889677, EPI_ISL_889678, EPI_ISL_889679, EPI_ISL_889680, EPI_ISL_889681, EPI_ISL_889682, EPI_ISL_889683, EPI_ISL_889684, EPI_ISL_889685, EPI_ISL_889686, EPI_ISL_889687, EPI_ISL_889688, EPI_ISL_889689, EPI_ISL_889690, EPI_ISL_889691, EPI_ISL_889692, EPI_ISL_889693, EPI_ISL_889694, EPI_ISL_889695, EPI_ISL_889696, EPI_ISL_889697, EPI_ISL_889698, EPI_ISL_889699, EPI_ISL_889700, EPI_ISL_889701, EPI_ISL_889702, EPI_ISL_889703, EPI_ISL_889704, EPI_ISL_889705, EPI_ISL_889706, EPI_ISL_889707, EPI_ISL_889708, EPI_ISL_889709, EPI_ISL_889710, EPI_ISL_889711, EPI_ISL_889712, EPI_ISL_889713, EPI_ISL_889714, EPI_ISL_889715, EPI_ISL_889716, EPI_ISL_889717, EPI_ISL_889718, EPI_ISL_889719, EPI_ISL_889720, EPI_ISL_889721, EPI_ISL_889722, EPI_ISL_889723, EPI_ISL_889724, EPI_ISL_889725, EPI_ISL_889726, EPI_ISL_889727, EPI_ISL_889728, EPI_ISL_889729, EPI_ISL_889730, EPI_ISL_889731, EPI_ISL_889732, EPI_ISL_889733, EPI_ISL_889734, EPI_ISL_889735, EPI_ISL_889736, EPI_ISL_889737, EPI_ISL_889738, EPI_ISL_889739, EPI_ISL_889740, EPI_ISL_889741, EPI_ISL_889742, EPI_ISL_889743, EPI_ISL_889744, EPI_ISL_889745, EPI_ISL_889746, EPI_ISL_889747, EPI_ISL_889748, EPI_ISL_889749, EPI_ISL_889750, EPI_ISL_889751, EPI_ISL_889752, EPI_ISL_889753, EPI_ISL_889754, EPI_ISL_889755, EPI_ISL_889756, EPI_ISL_889757, EPI_ISL_889758, EPI_ISL_889759, EPI_ISL_889760, EPI_ISL_889761, EPI_ISL_889762, EPI_ISL_889763, EPI_ISL_889764, EPI_ISL_889765, EPI_ISL_889766, EPI_ISL_889767, EPI_ISL_889768, EPI_ISL_889769, EPI_ISL_889770, EPI_ISL_889771, EPI_ISL_889772 | | | |
| see above | LSUHS Emerging Viral Threat Laboratory | Microbial Genome Sequencing Center | Jeremy P. Kamil, Jennifer L. Carroll, Camille F. Abshire, Maarten Van Diest, Mohammed N.A. Siddiquey, Andrew D. Yurochko, Martin J. Sapp, Rona S. Scott, Christopher G. Kevil, Daniel J. Snyder, Vaughn S. Cooper, John A. Vanchiere |
| EPI_ISL_889773, EPI_ISL_889774, EPI_ISL_889775 | LSUHS Emerging Viral Threat Laboratory | Microbial Genome Sequencing Center | Mohammed N.A. Siddiquey, Jennifer L. Carroll, Camille F. Abshire, Maarten Van Diest, Andrew D. Yurochko, Martin J. Sapp, Rona S. Scott, Christopher G. Kevil, Daniel J. Snyder, Vaughn S. Cooper, John A. Vanchiere, Jeremy P. Kamil, |
| EPI_ISL_889776, EPI_ISL_889777, EPI_ISL_889778, EPI_ISL_889779, EPI_ISL_889780, EPI_ISL_889781, EPI_ISL_889782, EPI_ISL_889783, EPI_ISL_889784, EPI_ISL_889785 | LSUHS Emerging Viral Threat Laboratory | Microbial Genome Sequencing Center | Maarten Van Diest, Jennifer L. Carroll, Camille F. Abshire, Mohammed N.A. Siddiquey, Andrew D. Yurochko, Martin J. Sapp, Rona S. Scott, Christopher G. Kevil, Daniel J. Snyder, Vaughn S. Cooper, John A. Vanchiere, Jeremy P. Kamil |
| EPI_ISL_889786, EPI_ISL_889787, EPI_ISL_889788, EPI_ISL_889789, EPI_ISL_889790, EPI_ISL_889791 | LSUHS Emerging Viral Threat Laboratory | Microbial Genome Sequencing Center | Jennifer L. Carroll, Rona S. Scott, Camille F. Abshire, Maarten Van Diest, Mohammed N.A. Siddiquey, Andrew D. Yurochko, Martin J. Sapp, Christopher G. Kevil, Daniel J. Snyder, Vaughn S. Cooper, John A. Vanchiere, Jeremy P. Kamil, |
| EPI_ISL_889792, EPI_ISL_889793, EPI_ISL_889794, EPI_ISL_889795, EPI_ISL_889796, EPI_ISL_889797, EPI_ISL_889798, EPI_ISL_889799, EPI_ISL_889800, EPI_ISL_889801 | LSUHS Emerging Viral Threat Laboratory | Microbial Genome Sequencing Center | Rona S. Scott, Jennifer L. Carroll, Camille F. Abshire, Maarten Van Diest, Mohammed N.A. Siddiquey, Andrew D. Yurochko, Martin J. Sapp, Christopher G. Kevil, Daniel J. Snyder, Vaughn S. Cooper, John A. Vanchiere, Jeremy P. Kamil, |
| EPI_ISL_890174, EPI_ISL_890175, EPI_ISL_890176, EPI_ISL_890178, EPI_ISL_890179, EPI_ISL_890180, EPI_ISL_890181, EPI_ISL_890182 | Ohio Department of Health Laboratory | Ohio Department of Health Laboratory | Holmes, Jennifer; Eric Brandt, Keoni Omura, Glen McGillivary, Caitlin McDonnell, Jade Mowery, Stephanie Mccracken, Tyler Payne, Kirtana Ramadugu, Erica Leasure, Brent Lee, Kelsey Florek, Heather Blankenship, Quanta Brown, and Tammy Bannerman |
| EPI_ISL_890184 | Seattle Flu Study | Seattle Flu Study | Deborah A. Nickerson, Chris D. Frazar, Jover Lee, Benjamin Pelle, Erica Ryke, Matthew Richardson, Amanda Adler, Elisabeth Brandstetter, Peter D. Han, Kairsten Fay, Misja Ilcisin, Kirsten Lacombe, Thomas R. Sibley, Melissa Truong, Caitlin R. Wolf, Karen Cowgill, Stephanie Schrag, Jeff Duchin, Michael Boeckh, Janet A. Englund, Michael Famulare, Barry R. Lutz, Mark J. Rieder, Lea M. Starita, Matthew Thompson, Helen Y. Chu, Trevor Bedford, Jay Shendure |
| EPI_ISL_890233 | School of Pharmacy, Shenandoah University | School of Pharmacy, Shenandoah University | Adams,S.M., Harralson,A.F., Kidd,R.S., Sawyer,G.W. |
| EPI_ISL_890235 | Respiratory Viruses Branch, Centers for Disease Control and Prevention | Centers for Disease Control and Prevention | Tao,Y., Li,Y., Zhang,J., Queen,K., Uehara,A., Cook,P., Paden,C.R., Wang,H. and Tong,S. |
| EPI_ISL_890236 | Center for Genome Sciences, US Army Medical Research Institute of Infectious Disease (USAMRIID) | Center for Genome Sciences, US Army Medical Research Institute of Infectious Disease (USAMRIID) | Kugelman,J., Di Paola,N., Richardson,J., Gibson,W., Bateman,S. and Jerke,K. |
| EPI_ISL_890360 | Johns Hopkins Hospital Department of Pathology | Johns Hopkins Hospital Department of Pathology | C. Paul Morris, Chun Huai Luo, Adannaya Amadi, Matthew Schwartz, Nicholas Gallagher, Heba H. Mostafa |
| EPI_ISL_890361, EPI_ISL_890362, EPI_ISL_890363, EPI_ISL_890364, EPI_ISL_890365, EPI_ISL_890366 | LSUHS Emerging Viral Threat Laboratory | Microbial Genome Sequencing Center | Jeremy P. Kamil, Jennifer L. Carroll, Camille F. Abshire, Maarten Van Diest, Mohammed N.A. Siddiquey, Andrew D. Yurochko, Martin J. Sapp, Rona S. Scott, Christopher G. Kevil, Daniel J. Snyder, Vaughn S. Cooper, John A. Vanchiere |
| EPI_ISL_890367, EPI_ISL_890368, EPI_ISL_890369, EPI_ISL_890370 | LSUHS Emerging Viral Threat Laboratory | Microbial Genome Sequencing Center | Jennifer L. Carroll, Rona S. Scott, Camille F. Abshire, Maarten Van Diest, Mohammed N.A. Siddiquey, Andrew D. Yurochko, Martin J. Sapp, Christopher G. Kevil, Daniel J. Snyder, Vaughn S. Cooper, John A. Vanchiere, Jeremy P. Kamil, |

EPI_ISL_890896, EPI_ISL_890898, EPI_ISL_890899, EPI_ISL_890900, EPI_ISL_890902, EPI_ISL_890903, EPI_ISL_890906, EPI_ISL_890907, EPI_ISL_890908, EPI_ISL_890909, EPI_ISL_890910, EPI_ISL_890911, EPI_ISL_890912, EPI_ISL_890913, EPI_ISL_890914, EPI_ISL_890915, EPI_ISL_890917, EPI_ISL_890918

| | | | |
|---|---|---|---|
| see above | Seattle Flu Study | Seattle Flu Study | Deborah A. Nickerson, Chris D. Frazar, Jover Lee, Benjamin Pelle, Erica Ryke, Matthew Richardson, Amanda Adler, Elisabeth Brandstetter, Peter D. Han, Kairsten Fay, Misja Ilcisin, Kirsten Lacombe, Thomas R. Sibley, Melissa Truong, Caitlin R. Wolf, Karen Cowgill, Stephanie Schrag, Jeff Duchin, Michael Boeckh, Janet A. Englund, Michael Famulare, Barry R. Lutz, Mark J. Rieder, Lea M. Starita, Matthew Thompson, Helen Y. Chu, Trevor Bedford, Jay Shendure |
| EPI_ISL_890919 | Seattle Flu Study | Seattle Flu Study | Deborah A. Nickerson, Chris D. Frazar, Jover Lee, Benjamin Pelle, Erica Ryke, Matthew Richardson, Amanda Adler, Elisabeth Brandstetter, Peter D. Han, Kairsten Fay, Misja Ilcisin, Kirsten Lacombe, Thomas R. Sibley, Melissa Truong, Caitlin R. Wolf, Michael Boeckh, Janet A. Englund, Michael Famulare, Barry R. Lutz, Mark J. Rieder, Lea M. Starita, Matthew Thompson, Jay Shendure, Trevor Bedford, Helen Y. Chu |
| EPI_ISL_890923, EPI_ISL_890924, EPI_ISL_890925, EPI_ISL_890926, EPI_ISL_890927, EPI_ISL_890929, EPI_ISL_890930, EPI_ISL_890937 | Seattle Flu Study | Seattle Flu Study | Deborah A. Nickerson, Chris D. Frazar, Jover Lee, Benjamin Pelle, Erica Ryke, Matthew Richardson, Amanda Adler, Elisabeth Brandstetter, Peter D. Han, Kairsten Fay, Misja Ilcisin, Kirsten Lacombe, Thomas R. Sibley, Melissa Truong, Caitlin R. Wolf, Karen Cowgill, Stephanie Schrag, Jeff Duchin, Michael Boeckh, Janet A. Englund, Michael Famulare, Barry R. Lutz, Mark J. Rieder, Lea M. Starita, Matthew Thompson, Helen Y. Chu, Trevor Bedford, Jay Shendure |
| EPI_ISL_890938 | Seattle Flu Study | Seattle Flu Study | Deborah A. Nickerson, Chris D. Frazar, Jover Lee, Benjamin Pelle, Erica Ryke, Matthew Richardson, Amanda Adler, Elisabeth Brandstetter, Peter D. Han, Kairsten Fay, Misja Ilcisin, Kirsten Lacombe, Thomas R. Sibley, Melissa Truong, Caitlin R. Wolf, Michael Boeckh, Janet A. Englund, Michael Famulare, Barry R. Lutz, Mark J. Rieder, Lea M. Starita, Matthew Thompson, Jay Shendure, Trevor Bedford, Helen Y. Chu |
| EPI_ISL_890939, EPI_ISL_890940, EPI_ISL_890941, EPI_ISL_890942, EPI_ISL_890943, EPI_ISL_890944, EPI_ISL_890946, EPI_ISL_890949 | Seattle Flu Study | Seattle Flu Study | Deborah A. Nickerson, Chris D. Frazar, Jover Lee, Benjamin Pelle, Erica Ryke, Matthew Richardson, Amanda Adler, Elisabeth Brandstetter, Peter D. Han, Kairsten Fay, Misja Ilcisin, Kirsten Lacombe, Thomas R. Sibley, Melissa Truong, Caitlin R. Wolf, Karen Cowgill, Stephanie Schrag, Jeff Duchin, Michael Boeckh, Janet A. Englund, Michael Famulare, Barry R. Lutz, Mark J. Rieder, Lea M. Starita, Matthew Thompson, Helen Y. Chu, Trevor Bedford, Jay Shendure |
| EPI_ISL_890950 | Seattle Flu Study | Seattle Flu Study | Deborah A. Nickerson, Chris D. Frazar, Jover Lee, Benjamin Pelle, Erica Ryke, Matthew Richardson, Amanda Adler, Elisabeth Brandstetter, Peter D. Han, Kairsten Fay, Misja Ilcisin, Kirsten Lacombe, Thomas R. Sibley, Melissa Truong, Caitlin R. Wolf, Michael Boeckh, Janet A. Englund, Michael Famulare, Barry R. Lutz, Mark J. Rieder, Lea M. Starita, Matthew Thompson, Jay Shendure, Trevor Bedford, Helen Y. Chu |

EPI_ISL_890951, EPI_ISL_890952, EPI_ISL_890953, EPI_ISL_890954, EPI_ISL_890955, EPI_ISL_890957, EPI_ISL_890958, EPI_ISL_890961, EPI_ISL_890962, EPI_ISL_890963, EPI_ISL_890964, EPI_ISL_890965, EPI_ISL_890966

| | | | |
|---|---|---|---|
| see above | Seattle Flu Study | Seattle Flu Study | Deborah A. Nickerson, Chris D. Frazar, Jover Lee, Benjamin Pelle, Erica Ryke, Matthew Richardson, Amanda Adler, Elisabeth Brandstetter, Peter D. Han, Kairsten Fay, Misja Ilcisin, Kirsten Lacombe, Thomas R. Sibley, Melissa Truong, Caitlin R. Wolf, Karen Cowgill, Stephanie Schrag, Jeff Duchin, Michael Boeckh, Janet A. Englund, Michael Famulare, Barry R. Lutz, Mark J. Rieder, Lea M. Starita, Matthew Thompson, Helen Y. Chu, Trevor Bedford, Jay Shendure |
| EPI_ISL_890967, EPI_ISL_890969 | Seattle Flu Study | Seattle Flu Study | Deborah A. Nickerson, Chris D. Frazar, Jover Lee, Benjamin Pelle, Erica Ryke, Matthew Richardson, Amanda Adler, Elisabeth Brandstetter, Peter D. Han, Kairsten Fay, Misja Ilcisin, Kirsten Lacombe, Thomas R. Sibley, Melissa Truong, Caitlin R. Wolf, Michael Boeckh, Janet A. Englund, Michael Famulare, Barry R. Lutz, Mark J. Rieder, Lea M. Starita, Matthew Thompson, Jay Shendure, Trevor Bedford, Helen Y. Chu |
| EPI_ISL_890970, EPI_ISL_890971, EPI_ISL_890973, EPI_ISL_890975, EPI_ISL_890977, EPI_ISL_890978, EPI_ISL_890980, EPI_ISL_890981, EPI_ISL_890982, EPI_ISL_890983 | Seattle Flu Study | Seattle Flu Study | Deborah A. Nickerson, Chris D. Frazar, Jover Lee, Benjamin Pelle, Erica Ryke, Matthew Richardson, Amanda Adler, Elisabeth Brandstetter, Peter D. Han, Kairsten Fay, Misja Ilcisin, Kirsten Lacombe, Thomas R. Sibley, Melissa Truong, Caitlin R. Wolf, Karen Cowgill, Stephanie Schrag, Jeff Duchin, Michael Boeckh, Janet A. Englund, Michael Famulare, Barry R. Lutz, Mark J. Rieder, Lea M. Starita, Matthew Thompson, Helen Y. Chu, Trevor Bedford, Jay Shendure |
| EPI_ISL_890985, EPI_ISL_890986, EPI_ISL_890987, EPI_ISL_890988 | Seattle Flu Study | Seattle Flu Study | Deborah A. Nickerson, Chris D. Frazar, Jover Lee, Benjamin Pelle, Erica Ryke, Matthew Richardson, Amanda Adler, Elisabeth Brandstetter, Peter D. Han, Kairsten Fay, Misja Ilcisin, Kirsten Lacombe, Thomas R. Sibley, Melissa Truong, Caitlin R. Wolf, Michael Boeckh, Janet A. Englund, Michael Famulare, Barry R. Lutz, Mark J. Rieder, Lea M. Starita, Matthew Thompson, Jay Shendure, Trevor Bedford, Helen Y. Chu |
| EPI_ISL_890989, EPI_ISL_890990 | Seattle Flu Study | Seattle Flu Study | Deborah A. Nickerson, Chris D. Frazar, Jover Lee, Benjamin Pelle, Erica Ryke, Matthew Richardson, Amanda Adler, Elisabeth Brandstetter, Peter D. Han, Kairsten Fay, Misja Ilcisin, Kirsten Lacombe, Thomas R. Sibley, Melissa Truong, Caitlin R. Wolf, Karen Cowgill, Stephanie Schrag, Jeff Duchin, Michael Boeckh, Janet A. Englund, Michael Famulare, Barry R. Lutz, Mark J. Rieder, Lea M. Starita, Matthew Thompson, Helen Y. Chu, Trevor Bedford, Jay Shendure |
| EPI_ISL_890991, EPI_ISL_890992 | Seattle Flu Study | Seattle Flu Study | Deborah A. Nickerson, Chris D. Frazar, Jover Lee, Benjamin Pelle, Erica Ryke, Matthew Richardson, Amanda Adler, Elisabeth Brandstetter, Peter D. Han, Kairsten Fay, Misja Ilcisin, Kirsten Lacombe, Thomas R. Sibley, Melissa Truong, Caitlin R. Wolf, Michael Boeckh, Janet A. Englund, Michael Famulare, Barry R. Lutz, Mark J. Rieder, Lea M. Starita, Matthew Thompson, Jay Shendure, Trevor Bedford, Helen Y. Chu |
| EPI_ISL_890993 | Seattle Flu Study | Seattle Flu Study | Deborah A. Nickerson, Chris D. Frazar, Jover Lee, Benjamin Pelle, Erica Ryke, Matthew Richardson, Amanda Adler, Elisabeth Brandstetter, Peter D. Han, Kairsten Fay, Misja Ilcisin, Kirsten Lacombe, Thomas R. Sibley, Melissa Truong, Caitlin R. Wolf, Karen Cowgill, Stephanie Schrag, Jeff Duchin, Michael Boeckh, Janet A. Englund, Michael Famulare, Barry R. Lutz, Mark J. Rieder, Lea M. Starita, Matthew Thompson, Helen Y. Chu, Trevor Bedford, Jay Shendure |
| EPI_ISL_890994, EPI_ISL_890995, EPI_ISL_890996, EPI_ISL_890997 | Seattle Flu Study | Seattle Flu Study | Deborah A. Nickerson, Chris D. Frazar, Jover Lee, Benjamin Pelle, Erica Ryke, Matthew Richardson, Amanda Adler, Elisabeth Brandstetter, Peter D. Han, Kairsten Fay, Misja Ilcisin, Kirsten Lacombe, Thomas R. Sibley, Melissa Truong, Caitlin R. Wolf, Michael Boeckh, Janet A. Englund, Michael Famulare, Barry R. Lutz, Mark J. Rieder, Lea M. Starita, Matthew Thompson, Jay Shendure, Trevor Bedford, Helen Y. Chu |

EPI_ISL_890999, EPI_ISL_891000, EPI_ISL_891001, EPI_ISL_891003, EPI_ISL_891005, EPI_ISL_891007, EPI_ISL_891008, EPI_ISL_891009, EPI_ISL_891010, EPI_ISL_891011, EPI_ISL_891012, EPI_ISL_891013, EPI_ISL_891014, EPI_ISL_891015, EPI_ISL_891016, EPI_ISL_891017, EPI_ISL_891018, EPI_ISL_891019, EPI_ISL_891020, EPI_ISL_891022, EPI_ISL_891023, EPI_ISL_891024, EPI_ISL_891025, EPI_ISL_891027, EPI_ISL_891028, EPI_ISL_891029, EPI_ISL_891030, EPI_ISL_891031, EPI_ISL_891032, EPI_ISL_891033, EPI_ISL_891036, EPI_ISL_891037, EPI_ISL_891038, EPI_ISL_891039, EPI_ISL_891040, EPI_ISL_891041, EPI_ISL_891043, EPI_ISL_891044, EPI_ISL_891045, EPI_ISL_891046, EPI_ISL_891047, EPI_ISL_891048, EPI_ISL_891049, EPI_ISL_891050, EPI_ISL_891051, EPI_ISL_891052, EPI_ISL_891053, EPI_ISL_891054, EPI_ISL_891055, EPI_ISL_891056

| | | | |
|---|---|---|---|
| see above | Washington State Department of Health | Seattle Flu Study | Deborah A. Nickerson, Chris D. Frazar, Jover Lee, Benjamin Pelle, Erica Ryke, Matthew Richardson, Amanda Adler, Elisabeth Brandstetter, Peter D. Han, Kairsten Fay, Misja Ilcisin, Kirsten Lacombe, Thomas R. Sibley, Melissa Truong, Caitlin R. Wolf, Romesh Gautom, Geoff Melly, Brian Hiatt, Philip Dykema, Scott Lindquist, Michael Boeckh, Janet A. Englund, Michael Famulare, Barry R. Lutz, Mark J. Rieder, Lea M. Starita, Matthew Thompson, Helen Y. Chu, Jay Shendure, Trevor Bedford |
| EPI_ISL_891058 | Seattle Flu Study | Seattle Flu Study | Deborah A. Nickerson, Chris D. Frazar, Jover Lee, Benjamin Pelle, Erica Ryke, Matthew Richardson, Amanda Adler, Elisabeth Brandstetter, Peter D. Han, Kairsten Fay, Misja Ilcisin, Kirsten Lacombe, Thomas R. Sibley, Melissa Truong, Caitlin R. Wolf, Karen Cowgill, Stephanie Schrag, Jeff Duchin, Michael Boeckh, Janet A. Englund, Michael Famulare, Barry R. Lutz, Mark J. Rieder, Lea M. Starita, Matthew Thompson, Helen Y. Chu, Trevor Bedford, Jay Shendure |
| EPI_ISL_891059 | FidaLab | Seattle Flu Study | Deborah A. Nickerson, Chris D. Frazar, Jover Lee, Benjamin Pelle, Erica Ryke, Matthew Richardson, Amanda Adler, Elisabeth Brandstetter, Peter D. Han, Kairsten Fay, Misja Ilcisin, Kirsten Lacombe, Thomas R. Sibley, Melissa Truong, Caitlin R. Wolf, Romesh Gautom, Geoff Melly, Brian Hiatt, Philip Dykema, Scott Lindquist, Michael Boeckh, Janet A. Englund, Michael Famulare, Barry R. Lutz, Mark J. Rieder, Lea M. Starita, Matthew Thompson, Helen Y. Chu, Jay Shendure, Trevor Bedford |
| EPI_ISL_891060, EPI_ISL_891061, EPI_ISL_891062, EPI_ISL_891063 | Seattle Flu Study | Seattle Flu Study | Deborah A. Nickerson, Chris D. Frazar, Jover Lee, Benjamin Pelle, Erica Ryke, Matthew Richardson, Amanda Adler, Elisabeth Brandstetter, Peter D. Han, Kairsten Fay, Misja Ilcisin, Kirsten Lacombe, Thomas R. Sibley, Melissa Truong, Caitlin R. Wolf, Michael Boeckh, Janet A. Englund, Michael Famulare, Barry R. Lutz, Mark J. Rieder, Lea M. Starita, Matthew Thompson, Jay Shendure, Trevor Bedford, Helen Y. Chu |
| EPI_ISL_891064, EPI_ISL_891065, EPI_ISL_891066, EPI_ISL_891067, EPI_ISL_891068, EPI_ISL_891069 | Seattle Flu Study | Seattle Flu Study | Deborah A. Nickerson, Chris D. Frazar, Jover Lee, Benjamin Pelle, Erica Ryke, Matthew Richardson, Amanda Adler, Elisabeth Brandstetter, Peter D. Han, Kairsten Fay, Misja Ilcisin, Kirsten Lacombe, Thomas R. Sibley, Melissa Truong, Caitlin R. Wolf, Karen Cowgill, Stephanie Schrag, Jeff Duchin, Michael Boeckh, Janet A. Englund, Michael Famulare, Barry R. Lutz, Mark J. Rieder, Lea M. Starita, Matthew Thompson, Helen Y. Chu, Trevor Bedford, Jay Shendure |
| EPI_ISL_891071 | Seattle Flu Study | Seattle Flu Study | Deborah A. Nickerson, Chris D. Frazar, Jover Lee, Benjamin Pelle, Erica Ryke, Matthew Richardson, Amanda Adler, Elisabeth Brandstetter, Peter D. Han, Kairsten Fay, Misja Ilcisin, Kirsten Lacombe, Thomas R. Sibley, Melissa Truong, Caitlin R. Wolf, Michael Boeckh, Janet A. Englund, Michael Famulare, Barry R. Lutz, Mark J. Rieder, Lea M. Starita, Matthew Thompson, Jay Shendure, Trevor Bedford, Helen Y. Chu |
| EPI_ISL_891073, EPI_ISL_891074 | Seattle Flu Study | Seattle Flu Study | Deborah A. Nickerson, Chris D. Frazar, Jover Lee, Benjamin Pelle, Erica Ryke, Matthew Richardson, Amanda Adler, Elisabeth Brandstetter, Peter D. Han, Kairsten Fay, Misja Ilcisin, Kirsten Lacombe, Thomas R. Sibley, Melissa Truong, Caitlin R. Wolf, Karen Cowgill, Stephanie Schrag, Jeff Duchin, Michael Boeckh, Janet A. Englund, Michael Famulare, Barry R. Lutz, Mark J. Rieder, Lea M. Starita, Matthew Thompson, Helen Y. Chu, Trevor Bedford, Jay Shendure |
| EPI_ISL_891075, EPI_ISL_891076 | Seattle Flu Study | Seattle Flu Study | Deborah A. Nickerson, Chris D. Frazar, Jover Lee, Benjamin Pelle, Erica Ryke, Matthew Richardson, Amanda Adler, Elisabeth Brandstetter, Peter D. Han, Kairsten Fay, Misja Ilcisin, Kirsten Lacombe, Thomas R. Sibley, Melissa Truong, Caitlin R. Wolf, Michael Boeckh, Janet A. Englund, Michael Famulare, Barry R. Lutz, Mark J. Rieder, Lea M. Starita, Matthew Thompson, Jay Shendure, Trevor Bedford, Helen Y. Chu |
| EPI_ISL_891077, EPI_ISL_891079, EPI_ISL_891080 | Seattle Flu Study | Seattle Flu Study | Deborah A. Nickerson, Chris D. Frazar, Jover Lee, Benjamin Pelle, Erica Ryke, Matthew Richardson, Amanda Adler, Elisabeth Brandstetter, Peter D. Han, Kairsten Fay, Misja Ilcisin, Kirsten Lacombe, Thomas R. Sibley, Melissa Truong, Caitlin R. Wolf, Karen Cowgill, Stephanie Schrag, Jeff Duchin, Michael Boeckh, Janet A. Englund, Michael Famulare, Barry R. Lutz, Mark J. Rieder, Lea M. Starita, Matthew Thompson, Helen Y. Chu, Trevor Bedford, Jay Shendure |
| EPI_ISL_891082, EPI_ISL_891083 | Seattle Flu Study | Seattle Flu Study | Deborah A. Nickerson, Chris D. Frazar, Jover Lee, Benjamin Pelle, Erica Ryke, Matthew Richardson, Amanda Adler, Elisabeth Brandstetter, Peter D. Han, Kairsten Fay, Misja Ilcisin, Kirsten Lacombe, Thomas R. Sibley, Melissa Truong, Caitlin R. Wolf, Michael Boeckh, Janet A. Englund, Michael Famulare, Barry R. Lutz, Mark J. Rieder, Lea M. Starita, Matthew Thompson, Jay Shendure, Trevor Bedford, Helen Y. Chu |
| EPI_ISL_891084, EPI_ISL_891085, EPI_ISL_891086, EPI_ISL_891087, EPI_ISL_891088, EPI_ISL_891089, EPI_ISL_891090, EPI_ISL_891091 | Seattle Flu Study | Seattle Flu Study | Deborah A. Nickerson, Chris D. Frazar, Jover Lee, Benjamin Pelle, Erica Ryke, Matthew Richardson, Amanda Adler, Elisabeth Brandstetter, Peter D. Han, Kairsten Fay, Misja Ilcisin, Kirsten Lacombe, Thomas R. Sibley, Melissa Truong, Caitlin R. Wolf, Karen Cowgill, Stephanie Schrag, Jeff Duchin, Michael Boeckh, Janet A. Englund, Michael Famulare, Barry R. Lutz, Mark J. Rieder, Lea M. Starita, Matthew Thompson, Helen Y. Chu, Trevor Bedford, Jay Shendure |

EPI_ISL_891094, EPI_ISL_891095, EPI_ISL_891097, EPI_ISL_891098, EPI_ISL_891100, EPI_ISL_891101, EPI_ISL_891102, EPI_ISL_891104, EPI_ISL_891105, EPI_ISL_891106, EPI_ISL_891107, EPI_ISL_891108, EPI_ISL_891110, EPI_ISL_891111, EPI_ISL_891112, EPI_ISL_891113, EPI_ISL_891114, EPI_ISL_891115, EPI_ISL_891116, EPI_ISL_891117, EPI_ISL_891118, EPI_ISL_891119, EPI_ISL_891120, EPI_ISL_891121, EPI_ISL_891122, EPI_ISL_891123, EPI_ISL_891125, EPI_ISL_891126, EPI_ISL_891127, EPI_ISL_891128, EPI_ISL_891129, EPI_ISL_891130, EPI_ISL_891131, EPI_ISL_891133, EPI_ISL_891135

| | | | |
|---|---|---|---|
| see above | Altius Institute for Biomedical Sciences | Seattle Flu Study | Deborah A. Nickerson, Chris D. Frazar, Jover Lee, Benjamin Pelle, Erica Ryke, Matthew Richardson, Amanda Adler, Elisabeth Brandstetter, Peter D. Han, Kairsten Fay, Misja Ilcisin, Kirsten Lacombe, Thomas R. Sibley, Melissa Truong, Caitlin R. Wolf, Ryan Alexander, Daniel Bates, Rebecca Bruders, Stephanie DeBaun, Clem Green, Muhammad Halimun, Kneshay Harper, Matt Hartman, Andrew Meuser, Alex Nguyen, Truong Nguyen, Sofia Olsson, Sadie Patraw, Hannah Petersen, Tobias Ragoczy, Joshua Richards, Jacob Rodriguez, John Stamatoyannopoulos, Julia Wald, Olivia Waltner, Michael Boeckh, Janet A. Englund, Michael Famulare, Barry R. Lutz, Mark J. Rieder, Lea M. Starita, Matthew Thompson, Helen Y. Chu, Jay Shendure, Trevor Bedford |

| | | | |
|---|---|---|---|
| EPI_ISL_891136, EPI_ISL_891138, EPI_ISL_891139, EPI_ISL_891140 | The Jackson Laboratory | The Jackson Laboratory | Lloyd M, Sanderson B, Srivastava A, Maurya R, Renzette N, Omerza G, Kelly K, Li L, Wei C L, Adams M |
| EPI_ISL_891141, EPI_ISL_891142, EPI_ISL_891143, EPI_ISL_891144 | Indiana Animal Disease Diagnostic Laboratory | Carpi Laboratory - Purdue University | Jack Dorman, Ilinca I Ciubotariu, Lev Gorenstein, Abebe A Fola, G Kenitra Hendrix, Rebecca P Wilkes, Giovanna Carpi |
| EPI_ISL_891145, EPI_ISL_891146, EPI_ISL_891147, EPI_ISL_891148, EPI_ISL_891149, EPI_ISL_891150 | University of Wisconsin-Madison AIDS Vaccine Research Laboratories | University of Wisconsin-Madison AIDS Vaccine Research Laboratories | Gage Moreno, Katarina Braun, et al. AIDS Vaccine Research Laboratories |
| EPI_ISL_891212 | Population Medicine and Diagnostic Sciences, Cornell University | Population Medicine and Diagnostic Sciences, Cornell University | Cardia Caserta,L., Mitchel,P.K., Plachorzyk,E. and Diel,D.G. |
| EPI_ISL_892248 | Ohio Department of Health Laboratory | Ohio Department of Health Laboratory | Holmes, Jennifer; Eric Brandt, Keoni Omura, Glen McGillivary, Caitlin McDonnell, Jade Mowery, Stephanie Mccracken, Tyler Payne, Kirtana Ramadugu, Erica Leasure, Brent Lee, Kelsey Florek, Heather Blankenship, Quanta Brown, and Tammy Bannerman |
| EPI_ISL_892249, EPI_ISL_892250, EPI_ISL_892251, EPI_ISL_892252, EPI_ISL_892253, EPI_ISL_892254, EPI_ISL_892255, EPI_ISL_892256, EPI_ISL_892257, EPI_ISL_892258, EPI_ISL_892259, EPI_ISL_892260, EPI_ISL_892261, EPI_ISL_892262, EPI_ISL_892263, EPI_ISL_892264, EPI_ISL_892265, EPI_ISL_892266 | | | |
| see above | MD Laboratories | Los Angeles County Public Health Laboratories | P. Hemarajata et al. |
| EPI_ISL_892267, EPI_ISL_892268 | UCLA Clinical Micro Lab | Los Angeles County Public Health Laboratories | P. Hemarajata et al. |
| EPI_ISL_892269, EPI_ISL_892364, EPI_ISL_892368 | Ohio Department of Health Laboratory | Ohio Department of Health Laboratory | Holmes, Jennifer; Eric Brandt, Keoni Omura, Glen McGillivary, Caitlin McDonnell, Jade Mowery, Stephanie Mccracken, Tyler Payne, Kirtana Ramadugu, Erica Leasure, Brent Lee, Kelsey Florek, Heather Blankenship, Quanta Brown, and Tammy Bannerman |
| EPI_ISL_893214 | Indiana Animal Disease Diagnostic Laboratory | Carpi Laboratory - Purdue University | Jack Dorman, Ilinca I Ciubotariu, Lev Gorenstein, Abebe A Fola, G Kenitra Hendrix, Rebecca P Wilkes, Giovanna Carpi |
| EPI_ISL_893367 | Ohio Department of Health Laboratory | Ohio Department of Health Laboratory | Holmes, Jennifer; Eric Brandt, Keoni Omura, Glen McGillivary, Caitlin McDonnell, Jade Mowery, Stephanie Mccracken, Tyler Payne, Kirtana Ramadugu, Erica Leasure, Brent Lee, Kelsey Florek, Heather Blankenship, Quanta Brown, and Tammy Bannerman |
| EPI_ISL_893721, EPI_ISL_893722, EPI_ISL_893723 | Oregon State Public Health Laboratory | Pathogen Discovery, Respiratory Viruses Branch, Division of Viral Diseases, Centers for Disease Control and Prevention | Ying Tao, Yan Li, Jing Zhang, Krista Queen, Anna Uehara, Peter Cook, Clinton R. Paden, Haibin Wang, Suxiang Tong |
| EPI_ISL_893724, EPI_ISL_893725, EPI_ISL_893726, EPI_ISL_893727, EPI_ISL_893728, EPI_ISL_893729, EPI_ISL_893730, EPI_ISL_893731, EPI_ISL_893732, EPI_ISL_893733, EPI_ISL_893734, EPI_ISL_893735 | | | |
| see above | Pima County Health Department | Pathogen Discovery, Respiratory Viruses Branch, Division of Viral Diseases, Centers for Disease Control and Prevention | Yan Li, Anna Montmayeur, Ying Tao, Jing Zhang, Krista Queen, Anna Uehara, Brian Lynch, Rachel Marine, Peter Cook, Clinton R. Paden, Haibin Wang, Suxiang Tong |
| EPI_ISL_893736, EPI_ISL_893737, EPI_ISL_893738 | Massachusetts Department of Public Health | Pathogen Discovery, Respiratory Viruses Branch, Division of Viral Diseases, Centers for Disease Control and Prevention | Yan Li, Anna Montmayeur, Ying Tao, Jing Zhang, Krista Queen, Anna Uehara, Brian Lynch, Rachel Marine, Peter Cook, Clinton R. Paden, Haibin Wang, Suxiang Tong |
| EPI_ISL_893739 | Florida Department of Public Health | Pathogen Discovery, Respiratory Viruses Branch, Division of Viral Diseases, Centers for Disease Control and Prevention | Yan Li, Anna Montmayeur, Ying Tao, Jing Zhang, Krista Queen, Anna Uehara, Brian Lynch, Rachel Marine, Peter Cook, Clinton R. Paden, Haibin Wang, Suxiang Tong |
| EPI_ISL_893740, EPI_ISL_893741, EPI_ISL_893742, EPI_ISL_894170, EPI_ISL_894947, EPI_ISL_895289, EPI_ISL_895727 | Ohio Department of Health Laboratory | Ohio Department of Health Laboratory | Holmes, Jennifer; Eric Brandt, Keoni Omura, Glen McGillivary, Caitlin McDonnell, Jade Mowery, Stephanie Mccracken, Tyler Payne, Kirtana Ramadugu, Erica Leasure, Brent Lee, Kelsey Florek, Heather Blankenship, Quanta Brown, and Tammy Bannerman |
| EPI_ISL_896216, EPI_ISL_896217 | Columbia University Irving Medical Center | Wadsworth Center, New York State Department of Health | Kirsten St. George, Daryl M. Lamson, Alexis Russel, Matthew Shudt, Melissa A Leisner, Jonathan Plitnick, Navjot Singh, John Kelly, Erasmus Schneider, Erica Lasek-Nesselquist |
| EPI_ISL_896218, EPI_ISL_896219, EPI_ISL_896220, EPI_ISL_896221, EPI_ISL_896222, EPI_ISL_896223, EPI_ISL_896224, EPI_ISL_896225 | SUNY UPSTATE MEDICAL UNIVERSITY | Wadsworth Center, New York State Department of Health | Kirsten St. George, Daryl M. Lamson, Alexis Russel, Matthew Shudt, Melissa A Leisner, Jonathan Plitnick, Navjot Singh, John Kelly, Erasmus Schneider, Erica Lasek-Nesselquist |
| EPI_ISL_896226 | TEMPUS LABS INC | Wadsworth Center, New York State Department of Health | Kirsten St. George, Daryl M. Lamson, Alexis Russel, Matthew Shudt, Melissa A Leisner, Jonathan Plitnick, Navjot Singh, John Kelly, Erasmus Schneider, Erica Lasek-Nesselquist |
| EPI_ISL_896227 | Columbia University Irving Medical Center | Wadsworth Center, New York State Department of Health | Kirsten St. George, Daryl M. Lamson, Alexis Russel, Matthew Shudt, Melissa A Leisner, Jonathan Plitnick, Navjot Singh, John Kelly, Erasmus Schneider, Erica Lasek-Nesselquist |
| EPI_ISL_896228, EPI_ISL_896229, EPI_ISL_896230 | SUNY UPSTATE MEDICAL UNIVERSITY | Wadsworth Center, New York State Department of Health | Kirsten St. George, Daryl M. Lamson, Alexis Russel, Matthew Shudt, Melissa A Leisner, Jonathan Plitnick, Navjot Singh, John Kelly, Erasmus Schneider, Erica Lasek-Nesselquist |
| EPI_ISL_896231, EPI_ISL_896232, EPI_ISL_896233, EPI_ISL_896234 | Columbia University Irving Medical Center | Wadsworth Center, New York State Department of Health | Kirsten St. George, Daryl M. Lamson, Alexis Russel, Matthew Shudt, Melissa A Leisner, Jonathan Plitnick, Navjot Singh, John Kelly, Erasmus Schneider, Erica Lasek-Nesselquist |
| EPI_ISL_896235 | SUNY UPSTATE MEDICAL UNIVERSITY | Wadsworth Center, New York State Department of Health | Kirsten St. George, Daryl M. Lamson, Alexis Russel, Matthew Shudt, Melissa A Leisner, Jonathan Plitnick, Navjot Singh, John Kelly, Erasmus Schneider, Erica Lasek-Nesselquist |
| EPI_ISL_896236, EPI_ISL_896237, EPI_ISL_896238, EPI_ISL_896239, EPI_ISL_896240, EPI_ISL_896241, EPI_ISL_896242, EPI_ISL_896243, EPI_ISL_896244, EPI_ISL_896245, EPI_ISL_896246 | | | |
| see above | Columbia University Irving Medical Center | Wadsworth Center, New York State Department of Health | Kirsten St. George, Daryl M. Lamson, Alexis Russel, Matthew Shudt, Melissa A Leisner, Jonathan Plitnick, Navjot Singh, John Kelly, Erasmus Schneider, Erica Lasek-Nesselquist |
| EPI_ISL_896247, EPI_ISL_896248, EPI_ISL_896249, EPI_ISL_896250, EPI_ISL_896251, EPI_ISL_896252, EPI_ISL_896253, EPI_ISL_896254, EPI_ISL_896255, EPI_ISL_896256, EPI_ISL_896257, EPI_ISL_896258, EPI_ISL_896259, EPI_ISL_896260, EPI_ISL_896261, EPI_ISL_896262, EPI_ISL_896263, EPI_ISL_896264, EPI_ISL_896265, EPI_ISL_896266, EPI_ISL_896267, EPI_ISL_896268, EPI_ISL_896269, EPI_ISL_896270, EPI_ISL_896271, EPI_ISL_896272, EPI_ISL_896273, EPI_ISL_896274, EPI_ISL_896275, EPI_ISL_896276, EPI_ISL_896277, EPI_ISL_896278, EPI_ISL_896279, EPI_ISL_896280, EPI_ISL_896281, EPI_ISL_896282, EPI_ISL_896283, EPI_ISL_896284, EPI_ISL_896285, EPI_ISL_896286, EPI_ISL_896287, EPI_ISL_896288, EPI_ISL_896289, EPI_ISL_896290, EPI_ISL_896291, EPI_ISL_896292, EPI_ISL_896293 | | | |
| see above | SUNY UPSTATE MEDICAL UNIVERSITY | Wadsworth Center, New York State Department of Health | Kirsten St. George, Daryl M. Lamson, Alexis Russel, Matthew Shudt, Melissa A Leisner, Jonathan Plitnick, Navjot Singh, John Kelly, Erasmus Schneider, Erica Lasek-Nesselquist |
| EPI_ISL_896294 | New York Presbyterian Hospital | Wadsworth Center, New York State Department of Health | Kirsten St. George, Daryl M. Lamson, Alexis Russel, Matthew Shudt, Melissa A Leisner, Jonathan Plitnick, Navjot Singh, John Kelly, Erasmus Schneider, Erica Lasek-Nesselquist |
| EPI_ISL_896295 | MEMORIAL SLOAN KETTERING CANCER CENTER | Wadsworth Center, New York State Department of Health | Kirsten St. George, Daryl M. Lamson, Alexis Russel, Matthew Shudt, Melissa A Leisner, Jonathan Plitnick, Navjot Singh, John Kelly, Erasmus Schneider, Erica Lasek-Nesselquist |
| EPI_ISL_896296 | New York Presbyterian Hospital | Wadsworth Center, New York State Department of Health | Kirsten St. George, Daryl M. Lamson, Alexis Russel, Matthew Shudt, Melissa A Leisner, Jonathan Plitnick, Navjot Singh, John Kelly, Erasmus Schneider, Erica Lasek-Nesselquist |
| EPI_ISL_896297 | BIO-REFERENCE LABORATORIES | Wadsworth Center, New York State Department of Health | Kirsten St. George, Daryl M. Lamson, Alexis Russel, Matthew Shudt, Melissa A Leisner, Jonathan Plitnick, Navjot Singh, John Kelly, Erasmus Schneider, Erica Lasek-Nesselquist |
| EPI_ISL_896298 | New York Presbyterian Hospital | Wadsworth Center, New York State Department of Health | Kirsten St. George, Daryl M. Lamson, Alexis Russel, Matthew Shudt, Melissa A Leisner, Jonathan Plitnick, Navjot Singh, John Kelly, Erasmus Schneider, Erica Lasek-Nesselquist |
| EPI_ISL_896299 | MEMORIAL SLOAN KETTERING CANCER CENTER | Wadsworth Center, New York State Department of Health | Kirsten St. George, Daryl M. Lamson, Alexis Russel, Matthew Shudt, Melissa A Leisner, Jonathan Plitnick, Navjot Singh, John Kelly, Erasmus Schneider, Erica Lasek-Nesselquist |
| EPI_ISL_896300 | BIO-REFERENCE LABORATORIES | Wadsworth Center, New York State Department of Health | Kirsten St. George, Daryl M. Lamson, Alexis Russel, Matthew Shudt, Melissa A Leisner, Jonathan Plitnick, Navjot Singh, John Kelly, Erasmus Schneider, Erica Lasek-Nesselquist |
| EPI_ISL_896301 | STONY BROOK UNIVERSITY HOSPITAL | Wadsworth Center, New York State Department of Health | Kirsten St. George, Daryl M. Lamson, Alexis Russel, Matthew Shudt, Melissa A Leisner, Jonathan Plitnick, Navjot Singh, John Kelly, Erasmus Schneider, Erica Lasek-Nesselquist |

| | | | |
|---|---|---|---|
| EPI_ISL_896302, EPI_ISL_896303, EPI_ISL_896304, EPI_ISL_896305, EPI_ISL_896306 | New York Presbyterian Hospital | Wadsworth Center, New York State Department of Health | Kirsten St. George, Daryl M. Lamson, Alexis Russel, Matthew Shudt, Melissa A Leisner, Jonathan Plitnick, Navjot Singh, John Kelly, Erasmus Schneider, Erica Lasek-Nesselquist |
| EPI_ISL_896307, EPI_ISL_896308, EPI_ISL_896309 | Wadsworth Center, New York State Department of Health | Wadsworth Center, New York State Department of Health | Kirsten St. George, Daryl M. Lamson, Alexis Russel, Matthew Shudt, Melissa A Leisner, Jonathan Plitnick, Navjot Singh, John Kelly, Erasmus Schneider, Erica Lasek-Nesselquist |
| EPI_ISL_896310, EPI_ISL_896311, EPI_ISL_896312, EPI_ISL_896313, EPI_ISL_896314, EPI_ISL_896315, EPI_ISL_896316, EPI_ISL_896317, EPI_ISL_896318, EPI_ISL_896319, EPI_ISL_896320, EPI_ISL_896321, EPI_ISL_896322, EPI_ISL_896323, EPI_ISL_896324, EPI_ISL_896325, EPI_ISL_896326, EPI_ISL_896327, EPI_ISL_896328, EPI_ISL_896329, EPI_ISL_896330, EPI_ISL_896331, EPI_ISL_896332, EPI_ISL_896333, EPI_ISL_896334, EPI_ISL_896335, EPI_ISL_896336, EPI_ISL_896337 | | | |
| see above | MEMORIAL SLOAN KETTERING CANCER CENTER | Wadsworth Center, New York State Department of Health | Kirsten St. George, Daryl M. Lamson, Alexis Russel, Matthew Shudt, Melissa A Leisner, Jonathan Plitnick, Navjot Singh, John Kelly, Erasmus Schneider, Erica Lasek-Nesselquist |
| EPI_ISL_896338, EPI_ISL_896339, EPI_ISL_896340, EPI_ISL_896341, EPI_ISL_896342 | New York Presbyterian Hospital | Wadsworth Center, New York State Department of Health | Kirsten St. George, Daryl M. Lamson, Alexis Russel, Matthew Shudt, Melissa A Leisner, Jonathan Plitnick, Navjot Singh, John Kelly, Erasmus Schneider, Erica Lasek-Nesselquist |
| EPI_ISL_896343, EPI_ISL_896344, EPI_ISL_896345, EPI_ISL_896346 | MEMORIAL SLOAN KETTERING CANCER CENTER | Wadsworth Center, New York State Department of Health | Kirsten St. George, Daryl M. Lamson, Alexis Russel, Matthew Shudt, Melissa A Leisner, Jonathan Plitnick, Navjot Singh, John Kelly, Erasmus Schneider, Erica Lasek-Nesselquist |
| EPI_ISL_896347, EPI_ISL_896348 | New York Presbyterian Hospital | Wadsworth Center, New York State Department of Health | Kirsten St. George, Daryl M. Lamson, Alexis Russel, Matthew Shudt, Melissa A Leisner, Jonathan Plitnick, Navjot Singh, John Kelly, Erasmus Schneider, Erica Lasek-Nesselquist |
| EPI_ISL_896349 | MEMORIAL SLOAN KETTERING CANCER CENTER | Wadsworth Center, New York State Department of Health | Kirsten St. George, Daryl M. Lamson, Alexis Russel, Matthew Shudt, Melissa A Leisner, Jonathan Plitnick, Navjot Singh, John Kelly, Erasmus Schneider, Erica Lasek-Nesselquist |
| EPI_ISL_896350, EPI_ISL_896351, EPI_ISL_896352, EPI_ISL_896353, EPI_ISL_896354, EPI_ISL_896355, EPI_ISL_896356, EPI_ISL_896357, EPI_ISL_896358, EPI_ISL_896359, EPI_ISL_896360, EPI_ISL_896361, EPI_ISL_896362, EPI_ISL_896363, EPI_ISL_896364, EPI_ISL_896365, EPI_ISL_896366, EPI_ISL_896367, EPI_ISL_896368, EPI_ISL_896369, EPI_ISL_896370, EPI_ISL_896371, EPI_ISL_896372 | | | |
| see above | New York Presbyterian Hospital | Wadsworth Center, New York State Department of Health | Kirsten St. George, Daryl M. Lamson, Alexis Russel, Matthew Shudt, Melissa A Leisner, Jonathan Plitnick, Navjot Singh, John Kelly, Erasmus Schneider, Erica Lasek-Nesselquist |
| EPI_ISL_896382, EPI_ISL_896383, EPI_ISL_896384 | Columbia University Irving Medical Center | Wadsworth Center, New York State Department of Health | Kirsten St. George, Daryl M. Lamson, Alexis Russel, Matthew Shudt, Melissa A Leisner, Jonathan Plitnick, Navjot Singh, John Kelly, Erasmus Schneider, Erica Lasek-Nesselquist |
| EPI_ISL_896385 | URMC LABS | Wadsworth Center, New York State Department of Health | Kirsten St. George, Daryl M. Lamson, Alexis Russel, Matthew Shudt, Melissa A Leisner, Jonathan Plitnick, Navjot Singh, John Kelly, Erasmus Schneider, Erica Lasek-Nesselquist |
| EPI_ISL_896386, EPI_ISL_896389 | Columbia University Irving Medical Center | Wadsworth Center, New York State Department of Health | Kirsten St. George, Daryl M. Lamson, Alexis Russel, Matthew Shudt, Melissa A Leisner, Jonathan Plitnick, Navjot Singh, John Kelly, Erasmus Schneider, Erica Lasek-Nesselquist |
| EPI_ISL_896390 | URMC LABS | Wadsworth Center, New York State Department of Health | Kirsten St. George, Daryl M. Lamson, Alexis Russel, Matthew Shudt, Melissa A Leisner, Jonathan Plitnick, Navjot Singh, John Kelly, Erasmus Schneider, Erica Lasek-Nesselquist |
| EPI_ISL_896391, EPI_ISL_896392, EPI_ISL_896393, EPI_ISL_896394, EPI_ISL_896395 | Columbia University Irving Medical Center | Wadsworth Center, New York State Department of Health | Kirsten St. George, Daryl M. Lamson, Alexis Russel, Matthew Shudt, Melissa A Leisner, Jonathan Plitnick, Navjot Singh, John Kelly, Erasmus Schneider, Erica Lasek-Nesselquist |
| EPI_ISL_896396, EPI_ISL_896397, EPI_ISL_896398, EPI_ISL_896399, EPI_ISL_896400, EPI_ISL_896401, EPI_ISL_896402, EPI_ISL_896403, EPI_ISL_896404, EPI_ISL_896405, EPI_ISL_896406, EPI_ISL_896407, EPI_ISL_896408, EPI_ISL_896409, EPI_ISL_896410, EPI_ISL_896411, EPI_ISL_896412, EPI_ISL_896413, EPI_ISL_896414, EPI_ISL_896415, EPI_ISL_896416, EPI_ISL_896417, EPI_ISL_896418, EPI_ISL_896419, EPI_ISL_896420, EPI_ISL_896421 | | | |
| see above | URMC LABS | Wadsworth Center, New York State Department of Health | Kirsten St. George, Daryl M. Lamson, Alexis Russel, Matthew Shudt, Melissa A Leisner, Jonathan Plitnick, Navjot Singh, John Kelly, Erasmus Schneider, Erica Lasek-Nesselquist |
| EPI_ISL_896422, EPI_ISL_896423, EPI_ISL_896424, EPI_ISL_896425, EPI_ISL_896426, EPI_ISL_896427, EPI_ISL_896428, EPI_ISL_896429, EPI_ISL_896430, EPI_ISL_896431, EPI_ISL_896432, EPI_ISL_896433, EPI_ISL_896434, EPI_ISL_896435, EPI_ISL_896436, EPI_ISL_896437, EPI_ISL_896438, EPI_ISL_896439, EPI_ISL_896440, EPI_ISL_896441, EPI_ISL_896442, EPI_ISL_896443, EPI_ISL_896444, EPI_ISL_896445, EPI_ISL_896446 | | | |
| see above | Columbia University Irving Medical Center | Wadsworth Center, New York State Department of Health | Kirsten St. George, Daryl M. Lamson, Alexis Russel, Matthew Shudt, Melissa A Leisner, Jonathan Plitnick, Navjot Singh, John Kelly, Erasmus Schneider, Erica Lasek-Nesselquist |
| EPI_ISL_896495, EPI_ISL_896496, EPI_ISL_896497 | URMC LABS | Wadsworth Center, New York State Department of Health | Kirsten St. George, Daryl M. Lamson, Alexis Russel, Matthew Shudt, Melissa A Leisner, Jonathan Plitnick, Navjot Singh, John Kelly, Erasmus Schneider, Erica Lasek-Nesselquist |
| EPI_ISL_896498, EPI_ISL_896499, EPI_ISL_896500, EPI_ISL_896501 | MEMORIAL SLOAN KETTERING CANCER CENTER | Wadsworth Center, New York State Department of Health | Kirsten St. George, Daryl M. Lamson, Alexis Russel, Matthew Shudt, Melissa A Leisner, Jonathan Plitnick, Navjot Singh, John Kelly, Erasmus Schneider, Erica Lasek-Nesselquist |
| EPI_ISL_896502, EPI_ISL_896503 | URMC LABS | Wadsworth Center, New York State Department of Health | Kirsten St. George, Daryl M. Lamson, Alexis Russel, Matthew Shudt, Melissa A Leisner, Jonathan Plitnick, Navjot Singh, John Kelly, Erasmus Schneider, Erica Lasek-Nesselquist |
| EPI_ISL_896504, EPI_ISL_896505, EPI_ISL_896506, EPI_ISL_896507 | New York Presbyterian Hospital | Wadsworth Center, New York State Department of Health | Kirsten St. George, Daryl M. Lamson, Alexis Russel, Matthew Shudt, Melissa A Leisner, Jonathan Plitnick, Navjot Singh, John Kelly, Erasmus Schneider, Erica Lasek-Nesselquist |
| EPI_ISL_896508, EPI_ISL_896509, EPI_ISL_896510, EPI_ISL_896511, EPI_ISL_896512, EPI_ISL_896513, EPI_ISL_896514, EPI_ISL_896515, EPI_ISL_896516, EPI_ISL_896517, EPI_ISL_896518, EPI_ISL_896519 | | | |
| see above | URMC LABS | Wadsworth Center, New York State Department of Health | Kirsten St. George, Daryl M. Lamson, Alexis Russel, Matthew Shudt, Melissa A Leisner, Jonathan Plitnick, Navjot Singh, John Kelly, Erasmus Schneider, Erica Lasek-Nesselquist |
| EPI_ISL_896520, EPI_ISL_896521, EPI_ISL_896522, EPI_ISL_896523, EPI_ISL_896524 | MEMORIAL SLOAN KETTERING CANCER CENTER | Wadsworth Center, New York State Department of Health | Kirsten St. George, Daryl M. Lamson, Alexis Russel, Matthew Shudt, Melissa A Leisner, Jonathan Plitnick, Navjot Singh, John Kelly, Erasmus Schneider, Erica Lasek-Nesselquist |
| EPI_ISL_896525, EPI_ISL_896526, EPI_ISL_896527, EPI_ISL_896528, EPI_ISL_896529 | New York Presbyterian Hospital | Wadsworth Center, New York State Department of Health | Kirsten St. George, Daryl M. Lamson, Alexis Russel, Matthew Shudt, Melissa A Leisner, Jonathan Plitnick, Navjot Singh, John Kelly, Erasmus Schneider, Erica Lasek-Nesselquist |
| EPI_ISL_896530 | MEMORIAL SLOAN KETTERING CANCER CENTER | Wadsworth Center, New York State Department of Health | Kirsten St. George, Daryl M. Lamson, Alexis Russel, Matthew Shudt, Melissa A Leisner, Jonathan Plitnick, Navjot Singh, John Kelly, Erasmus Schneider, Erica Lasek-Nesselquist |
| EPI_ISL_896531, EPI_ISL_896532 | New York Presbyterian Hospital | Wadsworth Center, New York State Department of Health | Kirsten St. George, Daryl M. Lamson, Alexis Russel, Matthew Shudt, Melissa A Leisner, Jonathan Plitnick, Navjot Singh, John Kelly, Erasmus Schneider, Erica Lasek-Nesselquist |
| EPI_ISL_896533 | MEMORIAL SLOAN KETTERING CANCER CENTER | Wadsworth Center, New York State Department of Health | Kirsten St. George, Daryl M. Lamson, Alexis Russel, Matthew Shudt, Melissa A Leisner, Jonathan Plitnick, Navjot Singh, John Kelly, Erasmus Schneider, Erica Lasek-Nesselquist |
| EPI_ISL_896534 | New York Presbyterian Hospital | Wadsworth Center, New York State Department of Health | Kirsten St. George, Daryl M. Lamson, Alexis Russel, Matthew Shudt, Melissa A Leisner, Jonathan Plitnick, Navjot Singh, John Kelly, Erasmus Schneider, Erica Lasek-Nesselquist |
| EPI_ISL_896535, EPI_ISL_896536 | MEMORIAL SLOAN KETTERING CANCER CENTER | Wadsworth Center, New York State Department of Health | Kirsten St. George, Daryl M. Lamson, Alexis Russel, Matthew Shudt, Melissa A Leisner, Jonathan Plitnick, Navjot Singh, John Kelly, Erasmus Schneider, Erica Lasek-Nesselquist |
| EPI_ISL_896537, EPI_ISL_896538, EPI_ISL_896539, EPI_ISL_896540 | New York Presbyterian Hospital | Wadsworth Center, New York State Department of Health | Kirsten St. George, Daryl M. Lamson, Alexis Russel, Matthew Shudt, Melissa A Leisner, Jonathan Plitnick, Navjot Singh, John Kelly, Erasmus Schneider, Erica Lasek-Nesselquist |
| EPI_ISL_896541, EPI_ISL_896542, EPI_ISL_896543, EPI_ISL_896544, EPI_ISL_896545, | MEMORIAL SLOAN KETTERING CANCER CENTER | Wadsworth Center, New York State Department of Health | Kirsten St. George, Daryl M. Lamson, Alexis Russel, Matthew Shudt, Melissa A Leisner, Jonathan Plitnick, Navjot Singh, John Kelly, Erasmus Schneider, Erica Lasek-Nesselquist |

| | | | |
|---|---|---|---|
| EPI_ISL_896546, EPI_ISL_896547, EPI_ISL_896548 | | | |
| EPI_ISL_896549, EPI_ISL_896550, EPI_ISL_896551 | New York Presbyterian Hospital | Wadsworth Center, New York State Department of Health | Kirsten St. George, Daryl M. Lamson, Alexis Russel, Matthew Shudt, Melissa A Leisner, Jonathan Plitnick, Navjot Singh, John Kelly, Erasmus Schneider, Erica Lasek-Nesselquist |
| EPI_ISL_896552 | MEMORIAL SLOAN KETTERING CANCER CENTER | Wadsworth Center, New York State Department of Health | Kirsten St. George, Daryl M. Lamson, Alexis Russel, Matthew Shudt, Melissa A Leisner, Jonathan Plitnick, Navjot Singh, John Kelly, Erasmus Schneider, Erica Lasek-Nesselquist |
| EPI_ISL_896553, EPI_ISL_896554 | New York Presbyterian Hospital | Wadsworth Center, New York State Department of Health | Kirsten St. George, Daryl M. Lamson, Alexis Russel, Matthew Shudt, Melissa A Leisner, Jonathan Plitnick, Navjot Singh, John Kelly, Erasmus Schneider, Erica Lasek-Nesselquist |
| EPI_ISL_896555 | MEMORIAL SLOAN KETTERING CANCER CENTER | Wadsworth Center, New York State Department of Health | Kirsten St. George, Daryl M. Lamson, Alexis Russel, Matthew Shudt, Melissa A Leisner, Jonathan Plitnick, Navjot Singh, John Kelly, Erasmus Schneider, Erica Lasek-Nesselquist |
| EPI_ISL_896556, EPI_ISL_896557, EPI_ISL_896558, EPI_ISL_896559, EPI_ISL_896560, EPI_ISL_896561 | New York Presbyterian Hospital | Wadsworth Center, New York State Department of Health | Kirsten St. George, Daryl M. Lamson, Alexis Russel, Matthew Shudt, Melissa A Leisner, Jonathan Plitnick, Navjot Singh, John Kelly, Erasmus Schneider, Erica Lasek-Nesselquist |
| EPI_ISL_896562 | MEMORIAL SLOAN KETTERING CANCER CENTER | Wadsworth Center, New York State Department of Health | Kirsten St. George, Daryl M. Lamson, Alexis Russel, Matthew Shudt, Melissa A Leisner, Jonathan Plitnick, Navjot Singh, John Kelly, Erasmus Schneider, Erica Lasek-Nesselquist |
| EPI_ISL_896563, EPI_ISL_896564, EPI_ISL_896565, EPI_ISL_896566, EPI_ISL_896567, EPI_ISL_896568, EPI_ISL_896569 | URMC LABS | Wadsworth Center, New York State Department of Health | Kirsten St. George, Daryl M. Lamson, Alexis Russel, Matthew Shudt, Melissa A Leisner, Jonathan Plitnick, Navjot Singh, John Kelly, Erasmus Schneider, Erica Lasek-Nesselquist |
| EPI_ISL_896570, EPI_ISL_896571 | MEMORIAL SLOAN KETTERING CANCER CENTER | Wadsworth Center, New York State Department of Health | Kirsten St. George, Daryl M. Lamson, Alexis Russel, Matthew Shudt, Melissa A Leisner, Jonathan Plitnick, Navjot Singh, John Kelly, Erasmus Schneider, Erica Lasek-Nesselquist |
| EPI_ISL_900054 | Houston Health Dept. | Houston Health Dept. | Ryker Penn, Pamela Brown |
| EPI_ISL_900691, EPI_ISL_900692, EPI_ISL_900693, EPI_ISL_900694, EPI_ISL_900695, EPI_ISL_900696, EPI_ISL_900697, EPI_ISL_900698, EPI_ISL_900699, EPI_ISL_900700, EPI_ISL_900701, EPI_ISL_900702, EPI_ISL_900703, EPI_ISL_900704, EPI_ISL_900705, EPI_ISL_900706, EPI_ISL_900707, EPI_ISL_900708, EPI_ISL_900709, EPI_ISL_900710, EPI_ISL_900711, EPI_ISL_900712, EPI_ISL_900713, EPI_ISL_900714, EPI_ISL_900715, EPI_ISL_900716, EPI_ISL_900717, EPI_ISL_900718, EPI_ISL_900719, EPI_ISL_900720, EPI_ISL_900721, EPI_ISL_900722, EPI_ISL_900723, EPI_ISL_900724, EPI_ISL_900725, EPI_ISL_900726, EPI_ISL_900727, EPI_ISL_900728, EPI_ISL_900729, EPI_ISL_900730, EPI_ISL_900731, EPI_ISL_900732, EPI_ISL_900733, EPI_ISL_900734, EPI_ISL_900735, EPI_ISL_900736, EPI_ISL_900737, EPI_ISL_900738, EPI_ISL_900739, EPI_ISL_900740, EPI_ISL_900741 | | | |
| see above | Bozeman Health Deaconess Hospital | Wiedenheft lab, Montana State University | Artem Nemudryi, Anna Nemudraia, Tanner Wiegand, Joseph Nichols, Deann T. Snyder, Jodi F. Hedges, Calvin Cicha, Helen Lee, Karl K. Vanderwood, Diane Bimczok, Mark A. Jutila and Blake Wiedenheft |
| EPI_ISL_900742 | Ohio Department of Health Laboratory | Ohio Department of Health Laboratory | Holmes, Jennifer; Eric Brandt, Keoni Omura, Glen McGillivary, Caitlin McDonnell, Jade Mowery, Stephanie Mccracken, Tyler Payne, Kirtana Ramadugu, Erica Leasure, Brent Lee, Kelsey Florek, Heather Blankenship, Quanta Brown, and Tammy Bannerman |
| EPI_ISL_902730, EPI_ISL_902731, EPI_ISL_902732, EPI_ISL_902733, EPI_ISL_902734 | Santa Clara County Public Health Laboratory | Santa Clara County Public Health Laboratory | Santa Clara County Public Health Department |
| EPI_ISL_902925 | MD Laboratories | Los Angeles County Public Health Laboratories | P. Hemarajata et al. |
| EPI_ISL_902927, EPI_ISL_902928, EPI_ISL_902929, EPI_ISL_902930, EPI_ISL_902931, EPI_ISL_902932, EPI_ISL_902933, EPI_ISL_902934, EPI_ISL_902935, EPI_ISL_902936, EPI_ISL_902937, EPI_ISL_902938, EPI_ISL_902939, EPI_ISL_902940, EPI_ISL_902941, EPI_ISL_902942, EPI_ISL_902943, EPI_ISL_902944, EPI_ISL_902945, EPI_ISL_902946, EPI_ISL_902947, EPI_ISL_902948, EPI_ISL_902949 | | | |
| see above | Maryland Public Health Laboratory | Maryland Public Health Laboratory | Maryland Department of Health Laboratories Administration |
| EPI_ISL_902950, EPI_ISL_902951, EPI_ISL_902952, EPI_ISL_902953, EPI_ISL_902954, EPI_ISL_902955, EPI_ISL_902956, EPI_ISL_902957, EPI_ISL_902958, EPI_ISL_902959, EPI_ISL_902960, EPI_ISL_902961, EPI_ISL_902962, EPI_ISL_902963, EPI_ISL_902964, EPI_ISL_902965, EPI_ISL_902966, EPI_ISL_902967, EPI_ISL_902968 | | | |
| see above | Clinical Diagnostics Laboratory, Diagnostic & Experimental Pathology, Lilly Research Laboratories | Clinical Diagnostics Laboratory, Diagnostic & Experimental Pathology, Lilly Research Laboratories | Tim Holzer, Mayuri Vaidya, Angie Fulford, Sam McNeely, Rachael Redmond, Phil Ebert, John Calley, Leslie ONeill Reising, Pat Finnegan, Erin Wray, John McElwee, Jeff Fill, Joe Oakley, Andrew Schade |
| EPI_ISL_902969 | Atlas Genomics - UW Virology Lab | UW Virology Lab | Pavitra Roychoudhury, Hong Xie, Lasata Shrestha, Michelle Lin, Meei-Li Huang, Keith R Jerome, Alexander Greninger |
| EPI_ISL_902970, EPI_ISL_902971, EPI_ISL_902972, EPI_ISL_902973, EPI_ISL_902974, EPI_ISL_902975, EPI_ISL_902976, EPI_ISL_902977, EPI_ISL_902978, EPI_ISL_902979, EPI_ISL_902980, EPI_ISL_902981, EPI_ISL_902982, EPI_ISL_902983, EPI_ISL_902984, EPI_ISL_902985, EPI_ISL_902986, EPI_ISL_902987, EPI_ISL_902988, EPI_ISL_902989, EPI_ISL_902990, EPI_ISL_902991, EPI_ISL_902992, EPI_ISL_902993, EPI_ISL_902994, EPI_ISL_902995 | | | |
| see above | Clinical Diagnostics Laboratory, Diagnostic & Experimental Pathology, Lilly Research Laboratories | Clinical Diagnostics Laboratory, Diagnostic & Experimental Pathology, Lilly Research Laboratories | Tim Holzer, Mayuri Vaidya, Angie Fulford, Sam McNeely, Rachael Redmond, Phil Ebert, John Calley, Leslie ONeill Reising, Pat Finnegan, Erin Wray, John McElwee, Jeff Fill, Joe Oakley, Andrew Schade |
| EPI_ISL_902996, EPI_ISL_902997, EPI_ISL_902998, EPI_ISL_903000, EPI_ISL_903001, EPI_ISL_903002, EPI_ISL_903003, EPI_ISL_903004, EPI_ISL_903005, EPI_ISL_903006, EPI_ISL_903007, EPI_ISL_903008, EPI_ISL_903009, EPI_ISL_903010, EPI_ISL_903012, EPI_ISL_903013, EPI_ISL_903014, EPI_ISL_903015, EPI_ISL_903016, EPI_ISL_903017, EPI_ISL_903018 | | | |
| see above | Seattle Flu Study | Seattle Flu Study | Deborah A. Nickerson, Chris D. Frazar, Jover Lee, Benjamin Pelle, Erica Ryke, Matthew Richardson, Amanda Adler, Elisabeth Brandstetter, Peter D. Han, Kairsten Fay, Misja Ilcisin, Kirsten Lacombe, Thomas R. Sibley, Melissa Truong, Caitlin R. Wolf, Michael Boeckh, Janet A. Englund, Michael Famulare, Barry R. Lutz, Mark J. Rieder, Lea M. Starita, Matthew Thompson, Jay Shendure, Trevor Bedford, Helen Y. Chu |
| EPI_ISL_903020, EPI_ISL_903021, EPI_ISL_903022, EPI_ISL_903024, EPI_ISL_903025 | Seattle Flu Study | Seattle Flu Study | Deborah A. Nickerson, Chris D. Frazar, Jover Lee, Benjamin Pelle, Erica Ryke, Matthew Richardson, Amanda Adler, Elisabeth Brandstetter, Peter D. Han, Kairsten Fay, Misja Ilcisin, Kirsten Lacombe, Thomas R. Sibley, Melissa Truong, Caitlin R. Wolf, Karen Cowgill, Stephanie Schrag, Jeff Duchin, Michael Boeckh, Janet A. Englund, Michael Famulare, Barry R. Lutz, Mark J. Rieder, Lea M. Starita, Matthew Thompson, Helen Y. Chu, Trevor Bedford, Jay Shendure |
| EPI_ISL_903027 | Seattle Flu Study | Seattle Flu Study | Deborah A. Nickerson, Chris D. Frazar, Jover Lee, Benjamin Pelle, Erica Ryke, Matthew Richardson, Amanda Adler, Elisabeth Brandstetter, Peter D. Han, Kairsten Fay, Misja Ilcisin, Kirsten Lacombe, Thomas R. Sibley, Melissa Truong, Caitlin R. Wolf, Michael Boeckh, Janet A. Englund, Michael Famulare, Barry R. Lutz, Mark J. Rieder, Lea M. Starita, Matthew Thompson, Jay Shendure, Trevor Bedford, Helen Y. Chu |
| EPI_ISL_903028, EPI_ISL_903029, EPI_ISL_903033, EPI_ISL_903035, EPI_ISL_903036, EPI_ISL_903039, EPI_ISL_903040, EPI_ISL_903041 | Seattle Flu Study | Seattle Flu Study | Deborah A. Nickerson, Chris D. Frazar, Jover Lee, Benjamin Pelle, Erica Ryke, Matthew Richardson, Amanda Adler, Elisabeth Brandstetter, Peter D. Han, Kairsten Fay, Misja Ilcisin, Kirsten Lacombe, Thomas R. Sibley, Melissa Truong, Caitlin R. Wolf, Karen Cowgill, Stephanie Schrag, Jeff Duchin, Michael Boeckh, Janet A. Englund, Michael Famulare, Barry R. Lutz, Mark J. Rieder, Lea M. Starita, Matthew Thompson, Helen Y. Chu, Trevor Bedford, Jay Shendure |
| EPI_ISL_903043, EPI_ISL_903044 | Seattle Flu Study | Seattle Flu Study | Deborah A. Nickerson, Chris D. Frazar, Jover Lee, Benjamin Pelle, Erica Ryke, Matthew Richardson, Amanda Adler, Elisabeth Brandstetter, Peter D. Han, Kairsten Fay, Misja Ilcisin, Kirsten Lacombe, Thomas R. Sibley, Melissa Truong, Caitlin R. Wolf, Michael Boeckh, Janet A. Englund, Michael Famulare, Barry R. Lutz, Mark J. Rieder, Lea M. Starita, Matthew Thompson, Jay Shendure, Trevor Bedford, Helen Y. Chu |
| EPI_ISL_903045 | Seattle Flu Study | Seattle Flu Study | Deborah A. Nickerson, Chris D. Frazar, Jover Lee, Benjamin Pelle, Erica Ryke, Matthew Richardson, Amanda Adler, Elisabeth Brandstetter, Peter D. Han, Kairsten Fay, Misja Ilcisin, Kirsten Lacombe, Thomas R. Sibley, Melissa Truong, Caitlin R. Wolf, Karen Cowgill, Stephanie Schrag, Jeff Duchin, Michael Boeckh, Janet A. Englund, Michael Famulare, Barry R. Lutz, Mark J. Rieder, Lea M. Starita, Matthew Thompson, Helen Y. Chu, Trevor Bedford, Jay Shendure |
| EPI_ISL_903048 | Seattle Flu Study | Seattle Flu Study | Deborah A. Nickerson, Chris D. Frazar, Jover Lee, Benjamin Pelle, Erica Ryke, Matthew Richardson, Amanda Adler, Elisabeth Brandstetter, Peter D. Han, Kairsten Fay, Misja Ilcisin, Kirsten Lacombe, Thomas R. Sibley, Melissa Truong, Caitlin R. Wolf, Michael Boeckh, Janet A. Englund, Michael Famulare, Barry R. Lutz, Mark J. Rieder, Lea M. Starita, Matthew Thompson, Jay Shendure, Trevor Bedford, Helen Y. Chu |
| EPI_ISL_903051, EPI_ISL_903053 | Seattle Flu Study | Seattle Flu Study | Deborah A. Nickerson, Chris D. Frazar, Jover Lee, Benjamin Pelle, Erica Ryke, Matthew Richardson, Amanda Adler, Elisabeth Brandstetter, Peter D. Han, Kairsten Fay, Misja Ilcisin, Kirsten Lacombe, Thomas R. Sibley, Melissa Truong, Caitlin R. Wolf, Karen Cowgill, Stephanie Schrag, Jeff Duchin, Michael Boeckh, Janet A. Englund, Michael Famulare, Barry R. Lutz, Mark J. Rieder, Lea M. Starita, Matthew Thompson, Helen Y. Chu, Trevor Bedford, Jay Shendure |
| EPI_ISL_903054 | Seattle Flu Study | Seattle Flu Study | Deborah A. Nickerson, Chris D. Frazar, Jover Lee, Benjamin Pelle, Erica Ryke, Matthew Richardson, Amanda Adler, Elisabeth Brandstetter, Peter D. Han, Kairsten Fay, Misja Ilcisin, Kirsten Lacombe, Thomas R. Sibley, Melissa Truong, Caitlin R. Wolf, Michael Boeckh, Janet A. Englund, Michael Famulare, Barry R. Lutz, Mark J. Rieder, Lea M. Starita, Matthew Thompson, Jay Shendure, Trevor Bedford, Helen Y. Chu |
| EPI_ISL_903055 | Seattle Flu Study | Seattle Flu Study | Deborah A. Nickerson, Chris D. Frazar, Jover Lee, Benjamin Pelle, Erica Ryke, Matthew Richardson, Amanda Adler, Elisabeth Brandstetter, Peter D. Han, Kairsten Fay, Misja Ilcisin, Kirsten Lacombe, Thomas R. Sibley, Melissa Truong, Caitlin R. Wolf, Karen Cowgill, Stephanie Schrag, Jeff Duchin, Michael Boeckh, Janet A. Englund, Michael Famulare, Barry R. Lutz, Mark J. Rieder, Lea M. Starita, Matthew Thompson, Helen Y. Chu, Trevor Bedford, Jay Shendure |
| EPI_ISL_903056 | Seattle Flu Study | Seattle Flu Study | Deborah A. Nickerson, Chris D. Frazar, Jover Lee, Benjamin Pelle, Erica Ryke, Matthew Richardson, Amanda Adler, Elisabeth Brandstetter, Peter D. Han, Kairsten Fay, Misja Ilcisin, Kirsten Lacombe, Thomas R. Sibley, Melissa Truong, Caitlin R. Wolf, Michael Boeckh, Janet A. Englund, Michael Famulare, Barry R. Lutz, Mark J. Rieder, Lea M. Starita, Matthew Thompson, Jay Shendure, Trevor Bedford, Helen Y. Chu |
| EPI_ISL_903057, EPI_ISL_903058, EPI_ISL_903060, EPI_ISL_903061, EPI_ISL_903062, EPI_ISL_903063 | Seattle Flu Study | Seattle Flu Study | Deborah A. Nickerson, Chris D. Frazar, Jover Lee, Benjamin Pelle, Erica Ryke, Matthew Richardson, Amanda Adler, Elisabeth Brandstetter, Peter D. Han, Kairsten Fay, Misja Ilcisin, Kirsten Lacombe, Thomas R. Sibley, Melissa Truong, Caitlin R. Wolf, Karen Cowgill, Stephanie Schrag, Jeff Duchin, Michael Boeckh, Janet A. Englund, Michael Famulare, Barry R. Lutz, Mark J. Rieder, Lea M. Starita, Matthew Thompson, Helen Y. Chu, Trevor Bedford, Jay Shendure |
| EPI_ISL_903064, EPI_ISL_903065, EPI_ISL_903066, | Seattle Flu Study | Seattle Flu Study | Deborah A. Nickerson, Chris D. Frazar, Jover Lee, Benjamin Pelle, Erica Ryke, Matthew Richardson, Amanda Adler, Elisabeth Brandstetter, Peter D. Han, Kairsten Fay, Misja Ilcisin, Kirsten Lacombe, Thomas R. Sibley, Melissa Truong, Caitlin R. Wolf, Michael Boeckh, Janet A. Englund, Michael Famulare, Barry R. Lutz, Mark J. Rieder, Lea M. Starita, Matthew Thompson, Jay Shendure, Trevor Bedford, Helen Y. Chu |

| | | | |
|---|---|---|---|
| EPI_ISL_903067, EPI_ISL_903068 | | | |
| EPI_ISL_903070, EPI_ISL_903071, EPI_ISL_903072 | Washington State Department of Health | Seattle Flu Study | Deborah A. Nickerson, Chris D. Frazar, Jover Lee, Benjamin Pelle, Erica Ryke, Matthew Richardson, Amanda Adler, Elisabeth Brandstetter, Peter D. Han, Kairsten Fay, Misja Ilcisin, Kirsten Lacombe, Thomas R. Sibley, Melissa Truong, Caitlin R. Wolf, Romesh Gautom, Geoff Melly, Brian Hiatt, Philip Dykema, Scott Lindquist, Michael Boeckh, Janet A. Englund, Michael Famulare, Barry R. Lutz, Mark J. Rieder, Lea M. Starita, Matthew Thompson, Helen Y. Chu, Jay Shendure, Trevor Bedford |
| EPI_ISL_903073 | Seattle Flu Study | Seattle Flu Study | Deborah A. Nickerson, Chris D. Frazar, Jover Lee, Benjamin Pelle, Erica Ryke, Matthew Richardson, Amanda Adler, Elisabeth Brandstetter, Peter D. Han, Kairsten Fay, Misja Ilcisin, Kirsten Lacombe, Thomas R. Sibley, Melissa Truong, Caitlin R. Wolf, Karen Cowgill, Stephanie Schrag, Jeff Duchin, Michael Boeckh, Janet A. Englund, Michael Famulare, Barry R. Lutz, Mark J. Rieder, Lea M. Starita, Matthew Thompson, Helen Y. Chu, Trevor Bedford, Jay Shendure |
| EPI_ISL_903074, EPI_ISL_903075, EPI_ISL_903077 | Seattle Flu Study | Seattle Flu Study | Deborah A. Nickerson, Chris D. Frazar, Jover Lee, Benjamin Pelle, Erica Ryke, Matthew Richardson, Amanda Adler, Elisabeth Brandstetter, Peter D. Han, Kairsten Fay, Misja Ilcisin, Kirsten Lacombe, Thomas R. Sibley, Melissa Truong, Caitlin R. Wolf, Michael Boeckh, Janet A. Englund, Michael Famulare, Barry R. Lutz, Mark J. Rieder, Lea M. Starita, Matthew Thompson, Jay Shendure, Trevor Bedford, Helen Y. Chu |
| EPI_ISL_903078, EPI_ISL_903079 | Seattle Flu Study | Seattle Flu Study | Deborah A. Nickerson, Chris D. Frazar, Jover Lee, Benjamin Pelle, Erica Ryke, Matthew Richardson, Amanda Adler, Elisabeth Brandstetter, Peter D. Han, Kairsten Fay, Misja Ilcisin, Kirsten Lacombe, Thomas R. Sibley, Melissa Truong, Caitlin R. Wolf, Karen Cowgill, Stephanie Schrag, Jeff Duchin, Michael Boeckh, Janet A. Englund, Michael Famulare, Barry R. Lutz, Mark J. Rieder, Lea M. Starita, Matthew Thompson, Helen Y. Chu, Trevor Bedford, Jay Shendure |
| EPI_ISL_903081 | Seattle Flu Study | Seattle Flu Study | Deborah A. Nickerson, Chris D. Frazar, Jover Lee, Benjamin Pelle, Erica Ryke, Matthew Richardson, Amanda Adler, Elisabeth Brandstetter, Peter D. Han, Kairsten Fay, Misja Ilcisin, Kirsten Lacombe, Thomas R. Sibley, Melissa Truong, Caitlin R. Wolf, Michael Boeckh, Janet A. Englund, Michael Famulare, Barry R. Lutz, Mark J. Rieder, Lea M. Starita, Matthew Thompson, Jay Shendure, Trevor Bedford, Helen Y. Chu |
| EPI_ISL_903082, EPI_ISL_903083 | Seattle Flu Study | Seattle Flu Study | Deborah A. Nickerson, Chris D. Frazar, Jover Lee, Benjamin Pelle, Erica Ryke, Matthew Richardson, Amanda Adler, Elisabeth Brandstetter, Peter D. Han, Kairsten Fay, Misja Ilcisin, Kirsten Lacombe, Thomas R. Sibley, Melissa Truong, Caitlin R. Wolf, Karen Cowgill, Stephanie Schrag, Jeff Duchin, Michael Boeckh, Janet A. Englund, Michael Famulare, Barry R. Lutz, Mark J. Rieder, Lea M. Starita, Matthew Thompson, Helen Y. Chu, Trevor Bedford, Jay Shendure |
| EPI_ISL_903084 | Seattle Flu Study | Seattle Flu Study | Deborah A. Nickerson, Chris D. Frazar, Jover Lee, Benjamin Pelle, Erica Ryke, Matthew Richardson, Amanda Adler, Elisabeth Brandstetter, Peter D. Han, Kairsten Fay, Misja Ilcisin, Kirsten Lacombe, Thomas R. Sibley, Melissa Truong, Caitlin R. Wolf, Michael Boeckh, Janet A. Englund, Michael Famulare, Barry R. Lutz, Mark J. Rieder, Lea M. Starita, Matthew Thompson, Jay Shendure, Trevor Bedford, Helen Y. Chu |
| EPI_ISL_903085, EPI_ISL_903086, EPI_ISL_903087, EPI_ISL_903090, EPI_ISL_903092, EPI_ISL_903093, EPI_ISL_903096, EPI_ISL_903097, EPI_ISL_903098, EPI_ISL_903099, EPI_ISL_903100, EPI_ISL_903101, EPI_ISL_903102, EPI_ISL_903104 | | | |
| see above | Seattle Flu Study | Seattle Flu Study | Deborah A. Nickerson, Chris D. Frazar, Jover Lee, Benjamin Pelle, Erica Ryke, Matthew Richardson, Amanda Adler, Elisabeth Brandstetter, Peter D. Han, Kairsten Fay, Misja Ilcisin, Kirsten Lacombe, Thomas R. Sibley, Melissa Truong, Caitlin R. Wolf, Karen Cowgill, Stephanie Schrag, Jeff Duchin, Michael Boeckh, Janet A. Englund, Michael Famulare, Barry R. Lutz, Mark J. Rieder, Lea M. Starita, Matthew Thompson, Helen Y. Chu, Trevor Bedford, Jay Shendure |
| EPI_ISL_903105, EPI_ISL_903106, EPI_ISL_903107, EPI_ISL_903108, EPI_ISL_903109, EPI_ISL_903110, EPI_ISL_903111, EPI_ISL_903112, EPI_ISL_903113, EPI_ISL_903114, EPI_ISL_903115, EPI_ISL_903116, EPI_ISL_903117, EPI_ISL_903118, EPI_ISL_903119, EPI_ISL_903120, EPI_ISL_903121, EPI_ISL_903122, EPI_ISL_903123, EPI_ISL_903124, EPI_ISL_903125, EPI_ISL_903126, EPI_ISL_903127, EPI_ISL_903128, EPI_ISL_903129, EPI_ISL_903130, EPI_ISL_903131, EPI_ISL_903132, EPI_ISL_903135, EPI_ISL_903136, EPI_ISL_903137, EPI_ISL_903138, EPI_ISL_903139, EPI_ISL_903141, EPI_ISL_903142, EPI_ISL_903143, EPI_ISL_903144, EPI_ISL_903145, EPI_ISL_903146, EPI_ISL_903147, EPI_ISL_903148, EPI_ISL_903149, EPI_ISL_903150, EPI_ISL_903151, EPI_ISL_903152, EPI_ISL_903154, EPI_ISL_903155, EPI_ISL_903156, EPI_ISL_903157, EPI_ISL_903158, EPI_ISL_903159, EPI_ISL_903160, EPI_ISL_903161, EPI_ISL_903163, EPI_ISL_903164, EPI_ISL_903165, EPI_ISL_903167, EPI_ISL_903168, EPI_ISL_903171, EPI_ISL_903172, EPI_ISL_903173, EPI_ISL_903174, EPI_ISL_903175, EPI_ISL_903176, EPI_ISL_903177, EPI_ISL_903178, EPI_ISL_903179 | | | |
| see above | Washington State Department of Health | Seattle Flu Study | Deborah A. Nickerson, Chris D. Frazar, Jover Lee, Benjamin Pelle, Erica Ryke, Matthew Richardson, Amanda Adler, Elisabeth Brandstetter, Peter D. Han, Kairsten Fay, Misja Ilcisin, Kirsten Lacombe, Thomas R. Sibley, Melissa Truong, Caitlin R. Wolf, Romesh Gautom, Geoff Melly, Brian Hiatt, Philip Dykema, Scott Lindquist, Michael Boeckh, Janet A. Englund, Michael Famulare, Barry R. Lutz, Mark J. Rieder, Lea M. Starita, Matthew Thompson, Helen Y. Chu, Jay Shendure, Trevor Bedford |
| EPI_ISL_903180, EPI_ISL_903181, EPI_ISL_903183, EPI_ISL_903184, EPI_ISL_903185, EPI_ISL_903187, EPI_ISL_903188, EPI_ISL_903189, EPI_ISL_903190, EPI_ISL_903191, EPI_ISL_903192, EPI_ISL_903193, EPI_ISL_903194, EPI_ISL_903195, EPI_ISL_903197, EPI_ISL_903198, EPI_ISL_903201, EPI_ISL_903202, EPI_ISL_903203, EPI_ISL_903204, EPI_ISL_903206, EPI_ISL_903207 | | | |
| see above | Seattle Flu Study | Seattle Flu Study | Deborah A. Nickerson, Chris D. Frazar, Jover Lee, Benjamin Pelle, Erica Ryke, Matthew Richardson, Amanda Adler, Elisabeth Brandstetter, Peter D. Han, Kairsten Fay, Misja Ilcisin, Kirsten Lacombe, Thomas R. Sibley, Melissa Truong, Caitlin R. Wolf, Karen Cowgill, Stephanie Schrag, Jeff Duchin, Michael Boeckh, Janet A. Englund, Michael Famulare, Barry R. Lutz, Mark J. Rieder, Lea M. Starita, Matthew Thompson, Helen Y. Chu, Trevor Bedford, Jay Shendure |
| EPI_ISL_903208, EPI_ISL_903209, EPI_ISL_903210, EPI_ISL_903211, EPI_ISL_903212, EPI_ISL_903213, EPI_ISL_903214, EPI_ISL_903215, EPI_ISL_903216, EPI_ISL_903217 | University of Michigan Clinical Microbiology Laboratory | Lauring Lab, University of Michigan, Department of Microbiology and Immunology | Valesano |
| EPI_ISL_903218, EPI_ISL_903219, EPI_ISL_903220, EPI_ISL_903221, EPI_ISL_903222, EPI_ISL_903223, EPI_ISL_903224, EPI_ISL_903225, EPI_ISL_903226, EPI_ISL_903227, EPI_ISL_903228 | | | |
| see above | Clinical Diagnostics Laboratory, Diagnostic & Experimental Pathology, Lilly Research Laboratories | Clinical Diagnostics Laboratory, Diagnostic & Experimental Pathology, Lilly Research Laboratories | Tim Holzer, Mayuri Vaidya, Angie Fulford, Sam McNeely, Rachael Redmond, Phil Ebert, John Calley, Leslie ONeill Reising, Pat Finnegan, Erin Wray, John McElwee, Jeff Fill, Joe Oakley, Andrew Schade |
| EPI_ISL_903229, EPI_ISL_903230, EPI_ISL_903231, EPI_ISL_903232, EPI_ISL_903233, EPI_ISL_903234, EPI_ISL_903235, EPI_ISL_903236, EPI_ISL_903237, EPI_ISL_903238, EPI_ISL_903239, EPI_ISL_903240, EPI_ISL_903241, EPI_ISL_903242, EPI_ISL_903243, EPI_ISL_903244, EPI_ISL_903245, EPI_ISL_903246, EPI_ISL_903247, EPI_ISL_903248, EPI_ISL_903249, EPI_ISL_903250, EPI_ISL_903251, EPI_ISL_903252, EPI_ISL_903253, EPI_ISL_903254, EPI_ISL_903255, EPI_ISL_903256, EPI_ISL_903257, EPI_ISL_903258, EPI_ISL_903259, EPI_ISL_903260, EPI_ISL_903261, EPI_ISL_903262, EPI_ISL_903263, EPI_ISL_903264, EPI_ISL_903265, EPI_ISL_903266, EPI_ISL_903267, EPI_ISL_903268, EPI_ISL_903269, EPI_ISL_903270, EPI_ISL_903271, EPI_ISL_903272, EPI_ISL_903273, EPI_ISL_903274, EPI_ISL_903275, EPI_ISL_903276, EPI_ISL_903277, EPI_ISL_903278, EPI_ISL_903279, EPI_ISL_903280, EPI_ISL_903281, EPI_ISL_903282, EPI_ISL_903283, EPI_ISL_903284, EPI_ISL_903285, EPI_ISL_903286, EPI_ISL_903287, EPI_ISL_903288, EPI_ISL_903289, EPI_ISL_903290, EPI_ISL_903291, EPI_ISL_903292, EPI_ISL_903293, EPI_ISL_903294, EPI_ISL_903295, EPI_ISL_903296, EPI_ISL_903297, EPI_ISL_903298, EPI_ISL_903299, EPI_ISL_903300, EPI_ISL_903301, EPI_ISL_903302, EPI_ISL_903303, EPI_ISL_903304, EPI_ISL_903305, EPI_ISL_903306, EPI_ISL_903307, EPI_ISL_903308, EPI_ISL_903309, EPI_ISL_903310, EPI_ISL_903311 | | | |
| see above | M Health Fairview | Minnesota Department of Health, Public Health Laboratory | Alexandra Lorentz, Jacob Garfin, Matt Plumb, and Xiong Wang |
| EPI_ISL_903312, EPI_ISL_903313, EPI_ISL_903314, EPI_ISL_903315, EPI_ISL_903316, EPI_ISL_903317, EPI_ISL_903318, EPI_ISL_903319, EPI_ISL_903320, EPI_ISL_903321, EPI_ISL_903322, EPI_ISL_903323, EPI_ISL_903324, EPI_ISL_903325, EPI_ISL_903326, EPI_ISL_903327, EPI_ISL_903328, EPI_ISL_903329, EPI_ISL_903330, EPI_ISL_903331, EPI_ISL_903332, EPI_ISL_903333, EPI_ISL_903334, EPI_ISL_903335, EPI_ISL_903336, EPI_ISL_903337, EPI_ISL_903338 | | | |
| see above | University of Iowa Hospitals & Clinics, Microbiology Laboratory | University or Iowa, Lung Biology and Cystic Fibrosis Research Center, Pezzulo Lab | Miguel E Ortiz, Alejandro A Pezzulo |
| EPI_ISL_903339, EPI_ISL_903340, EPI_ISL_903341, EPI_ISL_903342, EPI_ISL_903343, EPI_ISL_903344, EPI_ISL_903345, EPI_ISL_903346 | Bozeman Health Deaconess Hospital | Wiedenheft lab, Montana State University | Artem Nemudryi, Anna Nemudraia, Tanner Wiegand, Joseph Nichols, Deann T. Snyder, Jodi F. Hedges, Calvin Cicha, Helen Lee, Karl K. Vanderwood, Diane Bimczok, Mark A. Jutila and Blake Wiedenheft |
| EPI_ISL_903348, EPI_ISL_903349, EPI_ISL_903350, EPI_ISL_903356, EPI_ISL_903357, EPI_ISL_903358, EPI_ISL_903359, EPI_ISL_903360 | Wyoming Public Health Laboratory | Wyoming Public Health Laboratory | Noah Hull, Taylor Fearing, Lynette Gumbleton, Channing Weber, Ashley Norberg, Bailey Bowcutt, and Wanda Manley |
| EPI_ISL_903361, EPI_ISL_903362, EPI_ISL_903363 | Arizona State University | Arizona State University | Emily A. Kaelin, LaRinda A. Holland, Peter T. Skidmore, Nicholas J. Mellor, Kristina Buss, Joy M. Blain, Valerie Harris, Joshua LaBaer, Vel Murugan, Efrem S. Lim |
| EPI_ISL_903367, EPI_ISL_903368, EPI_ISL_903369, EPI_ISL_903370, EPI_ISL_903371, EPI_ISL_903372 | Los Angeles County Public Health Laboratories | Los Angeles County Public Health Laboratories | P. Hemarajata et al. |
| EPI_ISL_903373, EPI_ISL_903374 | MD Laboratories | Los Angeles County Public Health Laboratories | P. Hemarajata et al. |
| EPI_ISL_903390, EPI_ISL_903391, EPI_ISL_903392, EPI_ISL_903393, EPI_ISL_903394, EPI_ISL_903395, EPI_ISL_903396, EPI_ISL_903397, EPI_ISL_903398, EPI_ISL_903399, EPI_ISL_903400, EPI_ISL_903401, EPI_ISL_903402, EPI_ISL_903403, EPI_ISL_903404, EPI_ISL_903405, EPI_ISL_903406, EPI_ISL_903407, EPI_ISL_903408, EPI_ISL_903409, EPI_ISL_903410, EPI_ISL_903411, EPI_ISL_903412, EPI_ISL_903413, EPI_ISL_903414, EPI_ISL_903415, EPI_ISL_903416, EPI_ISL_903417, EPI_ISL_903418, EPI_ISL_903419, EPI_ISL_903420, EPI_ISL_903421, EPI_ISL_903422, EPI_ISL_903423, EPI_ISL_903424, EPI_ISL_903425, EPI_ISL_903426, EPI_ISL_903427, EPI_ISL_903428, EPI_ISL_903429, EPI_ISL_903430, EPI_ISL_903431, EPI_ISL_903432, EPI_ISL_903433, EPI_ISL_903434, EPI_ISL_903435, EPI_ISL_903436, EPI_ISL_903437, EPI_ISL_903438, EPI_ISL_903439, EPI_ISL_903440, EPI_ISL_903441, EPI_ISL_903442, EPI_ISL_903443, EPI_ISL_903444, EPI_ISL_903445, EPI_ISL_903446, EPI_ISL_903447, EPI_ISL_903448, EPI_ISL_903449, EPI_ISL_903450, EPI_ISL_903451, EPI_ISL_903452, EPI_ISL_903453, EPI_ISL_903454, EPI_ISL_903455, EPI_ISL_903456, EPI_ISL_903457, EPI_ISL_903458, EPI_ISL_903459, EPI_ISL_903460, EPI_ISL_903461, EPI_ISL_903462, EPI_ISL_903463, EPI_ISL_903464, EPI_ISL_903465, EPI_ISL_903466, EPI_ISL_903467, EPI_ISL_903468, EPI_ISL_903469, EPI_ISL_903470, EPI_ISL_903471, EPI_ISL_903472, EPI_ISL_903473, EPI_ISL_903474, EPI_ISL_903475, EPI_ISL_903476, EPI_ISL_903477, EPI_ISL_903478, EPI_ISL_903479, EPI_ISL_903480, EPI_ISL_903481, EPI_ISL_903482, EPI_ISL_903483, EPI_ISL_903484, EPI_ISL_903485, EPI_ISL_903486, EPI_ISL_903487, EPI_ISL_903488, EPI_ISL_903489, EPI_ISL_903490, EPI_ISL_903491, EPI_ISL_903492, EPI_ISL_903493, EPI_ISL_903494, EPI_ISL_903495, EPI_ISL_903496, EPI_ISL_903497, EPI_ISL_903498, EPI_ISL_903499, EPI_ISL_903500, EPI_ISL_903501, EPI_ISL_903502, EPI_ISL_903503, EPI_ISL_903504, EPI_ISL_903505, EPI_ISL_903506, EPI_ISL_903507, EPI_ISL_903508, EPI_ISL_903509, EPI_ISL_903510, EPI_ISL_903511, EPI_ISL_903512, EPI_ISL_903513, EPI_ISL_903514, EPI_ISL_903515, EPI_ISL_903516, EPI_ISL_903517, EPI_ISL_903518, EPI_ISL_903519, EPI_ISL_903520, EPI_ISL_903521, EPI_ISL_903522, EPI_ISL_903523, EPI_ISL_903524, EPI_ISL_903525, EPI_ISL_903526, EPI_ISL_903527, EPI_ISL_903528, EPI_ISL_903529, EPI_ISL_903530, EPI_ISL_903531, EPI_ISL_903532, EPI_ISL_903533, EPI_ISL_903534, EPI_ISL_903535, EPI_ISL_903536, EPI_ISL_903537, EPI_ISL_903538, EPI_ISL_903539, EPI_ISL_903540, EPI_ISL_903541, EPI_ISL_903542, EPI_ISL_903543, EPI_ISL_903544, EPI_ISL_903545, EPI_ISL_903546, EPI_ISL_903547, EPI_ISL_903548, EPI_ISL_903549, EPI_ISL_903550, EPI_ISL_903551, EPI_ISL_903552, EPI_ISL_903553, EPI_ISL_903554, EPI_ISL_903555, EPI_ISL_903556, EPI_ISL_903557, EPI_ISL_903558, EPI_ISL_903559, EPI_ISL_903560, EPI_ISL_903561, EPI_ISL_903562, EPI_ISL_903563, EPI_ISL_903564 | | | |
| see above | Quest Diagnostics | Quest Diagnostics | Rosenthal,S.H., Gerasimova,A., Kagan,R.M., Anderson, B., Hua, M., Liu Y., Bernstein, L.E., Livingston, K.E., Perez, A., Shalhout, D.F., Shlyakhter, I.A., Owen, R., Tanpaiboon, P., Lacbawan, F. |
| EPI_ISL_903565 | AR Dept. of Health-Public Health Lab | Genomics and Discovery, Respiratory Viruses Branch, Division of Viral Diseases, Centers for Disease Control and Prevention | Krista Queen, Yan Li, Ying Tao, Jing Zhang, Anna Uehara, Anna Montmayeur, Clinton R. Paden, Peter W. Cook, Rachel Marine, Mili Sheth, Jasmine Padilla, Sarah Nobles, Mark Burroughs, Lori Rowe, Haibin Wang, Ben L. Rambo-Martin, Dhwani Batra, Justin Lee, Suxiang Tong |
| EPI_ISL_903568 | NC State Laboratory of Public Health | Genomics and Discovery, Respiratory Viruses Branch, Division of Viral Diseases, Centers for Disease Control and Prevention | Krista Queen, Yan Li, Ying Tao, Jing Zhang, Anna Uehara, Anna Montmayeur, Clinton R. Paden, Peter W. Cook, Rachel Marine, Mili Sheth, Jasmine Padilla, Sarah Nobles, Mark Burroughs, Lori Rowe, Haibin Wang, Ben L. Rambo-Martin, Dhwani Batra, Justin Lee, Suxiang Tong |
| EPI_ISL_903571 | ID Bureau of Laboratories | Genomics and Discovery, Respiratory | Krista Queen, Yan Li, Ying Tao, Jing Zhang, Anna Uehara, Anna Montmayeur, Clinton R. Paden, Peter W. Cook, Rachel Marine, Mili Sheth, Jasmine Padilla, Sarah Nobles, Mark Burroughs, Lori Rowe, Haibin Wang, Ben L. Rambo-Martin, Dhwani Batra, Justin Lee, Suxiang Tong |

| | | | |
|---|---|---|---|
| | | Viruses Branch, Division of Viral Diseases, Centers for Disease Control and Prevention | |
| EPI_ISL_903578 | WA State Department of Health | Genomics and Discovery, Respiratory Viruses Branch, Division of Viral Diseases, Centers for Disease Control and Prevention | Krista Queen, Yan Li, Ying Tao, Jing Zhang, Anna Uehara, Anna Montmayeur, Clinton R. Paden, Peter W. Cook, Rachel Marine, Mili Sheth, Jasmine Padilla, Sarah Nobles, Mark Burroughs, Lori Rowe, Haibin Wang, Ben L. Rambo-Martin, Dhwani Batra, Justin Lee, Suxiang Tong |
| EPI_ISL_903582 | TN Division of Laboratory Services | Genomics and Discovery, Respiratory Viruses Branch, Division of Viral Diseases, Centers for Disease Control and Prevention | Krista Queen, Yan Li, Ying Tao, Jing Zhang, Anna Uehara, Anna Montmayeur, Clinton R. Paden, Peter W. Cook, Rachel Marine, Mili Sheth, Jasmine Padilla, Sarah Nobles, Mark Burroughs, Lori Rowe, Haibin Wang, Ben L. Rambo-Martin, Dhwani Batra, Justin Lee, Suxiang Tong |
| EPI_ISL_903583 | HI Dept. of Health, State Laboratories Division | Genomics and Discovery, Respiratory Viruses Branch, Division of Viral Diseases, Centers for Disease Control and Prevention | Krista Queen, Yan Li, Ying Tao, Jing Zhang, Anna Uehara, Anna Montmayeur, Clinton R. Paden, Peter W. Cook, Rachel Marine, Mili Sheth, Jasmine Padilla, Sarah Nobles, Mark Burroughs, Lori Rowe, Haibin Wang, Ben L. Rambo-Martin, Dhwani Batra, Justin Lee, Suxiang Tong |
| EPI_ISL_903587, EPI_ISL_903588 | IL Dept. of Public Health Springfield Laboratory | Genomics and Discovery, Respiratory Viruses Branch, Division of Viral Diseases, Centers for Disease Control and Prevention | Krista Queen, Yan Li, Ying Tao, Jing Zhang, Anna Uehara, Anna Montmayeur, Clinton R. Paden, Peter W. Cook, Rachel Marine, Mili Sheth, Jasmine Padilla, Sarah Nobles, Mark Burroughs, Lori Rowe, Haibin Wang, Ben L. Rambo-Martin, Dhwani Batra, Justin Lee, Suxiang Tong |
| EPI_ISL_903589 | MI - Michigan Department of Health and Human Services - Bureau of Laboratories | Genomics and Discovery, Respiratory Viruses Branch, Division of Viral Diseases, Centers for Disease Control and Prevention | Krista Queen, Yan Li, Ying Tao, Jing Zhang, Anna Uehara, Anna Montmayeur, Clinton R. Paden, Peter W. Cook, Rachel Marine, Mili Sheth, Jasmine Padilla, Sarah Nobles, Mark Burroughs, Lori Rowe, Haibin Wang, Ben L. Rambo-Martin, Dhwani Batra, Justin Lee, Suxiang Tong |
| EPI_ISL_903592 | PA Department of Health, Bureau of Laboratories | Genomics and Discovery, Respiratory Viruses Branch, Division of Viral Diseases, Centers for Disease Control and Prevention | Krista Queen, Yan Li, Ying Tao, Jing Zhang, Anna Uehara, Anna Montmayeur, Clinton R. Paden, Peter W. Cook, Rachel Marine, Mili Sheth, Jasmine Padilla, Sarah Nobles, Mark Burroughs, Lori Rowe, Haibin Wang, Ben L. Rambo-Martin, Dhwani Batra, Justin Lee, Suxiang Tong |
| EPI_ISL_903594, EPI_ISL_903595 | NJ Public Health and Environmental Laboratories | Genomics and Discovery, Respiratory Viruses Branch, Division of Viral Diseases, Centers for Disease Control and Prevention | Krista Queen, Yan Li, Ying Tao, Jing Zhang, Anna Uehara, Anna Montmayeur, Clinton R. Paden, Peter W. Cook, Rachel Marine, Mili Sheth, Jasmine Padilla, Sarah Nobles, Mark Burroughs, Lori Rowe, Haibin Wang, Ben L. Rambo-Martin, Dhwani Batra, Justin Lee, Suxiang Tong |
| EPI_ISL_903596 | OK Public Health Laboratory, Oklahoma State DOH | Genomics and Discovery, Respiratory Viruses Branch, Division of Viral Diseases, Centers for Disease Control and Prevention | Krista Queen, Yan Li, Ying Tao, Jing Zhang, Anna Uehara, Anna Montmayeur, Clinton R. Paden, Peter W. Cook, Rachel Marine, Mili Sheth, Jasmine Padilla, Sarah Nobles, Mark Burroughs, Lori Rowe, Haibin Wang, Ben L. Rambo-Martin, Dhwani Batra, Justin Lee, Suxiang Tong |
| EPI_ISL_903598 | UT-Unified State Labs: Public Health Utah DOH | Genomics and Discovery, Respiratory Viruses Branch, Division of Viral Diseases, Centers for Disease Control and Prevention | Krista Queen, Yan Li, Ying Tao, Jing Zhang, Anna Uehara, Anna Montmayeur, Clinton R. Paden, Peter W. Cook, Rachel Marine, Mili Sheth, Jasmine Padilla, Sarah Nobles, Mark Burroughs, Lori Rowe, Haibin Wang, Ben L. Rambo-Martin, Dhwani Batra, Justin Lee, Suxiang Tong |
| EPI_ISL_903599 | WY Public Health Laboratory | Genomics and Discovery, Respiratory Viruses Branch, Division of Viral Diseases, Centers for Disease Control and Prevention | Krista Queen, Yan Li, Ying Tao, Jing Zhang, Anna Uehara, Anna Montmayeur, Clinton R. Paden, Peter W. Cook, Rachel Marine, Mili Sheth, Jasmine Padilla, Sarah Nobles, Mark Burroughs, Lori Rowe, Haibin Wang, Ben L. Rambo-Martin, Dhwani Batra, Justin Lee, Suxiang Tong |
| EPI_ISL_903601, EPI_ISL_903602 | OR State PHL-Virology/Immunology Section | Genomics and Discovery, Respiratory Viruses Branch, Division of Viral Diseases, Centers for Disease Control and Prevention | Krista Queen, Yan Li, Ying Tao, Jing Zhang, Anna Uehara, Anna Montmayeur, Clinton R. Paden, Peter W. Cook, Rachel Marine, Mili Sheth, Jasmine Padilla, Sarah Nobles, Mark Burroughs, Lori Rowe, Haibin Wang, Ben L. Rambo-Martin, Dhwani Batra, Justin Lee, Suxiang Tong |
| EPI_ISL_903603 | ID Bureau of Laboratories | Genomics and Discovery, Respiratory Viruses Branch, Division of Viral Diseases, Centers for Disease Control and Prevention | Krista Queen, Yan Li, Ying Tao, Jing Zhang, Anna Uehara, Anna Montmayeur, Clinton R. Paden, Peter W. Cook, Rachel Marine, Mili Sheth, Jasmine Padilla, Sarah Nobles, Mark Burroughs, Lori Rowe, Haibin Wang, Ben L. Rambo-Martin, Dhwani Batra, Justin Lee, Suxiang Tong |
| EPI_ISL_903604 | VA-Division of Consolidated Laboratory Services | Genomics and Discovery, Respiratory Viruses Branch, Division of Viral Diseases, Centers for Disease Control and Prevention | Krista Queen, Yan Li, Ying Tao, Jing Zhang, Anna Uehara, Anna Montmayeur, Clinton R. Paden, Peter W. Cook, Rachel Marine, Mili Sheth, Jasmine Padilla, Sarah Nobles, Mark Burroughs, Lori Rowe, Haibin Wang, Ben L. Rambo-Martin, Dhwani Batra, Justin Lee, Suxiang Tong |
| EPI_ISL_903607 | IL Dept. of Public Health Springfield Laboratory | Genomics and Discovery, Respiratory Viruses Branch, Division of Viral Diseases, Centers for Disease Control and Prevention | Krista Queen, Yan Li, Ying Tao, Jing Zhang, Anna Uehara, Anna Montmayeur, Clinton R. Paden, Peter W. Cook, Rachel Marine, Mili Sheth, Jasmine Padilla, Sarah Nobles, Mark Burroughs, Lori Rowe, Haibin Wang, Ben L. Rambo-Martin, Dhwani Batra, Justin Lee, Suxiang Tong |
| EPI_ISL_903608 | MN PHL Division, Minnesota Department of Health | Genomics and Discovery, Respiratory Viruses Branch, Division of Viral Diseases, Centers for Disease Control and Prevention | Krista Queen, Yan Li, Ying Tao, Jing Zhang, Anna Uehara, Anna Montmayeur, Clinton R. Paden, Peter W. Cook, Rachel Marine, Mili Sheth, Jasmine Padilla, Sarah Nobles, Mark Burroughs, Lori Rowe, Haibin Wang, Ben L. Rambo-Martin, Dhwani Batra, Justin Lee, Suxiang Tong |
| EPI_ISL_903609 | NJ Public Health and Environmental Laboratories | Genomics and Discovery, Respiratory Viruses Branch, Division of Viral Diseases, Centers for Disease Control and Prevention | Krista Queen, Yan Li, Ying Tao, Jing Zhang, Anna Uehara, Anna Montmayeur, Clinton R. Paden, Peter W. Cook, Rachel Marine, Mili Sheth, Jasmine Padilla, Sarah Nobles, Mark Burroughs, Lori Rowe, Haibin Wang, Ben L. Rambo-Martin, Dhwani Batra, Justin Lee, Suxiang Tong |
| EPI_ISL_903610 | NV State Public Health Laboratory | Genomics and Discovery, Respiratory Viruses Branch, Division of Viral Diseases, Centers for Disease Control and Prevention | Krista Queen, Yan Li, Ying Tao, Jing Zhang, Anna Uehara, Anna Montmayeur, Clinton R. Paden, Peter W. Cook, Rachel Marine, Mili Sheth, Jasmine Padilla, Sarah Nobles, Mark Burroughs, Lori Rowe, Haibin Wang, Ben L. Rambo-Martin, Dhwani Batra, Justin Lee, Suxiang Tong |
| EPI_ISL_903611, EPI_ISL_903612 | OR State PHL-Virology/Immunology Section | Genomics and Discovery, Respiratory Viruses Branch, Division of Viral Diseases, Centers for Disease Control and Prevention | Krista Queen, Yan Li, Ying Tao, Jing Zhang, Anna Uehara, Anna Montmayeur, Clinton R. Paden, Peter W. Cook, Rachel Marine, Mili Sheth, Jasmine Padilla, Sarah Nobles, Mark Burroughs, Lori Rowe, Haibin Wang, Ben L. Rambo-Martin, Dhwani Batra, Justin Lee, Suxiang Tong |
| EPI_ISL_903613 | LA Office of Public Health Laboratories | Genomics and Discovery, Respiratory Viruses Branch, Division of Viral Diseases, Centers for Disease Control and Prevention | Krista Queen, Yan Li, Ying Tao, Jing Zhang, Anna Uehara, Anna Montmayeur, Clinton R. Paden, Peter W. Cook, Rachel Marine, Mili Sheth, Jasmine Padilla, Sarah Nobles, Mark Burroughs, Lori Rowe, Haibin Wang, Ben L. Rambo-Martin, Dhwani Batra, Justin Lee, Suxiang Tong |
| EPI_ISL_903616 | IN State Department of Health Laboratory Services | Genomics and Discovery, Respiratory Viruses Branch, Division of Viral Diseases, Centers for Disease Control and Prevention | Krista Queen, Yan Li, Ying Tao, Jing Zhang, Anna Uehara, Anna Montmayeur, Clinton R. Paden, Peter W. Cook, Rachel Marine, Mili Sheth, Jasmine Padilla, Sarah Nobles, Mark Burroughs, Lori Rowe, Haibin Wang, Ben L. Rambo-Martin, Dhwani Batra, Justin Lee, Suxiang Tong |
| EPI_ISL_903617 | UT-Unified State Labs: Public Health Utah DOH | Genomics and Discovery, Respiratory Viruses Branch, Division of Viral Diseases, Centers for Disease Control and Prevention | Krista Queen, Yan Li, Ying Tao, Jing Zhang, Anna Uehara, Anna Montmayeur, Clinton R. Paden, Peter W. Cook, Rachel Marine, Mili Sheth, Jasmine Padilla, Sarah Nobles, Mark Burroughs, Lori Rowe, Haibin Wang, Ben L. Rambo-Martin, Dhwani Batra, Justin Lee, Suxiang Tong |
| EPI_ISL_903618 | MN PHL Division, Minnesota Department of Health | Genomics and Discovery, Respiratory Viruses Branch, Division of Viral Diseases, Centers for Disease Control and Prevention | Krista Queen, Yan Li, Ying Tao, Jing Zhang, Anna Uehara, Anna Montmayeur, Clinton R. Paden, Peter W. Cook, Rachel Marine, Mili Sheth, Jasmine Padilla, Sarah Nobles, Mark Burroughs, Lori Rowe, Haibin Wang, Ben L. Rambo-Martin, Dhwani Batra, Justin Lee, Suxiang Tong |
| EPI_ISL_903619 | ID Bureau of Laboratories | Genomics and Discovery, Respiratory Viruses Branch, Division of Viral Diseases, Centers for Disease Control and Prevention | Krista Queen, Yan Li, Ying Tao, Jing Zhang, Anna Uehara, Anna Montmayeur, Clinton R. Paden, Peter W. Cook, Rachel Marine, Mili Sheth, Jasmine Padilla, Sarah Nobles, Mark Burroughs, Lori Rowe, Haibin Wang, Ben L. Rambo-Martin, Dhwani Batra, Justin Lee, Suxiang Tong |
| EPI_ISL_903621 | IA State Hygienic Laboratory | Genomics and Discovery, Respiratory Viruses Branch, Division of Viral Diseases, Centers for Disease Control and Prevention | Krista Queen, Yan Li, Ying Tao, Jing Zhang, Anna Uehara, Anna Montmayeur, Clinton R. Paden, Peter W. Cook, Rachel Marine, Mili Sheth, Jasmine Padilla, Sarah Nobles, Mark Burroughs, Lori Rowe, Haibin Wang, Ben L. Rambo-Martin, Dhwani Batra, Justin Lee, Suxiang Tong |
| EPI_ISL_903623 | ND Dept. of Health Laboratory Services-Microbiology | Genomics and Discovery, Respiratory Viruses Branch, Division of Viral Diseases, Centers for Disease Control and Prevention | Krista Queen, Yan Li, Ying Tao, Jing Zhang, Anna Uehara, Anna Montmayeur, Clinton R. Paden, Peter W. Cook, Rachel Marine, Mili Sheth, Jasmine Padilla, Sarah Nobles, Mark Burroughs, Lori Rowe, Haibin Wang, Ben L. Rambo-Martin, Dhwani Batra, Justin Lee, Suxiang Tong |
| EPI_ISL_903624 | ID Bureau of Laboratories | Genomics and Discovery, Respiratory Viruses Branch, Division of Viral Diseases, Centers for Disease Control and Prevention | Krista Queen, Yan Li, Ying Tao, Jing Zhang, Anna Uehara, Anna Montmayeur, Clinton R. Paden, Peter W. Cook, Rachel Marine, Mili Sheth, Jasmine Padilla, Sarah Nobles, Mark Burroughs, Lori Rowe, Haibin Wang, Ben L. Rambo-Martin, Dhwani Batra, Justin Lee, Suxiang Tong |
| EPI_ISL_903625 | PA Department of Health, Bureau of Laboratories | Genomics and Discovery, Respiratory Viruses Branch, Division of Viral Diseases, Centers for Disease Control and Prevention | Krista Queen, Yan Li, Ying Tao, Jing Zhang, Anna Uehara, Anna Montmayeur, Clinton R. Paden, Peter W. Cook, Rachel Marine, Mili Sheth, Jasmine Padilla, Sarah Nobles, Mark Burroughs, Lori Rowe, Haibin Wang, Ben L. Rambo-Martin, Dhwani Batra, Justin Lee, Suxiang Tong |
| EPI_ISL_903627 | LA Office of Public Health Laboratories | Genomics and Discovery, Respiratory Viruses Branch, Division of Viral Diseases, Centers for Disease Control and Prevention | Krista Queen, Yan Li, Ying Tao, Jing Zhang, Anna Uehara, Anna Montmayeur, Clinton R. Paden, Peter W. Cook, Rachel Marine, Mili Sheth, Jasmine Padilla, Sarah Nobles, Mark Burroughs, Lori Rowe, Haibin Wang, Ben L. Rambo-Martin, Dhwani Batra, Justin Lee, Suxiang Tong |
| EPI_ISL_903628 | MS Public Health Laboratory | Genomics and Discovery, Respiratory Viruses Branch, Division of Viral Diseases, Centers for Disease Control and Prevention | Krista Queen, Yan Li, Ying Tao, Jing Zhang, Anna Uehara, Anna Montmayeur, Clinton R. Paden, Peter W. Cook, Rachel Marine, Mili Sheth, Jasmine Padilla, Sarah Nobles, Mark Burroughs, Lori Rowe, Haibin Wang, Ben L. Rambo-Martin, Dhwani Batra, Justin Lee, Suxiang Tong |
| EPI_ISL_903629 | IA State Hygienic Laboratory | Genomics and Discovery, Respiratory Viruses Branch, Division of Viral Diseases, Centers for Disease Control and Prevention | Krista Queen, Yan Li, Ying Tao, Jing Zhang, Anna Uehara, Anna Montmayeur, Clinton R. Paden, Peter W. Cook, Rachel Marine, Mili Sheth, Jasmine Padilla, Sarah Nobles, Mark Burroughs, Lori Rowe, Haibin Wang, Ben L. Rambo-Martin, Dhwani Batra, Justin Lee, Suxiang Tong |
| EPI_ISL_903630 | MS Public Health Laboratory | Genomics and Discovery, Respiratory Viruses Branch, Division of Viral Diseases, Centers for Disease Control and Prevention | Krista Queen, Yan Li, Ying Tao, Jing Zhang, Anna Uehara, Anna Montmayeur, Clinton R. Paden, Peter W. Cook, Rachel Marine, Mili Sheth, Jasmine Padilla, Sarah Nobles, Mark Burroughs, Lori Rowe, Haibin Wang, Ben L. Rambo-Martin, Dhwani Batra, Justin Lee, Suxiang Tong |

| | | | |
|---|---|---|---|
| EPI_ISL_903631 | WA State Department of Health | Genomics and Discovery, Respiratory Viruses Branch, Division of Viral Diseases, Centers for Disease Control and Prevention | Krista Queen, Yan Li, Ying Tao, Jing Zhang, Anna Uehara, Anna Montmayeur, Clinton R. Paden, Peter W. Cook, Rachel Marine, Mili Sheth, Jasmine Padilla, Sarah Nobles, Mark Burroughs, Lori Rowe, Haibin Wang, Ben L. Rambo-Martin, Dhwani Batra, Justin Lee, Suxiang Tong |
| EPI_ISL_903632 | NC State Laboratory of Public Health | Genomics and Discovery, Respiratory Viruses Branch, Division of Viral Diseases, Centers for Disease Control and Prevention | Krista Queen, Yan Li, Ying Tao, Jing Zhang, Anna Uehara, Anna Montmayeur, Clinton R. Paden, Peter W. Cook, Rachel Marine, Mili Sheth, Jasmine Padilla, Sarah Nobles, Mark Burroughs, Lori Rowe, Haibin Wang, Ben L. Rambo-Martin, Dhwani Batra, Justin Lee, Suxiang Tong |
| EPI_ISL_903633 | WA State Department of Health | Genomics and Discovery, Respiratory Viruses Branch, Division of Viral Diseases, Centers for Disease Control and Prevention | Krista Queen, Yan Li, Ying Tao, Jing Zhang, Anna Uehara, Anna Montmayeur, Clinton R. Paden, Peter W. Cook, Rachel Marine, Mili Sheth, Jasmine Padilla, Sarah Nobles, Mark Burroughs, Lori Rowe, Haibin Wang, Ben L. Rambo-Martin, Dhwani Batra, Justin Lee, Suxiang Tong |
| EPI_ISL_903635, EPI_ISL_903636 | VT Dept. of Health Laboratory | Genomics and Discovery, Respiratory Viruses Branch, Division of Viral Diseases, Centers for Disease Control and Prevention | Krista Queen, Yan Li, Ying Tao, Jing Zhang, Anna Uehara, Anna Montmayeur, Clinton R. Paden, Peter W. Cook, Rachel Marine, Mili Sheth, Jasmine Padilla, Sarah Nobles, Mark Burroughs, Lori Rowe, Haibin Wang, Ben L. Rambo-Martin, Dhwani Batra, Justin Lee, Suxiang Tong |
| EPI_ISL_903638 | NJ Public Health and Environmental Laboratories | Genomics and Discovery, Respiratory Viruses Branch, Division of Viral Diseases, Centers for Disease Control and Prevention | Krista Queen, Yan Li, Ying Tao, Jing Zhang, Anna Uehara, Anna Montmayeur, Clinton R. Paden, Peter W. Cook, Rachel Marine, Mili Sheth, Jasmine Padilla, Sarah Nobles, Mark Burroughs, Lori Rowe, Haibin Wang, Ben L. Rambo-Martin, Dhwani Batra, Justin Lee, Suxiang Tong |
| EPI_ISL_903641, EPI_ISL_903643, EPI_ISL_903644, EPI_ISL_903646, EPI_ISL_903647 | RI State Health Laboratories | Genomics and Discovery, Respiratory Viruses Branch, Division of Viral Diseases, Centers for Disease Control and Prevention | Krista Queen, Yan Li, Ying Tao, Jing Zhang, Anna Uehara, Anna Montmayeur, Clinton R. Paden, Peter W. Cook, Rachel Marine, Mili Sheth, Jasmine Padilla, Sarah Nobles, Mark Burroughs, Lori Rowe, Haibin Wang, Ben L. Rambo-Martin, Dhwani Batra, Justin Lee, Suxiang Tong |
| EPI_ISL_903648 | CO Dept. of Public Health and Environment, Lab Services Division | Genomics and Discovery, Respiratory Viruses Branch, Division of Viral Diseases, Centers for Disease Control and Prevention | Krista Queen, Yan Li, Ying Tao, Jing Zhang, Anna Uehara, Anna Montmayeur, Clinton R. Paden, Peter W. Cook, Rachel Marine, Mili Sheth, Jasmine Padilla, Sarah Nobles, Mark Burroughs, Lori Rowe, Haibin Wang, Ben L. Rambo-Martin, Dhwani Batra, Justin Lee, Suxiang Tong |
| EPI_ISL_903650 | VT Dept. of Health Laboratory | Genomics and Discovery, Respiratory Viruses Branch, Division of Viral Diseases, Centers for Disease Control and Prevention | Krista Queen, Yan Li, Ying Tao, Jing Zhang, Anna Uehara, Anna Montmayeur, Clinton R. Paden, Peter W. Cook, Rachel Marine, Mili Sheth, Jasmine Padilla, Sarah Nobles, Mark Burroughs, Lori Rowe, Haibin Wang, Ben L. Rambo-Martin, Dhwani Batra, Justin Lee, Suxiang Tong |
| EPI_ISL_903651 | LA Office of Public Health Laboratories | Genomics and Discovery, Respiratory Viruses Branch, Division of Viral Diseases, Centers for Disease Control and Prevention | Krista Queen, Yan Li, Ying Tao, Jing Zhang, Anna Uehara, Anna Montmayeur, Clinton R. Paden, Peter W. Cook, Rachel Marine, Mili Sheth, Jasmine Padilla, Sarah Nobles, Mark Burroughs, Lori Rowe, Haibin Wang, Ben L. Rambo-Martin, Dhwani Batra, Justin Lee, Suxiang Tong |
| EPI_ISL_903652 | NC State Laboratory of Public Health | Genomics and Discovery, Respiratory Viruses Branch, Division of Viral Diseases, Centers for Disease Control and Prevention | Krista Queen, Yan Li, Ying Tao, Jing Zhang, Anna Uehara, Anna Montmayeur, Clinton R. Paden, Peter W. Cook, Rachel Marine, Mili Sheth, Jasmine Padilla, Sarah Nobles, Mark Burroughs, Lori Rowe, Haibin Wang, Ben L. Rambo-Martin, Dhwani Batra, Justin Lee, Suxiang Tong |
| EPI_ISL_903653 | WA State Department of Health | Genomics and Discovery, Respiratory Viruses Branch, Division of Viral Diseases, Centers for Disease Control and Prevention | Krista Queen, Yan Li, Ying Tao, Jing Zhang, Anna Uehara, Anna Montmayeur, Clinton R. Paden, Peter W. Cook, Rachel Marine, Mili Sheth, Jasmine Padilla, Sarah Nobles, Mark Burroughs, Lori Rowe, Haibin Wang, Ben L. Rambo-Martin, Dhwani Batra, Justin Lee, Suxiang Tong |
| EPI_ISL_903654 | NJ Public Health and Environmental Laboratories | Genomics and Discovery, Respiratory Viruses Branch, Division of Viral Diseases, Centers for Disease Control and Prevention | Krista Queen, Yan Li, Ying Tao, Jing Zhang, Anna Uehara, Anna Montmayeur, Clinton R. Paden, Peter W. Cook, Rachel Marine, Mili Sheth, Jasmine Padilla, Sarah Nobles, Mark Burroughs, Lori Rowe, Haibin Wang, Ben L. Rambo-Martin, Dhwani Batra, Justin Lee, Suxiang Tong |
| EPI_ISL_903656 | RI State Health Laboratories | Genomics and Discovery, Respiratory Viruses Branch, Division of Viral Diseases, Centers for Disease Control and Prevention | Krista Queen, Yan Li, Ying Tao, Jing Zhang, Anna Uehara, Anna Montmayeur, Clinton R. Paden, Peter W. Cook, Rachel Marine, Mili Sheth, Jasmine Padilla, Sarah Nobles, Mark Burroughs, Lori Rowe, Haibin Wang, Ben L. Rambo-Martin, Dhwani Batra, Justin Lee, Suxiang Tong |
| EPI_ISL_903657 | VT Dept. of Health Laboratory | Genomics and Discovery, Respiratory Viruses Branch, Division of Viral Diseases, Centers for Disease Control and Prevention | Krista Queen, Yan Li, Ying Tao, Jing Zhang, Anna Uehara, Anna Montmayeur, Clinton R. Paden, Peter W. Cook, Rachel Marine, Mili Sheth, Jasmine Padilla, Sarah Nobles, Mark Burroughs, Lori Rowe, Haibin Wang, Ben L. Rambo-Martin, Dhwani Batra, Justin Lee, Suxiang Tong |
| EPI_ISL_903659, EPI_ISL_903660 | MN PHL Division, Minnesota Department of Health | Genomics and Discovery, Respiratory Viruses Branch, Division of Viral Diseases, Centers for Disease Control and Prevention | Krista Queen, Yan Li, Ying Tao, Jing Zhang, Anna Uehara, Anna Montmayeur, Clinton R. Paden, Peter W. Cook, Rachel Marine, Mili Sheth, Jasmine Padilla, Sarah Nobles, Mark Burroughs, Lori Rowe, Haibin Wang, Ben L. Rambo-Martin, Dhwani Batra, Justin Lee, Suxiang Tong |
| EPI_ISL_903661, EPI_ISL_903662 | PA Department of Health, Bureau of Laboratories | Genomics and Discovery, Respiratory Viruses Branch, Division of Viral Diseases, Centers for Disease Control and Prevention | Krista Queen, Yan Li, Ying Tao, Jing Zhang, Anna Uehara, Anna Montmayeur, Clinton R. Paden, Peter W. Cook, Rachel Marine, Mili Sheth, Jasmine Padilla, Sarah Nobles, Mark Burroughs, Lori Rowe, Haibin Wang, Ben L. Rambo-Martin, Dhwani Batra, Justin Lee, Suxiang Tong |
| EPI_ISL_903663 | IL Dept. of Public Health Springfield Laboratory | Genomics and Discovery, Respiratory Viruses Branch, Division of Viral Diseases, Centers for Disease Control and Prevention | Krista Queen, Yan Li, Ying Tao, Jing Zhang, Anna Uehara, Anna Montmayeur, Clinton R. Paden, Peter W. Cook, Rachel Marine, Mili Sheth, Jasmine Padilla, Sarah Nobles, Mark Burroughs, Lori Rowe, Haibin Wang, Ben L. Rambo-Martin, Dhwani Batra, Justin Lee, Suxiang Tong |
| EPI_ISL_903664 | NV State Public Health Laboratory | Genomics and Discovery, Respiratory Viruses Branch, Division of Viral Diseases, Centers for Disease Control and Prevention | Krista Queen, Yan Li, Ying Tao, Jing Zhang, Anna Uehara, Anna Montmayeur, Clinton R. Paden, Peter W. Cook, Rachel Marine, Mili Sheth, Jasmine Padilla, Sarah Nobles, Mark Burroughs, Lori Rowe, Haibin Wang, Ben L. Rambo-Martin, Dhwani Batra, Justin Lee, Suxiang Tong |
| EPI_ISL_903666 | OR State PHL-Virology/Immunology Section | Genomics and Discovery, Respiratory Viruses Branch, Division of Viral Diseases, Centers for Disease Control and Prevention | Krista Queen, Yan Li, Ying Tao, Jing Zhang, Anna Uehara, Anna Montmayeur, Clinton R. Paden, Peter W. Cook, Rachel Marine, Mili Sheth, Jasmine Padilla, Sarah Nobles, Mark Burroughs, Lori Rowe, Haibin Wang, Ben L. Rambo-Martin, Dhwani Batra, Justin Lee, Suxiang Tong |
| EPI_ISL_903667 | NH Dept. of Health and Human Services Public Health Labs | Genomics and Discovery, Respiratory Viruses Branch, Division of Viral Diseases, Centers for Disease Control and Prevention | Krista Queen, Yan Li, Ying Tao, Jing Zhang, Anna Uehara, Anna Montmayeur, Clinton R. Paden, Peter W. Cook, Rachel Marine, Mili Sheth, Jasmine Padilla, Sarah Nobles, Mark Burroughs, Lori Rowe, Haibin Wang, Ben L. Rambo-Martin, Dhwani Batra, Justin Lee, Suxiang Tong |
| EPI_ISL_903668 | SC Dept of Health and Env. Control-Bureau of Laboratories | Genomics and Discovery, Respiratory Viruses Branch, Division of Viral Diseases, Centers for Disease Control and Prevention | Krista Queen, Yan Li, Ying Tao, Jing Zhang, Anna Uehara, Anna Montmayeur, Clinton R. Paden, Peter W. Cook, Rachel Marine, Mili Sheth, Jasmine Padilla, Sarah Nobles, Mark Burroughs, Lori Rowe, Haibin Wang, Ben L. Rambo-Martin, Dhwani Batra, Justin Lee, Suxiang Tong |
| EPI_ISL_903669 | IA State Hygienic Laboratory | Genomics and Discovery, Respiratory Viruses Branch, Division of Viral Diseases, Centers for Disease Control and Prevention | Krista Queen, Yan Li, Ying Tao, Jing Zhang, Anna Uehara, Anna Montmayeur, Clinton R. Paden, Peter W. Cook, Rachel Marine, Mili Sheth, Jasmine Padilla, Sarah Nobles, Mark Burroughs, Lori Rowe, Haibin Wang, Ben L. Rambo-Martin, Dhwani Batra, Justin Lee, Suxiang Tong |
| EPI_ISL_903671, EPI_ISL_903672 | SC Dept of Health and Env. Control-Bureau of Laboratories | Genomics and Discovery, Respiratory Viruses Branch, Division of Viral Diseases, Centers for Disease Control and Prevention | Krista Queen, Yan Li, Ying Tao, Jing Zhang, Anna Uehara, Anna Montmayeur, Clinton R. Paden, Peter W. Cook, Rachel Marine, Mili Sheth, Jasmine Padilla, Sarah Nobles, Mark Burroughs, Lori Rowe, Haibin Wang, Ben L. Rambo-Martin, Dhwani Batra, Justin Lee, Suxiang Tong |
| EPI_ISL_903675 | MI - Michigan Department of Health and Human Services - Bureau of Laboratories | Genomics and Discovery, Respiratory Viruses Branch, Division of Viral Diseases, Centers for Disease Control and Prevention | Krista Queen, Yan Li, Ying Tao, Jing Zhang, Anna Uehara, Anna Montmayeur, Clinton R. Paden, Peter W. Cook, Rachel Marine, Mili Sheth, Jasmine Padilla, Sarah Nobles, Mark Burroughs, Lori Rowe, Haibin Wang, Ben L. Rambo-Martin, Dhwani Batra, Justin Lee, Suxiang Tong |
| EPI_ISL_903677 | MN PHL Division, Minnesota Department of Health | Genomics and Discovery, Respiratory Viruses Branch, Division of Viral Diseases, Centers for Disease Control and Prevention | Krista Queen, Yan Li, Ying Tao, Jing Zhang, Anna Uehara, Anna Montmayeur, Clinton R. Paden, Peter W. Cook, Rachel Marine, Mili Sheth, Jasmine Padilla, Sarah Nobles, Mark Burroughs, Lori Rowe, Haibin Wang, Ben L. Rambo-Martin, Dhwani Batra, Justin Lee, Suxiang Tong |
| EPI_ISL_903680 | NJ Public Health and Environmental Laboratories | Genomics and Discovery, Respiratory Viruses Branch, Division of Viral Diseases, Centers for Disease Control and Prevention | Krista Queen, Yan Li, Ying Tao, Jing Zhang, Anna Uehara, Anna Montmayeur, Clinton R. Paden, Peter W. Cook, Rachel Marine, Mili Sheth, Jasmine Padilla, Sarah Nobles, Mark Burroughs, Lori Rowe, Haibin Wang, Ben L. Rambo-Martin, Dhwani Batra, Justin Lee, Suxiang Tong |
| EPI_ISL_903682 | IL Dept. of Public Health Springfield Laboratory | Genomics and Discovery, Respiratory Viruses Branch, Division of Viral Diseases, Centers for Disease Control and Prevention | Krista Queen, Yan Li, Ying Tao, Jing Zhang, Anna Uehara, Anna Montmayeur, Clinton R. Paden, Peter W. Cook, Rachel Marine, Mili Sheth, Jasmine Padilla, Sarah Nobles, Mark Burroughs, Lori Rowe, Haibin Wang, Ben L. Rambo-Martin, Dhwani Batra, Justin Lee, Suxiang Tong |
| EPI_ISL_903683 | NJ Public Health and Environmental Laboratories | Genomics and Discovery, Respiratory Viruses Branch, Division of Viral Diseases, Centers for Disease Control and Prevention | Krista Queen, Yan Li, Ying Tao, Jing Zhang, Anna Uehara, Anna Montmayeur, Clinton R. Paden, Peter W. Cook, Rachel Marine, Mili Sheth, Jasmine Padilla, Sarah Nobles, Mark Burroughs, Lori Rowe, Haibin Wang, Ben L. Rambo-Martin, Dhwani Batra, Justin Lee, Suxiang Tong |
| EPI_ISL_903684 | WI State Laboratory of Hygiene | Genomics and Discovery, Respiratory Viruses Branch, Division of Viral Diseases, Centers for Disease Control and Prevention | Krista Queen, Yan Li, Ying Tao, Jing Zhang, Anna Uehara, Anna Montmayeur, Clinton R. Paden, Peter W. Cook, Rachel Marine, Mili Sheth, Jasmine Padilla, Sarah Nobles, Mark Burroughs, Lori Rowe, Haibin Wang, Ben L. Rambo-Martin, Dhwani Batra, Justin Lee, Suxiang Tong |
| EPI_ISL_903686 | NJ Public Health and Environmental Laboratories | Genomics and Discovery, Respiratory Viruses Branch, Division of Viral Diseases, Centers for Disease Control and Prevention | Krista Queen, Yan Li, Ying Tao, Jing Zhang, Anna Uehara, Anna Montmayeur, Clinton R. Paden, Peter W. Cook, Rachel Marine, Mili Sheth, Jasmine Padilla, Sarah Nobles, Mark Burroughs, Lori Rowe, Haibin Wang, Ben L. Rambo-Martin, Dhwani Batra, Justin Lee, Suxiang Tong |
| EPI_ISL_903688 | SC Dept of Health and Env. Control-Bureau of Laboratories | Genomics and Discovery, Respiratory Viruses Branch, Division of Viral Diseases, Centers for Disease Control and Prevention | Krista Queen, Yan Li, Ying Tao, Jing Zhang, Anna Uehara, Anna Montmayeur, Clinton R. Paden, Peter W. Cook, Rachel Marine, Mili Sheth, Jasmine Padilla, Sarah Nobles, Mark Burroughs, Lori Rowe, Haibin Wang, Ben L. Rambo-Martin, Dhwani Batra, Justin Lee, Suxiang Tong |
| EPI_ISL_903689 | LA Office of Public Health Laboratories | Genomics and Discovery, Respiratory | Krista Queen, Yan Li, Ying Tao, Jing Zhang, Anna Uehara, Anna Montmayeur, Clinton R. Paden, Peter W. Cook, Rachel Marine, Mili Sheth, Jasmine Padilla, Sarah Nobles, Mark Burroughs, Lori Rowe, Haibin Wang, Ben L. Rambo-Martin, Dhwani Batra, Justin Lee, Suxiang Tong |

| | | | |
|---|---|---|---|
| | | Viruses Branch, Division of Viral Diseases, Centers for Disease Control and Prevention | |
| EPI_ISL_903690 | NV State Public Health Laboratory | Genomics and Discovery, Respiratory Viruses Branch, Division of Viral Diseases, Centers for Disease Control and Prevention | Krista Queen, Yan Li, Ying Tao, Jing Zhang, Anna Uehara, Anna Montmayeur, Clinton R. Paden, Peter W. Cook, Rachel Marine, Mili Sheth, Jasmine Padilla, Sarah Nobles, Mark Burroughs, Lori Rowe, Haibin Wang, Ben L. Rambo-Martin, Dhwani Batra, Justin Lee, Suxiang Tong |
| EPI_ISL_903691 | MN PHL Division, Minnesota Department of Health | Genomics and Discovery, Respiratory Viruses Branch, Division of Viral Diseases, Centers for Disease Control and Prevention | Krista Queen, Yan Li, Ying Tao, Jing Zhang, Anna Uehara, Anna Montmayeur, Clinton R. Paden, Peter W. Cook, Rachel Marine, Mili Sheth, Jasmine Padilla, Sarah Nobles, Mark Burroughs, Lori Rowe, Haibin Wang, Ben L. Rambo-Martin, Dhwani Batra, Justin Lee, Suxiang Tong |
| EPI_ISL_903692 | NJ Public Health and Environmental Laboratories | Genomics and Discovery, Respiratory Viruses Branch, Division of Viral Diseases, Centers for Disease Control and Prevention | Krista Queen, Yan Li, Ying Tao, Jing Zhang, Anna Uehara, Anna Montmayeur, Clinton R. Paden, Peter W. Cook, Rachel Marine, Mili Sheth, Jasmine Padilla, Sarah Nobles, Mark Burroughs, Lori Rowe, Haibin Wang, Ben L. Rambo-Martin, Dhwani Batra, Justin Lee, Suxiang Tong |
| EPI_ISL_903693 | LA Office of Public Health Laboratories | Genomics and Discovery, Respiratory Viruses Branch, Division of Viral Diseases, Centers for Disease Control and Prevention | Krista Queen, Yan Li, Ying Tao, Jing Zhang, Anna Uehara, Anna Montmayeur, Clinton R. Paden, Peter W. Cook, Rachel Marine, Mili Sheth, Jasmine Padilla, Sarah Nobles, Mark Burroughs, Lori Rowe, Haibin Wang, Ben L. Rambo-Martin, Dhwani Batra, Justin Lee, Suxiang Tong |
| EPI_ISL_903694, EPI_ISL_903696 | VT Dept. of Health Laboratory | Genomics and Discovery, Respiratory Viruses Branch, Division of Viral Diseases, Centers for Disease Control and Prevention | Krista Queen, Yan Li, Ying Tao, Jing Zhang, Anna Uehara, Anna Montmayeur, Clinton R. Paden, Peter W. Cook, Rachel Marine, Mili Sheth, Jasmine Padilla, Sarah Nobles, Mark Burroughs, Lori Rowe, Haibin Wang, Ben L. Rambo-Martin, Dhwani Batra, Justin Lee, Suxiang Tong |
| EPI_ISL_903697 | AR Dept. of Health-Public Health Lab | Genomics and Discovery, Respiratory Viruses Branch, Division of Viral Diseases, Centers for Disease Control and Prevention | Krista Queen, Yan Li, Ying Tao, Jing Zhang, Anna Uehara, Anna Montmayeur, Clinton R. Paden, Peter W. Cook, Rachel Marine, Mili Sheth, Jasmine Padilla, Sarah Nobles, Mark Burroughs, Lori Rowe, Haibin Wang, Ben L. Rambo-Martin, Dhwani Batra, Justin Lee, Suxiang Tong |
| EPI_ISL_903699 | NJ Public Health and Environmental Laboratories | Genomics and Discovery, Respiratory Viruses Branch, Division of Viral Diseases, Centers for Disease Control and Prevention | Krista Queen, Yan Li, Ying Tao, Jing Zhang, Anna Uehara, Anna Montmayeur, Clinton R. Paden, Peter W. Cook, Rachel Marine, Mili Sheth, Jasmine Padilla, Sarah Nobles, Mark Burroughs, Lori Rowe, Haibin Wang, Ben L. Rambo-Martin, Dhwani Batra, Justin Lee, Suxiang Tong |
| EPI_ISL_903700 | NV State Public Health Laboratory | Genomics and Discovery, Respiratory Viruses Branch, Division of Viral Diseases, Centers for Disease Control and Prevention | Krista Queen, Yan Li, Ying Tao, Jing Zhang, Anna Uehara, Anna Montmayeur, Clinton R. Paden, Peter W. Cook, Rachel Marine, Mili Sheth, Jasmine Padilla, Sarah Nobles, Mark Burroughs, Lori Rowe, Haibin Wang, Ben L. Rambo-Martin, Dhwani Batra, Justin Lee, Suxiang Tong |
| EPI_ISL_903701 | MN PHL Division, Minnesota Department of Health | Genomics and Discovery, Respiratory Viruses Branch, Division of Viral Diseases, Centers for Disease Control and Prevention | Krista Queen, Yan Li, Ying Tao, Jing Zhang, Anna Uehara, Anna Montmayeur, Clinton R. Paden, Peter W. Cook, Rachel Marine, Mili Sheth, Jasmine Padilla, Sarah Nobles, Mark Burroughs, Lori Rowe, Haibin Wang, Ben L. Rambo-Martin, Dhwani Batra, Justin Lee, Suxiang Tong |
| EPI_ISL_903702, EPI_ISL_903703 | NV State Public Health Laboratory | Genomics and Discovery, Respiratory Viruses Branch, Division of Viral Diseases, Centers for Disease Control and Prevention | Krista Queen, Yan Li, Ying Tao, Jing Zhang, Anna Uehara, Anna Montmayeur, Clinton R. Paden, Peter W. Cook, Rachel Marine, Mili Sheth, Jasmine Padilla, Sarah Nobles, Mark Burroughs, Lori Rowe, Haibin Wang, Ben L. Rambo-Martin, Dhwani Batra, Justin Lee, Suxiang Tong |
| EPI_ISL_903705 | MI - Michigan Department of Health and Human Services - Bureau of Laboratories | Genomics and Discovery, Respiratory Viruses Branch, Division of Viral Diseases, Centers for Disease Control and Prevention | Krista Queen, Yan Li, Ying Tao, Jing Zhang, Anna Uehara, Anna Montmayeur, Clinton R. Paden, Peter W. Cook, Rachel Marine, Mili Sheth, Jasmine Padilla, Sarah Nobles, Mark Burroughs, Lori Rowe, Haibin Wang, Ben L. Rambo-Martin, Dhwani Batra, Justin Lee, Suxiang Tong |
| EPI_ISL_903706 | MN PHL Division, Minnesota Department of Health | Genomics and Discovery, Respiratory Viruses Branch, Division of Viral Diseases, Centers for Disease Control and Prevention | Krista Queen, Yan Li, Ying Tao, Jing Zhang, Anna Uehara, Anna Montmayeur, Clinton R. Paden, Peter W. Cook, Rachel Marine, Mili Sheth, Jasmine Padilla, Sarah Nobles, Mark Burroughs, Lori Rowe, Haibin Wang, Ben L. Rambo-Martin, Dhwani Batra, Justin Lee, Suxiang Tong |
| EPI_ISL_903708 | MI - Michigan Department of Health and Human Services - Bureau of Laboratories | Genomics and Discovery, Respiratory Viruses Branch, Division of Viral Diseases, Centers for Disease Control and Prevention | Krista Queen, Yan Li, Ying Tao, Jing Zhang, Anna Uehara, Anna Montmayeur, Clinton R. Paden, Peter W. Cook, Rachel Marine, Mili Sheth, Jasmine Padilla, Sarah Nobles, Mark Burroughs, Lori Rowe, Haibin Wang, Ben L. Rambo-Martin, Dhwani Batra, Justin Lee, Suxiang Tong |
| EPI_ISL_903709 | LA Office of Public Health Laboratories | Genomics and Discovery, Respiratory Viruses Branch, Division of Viral Diseases, Centers for Disease Control and Prevention | Krista Queen, Yan Li, Ying Tao, Jing Zhang, Anna Uehara, Anna Montmayeur, Clinton R. Paden, Peter W. Cook, Rachel Marine, Mili Sheth, Jasmine Padilla, Sarah Nobles, Mark Burroughs, Lori Rowe, Haibin Wang, Ben L. Rambo-Martin, Dhwani Batra, Justin Lee, Suxiang Tong |
| EPI_ISL_903710 | MI - Michigan Department of Health and Human Services - Bureau of Laboratories | Genomics and Discovery, Respiratory Viruses Branch, Division of Viral Diseases, Centers for Disease Control and Prevention | Krista Queen, Yan Li, Ying Tao, Jing Zhang, Anna Uehara, Anna Montmayeur, Clinton R. Paden, Peter W. Cook, Rachel Marine, Mili Sheth, Jasmine Padilla, Sarah Nobles, Mark Burroughs, Lori Rowe, Haibin Wang, Ben L. Rambo-Martin, Dhwani Batra, Justin Lee, Suxiang Tong |
| EPI_ISL_903711 | NJ Public Health and Environmental Laboratories | Genomics and Discovery, Respiratory Viruses Branch, Division of Viral Diseases, Centers for Disease Control and Prevention | Krista Queen, Yan Li, Ying Tao, Jing Zhang, Anna Uehara, Anna Montmayeur, Clinton R. Paden, Peter W. Cook, Rachel Marine, Mili Sheth, Jasmine Padilla, Sarah Nobles, Mark Burroughs, Lori Rowe, Haibin Wang, Ben L. Rambo-Martin, Dhwani Batra, Justin Lee, Suxiang Tong |
| EPI_ISL_903713 | MN PHL Division, Minnesota Department of Health | Genomics and Discovery, Respiratory Viruses Branch, Division of Viral Diseases, Centers for Disease Control and Prevention | Krista Queen, Yan Li, Ying Tao, Jing Zhang, Anna Uehara, Anna Montmayeur, Clinton R. Paden, Peter W. Cook, Rachel Marine, Mili Sheth, Jasmine Padilla, Sarah Nobles, Mark Burroughs, Lori Rowe, Haibin Wang, Ben L. Rambo-Martin, Dhwani Batra, Justin Lee, Suxiang Tong |
| EPI_ISL_903714 | NJ Public Health and Environmental Laboratories | Genomics and Discovery, Respiratory Viruses Branch, Division of Viral Diseases, Centers for Disease Control and Prevention | Krista Queen, Yan Li, Ying Tao, Jing Zhang, Anna Uehara, Anna Montmayeur, Clinton R. Paden, Peter W. Cook, Rachel Marine, Mili Sheth, Jasmine Padilla, Sarah Nobles, Mark Burroughs, Lori Rowe, Haibin Wang, Ben L. Rambo-Martin, Dhwani Batra, Justin Lee, Suxiang Tong |
| EPI_ISL_903715 | VT Dept. of Health Laboratory | Genomics and Discovery, Respiratory Viruses Branch, Division of Viral Diseases, Centers for Disease Control and Prevention | Krista Queen, Yan Li, Ying Tao, Jing Zhang, Anna Uehara, Anna Montmayeur, Clinton R. Paden, Peter W. Cook, Rachel Marine, Mili Sheth, Jasmine Padilla, Sarah Nobles, Mark Burroughs, Lori Rowe, Haibin Wang, Ben L. Rambo-Martin, Dhwani Batra, Justin Lee, Suxiang Tong |
| EPI_ISL_903717 | SC Dept of Health and Env. Control-Bureau of Laboratories | Genomics and Discovery, Respiratory Viruses Branch, Division of Viral Diseases, Centers for Disease Control and Prevention | Krista Queen, Yan Li, Ying Tao, Jing Zhang, Anna Uehara, Anna Montmayeur, Clinton R. Paden, Peter W. Cook, Rachel Marine, Mili Sheth, Jasmine Padilla, Sarah Nobles, Mark Burroughs, Lori Rowe, Haibin Wang, Ben L. Rambo-Martin, Dhwani Batra, Justin Lee, Suxiang Tong |
| EPI_ISL_903718 | VT Dept. of Health Laboratory | Genomics and Discovery, Respiratory Viruses Branch, Division of Viral Diseases, Centers for Disease Control and Prevention | Krista Queen, Yan Li, Ying Tao, Jing Zhang, Anna Uehara, Anna Montmayeur, Clinton R. Paden, Peter W. Cook, Rachel Marine, Mili Sheth, Jasmine Padilla, Sarah Nobles, Mark Burroughs, Lori Rowe, Haibin Wang, Ben L. Rambo-Martin, Dhwani Batra, Justin Lee, Suxiang Tong |
| EPI_ISL_903719 | NV State Public Health Laboratory | Genomics and Discovery, Respiratory Viruses Branch, Division of Viral Diseases, Centers for Disease Control and Prevention | Krista Queen, Yan Li, Ying Tao, Jing Zhang, Anna Uehara, Anna Montmayeur, Clinton R. Paden, Peter W. Cook, Rachel Marine, Mili Sheth, Jasmine Padilla, Sarah Nobles, Mark Burroughs, Lori Rowe, Haibin Wang, Ben L. Rambo-Martin, Dhwani Batra, Justin Lee, Suxiang Tong |
| EPI_ISL_903720 | LA Office of Public Health Laboratories | Genomics and Discovery, Respiratory Viruses Branch, Division of Viral Diseases, Centers for Disease Control and Prevention | Krista Queen, Yan Li, Ying Tao, Jing Zhang, Anna Uehara, Anna Montmayeur, Clinton R. Paden, Peter W. Cook, Rachel Marine, Mili Sheth, Jasmine Padilla, Sarah Nobles, Mark Burroughs, Lori Rowe, Haibin Wang, Ben L. Rambo-Martin, Dhwani Batra, Justin Lee, Suxiang Tong |
| EPI_ISL_903721 | NV State Public Health Laboratory | Genomics and Discovery, Respiratory Viruses Branch, Division of Viral Diseases, Centers for Disease Control and Prevention | Krista Queen, Yan Li, Ying Tao, Jing Zhang, Anna Uehara, Anna Montmayeur, Clinton R. Paden, Peter W. Cook, Rachel Marine, Mili Sheth, Jasmine Padilla, Sarah Nobles, Mark Burroughs, Lori Rowe, Haibin Wang, Ben L. Rambo-Martin, Dhwani Batra, Justin Lee, Suxiang Tong |
| EPI_ISL_903724 | MI - Michigan Department of Health and Human Services - Bureau of Laboratories | Genomics and Discovery, Respiratory Viruses Branch, Division of Viral Diseases, Centers for Disease Control and Prevention | Krista Queen, Yan Li, Ying Tao, Jing Zhang, Anna Uehara, Anna Montmayeur, Clinton R. Paden, Peter W. Cook, Rachel Marine, Mili Sheth, Jasmine Padilla, Sarah Nobles, Mark Burroughs, Lori Rowe, Haibin Wang, Ben L. Rambo-Martin, Dhwani Batra, Justin Lee, Suxiang Tong |
| EPI_ISL_903725 | SD Public Health Laboratory | Genomics and Discovery, Respiratory Viruses Branch, Division of Viral Diseases, Centers for Disease Control and Prevention | Krista Queen, Yan Li, Ying Tao, Jing Zhang, Anna Uehara, Anna Montmayeur, Clinton R. Paden, Peter W. Cook, Rachel Marine, Mili Sheth, Jasmine Padilla, Sarah Nobles, Mark Burroughs, Lori Rowe, Haibin Wang, Ben L. Rambo-Martin, Dhwani Batra, Justin Lee, Suxiang Tong |
| EPI_ISL_903727 | NV State Public Health Laboratory | Genomics and Discovery, Respiratory Viruses Branch, Division of Viral Diseases, Centers for Disease Control and Prevention | Krista Queen, Yan Li, Ying Tao, Jing Zhang, Anna Uehara, Anna Montmayeur, Clinton R. Paden, Peter W. Cook, Rachel Marine, Mili Sheth, Jasmine Padilla, Sarah Nobles, Mark Burroughs, Lori Rowe, Haibin Wang, Ben L. Rambo-Martin, Dhwani Batra, Justin Lee, Suxiang Tong |
| EPI_ISL_903728 | LA Office of Public Health Laboratories | Genomics and Discovery, Respiratory Viruses Branch, Division of Viral Diseases, Centers for Disease Control and Prevention | Krista Queen, Yan Li, Ying Tao, Jing Zhang, Anna Uehara, Anna Montmayeur, Clinton R. Paden, Peter W. Cook, Rachel Marine, Mili Sheth, Jasmine Padilla, Sarah Nobles, Mark Burroughs, Lori Rowe, Haibin Wang, Ben L. Rambo-Martin, Dhwani Batra, Justin Lee, Suxiang Tong |
| EPI_ISL_903730 | VT Dept. of Health Laboratory | Genomics and Discovery, Respiratory Viruses Branch, Division of Viral Diseases, Centers for Disease Control and Prevention | Krista Queen, Yan Li, Ying Tao, Jing Zhang, Anna Uehara, Anna Montmayeur, Clinton R. Paden, Peter W. Cook, Rachel Marine, Mili Sheth, Jasmine Padilla, Sarah Nobles, Mark Burroughs, Lori Rowe, Haibin Wang, Ben L. Rambo-Martin, Dhwani Batra, Justin Lee, Suxiang Tong |
| EPI_ISL_903731, EPI_ISL_903732, EPI_ISL_903733 | NV State Public Health Laboratory | Genomics and Discovery, Respiratory Viruses Branch, Division of Viral Diseases, Centers for Disease Control and Prevention | Krista Queen, Yan Li, Ying Tao, Jing Zhang, Anna Uehara, Anna Montmayeur, Clinton R. Paden, Peter W. Cook, Rachel Marine, Mili Sheth, Jasmine Padilla, Sarah Nobles, Mark Burroughs, Lori Rowe, Haibin Wang, Ben L. Rambo-Martin, Dhwani Batra, Justin Lee, Suxiang Tong |
| EPI_ISL_903734 | NJ Public Health and Environmental Laboratories | Genomics and Discovery, Respiratory Viruses Branch, Division of Viral Diseases, Centers for Disease Control and Prevention | Krista Queen, Yan Li, Ying Tao, Jing Zhang, Anna Uehara, Anna Montmayeur, Clinton R. Paden, Peter W. Cook, Rachel Marine, Mili Sheth, Jasmine Padilla, Sarah Nobles, Mark Burroughs, Lori Rowe, Haibin Wang, Ben L. Rambo-Martin, Dhwani Batra, Justin Lee, Suxiang Tong |
| EPI_ISL_903735 | NV State Public Health Laboratory | Genomics and Discovery, Respiratory | Krista Queen, Yan Li, Ying Tao, Jing Zhang, Anna Uehara, Anna Montmayeur, Clinton R. Paden, Peter W. Cook, Rachel Marine, Mili Sheth, Jasmine Padilla, Sarah Nobles, Mark Burroughs, Lori Rowe, Haibin Wang, Ben L. Rambo-Martin, Dhwani Batra, Justin Lee, Suxiang Tong |

| | | | |
|---|---|---|---|
| | | Viruses Branch, Division of Viral Diseases, Centers for Disease Control and Prevention | |
| EPI_ISL_903736 | PA Department of Health, Bureau of Laboratories | Genomics and Discovery, Respiratory Viruses Branch, Division of Viral Diseases, Centers for Disease Control and Prevention | Krista Queen, Yan Li, Ying Tao, Jing Zhang, Anna Uehara, Anna Montmayeur, Clinton R. Paden, Peter W. Cook, Rachel Marine, Mili Sheth, Jasmine Padilla, Sarah Nobles, Mark Burroughs, Lori Rowe, Haibin Wang, Ben L. Rambo-Martin, Dhwani Batra, Justin Lee, Suxiang Tong |
| EPI_ISL_903737, EPI_ISL_903738 | MN PHL Division, Minnesota Department of Health | Genomics and Discovery, Respiratory Viruses Branch, Division of Viral Diseases, Centers for Disease Control and Prevention | Krista Queen, Yan Li, Ying Tao, Jing Zhang, Anna Uehara, Anna Montmayeur, Clinton R. Paden, Peter W. Cook, Rachel Marine, Mili Sheth, Jasmine Padilla, Sarah Nobles, Mark Burroughs, Lori Rowe, Haibin Wang, Ben L. Rambo-Martin, Dhwani Batra, Justin Lee, Suxiang Tong |
| EPI_ISL_903739 | LA Office of Public Health Laboratories | Genomics and Discovery, Respiratory Viruses Branch, Division of Viral Diseases, Centers for Disease Control and Prevention | Krista Queen, Yan Li, Ying Tao, Jing Zhang, Anna Uehara, Anna Montmayeur, Clinton R. Paden, Peter W. Cook, Rachel Marine, Mili Sheth, Jasmine Padilla, Sarah Nobles, Mark Burroughs, Lori Rowe, Haibin Wang, Ben L. Rambo-Martin, Dhwani Batra, Justin Lee, Suxiang Tong |
| EPI_ISL_903741 | VA-Division of Consolidated Laboratory Services | Genomics and Discovery, Respiratory Viruses Branch, Division of Viral Diseases, Centers for Disease Control and Prevention | Krista Queen, Yan Li, Ying Tao, Jing Zhang, Anna Uehara, Anna Montmayeur, Clinton R. Paden, Peter W. Cook, Rachel Marine, Mili Sheth, Jasmine Padilla, Sarah Nobles, Mark Burroughs, Lori Rowe, Haibin Wang, Ben L. Rambo-Martin, Dhwani Batra, Justin Lee, Suxiang Tong |
| EPI_ISL_903742 | IL Dept. of Public Health Springfield Laboratory | Genomics and Discovery, Respiratory Viruses Branch, Division of Viral Diseases, Centers for Disease Control and Prevention | Krista Queen, Yan Li, Ying Tao, Jing Zhang, Anna Uehara, Anna Montmayeur, Clinton R. Paden, Peter W. Cook, Rachel Marine, Mili Sheth, Jasmine Padilla, Sarah Nobles, Mark Burroughs, Lori Rowe, Haibin Wang, Ben L. Rambo-Martin, Dhwani Batra, Justin Lee, Suxiang Tong |
| EPI_ISL_903743 | DC Public Health Lab/ Dept. of Forensic Sciences | Genomics and Discovery, Respiratory Viruses Branch, Division of Viral Diseases, Centers for Disease Control and Prevention | Krista Queen, Yan Li, Ying Tao, Jing Zhang, Anna Uehara, Anna Montmayeur, Clinton R. Paden, Peter W. Cook, Rachel Marine, Mili Sheth, Jasmine Padilla, Sarah Nobles, Mark Burroughs, Lori Rowe, Haibin Wang, Ben L. Rambo-Martin, Dhwani Batra, Justin Lee, Suxiang Tong |
| EPI_ISL_903744 | SD Public Health Laboratory | Genomics and Discovery, Respiratory Viruses Branch, Division of Viral Diseases, Centers for Disease Control and Prevention | Krista Queen, Yan Li, Ying Tao, Jing Zhang, Anna Uehara, Anna Montmayeur, Clinton R. Paden, Peter W. Cook, Rachel Marine, Mili Sheth, Jasmine Padilla, Sarah Nobles, Mark Burroughs, Lori Rowe, Haibin Wang, Ben L. Rambo-Martin, Dhwani Batra, Justin Lee, Suxiang Tong |
| EPI_ISL_903745 | MS Public Health Laboratory | Genomics and Discovery, Respiratory Viruses Branch, Division of Viral Diseases, Centers for Disease Control and Prevention | Krista Queen, Yan Li, Ying Tao, Jing Zhang, Anna Uehara, Anna Montmayeur, Clinton R. Paden, Peter W. Cook, Rachel Marine, Mili Sheth, Jasmine Padilla, Sarah Nobles, Mark Burroughs, Lori Rowe, Haibin Wang, Ben L. Rambo-Martin, Dhwani Batra, Justin Lee, Suxiang Tong |
| EPI_ISL_903746 | MN PHL Division, Minnesota Department of Health | Genomics and Discovery, Respiratory Viruses Branch, Division of Viral Diseases, Centers for Disease Control and Prevention | Krista Queen, Yan Li, Ying Tao, Jing Zhang, Anna Uehara, Anna Montmayeur, Clinton R. Paden, Peter W. Cook, Rachel Marine, Mili Sheth, Jasmine Padilla, Sarah Nobles, Mark Burroughs, Lori Rowe, Haibin Wang, Ben L. Rambo-Martin, Dhwani Batra, Justin Lee, Suxiang Tong |
| EPI_ISL_903747 | NE Public Health Laboratory | Genomics and Discovery, Respiratory Viruses Branch, Division of Viral Diseases, Centers for Disease Control and Prevention | Krista Queen, Yan Li, Ying Tao, Jing Zhang, Anna Uehara, Anna Montmayeur, Clinton R. Paden, Peter W. Cook, Rachel Marine, Mili Sheth, Jasmine Padilla, Sarah Nobles, Mark Burroughs, Lori Rowe, Haibin Wang, Ben L. Rambo-Martin, Dhwani Batra, Justin Lee, Suxiang Tong |
| EPI_ISL_903748 | PA Department of Health, Bureau of Laboratories | Genomics and Discovery, Respiratory Viruses Branch, Division of Viral Diseases, Centers for Disease Control and Prevention | Krista Queen, Yan Li, Ying Tao, Jing Zhang, Anna Uehara, Anna Montmayeur, Clinton R. Paden, Peter W. Cook, Rachel Marine, Mili Sheth, Jasmine Padilla, Sarah Nobles, Mark Burroughs, Lori Rowe, Haibin Wang, Ben L. Rambo-Martin, Dhwani Batra, Justin Lee, Suxiang Tong |
| EPI_ISL_903749 | WY Public Health Laboratory | Genomics and Discovery, Respiratory Viruses Branch, Division of Viral Diseases, Centers for Disease Control and Prevention | Krista Queen, Yan Li, Ying Tao, Jing Zhang, Anna Uehara, Anna Montmayeur, Clinton R. Paden, Peter W. Cook, Rachel Marine, Mili Sheth, Jasmine Padilla, Sarah Nobles, Mark Burroughs, Lori Rowe, Haibin Wang, Ben L. Rambo-Martin, Dhwani Batra, Justin Lee, Suxiang Tong |
| EPI_ISL_903751 | PA Department of Health, Bureau of Laboratories | Genomics and Discovery, Respiratory Viruses Branch, Division of Viral Diseases, Centers for Disease Control and Prevention | Krista Queen, Yan Li, Ying Tao, Jing Zhang, Anna Uehara, Anna Montmayeur, Clinton R. Paden, Peter W. Cook, Rachel Marine, Mili Sheth, Jasmine Padilla, Sarah Nobles, Mark Burroughs, Lori Rowe, Haibin Wang, Ben L. Rambo-Martin, Dhwani Batra, Justin Lee, Suxiang Tong |
| EPI_ISL_903752 | LA Office of Public Health Laboratories | Genomics and Discovery, Respiratory Viruses Branch, Division of Viral Diseases, Centers for Disease Control and Prevention | Krista Queen, Yan Li, Ying Tao, Jing Zhang, Anna Uehara, Anna Montmayeur, Clinton R. Paden, Peter W. Cook, Rachel Marine, Mili Sheth, Jasmine Padilla, Sarah Nobles, Mark Burroughs, Lori Rowe, Haibin Wang, Ben L. Rambo-Martin, Dhwani Batra, Justin Lee, Suxiang Tong |
| EPI_ISL_903753 | ID Bureau of Laboratories | Genomics and Discovery, Respiratory Viruses Branch, Division of Viral Diseases, Centers for Disease Control and Prevention | Krista Queen, Yan Li, Ying Tao, Jing Zhang, Anna Uehara, Anna Montmayeur, Clinton R. Paden, Peter W. Cook, Rachel Marine, Mili Sheth, Jasmine Padilla, Sarah Nobles, Mark Burroughs, Lori Rowe, Haibin Wang, Ben L. Rambo-Martin, Dhwani Batra, Justin Lee, Suxiang Tong |
| EPI_ISL_903754 | IN State Department of Health Laboratory Services | Genomics and Discovery, Respiratory Viruses Branch, Division of Viral Diseases, Centers for Disease Control and Prevention | Krista Queen, Yan Li, Ying Tao, Jing Zhang, Anna Uehara, Anna Montmayeur, Clinton R. Paden, Peter W. Cook, Rachel Marine, Mili Sheth, Jasmine Padilla, Sarah Nobles, Mark Burroughs, Lori Rowe, Haibin Wang, Ben L. Rambo-Martin, Dhwani Batra, Justin Lee, Suxiang Tong |
| EPI_ISL_903757 | TN Division of Laboratory Services | Genomics and Discovery, Respiratory Viruses Branch, Division of Viral Diseases, Centers for Disease Control and Prevention | Krista Queen, Yan Li, Ying Tao, Jing Zhang, Anna Uehara, Anna Montmayeur, Clinton R. Paden, Peter W. Cook, Rachel Marine, Mili Sheth, Jasmine Padilla, Sarah Nobles, Mark Burroughs, Lori Rowe, Haibin Wang, Ben L. Rambo-Martin, Dhwani Batra, Justin Lee, Suxiang Tong |
| EPI_ISL_903760 | MN PHL Division, Minnesota Department of Health | Genomics and Discovery, Respiratory Viruses Branch, Division of Viral Diseases, Centers for Disease Control and Prevention | Krista Queen, Yan Li, Ying Tao, Jing Zhang, Anna Uehara, Anna Montmayeur, Clinton R. Paden, Peter W. Cook, Rachel Marine, Mili Sheth, Jasmine Padilla, Sarah Nobles, Mark Burroughs, Lori Rowe, Haibin Wang, Ben L. Rambo-Martin, Dhwani Batra, Justin Lee, Suxiang Tong |
| EPI_ISL_903763 | WY Public Health Laboratory | Genomics and Discovery, Respiratory Viruses Branch, Division of Viral Diseases, Centers for Disease Control and Prevention | Krista Queen, Yan Li, Ying Tao, Jing Zhang, Anna Uehara, Anna Montmayeur, Clinton R. Paden, Peter W. Cook, Rachel Marine, Mili Sheth, Jasmine Padilla, Sarah Nobles, Mark Burroughs, Lori Rowe, Haibin Wang, Ben L. Rambo-Martin, Dhwani Batra, Justin Lee, Suxiang Tong |
| EPI_ISL_903765 | LA Office of Public Health Laboratories | Genomics and Discovery, Respiratory Viruses Branch, Division of Viral Diseases, Centers for Disease Control and Prevention | Krista Queen, Yan Li, Ying Tao, Jing Zhang, Anna Uehara, Anna Montmayeur, Clinton R. Paden, Peter W. Cook, Rachel Marine, Mili Sheth, Jasmine Padilla, Sarah Nobles, Mark Burroughs, Lori Rowe, Haibin Wang, Ben L. Rambo-Martin, Dhwani Batra, Justin Lee, Suxiang Tong |
| EPI_ISL_903766 | UT-Unified State Labs: Public Health Utah DOH | Genomics and Discovery, Respiratory Viruses Branch, Division of Viral Diseases, Centers for Disease Control and Prevention | Krista Queen, Yan Li, Ying Tao, Jing Zhang, Anna Uehara, Anna Montmayeur, Clinton R. Paden, Peter W. Cook, Rachel Marine, Mili Sheth, Jasmine Padilla, Sarah Nobles, Mark Burroughs, Lori Rowe, Haibin Wang, Ben L. Rambo-Martin, Dhwani Batra, Justin Lee, Suxiang Tong |
| EPI_ISL_903767, EPI_ISL_903768 | TX DSHS, Lab Services Section MC 1947 | Genomics and Discovery, Respiratory Viruses Branch, Division of Viral Diseases, Centers for Disease Control and Prevention | Krista Queen, Yan Li, Ying Tao, Jing Zhang, Anna Uehara, Anna Montmayeur, Clinton R. Paden, Peter W. Cook, Rachel Marine, Mili Sheth, Jasmine Padilla, Sarah Nobles, Mark Burroughs, Lori Rowe, Haibin Wang, Ben L. Rambo-Martin, Dhwani Batra, Justin Lee, Suxiang Tong |
| EPI_ISL_903771 | VA-Division of Consolidated Laboratory Services | Genomics and Discovery, Respiratory Viruses Branch, Division of Viral Diseases, Centers for Disease Control and Prevention | Krista Queen, Yan Li, Ying Tao, Jing Zhang, Anna Uehara, Anna Montmayeur, Clinton R. Paden, Peter W. Cook, Rachel Marine, Mili Sheth, Jasmine Padilla, Sarah Nobles, Mark Burroughs, Lori Rowe, Haibin Wang, Ben L. Rambo-Martin, Dhwani Batra, Justin Lee, Suxiang Tong |
| EPI_ISL_903772 | IA State Hygienic Laboratory | Genomics and Discovery, Respiratory Viruses Branch, Division of Viral Diseases, Centers for Disease Control and Prevention | Krista Queen, Yan Li, Ying Tao, Jing Zhang, Anna Uehara, Anna Montmayeur, Clinton R. Paden, Peter W. Cook, Rachel Marine, Mili Sheth, Jasmine Padilla, Sarah Nobles, Mark Burroughs, Lori Rowe, Haibin Wang, Ben L. Rambo-Martin, Dhwani Batra, Justin Lee, Suxiang Tong |
| EPI_ISL_903773 | IN State Department of Health Laboratory Services | Genomics and Discovery, Respiratory Viruses Branch, Division of Viral Diseases, Centers for Disease Control and Prevention | Krista Queen, Yan Li, Ying Tao, Jing Zhang, Anna Uehara, Anna Montmayeur, Clinton R. Paden, Peter W. Cook, Rachel Marine, Mili Sheth, Jasmine Padilla, Sarah Nobles, Mark Burroughs, Lori Rowe, Haibin Wang, Ben L. Rambo-Martin, Dhwani Batra, Justin Lee, Suxiang Tong |
| EPI_ISL_903774 | NE Public Health Laboratory | Genomics and Discovery, Respiratory Viruses Branch, Division of Viral Diseases, Centers for Disease Control and Prevention | Krista Queen, Yan Li, Ying Tao, Jing Zhang, Anna Uehara, Anna Montmayeur, Clinton R. Paden, Peter W. Cook, Rachel Marine, Mili Sheth, Jasmine Padilla, Sarah Nobles, Mark Burroughs, Lori Rowe, Haibin Wang, Ben L. Rambo-Martin, Dhwani Batra, Justin Lee, Suxiang Tong |
| EPI_ISL_903776 | LA Office of Public Health Laboratories | Genomics and Discovery, Respiratory Viruses Branch, Division of Viral Diseases, Centers for Disease Control and Prevention | Krista Queen, Yan Li, Ying Tao, Jing Zhang, Anna Uehara, Anna Montmayeur, Clinton R. Paden, Peter W. Cook, Rachel Marine, Mili Sheth, Jasmine Padilla, Sarah Nobles, Mark Burroughs, Lori Rowe, Haibin Wang, Ben L. Rambo-Martin, Dhwani Batra, Justin Lee, Suxiang Tong |
| EPI_ISL_903777 | NC State Laboratory of Public Health | Genomics and Discovery, Respiratory Viruses Branch, Division of Viral Diseases, Centers for Disease Control and Prevention | Krista Queen, Yan Li, Ying Tao, Jing Zhang, Anna Uehara, Anna Montmayeur, Clinton R. Paden, Peter W. Cook, Rachel Marine, Mili Sheth, Jasmine Padilla, Sarah Nobles, Mark Burroughs, Lori Rowe, Haibin Wang, Ben L. Rambo-Martin, Dhwani Batra, Justin Lee, Suxiang Tong |
| EPI_ISL_903778 | PA Department of Health, Bureau of Laboratories | Genomics and Discovery, Respiratory Viruses Branch, Division of Viral Diseases, Centers for Disease Control and Prevention | Krista Queen, Yan Li, Ying Tao, Jing Zhang, Anna Uehara, Anna Montmayeur, Clinton R. Paden, Peter W. Cook, Rachel Marine, Mili Sheth, Jasmine Padilla, Sarah Nobles, Mark Burroughs, Lori Rowe, Haibin Wang, Ben L. Rambo-Martin, Dhwani Batra, Justin Lee, Suxiang Tong |
| EPI_ISL_903779 | IL Dept. of Public Health Springfield Laboratory | Genomics and Discovery, Respiratory Viruses Branch, Division of Viral Diseases, Centers for Disease Control and Prevention | Krista Queen, Yan Li, Ying Tao, Jing Zhang, Anna Uehara, Anna Montmayeur, Clinton R. Paden, Peter W. Cook, Rachel Marine, Mili Sheth, Jasmine Padilla, Sarah Nobles, Mark Burroughs, Lori Rowe, Haibin Wang, Ben L. Rambo-Martin, Dhwani Batra, Justin Lee, Suxiang Tong |
| EPI_ISL_903781 | CO Dept. of Public Health and Environment, Lab Services Division | Genomics and Discovery, Respiratory Viruses Branch, Division of Viral Diseases, Centers for Disease Control and Prevention | Krista Queen, Yan Li, Ying Tao, Jing Zhang, Anna Uehara, Anna Montmayeur, Clinton R. Paden, Peter W. Cook, Rachel Marine, Mili Sheth, Jasmine Padilla, Sarah Nobles, Mark Burroughs, Lori Rowe, Haibin Wang, Ben L. Rambo-Martin, Dhwani Batra, Justin Lee, Suxiang Tong |

| | | | |
|---|---|---|---|
| EPI_ISL_903782 | PA Department of Health, Bureau of Laboratories | Genomics and Discovery, Respiratory Viruses Branch, Division of Viral Diseases, Centers for Disease Control and Prevention | Krista Queen, Yan Li, Ying Tao, Jing Zhang, Anna Uehara, Anna Montmayeur, Clinton R. Paden, Peter W. Cook, Rachel Marine, Mili Sheth, Jasmine Padilla, Sarah Nobles, Mark Burroughs, Lori Rowe, Haibin Wang, Ben L. Rambo-Martin, Dhwani Batra, Justin Lee, Suxiang Tong |
| EPI_ISL_903784 | WVDHHR - Office of Laboratory Services | Genomics and Discovery, Respiratory Viruses Branch, Division of Viral Diseases, Centers for Disease Control and Prevention | Krista Queen, Yan Li, Ying Tao, Jing Zhang, Anna Uehara, Anna Montmayeur, Clinton R. Paden, Peter W. Cook, Rachel Marine, Mili Sheth, Jasmine Padilla, Sarah Nobles, Mark Burroughs, Lori Rowe, Haibin Wang, Ben L. Rambo-Martin, Dhwani Batra, Justin Lee, Suxiang Tong |
| EPI_ISL_903785 | PA Department of Health, Bureau of Laboratories | Genomics and Discovery, Respiratory Viruses Branch, Division of Viral Diseases, Centers for Disease Control and Prevention | Krista Queen, Yan Li, Ying Tao, Jing Zhang, Anna Uehara, Anna Montmayeur, Clinton R. Paden, Peter W. Cook, Rachel Marine, Mili Sheth, Jasmine Padilla, Sarah Nobles, Mark Burroughs, Lori Rowe, Haibin Wang, Ben L. Rambo-Martin, Dhwani Batra, Justin Lee, Suxiang Tong |
| EPI_ISL_903786 | WVDHHR - Office of Laboratory Services | Genomics and Discovery, Respiratory Viruses Branch, Division of Viral Diseases, Centers for Disease Control and Prevention | Krista Queen, Yan Li, Ying Tao, Jing Zhang, Anna Uehara, Anna Montmayeur, Clinton R. Paden, Peter W. Cook, Rachel Marine, Mili Sheth, Jasmine Padilla, Sarah Nobles, Mark Burroughs, Lori Rowe, Haibin Wang, Ben L. Rambo-Martin, Dhwani Batra, Justin Lee, Suxiang Tong |
| EPI_ISL_903787 | MO State Public Health Laboratory | Genomics and Discovery, Respiratory Viruses Branch, Division of Viral Diseases, Centers for Disease Control and Prevention | Krista Queen, Yan Li, Ying Tao, Jing Zhang, Anna Uehara, Anna Montmayeur, Clinton R. Paden, Peter W. Cook, Rachel Marine, Mili Sheth, Jasmine Padilla, Sarah Nobles, Mark Burroughs, Lori Rowe, Haibin Wang, Ben L. Rambo-Martin, Dhwani Batra, Justin Lee, Suxiang Tong |
| EPI_ISL_903788 | TX DSHS, Lab Services Section MC 1947 | Genomics and Discovery, Respiratory Viruses Branch, Division of Viral Diseases, Centers for Disease Control and Prevention | Krista Queen, Yan Li, Ying Tao, Jing Zhang, Anna Uehara, Anna Montmayeur, Clinton R. Paden, Peter W. Cook, Rachel Marine, Mili Sheth, Jasmine Padilla, Sarah Nobles, Mark Burroughs, Lori Rowe, Haibin Wang, Ben L. Rambo-Martin, Dhwani Batra, Justin Lee, Suxiang Tong |
| EPI_ISL_903789 | IL Dept. of Public Health Springfield Laboratory | Genomics and Discovery, Respiratory Viruses Branch, Division of Viral Diseases, Centers for Disease Control and Prevention | Krista Queen, Yan Li, Ying Tao, Jing Zhang, Anna Uehara, Anna Montmayeur, Clinton R. Paden, Peter W. Cook, Rachel Marine, Mili Sheth, Jasmine Padilla, Sarah Nobles, Mark Burroughs, Lori Rowe, Haibin Wang, Ben L. Rambo-Martin, Dhwani Batra, Justin Lee, Suxiang Tong |
| EPI_ISL_903790 | PA Department of Health, Bureau of Laboratories | Genomics and Discovery, Respiratory Viruses Branch, Division of Viral Diseases, Centers for Disease Control and Prevention | Krista Queen, Yan Li, Ying Tao, Jing Zhang, Anna Uehara, Anna Montmayeur, Clinton R. Paden, Peter W. Cook, Rachel Marine, Mili Sheth, Jasmine Padilla, Sarah Nobles, Mark Burroughs, Lori Rowe, Haibin Wang, Ben L. Rambo-Martin, Dhwani Batra, Justin Lee, Suxiang Tong |
| EPI_ISL_903791 | NE Public Health Laboratory | Genomics and Discovery, Respiratory Viruses Branch, Division of Viral Diseases, Centers for Disease Control and Prevention | Krista Queen, Yan Li, Ying Tao, Jing Zhang, Anna Uehara, Anna Montmayeur, Clinton R. Paden, Peter W. Cook, Rachel Marine, Mili Sheth, Jasmine Padilla, Sarah Nobles, Mark Burroughs, Lori Rowe, Haibin Wang, Ben L. Rambo-Martin, Dhwani Batra, Justin Lee, Suxiang Tong |
| EPI_ISL_903792 | VA-Division of Consolidated Laboratory Services | Genomics and Discovery, Respiratory Viruses Branch, Division of Viral Diseases, Centers for Disease Control and Prevention | Krista Queen, Yan Li, Ying Tao, Jing Zhang, Anna Uehara, Anna Montmayeur, Clinton R. Paden, Peter W. Cook, Rachel Marine, Mili Sheth, Jasmine Padilla, Sarah Nobles, Mark Burroughs, Lori Rowe, Haibin Wang, Ben L. Rambo-Martin, Dhwani Batra, Justin Lee, Suxiang Tong |
| EPI_ISL_903793 | DC Public Health Lab/ Dept. of Forensic Sciences | Genomics and Discovery, Respiratory Viruses Branch, Division of Viral Diseases, Centers for Disease Control and Prevention | Krista Queen, Yan Li, Ying Tao, Jing Zhang, Anna Uehara, Anna Montmayeur, Clinton R. Paden, Peter W. Cook, Rachel Marine, Mili Sheth, Jasmine Padilla, Sarah Nobles, Mark Burroughs, Lori Rowe, Haibin Wang, Ben L. Rambo-Martin, Dhwani Batra, Justin Lee, Suxiang Tong |
| EPI_ISL_903795 | WVDHHR - Office of Laboratory Services | Genomics and Discovery, Respiratory Viruses Branch, Division of Viral Diseases, Centers for Disease Control and Prevention | Krista Queen, Yan Li, Ying Tao, Jing Zhang, Anna Uehara, Anna Montmayeur, Clinton R. Paden, Peter W. Cook, Rachel Marine, Mili Sheth, Jasmine Padilla, Sarah Nobles, Mark Burroughs, Lori Rowe, Haibin Wang, Ben L. Rambo-Martin, Dhwani Batra, Justin Lee, Suxiang Tong |
| EPI_ISL_903798 | CO Dept. of Public Health and Environment, Lab Services Division | Genomics and Discovery, Respiratory Viruses Branch, Division of Viral Diseases, Centers for Disease Control and Prevention | Krista Queen, Yan Li, Ying Tao, Jing Zhang, Anna Uehara, Anna Montmayeur, Clinton R. Paden, Peter W. Cook, Rachel Marine, Mili Sheth, Jasmine Padilla, Sarah Nobles, Mark Burroughs, Lori Rowe, Haibin Wang, Ben L. Rambo-Martin, Dhwani Batra, Justin Lee, Suxiang Tong |
| EPI_ISL_903801 | WI State Laboratory of Hygiene | Genomics and Discovery, Respiratory Viruses Branch, Division of Viral Diseases, Centers for Disease Control and Prevention | Krista Queen, Yan Li, Ying Tao, Jing Zhang, Anna Uehara, Anna Montmayeur, Clinton R. Paden, Peter W. Cook, Rachel Marine, Mili Sheth, Jasmine Padilla, Sarah Nobles, Mark Burroughs, Lori Rowe, Haibin Wang, Ben L. Rambo-Martin, Dhwani Batra, Justin Lee, Suxiang Tong |
| EPI_ISL_903802 | TN Division of Laboratory Services | Genomics and Discovery, Respiratory Viruses Branch, Division of Viral Diseases, Centers for Disease Control and Prevention | Krista Queen, Yan Li, Ying Tao, Jing Zhang, Anna Uehara, Anna Montmayeur, Clinton R. Paden, Peter W. Cook, Rachel Marine, Mili Sheth, Jasmine Padilla, Sarah Nobles, Mark Burroughs, Lori Rowe, Haibin Wang, Ben L. Rambo-Martin, Dhwani Batra, Justin Lee, Suxiang Tong |
| EPI_ISL_903803 | WY Public Health Laboratory | Genomics and Discovery, Respiratory Viruses Branch, Division of Viral Diseases, Centers for Disease Control and Prevention | Krista Queen, Yan Li, Ying Tao, Jing Zhang, Anna Uehara, Anna Montmayeur, Clinton R. Paden, Peter W. Cook, Rachel Marine, Mili Sheth, Jasmine Padilla, Sarah Nobles, Mark Burroughs, Lori Rowe, Haibin Wang, Ben L. Rambo-Martin, Dhwani Batra, Justin Lee, Suxiang Tong |
| EPI_ISL_903805 | TN Division of Laboratory Services | Genomics and Discovery, Respiratory Viruses Branch, Division of Viral Diseases, Centers for Disease Control and Prevention | Krista Queen, Yan Li, Ying Tao, Jing Zhang, Anna Uehara, Anna Montmayeur, Clinton R. Paden, Peter W. Cook, Rachel Marine, Mili Sheth, Jasmine Padilla, Sarah Nobles, Mark Burroughs, Lori Rowe, Haibin Wang, Ben L. Rambo-Martin, Dhwani Batra, Justin Lee, Suxiang Tong |
| EPI_ISL_903806 | WY Public Health Laboratory | Genomics and Discovery, Respiratory Viruses Branch, Division of Viral Diseases, Centers for Disease Control and Prevention | Krista Queen, Yan Li, Ying Tao, Jing Zhang, Anna Uehara, Anna Montmayeur, Clinton R. Paden, Peter W. Cook, Rachel Marine, Mili Sheth, Jasmine Padilla, Sarah Nobles, Mark Burroughs, Lori Rowe, Haibin Wang, Ben L. Rambo-Martin, Dhwani Batra, Justin Lee, Suxiang Tong |
| EPI_ISL_903807 | IL Dept. of Public Health Springfield Laboratory | Genomics and Discovery, Respiratory Viruses Branch, Division of Viral Diseases, Centers for Disease Control and Prevention | Krista Queen, Yan Li, Ying Tao, Jing Zhang, Anna Uehara, Anna Montmayeur, Clinton R. Paden, Peter W. Cook, Rachel Marine, Mili Sheth, Jasmine Padilla, Sarah Nobles, Mark Burroughs, Lori Rowe, Haibin Wang, Ben L. Rambo-Martin, Dhwani Batra, Justin Lee, Suxiang Tong |
| EPI_ISL_903809 | WY Public Health Laboratory | Genomics and Discovery, Respiratory Viruses Branch, Division of Viral Diseases, Centers for Disease Control and Prevention | Krista Queen, Yan Li, Ying Tao, Jing Zhang, Anna Uehara, Anna Montmayeur, Clinton R. Paden, Peter W. Cook, Rachel Marine, Mili Sheth, Jasmine Padilla, Sarah Nobles, Mark Burroughs, Lori Rowe, Haibin Wang, Ben L. Rambo-Martin, Dhwani Batra, Justin Lee, Suxiang Tong |
| EPI_ISL_903812 | IL Dept. of Public Health Springfield Laboratory | Genomics and Discovery, Respiratory Viruses Branch, Division of Viral Diseases, Centers for Disease Control and Prevention | Krista Queen, Yan Li, Ying Tao, Jing Zhang, Anna Uehara, Anna Montmayeur, Clinton R. Paden, Peter W. Cook, Rachel Marine, Mili Sheth, Jasmine Padilla, Sarah Nobles, Mark Burroughs, Lori Rowe, Haibin Wang, Ben L. Rambo-Martin, Dhwani Batra, Justin Lee, Suxiang Tong |
| EPI_ISL_903813 | WVDHHR - Office of Laboratory Services | Genomics and Discovery, Respiratory Viruses Branch, Division of Viral Diseases, Centers for Disease Control and Prevention | Krista Queen, Yan Li, Ying Tao, Jing Zhang, Anna Uehara, Anna Montmayeur, Clinton R. Paden, Peter W. Cook, Rachel Marine, Mili Sheth, Jasmine Padilla, Sarah Nobles, Mark Burroughs, Lori Rowe, Haibin Wang, Ben L. Rambo-Martin, Dhwani Batra, Justin Lee, Suxiang Tong |
| EPI_ISL_903814 | ND Dept. of Health Laboratory Services-Microbiology | Genomics and Discovery, Respiratory Viruses Branch, Division of Viral Diseases, Centers for Disease Control and Prevention | Krista Queen, Yan Li, Ying Tao, Jing Zhang, Anna Uehara, Anna Montmayeur, Clinton R. Paden, Peter W. Cook, Rachel Marine, Mili Sheth, Jasmine Padilla, Sarah Nobles, Mark Burroughs, Lori Rowe, Haibin Wang, Ben L. Rambo-Martin, Dhwani Batra, Justin Lee, Suxiang Tong |
| EPI_ISL_903815 | IA State Hygienic Laboratory | Genomics and Discovery, Respiratory Viruses Branch, Division of Viral Diseases, Centers for Disease Control and Prevention | Krista Queen, Yan Li, Ying Tao, Jing Zhang, Anna Uehara, Anna Montmayeur, Clinton R. Paden, Peter W. Cook, Rachel Marine, Mili Sheth, Jasmine Padilla, Sarah Nobles, Mark Burroughs, Lori Rowe, Haibin Wang, Ben L. Rambo-Martin, Dhwani Batra, Justin Lee, Suxiang Tong |
| EPI_ISL_903817 | NE Public Health Laboratory | Genomics and Discovery, Respiratory Viruses Branch, Division of Viral Diseases, Centers for Disease Control and Prevention | Krista Queen, Yan Li, Ying Tao, Jing Zhang, Anna Uehara, Anna Montmayeur, Clinton R. Paden, Peter W. Cook, Rachel Marine, Mili Sheth, Jasmine Padilla, Sarah Nobles, Mark Burroughs, Lori Rowe, Haibin Wang, Ben L. Rambo-Martin, Dhwani Batra, Justin Lee, Suxiang Tong |
| EPI_ISL_903819 | SC Dept of Health and Env. Control-Bureau of Laboratories | Genomics and Discovery, Respiratory Viruses Branch, Division of Viral Diseases, Centers for Disease Control and Prevention | Krista Queen, Yan Li, Ying Tao, Jing Zhang, Anna Uehara, Anna Montmayeur, Clinton R. Paden, Peter W. Cook, Rachel Marine, Mili Sheth, Jasmine Padilla, Sarah Nobles, Mark Burroughs, Lori Rowe, Haibin Wang, Ben L. Rambo-Martin, Dhwani Batra, Justin Lee, Suxiang Tong |
| EPI_ISL_903820, EPI_ISL_903821 | PA Department of Health, Bureau of Laboratories | Genomics and Discovery, Respiratory Viruses Branch, Division of Viral Diseases, Centers for Disease Control and Prevention | Krista Queen, Yan Li, Ying Tao, Jing Zhang, Anna Uehara, Anna Montmayeur, Clinton R. Paden, Peter W. Cook, Rachel Marine, Mili Sheth, Jasmine Padilla, Sarah Nobles, Mark Burroughs, Lori Rowe, Haibin Wang, Ben L. Rambo-Martin, Dhwani Batra, Justin Lee, Suxiang Tong |
| EPI_ISL_903822 | WI State Laboratory of Hygiene | Genomics and Discovery, Respiratory Viruses Branch, Division of Viral Diseases, Centers for Disease Control and Prevention | Krista Queen, Yan Li, Ying Tao, Jing Zhang, Anna Uehara, Anna Montmayeur, Clinton R. Paden, Peter W. Cook, Rachel Marine, Mili Sheth, Jasmine Padilla, Sarah Nobles, Mark Burroughs, Lori Rowe, Haibin Wang, Ben L. Rambo-Martin, Dhwani Batra, Justin Lee, Suxiang Tong |
| EPI_ISL_903824 | VA-Division of Consolidated Laboratory Services | Genomics and Discovery, Respiratory Viruses Branch, Division of Viral Diseases, Centers for Disease Control and Prevention | Krista Queen, Yan Li, Ying Tao, Jing Zhang, Anna Uehara, Anna Montmayeur, Clinton R. Paden, Peter W. Cook, Rachel Marine, Mili Sheth, Jasmine Padilla, Sarah Nobles, Mark Burroughs, Lori Rowe, Haibin Wang, Ben L. Rambo-Martin, Dhwani Batra, Justin Lee, Suxiang Tong |
| EPI_ISL_903825 | PA Department of Health, Bureau of Laboratories | Genomics and Discovery, Respiratory Viruses Branch, Division of Viral Diseases, Centers for Disease Control and Prevention | Krista Queen, Yan Li, Ying Tao, Jing Zhang, Anna Uehara, Anna Montmayeur, Clinton R. Paden, Peter W. Cook, Rachel Marine, Mili Sheth, Jasmine Padilla, Sarah Nobles, Mark Burroughs, Lori Rowe, Haibin Wang, Ben L. Rambo-Martin, Dhwani Batra, Justin Lee, Suxiang Tong |
| EPI_ISL_903826 | VA-Division of Consolidated Laboratory Services | Genomics and Discovery, Respiratory Viruses Branch, Division of Viral Diseases, Centers for Disease Control and Prevention | Krista Queen, Yan Li, Ying Tao, Jing Zhang, Anna Uehara, Anna Montmayeur, Clinton R. Paden, Peter W. Cook, Rachel Marine, Mili Sheth, Jasmine Padilla, Sarah Nobles, Mark Burroughs, Lori Rowe, Haibin Wang, Ben L. Rambo-Martin, Dhwani Batra, Justin Lee, Suxiang Tong |
| EPI_ISL_903828 | IL Dept. of Public Health Springfield Laboratory | Genomics and Discovery, Respiratory Viruses Branch, Division of Viral Diseases, Centers for Disease Control and Prevention | Krista Queen, Yan Li, Ying Tao, Jing Zhang, Anna Uehara, Anna Montmayeur, Clinton R. Paden, Peter W. Cook, Rachel Marine, Mili Sheth, Jasmine Padilla, Sarah Nobles, Mark Burroughs, Lori Rowe, Haibin Wang, Ben L. Rambo-Martin, Dhwani Batra, Justin Lee, Suxiang Tong |

| | | | |
|---|---|---|---|
| EPI_ISL_903829 | MO State Public Health Laboratory | Genomics and Discovery, Respiratory Viruses Branch, Division of Viral Diseases, Centers for Disease Control and Prevention | Krista Queen, Yan Li, Ying Tao, Jing Zhang, Anna Uehara, Anna Montmayeur, Clinton R. Paden, Peter W. Cook, Rachel Marine, Mili Sheth, Jasmine Padilla, Sarah Nobles, Mark Burroughs, Lori Rowe, Haibin Wang, Ben L. Rambo-Martin, Dhwani Batra, Justin Lee, Suxiang Tong |
| EPI_ISL_903830 | VA-Division of Consolidated Laboratory Services | Genomics and Discovery, Respiratory Viruses Branch, Division of Viral Diseases, Centers for Disease Control and Prevention | Krista Queen, Yan Li, Ying Tao, Jing Zhang, Anna Uehara, Anna Montmayeur, Clinton R. Paden, Peter W. Cook, Rachel Marine, Mili Sheth, Jasmine Padilla, Sarah Nobles, Mark Burroughs, Lori Rowe, Haibin Wang, Ben L. Rambo-Martin, Dhwani Batra, Justin Lee, Suxiang Tong |
| EPI_ISL_903831 | MO State Public Health Laboratory | Genomics and Discovery, Respiratory Viruses Branch, Division of Viral Diseases, Centers for Disease Control and Prevention | Krista Queen, Yan Li, Ying Tao, Jing Zhang, Anna Uehara, Anna Montmayeur, Clinton R. Paden, Peter W. Cook, Rachel Marine, Mili Sheth, Jasmine Padilla, Sarah Nobles, Mark Burroughs, Lori Rowe, Haibin Wang, Ben L. Rambo-Martin, Dhwani Batra, Justin Lee, Suxiang Tong |
| EPI_ISL_903832 | LA Office of Public Health Laboratories | Genomics and Discovery, Respiratory Viruses Branch, Division of Viral Diseases, Centers for Disease Control and Prevention | Krista Queen, Yan Li, Ying Tao, Jing Zhang, Anna Uehara, Anna Montmayeur, Clinton R. Paden, Peter W. Cook, Rachel Marine, Mili Sheth, Jasmine Padilla, Sarah Nobles, Mark Burroughs, Lori Rowe, Haibin Wang, Ben L. Rambo-Martin, Dhwani Batra, Justin Lee, Suxiang Tong |
| EPI_ISL_903834 | PA Department of Health, Bureau of Laboratories | Genomics and Discovery, Respiratory Viruses Branch, Division of Viral Diseases, Centers for Disease Control and Prevention | Krista Queen, Yan Li, Ying Tao, Jing Zhang, Anna Uehara, Anna Montmayeur, Clinton R. Paden, Peter W. Cook, Rachel Marine, Mili Sheth, Jasmine Padilla, Sarah Nobles, Mark Burroughs, Lori Rowe, Haibin Wang, Ben L. Rambo-Martin, Dhwani Batra, Justin Lee, Suxiang Tong |
| EPI_ISL_903836 | SD Public Health Laboratory | Genomics and Discovery, Respiratory Viruses Branch, Division of Viral Diseases, Centers for Disease Control and Prevention | Krista Queen, Yan Li, Ying Tao, Jing Zhang, Anna Uehara, Anna Montmayeur, Clinton R. Paden, Peter W. Cook, Rachel Marine, Mili Sheth, Jasmine Padilla, Sarah Nobles, Mark Burroughs, Lori Rowe, Haibin Wang, Ben L. Rambo-Martin, Dhwani Batra, Justin Lee, Suxiang Tong |
| EPI_ISL_903837 | PA Department of Health, Bureau of Laboratories | Genomics and Discovery, Respiratory Viruses Branch, Division of Viral Diseases, Centers for Disease Control and Prevention | Krista Queen, Yan Li, Ying Tao, Jing Zhang, Anna Uehara, Anna Montmayeur, Clinton R. Paden, Peter W. Cook, Rachel Marine, Mili Sheth, Jasmine Padilla, Sarah Nobles, Mark Burroughs, Lori Rowe, Haibin Wang, Ben L. Rambo-Martin, Dhwani Batra, Justin Lee, Suxiang Tong |
| EPI_ISL_903840 | IA State Hygienic Laboratory | Genomics and Discovery, Respiratory Viruses Branch, Division of Viral Diseases, Centers for Disease Control and Prevention | Krista Queen, Yan Li, Ying Tao, Jing Zhang, Anna Uehara, Anna Montmayeur, Clinton R. Paden, Peter W. Cook, Rachel Marine, Mili Sheth, Jasmine Padilla, Sarah Nobles, Mark Burroughs, Lori Rowe, Haibin Wang, Ben L. Rambo-Martin, Dhwani Batra, Justin Lee, Suxiang Tong |
| EPI_ISL_903843 | OR State PHL-Virology/Immunology Section | Genomics and Discovery, Respiratory Viruses Branch, Division of Viral Diseases, Centers for Disease Control and Prevention | Krista Queen, Yan Li, Ying Tao, Jing Zhang, Anna Uehara, Anna Montmayeur, Clinton R. Paden, Peter W. Cook, Rachel Marine, Mili Sheth, Jasmine Padilla, Sarah Nobles, Mark Burroughs, Lori Rowe, Haibin Wang, Ben L. Rambo-Martin, Dhwani Batra, Justin Lee, Suxiang Tong |
| EPI_ISL_903845 | NE Public Health Laboratory | Genomics and Discovery, Respiratory Viruses Branch, Division of Viral Diseases, Centers for Disease Control and Prevention | Krista Queen, Yan Li, Ying Tao, Jing Zhang, Anna Uehara, Anna Montmayeur, Clinton R. Paden, Peter W. Cook, Rachel Marine, Mili Sheth, Jasmine Padilla, Sarah Nobles, Mark Burroughs, Lori Rowe, Haibin Wang, Ben L. Rambo-Martin, Dhwani Batra, Justin Lee, Suxiang Tong |
| EPI_ISL_903846 | WI State Laboratory of Hygiene | Genomics and Discovery, Respiratory Viruses Branch, Division of Viral Diseases, Centers for Disease Control and Prevention | Krista Queen, Yan Li, Ying Tao, Jing Zhang, Anna Uehara, Anna Montmayeur, Clinton R. Paden, Peter W. Cook, Rachel Marine, Mili Sheth, Jasmine Padilla, Sarah Nobles, Mark Burroughs, Lori Rowe, Haibin Wang, Ben L. Rambo-Martin, Dhwani Batra, Justin Lee, Suxiang Tong |
| EPI_ISL_903847 | IL Dept. of Public Health Springfield Laboratory | Genomics and Discovery, Respiratory Viruses Branch, Division of Viral Diseases, Centers for Disease Control and Prevention | Krista Queen, Yan Li, Ying Tao, Jing Zhang, Anna Uehara, Anna Montmayeur, Clinton R. Paden, Peter W. Cook, Rachel Marine, Mili Sheth, Jasmine Padilla, Sarah Nobles, Mark Burroughs, Lori Rowe, Haibin Wang, Ben L. Rambo-Martin, Dhwani Batra, Justin Lee, Suxiang Tong |
| EPI_ISL_903850 | MS Public Health Laboratory | Genomics and Discovery, Respiratory Viruses Branch, Division of Viral Diseases, Centers for Disease Control and Prevention | Krista Queen, Yan Li, Ying Tao, Jing Zhang, Anna Uehara, Anna Montmayeur, Clinton R. Paden, Peter W. Cook, Rachel Marine, Mili Sheth, Jasmine Padilla, Sarah Nobles, Mark Burroughs, Lori Rowe, Haibin Wang, Ben L. Rambo-Martin, Dhwani Batra, Justin Lee, Suxiang Tong |
| EPI_ISL_903851 | CT-Dr. Katherine A. Kelley State Public Health Lab | Genomics and Discovery, Respiratory Viruses Branch, Division of Viral Diseases, Centers for Disease Control and Prevention | Krista Queen, Yan Li, Ying Tao, Jing Zhang, Anna Uehara, Anna Montmayeur, Clinton R. Paden, Peter W. Cook, Rachel Marine, Mili Sheth, Jasmine Padilla, Sarah Nobles, Mark Burroughs, Lori Rowe, Haibin Wang, Ben L. Rambo-Martin, Dhwani Batra, Justin Lee, Suxiang Tong |
| EPI_ISL_903852 | PA Department of Health, Bureau of Laboratories | Genomics and Discovery, Respiratory Viruses Branch, Division of Viral Diseases, Centers for Disease Control and Prevention | Krista Queen, Yan Li, Ying Tao, Jing Zhang, Anna Uehara, Anna Montmayeur, Clinton R. Paden, Peter W. Cook, Rachel Marine, Mili Sheth, Jasmine Padilla, Sarah Nobles, Mark Burroughs, Lori Rowe, Haibin Wang, Ben L. Rambo-Martin, Dhwani Batra, Justin Lee, Suxiang Tong |
| EPI_ISL_903853 | MT Public Health Laboratory | Genomics and Discovery, Respiratory Viruses Branch, Division of Viral Diseases, Centers for Disease Control and Prevention | Krista Queen, Yan Li, Ying Tao, Jing Zhang, Anna Uehara, Anna Montmayeur, Clinton R. Paden, Peter W. Cook, Rachel Marine, Mili Sheth, Jasmine Padilla, Sarah Nobles, Mark Burroughs, Lori Rowe, Haibin Wang, Ben L. Rambo-Martin, Dhwani Batra, Justin Lee, Suxiang Tong |
| EPI_ISL_903854 | PA Department of Health, Bureau of Laboratories | Genomics and Discovery, Respiratory Viruses Branch, Division of Viral Diseases, Centers for Disease Control and Prevention | Krista Queen, Yan Li, Ying Tao, Jing Zhang, Anna Uehara, Anna Montmayeur, Clinton R. Paden, Peter W. Cook, Rachel Marine, Mili Sheth, Jasmine Padilla, Sarah Nobles, Mark Burroughs, Lori Rowe, Haibin Wang, Ben L. Rambo-Martin, Dhwani Batra, Justin Lee, Suxiang Tong |
| EPI_ISL_903857 | MN PHL Division, Minnesota Department of Health | Genomics and Discovery, Respiratory Viruses Branch, Division of Viral Diseases, Centers for Disease Control and Prevention | Krista Queen, Yan Li, Ying Tao, Jing Zhang, Anna Uehara, Anna Montmayeur, Clinton R. Paden, Peter W. Cook, Rachel Marine, Mili Sheth, Jasmine Padilla, Sarah Nobles, Mark Burroughs, Lori Rowe, Haibin Wang, Ben L. Rambo-Martin, Dhwani Batra, Justin Lee, Suxiang Tong |
| EPI_ISL_903858 | ND Dept. of Health Laboratory Services-Microbiology | Genomics and Discovery, Respiratory Viruses Branch, Division of Viral Diseases, Centers for Disease Control and Prevention | Krista Queen, Yan Li, Ying Tao, Jing Zhang, Anna Uehara, Anna Montmayeur, Clinton R. Paden, Peter W. Cook, Rachel Marine, Mili Sheth, Jasmine Padilla, Sarah Nobles, Mark Burroughs, Lori Rowe, Haibin Wang, Ben L. Rambo-Martin, Dhwani Batra, Justin Lee, Suxiang Tong |
| EPI_ISL_903859 | PA Department of Health, Bureau of Laboratories | Genomics and Discovery, Respiratory Viruses Branch, Division of Viral Diseases, Centers for Disease Control and Prevention | Krista Queen, Yan Li, Ying Tao, Jing Zhang, Anna Uehara, Anna Montmayeur, Clinton R. Paden, Peter W. Cook, Rachel Marine, Mili Sheth, Jasmine Padilla, Sarah Nobles, Mark Burroughs, Lori Rowe, Haibin Wang, Ben L. Rambo-Martin, Dhwani Batra, Justin Lee, Suxiang Tong |
| EPI_ISL_903863 | WY Public Health Laboratory | Genomics and Discovery, Respiratory Viruses Branch, Division of Viral Diseases, Centers for Disease Control and Prevention | Krista Queen, Yan Li, Ying Tao, Jing Zhang, Anna Uehara, Anna Montmayeur, Clinton R. Paden, Peter W. Cook, Rachel Marine, Mili Sheth, Jasmine Padilla, Sarah Nobles, Mark Burroughs, Lori Rowe, Haibin Wang, Ben L. Rambo-Martin, Dhwani Batra, Justin Lee, Suxiang Tong |
| EPI_ISL_903864 | DC Public Health Lab/ Dept. of Forensic Sciences | Genomics and Discovery, Respiratory Viruses Branch, Division of Viral Diseases, Centers for Disease Control and Prevention | Krista Queen, Yan Li, Ying Tao, Jing Zhang, Anna Uehara, Anna Montmayeur, Clinton R. Paden, Peter W. Cook, Rachel Marine, Mili Sheth, Jasmine Padilla, Sarah Nobles, Mark Burroughs, Lori Rowe, Haibin Wang, Ben L. Rambo-Martin, Dhwani Batra, Justin Lee, Suxiang Tong |
| EPI_ISL_903865 | WI State Laboratory of Hygiene | Genomics and Discovery, Respiratory Viruses Branch, Division of Viral Diseases, Centers for Disease Control and Prevention | Krista Queen, Yan Li, Ying Tao, Jing Zhang, Anna Uehara, Anna Montmayeur, Clinton R. Paden, Peter W. Cook, Rachel Marine, Mili Sheth, Jasmine Padilla, Sarah Nobles, Mark Burroughs, Lori Rowe, Haibin Wang, Ben L. Rambo-Martin, Dhwani Batra, Justin Lee, Suxiang Tong |
| EPI_ISL_903866 | MD DOH Laboratories Administration | Genomics and Discovery, Respiratory Viruses Branch, Division of Viral Diseases, Centers for Disease Control and Prevention | Krista Queen, Yan Li, Ying Tao, Jing Zhang, Anna Uehara, Anna Montmayeur, Clinton R. Paden, Peter W. Cook, Rachel Marine, Mili Sheth, Jasmine Padilla, Sarah Nobles, Mark Burroughs, Lori Rowe, Haibin Wang, Ben L. Rambo-Martin, Dhwani Batra, Justin Lee, Suxiang Tong |
| EPI_ISL_903867 | NC State Laboratory of Public Health | Genomics and Discovery, Respiratory Viruses Branch, Division of Viral Diseases, Centers for Disease Control and Prevention | Krista Queen, Yan Li, Ying Tao, Jing Zhang, Anna Uehara, Anna Montmayeur, Clinton R. Paden, Peter W. Cook, Rachel Marine, Mili Sheth, Jasmine Padilla, Sarah Nobles, Mark Burroughs, Lori Rowe, Haibin Wang, Ben L. Rambo-Martin, Dhwani Batra, Justin Lee, Suxiang Tong |
| EPI_ISL_903868 | MN PHL Division, Minnesota Department of Health | Genomics and Discovery, Respiratory Viruses Branch, Division of Viral Diseases, Centers for Disease Control and Prevention | Krista Queen, Yan Li, Ying Tao, Jing Zhang, Anna Uehara, Anna Montmayeur, Clinton R. Paden, Peter W. Cook, Rachel Marine, Mili Sheth, Jasmine Padilla, Sarah Nobles, Mark Burroughs, Lori Rowe, Haibin Wang, Ben L. Rambo-Martin, Dhwani Batra, Justin Lee, Suxiang Tong |
| EPI_ISL_903871 | NC State Laboratory of Public Health | Genomics and Discovery, Respiratory Viruses Branch, Division of Viral Diseases, Centers for Disease Control and Prevention | Krista Queen, Yan Li, Ying Tao, Jing Zhang, Anna Uehara, Anna Montmayeur, Clinton R. Paden, Peter W. Cook, Rachel Marine, Mili Sheth, Jasmine Padilla, Sarah Nobles, Mark Burroughs, Lori Rowe, Haibin Wang, Ben L. Rambo-Martin, Dhwani Batra, Justin Lee, Suxiang Tong |
| EPI_ISL_903872 | SC Dept of Health and Env. Control-Bureau of Laboratories | Genomics and Discovery, Respiratory Viruses Branch, Division of Viral Diseases, Centers for Disease Control and Prevention | Krista Queen, Yan Li, Ying Tao, Jing Zhang, Anna Uehara, Anna Montmayeur, Clinton R. Paden, Peter W. Cook, Rachel Marine, Mili Sheth, Jasmine Padilla, Sarah Nobles, Mark Burroughs, Lori Rowe, Haibin Wang, Ben L. Rambo-Martin, Dhwani Batra, Justin Lee, Suxiang Tong |
| EPI_ISL_903874 | MO State Public Health Laboratory | Genomics and Discovery, Respiratory Viruses Branch, Division of Viral Diseases, Centers for Disease Control and Prevention | Krista Queen, Yan Li, Ying Tao, Jing Zhang, Anna Uehara, Anna Montmayeur, Clinton R. Paden, Peter W. Cook, Rachel Marine, Mili Sheth, Jasmine Padilla, Sarah Nobles, Mark Burroughs, Lori Rowe, Haibin Wang, Ben L. Rambo-Martin, Dhwani Batra, Justin Lee, Suxiang Tong |
| EPI_ISL_903875 | IN State Department of Health Laboratory Services | Genomics and Discovery, Respiratory Viruses Branch, Division of Viral Diseases, Centers for Disease Control and Prevention | Krista Queen, Yan Li, Ying Tao, Jing Zhang, Anna Uehara, Anna Montmayeur, Clinton R. Paden, Peter W. Cook, Rachel Marine, Mili Sheth, Jasmine Padilla, Sarah Nobles, Mark Burroughs, Lori Rowe, Haibin Wang, Ben L. Rambo-Martin, Dhwani Batra, Justin Lee, Suxiang Tong |
| EPI_ISL_903876 | PA Department of Health, Bureau of Laboratories | Genomics and Discovery, Respiratory Viruses Branch, Division of Viral Diseases, Centers for Disease Control and Prevention | Krista Queen, Yan Li, Ying Tao, Jing Zhang, Anna Uehara, Anna Montmayeur, Clinton R. Paden, Peter W. Cook, Rachel Marine, Mili Sheth, Jasmine Padilla, Sarah Nobles, Mark Burroughs, Lori Rowe, Haibin Wang, Ben L. Rambo-Martin, Dhwani Batra, Justin Lee, Suxiang Tong |
| EPI_ISL_903877 | LA Office of Public Health Laboratories | Genomics and Discovery, Respiratory Viruses Branch, Division of Viral Diseases, Centers for Disease Control and Prevention | Krista Queen, Yan Li, Ying Tao, Jing Zhang, Anna Uehara, Anna Montmayeur, Clinton R. Paden, Peter W. Cook, Rachel Marine, Mili Sheth, Jasmine Padilla, Sarah Nobles, Mark Burroughs, Lori Rowe, Haibin Wang, Ben L. Rambo-Martin, Dhwani Batra, Justin Lee, Suxiang Tong |

| | | | |
|---|---|---|---|
| EPI_ISL_903879 | UT-Unified State Labs: Public Health Utah DOH | Genomics and Discovery, Respiratory Viruses Branch, Division of Viral Diseases, Centers for Disease Control and Prevention | Krista Queen, Yan Li, Ying Tao, Jing Zhang, Anna Uehara, Anna Montmayeur, Clinton R. Paden, Peter W. Cook, Rachel Marine, Mili Sheth, Jasmine Padilla, Sarah Nobles, Mark Burroughs, Lori Rowe, Haibin Wang, Ben L. Rambo-Martin, Dhwani Batra, Justin Lee, Suxiang Tong |
| EPI_ISL_903881 | WVDHHR - Office of Laboratory Services | Genomics and Discovery, Respiratory Viruses Branch, Division of Viral Diseases, Centers for Disease Control and Prevention | Krista Queen, Yan Li, Ying Tao, Jing Zhang, Anna Uehara, Anna Montmayeur, Clinton R. Paden, Peter W. Cook, Rachel Marine, Mili Sheth, Jasmine Padilla, Sarah Nobles, Mark Burroughs, Lori Rowe, Haibin Wang, Ben L. Rambo-Martin, Dhwani Batra, Justin Lee, Suxiang Tong |
| EPI_ISL_903882 | TN Division of Laboratory Services | Genomics and Discovery, Respiratory Viruses Branch, Division of Viral Diseases, Centers for Disease Control and Prevention | Krista Queen, Yan Li, Ying Tao, Jing Zhang, Anna Uehara, Anna Montmayeur, Clinton R. Paden, Peter W. Cook, Rachel Marine, Mili Sheth, Jasmine Padilla, Sarah Nobles, Mark Burroughs, Lori Rowe, Haibin Wang, Ben L. Rambo-Martin, Dhwani Batra, Justin Lee, Suxiang Tong |
| EPI_ISL_903883 | IN State Department of Health Laboratory Services | Genomics and Discovery, Respiratory Viruses Branch, Division of Viral Diseases, Centers for Disease Control and Prevention | Krista Queen, Yan Li, Ying Tao, Jing Zhang, Anna Uehara, Anna Montmayeur, Clinton R. Paden, Peter W. Cook, Rachel Marine, Mili Sheth, Jasmine Padilla, Sarah Nobles, Mark Burroughs, Lori Rowe, Haibin Wang, Ben L. Rambo-Martin, Dhwani Batra, Justin Lee, Suxiang Tong |
| EPI_ISL_903884 | TX DSHS, Lab Services Section MC 1947 | Genomics and Discovery, Respiratory Viruses Branch, Division of Viral Diseases, Centers for Disease Control and Prevention | Krista Queen, Yan Li, Ying Tao, Jing Zhang, Anna Uehara, Anna Montmayeur, Clinton R. Paden, Peter W. Cook, Rachel Marine, Mili Sheth, Jasmine Padilla, Sarah Nobles, Mark Burroughs, Lori Rowe, Haibin Wang, Ben L. Rambo-Martin, Dhwani Batra, Justin Lee, Suxiang Tong |
| EPI_ISL_903885 | NE Public Health Laboratory | Genomics and Discovery, Respiratory Viruses Branch, Division of Viral Diseases, Centers for Disease Control and Prevention | Krista Queen, Yan Li, Ying Tao, Jing Zhang, Anna Uehara, Anna Montmayeur, Clinton R. Paden, Peter W. Cook, Rachel Marine, Mili Sheth, Jasmine Padilla, Sarah Nobles, Mark Burroughs, Lori Rowe, Haibin Wang, Ben L. Rambo-Martin, Dhwani Batra, Justin Lee, Suxiang Tong |
| EPI_ISL_903886 | IN State Department of Health Laboratory Services | Genomics and Discovery, Respiratory Viruses Branch, Division of Viral Diseases, Centers for Disease Control and Prevention | Krista Queen, Yan Li, Ying Tao, Jing Zhang, Anna Uehara, Anna Montmayeur, Clinton R. Paden, Peter W. Cook, Rachel Marine, Mili Sheth, Jasmine Padilla, Sarah Nobles, Mark Burroughs, Lori Rowe, Haibin Wang, Ben L. Rambo-Martin, Dhwani Batra, Justin Lee, Suxiang Tong |
| EPI_ISL_903887 | ID Bureau of Laboratories | Genomics and Discovery, Respiratory Viruses Branch, Division of Viral Diseases, Centers for Disease Control and Prevention | Krista Queen, Yan Li, Ying Tao, Jing Zhang, Anna Uehara, Anna Montmayeur, Clinton R. Paden, Peter W. Cook, Rachel Marine, Mili Sheth, Jasmine Padilla, Sarah Nobles, Mark Burroughs, Lori Rowe, Haibin Wang, Ben L. Rambo-Martin, Dhwani Batra, Justin Lee, Suxiang Tong |
| EPI_ISL_903888 | MS Public Health Laboratory | Genomics and Discovery, Respiratory Viruses Branch, Division of Viral Diseases, Centers for Disease Control and Prevention | Krista Queen, Yan Li, Ying Tao, Jing Zhang, Anna Uehara, Anna Montmayeur, Clinton R. Paden, Peter W. Cook, Rachel Marine, Mili Sheth, Jasmine Padilla, Sarah Nobles, Mark Burroughs, Lori Rowe, Haibin Wang, Ben L. Rambo-Martin, Dhwani Batra, Justin Lee, Suxiang Tong |
| EPI_ISL_903889 | TX DSHS, Lab Services Section MC 1947 | Genomics and Discovery, Respiratory Viruses Branch, Division of Viral Diseases, Centers for Disease Control and Prevention | Krista Queen, Yan Li, Ying Tao, Jing Zhang, Anna Uehara, Anna Montmayeur, Clinton R. Paden, Peter W. Cook, Rachel Marine, Mili Sheth, Jasmine Padilla, Sarah Nobles, Mark Burroughs, Lori Rowe, Haibin Wang, Ben L. Rambo-Martin, Dhwani Batra, Justin Lee, Suxiang Tong |
| EPI_ISL_903890 | RI State Health Laboratories | Genomics and Discovery, Respiratory Viruses Branch, Division of Viral Diseases, Centers for Disease Control and Prevention | Krista Queen, Yan Li, Ying Tao, Jing Zhang, Anna Uehara, Anna Montmayeur, Clinton R. Paden, Peter W. Cook, Rachel Marine, Mili Sheth, Jasmine Padilla, Sarah Nobles, Mark Burroughs, Lori Rowe, Haibin Wang, Ben L. Rambo-Martin, Dhwani Batra, Justin Lee, Suxiang Tong |
| EPI_ISL_903891 | UT-Unified State Labs: Public Health Utah DOH | Genomics and Discovery, Respiratory Viruses Branch, Division of Viral Diseases, Centers for Disease Control and Prevention | Krista Queen, Yan Li, Ying Tao, Jing Zhang, Anna Uehara, Anna Montmayeur, Clinton R. Paden, Peter W. Cook, Rachel Marine, Mili Sheth, Jasmine Padilla, Sarah Nobles, Mark Burroughs, Lori Rowe, Haibin Wang, Ben L. Rambo-Martin, Dhwani Batra, Justin Lee, Suxiang Tong |
| EPI_ISL_903893 | TX DSHS, Lab Services Section MC 1947 | Genomics and Discovery, Respiratory Viruses Branch, Division of Viral Diseases, Centers for Disease Control and Prevention | Krista Queen, Yan Li, Ying Tao, Jing Zhang, Anna Uehara, Anna Montmayeur, Clinton R. Paden, Peter W. Cook, Rachel Marine, Mili Sheth, Jasmine Padilla, Sarah Nobles, Mark Burroughs, Lori Rowe, Haibin Wang, Ben L. Rambo-Martin, Dhwani Batra, Justin Lee, Suxiang Tong |
| EPI_ISL_903894 | SD Public Health Laboratory | Genomics and Discovery, Respiratory Viruses Branch, Division of Viral Diseases, Centers for Disease Control and Prevention | Krista Queen, Yan Li, Ying Tao, Jing Zhang, Anna Uehara, Anna Montmayeur, Clinton R. Paden, Peter W. Cook, Rachel Marine, Mili Sheth, Jasmine Padilla, Sarah Nobles, Mark Burroughs, Lori Rowe, Haibin Wang, Ben L. Rambo-Martin, Dhwani Batra, Justin Lee, Suxiang Tong |
| EPI_ISL_903895 | WY Public Health Laboratory | Genomics and Discovery, Respiratory Viruses Branch, Division of Viral Diseases, Centers for Disease Control and Prevention | Krista Queen, Yan Li, Ying Tao, Jing Zhang, Anna Uehara, Anna Montmayeur, Clinton R. Paden, Peter W. Cook, Rachel Marine, Mili Sheth, Jasmine Padilla, Sarah Nobles, Mark Burroughs, Lori Rowe, Haibin Wang, Ben L. Rambo-Martin, Dhwani Batra, Justin Lee, Suxiang Tong |
| EPI_ISL_903896 | IL Dept. of Public Health Springfield Laboratory | Genomics and Discovery, Respiratory Viruses Branch, Division of Viral Diseases, Centers for Disease Control and Prevention | Krista Queen, Yan Li, Ying Tao, Jing Zhang, Anna Uehara, Anna Montmayeur, Clinton R. Paden, Peter W. Cook, Rachel Marine, Mili Sheth, Jasmine Padilla, Sarah Nobles, Mark Burroughs, Lori Rowe, Haibin Wang, Ben L. Rambo-Martin, Dhwani Batra, Justin Lee, Suxiang Tong |
| EPI_ISL_903897 | TX DSHS, Lab Services Section MC 1947 | Genomics and Discovery, Respiratory Viruses Branch, Division of Viral Diseases, Centers for Disease Control and Prevention | Krista Queen, Yan Li, Ying Tao, Jing Zhang, Anna Uehara, Anna Montmayeur, Clinton R. Paden, Peter W. Cook, Rachel Marine, Mili Sheth, Jasmine Padilla, Sarah Nobles, Mark Burroughs, Lori Rowe, Haibin Wang, Ben L. Rambo-Martin, Dhwani Batra, Justin Lee, Suxiang Tong |
| EPI_ISL_903898 | MO State Public Health Laboratory | Genomics and Discovery, Respiratory Viruses Branch, Division of Viral Diseases, Centers for Disease Control and Prevention | Krista Queen, Yan Li, Ying Tao, Jing Zhang, Anna Uehara, Anna Montmayeur, Clinton R. Paden, Peter W. Cook, Rachel Marine, Mili Sheth, Jasmine Padilla, Sarah Nobles, Mark Burroughs, Lori Rowe, Haibin Wang, Ben L. Rambo-Martin, Dhwani Batra, Justin Lee, Suxiang Tong |
| EPI_ISL_903900 | IA State Hygienic Laboratory | Genomics and Discovery, Respiratory Viruses Branch, Division of Viral Diseases, Centers for Disease Control and Prevention | Krista Queen, Yan Li, Ying Tao, Jing Zhang, Anna Uehara, Anna Montmayeur, Clinton R. Paden, Peter W. Cook, Rachel Marine, Mili Sheth, Jasmine Padilla, Sarah Nobles, Mark Burroughs, Lori Rowe, Haibin Wang, Ben L. Rambo-Martin, Dhwani Batra, Justin Lee, Suxiang Tong |
| EPI_ISL_903901 | NE Public Health Laboratory | Genomics and Discovery, Respiratory Viruses Branch, Division of Viral Diseases, Centers for Disease Control and Prevention | Krista Queen, Yan Li, Ying Tao, Jing Zhang, Anna Uehara, Anna Montmayeur, Clinton R. Paden, Peter W. Cook, Rachel Marine, Mili Sheth, Jasmine Padilla, Sarah Nobles, Mark Burroughs, Lori Rowe, Haibin Wang, Ben L. Rambo-Martin, Dhwani Batra, Justin Lee, Suxiang Tong |
| EPI_ISL_903903 | WI State Laboratory of Hygiene | Genomics and Discovery, Respiratory Viruses Branch, Division of Viral Diseases, Centers for Disease Control and Prevention | Krista Queen, Yan Li, Ying Tao, Jing Zhang, Anna Uehara, Anna Montmayeur, Clinton R. Paden, Peter W. Cook, Rachel Marine, Mili Sheth, Jasmine Padilla, Sarah Nobles, Mark Burroughs, Lori Rowe, Haibin Wang, Ben L. Rambo-Martin, Dhwani Batra, Justin Lee, Suxiang Tong |
| EPI_ISL_903904 | IN State Department of Health Laboratory Services | Genomics and Discovery, Respiratory Viruses Branch, Division of Viral Diseases, Centers for Disease Control and Prevention | Krista Queen, Yan Li, Ying Tao, Jing Zhang, Anna Uehara, Anna Montmayeur, Clinton R. Paden, Peter W. Cook, Rachel Marine, Mili Sheth, Jasmine Padilla, Sarah Nobles, Mark Burroughs, Lori Rowe, Haibin Wang, Ben L. Rambo-Martin, Dhwani Batra, Justin Lee, Suxiang Tong |
| EPI_ISL_903905 | MO State Public Health Laboratory | Genomics and Discovery, Respiratory Viruses Branch, Division of Viral Diseases, Centers for Disease Control and Prevention | Krista Queen, Yan Li, Ying Tao, Jing Zhang, Anna Uehara, Anna Montmayeur, Clinton R. Paden, Peter W. Cook, Rachel Marine, Mili Sheth, Jasmine Padilla, Sarah Nobles, Mark Burroughs, Lori Rowe, Haibin Wang, Ben L. Rambo-Martin, Dhwani Batra, Justin Lee, Suxiang Tong |
| EPI_ISL_903906 | LA Office of Public Health Laboratories | Genomics and Discovery, Respiratory Viruses Branch, Division of Viral Diseases, Centers for Disease Control and Prevention | Krista Queen, Yan Li, Ying Tao, Jing Zhang, Anna Uehara, Anna Montmayeur, Clinton R. Paden, Peter W. Cook, Rachel Marine, Mili Sheth, Jasmine Padilla, Sarah Nobles, Mark Burroughs, Lori Rowe, Haibin Wang, Ben L. Rambo-Martin, Dhwani Batra, Justin Lee, Suxiang Tong |
| EPI_ISL_903908 | ID Bureau of Laboratories | Genomics and Discovery, Respiratory Viruses Branch, Division of Viral Diseases, Centers for Disease Control and Prevention | Krista Queen, Yan Li, Ying Tao, Jing Zhang, Anna Uehara, Anna Montmayeur, Clinton R. Paden, Peter W. Cook, Rachel Marine, Mili Sheth, Jasmine Padilla, Sarah Nobles, Mark Burroughs, Lori Rowe, Haibin Wang, Ben L. Rambo-Martin, Dhwani Batra, Justin Lee, Suxiang Tong |
| EPI_ISL_903909 | PA Department of Health, Bureau of Laboratories | Genomics and Discovery, Respiratory Viruses Branch, Division of Viral Diseases, Centers for Disease Control and Prevention | Krista Queen, Yan Li, Ying Tao, Jing Zhang, Anna Uehara, Anna Montmayeur, Clinton R. Paden, Peter W. Cook, Rachel Marine, Mili Sheth, Jasmine Padilla, Sarah Nobles, Mark Burroughs, Lori Rowe, Haibin Wang, Ben L. Rambo-Martin, Dhwani Batra, Justin Lee, Suxiang Tong |
| EPI_ISL_903910 | UT-Unified State Labs: Public Health Utah DOH | Genomics and Discovery, Respiratory Viruses Branch, Division of Viral Diseases, Centers for Disease Control and Prevention | Krista Queen, Yan Li, Ying Tao, Jing Zhang, Anna Uehara, Anna Montmayeur, Clinton R. Paden, Peter W. Cook, Rachel Marine, Mili Sheth, Jasmine Padilla, Sarah Nobles, Mark Burroughs, Lori Rowe, Haibin Wang, Ben L. Rambo-Martin, Dhwani Batra, Justin Lee, Suxiang Tong |
| EPI_ISL_903911 | IL Dept. of Public Health Springfield Laboratory | Genomics and Discovery, Respiratory Viruses Branch, Division of Viral Diseases, Centers for Disease Control and Prevention | Krista Queen, Yan Li, Ying Tao, Jing Zhang, Anna Uehara, Anna Montmayeur, Clinton R. Paden, Peter W. Cook, Rachel Marine, Mili Sheth, Jasmine Padilla, Sarah Nobles, Mark Burroughs, Lori Rowe, Haibin Wang, Ben L. Rambo-Martin, Dhwani Batra, Justin Lee, Suxiang Tong |
| EPI_ISL_903912 | TN Division of Laboratory Services | Genomics and Discovery, Respiratory Viruses Branch, Division of Viral Diseases, Centers for Disease Control and Prevention | Krista Queen, Yan Li, Ying Tao, Jing Zhang, Anna Uehara, Anna Montmayeur, Clinton R. Paden, Peter W. Cook, Rachel Marine, Mili Sheth, Jasmine Padilla, Sarah Nobles, Mark Burroughs, Lori Rowe, Haibin Wang, Ben L. Rambo-Martin, Dhwani Batra, Justin Lee, Suxiang Tong |
| EPI_ISL_903914 | RI State Health Laboratories | Genomics and Discovery, Respiratory Viruses Branch, Division of Viral Diseases, Centers for Disease Control and Prevention | Krista Queen, Yan Li, Ying Tao, Jing Zhang, Anna Uehara, Anna Montmayeur, Clinton R. Paden, Peter W. Cook, Rachel Marine, Mili Sheth, Jasmine Padilla, Sarah Nobles, Mark Burroughs, Lori Rowe, Haibin Wang, Ben L. Rambo-Martin, Dhwani Batra, Justin Lee, Suxiang Tong |
| EPI_ISL_903916 | MO State Public Health Laboratory | Genomics and Discovery, Respiratory Viruses Branch, Division of Viral Diseases, Centers for Disease Control and Prevention | Krista Queen, Yan Li, Ying Tao, Jing Zhang, Anna Uehara, Anna Montmayeur, Clinton R. Paden, Peter W. Cook, Rachel Marine, Mili Sheth, Jasmine Padilla, Sarah Nobles, Mark Burroughs, Lori Rowe, Haibin Wang, Ben L. Rambo-Martin, Dhwani Batra, Justin Lee, Suxiang Tong |
| EPI_ISL_903917 | PA Department of Health, Bureau of Laboratories | Genomics and Discovery, Respiratory Viruses Branch, Division of Viral Diseases, Centers for Disease Control and Prevention | Krista Queen, Yan Li, Ying Tao, Jing Zhang, Anna Uehara, Anna Montmayeur, Clinton R. Paden, Peter W. Cook, Rachel Marine, Mili Sheth, Jasmine Padilla, Sarah Nobles, Mark Burroughs, Lori Rowe, Haibin Wang, Ben L. Rambo-Martin, Dhwani Batra, Justin Lee, Suxiang Tong |

| | | | |
|---|---|---|---|
| EPI_ISL_903920 | IA State Hygienic Laboratory | Genomics and Discovery, Respiratory Viruses Branch, Division of Viral Diseases, Centers for Disease Control and Prevention | Krista Queen, Yan Li, Ying Tao, Jing Zhang, Anna Uehara, Anna Montmayeur, Clinton R. Paden, Peter W. Cook, Rachel Marine, Mili Sheth, Jasmine Padilla, Sarah Nobles, Mark Burroughs, Lori Rowe, Haibin Wang, Ben L. Rambo-Martin, Dhwani Batra, Justin Lee, Suxiang Tong |
| EPI_ISL_903921 | CT-Dr. Katherine A. Kelley State Public Health Lab | Genomics and Discovery, Respiratory Viruses Branch, Division of Viral Diseases, Centers for Disease Control and Prevention | Krista Queen, Yan Li, Ying Tao, Jing Zhang, Anna Uehara, Anna Montmayeur, Clinton R. Paden, Peter W. Cook, Rachel Marine, Mili Sheth, Jasmine Padilla, Sarah Nobles, Mark Burroughs, Lori Rowe, Haibin Wang, Ben L. Rambo-Martin, Dhwani Batra, Justin Lee, Suxiang Tong |
| EPI_ISL_903922 | DC Public Health Lab/ Dept. of Forensic Sciences | Genomics and Discovery, Respiratory Viruses Branch, Division of Viral Diseases, Centers for Disease Control and Prevention | Krista Queen, Yan Li, Ying Tao, Jing Zhang, Anna Uehara, Anna Montmayeur, Clinton R. Paden, Peter W. Cook, Rachel Marine, Mili Sheth, Jasmine Padilla, Sarah Nobles, Mark Burroughs, Lori Rowe, Haibin Wang, Ben L. Rambo-Martin, Dhwani Batra, Justin Lee, Suxiang Tong |
| EPI_ISL_903923 | ID Bureau of Laboratories | Genomics and Discovery, Respiratory Viruses Branch, Division of Viral Diseases, Centers for Disease Control and Prevention | Krista Queen, Yan Li, Ying Tao, Jing Zhang, Anna Uehara, Anna Montmayeur, Clinton R. Paden, Peter W. Cook, Rachel Marine, Mili Sheth, Jasmine Padilla, Sarah Nobles, Mark Burroughs, Lori Rowe, Haibin Wang, Ben L. Rambo-Martin, Dhwani Batra, Justin Lee, Suxiang Tong |
| EPI_ISL_903924 | TN Division of Laboratory Services | Genomics and Discovery, Respiratory Viruses Branch, Division of Viral Diseases, Centers for Disease Control and Prevention | Krista Queen, Yan Li, Ying Tao, Jing Zhang, Anna Uehara, Anna Montmayeur, Clinton R. Paden, Peter W. Cook, Rachel Marine, Mili Sheth, Jasmine Padilla, Sarah Nobles, Mark Burroughs, Lori Rowe, Haibin Wang, Ben L. Rambo-Martin, Dhwani Batra, Justin Lee, Suxiang Tong |
| EPI_ISL_903925 | RI State Health Laboratories | Genomics and Discovery, Respiratory Viruses Branch, Division of Viral Diseases, Centers for Disease Control and Prevention | Krista Queen, Yan Li, Ying Tao, Jing Zhang, Anna Uehara, Anna Montmayeur, Clinton R. Paden, Peter W. Cook, Rachel Marine, Mili Sheth, Jasmine Padilla, Sarah Nobles, Mark Burroughs, Lori Rowe, Haibin Wang, Ben L. Rambo-Martin, Dhwani Batra, Justin Lee, Suxiang Tong |
| EPI_ISL_903926 | PA Department of Health, Bureau of Laboratories | Genomics and Discovery, Respiratory Viruses Branch, Division of Viral Diseases, Centers for Disease Control and Prevention | Krista Queen, Yan Li, Ying Tao, Jing Zhang, Anna Uehara, Anna Montmayeur, Clinton R. Paden, Peter W. Cook, Rachel Marine, Mili Sheth, Jasmine Padilla, Sarah Nobles, Mark Burroughs, Lori Rowe, Haibin Wang, Ben L. Rambo-Martin, Dhwani Batra, Justin Lee, Suxiang Tong |
| EPI_ISL_903927 | IA State Hygienic Laboratory | Genomics and Discovery, Respiratory Viruses Branch, Division of Viral Diseases, Centers for Disease Control and Prevention | Krista Queen, Yan Li, Ying Tao, Jing Zhang, Anna Uehara, Anna Montmayeur, Clinton R. Paden, Peter W. Cook, Rachel Marine, Mili Sheth, Jasmine Padilla, Sarah Nobles, Mark Burroughs, Lori Rowe, Haibin Wang, Ben L. Rambo-Martin, Dhwani Batra, Justin Lee, Suxiang Tong |
| EPI_ISL_903929 | VT Dept. of Health Laboratory | Genomics and Discovery, Respiratory Viruses Branch, Division of Viral Diseases, Centers for Disease Control and Prevention | Krista Queen, Yan Li, Ying Tao, Jing Zhang, Anna Uehara, Anna Montmayeur, Clinton R. Paden, Peter W. Cook, Rachel Marine, Mili Sheth, Jasmine Padilla, Sarah Nobles, Mark Burroughs, Lori Rowe, Haibin Wang, Ben L. Rambo-Martin, Dhwani Batra, Justin Lee, Suxiang Tong |
| EPI_ISL_903930 | WY Public Health Laboratory | Genomics and Discovery, Respiratory Viruses Branch, Division of Viral Diseases, Centers for Disease Control and Prevention | Krista Queen, Yan Li, Ying Tao, Jing Zhang, Anna Uehara, Anna Montmayeur, Clinton R. Paden, Peter W. Cook, Rachel Marine, Mili Sheth, Jasmine Padilla, Sarah Nobles, Mark Burroughs, Lori Rowe, Haibin Wang, Ben L. Rambo-Martin, Dhwani Batra, Justin Lee, Suxiang Tong |
| EPI_ISL_903931 | TX DSHS, Lab Services Section MC 1947 | Genomics and Discovery, Respiratory Viruses Branch, Division of Viral Diseases, Centers for Disease Control and Prevention | Krista Queen, Yan Li, Ying Tao, Jing Zhang, Anna Uehara, Anna Montmayeur, Clinton R. Paden, Peter W. Cook, Rachel Marine, Mili Sheth, Jasmine Padilla, Sarah Nobles, Mark Burroughs, Lori Rowe, Haibin Wang, Ben L. Rambo-Martin, Dhwani Batra, Justin Lee, Suxiang Tong |
| EPI_ISL_903933 | PA Department of Health, Bureau of Laboratories | Genomics and Discovery, Respiratory Viruses Branch, Division of Viral Diseases, Centers for Disease Control and Prevention | Krista Queen, Yan Li, Ying Tao, Jing Zhang, Anna Uehara, Anna Montmayeur, Clinton R. Paden, Peter W. Cook, Rachel Marine, Mili Sheth, Jasmine Padilla, Sarah Nobles, Mark Burroughs, Lori Rowe, Haibin Wang, Ben L. Rambo-Martin, Dhwani Batra, Justin Lee, Suxiang Tong |
| EPI_ISL_903934 | SD Public Health Laboratory | Genomics and Discovery, Respiratory Viruses Branch, Division of Viral Diseases, Centers for Disease Control and Prevention | Krista Queen, Yan Li, Ying Tao, Jing Zhang, Anna Uehara, Anna Montmayeur, Clinton R. Paden, Peter W. Cook, Rachel Marine, Mili Sheth, Jasmine Padilla, Sarah Nobles, Mark Burroughs, Lori Rowe, Haibin Wang, Ben L. Rambo-Martin, Dhwani Batra, Justin Lee, Suxiang Tong |
| EPI_ISL_903936 | CO Dept. of Public Health and Environment, Lab Services Division | Genomics and Discovery, Respiratory Viruses Branch, Division of Viral Diseases, Centers for Disease Control and Prevention | Krista Queen, Yan Li, Ying Tao, Jing Zhang, Anna Uehara, Anna Montmayeur, Clinton R. Paden, Peter W. Cook, Rachel Marine, Mili Sheth, Jasmine Padilla, Sarah Nobles, Mark Burroughs, Lori Rowe, Haibin Wang, Ben L. Rambo-Martin, Dhwani Batra, Justin Lee, Suxiang Tong |
| EPI_ISL_903937 | MS Public Health Laboratory | Genomics and Discovery, Respiratory Viruses Branch, Division of Viral Diseases, Centers for Disease Control and Prevention | Krista Queen, Yan Li, Ying Tao, Jing Zhang, Anna Uehara, Anna Montmayeur, Clinton R. Paden, Peter W. Cook, Rachel Marine, Mili Sheth, Jasmine Padilla, Sarah Nobles, Mark Burroughs, Lori Rowe, Haibin Wang, Ben L. Rambo-Martin, Dhwani Batra, Justin Lee, Suxiang Tong |
| EPI_ISL_903938 | UT-Unified State Labs: Public Health Utah DOH | Genomics and Discovery, Respiratory Viruses Branch, Division of Viral Diseases, Centers for Disease Control and Prevention | Krista Queen, Yan Li, Ying Tao, Jing Zhang, Anna Uehara, Anna Montmayeur, Clinton R. Paden, Peter W. Cook, Rachel Marine, Mili Sheth, Jasmine Padilla, Sarah Nobles, Mark Burroughs, Lori Rowe, Haibin Wang, Ben L. Rambo-Martin, Dhwani Batra, Justin Lee, Suxiang Tong |
| EPI_ISL_903939 | TN Division of Laboratory Services | Genomics and Discovery, Respiratory Viruses Branch, Division of Viral Diseases, Centers for Disease Control and Prevention | Krista Queen, Yan Li, Ying Tao, Jing Zhang, Anna Uehara, Anna Montmayeur, Clinton R. Paden, Peter W. Cook, Rachel Marine, Mili Sheth, Jasmine Padilla, Sarah Nobles, Mark Burroughs, Lori Rowe, Haibin Wang, Ben L. Rambo-Martin, Dhwani Batra, Justin Lee, Suxiang Tong |
| EPI_ISL_903940 | WI State Laboratory of Hygiene | Genomics and Discovery, Respiratory Viruses Branch, Division of Viral Diseases, Centers for Disease Control and Prevention | Krista Queen, Yan Li, Ying Tao, Jing Zhang, Anna Uehara, Anna Montmayeur, Clinton R. Paden, Peter W. Cook, Rachel Marine, Mili Sheth, Jasmine Padilla, Sarah Nobles, Mark Burroughs, Lori Rowe, Haibin Wang, Ben L. Rambo-Martin, Dhwani Batra, Justin Lee, Suxiang Tong |
| EPI_ISL_903942, EPI_ISL_903943, EPI_ISL_903944, EPI_ISL_903946, EPI_ISL_903947 | ND Dept. of Health Laboratory Services-Microbiology | Genomics and Discovery, Respiratory Viruses Branch, Division of Viral Diseases, Centers for Disease Control and Prevention | Krista Queen, Yan Li, Ying Tao, Jing Zhang, Anna Uehara, Anna Montmayeur, Clinton R. Paden, Peter W. Cook, Rachel Marine, Mili Sheth, Jasmine Padilla, Sarah Nobles, Mark Burroughs, Lori Rowe, Haibin Wang, Ben L. Rambo-Martin, Dhwani Batra, Justin Lee, Suxiang Tong |
| EPI_ISL_903948, EPI_ISL_903949 | WA State Department of Health | Genomics and Discovery, Respiratory Viruses Branch, Division of Viral Diseases, Centers for Disease Control and Prevention | Krista Queen, Yan Li, Ying Tao, Jing Zhang, Anna Uehara, Anna Montmayeur, Clinton R. Paden, Peter W. Cook, Rachel Marine, Mili Sheth, Jasmine Padilla, Sarah Nobles, Mark Burroughs, Lori Rowe, Haibin Wang, Ben L. Rambo-Martin, Dhwani Batra, Justin Lee, Suxiang Tong |
| EPI_ISL_903951, EPI_ISL_903953 | MN PHL Division, Minnesota Department of Health | Genomics and Discovery, Respiratory Viruses Branch, Division of Viral Diseases, Centers for Disease Control and Prevention | Krista Queen, Yan Li, Ying Tao, Jing Zhang, Anna Uehara, Anna Montmayeur, Clinton R. Paden, Peter W. Cook, Rachel Marine, Mili Sheth, Jasmine Padilla, Sarah Nobles, Mark Burroughs, Lori Rowe, Haibin Wang, Ben L. Rambo-Martin, Dhwani Batra, Justin Lee, Suxiang Tong |
| EPI_ISL_903955, EPI_ISL_903956 | WA State Department of Health | Genomics and Discovery, Respiratory Viruses Branch, Division of Viral Diseases, Centers for Disease Control and Prevention | Krista Queen, Yan Li, Ying Tao, Jing Zhang, Anna Uehara, Anna Montmayeur, Clinton R. Paden, Peter W. Cook, Rachel Marine, Mili Sheth, Jasmine Padilla, Sarah Nobles, Mark Burroughs, Lori Rowe, Haibin Wang, Ben L. Rambo-Martin, Dhwani Batra, Justin Lee, Suxiang Tong |
| EPI_ISL_903958 | MD DOH Laboratories Administration | Genomics and Discovery, Respiratory Viruses Branch, Division of Viral Diseases, Centers for Disease Control and Prevention | Krista Queen, Yan Li, Ying Tao, Jing Zhang, Anna Uehara, Anna Montmayeur, Clinton R. Paden, Peter W. Cook, Rachel Marine, Mili Sheth, Jasmine Padilla, Sarah Nobles, Mark Burroughs, Lori Rowe, Haibin Wang, Ben L. Rambo-Martin, Dhwani Batra, Justin Lee, Suxiang Tong |
| EPI_ISL_903959 | DC Public Health Lab/ Dept. of Forensic Sciences | Genomics and Discovery, Respiratory Viruses Branch, Division of Viral Diseases, Centers for Disease Control and Prevention | Krista Queen, Yan Li, Ying Tao, Jing Zhang, Anna Uehara, Anna Montmayeur, Clinton R. Paden, Peter W. Cook, Rachel Marine, Mili Sheth, Jasmine Padilla, Sarah Nobles, Mark Burroughs, Lori Rowe, Haibin Wang, Ben L. Rambo-Martin, Dhwani Batra, Justin Lee, Suxiang Tong |
| EPI_ISL_903960, EPI_ISL_903961, EPI_ISL_903962 | WA State Department of Health | Genomics and Discovery, Respiratory Viruses Branch, Division of Viral Diseases, Centers for Disease Control and Prevention | Krista Queen, Yan Li, Ying Tao, Jing Zhang, Anna Uehara, Anna Montmayeur, Clinton R. Paden, Peter W. Cook, Rachel Marine, Mili Sheth, Jasmine Padilla, Sarah Nobles, Mark Burroughs, Lori Rowe, Haibin Wang, Ben L. Rambo-Martin, Dhwani Batra, Justin Lee, Suxiang Tong |
| EPI_ISL_903964 | DC Public Health Lab/ Dept. of Forensic Sciences | Genomics and Discovery, Respiratory Viruses Branch, Division of Viral Diseases, Centers for Disease Control and Prevention | Krista Queen, Yan Li, Ying Tao, Jing Zhang, Anna Uehara, Anna Montmayeur, Clinton R. Paden, Peter W. Cook, Rachel Marine, Mili Sheth, Jasmine Padilla, Sarah Nobles, Mark Burroughs, Lori Rowe, Haibin Wang, Ben L. Rambo-Martin, Dhwani Batra, Justin Lee, Suxiang Tong |
| EPI_ISL_903965, EPI_ISL_903966 | WA State Department of Health | Genomics and Discovery, Respiratory Viruses Branch, Division of Viral Diseases, Centers for Disease Control and Prevention | Krista Queen, Yan Li, Ying Tao, Jing Zhang, Anna Uehara, Anna Montmayeur, Clinton R. Paden, Peter W. Cook, Rachel Marine, Mili Sheth, Jasmine Padilla, Sarah Nobles, Mark Burroughs, Lori Rowe, Haibin Wang, Ben L. Rambo-Martin, Dhwani Batra, Justin Lee, Suxiang Tong |
| EPI_ISL_903967 | MD DOH Laboratories Administration | Genomics and Discovery, Respiratory Viruses Branch, Division of Viral Diseases, Centers for Disease Control and Prevention | Krista Queen, Yan Li, Ying Tao, Jing Zhang, Anna Uehara, Anna Montmayeur, Clinton R. Paden, Peter W. Cook, Rachel Marine, Mili Sheth, Jasmine Padilla, Sarah Nobles, Mark Burroughs, Lori Rowe, Haibin Wang, Ben L. Rambo-Martin, Dhwani Batra, Justin Lee, Suxiang Tong |
| EPI_ISL_903968 | WA State Department of Health | Genomics and Discovery, Respiratory Viruses Branch, Division of Viral Diseases, Centers for Disease Control and Prevention | Krista Queen, Yan Li, Ying Tao, Jing Zhang, Anna Uehara, Anna Montmayeur, Clinton R. Paden, Peter W. Cook, Rachel Marine, Mili Sheth, Jasmine Padilla, Sarah Nobles, Mark Burroughs, Lori Rowe, Haibin Wang, Ben L. Rambo-Martin, Dhwani Batra, Justin Lee, Suxiang Tong |
| EPI_ISL_903969 | DC Public Health Lab/ Dept. of Forensic Sciences | Genomics and Discovery, Respiratory Viruses Branch, Division of Viral Diseases, Centers for Disease Control and Prevention | Krista Queen, Yan Li, Ying Tao, Jing Zhang, Anna Uehara, Anna Montmayeur, Clinton R. Paden, Peter W. Cook, Rachel Marine, Mili Sheth, Jasmine Padilla, Sarah Nobles, Mark Burroughs, Lori Rowe, Haibin Wang, Ben L. Rambo-Martin, Dhwani Batra, Justin Lee, Suxiang Tong |
| EPI_ISL_903970, EPI_ISL_903971, EPI_ISL_903972 | WA State Department of Health | Genomics and Discovery, Respiratory Viruses Branch, Division of Viral Diseases, Centers for Disease Control and Prevention | Krista Queen, Yan Li, Ying Tao, Jing Zhang, Anna Uehara, Anna Montmayeur, Clinton R. Paden, Peter W. Cook, Rachel Marine, Mili Sheth, Jasmine Padilla, Sarah Nobles, Mark Burroughs, Lori Rowe, Haibin Wang, Ben L. Rambo-Martin, Dhwani Batra, Justin Lee, Suxiang Tong |
| EPI_ISL_903973 | DC Public Health Lab/ Dept. of Forensic Sciences | Genomics and Discovery, Respiratory Viruses Branch, Division of Viral Diseases, | Krista Queen, Yan Li, Ying Tao, Jing Zhang, Anna Uehara, Anna Montmayeur, Clinton R. Paden, Peter W. Cook, Rachel Marine, Mili Sheth, Jasmine Padilla, Sarah Nobles, Mark Burroughs, Lori Rowe, Haibin Wang, Ben L. Rambo-Martin, Dhwani Batra, Justin Lee, Suxiang Tong |

| | | | |
|---|---|---|---|
| | | Centers for Disease Control and Prevention | |
| EPI_ISL_903974, EPI_ISL_903975 | WA State Department of Health | Genomics and Discovery, Respiratory Viruses Branch, Division of Viral Diseases, Centers for Disease Control and Prevention | Krista Queen, Yan Li, Ying Tao, Jing Zhang, Anna Uehara, Anna Montmayeur, Clinton R. Paden, Peter W. Cook, Rachel Marine, Mili Sheth, Jasmine Padilla, Sarah Nobles, Mark Burroughs, Lori Rowe, Haibin Wang, Ben L. Rambo-Martin, Dhwani Batra, Justin Lee, Suxiang Tong |
| EPI_ISL_903976 | DC Public Health Lab/ Dept. of Forensic Sciences | Genomics and Discovery, Respiratory Viruses Branch, Division of Viral Diseases, Centers for Disease Control and Prevention | Krista Queen, Yan Li, Ying Tao, Jing Zhang, Anna Uehara, Anna Montmayeur, Clinton R. Paden, Peter W. Cook, Rachel Marine, Mili Sheth, Jasmine Padilla, Sarah Nobles, Mark Burroughs, Lori Rowe, Haibin Wang, Ben L. Rambo-Martin, Dhwani Batra, Justin Lee, Suxiang Tong |
| EPI_ISL_903978 | WA State Department of Health | Genomics and Discovery, Respiratory Viruses Branch, Division of Viral Diseases, Centers for Disease Control and Prevention | Krista Queen, Yan Li, Ying Tao, Jing Zhang, Anna Uehara, Anna Montmayeur, Clinton R. Paden, Peter W. Cook, Rachel Marine, Mili Sheth, Jasmine Padilla, Sarah Nobles, Mark Burroughs, Lori Rowe, Haibin Wang, Ben L. Rambo-Martin, Dhwani Batra, Justin Lee, Suxiang Tong |
| EPI_ISL_904017 | NCSLPH | NCSLPH | Kitty Chase, Michael C Miller, Shermalyn Greene, William Glover |
| EPI_ISL_904035, EPI_ISL_904036, EPI_ISL_904037, EPI_ISL_904038, EPI_ISL_904039, EPI_ISL_904040, EPI_ISL_904041, EPI_ISL_904042, EPI_ISL_904043, EPI_ISL_904044, EPI_ISL_904045, EPI_ISL_904046, EPI_ISL_904047, EPI_ISL_904048, EPI_ISL_904049, EPI_ISL_904050, EPI_ISL_904051, EPI_ISL_904052, EPI_ISL_904053, EPI_ISL_904054, EPI_ISL_904055, EPI_ISL_904056, EPI_ISL_904057, EPI_ISL_904058, EPI_ISL_904059, EPI_ISL_904060, EPI_ISL_904061, EPI_ISL_904062, EPI_ISL_904063, EPI_ISL_904064, EPI_ISL_904065, EPI_ISL_904066, EPI_ISL_904067, EPI_ISL_904068, EPI_ISL_904069, EPI_ISL_904070, EPI_ISL_904071, EPI_ISL_904072, EPI_ISL_904073, EPI_ISL_904074, EPI_ISL_904075, EPI_ISL_904076, EPI_ISL_904077, EPI_ISL_904078, EPI_ISL_904079, EPI_ISL_904080, EPI_ISL_904081, EPI_ISL_904082, EPI_ISL_904083, EPI_ISL_904084, EPI_ISL_904085, EPI_ISL_904086, EPI_ISL_904087, EPI_ISL_904089, EPI_ISL_904090, EPI_ISL_904091, EPI_ISL_904092, EPI_ISL_904093, EPI_ISL_904094, EPI_ISL_904095, EPI_ISL_904096, EPI_ISL_904097, EPI_ISL_904098, EPI_ISL_904099, EPI_ISL_904100, EPI_ISL_904101, EPI_ISL_904102, EPI_ISL_904103, EPI_ISL_904104, EPI_ISL_904105, EPI_ISL_904106, EPI_ISL_904107, EPI_ISL_904108, EPI_ISL_904109, EPI_ISL_904110, EPI_ISL_904111, EPI_ISL_904112, EPI_ISL_904115, EPI_ISL_904116, EPI_ISL_904117, EPI_ISL_904118, EPI_ISL_904119 | | | |
| see above | New Mexico Department of Health Scientific Laboratory | New Mexico Department of Health Scientific Laboratory | Ellie Johnson, Anastacia Griego-Fisher, D'eldra Malone |
| EPI_ISL_904127, EPI_ISL_904128, EPI_ISL_904129, EPI_ISL_904130, EPI_ISL_904131, EPI_ISL_904132, EPI_ISL_904133, EPI_ISL_904134 | Quest Diagnostics | Quest Diagnostics | Rosenthal,S.H., Gerasimova,A., Kagan,R.M., Anderson, B., Hua, M., Liu Y., Bernstein, L.E., Livingston, K.E., Perez, A., Shalhout, D.F., Shlyakhter, I.A., Owen, R., Tanpaiboon, P., Lacbawan, F. |
| EPI_ISL_905765, EPI_ISL_905766, EPI_ISL_905767, EPI_ISL_905768, EPI_ISL_905769, EPI_ISL_905770, EPI_ISL_905771, EPI_ISL_905772 | MD Laboratories | Los Angeles County Public Health Laboratories | P. Hemarajata et al. |
| EPI_ISL_905773, EPI_ISL_905774, EPI_ISL_905775, EPI_ISL_905776, EPI_ISL_905777, EPI_ISL_905778, EPI_ISL_905779, EPI_ISL_905780, EPI_ISL_905781, EPI_ISL_905782, EPI_ISL_905783, EPI_ISL_905784, EPI_ISL_905785, EPI_ISL_905786, EPI_ISL_905787, EPI_ISL_905788, EPI_ISL_905789, EPI_ISL_905790, EPI_ISL_905791 | | | |
| see above | UCLA Clinical Micro Lab | Los Angeles County Public Health Laboratories | P. Hemarajata et al. |
| EPI_ISL_905792, EPI_ISL_905793, EPI_ISL_905794, EPI_ISL_905795, EPI_ISL_905797, EPI_ISL_905798, EPI_ISL_905800, EPI_ISL_905801, EPI_ISL_905802, EPI_ISL_905803, EPI_ISL_905804, EPI_ISL_905805, EPI_ISL_905806, EPI_ISL_905807, EPI_ISL_905808, EPI_ISL_905809, EPI_ISL_905810, EPI_ISL_905811, EPI_ISL_905812, EPI_ISL_905813, EPI_ISL_905814, EPI_ISL_905815, EPI_ISL_905816, EPI_ISL_905817, EPI_ISL_905818, EPI_ISL_905819, EPI_ISL_905820, EPI_ISL_905821, EPI_ISL_905822, EPI_ISL_905823, EPI_ISL_905824, EPI_ISL_905825, EPI_ISL_905826, EPI_ISL_905827, EPI_ISL_905828, EPI_ISL_905832, EPI_ISL_905833, EPI_ISL_905834, EPI_ISL_905835, EPI_ISL_905836, EPI_ISL_905837, EPI_ISL_905839, EPI_ISL_905840, EPI_ISL_905841, EPI_ISL_905842, EPI_ISL_905843, EPI_ISL_905844, EPI_ISL_905845, EPI_ISL_905846, EPI_ISL_905847, EPI_ISL_905848, EPI_ISL_905849, EPI_ISL_905850, EPI_ISL_905851, EPI_ISL_905852, EPI_ISL_905853, EPI_ISL_905854, EPI_ISL_905855, EPI_ISL_905856, EPI_ISL_905857, EPI_ISL_905859, EPI_ISL_905860, EPI_ISL_905861, EPI_ISL_905862, EPI_ISL_905863, EPI_ISL_905864, EPI_ISL_905865, EPI_ISL_905866, EPI_ISL_905867, EPI_ISL_905868, EPI_ISL_905869, EPI_ISL_905870, EPI_ISL_905871, EPI_ISL_905872, EPI_ISL_905873, EPI_ISL_905874, EPI_ISL_905875, EPI_ISL_905876, EPI_ISL_905877, EPI_ISL_905878, EPI_ISL_905879, EPI_ISL_905880, EPI_ISL_905881, EPI_ISL_905883, EPI_ISL_905884, EPI_ISL_905885, EPI_ISL_905886, EPI_ISL_905887, EPI_ISL_905888, EPI_ISL_905889, EPI_ISL_905890, EPI_ISL_905891, EPI_ISL_905893, EPI_ISL_905894, EPI_ISL_905895, EPI_ISL_905896, EPI_ISL_905897, EPI_ISL_905898, EPI_ISL_905899, EPI_ISL_905900, EPI_ISL_905901, EPI_ISL_905903, EPI_ISL_905904, EPI_ISL_905905, EPI_ISL_905906, EPI_ISL_905908, EPI_ISL_905909, EPI_ISL_905910, EPI_ISL_905911, EPI_ISL_905912, EPI_ISL_905913, EPI_ISL_905915, EPI_ISL_905916, EPI_ISL_905917, EPI_ISL_905918, EPI_ISL_905919, EPI_ISL_905920, EPI_ISL_905921, EPI_ISL_905922, EPI_ISL_905923, EPI_ISL_905924, EPI_ISL_905925, EPI_ISL_905926, EPI_ISL_905927, EPI_ISL_905928, EPI_ISL_905929, EPI_ISL_905930, EPI_ISL_905931, EPI_ISL_905933, EPI_ISL_905935, EPI_ISL_905937, EPI_ISL_905938, EPI_ISL_905939, EPI_ISL_905940, EPI_ISL_905941, EPI_ISL_905942, EPI_ISL_905944, EPI_ISL_905945, EPI_ISL_905946, EPI_ISL_905947, EPI_ISL_905948, EPI_ISL_905949, EPI_ISL_905950, EPI_ISL_905951, EPI_ISL_905952, EPI_ISL_905953, EPI_ISL_905954, EPI_ISL_905955, EPI_ISL_905957, EPI_ISL_905958, EPI_ISL_905960, EPI_ISL_905961, EPI_ISL_905962, EPI_ISL_905963, EPI_ISL_905964, EPI_ISL_905965, EPI_ISL_905966, EPI_ISL_905967, EPI_ISL_905969, EPI_ISL_905970, EPI_ISL_905971, EPI_ISL_905972, EPI_ISL_905973, EPI_ISL_905974, EPI_ISL_905975, EPI_ISL_905976, EPI_ISL_905977, EPI_ISL_905979, EPI_ISL_905980, EPI_ISL_905981, EPI_ISL_905982, EPI_ISL_905983, EPI_ISL_905984, EPI_ISL_905985, EPI_ISL_905986, EPI_ISL_905987, EPI_ISL_905989, EPI_ISL_905992, EPI_ISL_905993, EPI_ISL_905994, EPI_ISL_905995, EPI_ISL_905996, EPI_ISL_905997, EPI_ISL_905998, EPI_ISL_906000, EPI_ISL_906001, EPI_ISL_906002, EPI_ISL_906003, EPI_ISL_906005, EPI_ISL_906006, EPI_ISL_906007, EPI_ISL_906008, EPI_ISL_906009, EPI_ISL_906010, EPI_ISL_906011, EPI_ISL_906012, EPI_ISL_906013, EPI_ISL_906014, EPI_ISL_906015, EPI_ISL_906016, EPI_ISL_906017, EPI_ISL_906018, EPI_ISL_906019, EPI_ISL_906020, EPI_ISL_906023, EPI_ISL_906025, EPI_ISL_906026, EPI_ISL_906027, EPI_ISL_906029, EPI_ISL_906030, EPI_ISL_906031, EPI_ISL_906032, EPI_ISL_906033, EPI_ISL_906034, EPI_ISL_906035, EPI_ISL_906036, EPI_ISL_906037, EPI_ISL_906038, EPI_ISL_906039, EPI_ISL_906040, EPI_ISL_906041, EPI_ISL_906042, EPI_ISL_906043, EPI_ISL_906044, EPI_ISL_906045, EPI_ISL_906046 | | | |
| see above | OHSU Lab Services Molecular Microbiology Lab | Oregon SARS-CoV-2 Genome Sequencing Center | Brendan L. O'Connell, Sally Grindstaff, Kayla Carter, Ruth V. Nichols, Alec J. Hirsch, Donna Hansel, Guang Fan, Xuan, Qin, Daniel N. Streblow, William B. Messer, Andrew C. Adey, Benjamin N. Bimber, Brian J. O'Roak |
| EPI_ISL_906047, EPI_ISL_906048 | MD Laboratories | Los Angeles County Public Health Laboratories | P. Hemarajata et al. |
| EPI_ISL_906146 | Gundersen Molecular Diagnostics Laboratory | Kabara Cancer Research Institute | Craig S. Richmond, Paraic A. Kenny |
| EPI_ISL_906147, EPI_ISL_906148, EPI_ISL_906149, EPI_ISL_906150, EPI_ISL_906151, EPI_ISL_906152, EPI_ISL_906153, EPI_ISL_906154, EPI_ISL_906155, EPI_ISL_906156, EPI_ISL_906157, EPI_ISL_906158, EPI_ISL_906159, EPI_ISL_906160, EPI_ISL_906161, EPI_ISL_906162, EPI_ISL_906163, EPI_ISL_906164, EPI_ISL_906165, EPI_ISL_906166, EPI_ISL_906167, EPI_ISL_906168, EPI_ISL_906169, EPI_ISL_906170, EPI_ISL_906171, EPI_ISL_906172, EPI_ISL_906173, EPI_ISL_906174, EPI_ISL_906175, EPI_ISL_906176, EPI_ISL_906177, EPI_ISL_906178, EPI_ISL_906179, EPI_ISL_906180, EPI_ISL_906181, EPI_ISL_906182, EPI_ISL_906183, EPI_ISL_906185, EPI_ISL_906186, EPI_ISL_906187, EPI_ISL_906188, EPI_ISL_906189, EPI_ISL_906190, EPI_ISL_906191, EPI_ISL_906192, EPI_ISL_906193, EPI_ISL_906194, EPI_ISL_906195, EPI_ISL_906196, EPI_ISL_906197, EPI_ISL_906198, EPI_ISL_906199, EPI_ISL_906200, EPI_ISL_906201, EPI_ISL_906202, EPI_ISL_906203, EPI_ISL_906204, EPI_ISL_906205, EPI_ISL_906206, EPI_ISL_906207, EPI_ISL_906208, EPI_ISL_906209, EPI_ISL_906210, EPI_ISL_906211, EPI_ISL_906212, EPI_ISL_906213, EPI_ISL_906214, EPI_ISL_906215, EPI_ISL_906216, EPI_ISL_906217, EPI_ISL_906218, EPI_ISL_906219, EPI_ISL_906220, EPI_ISL_906221, EPI_ISL_906222, EPI_ISL_906223, EPI_ISL_906224, EPI_ISL_906225, EPI_ISL_906226, EPI_ISL_906227, EPI_ISL_906228, EPI_ISL_906229, EPI_ISL_906230, EPI_ISL_906231, EPI_ISL_906232, EPI_ISL_906233, EPI_ISL_906234, EPI_ISL_906235, EPI_ISL_906236, EPI_ISL_906237, EPI_ISL_906238, EPI_ISL_906239, EPI_ISL_906240, EPI_ISL_906241, EPI_ISL_906242, EPI_ISL_906243, EPI_ISL_906244, EPI_ISL_906245, EPI_ISL_906246, EPI_ISL_906247, EPI_ISL_906248, EPI_ISL_906249, EPI_ISL_906250, EPI_ISL_906251, EPI_ISL_906252, EPI_ISL_906253, EPI_ISL_906254, EPI_ISL_906255, EPI_ISL_906256, EPI_ISL_906257, EPI_ISL_906258, EPI_ISL_906259, EPI_ISL_906260, EPI_ISL_906261, EPI_ISL_906262, EPI_ISL_906263, EPI_ISL_906264, EPI_ISL_906265, EPI_ISL_906266, EPI_ISL_906267, EPI_ISL_906268, EPI_ISL_906269, EPI_ISL_906270, EPI_ISL_906271, EPI_ISL_906272, EPI_ISL_906273, EPI_ISL_906274 | | | |
| see above | University of Wisconsin-Madison AIDS Vaccine Research Laboratories | University of Wisconsin-Madison AIDS Vaccine Research Laboratories | Gage Moreno, Katarina Braun, et al. AIDS Vaccine Research Laboratories |
| EPI_ISL_906307, EPI_ISL_906308, EPI_ISL_906309, EPI_ISL_906310, EPI_ISL_906311, EPI_ISL_906312, EPI_ISL_906313, EPI_ISL_906314, EPI_ISL_906315, EPI_ISL_906316, EPI_ISL_906317, EPI_ISL_906318, EPI_ISL_906319, EPI_ISL_906320, EPI_ISL_906321, EPI_ISL_906322, EPI_ISL_906323, EPI_ISL_906324, EPI_ISL_906325, EPI_ISL_906326, EPI_ISL_906327, EPI_ISL_906328, EPI_ISL_906329, EPI_ISL_906330, EPI_ISL_906331, EPI_ISL_906332, EPI_ISL_906333, EPI_ISL_906334, EPI_ISL_906335, EPI_ISL_906336, EPI_ISL_906337, EPI_ISL_906338, EPI_ISL_906339, EPI_ISL_906340, EPI_ISL_906341, EPI_ISL_906342, EPI_ISL_906343, EPI_ISL_906344, EPI_ISL_906345, EPI_ISL_906346, EPI_ISL_906347, EPI_ISL_906348, EPI_ISL_906349, EPI_ISL_906350, EPI_ISL_906351, EPI_ISL_906352, EPI_ISL_906353, EPI_ISL_906354, EPI_ISL_906355, EPI_ISL_906356, EPI_ISL_906357, EPI_ISL_906358, EPI_ISL_906359, EPI_ISL_906360, EPI_ISL_906361, EPI_ISL_906362, EPI_ISL_906363, EPI_ISL_906364, EPI_ISL_906365, EPI_ISL_906366, EPI_ISL_906367, EPI_ISL_906368, EPI_ISL_906369, EPI_ISL_906370, EPI_ISL_906371, EPI_ISL_906372, EPI_ISL_906373, EPI_ISL_906374, EPI_ISL_906375, EPI_ISL_906376, EPI_ISL_906377, EPI_ISL_906378, EPI_ISL_906379, EPI_ISL_906380, EPI_ISL_906381, EPI_ISL_906382, EPI_ISL_906383, EPI_ISL_906384, EPI_ISL_906385, EPI_ISL_906386, EPI_ISL_906387, EPI_ISL_906388, EPI_ISL_906389, EPI_ISL_906390, EPI_ISL_906391, EPI_ISL_906392, EPI_ISL_906393, EPI_ISL_906394, EPI_ISL_906395, EPI_ISL_906396, EPI_ISL_906397, EPI_ISL_906398, EPI_ISL_906399, EPI_ISL_906400, EPI_ISL_906401, EPI_ISL_906402, EPI_ISL_906403, EPI_ISL_906404, EPI_ISL_906405, EPI_ISL_906406, EPI_ISL_906407, EPI_ISL_906408, EPI_ISL_906409, EPI_ISL_906410, EPI_ISL_906411, EPI_ISL_906412, EPI_ISL_906413, EPI_ISL_906414, EPI_ISL_906415, EPI_ISL_906416, EPI_ISL_906417, EPI_ISL_906418, EPI_ISL_906419, EPI_ISL_906420, EPI_ISL_906421, EPI_ISL_906422, EPI_ISL_906423, EPI_ISL_906424, EPI_ISL_906425, EPI_ISL_906426, EPI_ISL_906427, EPI_ISL_906428, EPI_ISL_906429, EPI_ISL_906430, EPI_ISL_906431, EPI_ISL_906432, EPI_ISL_906433, EPI_ISL_906434, EPI_ISL_906435, EPI_ISL_906436, EPI_ISL_906437, EPI_ISL_906438, EPI_ISL_906439, EPI_ISL_906440, EPI_ISL_906441, EPI_ISL_906442, EPI_ISL_906443, EPI_ISL_906444, EPI_ISL_906445, EPI_ISL_906446, EPI_ISL_906447, EPI_ISL_906448, EPI_ISL_906449, EPI_ISL_906450, EPI_ISL_906451, EPI_ISL_906452, EPI_ISL_906453, EPI_ISL_906454, EPI_ISL_906455, EPI_ISL_906456, EPI_ISL_906457, EPI_ISL_906458, EPI_ISL_906459, EPI_ISL_906460, EPI_ISL_906461, EPI_ISL_906462, EPI_ISL_906463, EPI_ISL_906464 | | | |
| see above | Quest Diagnostics | Quest Diagnostics | Rosenthal,S.H., Gerasimova,A., Kagan,R.M., Anderson, B., Hua, M., Liu Y., Bernstein, L.E., Livingston, K.E., Perez, A., Shalhout, D.F., Shlyakhter, I.A., Owen, R., Tanpaiboon, P., Lacbawan, F. |
| EPI_ISL_906562, EPI_ISL_906563, EPI_ISL_906564, EPI_ISL_906565, EPI_ISL_906566, EPI_ISL_906567, EPI_ISL_906568, EPI_ISL_906569, EPI_ISL_906570, EPI_ISL_906571, EPI_ISL_906572, EPI_ISL_906573, EPI_ISL_906574, EPI_ISL_906575, EPI_ISL_906576, EPI_ISL_906577, EPI_ISL_906578, EPI_ISL_906579, EPI_ISL_906581, EPI_ISL_906582, EPI_ISL_906583, EPI_ISL_906584, EPI_ISL_906585, EPI_ISL_906586, EPI_ISL_906587, EPI_ISL_906588, EPI_ISL_906589, EPI_ISL_906590, EPI_ISL_906591, EPI_ISL_906592, EPI_ISL_906593, EPI_ISL_906594, EPI_ISL_906595, EPI_ISL_906596, EPI_ISL_906597, EPI_ISL_906598, EPI_ISL_906599, EPI_ISL_906600, EPI_ISL_906601, EPI_ISL_906602, EPI_ISL_906603, EPI_ISL_906604, EPI_ISL_906605, EPI_ISL_906606, EPI_ISL_906607, EPI_ISL_906608, EPI_ISL_906609, EPI_ISL_906610, EPI_ISL_906611, EPI_ISL_906612, EPI_ISL_906613, EPI_ISL_906614, EPI_ISL_906616, EPI_ISL_906617, EPI_ISL_906618, EPI_ISL_906619, EPI_ISL_906620, EPI_ISL_906622, EPI_ISL_906624, EPI_ISL_906625, EPI_ISL_906626, EPI_ISL_906627, EPI_ISL_906628, EPI_ISL_906629, EPI_ISL_906631, EPI_ISL_906632, EPI_ISL_906633, EPI_ISL_906634, EPI_ISL_906635, EPI_ISL_906636, EPI_ISL_906637, EPI_ISL_906638, EPI_ISL_906639, EPI_ISL_906640, EPI_ISL_906641, EPI_ISL_906642, EPI_ISL_906643, EPI_ISL_906644, EPI_ISL_906645, EPI_ISL_906646, EPI_ISL_906647, EPI_ISL_906648, EPI_ISL_906649, EPI_ISL_906650, EPI_ISL_906651, EPI_ISL_906652, EPI_ISL_906653, EPI_ISL_906654, EPI_ISL_906655, EPI_ISL_906656, EPI_ISL_906657, EPI_ISL_906658, EPI_ISL_906659, EPI_ISL_906660, EPI_ISL_906661, EPI_ISL_906662, EPI_ISL_906663, EPI_ISL_906664, EPI_ISL_906665, EPI_ISL_906666, EPI_ISL_906667, EPI_ISL_906668, EPI_ISL_906669, EPI_ISL_906670, EPI_ISL_906671, EPI_ISL_906672, EPI_ISL_906673, EPI_ISL_906674, EPI_ISL_906675, EPI_ISL_906676, EPI_ISL_906677, EPI_ISL_906678, EPI_ISL_906679, EPI_ISL_906681, EPI_ISL_906682, EPI_ISL_906683, EPI_ISL_906684, EPI_ISL_906685, EPI_ISL_906686, EPI_ISL_906687, EPI_ISL_906688, EPI_ISL_906690, EPI_ISL_906691, EPI_ISL_906692, EPI_ISL_906693, EPI_ISL_906694, EPI_ISL_906696, EPI_ISL_906697, EPI_ISL_906698, EPI_ISL_906699, EPI_ISL_906700, EPI_ISL_906701, EPI_ISL_906702, EPI_ISL_906703, EPI_ISL_906704, EPI_ISL_906705, EPI_ISL_906707, EPI_ISL_906708, EPI_ISL_906709, EPI_ISL_906710 | | | |
| see above | Maine Health and Environmental Testing Laboratory (Maine HETL) | Tewhey Lab, The Jackson Laboratory | Matluk,N., Dewey,H., Iosue,F., Barter,M., Lynch,R., Munger,H. and Tewhey,R. |
| EPI_ISL_906777, EPI_ISL_906778, EPI_ISL_906779 | New Mexico Department of Health Scientific Laboratory | New Mexico Department of Health Scientific Laboratory | Ellie Johnson, Anastacia Griego-Fisher, D'eldra Malone |
| EPI_ISL_906780, EPI_ISL_906781, EPI_ISL_906790, EPI_ISL_906792 | Center for Genome Research and Biocomputing | Center for Genome Research and Biocomputing | Oregon State University TRACE Project |
| EPI_ISL_906848, EPI_ISL_906849, EPI_ISL_906850, EPI_ISL_906852 | Respiratory Viruses Branch, Centers for Disease Control and Prevention | Respiratory Viruses Branch, Centers for Disease Control and Prevention | Tao,Y., Li,Y., Zhang,J., Queen,K., Uehara,A., Cook,P., Paden,C.R., Wang,H., Tong,S. |
| EPI_ISL_906885, EPI_ISL_906886, EPI_ISL_906887, EPI_ISL_906889, EPI_ISL_906891, EPI_ISL_906892, EPI_ISL_906894, EPI_ISL_906895, EPI_ISL_906897, EPI_ISL_906898, EPI_ISL_906899, EPI_ISL_906900, EPI_ISL_906903, EPI_ISL_906906, EPI_ISL_906908, EPI_ISL_906909, EPI_ISL_906913 | | | |
| see above | Bureau of Public Health Laboratories, Florida Department of Health (BPHL, FLDOH) | Bureau of Public Health Laboratories, Florida Department of Health (BPHL, FLDOH) | Schmedes,S., Blanton,J. |
| EPI_ISL_906920, EPI_ISL_906921, EPI_ISL_906922, EPI_ISL_906923, EPI_ISL_906924, EPI_ISL_906925, EPI_ISL_906926, EPI_ISL_906927, EPI_ISL_906928, EPI_ISL_906929, EPI_ISL_906930, EPI_ISL_906931, EPI_ISL_906932, EPI_ISL_906933, EPI_ISL_906934, EPI_ISL_906935, EPI_ISL_906936, EPI_ISL_906937, EPI_ISL_906938, EPI_ISL_906939, EPI_ISL_906940, EPI_ISL_906941, EPI_ISL_906942, EPI_ISL_906943, EPI_ISL_906944, EPI_ISL_906945, EPI_ISL_906946, EPI_ISL_906947, EPI_ISL_906948, EPI_ISL_906949, EPI_ISL_906950, EPI_ISL_906951, EPI_ISL_906952, EPI_ISL_906953, EPI_ISL_906954, EPI_ISL_906955, EPI_ISL_906956, EPI_ISL_906957, EPI_ISL_906958, EPI_ISL_906959, EPI_ISL_906960, EPI_ISL_906961, EPI_ISL_906962, EPI_ISL_906963, EPI_ISL_906964, EPI_ISL_906965, EPI_ISL_906966, EPI_ISL_906967, EPI_ISL_906968, EPI_ISL_906969, EPI_ISL_906970, EPI_ISL_906971, EPI_ISL_906972, EPI_ISL_906973, EPI_ISL_906974, EPI_ISL_906976, EPI_ISL_906977, EPI_ISL_906978, EPI_ISL_906979, EPI_ISL_906980, EPI_ISL_906981, EPI_ISL_906982, EPI_ISL_906983, EPI_ISL_906984, EPI_ISL_906985, EPI_ISL_906986, EPI_ISL_906987, EPI_ISL_906988, EPI_ISL_906989, EPI_ISL_906990, EPI_ISL_906991, EPI_ISL_906992, EPI_ISL_906993, EPI_ISL_906994, EPI_ISL_906995, EPI_ISL_906996, EPI_ISL_906997, EPI_ISL_906998, EPI_ISL_906999, EPI_ISL_907000, EPI_ISL_907001, EPI_ISL_907002, EPI_ISL_907003, EPI_ISL_907004, EPI_ISL_907005, EPI_ISL_907006, EPI_ISL_907007, EPI_ISL_907008, EPI_ISL_907009, EPI_ISL_907010, EPI_ISL_907011, EPI_ISL_907012, EPI_ISL_907013, EPI_ISL_907014, EPI_ISL_907015, EPI_ISL_907016, EPI_ISL_907017, EPI_ISL_907018, EPI_ISL_907019, EPI_ISL_907020, EPI_ISL_907021, EPI_ISL_907022, EPI_ISL_907023, EPI_ISL_907024, EPI_ISL_907025, EPI_ISL_907026, EPI_ISL_907027, EPI_ISL_907028, EPI_ISL_907029, EPI_ISL_907030, EPI_ISL_907031, EPI_ISL_907032, EPI_ISL_907033, EPI_ISL_907034, EPI_ISL_907035, EPI_ISL_907036, EPI_ISL_907037, EPI_ISL_907038, EPI_ISL_907039, EPI_ISL_907040, EPI_ISL_907041, EPI_ISL_907042, EPI_ISL_907043, EPI_ISL_907044, EPI_ISL_907045, EPI_ISL_907046, EPI_ISL_907047, EPI_ISL_907048, EPI_ISL_907049, EPI_ISL_907050, EPI_ISL_907051, EPI_ISL_907052, EPI_ISL_907053, EPI_ISL_907054, EPI_ISL_907055, EPI_ISL_907056, EPI_ISL_907057, EPI_ISL_907058, EPI_ISL_907059, EPI_ISL_907060, EPI_ISL_907061, EPI_ISL_907062, EPI_ISL_907063, EPI_ISL_907064, EPI_ISL_907065, EPI_ISL_907066, EPI_ISL_907067, EPI_ISL_907068, EPI_ISL_907069, EPI_ISL_907070, EPI_ISL_907071, EPI_ISL_907072, EPI_ISL_907073, EPI_ISL_907074 | | | |

| | | | |
|---|---|---|---|
| see above | Infectious Diseases, Quest Diagnostics | Infectious Diseases, Quest Diagnostics | Rosenthal,S.H., Gerasimova,A., Kagan,R.M., Anderson,B., Bernstein,L.E., Livingston,K.E., Hua,M., Liu,Y., Shalhout,D.F., Owen,R., Lacbawan,F. |
| EPI_ISL_909741, EPI_ISL_909743 | Ohio Department of Health Laboratory | Ohio Department of Health Laboratory | Holmes, Jennifer; Eric Brandt, Keoni Omura, Glen McGillivary, Caitlin McDonnell, Jade Mowery, Stephanie Mccracken, Tyler Payne, Kirtana Ramadugu, Erica Leasure, Brent Lee, Kelsey Florek, Heather Blankenship, Quanta Brown, and Tammy Bannerman |
| EPI_ISL_911353, EPI_ISL_911354, EPI_ISL_911355, EPI_ISL_911356, EPI_ISL_911357, EPI_ISL_911358, EPI_ISL_911359, EPI_ISL_911360, EPI_ISL_911361, EPI_ISL_911362, EPI_ISL_911363, EPI_ISL_911364, EPI_ISL_911365, EPI_ISL_911366, EPI_ISL_911367, EPI_ISL_911368, EPI_ISL_911369, EPI_ISL_911370, EPI_ISL_911371 | | | |
| see above | Clinical Diagnostics Laboratory, Diagnostic & Experimental Pathology, Lilly Research Laboratories | Clinical Diagnostics Laboratory, Diagnostic & Experimental Pathology, Lilly Research Laboratories | Tim Holzer, Mayuri Vaidya, Angie Fulford, Sam McNeely, Rachael Redmond, Phil Ebert, John Calley, Leslie ONeill Reising, Pat Finnegan, Erin Wray, John McElwee, Jeff Fill, Joe Oakley, Andrew Schade |
| EPI_ISL_911372, EPI_ISL_911373, EPI_ISL_911374, EPI_ISL_911375, EPI_ISL_911376, EPI_ISL_911377, EPI_ISL_911378, EPI_ISL_911379, EPI_ISL_911380, EPI_ISL_911381, EPI_ISL_911382, EPI_ISL_911383, EPI_ISL_911384, EPI_ISL_911385 | | | |
| see above | University of Michigan Clinical Microbiology Laboratory | Lauring Lab, University of Michigan, Department of Microbiology and Immunology | Valesano |
| EPI_ISL_911402, EPI_ISL_911403, EPI_ISL_911404, EPI_ISL_911405, EPI_ISL_911406, EPI_ISL_911407, EPI_ISL_911408, EPI_ISL_911409, EPI_ISL_911410, EPI_ISL_911411, EPI_ISL_911412, EPI_ISL_911413, EPI_ISL_911414, EPI_ISL_911415, EPI_ISL_911416, EPI_ISL_911417, EPI_ISL_911418, EPI_ISL_911419, EPI_ISL_911420, EPI_ISL_911421, EPI_ISL_911422, EPI_ISL_911423, EPI_ISL_911424, EPI_ISL_911425, EPI_ISL_911426, EPI_ISL_911427, EPI_ISL_911428, EPI_ISL_911429, EPI_ISL_911443, EPI_ISL_911444, EPI_ISL_911445, EPI_ISL_911446, EPI_ISL_911447, EPI_ISL_911448, EPI_ISL_911449, EPI_ISL_911450, EPI_ISL_911451, EPI_ISL_911452, EPI_ISL_911453, EPI_ISL_911454, EPI_ISL_911455, EPI_ISL_911456, EPI_ISL_911457, EPI_ISL_911458, EPI_ISL_911459, EPI_ISL_911460, EPI_ISL_911461, EPI_ISL_911462, EPI_ISL_911463, EPI_ISL_911464, EPI_ISL_911465, EPI_ISL_911466, EPI_ISL_911467, EPI_ISL_911468, EPI_ISL_911469, EPI_ISL_911470 | | | |
| see above | Clinical Diagnostics Laboratory, Diagnostic & Experimental Pathology, Lilly Research Laboratories | Clinical Diagnostics Laboratory, Diagnostic & Experimental Pathology, Lilly Research Laboratories | Tim Holzer, Mayuri Vaidya, Angie Fulford, Sam McNeely, Rachael Redmond, Phil Ebert, John Calley, Leslie ONeill Reising, Pat Finnegan, Erin Wray, John McElwee, Jeff Fill, Joe Oakley, Andrew Schade |
| EPI_ISL_911471, EPI_ISL_911472, EPI_ISL_911473, EPI_ISL_911474, EPI_ISL_911475, EPI_ISL_911476, EPI_ISL_911477, EPI_ISL_911478, EPI_ISL_911479, EPI_ISL_911480, EPI_ISL_911481, EPI_ISL_911482, EPI_ISL_911483, EPI_ISL_911484, EPI_ISL_911485, EPI_ISL_911486, EPI_ISL_911487, EPI_ISL_911488, EPI_ISL_911489, EPI_ISL_911490, EPI_ISL_911491, EPI_ISL_911492, EPI_ISL_911493 | | | |
| see above | Houston Health Dept. | Houston Health Dept. | Ryker Penn, Pamela Brown, Adolpho Lara |
| EPI_ISL_911514, EPI_ISL_911516, EPI_ISL_911517 | Ohio Department of Health Laboratory | Ohio Department of Health Laboratory | Holmes, Jennifer; Eric Brandt, Keoni Omura, Glen McGillivary, Caitlin McDonnell, Jade Mowery, Stephanie Mccracken, Tyler Payne, Kirtana Ramadugu, Erica Leasure, Brent Lee, Kelsey Florek, Heather Blankenship, Quanta Brown, and Tammy Bannerman |
| EPI_ISL_911520, EPI_ISL_911521 | Florida Bureau of Public Health Laboratories | Florida Bureau of Public Health Laboratories | Sarah Schmedes, Jason Blanton |
| EPI_ISL_911528, EPI_ISL_911529, EPI_ISL_911530, EPI_ISL_911531 | M Health Fairview | Minnesota Department of Health, Public Health Laboratory | Alexandra Lorentz, Jacob Garfin, Matt Plumb, and Xiong Wang |
| EPI_ISL_911548 | ARUP laboratories | ARUP Laboratories | Hymas W, Slechta ES, Pyne MT, Mallory MA, Simmon KE, Shakir SM, Hillyard DR, Barker AP |
| EPI_ISL_911549, EPI_ISL_911550, EPI_ISL_911551, EPI_ISL_911552, EPI_ISL_911553, EPI_ISL_911554, EPI_ISL_911555, EPI_ISL_911556, EPI_ISL_911557, EPI_ISL_911558, EPI_ISL_911559, EPI_ISL_911560, EPI_ISL_911561, EPI_ISL_911562, EPI_ISL_911563, EPI_ISL_911564, EPI_ISL_911565, EPI_ISL_911566, EPI_ISL_911567, EPI_ISL_911568, EPI_ISL_911569, EPI_ISL_911570, EPI_ISL_911571, EPI_ISL_911572, EPI_ISL_911573, EPI_ISL_911574, EPI_ISL_911575, EPI_ISL_911576, EPI_ISL_911577, EPI_ISL_911578, EPI_ISL_911579 | | | |
| see above | Clinical Diagnostics Laboratory, Diagnostic & Experimental Pathology, Lilly Research Laboratories | Clinical Diagnostics Laboratory, Diagnostic & Experimental Pathology, Lilly Research Laboratories | Tim Holzer, Mayuri Vaidya, Angie Fulford, Sam McNeely, Rachael Redmond, Phil Ebert, John Calley, Leslie ONeill Reising, Pat Finnegan, Erin Wray, John McElwee, Jeff Fill, Joe Oakley, Andrew Schade |
| EPI_ISL_911580, EPI_ISL_911581, EPI_ISL_911582, EPI_ISL_911583, EPI_ISL_911584, EPI_ISL_911585, EPI_ISL_911586, EPI_ISL_911587, EPI_ISL_911588, EPI_ISL_911589, EPI_ISL_911591, EPI_ISL_911592, EPI_ISL_911593, EPI_ISL_911594, EPI_ISL_911595, EPI_ISL_911596, EPI_ISL_911597, EPI_ISL_911598, EPI_ISL_911599, EPI_ISL_911600, EPI_ISL_911601, EPI_ISL_911602, EPI_ISL_911603 | | | |
| see above | Clinical Molecular Microbiology Laboratory, UNC Hospitals | Jeremy Wang | Jeremy Wang, Alexander Rubinsteyn, Colleen Rice, Jason Smedberg, Melissa Miller, Corbin Jones, Robert Hagan |
| EPI_ISL_911604, EPI_ISL_911605, EPI_ISL_911606, EPI_ISL_911607, EPI_ISL_911608, EPI_ISL_911609, EPI_ISL_911610, EPI_ISL_911611, EPI_ISL_911612, EPI_ISL_911613, EPI_ISL_911614, EPI_ISL_911615, EPI_ISL_911616, EPI_ISL_911617, EPI_ISL_911618, EPI_ISL_911619, EPI_ISL_911620, EPI_ISL_911621, EPI_ISL_911622, EPI_ISL_911623, EPI_ISL_911624, EPI_ISL_911625, EPI_ISL_911626, EPI_ISL_911627, EPI_ISL_911628, EPI_ISL_911629, EPI_ISL_911630, EPI_ISL_911631, EPI_ISL_911632 | | | |
| see above | Clinical Diagnostics Laboratory, Diagnostic & Experimental Pathology, Lilly Research Laboratories | Clinical Diagnostics Laboratory, Diagnostic & Experimental Pathology, Lilly Research Laboratories | Tim Holzer, Mayuri Vaidya, Angie Fulford, Sam McNeely, Rachael Redmond, Phil Ebert, John Calley, Leslie ONeill Reising, Pat Finnegan, Erin Wray, John McElwee, Jeff Fill, Joe Oakley, Andrew Schade |
| EPI_ISL_911635, EPI_ISL_911637, EPI_ISL_911638, EPI_ISL_911639, EPI_ISL_911642, EPI_ISL_911643, EPI_ISL_911645, EPI_ISL_911647, EPI_ISL_911648, EPI_ISL_911649, EPI_ISL_911653, EPI_ISL_911654, EPI_ISL_911655, EPI_ISL_911656, EPI_ISL_911657, EPI_ISL_911659, EPI_ISL_911660, EPI_ISL_911661, EPI_ISL_911662, EPI_ISL_911667, EPI_ISL_911668, EPI_ISL_911669, EPI_ISL_911670, EPI_ISL_911672, EPI_ISL_911673 | | | |
| see above | Texas Department of State Health Services (TXDSHS) | Texas Department of State Health Services (TXDSHS) | Bonnie Oh, Anita Pokharel, James Daniel Bonser, Myong Koag, Chung Wang, Rachel Lee, Grace Kubin, Rashmi Tuladhar, Mayela Pedrueza, Maliha Rahman, Jenny Zhang |
| EPI_ISL_911674 | Clinical Molecular Microbiology Laboratory, UNC Hospitals | Jeremy Wang | Jeremy Wang, Alexander Rubinsteyn, Colleen Rice, Jason Smedberg, Melissa Miller, Corbin Jones, Robert Hagan |
| EPI_ISL_911676, EPI_ISL_911677, EPI_ISL_911678, EPI_ISL_911679, EPI_ISL_911680, EPI_ISL_911681, EPI_ISL_911682, EPI_ISL_911683, EPI_ISL_911684, EPI_ISL_911685, EPI_ISL_911686, EPI_ISL_911687, EPI_ISL_911688, EPI_ISL_911689, EPI_ISL_911690, EPI_ISL_911691, EPI_ISL_911692, EPI_ISL_911693, EPI_ISL_911694, EPI_ISL_911695, EPI_ISL_911696, EPI_ISL_911697, EPI_ISL_911698, EPI_ISL_911699, EPI_ISL_911700, EPI_ISL_911701, EPI_ISL_911702, EPI_ISL_911703 | | | |
| see above | Alaska State Virology Laboratory | Alaska State Virology Laboratory | Stephanie DeRonde, Lisa Smith, Ph.D., Jack Chen, Ph.D. |
| EPI_ISL_911704, EPI_ISL_911705, EPI_ISL_911706 | Quest Diagnostics | Quest Diagnostics | Rosenthal,S.H., Gerasimova,A., Kagan,R.M., Anderson, B., Hua, M., Liu Y., Bernstein, L.E., Livingston, K.E., Perez, A., Shalhout, D.F., Shlyakhter, I.A., Owen, R., Tanpaiboon, P., Lacbawan, F. |
| EPI_ISL_911710, EPI_ISL_911711, EPI_ISL_911712, EPI_ISL_911713, EPI_ISL_911714, EPI_ISL_911715, EPI_ISL_911716, EPI_ISL_911717, EPI_ISL_911718, EPI_ISL_911719, EPI_ISL_911720, EPI_ISL_911721, EPI_ISL_911722, EPI_ISL_911723, EPI_ISL_911724, EPI_ISL_911725, EPI_ISL_911726, EPI_ISL_911727, EPI_ISL_911728, EPI_ISL_911729, EPI_ISL_911730, EPI_ISL_911731, EPI_ISL_911733, EPI_ISL_911734, EPI_ISL_911735, EPI_ISL_911736, EPI_ISL_911737, EPI_ISL_911738, EPI_ISL_911739, EPI_ISL_911740, EPI_ISL_911741, EPI_ISL_911742, EPI_ISL_911743, EPI_ISL_911744, EPI_ISL_911745, EPI_ISL_911751, EPI_ISL_911752, EPI_ISL_911753, EPI_ISL_911754 | | | |
| see above | Clinical Diagnostics Laboratory, Diagnostic & Experimental Pathology, Lilly Research Laboratories | Clinical Diagnostics Laboratory, Diagnostic & Experimental Pathology, Lilly Research Laboratories | Tim Holzer, Mayuri Vaidya, Angie Fulford, Sam McNeely, Rachael Redmond, Phil Ebert, John Calley, Leslie ONeill Reising, Pat Finnegan, Erin Wray, John McElwee, Jeff Fill, Joe Oakley, Andrew Schade |
| EPI_ISL_911755, EPI_ISL_911756, EPI_ISL_911757, EPI_ISL_911758, EPI_ISL_911759, EPI_ISL_911760, EPI_ISL_911762, EPI_ISL_911763, EPI_ISL_911764, EPI_ISL_911765, EPI_ISL_911766, EPI_ISL_911768, EPI_ISL_911769, EPI_ISL_911770, EPI_ISL_911771, EPI_ISL_911772, EPI_ISL_911773, EPI_ISL_911774, EPI_ISL_911775, EPI_ISL_911776, EPI_ISL_911777, EPI_ISL_911778, EPI_ISL_911779, EPI_ISL_911780, EPI_ISL_911781, EPI_ISL_911782, EPI_ISL_911783, EPI_ISL_911784, EPI_ISL_911785, EPI_ISL_911786, EPI_ISL_911787, EPI_ISL_911788, EPI_ISL_911789, EPI_ISL_911790, EPI_ISL_911791, EPI_ISL_911793, EPI_ISL_911794, EPI_ISL_911795, EPI_ISL_911796, EPI_ISL_911797, EPI_ISL_911798, EPI_ISL_911799, EPI_ISL_911800, EPI_ISL_911801, EPI_ISL_911802, EPI_ISL_911803, EPI_ISL_911804, EPI_ISL_911805, EPI_ISL_911806, EPI_ISL_911807, EPI_ISL_911808, EPI_ISL_911809, EPI_ISL_911810, EPI_ISL_911811, EPI_ISL_911812, EPI_ISL_911813, EPI_ISL_911814, EPI_ISL_911815, EPI_ISL_911816, EPI_ISL_911817, EPI_ISL_911818, EPI_ISL_911819, EPI_ISL_911820, EPI_ISL_911821, EPI_ISL_911822, EPI_ISL_911823, EPI_ISL_911824, EPI_ISL_911825, EPI_ISL_911826, EPI_ISL_911827, EPI_ISL_911828, EPI_ISL_911829, EPI_ISL_911830, EPI_ISL_911831, EPI_ISL_911832, EPI_ISL_911834, EPI_ISL_911835, EPI_ISL_911836, EPI_ISL_911837, EPI_ISL_911838, EPI_ISL_911839, EPI_ISL_911840, EPI_ISL_911841, EPI_ISL_911842, EPI_ISL_911843, EPI_ISL_911844, EPI_ISL_911845, EPI_ISL_911846, EPI_ISL_911847, EPI_ISL_911848, EPI_ISL_911849, EPI_ISL_911850, EPI_ISL_911851, EPI_ISL_911852, EPI_ISL_911853, EPI_ISL_911854, EPI_ISL_911855, EPI_ISL_911856, EPI_ISL_911857, EPI_ISL_911858, EPI_ISL_911859, EPI_ISL_911860, EPI_ISL_911861, EPI_ISL_911862, EPI_ISL_911863, EPI_ISL_911864, EPI_ISL_911865, EPI_ISL_911866, EPI_ISL_911867, EPI_ISL_911868, EPI_ISL_911869, EPI_ISL_911870, EPI_ISL_911871, EPI_ISL_911872, EPI_ISL_911873, EPI_ISL_911874, EPI_ISL_911875, EPI_ISL_911876, EPI_ISL_911877, EPI_ISL_911878, EPI_ISL_911879, EPI_ISL_911880, EPI_ISL_911881, EPI_ISL_911882, EPI_ISL_911883, EPI_ISL_911884, EPI_ISL_911885, EPI_ISL_911886, EPI_ISL_911887, EPI_ISL_911888, EPI_ISL_911889, EPI_ISL_911890, EPI_ISL_911891, EPI_ISL_911892, EPI_ISL_911893, EPI_ISL_911894, EPI_ISL_911895, EPI_ISL_911896, EPI_ISL_911897, EPI_ISL_911898, EPI_ISL_911899, EPI_ISL_911900, EPI_ISL_911901, EPI_ISL_911902, EPI_ISL_911903, EPI_ISL_911904, EPI_ISL_911905, EPI_ISL_911906, EPI_ISL_911907, EPI_ISL_911908, EPI_ISL_911909, EPI_ISL_911910, EPI_ISL_911911, EPI_ISL_911912, EPI_ISL_911913, EPI_ISL_911914, EPI_ISL_911915, EPI_ISL_911916, EPI_ISL_911917 | | | |
| see above | Johns Hopkins Hospital Department of Pathology | Johns Hopkins Hospital Department of Pathology | C. Paul Morris, Chun Huai Luo, Adannaya Amadi, Matthew Schwartz, Nicholas Gallagher, Heba H. Mostafa |
| EPI_ISL_911918, EPI_ISL_911920, EPI_ISL_911921, EPI_ISL_911922 | SRL (Sonic Reference Laboratory - Sonic Healthcare) | Pathogen Discovery, Respiratory Viruses Branch, Division of Viral Diseases, Centers for Disease Control and Prevention | Ying Tao, Yan Li, Jing Zhang, Krista Queen, Anna Uehara, Peter Cook, Clinton R. Paden, Haibin Wang, Suxiang Tong |
| EPI_ISL_911923 | IL Department of Public Health Chicago Laboratory/Tempus Labs | Pathogen Discovery, Respiratory Viruses Branch, Division of Viral Diseases, Centers for Disease Control and Prevention | Ying Tao, Yan Li, Jing Zhang, Krista Queen, Anna Uehara, Peter Cook, Clinton R. Paden, Haibin Wang, Suxiang Tong |
| EPI_ISL_911924, EPI_ISL_911925, EPI_ISL_911926, EPI_ISL_911927, EPI_ISL_911928 | Clinical Diagnostics Laboratory, Diagnostic & Experimental Pathology, Lilly Research Laboratories | Clinical Diagnostics Laboratory, Diagnostic & Experimental Pathology, Lilly Research Laboratories | Tim Holzer, Mayuri Vaidya, Angie Fulford, Sam McNeely, Rachael Redmond, Phil Ebert, John Calley, Leslie ONeill Reising, Pat Finnegan, Erin Wray, John McElwee, Jeff Fill, Joe Oakley, Andrew Schade |
| EPI_ISL_911929, EPI_ISL_911930, EPI_ISL_911931, EPI_ISL_911932, EPI_ISL_911933, EPI_ISL_911934, EPI_ISL_911935, EPI_ISL_911936, EPI_ISL_911937, EPI_ISL_911938, EPI_ISL_911939, EPI_ISL_911940, EPI_ISL_911941, EPI_ISL_911942, EPI_ISL_911943, EPI_ISL_911944, EPI_ISL_911945, EPI_ISL_911947, EPI_ISL_911948, EPI_ISL_911949, EPI_ISL_911950, EPI_ISL_911951, EPI_ISL_911952, EPI_ISL_911953, EPI_ISL_911954, EPI_ISL_911955, EPI_ISL_911956, EPI_ISL_911957, EPI_ISL_911958, EPI_ISL_911959, EPI_ISL_911960, EPI_ISL_911961, EPI_ISL_911962, EPI_ISL_911963, EPI_ISL_911964, EPI_ISL_911965, EPI_ISL_911966 | | | |
| see above | Wyoming Public Health Laboratory | Wyoming Public Health Laboratory | Noah Hull, Taylor Fearing, Lynette Gumbleton, Channing Weber, Ashley Norberg, Bailey Bowcutt, and Wanda Manley |
| EPI_ISL_911967 | LabCorp | Wyoming Public Health Laboratory | Noah Hull, Taylor Fearing, Lynette Gumbleton, Channing Weber, Ashley Norberg, Bailey Bowcutt, and Wanda Manley |
| EPI_ISL_911970 | Seattle Flu Study | Seattle Flu Study | Deborah A. Nickerson, Chris D. Frazar, Jover Lee, Benjamin Pelle, Erica Ryke, Matthew Richardson, Amanda Adler, Elisabeth Brandstetter, Peter D. Han, Kairsten Fay, Misja Ilcisin, Kirsten Lacombe, Thomas R. Sibley, Melissa Truong, Caitlin R. Wolf, Karen Cowgill, Stephanie Schrag, Jeff Duchin, Michael Boeckh, Janet A. Englund, Michael Famulare, Barry R. Lutz, Mark J. Rieder, Lea M. Starita, Matthew Thompson, Helen Y. Chu, Trevor Bedford, Jay Shendure |
| EPI_ISL_911972, EPI_ISL_911973, EPI_ISL_911974, EPI_ISL_911975, EPI_ISL_911976, EPI_ISL_911977, EPI_ISL_911978, EPI_ISL_911979, EPI_ISL_911980, EPI_ISL_911982, EPI_ISL_911983, EPI_ISL_911985, EPI_ISL_911986, EPI_ISL_911987, EPI_ISL_911990, EPI_ISL_911992, EPI_ISL_911993, EPI_ISL_911994, EPI_ISL_911996, EPI_ISL_911998, EPI_ISL_911999, EPI_ISL_912000, EPI_ISL_912001, EPI_ISL_912002, EPI_ISL_912003, EPI_ISL_912004, EPI_ISL_912005, EPI_ISL_912006, EPI_ISL_912007, EPI_ISL_912008, EPI_ISL_912009, EPI_ISL_912010, EPI_ISL_912012, EPI_ISL_912013, EPI_ISL_912014, EPI_ISL_912015, EPI_ISL_912016, EPI_ISL_912017, EPI_ISL_912019, EPI_ISL_912020, EPI_ISL_912022, EPI_ISL_912024, EPI_ISL_912025, EPI_ISL_912026, EPI_ISL_912027, EPI_ISL_912028, EPI_ISL_912030, EPI_ISL_912031, EPI_ISL_912032, EPI_ISL_912033, EPI_ISL_912034 | | | |
| see above | Altius Institute for Biomedical Sciences | Seattle Flu Study | Deborah A. Nickerson, Chris D. Frazar, Jover Lee, Benjamin Pelle, Erica Ryke, Matthew Richardson, Amanda Adler, Elisabeth Brandstetter, Peter D. Han, Kairsten Fay, Misja Ilcisin, Kirsten Lacombe, Thomas R. Sibley, Melissa Truong, Caitlin R. Wolf, Ryan Alexander, Daniel Bates, Rebecca Bruders, Stephanie DeBaun, Clem Green, Muhammad Halimun, Jessica Halow, Kneshay Harper, Matt Hartman, Andrew Meuser, Alex Nguyen, Truong Nguyen, Sofia Olsson, Sadie Patraw, Hannah Petersen, Tobias Ragoczy, Joshua Richards, Jacob Rodriguez, John Stamatoyannopoulos, Julia Wald, Olivia Waltner, Michael Boeckh, Janet A. Englund, Michael Famulare, Barry R. Lutz, Mark J. Rieder, Lea M. Starita, Matthew Thompson, Helen Y. Chu, Jay Shendure, Trevor Bedford |
| EPI_ISL_912035, EPI_ISL_912037, EPI_ISL_912038, EPI_ISL_912039, EPI_ISL_912041, EPI_ISL_912042, EPI_ISL_912043, EPI_ISL_912044, EPI_ISL_912045, EPI_ISL_912046, EPI_ISL_912047, EPI_ISL_912048, EPI_ISL_912049, EPI_ISL_912051, EPI_ISL_912052, EPI_ISL_912053, EPI_ISL_912054, EPI_ISL_912056, EPI_ISL_912058, EPI_ISL_912060, EPI_ISL_912063, EPI_ISL_912064, EPI_ISL_912066, EPI_ISL_912068, EPI_ISL_912069, EPI_ISL_912070, EPI_ISL_912071, EPI_ISL_912072, EPI_ISL_912073, EPI_ISL_912074, EPI_ISL_912075, EPI_ISL_912076, EPI_ISL_912077, EPI_ISL_912078, EPI_ISL_912079, EPI_ISL_912080, EPI_ISL_912081, EPI_ISL_912082, EPI_ISL_912084, EPI_ISL_912085, EPI_ISL_912086, EPI_ISL_912088, EPI_ISL_912089, EPI_ISL_912091, EPI_ISL_912092, EPI_ISL_912093, EPI_ISL_912094, EPI_ISL_912095, | | | |

| | | | |
|---|---|---|---|
| EPI_ISL_912096, EPI_ISL_912097, EPI_ISL_912099, EPI_ISL_912101, EPI_ISL_912102, EPI_ISL_912103, EPI_ISL_912105, EPI_ISL_912106, EPI_ISL_912107, EPI_ISL_912108 | | | |
| see above | Washington State Department of Health | Seattle Flu Study | Deborah A. Nickerson, Chris D. Frazar, Jover Lee, Benjamin Pelle, Erica Ryke, Matthew Richardson, Amanda Adler, Elisabeth Brandstetter, Peter D. Han, Kairsten Fay, Misja Ilcisin, Kirsten Lacombe, Thomas R. Sibley, Melissa Truong, Caitlin R. Wolf, Romesh Gautom, Geoff Melly, Brian Hiatt, Philip Dykema, Scott Lindquist, Michael Boeckh, Janet A. Englund, Michael Famulare, Barry R. Lutz, Mark J. Rieder, Lea M. Starita, Matthew Thompson, Helen Y. Chu, Jay Shendure, Trevor Bedford |
| EPI_ISL_912109, EPI_ISL_912110, EPI_ISL_912112, EPI_ISL_912114, EPI_ISL_912115, EPI_ISL_912116, EPI_ISL_912117, EPI_ISL_912118, EPI_ISL_912119, EPI_ISL_912120, EPI_ISL_912121, EPI_ISL_912122, EPI_ISL_912123, EPI_ISL_912124, EPI_ISL_912125, EPI_ISL_912127, EPI_ISL_912131, EPI_ISL_912132, EPI_ISL_912133, EPI_ISL_912135, EPI_ISL_912136, EPI_ISL_912137, EPI_ISL_912138, EPI_ISL_912140, EPI_ISL_912142, EPI_ISL_912143, EPI_ISL_912145, EPI_ISL_912146, EPI_ISL_912150, EPI_ISL_912151, EPI_ISL_912152, EPI_ISL_912153, EPI_ISL_912154, EPI_ISL_912155, EPI_ISL_912156, EPI_ISL_912158, EPI_ISL_912159 | | | |
| see above | Altius Institute for Biomedical Sciences | Seattle Flu Study | Deborah A. Nickerson, Chris D. Frazar, Jover Lee, Benjamin Pelle, Erica Ryke, Matthew Richardson, Amanda Adler, Elisabeth Brandstetter, Peter D. Han, Kairsten Fay, Misja Ilcisin, Kirsten Lacombe, Thomas R. Sibley, Melissa Truong, Caitlin R. Wolf, Ryan Alexander, Daniel Bates, Rebecca Bruders, Stephanie DeBaun, Clem Green, Muhammad Halimun, Jessica Halow, Kneshay Harper, Matt Hartman, Andrew Meuser, Alex Nguyen, Truong Nguyen, Sofia Olsson, Sadie Patraw, Hannah Petersen, Tobias Ragoczy, Joshua Richards, Jacob Rodriguez, John Stamatoyannopoulos, Julia Wald, Olivia Waltner, Michael Boeckh, Janet A. Englund, Michael Famulare, Barry R. Lutz, Mark J. Rieder, Lea M. Starita, Matthew Thompson, Helen Y. Chu, Jay Shendure, Trevor Bedford |
| EPI_ISL_912161 | Tempus | Grubaugh Lab - Yale School of Public Health | Tara Alpert, Joseph Fauver, Anderson Brito, Mallery Breban, Anne Wyllie, Chantal Vogels, Mary Petrone, Annie Watkins, Chaney Kalinich, Isabel Ott, Nathan Grubaugh |
| EPI_ISL_912163, EPI_ISL_912164 | Connecticut Department of Health | Grubaugh Lab - Yale School of Public Health | Tara Alpert, Joseph Fauver, Anderson Brito, Mallery Breban, Anne Wyllie, Chantal Vogels, Mary Petrone, Annie Watkins, Chaney Kalinich, Isabel Ott, Nathan Grubaugh |
| EPI_ISL_912168 | Yale Pathology Lab | Grubaugh Lab - Yale School of Public Health | Tara Alpert, Joseph Fauver, Chen Liu, Pei Hui, Jianhui Wang, Susan Bell and Han Zhou, Anderson Brito, Mallery Breban, Anne Wyllie, Chantal Vogels, Mary Petrone, Chaney Kalinich, Isabel Ott, Arnau Casanovas, Catherine Muenker, Adam Moore, Alice Lu, Maria Tokuyama, Patrick Wong, Peiwen Lu, Saad Omer, Richard Martinello, Allison Nelson, Shelli Farhadian, Akiko Iwasaki, Charlese Dela Cruz, Albert Ko, Nathan Grubaugh |
| EPI_ISL_912172, EPI_ISL_912176, EPI_ISL_912177 | Tempus | Grubaugh Lab - Yale School of Public Health | Tara Alpert, Joseph Fauver, Anderson Brito, Mallery Breban, Anne Wyllie, Chantal Vogels, Mary Petrone, Annie Watkins, Chaney Kalinich, Isabel Ott, Nathan Grubaugh |
| EPI_ISL_912187, EPI_ISL_912188, EPI_ISL_912189, EPI_ISL_912190, EPI_ISL_912191, EPI_ISL_912192, EPI_ISL_912193, EPI_ISL_912194 | California Institute of Technology | Chan-Zuckerberg Biohub | CZB Cliahub Consortium |
| EPI_ISL_913116, EPI_ISL_913117, EPI_ISL_913118, EPI_ISL_913119, EPI_ISL_913120, EPI_ISL_913121, EPI_ISL_913122, EPI_ISL_913123, EPI_ISL_913124, EPI_ISL_913125, EPI_ISL_913126, EPI_ISL_913127, EPI_ISL_913128, EPI_ISL_913129, EPI_ISL_913130, EPI_ISL_913131, EPI_ISL_913132, EPI_ISL_913133, EPI_ISL_913134, EPI_ISL_913135, EPI_ISL_913136, EPI_ISL_913137, EPI_ISL_913138, EPI_ISL_913139, EPI_ISL_913140, EPI_ISL_913141, EPI_ISL_913142, EPI_ISL_913143, EPI_ISL_913144, EPI_ISL_913145, EPI_ISL_913146, EPI_ISL_913147, EPI_ISL_913148, EPI_ISL_913149, EPI_ISL_913150, EPI_ISL_913151, EPI_ISL_913152, EPI_ISL_913153, EPI_ISL_913154, EPI_ISL_913155, EPI_ISL_913156, EPI_ISL_913157, EPI_ISL_913158, EPI_ISL_913159, EPI_ISL_913160, EPI_ISL_913161, EPI_ISL_913162, EPI_ISL_913163, EPI_ISL_913164, EPI_ISL_913165, EPI_ISL_913166, EPI_ISL_913167, EPI_ISL_913168, EPI_ISL_913169, EPI_ISL_913170, EPI_ISL_913171, EPI_ISL_913172, EPI_ISL_913173, EPI_ISL_913174, EPI_ISL_913175, EPI_ISL_913176, EPI_ISL_913177, EPI_ISL_913178, EPI_ISL_913179, EPI_ISL_913180, EPI_ISL_913181, EPI_ISL_913182, EPI_ISL_913183, EPI_ISL_913184, EPI_ISL_913185, EPI_ISL_913186, EPI_ISL_913187, EPI_ISL_913188, EPI_ISL_913189, EPI_ISL_913190, EPI_ISL_913191, EPI_ISL_913192, EPI_ISL_913193, EPI_ISL_913194, EPI_ISL_913195, EPI_ISL_913196, EPI_ISL_913197, EPI_ISL_913198, EPI_ISL_913199, EPI_ISL_913200, EPI_ISL_913201, EPI_ISL_913202, EPI_ISL_913203, EPI_ISL_913204, EPI_ISL_913205, EPI_ISL_913206, EPI_ISL_913207, EPI_ISL_913208, EPI_ISL_913209, EPI_ISL_913210, EPI_ISL_913211, EPI_ISL_913212, EPI_ISL_913213, EPI_ISL_913214, EPI_ISL_913215, EPI_ISL_913216, EPI_ISL_913217, EPI_ISL_913218, EPI_ISL_913219, EPI_ISL_913220, EPI_ISL_913221, EPI_ISL_913222, EPI_ISL_913223, EPI_ISL_913224, EPI_ISL_913225, EPI_ISL_913226, EPI_ISL_913227, EPI_ISL_913228, EPI_ISL_913229, EPI_ISL_913230, EPI_ISL_913231, EPI_ISL_913232, EPI_ISL_913233, EPI_ISL_913234, EPI_ISL_913235, EPI_ISL_913236, EPI_ISL_913237, EPI_ISL_913238, EPI_ISL_913239, EPI_ISL_913240, EPI_ISL_913241, EPI_ISL_913242, EPI_ISL_913243, EPI_ISL_913244, EPI_ISL_913245, EPI_ISL_913246, EPI_ISL_913247, EPI_ISL_913248, EPI_ISL_913249, EPI_ISL_913250, EPI_ISL_913251, EPI_ISL_913252, EPI_ISL_913253, EPI_ISL_913254, EPI_ISL_913255 | | | |
| see above | University of Michigan Clinical Microbiology Laboratory | Lauring Lab, University of Michigan, Department of Microbiology and Immunology | Valesano |
| EPI_ISL_913393, EPI_ISL_913394, EPI_ISL_913395, EPI_ISL_913396, EPI_ISL_913397, EPI_ISL_913398, EPI_ISL_913399, EPI_ISL_913400, EPI_ISL_913401, EPI_ISL_913402, EPI_ISL_913403, EPI_ISL_913404, EPI_ISL_913405, EPI_ISL_913406, EPI_ISL_913407, EPI_ISL_913408, EPI_ISL_913409, EPI_ISL_913410, EPI_ISL_913411, EPI_ISL_913412, EPI_ISL_913433, EPI_ISL_913434, EPI_ISL_913435 | | | |
| see above | Massachusetts State Public Health Laboratory | Massachusetts State Public Health Laboratory | Andrew Lang, Timelia Fink, Glen Gallagher, Sandra Smole |
| EPI_ISL_913514, EPI_ISL_913515, EPI_ISL_913516, EPI_ISL_913517, EPI_ISL_913518 | University of Michigan Clinical Microbiology Laboratory | Lauring Lab, University of Michigan, Department of Microbiology and Immunology | Valesano |
| EPI_ISL_913519, EPI_ISL_913520, EPI_ISL_913521, EPI_ISL_913522, EPI_ISL_913523, EPI_ISL_913524, EPI_ISL_913525, EPI_ISL_913526, EPI_ISL_913527, EPI_ISL_913528, EPI_ISL_913529, EPI_ISL_913530, EPI_ISL_913531, EPI_ISL_913532, EPI_ISL_913533, EPI_ISL_913534, EPI_ISL_913535, EPI_ISL_913536, EPI_ISL_913537, EPI_ISL_913538, EPI_ISL_913539, EPI_ISL_913540, EPI_ISL_913541, EPI_ISL_913542, EPI_ISL_913543, EPI_ISL_913544, EPI_ISL_913545, EPI_ISL_913546, EPI_ISL_913547, EPI_ISL_913548, EPI_ISL_913549, EPI_ISL_913550, EPI_ISL_913551, EPI_ISL_913552, EPI_ISL_913553, EPI_ISL_913554, EPI_ISL_913555, EPI_ISL_913556, EPI_ISL_913557, EPI_ISL_913558, EPI_ISL_913559, EPI_ISL_913560, EPI_ISL_913561, EPI_ISL_913562, EPI_ISL_913563, EPI_ISL_913564, EPI_ISL_913565, EPI_ISL_913566, EPI_ISL_913567, EPI_ISL_913568, EPI_ISL_913569, EPI_ISL_913570, EPI_ISL_913571, EPI_ISL_913572, EPI_ISL_913573, EPI_ISL_913574, EPI_ISL_913575, EPI_ISL_913576, EPI_ISL_913577, EPI_ISL_913578, EPI_ISL_913579, EPI_ISL_913580, EPI_ISL_913581, EPI_ISL_913582, EPI_ISL_913583, EPI_ISL_913584, EPI_ISL_913585 | | | |
| see above | M Health Fairview | Minnesota Department of Health, Public Health Laboratory | Alexandra Lorentz, Jacob Garfin, Matt Plumb, and Xiong Wang |
| EPI_ISL_913586, EPI_ISL_913587, EPI_ISL_913588, EPI_ISL_913589, EPI_ISL_913590, EPI_ISL_913591 | Vault Health | Minnesota Department of Health, Public Health Laboratory | Alexandra Lorentz, Jacob Garfin, Matt Plumb, and Xiong Wang |
| EPI_ISL_913611, EPI_ISL_913612, EPI_ISL_913613, EPI_ISL_913614, EPI_ISL_913615, EPI_ISL_913616, EPI_ISL_913617, EPI_ISL_913619, EPI_ISL_913620, EPI_ISL_913621, EPI_ISL_913623, EPI_ISL_913624, EPI_ISL_913626, EPI_ISL_913627, EPI_ISL_913628, EPI_ISL_913629, EPI_ISL_913630, EPI_ISL_913631, EPI_ISL_913632, EPI_ISL_913633, EPI_ISL_913634, EPI_ISL_913635, EPI_ISL_913636, EPI_ISL_913637, EPI_ISL_913639, EPI_ISL_913640, EPI_ISL_913641, EPI_ISL_913642, EPI_ISL_913643, EPI_ISL_913644, EPI_ISL_913645, EPI_ISL_913646, EPI_ISL_913648, EPI_ISL_913651, EPI_ISL_913652, EPI_ISL_913653, EPI_ISL_913654, EPI_ISL_913655, EPI_ISL_913656, EPI_ISL_913657, EPI_ISL_913658, EPI_ISL_913659, EPI_ISL_913660, EPI_ISL_913661, EPI_ISL_913662, EPI_ISL_913663, EPI_ISL_913664, EPI_ISL_913665 | | | |
| see above | Michigan Department of Health and Human Services, Bureau of Laboratories | Michigan Department of Health and Human Services, Bureau of Laboratories | Blankenship HM, Riner D, Soehnlen MK |
| EPI_ISL_913667 | LA Office of Public Health Laboratories | Centers for Disease Control and Prevention Division of Viral Diseases, Pathogen Discovery | Ying Tao, Yan Li, Jing Zhang, Krista Queen, Anna Uehara, Peter Cook, Clinton R. Paden, Haibin Wang, Suxiang Tong |
| EPI_ISL_913668, EPI_ISL_913669, EPI_ISL_913670, EPI_ISL_913671, EPI_ISL_913672, EPI_ISL_913673, EPI_ISL_913674, EPI_ISL_913675 | Vault Health | Minnesota Department of Health, Public Health Laboratory | Alexandra Lorentz, Jacob Garfin, Matt Plumb, and Xiong Wang |
| EPI_ISL_913676, EPI_ISL_913677, EPI_ISL_913678, EPI_ISL_913679, EPI_ISL_913680, EPI_ISL_913681, EPI_ISL_913682, EPI_ISL_913683, EPI_ISL_913684, EPI_ISL_913685, EPI_ISL_913686, EPI_ISL_913687, EPI_ISL_913688, EPI_ISL_913689, EPI_ISL_913690, EPI_ISL_913691, EPI_ISL_913692, EPI_ISL_913693, EPI_ISL_913694, EPI_ISL_913695, EPI_ISL_913696, EPI_ISL_913697, EPI_ISL_913698, EPI_ISL_913699, EPI_ISL_913700, EPI_ISL_913701, EPI_ISL_913702, EPI_ISL_913703, EPI_ISL_913704, EPI_ISL_913705, EPI_ISL_913706, EPI_ISL_913707, EPI_ISL_913708, EPI_ISL_913709, EPI_ISL_913710, EPI_ISL_913711, EPI_ISL_913712, EPI_ISL_913713, EPI_ISL_913714, EPI_ISL_913715, EPI_ISL_913716, EPI_ISL_913717, EPI_ISL_913718, EPI_ISL_913719, EPI_ISL_913720, EPI_ISL_913721, EPI_ISL_913722, EPI_ISL_913723, EPI_ISL_913724, EPI_ISL_913725 | | | |
| see above | Minnesota Department of Health, Public Health Laboratory | Minnesota Department of Health, Public Health Laboratory | Alexandra Lorentz, Jacob Garfin, Matt Plumb, and Xiong Wang |
| EPI_ISL_913726 | Missouri State Public Health Laboratory | Minnesota Department of Health, Public Health Laboratory | Alexandra Lorentz, Jacob Garfin, Matt Plumb, and Xiong Wang |
| EPI_ISL_913727, EPI_ISL_913728, EPI_ISL_913729, EPI_ISL_913730, EPI_ISL_913731, EPI_ISL_913732, EPI_ISL_913733, EPI_ISL_913734, EPI_ISL_913735, EPI_ISL_913736 | M Health Fairview | Minnesota Department of Health, Public Health Laboratory | Alexandra Lorentz, Jacob Garfin, Matt Plumb, and Xiong Wang |
| EPI_ISL_913737, EPI_ISL_913738, EPI_ISL_913739 | Minnesota Department of Health, Public Health Laboratory | Minnesota Department of Health, Public Health Laboratory | Alexandra Lorentz, Jacob Garfin, Matt Plumb, and Xiong Wang |
| EPI_ISL_913740, EPI_ISL_913741, EPI_ISL_913742, EPI_ISL_913744, EPI_ISL_913745, EPI_ISL_913746, EPI_ISL_913747 | DC Public Health Lab/ Dept. of Forensic Sciences | DC Public Health Lab/ Dept. of Forensic Sciences | Scott Nguyen, Elizabeth Zelaya, Connie Maza, Monica Mann, Brittany Hamilton, David Payne, Jocelyn Hauser |
| EPI_ISL_913771, EPI_ISL_913774, EPI_ISL_913778, EPI_ISL_913782, EPI_ISL_913788, EPI_ISL_913791, EPI_ISL_913795, EPI_ISL_913798, EPI_ISL_913802, EPI_ISL_913805, EPI_ISL_913808, EPI_ISL_913811, EPI_ISL_913815, EPI_ISL_913818, EPI_ISL_913822, EPI_ISL_913825, EPI_ISL_913829, EPI_ISL_913832, EPI_ISL_913836, EPI_ISL_913839, EPI_ISL_913842, EPI_ISL_913846, EPI_ISL_913849, EPI_ISL_913852, EPI_ISL_913856, EPI_ISL_913859, EPI_ISL_913862, EPI_ISL_913866, EPI_ISL_913870, EPI_ISL_913873, EPI_ISL_913877, EPI_ISL_913879, EPI_ISL_913880, EPI_ISL_913881, EPI_ISL_913882, EPI_ISL_913883, EPI_ISL_913884, EPI_ISL_913885, EPI_ISL_913886, EPI_ISL_913887, EPI_ISL_913888, EPI_ISL_913890, EPI_ISL_913891, EPI_ISL_913892, EPI_ISL_913893, EPI_ISL_913894, EPI_ISL_913895, EPI_ISL_913896, EPI_ISL_913897, EPI_ISL_913899, EPI_ISL_913900, EPI_ISL_913901, EPI_ISL_913902, EPI_ISL_913903, EPI_ISL_913904, EPI_ISL_913905, EPI_ISL_913906, EPI_ISL_913907, EPI_ISL_913908, EPI_ISL_913909, EPI_ISL_913910 | | | |
| see above | TGen North | TGen North | "Jolene Bowers, Megan Folkerts, Chris French, Hayley Yaglom, Ashlyn Pfeiffer, Darrin Lemmer, Dave Engelthaler, The Arizona COVID Genomics Union (ACGU)" |

| | | | |
|---|---|---|---|
| EPI_ISL_913911, EPI_ISL_913912, EPI_ISL_913913, EPI_ISL_913914, EPI_ISL_913915, EPI_ISL_913916, EPI_ISL_913917, EPI_ISL_913918, EPI_ISL_913919, EPI_ISL_913920, EPI_ISL_913921, EPI_ISL_913922, EPI_ISL_913923, EPI_ISL_913924, EPI_ISL_913925, EPI_ISL_913926, EPI_ISL_913927, EPI_ISL_913928, EPI_ISL_913929, EPI_ISL_913930, EPI_ISL_913931, EPI_ISL_913932, EPI_ISL_913933, EPI_ISL_913934, EPI_ISL_913935, EPI_ISL_913936, EPI_ISL_913937, EPI_ISL_913938, EPI_ISL_913939, EPI_ISL_913940, EPI_ISL_913941, EPI_ISL_913942, EPI_ISL_913943, EPI_ISL_913944, EPI_ISL_913945, EPI_ISL_913946, EPI_ISL_913947, EPI_ISL_913948, EPI_ISL_913949, EPI_ISL_913950, EPI_ISL_913951, EPI_ISL_913952, EPI_ISL_913953, EPI_ISL_913954, EPI_ISL_913955, EPI_ISL_913956, EPI_ISL_913957, EPI_ISL_913958, EPI_ISL_913959, EPI_ISL_913960, EPI_ISL_913961, EPI_ISL_913962, EPI_ISL_913963, EPI_ISL_913964, EPI_ISL_913965, EPI_ISL_913966, EPI_ISL_913967, EPI_ISL_913969, EPI_ISL_913970, EPI_ISL_913971, EPI_ISL_913972, EPI_ISL_913973, EPI_ISL_913974, EPI_ISL_913975, EPI_ISL_913976, EPI_ISL_913977, EPI_ISL_913978, EPI_ISL_913979, EPI_ISL_913980, EPI_ISL_913981, EPI_ISL_913982, EPI_ISL_913983, EPI_ISL_913984 | | | |
| see above | Instituto de Diagnostico y Referencia Epidemiologicos INDRE_RNLSP | Instituto de Diagnostico y Referencia Epidemiologicos (INDRE) | Claudia Wong-Arambula, Abril Rodriguez-Maldonado, Fabiola Garces-Ayala, Adnan Araiza-Rodriguez, David Fragoso-Fonseca, Sergio Rangel-Guerrero, Mayra Jimenez-Morales, Nancy Munoz-Hernandez, Natividad Cruz-Ortiz, Tatiana Nunez-Garcia, Gisela Barrera-Badillo, Lucia Hernandez-Rivas, Irma Lopez-Martinez, Ernesto Ramirez-Gonzalez. |
| EPI_ISL_913985, EPI_ISL_913987, EPI_ISL_913988, EPI_ISL_913989 | TX DSHS, Lab Services Section MC 1947 | Pathogen Discovery, Respiratory Viruses Branch, Division of Viral Diseases, Centers for Disease Control and Prevention | Ying Tao, Yan Li, Jing Zhang, Krista Queen, Anna Uehara, Peter Cook, Clinton R. Paden, Haibin Wang, Suxiang Tong |
| EPI_ISL_913990 | IL Department of Public Health Chicago Laboratory | Pathogen Discovery, Respiratory Viruses Branch, Division of Viral Diseases, Centers for Disease Control and Prevention | Ying Tao, Yan Li, Jing Zhang, Krista Queen, Anna Uehara, Peter Cook, Clinton R. Paden, Haibin Wang, Suxiang Tong |
| EPI_ISL_913991 | IL Dept. of Public Health Springfield Laboratory | Pathogen Discovery, Respiratory Viruses Branch, Division of Viral Diseases, Centers for Disease Control and Prevention | Ying Tao, Yan Li, Jing Zhang, Krista Queen, Anna Uehara, Peter Cook, Clinton R. Paden, Haibin Wang, Suxiang Tong |
| EPI_ISL_913992, EPI_ISL_913993, EPI_ISL_913994 | TN Division of Laboratory Services | Pathogen Discovery, Respiratory Viruses Branch, Division of Viral Diseases, Centers for Disease Control and Prevention | Ying Tao, Yan Li, Jing Zhang, Krista Queen, Anna Uehara, Peter Cook, Clinton R. Paden, Haibin Wang, Suxiang Tong |
| EPI_ISL_913995, EPI_ISL_913996 | IL Dept. of Public Health Springfield Laboratory | Pathogen Discovery, Respiratory Viruses Branch, Division of Viral Diseases, Centers for Disease Control and Prevention | Ying Tao, Yan Li, Jing Zhang, Krista Queen, Anna Uehara, Peter Cook, Clinton R. Paden, Haibin Wang, Suxiang Tong |
| EPI_ISL_913997, EPI_ISL_913998 | LA Office of Public Health Laboratories | Pathogen Discovery, Respiratory Viruses Branch, Division of Viral Diseases, Centers for Disease Control and Prevention | Ying Tao, Yan Li, Jing Zhang, Krista Queen, Anna Uehara, Peter Cook, Clinton R. Paden, Haibin Wang, Suxiang Tong |
| EPI_ISL_913999 | TX DSHS, Lab Services Section MC 1947 | Pathogen Discovery, Respiratory Viruses Branch, Division of Viral Diseases, Centers for Disease Control and Prevention | Ying Tao, Yan Li, Jing Zhang, Krista Queen, Anna Uehara, Peter Cook, Clinton R. Paden, Haibin Wang, Suxiang Tong |
| EPI_ISL_914000, EPI_ISL_914001, EPI_ISL_914002 | Quest Diagnostics Tucker GA | Pathogen Discovery, Respiratory Viruses Branch, Division of Viral Diseases, Centers for Disease Control and Prevention | Ying Tao, Yan Li, Jing Zhang, Krista Queen, Anna Uehara, Peter Cook, Clinton R. Paden, Haibin Wang, Suxiang Tong |
| EPI_ISL_914003, EPI_ISL_914004, EPI_ISL_914005, EPI_ISL_914006, EPI_ISL_914007, EPI_ISL_914008, EPI_ISL_914009 | Clinical Pathology Lab | Pathogen Discovery, Respiratory Viruses Branch, Division of Viral Diseases, Centers for Disease Control and Prevention | Ying Tao, Yan Li, Jing Zhang, Krista Queen, Anna Uehara, Peter Cook, Clinton R. Paden, Haibin Wang, Suxiang Tong |
| EPI_ISL_914010, EPI_ISL_914011 | TMC/Pathology | Pathogen Discovery, Respiratory Viruses Branch, Division of Viral Diseases, Centers for Disease Control and Prevention | Ying Tao, Yan Li, Jing Zhang, Krista Queen, Anna Uehara, Peter Cook, Clinton R. Paden, Haibin Wang, Suxiang Tong |
| EPI_ISL_914012 | LA Office of Public Health Laboratories | Pathogen Discovery, Respiratory Viruses Branch, Division of Viral Diseases, Centers for Disease Control and Prevention | Ying Tao, Yan Li, Jing Zhang, Krista Queen, Anna Uehara, Peter Cook, Clinton R. Paden, Haibin Wang, Suxiang Tong |
| EPI_ISL_914013, EPI_ISL_914014, EPI_ISL_914015, EPI_ISL_914016, EPI_ISL_914017, EPI_ISL_914018, EPI_ISL_914019, EPI_ISL_914020, EPI_ISL_914021, EPI_ISL_914022, EPI_ISL_914023, EPI_ISL_914024, EPI_ISL_914025, EPI_ISL_914026, EPI_ISL_914027, EPI_ISL_914028, EPI_ISL_914029 | | | |
| see above | IL Department of Public Health Chicago Laboratory | Pathogen Discovery, Respiratory Viruses Branch, Division of Viral Diseases, Centers for Disease Control and Prevention | Ying Tao, Yan Li, Jing Zhang, Krista Queen, Anna Uehara, Peter Cook, Clinton R. Paden, Haibin Wang, Suxiang Tong |
| EPI_ISL_914030, EPI_ISL_914031, EPI_ISL_914032, EPI_ISL_914033, EPI_ISL_914034 | IL Department of Public Health Springfield Laboratory | Pathogen Discovery, Respiratory Viruses Branch, Division of Viral Diseases, Centers for Disease Control and Prevention | Ying Tao, Yan Li, Jing Zhang, Krista Queen, Anna Uehara, Peter Cook, Clinton R. Paden, Haibin Wang, Suxiang Tong |
| EPI_ISL_914040, EPI_ISL_914041, EPI_ISL_914042, EPI_ISL_914044, EPI_ISL_914045, EPI_ISL_914046, EPI_ISL_914047, EPI_ISL_914048, EPI_ISL_914049, EPI_ISL_914050, EPI_ISL_914051, EPI_ISL_914052, EPI_ISL_914055, EPI_ISL_914056, EPI_ISL_914057, EPI_ISL_914058, EPI_ISL_914059, EPI_ISL_914061, EPI_ISL_914062, EPI_ISL_914063, EPI_ISL_914064, EPI_ISL_914065, EPI_ISL_914066, EPI_ISL_914067, EPI_ISL_914068, EPI_ISL_914069, EPI_ISL_914070, EPI_ISL_914072, EPI_ISL_914073, EPI_ISL_914074, EPI_ISL_914076, EPI_ISL_914077, EPI_ISL_914078, EPI_ISL_914079, EPI_ISL_914080, EPI_ISL_914081, EPI_ISL_914084, EPI_ISL_914085, EPI_ISL_914087, EPI_ISL_914088, EPI_ISL_914089, EPI_ISL_914090, EPI_ISL_914091, EPI_ISL_914092, EPI_ISL_914094, EPI_ISL_914095, EPI_ISL_914096, EPI_ISL_914097, EPI_ISL_914098, EPI_ISL_914099, EPI_ISL_914100, EPI_ISL_914101, EPI_ISL_914102, EPI_ISL_914103, EPI_ISL_914104, EPI_ISL_914105, EPI_ISL_914106, EPI_ISL_914107, EPI_ISL_914108, EPI_ISL_914110, EPI_ISL_914111, EPI_ISL_914112, EPI_ISL_914113, EPI_ISL_914114, EPI_ISL_914115, EPI_ISL_914116, EPI_ISL_914117, EPI_ISL_914119, EPI_ISL_914120, EPI_ISL_914121, EPI_ISL_914122, EPI_ISL_914123, EPI_ISL_914124, EPI_ISL_914125, EPI_ISL_914126, EPI_ISL_914127, EPI_ISL_914128, EPI_ISL_914129, EPI_ISL_914130, EPI_ISL_914131, EPI_ISL_914132, EPI_ISL_914133, EPI_ISL_914134, EPI_ISL_914135, EPI_ISL_914136, EPI_ISL_914137, EPI_ISL_914138, EPI_ISL_914139, EPI_ISL_914140, EPI_ISL_914141, EPI_ISL_914142, EPI_ISL_914143, EPI_ISL_914144, EPI_ISL_914145, EPI_ISL_914146, EPI_ISL_914147, EPI_ISL_914148, EPI_ISL_914149, EPI_ISL_914150, EPI_ISL_914151, EPI_ISL_914152, EPI_ISL_914153, EPI_ISL_914154, EPI_ISL_914155, EPI_ISL_914156, EPI_ISL_914157, EPI_ISL_914158, EPI_ISL_914159, EPI_ISL_914160, EPI_ISL_914161, EPI_ISL_914163, EPI_ISL_914164, EPI_ISL_914165, EPI_ISL_914166, EPI_ISL_914167, EPI_ISL_914168, EPI_ISL_914169, EPI_ISL_914170, EPI_ISL_914171, EPI_ISL_914172, EPI_ISL_914173, EPI_ISL_914174, EPI_ISL_914175, EPI_ISL_914177, EPI_ISL_914178, EPI_ISL_914179, EPI_ISL_914180, EPI_ISL_914181, EPI_ISL_914182, EPI_ISL_914184, EPI_ISL_914185, EPI_ISL_914186, EPI_ISL_914187, EPI_ISL_914188, EPI_ISL_914189, EPI_ISL_914190, EPI_ISL_914191, EPI_ISL_914192, EPI_ISL_914193, EPI_ISL_914194, EPI_ISL_914195, EPI_ISL_914196, EPI_ISL_914198, EPI_ISL_914199, EPI_ISL_914200, EPI_ISL_914201, EPI_ISL_914202, EPI_ISL_914203, EPI_ISL_914204, EPI_ISL_914205, EPI_ISL_914206, EPI_ISL_914207, EPI_ISL_914208, EPI_ISL_914209, EPI_ISL_914210, EPI_ISL_914211, EPI_ISL_914212, EPI_ISL_914213, EPI_ISL_914214, EPI_ISL_914215, EPI_ISL_914216, EPI_ISL_914218, EPI_ISL_914219, EPI_ISL_914220, EPI_ISL_914221, EPI_ISL_914222, EPI_ISL_914223, EPI_ISL_914224, EPI_ISL_914225, EPI_ISL_914226, EPI_ISL_914227, EPI_ISL_914228, EPI_ISL_914229, EPI_ISL_914230, EPI_ISL_914231, EPI_ISL_914232, EPI_ISL_914233, EPI_ISL_914234, EPI_ISL_914235, EPI_ISL_914236, EPI_ISL_914237, EPI_ISL_914238, EPI_ISL_914239, EPI_ISL_914240, EPI_ISL_914241, EPI_ISL_914243, EPI_ISL_914244, EPI_ISL_914245, EPI_ISL_914246, EPI_ISL_914250, EPI_ISL_914251, EPI_ISL_914252, EPI_ISL_914253, EPI_ISL_914254, EPI_ISL_914255, EPI_ISL_914256, EPI_ISL_914257, EPI_ISL_914258, EPI_ISL_914259, EPI_ISL_914261, EPI_ISL_914262, EPI_ISL_914263, EPI_ISL_914264, EPI_ISL_914265, EPI_ISL_914266, EPI_ISL_914267, EPI_ISL_914268, EPI_ISL_914269, EPI_ISL_914270, EPI_ISL_914271, EPI_ISL_914272, EPI_ISL_914273, EPI_ISL_914274, EPI_ISL_914275, EPI_ISL_914276, EPI_ISL_914277, EPI_ISL_914278, EPI_ISL_914279, EPI_ISL_914281, EPI_ISL_914282, EPI_ISL_914283, EPI_ISL_914284, EPI_ISL_914285, EPI_ISL_914286, EPI_ISL_914288, EPI_ISL_914289, EPI_ISL_914290, EPI_ISL_914291, EPI_ISL_914292, EPI_ISL_914293, EPI_ISL_914294, EPI_ISL_914295, EPI_ISL_914296, EPI_ISL_914297, EPI_ISL_914298, EPI_ISL_914299, EPI_ISL_914300, EPI_ISL_914301, EPI_ISL_914302, EPI_ISL_914303, EPI_ISL_914304, EPI_ISL_914305, EPI_ISL_914306, EPI_ISL_914307, EPI_ISL_914308, EPI_ISL_914309, EPI_ISL_914310, EPI_ISL_914311, EPI_ISL_914313, EPI_ISL_914314, EPI_ISL_914315, EPI_ISL_914316, EPI_ISL_914317, EPI_ISL_914318, EPI_ISL_914319, EPI_ISL_914320, EPI_ISL_914321, EPI_ISL_914322, EPI_ISL_914323, EPI_ISL_914324, EPI_ISL_914325, EPI_ISL_914326, EPI_ISL_914327, EPI_ISL_914328, EPI_ISL_914329, EPI_ISL_914330, EPI_ISL_914331, EPI_ISL_914332, EPI_ISL_914333, EPI_ISL_914334, EPI_ISL_914335, EPI_ISL_914336, EPI_ISL_914337, EPI_ISL_914338, EPI_ISL_914339, EPI_ISL_914340, EPI_ISL_914341, EPI_ISL_914342, EPI_ISL_914343, EPI_ISL_914344, EPI_ISL_914346, EPI_ISL_914347, EPI_ISL_914348, EPI_ISL_914349, EPI_ISL_914350, EPI_ISL_914351, EPI_ISL_914352, EPI_ISL_914353, EPI_ISL_914354, EPI_ISL_914356, EPI_ISL_914357, EPI_ISL_914358, EPI_ISL_914359, EPI_ISL_914360, EPI_ISL_914361, EPI_ISL_914362, EPI_ISL_914363, EPI_ISL_914364, EPI_ISL_914365, EPI_ISL_914366, EPI_ISL_914367, EPI_ISL_914368, EPI_ISL_914369, EPI_ISL_914371, EPI_ISL_914372, EPI_ISL_914373, EPI_ISL_914374, EPI_ISL_914375, EPI_ISL_914377, EPI_ISL_914378, EPI_ISL_914379, EPI_ISL_914380, EPI_ISL_914382, EPI_ISL_914383, EPI_ISL_914384, EPI_ISL_914385, EPI_ISL_914388, EPI_ISL_914389, EPI_ISL_914390, EPI_ISL_914391, EPI_ISL_914392, EPI_ISL_914393, EPI_ISL_914394, EPI_ISL_914395, EPI_ISL_914396, EPI_ISL_914397, EPI_ISL_914400, EPI_ISL_914401, EPI_ISL_914402, EPI_ISL_914403, EPI_ISL_914404, EPI_ISL_914406, EPI_ISL_914408, EPI_ISL_914409, EPI_ISL_914410, EPI_ISL_914411, EPI_ISL_914412, EPI_ISL_914414, EPI_ISL_914415, EPI_ISL_914416, EPI_ISL_914418, EPI_ISL_914419, EPI_ISL_914421, EPI_ISL_914422, EPI_ISL_914424, EPI_ISL_914426, EPI_ISL_914429, EPI_ISL_914430, EPI_ISL_914431, EPI_ISL_914432, EPI_ISL_914433, EPI_ISL_914435, EPI_ISL_914436, EPI_ISL_914437, EPI_ISL_914438, EPI_ISL_914439, EPI_ISL_914440, EPI_ISL_914442, EPI_ISL_914443, EPI_ISL_914444, EPI_ISL_914445, EPI_ISL_914447, EPI_ISL_914448, EPI_ISL_914449, EPI_ISL_914450, EPI_ISL_914451, EPI_ISL_914452, EPI_ISL_914453, EPI_ISL_914454, EPI_ISL_914455, EPI_ISL_914456, EPI_ISL_914457, EPI_ISL_914458, EPI_ISL_914459, EPI_ISL_914460, EPI_ISL_914461, EPI_ISL_914462, EPI_ISL_914463, EPI_ISL_914464, EPI_ISL_914465, EPI_ISL_914466, EPI_ISL_914467, EPI_ISL_914468, EPI_ISL_914469, EPI_ISL_914470, EPI_ISL_914471, EPI_ISL_914472, EPI_ISL_914473, EPI_ISL_914474, EPI_ISL_914475, EPI_ISL_914476, EPI_ISL_914477, EPI_ISL_914478, EPI_ISL_914479, EPI_ISL_914481, EPI_ISL_914482, EPI_ISL_914483, EPI_ISL_914484, EPI_ISL_914485, EPI_ISL_914486, EPI_ISL_914487, EPI_ISL_914488, EPI_ISL_914489, EPI_ISL_914490, EPI_ISL_914491, EPI_ISL_914492, EPI_ISL_914493, EPI_ISL_914494, EPI_ISL_914495, EPI_ISL_914496, EPI_ISL_914498, EPI_ISL_914500, EPI_ISL_914501, EPI_ISL_914502, EPI_ISL_914503, EPI_ISL_914504, EPI_ISL_914505, EPI_ISL_914506, EPI_ISL_914507, EPI_ISL_914508, EPI_ISL_914509, EPI_ISL_914510, EPI_ISL_914511, EPI_ISL_914512, EPI_ISL_914513, EPI_ISL_914514, EPI_ISL_914515, EPI_ISL_914516, EPI_ISL_914518, EPI_ISL_914519, EPI_ISL_914520, EPI_ISL_914521, EPI_ISL_914522, EPI_ISL_914523, EPI_ISL_914524, EPI_ISL_914525, EPI_ISL_914526, EPI_ISL_914527, EPI_ISL_914528, EPI_ISL_914529, EPI_ISL_914534, EPI_ISL_914535, EPI_ISL_914536, EPI_ISL_914537, EPI_ISL_914538, EPI_ISL_914540, EPI_ISL_914543, EPI_ISL_914544, EPI_ISL_914547, EPI_ISL_914548, EPI_ISL_914550, EPI_ISL_914552, EPI_ISL_914553, EPI_ISL_914555, EPI_ISL_914556, EPI_ISL_914557, EPI_ISL_914558, EPI_ISL_914559, EPI_ISL_914560, EPI_ISL_914562, EPI_ISL_914563, EPI_ISL_914564, EPI_ISL_914565, EPI_ISL_914566, EPI_ISL_914568, EPI_ISL_914569, EPI_ISL_914570, EPI_ISL_914571, EPI_ISL_914572, EPI_ISL_914575, EPI_ISL_914576, EPI_ISL_914578, EPI_ISL_914579, EPI_ISL_914580, EPI_ISL_914581, EPI_ISL_914584, EPI_ISL_914585, EPI_ISL_914586, EPI_ISL_914587, EPI_ISL_914588, EPI_ISL_914589, EPI_ISL_914590, EPI_ISL_914591, EPI_ISL_914593, EPI_ISL_914594, EPI_ISL_914595, EPI_ISL_914596, EPI_ISL_914599, EPI_ISL_914600, EPI_ISL_914601, EPI_ISL_914602, EPI_ISL_914603, EPI_ISL_914604, EPI_ISL_914605, EPI_ISL_914606, EPI_ISL_914608, EPI_ISL_914609, EPI_ISL_914610, EPI_ISL_914611, EPI_ISL_914613, EPI_ISL_914614, EPI_ISL_914615, EPI_ISL_914616, EPI_ISL_914617, EPI_ISL_914618, EPI_ISL_914619, EPI_ISL_914620, EPI_ISL_914623, EPI_ISL_914624, EPI_ISL_914625, EPI_ISL_914627, EPI_ISL_914628, EPI_ISL_914631, EPI_ISL_914632, EPI_ISL_914633, EPI_ISL_914635, EPI_ISL_914636, EPI_ISL_914637 | | | |
| see above | TGen North | TGen North | "Jolene Bowers, Megan Folkerts, Chris French, Hayley Yaglom, Ashlyn Pfeiffer, Darrin Lemmer, Dave Engelthaler, The Arizona COVID Genomics Union (ACGU)" |
| EPI_ISL_914638, EPI_ISL_914639, EPI_ISL_914641, EPI_ISL_914642, EPI_ISL_914643, EPI_ISL_914644, EPI_ISL_914646 | Santa Clara County Public Health Laboratory | Santa Clara County Public Health Laboratory | Santa Clara County Public Health Department |
| EPI_ISL_914647, EPI_ISL_914648, EPI_ISL_914649, EPI_ISL_914650, EPI_ISL_914651, EPI_ISL_914652, EPI_ISL_914653, EPI_ISL_914654, EPI_ISL_914655, EPI_ISL_914656, EPI_ISL_914657, EPI_ISL_914658, EPI_ISL_914659, EPI_ISL_914660, EPI_ISL_914661, EPI_ISL_914662, EPI_ISL_914663, EPI_ISL_914664, EPI_ISL_914665, EPI_ISL_914666, EPI_ISL_914667, EPI_ISL_914668, EPI_ISL_914669, EPI_ISL_914670, EPI_ISL_914671, EPI_ISL_914672, EPI_ISL_914673, EPI_ISL_914674, EPI_ISL_914675, EPI_ISL_914676, EPI_ISL_914677, EPI_ISL_914678, EPI_ISL_914679, EPI_ISL_914680, EPI_ISL_914681, EPI_ISL_914682, EPI_ISL_914683, EPI_ISL_914684, EPI_ISL_914685, EPI_ISL_914686, EPI_ISL_914687, EPI_ISL_914688, EPI_ISL_914689, EPI_ISL_914690, EPI_ISL_914691, EPI_ISL_914692, EPI_ISL_914693, EPI_ISL_914694, EPI_ISL_914695, EPI_ISL_914696, EPI_ISL_914697, EPI_ISL_914698, EPI_ISL_914699, EPI_ISL_914700, EPI_ISL_914701 | | | |
| see above | Utah Public Health Laboratory | Utah Public Health Laboratory | Erin L. Young, Kelly F. Oakeson, Tara Gallagher |
| EPI_ISL_914702, EPI_ISL_914703, EPI_ISL_914704, EPI_ISL_914705, EPI_ISL_914706, EPI_ISL_914707, EPI_ISL_914708, EPI_ISL_914709, EPI_ISL_914710, EPI_ISL_914711, EPI_ISL_914712, EPI_ISL_914713, EPI_ISL_914714, EPI_ISL_914715, EPI_ISL_914716, EPI_ISL_914717, EPI_ISL_914718, EPI_ISL_914719, EPI_ISL_914720, EPI_ISL_914721, EPI_ISL_914722, EPI_ISL_914723, EPI_ISL_914724 | | | |
| see above | Maryland Public Health Laboratory (MD PHL) | Maryland Public Health Laboratory (MD PHL) | Maryland Department of Health Laboratories Administration |
| EPI_ISL_914725, EPI_ISL_914726, EPI_ISL_914727, EPI_ISL_914730, EPI_ISL_914731, EPI_ISL_914732, EPI_ISL_914733, EPI_ISL_914734, EPI_ISL_914735, EPI_ISL_914736, EPI_ISL_914737, EPI_ISL_914738, EPI_ISL_914739, EPI_ISL_914740, EPI_ISL_914741, EPI_ISL_914752, EPI_ISL_914753, EPI_ISL_914754, EPI_ISL_914755, EPI_ISL_914756, EPI_ISL_914757, EPI_ISL_914758, EPI_ISL_914759, EPI_ISL_914760, EPI_ISL_914761, EPI_ISL_914762, EPI_ISL_914763, EPI_ISL_914765, EPI_ISL_914766, EPI_ISL_914767, EPI_ISL_914768, EPI_ISL_914769, EPI_ISL_914770, EPI_ISL_914772, EPI_ISL_914773, EPI_ISL_914774, EPI_ISL_914775, EPI_ISL_914776, EPI_ISL_914777, EPI_ISL_914778, EPI_ISL_914779, EPI_ISL_914780, EPI_ISL_914781, EPI_ISL_914782, EPI_ISL_914784, EPI_ISL_914785, EPI_ISL_914786, EPI_ISL_914787, EPI_ISL_914788, EPI_ISL_914789, EPI_ISL_914790, EPI_ISL_914791, EPI_ISL_914792, EPI_ISL_914793 | | | |
| see above | Wyoming Public Health Laboratory | Wyoming Public Health Laboratory | Noah Hull, Taylor Fearing, Lynette Gumbleton, Channing Weber, Ashley Norberg, Bailey Bowcutt, and Wanda Manley |
| EPI_ISL_914841, EPI_ISL_914842, EPI_ISL_914843, EPI_ISL_914844, EPI_ISL_914845, EPI_ISL_914846, EPI_ISL_914847, EPI_ISL_914848, EPI_ISL_914849, EPI_ISL_914850, EPI_ISL_914851, EPI_ISL_914852, EPI_ISL_914853, EPI_ISL_914854, EPI_ISL_914855, EPI_ISL_914856, EPI_ISL_914857, EPI_ISL_914858, EPI_ISL_914859, EPI_ISL_914860, EPI_ISL_914861, EPI_ISL_914862, EPI_ISL_914863, EPI_ISL_914864, EPI_ISL_914865 | | | |
| see above | Virginia Division of Consolidated Laboratory Services (DCLS) | Virginia Division of Consolidated Laboratory Services (DCLS) | Virginia DCLS |

| | | | |
|---|---|---|---|
| EPI_ISL_914866, EPI_ISL_914867, EPI_ISL_914868, EPI_ISL_914869, EPI_ISL_914870, EPI_ISL_914871, EPI_ISL_914872, EPI_ISL_914873, EPI_ISL_914874 | Kansas Health and Environmental Lab | Kansas Health and Environmental Lab | Mike Grose, Paige Drury, Carissa Robertson, Ben Olsen, and Phil Adam |
| EPI_ISL_914878, EPI_ISL_914879, EPI_ISL_914880 | Instituto de Diagnostico y Referencia Epidemiologicos INDRE_RNLSP | Instituto de Diagnostico y Referencia Epidemiologicos (INDRE) | Claudia Wong-Arambula, Abril Rodriguez-Maldonado, Fabiola Garces-Ayala, Adnan Araiza-Rodriguez, David Fragoso-Fonseca, Sergio Rangel-Guerrero, Mayra Jimenez-Morales, Nancy Munoz-Hernandez, Natividad Cruz-Ortiz, Tatiana Nunez-Garcia, Gisela Barrera-Badillo, Lucia Hernandez-Rivas, Irma Lopez-Martinez, Ernesto Ramirez-Gonzalez. |
| EPI_ISL_914890, EPI_ISL_914891, EPI_ISL_914892, EPI_ISL_914893, EPI_ISL_914894, EPI_ISL_914895, EPI_ISL_914896, EPI_ISL_914897, EPI_ISL_914898, EPI_ISL_914899, EPI_ISL_914900, EPI_ISL_914901, EPI_ISL_914902, EPI_ISL_914903, EPI_ISL_914904, EPI_ISL_914905, EPI_ISL_914906, EPI_ISL_914907, EPI_ISL_914908, EPI_ISL_914909, EPI_ISL_914910, EPI_ISL_914911, EPI_ISL_914912, EPI_ISL_914913, EPI_ISL_914914, EPI_ISL_914915, EPI_ISL_914916, EPI_ISL_914917, EPI_ISL_914918, EPI_ISL_914919, EPI_ISL_914920, EPI_ISL_914921, EPI_ISL_914922, EPI_ISL_914923, EPI_ISL_914924, EPI_ISL_914925, EPI_ISL_914926, EPI_ISL_914927, EPI_ISL_914928, EPI_ISL_914929, EPI_ISL_914930, EPI_ISL_914931, EPI_ISL_914932, EPI_ISL_914933, EPI_ISL_914934, EPI_ISL_914935, EPI_ISL_914936, EPI_ISL_914937, EPI_ISL_914938, EPI_ISL_914939, EPI_ISL_914940, EPI_ISL_914941, EPI_ISL_914942, EPI_ISL_914943, EPI_ISL_914944, EPI_ISL_914945, EPI_ISL_914946, EPI_ISL_914947, EPI_ISL_914948, EPI_ISL_914949, EPI_ISL_914950, EPI_ISL_914951, EPI_ISL_914952, EPI_ISL_914953, EPI_ISL_914954, EPI_ISL_914955, EPI_ISL_914956, EPI_ISL_914957, EPI_ISL_914958, EPI_ISL_914959, EPI_ISL_914960, EPI_ISL_914961, EPI_ISL_914962, EPI_ISL_914963, EPI_ISL_914964, EPI_ISL_914965, EPI_ISL_914966, EPI_ISL_914967, EPI_ISL_914968, EPI_ISL_914969, EPI_ISL_915193, EPI_ISL_915194, EPI_ISL_915195, EPI_ISL_915196, EPI_ISL_915197, EPI_ISL_915198, EPI_ISL_915199, EPI_ISL_915200, EPI_ISL_915201, EPI_ISL_915202, EPI_ISL_915203, EPI_ISL_915204, EPI_ISL_915205, EPI_ISL_915206, EPI_ISL_915207, EPI_ISL_915208, EPI_ISL_915209, EPI_ISL_915210, EPI_ISL_915211, EPI_ISL_915212, EPI_ISL_915213, EPI_ISL_915214, EPI_ISL_915215, EPI_ISL_915216, EPI_ISL_915217, EPI_ISL_915218, EPI_ISL_915219, EPI_ISL_915220, EPI_ISL_915221, EPI_ISL_915222, EPI_ISL_915223, EPI_ISL_915224, EPI_ISL_915225, EPI_ISL_915226, EPI_ISL_915227, EPI_ISL_915228, EPI_ISL_915229, EPI_ISL_915230, EPI_ISL_915231, EPI_ISL_915232, EPI_ISL_915233, EPI_ISL_915234, EPI_ISL_915235, EPI_ISL_915236, EPI_ISL_915237, EPI_ISL_915238, EPI_ISL_915239, EPI_ISL_915240, EPI_ISL_915241, EPI_ISL_915242, EPI_ISL_915243, EPI_ISL_915244, EPI_ISL_915245, EPI_ISL_915246, EPI_ISL_915247, EPI_ISL_915248, EPI_ISL_915249, EPI_ISL_915250, EPI_ISL_915251, EPI_ISL_915252, EPI_ISL_915253, EPI_ISL_915254, EPI_ISL_915255, EPI_ISL_915256, EPI_ISL_915257, EPI_ISL_915258, EPI_ISL_915259, EPI_ISL_915260, EPI_ISL_915261, EPI_ISL_915262, EPI_ISL_915263, EPI_ISL_915264, EPI_ISL_915265, EPI_ISL_915266, EPI_ISL_915267, EPI_ISL_915268, EPI_ISL_915269, EPI_ISL_915270, EPI_ISL_915271, EPI_ISL_915272, EPI_ISL_915273, EPI_ISL_915274, EPI_ISL_915275, EPI_ISL_915276, EPI_ISL_915277, EPI_ISL_915278, EPI_ISL_915279, EPI_ISL_915280, EPI_ISL_915281, EPI_ISL_915282, EPI_ISL_915283, EPI_ISL_915284, EPI_ISL_915285, EPI_ISL_915286, EPI_ISL_915287, EPI_ISL_915288, EPI_ISL_915289, EPI_ISL_915290, EPI_ISL_915291, EPI_ISL_915292, EPI_ISL_915293, EPI_ISL_915294, EPI_ISL_915295, EPI_ISL_915296, EPI_ISL_915297, EPI_ISL_915298, EPI_ISL_915299, EPI_ISL_915300, EPI_ISL_915301, EPI_ISL_915302, EPI_ISL_915303, EPI_ISL_915304, EPI_ISL_915305, EPI_ISL_915306, EPI_ISL_915307, EPI_ISL_915308, EPI_ISL_915309, EPI_ISL_915310, EPI_ISL_915311, EPI_ISL_915312, EPI_ISL_915313, EPI_ISL_915314, EPI_ISL_915315, EPI_ISL_915316, EPI_ISL_915317, EPI_ISL_915318, EPI_ISL_915319, EPI_ISL_915320, EPI_ISL_915321, EPI_ISL_915322, EPI_ISL_915323, EPI_ISL_915324, EPI_ISL_915325, EPI_ISL_915326, EPI_ISL_915327, EPI_ISL_915328, EPI_ISL_915329, EPI_ISL_915330, EPI_ISL_915331, EPI_ISL_915332, EPI_ISL_915333, EPI_ISL_915334, EPI_ISL_915335, EPI_ISL_915336, EPI_ISL_915337, EPI_ISL_915338, EPI_ISL_915339, EPI_ISL_915340, EPI_ISL_915341, EPI_ISL_915342, EPI_ISL_915343, EPI_ISL_915344, EPI_ISL_915345, EPI_ISL_915346, EPI_ISL_915347, EPI_ISL_915348, EPI_ISL_915349, EPI_ISL_915350, EPI_ISL_915351, EPI_ISL_915352, EPI_ISL_915353, EPI_ISL_915354, EPI_ISL_915355, EPI_ISL_915356, EPI_ISL_915357 | | | |
| see above | Quest Diagnostics | Quest Diagnostics | Rosenthal,S.H., Gerasimova,A., Kagan,R.M., Anderson, B., Hua, M., Liu Y., Bernstein, L.E., Livingston, K.E., Perez, A., Shalhout, D.F., Shlyakhter, I.A., Owen, R., Tanpaiboon, P., Lacbawan, F. |
| EPI_ISL_915434, EPI_ISL_915435, EPI_ISL_915436, EPI_ISL_915437 | MD Laboratories | Los Angeles County Public Health Laboratories | P. Hemarajata et al. |
| EPI_ISL_915438 | Los Angeles County Public Health Laboratories | Los Angeles County Public Health Laboratories | P. Hemarajata et al. |
| EPI_ISL_915439 | MD Laboratories | Los Angeles County Public Health Laboratories | P. Hemarajata et al. |
| EPI_ISL_917936, EPI_ISL_917938, EPI_ISL_917941 | Los Angeles County Public Health Laboratories | Los Angeles County Public Health Laboratories | P. Hemarajata et al. |
| EPI_ISL_917943 | Flow Diagnostics Lab | Los Angeles County Public Health Laboratories | P. Hemarajata et al. |
| EPI_ISL_917944, EPI_ISL_917945, EPI_ISL_917946, EPI_ISL_917947, EPI_ISL_917948, EPI_ISL_917949, EPI_ISL_917950 | MD Laboratories | Los Angeles County Public Health Laboratories | P. Hemarajata et al. |
| EPI_ISL_917951, EPI_ISL_917952, EPI_ISL_917953 | Los Angeles County Public Health Laboratories | Los Angeles County Public Health Laboratories | P. Hemarajata et al. |
| EPI_ISL_917954 | Pomona Valley Hospital Medical Center | Los Angeles County Public Health Laboratories | P. Hemarajata et al. |
| EPI_ISL_917955 | Los Angeles County Public Health Laboratories | Los Angeles County Public Health Laboratories | P. Hemarajata et al. |
| EPI_ISL_918177, EPI_ISL_918178, EPI_ISL_918179, EPI_ISL_918180, EPI_ISL_918181, EPI_ISL_918182, EPI_ISL_918183, EPI_ISL_918184, EPI_ISL_918185, EPI_ISL_918186, EPI_ISL_918187, EPI_ISL_918188, EPI_ISL_918189, EPI_ISL_918190, EPI_ISL_918191, EPI_ISL_918192, EPI_ISL_918201, EPI_ISL_918202, EPI_ISL_918203, EPI_ISL_918204, EPI_ISL_918205, EPI_ISL_918206, EPI_ISL_918207, EPI_ISL_918208, EPI_ISL_918209, EPI_ISL_918210, EPI_ISL_918211, EPI_ISL_918212, EPI_ISL_918213, EPI_ISL_918214, EPI_ISL_918215, EPI_ISL_918216, EPI_ISL_918217, EPI_ISL_918218, EPI_ISL_918219, EPI_ISL_918220, EPI_ISL_918221, EPI_ISL_918222, EPI_ISL_918223, EPI_ISL_918224, EPI_ISL_918225, EPI_ISL_918226, EPI_ISL_918227, EPI_ISL_918228, EPI_ISL_918229, EPI_ISL_918230, EPI_ISL_918231, EPI_ISL_918233, EPI_ISL_918234, EPI_ISL_918235, EPI_ISL_918236, EPI_ISL_918237, EPI_ISL_918238, EPI_ISL_918239, EPI_ISL_918240, EPI_ISL_918241, EPI_ISL_918242, EPI_ISL_918243, EPI_ISL_918244, EPI_ISL_918245, EPI_ISL_918246, EPI_ISL_918247, EPI_ISL_918248, EPI_ISL_918249, EPI_ISL_918250, EPI_ISL_918251, EPI_ISL_918252, EPI_ISL_918253, EPI_ISL_918254, EPI_ISL_918255, EPI_ISL_918256, EPI_ISL_918257, EPI_ISL_918258, EPI_ISL_918259, EPI_ISL_918260, EPI_ISL_918261, EPI_ISL_918262 | | | |
| see above | Innovative Genomics Institute, UC Berkeley | Innovative Genomics Institute, UC Berkeley | Stacia Wyman, Haridha Shivram, Phil Frankino, Liana Lareau, Shana McDevitt, Justin Choi |
| EPI_ISL_918485, EPI_ISL_918491, EPI_ISL_918494 | Ohio Department of Health Laboratory | Ohio Department of Health Laboratory | Holmes, Jennifer; Eric Brandt, Keoni Omura, Glen McGillivary, Caitlin McDonnell, Jade Mowery, Stephanie Mccracken, Tyler Payne, Kirtana Ramadugu, Erica Leasure, Brent Lee, Kelsey Florek, Heather Blankenship, Quanta Brown, and Tammy Bannerman |
| EPI_ISL_918495 | Florida Bureau of Public Health Laboratories | Florida Bureau of Public Health Laboratories | Sarah Schmedes, Jason Blanton |
| EPI_ISL_918496 | BSWMC-Marble Falls | BSWMC-Temple Molecular | Ari Rao, Linden Morales, Kimberly Walker, Shelby Hendrickson |
| EPI_ISL_918498, EPI_ISL_918562 | Ohio Department of Health Laboratory | Ohio Department of Health Laboratory | Holmes, Jennifer; Eric Brandt, Keoni Omura, Glen McGillivary, Caitlin McDonnell, Jade Mowery, Stephanie Mccracken, Tyler Payne, Kirtana Ramadugu, Erica Leasure, Brent Lee, Kelsey Florek, Heather Blankenship, Quanta Brown, and Tammy Bannerman |
| EPI_ISL_925048, EPI_ISL_925049, EPI_ISL_925050, EPI_ISL_925051, EPI_ISL_925052, EPI_ISL_925053, EPI_ISL_925054, EPI_ISL_925055, EPI_ISL_925056, EPI_ISL_925057, EPI_ISL_925058, EPI_ISL_925059, EPI_ISL_925060, EPI_ISL_925061, EPI_ISL_925062, EPI_ISL_925063, EPI_ISL_925064, EPI_ISL_925065, EPI_ISL_925066, EPI_ISL_925068, EPI_ISL_925075, EPI_ISL_925078, EPI_ISL_925079, EPI_ISL_925080 | | | |
| see above | TXDSHS | TXDSHS | Bonnie Oh, Anita Pokharel, James Daniel Bonser, Myong Koag, Chung Wang, Rachel Lee, Grace Kubin, Rashmi Tuladhar, Mayela Pedrueza, Maliha Rahman, Jenny Zhang |
| EPI_ISL_925081, EPI_ISL_925082, EPI_ISL_925083, EPI_ISL_925084, EPI_ISL_925085, EPI_ISL_925091, EPI_ISL_925092 | Wyoming Public Health Laboratory | Wyoming Public Health Laboratory | Noah Hull, Taylor Fearing, Lynette Gumbleton, Channing Weber, Ashley Norberg, Bailey Bowcutt, and Wanda Manley |
| EPI_ISL_925137, EPI_ISL_925138, EPI_ISL_925139, EPI_ISL_925140 | Quest Diagnostics | Quest Diagnostics | Rosenthal,S.H., Gerasimova,A., Kagan,R.M., Anderson, B., Hua, M., Liu Y., Bernstein, L.E., Livingston, K.E., Perez, A., Shalhout, D.F., Shlyakhter, I.A., Owen, R., Tanpaiboon, P., Lacbawan, F. |
| EPI_ISL_925141, EPI_ISL_925142, EPI_ISL_925143, EPI_ISL_925144, EPI_ISL_925145, EPI_ISL_925146, EPI_ISL_925147, EPI_ISL_925148, EPI_ISL_925149, EPI_ISL_925150, EPI_ISL_925151, EPI_ISL_925152, EPI_ISL_925153, EPI_ISL_925154, EPI_ISL_925155, EPI_ISL_925156, EPI_ISL_925157, EPI_ISL_925158, EPI_ISL_925159, EPI_ISL_925160, EPI_ISL_925161, EPI_ISL_925162, EPI_ISL_925163, EPI_ISL_925164, EPI_ISL_925165, EPI_ISL_925166, EPI_ISL_925167, EPI_ISL_925168, EPI_ISL_925169, EPI_ISL_925170, EPI_ISL_925171, EPI_ISL_925172, EPI_ISL_925174, EPI_ISL_925175, EPI_ISL_925176, EPI_ISL_925177, EPI_ISL_925178, EPI_ISL_925179, EPI_ISL_925180, EPI_ISL_925181, EPI_ISL_925182, EPI_ISL_925183, EPI_ISL_925184, EPI_ISL_925185, EPI_ISL_925186, EPI_ISL_925187, EPI_ISL_925188, EPI_ISL_925189, EPI_ISL_925190, EPI_ISL_925191, EPI_ISL_925192, EPI_ISL_925193, EPI_ISL_925194, EPI_ISL_925195, EPI_ISL_925196, EPI_ISL_925197, EPI_ISL_925198, EPI_ISL_925199, EPI_ISL_925200, EPI_ISL_925201, EPI_ISL_925202, EPI_ISL_925203, EPI_ISL_925204, EPI_ISL_925205, EPI_ISL_925206, EPI_ISL_925207 | | | |
| see above | Virginia DCLS | Virginia DCLS | Virginia DCLS |
| EPI_ISL_925208 | Baylor Scott & White-Temple | Baylor Scott & White-Temple | Ari Rao, Linden Morales, Kimberly Walker, Marcus Volz, Shelby Hendrickson |
| EPI_ISL_925209, EPI_ISL_925210, EPI_ISL_925211, EPI_ISL_925212, EPI_ISL_925213, EPI_ISL_925214, EPI_ISL_925215, EPI_ISL_925216, EPI_ISL_925217, EPI_ISL_925218, EPI_ISL_925219, EPI_ISL_925220, EPI_ISL_925221, EPI_ISL_925222, EPI_ISL_925223, EPI_ISL_925224, EPI_ISL_925225, EPI_ISL_925226, EPI_ISL_925227, EPI_ISL_925228, EPI_ISL_925229, EPI_ISL_925230, EPI_ISL_925231, EPI_ISL_925232, EPI_ISL_925233, EPI_ISL_925234, EPI_ISL_925235, EPI_ISL_925236, EPI_ISL_925237, EPI_ISL_925238, EPI_ISL_925239, EPI_ISL_925240, EPI_ISL_925241, EPI_ISL_925242, EPI_ISL_925243, EPI_ISL_925244, EPI_ISL_925245, EPI_ISL_925246, EPI_ISL_925247, EPI_ISL_925248, EPI_ISL_925249, EPI_ISL_925250, EPI_ISL_925251, EPI_ISL_925252, EPI_ISL_925253, EPI_ISL_925254, EPI_ISL_925255, EPI_ISL_925256, EPI_ISL_925257 | | | |
| see above | New Mexico Department of Health Scientific Laboratory | New Mexico Department of Health Scientific Laboratory | Ellie Johnson, Anastacia Griego-Fisher, D'eldra Malone, Jennifer Benoit |
| EPI_ISL_925259, EPI_ISL_925265, EPI_ISL_925266, EPI_ISL_925270, EPI_ISL_925282, EPI_ISL_925288, EPI_ISL_925292, EPI_ISL_925294, EPI_ISL_925298, | Wyoming Public Health Laboratory | Wyoming Public Health Laboratory | Noah Hull, Taylor Fearing, Lynette Gumbleton, Channing Weber, Ashley Norberg, Bailey Bowcutt, and Wanda Manley |

| | | | |
|---|---|---|---|
| EPI_ISL_925300 | | | |
| EPI_ISL_925361 | NCSLPH | NCSLPH | Chase K, Miller MC, Greene S, Glover W |
| EPI_ISL_925509, EPI_ISL_925510, EPI_ISL_925511, EPI_ISL_925512, EPI_ISL_925513, EPI_ISL_925514, EPI_ISL_925515, EPI_ISL_925516, EPI_ISL_925517, EPI_ISL_925518, EPI_ISL_925519, EPI_ISL_925520, EPI_ISL_925521, EPI_ISL_925522, EPI_ISL_925523, EPI_ISL_925525, EPI_ISL_925526, EPI_ISL_925527, EPI_ISL_925528, EPI_ISL_925529, EPI_ISL_925530, EPI_ISL_925531, EPI_ISL_925532, EPI_ISL_925533, EPI_ISL_925534, EPI_ISL_925824, EPI_ISL_925825, EPI_ISL_925826, EPI_ISL_925827, EPI_ISL_925828, EPI_ISL_925829, EPI_ISL_925830, EPI_ISL_925832, EPI_ISL_925833, EPI_ISL_925834, EPI_ISL_925835, EPI_ISL_925836, EPI_ISL_925837, EPI_ISL_925838, EPI_ISL_925839, EPI_ISL_925841, EPI_ISL_925842, EPI_ISL_925843, EPI_ISL_925844, EPI_ISL_925845 | | | |
| see above | Arizona State Public Health Laboratory | Arizona State Public Health Laboratory | Trung Huynh, Jessica Escobar, Katherine Fullerton, Nobuko Fukushima, Stacy White, Linda Getsinger, Victor Waddell |
| EPI_ISL_929763 | NCSLPH | NCSLPH | Chase K, Miller MC, Greene S, Glover W |
| EPI_ISL_930635, EPI_ISL_930638, EPI_ISL_930639, EPI_ISL_930641, EPI_ISL_930642, EPI_ISL_930643, EPI_ISL_930645, EPI_ISL_930652, EPI_ISL_930653, EPI_ISL_930654, EPI_ISL_930655, EPI_ISL_930657, EPI_ISL_930658, EPI_ISL_930659, EPI_ISL_930660, EPI_ISL_930661, EPI_ISL_930663 | | | |
| see above | Arizona State Public Health Laboratory | Arizona State Public Health Laboratory | Trung Huynh, Jessica Escobar, Katherine Fullerton, Nobuko Fukushima, Stacy White, Linda Getsinger, Victor Waddell |
| EPI_ISL_930665, EPI_ISL_930666, EPI_ISL_930667, EPI_ISL_930668, EPI_ISL_930669, EPI_ISL_930670, EPI_ISL_930671, EPI_ISL_930672, EPI_ISL_930673, EPI_ISL_930674, EPI_ISL_930675, EPI_ISL_930676, EPI_ISL_930677, EPI_ISL_930678, EPI_ISL_930679, EPI_ISL_930680, EPI_ISL_930681, EPI_ISL_930682, EPI_ISL_930683, EPI_ISL_930684, EPI_ISL_930685, EPI_ISL_930686, EPI_ISL_930687, EPI_ISL_930688, EPI_ISL_930689, EPI_ISL_930690, EPI_ISL_930691, EPI_ISL_930692, EPI_ISL_930693, EPI_ISL_930694, EPI_ISL_930695, EPI_ISL_930696, EPI_ISL_930697, EPI_ISL_930698, EPI_ISL_930699, EPI_ISL_930700, EPI_ISL_930701, EPI_ISL_930702, EPI_ISL_930703, EPI_ISL_930704, EPI_ISL_930705, EPI_ISL_930706, EPI_ISL_930707, EPI_ISL_930708, EPI_ISL_930709, EPI_ISL_930710, EPI_ISL_930711, EPI_ISL_930712, EPI_ISL_930713, EPI_ISL_930714, EPI_ISL_930715, EPI_ISL_930716, EPI_ISL_930717, EPI_ISL_930718, EPI_ISL_930719, EPI_ISL_930720, EPI_ISL_930721, EPI_ISL_930722, EPI_ISL_930723, EPI_ISL_930724, EPI_ISL_930725, EPI_ISL_930726, EPI_ISL_930727, EPI_ISL_930728, EPI_ISL_930729, EPI_ISL_930730, EPI_ISL_930731, EPI_ISL_930732, EPI_ISL_930733, EPI_ISL_930734, EPI_ISL_930735, EPI_ISL_930736, EPI_ISL_930737, EPI_ISL_930738, EPI_ISL_930739, EPI_ISL_930740, EPI_ISL_930741, EPI_ISL_930742, EPI_ISL_930743, EPI_ISL_930744, EPI_ISL_930745, EPI_ISL_930746, EPI_ISL_930747, EPI_ISL_930748, EPI_ISL_930749, EPI_ISL_930750, EPI_ISL_930751, EPI_ISL_930752, EPI_ISL_930753, EPI_ISL_930754, EPI_ISL_930755, EPI_ISL_930756, EPI_ISL_930757, EPI_ISL_930758, EPI_ISL_930759, EPI_ISL_930760, EPI_ISL_930761, EPI_ISL_930762, EPI_ISL_930763, EPI_ISL_930764, EPI_ISL_930765, EPI_ISL_930766, EPI_ISL_930767, EPI_ISL_930768, EPI_ISL_930769, EPI_ISL_930770, EPI_ISL_930771, EPI_ISL_930772, EPI_ISL_930773, EPI_ISL_930774, EPI_ISL_930775, EPI_ISL_930776, EPI_ISL_930777, EPI_ISL_930778, EPI_ISL_930779, EPI_ISL_930780, EPI_ISL_930781, EPI_ISL_930782 | | | |
| see above | Utah Public Health Laboratory | Utah Public Health Laboratory | Erin L. Young, Kelly F. Oakeson, Tara Gallagher |
| EPI_ISL_930783 | Instituto Nacional de Medicina Genomica | Instituto Nacional de Medicina Genomica | Hidalgo-Miranda A, Mendoza-Vargas A, Reyes-Grajeda JP, Cisneros-Villanueva M, Cedro-Tanda A,Peñaloza-Figueroa F, Herrera-Montalvo LA |
| EPI_ISL_930789, EPI_ISL_930791, EPI_ISL_930792 | Monterey County Public Health Laboratory | Monterey County Public Health Laboratory | C. Anaya et al., |
| EPI_ISL_930794, EPI_ISL_930796 | Monterey County Public Health Laboratory | Monterey County Public Health Laboratory | C. Anaya et al. |
| EPI_ISL_930797 | Monterey County Public Health Laboratory | Monterey County Public Health Laboratory | C. Anaya et al., |
| EPI_ISL_930800 | Monterey County Public Health Laboratory | Monterey County Public Health Laboratory | C. Anaya et al. |
| EPI_ISL_930804 | Monterey County Public Health Laboratory | Monterey County Public Health Laboratory | C. Anaya et al., |
| EPI_ISL_930805 | Monterey County Public Health Laboratory | Monterey County Public Health Laboratory | C. Anaya et al. |
| EPI_ISL_930830, EPI_ISL_930833, EPI_ISL_930845, EPI_ISL_930847, EPI_ISL_930848, EPI_ISL_930851 | Monterey County Public Health Laboratory | Monterey County Public Health Laboratory | C. Anaya et al., |
| EPI_ISL_930852 | Monterey County Public Health Laboratory | Monterey County Public Health Laboratory | C. Anaya et al. |
| EPI_ISL_931451, EPI_ISL_931452, EPI_ISL_931453, EPI_ISL_931454, EPI_ISL_931455, EPI_ISL_931456, EPI_ISL_931457, EPI_ISL_931458, EPI_ISL_931459, EPI_ISL_931460, EPI_ISL_931461, EPI_ISL_931462, EPI_ISL_931463, EPI_ISL_931464, EPI_ISL_931465, EPI_ISL_931466, EPI_ISL_931467, EPI_ISL_931468, EPI_ISL_931469, EPI_ISL_931470, EPI_ISL_931471, EPI_ISL_931486, EPI_ISL_931487, EPI_ISL_931488, EPI_ISL_931489, EPI_ISL_931490, EPI_ISL_931491, EPI_ISL_931492, EPI_ISL_931493, EPI_ISL_931494, EPI_ISL_931495, EPI_ISL_931496, EPI_ISL_931497, EPI_ISL_931498, EPI_ISL_931499, EPI_ISL_931500, EPI_ISL_931501, EPI_ISL_931502, EPI_ISL_931503, EPI_ISL_931504, EPI_ISL_931505, EPI_ISL_931506 | | | |
| see above | Maryland Public Health Laboratory (MD PHL) | Maryland Public Health Laboratory (MD PHL) | Maryland Department of Health Laboratories Administration |
| EPI_ISL_931507, EPI_ISL_931508, EPI_ISL_931509, EPI_ISL_931510, EPI_ISL_931511, EPI_ISL_931512, EPI_ISL_931513, EPI_ISL_931514, EPI_ISL_931515, EPI_ISL_931516, EPI_ISL_931517, EPI_ISL_931518, EPI_ISL_931519, EPI_ISL_931520, EPI_ISL_931521, EPI_ISL_931522, EPI_ISL_931523, EPI_ISL_931524, EPI_ISL_931525, EPI_ISL_931526, EPI_ISL_931527, EPI_ISL_931528, EPI_ISL_931529, EPI_ISL_931530, EPI_ISL_931531, EPI_ISL_931532, EPI_ISL_931533, EPI_ISL_931534, EPI_ISL_931535, EPI_ISL_931536, EPI_ISL_931537, EPI_ISL_931538, EPI_ISL_931539, EPI_ISL_931540, EPI_ISL_931541, EPI_ISL_931542, EPI_ISL_931543, EPI_ISL_931544, EPI_ISL_931545, EPI_ISL_931546, EPI_ISL_931547, EPI_ISL_931548, EPI_ISL_931549, EPI_ISL_931550, EPI_ISL_931551, EPI_ISL_931552, EPI_ISL_931553, EPI_ISL_931554, EPI_ISL_931555, EPI_ISL_931556, EPI_ISL_931557, EPI_ISL_931558, EPI_ISL_931559, EPI_ISL_931560, EPI_ISL_931561, EPI_ISL_931562, EPI_ISL_931563, EPI_ISL_931564, EPI_ISL_931565, EPI_ISL_931566, EPI_ISL_931567, EPI_ISL_931568, EPI_ISL_931569, EPI_ISL_931570, EPI_ISL_931571, EPI_ISL_931572, EPI_ISL_931573, EPI_ISL_931574, EPI_ISL_931575, EPI_ISL_931576, EPI_ISL_931577, EPI_ISL_931578, EPI_ISL_931579, EPI_ISL_931580, EPI_ISL_931581, EPI_ISL_931582, EPI_ISL_931583 | | | |
| see above | Utah Public Health Laboratory | Utah Public Health Laboratory | Erin L. Young, Kelly F. Oakeson, Tara Gallagher |
| EPI_ISL_933512, EPI_ISL_933513, EPI_ISL_933514, EPI_ISL_933515, EPI_ISL_933516, EPI_ISL_933517, EPI_ISL_933518, EPI_ISL_933519, EPI_ISL_933520, EPI_ISL_933521, EPI_ISL_933522, EPI_ISL_933523, EPI_ISL_933524, EPI_ISL_933525, EPI_ISL_933526, EPI_ISL_933527, EPI_ISL_933528, EPI_ISL_933529, EPI_ISL_933530, EPI_ISL_933531 | | | |
| see above | Instituto Nacional de Medicina Genomica | Instituto Nacional de Medicina Genomica | Hidalgo-Miranda A, Mendoza-Vargas A, Reyes-Grajeda JP, Cisneros-Villanueva M, Cedro-Tanda A,Peñaloza-Figueroa F, Herrera-Montalvo LA |
| EPI_ISL_933540, EPI_ISL_933541, EPI_ISL_933542, EPI_ISL_933543, EPI_ISL_933544, EPI_ISL_933545, EPI_ISL_933546, EPI_ISL_933547, EPI_ISL_933548, EPI_ISL_933549, EPI_ISL_933550, EPI_ISL_933551, EPI_ISL_933552, EPI_ISL_933553, EPI_ISL_933554, EPI_ISL_933555, EPI_ISL_933556, EPI_ISL_933557, EPI_ISL_933558, EPI_ISL_933559, EPI_ISL_933560, EPI_ISL_933561, EPI_ISL_933562, EPI_ISL_933563, EPI_ISL_933580, EPI_ISL_933581, EPI_ISL_933582, EPI_ISL_933583, EPI_ISL_933584, EPI_ISL_933585, EPI_ISL_933587, EPI_ISL_933588, EPI_ISL_933589, EPI_ISL_933590, EPI_ISL_933591, EPI_ISL_933592, EPI_ISL_933593, EPI_ISL_933594, EPI_ISL_933595, EPI_ISL_933596, EPI_ISL_933597, EPI_ISL_933598, EPI_ISL_933599, EPI_ISL_933600, EPI_ISL_933601 | | | |
| see above | Arizona State Public Health Laboratory | Arizona State Public Health Laboratory | Trung Huynh, Jessica Escobar, Katherine Fullerton, Nobuko Fukushima, Stacy White, Linda Getsinger, Victor Waddell |
| EPI_ISL_933661, EPI_ISL_933662, EPI_ISL_933663, EPI_ISL_933664, EPI_ISL_933665, EPI_ISL_933666, EPI_ISL_933667, EPI_ISL_933668, EPI_ISL_933669, EPI_ISL_933670, EPI_ISL_933671, EPI_ISL_933672, EPI_ISL_933673, EPI_ISL_933674, EPI_ISL_933675, EPI_ISL_933676, EPI_ISL_933677, EPI_ISL_933678, EPI_ISL_933679, EPI_ISL_933680, EPI_ISL_933681, EPI_ISL_933682, EPI_ISL_933683, EPI_ISL_933684, EPI_ISL_933685, EPI_ISL_933686, EPI_ISL_933687, EPI_ISL_933688, EPI_ISL_933689, EPI_ISL_933690, EPI_ISL_933691, EPI_ISL_933692, EPI_ISL_933693, EPI_ISL_933694, EPI_ISL_933695, EPI_ISL_933696, EPI_ISL_933697, EPI_ISL_933698, EPI_ISL_933699, EPI_ISL_933700, EPI_ISL_933701, EPI_ISL_933702, EPI_ISL_933703, EPI_ISL_933704, EPI_ISL_933705, EPI_ISL_933706, EPI_ISL_933707, EPI_ISL_933708, EPI_ISL_933709, EPI_ISL_933710, EPI_ISL_933711, EPI_ISL_933712 | | | |
| see above | Instituto de Diagnostico y Referencia Epidemiologicos INDRE_RNLSP | Instituto de Diagnostico y Referencia Epidemiologicos (INDRE) | Claudia Wong-Arambula, Abril Rodriguez-Maldonado, Fabiola Garces-Ayala, Adnan Araiza-Rodriguez, David Fragoso-Fonseca, Sergio Rangel-Guerrero, Mayra Jimenez-Morales, Nancy Munoz-Hernandez, Natividad Cruz-Ortiz, Tatiana Nunez-Garcia, Gisela Barrera-Badillo, Lucia Hernandez-Rivas, Irma Lopez-Martinez, Ernesto Ramirez-Gonzalez. |
| EPI_ISL_933735, EPI_ISL_933736, EPI_ISL_933737, EPI_ISL_933738, EPI_ISL_933739, EPI_ISL_933741, EPI_ISL_933742, EPI_ISL_933743, EPI_ISL_933744, EPI_ISL_933745, EPI_ISL_933746, EPI_ISL_933747, EPI_ISL_933748, EPI_ISL_933749, EPI_ISL_933750, EPI_ISL_933751, EPI_ISL_933752, EPI_ISL_933753, EPI_ISL_933754, EPI_ISL_933755, EPI_ISL_933756, EPI_ISL_933757, EPI_ISL_933758, EPI_ISL_933759, EPI_ISL_933760, EPI_ISL_933761, EPI_ISL_933762, EPI_ISL_933763, EPI_ISL_933764, EPI_ISL_933765 | | | |
| see above | DPHL | Delaware Public Health Lab | Gregory Hovan |
| EPI_ISL_934414, EPI_ISL_934417, EPI_ISL_934973, EPI_ISL_934975 | Ohio Department of Health Laboratory | Ohio Department of Health Laboratory | Holmes, Jennifer; Eric Brandt, Keoni Omura, Glen McGillivary, Caitlin McDonnell, Jade Mowery, Stephanie Mccracken, Tyler Payne, Kirtana Ramadugu, Erica Leasure, Brent Lee, Kelsey Florek, Heather Blankenship, Quanta Brown, and Tammy Bannerman |
| EPI_ISL_934977 | Northwest Pathology | UW Virology Lab | Pavitra Roychoudhury, Hong Xie, Lasata Shrestha, Michelle Lin, Meei-Li Huang, Keith R Jerome, Alexander Greninger |
| EPI_ISL_935012, EPI_ISL_935013, EPI_ISL_935014, EPI_ISL_935015, EPI_ISL_935016 | University of Michigan Clinical Microbiology Laboratory | Lauring Lab, University of Michigan, Department of Microbiology and Immunology | Valesano |
| EPI_ISL_935020 | Ohio Department of Health Laboratory | Ohio Department of Health Laboratory | Holmes, Jennifer; Eric Brandt, Keoni Omura, Glen McGillivary, Caitlin McDonnell, Jade Mowery, Stephanie Mccracken, Tyler Payne, Kirtana Ramadugu, Erica Leasure, Brent Lee, Kelsey Florek, Heather Blankenship, Quanta Brown, and Tammy Bannerman |
| EPI_ISL_935022 | Northwest Pathology | UW Virology Lab | Pavitra Roychoudhury, Hong Xie, Lasata Shrestha, Michelle Lin, Meei-Li Huang, Keith R Jerome, Alexander Greninger |
| EPI_ISL_935023, EPI_ISL_935024, EPI_ISL_935027, EPI_ISL_935031, EPI_ISL_935036 | Ohio Department of Health Laboratory | Ohio Department of Health Laboratory | Holmes, Jennifer; Eric Brandt, Keoni Omura, Glen McGillivary, Caitlin McDonnell, Jade Mowery, Stephanie Mccracken, Tyler Payne, Kirtana Ramadugu, Erica Leasure, Brent Lee, Kelsey Florek, Heather Blankenship, Quanta Brown, and Tammy Bannerman |
| EPI_ISL_935047 | MD PHL | MD PHL | Maryland Department of Health Laboratories Administration |
| EPI_ISL_935155 | Cytocheck Laboratory | Kansas Health and Environmental Lab | Mike Grose, Paige Drury, Carissa Robertson, Ben Olsen, and Phil Adam |
| EPI_ISL_935171, EPI_ISL_935172, EPI_ISL_935173, EPI_ISL_935175, EPI_ISL_935177, EPI_ISL_935179, EPI_ISL_935180, EPI_ISL_935181, EPI_ISL_935182, EPI_ISL_935184, EPI_ISL_935185, EPI_ISL_935186 | | | |
| see above | Wyoming Public Health Laboratory | Wyoming Public Health Laboratory | Noah Hull, Taylor Fearing, Lynette Gumbleton, Channing Weber, Ashley Norberg, Bailey Bowcutt, and Wanda Manley |
| EPI_ISL_935359, EPI_ISL_935360, EPI_ISL_935361, EPI_ISL_935362, EPI_ISL_935363, EPI_ISL_935364, EPI_ISL_935365, EPI_ISL_935366, EPI_ISL_935367, EPI_ISL_935368, EPI_ISL_935369, EPI_ISL_935370, EPI_ISL_935371, EPI_ISL_935373, EPI_ISL_935374, EPI_ISL_935375, EPI_ISL_935376, EPI_ISL_935377, EPI_ISL_935378, EPI_ISL_935379, EPI_ISL_935380, EPI_ISL_935381, EPI_ISL_935382, EPI_ISL_935383, EPI_ISL_935384, EPI_ISL_935385, EPI_ISL_935386, EPI_ISL_935387, EPI_ISL_935388, EPI_ISL_935389, EPI_ISL_935390, EPI_ISL_935391, EPI_ISL_935393, EPI_ISL_935394, EPI_ISL_935395, EPI_ISL_935396, EPI_ISL_935397, EPI_ISL_935398, EPI_ISL_935399, EPI_ISL_935400, EPI_ISL_935401, EPI_ISL_935402, EPI_ISL_935403, EPI_ISL_935404, EPI_ISL_935405, EPI_ISL_935406, EPI_ISL_935407, EPI_ISL_935408, EPI_ISL_935409, EPI_ISL_935410, EPI_ISL_935411, EPI_ISL_935412, EPI_ISL_935413, EPI_ISL_935414, EPI_ISL_935415, EPI_ISL_935416, EPI_ISL_935417, EPI_ISL_935420, EPI_ISL_935421, EPI_ISL_935422, EPI_ISL_935423, EPI_ISL_935424, EPI_ISL_935425, EPI_ISL_935426, EPI_ISL_935428, EPI_ISL_935429, EPI_ISL_935430, EPI_ISL_935431, EPI_ISL_935432, EPI_ISL_935433, EPI_ISL_935434, EPI_ISL_935435, EPI_ISL_935436, EPI_ISL_935437, EPI_ISL_935438, EPI_ISL_935439, EPI_ISL_935441, EPI_ISL_935442, EPI_ISL_935443, EPI_ISL_935444, EPI_ISL_935445, EPI_ISL_935446, EPI_ISL_935447, EPI_ISL_935448, EPI_ISL_935449, EPI_ISL_935450, EPI_ISL_935451, EPI_ISL_935452, EPI_ISL_935453, EPI_ISL_935454, EPI_ISL_935455, EPI_ISL_935456, EPI_ISL_935457, EPI_ISL_935458, EPI_ISL_935459, EPI_ISL_935460, EPI_ISL_935461, EPI_ISL_935462, EPI_ISL_935463, EPI_ISL_935464, EPI_ISL_935466, EPI_ISL_935467, EPI_ISL_935468, EPI_ISL_935469, EPI_ISL_935470, EPI_ISL_935471, EPI_ISL_935472, EPI_ISL_935473, EPI_ISL_935474, EPI_ISL_935475, EPI_ISL_935476, EPI_ISL_935477, EPI_ISL_935478, EPI_ISL_935479, EPI_ISL_935480, EPI_ISL_935481, EPI_ISL_935482, EPI_ISL_935484, EPI_ISL_935485, EPI_ISL_935487, EPI_ISL_935488, EPI_ISL_935489, EPI_ISL_935490, EPI_ISL_935491, EPI_ISL_935492, EPI_ISL_935493, EPI_ISL_935494, EPI_ISL_935495, EPI_ISL_935496, EPI_ISL_935498, EPI_ISL_935499, EPI_ISL_935500, EPI_ISL_935501, EPI_ISL_935502, EPI_ISL_935503, EPI_ISL_935504, EPI_ISL_935505, EPI_ISL_935506, EPI_ISL_935507, EPI_ISL_935508, EPI_ISL_935510, EPI_ISL_935511, EPI_ISL_935512, EPI_ISL_935513, EPI_ISL_935514, EPI_ISL_935515, EPI_ISL_935516, EPI_ISL_935517, EPI_ISL_935518, EPI_ISL_935519 | | | |
| see above | Florida Bureau of Public Health Laboratories | Florida Bureau of Public Health Laboratories | Sarah Schmedes, Jason Blanton |
| EPI_ISL_935520, EPI_ISL_935521 | Ohio Department of Health Laboratory | Ohio Department of Health Laboratory | Holmes, Jennifer; Eric Brandt, Keoni Omura, Glen McGillivary, Caitlin McDonnell, Jade Mowery, Stephanie Mccracken, Tyler Payne, Kirtana Ramadugu, Erica Leasure, Brent Lee, Kelsey Florek, Heather Blankenship, Quanta Brown, and Tammy Bannerman |

| | | | |
|---|---|---|---|
| EPI_ISL_935522, EPI_ISL_935523, EPI_ISL_935524 | University of Massachusetts Medical School | Infectious Disease Program, Broad Institute of Harvard and MIT | Tomkins-Tinch, Christopher H.; Daly, Jennifer S.; Gladden-Young, Adrianne; Theodoropoulos, Nicole M.; Madaio, Michael; Yu, Neng; Vanguri, Vijay K; Siddle, Katherine J.; Adams, Gordon; Krasilnikova, Lydia A.; Movahedi, Babak; Bozorgzadeh, Adel; Simin, Karl; Lemieux, Jacob E.; Luban, Jeremy; Park, Daniel J.; MacInnis, Bronwyn L.; Sabeti, Pardis C.; Levitz, Stuart M. |
| EPI_ISL_935525 | Ohio Department of Health Laboratory | Ohio Department of Health Laboratory | Holmes, Jennifer; Eric Brandt, Keoni Omura, Glen McGillivary, Caitlin McDonnell, Jade Mowery, Stephanie Mccracken, Tyler Payne, Kirtana Ramadugu, Erica Leasure, Brent Lee, Kelsey Florek, Heather Blankenship, Quanta Brown, and Tammy Bannerman |
| EPI_ISL_935526 | University of Massachusetts Medical School | Infectious Disease Program, Broad Institute of Harvard and MIT | Tomkins-Tinch, Christopher H.; Daly, Jennifer S.; Gladden-Young, Adrianne; Theodoropoulos, Nicole M.; Madaio, Michael; Yu, Neng; Vanguri, Vijay K; Siddle, Katherine J.; Adams, Gordon; Krasilnikova, Lydia A.; Movahedi, Babak; Bozorgzadeh, Adel; Simin, Karl; Lemieux, Jacob E.; Luban, Jeremy; Park, Daniel J.; MacInnis, Bronwyn L.; Sabeti, Pardis C.; Levitz, Stuart M. |
| EPI_ISL_935696, EPI_ISL_935697, EPI_ISL_935698, EPI_ISL_935699, EPI_ISL_935700, EPI_ISL_935701, EPI_ISL_935702, EPI_ISL_935703, EPI_ISL_935704, EPI_ISL_935705, EPI_ISL_935706, EPI_ISL_935707, EPI_ISL_935708, EPI_ISL_935709, EPI_ISL_935710, EPI_ISL_935711, EPI_ISL_935712, EPI_ISL_935713, EPI_ISL_935714, EPI_ISL_935715, EPI_ISL_935716, EPI_ISL_935717, EPI_ISL_935718, EPI_ISL_935719, EPI_ISL_935720, EPI_ISL_935721, EPI_ISL_935722, EPI_ISL_935723, EPI_ISL_935724, EPI_ISL_935725, EPI_ISL_935726, EPI_ISL_935727, EPI_ISL_935728, EPI_ISL_935729, EPI_ISL_935730, EPI_ISL_935731, EPI_ISL_935732, EPI_ISL_935733, EPI_ISL_935734, EPI_ISL_935735, EPI_ISL_935736, EPI_ISL_935737, EPI_ISL_935738, EPI_ISL_935739, EPI_ISL_935740, EPI_ISL_935741, EPI_ISL_935742, EPI_ISL_935743, EPI_ISL_935744, EPI_ISL_935745, EPI_ISL_935746, EPI_ISL_935747, EPI_ISL_935748, EPI_ISL_935749, EPI_ISL_935750, EPI_ISL_935751, EPI_ISL_935752 | | | |
| see above | Houston Health Dept. | Houston Health Dept. | Ryker Penn, Pamela Brown, Adolpho Lara |
| EPI_ISL_935973, EPI_ISL_935974, EPI_ISL_935975, EPI_ISL_935976, EPI_ISL_935977, EPI_ISL_935978, EPI_ISL_935979, EPI_ISL_935980, EPI_ISL_935981, EPI_ISL_935982, EPI_ISL_935983, EPI_ISL_935984, EPI_ISL_935985, EPI_ISL_935986, EPI_ISL_935987, EPI_ISL_935988, EPI_ISL_935989, EPI_ISL_935990, EPI_ISL_935991, EPI_ISL_935992 | | | |
| see above | GLENS FALLS HOSPITAL LABORATORY | Wadsworth Center, New York State Department of Health | Kirsten St. George, Daryl M. Lamson, Alexis Russel, Matthew Shudt, Melissa A Leisner, Jonathan Plitnick, Navjot Singh, John Kelly, Erasmus Schneider, Erica Lasek-Nesselquist |
| EPI_ISL_935993, EPI_ISL_935994, EPI_ISL_935995, EPI_ISL_935996, EPI_ISL_935997, EPI_ISL_935998, EPI_ISL_935999, EPI_ISL_936000, EPI_ISL_936001, EPI_ISL_936002, EPI_ISL_936003, EPI_ISL_936004, EPI_ISL_936005, EPI_ISL_936006, EPI_ISL_936007, EPI_ISL_936008, EPI_ISL_936009, EPI_ISL_936010, EPI_ISL_936011, EPI_ISL_936012, EPI_ISL_936013, EPI_ISL_936014, EPI_ISL_936015, EPI_ISL_936016, EPI_ISL_936017, EPI_ISL_936018, EPI_ISL_936019, EPI_ISL_936020, EPI_ISL_936021, EPI_ISL_936022 | | | |
| see above | SUNY UPSTATE MEDICAL UNIVERSITY | Wadsworth Center, New York State Department of Health | Kirsten St. George, Daryl M. Lamson, Alexis Russel, Matthew Shudt, Melissa A Leisner, Jonathan Plitnick, Navjot Singh, John Kelly, Erasmus Schneider, Erica Lasek-Nesselquist |
| EPI_ISL_936023, EPI_ISL_936024, EPI_ISL_936025, EPI_ISL_936026, EPI_ISL_936027, EPI_ISL_936028, EPI_ISL_936029, EPI_ISL_936030, EPI_ISL_936031, EPI_ISL_936032, EPI_ISL_936033, EPI_ISL_936034, EPI_ISL_936035 | | | |
| see above | THE MARY IMOGENE BASSETT HOSPITAL | Wadsworth Center, New York State Department of Health | Kirsten St. George, Daryl M. Lamson, Alexis Russel, Matthew Shudt, Melissa A Leisner, Jonathan Plitnick, Navjot Singh, John Kelly, Erasmus Schneider, Erica Lasek-Nesselquist |
| EPI_ISL_936036, EPI_ISL_936037, EPI_ISL_936038, EPI_ISL_936039, EPI_ISL_936040, EPI_ISL_936041, EPI_ISL_936042, EPI_ISL_936043, EPI_ISL_936044, EPI_ISL_936045 | Columbia University Irving Medical Center | Wadsworth Center, New York State Department of Health | Kirsten St. George, Daryl M. Lamson, Alexis Russel, Matthew Shudt, Melissa A Leisner, Jonathan Plitnick, Navjot Singh, John Kelly, Erasmus Schneider, Erica Lasek-Nesselquist |
| EPI_ISL_936046, EPI_ISL_936047, EPI_ISL_936048, EPI_ISL_936049, EPI_ISL_936050, EPI_ISL_936051, EPI_ISL_936052, EPI_ISL_936053, EPI_ISL_936054, EPI_ISL_936055, EPI_ISL_936056, EPI_ISL_936057, EPI_ISL_936058, EPI_ISL_936059 | | | |
| see above | ADIRONDACK MEDICAL CENTER | Wadsworth Center, New York State Department of Health | Kirsten St. George, Daryl M. Lamson, Alexis Russel, Matthew Shudt, Melissa A Leisner, Jonathan Plitnick, Navjot Singh, John Kelly, Erasmus Schneider, Erica Lasek-Nesselquist |
| EPI_ISL_936060, EPI_ISL_936061, EPI_ISL_936062 | KALEIDA CENTER FOR LABORATORY MEDICINE | Wadsworth Center, New York State Department of Health | Kirsten St. George, Daryl M. Lamson, Alexis Russel, Matthew Shudt, Melissa A Leisner, Jonathan Plitnick, Navjot Singh, John Kelly, Erasmus Schneider, Erica Lasek-Nesselquist |
| EPI_ISL_936063, EPI_ISL_936064, EPI_ISL_936065, EPI_ISL_936066 | Columbia University Irving Medical Center | Wadsworth Center, New York State Department of Health | Kirsten St. George, Daryl M. Lamson, Alexis Russel, Matthew Shudt, Melissa A Leisner, Jonathan Plitnick, Navjot Singh, John Kelly, Erasmus Schneider, Erica Lasek-Nesselquist |
| EPI_ISL_936067, EPI_ISL_936068 | KALEIDA CENTER FOR LABORATORY MEDICINE | Wadsworth Center, New York State Department of Health | Kirsten St. George, Daryl M. Lamson, Alexis Russel, Matthew Shudt, Melissa A Leisner, Jonathan Plitnick, Navjot Singh, John Kelly, Erasmus Schneider, Erica Lasek-Nesselquist |
| EPI_ISL_936069 | Columbia University Irving Medical Center | Wadsworth Center, New York State Department of Health | Kirsten St. George, Daryl M. Lamson, Alexis Russel, Matthew Shudt, Melissa A Leisner, Jonathan Plitnick, Navjot Singh, John Kelly, Erasmus Schneider, Erica Lasek-Nesselquist |
| EPI_ISL_936070, EPI_ISL_936071, EPI_ISL_936072, EPI_ISL_936073, EPI_ISL_936074, EPI_ISL_936075, EPI_ISL_936076, EPI_ISL_936077, EPI_ISL_936078, EPI_ISL_936079, EPI_ISL_936080, EPI_ISL_936081 | | | |
| see above | KALEIDA CENTER FOR LABORATORY MEDICINE | Wadsworth Center, New York State Department of Health | Kirsten St. George, Daryl M. Lamson, Alexis Russel, Matthew Shudt, Melissa A Leisner, Jonathan Plitnick, Navjot Singh, John Kelly, Erasmus Schneider, Erica Lasek-Nesselquist |
| EPI_ISL_936082 | Columbia University Irving Medical Center | Wadsworth Center, New York State Department of Health | Kirsten St. George, Daryl M. Lamson, Alexis Russel, Matthew Shudt, Melissa A Leisner, Jonathan Plitnick, Navjot Singh, John Kelly, Erasmus Schneider, Erica Lasek-Nesselquist |
| EPI_ISL_936083, EPI_ISL_936084, EPI_ISL_936085 | KALEIDA CENTER FOR LABORATORY MEDICINE | Wadsworth Center, New York State Department of Health | Kirsten St. George, Daryl M. Lamson, Alexis Russel, Matthew Shudt, Melissa A Leisner, Jonathan Plitnick, Navjot Singh, John Kelly, Erasmus Schneider, Erica Lasek-Nesselquist |
| EPI_ISL_936086 | Columbia University Irving Medical Center | Wadsworth Center, New York State Department of Health | Kirsten St. George, Daryl M. Lamson, Alexis Russel, Matthew Shudt, Melissa A Leisner, Jonathan Plitnick, Navjot Singh, John Kelly, Erasmus Schneider, Erica Lasek-Nesselquist |
| EPI_ISL_936087, EPI_ISL_936088 | KALEIDA CENTER FOR LABORATORY MEDICINE | Wadsworth Center, New York State Department of Health | Kirsten St. George, Daryl M. Lamson, Alexis Russel, Matthew Shudt, Melissa A Leisner, Jonathan Plitnick, Navjot Singh, John Kelly, Erasmus Schneider, Erica Lasek-Nesselquist |
| EPI_ISL_936089 | Columbia University Irving Medical Center | Wadsworth Center, New York State Department of Health | Kirsten St. George, Daryl M. Lamson, Alexis Russel, Matthew Shudt, Melissa A Leisner, Jonathan Plitnick, Navjot Singh, John Kelly, Erasmus Schneider, Erica Lasek-Nesselquist |
| EPI_ISL_936090, EPI_ISL_936091 | KALEIDA CENTER FOR LABORATORY MEDICINE | Wadsworth Center, New York State Department of Health | Kirsten St. George, Daryl M. Lamson, Alexis Russel, Matthew Shudt, Melissa A Leisner, Jonathan Plitnick, Navjot Singh, John Kelly, Erasmus Schneider, Erica Lasek-Nesselquist |
| EPI_ISL_936092, EPI_ISL_936093 | Columbia University Irving Medical Center | Wadsworth Center, New York State Department of Health | Kirsten St. George, Daryl M. Lamson, Alexis Russel, Matthew Shudt, Melissa A Leisner, Jonathan Plitnick, Navjot Singh, John Kelly, Erasmus Schneider, Erica Lasek-Nesselquist |
| EPI_ISL_936094, EPI_ISL_936095, EPI_ISL_936096, EPI_ISL_936097, EPI_ISL_936098, EPI_ISL_936099, EPI_ISL_936100, EPI_ISL_936101, EPI_ISL_936102 | KALEIDA CENTER FOR LABORATORY MEDICINE | Wadsworth Center, New York State Department of Health | Kirsten St. George, Daryl M. Lamson, Alexis Russel, Matthew Shudt, Melissa A Leisner, Jonathan Plitnick, Navjot Singh, John Kelly, Erasmus Schneider, Erica Lasek-Nesselquist |
| EPI_ISL_936103, EPI_ISL_936104, EPI_ISL_936105, EPI_ISL_936106, EPI_ISL_936107, EPI_ISL_936108, EPI_ISL_936109, EPI_ISL_936110, EPI_ISL_936111, EPI_ISL_936112, EPI_ISL_936113, EPI_ISL_936114, EPI_ISL_936115, EPI_ISL_936116, EPI_ISL_936117, EPI_ISL_936118, EPI_ISL_936119, EPI_ISL_936120, EPI_ISL_936121, EPI_ISL_936122, EPI_ISL_936123, EPI_ISL_936124, EPI_ISL_936125, EPI_ISL_936126, EPI_ISL_936127, EPI_ISL_936128, EPI_ISL_936129 | | | |
| see above | WESTCHESTER MEDICAL CENTER | Wadsworth Center, New York State Department of Health | Kirsten St. George, Daryl M. Lamson, Alexis Russel, Matthew Shudt, Melissa A Leisner, Jonathan Plitnick, Navjot Singh, John Kelly, Erasmus Schneider, Erica Lasek-Nesselquist |
| EPI_ISL_936130, EPI_ISL_936131, EPI_ISL_936132, EPI_ISL_936133, EPI_ISL_936134, EPI_ISL_936135, EPI_ISL_936136, EPI_ISL_936137 | New York Presbyterian Hospital | Wadsworth Center, New York State Department of Health | Kirsten St. George, Daryl M. Lamson, Alexis Russel, Matthew Shudt, Melissa A Leisner, Jonathan Plitnick, Navjot Singh, John Kelly, Erasmus Schneider, Erica Lasek-Nesselquist |
| EPI_ISL_936138, EPI_ISL_936139, EPI_ISL_936140, EPI_ISL_936141 | THE MARY IMOGENE BASSETT HOSPITAL | Wadsworth Center, New York State Department of Health | Kirsten St. George, Daryl M. Lamson, Alexis Russel, Matthew Shudt, Melissa A Leisner, Jonathan Plitnick, Navjot Singh, John Kelly, Erasmus Schneider, Erica Lasek-Nesselquist |
| EPI_ISL_936142, EPI_ISL_936143 | SUNY UPSTATE MEDICAL UNIVERSITY | Wadsworth Center, New York State Department of Health | Kirsten St. George, Daryl M. Lamson, Alexis Russel, Matthew Shudt, Melissa A Leisner, Jonathan Plitnick, Navjot Singh, John Kelly, Erasmus Schneider, Erica Lasek-Nesselquist |
| EPI_ISL_936144 | KALEIDA CENTER FOR LABORATORY MEDICINE | Wadsworth Center, New York State Department of Health | Kirsten St. George, Daryl M. Lamson, Alexis Russel, Matthew Shudt, Melissa A Leisner, Jonathan Plitnick, Navjot Singh, John Kelly, Erasmus Schneider, Erica Lasek-Nesselquist |
| EPI_ISL_936145, EPI_ISL_936146, EPI_ISL_936147 | New York Presbyterian Hospital | Wadsworth Center, New York State Department of Health | Kirsten St. George, Daryl M. Lamson, Alexis Russel, Matthew Shudt, Melissa A Leisner, Jonathan Plitnick, Navjot Singh, John Kelly, Erasmus Schneider, Erica Lasek-Nesselquist |

| | | | |
|---|---|---|---|
| EPI_ISL_936148 | Wadsworth Center, New York State Department of Health | Wadsworth Center, New York State Department of Health | Kirsten St. George, Daryl M. Lamson, Alexis Russel, Matthew Shudt, Melissa A Leisner, Jonathan Plitnick, Navjot Singh, John Kelly, Erasmus Schneider, Erica Lasek-Nesselquist |
| EPI_ISL_936149, EPI_ISL_936150, EPI_ISL_936151, EPI_ISL_936152 | MEMORIAL SLOAN KETTERING CANCER CENTER | Wadsworth Center, New York State Department of Health | Kirsten St. George, Daryl M. Lamson, Alexis Russel, Matthew Shudt, Melissa A Leisner, Jonathan Plitnick, Navjot Singh, John Kelly, Erasmus Schneider, Erica Lasek-Nesselquist |
| EPI_ISL_936153 | THE MARY IMOGENE BASSETT HOSPITAL | Wadsworth Center, New York State Department of Health | Kirsten St. George, Daryl M. Lamson, Alexis Russel, Matthew Shudt, Melissa A Leisner, Jonathan Plitnick, Navjot Singh, John Kelly, Erasmus Schneider, Erica Lasek-Nesselquist |
| EPI_ISL_936154, EPI_ISL_936155, EPI_ISL_936156, EPI_ISL_936157, EPI_ISL_936158, EPI_ISL_936159, EPI_ISL_936160, EPI_ISL_936161, EPI_ISL_936162, EPI_ISL_936163, EPI_ISL_936164, EPI_ISL_936165, EPI_ISL_936166, EPI_ISL_936167, EPI_ISL_936168, EPI_ISL_936169, EPI_ISL_936170, EPI_ISL_936171, EPI_ISL_936172 | | | |
| see above | MEMORIAL SLOAN KETTERING CANCER CENTER | Wadsworth Center, New York State Department of Health | Kirsten St. George, Daryl M. Lamson, Alexis Russel, Matthew Shudt, Melissa A Leisner, Jonathan Plitnick, Navjot Singh, John Kelly, Erasmus Schneider, Erica Lasek-Nesselquist |
| EPI_ISL_936173 | TEMPUS LABS INC | Wadsworth Center, New York State Department of Health | Kirsten St. George, Daryl M. Lamson, Alexis Russel, Matthew Shudt, Melissa A Leisner, Jonathan Plitnick, Navjot Singh, John Kelly, Erasmus Schneider, Erica Lasek-Nesselquist |
| EPI_ISL_936174, EPI_ISL_936175, EPI_ISL_936176, EPI_ISL_936177, EPI_ISL_936178, EPI_ISL_936179 | MEMORIAL SLOAN KETTERING CANCER CENTER | Wadsworth Center, New York State Department of Health | Kirsten St. George, Daryl M. Lamson, Alexis Russel, Matthew Shudt, Melissa A Leisner, Jonathan Plitnick, Navjot Singh, John Kelly, Erasmus Schneider, Erica Lasek-Nesselquist |
| EPI_ISL_936180, EPI_ISL_936181 | ALBANY MEDICAL CENTER | Wadsworth Center, New York State Department of Health | Kirsten St. George, Daryl M. Lamson, Alexis Russel, Matthew Shudt, Melissa A Leisner, Jonathan Plitnick, Navjot Singh, John Kelly, Erasmus Schneider, Erica Lasek-Nesselquist |
| EPI_ISL_936182, EPI_ISL_936183, EPI_ISL_936184, EPI_ISL_936185, EPI_ISL_936186, EPI_ISL_936187, EPI_ISL_936188, EPI_ISL_936189, EPI_ISL_936190, EPI_ISL_936191, EPI_ISL_936192, EPI_ISL_936193, EPI_ISL_936194, EPI_ISL_936195, EPI_ISL_936196, EPI_ISL_936197, EPI_ISL_936198, EPI_ISL_936199, EPI_ISL_936200, EPI_ISL_936201, EPI_ISL_936202, EPI_ISL_936203, EPI_ISL_936204, EPI_ISL_936205, EPI_ISL_936206, EPI_ISL_936207, EPI_ISL_936208, EPI_ISL_936209, EPI_ISL_936210, EPI_ISL_936211 | | | |
| see above | Houston Health Dept. | Houston Health Dept. | Ryker Penn, Pamela Brown, Adolpho Lara |
| EPI_ISL_936212, EPI_ISL_936213, EPI_ISL_936214, EPI_ISL_936215, EPI_ISL_936216, EPI_ISL_936217, EPI_ISL_936218, EPI_ISL_936219, EPI_ISL_936220, EPI_ISL_936221, EPI_ISL_936222, EPI_ISL_936223, EPI_ISL_936224, EPI_ISL_936225, EPI_ISL_936226, EPI_ISL_936227, EPI_ISL_936228, EPI_ISL_936229, EPI_ISL_936230, EPI_ISL_936231, EPI_ISL_936232, EPI_ISL_936233, EPI_ISL_936234, EPI_ISL_936235, EPI_ISL_936236, EPI_ISL_936237, EPI_ISL_936238, EPI_ISL_936239, EPI_ISL_936240, EPI_ISL_936241, EPI_ISL_936242, EPI_ISL_936243, EPI_ISL_936244, EPI_ISL_936245, EPI_ISL_936246, EPI_ISL_936247 | | | |
| see above | Wadsworth Center, New York State Department of Health | Wadsworth Center, New York State Department of Health | Kirsten St. George, Daryl M. Lamson, Alexis Russel, Matthew Shudt, Melissa A Leisner, Jonathan Plitnick, Navjot Singh, John Kelly, Erasmus Schneider, Erica Lasek-Nesselquist |
| EPI_ISL_936248, EPI_ISL_936249, EPI_ISL_936250, EPI_ISL_936251, EPI_ISL_936252, EPI_ISL_936253, EPI_ISL_936254, EPI_ISL_936255, EPI_ISL_936256, EPI_ISL_936257, EPI_ISL_936258, EPI_ISL_936259, EPI_ISL_936260, EPI_ISL_936261, EPI_ISL_936262, EPI_ISL_936263, EPI_ISL_936264, EPI_ISL_936265, EPI_ISL_936266, EPI_ISL_936267, EPI_ISL_936268, EPI_ISL_936269, EPI_ISL_936270, EPI_ISL_936271, EPI_ISL_936272, EPI_ISL_936273, EPI_ISL_936274, EPI_ISL_936275, EPI_ISL_936276, EPI_ISL_936277, EPI_ISL_936278, EPI_ISL_936279, EPI_ISL_936280, EPI_ISL_936281, EPI_ISL_936282, EPI_ISL_936283, EPI_ISL_936284, EPI_ISL_936285, EPI_ISL_936286, EPI_ISL_936287, EPI_ISL_936288, EPI_ISL_936289, EPI_ISL_936290, EPI_ISL_936291, EPI_ISL_936292, EPI_ISL_936293, EPI_ISL_936294, EPI_ISL_936295, EPI_ISL_936296, EPI_ISL_936297, EPI_ISL_936298, EPI_ISL_936299 | | | |
| see above | MONTEFIORE MEDICAL CENTER LABORATORIES | Wadsworth Center, New York State Department of Health | Kirsten St. George, Daryl M. Lamson, Alexis Russel, Matthew Shudt, Melissa A Leisner, Jonathan Plitnick, Navjot Singh, John Kelly, Erasmus Schneider, Erica Lasek-Nesselquist |
| EPI_ISL_936300, EPI_ISL_936301, EPI_ISL_936302 | Wadsworth Center, New York State Department of Health | Wadsworth Center, New York State Department of Health | Kirsten St. George, Daryl M. Lamson, Alexis Russel, Matthew Shudt, Melissa A Leisner, Jonathan Plitnick, Navjot Singh, John Kelly, Erasmus Schneider, Erica Lasek-Nesselquist |
| EPI_ISL_936387, EPI_ISL_936388, EPI_ISL_936389, EPI_ISL_936390, EPI_ISL_936391, EPI_ISL_936393, EPI_ISL_936394, EPI_ISL_936395, EPI_ISL_936396, EPI_ISL_936397, EPI_ISL_936398, EPI_ISL_936399, EPI_ISL_936400, EPI_ISL_936401, EPI_ISL_936402, EPI_ISL_936404, EPI_ISL_936405, EPI_ISL_936406, EPI_ISL_936407, EPI_ISL_936408, EPI_ISL_936409, EPI_ISL_936410, EPI_ISL_936411, EPI_ISL_936412, EPI_ISL_936413, EPI_ISL_936414, EPI_ISL_936415, EPI_ISL_936416, EPI_ISL_936417, EPI_ISL_936418, EPI_ISL_936419, EPI_ISL_936420, EPI_ISL_936422, EPI_ISL_936423, EPI_ISL_936425, EPI_ISL_936426, EPI_ISL_936427, EPI_ISL_936428, EPI_ISL_936429, EPI_ISL_936430, EPI_ISL_936431, EPI_ISL_936433, EPI_ISL_936434, EPI_ISL_936435, EPI_ISL_936436, EPI_ISL_936437, EPI_ISL_936438, EPI_ISL_936439, EPI_ISL_936440, EPI_ISL_936441, EPI_ISL_936442, EPI_ISL_936443, EPI_ISL_936444, EPI_ISL_936445, EPI_ISL_936446, EPI_ISL_936447, EPI_ISL_936448, EPI_ISL_936449, EPI_ISL_936450, EPI_ISL_936451, EPI_ISL_936452, EPI_ISL_936453, EPI_ISL_936454, EPI_ISL_936455, EPI_ISL_936456, EPI_ISL_936457, EPI_ISL_936458, EPI_ISL_936459, EPI_ISL_936460, EPI_ISL_936461, EPI_ISL_936462, EPI_ISL_936463 | | | |
| see above | TGen North | TGen North | Jolene Bowers, Megan Folkerts, Chris French, Hayley Yaglom, Ashlyn Pfeiffer, Darrin Lemmer, Dave Engelthaler, The Arizona COVID Genomics Union (ACGU) |
| EPI_ISL_936493 | Santa Clara County Public Health Laboratory | Santa Clara County Public Health Laboratory | Santa Clara County Public Health Department |
| EPI_ISL_936497, EPI_ISL_936499, EPI_ISL_936500, EPI_ISL_936501, EPI_ISL_936502, EPI_ISL_936504, EPI_ISL_936505, EPI_ISL_936506, EPI_ISL_936507, EPI_ISL_936508, EPI_ISL_936509, EPI_ISL_936510, EPI_ISL_936511, EPI_ISL_936512, EPI_ISL_936513, EPI_ISL_936514, EPI_ISL_936516, EPI_ISL_936517, EPI_ISL_936518, EPI_ISL_936519, EPI_ISL_936520, EPI_ISL_936521, EPI_ISL_936522, EPI_ISL_936523, EPI_ISL_936524, EPI_ISL_936525, EPI_ISL_936526, EPI_ISL_936527, EPI_ISL_936528, EPI_ISL_936529, EPI_ISL_936530, EPI_ISL_936531, EPI_ISL_936532, EPI_ISL_936533, EPI_ISL_936534, EPI_ISL_936535, EPI_ISL_936536, EPI_ISL_936537, EPI_ISL_936538, EPI_ISL_936539, EPI_ISL_936540, EPI_ISL_936541, EPI_ISL_936542, EPI_ISL_936543, EPI_ISL_936544, EPI_ISL_936545, EPI_ISL_936546, EPI_ISL_936547, EPI_ISL_936548, EPI_ISL_936549, EPI_ISL_936550, EPI_ISL_936551, EPI_ISL_936552, EPI_ISL_936553, EPI_ISL_936556, EPI_ISL_936557, EPI_ISL_936558, EPI_ISL_936559, EPI_ISL_936560, EPI_ISL_936561, EPI_ISL_936562, EPI_ISL_936563, EPI_ISL_936564, EPI_ISL_936565, EPI_ISL_936566, EPI_ISL_936567, EPI_ISL_936568, EPI_ISL_936569, EPI_ISL_936570, EPI_ISL_936571, EPI_ISL_936572, EPI_ISL_936573, EPI_ISL_936574, EPI_ISL_936575, EPI_ISL_936577, EPI_ISL_936578, EPI_ISL_936579, EPI_ISL_936580, EPI_ISL_936581, EPI_ISL_936583, EPI_ISL_936584, EPI_ISL_936585, EPI_ISL_936586, EPI_ISL_936587, EPI_ISL_936588, EPI_ISL_936589, EPI_ISL_936590, EPI_ISL_936591, EPI_ISL_936592, EPI_ISL_936593, EPI_ISL_936594, EPI_ISL_936595, EPI_ISL_936596, EPI_ISL_936597, EPI_ISL_936598, EPI_ISL_936599, EPI_ISL_936600, EPI_ISL_936601, EPI_ISL_936602, EPI_ISL_936603, EPI_ISL_936604, EPI_ISL_936605, EPI_ISL_936606, EPI_ISL_936607, EPI_ISL_936608, EPI_ISL_936609, EPI_ISL_936610, EPI_ISL_936611, EPI_ISL_936612, EPI_ISL_936613, EPI_ISL_936614, EPI_ISL_936615, EPI_ISL_936616, EPI_ISL_936617, EPI_ISL_936618, EPI_ISL_936619, EPI_ISL_936620, EPI_ISL_936621, EPI_ISL_936622, EPI_ISL_936623, EPI_ISL_936624, EPI_ISL_936625, EPI_ISL_936626, EPI_ISL_936627, EPI_ISL_936628, EPI_ISL_936629, EPI_ISL_936630, EPI_ISL_936631, EPI_ISL_936632, EPI_ISL_936633, EPI_ISL_936634, EPI_ISL_936635, EPI_ISL_936636, EPI_ISL_936637, EPI_ISL_936638, EPI_ISL_936639, EPI_ISL_936640, EPI_ISL_936641, EPI_ISL_936642, EPI_ISL_936643, EPI_ISL_936644, EPI_ISL_936645, EPI_ISL_936647, EPI_ISL_936648, EPI_ISL_936649, EPI_ISL_936650, EPI_ISL_936651, EPI_ISL_936652, EPI_ISL_936653, EPI_ISL_936654, EPI_ISL_936655, EPI_ISL_936656, EPI_ISL_936657, EPI_ISL_936658, EPI_ISL_936659, EPI_ISL_936660, EPI_ISL_936661, EPI_ISL_936662, EPI_ISL_936663, EPI_ISL_936664, EPI_ISL_936665, EPI_ISL_936666, EPI_ISL_936667, EPI_ISL_936668, EPI_ISL_936669, EPI_ISL_936670, EPI_ISL_936671, EPI_ISL_936672, EPI_ISL_936673, EPI_ISL_936675, EPI_ISL_936676, EPI_ISL_936677, EPI_ISL_936678, EPI_ISL_936679, EPI_ISL_936680, EPI_ISL_936681, EPI_ISL_936682, EPI_ISL_936683, EPI_ISL_936684, EPI_ISL_936685, EPI_ISL_936686, EPI_ISL_936687, EPI_ISL_936688, EPI_ISL_936689, EPI_ISL_936690, EPI_ISL_936691, EPI_ISL_936692, EPI_ISL_936693, EPI_ISL_936694, EPI_ISL_936695, EPI_ISL_936696, EPI_ISL_936697, EPI_ISL_936698, EPI_ISL_936699, EPI_ISL_936700, EPI_ISL_936701, EPI_ISL_936702, EPI_ISL_936703, EPI_ISL_936704, EPI_ISL_936705, EPI_ISL_936706, EPI_ISL_936707, EPI_ISL_936708, EPI_ISL_936709, EPI_ISL_936710, EPI_ISL_936711, EPI_ISL_936712, EPI_ISL_936713, EPI_ISL_936714, EPI_ISL_936715, EPI_ISL_936716, EPI_ISL_936717, EPI_ISL_936718, EPI_ISL_936719, EPI_ISL_936720, EPI_ISL_936721, EPI_ISL_936722, EPI_ISL_936723, EPI_ISL_936724, EPI_ISL_936725, EPI_ISL_936726, EPI_ISL_936727, EPI_ISL_936728, EPI_ISL_936729, EPI_ISL_936730, EPI_ISL_936731, EPI_ISL_936732, EPI_ISL_936733, EPI_ISL_936734, EPI_ISL_936735, EPI_ISL_936736, EPI_ISL_936737, EPI_ISL_936738, EPI_ISL_936739, EPI_ISL_936740, EPI_ISL_936741, EPI_ISL_936742, EPI_ISL_936743, EPI_ISL_936744, EPI_ISL_936745, EPI_ISL_936746, EPI_ISL_936747, EPI_ISL_936748, EPI_ISL_936749, EPI_ISL_936750, EPI_ISL_936751, EPI_ISL_936752, EPI_ISL_936753, EPI_ISL_936754, EPI_ISL_936755, EPI_ISL_936756, EPI_ISL_936757, EPI_ISL_936758, EPI_ISL_936759, EPI_ISL_936760, EPI_ISL_936761, EPI_ISL_936762, EPI_ISL_936763, EPI_ISL_936764, EPI_ISL_936765, EPI_ISL_936766, EPI_ISL_936767, EPI_ISL_936768, EPI_ISL_936769, EPI_ISL_936770, EPI_ISL_936771, EPI_ISL_936772, EPI_ISL_936773, EPI_ISL_936774, EPI_ISL_936775, EPI_ISL_936776, EPI_ISL_936777, EPI_ISL_936778, EPI_ISL_936779, EPI_ISL_936780, EPI_ISL_936781, EPI_ISL_936782, EPI_ISL_936783, EPI_ISL_936784, EPI_ISL_936785, EPI_ISL_936786, EPI_ISL_936787, EPI_ISL_936788, EPI_ISL_936789, EPI_ISL_936790, EPI_ISL_936791, EPI_ISL_936792, EPI_ISL_936793, EPI_ISL_936794, EPI_ISL_936795, EPI_ISL_936796, EPI_ISL_936797, EPI_ISL_936798, EPI_ISL_936799, EPI_ISL_936800, EPI_ISL_936801, EPI_ISL_936802, EPI_ISL_936803, EPI_ISL_936804, EPI_ISL_936805, EPI_ISL_936806, EPI_ISL_936807, EPI_ISL_936808, EPI_ISL_936809, EPI_ISL_936810, EPI_ISL_936811, EPI_ISL_936812, EPI_ISL_936813, EPI_ISL_936814, EPI_ISL_936815, EPI_ISL_936816, EPI_ISL_936817, EPI_ISL_936818, EPI_ISL_936819, EPI_ISL_936820, EPI_ISL_936821, EPI_ISL_936822, EPI_ISL_936823, EPI_ISL_936824, EPI_ISL_936825, EPI_ISL_936826, EPI_ISL_936827, EPI_ISL_936828, EPI_ISL_936829, EPI_ISL_936830, EPI_ISL_936831, EPI_ISL_936832, EPI_ISL_936833, EPI_ISL_936834, EPI_ISL_936835, EPI_ISL_936836, EPI_ISL_936837, EPI_ISL_936838, EPI_ISL_936839, EPI_ISL_936840, EPI_ISL_936841, EPI_ISL_936842, EPI_ISL_936843, EPI_ISL_936844, EPI_ISL_936845, EPI_ISL_936846, EPI_ISL_936847, EPI_ISL_936848, EPI_ISL_936849, EPI_ISL_936850, EPI_ISL_936851, EPI_ISL_936852, EPI_ISL_936853, EPI_ISL_936854, EPI_ISL_936855, EPI_ISL_936856, EPI_ISL_936857, EPI_ISL_936858, EPI_ISL_936859, EPI_ISL_936860, EPI_ISL_936861, EPI_ISL_936862, EPI_ISL_936863, EPI_ISL_936864, EPI_ISL_936865, EPI_ISL_936866, EPI_ISL_936867, EPI_ISL_936868, EPI_ISL_936869, EPI_ISL_936870, EPI_ISL_936871, EPI_ISL_936872, EPI_ISL_936873, EPI_ISL_936874, EPI_ISL_936875, EPI_ISL_936876, EPI_ISL_936877, EPI_ISL_936878, EPI_ISL_936879, EPI_ISL_936880, EPI_ISL_936881, EPI_ISL_936882, EPI_ISL_936883, EPI_ISL_936884, EPI_ISL_936885, EPI_ISL_936886, EPI_ISL_936887, EPI_ISL_936888, EPI_ISL_936889, EPI_ISL_936890, EPI_ISL_936891, EPI_ISL_936892, EPI_ISL_936893, EPI_ISL_936894, EPI_ISL_936895, EPI_ISL_936896, EPI_ISL_936897, EPI_ISL_936898, EPI_ISL_936899, EPI_ISL_936900, EPI_ISL_936901, EPI_ISL_936902, EPI_ISL_936903, EPI_ISL_936904, EPI_ISL_936905, EPI_ISL_936906, EPI_ISL_936907, EPI_ISL_936908, EPI_ISL_936909, EPI_ISL_936910, EPI_ISL_936911, EPI_ISL_936912, EPI_ISL_936913, EPI_ISL_936914, EPI_ISL_936915, EPI_ISL_936917, EPI_ISL_936918, EPI_ISL_936919, EPI_ISL_936920, EPI_ISL_936921, EPI_ISL_936922, EPI_ISL_936923, EPI_ISL_936924, EPI_ISL_936925, EPI_ISL_936926, EPI_ISL_936927, EPI_ISL_936928, EPI_ISL_936929, EPI_ISL_936930, EPI_ISL_936931, EPI_ISL_936932, EPI_ISL_936933, EPI_ISL_936934, EPI_ISL_936935, EPI_ISL_936936, EPI_ISL_936937, EPI_ISL_936938, EPI_ISL_936939, EPI_ISL_936940, EPI_ISL_936941, EPI_ISL_936942, EPI_ISL_936943, EPI_ISL_936944, EPI_ISL_936945, EPI_ISL_936946, EPI_ISL_936947, EPI_ISL_936948, EPI_ISL_936949, EPI_ISL_936950, EPI_ISL_936951, EPI_ISL_936952, EPI_ISL_936953, EPI_ISL_936954, EPI_ISL_936955, EPI_ISL_936956, EPI_ISL_936957, EPI_ISL_936958, EPI_ISL_936959, EPI_ISL_936960, EPI_ISL_936961, EPI_ISL_936962, EPI_ISL_936963, EPI_ISL_936964, EPI_ISL_936965, EPI_ISL_936966, EPI_ISL_936967, EPI_ISL_936968, EPI_ISL_936969, EPI_ISL_936970, EPI_ISL_936971, EPI_ISL_936972, EPI_ISL_936973, EPI_ISL_936974, EPI_ISL_936975, EPI_ISL_936976, EPI_ISL_936977, EPI_ISL_936978, EPI_ISL_936979, EPI_ISL_936980, EPI_ISL_936981, EPI_ISL_936982, EPI_ISL_936983, EPI_ISL_936984, EPI_ISL_936985, EPI_ISL_936986, EPI_ISL_936987, EPI_ISL_936988, EPI_ISL_936989, EPI_ISL_936990, EPI_ISL_936991, EPI_ISL_936992, EPI_ISL_936993, EPI_ISL_936994, EPI_ISL_936995, EPI_ISL_936996, EPI_ISL_936997, EPI_ISL_936998, EPI_ISL_936999, EPI_ISL_937000, EPI_ISL_937001, EPI_ISL_937002, EPI_ISL_937003, EPI_ISL_937004, EPI_ISL_937005, EPI_ISL_937006, EPI_ISL_937007, EPI_ISL_937008, EPI_ISL_937009, EPI_ISL_937010, EPI_ISL_937011, EPI_ISL_937012, EPI_ISL_937013, EPI_ISL_937014, EPI_ISL_937015, EPI_ISL_937016, EPI_ISL_937017, EPI_ISL_937018, EPI_ISL_937019, EPI_ISL_937020, EPI_ISL_937021, EPI_ISL_937022, EPI_ISL_937023 | | | |
| see above | Northwestern Memorial Hospital | Ozer Lab | Ramon Lorenzo-Redondo, Lacy M. Simons, Chad J. Achenbach, Lawrence J. Jennings, Michael G. Ison, Judd F. Hultquist, Egon A. Ozer |
| EPI_ISL_937024, EPI_ISL_937025, EPI_ISL_937026, EPI_ISL_937027, EPI_ISL_937028, EPI_ISL_937029, EPI_ISL_937030, EPI_ISL_937031, EPI_ISL_937032, EPI_ISL_937033, EPI_ISL_937034, EPI_ISL_937035, EPI_ISL_937036, EPI_ISL_937037, EPI_ISL_937038, EPI_ISL_937039, EPI_ISL_937040, EPI_ISL_937041, EPI_ISL_937042, EPI_ISL_937043, EPI_ISL_937044, EPI_ISL_937045, EPI_ISL_937046, EPI_ISL_937047, EPI_ISL_937048, EPI_ISL_937049, EPI_ISL_937050, EPI_ISL_937051, EPI_ISL_937052, EPI_ISL_937053, EPI_ISL_937054, EPI_ISL_937055, EPI_ISL_937056, EPI_ISL_937057, EPI_ISL_937058, EPI_ISL_937059, EPI_ISL_937060, EPI_ISL_937061, EPI_ISL_937062, EPI_ISL_937063, EPI_ISL_937064, EPI_ISL_937065, EPI_ISL_937066, EPI_ISL_937067, EPI_ISL_937068, EPI_ISL_937069, EPI_ISL_937070, EPI_ISL_937071, EPI_ISL_937072, EPI_ISL_937073, EPI_ISL_937074, EPI_ISL_937075, EPI_ISL_937076, EPI_ISL_937077, EPI_ISL_937078, EPI_ISL_937079, EPI_ISL_937080, EPI_ISL_937081, EPI_ISL_937082, EPI_ISL_937083, EPI_ISL_937084, EPI_ISL_937085, EPI_ISL_937086, EPI_ISL_937087, EPI_ISL_937088, EPI_ISL_937089, EPI_ISL_937090, EPI_ISL_937091, EPI_ISL_937092, EPI_ISL_937093, EPI_ISL_937094, EPI_ISL_937095, EPI_ISL_937096, EPI_ISL_937097, EPI_ISL_937098, EPI_ISL_937099, EPI_ISL_937100, EPI_ISL_937101, EPI_ISL_937102, EPI_ISL_937103, EPI_ISL_937104, EPI_ISL_937105, EPI_ISL_937106, EPI_ISL_937107, EPI_ISL_937108, EPI_ISL_937109, EPI_ISL_937110, EPI_ISL_937111, EPI_ISL_937112, EPI_ISL_937113, EPI_ISL_937114, EPI_ISL_937115, EPI_ISL_937116, EPI_ISL_937117, EPI_ISL_937118, EPI_ISL_937119, EPI_ISL_937120 | | | |
| see above | Quest Diagnostics | Quest Diagnostics | Rosenthal,S.H., Gerasimova,A., Kagan,R.M., Anderson, B., Livingston, K.E., Hua, M., Liu Y., Shalhout, D.F., Owen, R., Lacbawan, F. |
| EPI_ISL_937121, EPI_ISL_937122, EPI_ISL_937123, EPI_ISL_937124, EPI_ISL_937125 | Quest Diagnostics | Quest Diagnostics | Rosenthal,S.H., Gerasimova,A., Kagan,R.M., Anderson, B., Hua, M., Liu Y., Bernstein, L.E., Livingston, K.E., Perez, A., Shalhout, D.F., Shlyakhter, I.A., Owen, R., Tanpaiboon, P., Lacbawan, F. |
| EPI_ISL_937126, EPI_ISL_937127, EPI_ISL_937128 | DOHMH Riverside | New York City Public Health Laboratory | Jade Wang, et al. |
| EPI_ISL_937129, EPI_ISL_937130, EPI_ISL_937131 | DOHMH Morrisania | New York City Public Health Laboratory | Jade Wang, et al. |
| EPI_ISL_937132, EPI_ISL_937133 | DOHMH Central Harlem | New York City Public Health Laboratory | Jade Wang, et al. |
| EPI_ISL_937134, EPI_ISL_937135 | DOHMH Fort Greene | New York City Public Health Laboratory | Jade Wang, et al. |
| EPI_ISL_937136, EPI_ISL_937137 | DOHMH Chelsea | New York City Public Health Laboratory | Jade Wang, et al. |
| EPI_ISL_937138 | DOHMH Crown Heights | New York City Public Health Laboratory | Jade Wang, et al. |
| EPI_ISL_937139, EPI_ISL_937140, EPI_ISL_937141, EPI_ISL_937142, EPI_ISL_937143, EPI_ISL_937144 | DOHMH Corona | New York City Public Health Laboratory | Jade Wang, et al. |
| EPI_ISL_937145, | DOHMH Jamaica | New York City Public Health Laboratory | Jade Wang, et al. |

| | | | |
|---|---|---|---|
| EPI_ISL_937146, EPI_ISL_937147, EPI_ISL_937148, EPI_ISL_937149, EPI_ISL_937150, EPI_ISL_937151 | | | |
| EPI_ISL_937152, EPI_ISL_937153, EPI_ISL_937154, EPI_ISL_937155, EPI_ISL_937156 | DOHMH PHL | New York City Public Health Laboratory | Jade Wang, et al. |
| EPI_ISL_937157, EPI_ISL_937158, EPI_ISL_937159, EPI_ISL_937160, EPI_ISL_937161, EPI_ISL_937162, EPI_ISL_937163 | DOHMH Jamaica | New York City Public Health Laboratory | Jade Wang, et al. |
| EPI_ISL_937164, EPI_ISL_937165 | DOHMH Corona | New York City Public Health Laboratory | Jade Wang, et al. |
| EPI_ISL_937166 | DOHMH Riverside | New York City Public Health Laboratory | Jade Wang, et al. |
| EPI_ISL_937167 | DOHMH Central Harlem | New York City Public Health Laboratory | Jade Wang, et al. |
| EPI_ISL_937168 | DOHMH Crown Heights | New York City Public Health Laboratory | Jade Wang, et al. |
| EPI_ISL_937169, EPI_ISL_937170, EPI_ISL_937171 | DOHMH PHL | New York City Public Health Laboratory | Jade Wang, et al. |
| EPI_ISL_937172 | DOHMH Central Harlem | New York City Public Health Laboratory | Jade Wang, et al. |
| EPI_ISL_937173 | DOHMH Crown Heights | New York City Public Health Laboratory | Jade Wang, et al. |
| EPI_ISL_937174 | DOHMH Chelsea | New York City Public Health Laboratory | Jade Wang, et al. |
| EPI_ISL_937175, EPI_ISL_937176, EPI_ISL_937177 | DOHMH Jamaica | New York City Public Health Laboratory | Jade Wang, et al. |
| EPI_ISL_937178, EPI_ISL_937179, EPI_ISL_937180, EPI_ISL_937181, EPI_ISL_937182, EPI_ISL_937183, EPI_ISL_937184 | DOHMH Corona | New York City Public Health Laboratory | Jade Wang, et al. |
| EPI_ISL_937185 | DOHMH PHL | New York City Public Health Laboratory | Jade Wang, et al. |
| EPI_ISL_937186, EPI_ISL_937187, EPI_ISL_937188, EPI_ISL_937189, EPI_ISL_937190 | OCME Office Of Chief Medical Examiner | New York City Public Health Laboratory | Jade Wang, et al. |
| EPI_ISL_937191, EPI_ISL_937192 | Department of Homeless Services | New York City Public Health Laboratory | Jade Wang, et al. |
| EPI_ISL_937193, EPI_ISL_937194, EPI_ISL_937195, EPI_ISL_937196, EPI_ISL_937197, EPI_ISL_937198, EPI_ISL_937199 | OCME Office Of Chief Medical Examiner | New York City Public Health Laboratory | Jade Wang, et al. |
| EPI_ISL_937200, EPI_ISL_937201 | DOHMH Fort Greene | New York City Public Health Laboratory | Jade Wang, et al. |
| EPI_ISL_937202, EPI_ISL_937203 | DOHMH Chelsea | New York City Public Health Laboratory | Jade Wang, et al. |
| EPI_ISL_937204, EPI_ISL_937205 | DOHMH Central Harlem | New York City Public Health Laboratory | Jade Wang, et al. |
| EPI_ISL_937206, EPI_ISL_937207 | DOHMH Morrisania | New York City Public Health Laboratory | Jade Wang, et al. |
| EPI_ISL_937208 | DOHMH Corona | New York City Public Health Laboratory | Jade Wang, et al. |
| EPI_ISL_937209, EPI_ISL_937210, EPI_ISL_937211, EPI_ISL_937212 | DOHMH PHL | New York City Public Health Laboratory | Jade Wang, et al. |
| EPI_ISL_937213 | DOHMH Jamaica | New York City Public Health Laboratory | Jade Wang, et al. |
| EPI_ISL_937214 | DOHMH Corona | New York City Public Health Laboratory | Jade Wang, et al. |
| EPI_ISL_937215 | DOHMH PHL | New York City Public Health Laboratory | Jade Wang, et al. |
| EPI_ISL_937216 | DOHMH Central Harlem | New York City Public Health Laboratory | Jade Wang, et al. |
| EPI_ISL_937217, EPI_ISL_937218 | DOHMH Chelsea | New York City Public Health Laboratory | Jade Wang, et al. |
| EPI_ISL_937219, EPI_ISL_937220, EPI_ISL_937221, EPI_ISL_937222, EPI_ISL_937223, EPI_ISL_937224, EPI_ISL_937225, EPI_ISL_937226 | DOHMH Jamaica | New York City Public Health Laboratory | Jade Wang, et al. |
| EPI_ISL_937227, EPI_ISL_937228, EPI_ISL_937229, EPI_ISL_937230, EPI_ISL_937231 | DOHMH Corona | New York City Public Health Laboratory | Jade Wang, et al. |
| EPI_ISL_937232, EPI_ISL_937233, EPI_ISL_937234, EPI_ISL_937235, EPI_ISL_937236 | DOHMH Morrisania | New York City Public Health Laboratory | Jade Wang, et al. |
| EPI_ISL_937237 | DOHMH Riverside | New York City Public Health Laboratory | Jade Wang, et al. |
| EPI_ISL_937238, EPI_ISL_937239 | DOHMH Morrisania | New York City Public Health Laboratory | Jade Wang, et al. |
| EPI_ISL_937240, EPI_ISL_937241, | OCME Office Of Chief Medical Examiner | New York City Public Health Laboratory | Jade Wang, et al. |

| | | | |
|---|---|---|---|
| EPI_ISL_937242, EPI_ISL_937243, EPI_ISL_937244, EPI_ISL_937245, EPI_ISL_937246, EPI_ISL_937247 | | | |
| EPI_ISL_937248, EPI_ISL_937249, EPI_ISL_937250 | Department of Homeless Services | New York City Public Health Laboratory | Jade Wang, et al. |
| EPI_ISL_937251, EPI_ISL_937252, EPI_ISL_937253, EPI_ISL_937254, EPI_ISL_937255 | OCME Office Of Chief Medical Examiner | New York City Public Health Laboratory | Jade Wang, et al. |
| EPI_ISL_937256, EPI_ISL_937257, EPI_ISL_937258, EPI_ISL_937259, EPI_ISL_937260, EPI_ISL_937261, EPI_ISL_937262, EPI_ISL_937263, EPI_ISL_937264, EPI_ISL_937265, EPI_ISL_937266, EPI_ISL_937267, EPI_ISL_937268, EPI_ISL_937269, EPI_ISL_937270, EPI_ISL_937271, EPI_ISL_937272 | | | |
| see above | NYC HH Elmhurst Hospital Medical Center | New York City Public Health Laboratory | Jade Wang, et al. |
| EPI_ISL_937273, EPI_ISL_937274, EPI_ISL_937277, EPI_ISL_937278, EPI_ISL_937280, EPI_ISL_937281, EPI_ISL_937282, EPI_ISL_937284, EPI_ISL_937285, EPI_ISL_937286, EPI_ISL_937287, EPI_ISL_937289, EPI_ISL_937291, EPI_ISL_937292, EPI_ISL_937293, EPI_ISL_937294, EPI_ISL_937295, EPI_ISL_937296, EPI_ISL_937297, EPI_ISL_937298, EPI_ISL_937299, EPI_ISL_937300, EPI_ISL_937301, EPI_ISL_937302, EPI_ISL_937303, EPI_ISL_937304, EPI_ISL_937305, EPI_ISL_937306, EPI_ISL_937307, EPI_ISL_937308, EPI_ISL_937309, EPI_ISL_937310, EPI_ISL_937311, EPI_ISL_937313, EPI_ISL_937314, EPI_ISL_937315, EPI_ISL_937316, EPI_ISL_937317, EPI_ISL_937319, EPI_ISL_937320, EPI_ISL_937322, EPI_ISL_937323, EPI_ISL_937324, EPI_ISL_937325, EPI_ISL_937326, EPI_ISL_937327, EPI_ISL_937329, EPI_ISL_937330, EPI_ISL_937331, EPI_ISL_937332, EPI_ISL_937333, EPI_ISL_937334, EPI_ISL_937335, EPI_ISL_937336, EPI_ISL_937337, EPI_ISL_937338, EPI_ISL_937339, EPI_ISL_937340, EPI_ISL_937341, EPI_ISL_937343, EPI_ISL_937344, EPI_ISL_937345, EPI_ISL_937354, EPI_ISL_937356, EPI_ISL_937357, EPI_ISL_937358, EPI_ISL_937359, EPI_ISL_937360, EPI_ISL_937361, EPI_ISL_937362, EPI_ISL_937363, EPI_ISL_937364, EPI_ISL_937365, EPI_ISL_937366, EPI_ISL_937367, EPI_ISL_937368, EPI_ISL_937369, EPI_ISL_937370, EPI_ISL_937371, EPI_ISL_937372, EPI_ISL_937373, EPI_ISL_937375, EPI_ISL_937376, EPI_ISL_937377, EPI_ISL_937379, EPI_ISL_937381, EPI_ISL_937382 | | | |
| see above | Utah Public Health Laboratory | Utah Public Health Laboratory | Erin L. Young, Kelly F. Oakeson, Tara Gallagher |
| EPI_ISL_937383, EPI_ISL_937384, EPI_ISL_937385, EPI_ISL_937386, EPI_ISL_937387, EPI_ISL_937388, EPI_ISL_937390, EPI_ISL_937391, EPI_ISL_937392, EPI_ISL_937393, EPI_ISL_937394, EPI_ISL_937395, EPI_ISL_937396, EPI_ISL_937397, EPI_ISL_937398, EPI_ISL_937399, EPI_ISL_937400, EPI_ISL_937401, EPI_ISL_937402, EPI_ISL_937403, EPI_ISL_937404, EPI_ISL_937405, EPI_ISL_937406, EPI_ISL_937407, EPI_ISL_937408, EPI_ISL_937409, EPI_ISL_937410, EPI_ISL_937411, EPI_ISL_937412, EPI_ISL_937413, EPI_ISL_937414, EPI_ISL_937415, EPI_ISL_937416, EPI_ISL_937418, EPI_ISL_937419, EPI_ISL_937420, EPI_ISL_937421, EPI_ISL_937422, EPI_ISL_937423, EPI_ISL_937424, EPI_ISL_937425, EPI_ISL_937426, EPI_ISL_937427, EPI_ISL_937428, EPI_ISL_937429, EPI_ISL_937430, EPI_ISL_937431, EPI_ISL_937432, EPI_ISL_937433, EPI_ISL_937434, EPI_ISL_937435, EPI_ISL_937436, EPI_ISL_937437, EPI_ISL_937438, EPI_ISL_937439, EPI_ISL_937440, EPI_ISL_937441, EPI_ISL_937442, EPI_ISL_937443, EPI_ISL_937444, EPI_ISL_937445, EPI_ISL_937446, EPI_ISL_937447, EPI_ISL_937448, EPI_ISL_937449, EPI_ISL_937450, EPI_ISL_937451, EPI_ISL_937452, EPI_ISL_937453, EPI_ISL_937454, EPI_ISL_937455, EPI_ISL_937456, EPI_ISL_937457, EPI_ISL_937458, EPI_ISL_937459, EPI_ISL_937460, EPI_ISL_937461, EPI_ISL_937462, EPI_ISL_937463, EPI_ISL_937464, EPI_ISL_937465, EPI_ISL_937466, EPI_ISL_937467, EPI_ISL_937468, EPI_ISL_937469, EPI_ISL_937470, EPI_ISL_937471, EPI_ISL_937472, EPI_ISL_937473, EPI_ISL_937474, EPI_ISL_937475, EPI_ISL_937476, EPI_ISL_937477, EPI_ISL_937478, EPI_ISL_937479, EPI_ISL_937480, EPI_ISL_937481, EPI_ISL_937482, EPI_ISL_937483, EPI_ISL_937484, EPI_ISL_937485, EPI_ISL_937486, EPI_ISL_937487, EPI_ISL_937488, EPI_ISL_937489, EPI_ISL_937490, EPI_ISL_937491, EPI_ISL_937492, EPI_ISL_937493, EPI_ISL_937494, EPI_ISL_937495, EPI_ISL_937496, EPI_ISL_937497, EPI_ISL_937498, EPI_ISL_937499, EPI_ISL_937500, EPI_ISL_937501, EPI_ISL_937502, EPI_ISL_937503, EPI_ISL_937504, EPI_ISL_937505, EPI_ISL_937506, EPI_ISL_937507, EPI_ISL_937508, EPI_ISL_937509, EPI_ISL_937510, EPI_ISL_937511, EPI_ISL_937512, EPI_ISL_937513 | | | |
| see above | Maine Health and Environmental Testing Laboratory (Maine HETL) | Tewhey Lab, The Jackson Laboratory | Matluk,N., Dewey,H., Iosue,F., Barter,M., Lynch,R., Munger,H. and Tewhey,R. |
| EPI_ISL_939609, EPI_ISL_939610, EPI_ISL_939611, EPI_ISL_939612, EPI_ISL_939613, EPI_ISL_939614, EPI_ISL_939615, EPI_ISL_939616, EPI_ISL_939617, EPI_ISL_939618 | Los Angeles County Public Health Laboratories | Los Angeles County Public Health Laboratories | P. Hemarajata et al. |
| EPI_ISL_939619, EPI_ISL_939620, EPI_ISL_939621, EPI_ISL_939622, EPI_ISL_939623, EPI_ISL_939624, EPI_ISL_939625 | USC Clinical Lab | Los Angeles County Public Health Laboratories | P. Hemarajata et al. |
| EPI_ISL_939651 | MD Laboratories | Los Angeles County Public Health Laboratories | P. Hemarajata et al. |
| EPI_ISL_939652 | Los Angeles County Public Health Laboratories | Los Angeles County Public Health Laboratories | P. Hemarajata et al. |
| EPI_ISL_939653 | MD Laboratories | Los Angeles County Public Health Laboratories | P. Hemarajata et al. |
| EPI_ISL_940145, EPI_ISL_940158, EPI_ISL_940553, EPI_ISL_940556, EPI_ISL_940559, EPI_ISL_940563 | Ohio Department of Health Laboratory | Ohio Department of Health Laboratory | Holmes, Jennifer; Eric Brandt, Keoni Omura, Glen McGillivary, Caitlin McDonnell, Jade Mowery, Stephanie Mccracken, Tyler Payne, Kirtana Ramadugu, Erica Leasure, Brent Lee, Kelsey Florek, Heather Blankenship, Quanta Brown, and Tammy Bannerman |
| EPI_ISL_940715, EPI_ISL_940716, EPI_ISL_940717, EPI_ISL_940718, EPI_ISL_940721, EPI_ISL_940723, EPI_ISL_940724, EPI_ISL_940725, EPI_ISL_940726, EPI_ISL_940727, EPI_ISL_940729, EPI_ISL_940730, EPI_ISL_940732 | | | |
| see above | City of Milwaukee Health Department Laboratory | City of Milwaukee Health Department Laboratory | Sanjib Bhattacharyya |
| EPI_ISL_940745, EPI_ISL_940748, EPI_ISL_940751, EPI_ISL_940754 | Ohio Department of Health Laboratory | Ohio Department of Health Laboratory | Holmes, Jennifer; Eric Brandt, Keoni Omura, Glen McGillivary, Caitlin McDonnell, Jade Mowery, Stephanie Mccracken, Tyler Payne, Kirtana Ramadugu, Erica Leasure, Brent Lee, Kelsey Florek, Heather Blankenship, Quanta Brown, and Tammy Bannerman |
| EPI_ISL_940756 | Centers for Disease Control and Prevention, Dengue Branch | Centers for Disease Control and Prevention, Dengue Branch | Gilberto A. Santiago, Glenda Gonzalez, Betzabel Flores, Keyla Charriez, Gabriela Paz-Bailey, Jorge L. Munoz-Jordan |
| EPI_ISL_940759 | Ohio Department of Health Laboratory | Ohio Department of Health Laboratory | Holmes, Jennifer; Eric Brandt, Keoni Omura, Glen McGillivary, Caitlin McDonnell, Jade Mowery, Stephanie Mccracken, Tyler Payne, Kirtana Ramadugu, Erica Leasure, Brent Lee, Kelsey Florek, Heather Blankenship, Quanta Brown, and Tammy Bannerman |
| EPI_ISL_940784, EPI_ISL_940785, EPI_ISL_940786 | City of Milwaukee Health Department Laboratory | City of Milwaukee Health Department Laboratory | Sanjib Bhattacharyya |
| EPI_ISL_940793, EPI_ISL_940794, EPI_ISL_940795, EPI_ISL_940796, EPI_ISL_940797, EPI_ISL_940798, EPI_ISL_940799, EPI_ISL_940800, EPI_ISL_940801, EPI_ISL_940802, EPI_ISL_940803, EPI_ISL_940804, EPI_ISL_940805, EPI_ISL_940806, EPI_ISL_940807, EPI_ISL_940808, EPI_ISL_940809, EPI_ISL_940810, EPI_ISL_940811, EPI_ISL_940812, EPI_ISL_940813, EPI_ISL_940814, EPI_ISL_940815, EPI_ISL_940816, EPI_ISL_940817, EPI_ISL_940818, EPI_ISL_940819, EPI_ISL_940820 | | | |
| see above | Houston Health Dept. | Houston Health Dept. | Ryker Penn, Pamela Brown, Adolpho Lara |
| EPI_ISL_940823, EPI_ISL_940824, EPI_ISL_940825, EPI_ISL_940826, EPI_ISL_940827, EPI_ISL_940828, EPI_ISL_940829, EPI_ISL_940830, EPI_ISL_940831, EPI_ISL_940832, EPI_ISL_940833, EPI_ISL_940834, EPI_ISL_940835, EPI_ISL_940836, EPI_ISL_940837, EPI_ISL_940838, EPI_ISL_940839, EPI_ISL_940840, EPI_ISL_940841 | | | |
| see above | Virginia DCLS | Virginia DCLS | Virginia DCLS |
| EPI_ISL_940846, EPI_ISL_940848, EPI_ISL_940860 | Ohio Department of Health Laboratory | Ohio Department of Health Laboratory | Holmes, Jennifer; Eric Brandt, Keoni Omura, Glen McGillivary, Caitlin McDonnell, Jade Mowery, Stephanie Mccracken, Tyler Payne, Kirtana Ramadugu, Erica Leasure, Brent Lee, Kelsey Florek, Heather Blankenship, Quanta Brown, and Tammy Bannerman |
| EPI_ISL_940894 | NCSLPH | NCSLPH | Chase K, Miller MC, Greene S, Glover W |
| EPI_ISL_940895, EPI_ISL_940896, EPI_ISL_940897, EPI_ISL_940898, EPI_ISL_940899, EPI_ISL_940900, EPI_ISL_940902, EPI_ISL_940903, EPI_ISL_940904, EPI_ISL_940905, EPI_ISL_940906, EPI_ISL_940907, EPI_ISL_940908, EPI_ISL_940909, EPI_ISL_940910, EPI_ISL_940911, EPI_ISL_940912, EPI_ISL_940913, EPI_ISL_940914, EPI_ISL_940915, EPI_ISL_940916, EPI_ISL_940917, EPI_ISL_940918, EPI_ISL_940919, EPI_ISL_940920, EPI_ISL_940921, EPI_ISL_940922, EPI_ISL_940923, EPI_ISL_940924, EPI_ISL_940925, EPI_ISL_940927, EPI_ISL_940928, EPI_ISL_940929, EPI_ISL_940930, EPI_ISL_940931, EPI_ISL_940932, EPI_ISL_940933, EPI_ISL_940934, EPI_ISL_940935, EPI_ISL_940936, EPI_ISL_940937, EPI_ISL_940938, EPI_ISL_940939, EPI_ISL_940940, EPI_ISL_940941, EPI_ISL_940942, EPI_ISL_940943, EPI_ISL_940944, EPI_ISL_940945, EPI_ISL_940946, EPI_ISL_940947, EPI_ISL_940948, EPI_ISL_940949, EPI_ISL_940950, EPI_ISL_940951, EPI_ISL_940952, EPI_ISL_940953, EPI_ISL_940954, EPI_ISL_940955, EPI_ISL_940956, EPI_ISL_940957, EPI_ISL_940958, EPI_ISL_940959, EPI_ISL_940960, EPI_ISL_940961, EPI_ISL_940962, EPI_ISL_940963, EPI_ISL_940964, EPI_ISL_940965, EPI_ISL_940966, EPI_ISL_940967, EPI_ISL_940968, EPI_ISL_940969, EPI_ISL_940970, EPI_ISL_940971, EPI_ISL_940972, EPI_ISL_940973, EPI_ISL_940974, EPI_ISL_940975, EPI_ISL_940976, EPI_ISL_940977, EPI_ISL_940978, EPI_ISL_940979, EPI_ISL_940980, EPI_ISL_940981, EPI_ISL_940982, EPI_ISL_940983, EPI_ISL_940984, EPI_ISL_940985, EPI_ISL_940986, EPI_ISL_940987, EPI_ISL_940988, EPI_ISL_940989, EPI_ISL_940990, EPI_ISL_940991, EPI_ISL_940992, EPI_ISL_940993, EPI_ISL_940994 | | | |
| see above | Centers for Disease Control and Prevention, Dengue Branch | Centers for Disease Control and Prevention, Dengue Branch | Gilberto A. Santiago, Glenda Gonzalez, Betzabel Flores, Keyla Charriez, Gabriela Paz-Bailey, Jorge L. Munoz-Jordan |
| EPI_ISL_941240, EPI_ISL_941241, EPI_ISL_941242, EPI_ISL_941243, EPI_ISL_941244, EPI_ISL_941245, EPI_ISL_941246, EPI_ISL_941247, EPI_ISL_941248, EPI_ISL_941249, EPI_ISL_941250, EPI_ISL_941251, EPI_ISL_941252, EPI_ISL_941253, EPI_ISL_941254, EPI_ISL_941255, EPI_ISL_941256, EPI_ISL_941257, EPI_ISL_941258, EPI_ISL_941259, EPI_ISL_941260, EPI_ISL_941261, EPI_ISL_941262, EPI_ISL_941263, EPI_ISL_941264, EPI_ISL_941265, EPI_ISL_941266, EPI_ISL_941267, EPI_ISL_941268, EPI_ISL_941269 | | | |
| see above | Virginia DCLS | Virginia DCLS | Virginia DCLS |
| EPI_ISL_941272 | Ohio Department of Health Laboratory | Ohio Department of Health Laboratory | Holmes, Jennifer; Eric Brandt, Keoni Omura, Glen McGillivary, Caitlin McDonnell, Jade Mowery, Stephanie Mccracken, Tyler Payne, Kirtana Ramadugu, Erica Leasure, Brent Lee, Kelsey Florek, Heather Blankenship, Quanta Brown, and Tammy Bannerman |
| EPI_ISL_941273 | NCSLPH | NCSLPH | Chase K, Miller MC, Greene S, Glover W |
| EPI_ISL_941274, | Ohio Department of Health Laboratory | Ohio Department of Health Laboratory | Holmes, Jennifer; Eric Brandt, Keoni Omura, Glen McGillivary, Caitlin McDonnell, Jade Mowery, Stephanie Mccracken, Tyler Payne, Kirtana Ramadugu, Erica Leasure, Brent Lee, Kelsey Florek, Heather Blankenship, Quanta Brown, and Tammy Bannerman |

| | | | |
|---|---|---|---|
| EPI_ISL_941298, EPI_ISL_941299 | | | |
| EPI_ISL_941300, EPI_ISL_941301, EPI_ISL_941302, EPI_ISL_941303, EPI_ISL_941310, EPI_ISL_941311, EPI_ISL_941312, EPI_ISL_941313, EPI_ISL_941314, EPI_ISL_941315, EPI_ISL_941316, EPI_ISL_941317, EPI_ISL_941318, EPI_ISL_941319, EPI_ISL_941320, EPI_ISL_941321, EPI_ISL_941322, EPI_ISL_941323, EPI_ISL_941324, EPI_ISL_941325, EPI_ISL_941326, EPI_ISL_941327, EPI_ISL_941328, EPI_ISL_941329, EPI_ISL_941330, EPI_ISL_941331, EPI_ISL_941332, EPI_ISL_941333, EPI_ISL_941334, EPI_ISL_941335 | | | |
| see above | New Mexico Department of Health Scientific Laboratory | New Mexico Department of Health Scientific Laboratory | Ellie Johnson, Anastacia Griego-Fisher, D'eldra Malone, Jennifer Benoit |
| EPI_ISL_941902, EPI_ISL_941903, EPI_ISL_941904, EPI_ISL_941905, EPI_ISL_941906, EPI_ISL_941907, EPI_ISL_941908, EPI_ISL_941909, EPI_ISL_941910, EPI_ISL_941911, EPI_ISL_941912, EPI_ISL_941913, EPI_ISL_941914, EPI_ISL_941915, EPI_ISL_941916, EPI_ISL_941917, EPI_ISL_941918, EPI_ISL_941919, EPI_ISL_941920, EPI_ISL_941921, EPI_ISL_941922, EPI_ISL_941923, EPI_ISL_941924 | | | |
| see above | Virginia DCLS | Virginia DCLS | Virginia DCLS |
| EPI_ISL_941927, EPI_ISL_941928, EPI_ISL_941929, EPI_ISL_941930, EPI_ISL_941931, EPI_ISL_941932, EPI_ISL_941934, EPI_ISL_941935, EPI_ISL_941936, EPI_ISL_941937, EPI_ISL_941938, EPI_ISL_941939 | | | |
| see above | Florida Bureau of Public Health Laboratories | Florida Bureau of Public Health Laboratories | Sarah Schmedes, Jason Blanton |
| EPI_ISL_942007, EPI_ISL_942008, EPI_ISL_942009, EPI_ISL_942010, EPI_ISL_942011, EPI_ISL_942012 | Centers for Disease Control and Prevention, Dengue Branch | Centers for Disease Control and Prevention, Dengue Branch | Gilberto A. Santiago, Glenda Gonzalez, Betzabel Flores, Keyla Charriez, Gabriela Paz-Bailey, Jorge L. Munoz-Jordan |
| EPI_ISL_942014, EPI_ISL_942015, EPI_ISL_942016, EPI_ISL_942017, EPI_ISL_942018, EPI_ISL_942019, EPI_ISL_942020, EPI_ISL_942021, EPI_ISL_942022, EPI_ISL_942023, EPI_ISL_942024, EPI_ISL_942025, EPI_ISL_942026, EPI_ISL_942027, EPI_ISL_942028, EPI_ISL_942029, EPI_ISL_942030, EPI_ISL_942031, EPI_ISL_942032, EPI_ISL_942033, EPI_ISL_942034, EPI_ISL_942035, EPI_ISL_942036, EPI_ISL_942037, EPI_ISL_942038, EPI_ISL_942039, EPI_ISL_942040, EPI_ISL_942041, EPI_ISL_942042, EPI_ISL_942043, EPI_ISL_942044, EPI_ISL_942045, EPI_ISL_942046, EPI_ISL_942047, EPI_ISL_942048, EPI_ISL_942049, EPI_ISL_942050, EPI_ISL_942051, EPI_ISL_942052, EPI_ISL_942053, EPI_ISL_942054, EPI_ISL_942055, EPI_ISL_942056, EPI_ISL_942057, EPI_ISL_942058, EPI_ISL_942059, EPI_ISL_942060, EPI_ISL_942061, EPI_ISL_942062, EPI_ISL_942063, EPI_ISL_942064, EPI_ISL_942065, EPI_ISL_942066, EPI_ISL_942067, EPI_ISL_942068, EPI_ISL_942069, EPI_ISL_942070, EPI_ISL_942071, EPI_ISL_942072, EPI_ISL_942073, EPI_ISL_942074, EPI_ISL_942075, EPI_ISL_942076, EPI_ISL_942077, EPI_ISL_942078, EPI_ISL_942079, EPI_ISL_942080, EPI_ISL_942081, EPI_ISL_942082, EPI_ISL_942083, EPI_ISL_942084, EPI_ISL_942085, EPI_ISL_942086, EPI_ISL_942087, EPI_ISL_942088, EPI_ISL_942089, EPI_ISL_942090, EPI_ISL_942091, EPI_ISL_942092, EPI_ISL_942093, EPI_ISL_942094, EPI_ISL_942095, EPI_ISL_942096, EPI_ISL_942097, EPI_ISL_942098, EPI_ISL_942099, EPI_ISL_942100, EPI_ISL_942101, EPI_ISL_942102, EPI_ISL_942103, EPI_ISL_942104, EPI_ISL_942105, EPI_ISL_942106, EPI_ISL_942107, EPI_ISL_942108, EPI_ISL_942109, EPI_ISL_942110, EPI_ISL_942111, EPI_ISL_942112, EPI_ISL_942113, EPI_ISL_942114, EPI_ISL_942115, EPI_ISL_942116, EPI_ISL_942117, EPI_ISL_942118, EPI_ISL_942119, EPI_ISL_942120, EPI_ISL_942121, EPI_ISL_942122, EPI_ISL_942123, EPI_ISL_942124, EPI_ISL_942125, EPI_ISL_942126, EPI_ISL_942127, EPI_ISL_942128, EPI_ISL_942129, EPI_ISL_942130, EPI_ISL_942131, EPI_ISL_942132, EPI_ISL_942133, EPI_ISL_942134, EPI_ISL_942135, EPI_ISL_942136, EPI_ISL_942137, EPI_ISL_942138, EPI_ISL_942139, EPI_ISL_942140, EPI_ISL_942141, EPI_ISL_942142, EPI_ISL_942143, EPI_ISL_942144, EPI_ISL_942145, EPI_ISL_942146, EPI_ISL_942147, EPI_ISL_942148, EPI_ISL_942149, EPI_ISL_942150, EPI_ISL_942151, EPI_ISL_942152, EPI_ISL_942153, EPI_ISL_942154, EPI_ISL_942155, EPI_ISL_942156, EPI_ISL_942157, EPI_ISL_942158, EPI_ISL_942159, EPI_ISL_942160, EPI_ISL_942161, EPI_ISL_942162, EPI_ISL_942163, EPI_ISL_942164, EPI_ISL_942165, EPI_ISL_942166, EPI_ISL_942167, EPI_ISL_942168, EPI_ISL_942169, EPI_ISL_942170, EPI_ISL_942171, EPI_ISL_942172, EPI_ISL_942173, EPI_ISL_942174, EPI_ISL_942175, EPI_ISL_942176, EPI_ISL_942177, EPI_ISL_942178, EPI_ISL_942179, EPI_ISL_942180, EPI_ISL_942181, EPI_ISL_942182, EPI_ISL_942183, EPI_ISL_942184, EPI_ISL_942185, EPI_ISL_942186, EPI_ISL_942187, EPI_ISL_942188, EPI_ISL_942189, EPI_ISL_942190, EPI_ISL_942191, EPI_ISL_942192, EPI_ISL_942193, EPI_ISL_942194, EPI_ISL_942195, EPI_ISL_942196, EPI_ISL_942197, EPI_ISL_942198, EPI_ISL_942199, EPI_ISL_942200, EPI_ISL_942201, EPI_ISL_942202, EPI_ISL_942203, EPI_ISL_942204, EPI_ISL_942205, EPI_ISL_942206, EPI_ISL_942207, EPI_ISL_942208, EPI_ISL_942209, EPI_ISL_942210, EPI_ISL_942211, EPI_ISL_942212, EPI_ISL_942213, EPI_ISL_942214, EPI_ISL_942215, EPI_ISL_942216, EPI_ISL_942217, EPI_ISL_942218, EPI_ISL_942219, EPI_ISL_942220, EPI_ISL_942221, EPI_ISL_942222, EPI_ISL_942223, EPI_ISL_942224, EPI_ISL_942225, EPI_ISL_942226, EPI_ISL_942227, EPI_ISL_942228, EPI_ISL_942229, EPI_ISL_942230, EPI_ISL_942231, EPI_ISL_942232, EPI_ISL_942233, EPI_ISL_942234, EPI_ISL_942235, EPI_ISL_942236, EPI_ISL_942237, EPI_ISL_942238, EPI_ISL_942239, EPI_ISL_942240, EPI_ISL_942241, EPI_ISL_942242, EPI_ISL_942243, EPI_ISL_942244, EPI_ISL_942245, EPI_ISL_942246, EPI_ISL_942247, EPI_ISL_942248, EPI_ISL_942249, EPI_ISL_942250, EPI_ISL_942251, EPI_ISL_942252, EPI_ISL_942253, EPI_ISL_942254, EPI_ISL_942255, EPI_ISL_942256, EPI_ISL_942257, EPI_ISL_942258, EPI_ISL_942259, EPI_ISL_942260, EPI_ISL_942261, EPI_ISL_942262, EPI_ISL_942263, EPI_ISL_942264, EPI_ISL_942265, EPI_ISL_942266, EPI_ISL_942267, EPI_ISL_942268, EPI_ISL_942269, EPI_ISL_942270, EPI_ISL_942271, EPI_ISL_942272, EPI_ISL_942273, EPI_ISL_942274, EPI_ISL_942275, EPI_ISL_942276, EPI_ISL_942277, EPI_ISL_942278, EPI_ISL_942279, EPI_ISL_942280, EPI_ISL_942281, EPI_ISL_942282, EPI_ISL_942283, EPI_ISL_942284, EPI_ISL_942285, EPI_ISL_942286, EPI_ISL_942287, EPI_ISL_942288, EPI_ISL_942289, EPI_ISL_942290, EPI_ISL_942291, EPI_ISL_942292, EPI_ISL_942293, EPI_ISL_942294, EPI_ISL_942295, EPI_ISL_942296, EPI_ISL_942297, EPI_ISL_942298, EPI_ISL_942299, EPI_ISL_942300, EPI_ISL_942301, EPI_ISL_942302, EPI_ISL_942303, EPI_ISL_942304, EPI_ISL_942305, EPI_ISL_942306, EPI_ISL_942307, EPI_ISL_942308, EPI_ISL_942309, EPI_ISL_942310, EPI_ISL_942311, EPI_ISL_942312, EPI_ISL_942313, EPI_ISL_942314, EPI_ISL_942315, EPI_ISL_942316, EPI_ISL_942317, EPI_ISL_942318, EPI_ISL_942319, EPI_ISL_942320, EPI_ISL_942321, EPI_ISL_942322, EPI_ISL_942323, EPI_ISL_942324, EPI_ISL_942325 | | | |
| see above | Wisconsin State Laboratory of Hygiene Communicable Disease Division | Wisconsin State Laboratory of Hygiene Communicable Disease Division | Kelsey R. Florek, Abigail C. Shockey |
| EPI_ISL_942326, EPI_ISL_942327, EPI_ISL_942328, EPI_ISL_942329, EPI_ISL_942330, EPI_ISL_942331, EPI_ISL_942332, EPI_ISL_942333, EPI_ISL_942334, EPI_ISL_942335, EPI_ISL_942336, EPI_ISL_942337, EPI_ISL_942338, EPI_ISL_942339, EPI_ISL_942340, EPI_ISL_942341, EPI_ISL_942342, EPI_ISL_942343, EPI_ISL_942344, EPI_ISL_942345, EPI_ISL_942346, EPI_ISL_942347, EPI_ISL_942348, EPI_ISL_942349, EPI_ISL_942350, EPI_ISL_942351, EPI_ISL_942352, EPI_ISL_942353, EPI_ISL_942354, EPI_ISL_942355, EPI_ISL_942356, EPI_ISL_942357, EPI_ISL_942358, EPI_ISL_942359, EPI_ISL_942360, EPI_ISL_942361, EPI_ISL_942362, EPI_ISL_942363, EPI_ISL_942364, EPI_ISL_942365, EPI_ISL_942366, EPI_ISL_942367, EPI_ISL_942368, EPI_ISL_942369, EPI_ISL_942370, EPI_ISL_942371, EPI_ISL_942372, EPI_ISL_942373 | | | |
| see above | MD PHL | MD PHL | Maryland Department of Health Laboratories Administration |
| EPI_ISL_942377, EPI_ISL_942378, EPI_ISL_942379, EPI_ISL_942380, EPI_ISL_942381, EPI_ISL_942382, EPI_ISL_942383, EPI_ISL_942384, EPI_ISL_942385, EPI_ISL_942386, EPI_ISL_942387, EPI_ISL_942388, EPI_ISL_942389, EPI_ISL_942390, EPI_ISL_942391, EPI_ISL_942392, EPI_ISL_942393, EPI_ISL_942394, EPI_ISL_942395, EPI_ISL_942396, EPI_ISL_942397, EPI_ISL_942398, EPI_ISL_942399, EPI_ISL_942400, EPI_ISL_942401, EPI_ISL_942402, EPI_ISL_942403, EPI_ISL_942404, EPI_ISL_942405 | | | |
| see above | Virginia DCLS | Virginia DCLS | Virginia DCLS |
| EPI_ISL_942408, EPI_ISL_942409, EPI_ISL_942410, EPI_ISL_942411, EPI_ISL_942412, EPI_ISL_942413, EPI_ISL_942414 | Gundersen Molecular Diagnostics Laboratory | Kabara Cancer Research Institute | Craig S. Richmond, Paraic A. Kenny |
| EPI_ISL_942415, EPI_ISL_942416, EPI_ISL_942417, EPI_ISL_942418 | Gundersen Clinical Microbiology Laboratory | Kabara Cancer Research Institute | Craig S. Richmond, Paraic A. Kenny |
| EPI_ISL_942419, EPI_ISL_942420, EPI_ISL_942421, EPI_ISL_942422, EPI_ISL_942423, EPI_ISL_942424, EPI_ISL_942425, EPI_ISL_942426, EPI_ISL_942427, EPI_ISL_942428, EPI_ISL_942429, EPI_ISL_942430, EPI_ISL_942431, EPI_ISL_942432, EPI_ISL_942433, EPI_ISL_942434, EPI_ISL_942435, EPI_ISL_942436, EPI_ISL_942437, EPI_ISL_942438, EPI_ISL_942439, EPI_ISL_942440, EPI_ISL_942441, EPI_ISL_942442, EPI_ISL_942443, EPI_ISL_942444, EPI_ISL_942445, EPI_ISL_942446, EPI_ISL_942447, EPI_ISL_942448, EPI_ISL_942449, EPI_ISL_942450, EPI_ISL_942451, EPI_ISL_942452, EPI_ISL_942453, EPI_ISL_942454, EPI_ISL_942455, EPI_ISL_942456, EPI_ISL_942457, EPI_ISL_942458, EPI_ISL_942459, EPI_ISL_942460, EPI_ISL_942461, EPI_ISL_942462, EPI_ISL_942463, EPI_ISL_942464, EPI_ISL_942465, EPI_ISL_942466, EPI_ISL_942467, EPI_ISL_942468, EPI_ISL_942469, EPI_ISL_942470, EPI_ISL_942471, EPI_ISL_942472, EPI_ISL_942473, EPI_ISL_942474, EPI_ISL_942475, EPI_ISL_942476, EPI_ISL_942477, EPI_ISL_942478, EPI_ISL_942479, EPI_ISL_942480, EPI_ISL_942481 | | | |
| see above | Gundersen Molecular Diagnostics Laboratory | Kabara Cancer Research Institute | Craig S. Richmond, Paraic A. Kenny |
| EPI_ISL_942482, EPI_ISL_942483, EPI_ISL_942484, EPI_ISL_942485, EPI_ISL_942486, EPI_ISL_942487, EPI_ISL_942488, EPI_ISL_942489, EPI_ISL_942490, EPI_ISL_942491, EPI_ISL_942492, EPI_ISL_942493, EPI_ISL_942494 | | | |
| see above | Gundersen Clinical Microbiology Laboratory | Kabara Cancer Research Institute | Craig S. Richmond, Paraic A. Kenny |
| EPI_ISL_942495, EPI_ISL_942496, EPI_ISL_942497, EPI_ISL_942498, EPI_ISL_942499, EPI_ISL_942500, EPI_ISL_942501, EPI_ISL_942502, EPI_ISL_942503, EPI_ISL_942504 | Gundersen Molecular Diagnostics Laboratory | Kabara Cancer Research Institute | Craig S. Richmond, Paraic A. Kenny |
| EPI_ISL_942505, EPI_ISL_942506 | Gundersen Clinical Microbiology Laboratory | Kabara Cancer Research Institute | Craig S. Richmond, Paraic A. Kenny |
| EPI_ISL_942507, EPI_ISL_942508, EPI_ISL_942509, EPI_ISL_942510, EPI_ISL_942511, EPI_ISL_942512, EPI_ISL_942513, EPI_ISL_942514, EPI_ISL_942516, EPI_ISL_942517, EPI_ISL_942518, EPI_ISL_942519, EPI_ISL_942520, EPI_ISL_942521, EPI_ISL_942522, EPI_ISL_942523, EPI_ISL_942524, EPI_ISL_942525, EPI_ISL_942526, EPI_ISL_942527, EPI_ISL_942528, EPI_ISL_942529, EPI_ISL_942530, EPI_ISL_942531, EPI_ISL_942532, EPI_ISL_942533, EPI_ISL_942534, EPI_ISL_942535, EPI_ISL_942536, EPI_ISL_942537, EPI_ISL_942538, EPI_ISL_942539, EPI_ISL_942540, EPI_ISL_942541, EPI_ISL_942542, EPI_ISL_942543, EPI_ISL_942544, EPI_ISL_942545, EPI_ISL_942546, EPI_ISL_942547, EPI_ISL_942548, EPI_ISL_942549, EPI_ISL_942550, EPI_ISL_942551, EPI_ISL_942552, EPI_ISL_942553, EPI_ISL_942554, EPI_ISL_942555, EPI_ISL_942556, EPI_ISL_942557, EPI_ISL_942558, EPI_ISL_942559, EPI_ISL_942560, EPI_ISL_942561, EPI_ISL_942562, EPI_ISL_942563, EPI_ISL_942564, EPI_ISL_942565, EPI_ISL_942566, EPI_ISL_942567, EPI_ISL_942568, EPI_ISL_942569, EPI_ISL_942570, EPI_ISL_942571, EPI_ISL_942572, EPI_ISL_942573, EPI_ISL_942574, EPI_ISL_942575, EPI_ISL_942576, EPI_ISL_942577, EPI_ISL_942578, EPI_ISL_942579, EPI_ISL_942580, EPI_ISL_942581, EPI_ISL_942582, EPI_ISL_942583, EPI_ISL_942584, EPI_ISL_942585, EPI_ISL_942586, EPI_ISL_942587, EPI_ISL_942588, EPI_ISL_942589, EPI_ISL_942590, EPI_ISL_942591, EPI_ISL_942592, EPI_ISL_942593, EPI_ISL_942594, EPI_ISL_942595, EPI_ISL_942596, EPI_ISL_942597, EPI_ISL_942598, EPI_ISL_942599, EPI_ISL_942600, EPI_ISL_942601, EPI_ISL_942602, EPI_ISL_942603, EPI_ISL_942604, EPI_ISL_942605, EPI_ISL_942606, EPI_ISL_942607, EPI_ISL_942608, EPI_ISL_942609, EPI_ISL_942610, EPI_ISL_942611, EPI_ISL_942612, EPI_ISL_942613, EPI_ISL_942614, EPI_ISL_942615, EPI_ISL_942616, EPI_ISL_942617, EPI_ISL_942618, EPI_ISL_942619, EPI_ISL_942620, EPI_ISL_942621, EPI_ISL_942622, EPI_ISL_942623, EPI_ISL_942624, EPI_ISL_942625, EPI_ISL_942626, EPI_ISL_942627, EPI_ISL_942628, EPI_ISL_942629, EPI_ISL_942630, EPI_ISL_942631, EPI_ISL_942632, EPI_ISL_942633, EPI_ISL_942634, EPI_ISL_942635, EPI_ISL_942636, EPI_ISL_942637, EPI_ISL_942638, EPI_ISL_942639, EPI_ISL_942640, EPI_ISL_942641, EPI_ISL_942642, EPI_ISL_942643, EPI_ISL_942644, EPI_ISL_942645, EPI_ISL_942646, EPI_ISL_942647, EPI_ISL_942648, EPI_ISL_942649, EPI_ISL_942650, EPI_ISL_942651, EPI_ISL_942652, EPI_ISL_942653, EPI_ISL_942654, EPI_ISL_942655, EPI_ISL_942656, EPI_ISL_942657, EPI_ISL_942658, EPI_ISL_942659, EPI_ISL_942660, EPI_ISL_942661, EPI_ISL_942662, EPI_ISL_942663, EPI_ISL_942664, EPI_ISL_942665, EPI_ISL_942666, EPI_ISL_942667, EPI_ISL_942668, EPI_ISL_942669, EPI_ISL_942670, EPI_ISL_942671, EPI_ISL_942672, EPI_ISL_942673, EPI_ISL_942674, EPI_ISL_942675, EPI_ISL_942676, EPI_ISL_942677, EPI_ISL_942678, EPI_ISL_942679, EPI_ISL_942680, EPI_ISL_942681, EPI_ISL_942682, EPI_ISL_942683, EPI_ISL_942684, EPI_ISL_942685, EPI_ISL_942686, EPI_ISL_942687, EPI_ISL_942688, EPI_ISL_942689, EPI_ISL_942690, EPI_ISL_942691, EPI_ISL_942692, EPI_ISL_942693, EPI_ISL_942694, EPI_ISL_942695, EPI_ISL_942696, EPI_ISL_942697, EPI_ISL_942698, EPI_ISL_942699, EPI_ISL_942700, EPI_ISL_942701, EPI_ISL_942702, EPI_ISL_942703, EPI_ISL_942704, EPI_ISL_942705, EPI_ISL_942706, EPI_ISL_942707, EPI_ISL_942708, EPI_ISL_942709, EPI_ISL_942710, EPI_ISL_942711, EPI_ISL_942712, EPI_ISL_942713, EPI_ISL_942714, EPI_ISL_942715, EPI_ISL_942716, EPI_ISL_942717, EPI_ISL_942718, EPI_ISL_942719, EPI_ISL_942720, EPI_ISL_942721, EPI_ISL_942722, EPI_ISL_942723, EPI_ISL_942724, EPI_ISL_942725, EPI_ISL_942726, EPI_ISL_942727, EPI_ISL_942728, EPI_ISL_942729, EPI_ISL_942730, EPI_ISL_942731, EPI_ISL_942732, EPI_ISL_942733, EPI_ISL_942734, EPI_ISL_942735, EPI_ISL_942736, EPI_ISL_942737, EPI_ISL_942738, EPI_ISL_942739, EPI_ISL_942740, EPI_ISL_942741, EPI_ISL_942742, EPI_ISL_942743, EPI_ISL_942744, EPI_ISL_942745, EPI_ISL_942746, EPI_ISL_942747, EPI_ISL_942748, EPI_ISL_942749, EPI_ISL_942750, EPI_ISL_942751, EPI_ISL_942752, EPI_ISL_942753, EPI_ISL_942754, EPI_ISL_942755, EPI_ISL_942756, EPI_ISL_942757, EPI_ISL_942758, EPI_ISL_942759, EPI_ISL_942760, EPI_ISL_942761, EPI_ISL_942762, EPI_ISL_942763, EPI_ISL_942764, EPI_ISL_942765, EPI_ISL_942766, EPI_ISL_942767, EPI_ISL_942768, EPI_ISL_942769, EPI_ISL_942770, EPI_ISL_942771, EPI_ISL_942772, EPI_ISL_942773, EPI_ISL_942774, EPI_ISL_942775, EPI_ISL_942776, EPI_ISL_942777, EPI_ISL_942778, EPI_ISL_942779, EPI_ISL_942780, EPI_ISL_942781, EPI_ISL_942782, EPI_ISL_942783, EPI_ISL_942784, EPI_ISL_942785, EPI_ISL_942786, EPI_ISL_942787, EPI_ISL_942788, EPI_ISL_942789, EPI_ISL_942790, EPI_ISL_942791, EPI_ISL_942792, EPI_ISL_942793, EPI_ISL_942794, EPI_ISL_942795, EPI_ISL_942796, EPI_ISL_942797, EPI_ISL_942798, EPI_ISL_942799, EPI_ISL_942800, EPI_ISL_942801, EPI_ISL_942802, EPI_ISL_942803, EPI_ISL_942804, EPI_ISL_942805, EPI_ISL_942806, EPI_ISL_942807, EPI_ISL_942808, EPI_ISL_942809, EPI_ISL_942810, EPI_ISL_942811, EPI_ISL_942812, EPI_ISL_942813, EPI_ISL_942814, EPI_ISL_942815, EPI_ISL_942816, EPI_ISL_942817, EPI_ISL_942818, EPI_ISL_942819, EPI_ISL_942820, EPI_ISL_942821, EPI_ISL_942822, EPI_ISL_942823, EPI_ISL_942824, EPI_ISL_942825, EPI_ISL_942826, EPI_ISL_942827, EPI_ISL_942828, EPI_ISL_942829, EPI_ISL_942830, EPI_ISL_942831, EPI_ISL_942832, EPI_ISL_942833, EPI_ISL_942834, EPI_ISL_942835, EPI_ISL_942836, EPI_ISL_942837, EPI_ISL_942838, EPI_ISL_942839, EPI_ISL_942840, EPI_ISL_942841, EPI_ISL_942842, EPI_ISL_942843, EPI_ISL_942844, EPI_ISL_942845, EPI_ISL_942846, EPI_ISL_942847, EPI_ISL_942848, EPI_ISL_942849, EPI_ISL_942850, EPI_ISL_942851, EPI_ISL_942852, EPI_ISL_942853, EPI_ISL_942854, EPI_ISL_942855, EPI_ISL_942856, EPI_ISL_942857, EPI_ISL_942858, EPI_ISL_942859, EPI_ISL_942860, EPI_ISL_942861, EPI_ISL_942862, EPI_ISL_942863, EPI_ISL_942864, EPI_ISL_942865, EPI_ISL_942866, EPI_ISL_942867, EPI_ISL_942868, EPI_ISL_942869, EPI_ISL_942870, EPI_ISL_942871, EPI_ISL_942872, EPI_ISL_942873, EPI_ISL_942874, EPI_ISL_942875, EPI_ISL_942876, EPI_ISL_942877, EPI_ISL_942878, EPI_ISL_942879, EPI_ISL_942880, EPI_ISL_942881, EPI_ISL_942882, EPI_ISL_942883, EPI_ISL_942884, EPI_ISL_942885, EPI_ISL_942886, EPI_ISL_942887, EPI_ISL_942888, EPI_ISL_942889, EPI_ISL_942890, EPI_ISL_942891, EPI_ISL_942892, EPI_ISL_942893, EPI_ISL_942894 | | | |
| see above | Gundersen Molecular Diagnostics Laboratory | Kabara Cancer Research Institute | Craig S. Richmond, Paraic A. Kenny |
| EPI_ISL_942900, EPI_ISL_942901, EPI_ISL_942902, EPI_ISL_942903, EPI_ISL_942904, EPI_ISL_942905, EPI_ISL_942906, EPI_ISL_942907, EPI_ISL_942908, EPI_ISL_942909, EPI_ISL_942910, EPI_ISL_942911, EPI_ISL_942912, EPI_ISL_942913, EPI_ISL_942914, EPI_ISL_942915, EPI_ISL_942916, EPI_ISL_942917, EPI_ISL_942918, EPI_ISL_942919, EPI_ISL_942920, EPI_ISL_942921, EPI_ISL_942922, EPI_ISL_942923, EPI_ISL_942924, EPI_ISL_942925, EPI_ISL_942926 | | | |
| see above | Houston Health Dept. | Houston Health Dept. | Ryker Penn, Pamela Brown, Adolpho Lara |
| EPI_ISL_942927 | LESP Nuevo Leon/Grupo de Diagnostico ARIES | Instituto de Diagnostico y Referencia Epidemiologicos (INDRE) | Claudia Wong-Arambula, Abril Rodriguez-Maldonado, Fabiola Garces-Ayala, Natividad Cruz-Ortiz, Tatiana Nunez-Garcia, Gisela Barrera-Badillo, Lucia Hernandez-Rivas, Irma Lopez-Martinez, Ernesto Ramirez-Gonzalez. |
| EPI_ISL_942928, EPI_ISL_942929 | Instituto de Diagnostico y Referencia Epidemiologicos INDRE_RNLSP | Instituto de Diagnostico y Referencia Epidemiologicos (INDRE) | Claudia Wong-Arambula, Abril Rodriguez-Maldonado, Fabiola Garces-Ayala, Natividad Cruz-Ortiz, Tatiana Nunez-Garcia, Gisela Barrera-Badillo, Lucia Hernandez-Rivas, Irma Lopez-Martinez, Ernesto Ramirez-Gonzalez. |
| EPI_ISL_942935, EPI_ISL_942936, | Wyoming Public Health Laboratory | Wyoming Public Health Laboratory | Noah Hull, Taylor Fearing, Lynette Gumbleton, Channing Weber, Ashley Norberg, Bailey Bowcutt, and Wanda Manley |

| | | | |
|---|---|---|---|
| EPI_ISL_942938, EPI_ISL_942939 | | | |
| EPI_ISL_942940 | Gundersen Molecular Diagnostics Laboratory | Kabara Cancer Research Institute | Craig S. Richmond, Paraic A. Kenny |
| EPI_ISL_942941 | Gundersen Clinical Microbiology Laboratory | Kabara Cancer Research Institute | Craig S. Richmond, Paraic A. Kenny |
| EPI_ISL_942942, EPI_ISL_942943, EPI_ISL_942944, EPI_ISL_942945, EPI_ISL_942946, EPI_ISL_942947, EPI_ISL_942948, EPI_ISL_942949, EPI_ISL_942950 | Gundersen Molecular Diagnostics Laboratory | Kabara Cancer Research Institute | Craig S. Richmond, Paraic A. Kenny |
| EPI_ISL_943550 | Baylor Scott & White-Temple | Baylor Scott & White-Temple | Ari Rao, Linden Morales, Kimberly Walker, Marcus Volz, Shelby Hendrickson |
| EPI_ISL_943569 | Arizona State University | Arizona State University | Peter T. Skidmore, Emily A. Kaelin, LaRinda A. Holland, Nicholas J. Mellor, Kristina Buss, Joy M. Blain, Valerie Harris, Joshua LaBaer, Vel Murugan, Efrem S. Lim |
| EPI_ISL_943614, EPI_ISL_943615, EPI_ISL_943623, EPI_ISL_943624, EPI_ISL_943625, EPI_ISL_943627, EPI_ISL_943629, EPI_ISL_943631, EPI_ISL_943639, EPI_ISL_943641, EPI_ISL_943642, EPI_ISL_943644, EPI_ISL_943646, EPI_ISL_943648, EPI_ISL_943649, EPI_ISL_943650, EPI_ISL_943653, EPI_ISL_943654, EPI_ISL_943655, EPI_ISL_943656, EPI_ISL_943661, EPI_ISL_943665, EPI_ISL_943666, EPI_ISL_943670, EPI_ISL_943673, EPI_ISL_943674, EPI_ISL_943680, EPI_ISL_943681, EPI_ISL_943683, EPI_ISL_943684, EPI_ISL_943685, EPI_ISL_943686, EPI_ISL_943688, EPI_ISL_943690, EPI_ISL_943691, EPI_ISL_943693, EPI_ISL_943694, EPI_ISL_943695, EPI_ISL_943696, EPI_ISL_943697, EPI_ISL_943700, EPI_ISL_943701, EPI_ISL_943702, EPI_ISL_943703, EPI_ISL_943704, EPI_ISL_943705, EPI_ISL_943706, EPI_ISL_943707, EPI_ISL_943708, EPI_ISL_943709, EPI_ISL_943710, EPI_ISL_943711, EPI_ISL_943714, EPI_ISL_943716, EPI_ISL_943718, EPI_ISL_943721, EPI_ISL_943722, EPI_ISL_943726, EPI_ISL_943727, EPI_ISL_943728, EPI_ISL_943730, EPI_ISL_943735, EPI_ISL_943736, EPI_ISL_943737, EPI_ISL_943738, EPI_ISL_943739, EPI_ISL_943743, EPI_ISL_943746, EPI_ISL_943747, EPI_ISL_943748, EPI_ISL_943750, EPI_ISL_943751, EPI_ISL_943752, EPI_ISL_943755, EPI_ISL_943757, EPI_ISL_943758, EPI_ISL_943759, EPI_ISL_943760, EPI_ISL_943761, EPI_ISL_943763, EPI_ISL_943764, EPI_ISL_943766, EPI_ISL_943768, EPI_ISL_943771, EPI_ISL_943772, EPI_ISL_943773, EPI_ISL_943775, EPI_ISL_943781, EPI_ISL_943786, EPI_ISL_943787, EPI_ISL_943788, EPI_ISL_943791, EPI_ISL_943796, EPI_ISL_943798, EPI_ISL_943799, EPI_ISL_943800, EPI_ISL_943803 | | | |
| see above | Utah Public Health Laboratory | Utah Public Health Laboratory | Erin L. Young, Kelly F. Oakeson, Tara Gallagher |
| EPI_ISL_943808 | Molecular Pathology, Pathology and Laboratory Medicine Institute, Cleveland Clinic, Ohio, USA | Molecular Pathology, Pathology and Laboratory Medicine Institute, Cleveland Clinic, Ohio, USA | Frank P. Esper, Yu-Wei Cheng, Thamali M. Adhikari, Zheng Jin Tu, Daniel H. Farkas, Gary Procop, Jennifer Ko, Timothy A. Chan, Jing Li, Brian P. Rubin |
| EPI_ISL_943810, EPI_ISL_943811, EPI_ISL_943812, EPI_ISL_943813, EPI_ISL_943814, EPI_ISL_943817, EPI_ISL_943818, EPI_ISL_943820, EPI_ISL_943821, EPI_ISL_943824, EPI_ISL_943828, EPI_ISL_943829, EPI_ISL_943832, EPI_ISL_943834, EPI_ISL_943835, EPI_ISL_943839, EPI_ISL_943841, EPI_ISL_943843, EPI_ISL_943844, EPI_ISL_943847, EPI_ISL_943848, EPI_ISL_943849, EPI_ISL_943850, EPI_ISL_943851, EPI_ISL_943852, EPI_ISL_943857, EPI_ISL_943858, EPI_ISL_943859, EPI_ISL_943860, EPI_ISL_943861, EPI_ISL_943863, EPI_ISL_943864, EPI_ISL_943865, EPI_ISL_943866, EPI_ISL_943867, EPI_ISL_943868, EPI_ISL_943869, EPI_ISL_943871, EPI_ISL_943872, EPI_ISL_943873, EPI_ISL_943874, EPI_ISL_943876, EPI_ISL_943877, EPI_ISL_943879, EPI_ISL_943881, EPI_ISL_943882, EPI_ISL_943883, EPI_ISL_943884, EPI_ISL_943885, EPI_ISL_943886, EPI_ISL_943887, EPI_ISL_943889, EPI_ISL_943890, EPI_ISL_943891, EPI_ISL_943892, EPI_ISL_943894, EPI_ISL_943895, EPI_ISL_943896, EPI_ISL_943898, EPI_ISL_943899, EPI_ISL_943903, EPI_ISL_943904, EPI_ISL_943905, EPI_ISL_943906, EPI_ISL_943907, EPI_ISL_943908, EPI_ISL_943909, EPI_ISL_943911, EPI_ISL_943912, EPI_ISL_943914, EPI_ISL_943915, EPI_ISL_943916, EPI_ISL_943917, EPI_ISL_943918, EPI_ISL_943919, EPI_ISL_943920, EPI_ISL_943921, EPI_ISL_943922, EPI_ISL_943924, EPI_ISL_943925, EPI_ISL_943926, EPI_ISL_943928, EPI_ISL_943930, EPI_ISL_943931, EPI_ISL_943933, EPI_ISL_943934, EPI_ISL_943935, EPI_ISL_943938, EPI_ISL_943939, EPI_ISL_943941, EPI_ISL_943945, EPI_ISL_943946, EPI_ISL_943947, EPI_ISL_943948, EPI_ISL_943949, EPI_ISL_943950, EPI_ISL_943951, EPI_ISL_943952, EPI_ISL_943953, EPI_ISL_943954, EPI_ISL_943956, EPI_ISL_943958, EPI_ISL_943959, EPI_ISL_943960, EPI_ISL_943961, EPI_ISL_943962, EPI_ISL_943964, EPI_ISL_943965, EPI_ISL_943966 | | | |
| see above | Utah Public Health Laboratory | Utah Public Health Laboratory | Erin L. Young, Kelly F. Oakeson, Tara Gallagher |
| EPI_ISL_944595, EPI_ISL_944597, EPI_ISL_944598, EPI_ISL_944605 | Connecticut Department of Public Health | Grubaugh Lab - Yale School of Public Health | Joseph Fauver, Tara Alpert, Anderson Brito, Mallery Breban, Anne Wyllie, Chantal Vogels, Mary Petrone, Annie Watkins, Chaney Kalinich, Isabel Ott, Nathan Grubaugh |
| EPI_ISL_944611, EPI_ISL_944612, EPI_ISL_944613, EPI_ISL_944614, EPI_ISL_944615, EPI_ISL_944616, EPI_ISL_944617, EPI_ISL_944618, EPI_ISL_944619, EPI_ISL_944620, EPI_ISL_944621, EPI_ISL_944622, EPI_ISL_944623, EPI_ISL_944624, EPI_ISL_944625, EPI_ISL_944626, EPI_ISL_944627, EPI_ISL_944628, EPI_ISL_944629, EPI_ISL_944630, EPI_ISL_944631, EPI_ISL_944632, EPI_ISL_944633 | | | |
| see above | Instituto Nacional de Medicina Genomica | Instituto Nacional de Medicina Genomica | Hidalgo-Miranda A, Mendoza-Vargas A, Reyes-Grajeda JP, Cisneros-Villanueva M, Cedro-Tanda A,Peñaloza-Figueroa F, Herrera-Montalvo LA |
| EPI_ISL_954754 | City of Milwaukee Health Department Laboratory | City of Milwaukee Health Department Laboratory | Sanjib Bhattacharyya |
| EPI_ISL_954818, EPI_ISL_954819, EPI_ISL_954820, EPI_ISL_954821, EPI_ISL_954822, EPI_ISL_954823, EPI_ISL_954824, EPI_ISL_954825, EPI_ISL_954826, EPI_ISL_954827, EPI_ISL_954828, EPI_ISL_954829, EPI_ISL_954830, EPI_ISL_954831, EPI_ISL_954832, EPI_ISL_954833, EPI_ISL_954834, EPI_ISL_954835, EPI_ISL_954836, EPI_ISL_954837, EPI_ISL_954838, EPI_ISL_954839, EPI_ISL_954840, EPI_ISL_954841, EPI_ISL_954842, EPI_ISL_954843, EPI_ISL_954844, EPI_ISL_954845, EPI_ISL_954846, EPI_ISL_954847, EPI_ISL_954848, EPI_ISL_954849, EPI_ISL_954850, EPI_ISL_954851, EPI_ISL_954852, EPI_ISL_954853, EPI_ISL_954854, EPI_ISL_954855, EPI_ISL_954856, EPI_ISL_954857, EPI_ISL_954858, EPI_ISL_954859, EPI_ISL_954860, EPI_ISL_954861, EPI_ISL_954862, EPI_ISL_954863, EPI_ISL_954886, EPI_ISL_954887, EPI_ISL_954888, EPI_ISL_954889, EPI_ISL_954890, EPI_ISL_954891, EPI_ISL_954892, EPI_ISL_954893, EPI_ISL_954894, EPI_ISL_954895, EPI_ISL_954896, EPI_ISL_954897, EPI_ISL_954898, EPI_ISL_954899, EPI_ISL_954900, EPI_ISL_954901, EPI_ISL_954902, EPI_ISL_954903, EPI_ISL_954904, EPI_ISL_954905, EPI_ISL_954906, EPI_ISL_954907, EPI_ISL_954908, EPI_ISL_954909, EPI_ISL_954910, EPI_ISL_954911, EPI_ISL_954912, EPI_ISL_954913, EPI_ISL_954914, EPI_ISL_954915, EPI_ISL_954916, EPI_ISL_954917, EPI_ISL_954918, EPI_ISL_954919, EPI_ISL_954920, EPI_ISL_954921, EPI_ISL_954922, EPI_ISL_954923, EPI_ISL_954924, EPI_ISL_954925, EPI_ISL_954926, EPI_ISL_954927, EPI_ISL_954928, EPI_ISL_954929, EPI_ISL_954930, EPI_ISL_954931, EPI_ISL_954932, EPI_ISL_954933, EPI_ISL_954934, EPI_ISL_954935, EPI_ISL_954936, EPI_ISL_954937, EPI_ISL_954938, EPI_ISL_954939, EPI_ISL_954940, EPI_ISL_954941, EPI_ISL_954942, EPI_ISL_954944, EPI_ISL_954945, EPI_ISL_954946, EPI_ISL_954947, EPI_ISL_954948, EPI_ISL_954949, EPI_ISL_954950, EPI_ISL_954951, EPI_ISL_954952, EPI_ISL_954953, EPI_ISL_954954, EPI_ISL_954955, EPI_ISL_954956, EPI_ISL_954957, EPI_ISL_954958, EPI_ISL_954959, EPI_ISL_954960, EPI_ISL_954961, EPI_ISL_954962, EPI_ISL_954963, EPI_ISL_954964, EPI_ISL_954965, EPI_ISL_954966, EPI_ISL_954967, EPI_ISL_954968, EPI_ISL_954969, EPI_ISL_954970, EPI_ISL_954971, EPI_ISL_954972, EPI_ISL_954973, EPI_ISL_954974, EPI_ISL_954975, EPI_ISL_954976, EPI_ISL_954977, EPI_ISL_954978, EPI_ISL_954979, EPI_ISL_954980, EPI_ISL_954981, EPI_ISL_954982, EPI_ISL_954983, EPI_ISL_954984, EPI_ISL_954985, EPI_ISL_954986, EPI_ISL_954987, EPI_ISL_954988, EPI_ISL_954989, EPI_ISL_954990, EPI_ISL_954991, EPI_ISL_954992, EPI_ISL_954993, EPI_ISL_954994, EPI_ISL_954995, EPI_ISL_954996, EPI_ISL_954997, EPI_ISL_954998, EPI_ISL_954999, EPI_ISL_955000, EPI_ISL_955001, EPI_ISL_955002, EPI_ISL_955003, EPI_ISL_955004, EPI_ISL_955005, EPI_ISL_955006, EPI_ISL_955007, EPI_ISL_955008, EPI_ISL_955009, EPI_ISL_955010, EPI_ISL_955011, EPI_ISL_955012, EPI_ISL_955013, EPI_ISL_955014, EPI_ISL_955015, EPI_ISL_955016, EPI_ISL_955017, EPI_ISL_955018, EPI_ISL_955019, EPI_ISL_955020, EPI_ISL_955021, EPI_ISL_955022, EPI_ISL_955023, EPI_ISL_955024, EPI_ISL_955025, EPI_ISL_955026, EPI_ISL_955027, EPI_ISL_955028, EPI_ISL_955029, EPI_ISL_955030, EPI_ISL_955031, EPI_ISL_955032, EPI_ISL_955033, EPI_ISL_955034, EPI_ISL_955035, EPI_ISL_955036, EPI_ISL_955037, EPI_ISL_955038, EPI_ISL_955039, EPI_ISL_955040, EPI_ISL_955041, EPI_ISL_955042, EPI_ISL_955043, EPI_ISL_955044, EPI_ISL_955045, EPI_ISL_955046, EPI_ISL_955047, EPI_ISL_955048, EPI_ISL_955049, EPI_ISL_955050, EPI_ISL_955051, EPI_ISL_955052, EPI_ISL_955053, EPI_ISL_955054, EPI_ISL_955055, EPI_ISL_955056, EPI_ISL_955057, EPI_ISL_955058, EPI_ISL_955059, EPI_ISL_955060 | | | |
| see above | Colorado Department of Public Health and Environment | Colorado Department of Puplic Health and Environment | Laura Bankers, Molly C. Hetherington-Rauth, Diana Ir, Shannon Ely, Shannon R. Matzinger, Sarah Elizabeth Totten, Emily A. Travanty |
| EPI_ISL_955104, EPI_ISL_955105, EPI_ISL_955106, EPI_ISL_955107, EPI_ISL_955108, EPI_ISL_955109, EPI_ISL_955110, EPI_ISL_955111, EPI_ISL_955112, EPI_ISL_955113, EPI_ISL_955114, EPI_ISL_955115, EPI_ISL_955116, EPI_ISL_955117, EPI_ISL_955118, EPI_ISL_955119, EPI_ISL_955120, EPI_ISL_955121, EPI_ISL_955122, EPI_ISL_955123, EPI_ISL_955124, EPI_ISL_955125 | | | |
| see above | Maryland Public Health Laboratory | Maryland Public Health Laboratory | Maryland Department of Health Laboratories Administration |
| EPI_ISL_955126, EPI_ISL_955127, EPI_ISL_955128 | Innovative Genomics Institute, UC Berkeley | Innovative Genomics Institute, UC Berkeley | Stacia Wyman, Haridha Shivram, Phil Frankino, Liana Lareau, Shana McDevitt, Justin Choi |
| EPI_ISL_955131, EPI_ISL_955132, EPI_ISL_955133, EPI_ISL_955134, EPI_ISL_955135 | Colorado Department of Public Health and Environment | Colorado Department of Puplic Health and Environment | Laura Bankers, Molly C. Hetherington-Rauth, Diana Ir, Shannon Ely, Shannon R. Matzinger, Sarah Elizabeth Totten, Emily A. Travanty |
| EPI_ISL_955189, EPI_ISL_955190, EPI_ISL_955191, EPI_ISL_955192, EPI_ISL_955193, EPI_ISL_955194, EPI_ISL_955195, EPI_ISL_955196, EPI_ISL_955197, EPI_ISL_955198, EPI_ISL_955199, EPI_ISL_955200, EPI_ISL_955201, EPI_ISL_955202, EPI_ISL_955203, EPI_ISL_955204, EPI_ISL_955205, EPI_ISL_955206, EPI_ISL_955207, EPI_ISL_955208, EPI_ISL_955209 | | | |
| see above | Instituto Nacional de Medicina Genomica | Instituto Nacional de Medicina Genomica | Hidalgo-Miranda A, Mendoza-Vargas A, Reyes-Grajeda JP, Cisneros-Villanueva M, Cedro-Tanda A,Peñaloza-Figueroa F, Herrera-Montalvo LA |
| EPI_ISL_955212, EPI_ISL_955213, EPI_ISL_955214, EPI_ISL_955215, EPI_ISL_955216, EPI_ISL_955217, EPI_ISL_955218, EPI_ISL_955219, EPI_ISL_955220, EPI_ISL_955221, EPI_ISL_955222, EPI_ISL_955223, EPI_ISL_955224, EPI_ISL_955225, EPI_ISL_955226, EPI_ISL_955227, EPI_ISL_955228 | | | |
| see above | Indiana Animal Disease Diagnostic Laboratory | Carpi Laboratory - Purdue University | Jack Dorman, Ilinca I Ciubotariu, Lev Gorenstein, Abebe A Fola, G Kenitra Hendrix, Rebecca P Wilkes, Giovanna Carpi |
| EPI_ISL_955230 | Mary Eliza Mahoney HC | NJ Public Health and Environmental Laboratories | Lindsey Bodnar, Shiv Verma, Dana Woell, Byeong Jeong |
| EPI_ISL_955231, EPI_ISL_955232 | BioReference Lab | NJ Public Health and Environmental Laboratories | Lindsey Bodnar, Shiv Verma, Dana Woell, Byeong Jeong |
| EPI_ISL_955233 | HGZMF 1 LA PAZ | BIOBANCO / COCTI | Borja-Aburto VH, Grajales-Muñiz C, Santacruz Tinoco CE, Rojas-Mendoza T, Ochoa Carrera LA, Sánchez A, Grande R, Isa P, Taboada B, López S, Arias C, Muñoz-Medina JE |
| EPI_ISL_955234 | HE CMN LA RAZA | BIOBANCO / COCTI | Borja-Aburto VH, Grajales-Muñiz C, Santacruz Tinoco CE, Rojas-Mendoza T, Ochoa Carrera LA, Sánchez A, Grande R, Isa P, Taboada B, López S, Arias C, Muñoz-Medina JE |
| EPI_ISL_955235 | UMF 23 UNIDAD MORELOS | BIOBANCO / COCTI | Borja-Aburto VH, Grajales-Muñiz C, Santacruz Tinoco CE, Rojas-Mendoza T, Ochoa Carrera LA, Sánchez A, Grande R, Isa P, Taboada B, López S, Arias C, Muñoz-Medina JE |
| EPI_ISL_955236 | HGR 1 VICENTE GUERRERO | BIOBANCO / COCTI | Borja-Aburto VH, Grajales-Muñiz C, Santacruz Tinoco CE, Rojas-Mendoza T, Ochoa Carrera LA, Sánchez A, Grande R, Isa P, Taboada B, López S, Arias C, Muñoz-Medina JE |
| EPI_ISL_955237 | UMF 75 NETZAHUALCOYOTL | BIOBANCO / COCTI | Borja-Aburto VH, Grajales-Muñiz C, Santacruz Tinoco CE, Rojas-Mendoza T, Ochoa Carrera LA, Sánchez A, Grande R, Isa P, Taboada B, López S, Arias C, Muñoz-Medina JE |
| EPI_ISL_955238 | HI CMN LA RAZA | BIOBANCO / COCTI | Borja-Aburto VH, Grajales-Muñiz C, Santacruz Tinoco CE, Rojas-Mendoza T, Ochoa Carrera LA, Sánchez A, Grande R, Isa P, Taboada B, López S, Arias C, Muñoz-Medina JE |
| EPI_ISL_955239 | HE CMN LA RAZA | BIOBANCO / COCTI | Borja-Aburto VH, Grajales-Muñiz C, Santacruz Tinoco CE, Rojas-Mendoza T, Ochoa Carrera LA, Sánchez A, Grande R, Isa P, Taboada B, López S, Arias C, Muñoz-Medina JE |
| EPI_ISL_955240, EPI_ISL_955241 | HGZ 30 MEXICALI | BIOBANCO / COCTI | Borja-Aburto VH, Grajales-Muñiz C, Santacruz Tinoco CE, Rojas-Mendoza T, Ochoa Carrera LA, Sánchez A, Grande R, Isa P, Taboada B, López S, Arias C, Muñoz-Medina JE |
| EPI_ISL_955242, EPI_ISL_955243 | HGSMF 26 CABO SAN LUCAS | BIOBANCO / COCTI | Borja-Aburto VH, Grajales-Muñiz C, Santacruz Tinoco CE, Rojas-Mendoza T, Ochoa Carrera LA, Sánchez A, Grande R, Isa P, Taboada B, López S, Arias C, Muñoz-Medina JE |
| EPI_ISL_955244 | UMF 26 ACAPULCO | BIOBANCO / COCTI | Borja-Aburto VH, Grajales-Muñiz C, Santacruz Tinoco CE, Rojas-Mendoza T, Ochoa Carrera LA, Sánchez A, Grande R, Isa P, Taboada B, López S, Arias C, Muñoz-Medina JE |
| EPI_ISL_955245 | HGR 1 VICENTE GUERRERO | BIOBANCO / COCTI | Borja-Aburto VH, Grajales-Muñiz C, Santacruz Tinoco CE, Rojas-Mendoza T, Ochoa Carrera LA, Sánchez A, Grande R, Isa P, Taboada B, López S, Arias C, Muñoz-Medina JE |
| EPI_ISL_955246 | HGOMF 60 TLANEPANTLA | BIOBANCO / COCTI | Borja-Aburto VH, Grajales-Muñiz C, Santacruz Tinoco CE, Rojas-Mendoza T, Ochoa Carrera LA, Sánchez A, Grande R, Isa P, Taboada B, López S, Arias C, Muñoz-Medina JE |
| EPI_ISL_955247 | HGR 200 TECAMAC | BIOBANCO / COCTI | Borja-Aburto VH, Grajales-Muñiz C, Santacruz Tinoco CE, Rojas-Mendoza T, Ochoa Carrera LA, Sánchez A, Grande R, Isa P, Taboada B, López S, Arias C, Muñoz-Medina JE |
| EPI_ISL_955248 | BCO SANGRE CM LA RAZA | BIOBANCO / COCTI | Borja-Aburto VH, Grajales-Muñiz C, Santacruz Tinoco CE, Rojas-Mendoza T, Ochoa Carrera LA, Sánchez A, Grande R, Isa P, Taboada B, López S, Arias C, Muñoz-Medina JE |
| EPI_ISL_955249 | HGZ 2A PASO-TRONCOSO | BIOBANCO / COCTI | Borja-Aburto VH, Grajales-Muñiz C, Santacruz Tinoco CE, Rojas-Mendoza T, Ochoa Carrera LA, Sánchez A, Grande R, Isa P, Taboada B, López S, Arias C, Muñoz-Medina JE |
| EPI_ISL_955250, EPI_ISL_955251, | HGSMF 26 CABO SAN LUCAS | BIOBANCO / COCTI | Borja-Aburto VH, Grajales-Muñiz C, Santacruz Tinoco CE, Rojas-Mendoza T, Ochoa Carrera LA, Sánchez A, Grande R, Isa P, Taboada B, López S, Arias C, Muñoz-Medina JE |

| | | | |
|---|---|---|---|
| EPI_ISL_955252, EPI_ISL_955253 | | | |
| EPI_ISL_955254 | UMF 28 DEL VALLE | BIOBANCO / COCTI | Borja-Aburto VH, Grajales-Muñiz C, Santacruz Tinoco CE, Rojas-Mendoza T, Ochoa Carrera LA, Sánchez A, Grande R, Isa P, Taboada B, López S, Arias C, Muñoz-Medina JE |
| EPI_ISL_955255 | HGSMF 26 CABO SAN LUCAS | BIOBANCO / COCTI | Borja-Aburto VH, Grajales-Muñiz C, Santacruz Tinoco CE, Rojas-Mendoza T, Ochoa Carrera LA, Sánchez A, Grande R, Isa P, Taboada B, López S, Arias C, Muñoz-Medina JE |
| EPI_ISL_955256 | HGZ 1 CARLOS MC GREGOR | BIOBANCO / COCTI | Borja-Aburto VH, Grajales-Muñiz C, Santacruz Tinoco CE, Rojas-Mendoza T, Ochoa Carrera LA, Sánchez A, Grande R, Isa P, Taboada B, López S, Arias C, Muñoz-Medina JE |
| EPI_ISL_955257 | HGZ 32 VILLA COAPA | BIOBANCO / COCTI | Borja-Aburto VH, Grajales-Muñiz C, Santacruz Tinoco CE, Rojas-Mendoza T, Ochoa Carrera LA, Sánchez A, Grande R, Isa P, Taboada B, López S, Arias C, Muñoz-Medina JE |
| EPI_ISL_955258, EPI_ISL_955259, EPI_ISL_955260 | BioReference Lab | NJ Public Health and Environmental Laboratories | Lindsey Bodnar, Shiv Verma, Dana Woell, Byeong Jeong |
| EPI_ISL_955261, EPI_ISL_955263 | Fulgent Genetics | Fulgent Genetics | Harry Gao, Mickey Li, John Gao, Joseph Fierro, Benafsh Sapra, Becky Tsai, Yan Meng, Doreen Ng, James Xie |
| EPI_ISL_955264 | Clinical Pathology Lab | Pathogen Discovery, Respiratory Viruses Branch, Division of Viral Diseases, Centers for Disease Control and Prevention | Ying Tao, Jing Zhang, Yan Li, Krista Queen, Anna Uehara, Peter Cook, Clinton R. Paden, Haibin Wang, Suxiang Tong |
| EPI_ISL_955265 | Sunrise Medical Laboratory | Pathogen Discovery, Respiratory Viruses Branch, Division of Viral Diseases, Centers for Disease Control and Prevention | Ying Tao, Jing Zhang, Yan Li, Krista Queen, Anna Uehara, Peter Cook, Clinton R. Paden, Haibin Wang, Suxiang Tong |
| EPI_ISL_955266, EPI_ISL_955267, EPI_ISL_955268 | Clinical Pathology Lab | Pathogen Discovery, Respiratory Viruses Branch, Division of Viral Diseases, Centers for Disease Control and Prevention | Ying Tao, Jing Zhang, Yan Li, Krista Queen, Anna Uehara, Peter Cook, Clinton R. Paden, Haibin Wang, Suxiang Tong |
| EPI_ISL_955269 | LA Office of Public Health Laboratories | Pathogen Discovery, Respiratory Viruses Branch, Division of Viral Diseases, Centers for Disease Control and Prevention | Ying Tao, Jing Zhang, Yan Li, Krista Queen, Anna Uehara, Peter Cook, Clinton R. Paden, Haibin Wang, Suxiang Tong |
| EPI_ISL_955270 | Clinical Pathology Lab | Pathogen Discovery, Respiratory Viruses Branch, Division of Viral Diseases, Centers for Disease Control and Prevention | Ying Tao, Jing Zhang, Yan Li, Krista Queen, Anna Uehara, Peter Cook, Clinton R. Paden, Haibin Wang, Suxiang Tong |
| EPI_ISL_955271 | American Esoteric Laboratory | Pathogen Discovery, Respiratory Viruses Branch, Division of Viral Diseases, Centers for Disease Control and Prevention | Ying Tao, Jing Zhang, Yan Li, Krista Queen, Anna Uehara, Peter Cook, Clinton R. Paden, Haibin Wang, Suxiang Tong |
| EPI_ISL_955272 | AR Dept. of Health-PHL, Molecular Diagnostics | Pathogen Discovery, Respiratory Viruses Branch, Division of Viral Diseases, Centers for Disease Control and Prevention | Ying Tao, Jing Zhang, Yan Li, Krista Queen, Anna Uehara, Peter Cook, Clinton R. Paden, Haibin Wang, Suxiang Tong |
| EPI_ISL_955273, EPI_ISL_955274 | American Esoteric Laboratory | Pathogen Discovery, Respiratory Viruses Branch, Division of Viral Diseases, Centers for Disease Control and Prevention | Ying Tao, Jing Zhang, Yan Li, Krista Queen, Anna Uehara, Peter Cook, Clinton R. Paden, Haibin Wang, Suxiang Tong |
| EPI_ISL_955275 | LA Office of Public Health Laboratories | Pathogen Discovery, Respiratory Viruses Branch, Division of Viral Diseases, Centers for Disease Control and Prevention | Ying Tao, Jing Zhang, Yan Li, Krista Queen, Anna Uehara, Peter Cook, Clinton R. Paden, Haibin Wang, Suxiang Tong |
| EPI_ISL_955276 | Clinical Pathology Lab | Pathogen Discovery, Respiratory Viruses Branch, Division of Viral Diseases, Centers for Disease Control and Prevention | Ying Tao, Jing Zhang, Yan Li, Krista Queen, Anna Uehara, Peter Cook, Clinton R. Paden, Haibin Wang, Suxiang Tong |
| EPI_ISL_955277 | GA Department of Public Health Laboratory | Pathogen Discovery, Respiratory Viruses Branch, Division of Viral Diseases, Centers for Disease Control and Prevention | Ying Tao, Jing Zhang, Yan Li, Krista Queen, Anna Uehara, Peter Cook, Clinton R. Paden, Haibin Wang, Suxiang Tong |
| EPI_ISL_955278, EPI_ISL_955279, EPI_ISL_955280, EPI_ISL_955281, EPI_ISL_955282 | Clinical Pathology Lab | Pathogen Discovery, Respiratory Viruses Branch, Division of Viral Diseases, Centers for Disease Control and Prevention | Ying Tao, Jing Zhang, Yan Li, Krista Queen, Anna Uehara, Peter Cook, Clinton R. Paden, Haibin Wang, Suxiang Tong |
| EPI_ISL_955284 | NC State Laboratory of Public Health | Pathogen Discovery, Respiratory Viruses Branch, Division of Viral Diseases, Centers for Disease Control and Prevention | Ying Tao, Jing Zhang, Yan Li, Krista Queen, Anna Uehara, Peter Cook, Clinton R. Paden, Haibin Wang, Suxiang Tong |
| EPI_ISL_955285 | American Esoteric Laboratory | Pathogen Discovery, Respiratory Viruses Branch, Division of Viral Diseases, Centers for Disease Control and Prevention | Ying Tao, Jing Zhang, Yan Li, Krista Queen, Anna Uehara, Peter Cook, Clinton R. Paden, Haibin Wang, Suxiang Tong |
| EPI_ISL_955286, EPI_ISL_955287 | Clinical Pathology Lab | Pathogen Discovery, Respiratory Viruses Branch, Division of Viral Diseases, Centers for Disease Control and Prevention | Ying Tao, Jing Zhang, Yan Li, Krista Queen, Anna Uehara, Peter Cook, Clinton R. Paden, Haibin Wang, Suxiang Tong |
| EPI_ISL_955288 | GA Department of Public Health Laboratory | Pathogen Discovery, Respiratory Viruses Branch, Division of Viral Diseases, Centers for Disease Control and Prevention | Ying Tao, Jing Zhang, Yan Li, Krista Queen, Anna Uehara, Peter Cook, Clinton R. Paden, Haibin Wang, Suxiang Tong |
| EPI_ISL_955289 | Clinical Pathology Lab | Pathogen Discovery, Respiratory Viruses Branch, Division of Viral Diseases, Centers for Disease Control and Prevention | Ying Tao, Jing Zhang, Yan Li, Krista Queen, Anna Uehara, Peter Cook, Clinton R. Paden, Haibin Wang, Suxiang Tong |
| EPI_ISL_955290 | TMC/Pathology | Pathogen Discovery, Respiratory Viruses Branch, Division of Viral Diseases, Centers for Disease Control and Prevention | Ying Tao, Jing Zhang, Yan Li, Krista Queen, Anna Uehara, Peter Cook, Clinton R. Paden, Haibin Wang, Suxiang Tong |
| EPI_ISL_955291 | IL Dept. of Public Health Springfield Laboratory | Pathogen Discovery, Respiratory Viruses Branch, Division of Viral Diseases, Centers for Disease Control and Prevention | Ying Tao, Jing Zhang, Yan Li, Krista Queen, Anna Uehara, Peter Cook, Clinton R. Paden, Haibin Wang, Suxiang Tong |
| EPI_ISL_955292, EPI_ISL_955293 | TMC/Pathology | Pathogen Discovery, Respiratory Viruses Branch, Division of Viral Diseases, Centers for Disease Control and Prevention | Ying Tao, Jing Zhang, Yan Li, Krista Queen, Anna Uehara, Peter Cook, Clinton R. Paden, Haibin Wang, Suxiang Tong |
| EPI_ISL_955294, EPI_ISL_955295, EPI_ISL_955296, EPI_ISL_955297 | Clinical Pathology Lab | Pathogen Discovery, Respiratory Viruses Branch, Division of Viral Diseases, Centers for Disease Control and Prevention | Ying Tao, Jing Zhang, Yan Li, Krista Queen, Anna Uehara, Peter Cook, Clinton R. Paden, Haibin Wang, Suxiang Tong |
| EPI_ISL_955298 | NC State Laboratory of Public Health | Pathogen Discovery, Respiratory Viruses Branch, Division of Viral Diseases, Centers for Disease Control and Prevention | Ying Tao, Jing Zhang, Yan Li, Krista Queen, Anna Uehara, Peter Cook, Clinton R. Paden, Haibin Wang, Suxiang Tong |
| EPI_ISL_955300 | Sunrise Medical Laboratory | Pathogen Discovery, Respiratory Viruses Branch, Division of Viral Diseases, Centers for Disease Control and Prevention | Ying Tao, Jing Zhang, Yan Li, Krista Queen, Anna Uehara, Peter Cook, Clinton R. Paden, Haibin Wang, Suxiang Tong |
| EPI_ISL_955301 | Clinical Pathology Lab | Pathogen Discovery, Respiratory Viruses Branch, Division of Viral Diseases, Centers for Disease Control and Prevention | Ying Tao, Jing Zhang, Yan Li, Krista Queen, Anna Uehara, Peter Cook, Clinton R. Paden, Haibin Wang, Suxiang Tong |
| EPI_ISL_955302, EPI_ISL_955303 | GA Department of Public Health Laboratory | Pathogen Discovery, Respiratory Viruses Branch, Division of Viral Diseases, Centers for Disease Control and Prevention | Ying Tao, Jing Zhang, Yan Li, Krista Queen, Anna Uehara, Peter Cook, Clinton R. Paden, Haibin Wang, Suxiang Tong |
| EPI_ISL_955304, EPI_ISL_955305, EPI_ISL_955306 | Clinical Pathology Lab | Pathogen Discovery, Respiratory Viruses Branch, Division of Viral Diseases, Centers for Disease Control and Prevention | Ying Tao, Jing Zhang, Yan Li, Krista Queen, Anna Uehara, Peter Cook, Clinton R. Paden, Haibin Wang, Suxiang Tong |
| EPI_ISL_955307, EPI_ISL_955308 | OK Public Health Laboratory, Oklahoma State DOH | Pathogen Discovery, Respiratory Viruses Branch, Division of Viral Diseases, Centers for Disease Control and Prevention | Ying Tao, Jing Zhang, Yan Li, Krista Queen, Anna Uehara, Peter Cook, Clinton R. Paden, Haibin Wang, Suxiang Tong |

| | | | |
|---|---|---|---|
| EPI_ISL_955309 | IL Dept. of Public Health Springfield Laboratory | Pathogen Discovery, Respiratory Viruses Branch, Division of Viral Diseases, Centers for Disease Control and Prevention | Ying Tao, Jing Zhang, Yan Li, Krista Queen, Anna Uehara, Peter Cook, Clinton R. Paden, Haibin Wang, Suxiang Tong |
| EPI_ISL_955310 | TN Division of Laboratory Services | Pathogen Discovery, Respiratory Viruses Branch, Division of Viral Diseases, Centers for Disease Control and Prevention | Ying Tao, Jing Zhang, Yan Li, Krista Queen, Anna Uehara, Peter Cook, Clinton R. Paden, Haibin Wang, Suxiang Tong |
| EPI_ISL_955312 | Clinical Pathology Lab | Pathogen Discovery, Respiratory Viruses Branch, Division of Viral Diseases, Centers for Disease Control and Prevention | Ying Tao, Jing Zhang, Yan Li, Krista Queen, Anna Uehara, Peter Cook, Clinton R. Paden, Haibin Wang, Suxiang Tong |
| EPI_ISL_955313 | AR Dept. of Health-PHL, Molecular Diagnostics | Pathogen Discovery, Respiratory Viruses Branch, Division of Viral Diseases, Centers for Disease Control and Prevention | Ying Tao, Jing Zhang, Yan Li, Krista Queen, Anna Uehara, Peter Cook, Clinton R. Paden, Haibin Wang, Suxiang Tong |
| EPI_ISL_955321 | Clinical Pathology Lab | Pathogen Discovery, Respiratory Viruses Branch, Division of Viral Diseases, Centers for Disease Control and Prevention | Ying Tao, Jing Zhang, Yan Li, Krista Queen, Anna Uehara, Peter Cook, Clinton R. Paden, Haibin Wang, Suxiang Tong |
| EPI_ISL_955322, EPI_ISL_955323, EPI_ISL_955324, EPI_ISL_955325, EPI_ISL_955326, EPI_ISL_955327 | American Esoteric Laboratory | Pathogen Discovery, Respiratory Viruses Branch, Division of Viral Diseases, Centers for Disease Control and Prevention | Ying Tao, Jing Zhang, Yan Li, Krista Queen, Anna Uehara, Peter Cook, Clinton R. Paden, Haibin Wang, Suxiang Tong |
| EPI_ISL_955328 | IL Dept. of Public Health Springfield Laboratory | Pathogen Discovery, Respiratory Viruses Branch, Division of Viral Diseases, Centers for Disease Control and Prevention | Ying Tao, Jing Zhang, Yan Li, Krista Queen, Anna Uehara, Peter Cook, Clinton R. Paden, Haibin Wang, Suxiang Tong |
| EPI_ISL_955329, EPI_ISL_955330 | GA Department of Public Health Laboratory | Pathogen Discovery, Respiratory Viruses Branch, Division of Viral Diseases, Centers for Disease Control and Prevention | Ying Tao, Jing Zhang, Yan Li, Krista Queen, Anna Uehara, Peter Cook, Clinton R. Paden, Haibin Wang, Suxiang Tong |
| EPI_ISL_955331, EPI_ISL_955332, EPI_ISL_955333 | AR Dept. of Health-PHL, Molecular Diagnostics | Pathogen Discovery, Respiratory Viruses Branch, Division of Viral Diseases, Centers for Disease Control and Prevention | Ying Tao, Jing Zhang, Yan Li, Krista Queen, Anna Uehara, Peter Cook, Clinton R. Paden, Haibin Wang, Suxiang Tong |
| EPI_ISL_955334, EPI_ISL_955335, EPI_ISL_955336 | GA Department of Public Health Laboratory | Pathogen Discovery, Respiratory Viruses Branch, Division of Viral Diseases, Centers for Disease Control and Prevention | Ying Tao, Jing Zhang, Yan Li, Krista Queen, Anna Uehara, Peter Cook, Clinton R. Paden, Haibin Wang, Suxiang Tong |
| EPI_ISL_955337, EPI_ISL_955338, EPI_ISL_955339, EPI_ISL_955340, EPI_ISL_955341, EPI_ISL_955342, EPI_ISL_955343, EPI_ISL_955344, EPI_ISL_955345, EPI_ISL_955346, EPI_ISL_955347, EPI_ISL_955348, EPI_ISL_955349, EPI_ISL_955350, EPI_ISL_955351, EPI_ISL_955352 | | | |
| see above | UC Davis- Department of Pathology and Laboratory Medicine | Chan-Zuckerberg Biohub | CZB Cliahub Consortium |
| EPI_ISL_955353, EPI_ISL_955354, EPI_ISL_955355, EPI_ISL_955356, EPI_ISL_955357, EPI_ISL_955358, EPI_ISL_955361, EPI_ISL_955362, EPI_ISL_955363, EPI_ISL_955364, EPI_ISL_955365, EPI_ISL_955366, EPI_ISL_955367, EPI_ISL_955368, EPI_ISL_955369, EPI_ISL_955370, EPI_ISL_955371, EPI_ISL_955372, EPI_ISL_955373, EPI_ISL_955374, EPI_ISL_955375, EPI_ISL_955376, EPI_ISL_955377, EPI_ISL_955378, EPI_ISL_955379, EPI_ISL_955380, EPI_ISL_955381, EPI_ISL_955382, EPI_ISL_955383, EPI_ISL_955384, EPI_ISL_955385 | | | |
| see above | Santa Clara County Public Health Laboratory | Chan-Zuckerberg Biohub | CZB Cliahub Consortium |
| EPI_ISL_955386, EPI_ISL_955387, EPI_ISL_955388, EPI_ISL_955389, EPI_ISL_955390, EPI_ISL_955391, EPI_ISL_955392, EPI_ISL_955393, EPI_ISL_955394, EPI_ISL_955395, EPI_ISL_955396, EPI_ISL_955397, EPI_ISL_955398, EPI_ISL_955399, EPI_ISL_955400, EPI_ISL_955401, EPI_ISL_955402, EPI_ISL_955403, EPI_ISL_955404, EPI_ISL_955405, EPI_ISL_955406, EPI_ISL_955407, EPI_ISL_955408, EPI_ISL_955409, EPI_ISL_955410, EPI_ISL_955411, EPI_ISL_955412, EPI_ISL_955413, EPI_ISL_955414, EPI_ISL_955415, EPI_ISL_955416, EPI_ISL_955417, EPI_ISL_955418, EPI_ISL_955419, EPI_ISL_955420, EPI_ISL_955421, EPI_ISL_955422, EPI_ISL_955423, EPI_ISL_955424, EPI_ISL_955425, EPI_ISL_955426 | | | |
| see above | Alameda County Public Health Lab | Chan-Zuckerberg Biohub | CZB Cliahub Consortium |
| EPI_ISL_955427, EPI_ISL_955428, EPI_ISL_955429, EPI_ISL_955430, EPI_ISL_955431, EPI_ISL_955432, EPI_ISL_955433, EPI_ISL_955434, EPI_ISL_955435, EPI_ISL_955436, EPI_ISL_955437, EPI_ISL_955438, EPI_ISL_955439, EPI_ISL_955440, EPI_ISL_955441, EPI_ISL_955442, EPI_ISL_955443, EPI_ISL_955444, EPI_ISL_955445, EPI_ISL_955446, EPI_ISL_955447, EPI_ISL_955448, EPI_ISL_955449, EPI_ISL_955450, EPI_ISL_955451, EPI_ISL_955452, EPI_ISL_955453, EPI_ISL_955454, EPI_ISL_955455, EPI_ISL_955456, EPI_ISL_955457, EPI_ISL_955458, EPI_ISL_955459, EPI_ISL_955460, EPI_ISL_955461, EPI_ISL_955462, EPI_ISL_955463, EPI_ISL_955464, EPI_ISL_955465, EPI_ISL_955466, EPI_ISL_955467, EPI_ISL_955468, EPI_ISL_955469, EPI_ISL_955470, EPI_ISL_955471, EPI_ISL_955472, EPI_ISL_955473, EPI_ISL_955474, EPI_ISL_955475, EPI_ISL_955476, EPI_ISL_955477, EPI_ISL_955478, EPI_ISL_955479, EPI_ISL_955480, EPI_ISL_955481, EPI_ISL_955482, EPI_ISL_955483, EPI_ISL_955484, EPI_ISL_955485, EPI_ISL_955486, EPI_ISL_955487, EPI_ISL_955488, EPI_ISL_955489, EPI_ISL_955490, EPI_ISL_955491, EPI_ISL_955492, EPI_ISL_955493, EPI_ISL_955494, EPI_ISL_955495 | | | |
| see above | Orange County Public Health Lab | Chan-Zuckerberg Biohub | CZB Cliahub Consortium |
| EPI_ISL_955496, EPI_ISL_955497, EPI_ISL_955498, EPI_ISL_955499, EPI_ISL_955500, EPI_ISL_955501, EPI_ISL_955502, EPI_ISL_955503, EPI_ISL_955504, EPI_ISL_955505, EPI_ISL_955506, EPI_ISL_955507, EPI_ISL_955508, EPI_ISL_955509, EPI_ISL_955510, EPI_ISL_955511, EPI_ISL_955512, EPI_ISL_955513, EPI_ISL_955514, EPI_ISL_955515, EPI_ISL_955516, EPI_ISL_955517, EPI_ISL_955518, EPI_ISL_955519, EPI_ISL_955520, EPI_ISL_955521, EPI_ISL_955522, EPI_ISL_955523, EPI_ISL_955524, EPI_ISL_955525, EPI_ISL_955526, EPI_ISL_955527, EPI_ISL_955528, EPI_ISL_955529, EPI_ISL_955530, EPI_ISL_955531, EPI_ISL_955532, EPI_ISL_955533, EPI_ISL_955534, EPI_ISL_955535, EPI_ISL_955536, EPI_ISL_955537, EPI_ISL_955538, EPI_ISL_955539, EPI_ISL_955540, EPI_ISL_955541, EPI_ISL_955542, EPI_ISL_955543, EPI_ISL_955544, EPI_ISL_955545, EPI_ISL_955546, EPI_ISL_955547, EPI_ISL_955548, EPI_ISL_955549, EPI_ISL_955550, EPI_ISL_955551, EPI_ISL_955552, EPI_ISL_955553, EPI_ISL_955554, EPI_ISL_955555, EPI_ISL_955556, EPI_ISL_955557, EPI_ISL_955558, EPI_ISL_955559, EPI_ISL_955560 | | | |
| see above | Humboldt County Public Health Laboratory | Chan-Zuckerberg Biohub | CZB Cliahub Consortium |
| EPI_ISL_955562, EPI_ISL_955563, EPI_ISL_955564, EPI_ISL_955565, EPI_ISL_955566, EPI_ISL_955567, EPI_ISL_955568, EPI_ISL_955569, EPI_ISL_955570, EPI_ISL_955571, EPI_ISL_955572, EPI_ISL_955573, EPI_ISL_955574, EPI_ISL_955575, EPI_ISL_955576, EPI_ISL_955577, EPI_ISL_955578, EPI_ISL_955579, EPI_ISL_955580, EPI_ISL_955581, EPI_ISL_955582, EPI_ISL_955583, EPI_ISL_955584, EPI_ISL_955585, EPI_ISL_955586, EPI_ISL_955587, EPI_ISL_955588, EPI_ISL_955589, EPI_ISL_955590, EPI_ISL_955591, EPI_ISL_955592, EPI_ISL_955593, EPI_ISL_955594, EPI_ISL_955595, EPI_ISL_955596, EPI_ISL_955597, EPI_ISL_955598, EPI_ISL_955599, EPI_ISL_955600, EPI_ISL_955601, EPI_ISL_955602, EPI_ISL_955603, EPI_ISL_955604, EPI_ISL_955605, EPI_ISL_955606, EPI_ISL_955607, EPI_ISL_955608, EPI_ISL_955609, EPI_ISL_955610, EPI_ISL_955611, EPI_ISL_955612, EPI_ISL_955613, EPI_ISL_955614, EPI_ISL_955615, EPI_ISL_955616, EPI_ISL_955617, EPI_ISL_955618, EPI_ISL_955619, EPI_ISL_955620, EPI_ISL_955621, EPI_ISL_955622, EPI_ISL_955623, EPI_ISL_955624, EPI_ISL_955625, EPI_ISL_955626, EPI_ISL_955627 | | | |
| see above | Santa Clara County Public Health Laboratory | Chan-Zuckerberg Biohub | CZB Cliahub Consortium |
| EPI_ISL_955628, EPI_ISL_955629, EPI_ISL_955630, EPI_ISL_955631, EPI_ISL_955632, EPI_ISL_955633, EPI_ISL_955634, EPI_ISL_955635, EPI_ISL_955636, EPI_ISL_955637, EPI_ISL_955638, EPI_ISL_955639, EPI_ISL_955640, EPI_ISL_955641, EPI_ISL_955642, EPI_ISL_955643, EPI_ISL_955644, EPI_ISL_955645, EPI_ISL_955646, EPI_ISL_955647, EPI_ISL_955648, EPI_ISL_955649, EPI_ISL_955650, EPI_ISL_955651, EPI_ISL_955652, EPI_ISL_955653, EPI_ISL_955654, EPI_ISL_955655, EPI_ISL_955656, EPI_ISL_955657, EPI_ISL_955658, EPI_ISL_955659, EPI_ISL_955660, EPI_ISL_955661, EPI_ISL_955662, EPI_ISL_955663, EPI_ISL_955664, EPI_ISL_955665, EPI_ISL_955666, EPI_ISL_955667, EPI_ISL_955668, EPI_ISL_955669, EPI_ISL_955670, EPI_ISL_955671, EPI_ISL_955672, EPI_ISL_955673, EPI_ISL_955674, EPI_ISL_955675, EPI_ISL_955676, EPI_ISL_955677, EPI_ISL_955678, EPI_ISL_955679, EPI_ISL_955680, EPI_ISL_955681, EPI_ISL_955682, EPI_ISL_955683, EPI_ISL_955684, EPI_ISL_955685, EPI_ISL_955686, EPI_ISL_955687, EPI_ISL_955688, EPI_ISL_955689, EPI_ISL_955690, EPI_ISL_955691, EPI_ISL_955692 | | | |
| see above | Orange County Public Health Lab | Chan-Zuckerberg Biohub | CZB Cliahub Consortium |
| EPI_ISL_955693, EPI_ISL_955694, EPI_ISL_955695, EPI_ISL_955696, EPI_ISL_955697, EPI_ISL_955698, EPI_ISL_955699, EPI_ISL_955700, EPI_ISL_955701, EPI_ISL_955702, EPI_ISL_955703, EPI_ISL_955704, EPI_ISL_955705, EPI_ISL_955706, EPI_ISL_955707, EPI_ISL_955708, EPI_ISL_955709, EPI_ISL_955710, EPI_ISL_955711, EPI_ISL_955712, EPI_ISL_955713, EPI_ISL_955714, EPI_ISL_955715, EPI_ISL_955716, EPI_ISL_955717, EPI_ISL_955718, EPI_ISL_955719, EPI_ISL_955720, EPI_ISL_955721, EPI_ISL_955722, EPI_ISL_955723, EPI_ISL_955724, EPI_ISL_955725, EPI_ISL_955726, EPI_ISL_955727, EPI_ISL_955728, EPI_ISL_955729, EPI_ISL_955730, EPI_ISL_955731, EPI_ISL_955732, EPI_ISL_955733, EPI_ISL_955734, EPI_ISL_955735, EPI_ISL_955736, EPI_ISL_955737, EPI_ISL_955738, EPI_ISL_955739, EPI_ISL_955740, EPI_ISL_955741, EPI_ISL_955742, EPI_ISL_955743, EPI_ISL_955744, EPI_ISL_955745, EPI_ISL_955746, EPI_ISL_955747, EPI_ISL_955748, EPI_ISL_955749, EPI_ISL_955750, EPI_ISL_955751, EPI_ISL_955752, EPI_ISL_955753, EPI_ISL_955754, EPI_ISL_955755, EPI_ISL_955756, EPI_ISL_955757, EPI_ISL_955758, EPI_ISL_955759, EPI_ISL_955760, EPI_ISL_955761, EPI_ISL_955762, EPI_ISL_955763, EPI_ISL_955764, EPI_ISL_955765, EPI_ISL_955766, EPI_ISL_955767, EPI_ISL_955768, EPI_ISL_955769, EPI_ISL_955770, EPI_ISL_955771, EPI_ISL_955772, EPI_ISL_955773, EPI_ISL_955774, EPI_ISL_955775, EPI_ISL_955776, EPI_ISL_955777 | | | |
| see above | Humboldt County Public Health Laboratory | Chan-Zuckerberg Biohub | CZB Cliahub Consortium |
| EPI_ISL_955778, EPI_ISL_955779, EPI_ISL_955780, EPI_ISL_955781, EPI_ISL_955782, EPI_ISL_955783, EPI_ISL_955784, EPI_ISL_955785, EPI_ISL_955786, EPI_ISL_955787, EPI_ISL_955788, EPI_ISL_955789, EPI_ISL_955790, EPI_ISL_955791, EPI_ISL_955792, EPI_ISL_955793, EPI_ISL_955794, EPI_ISL_955795, EPI_ISL_955796, EPI_ISL_955797, EPI_ISL_955798, EPI_ISL_955799, EPI_ISL_955800, EPI_ISL_955801, EPI_ISL_955802, EPI_ISL_955803, EPI_ISL_955804, EPI_ISL_955805, EPI_ISL_955806, EPI_ISL_955807, EPI_ISL_955808, EPI_ISL_955809, EPI_ISL_955810, EPI_ISL_955811, EPI_ISL_955812, EPI_ISL_955813, EPI_ISL_955814, EPI_ISL_955815, EPI_ISL_955816, EPI_ISL_955817, EPI_ISL_955818, EPI_ISL_955819, EPI_ISL_955820, EPI_ISL_955821, EPI_ISL_955822, EPI_ISL_955823, EPI_ISL_955824, EPI_ISL_955825, EPI_ISL_955826, EPI_ISL_955827, EPI_ISL_955828, EPI_ISL_955829, EPI_ISL_955830, EPI_ISL_955831, EPI_ISL_955832, EPI_ISL_955833, EPI_ISL_955834, EPI_ISL_955835, EPI_ISL_955836, EPI_ISL_955837, EPI_ISL_955838, EPI_ISL_955839, EPI_ISL_955840, EPI_ISL_955841, EPI_ISL_955842, EPI_ISL_955843, EPI_ISL_955844, EPI_ISL_955845, EPI_ISL_955846, EPI_ISL_955847, EPI_ISL_955848, EPI_ISL_955849, EPI_ISL_955850, EPI_ISL_955851, EPI_ISL_955852, EPI_ISL_955853, EPI_ISL_955854, EPI_ISL_955855, EPI_ISL_955856, EPI_ISL_955857, EPI_ISL_955858, EPI_ISL_955859, EPI_ISL_955860, EPI_ISL_955861, EPI_ISL_955862, EPI_ISL_955863, EPI_ISL_955864, EPI_ISL_955865, EPI_ISL_955866, EPI_ISL_955867, EPI_ISL_955868, EPI_ISL_955869, EPI_ISL_955870, EPI_ISL_955871, EPI_ISL_955872, EPI_ISL_955873, EPI_ISL_955874, EPI_ISL_955875, EPI_ISL_955876, EPI_ISL_955877, EPI_ISL_955878, EPI_ISL_955879, EPI_ISL_955880, EPI_ISL_955881, EPI_ISL_955882, EPI_ISL_955883, EPI_ISL_955884, EPI_ISL_955885, EPI_ISL_955886, EPI_ISL_955887, EPI_ISL_955888, EPI_ISL_955889, EPI_ISL_955890, EPI_ISL_955891, EPI_ISL_955892, EPI_ISL_955893, EPI_ISL_955894, EPI_ISL_955895, EPI_ISL_955896, EPI_ISL_955897, EPI_ISL_955898, EPI_ISL_955899, EPI_ISL_955900, EPI_ISL_955901, EPI_ISL_955902, EPI_ISL_955903, EPI_ISL_955904, EPI_ISL_955906, EPI_ISL_955907, EPI_ISL_955908, EPI_ISL_955909, EPI_ISL_955910, EPI_ISL_955911, EPI_ISL_955912, EPI_ISL_955913, EPI_ISL_955914, EPI_ISL_955915, EPI_ISL_955916, EPI_ISL_955917, EPI_ISL_955918, EPI_ISL_955919, EPI_ISL_955920, EPI_ISL_955921, EPI_ISL_955922, EPI_ISL_955923, EPI_ISL_955924, EPI_ISL_955925, EPI_ISL_955926, EPI_ISL_955927, EPI_ISL_955928, EPI_ISL_955929, EPI_ISL_955930, EPI_ISL_955931, EPI_ISL_955932, EPI_ISL_955933, EPI_ISL_955934, EPI_ISL_955935, EPI_ISL_955936, EPI_ISL_955937, EPI_ISL_955938 | | | |
| see above | University of Michigan Clinical Microbiology Laboratory | Lauring Lab, University of Michigan, Department of Microbiology and Immunology | Valesano |
| EPI_ISL_955982, EPI_ISL_955983, EPI_ISL_955984, EPI_ISL_955985, EPI_ISL_955986, EPI_ISL_955987, EPI_ISL_955988, EPI_ISL_955989, EPI_ISL_955990, EPI_ISL_955991, EPI_ISL_955992, EPI_ISL_955993, EPI_ISL_955994, EPI_ISL_955995, EPI_ISL_955996, EPI_ISL_955997, EPI_ISL_955998, EPI_ISL_955999, EPI_ISL_956000, EPI_ISL_956001, EPI_ISL_956002, EPI_ISL_956003, EPI_ISL_956004, EPI_ISL_956005, EPI_ISL_956006, EPI_ISL_956007, EPI_ISL_956008, EPI_ISL_956009, EPI_ISL_956010, EPI_ISL_956011, EPI_ISL_956012, EPI_ISL_956013, EPI_ISL_956014, EPI_ISL_956015, EPI_ISL_956016, EPI_ISL_956017, EPI_ISL_956018, EPI_ISL_956019, EPI_ISL_956020, EPI_ISL_956021, EPI_ISL_956022, EPI_ISL_956023, EPI_ISL_956024, EPI_ISL_956025, EPI_ISL_956026, EPI_ISL_956027, EPI_ISL_956028, EPI_ISL_956029, EPI_ISL_956030, EPI_ISL_956031, EPI_ISL_956032, EPI_ISL_956033, EPI_ISL_956034, EPI_ISL_956035, EPI_ISL_956036, EPI_ISL_956037, EPI_ISL_956038, EPI_ISL_956039, EPI_ISL_956040, EPI_ISL_956041, EPI_ISL_956042, EPI_ISL_956043, EPI_ISL_956044, EPI_ISL_956045, EPI_ISL_956046, EPI_ISL_956047, EPI_ISL_956048, EPI_ISL_956049, EPI_ISL_956050, EPI_ISL_956051, EPI_ISL_956052, EPI_ISL_956053, EPI_ISL_956054, EPI_ISL_956055, EPI_ISL_956056, EPI_ISL_956057, EPI_ISL_956058, EPI_ISL_956059, EPI_ISL_956060, EPI_ISL_956061, EPI_ISL_956062, EPI_ISL_956063, EPI_ISL_956064, EPI_ISL_956065, EPI_ISL_956066, EPI_ISL_956067, EPI_ISL_956068, EPI_ISL_956069, EPI_ISL_956070, EPI_ISL_956071, EPI_ISL_956072, EPI_ISL_956073, EPI_ISL_956074, EPI_ISL_956075, EPI_ISL_956076, EPI_ISL_956077, EPI_ISL_956078, EPI_ISL_956079, EPI_ISL_956080, EPI_ISL_956081, EPI_ISL_956082, EPI_ISL_956083, EPI_ISL_956084, EPI_ISL_956085, EPI_ISL_956086, EPI_ISL_956087, EPI_ISL_956088, EPI_ISL_956089, EPI_ISL_956090, EPI_ISL_956091, EPI_ISL_956092, EPI_ISL_956093, EPI_ISL_956094, EPI_ISL_956095, EPI_ISL_956096, EPI_ISL_956097, EPI_ISL_956098, EPI_ISL_956099, EPI_ISL_956100, EPI_ISL_956101, EPI_ISL_956102, EPI_ISL_956103, EPI_ISL_956104, EPI_ISL_956105, EPI_ISL_956106, EPI_ISL_956107, EPI_ISL_956108, EPI_ISL_956109, EPI_ISL_956110, EPI_ISL_956111, EPI_ISL_956112, EPI_ISL_956113, EPI_ISL_956114, EPI_ISL_956115, EPI_ISL_956116, EPI_ISL_956117, EPI_ISL_956118, EPI_ISL_956119, EPI_ISL_956120, EPI_ISL_956121, EPI_ISL_956122, EPI_ISL_956123, EPI_ISL_956124, EPI_ISL_956125, EPI_ISL_956126, EPI_ISL_956127, EPI_ISL_956128, EPI_ISL_956129, EPI_ISL_956130, EPI_ISL_956131, EPI_ISL_956132, EPI_ISL_956133, EPI_ISL_956134, EPI_ISL_956135, EPI_ISL_956136, EPI_ISL_956137, EPI_ISL_956138, EPI_ISL_956139, EPI_ISL_956140, EPI_ISL_956141, EPI_ISL_956142, EPI_ISL_956143, EPI_ISL_956144, EPI_ISL_956145, EPI_ISL_956146, EPI_ISL_956147, EPI_ISL_956148, EPI_ISL_956149, EPI_ISL_956150, EPI_ISL_956151, EPI_ISL_956152, EPI_ISL_956153, EPI_ISL_956154, EPI_ISL_956155, EPI_ISL_956156, EPI_ISL_956157, EPI_ISL_956158, EPI_ISL_956159, EPI_ISL_956160, EPI_ISL_956161, EPI_ISL_956162, EPI_ISL_956163, EPI_ISL_956164, EPI_ISL_956165, EPI_ISL_956166, EPI_ISL_956167, EPI_ISL_956168, EPI_ISL_956169, EPI_ISL_956170, EPI_ISL_956171, EPI_ISL_956172, EPI_ISL_956173, EPI_ISL_956174, EPI_ISL_956175, EPI_ISL_956176, EPI_ISL_956177, EPI_ISL_956178, EPI_ISL_956179, EPI_ISL_956180, EPI_ISL_956181, EPI_ISL_956182, EPI_ISL_956183, EPI_ISL_956184, EPI_ISL_956185, EPI_ISL_956186, EPI_ISL_956187, EPI_ISL_956188, EPI_ISL_956189, EPI_ISL_956190, EPI_ISL_956191, EPI_ISL_956192, EPI_ISL_956193, EPI_ISL_956194, EPI_ISL_956195, EPI_ISL_956196, EPI_ISL_956197, EPI_ISL_956198, EPI_ISL_956199, EPI_ISL_956200, EPI_ISL_956201, EPI_ISL_956202, EPI_ISL_956203, EPI_ISL_956204, EPI_ISL_956205, EPI_ISL_956206, EPI_ISL_956207, EPI_ISL_956208, EPI_ISL_956209, EPI_ISL_956210, EPI_ISL_956211, EPI_ISL_956212, EPI_ISL_956213, EPI_ISL_956214, EPI_ISL_956215, EPI_ISL_956216, EPI_ISL_956217, EPI_ISL_956218, EPI_ISL_956219, EPI_ISL_956220, EPI_ISL_956221, EPI_ISL_956222, EPI_ISL_956223, EPI_ISL_956224, EPI_ISL_956225, EPI_ISL_956226, EPI_ISL_956227, EPI_ISL_956228, EPI_ISL_956229, EPI_ISL_956230, EPI_ISL_956231, EPI_ISL_956232, EPI_ISL_956233, EPI_ISL_956234, EPI_ISL_956235, EPI_ISL_956236, EPI_ISL_956237, EPI_ISL_956238, EPI_ISL_956239, EPI_ISL_956240, EPI_ISL_956241, EPI_ISL_956242, EPI_ISL_956243, EPI_ISL_956244, EPI_ISL_956245, EPI_ISL_956246, EPI_ISL_956247, EPI_ISL_956248, EPI_ISL_956249, EPI_ISL_956250, EPI_ISL_956251, EPI_ISL_956252, EPI_ISL_956253, EPI_ISL_956254, EPI_ISL_956255, EPI_ISL_956256, EPI_ISL_956257, EPI_ISL_956258, EPI_ISL_956259, EPI_ISL_956260, EPI_ISL_956261, EPI_ISL_956262, EPI_ISL_956263, EPI_ISL_956264, EPI_ISL_956265, EPI_ISL_956266, EPI_ISL_956267, EPI_ISL_956268, EPI_ISL_956269 | | | |
| see above | Pathology and Laboratory Medicine Institute, Cleveland Clinic, Ohio, USA | Pathology and Laboratory Medicine Institute, Cleveland Clinic, Ohio, USA | Frank P. Esper, Yu-Wei Cheng, Thamali M. Adhikari, Zheng Jin Tu, Daniel H. Farkas, Gary Procop, Jennifer Ko, Timothy A. Chan, Jing Li, Brian P. Rubin |
| EPI_ISL_956339 | University Hospitals Translational Laboratory (UHTL), University Hospitals | University Hospitals Translational Laboratory (UHTL), University Hospitals | Sadri,N., Alouani,D. and Song,X. |
| EPI_ISL_956340, EPI_ISL_956341, EPI_ISL_956342, EPI_ISL_956343, EPI_ISL_956344, EPI_ISL_956345, EPI_ISL_956346, EPI_ISL_956347, EPI_ISL_956348, EPI_ISL_956349, EPI_ISL_956350, EPI_ISL_956351, EPI_ISL_956352, EPI_ISL_956353, EPI_ISL_956354, EPI_ISL_956355, EPI_ISL_956356, EPI_ISL_956357, EPI_ISL_956358, EPI_ISL_956359, EPI_ISL_956360, EPI_ISL_956361, EPI_ISL_956362, EPI_ISL_956363, EPI_ISL_956364, EPI_ISL_956365, EPI_ISL_956366 | | | |
| see above | Houston Health Department, Disease Prevention and Control | Houston Health Department, Disease Prevention and Control | Penn,R., Brown,P., Lara,A. |

| | | | |
|---|---|---|---|
| EPI_ISL_959378, EPI_ISL_959379, EPI_ISL_959380, EPI_ISL_959381, EPI_ISL_959382, EPI_ISL_959383, EPI_ISL_959385, EPI_ISL_959387, EPI_ISL_959414, EPI_ISL_959415, EPI_ISL_959417, EPI_ISL_959472, EPI_ISL_959473, EPI_ISL_959474 | | | |
| see above | Ohio Department of Health Laboratory | Ohio Department of Health Laboratory | Holmes, Jennifer; Eric Brandt, Keoni Omura, Glen McGillivary, Caitlin McDonnell, Jade Mowery, Stephanie Mccracken, Tyler Payne, Kirtana Ramadugu, Erica Leasure, Brent Lee, Kelsey Florek, Heather Blankenship, Quanta Brown, and Tammy Bannerman |
| EPI_ISL_959539, EPI_ISL_959540, EPI_ISL_959541, EPI_ISL_959542, EPI_ISL_959543 | University of Michigan Clinical Microbiology Laboratory | Lauring Lab, University of Michigan, Department of Microbiology and Immunology | Valesano |
| EPI_ISL_960188, EPI_ISL_960189, EPI_ISL_960190, EPI_ISL_960191, EPI_ISL_960192, EPI_ISL_960193, EPI_ISL_960194, EPI_ISL_960195, EPI_ISL_960196, EPI_ISL_960197, EPI_ISL_960198, EPI_ISL_960199, EPI_ISL_960200, EPI_ISL_960201, EPI_ISL_960202, EPI_ISL_960203, EPI_ISL_960204, EPI_ISL_960205, EPI_ISL_960206, EPI_ISL_960207, EPI_ISL_960208, EPI_ISL_960209, EPI_ISL_960210, EPI_ISL_960211, EPI_ISL_960212, EPI_ISL_960213, EPI_ISL_960214, EPI_ISL_960215, EPI_ISL_960216, EPI_ISL_960217, EPI_ISL_960218, EPI_ISL_960219, EPI_ISL_960220, EPI_ISL_960221, EPI_ISL_960222 | | | |
| see above | Virginia Division of Consolidated Laboratory Services | Virginia Division of Consolidated Laboratory Services | Virginia DCLS |
| EPI_ISL_960313, EPI_ISL_960314, EPI_ISL_960315, EPI_ISL_960316, EPI_ISL_960317, EPI_ISL_960318, EPI_ISL_960319, EPI_ISL_960320, EPI_ISL_960321, EPI_ISL_960322, EPI_ISL_960323, EPI_ISL_960324, EPI_ISL_960325, EPI_ISL_960326, EPI_ISL_960327, EPI_ISL_960328, EPI_ISL_960329, EPI_ISL_960330, EPI_ISL_960331, EPI_ISL_960332, EPI_ISL_960333, EPI_ISL_960334, EPI_ISL_960335, EPI_ISL_960336, EPI_ISL_960337, EPI_ISL_960338, EPI_ISL_960339, EPI_ISL_960340, EPI_ISL_960341, EPI_ISL_960342, EPI_ISL_960343, EPI_ISL_960344, EPI_ISL_960345, EPI_ISL_960346, EPI_ISL_960347, EPI_ISL_960348, EPI_ISL_960349, EPI_ISL_960350, EPI_ISL_960351, EPI_ISL_960352, EPI_ISL_960353, EPI_ISL_960354, EPI_ISL_960355, EPI_ISL_960356, EPI_ISL_960357, EPI_ISL_960358, EPI_ISL_960359, EPI_ISL_960360, EPI_ISL_960361, EPI_ISL_960362, EPI_ISL_960363, EPI_ISL_960364, EPI_ISL_960365, EPI_ISL_960366, EPI_ISL_960367, EPI_ISL_960368, EPI_ISL_960369, EPI_ISL_960370, EPI_ISL_960371, EPI_ISL_960372, EPI_ISL_960373, EPI_ISL_960374, EPI_ISL_960375, EPI_ISL_960376, EPI_ISL_960377, EPI_ISL_960378, EPI_ISL_960379, EPI_ISL_960380, EPI_ISL_960381, EPI_ISL_960382, EPI_ISL_960383, EPI_ISL_960389, EPI_ISL_960390, EPI_ISL_960391, EPI_ISL_960392, EPI_ISL_960393, EPI_ISL_960394, EPI_ISL_960395, EPI_ISL_960396, EPI_ISL_960397, EPI_ISL_960398, EPI_ISL_960399, EPI_ISL_960400, EPI_ISL_960401, EPI_ISL_960402, EPI_ISL_960403 | | | |
| see above | University of Wisconsin-Madison AIDS Vaccine Research Laboratories | University of Wisconsin-Madison AIDS Vaccine Research Laboratories | Gage Moreno, Katarina Braun, et al. AIDS Vaccine Research Laboratories |
| EPI_ISL_961105, EPI_ISL_961106, EPI_ISL_961107, EPI_ISL_961109, EPI_ISL_961110, EPI_ISL_961111, EPI_ISL_961112, EPI_ISL_961113, EPI_ISL_961114, EPI_ISL_961115, EPI_ISL_961116, EPI_ISL_961117, EPI_ISL_961118, EPI_ISL_961119, EPI_ISL_961120, EPI_ISL_961121, EPI_ISL_961122, EPI_ISL_961123, EPI_ISL_961124, EPI_ISL_961125, EPI_ISL_961126, EPI_ISL_961128, EPI_ISL_961129, EPI_ISL_961130, EPI_ISL_961131 | | | |
| see above | Delaware Public Health Laboratory | Delaware Public Health Lab | Gregory Hovan |
| EPI_ISL_961134, EPI_ISL_961135, EPI_ISL_961136, EPI_ISL_961137, EPI_ISL_961138, EPI_ISL_961139, EPI_ISL_961140, EPI_ISL_961141, EPI_ISL_961142, EPI_ISL_961143, EPI_ISL_961144, EPI_ISL_961145, EPI_ISL_961146, EPI_ISL_961147, EPI_ISL_961148, EPI_ISL_961149, EPI_ISL_961150, EPI_ISL_961151, EPI_ISL_961152, EPI_ISL_961153, EPI_ISL_961154, EPI_ISL_961155, EPI_ISL_961156, EPI_ISL_961157, EPI_ISL_961158, EPI_ISL_961159, EPI_ISL_961160, EPI_ISL_961161, EPI_ISL_961162, EPI_ISL_961163, EPI_ISL_961180 | | | |
| see above | Texas Department of State Health Services | Texas Department of State Health Services | Bonnie Oh, Anita Pokharel, James Daniel Bonser, Myong Koag, Chung Wang, Rachel Lee, Grace Kubin, Rashmi Tuladhar, Mayela Pedrueza, Maliha Rahman, Jenny Zhang |
| EPI_ISL_961288, EPI_ISL_961289, EPI_ISL_961290, EPI_ISL_961291, EPI_ISL_961292, EPI_ISL_961293, EPI_ISL_961294, EPI_ISL_961295, EPI_ISL_961296, EPI_ISL_961297, EPI_ISL_961298, EPI_ISL_961299, EPI_ISL_961300, EPI_ISL_961301, EPI_ISL_961302, EPI_ISL_961303, EPI_ISL_961304, EPI_ISL_961305, EPI_ISL_961306, EPI_ISL_961307, EPI_ISL_961308, EPI_ISL_961309, EPI_ISL_961310, EPI_ISL_961311, EPI_ISL_961312, EPI_ISL_961313, EPI_ISL_961314, EPI_ISL_961315, EPI_ISL_961316, EPI_ISL_961317, EPI_ISL_961318, EPI_ISL_961319, EPI_ISL_961320, EPI_ISL_961321, EPI_ISL_961323, EPI_ISL_961324, EPI_ISL_961325, EPI_ISL_961326, EPI_ISL_961327, EPI_ISL_961328, EPI_ISL_961329, EPI_ISL_961330, EPI_ISL_961331, EPI_ISL_961332, EPI_ISL_961333, EPI_ISL_961334, EPI_ISL_961335, EPI_ISL_961336, EPI_ISL_961337, EPI_ISL_961338, EPI_ISL_961339, EPI_ISL_961340, EPI_ISL_961341, EPI_ISL_961342, EPI_ISL_961343, EPI_ISL_961344, EPI_ISL_961345, EPI_ISL_961346, EPI_ISL_961347, EPI_ISL_961348, EPI_ISL_961349, EPI_ISL_961350, EPI_ISL_961351 | | | |
| see above | Biotia | Biotia | Dorottya Nagy-Szakal, Mara Couto-Rodriguez, Heather Wells, Joseph Barrows, Marilyne Debieu, Kristin Butcher, Siyuan Chen, Agnes Berki, Courteny Hager, Robert Boorstein, Mariah Taylor, Colleen Jonsson, Christopher Mason, Niamh O'Hara |
| EPI_ISL_961466 | Laboratorios Lister | Instituto de Diagnostico y Referencia Epidemiologicos (INDRE) | Claudia Wong-Arambula, Abril Rodriguez-Maldonado, Fabiola Garces-Ayala, Natividad Cruz-Ortiz, Tatiana Nunez-Garcia, Gisela Barrera-Badillo, Lucia Hernandez-Rivas, Irma Lopez-Martinez, Ernesto Ramirez-Gonzalez. |
| EPI_ISL_961467, EPI_ISL_961468, EPI_ISL_961469 | LESP Nuevo Leon/Grupo de Diagnostico ARIES | Instituto de Diagnostico y Referencia Epidemiologicos (INDRE) | Claudia Wong-Arambula, Abril Rodriguez-Maldonado, Fabiola Garces-Ayala, Natividad Cruz-Ortiz, Tatiana Nunez-Garcia, Gisela Barrera-Badillo, Lucia Hernandez-Rivas, Irma Lopez-Martinez, Ernesto Ramirez-Gonzalez. |
| EPI_ISL_961470, EPI_ISL_961471, EPI_ISL_961472, EPI_ISL_961473, EPI_ISL_961474, EPI_ISL_961475, EPI_ISL_961476, EPI_ISL_961477, EPI_ISL_961478, EPI_ISL_961479, EPI_ISL_961480, EPI_ISL_961481, EPI_ISL_961482, EPI_ISL_961483, EPI_ISL_961484, EPI_ISL_961485, EPI_ISL_961486, EPI_ISL_961487, EPI_ISL_961488, EPI_ISL_961489, EPI_ISL_961490, EPI_ISL_961491, EPI_ISL_961492, EPI_ISL_961493, EPI_ISL_961494, EPI_ISL_961495, EPI_ISL_961496, EPI_ISL_961497, EPI_ISL_961498, EPI_ISL_961499, EPI_ISL_961500, EPI_ISL_961501, EPI_ISL_961502, EPI_ISL_961503, EPI_ISL_961504, EPI_ISL_961505, EPI_ISL_961506, EPI_ISL_961507, EPI_ISL_961508, EPI_ISL_961509, EPI_ISL_961510, EPI_ISL_961511, EPI_ISL_961512, EPI_ISL_961513, EPI_ISL_961514, EPI_ISL_961515, EPI_ISL_961516, EPI_ISL_961517, EPI_ISL_961518, EPI_ISL_961519, EPI_ISL_961520, EPI_ISL_961521, EPI_ISL_961528, EPI_ISL_961529, EPI_ISL_961530, EPI_ISL_961531, EPI_ISL_961532, EPI_ISL_961533, EPI_ISL_961534, EPI_ISL_961535, EPI_ISL_961536, EPI_ISL_961537, EPI_ISL_961538, EPI_ISL_961539, EPI_ISL_961540, EPI_ISL_961541, EPI_ISL_961542, EPI_ISL_961543, EPI_ISL_961544, EPI_ISL_961545, EPI_ISL_961546, EPI_ISL_961547, EPI_ISL_961548, EPI_ISL_961549, EPI_ISL_961550, EPI_ISL_961551, EPI_ISL_961552, EPI_ISL_961553, EPI_ISL_961554 | | | |
| see above | Michigan Department of Health and Human Services, Bureau of Laboratories | Michigan Department of Health and Human Services, Bureau of Laboratories | Blankenship HM, Riner D, Soehnlen MK |
| EPI_ISL_961760, EPI_ISL_961761, EPI_ISL_961762, EPI_ISL_961763, EPI_ISL_961765, EPI_ISL_961766, EPI_ISL_961767, EPI_ISL_961768, EPI_ISL_961771, EPI_ISL_961772, EPI_ISL_961773, EPI_ISL_961774, EPI_ISL_961777, EPI_ISL_961780, EPI_ISL_961781, EPI_ISL_961785, EPI_ISL_961791, EPI_ISL_961792 | | | |
| see above | Laboratorio de Infectología, Servicio de Infectología, Hospital Universitario Dr. José Eleuterio González - Universidad Autónoma de Nuevo León | Laboratorio de Infectología Molecular, Departamento de Bioquímica y Medicina Molecular,Facultad de Medicina - Universidad Autónoma de Nuevo León | Kame A. Galán-Huerta, María F. Herrera-Saldivar, Natalia Martínez-Acuña, Sonia A. Lozano-Sepúlveda, Daniel Arellanos-Soto, Ana M. Rivas-Estilla, Paola Bocanegra-Ibarias, Samantha M. Flores-Treviño, Elvira Garza-González, Eduardo Perez-Alba, Laura Nuzzolo-Shihadeh, Adrian Camacho-Ortiz |
| EPI_ISL_961883, EPI_ISL_961884, EPI_ISL_961885, EPI_ISL_961886, EPI_ISL_961887, EPI_ISL_961888, EPI_ISL_961889, EPI_ISL_961890, EPI_ISL_961891, EPI_ISL_961892, EPI_ISL_961893, EPI_ISL_961894, EPI_ISL_961895, EPI_ISL_961896, EPI_ISL_961897, EPI_ISL_961898, EPI_ISL_961899, EPI_ISL_961900, EPI_ISL_961901, EPI_ISL_961902, EPI_ISL_961903, EPI_ISL_961904, EPI_ISL_961905, EPI_ISL_961906, EPI_ISL_961907, EPI_ISL_961908, EPI_ISL_961909, EPI_ISL_961910, EPI_ISL_961911, EPI_ISL_961912, EPI_ISL_961913, EPI_ISL_961914, EPI_ISL_961915, EPI_ISL_961916, EPI_ISL_961917, EPI_ISL_961918, EPI_ISL_961919, EPI_ISL_961920, EPI_ISL_961921, EPI_ISL_961922, EPI_ISL_961923, EPI_ISL_961924, EPI_ISL_961925, EPI_ISL_961926, EPI_ISL_961927, EPI_ISL_961928, EPI_ISL_961929, EPI_ISL_961930, EPI_ISL_961931, EPI_ISL_961932, EPI_ISL_961933, EPI_ISL_961934, EPI_ISL_961935, EPI_ISL_961936, EPI_ISL_961937, EPI_ISL_961938, EPI_ISL_961939, EPI_ISL_961940, EPI_ISL_961941, EPI_ISL_961942, EPI_ISL_961943, EPI_ISL_961944, EPI_ISL_961945, EPI_ISL_961946, EPI_ISL_961947, EPI_ISL_961948, EPI_ISL_961949, EPI_ISL_961950, EPI_ISL_961951, EPI_ISL_961952, EPI_ISL_961953, EPI_ISL_961954, EPI_ISL_961955, EPI_ISL_961956, EPI_ISL_961957, EPI_ISL_961958, EPI_ISL_961959, EPI_ISL_961960, EPI_ISL_961961, EPI_ISL_961962, EPI_ISL_961963, EPI_ISL_961964, EPI_ISL_961965, EPI_ISL_961966, EPI_ISL_961967, EPI_ISL_961968, EPI_ISL_961969, EPI_ISL_961970, EPI_ISL_961971, EPI_ISL_961972, EPI_ISL_961973, EPI_ISL_961974, EPI_ISL_961975, EPI_ISL_961976, EPI_ISL_961977, EPI_ISL_961978, EPI_ISL_962009, EPI_ISL_962010, EPI_ISL_962011, EPI_ISL_962012, EPI_ISL_962013, EPI_ISL_962014, EPI_ISL_962015, EPI_ISL_962016, EPI_ISL_962017, EPI_ISL_962018, EPI_ISL_962019, EPI_ISL_962020, EPI_ISL_962021, EPI_ISL_962022, EPI_ISL_962023, EPI_ISL_962024, EPI_ISL_962025, EPI_ISL_962026, EPI_ISL_962027, EPI_ISL_962028, EPI_ISL_962029, EPI_ISL_962030, EPI_ISL_962031, EPI_ISL_962032, EPI_ISL_962033, EPI_ISL_962034, EPI_ISL_962035, EPI_ISL_962036, EPI_ISL_962037, EPI_ISL_962038, EPI_ISL_962039, EPI_ISL_962040, EPI_ISL_962041, EPI_ISL_962042, EPI_ISL_962043, EPI_ISL_962044, EPI_ISL_962045, EPI_ISL_962046, EPI_ISL_962047, EPI_ISL_962048, EPI_ISL_962049, EPI_ISL_962050, EPI_ISL_962051, EPI_ISL_962052, EPI_ISL_962053, EPI_ISL_962054, EPI_ISL_962055, EPI_ISL_962056, EPI_ISL_962057, EPI_ISL_962058, EPI_ISL_962059, EPI_ISL_962060, EPI_ISL_962061, EPI_ISL_962062, EPI_ISL_962063, EPI_ISL_962064, EPI_ISL_962065, EPI_ISL_962066, EPI_ISL_962067, EPI_ISL_962068, EPI_ISL_962069, EPI_ISL_962070, EPI_ISL_962071, EPI_ISL_962072, EPI_ISL_962073, EPI_ISL_962074, EPI_ISL_962075, EPI_ISL_962076, EPI_ISL_962077, EPI_ISL_962078, EPI_ISL_962079, EPI_ISL_962080, EPI_ISL_962081, EPI_ISL_962082, EPI_ISL_962083, EPI_ISL_962084, EPI_ISL_962085, EPI_ISL_962086, EPI_ISL_962087, EPI_ISL_962088, EPI_ISL_962089, EPI_ISL_962090, EPI_ISL_962091, EPI_ISL_962092, EPI_ISL_962093, EPI_ISL_962094, EPI_ISL_962095, EPI_ISL_962096, EPI_ISL_962097, EPI_ISL_962098, EPI_ISL_962099, EPI_ISL_962100, EPI_ISL_962101, EPI_ISL_962102, EPI_ISL_962103, EPI_ISL_962104, EPI_ISL_962105, EPI_ISL_962106, EPI_ISL_962107, EPI_ISL_962108, EPI_ISL_962109, EPI_ISL_962110, EPI_ISL_962111, EPI_ISL_962112, EPI_ISL_962113, EPI_ISL_962114, EPI_ISL_962115, EPI_ISL_962116, EPI_ISL_962117, EPI_ISL_962118, EPI_ISL_962119, EPI_ISL_962120, EPI_ISL_962121, EPI_ISL_962122, EPI_ISL_962123, EPI_ISL_962124, EPI_ISL_962125, EPI_ISL_962126, EPI_ISL_962127, EPI_ISL_962128, EPI_ISL_962129, EPI_ISL_962130, EPI_ISL_962131, EPI_ISL_962132, EPI_ISL_962133, EPI_ISL_962134, EPI_ISL_962135, EPI_ISL_962136, EPI_ISL_962137, EPI_ISL_962138, EPI_ISL_962139, EPI_ISL_962140, EPI_ISL_962141, EPI_ISL_962142, EPI_ISL_962143, EPI_ISL_962144, EPI_ISL_962145, EPI_ISL_962146, EPI_ISL_962147, EPI_ISL_962148, EPI_ISL_962149, EPI_ISL_962150, EPI_ISL_962151, EPI_ISL_962152, EPI_ISL_962153, EPI_ISL_962154, EPI_ISL_962155, EPI_ISL_962156, EPI_ISL_962157, EPI_ISL_962158, EPI_ISL_962159, EPI_ISL_962160, EPI_ISL_962161, EPI_ISL_962162, EPI_ISL_962163, EPI_ISL_962164, EPI_ISL_962165, EPI_ISL_962166, EPI_ISL_962167, EPI_ISL_962168, EPI_ISL_962169, EPI_ISL_962170, EPI_ISL_962171, EPI_ISL_962172, EPI_ISL_962173, EPI_ISL_962174, EPI_ISL_962175, EPI_ISL_962176, EPI_ISL_962177, EPI_ISL_962178 | | | |
| see above | Illinois Department of Public Health | Gagnon Lab, Southern Illinois University | Keith Gagnon |
| EPI_ISL_962182 | Ohio Department of Health Laboratory | Ohio Department of Health Laboratory | Holmes, Jennifer; Eric Brandt, Keoni Omura, Glen McGillivary, Caitlin McDonnell, Jade Mowery, Stephanie Mccracken, Tyler Payne, Kirtana Ramadugu, Erica Leasure, Brent Lee, Kelsey Florek, Heather Blankenship, Quanta Brown, and Tammy Bannerman |
| EPI_ISL_962208, EPI_ISL_962210, EPI_ISL_962212, EPI_ISL_962213, EPI_ISL_962214, EPI_ISL_962215, EPI_ISL_962216, EPI_ISL_962219, EPI_ISL_962221, EPI_ISL_962222, EPI_ISL_962225, EPI_ISL_962227, EPI_ISL_962228, EPI_ISL_962229, EPI_ISL_962231, EPI_ISL_962232, EPI_ISL_962234, EPI_ISL_962235, EPI_ISL_962236, EPI_ISL_962237, EPI_ISL_962239, EPI_ISL_962243, EPI_ISL_962244, EPI_ISL_962246, EPI_ISL_962247, EPI_ISL_962248, EPI_ISL_962249, EPI_ISL_962250, EPI_ISL_962251, EPI_ISL_962253, EPI_ISL_962254, EPI_ISL_962255, EPI_ISL_962256, EPI_ISL_962257, EPI_ISL_962259, EPI_ISL_962260, EPI_ISL_962261, EPI_ISL_962262, EPI_ISL_962263 | | | |
| see above | Seattle Flu Study | Seattle Flu Study | Deborah A. Nickerson, Chris D. Frazar, Jover Lee, Benjamin Pelle, Erica Ryke, Matthew Richardson, Amanda Adler, Elisabeth Brandstetter, Peter D. Han, Kairsten Fay, Misja Ilcisin, Kirsten Lacombe, Thomas R. Sibley, Melissa Truong, Caitlin R. Wolf, Karen Cowgill, Stephanie Schrag, Jeff Duchin, Michael Boeckh, Janet A. Englund, Michael Famulare, Barry R. Lutz, Mark J. Rieder, Lea M. Starita, Matthew Thompson, Helen Y. Chu, Trevor Bedford, Jay Shendure |
| EPI_ISL_962264, EPI_ISL_962265, EPI_ISL_962266, EPI_ISL_962267, EPI_ISL_962268, EPI_ISL_962269, EPI_ISL_962270, EPI_ISL_962271, EPI_ISL_962272, EPI_ISL_962273, EPI_ISL_962274 | | | |
| see above | Seattle Flu Study | Seattle Flu Study | Deborah A. Nickerson, Chris D. Frazar, Jover Lee, Benjamin Pelle, Erica Ryke, Matthew Richardson, Amanda Adler, Elisabeth Brandstetter, Peter D. Han, Kairsten Fay, Misja Ilcisin, Kirsten Lacombe, Thomas R. Sibley, Melissa Truong, Caitlin R. Wolf, Michael Boeckh, Janet A. Englund, Michael Famulare, Barry R. Lutz, Mark J. Rieder, Lea M. Starita, Matthew Thompson, Jay Shendure, Trevor Bedford, Helen Y. Chu |
| EPI_ISL_962275 | Seattle Flu Study | Seattle Flu Study | Deborah A. Nickerson, Chris D. Frazar, Jover Lee, Benjamin Pelle, Erica Ryke, Matthew Richardson, Amanda Adler, Elisabeth Brandstetter, Peter D. Han, Kairsten Fay, Misja Ilcisin, Kirsten Lacombe, Thomas R. Sibley, Melissa Truong, Caitlin R. Wolf, Karen Cowgill, Stephanie Schrag, Jeff Duchin, Michael Boeckh, Janet A. Englund, Michael Famulare, Barry R. Lutz, Mark J. Rieder, Lea M. Starita, Matthew Thompson, Helen Y. Chu, Trevor Bedford, Jay Shendure |
| EPI_ISL_962276 | Seattle Flu Study | Seattle Flu Study | Deborah A. Nickerson, Chris D. Frazar, Jover Lee, Benjamin Pelle, Erica Ryke, Matthew Richardson, Amanda Adler, Elisabeth Brandstetter, Peter D. Han, Kairsten Fay, Misja Ilcisin, Kirsten Lacombe, Thomas R. Sibley, Melissa Truong, Caitlin R. Wolf, Michael Boeckh, Janet A. Englund, Michael Famulare, Barry R. Lutz, Mark J. Rieder, Lea M. Starita, Matthew Thompson, Jay Shendure, Trevor Bedford, Helen Y. Chu |
| EPI_ISL_962277, EPI_ISL_962278, EPI_ISL_962280, EPI_ISL_962281, EPI_ISL_962282, EPI_ISL_962283, EPI_ISL_962284, EPI_ISL_962286, EPI_ISL_962288, EPI_ISL_962290, EPI_ISL_962291, EPI_ISL_962292, EPI_ISL_962293, EPI_ISL_962294, EPI_ISL_962295, EPI_ISL_962296, EPI_ISL_962297, EPI_ISL_962298, EPI_ISL_962299 | | | |
| see above | Seattle Flu Study | Seattle Flu Study | Deborah A. Nickerson, Chris D. Frazar, Jover Lee, Benjamin Pelle, Erica Ryke, Matthew Richardson, Amanda Adler, Elisabeth Brandstetter, Peter D. Han, Kairsten Fay, Misja Ilcisin, Kirsten Lacombe, Thomas R. Sibley, Melissa Truong, Caitlin R. Wolf, Karen Cowgill, Stephanie Schrag, Jeff Duchin, Michael Boeckh, Janet A. Englund, Michael Famulare, Barry R. Lutz, Mark J. Rieder, Lea M. Starita, Matthew Thompson, Helen Y. Chu, Trevor Bedford, Jay Shendure |
| EPI_ISL_962301 | Seattle Flu Study | Seattle Flu Study | Deborah A. Nickerson, Chris D. Frazar, Jover Lee, Benjamin Pelle, Erica Ryke, Matthew Richardson, Amanda Adler, Elisabeth Brandstetter, Peter D. Han, Kairsten Fay, Misja Ilcisin, Kirsten Lacombe, Thomas R. Sibley, Melissa Truong, Caitlin R. Wolf, Michael Boeckh, Janet A. Englund, Michael Famulare, Barry R. Lutz, Mark J. Rieder, Lea M. Starita, Matthew Thompson, Jay Shendure, Trevor Bedford, Helen Y. Chu |
| EPI_ISL_962302 | Seattle Flu Study | Seattle Flu Study | Deborah A. Nickerson, Chris D. Frazar, Jover Lee, Benjamin Pelle, Erica Ryke, Matthew Richardson, Amanda Adler, Elisabeth Brandstetter, Peter D. Han, Kairsten Fay, Misja Ilcisin, Kirsten Lacombe, Thomas R. Sibley, Melissa Truong, Caitlin R. Wolf, Karen Cowgill, Stephanie Schrag, Jeff Duchin, Michael Boeckh, Janet A. Englund, Michael Famulare, Barry R. Lutz, Mark J. Rieder, Lea M. Starita, Matthew Thompson, Helen Y. Chu, Trevor Bedford, Jay Shendure |
| EPI_ISL_962303, EPI_ISL_962304 | Seattle Flu Study | Seattle Flu Study | Deborah A. Nickerson, Chris D. Frazar, Jover Lee, Benjamin Pelle, Erica Ryke, Matthew Richardson, Amanda Adler, Elisabeth Brandstetter, Peter D. Han, Kairsten Fay, Misja Ilcisin, Kirsten Lacombe, Thomas R. Sibley, Melissa Truong, Caitlin R. Wolf, Michael Boeckh, Janet A. Englund, Michael Famulare, Barry R. Lutz, Mark J. Rieder, Lea M. Starita, Matthew Thompson, Jay Shendure, Trevor Bedford, Helen Y. Chu |
| EPI_ISL_962305, EPI_ISL_962306, EPI_ISL_962307, EPI_ISL_962308, EPI_ISL_962310, EPI_ISL_962312, EPI_ISL_962313, EPI_ISL_962315, EPI_ISL_962316, EPI_ISL_962317, EPI_ISL_962318, EPI_ISL_962319, EPI_ISL_962320, EPI_ISL_962321, EPI_ISL_962322, EPI_ISL_962323, EPI_ISL_962324, EPI_ISL_962325, EPI_ISL_962326, EPI_ISL_962328, EPI_ISL_962329, EPI_ISL_962330, EPI_ISL_962331, EPI_ISL_962332, EPI_ISL_962334, EPI_ISL_962335, EPI_ISL_962337, EPI_ISL_962338, EPI_ISL_962339, EPI_ISL_962340, EPI_ISL_962341, EPI_ISL_962343, EPI_ISL_962344, EPI_ISL_962346, EPI_ISL_962348, EPI_ISL_962349, EPI_ISL_962350, EPI_ISL_962351, EPI_ISL_962352, EPI_ISL_962353, EPI_ISL_962355, EPI_ISL_962356, EPI_ISL_962357, EPI_ISL_962358, EPI_ISL_962359, EPI_ISL_962360, EPI_ISL_962361, EPI_ISL_962362, EPI_ISL_962363, EPI_ISL_962364, EPI_ISL_962365, EPI_ISL_962367 | | | |
| see above | Altius Institute for Biomedical Sciences | Seattle Flu Study | Deborah A. Nickerson, Chris D. Frazar, Jover Lee, Benjamin Pelle, Erica Ryke, Matthew Richardson, Amanda Adler, Elisabeth Brandstetter, Peter D. Han, Kairsten Fay, Misja Ilcisin, Kirsten Lacombe, Thomas R. Sibley, Melissa Truong, Caitlin R. Wolf, Ryan Alexander, Daniel Bates, Rebecca Bruders, Stephanie DeBaun, Clem Green, Muhammad Halimun, Jessica Halow, Kneshay Harper, Matt Hartman, Andrew Meuser, Alex Nguyen, Truong Nguyen, Sofia Olsson, Sadie Patraw, Hannah Petersen, Tobias Ragoczy, Joshua Richards, Jacob Rodriguez, John Stamatoyannopoulos, Julia Wald, Olivia Waltner, Michael Boeckh, Janet A. Englund, Michael Famulare, Barry R. Lutz, Mark J. Rieder, Lea M. Starita, Matthew Thompson, Helen Y. Chu, Jay Shendure, Trevor Bedford |
| EPI_ISL_962368, EPI_ISL_962369, EPI_ISL_962370, EPI_ISL_962371, EPI_ISL_962372, EPI_ISL_962373, EPI_ISL_962376, EPI_ISL_962377 | Seattle Flu Study | Seattle Flu Study | Deborah A. Nickerson, Chris D. Frazar, Jover Lee, Benjamin Pelle, Erica Ryke, Matthew Richardson, Amanda Adler, Elisabeth Brandstetter, Peter D. Han, Kairsten Fay, Misja Ilcisin, Kirsten Lacombe, Thomas R. Sibley, Melissa Truong, Caitlin R. Wolf, Karen Cowgill, Stephanie Schrag, Jeff Duchin, Michael Boeckh, Janet A. Englund, Michael Famulare, Barry R. Lutz, Mark J. Rieder, Lea M. Starita, Matthew Thompson, Helen Y. Chu, Trevor Bedford, Jay Shendure |
| EPI_ISL_962378, EPI_ISL_962379, EPI_ISL_962381, EPI_ISL_962382, EPI_ISL_962386, EPI_ISL_962387, EPI_ISL_962388, EPI_ISL_962391, EPI_ISL_962392, EPI_ISL_962393, EPI_ISL_962398, EPI_ISL_962399, EPI_ISL_962401, EPI_ISL_962402, EPI_ISL_962403, EPI_ISL_962404, EPI_ISL_962405, EPI_ISL_962406, EPI_ISL_962407, EPI_ISL_962409, EPI_ISL_962410, EPI_ISL_962411, EPI_ISL_962412, EPI_ISL_962413, EPI_ISL_962417, EPI_ISL_962418, EPI_ISL_962419, EPI_ISL_962420, EPI_ISL_962421, EPI_ISL_962422, EPI_ISL_962423, EPI_ISL_962424, EPI_ISL_962425, EPI_ISL_962426, EPI_ISL_962427, EPI_ISL_962429, EPI_ISL_962430, EPI_ISL_962431, EPI_ISL_962432, EPI_ISL_962433, EPI_ISL_962434, EPI_ISL_962435, EPI_ISL_962436, EPI_ISL_962437, EPI_ISL_962439, EPI_ISL_962440, EPI_ISL_962441 | | | |

| | | | |
|---|---|---|---|
| see above | Washington State Department of Health | Seattle Flu Study | Deborah A. Nickerson, Chris D. Frazar, Jover Lee, Benjamin Pelle, Erica Ryke, Matthew Richardson, Amanda Adler, Elisabeth Brandstetter, Peter D. Han, Kairsten Fay, Misja Ilcisin, Kirsten Lacombe, Thomas R. Sibley, Melissa Truong, Caitlin R. Wolf, Romesh Gautom, Geoff Melly, Brian Hiatt, Philip Dykema, Scott Lindquist, Michael Boeckh, Janet A. Englund, Michael Famulare, Barry R. Lutz, Mark J. Rieder, Lea M. Starita, Matthew Thompson, Helen Y. Chu, Jay Shendure, Trevor Bedford |
| EPI_ISL_962442 | Evergreen Healthcare | Seattle Flu Study | Deborah A. Nickerson, Chris D. Frazar, Jover Lee, Benjamin Pelle, Erica Ryke, Matthew Richardson, Amanda Adler, Elisabeth Brandstetter, Peter D. Han, Kairsten Fay, Misja Ilcisin, Kirsten Lacombe, Thomas R. Sibley, Melissa Truong, Caitlin R. Wolf, Romesh Gautom, Geoff Melly, Brian Hiatt, Philip Dykema, Scott Lindquist, Michael Boeckh, Janet A. Englund, Michael Famulare, Barry R. Lutz, Mark J. Rieder, Lea M. Starita, Matthew Thompson, Helen Y. Chu, Jay Shendure, Trevor Bedford |
| EPI_ISL_962444 | Seattle Flu Study | Seattle Flu Study | Deborah A. Nickerson, Chris D. Frazar, Jover Lee, Benjamin Pelle, Erica Ryke, Matthew Richardson, Amanda Adler, Elisabeth Brandstetter, Peter D. Han, Kairsten Fay, Misja Ilcisin, Kirsten Lacombe, Thomas R. Sibley, Melissa Truong, Caitlin R. Wolf, Karen Cowgill, Stephanie Schrag, Jeff Duchin, Michael Boeckh, Janet A. Englund, Michael Famulare, Barry R. Lutz, Mark J. Rieder, Lea M. Starita, Matthew Thompson, Helen Y. Chu, Trevor Bedford, Jay Shendure |
| EPI_ISL_962445 | Seattle Flu Study | Seattle Flu Study | Deborah A. Nickerson, Chris D. Frazar, Jover Lee, Benjamin Pelle, Erica Ryke, Matthew Richardson, Amanda Adler, Elisabeth Brandstetter, Peter D. Han, Kairsten Fay, Misja Ilcisin, Kirsten Lacombe, Thomas R. Sibley, Melissa Truong, Caitlin R. Wolf, Michael Boeckh, Janet A. Englund, Michael Famulare, Barry R. Lutz, Mark J. Rieder, Lea M. Starita, Matthew Thompson, Jay Shendure, Trevor Bedford, Helen Y. Chu |
| EPI_ISL_962446, EPI_ISL_962447, EPI_ISL_962448, EPI_ISL_962449, EPI_ISL_962450, EPI_ISL_962452, EPI_ISL_962454, EPI_ISL_962455, EPI_ISL_962456 | Seattle Flu Study | Seattle Flu Study | Deborah A. Nickerson, Chris D. Frazar, Jover Lee, Benjamin Pelle, Erica Ryke, Matthew Richardson, Amanda Adler, Elisabeth Brandstetter, Peter D. Han, Kairsten Fay, Misja Ilcisin, Kirsten Lacombe, Thomas R. Sibley, Melissa Truong, Caitlin R. Wolf, Karen Cowgill, Stephanie Schrag, Jeff Duchin, Michael Boeckh, Janet A. Englund, Michael Famulare, Barry R. Lutz, Mark J. Rieder, Lea M. Starita, Matthew Thompson, Helen Y. Chu, Trevor Bedford, Jay Shendure |
| EPI_ISL_962457, EPI_ISL_962458 | Seattle Flu Study | Seattle Flu Study | Deborah A. Nickerson, Chris D. Frazar, Jover Lee, Benjamin Pelle, Erica Ryke, Matthew Richardson, Amanda Adler, Elisabeth Brandstetter, Peter D. Han, Kairsten Fay, Misja Ilcisin, Kirsten Lacombe, Thomas R. Sibley, Melissa Truong, Caitlin R. Wolf, Michael Boeckh, Janet A. Englund, Michael Famulare, Barry R. Lutz, Mark J. Rieder, Lea M. Starita, Matthew Thompson, Jay Shendure, Trevor Bedford, Helen Y. Chu |
| EPI_ISL_962459, EPI_ISL_962460, EPI_ISL_962461, EPI_ISL_962462, EPI_ISL_962463, EPI_ISL_962465, EPI_ISL_962466, EPI_ISL_962468, EPI_ISL_962470, EPI_ISL_962471, EPI_ISL_962472, EPI_ISL_962473, EPI_ISL_962474, EPI_ISL_962475, EPI_ISL_962476, EPI_ISL_962479, EPI_ISL_962480 | | | |
| see above | Seattle Flu Study | Seattle Flu Study | Deborah A. Nickerson, Chris D. Frazar, Jover Lee, Benjamin Pelle, Erica Ryke, Matthew Richardson, Amanda Adler, Elisabeth Brandstetter, Peter D. Han, Kairsten Fay, Misja Ilcisin, Kirsten Lacombe, Thomas R. Sibley, Melissa Truong, Caitlin R. Wolf, Karen Cowgill, Stephanie Schrag, Jeff Duchin, Michael Boeckh, Janet A. Englund, Michael Famulare, Barry R. Lutz, Mark J. Rieder, Lea M. Starita, Matthew Thompson, Helen Y. Chu, Trevor Bedford, Jay Shendure |
| EPI_ISL_962505, EPI_ISL_962506, EPI_ISL_962507, EPI_ISL_962508, EPI_ISL_962509, EPI_ISL_962510, EPI_ISL_962511, EPI_ISL_962512, EPI_ISL_962513, EPI_ISL_962514 | UCLA Clinical Micro Lab | Los Angeles County Public Health Laboratories | P. Hemarajata et al. |
| EPI_ISL_962515, EPI_ISL_962516 | MD Laboratories | Los Angeles County Public Health Laboratories | P. Hemarajata et al. |
| EPI_ISL_962517, EPI_ISL_962518, EPI_ISL_962519, EPI_ISL_962520 | VA Medical Center | Los Angeles County Public Health Laboratories | P. Hemarajata et al. |
| EPI_ISL_962521 | UCLA Clinical Micro Lab | Los Angeles County Public Health Laboratories | P. Hemarajata et al. |
| EPI_ISL_962585, EPI_ISL_962586, EPI_ISL_962587, EPI_ISL_962588, EPI_ISL_962589, EPI_ISL_962590, EPI_ISL_962591, EPI_ISL_962592, EPI_ISL_962593, EPI_ISL_962594, EPI_ISL_962595, EPI_ISL_962596, EPI_ISL_962597, EPI_ISL_962598, EPI_ISL_962599, EPI_ISL_962600, EPI_ISL_962601 | | | |
| see above | Scripps Medical Laboratory | Andersen lab at Scripps Research | SEARCH Alliance San Diego with Michael Quigley, Ellen Stefanski, Ian Mchardy |
| EPI_ISL_962602, EPI_ISL_962603, EPI_ISL_962604, EPI_ISL_962605, EPI_ISL_962606, EPI_ISL_962607, EPI_ISL_962608 | Sharp HealthCare Laboratory | Andersen lab at Scripps Research | SEARCH Alliance San Diego with Aaron Harding, Jacquelyn Berumen, Cathy Woerle, Liam McGinnis, Art Mendoza, Omid Bakhtar |
| EPI_ISL_962609, EPI_ISL_962610, EPI_ISL_962611, EPI_ISL_962612, EPI_ISL_962613, EPI_ISL_962614, EPI_ISL_962615, EPI_ISL_962616, EPI_ISL_962617 | San Diego County Public Health Laboratory | Andersen lab at Scripps Research | SEARCH Alliance San Diego with Tracy Basler, Jovan Shephard, Brett Austin |
| EPI_ISL_962618, EPI_ISL_962619, EPI_ISL_962620, EPI_ISL_962621, EPI_ISL_962622, EPI_ISL_962623, EPI_ISL_962624, EPI_ISL_962625, EPI_ISL_962626, EPI_ISL_962627, EPI_ISL_962628, EPI_ISL_962629, EPI_ISL_962630, EPI_ISL_962631, EPI_ISL_962632, EPI_ISL_962633, EPI_ISL_962634, EPI_ISL_962635, EPI_ISL_962636, EPI_ISL_962637, EPI_ISL_962638, EPI_ISL_962639, EPI_ISL_962640, EPI_ISL_962641, EPI_ISL_962642, EPI_ISL_962643, EPI_ISL_962644, EPI_ISL_962645, EPI_ISL_962646, EPI_ISL_962647, EPI_ISL_962648, EPI_ISL_962649 | | | |
| see above | Scripps Medical Laboratory | Andersen lab at Scripps Research | SEARCH Alliance San Diego with Michael Quigley, Ellen Stefanski, Ian Mchardy |
| EPI_ISL_962650, EPI_ISL_962651, EPI_ISL_962652, EPI_ISL_962653, EPI_ISL_962654, EPI_ISL_962655, EPI_ISL_962656, EPI_ISL_962657, EPI_ISL_962658, EPI_ISL_962659, EPI_ISL_962660, EPI_ISL_962661, EPI_ISL_962662, EPI_ISL_962663, EPI_ISL_962664, EPI_ISL_962665, EPI_ISL_962666, EPI_ISL_962667, EPI_ISL_962668, EPI_ISL_962669, EPI_ISL_962670, EPI_ISL_962671, EPI_ISL_962672, EPI_ISL_962673, EPI_ISL_962674, EPI_ISL_962675, EPI_ISL_962676, EPI_ISL_962677, EPI_ISL_962678, EPI_ISL_962679, EPI_ISL_962680, EPI_ISL_962681, EPI_ISL_962682, EPI_ISL_962683, EPI_ISL_962684, EPI_ISL_962685, EPI_ISL_962686, EPI_ISL_962687, EPI_ISL_962688, EPI_ISL_962689, EPI_ISL_962690, EPI_ISL_962691, EPI_ISL_962692, EPI_ISL_962693, EPI_ISL_962694, EPI_ISL_962695, EPI_ISL_962696, EPI_ISL_962697, EPI_ISL_962698, EPI_ISL_962699, EPI_ISL_962700, EPI_ISL_962701, EPI_ISL_962702, EPI_ISL_962703, EPI_ISL_962704, EPI_ISL_962705, EPI_ISL_962706, EPI_ISL_962707, EPI_ISL_962708, EPI_ISL_962709, EPI_ISL_962710, EPI_ISL_962711, EPI_ISL_962712, EPI_ISL_962713, EPI_ISL_962714, EPI_ISL_962715, EPI_ISL_962716, EPI_ISL_962717, EPI_ISL_962718, EPI_ISL_962719, EPI_ISL_962720, EPI_ISL_962721, EPI_ISL_962722, EPI_ISL_962723, EPI_ISL_962724, EPI_ISL_962725, EPI_ISL_962726, EPI_ISL_962727, EPI_ISL_962728, EPI_ISL_962729, EPI_ISL_962730, EPI_ISL_962731, EPI_ISL_962732, EPI_ISL_962733, EPI_ISL_962734, EPI_ISL_962735, EPI_ISL_962736, EPI_ISL_962737, EPI_ISL_962738, EPI_ISL_962739, EPI_ISL_962740, EPI_ISL_962741, EPI_ISL_962742, EPI_ISL_962743, EPI_ISL_962744, EPI_ISL_962745, EPI_ISL_962746, EPI_ISL_962747, EPI_ISL_962748, EPI_ISL_962749, EPI_ISL_962750, EPI_ISL_962751, EPI_ISL_962752, EPI_ISL_962753, EPI_ISL_962754, EPI_ISL_962755, EPI_ISL_962756, EPI_ISL_962757, EPI_ISL_962758, EPI_ISL_962759, EPI_ISL_962760, EPI_ISL_962761, EPI_ISL_962762, EPI_ISL_962763, EPI_ISL_962764, EPI_ISL_962765 | | | |
| see above | Sharp HealthCare Laboratory | Andersen lab at Scripps Research | SEARCH Alliance San Diego with Aaron Harding, Jacquelyn Berumen, Cathy Woerle, Liam McGinnis, Art Mendoza, Omid Bakhtar |
| EPI_ISL_962766, EPI_ISL_962767, EPI_ISL_962768, EPI_ISL_962769, EPI_ISL_962770, EPI_ISL_962771, EPI_ISL_962772, EPI_ISL_962773, EPI_ISL_962774, EPI_ISL_962775, EPI_ISL_962776, EPI_ISL_962777, EPI_ISL_962778, EPI_ISL_962779, EPI_ISL_962780, EPI_ISL_962781, EPI_ISL_962782, EPI_ISL_962783, EPI_ISL_962784, EPI_ISL_962785, EPI_ISL_962786, EPI_ISL_962787, EPI_ISL_962788, EPI_ISL_962789, EPI_ISL_962790, EPI_ISL_962791, EPI_ISL_962792, EPI_ISL_962793, EPI_ISL_962794, EPI_ISL_962795, EPI_ISL_962796, EPI_ISL_962797, EPI_ISL_962798, EPI_ISL_962799, EPI_ISL_962800, EPI_ISL_962801, EPI_ISL_962802, EPI_ISL_962803, EPI_ISL_962804, EPI_ISL_962805, EPI_ISL_962806, EPI_ISL_962807, EPI_ISL_962808, EPI_ISL_962809, EPI_ISL_962810, EPI_ISL_962811 | | | |
| see above | San Diego County Public Health Laboratory | Andersen lab at Scripps Research | SEARCH Alliance San Diego with Tracy Basler, Jovan Shephard, Brett Austin |
| EPI_ISL_962812, EPI_ISL_962813, EPI_ISL_962814, EPI_ISL_962815 | Robert Garry lab | Andersen lab at Scripps Research | Allison Smither, Gilberto Sabino-Santos, Patricia Snarski, Lilia Melnik, Antoinette Bell, Kaylynn Genemaras, Arnaud Drouin, Dahlene Fusco, Robert Garry with SEARCH Alliance San Diego |
| EPI_ISL_962833, EPI_ISL_962834, EPI_ISL_962835, EPI_ISL_962836, EPI_ISL_962837, EPI_ISL_962838, EPI_ISL_962839, EPI_ISL_962840, EPI_ISL_962841, EPI_ISL_962842, EPI_ISL_962843, EPI_ISL_962844, EPI_ISL_962845, EPI_ISL_962846 | | | |
| see above | UCLA Clinical Micro Lab | Los Angeles County Public Health Laboratories | P. Hemarajata et al. |
| EPI_ISL_962847 | MD Laboratories | Los Angeles County Public Health Laboratories | P. Hemarajata et al. |
| EPI_ISL_962848, EPI_ISL_962849, EPI_ISL_962850, EPI_ISL_962851, EPI_ISL_962852, EPI_ISL_962853, EPI_ISL_962854, EPI_ISL_962855, EPI_ISL_962856, EPI_ISL_962857, EPI_ISL_962858, EPI_ISL_962859, EPI_ISL_962860, EPI_ISL_962861, EPI_ISL_962862, EPI_ISL_962863, EPI_ISL_962864, EPI_ISL_962865, EPI_ISL_962866, EPI_ISL_962867 | | | |
| see above | Los Angeles County Public Health Laboratories | Los Angeles County Public Health Laboratories | P. Hemarajata et al. |
| EPI_ISL_962879 | UCLA Clinical Micro Lab | Los Angeles County Public Health Laboratories | P. Hemarajata et al. |
| EPI_ISL_965016, EPI_ISL_965017, EPI_ISL_965018, EPI_ISL_965019, EPI_ISL_965020, EPI_ISL_965021, EPI_ISL_965022, EPI_ISL_965023, EPI_ISL_965024 | Florida Bureau of Public Health Laboratories | Florida Bureau of Public Health Laboratories | Sarah Schmedes, Jason Blanton |
| EPI_ISL_965030, EPI_ISL_965032, | Ohio Department of Health Laboratory | Ohio Department of Health Laboratory | Holmes, Jennifer; Eric Brandt, Keoni Omura, Glen McGillivary, Caitlin McDonnell, Jade Mowery, Stephanie Mccracken, Tyler Payne, Kirtana Ramadugu, Erica Leasure, Brent Lee, Kelsey Florek, Heather Blankenship, Quanta Brown, and Tammy Bannerman |

| | | | |
|---|---|---|---|
| EPI_ISL_965034 | | | |
| EPI_ISL_965044, EPI_ISL_965045, EPI_ISL_965046, EPI_ISL_965047, EPI_ISL_965048, EPI_ISL_965049, EPI_ISL_965050, EPI_ISL_965051, EPI_ISL_965052, EPI_ISL_965053, EPI_ISL_965054, EPI_ISL_965056, EPI_ISL_965057, EPI_ISL_965058, EPI_ISL_965059, EPI_ISL_965060, EPI_ISL_965061, EPI_ISL_965062, EPI_ISL_965063, EPI_ISL_965064, EPI_ISL_965065, EPI_ISL_965066, EPI_ISL_965067, EPI_ISL_965068, EPI_ISL_965069, EPI_ISL_965070, EPI_ISL_965071, EPI_ISL_965077, EPI_ISL_965078, EPI_ISL_965079, EPI_ISL_965080, EPI_ISL_965081, EPI_ISL_965082, EPI_ISL_965083, EPI_ISL_965084, EPI_ISL_965085, EPI_ISL_965086, EPI_ISL_965087, EPI_ISL_965090, EPI_ISL_965091, EPI_ISL_965092, EPI_ISL_965093, EPI_ISL_965094, EPI_ISL_965095, EPI_ISL_965096, EPI_ISL_965097, EPI_ISL_965098, EPI_ISL_965099, EPI_ISL_965100, EPI_ISL_965102, EPI_ISL_965103, EPI_ISL_965104, EPI_ISL_965105, EPI_ISL_965106, EPI_ISL_965107, EPI_ISL_965108, EPI_ISL_965109, EPI_ISL_965110, EPI_ISL_965111, EPI_ISL_965112 | | | |
| see above | Wyoming Public Health Laboratory | Wyoming Public Health Laboratory | Noah Hull, Taylor Fearing, Lynette Gumbleton, Channing Weber, Ashley Norberg, Bailey Bowcutt, and Wanda Manley |
| EPI_ISL_965113, EPI_ISL_965121, EPI_ISL_965143, EPI_ISL_965149 | Ohio Department of Health Laboratory | Ohio Department of Health Laboratory | Holmes, Jennifer; Eric Brandt, Keoni Omura, Glen McGillivary, Caitlin McDonnell, Jade Mowery, Stephanie Mccracken, Tyler Payne, Kirtana Ramadugu, Erica Leasure, Brent Lee, Kelsey Florek, Heather Blankenship, Quanta Brown, and Tammy Bannerman |
| EPI_ISL_965155, EPI_ISL_965156 | Mary Eliza Mahoney HC | NJ_PHEL | Lindsey Bodnar, Shiv Verma, Dana Woell, Byeong Jeong |
| EPI_ISL_965157, EPI_ISL_965158, EPI_ISL_965159, EPI_ISL_965160, EPI_ISL_965161, EPI_ISL_965162, EPI_ISL_965163, EPI_ISL_965164, EPI_ISL_965165, EPI_ISL_965166, EPI_ISL_965167, EPI_ISL_965168, EPI_ISL_965169, EPI_ISL_965170, EPI_ISL_965171, EPI_ISL_965172, EPI_ISL_965173, EPI_ISL_965174, EPI_ISL_965175, EPI_ISL_965176, EPI_ISL_965177 | | | |
| see above | Clinical Molecular Microbiology Laboratory, UNC Hospitals | Jeremy Wang | Jeremy Wang, Alexander Rubinsteyn, Colleen Rice, Jason Smedberg, Shawn Hawken, Melissa Miller, Corbin Jones, Robert Hagan |
| EPI_ISL_965186, EPI_ISL_965187, EPI_ISL_965188, EPI_ISL_965189, EPI_ISL_965190, EPI_ISL_965191, EPI_ISL_965192, EPI_ISL_965193, EPI_ISL_965194, EPI_ISL_965195, EPI_ISL_965196, EPI_ISL_965197, EPI_ISL_965198, EPI_ISL_965199, EPI_ISL_965200, EPI_ISL_965201, EPI_ISL_965202, EPI_ISL_965203, EPI_ISL_965204, EPI_ISL_965205, EPI_ISL_965206, EPI_ISL_965207, EPI_ISL_965208, EPI_ISL_965209, EPI_ISL_965210, EPI_ISL_965211, EPI_ISL_965212, EPI_ISL_965213, EPI_ISL_965214, EPI_ISL_965215, EPI_ISL_965216, EPI_ISL_965217, EPI_ISL_965218, EPI_ISL_965219, EPI_ISL_965220, EPI_ISL_965221 | | | |
| see above | Virginia Division of Consolidated Laboratory Services | Virginia Division of Consolidated Laboratory Services | Virginia DCLS |
| EPI_ISL_965301, EPI_ISL_965302 | Ohio Department of Health Laboratory | Ohio Department of Health Laboratory | Holmes, Jennifer; Eric Brandt, Keoni Omura, Glen McGillivary, Caitlin McDonnell, Jade Mowery, Stephanie Mccracken, Tyler Payne, Kirtana Ramadugu, Erica Leasure, Brent Lee, Kelsey Florek, Heather Blankenship, Quanta Brown, and Tammy Bannerman |
| EPI_ISL_965303, EPI_ISL_965528, EPI_ISL_965770, EPI_ISL_965812 | NYU Langone Health | Departments of Pathology and Medicine, New York University School of Medicine | Adriana Heguy, Dacia Dimartino, Emily Guzman, Christian Marier, Peter Meyn, Sitharam Ramaswami, Gael Westby, Paul Zappile, Yutong Zhang, Paolo Cotzia, Guiqing Wang |
| EPI_ISL_965813 | Ohio Department of Health Laboratory | Ohio Department of Health Laboratory | Holmes, Jennifer; Eric Brandt, Keoni Omura, Glen McGillivary, Caitlin McDonnell, Jade Mowery, Stephanie Mccracken, Tyler Payne, Kirtana Ramadugu, Erica Leasure, Brent Lee, Kelsey Florek, Heather Blankenship, Quanta Brown, and Tammy Bannerman |
| EPI_ISL_965814 | NYU Langone Health | Departments of Pathology and Medicine, New York University School of Medicine | Adriana Heguy, Dacia Dimartino, Emily Guzman, Christian Marier, Peter Meyn, Sitharam Ramaswami, Gael Westby, Paul Zappile, Yutong Zhang, Paolo Cotzia, Guiqing Wang |
| EPI_ISL_965815, EPI_ISL_965816, EPI_ISL_965817, EPI_ISL_965818, EPI_ISL_965819, EPI_ISL_965820, EPI_ISL_965821, EPI_ISL_965822, EPI_ISL_965823, EPI_ISL_965824, EPI_ISL_965825 | | | |
| see above | Kentucky State Public Health Lab | Kentucky State Public Health Lab | Stephanie Lunn, Karim George, Joshua Tobias, William Grooms, Vaneet Arora, Matthew Johnson, Rachel Zinner, Rhonda Lucas |
| EPI_ISL_965826 | NYU Langone Health | Departments of Pathology and Medicine, New York University School of Medicine | Adriana Heguy, Dacia Dimartino, Emily Guzman, Christian Marier, Peter Meyn, Sitharam Ramaswami, Gael Westby, Paul Zappile, Yutong Zhang, Paolo Cotzia, Guiqing Wang |
| EPI_ISL_965827, EPI_ISL_965828, EPI_ISL_965829, EPI_ISL_965830, EPI_ISL_965831, EPI_ISL_965832, EPI_ISL_965833, EPI_ISL_965834, EPI_ISL_965835 | Kentucky State Public Health Lab | Kentucky State Public Health Lab | Stephanie Lunn, Karim George, Joshua Tobias, William Grooms, Vaneet Arora, Matthew Johnson, Rachel Zinner, Rhonda Lucas |
| EPI_ISL_965837, EPI_ISL_965838 | NYU Langone Health | Departments of Pathology and Medicine, New York University School of Medicine | Adriana Heguy, Dacia Dimartino, Emily Guzman, Christian Marier, Peter Meyn, Sitharam Ramaswami, Gael Westby, Paul Zappile, Yutong Zhang, Paolo Cotzia, Guiqing Wang |
| EPI_ISL_965840, EPI_ISL_965841, EPI_ISL_965842, EPI_ISL_965843, EPI_ISL_965844, EPI_ISL_965845, EPI_ISL_965846, EPI_ISL_965847, EPI_ISL_965848, EPI_ISL_965849, EPI_ISL_965850, EPI_ISL_965851, EPI_ISL_965852, EPI_ISL_965853, EPI_ISL_965854 | | | |
| see above | GA Department of Public Health | GA Department of Public Health | Stacy Reeves, Jonathan Edwards, Cynthia Dixey, Tonia Parrott |
| EPI_ISL_965855, EPI_ISL_965860, EPI_ISL_965861, EPI_ISL_965864, EPI_ISL_965865, EPI_ISL_965866, EPI_ISL_965867, EPI_ISL_965868, EPI_ISL_965871, EPI_ISL_965874, EPI_ISL_965875, EPI_ISL_965877, EPI_ISL_965878, EPI_ISL_965879, EPI_ISL_965880, EPI_ISL_965881, EPI_ISL_965882, EPI_ISL_965883, EPI_ISL_965884, EPI_ISL_965885, EPI_ISL_965886, EPI_ISL_965887, EPI_ISL_965888, EPI_ISL_965890, EPI_ISL_965891, EPI_ISL_965892, EPI_ISL_965893, EPI_ISL_965895, EPI_ISL_965896, EPI_ISL_965898, EPI_ISL_965899, EPI_ISL_965900, EPI_ISL_965901, EPI_ISL_965902, EPI_ISL_965903, EPI_ISL_965904, EPI_ISL_965905 | | | |
| see above | Massachusetts State Public Health Laboratory | Massachusetts State Public Health Laboratory | Andrew Lang, Timelia Fink, Glen Gallagher, Sandra Smole |
| EPI_ISL_965906, EPI_ISL_965907, EPI_ISL_965908 | Clinical Molecular Microbiology Laboratory, UNC Hospitals | Jeremy Wang | Jeremy Wang, Alexander Rubinsteyn, Colleen Rice, Jason Smedberg, Shawn Hawken, Melissa Miller, Corbin Jones, Robert Hagan |
| EPI_ISL_965910, EPI_ISL_965948, EPI_ISL_965962, EPI_ISL_965963, EPI_ISL_965964, EPI_ISL_965965, EPI_ISL_965967, EPI_ISL_965968, EPI_ISL_965969, EPI_ISL_965970, EPI_ISL_965971, EPI_ISL_965972, EPI_ISL_965973 | | | |
| see above | NYU Langone Health | Departments of Pathology and Medicine, New York University School of Medicine | Adriana Heguy, Dacia Dimartino, Emily Guzman, Christian Marier, Peter Meyn, Sitharam Ramaswami, Gael Westby, Paul Zappile, Yutong Zhang, Paolo Cotzia, Guiqing Wang |
| EPI_ISL_965976 | Seattle VA Medical Center | Seattle Flu Study | Deborah A. Nickerson, Chris D. Frazar, Jover Lee, Benjamin Pelle, Erica Ryke, Matthew Richardson, Amanda Adler, Elisabeth Brandstetter, Peter D. Han, Kairsten Fay, Misja Ilcisin, Kirsten Lacombe, Thomas R. Sibley, Melissa Truong, Caitlin R. Wolf, Romesh Gautom, Geoff Melly, Brian Hiatt, Philip Dykema, Scott Lindquist, Michael Boeckh, Janet A. Englund, Michael Famulare, Barry R. Lutz, Mark J. Rieder, Lea M. Starita, Matthew Thompson, Helen Y. Chu, Jay Shendure, Trevor Bedford |
| EPI_ISL_965977, EPI_ISL_965980, EPI_ISL_965989, EPI_ISL_966020, EPI_ISL_966044, EPI_ISL_966062, EPI_ISL_966084, EPI_ISL_966108, EPI_ISL_966160, EPI_ISL_966184, EPI_ISL_966204, EPI_ISL_966227, EPI_ISL_966245, EPI_ISL_966264 | | | |
| see above | NYU Langone Health | Departments of Pathology and Medicine, New York University School of Medicine | Adriana Heguy, Dacia Dimartino, Emily Guzman, Christian Marier, Peter Meyn, Sitharam Ramaswami, Gael Westby, Paul Zappile, Yutong Zhang, Paolo Cotzia, Guiqing Wang |
| EPI_ISL_966285 | Baylor Scott & White - Irving | Baylor Scott & White - Temple | Ari Rao, Linden Morales, Kimberly Walker, Marcus Volz, Shelby Hendrickson |
| EPI_ISL_966289, EPI_ISL_966308 | NYU Langone Health | Departments of Pathology and Medicine, New York University School of Medicine | Adriana Heguy, Dacia Dimartino, Emily Guzman, Christian Marier, Peter Meyn, Sitharam Ramaswami, Gael Westby, Paul Zappile, Yutong Zhang, Paolo Cotzia, Guiqing Wang |
| EPI_ISL_966310 | Baylor Scott & White - Temple | Baylor Scott & White - Temple | Ari Rao, Linden Morales, Kimberly Walker, Marcus Volz, Shelby Hendrickson |
| EPI_ISL_966311 | NYU Langone Health | Departments of Pathology and Medicine, New York University School of Medicine | Adriana Heguy, Dacia Dimartino, Emily Guzman, Christian Marier, Peter Meyn, Sitharam Ramaswami, Gael Westby, Paul Zappile, Yutong Zhang, Paolo Cotzia, Guiqing Wang |
| EPI_ISL_966313 | Baylor Scott & White - Temple | Baylor Scott & White - Temple | Ari Rao, Linden Morales, Kimberly Walker, Marcus Volz, Shelby Hendrickson |
| EPI_ISL_966314 | NYU Langone Health | Departments of Pathology and Medicine, New York University School of Medicine | Adriana Heguy, Dacia Dimartino, Emily Guzman, Christian Marier, Peter Meyn, Sitharam Ramaswami, Gael Westby, Paul Zappile, Yutong Zhang, Paolo Cotzia, Guiqing Wang |
| EPI_ISL_966315, EPI_ISL_966316, EPI_ISL_966317, EPI_ISL_966318, EPI_ISL_966319, EPI_ISL_966320, EPI_ISL_966321, EPI_ISL_966322, EPI_ISL_966323, EPI_ISL_966324, EPI_ISL_966325, EPI_ISL_966326, EPI_ISL_966327, EPI_ISL_966328, EPI_ISL_966329, EPI_ISL_966330, EPI_ISL_966331, EPI_ISL_966332 | | | |
| see above | Kentucky State Public Health Lab | Kentucky State Public Health Lab | Stephanie Lunn, Karim George, Joshua Tobias, William Grooms, Vaneet Arora, Matthew Johnson, Rachel Zinner, Rhonda Lucas |
| EPI_ISL_966333 | NYU Langone Health | Departments of Pathology and Medicine, New York University School of Medicine | Adriana Heguy, Dacia Dimartino, Emily Guzman, Christian Marier, Peter Meyn, Sitharam Ramaswami, Gael Westby, Paul Zappile, Yutong Zhang, Paolo Cotzia, Guiqing Wang |
| EPI_ISL_966334, EPI_ISL_966335, EPI_ISL_966336, EPI_ISL_966337 | Kansas Health and Environmental Lab | Kansas Health and Environmental Lab | Mike Grose, Paige Drury, Carissa Robertson, Ben Olsen, and Phil Adam |
| EPI_ISL_966338 | NYU Langone Health | Departments of Pathology and Medicine, New York University School of Medicine | Adriana Heguy, Dacia Dimartino, Emily Guzman, Christian Marier, Peter Meyn, Sitharam Ramaswami, Gael Westby, Paul Zappile, Yutong Zhang, Paolo Cotzia, Guiqing Wang |
| EPI_ISL_966339, EPI_ISL_966340, EPI_ISL_966341, EPI_ISL_966342, EPI_ISL_966343, EPI_ISL_966344, EPI_ISL_966345, EPI_ISL_966346, EPI_ISL_966347, EPI_ISL_966348, EPI_ISL_966349, EPI_ISL_966350, EPI_ISL_966351, EPI_ISL_966352, EPI_ISL_966353, EPI_ISL_966354 | | | |
| see above | Kansas Health and Environmental Lab | Kansas Health and Environmental Lab | Mike Grose, Paige Drury, Carissa Robertson, Ben Olsen, and Phil Adam |
| EPI_ISL_966355, EPI_ISL_966356 | Kentucky State Public Health Lab | Kentucky State Public Health Lab | Stephanie Lunn, Karim George, Joshua Tobias, William Grooms, Vaneet Arora, Matthew Johnson, Rachel Zinner, Rhonda Lucas |
| EPI_ISL_966357 | NYU Langone Health | Departments of Pathology and Medicine, New York University School of Medicine | Adriana Heguy, Dacia Dimartino, Emily Guzman, Christian Marier, Peter Meyn, Sitharam Ramaswami, Gael Westby, Paul Zappile, Yutong Zhang, Paolo Cotzia, Guiqing Wang |
| EPI_ISL_966358, EPI_ISL_966359, EPI_ISL_966360, EPI_ISL_966361, EPI_ISL_966362, EPI_ISL_966363, EPI_ISL_966364, EPI_ISL_966365, EPI_ISL_966366, EPI_ISL_966367, EPI_ISL_966368, EPI_ISL_966369, EPI_ISL_966370, EPI_ISL_966371, EPI_ISL_966372, EPI_ISL_966373, EPI_ISL_966374 | | | |
| see above | Kentucky State Public Health Lab | Kentucky State Public Health Lab | Stephanie Lunn, Karim George, Joshua Tobias, William Grooms, Vaneet Arora, Matthew Johnson, Rachel Zinner, Rhonda Lucas |
| EPI_ISL_966375, EPI_ISL_966376, EPI_ISL_966377, EPI_ISL_966378 | NYU Langone Health | Departments of Pathology and Medicine, New York University School of Medicine | Adriana Heguy, Dacia Dimartino, Emily Guzman, Christian Marier, Peter Meyn, Sitharam Ramaswami, Gael Westby, Paul Zappile, Yutong Zhang, Paolo Cotzia, Guiqing Wang |

| | | | |
|---|---|---|---|
| EPI_ISL_966379 | OCME Office Of Chief Medical Examiner | New York City Public Health Laboratory | Jade Wang, et al. |
| EPI_ISL_966380 | NYU Langone Health | Departments of Pathology and Medicine, New York University School of Medicine | Adriana Heguy, Dacia Dimartino, Emily Guzman, Christian Marier, Peter Meyn, Sitharam Ramaswami, Gael Westby, Paul Zappile, Yutong Zhang, Paolo Cotzia, Guiqing Wang |
| EPI_ISL_966381 | OCME Office Of Chief Medical Examiner | New York City Public Health Laboratory | Jade Wang, et al. |
| EPI_ISL_966382 | DOHMH Jamaica | New York City Public Health Laboratory | Jade Wang, et al. |
| EPI_ISL_966383 | OCME Office Of Chief Medical Examiner | New York City Public Health Laboratory | Jade Wang, et al. |
| EPI_ISL_966384 | DOHMH Jamaica | New York City Public Health Laboratory | Jade Wang, et al. |
| EPI_ISL_966385 | Department of Homeless Services | New York City Public Health Laboratory | Jade Wang, et al. |
| EPI_ISL_966386 | DOHMH Corona | New York City Public Health Laboratory | Jade Wang, et al. |
| EPI_ISL_966387, EPI_ISL_966388 | DOHMH Jamaica | New York City Public Health Laboratory | Jade Wang, et al. |
| EPI_ISL_966389 | DOHMH Central Harlem | New York City Public Health Laboratory | Jade Wang, et al. |
| EPI_ISL_966390, EPI_ISL_966391 | OCME Office Of Chief Medical Examiner | New York City Public Health Laboratory | Jade Wang, et al. |
| EPI_ISL_966392 | DOHMH Corona | New York City Public Health Laboratory | Jade Wang, et al. |
| EPI_ISL_966393 | OCME Office Of Chief Medical Examiner | New York City Public Health Laboratory | Jade Wang, et al. |
| EPI_ISL_966394 | DOHMH Corona | New York City Public Health Laboratory | Jade Wang, et al. |
| EPI_ISL_966395 | DOHMH Jamaica | New York City Public Health Laboratory | Jade Wang, et al. |
| EPI_ISL_966396, EPI_ISL_966397 | OCME Office Of Chief Medical Examiner | New York City Public Health Laboratory | Jade Wang, et al. |
| EPI_ISL_966398 | DOHMH Jamaica | New York City Public Health Laboratory | Jade Wang, et al. |
| EPI_ISL_966399 | DOHMH Corona | New York City Public Health Laboratory | Jade Wang, et al. |
| EPI_ISL_966400 | OCME Office Of Chief Medical Examiner | New York City Public Health Laboratory | Jade Wang, et al. |
| EPI_ISL_966401 | Department of Homeless Services | New York City Public Health Laboratory | Jade Wang, et al. |
| EPI_ISL_966402, EPI_ISL_966403 | OCME Office Of Chief Medical Examiner | New York City Public Health Laboratory | Jade Wang, et al. |
| EPI_ISL_966404 | DOHMH PHL | New York City Public Health Laboratory | Jade Wang, et al. |
| EPI_ISL_966405 | DOHMH Jamaica | New York City Public Health Laboratory | Jade Wang, et al. |
| EPI_ISL_966406 | NYU Langone Health | Departments of Pathology and Medicine, New York University School of Medicine | Adriana Heguy, Dacia Dimartino, Emily Guzman, Christian Marier, Peter Meyn, Sitharam Ramaswami, Gael Westby, Paul Zappile, Yutong Zhang, Paolo Cotzia, Guiqing Wang |
| EPI_ISL_966407, EPI_ISL_966408 | DOHMH Morrisania | New York City Public Health Laboratory | Jade Wang, et al. |
| EPI_ISL_966409, EPI_ISL_966410 | DOHMH Corona | New York City Public Health Laboratory | Jade Wang, et al. |
| EPI_ISL_966411 | OCME Office Of Chief Medical Examiner | New York City Public Health Laboratory | Jade Wang, et al. |
| EPI_ISL_966412 | DOHMH Jamaica | New York City Public Health Laboratory | Jade Wang, et al. |
| EPI_ISL_966413 | DOHMH Central Harlem | New York City Public Health Laboratory | Jade Wang, et al. |
| EPI_ISL_966414 | DOHMH Morrisania | New York City Public Health Laboratory | Jade Wang, et al. |
| EPI_ISL_966415 | DOHMH Corona | New York City Public Health Laboratory | Jade Wang, et al. |
| EPI_ISL_966416 | DOHMH Central Harlem | New York City Public Health Laboratory | Jade Wang, et al. |
| EPI_ISL_966417, EPI_ISL_966418, EPI_ISL_966419 | OCME Office Of Chief Medical Examiner | New York City Public Health Laboratory | Jade Wang, et al. |
| EPI_ISL_966420 | DOHMH Morrisania | New York City Public Health Laboratory | Jade Wang, et al. |
| EPI_ISL_966421, EPI_ISL_966422 | DOHMH Jamaica | New York City Public Health Laboratory | Jade Wang, et al. |
| EPI_ISL_966423 | DOHMH Morrisania | New York City Public Health Laboratory | Jade Wang, et al. |
| EPI_ISL_966424 | DOHMH Jamaica | New York City Public Health Laboratory | Jade Wang, et al. |
| EPI_ISL_966425 | DOHMH Corona | New York City Public Health Laboratory | Jade Wang, et al. |
| EPI_ISL_966426 | DOHMH PHL | New York City Public Health Laboratory | Jade Wang, et al. |
| EPI_ISL_966427 | OCME Office Of Chief Medical Examiner | New York City Public Health Laboratory | Jade Wang, et al. |
| EPI_ISL_966428 | NYU Langone Health | Departments of Pathology and Medicine, New York University School of Medicine | Adriana Heguy, Dacia Dimartino, Emily Guzman, Christian Marier, Peter Meyn, Sitharam Ramaswami, Gael Westby, Paul Zappile, Yutong Zhang, Paolo Cotzia, Guiqing Wang |
| EPI_ISL_966429, EPI_ISL_966430, EPI_ISL_966431 | OCME Office Of Chief Medical Examiner | New York City Public Health Laboratory | Jade Wang, et al. |
| EPI_ISL_966432, EPI_ISL_966433 | DOHMH Jamaica | New York City Public Health Laboratory | Jade Wang, et al. |
| EPI_ISL_966434, EPI_ISL_966435, EPI_ISL_966436, EPI_ISL_966437 | DOHMH Morrisania | New York City Public Health Laboratory | Jade Wang, et al. |
| EPI_ISL_966438, EPI_ISL_966439, EPI_ISL_966440, EPI_ISL_966441, EPI_ISL_966442 | OCME Office Of Chief Medical Examiner | New York City Public Health Laboratory | Jade Wang, et al. |
| EPI_ISL_966443 | DOHMH Fort Greene | New York City Public Health Laboratory | Jade Wang, et al. |
| EPI_ISL_966444 | DOHMH PHL | New York City Public Health Laboratory | Jade Wang, et al. |
| EPI_ISL_966445, EPI_ISL_966446 | DOHMH Corona | New York City Public Health Laboratory | Jade Wang, et al. |
| EPI_ISL_966447, EPI_ISL_966448, EPI_ISL_966449 | DOHMH Jamaica | New York City Public Health Laboratory | Jade Wang, et al. |
| EPI_ISL_966450 | DOHMH Central Harlem | New York City Public Health Laboratory | Jade Wang, et al. |
| EPI_ISL_966451 | DOHMH Morrisania | New York City Public Health Laboratory | Jade Wang, et al. |
| EPI_ISL_966453, EPI_ISL_966454, EPI_ISL_966455, EPI_ISL_966456, EPI_ISL_966457, EPI_ISL_966458 | OCME Office Of Chief Medical Examiner | New York City Public Health Laboratory | Jade Wang, et al. |
| EPI_ISL_966459 | DOHMH Corona | New York City Public Health Laboratory | Jade Wang, et al. |
| EPI_ISL_966460 | DOHMH Morrisania | New York City Public Health Laboratory | Jade Wang, et al. |
| EPI_ISL_966461, EPI_ISL_966462, | DOHMH Jamaica | New York City Public Health Laboratory | Jade Wang, et al. |

| | | | |
|---|---|---|---|
| EPI_ISL_966463 | | | |
| EPI_ISL_966464, EPI_ISL_966465, EPI_ISL_966466 | DOHMH PHL | New York City Public Health Laboratory | Jade Wang, et al. |
| EPI_ISL_966467 | DOHMH Corona | New York City Public Health Laboratory | Jade Wang, et al. |
| EPI_ISL_966468 | DOHMH Chelsea | New York City Public Health Laboratory | Jade Wang, et al. |
| EPI_ISL_966469 | Department of Homeless Services | New York City Public Health Laboratory | Jade Wang, et al. |
| EPI_ISL_966470, EPI_ISL_966471 | OCME Office Of Chief Medical Examiner | New York City Public Health Laboratory | Jade Wang, et al. |
| EPI_ISL_966472 | NYU Langone Health | Departments of Pathology and Medicine, New York University School of Medicine | Adriana Heguy, Dacia Dimartino, Emily Guzman, Christian Marier, Peter Meyn, Sitharam Ramaswami, Gael Westby, Paul Zappile, Yutong Zhang, Paolo Cotzia, Guiqing Wang |
| EPI_ISL_966473 | OCME Office Of Chief Medical Examiner | New York City Public Health Laboratory | Jade Wang, et al. |
| EPI_ISL_966474 | DOHMH Corona | New York City Public Health Laboratory | Jade Wang, et al. |
| EPI_ISL_966475 | DOHMH Central Harlem | New York City Public Health Laboratory | Jade Wang, et al. |
| EPI_ISL_966476, EPI_ISL_966477, EPI_ISL_966478 | DOHMH Corona | New York City Public Health Laboratory | Jade Wang, et al. |
| EPI_ISL_966479 | DOHMH Jamaica | New York City Public Health Laboratory | Jade Wang, et al. |
| EPI_ISL_966480 | DOHMH Corona | New York City Public Health Laboratory | Jade Wang, et al. |
| EPI_ISL_966481 | DOHMH Jamaica | New York City Public Health Laboratory | Jade Wang, et al. |
| EPI_ISL_966482 | DOHMH Corona | New York City Public Health Laboratory | Jade Wang, et al. |
| EPI_ISL_966483 | DOHMH Chelsea | New York City Public Health Laboratory | Jade Wang, et al. |
| EPI_ISL_966484 | DOHMH Crown Heights | New York City Public Health Laboratory | Jade Wang, et al. |
| EPI_ISL_966485, EPI_ISL_966486, EPI_ISL_966487, EPI_ISL_966488 | OCME Office Of Chief Medical Examiner | New York City Public Health Laboratory | Jade Wang, et al. |
| EPI_ISL_966489 | NYU Langone Health | Departments of Pathology and Medicine, New York University School of Medicine | Adriana Heguy, Dacia Dimartino, Emily Guzman, Christian Marier, Peter Meyn, Sitharam Ramaswami, Gael Westby, Paul Zappile, Yutong Zhang, Paolo Cotzia, Guiqing Wang |
| EPI_ISL_966490 | OCME Office Of Chief Medical Examiner | New York City Public Health Laboratory | Jade Wang, et al. |
| EPI_ISL_966491 | DOHMH Corona | New York City Public Health Laboratory | Jade Wang, et al. |
| EPI_ISL_966492, EPI_ISL_966493, EPI_ISL_966494, EPI_ISL_966495 | OCME Office Of Chief Medical Examiner | New York City Public Health Laboratory | Jade Wang, et al. |
| EPI_ISL_966496 | NYU Langone Health | Departments of Pathology and Medicine, New York University School of Medicine | Adriana Heguy, Dacia Dimartino, Emily Guzman, Christian Marier, Peter Meyn, Sitharam Ramaswami, Gael Westby, Paul Zappile, Yutong Zhang, Paolo Cotzia, Guiqing Wang |
| EPI_ISL_966497, EPI_ISL_966498, EPI_ISL_966499, EPI_ISL_966500, EPI_ISL_966501, EPI_ISL_966502, EPI_ISL_966503, EPI_ISL_966504, EPI_ISL_966505, EPI_ISL_966506, EPI_ISL_966507, EPI_ISL_966508, EPI_ISL_966509, EPI_ISL_966510, EPI_ISL_966511, EPI_ISL_966512, EPI_ISL_966514, EPI_ISL_966515, EPI_ISL_966516, EPI_ISL_966517, EPI_ISL_966518, EPI_ISL_966519, EPI_ISL_966520, EPI_ISL_966521, EPI_ISL_966522, EPI_ISL_966523, EPI_ISL_966524, EPI_ISL_966525, EPI_ISL_966526, EPI_ISL_966527, EPI_ISL_966528, EPI_ISL_966529, EPI_ISL_966530, EPI_ISL_966531, EPI_ISL_966532, EPI_ISL_966534, EPI_ISL_966535, EPI_ISL_966536, EPI_ISL_966537, EPI_ISL_966538, EPI_ISL_966539, EPI_ISL_966540, EPI_ISL_966541, EPI_ISL_966542, EPI_ISL_966543, EPI_ISL_966544, EPI_ISL_966545, EPI_ISL_966546, EPI_ISL_966547, EPI_ISL_966548 | | | |
| see above | Helix/Illumina | Respiratory Viruses Branch, Division of Viral Diseases, Centers for Disease Control and Prevention | Peter W. Cook,Dakota Howard,Dhwani Batra,Ben L. Rambo-Martin,Eileen de Feo,Jan Antico,Christine Tran,Matthew Tolentino,Shannon Wickline,Kim Gietzen,Brad Sickler,Jingtao Liu,Eric Allen,Phil Febbo,Summer Galloway,Nicole L. Washington,Simon White,Geraint Levan,Kelly Schiabor Barrett,Elizabeth Cirulli,Alexandre Bolze,Ary Ascencio,Charlotte Rivera-Garcia,Ryan Cho,Jason Nguyen,Sherry Wang,Jimmy Ramirez,Tyler Cassens,Efren Sandoval,Magnus Isaksson,William Lee,David Becker,Marc Laurent,James Lu,Clinton R. Paden,Suxiang Tong,Duncan MacCannell, |
| EPI_ISL_966549, EPI_ISL_966551, EPI_ISL_966552 | NYU Langone Health | Departments of Pathology and Medicine, New York University School of Medicine | Adriana Heguy, Dacia Dimartino, Emily Guzman, Christian Marier, Peter Meyn, Sitharam Ramaswami, Gael Westby, Paul Zappile, Yutong Zhang, Paolo Cotzia, Guiqing Wang |
| EPI_ISL_966553, EPI_ISL_966554, EPI_ISL_966555, EPI_ISL_966556, EPI_ISL_966557, EPI_ISL_966558, EPI_ISL_966559, EPI_ISL_966560, EPI_ISL_966561, EPI_ISL_966562, EPI_ISL_966563, EPI_ISL_966564 | | | |
| see above | Helix/Illumina | Respiratory Viruses Branch, Division of Viral Diseases, Centers for Disease Control and Prevention | Peter W. Cook,Dakota Howard,Dhwani Batra,Ben L. Rambo-Martin,Eileen de Feo,Jan Antico,Christine Tran,Matthew Tolentino,Shannon Wickline,Kim Gietzen,Brad Sickler,Jingtao Liu,Eric Allen,Phil Febbo,Summer Galloway,Nicole L. Washington,Simon White,Geraint Levan,Kelly Schiabor Barrett,Elizabeth Cirulli,Alexandre Bolze,Ary Ascencio,Charlotte Rivera-Garcia,Ryan Cho,Jason Nguyen,Sherry Wang,Jimmy Ramirez,Tyler Cassens,Efren Sandoval,Magnus Isaksson,William Lee,David Becker,Marc Laurent,James Lu,Clinton R. Paden,Suxiang Tong,Duncan MacCannell, |
| EPI_ISL_966565 | NYU Langone Health | Departments of Pathology and Medicine, New York University School of Medicine | Adriana Heguy, Dacia Dimartino, Emily Guzman, Christian Marier, Peter Meyn, Sitharam Ramaswami, Gael Westby, Paul Zappile, Yutong Zhang, Paolo Cotzia, Guiqing Wang |
| EPI_ISL_966566, EPI_ISL_966567, EPI_ISL_966568, EPI_ISL_966569, EPI_ISL_966570, EPI_ISL_966571, EPI_ISL_966572, EPI_ISL_966573, EPI_ISL_966574, EPI_ISL_966575, EPI_ISL_966576, EPI_ISL_966577, EPI_ISL_966578, EPI_ISL_966579, EPI_ISL_966580, EPI_ISL_966581, EPI_ISL_966582, EPI_ISL_966583, EPI_ISL_966584, EPI_ISL_966585, EPI_ISL_966586, EPI_ISL_966587, EPI_ISL_966588, EPI_ISL_966589, EPI_ISL_966590, EPI_ISL_966592, EPI_ISL_966593, EPI_ISL_966594, EPI_ISL_966595, EPI_ISL_966596, EPI_ISL_966597, EPI_ISL_966598, EPI_ISL_966599, EPI_ISL_966600, EPI_ISL_966601 | | | |
| see above | Helix/Illumina | Respiratory Viruses Branch, Division of Viral Diseases, Centers for Disease Control and Prevention | Peter W. Cook,Dakota Howard,Dhwani Batra,Ben L. Rambo-Martin,Eileen de Feo,Jan Antico,Christine Tran,Matthew Tolentino,Shannon Wickline,Kim Gietzen,Brad Sickler,Jingtao Liu,Eric Allen,Phil Febbo,Summer Galloway,Nicole L. Washington,Simon White,Geraint Levan,Kelly Schiabor Barrett,Elizabeth Cirulli,Alexandre Bolze,Ary Ascencio,Charlotte Rivera-Garcia,Ryan Cho,Jason Nguyen,Sherry Wang,Jimmy Ramirez,Tyler Cassens,Efren Sandoval,Magnus Isaksson,William Lee,David Becker,Marc Laurent,James Lu,Clinton R. Paden,Suxiang Tong,Duncan MacCannell, |
| EPI_ISL_966605 | NYU Langone Health | Departments of Pathology and Medicine, New York University School of Medicine | Adriana Heguy, Dacia Dimartino, Emily Guzman, Christian Marier, Peter Meyn, Sitharam Ramaswami, Gael Westby, Paul Zappile, Yutong Zhang, Paolo Cotzia, Guiqing Wang |
| EPI_ISL_966606, EPI_ISL_966607, EPI_ISL_966608, EPI_ISL_966609, EPI_ISL_966610, EPI_ISL_966612, EPI_ISL_966613, EPI_ISL_966614, EPI_ISL_966615, EPI_ISL_966616, EPI_ISL_966617, EPI_ISL_966618, EPI_ISL_966619, EPI_ISL_966620, EPI_ISL_966621, EPI_ISL_966622 | | | |
| see above | Helix/Illumina | Respiratory Viruses Branch, Division of Viral Diseases, Centers for Disease Control and Prevention | Peter W. Cook,Dakota Howard,Dhwani Batra,Ben L. Rambo-Martin,Eileen de Feo,Jan Antico,Christine Tran,Matthew Tolentino,Shannon Wickline,Kim Gietzen,Brad Sickler,Jingtao Liu,Eric Allen,Phil Febbo,Summer Galloway,Nicole L. Washington,Simon White,Geraint Levan,Kelly Schiabor Barrett,Elizabeth Cirulli,Alexandre Bolze,Ary Ascencio,Charlotte Rivera-Garcia,Ryan Cho,Jason Nguyen,Sherry Wang,Jimmy Ramirez,Tyler Cassens,Efren Sandoval,Magnus Isaksson,William Lee,David Becker,Marc Laurent,James Lu,Clinton R. Paden,Suxiang Tong,Duncan MacCannell, |
| EPI_ISL_966623 | NYU Langone Health | Departments of Pathology and Medicine, New York University School of Medicine | Adriana Heguy, Dacia Dimartino, Emily Guzman, Christian Marier, Peter Meyn, Sitharam Ramaswami, Gael Westby, Paul Zappile, Yutong Zhang, Paolo Cotzia, Guiqing Wang |
| EPI_ISL_966624, EPI_ISL_966625, EPI_ISL_966626, EPI_ISL_966627, EPI_ISL_966628, EPI_ISL_966629, EPI_ISL_966630, EPI_ISL_966631, EPI_ISL_966632, EPI_ISL_966633, EPI_ISL_966634, EPI_ISL_966635, EPI_ISL_966636, EPI_ISL_966637, EPI_ISL_966638, EPI_ISL_966639, EPI_ISL_966640, EPI_ISL_966641, EPI_ISL_966642, EPI_ISL_966643, EPI_ISL_966644, EPI_ISL_966645 | | | |
| see above | Helix/Illumina | Respiratory Viruses Branch, Division of Viral Diseases, Centers for Disease Control and Prevention | Peter W. Cook,Dakota Howard,Dhwani Batra,Ben L. Rambo-Martin,Eileen de Feo,Jan Antico,Christine Tran,Matthew Tolentino,Shannon Wickline,Kim Gietzen,Brad Sickler,Jingtao Liu,Eric Allen,Phil Febbo,Summer Galloway,Nicole L. Washington,Simon White,Geraint Levan,Kelly Schiabor Barrett,Elizabeth Cirulli,Alexandre Bolze,Ary Ascencio,Charlotte Rivera-Garcia,Ryan Cho,Jason Nguyen,Sherry Wang,Jimmy Ramirez,Tyler Cassens,Efren Sandoval,Magnus Isaksson,William Lee,David Becker,Marc Laurent,James Lu,Clinton R. Paden,Suxiang Tong,Duncan MacCannell, |
| EPI_ISL_966646 | NYU Langone Health | Departments of Pathology and Medicine, New York University School of Medicine | Adriana Heguy, Dacia Dimartino, Emily Guzman, Christian Marier, Peter Meyn, Sitharam Ramaswami, Gael Westby, Paul Zappile, Yutong Zhang, Paolo Cotzia, Guiqing Wang |
| EPI_ISL_966647, EPI_ISL_966648, EPI_ISL_966649, EPI_ISL_966650, EPI_ISL_966651, EPI_ISL_966652, EPI_ISL_966653, EPI_ISL_966654, EPI_ISL_966655, EPI_ISL_966656, EPI_ISL_966657, EPI_ISL_966658, EPI_ISL_966659, EPI_ISL_966660, EPI_ISL_966661, EPI_ISL_966662, EPI_ISL_966664, EPI_ISL_966665, EPI_ISL_966666, EPI_ISL_966667, EPI_ISL_966668, EPI_ISL_966669, EPI_ISL_966670, EPI_ISL_966671, EPI_ISL_966672, EPI_ISL_966673, EPI_ISL_966674, EPI_ISL_966675, EPI_ISL_966676, EPI_ISL_966677, EPI_ISL_966678, EPI_ISL_966679, EPI_ISL_966680, EPI_ISL_966681 | | | |
| see above | Helix/Illumina | Respiratory Viruses Branch, Division of Viral Diseases, Centers for Disease Control and Prevention | Peter W. Cook,Dakota Howard,Dhwani Batra,Ben L. Rambo-Martin,Eileen de Feo,Jan Antico,Christine Tran,Matthew Tolentino,Shannon Wickline,Kim Gietzen,Brad Sickler,Jingtao Liu,Eric Allen,Phil Febbo,Summer Galloway,Nicole L. Washington,Simon White,Geraint Levan,Kelly Schiabor Barrett,Elizabeth Cirulli,Alexandre Bolze,Ary Ascencio,Charlotte Rivera-Garcia,Ryan Cho,Jason Nguyen,Sherry Wang,Jimmy Ramirez,Tyler Cassens,Efren Sandoval,Magnus Isaksson,William Lee,David Becker,Marc Laurent,James Lu,Clinton R. Paden,Suxiang Tong,Duncan MacCannell, |
| EPI_ISL_966682 | NYU Langone Health | Departments of Pathology and Medicine, New York University School of Medicine | Adriana Heguy, Dacia Dimartino, Emily Guzman, Christian Marier, Peter Meyn, Sitharam Ramaswami, Gael Westby, Paul Zappile, Yutong Zhang, Paolo Cotzia, Guiqing Wang |
| EPI_ISL_966683, EPI_ISL_966684, EPI_ISL_966685, EPI_ISL_966686, EPI_ISL_966687, EPI_ISL_966688, EPI_ISL_966689, EPI_ISL_966690, EPI_ISL_966691, EPI_ISL_966692, EPI_ISL_966693, EPI_ISL_966694, EPI_ISL_966695, EPI_ISL_966696, EPI_ISL_966697, EPI_ISL_966698, EPI_ISL_966699, EPI_ISL_966700, EPI_ISL_966701, EPI_ISL_966702, EPI_ISL_966703, EPI_ISL_966704, EPI_ISL_966705, EPI_ISL_966706, EPI_ISL_966708, EPI_ISL_966709, EPI_ISL_966710, EPI_ISL_966711, EPI_ISL_966712, EPI_ISL_966713, EPI_ISL_966714, EPI_ISL_966715, EPI_ISL_966716, EPI_ISL_966717, EPI_ISL_966718, EPI_ISL_966719, EPI_ISL_966720, EPI_ISL_966721, EPI_ISL_966722, EPI_ISL_966723, EPI_ISL_966724, EPI_ISL_966725, EPI_ISL_966726, EPI_ISL_966727, EPI_ISL_966728, EPI_ISL_966729, EPI_ISL_966730, EPI_ISL_966731, EPI_ISL_966732, EPI_ISL_966733, EPI_ISL_966734, EPI_ISL_966735, EPI_ISL_966736, EPI_ISL_966737, EPI_ISL_966738, EPI_ISL_966739, EPI_ISL_966740, EPI_ISL_966741, EPI_ISL_966742, EPI_ISL_966743, EPI_ISL_966744, EPI_ISL_966745, EPI_ISL_966746, EPI_ISL_966747, EPI_ISL_966748, EPI_ISL_966749, EPI_ISL_966750, EPI_ISL_966751, EPI_ISL_966752, EPI_ISL_966753, EPI_ISL_966754, EPI_ISL_966755 | | | |
| see above | Helix/Illumina | Respiratory Viruses Branch, Division of Viral Diseases, Centers for Disease Control and Prevention | Peter W. Cook,Dakota Howard,Dhwani Batra,Ben L. Rambo-Martin,Eileen de Feo,Jan Antico,Christine Tran,Matthew Tolentino,Shannon Wickline,Kim Gietzen,Brad Sickler,Jingtao Liu,Eric Allen,Phil Febbo,Summer Galloway,Nicole L. Washington,Simon White,Geraint Levan,Kelly Schiabor Barrett,Elizabeth Cirulli,Alexandre Bolze,Ary Ascencio,Charlotte Rivera-Garcia,Ryan Cho,Jason Nguyen,Sherry Wang,Jimmy Ramirez,Tyler Cassens,Efren Sandoval,Magnus Isaksson,William Lee,David Becker,Marc Laurent,James Lu,Clinton R. Paden,Suxiang Tong,Duncan MacCannell, |
| EPI_ISL_966756, EPI_ISL_966757, EPI_ISL_966758, EPI_ISL_966759, EPI_ISL_966760, EPI_ISL_966761, EPI_ISL_966762, EPI_ISL_966763, EPI_ISL_966764, EPI_ISL_966765, EPI_ISL_966766, EPI_ISL_966767, EPI_ISL_966768, EPI_ISL_966769, EPI_ISL_966770, EPI_ISL_966771, EPI_ISL_966772, EPI_ISL_966773, EPI_ISL_966774, EPI_ISL_966775, EPI_ISL_966776, EPI_ISL_966777, EPI_ISL_966778, EPI_ISL_966779, EPI_ISL_966780, EPI_ISL_966781, EPI_ISL_966782, EPI_ISL_966783, EPI_ISL_966784, EPI_ISL_966785, EPI_ISL_966786, EPI_ISL_966787, EPI_ISL_966788, EPI_ISL_966789, EPI_ISL_966790, EPI_ISL_966791, EPI_ISL_966792, EPI_ISL_966793, EPI_ISL_966794, EPI_ISL_966795, EPI_ISL_966796, EPI_ISL_966797, EPI_ISL_966798, EPI_ISL_966799, EPI_ISL_966800, EPI_ISL_966801, EPI_ISL_966802, EPI_ISL_966803, EPI_ISL_966804, EPI_ISL_966805, EPI_ISL_966806, EPI_ISL_966807, EPI_ISL_966808, EPI_ISL_966809, EPI_ISL_966810, EPI_ISL_966811, EPI_ISL_966812, EPI_ISL_966813, EPI_ISL_966814, EPI_ISL_966815, EPI_ISL_966816, EPI_ISL_966817, EPI_ISL_966818, EPI_ISL_966819, EPI_ISL_966820, EPI_ISL_966821, EPI_ISL_966823, EPI_ISL_966824, EPI_ISL_966825, EPI_ISL_966826, EPI_ISL_966827, EPI_ISL_966828, EPI_ISL_966829, EPI_ISL_966830, EPI_ISL_966831, EPI_ISL_966832, EPI_ISL_966833, EPI_ISL_966834, EPI_ISL_966835, EPI_ISL_966836, EPI_ISL_966837, EPI_ISL_966838, EPI_ISL_966841, EPI_ISL_966842, EPI_ISL_966843, EPI_ISL_966844, EPI_ISL_966845, EPI_ISL_966846, EPI_ISL_966847, EPI_ISL_966848, EPI_ISL_966849, EPI_ISL_966850, EPI_ISL_966851 | | | |
| see above | Maine HETL | Tewhey Lab, The Jackson Laboratory | Matluk,N., Dewey,H., Iosue,F., Barter,M., Lynch,R., Munger,H. and Tewhey,R. |
| EPI_ISL_966852, EPI_ISL_966853, EPI_ISL_966854, EPI_ISL_966855, EPI_ISL_966856, EPI_ISL_966857, EPI_ISL_966858, EPI_ISL_966859, EPI_ISL_966860, EPI_ISL_966861, EPI_ISL_966862, EPI_ISL_966863, EPI_ISL_966864, EPI_ISL_966865, EPI_ISL_966867, EPI_ISL_966868, EPI_ISL_966869, EPI_ISL_966870, EPI_ISL_966871, EPI_ISL_966872, EPI_ISL_966873, EPI_ISL_966874, EPI_ISL_966875, EPI_ISL_966876, | | | |

EPI_ISL_966877, EPI_ISL_966878, EPI_ISL_966879, EPI_ISL_966880, EPI_ISL_966881, EPI_ISL_966882, EPI_ISL_966883, EPI_ISL_966884, EPI_ISL_966885, EPI_ISL_966886, EPI_ISL_966887, EPI_ISL_966888, EPI_ISL_966889, EPI_ISL_966890, EPI_ISL_966891, EPI_ISL_966892, EPI_ISL_966893, EPI_ISL_966894, EPI_ISL_966895, EPI_ISL_966896, EPI_ISL_966897, EPI_ISL_966898, EPI_ISL_966899, EPI_ISL_966900, EPI_ISL_966901, EPI_ISL_966902, EPI_ISL_966903, EPI_ISL_966904, EPI_ISL_966905, EPI_ISL_966906, EPI_ISL_966907, EPI_ISL_966908, EPI_ISL_966909, EPI_ISL_966910, EPI_ISL_966911, EPI_ISL_966912, EPI_ISL_966913, EPI_ISL_966914, EPI_ISL_966915, EPI_ISL_966916, EPI_ISL_966917, EPI_ISL_966918, EPI_ISL_966919, EPI_ISL_966920, EPI_ISL_966921, EPI_ISL_966922, EPI_ISL_966923, EPI_ISL_966924, EPI_ISL_966925, EPI_ISL_966926, EPI_ISL_966927, EPI_ISL_966928, EPI_ISL_966929, EPI_ISL_966930, EPI_ISL_966931, EPI_ISL_966932, EPI_ISL_966933, EPI_ISL_966934, EPI_ISL_966935, EPI_ISL_966936, EPI_ISL_966937, EPI_ISL_966938, EPI_ISL_966939, EPI_ISL_966941, EPI_ISL_966942, EPI_ISL_966943, EPI_ISL_966944, EPI_ISL_966945, EPI_ISL_966946, EPI_ISL_966947, EPI_ISL_966948, EPI_ISL_966949, EPI_ISL_966950, EPI_ISL_966951, EPI_ISL_966952, EPI_ISL_966953, EPI_ISL_966954, EPI_ISL_966955, EPI_ISL_966956, EPI_ISL_966957, EPI_ISL_966958, EPI_ISL_966959, EPI_ISL_966960, EPI_ISL_966961, EPI_ISL_966962, EPI_ISL_966963, EPI_ISL_966964, EPI_ISL_966965, EPI_ISL_966966, EPI_ISL_966967, EPI_ISL_966968, EPI_ISL_966969, EPI_ISL_966970, EPI_ISL_966971, EPI_ISL_966972, EPI_ISL_966973, EPI_ISL_966974, EPI_ISL_966975, EPI_ISL_966976, EPI_ISL_966977, EPI_ISL_966978, EPI_ISL_966979, EPI_ISL_966980, EPI_ISL_966981, EPI_ISL_966982, EPI_ISL_966983, EPI_ISL_966984, EPI_ISL_966985, EPI_ISL_966986, EPI_ISL_966987, EPI_ISL_966988, EPI_ISL_966989, EPI_ISL_966990, EPI_ISL_966991, EPI_ISL_966992, EPI_ISL_966993, EPI_ISL_966994, EPI_ISL_966995, EPI_ISL_966996, EPI_ISL_966997, EPI_ISL_966998, EPI_ISL_966999, EPI_ISL_967000, EPI_ISL_967002, EPI_ISL_967003, EPI_ISL_967004, EPI_ISL_967005, EPI_ISL_967006, EPI_ISL_967007, EPI_ISL_967008, EPI_ISL_967009, EPI_ISL_967010, EPI_ISL_967011, EPI_ISL_967012, EPI_ISL_967013, EPI_ISL_967014, EPI_ISL_967015, EPI_ISL_967016, EPI_ISL_967017, EPI_ISL_967018, EPI_ISL_967019, EPI_ISL_967020, EPI_ISL_967021, EPI_ISL_967022, EPI_ISL_967023, EPI_ISL_967024, EPI_ISL_967025, EPI_ISL_967026, EPI_ISL_967027, EPI_ISL_967028, EPI_ISL_967029, EPI_ISL_967030, EPI_ISL_967031, EPI_ISL_967032, EPI_ISL_967033, EPI_ISL_967034, EPI_ISL_967035, EPI_ISL_967036, EPI_ISL_967037, EPI_ISL_967038, EPI_ISL_967039, EPI_ISL_967040, EPI_ISL_967041, EPI_ISL_967042, EPI_ISL_967043, EPI_ISL_967044, EPI_ISL_967045, EPI_ISL_967046, EPI_ISL_967047, EPI_ISL_967048, EPI_ISL_967049, EPI_ISL_967050, EPI_ISL_967051, EPI_ISL_967052, EPI_ISL_967053, EPI_ISL_967054, EPI_ISL_967055, EPI_ISL_967056, EPI_ISL_967057, EPI_ISL_967058, EPI_ISL_967059, EPI_ISL_967060, EPI_ISL_967061, EPI_ISL_967062, EPI_ISL_967063, EPI_ISL_967064, EPI_ISL_967065, EPI_ISL_967066, EPI_ISL_967067, EPI_ISL_967068, EPI_ISL_967069, EPI_ISL_967070, EPI_ISL_967071, EPI_ISL_967072, EPI_ISL_967073, EPI_ISL_967074, EPI_ISL_967075, EPI_ISL_967076, EPI_ISL_967077, EPI_ISL_967078, EPI_ISL_967079, EPI_ISL_967080, EPI_ISL_967081, EPI_ISL_967082, EPI_ISL_967083, EPI_ISL_967084, EPI_ISL_967085, EPI_ISL_967086, EPI_ISL_967087, EPI_ISL_967088, EPI_ISL_967089, EPI_ISL_967090, EPI_ISL_967091, EPI_ISL_967092, EPI_ISL_967093, EPI_ISL_967094, EPI_ISL_967095, EPI_ISL_967096, EPI_ISL_967097, EPI_ISL_967098, EPI_ISL_967099, EPI_ISL_967100, EPI_ISL_967101, EPI_ISL_967102, EPI_ISL_967103, EPI_ISL_967104, EPI_ISL_967105, EPI_ISL_967106, EPI_ISL_967107, EPI_ISL_967108, EPI_ISL_967109, EPI_ISL_967110, EPI_ISL_967111, EPI_ISL_967112, EPI_ISL_967113, EPI_ISL_967114, EPI_ISL_967115, EPI_ISL_967116, EPI_ISL_967117, EPI_ISL_967118, EPI_ISL_967119, EPI_ISL_967120, EPI_ISL_967121, EPI_ISL_967122, EPI_ISL_967123, EPI_ISL_967124, EPI_ISL_967125, EPI_ISL_967126, EPI_ISL_967128, EPI_ISL_967129, EPI_ISL_967130, EPI_ISL_967131, EPI_ISL_967132, EPI_ISL_967133, EPI_ISL_967134, EPI_ISL_967135, EPI_ISL_967136, EPI_ISL_967137, EPI_ISL_967138, EPI_ISL_967139, EPI_ISL_967140, EPI_ISL_967141, EPI_ISL_967142, EPI_ISL_967143, EPI_ISL_967144, EPI_ISL_967145, EPI_ISL_967146, EPI_ISL_967147, EPI_ISL_967148, EPI_ISL_967149, EPI_ISL_967150, EPI_ISL_967151, EPI_ISL_967152, EPI_ISL_967153, EPI_ISL_967154, EPI_ISL_967155, EPI_ISL_967156, EPI_ISL_967157, EPI_ISL_967158, EPI_ISL_967159, EPI_ISL_967160, EPI_ISL_967161, EPI_ISL_967162, EPI_ISL_967163, EPI_ISL_967164, EPI_ISL_967165, EPI_ISL_967166, EPI_ISL_967167, EPI_ISL_967168, EPI_ISL_967169, EPI_ISL_967170, EPI_ISL_967171, EPI_ISL_967172, EPI_ISL_967173, EPI_ISL_967174, EPI_ISL_967175, EPI_ISL_967176, EPI_ISL_967177, EPI_ISL_967178, EPI_ISL_967179, EPI_ISL_967180, EPI_ISL_967181, EPI_ISL_967182, EPI_ISL_967185, EPI_ISL_967186, EPI_ISL_967187, EPI_ISL_967188, EPI_ISL_967189, EPI_ISL_967190, EPI_ISL_967191, EPI_ISL_967192, EPI_ISL_967193, EPI_ISL_967194, EPI_ISL_967195, EPI_ISL_967196, EPI_ISL_967197, EPI_ISL_967198, EPI_ISL_967199, EPI_ISL_967200, EPI_ISL_967201, EPI_ISL_967202, EPI_ISL_967203, EPI_ISL_967204, EPI_ISL_967205, EPI_ISL_967206, EPI_ISL_967207, EPI_ISL_967208, EPI_ISL_967209, EPI_ISL_967210, EPI_ISL_967211, EPI_ISL_967212, EPI_ISL_967213, EPI_ISL_967214, EPI_ISL_967215, EPI_ISL_967216, EPI_ISL_967217, EPI_ISL_967218, EPI_ISL_967219, EPI_ISL_967220, EPI_ISL_967221, EPI_ISL_967222, EPI_ISL_967223, EPI_ISL_967224, EPI_ISL_967225, EPI_ISL_967226, EPI_ISL_967227, EPI_ISL_967228, EPI_ISL_967229, EPI_ISL_967230, EPI_ISL_967231, EPI_ISL_967232, EPI_ISL_967233, EPI_ISL_967234, EPI_ISL_967235, EPI_ISL_967236, EPI_ISL_967237, EPI_ISL_967238, EPI_ISL_967239, EPI_ISL_967240, EPI_ISL_967241, EPI_ISL_967242, EPI_ISL_967243, EPI_ISL_967244, EPI_ISL_967245, EPI_ISL_967246, EPI_ISL_967247, EPI_ISL_967248, EPI_ISL_967249, EPI_ISL_967250, EPI_ISL_967251, EPI_ISL_967252, EPI_ISL_967253, EPI_ISL_967254, EPI_ISL_967255, EPI_ISL_967256, EPI_ISL_967257, EPI_ISL_967258, EPI_ISL_967259, EPI_ISL_967260, EPI_ISL_967261, EPI_ISL_967262, EPI_ISL_967263, EPI_ISL_967264, EPI_ISL_967265, EPI_ISL_967266, EPI_ISL_967267, EPI_ISL_967268, EPI_ISL_967269, EPI_ISL_967270, EPI_ISL_967271, EPI_ISL_967272, EPI_ISL_967273, EPI_ISL_967274, EPI_ISL_967275, EPI_ISL_967276, EPI_ISL_967277, EPI_ISL_967278, EPI_ISL_967279, EPI_ISL_967280, EPI_ISL_967281, EPI_ISL_967282, EPI_ISL_967283, EPI_ISL_967284, EPI_ISL_967285, EPI_ISL_967286, EPI_ISL_967287, EPI_ISL_967288, EPI_ISL_967289, EPI_ISL_967290, EPI_ISL_967291, EPI_ISL_967292, EPI_ISL_967293, EPI_ISL_967294, EPI_ISL_967295, EPI_ISL_967296, EPI_ISL_967297, EPI_ISL_967298, EPI_ISL_967299, EPI_ISL_967300, EPI_ISL_967301, EPI_ISL_967302, EPI_ISL_967303, EPI_ISL_967304, EPI_ISL_967305, EPI_ISL_967306, EPI_ISL_967307, EPI_ISL_967308, EPI_ISL_967309, EPI_ISL_967310, EPI_ISL_967311, EPI_ISL_967312, EPI_ISL_967313, EPI_ISL_967314, EPI_ISL_967315, EPI_ISL_967316, EPI_ISL_967317, EPI_ISL_967318, EPI_ISL_967319, EPI_ISL_967320, EPI_ISL_967321, EPI_ISL_967322, EPI_ISL_967323, EPI_ISL_967324, EPI_ISL_967325, EPI_ISL_967326, EPI_ISL_967327, EPI_ISL_967328, EPI_ISL_967329, EPI_ISL_967330, EPI_ISL_967331, EPI_ISL_967332, EPI_ISL_967333, EPI_ISL_967334, EPI_ISL_967335, EPI_ISL_967336, EPI_ISL_967337, EPI_ISL_967338, EPI_ISL_967339, EPI_ISL_967340, EPI_ISL_967341, EPI_ISL_967342, EPI_ISL_967343, EPI_ISL_967344, EPI_ISL_967345, EPI_ISL_967346, EPI_ISL_967347, EPI_ISL_967348, EPI_ISL_967349, EPI_ISL_967350, EPI_ISL_967351, EPI_ISL_967352, EPI_ISL_967353, EPI_ISL_967354, EPI_ISL_967355, EPI_ISL_967356, EPI_ISL_967357, EPI_ISL_967358, EPI_ISL_967359, EPI_ISL_967360, EPI_ISL_967361, EPI_ISL_967362, EPI_ISL_967363, EPI_ISL_967364, EPI_ISL_967365, EPI_ISL_967366, EPI_ISL_967367, EPI_ISL_967368, EPI_ISL_967369, EPI_ISL_967370, EPI_ISL_967371, EPI_ISL_967372, EPI_ISL_967373, EPI_ISL_967374, EPI_ISL_967375, EPI_ISL_967376, EPI_ISL_967377, EPI_ISL_967378, EPI_ISL_967379, EPI_ISL_967380, EPI_ISL_967381, EPI_ISL_967382, EPI_ISL_967383, EPI_ISL_967384, EPI_ISL_967385, EPI_ISL_967386, EPI_ISL_967387, EPI_ISL_967388, EPI_ISL_967389, EPI_ISL_967390, EPI_ISL_967391, EPI_ISL_967392, EPI_ISL_967393, EPI_ISL_967394, EPI_ISL_967395, EPI_ISL_967396, EPI_ISL_967397, EPI_ISL_967398, EPI_ISL_967399, EPI_ISL_967400, EPI_ISL_967401, EPI_ISL_967402, EPI_ISL_967403, EPI_ISL_967404, EPI_ISL_967405, EPI_ISL_967406, EPI_ISL_967407, EPI_ISL_967408, EPI_ISL_967409, EPI_ISL_967410, EPI_ISL_967411, EPI_ISL_967412, EPI_ISL_967413, EPI_ISL_967414, EPI_ISL_967415, EPI_ISL_967416, EPI_ISL_967417, EPI_ISL_967418, EPI_ISL_967419, EPI_ISL_967420, EPI_ISL_967421, EPI_ISL_967422, EPI_ISL_967423, EPI_ISL_967424, EPI_ISL_967425, EPI_ISL_967426, EPI_ISL_967427, EPI_ISL_967428, EPI_ISL_967429, EPI_ISL_967430, EPI_ISL_967431, EPI_ISL_967432, EPI_ISL_967433, EPI_ISL_967434, EPI_ISL_967436, EPI_ISL_967437, EPI_ISL_967438, EPI_ISL_967439, EPI_ISL_967440, EPI_ISL_967441, EPI_ISL_967442, EPI_ISL_967443, EPI_ISL_967444, EPI_ISL_967445, EPI_ISL_967446, EPI_ISL_967447, EPI_ISL_967448, EPI_ISL_967449, EPI_ISL_967450, EPI_ISL_967451, EPI_ISL_967452, EPI_ISL_967453, EPI_ISL_967454, EPI_ISL_967455, EPI_ISL_967456, EPI_ISL_967457, EPI_ISL_967458, EPI_ISL_967459, EPI_ISL_967460, EPI_ISL_967461, EPI_ISL_967465, EPI_ISL_967466, EPI_ISL_967467, EPI_ISL_967468, EPI_ISL_967469, EPI_ISL_967470, EPI_ISL_967471, EPI_ISL_967472, EPI_ISL_967473, EPI_ISL_967474, EPI_ISL_967475, EPI_ISL_967476, EPI_ISL_967477, EPI_ISL_967478, EPI_ISL_967479, EPI_ISL_967480, EPI_ISL_967481, EPI_ISL_967482, EPI_ISL_967483, EPI_ISL_967484, EPI_ISL_967485, EPI_ISL_967486, EPI_ISL_967487, EPI_ISL_967488, EPI_ISL_967489, EPI_ISL_967490, EPI_ISL_967491, EPI_ISL_967492, EPI_ISL_967493, EPI_ISL_967494, EPI_ISL_967495, EPI_ISL_967496, EPI_ISL_967497, EPI_ISL_967498, EPI_ISL_967499, EPI_ISL_967502, EPI_ISL_967503, EPI_ISL_967504, EPI_ISL_967505, EPI_ISL_967506, EPI_ISL_967507, EPI_ISL_967508, EPI_ISL_967509, EPI_ISL_967510, EPI_ISL_967511, EPI_ISL_967512, EPI_ISL_967513, EPI_ISL_967514, EPI_ISL_967515, EPI_ISL_967516, EPI_ISL_967517, EPI_ISL_967518, EPI_ISL_967519, EPI_ISL_967520, EPI_ISL_967521, EPI_ISL_967522

| | | | |
|---|---|---|---|
| see above | Helix/Illumina | Respiratory Viruses Branch, Division of Viral Diseases, Centers for Disease Control and Prevention | Peter W. Cook,Dakota Howard,Dhwani Batra,Ben L. Rambo-Martin,Eileen de Feo,Jan Antico,Christine Tran,Matthew Tolentino,Shannon Wickline,Kim Gietzen,Brad Sickler,Jingtao Liu,Eric Allen,Phil Febbo,Summer Galloway,Nicole L. Washington,Simon White,Geraint Levan,Kelly Schiabor Barrett,Elizabeth Cirulli,Alexandre Bolze,Ary Ascencio,Charlotte Rivera-Garcia,Ryan Cho,Jason Nguyen,Sherry Wang,Jimmy Ramirez,Tyler Cassens,Efren Sandoval,Magnus Isaksson,William Lee,David Becker,Marc Laurent,James Lu,Clinton R. Paden,Suxiang Tong,Duncan MacCannell, |
| EPI_ISL_967523 | State Laboratories Division, Hawaii State Department of Health | State Laboratories Division, Hawaii State Department of Health | Pamela O'Brien, Drew Kuwazaki, Ayana Garnet, Razvan Sultana, Edward Desmond |
| EPI_ISL_967524 | Helix/Illumina | Respiratory Viruses Branch, Division of Viral Diseases, Centers for Disease Control and Prevention | Peter W. Cook,Dakota Howard,Dhwani Batra,Ben L. Rambo-Martin,Eileen de Feo,Jan Antico,Christine Tran,Matthew Tolentino,Shannon Wickline,Kim Gietzen,Brad Sickler,Jingtao Liu,Eric Allen,Phil Febbo,Summer Galloway,Nicole L. Washington,Simon White,Geraint Levan,Kelly Schiabor Barrett,Elizabeth Cirulli,Alexandre Bolze,Ary Ascencio,Charlotte Rivera-Garcia,Ryan Cho,Jason Nguyen,Sherry Wang,Jimmy Ramirez,Tyler Cassens,Efren Sandoval,Magnus Isaksson,William Lee,David Becker,Marc Laurent,James Lu,Clinton R. Paden,Suxiang Tong,Duncan MacCannell, |
| EPI_ISL_967525, EPI_ISL_967526, EPI_ISL_967527, EPI_ISL_967528, EPI_ISL_967529, EPI_ISL_967530 | State Laboratories Division, Hawaii State Department of Health | State Laboratories Division, Hawaii State Department of Health | Pamela O'Brien, Drew Kuwazaki, Ayana Garnet, Razvan Sultana, Edward Desmond |
| EPI_ISL_967531 | Helix/Illumina | Respiratory Viruses Branch, Division of Viral Diseases, Centers for Disease Control and Prevention | Peter W. Cook,Dakota Howard,Dhwani Batra,Ben L. Rambo-Martin,Eileen de Feo,Jan Antico,Christine Tran,Matthew Tolentino,Shannon Wickline,Kim Gietzen,Brad Sickler,Jingtao Liu,Eric Allen,Phil Febbo,Summer Galloway,Nicole L. Washington,Simon White,Geraint Levan,Kelly Schiabor Barrett,Elizabeth Cirulli,Alexandre Bolze,Ary Ascencio,Charlotte Rivera-Garcia,Ryan Cho,Jason Nguyen,Sherry Wang,Jimmy Ramirez,Tyler Cassens,Efren Sandoval,Magnus Isaksson,William Lee,David Becker,Marc Laurent,James Lu,Clinton R. Paden,Suxiang Tong,Duncan MacCannell, |
| EPI_ISL_967532, EPI_ISL_967533, EPI_ISL_967534, EPI_ISL_967535, EPI_ISL_967536 | State Laboratories Division, Hawaii State Department of Health | State Laboratories Division, Hawaii State Department of Health | Pamela O'Brien, Drew Kuwazaki, Ayana Garnet, Razvan Sultana, Edward Desmond |
| EPI_ISL_967537 | Helix/Illumina | Respiratory Viruses Branch, Division of Viral Diseases, Centers for Disease Control and Prevention | Peter W. Cook,Dakota Howard,Dhwani Batra,Ben L. Rambo-Martin,Eileen de Feo,Jan Antico,Christine Tran,Matthew Tolentino,Shannon Wickline,Kim Gietzen,Brad Sickler,Jingtao Liu,Eric Allen,Phil Febbo,Summer Galloway,Nicole L. Washington,Simon White,Geraint Levan,Kelly Schiabor Barrett,Elizabeth Cirulli,Alexandre Bolze,Ary Ascencio,Charlotte Rivera-Garcia,Ryan Cho,Jason Nguyen,Sherry Wang,Jimmy Ramirez,Tyler Cassens,Efren Sandoval,Magnus Isaksson,William Lee,David Becker,Marc Laurent,James Lu,Clinton R. Paden,Suxiang Tong,Duncan MacCannell, |
| EPI_ISL_967538, EPI_ISL_967539, EPI_ISL_967540, EPI_ISL_967541, EPI_ISL_967542, EPI_ISL_967543, EPI_ISL_967544, EPI_ISL_967545, EPI_ISL_967546 | State Laboratories Division, Hawaii State Department of Health | State Laboratories Division, Hawaii State Department of Health | Pamela O'Brien, Drew Kuwazaki, Ayana Garnet, Razvan Sultana, Edward Desmond |
| EPI_ISL_967547 | Helix/Illumina | Respiratory Viruses Branch, Division of Viral Diseases, Centers for Disease Control and Prevention | Peter W. Cook,Dakota Howard,Dhwani Batra,Ben L. Rambo-Martin,Eileen de Feo,Jan Antico,Christine Tran,Matthew Tolentino,Shannon Wickline,Kim Gietzen,Brad Sickler,Jingtao Liu,Eric Allen,Phil Febbo,Summer Galloway,Nicole L. Washington,Simon White,Geraint Levan,Kelly Schiabor Barrett,Elizabeth Cirulli,Alexandre Bolze,Ary Ascencio,Charlotte Rivera-Garcia,Ryan Cho,Jason Nguyen,Sherry Wang,Jimmy Ramirez,Tyler Cassens,Efren Sandoval,Magnus Isaksson,William Lee,David Becker,Marc Laurent,James Lu,Clinton R. Paden,Suxiang Tong,Duncan MacCannell, |
| EPI_ISL_967548, EPI_ISL_967549, EPI_ISL_967550, EPI_ISL_967551, EPI_ISL_967552, EPI_ISL_967553 | State Laboratories Division, Hawaii State Department of Health | State Laboratories Division, Hawaii State Department of Health | Pamela O'Brien, Drew Kuwazaki, Ayana Garnet, Razvan Sultana, Edward Desmond |
| EPI_ISL_967554 | Helix/Illumina | Respiratory Viruses Branch, Division of Viral Diseases, Centers for Disease Control and Prevention | Peter W. Cook,Dakota Howard,Dhwani Batra,Ben L. Rambo-Martin,Eileen de Feo,Jan Antico,Christine Tran,Matthew Tolentino,Shannon Wickline,Kim Gietzen,Brad Sickler,Jingtao Liu,Eric Allen,Phil Febbo,Summer Galloway,Nicole L. Washington,Simon White,Geraint Levan,Kelly Schiabor Barrett,Elizabeth Cirulli,Alexandre Bolze,Ary Ascencio,Charlotte Rivera-Garcia,Ryan Cho,Jason Nguyen,Sherry Wang,Jimmy Ramirez,Tyler Cassens,Efren Sandoval,Magnus Isaksson,William Lee,David Becker,Marc Laurent,James Lu,Clinton R. Paden,Suxiang Tong,Duncan MacCannell, |

EPI_ISL_967555, EPI_ISL_967556, EPI_ISL_967557, EPI_ISL_967558, EPI_ISL_967559, EPI_ISL_967560, EPI_ISL_967561, EPI_ISL_967562, EPI_ISL_967563, EPI_ISL_967564, EPI_ISL_967565, EPI_ISL_967566

| | | | |
|---|---|---|---|
| see above | State Laboratories Division, Hawaii State Department of Health | State Laboratories Division, Hawaii State Department of Health | Pamela O'Brien, Drew Kuwazaki, Ayana Garnet, Razvan Sultana, Edward Desmond |
| EPI_ISL_967567 | Helix/Illumina | Respiratory Viruses Branch, Division of Viral Diseases, Centers for Disease Control and Prevention | Peter W. Cook,Dakota Howard,Dhwani Batra,Ben L. Rambo-Martin,Eileen de Feo,Jan Antico,Christine Tran,Matthew Tolentino,Shannon Wickline,Kim Gietzen,Brad Sickler,Jingtao Liu,Eric Allen,Phil Febbo,Summer Galloway,Nicole L. Washington,Simon White,Geraint Levan,Kelly Schiabor Barrett,Elizabeth Cirulli,Alexandre Bolze,Ary Ascencio,Charlotte Rivera-Garcia,Ryan Cho,Jason Nguyen,Sherry Wang,Jimmy Ramirez,Tyler Cassens,Efren Sandoval,Magnus Isaksson,William Lee,David Becker,Marc Laurent,James Lu,Clinton R. Paden,Suxiang Tong,Duncan MacCannell, |
| EPI_ISL_967568, EPI_ISL_967569, EPI_ISL_967570, EPI_ISL_967571, EPI_ISL_967572, EPI_ISL_967573, EPI_ISL_967574, EPI_ISL_967575 | State Laboratories Division, Hawaii State Department of Health | State Laboratories Division, Hawaii State Department of Health | Pamela O'Brien, Drew Kuwazaki, Ayana Garnet, Razvan Sultana, Edward Desmond |
| EPI_ISL_967576 | Helix/Illumina | Respiratory Viruses Branch, Division of Viral Diseases, Centers for Disease Control and Prevention | Peter W. Cook,Dakota Howard,Dhwani Batra,Ben L. Rambo-Martin,Eileen de Feo,Jan Antico,Christine Tran,Matthew Tolentino,Shannon Wickline,Kim Gietzen,Brad Sickler,Jingtao Liu,Eric Allen,Phil Febbo,Summer Galloway,Nicole L. Washington,Simon White,Geraint Levan,Kelly Schiabor Barrett,Elizabeth Cirulli,Alexandre Bolze,Ary Ascencio,Charlotte Rivera-Garcia,Ryan Cho,Jason Nguyen,Sherry Wang,Jimmy Ramirez,Tyler Cassens,Efren Sandoval,Magnus Isaksson,William Lee,David Becker,Marc Laurent,James Lu,Clinton R. Paden,Suxiang Tong,Duncan MacCannell, |
| EPI_ISL_967577, EPI_ISL_967578, EPI_ISL_967579, EPI_ISL_967580, EPI_ISL_967581, EPI_ISL_967582 | State Laboratories Division, Hawaii State Department of Health | State Laboratories Division, Hawaii State Department of Health | Pamela O'Brien, Drew Kuwazaki, Ayana Garnet, Razvan Sultana, Edward Desmond |
| EPI_ISL_967583 | Helix/Illumina | Respiratory Viruses Branch, Division of Viral Diseases, Centers for Disease Control and Prevention | Peter W. Cook,Dakota Howard,Dhwani Batra,Ben L. Rambo-Martin,Eileen de Feo,Jan Antico,Christine Tran,Matthew Tolentino,Shannon Wickline,Kim Gietzen,Brad Sickler,Jingtao Liu,Eric Allen,Phil Febbo,Summer Galloway,Nicole L. Washington,Simon White,Geraint Levan,Kelly Schiabor Barrett,Elizabeth Cirulli,Alexandre Bolze,Ary Ascencio,Charlotte Rivera-Garcia,Ryan Cho,Jason Nguyen,Sherry Wang,Jimmy Ramirez,Tyler Cassens,Efren Sandoval,Magnus Isaksson,William Lee,David Becker,Marc Laurent,James Lu,Clinton R. Paden,Suxiang Tong,Duncan MacCannell, |
| EPI_ISL_967584, EPI_ISL_967585, EPI_ISL_967586, | State Laboratories Division, Hawaii State Department of Health | State Laboratories Division, Hawaii State Department of Health | Pamela O'Brien, Drew Kuwazaki, Ayana Garnet, Razvan Sultana, Edward Desmond |

| | | | |
|---|---|---|---|
| EPI_ISL_967587, EPI_ISL_967588, EPI_ISL_967589 | | | |
| EPI_ISL_967590 | Helix/Illumina | Respiratory Viruses Branch, Division of Viral Diseases, Centers for Disease Control and Prevention | Peter W. Cook,Dakota Howard,Dhwani Batra,Ben L. Rambo-Martin,Eileen de Feo,Jan Antico,Christine Tran,Matthew Tolentino,Shannon Wickline,Kim Gietzen,Brad Sickler,Jingtao Liu,Eric Allen,Phil Febbo,Summer Galloway,Nicole L. Washington,Simon White,Geraint Levan,Kelly Schiabor Barrett,Elizabeth Cirulli,Alexandre Bolze,Ary Ascencio,Charlotte Rivera-Garcia,Ryan Cho,Jason Nguyen,Sherry Wang,Jimmy Ramirez,Tyler Cassens,Efren Sandoval,Magnus Isaksson,William Lee,David Becker,Marc Laurent,James Lu,Clinton R. Paden,Suxiang Tong,Duncan MacCannell, |
| EPI_ISL_967591, EPI_ISL_967592, EPI_ISL_967593, EPI_ISL_967594, EPI_ISL_967595, EPI_ISL_967596, EPI_ISL_967597, EPI_ISL_967598 | State Laboratories Division, Hawaii State Department of Health | State Laboratories Division, Hawaii State Department of Health | Pamela O'Brien, Drew Kuwazaki, Ayana Garnet, Razvan Sultana, Edward Desmond |
| EPI_ISL_967599 | Helix/Illumina | Respiratory Viruses Branch, Division of Viral Diseases, Centers for Disease Control and Prevention | Peter W. Cook,Dakota Howard,Dhwani Batra,Ben L. Rambo-Martin,Eileen de Feo,Jan Antico,Christine Tran,Matthew Tolentino,Shannon Wickline,Kim Gietzen,Brad Sickler,Jingtao Liu,Eric Allen,Phil Febbo,Summer Galloway,Nicole L. Washington,Simon White,Geraint Levan,Kelly Schiabor Barrett,Elizabeth Cirulli,Alexandre Bolze,Ary Ascencio,Charlotte Rivera-Garcia,Ryan Cho,Jason Nguyen,Sherry Wang,Jimmy Ramirez,Tyler Cassens,Efren Sandoval,Magnus Isaksson,William Lee,David Becker,Marc Laurent,James Lu,Clinton R. Paden,Suxiang Tong,Duncan MacCannell, |
| EPI_ISL_967600, EPI_ISL_967601, EPI_ISL_967602, EPI_ISL_967603, EPI_ISL_967604, EPI_ISL_967605, EPI_ISL_967606, EPI_ISL_967607, EPI_ISL_967608 | State Laboratories Division, Hawaii State Department of Health | State Laboratories Division, Hawaii State Department of Health | Pamela O'Brien, Drew Kuwazaki, Ayana Garnet, Razvan Sultana, Edward Desmond |
| EPI_ISL_967609 | Helix/Illumina | Respiratory Viruses Branch, Division of Viral Diseases, Centers for Disease Control and Prevention | Peter W. Cook,Dakota Howard,Dhwani Batra,Ben L. Rambo-Martin,Eileen de Feo,Jan Antico,Christine Tran,Matthew Tolentino,Shannon Wickline,Kim Gietzen,Brad Sickler,Jingtao Liu,Eric Allen,Phil Febbo,Summer Galloway,Nicole L. Washington,Simon White,Geraint Levan,Kelly Schiabor Barrett,Elizabeth Cirulli,Alexandre Bolze,Ary Ascencio,Charlotte Rivera-Garcia,Ryan Cho,Jason Nguyen,Sherry Wang,Jimmy Ramirez,Tyler Cassens,Efren Sandoval,Magnus Isaksson,William Lee,David Becker,Marc Laurent,James Lu,Clinton R. Paden,Suxiang Tong,Duncan MacCannell, |
| EPI_ISL_967610, EPI_ISL_967611, EPI_ISL_967612, EPI_ISL_967613, EPI_ISL_967614, EPI_ISL_967615, EPI_ISL_967616 | State Laboratories Division, Hawaii State Department of Health | State Laboratories Division, Hawaii State Department of Health | Pamela O'Brien, Drew Kuwazaki, Ayana Garnet, Razvan Sultana, Edward Desmond |
| EPI_ISL_967617 | Helix/Illumina | Respiratory Viruses Branch, Division of Viral Diseases, Centers for Disease Control and Prevention | Peter W. Cook,Dakota Howard,Dhwani Batra,Ben L. Rambo-Martin,Eileen de Feo,Jan Antico,Christine Tran,Matthew Tolentino,Shannon Wickline,Kim Gietzen,Brad Sickler,Jingtao Liu,Eric Allen,Phil Febbo,Summer Galloway,Nicole L. Washington,Simon White,Geraint Levan,Kelly Schiabor Barrett,Elizabeth Cirulli,Alexandre Bolze,Ary Ascencio,Charlotte Rivera-Garcia,Ryan Cho,Jason Nguyen,Sherry Wang,Jimmy Ramirez,Tyler Cassens,Efren Sandoval,Magnus Isaksson,William Lee,David Becker,Marc Laurent,James Lu,Clinton R. Paden,Suxiang Tong,Duncan MacCannell, |
| EPI_ISL_967618, EPI_ISL_967619, EPI_ISL_967620, EPI_ISL_967621, EPI_ISL_967622, EPI_ISL_967623, EPI_ISL_967624, EPI_ISL_967625, EPI_ISL_967626, EPI_ISL_967627, EPI_ISL_967628 | | | |
| see above | State Laboratories Division, Hawaii State Department of Health | State Laboratories Division, Hawaii State Department of Health | Pamela O'Brien, Drew Kuwazaki, Ayana Garnet, Razvan Sultana, Edward Desmond |
| EPI_ISL_967629 | Helix/Illumina | Respiratory Viruses Branch, Division of Viral Diseases, Centers for Disease Control and Prevention | Peter W. Cook,Dakota Howard,Dhwani Batra,Ben L. Rambo-Martin,Eileen de Feo,Jan Antico,Christine Tran,Matthew Tolentino,Shannon Wickline,Kim Gietzen,Brad Sickler,Jingtao Liu,Eric Allen,Phil Febbo,Summer Galloway,Nicole L. Washington,Simon White,Geraint Levan,Kelly Schiabor Barrett,Elizabeth Cirulli,Alexandre Bolze,Ary Ascencio,Charlotte Rivera-Garcia,Ryan Cho,Jason Nguyen,Sherry Wang,Jimmy Ramirez,Tyler Cassens,Efren Sandoval,Magnus Isaksson,William Lee,David Becker,Marc Laurent,James Lu,Clinton R. Paden,Suxiang Tong,Duncan MacCannell, |
| EPI_ISL_967630, EPI_ISL_967631, EPI_ISL_967632, EPI_ISL_967633, EPI_ISL_967634, EPI_ISL_967635, EPI_ISL_967636, EPI_ISL_967637, EPI_ISL_967638, EPI_ISL_967639, EPI_ISL_967640, EPI_ISL_967641 | | | |
| see above | State Laboratories Division, Hawaii State Department of Health | State Laboratories Division, Hawaii State Department of Health | Pamela O'Brien, Drew Kuwazaki, Ayana Garnet, Razvan Sultana, Edward Desmond |
| EPI_ISL_967642 | Helix/Illumina | Respiratory Viruses Branch, Division of Viral Diseases, Centers for Disease Control and Prevention | Peter W. Cook,Dakota Howard,Dhwani Batra,Ben L. Rambo-Martin,Eileen de Feo,Jan Antico,Christine Tran,Matthew Tolentino,Shannon Wickline,Kim Gietzen,Brad Sickler,Jingtao Liu,Eric Allen,Phil Febbo,Summer Galloway,Nicole L. Washington,Simon White,Geraint Levan,Kelly Schiabor Barrett,Elizabeth Cirulli,Alexandre Bolze,Ary Ascencio,Charlotte Rivera-Garcia,Ryan Cho,Jason Nguyen,Sherry Wang,Jimmy Ramirez,Tyler Cassens,Efren Sandoval,Magnus Isaksson,William Lee,David Becker,Marc Laurent,James Lu,Clinton R. Paden,Suxiang Tong,Duncan MacCannell, |
| EPI_ISL_967643, EPI_ISL_967644, EPI_ISL_967645, EPI_ISL_967646, EPI_ISL_967647, EPI_ISL_967648, EPI_ISL_967649, EPI_ISL_967650, EPI_ISL_967651, EPI_ISL_967652, EPI_ISL_967653, EPI_ISL_967654 | | | |
| see above | State Laboratories Division, Hawaii State Department of Health | State Laboratories Division, Hawaii State Department of Health | Pamela O'Brien, Drew Kuwazaki, Ayana Garnet, Razvan Sultana, Edward Desmond |
| EPI_ISL_967655 | Helix/Illumina | Respiratory Viruses Branch, Division of Viral Diseases, Centers for Disease Control and Prevention | Peter W. Cook,Dakota Howard,Dhwani Batra,Ben L. Rambo-Martin,Eileen de Feo,Jan Antico,Christine Tran,Matthew Tolentino,Shannon Wickline,Kim Gietzen,Brad Sickler,Jingtao Liu,Eric Allen,Phil Febbo,Summer Galloway,Nicole L. Washington,Simon White,Geraint Levan,Kelly Schiabor Barrett,Elizabeth Cirulli,Alexandre Bolze,Ary Ascencio,Charlotte Rivera-Garcia,Ryan Cho,Jason Nguyen,Sherry Wang,Jimmy Ramirez,Tyler Cassens,Efren Sandoval,Magnus Isaksson,William Lee,David Becker,Marc Laurent,James Lu,Clinton R. Paden,Suxiang Tong,Duncan MacCannell, |
| EPI_ISL_967656, EPI_ISL_967657, EPI_ISL_967658, EPI_ISL_967659, EPI_ISL_967660, EPI_ISL_967661 | State Laboratories Division, Hawaii State Department of Health | State Laboratories Division, Hawaii State Department of Health | Pamela O'Brien, Drew Kuwazaki, Ayana Garnet, Razvan Sultana, Edward Desmond |
| EPI_ISL_967662 | Helix/Illumina | Respiratory Viruses Branch, Division of Viral Diseases, Centers for Disease Control and Prevention | Peter W. Cook,Dakota Howard,Dhwani Batra,Ben L. Rambo-Martin,Eileen de Feo,Jan Antico,Christine Tran,Matthew Tolentino,Shannon Wickline,Kim Gietzen,Brad Sickler,Jingtao Liu,Eric Allen,Phil Febbo,Summer Galloway,Nicole L. Washington,Simon White,Geraint Levan,Kelly Schiabor Barrett,Elizabeth Cirulli,Alexandre Bolze,Ary Ascencio,Charlotte Rivera-Garcia,Ryan Cho,Jason Nguyen,Sherry Wang,Jimmy Ramirez,Tyler Cassens,Efren Sandoval,Magnus Isaksson,William Lee,David Becker,Marc Laurent,James Lu,Clinton R. Paden,Suxiang Tong,Duncan MacCannell, |
| EPI_ISL_967663, EPI_ISL_967665, EPI_ISL_967666, EPI_ISL_967667, EPI_ISL_967668, EPI_ISL_967669, EPI_ISL_967670 | State Laboratories Division, Hawaii State Department of Health | State Laboratories Division, Hawaii State Department of Health | Pamela O'Brien, Drew Kuwazaki, Ayana Garnet, Razvan Sultana, Edward Desmond |
| EPI_ISL_967671 | Helix/Illumina | Respiratory Viruses Branch, Division of Viral Diseases, Centers for Disease Control and Prevention | Peter W. Cook,Dakota Howard,Dhwani Batra,Ben L. Rambo-Martin,Eileen de Feo,Jan Antico,Christine Tran,Matthew Tolentino,Shannon Wickline,Kim Gietzen,Brad Sickler,Jingtao Liu,Eric Allen,Phil Febbo,Summer Galloway,Nicole L. Washington,Simon White,Geraint Levan,Kelly Schiabor Barrett,Elizabeth Cirulli,Alexandre Bolze,Ary Ascencio,Charlotte Rivera-Garcia,Ryan Cho,Jason Nguyen,Sherry Wang,Jimmy Ramirez,Tyler Cassens,Efren Sandoval,Magnus Isaksson,William Lee,David Becker,Marc Laurent,James Lu,Clinton R. Paden,Suxiang Tong,Duncan MacCannell, |
| EPI_ISL_967672, EPI_ISL_967673, EPI_ISL_967674, EPI_ISL_967675, EPI_ISL_967676 | State Laboratories Division, Hawaii State Department of Health | State Laboratories Division, Hawaii State Department of Health | Pamela O'Brien, Drew Kuwazaki, Ayana Garnet, Razvan Sultana, Edward Desmond |
| EPI_ISL_967678 | Helix/Illumina | Respiratory Viruses Branch, Division of Viral Diseases, Centers for Disease Control and Prevention | Peter W. Cook,Dakota Howard,Dhwani Batra,Ben L. Rambo-Martin,Eileen de Feo,Jan Antico,Christine Tran,Matthew Tolentino,Shannon Wickline,Kim Gietzen,Brad Sickler,Jingtao Liu,Eric Allen,Phil Febbo,Summer Galloway,Nicole L. Washington,Simon White,Geraint Levan,Kelly Schiabor Barrett,Elizabeth Cirulli,Alexandre Bolze,Ary Ascencio,Charlotte Rivera-Garcia,Ryan Cho,Jason Nguyen,Sherry Wang,Jimmy Ramirez,Tyler Cassens,Efren Sandoval,Magnus Isaksson,William Lee,David Becker,Marc Laurent,James Lu,Clinton R. Paden,Suxiang Tong,Duncan MacCannell, |
| EPI_ISL_967679, EPI_ISL_967680, EPI_ISL_967681, EPI_ISL_967682, EPI_ISL_967683 | State Laboratories Division, Hawaii State Department of Health | State Laboratories Division, Hawaii State Department of Health | Pamela O'Brien, Drew Kuwazaki, Ayana Garnet, Razvan Sultana, Edward Desmond |
| EPI_ISL_967684 | Helix/Illumina | Respiratory Viruses Branch, Division of Viral Diseases, Centers for Disease Control and Prevention | Peter W. Cook,Dakota Howard,Dhwani Batra,Ben L. Rambo-Martin,Eileen de Feo,Jan Antico,Christine Tran,Matthew Tolentino,Shannon Wickline,Kim Gietzen,Brad Sickler,Jingtao Liu,Eric Allen,Phil Febbo,Summer Galloway,Nicole L. Washington,Simon White,Geraint Levan,Kelly Schiabor Barrett,Elizabeth Cirulli,Alexandre Bolze,Ary Ascencio,Charlotte Rivera-Garcia,Ryan Cho,Jason Nguyen,Sherry Wang,Jimmy Ramirez,Tyler Cassens,Efren Sandoval,Magnus Isaksson,William Lee,David Becker,Marc Laurent,James Lu,Clinton R. Paden,Suxiang Tong,Duncan MacCannell, |
| EPI_ISL_967685, EPI_ISL_967686, EPI_ISL_967687, EPI_ISL_967688, EPI_ISL_967689, EPI_ISL_967690, | State Laboratories Division, Hawaii State Department of Health | State Laboratories Division, Hawaii State Department of Health | Pamela O'Brien, Drew Kuwazaki, Ayana Garnet, Razvan Sultana, Edward Desmond |

| | | | |
|---|---|---|---|
| EPI_ISL_967691, EPI_ISL_967692 | | | |
| EPI_ISL_967693 | Helix/Illumina | Respiratory Viruses Branch, Division of Viral Diseases, Centers for Disease Control and Prevention | Peter W. Cook,Dakota Howard,Dhwani Batra,Ben L. Rambo-Martin,Eileen de Feo,Jan Antico,Christine Tran,Matthew Tolentino,Shannon Wickline,Kim Gietzen,Brad Sickler,Jingtao Liu,Eric Allen,Phil Febbo,Summer Galloway,Nicole L. Washington,Simon White,Geraint Levan,Kelly Schiabor Barrett,Elizabeth Cirulli,Alexandre Bolze,Ary Ascencio,Charlotte Rivera-Garcia,Ryan Cho,Jason Nguyen,Sherry Wang,Jimmy Ramirez,Tyler Cassens,Efren Sandoval,Magnus Isaksson,William Lee,David Becker,Marc Laurent,James Lu,Clinton R. Paden,Suxiang Tong,Duncan MacCannell, |
| EPI_ISL_967694, EPI_ISL_967695, EPI_ISL_967696, EPI_ISL_967697, EPI_ISL_967698, EPI_ISL_967699 | State Laboratories Division, Hawaii State Department of Health | State Laboratories Division, Hawaii State Department of Health | Pamela O'Brien, Drew Kuwazaki, Ayana Garnet, Razvan Sultana, Edward Desmond |
| EPI_ISL_967700 | Helix/Illumina | Respiratory Viruses Branch, Division of Viral Diseases, Centers for Disease Control and Prevention | Peter W. Cook,Dakota Howard,Dhwani Batra,Ben L. Rambo-Martin,Eileen de Feo,Jan Antico,Christine Tran,Matthew Tolentino,Shannon Wickline,Kim Gietzen,Brad Sickler,Jingtao Liu,Eric Allen,Phil Febbo,Summer Galloway,Nicole L. Washington,Simon White,Geraint Levan,Kelly Schiabor Barrett,Elizabeth Cirulli,Alexandre Bolze,Ary Ascencio,Charlotte Rivera-Garcia,Ryan Cho,Jason Nguyen,Sherry Wang,Jimmy Ramirez,Tyler Cassens,Efren Sandoval,Magnus Isaksson,William Lee,David Becker,Marc Laurent,James Lu,Clinton R. Paden,Suxiang Tong,Duncan MacCannell, |
| EPI_ISL_967701, EPI_ISL_967702, EPI_ISL_967703, EPI_ISL_967704, EPI_ISL_967705, EPI_ISL_967706, EPI_ISL_967707, EPI_ISL_967708, EPI_ISL_967709 | State Laboratories Division, Hawaii State Department of Health | State Laboratories Division, Hawaii State Department of Health | Pamela O'Brien, Drew Kuwazaki, Ayana Garnet, Razvan Sultana, Edward Desmond |
| EPI_ISL_967710 | Helix/Illumina | Respiratory Viruses Branch, Division of Viral Diseases, Centers for Disease Control and Prevention | Peter W. Cook,Dakota Howard,Dhwani Batra,Ben L. Rambo-Martin,Eileen de Feo,Jan Antico,Christine Tran,Matthew Tolentino,Shannon Wickline,Kim Gietzen,Brad Sickler,Jingtao Liu,Eric Allen,Phil Febbo,Summer Galloway,Nicole L. Washington,Simon White,Geraint Levan,Kelly Schiabor Barrett,Elizabeth Cirulli,Alexandre Bolze,Ary Ascencio,Charlotte Rivera-Garcia,Ryan Cho,Jason Nguyen,Sherry Wang,Jimmy Ramirez,Tyler Cassens,Efren Sandoval,Magnus Isaksson,William Lee,David Becker,Marc Laurent,James Lu,Clinton R. Paden,Suxiang Tong,Duncan MacCannell, |
| EPI_ISL_967711, EPI_ISL_967712, EPI_ISL_967713, EPI_ISL_967714, EPI_ISL_967715, EPI_ISL_967717, EPI_ISL_967718 | State Laboratories Division, Hawaii State Department of Health | State Laboratories Division, Hawaii State Department of Health | Pamela O'Brien, Drew Kuwazaki, Ayana Garnet, Razvan Sultana, Edward Desmond |
| EPI_ISL_967719 | Helix/Illumina | Respiratory Viruses Branch, Division of Viral Diseases, Centers for Disease Control and Prevention | Peter W. Cook,Dakota Howard,Dhwani Batra,Ben L. Rambo-Martin,Eileen de Feo,Jan Antico,Christine Tran,Matthew Tolentino,Shannon Wickline,Kim Gietzen,Brad Sickler,Jingtao Liu,Eric Allen,Phil Febbo,Summer Galloway,Nicole L. Washington,Simon White,Geraint Levan,Kelly Schiabor Barrett,Elizabeth Cirulli,Alexandre Bolze,Ary Ascencio,Charlotte Rivera-Garcia,Ryan Cho,Jason Nguyen,Sherry Wang,Jimmy Ramirez,Tyler Cassens,Efren Sandoval,Magnus Isaksson,William Lee,David Becker,Marc Laurent,James Lu,Clinton R. Paden,Suxiang Tong,Duncan MacCannell, |
| EPI_ISL_967720, EPI_ISL_967721, EPI_ISL_967722, EPI_ISL_967723, EPI_ISL_967724, EPI_ISL_967725 | State Laboratories Division, Hawaii State Department of Health | State Laboratories Division, Hawaii State Department of Health | Pamela O'Brien, Drew Kuwazaki, Ayana Garnet, Razvan Sultana, Edward Desmond |
| EPI_ISL_967726 | Helix/Illumina | Respiratory Viruses Branch, Division of Viral Diseases, Centers for Disease Control and Prevention | Peter W. Cook,Dakota Howard,Dhwani Batra,Ben L. Rambo-Martin,Eileen de Feo,Jan Antico,Christine Tran,Matthew Tolentino,Shannon Wickline,Kim Gietzen,Brad Sickler,Jingtao Liu,Eric Allen,Phil Febbo,Summer Galloway,Nicole L. Washington,Simon White,Geraint Levan,Kelly Schiabor Barrett,Elizabeth Cirulli,Alexandre Bolze,Ary Ascencio,Charlotte Rivera-Garcia,Ryan Cho,Jason Nguyen,Sherry Wang,Jimmy Ramirez,Tyler Cassens,Efren Sandoval,Magnus Isaksson,William Lee,David Becker,Marc Laurent,James Lu,Clinton R. Paden,Suxiang Tong,Duncan MacCannell, |
| EPI_ISL_967727, EPI_ISL_967729, EPI_ISL_967730, EPI_ISL_967731, EPI_ISL_967732, EPI_ISL_967733, EPI_ISL_967734, EPI_ISL_967735 | State Laboratories Division, Hawaii State Department of Health | State Laboratories Division, Hawaii State Department of Health | Pamela O'Brien, Drew Kuwazaki, Ayana Garnet, Razvan Sultana, Edward Desmond |
| EPI_ISL_967736 | Helix/Illumina | Respiratory Viruses Branch, Division of Viral Diseases, Centers for Disease Control and Prevention | Peter W. Cook,Dakota Howard,Dhwani Batra,Ben L. Rambo-Martin,Eileen de Feo,Jan Antico,Christine Tran,Matthew Tolentino,Shannon Wickline,Kim Gietzen,Brad Sickler,Jingtao Liu,Eric Allen,Phil Febbo,Summer Galloway,Nicole L. Washington,Simon White,Geraint Levan,Kelly Schiabor Barrett,Elizabeth Cirulli,Alexandre Bolze,Ary Ascencio,Charlotte Rivera-Garcia,Ryan Cho,Jason Nguyen,Sherry Wang,Jimmy Ramirez,Tyler Cassens,Efren Sandoval,Magnus Isaksson,William Lee,David Becker,Marc Laurent,James Lu,Clinton R. Paden,Suxiang Tong,Duncan MacCannell, |
| EPI_ISL_967737, EPI_ISL_967738, EPI_ISL_967739, EPI_ISL_967740, EPI_ISL_967741, EPI_ISL_967742, EPI_ISL_967743, EPI_ISL_967744, EPI_ISL_967745, EPI_ISL_967746, EPI_ISL_967747 | | | |
| see above | State Laboratories Division, Hawaii State Department of Health | State Laboratories Division, Hawaii State Department of Health | Pamela O'Brien, Drew Kuwazaki, Ayana Garnet, Razvan Sultana, Edward Desmond |
| EPI_ISL_967748 | Helix/Illumina | Respiratory Viruses Branch, Division of Viral Diseases, Centers for Disease Control and Prevention | Peter W. Cook,Dakota Howard,Dhwani Batra,Ben L. Rambo-Martin,Eileen de Feo,Jan Antico,Christine Tran,Matthew Tolentino,Shannon Wickline,Kim Gietzen,Brad Sickler,Jingtao Liu,Eric Allen,Phil Febbo,Summer Galloway,Nicole L. Washington,Simon White,Geraint Levan,Kelly Schiabor Barrett,Elizabeth Cirulli,Alexandre Bolze,Ary Ascencio,Charlotte Rivera-Garcia,Ryan Cho,Jason Nguyen,Sherry Wang,Jimmy Ramirez,Tyler Cassens,Efren Sandoval,Magnus Isaksson,William Lee,David Becker,Marc Laurent,James Lu,Clinton R. Paden,Suxiang Tong,Duncan MacCannell, |
| EPI_ISL_967749, EPI_ISL_967750, EPI_ISL_967751, EPI_ISL_967752 | State Laboratories Division, Hawaii State Department of Health | State Laboratories Division, Hawaii State Department of Health | Pamela O'Brien, Drew Kuwazaki, Ayana Garnet, Razvan Sultana, Edward Desmond |
| EPI_ISL_967753 | Helix/Illumina | Respiratory Viruses Branch, Division of Viral Diseases, Centers for Disease Control and Prevention | Peter W. Cook,Dakota Howard,Dhwani Batra,Ben L. Rambo-Martin,Eileen de Feo,Jan Antico,Christine Tran,Matthew Tolentino,Shannon Wickline,Kim Gietzen,Brad Sickler,Jingtao Liu,Eric Allen,Phil Febbo,Summer Galloway,Nicole L. Washington,Simon White,Geraint Levan,Kelly Schiabor Barrett,Elizabeth Cirulli,Alexandre Bolze,Ary Ascencio,Charlotte Rivera-Garcia,Ryan Cho,Jason Nguyen,Sherry Wang,Jimmy Ramirez,Tyler Cassens,Efren Sandoval,Magnus Isaksson,William Lee,David Becker,Marc Laurent,James Lu,Clinton R. Paden,Suxiang Tong,Duncan MacCannell, |
| EPI_ISL_967754, EPI_ISL_967755, EPI_ISL_967756, EPI_ISL_967757, EPI_ISL_967758, EPI_ISL_967759, EPI_ISL_967760 | State Laboratories Division, Hawaii State Department of Health | State Laboratories Division, Hawaii State Department of Health | Pamela O'Brien, Drew Kuwazaki, Ayana Garnet, Razvan Sultana, Edward Desmond |
| EPI_ISL_967761 | Helix/Illumina | Respiratory Viruses Branch, Division of Viral Diseases, Centers for Disease Control and Prevention | Peter W. Cook,Dakota Howard,Dhwani Batra,Ben L. Rambo-Martin,Eileen de Feo,Jan Antico,Christine Tran,Matthew Tolentino,Shannon Wickline,Kim Gietzen,Brad Sickler,Jingtao Liu,Eric Allen,Phil Febbo,Summer Galloway,Nicole L. Washington,Simon White,Geraint Levan,Kelly Schiabor Barrett,Elizabeth Cirulli,Alexandre Bolze,Ary Ascencio,Charlotte Rivera-Garcia,Ryan Cho,Jason Nguyen,Sherry Wang,Jimmy Ramirez,Tyler Cassens,Efren Sandoval,Magnus Isaksson,William Lee,David Becker,Marc Laurent,James Lu,Clinton R. Paden,Suxiang Tong,Duncan MacCannell, |
| EPI_ISL_967762, EPI_ISL_967763, EPI_ISL_967764, EPI_ISL_967765, EPI_ISL_967766 | State Laboratories Division, Hawaii State Department of Health | State Laboratories Division, Hawaii State Department of Health | Pamela O'Brien, Drew Kuwazaki, Ayana Garnet, Razvan Sultana, Edward Desmond |
| EPI_ISL_967767 | Helix/Illumina | Respiratory Viruses Branch, Division of Viral Diseases, Centers for Disease Control and Prevention | Peter W. Cook,Dakota Howard,Dhwani Batra,Ben L. Rambo-Martin,Eileen de Feo,Jan Antico,Christine Tran,Matthew Tolentino,Shannon Wickline,Kim Gietzen,Brad Sickler,Jingtao Liu,Eric Allen,Phil Febbo,Summer Galloway,Nicole L. Washington,Simon White,Geraint Levan,Kelly Schiabor Barrett,Elizabeth Cirulli,Alexandre Bolze,Ary Ascencio,Charlotte Rivera-Garcia,Ryan Cho,Jason Nguyen,Sherry Wang,Jimmy Ramirez,Tyler Cassens,Efren Sandoval,Magnus Isaksson,William Lee,David Becker,Marc Laurent,James Lu,Clinton R. Paden,Suxiang Tong,Duncan MacCannell, |
| EPI_ISL_967768, EPI_ISL_967769, EPI_ISL_967770, EPI_ISL_967771 | State Laboratories Division, Hawaii State Department of Health | State Laboratories Division, Hawaii State Department of Health | Pamela O'Brien, Drew Kuwazaki, Ayana Garnet, Razvan Sultana, Edward Desmond |
| EPI_ISL_967772, EPI_ISL_967773, EPI_ISL_967774, EPI_ISL_967775, EPI_ISL_967776, EPI_ISL_967777, EPI_ISL_967778, EPI_ISL_967779, EPI_ISL_967780, EPI_ISL_967781, EPI_ISL_967783, EPI_ISL_967784, EPI_ISL_967785, EPI_ISL_967786, EPI_ISL_967787, EPI_ISL_967788, EPI_ISL_967789, EPI_ISL_967790, EPI_ISL_967791, EPI_ISL_967792, EPI_ISL_967793, EPI_ISL_967794, EPI_ISL_967795, EPI_ISL_967796, EPI_ISL_967797, EPI_ISL_967798, EPI_ISL_967799, EPI_ISL_967800, EPI_ISL_967801, EPI_ISL_967802, EPI_ISL_967803, EPI_ISL_967804, EPI_ISL_967805, EPI_ISL_967806, EPI_ISL_967807, EPI_ISL_967808, EPI_ISL_967809, EPI_ISL_967810, EPI_ISL_967811, EPI_ISL_967812, EPI_ISL_967813, EPI_ISL_967814, EPI_ISL_967815, EPI_ISL_967816, EPI_ISL_967817, EPI_ISL_967818, EPI_ISL_967819, EPI_ISL_967820, EPI_ISL_967821, EPI_ISL_967822, EPI_ISL_967823, EPI_ISL_967824, EPI_ISL_967825, EPI_ISL_967826, EPI_ISL_967827, EPI_ISL_967828, EPI_ISL_967829, EPI_ISL_967830, EPI_ISL_967831, EPI_ISL_967832, EPI_ISL_967833, EPI_ISL_967834, EPI_ISL_967835, EPI_ISL_967836, EPI_ISL_967837, EPI_ISL_967838, EPI_ISL_967839, EPI_ISL_967840, EPI_ISL_967841, EPI_ISL_967842, EPI_ISL_967843, EPI_ISL_967844, EPI_ISL_967845, EPI_ISL_967846, EPI_ISL_967847, EPI_ISL_967848, EPI_ISL_967849, EPI_ISL_967850, EPI_ISL_967851, EPI_ISL_967852, EPI_ISL_967853, EPI_ISL_967854, EPI_ISL_967855, EPI_ISL_967856, EPI_ISL_967857, EPI_ISL_967858, EPI_ISL_967859, EPI_ISL_967860, EPI_ISL_967861, EPI_ISL_967862, EPI_ISL_967863, EPI_ISL_967864, EPI_ISL_967865, EPI_ISL_967866, EPI_ISL_967867, EPI_ISL_967868, EPI_ISL_967869, EPI_ISL_967870, EPI_ISL_967871, EPI_ISL_967872, EPI_ISL_967873, EPI_ISL_967874, EPI_ISL_967875, EPI_ISL_967876, EPI_ISL_967877, EPI_ISL_967878, EPI_ISL_967879, EPI_ISL_967880, EPI_ISL_967881 | | | |
| see above | Helix/Illumina | Respiratory Viruses Branch, Division of Viral Diseases, Centers for Disease Control and Prevention | Peter W. Cook,Dakota Howard,Dhwani Batra,Ben L. Rambo-Martin,Eileen de Feo,Jan Antico,Christine Tran,Matthew Tolentino,Shannon Wickline,Kim Gietzen,Brad Sickler,Jingtao Liu,Eric Allen,Phil Febbo,Summer Galloway,Nicole L. Washington,Simon White,Geraint Levan,Kelly Schiabor Barrett,Elizabeth Cirulli,Alexandre Bolze,Ary Ascencio,Charlotte Rivera-Garcia,Ryan Cho,Jason Nguyen,Sherry Wang,Jimmy Ramirez,Tyler Cassens,Efren Sandoval,Magnus Isaksson,William Lee,David Becker,Marc Laurent,James Lu,Clinton R. Paden,Suxiang Tong,Duncan MacCannell, |
| EPI_ISL_967882, | Arizona State Public Health Laboratory | Arizona State Public Health Laboratory | Trung Huynh, Jessica Escobar, Katherine Fullerton, Nobuko Fukushima, Stacy White, Linda Getsinger, Victor Waddell |

| | | | |
|---|---|---|---|
| EPI_ISL_967883,<br>EPI_ISL_967884 | | | |
| EPI_ISL_967886,<br>EPI_ISL_967887,<br>EPI_ISL_967889 | TGen North | TGen North | "Jolene Bowers, Megan Folkerts, Chris French, Hayley Yaglom, Ashlyn Pfeiffer, Darrin Lemmer, Dave Engelthaler, The Arizona COVID Genomics Union (ACGU)" |
| EPI_ISL_967891,<br>EPI_ISL_967892,<br>EPI_ISL_967893 | Sonora Quest Laboratories | TGen North | "Jolene Bowers, Megan Folkerts, Chris French, Hayley Yaglom, Ashlyn Pfeiffer, Darrin Lemmer, Dave Engelthaler, The Arizona COVID Genomics Union (ACGU)" |
| EPI_ISL_967894 | TGen North | TGen North | "Jolene Bowers, Megan Folkerts, Chris French, Hayley Yaglom, Ashlyn Pfeiffer, Darrin Lemmer, Dave Engelthaler, The Arizona COVID Genomics Union (ACGU)" |
| EPI_ISL_967895,<br>EPI_ISL_967896,<br>EPI_ISL_967898,<br>EPI_ISL_967900 | Sonora Quest Laboratories | TGen North | "Jolene Bowers, Megan Folkerts, Chris French, Hayley Yaglom, Ashlyn Pfeiffer, Darrin Lemmer, Dave Engelthaler, The Arizona COVID Genomics Union (ACGU)" |
| EPI_ISL_967902 | TGen North | TGen North | "Jolene Bowers, Megan Folkerts, Chris French, Hayley Yaglom, Ashlyn Pfeiffer, Darrin Lemmer, Dave Engelthaler, The Arizona COVID Genomics Union (ACGU)" |
| EPI_ISL_967903 | Sonora Quest Laboratories | TGen North | "Jolene Bowers, Megan Folkerts, Chris French, Hayley Yaglom, Ashlyn Pfeiffer, Darrin Lemmer, Dave Engelthaler, The Arizona COVID Genomics Union (ACGU)" |
| EPI_ISL_967905,<br>EPI_ISL_967906 | TGen North | TGen North | "Jolene Bowers, Megan Folkerts, Chris French, Hayley Yaglom, Ashlyn Pfeiffer, Darrin Lemmer, Dave Engelthaler, The Arizona COVID Genomics Union (ACGU)" |
| EPI_ISL_967907 | Sonora Quest Laboratories | TGen North | "Jolene Bowers, Megan Folkerts, Chris French, Hayley Yaglom, Ashlyn Pfeiffer, Darrin Lemmer, Dave Engelthaler, The Arizona COVID Genomics Union (ACGU)" |
| EPI_ISL_967908 | TGen North | TGen North | "Jolene Bowers, Megan Folkerts, Chris French, Hayley Yaglom, Ashlyn Pfeiffer, Darrin Lemmer, Dave Engelthaler, The Arizona COVID Genomics Union (ACGU)" |
| EPI_ISL_967909 | Sonora Quest Laboratories | TGen North | "Jolene Bowers, Megan Folkerts, Chris French, Hayley Yaglom, Ashlyn Pfeiffer, Darrin Lemmer, Dave Engelthaler, The Arizona COVID Genomics Union (ACGU)" |
| EPI_ISL_967911,<br>EPI_ISL_967912,<br>EPI_ISL_967913,<br>EPI_ISL_967914 | TGen North | TGen North | "Jolene Bowers, Megan Folkerts, Chris French, Hayley Yaglom, Ashlyn Pfeiffer, Darrin Lemmer, Dave Engelthaler, The Arizona COVID Genomics Union (ACGU)" |
| EPI_ISL_967916,<br>EPI_ISL_967917,<br>EPI_ISL_967918 | Sonora Quest Laboratories | TGen North | "Jolene Bowers, Megan Folkerts, Chris French, Hayley Yaglom, Ashlyn Pfeiffer, Darrin Lemmer, Dave Engelthaler, The Arizona COVID Genomics Union (ACGU)" |
| EPI_ISL_967919,<br>EPI_ISL_967921 | TGen North | TGen North | "Jolene Bowers, Megan Folkerts, Chris French, Hayley Yaglom, Ashlyn Pfeiffer, Darrin Lemmer, Dave Engelthaler, The Arizona COVID Genomics Union (ACGU)" |
| EPI_ISL_967922 | Sonora Quest Laboratories | TGen North | "Jolene Bowers, Megan Folkerts, Chris French, Hayley Yaglom, Ashlyn Pfeiffer, Darrin Lemmer, Dave Engelthaler, The Arizona COVID Genomics Union (ACGU)" |
| EPI_ISL_967925 | TGen North | TGen North | "Jolene Bowers, Megan Folkerts, Chris French, Hayley Yaglom, Ashlyn Pfeiffer, Darrin Lemmer, Dave Engelthaler, The Arizona COVID Genomics Union (ACGU)" |
| EPI_ISL_967926 | Sonora Quest Laboratories | TGen North | "Jolene Bowers, Megan Folkerts, Chris French, Hayley Yaglom, Ashlyn Pfeiffer, Darrin Lemmer, Dave Engelthaler, The Arizona COVID Genomics Union (ACGU)" |
| EPI_ISL_967928 | TGen North | TGen North | "Jolene Bowers, Megan Folkerts, Chris French, Hayley Yaglom, Ashlyn Pfeiffer, Darrin Lemmer, Dave Engelthaler, The Arizona COVID Genomics Union (ACGU)" |
| EPI_ISL_967929 | Sonora Quest Laboratories | TGen North | "Jolene Bowers, Megan Folkerts, Chris French, Hayley Yaglom, Ashlyn Pfeiffer, Darrin Lemmer, Dave Engelthaler, The Arizona COVID Genomics Union (ACGU)" |
| EPI_ISL_967930,<br>EPI_ISL_967931 | TGen North | TGen North | "Jolene Bowers, Megan Folkerts, Chris French, Hayley Yaglom, Ashlyn Pfeiffer, Darrin Lemmer, Dave Engelthaler, The Arizona COVID Genomics Union (ACGU)" |
| EPI_ISL_967932,<br>EPI_ISL_967933 | Sonora Quest Laboratories | TGen North | "Jolene Bowers, Megan Folkerts, Chris French, Hayley Yaglom, Ashlyn Pfeiffer, Darrin Lemmer, Dave Engelthaler, The Arizona COVID Genomics Union (ACGU)" |
| EPI_ISL_967935,<br>EPI_ISL_967936,<br>EPI_ISL_967937,<br>EPI_ISL_967938,<br>EPI_ISL_967939 | TGen North | TGen North | "Jolene Bowers, Megan Folkerts, Chris French, Hayley Yaglom, Ashlyn Pfeiffer, Darrin Lemmer, Dave Engelthaler, The Arizona COVID Genomics Union (ACGU)" |
| EPI_ISL_967940 | Sonora Quest Laboratories | TGen North | "Jolene Bowers, Megan Folkerts, Chris French, Hayley Yaglom, Ashlyn Pfeiffer, Darrin Lemmer, Dave Engelthaler, The Arizona COVID Genomics Union (ACGU)" |
| EPI_ISL_967941 | TGen North | TGen North | "Jolene Bowers, Megan Folkerts, Chris French, Hayley Yaglom, Ashlyn Pfeiffer, Darrin Lemmer, Dave Engelthaler, The Arizona COVID Genomics Union (ACGU)" |
| EPI_ISL_967942 | Sonora Quest Laboratories | TGen North | "Jolene Bowers, Megan Folkerts, Chris French, Hayley Yaglom, Ashlyn Pfeiffer, Darrin Lemmer, Dave Engelthaler, The Arizona COVID Genomics Union (ACGU)" |
| EPI_ISL_967944 | TGen North | TGen North | "Jolene Bowers, Megan Folkerts, Chris French, Hayley Yaglom, Ashlyn Pfeiffer, Darrin Lemmer, Dave Engelthaler, The Arizona COVID Genomics Union (ACGU)" |
| EPI_ISL_967946 | Sonora Quest Laboratories | TGen North | "Jolene Bowers, Megan Folkerts, Chris French, Hayley Yaglom, Ashlyn Pfeiffer, Darrin Lemmer, Dave Engelthaler, The Arizona COVID Genomics Union (ACGU)" |
| EPI_ISL_967947,<br>EPI_ISL_967948,<br>EPI_ISL_967949,<br>EPI_ISL_967950 | TGen North | TGen North | "Jolene Bowers, Megan Folkerts, Chris French, Hayley Yaglom, Ashlyn Pfeiffer, Darrin Lemmer, Dave Engelthaler, The Arizona COVID Genomics Union (ACGU)" |
| EPI_ISL_967951 | Sonora Quest Laboratories | TGen North | "Jolene Bowers, Megan Folkerts, Chris French, Hayley Yaglom, Ashlyn Pfeiffer, Darrin Lemmer, Dave Engelthaler, The Arizona COVID Genomics Union (ACGU)" |
| EPI_ISL_967952,<br>EPI_ISL_967953,<br>EPI_ISL_967955,<br>EPI_ISL_967956 | TGen North | TGen North | "Jolene Bowers, Megan Folkerts, Chris French, Hayley Yaglom, Ashlyn Pfeiffer, Darrin Lemmer, Dave Engelthaler, The Arizona COVID Genomics Union (ACGU)" |
| EPI_ISL_967958 | Sonora Quest Laboratories | TGen North | "Jolene Bowers, Megan Folkerts, Chris French, Hayley Yaglom, Ashlyn Pfeiffer, Darrin Lemmer, Dave Engelthaler, The Arizona COVID Genomics Union (ACGU)" |
| EPI_ISL_967959,<br>EPI_ISL_967960,<br>EPI_ISL_967961 | TGen North | TGen North | "Jolene Bowers, Megan Folkerts, Chris French, Hayley Yaglom, Ashlyn Pfeiffer, Darrin Lemmer, Dave Engelthaler, The Arizona COVID Genomics Union (ACGU)" |
| EPI_ISL_967962,<br>EPI_ISL_967963,<br>EPI_ISL_967964,<br>EPI_ISL_967965 | Sonora Quest Laboratories | TGen North | "Jolene Bowers, Megan Folkerts, Chris French, Hayley Yaglom, Ashlyn Pfeiffer, Darrin Lemmer, Dave Engelthaler, The Arizona COVID Genomics Union (ACGU)" |
| EPI_ISL_967966,<br>EPI_ISL_967967 | TGen North | TGen North | "Jolene Bowers, Megan Folkerts, Chris French, Hayley Yaglom, Ashlyn Pfeiffer, Darrin Lemmer, Dave Engelthaler, The Arizona COVID Genomics Union (ACGU)" |
| EPI_ISL_967968 | Sonora Quest Laboratories | TGen North | "Jolene Bowers, Megan Folkerts, Chris French, Hayley Yaglom, Ashlyn Pfeiffer, Darrin Lemmer, Dave Engelthaler, The Arizona COVID Genomics Union (ACGU)" |
| EPI_ISL_967970,<br>EPI_ISL_967971 | TGen North | TGen North | "Jolene Bowers, Megan Folkerts, Chris French, Hayley Yaglom, Ashlyn Pfeiffer, Darrin Lemmer, Dave Engelthaler, The Arizona COVID Genomics Union (ACGU)" |
| EPI_ISL_967972 | Sonora Quest Laboratories | TGen North | "Jolene Bowers, Megan Folkerts, Chris French, Hayley Yaglom, Ashlyn Pfeiffer, Darrin Lemmer, Dave Engelthaler, The Arizona COVID Genomics Union (ACGU)" |
| EPI_ISL_967973,<br>EPI_ISL_967974,<br>EPI_ISL_967976,<br>EPI_ISL_967977 | TGen North | TGen North | "Jolene Bowers, Megan Folkerts, Chris French, Hayley Yaglom, Ashlyn Pfeiffer, Darrin Lemmer, Dave Engelthaler, The Arizona COVID Genomics Union (ACGU)" |
| EPI_ISL_967978 | Sonora Quest Laboratories | TGen North | "Jolene Bowers, Megan Folkerts, Chris French, Hayley Yaglom, Ashlyn Pfeiffer, Darrin Lemmer, Dave Engelthaler, The Arizona COVID Genomics Union (ACGU)" |
| EPI_ISL_967979 | TGen North | TGen North | "Jolene Bowers, Megan Folkerts, Chris French, Hayley Yaglom, Ashlyn Pfeiffer, Darrin Lemmer, Dave Engelthaler, The Arizona COVID Genomics Union (ACGU)" |
| EPI_ISL_967981 | Sonora Quest Laboratories | TGen North | "Jolene Bowers, Megan Folkerts, Chris French, Hayley Yaglom, Ashlyn Pfeiffer, Darrin Lemmer, Dave Engelthaler, The Arizona COVID Genomics Union (ACGU)" |
| EPI_ISL_967983 | TGen North | TGen North | "Jolene Bowers, Megan Folkerts, Chris French, Hayley Yaglom, Ashlyn Pfeiffer, Darrin Lemmer, Dave Engelthaler, The Arizona COVID Genomics Union (ACGU)" |
| EPI_ISL_967985 | Sonora Quest Laboratories | TGen North | "Jolene Bowers, Megan Folkerts, Chris French, Hayley Yaglom, Ashlyn Pfeiffer, Darrin Lemmer, Dave Engelthaler, The Arizona COVID Genomics Union (ACGU)" |
| EPI_ISL_967987,<br>EPI_ISL_967988,<br>EPI_ISL_967990 | TGen North | TGen North | "Jolene Bowers, Megan Folkerts, Chris French, Hayley Yaglom, Ashlyn Pfeiffer, Darrin Lemmer, Dave Engelthaler, The Arizona COVID Genomics Union (ACGU)" |
| EPI_ISL_967991 | Sonora Quest Laboratories | TGen North | "Jolene Bowers, Megan Folkerts, Chris French, Hayley Yaglom, Ashlyn Pfeiffer, Darrin Lemmer, Dave Engelthaler, The Arizona COVID Genomics Union (ACGU)" |
| EPI_ISL_967992 | TGen North | TGen North | "Jolene Bowers, Megan Folkerts, Chris French, Hayley Yaglom, Ashlyn Pfeiffer, Darrin Lemmer, Dave Engelthaler, The Arizona COVID Genomics Union (ACGU)" |
| EPI_ISL_967993,<br>EPI_ISL_967994,<br>EPI_ISL_967995,<br>EPI_ISL_967996 | Sonora Quest Laboratories | TGen North | "Jolene Bowers, Megan Folkerts, Chris French, Hayley Yaglom, Ashlyn Pfeiffer, Darrin Lemmer, Dave Engelthaler, The Arizona COVID Genomics Union (ACGU)" |

| | | | |
|---|---|---|---|
| EPI_ISL_967997, EPI_ISL_967999 | TGen North | TGen North | "Jolene Bowers, Megan Folkerts, Chris French, Hayley Yaglom, Ashlyn Pfeiffer, Darrin Lemmer, Dave Engelthaler, The Arizona COVID Genomics Union (ACGU)" |
| EPI_ISL_968000 | Sonora Quest Laboratories | TGen North | "Jolene Bowers, Megan Folkerts, Chris French, Hayley Yaglom, Ashlyn Pfeiffer, Darrin Lemmer, Dave Engelthaler, The Arizona COVID Genomics Union (ACGU)" |
| EPI_ISL_968001, EPI_ISL_968002, EPI_ISL_968003, EPI_ISL_968005 | TGen North | TGen North | "Jolene Bowers, Megan Folkerts, Chris French, Hayley Yaglom, Ashlyn Pfeiffer, Darrin Lemmer, Dave Engelthaler, The Arizona COVID Genomics Union (ACGU)" |
| EPI_ISL_968006, EPI_ISL_968008, EPI_ISL_968010 | Sonora Quest Laboratories | TGen North | "Jolene Bowers, Megan Folkerts, Chris French, Hayley Yaglom, Ashlyn Pfeiffer, Darrin Lemmer, Dave Engelthaler, The Arizona COVID Genomics Union (ACGU)" |
| EPI_ISL_968011, EPI_ISL_968012, EPI_ISL_968013 | TGen North | TGen North | "Jolene Bowers, Megan Folkerts, Chris French, Hayley Yaglom, Ashlyn Pfeiffer, Darrin Lemmer, Dave Engelthaler, The Arizona COVID Genomics Union (ACGU)" |
| EPI_ISL_968014 | Sonora Quest Laboratories | TGen North | "Jolene Bowers, Megan Folkerts, Chris French, Hayley Yaglom, Ashlyn Pfeiffer, Darrin Lemmer, Dave Engelthaler, The Arizona COVID Genomics Union (ACGU)" |
| EPI_ISL_968015, EPI_ISL_968016, EPI_ISL_968017 | TGen North | TGen North | "Jolene Bowers, Megan Folkerts, Chris French, Hayley Yaglom, Ashlyn Pfeiffer, Darrin Lemmer, Dave Engelthaler, The Arizona COVID Genomics Union (ACGU)" |
| EPI_ISL_968018 | Sonora Quest Laboratories | TGen North | "Jolene Bowers, Megan Folkerts, Chris French, Hayley Yaglom, Ashlyn Pfeiffer, Darrin Lemmer, Dave Engelthaler, The Arizona COVID Genomics Union (ACGU)" |
| EPI_ISL_968020 | TGen North | TGen North | "Jolene Bowers, Megan Folkerts, Chris French, Hayley Yaglom, Ashlyn Pfeiffer, Darrin Lemmer, Dave Engelthaler, The Arizona COVID Genomics Union (ACGU)" |
| EPI_ISL_968021, EPI_ISL_968023, EPI_ISL_968024, EPI_ISL_968025, EPI_ISL_968026, EPI_ISL_968028, EPI_ISL_968029, EPI_ISL_968031 | Sonora Quest Laboratories | TGen North | "Jolene Bowers, Megan Folkerts, Chris French, Hayley Yaglom, Ashlyn Pfeiffer, Darrin Lemmer, Dave Engelthaler, The Arizona COVID Genomics Union (ACGU)" |
| EPI_ISL_968032 | TGen North | TGen North | "Jolene Bowers, Megan Folkerts, Chris French, Hayley Yaglom, Ashlyn Pfeiffer, Darrin Lemmer, Dave Engelthaler, The Arizona COVID Genomics Union (ACGU)" |
| EPI_ISL_968033 | Sonora Quest Laboratories | TGen North | "Jolene Bowers, Megan Folkerts, Chris French, Hayley Yaglom, Ashlyn Pfeiffer, Darrin Lemmer, Dave Engelthaler, The Arizona COVID Genomics Union (ACGU)" |
| EPI_ISL_968034 | TGen North | TGen North | "Jolene Bowers, Megan Folkerts, Chris French, Hayley Yaglom, Ashlyn Pfeiffer, Darrin Lemmer, Dave Engelthaler, The Arizona COVID Genomics Union (ACGU)" |
| EPI_ISL_968035, EPI_ISL_968036 | Sonora Quest Laboratories | TGen North | "Jolene Bowers, Megan Folkerts, Chris French, Hayley Yaglom, Ashlyn Pfeiffer, Darrin Lemmer, Dave Engelthaler, The Arizona COVID Genomics Union (ACGU)" |
| EPI_ISL_968037, EPI_ISL_968038 | TGen North | TGen North | "Jolene Bowers, Megan Folkerts, Chris French, Hayley Yaglom, Ashlyn Pfeiffer, Darrin Lemmer, Dave Engelthaler, The Arizona COVID Genomics Union (ACGU)" |
| EPI_ISL_968039, EPI_ISL_968040 | Sonora Quest Laboratories | TGen North | "Jolene Bowers, Megan Folkerts, Chris French, Hayley Yaglom, Ashlyn Pfeiffer, Darrin Lemmer, Dave Engelthaler, The Arizona COVID Genomics Union (ACGU)" |
| EPI_ISL_968041 | TGen North | TGen North | "Jolene Bowers, Megan Folkerts, Chris French, Hayley Yaglom, Ashlyn Pfeiffer, Darrin Lemmer, Dave Engelthaler, The Arizona COVID Genomics Union (ACGU)" |
| EPI_ISL_968042, EPI_ISL_968043 | Sonora Quest Laboratories | TGen North | "Jolene Bowers, Megan Folkerts, Chris French, Hayley Yaglom, Ashlyn Pfeiffer, Darrin Lemmer, Dave Engelthaler, The Arizona COVID Genomics Union (ACGU)" |
| EPI_ISL_968044, EPI_ISL_968045 | TGen North | TGen North | "Jolene Bowers, Megan Folkerts, Chris French, Hayley Yaglom, Ashlyn Pfeiffer, Darrin Lemmer, Dave Engelthaler, The Arizona COVID Genomics Union (ACGU)" |
| EPI_ISL_968046, EPI_ISL_968047, EPI_ISL_968048, EPI_ISL_968049 | Sonora Quest Laboratories | TGen North | "Jolene Bowers, Megan Folkerts, Chris French, Hayley Yaglom, Ashlyn Pfeiffer, Darrin Lemmer, Dave Engelthaler, The Arizona COVID Genomics Union (ACGU)" |
| EPI_ISL_968050, EPI_ISL_968051 | TGen North | TGen North | "Jolene Bowers, Megan Folkerts, Chris French, Hayley Yaglom, Ashlyn Pfeiffer, Darrin Lemmer, Dave Engelthaler, The Arizona COVID Genomics Union (ACGU)" |
| EPI_ISL_968052 | Sonora Quest Laboratories | TGen North | "Jolene Bowers, Megan Folkerts, Chris French, Hayley Yaglom, Ashlyn Pfeiffer, Darrin Lemmer, Dave Engelthaler, The Arizona COVID Genomics Union (ACGU)" |
| EPI_ISL_968053, EPI_ISL_968055 | TGen North | TGen North | "Jolene Bowers, Megan Folkerts, Chris French, Hayley Yaglom, Ashlyn Pfeiffer, Darrin Lemmer, Dave Engelthaler, The Arizona COVID Genomics Union (ACGU)" |
| EPI_ISL_968056 | Sonora Quest Laboratories | TGen North | "Jolene Bowers, Megan Folkerts, Chris French, Hayley Yaglom, Ashlyn Pfeiffer, Darrin Lemmer, Dave Engelthaler, The Arizona COVID Genomics Union (ACGU)" |
| EPI_ISL_968057 | TGen North | TGen North | "Jolene Bowers, Megan Folkerts, Chris French, Hayley Yaglom, Ashlyn Pfeiffer, Darrin Lemmer, Dave Engelthaler, The Arizona COVID Genomics Union (ACGU)" |
| EPI_ISL_968058, EPI_ISL_968060, EPI_ISL_968061, EPI_ISL_968062 | Sonora Quest Laboratories | TGen North | "Jolene Bowers, Megan Folkerts, Chris French, Hayley Yaglom, Ashlyn Pfeiffer, Darrin Lemmer, Dave Engelthaler, The Arizona COVID Genomics Union (ACGU)" |
| EPI_ISL_968063, EPI_ISL_968064, EPI_ISL_968065 | TGen North | TGen North | "Jolene Bowers, Megan Folkerts, Chris French, Hayley Yaglom, Ashlyn Pfeiffer, Darrin Lemmer, Dave Engelthaler, The Arizona COVID Genomics Union (ACGU)" |
| EPI_ISL_968066 | Sonora Quest Laboratories | TGen North | "Jolene Bowers, Megan Folkerts, Chris French, Hayley Yaglom, Ashlyn Pfeiffer, Darrin Lemmer, Dave Engelthaler, The Arizona COVID Genomics Union (ACGU)" |
| EPI_ISL_968067, EPI_ISL_968068 | TGen North | TGen North | "Jolene Bowers, Megan Folkerts, Chris French, Hayley Yaglom, Ashlyn Pfeiffer, Darrin Lemmer, Dave Engelthaler, The Arizona COVID Genomics Union (ACGU)" |
| EPI_ISL_968069, EPI_ISL_968070, EPI_ISL_968071, EPI_ISL_968072, EPI_ISL_968073, EPI_ISL_968074, EPI_ISL_968075, EPI_ISL_968076, EPI_ISL_968077, EPI_ISL_968078, EPI_ISL_968079 | | | |
| see above | Monterey County Public Health Laboratory | Monterey County Public Health Laboratory | Monterey County Public Health Laboratory |
| EPI_ISL_968080 | Pandemic Response Lab - NYC | Pandemic Response Lab, R&D | Henry Lee, Michael Hammerling, Melissa Hopkins, Cybill del Castillo, William Ward, Pradeep Bugga, Haiping Hao, Jon Laurent |
| EPI_ISL_968082, EPI_ISL_968083, EPI_ISL_968084, EPI_ISL_968085, EPI_ISL_968086, EPI_ISL_968087, EPI_ISL_968088, EPI_ISL_968090, EPI_ISL_968091, EPI_ISL_968092, EPI_ISL_968093, EPI_ISL_968094, EPI_ISL_968095, EPI_ISL_968096, EPI_ISL_968097, EPI_ISL_968098, EPI_ISL_968099, EPI_ISL_968100, EPI_ISL_968101, EPI_ISL_968102, EPI_ISL_968103, EPI_ISL_968104, EPI_ISL_968105, EPI_ISL_968106, EPI_ISL_968107, EPI_ISL_968108, EPI_ISL_968109, EPI_ISL_968110, EPI_ISL_968111, EPI_ISL_968112, EPI_ISL_968113, EPI_ISL_968114, EPI_ISL_968115, EPI_ISL_968116, EPI_ISL_968117, EPI_ISL_968118, EPI_ISL_968119, EPI_ISL_968120, EPI_ISL_968121, EPI_ISL_968122, EPI_ISL_968123, EPI_ISL_968124, EPI_ISL_968125, EPI_ISL_968126, EPI_ISL_968127, EPI_ISL_968128, EPI_ISL_968129, EPI_ISL_968130, EPI_ISL_968131, EPI_ISL_968132, EPI_ISL_968133, EPI_ISL_968134, EPI_ISL_968135, EPI_ISL_968136, EPI_ISL_968137, EPI_ISL_968138, EPI_ISL_968139 | | | |
| see above | Houston Health Department, Disease Prevention and Control | Houston Health Department, Disease Prevention and Control | Penn,R., Brown,P., Lara,A. |
| EPI_ISL_968140, EPI_ISL_968141, EPI_ISL_968142, EPI_ISL_968143, EPI_ISL_968144, EPI_ISL_968145, EPI_ISL_968146, EPI_ISL_968147, EPI_ISL_968148, EPI_ISL_968149, EPI_ISL_968150, EPI_ISL_968151, EPI_ISL_968152, EPI_ISL_968153, EPI_ISL_968154, EPI_ISL_968155, EPI_ISL_968156, EPI_ISL_968157, EPI_ISL_968158, EPI_ISL_968159, EPI_ISL_968160, EPI_ISL_968161, EPI_ISL_968162, EPI_ISL_968163, EPI_ISL_968164, EPI_ISL_968165, EPI_ISL_968166, EPI_ISL_968167, EPI_ISL_968168, EPI_ISL_968169, EPI_ISL_968170, EPI_ISL_968171, EPI_ISL_968172, EPI_ISL_968173, EPI_ISL_968174, EPI_ISL_968175, EPI_ISL_968176, EPI_ISL_968177, EPI_ISL_968178, EPI_ISL_968179, EPI_ISL_968180, EPI_ISL_968181, EPI_ISL_968182, EPI_ISL_968183, EPI_ISL_968184, EPI_ISL_968185, EPI_ISL_968186, EPI_ISL_968187, EPI_ISL_968188 | | | |
| see above | Clinical Molecular Microbiology Laboratory, UNC Hospital | Dirk Dittmer | Justin T. Landis , Razia Moorad , Brent A. Eason, Melissa B. Miller, Linda Pluta, Dirk Dittmer, Angelica Juarez, Cecilia Thompson, Shawn Hawken, Cameroon Grant, Evelyn Hoffman, Patricio Cano, Jason Wong, Carolina Caro-Vegas, Ryan McNamara, Blossom Damania. |
| EPI_ISL_968189, EPI_ISL_968190, EPI_ISL_968191, EPI_ISL_968192, EPI_ISL_968193, EPI_ISL_968194, EPI_ISL_968195, EPI_ISL_968196, EPI_ISL_968197, EPI_ISL_968198, EPI_ISL_968199, EPI_ISL_968200, EPI_ISL_968201, EPI_ISL_968202, EPI_ISL_968203, EPI_ISL_968204, EPI_ISL_968205, EPI_ISL_968206, EPI_ISL_968207, EPI_ISL_968208, EPI_ISL_968209, EPI_ISL_968210, EPI_ISL_968211 | | | |
| see above | Arizona State Public Health Laboratory | Arizona State Public Health Laboratory | Trung Huynh, Jessica Escobar, Katherine Fullerton, Nobuko Fukushima, Stacy White, Linda Getsinger, Victor Waddell |
| EPI_ISL_976910, EPI_ISL_976911, EPI_ISL_976913, EPI_ISL_976915, EPI_ISL_976916, EPI_ISL_976917, EPI_ISL_976918, EPI_ISL_976919, EPI_ISL_976920, EPI_ISL_976921, EPI_ISL_976922, EPI_ISL_976923, EPI_ISL_976924, EPI_ISL_976925, EPI_ISL_976927, EPI_ISL_976928, EPI_ISL_976929, EPI_ISL_976930, EPI_ISL_976931, EPI_ISL_976933, EPI_ISL_976934, EPI_ISL_976935, EPI_ISL_976936, EPI_ISL_976937, EPI_ISL_976939 | | | |
| see above | University of Massachusetts Medical School | Infectious Disease Program, Broad Institute of Harvard and MIT | Lemieux,J.E., Siddle,K.J., Ward,D., Ellison,R., Adams,G., Gladden-Young,A., Lagerborg,K., Rudy,M., DeRuff,K., Carter,A., Normandin,E., Bauer,M., Reilly,S., Tomkins-Tinch,C., Loreth,C., Chaluvadi,S., Birren,B.W., Gallagher,G., Smole,S., Park,D.J., MacInnis,B.L., and Sabeti,P.C. |
| EPI_ISL_976941, EPI_ISL_976942, EPI_ISL_976943, EPI_ISL_976944, EPI_ISL_976945, EPI_ISL_976946, EPI_ISL_976947, EPI_ISL_976948, EPI_ISL_976949, EPI_ISL_976950, EPI_ISL_976951, EPI_ISL_976952, EPI_ISL_976953, EPI_ISL_976954, EPI_ISL_976955, EPI_ISL_976956, EPI_ISL_976957, EPI_ISL_976958, EPI_ISL_976959, EPI_ISL_976960, EPI_ISL_976961, EPI_ISL_976962, EPI_ISL_976963, EPI_ISL_976964, EPI_ISL_976965, EPI_ISL_976966, EPI_ISL_976967, EPI_ISL_976968, EPI_ISL_976969, EPI_ISL_976970, EPI_ISL_976971, EPI_ISL_976972, EPI_ISL_976973, EPI_ISL_976974, EPI_ISL_976975, EPI_ISL_976976, EPI_ISL_976977, EPI_ISL_976978, EPI_ISL_976979, EPI_ISL_976980, EPI_ISL_976983, EPI_ISL_976984, EPI_ISL_976985, EPI_ISL_976986, EPI_ISL_976987, EPI_ISL_976988, EPI_ISL_976989, EPI_ISL_976990, EPI_ISL_976991, EPI_ISL_976992, EPI_ISL_976993, EPI_ISL_976994, EPI_ISL_976995, EPI_ISL_976996, EPI_ISL_976997, EPI_ISL_976998, EPI_ISL_976999, EPI_ISL_977000, EPI_ISL_977001, EPI_ISL_977002, EPI_ISL_977003, EPI_ISL_977004, EPI_ISL_977005, EPI_ISL_977006, EPI_ISL_977007, EPI_ISL_977008, EPI_ISL_977009, EPI_ISL_977010, EPI_ISL_977011, EPI_ISL_977012, EPI_ISL_977013, EPI_ISL_977014, EPI_ISL_977015, EPI_ISL_977016, EPI_ISL_977017, EPI_ISL_977018, EPI_ISL_977019, EPI_ISL_977020, EPI_ISL_977021, EPI_ISL_977022, EPI_ISL_977023 | | | |
| see above | Broad Institute Clinical Research Sequencing Platform | Infectious Disease Program, Broad Institute of Harvard and MIT | Lemieux,J.E., Siddle,K.J., Adams,G., Gladden-Young,A., Lagerborg,K., Rudy,M., DeRuff,K., Carter,A., Normandin,E., Bauer,M., Reilly,S., Tomkins-Tinch,C., Loreth,C., Chaluvadi,S., Birren,B.W., Gallagher,G., Smole,S., Park,D.J., MacInnis,B.L., and Sabeti,P.C. |
| EPI_ISL_977026, EPI_ISL_977027, EPI_ISL_977029, EPI_ISL_977031, EPI_ISL_977032, EPI_ISL_977033, EPI_ISL_977034, EPI_ISL_977035, EPI_ISL_977036, EPI_ISL_977039, EPI_ISL_977042, EPI_ISL_977043, EPI_ISL_977044, EPI_ISL_977045, EPI_ISL_977048, EPI_ISL_977049, EPI_ISL_977050, EPI_ISL_977051, EPI_ISL_977052, EPI_ISL_977053, EPI_ISL_977054, EPI_ISL_977056, EPI_ISL_977057, EPI_ISL_977058, EPI_ISL_977061, EPI_ISL_977062, EPI_ISL_977065, EPI_ISL_977066, EPI_ISL_977067, EPI_ISL_977069, EPI_ISL_977070, EPI_ISL_977072, EPI_ISL_977073, EPI_ISL_977074, EPI_ISL_977075, EPI_ISL_977076, EPI_ISL_977077 | | | |

| | | | |
|---|---|---|---|
| see above | Rhode Island Department of Health | Infectious Disease Program, Broad Institute of Harvard and MIT | Lemieux,J.E., Siddle,K.J., Huard,R., King,E., Azevedo,K., Miller,A., Adams,G., Gladden-Young,A., Lagerborg,K., Rudy,M., DeRuff,K., Carter,A., Normandin,E., Bauer,M., Reilly,S., Tomkins-Tinch,C., Loreth,C., Chaluvadi,S., Birren,B.W., Gallagher,G., Smole,S., Park,D.J., MacInnis,B.L., and Sabeti,P.C. |
| EPI_ISL_977078, EPI_ISL_977079, EPI_ISL_977080, EPI_ISL_977081, EPI_ISL_977082, EPI_ISL_977083, EPI_ISL_977084, EPI_ISL_977088, EPI_ISL_977091, EPI_ISL_977094, EPI_ISL_977095, EPI_ISL_977097, EPI_ISL_977098, EPI_ISL_977099, EPI_ISL_977100, EPI_ISL_977101, EPI_ISL_977103, EPI_ISL_977104, EPI_ISL_977105, EPI_ISL_977106, EPI_ISL_977107, EPI_ISL_977108, EPI_ISL_977109, EPI_ISL_977112, EPI_ISL_977113, EPI_ISL_977114, EPI_ISL_977115, EPI_ISL_977119 | | | |
| see above | Massachusetts General Hospital | Infectious Disease Program, Broad Institute of Harvard and MIT | Lemieux,J.E., Siddle,K.J., Shaw,B., Adams,G., Pierce,V., Turbett,S., Anahtar,M., Branda,J., Slater,D., Harris,J., Lin,A.E., Gladden-Young,A., Lagerborg,K., Rudy,M., DeRuff,K., Carter,A., Normandin,E., Bauer,M., Reilly,S., Tomkins-Tinch,C., Loreth,C., Chaluvadi,S., Neumann,A., Cusick,C., Chapman,S.B., Gnirke,A., Flowers,K., Cerrato,F., Birren,B.W., Gallagher,G., Smole,S., Park,D.J., MacInnis,B.L., Ryan,E., LaRocque,R., Rosenberg,E. and Sabeti,P.C. |
| EPI_ISL_977120, EPI_ISL_977121, EPI_ISL_977127, EPI_ISL_977128, EPI_ISL_977129, EPI_ISL_977130, EPI_ISL_977132, EPI_ISL_977133, EPI_ISL_977134, EPI_ISL_977136, EPI_ISL_977139, EPI_ISL_977141, EPI_ISL_977142, EPI_ISL_977143, EPI_ISL_977144 | | | |
| see above | Flow Health | Infectious Disease Program, Broad Institute of Harvard and MIT | Lemieux,J.E., Siddle,K.J., Adams,G., Gladden-Young,A., Lagerborg,K., Rudy,M., DeRuff,K., Carter,A., Normandin,E., Bauer,M., Reilly,S., Tomkins-Tinch,C., Loreth,C., Chaluvadi,S., Birren,B.W., Gallagher,G., Smole,S., Park,D.J., MacInnis,B.L., and Sabeti,P.C. |
| EPI_ISL_977504, EPI_ISL_977508, EPI_ISL_977514, EPI_ISL_977515, EPI_ISL_977536 | NCSLPH | NCSLPH | Chase K, Miller MC, Greene S, Glover W |
| EPI_ISL_977566, EPI_ISL_977567, EPI_ISL_977568, EPI_ISL_977569, EPI_ISL_977570, EPI_ISL_977571, EPI_ISL_977572, EPI_ISL_977573, EPI_ISL_977574, EPI_ISL_977575, EPI_ISL_977576, EPI_ISL_977577, EPI_ISL_977578, EPI_ISL_977579, EPI_ISL_977580 | | | |
| see above | University of Michigan Clinical Microbiology Laboratory | Lauring Lab, University of Michigan, Department of Microbiology and Immunology | Valesano |
| EPI_ISL_977660, EPI_ISL_977665 | Auburn Sports Medicine | Synergy Laboratories | Megan Cornwell |
| EPI_ISL_977666 | NYU Langone Health | Departments of Pathology and Medicine, New York University School of Medicine | Adriana Heguy, Dacia Dimartino, Emily Guzman, Christian Marier, Peter Meyn, Sitharam Ramaswami, Gael Westby, Paul Zappile, Yutong Zhang, Paolo Cotzia, Guiqing Wang |
| EPI_ISL_977722, EPI_ISL_977723, EPI_ISL_977724, EPI_ISL_977725, EPI_ISL_977726, EPI_ISL_977727, EPI_ISL_977728, EPI_ISL_977729, EPI_ISL_977730, EPI_ISL_977731, EPI_ISL_977734, EPI_ISL_977738, EPI_ISL_977740, EPI_ISL_977741, EPI_ISL_977742, EPI_ISL_977745, EPI_ISL_977747, EPI_ISL_977748, EPI_ISL_977749, EPI_ISL_977750, EPI_ISL_977751, EPI_ISL_977752, EPI_ISL_977753, EPI_ISL_977754, EPI_ISL_977755, EPI_ISL_977757, EPI_ISL_977758, EPI_ISL_977759, EPI_ISL_977760, EPI_ISL_977761, EPI_ISL_977762, EPI_ISL_977763, EPI_ISL_977764, EPI_ISL_977765, EPI_ISL_977766, EPI_ISL_977767, EPI_ISL_977768, EPI_ISL_977769, EPI_ISL_977770, EPI_ISL_977771, EPI_ISL_977773, EPI_ISL_977775, EPI_ISL_977776, EPI_ISL_977777, EPI_ISL_977778, EPI_ISL_977779, EPI_ISL_977780, EPI_ISL_977781, EPI_ISL_977782, EPI_ISL_977783, EPI_ISL_977784, EPI_ISL_977785, EPI_ISL_977786, EPI_ISL_977787, EPI_ISL_977788, EPI_ISL_977789, EPI_ISL_977790, EPI_ISL_977792, EPI_ISL_977793, EPI_ISL_977794, EPI_ISL_977795, EPI_ISL_977796, EPI_ISL_977797, EPI_ISL_977798, EPI_ISL_977799, EPI_ISL_977801, EPI_ISL_977804, EPI_ISL_977805, EPI_ISL_977806, EPI_ISL_977807, EPI_ISL_977808, EPI_ISL_977809, EPI_ISL_977810, EPI_ISL_977811, EPI_ISL_977812, EPI_ISL_977813, EPI_ISL_977814, EPI_ISL_977815, EPI_ISL_977816, EPI_ISL_977817, EPI_ISL_977818, EPI_ISL_977819, EPI_ISL_977820, EPI_ISL_977821, EPI_ISL_977822, EPI_ISL_977823, EPI_ISL_977824, EPI_ISL_977825, EPI_ISL_977826, EPI_ISL_977827, EPI_ISL_977833, EPI_ISL_977837, EPI_ISL_977838, EPI_ISL_977839, EPI_ISL_977841, EPI_ISL_977842, EPI_ISL_977844, EPI_ISL_977845, EPI_ISL_977846, EPI_ISL_977847, EPI_ISL_977848, EPI_ISL_977849, EPI_ISL_977850, EPI_ISL_977851, EPI_ISL_977864, EPI_ISL_977865, EPI_ISL_977866, EPI_ISL_977867, EPI_ISL_977878 | | | |
| see above | California Department of Public Health | Chiu Laboratory, University of California, San Francisco | Charles Chiu, Xianding (Wayne) Deng, Candace Wang, Venice Servellita, Jill Hacker, Debra Wadford |
| EPI_ISL_977885, EPI_ISL_977886, EPI_ISL_977887, EPI_ISL_977888, EPI_ISL_977889, EPI_ISL_977890, EPI_ISL_977893, EPI_ISL_977894, EPI_ISL_977895, EPI_ISL_977897, EPI_ISL_977898, EPI_ISL_977899, EPI_ISL_977901, EPI_ISL_977902, EPI_ISL_977905, EPI_ISL_977906, EPI_ISL_977907, EPI_ISL_977908, EPI_ISL_977909, EPI_ISL_977912, EPI_ISL_977913, EPI_ISL_977914, EPI_ISL_977915, EPI_ISL_977916, EPI_ISL_977917, EPI_ISL_977918, EPI_ISL_977919, EPI_ISL_977920, EPI_ISL_977921, EPI_ISL_977922, EPI_ISL_977923, EPI_ISL_977924, EPI_ISL_977925, EPI_ISL_977926, EPI_ISL_977929, EPI_ISL_977930, EPI_ISL_977939, EPI_ISL_977945, EPI_ISL_977949, EPI_ISL_977950, EPI_ISL_977951, EPI_ISL_977957, EPI_ISL_977963, EPI_ISL_977964, EPI_ISL_977965, EPI_ISL_977967, EPI_ISL_977972, EPI_ISL_977974, EPI_ISL_977976, EPI_ISL_977977, EPI_ISL_977979, EPI_ISL_977982, EPI_ISL_977983 | | | |
| see above | Chiu Laboratory, University of California, San Francisco | Chiu Laboratory, University of California, San Francisco | Charles Chiu, Xianding (Wayne) Deng, Candace Wang, Venice Servellita, Jill Hacker, Debra Wadford |
| EPI_ISL_977984 | ` | Chiu Laboratory, University of California, San Francisco | Charles Chiu, Xianding (Wayne) Deng, Candace Wang, Venice Servellita, Jill Hacker, Debra Wadford |
| EPI_ISL_977985, EPI_ISL_977986, EPI_ISL_977988, EPI_ISL_977989, EPI_ISL_977991, EPI_ISL_977992, EPI_ISL_977993, EPI_ISL_977994, EPI_ISL_977996, EPI_ISL_977997, EPI_ISL_977999, EPI_ISL_978000, EPI_ISL_978001, EPI_ISL_978002, EPI_ISL_978003, EPI_ISL_978004, EPI_ISL_978006, EPI_ISL_978007, EPI_ISL_978008, EPI_ISL_978010, EPI_ISL_978012, EPI_ISL_978014, EPI_ISL_978015, EPI_ISL_978016, EPI_ISL_978017, EPI_ISL_978018, EPI_ISL_978019, EPI_ISL_978020, EPI_ISL_978021, EPI_ISL_978022, EPI_ISL_978023, EPI_ISL_978024, EPI_ISL_978025, EPI_ISL_978026, EPI_ISL_978027, EPI_ISL_978028, EPI_ISL_978030, EPI_ISL_978031, EPI_ISL_978033, EPI_ISL_978035, EPI_ISL_978036, EPI_ISL_978037, EPI_ISL_978038, EPI_ISL_978040, EPI_ISL_978041, EPI_ISL_978042, EPI_ISL_978043, EPI_ISL_978047, EPI_ISL_978048, EPI_ISL_978050, EPI_ISL_978053, EPI_ISL_978055, EPI_ISL_978057, EPI_ISL_978058, EPI_ISL_978059, EPI_ISL_978060, EPI_ISL_978061, EPI_ISL_978062, EPI_ISL_978063, EPI_ISL_978064, EPI_ISL_978065, EPI_ISL_978067, EPI_ISL_978068, EPI_ISL_978071, EPI_ISL_978073, EPI_ISL_978074, EPI_ISL_978075, EPI_ISL_978077, EPI_ISL_978078, EPI_ISL_978079, EPI_ISL_978081, EPI_ISL_978082, EPI_ISL_978083, EPI_ISL_978084, EPI_ISL_978087, EPI_ISL_978088, EPI_ISL_978089, EPI_ISL_978091, EPI_ISL_978092, EPI_ISL_978093, EPI_ISL_978095, EPI_ISL_978096, EPI_ISL_978097, EPI_ISL_978099, EPI_ISL_978100, EPI_ISL_978101, EPI_ISL_978102, EPI_ISL_978103, EPI_ISL_978105, EPI_ISL_978107, EPI_ISL_978108, EPI_ISL_978109, EPI_ISL_978110, EPI_ISL_978111, EPI_ISL_978112, EPI_ISL_978113, EPI_ISL_978115, EPI_ISL_978116, EPI_ISL_978117, EPI_ISL_978118, EPI_ISL_978119, EPI_ISL_978120, EPI_ISL_978122, EPI_ISL_978123, EPI_ISL_978124, EPI_ISL_978125, EPI_ISL_978126, EPI_ISL_978127, EPI_ISL_978128, EPI_ISL_978129, EPI_ISL_978130, EPI_ISL_978131, EPI_ISL_978132, EPI_ISL_978133, EPI_ISL_978134, EPI_ISL_978135, EPI_ISL_978136, EPI_ISL_978137, EPI_ISL_978138, EPI_ISL_978139, EPI_ISL_978140, EPI_ISL_978141, EPI_ISL_978142, EPI_ISL_978144, EPI_ISL_978147, EPI_ISL_978148, EPI_ISL_978150, EPI_ISL_978154, EPI_ISL_978155, EPI_ISL_978158, EPI_ISL_978159, EPI_ISL_978163, EPI_ISL_978164, EPI_ISL_978165, EPI_ISL_978166, EPI_ISL_978167, EPI_ISL_978168 | | | |
| see above | Chiu Laboratory, University of California, San Francisco | Chiu Laboratory, University of California, San Francisco | Charles Chiu, Xianding (Wayne) Deng, Candace Wang, Venice Servellita, Jill Hacker, Debra Wadford |
| EPI_ISL_978169, EPI_ISL_978170, EPI_ISL_978171, EPI_ISL_978172, EPI_ISL_978173, EPI_ISL_978174, EPI_ISL_978175, EPI_ISL_978176, EPI_ISL_978177, EPI_ISL_978178, EPI_ISL_978179, EPI_ISL_978180, EPI_ISL_978181, EPI_ISL_978182, EPI_ISL_978183, EPI_ISL_978184, EPI_ISL_978185, EPI_ISL_978186, EPI_ISL_978187, EPI_ISL_978188, EPI_ISL_978189, EPI_ISL_978190, EPI_ISL_978191, EPI_ISL_978192, EPI_ISL_978193, EPI_ISL_978194, EPI_ISL_978195, EPI_ISL_978196, EPI_ISL_978197, EPI_ISL_978198, EPI_ISL_978199, EPI_ISL_978200, EPI_ISL_978201, EPI_ISL_978202, EPI_ISL_978203, EPI_ISL_978204, EPI_ISL_978205, EPI_ISL_978206, EPI_ISL_978207, EPI_ISL_978208, EPI_ISL_978209, EPI_ISL_978210, EPI_ISL_978211, EPI_ISL_978212, EPI_ISL_978213, EPI_ISL_978214, EPI_ISL_978215, EPI_ISL_978216, EPI_ISL_978217, EPI_ISL_978218, EPI_ISL_978219, EPI_ISL_978220, EPI_ISL_978221, EPI_ISL_978222, EPI_ISL_978223, EPI_ISL_978224, EPI_ISL_978225, EPI_ISL_978226 | | | |
| see above | Virginia Division of Consolidated Laboratory Services | Virginia Division of Consolidated Laboratory Services | Virginia DCLS |
| EPI_ISL_978227, EPI_ISL_978229, EPI_ISL_978230, EPI_ISL_978232, EPI_ISL_978233, EPI_ISL_978234, EPI_ISL_978235, EPI_ISL_978236, EPI_ISL_978237, EPI_ISL_978238, EPI_ISL_978239, EPI_ISL_978240, EPI_ISL_978241, EPI_ISL_978242, EPI_ISL_978243, EPI_ISL_978244, EPI_ISL_978245, EPI_ISL_978246, EPI_ISL_978247, EPI_ISL_978248, EPI_ISL_978249, EPI_ISL_978251, EPI_ISL_978252, EPI_ISL_978255, EPI_ISL_978256, EPI_ISL_978257, EPI_ISL_978261, EPI_ISL_978262, EPI_ISL_978264, EPI_ISL_978265, EPI_ISL_978266, EPI_ISL_978267, EPI_ISL_978268, EPI_ISL_978269, EPI_ISL_978270, EPI_ISL_978271, EPI_ISL_978274, EPI_ISL_978275, EPI_ISL_978276, EPI_ISL_978277, EPI_ISL_978278, EPI_ISL_978279, EPI_ISL_978280, EPI_ISL_978283, EPI_ISL_978284, EPI_ISL_978286, EPI_ISL_978287, EPI_ISL_978288, EPI_ISL_978289, EPI_ISL_978290, EPI_ISL_978291, EPI_ISL_978292, EPI_ISL_978295, EPI_ISL_978296, EPI_ISL_978298, EPI_ISL_978299, EPI_ISL_978300, EPI_ISL_978301, EPI_ISL_978303, EPI_ISL_978304, EPI_ISL_978306, EPI_ISL_978307, EPI_ISL_978308, EPI_ISL_978309, EPI_ISL_978311, EPI_ISL_978312, EPI_ISL_978313, EPI_ISL_978314, EPI_ISL_978315, EPI_ISL_978316, EPI_ISL_978317, EPI_ISL_978318, EPI_ISL_978319, EPI_ISL_978320, EPI_ISL_978322, EPI_ISL_978323, EPI_ISL_978325, EPI_ISL_978326, EPI_ISL_978327, EPI_ISL_978329, EPI_ISL_978330, EPI_ISL_978331, EPI_ISL_978332, EPI_ISL_978333, EPI_ISL_978334, EPI_ISL_978335, EPI_ISL_978336, EPI_ISL_978337, EPI_ISL_978338, EPI_ISL_978339, EPI_ISL_978340, EPI_ISL_978341, EPI_ISL_978343, EPI_ISL_978344, EPI_ISL_978345, EPI_ISL_978346, EPI_ISL_978347, EPI_ISL_978350, EPI_ISL_978352 | | | |
| see above | Texas Department of State Health Services | Texas Department of State Health Services | Bonnie Oh, Anita Pokharel, James Daniel Bonser, Myong Koag, Chung Wang, Rachel Lee, Grace Kubin, Rashmi Tuladhar, Mayela Pedrueza, Maliha Rahman, Jenny Zhang |
| EPI_ISL_978353, EPI_ISL_978354, EPI_ISL_978355, EPI_ISL_978357, EPI_ISL_978358, EPI_ISL_978359, EPI_ISL_978360, EPI_ISL_978361, EPI_ISL_978362, EPI_ISL_978363, EPI_ISL_978364, EPI_ISL_978365, EPI_ISL_978366, EPI_ISL_978367, EPI_ISL_978368, EPI_ISL_978369, EPI_ISL_978370, EPI_ISL_978371, EPI_ISL_978372, EPI_ISL_978373, EPI_ISL_978374, EPI_ISL_978375, EPI_ISL_978376, EPI_ISL_978377, EPI_ISL_978378, EPI_ISL_978379, EPI_ISL_978380, EPI_ISL_978381, EPI_ISL_978382, EPI_ISL_978383, EPI_ISL_978384, EPI_ISL_978385, EPI_ISL_978386, EPI_ISL_978387, EPI_ISL_978388, EPI_ISL_978389, EPI_ISL_978391, EPI_ISL_978392, EPI_ISL_978393, EPI_ISL_978394, EPI_ISL_978395, EPI_ISL_978397, EPI_ISL_978398, EPI_ISL_978399, EPI_ISL_978400, EPI_ISL_978401, EPI_ISL_978402, EPI_ISL_978403, EPI_ISL_978404, EPI_ISL_978406, EPI_ISL_978407, EPI_ISL_978409, EPI_ISL_978410, EPI_ISL_978411, EPI_ISL_978414, EPI_ISL_978416, EPI_ISL_978417, EPI_ISL_978418, EPI_ISL_978423, EPI_ISL_978425, EPI_ISL_978427, EPI_ISL_978429, EPI_ISL_978431, EPI_ISL_978432, EPI_ISL_978433, EPI_ISL_978434, EPI_ISL_978435, EPI_ISL_978436, EPI_ISL_978437, EPI_ISL_978438, EPI_ISL_978439, EPI_ISL_978440, EPI_ISL_978441, EPI_ISL_978442, EPI_ISL_978443, EPI_ISL_978444, EPI_ISL_978445, EPI_ISL_978446, EPI_ISL_978447, EPI_ISL_978448, EPI_ISL_978449, EPI_ISL_978450, EPI_ISL_978451, EPI_ISL_978452, EPI_ISL_978453, EPI_ISL_978454, EPI_ISL_978455, EPI_ISL_978456, EPI_ISL_978457, EPI_ISL_978458, EPI_ISL_978459, EPI_ISL_978460, EPI_ISL_978461, EPI_ISL_978462, EPI_ISL_978463, EPI_ISL_978464, EPI_ISL_978465, EPI_ISL_978466, EPI_ISL_978467, EPI_ISL_978468, EPI_ISL_978469, EPI_ISL_978470, EPI_ISL_978471, EPI_ISL_978472, EPI_ISL_978473, EPI_ISL_978474, EPI_ISL_978475, EPI_ISL_978476, EPI_ISL_978477, EPI_ISL_978478, EPI_ISL_978479, EPI_ISL_978480, EPI_ISL_978481, EPI_ISL_978482, EPI_ISL_978483, EPI_ISL_978484, EPI_ISL_978485, EPI_ISL_978486, EPI_ISL_978487 | | | |
| see above | Arizona State Public Health Laboratory | Arizona State Public Health Laboratory | Trung Huynh, Jessica Escobar, Katherine Fullerton, Nobuko Fukushima, Stacy White, Linda Getsinger, Victor Waddell |
| EPI_ISL_978553 | Kansas Health and Environmental Lab | Kansas Health and Environmental Lab | Mike Grose, Paige Drury, Carissa Robertson, Ben Olsen, and Phil Adam |
| EPI_ISL_978555, EPI_ISL_978556, EPI_ISL_978557, EPI_ISL_978558, EPI_ISL_978559, EPI_ISL_978560, EPI_ISL_978561, EPI_ISL_978562, EPI_ISL_978563, EPI_ISL_978564, EPI_ISL_978565, EPI_ISL_978566, EPI_ISL_978567, EPI_ISL_978568, EPI_ISL_978569, EPI_ISL_978570, EPI_ISL_978571, EPI_ISL_978572, EPI_ISL_978573, EPI_ISL_978574, EPI_ISL_978575, EPI_ISL_978576, EPI_ISL_978577, EPI_ISL_978578, EPI_ISL_978579, EPI_ISL_978580, EPI_ISL_978581, EPI_ISL_978582, EPI_ISL_978583, EPI_ISL_978584, EPI_ISL_978585, EPI_ISL_978586, EPI_ISL_978587, EPI_ISL_978588, EPI_ISL_978589, EPI_ISL_978590, EPI_ISL_978591, EPI_ISL_978592, EPI_ISL_978593, EPI_ISL_978594, EPI_ISL_978595, EPI_ISL_978596, EPI_ISL_978597, EPI_ISL_978598, EPI_ISL_978599, EPI_ISL_978600, EPI_ISL_978601, EPI_ISL_978602, EPI_ISL_978603, EPI_ISL_978604, EPI_ISL_978605, EPI_ISL_978606, EPI_ISL_978607, EPI_ISL_978608, EPI_ISL_978609, EPI_ISL_978610, EPI_ISL_978611, EPI_ISL_978612, EPI_ISL_978613, EPI_ISL_978614, EPI_ISL_978615, EPI_ISL_978616, EPI_ISL_978617, EPI_ISL_978618, EPI_ISL_978619, EPI_ISL_978620, EPI_ISL_978621, EPI_ISL_978622, EPI_ISL_978623, EPI_ISL_978624, EPI_ISL_978625, EPI_ISL_978626, EPI_ISL_978627, EPI_ISL_978628, EPI_ISL_978629, EPI_ISL_978630, EPI_ISL_978631, EPI_ISL_978632, EPI_ISL_978633, EPI_ISL_978634, EPI_ISL_978635, EPI_ISL_978636, EPI_ISL_978637, EPI_ISL_978638, EPI_ISL_978639, EPI_ISL_978640, EPI_ISL_978641, EPI_ISL_978642, EPI_ISL_978643, EPI_ISL_978644, EPI_ISL_978645, EPI_ISL_978646, EPI_ISL_978647, EPI_ISL_978648, EPI_ISL_978649, EPI_ISL_978650, EPI_ISL_978651, EPI_ISL_978653, EPI_ISL_978654, EPI_ISL_978655, EPI_ISL_978656, EPI_ISL_978657, EPI_ISL_978658, EPI_ISL_978660, EPI_ISL_978661, EPI_ISL_978662, EPI_ISL_978663, EPI_ISL_978664, EPI_ISL_978665, EPI_ISL_978666, EPI_ISL_978667, EPI_ISL_978668, EPI_ISL_978669, EPI_ISL_978670, EPI_ISL_978671, EPI_ISL_978672, EPI_ISL_978673, EPI_ISL_978674, EPI_ISL_978675, EPI_ISL_978676, EPI_ISL_978677, EPI_ISL_978678, EPI_ISL_978679, EPI_ISL_978680, EPI_ISL_978681, EPI_ISL_978682, EPI_ISL_978683, EPI_ISL_978684, EPI_ISL_978685, EPI_ISL_978686, EPI_ISL_978687, EPI_ISL_978690, EPI_ISL_978691, EPI_ISL_978692, EPI_ISL_978693, EPI_ISL_978694, EPI_ISL_978695, EPI_ISL_978696, EPI_ISL_978697, EPI_ISL_978699, EPI_ISL_978700, EPI_ISL_978701, EPI_ISL_978702, EPI_ISL_978703, EPI_ISL_978704, EPI_ISL_978705, EPI_ISL_978706, EPI_ISL_978707, EPI_ISL_978708, EPI_ISL_978709, EPI_ISL_978710, EPI_ISL_978711, EPI_ISL_978713, EPI_ISL_978714, EPI_ISL_978715, EPI_ISL_978716, EPI_ISL_978717, EPI_ISL_978718, EPI_ISL_978719, EPI_ISL_978720, EPI_ISL_978721, EPI_ISL_978722, EPI_ISL_978723, EPI_ISL_978724, EPI_ISL_978725, EPI_ISL_978726, EPI_ISL_978727, EPI_ISL_978728, EPI_ISL_978729, EPI_ISL_978730, EPI_ISL_978731, EPI_ISL_978732, EPI_ISL_978733, EPI_ISL_978735, EPI_ISL_978736, EPI_ISL_978737, EPI_ISL_978739, EPI_ISL_978740, EPI_ISL_978741, EPI_ISL_978742, EPI_ISL_978743, EPI_ISL_978744, EPI_ISL_978745, EPI_ISL_978746, EPI_ISL_978747, EPI_ISL_978748, EPI_ISL_978749, EPI_ISL_978750, EPI_ISL_978751, EPI_ISL_978752, EPI_ISL_978753, EPI_ISL_978754, EPI_ISL_978755, EPI_ISL_978756, EPI_ISL_978757, EPI_ISL_978759, EPI_ISL_978760, EPI_ISL_978761, EPI_ISL_978762, EPI_ISL_978763, EPI_ISL_978764, EPI_ISL_978765, EPI_ISL_978766, EPI_ISL_978767, EPI_ISL_978768, EPI_ISL_978769, EPI_ISL_978770, EPI_ISL_978772, EPI_ISL_978773, EPI_ISL_978775, EPI_ISL_978776, EPI_ISL_978777, EPI_ISL_978778, EPI_ISL_978779, EPI_ISL_978780, EPI_ISL_978781, EPI_ISL_978782, EPI_ISL_978783, EPI_ISL_978784, EPI_ISL_978785, EPI_ISL_978786, EPI_ISL_978787 | | | |
| see above | Helix/Illumina | Respiratory Viruses Branch, Division of Viral Diseases, Centers for Disease Control and Prevention | Peter W. Cook,Dakota Howard,Dhwani Batra,Ben L. Rambo-Martin,Eileen de Feo,Jan Antico,Christine Tran,Matthew Tolentino,Shannon Wickline,Kim Gietzen,Brad Sickler,Jingtao Liu,Eric Allen,Phil Febbo,Summer Galloway,Nicole L. Washington,Simon White,Geraint Levan,Kelly Schiabor Barrett,Elizabeth Cirulli,Alexandre Bolze,Ary Ascencio,Charlotte Rivera-Garcia,Ryan Cho,Jason Nguyen,Sherry Wang,Jimmy Ramirez,Tyler Cassens,Efren Sandoval,Magnus Isaksson,William Lee,David Becker,Marc Laurent,James Lu,Clinton R. Paden,Suxiang Tong,Duncan MacCannell, |
| EPI_ISL_978955, EPI_ISL_978957, EPI_ISL_978958, EPI_ISL_978959, EPI_ISL_978962, EPI_ISL_978963, EPI_ISL_978964, EPI_ISL_978965, EPI_ISL_978967, EPI_ISL_978968, EPI_ISL_978970, EPI_ISL_978971, EPI_ISL_978972, EPI_ISL_978973 | | | |
| see above | Chiu Laboratory, University of California, San Francisco | Chiu Laboratory, University of California, San Francisco | Charles Chiu, Xianding (Wayne) Deng, Candace Wang, Venice Servellita, Jill Hacker, Debra Wadford |
| EPI_ISL_978977, EPI_ISL_978978, EPI_ISL_978979, EPI_ISL_978980, EPI_ISL_978981, EPI_ISL_978982 | California Department of Public Health | Chiu Laboratory, University of California, San Francisco | Charles Chiu, Xianding (Wayne) Deng, Candace Wang, Venice Servellita, Jill Hacker, Debra Wadford |
| EPI_ISL_978989, EPI_ISL_978990, EPI_ISL_978991, EPI_ISL_978992, EPI_ISL_978993, EPI_ISL_978994, EPI_ISL_978995, EPI_ISL_978996, EPI_ISL_978997, EPI_ISL_978998, EPI_ISL_978999, EPI_ISL_979000, EPI_ISL_979001, EPI_ISL_979002, EPI_ISL_979003, EPI_ISL_979004, EPI_ISL_979005, EPI_ISL_979006, EPI_ISL_979007, EPI_ISL_979008, EPI_ISL_979009, EPI_ISL_979010, EPI_ISL_979011, EPI_ISL_979012, EPI_ISL_979013, EPI_ISL_979014, EPI_ISL_979015, EPI_ISL_979016, EPI_ISL_979017, EPI_ISL_979018, EPI_ISL_979019, EPI_ISL_979020, EPI_ISL_979021, EPI_ISL_979022, EPI_ISL_979023, EPI_ISL_979024, EPI_ISL_979025, EPI_ISL_979026, EPI_ISL_979027, EPI_ISL_979028, EPI_ISL_979029, EPI_ISL_979030, EPI_ISL_979031, EPI_ISL_979032, EPI_ISL_979033, EPI_ISL_979034, EPI_ISL_979035, EPI_ISL_979036, EPI_ISL_979037, EPI_ISL_979038, EPI_ISL_979039, EPI_ISL_979040, EPI_ISL_979041, EPI_ISL_979042, EPI_ISL_979043, EPI_ISL_979044, EPI_ISL_979045, EPI_ISL_979046, EPI_ISL_979047, EPI_ISL_979048, EPI_ISL_979049, EPI_ISL_979050, EPI_ISL_979051, EPI_ISL_979052, EPI_ISL_979054, EPI_ISL_979055, EPI_ISL_979056, EPI_ISL_979057, EPI_ISL_979058, EPI_ISL_979059, EPI_ISL_979060, EPI_ISL_979061, EPI_ISL_979062, EPI_ISL_979063, EPI_ISL_979064, EPI_ISL_979065, EPI_ISL_979066, EPI_ISL_979067, EPI_ISL_979069, EPI_ISL_979070, EPI_ISL_979071, EPI_ISL_979072, EPI_ISL_979073, EPI_ISL_979074, EPI_ISL_979075, EPI_ISL_979076, EPI_ISL_979077, EPI_ISL_979078, EPI_ISL_979079, EPI_ISL_979080, EPI_ISL_979081, EPI_ISL_979082, EPI_ISL_979083, EPI_ISL_979084, EPI_ISL_979085, EPI_ISL_979086, EPI_ISL_979087, EPI_ISL_979088, EPI_ISL_979089, EPI_ISL_979090, EPI_ISL_979091, EPI_ISL_979092, EPI_ISL_979093, EPI_ISL_979094, EPI_ISL_979095, EPI_ISL_979096, EPI_ISL_979097, EPI_ISL_979098, EPI_ISL_979099, EPI_ISL_979100, EPI_ISL_979101, EPI_ISL_979102, EPI_ISL_979103, EPI_ISL_979104, EPI_ISL_979105, EPI_ISL_979106, EPI_ISL_979107, EPI_ISL_979108, EPI_ISL_979109, EPI_ISL_979110, EPI_ISL_979111, EPI_ISL_979112, EPI_ISL_979113, EPI_ISL_979114, EPI_ISL_979115, EPI_ISL_979116, EPI_ISL_979117, EPI_ISL_979118, EPI_ISL_979119, EPI_ISL_979120, EPI_ISL_979121, EPI_ISL_979122, EPI_ISL_979123, EPI_ISL_979124, EPI_ISL_979125, EPI_ISL_979126, EPI_ISL_979127, EPI_ISL_979128, EPI_ISL_979129, EPI_ISL_979130, EPI_ISL_979131, EPI_ISL_979132, EPI_ISL_979133, EPI_ISL_979134, EPI_ISL_979135, EPI_ISL_979136, EPI_ISL_979137, EPI_ISL_979138, EPI_ISL_979139, EPI_ISL_979140, EPI_ISL_979141, EPI_ISL_979142, EPI_ISL_979143, EPI_ISL_979144, EPI_ISL_979145, EPI_ISL_979146, EPI_ISL_979147, EPI_ISL_979148, EPI_ISL_979149, EPI_ISL_979150, EPI_ISL_979151, EPI_ISL_979152, EPI_ISL_979154, EPI_ISL_979155, EPI_ISL_979156, EPI_ISL_979157, EPI_ISL_979158 | | | |
| see above | Santa Clara County Public Health Laboratory | Chan-Zuckerberg Biohub | CZB Cliahub Consortium |
| EPI_ISL_979159, EPI_ISL_979160, EPI_ISL_979161 | County of San Luis Obispo Public Health Laboratory | Chan-Zuckerberg Biohub | CZB Cliahub Consortium |

| | | | |
|---|---|---|---|
| EPI_ISL_979162, EPI_ISL_979163, EPI_ISL_979164, EPI_ISL_979165, EPI_ISL_979166, EPI_ISL_979167, EPI_ISL_979168, EPI_ISL_979169, EPI_ISL_979170, EPI_ISL_979171, EPI_ISL_979172, EPI_ISL_979175, EPI_ISL_979176, EPI_ISL_979177, EPI_ISL_979178, EPI_ISL_979179, EPI_ISL_979180, EPI_ISL_979181, EPI_ISL_979182, EPI_ISL_979183, EPI_ISL_979184, EPI_ISL_979185, EPI_ISL_979186, EPI_ISL_979187, EPI_ISL_979188, EPI_ISL_979189, EPI_ISL_979190, EPI_ISL_979191, EPI_ISL_979192, EPI_ISL_979193, EPI_ISL_979194, EPI_ISL_979195, EPI_ISL_979196, EPI_ISL_979197, EPI_ISL_979198, EPI_ISL_979199, EPI_ISL_979200, EPI_ISL_979201, EPI_ISL_979202, EPI_ISL_979203, EPI_ISL_979204, EPI_ISL_979206, EPI_ISL_979207, EPI_ISL_979208, EPI_ISL_979209, EPI_ISL_979210, EPI_ISL_979211, EPI_ISL_979212, EPI_ISL_979213, EPI_ISL_979214, EPI_ISL_979215, EPI_ISL_979216, EPI_ISL_979217, EPI_ISL_979218, EPI_ISL_979219, EPI_ISL_979220, EPI_ISL_979221, EPI_ISL_979222, EPI_ISL_979223, EPI_ISL_979224, EPI_ISL_979225, EPI_ISL_979226, EPI_ISL_979227, EPI_ISL_979228, EPI_ISL_979229, EPI_ISL_979230, EPI_ISL_979231, EPI_ISL_979233, EPI_ISL_979234, EPI_ISL_979235, EPI_ISL_979236, EPI_ISL_979237, EPI_ISL_979238, EPI_ISL_979239, EPI_ISL_979240, EPI_ISL_979241, EPI_ISL_979242, EPI_ISL_979243, EPI_ISL_979244, EPI_ISL_979245 | | | |
| see above | Humboldt County Public Health Laboratory | Chan-Zuckerberg Biohub | CZB Cliahub Consortium |
| EPI_ISL_979327, EPI_ISL_979328, EPI_ISL_979329, EPI_ISL_979330, EPI_ISL_979331, EPI_ISL_979332, EPI_ISL_979333, EPI_ISL_979334, EPI_ISL_979335, EPI_ISL_979336, EPI_ISL_979337, EPI_ISL_979338, EPI_ISL_979339, EPI_ISL_979340, EPI_ISL_979341, EPI_ISL_979342, EPI_ISL_979343, EPI_ISL_979344, EPI_ISL_979345, EPI_ISL_979346, EPI_ISL_979347, EPI_ISL_979348, EPI_ISL_979349, EPI_ISL_979350, EPI_ISL_979351, EPI_ISL_979352 | | | |
| see above | Laboratorio Estatal de Salud Pública de Nuevo León | Laboratorio de Infectología Molecular, Departamento de Bioquímica y Medicina Molecular,Facultad de Medicina - Universidad Autónoma de Nuevo León | Kame A. Galán-Huerta, María F. Herrera-Saldivar, Natalia Martínez-Acuña, Sonia A. Lozano-Sepúlveda, Daniel Arellanos-Soto, Ana M. Rivas-Estilla, Samuel Buentello-Wong, Else del Carmen García-García, Gloria A. Jasso-de-la-Peña, Roberto Montes-de-Oca, Consuelo Treviño-Garza, Manuel E. de-la-O-Cavazos |
| EPI_ISL_979366, EPI_ISL_979367, EPI_ISL_979368, EPI_ISL_979369, EPI_ISL_979370, EPI_ISL_979371, EPI_ISL_979372, EPI_ISL_979373, EPI_ISL_979374, EPI_ISL_979375, EPI_ISL_979376, EPI_ISL_979377, EPI_ISL_979378 | | | |
| see above | The Jackson Laboratory | The Jackson Laboratory | Lloyd M, Sanderson B, Srivastava A, Maurya R, Renzette N, Omerza G, Kelly K, Li L, Wei C L, Adams M |
| EPI_ISL_979379, EPI_ISL_979380, EPI_ISL_979381, EPI_ISL_979382, EPI_ISL_979383, EPI_ISL_979384, EPI_ISL_979385, EPI_ISL_979386, EPI_ISL_979387, EPI_ISL_979388, EPI_ISL_979389, EPI_ISL_979390, EPI_ISL_979391, EPI_ISL_979392, EPI_ISL_979393, EPI_ISL_979394, EPI_ISL_979395, EPI_ISL_979396, EPI_ISL_979397, EPI_ISL_979398, EPI_ISL_979399, EPI_ISL_979400, EPI_ISL_979401, EPI_ISL_979402, EPI_ISL_979403, EPI_ISL_979404, EPI_ISL_979405, EPI_ISL_979406, EPI_ISL_979407, EPI_ISL_979408, EPI_ISL_979409 | | | |
| see above | Houston Health Dept. | Houston Health Dept. | Ryker Penn, Pamela Brown, Adolpho Lara |
| EPI_ISL_979410, EPI_ISL_979411, EPI_ISL_979412, EPI_ISL_979413, EPI_ISL_979414, EPI_ISL_979415, EPI_ISL_979416, EPI_ISL_979417, EPI_ISL_979418, EPI_ISL_979419, EPI_ISL_979420, EPI_ISL_979421, EPI_ISL_979423, EPI_ISL_979424, EPI_ISL_979425, EPI_ISL_979426, EPI_ISL_979427, EPI_ISL_979428, EPI_ISL_979429, EPI_ISL_979431, EPI_ISL_979432, EPI_ISL_979433, EPI_ISL_979434, EPI_ISL_979435, EPI_ISL_979436, EPI_ISL_979437, EPI_ISL_979438, EPI_ISL_979439, EPI_ISL_979440, EPI_ISL_979441, EPI_ISL_979442, EPI_ISL_979443, EPI_ISL_979444, EPI_ISL_979446, EPI_ISL_979447, EPI_ISL_979448, EPI_ISL_979449, EPI_ISL_979450, EPI_ISL_979451, EPI_ISL_979452 | | | |
| see above | New Mexico Department of Health Scientific Laboratory | New Mexico Department of Health Scientific Laboratory | Ellie Johnson, Anastacia Griego-Fisher, D'eldra Malone, Jennifer Benoit |
| EPI_ISL_979453, EPI_ISL_979454 | University of Iowa Hospitals and Clinics | State Hygienic Laboratory at the University of Iowa | Valerie Reeb, Wes Hottel, Alankar Kampoowale |
| EPI_ISL_979455 | Test Iowa | State Hygienic Laboratory at the University of Iowa | Valerie Reeb, Wes Hottel, Alankar Kampoowale |
| EPI_ISL_979456 | University of Iowa Hospitals and Clinics | State Hygienic Laboratory at the University of Iowa | Valerie Reeb, Wes Hottel, Alankar Kampoowale |
| EPI_ISL_979457 | State Hygienic Laboratory at the University of Iowa | State Hygienic Laboratory at the University of Iowa | Valerie Reeb, Erik Twait, Wes Hottel, Alankar Kampoowale |
| EPI_ISL_979458 | Test Iowa | State Hygienic Laboratory at the University of Iowa | Valerie Reeb, Erik Twait, Wes Hottel, Alankar Kampoowale |
| EPI_ISL_979459, EPI_ISL_979460, EPI_ISL_979461, EPI_ISL_979462, EPI_ISL_979463, EPI_ISL_979464, EPI_ISL_979465, EPI_ISL_979466, EPI_ISL_979467, EPI_ISL_979468, EPI_ISL_979469, EPI_ISL_979470, EPI_ISL_979471, EPI_ISL_979472, EPI_ISL_979473, EPI_ISL_979474, EPI_ISL_979475, EPI_ISL_979476, EPI_ISL_979477, EPI_ISL_979478, EPI_ISL_979479, EPI_ISL_979480, EPI_ISL_979481, EPI_ISL_979482, EPI_ISL_979483, EPI_ISL_979484, EPI_ISL_979485, EPI_ISL_979486, EPI_ISL_979488, EPI_ISL_979489, EPI_ISL_979490, EPI_ISL_979491, EPI_ISL_979492, EPI_ISL_979493, EPI_ISL_979494, EPI_ISL_979495, EPI_ISL_979496, EPI_ISL_979497, EPI_ISL_979498, EPI_ISL_979500, EPI_ISL_979501, EPI_ISL_979502, EPI_ISL_979503, EPI_ISL_979504, EPI_ISL_979505, EPI_ISL_979506, EPI_ISL_979507, EPI_ISL_979508, EPI_ISL_979509, EPI_ISL_979510, EPI_ISL_979511, EPI_ISL_979512, EPI_ISL_979513, EPI_ISL_979514, EPI_ISL_979515, EPI_ISL_979516, EPI_ISL_979517, EPI_ISL_979518, EPI_ISL_979519, EPI_ISL_979520, EPI_ISL_979521, EPI_ISL_979522, EPI_ISL_979523, EPI_ISL_979524, EPI_ISL_979525, EPI_ISL_979526, EPI_ISL_979527, EPI_ISL_979528, EPI_ISL_979529, EPI_ISL_979530, EPI_ISL_979531, EPI_ISL_979532, EPI_ISL_979533, EPI_ISL_979534, EPI_ISL_979535, EPI_ISL_979536, EPI_ISL_979537, EPI_ISL_979538, EPI_ISL_979539, EPI_ISL_979540, EPI_ISL_979541, EPI_ISL_979542, EPI_ISL_979543 | | | |
| see above | Eurofins Diatherix | Hudsonalpha Genome Sequencing Center | Jane Grimwood, Melissa Williams, Lori H. Handley, Joshua Stough, Leslie Malone, Stefan Brzezinski, Ada Stewart, Teresa Jones, Jenell Webber, John Lovell, Jennifer Cart, and Jeremy Schmutz |
| EPI_ISL_979545, EPI_ISL_979546, EPI_ISL_979547, EPI_ISL_979548, EPI_ISL_979549, EPI_ISL_979550, EPI_ISL_979551, EPI_ISL_979552, EPI_ISL_979553, EPI_ISL_979554, EPI_ISL_979555, EPI_ISL_979556, EPI_ISL_979557, EPI_ISL_979558, EPI_ISL_979559, EPI_ISL_979560, EPI_ISL_979561, EPI_ISL_979562, EPI_ISL_979563, EPI_ISL_979564, EPI_ISL_979565, EPI_ISL_979566, EPI_ISL_979567, EPI_ISL_979568, EPI_ISL_979569, EPI_ISL_979570, EPI_ISL_979571, EPI_ISL_979572, EPI_ISL_979573, EPI_ISL_979574, EPI_ISL_979575, EPI_ISL_979576, EPI_ISL_979577, EPI_ISL_979578, EPI_ISL_979579, EPI_ISL_979580, EPI_ISL_979581, EPI_ISL_979582, EPI_ISL_979583, EPI_ISL_979584, EPI_ISL_979585, EPI_ISL_979586, EPI_ISL_979587, EPI_ISL_979588, EPI_ISL_979589, EPI_ISL_979590, EPI_ISL_979592, EPI_ISL_979593, EPI_ISL_979594, EPI_ISL_979595, EPI_ISL_979596, EPI_ISL_979597, EPI_ISL_979598, EPI_ISL_979599, EPI_ISL_979600, EPI_ISL_979601, EPI_ISL_979602, EPI_ISL_979603, EPI_ISL_979604, EPI_ISL_979605, EPI_ISL_979606, EPI_ISL_979607, EPI_ISL_979608, EPI_ISL_979609, EPI_ISL_979610, EPI_ISL_979611, EPI_ISL_979612, EPI_ISL_979613, EPI_ISL_979614, EPI_ISL_979615, EPI_ISL_979616, EPI_ISL_979617, EPI_ISL_979618, EPI_ISL_979619, EPI_ISL_979620, EPI_ISL_979621, EPI_ISL_979622, EPI_ISL_979623, EPI_ISL_979624, EPI_ISL_979625, EPI_ISL_979626, EPI_ISL_979627, EPI_ISL_979628, EPI_ISL_979629, EPI_ISL_979630, EPI_ISL_979631, EPI_ISL_979632, EPI_ISL_979633, EPI_ISL_979634, EPI_ISL_979635, EPI_ISL_979636, EPI_ISL_979637, EPI_ISL_979638, EPI_ISL_979639, EPI_ISL_979640, EPI_ISL_979641, EPI_ISL_979642 | | | |
| see above | Santa Clara County Public Health Laboratory | Chan-Zuckerberg Biohub | CZB Cliahub Consortium |
| EPI_ISL_979643, EPI_ISL_979644, EPI_ISL_979645, EPI_ISL_979646, EPI_ISL_979647, EPI_ISL_979648, EPI_ISL_979649, EPI_ISL_979650, EPI_ISL_979651, EPI_ISL_979652, EPI_ISL_979653, EPI_ISL_979654, EPI_ISL_979655, EPI_ISL_979656, EPI_ISL_979657, EPI_ISL_979658, EPI_ISL_979659, EPI_ISL_979660, EPI_ISL_979661, EPI_ISL_979662, EPI_ISL_979663, EPI_ISL_979664, EPI_ISL_979665, EPI_ISL_979666, EPI_ISL_979667, EPI_ISL_979668, EPI_ISL_979669, EPI_ISL_979670, EPI_ISL_979671, EPI_ISL_979672, EPI_ISL_979673, EPI_ISL_979674, EPI_ISL_979675, EPI_ISL_979676, EPI_ISL_979677, EPI_ISL_979678, EPI_ISL_979679, EPI_ISL_979680, EPI_ISL_979681, EPI_ISL_979682, EPI_ISL_979683, EPI_ISL_979684, EPI_ISL_979685, EPI_ISL_979686, EPI_ISL_979687, EPI_ISL_979688, EPI_ISL_979689, EPI_ISL_979690, EPI_ISL_979691, EPI_ISL_979694, EPI_ISL_979695, EPI_ISL_979696, EPI_ISL_979697, EPI_ISL_979698, EPI_ISL_979699, EPI_ISL_979700, EPI_ISL_979701, EPI_ISL_979702, EPI_ISL_979703, EPI_ISL_979704, EPI_ISL_979705, EPI_ISL_979706, EPI_ISL_979707, EPI_ISL_979708, EPI_ISL_979709 | | | |
| see above | Orange County Public Health Lab | Chan-Zuckerberg Biohub | CZB Cliahub Consortium |
| EPI_ISL_979710, EPI_ISL_979711, EPI_ISL_979712, EPI_ISL_979713, EPI_ISL_979714, EPI_ISL_979715, EPI_ISL_979716, EPI_ISL_979717, EPI_ISL_979718, EPI_ISL_979719, EPI_ISL_979720, EPI_ISL_979721, EPI_ISL_979722, EPI_ISL_979723, EPI_ISL_979724, EPI_ISL_979725, EPI_ISL_979726, EPI_ISL_979727, EPI_ISL_979728, EPI_ISL_979729, EPI_ISL_979730, EPI_ISL_979731, EPI_ISL_979732, EPI_ISL_979733, EPI_ISL_979734, EPI_ISL_979735, EPI_ISL_979736, EPI_ISL_979737, EPI_ISL_979738, EPI_ISL_979739, EPI_ISL_979740, EPI_ISL_979741, EPI_ISL_979742, EPI_ISL_979743, EPI_ISL_979744, EPI_ISL_979745, EPI_ISL_979746, EPI_ISL_979747, EPI_ISL_979748, EPI_ISL_979749, EPI_ISL_979750, EPI_ISL_979751, EPI_ISL_979752, EPI_ISL_979753, EPI_ISL_979754, EPI_ISL_979755, EPI_ISL_979756, EPI_ISL_979757, EPI_ISL_979758, EPI_ISL_979759, EPI_ISL_979760, EPI_ISL_979761, EPI_ISL_979762, EPI_ISL_979763, EPI_ISL_979764, EPI_ISL_979765, EPI_ISL_979766, EPI_ISL_979767, EPI_ISL_979768, EPI_ISL_979769, EPI_ISL_979770, EPI_ISL_979771, EPI_ISL_979772, EPI_ISL_979773, EPI_ISL_979774, EPI_ISL_979775, EPI_ISL_979776, EPI_ISL_979777, EPI_ISL_979778, EPI_ISL_979779, EPI_ISL_979780, EPI_ISL_979781, EPI_ISL_979782, EPI_ISL_979783, EPI_ISL_979784, EPI_ISL_979785, EPI_ISL_979786, EPI_ISL_979787, EPI_ISL_979788, EPI_ISL_979789, EPI_ISL_979790, EPI_ISL_979791, EPI_ISL_979792, EPI_ISL_979793, EPI_ISL_979794, EPI_ISL_979795, EPI_ISL_979796, EPI_ISL_979797 | | | |
| see above | Humboldt County Public Health Laboratory | Chan-Zuckerberg Biohub | CZB Cliahub Consortium |
| EPI_ISL_980840, EPI_ISL_980841, EPI_ISL_980842, EPI_ISL_980843, EPI_ISL_980844, EPI_ISL_980845, EPI_ISL_980846, EPI_ISL_980847, EPI_ISL_980848, EPI_ISL_980849, EPI_ISL_980850, EPI_ISL_980851, EPI_ISL_980852, EPI_ISL_980853, EPI_ISL_980854, EPI_ISL_980855, EPI_ISL_980856, EPI_ISL_980857, EPI_ISL_980858, EPI_ISL_980859, EPI_ISL_980860, EPI_ISL_980861, EPI_ISL_980862, EPI_ISL_980863, EPI_ISL_980864, EPI_ISL_980865, EPI_ISL_980866, EPI_ISL_980867, EPI_ISL_980868, EPI_ISL_980869, EPI_ISL_980870, EPI_ISL_980871, EPI_ISL_980872, EPI_ISL_980873, EPI_ISL_980874, EPI_ISL_980875, EPI_ISL_980876, EPI_ISL_980877, EPI_ISL_980878, EPI_ISL_980879, EPI_ISL_980880, EPI_ISL_980881, EPI_ISL_980882, EPI_ISL_980883, EPI_ISL_980884, EPI_ISL_980885, EPI_ISL_980886, EPI_ISL_980887, EPI_ISL_980888, EPI_ISL_980889, EPI_ISL_980890, EPI_ISL_980891, EPI_ISL_980892, EPI_ISL_980893, EPI_ISL_980894, EPI_ISL_980895, EPI_ISL_980896, EPI_ISL_980897, EPI_ISL_980898, EPI_ISL_980899, EPI_ISL_980900, EPI_ISL_980901, EPI_ISL_980902, EPI_ISL_980903, EPI_ISL_980904, EPI_ISL_980905, EPI_ISL_980906, EPI_ISL_980907, EPI_ISL_980932, EPI_ISL_980933, EPI_ISL_980934, EPI_ISL_980935, EPI_ISL_980936, EPI_ISL_980937, EPI_ISL_980938, EPI_ISL_980939, EPI_ISL_980940, EPI_ISL_980941, EPI_ISL_980942, EPI_ISL_980943, EPI_ISL_980944, EPI_ISL_980945, EPI_ISL_980946, EPI_ISL_980947, EPI_ISL_980948, EPI_ISL_980949, EPI_ISL_980950, EPI_ISL_980951, EPI_ISL_980952, EPI_ISL_980953, EPI_ISL_980954, EPI_ISL_980955, EPI_ISL_980956, EPI_ISL_980957, EPI_ISL_980958, EPI_ISL_980959, EPI_ISL_980960, EPI_ISL_980961, EPI_ISL_980962, EPI_ISL_980963, EPI_ISL_980964, EPI_ISL_980965, EPI_ISL_980966, EPI_ISL_980967, EPI_ISL_980968, EPI_ISL_980969, EPI_ISL_980970, EPI_ISL_980971, EPI_ISL_980972, EPI_ISL_980973, EPI_ISL_980974, EPI_ISL_980975, EPI_ISL_980976, EPI_ISL_980977, EPI_ISL_980978, EPI_ISL_980979, EPI_ISL_980980, EPI_ISL_980981, EPI_ISL_980982, EPI_ISL_980983, EPI_ISL_980984, EPI_ISL_980985, EPI_ISL_980986, EPI_ISL_980987, EPI_ISL_980988, EPI_ISL_980989, EPI_ISL_980990, EPI_ISL_980991, EPI_ISL_980992, EPI_ISL_980993, EPI_ISL_980994, EPI_ISL_980995, EPI_ISL_980996, EPI_ISL_980997, EPI_ISL_980998, EPI_ISL_980999, EPI_ISL_981000, EPI_ISL_981001, EPI_ISL_981002, EPI_ISL_981003 | | | |
| see above | Innovative Genomics Institute, UC Berkeley | Innovative Genomics Institute, UC Berkeley | Stacia Wyman, Haridha Shivram, Phil Frankino, Liana Lareau |
| EPI_ISL_981057, EPI_ISL_981058, EPI_ISL_981059, EPI_ISL_981060, EPI_ISL_981061, EPI_ISL_981062, EPI_ISL_981063, EPI_ISL_981064, EPI_ISL_981065, EPI_ISL_981066, EPI_ISL_981067, EPI_ISL_981070, EPI_ISL_981071, EPI_ISL_981072, EPI_ISL_981073, EPI_ISL_981074, EPI_ISL_981075, EPI_ISL_981076, EPI_ISL_981077, EPI_ISL_981078, EPI_ISL_981079, EPI_ISL_981080, EPI_ISL_981081, EPI_ISL_981082, EPI_ISL_981083, EPI_ISL_981084, EPI_ISL_981085, EPI_ISL_981086, EPI_ISL_981087, EPI_ISL_981088, EPI_ISL_981089, EPI_ISL_981090, EPI_ISL_981091, EPI_ISL_981092, EPI_ISL_981093, EPI_ISL_981094, EPI_ISL_981095, EPI_ISL_981096, EPI_ISL_981097, EPI_ISL_981098, EPI_ISL_981099, EPI_ISL_981100, EPI_ISL_981101, EPI_ISL_981102, EPI_ISL_981103, EPI_ISL_981104, EPI_ISL_981105, EPI_ISL_981106, EPI_ISL_981107, EPI_ISL_981108, EPI_ISL_981109, EPI_ISL_981110, EPI_ISL_981111, EPI_ISL_981112, EPI_ISL_981113, EPI_ISL_981114, EPI_ISL_981115, EPI_ISL_981116, EPI_ISL_981117, EPI_ISL_981118, EPI_ISL_981119, EPI_ISL_981120, EPI_ISL_981121, EPI_ISL_981122, EPI_ISL_981123, EPI_ISL_981127, EPI_ISL_981128, EPI_ISL_981129, EPI_ISL_981130, EPI_ISL_981131, EPI_ISL_981132, EPI_ISL_981133, EPI_ISL_981134, EPI_ISL_981135, EPI_ISL_981136, EPI_ISL_981137, EPI_ISL_981138, EPI_ISL_981139, EPI_ISL_981140, EPI_ISL_981141, EPI_ISL_981142, EPI_ISL_981143, EPI_ISL_981144, EPI_ISL_981145, EPI_ISL_981146, EPI_ISL_981147, EPI_ISL_981148, EPI_ISL_981149, EPI_ISL_981150, EPI_ISL_981151, EPI_ISL_981152, EPI_ISL_981153, EPI_ISL_981154, EPI_ISL_981155, EPI_ISL_981156, EPI_ISL_981157, EPI_ISL_981158, EPI_ISL_981159, EPI_ISL_981160, EPI_ISL_981161, EPI_ISL_981162, EPI_ISL_981163, EPI_ISL_981164, EPI_ISL_981165, EPI_ISL_981166, EPI_ISL_981167, EPI_ISL_981168, EPI_ISL_981169, EPI_ISL_981170, EPI_ISL_981171, EPI_ISL_981172, EPI_ISL_981173, EPI_ISL_981174, EPI_ISL_981175, EPI_ISL_981176, EPI_ISL_981177, EPI_ISL_981178, EPI_ISL_981179, EPI_ISL_981180, EPI_ISL_981181, EPI_ISL_981182, EPI_ISL_981183, EPI_ISL_981184, EPI_ISL_981185, EPI_ISL_981186, EPI_ISL_981187, EPI_ISL_981188, EPI_ISL_981189, EPI_ISL_981190, EPI_ISL_981191, EPI_ISL_981192, EPI_ISL_981193, EPI_ISL_981194, EPI_ISL_981195, EPI_ISL_981196, EPI_ISL_981197, EPI_ISL_981198, EPI_ISL_981199, EPI_ISL_981200, EPI_ISL_981201 | | | |
| see above | Johns Hopkins Hospital Department of Pathology | Johns Hopkins Hospital Department of Pathology | C. Paul Morris, Chun Huai Luo, Adannaya Amadi, Matthew Schwartz, Nicholas Gallagher, Heba H. Mostafa |
| EPI_ISL_981202, EPI_ISL_981305, EPI_ISL_981306, EPI_ISL_981307, EPI_ISL_981368, EPI_ISL_981369 | Ohio Department of Health Laboratory | Ohio Department of Health Laboratory | Holmes, Jennifer; Eric Brandt, Keoni Omura, Glen McGillivary, Caitlin McDonnell, Jade Mowery, Stephanie Mccracken, Tyler Payne, Kirtana Ramadugu, Erica Leasure, Brent Lee, Kelsey Florek, Heather Blankenship, Quanta Brown, and Tammy Bannerman |
| EPI_ISL_981976 | Sonora Quest Laboratories | TGen North | "Jolene Bowers, Megan Folkerts, Chris French, Hayley Yaglom, Ashlyn Pfeiffer, Darrin Lemmer, Dave Engelthaler, The Arizona COVID Genomics Union (ACGU)" |
| EPI_ISL_981978 | TGen North | TGen North | "Jolene Bowers, Megan Folkerts, Chris French, Hayley Yaglom, Ashlyn Pfeiffer, Darrin Lemmer, Dave Engelthaler, The Arizona COVID Genomics Union (ACGU)" |
| EPI_ISL_981979, EPI_ISL_981980 | Sonora Quest Laboratories | TGen North | "Jolene Bowers, Megan Folkerts, Chris French, Hayley Yaglom, Ashlyn Pfeiffer, Darrin Lemmer, Dave Engelthaler, The Arizona COVID Genomics Union (ACGU)" |
| EPI_ISL_981982 | TGen North | TGen North | "Jolene Bowers, Megan Folkerts, Chris French, Hayley Yaglom, Ashlyn Pfeiffer, Darrin Lemmer, Dave Engelthaler, The Arizona COVID Genomics Union (ACGU)" |
| EPI_ISL_981983, EPI_ISL_981984, EPI_ISL_981986 | Sonora Quest Laboratories | TGen North | "Jolene Bowers, Megan Folkerts, Chris French, Hayley Yaglom, Ashlyn Pfeiffer, Darrin Lemmer, Dave Engelthaler, The Arizona COVID Genomics Union (ACGU)" |
| EPI_ISL_981987, EPI_ISL_981988 | TGen North | TGen North | "Jolene Bowers, Megan Folkerts, Chris French, Hayley Yaglom, Ashlyn Pfeiffer, Darrin Lemmer, Dave Engelthaler, The Arizona COVID Genomics Union (ACGU)" |
| EPI_ISL_981989 | Sonora Quest Laboratories | TGen North | "Jolene Bowers, Megan Folkerts, Chris French, Hayley Yaglom, Ashlyn Pfeiffer, Darrin Lemmer, Dave Engelthaler, The Arizona COVID Genomics Union (ACGU)" |
| EPI_ISL_981990 | TGen North | TGen North | "Jolene Bowers, Megan Folkerts, Chris French, Hayley Yaglom, Ashlyn Pfeiffer, Darrin Lemmer, Dave Engelthaler, The Arizona COVID Genomics Union (ACGU)" |
| EPI_ISL_981991, EPI_ISL_981992, EPI_ISL_981993, EPI_ISL_981994, EPI_ISL_981995 | Sonora Quest Laboratories | TGen North | "Jolene Bowers, Megan Folkerts, Chris French, Hayley Yaglom, Ashlyn Pfeiffer, Darrin Lemmer, Dave Engelthaler, The Arizona COVID Genomics Union (ACGU)" |
| EPI_ISL_981996 | TGen North | TGen North | "Jolene Bowers, Megan Folkerts, Chris French, Hayley Yaglom, Ashlyn Pfeiffer, Darrin Lemmer, Dave Engelthaler, The Arizona COVID Genomics Union (ACGU)" |

| | | | |
|---|---|---|---|
| EPI_ISL_981997 | Sonora Quest Laboratories | TGen North | "Jolene Bowers, Megan Folkerts, Chris French, Hayley Yaglom, Ashlyn Pfeiffer, Darrin Lemmer, Dave Engelthaler, The Arizona COVID Genomics Union (ACGU)" |
| EPI_ISL_981998, EPI_ISL_981999, EPI_ISL_982001, EPI_ISL_982003 | TGen North | TGen North | "Jolene Bowers, Megan Folkerts, Chris French, Hayley Yaglom, Ashlyn Pfeiffer, Darrin Lemmer, Dave Engelthaler, The Arizona COVID Genomics Union (ACGU)" |
| EPI_ISL_982005 | Sonora Quest Laboratories | TGen North | "Jolene Bowers, Megan Folkerts, Chris French, Hayley Yaglom, Ashlyn Pfeiffer, Darrin Lemmer, Dave Engelthaler, The Arizona COVID Genomics Union (ACGU)" |
| EPI_ISL_982007, EPI_ISL_982008 | TGen North | TGen North | "Jolene Bowers, Megan Folkerts, Chris French, Hayley Yaglom, Ashlyn Pfeiffer, Darrin Lemmer, Dave Engelthaler, The Arizona COVID Genomics Union (ACGU)" |
| EPI_ISL_982009 | Sonora Quest Laboratories | TGen North | "Jolene Bowers, Megan Folkerts, Chris French, Hayley Yaglom, Ashlyn Pfeiffer, Darrin Lemmer, Dave Engelthaler, The Arizona COVID Genomics Union (ACGU)" |
| EPI_ISL_982010, EPI_ISL_982011, EPI_ISL_982012 | TGen North | TGen North | "Jolene Bowers, Megan Folkerts, Chris French, Hayley Yaglom, Ashlyn Pfeiffer, Darrin Lemmer, Dave Engelthaler, The Arizona COVID Genomics Union (ACGU)" |
| EPI_ISL_982013, EPI_ISL_982014, EPI_ISL_982015 | Sonora Quest Laboratories | TGen North | "Jolene Bowers, Megan Folkerts, Chris French, Hayley Yaglom, Ashlyn Pfeiffer, Darrin Lemmer, Dave Engelthaler, The Arizona COVID Genomics Union (ACGU)" |
| EPI_ISL_982016 | TGen North | TGen North | "Jolene Bowers, Megan Folkerts, Chris French, Hayley Yaglom, Ashlyn Pfeiffer, Darrin Lemmer, Dave Engelthaler, The Arizona COVID Genomics Union (ACGU)" |
| EPI_ISL_982018 | Sonora Quest Laboratories | TGen North | "Jolene Bowers, Megan Folkerts, Chris French, Hayley Yaglom, Ashlyn Pfeiffer, Darrin Lemmer, Dave Engelthaler, The Arizona COVID Genomics Union (ACGU)" |
| EPI_ISL_982019, EPI_ISL_982020, EPI_ISL_982021 | TGen North | TGen North | "Jolene Bowers, Megan Folkerts, Chris French, Hayley Yaglom, Ashlyn Pfeiffer, Darrin Lemmer, Dave Engelthaler, The Arizona COVID Genomics Union (ACGU)" |
| EPI_ISL_982024, EPI_ISL_982025 | Sonora Quest Laboratories | TGen North | "Jolene Bowers, Megan Folkerts, Chris French, Hayley Yaglom, Ashlyn Pfeiffer, Darrin Lemmer, Dave Engelthaler, The Arizona COVID Genomics Union (ACGU)" |
| EPI_ISL_982026, EPI_ISL_982027 | TGen North | TGen North | "Jolene Bowers, Megan Folkerts, Chris French, Hayley Yaglom, Ashlyn Pfeiffer, Darrin Lemmer, Dave Engelthaler, The Arizona COVID Genomics Union (ACGU)" |
| EPI_ISL_982028 | Sonora Quest Laboratories | TGen North | "Jolene Bowers, Megan Folkerts, Chris French, Hayley Yaglom, Ashlyn Pfeiffer, Darrin Lemmer, Dave Engelthaler, The Arizona COVID Genomics Union (ACGU)" |
| EPI_ISL_982029 | TGen North | TGen North | "Jolene Bowers, Megan Folkerts, Chris French, Hayley Yaglom, Ashlyn Pfeiffer, Darrin Lemmer, Dave Engelthaler, The Arizona COVID Genomics Union (ACGU)" |
| EPI_ISL_982030, EPI_ISL_982031, EPI_ISL_982032, EPI_ISL_982033, EPI_ISL_982034, EPI_ISL_982035, EPI_ISL_982036, EPI_ISL_982037 | Sonora Quest Laboratories | TGen North | "Jolene Bowers, Megan Folkerts, Chris French, Hayley Yaglom, Ashlyn Pfeiffer, Darrin Lemmer, Dave Engelthaler, The Arizona COVID Genomics Union (ACGU)" |
| EPI_ISL_982038 | TGen North | TGen North | "Jolene Bowers, Megan Folkerts, Chris French, Hayley Yaglom, Ashlyn Pfeiffer, Darrin Lemmer, Dave Engelthaler, The Arizona COVID Genomics Union (ACGU)" |
| EPI_ISL_982039, EPI_ISL_982041, EPI_ISL_982042 | Sonora Quest Laboratories | TGen North | "Jolene Bowers, Megan Folkerts, Chris French, Hayley Yaglom, Ashlyn Pfeiffer, Darrin Lemmer, Dave Engelthaler, The Arizona COVID Genomics Union (ACGU)" |
| EPI_ISL_982043, EPI_ISL_982044 | TGen North | TGen North | "Jolene Bowers, Megan Folkerts, Chris French, Hayley Yaglom, Ashlyn Pfeiffer, Darrin Lemmer, Dave Engelthaler, The Arizona COVID Genomics Union (ACGU)" |
| EPI_ISL_982045, EPI_ISL_982046 | Sonora Quest Laboratories | TGen North | "Jolene Bowers, Megan Folkerts, Chris French, Hayley Yaglom, Ashlyn Pfeiffer, Darrin Lemmer, Dave Engelthaler, The Arizona COVID Genomics Union (ACGU)" |
| EPI_ISL_982047, EPI_ISL_982048 | TGen North | TGen North | "Jolene Bowers, Megan Folkerts, Chris French, Hayley Yaglom, Ashlyn Pfeiffer, Darrin Lemmer, Dave Engelthaler, The Arizona COVID Genomics Union (ACGU)" |
| EPI_ISL_982049, EPI_ISL_982050, EPI_ISL_982051 | Sonora Quest Laboratories | TGen North | "Jolene Bowers, Megan Folkerts, Chris French, Hayley Yaglom, Ashlyn Pfeiffer, Darrin Lemmer, Dave Engelthaler, The Arizona COVID Genomics Union (ACGU)" |
| EPI_ISL_982052, EPI_ISL_982053 | TGen North | TGen North | "Jolene Bowers, Megan Folkerts, Chris French, Hayley Yaglom, Ashlyn Pfeiffer, Darrin Lemmer, Dave Engelthaler, The Arizona COVID Genomics Union (ACGU)" |
| EPI_ISL_982054, EPI_ISL_982055 | Sonora Quest Laboratories | TGen North | "Jolene Bowers, Megan Folkerts, Chris French, Hayley Yaglom, Ashlyn Pfeiffer, Darrin Lemmer, Dave Engelthaler, The Arizona COVID Genomics Union (ACGU)" |
| EPI_ISL_982056 | TGen North | TGen North | "Jolene Bowers, Megan Folkerts, Chris French, Hayley Yaglom, Ashlyn Pfeiffer, Darrin Lemmer, Dave Engelthaler, The Arizona COVID Genomics Union (ACGU)" |
| EPI_ISL_982058 | Sonora Quest Laboratories | TGen North | "Jolene Bowers, Megan Folkerts, Chris French, Hayley Yaglom, Ashlyn Pfeiffer, Darrin Lemmer, Dave Engelthaler, The Arizona COVID Genomics Union (ACGU)" |
| EPI_ISL_982059 | TGen North | TGen North | "Jolene Bowers, Megan Folkerts, Chris French, Hayley Yaglom, Ashlyn Pfeiffer, Darrin Lemmer, Dave Engelthaler, The Arizona COVID Genomics Union (ACGU)" |
| EPI_ISL_982060, EPI_ISL_982062 | Sonora Quest Laboratories | TGen North | "Jolene Bowers, Megan Folkerts, Chris French, Hayley Yaglom, Ashlyn Pfeiffer, Darrin Lemmer, Dave Engelthaler, The Arizona COVID Genomics Union (ACGU)" |
| EPI_ISL_982063 | TGen North | TGen North | "Jolene Bowers, Megan Folkerts, Chris French, Hayley Yaglom, Ashlyn Pfeiffer, Darrin Lemmer, Dave Engelthaler, The Arizona COVID Genomics Union (ACGU)" |
| EPI_ISL_982064 | Sonora Quest Laboratories | TGen North | "Jolene Bowers, Megan Folkerts, Chris French, Hayley Yaglom, Ashlyn Pfeiffer, Darrin Lemmer, Dave Engelthaler, The Arizona COVID Genomics Union (ACGU)" |
| EPI_ISL_982065 | TGen North | TGen North | "Jolene Bowers, Megan Folkerts, Chris French, Hayley Yaglom, Ashlyn Pfeiffer, Darrin Lemmer, Dave Engelthaler, The Arizona COVID Genomics Union (ACGU)" |
| EPI_ISL_982066, EPI_ISL_982067 | Sonora Quest Laboratories | TGen North | "Jolene Bowers, Megan Folkerts, Chris French, Hayley Yaglom, Ashlyn Pfeiffer, Darrin Lemmer, Dave Engelthaler, The Arizona COVID Genomics Union (ACGU)" |
| EPI_ISL_982068 | TGen North | TGen North | "Jolene Bowers, Megan Folkerts, Chris French, Hayley Yaglom, Ashlyn Pfeiffer, Darrin Lemmer, Dave Engelthaler, The Arizona COVID Genomics Union (ACGU)" |
| EPI_ISL_982069 | Sonora Quest Laboratories | TGen North | "Jolene Bowers, Megan Folkerts, Chris French, Hayley Yaglom, Ashlyn Pfeiffer, Darrin Lemmer, Dave Engelthaler, The Arizona COVID Genomics Union (ACGU)" |
| EPI_ISL_982070 | TGen North | TGen North | "Jolene Bowers, Megan Folkerts, Chris French, Hayley Yaglom, Ashlyn Pfeiffer, Darrin Lemmer, Dave Engelthaler, The Arizona COVID Genomics Union (ACGU)" |
| EPI_ISL_982073, EPI_ISL_982074, EPI_ISL_982075, EPI_ISL_982076, EPI_ISL_982077 | Sonora Quest Laboratories | TGen North | "Jolene Bowers, Megan Folkerts, Chris French, Hayley Yaglom, Ashlyn Pfeiffer, Darrin Lemmer, Dave Engelthaler, The Arizona COVID Genomics Union (ACGU)" |
| EPI_ISL_982078 | TGen North | TGen North | "Jolene Bowers, Megan Folkerts, Chris French, Hayley Yaglom, Ashlyn Pfeiffer, Darrin Lemmer, Dave Engelthaler, The Arizona COVID Genomics Union (ACGU)" |
| EPI_ISL_982079, EPI_ISL_982081, EPI_ISL_982082, EPI_ISL_982083, EPI_ISL_982084 | Sonora Quest Laboratories | TGen North | "Jolene Bowers, Megan Folkerts, Chris French, Hayley Yaglom, Ashlyn Pfeiffer, Darrin Lemmer, Dave Engelthaler, The Arizona COVID Genomics Union (ACGU)" |
| EPI_ISL_982085, EPI_ISL_982086 | TGen North | TGen North | "Jolene Bowers, Megan Folkerts, Chris French, Hayley Yaglom, Ashlyn Pfeiffer, Darrin Lemmer, Dave Engelthaler, The Arizona COVID Genomics Union (ACGU)" |
| EPI_ISL_982087, EPI_ISL_982088, EPI_ISL_982089, EPI_ISL_982090, EPI_ISL_982091, EPI_ISL_982092, EPI_ISL_982093, EPI_ISL_982094, EPI_ISL_982095, EPI_ISL_982096 | Sonora Quest Laboratories | TGen North | "Jolene Bowers, Megan Folkerts, Chris French, Hayley Yaglom, Ashlyn Pfeiffer, Darrin Lemmer, Dave Engelthaler, The Arizona COVID Genomics Union (ACGU)" |
| EPI_ISL_982097 | TGen North | TGen North | "Jolene Bowers, Megan Folkerts, Chris French, Hayley Yaglom, Ashlyn Pfeiffer, Darrin Lemmer, Dave Engelthaler, The Arizona COVID Genomics Union (ACGU)" |
| EPI_ISL_982098, EPI_ISL_982099, EPI_ISL_982100, EPI_ISL_982101, EPI_ISL_982103 | Sonora Quest Laboratories | TGen North | "Jolene Bowers, Megan Folkerts, Chris French, Hayley Yaglom, Ashlyn Pfeiffer, Darrin Lemmer, Dave Engelthaler, The Arizona COVID Genomics Union (ACGU)" |

| | | | |
|---|---|---|---|
| EPI_ISL_982106 | TGen North | TGen North | "Jolene Bowers, Megan Folkerts, Chris French, Hayley Yaglom, Ashlyn Pfeiffer, Darrin Lemmer, Dave Engelthaler, The Arizona COVID Genomics Union (ACGU)" |
| EPI_ISL_982108 | Sonora Quest Laboratories | TGen North | "Jolene Bowers, Megan Folkerts, Chris French, Hayley Yaglom, Ashlyn Pfeiffer, Darrin Lemmer, Dave Engelthaler, The Arizona COVID Genomics Union (ACGU)" |
| EPI_ISL_982109 | TGen North | TGen North | "Jolene Bowers, Megan Folkerts, Chris French, Hayley Yaglom, Ashlyn Pfeiffer, Darrin Lemmer, Dave Engelthaler, The Arizona COVID Genomics Union (ACGU)" |
| EPI_ISL_982110 | Sonora Quest Laboratories | TGen North | "Jolene Bowers, Megan Folkerts, Chris French, Hayley Yaglom, Ashlyn Pfeiffer, Darrin Lemmer, Dave Engelthaler, The Arizona COVID Genomics Union (ACGU)" |
| EPI_ISL_982111, EPI_ISL_982115 | TGen North | TGen North | "Jolene Bowers, Megan Folkerts, Chris French, Hayley Yaglom, Ashlyn Pfeiffer, Darrin Lemmer, Dave Engelthaler, The Arizona COVID Genomics Union (ACGU)" |
| EPI_ISL_982344, EPI_ISL_982345, EPI_ISL_982346, EPI_ISL_982347, EPI_ISL_982348, EPI_ISL_982349, EPI_ISL_982350, EPI_ISL_982351, EPI_ISL_982352, EPI_ISL_982353, EPI_ISL_982354, EPI_ISL_982355, EPI_ISL_982356, EPI_ISL_982357, EPI_ISL_982358, EPI_ISL_982359, EPI_ISL_982360, EPI_ISL_982361, EPI_ISL_982362, EPI_ISL_982363, EPI_ISL_982364, EPI_ISL_982365, EPI_ISL_982366, EPI_ISL_982367, EPI_ISL_982368, EPI_ISL_982369, EPI_ISL_982370, EPI_ISL_982371, EPI_ISL_982372, EPI_ISL_982373, EPI_ISL_982374, EPI_ISL_982375, EPI_ISL_982376, EPI_ISL_982377, EPI_ISL_982378, EPI_ISL_982379, EPI_ISL_982380, EPI_ISL_982381, EPI_ISL_982382, EPI_ISL_982383, EPI_ISL_982384, EPI_ISL_982385, EPI_ISL_982386, EPI_ISL_982387, EPI_ISL_982388, EPI_ISL_982389, EPI_ISL_982390, EPI_ISL_982391, EPI_ISL_982392, EPI_ISL_982393, EPI_ISL_982394, EPI_ISL_982395, EPI_ISL_982396, EPI_ISL_982397, EPI_ISL_982398, EPI_ISL_982399, EPI_ISL_982400, EPI_ISL_982401, EPI_ISL_982402, EPI_ISL_982403, EPI_ISL_982404, EPI_ISL_982405, EPI_ISL_982406, EPI_ISL_982407, EPI_ISL_982408, EPI_ISL_982409, EPI_ISL_982410, EPI_ISL_982411, EPI_ISL_982412, EPI_ISL_982413 | | | |
| see above | M Health Fairview | Minnesota Department of Health, Public Health Laboratory | Alexandra Lorentz, Jacob Garfin, Matt Plumb, and Xiong Wang |
| EPI_ISL_1000967 | The Ohio State University Wexner Medical Center | James Molecular Lab | Huolin Tu, Matthew R Avenarius, Laura Kubatko, Matthew Hunt, Xiaokang Pan, Peng Ru, Jason Garee, Keelie Thomas, Peter Mohler, Preeti Pancholi, Dan Jones |
| EPI_ISL_1000971 | Kansas Health and Environmental Lab | Kansas Health and Environmental Lab | Mike Grose, Paige Drury, Carissa Robertson, Ben Olsen, and Phil Adam |
| EPI_ISL_1001005, EPI_ISL_1001006, EPI_ISL_1001007, EPI_ISL_1001008, EPI_ISL_1001009, EPI_ISL_1001010, EPI_ISL_1001012, EPI_ISL_1001013, EPI_ISL_1001014, EPI_ISL_1001017, EPI_ISL_1001018, EPI_ISL_1001019, EPI_ISL_1001020, EPI_ISL_1001021, EPI_ISL_1001022, EPI_ISL_1001023, EPI_ISL_1001024, EPI_ISL_1001025, EPI_ISL_1001026, EPI_ISL_1001027, EPI_ISL_1001028, EPI_ISL_1001029, EPI_ISL_1001030, EPI_ISL_1001031, EPI_ISL_1001033, EPI_ISL_1001034, EPI_ISL_1001035, EPI_ISL_1001036, EPI_ISL_1001037, EPI_ISL_1001038, EPI_ISL_1001039 | | | |
| see above | Santa Clara County Public Health Laboratory | Santa Clara County Public Health Laboratory | Santa Clara County Public Health Department |
| EPI_ISL_1001265, EPI_ISL_1001267, EPI_ISL_1001268, EPI_ISL_1001269, EPI_ISL_1001270, EPI_ISL_1001271, EPI_ISL_1001272, EPI_ISL_1001273, EPI_ISL_1001274, EPI_ISL_1001275, EPI_ISL_1001276, EPI_ISL_1001277, EPI_ISL_1001278, EPI_ISL_1001279, EPI_ISL_1001280, EPI_ISL_1001281, EPI_ISL_1001282, EPI_ISL_1001283, EPI_ISL_1001284, EPI_ISL_1001285, EPI_ISL_1001286, EPI_ISL_1001287, EPI_ISL_1001288, EPI_ISL_1001289, EPI_ISL_1001290, EPI_ISL_1001291, EPI_ISL_1001292, EPI_ISL_1001293, EPI_ISL_1001294, EPI_ISL_1001295, EPI_ISL_1001296, EPI_ISL_1001297, EPI_ISL_1001298 | | | |
| see above | University of Michigan Clinical Microbiology Laboratory | Lauring Lab, University of Michigan, Department of Microbiology and Immunology | Valesano |
| EPI_ISL_1001460 | Clinical Microbiology Laboratory, NewYork Presbyterian Hospital/Columbia University Irving Medical Center | Uhlemann Laboratory, Columbia University Irving Medical Center | Medini K. Annavajhala, Anne-Catrin Uhlemann, Hiroshi Mori, David Ho |
| EPI_ISL_1004178 | Oregon State University TRACE Project | Center for Genome Research and Biocomputing | Oregon State University TRACE Project |
| EPI_ISL_1004179, EPI_ISL_1004180, EPI_ISL_1004181, EPI_ISL_1004182, EPI_ISL_1004183, EPI_ISL_1004184, EPI_ISL_1004185, EPI_ISL_1004186, EPI_ISL_1004187, EPI_ISL_1004188, EPI_ISL_1004189, EPI_ISL_1004190, EPI_ISL_1004191, EPI_ISL_1004192, EPI_ISL_1004193, EPI_ISL_1004194, EPI_ISL_1004195, EPI_ISL_1004196, EPI_ISL_1004197, EPI_ISL_1004198, EPI_ISL_1004199, EPI_ISL_1004200, EPI_ISL_1004201, EPI_ISL_1004202, EPI_ISL_1004203, EPI_ISL_1004204, EPI_ISL_1004205, EPI_ISL_1004206, EPI_ISL_1004207, EPI_ISL_1004208, EPI_ISL_1004209, EPI_ISL_1004210, EPI_ISL_1004211, EPI_ISL_1004212 | | | |
| see above | Oregon State University TRACE Project | OSU Center for Genome Research and Biocomputing | Oregon State University TRACE Project |
| EPI_ISL_1004213, EPI_ISL_1004214, EPI_ISL_1004215, EPI_ISL_1004216, EPI_ISL_1004217 | Oregon State University TRACE Project | Center for Genome Research and Biocomputing | Oregon State University TRACE Project |
| EPI_ISL_1004218, EPI_ISL_1004219, EPI_ISL_1004220, EPI_ISL_1004221, EPI_ISL_1004222, EPI_ISL_1004223, EPI_ISL_1004224, EPI_ISL_1004225, EPI_ISL_1004226, EPI_ISL_1004227, EPI_ISL_1004228, EPI_ISL_1004229, EPI_ISL_1004230, EPI_ISL_1004231, EPI_ISL_1004232 | | | |
| see above | Kansas Health and Environmental Lab | Kansas Health and Environmental Lab | Mike Grose, Carissa Robertson, Ben Olsen, and Phil Adam |
| EPI_ISL_1005483, EPI_ISL_1005484, EPI_ISL_1005485, EPI_ISL_1005486, EPI_ISL_1005487, EPI_ISL_1005488, EPI_ISL_1005489, EPI_ISL_1005490 | Los Angeles County Public Health Laboratories | Los Angeles County Public Health Laboratories | P. Hemarajata et al. |
| EPI_ISL_1005491, EPI_ISL_1005492, EPI_ISL_1005494, EPI_ISL_1005495, EPI_ISL_1005496, EPI_ISL_1005497, EPI_ISL_1005498, EPI_ISL_1005499, EPI_ISL_1005500, EPI_ISL_1005501, EPI_ISL_1005502, EPI_ISL_1005503, EPI_ISL_1005505, EPI_ISL_1005506, EPI_ISL_1005507 | | | |
| see above | USC Clinical Lab | Los Angeles County Public Health Laboratories | P. Hemarajata et al. |
| EPI_ISL_1005508, EPI_ISL_1005509, EPI_ISL_1005510 | Los Angeles County Public Health Laboratories | Los Angeles County Public Health Laboratories | P. Hemarajata et al. |
| EPI_ISL_1005511, EPI_ISL_1005512, EPI_ISL_1005513, EPI_ISL_1005514, EPI_ISL_1005515, EPI_ISL_1005516, EPI_ISL_1005517, EPI_ISL_1005518, EPI_ISL_1005520, EPI_ISL_1005521 | UCLA Clinical Micro Lab | Los Angeles County Public Health Laboratories | P. Hemarajata et al. |
| EPI_ISL_1005523, EPI_ISL_1005524, EPI_ISL_1005528, EPI_ISL_1005529, EPI_ISL_1005530, EPI_ISL_1005531, EPI_ISL_1005532, EPI_ISL_1005534 | USC Clinical Lab | Los Angeles County Public Health Laboratories | P. Hemarajata et al. |
| EPI_ISL_1005542, EPI_ISL_1005543, EPI_ISL_1005544, EPI_ISL_1005545, EPI_ISL_1005546, EPI_ISL_1005547, EPI_ISL_1005548, EPI_ISL_1005549, EPI_ISL_1005550, EPI_ISL_1005551, EPI_ISL_1005552, EPI_ISL_1005553, EPI_ISL_1005554 | | | |
| see above | UCLA Clinical Micro Lab | Los Angeles County Public Health Laboratories | P. Hemarajata et al. |
| EPI_ISL_1005555 | MD Laboratories | Los Angeles County Public Health Laboratories | P. Hemarajata et al. |
| EPI_ISL_1005557 | Los Angeles Community Hospital | Los Angeles County Public Health Laboratories | P. Hemarajata et al. |
| EPI_ISL_1005558 | USC Clinical Lab | Los Angeles County Public Health Laboratories | P. Hemarajata et al. |
| EPI_ISL_1005559, EPI_ISL_1005560, EPI_ISL_1005561, EPI_ISL_1005562, EPI_ISL_1005563, EPI_ISL_1005564, EPI_ISL_1005565, EPI_ISL_1005566, EPI_ISL_1005567, EPI_ISL_1005568, EPI_ISL_1005569, EPI_ISL_1005570, EPI_ISL_1005571, EPI_ISL_1005572, EPI_ISL_1005573, EPI_ISL_1005574, EPI_ISL_1005575, EPI_ISL_1005576, EPI_ISL_1005577, EPI_ISL_1005578, EPI_ISL_1005579, EPI_ISL_1005580, EPI_ISL_1005581, EPI_ISL_1005582 | | | |
| see above | Instituto Nacional de Medicina Genomica | Instituto Nacional de Medicina Genomica | Hidalgo-Miranda A, Mendoza-Vargas A, Reyes-Grajeda JP, Cisneros-Villanueva M, Cedro-Tanda A,Peñaloza-Figueroa F, Herrera-Montalvo LA |
| EPI_ISL_1005583, EPI_ISL_1005584, EPI_ISL_1005585, EPI_ISL_1005586, EPI_ISL_1005587, EPI_ISL_1005588, EPI_ISL_1005589, EPI_ISL_1005590, EPI_ISL_1005591, EPI_ISL_1005592, EPI_ISL_1005593, EPI_ISL_1005594, EPI_ISL_1005595, EPI_ISL_1005596, EPI_ISL_1005597, EPI_ISL_1005598, EPI_ISL_1005599, EPI_ISL_1005600, EPI_ISL_1005601, EPI_ISL_1005602, EPI_ISL_1005603, EPI_ISL_1005604, EPI_ISL_1005605, EPI_ISL_1005606, EPI_ISL_1005607, EPI_ISL_1005608, EPI_ISL_1005609, EPI_ISL_1005610, EPI_ISL_1005611, EPI_ISL_1005612, EPI_ISL_1005613, EPI_ISL_1005614, EPI_ISL_1005615, EPI_ISL_1005616, EPI_ISL_1005617, EPI_ISL_1005618, EPI_ISL_1005619, EPI_ISL_1005620, EPI_ISL_1005621, EPI_ISL_1005622, EPI_ISL_1005623, EPI_ISL_1005624, EPI_ISL_1005625, EPI_ISL_1005626, EPI_ISL_1005627, EPI_ISL_1005628, EPI_ISL_1005629, EPI_ISL_1005630, EPI_ISL_1005631, EPI_ISL_1005632, EPI_ISL_1005633, EPI_ISL_1005634, EPI_ISL_1005635, EPI_ISL_1005636, EPI_ISL_1005637, EPI_ISL_1005638, EPI_ISL_1005639, EPI_ISL_1005640, EPI_ISL_1005641, EPI_ISL_1005642, EPI_ISL_1005643, EPI_ISL_1005644, EPI_ISL_1005645, EPI_ISL_1005646, EPI_ISL_1005647, EPI_ISL_1005648, EPI_ISL_1005649, EPI_ISL_1005650, EPI_ISL_1005651, EPI_ISL_1005652, EPI_ISL_1005653, EPI_ISL_1005654 | | | |
| see above | Fulgent Genetics | Fulgent Genetics | Harry Gao, Mickey Li, John Gao, Joseph Fierro, Benafsh Sapra, Becky Tsai, Yan Meng, Doreen Ng, James Xie |
| EPI_ISL_1005656, EPI_ISL_1005657, EPI_ISL_1005658, EPI_ISL_1005659, EPI_ISL_1005660, EPI_ISL_1005661, EPI_ISL_1005662, EPI_ISL_1005663, EPI_ISL_1005664, EPI_ISL_1005665, EPI_ISL_1005666, EPI_ISL_1005667, EPI_ISL_1005668, EPI_ISL_1005669, EPI_ISL_1005670, EPI_ISL_1005671, EPI_ISL_1005672, EPI_ISL_1005673 | | | |
| see above | Instituto Nacional de Medicina Genomica | Instituto Nacional de Medicina Genomica | Hidalgo-Miranda A, Mendoza-Vargas A, Reyes-Grajeda JP, Cisneros-Villanueva M, Cedro-Tanda A,Peñaloza-Figueroa F, Herrera-Montalvo LA |

| | | | |
|---|---|---|---|
| EPI_ISL_1007745, EPI_ISL_1007809 | Ohio Department of Health Laboratory | Ohio Department of Health Laboratory | Holmes, Jennifer; Eric Brandt, Keoni Omura, Glen McGillivary, Caitlin McDonnell, Jade Mowery, Stephanie Mccracken, Tyler Payne, Kirtana Ramadugu, Erica Leasure, Brent Lee, Kelsey Florek, Heather Blankenship, Quanta Brown, and Tammy Bannerman |
| EPI_ISL_1007992 | Wyoming Public Health Laboratory | Wyoming Public Health Laboratory | Noah Hull, Taylor Fearing, Lynette Gumbleton, Channing Weber, Ashley Norberg, Bailey Bowcutt, and Wanda Manley |
| EPI_ISL_1008389, EPI_ISL_1008390 | Ohio Department of Health Laboratory | Ohio Department of Health Laboratory | Holmes, Jennifer; Eric Brandt, Keoni Omura, Glen McGillivary, Caitlin McDonnell, Jade Mowery, Stephanie Mccracken, Tyler Payne, Kirtana Ramadugu, Erica Leasure, Brent Lee, Kelsey Florek, Heather Blankenship, Quanta Brown, and Tammy Bannerman |
| EPI_ISL_1008391, EPI_ISL_1008392, EPI_ISL_1008393 | Wyoming Public Health Laboratory | Wyoming Public Health Laboratory | Noah Hull, Taylor Fearing, Lynette Gumbleton, Channing Weber, Ashley Norberg, Bailey Bowcutt, and Wanda Manley |
| EPI_ISL_1008394 | Ohio Department of Health Laboratory | Ohio Department of Health Laboratory | Holmes, Jennifer; Eric Brandt, Keoni Omura, Glen McGillivary, Caitlin McDonnell, Jade Mowery, Stephanie Mccracken, Tyler Payne, Kirtana Ramadugu, Erica Leasure, Brent Lee, Kelsey Florek, Heather Blankenship, Quanta Brown, and Tammy Bannerman |
| EPI_ISL_1008395, EPI_ISL_1008604, EPI_ISL_1008605, EPI_ISL_1008606, EPI_ISL_1008607, EPI_ISL_1008608, EPI_ISL_1008609 | Wyoming Public Health Laboratory | Wyoming Public Health Laboratory | Noah Hull, Taylor Fearing, Lynette Gumbleton, Channing Weber, Ashley Norberg, Bailey Bowcutt, and Wanda Manley |
| EPI_ISL_1008610 | Ohio Department of Health Laboratory | Ohio Department of Health Laboratory | Holmes, Jennifer; Eric Brandt, Keoni Omura, Glen McGillivary, Caitlin McDonnell, Jade Mowery, Stephanie Mccracken, Tyler Payne, Kirtana Ramadugu, Erica Leasure, Brent Lee, Kelsey Florek, Heather Blankenship, Quanta Brown, and Tammy Bannerman |
| EPI_ISL_1008611, EPI_ISL_1008614, EPI_ISL_1008615, EPI_ISL_1008657, EPI_ISL_1008658, EPI_ISL_1008659, EPI_ISL_1008660, EPI_ISL_1008661, EPI_ISL_1008662 | Wyoming Public Health Laboratory | Wyoming Public Health Laboratory | Noah Hull, Taylor Fearing, Lynette Gumbleton, Channing Weber, Ashley Norberg, Bailey Bowcutt, and Wanda Manley |
| EPI_ISL_1008713 | Lister Laboratorio de Referencia S.A. de C.V. | Instituto de diagnóstico y Referencia Epidemiologicos (INDRE) Departamento de Virología | Claudia Wong-Arambula, Abril Rodriguez-Maldonado, Fabiola Garces-Ayala, Natividad Cruz-Ortiz, Tatiana Nunez-Garcia, Gisela Barrera-Badillo, Lucia Hernandez-Rivas, Irma Lopez-Martinez, Ernesto Ramirez-Gonzalez. |
| EPI_ISL_1008714 | Unidad de Patología Clínica | Instituto de diagnóstico y Referencia Epidemiologicos (INDRE) Departamento de Virología | Claudia Wong-Arambula, Abril Rodriguez-Maldonado, Fabiola Garces-Ayala, Natividad Cruz-Ortiz, Tatiana Nunez-Garcia, Gisela Barrera-Badillo, Lucia Hernandez-Rivas, Irma Lopez-Martinez, Ernesto Ramirez-Gonzalez. |
| EPI_ISL_1008761, EPI_ISL_1008762, EPI_ISL_1008763, EPI_ISL_1008764, EPI_ISL_1008765, EPI_ISL_1008766, EPI_ISL_1008767, EPI_ISL_1008768, EPI_ISL_1008770, EPI_ISL_1008771, EPI_ISL_1008772, EPI_ISL_1008773, EPI_ISL_1008774, EPI_ISL_1008775, EPI_ISL_1008776, EPI_ISL_1008777, EPI_ISL_1008778, EPI_ISL_1008779, EPI_ISL_1008780, EPI_ISL_1008781, EPI_ISL_1008782, EPI_ISL_1008783, EPI_ISL_1008784, EPI_ISL_1008785, EPI_ISL_1008786, EPI_ISL_1008787, EPI_ISL_1008788, EPI_ISL_1008789, EPI_ISL_1008790, EPI_ISL_1008791, EPI_ISL_1008792, EPI_ISL_1008793, EPI_ISL_1008794, EPI_ISL_1008795, EPI_ISL_1008796, EPI_ISL_1008797, EPI_ISL_1008799, EPI_ISL_1008800, EPI_ISL_1008801, EPI_ISL_1008802, EPI_ISL_1008803, EPI_ISL_1008804, EPI_ISL_1008805, EPI_ISL_1008806, EPI_ISL_1008807, EPI_ISL_1008808, EPI_ISL_1008809, EPI_ISL_1008810, EPI_ISL_1008811, EPI_ISL_1008812, EPI_ISL_1008813, EPI_ISL_1008814, EPI_ISL_1008815, EPI_ISL_1008816, EPI_ISL_1008817, EPI_ISL_1008818, EPI_ISL_1008819, EPI_ISL_1008820, EPI_ISL_1008821, EPI_ISL_1008823, EPI_ISL_1008825, EPI_ISL_1008826, EPI_ISL_1008827, EPI_ISL_1008828, EPI_ISL_1008829, EPI_ISL_1008830, EPI_ISL_1008831, EPI_ISL_1008832, EPI_ISL_1008833, EPI_ISL_1008834, EPI_ISL_1008835, EPI_ISL_1008836, EPI_ISL_1008837, EPI_ISL_1008838, EPI_ISL_1008839, EPI_ISL_1008840, EPI_ISL_1008842, EPI_ISL_1008843, EPI_ISL_1008844, EPI_ISL_1008845, EPI_ISL_1008846, EPI_ISL_1008847, EPI_ISL_1008848, EPI_ISL_1008849, EPI_ISL_1008850, EPI_ISL_1008851, EPI_ISL_1008852, EPI_ISL_1008853, EPI_ISL_1008854, EPI_ISL_1008855, EPI_ISL_1008856, EPI_ISL_1008857, EPI_ISL_1008858, EPI_ISL_1008860, EPI_ISL_1008861, EPI_ISL_1008862, EPI_ISL_1008863, EPI_ISL_1008864, EPI_ISL_1008865, EPI_ISL_1008866, EPI_ISL_1008867, EPI_ISL_1008868, EPI_ISL_1008869, EPI_ISL_1008870, EPI_ISL_1008871, EPI_ISL_1008872, EPI_ISL_1008873, EPI_ISL_1008875 | | | |
| see above | Michigan Department of Health and Human Services, Bureau of Laboratories | Michigan Department of Health and Human Services, Bureau of Laboratories | Blankenship HM, Riner D, Soehnlen MK |
| EPI_ISL_1008893, EPI_ISL_1008894, EPI_ISL_1008895, EPI_ISL_1008896 | IL Dept. of Public Health Springfield Laboratory | Pathogen Discovery, Respiratory Viruses Branch, Division of Viral Diseases, Centers for Disease Control and Prevention | Ying Tao, Jing Zhang, Yan Li, Krista Queen, Anna Uehara, Peter Cook, Clinton R. Paden, Haibin Wang, Suxiang Tong |
| EPI_ISL_1008897, EPI_ISL_1008898, EPI_ISL_1008899, EPI_ISL_1008900 | LA Office of Public Health Laboratories | Pathogen Discovery, Respiratory Viruses Branch, Division of Viral Diseases, Centers for Disease Control and Prevention | Ying Tao, Jing Zhang, Yan Li, Krista Queen, Anna Uehara, Peter Cook, Clinton R. Paden, Haibin Wang, Suxiang Tong |
| EPI_ISL_1008902, EPI_ISL_1008903 | PA Department of Health, Bureau of Laboratories | Pathogen Discovery, Respiratory Viruses Branch, Division of Viral Diseases, Centers for Disease Control and Prevention | Ying Tao, Jing Zhang, Yan Li, Krista Queen, Anna Uehara, Peter Cook, Clinton R. Paden, Haibin Wang, Suxiang Tong |
| EPI_ISL_1008904, EPI_ISL_1008905, EPI_ISL_1008906 | TN Division of Laboratory Services | Pathogen Discovery, Respiratory Viruses Branch, Division of Viral Diseases, Centers for Disease Control and Prevention | Ying Tao, Jing Zhang, Yan Li, Krista Queen, Anna Uehara, Peter Cook, Clinton R. Paden, Haibin Wang, Suxiang Tong |
| EPI_ISL_1008907, EPI_ISL_1008908, EPI_ISL_1008909 | Resolve Diagnostics | Pathogen Discovery, Respiratory Viruses Branch, Division of Viral Diseases, Centers for Disease Control and Prevention | Ying Tao, Jing Zhang, Yan Li, Krista Queen, Anna Uehara, Peter Cook, Clinton R. Paden, Haibin Wang, Suxiang Tong |
| EPI_ISL_1008910, EPI_ISL_1008911, EPI_ISL_1008912, EPI_ISL_1008913, EPI_ISL_1008914, EPI_ISL_1008915 | GA Department of Public Health Laboratory | Pathogen Discovery, Respiratory Viruses Branch, Division of Viral Diseases, Centers for Disease Control and Prevention | Ying Tao, Jing Zhang, Yan Li, Krista Queen, Anna Uehara, Peter Cook, Clinton R. Paden, Haibin Wang, Suxiang Tong |
| EPI_ISL_1008916 | CT-Dr. Katherine A. Kelley State Public Health Lab | Pathogen Discovery, Respiratory Viruses Branch, Division of Viral Diseases, Centers for Disease Control and Prevention | Ying Tao, Jing Zhang, Yan Li, Krista Queen, Anna Uehara, Peter Cook, Clinton R. Paden, Haibin Wang, Suxiang Tong |
| EPI_ISL_1008917, EPI_ISL_1008918 | IN State Department of Health Laboratory Services | Pathogen Discovery, Respiratory Viruses Branch, Division of Viral Diseases, Centers for Disease Control and Prevention | Ying Tao, Jing Zhang, Yan Li, Krista Queen, Anna Uehara, Peter Cook, Clinton R. Paden, Haibin Wang, Suxiang Tong |
| EPI_ISL_1008919, EPI_ISL_1008920 | IL Department of Public Health Chicago Laboratory | Pathogen Discovery, Respiratory Viruses Branch, Division of Viral Diseases, Centers for Disease Control and Prevention | Ying Tao, Jing Zhang, Yan Li, Krista Queen, Anna Uehara, Peter Cook, Clinton R. Paden, Haibin Wang, Suxiang Tong |
| EPI_ISL_1008921, EPI_ISL_1008922, EPI_ISL_1008923 | LA Office of Public Health Laboratories | Pathogen Discovery, Respiratory Viruses Branch, Division of Viral Diseases, Centers for Disease Control and Prevention | Ying Tao, Jing Zhang, Yan Li, Krista Queen, Anna Uehara, Peter Cook, Clinton R. Paden, Haibin Wang, Suxiang Tong |
| EPI_ISL_1008924, EPI_ISL_1008925, EPI_ISL_1008926 | IN State Department of Health Laboratory Services | Pathogen Discovery, Respiratory Viruses Branch, Division of Viral Diseases, Centers for Disease Control and Prevention | Ying Tao, Jing Zhang, Yan Li, Krista Queen, Anna Uehara, Peter Cook, Clinton R. Paden, Haibin Wang, Suxiang Tong |
| EPI_ISL_1008928, EPI_ISL_1008929 | MS Public Health Laboratory | Pathogen Discovery, Respiratory Viruses Branch, Division of Viral Diseases, Centers for Disease Control and Prevention | Ying Tao, Jing Zhang, Yan Li, Krista Queen, Anna Uehara, Peter Cook, Clinton R. Paden, Haibin Wang, Suxiang Tong |
| EPI_ISL_1009125, EPI_ISL_1009126 | Yale Pathology Lab | Grubaugh Lab - Yale School of Public Health | Tara Alpert, Joseph Fauver, Chen Liu, Pei Hui, Jianhui Wang, Susan Bell and Han Zhou, Anderson Brito, Mallery Breban, Anne Wyllie, Chantal Vogels, Mary Petrone, Chaney Kalinich, Isabel Ott, Arnau Casanovas, Catherine Muenker, Adam Moore, Alice Lu, Maria Tokuyama, Patrick Wong, Peiwen Lu, Saad Omer, Richard Martinello, Allison Nelson, Shelli Farhadian, Akiko Iwasaki, Charlese Dela Cruz, Albert Ko, Nathan Grubaugh |
| EPI_ISL_1009162, EPI_ISL_1009163, EPI_ISL_1009164, EPI_ISL_1009165, EPI_ISL_1009166, EPI_ISL_1009167, EPI_ISL_1009168, EPI_ISL_1009169, EPI_ISL_1009170, EPI_ISL_1009171, EPI_ISL_1009172, EPI_ISL_1009174, EPI_ISL_1009175, EPI_ISL_1009176, EPI_ISL_1009177, EPI_ISL_1009178, EPI_ISL_1009179, EPI_ISL_1009183, EPI_ISL_1009185, EPI_ISL_1009187, EPI_ISL_1009188, EPI_ISL_1009189, EPI_ISL_1009190, EPI_ISL_1009191, EPI_ISL_1009193, EPI_ISL_1009194, EPI_ISL_1009195, EPI_ISL_1009196, EPI_ISL_1009197, EPI_ISL_1009198, EPI_ISL_1009199, EPI_ISL_1009201, EPI_ISL_1009202, EPI_ISL_1009203, EPI_ISL_1009204, EPI_ISL_1009205, EPI_ISL_1009206, EPI_ISL_1009208, EPI_ISL_1009209, EPI_ISL_1009210, EPI_ISL_1009212 | | | |
| see above | American Esoteric Laboratories | Colleen B. Jonsson | Mariah K. Taylor, Walter Reichard, Jyothi Parvathareddy, Colleen B. Jonsson |
| EPI_ISL_1009215, EPI_ISL_1009216, EPI_ISL_1009217, EPI_ISL_1009218, EPI_ISL_1009220, EPI_ISL_1009221, EPI_ISL_1009222, EPI_ISL_1009223, EPI_ISL_1009224, EPI_ISL_1009227, EPI_ISL_1009229, EPI_ISL_1009230, EPI_ISL_1009231, EPI_ISL_1009232, EPI_ISL_1009233, EPI_ISL_1009235, EPI_ISL_1009236, EPI_ISL_1009237, EPI_ISL_1009238, EPI_ISL_1009239, EPI_ISL_1009240, EPI_ISL_1009241, EPI_ISL_1009242, EPI_ISL_1009243, EPI_ISL_1009244, EPI_ISL_1009245, EPI_ISL_1009247, EPI_ISL_1009248, EPI_ISL_1009249, EPI_ISL_1009250, EPI_ISL_1009251, EPI_ISL_1009252, EPI_ISL_1009253, EPI_ISL_1009254, EPI_ISL_1009255, EPI_ISL_1009256, EPI_ISL_1009257, EPI_ISL_1009258, EPI_ISL_1009259, EPI_ISL_1009260, EPI_ISL_1009261, EPI_ISL_1009262, EPI_ISL_1009263, EPI_ISL_1009264, EPI_ISL_1009265, EPI_ISL_1009266, EPI_ISL_1009267, EPI_ISL_1009268, EPI_ISL_1009269, EPI_ISL_1009270, EPI_ISL_1009271, EPI_ISL_1009274, EPI_ISL_1009275, EPI_ISL_1009277, EPI_ISL_1009280, EPI_ISL_1009281, EPI_ISL_1009282, EPI_ISL_1009283, EPI_ISL_1009284, EPI_ISL_1009286, EPI_ISL_1009287 | | | |
| see above | Wyoming Public Health Laboratory | Wyoming Public Health Laboratory | Noah Hull, Taylor Fearing, Lynette Gumbleton, Channing Weber, Ashley Norberg, Bailey Bowcutt, and Wanda Manley |
| EPI_ISL_1009289, EPI_ISL_1009290, EPI_ISL_1009291, EPI_ISL_1009292, EPI_ISL_1009293, EPI_ISL_1009294, EPI_ISL_1009295, EPI_ISL_1009296, EPI_ISL_1009297, EPI_ISL_1009298, EPI_ISL_1009299, EPI_ISL_1009300, EPI_ISL_1009301, EPI_ISL_1009302, EPI_ISL_1009303, EPI_ISL_1009304, EPI_ISL_1009305, EPI_ISL_1009306, EPI_ISL_1009307, EPI_ISL_1009308, EPI_ISL_1009309, EPI_ISL_1009310, EPI_ISL_1009311, EPI_ISL_1009312, EPI_ISL_1009313, EPI_ISL_1009314, EPI_ISL_1009315, EPI_ISL_1009316, EPI_ISL_1009317, EPI_ISL_1009318, EPI_ISL_1009319, EPI_ISL_1009320, EPI_ISL_1009321, EPI_ISL_1009322, EPI_ISL_1009323, EPI_ISL_1009324, EPI_ISL_1009325, EPI_ISL_1009326, EPI_ISL_1009327, EPI_ISL_1009328, EPI_ISL_1009329, EPI_ISL_1009330, EPI_ISL_1009331, EPI_ISL_1009332, EPI_ISL_1009333, EPI_ISL_1009334, EPI_ISL_1009335, EPI_ISL_1009336, EPI_ISL_1009337, EPI_ISL_1009338, EPI_ISL_1009339, EPI_ISL_1009340, EPI_ISL_1009341, EPI_ISL_1009342, EPI_ISL_1009343, EPI_ISL_1009344, EPI_ISL_1009345, EPI_ISL_1009346, EPI_ISL_1009347, EPI_ISL_1009348, EPI_ISL_1009349, EPI_ISL_1009350, EPI_ISL_1009351, EPI_ISL_1009352, EPI_ISL_1009353, EPI_ISL_1009354, EPI_ISL_1009355, EPI_ISL_1009356, EPI_ISL_1009357, EPI_ISL_1009358, EPI_ISL_1009359, EPI_ISL_1009360, EPI_ISL_1009361, EPI_ISL_1009362, EPI_ISL_1009363, EPI_ISL_1009364, EPI_ISL_1009365, EPI_ISL_1009366, EPI_ISL_1009367, EPI_ISL_1009368, EPI_ISL_1009369, EPI_ISL_1009370, EPI_ISL_1009371, EPI_ISL_1009372, EPI_ISL_1009373, EPI_ISL_1009374, EPI_ISL_1009375, EPI_ISL_1009376, EPI_ISL_1009377, EPI_ISL_1009378, EPI_ISL_1009379, EPI_ISL_1009380, EPI_ISL_1009381, EPI_ISL_1009382, EPI_ISL_1009383, EPI_ISL_1009384, EPI_ISL_1009385, EPI_ISL_1009386, EPI_ISL_1009387, EPI_ISL_1009388, EPI_ISL_1009389, EPI_ISL_1009390, EPI_ISL_1009391, EPI_ISL_1009392, EPI_ISL_1009393, EPI_ISL_1009394, EPI_ISL_1009395, EPI_ISL_1009396, EPI_ISL_1009397, EPI_ISL_1009398, EPI_ISL_1009399, EPI_ISL_1009400, EPI_ISL_1009401, EPI_ISL_1009402, EPI_ISL_1009403, EPI_ISL_1009404, EPI_ISL_1009405, EPI_ISL_1009406, EPI_ISL_1009407, EPI_ISL_1009408, EPI_ISL_1009409, EPI_ISL_1009410, EPI_ISL_1009411, EPI_ISL_1009412, EPI_ISL_1009413, EPI_ISL_1009414, EPI_ISL_1009415, EPI_ISL_1009416, EPI_ISL_1009417, EPI_ISL_1009418, EPI_ISL_1009419, EPI_ISL_1009420, EPI_ISL_1009421, EPI_ISL_1009422, EPI_ISL_1009423, EPI_ISL_1009424, EPI_ISL_1009425, EPI_ISL_1009426, EPI_ISL_1009427, EPI_ISL_1009428, EPI_ISL_1009429, EPI_ISL_1009430, EPI_ISL_1009431, EPI_ISL_1009432, EPI_ISL_1009433, EPI_ISL_1009434, EPI_ISL_1009435, EPI_ISL_1009436, EPI_ISL_1009437, EPI_ISL_1009438, EPI_ISL_1009439, EPI_ISL_1009440, EPI_ISL_1009441, EPI_ISL_1009442, EPI_ISL_1009443, EPI_ISL_1009444, EPI_ISL_1009445, EPI_ISL_1009446, EPI_ISL_1009447, EPI_ISL_1009448, EPI_ISL_1009449, EPI_ISL_1009450, EPI_ISL_1009451, EPI_ISL_1009452, EPI_ISL_1009453, EPI_ISL_1009454, EPI_ISL_1009455, EPI_ISL_1009456, EPI_ISL_1009457, EPI_ISL_1009458, EPI_ISL_1009459, EPI_ISL_1009460, EPI_ISL_1009461, EPI_ISL_1009462, EPI_ISL_1009463, EPI_ISL_1009464, | | | |

| | | | |
|---|---|---|---|
| EPI_ISL_1009465, EPI_ISL_1009466, EPI_ISL_1009467, EPI_ISL_1009468, EPI_ISL_1009469, EPI_ISL_1009470, EPI_ISL_1009471, EPI_ISL_1009472, EPI_ISL_1009473, EPI_ISL_1009474, EPI_ISL_1009475, EPI_ISL_1009476, EPI_ISL_1009477, EPI_ISL_1009478, EPI_ISL_1009479, EPI_ISL_1009480, EPI_ISL_1009481, EPI_ISL_1009482, EPI_ISL_1009483, EPI_ISL_1009484, EPI_ISL_1009485, EPI_ISL_1009486, EPI_ISL_1009487, EPI_ISL_1009488, EPI_ISL_1009489, EPI_ISL_1009490, EPI_ISL_1009491, EPI_ISL_1009492, EPI_ISL_1009493, EPI_ISL_1009494, EPI_ISL_1009495, EPI_ISL_1009496, EPI_ISL_1009497, EPI_ISL_1009498, EPI_ISL_1009499, EPI_ISL_1009500, EPI_ISL_1009501, EPI_ISL_1009502, EPI_ISL_1009503, EPI_ISL_1009504, EPI_ISL_1009505, EPI_ISL_1009506, EPI_ISL_1009507, EPI_ISL_1009508, EPI_ISL_1009509, EPI_ISL_1009510, EPI_ISL_1009511, EPI_ISL_1009512, EPI_ISL_1009513, EPI_ISL_1009514, EPI_ISL_1009515, EPI_ISL_1009516, EPI_ISL_1009517, EPI_ISL_1009518, EPI_ISL_1009519, EPI_ISL_1009520, EPI_ISL_1009521, EPI_ISL_1009522, EPI_ISL_1009523, EPI_ISL_1009524, EPI_ISL_1009525, EPI_ISL_1009526, EPI_ISL_1009527, EPI_ISL_1009528, EPI_ISL_1009529, EPI_ISL_1009530, EPI_ISL_1009531, EPI_ISL_1009532, EPI_ISL_1009533, EPI_ISL_1009534, EPI_ISL_1009535, EPI_ISL_1009536, EPI_ISL_1009537, EPI_ISL_1009538, EPI_ISL_1009539, EPI_ISL_1009540, EPI_ISL_1009541, EPI_ISL_1009542, EPI_ISL_1009543, EPI_ISL_1009544, EPI_ISL_1009545, EPI_ISL_1009546, EPI_ISL_1009547, EPI_ISL_1009548, EPI_ISL_1009549, EPI_ISL_1009550, EPI_ISL_1009551, EPI_ISL_1009552, EPI_ISL_1009553, EPI_ISL_1009554, EPI_ISL_1009555, EPI_ISL_1009556, EPI_ISL_1009557, EPI_ISL_1009558, EPI_ISL_1009559, EPI_ISL_1009560, EPI_ISL_1009561, EPI_ISL_1009562, EPI_ISL_1009563, EPI_ISL_1009564, EPI_ISL_1009565, EPI_ISL_1009566, EPI_ISL_1009567, EPI_ISL_1009568, EPI_ISL_1009569, EPI_ISL_1009570, EPI_ISL_1009571, EPI_ISL_1009572, EPI_ISL_1009573, EPI_ISL_1009574, EPI_ISL_1009575, EPI_ISL_1009576, EPI_ISL_1009577, EPI_ISL_1009578, EPI_ISL_1009579, EPI_ISL_1009580, EPI_ISL_1009581, EPI_ISL_1009582, EPI_ISL_1009583, EPI_ISL_1009584, EPI_ISL_1009585, EPI_ISL_1009586, EPI_ISL_1009587, EPI_ISL_1009588, EPI_ISL_1009589, EPI_ISL_1009590, EPI_ISL_1009591, EPI_ISL_1009592, EPI_ISL_1009593, EPI_ISL_1009594, EPI_ISL_1009595, EPI_ISL_1009596, EPI_ISL_1009597, EPI_ISL_1009598, EPI_ISL_1009599, EPI_ISL_1009600, EPI_ISL_1009601, EPI_ISL_1009602, EPI_ISL_1009603, EPI_ISL_1009604, EPI_ISL_1009605, EPI_ISL_1009606, EPI_ISL_1009607, EPI_ISL_1009608, EPI_ISL_1009609, EPI_ISL_1009610, EPI_ISL_1009611, EPI_ISL_1009612, EPI_ISL_1009613, EPI_ISL_1009614, EPI_ISL_1009615, EPI_ISL_1009616 | | | |
| see above | Erie County Public Health (ECPHL) | University at Buffalo Genomics and Bioinformatics Core | Jonathan Bard, Natalie Lamb, Alyssa Pohlman, Brandon Marzullo, Amanda Boccolucci, Norma Nowak, Donald Yergeau, Jennifer Surtees |
| EPI_ISL_1009620, EPI_ISL_1009621, EPI_ISL_1009622, EPI_ISL_1009624 | School of Pharmacy, Shenandoah University | School of Pharmacy, Shenandoah University | Adams,S.M., Harralson,A.F., Kidd,R.S., Sawyer,G.W. |
| EPI_ISL_1009632, EPI_ISL_1009633, EPI_ISL_1009634, EPI_ISL_1009635, EPI_ISL_1009636, EPI_ISL_1009637, EPI_ISL_1009638, EPI_ISL_1009639, EPI_ISL_1009640, EPI_ISL_1009641, EPI_ISL_1009642, EPI_ISL_1009643, EPI_ISL_1009644, EPI_ISL_1009645, EPI_ISL_1009646, EPI_ISL_1009647, EPI_ISL_1009648, EPI_ISL_1009649, EPI_ISL_1009650, EPI_ISL_1009651, EPI_ISL_1009652, EPI_ISL_1009653, EPI_ISL_1009654 | | | |
| see above | DPHL | Delaware Public Health Lab | Gregory Hovan |
| EPI_ISL_1009657, EPI_ISL_1009660, EPI_ISL_1009661, EPI_ISL_1009662, EPI_ISL_1009663, EPI_ISL_1009664 | Santa Clara County Public Health Laboratory | Santa Clara County Public Health Laboratory | Santa Clara County Public Health Department |
| EPI_ISL_1009666, EPI_ISL_1009668, EPI_ISL_1009670 | Auburn Sports Medicine | Synergy Laboratories | Megan Cornwell |
| EPI_ISL_1009712, EPI_ISL_1009713, EPI_ISL_1009715, EPI_ISL_1009716, EPI_ISL_1009717, EPI_ISL_1009718, EPI_ISL_1009719, EPI_ISL_1009720, EPI_ISL_1009721, EPI_ISL_1009722, EPI_ISL_1009723, EPI_ISL_1009724, EPI_ISL_1009725, EPI_ISL_1009729, EPI_ISL_1009732, EPI_ISL_1009733, EPI_ISL_1009735, EPI_ISL_1009747, EPI_ISL_1009748, EPI_ISL_1009749, EPI_ISL_1009751, EPI_ISL_1009754, EPI_ISL_1009755, EPI_ISL_1009756, EPI_ISL_1009757, EPI_ISL_1009759, EPI_ISL_1009760, EPI_ISL_1009761, EPI_ISL_1009762, EPI_ISL_1009764, EPI_ISL_1009765, EPI_ISL_1009766, EPI_ISL_1009767, EPI_ISL_1009768, EPI_ISL_1009769, EPI_ISL_1009770, EPI_ISL_1009774, EPI_ISL_1009775, EPI_ISL_1009776, EPI_ISL_1009777, EPI_ISL_1009780, EPI_ISL_1009781, EPI_ISL_1009783, EPI_ISL_1009784, EPI_ISL_1009785, EPI_ISL_1009786, EPI_ISL_1009787, EPI_ISL_1009788, EPI_ISL_1009795 | | | |
| see above | Massachusetts State Public Health Laboratory | Massachusetts State Public Health Laboratory | Andrew Lang, Timelia Fink, Glen Gallagher, Sandra Smole |
| EPI_ISL_1009796, EPI_ISL_1009797, EPI_ISL_1009800, EPI_ISL_1009801, EPI_ISL_1009802, EPI_ISL_1009803, EPI_ISL_1009804, EPI_ISL_1009805, EPI_ISL_1009806, EPI_ISL_1009807, EPI_ISL_1009808, EPI_ISL_1009809, EPI_ISL_1009810, EPI_ISL_1009811, EPI_ISL_1009812 | | | |
| see above | Curative Labs, Inc. | Wyoming Public Health Laboratory | Noah Hull, Taylor Fearing, Lynette Gumbleton, Channing Weber, Ashley Norberg, Bailey Bowcutt, and Wanda Manley |
| EPI_ISL_1009813 | UC Davis Genome Center and Healthy Davis Together | UC Davis Genome Center | Sam Diaz-Muñoz and AJ Campbell |
| EPI_ISL_1009815, EPI_ISL_1009816, EPI_ISL_1009817, EPI_ISL_1009818, EPI_ISL_1009820, EPI_ISL_1009821, EPI_ISL_1009823, EPI_ISL_1009824, EPI_ISL_1009826 | Curative Labs, Inc. | Wyoming Public Health Laboratory | Noah Hull, Taylor Fearing, Lynette Gumbleton, Channing Weber, Ashley Norberg, Bailey Bowcutt, and Wanda Manley |
| EPI_ISL_1009827 | UC Davis Genome Center and Healthy Davis Together | UC Davis Genome Center | Sam Diaz-Muñoz and AJ Campbell |
| EPI_ISL_1009828, EPI_ISL_1009829, EPI_ISL_1009830, EPI_ISL_1009831, EPI_ISL_1009832, EPI_ISL_1009833, EPI_ISL_1009834, EPI_ISL_1009835, EPI_ISL_1009836, EPI_ISL_1009837, EPI_ISL_1009838, EPI_ISL_1009839, EPI_ISL_1009840, EPI_ISL_1009841, EPI_ISL_1009842, EPI_ISL_1009843, EPI_ISL_1009846, EPI_ISL_1009847, EPI_ISL_1009848, EPI_ISL_1009849, EPI_ISL_1009850, EPI_ISL_1009851, EPI_ISL_1009852, EPI_ISL_1009853, EPI_ISL_1009854, EPI_ISL_1009855, EPI_ISL_1009856, EPI_ISL_1009857, EPI_ISL_1009858, EPI_ISL_1009859, EPI_ISL_1009860, EPI_ISL_1009861, EPI_ISL_1009862, EPI_ISL_1009863, EPI_ISL_1009864, EPI_ISL_1009865, EPI_ISL_1009866, EPI_ISL_1009867 | | | |
| see above | Curative Labs, Inc. | Wyoming Public Health Laboratory | Noah Hull, Taylor Fearing, Lynette Gumbleton, Channing Weber, Ashley Norberg, Bailey Bowcutt, and Wanda Manley |
| EPI_ISL_1009870, EPI_ISL_1009871, EPI_ISL_1009873, EPI_ISL_1009876, EPI_ISL_1009877, EPI_ISL_1009880, EPI_ISL_1009881, EPI_ISL_1009883, EPI_ISL_1009885, EPI_ISL_1009886, EPI_ISL_1009887, EPI_ISL_1009892, EPI_ISL_1009894, EPI_ISL_1009899, EPI_ISL_1009906, EPI_ISL_1009909, EPI_ISL_1009916, EPI_ISL_1009919, EPI_ISL_1009924, EPI_ISL_1009928, EPI_ISL_1009932, EPI_ISL_1009933, EPI_ISL_1009941, EPI_ISL_1009942, EPI_ISL_1009945, EPI_ISL_1009948, EPI_ISL_1009952, EPI_ISL_1009953, EPI_ISL_1009954, EPI_ISL_1009955, EPI_ISL_1009963, EPI_ISL_1009964, EPI_ISL_1009966, EPI_ISL_1009967, EPI_ISL_1009969, EPI_ISL_1009971, EPI_ISL_1009976, EPI_ISL_1009977, EPI_ISL_1009980, EPI_ISL_1009981, EPI_ISL_1009982, EPI_ISL_1009988, EPI_ISL_1009991, EPI_ISL_1009992, EPI_ISL_1009993, EPI_ISL_1009995, EPI_ISL_1009999, EPI_ISL_1010000, EPI_ISL_1010001, EPI_ISL_1010002, EPI_ISL_1010004, EPI_ISL_1010007, EPI_ISL_1010013, EPI_ISL_1010017, EPI_ISL_1010018, EPI_ISL_1010019, EPI_ISL_1010020, EPI_ISL_1010022, EPI_ISL_1010024, EPI_ISL_1010025, EPI_ISL_1010026, EPI_ISL_1010027, EPI_ISL_1010028, EPI_ISL_1010029, EPI_ISL_1010030, EPI_ISL_1010032, EPI_ISL_1010037, EPI_ISL_1010040, EPI_ISL_1010044, EPI_ISL_1010045, EPI_ISL_1010046, EPI_ISL_1010050, EPI_ISL_1010051, EPI_ISL_1010054, EPI_ISL_1010055, EPI_ISL_1010056, EPI_ISL_1010058, EPI_ISL_1010060, EPI_ISL_1010061, EPI_ISL_1010063, EPI_ISL_1010066, EPI_ISL_1010067, EPI_ISL_1010078, EPI_ISL_1010080, EPI_ISL_1010081, EPI_ISL_1010083, EPI_ISL_1010084, EPI_ISL_1010086, EPI_ISL_1010088, EPI_ISL_1010089, EPI_ISL_1010091, EPI_ISL_1010093, EPI_ISL_1010094, EPI_ISL_1010096, EPI_ISL_1010101, EPI_ISL_1010103, EPI_ISL_1010104, EPI_ISL_1010106, EPI_ISL_1010107, EPI_ISL_1010108, EPI_ISL_1010109, EPI_ISL_1010110, EPI_ISL_1010111, EPI_ISL_1010112, EPI_ISL_1010113, EPI_ISL_1010114, EPI_ISL_1010118, EPI_ISL_1010122, EPI_ISL_1010124, EPI_ISL_1010131, EPI_ISL_1010133, EPI_ISL_1010135, EPI_ISL_1010137, EPI_ISL_1010138, EPI_ISL_1010141, EPI_ISL_1010143, EPI_ISL_1010144, EPI_ISL_1010153, EPI_ISL_1010155, EPI_ISL_1010157, EPI_ISL_1010161, EPI_ISL_1010163, EPI_ISL_1010164, EPI_ISL_1010167, EPI_ISL_1010168, EPI_ISL_1010170, EPI_ISL_1010172, EPI_ISL_1010174, EPI_ISL_1010175, EPI_ISL_1010176, EPI_ISL_1010179, EPI_ISL_1010180, EPI_ISL_1010183, EPI_ISL_1010184, EPI_ISL_1010185, EPI_ISL_1010187, EPI_ISL_1010189, EPI_ISL_1010190, EPI_ISL_1010191, EPI_ISL_1010194, EPI_ISL_1010195, EPI_ISL_1010196, EPI_ISL_1010202, EPI_ISL_1010206, EPI_ISL_1010207, EPI_ISL_1010209, EPI_ISL_1010211, EPI_ISL_1010214, EPI_ISL_1010215, EPI_ISL_1010216, EPI_ISL_1010223, EPI_ISL_1010224, EPI_ISL_1010227, EPI_ISL_1010230, EPI_ISL_1010234, EPI_ISL_1010236, EPI_ISL_1010237, EPI_ISL_1010239, EPI_ISL_1010240, EPI_ISL_1010241, EPI_ISL_1010243, EPI_ISL_1010245, EPI_ISL_1010250, EPI_ISL_1010252, EPI_ISL_1010255, EPI_ISL_1010263, EPI_ISL_1010268, EPI_ISL_1010279, EPI_ISL_1010280, EPI_ISL_1010283, EPI_ISL_1010289, EPI_ISL_1010295, EPI_ISL_1010302, EPI_ISL_1010305, EPI_ISL_1010306, EPI_ISL_1010309, EPI_ISL_1010314, EPI_ISL_1010318, EPI_ISL_1010319, EPI_ISL_1010321, EPI_ISL_1010323, EPI_ISL_1010324, EPI_ISL_1010327, EPI_ISL_1010331, EPI_ISL_1010332, EPI_ISL_1010333, EPI_ISL_1010336, EPI_ISL_1010343, EPI_ISL_1010348, EPI_ISL_1010349, EPI_ISL_1010351, EPI_ISL_1010363, EPI_ISL_1010364, EPI_ISL_1010371, EPI_ISL_1010373, EPI_ISL_1010375, EPI_ISL_1010376, EPI_ISL_1010377, EPI_ISL_1010380, EPI_ISL_1010388, EPI_ISL_1010389, EPI_ISL_1010393, EPI_ISL_1010394, EPI_ISL_1010396, EPI_ISL_1010398, EPI_ISL_1010400, EPI_ISL_1010404, EPI_ISL_1010407, EPI_ISL_1010409, EPI_ISL_1010411, EPI_ISL_1010415, EPI_ISL_1010416, EPI_ISL_1010422, EPI_ISL_1010428, EPI_ISL_1010430, EPI_ISL_1010432, EPI_ISL_1010437, EPI_ISL_1010438, EPI_ISL_1010439, EPI_ISL_1010442, EPI_ISL_1010453, EPI_ISL_1010458, EPI_ISL_1010459, EPI_ISL_1010460, EPI_ISL_1010464, EPI_ISL_1010468, EPI_ISL_1010471, EPI_ISL_1010472, EPI_ISL_1010474, EPI_ISL_1010475, EPI_ISL_1010477, EPI_ISL_1010478, EPI_ISL_1010486, EPI_ISL_1010487, EPI_ISL_1010489, EPI_ISL_1010491, EPI_ISL_1010492, EPI_ISL_1010494, EPI_ISL_1010495, EPI_ISL_1010496, EPI_ISL_1010498, EPI_ISL_1010503, EPI_ISL_1010508, EPI_ISL_1010512, EPI_ISL_1010513, EPI_ISL_1010514, EPI_ISL_1010516, EPI_ISL_1010519, EPI_ISL_1010522, EPI_ISL_1010524, EPI_ISL_1010525, EPI_ISL_1010526, EPI_ISL_1010527, EPI_ISL_1010530, EPI_ISL_1010531, EPI_ISL_1010536, EPI_ISL_1010537, EPI_ISL_1010538, EPI_ISL_1010540, EPI_ISL_1010542, EPI_ISL_1010544, EPI_ISL_1010545, EPI_ISL_1010549, EPI_ISL_1010550, EPI_ISL_1010553, EPI_ISL_1010554, EPI_ISL_1010555, EPI_ISL_1010556, EPI_ISL_1010557, EPI_ISL_1010558, EPI_ISL_1010564, EPI_ISL_1010565, EPI_ISL_1010566, EPI_ISL_1010567, EPI_ISL_1010572, EPI_ISL_1010574, EPI_ISL_1010577, EPI_ISL_1010578, EPI_ISL_1010582, EPI_ISL_1010585, EPI_ISL_1010589, EPI_ISL_1010593, EPI_ISL_1010596, EPI_ISL_1010597, EPI_ISL_1010601, EPI_ISL_1010602, EPI_ISL_1010610, EPI_ISL_1010611, EPI_ISL_1010614, EPI_ISL_1010617, EPI_ISL_1010620, EPI_ISL_1010621, EPI_ISL_1010622, EPI_ISL_1010624, EPI_ISL_1010626, EPI_ISL_1010627, EPI_ISL_1010628, EPI_ISL_1010630, EPI_ISL_1010631, EPI_ISL_1010633, EPI_ISL_1010636, EPI_ISL_1010639, EPI_ISL_1010641, EPI_ISL_1010642, EPI_ISL_1010644, EPI_ISL_1010647, EPI_ISL_1010652, EPI_ISL_1010656, EPI_ISL_1010659, EPI_ISL_1010663, EPI_ISL_1010664, EPI_ISL_1010666, EPI_ISL_1010671, EPI_ISL_1010673, EPI_ISL_1010674, EPI_ISL_1010681, EPI_ISL_1010683, EPI_ISL_1010687, EPI_ISL_1010689, EPI_ISL_1010692, EPI_ISL_1010693, EPI_ISL_1010698 | | | |
| see above | Fulgent Genetics | Fulgent Genetics | Harry Gao, Mickey Li, John Gao, Joseph Fierro, Benafsh Sapra, Becky Tsai, Yan Meng, Doreen Ng, James Xie |
| EPI_ISL_1010732, EPI_ISL_1010733, EPI_ISL_1010734, EPI_ISL_1010735, EPI_ISL_1010736 | University of Iowa Hospitals & Clinics, Microbiology Laboratory | University or Iowa, Lung Biology and Cystic Fibrosis Research Center, Pezzulo Lab | Miguel E Ortiz, Alejandro A Pezzulo |
| EPI_ISL_1010737, EPI_ISL_1010738, EPI_ISL_1010739, EPI_ISL_1010741, EPI_ISL_1010744, EPI_ISL_1010746, EPI_ISL_1010747, EPI_ISL_1010748, EPI_ISL_1010749, EPI_ISL_1010750, EPI_ISL_1010751, EPI_ISL_1010753, EPI_ISL_1010755, EPI_ISL_1010756, EPI_ISL_1010758, EPI_ISL_1010759, EPI_ISL_1010760, EPI_ISL_1010761, EPI_ISL_1010765, EPI_ISL_1010767, EPI_ISL_1010768, EPI_ISL_1010770, EPI_ISL_1010771, EPI_ISL_1010772, EPI_ISL_1010775, EPI_ISL_1010776, EPI_ISL_1010777, EPI_ISL_1010780, EPI_ISL_1010782, EPI_ISL_1010783, EPI_ISL_1010784, EPI_ISL_1010790, EPI_ISL_1010795, EPI_ISL_1010796, EPI_ISL_1010797, EPI_ISL_1010798, EPI_ISL_1010799, EPI_ISL_1010800, EPI_ISL_1010804, EPI_ISL_1010805, EPI_ISL_1010806, EPI_ISL_1010808, EPI_ISL_1010810, EPI_ISL_1010813, EPI_ISL_1010817, EPI_ISL_1010818, EPI_ISL_1010820, EPI_ISL_1010821, EPI_ISL_1010824, EPI_ISL_1010825, EPI_ISL_1010826, EPI_ISL_1010827, EPI_ISL_1010828, EPI_ISL_1010829, EPI_ISL_1010830, EPI_ISL_1010833, EPI_ISL_1010834, EPI_ISL_1010836, EPI_ISL_1010837, EPI_ISL_1010838, EPI_ISL_1010839, EPI_ISL_1010844, EPI_ISL_1010846, EPI_ISL_1010848, EPI_ISL_1010851, EPI_ISL_1010855, EPI_ISL_1010858, EPI_ISL_1010862, EPI_ISL_1010863, EPI_ISL_1010865, EPI_ISL_1010866, EPI_ISL_1010868, EPI_ISL_1010869, EPI_ISL_1010870, EPI_ISL_1010871, EPI_ISL_1010872, EPI_ISL_1010873, EPI_ISL_1010874, EPI_ISL_1010876, EPI_ISL_1010880, EPI_ISL_1010881, EPI_ISL_1010882, EPI_ISL_1010885, EPI_ISL_1010887, EPI_ISL_1010890, EPI_ISL_1010891, EPI_ISL_1010892, EPI_ISL_1010893, EPI_ISL_1010894, EPI_ISL_1010895, EPI_ISL_1010896, EPI_ISL_1010898, EPI_ISL_1010899, EPI_ISL_1010901, EPI_ISL_1010902, EPI_ISL_1010904, EPI_ISL_1010905, EPI_ISL_1010906, EPI_ISL_1010907, EPI_ISL_1010908, EPI_ISL_1010910, EPI_ISL_1010911, EPI_ISL_1010912, EPI_ISL_1010915, EPI_ISL_1010917, EPI_ISL_1010918, EPI_ISL_1010919, EPI_ISL_1010920, EPI_ISL_1010921, EPI_ISL_1010923, EPI_ISL_1010926, EPI_ISL_1010929, EPI_ISL_1010931, EPI_ISL_1010934, EPI_ISL_1010935, EPI_ISL_1010936, EPI_ISL_1010939, EPI_ISL_1010940, EPI_ISL_1010941, EPI_ISL_1010942, EPI_ISL_1010943, EPI_ISL_1010946, EPI_ISL_1010949, EPI_ISL_1010952, EPI_ISL_1010954, EPI_ISL_1010955, EPI_ISL_1010956, EPI_ISL_1010957, EPI_ISL_1010958, EPI_ISL_1010959, EPI_ISL_1010962, EPI_ISL_1010963, EPI_ISL_1010965, EPI_ISL_1010968, EPI_ISL_1010969, EPI_ISL_1010970, EPI_ISL_1010974, EPI_ISL_1010975, EPI_ISL_1010976, EPI_ISL_1010978, EPI_ISL_1010979, EPI_ISL_1010983, EPI_ISL_1010984, EPI_ISL_1010986, EPI_ISL_1010987, EPI_ISL_1010988, EPI_ISL_1010992, EPI_ISL_1010993, EPI_ISL_1011004, EPI_ISL_1011006, EPI_ISL_1011008, EPI_ISL_1011009, EPI_ISL_1011011, EPI_ISL_1011015, EPI_ISL_1011016, EPI_ISL_1011017, EPI_ISL_1011018, EPI_ISL_1011023, EPI_ISL_1011024, EPI_ISL_1011025, EPI_ISL_1011029, EPI_ISL_1011031, EPI_ISL_1011032, EPI_ISL_1011035, EPI_ISL_1011036, EPI_ISL_1011037, EPI_ISL_1011039, EPI_ISL_1011040, EPI_ISL_1011042, EPI_ISL_1011043, EPI_ISL_1011047, EPI_ISL_1011048, EPI_ISL_1011049, EPI_ISL_1011052, EPI_ISL_1011054, EPI_ISL_1011056, EPI_ISL_1011059, EPI_ISL_1011061, EPI_ISL_1011062, EPI_ISL_1011066, EPI_ISL_1011067, EPI_ISL_1011072, EPI_ISL_1011073, EPI_ISL_1011075, EPI_ISL_1011076, EPI_ISL_1011077, EPI_ISL_1011078, EPI_ISL_1011079, EPI_ISL_1011081, EPI_ISL_1011083, EPI_ISL_1011084, EPI_ISL_1011088, EPI_ISL_1011090, EPI_ISL_1011092, EPI_ISL_1011093, EPI_ISL_1011095, EPI_ISL_1011096, EPI_ISL_1011097, EPI_ISL_1011098, EPI_ISL_1011100, EPI_ISL_1011101, EPI_ISL_1011102, EPI_ISL_1011104, EPI_ISL_1011105, EPI_ISL_1011106, EPI_ISL_1011108, EPI_ISL_1011110, EPI_ISL_1011112, EPI_ISL_1011113, EPI_ISL_1011115, EPI_ISL_1011117, EPI_ISL_1011119, EPI_ISL_1011121, EPI_ISL_1011126, EPI_ISL_1011129, EPI_ISL_1011130, EPI_ISL_1011131, EPI_ISL_1011134, EPI_ISL_1011138, EPI_ISL_1011140, EPI_ISL_1011143, EPI_ISL_1011144, EPI_ISL_1011145, EPI_ISL_1011146, EPI_ISL_1011147, EPI_ISL_1011148, EPI_ISL_1011152, EPI_ISL_1011154, EPI_ISL_1011158, EPI_ISL_1011159, EPI_ISL_1011163, EPI_ISL_1011164, EPI_ISL_1011166, EPI_ISL_1011169, EPI_ISL_1011170, EPI_ISL_1011171, EPI_ISL_1011173, EPI_ISL_1011175, EPI_ISL_1011176, EPI_ISL_1011178, EPI_ISL_1011182, EPI_ISL_1011184, EPI_ISL_1011185, EPI_ISL_1011186, EPI_ISL_1011189, EPI_ISL_1011190, EPI_ISL_1011192, EPI_ISL_1011195, EPI_ISL_1011201, EPI_ISL_1011204, EPI_ISL_1011206, EPI_ISL_1011208, EPI_ISL_1011209, EPI_ISL_1011213, EPI_ISL_1011214, EPI_ISL_1011218, EPI_ISL_1011219, EPI_ISL_1011222, EPI_ISL_1011223, EPI_ISL_1011227, EPI_ISL_1011230, EPI_ISL_1011233, EPI_ISL_1011236, EPI_ISL_1011238, EPI_ISL_1011239, EPI_ISL_1011240, EPI_ISL_1011241, EPI_ISL_1011242, EPI_ISL_1011243, EPI_ISL_1011244, EPI_ISL_1011247, EPI_ISL_1011248, EPI_ISL_1011250, EPI_ISL_1011251, EPI_ISL_1011252, EPI_ISL_1011253, EPI_ISL_1011254, EPI_ISL_1011256, EPI_ISL_1011258, EPI_ISL_1011259, EPI_ISL_1011260, EPI_ISL_1011264, EPI_ISL_1011268, EPI_ISL_1011270, EPI_ISL_1011272, EPI_ISL_1011273, EPI_ISL_1011274, EPI_ISL_1011279, EPI_ISL_1011280, EPI_ISL_1011284, EPI_ISL_1011285, EPI_ISL_1011286, EPI_ISL_1011288, EPI_ISL_1011291, EPI_ISL_1011294, EPI_ISL_1011299, EPI_ISL_1011302, EPI_ISL_1011305, EPI_ISL_1011306, EPI_ISL_1011308, EPI_ISL_1011309, EPI_ISL_1011310, EPI_ISL_1011311, EPI_ISL_1011315, EPI_ISL_1011316, EPI_ISL_1011319, EPI_ISL_1011320, EPI_ISL_1011323, EPI_ISL_1011326, EPI_ISL_1011327, EPI_ISL_1011329, EPI_ISL_1011332, EPI_ISL_1011339, EPI_ISL_1011340, EPI_ISL_1011342, EPI_ISL_1011345, EPI_ISL_1011350, EPI_ISL_1011356, EPI_ISL_1011358, EPI_ISL_1011361, EPI_ISL_1011364, EPI_ISL_1011365, EPI_ISL_1011366, EPI_ISL_1011370, EPI_ISL_1011371, EPI_ISL_1011374, EPI_ISL_1011376, EPI_ISL_1011378, EPI_ISL_1011382, EPI_ISL_1011383, EPI_ISL_1011386, EPI_ISL_1011395, EPI_ISL_1011397, EPI_ISL_1011398, EPI_ISL_1011401, EPI_ISL_1011402, EPI_ISL_1011404, EPI_ISL_1011405, EPI_ISL_1011406, EPI_ISL_1011408, EPI_ISL_1011410, EPI_ISL_1011411, EPI_ISL_1011415, EPI_ISL_1011418, EPI_ISL_1011419, EPI_ISL_1011421, EPI_ISL_1011422, EPI_ISL_1011424, EPI_ISL_1011428, EPI_ISL_1011431, EPI_ISL_1011433, EPI_ISL_1011437, EPI_ISL_1011439, EPI_ISL_1011440, EPI_ISL_1011444, EPI_ISL_1011446, EPI_ISL_1011451, EPI_ISL_1011452, EPI_ISL_1011456, EPI_ISL_1011457, EPI_ISL_1011458, EPI_ISL_1011462, EPI_ISL_1011468, EPI_ISL_1011470, EPI_ISL_1011473, EPI_ISL_1011474, EPI_ISL_1011476, EPI_ISL_1011477, EPI_ISL_1011478, EPI_ISL_1011479, EPI_ISL_1011480, EPI_ISL_1011481, EPI_ISL_1011482, EPI_ISL_1011484, EPI_ISL_1011487, EPI_ISL_1011488, EPI_ISL_1011489, EPI_ISL_1011490, EPI_ISL_1011491, EPI_ISL_1011494, EPI_ISL_1011497, EPI_ISL_1011498, EPI_ISL_1011501, EPI_ISL_1011503, EPI_ISL_1011505, EPI_ISL_1011506, EPI_ISL_1011508 | | | |
| see above | Fulgent Genetics | Fulgent Genetics | Harry Gao, Mickey Li, John Gao, Joseph Fierro, Benafsh Sapra, Becky Tsai, Yan Meng, Doreen Ng, James Xie |
| EPI_ISL_1011511, EPI_ISL_1011512, EPI_ISL_1011513, EPI_ISL_1011514, EPI_ISL_1011516, EPI_ISL_1011517, EPI_ISL_1011519, EPI_ISL_1011520, EPI_ISL_1011522, EPI_ISL_1011523, EPI_ISL_1011524, EPI_ISL_1011525, EPI_ISL_1011527, EPI_ISL_1011528, EPI_ISL_1011529, EPI_ISL_1011531, EPI_ISL_1011533, EPI_ISL_1011534, EPI_ISL_1011535, EPI_ISL_1011536, EPI_ISL_1011538, EPI_ISL_1011539, EPI_ISL_1011541, EPI_ISL_1011542, EPI_ISL_1011543, EPI_ISL_1011544, EPI_ISL_1011545, EPI_ISL_1011546, EPI_ISL_1011547, EPI_ISL_1011548, EPI_ISL_1011549, EPI_ISL_1011550, EPI_ISL_1011551, EPI_ISL_1011553, EPI_ISL_1011554, EPI_ISL_1011555, EPI_ISL_1011556, EPI_ISL_1011557, EPI_ISL_1011559, EPI_ISL_1011560, EPI_ISL_1011561, EPI_ISL_1011563, EPI_ISL_1011564, EPI_ISL_1011565, EPI_ISL_1011566, EPI_ISL_1011567, EPI_ISL_1011568, EPI_ISL_1011569, EPI_ISL_1011570, EPI_ISL_1011571, EPI_ISL_1011572, EPI_ISL_1011573, EPI_ISL_1011575, EPI_ISL_1011576, EPI_ISL_1011577, EPI_ISL_1011578, EPI_ISL_1011580, EPI_ISL_1011581, EPI_ISL_1011583, EPI_ISL_1011584, EPI_ISL_1011586, EPI_ISL_1011587, EPI_ISL_1011588, EPI_ISL_1011589, EPI_ISL_1011590, EPI_ISL_1011591, EPI_ISL_1011593, EPI_ISL_1011595, EPI_ISL_1011596, EPI_ISL_1011597 | | | |
| see above | Massachusetts General Hospital | Infectious Disease Program, Broad Institute of Harvard and MIT | Lemieux,J.E., Siddle,K.J., Shaw,B., Adams,G., Pierce,V., Turbett,S., Anahtar,M., Branda,J., Slater,D., Harris,J., Lin,A.E., Gladden-Young,A., Lagerborg,K., Rudy,M., DeRuff,K., Carter,A., Normandin,E., Bauer,M., Reilly,S., Tomkins-Tinch,C., Loreth,C., Chaluvadi,S., Neumann,A., Cusick,C., Chapman,S.B., Gnirke,A., Flowers,K., Cerrato,F., Birren,B.W., Gallagher,G., Smole,S., Park,D.J., MacInnis,B.L., Ryan,E., LaRocque,R., Rosenberg,E. and Sabeti,P.C. |
| EPI_ISL_1011599, EPI_ISL_1011600, EPI_ISL_1011601, EPI_ISL_1011602, EPI_ISL_1011603, EPI_ISL_1011606, EPI_ISL_1011607, EPI_ISL_1011608, EPI_ISL_1011609, EPI_ISL_1011610, EPI_ISL_1011612, EPI_ISL_1011613, EPI_ISL_1011614, EPI_ISL_1011615, EPI_ISL_1011616, EPI_ISL_1011617, EPI_ISL_1011618, EPI_ISL_1011620, EPI_ISL_1011621, EPI_ISL_1011622, EPI_ISL_1011623, EPI_ISL_1011624, EPI_ISL_1011626, EPI_ISL_1011627, EPI_ISL_1011628, EPI_ISL_1011629, EPI_ISL_1011630, EPI_ISL_1011631, EPI_ISL_1011632, EPI_ISL_1011634, EPI_ISL_1011635, EPI_ISL_1011636, EPI_ISL_1011637, EPI_ISL_1011638, EPI_ISL_1011641, EPI_ISL_1011642, EPI_ISL_1011643, EPI_ISL_1011645, EPI_ISL_1011646, EPI_ISL_1011647, EPI_ISL_1011649, EPI_ISL_1011650, EPI_ISL_1011651, EPI_ISL_1011652, EPI_ISL_1011653, EPI_ISL_1011654, EPI_ISL_1011655, EPI_ISL_1011656, EPI_ISL_1011657, EPI_ISL_1011658, EPI_ISL_1011659, EPI_ISL_1011660, EPI_ISL_1011661, EPI_ISL_1011662, EPI_ISL_1011663, EPI_ISL_1011664, EPI_ISL_1011666, EPI_ISL_1011667, EPI_ISL_1011668, EPI_ISL_1011669, EPI_ISL_1011670, EPI_ISL_1011671, EPI_ISL_1011674, EPI_ISL_1011675, EPI_ISL_1011676 | | | |
| see above | Warrior Diagnostics Laboratory | Infectious Disease Program, Broad Institute of Harvard and MIT | Lemieux,J.E., Siddle,K.J., Marshall,J., ONeill,M., Bronson,A., Adams,G., Gladden-Young,A., Lagerborg,K., Rudy,M., DeRuff,K., Carter,A., Normandin,E., Bauer,M., Reilly,S., Tomkins-Tinch,C., Loreth,C., Chaluvadi,S., Birren,B.W., Gallagher,G., Smole,S., Park,D.J., MacInnis,B.L., and Sabeti,P.C. |

| | | | |
|---|---|---|---|
| EPI_ISL_1011678, EPI_ISL_1011679, EPI_ISL_1011680, EPI_ISL_1011681, EPI_ISL_1011682, EPI_ISL_1011683, EPI_ISL_1011684, EPI_ISL_1011685, EPI_ISL_1011686, EPI_ISL_1011687, EPI_ISL_1011688, EPI_ISL_1011689, EPI_ISL_1011690, EPI_ISL_1011691, EPI_ISL_1011692, EPI_ISL_1011693, EPI_ISL_1011694, EPI_ISL_1011695, EPI_ISL_1011696, EPI_ISL_1011697, EPI_ISL_1011698, EPI_ISL_1011699, EPI_ISL_1011700, EPI_ISL_1011701, EPI_ISL_1011702, EPI_ISL_1011703, EPI_ISL_1011704, EPI_ISL_1011705 | | | |
| see above | Broad Institute Clinical Research Sequencing Platform | Infectious Disease Program, Broad Institute of Harvard and MIT | Lemieux,J.E., Siddle,K.J., Adams,G., Gladden-Young,A., Lagerborg,K., Rudy,M., DeRuff,K., Carter,A., Normandin,E., Bauer,M., Reilly,S., Tomkins-Tinch,C., Loreth,C., Chaluvadi,S., Birren,B.W., Gallagher,G., Smole,S., Park,D.J., MacInnis,B.L., and Sabeti,P.C. |
| EPI_ISL_1011706, EPI_ISL_1011707, EPI_ISL_1011708, EPI_ISL_1011709, EPI_ISL_1011710, EPI_ISL_1011711, EPI_ISL_1011712, EPI_ISL_1011713, EPI_ISL_1011714, EPI_ISL_1011715, EPI_ISL_1011717, EPI_ISL_1011718, EPI_ISL_1011719, EPI_ISL_1011720, EPI_ISL_1011721, EPI_ISL_1011722, EPI_ISL_1011723, EPI_ISL_1011724, EPI_ISL_1011725, EPI_ISL_1011726, EPI_ISL_1011727, EPI_ISL_1011728, EPI_ISL_1011729, EPI_ISL_1011730, EPI_ISL_1011731, EPI_ISL_1011732, EPI_ISL_1011733, EPI_ISL_1011734, EPI_ISL_1011735, EPI_ISL_1011736, EPI_ISL_1011737, EPI_ISL_1011738, EPI_ISL_1011739, EPI_ISL_1011740, EPI_ISL_1011741, EPI_ISL_1011742, EPI_ISL_1011743, EPI_ISL_1011744, EPI_ISL_1011745, EPI_ISL_1011746, EPI_ISL_1011747, EPI_ISL_1011748, EPI_ISL_1011749, EPI_ISL_1011750 | | | |
| see above | Rhode Island Department of Health | Infectious Disease Program, Broad Institute of Harvard and MIT | Lemieux,J.E., Siddle,K.J., Huard,R., King,E., Azevedo,K., Miller,A., Adams,G., Gladden-Young,A., Lagerborg,K., Rudy,M., DeRuff,K., Carter,A., Normandin,E., Bauer,M., Reilly,S., Tomkins-Tinch,C., Loreth,C., Chaluvadi,S., Birren,B.W., Gallagher,G., Smole,S., Park,D.J., MacInnis,B.L., and Sabeti,P.C. |
| EPI_ISL_1012883, EPI_ISL_1012884, EPI_ISL_1012885, EPI_ISL_1012886, EPI_ISL_1012887, EPI_ISL_1012888, EPI_ISL_1012889, EPI_ISL_1012890, EPI_ISL_1012891, EPI_ISL_1012892, EPI_ISL_1012893, EPI_ISL_1012894, EPI_ISL_1012895, EPI_ISL_1012896, EPI_ISL_1012897, EPI_ISL_1012898, EPI_ISL_1012899, EPI_ISL_1012900, EPI_ISL_1012901, EPI_ISL_1012902, EPI_ISL_1012904, EPI_ISL_1012905, EPI_ISL_1012906, EPI_ISL_1012907, EPI_ISL_1012908, EPI_ISL_1012909, EPI_ISL_1012910, EPI_ISL_1012911, EPI_ISL_1012912, EPI_ISL_1012913, EPI_ISL_1012914, EPI_ISL_1012915, EPI_ISL_1012916, EPI_ISL_1012917, EPI_ISL_1012918, EPI_ISL_1012919, EPI_ISL_1012920, EPI_ISL_1012921 | | | |
| see above | New Mexico Department of Health Scientific Laboratory | New Mexico Department of Health Scientific Laboratory | Ellie Johnson, Anastacia Griego-Fisher, D'eldra Malone, Jennifer Benoit |
| EPI_ISL_1014593 | Ohio Department of Health Laboratory | Ohio Department of Health Laboratory | Holmes, Jennifer; Eric Brandt, Keoni Omura, Glen McGillivary, Caitlin McDonnell, Jade Mowery, Stephanie Mccracken, Tyler Payne, Kirtana Ramadugu, Erica Leasure, Brent Lee, Kelsey Florek, Heather Blankenship, Quanta Brown, and Tammy Bannerman |
| EPI_ISL_1014688 | Ohio Department of Health Laboratory | Ohio Department of Health Laboratory | 8995 East Main Street Reynoldsburg, Ohio 43068 |
| EPI_ISL_1014689 | Ohio Department of Health Laboratory | Ohio Department of Health Laboratory | Holmes, Jennifer; Eric Brandt, Keoni Omura, Glen McGillivary, Caitlin McDonnell, Jade Mowery, Stephanie Mccracken, Tyler Payne, Kirtana Ramadugu, Erica Leasure, Brent Lee, Kelsey Florek, Heather Blankenship, Quanta Brown, and Tammy Bannerman |
| EPI_ISL_1014767, EPI_ISL_1014768, EPI_ISL_1014769, EPI_ISL_1014770, EPI_ISL_1014771, EPI_ISL_1014772, EPI_ISL_1014773, EPI_ISL_1014774, EPI_ISL_1014775, EPI_ISL_1014776, EPI_ISL_1014777, EPI_ISL_1014778, EPI_ISL_1014779, EPI_ISL_1014780, EPI_ISL_1014781, EPI_ISL_1014782, EPI_ISL_1014783, EPI_ISL_1014784, EPI_ISL_1014785, EPI_ISL_1014786, EPI_ISL_1014787, EPI_ISL_1014788, EPI_ISL_1014789, EPI_ISL_1014790, EPI_ISL_1014791, EPI_ISL_1014792, EPI_ISL_1014793, EPI_ISL_1014794, EPI_ISL_1014795, EPI_ISL_1014796, EPI_ISL_1014797, EPI_ISL_1014798, EPI_ISL_1014799, EPI_ISL_1014800, EPI_ISL_1014801, EPI_ISL_1014802, EPI_ISL_1014803, EPI_ISL_1014804, EPI_ISL_1014805, EPI_ISL_1014806, EPI_ISL_1014807, EPI_ISL_1014808, EPI_ISL_1014809, EPI_ISL_1014810, EPI_ISL_1014811, EPI_ISL_1014812, EPI_ISL_1014813, EPI_ISL_1014814, EPI_ISL_1014815, EPI_ISL_1014816, EPI_ISL_1014817, EPI_ISL_1014818, EPI_ISL_1014819, EPI_ISL_1014820, EPI_ISL_1014821, EPI_ISL_1014822, EPI_ISL_1014823, EPI_ISL_1014824, EPI_ISL_1014825, EPI_ISL_1014826, EPI_ISL_1014827, EPI_ISL_1014828, EPI_ISL_1014829, EPI_ISL_1014830, EPI_ISL_1014831, EPI_ISL_1014832, EPI_ISL_1014833, EPI_ISL_1014834, EPI_ISL_1014835, EPI_ISL_1014836, EPI_ISL_1014837, EPI_ISL_1014838, EPI_ISL_1014839, EPI_ISL_1014840, EPI_ISL_1014841, EPI_ISL_1014842, EPI_ISL_1014843, EPI_ISL_1014844, EPI_ISL_1014845, EPI_ISL_1014846, EPI_ISL_1014847, EPI_ISL_1014848, EPI_ISL_1014849, EPI_ISL_1014850, EPI_ISL_1014851, EPI_ISL_1014852, EPI_ISL_1014853, EPI_ISL_1014854, EPI_ISL_1014855, EPI_ISL_1014856, EPI_ISL_1014857, EPI_ISL_1014858, EPI_ISL_1014859, EPI_ISL_1014860, EPI_ISL_1014861, EPI_ISL_1014862, EPI_ISL_1014863, EPI_ISL_1014864, EPI_ISL_1014865, EPI_ISL_1014866, EPI_ISL_1014867, EPI_ISL_1014868, EPI_ISL_1014869, EPI_ISL_1014870, EPI_ISL_1014871, EPI_ISL_1014872, EPI_ISL_1014873, EPI_ISL_1014874, EPI_ISL_1014875, EPI_ISL_1014876, EPI_ISL_1014877, EPI_ISL_1014878, EPI_ISL_1014879, EPI_ISL_1014880, EPI_ISL_1014881, EPI_ISL_1014882, EPI_ISL_1014883, EPI_ISL_1014884, EPI_ISL_1014885, EPI_ISL_1014886, EPI_ISL_1014887, EPI_ISL_1014888, EPI_ISL_1014889, EPI_ISL_1014890, EPI_ISL_1014891, EPI_ISL_1014892, EPI_ISL_1014893, EPI_ISL_1014894, EPI_ISL_1014895, EPI_ISL_1014896, EPI_ISL_1014897, EPI_ISL_1014898, EPI_ISL_1014899, EPI_ISL_1014900, EPI_ISL_1014901, EPI_ISL_1014902, EPI_ISL_1014903 | | | |
| see above | University of Michigan Clinical Microbiology Laboratory | Lauring Lab, University of Michigan, Department of Microbiology and Immunology | Valesano |
| EPI_ISL_1014937 | Ohio Department of Health Laboratory | Ohio Department of Health Laboratory | Holmes, Jennifer; Eric Brandt, Keoni Omura, Glen McGillivary, Caitlin McDonnell, Jade Mowery, Stephanie Mccracken, Tyler Payne, Kirtana Ramadugu, Erica Leasure, Brent Lee, Kelsey Florek, Heather Blankenship, Quanta Brown, and Tammy Bannerman |
| EPI_ISL_1014988, EPI_ISL_1014989, EPI_ISL_1014990, EPI_ISL_1014991, EPI_ISL_1014992 | University of Michigan Clinical Microbiology Laboratory | Lauring Lab, University of Michigan, Department of Microbiology and Immunology | Valesano |
| EPI_ISL_1014993, EPI_ISL_1014995 | Arizona State Public Health Laboratory | Arizona State Public Health Laboratory | Trung Huynh, Jessica Escobar, Katherine Fullerton, Nobuko Fukushima, Stacy White, Linda Getsinger, Victor Waddell |
| EPI_ISL_1015335, EPI_ISL_1015336, EPI_ISL_1015338, EPI_ISL_1015339, EPI_ISL_1015340, EPI_ISL_1015341 | Univeristy of New Mexico Hospital | Center for Global Health, University of New Mexico Health Sciences Center | Daryl Domman, Kurt Schwalm, Justin Bacca, Jon Femling, Darrell Dinwiddie |
| EPI_ISL_1015342, EPI_ISL_1015343, EPI_ISL_1015344, EPI_ISL_1015345, EPI_ISL_1015346, EPI_ISL_1015347, EPI_ISL_1015348, EPI_ISL_1015349, EPI_ISL_1015350, EPI_ISL_1015351, EPI_ISL_1015352 | | | |
| see above | New Mexico Department of Health Scientific Laboratory | Center for Global Health, University of New Mexico Health Sciences Center | Daryl Domman, Kurt Schwalm, Twila Kunde, Joseph Hicks, Anastacia Griego, Michael Edwards, Darrell Dinwiddie |
| EPI_ISL_1015361 | Coast Diagnostics | Synergy Laboratories | Megan Cornwell |
| EPI_ISL_1015418, EPI_ISL_1015419, EPI_ISL_1015420, EPI_ISL_1015421, EPI_ISL_1015422, EPI_ISL_1015423, EPI_ISL_1015424, EPI_ISL_1015425, EPI_ISL_1015426, EPI_ISL_1015427, EPI_ISL_1015428, EPI_ISL_1015429, EPI_ISL_1015430, EPI_ISL_1015431, EPI_ISL_1015432, EPI_ISL_1015433, EPI_ISL_1015434, EPI_ISL_1015435, EPI_ISL_1015436, EPI_ISL_1015437, EPI_ISL_1015438, EPI_ISL_1015439, EPI_ISL_1015440, EPI_ISL_1015441, EPI_ISL_1015442, EPI_ISL_1015443, EPI_ISL_1015444, EPI_ISL_1015445, EPI_ISL_1015446, EPI_ISL_1015459 | | | |
| see above | Texas Department of State Health Services | Texas Department of State Health Services | Bonnie Oh, Anita Pokharel, James Daniel Bonser, Myong Koag, Chung Wang, Rachel Lee, Grace Kubin, Rashmi Tuladhar, Mayela Pedrueza, Maliha Rahman, Jenny Zhang |
| EPI_ISL_1015493, EPI_ISL_1015494, EPI_ISL_1015495, EPI_ISL_1015498, EPI_ISL_1015499, EPI_ISL_1015500, EPI_ISL_1015501, EPI_ISL_1015502, EPI_ISL_1015503, EPI_ISL_1015504, EPI_ISL_1015505, EPI_ISL_1015508, EPI_ISL_1015509, EPI_ISL_1015510, EPI_ISL_1015511, EPI_ISL_1015512, EPI_ISL_1015513, EPI_ISL_1015514 | | | |
| see above | City of Milwaukee Health Department Laboratory | City of Milwaukee Health Department Laboratory | Sanjib Bhattacharyya |
| EPI_ISL_1015561, EPI_ISL_1015562, EPI_ISL_1015563, EPI_ISL_1015564, EPI_ISL_1015565, EPI_ISL_1015566, EPI_ISL_1015567, EPI_ISL_1015568, EPI_ISL_1015571, EPI_ISL_1015572, EPI_ISL_1015573, EPI_ISL_1015574, EPI_ISL_1015575, EPI_ISL_1015576, EPI_ISL_1015577, EPI_ISL_1015578, EPI_ISL_1015579, EPI_ISL_1015580, EPI_ISL_1015581, EPI_ISL_1015582, EPI_ISL_1015583, EPI_ISL_1015584, EPI_ISL_1015585, EPI_ISL_1015586, EPI_ISL_1015587, EPI_ISL_1015588, EPI_ISL_1015589, EPI_ISL_1015590, EPI_ISL_1015591, EPI_ISL_1015592, EPI_ISL_1015593, EPI_ISL_1015594, EPI_ISL_1015595, EPI_ISL_1015596, EPI_ISL_1015597, EPI_ISL_1015598, EPI_ISL_1015599, EPI_ISL_1015600, EPI_ISL_1015601, EPI_ISL_1015602, EPI_ISL_1015603, EPI_ISL_1015604, EPI_ISL_1015605, EPI_ISL_1015606, EPI_ISL_1015607, EPI_ISL_1015608, EPI_ISL_1015609, EPI_ISL_1015610, EPI_ISL_1015611, EPI_ISL_1015612, EPI_ISL_1015613, EPI_ISL_1015614, EPI_ISL_1015615, EPI_ISL_1015616, EPI_ISL_1015617, EPI_ISL_1015618, EPI_ISL_1015619, EPI_ISL_1015620, EPI_ISL_1015621, EPI_ISL_1015622, EPI_ISL_1015623, EPI_ISL_1015624, EPI_ISL_1015625, EPI_ISL_1015626, EPI_ISL_1015627, EPI_ISL_1015628, EPI_ISL_1015629, EPI_ISL_1015630, EPI_ISL_1015631, EPI_ISL_1015632, EPI_ISL_1015633, EPI_ISL_1015634, EPI_ISL_1015635, EPI_ISL_1015636, EPI_ISL_1015637, EPI_ISL_1015638, EPI_ISL_1015639, EPI_ISL_1015640, EPI_ISL_1015641, EPI_ISL_1015642, EPI_ISL_1015643, EPI_ISL_1015644, EPI_ISL_1015645, EPI_ISL_1015646, EPI_ISL_1015647, EPI_ISL_1015648, EPI_ISL_1015649, EPI_ISL_1015650, EPI_ISL_1015651, EPI_ISL_1015652 | | | |
| see above | Maryland Public Health Laboratory | Maryland Public Health Laboratory | Maryland Department of Health Laboratories Administration |
| EPI_ISL_1015653, EPI_ISL_1015654, EPI_ISL_1015655, EPI_ISL_1015656, EPI_ISL_1015657, EPI_ISL_1015658, EPI_ISL_1015659, EPI_ISL_1015660, EPI_ISL_1015661, EPI_ISL_1015662, EPI_ISL_1015663, EPI_ISL_1015664, EPI_ISL_1015665, EPI_ISL_1015666, EPI_ISL_1015667, EPI_ISL_1015668, EPI_ISL_1015669, EPI_ISL_1015670, EPI_ISL_1015671, EPI_ISL_1015672, EPI_ISL_1015673, EPI_ISL_1015674, EPI_ISL_1015675, EPI_ISL_1015676, EPI_ISL_1015677, EPI_ISL_1015678, EPI_ISL_1015679, EPI_ISL_1015680, EPI_ISL_1015681, EPI_ISL_1015682, EPI_ISL_1015683, EPI_ISL_1015684, EPI_ISL_1015685, EPI_ISL_1015686, EPI_ISL_1015687, EPI_ISL_1015688, EPI_ISL_1015689, EPI_ISL_1015690, EPI_ISL_1015691, EPI_ISL_1015692, EPI_ISL_1015693, EPI_ISL_1015694, EPI_ISL_1015695, EPI_ISL_1015696, EPI_ISL_1015697, EPI_ISL_1015698, EPI_ISL_1015699, EPI_ISL_1015700, EPI_ISL_1015701, EPI_ISL_1015702, EPI_ISL_1015703, EPI_ISL_1015704, EPI_ISL_1015705, EPI_ISL_1015706, EPI_ISL_1015707, EPI_ISL_1015708, EPI_ISL_1015709, EPI_ISL_1015710, EPI_ISL_1015711, EPI_ISL_1015712 | | | |
| see above | Columbia University Irving Medical Center | Wadsworth Center, New York State Department of Health | Kirsten St. George, Daryl M. Lamson, Alexis Russel, Matthew Shudt, Melissa A Leisner, Jonathan Plitnick, Navjot Singh, John Kelly, Erasmus Schneider, Erica Lasek-Nesselquist |
| EPI_ISL_1015713, EPI_ISL_1015714, EPI_ISL_1015715, EPI_ISL_1015716, EPI_ISL_1015717, EPI_ISL_1015718, EPI_ISL_1015719, EPI_ISL_1015720, EPI_ISL_1015721, EPI_ISL_1015722, EPI_ISL_1015723, EPI_ISL_1015725, EPI_ISL_1015726, EPI_ISL_1015727, EPI_ISL_1015728, EPI_ISL_1015729, EPI_ISL_1015730, EPI_ISL_1015731 | | | |
| see above | Arizona State Public Health Laboratory | Arizona State Public Health Laboratory | Trung Huynh, Jessica Escobar, Katherine Fullerton, Nobuko Fukushima, Stacy White, Linda Getsinger, Victor Waddell |
| EPI_ISL_1015733 | MD Laboratories | Los Angeles County Public Health Laboratories | P. Hemarajata et al. |
| EPI_ISL_1015736, EPI_ISL_1015738, EPI_ISL_1015739, EPI_ISL_1015740, EPI_ISL_1015741, EPI_ISL_1015742, EPI_ISL_1015743, EPI_ISL_1015744, EPI_ISL_1015745, EPI_ISL_1015746, EPI_ISL_1015747, EPI_ISL_1015749, EPI_ISL_1015752, EPI_ISL_1015754, EPI_ISL_1015755 | | | |
| see above | Seattle Flu Study | Seattle Flu Study | Deborah A. Nickerson, Chris D. Frazar, Jover Lee, Benjamin Pelle, Erica Ryke, Matthew Richardson, Amanda Adler, Elisabeth Brandstetter, Peter D. Han, Kairsten Fay, Misja Ilcisin, Kirsten Lacombe, Thomas R. Sibley, Melissa Truong, Caitlin R. Wolf, Karen Cowgill, Stephanie Schrag, Jeff Duchin, Michael Boeckh, Janet A. Englund, Michael Famulare, Barry R. Lutz, Mark J. Rieder, Lea M. Starita, Matthew Thompson, Helen Y. Chu, Trevor Bedford, Jay Shendure |
| EPI_ISL_1015758, EPI_ISL_1015759, EPI_ISL_1015760, EPI_ISL_1015763, EPI_ISL_1015764, EPI_ISL_1015765, EPI_ISL_1015767, EPI_ISL_1015768, EPI_ISL_1015770, EPI_ISL_1015771, EPI_ISL_1015772, EPI_ISL_1015773, EPI_ISL_1015775, EPI_ISL_1015776, EPI_ISL_1015777, EPI_ISL_1015778, EPI_ISL_1015779, EPI_ISL_1015780, EPI_ISL_1015781, EPI_ISL_1015782, EPI_ISL_1015783, EPI_ISL_1015784, EPI_ISL_1015785, EPI_ISL_1015787, EPI_ISL_1015788, EPI_ISL_1015789, EPI_ISL_1015790, EPI_ISL_1015791, EPI_ISL_1015792, EPI_ISL_1015793, EPI_ISL_1015794, EPI_ISL_1015796, EPI_ISL_1015798, EPI_ISL_1015799, EPI_ISL_1015800, EPI_ISL_1015801, EPI_ISL_1015803, EPI_ISL_1015804, EPI_ISL_1015805, EPI_ISL_1015807, EPI_ISL_1015808, EPI_ISL_1015809, EPI_ISL_1015810, EPI_ISL_1015811, EPI_ISL_1015812, EPI_ISL_1015814, EPI_ISL_1015815, EPI_ISL_1015816, EPI_ISL_1015817, EPI_ISL_1015821, EPI_ISL_1015822, EPI_ISL_1015823, EPI_ISL_1015824, EPI_ISL_1015825, EPI_ISL_1015826, EPI_ISL_1015827, EPI_ISL_1015828, EPI_ISL_1015829, EPI_ISL_1015830, EPI_ISL_1015831, EPI_ISL_1015832, EPI_ISL_1015835, EPI_ISL_1015836, EPI_ISL_1015837, EPI_ISL_1015838, EPI_ISL_1015839, EPI_ISL_1015840, EPI_ISL_1015842, EPI_ISL_1015843, EPI_ISL_1015844, EPI_ISL_1015845, EPI_ISL_1015847, EPI_ISL_1015849, EPI_ISL_1015850, EPI_ISL_1015851, EPI_ISL_1015852, EPI_ISL_1015853, EPI_ISL_1015854, EPI_ISL_1015855, EPI_ISL_1015856, EPI_ISL_1015857, EPI_ISL_1015858, EPI_ISL_1015859, EPI_ISL_1015861, EPI_ISL_1015862, EPI_ISL_1015863, EPI_ISL_1015864, EPI_ISL_1015865, EPI_ISL_1015866, EPI_ISL_1015868, EPI_ISL_1015869, EPI_ISL_1015870, EPI_ISL_1015872, EPI_ISL_1015874, EPI_ISL_1015875, EPI_ISL_1015876 | | | |
| see above | Altius Institute for Biomedical Sciences | Seattle Flu Study | Deborah A. Nickerson, Chris D. Frazar, Jover Lee, Benjamin Pelle, Erica Ryke, Matthew Richardson, Amanda Adler, Elisabeth Brandstetter, Peter D. Han, Kairsten Fay, Misja Ilcisin, Kirsten Lacombe, Thomas R. Sibley, Melissa Truong, Caitlin R. Wolf, Ryan Alexander, Daniel Bates, Rebecca Bruders, Stephanie DeBaun, Clem Green, Muhammad Halimun, Jessica Halow, Kneshay Harper, Matt Hartman, Andrew Meuser, Alex Nguyen, Truong Nguyen, Sofia Olsson, Sadie Patraw, Hannah Petersen, Tobias Ragoczy, Joshua Richards, Jacob Rodriguez, John Stamatoyannopoulos, Julia Wald, Olivia Waltner, Michael Boeckh, Janet A. Englund, Michael Famulare, Barry R. Lutz, Mark J. Rieder, Lea M. Starita, Matthew Thompson, Helen Y. Chu, Jay Shendure, Trevor Bedford |
| EPI_ISL_1015878, EPI_ISL_1015881, EPI_ISL_1015882, EPI_ISL_1015884, EPI_ISL_1015885 | Seattle Flu Study | Seattle Flu Study | Deborah A. Nickerson, Chris D. Frazar, Jover Lee, Benjamin Pelle, Erica Ryke, Matthew Richardson, Amanda Adler, Elisabeth Brandstetter, Peter D. Han, Kairsten Fay, Misja Ilcisin, Kirsten Lacombe, Thomas R. Sibley, Melissa Truong, Caitlin R. Wolf, Karen Cowgill, Stephanie Schrag, Jeff Duchin, Michael Boeckh, Janet A. Englund, Michael Famulare, Barry R. Lutz, Mark J. Rieder, Lea M. Starita, Matthew Thompson, Helen Y. Chu, Trevor Bedford, Jay Shendure |
| EPI_ISL_1015887, EPI_ISL_1015888, EPI_ISL_1015889, EPI_ISL_1015890, EPI_ISL_1015892, EPI_ISL_1015893, EPI_ISL_1015894, EPI_ISL_1015895, EPI_ISL_1015897, EPI_ISL_1015898, EPI_ISL_1015899, EPI_ISL_1015900, EPI_ISL_1015901, EPI_ISL_1015902, EPI_ISL_1015904, EPI_ISL_1015906, EPI_ISL_1015908, EPI_ISL_1015909, EPI_ISL_1015910, EPI_ISL_1015911, EPI_ISL_1015912, EPI_ISL_1015916, EPI_ISL_1015917, EPI_ISL_1015918, EPI_ISL_1015919, EPI_ISL_1015921, EPI_ISL_1015923, EPI_ISL_1015926, EPI_ISL_1015927, EPI_ISL_1015928, EPI_ISL_1015932, EPI_ISL_1015933, EPI_ISL_1015934, EPI_ISL_1015935, EPI_ISL_1015938, EPI_ISL_1015939, EPI_ISL_1015940, EPI_ISL_1015941, EPI_ISL_1015943, EPI_ISL_1015944, EPI_ISL_1015945, EPI_ISL_1015946, EPI_ISL_1015947, EPI_ISL_1015948, EPI_ISL_1015949, EPI_ISL_1015950, EPI_ISL_1015951, EPI_ISL_1015952, EPI_ISL_1015953, EPI_ISL_1015954, EPI_ISL_1015955, EPI_ISL_1015956, EPI_ISL_1015957, EPI_ISL_1015959, EPI_ISL_1015960, EPI_ISL_1015962, EPI_ISL_1015963, EPI_ISL_1015964, EPI_ISL_1015965, EPI_ISL_1015966, EPI_ISL_1015967, EPI_ISL_1015968, EPI_ISL_1015969, EPI_ISL_1015971, EPI_ISL_1015972, EPI_ISL_1015975, EPI_ISL_1015976, EPI_ISL_1015977, EPI_ISL_1015978, EPI_ISL_1015979, EPI_ISL_1015980, EPI_ISL_1015981, EPI_ISL_1015982, EPI_ISL_1015983, EPI_ISL_1015984, EPI_ISL_1015985, EPI_ISL_1015986, EPI_ISL_1015988, EPI_ISL_1015989, EPI_ISL_1015992, EPI_ISL_1015993, EPI_ISL_1015995, EPI_ISL_1015997, EPI_ISL_1015999, EPI_ISL_1016000, EPI_ISL_1016001, EPI_ISL_1016003 | | | |
| see above | Altius Institute for Biomedical Sciences | Seattle Flu Study | Deborah A. Nickerson, Chris D. Frazar, Jover Lee, Benjamin Pelle, Erica Ryke, Matthew Richardson, Amanda Adler, Elisabeth Brandstetter, Peter D. Han, Kairsten Fay, Misja Ilcisin, Kirsten Lacombe, Thomas R. Sibley, Melissa Truong, Caitlin R. Wolf, Ryan Alexander, Daniel Bates, Rebecca Bruders, Stephanie DeBaun, Clem Green, Muhammad Halimun, Jessica Halow, Kneshay Harper, Matt Hartman, Andrew Meuser, Alex Nguyen, Truong Nguyen, Sofia Olsson, Sadie Patraw, Hannah Petersen, Tobias Ragoczy, Joshua Richards, Jacob Rodriguez, John Stamatoyannopoulos, Julia Wald, Olivia Waltner, Michael Boeckh, Janet A. Englund, Michael Famulare, Barry R. Lutz, Mark J. Rieder, Lea M. Starita, Matthew Thompson, Helen Y. Chu, Jay Shendure, Trevor Bedford |
| EPI_ISL_1016004, EPI_ISL_1016005, EPI_ISL_1016006 | Seattle Flu Study | Seattle Flu Study | Deborah A. Nickerson, Chris D. Frazar, Jover Lee, Benjamin Pelle, Erica Ryke, Matthew Richardson, Amanda Adler, Elisabeth Brandstetter, Peter D. Han, Kairsten Fay, Misja Ilcisin, Kirsten Lacombe, Thomas R. Sibley, Melissa Truong, Caitlin R. Wolf, Karen Cowgill, Stephanie Schrag, Jeff Duchin, Michael Boeckh, Janet A. Englund, Michael Famulare, Barry R. Lutz, Mark J. Rieder, Lea M. Starita, Matthew Thompson, Helen Y. Chu, Trevor Bedford, Jay Shendure |
| EPI_ISL_1016013, EPI_ISL_1016014, EPI_ISL_1016015, | NYU Langone Health | Departments of Pathology and Medicine, New York University School of Medicine | Adriana Heguy, Dacia Dimartino, Emily Guzman, Christian Marier, Peter Meyn, Sitharam Ramaswami, Gael Westby, Paul Zappile, Yutong Zhang, Paolo Cotzia, Guiqing Wang |

| | | | |
|---|---|---|---|
| EPI_ISL_1016016, EPI_ISL_1016017, EPI_ISL_1016018, EPI_ISL_1016075, EPI_ISL_1016100 | | | |
| EPI_ISL_1016101, EPI_ISL_1016102, EPI_ISL_1016103, EPI_ISL_1016104, EPI_ISL_1016105, EPI_ISL_1016106, EPI_ISL_1016107, EPI_ISL_1016108, EPI_ISL_1016109, EPI_ISL_1016110 | URMC LABS | Wadsworth Center, New York State Department of Health | Kirsten St. George, Daryl M. Lamson, Alexis Russel, Matthew Shudt, Melissa A Leisner, Jonathan Plitnick, Navjot Singh, John Kelly, Erasmus Schneider, Erica Lasek-Nesselquist |
| EPI_ISL_1016111, EPI_ISL_1016112, EPI_ISL_1016113, EPI_ISL_1016114, EPI_ISL_1016115, EPI_ISL_1016116, EPI_ISL_1016117 | Columbia University Irving Medical Center | Wadsworth Center, New York State Department of Health | Kirsten St. George, Daryl M. Lamson, Alexis Russel, Matthew Shudt, Melissa A Leisner, Jonathan Plitnick, Navjot Singh, John Kelly, Erasmus Schneider, Erica Lasek-Nesselquist |
| EPI_ISL_1016118, EPI_ISL_1016119, EPI_ISL_1016120, EPI_ISL_1016121, EPI_ISL_1016122, EPI_ISL_1016123, EPI_ISL_1016124, EPI_ISL_1016125, EPI_ISL_1016126 | URMC LABS | Wadsworth Center, New York State Department of Health | Kirsten St. George, Daryl M. Lamson, Alexis Russel, Matthew Shudt, Melissa A Leisner, Jonathan Plitnick, Navjot Singh, John Kelly, Erasmus Schneider, Erica Lasek-Nesselquist |
| EPI_ISL_1016127, EPI_ISL_1016128, EPI_ISL_1016129, EPI_ISL_1016130, EPI_ISL_1016131, EPI_ISL_1016132, EPI_ISL_1016133, EPI_ISL_1016134, EPI_ISL_1016135 | THE MARY IMOGENE BASSETT HOSPITAL | Wadsworth Center, New York State Department of Health | Kirsten St. George, Daryl M. Lamson, Alexis Russel, Matthew Shudt, Melissa A Leisner, Jonathan Plitnick, Navjot Singh, John Kelly, Erasmus Schneider, Erica Lasek-Nesselquist |
| EPI_ISL_1016136 | NORTHWELL HEALTH LABORATORIES | Wadsworth Center, New York State Department of Health | Kirsten St. George, Daryl M. Lamson, Alexis Russel, Matthew Shudt, Melissa A Leisner, Jonathan Plitnick, Navjot Singh, John Kelly, Erasmus Schneider, Erica Lasek-Nesselquist |
| EPI_ISL_1016137 | THE MARY IMOGENE BASSETT HOSPITAL | Wadsworth Center, New York State Department of Health | Kirsten St. George, Daryl M. Lamson, Alexis Russel, Matthew Shudt, Melissa A Leisner, Jonathan Plitnick, Navjot Singh, John Kelly, Erasmus Schneider, Erica Lasek-Nesselquist |
| EPI_ISL_1016138 | NORTHWELL HEALTH LABORATORIES | Wadsworth Center, New York State Department of Health | Kirsten St. George, Daryl M. Lamson, Alexis Russel, Matthew Shudt, Melissa A Leisner, Jonathan Plitnick, Navjot Singh, John Kelly, Erasmus Schneider, Erica Lasek-Nesselquist |
| EPI_ISL_1016139 | THE MARY IMOGENE BASSETT HOSPITAL | Wadsworth Center, New York State Department of Health | Kirsten St. George, Daryl M. Lamson, Alexis Russel, Matthew Shudt, Melissa A Leisner, Jonathan Plitnick, Navjot Singh, John Kelly, Erasmus Schneider, Erica Lasek-Nesselquist |
| EPI_ISL_1016140 | NORTHWELL HEALTH LABORATORIES | Wadsworth Center, New York State Department of Health | Kirsten St. George, Daryl M. Lamson, Alexis Russel, Matthew Shudt, Melissa A Leisner, Jonathan Plitnick, Navjot Singh, John Kelly, Erasmus Schneider, Erica Lasek-Nesselquist |
| EPI_ISL_1016141, EPI_ISL_1016142 | THE MARY IMOGENE BASSETT HOSPITAL | Wadsworth Center, New York State Department of Health | Kirsten St. George, Daryl M. Lamson, Alexis Russel, Matthew Shudt, Melissa A Leisner, Jonathan Plitnick, Navjot Singh, John Kelly, Erasmus Schneider, Erica Lasek-Nesselquist |
| EPI_ISL_1016143, EPI_ISL_1016144, EPI_ISL_1016145, EPI_ISL_1016146, EPI_ISL_1016147, EPI_ISL_1016148, EPI_ISL_1016149, EPI_ISL_1016150, EPI_ISL_1016151, EPI_ISL_1016152, EPI_ISL_1016153 | | | |
| see above | NORTHWELL HEALTH LABORATORIES | Wadsworth Center, New York State Department of Health | Kirsten St. George, Daryl M. Lamson, Alexis Russel, Matthew Shudt, Melissa A Leisner, Jonathan Plitnick, Navjot Singh, John Kelly, Erasmus Schneider, Erica Lasek-Nesselquist |
| EPI_ISL_1016154, EPI_ISL_1016155 | THE MARY IMOGENE BASSETT HOSPITAL | Wadsworth Center, New York State Department of Health | Kirsten St. George, Daryl M. Lamson, Alexis Russel, Matthew Shudt, Melissa A Leisner, Jonathan Plitnick, Navjot Singh, John Kelly, Erasmus Schneider, Erica Lasek-Nesselquist |
| EPI_ISL_1016156, EPI_ISL_1016157, EPI_ISL_1016158, EPI_ISL_1016159, EPI_ISL_1016160, EPI_ISL_1016161, EPI_ISL_1016162, EPI_ISL_1016163, EPI_ISL_1016164, EPI_ISL_1016165, EPI_ISL_1016166, EPI_ISL_1016167, EPI_ISL_1016168 | | | |
| see above | NORTHWELL HEALTH LABORATORIES | Wadsworth Center, New York State Department of Health | Kirsten St. George, Daryl M. Lamson, Alexis Russel, Matthew Shudt, Melissa A Leisner, Jonathan Plitnick, Navjot Singh, John Kelly, Erasmus Schneider, Erica Lasek-Nesselquist |
| EPI_ISL_1016169 | TEMPUS LABS INC | Wadsworth Center, New York State Department of Health | Kirsten St. George, Daryl M. Lamson, Alexis Russel, Matthew Shudt, Melissa A Leisner, Jonathan Plitnick, Navjot Singh, John Kelly, Erasmus Schneider, Erica Lasek-Nesselquist |
| EPI_ISL_1016170 | NYU Langone Health | Departments of Pathology and Medicine, New York University School of Medicine | Adriana Heguy, Dacia Dimartino, Emily Guzman, Christian Marier, Peter Meyn, Sitharam Ramaswami, Gael Westby, Paul Zappile, Yutong Zhang, Paolo Cotzia, Guiqing Wang |
| EPI_ISL_1016171, EPI_ISL_1016172, EPI_ISL_1016173, EPI_ISL_1016174, EPI_ISL_1016175, EPI_ISL_1016176, EPI_ISL_1016177, EPI_ISL_1016178, EPI_ISL_1016179, EPI_ISL_1016180, EPI_ISL_1016181, EPI_ISL_1016182, EPI_ISL_1016183, EPI_ISL_1016184, EPI_ISL_1016185, EPI_ISL_1016186, EPI_ISL_1016187, EPI_ISL_1016188, EPI_ISL_1016189, EPI_ISL_1016190, EPI_ISL_1016191, EPI_ISL_1016192, EPI_ISL_1016193, EPI_ISL_1016194, EPI_ISL_1016195, EPI_ISL_1016196, EPI_ISL_1016197, EPI_ISL_1016198, EPI_ISL_1016199, EPI_ISL_1016201, EPI_ISL_1016202, EPI_ISL_1016203, EPI_ISL_1016204, EPI_ISL_1016205, EPI_ISL_1016206, EPI_ISL_1016207, EPI_ISL_1016208, EPI_ISL_1016209, EPI_ISL_1016210, EPI_ISL_1016211, EPI_ISL_1016212, EPI_ISL_1016213, EPI_ISL_1016214, EPI_ISL_1016215, EPI_ISL_1016216, EPI_ISL_1016217, EPI_ISL_1016218, EPI_ISL_1016219 | | | |
| see above | MONTEFIORE MEDICAL CENTER LABORATORIES | Wadsworth Center, New York State Department of Health | Kirsten St. George, Daryl M. Lamson, Alexis Russel, Matthew Shudt, Melissa A Leisner, Jonathan Plitnick, Navjot Singh, John Kelly, Erasmus Schneider, Erica Lasek-Nesselquist |
| EPI_ISL_1016220, EPI_ISL_1016221, EPI_ISL_1016222, EPI_ISL_1016223, EPI_ISL_1016224, EPI_ISL_1016225, EPI_ISL_1016226, EPI_ISL_1016227, EPI_ISL_1016228, EPI_ISL_1016229, EPI_ISL_1016230, EPI_ISL_1016231, EPI_ISL_1016232, EPI_ISL_1016233, EPI_ISL_1016234, EPI_ISL_1016235, EPI_ISL_1016236, EPI_ISL_1016237, EPI_ISL_1016238, EPI_ISL_1016239, EPI_ISL_1016240, EPI_ISL_1016241, EPI_ISL_1016242, EPI_ISL_1016243, EPI_ISL_1016244, EPI_ISL_1016245, EPI_ISL_1016246, EPI_ISL_1016247, EPI_ISL_1016248 | | | |
| see above | New York Presbyterian Hospital | Wadsworth Center, New York State Department of Health | Kirsten St. George, Daryl M. Lamson, Alexis Russel, Matthew Shudt, Melissa A Leisner, Jonathan Plitnick, Navjot Singh, John Kelly, Erasmus Schneider, Erica Lasek-Nesselquist |
| EPI_ISL_1016249, EPI_ISL_1016250, EPI_ISL_1016251, EPI_ISL_1016252, EPI_ISL_1016253, EPI_ISL_1016254, EPI_ISL_1016255, EPI_ISL_1016256 | TEMPUS LABS INC | Wadsworth Center, New York State Department of Health | Kirsten St. George, Daryl M. Lamson, Alexis Russel, Matthew Shudt, Melissa A Leisner, Jonathan Plitnick, Navjot Singh, John Kelly, Erasmus Schneider, Erica Lasek-Nesselquist |
| EPI_ISL_1016257 | THE MARY IMOGENE BASSETT HOSPITAL | Wadsworth Center, New York State Department of Health | Kirsten St. George, Daryl M. Lamson, Alexis Russel, Matthew Shudt, Melissa A Leisner, Jonathan Plitnick, Navjot Singh, John Kelly, Erasmus Schneider, Erica Lasek-Nesselquist |
| EPI_ISL_1016258, EPI_ISL_1016259 | New York Presbyterian Hospital | Wadsworth Center, New York State Department of Health | Kirsten St. George, Daryl M. Lamson, Alexis Russel, Matthew Shudt, Melissa A Leisner, Jonathan Plitnick, Navjot Singh, John Kelly, Erasmus Schneider, Erica Lasek-Nesselquist |
| EPI_ISL_1016260, EPI_ISL_1016261 | Columbia University Irving Medical Center | Wadsworth Center, New York State Department of Health | Kirsten St. George, Daryl M. Lamson, Alexis Russel, Matthew Shudt, Melissa A Leisner, Jonathan Plitnick, Navjot Singh, John Kelly, Erasmus Schneider, Erica Lasek-Nesselquist |
| EPI_ISL_1016262, EPI_ISL_1016263, EPI_ISL_1016264, EPI_ISL_1016265, EPI_ISL_1016266 | New York Presbyterian Hospital | Wadsworth Center, New York State Department of Health | Kirsten St. George, Daryl M. Lamson, Alexis Russel, Matthew Shudt, Melissa A Leisner, Jonathan Plitnick, Navjot Singh, John Kelly, Erasmus Schneider, Erica Lasek-Nesselquist |
| EPI_ISL_1016267, EPI_ISL_1016268 | Columbia University Irving Medical Center | Wadsworth Center, New York State Department of Health | Kirsten St. George, Daryl M. Lamson, Alexis Russel, Matthew Shudt, Melissa A Leisner, Jonathan Plitnick, Navjot Singh, John Kelly, Erasmus Schneider, Erica Lasek-Nesselquist |
| EPI_ISL_1016269 | New York Presbyterian Hospital | Wadsworth Center, New York State Department of Health | Kirsten St. George, Daryl M. Lamson, Alexis Russel, Matthew Shudt, Melissa A Leisner, Jonathan Plitnick, Navjot Singh, John Kelly, Erasmus Schneider, Erica Lasek-Nesselquist |
| EPI_ISL_1016270 | Columbia University Irving Medical | Wadsworth Center, New York State | Kirsten St. George, Daryl M. Lamson, Alexis Russel, Matthew Shudt, Melissa A Leisner, Jonathan Plitnick, Navjot Singh, John Kelly, Erasmus Schneider, Erica Lasek-Nesselquist |

| | | | |
|---|---|---|---|
| | Center | Department of Health | |
| EPI_ISL_1016271, EPI_ISL_1016272, EPI_ISL_1016273, EPI_ISL_1016274, EPI_ISL_1016275, EPI_ISL_1016276, EPI_ISL_1016277, EPI_ISL_1016278, EPI_ISL_1016279, EPI_ISL_1016280 | New York Presbyterian Hospital | Wadsworth Center, New York State Department of Health | Kirsten St. George, Daryl M. Lamson, Alexis Russel, Matthew Shudt, Melissa A Leisner, Jonathan Plitnick, Navjot Singh, John Kelly, Erasmus Schneider, Erica Lasek-Nesselquist |
| EPI_ISL_1016281, EPI_ISL_1016282, EPI_ISL_1016283, EPI_ISL_1016284 | Columbia University Irving Medical Center | Wadsworth Center, New York State Department of Health | Kirsten St. George, Daryl M. Lamson, Alexis Russel, Matthew Shudt, Melissa A Leisner, Jonathan Plitnick, Navjot Singh, John Kelly, Erasmus Schneider, Erica Lasek-Nesselquist |
| EPI_ISL_1016285, EPI_ISL_1016286 | SUNY UPSTATE MEDICAL UNIVERSITY | Wadsworth Center, New York State Department of Health | Kirsten St. George, Daryl M. Lamson, Alexis Russel, Matthew Shudt, Melissa A Leisner, Jonathan Plitnick, Navjot Singh, John Kelly, Erasmus Schneider, Erica Lasek-Nesselquist |
| EPI_ISL_1016287, EPI_ISL_1016288, EPI_ISL_1016289, EPI_ISL_1016290, EPI_ISL_1016291, EPI_ISL_1016292, EPI_ISL_1016293, EPI_ISL_1016294, EPI_ISL_1016295, EPI_ISL_1016296, EPI_ISL_1016297 | | | |
| see above | Columbia University Irving Medical Center | Wadsworth Center, New York State Department of Health | Kirsten St. George, Daryl M. Lamson, Alexis Russel, Matthew Shudt, Melissa A Leisner, Jonathan Plitnick, Navjot Singh, John Kelly, Erasmus Schneider, Erica Lasek-Nesselquist |
| EPI_ISL_1016298, EPI_ISL_1016299, EPI_ISL_1016300, EPI_ISL_1016301, EPI_ISL_1016302, EPI_ISL_1016303 | SUNY UPSTATE MEDICAL UNIVERSITY | Wadsworth Center, New York State Department of Health | Kirsten St. George, Daryl M. Lamson, Alexis Russel, Matthew Shudt, Melissa A Leisner, Jonathan Plitnick, Navjot Singh, John Kelly, Erasmus Schneider, Erica Lasek-Nesselquist |
| EPI_ISL_1016304, EPI_ISL_1016305, EPI_ISL_1016306, EPI_ISL_1016307, EPI_ISL_1016308, EPI_ISL_1016309, EPI_ISL_1016310 | New York Presbyterian Hospital | Wadsworth Center, New York State Department of Health | Kirsten St. George, Daryl M. Lamson, Alexis Russel, Matthew Shudt, Melissa A Leisner, Jonathan Plitnick, Navjot Singh, John Kelly, Erasmus Schneider, Erica Lasek-Nesselquist |
| EPI_ISL_1016311, EPI_ISL_1016312, EPI_ISL_1016313, EPI_ISL_1016314, EPI_ISL_1016315, EPI_ISL_1016316, EPI_ISL_1016317, EPI_ISL_1016318, EPI_ISL_1016319, EPI_ISL_1016320, EPI_ISL_1016321, EPI_ISL_1016322 | | | |
| see above | SUNY UPSTATE MEDICAL UNIVERSITY | Wadsworth Center, New York State Department of Health | Kirsten St. George, Daryl M. Lamson, Alexis Russel, Matthew Shudt, Melissa A Leisner, Jonathan Plitnick, Navjot Singh, John Kelly, Erasmus Schneider, Erica Lasek-Nesselquist |
| EPI_ISL_1016323, EPI_ISL_1016324, EPI_ISL_1016325, EPI_ISL_1016326 | Columbia University Irving Medical Center | Wadsworth Center, New York State Department of Health | Kirsten St. George, Daryl M. Lamson, Alexis Russel, Matthew Shudt, Melissa A Leisner, Jonathan Plitnick, Navjot Singh, John Kelly, Erasmus Schneider, Erica Lasek-Nesselquist |
| EPI_ISL_1016327 | SUNY UPSTATE MEDICAL UNIVERSITY | Wadsworth Center, New York State Department of Health | Kirsten St. George, Daryl M. Lamson, Alexis Russel, Matthew Shudt, Melissa A Leisner, Jonathan Plitnick, Navjot Singh, John Kelly, Erasmus Schneider, Erica Lasek-Nesselquist |
| EPI_ISL_1016328, EPI_ISL_1016329, EPI_ISL_1016330, EPI_ISL_1016331 | Columbia University Irving Medical Center | Wadsworth Center, New York State Department of Health | Kirsten St. George, Daryl M. Lamson, Alexis Russel, Matthew Shudt, Melissa A Leisner, Jonathan Plitnick, Navjot Singh, John Kelly, Erasmus Schneider, Erica Lasek-Nesselquist |
| EPI_ISL_1016332 | WESTCHESTER MEDICAL CENTER | Wadsworth Center, New York State Department of Health | Kirsten St. George, Daryl M. Lamson, Alexis Russel, Matthew Shudt, Melissa A Leisner, Jonathan Plitnick, Navjot Singh, John Kelly, Erasmus Schneider, Erica Lasek-Nesselquist |
| EPI_ISL_1016333, EPI_ISL_1016334 | URMC LABS | Wadsworth Center, New York State Department of Health | Kirsten St. George, Daryl M. Lamson, Alexis Russel, Matthew Shudt, Melissa A Leisner, Jonathan Plitnick, Navjot Singh, John Kelly, Erasmus Schneider, Erica Lasek-Nesselquist |
| EPI_ISL_1016335, EPI_ISL_1016336 | New York Presbyterian Hospital | Wadsworth Center, New York State Department of Health | Kirsten St. George, Daryl M. Lamson, Alexis Russel, Matthew Shudt, Melissa A Leisner, Jonathan Plitnick, Navjot Singh, John Kelly, Erasmus Schneider, Erica Lasek-Nesselquist |
| EPI_ISL_1016337, EPI_ISL_1016338 | MEMORIAL SLOAN KETTERING CANCER CENTER | Wadsworth Center, New York State Department of Health | Kirsten St. George, Daryl M. Lamson, Alexis Russel, Matthew Shudt, Melissa A Leisner, Jonathan Plitnick, Navjot Singh, John Kelly, Erasmus Schneider, Erica Lasek-Nesselquist |
| EPI_ISL_1016339 | URMC LABS | Wadsworth Center, New York State Department of Health | Kirsten St. George, Daryl M. Lamson, Alexis Russel, Matthew Shudt, Melissa A Leisner, Jonathan Plitnick, Navjot Singh, John Kelly, Erasmus Schneider, Erica Lasek-Nesselquist |
| EPI_ISL_1016340 | WESTCHESTER MEDICAL CENTER | Wadsworth Center, New York State Department of Health | Kirsten St. George, Daryl M. Lamson, Alexis Russel, Matthew Shudt, Melissa A Leisner, Jonathan Plitnick, Navjot Singh, John Kelly, Erasmus Schneider, Erica Lasek-Nesselquist |
| EPI_ISL_1016341 | New York Presbyterian Hospital | Wadsworth Center, New York State Department of Health | Kirsten St. George, Daryl M. Lamson, Alexis Russel, Matthew Shudt, Melissa A Leisner, Jonathan Plitnick, Navjot Singh, John Kelly, Erasmus Schneider, Erica Lasek-Nesselquist |
| EPI_ISL_1016342, EPI_ISL_1016343 | MEMORIAL SLOAN KETTERING CANCER CENTER | Wadsworth Center, New York State Department of Health | Kirsten St. George, Daryl M. Lamson, Alexis Russel, Matthew Shudt, Melissa A Leisner, Jonathan Plitnick, Navjot Singh, John Kelly, Erasmus Schneider, Erica Lasek-Nesselquist |
| EPI_ISL_1016344, EPI_ISL_1016345, EPI_ISL_1016346 | WESTCHESTER MEDICAL CENTER | Wadsworth Center, New York State Department of Health | Kirsten St. George, Daryl M. Lamson, Alexis Russel, Matthew Shudt, Melissa A Leisner, Jonathan Plitnick, Navjot Singh, John Kelly, Erasmus Schneider, Erica Lasek-Nesselquist |
| EPI_ISL_1016347, EPI_ISL_1016348, EPI_ISL_1016349, EPI_ISL_1016350, EPI_ISL_1016351, EPI_ISL_1016352, EPI_ISL_1016353, EPI_ISL_1016354, EPI_ISL_1016355, EPI_ISL_1016356, EPI_ISL_1016357, EPI_ISL_1016358, EPI_ISL_1016359, EPI_ISL_1016360, EPI_ISL_1016361, EPI_ISL_1016362, EPI_ISL_1016363, EPI_ISL_1016364 | | | |
| see above | URMC LABS | Wadsworth Center, New York State Department of Health | Kirsten St. George, Daryl M. Lamson, Alexis Russel, Matthew Shudt, Melissa A Leisner, Jonathan Plitnick, Navjot Singh, John Kelly, Erasmus Schneider, Erica Lasek-Nesselquist |
| EPI_ISL_1016365 | WESTCHESTER MEDICAL CENTER | Wadsworth Center, New York State Department of Health | Kirsten St. George, Daryl M. Lamson, Alexis Russel, Matthew Shudt, Melissa A Leisner, Jonathan Plitnick, Navjot Singh, John Kelly, Erasmus Schneider, Erica Lasek-Nesselquist |
| EPI_ISL_1016366, EPI_ISL_1016367, EPI_ISL_1016368, EPI_ISL_1016369, EPI_ISL_1016370, EPI_ISL_1016371, EPI_ISL_1016372 | New York Presbyterian Hospital | Wadsworth Center, New York State Department of Health | Kirsten St. George, Daryl M. Lamson, Alexis Russel, Matthew Shudt, Melissa A Leisner, Jonathan Plitnick, Navjot Singh, John Kelly, Erasmus Schneider, Erica Lasek-Nesselquist |
| EPI_ISL_1016373, EPI_ISL_1016374, EPI_ISL_1016375, EPI_ISL_1016376, EPI_ISL_1016377, EPI_ISL_1016378, EPI_ISL_1016379, EPI_ISL_1016380 | WESTCHESTER MEDICAL CENTER | Wadsworth Center, New York State Department of Health | Kirsten St. George, Daryl M. Lamson, Alexis Russel, Matthew Shudt, Melissa A Leisner, Jonathan Plitnick, Navjot Singh, John Kelly, Erasmus Schneider, Erica Lasek-Nesselquist |
| EPI_ISL_1016381, EPI_ISL_1016382, EPI_ISL_1016383, EPI_ISL_1016384, EPI_ISL_1016385, EPI_ISL_1016386, EPI_ISL_1016387, EPI_ISL_1016388, EPI_ISL_1016389, EPI_ISL_1016390, EPI_ISL_1016391, EPI_ISL_1016392, EPI_ISL_1016393, EPI_ISL_1016394, EPI_ISL_1016395, EPI_ISL_1016396, EPI_ISL_1016397, EPI_ISL_1016398, EPI_ISL_1016399, EPI_ISL_1016400, EPI_ISL_1016401 | | | |
| see above | New York Presbyterian Hospital | Wadsworth Center, New York State Department of Health | Kirsten St. George, Daryl M. Lamson, Alexis Russel, Matthew Shudt, Melissa A Leisner, Jonathan Plitnick, Navjot Singh, John Kelly, Erasmus Schneider, Erica Lasek-Nesselquist |
| EPI_ISL_1016402, EPI_ISL_1016403, EPI_ISL_1016404, EPI_ISL_1016405, EPI_ISL_1016406, EPI_ISL_1016407, EPI_ISL_1016408, EPI_ISL_1016409, EPI_ISL_1016410, | MEMORIAL SLOAN KETTERING CANCER CENTER | Wadsworth Center, New York State Department of Health | Kirsten St. George, Daryl M. Lamson, Alexis Russel, Matthew Shudt, Melissa A Leisner, Jonathan Plitnick, Navjot Singh, John Kelly, Erasmus Schneider, Erica Lasek-Nesselquist |

| | | | |
|---|---|---|---|
| EPI_ISL_1016411 | | | |
| EPI_ISL_1016412 | STONY BROOK UNIVERSITY HOSPITAL | Wadsworth Center, New York State Department of Health | Kirsten St. George, Daryl M. Lamson, Alexis Russel, Matthew Shudt, Melissa A Leisner, Jonathan Plitnick, Navjot Singh, John Kelly, Erasmus Schneider, Erica Lasek-Nesselquist |
| EPI_ISL_1016413 | NORTHWELL HEALTH LABORATORIES | Wadsworth Center, New York State Department of Health | Kirsten St. George, Daryl M. Lamson, Alexis Russel, Matthew Shudt, Melissa A Leisner, Jonathan Plitnick, Navjot Singh, John Kelly, Erasmus Schneider, Erica Lasek-Nesselquist |
| EPI_ISL_1016414, EPI_ISL_1016415 | URMC LABS | Wadsworth Center, New York State Department of Health | Kirsten St. George, Daryl M. Lamson, Alexis Russel, Matthew Shudt, Melissa A Leisner, Jonathan Plitnick, Navjot Singh, John Kelly, Erasmus Schneider, Erica Lasek-Nesselquist |
| EPI_ISL_1016416 | STONY BROOK UNIVERSITY HOSPITAL | Wadsworth Center, New York State Department of Health | Kirsten St. George, Daryl M. Lamson, Alexis Russel, Matthew Shudt, Melissa A Leisner, Jonathan Plitnick, Navjot Singh, John Kelly, Erasmus Schneider, Erica Lasek-Nesselquist |
| EPI_ISL_1016417 | ALBANY MEDICAL CENTER HOSPITAL CLINICAL LABORATORIES | Wadsworth Center, New York State Department of Health | Kirsten St. George, Daryl M. Lamson, Alexis Russel, Matthew Shudt, Melissa A Leisner, Jonathan Plitnick, Navjot Singh, John Kelly, Erasmus Schneider, Erica Lasek-Nesselquist |
| EPI_ISL_1016418, EPI_ISL_1016419, EPI_ISL_1016420, EPI_ISL_1016421, EPI_ISL_1016422, EPI_ISL_1016423, EPI_ISL_1016424, EPI_ISL_1016425, EPI_ISL_1016426, EPI_ISL_1016427, EPI_ISL_1016428, EPI_ISL_1016429, EPI_ISL_1016430, EPI_ISL_1016431, EPI_ISL_1016432, EPI_ISL_1016433, EPI_ISL_1016434, EPI_ISL_1016435, EPI_ISL_1016436, EPI_ISL_1016437, EPI_ISL_1016438, EPI_ISL_1016439, EPI_ISL_1016440, EPI_ISL_1016441, EPI_ISL_1016442, EPI_ISL_1016443, EPI_ISL_1016444, EPI_ISL_1016445, EPI_ISL_1016446, EPI_ISL_1016447, EPI_ISL_1016448, EPI_ISL_1016449, EPI_ISL_1016450, EPI_ISL_1016451, EPI_ISL_1016452, EPI_ISL_1016453, EPI_ISL_1016454, EPI_ISL_1016455, EPI_ISL_1016456, EPI_ISL_1016457 | | | |
| see above | URMC LABS | Wadsworth Center, New York State Department of Health | Kirsten St. George, Daryl M. Lamson, Alexis Russel, Matthew Shudt, Melissa A Leisner, Jonathan Plitnick, Navjot Singh, John Kelly, Erasmus Schneider, Erica Lasek-Nesselquist |
| EPI_ISL_1016458 | NORTHWELL HEALTH LABORATORIES | Wadsworth Center, New York State Department of Health | Kirsten St. George, Daryl M. Lamson, Alexis Russel, Matthew Shudt, Melissa A Leisner, Jonathan Plitnick, Navjot Singh, John Kelly, Erasmus Schneider, Erica Lasek-Nesselquist |
| EPI_ISL_1016459, EPI_ISL_1016460, EPI_ISL_1016461, EPI_ISL_1016462, EPI_ISL_1016463, EPI_ISL_1016464, EPI_ISL_1016465, EPI_ISL_1016466, EPI_ISL_1016467 | STONY BROOK UNIVERSITY HOSPITAL | Wadsworth Center, New York State Department of Health | Kirsten St. George, Daryl M. Lamson, Alexis Russel, Matthew Shudt, Melissa A Leisner, Jonathan Plitnick, Navjot Singh, John Kelly, Erasmus Schneider, Erica Lasek-Nesselquist |
| EPI_ISL_1016468, EPI_ISL_1016469, EPI_ISL_1016470, EPI_ISL_1016471, EPI_ISL_1016472, EPI_ISL_1016473, EPI_ISL_1016474, EPI_ISL_1016475 | ALBANY MEDICAL CENTER HOSPITAL CLINICAL LABORATORIES | Wadsworth Center, New York State Department of Health | Kirsten St. George, Daryl M. Lamson, Alexis Russel, Matthew Shudt, Melissa A Leisner, Jonathan Plitnick, Navjot Singh, John Kelly, Erasmus Schneider, Erica Lasek-Nesselquist |
| EPI_ISL_1016476, EPI_ISL_1016477, EPI_ISL_1016478 | STONY BROOK UNIVERSITY HOSPITAL | Wadsworth Center, New York State Department of Health | Kirsten St. George, Daryl M. Lamson, Alexis Russel, Matthew Shudt, Melissa A Leisner, Jonathan Plitnick, Navjot Singh, John Kelly, Erasmus Schneider, Erica Lasek-Nesselquist |
| EPI_ISL_1016479, EPI_ISL_1016480, EPI_ISL_1016481, EPI_ISL_1016482, EPI_ISL_1016483, EPI_ISL_1016488, EPI_ISL_1016489, EPI_ISL_1016490, EPI_ISL_1016491, EPI_ISL_1016492, EPI_ISL_1016493, EPI_ISL_1016494, EPI_ISL_1016495, EPI_ISL_1016496, EPI_ISL_1016497, EPI_ISL_1016498, EPI_ISL_1016499, EPI_ISL_1016500, EPI_ISL_1016501, EPI_ISL_1016502, EPI_ISL_1016503, EPI_ISL_1016504, EPI_ISL_1016505, EPI_ISL_1016506, EPI_ISL_1016507 | | | |
| see above | University of Miami Immunology and Histocompatibility Laboratory | University of Miami Immunology and Histocompatibility Laboratory | Emilio Margolles-Clark, PhD and Phillip Ruiz, MD, PhD |
| EPI_ISL_1016509, EPI_ISL_1016510, EPI_ISL_1016511, EPI_ISL_1016512, EPI_ISL_1016513, EPI_ISL_1016514, EPI_ISL_1016515, EPI_ISL_1016516, EPI_ISL_1016517, EPI_ISL_1016518, EPI_ISL_1016519, EPI_ISL_1016520, EPI_ISL_1016521, EPI_ISL_1016522, EPI_ISL_1016523, EPI_ISL_1016524, EPI_ISL_1016525, EPI_ISL_1016526, EPI_ISL_1016527, EPI_ISL_1016528, EPI_ISL_1016529, EPI_ISL_1016530, EPI_ISL_1016531, EPI_ISL_1016532, EPI_ISL_1016533, EPI_ISL_1016534, EPI_ISL_1016535, EPI_ISL_1016536, EPI_ISL_1016537, EPI_ISL_1016538, EPI_ISL_1016539, EPI_ISL_1016540, EPI_ISL_1016541, EPI_ISL_1016542, EPI_ISL_1016543, EPI_ISL_1016544, EPI_ISL_1016545, EPI_ISL_1016546, EPI_ISL_1016547, EPI_ISL_1016548, EPI_ISL_1016549, EPI_ISL_1016550, EPI_ISL_1016551, EPI_ISL_1016552, EPI_ISL_1016553, EPI_ISL_1016554, EPI_ISL_1016555, EPI_ISL_1016556, EPI_ISL_1016557, EPI_ISL_1016558, EPI_ISL_1016559, EPI_ISL_1016560, EPI_ISL_1016561, EPI_ISL_1016562, EPI_ISL_1016563, EPI_ISL_1016564, EPI_ISL_1016565, EPI_ISL_1016566, EPI_ISL_1016567, EPI_ISL_1016568, EPI_ISL_1016569, EPI_ISL_1016570, EPI_ISL_1016571, EPI_ISL_1016572, EPI_ISL_1016573, EPI_ISL_1016574, EPI_ISL_1016594, EPI_ISL_1016595, EPI_ISL_1016596, EPI_ISL_1016597, EPI_ISL_1016598, EPI_ISL_1016599, EPI_ISL_1016600, EPI_ISL_1016601, EPI_ISL_1016602, EPI_ISL_1016603, EPI_ISL_1016604, EPI_ISL_1016605, EPI_ISL_1016606, EPI_ISL_1016607, EPI_ISL_1016608, EPI_ISL_1016609, EPI_ISL_1016610, EPI_ISL_1016611, EPI_ISL_1016612, EPI_ISL_1016613, EPI_ISL_1016614, EPI_ISL_1016615, EPI_ISL_1016616, EPI_ISL_1016617, EPI_ISL_1016618, EPI_ISL_1016619, EPI_ISL_1016620, EPI_ISL_1016621, EPI_ISL_1016622, EPI_ISL_1016623, EPI_ISL_1016624, EPI_ISL_1016625, EPI_ISL_1016626, EPI_ISL_1016627, EPI_ISL_1016628, EPI_ISL_1016629, EPI_ISL_1016630, EPI_ISL_1016631, EPI_ISL_1016632, EPI_ISL_1016633, EPI_ISL_1016634, EPI_ISL_1016635, EPI_ISL_1016636, EPI_ISL_1016637, EPI_ISL_1016638, EPI_ISL_1016639, EPI_ISL_1016640, EPI_ISL_1016641, EPI_ISL_1016642, EPI_ISL_1016643, EPI_ISL_1016644, EPI_ISL_1016645, EPI_ISL_1016646, EPI_ISL_1016647, EPI_ISL_1016648, EPI_ISL_1016649, EPI_ISL_1016650, EPI_ISL_1016651, EPI_ISL_1016652, EPI_ISL_1016653, EPI_ISL_1016654, EPI_ISL_1016655, EPI_ISL_1016656, EPI_ISL_1016657, EPI_ISL_1016658, EPI_ISL_1016659, EPI_ISL_1016660, EPI_ISL_1016661, EPI_ISL_1016662, EPI_ISL_1016663, EPI_ISL_1016664, EPI_ISL_1016665, EPI_ISL_1016666, EPI_ISL_1016667, EPI_ISL_1016668, EPI_ISL_1016669, EPI_ISL_1016670, EPI_ISL_1016671, EPI_ISL_1016672, EPI_ISL_1016673, EPI_ISL_1016674, EPI_ISL_1016675, EPI_ISL_1016676, EPI_ISL_1016677, EPI_ISL_1016678, EPI_ISL_1016679, EPI_ISL_1016680, EPI_ISL_1016681, EPI_ISL_1016682, EPI_ISL_1016683, EPI_ISL_1016684, EPI_ISL_1016685, EPI_ISL_1016686, EPI_ISL_1016687, EPI_ISL_1016688, EPI_ISL_1016689, EPI_ISL_1016691, EPI_ISL_1016692, EPI_ISL_1016693, EPI_ISL_1016694, EPI_ISL_1016695, EPI_ISL_1016696, EPI_ISL_1016697, EPI_ISL_1016698, EPI_ISL_1016699, EPI_ISL_1016700, EPI_ISL_1016701, EPI_ISL_1016702, EPI_ISL_1016703, EPI_ISL_1016704, EPI_ISL_1016705, EPI_ISL_1016706, EPI_ISL_1016707, EPI_ISL_1016708, EPI_ISL_1016709, EPI_ISL_1016710, EPI_ISL_1016711, EPI_ISL_1016712, EPI_ISL_1016713, EPI_ISL_1016714, EPI_ISL_1016715, EPI_ISL_1016716, EPI_ISL_1016717, EPI_ISL_1016718, EPI_ISL_1016719, EPI_ISL_1016720, EPI_ISL_1016721, EPI_ISL_1016722, EPI_ISL_1016723, EPI_ISL_1016724, EPI_ISL_1016725, EPI_ISL_1016726, EPI_ISL_1016727, EPI_ISL_1016728, EPI_ISL_1016729, EPI_ISL_1016730, EPI_ISL_1016731, EPI_ISL_1016732, EPI_ISL_1016733, EPI_ISL_1016734, EPI_ISL_1016735, EPI_ISL_1016736, EPI_ISL_1016737, EPI_ISL_1016738, EPI_ISL_1016739, EPI_ISL_1016740, EPI_ISL_1016741, EPI_ISL_1016742, EPI_ISL_1016743, EPI_ISL_1016744, EPI_ISL_1016745, EPI_ISL_1016746, EPI_ISL_1016747, EPI_ISL_1016748, EPI_ISL_1016749, EPI_ISL_1016750, EPI_ISL_1016751, EPI_ISL_1016752, EPI_ISL_1016753, EPI_ISL_1016754, EPI_ISL_1016755, EPI_ISL_1016756, EPI_ISL_1016757, EPI_ISL_1016758, EPI_ISL_1016759, EPI_ISL_1016760, EPI_ISL_1016761, EPI_ISL_1016762, EPI_ISL_1016763, EPI_ISL_1016764, EPI_ISL_1016765, EPI_ISL_1016766, EPI_ISL_1016767, EPI_ISL_1016768, EPI_ISL_1016769, EPI_ISL_1016770, EPI_ISL_1016771, EPI_ISL_1016772, EPI_ISL_1016773, EPI_ISL_1016774, EPI_ISL_1016775, EPI_ISL_1016776, EPI_ISL_1016777, EPI_ISL_1016778, EPI_ISL_1016779, EPI_ISL_1016780, EPI_ISL_1016781, EPI_ISL_1016782, EPI_ISL_1016783, EPI_ISL_1016784, EPI_ISL_1016785, EPI_ISL_1016786, EPI_ISL_1016787, EPI_ISL_1016788, EPI_ISL_1016789, EPI_ISL_1016790, EPI_ISL_1016791, EPI_ISL_1016792, EPI_ISL_1016793, EPI_ISL_1016794, EPI_ISL_1016795, EPI_ISL_1016796, EPI_ISL_1016797, EPI_ISL_1016798, EPI_ISL_1016799, EPI_ISL_1016800, EPI_ISL_1016801, EPI_ISL_1016802, EPI_ISL_1016803, EPI_ISL_1016804, EPI_ISL_1016805, EPI_ISL_1016806, EPI_ISL_1016807, EPI_ISL_1016808, EPI_ISL_1016809, EPI_ISL_1016810, EPI_ISL_1016811, EPI_ISL_1016812, EPI_ISL_1016813, EPI_ISL_1016814, EPI_ISL_1016815, EPI_ISL_1016816, EPI_ISL_1016817, EPI_ISL_1016818, EPI_ISL_1016819, EPI_ISL_1016820, EPI_ISL_1016823, EPI_ISL_1016824, EPI_ISL_1016825, EPI_ISL_1016826, EPI_ISL_1016827, EPI_ISL_1016828, EPI_ISL_1016829, EPI_ISL_1016830, EPI_ISL_1016831, EPI_ISL_1016832, EPI_ISL_1016833, EPI_ISL_1016834, EPI_ISL_1016835, EPI_ISL_1016836, EPI_ISL_1016837, EPI_ISL_1016838, EPI_ISL_1016839, EPI_ISL_1016840, EPI_ISL_1016841 | | | |
| see above | Helix/Illumina | Respiratory Viruses Branch, Division of Viral Diseases, Centers for Disease Control and Prevention | Peter W. Cook, Dakota Howard, Dhwani Batra, Ben L. Rambo-Martin, Eileen de Feo, Jan Antico, Christine Tran, Matthew Tolentino, Shannon Wickline, Kim Gietzen, Brad Sickler, Jingtao Liu, Eric Allen, Phil Febbo, Summer Galloway, Nicole L. Washington, Simon White, Geraint Levan, Kelly Schiabor Barrett, Elizabeth Cirulli, Alexandre Bolze, Ary Ascencio, Charlotte Rivera-Garcia, Ryan Cho, Jason Nguyen, Sherry Wang, Jimmy Ramirez, Tyler Cassens, Efren Sandoval, Magnus Isaksson, William Lee, David Becker, Marc Laurent, James Lu, Clinton R. Paden, Suxiang Tong, Duncan MacCannell |
| EPI_ISL_1016887, EPI_ISL_1016888, EPI_ISL_1016891, EPI_ISL_1016892, EPI_ISL_1016893, EPI_ISL_1016894, EPI_ISL_1016895, EPI_ISL_1016897, EPI_ISL_1016898, EPI_ISL_1016899, EPI_ISL_1016900, EPI_ISL_1016901, EPI_ISL_1016902, EPI_ISL_1016903, EPI_ISL_1016904, EPI_ISL_1016905, EPI_ISL_1016906, EPI_ISL_1016907, EPI_ISL_1016908, EPI_ISL_1016909, EPI_ISL_1016910, EPI_ISL_1016911, EPI_ISL_1016912, EPI_ISL_1016914, EPI_ISL_1016915, EPI_ISL_1016917, EPI_ISL_1016918, EPI_ISL_1016919, EPI_ISL_1016920, EPI_ISL_1016921, EPI_ISL_1016922, EPI_ISL_1016923, EPI_ISL_1016924, EPI_ISL_1016925, EPI_ISL_1016926, EPI_ISL_1016927, EPI_ISL_1016928, EPI_ISL_1016929, EPI_ISL_1016930, EPI_ISL_1016931, EPI_ISL_1016932, EPI_ISL_1016933, EPI_ISL_1016934, EPI_ISL_1016935, EPI_ISL_1016936, EPI_ISL_1016937, EPI_ISL_1016938, EPI_ISL_1016939, EPI_ISL_1016940, EPI_ISL_1016941, EPI_ISL_1016942, EPI_ISL_1016943, EPI_ISL_1016944, EPI_ISL_1016945, EPI_ISL_1016946 | | | |
| see above | Arizona State Public Health Laboratory | Arizona State Public Health Laboratory | Trung Huynh, Jessica Escobar, Katherine Fullerton, Nobuko Fukushima, Stacy White, Linda Getsinger, Victor Waddell |
| EPI_ISL_1016947, EPI_ISL_1016948, EPI_ISL_1016949, EPI_ISL_1016950, EPI_ISL_1016954, EPI_ISL_1016955, EPI_ISL_1016956, EPI_ISL_1016957, EPI_ISL_1016958, EPI_ISL_1016959, EPI_ISL_1016960, EPI_ISL_1016961, EPI_ISL_1016962, EPI_ISL_1016963, EPI_ISL_1016964, EPI_ISL_1016965, EPI_ISL_1016966, EPI_ISL_1016967 | | | |
| see above | UNMC Emerging Pathogens Laboratory | UNMC COVID-19 Response Team | UNMC COVID-19 Response Team |
| EPI_ISL_1016971, EPI_ISL_1016972, EPI_ISL_1016973, EPI_ISL_1016974 | South Dakota Public Health Laboratory | University of Minnesota Genomics Center | Daryl M. Gohl, Benjamin Auch, John Garbe, Jaquelyn Kuriger-Laber, Corbin Dirkx, and Chris Carlson |
| EPI_ISL_1016975 | M Health Fairview | University of Minnesota Genomics Center | Daryl M. Gohl, Benjamin Auch, John Garbe, Jaquelyn Kuriger-Laber, Corbin Dirkx, Andy Nelson, and Sean Wang |
| EPI_ISL_1016976, EPI_ISL_1016977, EPI_ISL_1016978 | South Dakota Public Health Laboratory | University of Minnesota Genomics Center | Daryl M. Gohl, Benjamin Auch, John Garbe, Jaquelyn Kuriger-Laber, Corbin Dirkx, and Chris Carlson |
| EPI_ISL_1016979 | M Health Fairview | University of Minnesota Genomics Center | Daryl M. Gohl, Benjamin Auch, John Garbe, Jaquelyn Kuriger-Laber, Corbin Dirkx, Andy Nelson, and Sean Wang |
| EPI_ISL_1016980, EPI_ISL_1016981, EPI_ISL_1016982, EPI_ISL_1016983, EPI_ISL_1016984 | South Dakota Public Health Laboratory | University of Minnesota Genomics Center | Daryl M. Gohl, Benjamin Auch, John Garbe, Jaquelyn Kuriger-Laber, Corbin Dirkx, and Chris Carlson |
| EPI_ISL_1016985, EPI_ISL_1016986 | M Health Fairview | University of Minnesota Genomics Center | Daryl M. Gohl, Benjamin Auch, John Garbe, Jaquelyn Kuriger-Laber, Corbin Dirkx, Andy Nelson, and Sean Wang |
| EPI_ISL_1016987 | South Dakota Public Health Laboratory | University of Minnesota Genomics Center | Daryl M. Gohl, Benjamin Auch, John Garbe, Jaquelyn Kuriger-Laber, Corbin Dirkx, and Chris Carlson |
| EPI_ISL_1016988 | M Health Fairview | University of Minnesota Genomics Center | Daryl M. Gohl, Benjamin Auch, John Garbe, Jaquelyn Kuriger-Laber, Corbin Dirkx, Andy Nelson, and Sean Wang |
| EPI_ISL_1016989, EPI_ISL_1016995, EPI_ISL_1016996, EPI_ISL_1016997, EPI_ISL_1016998, EPI_ISL_1016999, EPI_ISL_1017000, EPI_ISL_1017001, EPI_ISL_1017002, EPI_ISL_1017003, EPI_ISL_1017004, EPI_ISL_1017005, EPI_ISL_1017006, EPI_ISL_1017007, EPI_ISL_1017008, EPI_ISL_1017009, EPI_ISL_1017010, EPI_ISL_1017011, EPI_ISL_1017012, EPI_ISL_1017013, EPI_ISL_1017014, EPI_ISL_1017015, EPI_ISL_1017016, EPI_ISL_1017017, EPI_ISL_1017018, EPI_ISL_1017019, EPI_ISL_1017020, EPI_ISL_1017021, EPI_ISL_1017022, EPI_ISL_1017023 | | | |
| see above | South Dakota Public Health Laboratory | University of Minnesota Genomics Center | Daryl M. Gohl, Benjamin Auch, John Garbe, Jaquelyn Kuriger-Laber, Corbin Dirkx, and Chris Carlson |
| EPI_ISL_1017024, EPI_ISL_1017025, EPI_ISL_1017026, EPI_ISL_1017027, EPI_ISL_1017028, EPI_ISL_1017029, EPI_ISL_1017030, EPI_ISL_1017031, EPI_ISL_1017032 | M Health Fairview | University of Minnesota Genomics Center | Daryl M. Gohl, Benjamin Auch, John Garbe, Jaquelyn Kuriger-Laber, Corbin Dirkx, Andy Nelson, and Sean Wang |
| EPI_ISL_1017033, | South Dakota Public Health Laboratory | University of Minnesota Genomics Center | Daryl M. Gohl, Benjamin Auch, John Garbe, Jaquelyn Kuriger-Laber, Corbin Dirkx, and Chris Carlson |

| | | | |
|---|---|---|---|
| EPI_ISL_1017034, EPI_ISL_1017035, EPI_ISL_1017036, EPI_ISL_1017037, EPI_ISL_1017038, EPI_ISL_1017039, EPI_ISL_1017040 | | | |
| EPI_ISL_1017041, EPI_ISL_1017042, EPI_ISL_1017043, EPI_ISL_1017044, EPI_ISL_1017045, EPI_ISL_1017046, EPI_ISL_1017047, EPI_ISL_1017048, EPI_ISL_1017049, EPI_ISL_1017050 | Providence Oregon Regional Laboratories | Providence St. Joseph Health Molecular Genomics Laboratory | Alexa K Dowdell, Brian D Piening, Fred L Robinson, Carlo B Bifulco, Mary Campbell |
| EPI_ISL_1017051 | Providence Saint Joseph Medical Center - Burbank | Providence St. Joseph Health Molecular Genomics Laboratory | Alexa K Dowdell, Brian D Piening, Fred L Robinson, Carlo B Bifulco, Mary Campbell |
| EPI_ISL_1017052 | Providence St. Joseph Health Orange - Chapman | Providence St. Joseph Health Molecular Genomics Laboratory | Alexa K Dowdell, Brian D Piening, Fred L Robinson, Carlo B Bifulco, Mary Campbell |
| EPI_ISL_1017053 | Providence Oregon Regional Laboratories | Providence St. Joseph Health Molecular Genomics Laboratory | Alexa K Dowdell, Brian D Piening, Fred L Robinson, Carlo B Bifulco, Mary Campbell |
| EPI_ISL_1017054 | Providence Sacred Heart Medical Center | Providence St. Joseph Health Molecular Genomics Laboratory | Alexa K Dowdell, Brian D Piening, Fred L Robinson, Carlo B Bifulco, Mary Campbell |
| EPI_ISL_1017055, EPI_ISL_1017056, EPI_ISL_1017057 | Providence Regional Medical Center Everett | Providence St. Joseph Health Molecular Genomics Laboratory | Alexa K Dowdell, Brian D Piening, Fred L Robinson, Carlo B Bifulco, Mary Campbell |
| EPI_ISL_1017058 | Providence St. Joseph Health Orange - Chapman | Providence St. Joseph Health Molecular Genomics Laboratory | Alexa K Dowdell, Brian D Piening, Fred L Robinson, Carlo B Bifulco, Mary Campbell |
| EPI_ISL_1017059, EPI_ISL_1017060, EPI_ISL_1017061, EPI_ISL_1017062, EPI_ISL_1017063 | Providence Saint Joseph Medical Center - Burbank | Providence St. Joseph Health Molecular Genomics Laboratory | Alexa K Dowdell, Brian D Piening, Fred L Robinson, Carlo B Bifulco, Mary Campbell |
| EPI_ISL_1017064 | Providence Oregon Regional Laboratories | Providence St. Joseph Health Molecular Genomics Laboratory | Alexa K Dowdell, Brian D Piening, Fred L Robinson, Carlo B Bifulco, Mary Campbell |
| EPI_ISL_1017065 | Providence Saint Joseph Medical Center - Burbank | Providence St. Joseph Health Molecular Genomics Laboratory | Alexa K Dowdell, Brian D Piening, Fred L Robinson, Carlo B Bifulco, Mary Campbell |
| EPI_ISL_1017066 | Providence Alaska Medical Center | Providence St. Joseph Health Molecular Genomics Laboratory | Alexa K Dowdell, Brian D Piening, Fred L Robinson, Carlo B Bifulco, Mary Campbell |
| EPI_ISL_1017067, EPI_ISL_1017068, EPI_ISL_1017069, EPI_ISL_1017070, EPI_ISL_1017071, EPI_ISL_1017072, EPI_ISL_1017073, EPI_ISL_1017074 | Providence Oregon Regional Laboratories | Providence St. Joseph Health Molecular Genomics Laboratory | Alexa K Dowdell, Brian D Piening, Fred L Robinson, Carlo B Bifulco, Mary Campbell |
| EPI_ISL_1017075, EPI_ISL_1017076, EPI_ISL_1017077, EPI_ISL_1017078, EPI_ISL_1017079, EPI_ISL_1017080, EPI_ISL_1017081, EPI_ISL_1017082, EPI_ISL_1017083, EPI_ISL_1017084, EPI_ISL_1017085, EPI_ISL_1017086, EPI_ISL_1017087, EPI_ISL_1017088, EPI_ISL_1017089 | | | |
| see above | Providence St. Joseph Health Orange - Chapman | Providence St. Joseph Health Molecular Genomics Laboratory | Alexa K Dowdell, Brian D Piening, Fred L Robinson, Carlo B Bifulco, Mary Campbell |
| EPI_ISL_1017090 | Providence Oregon Regional Laboratories | Providence St. Joseph Health Molecular Genomics Laboratory | Alexa K Dowdell, Brian D Piening, Fred L Robinson, Carlo B Bifulco, Mary Campbell |
| EPI_ISL_1017091, EPI_ISL_1017092, EPI_ISL_1017093, EPI_ISL_1017094, EPI_ISL_1017095 | Providence St. Joseph Health Orange - Chapman | Providence St. Joseph Health Molecular Genomics Laboratory | Alexa K Dowdell, Brian D Piening, Fred L Robinson, Carlo B Bifulco, Mary Campbell |
| EPI_ISL_1017096 | Providence Oregon Regional Laboratories | Providence St. Joseph Health Molecular Genomics Laboratory | Alexa K Dowdell, Brian D Piening, Fred L Robinson, Carlo B Bifulco, Mary Campbell |
| EPI_ISL_1017097, EPI_ISL_1017098, EPI_ISL_1017099 | Providence Saint Joseph Medical Center - Burbank | Providence St. Joseph Health Molecular Genomics Laboratory | Alexa K Dowdell, Brian D Piening, Fred L Robinson, Carlo B Bifulco, Mary Campbell |
| EPI_ISL_1017100, EPI_ISL_1017101, EPI_ISL_1017102, EPI_ISL_1017103, EPI_ISL_1017104, EPI_ISL_1017105 | Providence Oregon Regional Laboratories | Providence St. Joseph Health Molecular Genomics Laboratory | Alexa K Dowdell, Brian D Piening, Fred L Robinson, Carlo B Bifulco, Mary Campbell |
| EPI_ISL_1017106 | Providence St. Joseph Health Orange - Chapman | Providence St. Joseph Health Molecular Genomics Laboratory | Alexa K Dowdell, Brian D Piening, Fred L Robinson, Carlo B Bifulco, Mary Campbell |
| EPI_ISL_1017107 | Providence Oregon Regional Laboratories | Providence St. Joseph Health Molecular Genomics Laboratory | Alexa K Dowdell, Brian D Piening, Fred L Robinson, Carlo B Bifulco, Mary Campbell |
| EPI_ISL_1017108, EPI_ISL_1017109, EPI_ISL_1017110, EPI_ISL_1017111 | Providence Saint Joseph Medical Center - Burbank | Providence St. Joseph Health Molecular Genomics Laboratory | Alexa K Dowdell, Brian D Piening, Fred L Robinson, Carlo B Bifulco, Mary Campbell |
| EPI_ISL_1017112, EPI_ISL_1017113, EPI_ISL_1017114, EPI_ISL_1017115 | Providence Oregon Regional Laboratories | Providence St. Joseph Health Molecular Genomics Laboratory | Alexa K Dowdell, Brian D Piening, Fred L Robinson, Carlo B Bifulco, Mary Campbell |
| EPI_ISL_1017116, EPI_ISL_1017117 | Providence Saint Joseph Medical Center - Burbank | Providence St. Joseph Health Molecular Genomics Laboratory | Alexa K Dowdell, Brian D Piening, Fred L Robinson, Carlo B Bifulco, Mary Campbell |
| EPI_ISL_1017118, EPI_ISL_1017119 | Providence St. Joseph Health Orange - Chapman | Providence St. Joseph Health Molecular Genomics Laboratory | Alexa K Dowdell, Brian D Piening, Fred L Robinson, Carlo B Bifulco, Mary Campbell |
| EPI_ISL_1017120, EPI_ISL_1017121, EPI_ISL_1017122, EPI_ISL_1017123, EPI_ISL_1017124, EPI_ISL_1017125 | Providence Oregon Regional Laboratories | Providence St. Joseph Health Molecular Genomics Laboratory | Alexa K Dowdell, Brian D Piening, Fred L Robinson, Carlo B Bifulco, Mary Campbell |
| EPI_ISL_1017126, EPI_ISL_1017127, EPI_ISL_1017128, EPI_ISL_1017129, EPI_ISL_1017130, EPI_ISL_1017131, | Providence Saint Joseph Medical Center - Burbank | Providence St. Joseph Health Molecular Genomics Laboratory | Alexa K Dowdell, Brian D Piening, Fred L Robinson, Carlo B Bifulco, Mary Campbell |

| | | | |
|---|---|---|---|
| EPI_ISL_1017132 | | | |
| EPI_ISL_1017133, EPI_ISL_1017134, EPI_ISL_1017135, EPI_ISL_1017136, EPI_ISL_1017137, EPI_ISL_1017138, EPI_ISL_1017139, EPI_ISL_1017140, EPI_ISL_1017141, EPI_ISL_1017142, EPI_ISL_1017143, EPI_ISL_1017144, EPI_ISL_1017145, EPI_ISL_1017146 | | | |
| see above | Providence St. Joseph Health Orange - Chapman | Providence St. Joseph Health Molecular Genomics Laboratory | Alexa K Dowdell, Brian D Piening, Fred L Robinson, Carlo B Bifulco, Mary Campbell |
| EPI_ISL_1017147, EPI_ISL_1017148, EPI_ISL_1017149, EPI_ISL_1017150, EPI_ISL_1017151, EPI_ISL_1017152, EPI_ISL_1017153, EPI_ISL_1017154, EPI_ISL_1017155, EPI_ISL_1017156, EPI_ISL_1017157, EPI_ISL_1017158, EPI_ISL_1017159, EPI_ISL_1017160 | | | |
| see above | Providence Saint Joseph Medical Center - Burbank | Providence St. Joseph Health Molecular Genomics Laboratory | Alexa K Dowdell, Brian D Piening, Fred L Robinson, Carlo B Bifulco, Mary Campbell |
| EPI_ISL_1017161, EPI_ISL_1017162, EPI_ISL_1017163, EPI_ISL_1017164, EPI_ISL_1017165, EPI_ISL_1017166, EPI_ISL_1017167, EPI_ISL_1017168, EPI_ISL_1017169, EPI_ISL_1017170, EPI_ISL_1017171 | | | |
| see above | Providence Regional Medical Center Everett | Providence St. Joseph Health Molecular Genomics Laboratory | Alexa K Dowdell, Brian D Piening, Fred L Robinson, Carlo B Bifulco, Mary Campbell |
| EPI_ISL_1017172, EPI_ISL_1017173, EPI_ISL_1017174, EPI_ISL_1017175, EPI_ISL_1017176, EPI_ISL_1017177, EPI_ISL_1017178, EPI_ISL_1017179 | Providence Oregon Regional Laboratories | Providence St. Joseph Health Molecular Genomics Laboratory | Alexa K Dowdell, Brian D Piening, Fred L Robinson, Carlo B Bifulco, Mary Campbell |
| EPI_ISL_1017180 | Providence Regional Medical Center Everett | Providence St. Joseph Health Molecular Genomics Laboratory | Alexa K Dowdell, Brian D Piening, Fred L Robinson, Carlo B Bifulco, Mary Campbell |
| EPI_ISL_1017181, EPI_ISL_1017182, EPI_ISL_1017183, EPI_ISL_1017184, EPI_ISL_1017185, EPI_ISL_1017186 | Providence Oregon Regional Laboratories | Providence St. Joseph Health Molecular Genomics Laboratory | Alexa K Dowdell, Brian D Piening, Fred L Robinson, Carlo B Bifulco, Mary Campbell |
| EPI_ISL_1017187, EPI_ISL_1017188, EPI_ISL_1017189, EPI_ISL_1017190, EPI_ISL_1017191, EPI_ISL_1017192, EPI_ISL_1017193, EPI_ISL_1017194, EPI_ISL_1017195, EPI_ISL_1017196, EPI_ISL_1017197, EPI_ISL_1017198, EPI_ISL_1017199, EPI_ISL_1017200, EPI_ISL_1017201, EPI_ISL_1017202, EPI_ISL_1017203, EPI_ISL_1017204, EPI_ISL_1017205, EPI_ISL_1017206, EPI_ISL_1017207, EPI_ISL_1017208, EPI_ISL_1017209, EPI_ISL_1017210, EPI_ISL_1017211, EPI_ISL_1017212, EPI_ISL_1017213, EPI_ISL_1017214, EPI_ISL_1017215 | | | |
| see above | Providence Alaska Medical Center | Providence St. Joseph Health Molecular Genomics Laboratory | Alexa K Dowdell, Brian D Piening, Fred L Robinson, Carlo B Bifulco, Mary Campbell |
| EPI_ISL_1017216, EPI_ISL_1017217, EPI_ISL_1017218, EPI_ISL_1017219, EPI_ISL_1017220, EPI_ISL_1017221, EPI_ISL_1017222, EPI_ISL_1017223, EPI_ISL_1017224, EPI_ISL_1017225, EPI_ISL_1017226, EPI_ISL_1017227, EPI_ISL_1017228, EPI_ISL_1017229, EPI_ISL_1017230, EPI_ISL_1017231, EPI_ISL_1017232, EPI_ISL_1017233, EPI_ISL_1017234, EPI_ISL_1017235, EPI_ISL_1017236 | | | |
| see above | Providence Sacred Heart Medical Center | Providence St. Joseph Health Molecular Genomics Laboratory | Alexa K Dowdell, Brian D Piening, Fred L Robinson, Carlo B Bifulco, Mary Campbell |
| EPI_ISL_1017237 | Providence Regional Medical Center Everett | Providence St. Joseph Health Molecular Genomics Laboratory | Alexa K Dowdell, Brian D Piening, Fred L Robinson, Carlo B Bifulco, Mary Campbell |
| EPI_ISL_1017238, EPI_ISL_1017239, EPI_ISL_1017240, EPI_ISL_1017241, EPI_ISL_1017242, EPI_ISL_1017243, EPI_ISL_1017244, EPI_ISL_1017245, EPI_ISL_1017246, EPI_ISL_1017247, EPI_ISL_1017248, EPI_ISL_1017249, EPI_ISL_1017250, EPI_ISL_1017251, EPI_ISL_1017252, EPI_ISL_1017253, EPI_ISL_1017254, EPI_ISL_1017255, EPI_ISL_1017256, EPI_ISL_1017257, EPI_ISL_1017258, EPI_ISL_1017259, EPI_ISL_1017260, EPI_ISL_1017261, EPI_ISL_1017262, EPI_ISL_1017263, EPI_ISL_1017264, EPI_ISL_1017265, EPI_ISL_1017266, EPI_ISL_1017267, EPI_ISL_1017268, EPI_ISL_1017269, EPI_ISL_1017270, EPI_ISL_1017271, EPI_ISL_1017272, EPI_ISL_1017273, EPI_ISL_1017274, EPI_ISL_1017275, EPI_ISL_1017276, EPI_ISL_1017277, EPI_ISL_1017278, EPI_ISL_1017279, EPI_ISL_1017280, EPI_ISL_1017281, EPI_ISL_1017282, EPI_ISL_1017283, EPI_ISL_1017284, EPI_ISL_1017285, EPI_ISL_1017286, EPI_ISL_1017287, EPI_ISL_1017288, EPI_ISL_1017289, EPI_ISL_1017290, EPI_ISL_1017291, EPI_ISL_1017292, EPI_ISL_1017293, EPI_ISL_1017294, EPI_ISL_1017295, EPI_ISL_1017296, EPI_ISL_1017297, EPI_ISL_1017298, EPI_ISL_1017299, EPI_ISL_1017300, EPI_ISL_1017301, EPI_ISL_1017302, EPI_ISL_1017303, EPI_ISL_1017304, EPI_ISL_1017305, EPI_ISL_1017306, EPI_ISL_1017307, EPI_ISL_1017308, EPI_ISL_1017309, EPI_ISL_1017310, EPI_ISL_1017311, EPI_ISL_1017312, EPI_ISL_1017313, EPI_ISL_1017314, EPI_ISL_1017315, EPI_ISL_1017316, EPI_ISL_1017317, EPI_ISL_1017318, EPI_ISL_1017319, EPI_ISL_1017320, EPI_ISL_1017321, EPI_ISL_1017322, EPI_ISL_1017323, EPI_ISL_1017324, EPI_ISL_1017325, EPI_ISL_1017326, EPI_ISL_1017327, EPI_ISL_1017328, EPI_ISL_1017329, EPI_ISL_1017330, EPI_ISL_1017331, EPI_ISL_1017332, EPI_ISL_1017333, EPI_ISL_1017334, EPI_ISL_1017335, EPI_ISL_1017336, EPI_ISL_1017337, EPI_ISL_1017338, EPI_ISL_1017339, EPI_ISL_1017340, EPI_ISL_1017341, EPI_ISL_1017342, EPI_ISL_1017343, EPI_ISL_1017344, EPI_ISL_1017345, EPI_ISL_1017346, EPI_ISL_1017347, EPI_ISL_1017348, EPI_ISL_1017349, EPI_ISL_1017350, EPI_ISL_1017351, EPI_ISL_1017352, EPI_ISL_1017353, EPI_ISL_1017354, EPI_ISL_1017355, EPI_ISL_1017356, EPI_ISL_1017357, EPI_ISL_1017358, EPI_ISL_1017359, EPI_ISL_1017360, EPI_ISL_1017361, EPI_ISL_1017362, EPI_ISL_1017363, EPI_ISL_1017364, EPI_ISL_1017365, EPI_ISL_1017366, EPI_ISL_1017367, EPI_ISL_1017368, EPI_ISL_1017369, EPI_ISL_1017370, EPI_ISL_1017371, EPI_ISL_1017372, EPI_ISL_1017373, EPI_ISL_1017374, EPI_ISL_1017375, EPI_ISL_1017376, EPI_ISL_1017377, EPI_ISL_1017378, EPI_ISL_1017379, EPI_ISL_1017380, EPI_ISL_1017381, EPI_ISL_1017382, EPI_ISL_1017383, EPI_ISL_1017384, EPI_ISL_1017385, EPI_ISL_1017386, EPI_ISL_1017387, EPI_ISL_1017388, EPI_ISL_1017389, EPI_ISL_1017390, EPI_ISL_1017391, EPI_ISL_1017392, EPI_ISL_1017393, EPI_ISL_1017394, EPI_ISL_1017395, EPI_ISL_1017396, EPI_ISL_1017397, EPI_ISL_1017398, EPI_ISL_1017399, EPI_ISL_1017400, EPI_ISL_1017401, EPI_ISL_1017402, EPI_ISL_1017403, EPI_ISL_1017404, EPI_ISL_1017405, EPI_ISL_1017406, EPI_ISL_1017407, EPI_ISL_1017408, EPI_ISL_1017409, EPI_ISL_1017410, EPI_ISL_1017411, EPI_ISL_1017412, EPI_ISL_1017413, EPI_ISL_1017414, EPI_ISL_1017415, EPI_ISL_1017416, EPI_ISL_1017417, EPI_ISL_1017418, EPI_ISL_1017419, EPI_ISL_1017420, EPI_ISL_1017421, EPI_ISL_1017422, EPI_ISL_1017423, EPI_ISL_1017424, EPI_ISL_1017425, EPI_ISL_1017426, EPI_ISL_1017427, EPI_ISL_1017428, EPI_ISL_1017429, EPI_ISL_1017430, EPI_ISL_1017431, EPI_ISL_1017432, EPI_ISL_1017433, EPI_ISL_1017434, EPI_ISL_1017435, EPI_ISL_1017436, EPI_ISL_1017437, EPI_ISL_1017438, EPI_ISL_1017439, EPI_ISL_1017440, EPI_ISL_1017441, EPI_ISL_1017442, EPI_ISL_1017443, EPI_ISL_1017444, EPI_ISL_1017445, EPI_ISL_1017446, EPI_ISL_1017447, EPI_ISL_1017448, EPI_ISL_1017449, EPI_ISL_1017450, EPI_ISL_1017451, EPI_ISL_1017452, EPI_ISL_1017453, EPI_ISL_1017454, EPI_ISL_1017455, EPI_ISL_1017456, EPI_ISL_1017457, EPI_ISL_1017458, EPI_ISL_1017459, EPI_ISL_1017460, EPI_ISL_1017461, EPI_ISL_1017462, EPI_ISL_1017463, EPI_ISL_1017464, EPI_ISL_1017465, EPI_ISL_1017466, EPI_ISL_1017467, EPI_ISL_1017468, EPI_ISL_1017469, EPI_ISL_1017470 | | | |
| see above | Providence Oregon Regional Laboratories | Providence St. Joseph Health Molecular Genomics Laboratory | Alexa K Dowdell, Brian D Piening, Fred L Robinson, Carlo B Bifulco, Mary Campbell |
| EPI_ISL_1017471, EPI_ISL_1017472, EPI_ISL_1017473, EPI_ISL_1017474, EPI_ISL_1017475, EPI_ISL_1017476, EPI_ISL_1017477, EPI_ISL_1017478, EPI_ISL_1017479, EPI_ISL_1017480, EPI_ISL_1017481, EPI_ISL_1017482, EPI_ISL_1017483, EPI_ISL_1017484, EPI_ISL_1017485, EPI_ISL_1017486, EPI_ISL_1017487, EPI_ISL_1017488, EPI_ISL_1017489, EPI_ISL_1017490, EPI_ISL_1017491, EPI_ISL_1017492, EPI_ISL_1017493, EPI_ISL_1017494, EPI_ISL_1017495, EPI_ISL_1017496, EPI_ISL_1017497, EPI_ISL_1017498, EPI_ISL_1017499, EPI_ISL_1017500, EPI_ISL_1017501, EPI_ISL_1017502, EPI_ISL_1017503, EPI_ISL_1017504, EPI_ISL_1017505, EPI_ISL_1017506, EPI_ISL_1017507, EPI_ISL_1017508, EPI_ISL_1017509, EPI_ISL_1017510, EPI_ISL_1017511, EPI_ISL_1017512, EPI_ISL_1017513, EPI_ISL_1017514, EPI_ISL_1017515, EPI_ISL_1017516, EPI_ISL_1017517, EPI_ISL_1017518, EPI_ISL_1017519, EPI_ISL_1017520, EPI_ISL_1017521, EPI_ISL_1017522, EPI_ISL_1017523, EPI_ISL_1017524, EPI_ISL_1017525, EPI_ISL_1017526, EPI_ISL_1017527, EPI_ISL_1017528, EPI_ISL_1017529, EPI_ISL_1017530, EPI_ISL_1017531, EPI_ISL_1017532, EPI_ISL_1017533, EPI_ISL_1017534, EPI_ISL_1017535, EPI_ISL_1017536, EPI_ISL_1017537, EPI_ISL_1017538, EPI_ISL_1017539, EPI_ISL_1017540, EPI_ISL_1017541, EPI_ISL_1017542, EPI_ISL_1017543, EPI_ISL_1017544, EPI_ISL_1017545, EPI_ISL_1017546, EPI_ISL_1017547, EPI_ISL_1017548, EPI_ISL_1017549, EPI_ISL_1017550, EPI_ISL_1017551, EPI_ISL_1017552, EPI_ISL_1017553, EPI_ISL_1017554, EPI_ISL_1017555, EPI_ISL_1017556, EPI_ISL_1017557, EPI_ISL_1017558, EPI_ISL_1017559, EPI_ISL_1017560, EPI_ISL_1017561, EPI_ISL_1017562, EPI_ISL_1017563, EPI_ISL_1017564, EPI_ISL_1017565, EPI_ISL_1017566, EPI_ISL_1017567, EPI_ISL_1017568, EPI_ISL_1017569, EPI_ISL_1017570, EPI_ISL_1017571, EPI_ISL_1017572, EPI_ISL_1017573, EPI_ISL_1017574, EPI_ISL_1017575, EPI_ISL_1017576, EPI_ISL_1017577, EPI_ISL_1017578, EPI_ISL_1017579, EPI_ISL_1017580, EPI_ISL_1017581, EPI_ISL_1017582, EPI_ISL_1017583, EPI_ISL_1017584 | | | |
| see above | Murphy Medical Associates | Grubaugh Lab - Yale School of Public Health | Mary Petrone, Joseph Fauver, Caleb Neal, Steven Murphy, Chantal Vogels, Mallery Breban, Annie Watkins, Tara Alpert, Nathan Grubaugh |
| EPI_ISL_1017585, EPI_ISL_1017586, EPI_ISL_1017587, EPI_ISL_1017588, EPI_ISL_1017589, EPI_ISL_1017590, EPI_ISL_1017591, EPI_ISL_1017592, EPI_ISL_1017593, EPI_ISL_1017594, EPI_ISL_1017596, EPI_ISL_1017597, EPI_ISL_1017598, EPI_ISL_1017599, EPI_ISL_1017600, EPI_ISL_1017601, EPI_ISL_1017602, EPI_ISL_1017603, EPI_ISL_1017604, EPI_ISL_1017605, EPI_ISL_1017606, EPI_ISL_1017607, EPI_ISL_1017608, EPI_ISL_1017609, EPI_ISL_1017610, EPI_ISL_1017611, EPI_ISL_1017612, EPI_ISL_1017613, EPI_ISL_1017614, EPI_ISL_1017615, EPI_ISL_1017616, EPI_ISL_1017617, EPI_ISL_1017618, EPI_ISL_1017619, EPI_ISL_1017620, EPI_ISL_1017621, EPI_ISL_1017622, EPI_ISL_1017623, EPI_ISL_1017625, EPI_ISL_1017626, EPI_ISL_1017627, EPI_ISL_1017628, EPI_ISL_1017629, EPI_ISL_1017630, EPI_ISL_1017631, EPI_ISL_1017632, EPI_ISL_1017633, EPI_ISL_1017634, EPI_ISL_1017635, EPI_ISL_1017636, EPI_ISL_1017637, EPI_ISL_1017638, EPI_ISL_1017639, EPI_ISL_1017640, EPI_ISL_1017641, EPI_ISL_1017642, EPI_ISL_1017643, EPI_ISL_1017644, EPI_ISL_1017645, EPI_ISL_1017646, EPI_ISL_1017647, EPI_ISL_1017648, EPI_ISL_1017649, EPI_ISL_1017650, EPI_ISL_1017651, EPI_ISL_1017652, EPI_ISL_1017653, EPI_ISL_1017654, EPI_ISL_1017655, EPI_ISL_1017656, EPI_ISL_1017657, EPI_ISL_1017658, EPI_ISL_1017659, EPI_ISL_1017660, EPI_ISL_1017661, EPI_ISL_1017662, EPI_ISL_1017663, EPI_ISL_1017664, EPI_ISL_1017665, EPI_ISL_1017666, EPI_ISL_1017667, EPI_ISL_1017668, EPI_ISL_1017669, EPI_ISL_1017670, EPI_ISL_1017671, EPI_ISL_1017672, EPI_ISL_1017673, EPI_ISL_1017674, EPI_ISL_1017675, EPI_ISL_1017676, EPI_ISL_1017677 | | | |
| see above | Connecticut Veterans' Affairs Hospital | Grubaugh Lab - Yale School of Public Health | Mary Petrone, Joseph Fauver, Tara Alpert, Chantal Vogels, Isabel Ott, Shaili Gupta, Danielle Plank, Nathan Grubaugh |
| EPI_ISL_1017713, EPI_ISL_1017718, EPI_ISL_1017719, EPI_ISL_1017721, EPI_ISL_1017722, EPI_ISL_1017723, EPI_ISL_1017724, EPI_ISL_1017725, EPI_ISL_1017727, EPI_ISL_1017729, EPI_ISL_1017730, EPI_ISL_1017731, EPI_ISL_1017732, EPI_ISL_1017733, EPI_ISL_1017735, EPI_ISL_1017737, EPI_ISL_1017738, EPI_ISL_1017739, EPI_ISL_1017743, EPI_ISL_1017744, EPI_ISL_1017745, EPI_ISL_1017746, EPI_ISL_1017747, EPI_ISL_1017748, EPI_ISL_1017750, EPI_ISL_1017751, EPI_ISL_1017753, EPI_ISL_1017754, EPI_ISL_1017755, EPI_ISL_1017758, EPI_ISL_1017759, EPI_ISL_1017760, EPI_ISL_1017761, EPI_ISL_1017762, EPI_ISL_1017763, EPI_ISL_1017764, EPI_ISL_1017765, EPI_ISL_1017766, EPI_ISL_1017767, EPI_ISL_1017768, EPI_ISL_1017769, EPI_ISL_1017771, EPI_ISL_1017772, EPI_ISL_1017773, EPI_ISL_1017774, EPI_ISL_1017775, EPI_ISL_1017777, EPI_ISL_1017778, EPI_ISL_1017779, EPI_ISL_1017780, EPI_ISL_1017781, EPI_ISL_1017782, EPI_ISL_1017784, EPI_ISL_1017785, EPI_ISL_1017786, EPI_ISL_1017787, EPI_ISL_1017788, EPI_ISL_1017789, EPI_ISL_1017790, EPI_ISL_1017791, EPI_ISL_1017792, EPI_ISL_1017793, EPI_ISL_1017794, EPI_ISL_1017795, EPI_ISL_1017797, EPI_ISL_1017798, EPI_ISL_1017799, EPI_ISL_1017800, EPI_ISL_1017801, EPI_ISL_1017802, EPI_ISL_1017803, EPI_ISL_1017804, EPI_ISL_1017805, EPI_ISL_1017806, EPI_ISL_1017807, EPI_ISL_1017810, EPI_ISL_1017811, EPI_ISL_1017812, EPI_ISL_1017813, EPI_ISL_1017814, EPI_ISL_1017815, EPI_ISL_1017816, EPI_ISL_1017817, EPI_ISL_1017818, EPI_ISL_1017819, EPI_ISL_1017820, EPI_ISL_1017824, EPI_ISL_1017825, EPI_ISL_1017826, EPI_ISL_1017827, EPI_ISL_1017828, EPI_ISL_1017829, EPI_ISL_1017830, EPI_ISL_1017831, EPI_ISL_1017832, EPI_ISL_1017833, EPI_ISL_1017834 | | | |
| see above | University of Wisconsin-Madison AIDS Vaccine Research Laboratories | University of Wisconsin-Madison AIDS Vaccine Research Laboratories | Gage Moreno, Katarina Braun, et al. AIDS Vaccine Research Laboratories |
| EPI_ISL_1017835, EPI_ISL_1017836, EPI_ISL_1017837, EPI_ISL_1017838, EPI_ISL_1017839, EPI_ISL_1017840, EPI_ISL_1017841, EPI_ISL_1017842, EPI_ISL_1017843, EPI_ISL_1017844, EPI_ISL_1017845, EPI_ISL_1017846, EPI_ISL_1017847, EPI_ISL_1017848, EPI_ISL_1017849, EPI_ISL_1017850, EPI_ISL_1017851, EPI_ISL_1017852, EPI_ISL_1017853, EPI_ISL_1017854, EPI_ISL_1017855, EPI_ISL_1017856, EPI_ISL_1017857, EPI_ISL_1017858, EPI_ISL_1017859, EPI_ISL_1017860, EPI_ISL_1017861, EPI_ISL_1017862, EPI_ISL_1017863, EPI_ISL_1017864, EPI_ISL_1017865, EPI_ISL_1017867, EPI_ISL_1017868, EPI_ISL_1017869, EPI_ISL_1017871, EPI_ISL_1017872, EPI_ISL_1017873, EPI_ISL_1017874, EPI_ISL_1017875, EPI_ISL_1017876, EPI_ISL_1017877, EPI_ISL_1017878, EPI_ISL_1017879, EPI_ISL_1017880, EPI_ISL_1017881, EPI_ISL_1017882, EPI_ISL_1017883, EPI_ISL_1017884, EPI_ISL_1017885, EPI_ISL_1017886, EPI_ISL_1017887, EPI_ISL_1017888, EPI_ISL_1017889, EPI_ISL_1017890, EPI_ISL_1017891, EPI_ISL_1017892, EPI_ISL_1017893, EPI_ISL_1017894, EPI_ISL_1017895, EPI_ISL_1017896, EPI_ISL_1017897, EPI_ISL_1017898, EPI_ISL_1017899, EPI_ISL_1017900, EPI_ISL_1017901, EPI_ISL_1017902, EPI_ISL_1017903, EPI_ISL_1017904, EPI_ISL_1017905, EPI_ISL_1017906, EPI_ISL_1017907, EPI_ISL_1017908, EPI_ISL_1017909, EPI_ISL_1017910, EPI_ISL_1017911, EPI_ISL_1017912, EPI_ISL_1017913, EPI_ISL_1017914, EPI_ISL_1017915, EPI_ISL_1017916, EPI_ISL_1017917, EPI_ISL_1017918, EPI_ISL_1017919, EPI_ISL_1017920, EPI_ISL_1017921, EPI_ISL_1017922, EPI_ISL_1017923, EPI_ISL_1017924, EPI_ISL_1017925, EPI_ISL_1017926, EPI_ISL_1017927, EPI_ISL_1017928, EPI_ISL_1017929, EPI_ISL_1017930, EPI_ISL_1017931, EPI_ISL_1017932, EPI_ISL_1017933, EPI_ISL_1017934, EPI_ISL_1017936, EPI_ISL_1017937, EPI_ISL_1017938, EPI_ISL_1017939, EPI_ISL_1017940, EPI_ISL_1017941, EPI_ISL_1017942, EPI_ISL_1017944, EPI_ISL_1017945, EPI_ISL_1017946, EPI_ISL_1017947, EPI_ISL_1017948, EPI_ISL_1017949, EPI_ISL_1017950, EPI_ISL_1017952, EPI_ISL_1017953, EPI_ISL_1017954, EPI_ISL_1017955, EPI_ISL_1017956, EPI_ISL_1017957, EPI_ISL_1017958, EPI_ISL_1017959, EPI_ISL_1017960, EPI_ISL_1017962, EPI_ISL_1017963, EPI_ISL_1017964, EPI_ISL_1017965, EPI_ISL_1017966, EPI_ISL_1017967, EPI_ISL_1017968 | | | |
| see above | Wyoming Public Health Laboratory | Wyoming Public Health Laboratory | Noah Hull, Taylor Fearing, Lynette Gumbleton, Channing Weber, Ashley Norberg, Bailey Bowcutt, and Wanda Manley |
| EPI_ISL_1017969, EPI_ISL_1017970 | DOHMH PHL | New York City Public Health Laboratory | Jade Wang, et al. |
| EPI_ISL_1017971, EPI_ISL_1017972, EPI_ISL_1017973 | DOHMH Riverside | New York City Public Health Laboratory | Jade Wang, et al. |
| EPI_ISL_1017974 | DOHMH Morrisania | New York City Public Health Laboratory | Jade Wang, et al. |
| EPI_ISL_1017975 | DOHMH Corona | New York City Public Health Laboratory | Jade Wang, et al. |
| EPI_ISL_1017976, EPI_ISL_1017977 | DOHMH Jamaica | New York City Public Health Laboratory | Jade Wang, et al. |
| EPI_ISL_1017978 | DOHMH Chelsea | New York City Public Health Laboratory | Jade Wang, et al. |
| EPI_ISL_1017979 | DOHMH PHL | New York City Public Health Laboratory | Jade Wang, et al. |
| EPI_ISL_1017980, EPI_ISL_1017981 | DOHMH Morrisania | New York City Public Health Laboratory | Jade Wang, et al. |
| EPI_ISL_1017982, EPI_ISL_1017983, EPI_ISL_1017984, EPI_ISL_1017985 | DOHMH Central Harlem | New York City Public Health Laboratory | Jade Wang, et al. |
| EPI_ISL_1017986, EPI_ISL_1017987, | DOHMH Corona | New York City Public Health Laboratory | Jade Wang, et al. |

| | | | |
|---|---|---|---|
| EPI_ISL_1017988 | | | |
| EPI_ISL_1017989, EPI_ISL_1017990 | Department of Homeless Services | New York City Public Health Laboratory | Jade Wang, et al. |
| EPI_ISL_1017991 | DOHMH PHL | New York City Public Health Laboratory | Jade Wang, et al. |
| EPI_ISL_1017992, EPI_ISL_1017993, EPI_ISL_1017994 | DOHMH Jamaica | New York City Public Health Laboratory | Jade Wang, et al. |
| EPI_ISL_1017995, EPI_ISL_1017996, EPI_ISL_1017997, EPI_ISL_1017998, EPI_ISL_1017999 | DOHMH Corona | New York City Public Health Laboratory | Jade Wang, et al. |
| EPI_ISL_1018000 | DOHMH Morrisania | New York City Public Health Laboratory | Jade Wang, et al. |
| EPI_ISL_1018001 | DOHMH Central Harlem | New York City Public Health Laboratory | Jade Wang, et al. |
| EPI_ISL_1018002, EPI_ISL_1018003, EPI_ISL_1018004 | DOHMH PHL | New York City Public Health Laboratory | Jade Wang, et al. |
| EPI_ISL_1018005 | DOHMH Corona | New York City Public Health Laboratory | Jade Wang, et al. |
| EPI_ISL_1018006, EPI_ISL_1018007, EPI_ISL_1018008, EPI_ISL_1018009, EPI_ISL_1018010 | DOHMH Morrisania | New York City Public Health Laboratory | Jade Wang, et al. |
| EPI_ISL_1018011 | DOHMH Chelsea | New York City Public Health Laboratory | Jade Wang, et al. |
| EPI_ISL_1018012, EPI_ISL_1018013, EPI_ISL_1018014, EPI_ISL_1018015, EPI_ISL_1018016, EPI_ISL_1018017, EPI_ISL_1018018, EPI_ISL_1018019, EPI_ISL_1018020 | DOHMH Jamaica | New York City Public Health Laboratory | Jade Wang, et al. |
| EPI_ISL_1018021 | DOHMH Crown Heights | New York City Public Health Laboratory | Jade Wang, et al. |
| EPI_ISL_1018022, EPI_ISL_1018023 | DOHMH Jamaica | New York City Public Health Laboratory | Jade Wang, et al. |
| EPI_ISL_1018024 | DOHMH Crown Heights | New York City Public Health Laboratory | Jade Wang, et al. |
| EPI_ISL_1018025, EPI_ISL_1018026 | DOHMH Morrisania | New York City Public Health Laboratory | Jade Wang, et al. |
| EPI_ISL_1018027, EPI_ISL_1018028, EPI_ISL_1018029, EPI_ISL_1018030, EPI_ISL_1018031 | DOHMH Corona | New York City Public Health Laboratory | Jade Wang, et al. |
| EPI_ISL_1018032, EPI_ISL_1018033, EPI_ISL_1018034, EPI_ISL_1018035, EPI_ISL_1018036, EPI_ISL_1018037, EPI_ISL_1018038, EPI_ISL_1018039, EPI_ISL_1018040, EPI_ISL_1018041, EPI_ISL_1018042, EPI_ISL_1018043, EPI_ISL_1018044, EPI_ISL_1018045, EPI_ISL_1018046, EPI_ISL_1018047, EPI_ISL_1018048, EPI_ISL_1018049 | | | |
| see above | OCME Office Of Chief Medical Examiner | New York City Public Health Laboratory | Jade Wang, et al. |
| EPI_ISL_1018050, EPI_ISL_1018051, EPI_ISL_1018052, EPI_ISL_1018053, EPI_ISL_1018054, EPI_ISL_1018055, EPI_ISL_1018056, EPI_ISL_1018057, EPI_ISL_1018058, EPI_ISL_1018059 | University Hospitals Translational Laboratory (UHTL), University Hospitals | University Hospitals Translational Laboratory (UHTL), University Hospitals | Sadri,N., Alouani,D. and Song,X. |
| EPI_ISL_1018061, EPI_ISL_1018062 | Los Angeles County Public Health Laboratories | Los Angeles County Public Health Laboratories | Hemarajata,P., Green,N. |
| EPI_ISL_1020332, EPI_ISL_1020338, EPI_ISL_1020340, EPI_ISL_1020341, EPI_ISL_1020409, EPI_ISL_1020410, EPI_ISL_1020486 | Ohio Department of Health Laboratory | Ohio Department of Health Laboratory | Holmes, Jennifer; Eric Brandt, Keoni Omura, Glen McGillivary, Caitlin McDonnell, Jade Mowery, Stephanie Mccracken, Tyler Payne, Kirtana Ramadugu, Erica Leasure, Brent Lee, Kelsey Florek, Heather Blankenship, Allison Black, Quanta Brown, and Tammy Bannerman |
| EPI_ISL_1020487, EPI_ISL_1020488, EPI_ISL_1020489, EPI_ISL_1020490, EPI_ISL_1020491, EPI_ISL_1020492, EPI_ISL_1020493, EPI_ISL_1020494, EPI_ISL_1020495, EPI_ISL_1020496, EPI_ISL_1020497, EPI_ISL_1020498, EPI_ISL_1020499, EPI_ISL_1020500, EPI_ISL_1020501, EPI_ISL_1020502, EPI_ISL_1020503, EPI_ISL_1020504, EPI_ISL_1020505, EPI_ISL_1020506, EPI_ISL_1020507, EPI_ISL_1020508, EPI_ISL_1020509, EPI_ISL_1020510, EPI_ISL_1020511, EPI_ISL_1020512, EPI_ISL_1020513, EPI_ISL_1020514, EPI_ISL_1020515, EPI_ISL_1020516, EPI_ISL_1020517, EPI_ISL_1020518, EPI_ISL_1020519, EPI_ISL_1020520, EPI_ISL_1020521, EPI_ISL_1020522, EPI_ISL_1020523, EPI_ISL_1020524, EPI_ISL_1020525, EPI_ISL_1020526, EPI_ISL_1020527, EPI_ISL_1020528, EPI_ISL_1020529, EPI_ISL_1020530, EPI_ISL_1020531, EPI_ISL_1020532, EPI_ISL_1020533, EPI_ISL_1020534, EPI_ISL_1020535, EPI_ISL_1020536, EPI_ISL_1020537, EPI_ISL_1020538, EPI_ISL_1020539, EPI_ISL_1020540, EPI_ISL_1020541, EPI_ISL_1020542, EPI_ISL_1020543, EPI_ISL_1020544, EPI_ISL_1020545, EPI_ISL_1020546, EPI_ISL_1020547, EPI_ISL_1020548, EPI_ISL_1020549, EPI_ISL_1020550 | | | |
| see above | Columbia University Irving Medical Center | Wadsworth Center, New York State Department of Health | Kirsten St. George, Daryl M. Lamson, Alexis Russel, Matthew Shudt, Melissa A Leisner, Jonathan Plitnick, Navjot Singh, John Kelly, Erasmus Schneider, Erica Lasek-Nesselquist |
| EPI_ISL_1020592, EPI_ISL_1020593, EPI_ISL_1020594, EPI_ISL_1020595, EPI_ISL_1020596, EPI_ISL_1020597, EPI_ISL_1020598, EPI_ISL_1020599, EPI_ISL_1020600, EPI_ISL_1020601, EPI_ISL_1020602, EPI_ISL_1020603, EPI_ISL_1020604, EPI_ISL_1020605, EPI_ISL_1020606, EPI_ISL_1020722, EPI_ISL_1020723, EPI_ISL_1020724, EPI_ISL_1020725, EPI_ISL_1020726, EPI_ISL_1020727, EPI_ISL_1020728, EPI_ISL_1020729, EPI_ISL_1020730, EPI_ISL_1020731, EPI_ISL_1020732, EPI_ISL_1020733, EPI_ISL_1020734, EPI_ISL_1020735, EPI_ISL_1020736, EPI_ISL_1020737, EPI_ISL_1020738, EPI_ISL_1020739, EPI_ISL_1020740, EPI_ISL_1020741, EPI_ISL_1020742, EPI_ISL_1020743, EPI_ISL_1020744, EPI_ISL_1020745, EPI_ISL_1020746, EPI_ISL_1020747, EPI_ISL_1020748, EPI_ISL_1020749, EPI_ISL_1020750, EPI_ISL_1020751, EPI_ISL_1020752, EPI_ISL_1020753, EPI_ISL_1020754, EPI_ISL_1020755, EPI_ISL_1020756, EPI_ISL_1020757, EPI_ISL_1020758, EPI_ISL_1020759, EPI_ISL_1020760, EPI_ISL_1020761, EPI_ISL_1020762, EPI_ISL_1020763, EPI_ISL_1020764, EPI_ISL_1020765, EPI_ISL_1020766, EPI_ISL_1020767, EPI_ISL_1020768, EPI_ISL_1020769, EPI_ISL_1020770, EPI_ISL_1020771, EPI_ISL_1020772, EPI_ISL_1020773, EPI_ISL_1020774, EPI_ISL_1020775, EPI_ISL_1020776, EPI_ISL_1020777, EPI_ISL_1020778, EPI_ISL_1020779, EPI_ISL_1020780, EPI_ISL_1020781, EPI_ISL_1020782, EPI_ISL_1020783, EPI_ISL_1020784, EPI_ISL_1020785, EPI_ISL_1020786, EPI_ISL_1020787, EPI_ISL_1020788, EPI_ISL_1020789, EPI_ISL_1020790, EPI_ISL_1020791, EPI_ISL_1020792, EPI_ISL_1020793, EPI_ISL_1020794, EPI_ISL_1020795, EPI_ISL_1020796, EPI_ISL_1020797, EPI_ISL_1020798, EPI_ISL_1020799, EPI_ISL_1020800, EPI_ISL_1020801, EPI_ISL_1020802, EPI_ISL_1020803, EPI_ISL_1020804, EPI_ISL_1020805, EPI_ISL_1020806, EPI_ISL_1020807, EPI_ISL_1020808, EPI_ISL_1020809, EPI_ISL_1020810, EPI_ISL_1020811, EPI_ISL_1020812, EPI_ISL_1020813, EPI_ISL_1020814, EPI_ISL_1020815, EPI_ISL_1020816, EPI_ISL_1020817, EPI_ISL_1020818, EPI_ISL_1020819, EPI_ISL_1020820, EPI_ISL_1020821, EPI_ISL_1020822, EPI_ISL_1020823, EPI_ISL_1020824, EPI_ISL_1020825, EPI_ISL_1020826, EPI_ISL_1020827, EPI_ISL_1020828, EPI_ISL_1020829, EPI_ISL_1020830, EPI_ISL_1020831, EPI_ISL_1020832, EPI_ISL_1020833, EPI_ISL_1020834, EPI_ISL_1021199, EPI_ISL_1021201, EPI_ISL_1021202, EPI_ISL_1021203, EPI_ISL_1021204, EPI_ISL_1021205, EPI_ISL_1021206, EPI_ISL_1021207, EPI_ISL_1021208, EPI_ISL_1021209, EPI_ISL_1021210, EPI_ISL_1021211, EPI_ISL_1021212, EPI_ISL_1021213, EPI_ISL_1021214, EPI_ISL_1021215, EPI_ISL_1021216, EPI_ISL_1021217, EPI_ISL_1021218, EPI_ISL_1021219, EPI_ISL_1021220, EPI_ISL_1021221, EPI_ISL_1021222, EPI_ISL_1021223, EPI_ISL_1021224, EPI_ISL_1021225, EPI_ISL_1021226, EPI_ISL_1021227, EPI_ISL_1021228, EPI_ISL_1021229, EPI_ISL_1021230, EPI_ISL_1021231, EPI_ISL_1021232, EPI_ISL_1021233, EPI_ISL_1021234, EPI_ISL_1021235, EPI_ISL_1021236, EPI_ISL_1021237, EPI_ISL_1021238, EPI_ISL_1021239, EPI_ISL_1021240, EPI_ISL_1021241, EPI_ISL_1021242, EPI_ISL_1021243, EPI_ISL_1021244, EPI_ISL_1021245, EPI_ISL_1021246, EPI_ISL_1021247, EPI_ISL_1021248, EPI_ISL_1021249, EPI_ISL_1021250, EPI_ISL_1021251, EPI_ISL_1021252, EPI_ISL_1021253, EPI_ISL_1021254, EPI_ISL_1021255, EPI_ISL_1021256, EPI_ISL_1021257, EPI_ISL_1021258, EPI_ISL_1021259, EPI_ISL_1021260, EPI_ISL_1021261, EPI_ISL_1021262, EPI_ISL_1021263, EPI_ISL_1021264, EPI_ISL_1021265, EPI_ISL_1021266, EPI_ISL_1021267, EPI_ISL_1021268, EPI_ISL_1021269, EPI_ISL_1021270, EPI_ISL_1021271, EPI_ISL_1021272, EPI_ISL_1021273, EPI_ISL_1021274, EPI_ISL_1021275, EPI_ISL_1021276, EPI_ISL_1021277, EPI_ISL_1021278, EPI_ISL_1021279, EPI_ISL_1021280, EPI_ISL_1021281, EPI_ISL_1021282, EPI_ISL_1021283, EPI_ISL_1021284, EPI_ISL_1021285, EPI_ISL_1021286, EPI_ISL_1021287, EPI_ISL_1021288, EPI_ISL_1021289, EPI_ISL_1021290, EPI_ISL_1021291, EPI_ISL_1021292, EPI_ISL_1021293, EPI_ISL_1021294, EPI_ISL_1021295, EPI_ISL_1021296, EPI_ISL_1021297, EPI_ISL_1021298, EPI_ISL_1021299, EPI_ISL_1021300, EPI_ISL_1021301, EPI_ISL_1021302, EPI_ISL_1021303, EPI_ISL_1021304, EPI_ISL_1021305, EPI_ISL_1021306, EPI_ISL_1021307, EPI_ISL_1021308, EPI_ISL_1021309, EPI_ISL_1021310, EPI_ISL_1021311, EPI_ISL_1021312, EPI_ISL_1021313, EPI_ISL_1021314, EPI_ISL_1021315, EPI_ISL_1021316, EPI_ISL_1021317, EPI_ISL_1021318, EPI_ISL_1021319, EPI_ISL_1021320, EPI_ISL_1021321, EPI_ISL_1021322, EPI_ISL_1021323, EPI_ISL_1021324, EPI_ISL_1021325, EPI_ISL_1021326, EPI_ISL_1021327, EPI_ISL_1021328, EPI_ISL_1021329, EPI_ISL_1021330, EPI_ISL_1021331, EPI_ISL_1021332, EPI_ISL_1021333, EPI_ISL_1021334, EPI_ISL_1021335, EPI_ISL_1021336, EPI_ISL_1021337, EPI_ISL_1021338, EPI_ISL_1021339, EPI_ISL_1021340, EPI_ISL_1021341, EPI_ISL_1021342, EPI_ISL_1021343, EPI_ISL_1021345, EPI_ISL_1021346, EPI_ISL_1021347, EPI_ISL_1021348, EPI_ISL_1021349, EPI_ISL_1021350, EPI_ISL_1021351, EPI_ISL_1021352, EPI_ISL_1021353, EPI_ISL_1021354, EPI_ISL_1021355, EPI_ISL_1021356, EPI_ISL_1021357, EPI_ISL_1021358, EPI_ISL_1021359, EPI_ISL_1021360, EPI_ISL_1021361, EPI_ISL_1021362, EPI_ISL_1021363, EPI_ISL_1021364, EPI_ISL_1021365, EPI_ISL_1021366, EPI_ISL_1021367, EPI_ISL_1021368, EPI_ISL_1021369, EPI_ISL_1021370, EPI_ISL_1021371, EPI_ISL_1021372, EPI_ISL_1021373, EPI_ISL_1021374, EPI_ISL_1021375, EPI_ISL_1021376, EPI_ISL_1021377, EPI_ISL_1021378, EPI_ISL_1021379, EPI_ISL_1021380, EPI_ISL_1021381, EPI_ISL_1021382, EPI_ISL_1021383, EPI_ISL_1021384, EPI_ISL_1021385, EPI_ISL_1021675, EPI_ISL_1021676, EPI_ISL_1021677, EPI_ISL_1021678, EPI_ISL_1021679, EPI_ISL_1021680, EPI_ISL_1021681, EPI_ISL_1021682, EPI_ISL_1021683, EPI_ISL_1021684, EPI_ISL_1021685, EPI_ISL_1021686, EPI_ISL_1021687, EPI_ISL_1021688, EPI_ISL_1021689, EPI_ISL_1021690, EPI_ISL_1021691, EPI_ISL_1021692, EPI_ISL_1021693, EPI_ISL_1021694, EPI_ISL_1021695, EPI_ISL_1021696, EPI_ISL_1021697, EPI_ISL_1021698, EPI_ISL_1021699, EPI_ISL_1021700, EPI_ISL_1021701, EPI_ISL_1021702, EPI_ISL_1021703, EPI_ISL_1021704, EPI_ISL_1021705, EPI_ISL_1021706, EPI_ISL_1021707, EPI_ISL_1021708, EPI_ISL_1021709, EPI_ISL_1021710, EPI_ISL_1021711, EPI_ISL_1021712, EPI_ISL_1021713, EPI_ISL_1021714, EPI_ISL_1021715, EPI_ISL_1021716, EPI_ISL_1021717, EPI_ISL_1021718, EPI_ISL_1021719, EPI_ISL_1021720, EPI_ISL_1021721, EPI_ISL_1021722, EPI_ISL_1021723, EPI_ISL_1021724, EPI_ISL_1021725, EPI_ISL_1021726, EPI_ISL_1021727, EPI_ISL_1021728, EPI_ISL_1021729, EPI_ISL_1021730, EPI_ISL_1021731, EPI_ISL_1021732, EPI_ISL_1021733, EPI_ISL_1021734, EPI_ISL_1021735, EPI_ISL_1021736, EPI_ISL_1021737, EPI_ISL_1021738, EPI_ISL_1021739, EPI_ISL_1021740, EPI_ISL_1021741, EPI_ISL_1021742, EPI_ISL_1021743, EPI_ISL_1021744, EPI_ISL_1021745, EPI_ISL_1021746, EPI_ISL_1021747, EPI_ISL_1021748, EPI_ISL_1021749, EPI_ISL_1021750, EPI_ISL_1021751, EPI_ISL_1021752, EPI_ISL_1021753, EPI_ISL_1021754, EPI_ISL_1021755, EPI_ISL_1021756, EPI_ISL_1021757, EPI_ISL_1021758, EPI_ISL_1021759, EPI_ISL_1021760, EPI_ISL_1021761, EPI_ISL_1021762, EPI_ISL_1021763, EPI_ISL_1021764, EPI_ISL_1021765, EPI_ISL_1021766, EPI_ISL_1021767, EPI_ISL_1021768, EPI_ISL_1021769, EPI_ISL_1021770, EPI_ISL_1021771, EPI_ISL_1021772, EPI_ISL_1021773, EPI_ISL_1021774, EPI_ISL_1021775, EPI_ISL_1021776, EPI_ISL_1021777, EPI_ISL_1021778, EPI_ISL_1021779, EPI_ISL_1021780, EPI_ISL_1021781, EPI_ISL_1021782, EPI_ISL_1021783, EPI_ISL_1021784, EPI_ISL_1021785, EPI_ISL_1021786, EPI_ISL_1021787, EPI_ISL_1021788, EPI_ISL_1021789, EPI_ISL_1021790, EPI_ISL_1021791, EPI_ISL_1021792, EPI_ISL_1021793, EPI_ISL_1021794, EPI_ISL_1021795, EPI_ISL_1021796, EPI_ISL_1021797, EPI_ISL_1021798, EPI_ISL_1021799, EPI_ISL_1021800, EPI_ISL_1021801, EPI_ISL_1021802, EPI_ISL_1021803, EPI_ISL_1021804, EPI_ISL_1021805, EPI_ISL_1021806, EPI_ISL_1021807, EPI_ISL_1021808, EPI_ISL_1021809, EPI_ISL_1021810, EPI_ISL_1021811, EPI_ISL_1021812, EPI_ISL_1021813, EPI_ISL_1021814, EPI_ISL_1021815, EPI_ISL_1021816, EPI_ISL_1021817, EPI_ISL_1021818, EPI_ISL_1021819, EPI_ISL_1021820, EPI_ISL_1021821, EPI_ISL_1021822, EPI_ISL_1021823, EPI_ISL_1021824, EPI_ISL_1021825, EPI_ISL_1021826, EPI_ISL_1021827, EPI_ISL_1021828, EPI_ISL_1021930, EPI_ISL_1021931, EPI_ISL_1021932, EPI_ISL_1021933, EPI_ISL_1021934, EPI_ISL_1021935, EPI_ISL_1021936, EPI_ISL_1021937, EPI_ISL_1021938, EPI_ISL_1021939, EPI_ISL_1021940, EPI_ISL_1021941, EPI_ISL_1021942, EPI_ISL_1021943, EPI_ISL_1021944, EPI_ISL_1021945, EPI_ISL_1021946, EPI_ISL_1021947, EPI_ISL_1021948, EPI_ISL_1021949, EPI_ISL_1021950, EPI_ISL_1021951, EPI_ISL_1021952, EPI_ISL_1021953, EPI_ISL_1021954, EPI_ISL_1021955, EPI_ISL_1021956, EPI_ISL_1021957, EPI_ISL_1021958, EPI_ISL_1021959, EPI_ISL_1021960, EPI_ISL_1021961, EPI_ISL_1021962, EPI_ISL_1021963, EPI_ISL_1021964, EPI_ISL_1021965, EPI_ISL_1021966, EPI_ISL_1021967, EPI_ISL_1021968, EPI_ISL_1021969, EPI_ISL_1021970, EPI_ISL_1021971, EPI_ISL_1021972, EPI_ISL_1021973, EPI_ISL_1021974, EPI_ISL_1021975, EPI_ISL_1021976, EPI_ISL_1021977, EPI_ISL_1021978, EPI_ISL_1021979, EPI_ISL_1021980, EPI_ISL_1021981, EPI_ISL_1021982, EPI_ISL_1021983, EPI_ISL_1021984, EPI_ISL_1021985, EPI_ISL_1021986, EPI_ISL_1021987, EPI_ISL_1021988, EPI_ISL_1021989, EPI_ISL_1022120, EPI_ISL_1022121, EPI_ISL_1022122, EPI_ISL_1022123, EPI_ISL_1022124, EPI_ISL_1022125, EPI_ISL_1022126, EPI_ISL_1022127, EPI_ISL_1022128, EPI_ISL_1022129, EPI_ISL_1022130, EPI_ISL_1022131, EPI_ISL_1022132, EPI_ISL_1022133, EPI_ISL_1022134, EPI_ISL_1022135, EPI_ISL_1022136, EPI_ISL_1022137, EPI_ISL_1022138, EPI_ISL_1022139, EPI_ISL_1022140, EPI_ISL_1022141, EPI_ISL_1022142, EPI_ISL_1022143, EPI_ISL_1022144, EPI_ISL_1022145, EPI_ISL_1022146, EPI_ISL_1022147, EPI_ISL_1022148, EPI_ISL_1022149, EPI_ISL_1022150, EPI_ISL_1022151, EPI_ISL_1022152, EPI_ISL_1022153, EPI_ISL_1022154, EPI_ISL_1022155, EPI_ISL_1022156, EPI_ISL_1022157, EPI_ISL_1022158, EPI_ISL_1022159, EPI_ISL_1022160, EPI_ISL_1022161, EPI_ISL_1022162, EPI_ISL_1022163, EPI_ISL_1022164, EPI_ISL_1022165, EPI_ISL_1022166, EPI_ISL_1022167, EPI_ISL_1022168, EPI_ISL_1022169, EPI_ISL_1022170, EPI_ISL_1022171, EPI_ISL_1022172, EPI_ISL_1022173, EPI_ISL_1022174, EPI_ISL_1022175, EPI_ISL_1022176, EPI_ISL_1022177, EPI_ISL_1022178, EPI_ISL_1022179, EPI_ISL_1022180, EPI_ISL_1022181, EPI_ISL_1022182, EPI_ISL_1022183, EPI_ISL_1022184, EPI_ISL_1022185, EPI_ISL_1022186, EPI_ISL_1022187, EPI_ISL_1022188, EPI_ISL_1022189, EPI_ISL_1022190, EPI_ISL_1022191, EPI_ISL_1022192, EPI_ISL_1022193, EPI_ISL_1022194, EPI_ISL_1022195, EPI_ISL_1022196, EPI_ISL_1022197, EPI_ISL_1022198, EPI_ISL_1022199, EPI_ISL_1022200, EPI_ISL_1022201, EPI_ISL_1022202, EPI_ISL_1022203, EPI_ISL_1022204, EPI_ISL_1022205, EPI_ISL_1022206, EPI_ISL_1022207, EPI_ISL_1022208, EPI_ISL_1022209, EPI_ISL_1022337, EPI_ISL_1022338, EPI_ISL_1022339, EPI_ISL_1022340, EPI_ISL_1022342, EPI_ISL_1022343, EPI_ISL_1022344, EPI_ISL_1022345, EPI_ISL_1022346, EPI_ISL_1022347, EPI_ISL_1022348, EPI_ISL_1022349, EPI_ISL_1022350, EPI_ISL_1022351, EPI_ISL_1022352, EPI_ISL_1022353, EPI_ISL_1022354, EPI_ISL_1022355, EPI_ISL_1022356, EPI_ISL_1022357, EPI_ISL_1022358, | | | |

| | | | |
|---|---|---|---|
| EPI_ISL_1022359, EPI_ISL_1022360, EPI_ISL_1022361, EPI_ISL_1022362, EPI_ISL_1022363, EPI_ISL_1022364, EPI_ISL_1022365, EPI_ISL_1022366, EPI_ISL_1022367, EPI_ISL_1022368, EPI_ISL_1022369, EPI_ISL_1022370, EPI_ISL_1022371, EPI_ISL_1022372, EPI_ISL_1022373, EPI_ISL_1022374, EPI_ISL_1022375, EPI_ISL_1022376, EPI_ISL_1022377, EPI_ISL_1022378, EPI_ISL_1022379, EPI_ISL_1022380, EPI_ISL_1022381, EPI_ISL_1022382, EPI_ISL_1022383, EPI_ISL_1022384, EPI_ISL_1022385, EPI_ISL_1022386, EPI_ISL_1022387, EPI_ISL_1022388, EPI_ISL_1022389, EPI_ISL_1022390, EPI_ISL_1022391, EPI_ISL_1022392, EPI_ISL_1022393, EPI_ISL_1022394, EPI_ISL_1022395, EPI_ISL_1022396, EPI_ISL_1022397, EPI_ISL_1022412, EPI_ISL_1022413, EPI_ISL_1022414, EPI_ISL_1022415, EPI_ISL_1022416, EPI_ISL_1022417 | | | |
| see above | Laboratory Corporation of America | Respiratory Viruses Branch, Division of Viral Diseases, Centers for Disease Control and Prevention | Peter W. Cook, Dakota Howard, Dhwani Batra, Ben L. Rambo-Martin, Clinton R. Paden, Suxiang Tong, Duncan MacCannell |
| EPI_ISL_1023582, EPI_ISL_1023584, EPI_ISL_1023588, EPI_ISL_1023590, EPI_ISL_1023592, EPI_ISL_1023594, EPI_ISL_1023595, EPI_ISL_1023597, EPI_ISL_1023599, EPI_ISL_1023601, EPI_ISL_1023603, EPI_ISL_1023605, EPI_ISL_1023606, EPI_ISL_1023608, EPI_ISL_1023610, EPI_ISL_1023612, EPI_ISL_1023614, EPI_ISL_1023615, EPI_ISL_1023617, EPI_ISL_1023671, EPI_ISL_1023673, EPI_ISL_1023675, EPI_ISL_1023677, EPI_ISL_1023679, EPI_ISL_1023681, EPI_ISL_1023683, EPI_ISL_1023686, EPI_ISL_1023688, EPI_ISL_1023690, EPI_ISL_1023692, EPI_ISL_1023693, EPI_ISL_1023695, EPI_ISL_1023697, EPI_ISL_1023699, EPI_ISL_1023701, EPI_ISL_1023703, EPI_ISL_1023704, EPI_ISL_1023706, EPI_ISL_1023708, EPI_ISL_1023710, EPI_ISL_1023712, EPI_ISL_1023714, EPI_ISL_1023716, EPI_ISL_1023718, EPI_ISL_1023720, EPI_ISL_1023721, EPI_ISL_1023723, EPI_ISL_1023725, EPI_ISL_1023727, EPI_ISL_1023729 | | | |
| see above | Wyoming Public Health Laboratory | Wyoming Public Health Laboratory | Noah Hull, Taylor Fearing, Lynette Gumbleton, Channing Weber, Ashley Norberg, Bailey Bowcutt, and Wanda Manley |
| EPI_ISL_1025963, EPI_ISL_1025964, EPI_ISL_1025965, EPI_ISL_1025966, EPI_ISL_1025967, EPI_ISL_1025968, EPI_ISL_1025969, EPI_ISL_1025970, EPI_ISL_1025971, EPI_ISL_1025972, EPI_ISL_1025973, EPI_ISL_1025974, EPI_ISL_1025975, EPI_ISL_1025976, EPI_ISL_1025977, EPI_ISL_1025978, EPI_ISL_1025979, EPI_ISL_1025980, EPI_ISL_1025981, EPI_ISL_1025982, EPI_ISL_1025983, EPI_ISL_1025984, EPI_ISL_1025985, EPI_ISL_1025986, EPI_ISL_1025987, EPI_ISL_1025988, EPI_ISL_1025989, EPI_ISL_1025990, EPI_ISL_1025991, EPI_ISL_1025992, EPI_ISL_1025993, EPI_ISL_1025994, EPI_ISL_1025995, EPI_ISL_1025996, EPI_ISL_1025997, EPI_ISL_1025998, EPI_ISL_1025999, EPI_ISL_1026000, EPI_ISL_1026001, EPI_ISL_1026002, EPI_ISL_1026003, EPI_ISL_1026005, EPI_ISL_1026006, EPI_ISL_1026007, EPI_ISL_1026008, EPI_ISL_1026009, EPI_ISL_1026010, EPI_ISL_1026011, EPI_ISL_1026012, EPI_ISL_1026013, EPI_ISL_1026014, EPI_ISL_1026015, EPI_ISL_1026016, EPI_ISL_1026017, EPI_ISL_1026018, EPI_ISL_1026019, EPI_ISL_1026020, EPI_ISL_1026021, EPI_ISL_1026022, EPI_ISL_1026023, EPI_ISL_1026024, EPI_ISL_1026025, EPI_ISL_1026026, EPI_ISL_1026027, EPI_ISL_1026028, EPI_ISL_1026029, EPI_ISL_1026847, EPI_ISL_1026848, EPI_ISL_1026849, EPI_ISL_1026850, EPI_ISL_1026851, EPI_ISL_1026852, EPI_ISL_1026853, EPI_ISL_1026854, EPI_ISL_1026855, EPI_ISL_1026856, EPI_ISL_1026857, EPI_ISL_1026858, EPI_ISL_1026859, EPI_ISL_1026860, EPI_ISL_1026861, EPI_ISL_1026862, EPI_ISL_1026863, EPI_ISL_1026864, EPI_ISL_1026865 | | | |
| see above | Laboratory Corporation of America | Respiratory Viruses Branch, Division of Viral Diseases, Centers for Disease Control and Prevention | Peter W. Cook, Dakota Howard, Dhwani Batra, Ben L. Rambo-Martin, Clinton R. Paden, Suxiang Tong, Duncan MacCannell |
| EPI_ISL_1026866, EPI_ISL_1026867, EPI_ISL_1026868, EPI_ISL_1026869, EPI_ISL_1026870, EPI_ISL_1026871, EPI_ISL_1026872, EPI_ISL_1026873, EPI_ISL_1026874, EPI_ISL_1026875, EPI_ISL_1026876, EPI_ISL_1026877, EPI_ISL_1026878, EPI_ISL_1026879, EPI_ISL_1026880, EPI_ISL_1026881, EPI_ISL_1026882, EPI_ISL_1026883, EPI_ISL_1026884, EPI_ISL_1026885, EPI_ISL_1026886, EPI_ISL_1026887, EPI_ISL_1026888, EPI_ISL_1026889, EPI_ISL_1026890, EPI_ISL_1026891, EPI_ISL_1026892, EPI_ISL_1026893, EPI_ISL_1026894, EPI_ISL_1026895, EPI_ISL_1026896, EPI_ISL_1026897, EPI_ISL_1026898, EPI_ISL_1026899, EPI_ISL_1026900, EPI_ISL_1026901, EPI_ISL_1026902, EPI_ISL_1026903, EPI_ISL_1026904, EPI_ISL_1026905, EPI_ISL_1026906, EPI_ISL_1026907, EPI_ISL_1026908, EPI_ISL_1026909, EPI_ISL_1026910, EPI_ISL_1026911, EPI_ISL_1026912, EPI_ISL_1026913, EPI_ISL_1026914, EPI_ISL_1026915, EPI_ISL_1026916, EPI_ISL_1026917, EPI_ISL_1026918, EPI_ISL_1026919, EPI_ISL_1026920, EPI_ISL_1026921, EPI_ISL_1026922, EPI_ISL_1026923, EPI_ISL_1026924, EPI_ISL_1026925, EPI_ISL_1026926, EPI_ISL_1026927, EPI_ISL_1026928, EPI_ISL_1026929, EPI_ISL_1026930, EPI_ISL_1026931, EPI_ISL_1026932, EPI_ISL_1026933, EPI_ISL_1026934, EPI_ISL_1026935, EPI_ISL_1026936, EPI_ISL_1026937, EPI_ISL_1026945, EPI_ISL_1026946, EPI_ISL_1026947, EPI_ISL_1026948, EPI_ISL_1026949, EPI_ISL_1026950, EPI_ISL_1026951, EPI_ISL_1026952, EPI_ISL_1026953, EPI_ISL_1026954, EPI_ISL_1026955, EPI_ISL_1026956, EPI_ISL_1026957, EPI_ISL_1026958, EPI_ISL_1026959, EPI_ISL_1026960, EPI_ISL_1026961, EPI_ISL_1026962, EPI_ISL_1026963, EPI_ISL_1026964, EPI_ISL_1026965, EPI_ISL_1026966, EPI_ISL_1026967, EPI_ISL_1026968, EPI_ISL_1026969, EPI_ISL_1026970, EPI_ISL_1026971, EPI_ISL_1026972, EPI_ISL_1026973, EPI_ISL_1026974, EPI_ISL_1026975, EPI_ISL_1026976, EPI_ISL_1026977, EPI_ISL_1026978, EPI_ISL_1026979, EPI_ISL_1026980, EPI_ISL_1026981, EPI_ISL_1026982, EPI_ISL_1026983, EPI_ISL_1026984, EPI_ISL_1026985, EPI_ISL_1026986, EPI_ISL_1026987, EPI_ISL_1026988, EPI_ISL_1026989, EPI_ISL_1026990, EPI_ISL_1026991, EPI_ISL_1026992, EPI_ISL_1026993, EPI_ISL_1026994, EPI_ISL_1026995, EPI_ISL_1026996, EPI_ISL_1026997, EPI_ISL_1026998, EPI_ISL_1026999, EPI_ISL_1027000, EPI_ISL_1027001, EPI_ISL_1027002, EPI_ISL_1027003, EPI_ISL_1027004, EPI_ISL_1027005, EPI_ISL_1027006, EPI_ISL_1027007, EPI_ISL_1027008, EPI_ISL_1027009, EPI_ISL_1027010, EPI_ISL_1027011, EPI_ISL_1027012, EPI_ISL_1027013, EPI_ISL_1027014, EPI_ISL_1027015, EPI_ISL_1027016, EPI_ISL_1027017, EPI_ISL_1027018, EPI_ISL_1027027, EPI_ISL_1027028, EPI_ISL_1027029, EPI_ISL_1027030, EPI_ISL_1027031, EPI_ISL_1027032, EPI_ISL_1027033, EPI_ISL_1027034, EPI_ISL_1027035, EPI_ISL_1027036, EPI_ISL_1027037, EPI_ISL_1027038, EPI_ISL_1027039, EPI_ISL_1027040, EPI_ISL_1027041, EPI_ISL_1027042, EPI_ISL_1027043, EPI_ISL_1027044, EPI_ISL_1027045, EPI_ISL_1027046, EPI_ISL_1027047, EPI_ISL_1027048, EPI_ISL_1027049, EPI_ISL_1027050, EPI_ISL_1027051, EPI_ISL_1027052, EPI_ISL_1027053, EPI_ISL_1027054, EPI_ISL_1027055, EPI_ISL_1027056, EPI_ISL_1027057, EPI_ISL_1027058, EPI_ISL_1027059, EPI_ISL_1027060, EPI_ISL_1027061, EPI_ISL_1027062, EPI_ISL_1027063, EPI_ISL_1027064, EPI_ISL_1027065, EPI_ISL_1027066, EPI_ISL_1027067, EPI_ISL_1027068, EPI_ISL_1027069, EPI_ISL_1027070, EPI_ISL_1027071, EPI_ISL_1027072, EPI_ISL_1027073, EPI_ISL_1027074, EPI_ISL_1027075, EPI_ISL_1027076, EPI_ISL_1027077, EPI_ISL_1027101, EPI_ISL_1027102, EPI_ISL_1027103, EPI_ISL_1027104, EPI_ISL_1027105, EPI_ISL_1027106, EPI_ISL_1027107, EPI_ISL_1027108, EPI_ISL_1027109, EPI_ISL_1027110, EPI_ISL_1027111, EPI_ISL_1027112, EPI_ISL_1027113, EPI_ISL_1027114, EPI_ISL_1027115, EPI_ISL_1027116, EPI_ISL_1027117, EPI_ISL_1027118, EPI_ISL_1027119, EPI_ISL_1027120, EPI_ISL_1027121, EPI_ISL_1027122, EPI_ISL_1027123, EPI_ISL_1027124, EPI_ISL_1027125, EPI_ISL_1027126, EPI_ISL_1027127, EPI_ISL_1027128, EPI_ISL_1027129, EPI_ISL_1027130, EPI_ISL_1027131, EPI_ISL_1027132, EPI_ISL_1027133, EPI_ISL_1027134, EPI_ISL_1027135, EPI_ISL_1027136, EPI_ISL_1027137, EPI_ISL_1027138, EPI_ISL_1027139, EPI_ISL_1027140, EPI_ISL_1027141, EPI_ISL_1027142, EPI_ISL_1027143, EPI_ISL_1027144, EPI_ISL_1027145, EPI_ISL_1027146, EPI_ISL_1027147, EPI_ISL_1027148, EPI_ISL_1027149, EPI_ISL_1027150, EPI_ISL_1027151, EPI_ISL_1027152, EPI_ISL_1027153, EPI_ISL_1027154, EPI_ISL_1027155, EPI_ISL_1027156, EPI_ISL_1027157, EPI_ISL_1027158, EPI_ISL_1027159, EPI_ISL_1027160, EPI_ISL_1027161, EPI_ISL_1027162, EPI_ISL_1027163, EPI_ISL_1027164, EPI_ISL_1027165, EPI_ISL_1027166, EPI_ISL_1027167, EPI_ISL_1027168, EPI_ISL_1027178, EPI_ISL_1027179, EPI_ISL_1027180, EPI_ISL_1027181, EPI_ISL_1027182, EPI_ISL_1027183, EPI_ISL_1027184, EPI_ISL_1027185, EPI_ISL_1027186, EPI_ISL_1027187, EPI_ISL_1027188, EPI_ISL_1027189, EPI_ISL_1027190, EPI_ISL_1027191, EPI_ISL_1027192, EPI_ISL_1027193, EPI_ISL_1027194, EPI_ISL_1027195, EPI_ISL_1027196, EPI_ISL_1027197, EPI_ISL_1027198, EPI_ISL_1027199, EPI_ISL_1027200, EPI_ISL_1027201, EPI_ISL_1027202, EPI_ISL_1027203, EPI_ISL_1027204, EPI_ISL_1027205, EPI_ISL_1027206, EPI_ISL_1027207, EPI_ISL_1027208, EPI_ISL_1027209, EPI_ISL_1027210, EPI_ISL_1027211, EPI_ISL_1027212, EPI_ISL_1027213, EPI_ISL_1027214, EPI_ISL_1027215, EPI_ISL_1027216, EPI_ISL_1027217, EPI_ISL_1027218, EPI_ISL_1027219, EPI_ISL_1027220, EPI_ISL_1027221, EPI_ISL_1027222, EPI_ISL_1027223, EPI_ISL_1027224, EPI_ISL_1027225, EPI_ISL_1027226, EPI_ISL_1027227, EPI_ISL_1027228, EPI_ISL_1027229, EPI_ISL_1027230, EPI_ISL_1027231, EPI_ISL_1027232, EPI_ISL_1027233, EPI_ISL_1027234, EPI_ISL_1027235, EPI_ISL_1027236, EPI_ISL_1027237, EPI_ISL_1027238, EPI_ISL_1027239, EPI_ISL_1027240, EPI_ISL_1027241, EPI_ISL_1027242, EPI_ISL_1027243, EPI_ISL_1027244, EPI_ISL_1027245, EPI_ISL_1027246 | | | |
| see above | Fulgent Genetics | Fulgent Genetics | Harry Gao, Mickey Li, John Gao, Joseph Fierro, Benafsh Sapra, Becky Tsai, Yan Meng, Doreen Ng, James Xie |
| EPI_ISL_1027249, EPI_ISL_1027250, EPI_ISL_1027251, EPI_ISL_1027252, EPI_ISL_1027253, EPI_ISL_1027254, EPI_ISL_1027255, EPI_ISL_1027256, EPI_ISL_1027257, EPI_ISL_1027258, EPI_ISL_1027259, EPI_ISL_1027260 | | | |
| see above | M Health Fairview | Minnesota Department of Health, Public Health Laboratory | Alexandra Lorentz, Jacob Garfin, Matt Plumb, and Xiong Wang |
| EPI_ISL_1027261, EPI_ISL_1027262, EPI_ISL_1027263, EPI_ISL_1027264 | Vault Health | Minnesota Department of Health, Public Health Laboratory | Alexandra Lorentz, Jacob Garfin, Matt Plumb, and Xiong Wang |
| EPI_ISL_1027265, EPI_ISL_1027266, EPI_ISL_1027267, EPI_ISL_1027268, EPI_ISL_1027269, EPI_ISL_1027270, EPI_ISL_1027271 | M Health Fairview | Minnesota Department of Health, Public Health Laboratory | Alexandra Lorentz, Jacob Garfin, Matt Plumb, and Xiong Wang |
| EPI_ISL_1027272, EPI_ISL_1027273, EPI_ISL_1027274, EPI_ISL_1027275, EPI_ISL_1027276, EPI_ISL_1027277, EPI_ISL_1027278, EPI_ISL_1027279, EPI_ISL_1027280, EPI_ISL_1027281 | Vault Health | Minnesota Department of Health, Public Health Laboratory | Alexandra Lorentz, Jacob Garfin, Matt Plumb, and Xiong Wang |
| EPI_ISL_1027282, EPI_ISL_1027283, EPI_ISL_1027284, EPI_ISL_1027285, EPI_ISL_1027286, EPI_ISL_1027287 | M Health Fairview | Minnesota Department of Health, Public Health Laboratory | Alexandra Lorentz, Jacob Garfin, Matt Plumb, and Xiong Wang |
| EPI_ISL_1027288, EPI_ISL_1027289, EPI_ISL_1027290, EPI_ISL_1027291, EPI_ISL_1027292 | Vault Health | Minnesota Department of Health, Public Health Laboratory | Alexandra Lorentz, Jacob Garfin, Matt Plumb, and Xiong Wang |
| EPI_ISL_1027293 | Santa Clara County Public Health Laboratory | Chan-Zuckerberg Biohub | CZB Cliahub Consortium |
| EPI_ISL_1027294 | Orange County Public Health Lab | Chan-Zuckerberg Biohub | CZB Cliahub Consortium |
| EPI_ISL_1027295 | Santa Clara County Public Health Laboratory | Chan-Zuckerberg Biohub | CZB Cliahub Consortium |
| EPI_ISL_1027296 | Alameda County Public Health Lab | Chan-Zuckerberg Biohub | CZB Cliahub Consortium |
| EPI_ISL_1027297, EPI_ISL_1027298, EPI_ISL_1027299, EPI_ISL_1027300, EPI_ISL_1027301, EPI_ISL_1027302 | Santa Clara County Public Health Laboratory | Chan-Zuckerberg Biohub | CZB Cliahub Consortium |
| EPI_ISL_1027303 | Alameda County Public Health Lab | Chan-Zuckerberg Biohub | CZB Cliahub Consortium |
| EPI_ISL_1027304, EPI_ISL_1027305, EPI_ISL_1027306 | Santa Clara County Public Health Laboratory | Chan-Zuckerberg Biohub | CZB Cliahub Consortium |
| EPI_ISL_1027307, EPI_ISL_1027308, EPI_ISL_1027309 | Alameda County Public Health Lab | Chan-Zuckerberg Biohub | CZB Cliahub Consortium |
| EPI_ISL_1027310, EPI_ISL_1027311, EPI_ISL_1027312, EPI_ISL_1027313, EPI_ISL_1027314, EPI_ISL_1027315, EPI_ISL_1027316, EPI_ISL_1027317, EPI_ISL_1027318, EPI_ISL_1027319, EPI_ISL_1027320 | | | |
| see above | Santa Clara County Public Health Laboratory | Chan-Zuckerberg Biohub | CZB Cliahub Consortium |
| EPI_ISL_1027321 | Orange County Public Health Lab | Chan-Zuckerberg Biohub | CZB Cliahub Consortium |
| EPI_ISL_1027322 | Santa Clara County Public Health Laboratory | Chan-Zuckerberg Biohub | CZB Cliahub Consortium |
| EPI_ISL_1027323, EPI_ISL_1027324 | Alameda County Public Health Lab | Chan-Zuckerberg Biohub | CZB Cliahub Consortium |
| EPI_ISL_1027325, | Santa Clara County Public Health | Chan-Zuckerberg Biohub | CZB Cliahub Consortium |

| | | | |
|---|---|---|---|
| EPI_ISL_1027326, EPI_ISL_1027327, EPI_ISL_1027328 | Laboratory | | |
| EPI_ISL_1027329 | Orange County Public Health Lab | Chan-Zuckerberg Biohub | CZB Cliahub Consortium |
| EPI_ISL_1027330 | Alameda County Public Health Lab | Chan-Zuckerberg Biohub | CZB Cliahub Consortium |
| EPI_ISL_1027331 | Orange County Public Health Lab | Chan-Zuckerberg Biohub | CZB Cliahub Consortium |
| EPI_ISL_1027332 | Santa Clara County Public Health Laboratory | Chan-Zuckerberg Biohub | CZB Cliahub Consortium |
| EPI_ISL_1027333 | Alameda County Public Health Lab | Chan-Zuckerberg Biohub | CZB Cliahub Consortium |
| EPI_ISL_1027334 | Santa Clara County Public Health Laboratory | Chan-Zuckerberg Biohub | CZB Cliahub Consortium |
| EPI_ISL_1027335 | Orange County Public Health Lab | Chan-Zuckerberg Biohub | CZB Cliahub Consortium |
| EPI_ISL_1027336 | Alameda County Public Health Lab | Chan-Zuckerberg Biohub | CZB Cliahub Consortium |
| EPI_ISL_1027337, EPI_ISL_1027338 | Santa Clara County Public Health Laboratory | Chan-Zuckerberg Biohub | CZB Cliahub Consortium |
| EPI_ISL_1027339, EPI_ISL_1027340 | Orange County Public Health Lab | Chan-Zuckerberg Biohub | CZB Cliahub Consortium |
| EPI_ISL_1027341 | Alameda County Public Health Lab | Chan-Zuckerberg Biohub | CZB Cliahub Consortium |
| EPI_ISL_1027342, EPI_ISL_1027343 | Santa Clara County Public Health Laboratory | Chan-Zuckerberg Biohub | CZB Cliahub Consortium |
| EPI_ISL_1027344 | Orange County Public Health Lab | Chan-Zuckerberg Biohub | CZB Cliahub Consortium |
| EPI_ISL_1027345 | Alameda County Public Health Lab | Chan-Zuckerberg Biohub | CZB Cliahub Consortium |
| EPI_ISL_1027346, EPI_ISL_1027347 | Santa Clara County Public Health Laboratory | Chan-Zuckerberg Biohub | CZB Cliahub Consortium |
| EPI_ISL_1027348 | Alameda County Public Health Lab | Chan-Zuckerberg Biohub | CZB Cliahub Consortium |
| EPI_ISL_1027349 | Santa Clara County Public Health Laboratory | Chan-Zuckerberg Biohub | CZB Cliahub Consortium |
| EPI_ISL_1027350, EPI_ISL_1027351 | Orange County Public Health Lab | Chan-Zuckerberg Biohub | CZB Cliahub Consortium |
| EPI_ISL_1027364 | Santa Clara County Public Health Laboratory | Chan-Zuckerberg Biohub | CZB Cliahub Consortium |
| EPI_ISL_1027365 | Alameda County Public Health Lab | Chan-Zuckerberg Biohub | CZB Cliahub Consortium |
| EPI_ISL_1027366, EPI_ISL_1027367 | County of San Luis Obispo Public Health Laboratory | Chan-Zuckerberg Biohub | CZB Cliahub Consortium |
| EPI_ISL_1027368, EPI_ISL_1027369, EPI_ISL_1027370, EPI_ISL_1027371, EPI_ISL_1027372, EPI_ISL_1027373, EPI_ISL_1027374, EPI_ISL_1027375, EPI_ISL_1027376, EPI_ISL_1027377 | Santa Clara County Public Health Laboratory | Chan-Zuckerberg Biohub | CZB Cliahub Consortium |
| EPI_ISL_1027378, EPI_ISL_1027379, EPI_ISL_1027380, EPI_ISL_1027381, EPI_ISL_1027382, EPI_ISL_1027383, EPI_ISL_1027384, EPI_ISL_1027385, EPI_ISL_1027386, EPI_ISL_1027387, EPI_ISL_1027388, EPI_ISL_1027389, EPI_ISL_1027390, EPI_ISL_1027391, EPI_ISL_1027392, EPI_ISL_1027393, EPI_ISL_1027394, EPI_ISL_1027395, EPI_ISL_1027396, EPI_ISL_1027397, EPI_ISL_1027398, EPI_ISL_1027399, EPI_ISL_1027400, EPI_ISL_1027401, EPI_ISL_1027402, EPI_ISL_1027403, EPI_ISL_1027404, EPI_ISL_1027405, EPI_ISL_1027406, EPI_ISL_1027407, EPI_ISL_1027408, EPI_ISL_1027409, EPI_ISL_1027410, EPI_ISL_1027411, EPI_ISL_1027412, EPI_ISL_1027413, EPI_ISL_1027414, EPI_ISL_1027415, EPI_ISL_1027416, EPI_ISL_1027417, EPI_ISL_1027418, EPI_ISL_1027419, EPI_ISL_1027420, EPI_ISL_1027421, EPI_ISL_1027422, EPI_ISL_1027423, EPI_ISL_1027424, EPI_ISL_1027425, EPI_ISL_1027426, EPI_ISL_1027427, EPI_ISL_1027428, EPI_ISL_1027429, EPI_ISL_1027430, EPI_ISL_1027431, EPI_ISL_1027432, EPI_ISL_1027433, EPI_ISL_1027434, EPI_ISL_1027435, EPI_ISL_1027436 | | | |
| see above | Orange County Public Health Lab | Chan-Zuckerberg Biohub | CZB Cliahub Consortium |
| EPI_ISL_1027437, EPI_ISL_1027438, EPI_ISL_1027439, EPI_ISL_1027440, EPI_ISL_1027441, EPI_ISL_1027442, EPI_ISL_1027443, EPI_ISL_1027444 | Santa Clara County Public Health Laboratory | Chan-Zuckerberg Biohub | CZB Cliahub Consortium |
| EPI_ISL_1027445, EPI_ISL_1027446, EPI_ISL_1027447, EPI_ISL_1027448, EPI_ISL_1027449, EPI_ISL_1027450, EPI_ISL_1027451, EPI_ISL_1027452, EPI_ISL_1027453, EPI_ISL_1027454, EPI_ISL_1027455, EPI_ISL_1027456, EPI_ISL_1027457, EPI_ISL_1027458, EPI_ISL_1027459, EPI_ISL_1027460, EPI_ISL_1027461, EPI_ISL_1027462, EPI_ISL_1027463, EPI_ISL_1027464, EPI_ISL_1027465, EPI_ISL_1027466, EPI_ISL_1027467, EPI_ISL_1027468, EPI_ISL_1027469, EPI_ISL_1027470, EPI_ISL_1027471, EPI_ISL_1027472, EPI_ISL_1027473 | | | |
| see above | Alameda County Public Health Lab | Chan-Zuckerberg Biohub | CZB Cliahub Consortium |
| EPI_ISL_1027474, EPI_ISL_1027475 | Santa Clara County Public Health Laboratory | Chan-Zuckerberg Biohub | CZB Cliahub Consortium |
| EPI_ISL_1027476 | Humboldt County Public Health Laboratory | Chan-Zuckerberg Biohub | CZB Cliahub Consortium |
| EPI_ISL_1027477, EPI_ISL_1027478 | Santa Clara County Public Health Laboratory | Chan-Zuckerberg Biohub | CZB Cliahub Consortium |
| EPI_ISL_1027479 | Alameda County Public Health Lab | Chan-Zuckerberg Biohub | CZB Cliahub Consortium |
| EPI_ISL_1027480, EPI_ISL_1027481, EPI_ISL_1027482, EPI_ISL_1027483, EPI_ISL_1027484, EPI_ISL_1027485, EPI_ISL_1027486 | Santa Clara County Public Health Laboratory | Chan-Zuckerberg Biohub | CZB Cliahub Consortium |
| EPI_ISL_1027487 | Orange County Public Health Lab | Chan-Zuckerberg Biohub | CZB Cliahub Consortium |
| EPI_ISL_1027488, EPI_ISL_1027489, EPI_ISL_1027490, EPI_ISL_1027491, EPI_ISL_1027492, EPI_ISL_1027493, EPI_ISL_1027494, EPI_ISL_1027495, EPI_ISL_1027496, EPI_ISL_1027497, EPI_ISL_1027498, EPI_ISL_1027499, EPI_ISL_1027500, EPI_ISL_1027501, EPI_ISL_1027502 | | | |
| see above | Santa Clara County Public Health Laboratory | Chan-Zuckerberg Biohub | CZB Cliahub Consortium |
| EPI_ISL_1027503, EPI_ISL_1027504 | Orange County Public Health Lab | Chan-Zuckerberg Biohub | CZB Cliahub Consortium |
| EPI_ISL_1027505, EPI_ISL_1027506 | Santa Clara County Public Health Laboratory | Chan-Zuckerberg Biohub | CZB Cliahub Consortium |
| EPI_ISL_1027507 | Orange County Public Health Lab | Chan-Zuckerberg Biohub | CZB Cliahub Consortium |
| EPI_ISL_1027508, EPI_ISL_1027509, EPI_ISL_1027510, EPI_ISL_1027511, EPI_ISL_1027528, EPI_ISL_1027529 | Santa Clara County Public Health Laboratory | Chan-Zuckerberg Biohub | CZB Cliahub Consortium |
| EPI_ISL_1027530, EPI_ISL_1027531, EPI_ISL_1027532 | Orange County Public Health Lab | Chan-Zuckerberg Biohub | CZB Cliahub Consortium |
| EPI_ISL_1027533, | Santa Clara County Public Health | Chan-Zuckerberg Biohub | CZB Cliahub Consortium |

| | | | |
|---|---|---|---|
| EPI_ISL_1027534 | Laboratory | | |
| EPI_ISL_1027535, EPI_ISL_1027536, EPI_ISL_1027537, EPI_ISL_1027538, EPI_ISL_1027539, EPI_ISL_1027540, EPI_ISL_1027541 | Humboldt County Public Health Laboratory | Chan-Zuckerberg Biohub | CZB Cliahub Consortium |
| EPI_ISL_1027542 | Santa Clara County Public Health Laboratory | Chan-Zuckerberg Biohub | CZB Cliahub Consortium |
| EPI_ISL_1027543 | Alameda County Public Health Lab | Chan-Zuckerberg Biohub | CZB Cliahub Consortium |
| EPI_ISL_1027544, EPI_ISL_1027545, EPI_ISL_1027546, EPI_ISL_1027547, EPI_ISL_1027548, EPI_ISL_1027549, EPI_ISL_1027550, EPI_ISL_1027551, EPI_ISL_1027552, EPI_ISL_1027553, EPI_ISL_1027554, EPI_ISL_1027555, EPI_ISL_1027556, EPI_ISL_1027557, EPI_ISL_1027558, EPI_ISL_1027559 | | | |
| see above | Santa Clara County Public Health Laboratory | Chan-Zuckerberg Biohub | CZB Cliahub Consortium |
| EPI_ISL_1027560 | Alameda County Public Health Lab | Chan-Zuckerberg Biohub | CZB Cliahub Consortium |
| EPI_ISL_1027561 | Humboldt County Public Health Laboratory | Chan-Zuckerberg Biohub | CZB Cliahub Consortium |
| EPI_ISL_1027562, EPI_ISL_1027563, EPI_ISL_1027564, EPI_ISL_1027565, EPI_ISL_1027566, EPI_ISL_1027567 | Santa Clara County Public Health Laboratory | Chan-Zuckerberg Biohub | CZB Cliahub Consortium |
| EPI_ISL_1027568 | Alameda County Public Health Lab | Chan-Zuckerberg Biohub | CZB Cliahub Consortium |
| EPI_ISL_1027569, EPI_ISL_1027570, EPI_ISL_1027571, EPI_ISL_1027572 | Santa Clara County Public Health Laboratory | Chan-Zuckerberg Biohub | CZB Cliahub Consortium |
| EPI_ISL_1027573 | Orange County Public Health Lab | Chan-Zuckerberg Biohub | CZB Cliahub Consortium |
| EPI_ISL_1027574, EPI_ISL_1027575, EPI_ISL_1027576, EPI_ISL_1027577 | Santa Clara County Public Health Laboratory | Chan-Zuckerberg Biohub | CZB Cliahub Consortium |
| EPI_ISL_1027578 | Alameda County Public Health Lab | Chan-Zuckerberg Biohub | CZB Cliahub Consortium |
| EPI_ISL_1027579, EPI_ISL_1027580, EPI_ISL_1027581 | Santa Clara County Public Health Laboratory | Chan-Zuckerberg Biohub | CZB Cliahub Consortium |
| EPI_ISL_1027582 | Alameda County Public Health Lab | Chan-Zuckerberg Biohub | CZB Cliahub Consortium |
| EPI_ISL_1027583, EPI_ISL_1027584 | Orange County Public Health Lab | Chan-Zuckerberg Biohub | CZB Cliahub Consortium |
| EPI_ISL_1027585, EPI_ISL_1027586 | Santa Clara County Public Health Laboratory | Chan-Zuckerberg Biohub | CZB Cliahub Consortium |
| EPI_ISL_1027587, EPI_ISL_1027588, EPI_ISL_1027589, EPI_ISL_1027590, EPI_ISL_1027591, EPI_ISL_1027592, EPI_ISL_1027593, EPI_ISL_1027594, EPI_ISL_1027595, EPI_ISL_1027596, EPI_ISL_1027597, EPI_ISL_1027598, EPI_ISL_1027599, EPI_ISL_1027600, EPI_ISL_1027601, EPI_ISL_1027602, EPI_ISL_1027603, EPI_ISL_1027604, EPI_ISL_1027605, EPI_ISL_1027606, EPI_ISL_1027607, EPI_ISL_1027608, EPI_ISL_1027609, EPI_ISL_1027610, EPI_ISL_1027611, EPI_ISL_1027612, EPI_ISL_1027613, EPI_ISL_1027614, EPI_ISL_1027615, EPI_ISL_1027616, EPI_ISL_1027617, EPI_ISL_1027618, EPI_ISL_1027619, EPI_ISL_1027620, EPI_ISL_1027621, EPI_ISL_1027622, EPI_ISL_1027623, EPI_ISL_1027624, EPI_ISL_1027625, EPI_ISL_1027626, EPI_ISL_1027627 | | | |
| see above | Orange County Public Health Lab | Chan-Zuckerberg Biohub | CZB Cliahub Consortium |
| EPI_ISL_1027629, EPI_ISL_1027630, EPI_ISL_1027631, EPI_ISL_1027632, EPI_ISL_1027633, EPI_ISL_1027634, EPI_ISL_1027635, EPI_ISL_1027636, EPI_ISL_1027658, EPI_ISL_1027659, EPI_ISL_1027660, EPI_ISL_1027661, EPI_ISL_1027662, EPI_ISL_1027663, EPI_ISL_1027664, EPI_ISL_1027665, EPI_ISL_1027666 | | | |
| see above | Mako Medical Laboratories | Mako Medical Laboratories | "Lauren Moon, Matthew Tugwell" |
| EPI_ISL_1027667 | Vault Health | Minnesota Department of Health, Public Health Laboratory | Alexandra Lorentz, Jacob Garfin, Matt Plumb, and Xiong Wang |
| EPI_ISL_1027668, EPI_ISL_1027669, EPI_ISL_1027670, EPI_ISL_1027671, EPI_ISL_1027672, EPI_ISL_1027673, EPI_ISL_1027674, EPI_ISL_1027675, EPI_ISL_1027676, EPI_ISL_1027677, EPI_ISL_1027678, EPI_ISL_1027679, EPI_ISL_1027680, EPI_ISL_1027681, EPI_ISL_1027682, EPI_ISL_1027683, EPI_ISL_1027684 | | | |
| see above | Laboratory Corporation of America | Respiratory Viruses Branch, Division of Viral Diseases, Centers for Disease Control and Prevention | Peter W. Cook, Dakota Howard, Dhwani Batra, Ben L. Rambo-Martin, Clinton R. Paden, Suxiang Tong, Duncan MacCannell |
| EPI_ISL_982414, EPI_ISL_982415, EPI_ISL_982416, EPI_ISL_982417, EPI_ISL_982418, EPI_ISL_982419, EPI_ISL_982420, EPI_ISL_982421, EPI_ISL_982422, EPI_ISL_982423, EPI_ISL_982424 | | | |
| see above | M Health Fairview | Minnesota Department of Health, Public Health Laboratory | Alexandra Lorentz, Jacob Garfin, Matt Plumb, and Xiong Wang |
| EPI_ISL_982425, EPI_ISL_982426, EPI_ISL_982427, EPI_ISL_982428, EPI_ISL_982429, EPI_ISL_982430, EPI_ISL_982431, EPI_ISL_982432, EPI_ISL_982433, EPI_ISL_982434, EPI_ISL_982435, EPI_ISL_982436, EPI_ISL_982437, EPI_ISL_982438, EPI_ISL_982439, EPI_ISL_982440, EPI_ISL_982441, EPI_ISL_982442, EPI_ISL_982443, EPI_ISL_982444, EPI_ISL_982445, EPI_ISL_982446, EPI_ISL_982447, EPI_ISL_982448, EPI_ISL_982449, EPI_ISL_982450, EPI_ISL_982451, EPI_ISL_982452, EPI_ISL_982453, EPI_ISL_982454, EPI_ISL_982455, EPI_ISL_982456, EPI_ISL_982457, EPI_ISL_982458, EPI_ISL_982459, EPI_ISL_982460, EPI_ISL_982461, EPI_ISL_982462, EPI_ISL_982463, EPI_ISL_982464, EPI_ISL_982465, EPI_ISL_982466, EPI_ISL_982467, EPI_ISL_982468, EPI_ISL_982469, EPI_ISL_982470, EPI_ISL_982471, EPI_ISL_982472, EPI_ISL_982473, EPI_ISL_982474, EPI_ISL_982475, EPI_ISL_982476, EPI_ISL_982477, EPI_ISL_982478, EPI_ISL_982479, EPI_ISL_982480, EPI_ISL_982481, EPI_ISL_982482, EPI_ISL_982483, EPI_ISL_982484, EPI_ISL_982485, EPI_ISL_982486, EPI_ISL_982487, EPI_ISL_982488, EPI_ISL_982489, EPI_ISL_982490, EPI_ISL_982491, EPI_ISL_982492, EPI_ISL_982493, EPI_ISL_982494, EPI_ISL_982495, EPI_ISL_982496, EPI_ISL_982497, EPI_ISL_982498 | | | |
| see above | MONTEFIORE MEDICAL CENTER LABORATORIES | Wadsworth Center, New York State Department of Health | Kirsten St. George, Daryl M. Lamson, Alexis Russel, Matthew Shudt, Melissa A Leisner, Jonathan Plitnick, Navjot Singh, John Kelly, Erasmus Schneider, Erica Lasek-Nesselquist |
| EPI_ISL_982499, EPI_ISL_982500, EPI_ISL_982501, EPI_ISL_982502, EPI_ISL_982503, EPI_ISL_982504 | ADIRONDACK MEDICAL CENTER | Wadsworth Center, New York State Department of Health | Kirsten St. George, Daryl M. Lamson, Alexis Russel, Matthew Shudt, Melissa A Leisner, Jonathan Plitnick, Navjot Singh, John Kelly, Erasmus Schneider, Erica Lasek-Nesselquist |
| EPI_ISL_982506, EPI_ISL_982507, EPI_ISL_982508, EPI_ISL_982509, EPI_ISL_982510, EPI_ISL_982511, EPI_ISL_982512, EPI_ISL_982513, EPI_ISL_982514, EPI_ISL_982515, EPI_ISL_982516, EPI_ISL_982517, EPI_ISL_982518, EPI_ISL_982519, EPI_ISL_982520, EPI_ISL_982521, EPI_ISL_982522, EPI_ISL_982523, EPI_ISL_982524, EPI_ISL_982525 | | | |
| see above | Kentucky State Public Health Lab | Kentucky State Public Health Lab | Stephanie Lunn, Karim George, Joshua Tobias, William Grooms, Vaneet Arora, Matthew Johnson, Rachel Zinner, Rhonda Lucas |
| EPI_ISL_982527, EPI_ISL_982528, EPI_ISL_982529, EPI_ISL_982530, EPI_ISL_982531, EPI_ISL_982532, EPI_ISL_982533, EPI_ISL_982534, EPI_ISL_982535, EPI_ISL_982536, EPI_ISL_982538, EPI_ISL_982540, EPI_ISL_982541, EPI_ISL_982542, EPI_ISL_982543, EPI_ISL_982544, EPI_ISL_982545, EPI_ISL_982546, EPI_ISL_982547, EPI_ISL_982548, EPI_ISL_982549, EPI_ISL_982550, EPI_ISL_982551, EPI_ISL_982552, EPI_ISL_982553, EPI_ISL_982554, EPI_ISL_982555, EPI_ISL_982556, EPI_ISL_982557, EPI_ISL_982558, EPI_ISL_982559, EPI_ISL_982560, EPI_ISL_982561, EPI_ISL_982562, EPI_ISL_982563, EPI_ISL_982564, EPI_ISL_982565, EPI_ISL_982566, EPI_ISL_982567, EPI_ISL_982568, EPI_ISL_982569, EPI_ISL_982570, EPI_ISL_982571, EPI_ISL_982572, EPI_ISL_982573, EPI_ISL_982574, EPI_ISL_982575, EPI_ISL_982576, EPI_ISL_982577, EPI_ISL_982578, EPI_ISL_982579, EPI_ISL_982580, EPI_ISL_982581, EPI_ISL_982582, EPI_ISL_982583, EPI_ISL_982584, EPI_ISL_982586, EPI_ISL_982587, EPI_ISL_982626, EPI_ISL_982631, EPI_ISL_982632, EPI_ISL_982633, EPI_ISL_982634, EPI_ISL_982635, EPI_ISL_982636, EPI_ISL_982637, EPI_ISL_982638, EPI_ISL_982639, EPI_ISL_982640, EPI_ISL_982641, EPI_ISL_982642, EPI_ISL_982643, EPI_ISL_982644, EPI_ISL_982645, EPI_ISL_982646, EPI_ISL_982647, EPI_ISL_982648, EPI_ISL_982649, EPI_ISL_982650, EPI_ISL_982651, EPI_ISL_982652, EPI_ISL_982653, EPI_ISL_982654, EPI_ISL_982655, EPI_ISL_982656, EPI_ISL_982657, EPI_ISL_982658, EPI_ISL_982659, EPI_ISL_982660, EPI_ISL_982661, EPI_ISL_982662, EPI_ISL_982663, EPI_ISL_982664, EPI_ISL_982665, EPI_ISL_982666, EPI_ISL_982667, EPI_ISL_982668, EPI_ISL_982669, EPI_ISL_982670, EPI_ISL_982671, EPI_ISL_982672, EPI_ISL_982673, EPI_ISL_982674, EPI_ISL_982675, EPI_ISL_982676, EPI_ISL_982677, EPI_ISL_982678, EPI_ISL_982679, EPI_ISL_982680, EPI_ISL_982681, EPI_ISL_982682, EPI_ISL_982683, EPI_ISL_982684, EPI_ISL_982685, EPI_ISL_982686, EPI_ISL_982687, EPI_ISL_982688, EPI_ISL_982689, EPI_ISL_982690, EPI_ISL_982691, EPI_ISL_982692, EPI_ISL_982693, EPI_ISL_982694, EPI_ISL_982695, EPI_ISL_982696, EPI_ISL_982697, EPI_ISL_982698, EPI_ISL_982699, EPI_ISL_982700, EPI_ISL_982701, EPI_ISL_982702, EPI_ISL_982703, EPI_ISL_982704, EPI_ISL_982705, EPI_ISL_982706, EPI_ISL_982707, EPI_ISL_982708, EPI_ISL_982709, EPI_ISL_982710, EPI_ISL_982711, EPI_ISL_982712, EPI_ISL_982713, EPI_ISL_982714, EPI_ISL_982715, EPI_ISL_982716, EPI_ISL_982717, EPI_ISL_982718, EPI_ISL_982719, EPI_ISL_982720, EPI_ISL_982721, EPI_ISL_982722, EPI_ISL_982723, EPI_ISL_982724, EPI_ISL_982725, EPI_ISL_982726, EPI_ISL_982727, EPI_ISL_982728, EPI_ISL_982729, EPI_ISL_982730, EPI_ISL_982731, EPI_ISL_982732, EPI_ISL_982733, EPI_ISL_982734, EPI_ISL_982735, EPI_ISL_982736, EPI_ISL_982737, EPI_ISL_982738, EPI_ISL_982739, EPI_ISL_982740, EPI_ISL_982741, EPI_ISL_982742, EPI_ISL_982743, EPI_ISL_982744, EPI_ISL_982745, EPI_ISL_982746, EPI_ISL_982747, EPI_ISL_982748, EPI_ISL_982749, EPI_ISL_982750, EPI_ISL_982751, EPI_ISL_982752, EPI_ISL_982753, EPI_ISL_982754, EPI_ISL_982755, EPI_ISL_982756, EPI_ISL_982757, EPI_ISL_982758, EPI_ISL_982759, EPI_ISL_982760, EPI_ISL_982761 | | | |
| see above | US Air Force School of Aerospace Medicine | US Air Force School of Aerospace Medicine | Anthony Fries, Jennifer Meyer, William Gruner, William Buggele, Amanda Javorina, Sarah Purves, Clarise Starr, Elizabeth Macias |
| EPI_ISL_982762, EPI_ISL_982763, EPI_ISL_982764, EPI_ISL_982765, EPI_ISL_982766, EPI_ISL_982767, EPI_ISL_982768, EPI_ISL_982769, EPI_ISL_982770, EPI_ISL_982771, EPI_ISL_982772, EPI_ISL_982773, EPI_ISL_982774, EPI_ISL_982775, EPI_ISL_982777, EPI_ISL_982778, EPI_ISL_982779, EPI_ISL_982780, EPI_ISL_982842, EPI_ISL_982843, EPI_ISL_982844 | | | |
| see above | Kentucky State Public Health Lab | Kentucky State Public Health Lab | Stephanie Lunn, Karim George, Joshua Tobias, William Grooms, Vaneet Arora, Matthew Johnson, Rachel Zinner, Rhonda Lucas |
| EPI_ISL_982845, EPI_ISL_982846, EPI_ISL_982847, EPI_ISL_982848, EPI_ISL_982849, EPI_ISL_982850, EPI_ISL_982851, EPI_ISL_982852 | UK Healthcare Clinical Microbiology | Kentucky State Public Health Lab | Stephanie Lunn, Karim George, Joshua Tobias, William Grooms, Vaneet Arora, Matthew Johnson, Rachel Zinner, Rhonda Lucas |
| EPI_ISL_982853, EPI_ISL_982854, EPI_ISL_982855, EPI_ISL_982856 | Kentucky State Public Health Lab | Kentucky State Public Health Lab | Stephanie Lunn, Karim George, Joshua Tobias, William Grooms, Vaneet Arora, Matthew Johnson, Rachel Zinner, Rhonda Lucas |

| | | | |
|---|---|---|---|
| EPI_ISL_982857, EPI_ISL_982858, EPI_ISL_982859, EPI_ISL_983063, EPI_ISL_983064, EPI_ISL_983065, EPI_ISL_983066, EPI_ISL_983067, EPI_ISL_983068, EPI_ISL_983069, EPI_ISL_983070, EPI_ISL_983071, EPI_ISL_983072 | | | |
| see above | Gravity Diagnostics | Kentucky State Public Health Lab | Stephanie Lunn, Karim George, Joshua Tobias, William Grooms, Vaneet Arora, Matthew Johnson, Rachel Zinner, Rhonda Lucas |
| EPI_ISL_983073, EPI_ISL_983074 | Kentucky State Public Health Lab | Kentucky State Public Health Lab | Stephanie Lunn, Karim George, Joshua Tobias, William Grooms, Vaneet Arora, Matthew Johnson, Rachel Zinner, Rhonda Lucas |
| EPI_ISL_983075, EPI_ISL_983076, EPI_ISL_983077, EPI_ISL_983078, EPI_ISL_983079, EPI_ISL_983080, EPI_ISL_983081 | Gravity Diagnostics | Kentucky State Public Health Lab | Stephanie Lunn, Karim George, Joshua Tobias, William Grooms, Vaneet Arora, Matthew Johnson, Rachel Zinner, Rhonda Lucas |
| EPI_ISL_983082 | Kentucky State Public Health Lab | Kentucky State Public Health Lab | Stephanie Lunn, Karim George, Joshua Tobias, William Grooms, Vaneet Arora, Matthew Johnson, Rachel Zinner, Rhonda Lucas |
| EPI_ISL_983083, EPI_ISL_983084 | Gravity Diagnostics | Kentucky State Public Health Lab | Stephanie Lunn, Karim George, Joshua Tobias, William Grooms, Vaneet Arora, Matthew Johnson, Rachel Zinner, Rhonda Lucas |
| EPI_ISL_983085 | Kentucky State Public Health Lab | Kentucky State Public Health Lab | Stephanie Lunn, Karim George, Joshua Tobias, William Grooms, Vaneet Arora, Matthew Johnson, Rachel Zinner, Rhonda Lucas |
| EPI_ISL_983086, EPI_ISL_983087, EPI_ISL_983088, EPI_ISL_983089, EPI_ISL_983090, EPI_ISL_983091 | Gravity Diagnostics | Kentucky State Public Health Lab | Stephanie Lunn, Karim George, Joshua Tobias, William Grooms, Vaneet Arora, Matthew Johnson, Rachel Zinner, Rhonda Lucas |
| EPI_ISL_983092, EPI_ISL_983093, EPI_ISL_983094, EPI_ISL_983095 | Kentucky State Public Health Lab | Kentucky State Public Health Lab | Stephanie Lunn, Karim George, Joshua Tobias, William Grooms, Vaneet Arora, Matthew Johnson, Rachel Zinner, Rhonda Lucas |
| EPI_ISL_983099, EPI_ISL_983100, EPI_ISL_983101, EPI_ISL_983102, EPI_ISL_983103, EPI_ISL_983104, EPI_ISL_983105, EPI_ISL_983106, EPI_ISL_983107, EPI_ISL_983108, EPI_ISL_983109, EPI_ISL_983110, EPI_ISL_983111, EPI_ISL_983112, EPI_ISL_983113, EPI_ISL_983114, EPI_ISL_983115 | | | |
| see above | MONTEFIORE MEDICAL CENTER LABORATORIES | Wadsworth Center, New York State Department of Health | Kirsten St. George, Daryl M. Lamson, Alexis Russel, Matthew Shudt, Melissa A Leisner, Jonathan Plitnick, Navjot Singh, John Kelly, Erasmus Schneider, Erica Lasek-Nesselquist |
| EPI_ISL_983116, EPI_ISL_983117, EPI_ISL_983118 | SUNY UPSTATE MEDICAL UNIVERSITY | Wadsworth Center, New York State Department of Health | Kirsten St. George, Daryl M. Lamson, Alexis Russel, Matthew Shudt, Melissa A Leisner, Jonathan Plitnick, Navjot Singh, John Kelly, Erasmus Schneider, Erica Lasek-Nesselquist |
| EPI_ISL_983119, EPI_ISL_983120, EPI_ISL_983121, EPI_ISL_983122, EPI_ISL_983123, EPI_ISL_983124, EPI_ISL_983125, EPI_ISL_983126, EPI_ISL_983127, EPI_ISL_983128, EPI_ISL_983129, EPI_ISL_983130, EPI_ISL_983131 | | | |
| see above | THE MARY IMOGENE BASSETT HOSPITAL | Wadsworth Center, New York State Department of Health | Kirsten St. George, Daryl M. Lamson, Alexis Russel, Matthew Shudt, Melissa A Leisner, Jonathan Plitnick, Navjot Singh, John Kelly, Erasmus Schneider, Erica Lasek-Nesselquist |
| EPI_ISL_983132, EPI_ISL_983133, EPI_ISL_983134, EPI_ISL_983135, EPI_ISL_983136, EPI_ISL_983137, EPI_ISL_983138, EPI_ISL_983139, EPI_ISL_983140, EPI_ISL_983141, EPI_ISL_983142, EPI_ISL_983143, EPI_ISL_983144, EPI_ISL_983145, EPI_ISL_983146, EPI_ISL_983147, EPI_ISL_983148, EPI_ISL_983149, EPI_ISL_983150, EPI_ISL_983151, EPI_ISL_983152, EPI_ISL_983153, EPI_ISL_983154, EPI_ISL_983155, EPI_ISL_983156, EPI_ISL_983157, EPI_ISL_983158, EPI_ISL_983159, EPI_ISL_983160, EPI_ISL_983161, EPI_ISL_983162, EPI_ISL_983163, EPI_ISL_983164, EPI_ISL_983165, EPI_ISL_983166, EPI_ISL_983167, EPI_ISL_983168, EPI_ISL_983169, EPI_ISL_983170 | | | |
| see above | MONTEFIORE MEDICAL CENTER LABORATORIES | Wadsworth Center, New York State Department of Health | Kirsten St. George, Daryl M. Lamson, Alexis Russel, Matthew Shudt, Melissa A Leisner, Jonathan Plitnick, Navjot Singh, John Kelly, Erasmus Schneider, Erica Lasek-Nesselquist |
| EPI_ISL_983240, EPI_ISL_983241, EPI_ISL_983242 | KALEIDA CENTER FOR LABORATORY MEDICINE | Wadsworth Center, New York State Department of Health | Kirsten St. George, Daryl M. Lamson, Alexis Russel, Matthew Shudt, Melissa A Leisner, Jonathan Plitnick, Navjot Singh, John Kelly, Erasmus Schneider, Erica Lasek-Nesselquist |
| EPI_ISL_983243 | SUNY UPSTATE MEDICAL UNIVERSITY | Wadsworth Center, New York State Department of Health | Kirsten St. George, Daryl M. Lamson, Alexis Russel, Matthew Shudt, Melissa A Leisner, Jonathan Plitnick, Navjot Singh, John Kelly, Erasmus Schneider, Erica Lasek-Nesselquist |
| EPI_ISL_983244 | KALEIDA CENTER FOR LABORATORY MEDICINE | Wadsworth Center, New York State Department of Health | Kirsten St. George, Daryl M. Lamson, Alexis Russel, Matthew Shudt, Melissa A Leisner, Jonathan Plitnick, Navjot Singh, John Kelly, Erasmus Schneider, Erica Lasek-Nesselquist |
| EPI_ISL_983245 | SUNY UPSTATE MEDICAL UNIVERSITY | Wadsworth Center, New York State Department of Health | Kirsten St. George, Daryl M. Lamson, Alexis Russel, Matthew Shudt, Melissa A Leisner, Jonathan Plitnick, Navjot Singh, John Kelly, Erasmus Schneider, Erica Lasek-Nesselquist |
| EPI_ISL_983246, EPI_ISL_983247 | KALEIDA CENTER FOR LABORATORY MEDICINE | Wadsworth Center, New York State Department of Health | Kirsten St. George, Daryl M. Lamson, Alexis Russel, Matthew Shudt, Melissa A Leisner, Jonathan Plitnick, Navjot Singh, John Kelly, Erasmus Schneider, Erica Lasek-Nesselquist |
| EPI_ISL_983248, EPI_ISL_983249 | SUNY UPSTATE MEDICAL UNIVERSITY | Wadsworth Center, New York State Department of Health | Kirsten St. George, Daryl M. Lamson, Alexis Russel, Matthew Shudt, Melissa A Leisner, Jonathan Plitnick, Navjot Singh, John Kelly, Erasmus Schneider, Erica Lasek-Nesselquist |
| EPI_ISL_983250, EPI_ISL_983251, EPI_ISL_983252, EPI_ISL_983253, EPI_ISL_983254, EPI_ISL_983255, EPI_ISL_983256, EPI_ISL_983257, EPI_ISL_983258, EPI_ISL_983259, EPI_ISL_983260, EPI_ISL_983261, EPI_ISL_983262, EPI_ISL_983263, EPI_ISL_983264, EPI_ISL_983265, EPI_ISL_983266, EPI_ISL_983267, EPI_ISL_983268, EPI_ISL_983269, EPI_ISL_983270, EPI_ISL_983271, EPI_ISL_983272, EPI_ISL_983273, EPI_ISL_983274, EPI_ISL_983275, EPI_ISL_983276, EPI_ISL_983277, EPI_ISL_983278, EPI_ISL_983279, EPI_ISL_983280, EPI_ISL_983281, EPI_ISL_983282 | | | |
| see above | KALEIDA CENTER FOR LABORATORY MEDICINE | Wadsworth Center, New York State Department of Health | Kirsten St. George, Daryl M. Lamson, Alexis Russel, Matthew Shudt, Melissa A Leisner, Jonathan Plitnick, Navjot Singh, John Kelly, Erasmus Schneider, Erica Lasek-Nesselquist |
| EPI_ISL_983283 | SUNY UPSTATE MEDICAL UNIVERSITY | Wadsworth Center, New York State Department of Health | Kirsten St. George, Daryl M. Lamson, Alexis Russel, Matthew Shudt, Melissa A Leisner, Jonathan Plitnick, Navjot Singh, John Kelly, Erasmus Schneider, Erica Lasek-Nesselquist |
| EPI_ISL_983284, EPI_ISL_983285, EPI_ISL_983286, EPI_ISL_983287, EPI_ISL_983288 | KALEIDA CENTER FOR LABORATORY MEDICINE | Wadsworth Center, New York State Department of Health | Kirsten St. George, Daryl M. Lamson, Alexis Russel, Matthew Shudt, Melissa A Leisner, Jonathan Plitnick, Navjot Singh, John Kelly, Erasmus Schneider, Erica Lasek-Nesselquist |
| EPI_ISL_983289, EPI_ISL_983290, EPI_ISL_983291, EPI_ISL_983292, EPI_ISL_983293, EPI_ISL_983294, EPI_ISL_983295 | SUNY UPSTATE MEDICAL UNIVERSITY | Wadsworth Center, New York State Department of Health | Kirsten St. George, Daryl M. Lamson, Alexis Russel, Matthew Shudt, Melissa A Leisner, Jonathan Plitnick, Navjot Singh, John Kelly, Erasmus Schneider, Erica Lasek-Nesselquist |
| EPI_ISL_983296, EPI_ISL_983297, EPI_ISL_983298, EPI_ISL_983299 | KALEIDA CENTER FOR LABORATORY MEDICINE | Wadsworth Center, New York State Department of Health | Kirsten St. George, Daryl M. Lamson, Alexis Russel, Matthew Shudt, Melissa A Leisner, Jonathan Plitnick, Navjot Singh, John Kelly, Erasmus Schneider, Erica Lasek-Nesselquist |
| EPI_ISL_983300, EPI_ISL_983301, EPI_ISL_983302, EPI_ISL_983303, EPI_ISL_983304, EPI_ISL_983305, EPI_ISL_983306, EPI_ISL_983307, EPI_ISL_983308, EPI_ISL_983309, EPI_ISL_983310, EPI_ISL_983311, EPI_ISL_983312, EPI_ISL_983313, EPI_ISL_983314, EPI_ISL_983315, EPI_ISL_983316, EPI_ISL_983317, EPI_ISL_983318, EPI_ISL_983319 | | | |
| see above | SUNY UPSTATE MEDICAL UNIVERSITY | Wadsworth Center, New York State Department of Health | Kirsten St. George, Daryl M. Lamson, Alexis Russel, Matthew Shudt, Melissa A Leisner, Jonathan Plitnick, Navjot Singh, John Kelly, Erasmus Schneider, Erica Lasek-Nesselquist |
| EPI_ISL_983331, EPI_ISL_983332, EPI_ISL_983333, EPI_ISL_983334, EPI_ISL_983335, EPI_ISL_983336, EPI_ISL_983337, EPI_ISL_983338, EPI_ISL_983339, EPI_ISL_983340, EPI_ISL_983341, EPI_ISL_983342, EPI_ISL_983343 | | | |
| see above | Pathology and Laboratory Medicine Institute, Cleveland Clinic, Ohio, USA | Pathology and Laboratory Medicine Institute, Cleveland Clinic, Ohio, USA | Frank P. Esper, Yu-Wei Cheng, Thamali M. Adhikari, Zheng Jin Tu, Daniel H. Farkas, Gary Procop, Jennifer Ko, Timothy A. Chan, Jing Li, Brian P. Rubin |
| EPI_ISL_983357, EPI_ISL_983358, EPI_ISL_983360, EPI_ISL_983361, EPI_ISL_983362, EPI_ISL_983363, EPI_ISL_983364, EPI_ISL_983365, EPI_ISL_983366, EPI_ISL_983367 | Kansas Health and Environmental Lab | Kansas Health and Environmental Lab | Mike Grose, Carissa Robertson, Ben Olsen, and Phil Adam |
| EPI_ISL_983371, EPI_ISL_983372, EPI_ISL_983373, EPI_ISL_983374, EPI_ISL_983375, EPI_ISL_983376, EPI_ISL_983377, EPI_ISL_983378, EPI_ISL_983379, EPI_ISL_983380, EPI_ISL_983381, EPI_ISL_983382, EPI_ISL_983383, EPI_ISL_983384, EPI_ISL_983385, EPI_ISL_983386, EPI_ISL_983387, EPI_ISL_983388, EPI_ISL_983389, EPI_ISL_983390, EPI_ISL_983391, EPI_ISL_983392, EPI_ISL_983393, EPI_ISL_983394, EPI_ISL_983395, EPI_ISL_983396, EPI_ISL_983397, EPI_ISL_983398, EPI_ISL_983399, EPI_ISL_983400, EPI_ISL_983401, EPI_ISL_983402, EPI_ISL_983403, EPI_ISL_983404, EPI_ISL_983405, EPI_ISL_983406, EPI_ISL_983407, EPI_ISL_983408 | | | |
| see above | Utah Public Health Laboratory | Utah Public Health Laboratory | Erin L. Young, Kelly F. Oakeson, Tara Gallagher |
| EPI_ISL_983438, EPI_ISL_983439, | URMC LABS | Wadsworth Center, New York State Department of Health | Kirsten St. George, Daryl M. Lamson, Alexis Russel, Matthew Shudt, Melissa A Leisner, Jonathan Plitnick, Navjot Singh, John Kelly, Erasmus Schneider, Erica Lasek-Nesselquist |

| | | | |
|---|---|---|---|
| EPI_ISL_983440, EPI_ISL_983441, EPI_ISL_983442, EPI_ISL_983443, EPI_ISL_983444 | | | |
| EPI_ISL_983445, EPI_ISL_983446 | THE MARY IMOGENE BASSETT HOSPITAL | Wadsworth Center, New York State Department of Health | Kirsten St. George, Daryl M. Lamson, Alexis Russel, Matthew Shudt, Melissa A Leisner, Jonathan Plitnick, Navjot Singh, John Kelly, Erasmus Schneider, Erica Lasek-Nesselquist |
| EPI_ISL_983447, EPI_ISL_983448, EPI_ISL_983449, EPI_ISL_983450, EPI_ISL_983451, EPI_ISL_983452, EPI_ISL_983453, EPI_ISL_983454, EPI_ISL_983455, EPI_ISL_983456, EPI_ISL_983457, EPI_ISL_983458, EPI_ISL_983459, EPI_ISL_983460, EPI_ISL_983461, EPI_ISL_983462, EPI_ISL_983463, EPI_ISL_983464, EPI_ISL_983465, EPI_ISL_983466, EPI_ISL_983467, EPI_ISL_983468, EPI_ISL_983469, EPI_ISL_983470, EPI_ISL_983471, EPI_ISL_983472, EPI_ISL_983473, EPI_ISL_983474, EPI_ISL_983475 | | | |
| see above | URMC LABS | Wadsworth Center, New York State Department of Health | Kirsten St. George, Daryl M. Lamson, Alexis Russel, Matthew Shudt, Melissa A Leisner, Jonathan Plitnick, Navjot Singh, John Kelly, Erasmus Schneider, Erica Lasek-Nesselquist |
| EPI_ISL_983476 | Wadsworth Center, New York State Department of Health | Wadsworth Center, New York State Department of Health | Kirsten St. George, Daryl M. Lamson, Alexis Russel, Matthew Shudt, Melissa A Leisner, Jonathan Plitnick, Navjot Singh, John Kelly, Erasmus Schneider, Erica Lasek-Nesselquist |
| EPI_ISL_983477 | THE MARY IMOGENE BASSETT HOSPITAL | Wadsworth Center, New York State Department of Health | Kirsten St. George, Daryl M. Lamson, Alexis Russel, Matthew Shudt, Melissa A Leisner, Jonathan Plitnick, Navjot Singh, John Kelly, Erasmus Schneider, Erica Lasek-Nesselquist |
| EPI_ISL_983478, EPI_ISL_983479, EPI_ISL_983480, EPI_ISL_983481, EPI_ISL_983482, EPI_ISL_983483, EPI_ISL_983484 | URMC LABS | Wadsworth Center, New York State Department of Health | Kirsten St. George, Daryl M. Lamson, Alexis Russel, Matthew Shudt, Melissa A Leisner, Jonathan Plitnick, Navjot Singh, John Kelly, Erasmus Schneider, Erica Lasek-Nesselquist |
| EPI_ISL_983485, EPI_ISL_983486, EPI_ISL_983487, EPI_ISL_983488, EPI_ISL_983489, EPI_ISL_983490, EPI_ISL_983491, EPI_ISL_983492 | THE MARY IMOGENE BASSETT HOSPITAL | Wadsworth Center, New York State Department of Health | Kirsten St. George, Daryl M. Lamson, Alexis Russel, Matthew Shudt, Melissa A Leisner, Jonathan Plitnick, Navjot Singh, John Kelly, Erasmus Schneider, Erica Lasek-Nesselquist |
| EPI_ISL_983494, EPI_ISL_983495, EPI_ISL_983496, EPI_ISL_983497, EPI_ISL_983498, EPI_ISL_983499 | Kansas Health and Environmental Lab | Kansas Health and Environmental Lab | Mike Grose, Paige Drury, Carissa Robertson, Ben Olsen, and Phil Adam |
| EPI_ISL_983588, EPI_ISL_983590, EPI_ISL_983591, EPI_ISL_983592, EPI_ISL_983593, EPI_ISL_983594, EPI_ISL_983595, EPI_ISL_983598, EPI_ISL_983600, EPI_ISL_983601, EPI_ISL_983603, EPI_ISL_983604, EPI_ISL_983605, EPI_ISL_983606, EPI_ISL_983607, EPI_ISL_983608, EPI_ISL_983609, EPI_ISL_983613, EPI_ISL_983614, EPI_ISL_983615, EPI_ISL_983617, EPI_ISL_983619, EPI_ISL_983620, EPI_ISL_983621, EPI_ISL_983622 | | | |
| see above | Texas Department of State Health Services | Texas Department of State Health Services | Bonnie Oh, Anita Pokharel, James Daniel Bonser, Myong Koag, Chung Wang, Rachel Lee, Grace Kubin, Rashmi Tuladhar, Mayela Pedrueza, Maliha Rahman, Jenny Zhang |
| EPI_ISL_983632, EPI_ISL_983633, EPI_ISL_983634, EPI_ISL_983635, EPI_ISL_983636, EPI_ISL_983637, EPI_ISL_983638, EPI_ISL_983639, EPI_ISL_983640, EPI_ISL_983641, EPI_ISL_983642, EPI_ISL_983643, EPI_ISL_983644, EPI_ISL_983645, EPI_ISL_983646, EPI_ISL_983647 | | | |
| see above | University of Michigan Clinical Microbiology Laboratory | Lauring Lab, University of Michigan, Department of Microbiology and Immunology | Valesano |
| EPI_ISL_983648 | North Carolina State Laboratory of Public Health | North Carolina State Laboratory of Public Health | Chase K, Miller MC, Greene S, Glover W |
| EPI_ISL_983649, EPI_ISL_983650, EPI_ISL_983651, EPI_ISL_983652, EPI_ISL_983653, EPI_ISL_983654, EPI_ISL_983655, EPI_ISL_983656, EPI_ISL_983657, EPI_ISL_983658, EPI_ISL_983659, EPI_ISL_983660, EPI_ISL_983661, EPI_ISL_983662, EPI_ISL_983663, EPI_ISL_983664, EPI_ISL_983665, EPI_ISL_983666, EPI_ISL_983667, EPI_ISL_983668, EPI_ISL_983669, EPI_ISL_983670, EPI_ISL_983671, EPI_ISL_983672, EPI_ISL_983673, EPI_ISL_983674, EPI_ISL_983675, EPI_ISL_983676, EPI_ISL_983677, EPI_ISL_983678, EPI_ISL_983679, EPI_ISL_983680, EPI_ISL_983681, EPI_ISL_983682, EPI_ISL_983683, EPI_ISL_983684, EPI_ISL_983685, EPI_ISL_983686, EPI_ISL_983687, EPI_ISL_983688, EPI_ISL_983689, EPI_ISL_983690, EPI_ISL_983691, EPI_ISL_983692, EPI_ISL_983693, EPI_ISL_983694, EPI_ISL_983695, EPI_ISL_983696, EPI_ISL_983697, EPI_ISL_983698 | | | |
| see above | Vault Health | Minnesota Department of Health, Public Health Laboratory | Alexandra Lorentz, Jacob Garfin, Matt Plumb, and Xiong Wang |
| EPI_ISL_983699, EPI_ISL_983700, EPI_ISL_983701, EPI_ISL_983702, EPI_ISL_983703, EPI_ISL_983704 | Essentia Health-St. Mary's Medical Center | Minnesota Department of Health, Public Health Laboratory | Alexandra Lorentz, Jacob Garfin, Matt Plumb, and Xiong Wang |
| EPI_ISL_983705, EPI_ISL_983706, EPI_ISL_983707, EPI_ISL_983708, EPI_ISL_983709, EPI_ISL_983710, EPI_ISL_983711, EPI_ISL_983712, EPI_ISL_983713, EPI_ISL_983714, EPI_ISL_983715, EPI_ISL_983716, EPI_ISL_983717, EPI_ISL_983718, EPI_ISL_983719, EPI_ISL_983720, EPI_ISL_983721, EPI_ISL_983722, EPI_ISL_983723, EPI_ISL_983724, EPI_ISL_983725, EPI_ISL_983726, EPI_ISL_983727, EPI_ISL_983728, EPI_ISL_983729, EPI_ISL_983730, EPI_ISL_983736, EPI_ISL_983737, EPI_ISL_983738, EPI_ISL_983739, EPI_ISL_983740, EPI_ISL_983741, EPI_ISL_983742, EPI_ISL_983743, EPI_ISL_983744, EPI_ISL_983745, EPI_ISL_983746, EPI_ISL_983747, EPI_ISL_983748, EPI_ISL_983749, EPI_ISL_983750, EPI_ISL_983751, EPI_ISL_983752, EPI_ISL_983753, EPI_ISL_983754, EPI_ISL_983755, EPI_ISL_983756, EPI_ISL_983757, EPI_ISL_983758, EPI_ISL_983759, EPI_ISL_983760, EPI_ISL_983761, EPI_ISL_983762, EPI_ISL_983763, EPI_ISL_983764, EPI_ISL_983765, EPI_ISL_983766, EPI_ISL_983767, EPI_ISL_983768, EPI_ISL_983769, EPI_ISL_983770, EPI_ISL_983771, EPI_ISL_983772, EPI_ISL_983773, EPI_ISL_983774, EPI_ISL_983775, EPI_ISL_983776, EPI_ISL_983777, EPI_ISL_983778, EPI_ISL_983779, EPI_ISL_983780, EPI_ISL_983781, EPI_ISL_983782, EPI_ISL_983783, EPI_ISL_983784, EPI_ISL_983785, EPI_ISL_983786, EPI_ISL_983787, EPI_ISL_983788, EPI_ISL_983789, EPI_ISL_983790, EPI_ISL_983791, EPI_ISL_983792, EPI_ISL_983793, EPI_ISL_983794, EPI_ISL_983795, EPI_ISL_983796, EPI_ISL_983797, EPI_ISL_983798, EPI_ISL_983799, EPI_ISL_983800, EPI_ISL_983801, EPI_ISL_983802, EPI_ISL_983803, EPI_ISL_983804, EPI_ISL_983805, EPI_ISL_983806, EPI_ISL_983807, EPI_ISL_983808, EPI_ISL_983809, EPI_ISL_983810, EPI_ISL_983811, EPI_ISL_983812, EPI_ISL_983813, EPI_ISL_983814, EPI_ISL_983815, EPI_ISL_983816, EPI_ISL_983817, EPI_ISL_983818, EPI_ISL_983819, EPI_ISL_983820, EPI_ISL_983821, EPI_ISL_983822, EPI_ISL_983823, EPI_ISL_983824, EPI_ISL_983825, EPI_ISL_983826, EPI_ISL_983827, EPI_ISL_983828, EPI_ISL_983829, EPI_ISL_983830, EPI_ISL_983831, EPI_ISL_983832, EPI_ISL_983833, EPI_ISL_983834, EPI_ISL_983835, EPI_ISL_983836, EPI_ISL_983837, EPI_ISL_983838, EPI_ISL_983839, EPI_ISL_983840, EPI_ISL_983841, EPI_ISL_983842, EPI_ISL_983843, EPI_ISL_983844, EPI_ISL_983845, EPI_ISL_983846, EPI_ISL_983847, EPI_ISL_983848, EPI_ISL_983849, EPI_ISL_983850, EPI_ISL_983851, EPI_ISL_983852, EPI_ISL_983853, EPI_ISL_983854, EPI_ISL_983855, EPI_ISL_983856, EPI_ISL_983857, EPI_ISL_983858, EPI_ISL_983859, EPI_ISL_983860, EPI_ISL_983861, EPI_ISL_983862 | | | |
| see above | Colorado Department of Public Health and Environment | Colorado Department of Puplic Health and Environment | Laura Bankers, Molly C. Hetherington-Rauth, Diana Ir, Shannon Ely, Shannon R. Matzinger, Sarah Elizabeth Totten, Emily A. Travanty |
| EPI_ISL_983870 | TwinStrand Biosciences, Inc. | TwinStrand Biosciences, Inc. | Tan Li, Alexandra Golubeva, Lindsey N Williams, Daniel D Sommer, Charles C Valentine III |
| EPI_ISL_983871, EPI_ISL_983872, EPI_ISL_983873, EPI_ISL_983874, EPI_ISL_983875, EPI_ISL_983876, EPI_ISL_983877, EPI_ISL_983878, EPI_ISL_983879, EPI_ISL_983880, EPI_ISL_983881, EPI_ISL_983882, EPI_ISL_983883, EPI_ISL_983884, EPI_ISL_983885, EPI_ISL_983886, EPI_ISL_983887, EPI_ISL_983888, EPI_ISL_983889, EPI_ISL_983890, EPI_ISL_983891, EPI_ISL_983892, EPI_ISL_983893, EPI_ISL_983894, EPI_ISL_983895, EPI_ISL_983896, EPI_ISL_983897, EPI_ISL_983898, EPI_ISL_983899, EPI_ISL_983900, EPI_ISL_983901, EPI_ISL_983902, EPI_ISL_983903, EPI_ISL_983904, EPI_ISL_983905, EPI_ISL_983906, EPI_ISL_983907, EPI_ISL_983908, EPI_ISL_983909, EPI_ISL_983910, EPI_ISL_983911, EPI_ISL_983912, EPI_ISL_983913, EPI_ISL_983914, EPI_ISL_983915, EPI_ISL_983916, EPI_ISL_983917, EPI_ISL_983918, EPI_ISL_983919, EPI_ISL_983920, EPI_ISL_983921, EPI_ISL_983922, EPI_ISL_983923, EPI_ISL_983924, EPI_ISL_983925, EPI_ISL_983926, EPI_ISL_983927, EPI_ISL_983928, EPI_ISL_983929, EPI_ISL_983930, EPI_ISL_983931, EPI_ISL_983932, EPI_ISL_983933, EPI_ISL_983934, EPI_ISL_983935, EPI_ISL_983936, EPI_ISL_983937, EPI_ISL_983938, EPI_ISL_983939, EPI_ISL_983940, EPI_ISL_983941, EPI_ISL_983942, EPI_ISL_983943, EPI_ISL_983944, EPI_ISL_983945, EPI_ISL_983946, EPI_ISL_983947, EPI_ISL_983948, EPI_ISL_983949, EPI_ISL_983950, EPI_ISL_983951, EPI_ISL_983952, EPI_ISL_983953, EPI_ISL_983954, EPI_ISL_983955, EPI_ISL_983956, EPI_ISL_983957, EPI_ISL_983958, EPI_ISL_983959, EPI_ISL_983960, EPI_ISL_983961, EPI_ISL_983962, EPI_ISL_983963, EPI_ISL_983964, EPI_ISL_983965, EPI_ISL_983966, EPI_ISL_983967, EPI_ISL_983968, EPI_ISL_983969, EPI_ISL_983970, EPI_ISL_983971, EPI_ISL_983972, EPI_ISL_983973, EPI_ISL_983974, EPI_ISL_983975, EPI_ISL_983976, EPI_ISL_983977, EPI_ISL_983978, EPI_ISL_983979, EPI_ISL_983980, EPI_ISL_983981, EPI_ISL_983982, EPI_ISL_983983, EPI_ISL_983984, EPI_ISL_983985, EPI_ISL_983986, EPI_ISL_983987, EPI_ISL_983988, EPI_ISL_983989, EPI_ISL_983990, EPI_ISL_983991, EPI_ISL_983992, EPI_ISL_983993, EPI_ISL_983994, EPI_ISL_983995, EPI_ISL_983996, EPI_ISL_983997, EPI_ISL_983998, EPI_ISL_983999, EPI_ISL_984000, EPI_ISL_984001, EPI_ISL_984002, EPI_ISL_984003, EPI_ISL_984004, EPI_ISL_984005, EPI_ISL_984006, EPI_ISL_984007, EPI_ISL_984008, EPI_ISL_984009, EPI_ISL_984010, EPI_ISL_984011, EPI_ISL_984012, EPI_ISL_984013, EPI_ISL_984014, EPI_ISL_984015, EPI_ISL_984016, EPI_ISL_984017, EPI_ISL_984018, EPI_ISL_984019, EPI_ISL_984020, EPI_ISL_984021, EPI_ISL_984022, EPI_ISL_984023, EPI_ISL_984024, EPI_ISL_984025, EPI_ISL_984026, EPI_ISL_984027, EPI_ISL_984028, EPI_ISL_984029, EPI_ISL_984030, EPI_ISL_984031, EPI_ISL_984032, EPI_ISL_984033, EPI_ISL_984034, EPI_ISL_984035, EPI_ISL_984036, EPI_ISL_984037, EPI_ISL_984038, EPI_ISL_984039, EPI_ISL_984040, EPI_ISL_984041, EPI_ISL_984042, EPI_ISL_984043, EPI_ISL_984044, EPI_ISL_984045, EPI_ISL_984046, EPI_ISL_984047, EPI_ISL_984048, EPI_ISL_984049, EPI_ISL_984050, EPI_ISL_984051, EPI_ISL_984052, EPI_ISL_984053, EPI_ISL_984054, EPI_ISL_984055, EPI_ISL_984056, EPI_ISL_984057, EPI_ISL_984058, EPI_ISL_984059, EPI_ISL_984060, EPI_ISL_984061, EPI_ISL_984062, EPI_ISL_984063, EPI_ISL_984064, EPI_ISL_984065, EPI_ISL_984066, EPI_ISL_984067, EPI_ISL_984068, EPI_ISL_984069, EPI_ISL_984070, EPI_ISL_984071, EPI_ISL_984072, EPI_ISL_984073, EPI_ISL_984074, EPI_ISL_984075, EPI_ISL_984076, EPI_ISL_984077, EPI_ISL_984078, EPI_ISL_984079, EPI_ISL_984080, EPI_ISL_984081, EPI_ISL_984082, EPI_ISL_984083, EPI_ISL_984084, EPI_ISL_984085, EPI_ISL_984086, EPI_ISL_984087, EPI_ISL_984088, EPI_ISL_984089, EPI_ISL_984090, EPI_ISL_984091, EPI_ISL_984092, EPI_ISL_984093, EPI_ISL_984094, EPI_ISL_984095, EPI_ISL_984096, EPI_ISL_984097, EPI_ISL_984098, EPI_ISL_984099, EPI_ISL_984100, EPI_ISL_984101, EPI_ISL_984102, EPI_ISL_984103, EPI_ISL_984104, EPI_ISL_984105, EPI_ISL_984106, EPI_ISL_984107, EPI_ISL_984108, EPI_ISL_984109, EPI_ISL_984110, EPI_ISL_984111, EPI_ISL_984112, EPI_ISL_984113, EPI_ISL_984114, EPI_ISL_984115, EPI_ISL_984116, EPI_ISL_984117, EPI_ISL_984118, EPI_ISL_984119, EPI_ISL_984120, EPI_ISL_984121, EPI_ISL_984122, EPI_ISL_984123, EPI_ISL_984124, EPI_ISL_984125, EPI_ISL_984126, EPI_ISL_984127, EPI_ISL_984128, EPI_ISL_984129, EPI_ISL_984130, EPI_ISL_984131, EPI_ISL_984132, EPI_ISL_984133, EPI_ISL_984134, EPI_ISL_984135, EPI_ISL_984136, EPI_ISL_984137, EPI_ISL_984138, EPI_ISL_984139, EPI_ISL_984140, EPI_ISL_984141, EPI_ISL_984142, EPI_ISL_984143, EPI_ISL_984144, EPI_ISL_984145, EPI_ISL_984146, EPI_ISL_984147, EPI_ISL_984148, EPI_ISL_984149, EPI_ISL_984150, EPI_ISL_984151, EPI_ISL_984152, EPI_ISL_984153, EPI_ISL_984154, EPI_ISL_984155, EPI_ISL_984156, EPI_ISL_984157, EPI_ISL_984158, EPI_ISL_984159, EPI_ISL_984160, EPI_ISL_984161, EPI_ISL_984162, EPI_ISL_984163, EPI_ISL_984164, EPI_ISL_984165, EPI_ISL_984166, EPI_ISL_984167, EPI_ISL_984168, EPI_ISL_984169, EPI_ISL_984170, EPI_ISL_984171, EPI_ISL_984172, EPI_ISL_984173, EPI_ISL_984174, EPI_ISL_984175, EPI_ISL_984176, EPI_ISL_984177, EPI_ISL_984178, EPI_ISL_984179, EPI_ISL_984180, EPI_ISL_984181, EPI_ISL_984182, EPI_ISL_984183, EPI_ISL_984184, EPI_ISL_984185, EPI_ISL_984186, EPI_ISL_984187, EPI_ISL_984188, EPI_ISL_984189, EPI_ISL_984190, EPI_ISL_984191, EPI_ISL_984192, EPI_ISL_984193, EPI_ISL_984194, EPI_ISL_984195, EPI_ISL_984196, EPI_ISL_984197, EPI_ISL_984198, EPI_ISL_984199, EPI_ISL_984200, EPI_ISL_984201, EPI_ISL_984202, EPI_ISL_984203, EPI_ISL_984204, EPI_ISL_984205, EPI_ISL_984206, EPI_ISL_984207, EPI_ISL_984208, EPI_ISL_984209, EPI_ISL_984210, EPI_ISL_984211, EPI_ISL_984212, EPI_ISL_984213, EPI_ISL_984214, EPI_ISL_984215, EPI_ISL_984216, EPI_ISL_984217, EPI_ISL_984218, EPI_ISL_984219, EPI_ISL_984220, EPI_ISL_984221, EPI_ISL_984222, EPI_ISL_984223, EPI_ISL_984224, EPI_ISL_984225, EPI_ISL_984226, EPI_ISL_984227, EPI_ISL_984228, EPI_ISL_984229, EPI_ISL_984230, EPI_ISL_984231, EPI_ISL_984232, EPI_ISL_984233, EPI_ISL_984234, EPI_ISL_984235, EPI_ISL_984236, EPI_ISL_984237, EPI_ISL_984238, EPI_ISL_984239, EPI_ISL_984240, EPI_ISL_984241 | | | |
| see above | Wisconsin State Laboratory of Hygiene Communicable Disease Division | Wisconsin State Laboratory of Hygiene Communicable Disease Division | Kelsey R. Florek, Abigail C. Shockey |
| EPI_ISL_984298, EPI_ISL_984299, EPI_ISL_984300 | Tampa General Hospital Esoteric Lab | Tampa General Hospital Esoteric Research & Development Lab | Grant Vestal, Deanna Becker, Dominic Uy, Amorce Lima, Suzane Silbert |
| EPI_ISL_984310, EPI_ISL_984311, EPI_ISL_984312, EPI_ISL_984314 | NCSLPH | NCSLPH | Chase K, Miller MC, Greene S, Glover W |
| EPI_ISL_984318, EPI_ISL_984319, EPI_ISL_984320, EPI_ISL_984321, EPI_ISL_984322, EPI_ISL_984323, EPI_ISL_984324, EPI_ISL_984325, EPI_ISL_984326, EPI_ISL_984327, EPI_ISL_984328, EPI_ISL_984329, EPI_ISL_984330, EPI_ISL_984331, EPI_ISL_984332, EPI_ISL_984333, EPI_ISL_984334, EPI_ISL_984335, EPI_ISL_984336, EPI_ISL_984337, EPI_ISL_984338, EPI_ISL_984339, EPI_ISL_984340, EPI_ISL_984341, EPI_ISL_984342, EPI_ISL_984343, EPI_ISL_984344, EPI_ISL_984345, EPI_ISL_984346, EPI_ISL_984347, EPI_ISL_984348, EPI_ISL_984349, EPI_ISL_984350, EPI_ISL_984351, EPI_ISL_984352, EPI_ISL_984353, EPI_ISL_984355, EPI_ISL_984356, EPI_ISL_984357, EPI_ISL_984358, EPI_ISL_984359, EPI_ISL_984360, EPI_ISL_984361, EPI_ISL_984362, EPI_ISL_984364, EPI_ISL_984365, EPI_ISL_984366, EPI_ISL_984367, EPI_ISL_984368, EPI_ISL_984369, EPI_ISL_984371, EPI_ISL_984372, EPI_ISL_984373, EPI_ISL_984374, EPI_ISL_984375, EPI_ISL_984376, EPI_ISL_984377, EPI_ISL_984378, EPI_ISL_984379, EPI_ISL_984380, EPI_ISL_984381, EPI_ISL_984382, EPI_ISL_984383, EPI_ISL_984384, EPI_ISL_984385, EPI_ISL_984386, EPI_ISL_984387, EPI_ISL_984388, EPI_ISL_984389, EPI_ISL_984390, EPI_ISL_984436, EPI_ISL_984437, EPI_ISL_984438, EPI_ISL_984439, EPI_ISL_984440, EPI_ISL_984441, EPI_ISL_984442, EPI_ISL_984443, EPI_ISL_984444, EPI_ISL_984445, EPI_ISL_984446, EPI_ISL_984447, EPI_ISL_984448, EPI_ISL_984449, EPI_ISL_984450, EPI_ISL_984451, EPI_ISL_984452, EPI_ISL_984453, EPI_ISL_984454, EPI_ISL_984455, EPI_ISL_984456, EPI_ISL_984457, EPI_ISL_984458, EPI_ISL_984459, EPI_ISL_984460, EPI_ISL_984461, EPI_ISL_984462, EPI_ISL_984463, EPI_ISL_984464, EPI_ISL_984465, EPI_ISL_984467, EPI_ISL_984468, EPI_ISL_984469, EPI_ISL_984470, EPI_ISL_984471, EPI_ISL_984472, EPI_ISL_984473, EPI_ISL_984474, EPI_ISL_984475, EPI_ISL_984476, EPI_ISL_984477, EPI_ISL_984478, EPI_ISL_984479, EPI_ISL_984480, EPI_ISL_984481, EPI_ISL_984482, EPI_ISL_984483, EPI_ISL_984484, EPI_ISL_984485, EPI_ISL_984486, EPI_ISL_984487, EPI_ISL_984488, EPI_ISL_984489, EPI_ISL_984490, EPI_ISL_984491, EPI_ISL_984492, EPI_ISL_984493, EPI_ISL_984494, EPI_ISL_984495, EPI_ISL_984496, EPI_ISL_984497, EPI_ISL_984498, EPI_ISL_984499, EPI_ISL_984500, EPI_ISL_984501, EPI_ISL_984502, EPI_ISL_984503, EPI_ISL_984504, EPI_ISL_984505, EPI_ISL_984506, EPI_ISL_984507, EPI_ISL_984508, EPI_ISL_984509, EPI_ISL_984510, EPI_ISL_984511, EPI_ISL_984512, EPI_ISL_984513, EPI_ISL_984514, EPI_ISL_984515, EPI_ISL_984516, EPI_ISL_984517, EPI_ISL_984518, EPI_ISL_984519, EPI_ISL_984520, EPI_ISL_984521, EPI_ISL_984522, EPI_ISL_984523, EPI_ISL_984524, EPI_ISL_984525, EPI_ISL_984526, EPI_ISL_984527, EPI_ISL_984528, EPI_ISL_984529, EPI_ISL_984530, EPI_ISL_984531, EPI_ISL_984532, EPI_ISL_984533, EPI_ISL_984534, | | | |

| | | | |
|---|---|---|---|
| EPI_ISL_984535, EPI_ISL_984536, EPI_ISL_984537, EPI_ISL_984538, EPI_ISL_984539, EPI_ISL_984540, EPI_ISL_984541, EPI_ISL_984542, EPI_ISL_984543, EPI_ISL_984544, EPI_ISL_984545, EPI_ISL_984546, EPI_ISL_984547, EPI_ISL_984548, EPI_ISL_984549, EPI_ISL_984550, EPI_ISL_984551, EPI_ISL_984552, EPI_ISL_984553, EPI_ISL_984554, EPI_ISL_984555, EPI_ISL_984556, EPI_ISL_984557, EPI_ISL_984558, EPI_ISL_984559, EPI_ISL_984560, EPI_ISL_984561, EPI_ISL_984562, EPI_ISL_984563, EPI_ISL_984564, EPI_ISL_984565, EPI_ISL_984566, EPI_ISL_984567, EPI_ISL_984568, EPI_ISL_984569, EPI_ISL_984570, EPI_ISL_984571, EPI_ISL_984572, EPI_ISL_984573, EPI_ISL_984574, EPI_ISL_984575, EPI_ISL_984576, EPI_ISL_984577, EPI_ISL_984578, EPI_ISL_984579, EPI_ISL_984580, EPI_ISL_984581, EPI_ISL_984582, EPI_ISL_984583, EPI_ISL_984584, EPI_ISL_984585, EPI_ISL_984586, EPI_ISL_984587, EPI_ISL_984588, EPI_ISL_984589, EPI_ISL_984590, EPI_ISL_984591, EPI_ISL_984592, EPI_ISL_984593, EPI_ISL_984594, EPI_ISL_984595, EPI_ISL_984596, EPI_ISL_984597, EPI_ISL_984598, EPI_ISL_984599, EPI_ISL_984600, EPI_ISL_984601, EPI_ISL_984602, EPI_ISL_984603, EPI_ISL_984604, EPI_ISL_984605, EPI_ISL_984606, EPI_ISL_984607, EPI_ISL_984608, EPI_ISL_984609, EPI_ISL_984610, EPI_ISL_984611, EPI_ISL_984612, EPI_ISL_984613 | | | |
| see above | California Department of Public Health | Chiu Laboratory, University of California, San Francisco | Charles Chiu, Xianding (Wayne) Deng, Candace Wang, Venice Servellita, Jill Hacker, Debra Wadford |
| EPI_ISL_984663, EPI_ISL_984664, EPI_ISL_984665, EPI_ISL_984666, EPI_ISL_984667, EPI_ISL_984668, EPI_ISL_984669 | University Hospitals Translational Laboratory (UHTL), University Hospitals | University Hospitals Translational Laboratory (UHTL), University Hospitals | Sadri,N., Alouani,D., Song,X. |
| EPI_ISL_984753 | Fulgent Genetics | Fulgent Genetics | Harry Gao, Mickey Li, John Gao, Joseph Fierro, Benafsh Sapra, Becky Tsai, Yan Meng, Doreen Ng, James Xie |
| EPI_ISL_984756, EPI_ISL_984757, EPI_ISL_984758, EPI_ISL_984759, EPI_ISL_984760, EPI_ISL_984761, EPI_ISL_984762, EPI_ISL_984763, EPI_ISL_984764, EPI_ISL_984765, EPI_ISL_984766, EPI_ISL_984767, EPI_ISL_984768, EPI_ISL_984769, EPI_ISL_984770, EPI_ISL_984771, EPI_ISL_984772, EPI_ISL_984773, EPI_ISL_984774, EPI_ISL_984775, EPI_ISL_984776, EPI_ISL_984777, EPI_ISL_984778, EPI_ISL_984779, EPI_ISL_984780, EPI_ISL_984781, EPI_ISL_984782, EPI_ISL_984783, EPI_ISL_984784, EPI_ISL_984785, EPI_ISL_984786, EPI_ISL_984787, EPI_ISL_984788, EPI_ISL_984789, EPI_ISL_984790, EPI_ISL_984791, EPI_ISL_984792, EPI_ISL_984793, EPI_ISL_984794, EPI_ISL_984795, EPI_ISL_984796, EPI_ISL_984797, EPI_ISL_984798, EPI_ISL_984799, EPI_ISL_984800, EPI_ISL_984801, EPI_ISL_984802, EPI_ISL_984803, EPI_ISL_984804, EPI_ISL_984805, EPI_ISL_984806, EPI_ISL_984807, EPI_ISL_984808, EPI_ISL_984809, EPI_ISL_984810, EPI_ISL_984811, EPI_ISL_984812, EPI_ISL_984813, EPI_ISL_984814, EPI_ISL_984815, EPI_ISL_984816, EPI_ISL_984817, EPI_ISL_984818, EPI_ISL_984819, EPI_ISL_984820, EPI_ISL_984821, EPI_ISL_984822, EPI_ISL_984823, EPI_ISL_984824, EPI_ISL_984825, EPI_ISL_984826, EPI_ISL_984827, EPI_ISL_984828, EPI_ISL_984829, EPI_ISL_984830, EPI_ISL_984831, EPI_ISL_984832, EPI_ISL_984833, EPI_ISL_984834, EPI_ISL_984835, EPI_ISL_984836, EPI_ISL_984837, EPI_ISL_984838, EPI_ISL_984839, EPI_ISL_984840, EPI_ISL_984841, EPI_ISL_984842, EPI_ISL_984843, EPI_ISL_984844, EPI_ISL_984845, EPI_ISL_984846, EPI_ISL_984847, EPI_ISL_984848, EPI_ISL_984849, EPI_ISL_984850, EPI_ISL_984851, EPI_ISL_984852, EPI_ISL_984853, EPI_ISL_984854, EPI_ISL_984855, EPI_ISL_984856, EPI_ISL_984857, EPI_ISL_984858, EPI_ISL_984859, EPI_ISL_984860, EPI_ISL_984861, EPI_ISL_984862, EPI_ISL_984863, EPI_ISL_984864, EPI_ISL_984865, EPI_ISL_984866, EPI_ISL_984867, EPI_ISL_984868, EPI_ISL_984869, EPI_ISL_984870, EPI_ISL_984871, EPI_ISL_984872, EPI_ISL_984873, EPI_ISL_984874, EPI_ISL_984875, EPI_ISL_984876, EPI_ISL_984877, EPI_ISL_984878, EPI_ISL_984879, EPI_ISL_984880, EPI_ISL_984881, EPI_ISL_984882, EPI_ISL_984883, EPI_ISL_984884, EPI_ISL_984885, EPI_ISL_984886, EPI_ISL_984887, EPI_ISL_984888, EPI_ISL_984889, EPI_ISL_984890, EPI_ISL_984891, EPI_ISL_984892, EPI_ISL_984893, EPI_ISL_984894, EPI_ISL_984895, EPI_ISL_984896, EPI_ISL_984897, EPI_ISL_984898, EPI_ISL_984899, EPI_ISL_984900, EPI_ISL_984901, EPI_ISL_984902, EPI_ISL_984903, EPI_ISL_984904, EPI_ISL_984905, EPI_ISL_984906, EPI_ISL_984907, EPI_ISL_984908, EPI_ISL_984909, EPI_ISL_984910, EPI_ISL_984911, EPI_ISL_984912, EPI_ISL_984913, EPI_ISL_984914, EPI_ISL_984915, EPI_ISL_984916, EPI_ISL_984917, EPI_ISL_984918, EPI_ISL_984919, EPI_ISL_984920, EPI_ISL_984921, EPI_ISL_984922, EPI_ISL_984923, EPI_ISL_984924, EPI_ISL_984925, EPI_ISL_984926, EPI_ISL_984927, EPI_ISL_984928, EPI_ISL_984929, EPI_ISL_984930, EPI_ISL_984931, EPI_ISL_984932, EPI_ISL_984933, EPI_ISL_984934, EPI_ISL_984935, EPI_ISL_984936, EPI_ISL_984937, EPI_ISL_984938, EPI_ISL_984939, EPI_ISL_984940, EPI_ISL_984941, EPI_ISL_984942, EPI_ISL_984943, EPI_ISL_984944, EPI_ISL_984945, EPI_ISL_984946, EPI_ISL_984947 | | | |
| see above | Pandemic Response Lab - NYC | Pandemic Response Lab, R&D | Henry Lee, Michael Hammerling, Melissa Hopkins, Cybill del Castillo, William Ward, Pradeep Bugga, Haiping Hao, Jon Laurent |
| EPI_ISL_984948 | Fulgent Genetics | Fulgent Genetics | Harry Gao, Mickey Li, John Gao, Joseph Fierro, Benafsh Sapra, Becky Tsai, Yan Meng, Doreen Ng, James Xie |
| EPI_ISL_985131, EPI_ISL_985134, EPI_ISL_985140 | School of Pharmacy, Shenandoah University | School of Pharmacy, Shenandoah University | Adams,S.M., Harralson,A.F., Kidd,R.S., Sawyer,G.W. |
| EPI_ISL_985149, EPI_ISL_985150, EPI_ISL_985151, EPI_ISL_985152, EPI_ISL_985153, EPI_ISL_985154, EPI_ISL_985155, EPI_ISL_985156, EPI_ISL_985157, EPI_ISL_985158, EPI_ISL_985159, EPI_ISL_985160, EPI_ISL_985161, EPI_ISL_985162, EPI_ISL_985163, EPI_ISL_985164, EPI_ISL_985165, EPI_ISL_985166, EPI_ISL_985167 | | | |
| see above | Instituto Nacional de Medicina Genomica | Instituto Nacional de Medicina Genomica | Hidalgo-Miranda A, Mendoza-Vargas A, Reyes-Grajeda JP, Cisneros-Villanueva M, Cedro-Tanda A,Peñaloza-Figueroa F, Herrera-Montalvo LA |
| EPI_ISL_985236 | University of Wisconsin-Madison AIDS Vaccine Research Laboratories | University of Wisconsin-Madison AIDS Vaccine Research Laboratories | Gage Moreno, Katarina Braun, et al. AIDS Vaccine Research Laboratories |
| EPI_ISL_985239, EPI_ISL_985240, EPI_ISL_985241, EPI_ISL_985242, EPI_ISL_985243, EPI_ISL_985244, EPI_ISL_985245, EPI_ISL_985246, EPI_ISL_985247, EPI_ISL_985248, EPI_ISL_985249, EPI_ISL_985250, EPI_ISL_985251, EPI_ISL_985252, EPI_ISL_985254, EPI_ISL_985255, EPI_ISL_985256, EPI_ISL_985257, EPI_ISL_985258, EPI_ISL_985259, EPI_ISL_985260, EPI_ISL_985261, EPI_ISL_985262, EPI_ISL_985263, EPI_ISL_985264, EPI_ISL_985265, EPI_ISL_985266, EPI_ISL_985267, EPI_ISL_985268, EPI_ISL_985269, EPI_ISL_985270, EPI_ISL_985271, EPI_ISL_985272, EPI_ISL_985273, EPI_ISL_985274, EPI_ISL_985275, EPI_ISL_985276, EPI_ISL_985277, EPI_ISL_985278, EPI_ISL_985279, EPI_ISL_985280, EPI_ISL_985281, EPI_ISL_985282, EPI_ISL_985283, EPI_ISL_985284, EPI_ISL_985285, EPI_ISL_985288, EPI_ISL_985289, EPI_ISL_985290, EPI_ISL_985291, EPI_ISL_985292, EPI_ISL_985293, EPI_ISL_985294, EPI_ISL_985296, EPI_ISL_985297, EPI_ISL_985298, EPI_ISL_985299, EPI_ISL_985300, EPI_ISL_985301, EPI_ISL_985302 | | | |
| see above | Delaware Public Health Laboratory (DPHL) | Delaware Public Health Lab | Gregory Hovan |
| EPI_ISL_994802, EPI_ISL_994803, EPI_ISL_994804, EPI_ISL_994805, EPI_ISL_994806, EPI_ISL_994807, EPI_ISL_994808, EPI_ISL_994809, EPI_ISL_994810, EPI_ISL_994811, EPI_ISL_994812, EPI_ISL_994813, EPI_ISL_994814, EPI_ISL_994815, EPI_ISL_994816, EPI_ISL_994817, EPI_ISL_994818, EPI_ISL_994819, EPI_ISL_994820, EPI_ISL_994821, EPI_ISL_994822, EPI_ISL_994823, EPI_ISL_994824, EPI_ISL_994825, EPI_ISL_994826, EPI_ISL_994827, EPI_ISL_994828, EPI_ISL_994829, EPI_ISL_994830, EPI_ISL_994831, EPI_ISL_994832, EPI_ISL_994833, EPI_ISL_994834, EPI_ISL_994835, EPI_ISL_994836, EPI_ISL_994837, EPI_ISL_994838, EPI_ISL_994839, EPI_ISL_994840, EPI_ISL_994841, EPI_ISL_994842, EPI_ISL_994843, EPI_ISL_994844, EPI_ISL_994845, EPI_ISL_994846, EPI_ISL_994847, EPI_ISL_994848, EPI_ISL_994849, EPI_ISL_994850, EPI_ISL_994851, EPI_ISL_994852, EPI_ISL_994853, EPI_ISL_994854, EPI_ISL_994855, EPI_ISL_994856, EPI_ISL_994857, EPI_ISL_994858, EPI_ISL_994859, EPI_ISL_994860, EPI_ISL_994861, EPI_ISL_994862, EPI_ISL_994863, EPI_ISL_994864, EPI_ISL_994865, EPI_ISL_994866, EPI_ISL_994867, EPI_ISL_994868, EPI_ISL_994869, EPI_ISL_994870, EPI_ISL_994871, EPI_ISL_994872, EPI_ISL_994873, EPI_ISL_994874, EPI_ISL_994875, EPI_ISL_994876, EPI_ISL_994877, EPI_ISL_994878, EPI_ISL_994879, EPI_ISL_994880, EPI_ISL_994881, EPI_ISL_994882, EPI_ISL_994883, EPI_ISL_994884, EPI_ISL_994885, EPI_ISL_994886, EPI_ISL_994887, EPI_ISL_994888, EPI_ISL_994889, EPI_ISL_994890, EPI_ISL_994891, EPI_ISL_994892, EPI_ISL_994893, EPI_ISL_994894, EPI_ISL_994895, EPI_ISL_994896, EPI_ISL_994897, EPI_ISL_994898, EPI_ISL_994899, EPI_ISL_994900, EPI_ISL_994901, EPI_ISL_994902, EPI_ISL_994903, EPI_ISL_994904, EPI_ISL_994905, EPI_ISL_994906, EPI_ISL_994907, EPI_ISL_994908, EPI_ISL_994909, EPI_ISL_994910, EPI_ISL_994911, EPI_ISL_994912, EPI_ISL_994913, EPI_ISL_994914, EPI_ISL_994915, EPI_ISL_994916, EPI_ISL_994917, EPI_ISL_994918, EPI_ISL_994919, EPI_ISL_994920, EPI_ISL_994921, EPI_ISL_994922, EPI_ISL_994923, EPI_ISL_994924, EPI_ISL_994925, EPI_ISL_994926, EPI_ISL_994927, EPI_ISL_994928, EPI_ISL_994929, EPI_ISL_994930, EPI_ISL_994931, EPI_ISL_994932, EPI_ISL_994933, EPI_ISL_994934, EPI_ISL_994935, EPI_ISL_994936, EPI_ISL_994937, EPI_ISL_994938, EPI_ISL_994939, EPI_ISL_994940, EPI_ISL_994941, EPI_ISL_994942, EPI_ISL_994943, EPI_ISL_994944, EPI_ISL_994945, EPI_ISL_994946, EPI_ISL_994947, EPI_ISL_994948, EPI_ISL_994949, EPI_ISL_994950, EPI_ISL_994951, EPI_ISL_994952, EPI_ISL_994953, EPI_ISL_994954, EPI_ISL_994955, EPI_ISL_994956, EPI_ISL_994957, EPI_ISL_994958, EPI_ISL_994959, EPI_ISL_994960, EPI_ISL_994961, EPI_ISL_994962, EPI_ISL_994963, EPI_ISL_994964, EPI_ISL_994965, EPI_ISL_994966, EPI_ISL_994967, EPI_ISL_994968, EPI_ISL_994969, EPI_ISL_994970, EPI_ISL_994971, EPI_ISL_994972, EPI_ISL_994973, EPI_ISL_994974, EPI_ISL_994975, EPI_ISL_994976, EPI_ISL_994977, EPI_ISL_994978, EPI_ISL_994979, EPI_ISL_994980, EPI_ISL_994981, EPI_ISL_994982, EPI_ISL_994983, EPI_ISL_994984, EPI_ISL_994985, EPI_ISL_994986, EPI_ISL_994987, EPI_ISL_994988, EPI_ISL_994989, EPI_ISL_994990, EPI_ISL_994991, EPI_ISL_994992, EPI_ISL_994993, EPI_ISL_994994, EPI_ISL_994995, EPI_ISL_994996, EPI_ISL_994997, EPI_ISL_994998, EPI_ISL_994999, EPI_ISL_995000, EPI_ISL_995001, EPI_ISL_995002, EPI_ISL_995003, EPI_ISL_995004, EPI_ISL_995005, EPI_ISL_995006, EPI_ISL_995007, EPI_ISL_995008, EPI_ISL_995009, EPI_ISL_995010, EPI_ISL_995011, EPI_ISL_995012, EPI_ISL_995013, EPI_ISL_995014, EPI_ISL_995015, EPI_ISL_995016, EPI_ISL_995017, EPI_ISL_995018, EPI_ISL_995019, EPI_ISL_995020, EPI_ISL_995021, EPI_ISL_995022, EPI_ISL_995023, EPI_ISL_995024, EPI_ISL_995025, EPI_ISL_995026, EPI_ISL_995027, EPI_ISL_995028, EPI_ISL_995029, EPI_ISL_995030, EPI_ISL_995031, EPI_ISL_995032, EPI_ISL_995033, EPI_ISL_995034, EPI_ISL_995035, EPI_ISL_995036, EPI_ISL_995037, EPI_ISL_995038, EPI_ISL_995039, EPI_ISL_995040, EPI_ISL_995041, EPI_ISL_995042, EPI_ISL_995043, EPI_ISL_995044, EPI_ISL_995045, EPI_ISL_995046, EPI_ISL_995047, EPI_ISL_995048, EPI_ISL_995049, EPI_ISL_995050, EPI_ISL_995051, EPI_ISL_995052, EPI_ISL_995053, EPI_ISL_995054, EPI_ISL_995055, EPI_ISL_995056, EPI_ISL_995057, EPI_ISL_995058, EPI_ISL_995059, EPI_ISL_995060, EPI_ISL_995061, EPI_ISL_995062, EPI_ISL_995063, EPI_ISL_995064, EPI_ISL_995065, EPI_ISL_995066, EPI_ISL_995067, EPI_ISL_995068, EPI_ISL_995069, EPI_ISL_995070, EPI_ISL_995071, EPI_ISL_995072, EPI_ISL_995073, EPI_ISL_995074, EPI_ISL_995075, EPI_ISL_995076, EPI_ISL_995077, EPI_ISL_995078, EPI_ISL_995079, EPI_ISL_995080, EPI_ISL_995081, EPI_ISL_995082, EPI_ISL_995083, EPI_ISL_995084, EPI_ISL_995085, EPI_ISL_995086, EPI_ISL_995087, EPI_ISL_995088, EPI_ISL_995089, EPI_ISL_995090, EPI_ISL_995091, EPI_ISL_995092, EPI_ISL_995093, EPI_ISL_995094, EPI_ISL_995095, EPI_ISL_995096, EPI_ISL_995097, EPI_ISL_995098, EPI_ISL_995099, EPI_ISL_995100, EPI_ISL_995101, EPI_ISL_995102, EPI_ISL_995103, EPI_ISL_995104, EPI_ISL_995105, EPI_ISL_995106, EPI_ISL_995107, EPI_ISL_995108, EPI_ISL_995109, EPI_ISL_995110, EPI_ISL_995111, EPI_ISL_995112, EPI_ISL_995113, EPI_ISL_995114, EPI_ISL_995115, EPI_ISL_995116, EPI_ISL_995117, EPI_ISL_995118, EPI_ISL_995119, EPI_ISL_995120, EPI_ISL_995121, EPI_ISL_995122, EPI_ISL_995123, EPI_ISL_995124, EPI_ISL_995125, EPI_ISL_995126, EPI_ISL_995127, EPI_ISL_995128, EPI_ISL_995129, EPI_ISL_995130, EPI_ISL_995131, EPI_ISL_995132, EPI_ISL_995133, EPI_ISL_995134, EPI_ISL_995135, EPI_ISL_995136, EPI_ISL_995137, EPI_ISL_995138, EPI_ISL_995139, EPI_ISL_995140, EPI_ISL_995141, EPI_ISL_995142, EPI_ISL_995143, EPI_ISL_995144, EPI_ISL_995145, EPI_ISL_995146, EPI_ISL_995147, EPI_ISL_995148, EPI_ISL_995149, EPI_ISL_995150, EPI_ISL_995151, EPI_ISL_995152, EPI_ISL_995153, EPI_ISL_995154, EPI_ISL_995155, EPI_ISL_995156, EPI_ISL_995157, EPI_ISL_995158, EPI_ISL_995159, EPI_ISL_995160, EPI_ISL_995161, EPI_ISL_995162, EPI_ISL_995163, EPI_ISL_995164, EPI_ISL_995165, EPI_ISL_995166, EPI_ISL_995167, EPI_ISL_995168, EPI_ISL_995169, EPI_ISL_995170, EPI_ISL_995171 | | | |
| see above | Pandemic Response Lab - NYC | Pandemic Response Lab, R&D | Henry Lee, Michael Hammerling, Melissa Hopkins, Cybill del Castillo, William Ward, Pradeep Bugga, Haiping Hao, Jon Laurent |
| EPI_ISL_1027686, EPI_ISL_1027687, EPI_ISL_1027688, EPI_ISL_1027689, EPI_ISL_1027690, EPI_ISL_1027691, EPI_ISL_1027692, EPI_ISL_1027693, EPI_ISL_1027694, EPI_ISL_1027695, EPI_ISL_1027696, EPI_ISL_1027697, EPI_ISL_1027698, EPI_ISL_1027699, EPI_ISL_1027700, EPI_ISL_1027701, EPI_ISL_1027702, EPI_ISL_1027703, EPI_ISL_1027704, EPI_ISL_1027705, EPI_ISL_1027706, EPI_ISL_1027707, EPI_ISL_1027708, EPI_ISL_1027709, EPI_ISL_1027710, EPI_ISL_1027711, EPI_ISL_1027712, EPI_ISL_1027713, EPI_ISL_1027714, EPI_ISL_1027715, EPI_ISL_1027716, EPI_ISL_1027717, EPI_ISL_1027718, EPI_ISL_1027719, EPI_ISL_1027720, EPI_ISL_1027721, EPI_ISL_1027722, EPI_ISL_1027723, EPI_ISL_1027724, EPI_ISL_1027725, EPI_ISL_1027726, EPI_ISL_1027727, EPI_ISL_1027728, EPI_ISL_1027729, EPI_ISL_1027730, EPI_ISL_1027731, EPI_ISL_1027732, EPI_ISL_1027733, EPI_ISL_1027734, EPI_ISL_1027735, EPI_ISL_1027737, EPI_ISL_1027738, EPI_ISL_1027739, EPI_ISL_1027740, EPI_ISL_1027741, EPI_ISL_1027742, EPI_ISL_1027743, EPI_ISL_1027744, EPI_ISL_1027745, EPI_ISL_1027746, EPI_ISL_1027747, EPI_ISL_1027748, EPI_ISL_1027749, EPI_ISL_1027750, EPI_ISL_1027751, EPI_ISL_1027752, EPI_ISL_1027753, EPI_ISL_1027754, EPI_ISL_1027755, EPI_ISL_1027756, EPI_ISL_1027757, EPI_ISL_1027758, EPI_ISL_1027759, EPI_ISL_1027760, EPI_ISL_1027761, EPI_ISL_1027762, EPI_ISL_1027763, EPI_ISL_1027764, EPI_ISL_1027765, EPI_ISL_1027766, EPI_ISL_1027767, EPI_ISL_1027768, EPI_ISL_1027769, EPI_ISL_1027770, EPI_ISL_1027771, EPI_ISL_1027772, EPI_ISL_1027773, EPI_ISL_1027774, EPI_ISL_1027775, EPI_ISL_1027776, EPI_ISL_1027777, EPI_ISL_1027778, EPI_ISL_1027779, EPI_ISL_1027780, EPI_ISL_1027781, EPI_ISL_1027782, EPI_ISL_1027783, EPI_ISL_1027784, EPI_ISL_1027785, EPI_ISL_1027786, EPI_ISL_1027787, EPI_ISL_1027788, EPI_ISL_1027789, EPI_ISL_1027790, EPI_ISL_1027791, EPI_ISL_1027792, EPI_ISL_1027793, EPI_ISL_1027794, EPI_ISL_1027795, EPI_ISL_1027796, EPI_ISL_1027797, EPI_ISL_1027798, EPI_ISL_1027799, EPI_ISL_1027800, EPI_ISL_1027801, EPI_ISL_1027802, EPI_ISL_1027803, EPI_ISL_1027804, EPI_ISL_1027805, EPI_ISL_1027806, EPI_ISL_1027807, EPI_ISL_1027808, EPI_ISL_1027809, EPI_ISL_1027810, EPI_ISL_1027811, EPI_ISL_1027812, EPI_ISL_1027813, EPI_ISL_1027814, EPI_ISL_1027815, EPI_ISL_1027816, EPI_ISL_1027817, EPI_ISL_1027818, EPI_ISL_1027819, EPI_ISL_1027820, EPI_ISL_1027821, EPI_ISL_1027822, EPI_ISL_1027823, EPI_ISL_1027824, EPI_ISL_1027825, EPI_ISL_1027826, EPI_ISL_1027827, EPI_ISL_1027828, EPI_ISL_1027829, EPI_ISL_1027830, EPI_ISL_1027831, EPI_ISL_1027832, EPI_ISL_1027833, EPI_ISL_1027834, EPI_ISL_1027835, EPI_ISL_1027836, EPI_ISL_1027837, EPI_ISL_1027838, EPI_ISL_1027839, EPI_ISL_1027840, EPI_ISL_1027841, EPI_ISL_1027842, EPI_ISL_1027843, EPI_ISL_1027844, EPI_ISL_1027845, EPI_ISL_1027846, EPI_ISL_1027847, EPI_ISL_1027848, EPI_ISL_1027849, EPI_ISL_1027850, EPI_ISL_1027851, EPI_ISL_1027852, EPI_ISL_1027853, EPI_ISL_1027854, EPI_ISL_1027855, EPI_ISL_1027856, EPI_ISL_1027857, EPI_ISL_1027858, EPI_ISL_1027859, EPI_ISL_1027860, EPI_ISL_1027861, EPI_ISL_1027862, EPI_ISL_1027863, EPI_ISL_1027864, EPI_ISL_1027865, EPI_ISL_1027866, EPI_ISL_1027867, EPI_ISL_1027868, EPI_ISL_1027869, EPI_ISL_1027870, EPI_ISL_1027871, EPI_ISL_1027872, EPI_ISL_1027873, EPI_ISL_1027874, EPI_ISL_1027875, EPI_ISL_1027876, EPI_ISL_1027877, EPI_ISL_1027878, EPI_ISL_1027879, EPI_ISL_1027880, EPI_ISL_1027881, EPI_ISL_1027882, EPI_ISL_1027883, EPI_ISL_1027884, EPI_ISL_1027885, EPI_ISL_1027886, EPI_ISL_1027887, EPI_ISL_1027888, EPI_ISL_1029050, EPI_ISL_1029051, EPI_ISL_1029052, EPI_ISL_1029053, EPI_ISL_1029054, EPI_ISL_1029055, EPI_ISL_1029056, EPI_ISL_1029057, EPI_ISL_1029058, EPI_ISL_1029059, EPI_ISL_1029060, EPI_ISL_1029061, EPI_ISL_1029062, EPI_ISL_1029063, EPI_ISL_1029064, EPI_ISL_1029065, EPI_ISL_1029066, EPI_ISL_1029067, EPI_ISL_1029068, EPI_ISL_1029069, EPI_ISL_1029070, EPI_ISL_1029071, EPI_ISL_1029072, EPI_ISL_1029073, EPI_ISL_1029074, EPI_ISL_1029075, EPI_ISL_1029076, EPI_ISL_1029077, EPI_ISL_1029078, EPI_ISL_1029079, EPI_ISL_1029080, EPI_ISL_1029081, EPI_ISL_1029082, EPI_ISL_1029083, EPI_ISL_1029084, EPI_ISL_1029085, EPI_ISL_1029086, EPI_ISL_1029087, EPI_ISL_1029088, EPI_ISL_1029089, EPI_ISL_1029090, EPI_ISL_1029091, EPI_ISL_1029092, EPI_ISL_1029093, EPI_ISL_1029094, EPI_ISL_1029095, EPI_ISL_1029096, EPI_ISL_1029097, EPI_ISL_1029098, EPI_ISL_1029099, EPI_ISL_1029100, EPI_ISL_1029101, EPI_ISL_1029102, EPI_ISL_1029103, EPI_ISL_1029104, EPI_ISL_1029105, EPI_ISL_1029106, EPI_ISL_1029107, EPI_ISL_1029108, EPI_ISL_1029109, EPI_ISL_1029110, EPI_ISL_1029111, EPI_ISL_1029112, EPI_ISL_1029113, EPI_ISL_1029114, EPI_ISL_1029115, EPI_ISL_1029116, EPI_ISL_1029117, EPI_ISL_1029118, EPI_ISL_1029119, EPI_ISL_1029120, EPI_ISL_1029121, EPI_ISL_1029122, EPI_ISL_1029123, EPI_ISL_1029124, EPI_ISL_1029125, EPI_ISL_1029126, EPI_ISL_1029127, EPI_ISL_1029128, EPI_ISL_1029129, EPI_ISL_1029130, EPI_ISL_1029131, EPI_ISL_1029132, EPI_ISL_1029133, EPI_ISL_1029134, EPI_ISL_1029135, EPI_ISL_1029136, EPI_ISL_1029137, EPI_ISL_1029138, EPI_ISL_1029139, EPI_ISL_1029140, EPI_ISL_1029141, EPI_ISL_1029142, EPI_ISL_1029143, EPI_ISL_1029144, EPI_ISL_1029145, EPI_ISL_1029146, EPI_ISL_1029147, EPI_ISL_1029148, EPI_ISL_1029149, EPI_ISL_1029150, EPI_ISL_1029151, EPI_ISL_1029152, EPI_ISL_1029153, EPI_ISL_1029154, EPI_ISL_1029155, EPI_ISL_1029156, EPI_ISL_1029157, EPI_ISL_1029158, EPI_ISL_1029159, EPI_ISL_1029160, EPI_ISL_1029161, EPI_ISL_1029162, EPI_ISL_1029163, EPI_ISL_1029164, EPI_ISL_1029165, EPI_ISL_1029166, EPI_ISL_1029167, EPI_ISL_1029168, EPI_ISL_1029169, EPI_ISL_1029170, EPI_ISL_1029171, EPI_ISL_1029172, EPI_ISL_1029173, EPI_ISL_1029174, EPI_ISL_1029175, EPI_ISL_1029176, EPI_ISL_1029177, EPI_ISL_1029178, EPI_ISL_1029179, EPI_ISL_1029180, EPI_ISL_1029181, EPI_ISL_1029182, EPI_ISL_1029183, EPI_ISL_1029184, EPI_ISL_1029185, EPI_ISL_1029186, EPI_ISL_1029187, EPI_ISL_1029188, EPI_ISL_1029189, EPI_ISL_1029190, EPI_ISL_1029191, EPI_ISL_1029192, EPI_ISL_1029193, EPI_ISL_1029194, EPI_ISL_1029195, EPI_ISL_1029196, EPI_ISL_1029197, EPI_ISL_1029198, EPI_ISL_1029199, EPI_ISL_1029200, EPI_ISL_1029201, EPI_ISL_1029202, EPI_ISL_1029203, EPI_ISL_1029204, EPI_ISL_1029205, EPI_ISL_1029206, EPI_ISL_1029207, EPI_ISL_1029208, EPI_ISL_1029209, EPI_ISL_1029210, EPI_ISL_1029211, EPI_ISL_1029212, EPI_ISL_1029213, EPI_ISL_1029214, EPI_ISL_1029215, EPI_ISL_1029216, EPI_ISL_1029217, EPI_ISL_1029218, EPI_ISL_1029219, EPI_ISL_1029220, EPI_ISL_1029221, EPI_ISL_1029222, EPI_ISL_1029223, EPI_ISL_1029224, EPI_ISL_1029225, EPI_ISL_1029226, EPI_ISL_1029227, EPI_ISL_1029228, EPI_ISL_1029229, EPI_ISL_1029230, EPI_ISL_1029231, EPI_ISL_1029232, EPI_ISL_1029233, EPI_ISL_1029234, EPI_ISL_1029235, EPI_ISL_1029236, EPI_ISL_1029237, EPI_ISL_1029238, EPI_ISL_1029239, EPI_ISL_1029240, EPI_ISL_1029241, EPI_ISL_1029242, EPI_ISL_1029243, EPI_ISL_1029244, EPI_ISL_1029245, EPI_ISL_1029246, EPI_ISL_1029247, EPI_ISL_1029248, EPI_ISL_1029249, EPI_ISL_1029250, EPI_ISL_1029251, EPI_ISL_1029252, EPI_ISL_1029253, EPI_ISL_1029254, EPI_ISL_1029255, EPI_ISL_1029256, EPI_ISL_1029257, EPI_ISL_1029258, EPI_ISL_1029259, EPI_ISL_1029260, EPI_ISL_1029261, EPI_ISL_1029262, EPI_ISL_1029263, EPI_ISL_1029264, EPI_ISL_1029265, EPI_ISL_1029266, EPI_ISL_1029267, EPI_ISL_1029268, EPI_ISL_1029269, EPI_ISL_1029270, EPI_ISL_1029271, EPI_ISL_1029272, EPI_ISL_1029273, EPI_ISL_1029274, EPI_ISL_1029275, EPI_ISL_1029276, EPI_ISL_1029277, EPI_ISL_1029278, EPI_ISL_1029279, EPI_ISL_1029280, EPI_ISL_1029281, EPI_ISL_1029282, EPI_ISL_1029283, EPI_ISL_1029284, EPI_ISL_1029285, EPI_ISL_1029286, EPI_ISL_1029287, EPI_ISL_1029288, EPI_ISL_1029289, EPI_ISL_1029290, EPI_ISL_1029291, EPI_ISL_1029292, EPI_ISL_1029293, EPI_ISL_1029294, EPI_ISL_1029295, EPI_ISL_1029296, EPI_ISL_1029297, EPI_ISL_1029298, EPI_ISL_1029299, EPI_ISL_1029300, EPI_ISL_1029301, EPI_ISL_1029303, EPI_ISL_1029304, EPI_ISL_1029305, EPI_ISL_1029306, EPI_ISL_1029307, EPI_ISL_1029308, EPI_ISL_1029309, EPI_ISL_1029310, EPI_ISL_1029311, EPI_ISL_1029312, EPI_ISL_1029313, EPI_ISL_1029314, EPI_ISL_1029315, EPI_ISL_1029316, EPI_ISL_1029317, EPI_ISL_1029318, EPI_ISL_1029319, EPI_ISL_1029320, EPI_ISL_1029321, EPI_ISL_1029322, EPI_ISL_1029323, EPI_ISL_1029324, EPI_ISL_1029325, EPI_ISL_1029326, EPI_ISL_1029327, EPI_ISL_1029328, EPI_ISL_1029329, EPI_ISL_1029330, EPI_ISL_1029331, EPI_ISL_1029332, EPI_ISL_1029333, EPI_ISL_1029334, EPI_ISL_1029336, EPI_ISL_1029337, EPI_ISL_1029338, EPI_ISL_1029339, EPI_ISL_1029340, EPI_ISL_1029341, EPI_ISL_1029342, EPI_ISL_1029343, EPI_ISL_1029344, EPI_ISL_1029345, EPI_ISL_1029346, EPI_ISL_1029347, EPI_ISL_1029348, EPI_ISL_1029349, EPI_ISL_1029350, EPI_ISL_1029351, EPI_ISL_1029352, EPI_ISL_1029353, EPI_ISL_1029354, EPI_ISL_1029355, EPI_ISL_1029356, EPI_ISL_1029357, EPI_ISL_1029358, EPI_ISL_1029359, EPI_ISL_1029360, EPI_ISL_1029361, EPI_ISL_1029362, EPI_ISL_1029363, EPI_ISL_1029364, EPI_ISL_1029365, EPI_ISL_1029366, EPI_ISL_1029367, EPI_ISL_1029368, EPI_ISL_1029369, EPI_ISL_1029370, EPI_ISL_1029371, EPI_ISL_1029372, EPI_ISL_1029373, EPI_ISL_1029374, EPI_ISL_1029375, EPI_ISL_1029376, EPI_ISL_1029377, EPI_ISL_1029378, EPI_ISL_1029379, EPI_ISL_1029380, EPI_ISL_1029381, EPI_ISL_1029382, EPI_ISL_1029383, EPI_ISL_1029384, EPI_ISL_1029385, EPI_ISL_1029386, EPI_ISL_1029387, EPI_ISL_1029388, EPI_ISL_1029389, EPI_ISL_1029390, EPI_ISL_1029391, EPI_ISL_1029392, EPI_ISL_1029393, EPI_ISL_1029394, EPI_ISL_1029395, EPI_ISL_1029396, EPI_ISL_1029397, EPI_ISL_1029398, EPI_ISL_1029399, EPI_ISL_1029400, EPI_ISL_1029401, EPI_ISL_1029402, EPI_ISL_1029403, EPI_ISL_1029404, EPI_ISL_1029405, EPI_ISL_1029406, EPI_ISL_1029407, EPI_ISL_1029408, EPI_ISL_1029409, EPI_ISL_1029410, EPI_ISL_1029411, EPI_ISL_1029412, EPI_ISL_1029413, EPI_ISL_1029414, EPI_ISL_1029415, EPI_ISL_1029416, EPI_ISL_1029417, EPI_ISL_1029418, EPI_ISL_1029419, EPI_ISL_1029420, EPI_ISL_1029421, EPI_ISL_1029422, EPI_ISL_1029423, EPI_ISL_1029424, EPI_ISL_1029425, EPI_ISL_1029426, EPI_ISL_1029427, EPI_ISL_1029428, EPI_ISL_1029429, EPI_ISL_1029430, EPI_ISL_1029431, EPI_ISL_1029432, EPI_ISL_1029433, EPI_ISL_1029434, EPI_ISL_1029435, EPI_ISL_1029436, EPI_ISL_1029437, EPI_ISL_1029438, EPI_ISL_1029439, EPI_ISL_1029440, EPI_ISL_1029441, EPI_ISL_1029442, EPI_ISL_1029443, EPI_ISL_1029444, EPI_ISL_1029445, EPI_ISL_1029446, EPI_ISL_1029447, EPI_ISL_1029448, EPI_ISL_1029449, EPI_ISL_1029450, EPI_ISL_1029451, EPI_ISL_1029452, EPI_ISL_1029453, EPI_ISL_1029454, EPI_ISL_1029455, EPI_ISL_1029456, EPI_ISL_1029457, EPI_ISL_1029458, EPI_ISL_1029459, EPI_ISL_1029460, EPI_ISL_1029461, EPI_ISL_1029462, EPI_ISL_1029463, EPI_ISL_1029464, EPI_ISL_1029465, EPI_ISL_1029466, EPI_ISL_1029467, EPI_ISL_1029468, EPI_ISL_1029469, EPI_ISL_1029470, EPI_ISL_1029471, EPI_ISL_1029472, EPI_ISL_1029473, EPI_ISL_1029474, EPI_ISL_1029475, EPI_ISL_1029476, EPI_ISL_1029477, EPI_ISL_1029478, EPI_ISL_1029479, EPI_ISL_1029480, EPI_ISL_1029481, EPI_ISL_1029482, EPI_ISL_1029483, EPI_ISL_1029484, EPI_ISL_1029485, EPI_ISL_1029486, EPI_ISL_1029487, EPI_ISL_1029488, EPI_ISL_1029489, EPI_ISL_1029490, EPI_ISL_1029491, EPI_ISL_1029492, EPI_ISL_1029493, EPI_ISL_1029494, EPI_ISL_1029495, EPI_ISL_1029496, EPI_ISL_1029497, EPI_ISL_1029498, EPI_ISL_1029499, EPI_ISL_1029500, EPI_ISL_1029501, EPI_ISL_1029502, EPI_ISL_1029503, EPI_ISL_1029504, EPI_ISL_1029505, EPI_ISL_1029506, EPI_ISL_1029507, EPI_ISL_1029508, EPI_ISL_1029509, EPI_ISL_1029510, EPI_ISL_1029511, EPI_ISL_1029512, EPI_ISL_1029513, EPI_ISL_1029514, EPI_ISL_1029515, EPI_ISL_1029516, EPI_ISL_1029517, EPI_ISL_1029518, EPI_ISL_1029519, EPI_ISL_1029520, EPI_ISL_1029521, EPI_ISL_1029522, EPI_ISL_1029523, EPI_ISL_1029524, EPI_ISL_1029525, EPI_ISL_1029526, EPI_ISL_1029527, EPI_ISL_1029528, EPI_ISL_1029529, EPI_ISL_1029530, EPI_ISL_1029531, EPI_ISL_1029532, EPI_ISL_1029533, EPI_ISL_1029534, EPI_ISL_1029535, EPI_ISL_1029536, EPI_ISL_1029537, EPI_ISL_1029538, EPI_ISL_1029539, EPI_ISL_1029540, EPI_ISL_1029541, EPI_ISL_1029542, EPI_ISL_1029543, EPI_ISL_1029544, EPI_ISL_1029545, EPI_ISL_1029546, EPI_ISL_1029547, EPI_ISL_1029548, EPI_ISL_1029549, EPI_ISL_1029550, EPI_ISL_1029551, EPI_ISL_1029552, EPI_ISL_1029553, EPI_ISL_1029554, EPI_ISL_1029555, EPI_ISL_1029556, EPI_ISL_1029557, EPI_ISL_1029558, EPI_ISL_1029559, EPI_ISL_1029560, EPI_ISL_1029561, EPI_ISL_1029562, EPI_ISL_1029563, EPI_ISL_1029564, EPI_ISL_1029565, EPI_ISL_1029566, EPI_ISL_1029567, EPI_ISL_1029568, EPI_ISL_1029569, EPI_ISL_1029570, EPI_ISL_1029571, EPI_ISL_1029572, EPI_ISL_1029573, EPI_ISL_1029574, EPI_ISL_1029575, EPI_ISL_1029576, EPI_ISL_1029577, EPI_ISL_1029578, EPI_ISL_1029579, EPI_ISL_1029580, EPI_ISL_1029581, EPI_ISL_1029582, EPI_ISL_1029583, EPI_ISL_1029584, EPI_ISL_1029585, EPI_ISL_1029586, EPI_ISL_1029587, EPI_ISL_1029588, EPI_ISL_1029589, EPI_ISL_1029590, EPI_ISL_1029591, EPI_ISL_1029592, EPI_ISL_1029593, EPI_ISL_1029594, EPI_ISL_1029595, EPI_ISL_1029596, EPI_ISL_1029597, EPI_ISL_1029598, EPI_ISL_1029599, EPI_ISL_1029600, EPI_ISL_1029601, EPI_ISL_1029602, EPI_ISL_1029603, EPI_ISL_1029604, EPI_ISL_1029605, EPI_ISL_1029606, EPI_ISL_1029607, EPI_ISL_1029608, EPI_ISL_1029609, EPI_ISL_1029610, EPI_ISL_1029611, EPI_ISL_1029612, EPI_ISL_1029613, EPI_ISL_1029614, EPI_ISL_1029615, EPI_ISL_1029616, EPI_ISL_1029617, EPI_ISL_1029618, EPI_ISL_1029619, EPI_ISL_1029620, EPI_ISL_1029621, EPI_ISL_1029622, EPI_ISL_1029623, EPI_ISL_1029624, EPI_ISL_1029625, EPI_ISL_1029626, EPI_ISL_1029627, EPI_ISL_1029628, EPI_ISL_1029629, EPI_ISL_1029630, EPI_ISL_1029631, EPI_ISL_1029632, EPI_ISL_1029633, EPI_ISL_1029634, EPI_ISL_1029635, EPI_ISL_1029636, EPI_ISL_1029637, EPI_ISL_1029638, EPI_ISL_1029639, EPI_ISL_1029640, EPI_ISL_1029641, EPI_ISL_1029642, EPI_ISL_1029643, EPI_ISL_1029644, EPI_ISL_1029645, EPI_ISL_1029646, EPI_ISL_1029647, EPI_ISL_1029648, EPI_ISL_1029649, EPI_ISL_1029650, EPI_ISL_1029651, EPI_ISL_1029652, EPI_ISL_1029653, EPI_ISL_1029654, EPI_ISL_1029655, EPI_ISL_1029656, EPI_ISL_1029657, EPI_ISL_1029658, EPI_ISL_1029659, EPI_ISL_1029660, EPI_ISL_1029661, EPI_ISL_1029662, EPI_ISL_1029663, EPI_ISL_1029664, EPI_ISL_1029665, EPI_ISL_1029666, EPI_ISL_1029667, EPI_ISL_1029668, EPI_ISL_1029669, EPI_ISL_1029670, EPI_ISL_1029671, EPI_ISL_1029672, EPI_ISL_1029673, EPI_ISL_1029674, EPI_ISL_1029675, EPI_ISL_1029676, EPI_ISL_1029677, EPI_ISL_1029678, EPI_ISL_1029679, EPI_ISL_1029680, EPI_ISL_1029681, EPI_ISL_1029682, EPI_ISL_1029683, EPI_ISL_1029684, EPI_ISL_1029685, EPI_ISL_1029686, EPI_ISL_1029687, EPI_ISL_1029688, EPI_ISL_1029689, EPI_ISL_1029690, EPI_ISL_1029691, EPI_ISL_1029692, EPI_ISL_1029693, EPI_ISL_1029694, EPI_ISL_1029695, EPI_ISL_1029696, EPI_ISL_1029697, EPI_ISL_1029698, EPI_ISL_1029699, EPI_ISL_1029700, EPI_ISL_1029701, EPI_ISL_1029702, EPI_ISL_1029703, EPI_ISL_1029704, EPI_ISL_1029705, EPI_ISL_1029706, EPI_ISL_1029707, EPI_ISL_1029708, EPI_ISL_1029709, EPI_ISL_1029710, EPI_ISL_1029711, EPI_ISL_1029712, EPI_ISL_1029713, EPI_ISL_1029714, EPI_ISL_1029715, EPI_ISL_1029716, EPI_ISL_1029717, EPI_ISL_1029718, EPI_ISL_1029719, EPI_ISL_1029720, EPI_ISL_1029721, EPI_ISL_1029722, EPI_ISL_1029723, EPI_ISL_1029724, EPI_ISL_1029725, EPI_ISL_1029726, EPI_ISL_1029727, EPI_ISL_1029728, EPI_ISL_1029729, EPI_ISL_1029730, EPI_ISL_1029731, EPI_ISL_1029732, EPI_ISL_1029733, EPI_ISL_1029734, EPI_ISL_1029735, EPI_ISL_1029736, EPI_ISL_1029737, EPI_ISL_1029738, EPI_ISL_1029739, EPI_ISL_1029740, EPI_ISL_1029741, EPI_ISL_1029742, EPI_ISL_1029743, EPI_ISL_1029744, EPI_ISL_1029745, EPI_ISL_1029746, EPI_ISL_1029747, EPI_ISL_1029748, EPI_ISL_1029749, EPI_ISL_1029750, EPI_ISL_1029751, EPI_ISL_1029752, EPI_ISL_1029753, EPI_ISL_1029754, EPI_ISL_1029755, EPI_ISL_1029756, EPI_ISL_1029757, EPI_ISL_1029758, EPI_ISL_1029759, EPI_ISL_1029760, EPI_ISL_1029761, EPI_ISL_1029762, EPI_ISL_1029763, EPI_ISL_1029764, EPI_ISL_1029765, EPI_ISL_1029766, EPI_ISL_1029767, EPI_ISL_1029768, EPI_ISL_1029769, EPI_ISL_1029770, EPI_ISL_1029771, EPI_ISL_1029772, EPI_ISL_1029773, EPI_ISL_1029774, EPI_ISL_1029775, EPI_ISL_1029776, EPI_ISL_1029777, EPI_ISL_1029778, EPI_ISL_1029779, EPI_ISL_1029780, EPI_ISL_1029781, EPI_ISL_1029782, EPI_ISL_1029783, EPI_ISL_1029784, EPI_ISL_1029785, EPI_ISL_1029786, EPI_ISL_1029787, EPI_ISL_1029788, EPI_ISL_1029789, EPI_ISL_1029790, EPI_ISL_1029791, EPI_ISL_1029792, EPI_ISL_1029793, EPI_ISL_1029794, EPI_ISL_1029795, EPI_ISL_1029796, EPI_ISL_1029797, EPI_ISL_1029798, EPI_ISL_1029799, EPI_ISL_1029800, EPI_ISL_1029801, EPI_ISL_1029802, EPI_ISL_1029803, EPI_ISL_1029804, EPI_ISL_1029805, EPI_ISL_1029806, EPI_ISL_1029807, EPI_ISL_1029808, EPI_ISL_1029809, EPI_ISL_1029810, EPI_ISL_1029811, EPI_ISL_1029812, EPI_ISL_1029813, EPI_ISL_1029814, EPI_ISL_1029815, EPI_ISL_1029816, EPI_ISL_1029817, EPI_ISL_1029818, EPI_ISL_1029819, EPI_ISL_1029820, EPI_ISL_1029821, EPI_ISL_1029822, EPI_ISL_1029823, EPI_ISL_1029824, EPI_ISL_1029825, EPI_ISL_1029826, EPI_ISL_1029827, EPI_ISL_1029828, EPI_ISL_1029829, EPI_ISL_1029830, EPI_ISL_1029831, EPI_ISL_1029832, EPI_ISL_1029833, EPI_ISL_1029834, EPI_ISL_1029835, EPI_ISL_1029836, EPI_ISL_1029837, EPI_ISL_1029838, EPI_ISL_1029839, EPI_ISL_1029840, EPI_ISL_1029841, EPI_ISL_1029842, EPI_ISL_1029843, EPI_ISL_1029844, EPI_ISL_1029845, EPI_ISL_1029846, EPI_ISL_1029847, EPI_ISL_1029848, EPI_ISL_1029849, EPI_ISL_1029850, EPI_ISL_1029851, EPI_ISL_1029852, EPI_ISL_1029853, EPI_ISL_1029854, EPI_ISL_1029855, EPI_ISL_1029856, EPI_ISL_1029857, EPI_ISL_1029858, EPI_ISL_1029859, EPI_ISL_1029860, EPI_ISL_1029861, EPI_ISL_1029862, EPI_ISL_1029863, EPI_ISL_1029864, EPI_ISL_1029865, EPI_ISL_1029866, EPI_ISL_1029867, EPI_ISL_1029868, EPI_ISL_1029869, EPI_ISL_1029870, EPI_ISL_1029871, EPI_ISL_1029872, EPI_ISL_1029873, EPI_ISL_1029874, EPI_ISL_1029875, EPI_ISL_1029876, EPI_ISL_1029877, EPI_ISL_1029878, EPI_ISL_1029879, EPI_ISL_1029880, EPI_ISL_1029881, EPI_ISL_1029882, EPI_ISL_1029883, EPI_ISL_1029884, EPI_ISL_1029885, EPI_ISL_1029886, EPI_ISL_1029887, EPI_ISL_1029888, EPI_ISL_1029889, EPI_ISL_1029890, EPI_ISL_1029891, EPI_ISL_1029892, EPI_ISL_1029893, EPI_ISL_1029894, EPI_ISL_1029895, EPI_ISL_1029896, EPI_ISL_1029897, EPI_ISL_1029898, EPI_ISL_1029899, EPI_ISL_1029900, EPI_ISL_1029901, EPI_ISL_1029902, EPI_ISL_1029903, EPI_ISL_1029904, EPI_ISL_1029905, EPI_ISL_1029906, EPI_ISL_1029907, EPI_ISL_1029908, EPI_ISL_1029909, EPI_ISL_1029910, EPI_ISL_1029911, EPI_ISL_1029912, EPI_ISL_1029913, EPI_ISL_1029914, EPI_ISL_1029915, EPI_ISL_1029916, EPI_ISL_1029917, EPI_ISL_1029918, EPI_ISL_1029919, EPI_ISL_1029920, EPI_ISL_1029921, EPI_ISL_1029922, EPI_ISL_1029923, EPI_ISL_1029924, EPI_ISL_1029925, EPI_ISL_1029927, EPI_ISL_1029928, EPI_ISL_1029929, EPI_ISL_1029930, EPI_ISL_1029931, EPI_ISL_1029932, EPI_ISL_1029933, EPI_ISL_1029934, EPI_ISL_1029935, EPI_ISL_1029936, EPI_ISL_1029937, EPI_ISL_1029938, EPI_ISL_1029939, EPI_ISL_1029940, EPI_ISL_1029941, EPI_ISL_1029942, EPI_ISL_1029943, EPI_ISL_1029944, EPI_ISL_1029945, EPI_ISL_1029946, EPI_ISL_1029947, EPI_ISL_1029948, EPI_ISL_1029949, EPI_ISL_1029950, | | | |

| | | | |
|---|---|---|---|
| EPI_ISL_1029951, EPI_ISL_1029952, EPI_ISL_1029953, EPI_ISL_1029954, EPI_ISL_1029955 | | | |
| see above | Laboratory Corporation of America | Respiratory Viruses Branch, Division of Viral Diseases, Centers for Disease Control and Prevention | Peter W. Cook, Dakota Howard, Dhwani Batra, Ben L. Rambo-Martin, Clinton R. Paden, Suxiang Tong, Duncan MacCannell |
| EPI_ISL_1029974, EPI_ISL_1029975, EPI_ISL_1029976, EPI_ISL_1029977, EPI_ISL_1029978, EPI_ISL_1029979, EPI_ISL_1029980, EPI_ISL_1029981, EPI_ISL_1029982, EPI_ISL_1029983, EPI_ISL_1029984, EPI_ISL_1029985, EPI_ISL_1029986, EPI_ISL_1029987, EPI_ISL_1029988, EPI_ISL_1029989, EPI_ISL_1029990, EPI_ISL_1029991, EPI_ISL_1029992, EPI_ISL_1029993, EPI_ISL_1029994, EPI_ISL_1029995, EPI_ISL_1029996, EPI_ISL_1029997, EPI_ISL_1029998, EPI_ISL_1029999, EPI_ISL_1030000, EPI_ISL_1030001, EPI_ISL_1030002, EPI_ISL_1030003, EPI_ISL_1030004, EPI_ISL_1030005, EPI_ISL_1030006, EPI_ISL_1030007, EPI_ISL_1030008, EPI_ISL_1030009, EPI_ISL_1030010, EPI_ISL_1030011, EPI_ISL_1030012, EPI_ISL_1030013, EPI_ISL_1030014, EPI_ISL_1030015, EPI_ISL_1030016, EPI_ISL_1030017, EPI_ISL_1030018, EPI_ISL_1030019, EPI_ISL_1030020, EPI_ISL_1030021, EPI_ISL_1030022, EPI_ISL_1030023, EPI_ISL_1030024, EPI_ISL_1030025, EPI_ISL_1030026, EPI_ISL_1030027, EPI_ISL_1030028, EPI_ISL_1030029, EPI_ISL_1030030, EPI_ISL_1030031, EPI_ISL_1030032, EPI_ISL_1030033, EPI_ISL_1030034, EPI_ISL_1030035, EPI_ISL_1030036, EPI_ISL_1030037, EPI_ISL_1030038, EPI_ISL_1030061, EPI_ISL_1030062, EPI_ISL_1030063, EPI_ISL_1030064, EPI_ISL_1030065, EPI_ISL_1030066, EPI_ISL_1030067, EPI_ISL_1030068, EPI_ISL_1030069, EPI_ISL_1030070, EPI_ISL_1030071, EPI_ISL_1030072, EPI_ISL_1030073, EPI_ISL_1030074, EPI_ISL_1030075, EPI_ISL_1030076, EPI_ISL_1030077, EPI_ISL_1030078, EPI_ISL_1030079, EPI_ISL_1030080, EPI_ISL_1030081, EPI_ISL_1030082, EPI_ISL_1030083, EPI_ISL_1030084, EPI_ISL_1030085, EPI_ISL_1030086, EPI_ISL_1030087, EPI_ISL_1030088, EPI_ISL_1030089, EPI_ISL_1030090, EPI_ISL_1030091, EPI_ISL_1030092, EPI_ISL_1030093, EPI_ISL_1030094, EPI_ISL_1030095, EPI_ISL_1030096, EPI_ISL_1030097, EPI_ISL_1030098, EPI_ISL_1030099, EPI_ISL_1030100, EPI_ISL_1030101, EPI_ISL_1030102, EPI_ISL_1030103, EPI_ISL_1030104, EPI_ISL_1030105, EPI_ISL_1030106, EPI_ISL_1030107, EPI_ISL_1030108, EPI_ISL_1030109, EPI_ISL_1030110, EPI_ISL_1030111, EPI_ISL_1030112, EPI_ISL_1030113, EPI_ISL_1030114, EPI_ISL_1030115, EPI_ISL_1030116, EPI_ISL_1030117, EPI_ISL_1030118, EPI_ISL_1030119, EPI_ISL_1030120, EPI_ISL_1030121, EPI_ISL_1030122, EPI_ISL_1030123, EPI_ISL_1030124, EPI_ISL_1030125, EPI_ISL_1030126, EPI_ISL_1030127, EPI_ISL_1030128, EPI_ISL_1030129, EPI_ISL_1030130, EPI_ISL_1030131, EPI_ISL_1030132, EPI_ISL_1030133, EPI_ISL_1030134, EPI_ISL_1030135, EPI_ISL_1030136, EPI_ISL_1030137, EPI_ISL_1030138, EPI_ISL_1030139, EPI_ISL_1030140, EPI_ISL_1030141, EPI_ISL_1030142, EPI_ISL_1030143, EPI_ISL_1030144, EPI_ISL_1030145, EPI_ISL_1030146, EPI_ISL_1030147, EPI_ISL_1030148, EPI_ISL_1030149, EPI_ISL_1030150, EPI_ISL_1030151, EPI_ISL_1030152, EPI_ISL_1030153, EPI_ISL_1030154, EPI_ISL_1030155, EPI_ISL_1030156, EPI_ISL_1030157, EPI_ISL_1030158, EPI_ISL_1030159, EPI_ISL_1030160, EPI_ISL_1030161, EPI_ISL_1030162, EPI_ISL_1030163, EPI_ISL_1030164, EPI_ISL_1030165, EPI_ISL_1030166, EPI_ISL_1030167, EPI_ISL_1030168, EPI_ISL_1030169, EPI_ISL_1030170, EPI_ISL_1030171, EPI_ISL_1030172, EPI_ISL_1030173, EPI_ISL_1030174, EPI_ISL_1030175, EPI_ISL_1030176, EPI_ISL_1030177, EPI_ISL_1030178, EPI_ISL_1030179, EPI_ISL_1030180, EPI_ISL_1030181, EPI_ISL_1030182, EPI_ISL_1030183, EPI_ISL_1030184, EPI_ISL_1030185, EPI_ISL_1030186, EPI_ISL_1030187, EPI_ISL_1030188, EPI_ISL_1030189, EPI_ISL_1030190, EPI_ISL_1030191, EPI_ISL_1030192, EPI_ISL_1030193, EPI_ISL_1030194, EPI_ISL_1030195, EPI_ISL_1030196, EPI_ISL_1030197, EPI_ISL_1030198, EPI_ISL_1030199, EPI_ISL_1030200, EPI_ISL_1030201, EPI_ISL_1030202, EPI_ISL_1030203, EPI_ISL_1030204, EPI_ISL_1030205, EPI_ISL_1030206, EPI_ISL_1030207, EPI_ISL_1030208, EPI_ISL_1030209, EPI_ISL_1030210, EPI_ISL_1030211, EPI_ISL_1030212, EPI_ISL_1030213, EPI_ISL_1030214, EPI_ISL_1030215, EPI_ISL_1030216, EPI_ISL_1030217, EPI_ISL_1030218, EPI_ISL_1030219, EPI_ISL_1030220, EPI_ISL_1030221, EPI_ISL_1030222, EPI_ISL_1030223, EPI_ISL_1030224, EPI_ISL_1030225, EPI_ISL_1030226, EPI_ISL_1030227, EPI_ISL_1030228, EPI_ISL_1030229, EPI_ISL_1030230, EPI_ISL_1030231, EPI_ISL_1030232, EPI_ISL_1030233, EPI_ISL_1030234, EPI_ISL_1030235, EPI_ISL_1030236, EPI_ISL_1030237, EPI_ISL_1030238, EPI_ISL_1030239, EPI_ISL_1030240, EPI_ISL_1030241, EPI_ISL_1030242, EPI_ISL_1030243, EPI_ISL_1030244, EPI_ISL_1030245, EPI_ISL_1030246, EPI_ISL_1030247, EPI_ISL_1030248, EPI_ISL_1030249, EPI_ISL_1030250, EPI_ISL_1030251 | | | |
| see above | Florida Bureau of Public Health Laboratories | Florida Bureau of Public Health Laboratories | Sarah Schmedes, Jason Blanton |
| EPI_ISL_1030252, EPI_ISL_1030253, EPI_ISL_1030254, EPI_ISL_1030255, EPI_ISL_1030256, EPI_ISL_1030257, EPI_ISL_1030258, EPI_ISL_1030259, EPI_ISL_1030260, EPI_ISL_1030261, EPI_ISL_1030262, EPI_ISL_1030263, EPI_ISL_1030264, EPI_ISL_1030265, EPI_ISL_1030266, EPI_ISL_1030267, EPI_ISL_1030268 | | | |
| see above | Santa Clara County Public Health Laboratory | Chan-Zuckerberg Biohub | CZB Cliahub Consortium |
| EPI_ISL_1030269 | Contra Costa County Public Health Lab | Chan-Zuckerberg Biohub | CZB Cliahub Consortium |
| EPI_ISL_1030270, EPI_ISL_1030271, EPI_ISL_1030272, EPI_ISL_1030273, EPI_ISL_1030274, EPI_ISL_1030275, EPI_ISL_1030276, EPI_ISL_1030277, EPI_ISL_1030278, EPI_ISL_1030279, EPI_ISL_1030280 | | | |
| see above | Santa Clara County Public Health Laboratory | Chan-Zuckerberg Biohub | CZB Cliahub Consortium |
| EPI_ISL_1030281, EPI_ISL_1030282 | Contra Costa County Public Health Lab | Chan-Zuckerberg Biohub | CZB Cliahub Consortium |
| EPI_ISL_1030283, EPI_ISL_1030284, EPI_ISL_1030285, EPI_ISL_1030286, EPI_ISL_1030287, EPI_ISL_1030288, EPI_ISL_1030289 | Santa Clara County Public Health Laboratory | Chan-Zuckerberg Biohub | CZB Cliahub Consortium |
| EPI_ISL_1030290, EPI_ISL_1030291, EPI_ISL_1030292, EPI_ISL_1030293, EPI_ISL_1030294, EPI_ISL_1030295, EPI_ISL_1030296, EPI_ISL_1030297, EPI_ISL_1030298, EPI_ISL_1030299 | Orange County Public Health Lab | Chan-Zuckerberg Biohub | CZB Cliahub Consortium |
| EPI_ISL_1030300, EPI_ISL_1030301 | Santa Clara County Public Health Laboratory | Chan-Zuckerberg Biohub | CZB Cliahub Consortium |
| EPI_ISL_1030302, EPI_ISL_1030303, EPI_ISL_1030306, EPI_ISL_1030307 | Contra Costa County Public Health Lab | Chan-Zuckerberg Biohub | CZB Cliahub Consortium |
| EPI_ISL_1030308 | Orange County Public Health Lab | Chan-Zuckerberg Biohub | CZB Cliahub Consortium |
| EPI_ISL_1030309, EPI_ISL_1030310, EPI_ISL_1030311 | Contra Costa County Public Health Lab | Chan-Zuckerberg Biohub | CZB Cliahub Consortium |
| EPI_ISL_1030312 | Orange County Public Health Lab | Chan-Zuckerberg Biohub | CZB Cliahub Consortium |
| EPI_ISL_1030313, EPI_ISL_1030314, EPI_ISL_1030315, EPI_ISL_1030316, EPI_ISL_1030317, EPI_ISL_1030318, EPI_ISL_1030319, EPI_ISL_1030320, EPI_ISL_1030321, EPI_ISL_1030322, EPI_ISL_1030323, EPI_ISL_1030324, EPI_ISL_1030325, EPI_ISL_1030326, EPI_ISL_1030327, EPI_ISL_1030328, EPI_ISL_1030329 | | | |
| see above | Contra Costa County Public Health Lab | Chan-Zuckerberg Biohub | CZB Cliahub Consortium |
| EPI_ISL_1030330 | Santa Clara County Public Health Laboratory | Chan-Zuckerberg Biohub | CZB Cliahub Consortium |
| EPI_ISL_1030331, EPI_ISL_1030332, EPI_ISL_1030333, EPI_ISL_1030334, EPI_ISL_1030335, EPI_ISL_1030336, EPI_ISL_1030337, EPI_ISL_1030338, EPI_ISL_1030339, EPI_ISL_1030340, EPI_ISL_1030341, EPI_ISL_1030342, EPI_ISL_1030343, EPI_ISL_1030344, EPI_ISL_1030345, EPI_ISL_1030346, EPI_ISL_1030347, EPI_ISL_1030348, EPI_ISL_1030349, EPI_ISL_1030350, EPI_ISL_1030351, EPI_ISL_1030352, EPI_ISL_1030353, EPI_ISL_1030354, EPI_ISL_1030355, EPI_ISL_1030356, EPI_ISL_1030357, EPI_ISL_1030358, EPI_ISL_1030359, EPI_ISL_1030360, EPI_ISL_1030361, EPI_ISL_1030362, EPI_ISL_1030363, EPI_ISL_1030364, EPI_ISL_1030365, EPI_ISL_1030366, EPI_ISL_1030367, EPI_ISL_1030368, EPI_ISL_1030369, EPI_ISL_1030370, EPI_ISL_1030371, EPI_ISL_1030372, EPI_ISL_1030373, EPI_ISL_1030374, EPI_ISL_1030375, EPI_ISL_1030376, EPI_ISL_1030377, EPI_ISL_1030378, EPI_ISL_1030379, EPI_ISL_1030380, EPI_ISL_1030381, EPI_ISL_1030382, EPI_ISL_1030383, EPI_ISL_1030384, EPI_ISL_1030385, EPI_ISL_1030386, EPI_ISL_1030387, EPI_ISL_1030388, EPI_ISL_1030389, EPI_ISL_1030390 | | | |
| see above | Contra Costa County Public Health Lab | Chan-Zuckerberg Biohub | CZB Cliahub Consortium |
| EPI_ISL_1030391, EPI_ISL_1030392, EPI_ISL_1030393, EPI_ISL_1030394, EPI_ISL_1030395, EPI_ISL_1030396, EPI_ISL_1030397, EPI_ISL_1030398, EPI_ISL_1030399, EPI_ISL_1030400, EPI_ISL_1030401, EPI_ISL_1030402, EPI_ISL_1030403, EPI_ISL_1030404, EPI_ISL_1030405, EPI_ISL_1030406, EPI_ISL_1030407, EPI_ISL_1030408, EPI_ISL_1030409, EPI_ISL_1030410, EPI_ISL_1030411, EPI_ISL_1030412, EPI_ISL_1030413, EPI_ISL_1030414, EPI_ISL_1030415, EPI_ISL_1030416, EPI_ISL_1030417, EPI_ISL_1030418, EPI_ISL_1030419, EPI_ISL_1030420, EPI_ISL_1030421, EPI_ISL_1030422, EPI_ISL_1030423, EPI_ISL_1030424, EPI_ISL_1030425, EPI_ISL_1030426, EPI_ISL_1030427, EPI_ISL_1030428, EPI_ISL_1030429, EPI_ISL_1030430, EPI_ISL_1030431, EPI_ISL_1030432, EPI_ISL_1030433, EPI_ISL_1030434, EPI_ISL_1030435, EPI_ISL_1030436, EPI_ISL_1030437, EPI_ISL_1030438, EPI_ISL_1030439, EPI_ISL_1030440, EPI_ISL_1030441, EPI_ISL_1030442, EPI_ISL_1030443 | | | |
| see above | Santa Clara County Public Health Laboratory | Chan-Zuckerberg Biohub | CZB Cliahub Consortium |
| EPI_ISL_1030444, EPI_ISL_1030445, EPI_ISL_1030446, EPI_ISL_1030447, EPI_ISL_1030448, EPI_ISL_1030449, EPI_ISL_1030450, EPI_ISL_1030451, EPI_ISL_1030452, EPI_ISL_1030453, EPI_ISL_1030454, EPI_ISL_1030455, EPI_ISL_1030456, EPI_ISL_1030457, EPI_ISL_1030458, EPI_ISL_1030459, EPI_ISL_1030460, EPI_ISL_1030461, EPI_ISL_1030462, EPI_ISL_1030463, EPI_ISL_1030464, EPI_ISL_1030465, EPI_ISL_1030466, EPI_ISL_1030467, EPI_ISL_1030468, EPI_ISL_1030469, EPI_ISL_1030470, EPI_ISL_1030471, EPI_ISL_1030472, EPI_ISL_1030473, EPI_ISL_1030474, EPI_ISL_1030475, EPI_ISL_1030476, EPI_ISL_1030477, EPI_ISL_1030478, EPI_ISL_1030479, EPI_ISL_1030480, EPI_ISL_1030481, EPI_ISL_1030482, EPI_ISL_1030483, EPI_ISL_1030484, EPI_ISL_1030485, EPI_ISL_1030486, EPI_ISL_1030487, EPI_ISL_1030488, EPI_ISL_1030489, EPI_ISL_1030490, EPI_ISL_1030491, EPI_ISL_1030492, EPI_ISL_1030493, EPI_ISL_1030494, EPI_ISL_1030495, EPI_ISL_1030496, EPI_ISL_1030497, EPI_ISL_1030498 | | | |
| see above | Orange County Public Health Lab | Chan-Zuckerberg Biohub | CZB Cliahub Consortium |
| EPI_ISL_1030499 | Humboldt County Public Health Laboratory | Chan-Zuckerberg Biohub | CZB Cliahub Consortium |
| EPI_ISL_1030500, EPI_ISL_1030501, EPI_ISL_1030502 | Santa Clara County Public Health Laboratory | Chan-Zuckerberg Biohub | CZB Cliahub Consortium |
| EPI_ISL_1030503, EPI_ISL_1030504, EPI_ISL_1030505, EPI_ISL_1030506, EPI_ISL_1030507, EPI_ISL_1030508, EPI_ISL_1030509, EPI_ISL_1030510, EPI_ISL_1030511, EPI_ISL_1030512, EPI_ISL_1030513, EPI_ISL_1030514, EPI_ISL_1030515, EPI_ISL_1030516, EPI_ISL_1030517, EPI_ISL_1030518, EPI_ISL_1030519, EPI_ISL_1030520, EPI_ISL_1030521, EPI_ISL_1030522, EPI_ISL_1030523, EPI_ISL_1030524, EPI_ISL_1030525, EPI_ISL_1030526, EPI_ISL_1030527, EPI_ISL_1030528, EPI_ISL_1030529, EPI_ISL_1030530, EPI_ISL_1030531, EPI_ISL_1030532, EPI_ISL_1030533, EPI_ISL_1030534, EPI_ISL_1030535, EPI_ISL_1030536, EPI_ISL_1030537, EPI_ISL_1030538, EPI_ISL_1030539, EPI_ISL_1030540, EPI_ISL_1030541, EPI_ISL_1030542, EPI_ISL_1030543, EPI_ISL_1030544, EPI_ISL_1030545, EPI_ISL_1030546, EPI_ISL_1030547, EPI_ISL_1030548, EPI_ISL_1030549, EPI_ISL_1030550, EPI_ISL_1030551, EPI_ISL_1030552, EPI_ISL_1030553, EPI_ISL_1030554, EPI_ISL_1030555, EPI_ISL_1030556, EPI_ISL_1030557, EPI_ISL_1030558, EPI_ISL_1030559, EPI_ISL_1030560, EPI_ISL_1030561, EPI_ISL_1030562, EPI_ISL_1030563, EPI_ISL_1030564, EPI_ISL_1030565, EPI_ISL_1030566, EPI_ISL_1030567, EPI_ISL_1030568, EPI_ISL_1030569, EPI_ISL_1030570, EPI_ISL_1030571, EPI_ISL_1030572, EPI_ISL_1030573, EPI_ISL_1030574, EPI_ISL_1030575, EPI_ISL_1030576, EPI_ISL_1030577, EPI_ISL_1030578, EPI_ISL_1030579, EPI_ISL_1030580, EPI_ISL_1030581, EPI_ISL_1030582, EPI_ISL_1030583, EPI_ISL_1030584, EPI_ISL_1030585, EPI_ISL_1030586, EPI_ISL_1030587, EPI_ISL_1030588, EPI_ISL_1030589, EPI_ISL_1030590, EPI_ISL_1030591, EPI_ISL_1030592, EPI_ISL_1030593, EPI_ISL_1030594, EPI_ISL_1030595, EPI_ISL_1030596, EPI_ISL_1030597, EPI_ISL_1030598, EPI_ISL_1030599, EPI_ISL_1030600, EPI_ISL_1030601, EPI_ISL_1030602, EPI_ISL_1030603, EPI_ISL_1030604, EPI_ISL_1030605, EPI_ISL_1030606, EPI_ISL_1030607, EPI_ISL_1030608, EPI_ISL_1030609, EPI_ISL_1030610, EPI_ISL_1030611, EPI_ISL_1030612, EPI_ISL_1030613, EPI_ISL_1030614, EPI_ISL_1030615, EPI_ISL_1030616, EPI_ISL_1030617, EPI_ISL_1030618, EPI_ISL_1030619, EPI_ISL_1030620, EPI_ISL_1030621, EPI_ISL_1030622, EPI_ISL_1030623, EPI_ISL_1030624, EPI_ISL_1030625, EPI_ISL_1030626, EPI_ISL_1030627, EPI_ISL_1030628, EPI_ISL_1030629, EPI_ISL_1030630, EPI_ISL_1030631, EPI_ISL_1030632, EPI_ISL_1030633, EPI_ISL_1030634, EPI_ISL_1030635, EPI_ISL_1030636, EPI_ISL_1030637, EPI_ISL_1030638, EPI_ISL_1030639, EPI_ISL_1030640, EPI_ISL_1030641, EPI_ISL_1030642, EPI_ISL_1030643, EPI_ISL_1030644, EPI_ISL_1030645, EPI_ISL_1030646, EPI_ISL_1030647, EPI_ISL_1030648, EPI_ISL_1030649, EPI_ISL_1030650, EPI_ISL_1030651, EPI_ISL_1030652, EPI_ISL_1030653, EPI_ISL_1030654, EPI_ISL_1030655, EPI_ISL_1030656, EPI_ISL_1030657, EPI_ISL_1030658, EPI_ISL_1030659, EPI_ISL_1030660, EPI_ISL_1030661, EPI_ISL_1030662, EPI_ISL_1030663, EPI_ISL_1030664, EPI_ISL_1030665, EPI_ISL_1030666, EPI_ISL_1030667, EPI_ISL_1030668, EPI_ISL_1030669, EPI_ISL_1030670, EPI_ISL_1030671, EPI_ISL_1030672, EPI_ISL_1030673, EPI_ISL_1030674, EPI_ISL_1030675, EPI_ISL_1030676, EPI_ISL_1030677, EPI_ISL_1030678, EPI_ISL_1030679, EPI_ISL_1030680, EPI_ISL_1030681, EPI_ISL_1030682, EPI_ISL_1030683, EPI_ISL_1030684, EPI_ISL_1030685, EPI_ISL_1030686, EPI_ISL_1030687, EPI_ISL_1030688, EPI_ISL_1030689, EPI_ISL_1030690, EPI_ISL_1030691, EPI_ISL_1030692, EPI_ISL_1030693, EPI_ISL_1030694, EPI_ISL_1030695, EPI_ISL_1030696, EPI_ISL_1030697, EPI_ISL_1030698, EPI_ISL_1030699, EPI_ISL_1030700, EPI_ISL_1030701, EPI_ISL_1030702, EPI_ISL_1030703, EPI_ISL_1030704, EPI_ISL_1030705, EPI_ISL_1030706, EPI_ISL_1030707, EPI_ISL_1030708, EPI_ISL_1030709, EPI_ISL_1030710, EPI_ISL_1030711, EPI_ISL_1030712, EPI_ISL_1030713, EPI_ISL_1030714, EPI_ISL_1030715, EPI_ISL_1030716, EPI_ISL_1030717, EPI_ISL_1030718, EPI_ISL_1030719, EPI_ISL_1030720, EPI_ISL_1030721, EPI_ISL_1030722, EPI_ISL_1030723, EPI_ISL_1030724, EPI_ISL_1030725, EPI_ISL_1030726, EPI_ISL_1030727, EPI_ISL_1030728, EPI_ISL_1030729, EPI_ISL_1030730, EPI_ISL_1030731, EPI_ISL_1030732, EPI_ISL_1030733, EPI_ISL_1030734, EPI_ISL_1030735, EPI_ISL_1030736, EPI_ISL_1030737, EPI_ISL_1030738, EPI_ISL_1030739, EPI_ISL_1030740, EPI_ISL_1030741, EPI_ISL_1030742, EPI_ISL_1030743, EPI_ISL_1030744, EPI_ISL_1030745, EPI_ISL_1030746, EPI_ISL_1031846, EPI_ISL_1031847, EPI_ISL_1031848, EPI_ISL_1031850, EPI_ISL_1031851, EPI_ISL_1031852, EPI_ISL_1031853, EPI_ISL_1031854, EPI_ISL_1031856, EPI_ISL_1031857, EPI_ISL_1031858, EPI_ISL_1031859, EPI_ISL_1031860, EPI_ISL_1031861, EPI_ISL_1031862, EPI_ISL_1031863, EPI_ISL_1031864, EPI_ISL_1031865, EPI_ISL_1031866, EPI_ISL_1031867, EPI_ISL_1031868, EPI_ISL_1031869, EPI_ISL_1031870, EPI_ISL_1031871, EPI_ISL_1031872, EPI_ISL_1031873, EPI_ISL_1031874, EPI_ISL_1031875, EPI_ISL_1031876, EPI_ISL_1031877, EPI_ISL_1031878, EPI_ISL_1031879, EPI_ISL_1031880, EPI_ISL_1031881, EPI_ISL_1031882, EPI_ISL_1031883, EPI_ISL_1031884, EPI_ISL_1031885, EPI_ISL_1031886, EPI_ISL_1031887, EPI_ISL_1031888, EPI_ISL_1031889, EPI_ISL_1031890, EPI_ISL_1031891, EPI_ISL_1031892, EPI_ISL_1031893, EPI_ISL_1031894, EPI_ISL_1031895, EPI_ISL_1031896, EPI_ISL_1031897, EPI_ISL_1031898, EPI_ISL_1031899, EPI_ISL_1031900, EPI_ISL_1031901, EPI_ISL_1031902, EPI_ISL_1031903, EPI_ISL_1031904, EPI_ISL_1031905, EPI_ISL_1031906, EPI_ISL_1031907, EPI_ISL_1031908, EPI_ISL_1031909, EPI_ISL_1031910, EPI_ISL_1031911, EPI_ISL_1031912, EPI_ISL_1031913, EPI_ISL_1031914, EPI_ISL_1031915, EPI_ISL_1031916, EPI_ISL_1031917, EPI_ISL_1031918, EPI_ISL_1031919, EPI_ISL_1031920, EPI_ISL_1031921, EPI_ISL_1031922, EPI_ISL_1031923, EPI_ISL_1031924, EPI_ISL_1031925, EPI_ISL_1031926, EPI_ISL_1031927, EPI_ISL_1031928, EPI_ISL_1031929, EPI_ISL_1031930, EPI_ISL_1031931, EPI_ISL_1031932, EPI_ISL_1031933, EPI_ISL_1031934, EPI_ISL_1031935, EPI_ISL_1031936, EPI_ISL_1031937, EPI_ISL_1031938, EPI_ISL_1031939, EPI_ISL_1031940, EPI_ISL_1031941, EPI_ISL_1031942, EPI_ISL_1031943, EPI_ISL_1031944, EPI_ISL_1031945, EPI_ISL_1031946, EPI_ISL_1031947, EPI_ISL_1031948, EPI_ISL_1031949, EPI_ISL_1031950, EPI_ISL_1031951, EPI_ISL_1031952, EPI_ISL_1031953, EPI_ISL_1031954, EPI_ISL_1031955, EPI_ISL_1031956, EPI_ISL_1031957, EPI_ISL_1031958, EPI_ISL_1031959, EPI_ISL_1031960, EPI_ISL_1031961, EPI_ISL_1031962, EPI_ISL_1031963, EPI_ISL_1031964, EPI_ISL_1031965, EPI_ISL_1031966, EPI_ISL_1031967, EPI_ISL_1031968, EPI_ISL_1031969, EPI_ISL_1031970, EPI_ISL_1031971, EPI_ISL_1031972, EPI_ISL_1031973, EPI_ISL_1031974, EPI_ISL_1031975, EPI_ISL_1031976, EPI_ISL_1031977, EPI_ISL_1031978, EPI_ISL_1031979, EPI_ISL_1031980, EPI_ISL_1031981, EPI_ISL_1031982, EPI_ISL_1031983, EPI_ISL_1031984, EPI_ISL_1031985, EPI_ISL_1031986, EPI_ISL_1031987, EPI_ISL_1031988, EPI_ISL_1031989, EPI_ISL_1031990, EPI_ISL_1031991, EPI_ISL_1031992, EPI_ISL_1031993, EPI_ISL_1031994, EPI_ISL_1031995, EPI_ISL_1031996, EPI_ISL_1031997, EPI_ISL_1031998, EPI_ISL_1031999, EPI_ISL_1032000, EPI_ISL_1032001, EPI_ISL_1032002, EPI_ISL_1032003, EPI_ISL_1032004, EPI_ISL_1032005, EPI_ISL_1032006, EPI_ISL_1032007, EPI_ISL_1032008, EPI_ISL_1032009, EPI_ISL_1032010, EPI_ISL_1032011, EPI_ISL_1032012, EPI_ISL_1032013, EPI_ISL_1032014, EPI_ISL_1032015, EPI_ISL_1032016, EPI_ISL_1032017, EPI_ISL_1032018, EPI_ISL_1032019, EPI_ISL_1032020, EPI_ISL_1032021, EPI_ISL_1032022, EPI_ISL_1032023, EPI_ISL_1032024, EPI_ISL_1032025, EPI_ISL_1032026, EPI_ISL_1032027, EPI_ISL_1032028, EPI_ISL_1032029, EPI_ISL_1032030, EPI_ISL_1032031, EPI_ISL_1032032, EPI_ISL_1032033, EPI_ISL_1032034, EPI_ISL_1032035, EPI_ISL_1032036, EPI_ISL_1032037, EPI_ISL_1032038, EPI_ISL_1032039, EPI_ISL_1032040, EPI_ISL_1032041, EPI_ISL_1032042, EPI_ISL_1032043, | | | |

| | | | |
|---|---|---|---|
| EPI_ISL_1032044, EPI_ISL_1032045, EPI_ISL_1032046, EPI_ISL_1032047, EPI_ISL_1032048, EPI_ISL_1032049, EPI_ISL_1032050, EPI_ISL_1032051, EPI_ISL_1032052, EPI_ISL_1032053, EPI_ISL_1032054, EPI_ISL_1032055, EPI_ISL_1032056, EPI_ISL_1032057, EPI_ISL_1032058, EPI_ISL_1032059, EPI_ISL_1032060, EPI_ISL_1032061, EPI_ISL_1032062, EPI_ISL_1032063, EPI_ISL_1032064, EPI_ISL_1032065, EPI_ISL_1032066, EPI_ISL_1032067, EPI_ISL_1032068, EPI_ISL_1032069, EPI_ISL_1032070, EPI_ISL_1032071, EPI_ISL_1032072, EPI_ISL_1032073, EPI_ISL_1032074, EPI_ISL_1032075, EPI_ISL_1032076, EPI_ISL_1032077, EPI_ISL_1032078, EPI_ISL_1032079, EPI_ISL_1032080, EPI_ISL_1032081, EPI_ISL_1032082, EPI_ISL_1032083, EPI_ISL_1032084, EPI_ISL_1032085, EPI_ISL_1032086, EPI_ISL_1032087, EPI_ISL_1032088, EPI_ISL_1032089, EPI_ISL_1032090, EPI_ISL_1032091, EPI_ISL_1032092, EPI_ISL_1032093, EPI_ISL_1032094, EPI_ISL_1032095, EPI_ISL_1032096, EPI_ISL_1032097, EPI_ISL_1032098, EPI_ISL_1032099, EPI_ISL_1032100, EPI_ISL_1032101, EPI_ISL_1032102, EPI_ISL_1032103, EPI_ISL_1032104, EPI_ISL_1032105, EPI_ISL_1032106, EPI_ISL_1032107, EPI_ISL_1032108, EPI_ISL_1032109, EPI_ISL_1032110, EPI_ISL_1032111, EPI_ISL_1032112, EPI_ISL_1032113, EPI_ISL_1032114, EPI_ISL_1032115, EPI_ISL_1032116, EPI_ISL_1032117, EPI_ISL_1032118, EPI_ISL_1032119, EPI_ISL_1032120, EPI_ISL_1032121, EPI_ISL_1032122, EPI_ISL_1032123, EPI_ISL_1032124, EPI_ISL_1032125, EPI_ISL_1032126, EPI_ISL_1032127, EPI_ISL_1032128, EPI_ISL_1032129, EPI_ISL_1032130, EPI_ISL_1032131, EPI_ISL_1032132, EPI_ISL_1032133, EPI_ISL_1032134, EPI_ISL_1032135, EPI_ISL_1032136, EPI_ISL_1032137, EPI_ISL_1032138, EPI_ISL_1032139, EPI_ISL_1032140, EPI_ISL_1032141, EPI_ISL_1032142, EPI_ISL_1032143, EPI_ISL_1032144, EPI_ISL_1032145, EPI_ISL_1032146, EPI_ISL_1032147, EPI_ISL_1032148, EPI_ISL_1032149, EPI_ISL_1032150, EPI_ISL_1032151, EPI_ISL_1032152, EPI_ISL_1032153, EPI_ISL_1032154, EPI_ISL_1032155, EPI_ISL_1032156, EPI_ISL_1032157, EPI_ISL_1032158, EPI_ISL_1032159, EPI_ISL_1032160, EPI_ISL_1032161, EPI_ISL_1032162, EPI_ISL_1032163, EPI_ISL_1032164, EPI_ISL_1032165, EPI_ISL_1032166, EPI_ISL_1032167, EPI_ISL_1032168, EPI_ISL_1032169, EPI_ISL_1032170, EPI_ISL_1032171, EPI_ISL_1032172, EPI_ISL_1032173, EPI_ISL_1032174, EPI_ISL_1032175, EPI_ISL_1032176, EPI_ISL_1032177, EPI_ISL_1032178, EPI_ISL_1032179, EPI_ISL_1032180, EPI_ISL_1032181, EPI_ISL_1032182, EPI_ISL_1032183, EPI_ISL_1032184, EPI_ISL_1032185, EPI_ISL_1032186, EPI_ISL_1032187, EPI_ISL_1032188, EPI_ISL_1032189, EPI_ISL_1032190, EPI_ISL_1032191, EPI_ISL_1032192, EPI_ISL_1032193, EPI_ISL_1032194, EPI_ISL_1032195, EPI_ISL_1032196, EPI_ISL_1032197, EPI_ISL_1032198, EPI_ISL_1032199, EPI_ISL_1032200, EPI_ISL_1032201, EPI_ISL_1032202, EPI_ISL_1032203, EPI_ISL_1032204, EPI_ISL_1032205, EPI_ISL_1032206, EPI_ISL_1032207, EPI_ISL_1032208, EPI_ISL_1032209, EPI_ISL_1032210, EPI_ISL_1032211, EPI_ISL_1032212, EPI_ISL_1032213, EPI_ISL_1032214, EPI_ISL_1032215, EPI_ISL_1032216, EPI_ISL_1032217, EPI_ISL_1032218, EPI_ISL_1032219, EPI_ISL_1032220, EPI_ISL_1032221, EPI_ISL_1032222, EPI_ISL_1032223, EPI_ISL_1032224, EPI_ISL_1032225, EPI_ISL_1032226, EPI_ISL_1032227, EPI_ISL_1032228, EPI_ISL_1032229, EPI_ISL_1032230, EPI_ISL_1032231, EPI_ISL_1032232, EPI_ISL_1032233, EPI_ISL_1032234, EPI_ISL_1032235, EPI_ISL_1032236, EPI_ISL_1032237, EPI_ISL_1032238, EPI_ISL_1032239, EPI_ISL_1032240, EPI_ISL_1032241, EPI_ISL_1032242, EPI_ISL_1032243, EPI_ISL_1032244, EPI_ISL_1032245, EPI_ISL_1032246, EPI_ISL_1032247, EPI_ISL_1032248, EPI_ISL_1032249, EPI_ISL_1032250, EPI_ISL_1032251, EPI_ISL_1032252, EPI_ISL_1032253, EPI_ISL_1032254, EPI_ISL_1032255, EPI_ISL_1032256, EPI_ISL_1032257, EPI_ISL_1032258, EPI_ISL_1032259, EPI_ISL_1032260, EPI_ISL_1032261, EPI_ISL_1032262, EPI_ISL_1032263, EPI_ISL_1032264, EPI_ISL_1032265, EPI_ISL_1032266, EPI_ISL_1032267, EPI_ISL_1032268, EPI_ISL_1032269, EPI_ISL_1032270, EPI_ISL_1032271, EPI_ISL_1032272, EPI_ISL_1032273, EPI_ISL_1032274, EPI_ISL_1032275, EPI_ISL_1032276, EPI_ISL_1032277, EPI_ISL_1032278, EPI_ISL_1032279, EPI_ISL_1032280, EPI_ISL_1032281, EPI_ISL_1032282, EPI_ISL_1032283, EPI_ISL_1032284, EPI_ISL_1032285, EPI_ISL_1032286, EPI_ISL_1032287, EPI_ISL_1032288, EPI_ISL_1032289, EPI_ISL_1032290, EPI_ISL_1032291, EPI_ISL_1032292, EPI_ISL_1032293, EPI_ISL_1032294, EPI_ISL_1032295, EPI_ISL_1032296, EPI_ISL_1032297, EPI_ISL_1032298, EPI_ISL_1032299, EPI_ISL_1032300, EPI_ISL_1032301, EPI_ISL_1032302, EPI_ISL_1032303, EPI_ISL_1032304, EPI_ISL_1032305, EPI_ISL_1032306, EPI_ISL_1032307, EPI_ISL_1032308, EPI_ISL_1032309, EPI_ISL_1032310, EPI_ISL_1032311, EPI_ISL_1032312, EPI_ISL_1032313, EPI_ISL_1032314, EPI_ISL_1032315, EPI_ISL_1032316, EPI_ISL_1032317, EPI_ISL_1032318, EPI_ISL_1032319, EPI_ISL_1032320, EPI_ISL_1032321, EPI_ISL_1032322, EPI_ISL_1032323, EPI_ISL_1032324, EPI_ISL_1032325, EPI_ISL_1032326, EPI_ISL_1032327, EPI_ISL_1032328, EPI_ISL_1032329, EPI_ISL_1032330, EPI_ISL_1032331, EPI_ISL_1032332, EPI_ISL_1032333, EPI_ISL_1032334, EPI_ISL_1032335, EPI_ISL_1032336, EPI_ISL_1032337, EPI_ISL_1032338, EPI_ISL_1032339, EPI_ISL_1032340, EPI_ISL_1032341, EPI_ISL_1032342, EPI_ISL_1032343, EPI_ISL_1032344, EPI_ISL_1032345, EPI_ISL_1032346, EPI_ISL_1032347, EPI_ISL_1032348, EPI_ISL_1032349, EPI_ISL_1032350, EPI_ISL_1032351, EPI_ISL_1032352, EPI_ISL_1032353, EPI_ISL_1032354, EPI_ISL_1032355, EPI_ISL_1032356, EPI_ISL_1032357, EPI_ISL_1032358, EPI_ISL_1032359, EPI_ISL_1032360, EPI_ISL_1032361, EPI_ISL_1032362, EPI_ISL_1032363, EPI_ISL_1032364, EPI_ISL_1032365, EPI_ISL_1032366, EPI_ISL_1032367, EPI_ISL_1032368, EPI_ISL_1032369, EPI_ISL_1032370, EPI_ISL_1032371, EPI_ISL_1032372, EPI_ISL_1032373, EPI_ISL_1032374, EPI_ISL_1032375, EPI_ISL_1032376, EPI_ISL_1032377, EPI_ISL_1032378, EPI_ISL_1032379, EPI_ISL_1032380, EPI_ISL_1032381, EPI_ISL_1032382, EPI_ISL_1032383, EPI_ISL_1032384, EPI_ISL_1032385, EPI_ISL_1032386, EPI_ISL_1032387, EPI_ISL_1032388, EPI_ISL_1032389, EPI_ISL_1032390, EPI_ISL_1032391, EPI_ISL_1032392, EPI_ISL_1032393, EPI_ISL_1032394, EPI_ISL_1032395, EPI_ISL_1032396, EPI_ISL_1032397, EPI_ISL_1032398, EPI_ISL_1032399, EPI_ISL_1032400, EPI_ISL_1032401, EPI_ISL_1032402, EPI_ISL_1032403, EPI_ISL_1032404, EPI_ISL_1032405, EPI_ISL_1032406, EPI_ISL_1032407, EPI_ISL_1032408, EPI_ISL_1032409, EPI_ISL_1032410, EPI_ISL_1032411, EPI_ISL_1032412, EPI_ISL_1032413, EPI_ISL_1032414, EPI_ISL_1032415, EPI_ISL_1032416, EPI_ISL_1032417, EPI_ISL_1032418, EPI_ISL_1032419, EPI_ISL_1032420, EPI_ISL_1032421, EPI_ISL_1032422, EPI_ISL_1032423, EPI_ISL_1032424, EPI_ISL_1032425, EPI_ISL_1032426, EPI_ISL_1032427, EPI_ISL_1032428, EPI_ISL_1032429, EPI_ISL_1032430, EPI_ISL_1032431, EPI_ISL_1032432, EPI_ISL_1032433, EPI_ISL_1032434, EPI_ISL_1032435, EPI_ISL_1032436, EPI_ISL_1032437, EPI_ISL_1032438, EPI_ISL_1032439, EPI_ISL_1032440, EPI_ISL_1032441, EPI_ISL_1032442, EPI_ISL_1032443, EPI_ISL_1032444, EPI_ISL_1032445, EPI_ISL_1032446, EPI_ISL_1032447, EPI_ISL_1032448, EPI_ISL_1032449, EPI_ISL_1032450, EPI_ISL_1032451, EPI_ISL_1032452, EPI_ISL_1032453, EPI_ISL_1032454, EPI_ISL_1032455, EPI_ISL_1032456, EPI_ISL_1032457, EPI_ISL_1032458, EPI_ISL_1032459, EPI_ISL_1032460, EPI_ISL_1032461, EPI_ISL_1032462, EPI_ISL_1032463, EPI_ISL_1032464, EPI_ISL_1032465, EPI_ISL_1032466, EPI_ISL_1032467, EPI_ISL_1032468, EPI_ISL_1032469, EPI_ISL_1032470, EPI_ISL_1032471, EPI_ISL_1032472, EPI_ISL_1032473, EPI_ISL_1032474, EPI_ISL_1032475, EPI_ISL_1032476, EPI_ISL_1032477, EPI_ISL_1032478, EPI_ISL_1032479, EPI_ISL_1032480, EPI_ISL_1032481, EPI_ISL_1032482, EPI_ISL_1032483, EPI_ISL_1032484, EPI_ISL_1032485, EPI_ISL_1032486, EPI_ISL_1032487, EPI_ISL_1032488, EPI_ISL_1032489, EPI_ISL_1032490, EPI_ISL_1032491, EPI_ISL_1032492, EPI_ISL_1032493, EPI_ISL_1032494, EPI_ISL_1032495, EPI_ISL_1032496, EPI_ISL_1032497, EPI_ISL_1032498, EPI_ISL_1032499, EPI_ISL_1032500, EPI_ISL_1032501, EPI_ISL_1032502, EPI_ISL_1032503, EPI_ISL_1032504, EPI_ISL_1032505, EPI_ISL_1032506, EPI_ISL_1032507, EPI_ISL_1032508, EPI_ISL_1032509, EPI_ISL_1032510, EPI_ISL_1032511, EPI_ISL_1032512, EPI_ISL_1032513, EPI_ISL_1032514, EPI_ISL_1032515, EPI_ISL_1032516, EPI_ISL_1032517, EPI_ISL_1032518, EPI_ISL_1032519, EPI_ISL_1032520, EPI_ISL_1032521, EPI_ISL_1032522, EPI_ISL_1032523, EPI_ISL_1032524, EPI_ISL_1032525, EPI_ISL_1032526, EPI_ISL_1032527, EPI_ISL_1032528, EPI_ISL_1032529, EPI_ISL_1032530, EPI_ISL_1032531, EPI_ISL_1032532, EPI_ISL_1032533, EPI_ISL_1032534, EPI_ISL_1032535, EPI_ISL_1032536, EPI_ISL_1032537, EPI_ISL_1032538, EPI_ISL_1032539, EPI_ISL_1032540, EPI_ISL_1032541, EPI_ISL_1032542, EPI_ISL_1032543, EPI_ISL_1032544, EPI_ISL_1032545, EPI_ISL_1032546, EPI_ISL_1032547, EPI_ISL_1032548, EPI_ISL_1032549, EPI_ISL_1032550, EPI_ISL_1032551, EPI_ISL_1032552, EPI_ISL_1032553, EPI_ISL_1032554, EPI_ISL_1032555, EPI_ISL_1032556, EPI_ISL_1032557, EPI_ISL_1032558, EPI_ISL_1032559, EPI_ISL_1032560, EPI_ISL_1032561, EPI_ISL_1032562, EPI_ISL_1032563, EPI_ISL_1032564, EPI_ISL_1032565, EPI_ISL_1032566, EPI_ISL_1032567, EPI_ISL_1032568, EPI_ISL_1032569, EPI_ISL_1032570, EPI_ISL_1032571, EPI_ISL_1032572, EPI_ISL_1032573, EPI_ISL_1032574, EPI_ISL_1032575, EPI_ISL_1032576, EPI_ISL_1032577, EPI_ISL_1032578, EPI_ISL_1032579, EPI_ISL_1032580, EPI_ISL_1032581, EPI_ISL_1032582 | | | |
| see above | Laboratory Corporation of America | Respiratory Viruses Branch, Division of Viral Diseases, Centers for Disease Control and Prevention | Peter W. Cook, Dakota Howard, Dhwani Batra, Ben L. Rambo-Martin, Clinton R. Paden, Suxiang Tong, Duncan MacCannell |
| EPI_ISL_1032583, EPI_ISL_1032584, EPI_ISL_1032590, EPI_ISL_1032591, EPI_ISL_1032592, EPI_ISL_1032593, EPI_ISL_1032594, EPI_ISL_1032595, EPI_ISL_1032596, EPI_ISL_1032597, EPI_ISL_1032598, EPI_ISL_1032599, EPI_ISL_1032600, EPI_ISL_1032601, EPI_ISL_1032602, EPI_ISL_1032603, EPI_ISL_1032604, EPI_ISL_1032605, EPI_ISL_1032606, EPI_ISL_1032607, EPI_ISL_1032608, EPI_ISL_1032609, EPI_ISL_1032610, EPI_ISL_1032611, EPI_ISL_1032612, EPI_ISL_1032613, EPI_ISL_1032614, EPI_ISL_1032615, EPI_ISL_1032616, EPI_ISL_1032617, EPI_ISL_1032631, EPI_ISL_1032632, EPI_ISL_1032633, EPI_ISL_1032634, EPI_ISL_1032654, EPI_ISL_1032655, EPI_ISL_1032658, EPI_ISL_1032660, EPI_ISL_1032661, EPI_ISL_1032664, EPI_ISL_1032665, EPI_ISL_1032666 | | | |
| see above | Wyoming Public Health Laboratory | Wyoming Public Health Laboratory | Noah Hull, Taylor Fearing, Lynette Gumbleton, Channing Weber, Ashley Norberg, Bailey Bowcutt, and Wanda Manley |
| EPI_ISL_1032667, EPI_ISL_1032668, EPI_ISL_1032669, EPI_ISL_1032670, EPI_ISL_1032672, EPI_ISL_1032675, EPI_ISL_1032677, EPI_ISL_1032678, EPI_ISL_1032679, EPI_ISL_1032680, EPI_ISL_1032681, EPI_ISL_1032682, EPI_ISL_1032683, EPI_ISL_1032684, EPI_ISL_1032685, EPI_ISL_1032687, EPI_ISL_1032689, EPI_ISL_1032690, EPI_ISL_1032691, EPI_ISL_1032693, EPI_ISL_1032695, EPI_ISL_1032696, EPI_ISL_1032697, EPI_ISL_1032698, EPI_ISL_1032699, EPI_ISL_1032700, EPI_ISL_1032701, EPI_ISL_1032702, EPI_ISL_1032703, EPI_ISL_1032704, EPI_ISL_1032705, EPI_ISL_1032707, EPI_ISL_1032708, EPI_ISL_1032709, EPI_ISL_1032710, EPI_ISL_1032711, EPI_ISL_1032712, EPI_ISL_1032713, EPI_ISL_1032714, EPI_ISL_1032715, EPI_ISL_1032716, EPI_ISL_1032717, EPI_ISL_1032718, EPI_ISL_1032719, EPI_ISL_1032720, EPI_ISL_1032721, EPI_ISL_1032722, EPI_ISL_1032723, EPI_ISL_1032724, EPI_ISL_1032725, EPI_ISL_1032726, EPI_ISL_1032727, EPI_ISL_1032728, EPI_ISL_1032729, EPI_ISL_1032730, EPI_ISL_1032731, EPI_ISL_1032732, EPI_ISL_1032733, EPI_ISL_1032734, EPI_ISL_1032736, EPI_ISL_1032737, EPI_ISL_1032738, EPI_ISL_1032739, EPI_ISL_1032740, EPI_ISL_1032741, EPI_ISL_1032742, EPI_ISL_1032743, EPI_ISL_1032744, EPI_ISL_1032745, EPI_ISL_1032746, EPI_ISL_1032747, EPI_ISL_1032749, EPI_ISL_1032750, EPI_ISL_1032751, EPI_ISL_1032752, EPI_ISL_1032753, EPI_ISL_1032754, EPI_ISL_1032755, EPI_ISL_1032756, EPI_ISL_1032757, EPI_ISL_1032758, EPI_ISL_1032759, EPI_ISL_1032760, EPI_ISL_1032761, EPI_ISL_1032762, EPI_ISL_1032763, EPI_ISL_1032764, EPI_ISL_1032766, EPI_ISL_1032767, EPI_ISL_1032768, EPI_ISL_1032770, EPI_ISL_1032771, EPI_ISL_1032772, EPI_ISL_1032773, EPI_ISL_1032774, EPI_ISL_1032776, EPI_ISL_1032777, EPI_ISL_1032778, EPI_ISL_1032779, EPI_ISL_1032780, EPI_ISL_1032781, EPI_ISL_1032782, EPI_ISL_1032783, EPI_ISL_1032784, EPI_ISL_1032785, EPI_ISL_1032786, EPI_ISL_1032787, EPI_ISL_1032788, EPI_ISL_1032789, EPI_ISL_1032790, EPI_ISL_1032791, EPI_ISL_1032792, EPI_ISL_1032794, EPI_ISL_1032795, EPI_ISL_1032796, EPI_ISL_1032797, EPI_ISL_1032798, EPI_ISL_1032800, EPI_ISL_1032801, EPI_ISL_1032802, EPI_ISL_1032804, EPI_ISL_1032805, EPI_ISL_1032807, EPI_ISL_1032808, EPI_ISL_1032809, EPI_ISL_1032810, EPI_ISL_1032811, EPI_ISL_1032812, EPI_ISL_1032813, EPI_ISL_1032814, EPI_ISL_1032815, EPI_ISL_1032816, EPI_ISL_1032817, EPI_ISL_1032818, EPI_ISL_1032819, EPI_ISL_1032820, EPI_ISL_1032821, EPI_ISL_1032822, EPI_ISL_1032823, EPI_ISL_1032824, EPI_ISL_1032825, EPI_ISL_1032826, EPI_ISL_1032827, EPI_ISL_1032828, EPI_ISL_1032829, EPI_ISL_1032830, EPI_ISL_1032831, EPI_ISL_1032832, EPI_ISL_1032833, EPI_ISL_1032834, EPI_ISL_1032835, EPI_ISL_1032836, EPI_ISL_1032837, EPI_ISL_1032838, EPI_ISL_1032839, EPI_ISL_1032840, EPI_ISL_1032841, EPI_ISL_1032842, EPI_ISL_1032843, EPI_ISL_1032844, EPI_ISL_1032845, EPI_ISL_1032846, EPI_ISL_1032847, EPI_ISL_1032848, EPI_ISL_1032850, EPI_ISL_1032851, EPI_ISL_1032852, EPI_ISL_1032853, EPI_ISL_1032854, EPI_ISL_1032855, EPI_ISL_1032856, EPI_ISL_1032857, EPI_ISL_1032858, EPI_ISL_1032859, EPI_ISL_1032860, EPI_ISL_1032861, EPI_ISL_1032862, EPI_ISL_1032863, EPI_ISL_1032864, EPI_ISL_1032865, EPI_ISL_1032866, EPI_ISL_1032867, EPI_ISL_1032868, EPI_ISL_1032869, EPI_ISL_1032870, EPI_ISL_1032871, EPI_ISL_1032872, EPI_ISL_1032873, EPI_ISL_1032874, EPI_ISL_1032875, EPI_ISL_1032876, EPI_ISL_1032877, EPI_ISL_1032878, EPI_ISL_1032881, EPI_ISL_1032882, EPI_ISL_1032883, EPI_ISL_1032884, EPI_ISL_1032885, EPI_ISL_1032886, EPI_ISL_1032887, EPI_ISL_1032889, EPI_ISL_1032890, EPI_ISL_1032891, EPI_ISL_1032892, EPI_ISL_1032894, EPI_ISL_1032895, EPI_ISL_1032896, EPI_ISL_1032897, EPI_ISL_1032898, EPI_ISL_1032899, EPI_ISL_1032900, EPI_ISL_1032901, EPI_ISL_1032902, EPI_ISL_1032903, EPI_ISL_1032905, EPI_ISL_1032907, EPI_ISL_1032908, EPI_ISL_1032909, EPI_ISL_1032910, EPI_ISL_1032911, EPI_ISL_1032913, EPI_ISL_1032914, EPI_ISL_1032915, EPI_ISL_1032916, EPI_ISL_1032917, EPI_ISL_1032918, EPI_ISL_1032919, EPI_ISL_1032921, EPI_ISL_1032922, EPI_ISL_1032923, EPI_ISL_1032924, EPI_ISL_1032925, EPI_ISL_1032926, EPI_ISL_1032927, EPI_ISL_1032928, EPI_ISL_1032929, EPI_ISL_1032930, EPI_ISL_1032931, EPI_ISL_1032932, EPI_ISL_1032933, EPI_ISL_1032934, EPI_ISL_1032935, EPI_ISL_1032936, EPI_ISL_1032937, EPI_ISL_1032938, EPI_ISL_1032939, EPI_ISL_1032941, EPI_ISL_1032942, EPI_ISL_1032943, EPI_ISL_1032944, EPI_ISL_1032945 | | | |
| see above | Illinois Department of Public Health | Gagnon Lab, Southern Illinois University | Keith Gagnon |
| EPI_ISL_1032946, EPI_ISL_1032947, EPI_ISL_1032949, EPI_ISL_1032952, EPI_ISL_1032953, EPI_ISL_1032955, EPI_ISL_1032957, EPI_ISL_1032959, EPI_ISL_1032960, EPI_ISL_1032962, EPI_ISL_1032963, EPI_ISL_1032964, EPI_ISL_1032967, EPI_ISL_1032969, EPI_ISL_1032972, EPI_ISL_1032973, EPI_ISL_1032975, EPI_ISL_1032979, EPI_ISL_1032980, EPI_ISL_1032981, EPI_ISL_1032982, EPI_ISL_1032983, EPI_ISL_1032984, EPI_ISL_1032986, EPI_ISL_1032987, EPI_ISL_1032989, EPI_ISL_1032990, EPI_ISL_1032991, EPI_ISL_1032993, EPI_ISL_1032995, EPI_ISL_1032999, EPI_ISL_1033002, EPI_ISL_1033003, EPI_ISL_1033004, EPI_ISL_1033006, EPI_ISL_1033009, EPI_ISL_1033010, EPI_ISL_1033011, EPI_ISL_1033012, EPI_ISL_1033013, EPI_ISL_1033014, EPI_ISL_1033017, EPI_ISL_1033018, EPI_ISL_1033020 | | | |
| see above | Fulgent Genetics | Fulgent Genetics | Harry Gao, Mickey Li, John Gao, Joseph Fierro, Benafsh Sapra, Becky Tsai, Yan Meng, Doreen Ng, James Xie |
| EPI_ISL_1034275, EPI_ISL_1034276, EPI_ISL_1034277, EPI_ISL_1034278, EPI_ISL_1034279 | Northwest Pathology | UW Virology Lab | Pavitra Roychoudhury, Lasata Shrestha, Shah A Mohamed Bakhash, Michelle Lin, Hong Xie, Meei-Li Huang, Keith R Jerome, Alexander Greninger |
| EPI_ISL_1035828, EPI_ISL_1035829, EPI_ISL_1035830, EPI_ISL_1035831, EPI_ISL_1035832, EPI_ISL_1035833, EPI_ISL_1035834, EPI_ISL_1035835, EPI_ISL_1035836, EPI_ISL_1035837, EPI_ISL_1035838, EPI_ISL_1035839, EPI_ISL_1035840, EPI_ISL_1035841, EPI_ISL_1035842, EPI_ISL_1035843, EPI_ISL_1035844, EPI_ISL_1035845, EPI_ISL_1035846, EPI_ISL_1035847, EPI_ISL_1035848, EPI_ISL_1035849, EPI_ISL_1035850, EPI_ISL_1035851, EPI_ISL_1035852, EPI_ISL_1035853, EPI_ISL_1035854, EPI_ISL_1035855, EPI_ISL_1035856, EPI_ISL_1035864, EPI_ISL_1035865, EPI_ISL_1035866, EPI_ISL_1035867, EPI_ISL_1035868, EPI_ISL_1035869, EPI_ISL_1035870 | | | |
| see above | North Dakota Department of Health, Public Health Laboratory | North Dakota Department of Health, Public Health Laboratory | Lisa Wingerter |
| EPI_ISL_1036056, EPI_ISL_1036057, EPI_ISL_1036234, EPI_ISL_1036241, EPI_ISL_1036242, EPI_ISL_1036243, EPI_ISL_1036244, EPI_ISL_1036245 | NCSLPH | NCSLPH | Chase K, Miller MC, Greene S, Glover W |
| EPI_ISL_1036246, EPI_ISL_1036247, EPI_ISL_1036248, EPI_ISL_1036249, EPI_ISL_1036250, EPI_ISL_1036251, EPI_ISL_1036252, EPI_ISL_1036253, EPI_ISL_1036254, EPI_ISL_1036255, EPI_ISL_1036256, EPI_ISL_1036257, EPI_ISL_1036258, EPI_ISL_1036259, EPI_ISL_1036260, EPI_ISL_1036261, EPI_ISL_1036262, EPI_ISL_1036263, EPI_ISL_1036264, EPI_ISL_1036265, EPI_ISL_1036266, EPI_ISL_1036267 | | | |
| see above | Center for Genome Research and Biocomputing | Center for Genome Research and Biocomputing | Oregon State University TRACE Project |
| EPI_ISL_1036268, EPI_ISL_1036270, EPI_ISL_1036271 | NCSLPH | NCSLPH | Chase K, Miller MC, Greene S, Glover W |
| EPI_ISL_1036273, EPI_ISL_1036274, EPI_ISL_1036275, EPI_ISL_1036276, EPI_ISL_1036277, EPI_ISL_1036278, EPI_ISL_1036279, EPI_ISL_1036280, EPI_ISL_1036281, EPI_ISL_1036282, EPI_ISL_1036283, EPI_ISL_1036284, EPI_ISL_1036285, EPI_ISL_1036286, EPI_ISL_1036287, EPI_ISL_1036288, EPI_ISL_1036289, EPI_ISL_1036290, EPI_ISL_1036291, EPI_ISL_1036292, EPI_ISL_1036293, EPI_ISL_1036294, EPI_ISL_1036295, EPI_ISL_1036296, EPI_ISL_1036297, EPI_ISL_1036298, EPI_ISL_1036299, EPI_ISL_1036300, EPI_ISL_1036301, EPI_ISL_1036302, EPI_ISL_1036303, EPI_ISL_1036304, EPI_ISL_1036305, EPI_ISL_1036306, EPI_ISL_1036307, EPI_ISL_1036308, EPI_ISL_1036309, EPI_ISL_1036310, EPI_ISL_1036311, EPI_ISL_1036312, EPI_ISL_1036313, EPI_ISL_1036314, EPI_ISL_1036315, EPI_ISL_1036316, EPI_ISL_1036317, EPI_ISL_1036318, EPI_ISL_1036319, EPI_ISL_1036320, EPI_ISL_1036321, EPI_ISL_1036322, EPI_ISL_1036323, EPI_ISL_1036324, EPI_ISL_1036325, EPI_ISL_1036326, EPI_ISL_1036327, EPI_ISL_1036328, EPI_ISL_1036329, EPI_ISL_1036330, EPI_ISL_1036331, EPI_ISL_1036332, EPI_ISL_1036333, EPI_ISL_1036334, EPI_ISL_1036335, EPI_ISL_1036336, EPI_ISL_1036337, EPI_ISL_1036338, EPI_ISL_1036339, EPI_ISL_1036340, EPI_ISL_1036341, EPI_ISL_1036342, EPI_ISL_1036343, EPI_ISL_1036344, EPI_ISL_1036345, EPI_ISL_1036346, EPI_ISL_1036347, EPI_ISL_1036348, EPI_ISL_1036349, EPI_ISL_1036350, EPI_ISL_1036351, EPI_ISL_1036352, EPI_ISL_1036353, EPI_ISL_1036354, EPI_ISL_1036355, EPI_ISL_1036356, EPI_ISL_1036357, EPI_ISL_1036358, EPI_ISL_1036359, EPI_ISL_1036360, EPI_ISL_1036361, EPI_ISL_1036362, EPI_ISL_1036363, EPI_ISL_1036364, EPI_ISL_1036365, EPI_ISL_1036366, EPI_ISL_1036367, EPI_ISL_1036368, EPI_ISL_1036369, EPI_ISL_1036370, EPI_ISL_1036371, EPI_ISL_1036372 | | | |
| see above | Johns Hopkins Hospital Department of Pathology | Johns Hopkins Hospital Department of Pathology | C. Paul Morris, Chun Huai Luo, Adannaya Amadi, Matthew Schwartz, Nicholas Gallagher, Heba H. Mostafa |
| EPI_ISL_1036433, EPI_ISL_1036434, EPI_ISL_1036435, EPI_ISL_1036436, EPI_ISL_1036437, EPI_ISL_1036438, EPI_ISL_1036439, EPI_ISL_1036440, EPI_ISL_1036441, EPI_ISL_1036442, EPI_ISL_1036443, EPI_ISL_1036444, EPI_ISL_1036445, EPI_ISL_1036446, EPI_ISL_1036447, EPI_ISL_1036448, EPI_ISL_1036449, EPI_ISL_1036450, EPI_ISL_1036452, EPI_ISL_1036453, EPI_ISL_1036454, EPI_ISL_1036455, EPI_ISL_1036456, EPI_ISL_1036457, EPI_ISL_1036458, EPI_ISL_1036459, EPI_ISL_1036460, EPI_ISL_1036461, EPI_ISL_1036462, EPI_ISL_1036463, EPI_ISL_1036464, EPI_ISL_1036465, EPI_ISL_1036467, EPI_ISL_1036468, EPI_ISL_1036469, EPI_ISL_1036472, EPI_ISL_1036473, EPI_ISL_1036476, EPI_ISL_1036477, EPI_ISL_1036478, EPI_ISL_1036479, EPI_ISL_1036480, EPI_ISL_1036481, EPI_ISL_1036483, EPI_ISL_1036485, EPI_ISL_1036486, EPI_ISL_1036487, EPI_ISL_1036488, EPI_ISL_1036489, EPI_ISL_1036490, EPI_ISL_1036491, EPI_ISL_1036492, EPI_ISL_1036494, EPI_ISL_1036495, EPI_ISL_1036496, EPI_ISL_1036497, EPI_ISL_1036498, EPI_ISL_1036499, EPI_ISL_1036500, EPI_ISL_1036501, EPI_ISL_1036502, EPI_ISL_1036503, EPI_ISL_1036504, EPI_ISL_1036505, EPI_ISL_1036506, EPI_ISL_1036507, EPI_ISL_1036508, EPI_ISL_1036509, EPI_ISL_1036511, EPI_ISL_1036512, EPI_ISL_1036513, EPI_ISL_1036516, EPI_ISL_1036517, EPI_ISL_1036518, EPI_ISL_1036520, EPI_ISL_1036521, EPI_ISL_1036522, EPI_ISL_1036523, EPI_ISL_1036524, EPI_ISL_1036525, EPI_ISL_1036526, EPI_ISL_1036527, EPI_ISL_1036528, EPI_ISL_1036529, EPI_ISL_1036530, EPI_ISL_1036531, EPI_ISL_1036532, EPI_ISL_1036533, EPI_ISL_1036534, EPI_ISL_1036535, EPI_ISL_1036536, EPI_ISL_1036538, EPI_ISL_1036539, EPI_ISL_1036540, EPI_ISL_1036541, EPI_ISL_1036542, EPI_ISL_1036543, EPI_ISL_1036544, EPI_ISL_1036545, EPI_ISL_1036546, EPI_ISL_1036547, EPI_ISL_1036548, EPI_ISL_1036550, EPI_ISL_1036551, EPI_ISL_1036553, EPI_ISL_1036554 | | | |
| see above | Sonora Quest Laboratories | TGen North | "Jolene Bowers, Megan Folkerts, Chris French, Hayley Yaglom, Ashlyn Pfeiffer, Darrin Lemmer, Dave Engelthaler, The Arizona COVID Genomics Union (ACGU)" |
| EPI_ISL_1036555, EPI_ISL_1036559, EPI_ISL_1036560, EPI_ISL_1036563, EPI_ISL_1036566, EPI_ISL_1036567, EPI_ISL_1036568, EPI_ISL_1036572, EPI_ISL_1036573, EPI_ISL_1036574, EPI_ISL_1036575, EPI_ISL_1036576, EPI_ISL_1036577, EPI_ISL_1036578, EPI_ISL_1036579, EPI_ISL_1036580, EPI_ISL_1036581, EPI_ISL_1036582, EPI_ISL_1036583, EPI_ISL_1036584, EPI_ISL_1036585, EPI_ISL_1036586, EPI_ISL_1036588, EPI_ISL_1036590 | | | |
| see above | TGen North | TGen North | "Jolene Bowers, Megan Folkerts, Chris French, Hayley Yaglom, Ashlyn Pfeiffer, Darrin Lemmer, Dave Engelthaler, The Arizona COVID Genomics Union (ACGU)" |
| EPI_ISL_1036624, EPI_ISL_1036625, EPI_ISL_1036626, EPI_ISL_1036627, EPI_ISL_1036628, EPI_ISL_1036630, EPI_ISL_1036631, EPI_ISL_1036632, EPI_ISL_1036633, EPI_ISL_1036634, EPI_ISL_1036635, EPI_ISL_1036636, EPI_ISL_1036637, EPI_ISL_1036638, EPI_ISL_1036639, EPI_ISL_1036640, EPI_ISL_1036641, EPI_ISL_1036642, EPI_ISL_1036643, EPI_ISL_1036644, EPI_ISL_1036645, EPI_ISL_1036646, EPI_ISL_1036648, EPI_ISL_1036649, EPI_ISL_1036650, EPI_ISL_1036651, EPI_ISL_1036652, EPI_ISL_1036653, EPI_ISL_1036654, EPI_ISL_1036655, EPI_ISL_1036656, EPI_ISL_1036657, EPI_ISL_1036661, EPI_ISL_1036662, EPI_ISL_1036663, EPI_ISL_1036664, EPI_ISL_1036665, EPI_ISL_1036666, EPI_ISL_1036667, EPI_ISL_1036668, EPI_ISL_1036669, EPI_ISL_1036670, EPI_ISL_1036671, EPI_ISL_1036672, EPI_ISL_1036673, EPI_ISL_1036674, EPI_ISL_1036675, EPI_ISL_1036676, EPI_ISL_1036677, EPI_ISL_1036678, EPI_ISL_1036679, EPI_ISL_1036680, EPI_ISL_1036681, EPI_ISL_1036682 | | | |
| see above | Wyoming Public Health Laboratory | Wyoming Public Health Laboratory | Noah Hull, Taylor Fearing, Lynette Gumbleton, Channing Weber, Ashley Norberg, Bailey Bowcutt, and Wanda Manley |
| EPI_ISL_1036684, EPI_ISL_1036685, EPI_ISL_1036686, EPI_ISL_1036687, EPI_ISL_1036688, EPI_ISL_1036689, EPI_ISL_1036690, EPI_ISL_1036691, EPI_ISL_1036692, EPI_ISL_1036693, EPI_ISL_1036694, EPI_ISL_1036695, EPI_ISL_1036696, EPI_ISL_1036697, EPI_ISL_1036698, EPI_ISL_1036699, EPI_ISL_1036700, EPI_ISL_1036701, EPI_ISL_1036702, EPI_ISL_1036703, EPI_ISL_1036704, EPI_ISL_1036705, EPI_ISL_1036706, EPI_ISL_1036707, EPI_ISL_1036708, EPI_ISL_1036709, EPI_ISL_1036710, EPI_ISL_1036711, EPI_ISL_1036712, EPI_ISL_1036713, EPI_ISL_1036714, EPI_ISL_1036715, EPI_ISL_1036716, EPI_ISL_1036717, EPI_ISL_1036718, EPI_ISL_1036719, EPI_ISL_1036720, EPI_ISL_1036721, EPI_ISL_1036722, EPI_ISL_1036723, EPI_ISL_1036724, EPI_ISL_1036725 | | | |
| see above | Helix/Illumina | Respiratory Viruses Branch, Division of Viral Diseases, Centers for Disease Control and Prevention | Peter W. Cook,Dakota Howard,Dhwani Batra,Ben L. Rambo-Martin,Eileen de Feo,Jan Antico,Christine Tran,Matthew Tolentino,Shannon Wickline,Kim Gietzen,Brad Sickler,Jingtao Liu,Eric Allen,Phil Febbo,Summer Galloway,Nicole L. Washington,Simon White,Geraint Levan,Kelly Schiabor Barrett,Elizabeth Cirulli,Alexandre Bolze,Ary Ascencio,Charlotte Rivera-Garcia,Ryan Cho,Jason Nguyen,Sherry Wang,Jimmy Ramirez,Tyler Cassens,Efren Sandoval,Magnus Isaksson,William Lee,David Becker,Marc Laurent,James Lu,Clinton R. Paden,Suxiang Tong,Duncan MacCannell |
| EPI_ISL_1036754 | Utah Public Health Laboratory | Utah Public Health Laboratory | Erin L. Young, Kelly F. Oakeson, Tara Gallagher |
| EPI_ISL_1036758, EPI_ISL_1036759, | Sonora Quest Laboratories | TGen North | Jolene Bowers, Megan Folkerts, Chris French, Hayley Yaglom, Ashlyn Pfeiffer, Darrin Lemmer, Dave Engelthaler, The Arizona COVID Genomics Union (ACGU) |

| | | | |
|---|---|---|---|
| EPI_ISL_1036760, EPI_ISL_1036761, EPI_ISL_1036762, EPI_ISL_1036763 | | | |
| EPI_ISL_1036764 | Sonora Quest Laboratories | TGen North | "Jolene Bowers, Megan Folkerts, Chris French, Hayley Yaglom, Ashlyn Pfeiffer, Darrin Lemmer, Dave Engelthaler, The Arizona COVID Genomics Union (ACGU)" |
| EPI_ISL_1036765, EPI_ISL_1036767, EPI_ISL_1036768, EPI_ISL_1036769, EPI_ISL_1036770, EPI_ISL_1036771, EPI_ISL_1036773, EPI_ISL_1036775, EPI_ISL_1036776, EPI_ISL_1036778, EPI_ISL_1036779, EPI_ISL_1036780 | | | |
| see above | Sonora Quest Laboratories | TGen North | Jolene Bowers, Megan Folkerts, Chris French, Hayley Yaglom, Ashlyn Pfeiffer, Darrin Lemmer, Dave Engelthaler, The Arizona COVID Genomics Union (ACGU) |
| EPI_ISL_1036781 | Sonora Quest Laboratories | TGen North | "Jolene Bowers, Megan Folkerts, Chris French, Hayley Yaglom, Ashlyn Pfeiffer, Darrin Lemmer, Dave Engelthaler, The Arizona COVID Genomics Union (ACGU)" |
| EPI_ISL_1036782, EPI_ISL_1036785, EPI_ISL_1036786, EPI_ISL_1036787, EPI_ISL_1036788, EPI_ISL_1036789, EPI_ISL_1036790, EPI_ISL_1036791 | Sonora Quest Laboratories | TGen North | Jolene Bowers, Megan Folkerts, Chris French, Hayley Yaglom, Ashlyn Pfeiffer, Darrin Lemmer, Dave Engelthaler, The Arizona COVID Genomics Union (ACGU) |
| EPI_ISL_1036792, EPI_ISL_1036793, EPI_ISL_1036795, EPI_ISL_1036796, EPI_ISL_1036797, EPI_ISL_1036798, EPI_ISL_1036800, EPI_ISL_1036801, EPI_ISL_1036802, EPI_ISL_1036803, EPI_ISL_1036804, EPI_ISL_1036805, EPI_ISL_1036806, EPI_ISL_1036808, EPI_ISL_1036809, EPI_ISL_1036810, EPI_ISL_1036811, EPI_ISL_1036812, EPI_ISL_1036813, EPI_ISL_1036814, EPI_ISL_1036820, EPI_ISL_1036821, EPI_ISL_1036822, EPI_ISL_1036823, EPI_ISL_1036824, EPI_ISL_1036825, EPI_ISL_1036826, EPI_ISL_1036827, EPI_ISL_1036828, EPI_ISL_1036829, EPI_ISL_1036830, EPI_ISL_1036831, EPI_ISL_1036832, EPI_ISL_1036833, EPI_ISL_1036834, EPI_ISL_1036835, EPI_ISL_1036836, EPI_ISL_1036837, EPI_ISL_1036838, EPI_ISL_1036839, EPI_ISL_1036840, EPI_ISL_1036841, EPI_ISL_1036842, EPI_ISL_1036843, EPI_ISL_1036844, EPI_ISL_1036845, EPI_ISL_1036846, EPI_ISL_1036847, EPI_ISL_1036848, EPI_ISL_1036855, EPI_ISL_1036856, EPI_ISL_1036857, EPI_ISL_1036858, EPI_ISL_1036859, EPI_ISL_1036860, EPI_ISL_1036861, EPI_ISL_1036862, EPI_ISL_1036863, EPI_ISL_1036864, EPI_ISL_1036865, EPI_ISL_1036866, EPI_ISL_1036867, EPI_ISL_1036868, EPI_ISL_1036869, EPI_ISL_1036870, EPI_ISL_1036871, EPI_ISL_1036872, EPI_ISL_1036873, EPI_ISL_1036874, EPI_ISL_1036875, EPI_ISL_1036876, EPI_ISL_1036877, EPI_ISL_1036878, EPI_ISL_1036882, EPI_ISL_1036883, EPI_ISL_1036884, EPI_ISL_1036885, EPI_ISL_1036886, EPI_ISL_1036887, EPI_ISL_1036888, EPI_ISL_1036889, EPI_ISL_1036890, EPI_ISL_1036891, EPI_ISL_1036892, EPI_ISL_1036893, EPI_ISL_1036894, EPI_ISL_1036895, EPI_ISL_1036896, EPI_ISL_1036897, EPI_ISL_1036898, EPI_ISL_1036899, EPI_ISL_1036900, EPI_ISL_1036901, EPI_ISL_1036902, EPI_ISL_1036903, EPI_ISL_1036904, EPI_ISL_1036905, EPI_ISL_1036906, EPI_ISL_1036907, EPI_ISL_1036908, EPI_ISL_1036909, EPI_ISL_1036910, EPI_ISL_1036911, EPI_ISL_1036912, EPI_ISL_1036913, EPI_ISL_1036914, EPI_ISL_1036915, EPI_ISL_1036916, EPI_ISL_1036917, EPI_ISL_1036918, EPI_ISL_1036919, EPI_ISL_1036920, EPI_ISL_1036921, EPI_ISL_1036922, EPI_ISL_1036923, EPI_ISL_1036924, EPI_ISL_1036925, EPI_ISL_1036926, EPI_ISL_1036927, EPI_ISL_1036928, EPI_ISL_1036929, EPI_ISL_1036930, EPI_ISL_1036931, EPI_ISL_1036932, EPI_ISL_1036933, EPI_ISL_1036934, EPI_ISL_1036936 | | | |
| see above | US Air Force School of Aerospace Medicine | US Air Force School of Aerospace Medicine | Anthony Fries, Jennifer Meyer, William Gruner, William Buggele, Amanda Javorina, Sarah Purves, Clarise Starr, Elizabeth Macias |
| EPI_ISL_1036937, EPI_ISL_1036938, EPI_ISL_1036940, EPI_ISL_1036941, EPI_ISL_1036942, EPI_ISL_1036943, EPI_ISL_1036944, EPI_ISL_1036945, EPI_ISL_1036946, EPI_ISL_1036947, EPI_ISL_1036948, EPI_ISL_1036949, EPI_ISL_1036950, EPI_ISL_1036951, EPI_ISL_1036952, EPI_ISL_1036953, EPI_ISL_1036954, EPI_ISL_1036955, EPI_ISL_1036956, EPI_ISL_1036957, EPI_ISL_1036958, EPI_ISL_1036959, EPI_ISL_1036960, EPI_ISL_1036961, EPI_ISL_1036962, EPI_ISL_1036963, EPI_ISL_1036964, EPI_ISL_1036965, EPI_ISL_1036966, EPI_ISL_1036967, EPI_ISL_1036968, EPI_ISL_1036969, EPI_ISL_1036970, EPI_ISL_1036971, EPI_ISL_1036972, EPI_ISL_1036973, EPI_ISL_1036974, EPI_ISL_1036975, EPI_ISL_1036976, EPI_ISL_1036977, EPI_ISL_1036978, EPI_ISL_1036979, EPI_ISL_1036980, EPI_ISL_1036981, EPI_ISL_1036982, EPI_ISL_1036983, EPI_ISL_1036984, EPI_ISL_1036985, EPI_ISL_1036986, EPI_ISL_1036987, EPI_ISL_1036988, EPI_ISL_1036989, EPI_ISL_1036990, EPI_ISL_1036991, EPI_ISL_1036992, EPI_ISL_1036993, EPI_ISL_1036994, EPI_ISL_1036995, EPI_ISL_1036996, EPI_ISL_1036997, EPI_ISL_1036998, EPI_ISL_1036999, EPI_ISL_1037000, EPI_ISL_1037001, EPI_ISL_1037002, EPI_ISL_1037003, EPI_ISL_1037004, EPI_ISL_1037005, EPI_ISL_1037006, EPI_ISL_1037007, EPI_ISL_1037008, EPI_ISL_1037009, EPI_ISL_1037010, EPI_ISL_1037011, EPI_ISL_1037012, EPI_ISL_1037013, EPI_ISL_1037014, EPI_ISL_1037015, EPI_ISL_1037016, EPI_ISL_1037017, EPI_ISL_1037018, EPI_ISL_1037019, EPI_ISL_1037020, EPI_ISL_1037021, EPI_ISL_1037022, EPI_ISL_1037023, EPI_ISL_1037024, EPI_ISL_1037025, EPI_ISL_1037026, EPI_ISL_1037027, EPI_ISL_1037028, EPI_ISL_1037029, EPI_ISL_1037030, EPI_ISL_1037031, EPI_ISL_1037032, EPI_ISL_1037033, EPI_ISL_1037034, EPI_ISL_1037035, EPI_ISL_1037036, EPI_ISL_1037037, EPI_ISL_1037038, EPI_ISL_1037039, EPI_ISL_1037040, EPI_ISL_1037041, EPI_ISL_1037042, EPI_ISL_1037043, EPI_ISL_1037044, EPI_ISL_1037045, EPI_ISL_1037046, EPI_ISL_1037047, EPI_ISL_1037048, EPI_ISL_1037049, EPI_ISL_1037050, EPI_ISL_1037051, EPI_ISL_1037052, EPI_ISL_1037053, EPI_ISL_1037054, EPI_ISL_1037055, EPI_ISL_1037056, EPI_ISL_1037057, EPI_ISL_1037058, EPI_ISL_1037059, EPI_ISL_1037060, EPI_ISL_1037061, EPI_ISL_1037062, EPI_ISL_1037063, EPI_ISL_1037064, EPI_ISL_1037065, EPI_ISL_1037066, EPI_ISL_1037067, EPI_ISL_1037068, EPI_ISL_1037069, EPI_ISL_1037070, EPI_ISL_1037071, EPI_ISL_1037072, EPI_ISL_1037073, EPI_ISL_1037074, EPI_ISL_1037075, EPI_ISL_1037076, EPI_ISL_1037077, EPI_ISL_1037078, EPI_ISL_1037079, EPI_ISL_1037080, EPI_ISL_1037081, EPI_ISL_1037082, EPI_ISL_1037083, EPI_ISL_1037084, EPI_ISL_1037085, EPI_ISL_1037086, EPI_ISL_1037087, EPI_ISL_1037088, EPI_ISL_1037089, EPI_ISL_1037090, EPI_ISL_1037091, EPI_ISL_1037092, EPI_ISL_1037093, EPI_ISL_1037094, EPI_ISL_1037095, EPI_ISL_1037096, EPI_ISL_1037097, EPI_ISL_1037098, EPI_ISL_1037099, EPI_ISL_1037100, EPI_ISL_1037101, EPI_ISL_1037102, EPI_ISL_1037103, EPI_ISL_1037104, EPI_ISL_1037105, EPI_ISL_1037106, EPI_ISL_1037107, EPI_ISL_1037108, EPI_ISL_1037109, EPI_ISL_1037110, EPI_ISL_1037111, EPI_ISL_1037112, EPI_ISL_1037113, EPI_ISL_1037114, EPI_ISL_1037115, EPI_ISL_1037116, EPI_ISL_1037117, EPI_ISL_1037118, EPI_ISL_1037119, EPI_ISL_1037120, EPI_ISL_1037121, EPI_ISL_1037122, EPI_ISL_1037123, EPI_ISL_1037124, EPI_ISL_1037125, EPI_ISL_1037126, EPI_ISL_1037127, EPI_ISL_1037128, EPI_ISL_1037129, EPI_ISL_1037130, EPI_ISL_1037131, EPI_ISL_1037132, EPI_ISL_1037133, EPI_ISL_1037134, EPI_ISL_1037135, EPI_ISL_1037136, EPI_ISL_1037137, EPI_ISL_1038282, EPI_ISL_1038283, EPI_ISL_1038284, EPI_ISL_1038285, EPI_ISL_1038286, EPI_ISL_1038287, EPI_ISL_1038288, EPI_ISL_1038289, EPI_ISL_1038290, EPI_ISL_1038291, EPI_ISL_1038292, EPI_ISL_1038293, EPI_ISL_1038294, EPI_ISL_1038295, EPI_ISL_1038296, EPI_ISL_1038297, EPI_ISL_1038298, EPI_ISL_1038299, EPI_ISL_1038300, EPI_ISL_1038301, EPI_ISL_1038302, EPI_ISL_1038303, EPI_ISL_1038304, EPI_ISL_1038305, EPI_ISL_1038306, EPI_ISL_1038307, EPI_ISL_1038308, EPI_ISL_1038309, EPI_ISL_1038310, EPI_ISL_1038311, EPI_ISL_1038312, EPI_ISL_1038313, EPI_ISL_1038314, EPI_ISL_1038315, EPI_ISL_1038316, EPI_ISL_1038317, EPI_ISL_1038318, EPI_ISL_1038319, EPI_ISL_1038320, EPI_ISL_1038321, EPI_ISL_1038322, EPI_ISL_1038323, EPI_ISL_1038324, EPI_ISL_1038325, EPI_ISL_1038326, EPI_ISL_1038327, EPI_ISL_1038328, EPI_ISL_1038329, EPI_ISL_1038330, EPI_ISL_1038331, EPI_ISL_1038332, EPI_ISL_1038333, EPI_ISL_1038334, EPI_ISL_1038335, EPI_ISL_1038336, EPI_ISL_1038337, EPI_ISL_1038338, EPI_ISL_1038339, EPI_ISL_1038340, EPI_ISL_1038341, EPI_ISL_1038342, EPI_ISL_1038343, EPI_ISL_1038344, EPI_ISL_1038345, EPI_ISL_1038346, EPI_ISL_1038347, EPI_ISL_1038348, EPI_ISL_1038349, EPI_ISL_1038350, EPI_ISL_1038351, EPI_ISL_1038352, EPI_ISL_1038353, EPI_ISL_1038354, EPI_ISL_1038355, EPI_ISL_1038356, EPI_ISL_1038357, EPI_ISL_1038358, EPI_ISL_1038359, EPI_ISL_1038360, EPI_ISL_1038361, EPI_ISL_1038362, EPI_ISL_1038363, EPI_ISL_1038364, EPI_ISL_1038365, EPI_ISL_1038366, EPI_ISL_1038367, EPI_ISL_1038368, EPI_ISL_1038369, EPI_ISL_1038370, EPI_ISL_1038371, EPI_ISL_1038372, EPI_ISL_1038373, EPI_ISL_1038374, EPI_ISL_1038375, EPI_ISL_1038376, EPI_ISL_1038377, EPI_ISL_1038378, EPI_ISL_1038379, EPI_ISL_1038380, EPI_ISL_1038381, EPI_ISL_1038382, EPI_ISL_1038383, EPI_ISL_1038384, EPI_ISL_1038385, EPI_ISL_1038386, EPI_ISL_1038387, EPI_ISL_1038388, EPI_ISL_1038389, EPI_ISL_1038390, EPI_ISL_1038391, EPI_ISL_1038392, EPI_ISL_1038393, EPI_ISL_1038394, EPI_ISL_1038395, EPI_ISL_1038396, EPI_ISL_1038397, EPI_ISL_1038398, EPI_ISL_1038399, EPI_ISL_1038400, EPI_ISL_1038401, EPI_ISL_1038402, EPI_ISL_1038403, EPI_ISL_1038404, EPI_ISL_1038405, EPI_ISL_1038406, EPI_ISL_1038407, EPI_ISL_1038408, EPI_ISL_1038409, EPI_ISL_1038410, EPI_ISL_1038411, EPI_ISL_1038412, EPI_ISL_1038413, EPI_ISL_1038414, EPI_ISL_1038415, EPI_ISL_1038416, EPI_ISL_1038417, EPI_ISL_1038418, EPI_ISL_1038419, EPI_ISL_1038420, EPI_ISL_1038421, EPI_ISL_1038422, EPI_ISL_1038423, EPI_ISL_1038424, EPI_ISL_1038425, EPI_ISL_1038426, EPI_ISL_1038427, EPI_ISL_1038428, EPI_ISL_1038429, EPI_ISL_1038430, EPI_ISL_1038431, EPI_ISL_1038432, EPI_ISL_1038433, EPI_ISL_1038434, EPI_ISL_1038435, EPI_ISL_1038436, EPI_ISL_1038437, EPI_ISL_1038438, EPI_ISL_1038439, EPI_ISL_1038440, EPI_ISL_1038441, EPI_ISL_1038442, EPI_ISL_1038443, EPI_ISL_1038444, EPI_ISL_1038445, EPI_ISL_1038446, EPI_ISL_1038447, EPI_ISL_1038448, EPI_ISL_1038449, EPI_ISL_1038450, EPI_ISL_1038451, EPI_ISL_1038452, EPI_ISL_1038453, EPI_ISL_1038454, EPI_ISL_1038455, EPI_ISL_1038456, EPI_ISL_1038457, EPI_ISL_1038458, EPI_ISL_1038459, EPI_ISL_1038460, EPI_ISL_1038461, EPI_ISL_1038462, EPI_ISL_1038463, EPI_ISL_1038464, EPI_ISL_1038465, EPI_ISL_1038466, EPI_ISL_1038467, EPI_ISL_1038468, EPI_ISL_1038469, EPI_ISL_1038470, EPI_ISL_1038471, EPI_ISL_1038472, EPI_ISL_1038473, EPI_ISL_1038474, EPI_ISL_1038475, EPI_ISL_1038476, EPI_ISL_1038477, EPI_ISL_1038478, EPI_ISL_1038479, EPI_ISL_1038480, EPI_ISL_1038481, EPI_ISL_1038482, EPI_ISL_1038483, EPI_ISL_1038484, EPI_ISL_1038485, EPI_ISL_1038486, EPI_ISL_1038487, EPI_ISL_1038488, EPI_ISL_1038489, EPI_ISL_1038490, EPI_ISL_1038491, EPI_ISL_1038492, EPI_ISL_1038493, EPI_ISL_1038494, EPI_ISL_1038495, EPI_ISL_1038496, EPI_ISL_1038497, EPI_ISL_1038498, EPI_ISL_1038499, EPI_ISL_1038500, EPI_ISL_1038501, EPI_ISL_1038502, EPI_ISL_1038503, EPI_ISL_1038504, EPI_ISL_1038505, EPI_ISL_1038506, EPI_ISL_1038507, EPI_ISL_1038508, EPI_ISL_1038509, EPI_ISL_1038510, EPI_ISL_1038511, EPI_ISL_1038512, EPI_ISL_1038513, EPI_ISL_1038514, EPI_ISL_1038515, EPI_ISL_1038516, EPI_ISL_1038517, EPI_ISL_1038518, EPI_ISL_1038519, EPI_ISL_1038520, EPI_ISL_1038521, EPI_ISL_1038522, EPI_ISL_1038523, EPI_ISL_1038524, EPI_ISL_1038525, EPI_ISL_1038526, EPI_ISL_1038527, EPI_ISL_1038528, EPI_ISL_1038529, EPI_ISL_1038530, EPI_ISL_1038531, EPI_ISL_1038532, EPI_ISL_1038533, EPI_ISL_1038534, EPI_ISL_1038535, EPI_ISL_1038536, EPI_ISL_1038537, EPI_ISL_1038538, EPI_ISL_1038539, EPI_ISL_1038540, EPI_ISL_1038541, EPI_ISL_1038542, EPI_ISL_1038543, EPI_ISL_1038544, EPI_ISL_1038545, EPI_ISL_1038546, EPI_ISL_1038547, EPI_ISL_1038548, EPI_ISL_1038549, EPI_ISL_1038550, EPI_ISL_1038551, EPI_ISL_1038552, EPI_ISL_1038553, EPI_ISL_1038554, EPI_ISL_1038555, EPI_ISL_1038556, EPI_ISL_1038557, EPI_ISL_1038558, EPI_ISL_1038559, EPI_ISL_1038560, EPI_ISL_1038561, EPI_ISL_1038562, EPI_ISL_1038563, EPI_ISL_1038564, EPI_ISL_1038565, EPI_ISL_1038566, EPI_ISL_1038567, EPI_ISL_1038568, EPI_ISL_1038569, EPI_ISL_1038570, EPI_ISL_1038571, EPI_ISL_1038572, EPI_ISL_1038573, EPI_ISL_1038574, EPI_ISL_1038575, EPI_ISL_1038576, EPI_ISL_1038577, EPI_ISL_1038578, EPI_ISL_1038579, EPI_ISL_1038580, EPI_ISL_1038581, EPI_ISL_1038582, EPI_ISL_1038583, EPI_ISL_1038584, EPI_ISL_1038585, EPI_ISL_1038586, EPI_ISL_1038587, EPI_ISL_1038588, EPI_ISL_1038589, EPI_ISL_1038590, EPI_ISL_1038591, EPI_ISL_1038592, EPI_ISL_1038593, EPI_ISL_1038594, EPI_ISL_1038595, EPI_ISL_1038596, EPI_ISL_1038597, EPI_ISL_1038598, EPI_ISL_1038599, EPI_ISL_1038600, EPI_ISL_1038601, EPI_ISL_1038602, EPI_ISL_1038603, EPI_ISL_1038604, EPI_ISL_1038605, EPI_ISL_1038606, EPI_ISL_1038607, EPI_ISL_1038608, EPI_ISL_1038609, EPI_ISL_1038610, EPI_ISL_1038611, EPI_ISL_1038612, EPI_ISL_1038613, EPI_ISL_1038614, EPI_ISL_1038615, EPI_ISL_1038616, EPI_ISL_1038617, EPI_ISL_1038618, EPI_ISL_1038619, EPI_ISL_1038620, EPI_ISL_1038621, EPI_ISL_1038622, EPI_ISL_1038623, EPI_ISL_1038624, EPI_ISL_1038625, EPI_ISL_1038626, EPI_ISL_1038627, EPI_ISL_1038628, EPI_ISL_1038629, EPI_ISL_1038630, EPI_ISL_1038631, EPI_ISL_1038632, EPI_ISL_1038633, EPI_ISL_1038634, EPI_ISL_1038635, EPI_ISL_1038636, EPI_ISL_1038637, EPI_ISL_1038638, EPI_ISL_1038639, EPI_ISL_1038640, EPI_ISL_1038641, EPI_ISL_1038642, EPI_ISL_1038643, EPI_ISL_1038644, EPI_ISL_1038645, EPI_ISL_1038646, EPI_ISL_1038647, EPI_ISL_1038648, EPI_ISL_1038649, EPI_ISL_1038650, EPI_ISL_1038651, EPI_ISL_1038652, EPI_ISL_1038653, EPI_ISL_1038654, EPI_ISL_1038655, EPI_ISL_1038656, EPI_ISL_1038657, EPI_ISL_1038658, EPI_ISL_1038659, EPI_ISL_1038660, EPI_ISL_1038661, EPI_ISL_1038662, EPI_ISL_1038663, EPI_ISL_1038664, EPI_ISL_1038665, EPI_ISL_1038666, EPI_ISL_1038667, EPI_ISL_1038668, EPI_ISL_1038669, EPI_ISL_1038670, EPI_ISL_1038671, EPI_ISL_1038672, EPI_ISL_1038673, EPI_ISL_1038674, EPI_ISL_1038675, EPI_ISL_1038676, EPI_ISL_1038677, EPI_ISL_1038678, EPI_ISL_1038679, EPI_ISL_1038680, EPI_ISL_1038681, EPI_ISL_1038682, EPI_ISL_1038683, EPI_ISL_1038684, EPI_ISL_1038685, EPI_ISL_1038686, EPI_ISL_1038687, EPI_ISL_1038688, EPI_ISL_1038689, EPI_ISL_1038690, EPI_ISL_1038691, EPI_ISL_1038692, EPI_ISL_1038693, EPI_ISL_1038694, EPI_ISL_1038695, EPI_ISL_1038696, EPI_ISL_1038697, EPI_ISL_1038698, EPI_ISL_1038699, EPI_ISL_1038700, EPI_ISL_1038701, EPI_ISL_1038702, EPI_ISL_1038703, EPI_ISL_1038704, EPI_ISL_1038705, EPI_ISL_1038706, EPI_ISL_1038707, EPI_ISL_1038708, EPI_ISL_1038709, EPI_ISL_1038710, EPI_ISL_1038711, EPI_ISL_1038712, EPI_ISL_1038713, EPI_ISL_1038714, EPI_ISL_1038715, EPI_ISL_1038716, EPI_ISL_1038717, EPI_ISL_1038718, EPI_ISL_1038719, EPI_ISL_1038720, EPI_ISL_1038721, EPI_ISL_1038722, EPI_ISL_1038723, EPI_ISL_1038724, EPI_ISL_1038725, EPI_ISL_1038726, EPI_ISL_1038727, EPI_ISL_1038728, EPI_ISL_1038729, EPI_ISL_1038730, EPI_ISL_1038731, EPI_ISL_1038732, EPI_ISL_1038733, EPI_ISL_1038734, EPI_ISL_1038735, EPI_ISL_1038736, EPI_ISL_1038737, EPI_ISL_1038738, EPI_ISL_1038739, EPI_ISL_1038740, EPI_ISL_1038741, EPI_ISL_1038742, EPI_ISL_1038743, EPI_ISL_1038744, EPI_ISL_1038745, EPI_ISL_1038746, EPI_ISL_1038747, EPI_ISL_1038748, EPI_ISL_1038749, EPI_ISL_1038750, EPI_ISL_1038751, EPI_ISL_1038752, EPI_ISL_1038753, EPI_ISL_1038754, EPI_ISL_1038755, EPI_ISL_1038756, EPI_ISL_1038757, EPI_ISL_1038758, EPI_ISL_1038759, EPI_ISL_1038760, EPI_ISL_1038761, EPI_ISL_1038762, EPI_ISL_1038763, EPI_ISL_1038764, EPI_ISL_1038765, EPI_ISL_1038766, EPI_ISL_1038767, EPI_ISL_1038768, EPI_ISL_1038769, EPI_ISL_1038770, EPI_ISL_1038771, EPI_ISL_1038772, EPI_ISL_1038773, EPI_ISL_1038774, EPI_ISL_1038775, EPI_ISL_1038776, EPI_ISL_1038777, EPI_ISL_1038778, EPI_ISL_1038779, EPI_ISL_1038780, EPI_ISL_1038781, EPI_ISL_1038782, EPI_ISL_1038783, EPI_ISL_1038784, EPI_ISL_1038785, EPI_ISL_1038786, EPI_ISL_1038787, EPI_ISL_1038788, EPI_ISL_1038789, EPI_ISL_1038790 | | | |
| see above | Laboratory Corporation of America | Respiratory Viruses Branch, Division of Viral Diseases, Centers for Disease Control and Prevention | Peter W. Cook, Dakota Howard, Dhwani Batra, Ben L. Rambo-Martin, Clinton R. Paden, Suxiang Tong, Duncan MacCannell |
| EPI_ISL_1038796 | US Air Force School of Aerospace Medicine | US Air Force School of Aerospace Medicine | Anthony Fries, Jennifer Meyer, William Gruner, William Buggele, Amanda Javorina, Sarah Purves, Clarise Starr, Elizabeth Macias |
| EPI_ISL_1038797, EPI_ISL_1038798, EPI_ISL_1038799, EPI_ISL_1038801, EPI_ISL_1038802, EPI_ISL_1038803, EPI_ISL_1038804, EPI_ISL_1038805, EPI_ISL_1038806, EPI_ISL_1038807, EPI_ISL_1038808, EPI_ISL_1038809, EPI_ISL_1038811, EPI_ISL_1038812, EPI_ISL_1038813, EPI_ISL_1038814, EPI_ISL_1038815, EPI_ISL_1038816, EPI_ISL_1038817, EPI_ISL_1038818, EPI_ISL_1038819, EPI_ISL_1038820, EPI_ISL_1038821, EPI_ISL_1038822, EPI_ISL_1038823, EPI_ISL_1038824, EPI_ISL_1038825, EPI_ISL_1038826, EPI_ISL_1038827, EPI_ISL_1038828, EPI_ISL_1038829, EPI_ISL_1038830, EPI_ISL_1038831, EPI_ISL_1038832, EPI_ISL_1038833, EPI_ISL_1038834, EPI_ISL_1038835, EPI_ISL_1038836, EPI_ISL_1038837, EPI_ISL_1038838, EPI_ISL_1038840, EPI_ISL_1038841 | | | |
| see above | Maryland Public Health Laboratory | Maryland Public Health Laboratory | Maryland Department of Health Laboratories Administration |
| EPI_ISL_1038842, EPI_ISL_1038843, EPI_ISL_1038845, EPI_ISL_1038846, EPI_ISL_1038847, EPI_ISL_1038848, EPI_ISL_1038849, EPI_ISL_1038850, EPI_ISL_1038851, EPI_ISL_1038852, EPI_ISL_1038853, EPI_ISL_1038854, EPI_ISL_1038855, EPI_ISL_1038859, EPI_ISL_1038860, EPI_ISL_1038861, EPI_ISL_1038862, EPI_ISL_1038863, EPI_ISL_1038864, EPI_ISL_1038865, EPI_ISL_1038866, EPI_ISL_1038867, EPI_ISL_1038868, EPI_ISL_1038869, EPI_ISL_1038870, EPI_ISL_1038871, EPI_ISL_1038872, EPI_ISL_1038873, EPI_ISL_1038874, EPI_ISL_1038875, EPI_ISL_1038876, EPI_ISL_1038877, EPI_ISL_1038878, EPI_ISL_1038879, EPI_ISL_1038880, EPI_ISL_1038881, EPI_ISL_1038882, EPI_ISL_1038883, EPI_ISL_1038884, EPI_ISL_1038885, EPI_ISL_1038886, EPI_ISL_1038887, EPI_ISL_1038888, EPI_ISL_1038889, EPI_ISL_1038890, EPI_ISL_1038891, EPI_ISL_1038892, EPI_ISL_1038893, EPI_ISL_1038894, EPI_ISL_1038895, EPI_ISL_1038896, EPI_ISL_1038897, EPI_ISL_1038898, EPI_ISL_1038899, EPI_ISL_1038900, EPI_ISL_1038901, EPI_ISL_1038902, EPI_ISL_1038903, EPI_ISL_1038904, EPI_ISL_1038905, EPI_ISL_1038906, EPI_ISL_1038907, EPI_ISL_1038908, EPI_ISL_1038909, EPI_ISL_1038910, EPI_ISL_1038911, EPI_ISL_1038912, EPI_ISL_1038913, EPI_ISL_1038914, EPI_ISL_1038915, EPI_ISL_1038916, EPI_ISL_1038917, EPI_ISL_1038918, EPI_ISL_1038919, EPI_ISL_1038920, EPI_ISL_1038921, EPI_ISL_1038922, EPI_ISL_1038923 | | | |
| see above | Colorado Department of Public Health and Environment | Colorado Department of Puplic Health and Environment | Laura Bankers, Molly C. Hetherington-Rauth, Diana Ir, Shannon Ely, Shannon R. Matzinger, Sarah Elizabeth Totten, Emily A. Travanty |
| EPI_ISL_1038925, EPI_ISL_1038926, EPI_ISL_1038927, EPI_ISL_1038928, EPI_ISL_1038929, EPI_ISL_1038930, EPI_ISL_1038931, EPI_ISL_1038932, EPI_ISL_1038933, EPI_ISL_1038934, EPI_ISL_1038935, EPI_ISL_1038936, EPI_ISL_1038937, EPI_ISL_1038938, EPI_ISL_1038939, EPI_ISL_1038940, EPI_ISL_1038941, EPI_ISL_1038942, EPI_ISL_1038943, EPI_ISL_1038944, EPI_ISL_1038945, EPI_ISL_1038946, EPI_ISL_1038948, EPI_ISL_1038949, EPI_ISL_1038950, EPI_ISL_1038951, EPI_ISL_1038952, EPI_ISL_1038953, EPI_ISL_1038954, EPI_ISL_1038955, EPI_ISL_1038956, EPI_ISL_1038957, EPI_ISL_1038958, EPI_ISL_1038959, EPI_ISL_1038960, EPI_ISL_1038961, EPI_ISL_1038962, EPI_ISL_1038963, EPI_ISL_1038964, EPI_ISL_1038965, EPI_ISL_1038966, EPI_ISL_1038967, EPI_ISL_1038968, EPI_ISL_1038969, EPI_ISL_1038970 | | | |
| see above | New Mexico Department of Health Scientific Laboratory | New Mexico Department of Health Scientific Laboratory | Ellie Johnson, Anastacia Griego-Fisher, D'eldra Malone, Jennifer Benoit |
| EPI_ISL_1038981 | Tempus | Grubaugh Lab - Yale School of Public Health | Joseph Fauver, Tara Alpert, Anderson Brito, Mallery Breban, Anne Wyllie, Chantal Vogels, Mary Petrone, Annie Watkins, Chaney Kalinich, Isabel Ott, Nathan Grubaugh |
| EPI_ISL_1038993 | Yale Clinical Virology Lab | Grubaugh Lab - Yale School of Public Health | Joseph Fauver, Tara Alpert, Anderson Brito, Mallery Breban, Anne Wyllie, Chantal Vogels, Mary Petrone, Annie Watkins, Chaney Kalinich, Isabel Ott, Nathan Grubaugh |
| EPI_ISL_1039003 | Grubaugh Lab - Yale School of Public Health | Grubaugh Lab - Yale School of Public Health | Joseph Fauver, Tara Alpert, Anderson Brito, Mallery Breban, Anne Wyllie, Chantal Vogels, Mary Petrone, Annie Watkins, Chaney Kalinich, Isabel Ott, Nathan Grubaugh |
| EPI_ISL_1039521, EPI_ISL_1039522, EPI_ISL_1039523 | LSUHS Emerging Viral Threat Laboratory | Microbial Genome Sequencing Center | Jeremy P. Kamil, Jennifer L. Carroll, Maarten Van Diest, Andrew D. Yurochko, Rona S. Scott, John A. Vanchiere, Christopher G. Kevil, Daniel J. Snyder, Vaughn S. Cooper |
| EPI_ISL_1039524 | LSUHS Emerging Viral Threat Laboratory | Microbial Genome Sequencing Center | Jennifer L. Carroll, Maarten Van Diest, Andrew D. Yurochko, Rona S. Scott, Christopher G. Kevil, John A. Vanchiere, Daniel J. Snyder, Vaughn S. Cooper, Jeremy P. Kamil |
| EPI_ISL_1039525 | LSUHS Emerging Viral Threat Laboratory | Microbial Genome Sequencing Center | Jeremy P. Kamil, Jennifer L. Carroll, Maarten Van Diest, Andrew D. Yurochko, Rona S. Scott, Christopher G. Kevil, Daniel J. Snyder, Vaughn S. Cooper, John A. Vanchiere |
| EPI_ISL_1039526 | LSUHS Emerging Viral Threat Laboratory | Microbial Genome Sequencing Center | Jeremy P. Kamil, Jennifer L. Carroll, Maarten Van Diest, Andrew D. Yurochko, Rona S. Scott, John A. Vanchiere, Christopher G. Kevil, Daniel J. Snyder, Vaughn S. Cooper |
| EPI_ISL_1039527, EPI_ISL_1039528, EPI_ISL_1039529 | LSUHS Emerging Viral Threat Laboratory | Microbial Genome Sequencing Center | Jeremy P. Kamil, Jennifer L. Carroll, Maarten Van Diest, Andrew D. Yurochko, Rona S. Scott, Christopher G. Kevil, Daniel J. Snyder, Vaughn S. Cooper, John A. Vanchiere |
| EPI_ISL_1039530 | LSUHS Emerging Viral Threat Laboratory | Microbial Genome Sequencing Center | Daniel J. Snyder, Rona S. Scott, Jennifer L. Carroll, Maarten Van Diest, Andrew D. Yurochko, Christopher G. Kevil, John A. Vanchiere, Vaughn S. Cooper, Jeremy P. Kamil |
| EPI_ISL_1039531, EPI_ISL_1039532, EPI_ISL_1039533, EPI_ISL_1039534, EPI_ISL_1039535, EPI_ISL_1039536, EPI_ISL_1039537, EPI_ISL_1039538, EPI_ISL_1039539, EPI_ISL_1039540, EPI_ISL_1039541, EPI_ISL_1039543, EPI_ISL_1039544, EPI_ISL_1039545, EPI_ISL_1039546, EPI_ISL_1039547, EPI_ISL_1039548, EPI_ISL_1039549, EPI_ISL_1039550, EPI_ISL_1039551, EPI_ISL_1039552, EPI_ISL_1039553, EPI_ISL_1039554, EPI_ISL_1039555, EPI_ISL_1039556, EPI_ISL_1039557, EPI_ISL_1039558, EPI_ISL_1039559, EPI_ISL_1039560, EPI_ISL_1039561, EPI_ISL_1039562, EPI_ISL_1039563, EPI_ISL_1039564, EPI_ISL_1039565, EPI_ISL_1039566, EPI_ISL_1039567, EPI_ISL_1039568, EPI_ISL_1039569, EPI_ISL_1039570, EPI_ISL_1039571, EPI_ISL_1039572, EPI_ISL_1039573, EPI_ISL_1039574, EPI_ISL_1039575, EPI_ISL_1039576, EPI_ISL_1039577, EPI_ISL_1039578, EPI_ISL_1039579, EPI_ISL_1039580, EPI_ISL_1039581, EPI_ISL_1039582, EPI_ISL_1039583, EPI_ISL_1039584, EPI_ISL_1039585, EPI_ISL_1039586, EPI_ISL_1039587, EPI_ISL_1039588, EPI_ISL_1039589, EPI_ISL_1039590, EPI_ISL_1039591, EPI_ISL_1039592, EPI_ISL_1039593 | | | |
| see above | LSUHS Emerging Viral Threat | Microbial Genome Sequencing Center | Jeremy P. Kamil, Jennifer L. Carroll, Maarten Van Diest, Andrew D. Yurochko, Rona S. Scott, John A. Vanchiere, Christopher G. Kevil, Daniel J. Snyder, Vaughn S. Cooper |

| | | | |
|---|---|---|---|
| | Laboratory | | |
| EPI_ISL_1039594, EPI_ISL_1039595, EPI_ISL_1039596, EPI_ISL_1039597, EPI_ISL_1039598, EPI_ISL_1039599, EPI_ISL_1039600, EPI_ISL_1039601, EPI_ISL_1039602, EPI_ISL_1039603, EPI_ISL_1039604, EPI_ISL_1039605, EPI_ISL_1039606, EPI_ISL_1039607, EPI_ISL_1039608, EPI_ISL_1039609, EPI_ISL_1039610, EPI_ISL_1039611, EPI_ISL_1039612, EPI_ISL_1039613, EPI_ISL_1039614, EPI_ISL_1039615, EPI_ISL_1039616, EPI_ISL_1039617, EPI_ISL_1039618, EPI_ISL_1039619, EPI_ISL_1039620, EPI_ISL_1039621, EPI_ISL_1039622, EPI_ISL_1039623, EPI_ISL_1039624, EPI_ISL_1039625, EPI_ISL_1039626, EPI_ISL_1039627, EPI_ISL_1039628, EPI_ISL_1039629, EPI_ISL_1039630, EPI_ISL_1039631, EPI_ISL_1039632, EPI_ISL_1039633, EPI_ISL_1039634, EPI_ISL_1039635, EPI_ISL_1039636, EPI_ISL_1039637, EPI_ISL_1039638, EPI_ISL_1039639, EPI_ISL_1039640, EPI_ISL_1039641, EPI_ISL_1039642, EPI_ISL_1039643, EPI_ISL_1039644, EPI_ISL_1039645, EPI_ISL_1039646, EPI_ISL_1039647, EPI_ISL_1039648 | | | |
| see above | LSUHS Emerging Viral Threat Laboratory | Microbial Genome Sequencing Center | Jeremy P. Kamil, Jennifer L. Carroll, Maarten Van Diest, Andrew D. Yurochko, Rona S. Scott, Christopher G. Kevil, Daniel J. Snyder, Vaughn S. Cooper, John A. Vanchiere |
| EPI_ISL_1039649, EPI_ISL_1039650, EPI_ISL_1039651, EPI_ISL_1039652, EPI_ISL_1039653, EPI_ISL_1039654, EPI_ISL_1039655, EPI_ISL_1039656, EPI_ISL_1039657, EPI_ISL_1039658, EPI_ISL_1039659 | | | |
| see above | LSUHS Emerging Viral Threat Laboratory | Microbial Genome Sequencing Center | Maarten Van Diest, Jennifer L. Carroll, Andrew D. Yurochko, Martin J. Sapp, Rona S. Scott, Christopher G. Kevil, Daniel J. Snyder, John A. Vanchiere, Vaughn S. Cooper, Jeremy P. Kamil |
| EPI_ISL_1039660, EPI_ISL_1039661, EPI_ISL_1039662, EPI_ISL_1039663, EPI_ISL_1039664, EPI_ISL_1039665, EPI_ISL_1039666, EPI_ISL_1039667 | LSUHS Emerging Viral Threat Laboratory | Microbial Genome Sequencing Center | Daniel J. Snyder, Rona S. Scott, Jennifer L. Carroll, Maarten Van Diest, Andrew D. Yurochko, Christopher G. Kevil, John A. Vanchiere, Vaughn S. Cooper, Jeremy P. Kamil |
| EPI_ISL_1039668, EPI_ISL_1039669, EPI_ISL_1039670, EPI_ISL_1039671, EPI_ISL_1039672, EPI_ISL_1039673, EPI_ISL_1039674, EPI_ISL_1039675, EPI_ISL_1039676, EPI_ISL_1039677, EPI_ISL_1039678 | | | |
| see above | LSUHS Emerging Viral Threat Laboratory | Microbial Genome Sequencing Center | Rona S. Scott, Jennifer L. Carroll, Maarten Van Diest, Andrew D. Yurochko, Christopher G. Kevil, John A. Vanchiere, Daniel J. Snyder, Vaughn S. Cooper, Jeremy P. Kamil |
| EPI_ISL_1039679, EPI_ISL_1039680, EPI_ISL_1039681, EPI_ISL_1039682, EPI_ISL_1039683, EPI_ISL_1039684, EPI_ISL_1039685, EPI_ISL_1039686, EPI_ISL_1039687, EPI_ISL_1039688, EPI_ISL_1039689 | | | |
| see above | LSUHS Emerging Viral Threat Laboratory | Microbial Genome Sequencing Center | Jennifer L. Carroll, Maarten Van Diest, Andrew D. Yurochko, Rona S. Scott, Christopher G. Kevil, John A. Vanchiere, Daniel J. Snyder, Vaughn S. Cooper, Jeremy P. Kamil |
| EPI_ISL_1039712, EPI_ISL_1039713, EPI_ISL_1039714, EPI_ISL_1039715, EPI_ISL_1039716, EPI_ISL_1039717, EPI_ISL_1039718, EPI_ISL_1039719, EPI_ISL_1039720, EPI_ISL_1039721, EPI_ISL_1039722, EPI_ISL_1039723, EPI_ISL_1039724, EPI_ISL_1039725, EPI_ISL_1039726, EPI_ISL_1039727, EPI_ISL_1039728, EPI_ISL_1039729, EPI_ISL_1039730, EPI_ISL_1039731, EPI_ISL_1039732, EPI_ISL_1039733, EPI_ISL_1039734, EPI_ISL_1039735, EPI_ISL_1039736 | | | |
| see above | Alaska State Virology Laboratory | Alaska State Virology Laboratory | Stephanie DeRonde, Lisa Smith, Ph.D., Jack Chen, Ph.D. |
| EPI_ISL_1039738, EPI_ISL_1039752, EPI_ISL_1039753, EPI_ISL_1039754, EPI_ISL_1039755, EPI_ISL_1039756, EPI_ISL_1039757, EPI_ISL_1039758, EPI_ISL_1039759, EPI_ISL_1039760, EPI_ISL_1039761, EPI_ISL_1039762, EPI_ISL_1039763, EPI_ISL_1039764, EPI_ISL_1039765, EPI_ISL_1039766, EPI_ISL_1039767, EPI_ISL_1039768, EPI_ISL_1039769, EPI_ISL_1039770, EPI_ISL_1039771, EPI_ISL_1039772, EPI_ISL_1039773, EPI_ISL_1039774, EPI_ISL_1039775, EPI_ISL_1039776, EPI_ISL_1039777, EPI_ISL_1039778, EPI_ISL_1039779, EPI_ISL_1039780, EPI_ISL_1039781 | | | |
| see above | Wyoming Public Health Laboratory | Wyoming Public Health Laboratory | Noah Hull, Taylor Fearing, Lynette Gumbleton, Channing Weber, Ashley Norberg, Bailey Bowcutt, and Wanda Manley |
| EPI_ISL_1039839, EPI_ISL_1039840, EPI_ISL_1039841, EPI_ISL_1039842, EPI_ISL_1039843, EPI_ISL_1039844, EPI_ISL_1039845, EPI_ISL_1039846, EPI_ISL_1039847, EPI_ISL_1039849, EPI_ISL_1039850, EPI_ISL_1039851, EPI_ISL_1039852, EPI_ISL_1039853, EPI_ISL_1039854, EPI_ISL_1039855, EPI_ISL_1039857, EPI_ISL_1039858, EPI_ISL_1039859, EPI_ISL_1039860, EPI_ISL_1039861, EPI_ISL_1039862, EPI_ISL_1039863, EPI_ISL_1039864, EPI_ISL_1039865, EPI_ISL_1039866, EPI_ISL_1039867, EPI_ISL_1039868, EPI_ISL_1039869, EPI_ISL_1039870, EPI_ISL_1039871, EPI_ISL_1039872, EPI_ISL_1039873, EPI_ISL_1039874, EPI_ISL_1039875, EPI_ISL_1039876, EPI_ISL_1039877, EPI_ISL_1039878, EPI_ISL_1039879, EPI_ISL_1039880, EPI_ISL_1039881, EPI_ISL_1039882, EPI_ISL_1039883, EPI_ISL_1039884, EPI_ISL_1039885, EPI_ISL_1039886, EPI_ISL_1039887, EPI_ISL_1039888, EPI_ISL_1039889, EPI_ISL_1039890, EPI_ISL_1039891, EPI_ISL_1039892, EPI_ISL_1039893, EPI_ISL_1039894, EPI_ISL_1039895, EPI_ISL_1039896, EPI_ISL_1039897, EPI_ISL_1039898, EPI_ISL_1039899, EPI_ISL_1039900, EPI_ISL_1039901, EPI_ISL_1039902, EPI_ISL_1039903, EPI_ISL_1039904, EPI_ISL_1039905, EPI_ISL_1039906, EPI_ISL_1039907, EPI_ISL_1039908, EPI_ISL_1039909, EPI_ISL_1039910, EPI_ISL_1039911, EPI_ISL_1039914, EPI_ISL_1039916, EPI_ISL_1039917, EPI_ISL_1039919, EPI_ISL_1039920, EPI_ISL_1039921, EPI_ISL_1039922, EPI_ISL_1039923, EPI_ISL_1039924, EPI_ISL_1039957, EPI_ISL_1039958 | | | |
| see above | University of Wisconsin-Madison AIDS Vaccine Research Laboratories | University of Wisconsin-Madison AIDS Vaccine Research Laboratories | Gage Moreno, Katarina Braun, et al. AIDS Vaccine Research Laboratories |
| EPI_ISL_1039999, EPI_ISL_1040000, EPI_ISL_1040001, EPI_ISL_1040002, EPI_ISL_1040003, EPI_ISL_1040004, EPI_ISL_1040005, EPI_ISL_1040006, EPI_ISL_1040007, EPI_ISL_1040008, EPI_ISL_1040009, EPI_ISL_1040010, EPI_ISL_1040011, EPI_ISL_1040012, EPI_ISL_1040013, EPI_ISL_1040014, EPI_ISL_1040015, EPI_ISL_1040016, EPI_ISL_1040017, EPI_ISL_1040018, EPI_ISL_1040019, EPI_ISL_1040020, EPI_ISL_1040021, EPI_ISL_1040022, EPI_ISL_1040023, EPI_ISL_1040024, EPI_ISL_1040025, EPI_ISL_1040026, EPI_ISL_1040027 | | | |
| see above | UW Virology Lab | UW Virology Lab | Pavitra Roychoudhury, Hong Xie, Lasata Shrestha, Shah Mohamed Bakhash, Robert Livingston, Michelle Lin, Noah Baker, Meei-Li Huang, Keith R Jerome, Alexander Greninger |
| EPI_ISL_1040051 | USC Clinical Lab | Los Angeles County Public Health Laboratories | P. Hemarajata et al. |
| EPI_ISL_1040052 | Los Angeles County Public Health Laboratories | Los Angeles County Public Health Laboratories | P. Hemarajata et al. |
| EPI_ISL_1040053, EPI_ISL_1040054, EPI_ISL_1040055, EPI_ISL_1040056, EPI_ISL_1040057, EPI_ISL_1040058, EPI_ISL_1040059, EPI_ISL_1040060, EPI_ISL_1040061, EPI_ISL_1040062, EPI_ISL_1040063, EPI_ISL_1040064, EPI_ISL_1040065, EPI_ISL_1040066, EPI_ISL_1040067, EPI_ISL_1040069, EPI_ISL_1040070, EPI_ISL_1040071 | | | |
| see above | UCLA Clinical Micro Lab | Los Angeles County Public Health Laboratories | P. Hemarajata et al. |
| EPI_ISL_1040072, EPI_ISL_1040073, EPI_ISL_1040074, EPI_ISL_1040075, EPI_ISL_1040076, EPI_ISL_1040077, EPI_ISL_1040078, EPI_ISL_1040079, EPI_ISL_1040080, EPI_ISL_1040081, EPI_ISL_1040082, EPI_ISL_1040083, EPI_ISL_1040084, EPI_ISL_1040085, EPI_ISL_1040086, EPI_ISL_1040087, EPI_ISL_1040088, EPI_ISL_1040089, EPI_ISL_1040090, EPI_ISL_1040091, EPI_ISL_1040092, EPI_ISL_1040093, EPI_ISL_1040094, EPI_ISL_1040095, EPI_ISL_1040096, EPI_ISL_1040097, EPI_ISL_1040098, EPI_ISL_1040099, EPI_ISL_1040100, EPI_ISL_1040101 | | | |
| see above | LSUHS Emerging Viral Threat Laboratory | Microbial Genome Sequencing Center | Jeremy P. Kamil, Jennifer L. Carroll, Maarten Van Diest, Andrew D. Yurochko, Rona S. Scott, John A. Vanchiere, Christopher G. Kevil, Daniel J. Snyder, Vaughn S. Cooper |
| EPI_ISL_1040102, EPI_ISL_1040103, EPI_ISL_1040104, EPI_ISL_1040105, EPI_ISL_1040106, EPI_ISL_1040107, EPI_ISL_1040108, EPI_ISL_1040109, EPI_ISL_1040110, EPI_ISL_1040111, EPI_ISL_1040112, EPI_ISL_1040113, EPI_ISL_1040114, EPI_ISL_1040115 | | | |
| see above | LSUHS Emerging Viral Threat Laboratory | Microbial Genome Sequencing Center | Jeremy P. Kamil, Jennifer L. Carroll, Maarten Van Diest, Andrew D. Yurochko, Rona S. Scott, Christopher G. Kevil, Daniel J. Snyder, Vaughn S. Cooper, John A. Vanchiere |
| EPI_ISL_1040116 | LSUHS Emerging Viral Threat Laboratory | Microbial Genome Sequencing Center | Maarten Van Diest, Jennifer L. Carroll, Andrew D. Yurochko, Rona S. Scott, Christopher G. Kevil, Daniel J. Snyder, John A. Vanchiere, Vaughn S. Cooper, Jeremy P. Kamil |
| EPI_ISL_1040117 | LSUHS Emerging Viral Threat Laboratory | Microbial Genome Sequencing Center | Daniel J. Snyder, Rona S. Scott, Jennifer L. Carroll, Maarten Van Diest, Andrew D. Yurochko, Christopher G. Kevil, John A. Vanchiere, Vaughn S. Cooper, Jeremy P. Kamil |
| EPI_ISL_1040118 | LSUHS Emerging Viral Threat Laboratory | Microbial Genome Sequencing Center | Rona S. Scott, Jennifer L. Carroll, Maarten Van Diest, Andrew D. Yurochko, Christopher G. Kevil, John A. Vanchiere, Daniel J. Snyder, Vaughn S. Cooper, Jeremy P. Kamil |
| EPI_ISL_1040167, EPI_ISL_1040168, EPI_ISL_1040169, EPI_ISL_1040170, EPI_ISL_1040171, EPI_ISL_1040172, EPI_ISL_1040173, EPI_ISL_1040174, EPI_ISL_1040176, EPI_ISL_1040177, EPI_ISL_1040178, EPI_ISL_1040179, EPI_ISL_1040180 | | | |
| see above | USC Clinical Lab | Los Angeles County Public Health Laboratories | P. Hemarajata et al. |
| EPI_ISL_1040425, EPI_ISL_1040426 | Los Angeles County Public Health Laboratories | Los Angeles County Public Health Laboratories | P. Hemarajata et al. |
| EPI_ISL_1040427, EPI_ISL_1040428, EPI_ISL_1040429, EPI_ISL_1040430, EPI_ISL_1040431, EPI_ISL_1040432, EPI_ISL_1040433, EPI_ISL_1040434, EPI_ISL_1040435, EPI_ISL_1040436, EPI_ISL_1040437 | | | |
| see above | USC Clinical Lab | Los Angeles County Public Health Laboratories | P. Hemarajata et al. |
| EPI_ISL_1040480 | Pandemic Response Lab - NYC | Pandemic Response Lab, R&D | Henry Lee, Michael Hammerling, Melissa Hopkins, Cybill del Castillo, William Ward, Pradeep Bugga, Haiping Hao, Jon Laurent |
| EPI_ISL_1040535, EPI_ISL_1040536, EPI_ISL_1040537, EPI_ISL_1040538, EPI_ISL_1040539, EPI_ISL_1040540, EPI_ISL_1040541, EPI_ISL_1040542, EPI_ISL_1040543, EPI_ISL_1040544, EPI_ISL_1040545, EPI_ISL_1040546, EPI_ISL_1040547, EPI_ISL_1040548, EPI_ISL_1040549, EPI_ISL_1040550, EPI_ISL_1040551, EPI_ISL_1040552, EPI_ISL_1040553, EPI_ISL_1040554, EPI_ISL_1040555, EPI_ISL_1040556, EPI_ISL_1040560, EPI_ISL_1040561, EPI_ISL_1040562, EPI_ISL_1040563, EPI_ISL_1040564, EPI_ISL_1040565, EPI_ISL_1040566, EPI_ISL_1040567, EPI_ISL_1040568, EPI_ISL_1040569, EPI_ISL_1040570, EPI_ISL_1040571, EPI_ISL_1040572, EPI_ISL_1040573, EPI_ISL_1040574, EPI_ISL_1040575, EPI_ISL_1040576, EPI_ISL_1040577, EPI_ISL_1040578, EPI_ISL_1040579, EPI_ISL_1040580, EPI_ISL_1040581, EPI_ISL_1040582, EPI_ISL_1040583, EPI_ISL_1040584, EPI_ISL_1040585, EPI_ISL_1040586, EPI_ISL_1040587, EPI_ISL_1040588, EPI_ISL_1040589, EPI_ISL_1040590, EPI_ISL_1040591, EPI_ISL_1040592, EPI_ISL_1040593, EPI_ISL_1040594, EPI_ISL_1040595, EPI_ISL_1040596, EPI_ISL_1040597, EPI_ISL_1040598, EPI_ISL_1040599, EPI_ISL_1040600, EPI_ISL_1040601, EPI_ISL_1040602, EPI_ISL_1040604, EPI_ISL_1040605, EPI_ISL_1040606, EPI_ISL_1040607, EPI_ISL_1040608, EPI_ISL_1040609, EPI_ISL_1040610, EPI_ISL_1040611, EPI_ISL_1040612, EPI_ISL_1040613, EPI_ISL_1040614, EPI_ISL_1040615, EPI_ISL_1040616, EPI_ISL_1040617, EPI_ISL_1040618, EPI_ISL_1040619, EPI_ISL_1040620, EPI_ISL_1040621, EPI_ISL_1040622, EPI_ISL_1040623, EPI_ISL_1040624, EPI_ISL_1040625, EPI_ISL_1040626, EPI_ISL_1040627, EPI_ISL_1040628, EPI_ISL_1040629, EPI_ISL_1040630, EPI_ISL_1040631, EPI_ISL_1040632, EPI_ISL_1040633, EPI_ISL_1040634, EPI_ISL_1040635, EPI_ISL_1040636, EPI_ISL_1040637, EPI_ISL_1040638, EPI_ISL_1040639, EPI_ISL_1040640, EPI_ISL_1040641, EPI_ISL_1040642, EPI_ISL_1040643 | | | |
| see above | Instituto Nacional de Medicina Genomica | Instituto Nacional de Medicina Genomica | Hidalgo-Miranda A, Mendoza-Vargas A, Reyes-Grajeda JP, Cisneros-Villanueva M, Cedro-Tanda A,Peñaloza-Figueroa F, Alcaraz-Millman M,Herrera-Montalvo LA |
| EPI_ISL_1040852, EPI_ISL_1040853, EPI_ISL_1040854, EPI_ISL_1040855, EPI_ISL_1040856, EPI_ISL_1040857, EPI_ISL_1040858, EPI_ISL_1040859, EPI_ISL_1040860, EPI_ISL_1040861, EPI_ISL_1040862, EPI_ISL_1040863, EPI_ISL_1040864, EPI_ISL_1040865, EPI_ISL_1040866, EPI_ISL_1040867, EPI_ISL_1040868, EPI_ISL_1040869, EPI_ISL_1040870, EPI_ISL_1040871, EPI_ISL_1040872, EPI_ISL_1040873, EPI_ISL_1040874, EPI_ISL_1040875, EPI_ISL_1040876, EPI_ISL_1040877, EPI_ISL_1040878, EPI_ISL_1040879, EPI_ISL_1040880, EPI_ISL_1040881, EPI_ISL_1040882, EPI_ISL_1040883, EPI_ISL_1040884, EPI_ISL_1040885, EPI_ISL_1040897, EPI_ISL_1040898, EPI_ISL_1040899, EPI_ISL_1040900, EPI_ISL_1040901, EPI_ISL_1040902, EPI_ISL_1040903, EPI_ISL_1040904, EPI_ISL_1040905, EPI_ISL_1040906, EPI_ISL_1040907, EPI_ISL_1040908, EPI_ISL_1040909, EPI_ISL_1040910, EPI_ISL_1040911, EPI_ISL_1040912, EPI_ISL_1040913, EPI_ISL_1040914 | | | |
| see above | NYU Langone Health | Departments of Pathology and Medicine, New York University School of Medicine | Adriana Heguy, Dacia Dimartino, Emily Guzman, Christian Marier, Peter Meyn, Sitharam Ramaswami, Gael Westby, Paul Zappile, Yutong Zhang, Paolo Cotzia, Guiqing Wang |
| EPI_ISL_1040927 | Biologia molecular de enfermedades emergentes y EPOC, Instituto Nacional de Enfermedades Respiratorias | Biologia molecular de enfermedades emergentes y EPOC, Instituto Nacional de Enfermedades Respiratorias | Vazquez-Perez Joel Armando, Mejía-Nepomuceno Fidencio, Hernández-Teran Alejandra, Serna Ricardo, Salas-Hernández Jorge, Wong-Arambula Claudia, Rodríguez-Maldonado Abril, Perez-Padilla Rogelio, Ramirez-Gonzalez Ernesto. |
| EPI_ISL_1041034, EPI_ISL_1041035, EPI_ISL_1041036, EPI_ISL_1041037, EPI_ISL_1041038, EPI_ISL_1041039, EPI_ISL_1041040, EPI_ISL_1041041, EPI_ISL_1041042, EPI_ISL_1041043, EPI_ISL_1041044, EPI_ISL_1041045, EPI_ISL_1041046, EPI_ISL_1041047, EPI_ISL_1041048, EPI_ISL_1041049, EPI_ISL_1041050, EPI_ISL_1041051, EPI_ISL_1041052, EPI_ISL_1041053 | | | |
| see above | Innovative Genomics Institute, UC Berkeley | Innovative Genomics Institute, UC Berkeley | Stacia Wyman, Haridha Shivram, Phil Frankino, Liana Lareau |
| EPI_ISL_1041054 | UW Virology Lab | UW Virology Lab | Pavitra Roychoudhury, Hong Xie, Lasata Shrestha, Shah Mohamed Bakhash, Robert Livingston, Michelle Lin, Noah Baker, Meei-Li Huang, Keith R Jerome, Alexander Greninger |
| EPI_ISL_1041055, EPI_ISL_1041056, EPI_ISL_1041057, EPI_ISL_1041058, EPI_ISL_1041059, EPI_ISL_1041060, EPI_ISL_1041061, EPI_ISL_1041062, EPI_ISL_1041063, EPI_ISL_1041064, EPI_ISL_1041065, EPI_ISL_1041066, EPI_ISL_1041067, EPI_ISL_1041068, EPI_ISL_1041069, EPI_ISL_1041070, EPI_ISL_1041071, EPI_ISL_1041072, EPI_ISL_1041073, EPI_ISL_1041074, EPI_ISL_1041076, EPI_ISL_1041077, EPI_ISL_1041078, EPI_ISL_1041079, EPI_ISL_1041080, EPI_ISL_1041081, EPI_ISL_1041082, EPI_ISL_1041084, EPI_ISL_1041085, EPI_ISL_1041086, EPI_ISL_1041087, EPI_ISL_1041088, EPI_ISL_1041089, EPI_ISL_1041090, EPI_ISL_1041091, EPI_ISL_1041092, EPI_ISL_1041093, EPI_ISL_1041094, EPI_ISL_1041095, EPI_ISL_1041096, EPI_ISL_1041097, EPI_ISL_1041098, EPI_ISL_1041099, EPI_ISL_1041100, EPI_ISL_1041101, EPI_ISL_1041102, EPI_ISL_1041103, EPI_ISL_1041104, EPI_ISL_1041105, EPI_ISL_1041106, EPI_ISL_1041107, EPI_ISL_1041181, EPI_ISL_1041182, EPI_ISL_1041183, EPI_ISL_1041184, EPI_ISL_1041185, EPI_ISL_1041186, EPI_ISL_1041187, EPI_ISL_1041188, EPI_ISL_1041189, EPI_ISL_1041190, EPI_ISL_1041191, EPI_ISL_1041192, EPI_ISL_1041193, EPI_ISL_1041194 | | | |
| see above | Innovative Genomics Institute, UC | Innovative Genomics Institute, UC Berkeley | Stacia Wyman, Haridha Shivram, Phil Frankino, Liana Lareau |

| | | | |
|---|---|---|---|
| | Berkeley | | |
| EPI_ISL_1041213, EPI_ISL_1041214, EPI_ISL_1041215, EPI_ISL_1041216, EPI_ISL_1041217, EPI_ISL_1041218, EPI_ISL_1041219, EPI_ISL_1041220, EPI_ISL_1041221, EPI_ISL_1041222, EPI_ISL_1041223, EPI_ISL_1041224, EPI_ISL_1041225, EPI_ISL_1041226, EPI_ISL_1041227, EPI_ISL_1041228, EPI_ISL_1041229, EPI_ISL_1041230, EPI_ISL_1041231, EPI_ISL_1041232, EPI_ISL_1041233, EPI_ISL_1041234, EPI_ISL_1041235, EPI_ISL_1041236, EPI_ISL_1041237, EPI_ISL_1041238, EPI_ISL_1041239, EPI_ISL_1041240, EPI_ISL_1041241, EPI_ISL_1041242, EPI_ISL_1041243, EPI_ISL_1041244, EPI_ISL_1041245, EPI_ISL_1041246, EPI_ISL_1041247, EPI_ISL_1041248, EPI_ISL_1041249, EPI_ISL_1041250, EPI_ISL_1041251, EPI_ISL_1041252, EPI_ISL_1041253, EPI_ISL_1041254, EPI_ISL_1041255, EPI_ISL_1041256, EPI_ISL_1041257, EPI_ISL_1041258, EPI_ISL_1041259, EPI_ISL_1041260, EPI_ISL_1041261, EPI_ISL_1041262, EPI_ISL_1041263, EPI_ISL_1041264, EPI_ISL_1041265, EPI_ISL_1041266, EPI_ISL_1041267, EPI_ISL_1041268, EPI_ISL_1041269, EPI_ISL_1041270, EPI_ISL_1041271, EPI_ISL_1041272, EPI_ISL_1041273, EPI_ISL_1041274, EPI_ISL_1041275, EPI_ISL_1041276, EPI_ISL_1041277, EPI_ISL_1041278, EPI_ISL_1041279, EPI_ISL_1041280, EPI_ISL_1041281, EPI_ISL_1041282, EPI_ISL_1041283, EPI_ISL_1041284, EPI_ISL_1041285, EPI_ISL_1041286, EPI_ISL_1041287, EPI_ISL_1041288, EPI_ISL_1041289, EPI_ISL_1041290, EPI_ISL_1041291, EPI_ISL_1041292, EPI_ISL_1041293, EPI_ISL_1041294, EPI_ISL_1041295, EPI_ISL_1041296, EPI_ISL_1041297, EPI_ISL_1041298, EPI_ISL_1041299, EPI_ISL_1041300, EPI_ISL_1041301, EPI_ISL_1041302, EPI_ISL_1041303, EPI_ISL_1041304, EPI_ISL_1041305, EPI_ISL_1041306, EPI_ISL_1041307, EPI_ISL_1041308, EPI_ISL_1041309, EPI_ISL_1041310, EPI_ISL_1041311, EPI_ISL_1041312, EPI_ISL_1041313, EPI_ISL_1041314, EPI_ISL_1041315, EPI_ISL_1041316, EPI_ISL_1041317, EPI_ISL_1041318, EPI_ISL_1041319, EPI_ISL_1041320, EPI_ISL_1041321, EPI_ISL_1041322, EPI_ISL_1041323, EPI_ISL_1041324, EPI_ISL_1041325, EPI_ISL_1041326, EPI_ISL_1041327, EPI_ISL_1041328, EPI_ISL_1041329, EPI_ISL_1041330, EPI_ISL_1041331, EPI_ISL_1041332, EPI_ISL_1041333, EPI_ISL_1041334, EPI_ISL_1041335, EPI_ISL_1041336, EPI_ISL_1041337, EPI_ISL_1041338, EPI_ISL_1041339, EPI_ISL_1041340, EPI_ISL_1041341, EPI_ISL_1041342, EPI_ISL_1041343, EPI_ISL_1041344, EPI_ISL_1041345, EPI_ISL_1041346, EPI_ISL_1041347, EPI_ISL_1041348, EPI_ISL_1041349, EPI_ISL_1041350, EPI_ISL_1041351, EPI_ISL_1041352, EPI_ISL_1041353, EPI_ISL_1041354, EPI_ISL_1041355, EPI_ISL_1041356, EPI_ISL_1041357, EPI_ISL_1041358, EPI_ISL_1041359, EPI_ISL_1041360, EPI_ISL_1041361, EPI_ISL_1041362, EPI_ISL_1041363, EPI_ISL_1041364, EPI_ISL_1041365, EPI_ISL_1041366, EPI_ISL_1041367, EPI_ISL_1041368, EPI_ISL_1041369, EPI_ISL_1041370, EPI_ISL_1041371, EPI_ISL_1041372, EPI_ISL_1041373, EPI_ISL_1041374, EPI_ISL_1041375, EPI_ISL_1041376, EPI_ISL_1041377, EPI_ISL_1041378, EPI_ISL_1041379, EPI_ISL_1041380, EPI_ISL_1041381, EPI_ISL_1041382, EPI_ISL_1041383, EPI_ISL_1041384, EPI_ISL_1041385, EPI_ISL_1041386, EPI_ISL_1041387, EPI_ISL_1041388, EPI_ISL_1041389, EPI_ISL_1041390, EPI_ISL_1041391, EPI_ISL_1041392, EPI_ISL_1041394, EPI_ISL_1041395, EPI_ISL_1041396, EPI_ISL_1041397, EPI_ISL_1041398, EPI_ISL_1041399, EPI_ISL_1041400, EPI_ISL_1041401, EPI_ISL_1041402, EPI_ISL_1041403, EPI_ISL_1041404, EPI_ISL_1041405, EPI_ISL_1041406, EPI_ISL_1041407, EPI_ISL_1041408, EPI_ISL_1041409, EPI_ISL_1041410, EPI_ISL_1041411, EPI_ISL_1041412, EPI_ISL_1041413, EPI_ISL_1041414, EPI_ISL_1041415, EPI_ISL_1041416, EPI_ISL_1041417, EPI_ISL_1041418, EPI_ISL_1041419, EPI_ISL_1041420, EPI_ISL_1041421, EPI_ISL_1041422, EPI_ISL_1041423, EPI_ISL_1041424, EPI_ISL_1041425, EPI_ISL_1041426, EPI_ISL_1041427, EPI_ISL_1041428, EPI_ISL_1041429, EPI_ISL_1041430, EPI_ISL_1041431, EPI_ISL_1041432, EPI_ISL_1041433, EPI_ISL_1041434, EPI_ISL_1041435, EPI_ISL_1041436, EPI_ISL_1041437, EPI_ISL_1041438, EPI_ISL_1041439, EPI_ISL_1041440, EPI_ISL_1041441, EPI_ISL_1041442, EPI_ISL_1041443, EPI_ISL_1041444, EPI_ISL_1041445, EPI_ISL_1041446, EPI_ISL_1041447, EPI_ISL_1041448, EPI_ISL_1041449, EPI_ISL_1041450, EPI_ISL_1041451, EPI_ISL_1041452, EPI_ISL_1041453, EPI_ISL_1041454, EPI_ISL_1041455, EPI_ISL_1041457, EPI_ISL_1041458, EPI_ISL_1041459, EPI_ISL_1041460, EPI_ISL_1041461, EPI_ISL_1041462, EPI_ISL_1041463, EPI_ISL_1041464, EPI_ISL_1041465, EPI_ISL_1041466, EPI_ISL_1041467, EPI_ISL_1041468, EPI_ISL_1041469, EPI_ISL_1041470, EPI_ISL_1041471, EPI_ISL_1041472, EPI_ISL_1041473, EPI_ISL_1041474, EPI_ISL_1041475, EPI_ISL_1041476, EPI_ISL_1041477, EPI_ISL_1041478, EPI_ISL_1041479, EPI_ISL_1041480, EPI_ISL_1041481, EPI_ISL_1041482, EPI_ISL_1041483, EPI_ISL_1041484, EPI_ISL_1041485, EPI_ISL_1041486, EPI_ISL_1041487, EPI_ISL_1041488, EPI_ISL_1041489, EPI_ISL_1041490, EPI_ISL_1041491, EPI_ISL_1041492, EPI_ISL_1041493, EPI_ISL_1041494, EPI_ISL_1041495, EPI_ISL_1041496, EPI_ISL_1041497, EPI_ISL_1041498, EPI_ISL_1041499, EPI_ISL_1041500, EPI_ISL_1041501, EPI_ISL_1041502, EPI_ISL_1041503, EPI_ISL_1041504, EPI_ISL_1041505, EPI_ISL_1041506, EPI_ISL_1041507, EPI_ISL_1041508, EPI_ISL_1041510, EPI_ISL_1041511, EPI_ISL_1041512, EPI_ISL_1041513, EPI_ISL_1041514, EPI_ISL_1041515, EPI_ISL_1041516, EPI_ISL_1041517, EPI_ISL_1041518, EPI_ISL_1041519, EPI_ISL_1041520, EPI_ISL_1041521, EPI_ISL_1041522, EPI_ISL_1041523, EPI_ISL_1041524, EPI_ISL_1041525, EPI_ISL_1041526, EPI_ISL_1041527, EPI_ISL_1041528, EPI_ISL_1041529, EPI_ISL_1041530, EPI_ISL_1041531, EPI_ISL_1041532, EPI_ISL_1041533, EPI_ISL_1041534, EPI_ISL_1041535, EPI_ISL_1041536, EPI_ISL_1041537, EPI_ISL_1041538, EPI_ISL_1041539, EPI_ISL_1041540, EPI_ISL_1041541, EPI_ISL_1041542, EPI_ISL_1041543, EPI_ISL_1041544, EPI_ISL_1041545, EPI_ISL_1041546, EPI_ISL_1041547, EPI_ISL_1041548, EPI_ISL_1041549, EPI_ISL_1041550, EPI_ISL_1041551, EPI_ISL_1041552, EPI_ISL_1041553, EPI_ISL_1041554, EPI_ISL_1041555, EPI_ISL_1041556, EPI_ISL_1041557, EPI_ISL_1041558, EPI_ISL_1041559, EPI_ISL_1041560, EPI_ISL_1041561, EPI_ISL_1041562, EPI_ISL_1041563, EPI_ISL_1041564, EPI_ISL_1041565, EPI_ISL_1041566, EPI_ISL_1041567, EPI_ISL_1041568, EPI_ISL_1041569, EPI_ISL_1041570, EPI_ISL_1041571, EPI_ISL_1041572, EPI_ISL_1041573, EPI_ISL_1041574, EPI_ISL_1041575, EPI_ISL_1041576, EPI_ISL_1041577, EPI_ISL_1041578, EPI_ISL_1041579, EPI_ISL_1041580, EPI_ISL_1041581, EPI_ISL_1041582, EPI_ISL_1041583, EPI_ISL_1041584, EPI_ISL_1041585, EPI_ISL_1041586, EPI_ISL_1041587, EPI_ISL_1041588, EPI_ISL_1041589, EPI_ISL_1041590, EPI_ISL_1041591, EPI_ISL_1041592, EPI_ISL_1041593, EPI_ISL_1041594, EPI_ISL_1041595, EPI_ISL_1041596, EPI_ISL_1041597, EPI_ISL_1041598, EPI_ISL_1041599, EPI_ISL_1041600, EPI_ISL_1041601, EPI_ISL_1041602, EPI_ISL_1041603, EPI_ISL_1041604, EPI_ISL_1041605, EPI_ISL_1041606, EPI_ISL_1041607, EPI_ISL_1041608, EPI_ISL_1041609, EPI_ISL_1041610, EPI_ISL_1041611, EPI_ISL_1041612, EPI_ISL_1041613, EPI_ISL_1041614, EPI_ISL_1041615, EPI_ISL_1041616, EPI_ISL_1041617, EPI_ISL_1041618, EPI_ISL_1041619, EPI_ISL_1041620, EPI_ISL_1041621, EPI_ISL_1041622, EPI_ISL_1041623, EPI_ISL_1041624, EPI_ISL_1041625, EPI_ISL_1041626, EPI_ISL_1041627, EPI_ISL_1041628, EPI_ISL_1041629, EPI_ISL_1041630, EPI_ISL_1041631, EPI_ISL_1041632, EPI_ISL_1041633, EPI_ISL_1041634, EPI_ISL_1041635, EPI_ISL_1041636, EPI_ISL_1041637, EPI_ISL_1041638, EPI_ISL_1041639, EPI_ISL_1041640, EPI_ISL_1041641, EPI_ISL_1041642, EPI_ISL_1041643, EPI_ISL_1041644, EPI_ISL_1041645, EPI_ISL_1041646, EPI_ISL_1041647, EPI_ISL_1041648, EPI_ISL_1041649, EPI_ISL_1041650, EPI_ISL_1041651, EPI_ISL_1041652, EPI_ISL_1041653, EPI_ISL_1041654, EPI_ISL_1041655, EPI_ISL_1041656, EPI_ISL_1041657, EPI_ISL_1041658, EPI_ISL_1041659, EPI_ISL_1041660, EPI_ISL_1041661, EPI_ISL_1041662, EPI_ISL_1041663, EPI_ISL_1041664, EPI_ISL_1041665, EPI_ISL_1041666, EPI_ISL_1041667, EPI_ISL_1041668, EPI_ISL_1041669, EPI_ISL_1041670, EPI_ISL_1041671, EPI_ISL_1041672, EPI_ISL_1041673, EPI_ISL_1041674, EPI_ISL_1041675, EPI_ISL_1041676, EPI_ISL_1041677, EPI_ISL_1041678, EPI_ISL_1041679, EPI_ISL_1041680, EPI_ISL_1041681, EPI_ISL_1041682, EPI_ISL_1041683, EPI_ISL_1041684, EPI_ISL_1041685, EPI_ISL_1041686, EPI_ISL_1041687, EPI_ISL_1041688, EPI_ISL_1041689, EPI_ISL_1041690, EPI_ISL_1041691, EPI_ISL_1041692, EPI_ISL_1041693, EPI_ISL_1041694, EPI_ISL_1041695, EPI_ISL_1041696, EPI_ISL_1041697, EPI_ISL_1041698, EPI_ISL_1041699, EPI_ISL_1041700, EPI_ISL_1041701, EPI_ISL_1041702, EPI_ISL_1041703, EPI_ISL_1041704, EPI_ISL_1041705, EPI_ISL_1041706, EPI_ISL_1041707, EPI_ISL_1041708, EPI_ISL_1041709, EPI_ISL_1041710, EPI_ISL_1041711, EPI_ISL_1041712, EPI_ISL_1041713, EPI_ISL_1041714, EPI_ISL_1041715, EPI_ISL_1041716, EPI_ISL_1041717, EPI_ISL_1041718, EPI_ISL_1041719, EPI_ISL_1041720, EPI_ISL_1041721, EPI_ISL_1041722, EPI_ISL_1041723, EPI_ISL_1041724, EPI_ISL_1041725, EPI_ISL_1041726, EPI_ISL_1041727, EPI_ISL_1041728, EPI_ISL_1041729, EPI_ISL_1041730, EPI_ISL_1041731, EPI_ISL_1041732, EPI_ISL_1041733, EPI_ISL_1041734, EPI_ISL_1041735, EPI_ISL_1041736, EPI_ISL_1041737, EPI_ISL_1041738, EPI_ISL_1041739, EPI_ISL_1041740, EPI_ISL_1041741, EPI_ISL_1041742, EPI_ISL_1041743, EPI_ISL_1041744, EPI_ISL_1041745, EPI_ISL_1041746, EPI_ISL_1041747, EPI_ISL_1041748, EPI_ISL_1041749, EPI_ISL_1041750, EPI_ISL_1041751, EPI_ISL_1041752, EPI_ISL_1041753, EPI_ISL_1041754, EPI_ISL_1041755, EPI_ISL_1041756, EPI_ISL_1041757, EPI_ISL_1041758, EPI_ISL_1041759, EPI_ISL_1041760, EPI_ISL_1041761, EPI_ISL_1041762, EPI_ISL_1041763, EPI_ISL_1041764, EPI_ISL_1041765, EPI_ISL_1041766, EPI_ISL_1041767, EPI_ISL_1041768, EPI_ISL_1041769, EPI_ISL_1041770, EPI_ISL_1041771, EPI_ISL_1041772, EPI_ISL_1041773, EPI_ISL_1041774, EPI_ISL_1041775, EPI_ISL_1041776, EPI_ISL_1041777, EPI_ISL_1041778, EPI_ISL_1041779, EPI_ISL_1041780, EPI_ISL_1041781, EPI_ISL_1041782, EPI_ISL_1041783, EPI_ISL_1041784, EPI_ISL_1041785, EPI_ISL_1041786, EPI_ISL_1041787, EPI_ISL_1041788, EPI_ISL_1041789, EPI_ISL_1041790, EPI_ISL_1041791, EPI_ISL_1041792, EPI_ISL_1041793, EPI_ISL_1041794, EPI_ISL_1041795, EPI_ISL_1041796, EPI_ISL_1041797, EPI_ISL_1041798, EPI_ISL_1041799, EPI_ISL_1041800, EPI_ISL_1041801, EPI_ISL_1041802, EPI_ISL_1041803, EPI_ISL_1041804, EPI_ISL_1041805, EPI_ISL_1041806, EPI_ISL_1041807, EPI_ISL_1041808, EPI_ISL_1041809, EPI_ISL_1041810, EPI_ISL_1041811, EPI_ISL_1041812, EPI_ISL_1041813, EPI_ISL_1041814, EPI_ISL_1041815, EPI_ISL_1041816, EPI_ISL_1041817, EPI_ISL_1041818, EPI_ISL_1041819, EPI_ISL_1041820, EPI_ISL_1041821, EPI_ISL_1041822, EPI_ISL_1041823, EPI_ISL_1041824, EPI_ISL_1041825, EPI_ISL_1041826, EPI_ISL_1041827, EPI_ISL_1041828, EPI_ISL_1041829, EPI_ISL_1041830, EPI_ISL_1041831, EPI_ISL_1041832, EPI_ISL_1041833, EPI_ISL_1041834, EPI_ISL_1041835, EPI_ISL_1041836, EPI_ISL_1041837, EPI_ISL_1041838, EPI_ISL_1041839, EPI_ISL_1041840, EPI_ISL_1041841, EPI_ISL_1041842, EPI_ISL_1041843, EPI_ISL_1041844, EPI_ISL_1041845, EPI_ISL_1041846, EPI_ISL_1041847, EPI_ISL_1041848, EPI_ISL_1041849, EPI_ISL_1041850, EPI_ISL_1041851, EPI_ISL_1041852, EPI_ISL_1041853, EPI_ISL_1041854, EPI_ISL_1041855, EPI_ISL_1041856, EPI_ISL_1041857, EPI_ISL_1041858, EPI_ISL_1041859, EPI_ISL_1041860, EPI_ISL_1041861, EPI_ISL_1041862, EPI_ISL_1041863, EPI_ISL_1041864, EPI_ISL_1041865, EPI_ISL_1041866, EPI_ISL_1041867, EPI_ISL_1041868, EPI_ISL_1041869, EPI_ISL_1041870, EPI_ISL_1041871, EPI_ISL_1041872, EPI_ISL_1041873, EPI_ISL_1041874, EPI_ISL_1041875, EPI_ISL_1041876, EPI_ISL_1041877, EPI_ISL_1041878, EPI_ISL_1041879, EPI_ISL_1041880, EPI_ISL_1041881, EPI_ISL_1041882, EPI_ISL_1041883, EPI_ISL_1041884, EPI_ISL_1041885, EPI_ISL_1041886, EPI_ISL_1041887, EPI_ISL_1041888, EPI_ISL_1041889, EPI_ISL_1041890, EPI_ISL_1041891, EPI_ISL_1041892, EPI_ISL_1041893, EPI_ISL_1041894, EPI_ISL_1041895, EPI_ISL_1041896, EPI_ISL_1041897, EPI_ISL_1041898, EPI_ISL_1041899, EPI_ISL_1041900, EPI_ISL_1041901, EPI_ISL_1041902, EPI_ISL_1041903, EPI_ISL_1041904, EPI_ISL_1041905, EPI_ISL_1041906, EPI_ISL_1041907, EPI_ISL_1041908, EPI_ISL_1041909, EPI_ISL_1041910, EPI_ISL_1041911, EPI_ISL_1041912, EPI_ISL_1041913, EPI_ISL_1041914, EPI_ISL_1041915, EPI_ISL_1041916, EPI_ISL_1041917, EPI_ISL_1041918, EPI_ISL_1041919, EPI_ISL_1041920, EPI_ISL_1041921, EPI_ISL_1041922, EPI_ISL_1041923, EPI_ISL_1041924, EPI_ISL_1041925, EPI_ISL_1041926, EPI_ISL_1041927, EPI_ISL_1041928, EPI_ISL_1041929, EPI_ISL_1041930 | | | |
| see above | Pandemic Response Lab - NYC | Pandemic Response Lab, R&D | Henry Lee, Michael Hammerling, Melissa Hopkins, Cybill del Castillo, William Ward, Pradeep Bugga, Haiping Hao, Jon Laurent |
| EPI_ISL_1048571, EPI_ISL_1048572, EPI_ISL_1048573, EPI_ISL_1048574, EPI_ISL_1048575, EPI_ISL_1048576, EPI_ISL_1048577, EPI_ISL_1048578, EPI_ISL_1048579, EPI_ISL_1048580, EPI_ISL_1048581, EPI_ISL_1048582, EPI_ISL_1048584, EPI_ISL_1048585, EPI_ISL_1048586, EPI_ISL_1048587, EPI_ISL_1048588, EPI_ISL_1048589, EPI_ISL_1048590, EPI_ISL_1048591, EPI_ISL_1048592, EPI_ISL_1048593, EPI_ISL_1048594, EPI_ISL_1048595, EPI_ISL_1048596, EPI_ISL_1048597, EPI_ISL_1048598, EPI_ISL_1048599, EPI_ISL_1048605, EPI_ISL_1048606, EPI_ISL_1048607, EPI_ISL_1048608, EPI_ISL_1048609, EPI_ISL_1048610, EPI_ISL_1048611, EPI_ISL_1048612, EPI_ISL_1048613, EPI_ISL_1048614, EPI_ISL_1048615, EPI_ISL_1048616, EPI_ISL_1048617, EPI_ISL_1048618, EPI_ISL_1048619, EPI_ISL_1048620, EPI_ISL_1048621, EPI_ISL_1048622, EPI_ISL_1048623, EPI_ISL_1048624, EPI_ISL_1048625, EPI_ISL_1048626, EPI_ISL_1048627, EPI_ISL_1048628, EPI_ISL_1048629, EPI_ISL_1048630, EPI_ISL_1048631, EPI_ISL_1048632, EPI_ISL_1048633, EPI_ISL_1048634, EPI_ISL_1048635, EPI_ISL_1048636, EPI_ISL_1048637, EPI_ISL_1048638, EPI_ISL_1048639, EPI_ISL_1048640, EPI_ISL_1048641, EPI_ISL_1048642, EPI_ISL_1048643, EPI_ISL_1048644, EPI_ISL_1048645, EPI_ISL_1048646, EPI_ISL_1048647, EPI_ISL_1048648, EPI_ISL_1048649, EPI_ISL_1048650, EPI_ISL_1048651, EPI_ISL_1048652, EPI_ISL_1048653, EPI_ISL_1048654, EPI_ISL_1048655, EPI_ISL_1048656, EPI_ISL_1048657, EPI_ISL_1048658, EPI_ISL_1048659, EPI_ISL_1048660, EPI_ISL_1048661, EPI_ISL_1048662, EPI_ISL_1048663, EPI_ISL_1048664, EPI_ISL_1048665, EPI_ISL_1048666, EPI_ISL_1048667, EPI_ISL_1048668, EPI_ISL_1048669, EPI_ISL_1048670, EPI_ISL_1048671, EPI_ISL_1048672, EPI_ISL_1048673, EPI_ISL_1048674, EPI_ISL_1048675, EPI_ISL_1048676, EPI_ISL_1048677, EPI_ISL_1048678, EPI_ISL_1048679, EPI_ISL_1048680, EPI_ISL_1048681, EPI_ISL_1048682, EPI_ISL_1048683, EPI_ISL_1048684, EPI_ISL_1048685, EPI_ISL_1048686, EPI_ISL_1048687, EPI_ISL_1048688, EPI_ISL_1048689, EPI_ISL_1048690, EPI_ISL_1048691, EPI_ISL_1048692, EPI_ISL_1048693, EPI_ISL_1048694, EPI_ISL_1048695, EPI_ISL_1048696, EPI_ISL_1048697, EPI_ISL_1048698, EPI_ISL_1048699, EPI_ISL_1048700, EPI_ISL_1048701, EPI_ISL_1048702, EPI_ISL_1048703, EPI_ISL_1048704, EPI_ISL_1048705, EPI_ISL_1048706, EPI_ISL_1048707, EPI_ISL_1048708, EPI_ISL_1048709, EPI_ISL_1048710, EPI_ISL_1048711, EPI_ISL_1048712, EPI_ISL_1048713, EPI_ISL_1048714, EPI_ISL_1048715, EPI_ISL_1048716, EPI_ISL_1048717, EPI_ISL_1048718, EPI_ISL_1048719, EPI_ISL_1048720, EPI_ISL_1048721, EPI_ISL_1048722, EPI_ISL_1048723, EPI_ISL_1048724, EPI_ISL_1048725, EPI_ISL_1048726, EPI_ISL_1048727, EPI_ISL_1048728, EPI_ISL_1048729, EPI_ISL_1048730, EPI_ISL_1048731, EPI_ISL_1048732, EPI_ISL_1048733, EPI_ISL_1048734, EPI_ISL_1048735, EPI_ISL_1048736, EPI_ISL_1048737, EPI_ISL_1048738, EPI_ISL_1048739, EPI_ISL_1048740, EPI_ISL_1048741, EPI_ISL_1048742, EPI_ISL_1048743, EPI_ISL_1048744, EPI_ISL_1048745, EPI_ISL_1048746, EPI_ISL_1048747, EPI_ISL_1048748, EPI_ISL_1048749, EPI_ISL_1048750, EPI_ISL_1048751, EPI_ISL_1048752, EPI_ISL_1048753, EPI_ISL_1048754, EPI_ISL_1048755, EPI_ISL_1048756, EPI_ISL_1048757, EPI_ISL_1048758, EPI_ISL_1048759, EPI_ISL_1048760, EPI_ISL_1048761, EPI_ISL_1048762, EPI_ISL_1048763, EPI_ISL_1048764, EPI_ISL_1048765, EPI_ISL_1048766, EPI_ISL_1048767, EPI_ISL_1048768, EPI_ISL_1048769, EPI_ISL_1048770, EPI_ISL_1048771, EPI_ISL_1048772, EPI_ISL_1048773, EPI_ISL_1048774, EPI_ISL_1048775, EPI_ISL_1048776, EPI_ISL_1048777, EPI_ISL_1048778, EPI_ISL_1048779, EPI_ISL_1048780, EPI_ISL_1048781, EPI_ISL_1048782, EPI_ISL_1048783, EPI_ISL_1048784, EPI_ISL_1048785, EPI_ISL_1048786, EPI_ISL_1048787, EPI_ISL_1048788, EPI_ISL_1048789, EPI_ISL_1048790, EPI_ISL_1048791, EPI_ISL_1048792, EPI_ISL_1048793, EPI_ISL_1048794, EPI_ISL_1048795, EPI_ISL_1048796 | | | |
| see above | Maine HETL | Tewhey Lab, The Jackson Laboratory | Matluk,N., Dewey,H., Iosue,F., Barter,M., Lynch,R., Munger,H. and Tewhey,R. |
| EPI_ISL_1048801, EPI_ISL_1048804 | Arizona State University | Arizona State University | Emily A. Kaelin, LaRinda A. Holland, Peter T. Skidmore, Nicholas J. Mellor, Kristina Buss, Joy M. Blain, Neal W Woodbury, Valerie Harris, Joshua LaBaer, Vel Murugan, Efrem S. Lim |
| EPI_ISL_1048830, EPI_ISL_1048831, EPI_ISL_1048832, EPI_ISL_1048833, EPI_ISL_1048834, EPI_ISL_1048835, EPI_ISL_1048836, EPI_ISL_1048837, EPI_ISL_1048838, EPI_ISL_1048839, EPI_ISL_1048840, EPI_ISL_1048841, EPI_ISL_1048842 | | | |
| see above | Kansas Health and Environmental Lab | Kansas Health and Environmental Lab | Mike Grose, Carissa Robertson, Ben Olsen, and Phil Adam |
| EPI_ISL_1049203 | NC State Laboratory of Public Health | Pathogen Discovery, Respiratory Viruses Branch, Division of Viral Diseases, Centers for Disease Control and Prevention | Ying Tao, Jing Zhang, Yan Li, Brian Lynch, Krista Queen, Anna Uehara, Peter Cook, Han Jia Ng, Clinton R. Paden, Haibin Wang, Suxiang Tong |
| EPI_ISL_1049204, EPI_ISL_1049205, EPI_ISL_1049206 | PA Department of Health, Bureau of Laboratories | Pathogen Discovery, Respiratory Viruses Branch, Division of Viral Diseases, Centers for Disease Control and Prevention | Ying Tao, Jing Zhang, Yan Li, Brian Lynch, Krista Queen, Anna Uehara, Peter Cook, Han Jia Ng, Clinton R. Paden, Haibin Wang, Suxiang Tong |
| EPI_ISL_1049207, EPI_ISL_1049208 | Blackhawk Genomics | Pathogen Discovery, Respiratory Viruses Branch, Division of Viral Diseases, Centers for Disease Control and Prevention | Ying Tao, Jing Zhang, Yan Li, Brian Lynch, Krista Queen, Anna Uehara, Peter Cook, Han Jia Ng, Clinton R. Paden, Haibin Wang, Suxiang Tong |
| EPI_ISL_1049209 | PA Department of Health, Bureau of Laboratories | Pathogen Discovery, Respiratory Viruses Branch, Division of Viral Diseases, Centers for Disease Control and Prevention | Ying Tao, Jing Zhang, Yan Li, Brian Lynch, Krista Queen, Anna Uehara, Peter Cook, Han Jia Ng, Clinton R. Paden, Haibin Wang, Suxiang Tong |
| EPI_ISL_1049210, EPI_ISL_1049211 | Blackhawk Genomics | Pathogen Discovery, Respiratory Viruses Branch, Division of Viral Diseases, Centers for Disease Control and Prevention | Ying Tao, Jing Zhang, Yan Li, Brian Lynch, Krista Queen, Anna Uehara, Peter Cook, Han Jia Ng, Clinton R. Paden, Haibin Wang, Suxiang Tong |
| EPI_ISL_1049212, EPI_ISL_1049213, EPI_ISL_1049214, EPI_ISL_1049215 | PA Department of Health, Bureau of Laboratories | Pathogen Discovery, Respiratory Viruses Branch, Division of Viral Diseases, Centers for Disease Control and Prevention | Ying Tao, Jing Zhang, Yan Li, Brian Lynch, Krista Queen, Anna Uehara, Peter Cook, Han Jia Ng, Clinton R. Paden, Haibin Wang, Suxiang Tong |
| EPI_ISL_1049220 | Blackhawk Genomics | Pathogen Discovery, Respiratory Viruses Branch, Division of Viral Diseases, Centers for Disease Control and Prevention | Ying Tao, Jing Zhang, Yan Li, Brian Lynch, Krista Queen, Anna Uehara, Peter Cook, Han Jia Ng, Clinton R. Paden, Haibin Wang, Suxiang Tong |
| EPI_ISL_1049221, EPI_ISL_1049222, EPI_ISL_1049223 | PA Department of Health, Bureau of Laboratories | Pathogen Discovery, Respiratory Viruses Branch, Division of Viral Diseases, Centers for Disease Control and Prevention | Ying Tao, Jing Zhang, Yan Li, Brian Lynch, Krista Queen, Anna Uehara, Peter Cook, Han Jia Ng, Clinton R. Paden, Haibin Wang, Suxiang Tong |
| EPI_ISL_1049224 | TN Division of Laboratory Services | Pathogen Discovery, Respiratory Viruses Branch, Division of Viral Diseases, Centers for Disease Control and Prevention | Ying Tao, Jing Zhang, Yan Li, Brian Lynch, Krista Queen, Anna Uehara, Peter Cook, Han Jia Ng, Clinton R. Paden, Haibin Wang, Suxiang Tong |
| EPI_ISL_1049226, EPI_ISL_1049227, EPI_ISL_1049228, EPI_ISL_1049229, EPI_ISL_1049230, EPI_ISL_1049231, EPI_ISL_1049232, EPI_ISL_1049233, EPI_ISL_1049234, EPI_ISL_1049235, EPI_ISL_1049236, EPI_ISL_1049237, EPI_ISL_1049238, EPI_ISL_1049239, EPI_ISL_1049240, EPI_ISL_1049241, EPI_ISL_1049242, EPI_ISL_1049243, EPI_ISL_1049244, EPI_ISL_1049245, EPI_ISL_1049246, EPI_ISL_1049247, EPI_ISL_1049248, EPI_ISL_1049249, EPI_ISL_1049250, EPI_ISL_1049251, EPI_ISL_1049252, EPI_ISL_1049253, EPI_ISL_1049254, EPI_ISL_1049255, EPI_ISL_1049256, EPI_ISL_1049257, EPI_ISL_1049258, EPI_ISL_1049259 | | | |
| see above | Utah Public Health Laboratory | Utah Public Health Laboratory | Erin L. Young, Kelly F. Oakeson, Tara Gallagher |
| EPI_ISL_1049262, EPI_ISL_1049263, EPI_ISL_1049264, EPI_ISL_1049265, EPI_ISL_1049266, EPI_ISL_1049267, EPI_ISL_1049268, EPI_ISL_1049269, EPI_ISL_1049270, EPI_ISL_1049271, EPI_ISL_1049272, EPI_ISL_1049273, EPI_ISL_1049274, EPI_ISL_1049275, EPI_ISL_1049276, EPI_ISL_1049277, EPI_ISL_1049278, EPI_ISL_1049279, EPI_ISL_1049280, EPI_ISL_1049281, EPI_ISL_1049282, EPI_ISL_1049295, EPI_ISL_1049296, EPI_ISL_1049297, EPI_ISL_1049298, EPI_ISL_1049299, EPI_ISL_1049300, EPI_ISL_1049301, EPI_ISL_1049302, EPI_ISL_1049303, EPI_ISL_1049304, EPI_ISL_1049305, EPI_ISL_1049306, EPI_ISL_1049307, EPI_ISL_1049308, EPI_ISL_1049309, EPI_ISL_1049310, EPI_ISL_1049311, EPI_ISL_1049312, EPI_ISL_1049313, EPI_ISL_1049314, EPI_ISL_1049315, EPI_ISL_1049316, EPI_ISL_1049317, EPI_ISL_1049318, EPI_ISL_1049319, EPI_ISL_1049320, EPI_ISL_1049321, EPI_ISL_1049322, EPI_ISL_1049323, EPI_ISL_1049324, EPI_ISL_1049325, EPI_ISL_1049326, EPI_ISL_1049327, EPI_ISL_1049328, EPI_ISL_1049329, EPI_ISL_1049330, EPI_ISL_1049331, EPI_ISL_1049332, EPI_ISL_1049333, EPI_ISL_1049334, EPI_ISL_1049335, EPI_ISL_1049336, EPI_ISL_1049337 | | | |
| see above | Innovative Genomics Institute, UC Berkeley | Innovative Genomics Institute, UC Berkeley | Stacia Wyman, Alison Ciling, Netravathi Krishnappa, Haridha Shivram, Phil Frankino, Liana Lareau |
| EPI_ISL_1049338, EPI_ISL_1049339 | Wadsworth Center, New York State Department.of Health | Wadsworth Center, New York State Department.of Health | Kirsten St. George, Daryl M. Lamson, Alexis Russel, Matthew Shudt, Melissa A Leisner, Jonathan Plitnick, Navjot Singh, John Kelly, Erasmus Schneider, Erica Lasek-Nesselquist |
| EPI_ISL_1049482 | MSHS Clinical Microbiology Laboratories | MSHS Pathogen Surveillance Program | Ana S. Gonzalez-Reiche, Hala Alshammary, Mitchell J. Sullivan, Brianne Ciferri, Ajay Obla, Angela Amoako, Mahmoud Awawda, Daniel Floda, Julia Matthews, Ashley Salimbangon, Levy Sominsky, Katherine Beach, Kayla Russo, Charles Gleason, Shelcie Fabre, Giulio Kleiner, Zenab Khan, Bremy Alburquerque, Adriana van de Guchte, Komal Srivastava, Matthew M. Hernandez, Jayeeta Dutta, Denise Jurczyszak, Nancy Francoeur, Betsaida Salom Melo, Irina Oussenko, Gintaras Deikus, Juan Soto, Shwetha Hara Sridhar, Ying-Chih Wang, Kathryn Twyman, Deena R. Altman, Robert Sebra, Adolfo García-Sastre, Marta Luksza, Gopi Patel, Sarah Schaefer, Melissa Gitman, Michael D. Nowak, Alberto Paniz-Mondolfi, Emilia Mia Sordillo, Viviana Simon, Harm van Bakel |
| EPI_ISL_1049483, EPI_ISL_1049484, EPI_ISL_1049485, EPI_ISL_1049486, EPI_ISL_1049487, EPI_ISL_1049488, EPI_ISL_1049489, EPI_ISL_1049490, EPI_ISL_1049491, EPI_ISL_1049492, EPI_ISL_1049493, EPI_ISL_1049494, EPI_ISL_1049495, EPI_ISL_1049496, EPI_ISL_1049497, EPI_ISL_1049498, EPI_ISL_1049499, EPI_ISL_1049500, EPI_ISL_1049501, EPI_ISL_1049502, EPI_ISL_1049503, EPI_ISL_1049504, EPI_ISL_1049505, EPI_ISL_1049506, EPI_ISL_1049507, EPI_ISL_1049508, EPI_ISL_1049509, EPI_ISL_1049510, EPI_ISL_1049511, EPI_ISL_1049512 | | | |
| see above | Alaska State Virology Laboratory | Alaska State Virology Laboratory | Stephanie DeRonde, Lisa Smith, Ph.D., Jack Chen, Ph.D. |
| EPI_ISL_1049513, EPI_ISL_1049514, EPI_ISL_1049515, EPI_ISL_1049516, EPI_ISL_1049517, EPI_ISL_1049518, EPI_ISL_1049520, EPI_ISL_1049521, EPI_ISL_1049522, EPI_ISL_1049523, EPI_ISL_1049525, EPI_ISL_1049527, EPI_ISL_1049528, EPI_ISL_1049530, EPI_ISL_1049531, EPI_ISL_1049532, EPI_ISL_1049533, EPI_ISL_1049534, EPI_ISL_1049536, EPI_ISL_1049537, EPI_ISL_1049539, EPI_ISL_1049540, EPI_ISL_1049541 | | | |
| see above | Seattle Flu Study | Seattle Flu Study | Deborah A. Nickerson, Chris D. Frazar, Jover Lee, Benjamin Pelle, Erica Ryke, Matthew Richardson, Amanda Adler, Elisabeth Brandstetter, Peter D. Han, Kairsten Fay, Misja Ilcisin, Kirsten Lacombe, Thomas R. Sibley, Melissa Truong, Caitlin R. Wolf, Karen Cowgill, Stephanie Schrag, Jeff Duchin, Michael Boeckh, Janet A. Englund, Michael Famulare, Barry R. Lutz, Mark J. Rieder, Lea M. Starita, Matthew Thompson, Helen Y. Chu, Trevor Bedford, Jay Shendure |

| | | | |
|---|---|---|---|
| EPI_ISL_1049542, EPI_ISL_1049543, EPI_ISL_1049544, EPI_ISL_1049545, EPI_ISL_1049546, EPI_ISL_1049547, EPI_ISL_1049548, EPI_ISL_1049549, EPI_ISL_1049550, EPI_ISL_1049551, EPI_ISL_1049552, EPI_ISL_1049553, EPI_ISL_1049554, EPI_ISL_1049555, EPI_ISL_1049556, EPI_ISL_1049557, EPI_ISL_1049558, EPI_ISL_1049559, EPI_ISL_1049560, EPI_ISL_1049561, EPI_ISL_1049562, EPI_ISL_1049563, EPI_ISL_1049566, EPI_ISL_1049567, EPI_ISL_1049569, EPI_ISL_1049570, EPI_ISL_1049571, EPI_ISL_1049572, EPI_ISL_1049573, EPI_ISL_1049574, EPI_ISL_1049575, EPI_ISL_1049577, EPI_ISL_1049578, EPI_ISL_1049582, EPI_ISL_1049583, EPI_ISL_1049584, EPI_ISL_1049585, EPI_ISL_1049587, EPI_ISL_1049590, EPI_ISL_1049591, EPI_ISL_1049592, EPI_ISL_1049593, EPI_ISL_1049594 | | | |
| see above | Washington State Department of Health | Seattle Flu Study | Deborah A. Nickerson, Chris D. Frazar, Jover Lee, Benjamin Pelle, Erica Ryke, Matthew Richardson, Amanda Adler, Elisabeth Brandstetter, Peter D. Han, Kairsten Fay, Misja Ilcisin, Kirsten Lacombe, Thomas R. Sibley, Melissa Truong, Caitlin R. Wolf, Romesh Gautom, Geoff Melly, Brian Hiatt, Philip Dykema, Scott Lindquist, Michael Boeckh, Janet A. Englund, Michael Famulare, Barry R. Lutz, Mark J. Rieder, Lea M. Starita, Matthew Thompson, Helen Y. Chu, Jay Shendure, Trevor Bedford |
| EPI_ISL_1049599, EPI_ISL_1049600 | Seattle Flu Study | Seattle Flu Study | Deborah A. Nickerson, Chris D. Frazar, Jover Lee, Benjamin Pelle, Erica Ryke, Matthew Richardson, Amanda Adler, Elisabeth Brandstetter, Peter D. Han, Kairsten Fay, Misja Ilcisin, Kirsten Lacombe, Thomas R. Sibley, Melissa Truong, Caitlin R. Wolf, Karen Cowgill, Stephanie Schrag, Jeff Duchin, Michael Boeckh, Janet A. Englund, Michael Famulare, Barry R. Lutz, Mark J. Rieder, Lea M. Starita, Matthew Thompson, Helen Y. Chu, Trevor Bedford, Jay Shendure |
| EPI_ISL_1049602 | Seattle Flu Study | Seattle Flu Study | Deborah A. Nickerson, Chris D. Frazar, Jover Lee, Benjamin Pelle, Erica Ryke, Matthew Richardson, Amanda Adler, Elisabeth Brandstetter, Peter D. Han, Kairsten Fay, Misja Ilcisin, Kirsten Lacombe, Thomas R. Sibley, Melissa Truong, Caitlin R. Wolf, Michael Boeckh, Janet A. Englund, Michael Famulare, Barry R. Lutz, Mark J. Rieder, Lea M. Starita, Matthew Thompson, Jay Shendure, Trevor Bedford, Helen Y. Chu |
| EPI_ISL_1049603 | Atlas Genomics | Seattle Flu Study | Deborah A. Nickerson, Chris D. Frazar, Jover Lee, Benjamin Pelle, Erica Ryke, Matthew Richardson, Amanda Adler, Elisabeth Brandstetter, Peter D. Han, Kairsten Fay, Misja Ilcisin, Kirsten Lacombe, Thomas R. Sibley, Melissa Truong, Caitlin R. Wolf, Romesh Gautom, Geoff Melly, Brian Hiatt, Philip Dykema, Scott Lindquist, Michael Boeckh, Janet A. Englund, Michael Famulare, Barry R. Lutz, Mark J. Rieder, Lea M. Starita, Matthew Thompson, Helen Y. Chu, Jay Shendure, Trevor Bedford |
| EPI_ISL_1049604 | Seattle Flu Study | Seattle Flu Study | Deborah A. Nickerson, Chris D. Frazar, Jover Lee, Benjamin Pelle, Erica Ryke, Matthew Richardson, Amanda Adler, Elisabeth Brandstetter, Peter D. Han, Kairsten Fay, Misja Ilcisin, Kirsten Lacombe, Thomas R. Sibley, Melissa Truong, Caitlin R. Wolf, Michael Boeckh, Janet A. Englund, Michael Famulare, Barry R. Lutz, Mark J. Rieder, Lea M. Starita, Matthew Thompson, Jay Shendure, Trevor Bedford, Helen Y. Chu |
| EPI_ISL_1049605, EPI_ISL_1049608, EPI_ISL_1049609 | Seattle Flu Study | Seattle Flu Study | Deborah A. Nickerson, Chris D. Frazar, Jover Lee, Benjamin Pelle, Erica Ryke, Matthew Richardson, Amanda Adler, Elisabeth Brandstetter, Peter D. Han, Kairsten Fay, Misja Ilcisin, Kirsten Lacombe, Thomas R. Sibley, Melissa Truong, Caitlin R. Wolf, Karen Cowgill, Stephanie Schrag, Jeff Duchin, Michael Boeckh, Janet A. Englund, Michael Famulare, Barry R. Lutz, Mark J. Rieder, Lea M. Starita, Matthew Thompson, Helen Y. Chu, Trevor Bedford, Jay Shendure |
| EPI_ISL_1049610, EPI_ISL_1049611, EPI_ISL_1049612, EPI_ISL_1049613, EPI_ISL_1049614, EPI_ISL_1049615, EPI_ISL_1049616, EPI_ISL_1049617, EPI_ISL_1049618, EPI_ISL_1049619, EPI_ISL_1049620, EPI_ISL_1049621, EPI_ISL_1049622, EPI_ISL_1049624, EPI_ISL_1049626, EPI_ISL_1049627, EPI_ISL_1049628, EPI_ISL_1049629, EPI_ISL_1049630, EPI_ISL_1049632, EPI_ISL_1049633, EPI_ISL_1049634, EPI_ISL_1049635, EPI_ISL_1049636, EPI_ISL_1049637, EPI_ISL_1049638, EPI_ISL_1049640, EPI_ISL_1049641, EPI_ISL_1049642, EPI_ISL_1049643, EPI_ISL_1049645, EPI_ISL_1049646 | | | |
| see above | Washington State Department of Health | Seattle Flu Study | Deborah A. Nickerson, Chris D. Frazar, Jover Lee, Benjamin Pelle, Erica Ryke, Matthew Richardson, Amanda Adler, Elisabeth Brandstetter, Peter D. Han, Kairsten Fay, Misja Ilcisin, Kirsten Lacombe, Thomas R. Sibley, Melissa Truong, Caitlin R. Wolf, Romesh Gautom, Geoff Melly, Brian Hiatt, Philip Dykema, Scott Lindquist, Michael Boeckh, Janet A. Englund, Michael Famulare, Barry R. Lutz, Mark J. Rieder, Lea M. Starita, Matthew Thompson, Helen Y. Chu, Jay Shendure, Trevor Bedford |
| EPI_ISL_1049647, EPI_ISL_1049648 | Seattle Flu Study | Seattle Flu Study | Deborah A. Nickerson, Chris D. Frazar, Jover Lee, Benjamin Pelle, Erica Ryke, Matthew Richardson, Amanda Adler, Elisabeth Brandstetter, Peter D. Han, Kairsten Fay, Misja Ilcisin, Kirsten Lacombe, Thomas R. Sibley, Melissa Truong, Caitlin R. Wolf, Karen Cowgill, Stephanie Schrag, Jeff Duchin, Michael Boeckh, Janet A. Englund, Michael Famulare, Barry R. Lutz, Mark J. Rieder, Lea M. Starita, Matthew Thompson, Helen Y. Chu, Trevor Bedford, Jay Shendure |
| EPI_ISL_1049649, EPI_ISL_1049650, EPI_ISL_1049651, EPI_ISL_1049652, EPI_ISL_1049653, EPI_ISL_1049654, EPI_ISL_1049655, EPI_ISL_1049656, EPI_ISL_1049657, EPI_ISL_1049659, EPI_ISL_1049661, EPI_ISL_1049662, EPI_ISL_1049663, EPI_ISL_1049664 | | | |
| see above | Washington State Department of Health | Seattle Flu Study | Deborah A. Nickerson, Chris D. Frazar, Jover Lee, Benjamin Pelle, Erica Ryke, Matthew Richardson, Amanda Adler, Elisabeth Brandstetter, Peter D. Han, Kairsten Fay, Misja Ilcisin, Kirsten Lacombe, Thomas R. Sibley, Melissa Truong, Caitlin R. Wolf, Romesh Gautom, Geoff Melly, Brian Hiatt, Philip Dykema, Scott Lindquist, Michael Boeckh, Janet A. Englund, Michael Famulare, Barry R. Lutz, Mark J. Rieder, Lea M. Starita, Matthew Thompson, Helen Y. Chu, Jay Shendure, Trevor Bedford |
| EPI_ISL_1049665 | Seattle Flu Study | Seattle Flu Study | Deborah A. Nickerson, Chris D. Frazar, Jover Lee, Benjamin Pelle, Erica Ryke, Matthew Richardson, Amanda Adler, Elisabeth Brandstetter, Peter D. Han, Kairsten Fay, Misja Ilcisin, Kirsten Lacombe, Thomas R. Sibley, Melissa Truong, Caitlin R. Wolf, Karen Cowgill, Stephanie Schrag, Jeff Duchin, Michael Boeckh, Janet A. Englund, Michael Famulare, Barry R. Lutz, Mark J. Rieder, Lea M. Starita, Matthew Thompson, Helen Y. Chu, Trevor Bedford, Jay Shendure |
| EPI_ISL_1049666, EPI_ISL_1049669, EPI_ISL_1049670, EPI_ISL_1049671, EPI_ISL_1049675, EPI_ISL_1049676, EPI_ISL_1049677, EPI_ISL_1049678, EPI_ISL_1049679, EPI_ISL_1049680, EPI_ISL_1049681, EPI_ISL_1049683, EPI_ISL_1049684, EPI_ISL_1049685, EPI_ISL_1049686, EPI_ISL_1049688, EPI_ISL_1049689, EPI_ISL_1049691, EPI_ISL_1049693, EPI_ISL_1049694, EPI_ISL_1049696, EPI_ISL_1049697, EPI_ISL_1049699, EPI_ISL_1049700, EPI_ISL_1049701, EPI_ISL_1049702, EPI_ISL_1049707, EPI_ISL_1049710, EPI_ISL_1049713, EPI_ISL_1049714, EPI_ISL_1049716, EPI_ISL_1049722, EPI_ISL_1049723, EPI_ISL_1049724, EPI_ISL_1049726, EPI_ISL_1049728, EPI_ISL_1049730, EPI_ISL_1049731, EPI_ISL_1049732, EPI_ISL_1049733, EPI_ISL_1049734, EPI_ISL_1049736, EPI_ISL_1049737, EPI_ISL_1049739, EPI_ISL_1049740, EPI_ISL_1049742, EPI_ISL_1049743, EPI_ISL_1049744, EPI_ISL_1049746, EPI_ISL_1049749, EPI_ISL_1049753, EPI_ISL_1049754, EPI_ISL_1049755, EPI_ISL_1049760, EPI_ISL_1049762, EPI_ISL_1049764, EPI_ISL_1049766, EPI_ISL_1049772, EPI_ISL_1049775, EPI_ISL_1049780, EPI_ISL_1049781, EPI_ISL_1049782, EPI_ISL_1049785, EPI_ISL_1049787, EPI_ISL_1049788, EPI_ISL_1049789, EPI_ISL_1049790, EPI_ISL_1049791, EPI_ISL_1049792, EPI_ISL_1049793, EPI_ISL_1049795, EPI_ISL_1049796, EPI_ISL_1049797, EPI_ISL_1049798, EPI_ISL_1049799, EPI_ISL_1049800, EPI_ISL_1049804, EPI_ISL_1049806, EPI_ISL_1049808, EPI_ISL_1049810, EPI_ISL_1049811, EPI_ISL_1049814, EPI_ISL_1049815, EPI_ISL_1049816, EPI_ISL_1049818, EPI_ISL_1049821, EPI_ISL_1049822, EPI_ISL_1049823, EPI_ISL_1049824, EPI_ISL_1049826, EPI_ISL_1049827, EPI_ISL_1049828, EPI_ISL_1049829, EPI_ISL_1049830, EPI_ISL_1049832, EPI_ISL_1049833, EPI_ISL_1049834, EPI_ISL_1049835, EPI_ISL_1049836, EPI_ISL_1049837, EPI_ISL_1049839, EPI_ISL_1049840, EPI_ISL_1049841 | | | |
| see above | Altius Institute for Biomedical Sciences | Seattle Flu Study | Deborah A. Nickerson, Chris D. Frazar, Jover Lee, Benjamin Pelle, Erica Ryke, Matthew Richardson, Amanda Adler, Elisabeth Brandstetter, Peter D. Han, Kairsten Fay, Misja Ilcisin, Kirsten Lacombe, Thomas R. Sibley, Melissa Truong, Caitlin R. Wolf, Ryan Alexander, Daniel Bates, Rebecca Bruders, Stephanie DeBaun, Clem Green, Muhammad Halimun, Jessica Halow, Kneshay Harper, Matt Hartman, Andrew Meuser, Alex Nguyen, Truong Nguyen, Sofia Olsson, Sadie Patraw, Hannah Petersen, Tobias Ragoczy, Joshua Richards, Jacob Rodriguez, John Stamatoyannopoulos, Julia Wald, Olivia Waltner, Michael Boeckh, Janet A. Englund, Michael Famulare, Barry R. Lutz, Mark J. Rieder, Lea M. Starita, Matthew Thompson, Helen Y. Chu, Jay Shendure, Trevor Bedford |
| EPI_ISL_1049843, EPI_ISL_1049844, EPI_ISL_1049845, EPI_ISL_1049846, EPI_ISL_1049847, EPI_ISL_1049848, EPI_ISL_1049849, EPI_ISL_1049850, EPI_ISL_1049851, EPI_ISL_1049852, EPI_ISL_1049853, EPI_ISL_1049854, EPI_ISL_1049855, EPI_ISL_1049856, EPI_ISL_1049857, EPI_ISL_1049858 | | | |
| see above | Kansas Health and Environmental Lab | Kansas Health and Environmental Lab | Mike Grose, Paige Drury, Carissa Robertson, Ben Olsen, and Phil Adam |
| EPI_ISL_1049859, EPI_ISL_1049860, EPI_ISL_1049861, EPI_ISL_1049862, EPI_ISL_1049863, EPI_ISL_1049864, EPI_ISL_1049865, EPI_ISL_1049866, EPI_ISL_1049867, EPI_ISL_1049868, EPI_ISL_1049869, EPI_ISL_1049870, EPI_ISL_1049871, EPI_ISL_1049872, EPI_ISL_1049873, EPI_ISL_1049874, EPI_ISL_1049875, EPI_ISL_1049876, EPI_ISL_1049877, EPI_ISL_1049878, EPI_ISL_1049879, EPI_ISL_1049880, EPI_ISL_1049881, EPI_ISL_1049882, EPI_ISL_1049883, EPI_ISL_1049884, EPI_ISL_1049922, EPI_ISL_1049923, EPI_ISL_1049924, EPI_ISL_1049925, EPI_ISL_1049926, EPI_ISL_1049927, EPI_ISL_1049928, EPI_ISL_1049929, EPI_ISL_1049930, EPI_ISL_1049931, EPI_ISL_1049932, EPI_ISL_1049933, EPI_ISL_1049934, EPI_ISL_1049935, EPI_ISL_1049936, EPI_ISL_1049937, EPI_ISL_1049938, EPI_ISL_1049939, EPI_ISL_1049940, EPI_ISL_1049941, EPI_ISL_1049942, EPI_ISL_1049943, EPI_ISL_1049944, EPI_ISL_1049945, EPI_ISL_1049946, EPI_ISL_1049947, EPI_ISL_1049948, EPI_ISL_1049949, EPI_ISL_1049950, EPI_ISL_1049951, EPI_ISL_1049952, EPI_ISL_1049953, EPI_ISL_1049954, EPI_ISL_1049955, EPI_ISL_1049956, EPI_ISL_1049957, EPI_ISL_1049958, EPI_ISL_1049959, EPI_ISL_1049960, EPI_ISL_1049961, EPI_ISL_1049962, EPI_ISL_1049963, EPI_ISL_1049964, EPI_ISL_1049965, EPI_ISL_1049966, EPI_ISL_1049967, EPI_ISL_1049968, EPI_ISL_1049969, EPI_ISL_1049970, EPI_ISL_1049971, EPI_ISL_1049972, EPI_ISL_1049973, EPI_ISL_1049974, EPI_ISL_1049975, EPI_ISL_1049976, EPI_ISL_1049977, EPI_ISL_1049978, EPI_ISL_1049979, EPI_ISL_1049980, EPI_ISL_1049981, EPI_ISL_1049982, EPI_ISL_1049983, EPI_ISL_1049984, EPI_ISL_1049985, EPI_ISL_1049986, EPI_ISL_1049987, EPI_ISL_1049988, EPI_ISL_1049989, EPI_ISL_1049990, EPI_ISL_1049991, EPI_ISL_1049992, EPI_ISL_1049993, EPI_ISL_1049994, EPI_ISL_1049995, EPI_ISL_1049996, EPI_ISL_1049997, EPI_ISL_1049998, EPI_ISL_1049999, EPI_ISL_1050000, EPI_ISL_1050001, EPI_ISL_1050002, EPI_ISL_1050003, EPI_ISL_1050004, EPI_ISL_1050005, EPI_ISL_1050006, EPI_ISL_1050007 | | | |
| see above | Florida Bureau of Public Health Laboratories | Florida Bureau of Public Health Laboratories | Sarah Schmedes, Jason Blanton |
| EPI_ISL_1050008, EPI_ISL_1050009, EPI_ISL_1050010, EPI_ISL_1050011, EPI_ISL_1050012, EPI_ISL_1050013, EPI_ISL_1050014, EPI_ISL_1050015, EPI_ISL_1050016, EPI_ISL_1050017, EPI_ISL_1050018, EPI_ISL_1050019, EPI_ISL_1050020, EPI_ISL_1050021, EPI_ISL_1050022, EPI_ISL_1050023, EPI_ISL_1050024, EPI_ISL_1050025, EPI_ISL_1050026, EPI_ISL_1050027, EPI_ISL_1050028, EPI_ISL_1050029, EPI_ISL_1050030, EPI_ISL_1050037, EPI_ISL_1050038, EPI_ISL_1050039, EPI_ISL_1050040, EPI_ISL_1050041, EPI_ISL_1050042, EPI_ISL_1050043, EPI_ISL_1050044, EPI_ISL_1050045, EPI_ISL_1050046, EPI_ISL_1050047, EPI_ISL_1050048, EPI_ISL_1050049, EPI_ISL_1050050, EPI_ISL_1050051, EPI_ISL_1050052, EPI_ISL_1050053, EPI_ISL_1050054, EPI_ISL_1050055, EPI_ISL_1050056, EPI_ISL_1050057, EPI_ISL_1050058, EPI_ISL_1050059, EPI_ISL_1050060, EPI_ISL_1050061, EPI_ISL_1050062, EPI_ISL_1050063, EPI_ISL_1050064 | | | |
| see above | NYU Langone Health | Departments of Pathology and Medicine, New York University School of Medicine | Adriana Heguy, Dacia Dimartino, Emily Guzman, Christian Marier, Peter Meyn, Sitharam Ramaswami, Gael Westby, Paul Zappile, Yutong Zhang, Paolo Cotzia, Guiqing Wang |
| EPI_ISL_1050502, EPI_ISL_1050703, EPI_ISL_1051176 | Arizona State University | Arizona State University | Peter T. Skidmore, LaRinda A. Holland, Emily A. Kaelin, Nicholas J. Mellor, Kristina Buss, Joy M. Blain, Neal W Woodbury, Valerie Harris, Joshua LaBaer, Vel Murugan, Efrem S. Lim |
| EPI_ISL_1053411, EPI_ISL_1053413, EPI_ISL_1053415, EPI_ISL_1053417, EPI_ISL_1053419, EPI_ISL_1053421, EPI_ISL_1053423, EPI_ISL_1053424, EPI_ISL_1053426, EPI_ISL_1053428, EPI_ISL_1053431, EPI_ISL_1053433, EPI_ISL_1053434, EPI_ISL_1053440, EPI_ISL_1053442, EPI_ISL_1053444, EPI_ISL_1053446, EPI_ISL_1053448, EPI_ISL_1053449, EPI_ISL_1054861, EPI_ISL_1054862, EPI_ISL_1054863, EPI_ISL_1054864, EPI_ISL_1054865, EPI_ISL_1054866, EPI_ISL_1054867, EPI_ISL_1054873, EPI_ISL_1054874, EPI_ISL_1054875, EPI_ISL_1054876, EPI_ISL_1054877, EPI_ISL_1054878, EPI_ISL_1054879, EPI_ISL_1054880, EPI_ISL_1054881, EPI_ISL_1054882, EPI_ISL_1054883, EPI_ISL_1054884, EPI_ISL_1054885 | | | |
| see above | Wyoming Public Health Laboratory | Wyoming Public Health Laboratory | Noah Hull, Taylor Fearing, Lynette Gumbleton, Channing Weber, Ashley Norberg, Bailey Bowcutt, and Wanda Manley |
| EPI_ISL_1054886, EPI_ISL_1054887, EPI_ISL_1054888, EPI_ISL_1054889, EPI_ISL_1054890, EPI_ISL_1054891 | Kansas Health and Environmental Lab | Kansas Health and Environmental Lab | Mike Grose, Paige Drury, Carissa Robertson, Ben Olsen, and Phil Adam |
| EPI_ISL_1054892, EPI_ISL_1054893, EPI_ISL_1054894, EPI_ISL_1054895, EPI_ISL_1054896, EPI_ISL_1054897, EPI_ISL_1054898, EPI_ISL_1054899, EPI_ISL_1054900, EPI_ISL_1054901, EPI_ISL_1054902, EPI_ISL_1054903, EPI_ISL_1054904, EPI_ISL_1054905, EPI_ISL_1054906, EPI_ISL_1054907, EPI_ISL_1054908, EPI_ISL_1054909, EPI_ISL_1054910, EPI_ISL_1054911, EPI_ISL_1054912, EPI_ISL_1054913, EPI_ISL_1054914, EPI_ISL_1054915, EPI_ISL_1054916, EPI_ISL_1054917, EPI_ISL_1054918, EPI_ISL_1054919, EPI_ISL_1054920, EPI_ISL_1054921, EPI_ISL_1054922, EPI_ISL_1054925, EPI_ISL_1054926, EPI_ISL_1054927, EPI_ISL_1054928, EPI_ISL_1054929, EPI_ISL_1054930, EPI_ISL_1054931, EPI_ISL_1054932, EPI_ISL_1054933, EPI_ISL_1054934, EPI_ISL_1054935, EPI_ISL_1054936, EPI_ISL_1054937, EPI_ISL_1054938, EPI_ISL_1054939 | | | |
| see above | North Dakota Department of Health, Public Health Laboratory | North Dakota Department of Health, Public Health Laboratory | Lisa Wingerter |
| EPI_ISL_1054940, EPI_ISL_1054941, EPI_ISL_1054942, EPI_ISL_1054943, EPI_ISL_1054944, EPI_ISL_1054945, EPI_ISL_1054946, EPI_ISL_1054947, EPI_ISL_1054948, EPI_ISL_1054949, EPI_ISL_1054950, EPI_ISL_1054951, EPI_ISL_1054952, EPI_ISL_1054953, EPI_ISL_1054954, EPI_ISL_1054955, EPI_ISL_1054956, EPI_ISL_1054957, EPI_ISL_1054958, EPI_ISL_1054959, EPI_ISL_1054960, EPI_ISL_1054961, EPI_ISL_1054962, EPI_ISL_1054963, EPI_ISL_1054964, EPI_ISL_1054965, EPI_ISL_1054966, EPI_ISL_1054967, EPI_ISL_1054968, EPI_ISL_1054969, EPI_ISL_1054970, EPI_ISL_1054971, EPI_ISL_1054972, EPI_ISL_1054973, EPI_ISL_1054974, EPI_ISL_1054975, EPI_ISL_1054976, EPI_ISL_1054977, EPI_ISL_1054978, EPI_ISL_1054979, EPI_ISL_1054980, EPI_ISL_1054981, EPI_ISL_1054982, EPI_ISL_1054983, EPI_ISL_1054984, EPI_ISL_1054985, EPI_ISL_1054986, EPI_ISL_1054987, EPI_ISL_1054988, EPI_ISL_1054989, EPI_ISL_1054990, EPI_ISL_1054991, EPI_ISL_1054992, EPI_ISL_1054993, EPI_ISL_1054994, EPI_ISL_1054995, EPI_ISL_1054996 | | | |
| see above | Instituto de Diagnostico y Referencia Epidemiologicos INDRE_RNLSP | Instituto de Diagnostico y Referencia Epidemiologicos (INDRE) | Claudia Wong-Arambula, Abril Rodriguez-Maldonado, Fabiola Garces-Ayala, Adnan Araiza-Rodriguez, David Fragoso-Fonseca, Sergio Rangel-Guerrero, Mayra Jimenez-Morales, Nancy Munoz-Hernandez, Natividad Cruz-Ortiz, Tatiana Nunez-Garcia, Gisela Barrera-Badillo, Lucia Hernandez-Rivas, Irma Lopez-Martinez, Ernesto Ramirez-Gonzalez. |
| EPI_ISL_1055005, EPI_ISL_1055006, EPI_ISL_1055007, EPI_ISL_1055008, EPI_ISL_1055009, EPI_ISL_1055010, EPI_ISL_1055011, EPI_ISL_1055012, EPI_ISL_1055013, EPI_ISL_1055014, EPI_ISL_1055015, EPI_ISL_1055016, EPI_ISL_1055017, EPI_ISL_1055018, EPI_ISL_1055019, EPI_ISL_1055020, EPI_ISL_1055021, EPI_ISL_1055022, EPI_ISL_1055023, EPI_ISL_1055024, EPI_ISL_1055025 | | | |
| see above | Instituto Nacional de Medicina Genomica | Instituto Nacional de Medicina Genomica | Hidalgo-Miranda A, Mendoza-Vargas A, Reyes-Grajeda JP, Cedro-Tanda A, Alcaraz N, Gonzalez-Barrera D, Rangel-DeLeon D, Ramirez-Vega O, Sifuentes-Rojas C, Cisneros-Villanueva M, Herrera-Montalvo LA |
| EPI_ISL_1055026, EPI_ISL_1055027, EPI_ISL_1055028, EPI_ISL_1055029, EPI_ISL_1055030, EPI_ISL_1055031, EPI_ISL_1055032, EPI_ISL_1055033, EPI_ISL_1055034, EPI_ISL_1055035, EPI_ISL_1055036 | | | |
| see above | New Mexico Department of Health Scientific Laboratory | New Mexico Department of Health Scientific Laboratory | Ellie Johnson, Anastacia Griego-Fisher, D'eldra Malone, Jennifer Benoit |
| EPI_ISL_1055366, EPI_ISL_1055367, EPI_ISL_1055368, EPI_ISL_1055369, EPI_ISL_1055370, EPI_ISL_1055371, EPI_ISL_1055372, EPI_ISL_1055373, EPI_ISL_1055374, EPI_ISL_1055375, EPI_ISL_1055376, EPI_ISL_1055377 | | | |
| see above | Oregon State Public Health Laboratory | Oregon SARS-CoV-2 Genome Sequencing Center | Brendan L. O'Connell, Sally Grindstaff, Kayla Carter, Sonia Acharya, John Fontana, LaDonna Grenz, Ruth V. Nichols, Alec J. Hirsch, Donna Hansel, Guang Fan, Xuan, Qin, Daniel N. Streblow, William B. Messer, Andrew C. Adey, Benjamin N. Bimber, Brian J. O'Roak |
| EPI_ISL_1055378, EPI_ISL_1055379, EPI_ISL_1055380 | Oregon State Public Health Laboratory | Oregon SARS-CoV-2 Genome Sequencing Center | Brendan L. O'Connell, Sally Grindstaff, Kayla Carter, Sonia Acharya, John Fontana, LaDonna Grenz, Ruth V. Nichols, Alec J. Hirsch, Donna Hansel, Guang Fan, Xuan Qin, Daniel N. Streblow, William B. Messer, Andrew C. Adey, Benjamin N. Bimber, Brian J. O'Roak |
| EPI_ISL_1055381, EPI_ISL_1055382 | Oregon State Public Health Laboratory | Oregon SARS-CoV-2 Genome Sequencing Center | Brendan L. O'Connell, Sally Grindstaff, Kayla Carter, Sonia Acharya, John Fontana, LaDonna Grenz, Ruth V. Nichols, Alec J. Hirsch, Donna Hansel, Guang Fan, Xuan, Qin, Daniel N. Streblow, William B. Messer, Andrew C. Adey, Benjamin N. Bimber, Brian J. O'Roak |
| EPI_ISL_1056953, EPI_ISL_1056954, EPI_ISL_1056955, EPI_ISL_1056958, EPI_ISL_1056960, EPI_ISL_1056969, EPI_ISL_1056973, EPI_ISL_1057037 | Ohio Department of Health Laboratory | Ohio Department of Health Laboratory | Holmes, Jennifer; Eric Brandt, Keoni Omura, Glen McGillivary, Caitlin McDonnell, Jade Mowery, Stephanie Mccracken, Tyler Payne, Kirtana Ramadugu, Erica Leasure, Brent Lee, Kelsey Florek, Heather Blankenship, Allison Black, Quanta Brown, and Tammy Bannerman |

| | | | |
|---|---|---|---|
| EPI_ISL_1058027, EPI_ISL_1058028 | School of Pharmacy, Shenandoah University | School of Pharmacy, Shenandoah University | Adams,S.M., Harralson,A.F., Kidd,R.S., Sawyer,G.W. |
| EPI_ISL_1058046 | Instituto Nacional de Medicina Genomica | Instituto Nacional de Medicina Genomica | Hidalgo-Miranda A, Mendoza-Vargas A, Reyes-Grajeda JP, Cedro-Tanda A, Alcaraz N, Gonzalez-Barrera D, Rangel-DeLeon D, Ramirez-Vega O, Sifuentes-Rojas C, Cisneros-Villanueva M, Herrera-Montalvo LA |
| EPI_ISL_1058061 | Baylor Scott & White - Southlake Family Medicine | Baylor Scott & White-Temple | Ari Rao, Linden Morales, Kimberly Walker, Marcus Volz, Shelby Hendrickson |
| EPI_ISL_1058066 | Baylor Scott & White - Irving | Baylor Scott & White-Temple | Ari Rao, Linden Morales, Kimberly Walker, Marcus Volz, Shelby Hendrickson |
| EPI_ISL_1058067, EPI_ISL_1058068, EPI_ISL_1058069, EPI_ISL_1058070, EPI_ISL_1058071, EPI_ISL_1058072, EPI_ISL_1058073, EPI_ISL_1058074, EPI_ISL_1058075 | Kansas Health and Environmental Lab | Kansas Health and Environmental Lab | Mike Grose, Paige Drury, Carissa Robertson, Ben Olsen, and Phil Adam |
| EPI_ISL_1058076, EPI_ISL_1058077, EPI_ISL_1058078 | Cytocheck Laboratory | Kansas Health and Environmental Lab | Mike Grose, Paige Drury, Carissa Robertson, Ben Olsen, and Phil Adam |
| EPI_ISL_1058688, EPI_ISL_1058689, EPI_ISL_1058690, EPI_ISL_1058691, EPI_ISL_1058692, EPI_ISL_1058693, EPI_ISL_1058694, EPI_ISL_1058695, EPI_ISL_1058696, EPI_ISL_1058697, EPI_ISL_1058698, EPI_ISL_1058699, EPI_ISL_1058700, EPI_ISL_1058701, EPI_ISL_1058702, EPI_ISL_1058703, EPI_ISL_1058704, EPI_ISL_1058705, EPI_ISL_1058707, EPI_ISL_1058709, EPI_ISL_1058710, EPI_ISL_1058711, EPI_ISL_1058712, EPI_ISL_1058713, EPI_ISL_1058714, EPI_ISL_1058715, EPI_ISL_1058716, EPI_ISL_1058717, EPI_ISL_1058718, EPI_ISL_1058719, EPI_ISL_1058720, EPI_ISL_1058721, EPI_ISL_1058722, EPI_ISL_1058723, EPI_ISL_1058724, EPI_ISL_1058725, EPI_ISL_1058726, EPI_ISL_1058727, EPI_ISL_1058728, EPI_ISL_1058729, EPI_ISL_1058730, EPI_ISL_1058732, EPI_ISL_1058734, EPI_ISL_1058735, EPI_ISL_1058736, EPI_ISL_1058737, EPI_ISL_1058738, EPI_ISL_1058739, EPI_ISL_1058740, EPI_ISL_1058741, EPI_ISL_1058742, EPI_ISL_1058743, EPI_ISL_1058744, EPI_ISL_1058745, EPI_ISL_1058746, EPI_ISL_1058747, EPI_ISL_1058748, EPI_ISL_1058749, EPI_ISL_1058750, EPI_ISL_1058752, EPI_ISL_1058753, EPI_ISL_1058754, EPI_ISL_1058755, EPI_ISL_1058756, EPI_ISL_1058757, EPI_ISL_1058759, EPI_ISL_1058760, EPI_ISL_1058761, EPI_ISL_1058762, EPI_ISL_1058763, EPI_ISL_1058764, EPI_ISL_1058765, EPI_ISL_1058766, EPI_ISL_1058767, EPI_ISL_1058778, EPI_ISL_1058779, EPI_ISL_1058780, EPI_ISL_1058781, EPI_ISL_1058782, EPI_ISL_1058783, EPI_ISL_1058784, EPI_ISL_1058785, EPI_ISL_1058786, EPI_ISL_1058787, EPI_ISL_1058788, EPI_ISL_1058789, EPI_ISL_1058790, EPI_ISL_1058792, EPI_ISL_1058793, EPI_ISL_1058794, EPI_ISL_1058795, EPI_ISL_1058796, EPI_ISL_1058797, EPI_ISL_1058798, EPI_ISL_1058799, EPI_ISL_1058800, EPI_ISL_1058801, EPI_ISL_1058802, EPI_ISL_1058804, EPI_ISL_1058805, EPI_ISL_1058806, EPI_ISL_1058807, EPI_ISL_1058808, EPI_ISL_1058809, EPI_ISL_1058810, EPI_ISL_1058811, EPI_ISL_1058814, EPI_ISL_1058815, EPI_ISL_1058816, EPI_ISL_1058817, EPI_ISL_1058818, EPI_ISL_1058819, EPI_ISL_1058820, EPI_ISL_1058821, EPI_ISL_1058822, EPI_ISL_1058823, EPI_ISL_1058824, EPI_ISL_1058825, EPI_ISL_1058826, EPI_ISL_1058827, EPI_ISL_1058828, EPI_ISL_1058829, EPI_ISL_1058830, EPI_ISL_1058831, EPI_ISL_1058832, EPI_ISL_1058833, EPI_ISL_1058834, EPI_ISL_1058835, EPI_ISL_1058837, EPI_ISL_1058838, EPI_ISL_1058839, EPI_ISL_1058840, EPI_ISL_1058841, EPI_ISL_1058842, EPI_ISL_1058843, EPI_ISL_1058844, EPI_ISL_1058845, EPI_ISL_1058846, EPI_ISL_1058847, EPI_ISL_1058848, EPI_ISL_1058849, EPI_ISL_1058850, EPI_ISL_1058851, EPI_ISL_1058852, EPI_ISL_1058854, EPI_ISL_1058855, EPI_ISL_1058856, EPI_ISL_1058857, EPI_ISL_1058858, EPI_ISL_1058859, EPI_ISL_1058860, EPI_ISL_1058861, EPI_ISL_1058862, EPI_ISL_1058863, EPI_ISL_1058864, EPI_ISL_1058865, EPI_ISL_1058866, EPI_ISL_1058867, EPI_ISL_1058868, EPI_ISL_1058869, EPI_ISL_1058870, EPI_ISL_1058871, EPI_ISL_1058872, EPI_ISL_1058873, EPI_ISL_1058874, EPI_ISL_1058875, EPI_ISL_1058876, EPI_ISL_1058877, EPI_ISL_1058878, EPI_ISL_1058879, EPI_ISL_1058880, EPI_ISL_1058881, EPI_ISL_1058882, EPI_ISL_1058883, EPI_ISL_1058884, EPI_ISL_1058885, EPI_ISL_1058886, EPI_ISL_1058887, EPI_ISL_1058888, EPI_ISL_1058889, EPI_ISL_1058890, EPI_ISL_1058891, EPI_ISL_1058892, EPI_ISL_1058893, EPI_ISL_1058894, EPI_ISL_1058895, EPI_ISL_1058896, EPI_ISL_1058897, EPI_ISL_1058898, EPI_ISL_1058899, EPI_ISL_1058900, EPI_ISL_1058901, EPI_ISL_1058902, EPI_ISL_1058903, EPI_ISL_1058904, EPI_ISL_1058905, EPI_ISL_1058906, EPI_ISL_1058908, EPI_ISL_1058909, EPI_ISL_1058910, EPI_ISL_1058911, EPI_ISL_1058912, EPI_ISL_1058913, EPI_ISL_1058914, EPI_ISL_1058915, EPI_ISL_1058916, EPI_ISL_1058917, EPI_ISL_1058918, EPI_ISL_1058919, EPI_ISL_1058920, EPI_ISL_1058921, EPI_ISL_1058922, EPI_ISL_1058923, EPI_ISL_1058924, EPI_ISL_1058925, EPI_ISL_1058926, EPI_ISL_1058927, EPI_ISL_1058928, EPI_ISL_1058929, EPI_ISL_1058930, EPI_ISL_1058931, EPI_ISL_1058934, EPI_ISL_1058935, EPI_ISL_1058936, EPI_ISL_1058937, EPI_ISL_1058938, EPI_ISL_1058939, EPI_ISL_1058940, EPI_ISL_1058941, EPI_ISL_1058942, EPI_ISL_1058943, EPI_ISL_1058944, EPI_ISL_1058945, EPI_ISL_1058946, EPI_ISL_1058947, EPI_ISL_1058948, EPI_ISL_1058949, EPI_ISL_1058950, EPI_ISL_1058954, EPI_ISL_1058955, EPI_ISL_1058956, EPI_ISL_1058957, EPI_ISL_1058958, EPI_ISL_1058959, EPI_ISL_1058960, EPI_ISL_1058961, EPI_ISL_1058962, EPI_ISL_1058963, EPI_ISL_1058964, EPI_ISL_1058965, EPI_ISL_1058966, EPI_ISL_1058967, EPI_ISL_1058968, EPI_ISL_1058970, EPI_ISL_1058972, EPI_ISL_1058973, EPI_ISL_1058974, EPI_ISL_1058976, EPI_ISL_1058977, EPI_ISL_1058978, EPI_ISL_1058979 | | | |
| see above | Michigan Department of Health and Human Services, Bureau of Laboratories | Michigan Department of Health and Human Services, Bureau of Laboratories | Blankenship HM, Riner D, Soehnlen MK |
| EPI_ISL_1058982, EPI_ISL_1058983, EPI_ISL_1058984, EPI_ISL_1058985, EPI_ISL_1058986, EPI_ISL_1058987, EPI_ISL_1058988, EPI_ISL_1058989, EPI_ISL_1058990, EPI_ISL_1058991, EPI_ISL_1058992, EPI_ISL_1058993, EPI_ISL_1058994, EPI_ISL_1058995, EPI_ISL_1058996, EPI_ISL_1058997, EPI_ISL_1058998, EPI_ISL_1058999, EPI_ISL_1059000, EPI_ISL_1059001, EPI_ISL_1059002, EPI_ISL_1059003, EPI_ISL_1059004, EPI_ISL_1059005, EPI_ISL_1059006, EPI_ISL_1059007, EPI_ISL_1059008, EPI_ISL_1059009, EPI_ISL_1059010, EPI_ISL_1059011, EPI_ISL_1059012, EPI_ISL_1059013, EPI_ISL_1059014, EPI_ISL_1059015, EPI_ISL_1059016, EPI_ISL_1059017, EPI_ISL_1059018, EPI_ISL_1059019, EPI_ISL_1059020, EPI_ISL_1059021, EPI_ISL_1059073, EPI_ISL_1059074, EPI_ISL_1059075, EPI_ISL_1059076, EPI_ISL_1059077, EPI_ISL_1059078, EPI_ISL_1059079, EPI_ISL_1059080, EPI_ISL_1059081, EPI_ISL_1059082, EPI_ISL_1059083, EPI_ISL_1059084, EPI_ISL_1059085, EPI_ISL_1059086, EPI_ISL_1059087, EPI_ISL_1059088, EPI_ISL_1059089, EPI_ISL_1059090, EPI_ISL_1059091, EPI_ISL_1059092, EPI_ISL_1059093, EPI_ISL_1059094, EPI_ISL_1059095, EPI_ISL_1059096, EPI_ISL_1059097, EPI_ISL_1059098, EPI_ISL_1059099, EPI_ISL_1059100, EPI_ISL_1059101, EPI_ISL_1059102, EPI_ISL_1059103, EPI_ISL_1059104, EPI_ISL_1059105, EPI_ISL_1059106, EPI_ISL_1059107, EPI_ISL_1059108, EPI_ISL_1059109, EPI_ISL_1059110, EPI_ISL_1059111, EPI_ISL_1059112, EPI_ISL_1059113, EPI_ISL_1059114, EPI_ISL_1059115, EPI_ISL_1059116, EPI_ISL_1059117, EPI_ISL_1059118, EPI_ISL_1059119, EPI_ISL_1059120, EPI_ISL_1059121, EPI_ISL_1059122, EPI_ISL_1059123, EPI_ISL_1059124, EPI_ISL_1059125, EPI_ISL_1059126, EPI_ISL_1059127, EPI_ISL_1059128, EPI_ISL_1059129, EPI_ISL_1059130, EPI_ISL_1059131, EPI_ISL_1059132, EPI_ISL_1059133, EPI_ISL_1059134, EPI_ISL_1059135, EPI_ISL_1059136, EPI_ISL_1059137, EPI_ISL_1059138, EPI_ISL_1059139, EPI_ISL_1059140, EPI_ISL_1059141, EPI_ISL_1059142, EPI_ISL_1059143, EPI_ISL_1059144, EPI_ISL_1059145, EPI_ISL_1059146, EPI_ISL_1059147, EPI_ISL_1059148, EPI_ISL_1059149, EPI_ISL_1059150, EPI_ISL_1059151, EPI_ISL_1059152, EPI_ISL_1059153, EPI_ISL_1059154, EPI_ISL_1059155, EPI_ISL_1059156, EPI_ISL_1059157, EPI_ISL_1059158, EPI_ISL_1059159, EPI_ISL_1059160, EPI_ISL_1059161, EPI_ISL_1059162, EPI_ISL_1059163, EPI_ISL_1059164, EPI_ISL_1059165, EPI_ISL_1059166, EPI_ISL_1059167, EPI_ISL_1059168, EPI_ISL_1059169, EPI_ISL_1059170, EPI_ISL_1059171, EPI_ISL_1059172, EPI_ISL_1059173, EPI_ISL_1059174, EPI_ISL_1059175, EPI_ISL_1059176, EPI_ISL_1059177, EPI_ISL_1059178, EPI_ISL_1059179, EPI_ISL_1059180, EPI_ISL_1059181, EPI_ISL_1059182, EPI_ISL_1059183, EPI_ISL_1059184, EPI_ISL_1059185, EPI_ISL_1059186, EPI_ISL_1059187, EPI_ISL_1059188, EPI_ISL_1059189, EPI_ISL_1059190, EPI_ISL_1059191, EPI_ISL_1059192, EPI_ISL_1059193, EPI_ISL_1059194, EPI_ISL_1059195, EPI_ISL_1059196, EPI_ISL_1059197, EPI_ISL_1059198, EPI_ISL_1059199, EPI_ISL_1059200, EPI_ISL_1059201, EPI_ISL_1059202, EPI_ISL_1059203, EPI_ISL_1059204, EPI_ISL_1059205, EPI_ISL_1059206, EPI_ISL_1059207, EPI_ISL_1059208, EPI_ISL_1059209, EPI_ISL_1059210, EPI_ISL_1059211, EPI_ISL_1059212, EPI_ISL_1059213, EPI_ISL_1059214, EPI_ISL_1059215, EPI_ISL_1059216, EPI_ISL_1059217, EPI_ISL_1059218, EPI_ISL_1059219, EPI_ISL_1059220, EPI_ISL_1059221, EPI_ISL_1059222, EPI_ISL_1059223, EPI_ISL_1059224, EPI_ISL_1059225, EPI_ISL_1059226, EPI_ISL_1059227, EPI_ISL_1059228, EPI_ISL_1059229, EPI_ISL_1059230, EPI_ISL_1059231, EPI_ISL_1059232, EPI_ISL_1059233, EPI_ISL_1059234, EPI_ISL_1059235, EPI_ISL_1059236, EPI_ISL_1059237, EPI_ISL_1059238 | | | |
| see above | University of Michigan Clinical Microbiology Laboratory | Lauring Lab, University of Michigan, Department of Microbiology and Immunology | Valesano |
| EPI_ISL_1059239, EPI_ISL_1059240, EPI_ISL_1059241, EPI_ISL_1059242, EPI_ISL_1059243, EPI_ISL_1059244, EPI_ISL_1059245, EPI_ISL_1059246, EPI_ISL_1059247, EPI_ISL_1059248, EPI_ISL_1059249, EPI_ISL_1059250, EPI_ISL_1059251, EPI_ISL_1059252, EPI_ISL_1059253, EPI_ISL_1059254, EPI_ISL_1059255, EPI_ISL_1059256, EPI_ISL_1059257, EPI_ISL_1059258, EPI_ISL_1059259, EPI_ISL_1059260, EPI_ISL_1059261, EPI_ISL_1059262, EPI_ISL_1059263, EPI_ISL_1059264, EPI_ISL_1059265, EPI_ISL_1059266, EPI_ISL_1059267, EPI_ISL_1059268, EPI_ISL_1059269, EPI_ISL_1059270, EPI_ISL_1059271, EPI_ISL_1059272, EPI_ISL_1059273, EPI_ISL_1059274, EPI_ISL_1059275, EPI_ISL_1059276, EPI_ISL_1059277, EPI_ISL_1059278, EPI_ISL_1059279, EPI_ISL_1059280, EPI_ISL_1059281, EPI_ISL_1059282, EPI_ISL_1059283, EPI_ISL_1059284, EPI_ISL_1059285, EPI_ISL_1059286, EPI_ISL_1059287, EPI_ISL_1059288, EPI_ISL_1059289, EPI_ISL_1059290, EPI_ISL_1059291, EPI_ISL_1059292, EPI_ISL_1059293, EPI_ISL_1059294, EPI_ISL_1059295 | | | |
| see above | Houston Health Dept. | Houston Health Dept. | Ryker Penn, Pamela Brown, Adolpho Lara |
| EPI_ISL_1059946, EPI_ISL_1059947, EPI_ISL_1060014, EPI_ISL_1060015, EPI_ISL_1060016, EPI_ISL_1060017, EPI_ISL_1060018, EPI_ISL_1060019, EPI_ISL_1060020, EPI_ISL_1060021, EPI_ISL_1060022, EPI_ISL_1060023, EPI_ISL_1060024, EPI_ISL_1060025, EPI_ISL_1060026, EPI_ISL_1060027, EPI_ISL_1060028, EPI_ISL_1060029, EPI_ISL_1060030, EPI_ISL_1060031, EPI_ISL_1060032, EPI_ISL_1060033, EPI_ISL_1060034, EPI_ISL_1060035, EPI_ISL_1060036, EPI_ISL_1060037, EPI_ISL_1060038, EPI_ISL_1060039 | | | |
| see above | Wyoming Public Health Laboratory | Wyoming Public Health Laboratory | Jim Mildenberger, Wanda Manley, Noah Hull, Taylor Fearing, Lynette Gumbleton, Channing Weber, Ashley Norberg, Chayse Rowley, Marley Goetz, Brian Dominguez, Elliot Thomasson, Cari Sloma, and Rob Christensen |
| EPI_ISL_1060040, EPI_ISL_1060041, EPI_ISL_1060042, EPI_ISL_1060043, EPI_ISL_1060044, EPI_ISL_1060045, EPI_ISL_1060046, EPI_ISL_1060047, EPI_ISL_1060048, EPI_ISL_1060049, EPI_ISL_1060050, EPI_ISL_1060051, EPI_ISL_1060052, EPI_ISL_1060053, EPI_ISL_1060054, EPI_ISL_1060055, EPI_ISL_1060056, EPI_ISL_1060057, EPI_ISL_1060058, EPI_ISL_1060059, EPI_ISL_1060060, EPI_ISL_1060061, EPI_ISL_1060062, EPI_ISL_1060063, EPI_ISL_1060064, EPI_ISL_1060065, EPI_ISL_1060066, EPI_ISL_1060067, EPI_ISL_1060068, EPI_ISL_1060069, EPI_ISL_1060070, EPI_ISL_1060071, EPI_ISL_1060072, EPI_ISL_1060073, EPI_ISL_1060074, EPI_ISL_1060075, EPI_ISL_1060076, EPI_ISL_1060077, EPI_ISL_1060078, EPI_ISL_1060079, EPI_ISL_1060080, EPI_ISL_1060081, EPI_ISL_1060082, EPI_ISL_1060083, EPI_ISL_1060084, EPI_ISL_1060085, EPI_ISL_1060086, EPI_ISL_1060087, EPI_ISL_1060088, EPI_ISL_1060089, EPI_ISL_1060090, EPI_ISL_1060091, EPI_ISL_1060092, EPI_ISL_1060093, EPI_ISL_1060094, EPI_ISL_1060095, EPI_ISL_1060096, EPI_ISL_1060097, EPI_ISL_1060098, EPI_ISL_1060099, EPI_ISL_1060161, EPI_ISL_1060162, EPI_ISL_1060163, EPI_ISL_1060164, EPI_ISL_1060165, EPI_ISL_1060166, EPI_ISL_1060167, EPI_ISL_1060168, EPI_ISL_1060169, EPI_ISL_1060170, EPI_ISL_1060171, EPI_ISL_1060172, EPI_ISL_1060173, EPI_ISL_1060174, EPI_ISL_1060175, EPI_ISL_1060176, EPI_ISL_1060177, EPI_ISL_1060178, EPI_ISL_1060179, EPI_ISL_1060180, EPI_ISL_1060181, EPI_ISL_1060182, EPI_ISL_1060183, EPI_ISL_1060184, EPI_ISL_1060185, EPI_ISL_1060186, EPI_ISL_1060187, EPI_ISL_1060188, EPI_ISL_1060189, EPI_ISL_1060190, EPI_ISL_1060191 | | | |
| see above | Innovative Genomics Institute, UC Berkeley | Innovative Genomics Institute, UC Berkeley | Stacia Wyman, Alison Ciling, Netravathi Krishnappa, Haridha Shivram, Phil Frankino, Liana Lareau |
| EPI_ISL_1060192, EPI_ISL_1060193, EPI_ISL_1060194, EPI_ISL_1060195, EPI_ISL_1060196, EPI_ISL_1060198, EPI_ISL_1060199, EPI_ISL_1060200, EPI_ISL_1060201, EPI_ISL_1060202, EPI_ISL_1060203, EPI_ISL_1060204, EPI_ISL_1060205, EPI_ISL_1060206, EPI_ISL_1060207, EPI_ISL_1060208, EPI_ISL_1060209, EPI_ISL_1060210, EPI_ISL_1060211, EPI_ISL_1060212, EPI_ISL_1060213, EPI_ISL_1060298, EPI_ISL_1060299, EPI_ISL_1060300, EPI_ISL_1060301, EPI_ISL_1060302, EPI_ISL_1060303, EPI_ISL_1060304, EPI_ISL_1060305, EPI_ISL_1060306, EPI_ISL_1060313, EPI_ISL_1060314, EPI_ISL_1060315, EPI_ISL_1060316, EPI_ISL_1060317, EPI_ISL_1060318, EPI_ISL_1060319, EPI_ISL_1060320, EPI_ISL_1060321, EPI_ISL_1060322, EPI_ISL_1060323, EPI_ISL_1060324, EPI_ISL_1060325, EPI_ISL_1060326, EPI_ISL_1060327, EPI_ISL_1060328, EPI_ISL_1060329, EPI_ISL_1060330, EPI_ISL_1060331, EPI_ISL_1060332 | | | |
| see above | Wyoming Public Health Laboratory | Wyoming Public Health Laboratory | Jim Mildenberger, Wanda Manley, Noah Hull, Taylor Fearing, Lynette Gumbleton, Channing Weber, Ashley Norberg, Chayse Rowley, Marley Goetz, Brian Dominguez, Elliot Thomasson, Cari Sloma, and Rob Christensen |
| EPI_ISL_1060543, EPI_ISL_1060544, EPI_ISL_1060545, EPI_ISL_1060550, EPI_ISL_1060551, EPI_ISL_1060552, EPI_ISL_1060553, EPI_ISL_1060554, EPI_ISL_1060555, EPI_ISL_1060556 | Santa Clara County Public Health Laboratory | Santa Clara County Public Health Laboratory | Santa Clara County Public Health Department |
| EPI_ISL_1060594, EPI_ISL_1060595, EPI_ISL_1060596 | Cytocheck Laboratory | Kansas Health and Environmental Lab | Mike Grose, Paige Drury, Carissa Robertson, Ben Olsen, and Phil Adam |
| EPI_ISL_1060598, EPI_ISL_1060599, EPI_ISL_1060600, EPI_ISL_1060601, EPI_ISL_1060602, EPI_ISL_1060603, EPI_ISL_1060604, EPI_ISL_1060605, EPI_ISL_1060606, EPI_ISL_1060607, EPI_ISL_1060608, EPI_ISL_1060609, EPI_ISL_1060610, EPI_ISL_1060611, EPI_ISL_1060612, EPI_ISL_1060613, EPI_ISL_1060614, EPI_ISL_1060615 | | | |
| see above | Instituto Nacional de Medicina Genomica | Instituto Nacional de Medicina Genomica | Hidalgo-Miranda A, Mendoza-Vargas A, Reyes-Grajeda JP, Cedro-Tanda A, Alcaraz N, Gonzalez-Barrera D, Rangel-DeLeon D, Ramirez-Vega O, Sifuentes-Rojas C, Cisneros-Villanueva M, Herrera-Montalvo LA |
| EPI_ISL_1060616, EPI_ISL_1060617, EPI_ISL_1060618, EPI_ISL_1060619, EPI_ISL_1060620, EPI_ISL_1060622, EPI_ISL_1060623, EPI_ISL_1060624, EPI_ISL_1060625, EPI_ISL_1060626, EPI_ISL_1060627, EPI_ISL_1060628, EPI_ISL_1060629, EPI_ISL_1060630, EPI_ISL_1060631, EPI_ISL_1060632, EPI_ISL_1060633, EPI_ISL_1060634, EPI_ISL_1060635, EPI_ISL_1060636, EPI_ISL_1060637, EPI_ISL_1060638, EPI_ISL_1060639, EPI_ISL_1060640, EPI_ISL_1060641, EPI_ISL_1060642, EPI_ISL_1060643, EPI_ISL_1060644, EPI_ISL_1060645, EPI_ISL_1060646, EPI_ISL_1060647, EPI_ISL_1060648, EPI_ISL_1060649, EPI_ISL_1060650, EPI_ISL_1060651, EPI_ISL_1060652, EPI_ISL_1060653, EPI_ISL_1060654, EPI_ISL_1060655, EPI_ISL_1060656, EPI_ISL_1060657, EPI_ISL_1060658, EPI_ISL_1060659, EPI_ISL_1060660, EPI_ISL_1060661, EPI_ISL_1060662, EPI_ISL_1060663, EPI_ISL_1060664, EPI_ISL_1060665, EPI_ISL_1060666, EPI_ISL_1060667, EPI_ISL_1060668, EPI_ISL_1060669, EPI_ISL_1060670, EPI_ISL_1060671, EPI_ISL_1060672, EPI_ISL_1060673, EPI_ISL_1060674, EPI_ISL_1060675 | | | |
| see above | NYU Langone Health | Departments of Pathology and Medicine, New York University School of Medicine | Adriana Heguy, Dacia Dimartino, Emily Guzman, Christian Marier, Peter Meyn, Sitharam Ramaswami, Gael Westby, Paul Zappile, Yutong Zhang, Paolo Cotzia, Guiqing Wang |
| EPI_ISL_1060676, EPI_ISL_1060677, EPI_ISL_1060678, EPI_ISL_1060679, EPI_ISL_1060680, EPI_ISL_1060681, EPI_ISL_1060682, EPI_ISL_1060683, EPI_ISL_1060684, EPI_ISL_1060685, EPI_ISL_1060686, EPI_ISL_1060687, EPI_ISL_1060688, EPI_ISL_1060689, EPI_ISL_1060690, EPI_ISL_1060691, EPI_ISL_1060692, EPI_ISL_1060693, EPI_ISL_1060694, EPI_ISL_1060695, EPI_ISL_1060696, EPI_ISL_1060697, EPI_ISL_1060698, EPI_ISL_1060699, EPI_ISL_1060700, EPI_ISL_1060701, EPI_ISL_1060702, EPI_ISL_1060703, EPI_ISL_1060704, EPI_ISL_1060705, EPI_ISL_1060706, EPI_ISL_1060707, EPI_ISL_1060714, EPI_ISL_1060715, EPI_ISL_1060716, EPI_ISL_1060717, EPI_ISL_1060718, EPI_ISL_1060719, EPI_ISL_1060720, EPI_ISL_1060721, EPI_ISL_1060722, EPI_ISL_1060723, EPI_ISL_1060724, EPI_ISL_1060725, EPI_ISL_1060726, EPI_ISL_1060727, EPI_ISL_1060728, EPI_ISL_1060729, EPI_ISL_1060730, EPI_ISL_1060731, EPI_ISL_1060732, EPI_ISL_1060733, EPI_ISL_1060734, EPI_ISL_1060735, EPI_ISL_1060736, EPI_ISL_1060737, EPI_ISL_1060738, EPI_ISL_1060739, EPI_ISL_1060740, EPI_ISL_1060741, EPI_ISL_1060742, EPI_ISL_1060743, EPI_ISL_1060744, EPI_ISL_1060745, EPI_ISL_1060746, EPI_ISL_1060747, EPI_ISL_1060748, EPI_ISL_1060749, EPI_ISL_1060750, EPI_ISL_1060751, EPI_ISL_1060752, EPI_ISL_1060753, EPI_ISL_1060754, EPI_ISL_1060755, EPI_ISL_1060756, EPI_ISL_1060757, EPI_ISL_1060758, EPI_ISL_1060759, EPI_ISL_1060760, EPI_ISL_1060761, EPI_ISL_1060762, EPI_ISL_1060763 | | | |
| see above | Instituto de Diagnostico y Referencia Epidemiologicos (INDRE)_RNLSP | Instituto de Diagnostico y Referencia Epidemiologicos (INDRE) | Claudia Wong-Arambula, Abril Rodriguez-Maldonado, Fabiola Garces-Ayala, Adnan Araiza-Rodriguez, David Fragoso-Fonseca, Sergio Rangel-Guerrero, Mayra Jimenez-Morales, Nancy Munoz-Hernandez, Natividad Cruz-Ortiz, Tatiana Nunez-Garcia, Gisela Barrera-Badillo, Lucia Hernandez-Rivas, Irma Lopez-Martinez, Ernesto Ramirez-Gonzalez. |
| EPI_ISL_1060764, EPI_ISL_1060765 | University of Michigan Clinical Microbiology Laboratory | Lauring Lab, University of Michigan, Department of Microbiology and Immunology | Valesano |
| EPI_ISL_1060766, EPI_ISL_1060767 | NJ Rapid Response Teams | New Jersey Public Health Environmental Laboratories (NJ_PHEL) | Lindsey Bodnar, Shiv Verma, Dana Woell, Byeong Jeong |
| EPI_ISL_1060768 | Mary Eliza Mahoney HC | New Jersey Public Health Environmental Laboratories (NJ_PHEL) | Lindsey Bodnar, Shiv Verma, Dana Woell, Byeong Jeong |
| EPI_ISL_1060769, EPI_ISL_1060770, EPI_ISL_1060771, EPI_ISL_1060772, EPI_ISL_1060773, EPI_ISL_1060774, EPI_ISL_1060775, EPI_ISL_1060776, EPI_ISL_1060777, EPI_ISL_1060778, EPI_ISL_1060779, EPI_ISL_1060780, EPI_ISL_1060781, EPI_ISL_1060782, EPI_ISL_1060783, EPI_ISL_1060784, EPI_ISL_1060785, EPI_ISL_1060786, EPI_ISL_1060787, EPI_ISL_1060788, EPI_ISL_1060789, EPI_ISL_1060790 | | | |

| | | | |
|---|---|---|---|
| see above | NJ Rapid Response Teams | New Jersey Public Health Environmental Laboratories (NJ_PHEL) | Lindsey Bodnar, Shiv Verma, Dana Woell, Byeong Jeong |
| EPI_ISL_1060791 | Virtua Health System-Voorhees | New Jersey Public Health Environmental Laboratories (NJ_PHEL) | Lindsey Bodnar, Shiv Verma, Dana Woell, Byeong Jeong |
| EPI_ISL_1060797 | Mary Eliza Mahoney HC | New Jersey Public Health Environmental Laboratories (NJ_PHEL) | Lindsey Bodnar, Shiv Verma, Dana Woell, Byeong Jeong |
| EPI_ISL_1060798, EPI_ISL_1060799, EPI_ISL_1060800, EPI_ISL_1060801, EPI_ISL_1060802, EPI_ISL_1060803, EPI_ISL_1060804, EPI_ISL_1060805 | NJ Rapid Response Teams | New Jersey Public Health Environmental Laboratories (NJ_PHEL) | Lindsey Bodnar, Shiv Verma, Dana Woell, Byeong Jeong |
| EPI_ISL_1060806 | New Jersey Department of Health, Public Health Laboratories | New Jersey Public Health Environmental Laboratories (NJ_PHEL) | Lindsey Bodnar, Shiv Verma, Dana Woell, Byeong Jeong |
| EPI_ISL_1060807, EPI_ISL_1060808, EPI_ISL_1060809, EPI_ISL_1060810, EPI_ISL_1060811, EPI_ISL_1060812 | NJ Rapid Response Teams | New Jersey Public Health Environmental Laboratories (NJ_PHEL) | Lindsey Bodnar, Shiv Verma, Dana Woell, Byeong Jeong |
| EPI_ISL_1060813, EPI_ISL_1060814, EPI_ISL_1060815, EPI_ISL_1060816, EPI_ISL_1060817, EPI_ISL_1060818, EPI_ISL_1060819, EPI_ISL_1060820, EPI_ISL_1060821, EPI_ISL_1060822, EPI_ISL_1060823, EPI_ISL_1060824, EPI_ISL_1060825, EPI_ISL_1060826, EPI_ISL_1060827, EPI_ISL_1060828, EPI_ISL_1060829, EPI_ISL_1060830, EPI_ISL_1060831, EPI_ISL_1060832, EPI_ISL_1060833, EPI_ISL_1060834, EPI_ISL_1060835, EPI_ISL_1060836, EPI_ISL_1060837, EPI_ISL_1060858, EPI_ISL_1060859, EPI_ISL_1060860, EPI_ISL_1060861, EPI_ISL_1060862, EPI_ISL_1060863, EPI_ISL_1060864, EPI_ISL_1060865, EPI_ISL_1060866, EPI_ISL_1060867, EPI_ISL_1060868, EPI_ISL_1060869, EPI_ISL_1060870 | | | |
| see above | Utah Public Health Laboratory | Utah Public Health Laboratory | Erin L. Young, Kelly F. Oakeson, Tara Gallagher |
| EPI_ISL_1060871, EPI_ISL_1060872, EPI_ISL_1060873, EPI_ISL_1060874, EPI_ISL_1060875 | Providence Oregon Regional Laboratories | Providence St. Joseph Health Molecular Genomics Laboratory | Alexa K Dowdell, Brian D Piening, Fred L Robinson, Carlo B Bifulco, Mary Campbell |
| EPI_ISL_1061279, EPI_ISL_1061280, EPI_ISL_1061281, EPI_ISL_1061282, EPI_ISL_1061283, EPI_ISL_1061284, EPI_ISL_1061285, EPI_ISL_1061286, EPI_ISL_1061287, EPI_ISL_1061288, EPI_ISL_1061289, EPI_ISL_1061290, EPI_ISL_1061291, EPI_ISL_1061292, EPI_ISL_1061293, EPI_ISL_1061294, EPI_ISL_1061295, EPI_ISL_1061296, EPI_ISL_1061297, EPI_ISL_1061298, EPI_ISL_1061299, EPI_ISL_1061300, EPI_ISL_1061301, EPI_ISL_1061302, EPI_ISL_1061303, EPI_ISL_1061304, EPI_ISL_1061305, EPI_ISL_1061306, EPI_ISL_1061307, EPI_ISL_1061308 | | | |
| see above | Alaska State Virology Laboratory | Alaska State Virology Laboratory | Stephanie DeRonde, Lisa Smith, Ph.D., Jack Chen, Ph.D. |
| EPI_ISL_1063904, EPI_ISL_1063907 | Tampa General Hospital Esoteric Lab | Tampa General Hospital Esoteric Research & Development Lab | Grant Vestal, Deanna Becker, Dominic Uy, Vicki Healer, Amorce Lima, Suzane Silbert |
| EPI_ISL_1063913, EPI_ISL_1063914, EPI_ISL_1063987, EPI_ISL_1063997, EPI_ISL_1064017 | Ohio Department of Health Laboratory | Ohio Department of Health Laboratory | Holmes, Jennifer; Eric Brandt, Keoni Omura, Glen McGillivary, Caitlin McDonnell, Jade Mowery, Stephanie Mccracken, Tyler Payne, Kirtana Ramadugu, Erica Leasure, Brent Lee, Kelsey Florek, Heather Blankenship, Allison Black, Quanta Brown, and Tammy Bannerman |
| EPI_ISL_1064172, EPI_ISL_1064173, EPI_ISL_1064174, EPI_ISL_1064175, EPI_ISL_1064177, EPI_ISL_1064178, EPI_ISL_1064179, EPI_ISL_1064180, EPI_ISL_1064184, EPI_ISL_1064185, EPI_ISL_1064186, EPI_ISL_1064187, EPI_ISL_1064188, EPI_ISL_1064189, EPI_ISL_1064190, EPI_ISL_1064191, EPI_ISL_1064193, EPI_ISL_1064194, EPI_ISL_1064195, EPI_ISL_1064196 | | | |
| see above | Arizona State Public Health Laboratory | Arizona State Public Health Laboratory | Trung Huynh, Jessica Escobar, Katherine Fullerton, Nobuko Fukushima, Stacy White, Linda Getsinger, Victor Waddell |
| EPI_ISL_1064200, EPI_ISL_1064201, EPI_ISL_1064202, EPI_ISL_1064203, EPI_ISL_1064204, EPI_ISL_1064205, EPI_ISL_1064206, EPI_ISL_1064207, EPI_ISL_1064208, EPI_ISL_1064210, EPI_ISL_1064211, EPI_ISL_1064212, EPI_ISL_1064213, EPI_ISL_1064214, EPI_ISL_1064215, EPI_ISL_1064216, EPI_ISL_1064217, EPI_ISL_1064218, EPI_ISL_1064219, EPI_ISL_1064220, EPI_ISL_1064221, EPI_ISL_1064222, EPI_ISL_1064223, EPI_ISL_1064224, EPI_ISL_1064225, EPI_ISL_1064226, EPI_ISL_1064227, EPI_ISL_1064228, EPI_ISL_1064230, EPI_ISL_1064231, EPI_ISL_1064232, EPI_ISL_1064233, EPI_ISL_1064234, EPI_ISL_1064235, EPI_ISL_1064236, EPI_ISL_1064237 | | | |
| see above | New Mexico Department of Health Scientific Laboratory | Center for Global Health, University of New Mexico Health Sciences Center | Daryl Domman, Kurt Schwalm, Twila Kunde, Joseph Hicks, Anastacia Griego, Michael Edwards, Darrell Dinwiddie |
| EPI_ISL_1064239, EPI_ISL_1064240, EPI_ISL_1064241, EPI_ISL_1064242, EPI_ISL_1064243, EPI_ISL_1064244, EPI_ISL_1064245, EPI_ISL_1064246, EPI_ISL_1064247, EPI_ISL_1064248, EPI_ISL_1064249, EPI_ISL_1064250, EPI_ISL_1064251, EPI_ISL_1064252, EPI_ISL_1064253, EPI_ISL_1064254, EPI_ISL_1064255, EPI_ISL_1064258, EPI_ISL_1064259, EPI_ISL_1064260, EPI_ISL_1064261, EPI_ISL_1064262, EPI_ISL_1064263, EPI_ISL_1064264, EPI_ISL_1064265, EPI_ISL_1064266, EPI_ISL_1064267, EPI_ISL_1064268, EPI_ISL_1064269, EPI_ISL_1064270, EPI_ISL_1064271, EPI_ISL_1064272, EPI_ISL_1064273, EPI_ISL_1064274, EPI_ISL_1064275, EPI_ISL_1064276, EPI_ISL_1064277, EPI_ISL_1064278, EPI_ISL_1064279, EPI_ISL_1064280, EPI_ISL_1064281, EPI_ISL_1064282, EPI_ISL_1064283, EPI_ISL_1064284, EPI_ISL_1064285, EPI_ISL_1064286, EPI_ISL_1064287, EPI_ISL_1064288, EPI_ISL_1064289, EPI_ISL_1064290, EPI_ISL_1064291, EPI_ISL_1064292, EPI_ISL_1064293, EPI_ISL_1064294, EPI_ISL_1064295, EPI_ISL_1064296, EPI_ISL_1064297, EPI_ISL_1064298, EPI_ISL_1064299, EPI_ISL_1064300, EPI_ISL_1064301, EPI_ISL_1064302, EPI_ISL_1064303, EPI_ISL_1064304, EPI_ISL_1064305, EPI_ISL_1064306, EPI_ISL_1064307, EPI_ISL_1064308, EPI_ISL_1064309, EPI_ISL_1064310, EPI_ISL_1064316, EPI_ISL_1064317, EPI_ISL_1064318, EPI_ISL_1064319, EPI_ISL_1064320, EPI_ISL_1064321, EPI_ISL_1064322, EPI_ISL_1064323 | | | |
| see above | University of Wisconsin-Madison AIDS Vaccine Research Laboratories | University of Wisconsin-Madison AIDS Vaccine Research Laboratories | Gage Moreno, Katarina Braun, et al. AIDS Vaccine Research Laboratories |
| EPI_ISL_1064324, EPI_ISL_1064325, EPI_ISL_1064326, EPI_ISL_1064327, EPI_ISL_1064328, EPI_ISL_1064329, EPI_ISL_1064330, EPI_ISL_1064331, EPI_ISL_1064332, EPI_ISL_1064333, EPI_ISL_1064334, EPI_ISL_1064335, EPI_ISL_1064336, EPI_ISL_1064337, EPI_ISL_1064338, EPI_ISL_1064339, EPI_ISL_1064340, EPI_ISL_1064341, EPI_ISL_1064345, EPI_ISL_1064346, EPI_ISL_1064347, EPI_ISL_1064348, EPI_ISL_1064349, EPI_ISL_1064350, EPI_ISL_1064351, EPI_ISL_1064352, EPI_ISL_1064353, EPI_ISL_1064354, EPI_ISL_1064355, EPI_ISL_1064356, EPI_ISL_1064357, EPI_ISL_1064358, EPI_ISL_1064359, EPI_ISL_1064360, EPI_ISL_1064361, EPI_ISL_1064362, EPI_ISL_1064363, EPI_ISL_1064364, EPI_ISL_1064365, EPI_ISL_1064366, EPI_ISL_1064367, EPI_ISL_1064368, EPI_ISL_1064369 | | | |
| see above | Maryland Public Health Laboratory | Maryland Public Health Laboratory | Maryland Department of Health Laboratories Administration |
| EPI_ISL_1064370, EPI_ISL_1064371, EPI_ISL_1064372, EPI_ISL_1064373, EPI_ISL_1064374, EPI_ISL_1064375, EPI_ISL_1064376, EPI_ISL_1064377 | Vault Health | Minnesota Department of Health, Public Health Laboratory | Alexandra Lorentz, Jacob Garfin, Matt Plumb, and Xiong Wang |
| EPI_ISL_1064378 | M Health Fairview | Minnesota Department of Health, Public Health Laboratory | Alexandra Lorentz, Jacob Garfin, Matt Plumb, and Xiong Wang |
| EPI_ISL_1064379, EPI_ISL_1064380, EPI_ISL_1064381, EPI_ISL_1064382, EPI_ISL_1064383, EPI_ISL_1064384 | Vault Health | Minnesota Department of Health, Public Health Laboratory | Alexandra Lorentz, Jacob Garfin, Matt Plumb, and Xiong Wang |
| EPI_ISL_1064385 | M Health Fairview | Minnesota Department of Health, Public Health Laboratory | Alexandra Lorentz, Jacob Garfin, Matt Plumb, and Xiong Wang |
| EPI_ISL_1064386, EPI_ISL_1064387, EPI_ISL_1064388, EPI_ISL_1064389, EPI_ISL_1064390, EPI_ISL_1064391, EPI_ISL_1064392, EPI_ISL_1064393, EPI_ISL_1064394, EPI_ISL_1064395, EPI_ISL_1064396, EPI_ISL_1064397, EPI_ISL_1064398, EPI_ISL_1064399, EPI_ISL_1064400, EPI_ISL_1064401, EPI_ISL_1064402 | | | |
| see above | Vault Health | Minnesota Department of Health, Public Health Laboratory | Alexandra Lorentz, Jacob Garfin, Matt Plumb, and Xiong Wang |
| EPI_ISL_1064403, EPI_ISL_1064404 | M Health Fairview | Minnesota Department of Health, Public Health Laboratory | Alexandra Lorentz, Jacob Garfin, Matt Plumb, and Xiong Wang |
| EPI_ISL_1064491 | Vault Health | Minnesota Department of Health, Public Health Laboratory | Alexandra Lorentz, Jacob Garfin, Matt Plumb, and Xiong Wang |
| EPI_ISL_1064492, EPI_ISL_1064493, EPI_ISL_1064494, EPI_ISL_1064495 | M Health Fairview | Minnesota Department of Health, Public Health Laboratory | Alexandra Lorentz, Jacob Garfin, Matt Plumb, and Xiong Wang |
| EPI_ISL_1064496, EPI_ISL_1064497, EPI_ISL_1064498 | Vault Health | Minnesota Department of Health, Public Health Laboratory | Alexandra Lorentz, Jacob Garfin, Matt Plumb, and Xiong Wang |
| EPI_ISL_1064499, EPI_ISL_1064500, EPI_ISL_1064501 | M Health Fairview | Minnesota Department of Health, Public Health Laboratory | Alexandra Lorentz, Jacob Garfin, Matt Plumb, and Xiong Wang |
| EPI_ISL_1064502 | Vault Health | Minnesota Department of Health, Public Health Laboratory | Alexandra Lorentz, Jacob Garfin, Matt Plumb, and Xiong Wang |
| EPI_ISL_1064503, EPI_ISL_1064504, EPI_ISL_1064505, EPI_ISL_1064506, EPI_ISL_1064507, EPI_ISL_1064508, EPI_ISL_1064509, EPI_ISL_1064510, EPI_ISL_1064511, EPI_ISL_1064512, EPI_ISL_1064513 | | | |
| see above | M Health Fairview | Minnesota Department of Health, Public Health Laboratory | Alexandra Lorentz, Jacob Garfin, Matt Plumb, and Xiong Wang |

| | | | |
|---|---|---|---|
| EPI_ISL_1064514 | Vault Health | Minnesota Department of Health, Public Health Laboratory | Alexandra Lorentz, Jacob Garfin, Matt Plumb, and Xiong Wang |
| EPI_ISL_1064515, EPI_ISL_1064516 | M Health Fairview | Minnesota Department of Health, Public Health Laboratory | Alexandra Lorentz, Jacob Garfin, Matt Plumb, and Xiong Wang |
| EPI_ISL_1064517, EPI_ISL_1064518, EPI_ISL_1064519, EPI_ISL_1064520 | Vault Health | Minnesota Department of Health, Public Health Laboratory | Alexandra Lorentz, Jacob Garfin, Matt Plumb, and Xiong Wang |
| EPI_ISL_1064521, EPI_ISL_1064522, EPI_ISL_1064523, EPI_ISL_1064524, EPI_ISL_1064525, EPI_ISL_1064526, EPI_ISL_1064527, EPI_ISL_1064528, EPI_ISL_1064529, EPI_ISL_1064530, EPI_ISL_1064531, EPI_ISL_1064532, EPI_ISL_1064533, EPI_ISL_1064534, EPI_ISL_1064535, EPI_ISL_1064536, EPI_ISL_1064537, EPI_ISL_1064538, EPI_ISL_1064539, EPI_ISL_1064540, EPI_ISL_1064541, EPI_ISL_1064542, EPI_ISL_1064543, EPI_ISL_1064544, EPI_ISL_1064545, EPI_ISL_1064546, EPI_ISL_1064547, EPI_ISL_1064548, EPI_ISL_1064549 | | | |
| see above | M Health Fairview | Minnesota Department of Health, Public Health Laboratory | Alexandra Lorentz, Jacob Garfin, Matt Plumb, and Xiong Wang |
| EPI_ISL_1064711, EPI_ISL_1064712, EPI_ISL_1064713, EPI_ISL_1064714, EPI_ISL_1064715, EPI_ISL_1064716, EPI_ISL_1064717, EPI_ISL_1064718, EPI_ISL_1064719, EPI_ISL_1064720, EPI_ISL_1064721, EPI_ISL_1064722, EPI_ISL_1064723, EPI_ISL_1064724, EPI_ISL_1064725, EPI_ISL_1064726, EPI_ISL_1064727, EPI_ISL_1064728, EPI_ISL_1064729, EPI_ISL_1064730, EPI_ISL_1064731, EPI_ISL_1064732, EPI_ISL_1064733, EPI_ISL_1064734, EPI_ISL_1064735 | | | |
| see above | Kansas Health and Environmental Lab | Kansas Health and Environmental Lab | Mike Grose, Paige Drury, Carissa Robertson, Ben Olsen, and Phil Adam |
| EPI_ISL_1064751, EPI_ISL_1064752, EPI_ISL_1064753, EPI_ISL_1064754 | OCME Office Of Chief Medical Examiner | New York City Public Health Laboratory | Jade Wang, et al. |
| EPI_ISL_1064755 | DOHMH Jamaica | New York City Public Health Laboratory | Jade Wang, et al. |
| EPI_ISL_1064756 | DOHMH PHL | New York City Public Health Laboratory | Jade Wang, et al. |
| EPI_ISL_1064757 | DOHMH Corona | New York City Public Health Laboratory | Jade Wang, et al. |
| EPI_ISL_1064758 | Department of Homeless Services | New York City Public Health Laboratory | Jade Wang, et al. |
| EPI_ISL_1064759 | DOHMH Jamaica | New York City Public Health Laboratory | Jade Wang, et al. |
| EPI_ISL_1064760, EPI_ISL_1064761 | DOHMH Corona | New York City Public Health Laboratory | Jade Wang, et al. |
| EPI_ISL_1064762 | DOHMH Fort Greene | New York City Public Health Laboratory | Jade Wang, et al. |
| EPI_ISL_1064763, EPI_ISL_1064764, EPI_ISL_1064765 | OCME Office Of Chief Medical Examiner | New York City Public Health Laboratory | Jade Wang, et al. |
| EPI_ISL_1064766 | DOHMH Chelsea | New York City Public Health Laboratory | Jade Wang, et al. |
| EPI_ISL_1064767 | DOHMH Corona | New York City Public Health Laboratory | Jade Wang, et al. |
| EPI_ISL_1064768 | Department of Homeless Services | New York City Public Health Laboratory | Jade Wang, et al. |
| EPI_ISL_1064769 | DOHMH PHL | New York City Public Health Laboratory | Jade Wang, et al. |
| EPI_ISL_1064770 | OCME Office Of Chief Medical Examiner | New York City Public Health Laboratory | Jade Wang, et al. |
| EPI_ISL_1064771 | DOHMH Fort Greene | New York City Public Health Laboratory | Jade Wang, et al. |
| EPI_ISL_1064772, EPI_ISL_1064773, EPI_ISL_1064774 | OCME Office Of Chief Medical Examiner | New York City Public Health Laboratory | Jade Wang, et al. |
| EPI_ISL_1064775 | DOHMH Central Harlem | New York City Public Health Laboratory | Jade Wang, et al. |
| EPI_ISL_1064776 | OCME Office Of Chief Medical Examiner | New York City Public Health Laboratory | Jade Wang, et al. |
| EPI_ISL_1064777 | DOHMH Crown Heights | New York City Public Health Laboratory | Jade Wang, et al. |
| EPI_ISL_1064778 | OCME Office Of Chief Medical Examiner | New York City Public Health Laboratory | Jade Wang, et al. |
| EPI_ISL_1064779 | DOHMH Chelsea | New York City Public Health Laboratory | Jade Wang, et al. |
| EPI_ISL_1064780 | DOHMH Fort Greene | New York City Public Health Laboratory | Jade Wang, et al. |
| EPI_ISL_1064781 | DOHMH Jamaica | New York City Public Health Laboratory | Jade Wang, et al. |
| EPI_ISL_1064782 | DOHMH Chelsea | New York City Public Health Laboratory | Jade Wang, et al. |
| EPI_ISL_1064783 | OCME Office Of Chief Medical Examiner | New York City Public Health Laboratory | Jade Wang, et al. |
| EPI_ISL_1064784 | DOHMH Corona | New York City Public Health Laboratory | Jade Wang, et al. |
| EPI_ISL_1064785 | DOHMH Fort Greene | New York City Public Health Laboratory | Jade Wang, et al. |
| EPI_ISL_1064786 | DOHMH Corona | New York City Public Health Laboratory | Jade Wang, et al. |
| EPI_ISL_1064787 | DOHMH Morrisania | New York City Public Health Laboratory | Jade Wang, et al. |
| EPI_ISL_1064788 | DOHMH Jamaica | New York City Public Health Laboratory | Jade Wang, et al. |
| EPI_ISL_1064789 | DOHMH Corona | New York City Public Health Laboratory | Jade Wang, et al. |
| EPI_ISL_1064790 | DOHMH Fort Greene | New York City Public Health Laboratory | Jade Wang, et al. |
| EPI_ISL_1064791 | DOHMH Crown Heights | New York City Public Health Laboratory | Jade Wang, et al. |
| EPI_ISL_1064792, EPI_ISL_1064793 | OCME Office Of Chief Medical Examiner | New York City Public Health Laboratory | Jade Wang, et al. |
| EPI_ISL_1064794 | DOHMH Fort Greene | New York City Public Health Laboratory | Jade Wang, et al. |
| EPI_ISL_1064795, EPI_ISL_1064796 | OCME Office Of Chief Medical Examiner | New York City Public Health Laboratory | Jade Wang, et al. |
| EPI_ISL_1064797 | DOHMH Chelsea | New York City Public Health Laboratory | Jade Wang, et al. |
| EPI_ISL_1064798 | DOHMH Crown Heights | New York City Public Health Laboratory | Jade Wang, et al. |
| EPI_ISL_1064799 | DOHMH PHL | New York City Public Health Laboratory | Jade Wang, et al. |
| EPI_ISL_1064800 | NYC HH Elmhurst Hospital Medical Center | New York City Public Health Laboratory | Jade Wang, et al. |
| EPI_ISL_1064801, EPI_ISL_1064802 | DOHMH Morrisania | New York City Public Health Laboratory | Jade Wang, et al. |
| EPI_ISL_1064803 | DOHMH Central Harlem | New York City Public Health Laboratory | Jade Wang, et al. |
| EPI_ISL_1064804 | OCME Office Of Chief Medical Examiner | New York City Public Health Laboratory | Jade Wang, et al. |
| EPI_ISL_1064805, EPI_ISL_1064806 | DOHMH PHL | New York City Public Health Laboratory | Jade Wang, et al. |
| EPI_ISL_1064807 | DOHMH Jamaica | New York City Public Health Laboratory | Jade Wang, et al. |
| EPI_ISL_1064808 | DOHMH Corona | New York City Public Health Laboratory | Jade Wang, et al. |
| EPI_ISL_1064809 | DOHMH Central Harlem | New York City Public Health Laboratory | Jade Wang, et al. |
| EPI_ISL_1064810 | Department of Homeless Services | New York City Public Health Laboratory | Jade Wang, et al. |
| EPI_ISL_1064811 | DOHMH Corona | New York City Public Health Laboratory | Jade Wang, et al. |
| EPI_ISL_1064812 | DOHMH Morrisania | New York City Public Health Laboratory | Jade Wang, et al. |
| EPI_ISL_1064813, EPI_ISL_1064814, EPI_ISL_1064815, EPI_ISL_1064816, EPI_ISL_1064817, EPI_ISL_1064818 | OCME Office Of Chief Medical Examiner | New York City Public Health Laboratory | Jade Wang, et al. |

| | | | |
|---|---|---|---|
| EPI_ISL_1064819 | DOHMH Jamaica | New York City Public Health Laboratory | Jade Wang, et al. |
| EPI_ISL_1064820 | DOHMH Central Harlem | New York City Public Health Laboratory | Jade Wang, et al. |
| EPI_ISL_1064821, EPI_ISL_1064822 | DOHMH PHL | New York City Public Health Laboratory | Jade Wang, et al. |
| EPI_ISL_1064823 | DOHMH Riverside | New York City Public Health Laboratory | Jade Wang, et al. |
| EPI_ISL_1064824 | DOHMH Crown Heights | New York City Public Health Laboratory | Jade Wang, et al. |
| EPI_ISL_1064825 | DOHMH Central Harlem | New York City Public Health Laboratory | Jade Wang, et al. |
| EPI_ISL_1064826 | DOHMH PHL | New York City Public Health Laboratory | Jade Wang, et al. |
| EPI_ISL_1064827, EPI_ISL_1064828 | Department of Homeless Services | New York City Public Health Laboratory | Jade Wang, et al. |
| EPI_ISL_1064829 | OCME Office Of Chief Medical Examiner | New York City Public Health Laboratory | Jade Wang, et al. |
| EPI_ISL_1064830 | DOHMH Jamaica | New York City Public Health Laboratory | Jade Wang, et al. |
| EPI_ISL_1064831 | DOHMH Central Harlem | New York City Public Health Laboratory | Jade Wang, et al. |
| EPI_ISL_1064832, EPI_ISL_1064833 | OCME Office Of Chief Medical Examiner | New York City Public Health Laboratory | Jade Wang, et al. |
| EPI_ISL_1064834 | DOHMH Morrisania | New York City Public Health Laboratory | Jade Wang, et al. |
| EPI_ISL_1064835 | DOHMH Crown Heights | New York City Public Health Laboratory | Jade Wang, et al. |
| EPI_ISL_1064836 | DOHMH Fort Greene | New York City Public Health Laboratory | Jade Wang, et al. |
| EPI_ISL_1064837, EPI_ISL_1064838, EPI_ISL_1064839, EPI_ISL_1064840 | DOHMH Jamaica | New York City Public Health Laboratory | Jade Wang, et al. |
| EPI_ISL_1064841, EPI_ISL_1064842 | DOHMH Morrisania | New York City Public Health Laboratory | Jade Wang, et al. |
| EPI_ISL_1064843, EPI_ISL_1064844, EPI_ISL_1064845, EPI_ISL_1064846 | OCME Office Of Chief Medical Examiner | New York City Public Health Laboratory | Jade Wang, et al. |
| EPI_ISL_1064847 | DOHMH Corona | New York City Public Health Laboratory | Jade Wang, et al. |
| EPI_ISL_1064848 | DOHMH Jamaica | New York City Public Health Laboratory | Jade Wang, et al. |
| EPI_ISL_1064849, EPI_ISL_1064850 | DOHMH Morrisania | New York City Public Health Laboratory | Jade Wang, et al. |
| EPI_ISL_1064851, EPI_ISL_1064852 | Department of Homeless Services | New York City Public Health Laboratory | Jade Wang, et al. |
| EPI_ISL_1064853, EPI_ISL_1064854, EPI_ISL_1064855, EPI_ISL_1064856, EPI_ISL_1064857 | OCME Office Of Chief Medical Examiner | New York City Public Health Laboratory | Jade Wang, et al. |
| EPI_ISL_1064858 | DOHMH Jamaica | New York City Public Health Laboratory | Jade Wang, et al. |
| EPI_ISL_1064859, EPI_ISL_1064860 | DOHMH PHL | New York City Public Health Laboratory | Jade Wang, et al. |
| EPI_ISL_1064861, EPI_ISL_1064862 | DOHMH Morrisania | New York City Public Health Laboratory | Jade Wang, et al. |
| EPI_ISL_1064863, EPI_ISL_1064864 | OCME Office Of Chief Medical Examiner | New York City Public Health Laboratory | Jade Wang, et al. |
| EPI_ISL_1064865, EPI_ISL_1064866, EPI_ISL_1064867, EPI_ISL_1064868, EPI_ISL_1064869 | DOHMH Jamaica | New York City Public Health Laboratory | Jade Wang, et al. |
| EPI_ISL_1064870, EPI_ISL_1064871 | DOHMH Corona | New York City Public Health Laboratory | Jade Wang, et al. |
| EPI_ISL_1064872 | DOHMH Chelsea | New York City Public Health Laboratory | Jade Wang, et al. |
| EPI_ISL_1064873, EPI_ISL_1064874 | DOHMH Corona | New York City Public Health Laboratory | Jade Wang, et al. |
| EPI_ISL_1064875 | DOHMH Morrisania | New York City Public Health Laboratory | Jade Wang, et al. |
| EPI_ISL_1064876 | DOHMH Central Harlem | New York City Public Health Laboratory | Jade Wang, et al. |
| EPI_ISL_1064877, EPI_ISL_1064878, EPI_ISL_1064879, EPI_ISL_1064880, EPI_ISL_1064881, EPI_ISL_1064882 | OCME Office Of Chief Medical Examiner | New York City Public Health Laboratory | Jade Wang, et al. |
| EPI_ISL_1064883 | DOHMH Crown Heights | New York City Public Health Laboratory | Jade Wang, et al. |
| EPI_ISL_1064884 | DOHMH Jamaica | New York City Public Health Laboratory | Jade Wang, et al. |
| EPI_ISL_1064885 | DOHMH Fort Greene | New York City Public Health Laboratory | Jade Wang, et al. |
| EPI_ISL_1064886 | DOHMH Jamaica | New York City Public Health Laboratory | Jade Wang, et al. |
| EPI_ISL_1064887 | DOHMH PHL | New York City Public Health Laboratory | Jade Wang, et al. |
| EPI_ISL_1064888 | DOHMH Corona | New York City Public Health Laboratory | Jade Wang, et al. |
| EPI_ISL_1064889 | Department of Homeless Services | New York City Public Health Laboratory | Jade Wang, et al. |
| EPI_ISL_1064890 | DOHMH Crown Heights | New York City Public Health Laboratory | Jade Wang, et al. |
| EPI_ISL_1064891 | DOHMH Fort Greene | New York City Public Health Laboratory | Jade Wang, et al. |
| EPI_ISL_1064892 | DOHMH Jamaica | New York City Public Health Laboratory | Jade Wang, et al. |
| EPI_ISL_1064893 | DOHMH Corona | New York City Public Health Laboratory | Jade Wang, et al. |
| EPI_ISL_1064894 | DOHMH PHL | New York City Public Health Laboratory | Jade Wang, et al. |
| EPI_ISL_1064895, EPI_ISL_1064896 | DOHMH Chelsea | New York City Public Health Laboratory | Jade Wang, et al. |
| EPI_ISL_1064897 | DOHMH Corona | New York City Public Health Laboratory | Jade Wang, et al. |
| EPI_ISL_1064898 | Department of Homeless Services | New York City Public Health Laboratory | Jade Wang, et al. |
| EPI_ISL_1064899, EPI_ISL_1064900, EPI_ISL_1064901, EPI_ISL_1064902, EPI_ISL_1064903 | OCME Office Of Chief Medical Examiner | New York City Public Health Laboratory | Jade Wang, et al. |
| EPI_ISL_1064904 | DOHMH Crown Heights | New York City Public Health Laboratory | Jade Wang, et al. |
| EPI_ISL_1064905 | DOHMH Corona | New York City Public Health Laboratory | Jade Wang, et al. |

| | | | |
|---|---|---|---|
| EPI_ISL_1064906, EPI_ISL_1064907 | DOHMH Jamaica | New York City Public Health Laboratory | Jade Wang, et al. |
| EPI_ISL_1064908 | DOHMH Riverside | New York City Public Health Laboratory | Jade Wang, et al. |
| EPI_ISL_1064909 | DOHMH Central Harlem | New York City Public Health Laboratory | Jade Wang, et al. |
| EPI_ISL_1064910, EPI_ISL_1064911, EPI_ISL_1064912, EPI_ISL_1064913, EPI_ISL_1064914 | OCME Office Of Chief Medical Examiner | New York City Public Health Laboratory | Jade Wang, et al. |
| EPI_ISL_1064915, EPI_ISL_1064916, EPI_ISL_1064917 | Univeristy of New Mexico Hospital | Center for Global Health, University of New Mexico Health Sciences Center | Daryl Domman, Kurt Schwalm, Justin Bacca, Jon Femling, Darrell Dinwiddie |
| EPI_ISL_1064918, EPI_ISL_1064919, EPI_ISL_1064920, EPI_ISL_1064921 | New Mexico Department of Health Scientific Laboratory | Center for Global Health, University of New Mexico Health Sciences Center | Daryl Domman, Kurt Schwalm, Twila Kunde, Joseph Hicks, Anastacia Griego, Michael Edwards, Darrell Dinwiddie |
| EPI_ISL_1064922 | Univeristy of New Mexico Hospital | Center for Global Health, University of New Mexico Health Sciences Center | Daryl Domman, Kurt Schwalm, Justin Bacca, Jon Femling, Darrell Dinwiddie |
| EPI_ISL_1064923, EPI_ISL_1064925 | New Mexico Department of Health Scientific Laboratory | Center for Global Health, University of New Mexico Health Sciences Center | Daryl Domman, Kurt Schwalm, Twila Kunde, Joseph Hicks, Anastacia Griego, Michael Edwards, Darrell Dinwiddie |
| EPI_ISL_1064926, EPI_ISL_1064927 | Univeristy of New Mexico Hospital | Center for Global Health, University of New Mexico Health Sciences Center | Daryl Domman, Kurt Schwalm, Justin Bacca, Jon Femling, Darrell Dinwiddie |
| EPI_ISL_1064928, EPI_ISL_1064929, EPI_ISL_1064930, EPI_ISL_1064931, EPI_ISL_1064932, EPI_ISL_1064933, EPI_ISL_1064934, EPI_ISL_1064935, EPI_ISL_1064936, EPI_ISL_1064937, EPI_ISL_1064938, EPI_ISL_1064939, EPI_ISL_1064940, EPI_ISL_1064941, EPI_ISL_1064942, EPI_ISL_1064943, EPI_ISL_1064944, EPI_ISL_1064945, EPI_ISL_1064946, EPI_ISL_1064947, EPI_ISL_1064948, EPI_ISL_1064949, EPI_ISL_1064950, EPI_ISL_1064951, EPI_ISL_1064952, EPI_ISL_1064953, EPI_ISL_1064954, EPI_ISL_1064955, EPI_ISL_1064956 | | | |
| see above | New Mexico Department of Health Scientific Laboratory | Center for Global Health, University of New Mexico Health Sciences Center | Daryl Domman, Kurt Schwalm, Twila Kunde, Joseph Hicks, Anastacia Griego, Michael Edwards, Darrell Dinwiddie |
| EPI_ISL_1064957, EPI_ISL_1064958, EPI_ISL_1064959, EPI_ISL_1064960, EPI_ISL_1064961, EPI_ISL_1064962, EPI_ISL_1064963, EPI_ISL_1064964, EPI_ISL_1064965, EPI_ISL_1064966, EPI_ISL_1064967, EPI_ISL_1064968, EPI_ISL_1064969, EPI_ISL_1064970, EPI_ISL_1064971, EPI_ISL_1064972, EPI_ISL_1064973, EPI_ISL_1064974, EPI_ISL_1064975, EPI_ISL_1064976, EPI_ISL_1064977, EPI_ISL_1064978, EPI_ISL_1064979, EPI_ISL_1064980, EPI_ISL_1064981, EPI_ISL_1064982, EPI_ISL_1064983 | | | |
| see above | Univeristy of New Mexico Hospital | Center for Global Health, University of New Mexico Health Sciences Center | Daryl Domman, Kurt Schwalm, Justin Bacca, Jon Femling, Darrell Dinwiddie |
| EPI_ISL_1066758, EPI_ISL_1066760, EPI_ISL_1066767, EPI_ISL_1066769, EPI_ISL_1066771, EPI_ISL_1066773, EPI_ISL_1066775, EPI_ISL_1066777, EPI_ISL_1066778, EPI_ISL_1066780, EPI_ISL_1066783, EPI_ISL_1066784, EPI_ISL_1066786, EPI_ISL_1066788, EPI_ISL_1066790, EPI_ISL_1066792, EPI_ISL_1066794, EPI_ISL_1066796 | | | |
| see above | Arizona State Public Health Laboratory | Arizona State Public Health Laboratory | Trung Huynh, Jessica Escobar, Katherine Fullerton, Nobuko Fukushima, Stacy White, Linda Getsinger, Victor Waddell |
| EPI_ISL_1067638 | Murphy Medical Associates | Grubaugh Lab - Yale School of Public Health | Mary Petrone, Joseph Fauver, Caleb Neal, Steven Murphy, Chantal Vogels, Mallery Breban, Annie Watkins, Tara Alpert, Nathan Grubaugh |
| EPI_ISL_1067639 | Connecticut Veterans' Affairs Hospital | Grubaugh Lab - Yale School of Public Health | Mary Petrone, Joseph Fauver, Tara Alpert, Chantal Vogels, Isabel Ott, Shaili Gupta, Danielle Plank, Nathan Grubaugh |
| EPI_ISL_1067640, EPI_ISL_1067641 | Murphy Medical Associates | Grubaugh Lab - Yale School of Public Health | Mary Petrone, Joseph Fauver, Caleb Neal, Steven Murphy, Chantal Vogels, Mallery Breban, Annie Watkins, Tara Alpert, Nathan Grubaugh |
| EPI_ISL_1067642, EPI_ISL_1067643, EPI_ISL_1067644 | Connecticut Veterans' Affairs Hospital | Grubaugh Lab - Yale School of Public Health | Mary Petrone, Joseph Fauver, Tara Alpert, Chantal Vogels, Isabel Ott, Shaili Gupta, Danielle Plank, Nathan Grubaugh |
| EPI_ISL_1067645 | Murphy Medical Associates | Grubaugh Lab - Yale School of Public Health | Mary Petrone, Joseph Fauver, Caleb Neal, Steven Murphy, Chantal Vogels, Mallery Breban, Annie Watkins, Tara Alpert, Nathan Grubaugh |
| EPI_ISL_1067646, EPI_ISL_1067647 | Connecticut Veterans' Affairs Hospital | Grubaugh Lab - Yale School of Public Health | Mary Petrone, Joseph Fauver, Tara Alpert, Chantal Vogels, Isabel Ott, Shaili Gupta, Danielle Plank, Nathan Grubaugh |
| EPI_ISL_1067648 | Murphy Medical Associates | Grubaugh Lab - Yale School of Public Health | Mary Petrone, Joseph Fauver, Caleb Neal, Steven Murphy, Chantal Vogels, Mallery Breban, Annie Watkins, Tara Alpert, Nathan Grubaugh |
| EPI_ISL_1067649 | Connecticut Veterans' Affairs Hospital | Grubaugh Lab - Yale School of Public Health | Mary Petrone, Joseph Fauver, Tara Alpert, Chantal Vogels, Isabel Ott, Shaili Gupta, Danielle Plank, Nathan Grubaugh |
| EPI_ISL_1067650, EPI_ISL_1067651, EPI_ISL_1067652, EPI_ISL_1067653, EPI_ISL_1067654 | Murphy Medical Associates | Grubaugh Lab - Yale School of Public Health | Mary Petrone, Joseph Fauver, Caleb Neal, Steven Murphy, Chantal Vogels, Mallery Breban, Annie Watkins, Tara Alpert, Nathan Grubaugh |
| EPI_ISL_1067655 | Connecticut Veterans' Affairs Hospital | Grubaugh Lab - Yale School of Public Health | Mary Petrone, Joseph Fauver, Tara Alpert, Chantal Vogels, Isabel Ott, Shaili Gupta, Danielle Plank, Nathan Grubaugh |
| EPI_ISL_1067656, EPI_ISL_1067657, EPI_ISL_1067658, EPI_ISL_1067659, EPI_ISL_1067692, EPI_ISL_1067693, EPI_ISL_1067694, EPI_ISL_1067695 | Murphy Medical Associates | Grubaugh Lab - Yale School of Public Health | Mary Petrone, Joseph Fauver, Caleb Neal, Steven Murphy, Chantal Vogels, Mallery Breban, Annie Watkins, Tara Alpert, Nathan Grubaugh |
| EPI_ISL_1067696, EPI_ISL_1067697, EPI_ISL_1067698, EPI_ISL_1067701, EPI_ISL_1067702, EPI_ISL_1067703, EPI_ISL_1067704, EPI_ISL_1067705, EPI_ISL_1067706, EPI_ISL_1067707, EPI_ISL_1067708, EPI_ISL_1067709, EPI_ISL_1067710, EPI_ISL_1067711, EPI_ISL_1067712, EPI_ISL_1067713, EPI_ISL_1067714, EPI_ISL_1067715, EPI_ISL_1067716 | | | |
| see above | Arizona State Public Health Laboratory | Arizona State Public Health Laboratory | Trung Huynh, Jessica Escobar, Katherine Fullerton, Nobuko Fukushima, Stacy White, Linda Getsinger, Victor Waddell |
| EPI_ISL_1067717, EPI_ISL_1067718, EPI_ISL_1067719, EPI_ISL_1067720, EPI_ISL_1067721, EPI_ISL_1067722, EPI_ISL_1067723, EPI_ISL_1067724, EPI_ISL_1067725 | Kansas Health and Environmental Lab | Kansas Health and Environmental Lab | Mike Grose, Paige Drury, Carissa Robertson, Ben Olsen, and Phil Adam |
| EPI_ISL_1067739, EPI_ISL_1067740, EPI_ISL_1067741, EPI_ISL_1067742, EPI_ISL_1067743, EPI_ISL_1067744, EPI_ISL_1067745, EPI_ISL_1067746, EPI_ISL_1067747, EPI_ISL_1067748, EPI_ISL_1067749, EPI_ISL_1067750, EPI_ISL_1067751, EPI_ISL_1067752, EPI_ISL_1067753, EPI_ISL_1067754, EPI_ISL_1067755, EPI_ISL_1067756, EPI_ISL_1067757, EPI_ISL_1067758, EPI_ISL_1067759, EPI_ISL_1067760, EPI_ISL_1067761, EPI_ISL_1067762, EPI_ISL_1067763, EPI_ISL_1067764, EPI_ISL_1067765, EPI_ISL_1067766, EPI_ISL_1067767, EPI_ISL_1067768, EPI_ISL_1067769, EPI_ISL_1067770, EPI_ISL_1067771, EPI_ISL_1067772, EPI_ISL_1067773, EPI_ISL_1067774, EPI_ISL_1067775, EPI_ISL_1067776, EPI_ISL_1067777, EPI_ISL_1067778, EPI_ISL_1067779, EPI_ISL_1067780, EPI_ISL_1067781, EPI_ISL_1067782, EPI_ISL_1067783, EPI_ISL_1067784, EPI_ISL_1067785, EPI_ISL_1067786, EPI_ISL_1067808, EPI_ISL_1067809, EPI_ISL_1067810, EPI_ISL_1067811, EPI_ISL_1067812, EPI_ISL_1067813, EPI_ISL_1067814, EPI_ISL_1067815, EPI_ISL_1067816, EPI_ISL_1067817 | | | |
| see above | Broad Institute Clinical Research Sequencing Platform | Infectious Disease Program, Broad Institute of Harvard and MIT | Lemieux,J.E., Siddle,K.J., Adams,G., Gladden-Young,A., Lagerborg,K., Rudy,M., DeRuff,K., Carter,A., Normandin,E., Bauer,M., Reilly,S., Tomkins-Tinch,C., Loreth,C., Chaluvadi,S., Birren,B.W., Gallagher,G., Smole,S., Park,D.J., MacInnis,B.L., and Sabeti,P.C. |
| EPI_ISL_1067818, EPI_ISL_1067819, EPI_ISL_1067820, EPI_ISL_1067821, EPI_ISL_1067822, EPI_ISL_1067823, EPI_ISL_1067824, EPI_ISL_1067825, EPI_ISL_1067826, EPI_ISL_1067827, EPI_ISL_1067828, EPI_ISL_1067829, EPI_ISL_1067830, EPI_ISL_1067831, EPI_ISL_1067832, EPI_ISL_1067833, EPI_ISL_1067834, EPI_ISL_1067835, EPI_ISL_1067836, EPI_ISL_1067837, EPI_ISL_1067838, EPI_ISL_1067839, EPI_ISL_1067840, EPI_ISL_1067841, EPI_ISL_1067842, EPI_ISL_1067843, EPI_ISL_1067844, EPI_ISL_1067845, EPI_ISL_1067846, EPI_ISL_1067847, EPI_ISL_1067848, EPI_ISL_1067849, EPI_ISL_1067850, EPI_ISL_1067851, EPI_ISL_1067852, EPI_ISL_1067853, EPI_ISL_1067854, EPI_ISL_1067855, EPI_ISL_1067856, EPI_ISL_1067857, EPI_ISL_1067858, EPI_ISL_1067859, EPI_ISL_1067860, EPI_ISL_1067861, EPI_ISL_1067862, EPI_ISL_1067863, EPI_ISL_1067864, EPI_ISL_1067865, EPI_ISL_1067866, EPI_ISL_1067867, EPI_ISL_1067868, EPI_ISL_1067869, EPI_ISL_1067870, EPI_ISL_1067871, EPI_ISL_1067872, EPI_ISL_1067873, EPI_ISL_1067874, EPI_ISL_1067875, EPI_ISL_1067876, EPI_ISL_1067877, EPI_ISL_1067878, EPI_ISL_1067879, EPI_ISL_1067880, EPI_ISL_1067881, EPI_ISL_1067882, EPI_ISL_1067883, EPI_ISL_1067884, EPI_ISL_1067885, EPI_ISL_1067886, EPI_ISL_1067887, EPI_ISL_1067888, EPI_ISL_1067889, EPI_ISL_1067890, EPI_ISL_1067891, EPI_ISL_1067892, EPI_ISL_1067893, EPI_ISL_1067894, EPI_ISL_1067895, EPI_ISL_1067896, EPI_ISL_1067897, EPI_ISL_1067898, EPI_ISL_1067899, EPI_ISL_1067900, EPI_ISL_1067901, EPI_ISL_1067902, EPI_ISL_1067903, EPI_ISL_1067904, EPI_ISL_1067905, EPI_ISL_1067906, EPI_ISL_1067907, EPI_ISL_1067908, EPI_ISL_1067909, EPI_ISL_1067910, EPI_ISL_1067911, EPI_ISL_1067912, EPI_ISL_1067913, EPI_ISL_1067914, EPI_ISL_1067915, EPI_ISL_1067916, EPI_ISL_1067917, EPI_ISL_1067918, EPI_ISL_1067919, EPI_ISL_1067920, EPI_ISL_1067921, EPI_ISL_1067922, EPI_ISL_1067923, EPI_ISL_1067924, EPI_ISL_1067925, EPI_ISL_1067926, EPI_ISL_1067927, EPI_ISL_1067928, EPI_ISL_1067929, EPI_ISL_1067930, EPI_ISL_1067931, EPI_ISL_1067932, EPI_ISL_1067933, EPI_ISL_1067934, EPI_ISL_1067935, EPI_ISL_1067936, EPI_ISL_1067937, EPI_ISL_1067938, EPI_ISL_1067939, EPI_ISL_1067940, EPI_ISL_1067941, EPI_ISL_1067942, EPI_ISL_1067943, EPI_ISL_1067944, EPI_ISL_1067945, EPI_ISL_1067946, EPI_ISL_1067947, EPI_ISL_1067948, EPI_ISL_1067949, EPI_ISL_1067950, EPI_ISL_1067951, EPI_ISL_1067952, EPI_ISL_1067953, EPI_ISL_1067954, EPI_ISL_1067955, EPI_ISL_1067956, EPI_ISL_1067957, EPI_ISL_1067958, EPI_ISL_1067959, EPI_ISL_1067960, EPI_ISL_1067961, EPI_ISL_1067962, EPI_ISL_1067963, EPI_ISL_1067964, EPI_ISL_1067965, EPI_ISL_1067966, EPI_ISL_1067967, EPI_ISL_1067968, EPI_ISL_1067969, EPI_ISL_1067970, EPI_ISL_1067971, EPI_ISL_1067972, EPI_ISL_1067973, EPI_ISL_1067974, EPI_ISL_1067975, EPI_ISL_1067976, EPI_ISL_1067977, EPI_ISL_1067978, EPI_ISL_1067979, EPI_ISL_1067980, EPI_ISL_1067981, EPI_ISL_1067982, EPI_ISL_1067983, EPI_ISL_1067984 | | | |
| see above | Wisconsin State Laboratory of Hygiene Communicable Disease Division | Wisconsin State Laboratory of Hygiene Communicable Disease Division | Kelsey R. Florek, Abigail C. Shockey |
| EPI_ISL_1067985, EPI_ISL_1067986, EPI_ISL_1067987, EPI_ISL_1067988, EPI_ISL_1067989, EPI_ISL_1067990, EPI_ISL_1067991, EPI_ISL_1067992, EPI_ISL_1067993, EPI_ISL_1067994, EPI_ISL_1067995, EPI_ISL_1067996, EPI_ISL_1067997, EPI_ISL_1067998, EPI_ISL_1067999, EPI_ISL_1068000, EPI_ISL_1068001, EPI_ISL_1068002, EPI_ISL_1068003, EPI_ISL_1068004, EPI_ISL_1068005, EPI_ISL_1068006, EPI_ISL_1068007, EPI_ISL_1068008, EPI_ISL_1068009, EPI_ISL_1068010, EPI_ISL_1068011, EPI_ISL_1068012, EPI_ISL_1068013, EPI_ISL_1068014, EPI_ISL_1068015, EPI_ISL_1068016, EPI_ISL_1068017, EPI_ISL_1068018, EPI_ISL_1068019, EPI_ISL_1068020, EPI_ISL_1068021, EPI_ISL_1068022, EPI_ISL_1068023, EPI_ISL_1068024 | | | |
| see above | Clinical Molecular Microbiology Laboratory, UNC Hospital | Dirk Dittmer | Justin T. Landis , Razia Moorad , Brent A. Eason, Melissa B. Miller, Linda Pluta, Dirk Dittmer, Angelica Juarez, Cecilia Thompson, Shawn Hawken, Cameroon Grant, Evelyn Hoffman, Patricio Cano, Jason Wong, Carolina Caro-Vegas, Ryan McNamara, Blossom Damania. |
| EPI_ISL_1068025, EPI_ISL_1068026, EPI_ISL_1068027, EPI_ISL_1068028, EPI_ISL_1068029, EPI_ISL_1068030, EPI_ISL_1068032, EPI_ISL_1068033, EPI_ISL_1068034, EPI_ISL_1068035, EPI_ISL_1068036, EPI_ISL_1068038, EPI_ISL_1068039, EPI_ISL_1068040, EPI_ISL_1068041, EPI_ISL_1068042, EPI_ISL_1068043, EPI_ISL_1068044, EPI_ISL_1068046, EPI_ISL_1068047, EPI_ISL_1068048, EPI_ISL_1068049, EPI_ISL_1068050, EPI_ISL_1068052, EPI_ISL_1068053, EPI_ISL_1068054, EPI_ISL_1068055, EPI_ISL_1068057, EPI_ISL_1068059, EPI_ISL_1068061, EPI_ISL_1068062, EPI_ISL_1068064, EPI_ISL_1068065, EPI_ISL_1068066, EPI_ISL_1068067, EPI_ISL_1068070, EPI_ISL_1068075, EPI_ISL_1068077 | | | |
| see above | Utah Public Health Laboratory | Utah Public Health Laboratory | Erin L. Young, Kelly F. Oakeson, Tara Gallagher |
| EPI_ISL_1068293, EPI_ISL_1068294, EPI_ISL_1068295, EPI_ISL_1068296, EPI_ISL_1068297, EPI_ISL_1068298, EPI_ISL_1068299, EPI_ISL_1068300, EPI_ISL_1068301, EPI_ISL_1068302, EPI_ISL_1068303, EPI_ISL_1068304, EPI_ISL_1068305, EPI_ISL_1068306, EPI_ISL_1068307, EPI_ISL_1068308, EPI_ISL_1068309, EPI_ISL_1068310, EPI_ISL_1068311, EPI_ISL_1068312, EPI_ISL_1068313, EPI_ISL_1068314 | | | |
| see above | Alaska State Virology Laboratory | Alaska State Virology Laboratory | Stephanie DeRonde, Lisa Smith, Ph.D., Jack Chen, Ph.D. |

| | | | |
|---|---|---|---|
| EPI_ISL_1068395, EPI_ISL_1068396, EPI_ISL_1068397, EPI_ISL_1068398, EPI_ISL_1068399, EPI_ISL_1068400, EPI_ISL_1068401, EPI_ISL_1068402, EPI_ISL_1068403, EPI_ISL_1068404, EPI_ISL_1068405, EPI_ISL_1068406, EPI_ISL_1068407, EPI_ISL_1068408, EPI_ISL_1068410, EPI_ISL_1068411, EPI_ISL_1068412, EPI_ISL_1068413, EPI_ISL_1068414, EPI_ISL_1068416, EPI_ISL_1068417, EPI_ISL_1068418, EPI_ISL_1068419, EPI_ISL_1068420, EPI_ISL_1068422, EPI_ISL_1068423, EPI_ISL_1068424, EPI_ISL_1068425, EPI_ISL_1068426, EPI_ISL_1068427, EPI_ISL_1068428, EPI_ISL_1068429, EPI_ISL_1068430, EPI_ISL_1068431, EPI_ISL_1068432, EPI_ISL_1068433, EPI_ISL_1068434, EPI_ISL_1068435, EPI_ISL_1068436, EPI_ISL_1068437, EPI_ISL_1068438, EPI_ISL_1068439, EPI_ISL_1068440, EPI_ISL_1068441, EPI_ISL_1068442, EPI_ISL_1068443, EPI_ISL_1068445, EPI_ISL_1068446, EPI_ISL_1068447, EPI_ISL_1068448, EPI_ISL_1068449, EPI_ISL_1068450, EPI_ISL_1068451, EPI_ISL_1068452, EPI_ISL_1068455, EPI_ISL_1068456, EPI_ISL_1068457, EPI_ISL_1068458, EPI_ISL_1068459, EPI_ISL_1068460, EPI_ISL_1068461, EPI_ISL_1068462, EPI_ISL_1068463, EPI_ISL_1068464, EPI_ISL_1068465, EPI_ISL_1068467, EPI_ISL_1068468, EPI_ISL_1068469, EPI_ISL_1068470, EPI_ISL_1068471, EPI_ISL_1068473, EPI_ISL_1068474, EPI_ISL_1068476, EPI_ISL_1068477, EPI_ISL_1068478, EPI_ISL_1068479, EPI_ISL_1068480, EPI_ISL_1068481, EPI_ISL_1068482, EPI_ISL_1068483, EPI_ISL_1068484, EPI_ISL_1068485, EPI_ISL_1068486, EPI_ISL_1068487, EPI_ISL_1068488, EPI_ISL_1068489, EPI_ISL_1068490, EPI_ISL_1068491, EPI_ISL_1068492, EPI_ISL_1068494, EPI_ISL_1068495, EPI_ISL_1068496, EPI_ISL_1068497, EPI_ISL_1068498, EPI_ISL_1068499, EPI_ISL_1068500, EPI_ISL_1068501, EPI_ISL_1068502, EPI_ISL_1068503, EPI_ISL_1068504, EPI_ISL_1068505, EPI_ISL_1068506, EPI_ISL_1068507, EPI_ISL_1068508, EPI_ISL_1068509, EPI_ISL_1068510, EPI_ISL_1068511, EPI_ISL_1068512, EPI_ISL_1068513, EPI_ISL_1068514, EPI_ISL_1068515, EPI_ISL_1068517, EPI_ISL_1068518, EPI_ISL_1068519, EPI_ISL_1068520, EPI_ISL_1068521, EPI_ISL_1068522, EPI_ISL_1068523, EPI_ISL_1068524, EPI_ISL_1068525, EPI_ISL_1068526, EPI_ISL_1068527, EPI_ISL_1068528, EPI_ISL_1068529, EPI_ISL_1068530, EPI_ISL_1068531, EPI_ISL_1068532, EPI_ISL_1068533, EPI_ISL_1068534, EPI_ISL_1068535, EPI_ISL_1068536, EPI_ISL_1068537, EPI_ISL_1068538, EPI_ISL_1068539, EPI_ISL_1068540, EPI_ISL_1068541, EPI_ISL_1068542, EPI_ISL_1068543, EPI_ISL_1068544, EPI_ISL_1068546, EPI_ISL_1068547, EPI_ISL_1068548, EPI_ISL_1068549, EPI_ISL_1068550, EPI_ISL_1068551, EPI_ISL_1068552, EPI_ISL_1068553, EPI_ISL_1068554, EPI_ISL_1068555, EPI_ISL_1068556, EPI_ISL_1068557, EPI_ISL_1068558, EPI_ISL_1068561, EPI_ISL_1068562, EPI_ISL_1068563, EPI_ISL_1068564, EPI_ISL_1068566, EPI_ISL_1068567, EPI_ISL_1068568, EPI_ISL_1068569, EPI_ISL_1068570, EPI_ISL_1068571, EPI_ISL_1068572, EPI_ISL_1068573, EPI_ISL_1068574, EPI_ISL_1068575, EPI_ISL_1068576, EPI_ISL_1068577, EPI_ISL_1068578, EPI_ISL_1068579, EPI_ISL_1068580, EPI_ISL_1068581, EPI_ISL_1068582, EPI_ISL_1068583, EPI_ISL_1068584, EPI_ISL_1068585, EPI_ISL_1068586, EPI_ISL_1068588, EPI_ISL_1068589, EPI_ISL_1068590, EPI_ISL_1068591, EPI_ISL_1068592, EPI_ISL_1068594, EPI_ISL_1068595, EPI_ISL_1068596, EPI_ISL_1068597, EPI_ISL_1068598, EPI_ISL_1068602, EPI_ISL_1068603, EPI_ISL_1068604, EPI_ISL_1068605, EPI_ISL_1068606, EPI_ISL_1068607, EPI_ISL_1068608, EPI_ISL_1068609, EPI_ISL_1068610, EPI_ISL_1068611, EPI_ISL_1068612, EPI_ISL_1068613, EPI_ISL_1068614, EPI_ISL_1068615, EPI_ISL_1068616, EPI_ISL_1068617, EPI_ISL_1068618, EPI_ISL_1068619, EPI_ISL_1068620, EPI_ISL_1068621, EPI_ISL_1068622, EPI_ISL_1068623, EPI_ISL_1068624, EPI_ISL_1068625, EPI_ISL_1068626, EPI_ISL_1068627, EPI_ISL_1068628, EPI_ISL_1068629, EPI_ISL_1068630, EPI_ISL_1068632, EPI_ISL_1068634, EPI_ISL_1068635, EPI_ISL_1068637, EPI_ISL_1068638, EPI_ISL_1068639, EPI_ISL_1068640, EPI_ISL_1068641, EPI_ISL_1068642, EPI_ISL_1068643, EPI_ISL_1068644, EPI_ISL_1068645, EPI_ISL_1068647, EPI_ISL_1068648, EPI_ISL_1068649, EPI_ISL_1068650, EPI_ISL_1068651, EPI_ISL_1068652, EPI_ISL_1068654, EPI_ISL_1068655, EPI_ISL_1068656, EPI_ISL_1068657, EPI_ISL_1068658, EPI_ISL_1068659, EPI_ISL_1068660, EPI_ISL_1068661, EPI_ISL_1068662, EPI_ISL_1068663, EPI_ISL_1068664, EPI_ISL_1068665, EPI_ISL_1068666, EPI_ISL_1068667, EPI_ISL_1068668, EPI_ISL_1068669, EPI_ISL_1068670, EPI_ISL_1068673, EPI_ISL_1068674, EPI_ISL_1068675, EPI_ISL_1068676, EPI_ISL_1068677, EPI_ISL_1068678, EPI_ISL_1068679, EPI_ISL_1068680, EPI_ISL_1068681, EPI_ISL_1068683, EPI_ISL_1068684, EPI_ISL_1068685, EPI_ISL_1068686, EPI_ISL_1068687, EPI_ISL_1068688, EPI_ISL_1068689, EPI_ISL_1068690, EPI_ISL_1068691, EPI_ISL_1068692, EPI_ISL_1068694, EPI_ISL_1068695, EPI_ISL_1068696, EPI_ISL_1068697, EPI_ISL_1068699, EPI_ISL_1068700, EPI_ISL_1068702, EPI_ISL_1068703, EPI_ISL_1068704, EPI_ISL_1068705, EPI_ISL_1068706, EPI_ISL_1068707, EPI_ISL_1068708, EPI_ISL_1068709, EPI_ISL_1068710, EPI_ISL_1068711, EPI_ISL_1068712, EPI_ISL_1068714, EPI_ISL_1068716, EPI_ISL_1068717, EPI_ISL_1068718, EPI_ISL_1068719, EPI_ISL_1068720, EPI_ISL_1068721, EPI_ISL_1068722, EPI_ISL_1068723, EPI_ISL_1068724, EPI_ISL_1068725, EPI_ISL_1068726, EPI_ISL_1068727, EPI_ISL_1068728, EPI_ISL_1068729, EPI_ISL_1068730, EPI_ISL_1068731, EPI_ISL_1068732, EPI_ISL_1068733, EPI_ISL_1068734, EPI_ISL_1068735, EPI_ISL_1068736, EPI_ISL_1068737, EPI_ISL_1068738, EPI_ISL_1068739, EPI_ISL_1068740, EPI_ISL_1068741, EPI_ISL_1068742, EPI_ISL_1068743, EPI_ISL_1068745, EPI_ISL_1068746, EPI_ISL_1068747, EPI_ISL_1068748, EPI_ISL_1068750, EPI_ISL_1068751, EPI_ISL_1068752, EPI_ISL_1068753, EPI_ISL_1068754, EPI_ISL_1068755, EPI_ISL_1068756, EPI_ISL_1068757, EPI_ISL_1068758, EPI_ISL_1068759, EPI_ISL_1068760, EPI_ISL_1068761, EPI_ISL_1068762, EPI_ISL_1068763, EPI_ISL_1068764, EPI_ISL_1068765, EPI_ISL_1068766, EPI_ISL_1068767, EPI_ISL_1068768, EPI_ISL_1068769, EPI_ISL_1068770, EPI_ISL_1068771, EPI_ISL_1068772, EPI_ISL_1068773, EPI_ISL_1068774, EPI_ISL_1068775, EPI_ISL_1068776, EPI_ISL_1068777, EPI_ISL_1068778, EPI_ISL_1068779, EPI_ISL_1068780, EPI_ISL_1068781, EPI_ISL_1068782, EPI_ISL_1068783, EPI_ISL_1068784, EPI_ISL_1068785, EPI_ISL_1068786, EPI_ISL_1068787, EPI_ISL_1068788, EPI_ISL_1068789, EPI_ISL_1068790, EPI_ISL_1068791, EPI_ISL_1068792, EPI_ISL_1068793, EPI_ISL_1068794, EPI_ISL_1068795, EPI_ISL_1068796, EPI_ISL_1068797, EPI_ISL_1068798, EPI_ISL_1068800, EPI_ISL_1068801, EPI_ISL_1068802, EPI_ISL_1068803, EPI_ISL_1068804, EPI_ISL_1068805, EPI_ISL_1068806, EPI_ISL_1068807, EPI_ISL_1068808, EPI_ISL_1068809, EPI_ISL_1068810, EPI_ISL_1068811, EPI_ISL_1068812, EPI_ISL_1068813, EPI_ISL_1068814, EPI_ISL_1068815, EPI_ISL_1068816, EPI_ISL_1068817, EPI_ISL_1068818, EPI_ISL_1068819, EPI_ISL_1068820, EPI_ISL_1068821, EPI_ISL_1068822, EPI_ISL_1068823, EPI_ISL_1068824, EPI_ISL_1068825, EPI_ISL_1068826, EPI_ISL_1068827, EPI_ISL_1068828, EPI_ISL_1068829, EPI_ISL_1068830, EPI_ISL_1068831, EPI_ISL_1068832, EPI_ISL_1068833, EPI_ISL_1068834, EPI_ISL_1068835, EPI_ISL_1068836, EPI_ISL_1068838, EPI_ISL_1068839, EPI_ISL_1068840, EPI_ISL_1068841, EPI_ISL_1068842, EPI_ISL_1068843, EPI_ISL_1068844, EPI_ISL_1068845, EPI_ISL_1068846, EPI_ISL_1068847, EPI_ISL_1068848, EPI_ISL_1068852, EPI_ISL_1068853, EPI_ISL_1068854, EPI_ISL_1068855, EPI_ISL_1068856, EPI_ISL_1068857, EPI_ISL_1068858, EPI_ISL_1068859, EPI_ISL_1068860, EPI_ISL_1068861, EPI_ISL_1068862, EPI_ISL_1068863, EPI_ISL_1068865, EPI_ISL_1068866, EPI_ISL_1068867, EPI_ISL_1068868, EPI_ISL_1068869, EPI_ISL_1068870, EPI_ISL_1068871, EPI_ISL_1068872, EPI_ISL_1068873, EPI_ISL_1068874, EPI_ISL_1068875, EPI_ISL_1068876, EPI_ISL_1068877, EPI_ISL_1068878, EPI_ISL_1068879, EPI_ISL_1068880, EPI_ISL_1068881, EPI_ISL_1068882, EPI_ISL_1068883, EPI_ISL_1068884, EPI_ISL_1068885, EPI_ISL_1068886, EPI_ISL_1068887, EPI_ISL_1068888, EPI_ISL_1068889, EPI_ISL_1068890, EPI_ISL_1068891, EPI_ISL_1068892, EPI_ISL_1068893, EPI_ISL_1068894, EPI_ISL_1068895, EPI_ISL_1068896, EPI_ISL_1068897, EPI_ISL_1068898, EPI_ISL_1068900, EPI_ISL_1068901, EPI_ISL_1068902, EPI_ISL_1068903, EPI_ISL_1068904, EPI_ISL_1068905, EPI_ISL_1068906, EPI_ISL_1068907, EPI_ISL_1068908, EPI_ISL_1068909, EPI_ISL_1068910, EPI_ISL_1068911, EPI_ISL_1068912, EPI_ISL_1068913, EPI_ISL_1068914, EPI_ISL_1068915, EPI_ISL_1068916, EPI_ISL_1068917, EPI_ISL_1068918, EPI_ISL_1068919, EPI_ISL_1068920, EPI_ISL_1068921, EPI_ISL_1068922, EPI_ISL_1068923, EPI_ISL_1068924, EPI_ISL_1068925, EPI_ISL_1068926, EPI_ISL_1068930, EPI_ISL_1068931, EPI_ISL_1068932, EPI_ISL_1068933, EPI_ISL_1068934, EPI_ISL_1068935, EPI_ISL_1068936, EPI_ISL_1068937, EPI_ISL_1068938, EPI_ISL_1068939, EPI_ISL_1068941, EPI_ISL_1068942, EPI_ISL_1068943, EPI_ISL_1068944, EPI_ISL_1068945, EPI_ISL_1068946, EPI_ISL_1068947, EPI_ISL_1068948, EPI_ISL_1068949, EPI_ISL_1068950, EPI_ISL_1068951, EPI_ISL_1068952, EPI_ISL_1068953, EPI_ISL_1068954, EPI_ISL_1068955, EPI_ISL_1068956, EPI_ISL_1068957, EPI_ISL_1068958, EPI_ISL_1068959, EPI_ISL_1068960, EPI_ISL_1068961, EPI_ISL_1068962, EPI_ISL_1068963, EPI_ISL_1068964, EPI_ISL_1068965, EPI_ISL_1068966, EPI_ISL_1068967, EPI_ISL_1068968, EPI_ISL_1068969, EPI_ISL_1068970, EPI_ISL_1068971, EPI_ISL_1068972, EPI_ISL_1068973, EPI_ISL_1068974, EPI_ISL_1068975, EPI_ISL_1068976, EPI_ISL_1068977, EPI_ISL_1068978, EPI_ISL_1068979, EPI_ISL_1068980, EPI_ISL_1068981, EPI_ISL_1068982, EPI_ISL_1068983, EPI_ISL_1068984, EPI_ISL_1068985, EPI_ISL_1068986, EPI_ISL_1068987, EPI_ISL_1068988, EPI_ISL_1068989, EPI_ISL_1068990, EPI_ISL_1068991, EPI_ISL_1068992, EPI_ISL_1068993, EPI_ISL_1068994, EPI_ISL_1068995, EPI_ISL_1069001, EPI_ISL_1069002, EPI_ISL_1069003, EPI_ISL_1069004, EPI_ISL_1069005, EPI_ISL_1069006, EPI_ISL_1069007, EPI_ISL_1069008, EPI_ISL_1069009, EPI_ISL_1069010, EPI_ISL_1069011, EPI_ISL_1069012, EPI_ISL_1069013, EPI_ISL_1069014, EPI_ISL_1069015, EPI_ISL_1069016, EPI_ISL_1069017, EPI_ISL_1069018, EPI_ISL_1069019, EPI_ISL_1069020, EPI_ISL_1069021, EPI_ISL_1069022, EPI_ISL_1069023, EPI_ISL_1069024, EPI_ISL_1069025, EPI_ISL_1069026, EPI_ISL_1069027, EPI_ISL_1069028, EPI_ISL_1069029, EPI_ISL_1069030, EPI_ISL_1069031, EPI_ISL_1069032, EPI_ISL_1069033, EPI_ISL_1069034, EPI_ISL_1069035, EPI_ISL_1069036, EPI_ISL_1069037, EPI_ISL_1069038, EPI_ISL_1069039, EPI_ISL_1069040, EPI_ISL_1069041, EPI_ISL_1069042, EPI_ISL_1069043, EPI_ISL_1069045, EPI_ISL_1069046, EPI_ISL_1069047, EPI_ISL_1069048, EPI_ISL_1069050, EPI_ISL_1069051, EPI_ISL_1069052, EPI_ISL_1069053, EPI_ISL_1069054, EPI_ISL_1069055, EPI_ISL_1069056, EPI_ISL_1069057, EPI_ISL_1069058, EPI_ISL_1069059, EPI_ISL_1069060, EPI_ISL_1069061, EPI_ISL_1069062, EPI_ISL_1069063, EPI_ISL_1069064, EPI_ISL_1069065, EPI_ISL_1069066, EPI_ISL_1069067, EPI_ISL_1069068, EPI_ISL_1069069, EPI_ISL_1069070, EPI_ISL_1069071, EPI_ISL_1069072, EPI_ISL_1069073, EPI_ISL_1069074, EPI_ISL_1069075, EPI_ISL_1069086, EPI_ISL_1069087, EPI_ISL_1069088, EPI_ISL_1069089, EPI_ISL_1069090, EPI_ISL_1069091, EPI_ISL_1069092, EPI_ISL_1069093, EPI_ISL_1069094, EPI_ISL_1069095, EPI_ISL_1069096, EPI_ISL_1069097, EPI_ISL_1069098, EPI_ISL_1069099, EPI_ISL_1069100, EPI_ISL_1069101, EPI_ISL_1069102, EPI_ISL_1069103, EPI_ISL_1069104, EPI_ISL_1069105, EPI_ISL_1069106, EPI_ISL_1069107, EPI_ISL_1069108, EPI_ISL_1069109, EPI_ISL_1069110, EPI_ISL_1069111, EPI_ISL_1069112, EPI_ISL_1069113, EPI_ISL_1069114, EPI_ISL_1069115, EPI_ISL_1069116, EPI_ISL_1069117, EPI_ISL_1069118, EPI_ISL_1069119, EPI_ISL_1069120, EPI_ISL_1069121, EPI_ISL_1069122, EPI_ISL_1069123, EPI_ISL_1069124, EPI_ISL_1069125, EPI_ISL_1069127, EPI_ISL_1069128, EPI_ISL_1069129, EPI_ISL_1069130, EPI_ISL_1069131, EPI_ISL_1069132, EPI_ISL_1069133, EPI_ISL_1069134, EPI_ISL_1069135, EPI_ISL_1069136, EPI_ISL_1069138, EPI_ISL_1069139, EPI_ISL_1069140, EPI_ISL_1069142, EPI_ISL_1069143, EPI_ISL_1069144, EPI_ISL_1069145, EPI_ISL_1069146, EPI_ISL_1069147, EPI_ISL_1069148, EPI_ISL_1069149, EPI_ISL_1069150, EPI_ISL_1069152 | | | |
| see above | Fulgent Genetics | Fulgent Genetics | Harry Gao, Mickey Li, John Gao, Joseph Fierro, Benafsh Sapra, Becky Tsai, Yan Meng, Doreen Ng, James Xie |
| EPI_ISL_1069221, EPI_ISL_1069222, EPI_ISL_1069223, EPI_ISL_1069224, EPI_ISL_1069225, EPI_ISL_1069226, EPI_ISL_1069227, EPI_ISL_1069228, EPI_ISL_1069229, EPI_ISL_1069230, EPI_ISL_1069231, EPI_ISL_1069232, EPI_ISL_1069233, EPI_ISL_1069234, EPI_ISL_1069235, EPI_ISL_1069236, EPI_ISL_1069237, EPI_ISL_1069238, EPI_ISL_1069239, EPI_ISL_1069240, EPI_ISL_1069241, EPI_ISL_1069242, EPI_ISL_1069243, EPI_ISL_1069244, EPI_ISL_1069245, EPI_ISL_1069246, EPI_ISL_1069247, EPI_ISL_1069248, EPI_ISL_1069249, EPI_ISL_1069250, EPI_ISL_1069251, EPI_ISL_1069252, EPI_ISL_1069253, EPI_ISL_1069254, EPI_ISL_1069255, EPI_ISL_1069256, EPI_ISL_1069257, EPI_ISL_1069258, EPI_ISL_1069259, EPI_ISL_1069260, EPI_ISL_1069261, EPI_ISL_1069262, EPI_ISL_1069263, EPI_ISL_1069264, EPI_ISL_1069265, EPI_ISL_1069266, EPI_ISL_1069267, EPI_ISL_1069268, EPI_ISL_1069269, EPI_ISL_1069270, EPI_ISL_1069271, EPI_ISL_1069272, EPI_ISL_1069273, EPI_ISL_1069274, EPI_ISL_1069275, EPI_ISL_1069276, EPI_ISL_1069277, EPI_ISL_1069278, EPI_ISL_1069279, EPI_ISL_1069280, EPI_ISL_1069281, EPI_ISL_1069282, EPI_ISL_1069283, EPI_ISL_1069284, EPI_ISL_1069285, EPI_ISL_1069286, EPI_ISL_1069287, EPI_ISL_1069288, EPI_ISL_1069289, EPI_ISL_1069290, EPI_ISL_1069291, EPI_ISL_1069292, EPI_ISL_1069293, EPI_ISL_1069294, EPI_ISL_1069295, EPI_ISL_1069296, EPI_ISL_1069297, EPI_ISL_1069298, EPI_ISL_1069299, EPI_ISL_1069300, EPI_ISL_1069301, EPI_ISL_1069302, EPI_ISL_1069303, EPI_ISL_1069304, EPI_ISL_1069305, EPI_ISL_1069306, EPI_ISL_1069307, EPI_ISL_1069308, EPI_ISL_1069309, EPI_ISL_1069310, EPI_ISL_1069311, EPI_ISL_1069312, EPI_ISL_1069313, EPI_ISL_1069314, EPI_ISL_1069315, EPI_ISL_1069316, EPI_ISL_1069317, EPI_ISL_1069318, EPI_ISL_1069319, EPI_ISL_1069320, EPI_ISL_1069321, EPI_ISL_1069322, EPI_ISL_1069323, EPI_ISL_1069324, EPI_ISL_1069325, EPI_ISL_1069326, EPI_ISL_1069327, EPI_ISL_1069328, EPI_ISL_1069329, EPI_ISL_1069330, EPI_ISL_1069331, EPI_ISL_1069332, EPI_ISL_1069333, EPI_ISL_1069334, EPI_ISL_1069335, EPI_ISL_1069336, EPI_ISL_1069337, EPI_ISL_1069338, EPI_ISL_1069339, EPI_ISL_1069340, EPI_ISL_1069341, EPI_ISL_1069342, EPI_ISL_1069343, EPI_ISL_1069344, EPI_ISL_1069345, EPI_ISL_1069346, EPI_ISL_1069347, EPI_ISL_1069348, EPI_ISL_1069349, EPI_ISL_1069350, EPI_ISL_1069351, EPI_ISL_1069352, EPI_ISL_1069353, EPI_ISL_1069354, EPI_ISL_1069355, EPI_ISL_1069356, EPI_ISL_1069357, EPI_ISL_1069358, EPI_ISL_1069359, EPI_ISL_1069360, EPI_ISL_1069361, EPI_ISL_1069362, EPI_ISL_1069363, EPI_ISL_1069364, EPI_ISL_1069365, EPI_ISL_1069366, EPI_ISL_1069367, EPI_ISL_1069368, EPI_ISL_1069369, EPI_ISL_1069370, EPI_ISL_1069371, EPI_ISL_1069372, EPI_ISL_1069373, EPI_ISL_1069374, EPI_ISL_1069375, EPI_ISL_1069376, EPI_ISL_1069377, EPI_ISL_1069378, EPI_ISL_1069379, EPI_ISL_1069380, EPI_ISL_1069381, EPI_ISL_1069382, EPI_ISL_1069383, EPI_ISL_1069384, EPI_ISL_1069385, EPI_ISL_1069386, EPI_ISL_1069387, EPI_ISL_1069388, EPI_ISL_1069389 | | | |
| see above | UW Virology Lab | UW Virology Lab | Pavitra Roychoudhury, Hong Xie, Lasata Shrestha, Shah Mohamed Bakhash, Robert Livingston, Michelle Lin, Noah Baker, Meei-Li Huang, Keith R Jerome, Alexander Greninger |
| EPI_ISL_1073813, EPI_ISL_1073814 | Kansas Health and Environmental Lab | Kansas Health and Environmental Lab | Mike Grose, Paige Drury, Carissa Robertson, Ben Olsen, and Phil Adam |
| EPI_ISL_1073815, EPI_ISL_1073816, EPI_ISL_1073817, EPI_ISL_1073818, EPI_ISL_1073819, EPI_ISL_1073820, EPI_ISL_1073821, EPI_ISL_1073822, EPI_ISL_1073823, EPI_ISL_1073824 | Kansas Health and Environmental Lab | Kansas Health and Environmental Lab | Mike Grose, Carissa Robertson, Ben Olsen, and Phil Adam |
| EPI_ISL_1074302, EPI_ISL_1074303, EPI_ISL_1074304, EPI_ISL_1074305, EPI_ISL_1074306, EPI_ISL_1074307, EPI_ISL_1074308, EPI_ISL_1074309, EPI_ISL_1074310, EPI_ISL_1074311, EPI_ISL_1074312, EPI_ISL_1074313, EPI_ISL_1074314, EPI_ISL_1074315, EPI_ISL_1074316, EPI_ISL_1074317, EPI_ISL_1074318, EPI_ISL_1074319, EPI_ISL_1074320, EPI_ISL_1074321, EPI_ISL_1074322, EPI_ISL_1074323, EPI_ISL_1074325, EPI_ISL_1074326, EPI_ISL_1074327, EPI_ISL_1074328, EPI_ISL_1074329, EPI_ISL_1074330, EPI_ISL_1074331, EPI_ISL_1074332, EPI_ISL_1074333, EPI_ISL_1074334, EPI_ISL_1074335, EPI_ISL_1074336, EPI_ISL_1074337, EPI_ISL_1074338, EPI_ISL_1074339, EPI_ISL_1074340, EPI_ISL_1074341, EPI_ISL_1074342, EPI_ISL_1074343, EPI_ISL_1074344, EPI_ISL_1074345, EPI_ISL_1074346, EPI_ISL_1074347, EPI_ISL_1074348, EPI_ISL_1074349, EPI_ISL_1074350, EPI_ISL_1074351, EPI_ISL_1074352, EPI_ISL_1074353, EPI_ISL_1074354, EPI_ISL_1074355, EPI_ISL_1074356, EPI_ISL_1074357, EPI_ISL_1074358, EPI_ISL_1074359, EPI_ISL_1074360, EPI_ISL_1074361, EPI_ISL_1074362, EPI_ISL_1074363, EPI_ISL_1074364, EPI_ISL_1074365, EPI_ISL_1074366, EPI_ISL_1074367, EPI_ISL_1074368, EPI_ISL_1074369, EPI_ISL_1074370, EPI_ISL_1074371, EPI_ISL_1074372, EPI_ISL_1074373, EPI_ISL_1074374, EPI_ISL_1074375, EPI_ISL_1074376, EPI_ISL_1074377, EPI_ISL_1074378, EPI_ISL_1074379, EPI_ISL_1074380, EPI_ISL_1074381, EPI_ISL_1074382, EPI_ISL_1074383, EPI_ISL_1074384, EPI_ISL_1074385, EPI_ISL_1074386, EPI_ISL_1074387, EPI_ISL_1074388, EPI_ISL_1074389, EPI_ISL_1074390, EPI_ISL_1074391, EPI_ISL_1074392, EPI_ISL_1074393, EPI_ISL_1074394, EPI_ISL_1074395, EPI_ISL_1074396, EPI_ISL_1074397, EPI_ISL_1074398, EPI_ISL_1074399, EPI_ISL_1074400, EPI_ISL_1074401, EPI_ISL_1074402, EPI_ISL_1074403, EPI_ISL_1074404, EPI_ISL_1074405, EPI_ISL_1074406, EPI_ISL_1074407, EPI_ISL_1074408, EPI_ISL_1074409, EPI_ISL_1074410, EPI_ISL_1074411, EPI_ISL_1074412, EPI_ISL_1074413, EPI_ISL_1074414, EPI_ISL_1074415, EPI_ISL_1074416, EPI_ISL_1074417, EPI_ISL_1074418, EPI_ISL_1074419, EPI_ISL_1074420, EPI_ISL_1074421, EPI_ISL_1074422, EPI_ISL_1074423, EPI_ISL_1074424, EPI_ISL_1074425, EPI_ISL_1074426, EPI_ISL_1074427, EPI_ISL_1074428, EPI_ISL_1074429, EPI_ISL_1074430, EPI_ISL_1074431, EPI_ISL_1074432, EPI_ISL_1074433, EPI_ISL_1074434, EPI_ISL_1074435, EPI_ISL_1074436, EPI_ISL_1074437, EPI_ISL_1074438, EPI_ISL_1074439, EPI_ISL_1074440, EPI_ISL_1074441, EPI_ISL_1074442, EPI_ISL_1074443, EPI_ISL_1074444, EPI_ISL_1074445, EPI_ISL_1074446, EPI_ISL_1074447, EPI_ISL_1074448, EPI_ISL_1074449, EPI_ISL_1074450, EPI_ISL_1074451, EPI_ISL_1074452, EPI_ISL_1074453, EPI_ISL_1074454, EPI_ISL_1074455, EPI_ISL_1074456, EPI_ISL_1074457, EPI_ISL_1074458, EPI_ISL_1074459, EPI_ISL_1074460, EPI_ISL_1074461, EPI_ISL_1074462, EPI_ISL_1074463, EPI_ISL_1074464, EPI_ISL_1074465, EPI_ISL_1074466, EPI_ISL_1074467, EPI_ISL_1074468, EPI_ISL_1074469, EPI_ISL_1074470, EPI_ISL_1074471, EPI_ISL_1074472, EPI_ISL_1074473, EPI_ISL_1074474, EPI_ISL_1074475, EPI_ISL_1074476, EPI_ISL_1074477, EPI_ISL_1074478, EPI_ISL_1074479, EPI_ISL_1074480, EPI_ISL_1074481, EPI_ISL_1074482, EPI_ISL_1074483, EPI_ISL_1074484, EPI_ISL_1074485, EPI_ISL_1074486, EPI_ISL_1074487, EPI_ISL_1074488, EPI_ISL_1074489, EPI_ISL_1074490, EPI_ISL_1074491, EPI_ISL_1074492, EPI_ISL_1074493, EPI_ISL_1074494, EPI_ISL_1074495, EPI_ISL_1074496, EPI_ISL_1074497, EPI_ISL_1074498, EPI_ISL_1074499, EPI_ISL_1074500, EPI_ISL_1074501, EPI_ISL_1074502, EPI_ISL_1074503, EPI_ISL_1074504, EPI_ISL_1074505, EPI_ISL_1074506, EPI_ISL_1074507, EPI_ISL_1074508, EPI_ISL_1074509, EPI_ISL_1074510, EPI_ISL_1074511, EPI_ISL_1074512, EPI_ISL_1074513, EPI_ISL_1074514, EPI_ISL_1074515, EPI_ISL_1074516, EPI_ISL_1074517, EPI_ISL_1074518, EPI_ISL_1074519, EPI_ISL_1074520, EPI_ISL_1074521, EPI_ISL_1074522, EPI_ISL_1074523, EPI_ISL_1074524, EPI_ISL_1074525, EPI_ISL_1074526, EPI_ISL_1074527, EPI_ISL_1074528, EPI_ISL_1074529, EPI_ISL_1074530, EPI_ISL_1074531, EPI_ISL_1074532, EPI_ISL_1074533, EPI_ISL_1074534, EPI_ISL_1074535, EPI_ISL_1074536, EPI_ISL_1074537, EPI_ISL_1074538, EPI_ISL_1074539, EPI_ISL_1074540, EPI_ISL_1074541, EPI_ISL_1074542, EPI_ISL_1074543, EPI_ISL_1074544, EPI_ISL_1074545, EPI_ISL_1074546, EPI_ISL_1074547, EPI_ISL_1074548, EPI_ISL_1074549, EPI_ISL_1074550, EPI_ISL_1074551, EPI_ISL_1074552, EPI_ISL_1074553, EPI_ISL_1074554, EPI_ISL_1074555, EPI_ISL_1074556, EPI_ISL_1074557, EPI_ISL_1074558, EPI_ISL_1074559, EPI_ISL_1074560, EPI_ISL_1074561, EPI_ISL_1074562, EPI_ISL_1074563, EPI_ISL_1074564, EPI_ISL_1074565, EPI_ISL_1074566, EPI_ISL_1074567, EPI_ISL_1074568, EPI_ISL_1074569, EPI_ISL_1074570, EPI_ISL_1074571, EPI_ISL_1074572, EPI_ISL_1074573, EPI_ISL_1074574, EPI_ISL_1074575, EPI_ISL_1074576, EPI_ISL_1074577, EPI_ISL_1074578, EPI_ISL_1074579, EPI_ISL_1074580, EPI_ISL_1074581, EPI_ISL_1074582, EPI_ISL_1074583, EPI_ISL_1074584, EPI_ISL_1074585, EPI_ISL_1074586, EPI_ISL_1074587, EPI_ISL_1074588, EPI_ISL_1074589, EPI_ISL_1074590, EPI_ISL_1074591, EPI_ISL_1074592, EPI_ISL_1074593, EPI_ISL_1074594, EPI_ISL_1074595, EPI_ISL_1074596, EPI_ISL_1074597, EPI_ISL_1074598, EPI_ISL_1074599, EPI_ISL_1074600, EPI_ISL_1074601, EPI_ISL_1074602, EPI_ISL_1074603, EPI_ISL_1074604, EPI_ISL_1074605, EPI_ISL_1074606, EPI_ISL_1074607, EPI_ISL_1074608, EPI_ISL_1074609, EPI_ISL_1074610, EPI_ISL_1074611, EPI_ISL_1074612, EPI_ISL_1074613, EPI_ISL_1074614, EPI_ISL_1074615, EPI_ISL_1074616, EPI_ISL_1074617, EPI_ISL_1074618, EPI_ISL_1074619, EPI_ISL_1074620, EPI_ISL_1074621, EPI_ISL_1074624, EPI_ISL_1074625, EPI_ISL_1074626, EPI_ISL_1074627, EPI_ISL_1074628, EPI_ISL_1074629, EPI_ISL_1074630, EPI_ISL_1074631, EPI_ISL_1074632, EPI_ISL_1074633, EPI_ISL_1074634, EPI_ISL_1074635, EPI_ISL_1074636, EPI_ISL_1074637, EPI_ISL_1074638, EPI_ISL_1074639, EPI_ISL_1074640, EPI_ISL_1074641, EPI_ISL_1074642, EPI_ISL_1074643, EPI_ISL_1074644, EPI_ISL_1074645, EPI_ISL_1074646, EPI_ISL_1074647, EPI_ISL_1074648, EPI_ISL_1074649, EPI_ISL_1074650, EPI_ISL_1074651, EPI_ISL_1074652, EPI_ISL_1074653, EPI_ISL_1074654, EPI_ISL_1074655, EPI_ISL_1074656, EPI_ISL_1074657, EPI_ISL_1074658, EPI_ISL_1074659, EPI_ISL_1074660, EPI_ISL_1074661, EPI_ISL_1074662, EPI_ISL_1074663, EPI_ISL_1074664, EPI_ISL_1074665, EPI_ISL_1074666, EPI_ISL_1074667, EPI_ISL_1074668, EPI_ISL_1074669, EPI_ISL_1074670, EPI_ISL_1074671, EPI_ISL_1074672, EPI_ISL_1074673, EPI_ISL_1074674, EPI_ISL_1074675, EPI_ISL_1074676, EPI_ISL_1074677, EPI_ISL_1074678, EPI_ISL_1074679, EPI_ISL_1074680, EPI_ISL_1074681, EPI_ISL_1074682, EPI_ISL_1074683, EPI_ISL_1074684, EPI_ISL_1074685, EPI_ISL_1074686, EPI_ISL_1074687, EPI_ISL_1074688, EPI_ISL_1074689, EPI_ISL_1074690, EPI_ISL_1074691, EPI_ISL_1074692, EPI_ISL_1074693, EPI_ISL_1074694, EPI_ISL_1074695, EPI_ISL_1074696, EPI_ISL_1074697, EPI_ISL_1074698, EPI_ISL_1074699, EPI_ISL_1074700, EPI_ISL_1074701, EPI_ISL_1074702, EPI_ISL_1074703, EPI_ISL_1074704, EPI_ISL_1074705, EPI_ISL_1074706, EPI_ISL_1074707, EPI_ISL_1074708, EPI_ISL_1074709, EPI_ISL_1074710, EPI_ISL_1074711, EPI_ISL_1074712, EPI_ISL_1074713, EPI_ISL_1074714, EPI_ISL_1074715, EPI_ISL_1074716, EPI_ISL_1074717, EPI_ISL_1074718, EPI_ISL_1074719, EPI_ISL_1074720, EPI_ISL_1074721, EPI_ISL_1074722, EPI_ISL_1074723, EPI_ISL_1074724, EPI_ISL_1074725, EPI_ISL_1074726, EPI_ISL_1074727, EPI_ISL_1074728, EPI_ISL_1074729, EPI_ISL_1074730, EPI_ISL_1074731, EPI_ISL_1074732, EPI_ISL_1074733, EPI_ISL_1074734, EPI_ISL_1074735, EPI_ISL_1074736, EPI_ISL_1074737, EPI_ISL_1074738, EPI_ISL_1074739, EPI_ISL_1074740, EPI_ISL_1074741, EPI_ISL_1074742, EPI_ISL_1074744, EPI_ISL_1074745, EPI_ISL_1074746, EPI_ISL_1074747, EPI_ISL_1074748, EPI_ISL_1074749, EPI_ISL_1074750, EPI_ISL_1074751, EPI_ISL_1074752, EPI_ISL_1074753, EPI_ISL_1074754, EPI_ISL_1074755, EPI_ISL_1074756, EPI_ISL_1074757, EPI_ISL_1074758, EPI_ISL_1074759, EPI_ISL_1074760, EPI_ISL_1074761, EPI_ISL_1074762, EPI_ISL_1074763, EPI_ISL_1074764, EPI_ISL_1074765, EPI_ISL_1074766, EPI_ISL_1074767, EPI_ISL_1074768, EPI_ISL_1074769, EPI_ISL_1074770, EPI_ISL_1074771, EPI_ISL_1074772, EPI_ISL_1074773, EPI_ISL_1074774, EPI_ISL_1074775, EPI_ISL_1074776, EPI_ISL_1074777, EPI_ISL_1074778, EPI_ISL_1074779, EPI_ISL_1074780, EPI_ISL_1074781, EPI_ISL_1074782, EPI_ISL_1074783, EPI_ISL_1074784, EPI_ISL_1074785, EPI_ISL_1074786, EPI_ISL_1074787, EPI_ISL_1074788, EPI_ISL_1074789, EPI_ISL_1074790, EPI_ISL_1074791, EPI_ISL_1074792, EPI_ISL_1074793, EPI_ISL_1074794, EPI_ISL_1074795, EPI_ISL_1074796, EPI_ISL_1074797, EPI_ISL_1074798, EPI_ISL_1074799, EPI_ISL_1074800, EPI_ISL_1074801, EPI_ISL_1074802, EPI_ISL_1074803, EPI_ISL_1074804, EPI_ISL_1074805, EPI_ISL_1074806, EPI_ISL_1074807, EPI_ISL_1074808, EPI_ISL_1074809, EPI_ISL_1074810, EPI_ISL_1074811, EPI_ISL_1074812, EPI_ISL_1074813, EPI_ISL_1074814, EPI_ISL_1074815, EPI_ISL_1074816, EPI_ISL_1074817, EPI_ISL_1074818, EPI_ISL_1074819, EPI_ISL_1074820, EPI_ISL_1074821, EPI_ISL_1074822, EPI_ISL_1074823, EPI_ISL_1074824, EPI_ISL_1074825, EPI_ISL_1074826, EPI_ISL_1074827, EPI_ISL_1074828, EPI_ISL_1074829, EPI_ISL_1074830, EPI_ISL_1074831, EPI_ISL_1074832, EPI_ISL_1074833, EPI_ISL_1074834, EPI_ISL_1074835, EPI_ISL_1074836, EPI_ISL_1074837, EPI_ISL_1074838, EPI_ISL_1074839, EPI_ISL_1074840, EPI_ISL_1074841, EPI_ISL_1074842, EPI_ISL_1074843, EPI_ISL_1074844, EPI_ISL_1074845, EPI_ISL_1074846, EPI_ISL_1074847, EPI_ISL_1074848, EPI_ISL_1074849, EPI_ISL_1074850, EPI_ISL_1074851, EPI_ISL_1074852, EPI_ISL_1074853, EPI_ISL_1074854, EPI_ISL_1074855, EPI_ISL_1074856, EPI_ISL_1074857, EPI_ISL_1074858, EPI_ISL_1074859, EPI_ISL_1074860, EPI_ISL_1074861, EPI_ISL_1074862, EPI_ISL_1074863, EPI_ISL_1074864, EPI_ISL_1074865, EPI_ISL_1074866, EPI_ISL_1074867, EPI_ISL_1074868, EPI_ISL_1074869, EPI_ISL_1074870, EPI_ISL_1074871, EPI_ISL_1074872, EPI_ISL_1074873, EPI_ISL_1074874, EPI_ISL_1074875, EPI_ISL_1074876, EPI_ISL_1074877, EPI_ISL_1074878, EPI_ISL_1074879, EPI_ISL_1074880, EPI_ISL_1074881, EPI_ISL_1074882, EPI_ISL_1074883, EPI_ISL_1074884, EPI_ISL_1074885, EPI_ISL_1074886, EPI_ISL_1074887, EPI_ISL_1074888, EPI_ISL_1074889, EPI_ISL_1074890, EPI_ISL_1074891, EPI_ISL_1074892, EPI_ISL_1074893, EPI_ISL_1074894, EPI_ISL_1074895, EPI_ISL_1074896, EPI_ISL_1074897, EPI_ISL_1074898, EPI_ISL_1074899, EPI_ISL_1074900, EPI_ISL_1074901, EPI_ISL_1074902, EPI_ISL_1074903, EPI_ISL_1074904, EPI_ISL_1074905, EPI_ISL_1074906, EPI_ISL_1074907, EPI_ISL_1074908, EPI_ISL_1074909, EPI_ISL_1074910, EPI_ISL_1074911, EPI_ISL_1074912, EPI_ISL_1074913, EPI_ISL_1074914, EPI_ISL_1074915, EPI_ISL_1074916, EPI_ISL_1074917, EPI_ISL_1074918, EPI_ISL_1074919, EPI_ISL_1074920, EPI_ISL_1074921, EPI_ISL_1074922, EPI_ISL_1074923, EPI_ISL_1074924, EPI_ISL_1074925, EPI_ISL_1074926, EPI_ISL_1074927, EPI_ISL_1074928, EPI_ISL_1074929, EPI_ISL_1074930, EPI_ISL_1074931, EPI_ISL_1074932, EPI_ISL_1074933, EPI_ISL_1074934, EPI_ISL_1074935, EPI_ISL_1074936, EPI_ISL_1074937, EPI_ISL_1074938, EPI_ISL_1074939, EPI_ISL_1074940, EPI_ISL_1074941, EPI_ISL_1074942, EPI_ISL_1074943, EPI_ISL_1074944, EPI_ISL_1074945, EPI_ISL_1074946, EPI_ISL_1074947, EPI_ISL_1074948, EPI_ISL_1074949, EPI_ISL_1074950, EPI_ISL_1074951, EPI_ISL_1074952, EPI_ISL_1074953, EPI_ISL_1074954, EPI_ISL_1074955, EPI_ISL_1074956, EPI_ISL_1074957, EPI_ISL_1074958, EPI_ISL_1074959, EPI_ISL_1074960, EPI_ISL_1074961, EPI_ISL_1074962, EPI_ISL_1074963, EPI_ISL_1074964, EPI_ISL_1074965, EPI_ISL_1074966, EPI_ISL_1074967, EPI_ISL_1074968, EPI_ISL_1074969, EPI_ISL_1074970, EPI_ISL_1074971, EPI_ISL_1074972, EPI_ISL_1074973, EPI_ISL_1074974, EPI_ISL_1074975, EPI_ISL_1074976, EPI_ISL_1074977, EPI_ISL_1074978, EPI_ISL_1074979, EPI_ISL_1074980, EPI_ISL_1074981, EPI_ISL_1074982, EPI_ISL_1074983, EPI_ISL_1074984, EPI_ISL_1074985, EPI_ISL_1074986, EPI_ISL_1074987, EPI_ISL_1074988, EPI_ISL_1074989, EPI_ISL_1074990, EPI_ISL_1074991, EPI_ISL_1074992, EPI_ISL_1074993, EPI_ISL_1074994, EPI_ISL_1074995, EPI_ISL_1074996, EPI_ISL_1074997, EPI_ISL_1074998, EPI_ISL_1074999, EPI_ISL_1075000, EPI_ISL_1075001, EPI_ISL_1075002, EPI_ISL_1075003, EPI_ISL_1075004, EPI_ISL_1075005, EPI_ISL_1075006, EPI_ISL_1075007, EPI_ISL_1075008, EPI_ISL_1075009, EPI_ISL_1075010, EPI_ISL_1075011, EPI_ISL_1075012, EPI_ISL_1075013, EPI_ISL_1075014, EPI_ISL_1075015, EPI_ISL_1075016, EPI_ISL_1075017, EPI_ISL_1075018, EPI_ISL_1075019, EPI_ISL_1075020, EPI_ISL_1075021, EPI_ISL_1075022, EPI_ISL_1075023, EPI_ISL_1075024, EPI_ISL_1075025, EPI_ISL_1075026, EPI_ISL_1075027, EPI_ISL_1075028, EPI_ISL_1075029, EPI_ISL_1075030, EPI_ISL_1075031, EPI_ISL_1075032, EPI_ISL_1075033, EPI_ISL_1075034, EPI_ISL_1075035, EPI_ISL_1075036, EPI_ISL_1075037, EPI_ISL_1075038, EPI_ISL_1075039, EPI_ISL_1075040, EPI_ISL_1075041, EPI_ISL_1075042, EPI_ISL_1075043, EPI_ISL_1075044, EPI_ISL_1075045, EPI_ISL_1075046, EPI_ISL_1075047, EPI_ISL_1075048, EPI_ISL_1075049, EPI_ISL_1075050, EPI_ISL_1075051, EPI_ISL_1075052, EPI_ISL_1075053, EPI_ISL_1075054, EPI_ISL_1075055, EPI_ISL_1075056, EPI_ISL_1075057, EPI_ISL_1075058, EPI_ISL_1075059, EPI_ISL_1075060, EPI_ISL_1075061, EPI_ISL_1075062, EPI_ISL_1075063, EPI_ISL_1075064, EPI_ISL_1075065, EPI_ISL_1075066, EPI_ISL_1075067, EPI_ISL_1075068, EPI_ISL_1075069, EPI_ISL_1075070, EPI_ISL_1075071, EPI_ISL_1075072, EPI_ISL_1075073, EPI_ISL_1075074, EPI_ISL_1075075, EPI_ISL_1075076, EPI_ISL_1075077, EPI_ISL_1075078, EPI_ISL_1075079, EPI_ISL_1075080, EPI_ISL_1075081, EPI_ISL_1075082, EPI_ISL_1075083, EPI_ISL_1075084, EPI_ISL_1075085, EPI_ISL_1075086, EPI_ISL_1075087, EPI_ISL_1075088, EPI_ISL_1075089, EPI_ISL_1075090, EPI_ISL_1075091, EPI_ISL_1075092, EPI_ISL_1075093, EPI_ISL_1075094, EPI_ISL_1075095, EPI_ISL_1075096, EPI_ISL_1075097, EPI_ISL_1075098, EPI_ISL_1075099, EPI_ISL_1075100, EPI_ISL_1075101, EPI_ISL_1075102, EPI_ISL_1075103, EPI_ISL_1075104, EPI_ISL_1075105, EPI_ISL_1075106, EPI_ISL_1075107, EPI_ISL_1075108, EPI_ISL_1075109, EPI_ISL_1075110, EPI_ISL_1075111, EPI_ISL_1075112, EPI_ISL_1075113, EPI_ISL_1075114, EPI_ISL_1075115, EPI_ISL_1075116, EPI_ISL_1075117, EPI_ISL_1075118, EPI_ISL_1075119, EPI_ISL_1075120, EPI_ISL_1075121, EPI_ISL_1075122, EPI_ISL_1075123, EPI_ISL_1075124, EPI_ISL_1075125, EPI_ISL_1075126, EPI_ISL_1075127, EPI_ISL_1075128, EPI_ISL_1075129, EPI_ISL_1075130, EPI_ISL_1075131, EPI_ISL_1075132, EPI_ISL_1075133, EPI_ISL_1075134, EPI_ISL_1075135, EPI_ISL_1075136, EPI_ISL_1075137, EPI_ISL_1075138, EPI_ISL_1075139, EPI_ISL_1075140, EPI_ISL_1075141, EPI_ISL_1075142, EPI_ISL_1075143, EPI_ISL_1075144, EPI_ISL_1075145, EPI_ISL_1075146, EPI_ISL_1075147, EPI_ISL_1075148, EPI_ISL_1075149, EPI_ISL_1075150, EPI_ISL_1075151, EPI_ISL_1075152, EPI_ISL_1075153, EPI_ISL_1075154, EPI_ISL_1075155, EPI_ISL_1075156, EPI_ISL_1075157, EPI_ISL_1075158, EPI_ISL_1075159, EPI_ISL_1075160, EPI_ISL_1075161, EPI_ISL_1075162, EPI_ISL_1075163, EPI_ISL_1075164, EPI_ISL_1075165, EPI_ISL_1075166, EPI_ISL_1075167, EPI_ISL_1075168, EPI_ISL_1075169, EPI_ISL_1075170, EPI_ISL_1075171, EPI_ISL_1075172, EPI_ISL_1075173, EPI_ISL_1075174, EPI_ISL_1075175, EPI_ISL_1075176, EPI_ISL_1075177, EPI_ISL_1075178, EPI_ISL_1075179, EPI_ISL_1075180, EPI_ISL_1075181, EPI_ISL_1075182, EPI_ISL_1075183, EPI_ISL_1075184, EPI_ISL_1075185, EPI_ISL_1075186, EPI_ISL_1075187, EPI_ISL_1075188, EPI_ISL_1075189, EPI_ISL_1075190, EPI_ISL_1075191, EPI_ISL_1075192, EPI_ISL_1075193, EPI_ISL_1075194, EPI_ISL_1075195, EPI_ISL_1075196, EPI_ISL_1075197, EPI_ISL_1075198, EPI_ISL_1075199, EPI_ISL_1075200, EPI_ISL_1075201, EPI_ISL_1075202, EPI_ISL_1075203, EPI_ISL_1075204, EPI_ISL_1075205, EPI_ISL_1075206, EPI_ISL_1075207, EPI_ISL_1075208, EPI_ISL_1075209, EPI_ISL_1075210, EPI_ISL_1075211, EPI_ISL_1075212, EPI_ISL_1075213, EPI_ISL_1075214, EPI_ISL_1075215, EPI_ISL_1075216, EPI_ISL_1075217, EPI_ISL_1075218, EPI_ISL_1075219, EPI_ISL_1075220, EPI_ISL_1075221, EPI_ISL_1075222, EPI_ISL_1075223, EPI_ISL_1075224, EPI_ISL_1075225, EPI_ISL_1075226, EPI_ISL_1075227, EPI_ISL_1075228, EPI_ISL_1075229, EPI_ISL_1075230, EPI_ISL_1075231, EPI_ISL_1075232, EPI_ISL_1075233, EPI_ISL_1075234, EPI_ISL_1075235, EPI_ISL_1075236, EPI_ISL_1075237, EPI_ISL_1075238, EPI_ISL_1075239, EPI_ISL_1075240, EPI_ISL_1075241, EPI_ISL_1075242, EPI_ISL_1075243, EPI_ISL_1075244, EPI_ISL_1075245, EPI_ISL_1075246, EPI_ISL_1075247, EPI_ISL_1075248, EPI_ISL_1075249, EPI_ISL_1075250, EPI_ISL_1075251, EPI_ISL_1075252, EPI_ISL_1075253, EPI_ISL_1075254, EPI_ISL_1075255, EPI_ISL_1075256, EPI_ISL_1075257, EPI_ISL_1075258, EPI_ISL_1075259, EPI_ISL_1075260, EPI_ISL_1075261, EPI_ISL_1075262, EPI_ISL_1075263, EPI_ISL_1075264, EPI_ISL_1075265, EPI_ISL_1075266 | | | |
| see above | Houston Methodist Hospital | Houston Methodist Hospital | S. Wesley Long, Randall J. Olsen, Paul A. Christensen, Sishir Subedi, Robert Olson, James J. Davis, Matthew Ojeda Saavedra, Prasanti Yerramilli, Layne Pruitt, Kristina Reppond, Madison N. Shyer, Jessica Cambric, Ilya J. Finkelstein, Jimmy Gollihar, and James M. Musser |
| EPI_ISL_1075267 | Instituto Nacional de Medicina Genomica | Instituto Nacional de Medicina Genomica | Hidalgo-Miranda A, Mendoza-Vargas A, Reyes-Grajeda JP, Cedro-Tanda A, Alcaraz N, Gonzalez-Barrera D, Rangel-DeLeon D, Ramirez-Vega O, Sifuentes-Rojas C, Cisneros-Villanueva M, Herrera-Montalvo LA |
| EPI_ISL_1075268, EPI_ISL_1075269, EPI_ISL_1075270, EPI_ISL_1075271, EPI_ISL_1075272, EPI_ISL_1075273, EPI_ISL_1075274, EPI_ISL_1075275, EPI_ISL_1075276, EPI_ISL_1075277, EPI_ISL_1075278, EPI_ISL_1075279, EPI_ISL_1075280, EPI_ISL_1075281, EPI_ISL_1075282, EPI_ISL_1075283, EPI_ISL_1075284, EPI_ISL_1075285, EPI_ISL_1075286, EPI_ISL_1075287, EPI_ISL_1075288, EPI_ISL_1075289, EPI_ISL_1075290, EPI_ISL_1075291, EPI_ISL_1075292, EPI_ISL_1075293, EPI_ISL_1075294, EPI_ISL_1075295, EPI_ISL_1075296, EPI_ISL_1075297, EPI_ISL_1075298, EPI_ISL_1075299, EPI_ISL_1075300, EPI_ISL_1075301, EPI_ISL_1075302, EPI_ISL_1075303, EPI_ISL_1075304, EPI_ISL_1075305, EPI_ISL_1075306, EPI_ISL_1075307, EPI_ISL_1075308, EPI_ISL_1075309, EPI_ISL_1075310, EPI_ISL_1075311, EPI_ISL_1075312, EPI_ISL_1075313, EPI_ISL_1075314, EPI_ISL_1075315, EPI_ISL_1075316, EPI_ISL_1075317, EPI_ISL_1075318, EPI_ISL_1075319, EPI_ISL_1075320, EPI_ISL_1075321, EPI_ISL_1075322, EPI_ISL_1075323, EPI_ISL_1075324, EPI_ISL_1075325, EPI_ISL_1075326, EPI_ISL_1075327, EPI_ISL_1075328, EPI_ISL_1075329, EPI_ISL_1075330, EPI_ISL_1075331, EPI_ISL_1075332, EPI_ISL_1075333, EPI_ISL_1075334, EPI_ISL_1075335, EPI_ISL_1075336, EPI_ISL_1075337, EPI_ISL_1075338, EPI_ISL_1075339, EPI_ISL_1075340, EPI_ISL_1075341, EPI_ISL_1075342, EPI_ISL_1075343, EPI_ISL_1075344, EPI_ISL_1075345, EPI_ISL_1075346, EPI_ISL_1075347, EPI_ISL_1075348, EPI_ISL_1075349, EPI_ISL_1075351, EPI_ISL_1075352, EPI_ISL_1075353, EPI_ISL_1075354, EPI_ISL_1075355, EPI_ISL_1075356, EPI_ISL_1075357, EPI_ISL_1075358, EPI_ISL_1075359, EPI_ISL_1075360, EPI_ISL_1075361, EPI_ISL_1075362, EPI_ISL_1075363, EPI_ISL_1075364, EPI_ISL_1075365, EPI_ISL_1075366, EPI_ISL_1075367, EPI_ISL_1075368, EPI_ISL_1075369, EPI_ISL_1075370, EPI_ISL_1075371, EPI_ISL_1075372, EPI_ISL_1075373, EPI_ISL_1075374, EPI_ISL_1075375, EPI_ISL_1075376, EPI_ISL_1075377, EPI_ISL_1075378, | | | |

EPI_ISL_1075379, EPI_ISL_1075380, EPI_ISL_1075381, EPI_ISL_1075382, EPI_ISL_1075383, EPI_ISL_1075384, EPI_ISL_1075385, EPI_ISL_1075386, EPI_ISL_1075387, EPI_ISL_1075388, EPI_ISL_1075389, EPI_ISL_1075390, EPI_ISL_1075391, EPI_ISL_1075392, EPI_ISL_1075393, EPI_ISL_1075394, EPI_ISL_1075395, EPI_ISL_1075396, EPI_ISL_1075397, EPI_ISL_1075398, EPI_ISL_1075399, EPI_ISL_1075400,
EPI_ISL_1075401, EPI_ISL_1075402, EPI_ISL_1075403, EPI_ISL_1075404, EPI_ISL_1075405, EPI_ISL_1075406, EPI_ISL_1075407, EPI_ISL_1075408, EPI_ISL_1075409, EPI_ISL_1075410, EPI_ISL_1075411, EPI_ISL_1075412, EPI_ISL_1075413, EPI_ISL_1075414, EPI_ISL_1075415, EPI_ISL_1075416, EPI_ISL_1075417, EPI_ISL_1075418, EPI_ISL_1075419, EPI_ISL_1075420, EPI_ISL_1075421, EPI_ISL_1075422,
EPI_ISL_1075423, EPI_ISL_1075424, EPI_ISL_1075425, EPI_ISL_1075426, EPI_ISL_1075427, EPI_ISL_1075428, EPI_ISL_1075429, EPI_ISL_1075430, EPI_ISL_1075431, EPI_ISL_1075432, EPI_ISL_1075433, EPI_ISL_1075434, EPI_ISL_1075435, EPI_ISL_1075436, EPI_ISL_1075437, EPI_ISL_1075438, EPI_ISL_1075439, EPI_ISL_1075440, EPI_ISL_1075441, EPI_ISL_1075442, EPI_ISL_1075443, EPI_ISL_1075444,
EPI_ISL_1075445, EPI_ISL_1075446, EPI_ISL_1075447, EPI_ISL_1075448, EPI_ISL_1075449, EPI_ISL_1075450, EPI_ISL_1075451, EPI_ISL_1075452, EPI_ISL_1075453, EPI_ISL_1075454, EPI_ISL_1075455, EPI_ISL_1075456, EPI_ISL_1075457, EPI_ISL_1075458, EPI_ISL_1075459, EPI_ISL_1075460, EPI_ISL_1075461, EPI_ISL_1075462, EPI_ISL_1075463, EPI_ISL_1075464, EPI_ISL_1075465, EPI_ISL_1075466,
EPI_ISL_1075467, EPI_ISL_1075468, EPI_ISL_1075469, EPI_ISL_1075470, EPI_ISL_1075471, EPI_ISL_1075472, EPI_ISL_1075473, EPI_ISL_1075474, EPI_ISL_1075475, EPI_ISL_1075476, EPI_ISL_1075477, EPI_ISL_1075478, EPI_ISL_1075479, EPI_ISL_1075480, EPI_ISL_1075481, EPI_ISL_1075482, EPI_ISL_1075483, EPI_ISL_1075484, EPI_ISL_1075485, EPI_ISL_1075486, EPI_ISL_1075487, EPI_ISL_1075488,
EPI_ISL_1075489, EPI_ISL_1075490, EPI_ISL_1075491, EPI_ISL_1075492, EPI_ISL_1075493, EPI_ISL_1075494, EPI_ISL_1075495, EPI_ISL_1075496, EPI_ISL_1075497, EPI_ISL_1075498, EPI_ISL_1075499, EPI_ISL_1075500, EPI_ISL_1075501, EPI_ISL_1075502, EPI_ISL_1075503, EPI_ISL_1075504, EPI_ISL_1075505, EPI_ISL_1075506, EPI_ISL_1075507, EPI_ISL_1075508, EPI_ISL_1075509, EPI_ISL_1075510,
EPI_ISL_1075511, EPI_ISL_1075512, EPI_ISL_1075513, EPI_ISL_1075514, EPI_ISL_1075515, EPI_ISL_1075516, EPI_ISL_1075517, EPI_ISL_1075518, EPI_ISL_1075519, EPI_ISL_1075520, EPI_ISL_1075521, EPI_ISL_1075522, EPI_ISL_1075523, EPI_ISL_1075524, EPI_ISL_1075525, EPI_ISL_1075526, EPI_ISL_1075527, EPI_ISL_1075528, EPI_ISL_1075529, EPI_ISL_1075530, EPI_ISL_1075531, EPI_ISL_1075532,
EPI_ISL_1075533, EPI_ISL_1075534, EPI_ISL_1075535, EPI_ISL_1075536, EPI_ISL_1075537, EPI_ISL_1075538, EPI_ISL_1075539, EPI_ISL_1075540, EPI_ISL_1075541, EPI_ISL_1075542, EPI_ISL_1075543, EPI_ISL_1075544, EPI_ISL_1075545, EPI_ISL_1075546, EPI_ISL_1075547, EPI_ISL_1075548, EPI_ISL_1075549, EPI_ISL_1075550, EPI_ISL_1075551, EPI_ISL_1075552, EPI_ISL_1075553, EPI_ISL_1075554,
EPI_ISL_1075555, EPI_ISL_1075556, EPI_ISL_1075557, EPI_ISL_1075558, EPI_ISL_1075559, EPI_ISL_1075560, EPI_ISL_1075561, EPI_ISL_1075562, EPI_ISL_1075563, EPI_ISL_1075564, EPI_ISL_1075565, EPI_ISL_1075566, EPI_ISL_1075567, EPI_ISL_1075568, EPI_ISL_1075569, EPI_ISL_1075570, EPI_ISL_1075571, EPI_ISL_1075572, EPI_ISL_1075573, EPI_ISL_1075574, EPI_ISL_1075575, EPI_ISL_1075576,
EPI_ISL_1075577, EPI_ISL_1075578, EPI_ISL_1075579, EPI_ISL_1075580, EPI_ISL_1075581, EPI_ISL_1075582, EPI_ISL_1075583, EPI_ISL_1075584, EPI_ISL_1075585, EPI_ISL_1075586, EPI_ISL_1075587, EPI_ISL_1075588, EPI_ISL_1075589, EPI_ISL_1075590, EPI_ISL_1075591, EPI_ISL_1075592, EPI_ISL_1075593, EPI_ISL_1075594, EPI_ISL_1075595, EPI_ISL_1075596, EPI_ISL_1075597, EPI_ISL_1075598,
EPI_ISL_1075599, EPI_ISL_1075600, EPI_ISL_1075601, EPI_ISL_1075602, EPI_ISL_1075603, EPI_ISL_1075604, EPI_ISL_1075605, EPI_ISL_1075606, EPI_ISL_1075607, EPI_ISL_1075608, EPI_ISL_1075609, EPI_ISL_1075610, EPI_ISL_1075611, EPI_ISL_1075612, EPI_ISL_1075613, EPI_ISL_1075614, EPI_ISL_1075615, EPI_ISL_1075616, EPI_ISL_1075617, EPI_ISL_1075618, EPI_ISL_1075619, EPI_ISL_1075620,
EPI_ISL_1075621, EPI_ISL_1075622, EPI_ISL_1075623, EPI_ISL_1075624, EPI_ISL_1075625, EPI_ISL_1075626, EPI_ISL_1075627, EPI_ISL_1075628, EPI_ISL_1075629, EPI_ISL_1075630, EPI_ISL_1075631, EPI_ISL_1075632, EPI_ISL_1075633, EPI_ISL_1075634, EPI_ISL_1075635, EPI_ISL_1075636, EPI_ISL_1075637, EPI_ISL_1075638, EPI_ISL_1075639, EPI_ISL_1075640, EPI_ISL_1075641, EPI_ISL_1075642,
EPI_ISL_1075643, EPI_ISL_1075644, EPI_ISL_1075645, EPI_ISL_1075646, EPI_ISL_1075647, EPI_ISL_1075648, EPI_ISL_1075649, EPI_ISL_1075650, EPI_ISL_1075651, EPI_ISL_1075652, EPI_ISL_1075653, EPI_ISL_1075654, EPI_ISL_1075655, EPI_ISL_1075656

| | | | |
|---|---|---|---|
| see above | Houston Methodist Hospital | Houston Methodist Hospital | S. Wesley Long, Randall J. Olsen, Paul A. Christensen, Sishir Subedi, Robert Olson, James J. Davis, Matthew Ojeda Saavedra, Prasanti Yerramilli, Layne Pruitt, Kristina Reppond, Madison N. Shyer, Jessica Cambric, Ilya J. Finkelstein, Jimmy Gollihar, and James M. Musser |
| EPI_ISL_1075657 | Instituto Nacional de Medicina Genomica | Instituto Nacional de Medicina Genomica | Hidalgo-Miranda A, Mendoza-Vargas A, Reyes-Grajeda JP, Cedro-Tanda A, Alcaraz N, Gonzalez-Barrera D, Rangel-DeLeon D, Ramirez-Vega O, Sifuentes-Rojas C, Cisneros-Villanueva M, Herrera-Montalvo LA. |

EPI_ISL_1075658, EPI_ISL_1075659, EPI_ISL_1075660, EPI_ISL_1075661, EPI_ISL_1075662, EPI_ISL_1075663, EPI_ISL_1075664, EPI_ISL_1075665, EPI_ISL_1075666, EPI_ISL_1075667, EPI_ISL_1075668, EPI_ISL_1075669, EPI_ISL_1075670, EPI_ISL_1075671, EPI_ISL_1075672, EPI_ISL_1075673, EPI_ISL_1075674, EPI_ISL_1075675, EPI_ISL_1075676, EPI_ISL_1075677, EPI_ISL_1075678, EPI_ISL_1075679,
EPI_ISL_1075680, EPI_ISL_1075681, EPI_ISL_1075682, EPI_ISL_1075683, EPI_ISL_1075684, EPI_ISL_1075685, EPI_ISL_1075686, EPI_ISL_1075687, EPI_ISL_1075688, EPI_ISL_1075689, EPI_ISL_1075690, EPI_ISL_1075691, EPI_ISL_1075692, EPI_ISL_1075693, EPI_ISL_1075694, EPI_ISL_1075695, EPI_ISL_1075696, EPI_ISL_1075697, EPI_ISL_1075698, EPI_ISL_1075699, EPI_ISL_1075700, EPI_ISL_1075701,
EPI_ISL_1075702, EPI_ISL_1075703, EPI_ISL_1075704, EPI_ISL_1075705, EPI_ISL_1075706, EPI_ISL_1075707, EPI_ISL_1075708, EPI_ISL_1075709, EPI_ISL_1075710, EPI_ISL_1075711, EPI_ISL_1075712, EPI_ISL_1075713, EPI_ISL_1075714, EPI_ISL_1075715, EPI_ISL_1075716, EPI_ISL_1075717, EPI_ISL_1075718, EPI_ISL_1075719, EPI_ISL_1075720, EPI_ISL_1075721, EPI_ISL_1075722, EPI_ISL_1075723,
EPI_ISL_1075724, EPI_ISL_1075725, EPI_ISL_1075726, EPI_ISL_1075727, EPI_ISL_1075728, EPI_ISL_1075729, EPI_ISL_1075730, EPI_ISL_1075731, EPI_ISL_1075732, EPI_ISL_1075733, EPI_ISL_1075734, EPI_ISL_1075735, EPI_ISL_1075736, EPI_ISL_1075737, EPI_ISL_1075738, EPI_ISL_1075739, EPI_ISL_1075740, EPI_ISL_1075741, EPI_ISL_1075742, EPI_ISL_1075743, EPI_ISL_1075744, EPI_ISL_1075745,
EPI_ISL_1075746, EPI_ISL_1075747, EPI_ISL_1075748, EPI_ISL_1075749, EPI_ISL_1075750, EPI_ISL_1075751, EPI_ISL_1075752, EPI_ISL_1075753, EPI_ISL_1075754, EPI_ISL_1075755, EPI_ISL_1075756, EPI_ISL_1075757, EPI_ISL_1075758, EPI_ISL_1075759, EPI_ISL_1075760, EPI_ISL_1075761, EPI_ISL_1075762, EPI_ISL_1075763, EPI_ISL_1075764, EPI_ISL_1075765, EPI_ISL_1075766, EPI_ISL_1075767,
EPI_ISL_1075768, EPI_ISL_1075769, EPI_ISL_1075770, EPI_ISL_1075771, EPI_ISL_1075772, EPI_ISL_1075773, EPI_ISL_1075774, EPI_ISL_1075775, EPI_ISL_1075776, EPI_ISL_1075777, EPI_ISL_1075778, EPI_ISL_1075779, EPI_ISL_1075780, EPI_ISL_1075781, EPI_ISL_1075782, EPI_ISL_1075783, EPI_ISL_1075784, EPI_ISL_1075785, EPI_ISL_1075786, EPI_ISL_1075787, EPI_ISL_1075788, EPI_ISL_1075789,
EPI_ISL_1075790, EPI_ISL_1075791, EPI_ISL_1075792, EPI_ISL_1075793, EPI_ISL_1075794, EPI_ISL_1075795, EPI_ISL_1075796, EPI_ISL_1075797, EPI_ISL_1075798, EPI_ISL_1075799, EPI_ISL_1075800, EPI_ISL_1075801, EPI_ISL_1075802, EPI_ISL_1075803, EPI_ISL_1075804, EPI_ISL_1075805, EPI_ISL_1075806, EPI_ISL_1075807, EPI_ISL_1075808, EPI_ISL_1075809, EPI_ISL_1075810, EPI_ISL_1075811,
EPI_ISL_1075812, EPI_ISL_1075813, EPI_ISL_1075814, EPI_ISL_1075815, EPI_ISL_1075816, EPI_ISL_1075817, EPI_ISL_1075818, EPI_ISL_1075819, EPI_ISL_1075820, EPI_ISL_1075821, EPI_ISL_1075822, EPI_ISL_1075823, EPI_ISL_1075824, EPI_ISL_1075825, EPI_ISL_1075826, EPI_ISL_1075827, EPI_ISL_1075828, EPI_ISL_1075829, EPI_ISL_1075830, EPI_ISL_1075831, EPI_ISL_1075832, EPI_ISL_1075833,
EPI_ISL_1075834, EPI_ISL_1075835, EPI_ISL_1075836, EPI_ISL_1075837, EPI_ISL_1075838, EPI_ISL_1075839, EPI_ISL_1075840, EPI_ISL_1075841, EPI_ISL_1075842, EPI_ISL_1075843, EPI_ISL_1075844, EPI_ISL_1075845, EPI_ISL_1075846, EPI_ISL_1075847, EPI_ISL_1075848, EPI_ISL_1075849, EPI_ISL_1075850, EPI_ISL_1075851, EPI_ISL_1075852, EPI_ISL_1075853, EPI_ISL_1075854, EPI_ISL_1075855,
EPI_ISL_1075856, EPI_ISL_1075857, EPI_ISL_1075858, EPI_ISL_1075859, EPI_ISL_1075860, EPI_ISL_1075861, EPI_ISL_1075862, EPI_ISL_1075863, EPI_ISL_1075864, EPI_ISL_1075865, EPI_ISL_1075866, EPI_ISL_1075867, EPI_ISL_1075868, EPI_ISL_1075869, EPI_ISL_1075870, EPI_ISL_1075871, EPI_ISL_1075872, EPI_ISL_1075873, EPI_ISL_1075874, EPI_ISL_1075875, EPI_ISL_1075876, EPI_ISL_1075877,
EPI_ISL_1075878, EPI_ISL_1075879, EPI_ISL_1075880, EPI_ISL_1075881, EPI_ISL_1075882, EPI_ISL_1075883, EPI_ISL_1075884, EPI_ISL_1075885, EPI_ISL_1075886, EPI_ISL_1075887, EPI_ISL_1075888, EPI_ISL_1075889, EPI_ISL_1075890, EPI_ISL_1075891, EPI_ISL_1075892, EPI_ISL_1075893, EPI_ISL_1075894, EPI_ISL_1075895, EPI_ISL_1075896, EPI_ISL_1075897, EPI_ISL_1075898, EPI_ISL_1075899,
EPI_ISL_1075900, EPI_ISL_1075901, EPI_ISL_1075902, EPI_ISL_1075903, EPI_ISL_1075904, EPI_ISL_1075905, EPI_ISL_1075906, EPI_ISL_1075907, EPI_ISL_1075908, EPI_ISL_1075909, EPI_ISL_1075910, EPI_ISL_1075911, EPI_ISL_1075912, EPI_ISL_1075913, EPI_ISL_1075914, EPI_ISL_1075915, EPI_ISL_1075916, EPI_ISL_1075917, EPI_ISL_1075918, EPI_ISL_1075919, EPI_ISL_1075920, EPI_ISL_1075921,
EPI_ISL_1075922, EPI_ISL_1075923, EPI_ISL_1075924, EPI_ISL_1075925, EPI_ISL_1075926, EPI_ISL_1075927, EPI_ISL_1075928, EPI_ISL_1075929, EPI_ISL_1075930, EPI_ISL_1075931, EPI_ISL_1075932, EPI_ISL_1075933, EPI_ISL_1075934, EPI_ISL_1075935, EPI_ISL_1075936, EPI_ISL_1075937, EPI_ISL_1075938, EPI_ISL_1075939, EPI_ISL_1075940, EPI_ISL_1075941, EPI_ISL_1075942, EPI_ISL_1075943,
EPI_ISL_1075944, EPI_ISL_1075945, EPI_ISL_1075946, EPI_ISL_1075947, EPI_ISL_1075948, EPI_ISL_1075949, EPI_ISL_1075950, EPI_ISL_1075951, EPI_ISL_1075952, EPI_ISL_1075953, EPI_ISL_1075954, EPI_ISL_1075955, EPI_ISL_1075956, EPI_ISL_1075957, EPI_ISL_1075958, EPI_ISL_1075959, EPI_ISL_1075960, EPI_ISL_1075961, EPI_ISL_1075962, EPI_ISL_1075963, EPI_ISL_1075964, EPI_ISL_1075965,
EPI_ISL_1075966, EPI_ISL_1075967, EPI_ISL_1075968, EPI_ISL_1075969, EPI_ISL_1075970, EPI_ISL_1075971, EPI_ISL_1075972, EPI_ISL_1075973, EPI_ISL_1075974, EPI_ISL_1075975, EPI_ISL_1075976, EPI_ISL_1075977, EPI_ISL_1075978, EPI_ISL_1075979, EPI_ISL_1075980, EPI_ISL_1075981, EPI_ISL_1075982, EPI_ISL_1075983, EPI_ISL_1075984, EPI_ISL_1075985, EPI_ISL_1075986, EPI_ISL_1075987,
EPI_ISL_1075988, EPI_ISL_1075989, EPI_ISL_1075990, EPI_ISL_1075991, EPI_ISL_1075992, EPI_ISL_1075993, EPI_ISL_1075994, EPI_ISL_1075995, EPI_ISL_1075996, EPI_ISL_1075997, EPI_ISL_1075998, EPI_ISL_1075999, EPI_ISL_1076000, EPI_ISL_1076001, EPI_ISL_1076002, EPI_ISL_1076003, EPI_ISL_1076004, EPI_ISL_1076005, EPI_ISL_1076006, EPI_ISL_1076007, EPI_ISL_1076008, EPI_ISL_1076009,
EPI_ISL_1076010, EPI_ISL_1076011, EPI_ISL_1076012, EPI_ISL_1076013, EPI_ISL_1076014, EPI_ISL_1076015, EPI_ISL_1076016, EPI_ISL_1076017, EPI_ISL_1076018, EPI_ISL_1076019, EPI_ISL_1076020, EPI_ISL_1076021, EPI_ISL_1076022, EPI_ISL_1076024, EPI_ISL_1076025, EPI_ISL_1076026, EPI_ISL_1076027, EPI_ISL_1076028, EPI_ISL_1076029, EPI_ISL_1076030, EPI_ISL_1076031, EPI_ISL_1076032,
EPI_ISL_1076033, EPI_ISL_1076034, EPI_ISL_1076035, EPI_ISL_1076036, EPI_ISL_1076037, EPI_ISL_1076038, EPI_ISL_1076039, EPI_ISL_1076040, EPI_ISL_1076041, EPI_ISL_1076042, EPI_ISL_1076043, EPI_ISL_1076044, EPI_ISL_1076045, EPI_ISL_1076046, EPI_ISL_1076047, EPI_ISL_1076048, EPI_ISL_1076049, EPI_ISL_1076050, EPI_ISL_1076051, EPI_ISL_1076052, EPI_ISL_1076053, EPI_ISL_1076054,
EPI_ISL_1076055, EPI_ISL_1076056, EPI_ISL_1076057, EPI_ISL_1076058, EPI_ISL_1076059, EPI_ISL_1076060, EPI_ISL_1076061, EPI_ISL_1076062, EPI_ISL_1076063, EPI_ISL_1076064, EPI_ISL_1076065, EPI_ISL_1076066, EPI_ISL_1076067, EPI_ISL_1076068, EPI_ISL_1076069, EPI_ISL_1076070, EPI_ISL_1076071, EPI_ISL_1076072, EPI_ISL_1076073, EPI_ISL_1076074, EPI_ISL_1076075, EPI_ISL_1076076,
EPI_ISL_1076077, EPI_ISL_1076078, EPI_ISL_1076079, EPI_ISL_1076080, EPI_ISL_1076081, EPI_ISL_1076082, EPI_ISL_1076083, EPI_ISL_1076084, EPI_ISL_1076085, EPI_ISL_1076086, EPI_ISL_1076087, EPI_ISL_1076088, EPI_ISL_1076089, EPI_ISL_1076090, EPI_ISL_1076091, EPI_ISL_1076092, EPI_ISL_1076093, EPI_ISL_1076094, EPI_ISL_1076095, EPI_ISL_1076096, EPI_ISL_1076097, EPI_ISL_1076098,
EPI_ISL_1076099, EPI_ISL_1076100, EPI_ISL_1076101, EPI_ISL_1076102, EPI_ISL_1076103, EPI_ISL_1076104, EPI_ISL_1076105, EPI_ISL_1076106, EPI_ISL_1076107, EPI_ISL_1076108, EPI_ISL_1076109, EPI_ISL_1076110, EPI_ISL_1076111, EPI_ISL_1076112, EPI_ISL_1076113, EPI_ISL_1076114, EPI_ISL_1076115, EPI_ISL_1076116, EPI_ISL_1076117, EPI_ISL_1076118, EPI_ISL_1076119, EPI_ISL_1076120,
EPI_ISL_1076121, EPI_ISL_1076122, EPI_ISL_1076123, EPI_ISL_1076124, EPI_ISL_1076125, EPI_ISL_1076126, EPI_ISL_1076127, EPI_ISL_1076128, EPI_ISL_1076129, EPI_ISL_1076130, EPI_ISL_1076131, EPI_ISL_1076132, EPI_ISL_1076133, EPI_ISL_1076134, EPI_ISL_1076135, EPI_ISL_1076136, EPI_ISL_1076137, EPI_ISL_1076138, EPI_ISL_1076139, EPI_ISL_1076140, EPI_ISL_1076141, EPI_ISL_1076142,
EPI_ISL_1076143, EPI_ISL_1076144, EPI_ISL_1076145, EPI_ISL_1076146, EPI_ISL_1076147, EPI_ISL_1076148, EPI_ISL_1076149, EPI_ISL_1076150, EPI_ISL_1076151, EPI_ISL_1076152, EPI_ISL_1076153, EPI_ISL_1076154, EPI_ISL_1076155, EPI_ISL_1076156, EPI_ISL_1076157, EPI_ISL_1076158, EPI_ISL_1076159, EPI_ISL_1076160, EPI_ISL_1076161, EPI_ISL_1076162, EPI_ISL_1076163, EPI_ISL_1076164,
EPI_ISL_1076165, EPI_ISL_1076166, EPI_ISL_1076167, EPI_ISL_1076168, EPI_ISL_1076169, EPI_ISL_1076170, EPI_ISL_1076171, EPI_ISL_1076172, EPI_ISL_1076173, EPI_ISL_1076174, EPI_ISL_1076175, EPI_ISL_1076176, EPI_ISL_1076177, EPI_ISL_1076178, EPI_ISL_1076179, EPI_ISL_1076180, EPI_ISL_1076181, EPI_ISL_1076182, EPI_ISL_1076183, EPI_ISL_1076184, EPI_ISL_1076185, EPI_ISL_1076186,
EPI_ISL_1076187, EPI_ISL_1076188, EPI_ISL_1076189, EPI_ISL_1076190, EPI_ISL_1076191, EPI_ISL_1076192, EPI_ISL_1076193, EPI_ISL_1076194, EPI_ISL_1076195, EPI_ISL_1076196, EPI_ISL_1076197, EPI_ISL_1076198, EPI_ISL_1076199, EPI_ISL_1076200, EPI_ISL_1076201, EPI_ISL_1076202, EPI_ISL_1076203, EPI_ISL_1076204, EPI_ISL_1076205, EPI_ISL_1076206, EPI_ISL_1076207, EPI_ISL_1076208,
EPI_ISL_1076209, EPI_ISL_1076210, EPI_ISL_1076211, EPI_ISL_1076212, EPI_ISL_1076213, EPI_ISL_1076214, EPI_ISL_1076215, EPI_ISL_1076216, EPI_ISL_1076217, EPI_ISL_1076218, EPI_ISL_1076219, EPI_ISL_1076220, EPI_ISL_1076221, EPI_ISL_1076222, EPI_ISL_1076223, EPI_ISL_1076224, EPI_ISL_1076225, EPI_ISL_1076226, EPI_ISL_1076227, EPI_ISL_1076228, EPI_ISL_1076229, EPI_ISL_1076230,
EPI_ISL_1076231, EPI_ISL_1076232, EPI_ISL_1076233, EPI_ISL_1076234, EPI_ISL_1076235, EPI_ISL_1076236, EPI_ISL_1076237, EPI_ISL_1076238, EPI_ISL_1076239, EPI_ISL_1076240, EPI_ISL_1076241, EPI_ISL_1076242, EPI_ISL_1076243, EPI_ISL_1076244, EPI_ISL_1076245, EPI_ISL_1076246, EPI_ISL_1076247, EPI_ISL_1076248, EPI_ISL_1076249, EPI_ISL_1076250, EPI_ISL_1076251, EPI_ISL_1076252,
EPI_ISL_1076253, EPI_ISL_1076254, EPI_ISL_1076255, EPI_ISL_1076256, EPI_ISL_1076257, EPI_ISL_1076258, EPI_ISL_1076259, EPI_ISL_1076260, EPI_ISL_1076261, EPI_ISL_1076262, EPI_ISL_1076263, EPI_ISL_1076264, EPI_ISL_1076265, EPI_ISL_1076266, EPI_ISL_1076267, EPI_ISL_1076268, EPI_ISL_1076269, EPI_ISL_1076270, EPI_ISL_1076271, EPI_ISL_1076272, EPI_ISL_1076273, EPI_ISL_1076274,
EPI_ISL_1076275, EPI_ISL_1076276, EPI_ISL_1076277, EPI_ISL_1076278, EPI_ISL_1076279, EPI_ISL_1076280, EPI_ISL_1076281, EPI_ISL_1076282, EPI_ISL_1076283, EPI_ISL_1076284, EPI_ISL_1076285, EPI_ISL_1076286, EPI_ISL_1076287, EPI_ISL_1076288, EPI_ISL_1076289, EPI_ISL_1076290, EPI_ISL_1076291, EPI_ISL_1076292, EPI_ISL_1076293, EPI_ISL_1076294, EPI_ISL_1076295, EPI_ISL_1076296,
EPI_ISL_1076297, EPI_ISL_1076298, EPI_ISL_1076299, EPI_ISL_1076300, EPI_ISL_1076301, EPI_ISL_1076302, EPI_ISL_1076303, EPI_ISL_1076304, EPI_ISL_1076305, EPI_ISL_1076307, EPI_ISL_1076308, EPI_ISL_1076309, EPI_ISL_1076310, EPI_ISL_1076311, EPI_ISL_1076312, EPI_ISL_1076313, EPI_ISL_1076314, EPI_ISL_1076315, EPI_ISL_1076316, EPI_ISL_1076317, EPI_ISL_1076318, EPI_ISL_1076319,
EPI_ISL_1076320, EPI_ISL_1076321, EPI_ISL_1076322, EPI_ISL_1076323, EPI_ISL_1076324, EPI_ISL_1076325, EPI_ISL_1076326, EPI_ISL_1076327, EPI_ISL_1076328, EPI_ISL_1076329, EPI_ISL_1076330, EPI_ISL_1076331, EPI_ISL_1076332, EPI_ISL_1076333, EPI_ISL_1076334, EPI_ISL_1076335, EPI_ISL_1076336, EPI_ISL_1076337, EPI_ISL_1076338, EPI_ISL_1076339, EPI_ISL_1076340, EPI_ISL_1076341,
EPI_ISL_1076342, EPI_ISL_1076343, EPI_ISL_1076344, EPI_ISL_1076345, EPI_ISL_1076346, EPI_ISL_1076347, EPI_ISL_1076348, EPI_ISL_1076349, EPI_ISL_1076350, EPI_ISL_1076351, EPI_ISL_1076352, EPI_ISL_1076357, EPI_ISL_1076360, EPI_ISL_1076365, EPI_ISL_1076368, EPI_ISL_1076372, EPI_ISL_1076375, EPI_ISL_1076378, EPI_ISL_1076382, EPI_ISL_1076386, EPI_ISL_1076389, EPI_ISL_1076393,
EPI_ISL_1076396, EPI_ISL_1076400, EPI_ISL_1076403, EPI_ISL_1076406, EPI_ISL_1076410, EPI_ISL_1076413, EPI_ISL_1076416, EPI_ISL_1076420, EPI_ISL_1076423, EPI_ISL_1076427, EPI_ISL_1076430, EPI_ISL_1076434, EPI_ISL_1076437, EPI_ISL_1076440, EPI_ISL_1076444, EPI_ISL_1076447, EPI_ISL_1076451, EPI_ISL_1076454, EPI_ISL_1076458, EPI_ISL_1076461, EPI_ISL_1076462, EPI_ISL_1076463,
EPI_ISL_1076464, EPI_ISL_1076465, EPI_ISL_1076466, EPI_ISL_1076467, EPI_ISL_1076468, EPI_ISL_1076469, EPI_ISL_1076470, EPI_ISL_1076471, EPI_ISL_1076472, EPI_ISL_1076473, EPI_ISL_1076474, EPI_ISL_1076475, EPI_ISL_1076476, EPI_ISL_1076477, EPI_ISL_1076478, EPI_ISL_1076479, EPI_ISL_1076480, EPI_ISL_1076481, EPI_ISL_1076482, EPI_ISL_1076483, EPI_ISL_1076484, EPI_ISL_1076485,
EPI_ISL_1076486, EPI_ISL_1076487, EPI_ISL_1076488, EPI_ISL_1076489, EPI_ISL_1076490, EPI_ISL_1076491, EPI_ISL_1076492, EPI_ISL_1076493, EPI_ISL_1076494, EPI_ISL_1076495, EPI_ISL_1076496, EPI_ISL_1076497, EPI_ISL_1076498, EPI_ISL_1076499, EPI_ISL_1076500, EPI_ISL_1076501, EPI_ISL_1076502, EPI_ISL_1076503, EPI_ISL_1076504, EPI_ISL_1076505, EPI_ISL_1076506, EPI_ISL_1076507,
EPI_ISL_1076508, EPI_ISL_1076509, EPI_ISL_1076510, EPI_ISL_1076511, EPI_ISL_1076512, EPI_ISL_1076513, EPI_ISL_1076514, EPI_ISL_1076515, EPI_ISL_1076516, EPI_ISL_1076517, EPI_ISL_1076518, EPI_ISL_1076519, EPI_ISL_1076520, EPI_ISL_1076521, EPI_ISL_1076522, EPI_ISL_1076523, EPI_ISL_1076524, EPI_ISL_1076525, EPI_ISL_1076526, EPI_ISL_1076527, EPI_ISL_1076528, EPI_ISL_1076529,
EPI_ISL_1076530, EPI_ISL_1076531, EPI_ISL_1076532, EPI_ISL_1076533, EPI_ISL_1076534, EPI_ISL_1076535, EPI_ISL_1076536, EPI_ISL_1076538, EPI_ISL_1076539, EPI_ISL_1076540, EPI_ISL_1076541, EPI_ISL_1076542, EPI_ISL_1076543, EPI_ISL_1076544, EPI_ISL_1076545, EPI_ISL_1076546, EPI_ISL_1076547, EPI_ISL_1076548, EPI_ISL_1076549, EPI_ISL_1076550, EPI_ISL_1076551, EPI_ISL_1076552,
EPI_ISL_1076553, EPI_ISL_1076554, EPI_ISL_1076555, EPI_ISL_1076556, EPI_ISL_1076557, EPI_ISL_1076558, EPI_ISL_1076559, EPI_ISL_1076560, EPI_ISL_1076561, EPI_ISL_1076562, EPI_ISL_1076563, EPI_ISL_1076564, EPI_ISL_1076565, EPI_ISL_1076566, EPI_ISL_1076567, EPI_ISL_1076568, EPI_ISL_1076569, EPI_ISL_1076570, EPI_ISL_1076571, EPI_ISL_1076572, EPI_ISL_1076573, EPI_ISL_1076574,
EPI_ISL_1076575, EPI_ISL_1076576, EPI_ISL_1076577, EPI_ISL_1076578, EPI_ISL_1076579, EPI_ISL_1076580, EPI_ISL_1076581, EPI_ISL_1076582, EPI_ISL_1076583, EPI_ISL_1076584, EPI_ISL_1076585, EPI_ISL_1076586, EPI_ISL_1076587, EPI_ISL_1076588, EPI_ISL_1076589, EPI_ISL_1076590, EPI_ISL_1076591, EPI_ISL_1076592, EPI_ISL_1076593, EPI_ISL_1076594, EPI_ISL_1076595, EPI_ISL_1076596,
EPI_ISL_1076597, EPI_ISL_1076598, EPI_ISL_1076599, EPI_ISL_1076600, EPI_ISL_1076601, EPI_ISL_1076602, EPI_ISL_1076603, EPI_ISL_1076604, EPI_ISL_1076605, EPI_ISL_1076606, EPI_ISL_1076607, EPI_ISL_1076608, EPI_ISL_1076609, EPI_ISL_1076610, EPI_ISL_1076611, EPI_ISL_1076612, EPI_ISL_1076613, EPI_ISL_1076614, EPI_ISL_1076615, EPI_ISL_1076616, EPI_ISL_1076617, EPI_ISL_1076618,
EPI_ISL_1076619, EPI_ISL_1076620, EPI_ISL_1076621, EPI_ISL_1076622, EPI_ISL_1076623, EPI_ISL_1076624, EPI_ISL_1076625, EPI_ISL_1076626, EPI_ISL_1076627, EPI_ISL_1076628, EPI_ISL_1076629, EPI_ISL_1076630, EPI_ISL_1076631, EPI_ISL_1076632, EPI_ISL_1076633, EPI_ISL_1076634, EPI_ISL_1076635, EPI_ISL_1076636, EPI_ISL_1076637, EPI_ISL_1076638, EPI_ISL_1076639, EPI_ISL_1076640,
EPI_ISL_1076641, EPI_ISL_1076642, EPI_ISL_1076643, EPI_ISL_1076644, EPI_ISL_1076645, EPI_ISL_1076646, EPI_ISL_1076647, EPI_ISL_1076648, EPI_ISL_1076649, EPI_ISL_1076650, EPI_ISL_1076651, EPI_ISL_1076652, EPI_ISL_1076653, EPI_ISL_1076654, EPI_ISL_1076655, EPI_ISL_1076656, EPI_ISL_1076657, EPI_ISL_1076658, EPI_ISL_1076659, EPI_ISL_1076660, EPI_ISL_1076661, EPI_ISL_1076662,
EPI_ISL_1076663, EPI_ISL_1076664, EPI_ISL_1076665, EPI_ISL_1076666, EPI_ISL_1076667, EPI_ISL_1076668, EPI_ISL_1076669, EPI_ISL_1076670, EPI_ISL_1076671, EPI_ISL_1076672, EPI_ISL_1076673, EPI_ISL_1076674, EPI_ISL_1076675, EPI_ISL_1076676, EPI_ISL_1076677, EPI_ISL_1076678, EPI_ISL_1076679, EPI_ISL_1076680, EPI_ISL_1076681, EPI_ISL_1076682, EPI_ISL_1076683, EPI_ISL_1076684,
EPI_ISL_1076685, EPI_ISL_1076686, EPI_ISL_1076687, EPI_ISL_1076688, EPI_ISL_1076690, EPI_ISL_1076691, EPI_ISL_1076692, EPI_ISL_1076693, EPI_ISL_1076694, EPI_ISL_1076695, EPI_ISL_1076696, EPI_ISL_1076697, EPI_ISL_1076698, EPI_ISL_1076699, EPI_ISL_1076700, EPI_ISL_1076701, EPI_ISL_1076702, EPI_ISL_1076703, EPI_ISL_1076704, EPI_ISL_1076705, EPI_ISL_1076706, EPI_ISL_1076707,
EPI_ISL_1076708, EPI_ISL_1076709, EPI_ISL_1076710, EPI_ISL_1076711, EPI_ISL_1076712, EPI_ISL_1076713, EPI_ISL_1076714, EPI_ISL_1076715, EPI_ISL_1076716, EPI_ISL_1076717, EPI_ISL_1076718, EPI_ISL_1076719, EPI_ISL_1076720, EPI_ISL_1076721, EPI_ISL_1076722, EPI_ISL_1076723, EPI_ISL_1076724, EPI_ISL_1076725, EPI_ISL_1076726, EPI_ISL_1076727, EPI_ISL_1076728, EPI_ISL_1076729,
EPI_ISL_1076730, EPI_ISL_1076731, EPI_ISL_1076732, EPI_ISL_1076733, EPI_ISL_1076734, EPI_ISL_1076735, EPI_ISL_1076736, EPI_ISL_1076737, EPI_ISL_1076738, EPI_ISL_1076739, EPI_ISL_1076740, EPI_ISL_1076741, EPI_ISL_1076742, EPI_ISL_1076743, EPI_ISL_1076744, EPI_ISL_1076745, EPI_ISL_1076746, EPI_ISL_1076747, EPI_ISL_1076748, EPI_ISL_1076749, EPI_ISL_1076750, EPI_ISL_1076751,
EPI_ISL_1076752, EPI_ISL_1076753, EPI_ISL_1076754, EPI_ISL_1076755, EPI_ISL_1076756, EPI_ISL_1076757, EPI_ISL_1076758, EPI_ISL_1076759, EPI_ISL_1076760, EPI_ISL_1076761, EPI_ISL_1076762, EPI_ISL_1076763, EPI_ISL_1076764, EPI_ISL_1076765, EPI_ISL_1076766, EPI_ISL_1076767, EPI_ISL_1076768, EPI_ISL_1076769, EPI_ISL_1076770, EPI_ISL_1076771, EPI_ISL_1076772, EPI_ISL_1076773,
EPI_ISL_1076774, EPI_ISL_1076775, EPI_ISL_1076776, EPI_ISL_1076777, EPI_ISL_1076778, EPI_ISL_1076779, EPI_ISL_1076780, EPI_ISL_1076781, EPI_ISL_1076782, EPI_ISL_1076783, EPI_ISL_1076784, EPI_ISL_1076785, EPI_ISL_1076786, EPI_ISL_1076787, EPI_ISL_1076788, EPI_ISL_1076789, EPI_ISL_1076790, EPI_ISL_1076791, EPI_ISL_1076792, EPI_ISL_1076793, EPI_ISL_1076794, EPI_ISL_1076795,
EPI_ISL_1076796, EPI_ISL_1076797, EPI_ISL_1076798, EPI_ISL_1076799, EPI_ISL_1076800, EPI_ISL_1076801, EPI_ISL_1076802, EPI_ISL_1076803, EPI_ISL_1076804, EPI_ISL_1076805, EPI_ISL_1076806, EPI_ISL_1076807, EPI_ISL_1076808, EPI_ISL_1076809, EPI_ISL_1076810, EPI_ISL_1076811, EPI_ISL_1076812, EPI_ISL_1076813, EPI_ISL_1076814, EPI_ISL_1076815, EPI_ISL_1076816, EPI_ISL_1076817,
EPI_ISL_1076818, EPI_ISL_1076819, EPI_ISL_1076820, EPI_ISL_1076821, EPI_ISL_1076822, EPI_ISL_1076823, EPI_ISL_1076824, EPI_ISL_1076825, EPI_ISL_1076826, EPI_ISL_1076827, EPI_ISL_1076828, EPI_ISL_1076829, EPI_ISL_1076830, EPI_ISL_1076831, EPI_ISL_1076832, EPI_ISL_1076833, EPI_ISL_1076834, EPI_ISL_1076835, EPI_ISL_1076836, EPI_ISL_1076837, EPI_ISL_1076838, EPI_ISL_1076839,
EPI_ISL_1076840, EPI_ISL_1076841, EPI_ISL_1076842, EPI_ISL_1076843, EPI_ISL_1076844, EPI_ISL_1076845, EPI_ISL_1076846, EPI_ISL_1076847, EPI_ISL_1076848, EPI_ISL_1076849, EPI_ISL_1076850, EPI_ISL_1076851, EPI_ISL_1076852, EPI_ISL_1076853, EPI_ISL_1076854, EPI_ISL_1076855, EPI_ISL_1076856, EPI_ISL_1076857, EPI_ISL_1076858, EPI_ISL_1076859, EPI_ISL_1076860, EPI_ISL_1076861,
EPI_ISL_1076862, EPI_ISL_1076863, EPI_ISL_1076864, EPI_ISL_1076865, EPI_ISL_1076866, EPI_ISL_1076867, EPI_ISL_1076868, EPI_ISL_1076869, EPI_ISL_1076870, EPI_ISL_1076871, EPI_ISL_1076872, EPI_ISL_1076873, EPI_ISL_1076874, EPI_ISL_1076875, EPI_ISL_1076876, EPI_ISL_1076877, EPI_ISL_1076878, EPI_ISL_1076879, EPI_ISL_1076880, EPI_ISL_1076881, EPI_ISL_1076882, EPI_ISL_1076883,
EPI_ISL_1076884, EPI_ISL_1076885, EPI_ISL_1076886, EPI_ISL_1076887, EPI_ISL_1076888, EPI_ISL_1076889, EPI_ISL_1076890, EPI_ISL_1076891, EPI_ISL_1076892, EPI_ISL_1076893, EPI_ISL_1076894, EPI_ISL_1076895, EPI_ISL_1076896, EPI_ISL_1076897, EPI_ISL_1076898, EPI_ISL_1076899, EPI_ISL_1076900, EPI_ISL_1076901, EPI_ISL_1076902, EPI_ISL_1076903, EPI_ISL_1076904, EPI_ISL_1076905,
EPI_ISL_1076906, EPI_ISL_1076907, EPI_ISL_1076908, EPI_ISL_1076909, EPI_ISL_1076910, EPI_ISL_1076911, EPI_ISL_1076912, EPI_ISL_1076913, EPI_ISL_1076914, EPI_ISL_1076915, EPI_ISL_1076916, EPI_ISL_1076917, EPI_ISL_1076918, EPI_ISL_1076919, EPI_ISL_1076920, EPI_ISL_1076921, EPI_ISL_1076922, EPI_ISL_1076923, EPI_ISL_1076924, EPI_ISL_1076925, EPI_ISL_1076926, EPI_ISL_1076927,
EPI_ISL_1076928, EPI_ISL_1076929, EPI_ISL_1076930, EPI_ISL_1076931, EPI_ISL_1076932, EPI_ISL_1076933, EPI_ISL_1076934, EPI_ISL_1076935, EPI_ISL_1076936, EPI_ISL_1076937, EPI_ISL_1076938, EPI_ISL_1076939, EPI_ISL_1076940, EPI_ISL_1076941, EPI_ISL_1076942, EPI_ISL_1076943, EPI_ISL_1076944, EPI_ISL_1076945, EPI_ISL_1076946, EPI_ISL_1076947, EPI_ISL_1076948, EPI_ISL_1076949,
EPI_ISL_1076950, EPI_ISL_1076951, EPI_ISL_1076952, EPI_ISL_1076953, EPI_ISL_1076954, EPI_ISL_1076955, EPI_ISL_1076956, EPI_ISL_1076957, EPI_ISL_1076958, EPI_ISL_1076959, EPI_ISL_1076960, EPI_ISL_1076961, EPI_ISL_1076962, EPI_ISL_1076963, EPI_ISL_1076964, EPI_ISL_1076965, EPI_ISL_1076966, EPI_ISL_1076967, EPI_ISL_1076968, EPI_ISL_1076969, EPI_ISL_1076970, EPI_ISL_1076971,
EPI_ISL_1076972, EPI_ISL_1076973, EPI_ISL_1076974, EPI_ISL_1076975, EPI_ISL_1076976, EPI_ISL_1076977, EPI_ISL_1076978, EPI_ISL_1076979, EPI_ISL_1076980, EPI_ISL_1076981, EPI_ISL_1076982, EPI_ISL_1076983, EPI_ISL_1076984, EPI_ISL_1076985, EPI_ISL_1076986, EPI_ISL_1076987, EPI_ISL_1076988, EPI_ISL_1076989, EPI_ISL_1076990, EPI_ISL_1076991, EPI_ISL_1076992, EPI_ISL_1076993,
EPI_ISL_1076994, EPI_ISL_1076995, EPI_ISL_1076996, EPI_ISL_1076997, EPI_ISL_1076998, EPI_ISL_1076999, EPI_ISL_1077000, EPI_ISL_1077001, EPI_ISL_1077002, EPI_ISL_1077003, EPI_ISL_1077004, EPI_ISL_1077005, EPI_ISL_1077006, EPI_ISL_1077007, EPI_ISL_1077008, EPI_ISL_1077009, EPI_ISL_1077010, EPI_ISL_1077011, EPI_ISL_1077012, EPI_ISL_1077013, EPI_ISL_1077014, EPI_ISL_1077015,
EPI_ISL_1077016, EPI_ISL_1077017, EPI_ISL_1077018, EPI_ISL_1077019, EPI_ISL_1077020, EPI_ISL_1077021, EPI_ISL_1077022, EPI_ISL_1077023, EPI_ISL_1077024, EPI_ISL_1077025, EPI_ISL_1077026, EPI_ISL_1077027, EPI_ISL_1077028, EPI_ISL_1077029, EPI_ISL_1077030, EPI_ISL_1077031, EPI_ISL_1077032, EPI_ISL_1077033, EPI_ISL_1077034, EPI_ISL_1077035, EPI_ISL_1077036, EPI_ISL_1077037,
EPI_ISL_1077038, EPI_ISL_1077039, EPI_ISL_1077040, EPI_ISL_1077041, EPI_ISL_1077042, EPI_ISL_1077043, EPI_ISL_1077044, EPI_ISL_1077045, EPI_ISL_1077046, EPI_ISL_1077047, EPI_ISL_1077048, EPI_ISL_1077049, EPI_ISL_1077050, EPI_ISL_1077051, EPI_ISL_1077052, EPI_ISL_1077053, EPI_ISL_1077054, EPI_ISL_1077055, EPI_ISL_1077056, EPI_ISL_1077057, EPI_ISL_1077058, EPI_ISL_1077059,
EPI_ISL_1077060, EPI_ISL_1077061, EPI_ISL_1077062, EPI_ISL_1077063, EPI_ISL_1077064, EPI_ISL_1077065, EPI_ISL_1077066, EPI_ISL_1077067, EPI_ISL_1077068, EPI_ISL_1077069, EPI_ISL_1077070, EPI_ISL_1077071, EPI_ISL_1077072, EPI_ISL_1077073, EPI_ISL_1077074, EPI_ISL_1077075, EPI_ISL_1077076, EPI_ISL_1077077, EPI_ISL_1077078, EPI_ISL_1077079, EPI_ISL_1077080, EPI_ISL_1077081,
EPI_ISL_1077082, EPI_ISL_1077083, EPI_ISL_1077084, EPI_ISL_1077085, EPI_ISL_1077086, EPI_ISL_1077087, EPI_ISL_1077088, EPI_ISL_1077089, EPI_ISL_1077090, EPI_ISL_1077091, EPI_ISL_1077092, EPI_ISL_1077093, EPI_ISL_1077094, EPI_ISL_1077095, EPI_ISL_1077096, EPI_ISL_1077097, EPI_ISL_1077098, EPI_ISL_1077099, EPI_ISL_1077100, EPI_ISL_1077101, EPI_ISL_1077102, EPI_ISL_1077103,
EPI_ISL_1077104, EPI_ISL_1077105, EPI_ISL_1077106, EPI_ISL_1077107, EPI_ISL_1077108, EPI_ISL_1077109, EPI_ISL_1077110, EPI_ISL_1077111, EPI_ISL_1077112, EPI_ISL_1077113, EPI_ISL_1077114, EPI_ISL_1077115, EPI_ISL_1077116, EPI_ISL_1077117, EPI_ISL_1077118, EPI_ISL_1077119, EPI_ISL_1077120, EPI_ISL_1077121, EPI_ISL_1077122, EPI_ISL_1077123, EPI_ISL_1077124, EPI_ISL_1077125,
EPI_ISL_1077126, EPI_ISL_1077127, EPI_ISL_1077128, EPI_ISL_1077129, EPI_ISL_1077130, EPI_ISL_1077131, EPI_ISL_1077132, EPI_ISL_1077133, EPI_ISL_1077134, EPI_ISL_1077135, EPI_ISL_1077136, EPI_ISL_1077137, EPI_ISL_1077138, EPI_ISL_1077139, EPI_ISL_1077140, EPI_ISL_1077141, EPI_ISL_1077142, EPI_ISL_1077143, EPI_ISL_1077144, EPI_ISL_1077145, EPI_ISL_1077146, EPI_ISL_1077147,
EPI_ISL_1077148, EPI_ISL_1077149, EPI_ISL_1077150, EPI_ISL_1077151, EPI_ISL_1077152, EPI_ISL_1077153, EPI_ISL_1077154, EPI_ISL_1077155, EPI_ISL_1077156, EPI_ISL_1077157, EPI_ISL_1077158, EPI_ISL_1077159, EPI_ISL_1077160, EPI_ISL_1077161, EPI_ISL_1077162, EPI_ISL_1077163, EPI_ISL_1077164, EPI_ISL_1077165, EPI_ISL_1077166, EPI_ISL_1077167, EPI_ISL_1077168, EPI_ISL_1077169,
EPI_ISL_1077170, EPI_ISL_1077171, EPI_ISL_1077172, EPI_ISL_1077173, EPI_ISL_1077174, EPI_ISL_1077175, EPI_ISL_1077176, EPI_ISL_1077177, EPI_ISL_1077178, EPI_ISL_1077179, EPI_ISL_1077180, EPI_ISL_1077181, EPI_ISL_1077182, EPI_ISL_1077183, EPI_ISL_1077184, EPI_ISL_1077185, EPI_ISL_1077186, EPI_ISL_1077187, EPI_ISL_1077188, EPI_ISL_1077189, EPI_ISL_1077190, EPI_ISL_1077191,
EPI_ISL_1077192, EPI_ISL_1077193, EPI_ISL_1077194, EPI_ISL_1077195, EPI_ISL_1077196, EPI_ISL_1077197, EPI_ISL_1077198, EPI_ISL_1077199, EPI_ISL_1077200, EPI_ISL_1077201, EPI_ISL_1077202, EPI_ISL_1077203, EPI_ISL_1077204, EPI_ISL_1077205, EPI_ISL_1077206, EPI_ISL_1077207, EPI_ISL_1077208, EPI_ISL_1077209, EPI_ISL_1077210, EPI_ISL_1077211, EPI_ISL_1077212, EPI_ISL_1077213,
EPI_ISL_1077214, EPI_ISL_1077215, EPI_ISL_1077216, EPI_ISL_1077217, EPI_ISL_1077218, EPI_ISL_1077219, EPI_ISL_1077220, EPI_ISL_1077221, EPI_ISL_1077222, EPI_ISL_1077223, EPI_ISL_1077224, EPI_ISL_1077225, EPI_ISL_1077226, EPI_ISL_1077227, EPI_ISL_1077228, EPI_ISL_1077229, EPI_ISL_1077230, EPI_ISL_1077231, EPI_ISL_1077232, EPI_ISL_1077233, EPI_ISL_1077234, EPI_ISL_1077235,
EPI_ISL_1077236, EPI_ISL_1077237, EPI_ISL_1077238, EPI_ISL_1077239, EPI_ISL_1077240, EPI_ISL_1077241, EPI_ISL_1077242, EPI_ISL_1077243, EPI_ISL_1077244, EPI_ISL_1077245, EPI_ISL_1077246, EPI_ISL_1077247, EPI_ISL_1077248, EPI_ISL_1077249, EPI_ISL_1077250, EPI_ISL_1077251, EPI_ISL_1077252, EPI_ISL_1077253, EPI_ISL_1077254, EPI_ISL_1077255, EPI_ISL_1077256, EPI_ISL_1077257,
EPI_ISL_1077258, EPI_ISL_1077259, EPI_ISL_1077260, EPI_ISL_1077261, EPI_ISL_1077262, EPI_ISL_1077263, EPI_ISL_1077264, EPI_ISL_1077265, EPI_ISL_1077266, EPI_ISL_1077267, EPI_ISL_1077268, EPI_ISL_1077269, EPI_ISL_1077270, EPI_ISL_1077271, EPI_ISL_1077272, EPI_ISL_1077273, EPI_ISL_1077274, EPI_ISL_1077275, EPI_ISL_1077276, EPI_ISL_1077277, EPI_ISL_1077278, EPI_ISL_1077279,
EPI_ISL_1077280, EPI_ISL_1077281, EPI_ISL_1077282, EPI_ISL_1077283, EPI_ISL_1077284, EPI_ISL_1077285, EPI_ISL_1077286, EPI_ISL_1077287, EPI_ISL_1077288, EPI_ISL_1077289, EPI_ISL_1077290, EPI_ISL_1077291, EPI_ISL_1077292, EPI_ISL_1077293, EPI_ISL_1077294, EPI_ISL_1077295, EPI_ISL_1077296, EPI_ISL_1077297, EPI_ISL_1077298, EPI_ISL_1077299, EPI_ISL_1077300, EPI_ISL_1077301,
EPI_ISL_1077302, EPI_ISL_1077303, EPI_ISL_1077304, EPI_ISL_1077305, EPI_ISL_1077306, EPI_ISL_1077307, EPI_ISL_1077308, EPI_ISL_1077309, EPI_ISL_1077310, EPI_ISL_1077311, EPI_ISL_1077312, EPI_ISL_1077313, EPI_ISL_1077314, EPI_ISL_1077315, EPI_ISL_1077316, EPI_ISL_1077317, EPI_ISL_1077318, EPI_ISL_1077319, EPI_ISL_1077320, EPI_ISL_1077321, EPI_ISL_1077322, EPI_ISL_1077323,
EPI_ISL_1077324, EPI_ISL_1077325, EPI_ISL_1077326, EPI_ISL_1077327, EPI_ISL_1077328, EPI_ISL_1077329, EPI_ISL_1077330, EPI_ISL_1077331, EPI_ISL_1077332, EPI_ISL_1077333, EPI_ISL_1077334, EPI_ISL_1077335, EPI_ISL_1077336, EPI_ISL_1077337, EPI_ISL_1077338, EPI_ISL_1077339, EPI_ISL_1077340, EPI_ISL_1077341, EPI_ISL_1077342, EPI_ISL_1077343, EPI_ISL_1077344, EPI_ISL_1077345,
EPI_ISL_1077346, EPI_ISL_1077347, EPI_ISL_1077350, EPI_ISL_1077351, EPI_ISL_1077352, EPI_ISL_1077353, EPI_ISL_1077354, EPI_ISL_1077355, EPI_ISL_1077356, EPI_ISL_1077357, EPI_ISL_1077358, EPI_ISL_1077359, EPI_ISL_1077360, EPI_ISL_1077361, EPI_ISL_1077362, EPI_ISL_1077363, EPI_ISL_1077364, EPI_ISL_1077365, EPI_ISL_1077366, EPI_ISL_1077367, EPI_ISL_1077368, EPI_ISL_1077369,
EPI_ISL_1077370, EPI_ISL_1077371, EPI_ISL_1077372, EPI_ISL_1077373, EPI_ISL_1077374, EPI_ISL_1077375, EPI_ISL_1077376, EPI_ISL_1077377, EPI_ISL_1077378, EPI_ISL_1077379, EPI_ISL_1077380, EPI_ISL_1077381, EPI_ISL_1077382, EPI_ISL_1077383, EPI_ISL_1077384, EPI_ISL_1077385, EPI_ISL_1077386, EPI_ISL_1077387, EPI_ISL_1077388, EPI_ISL_1077389, EPI_ISL_1077390, EPI_ISL_1077391,
EPI_ISL_1077392, EPI_ISL_1077393, EPI_ISL_1077394, EPI_ISL_1077395, EPI_ISL_1077396, EPI_ISL_1077397, EPI_ISL_1077398, EPI_ISL_1077399, EPI_ISL_1077400, EPI_ISL_1077401, EPI_ISL_1077402, EPI_ISL_1077403, EPI_ISL_1077404, EPI_ISL_1077405, EPI_ISL_1077406, EPI_ISL_1077407, EPI_ISL_1077408, EPI_ISL_1077409, EPI_ISL_1077410, EPI_ISL_1077411, EPI_ISL_1077412, EPI_ISL_1077413,
EPI_ISL_1077414, EPI_ISL_1077415, EPI_ISL_1077416, EPI_ISL_1077417, EPI_ISL_1077418, EPI_ISL_1077419, EPI_ISL_1077420, EPI_ISL_1077421, EPI_ISL_1077422, EPI_ISL_1077423, EPI_ISL_1077424, EPI_ISL_1077425, EPI_ISL_1077426, EPI_ISL_1077427, EPI_ISL_1077428, EPI_ISL_1077429, EPI_ISL_1077430, EPI_ISL_1077431, EPI_ISL_1077432, EPI_ISL_1077433, EPI_ISL_1077434, EPI_ISL_1077435,
EPI_ISL_1077436, EPI_ISL_1077437, EPI_ISL_1077438, EPI_ISL_1077439, EPI_ISL_1077440, EPI_ISL_1077441, EPI_ISL_1077442, EPI_ISL_1077443, EPI_ISL_1077444, EPI_ISL_1077445, EPI_ISL_1077446, EPI_ISL_1077447, EPI_ISL_1077448, EPI_ISL_1077449, EPI_ISL_1077450, EPI_ISL_1077451, EPI_ISL_1077452, EPI_ISL_1077453, EPI_ISL_1077454, EPI_ISL_1077455, EPI_ISL_1077456, EPI_ISL_1077457,
EPI_ISL_1077458, EPI_ISL_1077459, EPI_ISL_1077460, EPI_ISL_1077461, EPI_ISL_1077462, EPI_ISL_1077463, EPI_ISL_1077464, EPI_ISL_1077465, EPI_ISL_1077466, EPI_ISL_1077467, EPI_ISL_1077468, EPI_ISL_1077469, EPI_ISL_1077470, EPI_ISL_1077471, EPI_ISL_1077472, EPI_ISL_1077473, EPI_ISL_1077474, EPI_ISL_1077475, EPI_ISL_1077476, EPI_ISL_1077477, EPI_ISL_1077478, EPI_ISL_1077479,
EPI_ISL_1077480, EPI_ISL_1077481, EPI_ISL_1077482, EPI_ISL_1077483, EPI_ISL_1077484, EPI_ISL_1077485, EPI_ISL_1077486, EPI_ISL_1077487, EPI_ISL_1077488, EPI_ISL_1077489, EPI_ISL_1077490, EPI_ISL_1077491, EPI_ISL_1077492, EPI_ISL_1077493, EPI_ISL_1077494, EPI_ISL_1077495, EPI_ISL_1077496, EPI_ISL_1077497, EPI_ISL_1077498, EPI_ISL_1077499, EPI_ISL_1077500, EPI_ISL_1077501,
EPI_ISL_1077502, EPI_ISL_1077503, EPI_ISL_1077504, EPI_ISL_1077505, EPI_ISL_1077506, EPI_ISL_1077507, EPI_ISL_1077508, EPI_ISL_1077509, EPI_ISL_1077510, EPI_ISL_1077511, EPI_ISL_1077512, EPI_ISL_1077513, EPI_ISL_1077514, EPI_ISL_1077515, EPI_ISL_1077516, EPI_ISL_1077517, EPI_ISL_1077518, EPI_ISL_1077519, EPI_ISL_1077520, EPI_ISL_1077521, EPI_ISL_1077522, EPI_ISL_1077523,
EPI_ISL_1077524, EPI_ISL_1077525, EPI_ISL_1077526, EPI_ISL_1077527, EPI_ISL_1077528, EPI_ISL_1077529, EPI_ISL_1077530, EPI_ISL_1077531, EPI_ISL_1077532, EPI_ISL_1077533, EPI_ISL_1077534, EPI_ISL_1077535, EPI_ISL_1077536, EPI_ISL_1077537, EPI_ISL_1077538, EPI_ISL_1077539, EPI_ISL_1077540, EPI_ISL_1077541, EPI_ISL_1077542, EPI_ISL_1077543, EPI_ISL_1077544, EPI_ISL_1077545,
EPI_ISL_1077546, EPI_ISL_1077547, EPI_ISL_1077548, EPI_ISL_1077549, EPI_ISL_1077550, EPI_ISL_1077551, EPI_ISL_1077552, EPI_ISL_1077553, EPI_ISL_1077554, EPI_ISL_1077555, EPI_ISL_1077556, EPI_ISL_1077557, EPI_ISL_1077558, EPI_ISL_1077559, EPI_ISL_1077560, EPI_ISL_1077561, EPI_ISL_1077562, EPI_ISL_1077563, EPI_ISL_1077564, EPI_ISL_1077565, EPI_ISL_1077566, EPI_ISL_1077567,
EPI_ISL_1077568, EPI_ISL_1077569, EPI_ISL_1077570, EPI_ISL_1077571, EPI_ISL_1077572, EPI_ISL_1077573, EPI_ISL_1077574, EPI_ISL_1077575, EPI_ISL_1077576, EPI_ISL_1077577, EPI_ISL_1077578, EPI_ISL_1077579, EPI_ISL_1077580, EPI_ISL_1077581, EPI_ISL_1077582, EPI_ISL_1077583, EPI_ISL_1077584, EPI_ISL_1077585, EPI_ISL_1077586, EPI_ISL_1077587, EPI_ISL_1077588, EPI_ISL_1077589,
EPI_ISL_1077590, EPI_ISL_1077591, EPI_ISL_1077592, EPI_ISL_1077593, EPI_ISL_1077594, EPI_ISL_1077595, EPI_ISL_1077596, EPI_ISL_1077597, EPI_ISL_1077598, EPI_ISL_1077599, EPI_ISL_1077600, EPI_ISL_1077601, EPI_ISL_1077602, EPI_ISL_1077603, EPI_ISL_1077604, EPI_ISL_1077605, EPI_ISL_1077606, EPI_ISL_1077607, EPI_ISL_1077608, EPI_ISL_1077609, EPI_ISL_1077610, EPI_ISL_1077611,
EPI_ISL_1077612, EPI_ISL_1077613, EPI_ISL_1077614, EPI_ISL_1077615, EPI_ISL_1077616, EPI_ISL_1077617, EPI_ISL_1077618, EPI_ISL_1077619, EPI_ISL_1077620, EPI_ISL_1077621, EPI_ISL_1077622, EPI_ISL_1077623, EPI_ISL_1077624, EPI_ISL_1077625, EPI_ISL_1077626, EPI_ISL_1077627, EPI_ISL_1077628, EPI_ISL_1077629, EPI_ISL_1077630, EPI_ISL_1077631, EPI_ISL_1077632, EPI_ISL_1077633,
EPI_ISL_1077634, EPI_ISL_1077635, EPI_ISL_1077636, EPI_ISL_1077637, EPI_ISL_1077638, EPI_ISL_1077639, EPI_ISL_1077640, EPI_ISL_1077641, EPI_ISL_1077642, EPI_ISL_1077643, EPI_ISL_1077644, EPI_ISL_1077645, EPI_ISL_1077646, EPI_ISL_1077647, EPI_ISL_1077648, EPI_ISL_1077649, EPI_ISL_1077650, EPI_ISL_1077651, EPI_ISL_1077652, EPI_ISL_1077653, EPI_ISL_1077654, EPI_ISL_1077655,
EPI_ISL_1077656, EPI_ISL_1077657, EPI_ISL_1077658, EPI_ISL_1077659, EPI_ISL_1077660, EPI_ISL_1077661, EPI_ISL_1077662, EPI_ISL_1077663, EPI_ISL_1077664, EPI_ISL_1077665, EPI_ISL_1077666, EPI_ISL_1077667, EPI_ISL_1077668, EPI_ISL_1077669, EPI_ISL_1077670, EPI_ISL_1077671, EPI_ISL_1077672, EPI_ISL_1077673, EPI_ISL_1077674, EPI_ISL_1077675, EPI_ISL_1077676, EPI_ISL_1077677,
EPI_ISL_1077678, EPI_ISL_1077679, EPI_ISL_1077680, EPI_ISL_1077681, EPI_ISL_1077682, EPI_ISL_1077683, EPI_ISL_1077684, EPI_ISL_1077685, EPI_ISL_1077686, EPI_ISL_1077687, EPI_ISL_1077688, EPI_ISL_1077689, EPI_ISL_1077690, EPI_ISL_1077691, EPI_ISL_1077692, EPI_ISL_1077693, EPI_ISL_1077694, EPI_ISL_1077695, EPI_ISL_1077696, EPI_ISL_1077697, EPI_ISL_1077698, EPI_ISL_1077699,
EPI_ISL_1077700, EPI_ISL_1077701, EPI_ISL_1077702, EPI_ISL_1077703, EPI_ISL_1077704, EPI_ISL_1077705, EPI_ISL_1077706, EPI_ISL_1077707, EPI_ISL_1077708, EPI_ISL_1077709, EPI_ISL_1077710, EPI_ISL_1077711, EPI_ISL_1077712, EPI_ISL_1077713, EPI_ISL_1077714, EPI_ISL_1077715, EPI_ISL_1077716, EPI_ISL_1077717, EPI_ISL_1077718, EPI_ISL_1077719, EPI_ISL_1077720, EPI_ISL_1077721,
EPI_ISL_1077722, EPI_ISL_1077723, EPI_ISL_1077724, EPI_ISL_1077725, EPI_ISL_1077726, EPI_ISL_1077727, EPI_ISL_1077728, EPI_ISL_1077729, EPI_ISL_1077730, EPI_ISL_1077731, EPI_ISL_1077732, EPI_ISL_1077733, EPI_ISL_1077734, EPI_ISL_1077735, EPI_ISL_1077736, EPI_ISL_1077737, EPI_ISL_1077738, EPI_ISL_1077739, EPI_ISL_1077740, EPI_ISL_1077741, EPI_ISL_1077742, EPI_ISL_1077743,
EPI_ISL_1077744, EPI_ISL_1077745, EPI_ISL_1077746, EPI_ISL_1077747, EPI_ISL_1077748, EPI_ISL_1077749, EPI_ISL_1077750, EPI_ISL_1077751, EPI_ISL_1077752, EPI_ISL_1077753, EPI_ISL_1077754, EPI_ISL_1077755, EPI_ISL_1077756, EPI_ISL_1077757, EPI_ISL_1077758, EPI_ISL_1077759, EPI_ISL_1077760, EPI_ISL_1077761, EPI_ISL_1077762, EPI_ISL_1077763, EPI_ISL_1077764, EPI_ISL_1077765,
EPI_ISL_1077766, EPI_ISL_1077767, EPI_ISL_1077768, EPI_ISL_1077769, EPI_ISL_1077770, EPI_ISL_1077771, EPI_ISL_1077772, EPI_ISL_1077773, EPI_ISL_1077774, EPI_ISL_1077775, EPI_ISL_1077776, EPI_ISL_1077777, EPI_ISL_1077778, EPI_ISL_1077779, EPI_ISL_1077780, EPI_ISL_1077781, EPI_ISL_1077782, EPI_ISL_1077783, EPI_ISL_1077784, EPI_ISL_1077785, EPI_ISL_1077786, EPI_ISL_1077787,

EPI_ISL_1077788, EPI_ISL_1077789, EPI_ISL_1077790, EPI_ISL_1077791, EPI_ISL_1077792, EPI_ISL_1077793, EPI_ISL_1077794, EPI_ISL_1077795, EPI_ISL_1077796, EPI_ISL_1077797, EPI_ISL_1077798, EPI_ISL_1077799, EPI_ISL_1077800, EPI_ISL_1077801, EPI_ISL_1077802, EPI_ISL_1077803, EPI_ISL_1077804, EPI_ISL_1077805, EPI_ISL_1077806, EPI_ISL_1077807, EPI_ISL_1077808, EPI_ISL_1077809,
EPI_ISL_1077810, EPI_ISL_1077811, EPI_ISL_1077812, EPI_ISL_1077813, EPI_ISL_1077814, EPI_ISL_1077815, EPI_ISL_1077816, EPI_ISL_1077817, EPI_ISL_1077818, EPI_ISL_1077819, EPI_ISL_1077820, EPI_ISL_1077821, EPI_ISL_1077822, EPI_ISL_1077823, EPI_ISL_1077824, EPI_ISL_1077825, EPI_ISL_1077826, EPI_ISL_1077827, EPI_ISL_1077828, EPI_ISL_1077829, EPI_ISL_1077830, EPI_ISL_1077831,
EPI_ISL_1077832, EPI_ISL_1077833, EPI_ISL_1077834, EPI_ISL_1077835, EPI_ISL_1077836, EPI_ISL_1077837, EPI_ISL_1077838, EPI_ISL_1077839, EPI_ISL_1077840, EPI_ISL_1077841, EPI_ISL_1077842, EPI_ISL_1077843, EPI_ISL_1077844, EPI_ISL_1077845, EPI_ISL_1077846, EPI_ISL_1077847, EPI_ISL_1077848, EPI_ISL_1077849, EPI_ISL_1077850, EPI_ISL_1077851, EPI_ISL_1077852, EPI_ISL_1077853,
EPI_ISL_1077854, EPI_ISL_1077855, EPI_ISL_1077856, EPI_ISL_1077857, EPI_ISL_1077858, EPI_ISL_1077859, EPI_ISL_1077860, EPI_ISL_1077861, EPI_ISL_1077862, EPI_ISL_1077863, EPI_ISL_1077864, EPI_ISL_1077865, EPI_ISL_1077866, EPI_ISL_1077867, EPI_ISL_1077868, EPI_ISL_1077869, EPI_ISL_1077870, EPI_ISL_1077871, EPI_ISL_1077872, EPI_ISL_1077873, EPI_ISL_1077874, EPI_ISL_1077875,
EPI_ISL_1077876, EPI_ISL_1077877, EPI_ISL_1077878, EPI_ISL_1077879, EPI_ISL_1077880, EPI_ISL_1077881, EPI_ISL_1077882, EPI_ISL_1077883, EPI_ISL_1077884, EPI_ISL_1077885, EPI_ISL_1077886, EPI_ISL_1077887, EPI_ISL_1077888, EPI_ISL_1077889, EPI_ISL_1077890, EPI_ISL_1077891, EPI_ISL_1077892, EPI_ISL_1077893, EPI_ISL_1077894, EPI_ISL_1077895, EPI_ISL_1077896, EPI_ISL_1077897,
EPI_ISL_1077898, EPI_ISL_1077899, EPI_ISL_1077900, EPI_ISL_1077901, EPI_ISL_1077902, EPI_ISL_1077903, EPI_ISL_1077904, EPI_ISL_1077905, EPI_ISL_1077906, EPI_ISL_1077907, EPI_ISL_1077908, EPI_ISL_1077909, EPI_ISL_1077910, EPI_ISL_1077911, EPI_ISL_1077912, EPI_ISL_1077913, EPI_ISL_1077914, EPI_ISL_1077915, EPI_ISL_1077916, EPI_ISL_1077917, EPI_ISL_1077918, EPI_ISL_1077919,
EPI_ISL_1077920, EPI_ISL_1077921, EPI_ISL_1077922, EPI_ISL_1077923, EPI_ISL_1077924, EPI_ISL_1077925, EPI_ISL_1077926, EPI_ISL_1077927, EPI_ISL_1077928, EPI_ISL_1077929, EPI_ISL_1077930, EPI_ISL_1077931, EPI_ISL_1077932, EPI_ISL_1077933, EPI_ISL_1077934, EPI_ISL_1077935, EPI_ISL_1077936, EPI_ISL_1077937, EPI_ISL_1077938, EPI_ISL_1077939, EPI_ISL_1077940, EPI_ISL_1077941,
EPI_ISL_1077942, EPI_ISL_1077943, EPI_ISL_1077944, EPI_ISL_1077945, EPI_ISL_1077946, EPI_ISL_1077947, EPI_ISL_1077948, EPI_ISL_1077949, EPI_ISL_1077950, EPI_ISL_1077951, EPI_ISL_1077952, EPI_ISL_1077953, EPI_ISL_1077954, EPI_ISL_1077955, EPI_ISL_1077956, EPI_ISL_1077957, EPI_ISL_1077958, EPI_ISL_1077959, EPI_ISL_1077960, EPI_ISL_1077961, EPI_ISL_1077962, EPI_ISL_1077963,
EPI_ISL_1077964, EPI_ISL_1077965, EPI_ISL_1077966, EPI_ISL_1077967, EPI_ISL_1077968, EPI_ISL_1077969, EPI_ISL_1077970, EPI_ISL_1077971, EPI_ISL_1077972, EPI_ISL_1077973, EPI_ISL_1077974, EPI_ISL_1077975, EPI_ISL_1077976, EPI_ISL_1077977, EPI_ISL_1077978, EPI_ISL_1077979, EPI_ISL_1077980, EPI_ISL_1077981, EPI_ISL_1077982, EPI_ISL_1077983, EPI_ISL_1077984, EPI_ISL_1077985,
EPI_ISL_1077986, EPI_ISL_1077987, EPI_ISL_1077988, EPI_ISL_1077989, EPI_ISL_1077990, EPI_ISL_1077991, EPI_ISL_1077992, EPI_ISL_1077993, EPI_ISL_1077994, EPI_ISL_1077995, EPI_ISL_1077996, EPI_ISL_1077997, EPI_ISL_1077998, EPI_ISL_1077999, EPI_ISL_1078000, EPI_ISL_1078001, EPI_ISL_1078002, EPI_ISL_1078003, EPI_ISL_1078004, EPI_ISL_1078005, EPI_ISL_1078006, EPI_ISL_1078007,
EPI_ISL_1078008, EPI_ISL_1078009, EPI_ISL_1078010, EPI_ISL_1078011, EPI_ISL_1078012, EPI_ISL_1078013, EPI_ISL_1078014, EPI_ISL_1078015, EPI_ISL_1078016, EPI_ISL_1078017, EPI_ISL_1078018, EPI_ISL_1078019, EPI_ISL_1078020, EPI_ISL_1078021, EPI_ISL_1078022, EPI_ISL_1078023, EPI_ISL_1078024, EPI_ISL_1078025, EPI_ISL_1078026, EPI_ISL_1078027, EPI_ISL_1078028, EPI_ISL_1078029,
EPI_ISL_1078030, EPI_ISL_1078031, EPI_ISL_1078032, EPI_ISL_1078033, EPI_ISL_1078034, EPI_ISL_1078035, EPI_ISL_1078036, EPI_ISL_1078037, EPI_ISL_1078038, EPI_ISL_1078039, EPI_ISL_1078040, EPI_ISL_1078041, EPI_ISL_1078042, EPI_ISL_1078043, EPI_ISL_1078044, EPI_ISL_1078045, EPI_ISL_1078046, EPI_ISL_1078047, EPI_ISL_1078048, EPI_ISL_1078049, EPI_ISL_1078050, EPI_ISL_1078051,
EPI_ISL_1078052, EPI_ISL_1078053, EPI_ISL_1078054, EPI_ISL_1078055, EPI_ISL_1078056, EPI_ISL_1078057, EPI_ISL_1078058, EPI_ISL_1078059, EPI_ISL_1078060, EPI_ISL_1078061, EPI_ISL_1078062, EPI_ISL_1078063, EPI_ISL_1078064, EPI_ISL_1078065, EPI_ISL_1078066, EPI_ISL_1078067, EPI_ISL_1078068, EPI_ISL_1078069, EPI_ISL_1078070, EPI_ISL_1078071, EPI_ISL_1078072, EPI_ISL_1078073,
EPI_ISL_1078074, EPI_ISL_1078075, EPI_ISL_1078076, EPI_ISL_1078077, EPI_ISL_1078078, EPI_ISL_1078079, EPI_ISL_1078080, EPI_ISL_1078081, EPI_ISL_1078082, EPI_ISL_1078083, EPI_ISL_1078084, EPI_ISL_1078085, EPI_ISL_1078086, EPI_ISL_1078087, EPI_ISL_1078088, EPI_ISL_1078089, EPI_ISL_1078090, EPI_ISL_1078091, EPI_ISL_1078092, EPI_ISL_1078093, EPI_ISL_1078094, EPI_ISL_1078095,
EPI_ISL_1078096, EPI_ISL_1078097, EPI_ISL_1078098, EPI_ISL_1078099, EPI_ISL_1078100, EPI_ISL_1078101, EPI_ISL_1078102, EPI_ISL_1078103, EPI_ISL_1078104, EPI_ISL_1078105, EPI_ISL_1078106, EPI_ISL_1078107, EPI_ISL_1078108, EPI_ISL_1078109, EPI_ISL_1078110, EPI_ISL_1078111, EPI_ISL_1078112, EPI_ISL_1078113, EPI_ISL_1078114, EPI_ISL_1078115, EPI_ISL_1078116, EPI_ISL_1078117,
EPI_ISL_1078118, EPI_ISL_1078119, EPI_ISL_1078120, EPI_ISL_1078121, EPI_ISL_1078122, EPI_ISL_1078123, EPI_ISL_1078124, EPI_ISL_1078125, EPI_ISL_1078126, EPI_ISL_1078127, EPI_ISL_1078128, EPI_ISL_1078129, EPI_ISL_1078130, EPI_ISL_1078131, EPI_ISL_1078132, EPI_ISL_1078133, EPI_ISL_1078134, EPI_ISL_1078135, EPI_ISL_1078136, EPI_ISL_1078137, EPI_ISL_1078138, EPI_ISL_1078139,
EPI_ISL_1078140, EPI_ISL_1078141, EPI_ISL_1078142, EPI_ISL_1078143, EPI_ISL_1078144, EPI_ISL_1078145, EPI_ISL_1078146, EPI_ISL_1078147, EPI_ISL_1078148, EPI_ISL_1078149, EPI_ISL_1078150, EPI_ISL_1078151, EPI_ISL_1078152, EPI_ISL_1078153, EPI_ISL_1078154, EPI_ISL_1078155, EPI_ISL_1078156, EPI_ISL_1078157, EPI_ISL_1078158, EPI_ISL_1078159, EPI_ISL_1078160, EPI_ISL_1078161,
EPI_ISL_1078162, EPI_ISL_1078163, EPI_ISL_1078164, EPI_ISL_1078165, EPI_ISL_1078166, EPI_ISL_1078167, EPI_ISL_1078168, EPI_ISL_1078169, EPI_ISL_1078170, EPI_ISL_1078171, EPI_ISL_1078172, EPI_ISL_1078173, EPI_ISL_1078174, EPI_ISL_1078175, EPI_ISL_1078176, EPI_ISL_1078177, EPI_ISL_1078178, EPI_ISL_1078179, EPI_ISL_1078180, EPI_ISL_1078181, EPI_ISL_1078182, EPI_ISL_1078183,
EPI_ISL_1078184, EPI_ISL_1078185, EPI_ISL_1078186, EPI_ISL_1078187, EPI_ISL_1078188, EPI_ISL_1078189, EPI_ISL_1078190, EPI_ISL_1078191, EPI_ISL_1078192, EPI_ISL_1078193, EPI_ISL_1078194, EPI_ISL_1078195, EPI_ISL_1078196, EPI_ISL_1078197, EPI_ISL_1078198, EPI_ISL_1078199, EPI_ISL_1078200, EPI_ISL_1078201, EPI_ISL_1078202, EPI_ISL_1078203, EPI_ISL_1078204, EPI_ISL_1078205,
EPI_ISL_1078206, EPI_ISL_1078207, EPI_ISL_1078208, EPI_ISL_1078209, EPI_ISL_1078210, EPI_ISL_1078211, EPI_ISL_1078212, EPI_ISL_1078213, EPI_ISL_1078214, EPI_ISL_1078215, EPI_ISL_1078216, EPI_ISL_1078217, EPI_ISL_1078218, EPI_ISL_1078219, EPI_ISL_1078220, EPI_ISL_1078221, EPI_ISL_1078222, EPI_ISL_1078223, EPI_ISL_1078224, EPI_ISL_1078225, EPI_ISL_1078226, EPI_ISL_1078227,
EPI_ISL_1078228, EPI_ISL_1078229, EPI_ISL_1078230, EPI_ISL_1078231, EPI_ISL_1078232, EPI_ISL_1078233, EPI_ISL_1078234, EPI_ISL_1078235, EPI_ISL_1078236, EPI_ISL_1078237, EPI_ISL_1078238, EPI_ISL_1078239, EPI_ISL_1078240, EPI_ISL_1078241, EPI_ISL_1078242, EPI_ISL_1078243, EPI_ISL_1078244, EPI_ISL_1078245, EPI_ISL_1078246, EPI_ISL_1078247, EPI_ISL_1078248, EPI_ISL_1078249,
EPI_ISL_1078250, EPI_ISL_1078251, EPI_ISL_1078252, EPI_ISL_1078253, EPI_ISL_1078254, EPI_ISL_1078255, EPI_ISL_1078256, EPI_ISL_1078257, EPI_ISL_1078258, EPI_ISL_1078259, EPI_ISL_1078260, EPI_ISL_1078261, EPI_ISL_1078262, EPI_ISL_1078263, EPI_ISL_1078264, EPI_ISL_1078265, EPI_ISL_1078266, EPI_ISL_1078267, EPI_ISL_1078268, EPI_ISL_1078269, EPI_ISL_1078270, EPI_ISL_1078271,
EPI_ISL_1078272, EPI_ISL_1078273, EPI_ISL_1078274, EPI_ISL_1078275, EPI_ISL_1078276, EPI_ISL_1078277, EPI_ISL_1078278, EPI_ISL_1078279, EPI_ISL_1078280, EPI_ISL_1078281, EPI_ISL_1078282, EPI_ISL_1078283, EPI_ISL_1078284, EPI_ISL_1078285, EPI_ISL_1078286, EPI_ISL_1078287, EPI_ISL_1078288, EPI_ISL_1078289, EPI_ISL_1078290, EPI_ISL_1078291, EPI_ISL_1078292, EPI_ISL_1078293,
EPI_ISL_1078294, EPI_ISL_1078295, EPI_ISL_1078296, EPI_ISL_1078297, EPI_ISL_1078298, EPI_ISL_1078299, EPI_ISL_1078300, EPI_ISL_1078301, EPI_ISL_1078302, EPI_ISL_1078303, EPI_ISL_1078304, EPI_ISL_1078305, EPI_ISL_1078306, EPI_ISL_1078307, EPI_ISL_1078308, EPI_ISL_1078309, EPI_ISL_1078310, EPI_ISL_1078311, EPI_ISL_1078312, EPI_ISL_1078313, EPI_ISL_1078314, EPI_ISL_1078315,
EPI_ISL_1078316, EPI_ISL_1078317, EPI_ISL_1078318, EPI_ISL_1078319, EPI_ISL_1078320, EPI_ISL_1078321, EPI_ISL_1078322, EPI_ISL_1078323, EPI_ISL_1078324, EPI_ISL_1078325, EPI_ISL_1078326, EPI_ISL_1078327, EPI_ISL_1078328, EPI_ISL_1078329, EPI_ISL_1078330, EPI_ISL_1078331, EPI_ISL_1078332, EPI_ISL_1078333, EPI_ISL_1078334, EPI_ISL_1078335, EPI_ISL_1078336, EPI_ISL_1078337,
EPI_ISL_1078338, EPI_ISL_1078339, EPI_ISL_1078340, EPI_ISL_1078341, EPI_ISL_1078342, EPI_ISL_1078343, EPI_ISL_1078344, EPI_ISL_1078345, EPI_ISL_1078346, EPI_ISL_1078347, EPI_ISL_1078348, EPI_ISL_1078349, EPI_ISL_1078350, EPI_ISL_1078351, EPI_ISL_1078352, EPI_ISL_1078353, EPI_ISL_1078354, EPI_ISL_1078355, EPI_ISL_1078356, EPI_ISL_1078357, EPI_ISL_1078358, EPI_ISL_1078359,
EPI_ISL_1078360, EPI_ISL_1078361, EPI_ISL_1078362, EPI_ISL_1078363, EPI_ISL_1078364, EPI_ISL_1078365, EPI_ISL_1078366, EPI_ISL_1078367, EPI_ISL_1078368, EPI_ISL_1078369, EPI_ISL_1078370, EPI_ISL_1078371, EPI_ISL_1078372, EPI_ISL_1078373, EPI_ISL_1078374, EPI_ISL_1078375, EPI_ISL_1078376, EPI_ISL_1078377, EPI_ISL_1078378, EPI_ISL_1078379, EPI_ISL_1078380, EPI_ISL_1078381,
EPI_ISL_1078382, EPI_ISL_1078383, EPI_ISL_1078384, EPI_ISL_1078385, EPI_ISL_1078386, EPI_ISL_1078387, EPI_ISL_1078388, EPI_ISL_1078389, EPI_ISL_1078390, EPI_ISL_1078391, EPI_ISL_1078392, EPI_ISL_1078393, EPI_ISL_1078394, EPI_ISL_1078395, EPI_ISL_1078396, EPI_ISL_1078397, EPI_ISL_1078398, EPI_ISL_1078399, EPI_ISL_1078400, EPI_ISL_1078401, EPI_ISL_1078402, EPI_ISL_1078403,
EPI_ISL_1078404, EPI_ISL_1078405, EPI_ISL_1078406, EPI_ISL_1078407, EPI_ISL_1078408, EPI_ISL_1078409, EPI_ISL_1078410, EPI_ISL_1078411, EPI_ISL_1078412, EPI_ISL_1078413, EPI_ISL_1078414, EPI_ISL_1078415, EPI_ISL_1078416, EPI_ISL_1078417, EPI_ISL_1078418, EPI_ISL_1078419, EPI_ISL_1078420, EPI_ISL_1078421, EPI_ISL_1078422, EPI_ISL_1078423, EPI_ISL_1078424, EPI_ISL_1078425,
EPI_ISL_1078426, EPI_ISL_1078427, EPI_ISL_1078428, EPI_ISL_1078429, EPI_ISL_1078430, EPI_ISL_1078431, EPI_ISL_1078432, EPI_ISL_1078433, EPI_ISL_1078434, EPI_ISL_1078435, EPI_ISL_1078436, EPI_ISL_1078437, EPI_ISL_1078438, EPI_ISL_1078439, EPI_ISL_1078440, EPI_ISL_1078441, EPI_ISL_1078442, EPI_ISL_1078443, EPI_ISL_1078444, EPI_ISL_1078445, EPI_ISL_1078446, EPI_ISL_1078447,
EPI_ISL_1078448, EPI_ISL_1078449, EPI_ISL_1078450, EPI_ISL_1078451, EPI_ISL_1078452, EPI_ISL_1078453, EPI_ISL_1078454, EPI_ISL_1078455, EPI_ISL_1078456, EPI_ISL_1078457, EPI_ISL_1078458, EPI_ISL_1078459, EPI_ISL_1078460, EPI_ISL_1078461, EPI_ISL_1078462, EPI_ISL_1078463, EPI_ISL_1078464, EPI_ISL_1078465, EPI_ISL_1078466, EPI_ISL_1078467, EPI_ISL_1078468, EPI_ISL_1078469,
EPI_ISL_1078470, EPI_ISL_1078471, EPI_ISL_1078472, EPI_ISL_1078473, EPI_ISL_1078474, EPI_ISL_1078475, EPI_ISL_1078476, EPI_ISL_1078477, EPI_ISL_1078478, EPI_ISL_1078479, EPI_ISL_1078480, EPI_ISL_1078481, EPI_ISL_1078482, EPI_ISL_1078483, EPI_ISL_1078484, EPI_ISL_1078485, EPI_ISL_1078486, EPI_ISL_1078487, EPI_ISL_1078488, EPI_ISL_1078489, EPI_ISL_1078490, EPI_ISL_1078491,
EPI_ISL_1078492, EPI_ISL_1078493, EPI_ISL_1078494, EPI_ISL_1078495, EPI_ISL_1078496, EPI_ISL_1078497, EPI_ISL_1078498, EPI_ISL_1078499, EPI_ISL_1078500, EPI_ISL_1078501, EPI_ISL_1078502, EPI_ISL_1078503, EPI_ISL_1078504, EPI_ISL_1078505, EPI_ISL_1078506, EPI_ISL_1078507, EPI_ISL_1078508, EPI_ISL_1078509, EPI_ISL_1078510, EPI_ISL_1078511, EPI_ISL_1078512, EPI_ISL_1078513,
EPI_ISL_1078514, EPI_ISL_1078515, EPI_ISL_1078516, EPI_ISL_1078517, EPI_ISL_1078518, EPI_ISL_1078519, EPI_ISL_1078520, EPI_ISL_1078521, EPI_ISL_1078522, EPI_ISL_1078523, EPI_ISL_1078524, EPI_ISL_1078525, EPI_ISL_1078526, EPI_ISL_1078527, EPI_ISL_1078528, EPI_ISL_1078529, EPI_ISL_1078530, EPI_ISL_1078531, EPI_ISL_1078532, EPI_ISL_1078533, EPI_ISL_1078534, EPI_ISL_1078535,
EPI_ISL_1078536, EPI_ISL_1078537, EPI_ISL_1078538, EPI_ISL_1078539, EPI_ISL_1078540, EPI_ISL_1078541, EPI_ISL_1078542, EPI_ISL_1078543, EPI_ISL_1078544, EPI_ISL_1078545, EPI_ISL_1078546, EPI_ISL_1078547, EPI_ISL_1078548, EPI_ISL_1078549, EPI_ISL_1078550, EPI_ISL_1078551, EPI_ISL_1078552, EPI_ISL_1078553, EPI_ISL_1078554, EPI_ISL_1078555, EPI_ISL_1078556, EPI_ISL_1078557,
EPI_ISL_1078558, EPI_ISL_1078559, EPI_ISL_1078560, EPI_ISL_1078561, EPI_ISL_1078562, EPI_ISL_1078563, EPI_ISL_1078564, EPI_ISL_1078565, EPI_ISL_1078566, EPI_ISL_1078567, EPI_ISL_1078568, EPI_ISL_1078569, EPI_ISL_1078570, EPI_ISL_1078571, EPI_ISL_1078572, EPI_ISL_1078573, EPI_ISL_1078574, EPI_ISL_1078575, EPI_ISL_1078576, EPI_ISL_1078577, EPI_ISL_1078578, EPI_ISL_1078579,
EPI_ISL_1078580, EPI_ISL_1078581, EPI_ISL_1078582, EPI_ISL_1078583, EPI_ISL_1078584, EPI_ISL_1078585, EPI_ISL_1078586, EPI_ISL_1078587, EPI_ISL_1078588, EPI_ISL_1078591, EPI_ISL_1078595, EPI_ISL_1078598, EPI_ISL_1078602, EPI_ISL_1078605, EPI_ISL_1078609, EPI_ISL_1078612, EPI_ISL_1078615, EPI_ISL_1078619, EPI_ISL_1078622, EPI_ISL_1078623, EPI_ISL_1078624, EPI_ISL_1078625,
EPI_ISL_1078626, EPI_ISL_1078627, EPI_ISL_1078628, EPI_ISL_1078629, EPI_ISL_1078630, EPI_ISL_1078631, EPI_ISL_1078632, EPI_ISL_1078633, EPI_ISL_1078634, EPI_ISL_1078635, EPI_ISL_1078636, EPI_ISL_1078637, EPI_ISL_1078638, EPI_ISL_1078639, EPI_ISL_1078640, EPI_ISL_1078641, EPI_ISL_1078642, EPI_ISL_1078643, EPI_ISL_1078644, EPI_ISL_1078645, EPI_ISL_1078646, EPI_ISL_1078647,
EPI_ISL_1078648, EPI_ISL_1078649, EPI_ISL_1078650, EPI_ISL_1078651, EPI_ISL_1078652, EPI_ISL_1078653, EPI_ISL_1078654, EPI_ISL_1078655, EPI_ISL_1078656, EPI_ISL_1078657, EPI_ISL_1078658, EPI_ISL_1078659, EPI_ISL_1078660, EPI_ISL_1078661, EPI_ISL_1078662, EPI_ISL_1078663, EPI_ISL_1078664, EPI_ISL_1078665, EPI_ISL_1078666, EPI_ISL_1078667, EPI_ISL_1078668, EPI_ISL_1078669,
EPI_ISL_1078670, EPI_ISL_1078671, EPI_ISL_1078672, EPI_ISL_1078673, EPI_ISL_1078674, EPI_ISL_1078675, EPI_ISL_1078676, EPI_ISL_1078677, EPI_ISL_1078678, EPI_ISL_1078679, EPI_ISL_1078680, EPI_ISL_1078681, EPI_ISL_1078682, EPI_ISL_1078683, EPI_ISL_1078684, EPI_ISL_1078685, EPI_ISL_1078686, EPI_ISL_1078687, EPI_ISL_1078688, EPI_ISL_1078689, EPI_ISL_1078690, EPI_ISL_1078691,
EPI_ISL_1078692, EPI_ISL_1078693, EPI_ISL_1078694, EPI_ISL_1078695, EPI_ISL_1078696, EPI_ISL_1078697, EPI_ISL_1078698, EPI_ISL_1078699, EPI_ISL_1078700, EPI_ISL_1078701, EPI_ISL_1078702, EPI_ISL_1078703, EPI_ISL_1078704, EPI_ISL_1078705, EPI_ISL_1078706, EPI_ISL_1078707, EPI_ISL_1078708, EPI_ISL_1078709, EPI_ISL_1078710, EPI_ISL_1078711, EPI_ISL_1078712, EPI_ISL_1078713,
EPI_ISL_1078714, EPI_ISL_1078715, EPI_ISL_1078716, EPI_ISL_1078717, EPI_ISL_1078718, EPI_ISL_1078719, EPI_ISL_1078720, EPI_ISL_1078721, EPI_ISL_1078722, EPI_ISL_1078723, EPI_ISL_1078724, EPI_ISL_1078725, EPI_ISL_1078726, EPI_ISL_1078727, EPI_ISL_1078728, EPI_ISL_1078729, EPI_ISL_1078730, EPI_ISL_1078731, EPI_ISL_1078732, EPI_ISL_1078733, EPI_ISL_1078734, EPI_ISL_1078735,
EPI_ISL_1078736, EPI_ISL_1078737, EPI_ISL_1078738, EPI_ISL_1078739, EPI_ISL_1078740, EPI_ISL_1078741, EPI_ISL_1078742, EPI_ISL_1078743, EPI_ISL_1078744, EPI_ISL_1078745, EPI_ISL_1078746, EPI_ISL_1078747, EPI_ISL_1078748, EPI_ISL_1078749, EPI_ISL_1078750, EPI_ISL_1078751, EPI_ISL_1078752, EPI_ISL_1078753, EPI_ISL_1078754, EPI_ISL_1078755, EPI_ISL_1078756, EPI_ISL_1078757,
EPI_ISL_1078758, EPI_ISL_1078759, EPI_ISL_1078760, EPI_ISL_1078761, EPI_ISL_1078762, EPI_ISL_1078763, EPI_ISL_1078764, EPI_ISL_1078765, EPI_ISL_1078766, EPI_ISL_1078767, EPI_ISL_1078768, EPI_ISL_1078769, EPI_ISL_1078770, EPI_ISL_1078771, EPI_ISL_1078772, EPI_ISL_1078773, EPI_ISL_1078774, EPI_ISL_1078775, EPI_ISL_1078776, EPI_ISL_1078777, EPI_ISL_1078778, EPI_ISL_1078779,
EPI_ISL_1078780, EPI_ISL_1078781, EPI_ISL_1078782, EPI_ISL_1078783, EPI_ISL_1078784, EPI_ISL_1078785, EPI_ISL_1078786, EPI_ISL_1078787, EPI_ISL_1078788, EPI_ISL_1078789, EPI_ISL_1078790, EPI_ISL_1078791, EPI_ISL_1078792, EPI_ISL_1078793, EPI_ISL_1078794, EPI_ISL_1078795, EPI_ISL_1078796, EPI_ISL_1078797, EPI_ISL_1078798, EPI_ISL_1078799, EPI_ISL_1078800, EPI_ISL_1078801,
EPI_ISL_1078802, EPI_ISL_1078803, EPI_ISL_1078804, EPI_ISL_1078805, EPI_ISL_1078806, EPI_ISL_1078807, EPI_ISL_1078808, EPI_ISL_1078809, EPI_ISL_1078810, EPI_ISL_1078811, EPI_ISL_1078812, EPI_ISL_1078813, EPI_ISL_1078814, EPI_ISL_1078815, EPI_ISL_1078816, EPI_ISL_1078817, EPI_ISL_1078818, EPI_ISL_1078819, EPI_ISL_1078820, EPI_ISL_1078821, EPI_ISL_1078822, EPI_ISL_1078823,
EPI_ISL_1078824, EPI_ISL_1078825, EPI_ISL_1078826, EPI_ISL_1078827, EPI_ISL_1078828, EPI_ISL_1078829, EPI_ISL_1078830, EPI_ISL_1078831, EPI_ISL_1078832, EPI_ISL_1078833, EPI_ISL_1078834, EPI_ISL_1078835, EPI_ISL_1078836, EPI_ISL_1078837, EPI_ISL_1078838, EPI_ISL_1078839, EPI_ISL_1078840, EPI_ISL_1078841, EPI_ISL_1078842, EPI_ISL_1078843, EPI_ISL_1078844, EPI_ISL_1078845,
EPI_ISL_1078846, EPI_ISL_1078847, EPI_ISL_1078848, EPI_ISL_1078849, EPI_ISL_1078850, EPI_ISL_1078851, EPI_ISL_1078852, EPI_ISL_1078853, EPI_ISL_1078854, EPI_ISL_1078855, EPI_ISL_1078856, EPI_ISL_1078857, EPI_ISL_1078858, EPI_ISL_1078859, EPI_ISL_1078860, EPI_ISL_1078861, EPI_ISL_1078862, EPI_ISL_1078863, EPI_ISL_1078864, EPI_ISL_1078865, EPI_ISL_1078866, EPI_ISL_1078867,
EPI_ISL_1078868, EPI_ISL_1078869, EPI_ISL_1078870, EPI_ISL_1078871, EPI_ISL_1078872, EPI_ISL_1078873, EPI_ISL_1078874, EPI_ISL_1078875, EPI_ISL_1078876, EPI_ISL_1078877, EPI_ISL_1078878, EPI_ISL_1078879, EPI_ISL_1078880, EPI_ISL_1078881, EPI_ISL_1078882, EPI_ISL_1078883, EPI_ISL_1078884, EPI_ISL_1078885, EPI_ISL_1078886, EPI_ISL_1078887, EPI_ISL_1078888, EPI_ISL_1078889,
EPI_ISL_1078890, EPI_ISL_1078891, EPI_ISL_1078892, EPI_ISL_1078893, EPI_ISL_1078894, EPI_ISL_1078895, EPI_ISL_1078896, EPI_ISL_1078897, EPI_ISL_1078898, EPI_ISL_1078899, EPI_ISL_1078900, EPI_ISL_1078901, EPI_ISL_1078902, EPI_ISL_1078903, EPI_ISL_1078904, EPI_ISL_1078905, EPI_ISL_1078906, EPI_ISL_1078907, EPI_ISL_1078908, EPI_ISL_1078909, EPI_ISL_1078910, EPI_ISL_1078911,
EPI_ISL_1078912, EPI_ISL_1078913, EPI_ISL_1078914, EPI_ISL_1078915, EPI_ISL_1078916, EPI_ISL_1078917, EPI_ISL_1078918, EPI_ISL_1078919, EPI_ISL_1078920, EPI_ISL_1078921, EPI_ISL_1078922, EPI_ISL_1078923, EPI_ISL_1078924, EPI_ISL_1078925, EPI_ISL_1078926, EPI_ISL_1078927, EPI_ISL_1078928, EPI_ISL_1078929, EPI_ISL_1078930, EPI_ISL_1078931, EPI_ISL_1078932, EPI_ISL_1078933,
EPI_ISL_1078934, EPI_ISL_1078935, EPI_ISL_1078936, EPI_ISL_1078937, EPI_ISL_1078938, EPI_ISL_1078939, EPI_ISL_1078940, EPI_ISL_1078941, EPI_ISL_1078942, EPI_ISL_1078943, EPI_ISL_1078944, EPI_ISL_1078945, EPI_ISL_1078946, EPI_ISL_1078947, EPI_ISL_1078948, EPI_ISL_1078949, EPI_ISL_1078950, EPI_ISL_1078951, EPI_ISL_1078952, EPI_ISL_1078953, EPI_ISL_1078954, EPI_ISL_1078955,
EPI_ISL_1078956, EPI_ISL_1078957, EPI_ISL_1078958, EPI_ISL_1078959, EPI_ISL_1078960, EPI_ISL_1078961, EPI_ISL_1078962, EPI_ISL_1078963, EPI_ISL_1078964, EPI_ISL_1078965, EPI_ISL_1078966, EPI_ISL_1078967, EPI_ISL_1078968, EPI_ISL_1078969, EPI_ISL_1078970, EPI_ISL_1078971, EPI_ISL_1078972, EPI_ISL_1078973, EPI_ISL_1078974, EPI_ISL_1078975, EPI_ISL_1078976, EPI_ISL_1078977,
EPI_ISL_1078978, EPI_ISL_1078979, EPI_ISL_1078980, EPI_ISL_1078982, EPI_ISL_1078985, EPI_ISL_1078990, EPI_ISL_1078994, EPI_ISL_1078998, EPI_ISL_1079001, EPI_ISL_1079005, EPI_ISL_1079009, EPI_ISL_1079010, EPI_ISL_1079011, EPI_ISL_1079012, EPI_ISL_1079013, EPI_ISL_1079014, EPI_ISL_1079015, EPI_ISL_1079016, EPI_ISL_1079017, EPI_ISL_1079018, EPI_ISL_1079019, EPI_ISL_1079020,
EPI_ISL_1079021, EPI_ISL_1079022, EPI_ISL_1079023, EPI_ISL_1079024, EPI_ISL_1079025, EPI_ISL_1079026, EPI_ISL_1079027, EPI_ISL_1079028, EPI_ISL_1079029, EPI_ISL_1079030, EPI_ISL_1079031, EPI_ISL_1079032, EPI_ISL_1079033, EPI_ISL_1079034, EPI_ISL_1079035, EPI_ISL_1079036, EPI_ISL_1079037, EPI_ISL_1079038, EPI_ISL_1079039, EPI_ISL_1079040, EPI_ISL_1079041, EPI_ISL_1079042,
EPI_ISL_1079043, EPI_ISL_1079044, EPI_ISL_1079045, EPI_ISL_1079046, EPI_ISL_1079047, EPI_ISL_1079048, EPI_ISL_1079049, EPI_ISL_1079050, EPI_ISL_1079051, EPI_ISL_1079052, EPI_ISL_1079053, EPI_ISL_1079054, EPI_ISL_1079055, EPI_ISL_1079056, EPI_ISL_1079057, EPI_ISL_1079058, EPI_ISL_1079059, EPI_ISL_1079060, EPI_ISL_1079061, EPI_ISL_1079062, EPI_ISL_1079063, EPI_ISL_1079064,
EPI_ISL_1079065, EPI_ISL_1079066, EPI_ISL_1079067, EPI_ISL_1079068, EPI_ISL_1079069, EPI_ISL_1079070, EPI_ISL_1079071, EPI_ISL_1079072, EPI_ISL_1079073, EPI_ISL_1079074, EPI_ISL_1079075, EPI_ISL_1079076, EPI_ISL_1079077, EPI_ISL_1079078, EPI_ISL_1079079, EPI_ISL_1079080, EPI_ISL_1079081, EPI_ISL_1079082, EPI_ISL_1079083, EPI_ISL_1079084, EPI_ISL_1079085, EPI_ISL_1079086,
EPI_ISL_1079087, EPI_ISL_1079088, EPI_ISL_1079089, EPI_ISL_1079090, EPI_ISL_1079091, EPI_ISL_1079092, EPI_ISL_1079093, EPI_ISL_1079094, EPI_ISL_1079095, EPI_ISL_1079096, EPI_ISL_1079097, EPI_ISL_1079098, EPI_ISL_1079099, EPI_ISL_1079100, EPI_ISL_1079101, EPI_ISL_1079102, EPI_ISL_1079103, EPI_ISL_1079104, EPI_ISL_1079105, EPI_ISL_1079106, EPI_ISL_1079107, EPI_ISL_1079108,
EPI_ISL_1079109, EPI_ISL_1079110, EPI_ISL_1079111, EPI_ISL_1079112, EPI_ISL_1079113, EPI_ISL_1079114, EPI_ISL_1079115, EPI_ISL_1079116, EPI_ISL_1079117, EPI_ISL_1079118, EPI_ISL_1079119, EPI_ISL_1079120, EPI_ISL_1079121, EPI_ISL_1079122, EPI_ISL_1079123, EPI_ISL_1079124, EPI_ISL_1079125, EPI_ISL_1079126, EPI_ISL_1079127, EPI_ISL_1079128, EPI_ISL_1079129, EPI_ISL_1079130,
EPI_ISL_1079131, EPI_ISL_1079132, EPI_ISL_1079133, EPI_ISL_1079134, EPI_ISL_1079135, EPI_ISL_1079136, EPI_ISL_1079137, EPI_ISL_1079138, EPI_ISL_1079139, EPI_ISL_1079140, EPI_ISL_1079141, EPI_ISL_1079142, EPI_ISL_1079143, EPI_ISL_1079144, EPI_ISL_1079145, EPI_ISL_1079146, EPI_ISL_1079147, EPI_ISL_1079148, EPI_ISL_1079149, EPI_ISL_1079150, EPI_ISL_1079151, EPI_ISL_1079152,
EPI_ISL_1079153, EPI_ISL_1079154, EPI_ISL_1079155, EPI_ISL_1079156, EPI_ISL_1079157, EPI_ISL_1079160, EPI_ISL_1079164, EPI_ISL_1079167, EPI_ISL_1079168, EPI_ISL_1079169, EPI_ISL_1079170, EPI_ISL_1079171, EPI_ISL_1079172, EPI_ISL_1079173, EPI_ISL_1079174, EPI_ISL_1079175, EPI_ISL_1079176, EPI_ISL_1079177, EPI_ISL_1079178, EPI_ISL_1079179, EPI_ISL_1079180, EPI_ISL_1079181,
EPI_ISL_1079182, EPI_ISL_1079183, EPI_ISL_1079184, EPI_ISL_1079185, EPI_ISL_1079186, EPI_ISL_1079187, EPI_ISL_1079188, EPI_ISL_1079189, EPI_ISL_1079190, EPI_ISL_1079191, EPI_ISL_1079192, EPI_ISL_1079193, EPI_ISL_1079194, EPI_ISL_1079195, EPI_ISL_1079196, EPI_ISL_1079197, EPI_ISL_1079198, EPI_ISL_1079199, EPI_ISL_1079200, EPI_ISL_1079201, EPI_ISL_1079202, EPI_ISL_1079203,
EPI_ISL_1079204, EPI_ISL_1079205, EPI_ISL_1079206, EPI_ISL_1079207, EPI_ISL_1079208, EPI_ISL_1079209, EPI_ISL_1079210, EPI_ISL_1079211, EPI_ISL_1079212, EPI_ISL_1079213, EPI_ISL_1079214, EPI_ISL_1079215, EPI_ISL_1079216, EPI_ISL_1079217, EPI_ISL_1079218, EPI_ISL_1079219, EPI_ISL_1079220, EPI_ISL_1079221, EPI_ISL_1079222, EPI_ISL_1079223, EPI_ISL_1079224, EPI_ISL_1079225,
EPI_ISL_1079226, EPI_ISL_1079227, EPI_ISL_1079228, EPI_ISL_1079229, EPI_ISL_1079230, EPI_ISL_1079231, EPI_ISL_1079232, EPI_ISL_1079233, EPI_ISL_1079234, EPI_ISL_1079235, EPI_ISL_1079236, EPI_ISL_1079237, EPI_ISL_1079238, EPI_ISL_1079239, EPI_ISL_1079240, EPI_ISL_1079241, EPI_ISL_1079242, EPI_ISL_1079243, EPI_ISL_1079244, EPI_ISL_1079245, EPI_ISL_1079246, EPI_ISL_1079247,
EPI_ISL_1079248, EPI_ISL_1079249, EPI_ISL_1079250, EPI_ISL_1079251, EPI_ISL_1079252, EPI_ISL_1079253, EPI_ISL_1079254, EPI_ISL_1079255, EPI_ISL_1079256, EPI_ISL_1079257, EPI_ISL_1079258, EPI_ISL_1079259, EPI_ISL_1079260, EPI_ISL_1079261, EPI_ISL_1079262, EPI_ISL_1079263, EPI_ISL_1079264, EPI_ISL_1079265, EPI_ISL_1079266, EPI_ISL_1079267, EPI_ISL_1079268, EPI_ISL_1079269,
EPI_ISL_1079270, EPI_ISL_1079271, EPI_ISL_1079272, EPI_ISL_1079273, EPI_ISL_1079274, EPI_ISL_1079275, EPI_ISL_1079276, EPI_ISL_1079277, EPI_ISL_1079278, EPI_ISL_1079279, EPI_ISL_1079280, EPI_ISL_1079281, EPI_ISL_1079282, EPI_ISL_1079283, EPI_ISL_1079284, EPI_ISL_1079285, EPI_ISL_1079286, EPI_ISL_1079287, EPI_ISL_1079288, EPI_ISL_1079289, EPI_ISL_1079290, EPI_ISL_1079291,
EPI_ISL_1079292, EPI_ISL_1079293, EPI_ISL_1079294, EPI_ISL_1079295, EPI_ISL_1079296, EPI_ISL_1079297, EPI_ISL_1079298, EPI_ISL_1079299, EPI_ISL_1079300, EPI_ISL_1079301, EPI_ISL_1079302, EPI_ISL_1079303, EPI_ISL_1079304, EPI_ISL_1079305, EPI_ISL_1079306, EPI_ISL_1079307, EPI_ISL_1079308, EPI_ISL_1079309, EPI_ISL_1079310, EPI_ISL_1079311, EPI_ISL_1079312, EPI_ISL_1079313,
EPI_ISL_1079314, EPI_ISL_1079315, EPI_ISL_1079316, EPI_ISL_1079317, EPI_ISL_1079318, EPI_ISL_1079319, EPI_ISL_1079320, EPI_ISL_1079321, EPI_ISL_1079322, EPI_ISL_1079323, EPI_ISL_1079324, EPI_ISL_1079325, EPI_ISL_1079326, EPI_ISL_1079327, EPI_ISL_1079328, EPI_ISL_1079329, EPI_ISL_1079330, EPI_ISL_1079331, EPI_ISL_1079332, EPI_ISL_1079333, EPI_ISL_1079334, EPI_ISL_1079335,
EPI_ISL_1079336, EPI_ISL_1079337, EPI_ISL_1079338, EPI_ISL_1079340, EPI_ISL_1079341, EPI_ISL_1079343, EPI_ISL_1079344, EPI_ISL_1079345, EPI_ISL_1079346, EPI_ISL_1079347, EPI_ISL_1079348, EPI_ISL_1079349, EPI_ISL_1079350, EPI_ISL_1079351, EPI_ISL_1079354, EPI_ISL_1079357, EPI_ISL_1079361, EPI_ISL_1079365, EPI_ISL_1079367, EPI_ISL_1079369, EPI_ISL_1079372, EPI_ISL_1079374,
EPI_ISL_1079375, EPI_ISL_1079377, EPI_ISL_1079379, EPI_ISL_1079381, EPI_ISL_1079384, EPI_ISL_1079385, EPI_ISL_1079387, EPI_ISL_1079389, EPI_ISL_1079393, EPI_ISL_1079396, EPI_ISL_1079397, EPI_ISL_1079399, EPI_ISL_1079401, EPI_ISL_1079402, EPI_ISL_1079403, EPI_ISL_1079404, EPI_ISL_1079405, EPI_ISL_1079406, EPI_ISL_1079407, EPI_ISL_1079408, EPI_ISL_1079409, EPI_ISL_1079410,
EPI_ISL_1079411, EPI_ISL_1079412, EPI_ISL_1079413, EPI_ISL_1079414, EPI_ISL_1079415, EPI_ISL_1079416, EPI_ISL_1079417, EPI_ISL_1079418, EPI_ISL_1079419, EPI_ISL_1079420, EPI_ISL_1079421, EPI_ISL_1079422, EPI_ISL_1079423, EPI_ISL_1079424, EPI_ISL_1079425, EPI_ISL_1079426, EPI_ISL_1079427, EPI_ISL_1079428, EPI_ISL_1079429, EPI_ISL_1079430, EPI_ISL_1079431, EPI_ISL_1079432,
EPI_ISL_1079433, EPI_ISL_1079434, EPI_ISL_1079435, EPI_ISL_1079437, EPI_ISL_1079438, EPI_ISL_1079439, EPI_ISL_1079440, EPI_ISL_1079441, EPI_ISL_1079442, EPI_ISL_1079443, EPI_ISL_1079444, EPI_ISL_1079445, EPI_ISL_1079446, EPI_ISL_1079447, EPI_ISL_1079448, EPI_ISL_1079449, EPI_ISL_1079450, EPI_ISL_1079452, EPI_ISL_1079453, EPI_ISL_1079454, EPI_ISL_1079455, EPI_ISL_1079456,
EPI_ISL_1079457, EPI_ISL_1079458, EPI_ISL_1079459, EPI_ISL_1079460, EPI_ISL_1079461, EPI_ISL_1079462, EPI_ISL_1079463, EPI_ISL_1079464, EPI_ISL_1079465, EPI_ISL_1079466, EPI_ISL_1079467, EPI_ISL_1079468, EPI_ISL_1079469, EPI_ISL_1079470, EPI_ISL_1079471, EPI_ISL_1079472, EPI_ISL_1079473, EPI_ISL_1079474, EPI_ISL_1079475, EPI_ISL_1079476, EPI_ISL_1079477, EPI_ISL_1079478,
EPI_ISL_1079479, EPI_ISL_1079480, EPI_ISL_1079481, EPI_ISL_1079482, EPI_ISL_1079483, EPI_ISL_1079484, EPI_ISL_1079485, EPI_ISL_1079486, EPI_ISL_1079487, EPI_ISL_1079488, EPI_ISL_1079489, EPI_ISL_1079490, EPI_ISL_1079491, EPI_ISL_1079492, EPI_ISL_1079493, EPI_ISL_1079494, EPI_ISL_1079495, EPI_ISL_1079496, EPI_ISL_1079497, EPI_ISL_1079498, EPI_ISL_1079499, EPI_ISL_1079500,
EPI_ISL_1079501, EPI_ISL_1079502, EPI_ISL_1079503, EPI_ISL_1079504, EPI_ISL_1079505, EPI_ISL_1079506, EPI_ISL_1079507, EPI_ISL_1079508, EPI_ISL_1079509, EPI_ISL_1079510, EPI_ISL_1079511, EPI_ISL_1079512, EPI_ISL_1079513, EPI_ISL_1079514, EPI_ISL_1079515, EPI_ISL_1079516, EPI_ISL_1079517, EPI_ISL_1079518, EPI_ISL_1079519, EPI_ISL_1079520, EPI_ISL_1079521, EPI_ISL_1079522,
EPI_ISL_1079523, EPI_ISL_1079524, EPI_ISL_1079525, EPI_ISL_1079526, EPI_ISL_1079527, EPI_ISL_1079528, EPI_ISL_1079529, EPI_ISL_1079530, EPI_ISL_1079531, EPI_ISL_1079532, EPI_ISL_1079533, EPI_ISL_1079534, EPI_ISL_1079535, EPI_ISL_1079536, EPI_ISL_1079537, EPI_ISL_1079538, EPI_ISL_1079539, EPI_ISL_1079540, EPI_ISL_1079541, EPI_ISL_1079542, EPI_ISL_1079543, EPI_ISL_1079544,
EPI_ISL_1079545, EPI_ISL_1079546, EPI_ISL_1079547, EPI_ISL_1079548, EPI_ISL_1079549, EPI_ISL_1079550, EPI_ISL_1079551, EPI_ISL_1079552, EPI_ISL_1079553, EPI_ISL_1079554, EPI_ISL_1079555, EPI_ISL_1079556, EPI_ISL_1079557, EPI_ISL_1079558, EPI_ISL_1079559, EPI_ISL_1079560, EPI_ISL_1079561, EPI_ISL_1079562, EPI_ISL_1079563, EPI_ISL_1079564, EPI_ISL_1079565, EPI_ISL_1079566,
EPI_ISL_1079567, EPI_ISL_1079568, EPI_ISL_1079569, EPI_ISL_1079570, EPI_ISL_1079571, EPI_ISL_1079572, EPI_ISL_1079573, EPI_ISL_1079574, EPI_ISL_1079575, EPI_ISL_1079576, EPI_ISL_1079577, EPI_ISL_1079578, EPI_ISL_1079579, EPI_ISL_1079580, EPI_ISL_1079581, EPI_ISL_1079582, EPI_ISL_1079583, EPI_ISL_1079584, EPI_ISL_1079585, EPI_ISL_1079586, EPI_ISL_1079587, EPI_ISL_1079588,
EPI_ISL_1079589, EPI_ISL_1079590, EPI_ISL_1079591, EPI_ISL_1079592, EPI_ISL_1079593, EPI_ISL_1079594, EPI_ISL_1079595, EPI_ISL_1079596, EPI_ISL_1079597, EPI_ISL_1079598, EPI_ISL_1079599, EPI_ISL_1079600, EPI_ISL_1079601, EPI_ISL_1079602, EPI_ISL_1079603, EPI_ISL_1079604, EPI_ISL_1079605, EPI_ISL_1079606, EPI_ISL_1079607, EPI_ISL_1079608, EPI_ISL_1079609, EPI_ISL_1079610,
EPI_ISL_1079611, EPI_ISL_1079612, EPI_ISL_1079613, EPI_ISL_1079614, EPI_ISL_1079615, EPI_ISL_1079616, EPI_ISL_1079617, EPI_ISL_1079618, EPI_ISL_1079619, EPI_ISL_1079620, EPI_ISL_1079621, EPI_ISL_1079622, EPI_ISL_1079623, EPI_ISL_1079624, EPI_ISL_1079625, EPI_ISL_1079626, EPI_ISL_1079627, EPI_ISL_1079628, EPI_ISL_1079629, EPI_ISL_1079630, EPI_ISL_1079631, EPI_ISL_1079632,
EPI_ISL_1079633, EPI_ISL_1079634, EPI_ISL_1079635, EPI_ISL_1079636, EPI_ISL_1079637, EPI_ISL_1079638, EPI_ISL_1079639, EPI_ISL_1079640, EPI_ISL_1079641, EPI_ISL_1079642, EPI_ISL_1079643, EPI_ISL_1079644, EPI_ISL_1079645, EPI_ISL_1079646, EPI_ISL_1079647, EPI_ISL_1079648, EPI_ISL_1079649, EPI_ISL_1079650, EPI_ISL_1079651, EPI_ISL_1079652, EPI_ISL_1079653, EPI_ISL_1079654,
EPI_ISL_1079655, EPI_ISL_1079656, EPI_ISL_1079657, EPI_ISL_1079658, EPI_ISL_1079659, EPI_ISL_1079660, EPI_ISL_1079661, EPI_ISL_1079662, EPI_ISL_1079663, EPI_ISL_1079664, EPI_ISL_1079665, EPI_ISL_1079666, EPI_ISL_1079667, EPI_ISL_1079668, EPI_ISL_1079669, EPI_ISL_1079670, EPI_ISL_1079671, EPI_ISL_1079672, EPI_ISL_1079673, EPI_ISL_1079674, EPI_ISL_1079675, EPI_ISL_1079676,
EPI_ISL_1079677, EPI_ISL_1079678, EPI_ISL_1079679, EPI_ISL_1079680, EPI_ISL_1079681, EPI_ISL_1079682, EPI_ISL_1079683, EPI_ISL_1079684, EPI_ISL_1079685, EPI_ISL_1079686, EPI_ISL_1079687, EPI_ISL_1079688, EPI_ISL_1079691, EPI_ISL_1079694, EPI_ISL_1079698, EPI_ISL_1079702, EPI_ISL_1079704, EPI_ISL_1079705, EPI_ISL_1079706, EPI_ISL_1079707, EPI_ISL_1079708, EPI_ISL_1079709,
EPI_ISL_1079710, EPI_ISL_1079711, EPI_ISL_1079712, EPI_ISL_1079713, EPI_ISL_1079714, EPI_ISL_1079715, EPI_ISL_1079716, EPI_ISL_1079717, EPI_ISL_1079718, EPI_ISL_1079719, EPI_ISL_1079720, EPI_ISL_1079721, EPI_ISL_1079722, EPI_ISL_1079723, EPI_ISL_1079724, EPI_ISL_1079725, EPI_ISL_1079726, EPI_ISL_1079727, EPI_ISL_1079728, EPI_ISL_1079729, EPI_ISL_1079730, EPI_ISL_1079731,
EPI_ISL_1079732, EPI_ISL_1079733, EPI_ISL_1079734, EPI_ISL_1079735, EPI_ISL_1079736, EPI_ISL_1079737, EPI_ISL_1079738, EPI_ISL_1079739, EPI_ISL_1079740, EPI_ISL_1079741, EPI_ISL_1079742, EPI_ISL_1079743, EPI_ISL_1079744, EPI_ISL_1079745, EPI_ISL_1079746, EPI_ISL_1079747, EPI_ISL_1079748, EPI_ISL_1079749, EPI_ISL_1079750, EPI_ISL_1079751, EPI_ISL_1079752, EPI_ISL_1079753,
EPI_ISL_1079754, EPI_ISL_1079755, EPI_ISL_1079756, EPI_ISL_1079757, EPI_ISL_1079758, EPI_ISL_1079759, EPI_ISL_1079760, EPI_ISL_1079761, EPI_ISL_1079762, EPI_ISL_1079763, EPI_ISL_1079764, EPI_ISL_1079765, EPI_ISL_1079766, EPI_ISL_1079767, EPI_ISL_1079768, EPI_ISL_1079769, EPI_ISL_1079770, EPI_ISL_1079771, EPI_ISL_1079772, EPI_ISL_1079773, EPI_ISL_1079774, EPI_ISL_1079775,
EPI_ISL_1079776, EPI_ISL_1079777, EPI_ISL_1079778, EPI_ISL_1079779, EPI_ISL_1079780, EPI_ISL_1079781, EPI_ISL_1079782, EPI_ISL_1079783, EPI_ISL_1079784, EPI_ISL_1079785, EPI_ISL_1079786, EPI_ISL_1079787, EPI_ISL_1079788, EPI_ISL_1079789, EPI_ISL_1079790, EPI_ISL_1079791, EPI_ISL_1079792, EPI_ISL_1079793, EPI_ISL_1079794, EPI_ISL_1079795, EPI_ISL_1079796, EPI_ISL_1079797,
EPI_ISL_1079798, EPI_ISL_1079799, EPI_ISL_1079800, EPI_ISL_1079801, EPI_ISL_1079802, EPI_ISL_1079803, EPI_ISL_1079804, EPI_ISL_1079805, EPI_ISL_1079806, EPI_ISL_1079807, EPI_ISL_1079808, EPI_ISL_1079809, EPI_ISL_1079810, EPI_ISL_1079811, EPI_ISL_1079812, EPI_ISL_1079813, EPI_ISL_1079814, EPI_ISL_1079815, EPI_ISL_1079816, EPI_ISL_1079817, EPI_ISL_1079818, EPI_ISL_1079819,
EPI_ISL_1079820, EPI_ISL_1079821, EPI_ISL_1079822, EPI_ISL_1079823, EPI_ISL_1079824, EPI_ISL_1079825, EPI_ISL_1079826, EPI_ISL_1079827, EPI_ISL_1079828, EPI_ISL_1079829, EPI_ISL_1079830, EPI_ISL_1079831, EPI_ISL_1079832, EPI_ISL_1079833, EPI_ISL_1079834, EPI_ISL_1079835, EPI_ISL_1079836, EPI_ISL_1079837, EPI_ISL_1079838, EPI_ISL_1079839, EPI_ISL_1079840, EPI_ISL_1079841,
EPI_ISL_1079842, EPI_ISL_1079843, EPI_ISL_1079844, EPI_ISL_1079845, EPI_ISL_1079846, EPI_ISL_1079847, EPI_ISL_1079848, EPI_ISL_1079849, EPI_ISL_1079850, EPI_ISL_1079851, EPI_ISL_1079852, EPI_ISL_1079853, EPI_ISL_1079854, EPI_ISL_1079855, EPI_ISL_1079856, EPI_ISL_1079857, EPI_ISL_1079858, EPI_ISL_1079859, EPI_ISL_1079860, EPI_ISL_1079861, EPI_ISL_1079862, EPI_ISL_1079863,
EPI_ISL_1079864, EPI_ISL_1079865, EPI_ISL_1079866, EPI_ISL_1079867, EPI_ISL_1079868, EPI_ISL_1079869, EPI_ISL_1079870, EPI_ISL_1079871, EPI_ISL_1079872, EPI_ISL_1079873, EPI_ISL_1079874, EPI_ISL_1079875, EPI_ISL_1079876, EPI_ISL_1079877, EPI_ISL_1079878, EPI_ISL_1079879, EPI_ISL_1079880, EPI_ISL_1079881, EPI_ISL_1079882, EPI_ISL_1079883, EPI_ISL_1079884, EPI_ISL_1079885,
EPI_ISL_1079886, EPI_ISL_1079887, EPI_ISL_1079888, EPI_ISL_1079889

| | | | |
|---|---|---|---|
| see above | Houston Methodist Hospital | Houston Methodist Hospital | S. Wesley Long, Randall J. Olsen, Paul A. Christensen, Sishir Subedi, Robert Olson, James J. Davis, Matthew Ojeda Saavedra, Prasanti Yerramilli, Layne Pruitt, Kristina Reppond, Madison N. Shyer, Jessica Cambric, Ilya J. Finkelstein, Jimmy Gollihar, and James M. Musser |
| EPI_ISL_1079890, EPI_ISL_1079891 | Kansas Health and Environmental Lab | Kansas Health and Environmental Lab | Mike Grose, Paige Drury, Carissa Robertson, Ben Olsen, and Phil Adam |
| EPI_ISL_1079892 | Houston Methodist Hospital | Houston Methodist Hospital | S. Wesley Long, Randall J. Olsen, Paul A. Christensen, Sishir Subedi, Robert Olson, James J. Davis, Matthew Ojeda Saavedra, Prasanti Yerramilli, Layne Pruitt, Kristina Reppond, Madison N. Shyer, Jessica Cambric, Ilya J. Finkelstein, Jimmy Gollihar, and James M. Musser |
| EPI_ISL_1079893, EPI_ISL_1079894 | Kansas Health and Environmental Lab | Kansas Health and Environmental Lab | Mike Grose, Paige Drury, Carissa Robertson, Ben Olsen, and Phil Adam |

EPI_ISL_1079895, EPI_ISL_1079896, EPI_ISL_1079897, EPI_ISL_1079898, EPI_ISL_1079899, EPI_ISL_1079900, EPI_ISL_1079901, EPI_ISL_1079902, EPI_ISL_1079903, EPI_ISL_1079904, EPI_ISL_1079905, EPI_ISL_1079906, EPI_ISL_1079907, EPI_ISL_1079908, EPI_ISL_1079909, EPI_ISL_1079910, EPI_ISL_1079911, EPI_ISL_1079912, EPI_ISL_1079913, EPI_ISL_1079914, EPI_ISL_1079915, EPI_ISL_1079916,
EPI_ISL_1079917, EPI_ISL_1079918, EPI_ISL_1079919, EPI_ISL_1079920, EPI_ISL_1079921, EPI_ISL_1079922, EPI_ISL_1079923, EPI_ISL_1079924, EPI_ISL_1079925, EPI_ISL_1079926, EPI_ISL_1079927, EPI_ISL_1079928, EPI_ISL_1079929, EPI_ISL_1079930, EPI_ISL_1079931, EPI_ISL_1079932, EPI_ISL_1079933, EPI_ISL_1079934, EPI_ISL_1079935, EPI_ISL_1079936, EPI_ISL_1079937, EPI_ISL_1079938,
EPI_ISL_1079939, EPI_ISL_1079940, EPI_ISL_1079941, EPI_ISL_1079942, EPI_ISL_1079943, EPI_ISL_1079944, EPI_ISL_1079945, EPI_ISL_1079946, EPI_ISL_1079947, EPI_ISL_1079948, EPI_ISL_1079949, EPI_ISL_1079950, EPI_ISL_1079951, EPI_ISL_1079952, EPI_ISL_1079953, EPI_ISL_1079954, EPI_ISL_1079955, EPI_ISL_1079956, EPI_ISL_1079957, EPI_ISL_1079958, EPI_ISL_1079959, EPI_ISL_1079960,
EPI_ISL_1079961, EPI_ISL_1079962, EPI_ISL_1079963, EPI_ISL_1079964, EPI_ISL_1079965, EPI_ISL_1079966, EPI_ISL_1079967, EPI_ISL_1079968, EPI_ISL_1079969, EPI_ISL_1079970, EPI_ISL_1079971, EPI_ISL_1079972, EPI_ISL_1079973, EPI_ISL_1079974, EPI_ISL_1079975, EPI_ISL_1079976, EPI_ISL_1079977, EPI_ISL_1079978, EPI_ISL_1079979, EPI_ISL_1079980, EPI_ISL_1079981, EPI_ISL_1079982,
EPI_ISL_1079983, EPI_ISL_1079984, EPI_ISL_1079985, EPI_ISL_1079986, EPI_ISL_1079987, EPI_ISL_1079988, EPI_ISL_1079989, EPI_ISL_1079990, EPI_ISL_1079991, EPI_ISL_1079997, EPI_ISL_1080000, EPI_ISL_1080002, EPI_ISL_1080003, EPI_ISL_1080004, EPI_ISL_1080005, EPI_ISL_1080006, EPI_ISL_1080007, EPI_ISL_1080008, EPI_ISL_1080009, EPI_ISL_1080010, EPI_ISL_1080011, EPI_ISL_1080012,
EPI_ISL_1080013, EPI_ISL_1080014, EPI_ISL_1080015, EPI_ISL_1080016, EPI_ISL_1080017, EPI_ISL_1080018, EPI_ISL_1080019, EPI_ISL_1080020, EPI_ISL_1080021, EPI_ISL_1080022, EPI_ISL_1080023, EPI_ISL_1080024, EPI_ISL_1080025, EPI_ISL_1080026, EPI_ISL_1080027, EPI_ISL_1080028, EPI_ISL_1080029, EPI_ISL_1080030, EPI_ISL_1080031, EPI_ISL_1080032, EPI_ISL_1080033, EPI_ISL_1080034,
EPI_ISL_1080035, EPI_ISL_1080036, EPI_ISL_1080037, EPI_ISL_1080038, EPI_ISL_1080039, EPI_ISL_1080040, EPI_ISL_1080041, EPI_ISL_1080042, EPI_ISL_1080043, EPI_ISL_1080044, EPI_ISL_1080045

| | | | |
|---|---|---|---|
| see above | Houston Methodist Hospital | Houston Methodist Hospital | S. Wesley Long, Randall J. Olsen, Paul A. Christensen, Sishir Subedi, Robert Olson, James J. Davis, Matthew Ojeda Saavedra, Prasanti Yerramilli, Layne Pruitt, Kristina Reppond, Madison N. Shyer, Jessica Cambric, Ilya J. Finkelstein, Jimmy Gollihar, and James M. Musser |
| EPI_ISL_1080046 | Ohio Department of Health Laboratory | Ohio Department of Health Laboratory | Holmes, Jennifer; Eric Brandt, Keoni Omura, Glen McGillivary, Caitlin McDonnell, Jade Mowery, Stephanie Mccracken, Tyler Payne, Kirtana Ramadugu, Erica Leasure, Brent Lee, Kelsey Florek, Heather Blankenship, Allison Black, Quanta Brown, and Tammy Bannerman |

| | | | |
|---|---|---|---|
| EPI_ISL_1080047, EPI_ISL_1080048, EPI_ISL_1080049, EPI_ISL_1080050, EPI_ISL_1080051, EPI_ISL_1080052, EPI_ISL_1080053, EPI_ISL_1080054, EPI_ISL_1080055, EPI_ISL_1080056, EPI_ISL_1080057, EPI_ISL_1080058, EPI_ISL_1080059, EPI_ISL_1080060, EPI_ISL_1080061, EPI_ISL_1080062, EPI_ISL_1080063, EPI_ISL_1080064, EPI_ISL_1080065, EPI_ISL_1080066, EPI_ISL_1080067, EPI_ISL_1080068, EPI_ISL_1080069, EPI_ISL_1080070, EPI_ISL_1080071, EPI_ISL_1080072, EPI_ISL_1080073, EPI_ISL_1080074, EPI_ISL_1080075, EPI_ISL_1080076, EPI_ISL_1080077, EPI_ISL_1080078, EPI_ISL_1080079, EPI_ISL_1080080, EPI_ISL_1080081, EPI_ISL_1080082, EPI_ISL_1080083, EPI_ISL_1080084, EPI_ISL_1080085, EPI_ISL_1080086, EPI_ISL_1080087, EPI_ISL_1080088, EPI_ISL_1080089, EPI_ISL_1080090, EPI_ISL_1080091, EPI_ISL_1080092, EPI_ISL_1080093, EPI_ISL_1080094, EPI_ISL_1080095, EPI_ISL_1080096, EPI_ISL_1080097, EPI_ISL_1080098, EPI_ISL_1080099, EPI_ISL_1080100, EPI_ISL_1080101, EPI_ISL_1080102, EPI_ISL_1080103, EPI_ISL_1080104, EPI_ISL_1080105, EPI_ISL_1080106, EPI_ISL_1080107, EPI_ISL_1080108, EPI_ISL_1080109, EPI_ISL_1080110, EPI_ISL_1080111, EPI_ISL_1080112, EPI_ISL_1080113, EPI_ISL_1080114, EPI_ISL_1080115, EPI_ISL_1080116, EPI_ISL_1080117, EPI_ISL_1080118, EPI_ISL_1080119, EPI_ISL_1080120, EPI_ISL_1080121, EPI_ISL_1080122, EPI_ISL_1080123, EPI_ISL_1080124, EPI_ISL_1080125, EPI_ISL_1080126, EPI_ISL_1080127, EPI_ISL_1080128, EPI_ISL_1080129, EPI_ISL_1080130, EPI_ISL_1080131, EPI_ISL_1080132, EPI_ISL_1080133, EPI_ISL_1080134, EPI_ISL_1080135, EPI_ISL_1080136, EPI_ISL_1080137, EPI_ISL_1080138, EPI_ISL_1080139, EPI_ISL_1080140, EPI_ISL_1080141, EPI_ISL_1080142, EPI_ISL_1080143, EPI_ISL_1080144, EPI_ISL_1080145, EPI_ISL_1080146, EPI_ISL_1080147, EPI_ISL_1080148, EPI_ISL_1080149, EPI_ISL_1080150, EPI_ISL_1080151, EPI_ISL_1080152, EPI_ISL_1080153, EPI_ISL_1080154, EPI_ISL_1080155, EPI_ISL_1080156, EPI_ISL_1080157, EPI_ISL_1080158, EPI_ISL_1080159, EPI_ISL_1080160, EPI_ISL_1080161, EPI_ISL_1080162, EPI_ISL_1080163, EPI_ISL_1080164, EPI_ISL_1080165, EPI_ISL_1080166, EPI_ISL_1080167, EPI_ISL_1080168, EPI_ISL_1080169, EPI_ISL_1080170, EPI_ISL_1080171, EPI_ISL_1080172, EPI_ISL_1080173, EPI_ISL_1080174, EPI_ISL_1080175, EPI_ISL_1080176, EPI_ISL_1080177, EPI_ISL_1080178, EPI_ISL_1080179, EPI_ISL_1080180, EPI_ISL_1080181, EPI_ISL_1080182, EPI_ISL_1080183, EPI_ISL_1080184, EPI_ISL_1080185, EPI_ISL_1080186, EPI_ISL_1080187, EPI_ISL_1080188, EPI_ISL_1080189, EPI_ISL_1080190, EPI_ISL_1080191, EPI_ISL_1080192, EPI_ISL_1080193, EPI_ISL_1080194, EPI_ISL_1080195, EPI_ISL_1080196, EPI_ISL_1080197, EPI_ISL_1080198, EPI_ISL_1080199, EPI_ISL_1080200, EPI_ISL_1080201, EPI_ISL_1080202, EPI_ISL_1080203, EPI_ISL_1080204, EPI_ISL_1080205, EPI_ISL_1080206, EPI_ISL_1080207, EPI_ISL_1080208, EPI_ISL_1080209, EPI_ISL_1080210, EPI_ISL_1080211, EPI_ISL_1080212, EPI_ISL_1080213, EPI_ISL_1080214, EPI_ISL_1080215, EPI_ISL_1080216, EPI_ISL_1080217, EPI_ISL_1080218, EPI_ISL_1080219, EPI_ISL_1080220, EPI_ISL_1080221, EPI_ISL_1080222, EPI_ISL_1080223, EPI_ISL_1080224, EPI_ISL_1080225, EPI_ISL_1080226, EPI_ISL_1080227, EPI_ISL_1080228, EPI_ISL_1080229, EPI_ISL_1080230, EPI_ISL_1080231, EPI_ISL_1080232, EPI_ISL_1080233, EPI_ISL_1080234, EPI_ISL_1080235, EPI_ISL_1080236, EPI_ISL_1080237, EPI_ISL_1080238, EPI_ISL_1080239, EPI_ISL_1080240, EPI_ISL_1080241, EPI_ISL_1080242, EPI_ISL_1080243, EPI_ISL_1080244, EPI_ISL_1080245, EPI_ISL_1080246, EPI_ISL_1080247, EPI_ISL_1080248, EPI_ISL_1080249, EPI_ISL_1080250, EPI_ISL_1080251, EPI_ISL_1080252, EPI_ISL_1080253, EPI_ISL_1080254, EPI_ISL_1080255, EPI_ISL_1080256, EPI_ISL_1080257, EPI_ISL_1080258, EPI_ISL_1080259, EPI_ISL_1080260, EPI_ISL_1080261, EPI_ISL_1080262, EPI_ISL_1080263, EPI_ISL_1080264, EPI_ISL_1080265, EPI_ISL_1080266, EPI_ISL_1080267, EPI_ISL_1080268, EPI_ISL_1080269, EPI_ISL_1080270, EPI_ISL_1080271, EPI_ISL_1080272, EPI_ISL_1080273, EPI_ISL_1080274, EPI_ISL_1080275, EPI_ISL_1080276, EPI_ISL_1080277, EPI_ISL_1080278, EPI_ISL_1080279, EPI_ISL_1080280, EPI_ISL_1080281, EPI_ISL_1080282, EPI_ISL_1080283, EPI_ISL_1080284, EPI_ISL_1080285, EPI_ISL_1080286, EPI_ISL_1080287, EPI_ISL_1080288, EPI_ISL_1080289, EPI_ISL_1080290, EPI_ISL_1080291, EPI_ISL_1080292, EPI_ISL_1080293, EPI_ISL_1080294, EPI_ISL_1080295, EPI_ISL_1080296, EPI_ISL_1080297, EPI_ISL_1080298, EPI_ISL_1080299, EPI_ISL_1080300, EPI_ISL_1080301, EPI_ISL_1080302, EPI_ISL_1080303, EPI_ISL_1080304, EPI_ISL_1080305, EPI_ISL_1080306, EPI_ISL_1080307, EPI_ISL_1080308, EPI_ISL_1080309, EPI_ISL_1080310, EPI_ISL_1080311, EPI_ISL_1080312, EPI_ISL_1080313, EPI_ISL_1080314, EPI_ISL_1080315, EPI_ISL_1080316, EPI_ISL_1080317, EPI_ISL_1080318, EPI_ISL_1080319, EPI_ISL_1080320, EPI_ISL_1080321, EPI_ISL_1080322, EPI_ISL_1080323, EPI_ISL_1080324, EPI_ISL_1080325, EPI_ISL_1080326, EPI_ISL_1080327, EPI_ISL_1080328, EPI_ISL_1080329, EPI_ISL_1080330, EPI_ISL_1080331, EPI_ISL_1080332, EPI_ISL_1080333 | | | |
| see above | Houston Methodist Hospital | Houston Methodist Hospital | S. Wesley Long, Randall J. Olsen, Paul A. Christensen, Sishir Subedi, Robert Olson, James J. Davis, Matthew Ojeda Saavedra, Prasanti Yerramilli, Layne Pruitt, Kristina Reppond, Madison N. Shyer, Jessica Cambric, Ilya J. Finkelstein, Jimmy Gollihar, and James M. Musser |
| EPI_ISL_1080334 | Mary Eliza Mahoney HC | NJ_PHEL | Lindsey Bodnar, Shiv Verma, Dana Woell, Byeong Jeong |
| EPI_ISL_1080335 | Houston Methodist Hospital | Houston Methodist Hospital | S. Wesley Long, Randall J. Olsen, Paul A. Christensen, Sishir Subedi, Robert Olson, James J. Davis, Matthew Ojeda Saavedra, Prasanti Yerramilli, Layne Pruitt, Kristina Reppond, Madison N. Shyer, Jessica Cambric, Ilya J. Finkelstein, Jimmy Gollihar, and James M. Musser |
| EPI_ISL_1080336 | RWJ University Hospital-New Brunswick | NJ_PHEL | Lindsey Bodnar, Shiv Verma, Dana Woell, Byeong Jeong |
| EPI_ISL_1080337 | NJ Rapid Response Teams | NJ_PHEL | Lindsey Bodnar, Shiv Verma, Dana Woell, Byeong Jeong |
| EPI_ISL_1080338 | Qdx Path Alliance PC | NJ_PHEL | Lindsey Bodnar, Shiv Verma, Dana Woell, Byeong Jeong |
| EPI_ISL_1080339 | New Jersey Department of Health, Public Health Laboratories | NJ_PHEL | Lindsey Bodnar, Shiv Verma, Dana Woell, Byeong Jeong |
| EPI_ISL_1080340 | Houston Methodist Hospital | Houston Methodist Hospital | S. Wesley Long, Randall J. Olsen, Paul A. Christensen, Sishir Subedi, Robert Olson, James J. Davis, Matthew Ojeda Saavedra, Prasanti Yerramilli, Layne Pruitt, Kristina Reppond, Madison N. Shyer, Jessica Cambric, Ilya J. Finkelstein, Jimmy Gollihar, and James M. Musser |
| EPI_ISL_1080341 | NJ Rapid Response Teams | NJ_PHEL | Lindsey Bodnar, Shiv Verma, Dana Woell, Byeong Jeong |
| EPI_ISL_1080342 | New Jersey Department of Health, Public Health Laboratories | NJ_PHEL | Lindsey Bodnar, Shiv Verma, Dana Woell, Byeong Jeong |
| EPI_ISL_1080343, EPI_ISL_1080344 | NJ Rapid Response Teams | NJ_PHEL | Lindsey Bodnar, Shiv Verma, Dana Woell, Byeong Jeong |
| EPI_ISL_1080345 | Houston Methodist Hospital | Houston Methodist Hospital | S. Wesley Long, Randall J. Olsen, Paul A. Christensen, Sishir Subedi, Robert Olson, James J. Davis, Matthew Ojeda Saavedra, Prasanti Yerramilli, Layne Pruitt, Kristina Reppond, Madison N. Shyer, Jessica Cambric, Ilya J. Finkelstein, Jimmy Gollihar, and James M. Musser |
| EPI_ISL_1080346 | NJ Rapid Response Teams | NJ_PHEL | Lindsey Bodnar, Shiv Verma, Dana Woell, Byeong Jeong |
| EPI_ISL_1080349, EPI_ISL_1080352, EPI_ISL_1080353, EPI_ISL_1080354, EPI_ISL_1080355, EPI_ISL_1080356, EPI_ISL_1080357, EPI_ISL_1080358, EPI_ISL_1080359, EPI_ISL_1080360, EPI_ISL_1080361, EPI_ISL_1080362, EPI_ISL_1080363, EPI_ISL_1080364, EPI_ISL_1080365, EPI_ISL_1080366, EPI_ISL_1080367, EPI_ISL_1080368, EPI_ISL_1080369, EPI_ISL_1080370, EPI_ISL_1080371, EPI_ISL_1080372, EPI_ISL_1080373, EPI_ISL_1080374, EPI_ISL_1080377, EPI_ISL_1080383, EPI_ISL_1080392, EPI_ISL_1080393, EPI_ISL_1080394, EPI_ISL_1080395, EPI_ISL_1080396, EPI_ISL_1080397, EPI_ISL_1080398, EPI_ISL_1080399, EPI_ISL_1080400, EPI_ISL_1080401, EPI_ISL_1080402, EPI_ISL_1080403, EPI_ISL_1080404, EPI_ISL_1080405, EPI_ISL_1080406, EPI_ISL_1080407, EPI_ISL_1080408, EPI_ISL_1080409, EPI_ISL_1080410, EPI_ISL_1080411, EPI_ISL_1080412, EPI_ISL_1080413, EPI_ISL_1080414, EPI_ISL_1080415, EPI_ISL_1080416, EPI_ISL_1080417, EPI_ISL_1080418, EPI_ISL_1080419, EPI_ISL_1080420, EPI_ISL_1080421, EPI_ISL_1080422, EPI_ISL_1080423, EPI_ISL_1080424, EPI_ISL_1080425, EPI_ISL_1080426, EPI_ISL_1080427, EPI_ISL_1080428, EPI_ISL_1080429, EPI_ISL_1080430, EPI_ISL_1080431, EPI_ISL_1080432 | | | |
| see above | Houston Methodist Hospital | Houston Methodist Hospital | S. Wesley Long, Randall J. Olsen, Paul A. Christensen, Sishir Subedi, Robert Olson, James J. Davis, Matthew Ojeda Saavedra, Prasanti Yerramilli, Layne Pruitt, Kristina Reppond, Madison N. Shyer, Jessica Cambric, Ilya J. Finkelstein, Jimmy Gollihar, and James M. Musser |
| EPI_ISL_1080433, EPI_ISL_1080434, EPI_ISL_1080435, EPI_ISL_1080436, EPI_ISL_1080437, EPI_ISL_1080438, EPI_ISL_1080439, EPI_ISL_1080440, EPI_ISL_1080441, EPI_ISL_1080442, EPI_ISL_1080443, EPI_ISL_1080444, EPI_ISL_1080445, EPI_ISL_1080446, EPI_ISL_1080447, EPI_ISL_1080448, EPI_ISL_1080449, EPI_ISL_1080450, EPI_ISL_1080451, EPI_ISL_1080452, EPI_ISL_1080453 | | | |
| see above | Instituto Nacional de Medicina Genomica | Instituto Nacional de Medicina Genomica | Hidalgo-Miranda A, Mendoza-Vargas A, Reyes-Grajeda JP, Cedro-Tanda A, Alcaraz N, Gonzalez-Barrera D, Rangel-DeLeon D, Ramirez-Vega O, Sifuentes-Rojas C, Cisneros-Villanueva M, Herrera-Montalvo LA |
| EPI_ISL_1080454 | Murphy Medical Associates | Grubaugh Lab - Yale School of Public Health | Mary Petrone, Joseph Fauver, Caleb Neal, Steven Murphy, Chantal Vogels, Mallery Breban, Annie Watkins, Tara Alpert, Nathan Grubaugh |
| EPI_ISL_1080458, EPI_ISL_1080459 | Connecticut Veterans' Affairs Hospital | Grubaugh Lab - Yale School of Public Health | Mary Petrone, Joseph Fauver, Tara Alpert, Chantal Vogels, Isabel Ott, Shaili Gupta, Danielle Plank, Nathan Grubaugh |
| EPI_ISL_1080460, EPI_ISL_1080461 | Murphy Medical Associates | Grubaugh Lab - Yale School of Public Health | Mary Petrone, Joseph Fauver, Caleb Neal, Steven Murphy, Chantal Vogels, Mallery Breban, Annie Watkins, Tara Alpert, Nathan Grubaugh |
| EPI_ISL_1080462 | Connecticut Veterans' Affairs Hospital | Grubaugh Lab - Yale School of Public Health | Mary Petrone, Joseph Fauver, Tara Alpert, Chantal Vogels, Isabel Ott, Shaili Gupta, Danielle Plank, Nathan Grubaugh |
| EPI_ISL_1080463 | Murphy Medical Associates | Grubaugh Lab - Yale School of Public Health | Mary Petrone, Joseph Fauver, Caleb Neal, Steven Murphy, Chantal Vogels, Mallery Breban, Annie Watkins, Tara Alpert, Nathan Grubaugh |
| EPI_ISL_1080603, EPI_ISL_1080604, EPI_ISL_1080680, EPI_ISL_1080749, EPI_ISL_1080750 | Ohio Department of Health Laboratory | Ohio Department of Health Laboratory | Holmes, Jennifer; Eric Brandt, Keoni Omura, Glen McGillivary, Caitlin McDonnell, Jade Mowery, Stephanie Mccracken, Tyler Payne, Kirtana Ramadugu, Erica Leasure, Brent Lee, Kelsey Florek, Heather Blankenship, Allison Black, Quanta Brown, and Tammy Bannerman |
| EPI_ISL_1080751, EPI_ISL_1080752, EPI_ISL_1080753, EPI_ISL_1080754, EPI_ISL_1080755, EPI_ISL_1080756, EPI_ISL_1080757, EPI_ISL_1080758, EPI_ISL_1080759, EPI_ISL_1080760, EPI_ISL_1080761, EPI_ISL_1080762, EPI_ISL_1080763, EPI_ISL_1080764, EPI_ISL_1080765, EPI_ISL_1080766, EPI_ISL_1080767, EPI_ISL_1080768, EPI_ISL_1080769, EPI_ISL_1080770, EPI_ISL_1080771, EPI_ISL_1080772, EPI_ISL_1080773, EPI_ISL_1080774, EPI_ISL_1080775, EPI_ISL_1080776, EPI_ISL_1080777, EPI_ISL_1080778, EPI_ISL_1080784, EPI_ISL_1080785, EPI_ISL_1080786, EPI_ISL_1080787, EPI_ISL_1080788, EPI_ISL_1080789, EPI_ISL_1080790, EPI_ISL_1080791, EPI_ISL_1080792, EPI_ISL_1080793, EPI_ISL_1080794, EPI_ISL_1080795, EPI_ISL_1080796, EPI_ISL_1080797, EPI_ISL_1080798, EPI_ISL_1080799, EPI_ISL_1080800, EPI_ISL_1080801, EPI_ISL_1080802, EPI_ISL_1080803, EPI_ISL_1080804, EPI_ISL_1080805, EPI_ISL_1080806, EPI_ISL_1080807, EPI_ISL_1080808, EPI_ISL_1080809, EPI_ISL_1080810 | | | |
| see above | Clinical Microbiology Laboratory, NewYork Presbyterian Hospital/Columbia University Irving Medical Center | Uhlemann Laboratory, Columbia University Irving Medical Center | Medini K. Annavajhala, Anne-Catrin Uhlemann, Hiroshi Mohri, David Ho |
| EPI_ISL_1080853 | Baylor Scott & White-Temple | Baylor Scott & White-Temple | Ari Rao, Linden Morales, Kimberly Walker, Marcus Volz, Shelby Hendrickson |
| EPI_ISL_1080854 | Ohio Department of Health Laboratory | Ohio Department of Health Laboratory | Holmes, Jennifer; Eric Brandt, Keoni Omura, Glen McGillivary, Caitlin McDonnell, Jade Mowery, Stephanie Mccracken, Tyler Payne, Kirtana Ramadugu, Erica Leasure, Brent Lee, Kelsey Florek, Heather Blankenship, Allison Black, Quanta Brown, and Tammy Bannerman |
| EPI_ISL_1080855, EPI_ISL_1080857 | Baylor Scott & White-Temple | Baylor Scott & White-Temple | Ari Rao, Linden Morales, Kimberly Walker, Marcus Volz, Shelby Hendrickson |
| EPI_ISL_1080858, EPI_ISL_1080859, EPI_ISL_1080860, EPI_ISL_1080861, EPI_ISL_1080862, EPI_ISL_1080863, EPI_ISL_1080864, EPI_ISL_1080865, EPI_ISL_1080866, EPI_ISL_1080867, EPI_ISL_1080868, EPI_ISL_1080869, EPI_ISL_1080870, EPI_ISL_1080871, EPI_ISL_1080872, EPI_ISL_1080873, EPI_ISL_1080874, EPI_ISL_1080875, EPI_ISL_1080876, EPI_ISL_1080877, EPI_ISL_1080878, EPI_ISL_1080879, EPI_ISL_1080880, EPI_ISL_1080881, EPI_ISL_1080882, EPI_ISL_1080883, EPI_ISL_1080884, EPI_ISL_1080885, EPI_ISL_1080886, EPI_ISL_1080887, EPI_ISL_1080888, EPI_ISL_1080889, EPI_ISL_1080890, EPI_ISL_1080891, EPI_ISL_1080892, EPI_ISL_1080893, EPI_ISL_1080894, EPI_ISL_1080895, EPI_ISL_1080896, EPI_ISL_1080897, EPI_ISL_1080898, EPI_ISL_1080899, EPI_ISL_1080900, EPI_ISL_1080901, EPI_ISL_1080902, EPI_ISL_1080903, EPI_ISL_1080904, EPI_ISL_1080905, EPI_ISL_1080906, EPI_ISL_1080907, EPI_ISL_1080908, EPI_ISL_1080909, EPI_ISL_1080910, EPI_ISL_1080911, EPI_ISL_1080912, EPI_ISL_1080913, EPI_ISL_1080914, EPI_ISL_1080915, EPI_ISL_1080916, EPI_ISL_1080917, EPI_ISL_1080918, EPI_ISL_1080919, EPI_ISL_1080920, EPI_ISL_1080921, EPI_ISL_1080922, EPI_ISL_1080923, EPI_ISL_1080924, EPI_ISL_1080925, EPI_ISL_1080926, EPI_ISL_1080927, EPI_ISL_1080928, EPI_ISL_1080929, EPI_ISL_1080930, EPI_ISL_1080934, EPI_ISL_1080938, EPI_ISL_1080942, EPI_ISL_1080946, EPI_ISL_1080951, EPI_ISL_1080955, EPI_ISL_1080960, EPI_ISL_1080964, EPI_ISL_1080968, EPI_ISL_1080973, EPI_ISL_1080977, EPI_ISL_1080980, EPI_ISL_1080985, EPI_ISL_1080986, EPI_ISL_1080987, EPI_ISL_1080988, EPI_ISL_1080989, EPI_ISL_1080990, EPI_ISL_1080991, EPI_ISL_1080992, EPI_ISL_1080993, EPI_ISL_1080994, EPI_ISL_1080995, EPI_ISL_1080996, EPI_ISL_1080997, EPI_ISL_1080998, EPI_ISL_1080999, EPI_ISL_1081000, EPI_ISL_1081001, EPI_ISL_1081002, EPI_ISL_1081003, EPI_ISL_1081004, EPI_ISL_1081005, EPI_ISL_1081006, EPI_ISL_1081007, EPI_ISL_1081008, EPI_ISL_1081009, EPI_ISL_1081010, EPI_ISL_1081011, EPI_ISL_1081012, EPI_ISL_1081013, EPI_ISL_1081014, EPI_ISL_1081015, EPI_ISL_1081016, EPI_ISL_1081017, EPI_ISL_1081018, EPI_ISL_1081019, EPI_ISL_1081020, EPI_ISL_1081021, EPI_ISL_1081022, EPI_ISL_1081023, EPI_ISL_1081024, EPI_ISL_1081025, EPI_ISL_1081026, EPI_ISL_1081027, EPI_ISL_1081028, EPI_ISL_1081029, EPI_ISL_1081030, EPI_ISL_1081031, EPI_ISL_1081032, EPI_ISL_1081033, EPI_ISL_1081034, EPI_ISL_1081035, EPI_ISL_1081036, EPI_ISL_1081037, EPI_ISL_1081038, EPI_ISL_1081039, EPI_ISL_1081040, EPI_ISL_1081041, EPI_ISL_1081042, EPI_ISL_1081043, EPI_ISL_1081044 | | | |
| see above | Houston Methodist Hospital | Houston Methodist Hospital | S. Wesley Long, Randall J. Olsen, Paul A. Christensen, Sishir Subedi, Robert Olson, James J. Davis, Matthew Ojeda Saavedra, Prasanti Yerramilli, Layne Pruitt, Kristina Reppond, Madison N. Shyer, Jessica Cambric, Ilya J. Finkelstein, Jimmy Gollihar, and James M. Musser |
| EPI_ISL_1081045, EPI_ISL_1081046, EPI_ISL_1081047, EPI_ISL_1081048, EPI_ISL_1081049, EPI_ISL_1081050, EPI_ISL_1081051, EPI_ISL_1081052, EPI_ISL_1081053, EPI_ISL_1081054, EPI_ISL_1081055, EPI_ISL_1081056, EPI_ISL_1081057, EPI_ISL_1081059, EPI_ISL_1081060, EPI_ISL_1081061, EPI_ISL_1081064, EPI_ISL_1081065, EPI_ISL_1081066, EPI_ISL_1081067, EPI_ISL_1081068, EPI_ISL_1081069, EPI_ISL_1081070, EPI_ISL_1081071, EPI_ISL_1081072, EPI_ISL_1081073, EPI_ISL_1081074, EPI_ISL_1081075, EPI_ISL_1081076, EPI_ISL_1081077, EPI_ISL_1081078, EPI_ISL_1081079, EPI_ISL_1081080, EPI_ISL_1081081, EPI_ISL_1081082, EPI_ISL_1081083, EPI_ISL_1081084, EPI_ISL_1081085, EPI_ISL_1081086, EPI_ISL_1081087, EPI_ISL_1081088, EPI_ISL_1081089, EPI_ISL_1081090, EPI_ISL_1081091, EPI_ISL_1081092, EPI_ISL_1081093, EPI_ISL_1081094, EPI_ISL_1081095, EPI_ISL_1081096 | | | |
| see above | Helix/Illumina | Respiratory Viruses Branch, Division of Viral Diseases, Centers for Disease Control and Prevention | Peter W. Cook, Dakota Howard, Dhwani Batra, Ben L. Rambo-Martin, Eileen de Feo, Jan Antico, Christine Tran, Matthew Tolentino, Shannon Wickline, Kim Gietzen, Brad Sickler, Jingtao Liu, Eric Allen, Phil Febbo, Summer Galloway, Nicole L. Washington, Simon White, Geraint Levan, Kelly Schiabor Barrett, Elizabeth Cirulli, Alexandre Bolze, Ary Ascencio, Charlotte Rivera-Garcia, Ryan Cho, Jason Nguyen, Sherry Wang, Jimmy Ramirez, Tyler Cassens, Efren Sandoval, Magnus Isaksson, William Lee, David Becker, Marc Laurent, James Lu, Clinton R. Paden, Suxiang Tong, Duncan MacCannell |
| EPI_ISL_1081136, EPI_ISL_1081149, EPI_ISL_1081150, EPI_ISL_1081151 | Baylor Scott & White-Temple | Baylor Scott & White-Temple | Ari Rao, Linden Morales, Kimberly Walker, Marcus Volz, Shelby Hendrickson |
| EPI_ISL_1081152, EPI_ISL_1081153, EPI_ISL_1081154, EPI_ISL_1081155, EPI_ISL_1081156, EPI_ISL_1081158, EPI_ISL_1081159, EPI_ISL_1081160, EPI_ISL_1081161, EPI_ISL_1081162, EPI_ISL_1081163, EPI_ISL_1081164, EPI_ISL_1081165, EPI_ISL_1081166, EPI_ISL_1081167, EPI_ISL_1081168, EPI_ISL_1081169 | | | |
| see above | Instituto Nacional de Medicina Genomica | Instituto Nacional de Medicina Genomica | Hidalgo-Miranda A, Mendoza-Vargas A, Reyes-Grajeda JP, Cedro-Tanda A, Alcaraz N, Gonzalez-Barrera D, Rangel-DeLeon D, Ramirez-Vega O, Sifuentes-Rojas C, Cisneros-Villanueva M, Herrera-Montalvo LA |
| EPI_ISL_1081170 | IEH Laboratories & Consulting Group | ID Genomics | Veronika Tchesnokova, Kirthi Kutumbaka, Kai Deng, Hemantha Kulakesara, Lydia Larson, Hao Geng, Curtis Fowler, Mansour Samadpour, Evgeni Sokurenko |
| EPI_ISL_1081171, EPI_ISL_1081172, EPI_ISL_1081173, EPI_ISL_1081174, EPI_ISL_1081175, EPI_ISL_1081176, EPI_ISL_1081177, EPI_ISL_1081178, EPI_ISL_1081179, EPI_ISL_1081180, EPI_ISL_1081181, EPI_ISL_1081182, EPI_ISL_1081183, EPI_ISL_1081184, EPI_ISL_1081185, EPI_ISL_1081186, EPI_ISL_1081187, EPI_ISL_1081188, EPI_ISL_1081189, EPI_ISL_1081190, EPI_ISL_1081191 | | | |
| see above | Baylor Scott & White-Temple | Baylor Scott & White-Temple | Ari Rao, Linden Morales, Kimberly Walker, Marcus Volz, Shelby Hendrickson |
| EPI_ISL_1081192 | Santa Clara County Public Health Laboratory | Santa Clara County Public Health Laboratory | Santa Clara County Public Health Department |
| EPI_ISL_1081201, EPI_ISL_1081203, EPI_ISL_1081206, EPI_ISL_1081208, EPI_ISL_1081209, | Laboratory Corporation of America | Respiratory Viruses Branch, Division of Viral Diseases, Centers for Disease Control and Prevention | Peter W. Cook, Dakota Howard, Dhwani Batra, Ben L. Rambo-Martin, Eileen de Feo, Jan Antico, Christine Tran, Matthew Tolentino, Shannon Wickline, Kim Gietzen, Brad Sickler, Jingtao Liu, Eric Allen, Phil Febbo, Summer Galloway, Nicole L. Washington, Simon White, Geraint Levan, Kelly Schiabor Barrett, Elizabeth Cirulli, Alexandre Bolze, Ary Ascencio, Charlotte Rivera-Garcia, Ryan Cho, Jason Nguyen, Sherry Wang, Jimmy Ramirez, Tyler Cassens, Efren Sandoval, Magnus Isaksson, William Lee, David Becker, Marc Laurent, James Lu, Clinton R. Paden, Suxiang Tong, Duncan MacCannell |

| | | | |
|---|---|---|---|
| EPI_ISL_1081210, EPI_ISL_1081212, EPI_ISL_1081213, EPI_ISL_1081214 | | | |
| EPI_ISL_1081219, EPI_ISL_1081220, EPI_ISL_1081221, EPI_ISL_1081222, EPI_ISL_1081223, EPI_ISL_1081224, EPI_ISL_1081225, EPI_ISL_1081226, EPI_ISL_1081227, EPI_ISL_1081228, EPI_ISL_1081229, EPI_ISL_1081230, EPI_ISL_1081231, EPI_ISL_1081232, EPI_ISL_1081233, EPI_ISL_1081234, EPI_ISL_1081235, EPI_ISL_1081236, EPI_ISL_1081237, EPI_ISL_1081238, EPI_ISL_1081239, EPI_ISL_1081240, EPI_ISL_1081241, EPI_ISL_1081242, EPI_ISL_1081243, EPI_ISL_1081244, EPI_ISL_1081245, EPI_ISL_1081246, EPI_ISL_1081247, EPI_ISL_1081248, EPI_ISL_1081249, EPI_ISL_1081250, EPI_ISL_1081251, EPI_ISL_1081252, EPI_ISL_1081253, EPI_ISL_1081254, EPI_ISL_1081255, EPI_ISL_1081256, EPI_ISL_1081257, EPI_ISL_1081258, EPI_ISL_1081259, EPI_ISL_1081261, EPI_ISL_1081262, EPI_ISL_1081263, EPI_ISL_1081264, EPI_ISL_1081265, EPI_ISL_1081266 | | | |
| see above | Maryland Public Health Laboratory | Maryland Public Health Laboratory | Maryland Department of Health Laboratories Administration |
| EPI_ISL_1081267, EPI_ISL_1081268, EPI_ISL_1081269, EPI_ISL_1081270, EPI_ISL_1081271, EPI_ISL_1081272, EPI_ISL_1081273, EPI_ISL_1081274, EPI_ISL_1081275, EPI_ISL_1081277, EPI_ISL_1081278, EPI_ISL_1081279, EPI_ISL_1081280 | | | |
| see above | Laboratory Corporation of America | Respiratory Viruses Branch, Division of Viral Diseases, Centers for Disease Control and Prevention | Peter W. Cook, Dakota Howard, Dhwani Batra, Ben L. Rambo-Martin, Eileen de Feo, Jan Antico, Christine Tran, Matthew Tolentino, Shannon Wickline, Kim Gietzen, Brad Sickler, Jingtao Liu, Eric Allen, Phil Febbo, Summer Galloway, Nicole L. Washington, Simon White, Geraint Levan, Kelly Schiabor Barrett, Elizabeth Cirulli, Alexandre Bolze, Ary Ascencio, Charlotte Rivera-Garcia, Ryan Cho, Jason Nguyen, Sherry Wang, Jimmy Ramirez, Tyler Cassens, Efren Sandoval, Magnus Isaksson, William Lee, David Becker, Marc Laurent, James Lu, Clinton R. Paden, Suxiang Tong, Duncan MacCannell |
| EPI_ISL_1081287, EPI_ISL_1081288 | Innovative Genomics Institute, UC Berkeley | Innovative Genomics Institute, UC Berkeley | Stacia Wyman, Alison Ciling, Netravathi Krishnappa, Haridha Shivram, Phil Frankino, Liana Lareau |
| EPI_ISL_1081292, EPI_ISL_1081294, EPI_ISL_1081295, EPI_ISL_1081296, EPI_ISL_1081297, EPI_ISL_1081298, EPI_ISL_1081299, EPI_ISL_1081300, EPI_ISL_1081301, EPI_ISL_1081302, EPI_ISL_1081303, EPI_ISL_1081307, EPI_ISL_1081309 | | | |
| see above | Wyoming Public Health Laboratory | Wyoming Public Health Laboratory | Jim Mildenberger, Wanda Manley, Noah Hull, Taylor Fearing, Lynette Gumbleton, Channing Weber, Ashley Norberg, Chayse Rowley, Marley Goetz, Brian Dominguez, Elliot Thomasson, Cari Sloma, and Rob Christensen |
| EPI_ISL_1081312, EPI_ISL_1081313, EPI_ISL_1081314, EPI_ISL_1081315, EPI_ISL_1081316, EPI_ISL_1081317, EPI_ISL_1081318, EPI_ISL_1081319, EPI_ISL_1081320, EPI_ISL_1081321, EPI_ISL_1081322, EPI_ISL_1081323 | | | |
| see above | Bozeman Health Deaconess Hospital | Wiedenheft lab, Montana State University | Artem Nemudryi, Anna Nemudraia, Tanner Wiegand, Joseph Nichols, Deann T. Snyder, Jodi F. Hedges, Calvin Cicha, Helen Lee, Karl K. Vanderwood, Diane Bimczok, Mark A. Jutila and Blake Wiedenheft |
| EPI_ISL_1081332, EPI_ISL_1081337, EPI_ISL_1081339, EPI_ISL_1081341, EPI_ISL_1081346, EPI_ISL_1081347 | IEH Laboratories and Consulting Group | IEH and ID Genomics | Veronika Tchesnokova, Hemantha Kulakesara, Lydia Larson, Kai Deng, Kirthi Kutumbaka, Hao Geng, Curtis Fowler, Mansour Samadpour, Evgeni Sokurenko |
| EPI_ISL_1081373, EPI_ISL_1081374, EPI_ISL_1081375, EPI_ISL_1081376, EPI_ISL_1081377, EPI_ISL_1081378, EPI_ISL_1081379, EPI_ISL_1081380, EPI_ISL_1081381, EPI_ISL_1081382, EPI_ISL_1081383, EPI_ISL_1081384, EPI_ISL_1081385, EPI_ISL_1081386, EPI_ISL_1081387, EPI_ISL_1081388, EPI_ISL_1081389, EPI_ISL_1081390, EPI_ISL_1081391, EPI_ISL_1081392, EPI_ISL_1081393, EPI_ISL_1081394, EPI_ISL_1081395, EPI_ISL_1081396, EPI_ISL_1081397, EPI_ISL_1081398, EPI_ISL_1081399, EPI_ISL_1081400, EPI_ISL_1081401, EPI_ISL_1081402, EPI_ISL_1081403, EPI_ISL_1081404, EPI_ISL_1081405, EPI_ISL_1081406, EPI_ISL_1081407, EPI_ISL_1081408, EPI_ISL_1081409, EPI_ISL_1081410, EPI_ISL_1081411, EPI_ISL_1081412, EPI_ISL_1081413, EPI_ISL_1081414, EPI_ISL_1081415, EPI_ISL_1081416, EPI_ISL_1081417, EPI_ISL_1081418, EPI_ISL_1081419, EPI_ISL_1081420, EPI_ISL_1081421, EPI_ISL_1081422 | | | |
| see above | Sharp HealthCare Laboratory | Andersen lab at Scripps Research | SEARCH Alliance San Diego with Aaron Harding, Jacquelyn Berumen, Cathy Woerle, Liam McGinnis, Art Mendoza, Omid Bakhtar |
| EPI_ISL_1081423, EPI_ISL_1081424, EPI_ISL_1081425, EPI_ISL_1081426, EPI_ISL_1081427, EPI_ISL_1081428, EPI_ISL_1081429, EPI_ISL_1081430, EPI_ISL_1081431, EPI_ISL_1081432, EPI_ISL_1081433, EPI_ISL_1081434, EPI_ISL_1081435, EPI_ISL_1081436, EPI_ISL_1081437, EPI_ISL_1081438, EPI_ISL_1081439, EPI_ISL_1081440, EPI_ISL_1081441, EPI_ISL_1081442, EPI_ISL_1081443, EPI_ISL_1081444, EPI_ISL_1081445, EPI_ISL_1081446, EPI_ISL_1081447, EPI_ISL_1081448, EPI_ISL_1081449, EPI_ISL_1081450, EPI_ISL_1081451, EPI_ISL_1081452, EPI_ISL_1081453, EPI_ISL_1081454, EPI_ISL_1081455, EPI_ISL_1081456 | | | |
| see above | Genomica Lab Molecular, M√©xico | Andersen lab at Scripps Research | SEARCH Alliance San Diego with Jonathan Gonzalez Garcia, Jose Roman Chavez Mendez, Jose Horacio Reyna Verdugo, Martin Gonzalez Ibarra, Luis Alberto Rangel Gonzalez |
| EPI_ISL_1081457, EPI_ISL_1081458, EPI_ISL_1081459, EPI_ISL_1081460, EPI_ISL_1081461, EPI_ISL_1081462, EPI_ISL_1081463, EPI_ISL_1081464, EPI_ISL_1081465, EPI_ISL_1081466, EPI_ISL_1081467, EPI_ISL_1081468, EPI_ISL_1081469, EPI_ISL_1081470, EPI_ISL_1081471, EPI_ISL_1081472 | | | |
| see above | San Diego County Public Health Laboratory | Andersen lab at Scripps Research | SEARCH Alliance San Diego with Tracy Basler, Jovan Shephard, Brett Austin |
| EPI_ISL_1081473, EPI_ISL_1081474, EPI_ISL_1081475, EPI_ISL_1081476, EPI_ISL_1081477, EPI_ISL_1081478, EPI_ISL_1081479, EPI_ISL_1081480, EPI_ISL_1081481, EPI_ISL_1081482, EPI_ISL_1081483, EPI_ISL_1081484, EPI_ISL_1081485, EPI_ISL_1081486, EPI_ISL_1081487, EPI_ISL_1081488, EPI_ISL_1081489, EPI_ISL_1081490, EPI_ISL_1081491, EPI_ISL_1081492, EPI_ISL_1081493, EPI_ISL_1081494, EPI_ISL_1081495 | | | |
| see above | Genomica Lab Molecular, M√©xico | Andersen lab at Scripps Research | SEARCH Alliance San Diego with Jonathan Gonzalez Garcia, Jose Roman Chavez Mendez, Jose Horacio Reyna Verdugo, Martin Gonzalez Ibarra, Luis Alberto Rangel Gonzalez |
| EPI_ISL_1081496, EPI_ISL_1081497, EPI_ISL_1081498, EPI_ISL_1081499, EPI_ISL_1081500, EPI_ISL_1081501, EPI_ISL_1081502, EPI_ISL_1081503, EPI_ISL_1081504, EPI_ISL_1081505, EPI_ISL_1081506, EPI_ISL_1081507, EPI_ISL_1081508, EPI_ISL_1081509, EPI_ISL_1081510, EPI_ISL_1081511, EPI_ISL_1081512, EPI_ISL_1081513, EPI_ISL_1081514, EPI_ISL_1081515, EPI_ISL_1081516, EPI_ISL_1081517, EPI_ISL_1081518, EPI_ISL_1081519, EPI_ISL_1081520, EPI_ISL_1081521, EPI_ISL_1081522, EPI_ISL_1081523, EPI_ISL_1081524, EPI_ISL_1081525, EPI_ISL_1081526, EPI_ISL_1081527, EPI_ISL_1081528, EPI_ISL_1081529, EPI_ISL_1081530, EPI_ISL_1081531, EPI_ISL_1081532, EPI_ISL_1081533, EPI_ISL_1081534, EPI_ISL_1081535, EPI_ISL_1081536, EPI_ISL_1081537, EPI_ISL_1081538, EPI_ISL_1081539, EPI_ISL_1081540, EPI_ISL_1081541, EPI_ISL_1081542, EPI_ISL_1081543, EPI_ISL_1081544, EPI_ISL_1081545, EPI_ISL_1081546, EPI_ISL_1081547, EPI_ISL_1081548, EPI_ISL_1081549 | | | |
| see above | Sharp HealthCare Laboratory | Andersen lab at Scripps Research | SEARCH Alliance San Diego with Aaron Harding, Jacquelyn Berumen, Cathy Woerle, Liam McGinnis, Art Mendoza, Omid Bakhtar |
| EPI_ISL_1081550, EPI_ISL_1081551, EPI_ISL_1081552, EPI_ISL_1081553, EPI_ISL_1081554, EPI_ISL_1081555, EPI_ISL_1081556, EPI_ISL_1081557, EPI_ISL_1081558, EPI_ISL_1081559, EPI_ISL_1081560, EPI_ISL_1081561, EPI_ISL_1081562, EPI_ISL_1081563, EPI_ISL_1081564, EPI_ISL_1081565, EPI_ISL_1081566, EPI_ISL_1081567 | | | |
| see above | Scripps Medical Laboratory | Andersen lab at Scripps Research | SEARCH Alliance San Diego with Michael Quigley, Ellen Stefanski, Ian Mchardy |
| EPI_ISL_1081568, EPI_ISL_1081569, EPI_ISL_1081570 | Sharp HealthCare Laboratory | Andersen lab at Scripps Research | SEARCH Alliance San Diego with Aaron Harding, Jacquelyn Berumen, Cathy Woerle, Liam McGinnis, Art Mendoza, Omid Bakhtar |
| EPI_ISL_1081571, EPI_ISL_1081572, EPI_ISL_1081573, EPI_ISL_1081574, EPI_ISL_1081575, EPI_ISL_1081576, EPI_ISL_1081577, EPI_ISL_1081578, EPI_ISL_1081579, EPI_ISL_1081580, EPI_ISL_1081581, EPI_ISL_1081582, EPI_ISL_1081583, EPI_ISL_1081584, EPI_ISL_1081585, EPI_ISL_1081586, EPI_ISL_1081587, EPI_ISL_1081588, EPI_ISL_1081589, EPI_ISL_1081590, EPI_ISL_1081591, EPI_ISL_1081592, EPI_ISL_1081593, EPI_ISL_1081594, EPI_ISL_1081595, EPI_ISL_1081596, EPI_ISL_1081597 | | | |
| see above | Rady's Childrens Hospital | Andersen lab at Scripps Research | SEARCH Alliance San Diego with Nanda Radamchar, David Dimmock, Linda Luo, Christina Clarke, Kathryn Bouic, Teresa Mueller, Denise Malicki |
| EPI_ISL_1081598, EPI_ISL_1081599, EPI_ISL_1081600, EPI_ISL_1081601, EPI_ISL_1081602, EPI_ISL_1081603, EPI_ISL_1081604, EPI_ISL_1081605, EPI_ISL_1081606, EPI_ISL_1081607, EPI_ISL_1081608, EPI_ISL_1081609, EPI_ISL_1081610, EPI_ISL_1081611, EPI_ISL_1081612, EPI_ISL_1081613, EPI_ISL_1081614, EPI_ISL_1081615, EPI_ISL_1081616, EPI_ISL_1081617, EPI_ISL_1081618 | | | |
| see above | Scripps Medical Laboratory | Andersen lab at Scripps Research | SEARCH Alliance San Diego with Michael Quigley, Ellen Stefanski, Ian Mchardy |
| EPI_ISL_1081619, EPI_ISL_1081620, EPI_ISL_1081621, EPI_ISL_1081622, EPI_ISL_1081623, EPI_ISL_1081624, EPI_ISL_1081625, EPI_ISL_1081626, EPI_ISL_1081627 | San Diego County Public Health Laboratory | Andersen lab at Scripps Research | SEARCH Alliance San Diego with Tracy Basler, Jovan Shephard, Brett Austin |
| EPI_ISL_1081628, EPI_ISL_1081629, EPI_ISL_1081630, EPI_ISL_1081631, EPI_ISL_1081632, EPI_ISL_1081633, EPI_ISL_1081634, EPI_ISL_1081635, EPI_ISL_1081636, EPI_ISL_1081637, EPI_ISL_1081638, EPI_ISL_1081639, EPI_ISL_1081640, EPI_ISL_1081641, EPI_ISL_1081642, EPI_ISL_1081643, EPI_ISL_1081644, EPI_ISL_1081645, EPI_ISL_1081646, EPI_ISL_1081647, EPI_ISL_1081648, EPI_ISL_1081649, EPI_ISL_1081650, EPI_ISL_1081651, EPI_ISL_1081652, EPI_ISL_1081653, EPI_ISL_1081654, EPI_ISL_1081655, EPI_ISL_1081656, EPI_ISL_1081657, EPI_ISL_1081658, EPI_ISL_1081659, EPI_ISL_1081660, EPI_ISL_1081661, EPI_ISL_1081662, EPI_ISL_1081663, EPI_ISL_1081664, EPI_ISL_1081665, EPI_ISL_1081666, EPI_ISL_1081667, EPI_ISL_1081668, EPI_ISL_1081669, EPI_ISL_1081670, EPI_ISL_1081671, EPI_ISL_1081672, EPI_ISL_1081673, EPI_ISL_1081674, EPI_ISL_1081675, EPI_ISL_1081676, EPI_ISL_1081677, EPI_ISL_1081678, EPI_ISL_1081679, EPI_ISL_1081680, EPI_ISL_1081681, EPI_ISL_1081682, EPI_ISL_1081683, EPI_ISL_1081684, EPI_ISL_1081685, EPI_ISL_1081686, EPI_ISL_1081687, EPI_ISL_1081688, EPI_ISL_1081689, EPI_ISL_1081690, EPI_ISL_1081691, EPI_ISL_1081692, EPI_ISL_1081693, EPI_ISL_1081694, EPI_ISL_1081695, EPI_ISL_1081696, EPI_ISL_1081697, EPI_ISL_1081698, EPI_ISL_1081699, EPI_ISL_1081700, EPI_ISL_1081701, EPI_ISL_1081702, EPI_ISL_1081703, EPI_ISL_1081704, EPI_ISL_1081705, EPI_ISL_1081706, EPI_ISL_1081707, EPI_ISL_1081708, EPI_ISL_1081709, EPI_ISL_1081710, EPI_ISL_1081711, EPI_ISL_1081712, EPI_ISL_1081713, EPI_ISL_1081714, EPI_ISL_1081715, EPI_ISL_1081716, EPI_ISL_1081717, EPI_ISL_1081718, EPI_ISL_1081719, EPI_ISL_1081720, EPI_ISL_1081721, EPI_ISL_1081722, EPI_ISL_1081723, EPI_ISL_1081724, EPI_ISL_1081725, EPI_ISL_1081726, EPI_ISL_1081727, EPI_ISL_1081728, EPI_ISL_1081729, EPI_ISL_1081730, EPI_ISL_1081731, EPI_ISL_1081732, EPI_ISL_1081733, EPI_ISL_1081734, EPI_ISL_1081735, EPI_ISL_1081736, EPI_ISL_1081737, EPI_ISL_1081738, EPI_ISL_1081739, EPI_ISL_1081740, EPI_ISL_1081741 | | | |
| see above | Scripps Medical Laboratory | Andersen lab at Scripps Research | SEARCH Alliance San Diego with Michael Quigley, Ellen Stefanski, Ian Mchardy |
| EPI_ISL_1081742, EPI_ISL_1081743, EPI_ISL_1081744, EPI_ISL_1081745, EPI_ISL_1081746, EPI_ISL_1081747, EPI_ISL_1081748, EPI_ISL_1081749, EPI_ISL_1081750, EPI_ISL_1081751, EPI_ISL_1081752, EPI_ISL_1081753, EPI_ISL_1081754, EPI_ISL_1081755, EPI_ISL_1081756, EPI_ISL_1081757, EPI_ISL_1081758, EPI_ISL_1081759, EPI_ISL_1081760, EPI_ISL_1081761, EPI_ISL_1081762, EPI_ISL_1081763, EPI_ISL_1081764, EPI_ISL_1081765, EPI_ISL_1081766, EPI_ISL_1081767, EPI_ISL_1081768, EPI_ISL_1081769, EPI_ISL_1081770, EPI_ISL_1081771, EPI_ISL_1081772, EPI_ISL_1081773, EPI_ISL_1081774, EPI_ISL_1081775, EPI_ISL_1081776, EPI_ISL_1081777, EPI_ISL_1081778, EPI_ISL_1081779, EPI_ISL_1081780, EPI_ISL_1081781, EPI_ISL_1081782, EPI_ISL_1081783, EPI_ISL_1081784, EPI_ISL_1081785, EPI_ISL_1081786, EPI_ISL_1081787, EPI_ISL_1081788, EPI_ISL_1081789, EPI_ISL_1081790, EPI_ISL_1081791 | | | |
| see above | San Diego County Public Health Laboratory | Andersen lab at Scripps Research | SEARCH Alliance San Diego with Tracy Basler, Jovan Shephard, Brett Austin |
| EPI_ISL_1082452, EPI_ISL_1082453, EPI_ISL_1082526, EPI_ISL_1082888, EPI_ISL_1083590, EPI_ISL_1084605, EPI_ISL_1084722, EPI_ISL_1084823, EPI_ISL_1085007, EPI_ISL_1085008 | Ohio Department of Health Laboratory | Ohio Department of Health Laboratory | Holmes, Jennifer; Eric Brandt, Keoni Omura, Glen McGillivary, Caitlin McDonnell, Jade Mowery, Stephanie Mccracken, Tyler Payne, Kirtana Ramadugu, Erica Leasure, Brent Lee, Kelsey Florek, Heather Blankenship, Allison Black, Quanta Brown, and Tammy Bannerman |
| EPI_ISL_1085093, EPI_ISL_1085161, EPI_ISL_1085163, EPI_ISL_1085166, EPI_ISL_1085168, EPI_ISL_1085171 | NYU Langone Health | Departments of Pathology and Medicine, New York University School of Medicine | Adriana Heguy, Dacia Dimartino, Emily Guzman, Christian Marier, Peter Meyn, Sitharam Ramaswami, Gael Westby, Paul Zappile, Yutong Zhang, Paolo Cotzia, Guiqing Wang |
| EPI_ISL_1085172 | Fidalab | Hyde Lab | Jennifer L. Hyde, Roger Peck, Quinne Feng |
| EPI_ISL_1085175 | NYU Langone Health | Departments of Pathology and Medicine, New York University School of Medicine | Adriana Heguy, Dacia Dimartino, Emily Guzman, Christian Marier, Peter Meyn, Sitharam Ramaswami, Gael Westby, Paul Zappile, Yutong Zhang, Paolo Cotzia, Guiqing Wang |
| EPI_ISL_1085421 | Instituto Nacional de Medicina Genomica | Instituto Nacional de Medicina Genomica | Hidalgo-Miranda A, Mendoza-Vargas A, Reyes-Grajeda JP, Cedro-Tanda A, Alcaraz N, Gonzalez-Barrera D, Rangel-DeLeon D, Ramirez-Vega O, Sifuentes-Rojas C, Cisneros-Villanueva M, Herrera-Montalvo LA |

EPI_ISL_1085676 University of Oregon COVID-19 MAP Laboratory | University of Oregon Genomics and Cell Characterization Core Facility (GC3F) | Douglas Turnbull, Ariana White, Jeff Bishop, Jason Carriere

EPI_ISL_1085677 University of Oregon COVID-19 MAP Laboratory | University of Oregon Genomics and Cell Characterization Core Facility (GC3F) | Douglas Turnbull, Ariana White, Jason Carriere, Jeff Bishop

EPI_ISL_1086218, EPI_ISL_1086219, EPI_ISL_1086220, EPI_ISL_1086221, EPI_ISL_1086222, EPI_ISL_1086223, EPI_ISL_1086224, EPI_ISL_1086225, EPI_ISL_1086226, EPI_ISL_1086227, EPI_ISL_1086228, EPI_ISL_1086229, EPI_ISL_1086230, EPI_ISL_1086231, EPI_ISL_1086232, EPI_ISL_1086233, EPI_ISL_1086234, EPI_ISL_1086235, EPI_ISL_1086236, EPI_ISL_1086237, EPI_ISL_1086238, EPI_ISL_1086239,
EPI_ISL_1086240, EPI_ISL_1086241, EPI_ISL_1086242, EPI_ISL_1086243, EPI_ISL_1086244, EPI_ISL_1086245, EPI_ISL_1086246, EPI_ISL_1086247, EPI_ISL_1086248, EPI_ISL_1086249, EPI_ISL_1086250, EPI_ISL_1086251, EPI_ISL_1086252, EPI_ISL_1086253, EPI_ISL_1086254, EPI_ISL_1086255, EPI_ISL_1086256, EPI_ISL_1086257, EPI_ISL_1086258, EPI_ISL_1086259, EPI_ISL_1086260, EPI_ISL_1086261,
EPI_ISL_1086262, EPI_ISL_1086263, EPI_ISL_1086264, EPI_ISL_1086265, EPI_ISL_1086266, EPI_ISL_1086267, EPI_ISL_1086268, EPI_ISL_1086269, EPI_ISL_1086270, EPI_ISL_1086271, EPI_ISL_1086272, EPI_ISL_1086273, EPI_ISL_1086274, EPI_ISL_1086275, EPI_ISL_1086276, EPI_ISL_1086277, EPI_ISL_1086278, EPI_ISL_1086279, EPI_ISL_1086280, EPI_ISL_1086281, EPI_ISL_1086282, EPI_ISL_1086283,
EPI_ISL_1086284, EPI_ISL_1086285, EPI_ISL_1086286, EPI_ISL_1086287, EPI_ISL_1086288, EPI_ISL_1086289, EPI_ISL_1086290, EPI_ISL_1086291, EPI_ISL_1086292, EPI_ISL_1086293, EPI_ISL_1086294, EPI_ISL_1086295, EPI_ISL_1086296, EPI_ISL_1086297, EPI_ISL_1086298, EPI_ISL_1086299, EPI_ISL_1086300, EPI_ISL_1086301, EPI_ISL_1086302, EPI_ISL_1086303, EPI_ISL_1086304, EPI_ISL_1086305,
EPI_ISL_1086306, EPI_ISL_1086307, EPI_ISL_1086308, EPI_ISL_1086309, EPI_ISL_1086310, EPI_ISL_1086311, EPI_ISL_1086312, EPI_ISL_1086313, EPI_ISL_1086314, EPI_ISL_1086315, EPI_ISL_1086316, EPI_ISL_1086317, EPI_ISL_1086318, EPI_ISL_1086319, EPI_ISL_1086320, EPI_ISL_1086321, EPI_ISL_1086322, EPI_ISL_1086323, EPI_ISL_1086324, EPI_ISL_1086325, EPI_ISL_1086326, EPI_ISL_1086327,
EPI_ISL_1086328, EPI_ISL_1086329, EPI_ISL_1086330, EPI_ISL_1086331, EPI_ISL_1086332, EPI_ISL_1086333, EPI_ISL_1086334, EPI_ISL_1086335, EPI_ISL_1086336, EPI_ISL_1086337, EPI_ISL_1086338, EPI_ISL_1086339, EPI_ISL_1086340, EPI_ISL_1086341, EPI_ISL_1086342, EPI_ISL_1086343, EPI_ISL_1086344, EPI_ISL_1086345, EPI_ISL_1086346, EPI_ISL_1086347, EPI_ISL_1086348, EPI_ISL_1086349,
EPI_ISL_1086350, EPI_ISL_1086351, EPI_ISL_1086352, EPI_ISL_1086353, EPI_ISL_1086354, EPI_ISL_1086355, EPI_ISL_1086356, EPI_ISL_1086357, EPI_ISL_1086358, EPI_ISL_1086359, EPI_ISL_1086360, EPI_ISL_1086361, EPI_ISL_1086362, EPI_ISL_1086363, EPI_ISL_1086364, EPI_ISL_1086365, EPI_ISL_1086366, EPI_ISL_1086367, EPI_ISL_1086368, EPI_ISL_1086369, EPI_ISL_1086370, EPI_ISL_1086371,
EPI_ISL_1086372, EPI_ISL_1086378, EPI_ISL_1086379, EPI_ISL_1086380, EPI_ISL_1086381, EPI_ISL_1086382, EPI_ISL_1086383, EPI_ISL_1086384, EPI_ISL_1086385, EPI_ISL_1086386, EPI_ISL_1086387, EPI_ISL_1086388, EPI_ISL_1086389, EPI_ISL_1086390, EPI_ISL_1086391, EPI_ISL_1086392, EPI_ISL_1086393, EPI_ISL_1086394, EPI_ISL_1086395, EPI_ISL_1086396, EPI_ISL_1086397, EPI_ISL_1086398,
EPI_ISL_1086399, EPI_ISL_1086400, EPI_ISL_1086401, EPI_ISL_1086402, EPI_ISL_1086403, EPI_ISL_1086404, EPI_ISL_1086405, EPI_ISL_1086406, EPI_ISL_1086407, EPI_ISL_1086408, EPI_ISL_1086409, EPI_ISL_1086410, EPI_ISL_1086411, EPI_ISL_1086412, EPI_ISL_1086413, EPI_ISL_1086414, EPI_ISL_1086415, EPI_ISL_1086416, EPI_ISL_1086417, EPI_ISL_1086418, EPI_ISL_1086419, EPI_ISL_1086420,
EPI_ISL_1086421, EPI_ISL_1086422, EPI_ISL_1086423, EPI_ISL_1086424, EPI_ISL_1086425, EPI_ISL_1086426, EPI_ISL_1086427, EPI_ISL_1086428, EPI_ISL_1086429, EPI_ISL_1086430, EPI_ISL_1086431, EPI_ISL_1086432, EPI_ISL_1086433, EPI_ISL_1086434, EPI_ISL_1086435, EPI_ISL_1086436, EPI_ISL_1086437, EPI_ISL_1086438, EPI_ISL_1086439, EPI_ISL_1086440, EPI_ISL_1086441, EPI_ISL_1086442,
EPI_ISL_1086443, EPI_ISL_1086444, EPI_ISL_1086445, EPI_ISL_1086446, EPI_ISL_1086447, EPI_ISL_1086448, EPI_ISL_1086449, EPI_ISL_1086450, EPI_ISL_1086451, EPI_ISL_1086452, EPI_ISL_1086453, EPI_ISL_1086454, EPI_ISL_1086455, EPI_ISL_1086456, EPI_ISL_1086457, EPI_ISL_1086458, EPI_ISL_1086459, EPI_ISL_1086460, EPI_ISL_1086461, EPI_ISL_1086462, EPI_ISL_1086463, EPI_ISL_1086464,
EPI_ISL_1086465, EPI_ISL_1086466, EPI_ISL_1086467, EPI_ISL_1086468, EPI_ISL_1086469, EPI_ISL_1086470, EPI_ISL_1086471, EPI_ISL_1086472, EPI_ISL_1086473, EPI_ISL_1086474, EPI_ISL_1086475, EPI_ISL_1086476, EPI_ISL_1086477, EPI_ISL_1086478, EPI_ISL_1086479, EPI_ISL_1086480, EPI_ISL_1086481, EPI_ISL_1086482, EPI_ISL_1086483, EPI_ISL_1086484, EPI_ISL_1086485, EPI_ISL_1086486,
EPI_ISL_1086487, EPI_ISL_1086488, EPI_ISL_1086489, EPI_ISL_1086490, EPI_ISL_1086491, EPI_ISL_1086492, EPI_ISL_1086493, EPI_ISL_1086494, EPI_ISL_1086495, EPI_ISL_1086496, EPI_ISL_1086497, EPI_ISL_1086498, EPI_ISL_1086499, EPI_ISL_1086500, EPI_ISL_1086501, EPI_ISL_1086502, EPI_ISL_1086503, EPI_ISL_1086504, EPI_ISL_1086505, EPI_ISL_1086506, EPI_ISL_1086507, EPI_ISL_1086508,
EPI_ISL_1086509, EPI_ISL_1086510, EPI_ISL_1086511, EPI_ISL_1086512, EPI_ISL_1086513, EPI_ISL_1086514, EPI_ISL_1086515, EPI_ISL_1086516, EPI_ISL_1086517, EPI_ISL_1086518, EPI_ISL_1086519, EPI_ISL_1086520, EPI_ISL_1086521, EPI_ISL_1086522, EPI_ISL_1086523, EPI_ISL_1086524, EPI_ISL_1086525, EPI_ISL_1086526, EPI_ISL_1086527, EPI_ISL_1086528, EPI_ISL_1086529, EPI_ISL_1086530,
EPI_ISL_1086531, EPI_ISL_1086532, EPI_ISL_1086533, EPI_ISL_1086534, EPI_ISL_1086535, EPI_ISL_1086536, EPI_ISL_1086537, EPI_ISL_1086538, EPI_ISL_1086539, EPI_ISL_1086540, EPI_ISL_1086541, EPI_ISL_1086542, EPI_ISL_1086543, EPI_ISL_1086544, EPI_ISL_1086545, EPI_ISL_1086546, EPI_ISL_1086547, EPI_ISL_1086548, EPI_ISL_1086549, EPI_ISL_1086550, EPI_ISL_1086551, EPI_ISL_1086552,
EPI_ISL_1086553, EPI_ISL_1086554, EPI_ISL_1086555, EPI_ISL_1086556, EPI_ISL_1086557, EPI_ISL_1086558, EPI_ISL_1086559, EPI_ISL_1086560, EPI_ISL_1086561, EPI_ISL_1086562, EPI_ISL_1086563, EPI_ISL_1086564, EPI_ISL_1086565, EPI_ISL_1086566, EPI_ISL_1086567, EPI_ISL_1086568, EPI_ISL_1086569, EPI_ISL_1086570, EPI_ISL_1086571, EPI_ISL_1086572, EPI_ISL_1086573, EPI_ISL_1086574,
EPI_ISL_1086575, EPI_ISL_1086576, EPI_ISL_1086577, EPI_ISL_1086578, EPI_ISL_1086579, EPI_ISL_1086580, EPI_ISL_1086581, EPI_ISL_1086582, EPI_ISL_1086583, EPI_ISL_1086584, EPI_ISL_1086585, EPI_ISL_1086586, EPI_ISL_1086587, EPI_ISL_1086588, EPI_ISL_1086589, EPI_ISL_1086590, EPI_ISL_1086591, EPI_ISL_1086592, EPI_ISL_1086593, EPI_ISL_1086594, EPI_ISL_1086595, EPI_ISL_1086596,
EPI_ISL_1086597, EPI_ISL_1086598, EPI_ISL_1086599, EPI_ISL_1086600, EPI_ISL_1086601, EPI_ISL_1086602, EPI_ISL_1086603, EPI_ISL_1086604, EPI_ISL_1086605, EPI_ISL_1086606, EPI_ISL_1086607, EPI_ISL_1086608, EPI_ISL_1086609, EPI_ISL_1086610, EPI_ISL_1086611, EPI_ISL_1086612, EPI_ISL_1086613, EPI_ISL_1086614, EPI_ISL_1086615, EPI_ISL_1086616, EPI_ISL_1086617, EPI_ISL_1086618,
EPI_ISL_1086619, EPI_ISL_1086620, EPI_ISL_1086621, EPI_ISL_1086622, EPI_ISL_1086623, EPI_ISL_1086624, EPI_ISL_1086625, EPI_ISL_1086626, EPI_ISL_1086627, EPI_ISL_1086628, EPI_ISL_1086629, EPI_ISL_1086630, EPI_ISL_1086631, EPI_ISL_1086632, EPI_ISL_1086633, EPI_ISL_1086634, EPI_ISL_1086635, EPI_ISL_1086636, EPI_ISL_1086637, EPI_ISL_1086638, EPI_ISL_1086639, EPI_ISL_1086640,
EPI_ISL_1086641, EPI_ISL_1086642, EPI_ISL_1086643, EPI_ISL_1086644, EPI_ISL_1086645, EPI_ISL_1086646, EPI_ISL_1086647, EPI_ISL_1086648, EPI_ISL_1086649, EPI_ISL_1086650, EPI_ISL_1086651, EPI_ISL_1086652, EPI_ISL_1086653, EPI_ISL_1086654, EPI_ISL_1086655, EPI_ISL_1086656, EPI_ISL_1086657, EPI_ISL_1086658, EPI_ISL_1086659, EPI_ISL_1086660, EPI_ISL_1086661, EPI_ISL_1086662,
EPI_ISL_1086663, EPI_ISL_1086664, EPI_ISL_1086665, EPI_ISL_1086666, EPI_ISL_1086667, EPI_ISL_1086668, EPI_ISL_1086669, EPI_ISL_1086670, EPI_ISL_1086671, EPI_ISL_1086672, EPI_ISL_1086673, EPI_ISL_1086674, EPI_ISL_1086675, EPI_ISL_1086676, EPI_ISL_1086677, EPI_ISL_1086678, EPI_ISL_1086679, EPI_ISL_1086680, EPI_ISL_1086681, EPI_ISL_1086682, EPI_ISL_1086683, EPI_ISL_1086684,
EPI_ISL_1086685, EPI_ISL_1086686, EPI_ISL_1086687, EPI_ISL_1086688, EPI_ISL_1086689, EPI_ISL_1086690, EPI_ISL_1086691, EPI_ISL_1086692, EPI_ISL_1086693, EPI_ISL_1086694, EPI_ISL_1086695, EPI_ISL_1086696, EPI_ISL_1086697, EPI_ISL_1086698, EPI_ISL_1086699, EPI_ISL_1086700, EPI_ISL_1086701, EPI_ISL_1086702, EPI_ISL_1086703, EPI_ISL_1086704, EPI_ISL_1086705, EPI_ISL_1086706,
EPI_ISL_1086707, EPI_ISL_1086708, EPI_ISL_1086709, EPI_ISL_1086710, EPI_ISL_1086711, EPI_ISL_1086712, EPI_ISL_1086713, EPI_ISL_1086714, EPI_ISL_1086715, EPI_ISL_1086716, EPI_ISL_1086717, EPI_ISL_1086718, EPI_ISL_1086719, EPI_ISL_1086720, EPI_ISL_1086721, EPI_ISL_1086722, EPI_ISL_1086723, EPI_ISL_1086724, EPI_ISL_1086725, EPI_ISL_1086726, EPI_ISL_1086727, EPI_ISL_1086728,
EPI_ISL_1086729, EPI_ISL_1086730, EPI_ISL_1086731, EPI_ISL_1086732, EPI_ISL_1086733, EPI_ISL_1086734, EPI_ISL_1086735, EPI_ISL_1086736, EPI_ISL_1086737, EPI_ISL_1086738, EPI_ISL_1086739, EPI_ISL_1086740, EPI_ISL_1086741, EPI_ISL_1086742, EPI_ISL_1086743, EPI_ISL_1086744, EPI_ISL_1086745, EPI_ISL_1086746, EPI_ISL_1086747, EPI_ISL_1086748, EPI_ISL_1086749, EPI_ISL_1086750,
EPI_ISL_1086751, EPI_ISL_1086752, EPI_ISL_1086753, EPI_ISL_1086754, EPI_ISL_1086755, EPI_ISL_1086756, EPI_ISL_1086757, EPI_ISL_1086758, EPI_ISL_1086759, EPI_ISL_1086760, EPI_ISL_1086761, EPI_ISL_1086762, EPI_ISL_1086763, EPI_ISL_1086764, EPI_ISL_1086765, EPI_ISL_1086766, EPI_ISL_1086767, EPI_ISL_1086768, EPI_ISL_1086769, EPI_ISL_1086770, EPI_ISL_1086771, EPI_ISL_1086772,
EPI_ISL_1086773, EPI_ISL_1086774, EPI_ISL_1086775, EPI_ISL_1086776, EPI_ISL_1086777, EPI_ISL_1086778, EPI_ISL_1086779, EPI_ISL_1086780, EPI_ISL_1086781, EPI_ISL_1086782, EPI_ISL_1086783, EPI_ISL_1086784, EPI_ISL_1086785, EPI_ISL_1086786, EPI_ISL_1086787, EPI_ISL_1086788, EPI_ISL_1086789, EPI_ISL_1086790, EPI_ISL_1086791, EPI_ISL_1086792, EPI_ISL_1086793, EPI_ISL_1086794,
EPI_ISL_1086795, EPI_ISL_1086796, EPI_ISL_1086797, EPI_ISL_1086798, EPI_ISL_1086799, EPI_ISL_1086800, EPI_ISL_1086801, EPI_ISL_1086802, EPI_ISL_1086803, EPI_ISL_1086804, EPI_ISL_1086805, EPI_ISL_1086806, EPI_ISL_1086807, EPI_ISL_1086808, EPI_ISL_1086809, EPI_ISL_1086810, EPI_ISL_1086811, EPI_ISL_1086812, EPI_ISL_1086813, EPI_ISL_1086814, EPI_ISL_1086815, EPI_ISL_1086816,
EPI_ISL_1086817, EPI_ISL_1086818, EPI_ISL_1086819, EPI_ISL_1086820, EPI_ISL_1086821, EPI_ISL_1086822, EPI_ISL_1086823, EPI_ISL_1086824, EPI_ISL_1086825, EPI_ISL_1086826, EPI_ISL_1086827, EPI_ISL_1086828, EPI_ISL_1086829, EPI_ISL_1086830, EPI_ISL_1086831, EPI_ISL_1086832, EPI_ISL_1086833, EPI_ISL_1086834, EPI_ISL_1086835, EPI_ISL_1086836, EPI_ISL_1086837, EPI_ISL_1086838,
EPI_ISL_1086839, EPI_ISL_1086840, EPI_ISL_1086841, EPI_ISL_1086842, EPI_ISL_1086843, EPI_ISL_1086844, EPI_ISL_1086845, EPI_ISL_1086846, EPI_ISL_1086847, EPI_ISL_1086848, EPI_ISL_1086849, EPI_ISL_1086850, EPI_ISL_1086851, EPI_ISL_1086852, EPI_ISL_1086853, EPI_ISL_1086854, EPI_ISL_1086855, EPI_ISL_1086856, EPI_ISL_1086859, EPI_ISL_1086860, EPI_ISL_1086861, EPI_ISL_1086862,
EPI_ISL_1086863, EPI_ISL_1086864, EPI_ISL_1086865, EPI_ISL_1086866, EPI_ISL_1086867, EPI_ISL_1086868, EPI_ISL_1086869, EPI_ISL_1086870, EPI_ISL_1086871, EPI_ISL_1086872, EPI_ISL_1086873, EPI_ISL_1086874, EPI_ISL_1086875, EPI_ISL_1086876, EPI_ISL_1086877, EPI_ISL_1086878, EPI_ISL_1086879, EPI_ISL_1086880, EPI_ISL_1086881, EPI_ISL_1086882, EPI_ISL_1086883, EPI_ISL_1086884,
EPI_ISL_1086885, EPI_ISL_1086886, EPI_ISL_1086887, EPI_ISL_1086888, EPI_ISL_1086889, EPI_ISL_1086890, EPI_ISL_1086891, EPI_ISL_1086892, EPI_ISL_1086893, EPI_ISL_1086894, EPI_ISL_1086895, EPI_ISL_1086896, EPI_ISL_1086897, EPI_ISL_1086898, EPI_ISL_1086899, EPI_ISL_1086900, EPI_ISL_1086901, EPI_ISL_1086902, EPI_ISL_1086903, EPI_ISL_1086904, EPI_ISL_1086905, EPI_ISL_1086906,
EPI_ISL_1086907, EPI_ISL_1086908, EPI_ISL_1086909, EPI_ISL_1086910, EPI_ISL_1086911, EPI_ISL_1086912, EPI_ISL_1086913, EPI_ISL_1086914, EPI_ISL_1086915, EPI_ISL_1086916, EPI_ISL_1086917, EPI_ISL_1086918, EPI_ISL_1086919, EPI_ISL_1086920, EPI_ISL_1086921, EPI_ISL_1086922, EPI_ISL_1086923, EPI_ISL_1086924, EPI_ISL_1086925, EPI_ISL_1086926, EPI_ISL_1086927, EPI_ISL_1086928,
EPI_ISL_1086929, EPI_ISL_1086930, EPI_ISL_1086931, EPI_ISL_1086932, EPI_ISL_1086933, EPI_ISL_1086934, EPI_ISL_1086935, EPI_ISL_1086936, EPI_ISL_1086937, EPI_ISL_1086938, EPI_ISL_1086939, EPI_ISL_1086940, EPI_ISL_1086941, EPI_ISL_1086942, EPI_ISL_1086943, EPI_ISL_1086944, EPI_ISL_1086945, EPI_ISL_1086946, EPI_ISL_1086947, EPI_ISL_1086948, EPI_ISL_1086949, EPI_ISL_1086950,
EPI_ISL_1086951, EPI_ISL_1086952, EPI_ISL_1086953, EPI_ISL_1086954, EPI_ISL_1086955, EPI_ISL_1086956, EPI_ISL_1086957, EPI_ISL_1086958, EPI_ISL_1086959, EPI_ISL_1086960, EPI_ISL_1086961, EPI_ISL_1086962, EPI_ISL_1086963, EPI_ISL_1086964, EPI_ISL_1086965, EPI_ISL_1086966, EPI_ISL_1086967, EPI_ISL_1086968, EPI_ISL_1086969, EPI_ISL_1086970, EPI_ISL_1086971, EPI_ISL_1086972,
EPI_ISL_1086973, EPI_ISL_1086974, EPI_ISL_1086975, EPI_ISL_1086976, EPI_ISL_1086977, EPI_ISL_1086978, EPI_ISL_1086979, EPI_ISL_1086980, EPI_ISL_1086981, EPI_ISL_1086982, EPI_ISL_1086983, EPI_ISL_1086984, EPI_ISL_1086985, EPI_ISL_1086986, EPI_ISL_1086987, EPI_ISL_1086988, EPI_ISL_1086989, EPI_ISL_1086990, EPI_ISL_1086991, EPI_ISL_1086992, EPI_ISL_1086993, EPI_ISL_1086994,
EPI_ISL_1086995, EPI_ISL_1086996, EPI_ISL_1086997, EPI_ISL_1086998, EPI_ISL_1086999, EPI_ISL_1087000, EPI_ISL_1087001, EPI_ISL_1087002, EPI_ISL_1087003, EPI_ISL_1087004, EPI_ISL_1087005, EPI_ISL_1087006, EPI_ISL_1087007, EPI_ISL_1087008, EPI_ISL_1087009, EPI_ISL_1087010, EPI_ISL_1087011, EPI_ISL_1087012, EPI_ISL_1087013, EPI_ISL_1087014, EPI_ISL_1087015, EPI_ISL_1087016,
EPI_ISL_1087017, EPI_ISL_1087018, EPI_ISL_1087019, EPI_ISL_1087020, EPI_ISL_1087021, EPI_ISL_1087022, EPI_ISL_1087023, EPI_ISL_1087024, EPI_ISL_1087025, EPI_ISL_1087026, EPI_ISL_1087027, EPI_ISL_1087028, EPI_ISL_1087029, EPI_ISL_1087030, EPI_ISL_1087031, EPI_ISL_1087032, EPI_ISL_1087033, EPI_ISL_1087034, EPI_ISL_1087035, EPI_ISL_1087036, EPI_ISL_1087037, EPI_ISL_1087038,
EPI_ISL_1087039, EPI_ISL_1087040, EPI_ISL_1087041, EPI_ISL_1087042, EPI_ISL_1087043, EPI_ISL_1087044, EPI_ISL_1087045, EPI_ISL_1087046, EPI_ISL_1087047, EPI_ISL_1087048, EPI_ISL_1087049, EPI_ISL_1087050, EPI_ISL_1087051, EPI_ISL_1087052, EPI_ISL_1087053, EPI_ISL_1087054, EPI_ISL_1087055, EPI_ISL_1087056, EPI_ISL_1087057, EPI_ISL_1087058, EPI_ISL_1087059, EPI_ISL_1087060,
EPI_ISL_1087061, EPI_ISL_1087062, EPI_ISL_1087063, EPI_ISL_1087064, EPI_ISL_1087065, EPI_ISL_1087066, EPI_ISL_1087067, EPI_ISL_1087068, EPI_ISL_1087069, EPI_ISL_1087070, EPI_ISL_1087071, EPI_ISL_1087072, EPI_ISL_1087073, EPI_ISL_1087074, EPI_ISL_1087075, EPI_ISL_1087076, EPI_ISL_1087077, EPI_ISL_1087078, EPI_ISL_1087079, EPI_ISL_1087080, EPI_ISL_1087081, EPI_ISL_1087082,
EPI_ISL_1087083, EPI_ISL_1087084, EPI_ISL_1087085, EPI_ISL_1087086, EPI_ISL_1087087, EPI_ISL_1087088, EPI_ISL_1087089, EPI_ISL_1087090, EPI_ISL_1087091, EPI_ISL_1087092, EPI_ISL_1087093, EPI_ISL_1087094, EPI_ISL_1087095, EPI_ISL_1087096, EPI_ISL_1087097, EPI_ISL_1087098, EPI_ISL_1087099, EPI_ISL_1087100, EPI_ISL_1087101, EPI_ISL_1087102, EPI_ISL_1087103, EPI_ISL_1087104,
EPI_ISL_1087105, EPI_ISL_1087106, EPI_ISL_1087107, EPI_ISL_1087108, EPI_ISL_1087109, EPI_ISL_1087110, EPI_ISL_1087111, EPI_ISL_1087112, EPI_ISL_1087113, EPI_ISL_1087114, EPI_ISL_1087115, EPI_ISL_1087116, EPI_ISL_1087117, EPI_ISL_1087118, EPI_ISL_1087119, EPI_ISL_1087120, EPI_ISL_1087121, EPI_ISL_1087122, EPI_ISL_1087123, EPI_ISL_1087124, EPI_ISL_1087125, EPI_ISL_1087126,
EPI_ISL_1087127, EPI_ISL_1087128, EPI_ISL_1087129, EPI_ISL_1087130, EPI_ISL_1087131, EPI_ISL_1087132, EPI_ISL_1087133, EPI_ISL_1087134, EPI_ISL_1087135, EPI_ISL_1087136, EPI_ISL_1087137, EPI_ISL_1087138, EPI_ISL_1087139, EPI_ISL_1087140, EPI_ISL_1087141, EPI_ISL_1087142, EPI_ISL_1087143, EPI_ISL_1087144, EPI_ISL_1087145, EPI_ISL_1087146, EPI_ISL_1087147, EPI_ISL_1087148,
EPI_ISL_1087149, EPI_ISL_1087150, EPI_ISL_1087151, EPI_ISL_1087152, EPI_ISL_1087153, EPI_ISL_1087154, EPI_ISL_1087155, EPI_ISL_1087156, EPI_ISL_1087157, EPI_ISL_1087158, EPI_ISL_1087159, EPI_ISL_1087160, EPI_ISL_1087161, EPI_ISL_1087162, EPI_ISL_1087163, EPI_ISL_1087164, EPI_ISL_1087165, EPI_ISL_1087166, EPI_ISL_1087167, EPI_ISL_1087168, EPI_ISL_1087169, EPI_ISL_1087170,
EPI_ISL_1087171, EPI_ISL_1087172, EPI_ISL_1087173, EPI_ISL_1087174, EPI_ISL_1087175, EPI_ISL_1087176, EPI_ISL_1087177, EPI_ISL_1087178, EPI_ISL_1087179, EPI_ISL_1087180, EPI_ISL_1087181, EPI_ISL_1087182, EPI_ISL_1087183, EPI_ISL_1087184, EPI_ISL_1087185, EPI_ISL_1087186, EPI_ISL_1087187, EPI_ISL_1087188, EPI_ISL_1087189, EPI_ISL_1087190, EPI_ISL_1087191, EPI_ISL_1087192,
EPI_ISL_1087193, EPI_ISL_1087194, EPI_ISL_1087195, EPI_ISL_1087196, EPI_ISL_1087197, EPI_ISL_1087198, EPI_ISL_1087199, EPI_ISL_1087200, EPI_ISL_1087201, EPI_ISL_1087202, EPI_ISL_1087203, EPI_ISL_1087204, EPI_ISL_1087205, EPI_ISL_1087206, EPI_ISL_1087207, EPI_ISL_1087208, EPI_ISL_1087209, EPI_ISL_1087210, EPI_ISL_1087211, EPI_ISL_1087212, EPI_ISL_1087213, EPI_ISL_1087214,
EPI_ISL_1087215, EPI_ISL_1087216, EPI_ISL_1087217, EPI_ISL_1087218, EPI_ISL_1087219, EPI_ISL_1087220, EPI_ISL_1087221, EPI_ISL_1087222, EPI_ISL_1087223, EPI_ISL_1087224, EPI_ISL_1087225, EPI_ISL_1087226, EPI_ISL_1087227, EPI_ISL_1087228, EPI_ISL_1087229, EPI_ISL_1087230, EPI_ISL_1087231, EPI_ISL_1087232, EPI_ISL_1087233, EPI_ISL_1087234, EPI_ISL_1087235, EPI_ISL_1087236,
EPI_ISL_1087237, EPI_ISL_1087238, EPI_ISL_1087239, EPI_ISL_1087240, EPI_ISL_1087241, EPI_ISL_1087242, EPI_ISL_1087243, EPI_ISL_1087244, EPI_ISL_1087245, EPI_ISL_1087246, EPI_ISL_1087247, EPI_ISL_1087248, EPI_ISL_1087249, EPI_ISL_1087250, EPI_ISL_1087251, EPI_ISL_1087252, EPI_ISL_1087253, EPI_ISL_1087254, EPI_ISL_1087255, EPI_ISL_1087256, EPI_ISL_1087257, EPI_ISL_1087258,
EPI_ISL_1087259, EPI_ISL_1087260, EPI_ISL_1087261, EPI_ISL_1087262, EPI_ISL_1087263, EPI_ISL_1087264, EPI_ISL_1087265, EPI_ISL_1087266, EPI_ISL_1087267, EPI_ISL_1087268, EPI_ISL_1087269, EPI_ISL_1087270, EPI_ISL_1087271

see above | Quest Diagnostics Incorporated | Respiratory Viruses Branch, Division of Viral Diseases, Centers for Disease Control and Prevention | Peter W. Cook, Dakota Howard, Dhwani Batra, Ben L. Rambo-Martin, S. H. Rosenthal, A. Gerasimova, R. M. Kagan, B. Anderson, M. Hua, Y. Liu, L.E. Bernstein, K.E. Livingston, A. Perez, I. A. Shlyakhter, R. V. Rolando, R. Owen, P. Tanpaiboon, F. Lacbawan, Clinton R. Paden, Suxiang Tong, Duncan MacCannell

EPI_ISL_1087272, EPI_ISL_1087273, EPI_ISL_1087274, EPI_ISL_1087275, EPI_ISL_1087276, EPI_ISL_1087277, EPI_ISL_1087278, EPI_ISL_1087279, EPI_ISL_1087280, EPI_ISL_1087281, EPI_ISL_1087282, EPI_ISL_1087283, EPI_ISL_1087284, EPI_ISL_1087285, EPI_ISL_1087286, EPI_ISL_1087287, EPI_ISL_1087288, EPI_ISL_1087289, EPI_ISL_1087290, EPI_ISL_1087291, EPI_ISL_1087292, EPI_ISL_1087293,
EPI_ISL_1087294, EPI_ISL_1087295, EPI_ISL_1087296, EPI_ISL_1087297, EPI_ISL_1087298, EPI_ISL_1087299, EPI_ISL_1087300, EPI_ISL_1087301, EPI_ISL_1087302, EPI_ISL_1087303, EPI_ISL_1087304, EPI_ISL_1087305, EPI_ISL_1087306, EPI_ISL_1087307, EPI_ISL_1087308, EPI_ISL_1087309, EPI_ISL_1087310, EPI_ISL_1087311, EPI_ISL_1087312, EPI_ISL_1087313, EPI_ISL_1087314, EPI_ISL_1087315,
EPI_ISL_1087316, EPI_ISL_1087317, EPI_ISL_1087318, EPI_ISL_1087319, EPI_ISL_1087320, EPI_ISL_1087321, EPI_ISL_1087322, EPI_ISL_1087323, EPI_ISL_1087324, EPI_ISL_1087325, EPI_ISL_1087326, EPI_ISL_1087327, EPI_ISL_1087328, EPI_ISL_1087329, EPI_ISL_1087330, EPI_ISL_1087331, EPI_ISL_1087332, EPI_ISL_1087333, EPI_ISL_1087334, EPI_ISL_1087335, EPI_ISL_1087336, EPI_ISL_1087337,
EPI_ISL_1087338, EPI_ISL_1087339, EPI_ISL_1087340, EPI_ISL_1087341, EPI_ISL_1087342, EPI_ISL_1087343, EPI_ISL_1087344, EPI_ISL_1087345, EPI_ISL_1087346, EPI_ISL_1087347, EPI_ISL_1087348, EPI_ISL_1087349, EPI_ISL_1087350, EPI_ISL_1087351, EPI_ISL_1087352

see above | NYU Langone Health | Departments of Pathology and Medicine, New York University School of Medicine | Adriana Heguy, Dacia Dimartino, Emily Guzman, Christian Marier, Peter Meyn, Sitharam Ramaswami, Gael Westby, Paul Zappile, Yutong Zhang, Paolo Cotzia, Guiqing Wang

EPI_ISL_1087414, EPI_ISL_1087415, EPI_ISL_1087416, EPI_ISL_1087417, EPI_ISL_1087418, EPI_ISL_1087419, EPI_ISL_1087420, EPI_ISL_1087421, EPI_ISL_1087422, EPI_ISL_1087423, EPI_ISL_1087424, EPI_ISL_1087425, EPI_ISL_1087426, EPI_ISL_1087427, EPI_ISL_1087428, EPI_ISL_1087429, EPI_ISL_1087430, EPI_ISL_1087431, EPI_ISL_1087432, EPI_ISL_1087433, EPI_ISL_1087434, EPI_ISL_1087435,
EPI_ISL_1087436, EPI_ISL_1087437, EPI_ISL_1087438, EPI_ISL_1087439, EPI_ISL_1087440, EPI_ISL_1087441, EPI_ISL_1087442, EPI_ISL_1087443, EPI_ISL_1087444, EPI_ISL_1087445, EPI_ISL_1087446, EPI_ISL_1087447, EPI_ISL_1087448, EPI_ISL_1087449, EPI_ISL_1087450, EPI_ISL_1087451, EPI_ISL_1087452, EPI_ISL_1087453, EPI_ISL_1087454, EPI_ISL_1087455, EPI_ISL_1087456, EPI_ISL_1087457,
EPI_ISL_1087458, EPI_ISL_1087459, EPI_ISL_1087460, EPI_ISL_1087461, EPI_ISL_1087462, EPI_ISL_1087463, EPI_ISL_1087464, EPI_ISL_1087465, EPI_ISL_1087466, EPI_ISL_1087467, EPI_ISL_1087468, EPI_ISL_1087469, EPI_ISL_1087470, EPI_ISL_1087471, EPI_ISL_1087472, EPI_ISL_1087473, EPI_ISL_1087474, EPI_ISL_1087475, EPI_ISL_1087476, EPI_ISL_1087477, EPI_ISL_1087478, EPI_ISL_1087479,
EPI_ISL_1087480, EPI_ISL_1087481, EPI_ISL_1087482, EPI_ISL_1087483, EPI_ISL_1087484, EPI_ISL_1087485, EPI_ISL_1087486, EPI_ISL_1087487, EPI_ISL_1087488, EPI_ISL_1087489, EPI_ISL_1087490, EPI_ISL_1087491, EPI_ISL_1087492, EPI_ISL_1087493, EPI_ISL_1087494, EPI_ISL_1087495, EPI_ISL_1087496, EPI_ISL_1087497, EPI_ISL_1087498, EPI_ISL_1087499, EPI_ISL_1087500, EPI_ISL_1087501,
EPI_ISL_1087502, EPI_ISL_1087503, EPI_ISL_1087504, EPI_ISL_1087505, EPI_ISL_1087506, EPI_ISL_1087507, EPI_ISL_1087508, EPI_ISL_1087509, EPI_ISL_1087510, EPI_ISL_1087511, EPI_ISL_1087512, EPI_ISL_1087513, EPI_ISL_1087514, EPI_ISL_1087515, EPI_ISL_1087516, EPI_ISL_1087517, EPI_ISL_1087518, EPI_ISL_1087519, EPI_ISL_1087520, EPI_ISL_1087521, EPI_ISL_1087522, EPI_ISL_1087523,
EPI_ISL_1087524, EPI_ISL_1087525, EPI_ISL_1087526, EPI_ISL_1087527, EPI_ISL_1087528, EPI_ISL_1087529, EPI_ISL_1087530, EPI_ISL_1087531, EPI_ISL_1087532, EPI_ISL_1087533, EPI_ISL_1087534, EPI_ISL_1087535, EPI_ISL_1087536, EPI_ISL_1087537, EPI_ISL_1087538, EPI_ISL_1087539, EPI_ISL_1087540, EPI_ISL_1087541, EPI_ISL_1087542, EPI_ISL_1087543, EPI_ISL_1087544, EPI_ISL_1087545,
EPI_ISL_1087546, EPI_ISL_1087547, EPI_ISL_1087548, EPI_ISL_1087549, EPI_ISL_1087550, EPI_ISL_1087551, EPI_ISL_1087552, EPI_ISL_1087553, EPI_ISL_1087554, EPI_ISL_1087555, EPI_ISL_1087556, EPI_ISL_1087557, EPI_ISL_1087559, EPI_ISL_1087560, EPI_ISL_1087561, EPI_ISL_1087562, EPI_ISL_1087563, EPI_ISL_1087564, EPI_ISL_1087565, EPI_ISL_1087566, EPI_ISL_1087567, EPI_ISL_1087568,
EPI_ISL_1087569, EPI_ISL_1087570, EPI_ISL_1087571, EPI_ISL_1087572, EPI_ISL_1087573, EPI_ISL_1087574, EPI_ISL_1087575, EPI_ISL_1087576, EPI_ISL_1087577, EPI_ISL_1087578, EPI_ISL_1087579, EPI_ISL_1087580, EPI_ISL_1087581, EPI_ISL_1087582, EPI_ISL_1087583, EPI_ISL_1087584, EPI_ISL_1087585, EPI_ISL_1087586, EPI_ISL_1087587, EPI_ISL_1087588, EPI_ISL_1087589, EPI_ISL_1087590,
EPI_ISL_1087591, EPI_ISL_1087592, EPI_ISL_1087593, EPI_ISL_1087594, EPI_ISL_1087595, EPI_ISL_1087596, EPI_ISL_1087597, EPI_ISL_1087598, EPI_ISL_1087599, EPI_ISL_1087600, EPI_ISL_1087601, EPI_ISL_1087602, EPI_ISL_1087603, EPI_ISL_1087604, EPI_ISL_1087605, EPI_ISL_1087606, EPI_ISL_1087607, EPI_ISL_1087608, EPI_ISL_1087609, EPI_ISL_1087610, EPI_ISL_1087611, EPI_ISL_1087612,
EPI_ISL_1087613, EPI_ISL_1087614, EPI_ISL_1087615, EPI_ISL_1087616, EPI_ISL_1087617, EPI_ISL_1087618, EPI_ISL_1087619, EPI_ISL_1087620, EPI_ISL_1087621, EPI_ISL_1087622, EPI_ISL_1087623, EPI_ISL_1087624, EPI_ISL_1087625, EPI_ISL_1087626, EPI_ISL_1087627, EPI_ISL_1087628, EPI_ISL_1087629, EPI_ISL_1087630, EPI_ISL_1087631, EPI_ISL_1087632, EPI_ISL_1087633, EPI_ISL_1087634,
EPI_ISL_1087635, EPI_ISL_1087636, EPI_ISL_1087637, EPI_ISL_1087638, EPI_ISL_1087639, EPI_ISL_1087640, EPI_ISL_1087641, EPI_ISL_1087642, EPI_ISL_1087643, EPI_ISL_1087644, EPI_ISL_1087645, EPI_ISL_1087646, EPI_ISL_1087647, EPI_ISL_1087648, EPI_ISL_1087649, EPI_ISL_1087650, EPI_ISL_1087651, EPI_ISL_1087652, EPI_ISL_1087653, EPI_ISL_1087654, EPI_ISL_1087655, EPI_ISL_1087656,
EPI_ISL_1087657, EPI_ISL_1087658, EPI_ISL_1087659, EPI_ISL_1087660, EPI_ISL_1087661, EPI_ISL_1087662, EPI_ISL_1087663, EPI_ISL_1087664, EPI_ISL_1087665, EPI_ISL_1087666, EPI_ISL_1087667, EPI_ISL_1087668, EPI_ISL_1087669, EPI_ISL_1087670, EPI_ISL_1087671, EPI_ISL_1087672, EPI_ISL_1087673, EPI_ISL_1087674, EPI_ISL_1087675, EPI_ISL_1087676, EPI_ISL_1087677, EPI_ISL_1087678,
EPI_ISL_1087679, EPI_ISL_1087680, EPI_ISL_1087681, EPI_ISL_1087682, EPI_ISL_1087683, EPI_ISL_1087684, EPI_ISL_1087685, EPI_ISL_1087686, EPI_ISL_1087687, EPI_ISL_1087688, EPI_ISL_1087689, EPI_ISL_1087690, EPI_ISL_1087691, EPI_ISL_1087692, EPI_ISL_1087693, EPI_ISL_1087694, EPI_ISL_1087695, EPI_ISL_1087696, EPI_ISL_1087697, EPI_ISL_1087698, EPI_ISL_1087699, EPI_ISL_1087700,
EPI_ISL_1087701, EPI_ISL_1087702, EPI_ISL_1087703, EPI_ISL_1087704, EPI_ISL_1087705, EPI_ISL_1087706, EPI_ISL_1087707, EPI_ISL_1087708, EPI_ISL_1087709, EPI_ISL_1087710, EPI_ISL_1087711, EPI_ISL_1087712, EPI_ISL_1087713, EPI_ISL_1087714, EPI_ISL_1087715, EPI_ISL_1087716, EPI_ISL_1087717, EPI_ISL_1087718, EPI_ISL_1087719, EPI_ISL_1087720, EPI_ISL_1087721, EPI_ISL_1087722,
EPI_ISL_1087723, EPI_ISL_1087724, EPI_ISL_1087725, EPI_ISL_1087726, EPI_ISL_1087727, EPI_ISL_1087728, EPI_ISL_1087729, EPI_ISL_1087730, EPI_ISL_1087731, EPI_ISL_1087732, EPI_ISL_1087733, EPI_ISL_1087734, EPI_ISL_1087735, EPI_ISL_1087736, EPI_ISL_1087737, EPI_ISL_1087738, EPI_ISL_1087739, EPI_ISL_1087740, EPI_ISL_1087741, EPI_ISL_1087742, EPI_ISL_1087743, EPI_ISL_1087744,
EPI_ISL_1087745, EPI_ISL_1087746, EPI_ISL_1087747, EPI_ISL_1087748, EPI_ISL_1087749, EPI_ISL_1087750, EPI_ISL_1087751, EPI_ISL_1087752, EPI_ISL_1087753, EPI_ISL_1087754, EPI_ISL_1087755, EPI_ISL_1087756, EPI_ISL_1087757, EPI_ISL_1087758, EPI_ISL_1087759, EPI_ISL_1087760, EPI_ISL_1087761, EPI_ISL_1087762, EPI_ISL_1087763, EPI_ISL_1087764, EPI_ISL_1087765, EPI_ISL_1087766,
EPI_ISL_1087767, EPI_ISL_1087768, EPI_ISL_1087769, EPI_ISL_1087770, EPI_ISL_1087771, EPI_ISL_1087772, EPI_ISL_1087773, EPI_ISL_1087774, EPI_ISL_1087775, EPI_ISL_1087776, EPI_ISL_1087777, EPI_ISL_1087778, EPI_ISL_1087779, EPI_ISL_1087780, EPI_ISL_1087781, EPI_ISL_1087782, EPI_ISL_1087783, EPI_ISL_1087784, EPI_ISL_1087785, EPI_ISL_1087786, EPI_ISL_1087787, EPI_ISL_1087788,
EPI_ISL_1087789, EPI_ISL_1087790, EPI_ISL_1087791, EPI_ISL_1087792, EPI_ISL_1087793, EPI_ISL_1087794, EPI_ISL_1087795, EPI_ISL_1087796, EPI_ISL_1087797, EPI_ISL_1087798, EPI_ISL_1087799, EPI_ISL_1087800, EPI_ISL_1087801, EPI_ISL_1087802, EPI_ISL_1087803, EPI_ISL_1087804, EPI_ISL_1087805, EPI_ISL_1087806, EPI_ISL_1087807, EPI_ISL_1087808, EPI_ISL_1087809, EPI_ISL_1087810,
EPI_ISL_1087811, EPI_ISL_1087812, EPI_ISL_1087813, EPI_ISL_1087814, EPI_ISL_1087815, EPI_ISL_1087816, EPI_ISL_1087817, EPI_ISL_1087818, EPI_ISL_1087819, EPI_ISL_1087820, EPI_ISL_1087821, EPI_ISL_1087822, EPI_ISL_1087823, EPI_ISL_1087824, EPI_ISL_1087825, EPI_ISL_1087826, EPI_ISL_1087827, EPI_ISL_1087828, EPI_ISL_1087829, EPI_ISL_1087830, EPI_ISL_1087831, EPI_ISL_1087832,
EPI_ISL_1087833, EPI_ISL_1087834, EPI_ISL_1087835, EPI_ISL_1087836, EPI_ISL_1087837, EPI_ISL_1087838, EPI_ISL_1087839, EPI_ISL_1087840, EPI_ISL_1087841, EPI_ISL_1087842, EPI_ISL_1087843, EPI_ISL_1087844, EPI_ISL_1087845, EPI_ISL_1087846, EPI_ISL_1087847, EPI_ISL_1087848, EPI_ISL_1087849, EPI_ISL_1087850, EPI_ISL_1087851, EPI_ISL_1087852, EPI_ISL_1087853, EPI_ISL_1087854,
EPI_ISL_1087855, EPI_ISL_1087856, EPI_ISL_1087857, EPI_ISL_1087858, EPI_ISL_1087859, EPI_ISL_1087860, EPI_ISL_1087861, EPI_ISL_1087862, EPI_ISL_1087863, EPI_ISL_1087864, EPI_ISL_1087865, EPI_ISL_1087866, EPI_ISL_1087867, EPI_ISL_1087868, EPI_ISL_1087869, EPI_ISL_1087870, EPI_ISL_1087871, EPI_ISL_1087872, EPI_ISL_1087873, EPI_ISL_1087874, EPI_ISL_1087875, EPI_ISL_1087876,
EPI_ISL_1087877, EPI_ISL_1087878, EPI_ISL_1087879, EPI_ISL_1087880, EPI_ISL_1087881, EPI_ISL_1087882, EPI_ISL_1087883, EPI_ISL_1087884, EPI_ISL_1087885, EPI_ISL_1087886, EPI_ISL_1087887, EPI_ISL_1087888, EPI_ISL_1087889, EPI_ISL_1087890, EPI_ISL_1087891, EPI_ISL_1087892, EPI_ISL_1087893, EPI_ISL_1087894, EPI_ISL_1087895, EPI_ISL_1087896, EPI_ISL_1087897, EPI_ISL_1087898,
EPI_ISL_1087899, EPI_ISL_1087900, EPI_ISL_1087901, EPI_ISL_1087902, EPI_ISL_1087903, EPI_ISL_1087904, EPI_ISL_1087905, EPI_ISL_1087906, EPI_ISL_1087907, EPI_ISL_1087908, EPI_ISL_1087909, EPI_ISL_1087910, EPI_ISL_1087911, EPI_ISL_1087912, EPI_ISL_1087913, EPI_ISL_1087914, EPI_ISL_1087915, EPI_ISL_1087916, EPI_ISL_1087917, EPI_ISL_1087918, EPI_ISL_1087919, EPI_ISL_1087920,
EPI_ISL_1087921, EPI_ISL_1087922, EPI_ISL_1087923, EPI_ISL_1087924, EPI_ISL_1087925, EPI_ISL_1087926, EPI_ISL_1087927, EPI_ISL_1087928, EPI_ISL_1087929, EPI_ISL_1087930, EPI_ISL_1087931, EPI_ISL_1087932, EPI_ISL_1087933, EPI_ISL_1087934, EPI_ISL_1087935, EPI_ISL_1087936, EPI_ISL_1087937, EPI_ISL_1087938, EPI_ISL_1087939, EPI_ISL_1087940, EPI_ISL_1087941, EPI_ISL_1087942,
EPI_ISL_1087943, EPI_ISL_1087944, EPI_ISL_1087945, EPI_ISL_1087946, EPI_ISL_1087947, EPI_ISL_1087948, EPI_ISL_1087949, EPI_ISL_1087950, EPI_ISL_1087951, EPI_ISL_1087952, EPI_ISL_1087953, EPI_ISL_1087954, EPI_ISL_1087955, EPI_ISL_1087956, EPI_ISL_1087957, EPI_ISL_1087958, EPI_ISL_1087959, EPI_ISL_1087960, EPI_ISL_1087961, EPI_ISL_1087962, EPI_ISL_1087963, EPI_ISL_1087964,
EPI_ISL_1087965, EPI_ISL_1087966, EPI_ISL_1087967, EPI_ISL_1087968, EPI_ISL_1087969, EPI_ISL_1087970, EPI_ISL_1087971, EPI_ISL_1087972, EPI_ISL_1087973, EPI_ISL_1087974, EPI_ISL_1087975, EPI_ISL_1087976, EPI_ISL_1087977, EPI_ISL_1087978, EPI_ISL_1087979, EPI_ISL_1087980, EPI_ISL_1087981, EPI_ISL_1087982, EPI_ISL_1087983, EPI_ISL_1087984, EPI_ISL_1087985, EPI_ISL_1087986,
EPI_ISL_1087987, EPI_ISL_1087988, EPI_ISL_1087989, EPI_ISL_1087990, EPI_ISL_1087991, EPI_ISL_1087992, EPI_ISL_1087993, EPI_ISL_1087994, EPI_ISL_1087995, EPI_ISL_1087996, EPI_ISL_1087997, EPI_ISL_1087998, EPI_ISL_1087999, EPI_ISL_1088000, EPI_ISL_1088001, EPI_ISL_1088002, EPI_ISL_1088003, EPI_ISL_1088004, EPI_ISL_1088005, EPI_ISL_1088006, EPI_ISL_1088007, EPI_ISL_1088077,
EPI_ISL_1088078, EPI_ISL_1088079, EPI_ISL_1088080, EPI_ISL_1088081, EPI_ISL_1088082, EPI_ISL_1088083, EPI_ISL_1088084, EPI_ISL_1088085, EPI_ISL_1088086, EPI_ISL_1088087, EPI_ISL_1088088, EPI_ISL_1088089, EPI_ISL_1088090, EPI_ISL_1088091, EPI_ISL_1088092, EPI_ISL_1088093, EPI_ISL_1088094, EPI_ISL_1088095, EPI_ISL_1088096, EPI_ISL_1088097, EPI_ISL_1088098, EPI_ISL_1088099,
EPI_ISL_1088100, EPI_ISL_1088101, EPI_ISL_1088102, EPI_ISL_1088103, EPI_ISL_1088104, EPI_ISL_1088105, EPI_ISL_1088106, EPI_ISL_1088107, EPI_ISL_1088108, EPI_ISL_1088109, EPI_ISL_1088110, EPI_ISL_1088111, EPI_ISL_1088112, EPI_ISL_1088113, EPI_ISL_1088114, EPI_ISL_1088115, EPI_ISL_1088116, EPI_ISL_1088117, EPI_ISL_1088118, EPI_ISL_1088119, EPI_ISL_1088120, EPI_ISL_1088121,
EPI_ISL_1088122, EPI_ISL_1088123, EPI_ISL_1088124, EPI_ISL_1088125, EPI_ISL_1088126, EPI_ISL_1088127, EPI_ISL_1088128, EPI_ISL_1088129, EPI_ISL_1088130, EPI_ISL_1088131, EPI_ISL_1088132, EPI_ISL_1088133, EPI_ISL_1088134, EPI_ISL_1088135, EPI_ISL_1088136, EPI_ISL_1088137, EPI_ISL_1088138, EPI_ISL_1088139, EPI_ISL_1088140, EPI_ISL_1088141, EPI_ISL_1088142, EPI_ISL_1088143,
EPI_ISL_1088144, EPI_ISL_1088145, EPI_ISL_1088146, EPI_ISL_1088147, EPI_ISL_1088148, EPI_ISL_1088149, EPI_ISL_1088150, EPI_ISL_1088151, EPI_ISL_1088152, EPI_ISL_1088153, EPI_ISL_1088154, EPI_ISL_1088155, EPI_ISL_1088156, EPI_ISL_1088157, EPI_ISL_1088158, EPI_ISL_1088159, EPI_ISL_1088160, EPI_ISL_1088161, EPI_ISL_1088162, EPI_ISL_1088163, EPI_ISL_1088164, EPI_ISL_1088165,
EPI_ISL_1088166, EPI_ISL_1088167, EPI_ISL_1088168, EPI_ISL_1088169, EPI_ISL_1088170, EPI_ISL_1088171, EPI_ISL_1088172, EPI_ISL_1088173, EPI_ISL_1088174, EPI_ISL_1088175, EPI_ISL_1088176, EPI_ISL_1088177, EPI_ISL_1088178, EPI_ISL_1088179, EPI_ISL_1088180, EPI_ISL_1088181, EPI_ISL_1088182, EPI_ISL_1088183, EPI_ISL_1088184, EPI_ISL_1088185, EPI_ISL_1088186, EPI_ISL_1088187,
EPI_ISL_1088188, EPI_ISL_1088189, EPI_ISL_1088190, EPI_ISL_1088191, EPI_ISL_1088192, EPI_ISL_1088193, EPI_ISL_1088194, EPI_ISL_1088195, EPI_ISL_1088196, EPI_ISL_1088197, EPI_ISL_1088198, EPI_ISL_1088199, EPI_ISL_1088200, EPI_ISL_1088201, EPI_ISL_1088202, EPI_ISL_1088203, EPI_ISL_1088204, EPI_ISL_1088205, EPI_ISL_1088206, EPI_ISL_1088207, EPI_ISL_1088208, EPI_ISL_1088209,
EPI_ISL_1088210, EPI_ISL_1088211, EPI_ISL_1088212, EPI_ISL_1088213, EPI_ISL_1088214, EPI_ISL_1088215, EPI_ISL_1088216, EPI_ISL_1088217, EPI_ISL_1088218, EPI_ISL_1088219, EPI_ISL_1088220, EPI_ISL_1088221, EPI_ISL_1088222, EPI_ISL_1088223, EPI_ISL_1088224, EPI_ISL_1088225, EPI_ISL_1088226, EPI_ISL_1088227, EPI_ISL_1088228, EPI_ISL_1088229, EPI_ISL_1088230, EPI_ISL_1088231,
EPI_ISL_1088232, EPI_ISL_1088233, EPI_ISL_1088234, EPI_ISL_1088235, EPI_ISL_1088236, EPI_ISL_1088237, EPI_ISL_1088238, EPI_ISL_1088239, EPI_ISL_1088240, EPI_ISL_1088241, EPI_ISL_1088242, EPI_ISL_1088243, EPI_ISL_1088244, EPI_ISL_1088245, EPI_ISL_1088246, EPI_ISL_1088247, EPI_ISL_1088248, EPI_ISL_1088249, EPI_ISL_1088250, EPI_ISL_1088251, EPI_ISL_1088252, EPI_ISL_1088253,
EPI_ISL_1088254, EPI_ISL_1088255, EPI_ISL_1088256, EPI_ISL_1088257, EPI_ISL_1088258, EPI_ISL_1088259, EPI_ISL_1088260, EPI_ISL_1088261, EPI_ISL_1088262, EPI_ISL_1088263, EPI_ISL_1088264, EPI_ISL_1088265, EPI_ISL_1088266, EPI_ISL_1088267, EPI_ISL_1088268, EPI_ISL_1088269, EPI_ISL_1088270, EPI_ISL_1088271, EPI_ISL_1088272, EPI_ISL_1088273, EPI_ISL_1088274, EPI_ISL_1088275,
EPI_ISL_1088276, EPI_ISL_1088277, EPI_ISL_1088278, EPI_ISL_1088279, EPI_ISL_1088280, EPI_ISL_1088281, EPI_ISL_1088282, EPI_ISL_1088283, EPI_ISL_1088284, EPI_ISL_1088285, EPI_ISL_1088286, EPI_ISL_1088287, EPI_ISL_1088288, EPI_ISL_1088289, EPI_ISL_1088290, EPI_ISL_1088291, EPI_ISL_1088292, EPI_ISL_1088293, EPI_ISL_1088294, EPI_ISL_1088295, EPI_ISL_1088296, EPI_ISL_1088297,
EPI_ISL_1088298, EPI_ISL_1088299, EPI_ISL_1088300, EPI_ISL_1088301, EPI_ISL_1088302, EPI_ISL_1088303, EPI_ISL_1088304, EPI_ISL_1088305, EPI_ISL_1088306, EPI_ISL_1088307, EPI_ISL_1088308, EPI_ISL_1088309, EPI_ISL_1088310, EPI_ISL_1088311, EPI_ISL_1088312, EPI_ISL_1088313, EPI_ISL_1088314, EPI_ISL_1088315, EPI_ISL_1088316, EPI_ISL_1088317, EPI_ISL_1088318, EPI_ISL_1088319,
EPI_ISL_1088320, EPI_ISL_1088321, EPI_ISL_1088322, EPI_ISL_1088323, EPI_ISL_1088324, EPI_ISL_1088325, EPI_ISL_1088326, EPI_ISL_1088327, EPI_ISL_1088328, EPI_ISL_1088329, EPI_ISL_1088330, EPI_ISL_1088331, EPI_ISL_1088332, EPI_ISL_1088333, EPI_ISL_1088334, EPI_ISL_1088335, EPI_ISL_1088336, EPI_ISL_1088337, EPI_ISL_1088338, EPI_ISL_1088339, EPI_ISL_1088340, EPI_ISL_1088341,
EPI_ISL_1088342, EPI_ISL_1088343, EPI_ISL_1088344, EPI_ISL_1088345, EPI_ISL_1088346, EPI_ISL_1088347, EPI_ISL_1088348, EPI_ISL_1088349, EPI_ISL_1088350, EPI_ISL_1088351, EPI_ISL_1088352, EPI_ISL_1088353, EPI_ISL_1088354, EPI_ISL_1088355, EPI_ISL_1088356, EPI_ISL_1088357, EPI_ISL_1088358, EPI_ISL_1088359, EPI_ISL_1088360, EPI_ISL_1088361, EPI_ISL_1088362, EPI_ISL_1088363,
EPI_ISL_1088364, EPI_ISL_1088365, EPI_ISL_1088366, EPI_ISL_1088367, EPI_ISL_1088368, EPI_ISL_1088369, EPI_ISL_1088370, EPI_ISL_1088371, EPI_ISL_1088372, EPI_ISL_1088373, EPI_ISL_1088374, EPI_ISL_1088375, EPI_ISL_1088376, EPI_ISL_1088377, EPI_ISL_1088378, EPI_ISL_1088379, EPI_ISL_1088380, EPI_ISL_1088381, EPI_ISL_1088382, EPI_ISL_1088383, EPI_ISL_1088384, EPI_ISL_1088385,
EPI_ISL_1088386, EPI_ISL_1088387, EPI_ISL_1088388, EPI_ISL_1088389, EPI_ISL_1088390, EPI_ISL_1088391, EPI_ISL_1088392, EPI_ISL_1088393, EPI_ISL_1088394, EPI_ISL_1088395, EPI_ISL_1088396, EPI_ISL_1088397, EPI_ISL_1088398, EPI_ISL_1088399, EPI_ISL_1088400, EPI_ISL_1088401, EPI_ISL_1088402, EPI_ISL_1088403, EPI_ISL_1088404, EPI_ISL_1088405, EPI_ISL_1088406, EPI_ISL_1088407,
EPI_ISL_1088408, EPI_ISL_1088409, EPI_ISL_1088410, EPI_ISL_1088411, EPI_ISL_1088412, EPI_ISL_1088413, EPI_ISL_1088414, EPI_ISL_1088415, EPI_ISL_1088416, EPI_ISL_1088417, EPI_ISL_1088418, EPI_ISL_1088419, EPI_ISL_1088420, EPI_ISL_1088421, EPI_ISL_1088422, EPI_ISL_1088423, EPI_ISL_1088424, EPI_ISL_1088425, EPI_ISL_1088426, EPI_ISL_1088427, EPI_ISL_1088428, EPI_ISL_1088429,
EPI_ISL_1088430, EPI_ISL_1088431, EPI_ISL_1088432, EPI_ISL_1088433, EPI_ISL_1088434, EPI_ISL_1088435, EPI_ISL_1088436, EPI_ISL_1088437, EPI_ISL_1088438, EPI_ISL_1088439, EPI_ISL_1088440, EPI_ISL_1088441, EPI_ISL_1088442, EPI_ISL_1088443, EPI_ISL_1088444, EPI_ISL_1088445, EPI_ISL_1088446, EPI_ISL_1088447, EPI_ISL_1088448, EPI_ISL_1088449, EPI_ISL_1088450, EPI_ISL_1088451,
EPI_ISL_1088452, EPI_ISL_1088453, EPI_ISL_1088454, EPI_ISL_1088455, EPI_ISL_1088456, EPI_ISL_1088457, EPI_ISL_1088458, EPI_ISL_1088459, EPI_ISL_1088460, EPI_ISL_1088461, EPI_ISL_1088462, EPI_ISL_1088463, EPI_ISL_1088464, EPI_ISL_1088465, EPI_ISL_1088466, EPI_ISL_1088467, EPI_ISL_1088468, EPI_ISL_1088469, EPI_ISL_1088470, EPI_ISL_1088471, EPI_ISL_1088472, EPI_ISL_1088473,
EPI_ISL_1088474, EPI_ISL_1088475, EPI_ISL_1088476, EPI_ISL_1088477, EPI_ISL_1088478, EPI_ISL_1088479, EPI_ISL_1088480, EPI_ISL_1088481, EPI_ISL_1088482, EPI_ISL_1088483, EPI_ISL_1088484, EPI_ISL_1088485, EPI_ISL_1088486, EPI_ISL_1088487, EPI_ISL_1088488, EPI_ISL_1088489, EPI_ISL_1088490, EPI_ISL_1088491, EPI_ISL_1088492, EPI_ISL_1088493, EPI_ISL_1088494, EPI_ISL_1088495,

| | | | |
|---|---|---|---|
| EPI_ISL_1088496, EPI_ISL_1088497, EPI_ISL_1088498, EPI_ISL_1088499, EPI_ISL_1088500, EPI_ISL_1088501, EPI_ISL_1088502, EPI_ISL_1088503, EPI_ISL_1088504, EPI_ISL_1088505, EPI_ISL_1088506, EPI_ISL_1088507, EPI_ISL_1088508, EPI_ISL_1088509, EPI_ISL_1088510, EPI_ISL_1088511, EPI_ISL_1088512, EPI_ISL_1088513, EPI_ISL_1088514, EPI_ISL_1088515, EPI_ISL_1088516, EPI_ISL_1088517, EPI_ISL_1088518, EPI_ISL_1088519, EPI_ISL_1088520, EPI_ISL_1088521, EPI_ISL_1088522, EPI_ISL_1088523, EPI_ISL_1088524, EPI_ISL_1088525, EPI_ISL_1088526, EPI_ISL_1088527, EPI_ISL_1088528, EPI_ISL_1088529, EPI_ISL_1088530, EPI_ISL_1088531, EPI_ISL_1088532, EPI_ISL_1088533, EPI_ISL_1088534, EPI_ISL_1088535, EPI_ISL_1088536, EPI_ISL_1088537, EPI_ISL_1088538, EPI_ISL_1088539, EPI_ISL_1088540, EPI_ISL_1088541, EPI_ISL_1088542, EPI_ISL_1088543, EPI_ISL_1088544, EPI_ISL_1088545, EPI_ISL_1088546, EPI_ISL_1088547, EPI_ISL_1088548, EPI_ISL_1088549, EPI_ISL_1088550 | | | |
| see above | Quest Diagnostics Incorporated | Respiratory Viruses Branch, Division of Viral Diseases, Centers for Disease Control and Prevention | Peter W. Cook, Dakota Howard, Dhwani Batra, Ben L. Rambo-Martin, S. H. Rosenthal, A. Gerasimova, R. M. Kagan, B. Anderson, M. Hua, Y. Liu, L.E. Bernstein, K.E. Livingston, A. Perez, I. A. Shlyakhter, R. V. Rolando, R. Owen, P. Tanpaiboon, F. Lacbawan, Clinton R. Paden, Suxiang Tong, Duncan MacCannell |
| EPI_ISL_1088585, EPI_ISL_1088586, EPI_ISL_1088587, EPI_ISL_1088588, EPI_ISL_1088589, EPI_ISL_1088590, EPI_ISL_1088591, EPI_ISL_1088592, EPI_ISL_1088593, EPI_ISL_1088594, EPI_ISL_1088595, EPI_ISL_1088596, EPI_ISL_1088597, EPI_ISL_1088598, EPI_ISL_1088599, EPI_ISL_1088600, EPI_ISL_1088601, EPI_ISL_1088602, EPI_ISL_1088603, EPI_ISL_1088604, EPI_ISL_1088605, EPI_ISL_1088606, EPI_ISL_1088607, EPI_ISL_1088608, EPI_ISL_1088609, EPI_ISL_1088610, EPI_ISL_1088611, EPI_ISL_1088612, EPI_ISL_1088613, EPI_ISL_1088614, EPI_ISL_1088615, EPI_ISL_1088616, EPI_ISL_1088617, EPI_ISL_1088618, EPI_ISL_1088619, EPI_ISL_1088620, EPI_ISL_1088621, EPI_ISL_1088622, EPI_ISL_1088623, EPI_ISL_1088624, EPI_ISL_1088625, EPI_ISL_1088626, EPI_ISL_1088627, EPI_ISL_1088628, EPI_ISL_1088629, EPI_ISL_1088630, EPI_ISL_1088631, EPI_ISL_1088632, EPI_ISL_1088633, EPI_ISL_1088634, EPI_ISL_1088635, EPI_ISL_1088636, EPI_ISL_1088637, EPI_ISL_1088638, EPI_ISL_1088639, EPI_ISL_1088640, EPI_ISL_1088641, EPI_ISL_1088642, EPI_ISL_1088643, EPI_ISL_1088644, EPI_ISL_1088645, EPI_ISL_1088646, EPI_ISL_1088647, EPI_ISL_1088648, EPI_ISL_1088649, EPI_ISL_1088650, EPI_ISL_1088651, EPI_ISL_1088652, EPI_ISL_1088653, EPI_ISL_1088654, EPI_ISL_1088655, EPI_ISL_1088656, EPI_ISL_1088657, EPI_ISL_1088658, EPI_ISL_1088659, EPI_ISL_1088660, EPI_ISL_1088661, EPI_ISL_1088662, EPI_ISL_1088663, EPI_ISL_1088664, EPI_ISL_1088665, EPI_ISL_1088666, EPI_ISL_1088667, EPI_ISL_1088668, EPI_ISL_1088669, EPI_ISL_1088670, EPI_ISL_1088671, EPI_ISL_1088672, EPI_ISL_1088673, EPI_ISL_1088674, EPI_ISL_1088675, EPI_ISL_1088676, EPI_ISL_1088677, EPI_ISL_1088678, EPI_ISL_1088679, EPI_ISL_1088680, EPI_ISL_1088681, EPI_ISL_1088682, EPI_ISL_1088683, EPI_ISL_1088684, EPI_ISL_1088685, EPI_ISL_1088686, EPI_ISL_1088687, EPI_ISL_1088688, EPI_ISL_1088689, EPI_ISL_1088690, EPI_ISL_1088691, EPI_ISL_1088692, EPI_ISL_1088693, EPI_ISL_1088694, EPI_ISL_1088695, EPI_ISL_1088696, EPI_ISL_1088697, EPI_ISL_1088698, EPI_ISL_1088699, EPI_ISL_1088700, EPI_ISL_1088701, EPI_ISL_1088702, EPI_ISL_1088703, EPI_ISL_1088704, EPI_ISL_1088705, EPI_ISL_1088706, EPI_ISL_1088707, EPI_ISL_1088708, EPI_ISL_1088709, EPI_ISL_1088710, EPI_ISL_1088711, EPI_ISL_1088712, EPI_ISL_1088713, EPI_ISL_1088714, EPI_ISL_1088715, EPI_ISL_1088716, EPI_ISL_1088717, EPI_ISL_1088718, EPI_ISL_1088719, EPI_ISL_1088720, EPI_ISL_1088721, EPI_ISL_1088722, EPI_ISL_1088723, EPI_ISL_1088724, EPI_ISL_1088725, EPI_ISL_1088726, EPI_ISL_1088727, EPI_ISL_1088728, EPI_ISL_1088729, EPI_ISL_1088730, EPI_ISL_1088731, EPI_ISL_1088732, EPI_ISL_1088733, EPI_ISL_1088734, EPI_ISL_1088735, EPI_ISL_1088736, EPI_ISL_1088737, EPI_ISL_1088738, EPI_ISL_1088739, EPI_ISL_1088740, EPI_ISL_1088741, EPI_ISL_1088742, EPI_ISL_1088743, EPI_ISL_1088744, EPI_ISL_1088745, EPI_ISL_1088746, EPI_ISL_1088747, EPI_ISL_1088748, EPI_ISL_1088749, EPI_ISL_1088750, EPI_ISL_1088751, EPI_ISL_1088752, EPI_ISL_1088753, EPI_ISL_1088754, EPI_ISL_1088755, EPI_ISL_1088756, EPI_ISL_1088757, EPI_ISL_1088758, EPI_ISL_1088759, EPI_ISL_1088760, EPI_ISL_1088761, EPI_ISL_1088762, EPI_ISL_1088763, EPI_ISL_1088764, EPI_ISL_1088765, EPI_ISL_1088766, EPI_ISL_1088767, EPI_ISL_1088768, EPI_ISL_1088769, EPI_ISL_1088770, EPI_ISL_1088771, EPI_ISL_1088772, EPI_ISL_1088773, EPI_ISL_1088774, EPI_ISL_1088775, EPI_ISL_1088776, EPI_ISL_1088777, EPI_ISL_1088778, EPI_ISL_1088779, EPI_ISL_1088780, EPI_ISL_1088781, EPI_ISL_1088782, EPI_ISL_1088783, EPI_ISL_1088784, EPI_ISL_1088785, EPI_ISL_1088786, EPI_ISL_1088787, EPI_ISL_1088788, EPI_ISL_1088789, EPI_ISL_1088790, EPI_ISL_1088791, EPI_ISL_1088792, EPI_ISL_1088793, EPI_ISL_1088794, EPI_ISL_1088795, EPI_ISL_1088796, EPI_ISL_1088797, EPI_ISL_1088798, EPI_ISL_1088799, EPI_ISL_1088800, EPI_ISL_1088801, EPI_ISL_1088802, EPI_ISL_1088803, EPI_ISL_1088804, EPI_ISL_1088805, EPI_ISL_1088806, EPI_ISL_1088807, EPI_ISL_1088808, EPI_ISL_1088809, EPI_ISL_1088810, EPI_ISL_1088811, EPI_ISL_1088812, EPI_ISL_1088813, EPI_ISL_1088814, EPI_ISL_1088815, EPI_ISL_1088816, EPI_ISL_1088817, EPI_ISL_1088818, EPI_ISL_1088819, EPI_ISL_1088820, EPI_ISL_1088821, EPI_ISL_1088822, EPI_ISL_1088823, EPI_ISL_1088824, EPI_ISL_1088825, EPI_ISL_1088826, EPI_ISL_1088827, EPI_ISL_1088828, EPI_ISL_1088829, EPI_ISL_1088830, EPI_ISL_1088831, EPI_ISL_1088832, EPI_ISL_1088833, EPI_ISL_1088834, EPI_ISL_1088835, EPI_ISL_1088836, EPI_ISL_1088837, EPI_ISL_1088838, EPI_ISL_1088839, EPI_ISL_1088840, EPI_ISL_1088841, EPI_ISL_1088842, EPI_ISL_1088843, EPI_ISL_1088844, EPI_ISL_1088845, EPI_ISL_1088846, EPI_ISL_1088847, EPI_ISL_1088848, EPI_ISL_1088890, EPI_ISL_1088893, EPI_ISL_1088894, EPI_ISL_1088895, EPI_ISL_1088896, EPI_ISL_1088897, EPI_ISL_1088898, EPI_ISL_1088899, EPI_ISL_1088900, EPI_ISL_1088901, EPI_ISL_1088902, EPI_ISL_1088903, EPI_ISL_1088904, EPI_ISL_1088905, EPI_ISL_1088906, EPI_ISL_1088907, EPI_ISL_1088908, EPI_ISL_1088909, EPI_ISL_1088910, EPI_ISL_1088911, EPI_ISL_1088912, EPI_ISL_1088913, EPI_ISL_1088914, EPI_ISL_1088915, EPI_ISL_1088916, EPI_ISL_1088917, EPI_ISL_1088918, EPI_ISL_1088919, EPI_ISL_1088920, EPI_ISL_1088921, EPI_ISL_1088922, EPI_ISL_1088923, EPI_ISL_1088924, EPI_ISL_1088925, EPI_ISL_1088926, EPI_ISL_1088927, EPI_ISL_1088928, EPI_ISL_1088929, EPI_ISL_1088930, EPI_ISL_1088931, EPI_ISL_1088932, EPI_ISL_1088933, EPI_ISL_1088934, EPI_ISL_1088935, EPI_ISL_1088936, EPI_ISL_1088937, EPI_ISL_1088938, EPI_ISL_1088939, EPI_ISL_1088940, EPI_ISL_1088941, EPI_ISL_1088942, EPI_ISL_1088943, EPI_ISL_1088944, EPI_ISL_1088945, EPI_ISL_1088946, EPI_ISL_1088947, EPI_ISL_1088948, EPI_ISL_1088949, EPI_ISL_1088950, EPI_ISL_1088951, EPI_ISL_1088952, EPI_ISL_1088953, EPI_ISL_1088954, EPI_ISL_1088955, EPI_ISL_1088956, EPI_ISL_1088957, EPI_ISL_1088958, EPI_ISL_1088959, EPI_ISL_1088960, EPI_ISL_1088961, EPI_ISL_1088962, EPI_ISL_1088963, EPI_ISL_1088964, EPI_ISL_1088965, EPI_ISL_1088966, EPI_ISL_1088967, EPI_ISL_1088968, EPI_ISL_1088969, EPI_ISL_1088970, EPI_ISL_1088971, EPI_ISL_1088972, EPI_ISL_1088973, EPI_ISL_1088974, EPI_ISL_1088975, EPI_ISL_1088976, EPI_ISL_1088977, EPI_ISL_1088978, EPI_ISL_1088979, EPI_ISL_1088980, EPI_ISL_1088981, EPI_ISL_1088982, EPI_ISL_1088983, EPI_ISL_1088984, EPI_ISL_1088985, EPI_ISL_1088986, EPI_ISL_1088987, EPI_ISL_1088988, EPI_ISL_1088989, EPI_ISL_1088990, EPI_ISL_1088991, EPI_ISL_1088992, EPI_ISL_1088993, EPI_ISL_1088994, EPI_ISL_1088995, EPI_ISL_1088996, EPI_ISL_1088997, EPI_ISL_1088998, EPI_ISL_1088999, EPI_ISL_1089000, EPI_ISL_1089001, EPI_ISL_1089002, EPI_ISL_1089003, EPI_ISL_1089004, EPI_ISL_1089005, EPI_ISL_1089006, EPI_ISL_1089007, EPI_ISL_1089008, EPI_ISL_1089009, EPI_ISL_1089010, EPI_ISL_1089011, EPI_ISL_1089012, EPI_ISL_1089013, EPI_ISL_1089014, EPI_ISL_1089015, EPI_ISL_1089016, EPI_ISL_1089017, EPI_ISL_1089018, EPI_ISL_1089019, EPI_ISL_1089020, EPI_ISL_1089021, EPI_ISL_1089022, EPI_ISL_1089023, EPI_ISL_1089024, EPI_ISL_1089025, EPI_ISL_1089026, EPI_ISL_1089027, EPI_ISL_1089028, EPI_ISL_1089029, EPI_ISL_1089030, EPI_ISL_1089031, EPI_ISL_1089032, EPI_ISL_1089033, EPI_ISL_1089034, EPI_ISL_1089035, EPI_ISL_1089036, EPI_ISL_1089037, EPI_ISL_1089038, EPI_ISL_1089039, EPI_ISL_1089040, EPI_ISL_1089041, EPI_ISL_1089042, EPI_ISL_1089043, EPI_ISL_1089044, EPI_ISL_1089045, EPI_ISL_1089046, EPI_ISL_1089047, EPI_ISL_1089048, EPI_ISL_1089049, EPI_ISL_1089050, EPI_ISL_1089051, EPI_ISL_1089052, EPI_ISL_1089053, EPI_ISL_1089054, EPI_ISL_1089055, EPI_ISL_1089056, EPI_ISL_1089057, EPI_ISL_1089058, EPI_ISL_1089059, EPI_ISL_1089060, EPI_ISL_1089061, EPI_ISL_1089062, EPI_ISL_1089063, EPI_ISL_1089064, EPI_ISL_1089065, EPI_ISL_1089066, EPI_ISL_1089067, EPI_ISL_1089068, EPI_ISL_1089069, EPI_ISL_1089070, EPI_ISL_1089071, EPI_ISL_1089072, EPI_ISL_1089073, EPI_ISL_1089074, EPI_ISL_1089075, EPI_ISL_1089076, EPI_ISL_1089077, EPI_ISL_1089078, EPI_ISL_1089079, EPI_ISL_1089080, EPI_ISL_1089081, EPI_ISL_1089082, EPI_ISL_1089083, EPI_ISL_1089084, EPI_ISL_1089085, EPI_ISL_1089086, EPI_ISL_1089087, EPI_ISL_1089088, EPI_ISL_1089089, EPI_ISL_1089090, EPI_ISL_1089091, EPI_ISL_1089092, EPI_ISL_1089093, EPI_ISL_1089094, EPI_ISL_1089095, EPI_ISL_1089096, EPI_ISL_1089097, EPI_ISL_1089098, EPI_ISL_1089099, EPI_ISL_1089100, EPI_ISL_1089101, EPI_ISL_1089102, EPI_ISL_1089103, EPI_ISL_1089104, EPI_ISL_1089105, EPI_ISL_1089106, EPI_ISL_1089107, EPI_ISL_1089108, EPI_ISL_1089109, EPI_ISL_1089110, EPI_ISL_1089111, EPI_ISL_1089112, EPI_ISL_1089113, EPI_ISL_1089114, EPI_ISL_1089115, EPI_ISL_1089116, EPI_ISL_1089117, EPI_ISL_1089118, EPI_ISL_1089119, EPI_ISL_1089120, EPI_ISL_1089121, EPI_ISL_1089122, EPI_ISL_1089123, EPI_ISL_1089124, EPI_ISL_1089125, EPI_ISL_1089126, EPI_ISL_1089127, EPI_ISL_1089128, EPI_ISL_1089129, EPI_ISL_1089130, EPI_ISL_1089131, EPI_ISL_1089132, EPI_ISL_1089133, EPI_ISL_1089134, EPI_ISL_1089135, EPI_ISL_1089136, EPI_ISL_1089137, EPI_ISL_1089138, EPI_ISL_1089139, EPI_ISL_1089140, EPI_ISL_1089141, EPI_ISL_1089142, EPI_ISL_1089143, EPI_ISL_1089144, EPI_ISL_1089145, EPI_ISL_1089146, EPI_ISL_1089147, EPI_ISL_1089148, EPI_ISL_1089149, EPI_ISL_1089150, EPI_ISL_1089151, EPI_ISL_1089152, EPI_ISL_1089153, EPI_ISL_1089154, EPI_ISL_1089155, EPI_ISL_1089156, EPI_ISL_1089157, EPI_ISL_1089158, EPI_ISL_1089159, EPI_ISL_1089160, EPI_ISL_1089161, EPI_ISL_1089162, EPI_ISL_1089163, EPI_ISL_1089164, EPI_ISL_1089165, EPI_ISL_1089166, EPI_ISL_1089167, EPI_ISL_1089168, EPI_ISL_1089169, EPI_ISL_1089170, EPI_ISL_1089171, EPI_ISL_1089172, EPI_ISL_1089173, EPI_ISL_1089174, EPI_ISL_1089175, EPI_ISL_1089176, EPI_ISL_1089177, EPI_ISL_1089178, EPI_ISL_1089179, EPI_ISL_1089180, EPI_ISL_1089181, EPI_ISL_1089182, EPI_ISL_1089183, EPI_ISL_1089184, EPI_ISL_1089185, EPI_ISL_1089186, EPI_ISL_1089187, EPI_ISL_1089188, EPI_ISL_1089189, EPI_ISL_1089190, EPI_ISL_1089191, EPI_ISL_1089192, EPI_ISL_1089193, EPI_ISL_1089194, EPI_ISL_1089195, EPI_ISL_1089196, EPI_ISL_1089197, EPI_ISL_1089198, EPI_ISL_1089199, EPI_ISL_1089200, EPI_ISL_1089201, EPI_ISL_1089202, EPI_ISL_1089203, EPI_ISL_1089204, EPI_ISL_1089205, EPI_ISL_1089206, EPI_ISL_1089207, EPI_ISL_1089208, EPI_ISL_1089209, EPI_ISL_1089210, EPI_ISL_1089211, EPI_ISL_1089212, EPI_ISL_1089213, EPI_ISL_1089214, EPI_ISL_1089215, EPI_ISL_1089216, EPI_ISL_1089217, EPI_ISL_1089218, EPI_ISL_1089219, EPI_ISL_1089220, EPI_ISL_1089221, EPI_ISL_1089222, EPI_ISL_1089223, EPI_ISL_1089224, EPI_ISL_1089225, EPI_ISL_1089226, EPI_ISL_1089227, EPI_ISL_1089228, EPI_ISL_1089229, EPI_ISL_1089230, EPI_ISL_1089231, EPI_ISL_1089232, EPI_ISL_1089233, EPI_ISL_1089234, EPI_ISL_1089235, EPI_ISL_1089236, EPI_ISL_1089237, EPI_ISL_1089238, EPI_ISL_1089239, EPI_ISL_1089240, EPI_ISL_1089241, EPI_ISL_1089242, EPI_ISL_1089243, EPI_ISL_1089244, EPI_ISL_1089245, EPI_ISL_1089246, EPI_ISL_1089247, EPI_ISL_1089248, EPI_ISL_1089249, EPI_ISL_1089250, EPI_ISL_1089251, EPI_ISL_1089252, EPI_ISL_1089253, EPI_ISL_1089254, EPI_ISL_1089255, EPI_ISL_1089256, EPI_ISL_1089257, EPI_ISL_1089258, EPI_ISL_1089259, EPI_ISL_1089260, EPI_ISL_1089261, EPI_ISL_1089262, EPI_ISL_1089263, EPI_ISL_1089264, EPI_ISL_1089265, EPI_ISL_1089266, EPI_ISL_1089267, EPI_ISL_1089268, EPI_ISL_1089269, EPI_ISL_1089270, EPI_ISL_1089271, EPI_ISL_1089272, EPI_ISL_1089273, EPI_ISL_1089274, EPI_ISL_1089275, EPI_ISL_1089276, EPI_ISL_1089277, EPI_ISL_1089278, EPI_ISL_1089279, EPI_ISL_1089280, EPI_ISL_1089281, EPI_ISL_1089282, EPI_ISL_1089283, EPI_ISL_1089284, EPI_ISL_1089285, EPI_ISL_1089286, EPI_ISL_1089287, EPI_ISL_1089288, EPI_ISL_1089289, EPI_ISL_1089290, EPI_ISL_1089291, EPI_ISL_1089292, EPI_ISL_1089293, EPI_ISL_1089294, EPI_ISL_1089295, EPI_ISL_1089296, EPI_ISL_1089297, EPI_ISL_1089298, EPI_ISL_1089299, EPI_ISL_1089300, EPI_ISL_1089301, EPI_ISL_1089302, EPI_ISL_1089303, EPI_ISL_1089304, EPI_ISL_1089305, EPI_ISL_1089306, EPI_ISL_1089307, EPI_ISL_1089308, EPI_ISL_1089309, EPI_ISL_1089310, EPI_ISL_1089311, EPI_ISL_1089312, EPI_ISL_1089313, EPI_ISL_1089314, EPI_ISL_1089315, EPI_ISL_1089316, EPI_ISL_1089317, EPI_ISL_1089318, EPI_ISL_1089319, EPI_ISL_1089320, EPI_ISL_1089321, EPI_ISL_1089322, EPI_ISL_1089323, EPI_ISL_1089324, EPI_ISL_1089325, EPI_ISL_1089326, EPI_ISL_1089327, EPI_ISL_1089328, EPI_ISL_1089329, EPI_ISL_1089330, EPI_ISL_1089331, EPI_ISL_1089332, EPI_ISL_1089333, EPI_ISL_1089334, EPI_ISL_1089335, EPI_ISL_1089336, EPI_ISL_1089337, EPI_ISL_1089338, EPI_ISL_1089339, EPI_ISL_1089340, EPI_ISL_1089341, EPI_ISL_1089342, EPI_ISL_1089343, EPI_ISL_1089344, EPI_ISL_1089345, EPI_ISL_1089346, EPI_ISL_1089347, EPI_ISL_1089348, EPI_ISL_1089349, EPI_ISL_1089350, EPI_ISL_1089351, EPI_ISL_1089352, EPI_ISL_1089353, EPI_ISL_1089354, EPI_ISL_1089355, EPI_ISL_1089356, EPI_ISL_1089357, EPI_ISL_1089358, EPI_ISL_1089359, EPI_ISL_1089360, EPI_ISL_1089361, EPI_ISL_1089362, EPI_ISL_1089363, EPI_ISL_1089364, EPI_ISL_1089365, EPI_ISL_1089366, EPI_ISL_1089367, EPI_ISL_1089368, EPI_ISL_1089369, EPI_ISL_1089370, EPI_ISL_1089371, EPI_ISL_1089372, EPI_ISL_1089373, EPI_ISL_1089374, EPI_ISL_1089375, EPI_ISL_1089376, EPI_ISL_1089377, EPI_ISL_1089378, EPI_ISL_1089379, EPI_ISL_1089380, EPI_ISL_1089381, EPI_ISL_1089382, EPI_ISL_1089383, EPI_ISL_1089384, EPI_ISL_1089385, EPI_ISL_1089386, EPI_ISL_1089387, EPI_ISL_1089388, EPI_ISL_1089389, EPI_ISL_1089390, EPI_ISL_1089391, EPI_ISL_1089392, EPI_ISL_1089393, EPI_ISL_1089394, EPI_ISL_1089395, EPI_ISL_1089396, EPI_ISL_1089397, EPI_ISL_1089398, EPI_ISL_1089399, EPI_ISL_1089400, EPI_ISL_1089401, EPI_ISL_1089402, EPI_ISL_1089403, EPI_ISL_1089404, EPI_ISL_1089405, EPI_ISL_1089406, EPI_ISL_1089407, EPI_ISL_1089408, EPI_ISL_1089409, EPI_ISL_1089410, EPI_ISL_1089411, EPI_ISL_1089412, EPI_ISL_1089413, EPI_ISL_1089414, EPI_ISL_1089415, EPI_ISL_1089416, EPI_ISL_1089417, EPI_ISL_1089418, EPI_ISL_1089419, EPI_ISL_1089420, EPI_ISL_1089421, EPI_ISL_1089422, EPI_ISL_1089423, EPI_ISL_1089424, EPI_ISL_1089425, EPI_ISL_1089426, EPI_ISL_1089427, EPI_ISL_1089428, EPI_ISL_1089429, EPI_ISL_1089430, EPI_ISL_1089431, EPI_ISL_1089432, EPI_ISL_1089433 | | | |
| see above | Helix / Illumina | Respiratory Viruses Branch, Division of Viral Diseases, Centers for Disease Control and Prevention | Peter W. Cook, Dakota Howard, Dhwani Batra, Ben L. Rambo-Martin, Eileen de Feo, Jan Antico, Christine Tran, Matthew Tolentino, Shannon Wickline, Kim Gietzen, Brad Sickler, Jingtao Liu, Eric Allen, Phil Febbo, Summer Galloway, Nicole L. Washington, Simon White, Geraint Levan, Kelly Schiabor Barrett, Elizabeth Cirulli, Alexandre Bolze, Ary Ascencio, Charlotte Rivera-Garcia, Ryan Cho, Jason Nguyen, Sherry Wang, Jimmy Ramirez, Tyler Cassens, Efren Sandoval, Magnus Isaksson, William Lee, David Becker, Marc Laurent, James Lu, Clinton R. Paden, Suxiang Tong, Duncan MacCannell |
| EPI_ISL_1090274, EPI_ISL_1090275, EPI_ISL_1090276, EPI_ISL_1090277, EPI_ISL_1090278, EPI_ISL_1090279, EPI_ISL_1090280, EPI_ISL_1090281, EPI_ISL_1090282, EPI_ISL_1090283, EPI_ISL_1090284, EPI_ISL_1090285, EPI_ISL_1090286, EPI_ISL_1090287, EPI_ISL_1090288, EPI_ISL_1090289, EPI_ISL_1090290, EPI_ISL_1090291, EPI_ISL_1090292, EPI_ISL_1090293, EPI_ISL_1090294, EPI_ISL_1090295, EPI_ISL_1090296, EPI_ISL_1090297, EPI_ISL_1090298, EPI_ISL_1090299, EPI_ISL_1090301, EPI_ISL_1090302, EPI_ISL_1090303, EPI_ISL_1090304, EPI_ISL_1090305, EPI_ISL_1090306, EPI_ISL_1090307, EPI_ISL_1090308, EPI_ISL_1090309, EPI_ISL_1090310, EPI_ISL_1090311, EPI_ISL_1090312, EPI_ISL_1090313, EPI_ISL_1090314, EPI_ISL_1090315, EPI_ISL_1090316, EPI_ISL_1090317, EPI_ISL_1090318, EPI_ISL_1090319, EPI_ISL_1090320, EPI_ISL_1090321, EPI_ISL_1090322, EPI_ISL_1090323, EPI_ISL_1090324, EPI_ISL_1090325, EPI_ISL_1090326, EPI_ISL_1090327, EPI_ISL_1090328, EPI_ISL_1090329, EPI_ISL_1090330, EPI_ISL_1090331, EPI_ISL_1090332, EPI_ISL_1090333, EPI_ISL_1090334, EPI_ISL_1090335, EPI_ISL_1090336, EPI_ISL_1090337, EPI_ISL_1090338, EPI_ISL_1090339, EPI_ISL_1090340, EPI_ISL_1090341, EPI_ISL_1090342, EPI_ISL_1090343, EPI_ISL_1090344, EPI_ISL_1090345, EPI_ISL_1090346, EPI_ISL_1090347, EPI_ISL_1090348, EPI_ISL_1090349, EPI_ISL_1090350, EPI_ISL_1090351, EPI_ISL_1090352, EPI_ISL_1090353, EPI_ISL_1090354, EPI_ISL_1090355, EPI_ISL_1090356, EPI_ISL_1090357, EPI_ISL_1090358, EPI_ISL_1090359, EPI_ISL_1090360, EPI_ISL_1090361, EPI_ISL_1090362, EPI_ISL_1090363, EPI_ISL_1090364, EPI_ISL_1090365, EPI_ISL_1090366, EPI_ISL_1090367, EPI_ISL_1090368, EPI_ISL_1090369, EPI_ISL_1090370, EPI_ISL_1090371, EPI_ISL_1090372, EPI_ISL_1090373, EPI_ISL_1090374, EPI_ISL_1090375, EPI_ISL_1090376, EPI_ISL_1090377, EPI_ISL_1090378, EPI_ISL_1090379, EPI_ISL_1090380, EPI_ISL_1090381, EPI_ISL_1090382, EPI_ISL_1090383, EPI_ISL_1090384, EPI_ISL_1090385, EPI_ISL_1090386, EPI_ISL_1090387, EPI_ISL_1090388, EPI_ISL_1090389, EPI_ISL_1090390, EPI_ISL_1090391, EPI_ISL_1090392, EPI_ISL_1090393, EPI_ISL_1090394, EPI_ISL_1090395, EPI_ISL_1090396, EPI_ISL_1090397, EPI_ISL_1090398, EPI_ISL_1090399, EPI_ISL_1090400, EPI_ISL_1090401, EPI_ISL_1090402, EPI_ISL_1090403, EPI_ISL_1090404, EPI_ISL_1090405, EPI_ISL_1090406, EPI_ISL_1090407, EPI_ISL_1090408, EPI_ISL_1090409, EPI_ISL_1090410, EPI_ISL_1090411, EPI_ISL_1090412, EPI_ISL_1090413, EPI_ISL_1090414, EPI_ISL_1090415, EPI_ISL_1090416, EPI_ISL_1090417, EPI_ISL_1090418, EPI_ISL_1090419, EPI_ISL_1090420, EPI_ISL_1090421, EPI_ISL_1090422, EPI_ISL_1090423, EPI_ISL_1090424, EPI_ISL_1090425, EPI_ISL_1090426, EPI_ISL_1090427, EPI_ISL_1090428, EPI_ISL_1090429, EPI_ISL_1090430, EPI_ISL_1090431, EPI_ISL_1090432, EPI_ISL_1090433, EPI_ISL_1090434, EPI_ISL_1090435, EPI_ISL_1090436, EPI_ISL_1090437, EPI_ISL_1090438, EPI_ISL_1090439, EPI_ISL_1090440, EPI_ISL_1090441, EPI_ISL_1090442, EPI_ISL_1090443, EPI_ISL_1090444, EPI_ISL_1090445, EPI_ISL_1090446, EPI_ISL_1090447, EPI_ISL_1090448, EPI_ISL_1090449, EPI_ISL_1090450, EPI_ISL_1090451, EPI_ISL_1090452, EPI_ISL_1090453, EPI_ISL_1090454, EPI_ISL_1090455, EPI_ISL_1090456, EPI_ISL_1090457, EPI_ISL_1090458, EPI_ISL_1090459, EPI_ISL_1090460, EPI_ISL_1090461, EPI_ISL_1090462, EPI_ISL_1090463, EPI_ISL_1090464, EPI_ISL_1090465, EPI_ISL_1090466, EPI_ISL_1090467, EPI_ISL_1090468, EPI_ISL_1090469, EPI_ISL_1090470, EPI_ISL_1090471, EPI_ISL_1090472, EPI_ISL_1090473, EPI_ISL_1090474, EPI_ISL_1090475, EPI_ISL_1090476, EPI_ISL_1090477, EPI_ISL_1090478, EPI_ISL_1090479, EPI_ISL_1090480, EPI_ISL_1090481, EPI_ISL_1090482, EPI_ISL_1090483, EPI_ISL_1090484, EPI_ISL_1090485, EPI_ISL_1090486, EPI_ISL_1090487, EPI_ISL_1090488, EPI_ISL_1090489, EPI_ISL_1090490, EPI_ISL_1090491, EPI_ISL_1090492, EPI_ISL_1090493, EPI_ISL_1090494, EPI_ISL_1090495, EPI_ISL_1090496, EPI_ISL_1090497, EPI_ISL_1090498, EPI_ISL_1090499, EPI_ISL_1090500, EPI_ISL_1090501, EPI_ISL_1090502, EPI_ISL_1090503, EPI_ISL_1090504, EPI_ISL_1090505, EPI_ISL_1090506, EPI_ISL_1090507, EPI_ISL_1090508, EPI_ISL_1090509, EPI_ISL_1090510, EPI_ISL_1090511, EPI_ISL_1090512, EPI_ISL_1090513, EPI_ISL_1090514, EPI_ISL_1090515, EPI_ISL_1090516, EPI_ISL_1090517, EPI_ISL_1090518, EPI_ISL_1090519, EPI_ISL_1090520, EPI_ISL_1090521, EPI_ISL_1090522, EPI_ISL_1090523, EPI_ISL_1090524, EPI_ISL_1090525, EPI_ISL_1090526, EPI_ISL_1090527, EPI_ISL_1090528, EPI_ISL_1090529, EPI_ISL_1090530, EPI_ISL_1090531, EPI_ISL_1090532, EPI_ISL_1090533, EPI_ISL_1090534, EPI_ISL_1090535, EPI_ISL_1090536, EPI_ISL_1090537, EPI_ISL_1090538, EPI_ISL_1090539, EPI_ISL_1090540, EPI_ISL_1090541, EPI_ISL_1090542, EPI_ISL_1090543, EPI_ISL_1090544, EPI_ISL_1090545, EPI_ISL_1090546, EPI_ISL_1090547, EPI_ISL_1090548, EPI_ISL_1090549, EPI_ISL_1090550, EPI_ISL_1090551, EPI_ISL_1090552, EPI_ISL_1090553, EPI_ISL_1090554, EPI_ISL_1090555, EPI_ISL_1090556, EPI_ISL_1090557, EPI_ISL_1090558, EPI_ISL_1090559, EPI_ISL_1090560, EPI_ISL_1090561, EPI_ISL_1090562, EPI_ISL_1090563, EPI_ISL_1090564, EPI_ISL_1090565, EPI_ISL_1090566, EPI_ISL_1090567, EPI_ISL_1090568, EPI_ISL_1090569, EPI_ISL_1090570, EPI_ISL_1090571, EPI_ISL_1090572, EPI_ISL_1090573, EPI_ISL_1090574, EPI_ISL_1090575, EPI_ISL_1090576, EPI_ISL_1090577, EPI_ISL_1090578, EPI_ISL_1090579, EPI_ISL_1090580, EPI_ISL_1090581, EPI_ISL_1090582, EPI_ISL_1090583, EPI_ISL_1090584, EPI_ISL_1090585, EPI_ISL_1090586, EPI_ISL_1090587, EPI_ISL_1090588, EPI_ISL_1090589, EPI_ISL_1090590, EPI_ISL_1090591, EPI_ISL_1090592, EPI_ISL_1090593, EPI_ISL_1090594, EPI_ISL_1090595, EPI_ISL_1090596, EPI_ISL_1090597, EPI_ISL_1090598, EPI_ISL_1090599, EPI_ISL_1090600, EPI_ISL_1090601, EPI_ISL_1090602, EPI_ISL_1090603, EPI_ISL_1090604, EPI_ISL_1090605, EPI_ISL_1090606, EPI_ISL_1090607, EPI_ISL_1090608, EPI_ISL_1090609, EPI_ISL_1090610, EPI_ISL_1090611, EPI_ISL_1090612, EPI_ISL_1090613, EPI_ISL_1090614, EPI_ISL_1090615, EPI_ISL_1090616, EPI_ISL_1090617, EPI_ISL_1090618, EPI_ISL_1090619, EPI_ISL_1090620, EPI_ISL_1090621, EPI_ISL_1090622, EPI_ISL_1090623, EPI_ISL_1090624, EPI_ISL_1090625, EPI_ISL_1090626, EPI_ISL_1090627, EPI_ISL_1090628, EPI_ISL_1090629, EPI_ISL_1090630, EPI_ISL_1090631, EPI_ISL_1090632, EPI_ISL_1090633, EPI_ISL_1090634, EPI_ISL_1090635, EPI_ISL_1090636, EPI_ISL_1090637, EPI_ISL_1090638, EPI_ISL_1090639, EPI_ISL_1090640, EPI_ISL_1090641, EPI_ISL_1090642, EPI_ISL_1090643, EPI_ISL_1090644, EPI_ISL_1090645, EPI_ISL_1090646, EPI_ISL_1090647, EPI_ISL_1090648, EPI_ISL_1090649, EPI_ISL_1090650, EPI_ISL_1090651, EPI_ISL_1090652, EPI_ISL_1090653, EPI_ISL_1090654, EPI_ISL_1090655, EPI_ISL_1090656, EPI_ISL_1090657, EPI_ISL_1090658, EPI_ISL_1090659, EPI_ISL_1090660, EPI_ISL_1090661, EPI_ISL_1090662, EPI_ISL_1090663, EPI_ISL_1090664, EPI_ISL_1090665, EPI_ISL_1090666, EPI_ISL_1090667, EPI_ISL_1090668, EPI_ISL_1090669, EPI_ISL_1090670, EPI_ISL_1090671, EPI_ISL_1090672, EPI_ISL_1090673, EPI_ISL_1090674, EPI_ISL_1090675, EPI_ISL_1090676, EPI_ISL_1090677, EPI_ISL_1090678, EPI_ISL_1090679, EPI_ISL_1090680, EPI_ISL_1090681, EPI_ISL_1090682, EPI_ISL_1090683, EPI_ISL_1090684, EPI_ISL_1090685, EPI_ISL_1090686, EPI_ISL_1090687, EPI_ISL_1090688, EPI_ISL_1090689, EPI_ISL_1090690, EPI_ISL_1090691, EPI_ISL_1090692, EPI_ISL_1090693, EPI_ISL_1090694, EPI_ISL_1090695, EPI_ISL_1090696, EPI_ISL_1090697, EPI_ISL_1090698, EPI_ISL_1090699, EPI_ISL_1090700, EPI_ISL_1090701, EPI_ISL_1090702, EPI_ISL_1090703, EPI_ISL_1090704, EPI_ISL_1090705, EPI_ISL_1090706, EPI_ISL_1090707, EPI_ISL_1090708, EPI_ISL_1090709, EPI_ISL_1090710, EPI_ISL_1090711, EPI_ISL_1090712, EPI_ISL_1090713, EPI_ISL_1090714, EPI_ISL_1090715, EPI_ISL_1090716, EPI_ISL_1090717, EPI_ISL_1090718, EPI_ISL_1090719, EPI_ISL_1090720, EPI_ISL_1090721, EPI_ISL_1090722, EPI_ISL_1090723, EPI_ISL_1090724, EPI_ISL_1090725, EPI_ISL_1090726, EPI_ISL_1090727, EPI_ISL_1090728, EPI_ISL_1090729, EPI_ISL_1090730, EPI_ISL_1090731, EPI_ISL_1090732, EPI_ISL_1090733, EPI_ISL_1090734, EPI_ISL_1090735, EPI_ISL_1090736, EPI_ISL_1090737, EPI_ISL_1090738, EPI_ISL_1090739, EPI_ISL_1090740, EPI_ISL_1090741, EPI_ISL_1090742, EPI_ISL_1090743, EPI_ISL_1090744, EPI_ISL_1090745, EPI_ISL_1090746, EPI_ISL_1090747, EPI_ISL_1090748, EPI_ISL_1090749, EPI_ISL_1090750, EPI_ISL_1090751, EPI_ISL_1090752, EPI_ISL_1090753, EPI_ISL_1090754, EPI_ISL_1090755, EPI_ISL_1090756, EPI_ISL_1090757, EPI_ISL_1090758, EPI_ISL_1090759, EPI_ISL_1090760, EPI_ISL_1090761, EPI_ISL_1090762, EPI_ISL_1090763, EPI_ISL_1090764, EPI_ISL_1090765, EPI_ISL_1090766, EPI_ISL_1090767, EPI_ISL_1090768, EPI_ISL_1090769, EPI_ISL_1090770, EPI_ISL_1090771, EPI_ISL_1090772, EPI_ISL_1090773, EPI_ISL_1090774, EPI_ISL_1090775, EPI_ISL_1090776, EPI_ISL_1090777, EPI_ISL_1090778, EPI_ISL_1090779, EPI_ISL_1090780, EPI_ISL_1090781, EPI_ISL_1090782, EPI_ISL_1090783, EPI_ISL_1090784, EPI_ISL_1090785, EPI_ISL_1090786, EPI_ISL_1090787, EPI_ISL_1090788, EPI_ISL_1090789, EPI_ISL_1090790, EPI_ISL_1090791, EPI_ISL_1090792, EPI_ISL_1090793, EPI_ISL_1090794, EPI_ISL_1090795, EPI_ISL_1090796, EPI_ISL_1090797, EPI_ISL_1090798, EPI_ISL_1090799, EPI_ISL_1090800, EPI_ISL_1090801, EPI_ISL_1090802, EPI_ISL_1090803, EPI_ISL_1090804, EPI_ISL_1090805, EPI_ISL_1090806, EPI_ISL_1090807, EPI_ISL_1090808, EPI_ISL_1090809, EPI_ISL_1090810, EPI_ISL_1090811, EPI_ISL_1090812, EPI_ISL_1090813, EPI_ISL_1090814, EPI_ISL_1090815, EPI_ISL_1090816, EPI_ISL_1090817, EPI_ISL_1090818, EPI_ISL_1090819, EPI_ISL_1090820, EPI_ISL_1090821, EPI_ISL_1090822, EPI_ISL_1090823, EPI_ISL_1090824, EPI_ISL_1090825, EPI_ISL_1090826, EPI_ISL_1090827, EPI_ISL_1090828, EPI_ISL_1090829, EPI_ISL_1090830, EPI_ISL_1090831, EPI_ISL_1090832, EPI_ISL_1090833, EPI_ISL_1090834, EPI_ISL_1090835, EPI_ISL_1090836, EPI_ISL_1090837, EPI_ISL_1090838, EPI_ISL_1090839, EPI_ISL_1090840, EPI_ISL_1090841, EPI_ISL_1090842, EPI_ISL_1090843, EPI_ISL_1090844, EPI_ISL_1090845, EPI_ISL_1090846, EPI_ISL_1090847, EPI_ISL_1090848, EPI_ISL_1090849, EPI_ISL_1090850, EPI_ISL_1090851, EPI_ISL_1090852, EPI_ISL_1090853, EPI_ISL_1090855, EPI_ISL_1090856, EPI_ISL_1090857, EPI_ISL_1090858, EPI_ISL_1090859, EPI_ISL_1090860, EPI_ISL_1090861, EPI_ISL_1090862, EPI_ISL_1090863, EPI_ISL_1090864, EPI_ISL_1090865, EPI_ISL_1090866, EPI_ISL_1090867, EPI_ISL_1090868, EPI_ISL_1090869, EPI_ISL_1090870, EPI_ISL_1090871, EPI_ISL_1090872, EPI_ISL_1090873, EPI_ISL_1090874, EPI_ISL_1090875, EPI_ISL_1090876, EPI_ISL_1090877, EPI_ISL_1090878, EPI_ISL_1090879, EPI_ISL_1090880, EPI_ISL_1090881, EPI_ISL_1090882, EPI_ISL_1090883, EPI_ISL_1090884, EPI_ISL_1090885, EPI_ISL_1090886, EPI_ISL_1090887, EPI_ISL_1090888, EPI_ISL_1090889, EPI_ISL_1090890, EPI_ISL_1090891, EPI_ISL_1090892, EPI_ISL_1090893, EPI_ISL_1090894, EPI_ISL_1090895, EPI_ISL_1090896, EPI_ISL_1090897, EPI_ISL_1090898, EPI_ISL_1090899, EPI_ISL_1090900, EPI_ISL_1090901, EPI_ISL_1090902, EPI_ISL_1090903, EPI_ISL_1090904, EPI_ISL_1090905, EPI_ISL_1090906, EPI_ISL_1090907, EPI_ISL_1090908, EPI_ISL_1090909, EPI_ISL_1090910, EPI_ISL_1090911, EPI_ISL_1090912, EPI_ISL_1090913, EPI_ISL_1090914, EPI_ISL_1090915, EPI_ISL_1090916, EPI_ISL_1090917, EPI_ISL_1090918, EPI_ISL_1090919, EPI_ISL_1090920, EPI_ISL_1090921, EPI_ISL_1090922, EPI_ISL_1090923, EPI_ISL_1090924, EPI_ISL_1090925, EPI_ISL_1090926, EPI_ISL_1090927, EPI_ISL_1090928, EPI_ISL_1090929, EPI_ISL_1090930, EPI_ISL_1090931, EPI_ISL_1090932, EPI_ISL_1090933, EPI_ISL_1090934, EPI_ISL_1090935, EPI_ISL_1090936, EPI_ISL_1090937, EPI_ISL_1090938, EPI_ISL_1090939, EPI_ISL_1090940, EPI_ISL_1090941, EPI_ISL_1090942, EPI_ISL_1090943, EPI_ISL_1090944, EPI_ISL_1090945, EPI_ISL_1090946, EPI_ISL_1090947, EPI_ISL_1090948, EPI_ISL_1090949, EPI_ISL_1090950, EPI_ISL_1090951, EPI_ISL_1090952, EPI_ISL_1090953, EPI_ISL_1090954, EPI_ISL_1090955, EPI_ISL_1090956, EPI_ISL_1090957, EPI_ISL_1090958, EPI_ISL_1090959, EPI_ISL_1090960, EPI_ISL_1090961, EPI_ISL_1090962, EPI_ISL_1090963, EPI_ISL_1090964, EPI_ISL_1090965, EPI_ISL_1090966, EPI_ISL_1090967, EPI_ISL_1090968, EPI_ISL_1090969, EPI_ISL_1090970, EPI_ISL_1090971, EPI_ISL_1090972, EPI_ISL_1090973, EPI_ISL_1090974, EPI_ISL_1090975, EPI_ISL_1090976, EPI_ISL_1090977, EPI_ISL_1090978, EPI_ISL_1090979, EPI_ISL_1090980, EPI_ISL_1090981, EPI_ISL_1090982, EPI_ISL_1090983, EPI_ISL_1090984, EPI_ISL_1090985, EPI_ISL_1090986, EPI_ISL_1090987, EPI_ISL_1090988, EPI_ISL_1090989, EPI_ISL_1090990, EPI_ISL_1090991, EPI_ISL_1090992, EPI_ISL_1090993, EPI_ISL_1090994, EPI_ISL_1090995, EPI_ISL_1090996, EPI_ISL_1090997, EPI_ISL_1090998, EPI_ISL_1090999, EPI_ISL_1091000, EPI_ISL_1091001, EPI_ISL_1091002, EPI_ISL_1091003, EPI_ISL_1091004, EPI_ISL_1091005, EPI_ISL_1091006, EPI_ISL_1091007, EPI_ISL_1091008, EPI_ISL_1091009, EPI_ISL_1091010, EPI_ISL_1091011, EPI_ISL_1091012, EPI_ISL_1091013, EPI_ISL_1091014, EPI_ISL_1091015, EPI_ISL_1091016, EPI_ISL_1091017, EPI_ISL_1091018, EPI_ISL_1091019, EPI_ISL_1091020, EPI_ISL_1091021, EPI_ISL_1091022, EPI_ISL_1091023, EPI_ISL_1091024, EPI_ISL_1091025, EPI_ISL_1091026, EPI_ISL_1091027, EPI_ISL_1091028, EPI_ISL_1091029, EPI_ISL_1091030, EPI_ISL_1091031, EPI_ISL_1091032, EPI_ISL_1091033, EPI_ISL_1091034, EPI_ISL_1091035, EPI_ISL_1091036, EPI_ISL_1091037, EPI_ISL_1091038, EPI_ISL_1091039, EPI_ISL_1091040, EPI_ISL_1091041, EPI_ISL_1091042, EPI_ISL_1091043, EPI_ISL_1091044, EPI_ISL_1091045, EPI_ISL_1091046, EPI_ISL_1091047, EPI_ISL_1091048, EPI_ISL_1091049, EPI_ISL_1091050, EPI_ISL_1091051, EPI_ISL_1091052, EPI_ISL_1091053, EPI_ISL_1091054, EPI_ISL_1091055, EPI_ISL_1091056, EPI_ISL_1091057, EPI_ISL_1091058, EPI_ISL_1091059, EPI_ISL_1091060, EPI_ISL_1091061, EPI_ISL_1091062, EPI_ISL_1091063, EPI_ISL_1091064, EPI_ISL_1091065, EPI_ISL_1091066, EPI_ISL_1091067, EPI_ISL_1091068, EPI_ISL_1091069, EPI_ISL_1091070, EPI_ISL_1091071, EPI_ISL_1091072, EPI_ISL_1091073, EPI_ISL_1091074, EPI_ISL_1091075, EPI_ISL_1091076, EPI_ISL_1091077, EPI_ISL_1091078, EPI_ISL_1091079, EPI_ISL_1091080, EPI_ISL_1091081, EPI_ISL_1091082, EPI_ISL_1091083, EPI_ISL_1091084, EPI_ISL_1091085, EPI_ISL_1091086, EPI_ISL_1091087, EPI_ISL_1091088, EPI_ISL_1091089 | | | |
| see above | Quest Diagnostics Incorporated | Respiratory Viruses Branch, Division of Viral Diseases, Centers for Disease Control and Prevention | Peter W. Cook, Dakota Howard, Dhwani Batra, Ben L. Rambo-Martin, S. H. Rosenthal, A. Gerasimova, R. M. Kagan, B. Anderson, M. Hua, Y. Liu, L.E. Bernstein, K.E. Livingston, A. Perez, I. A. Shlyakhter, R. V. Rolando, R. Owen, P. Tanpaiboon, F. Lacbawan, Clinton R. Paden, Suxiang Tong, Duncan MacCannell |
| EPI_ISL_1091241, EPI_ISL_1091242, EPI_ISL_1091243, EPI_ISL_1091244, EPI_ISL_1091245, EPI_ISL_1091246 | Laboratorio Estatal de Salud Pública de Nuevo León | Laboratorio de Infectología Molecular, Departamento de Bioquímica y Medicina Molecular,Facultad de Medicina - Universidad Autónoma de Nuevo León | Kame A. Galán-Huerta, María F. Herrera-Saldivar, Natalia Martínez-Acuña, Sonia A. Lozano-Sepúlveda, Daniel Arellanos-Soto, Ana M. Rivas-Estilla, Samuel Buentello-Wong, Else del Carmen García-García, Gloria A. Jasso-de-la-Peña, Roberto Montes-de-Oca, Consuelo Treviño-Garza, Manuel E. de-la-O-Cavazos |
| EPI_ISL_1091247, EPI_ISL_1091248 | Laboratorio PGM | Laboratorio de Infectología Molecular, Departamento de Bioquímica y Medicina Molecular,Facultad de Medicina - Universidad Autónoma de Nuevo León | Kame A. Galán-Huerta, María F. Herrera-Saldivar, Natalia Martínez-Acuña, Sonia A. Lozano-Sepúlveda, Daniel Arellanos-Soto, Ana M. Rivas-Estilla, Javier Ramos-Jimenez, Gabriela Elizondo, Eduardo Garza-de-la-Peña |
| EPI_ISL_1091249 | Laboratorio Estatal de Salud Pública de Nuevo León | Laboratorio de Infectología Molecular, Departamento de Bioquímica y Medicina Molecular,Facultad de Medicina - Universidad Autónoma de Nuevo León | Kame A. Galán-Huerta, María F. Herrera-Saldivar, Natalia Martínez-Acuña, Sonia A. Lozano-Sepúlveda, Daniel Arellanos-Soto, Ana M. Rivas-Estilla, Samuel Buentello-Wong, Else del Carmen García-García, Gloria A. Jasso-de-la-Peña, Roberto Montes-de-Oca, Consuelo Treviño-Garza, Manuel E. de-la-O-Cavazos |
| EPI_ISL_1091252 | Laboratorio PGM | Laboratorio de Infectología Molecular, Departamento de Bioquímica y Medicina Molecular,Facultad de Medicina - Universidad Autónoma de Nuevo León | Kame A. Galán-Huerta, María F. Herrera-Saldivar, Natalia Martínez-Acuña, Sonia A. Lozano-Sepúlveda, Daniel Arellanos-Soto, Ana M. Rivas-Estilla, Javier Ramos-Jimenez, Gabriela Elizondo, Eduardo Garza-de-la-Peña |
| EPI_ISL_1091253, EPI_ISL_1091254, | Laboratorio Estatal de Salud Pública de Nuevo León | Laboratorio de Infectología Molecular, Departamento de Bioquímica y Medicina | Kame A. Galán-Huerta, María F. Herrera-Saldivar, Natalia Martínez-Acuña, Sonia A. Lozano-Sepúlveda, Daniel Arellanos-Soto, Ana M. Rivas-Estilla, Samuel Buentello-Wong, Else del Carmen García-García, Gloria A. Jasso-de-la-Peña, Roberto Montes-de-Oca, Consuelo Treviño-Garza, Manuel E. de-la-O-Cavazos |

| | | | |
|---|---|---|---|
| EPI_ISL_1091255, EPI_ISL_1091256, EPI_ISL_1091257 | | Molecular,Facultad de Medicina - Universidad Autónoma de Nuevo León | |
| EPI_ISL_1091258, EPI_ISL_1091259, EPI_ISL_1091260 | Laboratorio PGM | Laboratorio de Infectología Molecular, Departamento de Bioquímica y Medicina Molecular,Facultad de Medicina - Universidad Autónoma de Nuevo León | Kame A. Galán-Huerta, María F. Herrera-Saldivar, Natalia Martínez-Acuña, Sonia A. Lozano-Sepúlveda, Daniel Arellanos-Soto, Ana M. Rivas-Estilla, Javier Ramos-Jimenez, Gabriela Elizondo, Eduardo Garza-de-la-Peña |
| EPI_ISL_1091261 | Laboratorio de Apoyo a la Vigilancia e Investigación Epidemiológica - CIBO | Laboratorio de Infectología Molecular, Departamento de Bioquímica y Medicina Molecular,Facultad de Medicina - Universidad Autónoma de Nuevo León | Kame A. Galán-Huerta, María F. Herrera-Saldivar, Natalia Martínez-Acuña, Sonia A. Lozano-Sepúlveda, Daniel Arellanos-Soto, Ana M. Rivas-Estilla, Javier Ramos-Jimenez, Claudia L. Charles-Niño, Alma M. Perez-Rios, Rafael Soto-Alvarez, Diego A. Gonzalez-Altamirano, Jose J. Figueroa-Delgadillo, Blanca M. Torres, Hector Montoya-Fuentes |
| EPI_ISL_1091262 | Laboratorio Estatal de Salud Pública de Nuevo León | Laboratorio de Infectología Molecular, Departamento de Bioquímica y Medicina Molecular,Facultad de Medicina - Universidad Autónoma de Nuevo León | Kame A. Galán-Huerta, María F. Herrera-Saldivar, Natalia Martínez-Acuña, Sonia A. Lozano-Sepúlveda, Daniel Arellanos-Soto, Ana M. Rivas-Estilla, Samuel Buentello-Wong, Else del Carmen García-García, Gloria A. Jasso-de-la-Peña, Roberto Montes-de-Oca, Consuelo Treviño-Garza, Manuel E. de-la-O-Cavazos |
| EPI_ISL_1091263, EPI_ISL_1091264, EPI_ISL_1091265, EPI_ISL_1091266, EPI_ISL_1091267, EPI_ISL_1091268, EPI_ISL_1091269, EPI_ISL_1091270, EPI_ISL_1091271, EPI_ISL_1091272, EPI_ISL_1091273, EPI_ISL_1091274 | | | |
| see above | Laboratorio PGM | Laboratorio de Infectología Molecular, Departamento de Bioquímica y Medicina Molecular,Facultad de Medicina - Universidad Autónoma de Nuevo León | Kame A. Galán-Huerta, María F. Herrera-Saldivar, Natalia Martínez-Acuña, Sonia A. Lozano-Sepúlveda, Daniel Arellanos-Soto, Ana M. Rivas-Estilla, Javier Ramos-Jimenez, Gabriela Elizondo, Eduardo Garza-de-la-Peña |
| EPI_ISL_1091576 | Arizona State University | Arizona State University | Peter T. Skidmore, LaRinda A. Holland, Emily A. Kaelin, Nicholas J. Mellor, Kristina Buss, Joy M. Blain, Neal W Woodbury, Valerie Harris, Joshua LaBaer, Vel Murugan, Efrem S. Lim |
| EPI_ISL_1091578, EPI_ISL_1091579, EPI_ISL_1091580, EPI_ISL_1091581, EPI_ISL_1091582, EPI_ISL_1091583, EPI_ISL_1091584, EPI_ISL_1091585, EPI_ISL_1091586, EPI_ISL_1091587, EPI_ISL_1091588, EPI_ISL_1091589, EPI_ISL_1091590, EPI_ISL_1091591, EPI_ISL_1091592, EPI_ISL_1091593, EPI_ISL_1091594, EPI_ISL_1091595, EPI_ISL_1091596, EPI_ISL_1091597, EPI_ISL_1091598, EPI_ISL_1091599, EPI_ISL_1091600, EPI_ISL_1091601, EPI_ISL_1091602, EPI_ISL_1091603, EPI_ISL_1091604, EPI_ISL_1091606, EPI_ISL_1091607, EPI_ISL_1091611, EPI_ISL_1091619, EPI_ISL_1091620, EPI_ISL_1091621, EPI_ISL_1091622, EPI_ISL_1091623, EPI_ISL_1091669, EPI_ISL_1091670, EPI_ISL_1091672, EPI_ISL_1091673, EPI_ISL_1091674, EPI_ISL_1091675, EPI_ISL_1091676, EPI_ISL_1091677, EPI_ISL_1091678, EPI_ISL_1091679, EPI_ISL_1091680, EPI_ISL_1091681, EPI_ISL_1091682, EPI_ISL_1091683, EPI_ISL_1091684, EPI_ISL_1091685, EPI_ISL_1091686, EPI_ISL_1091687, EPI_ISL_1091688, EPI_ISL_1091689, EPI_ISL_1091690, EPI_ISL_1091691, EPI_ISL_1091692, EPI_ISL_1091693, EPI_ISL_1091694, EPI_ISL_1091695, EPI_ISL_1091696, EPI_ISL_1091697, EPI_ISL_1091698, EPI_ISL_1091699, EPI_ISL_1091700, EPI_ISL_1091701, EPI_ISL_1091702, EPI_ISL_1091703, EPI_ISL_1091704, EPI_ISL_1091705, EPI_ISL_1091706, EPI_ISL_1091707, EPI_ISL_1091708, EPI_ISL_1091709, EPI_ISL_1091710, EPI_ISL_1091711, EPI_ISL_1091712, EPI_ISL_1091713, EPI_ISL_1091714, EPI_ISL_1091715, EPI_ISL_1091716, EPI_ISL_1091720, EPI_ISL_1091721, EPI_ISL_1091722, EPI_ISL_1091723, EPI_ISL_1091724, EPI_ISL_1091725, EPI_ISL_1091726, EPI_ISL_1091727, EPI_ISL_1091728, EPI_ISL_1091729, EPI_ISL_1091730, EPI_ISL_1091731, EPI_ISL_1091732, EPI_ISL_1091733, EPI_ISL_1091734, EPI_ISL_1091735, EPI_ISL_1091736, EPI_ISL_1091737, EPI_ISL_1091738, EPI_ISL_1091739, EPI_ISL_1091740, EPI_ISL_1091741, EPI_ISL_1091742, EPI_ISL_1091743, EPI_ISL_1091744, EPI_ISL_1091745, EPI_ISL_1091746, EPI_ISL_1091747, EPI_ISL_1091748, EPI_ISL_1091749, EPI_ISL_1091750, EPI_ISL_1091751, EPI_ISL_1091752, EPI_ISL_1091753, EPI_ISL_1091754, EPI_ISL_1091755, EPI_ISL_1091756, EPI_ISL_1091757, EPI_ISL_1091758, EPI_ISL_1091759, EPI_ISL_1091760, EPI_ISL_1091761, EPI_ISL_1091762, EPI_ISL_1091763, EPI_ISL_1091764, EPI_ISL_1091765, EPI_ISL_1091766, EPI_ISL_1091767, EPI_ISL_1091768, EPI_ISL_1091769, EPI_ISL_1091770, EPI_ISL_1091771, EPI_ISL_1091772, EPI_ISL_1091773, EPI_ISL_1091774, EPI_ISL_1091775, EPI_ISL_1091776 | | | |
| see above | US Air Force School of Aerospace Medicine | US Air Force School of Aerospace Medicine | Anthony Fries, Jennifer Meyer, William Gruner, William Buggele, Amanda Javorina, Sarah Purves, Clarise Starr, Elizabeth Macias |
| EPI_ISL_1091800 | Yale Clinical Virology Laboratory | Grubaugh Lab - Yale School of Public Health | Mary Petrone, Joseph Fauver, Tara Alpert, Anderson Brito, Mallery Breban, Anne Wyllie, Chantal Vogels, Annie Watkins, Chaney Kalinich, Isabel Ott, Nathan Grubaugh |
| EPI_ISL_1091801, EPI_ISL_1091802, EPI_ISL_1091803 | Murphy Medical Associates | Grubaugh Lab - Yale School of Public Health | Mary Petrone, Joseph Fauver, Caleb Neal, Steven Murphy, Chantal Vogels, Mallery Breban, Annie Watkins, Tara Alpert, Nathan Grubaugh |
| EPI_ISL_1091804 | Yale Clinical Virology Laboratory | Grubaugh Lab - Yale School of Public Health | Mary Petrone, Joseph Fauver, Tara Alpert, Anderson Brito, Mallery Breban, Anne Wyllie, Chantal Vogels, Annie Watkins, Chaney Kalinich, Isabel Ott, Nathan Grubaugh |
| EPI_ISL_1091805 | Connecticut Department of Public Health | Grubaugh Lab - Yale School of Public Health | Mary Petrone, Joseph Fauver, Tara Alpert, Anderson Brito, Mallery Breban, Anne Wyllie, Chantal Vogels, Annie Watkins, Chaney Kalinich, Isabel Ott, Nathan Grubaugh |
| EPI_ISL_1091806, EPI_ISL_1091807 | Murphy Medical Associates | Grubaugh Lab - Yale School of Public Health | Mary Petrone, Joseph Fauver, Caleb Neal, Steven Murphy, Chantal Vogels, Mallery Breban, Annie Watkins, Tara Alpert, Nathan Grubaugh |
| EPI_ISL_1091808, EPI_ISL_1091809 | Connecticut Department of Public Health | Grubaugh Lab - Yale School of Public Health | Mary Petrone, Joseph Fauver, Tara Alpert, Anderson Brito, Mallery Breban, Anne Wyllie, Chantal Vogels, Annie Watkins, Chaney Kalinich, Isabel Ott, Nathan Grubaugh |
| EPI_ISL_1091810 | Murphy Medical Associates | Grubaugh Lab - Yale School of Public Health | Mary Petrone, Joseph Fauver, Caleb Neal, Steven Murphy, Chantal Vogels, Mallery Breban, Annie Watkins, Tara Alpert, Nathan Grubaugh |
| EPI_ISL_1091811 | Connecticut Department of Public Health | Grubaugh Lab - Yale School of Public Health | Mary Petrone, Joseph Fauver, Tara Alpert, Anderson Brito, Mallery Breban, Anne Wyllie, Chantal Vogels, Annie Watkins, Chaney Kalinich, Isabel Ott, Nathan Grubaugh |
| EPI_ISL_1091812, EPI_ISL_1091813 | Murphy Medical Associates | Grubaugh Lab - Yale School of Public Health | Mary Petrone, Joseph Fauver, Caleb Neal, Steven Murphy, Chantal Vogels, Mallery Breban, Annie Watkins, Tara Alpert, Nathan Grubaugh |
| EPI_ISL_1091814, EPI_ISL_1091815, EPI_ISL_1091816 | Yale Clinical Virology Laboratory | Grubaugh Lab - Yale School of Public Health | Mary Petrone, Joseph Fauver, Tara Alpert, Anderson Brito, Mallery Breban, Anne Wyllie, Chantal Vogels, Annie Watkins, Chaney Kalinich, Isabel Ott, Nathan Grubaugh |
| EPI_ISL_1091817, EPI_ISL_1091818, EPI_ISL_1091819, EPI_ISL_1091820 | Murphy Medical Associates | Grubaugh Lab - Yale School of Public Health | Mary Petrone, Joseph Fauver, Caleb Neal, Steven Murphy, Chantal Vogels, Mallery Breban, Annie Watkins, Tara Alpert, Nathan Grubaugh |
| EPI_ISL_1091821, EPI_ISL_1091822, EPI_ISL_1091823 | Connecticut Department of Public Health | Grubaugh Lab - Yale School of Public Health | Mary Petrone, Joseph Fauver, Tara Alpert, Anderson Brito, Mallery Breban, Anne Wyllie, Chantal Vogels, Annie Watkins, Chaney Kalinich, Isabel Ott, Nathan Grubaugh |
| EPI_ISL_1091824 | Murphy Medical Associates | Grubaugh Lab - Yale School of Public Health | Mary Petrone, Joseph Fauver, Caleb Neal, Steven Murphy, Chantal Vogels, Mallery Breban, Annie Watkins, Tara Alpert, Nathan Grubaugh |
| EPI_ISL_1091825 | Yale Clinical Virology Laboratory | Grubaugh Lab - Yale School of Public Health | Mary Petrone, Joseph Fauver, Tara Alpert, Anderson Brito, Mallery Breban, Anne Wyllie, Chantal Vogels, Annie Watkins, Chaney Kalinich, Isabel Ott, Nathan Grubaugh |
| EPI_ISL_1091860, EPI_ISL_1091861, EPI_ISL_1091862, EPI_ISL_1091863, EPI_ISL_1091864 | Murphy Medical Associates | Grubaugh Lab - Yale School of Public Health | Mary Petrone, Joseph Fauver, Caleb Neal, Steven Murphy, Chantal Vogels, Mallery Breban, Annie Watkins, Tara Alpert, Nathan Grubaugh |
| EPI_ISL_1091865 | Connecticut Department of Public Health | Grubaugh Lab - Yale School of Public Health | Mary Petrone, Joseph Fauver, Tara Alpert, Anderson Brito, Mallery Breban, Anne Wyllie, Chantal Vogels, Annie Watkins, Chaney Kalinich, Isabel Ott, Nathan Grubaugh |
| EPI_ISL_1091866, EPI_ISL_1091867, EPI_ISL_1091868 | Murphy Medical Associates | Grubaugh Lab - Yale School of Public Health | Mary Petrone, Joseph Fauver, Caleb Neal, Steven Murphy, Chantal Vogels, Mallery Breban, Annie Watkins, Tara Alpert, Nathan Grubaugh |
| EPI_ISL_1091869 | Connecticut Veterans' Affairs Hospital | Grubaugh Lab - Yale School of Public Health | Mary Petrone, Joseph Fauver, Tara Alpert, Chantal Vogels, Isabel Ott, Shaili Gupta, Danielle Plank, Nathan Grubaugh |
| EPI_ISL_1091870 | Murphy Medical Associates | Grubaugh Lab - Yale School of Public Health | Mary Petrone, Joseph Fauver, Caleb Neal, Steven Murphy, Chantal Vogels, Mallery Breban, Annie Watkins, Tara Alpert, Nathan Grubaugh |
| EPI_ISL_1091872, EPI_ISL_1091873, EPI_ISL_1091874, EPI_ISL_1091875, EPI_ISL_1091876, EPI_ISL_1091877, EPI_ISL_1091878, EPI_ISL_1091879 | Tempus | Grubaugh Lab - Yale School of Public Health | Mary Petrone, Joseph Fauver, Tara Alpert, Anderson Brito, Mallery Breban, Anne Wyllie, Chantal Vogels, Annie Watkins, Chaney Kalinich, Isabel Ott, Nathan Grubaugh |
| EPI_ISL_1091883, EPI_ISL_1091887, EPI_ISL_1091888, EPI_ISL_1091889, EPI_ISL_1091890 | Connecticut Veterans' Affairs Hospital | Grubaugh Lab - Yale School of Public Health | Mary Petrone, Joseph Fauver, Tara Alpert, Chantal Vogels, Isabel Ott, Shaili Gupta, Danielle Plank, Nathan Grubaugh |
| EPI_ISL_1091891, EPI_ISL_1091892, EPI_ISL_1091893, EPI_ISL_1091894, EPI_ISL_1091895, EPI_ISL_1091896 | Tempus | Grubaugh Lab - Yale School of Public Health | Mary Petrone, Joseph Fauver, Tara Alpert, Anderson Brito, Mallery Breban, Anne Wyllie, Chantal Vogels, Annie Watkins, Chaney Kalinich, Isabel Ott, Nathan Grubaugh |
| EPI_ISL_1091897, EPI_ISL_1091898, EPI_ISL_1091899, EPI_ISL_1091904, EPI_ISL_1091905, EPI_ISL_1091906, EPI_ISL_1091907, EPI_ISL_1091908, EPI_ISL_1091909, EPI_ISL_1091910, EPI_ISL_1091911, EPI_ISL_1091912, EPI_ISL_1091913, EPI_ISL_1091914, EPI_ISL_1091915 | | | |
| see above | New York Presbetyrian / Weill Cornell Medicine | Grubaugh Lab - Yale School of Public Health | Mary Petrone, Joseph Fauver, Tara Alpert, Anderson Brito, Mallery Breban, Anne Wyllie, Chantal Vogels, Annie Watkins, Chaney Kalinich, Isabel Ott, Nathan Grubaugh |
| EPI_ISL_1091916, EPI_ISL_1091917, EPI_ISL_1091918 | Murphy Medical Associates | Grubaugh Lab - Yale School of Public Health | Mary Petrone, Joseph Fauver, Caleb Neal, Steven Murphy, Chantal Vogels, Mallery Breban, Annie Watkins, Tara Alpert, Nathan Grubaugh |

| | | | |
|---|---|---|---|
| EPI_ISL_1091924, EPI_ISL_1091925, EPI_ISL_1091926, EPI_ISL_1091927, EPI_ISL_1091928, EPI_ISL_1091929, EPI_ISL_1091930, EPI_ISL_1091931, EPI_ISL_1091932, EPI_ISL_1091933, EPI_ISL_1091934, EPI_ISL_1091935, EPI_ISL_1091936, EPI_ISL_1091937, EPI_ISL_1091938, EPI_ISL_1091939, EPI_ISL_1091940, EPI_ISL_1091941, EPI_ISL_1091942, EPI_ISL_1091943, EPI_ISL_1091944, EPI_ISL_1091945, EPI_ISL_1091946, EPI_ISL_1091947, EPI_ISL_1091948, EPI_ISL_1091949, EPI_ISL_1091950, EPI_ISL_1091951, EPI_ISL_1091952, EPI_ISL_1091953, EPI_ISL_1091954, EPI_ISL_1091955, EPI_ISL_1091956, EPI_ISL_1091957, EPI_ISL_1091958, EPI_ISL_1091959, EPI_ISL_1091960, EPI_ISL_1091961, EPI_ISL_1091962, EPI_ISL_1091963, EPI_ISL_1091964, EPI_ISL_1091965, EPI_ISL_1091966, EPI_ISL_1091967, EPI_ISL_1091968, EPI_ISL_1091969, EPI_ISL_1091970, EPI_ISL_1091971, EPI_ISL_1091972, EPI_ISL_1091973, EPI_ISL_1091974, EPI_ISL_1091975, EPI_ISL_1091982, EPI_ISL_1091983, EPI_ISL_1091984, EPI_ISL_1091985, EPI_ISL_1091986, EPI_ISL_1091987, EPI_ISL_1091988, EPI_ISL_1091989, EPI_ISL_1091990, EPI_ISL_1091991, EPI_ISL_1091992, EPI_ISL_1091993, EPI_ISL_1091994, EPI_ISL_1091995, EPI_ISL_1091996, EPI_ISL_1091997, EPI_ISL_1091998, EPI_ISL_1091999, EPI_ISL_1092000, EPI_ISL_1092001, EPI_ISL_1092002, EPI_ISL_1092003, EPI_ISL_1092004 | | | |
| see above | Johns Hopkins Hospital Department of Pathology | Johns Hopkins Hospital Department of Pathology | C. Paul Morris, Chun Huai Luo, Adannaya Amadi, Matthew Schwartz, Nicholas Gallagher, Heba H. Mostafa |
| EPI_ISL_1092009 | Kansas Health and Environmental Lab | Kansas Health and Environmental Lab | Mike Grose, Paige Drury, Carissa Robertson, Ben Olsen, and Phil Adam |
| EPI_ISL_1092361 | Laboratorio de Apoyo a la Vigilancia e Investigación Epidemiológica - CIBO | Laboratorio de Infectología Molecular, Departamento de Bioquímica y Medicina Molecular,Facultad de Medicina - Universidad Autónoma de Nuevo León | Kame A. Galán-Huerta, María F. Herrera-Saldivar, Natalia Martínez-Acuña, Sonia A. Lozano-Sepúlveda, Daniel Arellanos-Soto, Ana M. Rivas-Estilla, Javier Ramos-Jimenez, Claudia L. Charles-Niño, Alma M. Perez-Rios, Rafael Soto-Alvarez, Diego A. Gonzalez-Altamirano, Jose J. Figueroa-Delgadillo, Blanca M. Torres, Hector Montoya-Fuentes |
| EPI_ISL_1092458 | Laboratorio Estatal de Salud Pública de Nuevo León | Laboratorio de Infectología Molecular, Departamento de Bioquímica y Medicina Molecular,Facultad de Medicina - Universidad Autónoma de Nuevo León | Kame A. Galán-Huerta, María F. Herrera-Saldivar, Natalia Martínez-Acuña, Sonia A. Lozano-Sepúlveda, Daniel Arellanos-Soto, Ana M. Rivas-Estilla, Samuel Buentello-Wong, Else del Carmen García-García, Gloria A. Jasso-de-la-Peña, Roberto Montes-de-Oca, Consuelo Treviño-Garza, Manuel E. de-la-O-Cavazos |
| EPI_ISL_1093053, EPI_ISL_1093058, EPI_ISL_1093059, EPI_ISL_1093060, EPI_ISL_1093061, EPI_ISL_1093062, EPI_ISL_1093063, EPI_ISL_1093064, EPI_ISL_1093065, EPI_ISL_1093066, EPI_ISL_1093067, EPI_ISL_1093068 | | | |
| see above | Santa Clara County Public Health Laboratory | Chan-Zuckerberg Biohub | CZB Cliahub Consortium |
| EPI_ISL_1093145, EPI_ISL_1093146, EPI_ISL_1093147 | Laboratorio de Diagnostico de Enfermedades Emergentes y Reemergentes | Instituto de Diagnostico y Referencia Epidemiologicos (INDRE) | Claudia Wong-Arambula, Abril Rodriguez-Maldonado, Fabiola Garces-Ayala, Natividad Cruz-Ortiz, Tatiana Nunez-Garcia, Natali Vega-Magana, Gisela Barrera-Badillo, Lucia Hernandez-Rivas, Irma Lopez-Martinez, Ernesto Ramirez-Gonzalez. |
| EPI_ISL_1093546, EPI_ISL_1093547, EPI_ISL_1093548, EPI_ISL_1093549, EPI_ISL_1093550, EPI_ISL_1093551, EPI_ISL_1093552, EPI_ISL_1093553, EPI_ISL_1093554, EPI_ISL_1093555, EPI_ISL_1093556, EPI_ISL_1093557, EPI_ISL_1093558, EPI_ISL_1093559, EPI_ISL_1093560, EPI_ISL_1093561, EPI_ISL_1093562, EPI_ISL_1093563, EPI_ISL_1093564, EPI_ISL_1093568, EPI_ISL_1093569, EPI_ISL_1093570, EPI_ISL_1093571, EPI_ISL_1093572, EPI_ISL_1093573, EPI_ISL_1093574, EPI_ISL_1093575, EPI_ISL_1093576, EPI_ISL_1093577, EPI_ISL_1093578, EPI_ISL_1093579, EPI_ISL_1093580, EPI_ISL_1093581, EPI_ISL_1093582, EPI_ISL_1093583, EPI_ISL_1093584, EPI_ISL_1093585, EPI_ISL_1093586, EPI_ISL_1093587, EPI_ISL_1093588, EPI_ISL_1093589, EPI_ISL_1093590, EPI_ISL_1093591, EPI_ISL_1093592, EPI_ISL_1093593, EPI_ISL_1093594, EPI_ISL_1093595, EPI_ISL_1093596, EPI_ISL_1093597, EPI_ISL_1093598, EPI_ISL_1093599, EPI_ISL_1093600, EPI_ISL_1093601 | | | |
| see above | Alameda County Public Health Department | Alameda County Public Health Department | Kristna Hsieh, Maria Elma, Kevin Libuit |
| EPI_ISL_1093602, EPI_ISL_1093603, EPI_ISL_1093604, EPI_ISL_1093605, EPI_ISL_1093606, EPI_ISL_1093607, EPI_ISL_1093608, EPI_ISL_1093609, EPI_ISL_1093610, EPI_ISL_1093611, EPI_ISL_1093612, EPI_ISL_1093613, EPI_ISL_1093614, EPI_ISL_1093615, EPI_ISL_1093616, EPI_ISL_1093617, EPI_ISL_1093618, EPI_ISL_1093619, EPI_ISL_1093620, EPI_ISL_1093621, EPI_ISL_1093622, EPI_ISL_1093623, EPI_ISL_1093624, EPI_ISL_1093625, EPI_ISL_1093626, EPI_ISL_1093627, EPI_ISL_1093628, EPI_ISL_1093629, EPI_ISL_1093630, EPI_ISL_1093631, EPI_ISL_1093632, EPI_ISL_1093633, EPI_ISL_1093634, EPI_ISL_1093635, EPI_ISL_1093636, EPI_ISL_1093637, EPI_ISL_1093638, EPI_ISL_1093639, EPI_ISL_1093640, EPI_ISL_1093641, EPI_ISL_1093642, EPI_ISL_1093643, EPI_ISL_1093644, EPI_ISL_1093645, EPI_ISL_1093646, EPI_ISL_1093647, EPI_ISL_1093648, EPI_ISL_1093649, EPI_ISL_1093650, EPI_ISL_1093651, EPI_ISL_1093652, EPI_ISL_1093653, EPI_ISL_1093654, EPI_ISL_1093655, EPI_ISL_1093656, EPI_ISL_1093657, EPI_ISL_1093658, EPI_ISL_1093659, EPI_ISL_1093660, EPI_ISL_1093661, EPI_ISL_1093662, EPI_ISL_1093663, EPI_ISL_1093664, EPI_ISL_1093665, EPI_ISL_1093666, EPI_ISL_1093667, EPI_ISL_1093668, EPI_ISL_1093669, EPI_ISL_1093670, EPI_ISL_1093671, EPI_ISL_1093672, EPI_ISL_1093673, EPI_ISL_1093674, EPI_ISL_1093675, EPI_ISL_1093676, EPI_ISL_1093677, EPI_ISL_1093678, EPI_ISL_1093679, EPI_ISL_1093680, EPI_ISL_1093681, EPI_ISL_1093682, EPI_ISL_1093683, EPI_ISL_1093684, EPI_ISL_1093685, EPI_ISL_1093686, EPI_ISL_1093687, EPI_ISL_1093688, EPI_ISL_1093689, EPI_ISL_1093690, EPI_ISL_1093691, EPI_ISL_1093692, EPI_ISL_1093693, EPI_ISL_1093694, EPI_ISL_1093695, EPI_ISL_1093696, EPI_ISL_1093697, EPI_ISL_1093698, EPI_ISL_1093699, EPI_ISL_1093700, EPI_ISL_1093701, EPI_ISL_1093702, EPI_ISL_1093703, EPI_ISL_1093704, EPI_ISL_1093705, EPI_ISL_1093706, EPI_ISL_1093707, EPI_ISL_1093708, EPI_ISL_1093709, EPI_ISL_1093710, EPI_ISL_1093711, EPI_ISL_1093712, EPI_ISL_1093713, EPI_ISL_1093714, EPI_ISL_1093715, EPI_ISL_1093716, EPI_ISL_1093717, EPI_ISL_1093718, EPI_ISL_1093719, EPI_ISL_1093720, EPI_ISL_1093721, EPI_ISL_1093722, EPI_ISL_1093723, EPI_ISL_1093724, EPI_ISL_1093725, EPI_ISL_1093726, EPI_ISL_1093727, EPI_ISL_1093728, EPI_ISL_1093729, EPI_ISL_1093730, EPI_ISL_1093731, EPI_ISL_1093732, EPI_ISL_1093733, EPI_ISL_1093734, EPI_ISL_1093735, EPI_ISL_1093736, EPI_ISL_1093737, EPI_ISL_1093738, EPI_ISL_1093739, EPI_ISL_1093740, EPI_ISL_1093741, EPI_ISL_1093742, EPI_ISL_1093743, EPI_ISL_1093744, EPI_ISL_1093745, EPI_ISL_1093746, EPI_ISL_1093747, EPI_ISL_1093748, EPI_ISL_1093749, EPI_ISL_1093750, EPI_ISL_1093751, EPI_ISL_1093752, EPI_ISL_1093753, EPI_ISL_1093754, EPI_ISL_1093755, EPI_ISL_1093756, EPI_ISL_1093757, EPI_ISL_1093758, EPI_ISL_1093759, EPI_ISL_1093760, EPI_ISL_1093761, EPI_ISL_1093762, EPI_ISL_1093763, EPI_ISL_1093764, EPI_ISL_1093765, EPI_ISL_1093766, EPI_ISL_1093767, EPI_ISL_1093768, EPI_ISL_1093769, EPI_ISL_1093770, EPI_ISL_1093771, EPI_ISL_1093772, EPI_ISL_1093773, EPI_ISL_1093774, EPI_ISL_1093775, EPI_ISL_1093776, EPI_ISL_1093777, EPI_ISL_1093778, EPI_ISL_1093779, EPI_ISL_1093780, EPI_ISL_1093781, EPI_ISL_1093782, EPI_ISL_1093783, EPI_ISL_1093784, EPI_ISL_1093785, EPI_ISL_1093786, EPI_ISL_1093787, EPI_ISL_1093788, EPI_ISL_1093789, EPI_ISL_1093790, EPI_ISL_1093791, EPI_ISL_1093792, EPI_ISL_1093793, EPI_ISL_1093794, EPI_ISL_1093795, EPI_ISL_1093796, EPI_ISL_1093797, EPI_ISL_1093798, EPI_ISL_1093799, EPI_ISL_1093800, EPI_ISL_1093801, EPI_ISL_1093802, EPI_ISL_1093803, EPI_ISL_1093804, EPI_ISL_1093805, EPI_ISL_1093806, EPI_ISL_1093807, EPI_ISL_1093808, EPI_ISL_1093809, EPI_ISL_1093810, EPI_ISL_1093811, EPI_ISL_1093837, EPI_ISL_1093838, EPI_ISL_1093839, EPI_ISL_1093840, EPI_ISL_1093841, EPI_ISL_1093842, EPI_ISL_1093843, EPI_ISL_1093844, EPI_ISL_1093845, EPI_ISL_1093846, EPI_ISL_1093847, EPI_ISL_1093848, EPI_ISL_1093849, EPI_ISL_1093850, EPI_ISL_1093851, EPI_ISL_1093852, EPI_ISL_1093853, EPI_ISL_1093854, EPI_ISL_1093855, EPI_ISL_1093856, EPI_ISL_1093857, EPI_ISL_1093858, EPI_ISL_1093859, EPI_ISL_1093860, EPI_ISL_1093861, EPI_ISL_1093862, EPI_ISL_1093863, EPI_ISL_1093864, EPI_ISL_1093865, EPI_ISL_1093866, EPI_ISL_1093867, EPI_ISL_1093868, EPI_ISL_1093869, EPI_ISL_1093870, EPI_ISL_1093871, EPI_ISL_1093872, EPI_ISL_1093873, EPI_ISL_1093874, EPI_ISL_1093875, EPI_ISL_1093876, EPI_ISL_1093877, EPI_ISL_1093878, EPI_ISL_1093879, EPI_ISL_1093880, EPI_ISL_1093881, EPI_ISL_1093882, EPI_ISL_1093883, EPI_ISL_1093884, EPI_ISL_1093885, EPI_ISL_1093886, EPI_ISL_1093887, EPI_ISL_1093888, EPI_ISL_1093889, EPI_ISL_1093890, EPI_ISL_1093891, EPI_ISL_1093892, EPI_ISL_1093893, EPI_ISL_1093894, EPI_ISL_1093895, EPI_ISL_1093896, EPI_ISL_1093897, EPI_ISL_1093898, EPI_ISL_1093899, EPI_ISL_1093900, EPI_ISL_1093901, EPI_ISL_1093902, EPI_ISL_1093903, EPI_ISL_1093904, EPI_ISL_1093905, EPI_ISL_1093906, EPI_ISL_1093907, EPI_ISL_1093908, EPI_ISL_1093909, EPI_ISL_1093910, EPI_ISL_1093911, EPI_ISL_1093912, EPI_ISL_1093913, EPI_ISL_1093914, EPI_ISL_1093915, EPI_ISL_1093916, EPI_ISL_1093917, EPI_ISL_1093918, EPI_ISL_1093919, EPI_ISL_1093920, EPI_ISL_1093921, EPI_ISL_1093922, EPI_ISL_1093923, EPI_ISL_1093924, EPI_ISL_1093925, EPI_ISL_1093926, EPI_ISL_1093927, EPI_ISL_1093928, EPI_ISL_1093929, EPI_ISL_1093930, EPI_ISL_1093931, EPI_ISL_1093932, EPI_ISL_1093933, EPI_ISL_1093934, EPI_ISL_1093935, EPI_ISL_1093936, EPI_ISL_1093937, EPI_ISL_1093938, EPI_ISL_1093939, EPI_ISL_1093940, EPI_ISL_1093941, EPI_ISL_1093942, EPI_ISL_1093943, EPI_ISL_1093944, EPI_ISL_1093945, EPI_ISL_1093946, EPI_ISL_1093947, EPI_ISL_1093948, EPI_ISL_1093949, EPI_ISL_1093950, EPI_ISL_1093951, EPI_ISL_1093952, EPI_ISL_1093953, EPI_ISL_1093954, EPI_ISL_1093955, EPI_ISL_1093956, EPI_ISL_1093957, EPI_ISL_1093958, EPI_ISL_1093959, EPI_ISL_1093960, EPI_ISL_1093961, EPI_ISL_1093962, EPI_ISL_1093963, EPI_ISL_1093964, EPI_ISL_1093965, EPI_ISL_1093966, EPI_ISL_1093967, EPI_ISL_1093968, EPI_ISL_1093969, EPI_ISL_1093970, EPI_ISL_1093971, EPI_ISL_1093972, EPI_ISL_1093973, EPI_ISL_1093974, EPI_ISL_1093975, EPI_ISL_1093976, EPI_ISL_1093977, EPI_ISL_1093978, EPI_ISL_1093979, EPI_ISL_1093980, EPI_ISL_1093981, EPI_ISL_1093982, EPI_ISL_1093983, EPI_ISL_1093984, EPI_ISL_1093985, EPI_ISL_1093986, EPI_ISL_1093987, EPI_ISL_1093988, EPI_ISL_1093989, EPI_ISL_1093990, EPI_ISL_1093991, EPI_ISL_1093992, EPI_ISL_1093993, EPI_ISL_1093994, EPI_ISL_1093995, EPI_ISL_1093996, EPI_ISL_1093997, EPI_ISL_1093998, EPI_ISL_1093999, EPI_ISL_1094000, EPI_ISL_1094001, EPI_ISL_1094002, EPI_ISL_1094003, EPI_ISL_1094004, EPI_ISL_1094005, EPI_ISL_1094006, EPI_ISL_1094007, EPI_ISL_1094008, EPI_ISL_1094009, EPI_ISL_1094010, EPI_ISL_1094011, EPI_ISL_1094012, EPI_ISL_1094013, EPI_ISL_1094014, EPI_ISL_1094015, EPI_ISL_1094016, EPI_ISL_1094017, EPI_ISL_1094018, EPI_ISL_1094019, EPI_ISL_1094020, EPI_ISL_1094021, EPI_ISL_1094022, EPI_ISL_1094023, EPI_ISL_1094024, EPI_ISL_1094025, EPI_ISL_1094026, EPI_ISL_1094027, EPI_ISL_1094028, EPI_ISL_1094029, EPI_ISL_1094030, EPI_ISL_1094031, EPI_ISL_1094032, EPI_ISL_1094033, EPI_ISL_1094034, EPI_ISL_1094035, EPI_ISL_1094036, EPI_ISL_1094037, EPI_ISL_1094038, EPI_ISL_1094039, EPI_ISL_1094040, EPI_ISL_1094041, EPI_ISL_1094042, EPI_ISL_1094043, EPI_ISL_1094044, EPI_ISL_1094045, EPI_ISL_1094046, EPI_ISL_1094047, EPI_ISL_1094048, EPI_ISL_1094049, EPI_ISL_1094050, EPI_ISL_1094051, EPI_ISL_1094052, EPI_ISL_1094053, EPI_ISL_1094054, EPI_ISL_1094055, EPI_ISL_1094056, EPI_ISL_1094057, EPI_ISL_1094058, EPI_ISL_1094059, EPI_ISL_1094060, EPI_ISL_1094061, EPI_ISL_1094062, EPI_ISL_1094063, EPI_ISL_1094064, EPI_ISL_1094065, EPI_ISL_1094066, EPI_ISL_1094067, EPI_ISL_1094068, EPI_ISL_1094069, EPI_ISL_1094070, EPI_ISL_1094071, EPI_ISL_1094072, EPI_ISL_1094073, EPI_ISL_1094074, EPI_ISL_1094075, EPI_ISL_1094076, EPI_ISL_1094077, EPI_ISL_1094078, EPI_ISL_1094079, EPI_ISL_1094080, EPI_ISL_1094081, EPI_ISL_1094082, EPI_ISL_1094083, EPI_ISL_1094084, EPI_ISL_1094085, EPI_ISL_1094086, EPI_ISL_1094087, EPI_ISL_1094088, EPI_ISL_1094089, EPI_ISL_1094090, EPI_ISL_1094091, EPI_ISL_1094092, EPI_ISL_1094093, EPI_ISL_1094094, EPI_ISL_1094095, EPI_ISL_1094096, EPI_ISL_1094097, EPI_ISL_1094098, EPI_ISL_1094099, EPI_ISL_1094100, EPI_ISL_1094101, EPI_ISL_1094102, EPI_ISL_1094103, EPI_ISL_1094104, EPI_ISL_1094105, EPI_ISL_1094106, EPI_ISL_1094107, EPI_ISL_1094108, EPI_ISL_1094109, EPI_ISL_1094110, EPI_ISL_1094111, EPI_ISL_1094112, EPI_ISL_1094113, EPI_ISL_1094114, EPI_ISL_1094115, EPI_ISL_1094116, EPI_ISL_1094117, EPI_ISL_1094118, EPI_ISL_1094119, EPI_ISL_1094120, EPI_ISL_1094121, EPI_ISL_1094122, EPI_ISL_1094123, EPI_ISL_1094124, EPI_ISL_1094125, EPI_ISL_1094126, EPI_ISL_1094127, EPI_ISL_1094128, EPI_ISL_1094129, EPI_ISL_1094130, EPI_ISL_1094131, EPI_ISL_1094132, EPI_ISL_1094133, EPI_ISL_1094134, EPI_ISL_1094135, EPI_ISL_1094136, EPI_ISL_1094137, EPI_ISL_1094138, EPI_ISL_1094139, EPI_ISL_1094140, EPI_ISL_1094141, EPI_ISL_1094142, EPI_ISL_1094143, EPI_ISL_1094144, EPI_ISL_1094145, EPI_ISL_1094146, EPI_ISL_1094147, EPI_ISL_1094148, EPI_ISL_1094149, EPI_ISL_1094150, EPI_ISL_1094151, EPI_ISL_1094152, EPI_ISL_1094153, EPI_ISL_1094154, EPI_ISL_1094155, EPI_ISL_1094156, EPI_ISL_1094157, EPI_ISL_1094158, EPI_ISL_1094159, EPI_ISL_1094160, EPI_ISL_1094161, EPI_ISL_1094162, EPI_ISL_1094163, EPI_ISL_1094164, EPI_ISL_1094165, EPI_ISL_1094166, EPI_ISL_1094167, EPI_ISL_1094168, EPI_ISL_1094169, EPI_ISL_1094170, EPI_ISL_1094171, EPI_ISL_1094172, EPI_ISL_1094173, EPI_ISL_1094174, EPI_ISL_1094175, EPI_ISL_1094176, EPI_ISL_1094177, EPI_ISL_1094178, EPI_ISL_1094179, EPI_ISL_1094180, EPI_ISL_1094181, EPI_ISL_1094182, EPI_ISL_1094183, EPI_ISL_1094184, EPI_ISL_1094185, EPI_ISL_1094186, EPI_ISL_1094187, EPI_ISL_1094188, EPI_ISL_1094189, EPI_ISL_1094190, EPI_ISL_1094191 | | | |
| see above | DeRisi Lab, University of California, San Francisco | DeRisi Lab, University of California, San Francisco | Sabrina Mann, Anthea Mitchell, Jamin Liu, Sara Sunshine, Matthew Laurie, Genay Pilarowski, Amy Kistler, Manu Vanaerschot, Patrick Ayscue, Lucy Li, Eric Chow, IDseq Team, James Peng, Carina Marquez, Luis Rubio, Gabriel Chamie, Diane Jones, Jon Jacobo, Susana Rojas, Susy Rojas, Valerie Tulier-Laiwa, Douglas Black, Jackie Martinez, Maya Petersen, Diane Havlir, and Joseph DeRisi |
| EPI_ISL_1094196 | NV State Public Health Laboratory | Respiratory Viruses Branch, Division of Viral Diseases, Centers for Disease Control and Prevention | Krista Queen, Yan Li, Ying Tao, Jing Zhang, Anna Uehara, Anna Montmayeur, Clinton R. Paden, Peter W. Cook, Rachel Marine, Mili Sheth, Jasmine Padilla, Sarah Nobles, Mark Burroughs, Lori Rowe, Haibin Wang, Ben L. Rambo-Martin, Dhwani Batra, Justin Lee, Suxiang Tong |
| EPI_ISL_1094197, EPI_ISL_1094199 | NC State Laboratory of Public Health | Respiratory Viruses Branch, Division of Viral Diseases, Centers for Disease Control and Prevention | Krista Queen, Yan Li, Ying Tao, Jing Zhang, Anna Uehara, Anna Montmayeur, Clinton R. Paden, Peter W. Cook, Rachel Marine, Mili Sheth, Jasmine Padilla, Sarah Nobles, Mark Burroughs, Lori Rowe, Haibin Wang, Ben L. Rambo-Martin, Dhwani Batra, Justin Lee, Suxiang Tong |
| EPI_ISL_1094200 | VT Dept. of Health Laboratory | Respiratory Viruses Branch, Division of Viral Diseases, Centers for Disease Control and Prevention | Krista Queen, Yan Li, Ying Tao, Jing Zhang, Anna Uehara, Anna Montmayeur, Clinton R. Paden, Peter W. Cook, Rachel Marine, Mili Sheth, Jasmine Padilla, Sarah Nobles, Mark Burroughs, Lori Rowe, Haibin Wang, Ben L. Rambo-Martin, Dhwani Batra, Justin Lee, Suxiang Tong |
| EPI_ISL_1094201 | NC State Laboratory of Public Health | Respiratory Viruses Branch, Division of Viral Diseases, Centers for Disease Control and Prevention | Krista Queen, Yan Li, Ying Tao, Jing Zhang, Anna Uehara, Anna Montmayeur, Clinton R. Paden, Peter W. Cook, Rachel Marine, Mili Sheth, Jasmine Padilla, Sarah Nobles, Mark Burroughs, Lori Rowe, Haibin Wang, Ben L. Rambo-Martin, Dhwani Batra, Justin Lee, Suxiang Tong |
| EPI_ISL_1094202 | IL Dept. of Public Health Springfield Laboratory | Respiratory Viruses Branch, Division of Viral Diseases, Centers for Disease Control and Prevention | Krista Queen, Yan Li, Ying Tao, Jing Zhang, Anna Uehara, Anna Montmayeur, Clinton R. Paden, Peter W. Cook, Rachel Marine, Mili Sheth, Jasmine Padilla, Sarah Nobles, Mark Burroughs, Lori Rowe, Haibin Wang, Ben L. Rambo-Martin, Dhwani Batra, Justin Lee, Suxiang Tong |
| EPI_ISL_1094203 | NC State Laboratory of Public Health | Respiratory Viruses Branch, Division of Viral Diseases, Centers for Disease Control and Prevention | Krista Queen, Yan Li, Ying Tao, Jing Zhang, Anna Uehara, Anna Montmayeur, Clinton R. Paden, Peter W. Cook, Rachel Marine, Mili Sheth, Jasmine Padilla, Sarah Nobles, Mark Burroughs, Lori Rowe, Haibin Wang, Ben L. Rambo-Martin, Dhwani Batra, Justin Lee, Suxiang Tong |
| EPI_ISL_1094204, EPI_ISL_1094206 | OH Department of Health Laboratory | Respiratory Viruses Branch, Division of Viral Diseases, Centers for Disease Control and Prevention | Krista Queen, Yan Li, Ying Tao, Jing Zhang, Anna Uehara, Anna Montmayeur, Clinton R. Paden, Peter W. Cook, Rachel Marine, Mili Sheth, Jasmine Padilla, Sarah Nobles, Mark Burroughs, Lori Rowe, Haibin Wang, Ben L. Rambo-Martin, Dhwani Batra, Justin Lee, Suxiang Tong |
| EPI_ISL_1094207 | MS Public Health Laboratory | Respiratory Viruses Branch, Division of Viral Diseases, Centers for Disease Control and Prevention | Krista Queen, Yan Li, Ying Tao, Jing Zhang, Anna Uehara, Anna Montmayeur, Clinton R. Paden, Peter W. Cook, Rachel Marine, Mili Sheth, Jasmine Padilla, Sarah Nobles, Mark Burroughs, Lori Rowe, Haibin Wang, Ben L. Rambo-Martin, Dhwani Batra, Justin Lee, Suxiang Tong |
| EPI_ISL_1094208 | AR Dept. of Health-Public Health Lab | Respiratory Viruses Branch, Division of Viral Diseases, Centers for Disease Control and Prevention | Krista Queen, Yan Li, Ying Tao, Jing Zhang, Anna Uehara, Anna Montmayeur, Clinton R. Paden, Peter W. Cook, Rachel Marine, Mili Sheth, Jasmine Padilla, Sarah Nobles, Mark Burroughs, Lori Rowe, Haibin Wang, Ben L. Rambo-Martin, Dhwani Batra, Justin Lee, Suxiang Tong |
| EPI_ISL_1094209 | HI Dept. of Health, State Laboratories Division | Respiratory Viruses Branch, Division of Viral Diseases, Centers for Disease Control and Prevention | Krista Queen, Yan Li, Ying Tao, Jing Zhang, Anna Uehara, Anna Montmayeur, Clinton R. Paden, Peter W. Cook, Rachel Marine, Mili Sheth, Jasmine Padilla, Sarah Nobles, Mark Burroughs, Lori Rowe, Haibin Wang, Ben L. Rambo-Martin, Dhwani Batra, Justin Lee, Suxiang Tong |
| EPI_ISL_1094212 | NC State Laboratory of Public Health | Respiratory Viruses Branch, Division of Viral Diseases, Centers for Disease Control and Prevention | Krista Queen, Yan Li, Ying Tao, Jing Zhang, Anna Uehara, Anna Montmayeur, Clinton R. Paden, Peter W. Cook, Rachel Marine, Mili Sheth, Jasmine Padilla, Sarah Nobles, Mark Burroughs, Lori Rowe, Haibin Wang, Ben L. Rambo-Martin, Dhwani Batra, Justin Lee, Suxiang Tong |
| EPI_ISL_1094213 | MT Public Health Laboratory | Respiratory Viruses Branch, Division of Viral Diseases, Centers for Disease Control and Prevention | Krista Queen, Yan Li, Ying Tao, Jing Zhang, Anna Uehara, Anna Montmayeur, Clinton R. Paden, Peter W. Cook, Rachel Marine, Mili Sheth, Jasmine Padilla, Sarah Nobles, Mark Burroughs, Lori Rowe, Haibin Wang, Ben L. Rambo-Martin, Dhwani Batra, Justin Lee, Suxiang Tong |
| EPI_ISL_1094215 | MS Public Health Laboratory | Respiratory Viruses Branch, Division of Viral Diseases, Centers for Disease Control and Prevention | Krista Queen, Yan Li, Ying Tao, Jing Zhang, Anna Uehara, Anna Montmayeur, Clinton R. Paden, Peter W. Cook, Rachel Marine, Mili Sheth, Jasmine Padilla, Sarah Nobles, Mark Burroughs, Lori Rowe, Haibin Wang, Ben L. Rambo-Martin, Dhwani Batra, Justin Lee, Suxiang Tong |
| EPI_ISL_1094216 | CDPH, Viral and Rickettsial Disease Laboratory | Respiratory Viruses Branch, Division of Viral Diseases, Centers for Disease Control and Prevention | Krista Queen, Yan Li, Ying Tao, Jing Zhang, Anna Uehara, Anna Montmayeur, Clinton R. Paden, Peter W. Cook, Rachel Marine, Mili Sheth, Jasmine Padilla, Sarah Nobles, Mark Burroughs, Lori Rowe, Haibin Wang, Ben L. Rambo-Martin, Dhwani Batra, Justin Lee, Suxiang Tong |
| EPI_ISL_1094217 | NYSDOH Wadsworth Center, Virology Lab | Respiratory Viruses Branch, Division of Viral Diseases, Centers for Disease Control and | Krista Queen, Yan Li, Ying Tao, Jing Zhang, Anna Uehara, Anna Montmayeur, Clinton R. Paden, Peter W. Cook, Rachel Marine, Mili Sheth, Jasmine Padilla, Sarah Nobles, Mark Burroughs, Lori Rowe, Haibin Wang, Ben L. Rambo-Martin, Dhwani Batra, Justin Lee, Suxiang Tong |

| | | | |
|---|---|---|---|
| | | Prevention | |
| EPI_ISL_1094218 | RI State Health Laboratories | Respiratory Viruses Branch, Division of Viral Diseases, Centers for Disease Control and Prevention | Krista Queen, Yan Li, Ying Tao, Jing Zhang, Anna Uehara, Anna Montmayeur, Clinton R. Paden, Peter W. Cook, Rachel Marine, Mili Sheth, Jasmine Padilla, Sarah Nobles, Mark Burroughs, Lori Rowe, Haibin Wang, Ben L. Rambo-Martin, Dhwani Batra, Justin Lee, Suxiang Tong |
| EPI_ISL_1094219 | NYSDOH Wadsworth Center, Virology Lab | Respiratory Viruses Branch, Division of Viral Diseases, Centers for Disease Control and Prevention | Krista Queen, Yan Li, Ying Tao, Jing Zhang, Anna Uehara, Anna Montmayeur, Clinton R. Paden, Peter W. Cook, Rachel Marine, Mili Sheth, Jasmine Padilla, Sarah Nobles, Mark Burroughs, Lori Rowe, Haibin Wang, Ben L. Rambo-Martin, Dhwani Batra, Justin Lee, Suxiang Tong |
| EPI_ISL_1094220 | MT Public Health Laboratory | Respiratory Viruses Branch, Division of Viral Diseases, Centers for Disease Control and Prevention | Krista Queen, Yan Li, Ying Tao, Jing Zhang, Anna Uehara, Anna Montmayeur, Clinton R. Paden, Peter W. Cook, Rachel Marine, Mili Sheth, Jasmine Padilla, Sarah Nobles, Mark Burroughs, Lori Rowe, Haibin Wang, Ben L. Rambo-Martin, Dhwani Batra, Justin Lee, Suxiang Tong |
| EPI_ISL_1094221 | HI Dept. of Health, State Laboratories Division | Respiratory Viruses Branch, Division of Viral Diseases, Centers for Disease Control and Prevention | Krista Queen, Yan Li, Ying Tao, Jing Zhang, Anna Uehara, Anna Montmayeur, Clinton R. Paden, Peter W. Cook, Rachel Marine, Mili Sheth, Jasmine Padilla, Sarah Nobles, Mark Burroughs, Lori Rowe, Haibin Wang, Ben L. Rambo-Martin, Dhwani Batra, Justin Lee, Suxiang Tong |
| EPI_ISL_1094222 | IL Dept. of Public Health Springfield Laboratory | Respiratory Viruses Branch, Division of Viral Diseases, Centers for Disease Control and Prevention | Krista Queen, Yan Li, Ying Tao, Jing Zhang, Anna Uehara, Anna Montmayeur, Clinton R. Paden, Peter W. Cook, Rachel Marine, Mili Sheth, Jasmine Padilla, Sarah Nobles, Mark Burroughs, Lori Rowe, Haibin Wang, Ben L. Rambo-Martin, Dhwani Batra, Justin Lee, Suxiang Tong |
| EPI_ISL_1094223 | TN Division of Laboratory Services | Respiratory Viruses Branch, Division of Viral Diseases, Centers for Disease Control and Prevention | Krista Queen, Yan Li, Ying Tao, Jing Zhang, Anna Uehara, Anna Montmayeur, Clinton R. Paden, Peter W. Cook, Rachel Marine, Mili Sheth, Jasmine Padilla, Sarah Nobles, Mark Burroughs, Lori Rowe, Haibin Wang, Ben L. Rambo-Martin, Dhwani Batra, Justin Lee, Suxiang Tong |
| EPI_ISL_1094224 | CDPH, Viral and Rickettsial Disease Laboratory | Respiratory Viruses Branch, Division of Viral Diseases, Centers for Disease Control and Prevention | Krista Queen, Yan Li, Ying Tao, Jing Zhang, Anna Uehara, Anna Montmayeur, Clinton R. Paden, Peter W. Cook, Rachel Marine, Mili Sheth, Jasmine Padilla, Sarah Nobles, Mark Burroughs, Lori Rowe, Haibin Wang, Ben L. Rambo-Martin, Dhwani Batra, Justin Lee, Suxiang Tong |
| EPI_ISL_1094226 | OK Public Health Laboratory, Oklahoma State DOH | Respiratory Viruses Branch, Division of Viral Diseases, Centers for Disease Control and Prevention | Krista Queen, Yan Li, Ying Tao, Jing Zhang, Anna Uehara, Anna Montmayeur, Clinton R. Paden, Peter W. Cook, Rachel Marine, Mili Sheth, Jasmine Padilla, Sarah Nobles, Mark Burroughs, Lori Rowe, Haibin Wang, Ben L. Rambo-Martin, Dhwani Batra, Justin Lee, Suxiang Tong |
| EPI_ISL_1094227 | NYSDOH Wadsworth Center, Virology Lab | Respiratory Viruses Branch, Division of Viral Diseases, Centers for Disease Control and Prevention | Krista Queen, Yan Li, Ying Tao, Jing Zhang, Anna Uehara, Anna Montmayeur, Clinton R. Paden, Peter W. Cook, Rachel Marine, Mili Sheth, Jasmine Padilla, Sarah Nobles, Mark Burroughs, Lori Rowe, Haibin Wang, Ben L. Rambo-Martin, Dhwani Batra, Justin Lee, Suxiang Tong |
| EPI_ISL_1094228 | ME Health and Environmental Testing Laboratory | Respiratory Viruses Branch, Division of Viral Diseases, Centers for Disease Control and Prevention | Krista Queen, Yan Li, Ying Tao, Jing Zhang, Anna Uehara, Anna Montmayeur, Clinton R. Paden, Peter W. Cook, Rachel Marine, Mili Sheth, Jasmine Padilla, Sarah Nobles, Mark Burroughs, Lori Rowe, Haibin Wang, Ben L. Rambo-Martin, Dhwani Batra, Justin Lee, Suxiang Tong |
| EPI_ISL_1094229 | NC State Laboratory of Public Health | Respiratory Viruses Branch, Division of Viral Diseases, Centers for Disease Control and Prevention | Krista Queen, Yan Li, Ying Tao, Jing Zhang, Anna Uehara, Anna Montmayeur, Clinton R. Paden, Peter W. Cook, Rachel Marine, Mili Sheth, Jasmine Padilla, Sarah Nobles, Mark Burroughs, Lori Rowe, Haibin Wang, Ben L. Rambo-Martin, Dhwani Batra, Justin Lee, Suxiang Tong |
| EPI_ISL_1094230 | OK Public Health Laboratory, Oklahoma State DOH | Respiratory Viruses Branch, Division of Viral Diseases, Centers for Disease Control and Prevention | Krista Queen, Yan Li, Ying Tao, Jing Zhang, Anna Uehara, Anna Montmayeur, Clinton R. Paden, Peter W. Cook, Rachel Marine, Mili Sheth, Jasmine Padilla, Sarah Nobles, Mark Burroughs, Lori Rowe, Haibin Wang, Ben L. Rambo-Martin, Dhwani Batra, Justin Lee, Suxiang Tong |
| EPI_ISL_1094231 | CO Dept. of Public Health and Environment, Lab Services Division | Respiratory Viruses Branch, Division of Viral Diseases, Centers for Disease Control and Prevention | Krista Queen, Yan Li, Ying Tao, Jing Zhang, Anna Uehara, Anna Montmayeur, Clinton R. Paden, Peter W. Cook, Rachel Marine, Mili Sheth, Jasmine Padilla, Sarah Nobles, Mark Burroughs, Lori Rowe, Haibin Wang, Ben L. Rambo-Martin, Dhwani Batra, Justin Lee, Suxiang Tong |
| EPI_ISL_1094232 | TN Division of Laboratory Services | Respiratory Viruses Branch, Division of Viral Diseases, Centers for Disease Control and Prevention | Krista Queen, Yan Li, Ying Tao, Jing Zhang, Anna Uehara, Anna Montmayeur, Clinton R. Paden, Peter W. Cook, Rachel Marine, Mili Sheth, Jasmine Padilla, Sarah Nobles, Mark Burroughs, Lori Rowe, Haibin Wang, Ben L. Rambo-Martin, Dhwani Batra, Justin Lee, Suxiang Tong |
| EPI_ISL_1094233 | CDPH, Viral and Rickettsial Disease Laboratory | Respiratory Viruses Branch, Division of Viral Diseases, Centers for Disease Control and Prevention | Krista Queen, Yan Li, Ying Tao, Jing Zhang, Anna Uehara, Anna Montmayeur, Clinton R. Paden, Peter W. Cook, Rachel Marine, Mili Sheth, Jasmine Padilla, Sarah Nobles, Mark Burroughs, Lori Rowe, Haibin Wang, Ben L. Rambo-Martin, Dhwani Batra, Justin Lee, Suxiang Tong |
| EPI_ISL_1094234 | CO Dept. of Public Health and Environment, Lab Services Division | Respiratory Viruses Branch, Division of Viral Diseases, Centers for Disease Control and Prevention | Krista Queen, Yan Li, Ying Tao, Jing Zhang, Anna Uehara, Anna Montmayeur, Clinton R. Paden, Peter W. Cook, Rachel Marine, Mili Sheth, Jasmine Padilla, Sarah Nobles, Mark Burroughs, Lori Rowe, Haibin Wang, Ben L. Rambo-Martin, Dhwani Batra, Justin Lee, Suxiang Tong |
| EPI_ISL_1094238 | AK State Public Health Lab, State Health Department | Respiratory Viruses Branch, Division of Viral Diseases, Centers for Disease Control and Prevention | Krista Queen, Yan Li, Ying Tao, Jing Zhang, Anna Uehara, Anna Montmayeur, Clinton R. Paden, Peter W. Cook, Rachel Marine, Mili Sheth, Jasmine Padilla, Sarah Nobles, Mark Burroughs, Lori Rowe, Haibin Wang, Ben L. Rambo-Martin, Dhwani Batra, Justin Lee, Suxiang Tong |
| EPI_ISL_1094239 | NC State Laboratory of Public Health | Respiratory Viruses Branch, Division of Viral Diseases, Centers for Disease Control and Prevention | Krista Queen, Yan Li, Ying Tao, Jing Zhang, Anna Uehara, Anna Montmayeur, Clinton R. Paden, Peter W. Cook, Rachel Marine, Mili Sheth, Jasmine Padilla, Sarah Nobles, Mark Burroughs, Lori Rowe, Haibin Wang, Ben L. Rambo-Martin, Dhwani Batra, Justin Lee, Suxiang Tong |
| EPI_ISL_1094241 | IN State Department of Health Laboratory Services | Respiratory Viruses Branch, Division of Viral Diseases, Centers for Disease Control and Prevention | Krista Queen, Yan Li, Ying Tao, Jing Zhang, Anna Uehara, Anna Montmayeur, Clinton R. Paden, Peter W. Cook, Rachel Marine, Mili Sheth, Jasmine Padilla, Sarah Nobles, Mark Burroughs, Lori Rowe, Haibin Wang, Ben L. Rambo-Martin, Dhwani Batra, Justin Lee, Suxiang Tong |
| EPI_ISL_1094242 | FL Bur. of Public Health Laboratories-Jacksonville | Respiratory Viruses Branch, Division of Viral Diseases, Centers for Disease Control and Prevention | Krista Queen, Yan Li, Ying Tao, Jing Zhang, Anna Uehara, Anna Montmayeur, Clinton R. Paden, Peter W. Cook, Rachel Marine, Mili Sheth, Jasmine Padilla, Sarah Nobles, Mark Burroughs, Lori Rowe, Haibin Wang, Ben L. Rambo-Martin, Dhwani Batra, Justin Lee, Suxiang Tong |
| EPI_ISL_1094243 | MD DOH Laboratories Administration | Respiratory Viruses Branch, Division of Viral Diseases, Centers for Disease Control and Prevention | Krista Queen, Yan Li, Ying Tao, Jing Zhang, Anna Uehara, Anna Montmayeur, Clinton R. Paden, Peter W. Cook, Rachel Marine, Mili Sheth, Jasmine Padilla, Sarah Nobles, Mark Burroughs, Lori Rowe, Haibin Wang, Ben L. Rambo-Martin, Dhwani Batra, Justin Lee, Suxiang Tong |
| EPI_ISL_1094244 | SC Dept of Health and Env. Control-Bureau of Laboratories | Respiratory Viruses Branch, Division of Viral Diseases, Centers for Disease Control and Prevention | Krista Queen, Yan Li, Ying Tao, Jing Zhang, Anna Uehara, Anna Montmayeur, Clinton R. Paden, Peter W. Cook, Rachel Marine, Mili Sheth, Jasmine Padilla, Sarah Nobles, Mark Burroughs, Lori Rowe, Haibin Wang, Ben L. Rambo-Martin, Dhwani Batra, Justin Lee, Suxiang Tong |
| EPI_ISL_1094245 | VT Dept. of Health Laboratory | Respiratory Viruses Branch, Division of Viral Diseases, Centers for Disease Control and Prevention | Krista Queen, Yan Li, Ying Tao, Jing Zhang, Anna Uehara, Anna Montmayeur, Clinton R. Paden, Peter W. Cook, Rachel Marine, Mili Sheth, Jasmine Padilla, Sarah Nobles, Mark Burroughs, Lori Rowe, Haibin Wang, Ben L. Rambo-Martin, Dhwani Batra, Justin Lee, Suxiang Tong |
| EPI_ISL_1094246 | KS Health and Environmental Laboratories | Respiratory Viruses Branch, Division of Viral Diseases, Centers for Disease Control and Prevention | Krista Queen, Yan Li, Ying Tao, Jing Zhang, Anna Uehara, Anna Montmayeur, Clinton R. Paden, Peter W. Cook, Rachel Marine, Mili Sheth, Jasmine Padilla, Sarah Nobles, Mark Burroughs, Lori Rowe, Haibin Wang, Ben L. Rambo-Martin, Dhwani Batra, Justin Lee, Suxiang Tong |
| EPI_ISL_1094247 | OK Public Health Laboratory, Oklahoma State DOH | Respiratory Viruses Branch, Division of Viral Diseases, Centers for Disease Control and Prevention | Krista Queen, Yan Li, Ying Tao, Jing Zhang, Anna Uehara, Anna Montmayeur, Clinton R. Paden, Peter W. Cook, Rachel Marine, Mili Sheth, Jasmine Padilla, Sarah Nobles, Mark Burroughs, Lori Rowe, Haibin Wang, Ben L. Rambo-Martin, Dhwani Batra, Justin Lee, Suxiang Tong |
| EPI_ISL_1094249, EPI_ISL_1094250 | IL Dept. of Public Health Springfield Laboratory | Respiratory Viruses Branch, Division of Viral Diseases, Centers for Disease Control and Prevention | Krista Queen, Yan Li, Ying Tao, Jing Zhang, Anna Uehara, Anna Montmayeur, Clinton R. Paden, Peter W. Cook, Rachel Marine, Mili Sheth, Jasmine Padilla, Sarah Nobles, Mark Burroughs, Lori Rowe, Haibin Wang, Ben L. Rambo-Martin, Dhwani Batra, Justin Lee, Suxiang Tong |
| EPI_ISL_1094251 | WA State Department of Health | Respiratory Viruses Branch, Division of Viral Diseases, Centers for Disease Control and Prevention | Krista Queen, Yan Li, Ying Tao, Jing Zhang, Anna Uehara, Anna Montmayeur, Clinton R. Paden, Peter W. Cook, Rachel Marine, Mili Sheth, Jasmine Padilla, Sarah Nobles, Mark Burroughs, Lori Rowe, Haibin Wang, Ben L. Rambo-Martin, Dhwani Batra, Justin Lee, Suxiang Tong |
| EPI_ISL_1094252 | OK Public Health Laboratory, Oklahoma State DOH | Respiratory Viruses Branch, Division of Viral Diseases, Centers for Disease Control and Prevention | Krista Queen, Yan Li, Ying Tao, Jing Zhang, Anna Uehara, Anna Montmayeur, Clinton R. Paden, Peter W. Cook, Rachel Marine, Mili Sheth, Jasmine Padilla, Sarah Nobles, Mark Burroughs, Lori Rowe, Haibin Wang, Ben L. Rambo-Martin, Dhwani Batra, Justin Lee, Suxiang Tong |
| EPI_ISL_1094253 | OH Department of Health Laboratory | Respiratory Viruses Branch, Division of Viral Diseases, Centers for Disease Control and Prevention | Krista Queen, Yan Li, Ying Tao, Jing Zhang, Anna Uehara, Anna Montmayeur, Clinton R. Paden, Peter W. Cook, Rachel Marine, Mili Sheth, Jasmine Padilla, Sarah Nobles, Mark Burroughs, Lori Rowe, Haibin Wang, Ben L. Rambo-Martin, Dhwani Batra, Justin Lee, Suxiang Tong |
| EPI_ISL_1094254 | KS Health and Environmental Laboratories | Respiratory Viruses Branch, Division of Viral Diseases, Centers for Disease Control and Prevention | Krista Queen, Yan Li, Ying Tao, Jing Zhang, Anna Uehara, Anna Montmayeur, Clinton R. Paden, Peter W. Cook, Rachel Marine, Mili Sheth, Jasmine Padilla, Sarah Nobles, Mark Burroughs, Lori Rowe, Haibin Wang, Ben L. Rambo-Martin, Dhwani Batra, Justin Lee, Suxiang Tong |
| EPI_ISL_1094255, EPI_ISL_1094256 | GA Department of Public Health Laboratory | Respiratory Viruses Branch, Division of Viral Diseases, Centers for Disease Control and Prevention | Krista Queen, Yan Li, Ying Tao, Jing Zhang, Anna Uehara, Anna Montmayeur, Clinton R. Paden, Peter W. Cook, Rachel Marine, Mili Sheth, Jasmine Padilla, Sarah Nobles, Mark Burroughs, Lori Rowe, Haibin Wang, Ben L. Rambo-Martin, Dhwani Batra, Justin Lee, Suxiang Tong |
| EPI_ISL_1094258 | IL Dept. of Public Health Springfield Laboratory | Respiratory Viruses Branch, Division of Viral Diseases, Centers for Disease Control and | Krista Queen, Yan Li, Ying Tao, Jing Zhang, Anna Uehara, Anna Montmayeur, Clinton R. Paden, Peter W. Cook, Rachel Marine, Mili Sheth, Jasmine Padilla, Sarah Nobles, Mark Burroughs, Lori Rowe, Haibin Wang, Ben L. Rambo-Martin, Dhwani Batra, Justin Lee, Suxiang Tong |

| | | | |
|---|---|---|---|
| | | Prevention | |
| EPI_ISL_1094259, EPI_ISL_1094260 | UT-Unified State Labs: Public Health Utah DOH | Respiratory Viruses Branch, Division of Viral Diseases, Centers for Disease Control and Prevention | Krista Queen, Yan Li, Ying Tao, Jing Zhang, Anna Uehara, Anna Montmayeur, Clinton R. Paden, Peter W. Cook, Rachel Marine, Mili Sheth, Jasmine Padilla, Sarah Nobles, Mark Burroughs, Lori Rowe, Haibin Wang, Ben L. Rambo-Martin, Dhwani Batra, Justin Lee, Suxiang Tong |
| EPI_ISL_1094261 | CO Dept. of Public Health and Environment, Lab Services Division | Respiratory Viruses Branch, Division of Viral Diseases, Centers for Disease Control and Prevention | Krista Queen, Yan Li, Ying Tao, Jing Zhang, Anna Uehara, Anna Montmayeur, Clinton R. Paden, Peter W. Cook, Rachel Marine, Mili Sheth, Jasmine Padilla, Sarah Nobles, Mark Burroughs, Lori Rowe, Haibin Wang, Ben L. Rambo-Martin, Dhwani Batra, Justin Lee, Suxiang Tong |
| EPI_ISL_1094262, EPI_ISL_1094263 | OH Department of Health Laboratory | Respiratory Viruses Branch, Division of Viral Diseases, Centers for Disease Control and Prevention | Krista Queen, Yan Li, Ying Tao, Jing Zhang, Anna Uehara, Anna Montmayeur, Clinton R. Paden, Peter W. Cook, Rachel Marine, Mili Sheth, Jasmine Padilla, Sarah Nobles, Mark Burroughs, Lori Rowe, Haibin Wang, Ben L. Rambo-Martin, Dhwani Batra, Justin Lee, Suxiang Tong |
| EPI_ISL_1094264 | NC State Laboratory of Public Health | Respiratory Viruses Branch, Division of Viral Diseases, Centers for Disease Control and Prevention | Krista Queen, Yan Li, Ying Tao, Jing Zhang, Anna Uehara, Anna Montmayeur, Clinton R. Paden, Peter W. Cook, Rachel Marine, Mili Sheth, Jasmine Padilla, Sarah Nobles, Mark Burroughs, Lori Rowe, Haibin Wang, Ben L. Rambo-Martin, Dhwani Batra, Justin Lee, Suxiang Tong |
| EPI_ISL_1094265 | FL Bur. of Public Health Laboratories-Jacksonville | Respiratory Viruses Branch, Division of Viral Diseases, Centers for Disease Control and Prevention | Krista Queen, Yan Li, Ying Tao, Jing Zhang, Anna Uehara, Anna Montmayeur, Clinton R. Paden, Peter W. Cook, Rachel Marine, Mili Sheth, Jasmine Padilla, Sarah Nobles, Mark Burroughs, Lori Rowe, Haibin Wang, Ben L. Rambo-Martin, Dhwani Batra, Justin Lee, Suxiang Tong |
| EPI_ISL_1094266, EPI_ISL_1094267 | VT Dept. of Health Laboratory | Respiratory Viruses Branch, Division of Viral Diseases, Centers for Disease Control and Prevention | Krista Queen, Yan Li, Ying Tao, Jing Zhang, Anna Uehara, Anna Montmayeur, Clinton R. Paden, Peter W. Cook, Rachel Marine, Mili Sheth, Jasmine Padilla, Sarah Nobles, Mark Burroughs, Lori Rowe, Haibin Wang, Ben L. Rambo-Martin, Dhwani Batra, Justin Lee, Suxiang Tong |
| EPI_ISL_1094268 | CDPH, Viral and Rickettsial Disease Laboratory | Respiratory Viruses Branch, Division of Viral Diseases, Centers for Disease Control and Prevention | Krista Queen, Yan Li, Ying Tao, Jing Zhang, Anna Uehara, Anna Montmayeur, Clinton R. Paden, Peter W. Cook, Rachel Marine, Mili Sheth, Jasmine Padilla, Sarah Nobles, Mark Burroughs, Lori Rowe, Haibin Wang, Ben L. Rambo-Martin, Dhwani Batra, Justin Lee, Suxiang Tong |
| EPI_ISL_1094269 | DE Public Health Laboratory | Respiratory Viruses Branch, Division of Viral Diseases, Centers for Disease Control and Prevention | Krista Queen, Yan Li, Ying Tao, Jing Zhang, Anna Uehara, Anna Montmayeur, Clinton R. Paden, Peter W. Cook, Rachel Marine, Mili Sheth, Jasmine Padilla, Sarah Nobles, Mark Burroughs, Lori Rowe, Haibin Wang, Ben L. Rambo-Martin, Dhwani Batra, Justin Lee, Suxiang Tong |
| EPI_ISL_1094270 | KS Health and Environmental Laboratories | Respiratory Viruses Branch, Division of Viral Diseases, Centers for Disease Control and Prevention | Krista Queen, Yan Li, Ying Tao, Jing Zhang, Anna Uehara, Anna Montmayeur, Clinton R. Paden, Peter W. Cook, Rachel Marine, Mili Sheth, Jasmine Padilla, Sarah Nobles, Mark Burroughs, Lori Rowe, Haibin Wang, Ben L. Rambo-Martin, Dhwani Batra, Justin Lee, Suxiang Tong |
| EPI_ISL_1094271 | ME Health and Environmental Testing Laboratory | Respiratory Viruses Branch, Division of Viral Diseases, Centers for Disease Control and Prevention | Krista Queen, Yan Li, Ying Tao, Jing Zhang, Anna Uehara, Anna Montmayeur, Clinton R. Paden, Peter W. Cook, Rachel Marine, Mili Sheth, Jasmine Padilla, Sarah Nobles, Mark Burroughs, Lori Rowe, Haibin Wang, Ben L. Rambo-Martin, Dhwani Batra, Justin Lee, Suxiang Tong |
| EPI_ISL_1094272 | AK State Public Health Lab, State Health Department | Respiratory Viruses Branch, Division of Viral Diseases, Centers for Disease Control and Prevention | Krista Queen, Yan Li, Ying Tao, Jing Zhang, Anna Uehara, Anna Montmayeur, Clinton R. Paden, Peter W. Cook, Rachel Marine, Mili Sheth, Jasmine Padilla, Sarah Nobles, Mark Burroughs, Lori Rowe, Haibin Wang, Ben L. Rambo-Martin, Dhwani Batra, Justin Lee, Suxiang Tong |
| EPI_ISL_1094273 | OR State PHL-Virology/Immunology Section | Respiratory Viruses Branch, Division of Viral Diseases, Centers for Disease Control and Prevention | Krista Queen, Yan Li, Ying Tao, Jing Zhang, Anna Uehara, Anna Montmayeur, Clinton R. Paden, Peter W. Cook, Rachel Marine, Mili Sheth, Jasmine Padilla, Sarah Nobles, Mark Burroughs, Lori Rowe, Haibin Wang, Ben L. Rambo-Martin, Dhwani Batra, Justin Lee, Suxiang Tong |
| EPI_ISL_1094274, EPI_ISL_1094275 | WA State Department of Health | Respiratory Viruses Branch, Division of Viral Diseases, Centers for Disease Control and Prevention | Krista Queen, Yan Li, Ying Tao, Jing Zhang, Anna Uehara, Anna Montmayeur, Clinton R. Paden, Peter W. Cook, Rachel Marine, Mili Sheth, Jasmine Padilla, Sarah Nobles, Mark Burroughs, Lori Rowe, Haibin Wang, Ben L. Rambo-Martin, Dhwani Batra, Justin Lee, Suxiang Tong |
| EPI_ISL_1094276 | HI Dept. of Health, State Laboratories Division | Respiratory Viruses Branch, Division of Viral Diseases, Centers for Disease Control and Prevention | Krista Queen, Yan Li, Ying Tao, Jing Zhang, Anna Uehara, Anna Montmayeur, Clinton R. Paden, Peter W. Cook, Rachel Marine, Mili Sheth, Jasmine Padilla, Sarah Nobles, Mark Burroughs, Lori Rowe, Haibin Wang, Ben L. Rambo-Martin, Dhwani Batra, Justin Lee, Suxiang Tong |
| EPI_ISL_1094277 | TX DSHS, Lab Services Section MC 1947 | Respiratory Viruses Branch, Division of Viral Diseases, Centers for Disease Control and Prevention | Krista Queen, Yan Li, Ying Tao, Jing Zhang, Anna Uehara, Anna Montmayeur, Clinton R. Paden, Peter W. Cook, Rachel Marine, Mili Sheth, Jasmine Padilla, Sarah Nobles, Mark Burroughs, Lori Rowe, Haibin Wang, Ben L. Rambo-Martin, Dhwani Batra, Justin Lee, Suxiang Tong |
| EPI_ISL_1094278, EPI_ISL_1094279, EPI_ISL_1094280, EPI_ISL_1094281 | MT Public Health Laboratory | Respiratory Viruses Branch, Division of Viral Diseases, Centers for Disease Control and Prevention | Krista Queen, Yan Li, Ying Tao, Jing Zhang, Anna Uehara, Anna Montmayeur, Clinton R. Paden, Peter W. Cook, Rachel Marine, Mili Sheth, Jasmine Padilla, Sarah Nobles, Mark Burroughs, Lori Rowe, Haibin Wang, Ben L. Rambo-Martin, Dhwani Batra, Justin Lee, Suxiang Tong |
| EPI_ISL_1094282 | NC State Laboratory of Public Health | Respiratory Viruses Branch, Division of Viral Diseases, Centers for Disease Control and Prevention | Krista Queen, Yan Li, Ying Tao, Jing Zhang, Anna Uehara, Anna Montmayeur, Clinton R. Paden, Peter W. Cook, Rachel Marine, Mili Sheth, Jasmine Padilla, Sarah Nobles, Mark Burroughs, Lori Rowe, Haibin Wang, Ben L. Rambo-Martin, Dhwani Batra, Justin Lee, Suxiang Tong |
| EPI_ISL_1094283 | MI - Michigan Department of Health and Human Services - Bureau of Laboratories | Respiratory Viruses Branch, Division of Viral Diseases, Centers for Disease Control and Prevention | Krista Queen, Yan Li, Ying Tao, Jing Zhang, Anna Uehara, Anna Montmayeur, Clinton R. Paden, Peter W. Cook, Rachel Marine, Mili Sheth, Jasmine Padilla, Sarah Nobles, Mark Burroughs, Lori Rowe, Haibin Wang, Ben L. Rambo-Martin, Dhwani Batra, Justin Lee, Suxiang Tong |
| EPI_ISL_1094284 | GA Department of Public Health Laboratory | Respiratory Viruses Branch, Division of Viral Diseases, Centers for Disease Control and Prevention | Krista Queen, Yan Li, Ying Tao, Jing Zhang, Anna Uehara, Anna Montmayeur, Clinton R. Paden, Peter W. Cook, Rachel Marine, Mili Sheth, Jasmine Padilla, Sarah Nobles, Mark Burroughs, Lori Rowe, Haibin Wang, Ben L. Rambo-Martin, Dhwani Batra, Justin Lee, Suxiang Tong |
| EPI_ISL_1094285, EPI_ISL_1094286 | NYSDOH Wadsworth Center, Virology Lab | Respiratory Viruses Branch, Division of Viral Diseases, Centers for Disease Control and Prevention | Krista Queen, Yan Li, Ying Tao, Jing Zhang, Anna Uehara, Anna Montmayeur, Clinton R. Paden, Peter W. Cook, Rachel Marine, Mili Sheth, Jasmine Padilla, Sarah Nobles, Mark Burroughs, Lori Rowe, Haibin Wang, Ben L. Rambo-Martin, Dhwani Batra, Justin Lee, Suxiang Tong |
| EPI_ISL_1094287 | RI State Health Laboratories | Respiratory Viruses Branch, Division of Viral Diseases, Centers for Disease Control and Prevention | Krista Queen, Yan Li, Ying Tao, Jing Zhang, Anna Uehara, Anna Montmayeur, Clinton R. Paden, Peter W. Cook, Rachel Marine, Mili Sheth, Jasmine Padilla, Sarah Nobles, Mark Burroughs, Lori Rowe, Haibin Wang, Ben L. Rambo-Martin, Dhwani Batra, Justin Lee, Suxiang Tong |
| EPI_ISL_1094288 | ME Health and Environmental Testing Laboratory | Respiratory Viruses Branch, Division of Viral Diseases, Centers for Disease Control and Prevention | Krista Queen, Yan Li, Ying Tao, Jing Zhang, Anna Uehara, Anna Montmayeur, Clinton R. Paden, Peter W. Cook, Rachel Marine, Mili Sheth, Jasmine Padilla, Sarah Nobles, Mark Burroughs, Lori Rowe, Haibin Wang, Ben L. Rambo-Martin, Dhwani Batra, Justin Lee, Suxiang Tong |
| EPI_ISL_1094289 | IL Dept. of Public Health Springfield Laboratory | Respiratory Viruses Branch, Division of Viral Diseases, Centers for Disease Control and Prevention | Krista Queen, Yan Li, Ying Tao, Jing Zhang, Anna Uehara, Anna Montmayeur, Clinton R. Paden, Peter W. Cook, Rachel Marine, Mili Sheth, Jasmine Padilla, Sarah Nobles, Mark Burroughs, Lori Rowe, Haibin Wang, Ben L. Rambo-Martin, Dhwani Batra, Justin Lee, Suxiang Tong |
| EPI_ISL_1094290 | OR State PHL-Virology/Immunology Section | Respiratory Viruses Branch, Division of Viral Diseases, Centers for Disease Control and Prevention | Krista Queen, Yan Li, Ying Tao, Jing Zhang, Anna Uehara, Anna Montmayeur, Clinton R. Paden, Peter W. Cook, Rachel Marine, Mili Sheth, Jasmine Padilla, Sarah Nobles, Mark Burroughs, Lori Rowe, Haibin Wang, Ben L. Rambo-Martin, Dhwani Batra, Justin Lee, Suxiang Tong |
| EPI_ISL_1094291 | MO State Public Health Laboratory | Respiratory Viruses Branch, Division of Viral Diseases, Centers for Disease Control and Prevention | Krista Queen, Yan Li, Ying Tao, Jing Zhang, Anna Uehara, Anna Montmayeur, Clinton R. Paden, Peter W. Cook, Rachel Marine, Mili Sheth, Jasmine Padilla, Sarah Nobles, Mark Burroughs, Lori Rowe, Haibin Wang, Ben L. Rambo-Martin, Dhwani Batra, Justin Lee, Suxiang Tong |
| EPI_ISL_1094292 | MI - Michigan Department of Health and Human Services - Bureau of Laboratories | Respiratory Viruses Branch, Division of Viral Diseases, Centers for Disease Control and Prevention | Krista Queen, Yan Li, Ying Tao, Jing Zhang, Anna Uehara, Anna Montmayeur, Clinton R. Paden, Peter W. Cook, Rachel Marine, Mili Sheth, Jasmine Padilla, Sarah Nobles, Mark Burroughs, Lori Rowe, Haibin Wang, Ben L. Rambo-Martin, Dhwani Batra, Justin Lee, Suxiang Tong |
| EPI_ISL_1094293 | FL Bur. of Public Health Laboratories-Jacksonville | Respiratory Viruses Branch, Division of Viral Diseases, Centers for Disease Control and Prevention | Krista Queen, Yan Li, Ying Tao, Jing Zhang, Anna Uehara, Anna Montmayeur, Clinton R. Paden, Peter W. Cook, Rachel Marine, Mili Sheth, Jasmine Padilla, Sarah Nobles, Mark Burroughs, Lori Rowe, Haibin Wang, Ben L. Rambo-Martin, Dhwani Batra, Justin Lee, Suxiang Tong |
| EPI_ISL_1094294 | NC State Laboratory of Public Health | Respiratory Viruses Branch, Division of Viral Diseases, Centers for Disease Control and Prevention | Krista Queen, Yan Li, Ying Tao, Jing Zhang, Anna Uehara, Anna Montmayeur, Clinton R. Paden, Peter W. Cook, Rachel Marine, Mili Sheth, Jasmine Padilla, Sarah Nobles, Mark Burroughs, Lori Rowe, Haibin Wang, Ben L. Rambo-Martin, Dhwani Batra, Justin Lee, Suxiang Tong |
| EPI_ISL_1094305 | PA Department of Health, Bureau of Laboratories | Respiratory Viruses Branch, Division of Viral Diseases, Centers for Disease Control and Prevention | Krista Queen, Yan Li, Ying Tao, Jing Zhang, Anna Uehara, Anna Montmayeur, Clinton R. Paden, Peter W. Cook, Rachel Marine, Mili Sheth, Jasmine Padilla, Sarah Nobles, Mark Burroughs, Lori Rowe, Haibin Wang, Ben L. Rambo-Martin, Dhwani Batra, Justin Lee, Suxiang Tong |
| EPI_ISL_1094306 | OH Department of Health Laboratory | Respiratory Viruses Branch, Division of Viral Diseases, Centers for Disease Control and Prevention | Krista Queen, Yan Li, Ying Tao, Jing Zhang, Anna Uehara, Anna Montmayeur, Clinton R. Paden, Peter W. Cook, Rachel Marine, Mili Sheth, Jasmine Padilla, Sarah Nobles, Mark Burroughs, Lori Rowe, Haibin Wang, Ben L. Rambo-Martin, Dhwani Batra, Justin Lee, Suxiang Tong |
| EPI_ISL_1094307 | NYSDOH Wadsworth Center, Virology Lab | Respiratory Viruses Branch, Division of Viral Diseases, Centers for Disease Control and Prevention | Krista Queen, Yan Li, Ying Tao, Jing Zhang, Anna Uehara, Anna Montmayeur, Clinton R. Paden, Peter W. Cook, Rachel Marine, Mili Sheth, Jasmine Padilla, Sarah Nobles, Mark Burroughs, Lori Rowe, Haibin Wang, Ben L. Rambo-Martin, Dhwani Batra, Justin Lee, Suxiang Tong |
| EPI_ISL_1094308 | AR Dept. of Health-Public Health Lab | Respiratory Viruses Branch, Division of Viral | Krista Queen, Yan Li, Ying Tao, Jing Zhang, Anna Uehara, Anna Montmayeur, Clinton R. Paden, Peter W. Cook, Rachel Marine, Mili Sheth, Jasmine Padilla, Sarah Nobles, Mark Burroughs, Lori Rowe, Haibin Wang, Ben L. Rambo-Martin, Dhwani Batra, Justin Lee, Suxiang Tong |

| | | | |
|---|---|---|---|
| | | Diseases, Centers for Disease Control and Prevention | |
| EPI_ISL_1094310 | MS Public Health Laboratory | Respiratory Viruses Branch, Division of Viral Diseases, Centers for Disease Control and Prevention | Krista Queen, Yan Li, Ying Tao, Jing Zhang, Anna Uehara, Anna Montmayeur, Clinton R. Paden, Peter W. Cook, Rachel Marine, Mili Sheth, Jasmine Padilla, Sarah Nobles, Mark Burroughs, Lori Rowe, Haibin Wang, Ben L. Rambo-Martin, Dhwani Batra, Justin Lee, Suxiang Tong |
| EPI_ISL_1094311 | DC Public Health Lab/ Dept. of Forensic Sciences | Respiratory Viruses Branch, Division of Viral Diseases, Centers for Disease Control and Prevention | Krista Queen, Yan Li, Ying Tao, Jing Zhang, Anna Uehara, Anna Montmayeur, Clinton R. Paden, Peter W. Cook, Rachel Marine, Mili Sheth, Jasmine Padilla, Sarah Nobles, Mark Burroughs, Lori Rowe, Haibin Wang, Ben L. Rambo-Martin, Dhwani Batra, Justin Lee, Suxiang Tong |
| EPI_ISL_1094312 | GA Department of Public Health Laboratory | Respiratory Viruses Branch, Division of Viral Diseases, Centers for Disease Control and Prevention | Krista Queen, Yan Li, Ying Tao, Jing Zhang, Anna Uehara, Anna Montmayeur, Clinton R. Paden, Peter W. Cook, Rachel Marine, Mili Sheth, Jasmine Padilla, Sarah Nobles, Mark Burroughs, Lori Rowe, Haibin Wang, Ben L. Rambo-Martin, Dhwani Batra, Justin Lee, Suxiang Tong |
| EPI_ISL_1094313 | KS Health and Environmental Laboratories | Respiratory Viruses Branch, Division of Viral Diseases, Centers for Disease Control and Prevention | Krista Queen, Yan Li, Ying Tao, Jing Zhang, Anna Uehara, Anna Montmayeur, Clinton R. Paden, Peter W. Cook, Rachel Marine, Mili Sheth, Jasmine Padilla, Sarah Nobles, Mark Burroughs, Lori Rowe, Haibin Wang, Ben L. Rambo-Martin, Dhwani Batra, Justin Lee, Suxiang Tong |
| EPI_ISL_1094314 | OR State PHL-Virology/Immunology Section | Respiratory Viruses Branch, Division of Viral Diseases, Centers for Disease Control and Prevention | Krista Queen, Yan Li, Ying Tao, Jing Zhang, Anna Uehara, Anna Montmayeur, Clinton R. Paden, Peter W. Cook, Rachel Marine, Mili Sheth, Jasmine Padilla, Sarah Nobles, Mark Burroughs, Lori Rowe, Haibin Wang, Ben L. Rambo-Martin, Dhwani Batra, Justin Lee, Suxiang Tong |
| EPI_ISL_1094315 | FL Bureau of Public Health Laboratories-Tampa | Respiratory Viruses Branch, Division of Viral Diseases, Centers for Disease Control and Prevention | Krista Queen, Yan Li, Ying Tao, Jing Zhang, Anna Uehara, Anna Montmayeur, Clinton R. Paden, Peter W. Cook, Rachel Marine, Mili Sheth, Jasmine Padilla, Sarah Nobles, Mark Burroughs, Lori Rowe, Haibin Wang, Ben L. Rambo-Martin, Dhwani Batra, Justin Lee, Suxiang Tong |
| EPI_ISL_1094316 | WA State Department of Health | Respiratory Viruses Branch, Division of Viral Diseases, Centers for Disease Control and Prevention | Krista Queen, Yan Li, Ying Tao, Jing Zhang, Anna Uehara, Anna Montmayeur, Clinton R. Paden, Peter W. Cook, Rachel Marine, Mili Sheth, Jasmine Padilla, Sarah Nobles, Mark Burroughs, Lori Rowe, Haibin Wang, Ben L. Rambo-Martin, Dhwani Batra, Justin Lee, Suxiang Tong |
| EPI_ISL_1094318 | AR Dept. of Health-Public Health Lab | Respiratory Viruses Branch, Division of Viral Diseases, Centers for Disease Control and Prevention | Krista Queen, Yan Li, Ying Tao, Jing Zhang, Anna Uehara, Anna Montmayeur, Clinton R. Paden, Peter W. Cook, Rachel Marine, Mili Sheth, Jasmine Padilla, Sarah Nobles, Mark Burroughs, Lori Rowe, Haibin Wang, Ben L. Rambo-Martin, Dhwani Batra, Justin Lee, Suxiang Tong |
| EPI_ISL_1094319 | AK State Public Health Lab, State Health Department | Respiratory Viruses Branch, Division of Viral Diseases, Centers for Disease Control and Prevention | Krista Queen, Yan Li, Ying Tao, Jing Zhang, Anna Uehara, Anna Montmayeur, Clinton R. Paden, Peter W. Cook, Rachel Marine, Mili Sheth, Jasmine Padilla, Sarah Nobles, Mark Burroughs, Lori Rowe, Haibin Wang, Ben L. Rambo-Martin, Dhwani Batra, Justin Lee, Suxiang Tong |
| EPI_ISL_1094320 | NC State Laboratory of Public Health | Respiratory Viruses Branch, Division of Viral Diseases, Centers for Disease Control and Prevention | Krista Queen, Yan Li, Ying Tao, Jing Zhang, Anna Uehara, Anna Montmayeur, Clinton R. Paden, Peter W. Cook, Rachel Marine, Mili Sheth, Jasmine Padilla, Sarah Nobles, Mark Burroughs, Lori Rowe, Haibin Wang, Ben L. Rambo-Martin, Dhwani Batra, Justin Lee, Suxiang Tong |
| EPI_ISL_1094322 | NYSDOH Wadsworth Center, Virology Lab | Respiratory Viruses Branch, Division of Viral Diseases, Centers for Disease Control and Prevention | Krista Queen, Yan Li, Ying Tao, Jing Zhang, Anna Uehara, Anna Montmayeur, Clinton R. Paden, Peter W. Cook, Rachel Marine, Mili Sheth, Jasmine Padilla, Sarah Nobles, Mark Burroughs, Lori Rowe, Haibin Wang, Ben L. Rambo-Martin, Dhwani Batra, Justin Lee, Suxiang Tong |
| EPI_ISL_1094323 | VA-Division of Consolidated Laboratory Services | Respiratory Viruses Branch, Division of Viral Diseases, Centers for Disease Control and Prevention | Krista Queen, Yan Li, Ying Tao, Jing Zhang, Anna Uehara, Anna Montmayeur, Clinton R. Paden, Peter W. Cook, Rachel Marine, Mili Sheth, Jasmine Padilla, Sarah Nobles, Mark Burroughs, Lori Rowe, Haibin Wang, Ben L. Rambo-Martin, Dhwani Batra, Justin Lee, Suxiang Tong |
| EPI_ISL_1094324 | MT Public Health Laboratory | Respiratory Viruses Branch, Division of Viral Diseases, Centers for Disease Control and Prevention | Krista Queen, Yan Li, Ying Tao, Jing Zhang, Anna Uehara, Anna Montmayeur, Clinton R. Paden, Peter W. Cook, Rachel Marine, Mili Sheth, Jasmine Padilla, Sarah Nobles, Mark Burroughs, Lori Rowe, Haibin Wang, Ben L. Rambo-Martin, Dhwani Batra, Justin Lee, Suxiang Tong |
| EPI_ISL_1094325 | CA-Los Angeles County Public Health Laboratory | Respiratory Viruses Branch, Division of Viral Diseases, Centers for Disease Control and Prevention | Krista Queen, Yan Li, Ying Tao, Jing Zhang, Anna Uehara, Anna Montmayeur, Clinton R. Paden, Peter W. Cook, Rachel Marine, Mili Sheth, Jasmine Padilla, Sarah Nobles, Mark Burroughs, Lori Rowe, Haibin Wang, Ben L. Rambo-Martin, Dhwani Batra, Justin Lee, Suxiang Tong |
| EPI_ISL_1094326 | IN State Department of Health Laboratory Services | Respiratory Viruses Branch, Division of Viral Diseases, Centers for Disease Control and Prevention | Krista Queen, Yan Li, Ying Tao, Jing Zhang, Anna Uehara, Anna Montmayeur, Clinton R. Paden, Peter W. Cook, Rachel Marine, Mili Sheth, Jasmine Padilla, Sarah Nobles, Mark Burroughs, Lori Rowe, Haibin Wang, Ben L. Rambo-Martin, Dhwani Batra, Justin Lee, Suxiang Tong |
| EPI_ISL_1094327 | AZ SPHL, Arizona Department of Health Services | Respiratory Viruses Branch, Division of Viral Diseases, Centers for Disease Control and Prevention | Krista Queen, Yan Li, Ying Tao, Jing Zhang, Anna Uehara, Anna Montmayeur, Clinton R. Paden, Peter W. Cook, Rachel Marine, Mili Sheth, Jasmine Padilla, Sarah Nobles, Mark Burroughs, Lori Rowe, Haibin Wang, Ben L. Rambo-Martin, Dhwani Batra, Justin Lee, Suxiang Tong |
| EPI_ISL_1094329 | IA State Hygienic Laboratory | Respiratory Viruses Branch, Division of Viral Diseases, Centers for Disease Control and Prevention | Krista Queen, Yan Li, Ying Tao, Jing Zhang, Anna Uehara, Anna Montmayeur, Clinton R. Paden, Peter W. Cook, Rachel Marine, Mili Sheth, Jasmine Padilla, Sarah Nobles, Mark Burroughs, Lori Rowe, Haibin Wang, Ben L. Rambo-Martin, Dhwani Batra, Justin Lee, Suxiang Tong |
| EPI_ISL_1094330, EPI_ISL_1094331, EPI_ISL_1094332 | DC Public Health Lab/ Dept. of Forensic Sciences | Respiratory Viruses Branch, Division of Viral Diseases, Centers for Disease Control and Prevention | Krista Queen, Yan Li, Ying Tao, Jing Zhang, Anna Uehara, Anna Montmayeur, Clinton R. Paden, Peter W. Cook, Rachel Marine, Mili Sheth, Jasmine Padilla, Sarah Nobles, Mark Burroughs, Lori Rowe, Haibin Wang, Ben L. Rambo-Martin, Dhwani Batra, Justin Lee, Suxiang Tong |
| EPI_ISL_1094333 | LA Office of Public Health Laboratories | Respiratory Viruses Branch, Division of Viral Diseases, Centers for Disease Control and Prevention | Krista Queen, Yan Li, Ying Tao, Jing Zhang, Anna Uehara, Anna Montmayeur, Clinton R. Paden, Peter W. Cook, Rachel Marine, Mili Sheth, Jasmine Padilla, Sarah Nobles, Mark Burroughs, Lori Rowe, Haibin Wang, Ben L. Rambo-Martin, Dhwani Batra, Justin Lee, Suxiang Tong |
| EPI_ISL_1094335, EPI_ISL_1094336 | NE Public Health Laboratory | Respiratory Viruses Branch, Division of Viral Diseases, Centers for Disease Control and Prevention | Krista Queen, Yan Li, Ying Tao, Jing Zhang, Anna Uehara, Anna Montmayeur, Clinton R. Paden, Peter W. Cook, Rachel Marine, Mili Sheth, Jasmine Padilla, Sarah Nobles, Mark Burroughs, Lori Rowe, Haibin Wang, Ben L. Rambo-Martin, Dhwani Batra, Justin Lee, Suxiang Tong |
| EPI_ISL_1094337 | OK Public Health Laboratory, Oklahoma State DOH | Respiratory Viruses Branch, Division of Viral Diseases, Centers for Disease Control and Prevention | Krista Queen, Yan Li, Ying Tao, Jing Zhang, Anna Uehara, Anna Montmayeur, Clinton R. Paden, Peter W. Cook, Rachel Marine, Mili Sheth, Jasmine Padilla, Sarah Nobles, Mark Burroughs, Lori Rowe, Haibin Wang, Ben L. Rambo-Martin, Dhwani Batra, Justin Lee, Suxiang Tong |
| EPI_ISL_1094338 | NC State Laboratory of Public Health | Respiratory Viruses Branch, Division of Viral Diseases, Centers for Disease Control and Prevention | Krista Queen, Yan Li, Ying Tao, Jing Zhang, Anna Uehara, Anna Montmayeur, Clinton R. Paden, Peter W. Cook, Rachel Marine, Mili Sheth, Jasmine Padilla, Sarah Nobles, Mark Burroughs, Lori Rowe, Haibin Wang, Ben L. Rambo-Martin, Dhwani Batra, Justin Lee, Suxiang Tong |
| EPI_ISL_1094339 | IN State Department of Health Laboratory Services | Respiratory Viruses Branch, Division of Viral Diseases, Centers for Disease Control and Prevention | Krista Queen, Yan Li, Ying Tao, Jing Zhang, Anna Uehara, Anna Montmayeur, Clinton R. Paden, Peter W. Cook, Rachel Marine, Mili Sheth, Jasmine Padilla, Sarah Nobles, Mark Burroughs, Lori Rowe, Haibin Wang, Ben L. Rambo-Martin, Dhwani Batra, Justin Lee, Suxiang Tong |
| EPI_ISL_1094340 | OK Public Health Laboratory, Oklahoma State DOH | Respiratory Viruses Branch, Division of Viral Diseases, Centers for Disease Control and Prevention | Krista Queen, Yan Li, Ying Tao, Jing Zhang, Anna Uehara, Anna Montmayeur, Clinton R. Paden, Peter W. Cook, Rachel Marine, Mili Sheth, Jasmine Padilla, Sarah Nobles, Mark Burroughs, Lori Rowe, Haibin Wang, Ben L. Rambo-Martin, Dhwani Batra, Justin Lee, Suxiang Tong |
| EPI_ISL_1094341, EPI_ISL_1094343, EPI_ISL_1094344 | DC Public Health Lab/ Dept. of Forensic Sciences | Respiratory Viruses Branch, Division of Viral Diseases, Centers for Disease Control and Prevention | Krista Queen, Yan Li, Ying Tao, Jing Zhang, Anna Uehara, Anna Montmayeur, Clinton R. Paden, Peter W. Cook, Rachel Marine, Mili Sheth, Jasmine Padilla, Sarah Nobles, Mark Burroughs, Lori Rowe, Haibin Wang, Ben L. Rambo-Martin, Dhwani Batra, Justin Lee, Suxiang Tong |
| EPI_ISL_1094345 | KY State Public Health Lab | Respiratory Viruses Branch, Division of Viral Diseases, Centers for Disease Control and Prevention | Krista Queen, Yan Li, Ying Tao, Jing Zhang, Anna Uehara, Anna Montmayeur, Clinton R. Paden, Peter W. Cook, Rachel Marine, Mili Sheth, Jasmine Padilla, Sarah Nobles, Mark Burroughs, Lori Rowe, Haibin Wang, Ben L. Rambo-Martin, Dhwani Batra, Justin Lee, Suxiang Tong |
| EPI_ISL_1094346 | AZ SPHL, Arizona Department of Health Services | Respiratory Viruses Branch, Division of Viral Diseases, Centers for Disease Control and Prevention | Krista Queen, Yan Li, Ying Tao, Jing Zhang, Anna Uehara, Anna Montmayeur, Clinton R. Paden, Peter W. Cook, Rachel Marine, Mili Sheth, Jasmine Padilla, Sarah Nobles, Mark Burroughs, Lori Rowe, Haibin Wang, Ben L. Rambo-Martin, Dhwani Batra, Justin Lee, Suxiang Tong |
| EPI_ISL_1094347 | MD DOH Laboratories Administration | Respiratory Viruses Branch, Division of Viral Diseases, Centers for Disease Control and Prevention | Krista Queen, Yan Li, Ying Tao, Jing Zhang, Anna Uehara, Anna Montmayeur, Clinton R. Paden, Peter W. Cook, Rachel Marine, Mili Sheth, Jasmine Padilla, Sarah Nobles, Mark Burroughs, Lori Rowe, Haibin Wang, Ben L. Rambo-Martin, Dhwani Batra, Justin Lee, Suxiang Tong |
| EPI_ISL_1094348 | CDPH, Viral and Rickettsial Disease Laboratory | Respiratory Viruses Branch, Division of Viral Diseases, Centers for Disease Control and Prevention | Krista Queen, Yan Li, Ying Tao, Jing Zhang, Anna Uehara, Anna Montmayeur, Clinton R. Paden, Peter W. Cook, Rachel Marine, Mili Sheth, Jasmine Padilla, Sarah Nobles, Mark Burroughs, Lori Rowe, Haibin Wang, Ben L. Rambo-Martin, Dhwani Batra, Justin Lee, Suxiang Tong |
| EPI_ISL_1094349 | KS Health and Environmental Laboratories | Respiratory Viruses Branch, Division of Viral Diseases, Centers for Disease Control and Prevention | Krista Queen, Yan Li, Ying Tao, Jing Zhang, Anna Uehara, Anna Montmayeur, Clinton R. Paden, Peter W. Cook, Rachel Marine, Mili Sheth, Jasmine Padilla, Sarah Nobles, Mark Burroughs, Lori Rowe, Haibin Wang, Ben L. Rambo-Martin, Dhwani Batra, Justin Lee, Suxiang Tong |
| EPI_ISL_1094350, EPI_ISL_1094351 | MI - Michigan Department of Health and Human Services - Bureau of Laboratories | Respiratory Viruses Branch, Division of Viral Diseases, Centers for Disease Control and Prevention | Krista Queen, Yan Li, Ying Tao, Jing Zhang, Anna Uehara, Anna Montmayeur, Clinton R. Paden, Peter W. Cook, Rachel Marine, Mili Sheth, Jasmine Padilla, Sarah Nobles, Mark Burroughs, Lori Rowe, Haibin Wang, Ben L. Rambo-Martin, Dhwani Batra, Justin Lee, Suxiang Tong |

| | | | |
|---|---|---|---|
| EPI_ISL_1094352 | NC State Laboratory of Public Health | Respiratory Viruses Branch, Division of Viral Diseases, Centers for Disease Control and Prevention | Krista Queen, Yan Li, Ying Tao, Jing Zhang, Anna Uehara, Anna Montmayeur, Clinton R. Paden, Peter W. Cook, Rachel Marine, Mili Sheth, Jasmine Padilla, Sarah Nobles, Mark Burroughs, Lori Rowe, Haibin Wang, Ben L. Rambo-Martin, Dhwani Batra, Justin Lee, Suxiang Tong |
| EPI_ISL_1094353 | AK State Public Health Lab, State Health Department | Respiratory Viruses Branch, Division of Viral Diseases, Centers for Disease Control and Prevention | Krista Queen, Yan Li, Ying Tao, Jing Zhang, Anna Uehara, Anna Montmayeur, Clinton R. Paden, Peter W. Cook, Rachel Marine, Mili Sheth, Jasmine Padilla, Sarah Nobles, Mark Burroughs, Lori Rowe, Haibin Wang, Ben L. Rambo-Martin, Dhwani Batra, Justin Lee, Suxiang Tong |
| EPI_ISL_1094354 | NC State Laboratory of Public Health | Respiratory Viruses Branch, Division of Viral Diseases, Centers for Disease Control and Prevention | Krista Queen, Yan Li, Ying Tao, Jing Zhang, Anna Uehara, Anna Montmayeur, Clinton R. Paden, Peter W. Cook, Rachel Marine, Mili Sheth, Jasmine Padilla, Sarah Nobles, Mark Burroughs, Lori Rowe, Haibin Wang, Ben L. Rambo-Martin, Dhwani Batra, Justin Lee, Suxiang Tong |
| EPI_ISL_1094355 | RI State Health Laboratories | Respiratory Viruses Branch, Division of Viral Diseases, Centers for Disease Control and Prevention | Krista Queen, Yan Li, Ying Tao, Jing Zhang, Anna Uehara, Anna Montmayeur, Clinton R. Paden, Peter W. Cook, Rachel Marine, Mili Sheth, Jasmine Padilla, Sarah Nobles, Mark Burroughs, Lori Rowe, Haibin Wang, Ben L. Rambo-Martin, Dhwani Batra, Justin Lee, Suxiang Tong |
| EPI_ISL_1094358 | MO State Public Health Laboratory | Respiratory Viruses Branch, Division of Viral Diseases, Centers for Disease Control and Prevention | Krista Queen, Yan Li, Ying Tao, Jing Zhang, Anna Uehara, Anna Montmayeur, Clinton R. Paden, Peter W. Cook, Rachel Marine, Mili Sheth, Jasmine Padilla, Sarah Nobles, Mark Burroughs, Lori Rowe, Haibin Wang, Ben L. Rambo-Martin, Dhwani Batra, Justin Lee, Suxiang Tong |
| EPI_ISL_1094359 | NYSDOH Wadsworth Center, Virology Lab | Respiratory Viruses Branch, Division of Viral Diseases, Centers for Disease Control and Prevention | Krista Queen, Yan Li, Ying Tao, Jing Zhang, Anna Uehara, Anna Montmayeur, Clinton R. Paden, Peter W. Cook, Rachel Marine, Mili Sheth, Jasmine Padilla, Sarah Nobles, Mark Burroughs, Lori Rowe, Haibin Wang, Ben L. Rambo-Martin, Dhwani Batra, Justin Lee, Suxiang Tong |
| EPI_ISL_1094361, EPI_ISL_1094362, EPI_ISL_1094363 | CA-Los Angeles County Public Health Laboratory | Respiratory Viruses Branch, Division of Viral Diseases, Centers for Disease Control and Prevention | Krista Queen, Yan Li, Ying Tao, Jing Zhang, Anna Uehara, Anna Montmayeur, Clinton R. Paden, Peter W. Cook, Rachel Marine, Mili Sheth, Jasmine Padilla, Sarah Nobles, Mark Burroughs, Lori Rowe, Haibin Wang, Ben L. Rambo-Martin, Dhwani Batra, Justin Lee, Suxiang Tong |
| EPI_ISL_1094364 | GA Department of Public Health Laboratory | Respiratory Viruses Branch, Division of Viral Diseases, Centers for Disease Control and Prevention | Krista Queen, Yan Li, Ying Tao, Jing Zhang, Anna Uehara, Anna Montmayeur, Clinton R. Paden, Peter W. Cook, Rachel Marine, Mili Sheth, Jasmine Padilla, Sarah Nobles, Mark Burroughs, Lori Rowe, Haibin Wang, Ben L. Rambo-Martin, Dhwani Batra, Justin Lee, Suxiang Tong |
| EPI_ISL_1094365 | MI - Michigan Department of Health and Human Services - Bureau of Laboratories | Respiratory Viruses Branch, Division of Viral Diseases, Centers for Disease Control and Prevention | Krista Queen, Yan Li, Ying Tao, Jing Zhang, Anna Uehara, Anna Montmayeur, Clinton R. Paden, Peter W. Cook, Rachel Marine, Mili Sheth, Jasmine Padilla, Sarah Nobles, Mark Burroughs, Lori Rowe, Haibin Wang, Ben L. Rambo-Martin, Dhwani Batra, Justin Lee, Suxiang Tong |
| EPI_ISL_1094366 | AK State Public Health Lab, State Health Department | Respiratory Viruses Branch, Division of Viral Diseases, Centers for Disease Control and Prevention | Krista Queen, Yan Li, Ying Tao, Jing Zhang, Anna Uehara, Anna Montmayeur, Clinton R. Paden, Peter W. Cook, Rachel Marine, Mili Sheth, Jasmine Padilla, Sarah Nobles, Mark Burroughs, Lori Rowe, Haibin Wang, Ben L. Rambo-Martin, Dhwani Batra, Justin Lee, Suxiang Tong |
| EPI_ISL_1094367 | OR State PHL-Virology/Immunology Section | Respiratory Viruses Branch, Division of Viral Diseases, Centers for Disease Control and Prevention | Krista Queen, Yan Li, Ying Tao, Jing Zhang, Anna Uehara, Anna Montmayeur, Clinton R. Paden, Peter W. Cook, Rachel Marine, Mili Sheth, Jasmine Padilla, Sarah Nobles, Mark Burroughs, Lori Rowe, Haibin Wang, Ben L. Rambo-Martin, Dhwani Batra, Justin Lee, Suxiang Tong |
| EPI_ISL_1094368, EPI_ISL_1094370 | NYSDOH Wadsworth Center, Virology Lab | Respiratory Viruses Branch, Division of Viral Diseases, Centers for Disease Control and Prevention | Krista Queen, Yan Li, Ying Tao, Jing Zhang, Anna Uehara, Anna Montmayeur, Clinton R. Paden, Peter W. Cook, Rachel Marine, Mili Sheth, Jasmine Padilla, Sarah Nobles, Mark Burroughs, Lori Rowe, Haibin Wang, Ben L. Rambo-Martin, Dhwani Batra, Justin Lee, Suxiang Tong |
| EPI_ISL_1094371 | WA State Department of Health | Respiratory Viruses Branch, Division of Viral Diseases, Centers for Disease Control and Prevention | Krista Queen, Yan Li, Ying Tao, Jing Zhang, Anna Uehara, Anna Montmayeur, Clinton R. Paden, Peter W. Cook, Rachel Marine, Mili Sheth, Jasmine Padilla, Sarah Nobles, Mark Burroughs, Lori Rowe, Haibin Wang, Ben L. Rambo-Martin, Dhwani Batra, Justin Lee, Suxiang Tong |
| EPI_ISL_1094374 | SD Public Health Laboratory | Respiratory Viruses Branch, Division of Viral Diseases, Centers for Disease Control and Prevention | Krista Queen, Yan Li, Ying Tao, Jing Zhang, Anna Uehara, Anna Montmayeur, Clinton R. Paden, Peter W. Cook, Rachel Marine, Mili Sheth, Jasmine Padilla, Sarah Nobles, Mark Burroughs, Lori Rowe, Haibin Wang, Ben L. Rambo-Martin, Dhwani Batra, Justin Lee, Suxiang Tong |
| EPI_ISL_1094375, EPI_ISL_1094376 | DE Public Health Laboratory | Respiratory Viruses Branch, Division of Viral Diseases, Centers for Disease Control and Prevention | Krista Queen, Yan Li, Ying Tao, Jing Zhang, Anna Uehara, Anna Montmayeur, Clinton R. Paden, Peter W. Cook, Rachel Marine, Mili Sheth, Jasmine Padilla, Sarah Nobles, Mark Burroughs, Lori Rowe, Haibin Wang, Ben L. Rambo-Martin, Dhwani Batra, Justin Lee, Suxiang Tong |
| EPI_ISL_1094377 | ND Dept. of Health Laboratory Services-Microbiology | Respiratory Viruses Branch, Division of Viral Diseases, Centers for Disease Control and Prevention | Krista Queen, Yan Li, Ying Tao, Jing Zhang, Anna Uehara, Anna Montmayeur, Clinton R. Paden, Peter W. Cook, Rachel Marine, Mili Sheth, Jasmine Padilla, Sarah Nobles, Mark Burroughs, Lori Rowe, Haibin Wang, Ben L. Rambo-Martin, Dhwani Batra, Justin Lee, Suxiang Tong |
| EPI_ISL_1094378 | NYSDOH Wadsworth Center, Virology Lab | Respiratory Viruses Branch, Division of Viral Diseases, Centers for Disease Control and Prevention | Krista Queen, Yan Li, Ying Tao, Jing Zhang, Anna Uehara, Anna Montmayeur, Clinton R. Paden, Peter W. Cook, Rachel Marine, Mili Sheth, Jasmine Padilla, Sarah Nobles, Mark Burroughs, Lori Rowe, Haibin Wang, Ben L. Rambo-Martin, Dhwani Batra, Justin Lee, Suxiang Tong |
| EPI_ISL_1094379 | PA Department of Health, Bureau of Laboratories | Respiratory Viruses Branch, Division of Viral Diseases, Centers for Disease Control and Prevention | Krista Queen, Yan Li, Ying Tao, Jing Zhang, Anna Uehara, Anna Montmayeur, Clinton R. Paden, Peter W. Cook, Rachel Marine, Mili Sheth, Jasmine Padilla, Sarah Nobles, Mark Burroughs, Lori Rowe, Haibin Wang, Ben L. Rambo-Martin, Dhwani Batra, Justin Lee, Suxiang Tong |
| EPI_ISL_1094380 | AK State Public Health Lab, State Health Department | Respiratory Viruses Branch, Division of Viral Diseases, Centers for Disease Control and Prevention | Krista Queen, Yan Li, Ying Tao, Jing Zhang, Anna Uehara, Anna Montmayeur, Clinton R. Paden, Peter W. Cook, Rachel Marine, Mili Sheth, Jasmine Padilla, Sarah Nobles, Mark Burroughs, Lori Rowe, Haibin Wang, Ben L. Rambo-Martin, Dhwani Batra, Justin Lee, Suxiang Tong |
| EPI_ISL_1094381 | VA-Division of Consolidated Laboratory Services | Respiratory Viruses Branch, Division of Viral Diseases, Centers for Disease Control and Prevention | Krista Queen, Yan Li, Ying Tao, Jing Zhang, Anna Uehara, Anna Montmayeur, Clinton R. Paden, Peter W. Cook, Rachel Marine, Mili Sheth, Jasmine Padilla, Sarah Nobles, Mark Burroughs, Lori Rowe, Haibin Wang, Ben L. Rambo-Martin, Dhwani Batra, Justin Lee, Suxiang Tong |
| EPI_ISL_1094382, EPI_ISL_1094383, EPI_ISL_1094385, EPI_ISL_1094386, EPI_ISL_1094387 | NM Dept. Health, Scientific Laboratory Division | Respiratory Viruses Branch, Division of Viral Diseases, Centers for Disease Control and Prevention | Krista Queen, Yan Li, Ying Tao, Jing Zhang, Anna Uehara, Anna Montmayeur, Clinton R. Paden, Peter W. Cook, Rachel Marine, Mili Sheth, Jasmine Padilla, Sarah Nobles, Mark Burroughs, Lori Rowe, Haibin Wang, Ben L. Rambo-Martin, Dhwani Batra, Justin Lee, Suxiang Tong |
| EPI_ISL_1094388 | IA State Hygienic Laboratory | Respiratory Viruses Branch, Division of Viral Diseases, Centers for Disease Control and Prevention | Krista Queen, Yan Li, Ying Tao, Jing Zhang, Anna Uehara, Anna Montmayeur, Clinton R. Paden, Peter W. Cook, Rachel Marine, Mili Sheth, Jasmine Padilla, Sarah Nobles, Mark Burroughs, Lori Rowe, Haibin Wang, Ben L. Rambo-Martin, Dhwani Batra, Justin Lee, Suxiang Tong |
| EPI_ISL_1094389, EPI_ISL_1094390 | NV State Public Health Laboratory | Respiratory Viruses Branch, Division of Viral Diseases, Centers for Disease Control and Prevention | Krista Queen, Yan Li, Ying Tao, Jing Zhang, Anna Uehara, Anna Montmayeur, Clinton R. Paden, Peter W. Cook, Rachel Marine, Mili Sheth, Jasmine Padilla, Sarah Nobles, Mark Burroughs, Lori Rowe, Haibin Wang, Ben L. Rambo-Martin, Dhwani Batra, Justin Lee, Suxiang Tong |
| EPI_ISL_1094391 | OR State PHL-Virology/Immunology Section | Respiratory Viruses Branch, Division of Viral Diseases, Centers for Disease Control and Prevention | Krista Queen, Yan Li, Ying Tao, Jing Zhang, Anna Uehara, Anna Montmayeur, Clinton R. Paden, Peter W. Cook, Rachel Marine, Mili Sheth, Jasmine Padilla, Sarah Nobles, Mark Burroughs, Lori Rowe, Haibin Wang, Ben L. Rambo-Martin, Dhwani Batra, Justin Lee, Suxiang Tong |
| EPI_ISL_1094392, EPI_ISL_1094393, EPI_ISL_1094394, EPI_ISL_1094395, EPI_ISL_1094396, EPI_ISL_1094398, EPI_ISL_1094399, EPI_ISL_1094400, EPI_ISL_1094401, EPI_ISL_1094402 | WA State Department of Health | Respiratory Viruses Branch, Division of Viral Diseases, Centers for Disease Control and Prevention | Krista Queen, Yan Li, Ying Tao, Jing Zhang, Anna Uehara, Anna Montmayeur, Clinton R. Paden, Peter W. Cook, Rachel Marine, Mili Sheth, Jasmine Padilla, Sarah Nobles, Mark Burroughs, Lori Rowe, Haibin Wang, Ben L. Rambo-Martin, Dhwani Batra, Justin Lee, Suxiang Tong |
| EPI_ISL_1094403, EPI_ISL_1094404, EPI_ISL_1094405, EPI_ISL_1094406, EPI_ISL_1094407, EPI_ISL_1094408, EPI_ISL_1094409 | DC Public Health Lab/ Dept. of Forensic Sciences | Respiratory Viruses Branch, Division of Viral Diseases, Centers for Disease Control and Prevention | Krista Queen, Yan Li, Ying Tao, Jing Zhang, Anna Uehara, Anna Montmayeur, Clinton R. Paden, Peter W. Cook, Rachel Marine, Mili Sheth, Jasmine Padilla, Sarah Nobles, Mark Burroughs, Lori Rowe, Haibin Wang, Ben L. Rambo-Martin, Dhwani Batra, Justin Lee, Suxiang Tong |
| EPI_ISL_1094410, EPI_ISL_1094412 | LA Office of Public Health Laboratories | Respiratory Viruses Branch, Division of Viral Diseases, Centers for Disease Control and Prevention | Krista Queen, Yan Li, Ying Tao, Jing Zhang, Anna Uehara, Anna Montmayeur, Clinton R. Paden, Peter W. Cook, Rachel Marine, Mili Sheth, Jasmine Padilla, Sarah Nobles, Mark Burroughs, Lori Rowe, Haibin Wang, Ben L. Rambo-Martin, Dhwani Batra, Justin Lee, Suxiang Tong |
| EPI_ISL_1094413, EPI_ISL_1094415 | MT Public Health Laboratory | Respiratory Viruses Branch, Division of Viral Diseases, Centers for Disease Control and Prevention | Krista Queen, Yan Li, Ying Tao, Jing Zhang, Anna Uehara, Anna Montmayeur, Clinton R. Paden, Peter W. Cook, Rachel Marine, Mili Sheth, Jasmine Padilla, Sarah Nobles, Mark Burroughs, Lori Rowe, Haibin Wang, Ben L. Rambo-Martin, Dhwani Batra, Justin Lee, Suxiang Tong |

| | | | |
|---|---|---|---|
| EPI_ISL_1094417, EPI_ISL_1094418, EPI_ISL_1094419, EPI_ISL_1094420, EPI_ISL_1094421, EPI_ISL_1094422, EPI_ISL_1094423, EPI_ISL_1094424 | MI - Michigan Department of Health and Human Services - Bureau of Laboratories | Respiratory Viruses Branch, Division of Viral Diseases, Centers for Disease Control and Prevention | Krista Queen, Yan Li, Ying Tao, Jing Zhang, Anna Uehara, Anna Montmayeur, Clinton R. Paden, Peter W. Cook, Rachel Marine, Mili Sheth, Jasmine Padilla, Sarah Nobles, Mark Burroughs, Lori Rowe, Haibin Wang, Ben L. Rambo-Martin, Dhwani Batra, Justin Lee, Suxiang Tong |
| EPI_ISL_1094425, EPI_ISL_1094426, EPI_ISL_1094427, EPI_ISL_1094429, EPI_ISL_1094430, EPI_ISL_1094431, EPI_ISL_1094432 | GA Department of Public Health Laboratory | Respiratory Viruses Branch, Division of Viral Diseases, Centers for Disease Control and Prevention | Krista Queen, Yan Li, Ying Tao, Jing Zhang, Anna Uehara, Anna Montmayeur, Clinton R. Paden, Peter W. Cook, Rachel Marine, Mili Sheth, Jasmine Padilla, Sarah Nobles, Mark Burroughs, Lori Rowe, Haibin Wang, Ben L. Rambo-Martin, Dhwani Batra, Justin Lee, Suxiang Tong |
| EPI_ISL_1094433, EPI_ISL_1094434, EPI_ISL_1094435, EPI_ISL_1094436, EPI_ISL_1094437, EPI_ISL_1094438, EPI_ISL_1094439, EPI_ISL_1094440, EPI_ISL_1094441, EPI_ISL_1094443, EPI_ISL_1094444 | | | |
| see above | NYSDOH Wadsworth Center, Virology Lab | Respiratory Viruses Branch, Division of Viral Diseases, Centers for Disease Control and Prevention | Krista Queen, Yan Li, Ying Tao, Jing Zhang, Anna Uehara, Anna Montmayeur, Clinton R. Paden, Peter W. Cook, Rachel Marine, Mili Sheth, Jasmine Padilla, Sarah Nobles, Mark Burroughs, Lori Rowe, Haibin Wang, Ben L. Rambo-Martin, Dhwani Batra, Justin Lee, Suxiang Tong |
| EPI_ISL_1094445, EPI_ISL_1094446, EPI_ISL_1094447, EPI_ISL_1094448, EPI_ISL_1094450, EPI_ISL_1094451, EPI_ISL_1094452, EPI_ISL_1094453, EPI_ISL_1094454, EPI_ISL_1094455, EPI_ISL_1094456, EPI_ISL_1094457 | | | |
| see above | PA Department of Health, Bureau of Laboratories | Respiratory Viruses Branch, Division of Viral Diseases, Centers for Disease Control and Prevention | Krista Queen, Yan Li, Ying Tao, Jing Zhang, Anna Uehara, Anna Montmayeur, Clinton R. Paden, Peter W. Cook, Rachel Marine, Mili Sheth, Jasmine Padilla, Sarah Nobles, Mark Burroughs, Lori Rowe, Haibin Wang, Ben L. Rambo-Martin, Dhwani Batra, Justin Lee, Suxiang Tong |
| EPI_ISL_1094467, EPI_ISL_1094468, EPI_ISL_1094469, EPI_ISL_1094470, EPI_ISL_1094471, EPI_ISL_1094472 | DE Public Health Laboratory | Respiratory Viruses Branch, Division of Viral Diseases, Centers for Disease Control and Prevention | Krista Queen, Yan Li, Ying Tao, Jing Zhang, Anna Uehara, Anna Montmayeur, Clinton R. Paden, Peter W. Cook, Rachel Marine, Mili Sheth, Jasmine Padilla, Sarah Nobles, Mark Burroughs, Lori Rowe, Haibin Wang, Ben L. Rambo-Martin, Dhwani Batra, Justin Lee, Suxiang Tong |
| EPI_ISL_1094488, EPI_ISL_1094489, EPI_ISL_1094490, EPI_ISL_1094491 | IL Dept. of Public Health Springfield Laboratory | Respiratory Viruses Branch, Division of Viral Diseases, Centers for Disease Control and Prevention | Krista Queen, Yan Li, Ying Tao, Jing Zhang, Anna Uehara, Anna Montmayeur, Clinton R. Paden, Peter W. Cook, Rachel Marine, Mili Sheth, Jasmine Padilla, Sarah Nobles, Mark Burroughs, Lori Rowe, Haibin Wang, Ben L. Rambo-Martin, Dhwani Batra, Justin Lee, Suxiang Tong |
| EPI_ISL_1094492, EPI_ISL_1094493, EPI_ISL_1094494 | NE Public Health Laboratory | Respiratory Viruses Branch, Division of Viral Diseases, Centers for Disease Control and Prevention | Krista Queen, Yan Li, Ying Tao, Jing Zhang, Anna Uehara, Anna Montmayeur, Clinton R. Paden, Peter W. Cook, Rachel Marine, Mili Sheth, Jasmine Padilla, Sarah Nobles, Mark Burroughs, Lori Rowe, Haibin Wang, Ben L. Rambo-Martin, Dhwani Batra, Justin Lee, Suxiang Tong |
| EPI_ISL_1094495, EPI_ISL_1094496 | WA State Department of Health | Respiratory Viruses Branch, Division of Viral Diseases, Centers for Disease Control and Prevention | Krista Queen, Yan Li, Ying Tao, Jing Zhang, Anna Uehara, Anna Montmayeur, Clinton R. Paden, Peter W. Cook, Rachel Marine, Mili Sheth, Jasmine Padilla, Sarah Nobles, Mark Burroughs, Lori Rowe, Haibin Wang, Ben L. Rambo-Martin, Dhwani Batra, Justin Lee, Suxiang Tong |
| EPI_ISL_1094497, EPI_ISL_1094498, EPI_ISL_1094499, EPI_ISL_1094500, EPI_ISL_1094501, EPI_ISL_1094502 | OK Public Health Laboratory, Oklahoma State DOH | Respiratory Viruses Branch, Division of Viral Diseases, Centers for Disease Control and Prevention | Krista Queen, Yan Li, Ying Tao, Jing Zhang, Anna Uehara, Anna Montmayeur, Clinton R. Paden, Peter W. Cook, Rachel Marine, Mili Sheth, Jasmine Padilla, Sarah Nobles, Mark Burroughs, Lori Rowe, Haibin Wang, Ben L. Rambo-Martin, Dhwani Batra, Justin Lee, Suxiang Tong |
| EPI_ISL_1094503, EPI_ISL_1094504, EPI_ISL_1094505, EPI_ISL_1094506, EPI_ISL_1094507, EPI_ISL_1094508, EPI_ISL_1094509, EPI_ISL_1094510 | UT-Unified State Labs: Public Health Utah DOH | Respiratory Viruses Branch, Division of Viral Diseases, Centers for Disease Control and Prevention | Krista Queen, Yan Li, Ying Tao, Jing Zhang, Anna Uehara, Anna Montmayeur, Clinton R. Paden, Peter W. Cook, Rachel Marine, Mili Sheth, Jasmine Padilla, Sarah Nobles, Mark Burroughs, Lori Rowe, Haibin Wang, Ben L. Rambo-Martin, Dhwani Batra, Justin Lee, Suxiang Tong |
| EPI_ISL_1094511, EPI_ISL_1094512, EPI_ISL_1094513, EPI_ISL_1094514, EPI_ISL_1094515, EPI_ISL_1094516 | CO Dept. of Public Health and Environment, Lab Services Division | Respiratory Viruses Branch, Division of Viral Diseases, Centers for Disease Control and Prevention | Krista Queen, Yan Li, Ying Tao, Jing Zhang, Anna Uehara, Anna Montmayeur, Clinton R. Paden, Peter W. Cook, Rachel Marine, Mili Sheth, Jasmine Padilla, Sarah Nobles, Mark Burroughs, Lori Rowe, Haibin Wang, Ben L. Rambo-Martin, Dhwani Batra, Justin Lee, Suxiang Tong |
| EPI_ISL_1094518, EPI_ISL_1094519 | MT Public Health Laboratory | Respiratory Viruses Branch, Division of Viral Diseases, Centers for Disease Control and Prevention | Krista Queen, Yan Li, Ying Tao, Jing Zhang, Anna Uehara, Anna Montmayeur, Clinton R. Paden, Peter W. Cook, Rachel Marine, Mili Sheth, Jasmine Padilla, Sarah Nobles, Mark Burroughs, Lori Rowe, Haibin Wang, Ben L. Rambo-Martin, Dhwani Batra, Justin Lee, Suxiang Tong |
| EPI_ISL_1094521, EPI_ISL_1094522, EPI_ISL_1094523, EPI_ISL_1094524, EPI_ISL_1094525, EPI_ISL_1094526, EPI_ISL_1094527, EPI_ISL_1094528, EPI_ISL_1094529, EPI_ISL_1094530, EPI_ISL_1094531, EPI_ISL_1094532, EPI_ISL_1094533, EPI_ISL_1094534 | | | |
| see above | OH Department of Health Laboratory | Respiratory Viruses Branch, Division of Viral Diseases, Centers for Disease Control and Prevention | Krista Queen, Yan Li, Ying Tao, Jing Zhang, Anna Uehara, Anna Montmayeur, Clinton R. Paden, Peter W. Cook, Rachel Marine, Mili Sheth, Jasmine Padilla, Sarah Nobles, Mark Burroughs, Lori Rowe, Haibin Wang, Ben L. Rambo-Martin, Dhwani Batra, Justin Lee, Suxiang Tong |
| EPI_ISL_1094535 | NC State Laboratory of Public Health | Respiratory Viruses Branch, Division of Viral Diseases, Centers for Disease Control and Prevention | Krista Queen, Yan Li, Ying Tao, Jing Zhang, Anna Uehara, Anna Montmayeur, Clinton R. Paden, Peter W. Cook, Rachel Marine, Mili Sheth, Jasmine Padilla, Sarah Nobles, Mark Burroughs, Lori Rowe, Haibin Wang, Ben L. Rambo-Martin, Dhwani Batra, Justin Lee, Suxiang Tong |
| EPI_ISL_1094536 | CDPH, Microbial Diseases Laboratory | Respiratory Viruses Branch, Division of Viral Diseases, Centers for Disease Control and Prevention | Krista Queen, Yan Li, Ying Tao, Jing Zhang, Anna Uehara, Anna Montmayeur, Clinton R. Paden, Peter W. Cook, Rachel Marine, Mili Sheth, Jasmine Padilla, Sarah Nobles, Mark Burroughs, Lori Rowe, Haibin Wang, Ben L. Rambo-Martin, Dhwani Batra, Justin Lee, Suxiang Tong |
| EPI_ISL_1094537, EPI_ISL_1094538, EPI_ISL_1094539 | FL Bur. of Public Health Laboratories-Jacksonville | Respiratory Viruses Branch, Division of Viral Diseases, Centers for Disease Control and Prevention | Krista Queen, Yan Li, Ying Tao, Jing Zhang, Anna Uehara, Anna Montmayeur, Clinton R. Paden, Peter W. Cook, Rachel Marine, Mili Sheth, Jasmine Padilla, Sarah Nobles, Mark Burroughs, Lori Rowe, Haibin Wang, Ben L. Rambo-Martin, Dhwani Batra, Justin Lee, Suxiang Tong |
| EPI_ISL_1094540, EPI_ISL_1094541 | FL Bureau of Public Health Laboratories-Tampa | Respiratory Viruses Branch, Division of Viral Diseases, Centers for Disease Control and Prevention | Krista Queen, Yan Li, Ying Tao, Jing Zhang, Anna Uehara, Anna Montmayeur, Clinton R. Paden, Peter W. Cook, Rachel Marine, Mili Sheth, Jasmine Padilla, Sarah Nobles, Mark Burroughs, Lori Rowe, Haibin Wang, Ben L. Rambo-Martin, Dhwani Batra, Justin Lee, Suxiang Tong |
| EPI_ISL_1094542, EPI_ISL_1094543 | SD Public Health Laboratory | Respiratory Viruses Branch, Division of Viral Diseases, Centers for Disease Control and Prevention | Krista Queen, Yan Li, Ying Tao, Jing Zhang, Anna Uehara, Anna Montmayeur, Clinton R. Paden, Peter W. Cook, Rachel Marine, Mili Sheth, Jasmine Padilla, Sarah Nobles, Mark Burroughs, Lori Rowe, Haibin Wang, Ben L. Rambo-Martin, Dhwani Batra, Justin Lee, Suxiang Tong |
| EPI_ISL_1094545, EPI_ISL_1094546 | FL Bureau of Public Health Laboratories-Miami | Respiratory Viruses Branch, Division of Viral Diseases, Centers for Disease Control and Prevention | Krista Queen, Yan Li, Ying Tao, Jing Zhang, Anna Uehara, Anna Montmayeur, Clinton R. Paden, Peter W. Cook, Rachel Marine, Mili Sheth, Jasmine Padilla, Sarah Nobles, Mark Burroughs, Lori Rowe, Haibin Wang, Ben L. Rambo-Martin, Dhwani Batra, Justin Lee, Suxiang Tong |
| EPI_ISL_1094547, EPI_ISL_1094548, EPI_ISL_1094549, EPI_ISL_1094550, EPI_ISL_1094551, EPI_ISL_1094552, EPI_ISL_1094553, EPI_ISL_1094554, EPI_ISL_1094555 | SC Dept of Health and Env. Control-Bureau of Laboratories | Respiratory Viruses Branch, Division of Viral Diseases, Centers for Disease Control and Prevention | Krista Queen, Yan Li, Ying Tao, Jing Zhang, Anna Uehara, Anna Montmayeur, Clinton R. Paden, Peter W. Cook, Rachel Marine, Mili Sheth, Jasmine Padilla, Sarah Nobles, Mark Burroughs, Lori Rowe, Haibin Wang, Ben L. Rambo-Martin, Dhwani Batra, Justin Lee, Suxiang Tong |
| EPI_ISL_1094556, EPI_ISL_1094557, EPI_ISL_1094558 | VT Dept. of Health Laboratory | Respiratory Viruses Branch, Division of Viral Diseases, Centers for Disease Control and Prevention | Krista Queen, Yan Li, Ying Tao, Jing Zhang, Anna Uehara, Anna Montmayeur, Clinton R. Paden, Peter W. Cook, Rachel Marine, Mili Sheth, Jasmine Padilla, Sarah Nobles, Mark Burroughs, Lori Rowe, Haibin Wang, Ben L. Rambo-Martin, Dhwani Batra, Justin Lee, Suxiang Tong |
| EPI_ISL_1094559, EPI_ISL_1094560, EPI_ISL_1094561, EPI_ISL_1094562, EPI_ISL_1094563, EPI_ISL_1094564, EPI_ISL_1094565, EPI_ISL_1094566, EPI_ISL_1094567, EPI_ISL_1094568, EPI_ISL_1094569, EPI_ISL_1094570, EPI_ISL_1094571 | | | |
| see above | CA-Los Angeles County Public Health Laboratory | Respiratory Viruses Branch, Division of Viral Diseases, Centers for Disease Control and | Krista Queen, Yan Li, Ying Tao, Jing Zhang, Anna Uehara, Anna Montmayeur, Clinton R. Paden, Peter W. Cook, Rachel Marine, Mili Sheth, Jasmine Padilla, Sarah Nobles, Mark Burroughs, Lori Rowe, Haibin Wang, Ben L. Rambo-Martin, Dhwani Batra, Justin Lee, Suxiang Tong |

| | | | |
|---|---|---|---|
| | | Prevention | |
| EPI_ISL_1094572, EPI_ISL_1094573, EPI_ISL_1094574, EPI_ISL_1094575, EPI_ISL_1094576 | VA-Division of Consolidated Laboratory Services | Respiratory Viruses Branch, Division of Viral Diseases, Centers for Disease Control and Prevention | Krista Queen, Yan Li, Ying Tao, Jing Zhang, Anna Uehara, Anna Montmayeur, Clinton R. Paden, Peter W. Cook, Rachel Marine, Mili Sheth, Jasmine Padilla, Sarah Nobles, Mark Burroughs, Lori Rowe, Haibin Wang, Ben L. Rambo-Martin, Dhwani Batra, Justin Lee, Suxiang Tong |
| EPI_ISL_1094577, EPI_ISL_1094579, EPI_ISL_1094580, EPI_ISL_1094581, EPI_ISL_1094583, EPI_ISL_1094584, EPI_ISL_1094585, EPI_ISL_1094586, EPI_ISL_1094587, EPI_ISL_1094588, EPI_ISL_1094589 | | | |
| see above | CDPH, Viral and Rickettsial Disease Laboratory | Respiratory Viruses Branch, Division of Viral Diseases, Centers for Disease Control and Prevention | Krista Queen, Yan Li, Ying Tao, Jing Zhang, Anna Uehara, Anna Montmayeur, Clinton R. Paden, Peter W. Cook, Rachel Marine, Mili Sheth, Jasmine Padilla, Sarah Nobles, Mark Burroughs, Lori Rowe, Haibin Wang, Ben L. Rambo-Martin, Dhwani Batra, Justin Lee, Suxiang Tong |
| EPI_ISL_1094590, EPI_ISL_1094591, EPI_ISL_1094592, EPI_ISL_1094593, EPI_ISL_1094594, EPI_ISL_1094595, EPI_ISL_1094596, EPI_ISL_1094597, EPI_ISL_1094598, EPI_ISL_1094599 | WI State Laboratory of Hygiene | Respiratory Viruses Branch, Division of Viral Diseases, Centers for Disease Control and Prevention | Krista Queen, Yan Li, Ying Tao, Jing Zhang, Anna Uehara, Anna Montmayeur, Clinton R. Paden, Peter W. Cook, Rachel Marine, Mili Sheth, Jasmine Padilla, Sarah Nobles, Mark Burroughs, Lori Rowe, Haibin Wang, Ben L. Rambo-Martin, Dhwani Batra, Justin Lee, Suxiang Tong |
| EPI_ISL_1094602, EPI_ISL_1094603, EPI_ISL_1094604, EPI_ISL_1094605, EPI_ISL_1094606, EPI_ISL_1094607, EPI_ISL_1094608, EPI_ISL_1094609, EPI_ISL_1094610, EPI_ISL_1094611, EPI_ISL_1094612, EPI_ISL_1094613, EPI_ISL_1094614, EPI_ISL_1094615, EPI_ISL_1094616, EPI_ISL_1094617, EPI_ISL_1094618, EPI_ISL_1094619, EPI_ISL_1094621 | | | |
| see above | TN Division of Laboratory Services | Respiratory Viruses Branch, Division of Viral Diseases, Centers for Disease Control and Prevention | Krista Queen, Yan Li, Ying Tao, Jing Zhang, Anna Uehara, Anna Montmayeur, Clinton R. Paden, Peter W. Cook, Rachel Marine, Mili Sheth, Jasmine Padilla, Sarah Nobles, Mark Burroughs, Lori Rowe, Haibin Wang, Ben L. Rambo-Martin, Dhwani Batra, Justin Lee, Suxiang Tong |
| EPI_ISL_1094622, EPI_ISL_1094623, EPI_ISL_1094624, EPI_ISL_1094625, EPI_ISL_1094626, EPI_ISL_1094627, EPI_ISL_1094628, EPI_ISL_1094629, EPI_ISL_1094630, EPI_ISL_1094631, EPI_ISL_1094632, EPI_ISL_1094633, EPI_ISL_1094634, EPI_ISL_1094635, EPI_ISL_1094636, EPI_ISL_1094637, EPI_ISL_1094638 | | | |
| see above | DE Public Health Laboratory | Respiratory Viruses Branch, Division of Viral Diseases, Centers for Disease Control and Prevention | Krista Queen, Yan Li, Ying Tao, Jing Zhang, Anna Uehara, Anna Montmayeur, Clinton R. Paden, Peter W. Cook, Rachel Marine, Mili Sheth, Jasmine Padilla, Sarah Nobles, Mark Burroughs, Lori Rowe, Haibin Wang, Ben L. Rambo-Martin, Dhwani Batra, Justin Lee, Suxiang Tong |
| EPI_ISL_1094639, EPI_ISL_1094640, EPI_ISL_1094641, EPI_ISL_1094642, EPI_ISL_1094643, EPI_ISL_1094644, EPI_ISL_1094645, EPI_ISL_1094646, EPI_ISL_1094647, EPI_ISL_1094649, EPI_ISL_1094650, EPI_ISL_1094653, EPI_ISL_1094654, EPI_ISL_1094655 | | | |
| see above | KS Health and Environmental Laboratories | Respiratory Viruses Branch, Division of Viral Diseases, Centers for Disease Control and Prevention | Krista Queen, Yan Li, Ying Tao, Jing Zhang, Anna Uehara, Anna Montmayeur, Clinton R. Paden, Peter W. Cook, Rachel Marine, Mili Sheth, Jasmine Padilla, Sarah Nobles, Mark Burroughs, Lori Rowe, Haibin Wang, Ben L. Rambo-Martin, Dhwani Batra, Justin Lee, Suxiang Tong |
| EPI_ISL_1094657, EPI_ISL_1094658, EPI_ISL_1094659, EPI_ISL_1094660 | RI State Health Laboratories | Respiratory Viruses Branch, Division of Viral Diseases, Centers for Disease Control and Prevention | Krista Queen, Yan Li, Ying Tao, Jing Zhang, Anna Uehara, Anna Montmayeur, Clinton R. Paden, Peter W. Cook, Rachel Marine, Mili Sheth, Jasmine Padilla, Sarah Nobles, Mark Burroughs, Lori Rowe, Haibin Wang, Ben L. Rambo-Martin, Dhwani Batra, Justin Lee, Suxiang Tong |
| EPI_ISL_1094661, EPI_ISL_1094662, EPI_ISL_1094663, EPI_ISL_1094664 | NM Dept. Health, Scientific Laboratory Division | Respiratory Viruses Branch, Division of Viral Diseases, Centers for Disease Control and Prevention | Krista Queen, Yan Li, Ying Tao, Jing Zhang, Anna Uehara, Anna Montmayeur, Clinton R. Paden, Peter W. Cook, Rachel Marine, Mili Sheth, Jasmine Padilla, Sarah Nobles, Mark Burroughs, Lori Rowe, Haibin Wang, Ben L. Rambo-Martin, Dhwani Batra, Justin Lee, Suxiang Tong |
| EPI_ISL_1094666, EPI_ISL_1094668, EPI_ISL_1094670, EPI_ISL_1094671 | WVDHHR - Office of Laboratory Services | Respiratory Viruses Branch, Division of Viral Diseases, Centers for Disease Control and Prevention | Krista Queen, Yan Li, Ying Tao, Jing Zhang, Anna Uehara, Anna Montmayeur, Clinton R. Paden, Peter W. Cook, Rachel Marine, Mili Sheth, Jasmine Padilla, Sarah Nobles, Mark Burroughs, Lori Rowe, Haibin Wang, Ben L. Rambo-Martin, Dhwani Batra, Justin Lee, Suxiang Tong |
| EPI_ISL_1094672, EPI_ISL_1094674, EPI_ISL_1094676, EPI_ISL_1094677, EPI_ISL_1094679, EPI_ISL_1094680 | ME Health and Environmental Testing Laboratory | Respiratory Viruses Branch, Division of Viral Diseases, Centers for Disease Control and Prevention | Krista Queen, Yan Li, Ying Tao, Jing Zhang, Anna Uehara, Anna Montmayeur, Clinton R. Paden, Peter W. Cook, Rachel Marine, Mili Sheth, Jasmine Padilla, Sarah Nobles, Mark Burroughs, Lori Rowe, Haibin Wang, Ben L. Rambo-Martin, Dhwani Batra, Justin Lee, Suxiang Tong |
| EPI_ISL_1094687, EPI_ISL_1094688, EPI_ISL_1094689, EPI_ISL_1094690, EPI_ISL_1094691, EPI_ISL_1094692, EPI_ISL_1094693, EPI_ISL_1094694, EPI_ISL_1094695, EPI_ISL_1094696, EPI_ISL_1094697, EPI_ISL_1094698, EPI_ISL_1094699, EPI_ISL_1094700, EPI_ISL_1094701, EPI_ISL_1094702, EPI_ISL_1094703, EPI_ISL_1094704, EPI_ISL_1094705, EPI_ISL_1094706 | | | |
| see above | IA State Hygienic Laboratory | Respiratory Viruses Branch, Division of Viral Diseases, Centers for Disease Control and Prevention | Krista Queen, Yan Li, Ying Tao, Jing Zhang, Anna Uehara, Anna Montmayeur, Clinton R. Paden, Peter W. Cook, Rachel Marine, Mili Sheth, Jasmine Padilla, Sarah Nobles, Mark Burroughs, Lori Rowe, Haibin Wang, Ben L. Rambo-Martin, Dhwani Batra, Justin Lee, Suxiang Tong |
| EPI_ISL_1094707, EPI_ISL_1094708, EPI_ISL_1094709, EPI_ISL_1094710, EPI_ISL_1094711, EPI_ISL_1094712, EPI_ISL_1094713, EPI_ISL_1094714, EPI_ISL_1094715, EPI_ISL_1094716, EPI_ISL_1094717, EPI_ISL_1094718, EPI_ISL_1094719 | | | |
| see above | AK State Public Health Lab, State Health Department | Respiratory Viruses Branch, Division of Viral Diseases, Centers for Disease Control and Prevention | Krista Queen, Yan Li, Ying Tao, Jing Zhang, Anna Uehara, Anna Montmayeur, Clinton R. Paden, Peter W. Cook, Rachel Marine, Mili Sheth, Jasmine Padilla, Sarah Nobles, Mark Burroughs, Lori Rowe, Haibin Wang, Ben L. Rambo-Martin, Dhwani Batra, Justin Lee, Suxiang Tong |
| EPI_ISL_1094720, EPI_ISL_1094721, EPI_ISL_1094722 | NV State Public Health Laboratory | Respiratory Viruses Branch, Division of Viral Diseases, Centers for Disease Control and Prevention | Krista Queen, Yan Li, Ying Tao, Jing Zhang, Anna Uehara, Anna Montmayeur, Clinton R. Paden, Peter W. Cook, Rachel Marine, Mili Sheth, Jasmine Padilla, Sarah Nobles, Mark Burroughs, Lori Rowe, Haibin Wang, Ben L. Rambo-Martin, Dhwani Batra, Justin Lee, Suxiang Tong |
| EPI_ISL_1094723, EPI_ISL_1094724, EPI_ISL_1094725 | IL Dept. of Public Health Springfield Laboratory | Respiratory Viruses Branch, Division of Viral Diseases, Centers for Disease Control and Prevention | Krista Queen, Yan Li, Ying Tao, Jing Zhang, Anna Uehara, Anna Montmayeur, Clinton R. Paden, Peter W. Cook, Rachel Marine, Mili Sheth, Jasmine Padilla, Sarah Nobles, Mark Burroughs, Lori Rowe, Haibin Wang, Ben L. Rambo-Martin, Dhwani Batra, Justin Lee, Suxiang Tong |
| EPI_ISL_1094726, EPI_ISL_1094727, EPI_ISL_1094728, EPI_ISL_1094729, EPI_ISL_1094730, EPI_ISL_1094731, EPI_ISL_1094732, EPI_ISL_1094733, EPI_ISL_1094734, EPI_ISL_1094735 | OR State PHL-Virology/Immunology Section | Respiratory Viruses Branch, Division of Viral Diseases, Centers for Disease Control and Prevention | Krista Queen, Yan Li, Ying Tao, Jing Zhang, Anna Uehara, Anna Montmayeur, Clinton R. Paden, Peter W. Cook, Rachel Marine, Mili Sheth, Jasmine Padilla, Sarah Nobles, Mark Burroughs, Lori Rowe, Haibin Wang, Ben L. Rambo-Martin, Dhwani Batra, Justin Lee, Suxiang Tong |
| EPI_ISL_1094736, EPI_ISL_1094737, EPI_ISL_1094738, EPI_ISL_1094739, EPI_ISL_1094740, EPI_ISL_1094741, EPI_ISL_1094742, EPI_ISL_1094743, EPI_ISL_1094744, EPI_ISL_1094745, EPI_ISL_1094746, EPI_ISL_1094747, EPI_ISL_1094748, EPI_ISL_1094749, EPI_ISL_1094750, EPI_ISL_1094751, EPI_ISL_1094752, EPI_ISL_1094753 | | | |
| see above | NE Public Health Laboratory | Respiratory Viruses Branch, Division of Viral Diseases, Centers for Disease Control and Prevention | Krista Queen, Yan Li, Ying Tao, Jing Zhang, Anna Uehara, Anna Montmayeur, Clinton R. Paden, Peter W. Cook, Rachel Marine, Mili Sheth, Jasmine Padilla, Sarah Nobles, Mark Burroughs, Lori Rowe, Haibin Wang, Ben L. Rambo-Martin, Dhwani Batra, Justin Lee, Suxiang Tong |
| EPI_ISL_1094754, EPI_ISL_1094755, EPI_ISL_1094756, EPI_ISL_1094757, EPI_ISL_1094758, EPI_ISL_1094759, EPI_ISL_1094760, EPI_ISL_1094761, EPI_ISL_1094762, EPI_ISL_1094763, EPI_ISL_1094764, EPI_ISL_1094765, EPI_ISL_1094766, EPI_ISL_1094767, EPI_ISL_1094768, EPI_ISL_1094769, EPI_ISL_1094770, EPI_ISL_1094771 | | | |
| see above | WA State Department of Health | Respiratory Viruses Branch, Division of Viral Diseases, Centers for Disease Control and Prevention | Krista Queen, Yan Li, Ying Tao, Jing Zhang, Anna Uehara, Anna Montmayeur, Clinton R. Paden, Peter W. Cook, Rachel Marine, Mili Sheth, Jasmine Padilla, Sarah Nobles, Mark Burroughs, Lori Rowe, Haibin Wang, Ben L. Rambo-Martin, Dhwani Batra, Justin Lee, Suxiang Tong |
| EPI_ISL_1094772, EPI_ISL_1094773, EPI_ISL_1094774, EPI_ISL_1094775 | HI Dept. of Health, State Laboratories Division | Respiratory Viruses Branch, Division of Viral Diseases, Centers for Disease Control and Prevention | Krista Queen, Yan Li, Ying Tao, Jing Zhang, Anna Uehara, Anna Montmayeur, Clinton R. Paden, Peter W. Cook, Rachel Marine, Mili Sheth, Jasmine Padilla, Sarah Nobles, Mark Burroughs, Lori Rowe, Haibin Wang, Ben L. Rambo-Martin, Dhwani Batra, Justin Lee, Suxiang Tong |
| EPI_ISL_1094776, EPI_ISL_1094778, EPI_ISL_1094779, EPI_ISL_1094780, EPI_ISL_1094781, EPI_ISL_1094782, EPI_ISL_1094783, EPI_ISL_1094784, EPI_ISL_1094785, EPI_ISL_1094786, EPI_ISL_1094787, EPI_ISL_1094788, EPI_ISL_1094789, EPI_ISL_1094790 | | | |
| see above | IN State Department of Health Laboratory Services | Respiratory Viruses Branch, Division of Viral Diseases, Centers for Disease Control and Prevention | Krista Queen, Yan Li, Ying Tao, Jing Zhang, Anna Uehara, Anna Montmayeur, Clinton R. Paden, Peter W. Cook, Rachel Marine, Mili Sheth, Jasmine Padilla, Sarah Nobles, Mark Burroughs, Lori Rowe, Haibin Wang, Ben L. Rambo-Martin, Dhwani Batra, Justin Lee, Suxiang Tong |
| EPI_ISL_1094791, EPI_ISL_1094792, EPI_ISL_1094793, EPI_ISL_1094794, EPI_ISL_1094795, EPI_ISL_1094796, EPI_ISL_1094797, EPI_ISL_1094798, EPI_ISL_1094799, EPI_ISL_1094800, EPI_ISL_1094801, EPI_ISL_1094802, EPI_ISL_1094803, EPI_ISL_1094804, EPI_ISL_1094805, EPI_ISL_1094806, EPI_ISL_1094807, EPI_ISL_1094808, EPI_ISL_1094809, EPI_ISL_1094810, EPI_ISL_1094811, EPI_ISL_1094812, EPI_ISL_1094813 | | | |
| see above | OK Public Health Laboratory, Oklahoma State DOH | Respiratory Viruses Branch, Division of Viral Diseases, Centers for Disease Control and Prevention | Krista Queen, Yan Li, Ying Tao, Jing Zhang, Anna Uehara, Anna Montmayeur, Clinton R. Paden, Peter W. Cook, Rachel Marine, Mili Sheth, Jasmine Padilla, Sarah Nobles, Mark Burroughs, Lori Rowe, Haibin Wang, Ben L. Rambo-Martin, Dhwani Batra, Justin Lee, Suxiang Tong |
| EPI_ISL_1094814, EPI_ISL_1094815, | AR Dept. of Health-Public Health Lab | Respiratory Viruses Branch, Division of Viral Diseases, Centers for Disease Control and | Krista Queen, Yan Li, Ying Tao, Jing Zhang, Anna Uehara, Anna Montmayeur, Clinton R. Paden, Peter W. Cook, Rachel Marine, Mili Sheth, Jasmine Padilla, Sarah Nobles, Mark Burroughs, Lori Rowe, Haibin Wang, Ben L. Rambo-Martin, Dhwani Batra, Justin Lee, Suxiang Tong |

| | | | |
|---|---|---|---|
| EPI_ISL_1094816, EPI_ISL_1094817, EPI_ISL_1094818, EPI_ISL_1094821, EPI_ISL_1094824, EPI_ISL_1094826, EPI_ISL_1094830 | | Prevention | |
| EPI_ISL_1094834, EPI_ISL_1094835, EPI_ISL_1094837, EPI_ISL_1094838, EPI_ISL_1094844, EPI_ISL_1094847, EPI_ISL_1094848, EPI_ISL_1094849 | UT-Unified State Labs: Public Health Utah DOH | Respiratory Viruses Branch, Division of Viral Diseases, Centers for Disease Control and Prevention | Krista Queen, Yan Li, Ying Tao, Jing Zhang, Anna Uehara, Anna Montmayeur, Clinton R. Paden, Peter W. Cook, Rachel Marine, Mili Sheth, Jasmine Padilla, Sarah Nobles, Mark Burroughs, Lori Rowe, Haibin Wang, Ben L. Rambo-Martin, Dhwani Batra, Justin Lee, Suxiang Tong |
| EPI_ISL_1094851, EPI_ISL_1094852, EPI_ISL_1094853, EPI_ISL_1094854 | DC Public Health Lab/ Dept. of Forensic Sciences | Respiratory Viruses Branch, Division of Viral Diseases, Centers for Disease Control and Prevention | Krista Queen, Yan Li, Ying Tao, Jing Zhang, Anna Uehara, Anna Montmayeur, Clinton R. Paden, Peter W. Cook, Rachel Marine, Mili Sheth, Jasmine Padilla, Sarah Nobles, Mark Burroughs, Lori Rowe, Haibin Wang, Ben L. Rambo-Martin, Dhwani Batra, Justin Lee, Suxiang Tong |
| EPI_ISL_1094855, EPI_ISL_1094857, EPI_ISL_1094858, EPI_ISL_1094860, EPI_ISL_1094863, EPI_ISL_1094867, EPI_ISL_1094868, EPI_ISL_1094870 | WY Public Health Laboratory | Respiratory Viruses Branch, Division of Viral Diseases, Centers for Disease Control and Prevention | Krista Queen, Yan Li, Ying Tao, Jing Zhang, Anna Uehara, Anna Montmayeur, Clinton R. Paden, Peter W. Cook, Rachel Marine, Mili Sheth, Jasmine Padilla, Sarah Nobles, Mark Burroughs, Lori Rowe, Haibin Wang, Ben L. Rambo-Martin, Dhwani Batra, Justin Lee, Suxiang Tong |
| EPI_ISL_1094871, EPI_ISL_1094872, EPI_ISL_1094873, EPI_ISL_1094875, EPI_ISL_1094876, EPI_ISL_1094877, EPI_ISL_1094881, EPI_ISL_1094882, EPI_ISL_1094883, EPI_ISL_1094884, EPI_ISL_1094886 | | | |
| see above | LA Office of Public Health Laboratories | Respiratory Viruses Branch, Division of Viral Diseases, Centers for Disease Control and Prevention | Krista Queen, Yan Li, Ying Tao, Jing Zhang, Anna Uehara, Anna Montmayeur, Clinton R. Paden, Peter W. Cook, Rachel Marine, Mili Sheth, Jasmine Padilla, Sarah Nobles, Mark Burroughs, Lori Rowe, Haibin Wang, Ben L. Rambo-Martin, Dhwani Batra, Justin Lee, Suxiang Tong |
| EPI_ISL_1094888, EPI_ISL_1094889, EPI_ISL_1094890, EPI_ISL_1094891, EPI_ISL_1094892, EPI_ISL_1094893, EPI_ISL_1094894, EPI_ISL_1094895, EPI_ISL_1094896, EPI_ISL_1094897, EPI_ISL_1094898 | | | |
| see above | AL Dept. of Public Health Bureau of Clinical Laboratories | Respiratory Viruses Branch, Division of Viral Diseases, Centers for Disease Control and Prevention | Krista Queen, Yan Li, Ying Tao, Jing Zhang, Anna Uehara, Anna Montmayeur, Clinton R. Paden, Peter W. Cook, Rachel Marine, Mili Sheth, Jasmine Padilla, Sarah Nobles, Mark Burroughs, Lori Rowe, Haibin Wang, Ben L. Rambo-Martin, Dhwani Batra, Justin Lee, Suxiang Tong |
| EPI_ISL_1094899, EPI_ISL_1094900, EPI_ISL_1094901, EPI_ISL_1094902, EPI_ISL_1094903, EPI_ISL_1094904, EPI_ISL_1094905, EPI_ISL_1094906 | CO Dept. of Public Health and Environment, Lab Services Division | Respiratory Viruses Branch, Division of Viral Diseases, Centers for Disease Control and Prevention | Krista Queen, Yan Li, Ying Tao, Jing Zhang, Anna Uehara, Anna Montmayeur, Clinton R. Paden, Peter W. Cook, Rachel Marine, Mili Sheth, Jasmine Padilla, Sarah Nobles, Mark Burroughs, Lori Rowe, Haibin Wang, Ben L. Rambo-Martin, Dhwani Batra, Justin Lee, Suxiang Tong |
| EPI_ISL_1094907, EPI_ISL_1094908, EPI_ISL_1094909, EPI_ISL_1094910, EPI_ISL_1094911, EPI_ISL_1094912, EPI_ISL_1094913, EPI_ISL_1094914, EPI_ISL_1094915, EPI_ISL_1094916, EPI_ISL_1094917, EPI_ISL_1094918, EPI_ISL_1094919, EPI_ISL_1094920 | | | |
| see above | ND Dept. of Health Laboratory Services-Microbiology | Respiratory Viruses Branch, Division of Viral Diseases, Centers for Disease Control and Prevention | Krista Queen, Yan Li, Ying Tao, Jing Zhang, Anna Uehara, Anna Montmayeur, Clinton R. Paden, Peter W. Cook, Rachel Marine, Mili Sheth, Jasmine Padilla, Sarah Nobles, Mark Burroughs, Lori Rowe, Haibin Wang, Ben L. Rambo-Martin, Dhwani Batra, Justin Lee, Suxiang Tong |
| EPI_ISL_1094921, EPI_ISL_1094922, EPI_ISL_1094923, EPI_ISL_1094924, EPI_ISL_1094925, EPI_ISL_1094926, EPI_ISL_1094927, EPI_ISL_1094928 | TX DSHS, Lab Services Section MC 1947 | Respiratory Viruses Branch, Division of Viral Diseases, Centers for Disease Control and Prevention | Krista Queen, Yan Li, Ying Tao, Jing Zhang, Anna Uehara, Anna Montmayeur, Clinton R. Paden, Peter W. Cook, Rachel Marine, Mili Sheth, Jasmine Padilla, Sarah Nobles, Mark Burroughs, Lori Rowe, Haibin Wang, Ben L. Rambo-Martin, Dhwani Batra, Justin Lee, Suxiang Tong |
| EPI_ISL_1094929, EPI_ISL_1094930, EPI_ISL_1094931, EPI_ISL_1094932, EPI_ISL_1094933, EPI_ISL_1094934, EPI_ISL_1094935, EPI_ISL_1094936, EPI_ISL_1094937, EPI_ISL_1094938, EPI_ISL_1094939, EPI_ISL_1094940, EPI_ISL_1094941, EPI_ISL_1094942, EPI_ISL_1094943, EPI_ISL_1094944 | | | |
| see above | MT Public Health Laboratory | Respiratory Viruses Branch, Division of Viral Diseases, Centers for Disease Control and Prevention | Krista Queen, Yan Li, Ying Tao, Jing Zhang, Anna Uehara, Anna Montmayeur, Clinton R. Paden, Peter W. Cook, Rachel Marine, Mili Sheth, Jasmine Padilla, Sarah Nobles, Mark Burroughs, Lori Rowe, Haibin Wang, Ben L. Rambo-Martin, Dhwani Batra, Justin Lee, Suxiang Tong |
| EPI_ISL_1094945, EPI_ISL_1094946, EPI_ISL_1094947, EPI_ISL_1094948, EPI_ISL_1094949, EPI_ISL_1094950, EPI_ISL_1094951, EPI_ISL_1094952, EPI_ISL_1094953, EPI_ISL_1094954, EPI_ISL_1094955, EPI_ISL_1094956, EPI_ISL_1094957, EPI_ISL_1094958, EPI_ISL_1094959, EPI_ISL_1094960, EPI_ISL_1094961 | | | |
| see above | KY State Public Health Lab | Respiratory Viruses Branch, Division of Viral Diseases, Centers for Disease Control and Prevention | Krista Queen, Yan Li, Ying Tao, Jing Zhang, Anna Uehara, Anna Montmayeur, Clinton R. Paden, Peter W. Cook, Rachel Marine, Mili Sheth, Jasmine Padilla, Sarah Nobles, Mark Burroughs, Lori Rowe, Haibin Wang, Ben L. Rambo-Martin, Dhwani Batra, Justin Lee, Suxiang Tong |
| EPI_ISL_1094962, EPI_ISL_1094963, EPI_ISL_1094964, EPI_ISL_1094965, EPI_ISL_1094966, EPI_ISL_1094967, EPI_ISL_1094968, EPI_ISL_1094969, EPI_ISL_1094970, EPI_ISL_1094971, EPI_ISL_1094972, EPI_ISL_1094973 | | | |
| see above | OH Department of Health Laboratory | Respiratory Viruses Branch, Division of Viral Diseases, Centers for Disease Control and Prevention | Krista Queen, Yan Li, Ying Tao, Jing Zhang, Anna Uehara, Anna Montmayeur, Clinton R. Paden, Peter W. Cook, Rachel Marine, Mili Sheth, Jasmine Padilla, Sarah Nobles, Mark Burroughs, Lori Rowe, Haibin Wang, Ben L. Rambo-Martin, Dhwani Batra, Justin Lee, Suxiang Tong |
| EPI_ISL_1094974, EPI_ISL_1094975, EPI_ISL_1094977, EPI_ISL_1094978, EPI_ISL_1094979, EPI_ISL_1094980, EPI_ISL_1094981, EPI_ISL_1094982, EPI_ISL_1094983 | MO State Public Health Laboratory | Respiratory Viruses Branch, Division of Viral Diseases, Centers for Disease Control and Prevention | Krista Queen, Yan Li, Ying Tao, Jing Zhang, Anna Uehara, Anna Montmayeur, Clinton R. Paden, Peter W. Cook, Rachel Marine, Mili Sheth, Jasmine Padilla, Sarah Nobles, Mark Burroughs, Lori Rowe, Haibin Wang, Ben L. Rambo-Martin, Dhwani Batra, Justin Lee, Suxiang Tong |
| EPI_ISL_1094984, EPI_ISL_1094985, EPI_ISL_1094986, EPI_ISL_1094987, EPI_ISL_1094988, EPI_ISL_1094989, EPI_ISL_1094990, EPI_ISL_1094991, EPI_ISL_1094992 | AZ SPHL, Arizona Department of Health Services | Respiratory Viruses Branch, Division of Viral Diseases, Centers for Disease Control and Prevention | Krista Queen, Yan Li, Ying Tao, Jing Zhang, Anna Uehara, Anna Montmayeur, Clinton R. Paden, Peter W. Cook, Rachel Marine, Mili Sheth, Jasmine Padilla, Sarah Nobles, Mark Burroughs, Lori Rowe, Haibin Wang, Ben L. Rambo-Martin, Dhwani Batra, Justin Lee, Suxiang Tong |
| EPI_ISL_1094995, EPI_ISL_1094996, EPI_ISL_1094997, EPI_ISL_1094999, EPI_ISL_1095000, EPI_ISL_1095004, EPI_ISL_1095006, EPI_ISL_1095008, EPI_ISL_1095009, EPI_ISL_1095011, EPI_ISL_1095012, EPI_ISL_1095013, EPI_ISL_1095014, EPI_ISL_1095016, EPI_ISL_1095020, EPI_ISL_1095021, EPI_ISL_1095022, EPI_ISL_1095023, EPI_ISL_1095024, EPI_ISL_1095025, EPI_ISL_1095028, EPI_ISL_1095029, EPI_ISL_1095030, EPI_ISL_1095033, EPI_ISL_1095034, EPI_ISL_1095035, EPI_ISL_1095038, EPI_ISL_1095039 | | | |
| see above | MD DOH Laboratories Administration | Respiratory Viruses Branch, Division of Viral Diseases, Centers for Disease Control and Prevention | Krista Queen, Yan Li, Ying Tao, Jing Zhang, Anna Uehara, Anna Montmayeur, Clinton R. Paden, Peter W. Cook, Rachel Marine, Mili Sheth, Jasmine Padilla, Sarah Nobles, Mark Burroughs, Lori Rowe, Haibin Wang, Ben L. Rambo-Martin, Dhwani Batra, Justin Lee, Suxiang Tong |
| EPI_ISL_1095040, EPI_ISL_1095041, EPI_ISL_1095042, EPI_ISL_1095043, EPI_ISL_1095044, EPI_ISL_1095045, EPI_ISL_1095046, EPI_ISL_1095047, EPI_ISL_1095048, EPI_ISL_1095049, EPI_ISL_1095050, EPI_ISL_1095051, EPI_ISL_1095052, EPI_ISL_1095053, EPI_ISL_1095054, EPI_ISL_1095055, EPI_ISL_1095056, EPI_ISL_1095057, EPI_ISL_1095058, EPI_ISL_1095059, EPI_ISL_1095060, EPI_ISL_1095061, EPI_ISL_1095062, EPI_ISL_1095063, EPI_ISL_1095064, EPI_ISL_1095065, EPI_ISL_1095066, EPI_ISL_1095067, EPI_ISL_1095068, EPI_ISL_1095069, EPI_ISL_1095070, EPI_ISL_1095071, EPI_ISL_1095072, EPI_ISL_1095073 | | | |
| see above | NC State Laboratory of Public Health | Respiratory Viruses Branch, Division of Viral Diseases, Centers for Disease Control and Prevention | Krista Queen, Yan Li, Ying Tao, Jing Zhang, Anna Uehara, Anna Montmayeur, Clinton R. Paden, Peter W. Cook, Rachel Marine, Mili Sheth, Jasmine Padilla, Sarah Nobles, Mark Burroughs, Lori Rowe, Haibin Wang, Ben L. Rambo-Martin, Dhwani Batra, Justin Lee, Suxiang Tong |
| EPI_ISL_1095074, EPI_ISL_1095075, EPI_ISL_1095076, EPI_ISL_1095077, EPI_ISL_1095078, EPI_ISL_1095079, EPI_ISL_1095080, EPI_ISL_1095081, EPI_ISL_1095082, EPI_ISL_1095083, EPI_ISL_1095084, EPI_ISL_1095085, EPI_ISL_1095086, EPI_ISL_1095087, EPI_ISL_1095088, EPI_ISL_1095089, EPI_ISL_1095091, EPI_ISL_1095092, EPI_ISL_1095093, EPI_ISL_1095094, EPI_ISL_1095095, EPI_ISL_1095096 | | | |
| see above | MI - Michigan Department of Health and Human Services - Bureau of Laboratories | Respiratory Viruses Branch, Division of Viral Diseases, Centers for Disease Control and Prevention | Krista Queen, Yan Li, Ying Tao, Jing Zhang, Anna Uehara, Anna Montmayeur, Clinton R. Paden, Peter W. Cook, Rachel Marine, Mili Sheth, Jasmine Padilla, Sarah Nobles, Mark Burroughs, Lori Rowe, Haibin Wang, Ben L. Rambo-Martin, Dhwani Batra, Justin Lee, Suxiang Tong |

| | | | |
|---|---|---|---|
| EPI_ISL_1095097, EPI_ISL_1095098, EPI_ISL_1095099, EPI_ISL_1095100, EPI_ISL_1095101, EPI_ISL_1095102, EPI_ISL_1095103, EPI_ISL_1095104, EPI_ISL_1095105, EPI_ISL_1095106, EPI_ISL_1095107, EPI_ISL_1095109, EPI_ISL_1095111, EPI_ISL_1095112, EPI_ISL_1095114, EPI_ISL_1095115, EPI_ISL_1095116, EPI_ISL_1095117, EPI_ISL_1095118, EPI_ISL_1095119 | | | |
| see above | GA Department of Public Health Laboratory | Respiratory Viruses Branch, Division of Viral Diseases, Centers for Disease Control and Prevention | Krista Queen, Yan Li, Ying Tao, Jing Zhang, Anna Uehara, Anna Montmayeur, Clinton R. Paden, Peter W. Cook, Rachel Marine, Mili Sheth, Jasmine Padilla, Sarah Nobles, Mark Burroughs, Lori Rowe, Haibin Wang, Ben L. Rambo-Martin, Dhwani Batra, Justin Lee, Suxiang Tong |
| EPI_ISL_1095120, EPI_ISL_1095121, EPI_ISL_1095122, EPI_ISL_1095123, EPI_ISL_1095124, EPI_ISL_1095125, EPI_ISL_1095126, EPI_ISL_1095127, EPI_ISL_1095128, EPI_ISL_1095129, EPI_ISL_1095130 | | | |
| see above | MS Public Health Laboratory | Respiratory Viruses Branch, Division of Viral Diseases, Centers for Disease Control and Prevention | Krista Queen, Yan Li, Ying Tao, Jing Zhang, Anna Uehara, Anna Montmayeur, Clinton R. Paden, Peter W. Cook, Rachel Marine, Mili Sheth, Jasmine Padilla, Sarah Nobles, Mark Burroughs, Lori Rowe, Haibin Wang, Ben L. Rambo-Martin, Dhwani Batra, Justin Lee, Suxiang Tong |
| EPI_ISL_1095131, EPI_ISL_1095132, EPI_ISL_1095134, EPI_ISL_1095135, EPI_ISL_1095136, EPI_ISL_1095138, EPI_ISL_1095139, EPI_ISL_1095140, EPI_ISL_1095141, EPI_ISL_1095143, EPI_ISL_1095144, EPI_ISL_1095145, EPI_ISL_1095146, EPI_ISL_1095147, EPI_ISL_1095148, EPI_ISL_1095150, EPI_ISL_1095151, EPI_ISL_1095152, EPI_ISL_1095153, EPI_ISL_1095154, EPI_ISL_1095155, EPI_ISL_1095156, EPI_ISL_1095157, EPI_ISL_1095158, EPI_ISL_1095159, EPI_ISL_1095160, EPI_ISL_1095161, EPI_ISL_1095162, EPI_ISL_1095163, EPI_ISL_1095164, EPI_ISL_1095165, EPI_ISL_1095166, EPI_ISL_1095167, EPI_ISL_1095168 | | | |
| see above | NYSDOH Wadsworth Center, Virology Lab | Respiratory Viruses Branch, Division of Viral Diseases, Centers for Disease Control and Prevention | Krista Queen, Yan Li, Ying Tao, Jing Zhang, Anna Uehara, Anna Montmayeur, Clinton R. Paden, Peter W. Cook, Rachel Marine, Mili Sheth, Jasmine Padilla, Sarah Nobles, Mark Burroughs, Lori Rowe, Haibin Wang, Ben L. Rambo-Martin, Dhwani Batra, Justin Lee, Suxiang Tong |
| EPI_ISL_1095169, EPI_ISL_1095170, EPI_ISL_1095171, EPI_ISL_1095172, EPI_ISL_1095173, EPI_ISL_1095174, EPI_ISL_1095175, EPI_ISL_1095176, EPI_ISL_1095177, EPI_ISL_1095178, EPI_ISL_1095179, EPI_ISL_1095180, EPI_ISL_1095181, EPI_ISL_1095182, EPI_ISL_1095183, EPI_ISL_1095185 | | | |
| see above | PA Department of Health, Bureau of Laboratories | Respiratory Viruses Branch, Division of Viral Diseases, Centers for Disease Control and Prevention | Krista Queen, Yan Li, Ying Tao, Jing Zhang, Anna Uehara, Anna Montmayeur, Clinton R. Paden, Peter W. Cook, Rachel Marine, Mili Sheth, Jasmine Padilla, Sarah Nobles, Mark Burroughs, Lori Rowe, Haibin Wang, Ben L. Rambo-Martin, Dhwani Batra, Justin Lee, Suxiang Tong |
| EPI_ISL_1095186 | FL Bur. of Public Health Laboratories-Jacksonville | Respiratory Viruses Branch, Division of Viral Diseases, Centers for Disease Control and Prevention | Krista Queen, Yan Li, Ying Tao, Jing Zhang, Anna Uehara, Anna Montmayeur, Clinton R. Paden, Peter W. Cook, Rachel Marine, Mili Sheth, Jasmine Padilla, Sarah Nobles, Mark Burroughs, Lori Rowe, Haibin Wang, Ben L. Rambo-Martin, Dhwani Batra, Justin Lee, Suxiang Tong |
| EPI_ISL_1095187, EPI_ISL_1095188, EPI_ISL_1095189 | VT Dept. of Health Laboratory | Respiratory Viruses Branch, Division of Viral Diseases, Centers for Disease Control and Prevention | Krista Queen, Yan Li, Ying Tao, Jing Zhang, Anna Uehara, Anna Montmayeur, Clinton R. Paden, Peter W. Cook, Rachel Marine, Mili Sheth, Jasmine Padilla, Sarah Nobles, Mark Burroughs, Lori Rowe, Haibin Wang, Ben L. Rambo-Martin, Dhwani Batra, Justin Lee, Suxiang Tong |
| EPI_ISL_1095190, EPI_ISL_1095191, EPI_ISL_1095192, EPI_ISL_1095193, EPI_ISL_1095194, EPI_ISL_1095195, EPI_ISL_1095196, EPI_ISL_1095197, EPI_ISL_1095198, EPI_ISL_1095199, EPI_ISL_1095200, EPI_ISL_1095201, EPI_ISL_1095203 | | | |
| see above | CDPH, Viral and Rickettsial Disease Laboratory | Respiratory Viruses Branch, Division of Viral Diseases, Centers for Disease Control and Prevention | Krista Queen, Yan Li, Ying Tao, Jing Zhang, Anna Uehara, Anna Montmayeur, Clinton R. Paden, Peter W. Cook, Rachel Marine, Mili Sheth, Jasmine Padilla, Sarah Nobles, Mark Burroughs, Lori Rowe, Haibin Wang, Ben L. Rambo-Martin, Dhwani Batra, Justin Lee, Suxiang Tong |
| EPI_ISL_1095204, EPI_ISL_1095205, EPI_ISL_1095206, EPI_ISL_1095208, EPI_ISL_1095209 | WI State Laboratory of Hygiene | Respiratory Viruses Branch, Division of Viral Diseases, Centers for Disease Control and Prevention | Krista Queen, Yan Li, Ying Tao, Jing Zhang, Anna Uehara, Anna Montmayeur, Clinton R. Paden, Peter W. Cook, Rachel Marine, Mili Sheth, Jasmine Padilla, Sarah Nobles, Mark Burroughs, Lori Rowe, Haibin Wang, Ben L. Rambo-Martin, Dhwani Batra, Justin Lee, Suxiang Tong |
| EPI_ISL_1095210, EPI_ISL_1095211, EPI_ISL_1095212, EPI_ISL_1095213, EPI_ISL_1095214 | KS Health and Environmental Laboratories | Respiratory Viruses Branch, Division of Viral Diseases, Centers for Disease Control and Prevention | Krista Queen, Yan Li, Ying Tao, Jing Zhang, Anna Uehara, Anna Montmayeur, Clinton R. Paden, Peter W. Cook, Rachel Marine, Mili Sheth, Jasmine Padilla, Sarah Nobles, Mark Burroughs, Lori Rowe, Haibin Wang, Ben L. Rambo-Martin, Dhwani Batra, Justin Lee, Suxiang Tong |
| EPI_ISL_1095216, EPI_ISL_1095217, EPI_ISL_1095219, EPI_ISL_1095220 | RI State Health Laboratories | Respiratory Viruses Branch, Division of Viral Diseases, Centers for Disease Control and Prevention | Krista Queen, Yan Li, Ying Tao, Jing Zhang, Anna Uehara, Anna Montmayeur, Clinton R. Paden, Peter W. Cook, Rachel Marine, Mili Sheth, Jasmine Padilla, Sarah Nobles, Mark Burroughs, Lori Rowe, Haibin Wang, Ben L. Rambo-Martin, Dhwani Batra, Justin Lee, Suxiang Tong |
| EPI_ISL_1095221 | WVDHHR - Office of Laboratory Services | Respiratory Viruses Branch, Division of Viral Diseases, Centers for Disease Control and Prevention | Krista Queen, Yan Li, Ying Tao, Jing Zhang, Anna Uehara, Anna Montmayeur, Clinton R. Paden, Peter W. Cook, Rachel Marine, Mili Sheth, Jasmine Padilla, Sarah Nobles, Mark Burroughs, Lori Rowe, Haibin Wang, Ben L. Rambo-Martin, Dhwani Batra, Justin Lee, Suxiang Tong |
| EPI_ISL_1095222, EPI_ISL_1095223, EPI_ISL_1095224 | ME Health and Environmental Testing Laboratory | Respiratory Viruses Branch, Division of Viral Diseases, Centers for Disease Control and Prevention | Krista Queen, Yan Li, Ying Tao, Jing Zhang, Anna Uehara, Anna Montmayeur, Clinton R. Paden, Peter W. Cook, Rachel Marine, Mili Sheth, Jasmine Padilla, Sarah Nobles, Mark Burroughs, Lori Rowe, Haibin Wang, Ben L. Rambo-Martin, Dhwani Batra, Justin Lee, Suxiang Tong |
| EPI_ISL_1095225, EPI_ISL_1095226 | IA State Hygienic Laboratory | Respiratory Viruses Branch, Division of Viral Diseases, Centers for Disease Control and Prevention | Krista Queen, Yan Li, Ying Tao, Jing Zhang, Anna Uehara, Anna Montmayeur, Clinton R. Paden, Peter W. Cook, Rachel Marine, Mili Sheth, Jasmine Padilla, Sarah Nobles, Mark Burroughs, Lori Rowe, Haibin Wang, Ben L. Rambo-Martin, Dhwani Batra, Justin Lee, Suxiang Tong |
| EPI_ISL_1095227, EPI_ISL_1095228, EPI_ISL_1095229, EPI_ISL_1095230, EPI_ISL_1095231, EPI_ISL_1095232 | AK State Public Health Lab, State Health Department | Respiratory Viruses Branch, Division of Viral Diseases, Centers for Disease Control and Prevention | Krista Queen, Yan Li, Ying Tao, Jing Zhang, Anna Uehara, Anna Montmayeur, Clinton R. Paden, Peter W. Cook, Rachel Marine, Mili Sheth, Jasmine Padilla, Sarah Nobles, Mark Burroughs, Lori Rowe, Haibin Wang, Ben L. Rambo-Martin, Dhwani Batra, Justin Lee, Suxiang Tong |
| EPI_ISL_1095233, EPI_ISL_1095234 | IL Dept. of Public Health Springfield Laboratory | Respiratory Viruses Branch, Division of Viral Diseases, Centers for Disease Control and Prevention | Krista Queen, Yan Li, Ying Tao, Jing Zhang, Anna Uehara, Anna Montmayeur, Clinton R. Paden, Peter W. Cook, Rachel Marine, Mili Sheth, Jasmine Padilla, Sarah Nobles, Mark Burroughs, Lori Rowe, Haibin Wang, Ben L. Rambo-Martin, Dhwani Batra, Justin Lee, Suxiang Tong |
| EPI_ISL_1095235, EPI_ISL_1095236, EPI_ISL_1095237, EPI_ISL_1095238, EPI_ISL_1095239, EPI_ISL_1095240, EPI_ISL_1095241, EPI_ISL_1095242 | OR State PHL-Virology/Immunology Section | Respiratory Viruses Branch, Division of Viral Diseases, Centers for Disease Control and Prevention | Krista Queen, Yan Li, Ying Tao, Jing Zhang, Anna Uehara, Anna Montmayeur, Clinton R. Paden, Peter W. Cook, Rachel Marine, Mili Sheth, Jasmine Padilla, Sarah Nobles, Mark Burroughs, Lori Rowe, Haibin Wang, Ben L. Rambo-Martin, Dhwani Batra, Justin Lee, Suxiang Tong |
| EPI_ISL_1095243, EPI_ISL_1095244, EPI_ISL_1095245, EPI_ISL_1095246, EPI_ISL_1095247, EPI_ISL_1095248 | IN State Department of Health Laboratory Services | Respiratory Viruses Branch, Division of Viral Diseases, Centers for Disease Control and Prevention | Krista Queen, Yan Li, Ying Tao, Jing Zhang, Anna Uehara, Anna Montmayeur, Clinton R. Paden, Peter W. Cook, Rachel Marine, Mili Sheth, Jasmine Padilla, Sarah Nobles, Mark Burroughs, Lori Rowe, Haibin Wang, Ben L. Rambo-Martin, Dhwani Batra, Justin Lee, Suxiang Tong |
| EPI_ISL_1095249, EPI_ISL_1095250, EPI_ISL_1095251, EPI_ISL_1095252, EPI_ISL_1095253 | AR Dept. of Health-Public Health Lab | Respiratory Viruses Branch, Division of Viral Diseases, Centers for Disease Control and Prevention | Krista Queen, Yan Li, Ying Tao, Jing Zhang, Anna Uehara, Anna Montmayeur, Clinton R. Paden, Peter W. Cook, Rachel Marine, Mili Sheth, Jasmine Padilla, Sarah Nobles, Mark Burroughs, Lori Rowe, Haibin Wang, Ben L. Rambo-Martin, Dhwani Batra, Justin Lee, Suxiang Tong |
| EPI_ISL_1095254, EPI_ISL_1095255, EPI_ISL_1095256, EPI_ISL_1095257, EPI_ISL_1095258, EPI_ISL_1095259 | WY Public Health Laboratory | Respiratory Viruses Branch, Division of Viral Diseases, Centers for Disease Control and Prevention | Krista Queen, Yan Li, Ying Tao, Jing Zhang, Anna Uehara, Anna Montmayeur, Clinton R. Paden, Peter W. Cook, Rachel Marine, Mili Sheth, Jasmine Padilla, Sarah Nobles, Mark Burroughs, Lori Rowe, Haibin Wang, Ben L. Rambo-Martin, Dhwani Batra, Justin Lee, Suxiang Tong |
| EPI_ISL_1095260, EPI_ISL_1095261 | LA Office of Public Health Laboratories | Respiratory Viruses Branch, Division of Viral Diseases, Centers for Disease Control and Prevention | Krista Queen, Yan Li, Ying Tao, Jing Zhang, Anna Uehara, Anna Montmayeur, Clinton R. Paden, Peter W. Cook, Rachel Marine, Mili Sheth, Jasmine Padilla, Sarah Nobles, Mark Burroughs, Lori Rowe, Haibin Wang, Ben L. Rambo-Martin, Dhwani Batra, Justin Lee, Suxiang Tong |
| EPI_ISL_1095262 | AL Dept. of Public Health Bureau of Clinical Laboratories | Respiratory Viruses Branch, Division of Viral Diseases, Centers for Disease Control and Prevention | Krista Queen, Yan Li, Ying Tao, Jing Zhang, Anna Uehara, Anna Montmayeur, Clinton R. Paden, Peter W. Cook, Rachel Marine, Mili Sheth, Jasmine Padilla, Sarah Nobles, Mark Burroughs, Lori Rowe, Haibin Wang, Ben L. Rambo-Martin, Dhwani Batra, Justin Lee, Suxiang Tong |
| EPI_ISL_1095263, EPI_ISL_1095264, EPI_ISL_1095265, EPI_ISL_1095266, EPI_ISL_1095267, EPI_ISL_1095268, EPI_ISL_1095269 | MO State Public Health Laboratory | Respiratory Viruses Branch, Division of Viral Diseases, Centers for Disease Control and Prevention | Krista Queen, Yan Li, Ying Tao, Jing Zhang, Anna Uehara, Anna Montmayeur, Clinton R. Paden, Peter W. Cook, Rachel Marine, Mili Sheth, Jasmine Padilla, Sarah Nobles, Mark Burroughs, Lori Rowe, Haibin Wang, Ben L. Rambo-Martin, Dhwani Batra, Justin Lee, Suxiang Tong |
| EPI_ISL_1095270, | AZ SPHL, Arizona Department of Health | Respiratory Viruses Branch, Division of Viral | Krista Queen, Yan Li, Ying Tao, Jing Zhang, Anna Uehara, Anna Montmayeur, Clinton R. Paden, Peter W. Cook, Rachel Marine, Mili Sheth, Jasmine Padilla, Sarah Nobles, Mark Burroughs, Lori Rowe, Haibin Wang, Ben L. Rambo-Martin, Dhwani Batra, Justin Lee, Suxiang Tong |

| | | | |
|---|---|---|---|
| EPI_ISL_1095271, EPI_ISL_1095272, EPI_ISL_1095273, EPI_ISL_1095274, EPI_ISL_1095275 | Services | Diseases, Centers for Disease Control and Prevention | |
| EPI_ISL_1095276, EPI_ISL_1095278, EPI_ISL_1095279, EPI_ISL_1095281, EPI_ISL_1095282, EPI_ISL_1095283, EPI_ISL_1095284, EPI_ISL_1095285, EPI_ISL_1095286 | MI - Michigan Department of Health and Human Services - Bureau of Laboratories | Respiratory Viruses Branch, Division of Viral Diseases, Centers for Disease Control and Prevention | Krista Queen, Yan Li, Ying Tao, Jing Zhang, Anna Uehara, Anna Montmayeur, Clinton R. Paden, Peter W. Cook, Rachel Marine, Mili Sheth, Jasmine Padilla, Sarah Nobles, Mark Burroughs, Lori Rowe, Haibin Wang, Ben L. Rambo-Martin, Dhwani Batra, Justin Lee, Suxiang Tong |
| EPI_ISL_1095287 | FL Bur. of Public Health Laboratories-Jacksonville | Respiratory Viruses Branch, Division of Viral Diseases, Centers for Disease Control and Prevention | Krista Queen, Yan Li, Ying Tao, Jing Zhang, Anna Uehara, Anna Montmayeur, Clinton R. Paden, Peter W. Cook, Rachel Marine, Mili Sheth, Jasmine Padilla, Sarah Nobles, Mark Burroughs, Lori Rowe, Haibin Wang, Ben L. Rambo-Martin, Dhwani Batra, Justin Lee, Suxiang Tong |
| EPI_ISL_1095288, EPI_ISL_1095289 | FL Bureau of Public Health Laboratories-Tampa | Respiratory Viruses Branch, Division of Viral Diseases, Centers for Disease Control and Prevention | Krista Queen, Yan Li, Ying Tao, Jing Zhang, Anna Uehara, Anna Montmayeur, Clinton R. Paden, Peter W. Cook, Rachel Marine, Mili Sheth, Jasmine Padilla, Sarah Nobles, Mark Burroughs, Lori Rowe, Haibin Wang, Ben L. Rambo-Martin, Dhwani Batra, Justin Lee, Suxiang Tong |
| EPI_ISL_1095290, EPI_ISL_1095291, EPI_ISL_1095292, EPI_ISL_1095293, EPI_ISL_1095294 | FL Bureau of Public Health Laboratories-Miami | Respiratory Viruses Branch, Division of Viral Diseases, Centers for Disease Control and Prevention | Krista Queen, Yan Li, Ying Tao, Jing Zhang, Anna Uehara, Anna Montmayeur, Clinton R. Paden, Peter W. Cook, Rachel Marine, Mili Sheth, Jasmine Padilla, Sarah Nobles, Mark Burroughs, Lori Rowe, Haibin Wang, Ben L. Rambo-Martin, Dhwani Batra, Justin Lee, Suxiang Tong |
| EPI_ISL_1095295, EPI_ISL_1095296 | VA-Division of Consolidated Laboratory Services | Respiratory Viruses Branch, Division of Viral Diseases, Centers for Disease Control and Prevention | Krista Queen, Yan Li, Ying Tao, Jing Zhang, Anna Uehara, Anna Montmayeur, Clinton R. Paden, Peter W. Cook, Rachel Marine, Mili Sheth, Jasmine Padilla, Sarah Nobles, Mark Burroughs, Lori Rowe, Haibin Wang, Ben L. Rambo-Martin, Dhwani Batra, Justin Lee, Suxiang Tong |
| EPI_ISL_1095297, EPI_ISL_1095298, EPI_ISL_1095299, EPI_ISL_1095300, EPI_ISL_1095301, EPI_ISL_1095302, EPI_ISL_1095303, EPI_ISL_1095304, EPI_ISL_1095305 | ND Dept. of Health Laboratory Services-Microbiology | Respiratory Viruses Branch, Division of Viral Diseases, Centers for Disease Control and Prevention | Krista Queen, Yan Li, Ying Tao, Jing Zhang, Anna Uehara, Anna Montmayeur, Clinton R. Paden, Peter W. Cook, Rachel Marine, Mili Sheth, Jasmine Padilla, Sarah Nobles, Mark Burroughs, Lori Rowe, Haibin Wang, Ben L. Rambo-Martin, Dhwani Batra, Justin Lee, Suxiang Tong |
| EPI_ISL_1095306 | MT Public Health Laboratory | Respiratory Viruses Branch, Division of Viral Diseases, Centers for Disease Control and Prevention | Krista Queen, Yan Li, Ying Tao, Jing Zhang, Anna Uehara, Anna Montmayeur, Clinton R. Paden, Peter W. Cook, Rachel Marine, Mili Sheth, Jasmine Padilla, Sarah Nobles, Mark Burroughs, Lori Rowe, Haibin Wang, Ben L. Rambo-Martin, Dhwani Batra, Justin Lee, Suxiang Tong |
| EPI_ISL_1095307, EPI_ISL_1095308, EPI_ISL_1095309, EPI_ISL_1095310, EPI_ISL_1095311, EPI_ISL_1095312, EPI_ISL_1095313, EPI_ISL_1095314, EPI_ISL_1095315, EPI_ISL_1095316 | SC Dept of Health and Env. Control-Bureau of Laboratories | Respiratory Viruses Branch, Division of Viral Diseases, Centers for Disease Control and Prevention | Krista Queen, Yan Li, Ying Tao, Jing Zhang, Anna Uehara, Anna Montmayeur, Clinton R. Paden, Peter W. Cook, Rachel Marine, Mili Sheth, Jasmine Padilla, Sarah Nobles, Mark Burroughs, Lori Rowe, Haibin Wang, Ben L. Rambo-Martin, Dhwani Batra, Justin Lee, Suxiang Tong |
| EPI_ISL_1095317 | TN Division of Laboratory Services | Respiratory Viruses Branch, Division of Viral Diseases, Centers for Disease Control and Prevention | Krista Queen, Yan Li, Ying Tao, Jing Zhang, Anna Uehara, Anna Montmayeur, Clinton R. Paden, Peter W. Cook, Rachel Marine, Mili Sheth, Jasmine Padilla, Sarah Nobles, Mark Burroughs, Lori Rowe, Haibin Wang, Ben L. Rambo-Martin, Dhwani Batra, Justin Lee, Suxiang Tong |
| EPI_ISL_1095318, EPI_ISL_1095319, EPI_ISL_1095320, EPI_ISL_1095321, EPI_ISL_1095322, EPI_ISL_1095323 | WA State Department of Health | Respiratory Viruses Branch, Division of Viral Diseases, Centers for Disease Control and Prevention | Krista Queen, Yan Li, Ying Tao, Jing Zhang, Anna Uehara, Anna Montmayeur, Clinton R. Paden, Peter W. Cook, Rachel Marine, Mili Sheth, Jasmine Padilla, Sarah Nobles, Mark Burroughs, Lori Rowe, Haibin Wang, Ben L. Rambo-Martin, Dhwani Batra, Justin Lee, Suxiang Tong |
| EPI_ISL_1095324, EPI_ISL_1095325, EPI_ISL_1095326, EPI_ISL_1095327, EPI_ISL_1095328, EPI_ISL_1095329, EPI_ISL_1095330, EPI_ISL_1095331, EPI_ISL_1095332 | MD DOH Laboratories Administration | Respiratory Viruses Branch, Division of Viral Diseases, Centers for Disease Control and Prevention | Krista Queen, Yan Li, Ying Tao, Jing Zhang, Anna Uehara, Anna Montmayeur, Clinton R. Paden, Peter W. Cook, Rachel Marine, Mili Sheth, Jasmine Padilla, Sarah Nobles, Mark Burroughs, Lori Rowe, Haibin Wang, Ben L. Rambo-Martin, Dhwani Batra, Justin Lee, Suxiang Tong |
| EPI_ISL_1095333, EPI_ISL_1095334, EPI_ISL_1095335, EPI_ISL_1095336, EPI_ISL_1095337, EPI_ISL_1095338, EPI_ISL_1095339, EPI_ISL_1095340, EPI_ISL_1095341, EPI_ISL_1095342, EPI_ISL_1095343, EPI_ISL_1095344, EPI_ISL_1095345, EPI_ISL_1095346, EPI_ISL_1095347, EPI_ISL_1095348, EPI_ISL_1095349, EPI_ISL_1095350, EPI_ISL_1095351, EPI_ISL_1095352, EPI_ISL_1095353, EPI_ISL_1095354, EPI_ISL_1095355, EPI_ISL_1095356, EPI_ISL_1095357, EPI_ISL_1095358, EPI_ISL_1095359, EPI_ISL_1095360, EPI_ISL_1095361, EPI_ISL_1095362 | | | |
| see above | NC State Laboratory of Public Health | Respiratory Viruses Branch, Division of Viral Diseases, Centers for Disease Control and Prevention | Krista Queen, Yan Li, Ying Tao, Jing Zhang, Anna Uehara, Anna Montmayeur, Clinton R. Paden, Peter W. Cook, Rachel Marine, Mili Sheth, Jasmine Padilla, Sarah Nobles, Mark Burroughs, Lori Rowe, Haibin Wang, Ben L. Rambo-Martin, Dhwani Batra, Justin Lee, Suxiang Tong |
| EPI_ISL_1095408, EPI_ISL_1095409, EPI_ISL_1095410, EPI_ISL_1095411, EPI_ISL_1095412, EPI_ISL_1095413, EPI_ISL_1095414, EPI_ISL_1095415, EPI_ISL_1095416, EPI_ISL_1095417, EPI_ISL_1095418, EPI_ISL_1095419, EPI_ISL_1095420, EPI_ISL_1095421, EPI_ISL_1095422, EPI_ISL_1095423, EPI_ISL_1095424, EPI_ISL_1095425, EPI_ISL_1095426, EPI_ISL_1095427, EPI_ISL_1095428, EPI_ISL_1095429, EPI_ISL_1095430, EPI_ISL_1095431, EPI_ISL_1095432, EPI_ISL_1095433, EPI_ISL_1095434, EPI_ISL_1095435, EPI_ISL_1095436, EPI_ISL_1095437, EPI_ISL_1095438, EPI_ISL_1095439, EPI_ISL_1095440, EPI_ISL_1095441, EPI_ISL_1095442, EPI_ISL_1095443, EPI_ISL_1095444, EPI_ISL_1095445, EPI_ISL_1095446, EPI_ISL_1095447, EPI_ISL_1095448 | | | |
| see above | Virginia Division of Consolidated Laboratory Services | Virginia Division of Consolidated Laboratory Services | Virginia DCLS |
| EPI_ISL_1095452, EPI_ISL_1095458, EPI_ISL_1095459, EPI_ISL_1095460, EPI_ISL_1095461, EPI_ISL_1095462, EPI_ISL_1095463, EPI_ISL_1095464, EPI_ISL_1095465, EPI_ISL_1095466, EPI_ISL_1095467, EPI_ISL_1095468, EPI_ISL_1095469, EPI_ISL_1095470, EPI_ISL_1095471, EPI_ISL_1095472, EPI_ISL_1095473, EPI_ISL_1095474, EPI_ISL_1095475, EPI_ISL_1095476, EPI_ISL_1095477, EPI_ISL_1095478, EPI_ISL_1095479, EPI_ISL_1095480, EPI_ISL_1095481, EPI_ISL_1095482, EPI_ISL_1095483, EPI_ISL_1095484 | | | |
| see above | University of Wisconsin-Madison AIDS Vaccine Research Laboratories | University of Wisconsin-Madison AIDS Vaccine Research Laboratories | Gage Moreno, Katarina Braun, et al. AIDS Vaccine Research Laboratories |
| EPI_ISL_1095493, EPI_ISL_1095494, EPI_ISL_1095495, EPI_ISL_1095496, EPI_ISL_1095497, EPI_ISL_1095498, EPI_ISL_1095499, EPI_ISL_1095500, EPI_ISL_1095501, EPI_ISL_1095502, EPI_ISL_1095503, EPI_ISL_1095504, EPI_ISL_1095505, EPI_ISL_1095506, EPI_ISL_1095507, EPI_ISL_1095508, EPI_ISL_1095509, EPI_ISL_1095510, EPI_ISL_1095511, EPI_ISL_1095512, EPI_ISL_1095513, EPI_ISL_1095514, EPI_ISL_1095515, EPI_ISL_1095516 | | | |
| see above | Instituto Nacional de Medicina Genomica | Instituto Nacional de Medicina Genomica | Hidalgo-Miranda A, Mendoza-Vargas A, Reyes-Grajeda JP, Cedro-Tanda A, Alcaraz N, Gonzalez-Barrera D, Rangel-DeLeon D, Ramirez-Vega O, Sifuentes-Rojas C, Cisneros-Villanueva M, Herrera-Montalvo LA |
| EPI_ISL_1095628, EPI_ISL_1095629, EPI_ISL_1095630, EPI_ISL_1095631, EPI_ISL_1095632 | Wyoming Public Health Laboratory | Wyoming Public Health Laboratory | Jim Mildenberger, Wanda Manley, Noah Hull, Taylor Fearing, Lynette Gumbleton, Channing Weber, Ashley Norberg, Chayse Rowley, Marley Goetz, Brian Dominguez, Elliot Thomasson, Cari Sloma, and Rob Christensen |
| EPI_ISL_1095645, EPI_ISL_1095646, EPI_ISL_1095647, EPI_ISL_1095648, EPI_ISL_1095649, EPI_ISL_1095650, EPI_ISL_1095651, EPI_ISL_1095652, EPI_ISL_1095653, EPI_ISL_1095654, EPI_ISL_1095655, EPI_ISL_1095656, EPI_ISL_1095657, EPI_ISL_1095658, EPI_ISL_1095659, EPI_ISL_1095660, EPI_ISL_1095661, EPI_ISL_1095662, EPI_ISL_1095663, EPI_ISL_1095664, EPI_ISL_1095665, EPI_ISL_1095666, EPI_ISL_1095667, EPI_ISL_1095668, EPI_ISL_1095669, EPI_ISL_1095670, EPI_ISL_1095671, EPI_ISL_1095672, EPI_ISL_1095673, EPI_ISL_1095674, EPI_ISL_1095675, EPI_ISL_1095676, EPI_ISL_1095677, EPI_ISL_1095678, EPI_ISL_1095679, EPI_ISL_1095680, EPI_ISL_1095681, EPI_ISL_1095682, EPI_ISL_1095683, EPI_ISL_1095684, EPI_ISL_1095685, EPI_ISL_1095686, EPI_ISL_1095687, EPI_ISL_1095688, EPI_ISL_1095689, EPI_ISL_1095690, EPI_ISL_1095691, EPI_ISL_1095692, EPI_ISL_1095693, EPI_ISL_1095694, EPI_ISL_1095695, EPI_ISL_1095696, EPI_ISL_1095697, EPI_ISL_1095698, EPI_ISL_1095699, EPI_ISL_1095700, EPI_ISL_1095701, EPI_ISL_1095702, EPI_ISL_1095703, EPI_ISL_1095704, EPI_ISL_1095709, EPI_ISL_1095710, EPI_ISL_1095711, EPI_ISL_1095712, EPI_ISL_1095713, EPI_ISL_1095714, EPI_ISL_1095715, EPI_ISL_1095716, EPI_ISL_1095717, EPI_ISL_1095718, EPI_ISL_1095719, EPI_ISL_1095720, EPI_ISL_1095721, EPI_ISL_1095722, EPI_ISL_1095723, EPI_ISL_1095724, EPI_ISL_1095725, EPI_ISL_1095726, EPI_ISL_1095727, EPI_ISL_1095728, EPI_ISL_1095729, EPI_ISL_1095730, EPI_ISL_1095731, EPI_ISL_1095732, EPI_ISL_1095733, EPI_ISL_1095734, EPI_ISL_1095735, EPI_ISL_1095736, EPI_ISL_1095737, EPI_ISL_1095738, EPI_ISL_1095739, EPI_ISL_1095740, EPI_ISL_1095741, EPI_ISL_1095742, EPI_ISL_1095743, EPI_ISL_1095744, EPI_ISL_1095745, EPI_ISL_1095746, EPI_ISL_1095747, EPI_ISL_1095748, EPI_ISL_1095749, EPI_ISL_1095750, EPI_ISL_1095751, EPI_ISL_1095752, EPI_ISL_1095753, EPI_ISL_1095754, EPI_ISL_1095755, EPI_ISL_1095756, EPI_ISL_1095757, EPI_ISL_1095758, EPI_ISL_1095759, EPI_ISL_1095760, EPI_ISL_1095761, EPI_ISL_1095762, EPI_ISL_1095763, EPI_ISL_1095764, EPI_ISL_1095765, EPI_ISL_1095766, EPI_ISL_1095767, EPI_ISL_1095768, EPI_ISL_1095769, EPI_ISL_1095770, EPI_ISL_1095771, EPI_ISL_1095772, EPI_ISL_1095773, EPI_ISL_1095777, EPI_ISL_1095778, EPI_ISL_1095779, EPI_ISL_1095780, EPI_ISL_1095781, EPI_ISL_1095782, EPI_ISL_1095783, EPI_ISL_1095784, EPI_ISL_1095785, EPI_ISL_1095786, EPI_ISL_1095787, EPI_ISL_1095788, EPI_ISL_1095789, EPI_ISL_1095790, EPI_ISL_1095791, EPI_ISL_1095792, EPI_ISL_1095793, EPI_ISL_1095794, EPI_ISL_1095795, EPI_ISL_1095796, EPI_ISL_1095797, EPI_ISL_1095798, EPI_ISL_1095799, EPI_ISL_1095800, EPI_ISL_1095801, EPI_ISL_1095802, EPI_ISL_1095803, EPI_ISL_1095804, EPI_ISL_1095805, EPI_ISL_1095806, EPI_ISL_1095807, EPI_ISL_1095808, EPI_ISL_1095809, EPI_ISL_1095810, EPI_ISL_1095811, EPI_ISL_1095812, EPI_ISL_1095813, EPI_ISL_1095814, EPI_ISL_1095815, EPI_ISL_1095816, EPI_ISL_1095817, EPI_ISL_1095818, EPI_ISL_1095819, EPI_ISL_1095820, EPI_ISL_1095821, EPI_ISL_1095822, EPI_ISL_1095823, EPI_ISL_1095824, EPI_ISL_1095825, EPI_ISL_1095826, EPI_ISL_1095827, EPI_ISL_1095828, EPI_ISL_1095829, EPI_ISL_1095830, EPI_ISL_1095831, EPI_ISL_1095832, EPI_ISL_1095833, EPI_ISL_1095834, EPI_ISL_1095835, EPI_ISL_1095836, EPI_ISL_1095837, EPI_ISL_1095838, EPI_ISL_1095839, EPI_ISL_1095840, EPI_ISL_1095841, EPI_ISL_1095842, EPI_ISL_1095843, EPI_ISL_1095844, EPI_ISL_1095845, EPI_ISL_1095846, EPI_ISL_1095847, EPI_ISL_1095848, EPI_ISL_1095849, EPI_ISL_1095850, EPI_ISL_1095851, EPI_ISL_1095852, EPI_ISL_1095853, EPI_ISL_1095854, EPI_ISL_1095857, EPI_ISL_1095858, EPI_ISL_1095859, EPI_ISL_1095860, EPI_ISL_1095861, EPI_ISL_1095862, EPI_ISL_1095863, EPI_ISL_1095864, EPI_ISL_1095865, EPI_ISL_1095866, EPI_ISL_1095867, EPI_ISL_1095868, EPI_ISL_1095869, EPI_ISL_1095870, EPI_ISL_1095871, EPI_ISL_1095872, EPI_ISL_1095873, EPI_ISL_1095874, EPI_ISL_1095875, EPI_ISL_1095876, EPI_ISL_1095877, EPI_ISL_1095878, EPI_ISL_1095879, EPI_ISL_1095880, EPI_ISL_1095881, EPI_ISL_1095882, EPI_ISL_1095883, EPI_ISL_1095884, EPI_ISL_1095885, EPI_ISL_1095886, EPI_ISL_1095887, EPI_ISL_1095888, EPI_ISL_1095889, EPI_ISL_1095890, EPI_ISL_1095891, EPI_ISL_1095892, EPI_ISL_1095893, EPI_ISL_1095894, EPI_ISL_1095895, | | | |

| | | | |
|---|---|---|---|
| EPI_ISL_1095896, EPI_ISL_1095897, EPI_ISL_1095898, EPI_ISL_1095899, EPI_ISL_1095900, EPI_ISL_1095901, EPI_ISL_1095902, EPI_ISL_1095903, EPI_ISL_1095904, EPI_ISL_1095905, EPI_ISL_1095906, EPI_ISL_1095907, EPI_ISL_1095908, EPI_ISL_1095909, EPI_ISL_1095910, EPI_ISL_1095914, EPI_ISL_1095915, EPI_ISL_1095916, EPI_ISL_1095917, EPI_ISL_1095918, EPI_ISL_1095919, EPI_ISL_1095920, EPI_ISL_1095921, EPI_ISL_1095922, EPI_ISL_1095923, EPI_ISL_1095924, EPI_ISL_1095925, EPI_ISL_1095926, EPI_ISL_1095927, EPI_ISL_1095928, EPI_ISL_1095929, EPI_ISL_1095930, EPI_ISL_1095931, EPI_ISL_1095932, EPI_ISL_1095933, EPI_ISL_1095934, EPI_ISL_1095935, EPI_ISL_1095936, EPI_ISL_1095937, EPI_ISL_1095938, EPI_ISL_1095939, EPI_ISL_1095940, EPI_ISL_1095941, EPI_ISL_1095942, EPI_ISL_1095943, EPI_ISL_1095944, EPI_ISL_1095945, EPI_ISL_1095946, EPI_ISL_1095947, EPI_ISL_1095948, EPI_ISL_1095949, EPI_ISL_1095950, EPI_ISL_1095951, EPI_ISL_1095952, EPI_ISL_1095953, EPI_ISL_1095954, EPI_ISL_1095955, EPI_ISL_1095956, EPI_ISL_1095957, EPI_ISL_1095958, EPI_ISL_1095959, EPI_ISL_1095960, EPI_ISL_1095961, EPI_ISL_1095962, EPI_ISL_1095963, EPI_ISL_1095964, EPI_ISL_1095965, EPI_ISL_1095966, EPI_ISL_1095967, EPI_ISL_1095968, EPI_ISL_1095969, EPI_ISL_1095970, EPI_ISL_1095971, EPI_ISL_1095972, EPI_ISL_1095973, EPI_ISL_1095974, EPI_ISL_1095975, EPI_ISL_1095976, EPI_ISL_1095977, EPI_ISL_1095978, EPI_ISL_1095979, EPI_ISL_1095980, EPI_ISL_1095981, EPI_ISL_1095982, EPI_ISL_1095983, EPI_ISL_1095984, EPI_ISL_1095985, EPI_ISL_1095986, EPI_ISL_1095987, EPI_ISL_1095988, EPI_ISL_1095989, EPI_ISL_1095990, EPI_ISL_1095991, EPI_ISL_1095992, EPI_ISL_1095993, EPI_ISL_1095994, EPI_ISL_1095995, EPI_ISL_1095996, EPI_ISL_1095997, EPI_ISL_1095998, EPI_ISL_1095999, EPI_ISL_1096000, EPI_ISL_1096001, EPI_ISL_1096002, EPI_ISL_1096003, EPI_ISL_1096004, EPI_ISL_1096005, EPI_ISL_1096006, EPI_ISL_1096007, EPI_ISL_1096008, EPI_ISL_1096009, EPI_ISL_1096010, EPI_ISL_1096011, EPI_ISL_1096012, EPI_ISL_1096013, EPI_ISL_1096014, EPI_ISL_1096015, EPI_ISL_1096016, EPI_ISL_1096017, EPI_ISL_1096018, EPI_ISL_1096019, EPI_ISL_1096020, EPI_ISL_1096021, EPI_ISL_1096022, EPI_ISL_1096023, EPI_ISL_1096024, EPI_ISL_1096025, EPI_ISL_1096026, EPI_ISL_1096027, EPI_ISL_1096028 | | | |
| see above | Fulgent Genetics | Fulgent Genetics | Harry Gao, Mickey Li, John Gao, Joseph Fierro, Benafsh Sapra, Becky Tsai, Yan Meng, Doreen Ng, James Xie |
| EPI_ISL_1096137, EPI_ISL_1096138 | Ohio State University Wexner Medical Center | James Molecular Laboratory | Huolin Tu, Pamela Snyder, Zachary Rudy, Sara Koenig, Saran Corcoran, Keelie Thomas, Preeti Pancholi, Dan Jones |
| EPI_ISL_1096141 | Departamento de Cuidados Intensivos-Unidad de Investigación Médica en Inmunoquímica, UMAE Hospital de Especialidades "Bernardo Sepúlveda Gutiérrez", Centro Médico Nacional Siglo XXI, Instituto Mexicano del Seguro Social (IMSS), México. | Laboratorio de Secuenciación-Centro de Instrumentos, IMSS México /Unidad de Investigación Médica en Inmunoquímica, UMAE Hospital de Especialidades "Bernardo Sepúlveda Gutiérrez", Centro Médico Nacional Siglo XXI, Instituto Mexicano del Seguro Social (IMSS), México. | Horacio Zamudio-Meza, Javier Gaytán-Cervantes, Carolina González-Torres, Alejandro Torres Flores, Lourdes Arriaga-Pizano, Eduardo Ferat-Osorio, Arturo Cérbulo-Vázquez, Luis Sánchez Hurtado, Elizabeth Rubí Duran Castañeda, Jesús Mena López, Juan Carlos Guerrero Escobar, Zuleima Maldonado Galeana, Aura Angélica López García, José Manuel Ruiz Rincón, Joaquín González-Ibarra, César Raúl González-Bonilla, Constantino López-Macías. |
| EPI_ISL_1096985 | Departamento de Cuidados Intensivos-Unidad de Investigación Médica en Inmunoquímica, UMAE Hospital de Especialidades "Bernardo Sepúlveda Gutiérrez", Centro Médico Nacional Siglo XXI, Instituto Mexicano del Seguro Social (IMSS) México. | Laboratorio de Secuenciación-Centro de Instrumentos, IMSS México /Unidad de Investigación Médica en Inmunoquímica, UMAE Hospital de Especialidades "Bernardo Sepúlveda Gutiérrez", Centro Médico Nacional Siglo XXI, Instituto Mexicano del Seguro Social (IMSS), México. | Javier Gaytán-Cervantes, Carolina González-Torres, Horacio Zamudio-Meza, Alejandro Torres Flores, Lourdes Arriaga-Pizano, Eduardo Ferat-Osorio, Arturo Cérbulo-Vázquez, Luis Sánchez Hurtado, Elizabeth Rubí Duran Castañeda, Jesús Mena López, Juan Carlos Guerrero Escobar, Zuleima Maldonado Galeana, Aura Angélica López García, José Manuel Ruiz Rincón, Joaquín González-Ibarra, César Raúl González-Bonilla, Constantino López-Macías. |
| EPI_ISL_1097018 | Baylor Scott & White-Temple | Baylor Scott & White-Temple | Ari Rao, Linden Morales, Kimberly Walker, Marcus Volz, Shelby Hendrickson |
| EPI_ISL_1097313, EPI_ISL_1097314, EPI_ISL_1097315, EPI_ISL_1097316, EPI_ISL_1097317, EPI_ISL_1097318, EPI_ISL_1097320, EPI_ISL_1097321, EPI_ISL_1097322, EPI_ISL_1097323, EPI_ISL_1097324, EPI_ISL_1097325, EPI_ISL_1097326, EPI_ISL_1097327, EPI_ISL_1097328, EPI_ISL_1097329, EPI_ISL_1097330, EPI_ISL_1097331, EPI_ISL_1097332, EPI_ISL_1097333, EPI_ISL_1097334, EPI_ISL_1097335, EPI_ISL_1097336, EPI_ISL_1097337, EPI_ISL_1097338, EPI_ISL_1097339, EPI_ISL_1097340, EPI_ISL_1097341, EPI_ISL_1097342, EPI_ISL_1097343, EPI_ISL_1097344, EPI_ISL_1097345, EPI_ISL_1097346 | | | |
| see above | University of Oregon COVID-19 MAP Laboratory | University of Oregon Genomics and Cell Characterization Core Facility (GC3F) | Douglas Turnbull, Ariana White, Jeff Bishop, Jason Carriere |
| EPI_ISL_1097347, EPI_ISL_1097348, EPI_ISL_1097349, EPI_ISL_1097350, EPI_ISL_1097351, EPI_ISL_1097352, EPI_ISL_1097353, EPI_ISL_1097354, EPI_ISL_1097355, EPI_ISL_1097356, EPI_ISL_1097357, EPI_ISL_1097358, EPI_ISL_1097359, EPI_ISL_1097360, EPI_ISL_1097361, EPI_ISL_1097362, EPI_ISL_1097363, EPI_ISL_1097364, EPI_ISL_1097365, EPI_ISL_1097366, EPI_ISL_1097367, EPI_ISL_1097368, EPI_ISL_1097369, EPI_ISL_1097370, EPI_ISL_1097371, EPI_ISL_1097386, EPI_ISL_1097387, EPI_ISL_1097388, EPI_ISL_1097389, EPI_ISL_1097390, EPI_ISL_1097391, EPI_ISL_1097392, EPI_ISL_1097393, EPI_ISL_1097394, EPI_ISL_1097395, EPI_ISL_1097396, EPI_ISL_1097397, EPI_ISL_1097398, EPI_ISL_1097399, EPI_ISL_1097400, EPI_ISL_1097401, EPI_ISL_1097402, EPI_ISL_1097403, EPI_ISL_1097404, EPI_ISL_1097405, EPI_ISL_1097406, EPI_ISL_1097407, EPI_ISL_1097408, EPI_ISL_1097409, EPI_ISL_1097410, EPI_ISL_1097411, EPI_ISL_1097412, EPI_ISL_1097413, EPI_ISL_1097414, EPI_ISL_1097415, EPI_ISL_1097416, EPI_ISL_1097417, EPI_ISL_1097418, EPI_ISL_1097419, EPI_ISL_1097420, EPI_ISL_1097421, EPI_ISL_1097422, EPI_ISL_1097423, EPI_ISL_1097424, EPI_ISL_1097425, EPI_ISL_1097426, EPI_ISL_1097427, EPI_ISL_1097428, EPI_ISL_1097429, EPI_ISL_1097430, EPI_ISL_1097431, EPI_ISL_1097432, EPI_ISL_1097433, EPI_ISL_1097434, EPI_ISL_1097435, EPI_ISL_1097436, EPI_ISL_1097437, EPI_ISL_1097438, EPI_ISL_1097439, EPI_ISL_1097440, EPI_ISL_1097441, EPI_ISL_1097442, EPI_ISL_1097443, EPI_ISL_1097444, EPI_ISL_1097445 | | | |
| see above | Florida Bureau of Public Health Laboratories | Florida Bureau of Public Health Laboratories | Sarah Schmedes, Jason Blanton |
| EPI_ISL_1097446, EPI_ISL_1097449, EPI_ISL_1097450, EPI_ISL_1097451, EPI_ISL_1097453, EPI_ISL_1097454, EPI_ISL_1097457, EPI_ISL_1097458, EPI_ISL_1097459, EPI_ISL_1097460, EPI_ISL_1097461, EPI_ISL_1097462, EPI_ISL_1097463, EPI_ISL_1097464, EPI_ISL_1097465, EPI_ISL_1097466, EPI_ISL_1097467, EPI_ISL_1097468, EPI_ISL_1097469, EPI_ISL_1097471, EPI_ISL_1097472, EPI_ISL_1097473, EPI_ISL_1097474, EPI_ISL_1097475, EPI_ISL_1097477, EPI_ISL_1097478, EPI_ISL_1097479, EPI_ISL_1097480, EPI_ISL_1097481, EPI_ISL_1097483, EPI_ISL_1097484, EPI_ISL_1097485, EPI_ISL_1097486, EPI_ISL_1097488, EPI_ISL_1097489, EPI_ISL_1097492, EPI_ISL_1097493, EPI_ISL_1097494, EPI_ISL_1097495, EPI_ISL_1097496, EPI_ISL_1097498, EPI_ISL_1097500, EPI_ISL_1097501, EPI_ISL_1097502, EPI_ISL_1097503, EPI_ISL_1097504, EPI_ISL_1097505, EPI_ISL_1097506, EPI_ISL_1097508, EPI_ISL_1097509, EPI_ISL_1097510, EPI_ISL_1097511, EPI_ISL_1097512, EPI_ISL_1097513, EPI_ISL_1097514, EPI_ISL_1097515, EPI_ISL_1097516, EPI_ISL_1097517, EPI_ISL_1097518, EPI_ISL_1097519, EPI_ISL_1097520, EPI_ISL_1097521, EPI_ISL_1097522, EPI_ISL_1097525, EPI_ISL_1097526, EPI_ISL_1097527, EPI_ISL_1097528, EPI_ISL_1097529, EPI_ISL_1097530, EPI_ISL_1097531, EPI_ISL_1097532, EPI_ISL_1097533, EPI_ISL_1097534, EPI_ISL_1097535, EPI_ISL_1097536, EPI_ISL_1097537, EPI_ISL_1097538, EPI_ISL_1097539, EPI_ISL_1097540, EPI_ISL_1097541, EPI_ISL_1097542, EPI_ISL_1097543, EPI_ISL_1097544, EPI_ISL_1097545, EPI_ISL_1097546, EPI_ISL_1097547, EPI_ISL_1097548, EPI_ISL_1097549, EPI_ISL_1097550, EPI_ISL_1097551, EPI_ISL_1097552, EPI_ISL_1097553, EPI_ISL_1097554, EPI_ISL_1097555, EPI_ISL_1097556, EPI_ISL_1097557, EPI_ISL_1097558, EPI_ISL_1097559, EPI_ISL_1097560, EPI_ISL_1097561, EPI_ISL_1097562, EPI_ISL_1097563, EPI_ISL_1097564, EPI_ISL_1097565, EPI_ISL_1097566, EPI_ISL_1097568, EPI_ISL_1097570, EPI_ISL_1097571, EPI_ISL_1097573, EPI_ISL_1097574, EPI_ISL_1097575, EPI_ISL_1097576, EPI_ISL_1097577, EPI_ISL_1097578, EPI_ISL_1097579, EPI_ISL_1097580, EPI_ISL_1097581, EPI_ISL_1097582, EPI_ISL_1097583, EPI_ISL_1097585, EPI_ISL_1097586, EPI_ISL_1097588, EPI_ISL_1097589, EPI_ISL_1097590, EPI_ISL_1097591, EPI_ISL_1097592, EPI_ISL_1097593, EPI_ISL_1097594, EPI_ISL_1097595, EPI_ISL_1097596, EPI_ISL_1097597, EPI_ISL_1097598, EPI_ISL_1097599, EPI_ISL_1097600, EPI_ISL_1097601, EPI_ISL_1097602, EPI_ISL_1097603, EPI_ISL_1097604, EPI_ISL_1097605, EPI_ISL_1097606, EPI_ISL_1097607, EPI_ISL_1097608, EPI_ISL_1097609, EPI_ISL_1097610, EPI_ISL_1097611, EPI_ISL_1097612, EPI_ISL_1097613, EPI_ISL_1097614, EPI_ISL_1097616, EPI_ISL_1097617, EPI_ISL_1097618, EPI_ISL_1097619, EPI_ISL_1097620, EPI_ISL_1097621, EPI_ISL_1097622, EPI_ISL_1097623, EPI_ISL_1097624, EPI_ISL_1097625, EPI_ISL_1097626, EPI_ISL_1097627, EPI_ISL_1097628, EPI_ISL_1097629, EPI_ISL_1097630, EPI_ISL_1097631, EPI_ISL_1097632, EPI_ISL_1097633, EPI_ISL_1097634, EPI_ISL_1097635, EPI_ISL_1097636, EPI_ISL_1097637, EPI_ISL_1097638, EPI_ISL_1097639, EPI_ISL_1097640, EPI_ISL_1097641, EPI_ISL_1097642, EPI_ISL_1097643 | | | |
| see above | Colorado Department of Public Health and Environment | Colorado Department of Puplic Health and Environment | Laura Bankers, Molly C. Hetherington-Rauth, Diana Ir, Shannon Ely, Shannon R. Matzinger, Sarah Elizabeth Totten, Emily A. Travanty |
| EPI_ISL_1097645, EPI_ISL_1097646, EPI_ISL_1097647, EPI_ISL_1097648, EPI_ISL_1097649, EPI_ISL_1097650, EPI_ISL_1097651, EPI_ISL_1097652, EPI_ISL_1097653, EPI_ISL_1097654, EPI_ISL_1097655, EPI_ISL_1097656, EPI_ISL_1097657, EPI_ISL_1097658, EPI_ISL_1097659, EPI_ISL_1097660, EPI_ISL_1097661, EPI_ISL_1097662, EPI_ISL_1097663, EPI_ISL_1097664, EPI_ISL_1097665, EPI_ISL_1097666, EPI_ISL_1097667, EPI_ISL_1097668, EPI_ISL_1097669, EPI_ISL_1097670, EPI_ISL_1097671, EPI_ISL_1097672, EPI_ISL_1097673, EPI_ISL_1097674, EPI_ISL_1097675, EPI_ISL_1097676, EPI_ISL_1097677, EPI_ISL_1097678, EPI_ISL_1097679, EPI_ISL_1097680, EPI_ISL_1097681, EPI_ISL_1097682, EPI_ISL_1097683, EPI_ISL_1097684, EPI_ISL_1097685, EPI_ISL_1097686, EPI_ISL_1097687, EPI_ISL_1097688, EPI_ISL_1097689, EPI_ISL_1097690, EPI_ISL_1097691, EPI_ISL_1097692, EPI_ISL_1097693, EPI_ISL_1097694, EPI_ISL_1097695, EPI_ISL_1097696, EPI_ISL_1097697, EPI_ISL_1097698, EPI_ISL_1097699, EPI_ISL_1097700, EPI_ISL_1097701, EPI_ISL_1097702, EPI_ISL_1097703, EPI_ISL_1097704, EPI_ISL_1097705, EPI_ISL_1097706, EPI_ISL_1097707, EPI_ISL_1097708, EPI_ISL_1097709, EPI_ISL_1097710, EPI_ISL_1097711, EPI_ISL_1097712, EPI_ISL_1097713, EPI_ISL_1097714, EPI_ISL_1097715, EPI_ISL_1097716, EPI_ISL_1097717, EPI_ISL_1097718, EPI_ISL_1097719, EPI_ISL_1097720, EPI_ISL_1097721, EPI_ISL_1097722, EPI_ISL_1097723 | | | |
| see above | Eurofins Diatherix | Hudsonalpha Genome Sequencing Center | Jane Grimwood, Melissa Williams, Lori H. Handley, Joshua Stough, Leslie Malone, Stefan Brzezinski, Ada Stewart, Teresa Jones, Jenell Webber, John Lovell, Jennifer Cart, and Jeremy Schmutz |
| EPI_ISL_1097724, EPI_ISL_1097725, EPI_ISL_1097726, EPI_ISL_1097727, EPI_ISL_1097728, EPI_ISL_1097729, EPI_ISL_1097730, EPI_ISL_1097731, EPI_ISL_1097732, EPI_ISL_1097733, EPI_ISL_1097734, EPI_ISL_1097735, EPI_ISL_1097736, EPI_ISL_1097737 | | | |
| see above | Los Angeles County Public Health Laboratories | Los Angeles County Public Health Laboratories | P. Hemarajata et al. |
| EPI_ISL_1097739 | Flow Diagnostics Lab | Los Angeles County Public Health Laboratories | P. Hemarajata et al. |
| EPI_ISL_1097740, EPI_ISL_1097741 | Los Angeles County Public Health Laboratories | Los Angeles County Public Health Laboratories | P. Hemarajata et al. |
| EPI_ISL_1097742 | UCLA Clinical Micro Lab | Los Angeles County Public Health Laboratories | P. Hemarajata et al. |
| EPI_ISL_1097743 | Los Angeles County Public Health Laboratories | Los Angeles County Public Health Laboratories | P. Hemarajata et al. |
| EPI_ISL_1097744, EPI_ISL_1097745, EPI_ISL_1097746, EPI_ISL_1097747, EPI_ISL_1097748, EPI_ISL_1097749, EPI_ISL_1097750, EPI_ISL_1097752, EPI_ISL_1097753, EPI_ISL_1097755, EPI_ISL_1097756, EPI_ISL_1097757, EPI_ISL_1097758, EPI_ISL_1097759, EPI_ISL_1097760, EPI_ISL_1097761, EPI_ISL_1097762, EPI_ISL_1097764, EPI_ISL_1097765, EPI_ISL_1097766, EPI_ISL_1097767, EPI_ISL_1097768, EPI_ISL_1097769, EPI_ISL_1097770, EPI_ISL_1097771, EPI_ISL_1097772, EPI_ISL_1097773, EPI_ISL_1097774, EPI_ISL_1097775, EPI_ISL_1097776, EPI_ISL_1097777, EPI_ISL_1097778, EPI_ISL_1097779, EPI_ISL_1097780 | | | |
| see above | New Mexico Department of Health Scientific Laboratory | New Mexico Department of Health Scientific Laboratory | Ellie Johnson, Anastacia Griego-Fisher, D'eldra Malone, Jennifer Benoit |
| EPI_ISL_1097781, EPI_ISL_1097782, EPI_ISL_1097783, EPI_ISL_1097784, EPI_ISL_1097785, EPI_ISL_1097786, EPI_ISL_1097787, EPI_ISL_1097788, EPI_ISL_1097789, EPI_ISL_1097790, EPI_ISL_1097791, EPI_ISL_1097792, EPI_ISL_1097793, EPI_ISL_1097794, EPI_ISL_1097795, EPI_ISL_1097796, EPI_ISL_1097797, EPI_ISL_1097798, EPI_ISL_1097799, EPI_ISL_1097800, EPI_ISL_1097801, EPI_ISL_1097802, EPI_ISL_1097803, EPI_ISL_1097804, EPI_ISL_1097805, EPI_ISL_1097806, EPI_ISL_1097807, EPI_ISL_1097808, EPI_ISL_1097809, EPI_ISL_1097810, EPI_ISL_1097811, EPI_ISL_1097812, EPI_ISL_1097813, EPI_ISL_1097814, EPI_ISL_1097815, EPI_ISL_1097816, EPI_ISL_1097817, EPI_ISL_1097818, EPI_ISL_1097819, EPI_ISL_1097820, EPI_ISL_1097821, EPI_ISL_1097822, EPI_ISL_1097823, EPI_ISL_1097824, EPI_ISL_1097825, EPI_ISL_1097826, EPI_ISL_1097827, EPI_ISL_1097828, EPI_ISL_1097829, EPI_ISL_1097830, EPI_ISL_1097831, EPI_ISL_1097832, EPI_ISL_1097833, EPI_ISL_1097834, EPI_ISL_1097835, EPI_ISL_1097836, EPI_ISL_1097837, EPI_ISL_1097838, EPI_ISL_1097839, EPI_ISL_1097840, EPI_ISL_1097841, EPI_ISL_1097842, EPI_ISL_1097843, EPI_ISL_1097844, EPI_ISL_1097845, EPI_ISL_1097846, EPI_ISL_1097847, EPI_ISL_1097848, EPI_ISL_1097849, EPI_ISL_1097850, EPI_ISL_1097851, EPI_ISL_1097852, EPI_ISL_1097853, EPI_ISL_1097854, EPI_ISL_1097855, EPI_ISL_1097856, EPI_ISL_1097857, EPI_ISL_1097858, EPI_ISL_1097859, EPI_ISL_1097860, EPI_ISL_1097861, EPI_ISL_1097862, EPI_ISL_1097863, EPI_ISL_1097864, EPI_ISL_1097865, EPI_ISL_1097866, EPI_ISL_1097867, EPI_ISL_1097868, EPI_ISL_1097869, EPI_ISL_1097870, EPI_ISL_1097871, EPI_ISL_1097872, EPI_ISL_1097873, EPI_ISL_1097874, EPI_ISL_1097875, EPI_ISL_1097876, EPI_ISL_1097877, EPI_ISL_1097878, EPI_ISL_1097879, EPI_ISL_1097880, EPI_ISL_1097881, EPI_ISL_1097882, EPI_ISL_1097883, EPI_ISL_1097884, EPI_ISL_1097885, EPI_ISL_1097886, EPI_ISL_1097887, EPI_ISL_1097888, EPI_ISL_1097889, EPI_ISL_1097890, EPI_ISL_1097891, EPI_ISL_1097892, EPI_ISL_1097893, EPI_ISL_1097894, EPI_ISL_1097895, EPI_ISL_1097896, EPI_ISL_1097897, EPI_ISL_1097898, EPI_ISL_1097899, EPI_ISL_1097900, EPI_ISL_1097901, EPI_ISL_1097902, EPI_ISL_1097903, EPI_ISL_1097904, EPI_ISL_1097905, EPI_ISL_1097906, EPI_ISL_1097907, EPI_ISL_1097908, EPI_ISL_1097909, EPI_ISL_1097910, EPI_ISL_1097911, EPI_ISL_1097912, EPI_ISL_1097913, EPI_ISL_1097914, EPI_ISL_1097915, EPI_ISL_1097916, EPI_ISL_1097917, EPI_ISL_1097918, EPI_ISL_1097919, EPI_ISL_1097920, EPI_ISL_1097921, EPI_ISL_1097922, EPI_ISL_1097923, EPI_ISL_1097924, EPI_ISL_1097925, EPI_ISL_1097926, EPI_ISL_1097927, EPI_ISL_1097928, EPI_ISL_1097929, EPI_ISL_1097930, EPI_ISL_1097931, EPI_ISL_1097932, EPI_ISL_1097933, EPI_ISL_1097934, EPI_ISL_1097935, EPI_ISL_1097936, EPI_ISL_1097937, EPI_ISL_1097938, EPI_ISL_1097939, EPI_ISL_1097940, EPI_ISL_1097941, EPI_ISL_1097942, EPI_ISL_1097943, EPI_ISL_1097944, EPI_ISL_1097945, EPI_ISL_1097946, EPI_ISL_1097947, EPI_ISL_1097948, EPI_ISL_1097949, EPI_ISL_1097950, EPI_ISL_1097951, EPI_ISL_1097952, EPI_ISL_1097953, EPI_ISL_1097954, EPI_ISL_1097955, EPI_ISL_1097956, EPI_ISL_1097957, EPI_ISL_1097958, EPI_ISL_1097959, EPI_ISL_1097960, EPI_ISL_1097961, EPI_ISL_1097962, EPI_ISL_1097963, EPI_ISL_1097964, EPI_ISL_1097965, EPI_ISL_1097966, EPI_ISL_1097967, EPI_ISL_1097968, EPI_ISL_1097969, EPI_ISL_1097970, EPI_ISL_1097971, EPI_ISL_1097972, EPI_ISL_1097973, EPI_ISL_1097974, EPI_ISL_1097975, EPI_ISL_1097976, EPI_ISL_1097977, EPI_ISL_1097978, EPI_ISL_1097979, EPI_ISL_1097980, EPI_ISL_1097981, EPI_ISL_1097982, EPI_ISL_1097983, EPI_ISL_1097984, EPI_ISL_1097985, EPI_ISL_1097986, EPI_ISL_1097987, EPI_ISL_1097988, EPI_ISL_1097989, EPI_ISL_1097990, EPI_ISL_1097991, EPI_ISL_1097992, EPI_ISL_1097993, EPI_ISL_1097994, EPI_ISL_1097995, EPI_ISL_1097996, EPI_ISL_1097997, EPI_ISL_1097998, EPI_ISL_1097999, EPI_ISL_1098000, EPI_ISL_1098001, EPI_ISL_1098002, EPI_ISL_1098003, EPI_ISL_1098004, EPI_ISL_1098005, EPI_ISL_1098006, EPI_ISL_1098007, EPI_ISL_1098008, EPI_ISL_1098009, EPI_ISL_1098010, EPI_ISL_1098011, EPI_ISL_1098012, EPI_ISL_1098013, EPI_ISL_1098014, EPI_ISL_1098015, EPI_ISL_1098016, EPI_ISL_1098017, EPI_ISL_1098018, EPI_ISL_1098019, EPI_ISL_1098020, EPI_ISL_1098021, EPI_ISL_1098022, EPI_ISL_1098023, EPI_ISL_1098024, EPI_ISL_1098025, EPI_ISL_1098026, EPI_ISL_1098027, EPI_ISL_1098028, EPI_ISL_1098029, EPI_ISL_1098030, EPI_ISL_1098031, EPI_ISL_1098032, EPI_ISL_1098033, EPI_ISL_1098034, EPI_ISL_1098035, EPI_ISL_1098036, EPI_ISL_1098037, EPI_ISL_1098038, EPI_ISL_1098039, EPI_ISL_1098040, EPI_ISL_1098041, EPI_ISL_1098042, EPI_ISL_1098043, EPI_ISL_1098044, EPI_ISL_1098045, EPI_ISL_1098046, EPI_ISL_1098047, EPI_ISL_1098048, EPI_ISL_1098049, EPI_ISL_1098050, EPI_ISL_1098051, EPI_ISL_1098052, EPI_ISL_1098053, EPI_ISL_1098054, EPI_ISL_1098055, EPI_ISL_1098056, EPI_ISL_1098057, EPI_ISL_1098058, EPI_ISL_1098059, EPI_ISL_1098060, EPI_ISL_1098061, EPI_ISL_1098062, EPI_ISL_1098063, EPI_ISL_1098064, EPI_ISL_1098065, EPI_ISL_1098066, EPI_ISL_1098067, EPI_ISL_1098068, EPI_ISL_1098069, EPI_ISL_1098070, EPI_ISL_1098071, EPI_ISL_1098072, EPI_ISL_1098073, EPI_ISL_1098074, EPI_ISL_1098075, EPI_ISL_1098076, EPI_ISL_1098077, EPI_ISL_1098078, EPI_ISL_1098079, EPI_ISL_1098080, EPI_ISL_1098081, EPI_ISL_1098082, EPI_ISL_1098083, EPI_ISL_1098084, EPI_ISL_1098085, EPI_ISL_1098086, EPI_ISL_1098087, EPI_ISL_1098088, EPI_ISL_1098089, EPI_ISL_1098090, EPI_ISL_1098091, EPI_ISL_1098092, EPI_ISL_1098093, EPI_ISL_1098094, EPI_ISL_1098095, EPI_ISL_1098096, EPI_ISL_1098097, EPI_ISL_1098098, EPI_ISL_1098099, EPI_ISL_1098100, EPI_ISL_1098101, EPI_ISL_1098102, EPI_ISL_1098103, EPI_ISL_1098104, EPI_ISL_1098105, EPI_ISL_1098106, EPI_ISL_1098107, EPI_ISL_1098108, EPI_ISL_1098109, EPI_ISL_1098110, EPI_ISL_1098111, EPI_ISL_1098112, EPI_ISL_1098113, EPI_ISL_1098114, EPI_ISL_1098115, EPI_ISL_1098116, EPI_ISL_1098117, EPI_ISL_1098118, EPI_ISL_1098119, EPI_ISL_1098120, EPI_ISL_1098121, EPI_ISL_1098122, EPI_ISL_1098123, EPI_ISL_1098124, EPI_ISL_1098125, EPI_ISL_1098126, EPI_ISL_1098127, EPI_ISL_1098128, EPI_ISL_1098129, EPI_ISL_1098130, EPI_ISL_1098131, EPI_ISL_1098132, EPI_ISL_1098133, EPI_ISL_1098134, EPI_ISL_1098135, EPI_ISL_1098136, EPI_ISL_1098137, EPI_ISL_1098138, EPI_ISL_1098139, EPI_ISL_1098140, EPI_ISL_1098141, EPI_ISL_1098142, EPI_ISL_1098143, EPI_ISL_1098144, EPI_ISL_1098145, EPI_ISL_1098146, EPI_ISL_1098147, EPI_ISL_1098148, EPI_ISL_1098149, EPI_ISL_1098150, EPI_ISL_1098151, EPI_ISL_1098152, EPI_ISL_1098153, EPI_ISL_1098154, EPI_ISL_1098155, EPI_ISL_1098156, EPI_ISL_1098157, EPI_ISL_1098158, EPI_ISL_1098159, EPI_ISL_1098160, EPI_ISL_1098161, EPI_ISL_1098162, EPI_ISL_1098163, EPI_ISL_1098164, EPI_ISL_1098165, EPI_ISL_1098166, EPI_ISL_1098167, EPI_ISL_1098168, EPI_ISL_1098169, EPI_ISL_1098170, EPI_ISL_1098171, EPI_ISL_1098172, EPI_ISL_1098173, EPI_ISL_1098174, EPI_ISL_1098175, EPI_ISL_1098176, EPI_ISL_1098177, EPI_ISL_1098178, EPI_ISL_1098179, EPI_ISL_1098180, EPI_ISL_1098181, EPI_ISL_1098182, EPI_ISL_1098183, EPI_ISL_1098184, EPI_ISL_1098185, EPI_ISL_1098186, EPI_ISL_1098187, EPI_ISL_1098188, EPI_ISL_1098189, EPI_ISL_1098190, EPI_ISL_1098191, EPI_ISL_1098192, EPI_ISL_1098193, EPI_ISL_1098194, EPI_ISL_1098195, EPI_ISL_1098196, EPI_ISL_1098197, EPI_ISL_1098198, EPI_ISL_1098199, EPI_ISL_1098200, EPI_ISL_1098201, EPI_ISL_1098202, EPI_ISL_1098203, EPI_ISL_1098204, EPI_ISL_1098205, EPI_ISL_1098206, EPI_ISL_1098207, EPI_ISL_1098208, EPI_ISL_1098209, EPI_ISL_1098210, EPI_ISL_1098211, EPI_ISL_1098212, EPI_ISL_1098213, EPI_ISL_1098214, EPI_ISL_1098215, EPI_ISL_1098216, EPI_ISL_1098217, EPI_ISL_1098218, EPI_ISL_1098219, EPI_ISL_1098220, EPI_ISL_1098221, EPI_ISL_1098222, EPI_ISL_1098223, EPI_ISL_1098224, EPI_ISL_1098225, EPI_ISL_1098226, EPI_ISL_1098227, EPI_ISL_1098228, EPI_ISL_1098229, EPI_ISL_1098230, EPI_ISL_1098231, EPI_ISL_1098232, EPI_ISL_1098233, EPI_ISL_1098234, EPI_ISL_1098235, EPI_ISL_1098236, EPI_ISL_1098237, EPI_ISL_1098238, EPI_ISL_1098239, EPI_ISL_1098240, EPI_ISL_1098241, EPI_ISL_1098242, EPI_ISL_1098243, EPI_ISL_1098244, EPI_ISL_1098245, EPI_ISL_1098246, EPI_ISL_1098247, EPI_ISL_1098248, EPI_ISL_1098249, EPI_ISL_1098250, EPI_ISL_1098251, EPI_ISL_1098252, EPI_ISL_1098253, EPI_ISL_1098254, EPI_ISL_1098255, EPI_ISL_1098256, EPI_ISL_1098257, EPI_ISL_1098258, EPI_ISL_1098259, EPI_ISL_1098260, EPI_ISL_1098261, EPI_ISL_1098262, EPI_ISL_1098263, EPI_ISL_1098264, EPI_ISL_1098265, EPI_ISL_1098266, EPI_ISL_1098267, EPI_ISL_1098268, EPI_ISL_1098269, EPI_ISL_1098270, EPI_ISL_1098271, EPI_ISL_1098272, EPI_ISL_1098273, EPI_ISL_1098274, EPI_ISL_1098275, EPI_ISL_1098276, EPI_ISL_1098277, EPI_ISL_1098278, EPI_ISL_1098279, EPI_ISL_1098280, EPI_ISL_1098281, EPI_ISL_1098282, EPI_ISL_1098283, EPI_ISL_1098284, EPI_ISL_1098285, EPI_ISL_1098286, EPI_ISL_1098287, EPI_ISL_1098288, EPI_ISL_1098289, EPI_ISL_1098290, EPI_ISL_1098291, EPI_ISL_1098292, EPI_ISL_1098293, EPI_ISL_1098294, EPI_ISL_1098295, EPI_ISL_1098296, EPI_ISL_1098297, EPI_ISL_1098298, EPI_ISL_1098299, EPI_ISL_1098300, EPI_ISL_1098301, EPI_ISL_1098302, EPI_ISL_1098303, EPI_ISL_1098304, EPI_ISL_1098305, EPI_ISL_1098306, EPI_ISL_1098307, EPI_ISL_1098308, EPI_ISL_1098309, EPI_ISL_1098310, EPI_ISL_1098311, EPI_ISL_1098312, EPI_ISL_1098313, EPI_ISL_1098314, EPI_ISL_1098315, EPI_ISL_1098316, EPI_ISL_1098317, EPI_ISL_1098318, EPI_ISL_1098319, EPI_ISL_1098320, EPI_ISL_1098321, EPI_ISL_1098322, EPI_ISL_1098323, EPI_ISL_1098324, EPI_ISL_1098325, EPI_ISL_1098326, EPI_ISL_1098327, EPI_ISL_1098328, EPI_ISL_1098329, EPI_ISL_1098330, EPI_ISL_1098331, EPI_ISL_1098332, EPI_ISL_1098333, EPI_ISL_1098334, EPI_ISL_1098335, EPI_ISL_1098336, EPI_ISL_1098337, EPI_ISL_1098338, EPI_ISL_1098339, EPI_ISL_1098340, EPI_ISL_1098341, EPI_ISL_1098342, EPI_ISL_1098343, EPI_ISL_1098344, EPI_ISL_1098345, EPI_ISL_1098346, EPI_ISL_1098347, EPI_ISL_1098348, EPI_ISL_1098349, EPI_ISL_1098350, EPI_ISL_1098351, EPI_ISL_1098352, EPI_ISL_1098353, EPI_ISL_1098354, EPI_ISL_1098355, EPI_ISL_1098356, EPI_ISL_1098357, EPI_ISL_1098358, EPI_ISL_1098359, EPI_ISL_1098360, EPI_ISL_1098361, EPI_ISL_1098362, EPI_ISL_1098363, EPI_ISL_1098364, EPI_ISL_1098365, EPI_ISL_1098366, EPI_ISL_1098367, EPI_ISL_1098368, EPI_ISL_1098369, EPI_ISL_1098370, EPI_ISL_1098371, EPI_ISL_1098372, EPI_ISL_1098373, EPI_ISL_1098374, EPI_ISL_1098375, EPI_ISL_1098376, EPI_ISL_1098377, EPI_ISL_1098378, EPI_ISL_1098379, EPI_ISL_1098380, EPI_ISL_1098381, EPI_ISL_1098382, EPI_ISL_1098383, EPI_ISL_1098384, EPI_ISL_1098385, EPI_ISL_1098386, EPI_ISL_1098387, EPI_ISL_1098388, EPI_ISL_1098389, EPI_ISL_1098390, EPI_ISL_1098391, EPI_ISL_1098392, EPI_ISL_1098393, EPI_ISL_1098394, EPI_ISL_1098395, EPI_ISL_1098396, EPI_ISL_1098397, EPI_ISL_1098398, EPI_ISL_1098399, EPI_ISL_1098400, EPI_ISL_1098401, EPI_ISL_1098402, EPI_ISL_1098403, EPI_ISL_1098404, EPI_ISL_1098405, EPI_ISL_1098406, EPI_ISL_1098407, EPI_ISL_1098408, EPI_ISL_1098409, EPI_ISL_1098410, EPI_ISL_1098411, EPI_ISL_1098412, EPI_ISL_1098413, EPI_ISL_1098414, EPI_ISL_1098415, EPI_ISL_1098416, EPI_ISL_1098417, EPI_ISL_1098418, EPI_ISL_1098419, EPI_ISL_1098420, EPI_ISL_1098421, EPI_ISL_1098422, EPI_ISL_1098423, EPI_ISL_1098424, EPI_ISL_1098425, EPI_ISL_1098426, EPI_ISL_1098427, EPI_ISL_1098428, EPI_ISL_1098429, EPI_ISL_1098430, EPI_ISL_1098431, EPI_ISL_1098432, EPI_ISL_1098433, EPI_ISL_1098434, EPI_ISL_1098435, EPI_ISL_1098436, EPI_ISL_1098437, EPI_ISL_1098438, EPI_ISL_1098439, EPI_ISL_1098440, EPI_ISL_1098441, EPI_ISL_1098442, EPI_ISL_1098443, EPI_ISL_1098444, EPI_ISL_1098445, EPI_ISL_1098446, EPI_ISL_1098447, EPI_ISL_1098448, EPI_ISL_1098449, EPI_ISL_1098450, EPI_ISL_1098451, EPI_ISL_1098452, EPI_ISL_1098453, EPI_ISL_1098454, EPI_ISL_1098455, EPI_ISL_1098456, EPI_ISL_1098457, EPI_ISL_1098458, EPI_ISL_1098459, EPI_ISL_1098460, EPI_ISL_1098461, EPI_ISL_1098462, EPI_ISL_1098463, EPI_ISL_1098464, EPI_ISL_1098465, EPI_ISL_1098466, EPI_ISL_1098467, EPI_ISL_1098468, EPI_ISL_1098469, EPI_ISL_1098470, EPI_ISL_1098471, EPI_ISL_1098472, EPI_ISL_1098473, EPI_ISL_1098474, EPI_ISL_1098475, EPI_ISL_1098476, EPI_ISL_1098477, EPI_ISL_1098478, EPI_ISL_1098479, EPI_ISL_1098480, EPI_ISL_1098481, EPI_ISL_1098482, EPI_ISL_1098483, EPI_ISL_1098484, EPI_ISL_1098485, EPI_ISL_1098486, EPI_ISL_1098487, EPI_ISL_1098488, EPI_ISL_1098489, EPI_ISL_1098490, EPI_ISL_1098491, EPI_ISL_1098492, EPI_ISL_1098493, EPI_ISL_1098494, EPI_ISL_1098495, EPI_ISL_1098496, EPI_ISL_1098497, EPI_ISL_1098498, EPI_ISL_1098499, EPI_ISL_1098500, EPI_ISL_1098501, EPI_ISL_1098502, EPI_ISL_1098503, EPI_ISL_1098504, EPI_ISL_1098505, EPI_ISL_1098506, EPI_ISL_1098507, EPI_ISL_1098508, EPI_ISL_1098509, EPI_ISL_1098510, EPI_ISL_1098511, EPI_ISL_1098512, EPI_ISL_1098513, EPI_ISL_1098514, EPI_ISL_1098515, EPI_ISL_1098516, EPI_ISL_1098517, EPI_ISL_1098518, EPI_ISL_1098519, EPI_ISL_1098520, EPI_ISL_1098521, EPI_ISL_1098522, EPI_ISL_1098523, EPI_ISL_1098524, EPI_ISL_1098525, EPI_ISL_1098526, EPI_ISL_1098527, EPI_ISL_1098528, EPI_ISL_1098529, EPI_ISL_1098530, EPI_ISL_1098531, EPI_ISL_1098532, EPI_ISL_1098533, EPI_ISL_1098534, EPI_ISL_1098535, EPI_ISL_1098536, EPI_ISL_1098537, EPI_ISL_1098538, EPI_ISL_1098539, EPI_ISL_1098540, EPI_ISL_1098541, EPI_ISL_1098542, EPI_ISL_1098543, EPI_ISL_1098544, EPI_ISL_1098545, EPI_ISL_1098546, EPI_ISL_1098547, EPI_ISL_1098548, EPI_ISL_1098549, EPI_ISL_1098550, EPI_ISL_1098551, EPI_ISL_1098552, EPI_ISL_1098553, EPI_ISL_1098554, EPI_ISL_1098555, EPI_ISL_1098556, EPI_ISL_1098557, EPI_ISL_1098558, EPI_ISL_1098559, EPI_ISL_1098560, EPI_ISL_1098561, EPI_ISL_1098562, EPI_ISL_1098563, EPI_ISL_1098564, EPI_ISL_1098565, EPI_ISL_1098566, EPI_ISL_1098567, EPI_ISL_1098568, EPI_ISL_1098569, EPI_ISL_1098570, EPI_ISL_1098571, EPI_ISL_1098572, EPI_ISL_1098573, EPI_ISL_1098574, EPI_ISL_1098575, EPI_ISL_1098576, EPI_ISL_1098577, EPI_ISL_1098578, EPI_ISL_1098579, EPI_ISL_1098580, EPI_ISL_1098581, EPI_ISL_1098582, EPI_ISL_1098583, EPI_ISL_1098584, EPI_ISL_1098585, EPI_ISL_1098586, EPI_ISL_1098587, EPI_ISL_1098588, EPI_ISL_1098589, EPI_ISL_1098590, EPI_ISL_1098591, EPI_ISL_1098592, EPI_ISL_1098593, EPI_ISL_1098594, | | | |

| | | | |
|---|---|---|---|
| EPI_ISL_1098595, EPI_ISL_1098596, EPI_ISL_1098597, EPI_ISL_1098598, EPI_ISL_1098599, EPI_ISL_1098600, EPI_ISL_1098601 | | | |
| see above | Pandemic Response Lab - NYC | Pandemic Response Lab, R&D | Henry Lee, Michael Hammerling, Melissa Hopkins, Cybill del Castillo, Shinyoung Clair Kang, William Ward, Pradeep Bugga, Haiping Hao, Jon Laurent |
| EPI_ISL_1098609, EPI_ISL_1098610, EPI_ISL_1098611, EPI_ISL_1098612, EPI_ISL_1098613, EPI_ISL_1098614, EPI_ISL_1098615, EPI_ISL_1098616, EPI_ISL_1098617, EPI_ISL_1098618, EPI_ISL_1098619, EPI_ISL_1098620, EPI_ISL_1098621, EPI_ISL_1098622, EPI_ISL_1098623, EPI_ISL_1098624, EPI_ISL_1098625, EPI_ISL_1098626, EPI_ISL_1098627 | | | |
| see above | UCLA Clinical Micro Lab | Los Angeles County Public Health Laboratories | P. Hemarajata et al. |
| EPI_ISL_1098628 | MD Laboratories | Los Angeles County Public Health Laboratories | P. Hemarajata et al. |
| EPI_ISL_1098631 | UCLA Clinical Micro Lab | Los Angeles County Public Health Laboratories | P. Hemarajata et al. |
| EPI_ISL_1098632, EPI_ISL_1098633 | Los Angeles County Public Health Laboratories | Los Angeles County Public Health Laboratories | P. Hemarajata et al. |
| EPI_ISL_1098636, EPI_ISL_1098637 | Flow Diagnostics Lab | Los Angeles County Public Health Laboratories | P. Hemarajata et al. |
| EPI_ISL_1098639, EPI_ISL_1098640 | Los Angeles County Public Health Laboratories | Los Angeles County Public Health Laboratories | P. Hemarajata et al. |
| EPI_ISL_1098800, EPI_ISL_1098801, EPI_ISL_1098802, EPI_ISL_1098803, EPI_ISL_1098804, EPI_ISL_1098805, EPI_ISL_1098806, EPI_ISL_1098807, EPI_ISL_1098808, EPI_ISL_1098809, EPI_ISL_1098810, EPI_ISL_1098811, EPI_ISL_1098812, EPI_ISL_1098813, EPI_ISL_1098814, EPI_ISL_1098815, EPI_ISL_1098816, EPI_ISL_1098817, EPI_ISL_1098818, EPI_ISL_1098819, EPI_ISL_1098820, EPI_ISL_1098821, EPI_ISL_1098823, EPI_ISL_1098824, EPI_ISL_1098825, EPI_ISL_1098826, EPI_ISL_1098827, EPI_ISL_1098829 | | | |
| see above | DeRisi Lab, University of California, San Francisco | DeRisi Lab, University of California, San Francisco | Sabrina Mann, Anthea Mitchell, Jamin Liu, Sara Sunshine, Matthew Laurie, Genay Pilarowski, Amy Kistler, Manu Vanaerschot, Patrick Ayscue, Lucy Li, Eric Chow, IDseq Team, James Peng, Carina Marquez, Luis Rubio, Gabriel Chamie, Diane Jones, Jon Jacobo, Susana Rojas, Susy Rojas, Valerie Tulier-Laiwa, Douglas Black, Jackie Martinez, Maya Petersen, Diane Havlir, and Joseph DeRisi |
| EPI_ISL_1109633 | INNO Diagnostics Reference Laboratory | RCMI-Center for Research Resources, Ponce Research Institute | Vanessa Rivera-Amill, Andrea Arias-García, Raphael Sánchez-Torres |
| EPI_ISL_1109990, EPI_ISL_1110024, EPI_ISL_1110026 | Baylor Scott & White-Irving | Baylor Scott & White-Temple | Ari Rao, Linden Morales, Kimberly Walker, Marcus Volz, Shelby Hendrickson |
| EPI_ISL_1110029, EPI_ISL_1110030, EPI_ISL_1110031, EPI_ISL_1110032, EPI_ISL_1110033, EPI_ISL_1110034, EPI_ISL_1110035, EPI_ISL_1110036, EPI_ISL_1110037, EPI_ISL_1110038, EPI_ISL_1110039, EPI_ISL_1110040, EPI_ISL_1110041, EPI_ISL_1110042, EPI_ISL_1110045, EPI_ISL_1110046, EPI_ISL_1110047, EPI_ISL_1110048, EPI_ISL_1110049, EPI_ISL_1110050, EPI_ISL_1110051, EPI_ISL_1110052, EPI_ISL_1110053, EPI_ISL_1110054, EPI_ISL_1110055, EPI_ISL_1110056, EPI_ISL_1110057, EPI_ISL_1110058, EPI_ISL_1110059, EPI_ISL_1110060, EPI_ISL_1110061, EPI_ISL_1110062, EPI_ISL_1110063, EPI_ISL_1110064, EPI_ISL_1110065, EPI_ISL_1110066, EPI_ISL_1110067, EPI_ISL_1110068, EPI_ISL_1110069, EPI_ISL_1110070, EPI_ISL_1110071, EPI_ISL_1110072, EPI_ISL_1110073, EPI_ISL_1110074, EPI_ISL_1110075, EPI_ISL_1110076, EPI_ISL_1110077, EPI_ISL_1110078, EPI_ISL_1110079, EPI_ISL_1110080, EPI_ISL_1110081, EPI_ISL_1110082, EPI_ISL_1110083, EPI_ISL_1110084, EPI_ISL_1110085, EPI_ISL_1110086, EPI_ISL_1110087, EPI_ISL_1110088, EPI_ISL_1110089, EPI_ISL_1110090, EPI_ISL_1110091, EPI_ISL_1110092, EPI_ISL_1110093, EPI_ISL_1110094, EPI_ISL_1110095, EPI_ISL_1110096, EPI_ISL_1110097, EPI_ISL_1110098, EPI_ISL_1110099, EPI_ISL_1110100, EPI_ISL_1110101, EPI_ISL_1110102, EPI_ISL_1110103, EPI_ISL_1110104, EPI_ISL_1110105, EPI_ISL_1110106, EPI_ISL_1110107, EPI_ISL_1110108, EPI_ISL_1110109, EPI_ISL_1110110, EPI_ISL_1110111, EPI_ISL_1110112, EPI_ISL_1110113, EPI_ISL_1110114, EPI_ISL_1110115, EPI_ISL_1110116, EPI_ISL_1110117, EPI_ISL_1110118, EPI_ISL_1110119, EPI_ISL_1110120, EPI_ISL_1110121, EPI_ISL_1110122, EPI_ISL_1110123, EPI_ISL_1110124, EPI_ISL_1110125, EPI_ISL_1110126, EPI_ISL_1110127, EPI_ISL_1110128, EPI_ISL_1110129, EPI_ISL_1110130, EPI_ISL_1110131, EPI_ISL_1110132, EPI_ISL_1110133, EPI_ISL_1110134, EPI_ISL_1110135, EPI_ISL_1110136, EPI_ISL_1110137 | | | |
| see above | UW Virology Lab | UW Virology Lab | Pavitra Roychoudhury, Hong Xie, Lasata Shrestha, Shah Mohamed Bakhash, Michelle Lin, Noah Baker, Meei-Li Huang, Keith R Jerome, Alexander Greninger |
| EPI_ISL_1110290, EPI_ISL_1110291, EPI_ISL_1110292, EPI_ISL_1110294 | Baylor Scott & White-Temple | Baylor Scott & White-Temple | Ari Rao, Linden Morales, Kimberly Walker, Marcus Volz, Shelby Hendrickson |
| EPI_ISL_1110553, EPI_ISL_1110554, EPI_ISL_1110555, EPI_ISL_1110556, EPI_ISL_1110557, EPI_ISL_1110558, EPI_ISL_1110559 | Kansas Health and Environmental Lab | Kansas Health and Environmental Lab | Mike Grose, Paige Drury, Carissa Robertson, Ben Olsen, and Phil Adam |
| EPI_ISL_1110560, EPI_ISL_1110561, EPI_ISL_1110562, EPI_ISL_1110563, EPI_ISL_1110564, EPI_ISL_1110565, EPI_ISL_1110566, EPI_ISL_1110567, EPI_ISL_1110568, EPI_ISL_1110569, EPI_ISL_1110570, EPI_ISL_1110571, EPI_ISL_1110572, EPI_ISL_1110573, EPI_ISL_1110574, EPI_ISL_1110575, EPI_ISL_1110576, EPI_ISL_1110577, EPI_ISL_1110578, EPI_ISL_1110579 | | | |
| see above | Kansas Health and Environmental Lab | Kansas Health and Environmental Lab | Mike Grose, Carissa Robertson, Ben Olsen, and Phil Adam |
| EPI_ISL_1110603, EPI_ISL_1110604, EPI_ISL_1110605, EPI_ISL_1110606, EPI_ISL_1110607, EPI_ISL_1110608, EPI_ISL_1110609, EPI_ISL_1110610, EPI_ISL_1110611, EPI_ISL_1110612, EPI_ISL_1110613, EPI_ISL_1110614, EPI_ISL_1110630, EPI_ISL_1110631, EPI_ISL_1110632, EPI_ISL_1110633, EPI_ISL_1110634, EPI_ISL_1110635, EPI_ISL_1110636, EPI_ISL_1110637, EPI_ISL_1110638, EPI_ISL_1110639, EPI_ISL_1110640, EPI_ISL_1110641, EPI_ISL_1110644, EPI_ISL_1110645, EPI_ISL_1110646, EPI_ISL_1110647, EPI_ISL_1110648, EPI_ISL_1110649 | | | |
| see above | University of Wisconsin-Madison AIDS Vaccine Research Laboratories | University of Wisconsin-Madison AIDS Vaccine Research Laboratories | Gage Moreno, Katarina Braun, et al. AIDS Vaccine Research Laboratories |
| EPI_ISL_1110650, EPI_ISL_1110651, EPI_ISL_1110652, EPI_ISL_1110653, EPI_ISL_1110654, EPI_ISL_1110655, EPI_ISL_1110656, EPI_ISL_1110657, EPI_ISL_1110658, EPI_ISL_1110659, EPI_ISL_1110660, EPI_ISL_1110661, EPI_ISL_1110662, EPI_ISL_1110663, EPI_ISL_1110664, EPI_ISL_1110665, EPI_ISL_1110666 | | | |
| see above | Helix/Illumina | Respiratory Viruses Branch, Division of Viral Diseases, Centers for Disease Control and Prevention | Peter W. Cook, Dakota Howard, Dhwani Batra, Ben L. Rambo-Martin, Eileen de Feo, Jan Antico, Christine Tran, Matthew Tolentino, Shannon Wickline, Kim Gietzen, Brad Sickler, Jingtao Liu, Eric Allen, Phil Febbo, Summer Galloway, Nicole L. Washington, Simon White, Geraint Levan, Kelly Schiabor Barrett, Elizabeth Cirulli, Alexandre Bolze, Ary Ascencio, Charlotte Rivera-Garcia, Ryan Cho, Jason Nguyen, Sherry Wang, Jimmy Ramirez, Tyler Cassens, Efren Sandoval, Magnus Isaksson, William Lee, David Becker, Marc Laurent, James Lu, Clinton R. Paden, Suxiang Tong, Duncan MacCannell |
| EPI_ISL_1110834, EPI_ISL_1110835, EPI_ISL_1110836, EPI_ISL_1110837, EPI_ISL_1110838, EPI_ISL_1110839, EPI_ISL_1110840, EPI_ISL_1110841, EPI_ISL_1110842, EPI_ISL_1110843, EPI_ISL_1110844, EPI_ISL_1110845, EPI_ISL_1110846, EPI_ISL_1110847, EPI_ISL_1110848, EPI_ISL_1110849, EPI_ISL_1110850, EPI_ISL_1110851, EPI_ISL_1110852, EPI_ISL_1110853, EPI_ISL_1110854, EPI_ISL_1110855, EPI_ISL_1110856, EPI_ISL_1110857, EPI_ISL_1110858, EPI_ISL_1110859, EPI_ISL_1110860, EPI_ISL_1110861, EPI_ISL_1110862, EPI_ISL_1110863, EPI_ISL_1110864, EPI_ISL_1110865, EPI_ISL_1110866, EPI_ISL_1110867, EPI_ISL_1110868, EPI_ISL_1110869, EPI_ISL_1110870, EPI_ISL_1110871, EPI_ISL_1110872, EPI_ISL_1110873, EPI_ISL_1110874 | | | |
| see above | Virginia Division of Consolidated Laboratory Services | Virginia Division of Consolidated Laboratory Services | Virginia DCLS |
| EPI_ISL_1111065 | IL Department of Public Health Chicago Laboratory | Pathogen Discovery, Respiratory Viruses Branch, Division of Viral Diseases, Centers for Disease Control and Prevention | Ying Tao, Jing Zhang, Yan Li, Brian Lynch, Anna Kelleher, Krista Queen, Anna Uehara, Peter Cook, Han Jia Ng, Clinton R. Paden, Haibin Wang, Suxiang Tong |
| EPI_ISL_1111066 | NC State Laboratory of Public Health | Pathogen Discovery, Respiratory Viruses Branch, Division of Viral Diseases, Centers for Disease Control and Prevention | Ying Tao, Jing Zhang, Yan Li, Brian Lynch, Anna Kelleher, Krista Queen, Anna Uehara, Peter Cook, Han Jia Ng, Clinton R. Paden, Haibin Wang, Suxiang Tong |
| EPI_ISL_1111067 | IL Department of Public Health Chicago Laboratory | Pathogen Discovery, Respiratory Viruses Branch, Division of Viral Diseases, Centers for Disease Control and Prevention | Ying Tao, Jing Zhang, Yan Li, Brian Lynch, Anna Kelleher, Krista Queen, Anna Uehara, Peter Cook, Han Jia Ng, Clinton R. Paden, Haibin Wang, Suxiang Tong |
| EPI_ISL_1111506, EPI_ISL_1111507, EPI_ISL_1111508, EPI_ISL_1111509, EPI_ISL_1111510, EPI_ISL_1111511, EPI_ISL_1111512, EPI_ISL_1111513, EPI_ISL_1111514, EPI_ISL_1111515, EPI_ISL_1111516, EPI_ISL_1111517, EPI_ISL_1111518, EPI_ISL_1111519, EPI_ISL_1111520, EPI_ISL_1111521, EPI_ISL_1111522, EPI_ISL_1111523, EPI_ISL_1111524, EPI_ISL_1111525, EPI_ISL_1111526, EPI_ISL_1111527, EPI_ISL_1111528, EPI_ISL_1111529, EPI_ISL_1111530 | | | |
| see above | Virginia Division of Consolidated Laboratory Services | Virginia Division of Consolidated Laboratory Services | Virginia DCLS |
| EPI_ISL_1111531, EPI_ISL_1111533, EPI_ISL_1111534, EPI_ISL_1111535, EPI_ISL_1111536, EPI_ISL_1111537, EPI_ISL_1111540, EPI_ISL_1111541, EPI_ISL_1111542, EPI_ISL_1111543, EPI_ISL_1111544, EPI_ISL_1111545, EPI_ISL_1111548 | | | |
| see above | Wyoming Public Health Laboratory | Wyoming Public Health Laboratory | Jim Mildenberger, Wanda Manley, Noah Hull, Taylor Fearing, Lynette Gumbleton, Channing Weber, Ashley Norberg, Chayse Rowley, Marley Goetz, Brian Dominguez, Elliot Thomasson, Cari Sloma, and Rob Christensen |
| EPI_ISL_1111549, EPI_ISL_1111550, EPI_ISL_1111551, EPI_ISL_1111552, EPI_ISL_1111553, EPI_ISL_1111554, EPI_ISL_1111555, EPI_ISL_1111556, EPI_ISL_1111557, EPI_ISL_1111558, EPI_ISL_1111559, EPI_ISL_1111560, EPI_ISL_1111561, EPI_ISL_1111562, EPI_ISL_1111563, EPI_ISL_1111564, EPI_ISL_1111565, EPI_ISL_1111566, EPI_ISL_1111567, EPI_ISL_1111568, EPI_ISL_1111569, EPI_ISL_1111570, EPI_ISL_1111571, EPI_ISL_1111572, EPI_ISL_1111573, EPI_ISL_1111574, EPI_ISL_1111575, EPI_ISL_1111576, EPI_ISL_1111577, EPI_ISL_1111578, EPI_ISL_1111579, EPI_ISL_1111580, EPI_ISL_1111581, EPI_ISL_1111582, EPI_ISL_1111583, EPI_ISL_1111584, EPI_ISL_1111585, EPI_ISL_1111586, EPI_ISL_1111587, EPI_ISL_1111588, EPI_ISL_1111589, EPI_ISL_1111590, EPI_ISL_1111591, EPI_ISL_1111592, EPI_ISL_1111593, EPI_ISL_1111594, EPI_ISL_1111595, EPI_ISL_1111596, EPI_ISL_1111597, EPI_ISL_1111598, EPI_ISL_1111599, EPI_ISL_1111600, EPI_ISL_1111601, EPI_ISL_1111602, EPI_ISL_1111603, EPI_ISL_1111604, EPI_ISL_1111605 | | | |
| see above | Virginia Division of Consolidated Laboratory Services | Virginia Division of Consolidated Laboratory Services | Virginia DCLS |
| EPI_ISL_1112215, EPI_ISL_1112216, EPI_ISL_1112217, EPI_ISL_1112218, EPI_ISL_1112219, EPI_ISL_1112220 | University of Wisconsin-Madison AIDS Vaccine Research Laboratories | University of Wisconsin-Madison AIDS Vaccine Research Laboratories | Gage Moreno, Katarina Braun, et al. AIDS Vaccine Research Laboratories |
| EPI_ISL_1112947, EPI_ISL_1112948, EPI_ISL_1112949, EPI_ISL_1112950, EPI_ISL_1112951, EPI_ISL_1112952, EPI_ISL_1112953, EPI_ISL_1112954, EPI_ISL_1112955, EPI_ISL_1112956, EPI_ISL_1112957, EPI_ISL_1112958, EPI_ISL_1112959, EPI_ISL_1112960, EPI_ISL_1112961, EPI_ISL_1112962, EPI_ISL_1112963, EPI_ISL_1112964, EPI_ISL_1112965, EPI_ISL_1112966, EPI_ISL_1112967, EPI_ISL_1112968, EPI_ISL_1112969, EPI_ISL_1112970, EPI_ISL_1112971, EPI_ISL_1112972, EPI_ISL_1112973, EPI_ISL_1112974, EPI_ISL_1112975, EPI_ISL_1112976, EPI_ISL_1112977, EPI_ISL_1112978, EPI_ISL_1112979, EPI_ISL_1112980, EPI_ISL_1112981, EPI_ISL_1112982, EPI_ISL_1112983, EPI_ISL_1112984, EPI_ISL_1112985, EPI_ISL_1112986, EPI_ISL_1112987, EPI_ISL_1112988, EPI_ISL_1112989, EPI_ISL_1112990, EPI_ISL_1112991, EPI_ISL_1112992, EPI_ISL_1112993, EPI_ISL_1112994, EPI_ISL_1112995, EPI_ISL_1112996, EPI_ISL_1112997, EPI_ISL_1112998, EPI_ISL_1112999, EPI_ISL_1113000, EPI_ISL_1113001, EPI_ISL_1113002, EPI_ISL_1113003, EPI_ISL_1113004, EPI_ISL_1113005, EPI_ISL_1113006, EPI_ISL_1113007, EPI_ISL_1113008, EPI_ISL_1113009, EPI_ISL_1113010, EPI_ISL_1113011, EPI_ISL_1113012, EPI_ISL_1113013, EPI_ISL_1113014, EPI_ISL_1113015, EPI_ISL_1113016, EPI_ISL_1113017, EPI_ISL_1113018, EPI_ISL_1113019, EPI_ISL_1113020, EPI_ISL_1113021, EPI_ISL_1113022, EPI_ISL_1113023, EPI_ISL_1113024, EPI_ISL_1113025, EPI_ISL_1113026, EPI_ISL_1113027, EPI_ISL_1113028, EPI_ISL_1113029, EPI_ISL_1113030, EPI_ISL_1113031, EPI_ISL_1113032, EPI_ISL_1113033, EPI_ISL_1113034, EPI_ISL_1113035, EPI_ISL_1113036, EPI_ISL_1113037, EPI_ISL_1113038, EPI_ISL_1113039, EPI_ISL_1113040, EPI_ISL_1113041 | | | |
| see above | Baylor College of Medicine/ GCID | Baylor College of Medicine/ GCID | Vasanthi Avadhanula, Erin Nicholson, David Henke, Pedro Piedra, Harsha Doddapaneni, Donna Muzny, Qingchang Meng, Hsu Chao, Zeineen Momin, Hua Shen, George Weissenberger, Kavya Kottapalli, Yimiti Meiheerguli, Sejal Salvi, Ginger Metcalf, Vipin Menon, Sara J.J. Cregeen, Matthew C. Ross, Tulin Ayvaz, Richard Sucgang, Kristi L. Hoffman, Matthew Wong, Joseph F. Petrosino |
| EPI_ISL_1113042, EPI_ISL_1113043, EPI_ISL_1113044, EPI_ISL_1113045, EPI_ISL_1113046, EPI_ISL_1113047, EPI_ISL_1113048, EPI_ISL_1113049, EPI_ISL_1113050, EPI_ISL_1113051, EPI_ISL_1113052, EPI_ISL_1113053, EPI_ISL_1113054, EPI_ISL_1113055 | | | |
| see above | Kansas Health and Environmental Lab | Kansas Health and Environmental Lab | Mike Grose, Carissa Robertson, Ben Olsen, and Phil Adam |
| EPI_ISL_1113137 | Rocky Mountain Laboratories | Rocky Mountain Laboratories | van Doremalen,N., Holbrook,M.G., Barbian,K.D., Bushmaker,C., Bushmaker,T., Martens,C.A. and Munster,V.J. |

| | | | |
|---|---|---|---|
| EPI_ISL_1113138 | Rocky Mountain Laboratories | Rocky Mountain Laboratories | van Doremalen,N., Holbrook,M.G., Barbian,K.D., Bushmaker,C.,Bushmaker,T., Martens,C.A. and Munster,V.J. |
| EPI_ISL_1113139, EPI_ISL_1113140 | NJ Rapid Response Teams | New Jersey Public Health and Environmental Laboratories (PHEL) | Lindsey Bodnar, Shiv Verma, Dana Woell, Byeong Jeong |
| EPI_ISL_1113141 | Mary Eliza Mahoney HC | New Jersey Public Health and Environmental Laboratories (PHEL) | Lindsey Bodnar, Shiv Verma, Dana Woell, Byeong Jeong |
| EPI_ISL_1113142, EPI_ISL_1113143 | NJ Rapid Response Teams | New Jersey Public Health and Environmental Laboratories (PHEL) | Lindsey Bodnar, Shiv Verma, Dana Woell, Byeong Jeong |
| EPI_ISL_1113144 | RWJ Univ. Hospital-New Brunswi | New Jersey Public Health and Environmental Laboratories (PHEL) | Lindsey Bodnar, Shiv Verma, Dana Woell, Byeong Jeong |
| EPI_ISL_1113145, EPI_ISL_1113146, EPI_ISL_1113147, EPI_ISL_1113148, EPI_ISL_1113149, EPI_ISL_1113150 | NJ Rapid Response Teams | New Jersey Public Health and Environmental Laboratories (PHEL) | Lindsey Bodnar, Shiv Verma, Dana Woell, Byeong Jeong |
| EPI_ISL_1113151 | Mary Eliza Mahoney HC | New Jersey Public Health and Environmental Laboratories (PHEL) | Lindsey Bodnar, Shiv Verma, Dana Woell, Byeong Jeong |
| EPI_ISL_1113152, EPI_ISL_1113153, EPI_ISL_1113154, EPI_ISL_1113155 | NJ Rapid Response Teams | New Jersey Public Health and Environmental Laboratories (PHEL) | Lindsey Bodnar, Shiv Verma, Dana Woell, Byeong Jeong |
| EPI_ISL_1113156 | New Jersey Department of Health, Public Health Laboratories | New Jersey Public Health and Environmental Laboratories (PHEL) | Lindsey Bodnar, Shiv Verma, Dana Woell, Byeong Jeong |
| EPI_ISL_1113157, EPI_ISL_1113158 | NJ Rapid Response Teams | New Jersey Public Health and Environmental Laboratories (PHEL) | Lindsey Bodnar, Shiv Verma, Dana Woell, Byeong Jeong |
| EPI_ISL_1113159 | Virtua Health System-Voorhees | New Jersey Public Health and Environmental Laboratories (PHEL) | Lindsey Bodnar, Shiv Verma, Dana Woell, Byeong Jeong |
| EPI_ISL_1113160, EPI_ISL_1113161, EPI_ISL_1113162, EPI_ISL_1113163, EPI_ISL_1113169 | NJ Rapid Response Teams | New Jersey Public Health and Environmental Laboratories (PHEL) | Lindsey Bodnar, Shiv Verma, Dana Woell, Byeong Jeong |
| EPI_ISL_1113170 | Mary Eliza Mahoney HC | New Jersey Public Health and Environmental Laboratories (PHEL) | Lindsey Bodnar, Shiv Verma, Dana Woell, Byeong Jeong |
| EPI_ISL_1113171 | New Jersey Department of Health, Public Health Laboratories | New Jersey Public Health and Environmental Laboratories (PHEL) | Lindsey Bodnar, Shiv Verma, Dana Woell, Byeong Jeong |
| EPI_ISL_1113172 | RWJ University Hospital-Rahway | New Jersey Public Health and Environmental Laboratories (PHEL) | Lindsey Bodnar, Shiv Verma, Dana Woell, Byeong Jeong |
| EPI_ISL_1113173, EPI_ISL_1113174, EPI_ISL_1113175 | NJ Rapid Response Teams | New Jersey Public Health and Environmental Laboratories (PHEL) | Lindsey Bodnar, Shiv Verma, Dana Woell, Byeong Jeong |
| EPI_ISL_1113176, EPI_ISL_1113177, EPI_ISL_1113178, EPI_ISL_1113179, EPI_ISL_1113180, EPI_ISL_1113181 | Gundersen Molecular Diagnostics Laboratory | Kabara Cancer Research Institute | Craig S. Richmond, Paraic A. Kenny |
| EPI_ISL_1113182, EPI_ISL_1113183, EPI_ISL_1113184, EPI_ISL_1113185 | Gundersen Clinical Microbiology Laboratory | Kabara Cancer Research Institute | Craig S. Richmond, Paraic A. Kenny |
| EPI_ISL_1113186, EPI_ISL_1113187, EPI_ISL_1113188, EPI_ISL_1113189, EPI_ISL_1113190, EPI_ISL_1113191, EPI_ISL_1113192, EPI_ISL_1113193, EPI_ISL_1113194, EPI_ISL_1113195, EPI_ISL_1113196, EPI_ISL_1113197, EPI_ISL_1113198, EPI_ISL_1113199, EPI_ISL_1113200, EPI_ISL_1113201, EPI_ISL_1113202, EPI_ISL_1113203, EPI_ISL_1113204, EPI_ISL_1113205, EPI_ISL_1113206, EPI_ISL_1113207, EPI_ISL_1113208, EPI_ISL_1113209, EPI_ISL_1113210, EPI_ISL_1113211, EPI_ISL_1113212, EPI_ISL_1113213, EPI_ISL_1113214, EPI_ISL_1113215, EPI_ISL_1113216, EPI_ISL_1113217, EPI_ISL_1113218, EPI_ISL_1113219, EPI_ISL_1113220, EPI_ISL_1113221, EPI_ISL_1113222, EPI_ISL_1113223, EPI_ISL_1113224, EPI_ISL_1113225 | | | |
| see above | Gundersen Molecular Diagnostics Laboratory | Kabara Cancer Research Institute | Craig S. Richmond, Paraic A. Kenny |
| EPI_ISL_1113226, EPI_ISL_1113227, EPI_ISL_1113228, EPI_ISL_1113229, EPI_ISL_1113230, EPI_ISL_1113231, EPI_ISL_1113232, EPI_ISL_1113233, EPI_ISL_1113234, EPI_ISL_1113236, EPI_ISL_1113237, EPI_ISL_1113238, EPI_ISL_1113240, EPI_ISL_1113241, EPI_ISL_1113242, EPI_ISL_1113244, EPI_ISL_1113245, EPI_ISL_1113246, EPI_ISL_1113247, EPI_ISL_1113249, EPI_ISL_1113250, EPI_ISL_1113251, EPI_ISL_1113252, EPI_ISL_1113253, EPI_ISL_1113254, EPI_ISL_1113255, EPI_ISL_1113256, EPI_ISL_1113257, EPI_ISL_1113258, EPI_ISL_1113259, EPI_ISL_1113260, EPI_ISL_1113261, EPI_ISL_1113262, EPI_ISL_1113263, EPI_ISL_1113265, EPI_ISL_1113266, EPI_ISL_1113267, EPI_ISL_1113268, EPI_ISL_1113269, EPI_ISL_1113270, EPI_ISL_1113272, EPI_ISL_1113273, EPI_ISL_1113274, EPI_ISL_1113275, EPI_ISL_1113276, EPI_ISL_1113277, EPI_ISL_1113279, EPI_ISL_1113280, EPI_ISL_1113281, EPI_ISL_1113282, EPI_ISL_1113285, EPI_ISL_1113286, EPI_ISL_1113288, EPI_ISL_1113289, EPI_ISL_1113291, EPI_ISL_1113292 | | | |
| see above | Sonora Quest Laboratories | TGen North | "Jolene Bowers, Megan Folkerts, Chris French, Hayley Yaglom, Ashlyn Pfeiffer, Darrin Lemmer, Dave Engelthaler, The Arizona COVID Genomics Union (ACGU)" |
| EPI_ISL_1113293, EPI_ISL_1113294, EPI_ISL_1113295, EPI_ISL_1113296, EPI_ISL_1113297, EPI_ISL_1113298, EPI_ISL_1113299, EPI_ISL_1113300, EPI_ISL_1113301, EPI_ISL_1113302, EPI_ISL_1113304, EPI_ISL_1113305, EPI_ISL_1113306, EPI_ISL_1113307, EPI_ISL_1113308, EPI_ISL_1113309, EPI_ISL_1113311, EPI_ISL_1113312, EPI_ISL_1113313, EPI_ISL_1113314, EPI_ISL_1113315, EPI_ISL_1113317, EPI_ISL_1113318, EPI_ISL_1113319, EPI_ISL_1113320, EPI_ISL_1113321, EPI_ISL_1113322, EPI_ISL_1113323, EPI_ISL_1113325, EPI_ISL_1113326, EPI_ISL_1113327, EPI_ISL_1113328 | | | |
| see above | Arizona Department of Health Services | TGen North | "Jolene Bowers, Megan Folkerts, Chris French, Hayley Yaglom, Ashlyn Pfeiffer, Darrin Lemmer, Dave Engelthaler, The Arizona COVID Genomics Union (ACGU)" |
| EPI_ISL_1113329, EPI_ISL_1113330, EPI_ISL_1113331, EPI_ISL_1113332, EPI_ISL_1113333, EPI_ISL_1113334, EPI_ISL_1113335, EPI_ISL_1113336, EPI_ISL_1113338, EPI_ISL_1113339, EPI_ISL_1113340, EPI_ISL_1113342, EPI_ISL_1113343, EPI_ISL_1113344, EPI_ISL_1113345, EPI_ISL_1113346, EPI_ISL_1113347, EPI_ISL_1113348, EPI_ISL_1113349, EPI_ISL_1113350, EPI_ISL_1113352, EPI_ISL_1113355, EPI_ISL_1113356, EPI_ISL_1113357, EPI_ISL_1113358, EPI_ISL_1113359, EPI_ISL_1113360, EPI_ISL_1113361, EPI_ISL_1113362, EPI_ISL_1113363, EPI_ISL_1113364, EPI_ISL_1113365, EPI_ISL_1113366, EPI_ISL_1113367, EPI_ISL_1113368, EPI_ISL_1113369, EPI_ISL_1113370, EPI_ISL_1113371, EPI_ISL_1113372, EPI_ISL_1113373, EPI_ISL_1113374, EPI_ISL_1113375, EPI_ISL_1113376, EPI_ISL_1113377, EPI_ISL_1113378, EPI_ISL_1113380, EPI_ISL_1113381, EPI_ISL_1113382, EPI_ISL_1113383, EPI_ISL_1113385, EPI_ISL_1113386 | | | |
| see above | Sonora Quest Laboratories | TGen North | "Jolene Bowers, Megan Folkerts, Chris French, Hayley Yaglom, Ashlyn Pfeiffer, Darrin Lemmer, Dave Engelthaler, The Arizona COVID Genomics Union (ACGU)" |
| EPI_ISL_1113387, EPI_ISL_1113388, EPI_ISL_1113390, EPI_ISL_1113391 | Arizona Department of Health Services | TGen North | "Jolene Bowers, Megan Folkerts, Chris French, Hayley Yaglom, Ashlyn Pfeiffer, Darrin Lemmer, Dave Engelthaler, The Arizona COVID Genomics Union (ACGU)" |
| EPI_ISL_1113392, EPI_ISL_1113393, EPI_ISL_1113394, EPI_ISL_1113395, EPI_ISL_1113396, EPI_ISL_1113397, EPI_ISL_1113398, EPI_ISL_1113399, EPI_ISL_1113400, EPI_ISL_1113401, EPI_ISL_1113402, EPI_ISL_1113403, EPI_ISL_1113404, EPI_ISL_1113405, EPI_ISL_1113406, EPI_ISL_1113407, EPI_ISL_1113408, EPI_ISL_1113409, EPI_ISL_1113410, EPI_ISL_1113411, EPI_ISL_1113412, EPI_ISL_1113413, EPI_ISL_1113414, EPI_ISL_1113415, EPI_ISL_1113416, EPI_ISL_1113417, EPI_ISL_1113418, EPI_ISL_1113419, EPI_ISL_1113420, EPI_ISL_1113421, EPI_ISL_1113422, EPI_ISL_1113426, EPI_ISL_1113427, EPI_ISL_1113428, EPI_ISL_1113429, EPI_ISL_1113430, EPI_ISL_1113431, EPI_ISL_1113432, EPI_ISL_1113433, EPI_ISL_1113434, EPI_ISL_1113435, EPI_ISL_1113436, EPI_ISL_1113437, EPI_ISL_1113438, EPI_ISL_1113440, EPI_ISL_1113441, EPI_ISL_1113442, EPI_ISL_1113443, EPI_ISL_1113444, EPI_ISL_1113445, EPI_ISL_1113446, EPI_ISL_1113447, EPI_ISL_1113448, EPI_ISL_1113449, EPI_ISL_1113451, EPI_ISL_1113453, EPI_ISL_1113455, EPI_ISL_1113456, EPI_ISL_1113457, EPI_ISL_1113458, EPI_ISL_1113460, EPI_ISL_1113461, EPI_ISL_1113462, EPI_ISL_1113463, EPI_ISL_1113464, EPI_ISL_1113465, EPI_ISL_1113467, EPI_ISL_1113468, EPI_ISL_1113469, EPI_ISL_1113470, EPI_ISL_1113471, EPI_ISL_1113472, EPI_ISL_1113473, EPI_ISL_1113474, EPI_ISL_1113475, EPI_ISL_1113476, EPI_ISL_1113477, EPI_ISL_1113478, EPI_ISL_1113479, EPI_ISL_1113480, EPI_ISL_1113481, EPI_ISL_1113482, EPI_ISL_1113483, EPI_ISL_1113484, EPI_ISL_1113485, EPI_ISL_1113486, EPI_ISL_1113488, EPI_ISL_1113489, EPI_ISL_1113490, EPI_ISL_1113491, EPI_ISL_1113492, EPI_ISL_1113493, EPI_ISL_1113494, EPI_ISL_1113495 | | | |
| see above | Sonora Quest Laboratories | TGen North | "Jolene Bowers, Megan Folkerts, Chris French, Hayley Yaglom, Ashlyn Pfeiffer, Darrin Lemmer, Dave Engelthaler, The Arizona COVID Genomics Union (ACGU)" |
| EPI_ISL_1113496, EPI_ISL_1113497, EPI_ISL_1113498, EPI_ISL_1113499, EPI_ISL_1113500, EPI_ISL_1113501, EPI_ISL_1113502, EPI_ISL_1113504, EPI_ISL_1113505, EPI_ISL_1113506, EPI_ISL_1113507, EPI_ISL_1113509, EPI_ISL_1113511, EPI_ISL_1113512, EPI_ISL_1113514, EPI_ISL_1113515, EPI_ISL_1113516, EPI_ISL_1113517, EPI_ISL_1113518, EPI_ISL_1113519, EPI_ISL_1113520, EPI_ISL_1113522, EPI_ISL_1113523, EPI_ISL_1113524, EPI_ISL_1113525, EPI_ISL_1113526, EPI_ISL_1113527, EPI_ISL_1113528, EPI_ISL_1113529, EPI_ISL_1113530, EPI_ISL_1113531, EPI_ISL_1113532, EPI_ISL_1113533, EPI_ISL_1113534, EPI_ISL_1113535, EPI_ISL_1113536, EPI_ISL_1113537, EPI_ISL_1113539, EPI_ISL_1113540, EPI_ISL_1113541, EPI_ISL_1113542, EPI_ISL_1113544, EPI_ISL_1113545, EPI_ISL_1113546, EPI_ISL_1113547, EPI_ISL_1113548, EPI_ISL_1113549, EPI_ISL_1113550, EPI_ISL_1113551, EPI_ISL_1113552, EPI_ISL_1113553, EPI_ISL_1113554, EPI_ISL_1113555, EPI_ISL_1113556, EPI_ISL_1113557, EPI_ISL_1113558, EPI_ISL_1113559, EPI_ISL_1113560, EPI_ISL_1113561, EPI_ISL_1113562, EPI_ISL_1113563, EPI_ISL_1113564, EPI_ISL_1113565 | | | |
| see above | Arizona Department of Health Services | TGen North | "Jolene Bowers, Megan Folkerts, Chris French, Hayley Yaglom, Ashlyn Pfeiffer, Darrin Lemmer, Dave Engelthaler, The Arizona COVID Genomics Union (ACGU)" |
| EPI_ISL_1113566, EPI_ISL_1113567, EPI_ISL_1113568 | Sonora Quest Laboratories | TGen North | "Jolene Bowers, Megan Folkerts, Chris French, Hayley Yaglom, Ashlyn Pfeiffer, Darrin Lemmer, Dave Engelthaler, The Arizona COVID Genomics Union (ACGU)" |
| EPI_ISL_1113569 | TGen North | TGen North | "Jolene Bowers, Megan Folkerts, Chris French, Hayley Yaglom, Ashlyn Pfeiffer, Darrin Lemmer, Dave Engelthaler, The Arizona COVID Genomics Union (ACGU)" |
| EPI_ISL_1113571, EPI_ISL_1113573, EPI_ISL_1113574, EPI_ISL_1113576, EPI_ISL_1113579, EPI_ISL_1113581, EPI_ISL_1113583, EPI_ISL_1113584, EPI_ISL_1113585, EPI_ISL_1113587, EPI_ISL_1113588, EPI_ISL_1113589, EPI_ISL_1113590, EPI_ISL_1113592, EPI_ISL_1113593, EPI_ISL_1113594, EPI_ISL_1113596, EPI_ISL_1113597, EPI_ISL_1113598, EPI_ISL_1113599, EPI_ISL_1113602, EPI_ISL_1113603, EPI_ISL_1113604, EPI_ISL_1113605, EPI_ISL_1113606, EPI_ISL_1113607, EPI_ISL_1113609, EPI_ISL_1113610, EPI_ISL_1113611, EPI_ISL_1113612, EPI_ISL_1113616, EPI_ISL_1113617, EPI_ISL_1113618, EPI_ISL_1113619, EPI_ISL_1113621, EPI_ISL_1113622, EPI_ISL_1113623, EPI_ISL_1113624, EPI_ISL_1113627, EPI_ISL_1113628, EPI_ISL_1113629, EPI_ISL_1113630, EPI_ISL_1113631, EPI_ISL_1113633, EPI_ISL_1113634, EPI_ISL_1113635, EPI_ISL_1113636, EPI_ISL_1113637, EPI_ISL_1113638, EPI_ISL_1113639, EPI_ISL_1113640, EPI_ISL_1113641, EPI_ISL_1113642, EPI_ISL_1113643, EPI_ISL_1113644, EPI_ISL_1113645, EPI_ISL_1113646, EPI_ISL_1113647, EPI_ISL_1113648, EPI_ISL_1113649, EPI_ISL_1113650, EPI_ISL_1113651, EPI_ISL_1113652, EPI_ISL_1113653, EPI_ISL_1113654, EPI_ISL_1113655, EPI_ISL_1113657, EPI_ISL_1113658, EPI_ISL_1113659, EPI_ISL_1113660, EPI_ISL_1113661, EPI_ISL_1113662, EPI_ISL_1113663, EPI_ISL_1113664, EPI_ISL_1113665, EPI_ISL_1113666, EPI_ISL_1113667, EPI_ISL_1113668, EPI_ISL_1113669, EPI_ISL_1113670, EPI_ISL_1113671, EPI_ISL_1113674, EPI_ISL_1113675, EPI_ISL_1113676, EPI_ISL_1113677, EPI_ISL_1113678, EPI_ISL_1113679, EPI_ISL_1113680, EPI_ISL_1113681, EPI_ISL_1113682, EPI_ISL_1113683, EPI_ISL_1113684, EPI_ISL_1113685, EPI_ISL_1113687, EPI_ISL_1113689, EPI_ISL_1113690, EPI_ISL_1113692, EPI_ISL_1113693, EPI_ISL_1113695, EPI_ISL_1113696, EPI_ISL_1113697, EPI_ISL_1113698, EPI_ISL_1113699, EPI_ISL_1113701, EPI_ISL_1113703 | | | |
| see above | Sonora Quest Laboratories | TGen North | "Jolene Bowers, Megan Folkerts, Chris French, Hayley Yaglom, Ashlyn Pfeiffer, Darrin Lemmer, Dave Engelthaler, The Arizona COVID Genomics Union (ACGU)" |
| EPI_ISL_1113704, EPI_ISL_1113705, EPI_ISL_1113706, EPI_ISL_1113707, | Kashi Clinical Laboratory | Kashi Clinical Laboratory | Zahra Kashi, Jenefer DeKoning |

| | | | |
|---|---|---|---|
| EPI_ISL_1113708 | | | |
| EPI_ISL_1113709, EPI_ISL_1113710, EPI_ISL_1113711, EPI_ISL_1113712, EPI_ISL_1113713, EPI_ISL_1113714, EPI_ISL_1113715, EPI_ISL_1113716, EPI_ISL_1113719, EPI_ISL_1113720, EPI_ISL_1113721, EPI_ISL_1113722, EPI_ISL_1113723, EPI_ISL_1113724, EPI_ISL_1113725, EPI_ISL_1113726, EPI_ISL_1113727, EPI_ISL_1113728, EPI_ISL_1113729, EPI_ISL_1113730, EPI_ISL_1113731, EPI_ISL_1113732, EPI_ISL_1113733, EPI_ISL_1113734, EPI_ISL_1113735, EPI_ISL_1113736, EPI_ISL_1113737, EPI_ISL_1113738, EPI_ISL_1113739, EPI_ISL_1113740, EPI_ISL_1113741, EPI_ISL_1113742, EPI_ISL_1113743, EPI_ISL_1113744, EPI_ISL_1113745, EPI_ISL_1113746, EPI_ISL_1113747, EPI_ISL_1113748, EPI_ISL_1113749, EPI_ISL_1113750, EPI_ISL_1113751, EPI_ISL_1113752, EPI_ISL_1113753 | | | |
| see above | New Mexico Department of Health Scientific Laboratory | New Mexico Department of Health Scientific Laboratory | Ellie Johnson, Anastacia Griego-Fisher, D'eldra Malone, Jennifer Benoit |
| EPI_ISL_1113754, EPI_ISL_1113759, EPI_ISL_1113760, EPI_ISL_1113761, EPI_ISL_1113762, EPI_ISL_1113763 | Kashi Clinical Laboratory | Kashi Clinical Laboratory | Zahra Kashi, Jenefer DeKoning |
| EPI_ISL_1113764, EPI_ISL_1113765, EPI_ISL_1113766, EPI_ISL_1113767, EPI_ISL_1113768, EPI_ISL_1113769, EPI_ISL_1113770, EPI_ISL_1113771, EPI_ISL_1113772, EPI_ISL_1113773, EPI_ISL_1113774, EPI_ISL_1113775, EPI_ISL_1113776, EPI_ISL_1113777, EPI_ISL_1113778, EPI_ISL_1113779, EPI_ISL_1113780, EPI_ISL_1113781, EPI_ISL_1113782, EPI_ISL_1113783, EPI_ISL_1113784, EPI_ISL_1113785, EPI_ISL_1113786, EPI_ISL_1113787, EPI_ISL_1113788, EPI_ISL_1113789, EPI_ISL_1113790, EPI_ISL_1113791, EPI_ISL_1113792, EPI_ISL_1113793, EPI_ISL_1113794, EPI_ISL_1113795, EPI_ISL_1113796, EPI_ISL_1113797, EPI_ISL_1113798, EPI_ISL_1113799, EPI_ISL_1113800, EPI_ISL_1113801, EPI_ISL_1113802, EPI_ISL_1113803, EPI_ISL_1113804, EPI_ISL_1113805, EPI_ISL_1113806, EPI_ISL_1113807, EPI_ISL_1113808, EPI_ISL_1113809, EPI_ISL_1113810, EPI_ISL_1113811, EPI_ISL_1113812, EPI_ISL_1113813, EPI_ISL_1113814, EPI_ISL_1113815, EPI_ISL_1113816, EPI_ISL_1113817, EPI_ISL_1113818, EPI_ISL_1113819, EPI_ISL_1113820, EPI_ISL_1113821, EPI_ISL_1113822, EPI_ISL_1113823, EPI_ISL_1113824, EPI_ISL_1113825, EPI_ISL_1113826, EPI_ISL_1113827, EPI_ISL_1113828, EPI_ISL_1113829, EPI_ISL_1113830, EPI_ISL_1113831, EPI_ISL_1113832, EPI_ISL_1113833, EPI_ISL_1113834, EPI_ISL_1113835, EPI_ISL_1113836, EPI_ISL_1113837, EPI_ISL_1113838, EPI_ISL_1113839, EPI_ISL_1113840, EPI_ISL_1113841, EPI_ISL_1113842 | | | |
| see above | Quest Diagnostics Incorporated | Respiratory Viruses Branch, Division of Viral Diseases, Centers for Disease Control and Prevention | Peter W. Cook, Dakota Howard, Dhwani Batra, Ben L. Rambo-Martin, S. H. Rosenthal, A. Gerasimova, R. M. Kagan, B. Anderson, M. Hua, Y. Liu, L.E. Bernstein, K.E. Livingston, A. Perez, I. A. Shlyakhter, R. V. Rolando, R. Owen, P. Tanpaiboon, F. Lacbawan, Clinton R. Paden, Suxiang Tong, Duncan MacCannell |
| EPI_ISL_1113843 | Kashi Clinical Laboratory | Kashi Clinical Laboratory | Zahra Kashi, Jenefer DeKoning |
| EPI_ISL_1113844, EPI_ISL_1113845, EPI_ISL_1113846, EPI_ISL_1113847, EPI_ISL_1113848, EPI_ISL_1113849, EPI_ISL_1113850, EPI_ISL_1113851, EPI_ISL_1113852, EPI_ISL_1113853, EPI_ISL_1113854, EPI_ISL_1113855, EPI_ISL_1113856, EPI_ISL_1113857, EPI_ISL_1113858, EPI_ISL_1113859, EPI_ISL_1113860, EPI_ISL_1113861, EPI_ISL_1113862, EPI_ISL_1113863, EPI_ISL_1113864, EPI_ISL_1113865, EPI_ISL_1113866, EPI_ISL_1113867, EPI_ISL_1113868, EPI_ISL_1113869, EPI_ISL_1113870, EPI_ISL_1113871, EPI_ISL_1113872, EPI_ISL_1113873, EPI_ISL_1113874, EPI_ISL_1113875, EPI_ISL_1113876, EPI_ISL_1113877, EPI_ISL_1113878, EPI_ISL_1113879, EPI_ISL_1113880, EPI_ISL_1113881, EPI_ISL_1113882, EPI_ISL_1113883, EPI_ISL_1113884, EPI_ISL_1113885, EPI_ISL_1113886, EPI_ISL_1113887, EPI_ISL_1113888, EPI_ISL_1113889, EPI_ISL_1113890, EPI_ISL_1113891, EPI_ISL_1113892, EPI_ISL_1113893, EPI_ISL_1113894, EPI_ISL_1113895, EPI_ISL_1113896, EPI_ISL_1113897, EPI_ISL_1113898, EPI_ISL_1113899, EPI_ISL_1113900, EPI_ISL_1113901, EPI_ISL_1113902, EPI_ISL_1113903, EPI_ISL_1113904, EPI_ISL_1113905, EPI_ISL_1113906, EPI_ISL_1113907, EPI_ISL_1113908, EPI_ISL_1113909, EPI_ISL_1113910, EPI_ISL_1113911, EPI_ISL_1113912, EPI_ISL_1113913, EPI_ISL_1113914, EPI_ISL_1113915, EPI_ISL_1113916, EPI_ISL_1113917, EPI_ISL_1113918, EPI_ISL_1113919, EPI_ISL_1113920, EPI_ISL_1113921, EPI_ISL_1113922, EPI_ISL_1113924, EPI_ISL_1113925, EPI_ISL_1113926, EPI_ISL_1113927, EPI_ISL_1113928, EPI_ISL_1113929, EPI_ISL_1113930, EPI_ISL_1113931, EPI_ISL_1113932, EPI_ISL_1113933, EPI_ISL_1113934, EPI_ISL_1113935, EPI_ISL_1113936, EPI_ISL_1113937, EPI_ISL_1113938, EPI_ISL_1113939, EPI_ISL_1113940, EPI_ISL_1113941, EPI_ISL_1113942, EPI_ISL_1113943, EPI_ISL_1113944, EPI_ISL_1113945, EPI_ISL_1113946, EPI_ISL_1113947, EPI_ISL_1113948, EPI_ISL_1113949, EPI_ISL_1113950, EPI_ISL_1113951, EPI_ISL_1113952, EPI_ISL_1113953, EPI_ISL_1113954, EPI_ISL_1113955, EPI_ISL_1113956, EPI_ISL_1113957, EPI_ISL_1113958, EPI_ISL_1113959, EPI_ISL_1113960, EPI_ISL_1113961, EPI_ISL_1113962, EPI_ISL_1113963, EPI_ISL_1113964, EPI_ISL_1113965, EPI_ISL_1113966, EPI_ISL_1113967, EPI_ISL_1113968, EPI_ISL_1113969, EPI_ISL_1113970, EPI_ISL_1113971, EPI_ISL_1113972, EPI_ISL_1113973, EPI_ISL_1113974, EPI_ISL_1113975, EPI_ISL_1113976, EPI_ISL_1113977, EPI_ISL_1113978, EPI_ISL_1113979, EPI_ISL_1113980, EPI_ISL_1113981, EPI_ISL_1113982, EPI_ISL_1113983, EPI_ISL_1113984, EPI_ISL_1113985, EPI_ISL_1113986, EPI_ISL_1113987, EPI_ISL_1113988, EPI_ISL_1113989, EPI_ISL_1113990, EPI_ISL_1113991, EPI_ISL_1113992, EPI_ISL_1113993, EPI_ISL_1113994, EPI_ISL_1113995, EPI_ISL_1113996, EPI_ISL_1113997, EPI_ISL_1113998, EPI_ISL_1113999, EPI_ISL_1114000, EPI_ISL_1114001, EPI_ISL_1114002, EPI_ISL_1114003, EPI_ISL_1114004, EPI_ISL_1114005, EPI_ISL_1114006, EPI_ISL_1114007, EPI_ISL_1114008, EPI_ISL_1114009, EPI_ISL_1114010, EPI_ISL_1114011, EPI_ISL_1114012, EPI_ISL_1114013, EPI_ISL_1114014, EPI_ISL_1114015, EPI_ISL_1114016, EPI_ISL_1114017, EPI_ISL_1114018, EPI_ISL_1114019, EPI_ISL_1114020, EPI_ISL_1114021, EPI_ISL_1114022, EPI_ISL_1114023, EPI_ISL_1114024, EPI_ISL_1114025, EPI_ISL_1114026, EPI_ISL_1114027, EPI_ISL_1114028, EPI_ISL_1114029, EPI_ISL_1114030, EPI_ISL_1114031, EPI_ISL_1114032, EPI_ISL_1114033, EPI_ISL_1114034, EPI_ISL_1114035, EPI_ISL_1114036, EPI_ISL_1114037, EPI_ISL_1114038, EPI_ISL_1114039, EPI_ISL_1114040, EPI_ISL_1114041, EPI_ISL_1114042, EPI_ISL_1114043, EPI_ISL_1114044, EPI_ISL_1114045, EPI_ISL_1114046, EPI_ISL_1114047, EPI_ISL_1114048, EPI_ISL_1114049, EPI_ISL_1114050, EPI_ISL_1114051, EPI_ISL_1114052, EPI_ISL_1114053, EPI_ISL_1114054, EPI_ISL_1114055, EPI_ISL_1114056, EPI_ISL_1114057, EPI_ISL_1114058, EPI_ISL_1114059, EPI_ISL_1114060, EPI_ISL_1114061, EPI_ISL_1114062, EPI_ISL_1114063, EPI_ISL_1114064, EPI_ISL_1114065 | | | |
| see above | Quest Diagnostics Incorporated | Respiratory Viruses Branch, Division of Viral Diseases, Centers for Disease Control and Prevention | Peter W. Cook, Dakota Howard, Dhwani Batra, Ben L. Rambo-Martin, S. H. Rosenthal, A. Gerasimova, R. M. Kagan, B. Anderson, M. Hua, Y. Liu, L.E. Bernstein, K.E. Livingston, A. Perez, I. A. Shlyakhter, R. V. Rolando, R. Owen, P. Tanpaiboon, F. Lacbawan, Clinton R. Paden, Suxiang Tong, Duncan MacCannell |
| EPI_ISL_1114066 | Kashi Clinical Laboratory | Kashi Clinical Laboratory | Zahra Kashi, Jenefer DeKoning |
| EPI_ISL_1114067, EPI_ISL_1114068, EPI_ISL_1114069, EPI_ISL_1114070, EPI_ISL_1114071, EPI_ISL_1114072, EPI_ISL_1114073, EPI_ISL_1114074, EPI_ISL_1114075, EPI_ISL_1114076, EPI_ISL_1114077, EPI_ISL_1114078, EPI_ISL_1114079, EPI_ISL_1114080, EPI_ISL_1114081, EPI_ISL_1114082, EPI_ISL_1114083, EPI_ISL_1114084, EPI_ISL_1114085, EPI_ISL_1114086, EPI_ISL_1114087, EPI_ISL_1114088, EPI_ISL_1114089, EPI_ISL_1114090, EPI_ISL_1114091, EPI_ISL_1114092, EPI_ISL_1114093, EPI_ISL_1114094, EPI_ISL_1114095, EPI_ISL_1114096, EPI_ISL_1114097, EPI_ISL_1114098, EPI_ISL_1114099, EPI_ISL_1114100, EPI_ISL_1114101, EPI_ISL_1114102, EPI_ISL_1114103, EPI_ISL_1114104, EPI_ISL_1114105, EPI_ISL_1114106, EPI_ISL_1114107, EPI_ISL_1114108, EPI_ISL_1114109, EPI_ISL_1114110, EPI_ISL_1114111, EPI_ISL_1114112, EPI_ISL_1114113, EPI_ISL_1114114, EPI_ISL_1114115, EPI_ISL_1114116, EPI_ISL_1114117, EPI_ISL_1114118, EPI_ISL_1114119, EPI_ISL_1114120, EPI_ISL_1114121, EPI_ISL_1114122, EPI_ISL_1114123, EPI_ISL_1114124, EPI_ISL_1114125, EPI_ISL_1114126, EPI_ISL_1114127, EPI_ISL_1114128, EPI_ISL_1114129, EPI_ISL_1114130, EPI_ISL_1114131, EPI_ISL_1114132, EPI_ISL_1114133, EPI_ISL_1114134, EPI_ISL_1114135, EPI_ISL_1114136, EPI_ISL_1114137, EPI_ISL_1114138, EPI_ISL_1114139, EPI_ISL_1114140, EPI_ISL_1114141, EPI_ISL_1114142, EPI_ISL_1114143, EPI_ISL_1114144, EPI_ISL_1114145, EPI_ISL_1114146, EPI_ISL_1114147, EPI_ISL_1114148, EPI_ISL_1114149, EPI_ISL_1114150 | | | |
| see above | Quest Diagnostics Incorporated | Respiratory Viruses Branch, Division of Viral Diseases, Centers for Disease Control and Prevention | Peter W. Cook, Dakota Howard, Dhwani Batra, Ben L. Rambo-Martin, S. H. Rosenthal, A. Gerasimova, R. M. Kagan, B. Anderson, M. Hua, Y. Liu, L.E. Bernstein, K.E. Livingston, A. Perez, I. A. Shlyakhter, R. V. Rolando, R. Owen, P. Tanpaiboon, F. Lacbawan, Clinton R. Paden, Suxiang Tong, Duncan MacCannell |
| EPI_ISL_1114155, EPI_ISL_1114156, EPI_ISL_1114158, EPI_ISL_1114161, EPI_ISL_1114162, EPI_ISL_1114163, EPI_ISL_1114164, EPI_ISL_1114166, EPI_ISL_1114167, EPI_ISL_1114169, EPI_ISL_1114171, EPI_ISL_1114174, EPI_ISL_1114175, EPI_ISL_1114177, EPI_ISL_1114178, EPI_ISL_1114179, EPI_ISL_1114180, EPI_ISL_1114182, EPI_ISL_1114183, EPI_ISL_1114184, EPI_ISL_1114185, EPI_ISL_1114186, EPI_ISL_1114187, EPI_ISL_1114191, EPI_ISL_1114192, EPI_ISL_1114193, EPI_ISL_1114194, EPI_ISL_1114195, EPI_ISL_1114197, EPI_ISL_1114198, EPI_ISL_1114199, EPI_ISL_1114200 | | | |
| see above | Seattle Flu Study | Seattle Flu Study | Deborah A. Nickerson, Chris D. Frazar, Jover Lee, Benjamin Pelle, Erica Ryke, Matthew Richardson, Amanda Adler, Elisabeth Brandstetter, Peter D. Han, Kairsten Fay, Misja Ilcisin, Kirsten Lacombe, Thomas R. Sibley, Melissa Truong, Caitlin R. Wolf, Karen Cowgill, Stephanie Schrag, Jeff Duchin, Michael Boeckh, Janet A. Englund, Michael Famulare, Barry R. Lutz, Mark J. Rieder, Lea M. Starita, Matthew Thompson, Helen Y. Chu, Trevor Bedford, Jay Shendure |
| EPI_ISL_1114201 | Seattle Flu Study | Seattle Flu Study | Deborah A. Nickerson, Chris D. Frazar, Jover Lee, Benjamin Pelle, Erica Ryke, Matthew Richardson, Amanda Adler, Elisabeth Brandstetter, Peter D. Han, Kairsten Fay, Misja Ilcisin, Kirsten Lacombe, Thomas R. Sibley, Melissa Truong, Caitlin R. Wolf, Michael Boeckh, Janet A. Englund, Michael Famulare, Barry R. Lutz, Mark J. Rieder, Lea M. Starita, Matthew Thompson, Jay Shendure, Trevor Bedford, Helen Y. Chu |
| EPI_ISL_1114203 | Seattle Flu Study | Seattle Flu Study | Deborah A. Nickerson, Chris D. Frazar, Jover Lee, Benjamin Pelle, Erica Ryke, Matthew Richardson, Amanda Adler, Elisabeth Brandstetter, Peter D. Han, Kairsten Fay, Misja Ilcisin, Kirsten Lacombe, Thomas R. Sibley, Melissa Truong, Caitlin R. Wolf, Karen Cowgill, Stephanie Schrag, Jeff Duchin, Michael Boeckh, Janet A. Englund, Michael Famulare, Barry R. Lutz, Mark J. Rieder, Lea M. Starita, Matthew Thompson, Helen Y. Chu, Trevor Bedford, Jay Shendure |
| EPI_ISL_1114204, EPI_ISL_1114205, EPI_ISL_1114206, EPI_ISL_1114210, EPI_ISL_1114211, EPI_ISL_1114212, EPI_ISL_1114214, EPI_ISL_1114215, EPI_ISL_1114216, EPI_ISL_1114217, EPI_ISL_1114218, EPI_ISL_1114219, EPI_ISL_1114220, EPI_ISL_1114222 | | | |
| see above | Washington State Department of Health | Seattle Flu Study | Deborah A. Nickerson, Chris D. Frazar, Jover Lee, Benjamin Pelle, Erica Ryke, Matthew Richardson, Amanda Adler, Elisabeth Brandstetter, Peter D. Han, Kairsten Fay, Misja Ilcisin, Kirsten Lacombe, Thomas R. Sibley, Melissa Truong, Caitlin R. Wolf, Romesh Gautom, Geoff Melly, Brian Hiatt, Philip Dykema, Scott Lindquist, Michael Boeckh, Janet A. Englund, Michael Famulare, Barry R. Lutz, Mark J. Rieder, Lea M. Starita, Matthew Thompson, Helen Y. Chu, Jay Shendure, Trevor Bedford |
| EPI_ISL_1114224 | Northwest Laboratory | Seattle Flu Study | Deborah A. Nickerson, Chris D. Frazar, Jover Lee, Benjamin Pelle, Erica Ryke, Matthew Richardson, Amanda Adler, Elisabeth Brandstetter, Peter D. Han, Kairsten Fay, Misja Ilcisin, Kirsten Lacombe, Thomas R. Sibley, Melissa Truong, Caitlin R. Wolf, Romesh Gautom, Geoff Melly, Brian Hiatt, Philip Dykema, Scott Lindquist, Michael Boeckh, Janet A. Englund, Michael Famulare, Barry R. Lutz, Mark J. Rieder, Lea M. Starita, Matthew Thompson, Helen Y. Chu, Jay Shendure, Trevor Bedford |
| EPI_ISL_1114225, EPI_ISL_1114226, EPI_ISL_1114227, EPI_ISL_1114228, EPI_ISL_1114229 | Seattle Flu Study | Seattle Flu Study | Deborah A. Nickerson, Chris D. Frazar, Jover Lee, Benjamin Pelle, Erica Ryke, Matthew Richardson, Amanda Adler, Elisabeth Brandstetter, Peter D. Han, Kairsten Fay, Misja Ilcisin, Kirsten Lacombe, Thomas R. Sibley, Melissa Truong, Caitlin R. Wolf, Karen Cowgill, Stephanie Schrag, Jeff Duchin, Michael Boeckh, Janet A. Englund, Michael Famulare, Barry R. Lutz, Mark J. Rieder, Lea M. Starita, Matthew Thompson, Helen Y. Chu, Trevor Bedford, Jay Shendure |
| EPI_ISL_1114231, EPI_ISL_1114233 | Seattle Flu Study | Seattle Flu Study | Deborah A. Nickerson, Chris D. Frazar, Jover Lee, Benjamin Pelle, Erica Ryke, Matthew Richardson, Amanda Adler, Elisabeth Brandstetter, Peter D. Han, Kairsten Fay, Misja Ilcisin, Kirsten Lacombe, Thomas R. Sibley, Melissa Truong, Caitlin R. Wolf, Michael Boeckh, Janet A. Englund, Michael Famulare, Barry R. Lutz, Mark J. Rieder, Lea M. Starita, Matthew Thompson, Jay Shendure, Trevor Bedford, Helen Y. Chu |
| EPI_ISL_1114234, EPI_ISL_1114235, EPI_ISL_1114236, EPI_ISL_1114237, EPI_ISL_1114238, EPI_ISL_1114239, EPI_ISL_1114240, EPI_ISL_1114241, EPI_ISL_1114242, EPI_ISL_1114243, EPI_ISL_1114244, EPI_ISL_1114245, EPI_ISL_1114246, EPI_ISL_1114247, EPI_ISL_1114248, EPI_ISL_1114249, EPI_ISL_1114250, EPI_ISL_1114251, EPI_ISL_1114252, EPI_ISL_1114253, EPI_ISL_1114254 | | | |
| see above | Washington State Department of Health | Seattle Flu Study | Deborah A. Nickerson, Chris D. Frazar, Jover Lee, Benjamin Pelle, Erica Ryke, Matthew Richardson, Amanda Adler, Elisabeth Brandstetter, Peter D. Han, Kairsten Fay, Misja Ilcisin, Kirsten Lacombe, Thomas R. Sibley, Melissa Truong, Caitlin R. Wolf, Romesh Gautom, Geoff Melly, Brian Hiatt, Philip Dykema, Scott Lindquist, Michael Boeckh, Janet A. Englund, Michael Famulare, Barry R. Lutz, Mark J. Rieder, Lea M. Starita, Matthew Thompson, Helen Y. Chu, Jay Shendure, Trevor Bedford |
| EPI_ISL_1114255, EPI_ISL_1114256 | Northwest Laboratory | Seattle Flu Study | Deborah A. Nickerson, Chris D. Frazar, Jover Lee, Benjamin Pelle, Erica Ryke, Matthew Richardson, Amanda Adler, Elisabeth Brandstetter, Peter D. Han, Kairsten Fay, Misja Ilcisin, Kirsten Lacombe, Thomas R. Sibley, Melissa Truong, Caitlin R. Wolf, Romesh Gautom, Geoff Melly, Brian Hiatt, Philip Dykema, Scott Lindquist, Michael Boeckh, Janet A. Englund, Michael Famulare, Barry R. Lutz, Mark J. Rieder, Lea M. Starita, Matthew Thompson, Helen Y. Chu, Jay Shendure, Trevor Bedford |
| EPI_ISL_1114257 | LabCorp Seattle | Seattle Flu Study | Deborah A. Nickerson, Chris D. Frazar, Jover Lee, Benjamin Pelle, Erica Ryke, Matthew Richardson, Amanda Adler, Elisabeth Brandstetter, Peter D. Han, Kairsten Fay, Misja Ilcisin, Kirsten Lacombe, Thomas R. Sibley, Melissa Truong, Caitlin R. Wolf, Romesh Gautom, Geoff Melly, Brian Hiatt, Philip Dykema, Scott Lindquist, Michael Boeckh, Janet A. Englund, Michael Famulare, Barry R. Lutz, Mark J. Rieder, Lea M. Starita, Matthew Thompson, Helen Y. Chu, Jay Shendure, Trevor Bedford |
| EPI_ISL_1114258 | Northwest Laboratory | Seattle Flu Study | Deborah A. Nickerson, Chris D. Frazar, Jover Lee, Benjamin Pelle, Erica Ryke, Matthew Richardson, Amanda Adler, Elisabeth Brandstetter, Peter D. Han, Kairsten Fay, Misja Ilcisin, Kirsten Lacombe, Thomas R. Sibley, Melissa Truong, Caitlin R. Wolf, Romesh Gautom, Geoff Melly, Brian Hiatt, Philip Dykema, Scott Lindquist, Michael Boeckh, Janet A. Englund, Michael Famulare, Barry R. Lutz, Mark J. Rieder, Lea M. Starita, Matthew Thompson, Helen Y. Chu, Jay Shendure, Trevor Bedford |
| EPI_ISL_1114259, EPI_ISL_1114260 | Seattle Flu Study | Seattle Flu Study | Deborah A. Nickerson, Chris D. Frazar, Jover Lee, Benjamin Pelle, Erica Ryke, Matthew Richardson, Amanda Adler, Elisabeth Brandstetter, Peter D. Han, Kairsten Fay, Misja Ilcisin, Kirsten Lacombe, Thomas R. Sibley, Melissa Truong, Caitlin R. Wolf, Karen Cowgill, Stephanie Schrag, Jeff Duchin, Michael Boeckh, Janet A. Englund, Michael Famulare, Barry R. Lutz, Mark J. Rieder, Lea M. Starita, Matthew Thompson, Helen Y. Chu, Trevor Bedford, Jay Shendure |
| EPI_ISL_1114261, EPI_ISL_1114263, EPI_ISL_1114264, EPI_ISL_1114266, EPI_ISL_1114267, EPI_ISL_1114268 | Seattle Flu Study | Seattle Flu Study | Deborah A. Nickerson, Chris D. Frazar, Jover Lee, Benjamin Pelle, Erica Ryke, Matthew Richardson, Amanda Adler, Elisabeth Brandstetter, Peter D. Han, Kairsten Fay, Misja Ilcisin, Kirsten Lacombe, Thomas R. Sibley, Melissa Truong, Caitlin R. Wolf, Michael Boeckh, Janet A. Englund, Michael Famulare, Barry R. Lutz, Mark J. Rieder, Lea M. Starita, Matthew Thompson, Jay Shendure, Trevor Bedford, Helen Y. Chu |
| EPI_ISL_1114270, EPI_ISL_1114271 | Seattle Flu Study | Seattle Flu Study | Deborah A. Nickerson, Chris D. Frazar, Jover Lee, Benjamin Pelle, Erica Ryke, Matthew Richardson, Amanda Adler, Elisabeth Brandstetter, Peter D. Han, Kairsten Fay, Misja Ilcisin, Kirsten Lacombe, Thomas R. Sibley, Melissa Truong, Caitlin R. Wolf, Karen Cowgill, Stephanie Schrag, Jeff Duchin, Michael Boeckh, Janet A. Englund, Michael Famulare, Barry R. Lutz, Mark J. Rieder, Lea M. Starita, Matthew Thompson, Helen Y. Chu, Trevor Bedford, Jay Shendure |
| EPI_ISL_1114276 | Seattle Flu Study | Seattle Flu Study | Deborah A. Nickerson, Chris D. Frazar, Jover Lee, Benjamin Pelle, Erica Ryke, Matthew Richardson, Amanda Adler, Elisabeth Brandstetter, Peter D. Han, Kairsten Fay, Misja Ilcisin, Kirsten Lacombe, Thomas R. Sibley, Melissa Truong, Caitlin R. Wolf, Michael Boeckh, Janet A. Englund, Michael Famulare, Barry R. Lutz, Mark J. Rieder, Lea M. Starita, Matthew Thompson, Jay Shendure, Trevor Bedford, Helen Y. Chu |
| EPI_ISL_1114277, EPI_ISL_1114278, EPI_ISL_1114279, EPI_ISL_1114280, EPI_ISL_1114281, EPI_ISL_1114282, EPI_ISL_1114283, EPI_ISL_1114284, EPI_ISL_1114285, EPI_ISL_1114287, EPI_ISL_1114288, EPI_ISL_1114289, EPI_ISL_1114290 | | | |
| see above | Seattle Flu Study | Seattle Flu Study | Deborah A. Nickerson, Chris D. Frazar, Jover Lee, Benjamin Pelle, Erica Ryke, Matthew Richardson, Amanda Adler, Elisabeth Brandstetter, Peter D. Han, Kairsten Fay, Misja Ilcisin, Kirsten Lacombe, Thomas R. Sibley, Melissa Truong, Caitlin R. Wolf, Karen Cowgill, Stephanie Schrag, Jeff Duchin, Michael Boeckh, Janet A. Englund, Michael Famulare, Barry R. Lutz, Mark J. Rieder, Lea M. Starita, Matthew Thompson, Helen Y. Chu, Trevor Bedford, Jay Shendure |
| EPI_ISL_1114291, EPI_ISL_1114292, EPI_ISL_1114293, EPI_ISL_1114295, EPI_ISL_1114300, EPI_ISL_1114301, EPI_ISL_1114302, EPI_ISL_1114305, EPI_ISL_1114307, EPI_ISL_1114309, EPI_ISL_1114315, EPI_ISL_1114316, EPI_ISL_1114317, EPI_ISL_1114319, EPI_ISL_1114321, EPI_ISL_1114322, EPI_ISL_1114323, EPI_ISL_1114326, EPI_ISL_1114331, EPI_ISL_1114332, EPI_ISL_1114333, EPI_ISL_1114335, EPI_ISL_1114336, EPI_ISL_1114337, EPI_ISL_1114338, EPI_ISL_1114341, EPI_ISL_1114342, EPI_ISL_1114344, EPI_ISL_1114345, EPI_ISL_1114347, EPI_ISL_1114348, EPI_ISL_1114349, EPI_ISL_1114350, EPI_ISL_1114351 | | | |
| see above | Altius Institute for Biomedical Sciences | Seattle Flu Study | Deborah A. Nickerson, Chris D. Frazar, Jover Lee, Benjamin Pelle, Erica Ryke, Matthew Richardson, Amanda Adler, Elisabeth Brandstetter, Peter D. Han, Kairsten Fay, Misja Ilcisin, Kirsten Lacombe, Thomas R. Sibley, Melissa Truong, Caitlin R. Wolf, Ryan Alexander, Daniel Bates, Rebecca Bruders, Stephanie DeBaun, Clem Green, Muhammad Halimun, Jessica Halow, Kneshay Harper, Matt Hartman, Andrew Meuser, Alex Nguyen, Truong Nguyen, Sofia Olsson, Sadie Patraw, Hannah Petersen, Tobias Ragoczy, Joshua Richards, Jacob Rodriguez, John Stamatoyannopoulos, Julia Wald, Olivia Waltner, Michael Boeckh, Janet A. Englund, Michael Famulare, Barry R. Lutz, Mark J. Rieder, Lea M. Starita, Matthew Thompson, Helen Y. Chu, Jay Shendure, Trevor Bedford |
| EPI_ISL_1114352 | Seattle Flu Study | Seattle Flu Study | Deborah A. Nickerson, Chris D. Frazar, Jover Lee, Benjamin Pelle, Erica Ryke, Matthew Richardson, Amanda Adler, Elisabeth Brandstetter, Peter D. Han, Kairsten Fay, Misja Ilcisin, Kirsten Lacombe, Thomas R. Sibley, Melissa Truong, Caitlin R. Wolf, Michael Boeckh, Janet A. Englund, Michael Famulare, Barry R. Lutz, Mark J. Rieder, Lea M. Starita, Matthew Thompson, Jay Shendure, Trevor Bedford, Helen Y. Chu |
| EPI_ISL_1114354, EPI_ISL_1114355, | Washington State Department of Health | Seattle Flu Study | Deborah A. Nickerson, Chris D. Frazar, Jover Lee, Benjamin Pelle, Erica Ryke, Matthew Richardson, Amanda Adler, Elisabeth Brandstetter, Peter D. Han, Kairsten Fay, Misja Ilcisin, Kirsten Lacombe, Thomas R. Sibley, Melissa Truong, Caitlin R. Wolf, Romesh Gautom, Geoff Melly, Brian Hiatt, Philip Dykema, Scott Lindquist, Michael Boeckh, Janet A. Englund, Michael Famulare, Barry R. Lutz, Mark J. Rieder, Lea M. Starita, Matthew Thompson, Helen Y. Chu, Jay Shendure, Trevor Bedford |

| | | | |
|---|---|---|---|
| EPI_ISL_1114357, EPI_ISL_1114358, EPI_ISL_1114360, EPI_ISL_1114361, EPI_ISL_1114363, EPI_ISL_1114364, EPI_ISL_1114366 | | | |
| EPI_ISL_1114369, EPI_ISL_1114370 | FidaLab | Seattle Flu Study | Deborah A. Nickerson, Chris D. Frazar, Jover Lee, Benjamin Pelle, Erica Ryke, Matthew Richardson, Amanda Adler, Elisabeth Brandstetter, Peter D. Han, Kairsten Fay, Misja Ilcisin, Kirsten Lacombe, Thomas R. Sibley, Melissa Truong, Caitlin R. Wolf, Romesh Gautom, Geoff Melly, Brian Hiatt, Philip Dykema, Scott Lindquist, Michael Boeckh, Janet A. Englund, Michael Famulare, Barry R. Lutz, Mark J. Rieder, Lea M. Starita, Matthew Thompson, Helen Y. Chu, Jay Shendure, Trevor Bedford |
| EPI_ISL_1114371 | Vancouver Clinic | Seattle Flu Study | Deborah A. Nickerson, Chris D. Frazar, Jover Lee, Benjamin Pelle, Erica Ryke, Matthew Richardson, Amanda Adler, Elisabeth Brandstetter, Peter D. Han, Kairsten Fay, Misja Ilcisin, Kirsten Lacombe, Thomas R. Sibley, Melissa Truong, Caitlin R. Wolf, Romesh Gautom, Geoff Melly, Brian Hiatt, Philip Dykema, Scott Lindquist, Michael Boeckh, Janet A. Englund, Michael Famulare, Barry R. Lutz, Mark J. Rieder, Lea M. Starita, Matthew Thompson, Helen Y. Chu, Jay Shendure, Trevor Bedford |
| EPI_ISL_1114373 | Northwest Laboratory | Seattle Flu Study | Deborah A. Nickerson, Chris D. Frazar, Jover Lee, Benjamin Pelle, Erica Ryke, Matthew Richardson, Amanda Adler, Elisabeth Brandstetter, Peter D. Han, Kairsten Fay, Misja Ilcisin, Kirsten Lacombe, Thomas R. Sibley, Melissa Truong, Caitlin R. Wolf, Romesh Gautom, Geoff Melly, Brian Hiatt, Philip Dykema, Scott Lindquist, Michael Boeckh, Janet A. Englund, Michael Famulare, Barry R. Lutz, Mark J. Rieder, Lea M. Starita, Matthew Thompson, Helen Y. Chu, Jay Shendure, Trevor Bedford |
| EPI_ISL_1114374 | Evergreen Health | Seattle Flu Study | Deborah A. Nickerson, Chris D. Frazar, Jover Lee, Benjamin Pelle, Erica Ryke, Matthew Richardson, Amanda Adler, Elisabeth Brandstetter, Peter D. Han, Kairsten Fay, Misja Ilcisin, Kirsten Lacombe, Thomas R. Sibley, Melissa Truong, Caitlin R. Wolf, Romesh Gautom, Geoff Melly, Brian Hiatt, Philip Dykema, Scott Lindquist, Michael Boeckh, Janet A. Englund, Michael Famulare, Barry R. Lutz, Mark J. Rieder, Lea M. Starita, Matthew Thompson, Helen Y. Chu, Jay Shendure, Trevor Bedford |
| EPI_ISL_1114376 | Providence Sacred Heart Medical Center-Spokane | Seattle Flu Study | Deborah A. Nickerson, Chris D. Frazar, Jover Lee, Benjamin Pelle, Erica Ryke, Matthew Richardson, Amanda Adler, Elisabeth Brandstetter, Peter D. Han, Kairsten Fay, Misja Ilcisin, Kirsten Lacombe, Thomas R. Sibley, Melissa Truong, Caitlin R. Wolf, Romesh Gautom, Geoff Melly, Brian Hiatt, Philip Dykema, Scott Lindquist, Michael Boeckh, Janet A. Englund, Michael Famulare, Barry R. Lutz, Mark J. Rieder, Lea M. Starita, Matthew Thompson, Helen Y. Chu, Jay Shendure, Trevor Bedford |
| EPI_ISL_1114377 | Northwest Laboratory | Seattle Flu Study | Deborah A. Nickerson, Chris D. Frazar, Jover Lee, Benjamin Pelle, Erica Ryke, Matthew Richardson, Amanda Adler, Elisabeth Brandstetter, Peter D. Han, Kairsten Fay, Misja Ilcisin, Kirsten Lacombe, Thomas R. Sibley, Melissa Truong, Caitlin R. Wolf, Romesh Gautom, Geoff Melly, Brian Hiatt, Philip Dykema, Scott Lindquist, Michael Boeckh, Janet A. Englund, Michael Famulare, Barry R. Lutz, Mark J. Rieder, Lea M. Starita, Matthew Thompson, Helen Y. Chu, Jay Shendure, Trevor Bedford |
| EPI_ISL_1114381, EPI_ISL_1114382, EPI_ISL_1114384 | Seattle Flu Study | Seattle Flu Study | Deborah A. Nickerson, Chris D. Frazar, Jover Lee, Benjamin Pelle, Erica Ryke, Matthew Richardson, Amanda Adler, Elisabeth Brandstetter, Peter D. Han, Kairsten Fay, Misja Ilcisin, Kirsten Lacombe, Thomas R. Sibley, Melissa Truong, Caitlin R. Wolf, Karen Cowgill, Stephanie Schrag, Jeff Duchin, Michael Boeckh, Janet A. Englund, Michael Famulare, Barry R. Lutz, Mark J. Rieder, Lea M. Starita, Matthew Thompson, Helen Y. Chu, Trevor Bedford, Jay Shendure |
| EPI_ISL_1114385 | Kaiser Permanente | Seattle Flu Study | Deborah A. Nickerson, Chris D. Frazar, Jover Lee, Benjamin Pelle, Erica Ryke, Matthew Richardson, Amanda Adler, Elisabeth Brandstetter, Peter D. Han, Kairsten Fay, Misja Ilcisin, Kirsten Lacombe, Thomas R. Sibley, Melissa Truong, Caitlin R. Wolf, Romesh Gautom, Geoff Melly, Brian Hiatt, Philip Dykema, Scott Lindquist, Michael Boeckh, Janet A. Englund, Michael Famulare, Barry R. Lutz, Mark J. Rieder, Lea M. Starita, Matthew Thompson, Helen Y. Chu, Jay Shendure, Trevor Bedford |
| EPI_ISL_1114388, EPI_ISL_1114389, EPI_ISL_1114390, EPI_ISL_1114391, EPI_ISL_1114392, EPI_ISL_1114393, EPI_ISL_1114394, EPI_ISL_1114398, EPI_ISL_1114405, EPI_ISL_1114406, EPI_ISL_1114407, EPI_ISL_1114408, EPI_ISL_1114409, EPI_ISL_1114410, EPI_ISL_1114411, EPI_ISL_1114412, EPI_ISL_1114413, EPI_ISL_1114414, EPI_ISL_1114415, EPI_ISL_1114417, EPI_ISL_1114418, EPI_ISL_1114419, EPI_ISL_1114420, EPI_ISL_1114421, EPI_ISL_1114422, EPI_ISL_1114423 | | | |
| see above | Wyoming Public Health Laboratory | Wyoming Public Health Laboratory | Jim Mildenberger, Wanda Manley, Noah Hull, Taylor Fearing, Lynette Gumbleton, Channing Weber, Ashley Norberg, Chayse Rowley, Marley Goetz, Brian Dominguez, Elliot Thomasson, Cari Sloma, and Rob Christensen |
| EPI_ISL_1114428, EPI_ISL_1114434, EPI_ISL_1114439, EPI_ISL_1114442, EPI_ISL_1114444, EPI_ISL_1114446, EPI_ISL_1114448, EPI_ISL_1114450, EPI_ISL_1114451, EPI_ISL_1114452, EPI_ISL_1114453, EPI_ISL_1114454, EPI_ISL_1114458, EPI_ISL_1114459, EPI_ISL_1114460, EPI_ISL_1114461, EPI_ISL_1114462, EPI_ISL_1114463, EPI_ISL_1114464, EPI_ISL_1114465, EPI_ISL_1114466, EPI_ISL_1114467, EPI_ISL_1114468 | | | |
| see above | Utah Public Health Laboratory | Utah Public Health Laboratory | Erin L. Young, Kelly F. Oakeson, Tara Gallagher |
| EPI_ISL_1114469 | Department of Health and Mental Hygiene Public Health Laboratory | New York City Public Health Laboratory | Jade Wang, et al. |
| EPI_ISL_1114470, EPI_ISL_1114471 | Department of Health and Mental Hygiene Central Harlem | New York City Public Health Laboratory | Jade Wang, et al. |
| EPI_ISL_1114472, EPI_ISL_1114473, EPI_ISL_1114474, EPI_ISL_1114475 | Department of Health and Mental Hygiene Corona | New York City Public Health Laboratory | Jade Wang, et al. |
| EPI_ISL_1114476 | Department of Health and Mental Hygiene Public Health Laboratory | New York City Public Health Laboratory | Jade Wang, et al. |
| EPI_ISL_1114477, EPI_ISL_1114478, EPI_ISL_1114479, EPI_ISL_1114480, EPI_ISL_1114481 | Department of Health and Mental Hygiene Jamaica | New York City Public Health Laboratory | Jade Wang, et al. |
| EPI_ISL_1114482, EPI_ISL_1114483, EPI_ISL_1114484 | Department of Health and Mental Hygiene Public Health Laboratory | New York City Public Health Laboratory | Jade Wang, et al. |
| EPI_ISL_1114485 | Department of Health and Mental Hygiene Crown Heights | New York City Public Health Laboratory | Jade Wang, et al. |
| EPI_ISL_1114486 | Department of Health and Mental Hygiene Public Health Laboratory | New York City Public Health Laboratory | Jade Wang, et al. |
| EPI_ISL_1114487, EPI_ISL_1114488, EPI_ISL_1114489, EPI_ISL_1114490, EPI_ISL_1114491 | Department of Health and Mental Hygiene Morrisania | New York City Public Health Laboratory | Jade Wang, et al. |
| EPI_ISL_1114492, EPI_ISL_1114493 | Department of Health and Mental Hygiene Central Harlem | New York City Public Health Laboratory | Jade Wang, et al. |
| EPI_ISL_1114494, EPI_ISL_1114495, EPI_ISL_1114496 | Department of Health and Mental Hygiene Fort Greene | New York City Public Health Laboratory | Jade Wang, et al. |
| EPI_ISL_1114497, EPI_ISL_1114498 | Department of Health and Mental Hygiene Riverside | New York City Public Health Laboratory | Jade Wang, et al. |
| EPI_ISL_1114499 | Department of Health and Mental Hygiene Crown Heights | New York City Public Health Laboratory | Jade Wang, et al. |
| EPI_ISL_1114500, EPI_ISL_1114501 | Department of Health and Mental Hygiene Chelsea | New York City Public Health Laboratory | Jade Wang, et al. |
| EPI_ISL_1114502 | Department of Health and Mental Hygiene Crown Heights | New York City Public Health Laboratory | Jade Wang, et al. |
| EPI_ISL_1114503 | Department of Health and Mental Hygiene Riverside | New York City Public Health Laboratory | Jade Wang, et al. |
| EPI_ISL_1114504, EPI_ISL_1114505, EPI_ISL_1114506, EPI_ISL_1114507 | Department of Health and Mental Hygiene Corona | New York City Public Health Laboratory | Jade Wang, et al. |
| EPI_ISL_1114508 | Department of Health and Mental Hygiene Jamaica | New York City Public Health Laboratory | Jade Wang, et al. |
| EPI_ISL_1114509 | Department of Health and Mental Hygiene Chelsea | New York City Public Health Laboratory | Jade Wang, et al. |
| EPI_ISL_1114510 | Department of Health and Mental Hygiene Public Health Laboratory | New York City Public Health Laboratory | Jade Wang, et al. |
| EPI_ISL_1114511, EPI_ISL_1114512 | Department of Homeless Services | New York City Public Health Laboratory | Jade Wang, et al. |
| EPI_ISL_1114513, EPI_ISL_1114514, EPI_ISL_1114515 | Department of Health and Mental Hygiene Corona | New York City Public Health Laboratory | Jade Wang, et al. |
| EPI_ISL_1114516, EPI_ISL_1114517 | Department of Health and Mental Hygiene Crown Heights | New York City Public Health Laboratory | Jade Wang, et al. |
| EPI_ISL_1114518 | Department of Health and Mental Hygiene Jamaica | New York City Public Health Laboratory | Jade Wang, et al. |
| EPI_ISL_1114519, EPI_ISL_1114520, EPI_ISL_1114521, | Department of Health and Mental Hygiene Public Health Laboratory | New York City Public Health Laboratory | Jade Wang, et al. |

| | | | |
|---|---|---|---|
| EPI_ISL_1114522, EPI_ISL_1114523 | | | |
| EPI_ISL_1114524 | Department of Health and Mental Hygiene Chelsea | New York City Public Health Laboratory | Jade Wang, et al. |
| EPI_ISL_1114525, EPI_ISL_1114526, EPI_ISL_1114527, EPI_ISL_1114528, EPI_ISL_1114529, EPI_ISL_1114530 | OCME Office Of Chief Medical Examiner | New York City Public Health Laboratory | Jade Wang, et al. |
| EPI_ISL_1114531, EPI_ISL_1114532, EPI_ISL_1114533, EPI_ISL_1114534, EPI_ISL_1114535, EPI_ISL_1114536, EPI_ISL_1114537, EPI_ISL_1114538, EPI_ISL_1114539, EPI_ISL_1114540, EPI_ISL_1114541, EPI_ISL_1114542, EPI_ISL_1114543, EPI_ISL_1114544, EPI_ISL_1114545, EPI_ISL_1114546, EPI_ISL_1114547, EPI_ISL_1114548, EPI_ISL_1114549, EPI_ISL_1114550, EPI_ISL_1114551, EPI_ISL_1114552, EPI_ISL_1114553, EPI_ISL_1114554, EPI_ISL_1114555, EPI_ISL_1114556, EPI_ISL_1114557, EPI_ISL_1114558, EPI_ISL_1114559, EPI_ISL_1114560, EPI_ISL_1114561, EPI_ISL_1114562, EPI_ISL_1114563, EPI_ISL_1114564, EPI_ISL_1114565, EPI_ISL_1114566, EPI_ISL_1114567, EPI_ISL_1114568, EPI_ISL_1114569, EPI_ISL_1114570, EPI_ISL_1114571, EPI_ISL_1114572, EPI_ISL_1114573, EPI_ISL_1114574, EPI_ISL_1114575, EPI_ISL_1114576, EPI_ISL_1114577, EPI_ISL_1114578, EPI_ISL_1114579, EPI_ISL_1114580, EPI_ISL_1114581, EPI_ISL_1114582, EPI_ISL_1114583, EPI_ISL_1114584, EPI_ISL_1114585, EPI_ISL_1114586, EPI_ISL_1114587, EPI_ISL_1114588, EPI_ISL_1114589, EPI_ISL_1114590, EPI_ISL_1114591, EPI_ISL_1114592, EPI_ISL_1114593, EPI_ISL_1114594, EPI_ISL_1114595, EPI_ISL_1114596, EPI_ISL_1114597, EPI_ISL_1114598, EPI_ISL_1114599, EPI_ISL_1114600, EPI_ISL_1114601, EPI_ISL_1114602, EPI_ISL_1114603, EPI_ISL_1114604, EPI_ISL_1114605, EPI_ISL_1114606, EPI_ISL_1114607, EPI_ISL_1114608, EPI_ISL_1114609, EPI_ISL_1114610, EPI_ISL_1114611, EPI_ISL_1114612, EPI_ISL_1114613, EPI_ISL_1114614, EPI_ISL_1114615, EPI_ISL_1114616, EPI_ISL_1114617, EPI_ISL_1114618, EPI_ISL_1114619, EPI_ISL_1114620, EPI_ISL_1114621, EPI_ISL_1114622, EPI_ISL_1114623, EPI_ISL_1114624, EPI_ISL_1114625, EPI_ISL_1114626, EPI_ISL_1114627, EPI_ISL_1114628, EPI_ISL_1114629, EPI_ISL_1114630, EPI_ISL_1114631, EPI_ISL_1114632, EPI_ISL_1114633, EPI_ISL_1114634, EPI_ISL_1114635, EPI_ISL_1114636, EPI_ISL_1114637, EPI_ISL_1114638, EPI_ISL_1114639, EPI_ISL_1114640, EPI_ISL_1114641, EPI_ISL_1114642, EPI_ISL_1114643, EPI_ISL_1114644, EPI_ISL_1114645, EPI_ISL_1114646, EPI_ISL_1114647, EPI_ISL_1114648, EPI_ISL_1114649, EPI_ISL_1114650, EPI_ISL_1114651, EPI_ISL_1114652, EPI_ISL_1114653, EPI_ISL_1114654, EPI_ISL_1114655, EPI_ISL_1114656, EPI_ISL_1114657, EPI_ISL_1114658, EPI_ISL_1114659, EPI_ISL_1114660, EPI_ISL_1114661, EPI_ISL_1114662, EPI_ISL_1114663, EPI_ISL_1114664, EPI_ISL_1114665, EPI_ISL_1114666, EPI_ISL_1114667, EPI_ISL_1114668, EPI_ISL_1114669, EPI_ISL_1114670, EPI_ISL_1114671 | | | |
| see above | Quest Diagnostics Incorporated | Respiratory Viruses Branch, Division of Viral Diseases, Centers for Disease Control and Prevention | Peter W. Cook, Dakota Howard, Dhwani Batra, Ben L. Rambo-Martin, S. H. Rosenthal, A. Gerasimova, R. M. Kagan, B. Anderson, M. Hua, Y. Liu, L.E. Bernstein, K.E. Livingston, A. Perez, I. A. Shlyakhter, R. V. Rolando, R. Owen, P. Tanpaiboon, F. Lacbawan, Clinton R. Paden, Suxiang Tong, Duncan MacCannell |
| EPI_ISL_1114672, EPI_ISL_1114673, EPI_ISL_1114674, EPI_ISL_1114675, EPI_ISL_1114676, EPI_ISL_1114677, EPI_ISL_1114678, EPI_ISL_1114679, EPI_ISL_1114680, EPI_ISL_1114681, EPI_ISL_1114682, EPI_ISL_1114683, EPI_ISL_1114684, EPI_ISL_1114685, EPI_ISL_1114686, EPI_ISL_1114687, EPI_ISL_1114688, EPI_ISL_1114689, EPI_ISL_1114690, EPI_ISL_1114691, EPI_ISL_1114692, EPI_ISL_1114693, EPI_ISL_1114694, EPI_ISL_1114695, EPI_ISL_1114696, EPI_ISL_1114697, EPI_ISL_1114698, EPI_ISL_1114699, EPI_ISL_1114700, EPI_ISL_1114701, EPI_ISL_1114702, EPI_ISL_1114703, EPI_ISL_1114705, EPI_ISL_1114706, EPI_ISL_1114707, EPI_ISL_1114708, EPI_ISL_1114709, EPI_ISL_1114710, EPI_ISL_1114711, EPI_ISL_1114712, EPI_ISL_1114713, EPI_ISL_1114714, EPI_ISL_1114715, EPI_ISL_1114716 | | | |
| see above | New Mexico Department of Health Scientific Laboratory | New Mexico Department of Health Scientific Laboratory | Ellie Johnson, Anastacia Griego-Fisher, D'eldra Malone, Jennifer Benoit |
| EPI_ISL_1116476, EPI_ISL_1116477, EPI_ISL_1116478, EPI_ISL_1116479, EPI_ISL_1116480, EPI_ISL_1116481, EPI_ISL_1116482, EPI_ISL_1116483, EPI_ISL_1116484, EPI_ISL_1116485, EPI_ISL_1116486, EPI_ISL_1116487, EPI_ISL_1116488, EPI_ISL_1116489, EPI_ISL_1116490, EPI_ISL_1116491, EPI_ISL_1116492, EPI_ISL_1116493, EPI_ISL_1116494, EPI_ISL_1116495, EPI_ISL_1116496, EPI_ISL_1116497, EPI_ISL_1116498, EPI_ISL_1116499, EPI_ISL_1116500, EPI_ISL_1116503, EPI_ISL_1116504, EPI_ISL_1116505, EPI_ISL_1116506, EPI_ISL_1116507, EPI_ISL_1116508, EPI_ISL_1116509, EPI_ISL_1116510, EPI_ISL_1116511, EPI_ISL_1116512, EPI_ISL_1116513, EPI_ISL_1116514, EPI_ISL_1116515, EPI_ISL_1116516, EPI_ISL_1116517, EPI_ISL_1116518, EPI_ISL_1116519, EPI_ISL_1116520, EPI_ISL_1116521, EPI_ISL_1116522, EPI_ISL_1116523, EPI_ISL_1116524, EPI_ISL_1116525, EPI_ISL_1116526, EPI_ISL_1116527, EPI_ISL_1116528, EPI_ISL_1116529, EPI_ISL_1116530, EPI_ISL_1116531, EPI_ISL_1116532, EPI_ISL_1116533, EPI_ISL_1116534, EPI_ISL_1116535, EPI_ISL_1116536, EPI_ISL_1116537, EPI_ISL_1116538, EPI_ISL_1116539, EPI_ISL_1116540, EPI_ISL_1116541, EPI_ISL_1116542, EPI_ISL_1116543, EPI_ISL_1116544, EPI_ISL_1116545, EPI_ISL_1116546, EPI_ISL_1116547, EPI_ISL_1116548, EPI_ISL_1116549, EPI_ISL_1116550, EPI_ISL_1116551, EPI_ISL_1116552, EPI_ISL_1116553, EPI_ISL_1116554 | | | |
| see above | NYU Langone Health | Departments of Pathology and Medicine, New York University School of Medicine | Adriana Heguy, Dacia Dimartino, Emily Guzman, Christian Marier, Peter Meyn, Sitharam Ramaswami, Gael Westby, Paul Zappile, Yutong Zhang, Paolo Cotzia, Guiqing Wang |
| EPI_ISL_1117353, EPI_ISL_1117354, EPI_ISL_1117355, EPI_ISL_1117356, EPI_ISL_1117357, EPI_ISL_1117358, EPI_ISL_1117359, EPI_ISL_1117360, EPI_ISL_1117361, EPI_ISL_1117362, EPI_ISL_1117363, EPI_ISL_1117364, EPI_ISL_1117365, EPI_ISL_1117366, EPI_ISL_1117367, EPI_ISL_1117368, EPI_ISL_1117369, EPI_ISL_1117370, EPI_ISL_1117371, EPI_ISL_1117372, EPI_ISL_1117373, EPI_ISL_1117374, EPI_ISL_1117375 | | | |
| see above | Instituto Nacional de Medicina Genomica | Instituto Nacional de Medicina Genomica | Hidalgo-Miranda A, Mendoza-Vargas A, Reyes-Grajeda JP, Cedro-Tanda A, Alcaraz N, Gonzalez-Barrera D, Rangel-DeLeon D, Ramirez-Vega O, Sifuentes-Rojas C, Cisneros-Villanueva M, Herrera-Montalvo LA |
| EPI_ISL_1117700, EPI_ISL_1117774, EPI_ISL_1118086, EPI_ISL_1118087, EPI_ISL_1118088, EPI_ISL_1118089, EPI_ISL_1118205, EPI_ISL_1118281, EPI_ISL_1118374 | Ohio Department of Health Laboratory | Ohio Department of Health Laboratory | Holmes, Jennifer; Eric Brandt, Keoni Omura, Glen McGillivary, Caitlin McDonnell, Jade Mowery, Stephanie Mccracken, Tyler Payne, Kirtana Ramadugu, Erica Leasure, Brent Lee, Kelsey Florek, Heather Blankenship, Allison Black, Quanta Brown, and Tammy Bannerman |
| EPI_ISL_1118690, EPI_ISL_1118691, EPI_ISL_1118692, EPI_ISL_1118693, EPI_ISL_1118694, EPI_ISL_1118695, EPI_ISL_1118696, EPI_ISL_1118697, EPI_ISL_1118698, EPI_ISL_1118699, EPI_ISL_1118700, EPI_ISL_1118701, EPI_ISL_1118702, EPI_ISL_1118703, EPI_ISL_1118704, EPI_ISL_1118705, EPI_ISL_1118706, EPI_ISL_1118707, EPI_ISL_1118708, EPI_ISL_1118709, EPI_ISL_1118710, EPI_ISL_1118711, EPI_ISL_1118712, EPI_ISL_1118713, EPI_ISL_1118714, EPI_ISL_1118715, EPI_ISL_1118716, EPI_ISL_1118717, EPI_ISL_1118718, EPI_ISL_1118719, EPI_ISL_1118720, EPI_ISL_1118721, EPI_ISL_1118722, EPI_ISL_1118723, EPI_ISL_1118724, EPI_ISL_1118725, EPI_ISL_1118726, EPI_ISL_1118727, EPI_ISL_1118728, EPI_ISL_1118729, EPI_ISL_1118730, EPI_ISL_1118731, EPI_ISL_1118732, EPI_ISL_1118733, EPI_ISL_1118734, EPI_ISL_1118735, EPI_ISL_1118736, EPI_ISL_1118737, EPI_ISL_1118738, EPI_ISL_1118739, EPI_ISL_1118740, EPI_ISL_1118741, EPI_ISL_1118742, EPI_ISL_1118743, EPI_ISL_1118744, EPI_ISL_1118745, EPI_ISL_1118746, EPI_ISL_1118747, EPI_ISL_1118748, EPI_ISL_1118749, EPI_ISL_1118750, EPI_ISL_1118751, EPI_ISL_1118752, EPI_ISL_1118753, EPI_ISL_1118754, EPI_ISL_1118755, EPI_ISL_1118756, EPI_ISL_1118757, EPI_ISL_1118758, EPI_ISL_1118759, EPI_ISL_1118760, EPI_ISL_1118761, EPI_ISL_1118762, EPI_ISL_1118763, EPI_ISL_1118764, EPI_ISL_1118765, EPI_ISL_1118766, EPI_ISL_1118767, EPI_ISL_1118768, EPI_ISL_1118769, EPI_ISL_1118770, EPI_ISL_1118771, EPI_ISL_1118772, EPI_ISL_1118773, EPI_ISL_1118774, EPI_ISL_1118775, EPI_ISL_1118776, EPI_ISL_1118777, EPI_ISL_1118778, EPI_ISL_1118779, EPI_ISL_1118780, EPI_ISL_1118781, EPI_ISL_1118782, EPI_ISL_1118783, EPI_ISL_1118784, EPI_ISL_1118785, EPI_ISL_1118786, EPI_ISL_1118787, EPI_ISL_1118788, EPI_ISL_1118789, EPI_ISL_1118790, EPI_ISL_1118791, EPI_ISL_1118792, EPI_ISL_1118793, EPI_ISL_1118794, EPI_ISL_1118795, EPI_ISL_1118796, EPI_ISL_1118797, EPI_ISL_1118798, EPI_ISL_1118799, EPI_ISL_1118800, EPI_ISL_1118801 | | | |
| see above | University of Michigan Clinical Microbiology Laboratory | Lauring Lab, University of Michigan, Department of Microbiology and Immunology | Valesano |
| EPI_ISL_1118802 | Ohio Department of Health Laboratory | Ohio Department of Health Laboratory | Holmes, Jennifer; Eric Brandt, Keoni Omura, Glen McGillivary, Caitlin McDonnell, Jade Mowery, Stephanie Mccracken, Tyler Payne, Kirtana Ramadugu, Erica Leasure, Brent Lee, Kelsey Florek, Heather Blankenship, Allison Black, Quanta Brown, and Tammy Bannerman |
| EPI_ISL_1118803, EPI_ISL_1118804, EPI_ISL_1118805, EPI_ISL_1118806, EPI_ISL_1118807, EPI_ISL_1118808, EPI_ISL_1118809, EPI_ISL_1118810, EPI_ISL_1118811, EPI_ISL_1118812, EPI_ISL_1118813, EPI_ISL_1118814, EPI_ISL_1118815, EPI_ISL_1118816, EPI_ISL_1118817 | | | |
| see above | University of Michigan Clinical Microbiology Laboratory | Lauring Lab, University of Michigan, Department of Microbiology and Immunology | Valesano |
| EPI_ISL_1118871 | Ohio Department of Health Laboratory | Ohio Department of Health Laboratory | Holmes, Jennifer; Eric Brandt, Keoni Omura, Glen McGillivary, Caitlin McDonnell, Jade Mowery, Stephanie Mccracken, Tyler Payne, Kirtana Ramadugu, Erica Leasure, Brent Lee, Kelsey Florek, Heather Blankenship, Allison Black, Quanta Brown, and Tammy Bannerman |
| EPI_ISL_1118875 | Virginia Division of Consolidated Laboratory Services | Virginia Division of Consolidated Laboratory Services | Virginia DCLS |
| EPI_ISL_1118885, EPI_ISL_1118886, EPI_ISL_1118887, EPI_ISL_1118888, EPI_ISL_1118889 | Kansas Health and Environmental Lab | Kansas Health and Environmental Lab | Mike Grose, Carissa Robertson, Ben Olsen, and Phil Adam |
| EPI_ISL_1118890 | Nebraska Public Health Laboratory (NPHL) | NPHL | NPHL COVID-19 Response Team |
| EPI_ISL_1118910, EPI_ISL_1118911, EPI_ISL_1118912, EPI_ISL_1118913, EPI_ISL_1118914 | Ohio Department of Health Laboratory | Ohio Department of Health Laboratory | Holmes, Jennifer; Eric Brandt, Keoni Omura, Glen McGillivary, Caitlin McDonnell, Jade Mowery, Stephanie Mccracken, Tyler Payne, Kirtana Ramadugu, Erica Leasure, Brent Lee, Kelsey Florek, Heather Blankenship, Allison Black, Quanta Brown, and Tammy Bannerman |
| EPI_ISL_1118915, EPI_ISL_1118916, EPI_ISL_1118917, EPI_ISL_1118918, EPI_ISL_1118919 | University of Michigan Clinical Microbiology Laboratory | Lauring Lab, University of Michigan, Department of Microbiology and Immunology | Valesano |
| EPI_ISL_1118920, EPI_ISL_1118921, EPI_ISL_1118922, EPI_ISL_1118924 | Ohio Department of Health Laboratory | Ohio Department of Health Laboratory | Holmes, Jennifer; Eric Brandt, Keoni Omura, Glen McGillivary, Caitlin McDonnell, Jade Mowery, Stephanie Mccracken, Tyler Payne, Kirtana Ramadugu, Erica Leasure, Brent Lee, Kelsey Florek, Heather Blankenship, Allison Black, Quanta Brown, and Tammy Bannerman |
| EPI_ISL_1119868 | TGen North | TGen North | "Jolene Bowers, Megan Folkerts, Chris French, Hayley Yaglom, Ashlyn Pfeiffer, Darrin Lemmer, Dave Engelthaler, The Arizona COVID Genomics Union (ACGU)" |
| EPI_ISL_1120108, EPI_ISL_1120109, EPI_ISL_1120110, EPI_ISL_1120111, EPI_ISL_1120112, EPI_ISL_1120113, EPI_ISL_1120114, EPI_ISL_1120115, EPI_ISL_1120116, EPI_ISL_1120117, EPI_ISL_1120118, EPI_ISL_1120119, EPI_ISL_1120121, EPI_ISL_1120122, EPI_ISL_1120123, EPI_ISL_1120124, EPI_ISL_1120125, EPI_ISL_1120126, EPI_ISL_1120127, EPI_ISL_1120128, EPI_ISL_1120129, EPI_ISL_1120130, EPI_ISL_1120131, EPI_ISL_1120132 | | | |
| see above | Kansas Health and Environmental Lab | Kansas Health and Environmental Lab | Mike Grose, Jonathan Barnell, Ben Olsen, and Phil Adam |
| EPI_ISL_1120614 | Hospital Margarita Maza de Juarez | CIAD LDM-LGM | Bruno Gome-Gil, Julissa Enciso-Ibarra |
| EPI_ISL_1120708 | Inno Diagnostics/Center for Research Resources, Ponce Medical School Foundation, Inc. | Centers for Disease Control and Prevention, Dengue Branch | Gilberto A. Santiago, Glenda Gonzalez, Betzabel Flores, Keyla Charriez, Andrea Arias-Garcia, Raphael Sanchez-Torres, Vanessa Rivera-Amill, Gabriela Paz-Bailey, Jorge L. Munoz-Jordan |
| EPI_ISL_1120878, EPI_ISL_1120879, EPI_ISL_1120880, EPI_ISL_1120881, EPI_ISL_1120882, EPI_ISL_1120883, EPI_ISL_1120884, EPI_ISL_1120885, EPI_ISL_1120886, EPI_ISL_1120887, EPI_ISL_1120888, EPI_ISL_1120889, EPI_ISL_1120890, EPI_ISL_1120891, EPI_ISL_1120892, EPI_ISL_1120893, EPI_ISL_1120894, EPI_ISL_1120895, EPI_ISL_1120896 | | | |
| see above | Instituto Nacional de Medicina Genomica | Instituto Nacional de Medicina Genomica | Hidalgo-Miranda A, Mendoza-Vargas A, Reyes-Grajeda JP, Cedro-Tanda A, Alcaraz N, Gonzalez-Barrera D, Rangel-DeLeon D, Ramirez-Vega O, Sifuentes-Rojas C, Cisneros-Villanueva M, Herrera-Montalvo LA |
| EPI_ISL_1120970, EPI_ISL_1121014, EPI_ISL_1121015, EPI_ISL_1121016, EPI_ISL_1121017, EPI_ISL_1121018, EPI_ISL_1121019, EPI_ISL_1121020, EPI_ISL_1121021, EPI_ISL_1121024, EPI_ISL_1121025, EPI_ISL_1121026, EPI_ISL_1121027, EPI_ISL_1121028, EPI_ISL_1121029, EPI_ISL_1121030 | | | |
| see above | Wyoming Public Health Laboratory | Wyoming Public Health Laboratory | Jim Mildenberger, Wanda Manley, Noah Hull, Taylor Fearing, Lynette Gumbleton, Channing Weber, Ashley Norberg, Chayse Rowley, Marley Goetz, Brian Dominguez, Elliot Thomasson, Cari Sloma, and Rob Christensen |
| EPI_ISL_1121041, EPI_ISL_1121042, EPI_ISL_1121043, EPI_ISL_1121044, EPI_ISL_1121045, EPI_ISL_1121047, EPI_ISL_1121048, EPI_ISL_1121050, EPI_ISL_1121051, EPI_ISL_1121052, EPI_ISL_1121053, EPI_ISL_1121054, EPI_ISL_1121055, EPI_ISL_1121056 | | | |
| see above | Kashi Clinical Laboratory | Kashi Clinical Laboratory | Zahra Kashi, Jenefer DeKoning |
| EPI_ISL_1121057, EPI_ISL_1121058, EPI_ISL_1121059, EPI_ISL_1121060, EPI_ISL_1121061, EPI_ISL_1121062, EPI_ISL_1121065, EPI_ISL_1121066, EPI_ISL_1121067, EPI_ISL_1121068, EPI_ISL_1121069, EPI_ISL_1121070, EPI_ISL_1121071, EPI_ISL_1121072, EPI_ISL_1121073, EPI_ISL_1121074, EPI_ISL_1121075, EPI_ISL_1121076, EPI_ISL_1121077, EPI_ISL_1121078, EPI_ISL_1121079, EPI_ISL_1121080, | | | |

| | | | |
|---|---|---|---|
| EPI_ISL_1121081, EPI_ISL_1121082, EPI_ISL_1121083, EPI_ISL_1121084, EPI_ISL_1121085, EPI_ISL_1121086, EPI_ISL_1121087, EPI_ISL_1121088, EPI_ISL_1121089, EPI_ISL_1121090, EPI_ISL_1121091, EPI_ISL_1121092, EPI_ISL_1121093, EPI_ISL_1121094, EPI_ISL_1121095, EPI_ISL_1121096, EPI_ISL_1121097, EPI_ISL_1121098, EPI_ISL_1121099, EPI_ISL_1121100, EPI_ISL_1121101, EPI_ISL_1121102, EPI_ISL_1121103, EPI_ISL_1121104, EPI_ISL_1121105, EPI_ISL_1121106, EPI_ISL_1121107, EPI_ISL_1121108, EPI_ISL_1121109, EPI_ISL_1121110, EPI_ISL_1121111, EPI_ISL_1121112, EPI_ISL_1121113, EPI_ISL_1121114, EPI_ISL_1121115, EPI_ISL_1121116, EPI_ISL_1121117, EPI_ISL_1121118, EPI_ISL_1121119, EPI_ISL_1121120, EPI_ISL_1121121, EPI_ISL_1121122, EPI_ISL_1121123, EPI_ISL_1121124, EPI_ISL_1121125, EPI_ISL_1121126, EPI_ISL_1121127, EPI_ISL_1121128, EPI_ISL_1121129, EPI_ISL_1121130, EPI_ISL_1121131, EPI_ISL_1121132, EPI_ISL_1121133, EPI_ISL_1121134, EPI_ISL_1121135, EPI_ISL_1121136, EPI_ISL_1121137, EPI_ISL_1121138, EPI_ISL_1121139, EPI_ISL_1121140, EPI_ISL_1121141 | | | |
| see above | Nebraska Public Health Laboratory | NPHL COVID-19 Response Team | NPHL COVID-19 Response Team |
| EPI_ISL_1121221 | Kashi Clinical Laboratory | Kashi Clinical Laboratory | Zahra Kashi, Jenefer DeKoning |
| EPI_ISL_1121222, EPI_ISL_1121223, EPI_ISL_1121224, EPI_ISL_1121225, EPI_ISL_1121226, EPI_ISL_1121227, EPI_ISL_1121228 | DC Public Health Lab/ Dept. of Forensic Sciences | DC Public Health Lab/ Dept. of Forensic Sciences | Scott Nguyen, Janis Doss, Elizabeth Zelaya, Connie Maza, Monica Mann, Brittany Hamilton, David Payne, Jocelyn Hauser |
| EPI_ISL_1121229, EPI_ISL_1121230, EPI_ISL_1121231, EPI_ISL_1121232 | Kashi Clinical Laboratory | Kashi Clinical Laboratory | Zahra Kashi, Jenefer DeKoning |
| EPI_ISL_1121234, EPI_ISL_1121235, EPI_ISL_1121236, EPI_ISL_1121237, EPI_ISL_1121238, EPI_ISL_1121239, EPI_ISL_1121240, EPI_ISL_1121241, EPI_ISL_1121242, EPI_ISL_1121243, EPI_ISL_1121244, EPI_ISL_1121245, EPI_ISL_1121246, EPI_ISL_1121247, EPI_ISL_1121248, EPI_ISL_1121249, EPI_ISL_1121250, EPI_ISL_1121251, EPI_ISL_1121252, EPI_ISL_1121253, EPI_ISL_1121254, EPI_ISL_1121257, EPI_ISL_1121258, EPI_ISL_1121259, EPI_ISL_1121260, EPI_ISL_1121261, EPI_ISL_1121262, EPI_ISL_1121263, EPI_ISL_1121264, EPI_ISL_1121265, EPI_ISL_1121266, EPI_ISL_1121267, EPI_ISL_1121268, EPI_ISL_1121269, EPI_ISL_1121270, EPI_ISL_1121271, EPI_ISL_1121272, EPI_ISL_1121273, EPI_ISL_1121274, EPI_ISL_1121275, EPI_ISL_1121276, EPI_ISL_1121277, EPI_ISL_1121278, EPI_ISL_1121279, EPI_ISL_1121280 | | | |
| see above | Maryland Public Health Laboratory | Maryland Public Health Laboratory | Maryland Department of Health Laboratories Administration |
| EPI_ISL_1121281, EPI_ISL_1121282, EPI_ISL_1121283, EPI_ISL_1121284, EPI_ISL_1121285, EPI_ISL_1121286, EPI_ISL_1121287, EPI_ISL_1121288, EPI_ISL_1121289, EPI_ISL_1121290, EPI_ISL_1121291, EPI_ISL_1121292, EPI_ISL_1121293, EPI_ISL_1121294, EPI_ISL_1121295, EPI_ISL_1121296, EPI_ISL_1121297, EPI_ISL_1121298, EPI_ISL_1121299, EPI_ISL_1121300, EPI_ISL_1121301, EPI_ISL_1121302 | | | |
| see above | DC Public Health Lab/ Dept. of Forensic Sciences | DC Public Health Lab/ Dept. of Forensic Sciences | Scott Nguyen, Janis Doss, Elizabeth Zelaya, Connie Maza, Monica Mann, Brittany Hamilton, David Payne, Jocelyn Hauser |
| EPI_ISL_1121304, EPI_ISL_1121331, EPI_ISL_1121332, EPI_ISL_1121333, EPI_ISL_1121334, EPI_ISL_1121335, EPI_ISL_1121337 | Kashi Clinical Laboratory | Kashi Clinical Laboratory | Zahra Kashi, Jenefer DeKoning |
| EPI_ISL_1121338, EPI_ISL_1121339, EPI_ISL_1121340, EPI_ISL_1121341, EPI_ISL_1121342, EPI_ISL_1121343, EPI_ISL_1121344, EPI_ISL_1121345, EPI_ISL_1121346, EPI_ISL_1121347, EPI_ISL_1121348, EPI_ISL_1121349, EPI_ISL_1121350, EPI_ISL_1121351, EPI_ISL_1121352, EPI_ISL_1121353, EPI_ISL_1121354 | | | |
| see above | Kansas Health and Environmental Lab | Kansas Health and Environmental Lab | Mike Grose, Jonathan Barnell, Ben Olsen, and Phil Adam |
| EPI_ISL_1121355 | Kashi Clinical Laboratory | Kashi Clinical Laboratory | Zahra Kashi, Jenefer DeKoning |
| EPI_ISL_1121356 | Mary Eliza Mahoney HC | New Jersey Public Health and Environmental Laboratories (NJ PHEL) | Lindsey Bodnar, Shiv K. Verma, Dana Woell, Byeong Jeong |
| EPI_ISL_1121357, EPI_ISL_1121360, EPI_ISL_1121362, EPI_ISL_1121363, EPI_ISL_1121364, EPI_ISL_1121365, EPI_ISL_1121366, EPI_ISL_1121367, EPI_ISL_1121368, EPI_ISL_1121369, EPI_ISL_1121370, EPI_ISL_1121371, EPI_ISL_1121372 | | | |
| see above | NJ Rapid Response Teams | New Jersey Public Health and Environmental Laboratories (NJ PHEL) | Lindsey Bodnar, Shiv K. Verma, Dana Woell, Byeong Jeong |
| EPI_ISL_1121488, EPI_ISL_1121489, EPI_ISL_1121490, EPI_ISL_1121491, EPI_ISL_1121492, EPI_ISL_1121493, EPI_ISL_1121494, EPI_ISL_1121495, EPI_ISL_1121496, EPI_ISL_1121497, EPI_ISL_1121498, EPI_ISL_1121499, EPI_ISL_1121500, EPI_ISL_1121501, EPI_ISL_1121502, EPI_ISL_1121504, EPI_ISL_1121505, EPI_ISL_1121506, EPI_ISL_1121507, EPI_ISL_1121508, EPI_ISL_1121509, EPI_ISL_1121510, EPI_ISL_1121511, EPI_ISL_1121512, EPI_ISL_1121513, EPI_ISL_1121514, EPI_ISL_1121515, EPI_ISL_1121516, EPI_ISL_1121517, EPI_ISL_1121518, EPI_ISL_1121519, EPI_ISL_1121520, EPI_ISL_1121521, EPI_ISL_1121522, EPI_ISL_1121523, EPI_ISL_1121524, EPI_ISL_1121525, EPI_ISL_1121526, EPI_ISL_1121527, EPI_ISL_1121528, EPI_ISL_1121529, EPI_ISL_1121530, EPI_ISL_1121531, EPI_ISL_1121532, EPI_ISL_1121533, EPI_ISL_1121534, EPI_ISL_1121535, EPI_ISL_1121536, EPI_ISL_1121537, EPI_ISL_1121538, EPI_ISL_1121539, EPI_ISL_1121540, EPI_ISL_1121542, EPI_ISL_1121543, EPI_ISL_1121544, EPI_ISL_1121545, EPI_ISL_1121546, EPI_ISL_1121547, EPI_ISL_1121548, EPI_ISL_1121549, EPI_ISL_1121550, EPI_ISL_1121551, EPI_ISL_1121552, EPI_ISL_1121553, EPI_ISL_1121554, EPI_ISL_1121555 | | | |
| see above | New Mexico Department of Health Scientific Laboratory | Center for Global Health, University of New Mexico Health Sciences Center | Daryl Domman, Kurt Schwalm, Twila Kunde, Joseph Hicks, Anastacia Griego, Michael Edwards, Darrell Dinwiddie |
| EPI_ISL_1121556 | M Health Fairview | Minnesota Department of Health, Public Health Laboratory | Alexandra Lorentz, Jacob Garfin, Matt Plumb, and Xiong Wang |
| EPI_ISL_1121557, EPI_ISL_1121558, EPI_ISL_1121559 | Minnesota Department of Health, Public Health Laboratory | Minnesota Department of Health, Public Health Laboratory | Alexandra Lorentz, Jacob Garfin, Matt Plumb, and Xiong Wang |
| EPI_ISL_1121560, EPI_ISL_1121561, EPI_ISL_1121562, EPI_ISL_1121563, EPI_ISL_1121564, EPI_ISL_1121565, EPI_ISL_1121566, EPI_ISL_1121567, EPI_ISL_1121568, EPI_ISL_1121569, EPI_ISL_1121570, EPI_ISL_1121571, EPI_ISL_1121572, EPI_ISL_1121573, EPI_ISL_1121574, EPI_ISL_1121575, EPI_ISL_1121576, EPI_ISL_1121577, EPI_ISL_1121578, EPI_ISL_1121579, EPI_ISL_1121580, EPI_ISL_1121581, EPI_ISL_1121582, EPI_ISL_1121583, EPI_ISL_1121584, EPI_ISL_1121585, EPI_ISL_1121586, EPI_ISL_1121587, EPI_ISL_1121588, EPI_ISL_1121589 | | | |
| see above | M Health Fairview | Minnesota Department of Health, Public Health Laboratory | Alexandra Lorentz, Jacob Garfin, Matt Plumb, and Xiong Wang |
| EPI_ISL_1121590 | Minnesota Department of Health, Public Health Laboratory | Minnesota Department of Health, Public Health Laboratory | Alexandra Lorentz, Jacob Garfin, Matt Plumb, and Xiong Wang |
| EPI_ISL_1121591, EPI_ISL_1121592 | M Health Fairview | Minnesota Department of Health, Public Health Laboratory | Alexandra Lorentz, Jacob Garfin, Matt Plumb, and Xiong Wang |
| EPI_ISL_1121593, EPI_ISL_1121594 | Vault Health | Minnesota Department of Health, Public Health Laboratory | Alexandra Lorentz, Jacob Garfin, Matt Plumb, and Xiong Wang |
| EPI_ISL_1121595 | M Health Fairview | Minnesota Department of Health, Public Health Laboratory | Alexandra Lorentz, Jacob Garfin, Matt Plumb, and Xiong Wang |
| EPI_ISL_1121597, EPI_ISL_1121598, EPI_ISL_1121599, EPI_ISL_1121600 | Vault Health | Minnesota Department of Health, Public Health Laboratory | Alexandra Lorentz, Jacob Garfin, Matt Plumb, and Xiong Wang |
| EPI_ISL_1121601, EPI_ISL_1121602 | M Health Fairview | Minnesota Department of Health, Public Health Laboratory | Alexandra Lorentz, Jacob Garfin, Matt Plumb, and Xiong Wang |
| EPI_ISL_1121603, EPI_ISL_1121604, EPI_ISL_1121605, EPI_ISL_1121606 | Vault Health | Minnesota Department of Health, Public Health Laboratory | Alexandra Lorentz, Jacob Garfin, Matt Plumb, and Xiong Wang |
| EPI_ISL_1121607 | M Health Fairview | Minnesota Department of Health, Public Health Laboratory | Alexandra Lorentz, Jacob Garfin, Matt Plumb, and Xiong Wang |
| EPI_ISL_1121608, EPI_ISL_1121609, EPI_ISL_1121610, EPI_ISL_1121611, EPI_ISL_1121612 | Vault Health | Minnesota Department of Health, Public Health Laboratory | Alexandra Lorentz, Jacob Garfin, Matt Plumb, and Xiong Wang |
| EPI_ISL_1121613 | M Health Fairview | Minnesota Department of Health, Public Health Laboratory | Alexandra Lorentz, Jacob Garfin, Matt Plumb, and Xiong Wang |
| EPI_ISL_1121614, EPI_ISL_1121615, EPI_ISL_1121617, EPI_ISL_1121618 | Vault Health | Minnesota Department of Health, Public Health Laboratory | Alexandra Lorentz, Jacob Garfin, Matt Plumb, and Xiong Wang |
| EPI_ISL_1121619 | M Health Fairview | Minnesota Department of Health, Public Health Laboratory | Alexandra Lorentz, Jacob Garfin, Matt Plumb, and Xiong Wang |
| EPI_ISL_1121620, EPI_ISL_1121621, EPI_ISL_1121622, EPI_ISL_1121623 | Vault Health | Minnesota Department of Health, Public Health Laboratory | Alexandra Lorentz, Jacob Garfin, Matt Plumb, and Xiong Wang |
| EPI_ISL_1121624 | Minnesota Department of Health, Public Health Laboratory | Minnesota Department of Health, Public Health Laboratory | Alexandra Lorentz, Jacob Garfin, Matt Plumb, and Xiong Wang |
| EPI_ISL_1121625, EPI_ISL_1121626, EPI_ISL_1121627 | Vault Health | Minnesota Department of Health, Public Health Laboratory | Alexandra Lorentz, Jacob Garfin, Matt Plumb, and Xiong Wang |

| | | | |
|---|---|---|---|
| EPI_ISL_1121628 | Minnesota Department of Health, Public Health Laboratory | Minnesota Department of Health, Public Health Laboratory | Alexandra Lorentz, Jacob Garfin, Matt Plumb, and Xiong Wang |
| EPI_ISL_1121629 | M Health Fairview | Minnesota Department of Health, Public Health Laboratory | Alexandra Lorentz, Jacob Garfin, Matt Plumb, and Xiong Wang |
| EPI_ISL_1121630, EPI_ISL_1121631, EPI_ISL_1121632 | Vault Health | Minnesota Department of Health, Public Health Laboratory | Alexandra Lorentz, Jacob Garfin, Matt Plumb, and Xiong Wang |
| EPI_ISL_1121633, EPI_ISL_1121634, EPI_ISL_1121635, EPI_ISL_1121636, EPI_ISL_1121637, EPI_ISL_1121638, EPI_ISL_1121639, EPI_ISL_1121640, EPI_ISL_1121641, EPI_ISL_1121642, EPI_ISL_1121643, EPI_ISL_1121644, EPI_ISL_1121645, EPI_ISL_1121646, EPI_ISL_1121647, EPI_ISL_1121648, EPI_ISL_1121649, EPI_ISL_1121650, EPI_ISL_1121651, EPI_ISL_1121652, EPI_ISL_1121653, EPI_ISL_1121654, EPI_ISL_1121655, EPI_ISL_1121656, EPI_ISL_1121657, EPI_ISL_1121658, EPI_ISL_1121659, EPI_ISL_1121660, EPI_ISL_1121661, EPI_ISL_1121662, EPI_ISL_1121663, EPI_ISL_1121664, EPI_ISL_1121665, EPI_ISL_1121666, EPI_ISL_1121667, EPI_ISL_1121668, EPI_ISL_1121669, EPI_ISL_1121670, EPI_ISL_1121671, EPI_ISL_1121672, EPI_ISL_1121673, EPI_ISL_1121674, EPI_ISL_1121675, EPI_ISL_1121676, EPI_ISL_1121677, EPI_ISL_1121678, EPI_ISL_1121679, EPI_ISL_1121680, EPI_ISL_1121681, EPI_ISL_1121682, EPI_ISL_1121683, EPI_ISL_1121684, EPI_ISL_1121685, EPI_ISL_1121686, EPI_ISL_1121687, EPI_ISL_1121688, EPI_ISL_1121689, EPI_ISL_1121690, EPI_ISL_1121691, EPI_ISL_1121692, EPI_ISL_1121693, EPI_ISL_1121694, EPI_ISL_1121695, EPI_ISL_1121696, EPI_ISL_1121697, EPI_ISL_1121698, EPI_ISL_1121699, EPI_ISL_1121700, EPI_ISL_1121701, EPI_ISL_1121702 | | | |
| see above | Broad Institute Clinical Research Sequencing Platform | Infectious Disease Program, Broad Institute of Harvard and MIT | Lemieux,J.E., Siddle,K.J., Adams,G., Gladden-Young,A., Lagerborg,K., Rudy,M., DeRuff,K., Carter,A., Normandin,E., Bauer,M., Reilly,S., Tomkins-Tinch,C., Loreth,C., Chaluvadi,S., Birren,B.W., Gallagher,G., Smole,S., Park,D.J., MacInnis,B.L., and Sabeti,P.C. |
| EPI_ISL_1121705, EPI_ISL_1121706, EPI_ISL_1121707, EPI_ISL_1121709, EPI_ISL_1121712, EPI_ISL_1121715, EPI_ISL_1121716, EPI_ISL_1121717, EPI_ISL_1121718, EPI_ISL_1121719, EPI_ISL_1121721, EPI_ISL_1121722, EPI_ISL_1121723, EPI_ISL_1121724, EPI_ISL_1121725, EPI_ISL_1121728, EPI_ISL_1121730, EPI_ISL_1121731, EPI_ISL_1121732, EPI_ISL_1121734, EPI_ISL_1121736, EPI_ISL_1121737, EPI_ISL_1121739, EPI_ISL_1121741, EPI_ISL_1121742, EPI_ISL_1121743, EPI_ISL_1121745, EPI_ISL_1121746, EPI_ISL_1121749, EPI_ISL_1121750, EPI_ISL_1121751, EPI_ISL_1121755, EPI_ISL_1121758, EPI_ISL_1121759, EPI_ISL_1121760, EPI_ISL_1121761, EPI_ISL_1121762, EPI_ISL_1121763, EPI_ISL_1121764, EPI_ISL_1121765, EPI_ISL_1121766, EPI_ISL_1121767, EPI_ISL_1121768, EPI_ISL_1121769, EPI_ISL_1121770, EPI_ISL_1121771, EPI_ISL_1121772, EPI_ISL_1121773, EPI_ISL_1121774, EPI_ISL_1121775, EPI_ISL_1121776, EPI_ISL_1121777, EPI_ISL_1121778, EPI_ISL_1121779, EPI_ISL_1121780, EPI_ISL_1121781, EPI_ISL_1121782, EPI_ISL_1121783, EPI_ISL_1121784, EPI_ISL_1121785, EPI_ISL_1121786, EPI_ISL_1121787, EPI_ISL_1121788, EPI_ISL_1121789, EPI_ISL_1121790, EPI_ISL_1121791, EPI_ISL_1121792, EPI_ISL_1121793, EPI_ISL_1121794, EPI_ISL_1121795, EPI_ISL_1121796, EPI_ISL_1121797, EPI_ISL_1121798, EPI_ISL_1121799, EPI_ISL_1121800, EPI_ISL_1121801, EPI_ISL_1121802, EPI_ISL_1121803, EPI_ISL_1121804, EPI_ISL_1121805, EPI_ISL_1121806, EPI_ISL_1121807, EPI_ISL_1121808, EPI_ISL_1121809, EPI_ISL_1121810, EPI_ISL_1121812 | | | |
| see above | Massachusetts General Hospital | Infectious Disease Program, Broad Institute of Harvard and MIT | Lemieux,J.E., Siddle,K.J., Shaw,B., Adams,G., Pierce,V., Turbett,S., Anahtar,M., Branda,J., Slater,D., Harris,J., Lin,A.E., Gladden-Young,A., Lagerborg,K., Rudy,M., DeRuff,K., Carter,A., Normandin,E., Bauer,M., Reilly,S., Tomkins-Tinch,C., Loreth,C., Chaluvadi,S., Neumann,A., Cusick,C., Chapman,S.B., Gnirke,A., Flowers,K., Cerrato,F., Birren,B.W., Gallagher,G., Smole,S., Park,D.J., MacInnis,B.L., Ryan,E., LaRocque,R., Rosenberg,E. and Sabeti,P.C. |
| EPI_ISL_1121813, EPI_ISL_1121814, EPI_ISL_1121816, EPI_ISL_1121817, EPI_ISL_1121818, EPI_ISL_1121821, EPI_ISL_1121822, EPI_ISL_1121823, EPI_ISL_1121824, EPI_ISL_1121825, EPI_ISL_1121826, EPI_ISL_1121829, EPI_ISL_1121830, EPI_ISL_1121831, EPI_ISL_1121832, EPI_ISL_1121833, EPI_ISL_1121834, EPI_ISL_1121835, EPI_ISL_1121836, EPI_ISL_1121837, EPI_ISL_1121838, EPI_ISL_1121839, EPI_ISL_1121840, EPI_ISL_1121841, EPI_ISL_1121842, EPI_ISL_1121843, EPI_ISL_1121844, EPI_ISL_1121846, EPI_ISL_1121847, EPI_ISL_1121848, EPI_ISL_1121849, EPI_ISL_1121850, EPI_ISL_1121851, EPI_ISL_1121852, EPI_ISL_1121853, EPI_ISL_1121854, EPI_ISL_1121855, EPI_ISL_1121856, EPI_ISL_1121857, EPI_ISL_1121858, EPI_ISL_1121859, EPI_ISL_1121860, EPI_ISL_1121861, EPI_ISL_1121862, EPI_ISL_1121863, EPI_ISL_1121864, EPI_ISL_1121865, EPI_ISL_1121866, EPI_ISL_1121867, EPI_ISL_1121868, EPI_ISL_1121869, EPI_ISL_1121870, EPI_ISL_1121872, EPI_ISL_1121873, EPI_ISL_1121874, EPI_ISL_1121875, EPI_ISL_1121876, EPI_ISL_1121877, EPI_ISL_1121878 | | | |
| see above | Rhode Island Department of Health | Infectious Disease Program, Broad Institute of Harvard and MIT | Lemieux,J.E., Siddle,K.J., Huard,R., King,E., Azevedo,K., Miller,A., Adams,G., Gladden-Young,A., Lagerborg,K., Rudy,M., DeRuff,K., Carter,A., Normandin,E., Bauer,M., Reilly,S., Tomkins-Tinch,C., Loreth,C., Chaluvadi,S., Birren,B.W., Gallagher,G., Smole,S., Park,D.J., MacInnis,B.L., and Sabeti,P.C. |
| EPI_ISL_1121879, EPI_ISL_1121880, EPI_ISL_1121881, EPI_ISL_1121882, EPI_ISL_1121883, EPI_ISL_1121884, EPI_ISL_1121885, EPI_ISL_1121886 | Broad Institute Clinical Research Sequencing Platform | Infectious Disease Program, Broad Institute of Harvard and MIT | Lemieux,J.E., Siddle,K.J., Adams,G., Gladden-Young,A., Lagerborg,K., Rudy,M., DeRuff,K., Carter,A., Normandin,E., Bauer,M., Reilly,S., Tomkins-Tinch,C., Loreth,C., Chaluvadi,S., Birren,B.W., Gallagher,G., Smole,S., Park,D.J., MacInnis,B.L., and Sabeti,P.C. |
| EPI_ISL_1121887 | Department of Health & Mental Hygiene Corona | New York City Public Health Laboratory | Jade Wang, et al. |
| EPI_ISL_1121888 | Department of Health & Mental Hygiene Morrisania | New York City Public Health Laboratory | Jade Wang, et al. |
| EPI_ISL_1121889 | Department of Health & Mental Hygiene Fort Greene | New York City Public Health Laboratory | Jade Wang, et al. |
| EPI_ISL_1121890, EPI_ISL_1121891, EPI_ISL_1121892 | Department of Health & Mental Hygiene Crown Heights | New York City Public Health Laboratory | Jade Wang, et al. |
| EPI_ISL_1121893 | Department of Health & Mental Hygiene Central Harlem | New York City Public Health Laboratory | Jade Wang, et al. |
| EPI_ISL_1121894 | Department of Health & Mental Hygiene PHL | New York City Public Health Laboratory | Jade Wang, et al. |
| EPI_ISL_1121895, EPI_ISL_1121896, EPI_ISL_1121897, EPI_ISL_1121898 | OCME Office Of Chief Medical Examiner | New York City Public Health Laboratory | Jade Wang, et al. |
| EPI_ISL_1121899 | Department of Health & Mental Hygiene Corona | New York City Public Health Laboratory | Jade Wang, et al. |
| EPI_ISL_1121900 | Department of Health & Mental Hygiene Central Harlem | New York City Public Health Laboratory | Jade Wang, et al. |
| EPI_ISL_1121901, EPI_ISL_1121902, EPI_ISL_1121903 | Department of Health & Mental Hygiene Fort Greene | New York City Public Health Laboratory | Jade Wang, et al. |
| EPI_ISL_1121904 | Department of Health & Mental Hygiene Crown Heights | New York City Public Health Laboratory | Jade Wang, et al. |
| EPI_ISL_1121905, EPI_ISL_1121906, EPI_ISL_1121907, EPI_ISL_1121908 | Department of Health & Mental Hygiene Corona | New York City Public Health Laboratory | Jade Wang, et al. |
| EPI_ISL_1121909, EPI_ISL_1121910, EPI_ISL_1121911, EPI_ISL_1121912 | Department of Health & Mental Hygiene PHL | New York City Public Health Laboratory | Jade Wang, et al. |
| EPI_ISL_1121913 | Department of Health & Mental Hygiene Morrisania | New York City Public Health Laboratory | Jade Wang, et al. |
| EPI_ISL_1121914 | Department of Health & Mental Hygiene Riverside | New York City Public Health Laboratory | Jade Wang, et al. |
| EPI_ISL_1121915, EPI_ISL_1121916, EPI_ISL_1121917, EPI_ISL_1121918 | Department of Health & Mental Hygiene Jamaica | New York City Public Health Laboratory | Jade Wang, et al. |
| EPI_ISL_1121919, EPI_ISL_1121920, EPI_ISL_1121921, EPI_ISL_1121922, EPI_ISL_1121923 | Department of Health & Mental Hygiene Morrisania | New York City Public Health Laboratory | Jade Wang, et al. |
| EPI_ISL_1121924 | Department of Health & Mental Hygiene Jamaica | New York City Public Health Laboratory | Jade Wang, et al. |
| EPI_ISL_1121925 | Department of Health & Mental Hygiene Chelsea | New York City Public Health Laboratory | Jade Wang, et al. |
| EPI_ISL_1121926 | Department of Health & Mental Hygiene Morrisania | New York City Public Health Laboratory | Jade Wang, et al. |
| EPI_ISL_1121927, EPI_ISL_1121928 | Department of Health & Mental Hygiene Chelsea | New York City Public Health Laboratory | Jade Wang, et al. |
| EPI_ISL_1121929, EPI_ISL_1121930, EPI_ISL_1121931 | Department of Health & Mental Hygiene PHL | New York City Public Health Laboratory | Jade Wang, et al. |
| EPI_ISL_1121932, EPI_ISL_1121933, EPI_ISL_1121934, EPI_ISL_1121935, EPI_ISL_1121936 | Department of Health & Mental Hygiene Corona | New York City Public Health Laboratory | Jade Wang, et al. |

| | | | |
|---|---|---|---|
| EPI_ISL_1121937 | Department of Health & Mental Hygiene Fort Greene | New York City Public Health Laboratory | Jade Wang, et al. |
| EPI_ISL_1121938, EPI_ISL_1121939 | Department of Health & Mental Hygiene Crown Heights | New York City Public Health Laboratory | Jade Wang, et al. |
| EPI_ISL_1121940, EPI_ISL_1121941, EPI_ISL_1121942 | Department of Health & Mental Hygiene Morrisania | New York City Public Health Laboratory | Jade Wang, et al. |
| EPI_ISL_1121943 | Department of Health & Mental Hygiene Riverside | New York City Public Health Laboratory | Jade Wang, et al. |
| EPI_ISL_1121944 | Department of Health & Mental Hygiene Morrisania | New York City Public Health Laboratory | Jade Wang, et al. |
| EPI_ISL_1121945, EPI_ISL_1121946 | Department of Health & Mental Hygiene Central Harlem | New York City Public Health Laboratory | Jade Wang, et al. |
| EPI_ISL_1121947, EPI_ISL_1121948, EPI_ISL_1121949 | Department of Health & Mental Hygiene Morrisania | New York City Public Health Laboratory | Jade Wang, et al. |
| EPI_ISL_1121950 | Department of Health & Mental Hygiene Central Harlem | New York City Public Health Laboratory | Jade Wang, et al. |
| EPI_ISL_1121951, EPI_ISL_1121952, EPI_ISL_1121953, EPI_ISL_1121954, EPI_ISL_1121955, EPI_ISL_1121956, EPI_ISL_1121957, EPI_ISL_1121958, EPI_ISL_1121959 | Department of Health & Mental Hygiene Jamaica | New York City Public Health Laboratory | Jade Wang, et al. |
| EPI_ISL_1121960 | Department of Health & Mental Hygiene Chelsea | New York City Public Health Laboratory | Jade Wang, et al. |
| EPI_ISL_1121963, EPI_ISL_1121964, EPI_ISL_1121965, EPI_ISL_1121966, EPI_ISL_1121967, EPI_ISL_1121968 | Broad Institute Clinical Research Sequencing Platform | Infectious Disease Program, Broad Institute of Harvard and MIT | Lemieux,J.E., Siddle,K.J., Adams,G., Gladden-Young,A., Lagerborg,K., Rudy,M., DeRuff,K., Carter,A., Normandin,E., Bauer,M., Reilly,S., Tomkins-Tinch,C., Loreth,C., Chaluvadi,S., Birren,B.W., Gallagher,G., Smole,S., Park,D.J., MacInnis,B.L., and Sabeti,P.C. |
| EPI_ISL_1121969, EPI_ISL_1121970, EPI_ISL_1121971 | Massachusetts General Hospital | Infectious Disease Program, Broad Institute of Harvard and MIT | Lemieux,J.E., Siddle,K.J., Shaw,B., Adams,G., Pierce,V., Turbett,S., Anahtar,M., Branda,J., Slater,D., Harris,J., Lin,A.E., Gladden-Young,A., Lagerborg,K., Rudy,M., DeRuff,K., Carter,A., Normandin,E., Bauer,M., Reilly,S., Tomkins-Tinch,C., Loreth,C., Chaluvadi,S., Neumann,A., Cusick,C., Chapman,S.B., Gnirke,A., Flowers,K., Cerrato,F., Birren,B.W., Gallagher,G., Smole,S., Park,D.J., MacInnis,B.L., Ryan,E., LaRocque,R., Rosenberg,E. and Sabeti,P.C. |
| EPI_ISL_1121972 | Rhode Island Department of Health | Infectious Disease Program, Broad Institute of Harvard and MIT | Lemieux,J.E., Siddle,K.J., Huard,R., King,E., Azevedo,K., Miller,A., Adams,G., Gladden-Young,A., Lagerborg,K., Rudy,M., DeRuff,K., Carter,A., Normandin,E., Bauer,M., Reilly,S., Tomkins-Tinch,C., Loreth,C., Chaluvadi,S., Birren,B.W., Gallagher,G., Smole,S., Park,D.J., MacInnis,B.L., and Sabeti,P.C. |
| EPI_ISL_1121973 | Broad Institute Clinical Research Sequencing Platform | Infectious Disease Program, Broad Institute of Harvard and MIT | Lemieux,J.E., Siddle,K.J., Adams,G., Gladden-Young,A., Lagerborg,K., Rudy,M., DeRuff,K., Carter,A., Normandin,E., Bauer,M., Reilly,S., Tomkins-Tinch,C., Loreth,C., Chaluvadi,S., Birren,B.W., Gallagher,G., Smole,S., Park,D.J., MacInnis,B.L., and Sabeti,P.C. |
| EPI_ISL_1121994, EPI_ISL_1121995, EPI_ISL_1121996, EPI_ISL_1121997, EPI_ISL_1121998, EPI_ISL_1122000, EPI_ISL_1122001, EPI_ISL_1122002, EPI_ISL_1122003, EPI_ISL_1122004, EPI_ISL_1122005, EPI_ISL_1122006, EPI_ISL_1122007, EPI_ISL_1122008, EPI_ISL_1122009, EPI_ISL_1122010, EPI_ISL_1122011, EPI_ISL_1122012, EPI_ISL_1122013 | | | |
| see above | Kentucky State Public Health Lab | Kentucky State Public Health Lab | Stephanie Lunn, Karim George, Joshua Tobias, William Grooms, Vaneet Arora, Matthew Johnson, Rachel Zinner, Rhonda Lucas |
| EPI_ISL_1122018, EPI_ISL_1122019, EPI_ISL_1122020, EPI_ISL_1122021, EPI_ISL_1122023, EPI_ISL_1122024, EPI_ISL_1122025, EPI_ISL_1122026, EPI_ISL_1122028, EPI_ISL_1122029, EPI_ISL_1122030, EPI_ISL_1122031, EPI_ISL_1122034, EPI_ISL_1122035, EPI_ISL_1122037, EPI_ISL_1122038, EPI_ISL_1122039, EPI_ISL_1122040, EPI_ISL_1122041, EPI_ISL_1122042, EPI_ISL_1122043, EPI_ISL_1122045, EPI_ISL_1122047, EPI_ISL_1122048, EPI_ISL_1122049, EPI_ISL_1122050, EPI_ISL_1122051, EPI_ISL_1122052, EPI_ISL_1122053, EPI_ISL_1122054, EPI_ISL_1122055, EPI_ISL_1122056, EPI_ISL_1122057, EPI_ISL_1122058, EPI_ISL_1122059, EPI_ISL_1122060, EPI_ISL_1122061, EPI_ISL_1122062, EPI_ISL_1122063, EPI_ISL_1122064, EPI_ISL_1122066, EPI_ISL_1122068, EPI_ISL_1122069, EPI_ISL_1122070, EPI_ISL_1122071, EPI_ISL_1122072, EPI_ISL_1122073, EPI_ISL_1122074, EPI_ISL_1122075, EPI_ISL_1122076, EPI_ISL_1122078, EPI_ISL_1122079, EPI_ISL_1122080, EPI_ISL_1122081, EPI_ISL_1122082, EPI_ISL_1122083, EPI_ISL_1122084, EPI_ISL_1122085, EPI_ISL_1122086, EPI_ISL_1122087, EPI_ISL_1122088, EPI_ISL_1122089, EPI_ISL_1122090, EPI_ISL_1122091, EPI_ISL_1122092, EPI_ISL_1122093, EPI_ISL_1122094, EPI_ISL_1122095, EPI_ISL_1122097, EPI_ISL_1122098, EPI_ISL_1122099, EPI_ISL_1122100, EPI_ISL_1122101, EPI_ISL_1122102, EPI_ISL_1122103, EPI_ISL_1122104, EPI_ISL_1122105, EPI_ISL_1122106, EPI_ISL_1122107, EPI_ISL_1122108, EPI_ISL_1122109, EPI_ISL_1122110, EPI_ISL_1122111, EPI_ISL_1122112, EPI_ISL_1122113, EPI_ISL_1122114, EPI_ISL_1122115, EPI_ISL_1122116, EPI_ISL_1122117, EPI_ISL_1122118, EPI_ISL_1122119, EPI_ISL_1122120, EPI_ISL_1122121, EPI_ISL_1122122, EPI_ISL_1122123, EPI_ISL_1122124, EPI_ISL_1122125, EPI_ISL_1122126, EPI_ISL_1122127, EPI_ISL_1122128, EPI_ISL_1122129, EPI_ISL_1122130, EPI_ISL_1122131, EPI_ISL_1122133, EPI_ISL_1122134, EPI_ISL_1122135, EPI_ISL_1122136, EPI_ISL_1122138, EPI_ISL_1122139, EPI_ISL_1122140, EPI_ISL_1122141, EPI_ISL_1122142, EPI_ISL_1122143, EPI_ISL_1122144, EPI_ISL_1122146, EPI_ISL_1122147, EPI_ISL_1122148, EPI_ISL_1122149, EPI_ISL_1122151, EPI_ISL_1122152, EPI_ISL_1122153, EPI_ISL_1122154, EPI_ISL_1122155, EPI_ISL_1122156, EPI_ISL_1122157, EPI_ISL_1122159, EPI_ISL_1122160, EPI_ISL_1122161, EPI_ISL_1122162, EPI_ISL_1122163, EPI_ISL_1122164, EPI_ISL_1122165, EPI_ISL_1122166, EPI_ISL_1122167, EPI_ISL_1122168, EPI_ISL_1122169, EPI_ISL_1122170, EPI_ISL_1122171, EPI_ISL_1122172, EPI_ISL_1122173, EPI_ISL_1122174, EPI_ISL_1122175, EPI_ISL_1122176, EPI_ISL_1122177, EPI_ISL_1122178, EPI_ISL_1122179, EPI_ISL_1122180, EPI_ISL_1122182, EPI_ISL_1122183, EPI_ISL_1122184, EPI_ISL_1122186, EPI_ISL_1122188, EPI_ISL_1122189, EPI_ISL_1122190, EPI_ISL_1122191, EPI_ISL_1122192, EPI_ISL_1122193, EPI_ISL_1122194, EPI_ISL_1122195, EPI_ISL_1122196, EPI_ISL_1122197, EPI_ISL_1122198, EPI_ISL_1122199, EPI_ISL_1122200, EPI_ISL_1122201, EPI_ISL_1122202, EPI_ISL_1122203, EPI_ISL_1122204, EPI_ISL_1122205, EPI_ISL_1122206, EPI_ISL_1122207, EPI_ISL_1122208, EPI_ISL_1122209, EPI_ISL_1122210, EPI_ISL_1122211, EPI_ISL_1122212, EPI_ISL_1122213, EPI_ISL_1122214, EPI_ISL_1122215, EPI_ISL_1122216, EPI_ISL_1122217, EPI_ISL_1122218, EPI_ISL_1122219, EPI_ISL_1122220, EPI_ISL_1122221, EPI_ISL_1122222, EPI_ISL_1122223, EPI_ISL_1122224, EPI_ISL_1122225, EPI_ISL_1122226, EPI_ISL_1122227, EPI_ISL_1122228, EPI_ISL_1122229, EPI_ISL_1122230, EPI_ISL_1122231, EPI_ISL_1122232, EPI_ISL_1122233, EPI_ISL_1122234, EPI_ISL_1122235, EPI_ISL_1122236, EPI_ISL_1122237, EPI_ISL_1122238, EPI_ISL_1122239, EPI_ISL_1122240, EPI_ISL_1122241, EPI_ISL_1122242, EPI_ISL_1122243, EPI_ISL_1122244, EPI_ISL_1122245, EPI_ISL_1122246, EPI_ISL_1122247, EPI_ISL_1122248, EPI_ISL_1122249, EPI_ISL_1122250, EPI_ISL_1122251, EPI_ISL_1122252, EPI_ISL_1122254, EPI_ISL_1122255, EPI_ISL_1122256, EPI_ISL_1122257, EPI_ISL_1122258, EPI_ISL_1122259, EPI_ISL_1122260, EPI_ISL_1122261, EPI_ISL_1122262, EPI_ISL_1122263, EPI_ISL_1122264, EPI_ISL_1122265, EPI_ISL_1122266, EPI_ISL_1122267, EPI_ISL_1122268, EPI_ISL_1122269, EPI_ISL_1122273, EPI_ISL_1122274, EPI_ISL_1122275, EPI_ISL_1122276, EPI_ISL_1122277, EPI_ISL_1122278, EPI_ISL_1122279, EPI_ISL_1122280, EPI_ISL_1122281, EPI_ISL_1122283, EPI_ISL_1122284, EPI_ISL_1122286, EPI_ISL_1122287, EPI_ISL_1122288, EPI_ISL_1122289, EPI_ISL_1122290, EPI_ISL_1122291, EPI_ISL_1122292, EPI_ISL_1122294, EPI_ISL_1122295, EPI_ISL_1122296, EPI_ISL_1122297, EPI_ISL_1122298, EPI_ISL_1122299, EPI_ISL_1122300, EPI_ISL_1122301, EPI_ISL_1122302, EPI_ISL_1122303, EPI_ISL_1122304, EPI_ISL_1122305, EPI_ISL_1122306, EPI_ISL_1122307, EPI_ISL_1122308, EPI_ISL_1122309, EPI_ISL_1122310, EPI_ISL_1122311, EPI_ISL_1122312, EPI_ISL_1122313, EPI_ISL_1122314, EPI_ISL_1122315, EPI_ISL_1122316, EPI_ISL_1122317, EPI_ISL_1122318, EPI_ISL_1122319, EPI_ISL_1122320, EPI_ISL_1122321, EPI_ISL_1122322, EPI_ISL_1122323, EPI_ISL_1122324, EPI_ISL_1122326, EPI_ISL_1122327, EPI_ISL_1122328, EPI_ISL_1122329, EPI_ISL_1122330, EPI_ISL_1122331, EPI_ISL_1122332, EPI_ISL_1122333, EPI_ISL_1122334, EPI_ISL_1122335, EPI_ISL_1122336, EPI_ISL_1122337, EPI_ISL_1122340, EPI_ISL_1122342, EPI_ISL_1122344, EPI_ISL_1122347, EPI_ISL_1122348 | | | |
| see above | OHSU Lab Services Molecular Microbiology Lab | Oregon SARS-CoV-2 Genome Sequencing Center | Brendan L. O'Connell, Sally Grindstaff, Kayla Carter, Ruth V. Nichols, Alec J. Hirsch, Donna Hansel, Guang Fan, Xuan, Qin, Daniel N. Streblow, William B. Messer, Andrew C. Adey, Benjamin N. Bimber, Brian J. O'Roak |
| EPI_ISL_1132133, EPI_ISL_1132134, EPI_ISL_1132135, EPI_ISL_1132136, EPI_ISL_1132137, EPI_ISL_1132138, EPI_ISL_1132139, EPI_ISL_1132140, EPI_ISL_1132141, EPI_ISL_1132142, EPI_ISL_1132143, EPI_ISL_1132144, EPI_ISL_1132145, EPI_ISL_1132146, EPI_ISL_1132147, EPI_ISL_1132148, EPI_ISL_1132149, EPI_ISL_1132150, EPI_ISL_1132151, EPI_ISL_1132152, EPI_ISL_1132153, EPI_ISL_1132154, EPI_ISL_1132155, EPI_ISL_1132156, EPI_ISL_1132157, EPI_ISL_1132158, EPI_ISL_1132159, EPI_ISL_1132160, EPI_ISL_1132161, EPI_ISL_1132162, EPI_ISL_1132163, EPI_ISL_1132164, EPI_ISL_1132165, EPI_ISL_1132166, EPI_ISL_1132167, EPI_ISL_1132168, EPI_ISL_1132169, EPI_ISL_1132170, EPI_ISL_1132171, EPI_ISL_1132172, EPI_ISL_1132173, EPI_ISL_1132174, EPI_ISL_1132175, EPI_ISL_1132176, EPI_ISL_1132177, EPI_ISL_1132178, EPI_ISL_1132179, EPI_ISL_1132180, EPI_ISL_1132181, EPI_ISL_1132182, EPI_ISL_1132183, EPI_ISL_1132184, EPI_ISL_1132185, EPI_ISL_1132186, EPI_ISL_1132187, EPI_ISL_1132188, EPI_ISL_1132189, EPI_ISL_1132190, EPI_ISL_1132191, EPI_ISL_1132192, EPI_ISL_1132193, EPI_ISL_1132194, EPI_ISL_1132195, EPI_ISL_1132196, EPI_ISL_1132197, EPI_ISL_1132198, EPI_ISL_1132199, EPI_ISL_1132200, EPI_ISL_1132201, EPI_ISL_1132202, EPI_ISL_1132203, EPI_ISL_1132204, EPI_ISL_1132205, EPI_ISL_1132206, EPI_ISL_1132207, EPI_ISL_1132208, EPI_ISL_1132209, EPI_ISL_1132210, EPI_ISL_1132211, EPI_ISL_1132212, EPI_ISL_1132213, EPI_ISL_1132214, EPI_ISL_1132215, EPI_ISL_1132216, EPI_ISL_1132217, EPI_ISL_1132218, EPI_ISL_1132219, EPI_ISL_1132220, EPI_ISL_1132221, EPI_ISL_1132222, EPI_ISL_1132223, EPI_ISL_1132224, EPI_ISL_1132225, EPI_ISL_1132226, EPI_ISL_1132227, EPI_ISL_1132228, EPI_ISL_1132229, EPI_ISL_1132230, EPI_ISL_1132231, EPI_ISL_1132232, EPI_ISL_1132250, EPI_ISL_1132251, EPI_ISL_1132278, EPI_ISL_1132279, EPI_ISL_1132280, EPI_ISL_1132281, EPI_ISL_1132282, EPI_ISL_1132283, EPI_ISL_1132284, EPI_ISL_1132285, EPI_ISL_1132286, EPI_ISL_1132287, EPI_ISL_1132288, EPI_ISL_1132289, EPI_ISL_1132290, EPI_ISL_1132291, EPI_ISL_1132292, EPI_ISL_1132293, EPI_ISL_1132294, EPI_ISL_1132295, EPI_ISL_1132296, EPI_ISL_1132297, EPI_ISL_1132298, EPI_ISL_1132299, EPI_ISL_1132300, EPI_ISL_1132301, EPI_ISL_1132302, EPI_ISL_1132303, EPI_ISL_1132304, EPI_ISL_1132305, EPI_ISL_1132306, EPI_ISL_1132307, EPI_ISL_1132308, EPI_ISL_1132309, EPI_ISL_1132310, EPI_ISL_1132311, EPI_ISL_1132312, EPI_ISL_1132313, EPI_ISL_1132314, EPI_ISL_1132315, EPI_ISL_1132316, EPI_ISL_1132317, EPI_ISL_1132318, EPI_ISL_1132319, EPI_ISL_1132320, EPI_ISL_1132321, EPI_ISL_1132322, EPI_ISL_1132323, EPI_ISL_1132324, EPI_ISL_1132325, EPI_ISL_1132326, EPI_ISL_1132327, EPI_ISL_1132328, EPI_ISL_1132329, EPI_ISL_1132330, EPI_ISL_1132331, EPI_ISL_1132332, EPI_ISL_1132333, EPI_ISL_1132334, EPI_ISL_1132335, EPI_ISL_1132336, EPI_ISL_1132337, EPI_ISL_1132338, EPI_ISL_1132339, EPI_ISL_1132340, EPI_ISL_1132341, EPI_ISL_1132342, EPI_ISL_1132343, EPI_ISL_1132344, EPI_ISL_1132345, EPI_ISL_1132346, EPI_ISL_1132347, EPI_ISL_1132348, EPI_ISL_1132349, EPI_ISL_1132350, EPI_ISL_1132351, EPI_ISL_1132352, EPI_ISL_1132353, EPI_ISL_1132354, EPI_ISL_1132355, EPI_ISL_1132356, EPI_ISL_1132357, EPI_ISL_1132358, EPI_ISL_1132359, EPI_ISL_1132360, EPI_ISL_1132361, EPI_ISL_1132362, EPI_ISL_1132363, EPI_ISL_1132364, EPI_ISL_1132365, EPI_ISL_1132366 | | | |
| see above | Florida Bureau of Public Health Laboratories | Florida Bureau of Public Health Laboratories | Sarah Schmedes, Jason Blanton |
| EPI_ISL_1132367 | University of Wisconsin-Madison AIDS Vaccine Research Laboratories | University of Wisconsin-Madison AIDS Vaccine Research Laboratories | Gage Moreno, Katarina Braun, et al. AIDS Vaccine Research Laboratories |
| EPI_ISL_1132643, EPI_ISL_1132644, EPI_ISL_1132645, EPI_ISL_1132646, EPI_ISL_1132647, EPI_ISL_1132648, EPI_ISL_1132649, EPI_ISL_1132650, EPI_ISL_1132651, EPI_ISL_1132652, EPI_ISL_1132653, EPI_ISL_1132654, EPI_ISL_1132655, EPI_ISL_1132656, EPI_ISL_1132657, EPI_ISL_1132658, EPI_ISL_1132659, EPI_ISL_1132661, EPI_ISL_1132662, EPI_ISL_1132663, EPI_ISL_1132664, EPI_ISL_1132665, EPI_ISL_1132666 | | | |
| see above | Arizona State Public Health Laboratory | Arizona State Public Health Laboratory | Trung Huynh, Jessica Escobar, Katherine Fullerton, Nobuko Fukushima, Stacy White, Linda Getsinger, Victor Waddell |
| EPI_ISL_1132729 | Arizona State University | Arizona State University | Emily A. Kaelin, LaRinda A. Holland, Peter T. Skidmore, Nicholas J. Mellor, Kristina Buss, Joy M. Blain, Neal W Woodbury, Valerie Harris, Joshua LaBaer, Vel Murugan, Efrem S. Lim |
| EPI_ISL_1132730, EPI_ISL_1132731 | Mayo Clinic & Mayo Clinic Laboratories | Minnesota Department of Health, Public Health Laboratory | Alexandra Lorentz, Jacob Garfin, Matt Plumb, and Xiong Wang |
| EPI_ISL_1132732, EPI_ISL_1132733 | Vault Health | Minnesota Department of Health, Public Health Laboratory | Alexandra Lorentz, Jacob Garfin, Matt Plumb, and Xiong Wang |
| EPI_ISL_1132734, EPI_ISL_1132735, EPI_ISL_1132736, EPI_ISL_1132737, EPI_ISL_1132738, EPI_ISL_1132739, EPI_ISL_1132740, EPI_ISL_1132741 | M Health Fairview | Minnesota Department of Health, Public Health Laboratory | Alexandra Lorentz, Jacob Garfin, Matt Plumb, and Xiong Wang |
| EPI_ISL_1132742 | Vault Health | Minnesota Department of Health, Public Health Laboratory | Alexandra Lorentz, Jacob Garfin, Matt Plumb, and Xiong Wang |
| EPI_ISL_1132743 | HealthPartners Central Lab | Minnesota Department of Health, Public Health Laboratory | Alexandra Lorentz, Jacob Garfin, Matt Plumb, and Xiong Wang |
| EPI_ISL_1132744, EPI_ISL_1132745, EPI_ISL_1132746, EPI_ISL_1132747, EPI_ISL_1132748, EPI_ISL_1132749, EPI_ISL_1132750, EPI_ISL_1132751, EPI_ISL_1132752, EPI_ISL_1132753, EPI_ISL_1132754, EPI_ISL_1132755, EPI_ISL_1132756, EPI_ISL_1132757, EPI_ISL_1132758, EPI_ISL_1132759, EPI_ISL_1132760, EPI_ISL_1132761, EPI_ISL_1132762, EPI_ISL_1132763, EPI_ISL_1132764, EPI_ISL_1132765, EPI_ISL_1132766, EPI_ISL_1132767, EPI_ISL_1132768, EPI_ISL_1132769, EPI_ISL_1132770, EPI_ISL_1132771, EPI_ISL_1132772, EPI_ISL_1132773, EPI_ISL_1132774, EPI_ISL_1132775, EPI_ISL_1132776, EPI_ISL_1132777, EPI_ISL_1132778, EPI_ISL_1132779, EPI_ISL_1132780, EPI_ISL_1132781, EPI_ISL_1132782, EPI_ISL_1132783, EPI_ISL_1132784, EPI_ISL_1132785, EPI_ISL_1132786, EPI_ISL_1132787, EPI_ISL_1132788, EPI_ISL_1132789, EPI_ISL_1132790 | | | |
| see above | M Health Fairview | Minnesota Department of Health, Public | Alexandra Lorentz, Jacob Garfin, Matt Plumb, and Xiong Wang |

| | | | |
|---|---|---|---|
| | | Health Laboratory | |
| EPI_ISL_1132791, EPI_ISL_1132792, EPI_ISL_1132793, EPI_ISL_1132794, EPI_ISL_1132795, EPI_ISL_1132796, EPI_ISL_1132797, EPI_ISL_1132798, EPI_ISL_1132799, EPI_ISL_1132800, EPI_ISL_1132801, EPI_ISL_1132802, EPI_ISL_1132803, EPI_ISL_1132804, EPI_ISL_1132805, EPI_ISL_1132806 | | | |
| see above | Vault Health | Minnesota Department of Health, Public Health Laboratory | Alexandra Lorentz, Jacob Garfin, Matt Plumb, and Xiong Wang |
| EPI_ISL_1132808, EPI_ISL_1132809, EPI_ISL_1132810, EPI_ISL_1132811, EPI_ISL_1132812 | M Health Fairview | Minnesota Department of Health, Public Health Laboratory | Alexandra Lorentz, Jacob Garfin, Matt Plumb, and Xiong Wang |
| EPI_ISL_1132813 | Allina Health Laboratory | Minnesota Department of Health, Public Health Laboratory | Alexandra Lorentz, Jacob Garfin, Matt Plumb, and Xiong Wang |
| EPI_ISL_1132845, EPI_ISL_1132846, EPI_ISL_1132847, EPI_ISL_1132848, EPI_ISL_1132849, EPI_ISL_1132850, EPI_ISL_1132851, EPI_ISL_1132852, EPI_ISL_1132853, EPI_ISL_1132854, EPI_ISL_1132855, EPI_ISL_1132856, EPI_ISL_1132857, EPI_ISL_1132858, EPI_ISL_1132859, EPI_ISL_1132860, EPI_ISL_1132861, EPI_ISL_1132862, EPI_ISL_1132863, EPI_ISL_1132864, EPI_ISL_1132865, EPI_ISL_1132866, EPI_ISL_1132867, EPI_ISL_1132868, EPI_ISL_1132869, EPI_ISL_1132870, EPI_ISL_1132871, EPI_ISL_1132872, EPI_ISL_1132873, EPI_ISL_1132874, EPI_ISL_1132875, EPI_ISL_1132876, EPI_ISL_1132877, EPI_ISL_1132878, EPI_ISL_1132879, EPI_ISL_1132880, EPI_ISL_1132881, EPI_ISL_1132882, EPI_ISL_1132883, EPI_ISL_1132884, EPI_ISL_1132885, EPI_ISL_1132886, EPI_ISL_1132887, EPI_ISL_1132888, EPI_ISL_1132889, EPI_ISL_1132890, EPI_ISL_1132891, EPI_ISL_1132892, EPI_ISL_1132893, EPI_ISL_1132894, EPI_ISL_1132895, EPI_ISL_1132896, EPI_ISL_1132897, EPI_ISL_1132898, EPI_ISL_1132899, EPI_ISL_1132900, EPI_ISL_1132901, EPI_ISL_1132902, EPI_ISL_1132903, EPI_ISL_1132904, EPI_ISL_1132905, EPI_ISL_1132906, EPI_ISL_1132907, EPI_ISL_1132908, EPI_ISL_1132909, EPI_ISL_1132910, EPI_ISL_1132917, EPI_ISL_1132918, EPI_ISL_1132919, EPI_ISL_1132920, EPI_ISL_1132921, EPI_ISL_1132922, EPI_ISL_1132923, EPI_ISL_1132924, EPI_ISL_1132925, EPI_ISL_1132926, EPI_ISL_1132927, EPI_ISL_1132928, EPI_ISL_1132929, EPI_ISL_1132930, EPI_ISL_1132931, EPI_ISL_1132932, EPI_ISL_1132933, EPI_ISL_1132934, EPI_ISL_1132935, EPI_ISL_1132936, EPI_ISL_1132938, EPI_ISL_1132939, EPI_ISL_1132940, EPI_ISL_1132941, EPI_ISL_1132942, EPI_ISL_1132943, EPI_ISL_1132944, EPI_ISL_1132945, EPI_ISL_1132946, EPI_ISL_1132947, EPI_ISL_1132948, EPI_ISL_1132949, EPI_ISL_1132950, EPI_ISL_1132951, EPI_ISL_1132953, EPI_ISL_1132954, EPI_ISL_1132955, EPI_ISL_1132956, EPI_ISL_1132957, EPI_ISL_1132958, EPI_ISL_1132959, EPI_ISL_1132960, EPI_ISL_1132961, EPI_ISL_1132962, EPI_ISL_1132963, EPI_ISL_1132964, EPI_ISL_1132965, EPI_ISL_1132966, EPI_ISL_1132967, EPI_ISL_1132968, EPI_ISL_1132969, EPI_ISL_1132970 | | | |
| see above | Fulgent Genetics | Fulgent Genetics | Harry Gao, Mickey Li, John Gao, Joseph Fierro, Benafsh Sapra, Becky Tsai, Yan Meng, Doreen Ng, James Xie |
| EPI_ISL_1132971, EPI_ISL_1132972, EPI_ISL_1132973, EPI_ISL_1132974, EPI_ISL_1132975, EPI_ISL_1132976, EPI_ISL_1132977, EPI_ISL_1132978, EPI_ISL_1132979, EPI_ISL_1132980, EPI_ISL_1132981, EPI_ISL_1132982, EPI_ISL_1132983, EPI_ISL_1132984, EPI_ISL_1132985, EPI_ISL_1132986, EPI_ISL_1132987, EPI_ISL_1132988, EPI_ISL_1132989, EPI_ISL_1132990, EPI_ISL_1132991, EPI_ISL_1132992, EPI_ISL_1132993, EPI_ISL_1132994, EPI_ISL_1132995, EPI_ISL_1132996, EPI_ISL_1132997, EPI_ISL_1132998, EPI_ISL_1132999, EPI_ISL_1133000, EPI_ISL_1133001, EPI_ISL_1133002, EPI_ISL_1133003, EPI_ISL_1133004, EPI_ISL_1133005, EPI_ISL_1133006, EPI_ISL_1133007, EPI_ISL_1133008, EPI_ISL_1133009, EPI_ISL_1133010, EPI_ISL_1133011, EPI_ISL_1133012, EPI_ISL_1133013, EPI_ISL_1133014, EPI_ISL_1133015, EPI_ISL_1133016, EPI_ISL_1133017, EPI_ISL_1133018, EPI_ISL_1133019, EPI_ISL_1133020, EPI_ISL_1133021, EPI_ISL_1133022, EPI_ISL_1133023, EPI_ISL_1133024, EPI_ISL_1133025, EPI_ISL_1133026, EPI_ISL_1133027, EPI_ISL_1133028, EPI_ISL_1133029, EPI_ISL_1133030, EPI_ISL_1133031, EPI_ISL_1133032, EPI_ISL_1133033, EPI_ISL_1133034, EPI_ISL_1133035, EPI_ISL_1133036, EPI_ISL_1133037, EPI_ISL_1133038, EPI_ISL_1133039, EPI_ISL_1133040, EPI_ISL_1133041, EPI_ISL_1133042, EPI_ISL_1133043, EPI_ISL_1133044, EPI_ISL_1133045, EPI_ISL_1133046, EPI_ISL_1133047, EPI_ISL_1133048, EPI_ISL_1133049, EPI_ISL_1133050, EPI_ISL_1133051, EPI_ISL_1133052, EPI_ISL_1133053, EPI_ISL_1133054, EPI_ISL_1133055, EPI_ISL_1133056, EPI_ISL_1133057, EPI_ISL_1133058, EPI_ISL_1133059, EPI_ISL_1133060, EPI_ISL_1133061, EPI_ISL_1133062, EPI_ISL_1133063, EPI_ISL_1133064, EPI_ISL_1133065, EPI_ISL_1133066, EPI_ISL_1133067, EPI_ISL_1133068, EPI_ISL_1133069, EPI_ISL_1133070, EPI_ISL_1133071, EPI_ISL_1133072, EPI_ISL_1133073, EPI_ISL_1133074, EPI_ISL_1133075, EPI_ISL_1133076, EPI_ISL_1133077, EPI_ISL_1133078, EPI_ISL_1133079, EPI_ISL_1133080, EPI_ISL_1133081, EPI_ISL_1133082, EPI_ISL_1133083, EPI_ISL_1133084, EPI_ISL_1133085, EPI_ISL_1133086, EPI_ISL_1133087, EPI_ISL_1133088, EPI_ISL_1133089, EPI_ISL_1133090, EPI_ISL_1133091, EPI_ISL_1133092, EPI_ISL_1133093, EPI_ISL_1133094, EPI_ISL_1133095, EPI_ISL_1133096, EPI_ISL_1133097, EPI_ISL_1133098, EPI_ISL_1133099 | | | |
| see above | University of Michigan Clinical Microbiology Laboratory | Lauring Lab, University of Michigan, Department of Microbiology and Immunology | Valesano |
| EPI_ISL_1133100, EPI_ISL_1133101, EPI_ISL_1133102 | Kansas Health and Environmental Lab | Kansas Health and Environmental Lab | Mike Grose, Jonathan Barnell, Ben Olsen, and Phil Adam |
| EPI_ISL_1133103 | Instituto Nacional de Medicina Genomica | Instituto Nacional de Medicina Genomica | Hidalgo-Miranda A, Mendoza-Vargas A, Reyes-Grajeda JP, Cedro-Tanda A, Alcaraz N, Gonzalez-Barrera D, Rangel-DeLeon D, Ramirez-Vega O, Sifuentes-Rojas C, Cisneros-Villanueva M, Herrera-Montalvo LA |
| EPI_ISL_1133106, EPI_ISL_1133107, EPI_ISL_1133108, EPI_ISL_1133109, EPI_ISL_1133110, EPI_ISL_1133111, EPI_ISL_1133112, EPI_ISL_1133113, EPI_ISL_1133114, EPI_ISL_1133115, EPI_ISL_1133116, EPI_ISL_1133117, EPI_ISL_1133118 | | | |
| see above | DC Public Health Lab/ Dept. of Forensic Sciences | DC Public Health Lab/ Dept. of Forensic Sciences | Scott Nguyen, Janis Doss, Elizabeth Zelaya, Sarah Scott, Connie Maza, Monica Mann, Brittany Hamilton, David Payne, Jocelyn Hauser |
| EPI_ISL_1133211, EPI_ISL_1133212, EPI_ISL_1133213, EPI_ISL_1133214, EPI_ISL_1133215, EPI_ISL_1133216, EPI_ISL_1133217, EPI_ISL_1133218, EPI_ISL_1133219, EPI_ISL_1133220, EPI_ISL_1133221, EPI_ISL_1133222 | | | |
| see above | Arizona State Public Health Laboratory | Arizona State Public Health Laboratory | Trung Huynh, Jessica Escobar, Katherine Fullerton, Nobuko Fukushima, Stacy White, Linda Getsinger, Victor Waddell |
| EPI_ISL_1133223, EPI_ISL_1133224, EPI_ISL_1133225, EPI_ISL_1133226 | Wyoming Public Health Laboratory | Wyoming Public Health Laboratory | Jim Mildenberger, Wanda Manley, Noah Hull, Taylor Fearing, Lynette Gumbleton, Channing Weber, Ashley Norberg, Chayse Rowley, Marley Goetz, Brian Dominguez, Elliot Thomasson, Cari Sloma, and Rob Christensen |
| EPI_ISL_1133792, EPI_ISL_1133793, EPI_ISL_1133794, EPI_ISL_1133796, EPI_ISL_1133797, EPI_ISL_1133798, EPI_ISL_1133799, EPI_ISL_1133800, EPI_ISL_1133801, EPI_ISL_1133802, EPI_ISL_1133803, EPI_ISL_1133804, EPI_ISL_1133805, EPI_ISL_1133806, EPI_ISL_1133807, EPI_ISL_1133808, EPI_ISL_1133809, EPI_ISL_1133811, EPI_ISL_1133812, EPI_ISL_1133813, EPI_ISL_1133814, EPI_ISL_1133815, EPI_ISL_1133816, EPI_ISL_1133817, EPI_ISL_1133818, EPI_ISL_1133819, EPI_ISL_1133820, EPI_ISL_1133821, EPI_ISL_1133822, EPI_ISL_1133824, EPI_ISL_1133825, EPI_ISL_1133826, EPI_ISL_1133827, EPI_ISL_1133828, EPI_ISL_1133829, EPI_ISL_1133830, EPI_ISL_1133832, EPI_ISL_1133833, EPI_ISL_1133835, EPI_ISL_1133836, EPI_ISL_1133837, EPI_ISL_1133839, EPI_ISL_1133840, EPI_ISL_1133841, EPI_ISL_1133842, EPI_ISL_1133843, EPI_ISL_1133845, EPI_ISL_1133846, EPI_ISL_1133847, EPI_ISL_1133849, EPI_ISL_1133850, EPI_ISL_1133851, EPI_ISL_1133852, EPI_ISL_1133853, EPI_ISL_1133854, EPI_ISL_1133855, EPI_ISL_1133856, EPI_ISL_1133858, EPI_ISL_1133859, EPI_ISL_1133860, EPI_ISL_1133861, EPI_ISL_1133862, EPI_ISL_1133863, EPI_ISL_1133864, EPI_ISL_1133865, EPI_ISL_1133866, EPI_ISL_1133867, EPI_ISL_1133868, EPI_ISL_1133869, EPI_ISL_1133870, EPI_ISL_1133871, EPI_ISL_1133873, EPI_ISL_1133874, EPI_ISL_1133875, EPI_ISL_1133876, EPI_ISL_1133877, EPI_ISL_1133878, EPI_ISL_1133879, EPI_ISL_1133880, EPI_ISL_1133881, EPI_ISL_1133882, EPI_ISL_1133883, EPI_ISL_1133884, EPI_ISL_1133886, EPI_ISL_1133887, EPI_ISL_1133888, EPI_ISL_1133889, EPI_ISL_1133890, EPI_ISL_1133891, EPI_ISL_1133892, EPI_ISL_1133894, EPI_ISL_1133895, EPI_ISL_1133896, EPI_ISL_1133897, EPI_ISL_1133898, EPI_ISL_1133899, EPI_ISL_1133900, EPI_ISL_1133901, EPI_ISL_1133902, EPI_ISL_1133903, EPI_ISL_1133904, EPI_ISL_1133905, EPI_ISL_1133906, EPI_ISL_1133907, EPI_ISL_1133908, EPI_ISL_1133909, EPI_ISL_1133910, EPI_ISL_1133911, EPI_ISL_1133912, EPI_ISL_1133913, EPI_ISL_1133914, EPI_ISL_1133915, EPI_ISL_1133916, EPI_ISL_1133917, EPI_ISL_1133918, EPI_ISL_1133919, EPI_ISL_1133920, EPI_ISL_1133921, EPI_ISL_1133922, EPI_ISL_1133923, EPI_ISL_1133924, EPI_ISL_1133925, EPI_ISL_1133926, EPI_ISL_1133928, EPI_ISL_1133931, EPI_ISL_1133932, EPI_ISL_1133933, EPI_ISL_1133934, EPI_ISL_1133935, EPI_ISL_1133936, EPI_ISL_1133937, EPI_ISL_1133939, EPI_ISL_1133941, EPI_ISL_1133942, EPI_ISL_1133944, EPI_ISL_1133945, EPI_ISL_1133946, EPI_ISL_1133948, EPI_ISL_1133949, EPI_ISL_1133951, EPI_ISL_1133953, EPI_ISL_1133954, EPI_ISL_1133955, EPI_ISL_1133956, EPI_ISL_1133957, EPI_ISL_1133958, EPI_ISL_1133959, EPI_ISL_1133960, EPI_ISL_1133961, EPI_ISL_1133962, EPI_ISL_1133963, EPI_ISL_1133964, EPI_ISL_1133965, EPI_ISL_1133966, EPI_ISL_1133967, EPI_ISL_1133969, EPI_ISL_1133970, EPI_ISL_1133971, EPI_ISL_1133972, EPI_ISL_1133973, EPI_ISL_1133974, EPI_ISL_1133975, EPI_ISL_1133976, EPI_ISL_1133977, EPI_ISL_1133978, EPI_ISL_1133980, EPI_ISL_1133981, EPI_ISL_1133982, EPI_ISL_1133983, EPI_ISL_1133984, EPI_ISL_1133985, EPI_ISL_1133987, EPI_ISL_1133988, EPI_ISL_1133989, EPI_ISL_1133990, EPI_ISL_1133991, EPI_ISL_1133992, EPI_ISL_1133993, EPI_ISL_1133994, EPI_ISL_1133995, EPI_ISL_1133996, EPI_ISL_1133998, EPI_ISL_1133999, EPI_ISL_1134000, EPI_ISL_1134001, EPI_ISL_1134002, EPI_ISL_1134003, EPI_ISL_1134004, EPI_ISL_1134005, EPI_ISL_1134006, EPI_ISL_1134008, EPI_ISL_1134009, EPI_ISL_1134011, EPI_ISL_1134012, EPI_ISL_1134013, EPI_ISL_1134014, EPI_ISL_1134015, EPI_ISL_1134016, EPI_ISL_1134017, EPI_ISL_1134021, EPI_ISL_1134022, EPI_ISL_1134023, EPI_ISL_1134024, EPI_ISL_1134025, EPI_ISL_1134026, EPI_ISL_1134027, EPI_ISL_1134028, EPI_ISL_1134029, EPI_ISL_1134030, EPI_ISL_1134031, EPI_ISL_1134032, EPI_ISL_1134033, EPI_ISL_1134034, EPI_ISL_1134035, EPI_ISL_1134036, EPI_ISL_1134037, EPI_ISL_1134039, EPI_ISL_1134041, EPI_ISL_1134042, EPI_ISL_1134044, EPI_ISL_1134045, EPI_ISL_1134046, EPI_ISL_1134047, EPI_ISL_1134048, EPI_ISL_1134049, EPI_ISL_1134050, EPI_ISL_1134051, EPI_ISL_1134052, EPI_ISL_1134053, EPI_ISL_1134054, EPI_ISL_1134055, EPI_ISL_1134056, EPI_ISL_1134057, EPI_ISL_1134058, EPI_ISL_1134059, EPI_ISL_1134060, EPI_ISL_1134061, EPI_ISL_1134062, EPI_ISL_1134063, EPI_ISL_1134064, EPI_ISL_1134065, EPI_ISL_1134066, EPI_ISL_1134067, EPI_ISL_1134068, EPI_ISL_1134069, EPI_ISL_1134070, EPI_ISL_1134071, EPI_ISL_1134072, EPI_ISL_1134073, EPI_ISL_1134074, EPI_ISL_1134075, EPI_ISL_1134076, EPI_ISL_1134077, EPI_ISL_1134078, EPI_ISL_1134079, EPI_ISL_1134080, EPI_ISL_1134081, EPI_ISL_1134082, EPI_ISL_1134083, EPI_ISL_1134084, EPI_ISL_1134085, EPI_ISL_1134086, EPI_ISL_1134087, EPI_ISL_1134088, EPI_ISL_1134089, EPI_ISL_1134090, EPI_ISL_1134091, EPI_ISL_1134092, EPI_ISL_1134093, EPI_ISL_1134094, EPI_ISL_1134095, EPI_ISL_1134096, EPI_ISL_1134097, EPI_ISL_1134098, EPI_ISL_1134099, EPI_ISL_1134100, EPI_ISL_1134101, EPI_ISL_1134102, EPI_ISL_1134103, EPI_ISL_1134104, EPI_ISL_1134105, EPI_ISL_1134106, EPI_ISL_1134107, EPI_ISL_1134108, EPI_ISL_1134109, EPI_ISL_1134110, EPI_ISL_1134111, EPI_ISL_1134112, EPI_ISL_1134113, EPI_ISL_1134114, EPI_ISL_1134115, EPI_ISL_1134116, EPI_ISL_1134117, EPI_ISL_1134118, EPI_ISL_1134119, EPI_ISL_1134120, EPI_ISL_1134122, EPI_ISL_1134123, EPI_ISL_1134124, EPI_ISL_1134125, EPI_ISL_1134126, EPI_ISL_1134127, EPI_ISL_1134128, EPI_ISL_1134129, EPI_ISL_1134130, EPI_ISL_1134131, EPI_ISL_1134132, EPI_ISL_1134133, EPI_ISL_1134134, EPI_ISL_1134135, EPI_ISL_1134136, EPI_ISL_1134137, EPI_ISL_1134138, EPI_ISL_1134139, EPI_ISL_1134140, EPI_ISL_1134141, EPI_ISL_1134142, EPI_ISL_1134143, EPI_ISL_1134144, EPI_ISL_1134145, EPI_ISL_1134146, EPI_ISL_1134147, EPI_ISL_1134148, EPI_ISL_1134149, EPI_ISL_1134150, EPI_ISL_1134151, EPI_ISL_1134152, EPI_ISL_1134153, EPI_ISL_1134154, EPI_ISL_1134155, EPI_ISL_1134156, EPI_ISL_1134157, EPI_ISL_1134158, EPI_ISL_1134160, EPI_ISL_1134161, EPI_ISL_1134162, EPI_ISL_1134163, EPI_ISL_1134164, EPI_ISL_1134165, EPI_ISL_1134166, EPI_ISL_1134167, EPI_ISL_1134168, EPI_ISL_1134169, EPI_ISL_1134170, EPI_ISL_1134171, EPI_ISL_1134172, EPI_ISL_1134173, EPI_ISL_1134174, EPI_ISL_1134175, EPI_ISL_1134177, EPI_ISL_1134178, EPI_ISL_1134179, EPI_ISL_1134180, EPI_ISL_1134181, EPI_ISL_1134182, EPI_ISL_1134183, EPI_ISL_1134184, EPI_ISL_1134185, EPI_ISL_1134186, EPI_ISL_1134187, EPI_ISL_1134188, EPI_ISL_1134189, EPI_ISL_1134190, EPI_ISL_1134191, EPI_ISL_1134192, EPI_ISL_1134193, EPI_ISL_1134194, EPI_ISL_1134195, EPI_ISL_1134196, EPI_ISL_1134197, EPI_ISL_1134198, EPI_ISL_1134199, EPI_ISL_1134200, EPI_ISL_1134201, EPI_ISL_1134202, EPI_ISL_1134203, EPI_ISL_1134204, EPI_ISL_1134205, EPI_ISL_1134206, EPI_ISL_1134207, EPI_ISL_1134208, EPI_ISL_1134209, EPI_ISL_1134210, EPI_ISL_1134212, EPI_ISL_1134213, EPI_ISL_1134214, EPI_ISL_1134215, EPI_ISL_1134216, EPI_ISL_1134217, EPI_ISL_1134218, EPI_ISL_1134219, EPI_ISL_1134220, EPI_ISL_1134221, EPI_ISL_1134222, EPI_ISL_1134223, EPI_ISL_1134224, EPI_ISL_1134225, EPI_ISL_1134226, EPI_ISL_1134227, EPI_ISL_1134228, EPI_ISL_1134229, EPI_ISL_1134230, EPI_ISL_1134231, EPI_ISL_1134232, EPI_ISL_1134233, EPI_ISL_1134234, EPI_ISL_1134235, EPI_ISL_1134236, EPI_ISL_1134237, EPI_ISL_1134238, EPI_ISL_1134239, EPI_ISL_1134240, EPI_ISL_1134241, EPI_ISL_1134243, EPI_ISL_1134244, EPI_ISL_1134245, EPI_ISL_1134246, EPI_ISL_1134247, EPI_ISL_1134248, EPI_ISL_1134249, EPI_ISL_1134250, EPI_ISL_1134251, EPI_ISL_1134252, EPI_ISL_1134253, EPI_ISL_1134254, EPI_ISL_1134255, EPI_ISL_1134256, EPI_ISL_1134257, EPI_ISL_1134258, EPI_ISL_1134259, EPI_ISL_1134261, EPI_ISL_1134262, EPI_ISL_1134264, EPI_ISL_1134265, EPI_ISL_1134266, EPI_ISL_1134267, EPI_ISL_1134268, EPI_ISL_1134269, EPI_ISL_1134270, EPI_ISL_1134271, EPI_ISL_1134272, EPI_ISL_1134273, EPI_ISL_1134274, EPI_ISL_1134275, EPI_ISL_1134276, EPI_ISL_1134277, EPI_ISL_1134279, EPI_ISL_1134280, EPI_ISL_1134281, EPI_ISL_1134282, EPI_ISL_1134283, EPI_ISL_1134284, EPI_ISL_1134285, EPI_ISL_1134286, EPI_ISL_1134287, EPI_ISL_1134288, EPI_ISL_1134289, EPI_ISL_1134290, EPI_ISL_1134291, EPI_ISL_1134292, EPI_ISL_1134293, EPI_ISL_1134294, EPI_ISL_1134295, EPI_ISL_1134296, EPI_ISL_1134297, EPI_ISL_1134298, EPI_ISL_1134299, EPI_ISL_1134300, EPI_ISL_1134301, EPI_ISL_1134302, EPI_ISL_1134303, EPI_ISL_1134304, EPI_ISL_1134305, EPI_ISL_1134306, EPI_ISL_1134307, EPI_ISL_1134309, EPI_ISL_1134310, EPI_ISL_1134311, EPI_ISL_1134312, EPI_ISL_1134313, EPI_ISL_1134314, EPI_ISL_1134315, EPI_ISL_1134316, EPI_ISL_1134317, EPI_ISL_1134318, EPI_ISL_1134319, EPI_ISL_1134320, EPI_ISL_1134321, EPI_ISL_1134322, EPI_ISL_1134324, EPI_ISL_1134325, EPI_ISL_1134326, EPI_ISL_1134327, EPI_ISL_1134328, EPI_ISL_1134329, EPI_ISL_1134331, EPI_ISL_1134332, EPI_ISL_1134333, EPI_ISL_1134334, EPI_ISL_1134335, EPI_ISL_1134336, EPI_ISL_1134337, EPI_ISL_1134338, EPI_ISL_1134339, EPI_ISL_1134340, EPI_ISL_1134341, EPI_ISL_1134342, EPI_ISL_1134343, EPI_ISL_1134344, EPI_ISL_1134345, EPI_ISL_1134346, EPI_ISL_1134347, EPI_ISL_1134348, EPI_ISL_1134349, EPI_ISL_1134350, EPI_ISL_1134351, EPI_ISL_1134352, EPI_ISL_1134353, EPI_ISL_1134354, EPI_ISL_1134355, EPI_ISL_1134356, EPI_ISL_1134357, EPI_ISL_1134358, EPI_ISL_1134359, EPI_ISL_1134360, EPI_ISL_1134361, EPI_ISL_1134362, EPI_ISL_1134363, EPI_ISL_1134364, EPI_ISL_1134365, EPI_ISL_1134366, EPI_ISL_1134368, EPI_ISL_1134369, EPI_ISL_1134370, EPI_ISL_1134371, EPI_ISL_1134372, EPI_ISL_1134373, EPI_ISL_1134374, EPI_ISL_1134376, EPI_ISL_1134377, EPI_ISL_1134378, EPI_ISL_1134379, EPI_ISL_1134380, EPI_ISL_1134381, EPI_ISL_1134383, EPI_ISL_1134384, EPI_ISL_1134386, EPI_ISL_1134387, EPI_ISL_1134388, EPI_ISL_1134389, EPI_ISL_1134390, EPI_ISL_1134391, EPI_ISL_1134392, EPI_ISL_1134393, EPI_ISL_1134395, EPI_ISL_1134396, EPI_ISL_1134397, EPI_ISL_1134398, EPI_ISL_1134399, EPI_ISL_1134400, EPI_ISL_1134401, EPI_ISL_1134402, EPI_ISL_1134403, EPI_ISL_1134404, EPI_ISL_1134405, EPI_ISL_1134406, EPI_ISL_1134407, EPI_ISL_1134408, EPI_ISL_1134409, EPI_ISL_1134410, EPI_ISL_1134412, EPI_ISL_1134413, EPI_ISL_1134414, EPI_ISL_1134415, EPI_ISL_1134416, EPI_ISL_1134417, EPI_ISL_1134418, EPI_ISL_1134419, EPI_ISL_1134420, EPI_ISL_1134421, EPI_ISL_1134422, EPI_ISL_1134423, EPI_ISL_1134424, EPI_ISL_1134425, EPI_ISL_1134426, EPI_ISL_1134427, EPI_ISL_1134428, EPI_ISL_1134429, EPI_ISL_1134430, EPI_ISL_1134431, EPI_ISL_1134432, EPI_ISL_1134433, EPI_ISL_1134434, EPI_ISL_1134435, EPI_ISL_1134436, EPI_ISL_1134437, EPI_ISL_1134438, EPI_ISL_1134439, EPI_ISL_1134440, EPI_ISL_1134441, EPI_ISL_1134442, EPI_ISL_1134443, EPI_ISL_1134444, EPI_ISL_1134445, EPI_ISL_1134446, EPI_ISL_1134447, EPI_ISL_1134448, EPI_ISL_1134449, EPI_ISL_1134450, EPI_ISL_1134451, EPI_ISL_1134452, EPI_ISL_1134453, EPI_ISL_1134454, EPI_ISL_1134455, EPI_ISL_1134456, EPI_ISL_1134457, EPI_ISL_1134458, EPI_ISL_1134459, EPI_ISL_1134460, EPI_ISL_1134461, EPI_ISL_1134462, EPI_ISL_1134463, EPI_ISL_1134464, EPI_ISL_1134465, EPI_ISL_1134466, EPI_ISL_1134467, EPI_ISL_1134468, EPI_ISL_1134469, EPI_ISL_1134470, EPI_ISL_1134471, EPI_ISL_1134472, EPI_ISL_1134473, EPI_ISL_1134474, EPI_ISL_1134475, EPI_ISL_1134476, EPI_ISL_1134477, EPI_ISL_1134478, EPI_ISL_1134479, EPI_ISL_1134480, EPI_ISL_1134481, EPI_ISL_1134482, EPI_ISL_1134484, EPI_ISL_1134485, EPI_ISL_1134486, EPI_ISL_1134487, EPI_ISL_1134488, EPI_ISL_1134489, EPI_ISL_1134490, EPI_ISL_1134491, EPI_ISL_1134492, EPI_ISL_1134493, EPI_ISL_1134494, EPI_ISL_1134495, EPI_ISL_1134496, EPI_ISL_1134497, EPI_ISL_1134498, EPI_ISL_1134499, EPI_ISL_1134500, EPI_ISL_1134501, EPI_ISL_1134502, EPI_ISL_1134503, EPI_ISL_1134504, EPI_ISL_1134505, EPI_ISL_1134506, EPI_ISL_1134508, EPI_ISL_1134509, EPI_ISL_1134510, EPI_ISL_1134511, EPI_ISL_1134513, EPI_ISL_1134515, EPI_ISL_1134516, EPI_ISL_1134517, EPI_ISL_1134518, EPI_ISL_1134519, EPI_ISL_1134520, EPI_ISL_1134521, EPI_ISL_1134522, EPI_ISL_1134523, EPI_ISL_1134524, EPI_ISL_1134525, EPI_ISL_1134526, EPI_ISL_1134528, EPI_ISL_1134529, EPI_ISL_1134530, EPI_ISL_1134531, EPI_ISL_1134532, EPI_ISL_1134533, EPI_ISL_1134534, EPI_ISL_1134535, EPI_ISL_1134537, EPI_ISL_1134538, EPI_ISL_1134539, EPI_ISL_1134540, EPI_ISL_1134541, EPI_ISL_1134542, EPI_ISL_1134543, EPI_ISL_1134544, EPI_ISL_1134545, EPI_ISL_1134546, EPI_ISL_1134547, EPI_ISL_1134549, EPI_ISL_1134550, EPI_ISL_1134551, EPI_ISL_1134552, EPI_ISL_1134553, EPI_ISL_1134554, EPI_ISL_1134555, EPI_ISL_1134556, EPI_ISL_1134557, EPI_ISL_1134558, EPI_ISL_1134559, EPI_ISL_1134560, EPI_ISL_1134562, EPI_ISL_1134563, EPI_ISL_1134564, EPI_ISL_1134565, EPI_ISL_1134566, EPI_ISL_1134567, EPI_ISL_1134568, EPI_ISL_1134569, EPI_ISL_1134570, EPI_ISL_1134571, EPI_ISL_1134572, EPI_ISL_1134573, EPI_ISL_1134574, EPI_ISL_1134575, EPI_ISL_1134577, EPI_ISL_1134578, EPI_ISL_1134579, EPI_ISL_1134580, EPI_ISL_1134581, EPI_ISL_1134582, EPI_ISL_1134583, EPI_ISL_1134585, EPI_ISL_1134586, EPI_ISL_1134587, EPI_ISL_1134589, EPI_ISL_1134590, EPI_ISL_1134591, EPI_ISL_1134592, EPI_ISL_1134593, EPI_ISL_1134594, EPI_ISL_1134595, EPI_ISL_1134596, EPI_ISL_1134597, EPI_ISL_1134598, EPI_ISL_1134599, EPI_ISL_1134600, EPI_ISL_1134601, EPI_ISL_1134602, EPI_ISL_1134603, EPI_ISL_1134604, EPI_ISL_1134605, EPI_ISL_1134606, EPI_ISL_1134607, EPI_ISL_1134608, EPI_ISL_1134609, EPI_ISL_1134611, EPI_ISL_1134612, EPI_ISL_1134613, EPI_ISL_1134615, EPI_ISL_1134616, EPI_ISL_1134617, EPI_ISL_1134618, EPI_ISL_1134619, EPI_ISL_1134620, EPI_ISL_1134621, EPI_ISL_1134622, EPI_ISL_1134623, EPI_ISL_1134624, EPI_ISL_1134625, EPI_ISL_1134626, EPI_ISL_1134627, EPI_ISL_1134628, EPI_ISL_1134629, EPI_ISL_1134630, EPI_ISL_1134631, EPI_ISL_1134632, EPI_ISL_1134633, EPI_ISL_1134634, EPI_ISL_1134635, EPI_ISL_1134636, EPI_ISL_1134637, EPI_ISL_1134638, EPI_ISL_1134639, EPI_ISL_1134640, EPI_ISL_1134641, EPI_ISL_1134642, EPI_ISL_1134643, EPI_ISL_1134644, EPI_ISL_1134645, EPI_ISL_1134646, EPI_ISL_1134647, EPI_ISL_1134648, EPI_ISL_1134649, EPI_ISL_1134650, EPI_ISL_1134651, EPI_ISL_1134652, EPI_ISL_1134653, EPI_ISL_1134654, EPI_ISL_1134656, EPI_ISL_1134657, EPI_ISL_1134658, EPI_ISL_1134659, EPI_ISL_1134660, EPI_ISL_1134661, EPI_ISL_1134662, EPI_ISL_1134663, EPI_ISL_1134664, EPI_ISL_1134665, EPI_ISL_1134666, EPI_ISL_1134667, EPI_ISL_1134669, EPI_ISL_1134670, EPI_ISL_1134671, EPI_ISL_1134673, EPI_ISL_1134674, EPI_ISL_1134675, EPI_ISL_1134676, EPI_ISL_1134677, EPI_ISL_1134678, EPI_ISL_1134679, EPI_ISL_1134680, EPI_ISL_1134681, EPI_ISL_1134683, EPI_ISL_1134684, EPI_ISL_1134685, EPI_ISL_1134686, EPI_ISL_1134687, EPI_ISL_1134688, EPI_ISL_1134689, EPI_ISL_1134690, EPI_ISL_1134692, EPI_ISL_1134693, EPI_ISL_1134694, EPI_ISL_1134695, EPI_ISL_1134696, EPI_ISL_1134697, EPI_ISL_1134699, EPI_ISL_1134700, EPI_ISL_1134701, EPI_ISL_1134702, EPI_ISL_1134704, EPI_ISL_1134705, EPI_ISL_1134706, EPI_ISL_1134707, EPI_ISL_1134708, EPI_ISL_1134709, EPI_ISL_1134710, EPI_ISL_1134711, EPI_ISL_1134712, EPI_ISL_1134713, EPI_ISL_1134714, EPI_ISL_1134715, EPI_ISL_1134716, EPI_ISL_1134717, EPI_ISL_1134718, EPI_ISL_1134719, EPI_ISL_1134720, EPI_ISL_1134721, EPI_ISL_1134723, EPI_ISL_1134724, EPI_ISL_1134725, EPI_ISL_1134726, EPI_ISL_1134728, EPI_ISL_1134729, EPI_ISL_1134730, EPI_ISL_1134733, EPI_ISL_1134734, EPI_ISL_1134735, EPI_ISL_1134736, EPI_ISL_1134737, EPI_ISL_1134738, EPI_ISL_1134739, EPI_ISL_1134740, EPI_ISL_1134742, EPI_ISL_1134743, EPI_ISL_1134745, EPI_ISL_1134747, EPI_ISL_1134748, EPI_ISL_1134749, EPI_ISL_1134750, EPI_ISL_1134751, EPI_ISL_1134752, EPI_ISL_1134753, EPI_ISL_1134754, EPI_ISL_1134755, EPI_ISL_1134756, EPI_ISL_1134757, EPI_ISL_1134758, EPI_ISL_1134759, EPI_ISL_1134761, EPI_ISL_1134763, EPI_ISL_1134765, EPI_ISL_1134766, EPI_ISL_1134767, EPI_ISL_1134768, EPI_ISL_1134769, EPI_ISL_1134770, EPI_ISL_1134771, EPI_ISL_1134772, EPI_ISL_1134774, EPI_ISL_1134776, EPI_ISL_1134777, EPI_ISL_1134778, EPI_ISL_1134779, EPI_ISL_1134780, EPI_ISL_1134781, EPI_ISL_1134782, EPI_ISL_1134783, EPI_ISL_1134784, EPI_ISL_1134786, EPI_ISL_1134787, EPI_ISL_1134789, EPI_ISL_1134790, EPI_ISL_1134791, EPI_ISL_1134792, EPI_ISL_1134793, EPI_ISL_1134794, EPI_ISL_1134795, EPI_ISL_1134796, EPI_ISL_1134797, EPI_ISL_1134798, EPI_ISL_1134799, EPI_ISL_1134800, EPI_ISL_1134801, EPI_ISL_1134803, EPI_ISL_1134804, EPI_ISL_1134805, EPI_ISL_1134806, EPI_ISL_1134807, EPI_ISL_1134808, EPI_ISL_1134809, EPI_ISL_1134810, EPI_ISL_1134811, EPI_ISL_1134812, EPI_ISL_1134813, EPI_ISL_1134814, EPI_ISL_1134815, EPI_ISL_1134816, EPI_ISL_1134817, EPI_ISL_1134818, EPI_ISL_1134819, EPI_ISL_1134820, EPI_ISL_1134821, EPI_ISL_1134822, EPI_ISL_1134823, EPI_ISL_1134826, EPI_ISL_1134827, EPI_ISL_1134828, EPI_ISL_1134829, EPI_ISL_1134830, EPI_ISL_1134832, EPI_ISL_1134833, EPI_ISL_1134834, EPI_ISL_1134836, EPI_ISL_1134837, EPI_ISL_1134838, EPI_ISL_1134839 | | | |
| see above | Helix/Illumina | Respiratory Viruses Branch, Division of Viral Diseases, Centers for Disease Control and Prevention | Peter W. Cook, Dakota Howard, Dhwani Batra, Ben L. Rambo-Martin, Eileen de Feo, Jan Antico, Christine Tran, Matthew Tolentino, Shannon Wickline, Kim Gietzen, Brad Sickler, Jingtao Liu, Eric Allen, Phil Febbo, Summer Galloway, Nicole L. Washington, Simon White, Geraint Levan, Kelly Schiabor Barrett, Elizabeth Cirulli, Alexandre Bolze, Ary Ascencio, Charlotte Rivera-Garcia, Ryan Cho, Jason Nguyen, Sherry Wang, Jimmy Ramirez, Tyler Cassens, Efren Sandoval, Magnus Isaksson, William Lee, David Becker, Marc Laurent, James Lu, Clinton R. Paden, Suxiang Tong, Duncan MacCannell |
| EPI_ISL_1134840, EPI_ISL_1134841, EPI_ISL_1134842, EPI_ISL_1134843, EPI_ISL_1134844, EPI_ISL_1134845, EPI_ISL_1134846, EPI_ISL_1134847, EPI_ISL_1134848, EPI_ISL_1134850, EPI_ISL_1134851, EPI_ISL_1134853, EPI_ISL_1134854, EPI_ISL_1134855, EPI_ISL_1134856, EPI_ISL_1134857, EPI_ISL_1134858, EPI_ISL_1134861, EPI_ISL_1134862, EPI_ISL_1134863, EPI_ISL_1134864, EPI_ISL_1134865, EPI_ISL_1134866, EPI_ISL_1134867, EPI_ISL_1134868, EPI_ISL_1134869, EPI_ISL_1134870, EPI_ISL_1134871, EPI_ISL_1134872, EPI_ISL_1134874, EPI_ISL_1134875, EPI_ISL_1134876, EPI_ISL_1134877, EPI_ISL_1134878, EPI_ISL_1134879, EPI_ISL_1134880, EPI_ISL_1134881, EPI_ISL_1134882, EPI_ISL_1134883, EPI_ISL_1134885, EPI_ISL_1134886, EPI_ISL_1134887, EPI_ISL_1134888, EPI_ISL_1134889, EPI_ISL_1134890, EPI_ISL_1134891, EPI_ISL_1134893, EPI_ISL_1134894, EPI_ISL_1134895, EPI_ISL_1134896, EPI_ISL_1134897, EPI_ISL_1134898, EPI_ISL_1134899, EPI_ISL_1134901, EPI_ISL_1134902, EPI_ISL_1134903 | | | |
| see above | Fulgent Genetics | Fulgent Genetics | Harry Gao, Mickey Li, John Gao, Joseph Fierro, Benafsh Sapra, Becky Tsai, Yan Meng, Doreen Ng, James Xie |
| EPI_ISL_1135078, EPI_ISL_1135079, EPI_ISL_1135080, EPI_ISL_1135081, EPI_ISL_1135082, EPI_ISL_1135083, EPI_ISL_1135084, EPI_ISL_1135085, EPI_ISL_1135086, EPI_ISL_1135087, EPI_ISL_1135088, EPI_ISL_1135089, EPI_ISL_1135090, EPI_ISL_1135091, EPI_ISL_1135092, EPI_ISL_1135093, EPI_ISL_1135094, EPI_ISL_1135095, EPI_ISL_1135096, EPI_ISL_1135097, EPI_ISL_1135098, EPI_ISL_1135099, EPI_ISL_1135100, EPI_ISL_1135101, EPI_ISL_1135102, EPI_ISL_1135103, EPI_ISL_1135104, EPI_ISL_1135105, EPI_ISL_1135106, EPI_ISL_1135107, EPI_ISL_1135108, EPI_ISL_1135109, EPI_ISL_1135110, EPI_ISL_1135111, EPI_ISL_1135112 | | | |
| see above | Arizona State Public Health Laboratory | Arizona State Public Health Laboratory | Trung Huynh, Jessica Escobar, Katherine Fullerton, Nobuko Fukushima, Stacy White, Linda Getsinger, Victor Waddell |
| EPI_ISL_1135292, EPI_ISL_1135294, EPI_ISL_1135296, EPI_ISL_1135297, EPI_ISL_1135299, EPI_ISL_1135300 | Quest Diagnostics Incorporated | Respiratory Viruses Branch, Division of Viral Diseases, Centers for Disease Control and Prevention | Peter W. Cook, Dakota Howard, Dhwani Batra, Ben L. Rambo-Martin, S. H. Rosenthal, A. Gerasimova, R. M. Kagan, B. Anderson, M. Hua, Y. Liu, L.E. Bernstein, K.E. Livingston, A. Perez, I. A. Shlyakhter, R. V. Rolando, R. Owen, P. Tanpaiboon, F. Lacbawan, Clinton R. Paden, Suxiang Tong, Duncan MacCannell |

| | | | |
|---|---|---|---|
| EPI_ISL_1135856, EPI_ISL_1135857, EPI_ISL_1135858, EPI_ISL_1135859, EPI_ISL_1135860, EPI_ISL_1135861, EPI_ISL_1135862, EPI_ISL_1135863, EPI_ISL_1135864, EPI_ISL_1135865, EPI_ISL_1135866, EPI_ISL_1135867, EPI_ISL_1135868, EPI_ISL_1135869, EPI_ISL_1135870, EPI_ISL_1135871, EPI_ISL_1135872, EPI_ISL_1135873, EPI_ISL_1135874, EPI_ISL_1135875, EPI_ISL_1135876, EPI_ISL_1135877, EPI_ISL_1135878, EPI_ISL_1135879, EPI_ISL_1135880, EPI_ISL_1135881, EPI_ISL_1135882, EPI_ISL_1135883, EPI_ISL_1135884, EPI_ISL_1135885, EPI_ISL_1135886, EPI_ISL_1135887, EPI_ISL_1135888, EPI_ISL_1135889, EPI_ISL_1135890, EPI_ISL_1135891, EPI_ISL_1135892, EPI_ISL_1135893, EPI_ISL_1135894, EPI_ISL_1135895, EPI_ISL_1135896, EPI_ISL_1135897, EPI_ISL_1135898, EPI_ISL_1135899, EPI_ISL_1135900, EPI_ISL_1135901, EPI_ISL_1135902, EPI_ISL_1135903, EPI_ISL_1135904, EPI_ISL_1135905, EPI_ISL_1135906, EPI_ISL_1135907, EPI_ISL_1135908, EPI_ISL_1135909, EPI_ISL_1135910, EPI_ISL_1135911, EPI_ISL_1135912, EPI_ISL_1135913, EPI_ISL_1135914, EPI_ISL_1135915, EPI_ISL_1135916, EPI_ISL_1135917, EPI_ISL_1135918, EPI_ISL_1135919, EPI_ISL_1135920, EPI_ISL_1135921, EPI_ISL_1135922, EPI_ISL_1135923, EPI_ISL_1135924, EPI_ISL_1135925, EPI_ISL_1135926, EPI_ISL_1135927, EPI_ISL_1135928, EPI_ISL_1135929, EPI_ISL_1135930, EPI_ISL_1135931, EPI_ISL_1135932, EPI_ISL_1135933, EPI_ISL_1135934, EPI_ISL_1135935, EPI_ISL_1135936, EPI_ISL_1135937, EPI_ISL_1135938, EPI_ISL_1135939, EPI_ISL_1135940, EPI_ISL_1135941, EPI_ISL_1135942, EPI_ISL_1135943, EPI_ISL_1135944, EPI_ISL_1135945, EPI_ISL_1135946, EPI_ISL_1135947, EPI_ISL_1135948, EPI_ISL_1135949, EPI_ISL_1135950, EPI_ISL_1135951, EPI_ISL_1135952, EPI_ISL_1135953, EPI_ISL_1135954, EPI_ISL_1135955, EPI_ISL_1135956, EPI_ISL_1135957, EPI_ISL_1136052, EPI_ISL_1136053, EPI_ISL_1136054, EPI_ISL_1136055, EPI_ISL_1136056, EPI_ISL_1136057, EPI_ISL_1136058, EPI_ISL_1136059, EPI_ISL_1136060, EPI_ISL_1136061, EPI_ISL_1136062, EPI_ISL_1136063, EPI_ISL_1136064, EPI_ISL_1136065, EPI_ISL_1136066, EPI_ISL_1136067, EPI_ISL_1136068, EPI_ISL_1136069, EPI_ISL_1136070, EPI_ISL_1136071, EPI_ISL_1136072, EPI_ISL_1136073, EPI_ISL_1136074, EPI_ISL_1136075, EPI_ISL_1136076, EPI_ISL_1136077, EPI_ISL_1136078, EPI_ISL_1136079, EPI_ISL_1136080, EPI_ISL_1136081, EPI_ISL_1136082, EPI_ISL_1136083, EPI_ISL_1136084, EPI_ISL_1136085, EPI_ISL_1136086, EPI_ISL_1136087, EPI_ISL_1136088, EPI_ISL_1136089, EPI_ISL_1136090, EPI_ISL_1136091, EPI_ISL_1136092, EPI_ISL_1136093, EPI_ISL_1136094, EPI_ISL_1136095, EPI_ISL_1136096, EPI_ISL_1136097, EPI_ISL_1136098, EPI_ISL_1136099, EPI_ISL_1136100, EPI_ISL_1136101, EPI_ISL_1136102, EPI_ISL_1136103, EPI_ISL_1136104, EPI_ISL_1136105, EPI_ISL_1136106, EPI_ISL_1136107, EPI_ISL_1136108, EPI_ISL_1136109, EPI_ISL_1136110, EPI_ISL_1136111, EPI_ISL_1136112, EPI_ISL_1136113, EPI_ISL_1136114, EPI_ISL_1136115, EPI_ISL_1136116, EPI_ISL_1136117, EPI_ISL_1136118, EPI_ISL_1136119, EPI_ISL_1136120, EPI_ISL_1136121, EPI_ISL_1136122, EPI_ISL_1136123, EPI_ISL_1136124, EPI_ISL_1136125, EPI_ISL_1136126, EPI_ISL_1136127, EPI_ISL_1136128, EPI_ISL_1136129, EPI_ISL_1136130, EPI_ISL_1136131, EPI_ISL_1136132, EPI_ISL_1136133, EPI_ISL_1136134, EPI_ISL_1136135, EPI_ISL_1136136, EPI_ISL_1136137, EPI_ISL_1136138, EPI_ISL_1136139, EPI_ISL_1136140, EPI_ISL_1136141, EPI_ISL_1136142, EPI_ISL_1136143, EPI_ISL_1136144, EPI_ISL_1136145, EPI_ISL_1136146, EPI_ISL_1136147, EPI_ISL_1136148, EPI_ISL_1136149, EPI_ISL_1136150, EPI_ISL_1136151, EPI_ISL_1136152, EPI_ISL_1136153, EPI_ISL_1136154, EPI_ISL_1136155, EPI_ISL_1136156, EPI_ISL_1136157, EPI_ISL_1136158, EPI_ISL_1136159, EPI_ISL_1136160, EPI_ISL_1136161, EPI_ISL_1136162, EPI_ISL_1136163, EPI_ISL_1136164, EPI_ISL_1136165, EPI_ISL_1136166, EPI_ISL_1136167, EPI_ISL_1136168, EPI_ISL_1136169, EPI_ISL_1136170, EPI_ISL_1136171, EPI_ISL_1136172, EPI_ISL_1136173, EPI_ISL_1136174, EPI_ISL_1136175, EPI_ISL_1136176, EPI_ISL_1136177, EPI_ISL_1136178, EPI_ISL_1136179, EPI_ISL_1136180, EPI_ISL_1136181, EPI_ISL_1136182, EPI_ISL_1136183, EPI_ISL_1136184, EPI_ISL_1136185, EPI_ISL_1136186, EPI_ISL_1136187, EPI_ISL_1136188, EPI_ISL_1136189, EPI_ISL_1136190, EPI_ISL_1136191, EPI_ISL_1136192, EPI_ISL_1136193, EPI_ISL_1136194, EPI_ISL_1136195, EPI_ISL_1136196, EPI_ISL_1136197, EPI_ISL_1136198, EPI_ISL_1136199, EPI_ISL_1136200, EPI_ISL_1136201, EPI_ISL_1136202, EPI_ISL_1136203, EPI_ISL_1136204, EPI_ISL_1136205, EPI_ISL_1136206, EPI_ISL_1136207, EPI_ISL_1136208, EPI_ISL_1136209, EPI_ISL_1136210, EPI_ISL_1136211, EPI_ISL_1136212, EPI_ISL_1136213, EPI_ISL_1136214, EPI_ISL_1136215, EPI_ISL_1136216, EPI_ISL_1136217, EPI_ISL_1136218, EPI_ISL_1136219, EPI_ISL_1136220, EPI_ISL_1136221, EPI_ISL_1136222, EPI_ISL_1136223, EPI_ISL_1136224, EPI_ISL_1136225, EPI_ISL_1136226, EPI_ISL_1136227, EPI_ISL_1136228, EPI_ISL_1136229, EPI_ISL_1136230, EPI_ISL_1136231, EPI_ISL_1136232, EPI_ISL_1136233, EPI_ISL_1136234, EPI_ISL_1136235, EPI_ISL_1136236, EPI_ISL_1136237, EPI_ISL_1136238, EPI_ISL_1136239, EPI_ISL_1136240, EPI_ISL_1136241, EPI_ISL_1136242, EPI_ISL_1136243, EPI_ISL_1136244, EPI_ISL_1136245, EPI_ISL_1136246, EPI_ISL_1136247, EPI_ISL_1136248, EPI_ISL_1136249, EPI_ISL_1136250, EPI_ISL_1136251, EPI_ISL_1136252, EPI_ISL_1136253, EPI_ISL_1136254, EPI_ISL_1136255, EPI_ISL_1136256, EPI_ISL_1136257, EPI_ISL_1136258, EPI_ISL_1136259, EPI_ISL_1136260, EPI_ISL_1136261, EPI_ISL_1136262, EPI_ISL_1136263, EPI_ISL_1136264, EPI_ISL_1136265, EPI_ISL_1136266, EPI_ISL_1136267, EPI_ISL_1136268, EPI_ISL_1136269, EPI_ISL_1136270, EPI_ISL_1136271, EPI_ISL_1136272, EPI_ISL_1136273, EPI_ISL_1136274, EPI_ISL_1136275, EPI_ISL_1136276, EPI_ISL_1136277, EPI_ISL_1136278, EPI_ISL_1136279, EPI_ISL_1136280, EPI_ISL_1136281, EPI_ISL_1136282, EPI_ISL_1136283, EPI_ISL_1136284, EPI_ISL_1136285, EPI_ISL_1136286, EPI_ISL_1136287, EPI_ISL_1136288, EPI_ISL_1136289, EPI_ISL_1136290, EPI_ISL_1136291, EPI_ISL_1136292, EPI_ISL_1136293, EPI_ISL_1136294, EPI_ISL_1136295, EPI_ISL_1136296, EPI_ISL_1136297, EPI_ISL_1136298, EPI_ISL_1136299, EPI_ISL_1136300, EPI_ISL_1136301, EPI_ISL_1136302, EPI_ISL_1136303, EPI_ISL_1136304, EPI_ISL_1136305, EPI_ISL_1136306, EPI_ISL_1136307, EPI_ISL_1136308, EPI_ISL_1136309, EPI_ISL_1136310, EPI_ISL_1136311, EPI_ISL_1136312, EPI_ISL_1136313, EPI_ISL_1136314, EPI_ISL_1136315, EPI_ISL_1136316, EPI_ISL_1136317, EPI_ISL_1136318, EPI_ISL_1136319, EPI_ISL_1136320, EPI_ISL_1136321, EPI_ISL_1136322, EPI_ISL_1136323, EPI_ISL_1136324, EPI_ISL_1136325, EPI_ISL_1136326, EPI_ISL_1136327, EPI_ISL_1136328, EPI_ISL_1136329, EPI_ISL_1136330, EPI_ISL_1136331, EPI_ISL_1136332, EPI_ISL_1136333, EPI_ISL_1136334, EPI_ISL_1136335, EPI_ISL_1136336, EPI_ISL_1136337, EPI_ISL_1136338, EPI_ISL_1136339, EPI_ISL_1136340, EPI_ISL_1136341, EPI_ISL_1136342, EPI_ISL_1136343, EPI_ISL_1136344, EPI_ISL_1136345, EPI_ISL_1136346, EPI_ISL_1136347, EPI_ISL_1136348, EPI_ISL_1136349, EPI_ISL_1136350, EPI_ISL_1136351, EPI_ISL_1136352, EPI_ISL_1136353, EPI_ISL_1136354, EPI_ISL_1136355, EPI_ISL_1136356, EPI_ISL_1136357, EPI_ISL_1136358, EPI_ISL_1136359, EPI_ISL_1136360, EPI_ISL_1136361, EPI_ISL_1136362, EPI_ISL_1136363, EPI_ISL_1136364, EPI_ISL_1136365, EPI_ISL_1136366, EPI_ISL_1136367, EPI_ISL_1136368, EPI_ISL_1136369, EPI_ISL_1136370, EPI_ISL_1136371, EPI_ISL_1136372, EPI_ISL_1136373, EPI_ISL_1136374, EPI_ISL_1136375, EPI_ISL_1136376, EPI_ISL_1136377, EPI_ISL_1136378, EPI_ISL_1136379, EPI_ISL_1136380, EPI_ISL_1136381, EPI_ISL_1136382, EPI_ISL_1136383, EPI_ISL_1136384, EPI_ISL_1136385, EPI_ISL_1136386, EPI_ISL_1136387, EPI_ISL_1136388, EPI_ISL_1136389, EPI_ISL_1136390, EPI_ISL_1136391, EPI_ISL_1136393, EPI_ISL_1136394, EPI_ISL_1136395, EPI_ISL_1136396, EPI_ISL_1136397, EPI_ISL_1136398, EPI_ISL_1136400, EPI_ISL_1136401, EPI_ISL_1136402, EPI_ISL_1136405, EPI_ISL_1136406, EPI_ISL_1136407, EPI_ISL_1136408, EPI_ISL_1136409, EPI_ISL_1136410, EPI_ISL_1136411, EPI_ISL_1136412, EPI_ISL_1136413, EPI_ISL_1136414, EPI_ISL_1136415, EPI_ISL_1136416, EPI_ISL_1136417, EPI_ISL_1136418, EPI_ISL_1136420, EPI_ISL_1136421, EPI_ISL_1136422, EPI_ISL_1136423, EPI_ISL_1136424, EPI_ISL_1136425, EPI_ISL_1136426, EPI_ISL_1136427, EPI_ISL_1136428, EPI_ISL_1136429, EPI_ISL_1136430, EPI_ISL_1136432, EPI_ISL_1136434, EPI_ISL_1136435, EPI_ISL_1136438, EPI_ISL_1136439, EPI_ISL_1136440, EPI_ISL_1136441, EPI_ISL_1136448, EPI_ISL_1136449, EPI_ISL_1136450, EPI_ISL_1136451, EPI_ISL_1136452, EPI_ISL_1136453, EPI_ISL_1136454, EPI_ISL_1136455, EPI_ISL_1136456, EPI_ISL_1136459, EPI_ISL_1136460, EPI_ISL_1136461, EPI_ISL_1136462, EPI_ISL_1136469, EPI_ISL_1136470, EPI_ISL_1136471, EPI_ISL_1136472, EPI_ISL_1136473, EPI_ISL_1136474, EPI_ISL_1136475, EPI_ISL_1136476, EPI_ISL_1136477, EPI_ISL_1136478, EPI_ISL_1136479, EPI_ISL_1136480, EPI_ISL_1136484, EPI_ISL_1136485, EPI_ISL_1136486, EPI_ISL_1136487, EPI_ISL_1136489, EPI_ISL_1136490, EPI_ISL_1136491, EPI_ISL_1136492, EPI_ISL_1136498, EPI_ISL_1136499, EPI_ISL_1136500, EPI_ISL_1136501, EPI_ISL_1136502, EPI_ISL_1136503, EPI_ISL_1136504, EPI_ISL_1136505, EPI_ISL_1136506, EPI_ISL_1136507, EPI_ISL_1136508, EPI_ISL_1136509, EPI_ISL_1136510, EPI_ISL_1136511, EPI_ISL_1136512, EPI_ISL_1136513, EPI_ISL_1136514, EPI_ISL_1136515, EPI_ISL_1136516, EPI_ISL_1136517, EPI_ISL_1136518, EPI_ISL_1136519, EPI_ISL_1136524, EPI_ISL_1136525, EPI_ISL_1136526, EPI_ISL_1136527, EPI_ISL_1136528, EPI_ISL_1136529, EPI_ISL_1136530, EPI_ISL_1136531, EPI_ISL_1136532, EPI_ISL_1136533, EPI_ISL_1136534, EPI_ISL_1136535, EPI_ISL_1136536, EPI_ISL_1136537, EPI_ISL_1136538, EPI_ISL_1136539, EPI_ISL_1136540, EPI_ISL_1136541, EPI_ISL_1136542, EPI_ISL_1136543, EPI_ISL_1136544, EPI_ISL_1136545, EPI_ISL_1136546, EPI_ISL_1136548, EPI_ISL_1136549, EPI_ISL_1136550, EPI_ISL_1136551, EPI_ISL_1136552, EPI_ISL_1136553, EPI_ISL_1136556, EPI_ISL_1136557, EPI_ISL_1136558, EPI_ISL_1136559, EPI_ISL_1136560, EPI_ISL_1136561, EPI_ISL_1136562, EPI_ISL_1136563, EPI_ISL_1136564, EPI_ISL_1136565, EPI_ISL_1136566, EPI_ISL_1136567, EPI_ISL_1136568, EPI_ISL_1136569, EPI_ISL_1136570, EPI_ISL_1136571, EPI_ISL_1136572, EPI_ISL_1136573, EPI_ISL_1136574, EPI_ISL_1136575, EPI_ISL_1136576, EPI_ISL_1136577, EPI_ISL_1136578, EPI_ISL_1136579, EPI_ISL_1136580, EPI_ISL_1136581, EPI_ISL_1136582, EPI_ISL_1136583, EPI_ISL_1136584, EPI_ISL_1136585, EPI_ISL_1136586, EPI_ISL_1136587, EPI_ISL_1136588, EPI_ISL_1136589, EPI_ISL_1136590, EPI_ISL_1136591, EPI_ISL_1136592, EPI_ISL_1136593, EPI_ISL_1136594, EPI_ISL_1136595, EPI_ISL_1136596, EPI_ISL_1136597, EPI_ISL_1136598, EPI_ISL_1136599, EPI_ISL_1136600, EPI_ISL_1136601, EPI_ISL_1136602, EPI_ISL_1136603, EPI_ISL_1136604, EPI_ISL_1136605, EPI_ISL_1136606, EPI_ISL_1136607, EPI_ISL_1136608, EPI_ISL_1136609, EPI_ISL_1136616, EPI_ISL_1136617, EPI_ISL_1136618, EPI_ISL_1136619, EPI_ISL_1136620, EPI_ISL_1136621, EPI_ISL_1136622, EPI_ISL_1136623, EPI_ISL_1136624, EPI_ISL_1136625, EPI_ISL_1136626, EPI_ISL_1136627, EPI_ISL_1136628, EPI_ISL_1136629, EPI_ISL_1136630, EPI_ISL_1136631, EPI_ISL_1136632, EPI_ISL_1136633, EPI_ISL_1136634, EPI_ISL_1136635, EPI_ISL_1136636, EPI_ISL_1136637, EPI_ISL_1136638, EPI_ISL_1136639, EPI_ISL_1136644, EPI_ISL_1136645, EPI_ISL_1136646, EPI_ISL_1136647, EPI_ISL_1136648, EPI_ISL_1136649, EPI_ISL_1136650, EPI_ISL_1136651, EPI_ISL_1136652, EPI_ISL_1136654, EPI_ISL_1136655, EPI_ISL_1136656, EPI_ISL_1136657, EPI_ISL_1136658, EPI_ISL_1136659, EPI_ISL_1136660, EPI_ISL_1136661, EPI_ISL_1136662, EPI_ISL_1136665, EPI_ISL_1136666, EPI_ISL_1136667, EPI_ISL_1136668, EPI_ISL_1136669, EPI_ISL_1136670, EPI_ISL_1136671, EPI_ISL_1136672, EPI_ISL_1136673, EPI_ISL_1136674, EPI_ISL_1136675, EPI_ISL_1136676, EPI_ISL_1136677, EPI_ISL_1136678, EPI_ISL_1136679, EPI_ISL_1136680, EPI_ISL_1136681, EPI_ISL_1136682, EPI_ISL_1136683, EPI_ISL_1136684, EPI_ISL_1136685, EPI_ISL_1136686, EPI_ISL_1136695, EPI_ISL_1136696, EPI_ISL_1136697, EPI_ISL_1136698, EPI_ISL_1136699, EPI_ISL_1136700, EPI_ISL_1136701, EPI_ISL_1136702, EPI_ISL_1136703, EPI_ISL_1136704, EPI_ISL_1136705, EPI_ISL_1136706, EPI_ISL_1136707, EPI_ISL_1136708, EPI_ISL_1136709, EPI_ISL_1136710, EPI_ISL_1136711, EPI_ISL_1136712, EPI_ISL_1136713, EPI_ISL_1136714, EPI_ISL_1136715, EPI_ISL_1136716, EPI_ISL_1136717, EPI_ISL_1136718, EPI_ISL_1136719, EPI_ISL_1136720, EPI_ISL_1136721, EPI_ISL_1136722, EPI_ISL_1136723, EPI_ISL_1136724, EPI_ISL_1136725, EPI_ISL_1136726, EPI_ISL_1136727, EPI_ISL_1136728, EPI_ISL_1136729, EPI_ISL_1136730, EPI_ISL_1136731, EPI_ISL_1136733, EPI_ISL_1136734, EPI_ISL_1136735, EPI_ISL_1136736, EPI_ISL_1136737, EPI_ISL_1136738, EPI_ISL_1136739, EPI_ISL_1136740, EPI_ISL_1136741, EPI_ISL_1136742, EPI_ISL_1136743, EPI_ISL_1136744, EPI_ISL_1136751, EPI_ISL_1136752, EPI_ISL_1136753, EPI_ISL_1136754, EPI_ISL_1136755, EPI_ISL_1136756, EPI_ISL_1136757, EPI_ISL_1136758, EPI_ISL_1136759, EPI_ISL_1136760, EPI_ISL_1136761, EPI_ISL_1136762, EPI_ISL_1136763, EPI_ISL_1136764, EPI_ISL_1136765, EPI_ISL_1136766, EPI_ISL_1136768, EPI_ISL_1136769, EPI_ISL_1136770, EPI_ISL_1136771, EPI_ISL_1136772, EPI_ISL_1136773, EPI_ISL_1136774, EPI_ISL_1136775, EPI_ISL_1136776, EPI_ISL_1136777, EPI_ISL_1136783, EPI_ISL_1136784, EPI_ISL_1136785, EPI_ISL_1136786, EPI_ISL_1136787, EPI_ISL_1136788, EPI_ISL_1136789, EPI_ISL_1136790, EPI_ISL_1136791, EPI_ISL_1136792, EPI_ISL_1136793, EPI_ISL_1136794, EPI_ISL_1136795, EPI_ISL_1136796, EPI_ISL_1136799, EPI_ISL_1136800, EPI_ISL_1136801, EPI_ISL_1136802, EPI_ISL_1136803, EPI_ISL_1136804, EPI_ISL_1136805, EPI_ISL_1136806, EPI_ISL_1136807, EPI_ISL_1136808, EPI_ISL_1136809, EPI_ISL_1136810, EPI_ISL_1136811, EPI_ISL_1136812, EPI_ISL_1136813, EPI_ISL_1136814, EPI_ISL_1136815, EPI_ISL_1136816, EPI_ISL_1136817, EPI_ISL_1136818, EPI_ISL_1136819, EPI_ISL_1136820, EPI_ISL_1136829, EPI_ISL_1136830, EPI_ISL_1136831, EPI_ISL_1136832, EPI_ISL_1136833, EPI_ISL_1136834, EPI_ISL_1136835, EPI_ISL_1136836, EPI_ISL_1136837, EPI_ISL_1136838, EPI_ISL_1136839, EPI_ISL_1136840, EPI_ISL_1136841, EPI_ISL_1136842, EPI_ISL_1136843, EPI_ISL_1136844, EPI_ISL_1136845, EPI_ISL_1136846, EPI_ISL_1136847, EPI_ISL_1136848, EPI_ISL_1136849, EPI_ISL_1136854, EPI_ISL_1136855, EPI_ISL_1136856, EPI_ISL_1136857, EPI_ISL_1136858, EPI_ISL_1136859, EPI_ISL_1136860, EPI_ISL_1136861, EPI_ISL_1136862, EPI_ISL_1136863, EPI_ISL_1136864, EPI_ISL_1136865, EPI_ISL_1136866, EPI_ISL_1136867, EPI_ISL_1136868, EPI_ISL_1136869, EPI_ISL_1136870, EPI_ISL_1136871, EPI_ISL_1136872, EPI_ISL_1136873, EPI_ISL_1136874, EPI_ISL_1136875, EPI_ISL_1136876, EPI_ISL_1136877, EPI_ISL_1136878, EPI_ISL_1136880, EPI_ISL_1136881, EPI_ISL_1136882, EPI_ISL_1136883, EPI_ISL_1136884, EPI_ISL_1136885, EPI_ISL_1136889, EPI_ISL_1136890, EPI_ISL_1136891, EPI_ISL_1136892, EPI_ISL_1136895, EPI_ISL_1136896, EPI_ISL_1136897, EPI_ISL_1136898, EPI_ISL_1136899, EPI_ISL_1136900, EPI_ISL_1136901, EPI_ISL_1136902, EPI_ISL_1136903, EPI_ISL_1136904, EPI_ISL_1136905, EPI_ISL_1136906, EPI_ISL_1136907, EPI_ISL_1136908, EPI_ISL_1136909, EPI_ISL_1136910, EPI_ISL_1136911, EPI_ISL_1136912, EPI_ISL_1136913, EPI_ISL_1136914, EPI_ISL_1136915, EPI_ISL_1136916, EPI_ISL_1136917, EPI_ISL_1136918, EPI_ISL_1136919, EPI_ISL_1136920, EPI_ISL_1136921, EPI_ISL_1136922, EPI_ISL_1136923, EPI_ISL_1136924, EPI_ISL_1136925, EPI_ISL_1136926, EPI_ISL_1136930, EPI_ISL_1136931, EPI_ISL_1136932, EPI_ISL_1136933, EPI_ISL_1136934, EPI_ISL_1136935, EPI_ISL_1136936, EPI_ISL_1136937, EPI_ISL_1136940, EPI_ISL_1136941, EPI_ISL_1136942, EPI_ISL_1136943, EPI_ISL_1136944, EPI_ISL_1136945, EPI_ISL_1136946, EPI_ISL_1136947, EPI_ISL_1136948, EPI_ISL_1136949, EPI_ISL_1136950, EPI_ISL_1136951, EPI_ISL_1136952, EPI_ISL_1136953, EPI_ISL_1136954, EPI_ISL_1136955, EPI_ISL_1136956, EPI_ISL_1136957, EPI_ISL_1136958, EPI_ISL_1136959, EPI_ISL_1136960, EPI_ISL_1136961, EPI_ISL_1136962, EPI_ISL_1136963, EPI_ISL_1136964, EPI_ISL_1136965, EPI_ISL_1136966, EPI_ISL_1136967, EPI_ISL_1136968, EPI_ISL_1136969, EPI_ISL_1136970, EPI_ISL_1136971, EPI_ISL_1136972, EPI_ISL_1136973, EPI_ISL_1136974 | | | |
| see above | Nevada State Public Health Laboratory | Nevada State Public Health Laboratory | Andrew Gorzalski, Mark Pandori |
| EPI_ISL_1136975, EPI_ISL_1136976, EPI_ISL_1136977, EPI_ISL_1136978, EPI_ISL_1136979, EPI_ISL_1136980, EPI_ISL_1136981, EPI_ISL_1136982, EPI_ISL_1136983, EPI_ISL_1136995, EPI_ISL_1136996, EPI_ISL_1136997, EPI_ISL_1136998, EPI_ISL_1136999, EPI_ISL_1137000, EPI_ISL_1137001, EPI_ISL_1137002, EPI_ISL_1137003, EPI_ISL_1137004, EPI_ISL_1137005, EPI_ISL_1137006, EPI_ISL_1137007, EPI_ISL_1137008, EPI_ISL_1137009, EPI_ISL_1137010, EPI_ISL_1137011, EPI_ISL_1137012, EPI_ISL_1137013, EPI_ISL_1137014, EPI_ISL_1137015, EPI_ISL_1137016, EPI_ISL_1137017, EPI_ISL_1137018, EPI_ISL_1137019, EPI_ISL_1137020, EPI_ISL_1137021, EPI_ISL_1137022, EPI_ISL_1137023, EPI_ISL_1137024, EPI_ISL_1137025, EPI_ISL_1137026, EPI_ISL_1137027, EPI_ISL_1137028, EPI_ISL_1137029, EPI_ISL_1137030, EPI_ISL_1137031, EPI_ISL_1137032, EPI_ISL_1137033, EPI_ISL_1137034, EPI_ISL_1137035, EPI_ISL_1137036, EPI_ISL_1137037, EPI_ISL_1137038, EPI_ISL_1137039, EPI_ISL_1137040, EPI_ISL_1137041, EPI_ISL_1137042, EPI_ISL_1137043, EPI_ISL_1137044, EPI_ISL_1137045, EPI_ISL_1137046, EPI_ISL_1137047 | | | |
| see above | Delaware Public Health Lab | Delaware Public Health Lab | Gregory Hovan |
| EPI_ISL_1137048, EPI_ISL_1137049, EPI_ISL_1137050, EPI_ISL_1137051, EPI_ISL_1137052, EPI_ISL_1137053, EPI_ISL_1137054, EPI_ISL_1137055, EPI_ISL_1137056, EPI_ISL_1137057, EPI_ISL_1137058, EPI_ISL_1137059, EPI_ISL_1137060, EPI_ISL_1137061, EPI_ISL_1137062, EPI_ISL_1137063, EPI_ISL_1137064, EPI_ISL_1137065, EPI_ISL_1137066, EPI_ISL_1137067, EPI_ISL_1137068, EPI_ISL_1137069, EPI_ISL_1137070, EPI_ISL_1137071, EPI_ISL_1137072, EPI_ISL_1137073, EPI_ISL_1137074, EPI_ISL_1137075, EPI_ISL_1137076, EPI_ISL_1137077, EPI_ISL_1137078, EPI_ISL_1137079, EPI_ISL_1137080 | | | |
| see above | Kansas Health and Environmental Lab | Kansas Health and Environmental Lab | Mike Grose, Jonathan Barnell, Ben Olsen, and Phil Adam |
| EPI_ISL_1137082, EPI_ISL_1137083, EPI_ISL_1137084, EPI_ISL_1137085, EPI_ISL_1137086, EPI_ISL_1137087, EPI_ISL_1137088, EPI_ISL_1137089, EPI_ISL_1137090, EPI_ISL_1137091, EPI_ISL_1137092, EPI_ISL_1137093, EPI_ISL_1137094, EPI_ISL_1137095, EPI_ISL_1137096, EPI_ISL_1137097, EPI_ISL_1137098, EPI_ISL_1137099, EPI_ISL_1137100, EPI_ISL_1137101, EPI_ISL_1137102, EPI_ISL_1137103, EPI_ISL_1137104, EPI_ISL_1137105, EPI_ISL_1137106, EPI_ISL_1137107, EPI_ISL_1137108, EPI_ISL_1137109, EPI_ISL_1137110, EPI_ISL_1137111, EPI_ISL_1137112, EPI_ISL_1137113, EPI_ISL_1137114, EPI_ISL_1137115, EPI_ISL_1137116, EPI_ISL_1137117, EPI_ISL_1137118, EPI_ISL_1137119, EPI_ISL_1137120, EPI_ISL_1137121, EPI_ISL_1137122, EPI_ISL_1137123, EPI_ISL_1137124, EPI_ISL_1137125, EPI_ISL_1137126, EPI_ISL_1137127, EPI_ISL_1137128, EPI_ISL_1137129, EPI_ISL_1137130, EPI_ISL_1137131, EPI_ISL_1137132, EPI_ISL_1137133, EPI_ISL_1137134, EPI_ISL_1137135, EPI_ISL_1137136, EPI_ISL_1137137, EPI_ISL_1137138, EPI_ISL_1137139, EPI_ISL_1137140, EPI_ISL_1137141, EPI_ISL_1137142, EPI_ISL_1137143, EPI_ISL_1137144, EPI_ISL_1137145, EPI_ISL_1137146, EPI_ISL_1137147, EPI_ISL_1137148, EPI_ISL_1137149, EPI_ISL_1137150, EPI_ISL_1137151, EPI_ISL_1137152, EPI_ISL_1137153, EPI_ISL_1137154, EPI_ISL_1137155, EPI_ISL_1137156, EPI_ISL_1137157, EPI_ISL_1137158, EPI_ISL_1137159, EPI_ISL_1137160, EPI_ISL_1137161, EPI_ISL_1137162, EPI_ISL_1137163, EPI_ISL_1137164, EPI_ISL_1137165, EPI_ISL_1137166, EPI_ISL_1137167, EPI_ISL_1137168, EPI_ISL_1137169 | | | |
| see above | Alameda County Public Health Lab | Chan-Zuckerberg Biohub | CZB Cliahub Consortium |
| EPI_ISL_1137171, EPI_ISL_1137172, EPI_ISL_1137173, EPI_ISL_1137174, EPI_ISL_1137175, EPI_ISL_1137176, EPI_ISL_1137177, EPI_ISL_1137178, EPI_ISL_1137179, EPI_ISL_1137180, EPI_ISL_1137181, EPI_ISL_1137182, EPI_ISL_1137183, EPI_ISL_1137184, EPI_ISL_1137185, EPI_ISL_1137186, EPI_ISL_1137187, EPI_ISL_1137188, EPI_ISL_1137189, EPI_ISL_1137190, EPI_ISL_1137191, EPI_ISL_1137192, EPI_ISL_1137193, EPI_ISL_1137194, EPI_ISL_1137195, EPI_ISL_1137196, EPI_ISL_1137197, EPI_ISL_1137198, EPI_ISL_1137199, EPI_ISL_1137200, EPI_ISL_1137201, EPI_ISL_1137202, EPI_ISL_1137203, EPI_ISL_1137204, EPI_ISL_1137205, EPI_ISL_1137206, EPI_ISL_1137207, EPI_ISL_1137208, EPI_ISL_1137209, EPI_ISL_1137210, EPI_ISL_1137211, EPI_ISL_1137212, EPI_ISL_1137213, EPI_ISL_1137214, EPI_ISL_1137215, EPI_ISL_1137216, EPI_ISL_1137217, EPI_ISL_1137218, EPI_ISL_1137219, EPI_ISL_1137220, EPI_ISL_1137221, EPI_ISL_1137222, EPI_ISL_1137223, EPI_ISL_1137224, EPI_ISL_1137225, EPI_ISL_1137226, EPI_ISL_1137227, EPI_ISL_1137228, EPI_ISL_1137229, EPI_ISL_1137230, EPI_ISL_1137231, EPI_ISL_1137232, EPI_ISL_1137233, EPI_ISL_1137234, EPI_ISL_1137235, EPI_ISL_1137236, EPI_ISL_1137237, EPI_ISL_1137238, EPI_ISL_1137239, EPI_ISL_1137240, EPI_ISL_1137241, EPI_ISL_1137242, EPI_ISL_1137243, EPI_ISL_1137244, EPI_ISL_1137245, EPI_ISL_1137246, EPI_ISL_1137247, EPI_ISL_1137248, EPI_ISL_1137249, EPI_ISL_1137250, EPI_ISL_1137251, EPI_ISL_1137252, EPI_ISL_1137253, EPI_ISL_1137254 | | | |
| see above | UPMC Clinical Microbiology Laboratory | Microbial Genome Sequencing Center; Microbial Genomic Epidemiology Laboratory, University of Pittsburgh | Lee H. Harrison, Jane W. Marsh, Marissa P. Griffith, Stephanie L. Mitchell, Vatsala R. Srinivasa, Kady D. Waggle, Daniel J. Snyder, Vaughn S. Cooper |
| EPI_ISL_1137255, EPI_ISL_1137256, EPI_ISL_1137257 | M Health Fairview | Minnesota Department of Health, Public Health Laboratory | Alexandra Lorentz, Jacob Garfin, Matt Plumb, and Xiong Wang |
| EPI_ISL_1137258, EPI_ISL_1137259, EPI_ISL_1137260, EPI_ISL_1137265, EPI_ISL_1137266, EPI_ISL_1137267, EPI_ISL_1137268, EPI_ISL_1137269, EPI_ISL_1137270, EPI_ISL_1137271, EPI_ISL_1137272, EPI_ISL_1137273, EPI_ISL_1137274, EPI_ISL_1137275, EPI_ISL_1137276, EPI_ISL_1137277, EPI_ISL_1137278, EPI_ISL_1137279, EPI_ISL_1137280, EPI_ISL_1137281, EPI_ISL_1137282, EPI_ISL_1137283, EPI_ISL_1137284, EPI_ISL_1137285, EPI_ISL_1137286, EPI_ISL_1137287, EPI_ISL_1137288, EPI_ISL_1137289, EPI_ISL_1137290, EPI_ISL_1137291, EPI_ISL_1137292, EPI_ISL_1137293, EPI_ISL_1137294, EPI_ISL_1137295, EPI_ISL_1137296, EPI_ISL_1137297, EPI_ISL_1137298, EPI_ISL_1137299, EPI_ISL_1137300, EPI_ISL_1137301, EPI_ISL_1137302, EPI_ISL_1137303 | | | |
| see above | Nevada State Public Health Laboratory | Nevada State Public Health Laboratory | Andrew Gorzalski, Mark Pandori |
| EPI_ISL_1137459 | LABDIS-UAGro | Instituto Nacional de Medicina Genomica | Hidalgo-Miranda A, Mendoza-Vargas A, Reyes-Grajeda JP, Cedro-Tanda A, Alcaraz N, Gonzalez-Barrera D, Rangel-DeLeon D, Ramirez-Vega O, Sifuentes-Rojas C, Cisneros-Villanueva M, Herrera-Montalvo LA, Berenice Illades-Aguiar, Oscar del Moral-Hernandez, Adolfo Roman-Roman, Marco Antonio-Leyva, Daniel Hernandez-Sotelo, Hugo A. Rodriguez-Ruiz, Fredy O Beltran-Anaya, Roberto Dircio-Maldonado, Julio C. Azucar-Hesiquio, Azucena D. Alvarez-Diaz, Ciresthel Bello-Rios, Gladys W. Valente-Nio, Jaqueline Loaeza-Loaeza, Madian Valadez-Quinones, Luis I. Basilio-Leyva. |
| EPI_ISL_1137460, EPI_ISL_1137461, EPI_ISL_1137462, EPI_ISL_1137463, EPI_ISL_1137464, EPI_ISL_1137465, EPI_ISL_1137466, EPI_ISL_1137467, EPI_ISL_1137468, EPI_ISL_1137469, EPI_ISL_1137470, EPI_ISL_1137471, EPI_ISL_1137472, EPI_ISL_1137473, EPI_ISL_1137474 | | | |
| see above | Instituto Nacional de Medicina Genomica | Instituto Nacional de Medicina Genomica | Hidalgo-Miranda A, Mendoza-Vargas A, Reyes-Grajeda JP, Cedro-Tanda A, Alcaraz N, Gonzalez-Barrera D, Rangel-DeLeon D, Ramirez-Vega O, Sifuentes-Rojas C, Cisneros-Villanueva M, Herrera-Montalvo LA |
| EPI_ISL_1137475, EPI_ISL_1137476, EPI_ISL_1137477 | LABDIS-UAGro | Instituto Nacional de Medicina Genomica | Hidalgo-Miranda A, Mendoza-Vargas A, Reyes-Grajeda JP, Cedro-Tanda A, Alcaraz N, Gonzalez-Barrera D, Rangel-DeLeon D, Ramirez-Vega O, Sifuentes-Rojas C, Cisneros-Villanueva M, Herrera-Montalvo LA, Berenice Illades-Aguiar, Oscar del Moral-Hernandez, Adolfo Roman-Roman, Marco Antonio-Leyva, Daniel Hernandez-Sotelo, Hugo A. Rodriguez-Ruiz, Fredy O Beltran-Anaya, Roberto Dircio-Maldonado, Julio C. Azucar-Hesiquio, Azucena D. Alvarez-Diaz, Ciresthel Bello-Rios, Gladys W. Valente-Nio, Jaqueline Loaeza-Loaeza, Madian Valadez-Quinones, Luis I. Basilio-Leyva. |
| EPI_ISL_1137492, EPI_ISL_1137493, EPI_ISL_1137494, EPI_ISL_1137495, EPI_ISL_1137496, EPI_ISL_1137497, EPI_ISL_1137498, EPI_ISL_1137499, EPI_ISL_1137500, EPI_ISL_1137501, EPI_ISL_1137502 | | | |
| see above | New Jersey Department of Health, Public Health and Environmental Laboratories | New Jersey Public Health and Environmental Laboratories | Lindsey Bodnar, Shiv K. Verma, Dana Woell, Byeong Jeong |
| EPI_ISL_1137506, EPI_ISL_1137507, EPI_ISL_1137509, EPI_ISL_1137510, EPI_ISL_1137511, EPI_ISL_1137513, EPI_ISL_1137514, EPI_ISL_1137515, EPI_ISL_1137516, EPI_ISL_1137517, EPI_ISL_1137519, EPI_ISL_1137520, EPI_ISL_1137522, EPI_ISL_1137523, EPI_ISL_1137526, EPI_ISL_1137529, EPI_ISL_1137532, EPI_ISL_1137534, EPI_ISL_1137537, EPI_ISL_1137538, EPI_ISL_1137539, EPI_ISL_1137546, EPI_ISL_1137547, EPI_ISL_1137550, EPI_ISL_1137551, EPI_ISL_1137553, EPI_ISL_1137554, EPI_ISL_1137556, EPI_ISL_1137557, EPI_ISL_1137558, EPI_ISL_1137559, EPI_ISL_1137560, EPI_ISL_1137561, EPI_ISL_1137562, EPI_ISL_1137565, EPI_ISL_1137566, EPI_ISL_1137567, EPI_ISL_1137568, EPI_ISL_1137573, EPI_ISL_1137574, EPI_ISL_1137582, EPI_ISL_1137587, EPI_ISL_1137588, EPI_ISL_1137589, EPI_ISL_1137590, EPI_ISL_1137592, EPI_ISL_1137593, EPI_ISL_1137594, EPI_ISL_1137596, EPI_ISL_1137598 | | | |
| see above | Clinical Molecular Microbiology Laboratory, UNC Hospitals | Jeremy Wang | Jeremy Wang, Alexander Rubinsteyn, Colleen Rice, Jason Smedberg, Shawn Hawken, Melissa Miller, Corbin Jones, Robert Hagan |
| EPI_ISL_1137604, EPI_ISL_1137605, EPI_ISL_1137606, EPI_ISL_1137607 | Wyoming Public Health Laboratory | Wyoming Public Health Laboratory | Jim Mildenberger, Wanda Manley, Noah Hull, Taylor Fearing, Lynette Gumbleton, Channing Weber, Ashley Norberg, Chayse Rowley, Marley Goetz, Brian Dominguez, Elliot Thomasson, Cari Sloma, and Rob Christensen |
| EPI_ISL_1137631, EPI_ISL_1137632, EPI_ISL_1137633, EPI_ISL_1137634, EPI_ISL_1137635, EPI_ISL_1137636, EPI_ISL_1137637, EPI_ISL_1137638, EPI_ISL_1137639, EPI_ISL_1137640, EPI_ISL_1137641, EPI_ISL_1137644, EPI_ISL_1137646, EPI_ISL_1137647, EPI_ISL_1137648 | | | |
| see above | Santa Clara County Public Health Laboratory | Santa Clara County Public Health Laboratory | Santa Clara County Public Health Department |
| EPI_ISL_1138425, EPI_ISL_1138426, EPI_ISL_1138427, EPI_ISL_1138428, EPI_ISL_1138429, EPI_ISL_1138431, EPI_ISL_1138432, EPI_ISL_1138435, EPI_ISL_1138436, EPI_ISL_1138437, EPI_ISL_1138438, EPI_ISL_1138440, EPI_ISL_1138442, EPI_ISL_1138443, EPI_ISL_1138445, EPI_ISL_1138446, EPI_ISL_1138447, EPI_ISL_1138448, EPI_ISL_1138450, EPI_ISL_1138451, EPI_ISL_1138452, EPI_ISL_1138454, EPI_ISL_1138455, EPI_ISL_1138456, EPI_ISL_1138458, EPI_ISL_1138459, EPI_ISL_1138460, EPI_ISL_1138461, EPI_ISL_1138462, EPI_ISL_1138464, EPI_ISL_1138465, EPI_ISL_1138466, EPI_ISL_1138467, EPI_ISL_1138468, EPI_ISL_1138469, EPI_ISL_1138470, EPI_ISL_1138472, EPI_ISL_1138473, EPI_ISL_1138474, EPI_ISL_1138475, EPI_ISL_1138476, EPI_ISL_1138478, EPI_ISL_1138479, EPI_ISL_1138480, EPI_ISL_1138481, EPI_ISL_1138483, EPI_ISL_1138484, EPI_ISL_1138485, EPI_ISL_1138486, EPI_ISL_1138487, EPI_ISL_1138489, EPI_ISL_1138490, EPI_ISL_1138491, EPI_ISL_1138492, EPI_ISL_1138493, EPI_ISL_1138494, EPI_ISL_1138495, EPI_ISL_1138496, EPI_ISL_1138497, EPI_ISL_1138498, EPI_ISL_1138499, EPI_ISL_1138500, EPI_ISL_1138502, EPI_ISL_1138503, EPI_ISL_1138504, EPI_ISL_1138505, EPI_ISL_1138506, EPI_ISL_1138507, EPI_ISL_1138508, EPI_ISL_1138509, EPI_ISL_1138510, EPI_ISL_1138511 | | | |
| see above | Illinois Department of Public Health | Gagnon Lab, Southern Illinois University | Keith Gagnon |

| | | | |
|---|---|---|---|
| EPI_ISL_1139158 | Ohio State University Wexner Medical Center | James Molecular Lab | D Jones, P Pancholi, P Snyder, Z Rudy |
| EPI_ISL_1139174, EPI_ISL_1139175, EPI_ISL_1139176, EPI_ISL_1139177, EPI_ISL_1139178, EPI_ISL_1139179, EPI_ISL_1139180, EPI_ISL_1139181, EPI_ISL_1139182, EPI_ISL_1139183, EPI_ISL_1139184, EPI_ISL_1139185, EPI_ISL_1139186, EPI_ISL_1139187, EPI_ISL_1139188, EPI_ISL_1139189, EPI_ISL_1139190, EPI_ISL_1139191, EPI_ISL_1139192, EPI_ISL_1139193, EPI_ISL_1139194, EPI_ISL_1139195, EPI_ISL_1139196, EPI_ISL_1139197, EPI_ISL_1139198, EPI_ISL_1139199, EPI_ISL_1139200, EPI_ISL_1139201, EPI_ISL_1139202, EPI_ISL_1139203, EPI_ISL_1139204, EPI_ISL_1139205, EPI_ISL_1139206, EPI_ISL_1139207, EPI_ISL_1139208, EPI_ISL_1139209, EPI_ISL_1139210, EPI_ISL_1139211, EPI_ISL_1139212, EPI_ISL_1139213, EPI_ISL_1139214, EPI_ISL_1139215, EPI_ISL_1139216, EPI_ISL_1139217, EPI_ISL_1139218, EPI_ISL_1139219, EPI_ISL_1139220, EPI_ISL_1139221, EPI_ISL_1139222, EPI_ISL_1139223, EPI_ISL_1139224, EPI_ISL_1139225, EPI_ISL_1139226, EPI_ISL_1139227, EPI_ISL_1139228, EPI_ISL_1139229, EPI_ISL_1139230, EPI_ISL_1139231, EPI_ISL_1139232, EPI_ISL_1139233, EPI_ISL_1139234, EPI_ISL_1139235, EPI_ISL_1139236, EPI_ISL_1139237, EPI_ISL_1139238, EPI_ISL_1139239, EPI_ISL_1139240, EPI_ISL_1139241, EPI_ISL_1139242, EPI_ISL_1139243, EPI_ISL_1139244, EPI_ISL_1139245, EPI_ISL_1139246, EPI_ISL_1139247, EPI_ISL_1139248, EPI_ISL_1139249, EPI_ISL_1139250, EPI_ISL_1139251, EPI_ISL_1139252, EPI_ISL_1139253, EPI_ISL_1139254, EPI_ISL_1139255, EPI_ISL_1139256, EPI_ISL_1139257, EPI_ISL_1139258, EPI_ISL_1139259, EPI_ISL_1139260, EPI_ISL_1139261, EPI_ISL_1139262, EPI_ISL_1139263, EPI_ISL_1139264, EPI_ISL_1139265, EPI_ISL_1139266, EPI_ISL_1139267, EPI_ISL_1139268, EPI_ISL_1139269, EPI_ISL_1139270, EPI_ISL_1139271, EPI_ISL_1139272, EPI_ISL_1139273, EPI_ISL_1139274, EPI_ISL_1139275, EPI_ISL_1139276, EPI_ISL_1139277, EPI_ISL_1139278, EPI_ISL_1139279, EPI_ISL_1139280, EPI_ISL_1139281, EPI_ISL_1139282, EPI_ISL_1139283, EPI_ISL_1139284, EPI_ISL_1139285, EPI_ISL_1139286, EPI_ISL_1139287, EPI_ISL_1139288, EPI_ISL_1139289, EPI_ISL_1139290, EPI_ISL_1139291, EPI_ISL_1139292, EPI_ISL_1139293, EPI_ISL_1139294, EPI_ISL_1139295, EPI_ISL_1139296, EPI_ISL_1139297, EPI_ISL_1139298, EPI_ISL_1139299, EPI_ISL_1139300, EPI_ISL_1139301, EPI_ISL_1139302, EPI_ISL_1139303, EPI_ISL_1139304, EPI_ISL_1139305, EPI_ISL_1139306, EPI_ISL_1139307, EPI_ISL_1139308, EPI_ISL_1139309, EPI_ISL_1139310, EPI_ISL_1139311, EPI_ISL_1139312, EPI_ISL_1139313, EPI_ISL_1139314, EPI_ISL_1139315, EPI_ISL_1139316, EPI_ISL_1139317, EPI_ISL_1139318, EPI_ISL_1139319, EPI_ISL_1139320, EPI_ISL_1139321, EPI_ISL_1139322, EPI_ISL_1139323, EPI_ISL_1139324, EPI_ISL_1139325, EPI_ISL_1139326, EPI_ISL_1139327, EPI_ISL_1139328, EPI_ISL_1139329, EPI_ISL_1139330, EPI_ISL_1139331, EPI_ISL_1139332, EPI_ISL_1139333, EPI_ISL_1139334, EPI_ISL_1139335, EPI_ISL_1139336, EPI_ISL_1139337, EPI_ISL_1139338, EPI_ISL_1139339, EPI_ISL_1139340, EPI_ISL_1139341, EPI_ISL_1139342, EPI_ISL_1139343, EPI_ISL_1139344, EPI_ISL_1139345, EPI_ISL_1139346, EPI_ISL_1139347, EPI_ISL_1139348, EPI_ISL_1139349, EPI_ISL_1139350, EPI_ISL_1139351, EPI_ISL_1139352, EPI_ISL_1139353, EPI_ISL_1139354, EPI_ISL_1139355, EPI_ISL_1139356, EPI_ISL_1139357, EPI_ISL_1139358, EPI_ISL_1139359, EPI_ISL_1139360, EPI_ISL_1139361, EPI_ISL_1139362, EPI_ISL_1139363, EPI_ISL_1139364, EPI_ISL_1139365, EPI_ISL_1139366, EPI_ISL_1139367, EPI_ISL_1139368, EPI_ISL_1139369, EPI_ISL_1139370, EPI_ISL_1139371, EPI_ISL_1139372, EPI_ISL_1139373, EPI_ISL_1139374, EPI_ISL_1139375, EPI_ISL_1139376, EPI_ISL_1139377, EPI_ISL_1139378, EPI_ISL_1139379, EPI_ISL_1139380, EPI_ISL_1139381, EPI_ISL_1139382, EPI_ISL_1139383, EPI_ISL_1139384, EPI_ISL_1139385, EPI_ISL_1139386, EPI_ISL_1139387, EPI_ISL_1139388, EPI_ISL_1139389, EPI_ISL_1139390, EPI_ISL_1139391, EPI_ISL_1139392, EPI_ISL_1139393, EPI_ISL_1139394, EPI_ISL_1139395, EPI_ISL_1139396, EPI_ISL_1139397, EPI_ISL_1139398, EPI_ISL_1139399, EPI_ISL_1139400, EPI_ISL_1139401, EPI_ISL_1139402, EPI_ISL_1139403, EPI_ISL_1139404, EPI_ISL_1139405, EPI_ISL_1139406, EPI_ISL_1139407, EPI_ISL_1139408, EPI_ISL_1139409, EPI_ISL_1139410, EPI_ISL_1139411, EPI_ISL_1139412, EPI_ISL_1139413, EPI_ISL_1139414, EPI_ISL_1139415, EPI_ISL_1139416, EPI_ISL_1139417, EPI_ISL_1139418, EPI_ISL_1139419, EPI_ISL_1139420, EPI_ISL_1139421, EPI_ISL_1139422, EPI_ISL_1139423, EPI_ISL_1139424, EPI_ISL_1139425, EPI_ISL_1139426, EPI_ISL_1139427, EPI_ISL_1139428, EPI_ISL_1139429, EPI_ISL_1139430, EPI_ISL_1139431, EPI_ISL_1139480, EPI_ISL_1139481, EPI_ISL_1139482, EPI_ISL_1139483, EPI_ISL_1139484, EPI_ISL_1139485, EPI_ISL_1139486, EPI_ISL_1139487, EPI_ISL_1139488, EPI_ISL_1139489, EPI_ISL_1139490, EPI_ISL_1139491, EPI_ISL_1139492, EPI_ISL_1139493, EPI_ISL_1139494, EPI_ISL_1139495, EPI_ISL_1139496, EPI_ISL_1139497, EPI_ISL_1139498, EPI_ISL_1139499, EPI_ISL_1139500, EPI_ISL_1139501, EPI_ISL_1139502, EPI_ISL_1139503, EPI_ISL_1139504, EPI_ISL_1139505, EPI_ISL_1139506, EPI_ISL_1139507, EPI_ISL_1139508, EPI_ISL_1139509, EPI_ISL_1139510, EPI_ISL_1139511, EPI_ISL_1139512, EPI_ISL_1139513, EPI_ISL_1139514, EPI_ISL_1139515, EPI_ISL_1139516, EPI_ISL_1139517, EPI_ISL_1139518, EPI_ISL_1139519, EPI_ISL_1139520, EPI_ISL_1139521, EPI_ISL_1139522, EPI_ISL_1139523, EPI_ISL_1139524, EPI_ISL_1139525, EPI_ISL_1139526, EPI_ISL_1139527, EPI_ISL_1139528, EPI_ISL_1139529, EPI_ISL_1139530, EPI_ISL_1139531, EPI_ISL_1139532, EPI_ISL_1139533, EPI_ISL_1139534, EPI_ISL_1139535, EPI_ISL_1139536, EPI_ISL_1139537, EPI_ISL_1139538, EPI_ISL_1139539, EPI_ISL_1139540, EPI_ISL_1139541, EPI_ISL_1139542, EPI_ISL_1139543, EPI_ISL_1139544, EPI_ISL_1139545, EPI_ISL_1139546, EPI_ISL_1139547, EPI_ISL_1139548, EPI_ISL_1139549, EPI_ISL_1139550, EPI_ISL_1139551, EPI_ISL_1139552, EPI_ISL_1139553, EPI_ISL_1139554, EPI_ISL_1139555, EPI_ISL_1139556, EPI_ISL_1139557, EPI_ISL_1139558, EPI_ISL_1139559, EPI_ISL_1139560, EPI_ISL_1139561, EPI_ISL_1139562, EPI_ISL_1139563, EPI_ISL_1139564, EPI_ISL_1139565, EPI_ISL_1139566, EPI_ISL_1139567, EPI_ISL_1139568, EPI_ISL_1139569, EPI_ISL_1139570, EPI_ISL_1139571, EPI_ISL_1139572, EPI_ISL_1139573, EPI_ISL_1139574, EPI_ISL_1139575, EPI_ISL_1139576, EPI_ISL_1139577, EPI_ISL_1139578, EPI_ISL_1139579, EPI_ISL_1139580, EPI_ISL_1139581, EPI_ISL_1139582, EPI_ISL_1139583, EPI_ISL_1139584, EPI_ISL_1139585, EPI_ISL_1139586, EPI_ISL_1139587, EPI_ISL_1139588, EPI_ISL_1139589, EPI_ISL_1139590, EPI_ISL_1139591, EPI_ISL_1139592, EPI_ISL_1139593, EPI_ISL_1139594, EPI_ISL_1139595, EPI_ISL_1139596, EPI_ISL_1139597, EPI_ISL_1139598, EPI_ISL_1139599, EPI_ISL_1139600, EPI_ISL_1139601, EPI_ISL_1139602, EPI_ISL_1139603, EPI_ISL_1139604, EPI_ISL_1139605, EPI_ISL_1139606, EPI_ISL_1139607, EPI_ISL_1139608, EPI_ISL_1139609, EPI_ISL_1139610, EPI_ISL_1139611, EPI_ISL_1139612, EPI_ISL_1139613, EPI_ISL_1139614, EPI_ISL_1139615, EPI_ISL_1139616, EPI_ISL_1139617, EPI_ISL_1139618, EPI_ISL_1139619, EPI_ISL_1139620, EPI_ISL_1139621, EPI_ISL_1139622, EPI_ISL_1139623, EPI_ISL_1139624, EPI_ISL_1139625, EPI_ISL_1139626, EPI_ISL_1139627, EPI_ISL_1139628, EPI_ISL_1139629, EPI_ISL_1139630, EPI_ISL_1139631, EPI_ISL_1139632, EPI_ISL_1139633, EPI_ISL_1139634, EPI_ISL_1139635, EPI_ISL_1139636, EPI_ISL_1139637, EPI_ISL_1139638, EPI_ISL_1139639, EPI_ISL_1139640, EPI_ISL_1139641, EPI_ISL_1139642, EPI_ISL_1139643, EPI_ISL_1139644, EPI_ISL_1139645, EPI_ISL_1139646, EPI_ISL_1139647, EPI_ISL_1139648, EPI_ISL_1139649, EPI_ISL_1139650, EPI_ISL_1139651, EPI_ISL_1139652, EPI_ISL_1139653, EPI_ISL_1139654, EPI_ISL_1139655, EPI_ISL_1139656, EPI_ISL_1139657, EPI_ISL_1139658, EPI_ISL_1139659, EPI_ISL_1139660, EPI_ISL_1139661, EPI_ISL_1139662, EPI_ISL_1139663, EPI_ISL_1139664, EPI_ISL_1139665, EPI_ISL_1139666, EPI_ISL_1139667, EPI_ISL_1139668, EPI_ISL_1139669, EPI_ISL_1139670, EPI_ISL_1139671, EPI_ISL_1139672, EPI_ISL_1139673, EPI_ISL_1139674, EPI_ISL_1139675, EPI_ISL_1139676, EPI_ISL_1139677, EPI_ISL_1139678, EPI_ISL_1139679, EPI_ISL_1139680, EPI_ISL_1139681, EPI_ISL_1139682, EPI_ISL_1139683, EPI_ISL_1139684, EPI_ISL_1139685, EPI_ISL_1139686, EPI_ISL_1139687, EPI_ISL_1139688, EPI_ISL_1139689, EPI_ISL_1139690, EPI_ISL_1139691, EPI_ISL_1139692, EPI_ISL_1139693, EPI_ISL_1139694, EPI_ISL_1139695, EPI_ISL_1139696, EPI_ISL_1139697, EPI_ISL_1139698, EPI_ISL_1139699, EPI_ISL_1139700, EPI_ISL_1139701, EPI_ISL_1139702, EPI_ISL_1139703, EPI_ISL_1139704, EPI_ISL_1139705, EPI_ISL_1139706, EPI_ISL_1139707, EPI_ISL_1139708, EPI_ISL_1139709, EPI_ISL_1139710, EPI_ISL_1139711, EPI_ISL_1139712, EPI_ISL_1139713, EPI_ISL_1139714, EPI_ISL_1139715, EPI_ISL_1139716, EPI_ISL_1139717, EPI_ISL_1139718, EPI_ISL_1139719, EPI_ISL_1139720, EPI_ISL_1139721, EPI_ISL_1139722, EPI_ISL_1139723, EPI_ISL_1139724, EPI_ISL_1139725, EPI_ISL_1139726, EPI_ISL_1139727, EPI_ISL_1139728, EPI_ISL_1139729, EPI_ISL_1139730, EPI_ISL_1139731, EPI_ISL_1139732, EPI_ISL_1139733, EPI_ISL_1139734, EPI_ISL_1139735, EPI_ISL_1139736, EPI_ISL_1139737, EPI_ISL_1139738, EPI_ISL_1139739, EPI_ISL_1139740, EPI_ISL_1139741, EPI_ISL_1139742, EPI_ISL_1139743, EPI_ISL_1139744, EPI_ISL_1139745, EPI_ISL_1139746, EPI_ISL_1139747, EPI_ISL_1139748, EPI_ISL_1139749, EPI_ISL_1139750, EPI_ISL_1139751, EPI_ISL_1139752, EPI_ISL_1139753, EPI_ISL_1139754, EPI_ISL_1139755, EPI_ISL_1139756, EPI_ISL_1139757, EPI_ISL_1139758, EPI_ISL_1139759, EPI_ISL_1139760, EPI_ISL_1139761, EPI_ISL_1139762, EPI_ISL_1139763, EPI_ISL_1139764, EPI_ISL_1139765, EPI_ISL_1139766, EPI_ISL_1139767, EPI_ISL_1139768, EPI_ISL_1139769, EPI_ISL_1139770, EPI_ISL_1139771, EPI_ISL_1139772, EPI_ISL_1139773, EPI_ISL_1139774, EPI_ISL_1139775, EPI_ISL_1139776, EPI_ISL_1139777, EPI_ISL_1139778, EPI_ISL_1139779, EPI_ISL_1139780, EPI_ISL_1139781, EPI_ISL_1139782, EPI_ISL_1139783, EPI_ISL_1139784, EPI_ISL_1139785, EPI_ISL_1139786, EPI_ISL_1139787, EPI_ISL_1139788, EPI_ISL_1139789, EPI_ISL_1139790, EPI_ISL_1139791, EPI_ISL_1139792, EPI_ISL_1139793, EPI_ISL_1139794, EPI_ISL_1139795, EPI_ISL_1139796, EPI_ISL_1139797, EPI_ISL_1139798, EPI_ISL_1139799, EPI_ISL_1139800, EPI_ISL_1139801, EPI_ISL_1139802, EPI_ISL_1139803, EPI_ISL_1139804, EPI_ISL_1139805, EPI_ISL_1139806, EPI_ISL_1139807, EPI_ISL_1139808, EPI_ISL_1139809, EPI_ISL_1139810, EPI_ISL_1139811, EPI_ISL_1139812, EPI_ISL_1139813, EPI_ISL_1139814, EPI_ISL_1139815, EPI_ISL_1139816, EPI_ISL_1139817, EPI_ISL_1139818, EPI_ISL_1139819, EPI_ISL_1139820, EPI_ISL_1139821, EPI_ISL_1139822, EPI_ISL_1139823, EPI_ISL_1139824, EPI_ISL_1139825, EPI_ISL_1139826, EPI_ISL_1139827, EPI_ISL_1139828, EPI_ISL_1139829, EPI_ISL_1139830, EPI_ISL_1139831, EPI_ISL_1139832, EPI_ISL_1139833, EPI_ISL_1139834, EPI_ISL_1139835, EPI_ISL_1139836, EPI_ISL_1139837, EPI_ISL_1139838, EPI_ISL_1139839, EPI_ISL_1139840, EPI_ISL_1139841, EPI_ISL_1139842, EPI_ISL_1139843, EPI_ISL_1139844, EPI_ISL_1139845, EPI_ISL_1139846, EPI_ISL_1139847, EPI_ISL_1139848, EPI_ISL_1139849, EPI_ISL_1139850, EPI_ISL_1139851, EPI_ISL_1139852, EPI_ISL_1139853, EPI_ISL_1139854, EPI_ISL_1139855, EPI_ISL_1139856, EPI_ISL_1139857, EPI_ISL_1139858, EPI_ISL_1139859, EPI_ISL_1139860, EPI_ISL_1139861, EPI_ISL_1139862, EPI_ISL_1139863, EPI_ISL_1139864, EPI_ISL_1139865, EPI_ISL_1139866, EPI_ISL_1139867, EPI_ISL_1139868, EPI_ISL_1139869, EPI_ISL_1139870, EPI_ISL_1139871, EPI_ISL_1139872, EPI_ISL_1139873, EPI_ISL_1139874, EPI_ISL_1139875, EPI_ISL_1139876, EPI_ISL_1139877, EPI_ISL_1139878, EPI_ISL_1139879, EPI_ISL_1139880, EPI_ISL_1139881, EPI_ISL_1139882, EPI_ISL_1139883, EPI_ISL_1139884, EPI_ISL_1139885, EPI_ISL_1139886, EPI_ISL_1139887, EPI_ISL_1139888, EPI_ISL_1139889, EPI_ISL_1139890, EPI_ISL_1139891, EPI_ISL_1139892, EPI_ISL_1139893, EPI_ISL_1139894, EPI_ISL_1139895, EPI_ISL_1139896, EPI_ISL_1139897, EPI_ISL_1139898, EPI_ISL_1139899, EPI_ISL_1139900, EPI_ISL_1139901, EPI_ISL_1139902, EPI_ISL_1139903, EPI_ISL_1139904, EPI_ISL_1139905, EPI_ISL_1139906, EPI_ISL_1139907, EPI_ISL_1139908, EPI_ISL_1139909, EPI_ISL_1139910, EPI_ISL_1139911, EPI_ISL_1139912, EPI_ISL_1139913, EPI_ISL_1139914, EPI_ISL_1139915, EPI_ISL_1139916, EPI_ISL_1139917, EPI_ISL_1139918, EPI_ISL_1139919, EPI_ISL_1139920, EPI_ISL_1139921, EPI_ISL_1139922, EPI_ISL_1139923, EPI_ISL_1139924, EPI_ISL_1139925, EPI_ISL_1139926, EPI_ISL_1139927, EPI_ISL_1139928, EPI_ISL_1139929, EPI_ISL_1139930, EPI_ISL_1139931, EPI_ISL_1139932, EPI_ISL_1139933, EPI_ISL_1139934, EPI_ISL_1139935, EPI_ISL_1139936, EPI_ISL_1139937, EPI_ISL_1139938, EPI_ISL_1139939, EPI_ISL_1139940, EPI_ISL_1139941, EPI_ISL_1139942, EPI_ISL_1139943, EPI_ISL_1139944, EPI_ISL_1139945, EPI_ISL_1139946, EPI_ISL_1139947, EPI_ISL_1139948, EPI_ISL_1139949, EPI_ISL_1139950, EPI_ISL_1139951, EPI_ISL_1139952, EPI_ISL_1139953, EPI_ISL_1139954, EPI_ISL_1139955, EPI_ISL_1139956, EPI_ISL_1139957, EPI_ISL_1139958, EPI_ISL_1139959, EPI_ISL_1139960, EPI_ISL_1139961, EPI_ISL_1139962, EPI_ISL_1139963, EPI_ISL_1139964, EPI_ISL_1139965, EPI_ISL_1139966, EPI_ISL_1139967, EPI_ISL_1139968, EPI_ISL_1139969, EPI_ISL_1139970, EPI_ISL_1139971, EPI_ISL_1139972, EPI_ISL_1139973, EPI_ISL_1139974, EPI_ISL_1139975, EPI_ISL_1139976, EPI_ISL_1139977, EPI_ISL_1139978, EPI_ISL_1139979, EPI_ISL_1139980, EPI_ISL_1139981, EPI_ISL_1139982, EPI_ISL_1139983, EPI_ISL_1139984, EPI_ISL_1139985, EPI_ISL_1139986, EPI_ISL_1139987, EPI_ISL_1139988, EPI_ISL_1139989, EPI_ISL_1139990, EPI_ISL_1139991, EPI_ISL_1139992, EPI_ISL_1139993, EPI_ISL_1139994, EPI_ISL_1139995, EPI_ISL_1139996, EPI_ISL_1139997, EPI_ISL_1139998, EPI_ISL_1139999, EPI_ISL_1140000, EPI_ISL_1140001, EPI_ISL_1140002, EPI_ISL_1140003, EPI_ISL_1140004, EPI_ISL_1140005, EPI_ISL_1140006, EPI_ISL_1140007, EPI_ISL_1140008, EPI_ISL_1140009, EPI_ISL_1140010, EPI_ISL_1140011, EPI_ISL_1140012, EPI_ISL_1140013, EPI_ISL_1140014, EPI_ISL_1140015, EPI_ISL_1140016, EPI_ISL_1140017, EPI_ISL_1140018, EPI_ISL_1140019, EPI_ISL_1140020, EPI_ISL_1140021, EPI_ISL_1140022, EPI_ISL_1140023, EPI_ISL_1140024, EPI_ISL_1140025, EPI_ISL_1140026, EPI_ISL_1140027, EPI_ISL_1140028, EPI_ISL_1140029, EPI_ISL_1140030, EPI_ISL_1140031, EPI_ISL_1140032, EPI_ISL_1140033, EPI_ISL_1140034, EPI_ISL_1140035, EPI_ISL_1140036, EPI_ISL_1140037, EPI_ISL_1140038, EPI_ISL_1140039, EPI_ISL_1140040, EPI_ISL_1140041, EPI_ISL_1140042, EPI_ISL_1140043, EPI_ISL_1140044, EPI_ISL_1140045, EPI_ISL_1140046 | | | |
| see above | Quest Diagnostics Incorporated | Respiratory Viruses Branch, Division of Viral Diseases, Centers for Disease Control and Prevention | Peter W. Cook, Dakota Howard, Dhwani Batra, Ben L. Rambo-Martin, S. H. Rosenthal, A. Gerasimova, R. M. Kagan, B. Anderson, M. Hua, Y. Liu, L.E. Bernstein, K.E. Livingston, A. Perez, I. A. Shlyakhter, R. V. Rolando, R. Owen, P. Tanpaiboon, F. Lacbawan, Clinton R. Paden, Suxiang Tong, Duncan MacCannell |
| EPI_ISL_1140055, EPI_ISL_1140056, EPI_ISL_1140057, EPI_ISL_1140058, EPI_ISL_1140059, EPI_ISL_1140062, EPI_ISL_1140063, EPI_ISL_1140064, EPI_ISL_1140065, EPI_ISL_1140066, EPI_ISL_1140067, EPI_ISL_1140068, EPI_ISL_1140069, EPI_ISL_1140071, EPI_ISL_1140072, EPI_ISL_1140073, EPI_ISL_1140074, EPI_ISL_1140075, EPI_ISL_1140076, EPI_ISL_1140077, EPI_ISL_1140078, EPI_ISL_1140079, EPI_ISL_1140080, EPI_ISL_1140081, EPI_ISL_1140082, EPI_ISL_1140083, EPI_ISL_1140084, EPI_ISL_1140085, EPI_ISL_1140086, EPI_ISL_1140087, EPI_ISL_1140088, EPI_ISL_1140089, EPI_ISL_1140090, EPI_ISL_1140091, EPI_ISL_1140092, EPI_ISL_1140093 | | | |
| see above | Wyoming Public Health Laboratory | Wyoming Public Health Laboratory | Jim Mildenberger, Wanda Manley, Noah Hull, Taylor Fearing, Lynette Gumbleton, Channing Weber, Ashley Norberg, Chayse Rowley, Marley Goetz, Brian Dominguez, Elliot Thomasson, Cari Sloma, and Rob Christensen |
| EPI_ISL_1157002, EPI_ISL_1157004, EPI_ISL_1157006, EPI_ISL_1157008, EPI_ISL_1157009, EPI_ISL_1157011, EPI_ISL_1157013, EPI_ISL_1157015, EPI_ISL_1157017, EPI_ISL_1157018, EPI_ISL_1157020, EPI_ISL_1157022, EPI_ISL_1157024, EPI_ISL_1157025, EPI_ISL_1157027, EPI_ISL_1157029, EPI_ISL_1157031, EPI_ISL_1157033, EPI_ISL_1157035, EPI_ISL_1157036, EPI_ISL_1157038, EPI_ISL_1157040, EPI_ISL_1157042, EPI_ISL_1157043, EPI_ISL_1157045, EPI_ISL_1157047 | | | |
| see above | Virginia Division of Consolidated Laboratory Services | Virginia Division of Consolidated Laboratory Services | Virginia DCLS |
| EPI_ISL_1157399, EPI_ISL_1157401, EPI_ISL_1157402, EPI_ISL_1157404, EPI_ISL_1157406, EPI_ISL_1157408, EPI_ISL_1157413, EPI_ISL_1157415, EPI_ISL_1157416, EPI_ISL_1157418, EPI_ISL_1157420, EPI_ISL_1157422, EPI_ISL_1157423, EPI_ISL_1157425, EPI_ISL_1157427, EPI_ISL_1157429, EPI_ISL_1157430, EPI_ISL_1157432, EPI_ISL_1157434, EPI_ISL_1157436, EPI_ISL_1157437, EPI_ISL_1157439, EPI_ISL_1157441, EPI_ISL_1157443, EPI_ISL_1157444, EPI_ISL_1157448, EPI_ISL_1157450, EPI_ISL_1157452, EPI_ISL_1157454, EPI_ISL_1157455, EPI_ISL_1157457, EPI_ISL_1157459, EPI_ISL_1157461, EPI_ISL_1157462, EPI_ISL_1157464, EPI_ISL_1157466, EPI_ISL_1157470, EPI_ISL_1157472, EPI_ISL_1157474, EPI_ISL_1157476, EPI_ISL_1157477, EPI_ISL_1157479, EPI_ISL_1157481, EPI_ISL_1157482, EPI_ISL_1157484, EPI_ISL_1157486, EPI_ISL_1157487, EPI_ISL_1157489, EPI_ISL_1157491, EPI_ISL_1157493, EPI_ISL_1157494, EPI_ISL_1157496, EPI_ISL_1157498, EPI_ISL_1157500, EPI_ISL_1157502, EPI_ISL_1157503, EPI_ISL_1157505, EPI_ISL_1157507, EPI_ISL_1157509, EPI_ISL_1157510, EPI_ISL_1157515, EPI_ISL_1157517, EPI_ISL_1157519, EPI_ISL_1157520, EPI_ISL_1157522, EPI_ISL_1157524, EPI_ISL_1157525, EPI_ISL_1157527, EPI_ISL_1157529, EPI_ISL_1157530, EPI_ISL_1157532, EPI_ISL_1157534, EPI_ISL_1157536, EPI_ISL_1157537, EPI_ISL_1157539, EPI_ISL_1157541, EPI_ISL_1157543, EPI_ISL_1157544, EPI_ISL_1157546, EPI_ISL_1157549, EPI_ISL_1157551, EPI_ISL_1157553, EPI_ISL_1157555, EPI_ISL_1157556, EPI_ISL_1157558, EPI_ISL_1157560, EPI_ISL_1157562, EPI_ISL_1157563, EPI_ISL_1157565, EPI_ISL_1157567, EPI_ISL_1157569, EPI_ISL_1157570, EPI_ISL_1157572, EPI_ISL_1157574, EPI_ISL_1157576, EPI_ISL_1157577, EPI_ISL_1157579, EPI_ISL_1157581, EPI_ISL_1157583, EPI_ISL_1157585 | | | |
| see above | Northwestern Memorial Hospital | Northwestern University - Ozer Lab | Ramon Lorenzo-Redondo, Lacy M. Simons, Taylor J. Dean, Chad J. Achenbach, Lawrence J. Jennings, Chao Qi, Michael G. Ison, Judd F. Hultquist, Egon A. Ozer |
| EPI_ISL_1158033, EPI_ISL_1158034, EPI_ISL_1158035, EPI_ISL_1158036, EPI_ISL_1158038, EPI_ISL_1158039, EPI_ISL_1158040, EPI_ISL_1158041, EPI_ISL_1158042, EPI_ISL_1158043, EPI_ISL_1158044, EPI_ISL_1158045, EPI_ISL_1158046, EPI_ISL_1158047, EPI_ISL_1158049, EPI_ISL_1158050, EPI_ISL_1158051, EPI_ISL_1158052, EPI_ISL_1158053, EPI_ISL_1158054, EPI_ISL_1158056, EPI_ISL_1158057, EPI_ISL_1158058, EPI_ISL_1158059 | | | |
| see above | Michigan Department of Health and Human Services, Bureau of Laboratories | Michigan Department of Health and Human Services, Bureau of Laboratories | Blankenship HM, Riner D, Soehnlen MK |
| EPI_ISL_1158060, EPI_ISL_1158061, EPI_ISL_1158062 | LABDIS-UAGro | Instituto Nacional de Medicina Genomica | Hidalgo-Miranda A, Mendoza-Vargas A, Reyes-Grajeda JP, Cedro-Tanda A, Alcaraz N, Gonzalez-Barrera D, Rangel-DeLeon D, Ramirez-Vega O, Sifuentes-Rojas C, Cisneros-Villanueva M, Herrera-Montalvo LA, Berenice Illades-Aguiar, Oscar del Moral-Hernandez, Adolfo Roman-Roman, Marco Antonio-Leyva, Daniel Hernandez-Sotelo, Hugo A. Rodriguez-Ruiz, Fredy O Beltran-Anaya, Roberto Dircio-Maldonado, Julio C. Azucar-Hesiquio, Azucena D. Alvarez-Diaz, Ciresthel Bello-Rios, Gladys W. Valente-Nio, Jaqueline Loaeza-Loaeza, Madian Valadez-Quinones, Luis I. Basilio-Leyva. |
| EPI_ISL_1158063, EPI_ISL_1158064, EPI_ISL_1158066, EPI_ISL_1158067, EPI_ISL_1158068, EPI_ISL_1158069, EPI_ISL_1158070, EPI_ISL_1158071, EPI_ISL_1158072, EPI_ISL_1158073, EPI_ISL_1158074, EPI_ISL_1158075, EPI_ISL_1158076, EPI_ISL_1158078, EPI_ISL_1158079, EPI_ISL_1158080, EPI_ISL_1158083, EPI_ISL_1158084, EPI_ISL_1158085, EPI_ISL_1158086, EPI_ISL_1158087, EPI_ISL_1158089, EPI_ISL_1158090, EPI_ISL_1158091, EPI_ISL_1158092, EPI_ISL_1158093, EPI_ISL_1158094, EPI_ISL_1158095, EPI_ISL_1158097, EPI_ISL_1158098, EPI_ISL_1158100, EPI_ISL_1158101, EPI_ISL_1158102, EPI_ISL_1158103, EPI_ISL_1158104, EPI_ISL_1158105, EPI_ISL_1158106, EPI_ISL_1158107, EPI_ISL_1158108, EPI_ISL_1158109, EPI_ISL_1158110, EPI_ISL_1158111, EPI_ISL_1158113, EPI_ISL_1158114, EPI_ISL_1158115, EPI_ISL_1158116, EPI_ISL_1158117, EPI_ISL_1158118, EPI_ISL_1158120, EPI_ISL_1158121, EPI_ISL_1158122, EPI_ISL_1158123, EPI_ISL_1158124, EPI_ISL_1158125, EPI_ISL_1158126, EPI_ISL_1158127, EPI_ISL_1158128, EPI_ISL_1158129, EPI_ISL_1158131, EPI_ISL_1158132, EPI_ISL_1158133, EPI_ISL_1158134, EPI_ISL_1158135, EPI_ISL_1158136, EPI_ISL_1158137, EPI_ISL_1158138, EPI_ISL_1158139, EPI_ISL_1158140, EPI_ISL_1158141, EPI_ISL_1158142, EPI_ISL_1158143, EPI_ISL_1158144, EPI_ISL_1158145, EPI_ISL_1158146, EPI_ISL_1158147, EPI_ISL_1158148, EPI_ISL_1158149, EPI_ISL_1158167, EPI_ISL_1158168, EPI_ISL_1158170, EPI_ISL_1158171, EPI_ISL_1158172, EPI_ISL_1158173, EPI_ISL_1158174, EPI_ISL_1158175, EPI_ISL_1158176, EPI_ISL_1158177, EPI_ISL_1158178, EPI_ISL_1158179, EPI_ISL_1158180, EPI_ISL_1158181, EPI_ISL_1158182, EPI_ISL_1158183, EPI_ISL_1158184, EPI_ISL_1158185, EPI_ISL_1158186, EPI_ISL_1158187, EPI_ISL_1158188, EPI_ISL_1158189, EPI_ISL_1158190, EPI_ISL_1158191, EPI_ISL_1158192, EPI_ISL_1158193, EPI_ISL_1158194, EPI_ISL_1158195, EPI_ISL_1158196, EPI_ISL_1158197, EPI_ISL_1158198, EPI_ISL_1158200, EPI_ISL_1158201, EPI_ISL_1158202, EPI_ISL_1158203, EPI_ISL_1158205, EPI_ISL_1158207, EPI_ISL_1158208, EPI_ISL_1158209, EPI_ISL_1158210, EPI_ISL_1158211, EPI_ISL_1158212, EPI_ISL_1158213, EPI_ISL_1158214, EPI_ISL_1158215, EPI_ISL_1158216, EPI_ISL_1158217, EPI_ISL_1158218, EPI_ISL_1158219, EPI_ISL_1158220, EPI_ISL_1158221, EPI_ISL_1158222, EPI_ISL_1158223, EPI_ISL_1158224, EPI_ISL_1158225, EPI_ISL_1158226, EPI_ISL_1158227, EPI_ISL_1158277, EPI_ISL_1158278, EPI_ISL_1158279, EPI_ISL_1158280, EPI_ISL_1158281, EPI_ISL_1158282, EPI_ISL_1158283, EPI_ISL_1158284, EPI_ISL_1158285, EPI_ISL_1158286, EPI_ISL_1158287, EPI_ISL_1158288, EPI_ISL_1158289, EPI_ISL_1158290, EPI_ISL_1158291, EPI_ISL_1158292, EPI_ISL_1158293, EPI_ISL_1158294, EPI_ISL_1158295, EPI_ISL_1158296, EPI_ISL_1158297, EPI_ISL_1158298, EPI_ISL_1158299, EPI_ISL_1158300, EPI_ISL_1158301, EPI_ISL_1158302, EPI_ISL_1158303, EPI_ISL_1158304, EPI_ISL_1158305, EPI_ISL_1158306, EPI_ISL_1158307, EPI_ISL_1158308, EPI_ISL_1158309, EPI_ISL_1158310, EPI_ISL_1158311, EPI_ISL_1158312, EPI_ISL_1158313, EPI_ISL_1158314, EPI_ISL_1158316, EPI_ISL_1158317, EPI_ISL_1158318, EPI_ISL_1158320, EPI_ISL_1158321, EPI_ISL_1158323, EPI_ISL_1158324, EPI_ISL_1158326, EPI_ISL_1158327, EPI_ISL_1158328, EPI_ISL_1158329, EPI_ISL_1158330, EPI_ISL_1158331, EPI_ISL_1158332, EPI_ISL_1158333, EPI_ISL_1158334, EPI_ISL_1158337, EPI_ISL_1158338, EPI_ISL_1158340, EPI_ISL_1158341, EPI_ISL_1158343, EPI_ISL_1158345, EPI_ISL_1158346, EPI_ISL_1158348, EPI_ISL_1158349, EPI_ISL_1158350, EPI_ISL_1158352, EPI_ISL_1158353, EPI_ISL_1158354, EPI_ISL_1158357, EPI_ISL_1158358, EPI_ISL_1158359, EPI_ISL_1158360, EPI_ISL_1158361, EPI_ISL_1158362, EPI_ISL_1158363, EPI_ISL_1158364, EPI_ISL_1158365, EPI_ISL_1158366, EPI_ISL_1158367, EPI_ISL_1158368, EPI_ISL_1158369, EPI_ISL_1158370, EPI_ISL_1158371, EPI_ISL_1158372, EPI_ISL_1158373, EPI_ISL_1158374, EPI_ISL_1158375, EPI_ISL_1158376, EPI_ISL_1158377, EPI_ISL_1158378, EPI_ISL_1158379, EPI_ISL_1158380, EPI_ISL_1158381, EPI_ISL_1158382, EPI_ISL_1158383, EPI_ISL_1158384, EPI_ISL_1158385, EPI_ISL_1158387, EPI_ISL_1158388, EPI_ISL_1158389, EPI_ISL_1158390, EPI_ISL_1158391, EPI_ISL_1158392, EPI_ISL_1158393, EPI_ISL_1158394, EPI_ISL_1158395, EPI_ISL_1158396, EPI_ISL_1158397, EPI_ISL_1158398, EPI_ISL_1158399, EPI_ISL_1158400, EPI_ISL_1158403, EPI_ISL_1158404, EPI_ISL_1158405, EPI_ISL_1158406, EPI_ISL_1158407, EPI_ISL_1158408, EPI_ISL_1158409, EPI_ISL_1158410, EPI_ISL_1158411, EPI_ISL_1158412, EPI_ISL_1158413, EPI_ISL_1158414, EPI_ISL_1158415, EPI_ISL_1158416, EPI_ISL_1158417, EPI_ISL_1158418, EPI_ISL_1158419, EPI_ISL_1158420, EPI_ISL_1158421, EPI_ISL_1158422, EPI_ISL_1158423, EPI_ISL_1158424, EPI_ISL_1158425, EPI_ISL_1158426, EPI_ISL_1158427, EPI_ISL_1158428, EPI_ISL_1158429, EPI_ISL_1158430, EPI_ISL_1158431, EPI_ISL_1158432, EPI_ISL_1158433, EPI_ISL_1158434, EPI_ISL_1158435, EPI_ISL_1158436, EPI_ISL_1158437, EPI_ISL_1158438, EPI_ISL_1158439, EPI_ISL_1158440, EPI_ISL_1158441, EPI_ISL_1158442, EPI_ISL_1158443, EPI_ISL_1158444, EPI_ISL_1158446, EPI_ISL_1158447, EPI_ISL_1158448, EPI_ISL_1158449, EPI_ISL_1158450, EPI_ISL_1158451, EPI_ISL_1158452, EPI_ISL_1158453, EPI_ISL_1158454, EPI_ISL_1158455, EPI_ISL_1158456, EPI_ISL_1158457, EPI_ISL_1158458, EPI_ISL_1158459, EPI_ISL_1158460, EPI_ISL_1158461, EPI_ISL_1158462, EPI_ISL_1158463, EPI_ISL_1158464, EPI_ISL_1158465, EPI_ISL_1158466, EPI_ISL_1158467, EPI_ISL_1158468, EPI_ISL_1158469, EPI_ISL_1158470, EPI_ISL_1158471, EPI_ISL_1158472, EPI_ISL_1158473, EPI_ISL_1158474, EPI_ISL_1158475, EPI_ISL_1158476, EPI_ISL_1158477, EPI_ISL_1158478, EPI_ISL_1158479, EPI_ISL_1158480, EPI_ISL_1158481, EPI_ISL_1158482, EPI_ISL_1158483, EPI_ISL_1158484, EPI_ISL_1158485, EPI_ISL_1158486, EPI_ISL_1158487, EPI_ISL_1158488, EPI_ISL_1158489, EPI_ISL_1158490, EPI_ISL_1158491, EPI_ISL_1158492, EPI_ISL_1158493, EPI_ISL_1158494, EPI_ISL_1158495, EPI_ISL_1158496, EPI_ISL_1158497, EPI_ISL_1158498, EPI_ISL_1158499, EPI_ISL_1158500, EPI_ISL_1158502, EPI_ISL_1158503, EPI_ISL_1158504, EPI_ISL_1158505, EPI_ISL_1158506, EPI_ISL_1158507, EPI_ISL_1158508, EPI_ISL_1158509, EPI_ISL_1158510, EPI_ISL_1158511, EPI_ISL_1158513, EPI_ISL_1158514, EPI_ISL_1158515, EPI_ISL_1158516, EPI_ISL_1158517, EPI_ISL_1158518, EPI_ISL_1158519 | | | |
| see above | Michigan Department of Health and Human Services, Bureau of Laboratories | Michigan Department of Health and Human Services, Bureau of Laboratories | Blankenship HM, Riner D, Soehnlen MK |
| EPI_ISL_1158929, EPI_ISL_1158930, EPI_ISL_1158931, EPI_ISL_1158932, EPI_ISL_1158933, EPI_ISL_1158934, EPI_ISL_1158935, EPI_ISL_1158936, EPI_ISL_1158937, EPI_ISL_1158938, EPI_ISL_1158939, EPI_ISL_1158940, EPI_ISL_1158941, EPI_ISL_1158942, EPI_ISL_1158943, EPI_ISL_1158944, EPI_ISL_1158945, EPI_ISL_1158946, EPI_ISL_1158947, EPI_ISL_1158948, EPI_ISL_1158949, EPI_ISL_1158950, EPI_ISL_1158951, EPI_ISL_1158952, EPI_ISL_1158953, EPI_ISL_1158954, EPI_ISL_1158955, EPI_ISL_1158956, EPI_ISL_1158957, EPI_ISL_1158958, EPI_ISL_1158959, EPI_ISL_1158960, EPI_ISL_1158961, EPI_ISL_1158962, EPI_ISL_1158963, EPI_ISL_1158964, EPI_ISL_1158965, EPI_ISL_1158966, EPI_ISL_1158967, EPI_ISL_1158968, EPI_ISL_1158969, EPI_ISL_1158970, EPI_ISL_1158971, EPI_ISL_1158972, EPI_ISL_1158973, EPI_ISL_1158974, EPI_ISL_1158975, EPI_ISL_1158976, EPI_ISL_1158977, EPI_ISL_1158978, EPI_ISL_1158979, EPI_ISL_1158980, EPI_ISL_1158981, EPI_ISL_1158982, EPI_ISL_1158983, EPI_ISL_1158984, EPI_ISL_1158985, EPI_ISL_1158986, EPI_ISL_1158987, EPI_ISL_1158988, EPI_ISL_1158989, EPI_ISL_1158990, EPI_ISL_1158991, EPI_ISL_1158992, EPI_ISL_1158993, EPI_ISL_1158994, EPI_ISL_1158995, EPI_ISL_1158996, EPI_ISL_1158997, EPI_ISL_1158998, EPI_ISL_1158999, EPI_ISL_1159000, EPI_ISL_1159001, EPI_ISL_1159002, EPI_ISL_1159003, EPI_ISL_1159004, EPI_ISL_1159005, EPI_ISL_1159006, EPI_ISL_1159007, EPI_ISL_1159008, EPI_ISL_1159009, EPI_ISL_1159010, EPI_ISL_1159011, EPI_ISL_1159012, EPI_ISL_1159013, EPI_ISL_1159014, EPI_ISL_1159015, EPI_ISL_1159016, EPI_ISL_1159017, EPI_ISL_1159018, EPI_ISL_1159019, EPI_ISL_1159020, EPI_ISL_1159021, EPI_ISL_1159022, EPI_ISL_1159023, EPI_ISL_1159024, EPI_ISL_1159025, EPI_ISL_1159026, EPI_ISL_1159027, EPI_ISL_1159028, EPI_ISL_1159029, EPI_ISL_1159030, EPI_ISL_1159031, EPI_ISL_1159032, EPI_ISL_1159033, EPI_ISL_1159034, EPI_ISL_1159035, EPI_ISL_1159036, EPI_ISL_1159037, EPI_ISL_1159038, EPI_ISL_1159039, EPI_ISL_1159040, EPI_ISL_1159041, EPI_ISL_1159042, EPI_ISL_1159043, EPI_ISL_1159044, EPI_ISL_1159045, EPI_ISL_1159046, EPI_ISL_1159047, EPI_ISL_1159048, EPI_ISL_1159049, EPI_ISL_1159050, EPI_ISL_1159051, EPI_ISL_1159052, EPI_ISL_1159053, EPI_ISL_1159054, EPI_ISL_1159055, EPI_ISL_1159056, EPI_ISL_1159057, EPI_ISL_1159058, EPI_ISL_1159059, EPI_ISL_1159060, EPI_ISL_1159061, EPI_ISL_1159062, EPI_ISL_1159063, EPI_ISL_1159064, EPI_ISL_1159065, EPI_ISL_1159066, EPI_ISL_1159067, EPI_ISL_1159068 | | | |
| see above | Quest Diagnostics Incorporated | Respiratory Viruses Branch, Division of Viral Diseases, Centers for Disease Control and Prevention | Peter W. Cook, Dakota Howard, Dhwani Batra, Ben L. Rambo-Martin, S. H. Rosenthal, A. Gerasimova, R. M. Kagan, B. Anderson, M. Hua, Y. Liu, L.E. Bernstein, K.E. Livingston, A. Perez, I. A. Shlyakhter, R. V. Rolando, R. Owen, P. Tanpaiboon, F. Lacbawan, Clinton R. Paden, Suxiang Tong, Duncan MacCannell |
| EPI_ISL_1159108, EPI_ISL_1159109, EPI_ISL_1159110, EPI_ISL_1159111, EPI_ISL_1159112, EPI_ISL_1159113, EPI_ISL_1159114, EPI_ISL_1159115, EPI_ISL_1159116, EPI_ISL_1159117, EPI_ISL_1159118, EPI_ISL_1159119, EPI_ISL_1159120, EPI_ISL_1159121, EPI_ISL_1159122, EPI_ISL_1159123, EPI_ISL_1159124 | | | |
| see above | DC Public Health Lab/ Dept. of Forensic Sciences | DC Public Health Lab/ Dept. of Forensic Sciences | Scott Nguyen, Janis Doss, Elizabeth Zelaya, Sarah Scott, Connie Maza, Monica Mann, Brittany Hamilton, David Payne, Jocelyn Hauser |
| EPI_ISL_1159230 | Synergy Laboratories | Synergy Laboratories | Megan Cornwell |
| EPI_ISL_1159233, EPI_ISL_1159234, EPI_ISL_1159235, EPI_ISL_1159236, EPI_ISL_1159237, EPI_ISL_1159238, EPI_ISL_1159239, EPI_ISL_1159240, EPI_ISL_1159241, EPI_ISL_1159242, EPI_ISL_1159243, EPI_ISL_1159244, EPI_ISL_1159245, EPI_ISL_1159246, EPI_ISL_1159247, EPI_ISL_1159248, EPI_ISL_1159249, EPI_ISL_1159250, EPI_ISL_1159251, EPI_ISL_1159252, EPI_ISL_1159253, EPI_ISL_1159254, EPI_ISL_1159255, EPI_ISL_1159256, EPI_ISL_1159257, EPI_ISL_1159258, EPI_ISL_1159259, EPI_ISL_1159260, EPI_ISL_1159261, EPI_ISL_1159262, EPI_ISL_1159263, EPI_ISL_1159264, EPI_ISL_1159265, EPI_ISL_1159266, EPI_ISL_1159267, EPI_ISL_1159268, EPI_ISL_1159269, EPI_ISL_1159270, EPI_ISL_1159271, EPI_ISL_1159272, EPI_ISL_1159273, EPI_ISL_1159274, EPI_ISL_1159275, EPI_ISL_1159276, EPI_ISL_1159277, EPI_ISL_1159278, EPI_ISL_1159279, EPI_ISL_1159280, EPI_ISL_1159281, EPI_ISL_1159282, EPI_ISL_1159283, EPI_ISL_1159284, EPI_ISL_1159285, EPI_ISL_1159286, EPI_ISL_1159287, EPI_ISL_1159288, EPI_ISL_1159289, EPI_ISL_1159290, EPI_ISL_1159291, EPI_ISL_1159292, EPI_ISL_1159293, EPI_ISL_1159294, EPI_ISL_1159295, EPI_ISL_1159296, EPI_ISL_1159297, EPI_ISL_1159298, EPI_ISL_1159299, EPI_ISL_1159300, EPI_ISL_1159301, EPI_ISL_1159302, EPI_ISL_1159303, EPI_ISL_1159304, EPI_ISL_1159305, EPI_ISL_1159306, EPI_ISL_1159307, EPI_ISL_1159308, EPI_ISL_1159309, EPI_ISL_1159310, EPI_ISL_1159311, EPI_ISL_1159312, EPI_ISL_1159313, EPI_ISL_1159314, EPI_ISL_1159315, EPI_ISL_1159316, EPI_ISL_1159317, EPI_ISL_1159318, EPI_ISL_1159319, EPI_ISL_1159320, EPI_ISL_1159321, EPI_ISL_1159322, EPI_ISL_1159323, EPI_ISL_1159324, EPI_ISL_1159325, EPI_ISL_1159326, EPI_ISL_1159327, EPI_ISL_1159328, EPI_ISL_1159329, EPI_ISL_1159330, EPI_ISL_1159331, EPI_ISL_1159332, EPI_ISL_1159333, EPI_ISL_1159334, EPI_ISL_1159335, EPI_ISL_1159336, EPI_ISL_1159337, EPI_ISL_1159338, EPI_ISL_1159339, EPI_ISL_1159340, EPI_ISL_1159341, EPI_ISL_1159342, EPI_ISL_1159343, EPI_ISL_1159344, EPI_ISL_1159345 | | | |

see above

Quest Diagnostics Incorporated

Centers for Disease Control and Prevention Division of Viral Diseases, Pathogen Discovery

Peter W. Cook, Dakota Howard, Dhwani Batra, Ben L. Rambo-Martin, S. H. Rosenthal, A. Gerasimova, R. M. Kagan, B. Anderson, M. Hua, Y. Liu, L.E. Bernstein, K.E. Livingston, A. Perez, I. A. Shlyakhter, R. V. Rolando, R. Owen, P. Tanpaiboon, F. Lacbawan, Clinton R. Paden, Suxiang Tong, Duncan MacCannell

EPI_ISL_1159392, EPI_ISL_1159393, EPI_ISL_1159394, EPI_ISL_1159395, EPI_ISL_1159396, EPI_ISL_1159397, EPI_ISL_1159399

University of Wisconsin-Madison AIDS Vaccine Research Laboratories

University of Wisconsin-Madison AIDS Vaccine Research Laboratories

Gage Moreno, Katarina Braun, et al. AIDS Vaccine Research Laboratories

EPI_ISL_1159400, EPI_ISL_1159401, EPI_ISL_1159402, EPI_ISL_1159403, EPI_ISL_1159404, EPI_ISL_1159405, EPI_ISL_1159406, EPI_ISL_1159407, EPI_ISL_1159408, EPI_ISL_1159409, EPI_ISL_1159410, EPI_ISL_1159411, EPI_ISL_1159412, EPI_ISL_1159413, EPI_ISL_1159414, EPI_ISL_1159415, EPI_ISL_1159416, EPI_ISL_1159417, EPI_ISL_1159418, EPI_ISL_1159419, EPI_ISL_1159420, EPI_ISL_1159421,
EPI_ISL_1159422, EPI_ISL_1159423, EPI_ISL_1159424, EPI_ISL_1159425, EPI_ISL_1159426, EPI_ISL_1159427, EPI_ISL_1159428, EPI_ISL_1159429, EPI_ISL_1159430, EPI_ISL_1159431, EPI_ISL_1159432, EPI_ISL_1159433, EPI_ISL_1159434, EPI_ISL_1159435, EPI_ISL_1159436, EPI_ISL_1159437, EPI_ISL_1159438, EPI_ISL_1159439, EPI_ISL_1159440, EPI_ISL_1159441, EPI_ISL_1159442, EPI_ISL_1159443,
EPI_ISL_1159444, EPI_ISL_1159445, EPI_ISL_1159446, EPI_ISL_1159447, EPI_ISL_1159448, EPI_ISL_1159449, EPI_ISL_1159450, EPI_ISL_1159451, EPI_ISL_1159452, EPI_ISL_1159453, EPI_ISL_1159454, EPI_ISL_1159455, EPI_ISL_1159456, EPI_ISL_1159457, EPI_ISL_1159458, EPI_ISL_1159459, EPI_ISL_1159460, EPI_ISL_1159461, EPI_ISL_1159462, EPI_ISL_1159463, EPI_ISL_1159464, EPI_ISL_1159465,
EPI_ISL_1159466, EPI_ISL_1159467, EPI_ISL_1159468, EPI_ISL_1159469, EPI_ISL_1159470, EPI_ISL_1159471, EPI_ISL_1159472, EPI_ISL_1159473, EPI_ISL_1159474, EPI_ISL_1159475, EPI_ISL_1159476, EPI_ISL_1159477, EPI_ISL_1159478, EPI_ISL_1159479, EPI_ISL_1159480, EPI_ISL_1159481, EPI_ISL_1159482, EPI_ISL_1159483, EPI_ISL_1159484, EPI_ISL_1159485, EPI_ISL_1159486, EPI_ISL_1159487,
EPI_ISL_1159488, EPI_ISL_1159489, EPI_ISL_1159490, EPI_ISL_1159491, EPI_ISL_1159492, EPI_ISL_1159493, EPI_ISL_1159494, EPI_ISL_1159495, EPI_ISL_1159496, EPI_ISL_1159497, EPI_ISL_1159498, EPI_ISL_1159499, EPI_ISL_1159500, EPI_ISL_1159501, EPI_ISL_1159502, EPI_ISL_1159503, EPI_ISL_1159504, EPI_ISL_1159505, EPI_ISL_1159506, EPI_ISL_1159507, EPI_ISL_1159508, EPI_ISL_1159509,
EPI_ISL_1159510, EPI_ISL_1159511, EPI_ISL_1159512, EPI_ISL_1159514, EPI_ISL_1159515, EPI_ISL_1159516, EPI_ISL_1159517, EPI_ISL_1159518, EPI_ISL_1159519, EPI_ISL_1159520, EPI_ISL_1159521, EPI_ISL_1159522, EPI_ISL_1159523, EPI_ISL_1159524, EPI_ISL_1159525, EPI_ISL_1159526, EPI_ISL_1159527, EPI_ISL_1159528, EPI_ISL_1159529, EPI_ISL_1159530, EPI_ISL_1159531, EPI_ISL_1159532,
EPI_ISL_1159533, EPI_ISL_1159534, EPI_ISL_1159535, EPI_ISL_1159536, EPI_ISL_1159537, EPI_ISL_1159538, EPI_ISL_1159539, EPI_ISL_1159540, EPI_ISL_1159541, EPI_ISL_1159542, EPI_ISL_1159543, EPI_ISL_1159544, EPI_ISL_1159545, EPI_ISL_1159546, EPI_ISL_1159547, EPI_ISL_1159548, EPI_ISL_1159549, EPI_ISL_1159550, EPI_ISL_1159551, EPI_ISL_1159552, EPI_ISL_1159553, EPI_ISL_1159554,
EPI_ISL_1159555, EPI_ISL_1159556, EPI_ISL_1159557, EPI_ISL_1159558, EPI_ISL_1159559, EPI_ISL_1159560, EPI_ISL_1159561, EPI_ISL_1159562, EPI_ISL_1159563, EPI_ISL_1159564, EPI_ISL_1159565, EPI_ISL_1159566, EPI_ISL_1159567, EPI_ISL_1159568, EPI_ISL_1159569, EPI_ISL_1159570, EPI_ISL_1159571, EPI_ISL_1159572, EPI_ISL_1159573, EPI_ISL_1159574, EPI_ISL_1159575, EPI_ISL_1159576,
EPI_ISL_1159577, EPI_ISL_1159578, EPI_ISL_1159579, EPI_ISL_1159580, EPI_ISL_1159581, EPI_ISL_1159582, EPI_ISL_1159583, EPI_ISL_1159584, EPI_ISL_1159585, EPI_ISL_1159586, EPI_ISL_1159587, EPI_ISL_1159588, EPI_ISL_1159589, EPI_ISL_1159590, EPI_ISL_1159591, EPI_ISL_1159592, EPI_ISL_1159593, EPI_ISL_1159594, EPI_ISL_1159595, EPI_ISL_1159596, EPI_ISL_1159597, EPI_ISL_1159598,
EPI_ISL_1159599, EPI_ISL_1159600, EPI_ISL_1159601, EPI_ISL_1159602, EPI_ISL_1159603, EPI_ISL_1159604, EPI_ISL_1159605, EPI_ISL_1159606, EPI_ISL_1159607, EPI_ISL_1159608, EPI_ISL_1159609, EPI_ISL_1159610, EPI_ISL_1159611, EPI_ISL_1159612, EPI_ISL_1159613, EPI_ISL_1159614, EPI_ISL_1159615, EPI_ISL_1159616, EPI_ISL_1159617, EPI_ISL_1159618, EPI_ISL_1159619, EPI_ISL_1159620,
EPI_ISL_1159621, EPI_ISL_1159622, EPI_ISL_1159623, EPI_ISL_1159624, EPI_ISL_1159625, EPI_ISL_1159626, EPI_ISL_1159627, EPI_ISL_1159628, EPI_ISL_1159629, EPI_ISL_1159630, EPI_ISL_1159631, EPI_ISL_1159632, EPI_ISL_1159633, EPI_ISL_1159634, EPI_ISL_1159635, EPI_ISL_1159636, EPI_ISL_1159637, EPI_ISL_1159638, EPI_ISL_1159639, EPI_ISL_1159640, EPI_ISL_1159641, EPI_ISL_1159642,
EPI_ISL_1159643, EPI_ISL_1159644, EPI_ISL_1159645, EPI_ISL_1159646, EPI_ISL_1159647, EPI_ISL_1159648, EPI_ISL_1159649, EPI_ISL_1159650, EPI_ISL_1159651, EPI_ISL_1159652, EPI_ISL_1159653, EPI_ISL_1159654, EPI_ISL_1159655, EPI_ISL_1159656, EPI_ISL_1159657, EPI_ISL_1159658, EPI_ISL_1159659, EPI_ISL_1159660, EPI_ISL_1159661, EPI_ISL_1159662, EPI_ISL_1159663, EPI_ISL_1159664,
EPI_ISL_1159665, EPI_ISL_1159666, EPI_ISL_1159667, EPI_ISL_1159668, EPI_ISL_1159669, EPI_ISL_1159670, EPI_ISL_1159671, EPI_ISL_1159672, EPI_ISL_1159673, EPI_ISL_1159674, EPI_ISL_1159675, EPI_ISL_1159676, EPI_ISL_1159677, EPI_ISL_1159678, EPI_ISL_1159679, EPI_ISL_1159680, EPI_ISL_1159681

see above

Quest Diagnostics Incorporated

Respiratory Viruses Branch, Division of Viral Diseases, Centers for Disease Control and Prevention

Peter W. Cook, Dakota Howard, Dhwani Batra, Ben L. Rambo-Martin, S. H. Rosenthal, A. Gerasimova, R. M. Kagan, B. Anderson, M. Hua, Y. Liu, L.E. Bernstein, K.E. Livingston, A. Perez, I. A. Shlyakhter, R. V. Rolando, R. Owen, P. Tanpaiboon, F. Lacbawan, Clinton R. Paden, Suxiang Tong, Duncan MacCannell

EPI_ISL_1159682, EPI_ISL_1159683, EPI_ISL_1159684, EPI_ISL_1159685, EPI_ISL_1159686, EPI_ISL_1159687, EPI_ISL_1159688, EPI_ISL_1159689, EPI_ISL_1159690, EPI_ISL_1159691, EPI_ISL_1159692, EPI_ISL_1159693, EPI_ISL_1159694, EPI_ISL_1159695, EPI_ISL_1159696

see above

Kansas Health and Environmental Lab

Kansas Health and Environmental Lab

Mike Grose, Jonathan Barnell, Ben Olsen, and Phil Adam

EPI_ISL_1159701, EPI_ISL_1159702, EPI_ISL_1159703, EPI_ISL_1159704, EPI_ISL_1159705, EPI_ISL_1159706, EPI_ISL_1159707, EPI_ISL_1159708, EPI_ISL_1159709, EPI_ISL_1159710, EPI_ISL_1159711, EPI_ISL_1159712, EPI_ISL_1159714, EPI_ISL_1159715, EPI_ISL_1159716, EPI_ISL_1159717, EPI_ISL_1159719, EPI_ISL_1159720, EPI_ISL_1159721, EPI_ISL_1159722, EPI_ISL_1159723, EPI_ISL_1159725,
EPI_ISL_1159728

see above

Atlas Genomics

Atlas Genomics

Michael Kalnoski, MD

EPI_ISL_1159730, EPI_ISL_1159731, EPI_ISL_1159732, EPI_ISL_1159733, EPI_ISL_1159734, EPI_ISL_1159735, EPI_ISL_1159736, EPI_ISL_1159737, EPI_ISL_1159738, EPI_ISL_1159739, EPI_ISL_1159740, EPI_ISL_1159741, EPI_ISL_1159744, EPI_ISL_1159748, EPI_ISL_1159750, EPI_ISL_1159758, EPI_ISL_1159759, EPI_ISL_1159760, EPI_ISL_1159761, EPI_ISL_1159762, EPI_ISL_1159763, EPI_ISL_1159764,
EPI_ISL_1159765, EPI_ISL_1159766, EPI_ISL_1159767, EPI_ISL_1159769, EPI_ISL_1159770, EPI_ISL_1159771, EPI_ISL_1159772, EPI_ISL_1159773, EPI_ISL_1159774, EPI_ISL_1159775, EPI_ISL_1159776, EPI_ISL_1159777, EPI_ISL_1159778, EPI_ISL_1159779, EPI_ISL_1159780, EPI_ISL_1159781, EPI_ISL_1159782, EPI_ISL_1159783, EPI_ISL_1159784, EPI_ISL_1159785, EPI_ISL_1159786, EPI_ISL_1159787,
EPI_ISL_1159788, EPI_ISL_1159789, EPI_ISL_1159790, EPI_ISL_1159791, EPI_ISL_1159792, EPI_ISL_1159793, EPI_ISL_1159794, EPI_ISL_1159795, EPI_ISL_1159796, EPI_ISL_1159797, EPI_ISL_1159798, EPI_ISL_1159799, EPI_ISL_1159800, EPI_ISL_1159801, EPI_ISL_1159802, EPI_ISL_1159803, EPI_ISL_1159804, EPI_ISL_1159805, EPI_ISL_1159806, EPI_ISL_1159807, EPI_ISL_1159808, EPI_ISL_1159809,
EPI_ISL_1159810, EPI_ISL_1159811, EPI_ISL_1159812, EPI_ISL_1159813, EPI_ISL_1159814, EPI_ISL_1159815, EPI_ISL_1159816, EPI_ISL_1159817, EPI_ISL_1159818, EPI_ISL_1159819, EPI_ISL_1159820, EPI_ISL_1159821, EPI_ISL_1159822, EPI_ISL_1159823, EPI_ISL_1159824, EPI_ISL_1159825, EPI_ISL_1159826, EPI_ISL_1159827, EPI_ISL_1159828, EPI_ISL_1159829, EPI_ISL_1159830, EPI_ISL_1159831,
EPI_ISL_1159832, EPI_ISL_1159833, EPI_ISL_1159834, EPI_ISL_1159835, EPI_ISL_1159836, EPI_ISL_1159837, EPI_ISL_1159838, EPI_ISL_1159839, EPI_ISL_1159840, EPI_ISL_1159841, EPI_ISL_1159842, EPI_ISL_1159843, EPI_ISL_1159844, EPI_ISL_1159845, EPI_ISL_1159846, EPI_ISL_1159847, EPI_ISL_1159848, EPI_ISL_1159849, EPI_ISL_1159850, EPI_ISL_1159851, EPI_ISL_1159852, EPI_ISL_1159853,
EPI_ISL_1159854, EPI_ISL_1159855, EPI_ISL_1159857, EPI_ISL_1159858, EPI_ISL_1159859, EPI_ISL_1159860, EPI_ISL_1159861, EPI_ISL_1159862, EPI_ISL_1159863, EPI_ISL_1159864, EPI_ISL_1159865, EPI_ISL_1159866, EPI_ISL_1159868, EPI_ISL_1159869, EPI_ISL_1159871, EPI_ISL_1159872, EPI_ISL_1159873, EPI_ISL_1159874, EPI_ISL_1159875, EPI_ISL_1159876, EPI_ISL_1159877, EPI_ISL_1159878,
EPI_ISL_1159879, EPI_ISL_1159880, EPI_ISL_1159881, EPI_ISL_1159882, EPI_ISL_1159883

see above

Wisconsin State Laboratory of Hygiene Communicable Disease Division

Wisconsin State Laboratory of Hygiene Communicable Disease Division

Kelsey R. Florek, Abigail C. Shockey

EPI_ISL_1159885, EPI_ISL_1159886, EPI_ISL_1159887, EPI_ISL_1159888, EPI_ISL_1159890, EPI_ISL_1159891, EPI_ISL_1159892, EPI_ISL_1159893, EPI_ISL_1159894, EPI_ISL_1159895, EPI_ISL_1159896, EPI_ISL_1159897, EPI_ISL_1159898, EPI_ISL_1159899, EPI_ISL_1159900, EPI_ISL_1159901, EPI_ISL_1159902, EPI_ISL_1159903, EPI_ISL_1159904, EPI_ISL_1159905, EPI_ISL_1159906, EPI_ISL_1159909,
EPI_ISL_1159910, EPI_ISL_1159911, EPI_ISL_1159912, EPI_ISL_1159913, EPI_ISL_1159914, EPI_ISL_1159915, EPI_ISL_1159916, EPI_ISL_1159917, EPI_ISL_1159918, EPI_ISL_1159919, EPI_ISL_1159920, EPI_ISL_1159921, EPI_ISL_1159922, EPI_ISL_1159923, EPI_ISL_1159924, EPI_ISL_1159925, EPI_ISL_1159926, EPI_ISL_1159927, EPI_ISL_1159928, EPI_ISL_1159929, EPI_ISL_1159930, EPI_ISL_1159931,
EPI_ISL_1159932, EPI_ISL_1159933, EPI_ISL_1159934, EPI_ISL_1159935, EPI_ISL_1159936, EPI_ISL_1159937, EPI_ISL_1159938, EPI_ISL_1159939, EPI_ISL_1159940, EPI_ISL_1159941, EPI_ISL_1159942, EPI_ISL_1159943, EPI_ISL_1159944, EPI_ISL_1159945, EPI_ISL_1159946, EPI_ISL_1159947, EPI_ISL_1159948, EPI_ISL_1159949, EPI_ISL_1159950, EPI_ISL_1159951, EPI_ISL_1159952, EPI_ISL_1159953,
EPI_ISL_1159954, EPI_ISL_1159955, EPI_ISL_1159956, EPI_ISL_1159957, EPI_ISL_1159958, EPI_ISL_1159959, EPI_ISL_1159960, EPI_ISL_1159961, EPI_ISL_1159962, EPI_ISL_1159963, EPI_ISL_1159964, EPI_ISL_1159965, EPI_ISL_1159966, EPI_ISL_1159967, EPI_ISL_1159968, EPI_ISL_1159969, EPI_ISL_1159970, EPI_ISL_1159971, EPI_ISL_1159972, EPI_ISL_1159973, EPI_ISL_1159974, EPI_ISL_1159975,
EPI_ISL_1159976, EPI_ISL_1159977, EPI_ISL_1159978, EPI_ISL_1159979, EPI_ISL_1159980, EPI_ISL_1159981, EPI_ISL_1159982, EPI_ISL_1159983, EPI_ISL_1159984, EPI_ISL_1159985, EPI_ISL_1159986, EPI_ISL_1159987, EPI_ISL_1159988, EPI_ISL_1159989, EPI_ISL_1159990, EPI_ISL_1159991, EPI_ISL_1159992, EPI_ISL_1159993, EPI_ISL_1159994, EPI_ISL_1159995, EPI_ISL_1159996, EPI_ISL_1159997,
EPI_ISL_1159998, EPI_ISL_1159999, EPI_ISL_1160000, EPI_ISL_1160001, EPI_ISL_1160002, EPI_ISL_1160003, EPI_ISL_1160004, EPI_ISL_1160005, EPI_ISL_1160006, EPI_ISL_1160007, EPI_ISL_1160008, EPI_ISL_1160009, EPI_ISL_1160010, EPI_ISL_1160011, EPI_ISL_1160012, EPI_ISL_1160013, EPI_ISL_1160014, EPI_ISL_1160015, EPI_ISL_1160016, EPI_ISL_1160017, EPI_ISL_1160018, EPI_ISL_1160019,
EPI_ISL_1160020, EPI_ISL_1160021, EPI_ISL_1160022, EPI_ISL_1160023, EPI_ISL_1160024, EPI_ISL_1160025, EPI_ISL_1160026, EPI_ISL_1160027, EPI_ISL_1160028, EPI_ISL_1160029, EPI_ISL_1160030, EPI_ISL_1160031, EPI_ISL_1160032, EPI_ISL_1160033, EPI_ISL_1160034, EPI_ISL_1160035, EPI_ISL_1160036, EPI_ISL_1160037, EPI_ISL_1160038, EPI_ISL_1160039, EPI_ISL_1160040, EPI_ISL_1160041,
EPI_ISL_1160042, EPI_ISL_1160043, EPI_ISL_1160044, EPI_ISL_1160045, EPI_ISL_1160046, EPI_ISL_1160047, EPI_ISL_1160048, EPI_ISL_1160049, EPI_ISL_1160050, EPI_ISL_1160051, EPI_ISL_1160052, EPI_ISL_1160053, EPI_ISL_1160054, EPI_ISL_1160055, EPI_ISL_1160056, EPI_ISL_1160057, EPI_ISL_1160058, EPI_ISL_1160059, EPI_ISL_1160060, EPI_ISL_1160061, EPI_ISL_1160062, EPI_ISL_1160063,
EPI_ISL_1160064, EPI_ISL_1160065, EPI_ISL_1160066, EPI_ISL_1160067, EPI_ISL_1160068, EPI_ISL_1160069, EPI_ISL_1160070, EPI_ISL_1160071, EPI_ISL_1160072, EPI_ISL_1160073, EPI_ISL_1160074, EPI_ISL_1160075, EPI_ISL_1160076, EPI_ISL_1160077, EPI_ISL_1160078, EPI_ISL_1160079, EPI_ISL_1160080, EPI_ISL_1160081, EPI_ISL_1160082, EPI_ISL_1160083, EPI_ISL_1160084, EPI_ISL_1160085,
EPI_ISL_1160086, EPI_ISL_1160087, EPI_ISL_1160088, EPI_ISL_1160089, EPI_ISL_1160090, EPI_ISL_1160091, EPI_ISL_1160092, EPI_ISL_1160913, EPI_ISL_1160914, EPI_ISL_1160915, EPI_ISL_1160916, EPI_ISL_1160917, EPI_ISL_1160918, EPI_ISL_1160919, EPI_ISL_1160920, EPI_ISL_1160921, EPI_ISL_1160922, EPI_ISL_1160923, EPI_ISL_1160924, EPI_ISL_1160925, EPI_ISL_1160926, EPI_ISL_1160927,
EPI_ISL_1160928, EPI_ISL_1160929, EPI_ISL_1160930, EPI_ISL_1160931, EPI_ISL_1160932, EPI_ISL_1160933, EPI_ISL_1160934, EPI_ISL_1160935, EPI_ISL_1160936, EPI_ISL_1160937, EPI_ISL_1160938, EPI_ISL_1160939, EPI_ISL_1160940, EPI_ISL_1160941, EPI_ISL_1160942, EPI_ISL_1160943, EPI_ISL_1160944, EPI_ISL_1160945, EPI_ISL_1160946, EPI_ISL_1160947, EPI_ISL_1160948, EPI_ISL_1160949,
EPI_ISL_1160950, EPI_ISL_1160951, EPI_ISL_1160952, EPI_ISL_1160953, EPI_ISL_1160954, EPI_ISL_1160955, EPI_ISL_1160956, EPI_ISL_1160957, EPI_ISL_1160958, EPI_ISL_1160959, EPI_ISL_1160960, EPI_ISL_1160961, EPI_ISL_1160962, EPI_ISL_1160963, EPI_ISL_1160964, EPI_ISL_1160965, EPI_ISL_1160966, EPI_ISL_1160967, EPI_ISL_1160968, EPI_ISL_1160969, EPI_ISL_1160970, EPI_ISL_1160971,
EPI_ISL_1160972, EPI_ISL_1160973, EPI_ISL_1160974, EPI_ISL_1160975, EPI_ISL_1160976, EPI_ISL_1160977, EPI_ISL_1160978, EPI_ISL_1160979, EPI_ISL_1160980, EPI_ISL_1160981, EPI_ISL_1160982, EPI_ISL_1160983, EPI_ISL_1160984, EPI_ISL_1160985, EPI_ISL_1160986, EPI_ISL_1160987, EPI_ISL_1160988, EPI_ISL_1160989, EPI_ISL_1160990, EPI_ISL_1160991, EPI_ISL_1160992, EPI_ISL_1160993,
EPI_ISL_1160994, EPI_ISL_1160995, EPI_ISL_1160996, EPI_ISL_1160997, EPI_ISL_1160998, EPI_ISL_1160999, EPI_ISL_1161000, EPI_ISL_1161001, EPI_ISL_1161002, EPI_ISL_1161003, EPI_ISL_1161004, EPI_ISL_1161005, EPI_ISL_1161007, EPI_ISL_1161008, EPI_ISL_1161010, EPI_ISL_1161012, EPI_ISL_1161015, EPI_ISL_1161017, EPI_ISL_1161019, EPI_ISL_1161021, EPI_ISL_1161023, EPI_ISL_1161025,
EPI_ISL_1161027, EPI_ISL_1161029, EPI_ISL_1161031, EPI_ISL_1161033, EPI_ISL_1161035, EPI_ISL_1161037, EPI_ISL_1161039, EPI_ISL_1161042, EPI_ISL_1161044, EPI_ISL_1161046, EPI_ISL_1161048, EPI_ISL_1161051, EPI_ISL_1161053, EPI_ISL_1161057, EPI_ISL_1161059, EPI_ISL_1161061, EPI_ISL_1161063, EPI_ISL_1161066, EPI_ISL_1161068, EPI_ISL_1161070, EPI_ISL_1161072, EPI_ISL_1161074,
EPI_ISL_1161076, EPI_ISL_1161078, EPI_ISL_1161079, EPI_ISL_1161080, EPI_ISL_1161081, EPI_ISL_1161082, EPI_ISL_1161083, EPI_ISL_1161084, EPI_ISL_1161085, EPI_ISL_1161086, EPI_ISL_1161087, EPI_ISL_1161088, EPI_ISL_1161089, EPI_ISL_1161090, EPI_ISL_1161091, EPI_ISL_1161092, EPI_ISL_1161093, EPI_ISL_1161094, EPI_ISL_1161095, EPI_ISL_1161096, EPI_ISL_1161097, EPI_ISL_1161098,
EPI_ISL_1161099, EPI_ISL_1161100, EPI_ISL_1161101, EPI_ISL_1161102, EPI_ISL_1161103, EPI_ISL_1161104, EPI_ISL_1161105, EPI_ISL_1161106, EPI_ISL_1161107, EPI_ISL_1161108, EPI_ISL_1161109, EPI_ISL_1161110, EPI_ISL_1161111, EPI_ISL_1161112, EPI_ISL_1161113, EPI_ISL_1161114, EPI_ISL_1161115, EPI_ISL_1161116, EPI_ISL_1161117, EPI_ISL_1161118, EPI_ISL_1161119, EPI_ISL_1161120,
EPI_ISL_1161121, EPI_ISL_1161122, EPI_ISL_1161123, EPI_ISL_1161124, EPI_ISL_1161125, EPI_ISL_1161126, EPI_ISL_1161127, EPI_ISL_1161128, EPI_ISL_1161129, EPI_ISL_1161130, EPI_ISL_1161131, EPI_ISL_1161132, EPI_ISL_1161133, EPI_ISL_1161134, EPI_ISL_1161135, EPI_ISL_1161136, EPI_ISL_1161137, EPI_ISL_1161138, EPI_ISL_1161139, EPI_ISL_1161140, EPI_ISL_1161141, EPI_ISL_1161142,
EPI_ISL_1161143, EPI_ISL_1161144, EPI_ISL_1161145, EPI_ISL_1161146, EPI_ISL_1161147, EPI_ISL_1161148, EPI_ISL_1161149, EPI_ISL_1161150, EPI_ISL_1161151, EPI_ISL_1161152, EPI_ISL_1161153, EPI_ISL_1161154, EPI_ISL_1161155, EPI_ISL_1161156, EPI_ISL_1161157, EPI_ISL_1161158, EPI_ISL_1161159, EPI_ISL_1161160, EPI_ISL_1161161, EPI_ISL_1161163, EPI_ISL_1161164, EPI_ISL_1161165,
EPI_ISL_1161166, EPI_ISL_1161167, EPI_ISL_1161168, EPI_ISL_1161169, EPI_ISL_1161170, EPI_ISL_1161171, EPI_ISL_1161172, EPI_ISL_1161174, EPI_ISL_1161175, EPI_ISL_1161176, EPI_ISL_1161177, EPI_ISL_1161178, EPI_ISL_1161179, EPI_ISL_1161180, EPI_ISL_1161181, EPI_ISL_1161182, EPI_ISL_1161183, EPI_ISL_1161184, EPI_ISL_1161185, EPI_ISL_1161186, EPI_ISL_1161187, EPI_ISL_1161188,
EPI_ISL_1161189, EPI_ISL_1161190, EPI_ISL_1161191, EPI_ISL_1161192, EPI_ISL_1161193, EPI_ISL_1161194, EPI_ISL_1161195, EPI_ISL_1161196, EPI_ISL_1161197, EPI_ISL_1161198, EPI_ISL_1161199, EPI_ISL_1161200, EPI_ISL_1161201, EPI_ISL_1161202, EPI_ISL_1161203, EPI_ISL_1161204, EPI_ISL_1161205, EPI_ISL_1161206, EPI_ISL_1161207, EPI_ISL_1161208, EPI_ISL_1161209, EPI_ISL_1161210,
EPI_ISL_1161211, EPI_ISL_1161212, EPI_ISL_1161213, EPI_ISL_1161214, EPI_ISL_1161215, EPI_ISL_1161216, EPI_ISL_1161217, EPI_ISL_1161218, EPI_ISL_1161219, EPI_ISL_1161220, EPI_ISL_1161221, EPI_ISL_1161222, EPI_ISL_1161223, EPI_ISL_1161224, EPI_ISL_1161225, EPI_ISL_1161226, EPI_ISL_1161227, EPI_ISL_1161228, EPI_ISL_1161229, EPI_ISL_1161230, EPI_ISL_1161231, EPI_ISL_1161232,
EPI_ISL_1161233, EPI_ISL_1161234, EPI_ISL_1161235, EPI_ISL_1161236, EPI_ISL_1161237, EPI_ISL_1161238, EPI_ISL_1161239, EPI_ISL_1161240, EPI_ISL_1161241, EPI_ISL_1161242, EPI_ISL_1161243, EPI_ISL_1161244, EPI_ISL_1161245, EPI_ISL_1161246, EPI_ISL_1161247, EPI_ISL_1161248, EPI_ISL_1161249, EPI_ISL_1161250, EPI_ISL_1161251, EPI_ISL_1161252, EPI_ISL_1161253, EPI_ISL_1161254,
EPI_ISL_1161255, EPI_ISL_1161256, EPI_ISL_1161257, EPI_ISL_1161258, EPI_ISL_1161259, EPI_ISL_1161260, EPI_ISL_1161261, EPI_ISL_1161262, EPI_ISL_1161263, EPI_ISL_1161264, EPI_ISL_1161265, EPI_ISL_1161266, EPI_ISL_1161267, EPI_ISL_1161268, EPI_ISL_1161269, EPI_ISL_1161270, EPI_ISL_1161271, EPI_ISL_1161272, EPI_ISL_1161273, EPI_ISL_1161274, EPI_ISL_1161275, EPI_ISL_1161276,
EPI_ISL_1161277, EPI_ISL_1161278, EPI_ISL_1161279, EPI_ISL_1161280, EPI_ISL_1161281, EPI_ISL_1161282, EPI_ISL_1161283, EPI_ISL_1161284, EPI_ISL_1161285, EPI_ISL_1161286, EPI_ISL_1161287, EPI_ISL_1161288, EPI_ISL_1161289, EPI_ISL_1161290, EPI_ISL_1161291, EPI_ISL_1161292, EPI_ISL_1161293, EPI_ISL_1161294, EPI_ISL_1161295, EPI_ISL_1161296, EPI_ISL_1161297, EPI_ISL_1161298,
EPI_ISL_1161299, EPI_ISL_1161300, EPI_ISL_1161301, EPI_ISL_1161302, EPI_ISL_1161303, EPI_ISL_1161304, EPI_ISL_1161305, EPI_ISL_1161306, EPI_ISL_1161307, EPI_ISL_1161308, EPI_ISL_1161309, EPI_ISL_1161310, EPI_ISL_1161311, EPI_ISL_1161312, EPI_ISL_1161313, EPI_ISL_1161314, EPI_ISL_1161315, EPI_ISL_1161316, EPI_ISL_1161317, EPI_ISL_1161318, EPI_ISL_1161319, EPI_ISL_1161320,
EPI_ISL_1161321, EPI_ISL_1161322, EPI_ISL_1161323, EPI_ISL_1161324, EPI_ISL_1161325, EPI_ISL_1161326, EPI_ISL_1161327, EPI_ISL_1161328, EPI_ISL_1161329, EPI_ISL_1161330, EPI_ISL_1161331, EPI_ISL_1161332, EPI_ISL_1161333, EPI_ISL_1161334, EPI_ISL_1161335, EPI_ISL_1161336, EPI_ISL_1161337, EPI_ISL_1161338, EPI_ISL_1161339, EPI_ISL_1161340, EPI_ISL_1161341, EPI_ISL_1161342,
EPI_ISL_1161343, EPI_ISL_1161344, EPI_ISL_1161345, EPI_ISL_1161346, EPI_ISL_1161347, EPI_ISL_1161348, EPI_ISL_1161349, EPI_ISL_1161350, EPI_ISL_1161351, EPI_ISL_1161352, EPI_ISL_1161353, EPI_ISL_1161354, EPI_ISL_1161355, EPI_ISL_1161356, EPI_ISL_1161357, EPI_ISL_1161358, EPI_ISL_1161359, EPI_ISL_1161360, EPI_ISL_1161361, EPI_ISL_1161362, EPI_ISL_1161363, EPI_ISL_1161364,
EPI_ISL_1161365, EPI_ISL_1161366, EPI_ISL_1161367, EPI_ISL_1161368, EPI_ISL_1161369, EPI_ISL_1161370, EPI_ISL_1161371, EPI_ISL_1161372, EPI_ISL_1161373, EPI_ISL_1161374, EPI_ISL_1161375, EPI_ISL_1161376, EPI_ISL_1161377, EPI_ISL_1161378, EPI_ISL_1161379, EPI_ISL_1161380, EPI_ISL_1161381, EPI_ISL_1161382, EPI_ISL_1161383, EPI_ISL_1161384, EPI_ISL_1161385, EPI_ISL_1161386,
EPI_ISL_1161387, EPI_ISL_1161388, EPI_ISL_1161389, EPI_ISL_1161390, EPI_ISL_1161391, EPI_ISL_1161392, EPI_ISL_1161393, EPI_ISL_1161394, EPI_ISL_1161395, EPI_ISL_1161396, EPI_ISL_1161397, EPI_ISL_1161398, EPI_ISL_1161399, EPI_ISL_1161400, EPI_ISL_1161416, EPI_ISL_1161417, EPI_ISL_1161418, EPI_ISL_1161419, EPI_ISL_1161420, EPI_ISL_1161421, EPI_ISL_1161422, EPI_ISL_1161423,
EPI_ISL_1161424, EPI_ISL_1161425, EPI_ISL_1161427, EPI_ISL_1161428, EPI_ISL_1161429, EPI_ISL_1161430, EPI_ISL_1161431, EPI_ISL_1161432, EPI_ISL_1161433, EPI_ISL_1161434, EPI_ISL_1161435, EPI_ISL_1161436, EPI_ISL_1161437, EPI_ISL_1161438, EPI_ISL_1161439, EPI_ISL_1161440, EPI_ISL_1161441, EPI_ISL_1161442, EPI_ISL_1161443, EPI_ISL_1161444, EPI_ISL_1161445, EPI_ISL_1161446,
EPI_ISL_1161447, EPI_ISL_1161448, EPI_ISL_1161449, EPI_ISL_1161450, EPI_ISL_1161451, EPI_ISL_1161452, EPI_ISL_1161453, EPI_ISL_1161454, EPI_ISL_1161455, EPI_ISL_1161456, EPI_ISL_1161457, EPI_ISL_1161458, EPI_ISL_1161459, EPI_ISL_1161460, EPI_ISL_1161461, EPI_ISL_1161462, EPI_ISL_1161463, EPI_ISL_1161464, EPI_ISL_1161465, EPI_ISL_1161466, EPI_ISL_1161467, EPI_ISL_1161468,
EPI_ISL_1161469, EPI_ISL_1161470, EPI_ISL_1161471, EPI_ISL_1161472, EPI_ISL_1161473, EPI_ISL_1161474, EPI_ISL_1161475, EPI_ISL_1161476, EPI_ISL_1161477, EPI_ISL_1161478, EPI_ISL_1161479, EPI_ISL_1161480, EPI_ISL_1161481, EPI_ISL_1161482, EPI_ISL_1161483, EPI_ISL_1161484, EPI_ISL_1161485, EPI_ISL_1161486, EPI_ISL_1161487, EPI_ISL_1161488, EPI_ISL_1161489, EPI_ISL_1161490,
EPI_ISL_1161491, EPI_ISL_1161492, EPI_ISL_1161493, EPI_ISL_1161494, EPI_ISL_1161495, EPI_ISL_1161496, EPI_ISL_1161497, EPI_ISL_1161498, EPI_ISL_1161499, EPI_ISL_1161500, EPI_ISL_1161501, EPI_ISL_1161502, EPI_ISL_1161503, EPI_ISL_1161504, EPI_ISL_1161505, EPI_ISL_1161506, EPI_ISL_1161507, EPI_ISL_1161508, EPI_ISL_1161509, EPI_ISL_1161510, EPI_ISL_1161511, EPI_ISL_1161512,
EPI_ISL_1161513, EPI_ISL_1161514, EPI_ISL_1161515, EPI_ISL_1161516, EPI_ISL_1161517, EPI_ISL_1161518, EPI_ISL_1161519, EPI_ISL_1161520, EPI_ISL_1161521, EPI_ISL_1161522, EPI_ISL_1161523, EPI_ISL_1161524, EPI_ISL_1161525, EPI_ISL_1161526, EPI_ISL_1161527, EPI_ISL_1161528, EPI_ISL_1161529, EPI_ISL_1161530, EPI_ISL_1161531, EPI_ISL_1161532, EPI_ISL_1161533, EPI_ISL_1161534,
EPI_ISL_1161535, EPI_ISL_1161536, EPI_ISL_1161537, EPI_ISL_1161538, EPI_ISL_1161539, EPI_ISL_1161540, EPI_ISL_1161541, EPI_ISL_1161542, EPI_ISL_1161543, EPI_ISL_1161544, EPI_ISL_1161545, EPI_ISL_1161546, EPI_ISL_1161547, EPI_ISL_1161548, EPI_ISL_1161549, EPI_ISL_1161550, EPI_ISL_1161551, EPI_ISL_1161552, EPI_ISL_1161553, EPI_ISL_1161554, EPI_ISL_1161555, EPI_ISL_1161556,
EPI_ISL_1161557, EPI_ISL_1161558, EPI_ISL_1161559, EPI_ISL_1161560, EPI_ISL_1161561, EPI_ISL_1161562, EPI_ISL_1161563, EPI_ISL_1161564, EPI_ISL_1161565, EPI_ISL_1161566, EPI_ISL_1161567, EPI_ISL_1161568, EPI_ISL_1161569, EPI_ISL_1161570, EPI_ISL_1161571, EPI_ISL_1161572, EPI_ISL_1161573, EPI_ISL_1161574, EPI_ISL_1161575, EPI_ISL_1161576, EPI_ISL_1161577, EPI_ISL_1161578,
EPI_ISL_1161579, EPI_ISL_1161580, EPI_ISL_1161581, EPI_ISL_1161582, EPI_ISL_1161583, EPI_ISL_1161584, EPI_ISL_1161585, EPI_ISL_1161586, EPI_ISL_1161587, EPI_ISL_1161588, EPI_ISL_1161589, EPI_ISL_1161590, EPI_ISL_1161591, EPI_ISL_1161592, EPI_ISL_1161593, EPI_ISL_1161594, EPI_ISL_1161595, EPI_ISL_1161596, EPI_ISL_1161597, EPI_ISL_1161598, EPI_ISL_1161599, EPI_ISL_1161600,
EPI_ISL_1161601, EPI_ISL_1161602, EPI_ISL_1161603, EPI_ISL_1161604, EPI_ISL_1161605, EPI_ISL_1161606, EPI_ISL_1161607, EPI_ISL_1161608, EPI_ISL_1161609, EPI_ISL_1161610, EPI_ISL_1161611, EPI_ISL_1161612, EPI_ISL_1161613, EPI_ISL_1161614, EPI_ISL_1161615, EPI_ISL_1161616, EPI_ISL_1161617, EPI_ISL_1161618, EPI_ISL_1161619, EPI_ISL_1161620, EPI_ISL_1161621, EPI_ISL_1161622,
EPI_ISL_1161623, EPI_ISL_1161624, EPI_ISL_1161625, EPI_ISL_1161626, EPI_ISL_1161627, EPI_ISL_1161628, EPI_ISL_1161629, EPI_ISL_1161630, EPI_ISL_1161631, EPI_ISL_1161632, EPI_ISL_1161633, EPI_ISL_1161634, EPI_ISL_1161635, EPI_ISL_1161636, EPI_ISL_1161637, EPI_ISL_1161638, EPI_ISL_1161639, EPI_ISL_1161640, EPI_ISL_1161641, EPI_ISL_1161642, EPI_ISL_1161643, EPI_ISL_1161644,
EPI_ISL_1161645, EPI_ISL_1161646, EPI_ISL_1161647, EPI_ISL_1161648, EPI_ISL_1161649, EPI_ISL_1161650, EPI_ISL_1161651, EPI_ISL_1161652, EPI_ISL_1161653, EPI_ISL_1161654, EPI_ISL_1161655, EPI_ISL_1161656, EPI_ISL_1161657, EPI_ISL_1161658, EPI_ISL_1161659, EPI_ISL_1161660, EPI_ISL_1161661, EPI_ISL_1161662, EPI_ISL_1161663, EPI_ISL_1161664, EPI_ISL_1161665, EPI_ISL_1161666,
EPI_ISL_1161667, EPI_ISL_1161668, EPI_ISL_1161669, EPI_ISL_1161670, EPI_ISL_1161671, EPI_ISL_1161672, EPI_ISL_1161673, EPI_ISL_1161674, EPI_ISL_1161675, EPI_ISL_1161676, EPI_ISL_1161677, EPI_ISL_1161678, EPI_ISL_1161679, EPI_ISL_1161680, EPI_ISL_1161681, EPI_ISL_1161682, EPI_ISL_1161683, EPI_ISL_1161684, EPI_ISL_1161685, EPI_ISL_1161686, EPI_ISL_1161687, EPI_ISL_1161688,
EPI_ISL_1161689, EPI_ISL_1161690, EPI_ISL_1161691, EPI_ISL_1161692, EPI_ISL_1161693, EPI_ISL_1161694, EPI_ISL_1161695, EPI_ISL_1161696, EPI_ISL_1161697, EPI_ISL_1161698, EPI_ISL_1161699, EPI_ISL_1161700, EPI_ISL_1161701, EPI_ISL_1161702, EPI_ISL_1161703, EPI_ISL_1161704, EPI_ISL_1161705, EPI_ISL_1161706, EPI_ISL_1161707, EPI_ISL_1161708, EPI_ISL_1161709, EPI_ISL_1161710,
EPI_ISL_1161711, EPI_ISL_1161712, EPI_ISL_1161713, EPI_ISL_1161714, EPI_ISL_1161715, EPI_ISL_1161716, EPI_ISL_1161717, EPI_ISL_1162988, EPI_ISL_1162989, EPI_ISL_1162990, EPI_ISL_1162991, EPI_ISL_1162992, EPI_ISL_1162993, EPI_ISL_1162994, EPI_ISL_1162995, EPI_ISL_1162996, EPI_ISL_1162997, EPI_ISL_1162998, EPI_ISL_1162999, EPI_ISL_1163000, EPI_ISL_1163001, EPI_ISL_1163002,
EPI_ISL_1163003, EPI_ISL_1163004, EPI_ISL_1163005, EPI_ISL_1163006, EPI_ISL_1163007, EPI_ISL_1163008, EPI_ISL_1163009, EPI_ISL_1163010, EPI_ISL_1163011, EPI_ISL_1163012, EPI_ISL_1163013, EPI_ISL_1163014, EPI_ISL_1163015, EPI_ISL_1163016, EPI_ISL_1163017, EPI_ISL_1163018, EPI_ISL_1163019, EPI_ISL_1163020, EPI_ISL_1163021, EPI_ISL_1163022, EPI_ISL_1163023, EPI_ISL_1163024,
EPI_ISL_1163025, EPI_ISL_1163026, EPI_ISL_1163027, EPI_ISL_1163028, EPI_ISL_1163029, EPI_ISL_1163030, EPI_ISL_1163031, EPI_ISL_1163032, EPI_ISL_1163033, EPI_ISL_1163034, EPI_ISL_1163035, EPI_ISL_1163036, EPI_ISL_1163037, EPI_ISL_1163038, EPI_ISL_1163039, EPI_ISL_1163040, EPI_ISL_1163041, EPI_ISL_1163042, EPI_ISL_1163043, EPI_ISL_1163044, EPI_ISL_1163045, EPI_ISL_1163046,
EPI_ISL_1163047, EPI_ISL_1163048, EPI_ISL_1163049, EPI_ISL_1163051, EPI_ISL_1163052, EPI_ISL_1163053, EPI_ISL_1163054, EPI_ISL_1163055, EPI_ISL_1163056, EPI_ISL_1163057, EPI_ISL_1163058, EPI_ISL_1163059, EPI_ISL_1163060, EPI_ISL_1163061, EPI_ISL_1163062, EPI_ISL_1163063, EPI_ISL_1163064, EPI_ISL_1163065, EPI_ISL_1163066, EPI_ISL_1163067, EPI_ISL_1163068, EPI_ISL_1163069,
EPI_ISL_1163070, EPI_ISL_1163071, EPI_ISL_1163072, EPI_ISL_1163073, EPI_ISL_1163074, EPI_ISL_1163075, EPI_ISL_1163076, EPI_ISL_1163077, EPI_ISL_1163078, EPI_ISL_1163079, EPI_ISL_1163080, EPI_ISL_1163081, EPI_ISL_1163082, EPI_ISL_1163083, EPI_ISL_1163084, EPI_ISL_1163085, EPI_ISL_1163086, EPI_ISL_1163087, EPI_ISL_1163088, EPI_ISL_1163089, EPI_ISL_1163090, EPI_ISL_1163091,
EPI_ISL_1163092, EPI_ISL_1163093, EPI_ISL_1163094, EPI_ISL_1163095, EPI_ISL_1163096, EPI_ISL_1163097, EPI_ISL_1163098, EPI_ISL_1163099, EPI_ISL_1163100, EPI_ISL_1163101, EPI_ISL_1163102, EPI_ISL_1163103, EPI_ISL_1163104, EPI_ISL_1163105, EPI_ISL_1163106, EPI_ISL_1163107, EPI_ISL_1163108, EPI_ISL_1163109, EPI_ISL_1163110, EPI_ISL_1163111, EPI_ISL_1163112, EPI_ISL_1163113,
EPI_ISL_1163114, EPI_ISL_1163115, EPI_ISL_1163116, EPI_ISL_1163117, EPI_ISL_1163118, EPI_ISL_1163119, EPI_ISL_1163120, EPI_ISL_1163121, EPI_ISL_1163122, EPI_ISL_1163123, EPI_ISL_1163124, EPI_ISL_1163125, EPI_ISL_1163126, EPI_ISL_1163127, EPI_ISL_1163128, EPI_ISL_1163129, EPI_ISL_1163130, EPI_ISL_1163131, EPI_ISL_1163132, EPI_ISL_1163133, EPI_ISL_1163134, EPI_ISL_1163135,
EPI_ISL_1163136, EPI_ISL_1163137, EPI_ISL_1163138, EPI_ISL_1163139, EPI_ISL_1163140, EPI_ISL_1163141, EPI_ISL_1163142, EPI_ISL_1163143, EPI_ISL_1163144, EPI_ISL_1163145, EPI_ISL_1163146, EPI_ISL_1163147, EPI_ISL_1163148, EPI_ISL_1163149, EPI_ISL_1163150, EPI_ISL_1163151, EPI_ISL_1163152, EPI_ISL_1163153, EPI_ISL_1163154, EPI_ISL_1163155, EPI_ISL_1163156, EPI_ISL_1163157,
EPI_ISL_1163158, EPI_ISL_1163159, EPI_ISL_1163160, EPI_ISL_1163161, EPI_ISL_1163163, EPI_ISL_1163164, EPI_ISL_1163166, EPI_ISL_1163167, EPI_ISL_1163168, EPI_ISL_1163169, EPI_ISL_1163170, EPI_ISL_1163171, EPI_ISL_1163172, EPI_ISL_1163173, EPI_ISL_1163175, EPI_ISL_1163176, EPI_ISL_1163177, EPI_ISL_1163178, EPI_ISL_1163179, EPI_ISL_1163180, EPI_ISL_1163181, EPI_ISL_1163182,
EPI_ISL_1163183, EPI_ISL_1163184, EPI_ISL_1163185, EPI_ISL_1163186, EPI_ISL_1163187, EPI_ISL_1163188, EPI_ISL_1163189, EPI_ISL_1163190, EPI_ISL_1163191, EPI_ISL_1163192, EPI_ISL_1163193, EPI_ISL_1163194, EPI_ISL_1163195, EPI_ISL_1163196, EPI_ISL_1163197, EPI_ISL_1163198, EPI_ISL_1163199, EPI_ISL_1163200, EPI_ISL_1163201, EPI_ISL_1163202, EPI_ISL_1163203, EPI_ISL_1163204,
EPI_ISL_1163205, EPI_ISL_1163206, EPI_ISL_1163207, EPI_ISL_1163208, EPI_ISL_1163209, EPI_ISL_1163210, EPI_ISL_1163211, EPI_ISL_1163212, EPI_ISL_1163213, EPI_ISL_1163214, EPI_ISL_1163215, EPI_ISL_1163216, EPI_ISL_1163217, EPI_ISL_1163218, EPI_ISL_1163219, EPI_ISL_1163220, EPI_ISL_1163221, EPI_ISL_1163222, EPI_ISL_1163223, EPI_ISL_1163224, EPI_ISL_1163225, EPI_ISL_1163226,
EPI_ISL_1163227, EPI_ISL_1163228, EPI_ISL_1163229, EPI_ISL_1163230, EPI_ISL_1163231, EPI_ISL_1163232, EPI_ISL_1163233, EPI_ISL_1163234, EPI_ISL_1163235, EPI_ISL_1163236, EPI_ISL_1163237, EPI_ISL_1163238, EPI_ISL_1163239, EPI_ISL_1163240, EPI_ISL_1163241, EPI_ISL_1163242, EPI_ISL_1163243, EPI_ISL_1163244, EPI_ISL_1163245, EPI_ISL_1163246, EPI_ISL_1163247, EPI_ISL_1163248,
EPI_ISL_1163249, EPI_ISL_1163250, EPI_ISL_1163251, EPI_ISL_1163252, EPI_ISL_1163253, EPI_ISL_1163254, EPI_ISL_1163255, EPI_ISL_1163256, EPI_ISL_1163257, EPI_ISL_1163258, EPI_ISL_1163259, EPI_ISL_1163260, EPI_ISL_1163261, EPI_ISL_1163262, EPI_ISL_1163263, EPI_ISL_1163264, EPI_ISL_1163265, EPI_ISL_1163266, EPI_ISL_1163267, EPI_ISL_1163268, EPI_ISL_1163269, EPI_ISL_1163270,
EPI_ISL_1163271, EPI_ISL_1163272, EPI_ISL_1163273, EPI_ISL_1163274, EPI_ISL_1163275, EPI_ISL_1163276, EPI_ISL_1163277, EPI_ISL_1163278, EPI_ISL_1163279, EPI_ISL_1163280, EPI_ISL_1163281, EPI_ISL_1163282, EPI_ISL_1163283, EPI_ISL_1163284, EPI_ISL_1163285, EPI_ISL_1163286, EPI_ISL_1163287, EPI_ISL_1163288, EPI_ISL_1163289, EPI_ISL_1163290, EPI_ISL_1163291, EPI_ISL_1163292,
EPI_ISL_1163293, EPI_ISL_1163294, EPI_ISL_1163295, EPI_ISL_1163296, EPI_ISL_1163297, EPI_ISL_1163298, EPI_ISL_1163299, EPI_ISL_1163300, EPI_ISL_1163301, EPI_ISL_1163302, EPI_ISL_1163303, EPI_ISL_1163304, EPI_ISL_1163305, EPI_ISL_1163306, EPI_ISL_1163307, EPI_ISL_1163308, EPI_ISL_1163309, EPI_ISL_1163310, EPI_ISL_1163311, EPI_ISL_1163312, EPI_ISL_1163313, EPI_ISL_1163314,
EPI_ISL_1163315, EPI_ISL_1163316, EPI_ISL_1163317, EPI_ISL_1163318, EPI_ISL_1163319, EPI_ISL_1163320, EPI_ISL_1163321, EPI_ISL_1163322, EPI_ISL_1163323, EPI_ISL_1163324, EPI_ISL_1163325, EPI_ISL_1163326, EPI_ISL_1163327, EPI_ISL_1163328, EPI_ISL_1163329, EPI_ISL_1163330, EPI_ISL_1163331, EPI_ISL_1163332, EPI_ISL_1163333, EPI_ISL_1163334, EPI_ISL_1163335, EPI_ISL_1163336,
EPI_ISL_1163337, EPI_ISL_1163338, EPI_ISL_1163339, EPI_ISL_1163340, EPI_ISL_1163341, EPI_ISL_1163342, EPI_ISL_1163343, EPI_ISL_1163344, EPI_ISL_1163345, EPI_ISL_1163346, EPI_ISL_1163347, EPI_ISL_1163348, EPI_ISL_1163349, EPI_ISL_1163350, EPI_ISL_1163351, EPI_ISL_1163352, EPI_ISL_1163353, EPI_ISL_1163354, EPI_ISL_1163355, EPI_ISL_1163356, EPI_ISL_1163357, EPI_ISL_1163358,
EPI_ISL_1163359, EPI_ISL_1163360, EPI_ISL_1163361, EPI_ISL_1163362, EPI_ISL_1163363, EPI_ISL_1163364, EPI_ISL_1163365, EPI_ISL_1163366, EPI_ISL_1163367, EPI_ISL_1163368, EPI_ISL_1163369, EPI_ISL_1163370, EPI_ISL_1163371, EPI_ISL_1163372, EPI_ISL_1163373, EPI_ISL_1163374, EPI_ISL_1163375, EPI_ISL_1163376, EPI_ISL_1163377, EPI_ISL_1163378, EPI_ISL_1163379, EPI_ISL_1163380,
EPI_ISL_1163381, EPI_ISL_1163382, EPI_ISL_1163383, EPI_ISL_1163384, EPI_ISL_1163385, EPI_ISL_1163386, EPI_ISL_1163387, EPI_ISL_1163388, EPI_ISL_1163389, EPI_ISL_1163390, EPI_ISL_1163391, EPI_ISL_1163392, EPI_ISL_1163393, EPI_ISL_1163394, EPI_ISL_1163395, EPI_ISL_1163396, EPI_ISL_1163397, EPI_ISL_1163398, EPI_ISL_1163399, EPI_ISL_1163400, EPI_ISL_1163401, EPI_ISL_1163402,
EPI_ISL_1163403, EPI_ISL_1163404, EPI_ISL_1163405, EPI_ISL_1163406, EPI_ISL_1163407, EPI_ISL_1163408, EPI_ISL_1163409, EPI_ISL_1163410, EPI_ISL_1163411, EPI_ISL_1163412, EPI_ISL_1163413, EPI_ISL_1163414, EPI_ISL_1163415, EPI_ISL_1163416, EPI_ISL_1163417, EPI_ISL_1163418, EPI_ISL_1163419, EPI_ISL_1163420, EPI_ISL_1163421, EPI_ISL_1163422, EPI_ISL_1163423, EPI_ISL_1163424,
EPI_ISL_1163425, EPI_ISL_1163426, EPI_ISL_1163427, EPI_ISL_1163428, EPI_ISL_1163429, EPI_ISL_1163430, EPI_ISL_1163431, EPI_ISL_1163432, EPI_ISL_1163433, EPI_ISL_1163434, EPI_ISL_1163435, EPI_ISL_1163436, EPI_ISL_1163437, EPI_ISL_1163438, EPI_ISL_1163439, EPI_ISL_1163440, EPI_ISL_1163441, EPI_ISL_1163442, EPI_ISL_1163443, EPI_ISL_1163445, EPI_ISL_1163446, EPI_ISL_1163447,
EPI_ISL_1163448, EPI_ISL_1163449, EPI_ISL_1163450, EPI_ISL_1163451, EPI_ISL_1163452, EPI_ISL_1163453, EPI_ISL_1163454, EPI_ISL_1163455, EPI_ISL_1163456, EPI_ISL_1163457, EPI_ISL_1163458, EPI_ISL_1163459, EPI_ISL_1163460, EPI_ISL_1163461, EPI_ISL_1163462, EPI_ISL_1163463, EPI_ISL_1163464, EPI_ISL_1163465, EPI_ISL_1163466, EPI_ISL_1163467, EPI_ISL_1163468, EPI_ISL_1163469,

| | | | |
|---|---|---|---|
| EPI_ISL_1163470, EPI_ISL_1163471, EPI_ISL_1163472, EPI_ISL_1163473, EPI_ISL_1163474, EPI_ISL_1163475, EPI_ISL_1163476, EPI_ISL_1163477, EPI_ISL_1163478, EPI_ISL_1163479, EPI_ISL_1163480, EPI_ISL_1163481, EPI_ISL_1163482, EPI_ISL_1163483, EPI_ISL_1163484, EPI_ISL_1163485, EPI_ISL_1163486, EPI_ISL_1163487, EPI_ISL_1163488, EPI_ISL_1163489, EPI_ISL_1163490, EPI_ISL_1163491, EPI_ISL_1163492, EPI_ISL_1163493, EPI_ISL_1163494, EPI_ISL_1163495, EPI_ISL_1163496, EPI_ISL_1163497, EPI_ISL_1163498, EPI_ISL_1163499, EPI_ISL_1163500, EPI_ISL_1163501, EPI_ISL_1163502, EPI_ISL_1163503, EPI_ISL_1163504, EPI_ISL_1163505, EPI_ISL_1163506, EPI_ISL_1163507, EPI_ISL_1163508, EPI_ISL_1163510, EPI_ISL_1163511, EPI_ISL_1163512, EPI_ISL_1163513, EPI_ISL_1163514, EPI_ISL_1163515, EPI_ISL_1163516, EPI_ISL_1163517, EPI_ISL_1163518, EPI_ISL_1163519, EPI_ISL_1163520, EPI_ISL_1163521, EPI_ISL_1163522, EPI_ISL_1163523, EPI_ISL_1163524, EPI_ISL_1163525, EPI_ISL_1163526, EPI_ISL_1163527, EPI_ISL_1163528, EPI_ISL_1163529 | | | |
| see above | Laboratory Corporation of America | Respiratory Viruses Branch, Division of Viral Diseases, Centers for Disease Control and Prevention | Peter W. Cook, Dakota Howard, Dhwani Batra, Ben L. Rambo-Martin, Minoo Agarwal, Eyad Almasri Debbie Boles, Ayla Burns, Nuthawin Charoensri, Oren Cohen, Susan Countryman, Mary Ann Cristobal, Bobbi Croy, Suzanne Dale, Hrushikesh Deshmukh, Amanda Douglas, Vincent Drouillon, Marcia Eisenberg, Howard Engler, Rama Ghatti, Prashant Gupta, Susan Hicks, Jake Humphrey, Lax Iyer, Manoj Jain, Mohan Kolli, Brian Krueger, Tim Kuphal, Stanley Letovsky, Michael Levandoski, Craig Lukasik, Jonathan Meltzer, Brian Norvell, Mindy Nye, Scott Parker, Christos Petropoulos, John Pruitt, Steven Ragan, Scott Ryan, Mike Sapeta, Jana Schroth, Suresh Babu Selvaraju, Goran Stevovic, Amanda Suchanek, Andrea Throop, Lyndon Tilson, Thomas Urban, Joe Voshell, Kimberly Wagner, Jonathan Williams, Mary Williamson, Qian Zeng, Tricia Zwiefelhofer, Clinton R. Paden, Suxiang Tong, Duncan MacCannell |
| EPI_ISL_1163533, EPI_ISL_1163534, EPI_ISL_1163535, EPI_ISL_1163536, EPI_ISL_1163537, EPI_ISL_1163538, EPI_ISL_1163539, EPI_ISL_1163540, EPI_ISL_1163541, EPI_ISL_1163542, EPI_ISL_1163543, EPI_ISL_1163544, EPI_ISL_1163545, EPI_ISL_1163546, EPI_ISL_1163547, EPI_ISL_1163548, EPI_ISL_1163549, EPI_ISL_1163550, EPI_ISL_1163551, EPI_ISL_1163552, EPI_ISL_1163553, EPI_ISL_1163554, EPI_ISL_1163555, EPI_ISL_1163556, EPI_ISL_1163557, EPI_ISL_1163558, EPI_ISL_1163559, EPI_ISL_1163560, EPI_ISL_1163561, EPI_ISL_1163562, EPI_ISL_1163563, EPI_ISL_1163564, EPI_ISL_1163565, EPI_ISL_1163566, EPI_ISL_1163567, EPI_ISL_1163568, EPI_ISL_1163569, EPI_ISL_1163570, EPI_ISL_1163571, EPI_ISL_1163572, EPI_ISL_1163573, EPI_ISL_1163574, EPI_ISL_1163575, EPI_ISL_1163576, EPI_ISL_1163577, EPI_ISL_1163578, EPI_ISL_1163579, EPI_ISL_1163580, EPI_ISL_1163581, EPI_ISL_1163582, EPI_ISL_1163583, EPI_ISL_1163584, EPI_ISL_1163585, EPI_ISL_1163586, EPI_ISL_1163587, EPI_ISL_1163588, EPI_ISL_1163589, EPI_ISL_1163590, EPI_ISL_1163591, EPI_ISL_1163592, EPI_ISL_1163593, EPI_ISL_1163594, EPI_ISL_1163595, EPI_ISL_1163596, EPI_ISL_1163597, EPI_ISL_1163598, EPI_ISL_1163599, EPI_ISL_1163600, EPI_ISL_1163601, EPI_ISL_1163602, EPI_ISL_1163603, EPI_ISL_1163604, EPI_ISL_1163605, EPI_ISL_1163606, EPI_ISL_1163608, EPI_ISL_1163609, EPI_ISL_1163610, EPI_ISL_1163611, EPI_ISL_1163612, EPI_ISL_1163613, EPI_ISL_1163614, EPI_ISL_1163615, EPI_ISL_1163616, EPI_ISL_1163617, EPI_ISL_1163618, EPI_ISL_1163619, EPI_ISL_1163620, EPI_ISL_1163621, EPI_ISL_1163622, EPI_ISL_1163623, EPI_ISL_1163624, EPI_ISL_1163625, EPI_ISL_1163626, EPI_ISL_1163627, EPI_ISL_1163628, EPI_ISL_1163629, EPI_ISL_1163630, EPI_ISL_1163631, EPI_ISL_1163632, EPI_ISL_1163633, EPI_ISL_1163634, EPI_ISL_1163635, EPI_ISL_1163636, EPI_ISL_1163637, EPI_ISL_1163638, EPI_ISL_1163639, EPI_ISL_1163640, EPI_ISL_1163641, EPI_ISL_1163642, EPI_ISL_1163643, EPI_ISL_1163644, EPI_ISL_1163645, EPI_ISL_1163646, EPI_ISL_1163647, EPI_ISL_1163648, EPI_ISL_1163649, EPI_ISL_1163650, EPI_ISL_1163651, EPI_ISL_1163652, EPI_ISL_1163653, EPI_ISL_1163654, EPI_ISL_1163655, EPI_ISL_1163656, EPI_ISL_1163657, EPI_ISL_1163658, EPI_ISL_1163659, EPI_ISL_1163660, EPI_ISL_1163661, EPI_ISL_1163662 | | | |
| see above | Maine HETL | Tewhey Lab, The Jackson Laboratory | Matluk,N., Dewey,H., Iosue,F., Barter,M., Lynch,R., Munger,H. and Tewhey,R. |
| EPI_ISL_1163664 | Tempus | Grubaugh Lab - Yale School of Public Health | Joseph Fauver, Tara Alpert, Anderson Brito, Mallery Breban, Anne Wyllie, Chantal Vogels, Mary Petrone, Annie Watkins, Chaney Kalinich, Isabel Ott, Nathan Grubaugh |
| EPI_ISL_1163668, EPI_ISL_1163669, EPI_ISL_1163670, EPI_ISL_1163671, EPI_ISL_1163672, EPI_ISL_1163673, EPI_ISL_1163674, EPI_ISL_1163675, EPI_ISL_1163677, EPI_ISL_1163678, EPI_ISL_1163679, EPI_ISL_1163680, EPI_ISL_1163681, EPI_ISL_1163682, EPI_ISL_1163683, EPI_ISL_1163684, EPI_ISL_1163685, EPI_ISL_1163686, EPI_ISL_1163687 | | | |
| see above | Yale Clinical Virology Lab | Grubaugh Lab - Yale School of Public Health | Joseph Fauver, Tara Alpert, Anderson Brito, Mallery Breban, Anne Wyllie, Chantal Vogels, Mary Petrone, Annie Watkins, Chaney Kalinich, Isabel Ott, Nathan Grubaugh |
| EPI_ISL_1163717, EPI_ISL_1163718, EPI_ISL_1163719, EPI_ISL_1163720, EPI_ISL_1163721, EPI_ISL_1163722, EPI_ISL_1163723, EPI_ISL_1163724, EPI_ISL_1163725, EPI_ISL_1163726, EPI_ISL_1163727, EPI_ISL_1163728, EPI_ISL_1163729, EPI_ISL_1163730, EPI_ISL_1163731, EPI_ISL_1163732, EPI_ISL_1163733, EPI_ISL_1163734, EPI_ISL_1163735 | | | |
| see above | Houston Health Dept. | Houston Health Dept. | Ryker Penn, Pamela Brown, Adolpho Lara |
| EPI_ISL_1163738, EPI_ISL_1163739, EPI_ISL_1163740, EPI_ISL_1163741, EPI_ISL_1163742, EPI_ISL_1163743, EPI_ISL_1163744, EPI_ISL_1163745, EPI_ISL_1163746, EPI_ISL_1163747, EPI_ISL_1163748, EPI_ISL_1163749, EPI_ISL_1163750, EPI_ISL_1163751, EPI_ISL_1163752, EPI_ISL_1163753, EPI_ISL_1163754, EPI_ISL_1163755, EPI_ISL_1163756, EPI_ISL_1163757, EPI_ISL_1163758, EPI_ISL_1163759, EPI_ISL_1163760, EPI_ISL_1163761, EPI_ISL_1163762, EPI_ISL_1163763, EPI_ISL_1163764, EPI_ISL_1163765, EPI_ISL_1163766, EPI_ISL_1163767, EPI_ISL_1163768, EPI_ISL_1163769, EPI_ISL_1163770, EPI_ISL_1163771, EPI_ISL_1163772, EPI_ISL_1163773, EPI_ISL_1163774, EPI_ISL_1163775, EPI_ISL_1163776, EPI_ISL_1163777, EPI_ISL_1163778, EPI_ISL_1163779, EPI_ISL_1163780, EPI_ISL_1163781, EPI_ISL_1163782, EPI_ISL_1163783, EPI_ISL_1163784, EPI_ISL_1163785, EPI_ISL_1163786, EPI_ISL_1163787, EPI_ISL_1163788, EPI_ISL_1163789, EPI_ISL_1163790, EPI_ISL_1163791, EPI_ISL_1163792, EPI_ISL_1163793 | | | |
| see above | Clinical Molecular Microbiology Laboratory, UNC Hospital | Dirk Dittmer | Justin T. Landis , Razia Moorad , Brent A. Eason, Melissa B. Miller, Linda Pluta, Dirk Dittmer, Angelica Juarez, Cecilia Thompson, Shawn Hawken, Cameroon Grant, Evelyn Hoffman, Patricio Cano, Jason Wong, Carolina Caro-Vegas, Ryan McNamara, Blossom Damania. |
| EPI_ISL_1163794 | Kansas Health and Environmental Lab | Kansas Health and Environmental Lab | Mike Grose, Jonathan Barnell, Ben Olsen, and Phil Adam |
| EPI_ISL_1163846, EPI_ISL_1163847, EPI_ISL_1163848, EPI_ISL_1163850, EPI_ISL_1163853, EPI_ISL_1163854, EPI_ISL_1163855, EPI_ISL_1163856, EPI_ISL_1163858, EPI_ISL_1163859, EPI_ISL_1163860, EPI_ISL_1163861, EPI_ISL_1163862, EPI_ISL_1163863, EPI_ISL_1163864, EPI_ISL_1163865, EPI_ISL_1163866, EPI_ISL_1163867, EPI_ISL_1163868, EPI_ISL_1163869, EPI_ISL_1163870, EPI_ISL_1163871, EPI_ISL_1163872, EPI_ISL_1163873, EPI_ISL_1163875, EPI_ISL_1163877, EPI_ISL_1163878, EPI_ISL_1163879, EPI_ISL_1163880, EPI_ISL_1163882, EPI_ISL_1163883, EPI_ISL_1163884, EPI_ISL_1163886, EPI_ISL_1163887, EPI_ISL_1163888, EPI_ISL_1164179, EPI_ISL_1164180, EPI_ISL_1164181, EPI_ISL_1164182 | | | |
| see above | Clinical Molecular Microbiology Laboratory, UNC Hospitals | Jeremy Wang | Jeremy Wang, Alexander Rubinsteyn, Colleen Rice, Jason Smedberg, Shawn Hawken, Melissa Miller, Corbin Jones, Robert Hagan |
| EPI_ISL_1164183 | Clinical Microbiology, Nationwide Children's Hospital | James Molecular Laboratory | Huanyu Wang, Pam Snyder, Zachary Rudy, Dan Jones, Amy Leber |
| EPI_ISL_1164360, EPI_ISL_1164361, EPI_ISL_1164362, EPI_ISL_1164363, EPI_ISL_1164364, EPI_ISL_1164365, EPI_ISL_1164366, EPI_ISL_1164367, EPI_ISL_1164368, EPI_ISL_1164370, EPI_ISL_1164371, EPI_ISL_1164372, EPI_ISL_1164373, EPI_ISL_1164374, EPI_ISL_1164375, EPI_ISL_1164376, EPI_ISL_1164377, EPI_ISL_1164378, EPI_ISL_1164379, EPI_ISL_1164380, EPI_ISL_1164381, EPI_ISL_1164382, EPI_ISL_1164383, EPI_ISL_1164384, EPI_ISL_1164385, EPI_ISL_1164386, EPI_ISL_1164387, EPI_ISL_1164388, EPI_ISL_1164389, EPI_ISL_1164390, EPI_ISL_1164391, EPI_ISL_1164392, EPI_ISL_1164393, EPI_ISL_1164394, EPI_ISL_1164395, EPI_ISL_1164396, EPI_ISL_1164397, EPI_ISL_1164398, EPI_ISL_1164399, EPI_ISL_1164400, EPI_ISL_1164401, EPI_ISL_1164402, EPI_ISL_1164403, EPI_ISL_1164404, EPI_ISL_1164405, EPI_ISL_1164406, EPI_ISL_1164407, EPI_ISL_1164408, EPI_ISL_1164409, EPI_ISL_1164410, EPI_ISL_1164411, EPI_ISL_1164412, EPI_ISL_1164413, EPI_ISL_1164414, EPI_ISL_1164415, EPI_ISL_1164416, EPI_ISL_1164417, EPI_ISL_1164418, EPI_ISL_1164419, EPI_ISL_1164420, EPI_ISL_1164421, EPI_ISL_1164422, EPI_ISL_1164423, EPI_ISL_1164424, EPI_ISL_1164425, EPI_ISL_1164426, EPI_ISL_1164427, EPI_ISL_1164428, EPI_ISL_1164429, EPI_ISL_1164430, EPI_ISL_1164431, EPI_ISL_1164432, EPI_ISL_1164433, EPI_ISL_1164434, EPI_ISL_1164435, EPI_ISL_1164436, EPI_ISL_1164437, EPI_ISL_1164438 | | | |
| see above | Fulgent Genetics | Fulgent Genetics | Harry Gao, Mickey Li, John Gao, Joseph Fierro, Benafsh Sapra, Becky Tsai, Yan Meng, Doreen Ng, James Xie |
| EPI_ISL_1164484, EPI_ISL_1164485 | Vault Health | Minnesota Department of Health, Public Health Laboratory | Alexandra Lorentz, Jacob Garfin, Matt Plumb, and Xiong Wang |
| EPI_ISL_1164486 | Sanford South University Medical Center | Minnesota Department of Health, Public Health Laboratory | Alexandra Lorentz, Jacob Garfin, Matt Plumb, and Xiong Wang |
| EPI_ISL_1164487, EPI_ISL_1164488, EPI_ISL_1164489 | Vault Health | Minnesota Department of Health, Public Health Laboratory | Alexandra Lorentz, Jacob Garfin, Matt Plumb, and Xiong Wang |
| EPI_ISL_1164490, EPI_ISL_1164491 | Sanford South University Medical Center | Minnesota Department of Health, Public Health Laboratory | Alexandra Lorentz, Jacob Garfin, Matt Plumb, and Xiong Wang |
| EPI_ISL_1164492, EPI_ISL_1164493, EPI_ISL_1164494, EPI_ISL_1164495, EPI_ISL_1164496 | Vault Health | Minnesota Department of Health, Public Health Laboratory | Alexandra Lorentz, Jacob Garfin, Matt Plumb, and Xiong Wang |
| EPI_ISL_1164497 | Sanford South University Medical Center | Minnesota Department of Health, Public Health Laboratory | Alexandra Lorentz, Jacob Garfin, Matt Plumb, and Xiong Wang |
| EPI_ISL_1164498, EPI_ISL_1164499, EPI_ISL_1164500, EPI_ISL_1164501, EPI_ISL_1164502, EPI_ISL_1164503, EPI_ISL_1164504, EPI_ISL_1164505, EPI_ISL_1164506, EPI_ISL_1164507 | Vault Health | Minnesota Department of Health, Public Health Laboratory | Alexandra Lorentz, Jacob Garfin, Matt Plumb, and Xiong Wang |
| EPI_ISL_1164757, EPI_ISL_1164758, EPI_ISL_1164759, EPI_ISL_1164760, EPI_ISL_1164761, EPI_ISL_1164762, EPI_ISL_1164763, EPI_ISL_1164764, EPI_ISL_1164765, EPI_ISL_1164768, EPI_ISL_1164769, EPI_ISL_1164770, EPI_ISL_1164771, EPI_ISL_1164772, EPI_ISL_1164773 | | | |
| see above | NJDOH, Public Health and Environmental Laboratories | NJ_PHEL | Lindsey Bodnar, Shiv K. Verma, Dana Woell, Byeong Jeong |
| EPI_ISL_1164774, EPI_ISL_1164775, EPI_ISL_1164777 | School of Pharmacy, Shenandoah University | School of Pharmacy, Shenandoah University | Adams,S.M., Harralson,A.F., Kidd,R.S., Sawyer,G.W. |
| EPI_ISL_1164782, EPI_ISL_1164783, EPI_ISL_1164784, EPI_ISL_1164785, EPI_ISL_1164786, EPI_ISL_1164787, EPI_ISL_1164788, EPI_ISL_1164790, EPI_ISL_1164791, EPI_ISL_1164792, EPI_ISL_1164793, EPI_ISL_1164795, EPI_ISL_1164796, EPI_ISL_1164797, EPI_ISL_1164798, EPI_ISL_1164799, EPI_ISL_1164800, EPI_ISL_1164801, EPI_ISL_1164802, EPI_ISL_1164803, EPI_ISL_1164804, EPI_ISL_1164805, EPI_ISL_1164806, EPI_ISL_1164807, EPI_ISL_1164808, EPI_ISL_1164809, EPI_ISL_1164810, EPI_ISL_1164811, EPI_ISL_1164812, EPI_ISL_1164813, EPI_ISL_1164814, EPI_ISL_1164815 | | | |
| see above | Idaho Bureau of Laboratories | IBL | "R. Beukelman, Matthew C. Burns, Aimee Ceniseros, Robert L. Voermans, Christopher Ball" |
| EPI_ISL_1164817, EPI_ISL_1164820, EPI_ISL_1164821, EPI_ISL_1164822, EPI_ISL_1164823, EPI_ISL_1164824, EPI_ISL_1164826, EPI_ISL_1164828, EPI_ISL_1164829, EPI_ISL_1164830, EPI_ISL_1164831, EPI_ISL_1164833, EPI_ISL_1164834, EPI_ISL_1164835, EPI_ISL_1164836, EPI_ISL_1164839, EPI_ISL_1164842, EPI_ISL_1164843, EPI_ISL_1164844 | | | |
| see above | Nevada State Public Health Laboratory | Nevada State Public Health Laboratory | Andrew Gorzalski, Mark Pandori |
| EPI_ISL_1164855 | Diagnostic Genomics Lab and Functional Genomics Core University of South Carolina | Diagnostic Genomics Lab and Functional Genomics Core, University of South Carolina | Carolyn Banister, Hao Ji, Diego Altomare, Herr Doktor Professor Genomiklaborsdirektor Universitätkovidenpandemieretter Phillip Buckhaults, Michael Shtutman, Helmut Albrecht |
| EPI_ISL_1164856, EPI_ISL_1164857, EPI_ISL_1164859, EPI_ISL_1164860, EPI_ISL_1164861 | LSUHS Emerging Viral Threat Laboratory | Microbial Genome Sequencing Center | Jeremy P. Kamil, Jennifer L. Carroll, Rona S. Scott, Maarten Van Diest, Andrew D. Yurochko, Christopher G. Kevil, Daniel J. Snyder, Vaughn S. Cooper, John A. Vanchiere |
| EPI_ISL_1164862, EPI_ISL_1164863, EPI_ISL_1164864, EPI_ISL_1164865 | LSUHS Emerging Viral Threat Laboratory | Microbial Genome Sequencing Center | Maarten Van Diest, Jennifer L. Carroll, Rona S. Scott, Andrew D. Yurochko, Christopher G. Kevil, Daniel J. Snyder, Vaughn S. Cooper, John A. Vanchiere, Jeremy P. Kamil |
| EPI_ISL_1164866, EPI_ISL_1164867, EPI_ISL_1164868, EPI_ISL_1164869, EPI_ISL_1164870 | LSUHS Emerging Viral Threat Laboratory | Microbial Genome Sequencing Center | Jennifer L. Carroll, Rona S. Scott, Maarten Van Diest, Andrew D. Yurochko, Christopher G. Kevil, Daniel J. Snyder, Vaughn S. Cooper, John A. Vanchiere, Jeremy P. Kamil |
| EPI_ISL_1164871, EPI_ISL_1164872, EPI_ISL_1164873 | LSUHS Emerging Viral Threat Laboratory | Microbial Genome Sequencing Center | Rona S. Scott, Jennifer L. Carroll, Maarten Van Diest, Andrew D. Yurochko, Christopher G. Kevil, Daniel J. Snyder, Vaughn S. Cooper, John A. Vanchiere, Jeremy P. Kamil |
| EPI_ISL_1164874, | LSUHS Emerging Viral Threat | Microbial Genome Sequencing Center | Daniel J. Snyder, Jennifer L. Carroll, Rona S. Scott, Maarten Van Diest, Andrew D. Yurochko, Christopher G. Kevil, Vaughn S. Cooper, John A. Vanchiere, Jeremy P. Kamil |

| | | | |
|---|---|---|---|
| EPI_ISL_1164875, EPI_ISL_1164876, EPI_ISL_1164877, EPI_ISL_1164878 | Laboratory | | |
| EPI_ISL_1164879, EPI_ISL_1164880, EPI_ISL_1164881, EPI_ISL_1164882, EPI_ISL_1164883, EPI_ISL_1164884, EPI_ISL_1164885, EPI_ISL_1164886, EPI_ISL_1164887, EPI_ISL_1164888, EPI_ISL_1164889, EPI_ISL_1164890, EPI_ISL_1164891, EPI_ISL_1164892, EPI_ISL_1164893, EPI_ISL_1164894, EPI_ISL_1164895, EPI_ISL_1164896, EPI_ISL_1164897, EPI_ISL_1164898, EPI_ISL_1164899, EPI_ISL_1164900, EPI_ISL_1164901, EPI_ISL_1164902, EPI_ISL_1164903, EPI_ISL_1164904, EPI_ISL_1164905, EPI_ISL_1164906, EPI_ISL_1164907, EPI_ISL_1164908, EPI_ISL_1164909, EPI_ISL_1164910, EPI_ISL_1164911, EPI_ISL_1164912, EPI_ISL_1164913, EPI_ISL_1164914, EPI_ISL_1164915, EPI_ISL_1164916, EPI_ISL_1164917, EPI_ISL_1164918 | | | |
| see above | LSUHS Emerging Viral Threat Laboratory | Microbial Genome Sequencing Center | Jeremy P. Kamil, Jennifer L. Carroll, Rona S. Scott, Maarten Van Diest, Andrew D. Yurochko, Christopher G. Kevil, Daniel J. Snyder, Vaughn S. Cooper, John A. Vanchiere |
| EPI_ISL_1164919 | LSUHS Emerging Viral Threat Laboratory | Microbial Genome Sequencing Center | Maarten Van Diest, Jennifer L. Carroll, Rona S. Scott, Andrew D. Yurochko, Christopher G. Kevil, Daniel J. Snyder, Vaughn S. Cooper, John A. Vanchiere, Jeremy P. Kamil |
| EPI_ISL_1164920 | LSUHS Emerging Viral Threat Laboratory | Microbial Genome Sequencing Center | Jeremy P. Kamil, Jennifer L. Carroll, Rona S. Scott, Maarten Van Diest, Andrew D. Yurochko, Christopher G. Kevil, Daniel J. Snyder, Vaughn S. Cooper, John A. Vanchiere |
| EPI_ISL_1164921 | LSUHS Emerging Viral Threat Laboratory | Microbial Genome Sequencing Center | Rona S. Scott, Jennifer L. Carroll, Maarten Van Diest, Andrew D. Yurochko, Christopher G. Kevil, Daniel J. Snyder, Vaughn S. Cooper, John A. Vanchiere, Jeremy P. Kamil |
| EPI_ISL_1164922, EPI_ISL_1164923, EPI_ISL_1164924, EPI_ISL_1164925, EPI_ISL_1164926 | LSUHS Emerging Viral Threat Laboratory | Microbial Genome Sequencing Center | Jeremy P. Kamil, Jennifer L. Carroll, Rona S. Scott, Maarten Van Diest, Andrew D. Yurochko, Christopher G. Kevil, Daniel J. Snyder, Vaughn S. Cooper, John A. Vanchiere |
| EPI_ISL_1164931, EPI_ISL_1164946, EPI_ISL_1165135, EPI_ISL_1165442, EPI_ISL_1165770 | Ohio Department of Health Laboratory | Ohio Department of Health Laboratory | Holmes, Jennifer; Eric Brandt, Keoni Omura, Glen McGillivary, Caitlin McDonnell, Jade Mowery, Stephanie Mccracken, Tyler Payne, Kirtana Ramadugu, Erica Leasure, Brent Lee, Kelsey Florek, Heather Blankenship, Allison Black, Quanta Brown, and Tammy Bannerman |
| EPI_ISL_1166109, EPI_ISL_1166110, EPI_ISL_1166111, EPI_ISL_1166112, EPI_ISL_1166113, EPI_ISL_1166114, EPI_ISL_1166115, EPI_ISL_1166116, EPI_ISL_1166117, EPI_ISL_1166118, EPI_ISL_1166119, EPI_ISL_1166120, EPI_ISL_1166121, EPI_ISL_1166122, EPI_ISL_1166123, EPI_ISL_1166145, EPI_ISL_1166146, EPI_ISL_1166189, EPI_ISL_1166190, EPI_ISL_1166191, EPI_ISL_1166192, EPI_ISL_1166193, EPI_ISL_1166194, EPI_ISL_1166195, EPI_ISL_1166196, EPI_ISL_1166197, EPI_ISL_1166198, EPI_ISL_1166199, EPI_ISL_1166200, EPI_ISL_1166201, EPI_ISL_1166202, EPI_ISL_1166203, EPI_ISL_1166204, EPI_ISL_1166205, EPI_ISL_1166206, EPI_ISL_1166207, EPI_ISL_1166208, EPI_ISL_1166209, EPI_ISL_1166210, EPI_ISL_1166217, EPI_ISL_1166218, EPI_ISL_1166219, EPI_ISL_1166220 | | | |
| see above | University of Wisconsin-Madison AIDS Vaccine Research Laboratories | University of Wisconsin-Madison AIDS Vaccine Research Laboratories | Gage Moreno, Katarina Braun, et al. AIDS Vaccine Research Laboratories |
| EPI_ISL_1166616, EPI_ISL_1166617, EPI_ISL_1166618, EPI_ISL_1166862, EPI_ISL_1166863, EPI_ISL_1166864, EPI_ISL_1166865, EPI_ISL_1166866, EPI_ISL_1166867, EPI_ISL_1166869, EPI_ISL_1166870, EPI_ISL_1166871, EPI_ISL_1166872, EPI_ISL_1166873, EPI_ISL_1166874, EPI_ISL_1166875, EPI_ISL_1166876, EPI_ISL_1167088, EPI_ISL_1167102, EPI_ISL_1167108, EPI_ISL_1167109, EPI_ISL_1167110, EPI_ISL_1167111, EPI_ISL_1167112, EPI_ISL_1167113, EPI_ISL_1167114, EPI_ISL_1167115, EPI_ISL_1167116, EPI_ISL_1167117, EPI_ISL_1167118, EPI_ISL_1167119, EPI_ISL_1167120, EPI_ISL_1167121, EPI_ISL_1167122, EPI_ISL_1167123, EPI_ISL_1167124 | | | |
| see above | Wyoming Public Health Laboratory | Wyoming Public Health Laboratory | Jim Mildenberger, Wanda Manley, Noah Hull, Taylor Fearing, Lynette Gumbleton, Channing Weber, Ashley Norberg, Chayse Rowley, Marley Goetz, Brian Dominguez, Elliot Thomasson, Cari Sloma, and Rob Christensen |
| EPI_ISL_1167200, EPI_ISL_1167201, EPI_ISL_1167202, EPI_ISL_1167203, EPI_ISL_1167204, EPI_ISL_1167205, EPI_ISL_1167206, EPI_ISL_1167207, EPI_ISL_1167208, EPI_ISL_1167209, EPI_ISL_1167210, EPI_ISL_1167211, EPI_ISL_1167212, EPI_ISL_1167213, EPI_ISL_1167214, EPI_ISL_1167215, EPI_ISL_1167216, EPI_ISL_1167217, EPI_ISL_1167218, EPI_ISL_1167219, EPI_ISL_1167233, EPI_ISL_1167234, EPI_ISL_1167235, EPI_ISL_1167236, EPI_ISL_1167237, EPI_ISL_1167238, EPI_ISL_1167239, EPI_ISL_1167240, EPI_ISL_1167241, EPI_ISL_1167242, EPI_ISL_1167243, EPI_ISL_1167244, EPI_ISL_1167245, EPI_ISL_1167246, EPI_ISL_1167247, EPI_ISL_1167248, EPI_ISL_1167249, EPI_ISL_1167250, EPI_ISL_1167251, EPI_ISL_1167252, EPI_ISL_1167253, EPI_ISL_1167254 | | | |
| see above | UW Virology Lab | UW Virology Lab | Pavitra Roychoudhury, Hong Xie, Lasata Shrestha, Shah Mohamed Bakhash, Michelle Lin, Noah Baker, Meei-Li Huang, Keith R Jerome, Alexander Greninger |
| EPI_ISL_1167255, EPI_ISL_1167256, EPI_ISL_1167257, EPI_ISL_1167258, EPI_ISL_1167259, EPI_ISL_1167260, EPI_ISL_1167261, EPI_ISL_1167262, EPI_ISL_1167263, EPI_ISL_1167264, EPI_ISL_1167265, EPI_ISL_1167266, EPI_ISL_1167267, EPI_ISL_1167268, EPI_ISL_1167269, EPI_ISL_1167270, EPI_ISL_1167271, EPI_ISL_1167272, EPI_ISL_1167274, EPI_ISL_1167275, EPI_ISL_1167276, EPI_ISL_1167277, EPI_ISL_1167278, EPI_ISL_1167279, EPI_ISL_1167280, EPI_ISL_1167281, EPI_ISL_1167282, EPI_ISL_1167283, EPI_ISL_1167285, EPI_ISL_1167286, EPI_ISL_1167287, EPI_ISL_1167288, EPI_ISL_1167289, EPI_ISL_1167290, EPI_ISL_1167291, EPI_ISL_1167292, EPI_ISL_1167293, EPI_ISL_1167294, EPI_ISL_1167295, EPI_ISL_1167296, EPI_ISL_1167297 | | | |
| see above | Maryland Public Health Laboratory | Maryland Public Health Laboratory | Maryland Department of Health Laboratories Administration |
| EPI_ISL_1167547, EPI_ISL_1167548, EPI_ISL_1167549, EPI_ISL_1167550, EPI_ISL_1167551, EPI_ISL_1167552, EPI_ISL_1167553, EPI_ISL_1167554, EPI_ISL_1167555, EPI_ISL_1167556, EPI_ISL_1167557, EPI_ISL_1167558, EPI_ISL_1167559, EPI_ISL_1167560, EPI_ISL_1167561, EPI_ISL_1167562, EPI_ISL_1167563, EPI_ISL_1167564, EPI_ISL_1167565, EPI_ISL_1167566, EPI_ISL_1167567, EPI_ISL_1167664, EPI_ISL_1167665 | | | |
| see above | Kansas Health and Environmental Lab | Kansas Health and Environmental Lab | Mike Grose, Jonathan Barnell, Ben Olsen, and Phil Adam |
| EPI_ISL_1167666 | Cytocheck Laboratory | Kansas Health and Environmental Lab | Mike Grose, Jonathan Barnell, Ben Olsen, and Phil Adam |
| EPI_ISL_1167940, EPI_ISL_1167944, EPI_ISL_1167946, EPI_ISL_1167950, EPI_ISL_1167954, EPI_ISL_1167957, EPI_ISL_1167960, EPI_ISL_1167962, EPI_ISL_1167963, EPI_ISL_1167971, EPI_ISL_1167974, EPI_ISL_1167975, EPI_ISL_1167978, EPI_ISL_1167980, EPI_ISL_1167987 | | | |
| see above | GA Department of Public Health Laboratory | Genomics and Discovery, Respiratory Viruses Branch, Division of Viral Diseases, Centers for Disease Control and Prevention | Brian Lynch, Krista Queen, Yan Li, Ying Tao, Jing Zhang, Anna Uehara, Anna Montmayeur, Clinton R. Paden, Peter W. Cook, Rachel Marine, Haibin Wang,, Suxiang Tong |
| EPI_ISL_1167989, EPI_ISL_1167991, EPI_ISL_1167992, EPI_ISL_1167993, EPI_ISL_1167994, EPI_ISL_1167995, EPI_ISL_1167996, EPI_ISL_1167997, EPI_ISL_1167998 | MO State Public Health Laboratory | Genomics and Discovery, Respiratory Viruses Branch, Division of Viral Diseases, Centers for Disease Control and Prevention | Brian Lynch, Krista Queen, Yan Li, Ying Tao, Jing Zhang, Anna Uehara, Anna Montmayeur, Clinton R. Paden, Peter W. Cook, Rachel Marine, Haibin Wang,, Suxiang Tong |
| EPI_ISL_1168003 | GA Department of Public Health Laboratory | Genomics and Discovery, Respiratory Viruses Branch, Division of Viral Diseases, Centers for Disease Control and Prevention | Brian Lynch, Krista Queen, Yan Li, Ying Tao, Jing Zhang, Anna Uehara, Anna Montmayeur, Clinton R. Paden, Peter W. Cook, Rachel Marine, Haibin Wang,, Suxiang Tong |
| EPI_ISL_1168221, EPI_ISL_1168223, EPI_ISL_1168225, EPI_ISL_1168226, EPI_ISL_1168230, EPI_ISL_1168231, EPI_ISL_1168232, EPI_ISL_1168233, EPI_ISL_1168234, EPI_ISL_1168236, EPI_ISL_1168241, EPI_ISL_1168242, EPI_ISL_1168243, EPI_ISL_1168245, EPI_ISL_1168246, EPI_ISL_1168248, EPI_ISL_1168249, EPI_ISL_1168250, EPI_ISL_1168251, EPI_ISL_1168252, EPI_ISL_1168253, EPI_ISL_1168254, EPI_ISL_1168256, EPI_ISL_1168257, EPI_ISL_1168261, EPI_ISL_1168262, EPI_ISL_1168264, EPI_ISL_1168265, EPI_ISL_1168266, EPI_ISL_1168267, EPI_ISL_1168268, EPI_ISL_1168269, EPI_ISL_1168270, EPI_ISL_1168271, EPI_ISL_1168272, EPI_ISL_1168274, EPI_ISL_1168279, EPI_ISL_1168284, EPI_ISL_1168286 | | | |
| see above | TXDSHS | TXDSHS | Rashmi Tuladhar, Bonnie Oh, Jenny Zhang, Maliha Rahman, Mayela Pedrueza, Anita Pokharel, Myong Koag, Chun Wang, Rachel Lee, Grace Kubin |
| EPI_ISL_1168348, EPI_ISL_1168349, EPI_ISL_1168350, EPI_ISL_1168351, EPI_ISL_1168352, EPI_ISL_1168353 | LSUHS Emerging Viral Threat Laboratory | Microbial Genome Sequencing Center | Jeremy P. Kamil, Jennifer L. Carroll, Rona S. Scott, Maarten Van Diest, Andrew D. Yurochko, Christopher G. Kevil, Daniel J. Snyder, Vaughn S. Cooper, John A. Vanchiere |
| EPI_ISL_1168354, EPI_ISL_1168355, EPI_ISL_1168356, EPI_ISL_1168357, EPI_ISL_1168358, EPI_ISL_1168359, EPI_ISL_1168360, EPI_ISL_1168361, EPI_ISL_1168362, EPI_ISL_1168363, EPI_ISL_1168364, EPI_ISL_1168365, EPI_ISL_1168366, EPI_ISL_1168367, EPI_ISL_1168368, EPI_ISL_1168369, EPI_ISL_1168370, EPI_ISL_1168371, EPI_ISL_1168372, EPI_ISL_1168373, EPI_ISL_1168374, EPI_ISL_1168375, EPI_ISL_1168376, EPI_ISL_1168377, EPI_ISL_1168378, EPI_ISL_1168379, EPI_ISL_1168380, EPI_ISL_1168383, EPI_ISL_1168384, EPI_ISL_1168385, EPI_ISL_1168386, EPI_ISL_1168387, EPI_ISL_1168388, EPI_ISL_1168389, EPI_ISL_1168390, EPI_ISL_1168391, EPI_ISL_1168392, EPI_ISL_1168393, EPI_ISL_1168394, EPI_ISL_1168395, EPI_ISL_1168396, EPI_ISL_1168397, EPI_ISL_1168398, EPI_ISL_1168399, EPI_ISL_1168400, EPI_ISL_1168401, EPI_ISL_1168402, EPI_ISL_1168403, EPI_ISL_1168404, EPI_ISL_1168405, EPI_ISL_1168406, EPI_ISL_1168407, EPI_ISL_1168408, EPI_ISL_1168409, EPI_ISL_1168410, EPI_ISL_1168411, EPI_ISL_1168412, EPI_ISL_1168413, EPI_ISL_1168414, EPI_ISL_1168415, EPI_ISL_1168416, EPI_ISL_1168417, EPI_ISL_1168418, EPI_ISL_1168420, EPI_ISL_1168421, EPI_ISL_1168422, EPI_ISL_1168423, EPI_ISL_1168424, EPI_ISL_1168425, EPI_ISL_1168426, EPI_ISL_1168427, EPI_ISL_1168428, EPI_ISL_1168429, EPI_ISL_1168430, EPI_ISL_1168431, EPI_ISL_1168432, EPI_ISL_1168433, EPI_ISL_1168434, EPI_ISL_1168435, EPI_ISL_1168436, EPI_ISL_1168437, EPI_ISL_1168438, EPI_ISL_1168439, EPI_ISL_1168440, EPI_ISL_1168441 | | | |
| see above | VA Connecticut Healthcare System | Grubaugh Lab - Yale School of Public Health | Mary Petrone, Joseph Fauver, Tara Alpert, Chantal Vogels, Isabel Ott, Shaili Gupta, Danielle Plank, Nathan Grubaugh |
| EPI_ISL_1168442, EPI_ISL_1168443, EPI_ISL_1168444, EPI_ISL_1168445, EPI_ISL_1168446, EPI_ISL_1168447, EPI_ISL_1168448, EPI_ISL_1168449, EPI_ISL_1168450, EPI_ISL_1168451 | DC Public Health Lab/ Dept. of Forensic Sciences | DC Public Health Lab/ Dept. of Forensic Sciences | Scott Nguyen, Janis Doss, Elizabeth Zelaya, Sarah Scott, Connie Maza, Monica Mann, Brittany Hamilton, David Payne, Jocelyn Hauser |
| EPI_ISL_1168452, EPI_ISL_1168453, EPI_ISL_1168454, EPI_ISL_1168455, EPI_ISL_1168456, EPI_ISL_1168457, EPI_ISL_1168458, EPI_ISL_1168459, EPI_ISL_1168460, EPI_ISL_1168461, EPI_ISL_1168462, EPI_ISL_1168463, EPI_ISL_1168464, EPI_ISL_1168465, EPI_ISL_1168466, EPI_ISL_1168467, EPI_ISL_1168468, EPI_ISL_1168469, EPI_ISL_1168470, EPI_ISL_1168471, EPI_ISL_1168472, EPI_ISL_1168473, EPI_ISL_1168474, EPI_ISL_1168475, EPI_ISL_1168476, EPI_ISL_1168477, EPI_ISL_1168478, EPI_ISL_1168479, EPI_ISL_1168480, EPI_ISL_1168481, EPI_ISL_1168482, EPI_ISL_1168483, EPI_ISL_1168484, EPI_ISL_1168485, EPI_ISL_1168486, EPI_ISL_1168487, EPI_ISL_1168488, EPI_ISL_1168489, EPI_ISL_1168490, EPI_ISL_1168491, EPI_ISL_1168492, EPI_ISL_1168493, EPI_ISL_1168494, EPI_ISL_1168495, EPI_ISL_1168496, EPI_ISL_1168497, EPI_ISL_1168498, EPI_ISL_1168499, EPI_ISL_1168500, EPI_ISL_1168501, EPI_ISL_1168502, EPI_ISL_1168503, EPI_ISL_1168504, EPI_ISL_1168505, EPI_ISL_1168506, EPI_ISL_1168507, EPI_ISL_1168508, EPI_ISL_1168509, EPI_ISL_1168510, EPI_ISL_1168511, EPI_ISL_1168512, EPI_ISL_1168513, EPI_ISL_1168514, EPI_ISL_1168515, EPI_ISL_1168516, EPI_ISL_1168517, EPI_ISL_1168518, EPI_ISL_1168519, EPI_ISL_1168520, EPI_ISL_1168521, EPI_ISL_1168522, EPI_ISL_1168523, EPI_ISL_1168524, EPI_ISL_1168525, EPI_ISL_1168526, EPI_ISL_1168527, EPI_ISL_1168528, EPI_ISL_1168529, EPI_ISL_1168530, EPI_ISL_1168531, EPI_ISL_1168532, EPI_ISL_1168533, EPI_ISL_1168534, EPI_ISL_1168535, EPI_ISL_1168536, EPI_ISL_1168537, EPI_ISL_1168538, EPI_ISL_1168539, EPI_ISL_1168540, EPI_ISL_1168541, EPI_ISL_1168542, EPI_ISL_1168543, EPI_ISL_1168544, EPI_ISL_1168545, EPI_ISL_1168546, EPI_ISL_1168547, EPI_ISL_1168548, EPI_ISL_1168549, EPI_ISL_1168550, EPI_ISL_1168551, EPI_ISL_1168552, EPI_ISL_1168553, EPI_ISL_1168554, EPI_ISL_1168555, EPI_ISL_1168556, EPI_ISL_1168557, EPI_ISL_1168558, EPI_ISL_1168559, EPI_ISL_1168560, EPI_ISL_1168561, EPI_ISL_1168562, EPI_ISL_1168563, EPI_ISL_1168564, EPI_ISL_1168565, EPI_ISL_1168566, EPI_ISL_1168567, EPI_ISL_1168568, EPI_ISL_1168569, EPI_ISL_1168570, EPI_ISL_1168571, EPI_ISL_1168572, EPI_ISL_1168573, EPI_ISL_1168574, EPI_ISL_1168575, EPI_ISL_1168576, EPI_ISL_1168577, EPI_ISL_1168578, EPI_ISL_1168579, EPI_ISL_1168580, EPI_ISL_1168581, EPI_ISL_1168582, EPI_ISL_1168583, EPI_ISL_1168584, EPI_ISL_1168585, EPI_ISL_1168586, EPI_ISL_1168587, EPI_ISL_1168588, EPI_ISL_1168589, EPI_ISL_1168590, EPI_ISL_1168591, EPI_ISL_1168592, EPI_ISL_1168593, EPI_ISL_1168594, EPI_ISL_1168595, EPI_ISL_1168596, EPI_ISL_1168597, EPI_ISL_1168598, EPI_ISL_1168599, EPI_ISL_1168600, EPI_ISL_1168601, EPI_ISL_1168602, EPI_ISL_1168603, EPI_ISL_1168604, EPI_ISL_1168606, EPI_ISL_1168607, EPI_ISL_1168608, EPI_ISL_1168609, EPI_ISL_1168610, EPI_ISL_1168611, EPI_ISL_1168612, EPI_ISL_1168613, EPI_ISL_1168614, EPI_ISL_1168615, EPI_ISL_1168616, EPI_ISL_1168617, EPI_ISL_1168618, EPI_ISL_1168619, EPI_ISL_1168620, EPI_ISL_1168621, EPI_ISL_1168622, EPI_ISL_1168623, EPI_ISL_1168624, EPI_ISL_1168625, EPI_ISL_1168626, EPI_ISL_1168627, EPI_ISL_1168628, EPI_ISL_1168629, EPI_ISL_1168630, EPI_ISL_1168631, EPI_ISL_1168632, EPI_ISL_1168633 | | | |
| see above | Instituto de Diagnostico y Referencia Epidemiologicos INDRE_RNLSP | Instituto de Diagnostico y Referencia Epidemiologicos (INDRE) | Claudia Wong-Arambula, Abril Rodriguez-Maldonado, Vanessa Rivero-Arredondo, Ariadna Medina-Benitez, Joaquin Quiroz-Mercado,David Fragoso-Fonseca, Sergio Rangel-Guerrero, Natividad Cruz-Ortiz, Tatiana Nunez-Garcia, Gisela Barrera-Badillo, Lucia Hernandez-Rivas, Irma Lopez-Martinez, Ernesto Ramirez-Gonzalez. |
| EPI_ISL_1168634, EPI_ISL_1168635, EPI_ISL_1168636, EPI_ISL_1168637, EPI_ISL_1168638, EPI_ISL_1168639, EPI_ISL_1168640, EPI_ISL_1168641, EPI_ISL_1168642, EPI_ISL_1168643, EPI_ISL_1168644, EPI_ISL_1168645, EPI_ISL_1168646, EPI_ISL_1168647, EPI_ISL_1168648, EPI_ISL_1168649, EPI_ISL_1168650, EPI_ISL_1168651, EPI_ISL_1168652 | | | |
| see above | Instituto Nacional de Medicina Genomica | Instituto Nacional de Medicina Genomica | Hidalgo-Miranda A, Mendoza-Vargas A, Reyes-Grajeda JP, Cedro-Tanda A, Alcaraz N, Gonzalez-Barrera D, Rangel-DeLeon D, Ramirez-Vega O, Sifuentes-Rojas C, Cisneros-Villanueva M, Herrera-Montalvo LA |

| | | | |
|---|---|---|---|
| EPI_ISL_1168653, EPI_ISL_1168654, EPI_ISL_1168655, EPI_ISL_1168656, EPI_ISL_1168657, EPI_ISL_1168658, EPI_ISL_1168659, EPI_ISL_1168660, EPI_ISL_1168661, EPI_ISL_1168662, EPI_ISL_1168663, EPI_ISL_1168664, EPI_ISL_1168665, EPI_ISL_1168666, EPI_ISL_1168667, EPI_ISL_1168668, EPI_ISL_1168669, EPI_ISL_1168670, EPI_ISL_1168671, EPI_ISL_1168672, EPI_ISL_1168673, EPI_ISL_1168674, EPI_ISL_1168675, EPI_ISL_1168676, EPI_ISL_1168677, EPI_ISL_1168678, EPI_ISL_1168679, EPI_ISL_1168680, EPI_ISL_1168681 | | | |
| see above | Centers for Disease Control and Prevention, Dengue Branch | Centers for Disease Control and Prevention, Dengue Branch | Gilberto A. Santiago, Glenda Gonzalez, Betzabel Flores, Keyla Charriez, Gabriela Paz-Bailey, Jorge L. Munoz-Jordan |
| EPI_ISL_1168682, EPI_ISL_1168683, EPI_ISL_1168684, EPI_ISL_1168685 | Inno Diagnostics/Center for Research Resources, Ponce Medical School Foundation, Inc. | Centers for Disease Control and Prevention, Dengue Branch | Gilberto A. Santiago, Glenda Gonzalez, Betzabel Flores, Keyla Charriez, Andrea Arias-Garcia, Raphael Sanchez-Torres, Vanessa Rivera-Amill, Gabriela Paz-Bailey, Jorge L. Munoz-Jordan |
| EPI_ISL_1168686, EPI_ISL_1168687, EPI_ISL_1168688, EPI_ISL_1168689, EPI_ISL_1168690, EPI_ISL_1168691, EPI_ISL_1168692, EPI_ISL_1168693, EPI_ISL_1168694, EPI_ISL_1168695, EPI_ISL_1168696 | | | |
| see above | Puerto Rico Department of Health | Centers for Disease Control and Prevention, Dengue Branch | Gilberto A. Santiago, Glenda Gonzalez, Betzabel Flores, Keyla Charriez, Gabriela Paz-Bailey, Jorge L. Munoz-Jordan |
| EPI_ISL_1168697 | Clinical Molecular Microbiology Laboratory, UNC Hospital | Dirk Dittmer | Justin T. Landis , Razia Moorad , Brent A. Eason, Melissa B. Miller, Linda Pluta, Dirk Dittmer, Angelica Juarez, Cecilia Thompson, Shawn Hawken, Cameroon Grant, Evelyn Hoffman, Patricio Cano, Jason Wong, Carolina Caro-Vegas, Ryan McNamara, Blossom Damania. |
| EPI_ISL_1168698, EPI_ISL_1168699, EPI_ISL_1168700, EPI_ISL_1168701, EPI_ISL_1168702, EPI_ISL_1168703, EPI_ISL_1168704, EPI_ISL_1168705, EPI_ISL_1168706, EPI_ISL_1168707, EPI_ISL_1168708, EPI_ISL_1168709, EPI_ISL_1168710, EPI_ISL_1168711, EPI_ISL_1168712, EPI_ISL_1168713, EPI_ISL_1168714, EPI_ISL_1168715, EPI_ISL_1168716, EPI_ISL_1168717, EPI_ISL_1168718, EPI_ISL_1168719, EPI_ISL_1168720, EPI_ISL_1168721, EPI_ISL_1168722, EPI_ISL_1168723, EPI_ISL_1168724, EPI_ISL_1168725, EPI_ISL_1168726, EPI_ISL_1168727 | | | |
| see above | Houston Health Dept. | Houston Health Dept. | Ryker Penn, Pamela Brown, Adolpho Lara |
| EPI_ISL_1168728, EPI_ISL_1168729, EPI_ISL_1168730, EPI_ISL_1168731, EPI_ISL_1168732, EPI_ISL_1168733, EPI_ISL_1168734, EPI_ISL_1168735 | Biotia | Biotia | Dorottya Nagy-Szakal, Mara Couto-Rodriguez, Heather Wells, Joseph Barrows, Marilyne Debieu, Kristin Butcher, Siyuan Chen, Agnes Berki, Courteny Hager, Robert Boorstein, Mariah Taylor, Colleen Jonsson, Christopher Mason, Niamh O'Hara |
| EPI_ISL_1168737 | Connecticut Department of Health | Grubaugh Lab - Yale School of Public Health | Tara Alpert, Joseph Fauver, Anderson Brito, Mallery Breban, Anne Wyllie, Chantal Vogels, Mary Petrone, Annie Watkins, Chaney Kalinich, Isabel Ott, Nathan Grubaugh |
| EPI_ISL_1168739, EPI_ISL_1168742, EPI_ISL_1168743, EPI_ISL_1168744, EPI_ISL_1168748, EPI_ISL_1168749, EPI_ISL_1168750, EPI_ISL_1168753, EPI_ISL_1168755, EPI_ISL_1168757, EPI_ISL_1168758, EPI_ISL_1168760, EPI_ISL_1168761 | | | |
| see above | Tempus | Grubaugh Lab - Yale School of Public Health | Tara Alpert, Joseph Fauver, Anderson Brito, Mallery Breban, Anne Wyllie, Chantal Vogels, Mary Petrone, Annie Watkins, Chaney Kalinich, Isabel Ott, Nathan Grubaugh |
| EPI_ISL_1168766, EPI_ISL_1168767, EPI_ISL_1168768 | School of Pharmacy, Shenandoah University | School of Pharmacy, Shenandoah University | Adams,S.M., Harralson,A.F., Kidd,R.S., Sawyer,G.W. |
| EPI_ISL_1169487 | MN PHL Division, Minnesota Department of Health | Genomics and Discovery, Respiratory Viruses Branch, Division of Viral Diseases, Centers for Disease Control and Prevention | Krista Queen, Yan Li, Ying Tao, Jing Zhang, Anna Uehara, Anna Montmayeur, Clinton R. Paden, Peter W. Cook, Rachel Marine, Mili Sheth, Jasmine Padilla, Sarah Nobles, Mark Burroughs, Lori Rowe, Haibin Wang, Ben L. Rambo-Martin, Dhwani Batra, Justin Lee, Suxiang Tong |
| EPI_ISL_1169488 | MI - Michigan Department of Health and Human Services - Bureau of Laboratories | Genomics and Discovery, Respiratory Viruses Branch, Division of Viral Diseases, Centers for Disease Control and Prevention | Krista Queen, Yan Li, Ying Tao, Jing Zhang, Anna Uehara, Anna Montmayeur, Clinton R. Paden, Peter W. Cook, Rachel Marine, Mili Sheth, Jasmine Padilla, Sarah Nobles, Mark Burroughs, Lori Rowe, Haibin Wang, Ben L. Rambo-Martin, Dhwani Batra, Justin Lee, Suxiang Tong |
| EPI_ISL_1169489 | FL Bureau of Public Health Laboratories-Miami | Genomics and Discovery, Respiratory Viruses Branch, Division of Viral Diseases, Centers for Disease Control and Prevention | Krista Queen, Yan Li, Ying Tao, Jing Zhang, Anna Uehara, Anna Montmayeur, Clinton R. Paden, Peter W. Cook, Rachel Marine, Mili Sheth, Jasmine Padilla, Sarah Nobles, Mark Burroughs, Lori Rowe, Haibin Wang, Ben L. Rambo-Martin, Dhwani Batra, Justin Lee, Suxiang Tong |
| EPI_ISL_1169490 | OR State PHL-Virology/Immunology Section | Genomics and Discovery, Respiratory Viruses Branch, Division of Viral Diseases, Centers for Disease Control and Prevention | Krista Queen, Yan Li, Ying Tao, Jing Zhang, Anna Uehara, Anna Montmayeur, Clinton R. Paden, Peter W. Cook, Rachel Marine, Mili Sheth, Jasmine Padilla, Sarah Nobles, Mark Burroughs, Lori Rowe, Haibin Wang, Ben L. Rambo-Martin, Dhwani Batra, Justin Lee, Suxiang Tong |
| EPI_ISL_1169491, EPI_ISL_1169492 | MN PHL Division, Minnesota Department of Health | Genomics and Discovery, Respiratory Viruses Branch, Division of Viral Diseases, Centers for Disease Control and Prevention | Krista Queen, Yan Li, Ying Tao, Jing Zhang, Anna Uehara, Anna Montmayeur, Clinton R. Paden, Peter W. Cook, Rachel Marine, Mili Sheth, Jasmine Padilla, Sarah Nobles, Mark Burroughs, Lori Rowe, Haibin Wang, Ben L. Rambo-Martin, Dhwani Batra, Justin Lee, Suxiang Tong |
| EPI_ISL_1169498 | OR State PHL-Virology/Immunology Section | Genomics and Discovery, Respiratory Viruses Branch, Division of Viral Diseases, Centers for Disease Control and Prevention | Krista Queen, Yan Li, Ying Tao, Jing Zhang, Anna Uehara, Anna Montmayeur, Clinton R. Paden, Peter W. Cook, Rachel Marine, Mili Sheth, Jasmine Padilla, Sarah Nobles, Mark Burroughs, Lori Rowe, Haibin Wang, Ben L. Rambo-Martin, Dhwani Batra, Justin Lee, Suxiang Tong |
| EPI_ISL_1169500 | WY Public Health Laboratory | Genomics and Discovery, Respiratory Viruses Branch, Division of Viral Diseases, Centers for Disease Control and Prevention | Krista Queen, Yan Li, Ying Tao, Jing Zhang, Anna Uehara, Anna Montmayeur, Clinton R. Paden, Peter W. Cook, Rachel Marine, Mili Sheth, Jasmine Padilla, Sarah Nobles, Mark Burroughs, Lori Rowe, Haibin Wang, Ben L. Rambo-Martin, Dhwani Batra, Justin Lee, Suxiang Tong |
| EPI_ISL_1169501 | SC Dept of Health and Env. Control-Bureau of Laboratories | Genomics and Discovery, Respiratory Viruses Branch, Division of Viral Diseases, Centers for Disease Control and Prevention | Krista Queen, Yan Li, Ying Tao, Jing Zhang, Anna Uehara, Anna Montmayeur, Clinton R. Paden, Peter W. Cook, Rachel Marine, Mili Sheth, Jasmine Padilla, Sarah Nobles, Mark Burroughs, Lori Rowe, Haibin Wang, Ben L. Rambo-Martin, Dhwani Batra, Justin Lee, Suxiang Tong |
| EPI_ISL_1169503 | MT Public Health Laboratory | Genomics and Discovery, Respiratory Viruses Branch, Division of Viral Diseases, Centers for Disease Control and Prevention | Krista Queen, Yan Li, Ying Tao, Jing Zhang, Anna Uehara, Anna Montmayeur, Clinton R. Paden, Peter W. Cook, Rachel Marine, Mili Sheth, Jasmine Padilla, Sarah Nobles, Mark Burroughs, Lori Rowe, Haibin Wang, Ben L. Rambo-Martin, Dhwani Batra, Justin Lee, Suxiang Tong |
| EPI_ISL_1169545, EPI_ISL_1169546, EPI_ISL_1169547, EPI_ISL_1169548, EPI_ISL_1169550, EPI_ISL_1169551, EPI_ISL_1169552, EPI_ISL_1169553, EPI_ISL_1169554, EPI_ISL_1169555, EPI_ISL_1169556, EPI_ISL_1169557, EPI_ISL_1169558, EPI_ISL_1169559, EPI_ISL_1169560, EPI_ISL_1169561 | | | |
| see above | MN PHL Division, Minnesota Department of Health | Genomics and Discovery, Respiratory Viruses Branch, Division of Viral Diseases, Centers for Disease Control and Prevention | Krista Queen, Yan Li, Ying Tao, Jing Zhang, Anna Uehara, Anna Montmayeur, Clinton R. Paden, Peter W. Cook, Rachel Marine, Mili Sheth, Jasmine Padilla, Sarah Nobles, Mark Burroughs, Lori Rowe, Haibin Wang, Ben L. Rambo-Martin, Dhwani Batra, Justin Lee, Suxiang Tong |
| EPI_ISL_1169562, EPI_ISL_1169563, EPI_ISL_1169564, EPI_ISL_1169565, EPI_ISL_1169566, EPI_ISL_1169567, EPI_ISL_1169568, EPI_ISL_1169569, EPI_ISL_1169570, EPI_ISL_1169571, EPI_ISL_1169572, EPI_ISL_1169573, EPI_ISL_1169574, EPI_ISL_1169575, EPI_ISL_1169576, EPI_ISL_1169577, EPI_ISL_1169578 | | | |
| see above | Swedish Medical Center, Seattle, WA, USA | Institute for Systems Biology | Jason D. Goldman, Chris Lausted, Kai Wang, Jared C. Roach, James R. Heath, Julie A. Wallick |
| EPI_ISL_1169579, EPI_ISL_1169580, EPI_ISL_1169581, EPI_ISL_1169582, EPI_ISL_1169583, EPI_ISL_1169584, EPI_ISL_1169585, EPI_ISL_1169586, EPI_ISL_1169587, EPI_ISL_1169590, EPI_ISL_1169591, EPI_ISL_1169592, EPI_ISL_1169593, EPI_ISL_1169594, EPI_ISL_1169595, EPI_ISL_1169596, EPI_ISL_1169597, EPI_ISL_1169598, EPI_ISL_1169599, EPI_ISL_1169600 | | | |
| see above | Santa Clara County Public Health Laboratory | Santa Clara County Public Health Laboratory | Santa Clara County Public Health Department |
| EPI_ISL_1169601, EPI_ISL_1169602, EPI_ISL_1169603, EPI_ISL_1169604, EPI_ISL_1169605 | Wyoming Public Health Laboratory | Wyoming Public Health Laboratory | Jim Mildenberger, Wanda Manley, Noah Hull, Taylor Fearing, Lynette Gumbleton, Channing Weber, Ashley Norberg, Chayse Rowley, Marley Goetz, Brian Dominguez, Elliot Thomasson, Cari Sloma, and Rob Christensen |
| EPI_ISL_1169609, EPI_ISL_1169610, EPI_ISL_1169611, EPI_ISL_1169612, EPI_ISL_1169613, EPI_ISL_1169614, EPI_ISL_1169615, EPI_ISL_1169616, EPI_ISL_1169617, EPI_ISL_1169618, EPI_ISL_1169619, EPI_ISL_1169620, EPI_ISL_1169621, EPI_ISL_1169622, EPI_ISL_1169623, EPI_ISL_1169624, EPI_ISL_1169625, EPI_ISL_1169626, EPI_ISL_1169627, EPI_ISL_1169628, EPI_ISL_1169629, EPI_ISL_1169630, EPI_ISL_1169631, EPI_ISL_1169632, EPI_ISL_1169633, EPI_ISL_1169634, EPI_ISL_1169635, EPI_ISL_1169636, EPI_ISL_1169637, EPI_ISL_1169638, EPI_ISL_1169639, EPI_ISL_1169640, EPI_ISL_1169641, EPI_ISL_1169642, EPI_ISL_1169643, EPI_ISL_1169644, EPI_ISL_1169645, EPI_ISL_1169646, EPI_ISL_1169647, EPI_ISL_1169648, EPI_ISL_1169649, EPI_ISL_1169650, EPI_ISL_1169651, EPI_ISL_1169652, EPI_ISL_1169653, EPI_ISL_1169654, EPI_ISL_1169655, EPI_ISL_1169656, EPI_ISL_1169657, EPI_ISL_1169658, EPI_ISL_1169659, EPI_ISL_1169660, EPI_ISL_1169661, EPI_ISL_1169662, EPI_ISL_1169663, EPI_ISL_1169664, EPI_ISL_1169665, EPI_ISL_1169666, EPI_ISL_1169667, EPI_ISL_1169668, EPI_ISL_1169669, EPI_ISL_1169670, EPI_ISL_1169671, EPI_ISL_1169672, EPI_ISL_1169673, EPI_ISL_1169674, EPI_ISL_1169687, EPI_ISL_1169688, EPI_ISL_1169689, EPI_ISL_1169690, EPI_ISL_1169691, EPI_ISL_1169692, EPI_ISL_1169693, EPI_ISL_1169694, EPI_ISL_1169695, EPI_ISL_1169696, EPI_ISL_1169697, EPI_ISL_1169698, EPI_ISL_1169699, EPI_ISL_1169700, EPI_ISL_1169701, EPI_ISL_1169702, EPI_ISL_1169703, EPI_ISL_1169704, EPI_ISL_1169705, EPI_ISL_1169706, EPI_ISL_1169707, EPI_ISL_1169708, EPI_ISL_1169709, EPI_ISL_1169710, EPI_ISL_1169711, EPI_ISL_1169712, EPI_ISL_1169713, EPI_ISL_1169714, EPI_ISL_1169715, EPI_ISL_1169716, EPI_ISL_1169717, EPI_ISL_1169718, EPI_ISL_1169719, EPI_ISL_1169720, EPI_ISL_1169721, EPI_ISL_1169722, EPI_ISL_1169723, EPI_ISL_1169724, EPI_ISL_1169725, EPI_ISL_1169726, EPI_ISL_1169727, EPI_ISL_1169728, EPI_ISL_1169729, EPI_ISL_1169730, EPI_ISL_1169731, EPI_ISL_1169732, EPI_ISL_1169733, EPI_ISL_1169734, EPI_ISL_1169735, EPI_ISL_1169736, EPI_ISL_1169737, EPI_ISL_1169738, EPI_ISL_1169739, EPI_ISL_1169740, EPI_ISL_1169741, EPI_ISL_1169742, EPI_ISL_1169743, EPI_ISL_1169744, EPI_ISL_1169745, EPI_ISL_1169746, EPI_ISL_1169747, EPI_ISL_1169748, EPI_ISL_1169749, EPI_ISL_1169750, EPI_ISL_1169751, EPI_ISL_1169752, EPI_ISL_1169753, EPI_ISL_1169754, EPI_ISL_1169755, EPI_ISL_1169756, EPI_ISL_1169757, EPI_ISL_1169758, EPI_ISL_1169759, EPI_ISL_1169760, EPI_ISL_1169761, EPI_ISL_1169762, EPI_ISL_1169763, EPI_ISL_1169764, EPI_ISL_1169765, EPI_ISL_1169766, EPI_ISL_1169767, EPI_ISL_1169768, EPI_ISL_1169769, EPI_ISL_1169770, EPI_ISL_1169771, EPI_ISL_1169772, EPI_ISL_1169773, EPI_ISL_1169774, EPI_ISL_1169775, EPI_ISL_1169776, EPI_ISL_1169777, EPI_ISL_1169778, EPI_ISL_1169779, EPI_ISL_1169780, EPI_ISL_1169781, EPI_ISL_1169782, EPI_ISL_1169783, EPI_ISL_1169784, EPI_ISL_1169785, EPI_ISL_1169786, EPI_ISL_1169787, EPI_ISL_1169788, EPI_ISL_1169789, EPI_ISL_1169790, EPI_ISL_1169791, EPI_ISL_1169792, EPI_ISL_1169793, EPI_ISL_1169794, EPI_ISL_1169795, EPI_ISL_1169796, EPI_ISL_1169797, EPI_ISL_1169798, EPI_ISL_1169799, EPI_ISL_1169800, EPI_ISL_1169801, EPI_ISL_1169802, EPI_ISL_1169803, EPI_ISL_1169804, EPI_ISL_1169805, EPI_ISL_1169806, EPI_ISL_1169807, EPI_ISL_1169808, EPI_ISL_1169809, EPI_ISL_1169810, EPI_ISL_1169811, EPI_ISL_1169812, EPI_ISL_1169813, EPI_ISL_1169814, EPI_ISL_1169815, EPI_ISL_1169816, EPI_ISL_1169817 | | | |
| see above | Colorado Department of Public Health and Environment | Colorado Department of Public Health and Environment | Laura Bankers, Molly C. Hetherington-Rauth, Diana Ir, Shannon Ely, Shannon R. Matzinger, Sarah Elizabeth Totten, Emily A. Travanty |
| EPI_ISL_1169818, EPI_ISL_1169819, EPI_ISL_1169820, EPI_ISL_1169821, EPI_ISL_1169822, EPI_ISL_1169823, EPI_ISL_1169824, EPI_ISL_1169826, EPI_ISL_1169827, EPI_ISL_1169828, EPI_ISL_1169829, EPI_ISL_1169830, EPI_ISL_1169831, EPI_ISL_1169832, EPI_ISL_1169833, EPI_ISL_1169834, EPI_ISL_1169835, EPI_ISL_1169836, EPI_ISL_1169837, EPI_ISL_1169838, EPI_ISL_1169839, EPI_ISL_1169840, EPI_ISL_1169841, EPI_ISL_1169842, EPI_ISL_1169843, EPI_ISL_1169844, EPI_ISL_1169845, EPI_ISL_1169846, EPI_ISL_1169847, EPI_ISL_1169848, EPI_ISL_1169849, EPI_ISL_1169850, EPI_ISL_1169851, EPI_ISL_1169852 | | | |
| see above | Nevada State Public Health Laboratory | Nevada State Public Health Laboratory | Andrew Gorzalski, Mark Pandori |
| EPI_ISL_1169912, EPI_ISL_1169913, EPI_ISL_1169914, EPI_ISL_1169915, EPI_ISL_1169916, EPI_ISL_1169917, EPI_ISL_1169918 | Nebraska Public Health Laboratory | NPHL COVID-19 Response Team | NPHL COVID-19 Response Team |
| EPI_ISL_1169937, EPI_ISL_1169938, EPI_ISL_1169939, EPI_ISL_1169940, EPI_ISL_1169941, EPI_ISL_1169942, EPI_ISL_1169960, EPI_ISL_1169961, EPI_ISL_1169962, EPI_ISL_1169963, EPI_ISL_1169964, EPI_ISL_1169965, EPI_ISL_1169966, EPI_ISL_1169967, EPI_ISL_1169968, EPI_ISL_1169969, EPI_ISL_1169970, EPI_ISL_1169971, EPI_ISL_1169972, EPI_ISL_1169973, EPI_ISL_1169974 | | | |
| see above | Wyoming Public Health Laboratory | Wyoming Public Health Laboratory | Jim Mildenberger, Wanda Manley, Noah Hull, Taylor Fearing, Lynette Gumbleton, Channing Weber, Ashley Norberg, Chayse Rowley, Marley Goetz, Brian Dominguez, Elliot Thomasson, Cari Sloma, and Rob Christensen |

| | | | |
|---|---|---|---|
| EPI_ISL_1170959, EPI_ISL_1170960, EPI_ISL_1170961, EPI_ISL_1170962, EPI_ISL_1170963, EPI_ISL_1170964, EPI_ISL_1170965, EPI_ISL_1170966, EPI_ISL_1170967, EPI_ISL_1170968, EPI_ISL_1170969, EPI_ISL_1170971, EPI_ISL_1170972, EPI_ISL_1170973, EPI_ISL_1170974, EPI_ISL_1170975, EPI_ISL_1170976, EPI_ISL_1170977, EPI_ISL_1170978, EPI_ISL_1170979, EPI_ISL_1170980, EPI_ISL_1170981, EPI_ISL_1170982, EPI_ISL_1170983, EPI_ISL_1170984, EPI_ISL_1170985, EPI_ISL_1170986, EPI_ISL_1170987, EPI_ISL_1170988, EPI_ISL_1170989, EPI_ISL_1170990, EPI_ISL_1170991, EPI_ISL_1170993, EPI_ISL_1170994, EPI_ISL_1170995, EPI_ISL_1170996, EPI_ISL_1170997, EPI_ISL_1170998, EPI_ISL_1170999, EPI_ISL_1171000, EPI_ISL_1171001, EPI_ISL_1171002, EPI_ISL_1171003, EPI_ISL_1171004, EPI_ISL_1171005, EPI_ISL_1171006, EPI_ISL_1171007, EPI_ISL_1171008, EPI_ISL_1171009, EPI_ISL_1171010, EPI_ISL_1171011, EPI_ISL_1171013, EPI_ISL_1171014, EPI_ISL_1171015, EPI_ISL_1171016, EPI_ISL_1171017, EPI_ISL_1171018, EPI_ISL_1171019, EPI_ISL_1171020, EPI_ISL_1171021, EPI_ISL_1171022, EPI_ISL_1171023, EPI_ISL_1171024, EPI_ISL_1171025, EPI_ISL_1171026, EPI_ISL_1171027, EPI_ISL_1171028, EPI_ISL_1171029, EPI_ISL_1171030, EPI_ISL_1171031, EPI_ISL_1171032, EPI_ISL_1171033, EPI_ISL_1171034, EPI_ISL_1171035, EPI_ISL_1171036, EPI_ISL_1171037, EPI_ISL_1171038, EPI_ISL_1171039, EPI_ISL_1171040, EPI_ISL_1171041, EPI_ISL_1171042, EPI_ISL_1171043, EPI_ISL_1171044, EPI_ISL_1171045, EPI_ISL_1171046, EPI_ISL_1171047, EPI_ISL_1171048, EPI_ISL_1171049, EPI_ISL_1171050, EPI_ISL_1171051, EPI_ISL_1171052, EPI_ISL_1171053, EPI_ISL_1171054, EPI_ISL_1171055, EPI_ISL_1171056, EPI_ISL_1171057, EPI_ISL_1171058, EPI_ISL_1171059, EPI_ISL_1171060, EPI_ISL_1171061, EPI_ISL_1171062, EPI_ISL_1171063, EPI_ISL_1171064, EPI_ISL_1171065, EPI_ISL_1171066, EPI_ISL_1171067, EPI_ISL_1171068, EPI_ISL_1171069, EPI_ISL_1171070, EPI_ISL_1171071, EPI_ISL_1171072, EPI_ISL_1171073, EPI_ISL_1171074, EPI_ISL_1171075, EPI_ISL_1171076, EPI_ISL_1171077, EPI_ISL_1171078, EPI_ISL_1171079, EPI_ISL_1171080, EPI_ISL_1171081, EPI_ISL_1171082, EPI_ISL_1171083, EPI_ISL_1171084, EPI_ISL_1171085, EPI_ISL_1171086, EPI_ISL_1171087, EPI_ISL_1171088, EPI_ISL_1171089, EPI_ISL_1171090, EPI_ISL_1171091, EPI_ISL_1171092, EPI_ISL_1171093, EPI_ISL_1171094, EPI_ISL_1171095, EPI_ISL_1171096, EPI_ISL_1171097, EPI_ISL_1171098, EPI_ISL_1171099, EPI_ISL_1171100, EPI_ISL_1171101, EPI_ISL_1171102, EPI_ISL_1171103, EPI_ISL_1171104, EPI_ISL_1171105, EPI_ISL_1171106, EPI_ISL_1171107, EPI_ISL_1171108, EPI_ISL_1171109, EPI_ISL_1171110, EPI_ISL_1171111, EPI_ISL_1171112, EPI_ISL_1171113, EPI_ISL_1171114, EPI_ISL_1171115, EPI_ISL_1171116, EPI_ISL_1171117, EPI_ISL_1171118, EPI_ISL_1171119, EPI_ISL_1171120, EPI_ISL_1171121, EPI_ISL_1171122, EPI_ISL_1171123, EPI_ISL_1171124, EPI_ISL_1171125, EPI_ISL_1171126, EPI_ISL_1171127, EPI_ISL_1171128, EPI_ISL_1171129, EPI_ISL_1171130, EPI_ISL_1171131, EPI_ISL_1171132, EPI_ISL_1171133, EPI_ISL_1171134, EPI_ISL_1171135, EPI_ISL_1171136, EPI_ISL_1171137, EPI_ISL_1171138, EPI_ISL_1171139, EPI_ISL_1171140, EPI_ISL_1171141, EPI_ISL_1171142, EPI_ISL_1171143, EPI_ISL_1171144, EPI_ISL_1171145, EPI_ISL_1171146, EPI_ISL_1171147, EPI_ISL_1171148, EPI_ISL_1171149, EPI_ISL_1171150, EPI_ISL_1171151, EPI_ISL_1171152, EPI_ISL_1171153, EPI_ISL_1171154, EPI_ISL_1171155, EPI_ISL_1171156, EPI_ISL_1171157, EPI_ISL_1171158, EPI_ISL_1171159, EPI_ISL_1171160, EPI_ISL_1171161, EPI_ISL_1171162, EPI_ISL_1171163, EPI_ISL_1171164, EPI_ISL_1171165, EPI_ISL_1171166, EPI_ISL_1171167, EPI_ISL_1171168, EPI_ISL_1171169, EPI_ISL_1171170, EPI_ISL_1171171, EPI_ISL_1171172, EPI_ISL_1171173, EPI_ISL_1171174, EPI_ISL_1171175, EPI_ISL_1171176, EPI_ISL_1171177, EPI_ISL_1171178, EPI_ISL_1171179, EPI_ISL_1171180, EPI_ISL_1171181, EPI_ISL_1171182, EPI_ISL_1171183, EPI_ISL_1171184, EPI_ISL_1171185, EPI_ISL_1171186, EPI_ISL_1171187, EPI_ISL_1171188, EPI_ISL_1171189, EPI_ISL_1171190, EPI_ISL_1171191, EPI_ISL_1171192, EPI_ISL_1171193, EPI_ISL_1171194, EPI_ISL_1171195, EPI_ISL_1171196, EPI_ISL_1171197, EPI_ISL_1171198, EPI_ISL_1171199, EPI_ISL_1171200, EPI_ISL_1171201, EPI_ISL_1171202, EPI_ISL_1171203, EPI_ISL_1171204, EPI_ISL_1171205, EPI_ISL_1171206, EPI_ISL_1171207, EPI_ISL_1171208, EPI_ISL_1171209, EPI_ISL_1171210, EPI_ISL_1171211, EPI_ISL_1171212, EPI_ISL_1171213, EPI_ISL_1171214, EPI_ISL_1171215, EPI_ISL_1171216, EPI_ISL_1171217, EPI_ISL_1171218, EPI_ISL_1171219, EPI_ISL_1171220, EPI_ISL_1171221, EPI_ISL_1171222, EPI_ISL_1171223, EPI_ISL_1171224, EPI_ISL_1171225, EPI_ISL_1171226, EPI_ISL_1171227, EPI_ISL_1171228, EPI_ISL_1171229, EPI_ISL_1171230, EPI_ISL_1171231, EPI_ISL_1171232, EPI_ISL_1171233, EPI_ISL_1171234, EPI_ISL_1171235, EPI_ISL_1171236, EPI_ISL_1171237, EPI_ISL_1171238, EPI_ISL_1171240, EPI_ISL_1171241, EPI_ISL_1171242, EPI_ISL_1171243, EPI_ISL_1171244, EPI_ISL_1171245, EPI_ISL_1171246, EPI_ISL_1171247, EPI_ISL_1171248, EPI_ISL_1171249, EPI_ISL_1171250, EPI_ISL_1171251, EPI_ISL_1171252, EPI_ISL_1171253, EPI_ISL_1171254, EPI_ISL_1171255, EPI_ISL_1171256, EPI_ISL_1171257, EPI_ISL_1171258, EPI_ISL_1171259, EPI_ISL_1171260, EPI_ISL_1171261, EPI_ISL_1171262, EPI_ISL_1171263, EPI_ISL_1171264, EPI_ISL_1171265, EPI_ISL_1171266, EPI_ISL_1171267, EPI_ISL_1171268, EPI_ISL_1171269, EPI_ISL_1171270, EPI_ISL_1171271, EPI_ISL_1171272, EPI_ISL_1171273, EPI_ISL_1171274, EPI_ISL_1171275, EPI_ISL_1171276, EPI_ISL_1171277, EPI_ISL_1171279, EPI_ISL_1171280, EPI_ISL_1171281, EPI_ISL_1171282, EPI_ISL_1171283, EPI_ISL_1171284, EPI_ISL_1171285, EPI_ISL_1171286, EPI_ISL_1171287, EPI_ISL_1171288, EPI_ISL_1171289, EPI_ISL_1171290, EPI_ISL_1171291, EPI_ISL_1171292, EPI_ISL_1171294, EPI_ISL_1171295, EPI_ISL_1171297, EPI_ISL_1171298, EPI_ISL_1171299, EPI_ISL_1171300, EPI_ISL_1171301, EPI_ISL_1171303, EPI_ISL_1171304, EPI_ISL_1171305, EPI_ISL_1171306, EPI_ISL_1171307, EPI_ISL_1171308, EPI_ISL_1171309, EPI_ISL_1171310, EPI_ISL_1171311, EPI_ISL_1171312, EPI_ISL_1171313, EPI_ISL_1171314, EPI_ISL_1171315, EPI_ISL_1171316, EPI_ISL_1171317, EPI_ISL_1171318, EPI_ISL_1171319, EPI_ISL_1171320, EPI_ISL_1171321, EPI_ISL_1171322, EPI_ISL_1171323, EPI_ISL_1171324, EPI_ISL_1171325, EPI_ISL_1171326, EPI_ISL_1171327, EPI_ISL_1171328, EPI_ISL_1171329, EPI_ISL_1171330, EPI_ISL_1171331, EPI_ISL_1171333, EPI_ISL_1171334, EPI_ISL_1171336, EPI_ISL_1171338, EPI_ISL_1171339, EPI_ISL_1171340, EPI_ISL_1171342, EPI_ISL_1171343, EPI_ISL_1171344, EPI_ISL_1171346, EPI_ISL_1171348, EPI_ISL_1171349, EPI_ISL_1171350, EPI_ISL_1171351, EPI_ISL_1171352, EPI_ISL_1171353, EPI_ISL_1171356, EPI_ISL_1171357, EPI_ISL_1171358, EPI_ISL_1171359, EPI_ISL_1171360, EPI_ISL_1171361, EPI_ISL_1171362, EPI_ISL_1171363, EPI_ISL_1171364, EPI_ISL_1171365, EPI_ISL_1171366, EPI_ISL_1171367, EPI_ISL_1171369, EPI_ISL_1171370, EPI_ISL_1171372, EPI_ISL_1171373, EPI_ISL_1171374, EPI_ISL_1171375, EPI_ISL_1171381, EPI_ISL_1171382, EPI_ISL_1171384, EPI_ISL_1171385, EPI_ISL_1171386, EPI_ISL_1171387, EPI_ISL_1171388, EPI_ISL_1171389, EPI_ISL_1171390, EPI_ISL_1171391, EPI_ISL_1171392, EPI_ISL_1171393, EPI_ISL_1171394, EPI_ISL_1171395, EPI_ISL_1171396, EPI_ISL_1171398, EPI_ISL_1171400, EPI_ISL_1171401, EPI_ISL_1171402, EPI_ISL_1171403, EPI_ISL_1171404, EPI_ISL_1171405, EPI_ISL_1171406, EPI_ISL_1171407, EPI_ISL_1171408, EPI_ISL_1171409, EPI_ISL_1171410, EPI_ISL_1171411, EPI_ISL_1171412, EPI_ISL_1171417, EPI_ISL_1171418, EPI_ISL_1171420, EPI_ISL_1171421, EPI_ISL_1171422, EPI_ISL_1171423, EPI_ISL_1171424, EPI_ISL_1171426, EPI_ISL_1171427, EPI_ISL_1171428, EPI_ISL_1171430, EPI_ISL_1171431, EPI_ISL_1171433, EPI_ISL_1171434, EPI_ISL_1171435, EPI_ISL_1171437, EPI_ISL_1171440, EPI_ISL_1171442, EPI_ISL_1171443, EPI_ISL_1171446, EPI_ISL_1171448, EPI_ISL_1171449, EPI_ISL_1171451, EPI_ISL_1171454, EPI_ISL_1171455, EPI_ISL_1171456, EPI_ISL_1171457, EPI_ISL_1171458, EPI_ISL_1171459, EPI_ISL_1171461, EPI_ISL_1171462, EPI_ISL_1171463, EPI_ISL_1171464 | | | |
| see above | OHSU Lab Services Molecular Microbiology Lab | Ginkgo Bioworks Clinical Laboratory | Brendan L. O'Connell, Sally Grindstaff, Kayla Carter, Ruth V. Nichols, Malaika Mckenzie-Bennett, James McGann, Jim Griffin, Keith Robison, Alex Plocik, Becky Schilling, Martha Pierson, Rebecca Littlefield, Michelle Spencer, Amanda Riccardi, Birgitte Simen, Alec J. Hirsch, Donna Hansel, Guang Fan, Xuan Qin, Daniel N. Streblow, William B. Messer, Andrew C. Adey, Benjamin N. Bimber, Brian J. O'Roak |
| EPI_ISL_1171707 | Baylor Scott & White-Temple | Baylor Scott & White-Temple | ri Rao, Linden Morales, Kimberly Walker, Marcus Volz, Shelby Hendrickson, Caitlin Maloney |
| EPI_ISL_1171710, EPI_ISL_1171711 | Baylor Scott & White-Temple | Baylor Scott & White-Temple | Ari Rao, Linden Morales, Kimberly Walker, Marcus Volz, Shelby Hendrickson, Caitlin Maloney |
| EPI_ISL_1171713, EPI_ISL_1171714, EPI_ISL_1171715, EPI_ISL_1171716, EPI_ISL_1171719, EPI_ISL_1171720, EPI_ISL_1171721, EPI_ISL_1171722, EPI_ISL_1171723, EPI_ISL_1171724, EPI_ISL_1171725, EPI_ISL_1171726, EPI_ISL_1171728, EPI_ISL_1171729, EPI_ISL_1171730, EPI_ISL_1171732, EPI_ISL_1171733, EPI_ISL_1171734, EPI_ISL_1171735, EPI_ISL_1171736, EPI_ISL_1171738, EPI_ISL_1171739, EPI_ISL_1171740, EPI_ISL_1171741, EPI_ISL_1171742, EPI_ISL_1171743, EPI_ISL_1171744, EPI_ISL_1171745, EPI_ISL_1171746, EPI_ISL_1171747, EPI_ISL_1171748, EPI_ISL_1171749, EPI_ISL_1171750, EPI_ISL_1171751, EPI_ISL_1171752, EPI_ISL_1171753, EPI_ISL_1171754, EPI_ISL_1171755, EPI_ISL_1171756, EPI_ISL_1171757, EPI_ISL_1171758, EPI_ISL_1171759, EPI_ISL_1171760, EPI_ISL_1171761, EPI_ISL_1171762, EPI_ISL_1171763, EPI_ISL_1171764, EPI_ISL_1171765, EPI_ISL_1171766, EPI_ISL_1171767, EPI_ISL_1171768, EPI_ISL_1171769 | | | |
| see above | Wyoming Public Health Laboratory | Wyoming Public Health Laboratory | Jim Mildenberger, Wanda Manley, Noah Hull, Taylor Fearing, Lynette Gumbleton, Channing Weber, Ashley Norberg, Chayse Rowley, Marley Goetz, Brian Dominguez, Elliot Thomasson, Cari Sloma, and Rob Christensen |
| EPI_ISL_1171770 | Baylor Scott & White-Temple | Baylor Scott & White-Temple | Ari Rao, Linden Morales, Kimberly Walker, Marcus Volz, Shelby Hendrickson, Caitlin Maloney |
| EPI_ISL_1171771, EPI_ISL_1171772, EPI_ISL_1171773, EPI_ISL_1171774, EPI_ISL_1171775, EPI_ISL_1171776, EPI_ISL_1171777, EPI_ISL_1171778, EPI_ISL_1171779, EPI_ISL_1171780, EPI_ISL_1171781, EPI_ISL_1171782, EPI_ISL_1171783, EPI_ISL_1171784, EPI_ISL_1171785, EPI_ISL_1171786, EPI_ISL_1171787, EPI_ISL_1171789, EPI_ISL_1171790, EPI_ISL_1171791, EPI_ISL_1171792, EPI_ISL_1171793, EPI_ISL_1171794, EPI_ISL_1171796, EPI_ISL_1171797, EPI_ISL_1171798, EPI_ISL_1171799, EPI_ISL_1171800, EPI_ISL_1171801, EPI_ISL_1171802, EPI_ISL_1171803, EPI_ISL_1171804, EPI_ISL_1171805, EPI_ISL_1171806, EPI_ISL_1171807, EPI_ISL_1171808, EPI_ISL_1171809, EPI_ISL_1171810, EPI_ISL_1171811, EPI_ISL_1171812, EPI_ISL_1171813, EPI_ISL_1171814, EPI_ISL_1171815, EPI_ISL_1171816, EPI_ISL_1171817, EPI_ISL_1171818, EPI_ISL_1171819, EPI_ISL_1171820, EPI_ISL_1171821, EPI_ISL_1171822, EPI_ISL_1171823, EPI_ISL_1171824, EPI_ISL_1171825, EPI_ISL_1171826, EPI_ISL_1171827, EPI_ISL_1171828, EPI_ISL_1171829, EPI_ISL_1171830, EPI_ISL_1171831, EPI_ISL_1171832, EPI_ISL_1171833, EPI_ISL_1171834, EPI_ISL_1171836, EPI_ISL_1171837, EPI_ISL_1171838, EPI_ISL_1171839, EPI_ISL_1171840, EPI_ISL_1171841, EPI_ISL_1171842, EPI_ISL_1171843, EPI_ISL_1171844, EPI_ISL_1171845, EPI_ISL_1171846, EPI_ISL_1171847, EPI_ISL_1171848, EPI_ISL_1171849, EPI_ISL_1171850, EPI_ISL_1171851, EPI_ISL_1171852, EPI_ISL_1171853, EPI_ISL_1171854, EPI_ISL_1171855 | | | |
| see above | Johns Hopkins Hospital Department of Pathology | Johns Hopkins Hospital Department of Pathology | C. Paul Morris, Chun Huai Luo, Adannaya Amadi, Matthew Schwartz, Nicholas Gallagher, Heba H. Mostafa |
| EPI_ISL_1171856 | MSHS Clinical Microbiology Laboratories | MSHS Pathogen Surveillance Program | Fatima Amanat, Mahima Thapa, Tinting Lei, Shaza M. Sayed Ahmed, Daniel C. Adelsberg, Juan Manuel Carreno, Shirin Strohmeier, Aaron J. Schmitz, Sarah Zafar, Julian Q Zhou, Willemijn Rijnink, Hala Alshammary, Nicholas Borcherding, Ana Gonzalez Reiche, Komal Srivastava, Emilia Mia Sordillo, Harm van Bakel, The Personalized Virology Initiative, Jackson S. Turner, Goran Bajic, Viviana Simon, Ali H. Ellebedy, Florian Krammer |
| EPI_ISL_1171857 | Department of Microbiology, Icahn School of Medicine at Mount Sinai | MSHS Pathogen Surveillance Program | Fatima Amanat, Mahima Thapa, Tinting Lei, Shaza M. Sayed Ahmed, Daniel C. Adelsberg, Juan Manuel Carreno, Shirin Strohmeier, Aaron J. Schmitz, Sarah Zafar, Julian Q Zhou, Willemijn Rijnink, Hala Alshammary, Nicholas Borcherding, Ana Gonzalez Reiche, Komal Srivastava, Emilia Mia Sordillo, Harm van Bakel, The Personalized Virology Initiative, Jackson S. Turner, Goran Bajic, Viviana Simon, Ali H. Ellebedy, Florian Krammer |
| EPI_ISL_1171858, EPI_ISL_1171859, EPI_ISL_1171860, EPI_ISL_1171861, EPI_ISL_1171862, EPI_ISL_1171864, EPI_ISL_1171865, EPI_ISL_1171866, EPI_ISL_1171867, EPI_ISL_1171868, EPI_ISL_1171869, EPI_ISL_1171870 | | | |
| see above | Kentucky State Public Health Lab | Kentucky State Public Health Lab | Stephanie Lunn, Karim George, Joshua Tobias, William Grooms, Vaneet Arora, Matthew Johnson, Rachel Zinner, Rhonda Lucas |
| EPI_ISL_1172018, EPI_ISL_1172019 | Baylor Scott & White-Temple | Baylor Scott & White-Temple | Ari Rao, Linden Morales, Kimberly Walker, Marcus Volz, Shelby Hendrickson, Caitlin Maloney |
| EPI_ISL_1172020 | Baylor Scott & White-Irving | Baylor Scott & White-Temple | Ari Rao, Linden Morales, Kimberly Walker, Marcus Volz, Shelby Hendrickson, Caitlin Maloney |
| EPI_ISL_1172021 | Baylor Scott & White-Temple | Baylor Scott & White-Temple | Ari Rao, Linden Morales, Kimberly Walker, Marcus Volz, Shelby Hendrickson, Caitlin Maloney |
| EPI_ISL_1172046, EPI_ISL_1172047, EPI_ISL_1172048, EPI_ISL_1172049, EPI_ISL_1172050, EPI_ISL_1172051, EPI_ISL_1172052, EPI_ISL_1172053, EPI_ISL_1172054, EPI_ISL_1172055, EPI_ISL_1172056, EPI_ISL_1172057, EPI_ISL_1172058, EPI_ISL_1172059, EPI_ISL_1172060, EPI_ISL_1172061, EPI_ISL_1172062, EPI_ISL_1172063, EPI_ISL_1172064, EPI_ISL_1172066, EPI_ISL_1172067, EPI_ISL_1172068, EPI_ISL_1172069, EPI_ISL_1172070, EPI_ISL_1172071, EPI_ISL_1172072, EPI_ISL_1172073, EPI_ISL_1172074, EPI_ISL_1172075, EPI_ISL_1172076, EPI_ISL_1172077, EPI_ISL_1172078, EPI_ISL_1172079, EPI_ISL_1172080, EPI_ISL_1172081, EPI_ISL_1172082, EPI_ISL_1172083, EPI_ISL_1172084, EPI_ISL_1172085, EPI_ISL_1172086, EPI_ISL_1172087, EPI_ISL_1172088, EPI_ISL_1172089, EPI_ISL_1172090, EPI_ISL_1172091, EPI_ISL_1172092, EPI_ISL_1172093, EPI_ISL_1172094, EPI_ISL_1172095, EPI_ISL_1172096, EPI_ISL_1172097, EPI_ISL_1172098, EPI_ISL_1172099, EPI_ISL_1172100, EPI_ISL_1172101, EPI_ISL_1172102, EPI_ISL_1172103, EPI_ISL_1172104, EPI_ISL_1172105, EPI_ISL_1172106, EPI_ISL_1172107, EPI_ISL_1172108, EPI_ISL_1172109 | | | |
| see above | Fulgent Genetics | Fulgent Genetics | Harry Gao, Mickey Li, John Gao, Joseph Fierro, Benafsh Sapra, Becky Tsai, Yan Meng, Doreen Ng, James Xie |
| EPI_ISL_1172110, EPI_ISL_1172111, EPI_ISL_1172112, EPI_ISL_1172113, EPI_ISL_1172114, EPI_ISL_1172115, EPI_ISL_1172117, EPI_ISL_1172118, EPI_ISL_1172119, EPI_ISL_1172120, EPI_ISL_1172121, EPI_ISL_1172122, EPI_ISL_1172123, EPI_ISL_1172124, EPI_ISL_1172125, EPI_ISL_1172126, EPI_ISL_1172127, EPI_ISL_1172128, EPI_ISL_1172129, EPI_ISL_1172130, EPI_ISL_1172131, EPI_ISL_1172132, EPI_ISL_1172133, EPI_ISL_1172134, EPI_ISL_1172135, EPI_ISL_1172137, EPI_ISL_1172138, EPI_ISL_1172139, EPI_ISL_1172140, EPI_ISL_1172141, EPI_ISL_1172142, EPI_ISL_1172143, EPI_ISL_1172144, EPI_ISL_1172145, EPI_ISL_1172146, EPI_ISL_1172147, EPI_ISL_1172150, EPI_ISL_1172151, EPI_ISL_1172152, EPI_ISL_1172153, EPI_ISL_1172154, EPI_ISL_1172155, EPI_ISL_1172156, EPI_ISL_1172157, EPI_ISL_1172158, EPI_ISL_1172159, EPI_ISL_1172160, EPI_ISL_1172162, EPI_ISL_1172163, EPI_ISL_1172164, EPI_ISL_1172165, EPI_ISL_1172166, EPI_ISL_1172167, EPI_ISL_1172168, EPI_ISL_1172169, EPI_ISL_1172170, EPI_ISL_1172171, EPI_ISL_1172172, EPI_ISL_1172173, EPI_ISL_1172174, EPI_ISL_1172175, EPI_ISL_1172176, EPI_ISL_1172177, EPI_ISL_1172178, EPI_ISL_1172179, EPI_ISL_1172180, EPI_ISL_1172181, EPI_ISL_1172183, EPI_ISL_1172184, EPI_ISL_1172185, EPI_ISL_1172186, EPI_ISL_1172187, EPI_ISL_1172188, EPI_ISL_1172189, EPI_ISL_1172190, EPI_ISL_1172191, EPI_ISL_1172193, EPI_ISL_1172194, EPI_ISL_1172195, EPI_ISL_1172196, EPI_ISL_1172197, EPI_ISL_1172198, EPI_ISL_1172199, EPI_ISL_1172200, EPI_ISL_1172201, EPI_ISL_1172202, EPI_ISL_1172203, EPI_ISL_1172204, EPI_ISL_1172205, EPI_ISL_1172206, EPI_ISL_1172207, EPI_ISL_1172208, EPI_ISL_1172209, EPI_ISL_1172210, EPI_ISL_1172211, EPI_ISL_1172212, EPI_ISL_1172213, EPI_ISL_1172214, EPI_ISL_1172215, EPI_ISL_1172216, EPI_ISL_1172217, EPI_ISL_1172218, EPI_ISL_1172220, EPI_ISL_1172221, EPI_ISL_1172222, EPI_ISL_1172223, EPI_ISL_1172224, EPI_ISL_1172225, EPI_ISL_1172226, EPI_ISL_1172227, EPI_ISL_1172228, EPI_ISL_1172230, EPI_ISL_1172231, EPI_ISL_1172232, EPI_ISL_1172233, EPI_ISL_1172234, EPI_ISL_1172235, EPI_ISL_1172236, EPI_ISL_1172237, EPI_ISL_1172238, EPI_ISL_1172239, EPI_ISL_1172241, EPI_ISL_1172242, EPI_ISL_1172243, EPI_ISL_1172244, EPI_ISL_1172245, EPI_ISL_1172247, EPI_ISL_1172248, EPI_ISL_1172249, EPI_ISL_1172250, EPI_ISL_1172251, EPI_ISL_1172252, EPI_ISL_1172254, EPI_ISL_1172255, EPI_ISL_1172256, EPI_ISL_1172257, EPI_ISL_1172258, EPI_ISL_1172259, EPI_ISL_1172260, EPI_ISL_1172261, EPI_ISL_1172263, EPI_ISL_1172264, EPI_ISL_1172265, EPI_ISL_1172266, EPI_ISL_1172267, EPI_ISL_1172268, EPI_ISL_1172269, EPI_ISL_1172270, EPI_ISL_1172272, EPI_ISL_1172273, EPI_ISL_1172274, EPI_ISL_1172275, EPI_ISL_1172276, EPI_ISL_1172277, EPI_ISL_1172278, EPI_ISL_1172279, EPI_ISL_1172280, EPI_ISL_1172281, EPI_ISL_1172283, EPI_ISL_1172285, EPI_ISL_1172286, EPI_ISL_1172287, EPI_ISL_1172288, EPI_ISL_1172290, EPI_ISL_1172291, EPI_ISL_1172292, EPI_ISL_1172293, EPI_ISL_1172294, EPI_ISL_1172296, EPI_ISL_1172297, EPI_ISL_1172298, EPI_ISL_1172299, EPI_ISL_1172301, EPI_ISL_1172302, EPI_ISL_1172303, EPI_ISL_1172304, EPI_ISL_1172305, EPI_ISL_1172306, EPI_ISL_1172307, EPI_ISL_1172308, EPI_ISL_1172309, EPI_ISL_1172310, EPI_ISL_1172311, EPI_ISL_1172312, EPI_ISL_1172313, EPI_ISL_1172314, EPI_ISL_1172316, EPI_ISL_1172318, EPI_ISL_1172319, EPI_ISL_1172320, EPI_ISL_1172321, EPI_ISL_1172323, EPI_ISL_1172324, EPI_ISL_1172325, EPI_ISL_1172326, EPI_ISL_1172327, EPI_ISL_1172328, EPI_ISL_1172329, EPI_ISL_1172330, EPI_ISL_1172331, EPI_ISL_1172333, EPI_ISL_1172334, EPI_ISL_1172335, EPI_ISL_1172336, EPI_ISL_1172337, EPI_ISL_1172338, EPI_ISL_1172339, EPI_ISL_1172340, EPI_ISL_1172341, EPI_ISL_1172342, EPI_ISL_1172343, EPI_ISL_1172344, EPI_ISL_1172345, EPI_ISL_1172346, EPI_ISL_1172348, EPI_ISL_1172349, EPI_ISL_1172350, EPI_ISL_1172351, EPI_ISL_1172352, EPI_ISL_1172353, EPI_ISL_1172354, EPI_ISL_1172356, EPI_ISL_1172357, EPI_ISL_1172358, EPI_ISL_1172359, EPI_ISL_1172360, EPI_ISL_1172361, EPI_ISL_1172362, EPI_ISL_1172363, EPI_ISL_1172364, EPI_ISL_1172365, EPI_ISL_1172366, EPI_ISL_1172367, EPI_ISL_1172368, EPI_ISL_1172370, EPI_ISL_1172371, EPI_ISL_1172372, EPI_ISL_1172373, EPI_ISL_1172374, EPI_ISL_1172375, EPI_ISL_1172376, EPI_ISL_1172377, EPI_ISL_1172378, EPI_ISL_1172379, EPI_ISL_1172380, EPI_ISL_1172381, EPI_ISL_1172382, EPI_ISL_1172383, EPI_ISL_1172384, EPI_ISL_1172385, EPI_ISL_1172386, EPI_ISL_1172387, EPI_ISL_1172388, EPI_ISL_1172389, EPI_ISL_1172390, EPI_ISL_1172391, EPI_ISL_1172392, EPI_ISL_1172393, EPI_ISL_1172394, EPI_ISL_1172395, EPI_ISL_1172396, EPI_ISL_1172397, EPI_ISL_1172398, EPI_ISL_1172399, EPI_ISL_1172400, EPI_ISL_1172401, EPI_ISL_1172402, EPI_ISL_1172405, EPI_ISL_1172406, EPI_ISL_1172407, EPI_ISL_1172409, EPI_ISL_1172410, EPI_ISL_1172411, EPI_ISL_1172412, EPI_ISL_1172413, EPI_ISL_1172415, EPI_ISL_1172416, EPI_ISL_1172417, EPI_ISL_1172418, EPI_ISL_1172419, EPI_ISL_1172420, EPI_ISL_1172421, EPI_ISL_1172422, EPI_ISL_1172423, EPI_ISL_1172424, EPI_ISL_1172425, EPI_ISL_1172426, EPI_ISL_1172427, EPI_ISL_1172428, EPI_ISL_1172429, EPI_ISL_1172430, EPI_ISL_1172431, EPI_ISL_1172432, EPI_ISL_1172433, EPI_ISL_1172434, EPI_ISL_1172435, EPI_ISL_1172436, EPI_ISL_1172438, EPI_ISL_1172439, EPI_ISL_1172440, EPI_ISL_1172441, EPI_ISL_1172442, EPI_ISL_1172443, EPI_ISL_1172444, EPI_ISL_1172445, EPI_ISL_1172446, EPI_ISL_1172447, EPI_ISL_1172448, EPI_ISL_1172449, EPI_ISL_1172450, EPI_ISL_1172451, EPI_ISL_1172452, EPI_ISL_1172453, EPI_ISL_1172454, EPI_ISL_1172455, EPI_ISL_1172456, EPI_ISL_1172457, EPI_ISL_1172458, EPI_ISL_1172460, EPI_ISL_1172461, EPI_ISL_1172463, EPI_ISL_1172464, EPI_ISL_1172465, EPI_ISL_1172466, EPI_ISL_1172467, EPI_ISL_1172468, EPI_ISL_1172469, EPI_ISL_1172470, EPI_ISL_1172471, EPI_ISL_1172473, EPI_ISL_1172474, EPI_ISL_1172475, EPI_ISL_1172476, EPI_ISL_1172477, EPI_ISL_1172478, EPI_ISL_1172479, EPI_ISL_1172480, EPI_ISL_1172481, EPI_ISL_1172483, EPI_ISL_1172484, EPI_ISL_1172485, EPI_ISL_1172486, EPI_ISL_1172487, EPI_ISL_1172488, EPI_ISL_1172489, EPI_ISL_1172490, EPI_ISL_1172491, EPI_ISL_1172492, EPI_ISL_1172493, EPI_ISL_1172494, EPI_ISL_1172495, EPI_ISL_1172496, EPI_ISL_1172497, EPI_ISL_1172498, EPI_ISL_1172499, EPI_ISL_1172500, EPI_ISL_1172501, EPI_ISL_1172502, EPI_ISL_1172503, EPI_ISL_1172504, EPI_ISL_1172505, EPI_ISL_1172506, EPI_ISL_1172507, EPI_ISL_1172508, EPI_ISL_1172509, EPI_ISL_1172510, EPI_ISL_1172511, EPI_ISL_1172512, EPI_ISL_1172513, EPI_ISL_1172514, EPI_ISL_1172515, EPI_ISL_1172516, EPI_ISL_1172517, EPI_ISL_1172518, EPI_ISL_1172519, EPI_ISL_1172520, EPI_ISL_1172521, EPI_ISL_1172522, EPI_ISL_1172523, EPI_ISL_1172524, EPI_ISL_1172525, EPI_ISL_1172526, EPI_ISL_1172527, EPI_ISL_1172528, EPI_ISL_1172531, EPI_ISL_1172532, EPI_ISL_1172533, EPI_ISL_1172534, EPI_ISL_1172535, EPI_ISL_1172536, EPI_ISL_1172537, EPI_ISL_1172538, EPI_ISL_1172539, EPI_ISL_1172541, EPI_ISL_1172542, EPI_ISL_1172544, EPI_ISL_1172545, EPI_ISL_1172546, EPI_ISL_1172547, EPI_ISL_1172548, EPI_ISL_1172549, EPI_ISL_1172550, EPI_ISL_1172551, EPI_ISL_1172552, EPI_ISL_1172553, EPI_ISL_1172554, EPI_ISL_1172555, EPI_ISL_1172556, EPI_ISL_1172557, EPI_ISL_1172558, EPI_ISL_1172559, EPI_ISL_1172560, EPI_ISL_1172561, EPI_ISL_1172562, EPI_ISL_1172563, EPI_ISL_1172564, EPI_ISL_1172566, EPI_ISL_1172568, EPI_ISL_1172570, EPI_ISL_1172571, EPI_ISL_1172572, EPI_ISL_1172573, EPI_ISL_1172574, EPI_ISL_1172575, EPI_ISL_1172576, EPI_ISL_1172577, EPI_ISL_1172579, EPI_ISL_1172580, EPI_ISL_1172581, EPI_ISL_1172582, EPI_ISL_1172583, EPI_ISL_1172584, EPI_ISL_1172585, EPI_ISL_1172586, EPI_ISL_1172587, EPI_ISL_1172588, EPI_ISL_1172589, EPI_ISL_1172590, EPI_ISL_1172591, EPI_ISL_1172592, EPI_ISL_1172593, EPI_ISL_1172594, EPI_ISL_1172595, EPI_ISL_1172596, EPI_ISL_1172597, EPI_ISL_1172598, EPI_ISL_1172599, EPI_ISL_1172600, EPI_ISL_1172602, EPI_ISL_1172603, EPI_ISL_1172604, EPI_ISL_1172605, EPI_ISL_1172606, EPI_ISL_1172607, EPI_ISL_1172608, EPI_ISL_1172609, EPI_ISL_1172610, EPI_ISL_1172611, EPI_ISL_1172612, EPI_ISL_1172613, EPI_ISL_1172614, EPI_ISL_1172616, EPI_ISL_1172617, EPI_ISL_1172618, EPI_ISL_1172619, EPI_ISL_1172620, EPI_ISL_1172621, EPI_ISL_1172622, EPI_ISL_1172623, EPI_ISL_1172624, EPI_ISL_1172625, EPI_ISL_1172626, EPI_ISL_1172627, EPI_ISL_1172628, EPI_ISL_1172629, EPI_ISL_1172630, EPI_ISL_1172631, EPI_ISL_1172632, EPI_ISL_1172633, EPI_ISL_1172634, EPI_ISL_1172635, EPI_ISL_1172636, EPI_ISL_1172637, EPI_ISL_1172639, EPI_ISL_1172640, EPI_ISL_1172641, EPI_ISL_1172643, EPI_ISL_1172644, EPI_ISL_1172645, EPI_ISL_1172646, EPI_ISL_1172647, EPI_ISL_1172648, EPI_ISL_1172649, EPI_ISL_1172650, EPI_ISL_1172651, EPI_ISL_1172652, EPI_ISL_1172653, EPI_ISL_1172654, EPI_ISL_1172655, EPI_ISL_1172656, EPI_ISL_1172658, EPI_ISL_1172659, EPI_ISL_1172660, EPI_ISL_1172661, EPI_ISL_1172662, EPI_ISL_1172663, EPI_ISL_1172664, EPI_ISL_1172665, EPI_ISL_1172666, EPI_ISL_1172667, EPI_ISL_1172669, EPI_ISL_1172670, EPI_ISL_1172671, EPI_ISL_1172673, EPI_ISL_1172674, EPI_ISL_1172675, EPI_ISL_1172676, EPI_ISL_1172677, EPI_ISL_1172678, EPI_ISL_1172679, EPI_ISL_1172681, EPI_ISL_1172682, EPI_ISL_1172683, EPI_ISL_1172684, EPI_ISL_1172685, EPI_ISL_1172686, EPI_ISL_1172687, EPI_ISL_1172688, EPI_ISL_1172689, EPI_ISL_1172690, EPI_ISL_1172691, EPI_ISL_1172692, EPI_ISL_1172693, EPI_ISL_1172694, EPI_ISL_1172695, EPI_ISL_1172696, EPI_ISL_1172697, EPI_ISL_1172698, EPI_ISL_1172699, EPI_ISL_1172700, EPI_ISL_1172702, EPI_ISL_1172703, EPI_ISL_1172704, EPI_ISL_1172705, EPI_ISL_1172706, EPI_ISL_1172707, EPI_ISL_1172708, EPI_ISL_1172709, EPI_ISL_1172710, EPI_ISL_1172711, EPI_ISL_1172712, EPI_ISL_1172713, EPI_ISL_1172715, EPI_ISL_1172716, EPI_ISL_1172717, EPI_ISL_1172718, EPI_ISL_1172719, EPI_ISL_1172720, EPI_ISL_1172721, EPI_ISL_1172722, EPI_ISL_1172723, EPI_ISL_1172724, EPI_ISL_1172725, EPI_ISL_1172727, EPI_ISL_1172728, EPI_ISL_1172729, EPI_ISL_1172730, EPI_ISL_1172731, EPI_ISL_1172732, EPI_ISL_1172733, EPI_ISL_1172734, EPI_ISL_1172735, EPI_ISL_1172736, EPI_ISL_1172737, EPI_ISL_1172738, EPI_ISL_1172739, EPI_ISL_1172740, EPI_ISL_1172741, EPI_ISL_1172742, EPI_ISL_1172743, EPI_ISL_1172745, EPI_ISL_1172746, EPI_ISL_1172747, EPI_ISL_1172749, EPI_ISL_1172750, EPI_ISL_1172751, EPI_ISL_1172752, EPI_ISL_1172753, EPI_ISL_1172756, EPI_ISL_1172757, EPI_ISL_1172758, EPI_ISL_1172759, EPI_ISL_1172760, EPI_ISL_1172761, EPI_ISL_1172762, EPI_ISL_1172763, EPI_ISL_1172764, EPI_ISL_1172765, EPI_ISL_1172766, EPI_ISL_1172767, EPI_ISL_1172769, EPI_ISL_1172770, EPI_ISL_1172771, EPI_ISL_1172772, EPI_ISL_1172773, EPI_ISL_1172774, EPI_ISL_1172775, EPI_ISL_1172776, EPI_ISL_1172778, EPI_ISL_1172779, EPI_ISL_1172780, EPI_ISL_1172781, EPI_ISL_1172782, EPI_ISL_1172783, EPI_ISL_1172784, EPI_ISL_1172786, EPI_ISL_1172787, EPI_ISL_1172788, EPI_ISL_1172789, EPI_ISL_1172790, EPI_ISL_1172791, EPI_ISL_1172792, EPI_ISL_1172793, EPI_ISL_1172794, EPI_ISL_1172795, EPI_ISL_1172796, EPI_ISL_1172797, EPI_ISL_1172798, EPI_ISL_1172799, EPI_ISL_1172800, EPI_ISL_1172801, EPI_ISL_1172802, EPI_ISL_1172803, EPI_ISL_1172804, EPI_ISL_1172805, EPI_ISL_1172806, EPI_ISL_1172807, EPI_ISL_1172808, EPI_ISL_1172809, EPI_ISL_1172810, EPI_ISL_1172811, EPI_ISL_1172812, EPI_ISL_1172813, EPI_ISL_1172814, EPI_ISL_1172815, EPI_ISL_1172816, EPI_ISL_1172817, EPI_ISL_1172818, EPI_ISL_1172819, EPI_ISL_1172820, EPI_ISL_1172821, EPI_ISL_1172822, EPI_ISL_1172823, EPI_ISL_1172824, EPI_ISL_1172825, EPI_ISL_1172827, EPI_ISL_1172828, EPI_ISL_1172830, EPI_ISL_1172831, EPI_ISL_1172832, EPI_ISL_1172833, EPI_ISL_1172834, EPI_ISL_1172835, EPI_ISL_1172837, EPI_ISL_1172838, EPI_ISL_1172839, EPI_ISL_1172840, EPI_ISL_1172841, EPI_ISL_1172842, EPI_ISL_1172843, EPI_ISL_1172844, EPI_ISL_1172845, EPI_ISL_1172846, EPI_ISL_1172847, EPI_ISL_1172848, EPI_ISL_1172849, EPI_ISL_1172850, EPI_ISL_1172851, EPI_ISL_1172852, EPI_ISL_1172853, EPI_ISL_1172855, EPI_ISL_1172856, EPI_ISL_1172857, EPI_ISL_1172858, EPI_ISL_1172859, EPI_ISL_1172860, EPI_ISL_1172861, EPI_ISL_1172862, EPI_ISL_1172863, EPI_ISL_1172864, EPI_ISL_1172866, EPI_ISL_1172867, EPI_ISL_1172868, EPI_ISL_1172869, EPI_ISL_1172870, EPI_ISL_1172871, EPI_ISL_1172872, EPI_ISL_1172873, EPI_ISL_1172874, EPI_ISL_1172875, EPI_ISL_1172876, EPI_ISL_1172877, EPI_ISL_1172878, EPI_ISL_1172879, EPI_ISL_1172880, EPI_ISL_1172881, EPI_ISL_1172882, EPI_ISL_1172883, EPI_ISL_1172884, EPI_ISL_1172885, EPI_ISL_1172886, EPI_ISL_1172887, EPI_ISL_1172888, EPI_ISL_1172889, EPI_ISL_1172890, EPI_ISL_1172891, EPI_ISL_1172892, EPI_ISL_1172893, EPI_ISL_1172895, EPI_ISL_1172896, EPI_ISL_1172897, EPI_ISL_1172898, EPI_ISL_1172899, EPI_ISL_1172900, EPI_ISL_1172901, EPI_ISL_1172902, EPI_ISL_1172903, EPI_ISL_1172904, EPI_ISL_1172905, EPI_ISL_1172906, EPI_ISL_1172907, EPI_ISL_1172909, EPI_ISL_1172910, EPI_ISL_1172911, EPI_ISL_1172912, EPI_ISL_1172913, EPI_ISL_1172914, EPI_ISL_1172915, EPI_ISL_1172917, EPI_ISL_1172918, EPI_ISL_1172919, EPI_ISL_1172920, EPI_ISL_1172921, EPI_ISL_1172922, EPI_ISL_1172923, EPI_ISL_1172924, EPI_ISL_1172925, EPI_ISL_1172926, EPI_ISL_1172927, EPI_ISL_1172928, EPI_ISL_1172929, EPI_ISL_1172930, EPI_ISL_1172931, EPI_ISL_1172932, EPI_ISL_1172933, EPI_ISL_1172934, EPI_ISL_1172935, EPI_ISL_1172936, EPI_ISL_1172937, EPI_ISL_1172939, EPI_ISL_1172940, EPI_ISL_1172941, EPI_ISL_1172944, EPI_ISL_1172945, EPI_ISL_1172946, EPI_ISL_1172947, EPI_ISL_1172948, EPI_ISL_1172949, EPI_ISL_1172950, EPI_ISL_1172951, EPI_ISL_1172952, EPI_ISL_1172953, EPI_ISL_1172954, EPI_ISL_1172955, EPI_ISL_1172956, EPI_ISL_1172957, EPI_ISL_1172958, EPI_ISL_1172959, EPI_ISL_1172960, EPI_ISL_1172962, EPI_ISL_1172963, EPI_ISL_1172964, EPI_ISL_1172965, EPI_ISL_1172966, EPI_ISL_1172967, EPI_ISL_1172968, EPI_ISL_1172969, EPI_ISL_1172970, EPI_ISL_1172971, EPI_ISL_1172972, EPI_ISL_1172973, EPI_ISL_1172974, EPI_ISL_1172976, EPI_ISL_1172977, EPI_ISL_1172978, EPI_ISL_1172979, EPI_ISL_1172980, EPI_ISL_1172981, EPI_ISL_1172982, EPI_ISL_1172983, EPI_ISL_1172984, EPI_ISL_1172985, EPI_ISL_1172986, EPI_ISL_1172987, EPI_ISL_1172988, EPI_ISL_1172989, EPI_ISL_1172991, EPI_ISL_1172993, EPI_ISL_1172994, EPI_ISL_1172995, EPI_ISL_1172996, EPI_ISL_1172998, EPI_ISL_1172999, EPI_ISL_1173000, EPI_ISL_1173001, EPI_ISL_1173002, EPI_ISL_1173003, EPI_ISL_1173004, EPI_ISL_1173005, EPI_ISL_1173006, EPI_ISL_1173007, EPI_ISL_1173008, EPI_ISL_1173009, EPI_ISL_1173010, EPI_ISL_1173011, EPI_ISL_1173012, EPI_ISL_1173013, EPI_ISL_1173014, EPI_ISL_1173015, EPI_ISL_1173016, EPI_ISL_1173017, EPI_ISL_1173018, EPI_ISL_1173019, EPI_ISL_1173021, EPI_ISL_1173022, EPI_ISL_1173023, EPI_ISL_1173024, EPI_ISL_1173025, EPI_ISL_1173026, EPI_ISL_1173027, EPI_ISL_1173029, EPI_ISL_1173030, EPI_ISL_1173031, EPI_ISL_1173032, EPI_ISL_1173033, EPI_ISL_1173034, EPI_ISL_1173035, EPI_ISL_1173036, EPI_ISL_1173037, EPI_ISL_1173038, EPI_ISL_1173039, EPI_ISL_1173040, EPI_ISL_1173041, EPI_ISL_1173042, EPI_ISL_1173043, EPI_ISL_1173044, EPI_ISL_1173045, EPI_ISL_1173046, EPI_ISL_1173047, EPI_ISL_1173048, EPI_ISL_1173049, EPI_ISL_1173050, EPI_ISL_1173051, EPI_ISL_1173052, EPI_ISL_1173053, EPI_ISL_1173054, EPI_ISL_1173056, EPI_ISL_1173057, EPI_ISL_1173058, EPI_ISL_1173059, EPI_ISL_1173060, EPI_ISL_1173061, EPI_ISL_1173062, EPI_ISL_1173063, EPI_ISL_1173064, EPI_ISL_1173065, EPI_ISL_1173066, EPI_ISL_1173067, EPI_ISL_1173069, EPI_ISL_1173070, EPI_ISL_1173071, EPI_ISL_1173072, EPI_ISL_1173073, EPI_ISL_1173075, EPI_ISL_1173076, EPI_ISL_1173077, EPI_ISL_1173078, EPI_ISL_1173079, EPI_ISL_1173080, EPI_ISL_1173081, EPI_ISL_1173082, EPI_ISL_1173083, EPI_ISL_1173084, EPI_ISL_1173085, EPI_ISL_1173086, EPI_ISL_1173087, EPI_ISL_1173088, EPI_ISL_1173089, EPI_ISL_1173090, EPI_ISL_1173092, EPI_ISL_1173093, EPI_ISL_1173094, EPI_ISL_1173095, EPI_ISL_1173096, EPI_ISL_1173097, EPI_ISL_1173098, EPI_ISL_1173101, EPI_ISL_1173103, EPI_ISL_1173104, EPI_ISL_1173105, EPI_ISL_1173106, EPI_ISL_1173107, EPI_ISL_1173108, EPI_ISL_1173109, EPI_ISL_1173110, EPI_ISL_1173111, EPI_ISL_1173112, EPI_ISL_1173113, EPI_ISL_1173114, EPI_ISL_1173116, EPI_ISL_1173117, EPI_ISL_1173118, EPI_ISL_1173119, EPI_ISL_1173121, EPI_ISL_1173122, EPI_ISL_1173123, EPI_ISL_1173124, EPI_ISL_1173126, EPI_ISL_1173127, EPI_ISL_1173128, EPI_ISL_1173129, EPI_ISL_1173130, EPI_ISL_1173131, EPI_ISL_1173132, EPI_ISL_1173133, EPI_ISL_1173134, EPI_ISL_1173135, EPI_ISL_1173136, EPI_ISL_1173137, EPI_ISL_1173138, EPI_ISL_1173139, EPI_ISL_1173141, EPI_ISL_1173142, EPI_ISL_1173143, EPI_ISL_1173144, EPI_ISL_1173145, EPI_ISL_1173146, EPI_ISL_1173147, EPI_ISL_1173148, EPI_ISL_1173149, EPI_ISL_1173151, EPI_ISL_1173152, EPI_ISL_1173153, EPI_ISL_1173154, EPI_ISL_1173155, EPI_ISL_1173156, EPI_ISL_1173157, EPI_ISL_1173158 | | | |
| see above | Pandemic Response Lab - NYC | Pandemic Response Lab, R&D | Henry Lee, Michael Hammerling, Melissa Hopkins, Cybill del Castillo, Shinyoung Clair Kang, William Ward, Pradeep Bugga, Haiping Hao, Jon Laurent |
| EPI_ISL_1180229, EPI_ISL_1180231 | Baylor Scott & White-Temple | Baylor Scott & White-Temple | Ari Rao, Linden Morales, Kimberly Walker, Marcus Volz, Shelby Hendrickson, Caitlin Maloney |
| EPI_ISL_1180233 | Ohio Department of Health Laboratory | Ohio Department of Health Laboratory | Holmes, Jennifer; Eric Brandt, Keoni Omura, Glen McGillivary, Caitlin McDonnell, Jade Mowery, Stephanie Mccracken, Tyler Payne, Kirtana Ramadugu, Erica Leasure, Brent Lee, Kelsey Florek, Heather Blankenship, Allison Black, Quanta Brown, and Tammy Bannerman |

| | | | |
|---|---|---|---|
| EPI_ISL_1180235 | Baylor Scott & White-Temple | Baylor Scott & White-Temple | Ari Rao, Linden Morales, Kimberly Walker, Marcus Volz, Shelby Hendrickson, Caitlin Maloney |
| EPI_ISL_1180236 | Ohio Department of Health Laboratory | Ohio Department of Health Laboratory | Holmes, Jennifer; Eric Brandt, Keoni Omura, Glen McGillivary, Caitlin McDonnell, Jade Mowery, Stephanie Mccracken, Tyler Payne, Kirtana Ramadugu, Erica Leasure, Brent Lee, Kelsey Florek, Heather Blankenship, Allison Black, Quanta Brown, and Tammy Bannerman |
| EPI_ISL_1180237, EPI_ISL_1180238 | Baylor Scott & White-Temple | Baylor Scott & White-Temple | Ari Rao, Linden Morales, Kimberly Walker, Marcus Volz, Shelby Hendrickson, Caitlin Maloney |
| EPI_ISL_1180239, EPI_ISL_1180240, EPI_ISL_1180241, EPI_ISL_1180242, EPI_ISL_1180243, EPI_ISL_1180244, EPI_ISL_1180245, EPI_ISL_1180246, EPI_ISL_1180247, EPI_ISL_1180248, EPI_ISL_1180249 | | | |
| see above | DC Public Health Lab/ Dept. of Forensic Sciences | DC Public Health Lab/ Dept. of Forensic Sciences | Scott Nguyen, Janis Doss, Elizabeth Zelaya, Sarah Scott, Connie Maza, Monica Mann, Brittany Hamilton, David Payne, Jocelyn Hauser |
| EPI_ISL_1180254 | Ohio Department of Health Laboratory | Ohio Department of Health Laboratory | Holmes, Jennifer; Eric Brandt, Keoni Omura, Glen McGillivary, Caitlin McDonnell, Jade Mowery, Stephanie Mccracken, Tyler Payne, Kirtana Ramadugu, Erica Leasure, Brent Lee, Kelsey Florek, Heather Blankenship, Allison Black, Quanta Brown, and Tammy Bannerman |
| EPI_ISL_1180255, EPI_ISL_1180256 | Baylor Scott & White-Temple | Baylor Scott & White-Temple | Ari Rao, Linden Morales, Kimberly Walker, Marcus Volz, Shelby Hendrickson, Caitlin Maloney |
| EPI_ISL_1180257 | Ohio Department of Health Laboratory | Ohio Department of Health Laboratory | Holmes, Jennifer; Eric Brandt, Keoni Omura, Glen McGillivary, Caitlin McDonnell, Jade Mowery, Stephanie Mccracken, Tyler Payne, Kirtana Ramadugu, Erica Leasure, Brent Lee, Kelsey Florek, Heather Blankenship, Allison Black, Quanta Brown, and Tammy Bannerman |
| EPI_ISL_1180258 | Baylor Scott & White-Temple | Baylor Scott & White-Temple | Ari Rao, Linden Morales, Kimberly Walker, Marcus Volz, Shelby Hendrickson, Caitlin Maloney |
| EPI_ISL_1180259 | Ohio Department of Health Laboratory | Ohio Department of Health Laboratory | Holmes, Jennifer; Eric Brandt, Keoni Omura, Glen McGillivary, Caitlin McDonnell, Jade Mowery, Stephanie Mccracken, Tyler Payne, Kirtana Ramadugu, Erica Leasure, Brent Lee, Kelsey Florek, Heather Blankenship, Allison Black, Quanta Brown, and Tammy Bannerman |
| EPI_ISL_1180279, EPI_ISL_1180669, EPI_ISL_1180670 | Baylor Scott & White-Temple | Baylor Scott & White-Temple | Ari Rao, Linden Morales, Kimberly Walker, Marcus Volz, Shelby Hendrickson, Caitlin Maloney |
| EPI_ISL_1180936, EPI_ISL_1180937, EPI_ISL_1180938, EPI_ISL_1180939, EPI_ISL_1180940, EPI_ISL_1180941, EPI_ISL_1180942, EPI_ISL_1180943, EPI_ISL_1180944, EPI_ISL_1180945, EPI_ISL_1180946, EPI_ISL_1180947, EPI_ISL_1180948, EPI_ISL_1180949, EPI_ISL_1180950, EPI_ISL_1180951, EPI_ISL_1180952, EPI_ISL_1180953, EPI_ISL_1180954, EPI_ISL_1180955, EPI_ISL_1180956, EPI_ISL_1180957 | | | |
| see above | UW Virology Lab | UW Virology Lab | Pavitra Roychoudhury, Hong Xie, Lasata Shrestha, Shah Mohamed Bakhash, Michelle Lin, Noah Baker, Sean Ellis, Saraswathi Sathees, Meei-Li Huang, Keith R Jerome, Alexander Greninger |
| EPI_ISL_1180958 | Baylor Scott & White-Temple | Baylor Scott & White-Temple | Ari Rao, Linden Morales, Kimberly Walker, Marcus Volz, Shelby Hendrickson, Caitlin Maloney |
| EPI_ISL_1180959, EPI_ISL_1180960, EPI_ISL_1180961, EPI_ISL_1180962, EPI_ISL_1180963, EPI_ISL_1180964, EPI_ISL_1180965, EPI_ISL_1180966, EPI_ISL_1180967, EPI_ISL_1180968, EPI_ISL_1180969, EPI_ISL_1180970, EPI_ISL_1180971, EPI_ISL_1180972, EPI_ISL_1180973, EPI_ISL_1180974, EPI_ISL_1180975, EPI_ISL_1180976, EPI_ISL_1180977, EPI_ISL_1180978, EPI_ISL_1180979, EPI_ISL_1180980, EPI_ISL_1180981, EPI_ISL_1180982, EPI_ISL_1180983, EPI_ISL_1180984, EPI_ISL_1180985, EPI_ISL_1180986, EPI_ISL_1180987, EPI_ISL_1180988, EPI_ISL_1180989, EPI_ISL_1180990, EPI_ISL_1180991, EPI_ISL_1180992, EPI_ISL_1180993, EPI_ISL_1180994, EPI_ISL_1180995, EPI_ISL_1180996, EPI_ISL_1180997, EPI_ISL_1180998, EPI_ISL_1180999, EPI_ISL_1181000, EPI_ISL_1181001, EPI_ISL_1181002, EPI_ISL_1181003, EPI_ISL_1181004, EPI_ISL_1181005, EPI_ISL_1181006, EPI_ISL_1181007, EPI_ISL_1181008, EPI_ISL_1181009, EPI_ISL_1181010, EPI_ISL_1181011, EPI_ISL_1181012, EPI_ISL_1181013, EPI_ISL_1181014, EPI_ISL_1181015, EPI_ISL_1181016, EPI_ISL_1181017, EPI_ISL_1181018, EPI_ISL_1181019, EPI_ISL_1181020, EPI_ISL_1181021, EPI_ISL_1181022, EPI_ISL_1181023, EPI_ISL_1181024, EPI_ISL_1181025, EPI_ISL_1181026, EPI_ISL_1181027, EPI_ISL_1181028, EPI_ISL_1181029, EPI_ISL_1181030, EPI_ISL_1181031, EPI_ISL_1181032, EPI_ISL_1181033, EPI_ISL_1181034, EPI_ISL_1181035, EPI_ISL_1181036, EPI_ISL_1181037, EPI_ISL_1181038, EPI_ISL_1181039, EPI_ISL_1181040, EPI_ISL_1181041, EPI_ISL_1181042, EPI_ISL_1181043, EPI_ISL_1181044, EPI_ISL_1181045, EPI_ISL_1181046, EPI_ISL_1181047, EPI_ISL_1181048, EPI_ISL_1181049, EPI_ISL_1181050, EPI_ISL_1181051, EPI_ISL_1181052, EPI_ISL_1181053, EPI_ISL_1181054, EPI_ISL_1181055, EPI_ISL_1181056, EPI_ISL_1181057, EPI_ISL_1181058, EPI_ISL_1181059, EPI_ISL_1181060, EPI_ISL_1181061, EPI_ISL_1181062, EPI_ISL_1181063, EPI_ISL_1181064, EPI_ISL_1181065, EPI_ISL_1181066, EPI_ISL_1181067, EPI_ISL_1181068, EPI_ISL_1181069, EPI_ISL_1181070, EPI_ISL_1181071, EPI_ISL_1181072, EPI_ISL_1181073, EPI_ISL_1181074, EPI_ISL_1181075, EPI_ISL_1181076, EPI_ISL_1181077, EPI_ISL_1181078, EPI_ISL_1181079, EPI_ISL_1181080, EPI_ISL_1181081, EPI_ISL_1181082, EPI_ISL_1181083, EPI_ISL_1181084, EPI_ISL_1181085, EPI_ISL_1181086, EPI_ISL_1181087, EPI_ISL_1181088, EPI_ISL_1181089, EPI_ISL_1181090, EPI_ISL_1181091, EPI_ISL_1181092, EPI_ISL_1181093, EPI_ISL_1181094, EPI_ISL_1181095, EPI_ISL_1181096, EPI_ISL_1181097, EPI_ISL_1181098, EPI_ISL_1181099, EPI_ISL_1181100, EPI_ISL_1181101, EPI_ISL_1181102, EPI_ISL_1181103, EPI_ISL_1181104, EPI_ISL_1181105, EPI_ISL_1181106, EPI_ISL_1181107, EPI_ISL_1181108, EPI_ISL_1181109, EPI_ISL_1181110, EPI_ISL_1181111, EPI_ISL_1181112, EPI_ISL_1181113, EPI_ISL_1181114, EPI_ISL_1181115 | | | |
| see above | UW Virology Lab | UW Virology Lab | Pavitra Roychoudhury, Hong Xie, Lasata Shrestha, Shah Mohamed Bakhash, Michelle Lin, Noah Baker, Sean Ellis, Saraswathi Sathees, Meei-Li Huang, Keith R Jerome, Alexander Greninger |
| EPI_ISL_1181629, EPI_ISL_1181630, EPI_ISL_1181631, EPI_ISL_1181632, EPI_ISL_1181633, EPI_ISL_1181634, EPI_ISL_1181635, EPI_ISL_1181636, EPI_ISL_1181637, EPI_ISL_1181638, EPI_ISL_1181639, EPI_ISL_1181640, EPI_ISL_1181641, EPI_ISL_1181642, EPI_ISL_1181643, EPI_ISL_1181644, EPI_ISL_1181645, EPI_ISL_1181646, EPI_ISL_1181648, EPI_ISL_1181649, EPI_ISL_1181650, EPI_ISL_1181651, EPI_ISL_1181654, EPI_ISL_1181657, EPI_ISL_1181661, EPI_ISL_1181662, EPI_ISL_1181663, EPI_ISL_1181664, EPI_ISL_1181665, EPI_ISL_1181667, EPI_ISL_1181669, EPI_ISL_1181670, EPI_ISL_1181671, EPI_ISL_1181672, EPI_ISL_1181673, EPI_ISL_1181674, EPI_ISL_1181675, EPI_ISL_1181676, EPI_ISL_1181677, EPI_ISL_1181678, EPI_ISL_1181680, EPI_ISL_1181681, EPI_ISL_1181682, EPI_ISL_1181685, EPI_ISL_1181686, EPI_ISL_1181687, EPI_ISL_1181688, EPI_ISL_1181690, EPI_ISL_1181691, EPI_ISL_1181692, EPI_ISL_1181694 | | | |
| see above | DPHL | Delaware Public Health Lab | Gregory Hovan |
| EPI_ISL_1181696, EPI_ISL_1181698, EPI_ISL_1181699 | Ohio Department of Health Laboratory | Ohio Department of Health Laboratory | Holmes, Jennifer; Eric Brandt, Keoni Omura, Glen McGillivary, Caitlin McDonnell, Jade Mowery, Stephanie Mccracken, Tyler Payne, Kirtana Ramadugu, Erica Leasure, Brent Lee, Kelsey Florek, Heather Blankenship, Allison Black, Quanta Brown, and Tammy Bannerman |
| EPI_ISL_1181700, EPI_ISL_1181701, EPI_ISL_1181702, EPI_ISL_1181703, EPI_ISL_1181704, EPI_ISL_1181705, EPI_ISL_1181706, EPI_ISL_1181707, EPI_ISL_1181708, EPI_ISL_1181709, EPI_ISL_1181710, EPI_ISL_1181711, EPI_ISL_1181712, EPI_ISL_1181713, EPI_ISL_1181714, EPI_ISL_1181715, EPI_ISL_1181716, EPI_ISL_1181717, EPI_ISL_1181718, EPI_ISL_1181719 | | | |
| see above | Instituto Nacional de Medicina Genomica | Instituto Nacional de Medicina Genomica | Hidalgo-Miranda A, Mendoza-Vargas A, Reyes-Grajeda JP, Cedro-Tanda A, Alcaraz N, Gonzalez-Barrera D, Rangel-DeLeon D, Ramirez-Vega O, Sifuentes-Rojas C, Cisneros-Villanueva M, Herrera-Montalvo LA |
| EPI_ISL_1181720 | NCSLPH | NCSLPH | Chase K, Miller MC, Greene S, Glover W |
| EPI_ISL_1181721, EPI_ISL_1181722 | Ohio Department of Health Laboratory | Ohio Department of Health Laboratory | Holmes, Jennifer; Eric Brandt, Keoni Omura, Glen McGillivary, Caitlin McDonnell, Jade Mowery, Stephanie Mccracken, Tyler Payne, Kirtana Ramadugu, Erica Leasure, Brent Lee, Kelsey Florek, Heather Blankenship, Allison Black, Quanta Brown, and Tammy Bannerman |
| EPI_ISL_1181723, EPI_ISL_1181738 | NCSLPH | NCSLPH | Chase K, Miller MC, Greene S, Glover W |
| EPI_ISL_1181819, EPI_ISL_1181820 | Ohio Department of Health Laboratory | Ohio Department of Health Laboratory | Holmes, Jennifer; Eric Brandt, Keoni Omura, Glen McGillivary, Caitlin McDonnell, Jade Mowery, Stephanie Mccracken, Tyler Payne, Kirtana Ramadugu, Erica Leasure, Brent Lee, Kelsey Florek, Heather Blankenship, Allison Black, Quanta Brown, and Tammy Bannerman |
| EPI_ISL_1181821 | NCSLPH | NCSLPH | Chase K, Miller MC, Greene S, Glover W |
| EPI_ISL_1181865, EPI_ISL_1181881 | Ohio Department of Health Laboratory | Ohio Department of Health Laboratory | Holmes, Jennifer; Eric Brandt, Keoni Omura, Glen McGillivary, Caitlin McDonnell, Jade Mowery, Stephanie Mccracken, Tyler Payne, Kirtana Ramadugu, Erica Leasure, Brent Lee, Kelsey Florek, Heather Blankenship, Allison Black, Quanta Brown, and Tammy Bannerman |
| EPI_ISL_1181888, EPI_ISL_1181889, EPI_ISL_1181890, EPI_ISL_1181891, EPI_ISL_1181893, EPI_ISL_1181895, EPI_ISL_1181896, EPI_ISL_1181898, EPI_ISL_1181900, EPI_ISL_1181901, EPI_ISL_1181902, EPI_ISL_1181903, EPI_ISL_1181905, EPI_ISL_1181906, EPI_ISL_1181907, EPI_ISL_1181908, EPI_ISL_1181909, EPI_ISL_1181910, EPI_ISL_1181911, EPI_ISL_1181912, EPI_ISL_1181913, EPI_ISL_1181914, EPI_ISL_1181915, EPI_ISL_1181916, EPI_ISL_1181917, EPI_ISL_1181918, EPI_ISL_1181919, EPI_ISL_1181920, EPI_ISL_1181921, EPI_ISL_1181922, EPI_ISL_1181923, EPI_ISL_1181924, EPI_ISL_1181925, EPI_ISL_1181926, EPI_ISL_1181927, EPI_ISL_1181928, EPI_ISL_1181929, EPI_ISL_1181930, EPI_ISL_1181931, EPI_ISL_1181932 | | | |
| see above | Wyoming Public Health Laboratory | Wyoming Public Health Laboratory | Jim Mildenberger, Wanda Manley, Noah Hull, Taylor Fearing, Lynette Gumbleton, Channing Weber, Ashley Norberg, Chayse Rowley, Marley Goetz, Brian Dominguez, Elliot Thomasson, Cari Sloma, and Rob Christensen |
| EPI_ISL_1181948, EPI_ISL_1181951, EPI_ISL_1181954, EPI_ISL_1181955, EPI_ISL_1181956, EPI_ISL_1181957 | DC Public Health Lab/ Dept. of Forensic Sciences | DC Public Health Lab/ Dept. of Forensic Sciences | Scott Nguyen, Janis Doss, Elizabeth Zelaya, Sarah Scott, Connie Maza, Monica Mann, Brittany Hamilton, David Payne, Jocelyn Hauser |
| EPI_ISL_1181968, EPI_ISL_1181969, EPI_ISL_1181971, EPI_ISL_1181972, EPI_ISL_1181973, EPI_ISL_1181974, EPI_ISL_1181975, EPI_ISL_1181976, EPI_ISL_1181977, EPI_ISL_1181978, EPI_ISL_1181979, EPI_ISL_1181980, EPI_ISL_1181981, EPI_ISL_1181982, EPI_ISL_1181983, EPI_ISL_1181984, EPI_ISL_1181985, EPI_ISL_1181986, EPI_ISL_1181987, EPI_ISL_1181988, EPI_ISL_1181989, EPI_ISL_1181990, EPI_ISL_1181991, EPI_ISL_1181992, EPI_ISL_1181993, EPI_ISL_1181994, EPI_ISL_1181995, EPI_ISL_1181996, EPI_ISL_1181997, EPI_ISL_1181998, EPI_ISL_1182000, EPI_ISL_1182001, EPI_ISL_1182002, EPI_ISL_1182003, EPI_ISL_1182004, EPI_ISL_1182005, EPI_ISL_1182006, EPI_ISL_1182007, EPI_ISL_1182008, EPI_ISL_1182009, EPI_ISL_1182010, EPI_ISL_1182011, EPI_ISL_1182012, EPI_ISL_1182013, EPI_ISL_1182014 | | | |
| see above | Maryland Public Health Laboratory | Maryland Public Health Laboratory | Maryland Department of Health Laboratories Administration |
| EPI_ISL_1182016, EPI_ISL_1182019, EPI_ISL_1182021, EPI_ISL_1182023, EPI_ISL_1182024, EPI_ISL_1182025, EPI_ISL_1182026 | Yale Clinical Virology Lab | Grubaugh Lab - Yale School of Public Health | Joseph Fauver, Mallery Breban, Isabell Ott, Tara Alpert, Mary Petrone, Anderson Brito, Chantal Vogels, Annie Watkins, Chaney Kalinich, Marie L. Landry, Nathan Grubaugh |
| EPI_ISL_1182027, EPI_ISL_1182029 | Connecticut DPH | Grubaugh Lab - Yale School of Public Health | Joseph Fauver, Mallery Breban, Isabell Ott, Tara Alpert, Mary Petrone, Anderson Brito, Chantal Vogels, Annie Watkins, Chaney Kalinich, Anthony Muyombwe, Randy Downing, Jafar Razeq, Stephen M. Bart, Nathan Grubaugh |
| EPI_ISL_1182032, EPI_ISL_1182033, EPI_ISL_1182034, EPI_ISL_1182035, EPI_ISL_1182036, EPI_ISL_1182038, EPI_ISL_1182039, EPI_ISL_1182040, EPI_ISL_1182044, EPI_ISL_1182045, EPI_ISL_1182046, EPI_ISL_1182047, EPI_ISL_1182048, EPI_ISL_1182051, EPI_ISL_1182052, EPI_ISL_1182053, EPI_ISL_1182054, EPI_ISL_1182055, EPI_ISL_1182056, EPI_ISL_1182057, EPI_ISL_1182058, EPI_ISL_1182059, EPI_ISL_1182060, EPI_ISL_1182061, EPI_ISL_1182062, EPI_ISL_1182063, EPI_ISL_1182064, EPI_ISL_1182065, EPI_ISL_1182066, EPI_ISL_1182067, EPI_ISL_1182068, EPI_ISL_1182069, EPI_ISL_1182070, EPI_ISL_1182071, EPI_ISL_1182072, EPI_ISL_1182073, EPI_ISL_1182075, EPI_ISL_1182076, EPI_ISL_1182077, EPI_ISL_1182078, EPI_ISL_1182079, EPI_ISL_1182080, EPI_ISL_1182081, EPI_ISL_1182082, EPI_ISL_1182083, EPI_ISL_1182084, EPI_ISL_1182085, EPI_ISL_1182087 | | | |
| see above | Tempus | Grubaugh Lab - Yale School of Public Health | Joseph Fauver, Mallery Breban, Isabell Ott, Tara Alpert, Mary Petrone, Anderson Brito, Chantal Vogels, Annie Watkins, Chaney Kalinich, Matthew J. MacKay, Gaurav Khullar, Jessica Metti, Joel T. Dudley, Megan Nash, Nike Beaubier, Christopher E. Mason, Nathan Grubaugh |
| EPI_ISL_1182088, EPI_ISL_1182091, EPI_ISL_1182092 | Murphy Medical Association | Grubaugh Lab - Yale School of Public Health | Joseph Fauver, Mallery Breban, Isabell Ott, Tara Alpert, Mary Petrone, Anderson Brito, Chantal Vogels, Annie Watkins, Chaney Kalinich, Caleb Neal, Eva Laszlo, Steven Murphy, Nathan Grubaugh |
| EPI_ISL_1182105, EPI_ISL_1182106, EPI_ISL_1182107, EPI_ISL_1182108, EPI_ISL_1182109, EPI_ISL_1182110, EPI_ISL_1182111, EPI_ISL_1182114, EPI_ISL_1182115, EPI_ISL_1182116, EPI_ISL_1182117, EPI_ISL_1182118, EPI_ISL_1182119, EPI_ISL_1182120, EPI_ISL_1182121, EPI_ISL_1182123, EPI_ISL_1182124, EPI_ISL_1182125, EPI_ISL_1182126, EPI_ISL_1182127, EPI_ISL_1182128, EPI_ISL_1182129, EPI_ISL_1182130, EPI_ISL_1182131, EPI_ISL_1182132, EPI_ISL_1182133, EPI_ISL_1182134, EPI_ISL_1182135, EPI_ISL_1182136, EPI_ISL_1182137, EPI_ISL_1182138, EPI_ISL_1182139, EPI_ISL_1182140, EPI_ISL_1182141, EPI_ISL_1182142, EPI_ISL_1182143, EPI_ISL_1182144, EPI_ISL_1182145, EPI_ISL_1182148, EPI_ISL_1182149, EPI_ISL_1182150, EPI_ISL_1182151, EPI_ISL_1182152, EPI_ISL_1182153, EPI_ISL_1182156, EPI_ISL_1182157, EPI_ISL_1182158, EPI_ISL_1182159, EPI_ISL_1182160, EPI_ISL_1182161, EPI_ISL_1182162, EPI_ISL_1182163, EPI_ISL_1182164, EPI_ISL_1182165, EPI_ISL_1182167, EPI_ISL_1182168, EPI_ISL_1182169, EPI_ISL_1182170, EPI_ISL_1182171, EPI_ISL_1182172, EPI_ISL_1182173, EPI_ISL_1182174, EPI_ISL_1182175, EPI_ISL_1182176, EPI_ISL_1182177, EPI_ISL_1182178, EPI_ISL_1182179, EPI_ISL_1182180, EPI_ISL_1182181, EPI_ISL_1182182, EPI_ISL_1182183, EPI_ISL_1182184, EPI_ISL_1182185, EPI_ISL_1182186, EPI_ISL_1182188, EPI_ISL_1182189, EPI_ISL_1182190, EPI_ISL_1182191, EPI_ISL_1182192, EPI_ISL_1182193, EPI_ISL_1182195, EPI_ISL_1182196, EPI_ISL_1182197, EPI_ISL_1182198, EPI_ISL_1182199, EPI_ISL_1182200, EPI_ISL_1182201, EPI_ISL_1182202, EPI_ISL_1182203, EPI_ISL_1182204, EPI_ISL_1182205, EPI_ISL_1182206, EPI_ISL_1182207, EPI_ISL_1182210, EPI_ISL_1182211, EPI_ISL_1182212, EPI_ISL_1182213, EPI_ISL_1182214, EPI_ISL_1182215, EPI_ISL_1182216, EPI_ISL_1182217, EPI_ISL_1182218, EPI_ISL_1182219, EPI_ISL_1182220, EPI_ISL_1182221, EPI_ISL_1182222, EPI_ISL_1182223, EPI_ISL_1182224, EPI_ISL_1182225, EPI_ISL_1182226, EPI_ISL_1182227, EPI_ISL_1182228, EPI_ISL_1182229, EPI_ISL_1182230, EPI_ISL_1182231, EPI_ISL_1182232, EPI_ISL_1182233, EPI_ISL_1182234, EPI_ISL_1182235, EPI_ISL_1182236, EPI_ISL_1182237, EPI_ISL_1182238, EPI_ISL_1182240, EPI_ISL_1182241, EPI_ISL_1182242, EPI_ISL_1182243, EPI_ISL_1182244, EPI_ISL_1182245, EPI_ISL_1182246, EPI_ISL_1182247, EPI_ISL_1182248, EPI_ISL_1182249, EPI_ISL_1182250, EPI_ISL_1182251, EPI_ISL_1182252, EPI_ISL_1182253, EPI_ISL_1182254, EPI_ISL_1182255, EPI_ISL_1182256, EPI_ISL_1182257, EPI_ISL_1182258, EPI_ISL_1182259, EPI_ISL_1182260, EPI_ISL_1182261, EPI_ISL_1182262, EPI_ISL_1182263, EPI_ISL_1182264, EPI_ISL_1182265, EPI_ISL_1182266, EPI_ISL_1182267, EPI_ISL_1182268, EPI_ISL_1182270, EPI_ISL_1182271, EPI_ISL_1182273, EPI_ISL_1182274, EPI_ISL_1182275, EPI_ISL_1182276, EPI_ISL_1182277, EPI_ISL_1182278, EPI_ISL_1182279, EPI_ISL_1182280, EPI_ISL_1182281, EPI_ISL_1182282, EPI_ISL_1182283, EPI_ISL_1182284, EPI_ISL_1182287, EPI_ISL_1182288, EPI_ISL_1182289, EPI_ISL_1182291, EPI_ISL_1182293, EPI_ISL_1182294, EPI_ISL_1182295, EPI_ISL_1182297, EPI_ISL_1182298, EPI_ISL_1182299, EPI_ISL_1182300, EPI_ISL_1182301, EPI_ISL_1182303, EPI_ISL_1182304, EPI_ISL_1182305, EPI_ISL_1182306, EPI_ISL_1182307, EPI_ISL_1182308, EPI_ISL_1182309, EPI_ISL_1182310, EPI_ISL_1182311, EPI_ISL_1182312, EPI_ISL_1182313, EPI_ISL_1182314, EPI_ISL_1182315, EPI_ISL_1182316, EPI_ISL_1182317, EPI_ISL_1182318, EPI_ISL_1182319, EPI_ISL_1182320, EPI_ISL_1182321, EPI_ISL_1182322, EPI_ISL_1182323, EPI_ISL_1182324, EPI_ISL_1182325, EPI_ISL_1182326, EPI_ISL_1182327, EPI_ISL_1182328, EPI_ISL_1182329, EPI_ISL_1182330, EPI_ISL_1182331, EPI_ISL_1182332, EPI_ISL_1182333, EPI_ISL_1182334, EPI_ISL_1182335, EPI_ISL_1182336, EPI_ISL_1182337, EPI_ISL_1182338, EPI_ISL_1182339, EPI_ISL_1182340, EPI_ISL_1182342, EPI_ISL_1182343, EPI_ISL_1182344, EPI_ISL_1182345, EPI_ISL_1182346, EPI_ISL_1182347, EPI_ISL_1182348, EPI_ISL_1182349, EPI_ISL_1182350, EPI_ISL_1182351, EPI_ISL_1182352, EPI_ISL_1182353, EPI_ISL_1182354, EPI_ISL_1182355, EPI_ISL_1182356, EPI_ISL_1182357, EPI_ISL_1182358, EPI_ISL_1182359, EPI_ISL_1182360, EPI_ISL_1182361, EPI_ISL_1182363, EPI_ISL_1182364, EPI_ISL_1182365, EPI_ISL_1182366, EPI_ISL_1182367, EPI_ISL_1182368, EPI_ISL_1182369, EPI_ISL_1182370, EPI_ISL_1182371, EPI_ISL_1182372, EPI_ISL_1182373, EPI_ISL_1182375, EPI_ISL_1182376, EPI_ISL_1182377, EPI_ISL_1182378, EPI_ISL_1182379, EPI_ISL_1182380, EPI_ISL_1182381, EPI_ISL_1182382, EPI_ISL_1182383, EPI_ISL_1182384, EPI_ISL_1182385, EPI_ISL_1182386, EPI_ISL_1182387, EPI_ISL_1182388, EPI_ISL_1182389, EPI_ISL_1182390, EPI_ISL_1182391, EPI_ISL_1182392, EPI_ISL_1182393, EPI_ISL_1182394, EPI_ISL_1182395, EPI_ISL_1182396, EPI_ISL_1182398, EPI_ISL_1182399, EPI_ISL_1182400, EPI_ISL_1182401, EPI_ISL_1182402, EPI_ISL_1182403, EPI_ISL_1182404, EPI_ISL_1182406, EPI_ISL_1182407, EPI_ISL_1182408, EPI_ISL_1182409, EPI_ISL_1182410, EPI_ISL_1182411, EPI_ISL_1182413, EPI_ISL_1182414, EPI_ISL_1182415, EPI_ISL_1182416, EPI_ISL_1182417, EPI_ISL_1182418, EPI_ISL_1182419, EPI_ISL_1182420, EPI_ISL_1182421, EPI_ISL_1182422, EPI_ISL_1182423, EPI_ISL_1182424, EPI_ISL_1182425, EPI_ISL_1182426, EPI_ISL_1182429, EPI_ISL_1182430, EPI_ISL_1182431, EPI_ISL_1182432, EPI_ISL_1182433, EPI_ISL_1182434, EPI_ISL_1182435, EPI_ISL_1182436, EPI_ISL_1182438, EPI_ISL_1182439, EPI_ISL_1182440, EPI_ISL_1182441, EPI_ISL_1182442, EPI_ISL_1182443 | | | |
| see above | Sonora Quest Laboratories | TGen North | Jolene Bowers, Heather Centner, Chris French, Hayley Yaglom, Ashlyn Pfeiffer, Darrin Lemmer, Dave Engelthaler, The Arizona COVID Genomics Union (ACGU) |

| | | | |
|---|---|---|---|
| EPI_ISL_1182445, EPI_ISL_1182446, EPI_ISL_1182448, EPI_ISL_1182449, EPI_ISL_1182451, EPI_ISL_1182453, EPI_ISL_1182455, EPI_ISL_1182456, EPI_ISL_1182457, EPI_ISL_1182459, EPI_ISL_1182460, EPI_ISL_1182461, EPI_ISL_1182462, EPI_ISL_1182463, EPI_ISL_1182464, EPI_ISL_1182465 | | | |
| see above | TGen North | TGen North | Jolene Bowers, Heather Centner, Chris French, Hayley Yaglom, Ashlyn Pfeiffer, Darrin Lemmer, Dave Engelthaler, The Arizona COVID Genomics Union (ACGU) |
| EPI_ISL_1182466, EPI_ISL_1182467, EPI_ISL_1182469, EPI_ISL_1182470, EPI_ISL_1182471, EPI_ISL_1182472, EPI_ISL_1182473, EPI_ISL_1182474, EPI_ISL_1182475, EPI_ISL_1182476, EPI_ISL_1182477, EPI_ISL_1182478, EPI_ISL_1182479, EPI_ISL_1182480, EPI_ISL_1182481, EPI_ISL_1182482, EPI_ISL_1182483, EPI_ISL_1182484, EPI_ISL_1182485, EPI_ISL_1182486, EPI_ISL_1182487, EPI_ISL_1182488, EPI_ISL_1182489, EPI_ISL_1182491, EPI_ISL_1182492, EPI_ISL_1182493, EPI_ISL_1182494, EPI_ISL_1182495, EPI_ISL_1182497, EPI_ISL_1182498, EPI_ISL_1182499, EPI_ISL_1182500, EPI_ISL_1182502, EPI_ISL_1182503, EPI_ISL_1182504, EPI_ISL_1182505, EPI_ISL_1182506, EPI_ISL_1182507, EPI_ISL_1182508, EPI_ISL_1182509, EPI_ISL_1182510, EPI_ISL_1182511, EPI_ISL_1182513, EPI_ISL_1182514, EPI_ISL_1182515, EPI_ISL_1182516, EPI_ISL_1182517, EPI_ISL_1182518, EPI_ISL_1182519, EPI_ISL_1182520, EPI_ISL_1182521, EPI_ISL_1182523, EPI_ISL_1182525, EPI_ISL_1182526, EPI_ISL_1182527, EPI_ISL_1182528, EPI_ISL_1182529, EPI_ISL_1182530, EPI_ISL_1182531 | | | |
| see above | Sonora Quest Laboratories | TGen North | Jolene Bowers, Heather Centner, Chris French, Hayley Yaglom, Ashlyn Pfeiffer, Darrin Lemmer, Dave Engelthaler, The Arizona COVID Genomics Union (ACGU) |
| EPI_ISL_1182535, EPI_ISL_1182537, EPI_ISL_1182539, EPI_ISL_1182540 | TGen North | TGen North | Jolene Bowers, Heather Centner, Chris French, Hayley Yaglom, Ashlyn Pfeiffer, Darrin Lemmer, Dave Engelthaler, The Arizona COVID Genomics Union (ACGU) |
| EPI_ISL_1182626, EPI_ISL_1182627, EPI_ISL_1182628, EPI_ISL_1182629, EPI_ISL_1182630, EPI_ISL_1182631, EPI_ISL_1182633, EPI_ISL_1182634, EPI_ISL_1182635, EPI_ISL_1182636, EPI_ISL_1182637, EPI_ISL_1182638, EPI_ISL_1182639, EPI_ISL_1182641, EPI_ISL_1182642, EPI_ISL_1182643, EPI_ISL_1182644, EPI_ISL_1182645, EPI_ISL_1182646, EPI_ISL_1182649, EPI_ISL_1182650, EPI_ISL_1182651, EPI_ISL_1182653, EPI_ISL_1182654, EPI_ISL_1182655, EPI_ISL_1182656 | | | |
| see above | Wyoming Public Health Laboratory | Wyoming Public Health Laboratory | Jim Mildenberger, Wanda Manley, Noah Hull, Taylor Fearing, Lynette Gumbleton, Channing Weber, Ashley Norberg, Chayse Rowley, Marley Goetz, Brian Dominguez, Elliot Thomasson, Cari Sloma, and Rob Christensen |
| EPI_ISL_1182657, EPI_ISL_1182658, EPI_ISL_1182659, EPI_ISL_1182660, EPI_ISL_1182661, EPI_ISL_1182662, EPI_ISL_1182663, EPI_ISL_1182664, EPI_ISL_1182665, EPI_ISL_1182666, EPI_ISL_1182667, EPI_ISL_1182668, EPI_ISL_1182669, EPI_ISL_1182671, EPI_ISL_1182672, EPI_ISL_1182673, EPI_ISL_1182674, EPI_ISL_1182676, EPI_ISL_1182677, EPI_ISL_1182678, EPI_ISL_1182679, EPI_ISL_1182681, EPI_ISL_1182682, EPI_ISL_1182683, EPI_ISL_1182684, EPI_ISL_1182685, EPI_ISL_1182686, EPI_ISL_1182687, EPI_ISL_1182688, EPI_ISL_1182689, EPI_ISL_1182690, EPI_ISL_1182691 | | | |
| see above | University of Wisconsin-Madison AIDS Vaccine Research Laboratories | University of Wisconsin-Madison AIDS Vaccine Research Laboratories | Gage Moreno, Katarina Braun, et al. AIDS Vaccine Research Laboratories |
| EPI_ISL_1182692 | Baylor Esoteric + Molecular Lab | Baylor Esoteric + Molecular Lab | Elizabeth Forrester, Dawn Richards |
| EPI_ISL_1182693, EPI_ISL_1182694, EPI_ISL_1182695, EPI_ISL_1182696, EPI_ISL_1182697, EPI_ISL_1182698, EPI_ISL_1182699, EPI_ISL_1182700, EPI_ISL_1182701, EPI_ISL_1182702, EPI_ISL_1182703, EPI_ISL_1182704, EPI_ISL_1182705, EPI_ISL_1182706, EPI_ISL_1182707, EPI_ISL_1182708, EPI_ISL_1182709, EPI_ISL_1182710, EPI_ISL_1182711, EPI_ISL_1182712, EPI_ISL_1182713, EPI_ISL_1182714, EPI_ISL_1182715, EPI_ISL_1182716, EPI_ISL_1182717, EPI_ISL_1182718, EPI_ISL_1182719, EPI_ISL_1182720, EPI_ISL_1182721, EPI_ISL_1182722, EPI_ISL_1182723, EPI_ISL_1182724, EPI_ISL_1182725, EPI_ISL_1182726, EPI_ISL_1182727, EPI_ISL_1182728, EPI_ISL_1182729, EPI_ISL_1182730, EPI_ISL_1182731, EPI_ISL_1182732, EPI_ISL_1182733, EPI_ISL_1182734, EPI_ISL_1182735, EPI_ISL_1182736, EPI_ISL_1182737, EPI_ISL_1182738, EPI_ISL_1182739, EPI_ISL_1182740, EPI_ISL_1182741, EPI_ISL_1182742, EPI_ISL_1182743, EPI_ISL_1182744, EPI_ISL_1182745, EPI_ISL_1182746, EPI_ISL_1182747, EPI_ISL_1182748, EPI_ISL_1182749, EPI_ISL_1182750, EPI_ISL_1182751, EPI_ISL_1182752, EPI_ISL_1182753, EPI_ISL_1182754, EPI_ISL_1182755, EPI_ISL_1182756, EPI_ISL_1182757, EPI_ISL_1182758, EPI_ISL_1182759, EPI_ISL_1182760, EPI_ISL_1182761, EPI_ISL_1182762, EPI_ISL_1182763, EPI_ISL_1182764, EPI_ISL_1182765, EPI_ISL_1182766, EPI_ISL_1182767, EPI_ISL_1182768, EPI_ISL_1182769, EPI_ISL_1182770, EPI_ISL_1182771, EPI_ISL_1182772, EPI_ISL_1182773, EPI_ISL_1182774, EPI_ISL_1182775, EPI_ISL_1182776, EPI_ISL_1182777, EPI_ISL_1182778, EPI_ISL_1182779, EPI_ISL_1182780, EPI_ISL_1182781, EPI_ISL_1182782, EPI_ISL_1182783, EPI_ISL_1182784, EPI_ISL_1182785, EPI_ISL_1182786, EPI_ISL_1182787, EPI_ISL_1182788, EPI_ISL_1182789, EPI_ISL_1182790, EPI_ISL_1182791, EPI_ISL_1182792, EPI_ISL_1182793, EPI_ISL_1182794, EPI_ISL_1182795, EPI_ISL_1182796, EPI_ISL_1182797, EPI_ISL_1182798, EPI_ISL_1182799, EPI_ISL_1182800, EPI_ISL_1182801, EPI_ISL_1182802, EPI_ISL_1182803, EPI_ISL_1182804, EPI_ISL_1182805, EPI_ISL_1182806, EPI_ISL_1182807, EPI_ISL_1182808, EPI_ISL_1182809, EPI_ISL_1182810, EPI_ISL_1182811 | | | |
| see above | Alaska State Virology Laboratory | Alaska State Virology Laboratory | Stephanie DeRonde, Lisa Smith, Ph.D., Jack Chen, Ph.D. |
| EPI_ISL_1183692, EPI_ISL_1183693, EPI_ISL_1183694, EPI_ISL_1183695, EPI_ISL_1183696, EPI_ISL_1183697, EPI_ISL_1183698, EPI_ISL_1183699, EPI_ISL_1183700, EPI_ISL_1183701, EPI_ISL_1183702, EPI_ISL_1183703, EPI_ISL_1183704, EPI_ISL_1183705, EPI_ISL_1183706, EPI_ISL_1183707, EPI_ISL_1183708, EPI_ISL_1183709, EPI_ISL_1183710, EPI_ISL_1183711, EPI_ISL_1183722, EPI_ISL_1183723, EPI_ISL_1183724, EPI_ISL_1183725, EPI_ISL_1183726, EPI_ISL_1183727, EPI_ISL_1183728, EPI_ISL_1183729, EPI_ISL_1183730, EPI_ISL_1183731, EPI_ISL_1183732, EPI_ISL_1183733, EPI_ISL_1183734, EPI_ISL_1183735, EPI_ISL_1183736, EPI_ISL_1183737, EPI_ISL_1183738, EPI_ISL_1183739, EPI_ISL_1183740, EPI_ISL_1183741, EPI_ISL_1183742, EPI_ISL_1183743, EPI_ISL_1183744, EPI_ISL_1183745, EPI_ISL_1183746, EPI_ISL_1183747, EPI_ISL_1183748, EPI_ISL_1183749, EPI_ISL_1183750, EPI_ISL_1183751, EPI_ISL_1183752, EPI_ISL_1183753, EPI_ISL_1183754, EPI_ISL_1183755, EPI_ISL_1183756, EPI_ISL_1183757, EPI_ISL_1183758, EPI_ISL_1183759, EPI_ISL_1183760, EPI_ISL_1183761, EPI_ISL_1183762, EPI_ISL_1183763, EPI_ISL_1183764, EPI_ISL_1183765 | | | |
| see above | Utah Public Health Laboratory | Utah Public Health Laboratory | Erin L. Young, Kelly F. Oakeson, Tara Gallagher |
| EPI_ISL_1183766, EPI_ISL_1183767, EPI_ISL_1183768, EPI_ISL_1183769, EPI_ISL_1183770, EPI_ISL_1183771, EPI_ISL_1183772, EPI_ISL_1183773, EPI_ISL_1183774, EPI_ISL_1183775, EPI_ISL_1183776, EPI_ISL_1183777, EPI_ISL_1183778, EPI_ISL_1183779, EPI_ISL_1183780, EPI_ISL_1183781, EPI_ISL_1183782, EPI_ISL_1183783, EPI_ISL_1183784, EPI_ISL_1183786, EPI_ISL_1183787, EPI_ISL_1183788, EPI_ISL_1183789, EPI_ISL_1183790, EPI_ISL_1183791, EPI_ISL_1183792, EPI_ISL_1183793, EPI_ISL_1183796, EPI_ISL_1183798, EPI_ISL_1183799, EPI_ISL_1183800, EPI_ISL_1183801, EPI_ISL_1183802, EPI_ISL_1183804, EPI_ISL_1183806, EPI_ISL_1183807, EPI_ISL_1183808, EPI_ISL_1183809, EPI_ISL_1183810, EPI_ISL_1183811, EPI_ISL_1183812, EPI_ISL_1183813, EPI_ISL_1183814, EPI_ISL_1183816, EPI_ISL_1183817, EPI_ISL_1183818, EPI_ISL_1183819, EPI_ISL_1183820, EPI_ISL_1183821, EPI_ISL_1183822, EPI_ISL_1183823, EPI_ISL_1183824, EPI_ISL_1183825, EPI_ISL_1183826, EPI_ISL_1183827, EPI_ISL_1183828, EPI_ISL_1183830, EPI_ISL_1183832, EPI_ISL_1183833 | | | |
| see above | Sonora Quest Laboratories | TGen North | Jolene Bowers, Heather Centner, Chris French, Hayley Yaglom, Ashlyn Pfeiffer, Darrin Lemmer, Dave Engelthaler, The Arizona COVID Genomics Union (ACGU) |
| EPI_ISL_1183834, EPI_ISL_1183835, EPI_ISL_1183836, EPI_ISL_1183837, EPI_ISL_1183838, EPI_ISL_1183839, EPI_ISL_1183840, EPI_ISL_1183841, EPI_ISL_1183842, EPI_ISL_1183843, EPI_ISL_1183844, EPI_ISL_1183845, EPI_ISL_1183846, EPI_ISL_1183847, EPI_ISL_1183848, EPI_ISL_1183849, EPI_ISL_1183850, EPI_ISL_1183852, EPI_ISL_1183853, EPI_ISL_1183854, EPI_ISL_1183855, EPI_ISL_1183856, EPI_ISL_1183858, EPI_ISL_1183859, EPI_ISL_1183860, EPI_ISL_1183861, EPI_ISL_1183862, EPI_ISL_1183863, EPI_ISL_1183864, EPI_ISL_1183865, EPI_ISL_1183866, EPI_ISL_1183867, EPI_ISL_1183868, EPI_ISL_1183869, EPI_ISL_1183870 | | | |
| see above | Arizona Department of Health Services | TGen North | Jolene Bowers, Heather Centner, Chris French, Hayley Yaglom, Ashlyn Pfeiffer, Darrin Lemmer, Dave Engelthaler, The Arizona COVID Genomics Union (ACGU) |
| EPI_ISL_1183871, EPI_ISL_1183872, EPI_ISL_1183873, EPI_ISL_1183876, EPI_ISL_1183877, EPI_ISL_1183878, EPI_ISL_1183879, EPI_ISL_1183880, EPI_ISL_1183881, EPI_ISL_1183882, EPI_ISL_1183883, EPI_ISL_1183884, EPI_ISL_1183885, EPI_ISL_1183886, EPI_ISL_1183887, EPI_ISL_1183889, EPI_ISL_1183890, EPI_ISL_1183891, EPI_ISL_1183892, EPI_ISL_1183894, EPI_ISL_1183895, EPI_ISL_1183896, EPI_ISL_1183897, EPI_ISL_1183898, EPI_ISL_1183899, EPI_ISL_1183900, EPI_ISL_1183901, EPI_ISL_1183902, EPI_ISL_1183903, EPI_ISL_1183904, EPI_ISL_1183905, EPI_ISL_1183907, EPI_ISL_1183908, EPI_ISL_1183909, EPI_ISL_1183911, EPI_ISL_1183913, EPI_ISL_1183914, EPI_ISL_1183915, EPI_ISL_1183916, EPI_ISL_1183917, EPI_ISL_1183918, EPI_ISL_1183919, EPI_ISL_1183920, EPI_ISL_1183921, EPI_ISL_1183922, EPI_ISL_1183923, EPI_ISL_1183924, EPI_ISL_1183925, EPI_ISL_1183926, EPI_ISL_1183927, EPI_ISL_1183928, EPI_ISL_1183929, EPI_ISL_1183930, EPI_ISL_1183931, EPI_ISL_1183933, EPI_ISL_1183934, EPI_ISL_1183935, EPI_ISL_1183936, EPI_ISL_1183937, EPI_ISL_1183938, EPI_ISL_1183939, EPI_ISL_1183940, EPI_ISL_1183941, EPI_ISL_1183942, EPI_ISL_1183943, EPI_ISL_1183944, EPI_ISL_1183946, EPI_ISL_1183947, EPI_ISL_1183948, EPI_ISL_1183949, EPI_ISL_1183950, EPI_ISL_1183951, EPI_ISL_1183952, EPI_ISL_1183953, EPI_ISL_1183955, EPI_ISL_1183956, EPI_ISL_1183957, EPI_ISL_1183958, EPI_ISL_1183959, EPI_ISL_1183960, EPI_ISL_1183961, EPI_ISL_1183962, EPI_ISL_1183963, EPI_ISL_1183964, EPI_ISL_1183965, EPI_ISL_1183966, EPI_ISL_1183967, EPI_ISL_1183969, EPI_ISL_1183970, EPI_ISL_1183972, EPI_ISL_1183973, EPI_ISL_1183974, EPI_ISL_1183975, EPI_ISL_1183976, EPI_ISL_1183977, EPI_ISL_1183979, EPI_ISL_1183980, EPI_ISL_1183981, EPI_ISL_1183983, EPI_ISL_1183984, EPI_ISL_1183985, EPI_ISL_1183986, EPI_ISL_1183987, EPI_ISL_1183988, EPI_ISL_1183990, EPI_ISL_1183991, EPI_ISL_1183992, EPI_ISL_1183993, EPI_ISL_1183994, EPI_ISL_1183995, EPI_ISL_1183996, EPI_ISL_1183997, EPI_ISL_1183998, EPI_ISL_1183999, EPI_ISL_1184000, EPI_ISL_1184003, EPI_ISL_1184004, EPI_ISL_1184005, EPI_ISL_1184006, EPI_ISL_1184007, EPI_ISL_1184008, EPI_ISL_1184009, EPI_ISL_1184010, EPI_ISL_1184011, EPI_ISL_1184012, EPI_ISL_1184013, EPI_ISL_1184014, EPI_ISL_1184015, EPI_ISL_1184016, EPI_ISL_1184017, EPI_ISL_1184020, EPI_ISL_1184021, EPI_ISL_1184022, EPI_ISL_1184023, EPI_ISL_1184025, EPI_ISL_1184026, EPI_ISL_1184031, EPI_ISL_1184032, EPI_ISL_1184033, EPI_ISL_1184034, EPI_ISL_1184035, EPI_ISL_1184036, EPI_ISL_1184037, EPI_ISL_1184038, EPI_ISL_1184039, EPI_ISL_1184040, EPI_ISL_1184041, EPI_ISL_1184042, EPI_ISL_1184043, EPI_ISL_1184044, EPI_ISL_1184047, EPI_ISL_1184048, EPI_ISL_1184049, EPI_ISL_1184050, EPI_ISL_1184051, EPI_ISL_1184052, EPI_ISL_1184053, EPI_ISL_1184054, EPI_ISL_1184055, EPI_ISL_1184057, EPI_ISL_1184058, EPI_ISL_1184059, EPI_ISL_1184060, EPI_ISL_1184061, EPI_ISL_1184063, EPI_ISL_1184064, EPI_ISL_1184065, EPI_ISL_1184066, EPI_ISL_1184067, EPI_ISL_1184068, EPI_ISL_1184069, EPI_ISL_1184070, EPI_ISL_1184072, EPI_ISL_1184073, EPI_ISL_1184074, EPI_ISL_1184075, EPI_ISL_1184078 | | | |
| see above | Sonora Quest Laboratories | TGen North | Jolene Bowers, Heather Centner, Chris French, Hayley Yaglom, Ashlyn Pfeiffer, Darrin Lemmer, Dave Engelthaler, The Arizona COVID Genomics Union (ACGU) |
| EPI_ISL_1184079, EPI_ISL_1184080, EPI_ISL_1184081, EPI_ISL_1184082, EPI_ISL_1184083, EPI_ISL_1184084, EPI_ISL_1184085, EPI_ISL_1184086, EPI_ISL_1184087, EPI_ISL_1184088, EPI_ISL_1184089, EPI_ISL_1184090, EPI_ISL_1184093 | | | |
| see above | Arizona State University | Arizona State University | Rabia Maqsood, LaRinda A. Holland, Peter T. Skidmore, Emily A. Kaelin, Nicholas J. Mellor, Kristina Buss, Joy M. Blain, Neal W Woodbury, Valerie Harris, Joshua LaBaer, Vel Murugan, Efrem S. Lim |
| EPI_ISL_1184099, EPI_ISL_1184100, EPI_ISL_1184101, EPI_ISL_1184102, EPI_ISL_1184103, EPI_ISL_1184104, EPI_ISL_1184105, EPI_ISL_1184107, EPI_ISL_1184108, EPI_ISL_1184110, EPI_ISL_1184112 | | | |
| see above | New Jersey Department of Health, Public Health and Environmental Laboratories | New Jersey Public Health and Environmental Laboratories | Lindsey Bodnar, Shiv K. Verma, Dana Woell, Byeong Jeong |
| EPI_ISL_1184113, EPI_ISL_1184114 | Rochester Regional Hospital | Multidrug-Resistant Organism Repository and Surveillance Network (MRSN) | Emil Lesho, Brendan Corey, Francois Lebreton, Ana C. Ong, Brett E. Swierczewski, Jason W. Bennett, Edward E. Walsh, and Patrick Mc Gann |
| EPI_ISL_1184115, EPI_ISL_1184116, EPI_ISL_1184117, EPI_ISL_1184118, EPI_ISL_1184119, EPI_ISL_1184120, EPI_ISL_1184121, EPI_ISL_1184122, EPI_ISL_1184123, EPI_ISL_1184124, EPI_ISL_1184125, EPI_ISL_1184126, EPI_ISL_1184127, EPI_ISL_1184128, EPI_ISL_1184129, EPI_ISL_1184130, EPI_ISL_1184131, EPI_ISL_1184132, EPI_ISL_1184133, EPI_ISL_1184134 | | | |
| see above | Kentucky State Public Health Lab | Kentucky State Public Health Lab | Stephanie Lunn, Karim George, Joshua Tobias, William Grooms, Vaneet Arora, Matthew Johnson, Rachel Zinner, Rhonda Lucas |
| EPI_ISL_1184135, EPI_ISL_1184136, EPI_ISL_1184137 | Rochester Regional Hospital | Multidrug-Resistant Organism Repository and Surveillance Network (MRSN) | Emil Lesho, Brendan Corey, Francois Lebreton, Ana C. Ong, Brett E. Swierczewski, Jason W. Bennett, Edward E. Walsh, and Patrick Mc Gann |
| EPI_ISL_1184138, EPI_ISL_1184139, EPI_ISL_1184140, EPI_ISL_1184141, EPI_ISL_1184142, EPI_ISL_1184143, EPI_ISL_1184144, EPI_ISL_1184145, EPI_ISL_1184146, EPI_ISL_1184147, EPI_ISL_1184148, EPI_ISL_1184149, EPI_ISL_1184150, EPI_ISL_1184151, EPI_ISL_1184152, EPI_ISL_1184153, EPI_ISL_1184154, EPI_ISL_1184155, EPI_ISL_1184156, EPI_ISL_1184157, EPI_ISL_1184158, EPI_ISL_1184159, EPI_ISL_1184160, EPI_ISL_1184161, EPI_ISL_1184162, EPI_ISL_1184163, EPI_ISL_1184164, EPI_ISL_1184165, EPI_ISL_1184166, EPI_ISL_1184167, EPI_ISL_1184168 | | | |
| see above | Virginia Division of Consolidated Laboratory Services | Virginia Division of Consolidated Laboratory Services | Virginia DCLS |
| EPI_ISL_1184169, EPI_ISL_1184170, EPI_ISL_1184171, EPI_ISL_1184172, EPI_ISL_1184173, EPI_ISL_1184174, EPI_ISL_1184175, EPI_ISL_1184176, EPI_ISL_1184177, EPI_ISL_1184178, EPI_ISL_1184179, EPI_ISL_1184180, EPI_ISL_1184181, EPI_ISL_1184182, EPI_ISL_1184183, EPI_ISL_1184184, EPI_ISL_1184185, EPI_ISL_1184186, EPI_ISL_1184187, EPI_ISL_1184188, EPI_ISL_1184189, EPI_ISL_1184190, EPI_ISL_1184191, EPI_ISL_1184192, EPI_ISL_1184193, EPI_ISL_1184194, EPI_ISL_1184195, EPI_ISL_1184196, EPI_ISL_1184197, EPI_ISL_1184198, EPI_ISL_1184199, EPI_ISL_1184200, EPI_ISL_1184201, EPI_ISL_1184202, EPI_ISL_1184203, EPI_ISL_1184204, EPI_ISL_1184205, EPI_ISL_1184206, EPI_ISL_1184207, EPI_ISL_1184208, EPI_ISL_1184209, EPI_ISL_1184210, EPI_ISL_1184211, EPI_ISL_1184212, EPI_ISL_1184213, EPI_ISL_1184214, EPI_ISL_1184215, EPI_ISL_1184216, EPI_ISL_1184217, EPI_ISL_1184218, EPI_ISL_1184219 | | | |
| see above | Indiana Animal Disease Diagnostic Laboratory | Carpi Laboratory - Purdue University | Jack Dorman, Ilinca I Ciubotariu, Lev Gorenstein, Abebe A Fola, G Kenitra Hendrix, Rebecca P Wilkes, Giovanna Carpi |
| EPI_ISL_1184220, EPI_ISL_1184221, EPI_ISL_1184222, EPI_ISL_1184223, EPI_ISL_1184224, EPI_ISL_1184225, EPI_ISL_1184226, EPI_ISL_1184227, EPI_ISL_1184228, EPI_ISL_1184230 | Vault Health | Minnesota Department of Health, Public Health Laboratory | Alexandra Lorentz, Jacob Garfin, Matt Plumb, and Xiong Wang |
| EPI_ISL_1184231, EPI_ISL_1184232, EPI_ISL_1184233, EPI_ISL_1184234, EPI_ISL_1184235, EPI_ISL_1184236 | M Health Fairview | Minnesota Department of Health, Public Health Laboratory | Alexandra Lorentz, Jacob Garfin, Matt Plumb, and Xiong Wang |
| EPI_ISL_1184237 | Vault Health | Minnesota Department of Health, Public Health Laboratory | Alexandra Lorentz, Jacob Garfin, Matt Plumb, and Xiong Wang |
| EPI_ISL_1184238, EPI_ISL_1184239, EPI_ISL_1184240, EPI_ISL_1184241, EPI_ISL_1184242, EPI_ISL_1184243, EPI_ISL_1184244, EPI_ISL_1184245, EPI_ISL_1184246, EPI_ISL_1184247, EPI_ISL_1184248, EPI_ISL_1184249, EPI_ISL_1184250, EPI_ISL_1184251, EPI_ISL_1184252, EPI_ISL_1184253, EPI_ISL_1184254, EPI_ISL_1184255, EPI_ISL_1184256, EPI_ISL_1184257, EPI_ISL_1184258, EPI_ISL_1184259, EPI_ISL_1184260, EPI_ISL_1184261, EPI_ISL_1184262, EPI_ISL_1184263, EPI_ISL_1184264, EPI_ISL_1184265, EPI_ISL_1184266, EPI_ISL_1184267, EPI_ISL_1184268, EPI_ISL_1184269, EPI_ISL_1184270, EPI_ISL_1184271, EPI_ISL_1184272, EPI_ISL_1184273, EPI_ISL_1184274, EPI_ISL_1184275, EPI_ISL_1184276, EPI_ISL_1184277, EPI_ISL_1184278, EPI_ISL_1184279, EPI_ISL_1184280, EPI_ISL_1184281, EPI_ISL_1184282, EPI_ISL_1184283, EPI_ISL_1184284, EPI_ISL_1184285, EPI_ISL_1184286 | | | |
| see above | Mayo Clinic & Mayo Clinic Laboratories | Minnesota Department of Health, Public Health Laboratory | Alexandra Lorentz, Jacob Garfin, Matt Plumb, and Xiong Wang |
| EPI_ISL_1184287, EPI_ISL_1184288, EPI_ISL_1184289, EPI_ISL_1184290, EPI_ISL_1184291, EPI_ISL_1184292, EPI_ISL_1184293, EPI_ISL_1184294, EPI_ISL_1184295, EPI_ISL_1184296, EPI_ISL_1184297, EPI_ISL_1184298, EPI_ISL_1184299, EPI_ISL_1184300, EPI_ISL_1184301, EPI_ISL_1184302 | | | |
| see above | M Health Fairview | Minnesota Department of Health, Public | Alexandra Lorentz, Jacob Garfin, Matt Plumb, and Xiong Wang |

| | | | |
|---|---|---|---|
| | | Health Laboratory | |
| EPI_ISL_1184304 | Mayo Clinic & Mayo Clinic Laboratories | Minnesota Department of Health, Public Health Laboratory | Alexandra Lorentz, Jacob Garfin, Matt Plumb, and Xiong Wang |
| EPI_ISL_1184305 | North Dakota Department of Health, Public Health Laboratory | North Dakota Department of Health, Public Health Laboratory | Lisa Wingerter |
| EPI_ISL_1184307, EPI_ISL_1184310, EPI_ISL_1184317, EPI_ISL_1184318, EPI_ISL_1184319, EPI_ISL_1184325, EPI_ISL_1184327, EPI_ISL_1184328, EPI_ISL_1184329, EPI_ISL_1184330, EPI_ISL_1184331, EPI_ISL_1184332, EPI_ISL_1184336, EPI_ISL_1184337, EPI_ISL_1184338, EPI_ISL_1184339, EPI_ISL_1184340, EPI_ISL_1184341, EPI_ISL_1184342, EPI_ISL_1184343, EPI_ISL_1184344, EPI_ISL_1184345, EPI_ISL_1184346, EPI_ISL_1184347, EPI_ISL_1184351, EPI_ISL_1184353, EPI_ISL_1184354, EPI_ISL_1184356, EPI_ISL_1184357, EPI_ISL_1184358, EPI_ISL_1184359, EPI_ISL_1184362, EPI_ISL_1184366, EPI_ISL_1184370, EPI_ISL_1184374, EPI_ISL_1184376, EPI_ISL_1184377, EPI_ISL_1184379, EPI_ISL_1184380, EPI_ISL_1184381, EPI_ISL_1184382, EPI_ISL_1184383, EPI_ISL_1184386, EPI_ISL_1184387, EPI_ISL_1184388, EPI_ISL_1184390, EPI_ISL_1184392, EPI_ISL_1184393, EPI_ISL_1184394, EPI_ISL_1184396, EPI_ISL_1184397, EPI_ISL_1184398, EPI_ISL_1184399, EPI_ISL_1184400, EPI_ISL_1184403, EPI_ISL_1184405, EPI_ISL_1184408, EPI_ISL_1184409, EPI_ISL_1184413, EPI_ISL_1184414, EPI_ISL_1184415, EPI_ISL_1184416, EPI_ISL_1184418, EPI_ISL_1184419, EPI_ISL_1184420, EPI_ISL_1184421, EPI_ISL_1184423, EPI_ISL_1184424, EPI_ISL_1184425, EPI_ISL_1184426, EPI_ISL_1184427, EPI_ISL_1184428, EPI_ISL_1184429, EPI_ISL_1184430, EPI_ISL_1184431, EPI_ISL_1184432, EPI_ISL_1184435, EPI_ISL_1184438, EPI_ISL_1184439, EPI_ISL_1184440, EPI_ISL_1184441, EPI_ISL_1184442, EPI_ISL_1184445, EPI_ISL_1184447, EPI_ISL_1184448, EPI_ISL_1184449, EPI_ISL_1184450, EPI_ISL_1184451, EPI_ISL_1184452, EPI_ISL_1184453, EPI_ISL_1184454, EPI_ISL_1184456, EPI_ISL_1184458, EPI_ISL_1184459, EPI_ISL_1184464, EPI_ISL_1184465, EPI_ISL_1184466, EPI_ISL_1184467, EPI_ISL_1184468 | | | |
| see above | Texas Department of State Health Services | Texas Department of State Health Services | Rashmi Tuladhar, Bonnie Oh, Jenny Zhang, Maliha Rahman, Mayela Pedrueza, Anita Pokharel, Myong Koag, Chun Wang, Rachel Lee, Grace Kubin |
| EPI_ISL_1184559, EPI_ISL_1184560, EPI_ISL_1184561, EPI_ISL_1184562, EPI_ISL_1184563, EPI_ISL_1184564, EPI_ISL_1184565, EPI_ISL_1184566, EPI_ISL_1184567, EPI_ISL_1184569, EPI_ISL_1184570, EPI_ISL_1184571, EPI_ISL_1184573, EPI_ISL_1184574, EPI_ISL_1184575, EPI_ISL_1184576, EPI_ISL_1184577, EPI_ISL_1184578, EPI_ISL_1184579, EPI_ISL_1184580, EPI_ISL_1184581 | | | |
| see above | Arizona State Public Health Laboratory | Arizona State Public Health Laboratory | Trung Huynh, Jessica Escobar, Katherine Fullerton, Nobuko Fukushima, Stacy White, Linda Getsinger, Victor Waddell |
| EPI_ISL_1184583, EPI_ISL_1184584, EPI_ISL_1184585, EPI_ISL_1184586, EPI_ISL_1184587, EPI_ISL_1184588, EPI_ISL_1184589, EPI_ISL_1184590, EPI_ISL_1184591, EPI_ISL_1184592, EPI_ISL_1184593, EPI_ISL_1184594, EPI_ISL_1184595, EPI_ISL_1184596, EPI_ISL_1184597, EPI_ISL_1184598, EPI_ISL_1184599, EPI_ISL_1184600, EPI_ISL_1184601, EPI_ISL_1184602, EPI_ISL_1184603, EPI_ISL_1184604, EPI_ISL_1184605, EPI_ISL_1184606, EPI_ISL_1184607, EPI_ISL_1184608, EPI_ISL_1184609, EPI_ISL_1184610, EPI_ISL_1184611, EPI_ISL_1184612, EPI_ISL_1184613, EPI_ISL_1184614, EPI_ISL_1184615, EPI_ISL_1184616, EPI_ISL_1184617, EPI_ISL_1184619, EPI_ISL_1184620, EPI_ISL_1184622, EPI_ISL_1184623 | | | |
| see above | Santa Clara County Public Health Laboratory | Chan-Zuckerberg Biohub | CZB Cliahub Consortium |
| EPI_ISL_1184624, EPI_ISL_1184625, EPI_ISL_1184626, EPI_ISL_1184627, EPI_ISL_1184628, EPI_ISL_1184629, EPI_ISL_1184630, EPI_ISL_1184631, EPI_ISL_1184632, EPI_ISL_1184633, EPI_ISL_1184634, EPI_ISL_1184635, EPI_ISL_1184636, EPI_ISL_1184637, EPI_ISL_1184638, EPI_ISL_1184639, EPI_ISL_1184640, EPI_ISL_1184641, EPI_ISL_1184642, EPI_ISL_1184643, EPI_ISL_1184644, EPI_ISL_1184645, EPI_ISL_1184646, EPI_ISL_1184647, EPI_ISL_1184648, EPI_ISL_1184649, EPI_ISL_1184650, EPI_ISL_1184651, EPI_ISL_1184652, EPI_ISL_1184653, EPI_ISL_1184654, EPI_ISL_1184655, EPI_ISL_1184656, EPI_ISL_1184657, EPI_ISL_1184658, EPI_ISL_1184659, EPI_ISL_1184660, EPI_ISL_1184661, EPI_ISL_1184662, EPI_ISL_1184663, EPI_ISL_1184664, EPI_ISL_1184665, EPI_ISL_1184666, EPI_ISL_1184667, EPI_ISL_1184668, EPI_ISL_1184669, EPI_ISL_1184670, EPI_ISL_1184671, EPI_ISL_1184672, EPI_ISL_1184673, EPI_ISL_1184674, EPI_ISL_1184675, EPI_ISL_1184676, EPI_ISL_1184677, EPI_ISL_1184678, EPI_ISL_1184679, EPI_ISL_1184680 | | | |
| see above | Alameda County Public Health Lab | Chan-Zuckerberg Biohub | CZB Cliahub Consortium |
| EPI_ISL_1184681, EPI_ISL_1184682, EPI_ISL_1184683, EPI_ISL_1184684, EPI_ISL_1184685, EPI_ISL_1184686, EPI_ISL_1184687, EPI_ISL_1184688, EPI_ISL_1184689, EPI_ISL_1184690, EPI_ISL_1184691, EPI_ISL_1184692, EPI_ISL_1184693, EPI_ISL_1184694, EPI_ISL_1184695, EPI_ISL_1184696, EPI_ISL_1184697, EPI_ISL_1184698, EPI_ISL_1184699, EPI_ISL_1184700, EPI_ISL_1184701, EPI_ISL_1184702, EPI_ISL_1184703, EPI_ISL_1184704, EPI_ISL_1184705, EPI_ISL_1184706, EPI_ISL_1184707, EPI_ISL_1184708, EPI_ISL_1184709, EPI_ISL_1184710, EPI_ISL_1184711, EPI_ISL_1184712, EPI_ISL_1184713, EPI_ISL_1184714, EPI_ISL_1184715, EPI_ISL_1184716, EPI_ISL_1184717, EPI_ISL_1184718, EPI_ISL_1184719, EPI_ISL_1184720, EPI_ISL_1184721, EPI_ISL_1184722, EPI_ISL_1184723, EPI_ISL_1184724, EPI_ISL_1184725, EPI_ISL_1184726, EPI_ISL_1184727, EPI_ISL_1184728, EPI_ISL_1184729, EPI_ISL_1184730, EPI_ISL_1184731, EPI_ISL_1184732, EPI_ISL_1184733, EPI_ISL_1184734, EPI_ISL_1184735, EPI_ISL_1184736, EPI_ISL_1184737, EPI_ISL_1184738, EPI_ISL_1184739, EPI_ISL_1184740, EPI_ISL_1184741, EPI_ISL_1184742, EPI_ISL_1184743, EPI_ISL_1184744, EPI_ISL_1184745, EPI_ISL_1184746, EPI_ISL_1184747, EPI_ISL_1184748, EPI_ISL_1184749, EPI_ISL_1184750, EPI_ISL_1184751, EPI_ISL_1184752, EPI_ISL_1184753, EPI_ISL_1184754, EPI_ISL_1184755, EPI_ISL_1184756, EPI_ISL_1184757, EPI_ISL_1184758, EPI_ISL_1184759, EPI_ISL_1184760, EPI_ISL_1184761, EPI_ISL_1184762, EPI_ISL_1184763, EPI_ISL_1184764, EPI_ISL_1184765, EPI_ISL_1184766, EPI_ISL_1184767, EPI_ISL_1184768, EPI_ISL_1184769, EPI_ISL_1184770, EPI_ISL_1184771, EPI_ISL_1184772, EPI_ISL_1184773, EPI_ISL_1184774, EPI_ISL_1184775, EPI_ISL_1184776, EPI_ISL_1184777, EPI_ISL_1184778, EPI_ISL_1184779, EPI_ISL_1184780, EPI_ISL_1184781, EPI_ISL_1184782, EPI_ISL_1184783, EPI_ISL_1184784, EPI_ISL_1184785, EPI_ISL_1184786, EPI_ISL_1184787, EPI_ISL_1184788, EPI_ISL_1184789, EPI_ISL_1184790, EPI_ISL_1184791, EPI_ISL_1184792, EPI_ISL_1184793, EPI_ISL_1184794, EPI_ISL_1184795, EPI_ISL_1184796, EPI_ISL_1184797, EPI_ISL_1184798, EPI_ISL_1184799, EPI_ISL_1184800, EPI_ISL_1184801, EPI_ISL_1184802, EPI_ISL_1184803, EPI_ISL_1184804, EPI_ISL_1184805, EPI_ISL_1184806, EPI_ISL_1184807, EPI_ISL_1184808 | | | |
| see above | Orange County Public Health Lab | Chan-Zuckerberg Biohub | CZB Cliahub Consortium |
| EPI_ISL_1184809, EPI_ISL_1184810, EPI_ISL_1184811, EPI_ISL_1184812, EPI_ISL_1184813, EPI_ISL_1184814, EPI_ISL_1184815, EPI_ISL_1184816, EPI_ISL_1184817, EPI_ISL_1184818, EPI_ISL_1184819, EPI_ISL_1184820, EPI_ISL_1184821, EPI_ISL_1184822, EPI_ISL_1184823, EPI_ISL_1184824 | | | |
| see above | CA DPH Viral and Rickettsial Disease Laboratory | Chan-Zuckerberg Biohub | CZB Cliahub Consortium |
| EPI_ISL_1184828, EPI_ISL_1184829, EPI_ISL_1184830, EPI_ISL_1184831, EPI_ISL_1184832, EPI_ISL_1184833, EPI_ISL_1184834, EPI_ISL_1184835, EPI_ISL_1184836, EPI_ISL_1184837, EPI_ISL_1184838, EPI_ISL_1184839, EPI_ISL_1184840, EPI_ISL_1184841, EPI_ISL_1184842, EPI_ISL_1184843, EPI_ISL_1184844, EPI_ISL_1184845, EPI_ISL_1184846, EPI_ISL_1184847, EPI_ISL_1184848, EPI_ISL_1184849, EPI_ISL_1184850, EPI_ISL_1184852, EPI_ISL_1184853, EPI_ISL_1184854, EPI_ISL_1184855, EPI_ISL_1184856, EPI_ISL_1184857, EPI_ISL_1184858, EPI_ISL_1184859, EPI_ISL_1184860 | | | |
| see above | Santa Clara County Public Health Laboratory | Chan-Zuckerberg Biohub | CZB Cliahub Consortium |
| EPI_ISL_1184861, EPI_ISL_1184862, EPI_ISL_1184863, EPI_ISL_1184864, EPI_ISL_1184865, EPI_ISL_1184866, EPI_ISL_1184867, EPI_ISL_1184868, EPI_ISL_1184869, EPI_ISL_1184870, EPI_ISL_1184871, EPI_ISL_1184872, EPI_ISL_1184873, EPI_ISL_1184874, EPI_ISL_1184875, EPI_ISL_1184876, EPI_ISL_1184877, EPI_ISL_1184878, EPI_ISL_1184879, EPI_ISL_1184880, EPI_ISL_1184881, EPI_ISL_1184882, EPI_ISL_1184883, EPI_ISL_1184884, EPI_ISL_1184885, EPI_ISL_1184886, EPI_ISL_1184887, EPI_ISL_1184888, EPI_ISL_1184889, EPI_ISL_1184890, EPI_ISL_1184891, EPI_ISL_1184892, EPI_ISL_1184893, EPI_ISL_1184894, EPI_ISL_1184895, EPI_ISL_1184897, EPI_ISL_1184898, EPI_ISL_1184899, EPI_ISL_1184900, EPI_ISL_1184901, EPI_ISL_1184902, EPI_ISL_1184903, EPI_ISL_1184904, EPI_ISL_1184905, EPI_ISL_1184906, EPI_ISL_1184907, EPI_ISL_1184908, EPI_ISL_1184909, EPI_ISL_1184910, EPI_ISL_1184911, EPI_ISL_1184912, EPI_ISL_1184913, EPI_ISL_1184914, EPI_ISL_1184915, EPI_ISL_1184916, EPI_ISL_1184917, EPI_ISL_1184918, EPI_ISL_1184919, EPI_ISL_1184920, EPI_ISL_1184921, EPI_ISL_1184922, EPI_ISL_1184923, EPI_ISL_1184924, EPI_ISL_1184925, EPI_ISL_1184926, EPI_ISL_1184927, EPI_ISL_1184928, EPI_ISL_1184929, EPI_ISL_1184930, EPI_ISL_1184931, EPI_ISL_1184932, EPI_ISL_1184933, EPI_ISL_1184935, EPI_ISL_1184936, EPI_ISL_1184937, EPI_ISL_1184938, EPI_ISL_1184939, EPI_ISL_1184940, EPI_ISL_1184941, EPI_ISL_1184942, EPI_ISL_1184943, EPI_ISL_1184944, EPI_ISL_1184945, EPI_ISL_1184946, EPI_ISL_1184947, EPI_ISL_1184948, EPI_ISL_1184949, EPI_ISL_1184950, EPI_ISL_1184951, EPI_ISL_1184952, EPI_ISL_1184953, EPI_ISL_1184954, EPI_ISL_1184955, EPI_ISL_1184956, EPI_ISL_1184957, EPI_ISL_1184958, EPI_ISL_1184959, EPI_ISL_1184960, EPI_ISL_1184961, EPI_ISL_1184962, EPI_ISL_1184963, EPI_ISL_1184965, EPI_ISL_1184966 | | | |
| see above | Alameda County Public Health Lab | Chan-Zuckerberg Biohub | CZB Cliahub Consortium |
| EPI_ISL_1184967, EPI_ISL_1184968, EPI_ISL_1184969, EPI_ISL_1184970, EPI_ISL_1184971, EPI_ISL_1184972, EPI_ISL_1184973, EPI_ISL_1184974, EPI_ISL_1184975, EPI_ISL_1184976, EPI_ISL_1184977, EPI_ISL_1184978, EPI_ISL_1184979, EPI_ISL_1184980, EPI_ISL_1184981, EPI_ISL_1184982, EPI_ISL_1184983, EPI_ISL_1184984, EPI_ISL_1184985, EPI_ISL_1184986, EPI_ISL_1184987, EPI_ISL_1184988, EPI_ISL_1184989, EPI_ISL_1184990, EPI_ISL_1184991, EPI_ISL_1184992, EPI_ISL_1184993, EPI_ISL_1184994, EPI_ISL_1184995, EPI_ISL_1184996, EPI_ISL_1184997, EPI_ISL_1184998, EPI_ISL_1184999, EPI_ISL_1185000, EPI_ISL_1185001, EPI_ISL_1185002, EPI_ISL_1185003, EPI_ISL_1185004, EPI_ISL_1185005, EPI_ISL_1185006, EPI_ISL_1185007, EPI_ISL_1185008, EPI_ISL_1185009, EPI_ISL_1185010, EPI_ISL_1185011, EPI_ISL_1185012, EPI_ISL_1185013, EPI_ISL_1185014, EPI_ISL_1185015 | | | |
| see above | Orange County Public Health Lab | Chan-Zuckerberg Biohub | CZB Cliahub Consortium |
| EPI_ISL_1185016, EPI_ISL_1185017, EPI_ISL_1185018, EPI_ISL_1185019, EPI_ISL_1185020, EPI_ISL_1185021, EPI_ISL_1185022, EPI_ISL_1185023, EPI_ISL_1185024, EPI_ISL_1185025 | CA DPH Viral and Rickettsial Disease Laboratory | Chan-Zuckerberg Biohub | CZB Cliahub Consortium |
| EPI_ISL_1185027, EPI_ISL_1185028 | Alameda County Public Health Lab | Chan-Zuckerberg Biohub | CZB Cliahub Consortium |
| EPI_ISL_1185029 | Santa Clara County Public Health Laboratory | Chan-Zuckerberg Biohub | CZB Cliahub Consortium |
| EPI_ISL_1185030 | CA DPH Viral and Rickettsial Disease Laboratory | Chan-Zuckerberg Biohub | CZB Cliahub Consortium |
| EPI_ISL_1185032 | Orange County Public Health Lab | Chan-Zuckerberg Biohub | CZB Cliahub Consortium |
| EPI_ISL_1185033, EPI_ISL_1185034, EPI_ISL_1185035, EPI_ISL_1185036, EPI_ISL_1185037, EPI_ISL_1185038 | Alameda County Public Health Lab | Chan-Zuckerberg Biohub | CZB Cliahub Consortium |
| EPI_ISL_1185039, EPI_ISL_1185040 | Orange County Public Health Lab | Chan-Zuckerberg Biohub | CZB Cliahub Consortium |
| EPI_ISL_1185041 | Alameda County Public Health Lab | Chan-Zuckerberg Biohub | CZB Cliahub Consortium |
| EPI_ISL_1185042 | CA DPH Viral and Rickettsial Disease Laboratory | Chan-Zuckerberg Biohub | CZB Cliahub Consortium |
| EPI_ISL_1185043 | Orange County Public Health Lab | Chan-Zuckerberg Biohub | CZB Cliahub Consortium |
| EPI_ISL_1185044 | Santa Clara County Public Health Laboratory | Chan-Zuckerberg Biohub | CZB Cliahub Consortium |
| EPI_ISL_1185045 | Orange County Public Health Lab | Chan-Zuckerberg Biohub | CZB Cliahub Consortium |
| EPI_ISL_1185046 | Santa Clara County Public Health Laboratory | Chan-Zuckerberg Biohub | CZB Cliahub Consortium |
| EPI_ISL_1185047 | Alameda County Public Health Lab | Chan-Zuckerberg Biohub | CZB Cliahub Consortium |
| EPI_ISL_1185048, EPI_ISL_1185049 | Orange County Public Health Lab | Chan-Zuckerberg Biohub | CZB Cliahub Consortium |
| EPI_ISL_1185050 | CA DPH Viral and Rickettsial Disease Laboratory | Chan-Zuckerberg Biohub | CZB Cliahub Consortium |
| EPI_ISL_1185051 | Santa Clara County Public Health Laboratory | Chan-Zuckerberg Biohub | CZB Cliahub Consortium |
| EPI_ISL_1185052 | CA DPH Viral and Rickettsial Disease Laboratory | Chan-Zuckerberg Biohub | CZB Cliahub Consortium |
| EPI_ISL_1185053 | Santa Clara County Public Health Laboratory | Chan-Zuckerberg Biohub | CZB Cliahub Consortium |
| EPI_ISL_1185054 | Alameda County Public Health Lab | Chan-Zuckerberg Biohub | CZB Cliahub Consortium |

| | | | |
|---|---|---|---|
| EPI_ISL_1185055 | Orange County Public Health Lab | Chan-Zuckerberg Biohub | CZB Cliahub Consortium |
| EPI_ISL_1185056, EPI_ISL_1185057 | Alameda County Public Health Lab | Chan-Zuckerberg Biohub | CZB Cliahub Consortium |
| EPI_ISL_1185058 | Orange County Public Health Lab | Chan-Zuckerberg Biohub | CZB Cliahub Consortium |
| EPI_ISL_1185059, EPI_ISL_1185060, EPI_ISL_1185061 | CA DPH Viral and Rickettsial Disease Laboratory | Chan-Zuckerberg Biohub | CZB Cliahub Consortium |
| EPI_ISL_1185062 | Orange County Public Health Lab | Chan-Zuckerberg Biohub | CZB Cliahub Consortium |
| EPI_ISL_1185063, EPI_ISL_1185064 | CA DPH Viral and Rickettsial Disease Laboratory | Chan-Zuckerberg Biohub | CZB Cliahub Consortium |
| EPI_ISL_1185065 | Santa Clara County Public Health Laboratory | Chan-Zuckerberg Biohub | CZB Cliahub Consortium |
| EPI_ISL_1185066 | Alameda County Public Health Lab | Chan-Zuckerberg Biohub | CZB Cliahub Consortium |
| EPI_ISL_1185067, EPI_ISL_1185068, EPI_ISL_1185069 | CA DPH Viral and Rickettsial Disease Laboratory | Chan-Zuckerberg Biohub | CZB Cliahub Consortium |
| EPI_ISL_1185070 | Orange County Public Health Lab | Chan-Zuckerberg Biohub | CZB Cliahub Consortium |
| EPI_ISL_1185071 | Alameda County Public Health Lab | Chan-Zuckerberg Biohub | CZB Cliahub Consortium |
| EPI_ISL_1185072 | Santa Clara County Public Health Laboratory | Chan-Zuckerberg Biohub | CZB Cliahub Consortium |
| EPI_ISL_1185073 | Alameda County Public Health Lab | Chan-Zuckerberg Biohub | CZB Cliahub Consortium |
| EPI_ISL_1185074 | Santa Clara County Public Health Laboratory | Chan-Zuckerberg Biohub | CZB Cliahub Consortium |
| EPI_ISL_1185075, EPI_ISL_1185076 | Orange County Public Health Lab | Chan-Zuckerberg Biohub | CZB Cliahub Consortium |
| EPI_ISL_1185077, EPI_ISL_1185078, EPI_ISL_1185079, EPI_ISL_1185080 | CA DPH Viral and Rickettsial Disease Laboratory | Chan-Zuckerberg Biohub | CZB Cliahub Consortium |
| EPI_ISL_1185081 | Santa Clara County Public Health Laboratory | Chan-Zuckerberg Biohub | CZB Cliahub Consortium |
| EPI_ISL_1185082 | Alameda County Public Health Lab | Chan-Zuckerberg Biohub | CZB Cliahub Consortium |
| EPI_ISL_1185083 | Santa Clara County Public Health Laboratory | Chan-Zuckerberg Biohub | CZB Cliahub Consortium |
| EPI_ISL_1185084 | CA DPH Viral and Rickettsial Disease Laboratory | Chan-Zuckerberg Biohub | CZB Cliahub Consortium |
| EPI_ISL_1185085 | Alameda County Public Health Lab | Chan-Zuckerberg Biohub | CZB Cliahub Consortium |
| EPI_ISL_1185086 | Santa Clara County Public Health Laboratory | Chan-Zuckerberg Biohub | CZB Cliahub Consortium |
| EPI_ISL_1185087 | Alameda County Public Health Lab | Chan-Zuckerberg Biohub | CZB Cliahub Consortium |
| EPI_ISL_1185089, EPI_ISL_1185090 | CA DPH Viral and Rickettsial Disease Laboratory | Chan-Zuckerberg Biohub | CZB Cliahub Consortium |
| EPI_ISL_1185091, EPI_ISL_1185092 | Santa Clara County Public Health Laboratory | Chan-Zuckerberg Biohub | CZB Cliahub Consortium |
| EPI_ISL_1185093 | Orange County Public Health Lab | Chan-Zuckerberg Biohub | CZB Cliahub Consortium |
| EPI_ISL_1185094, EPI_ISL_1185095 | Alameda County Public Health Lab | Chan-Zuckerberg Biohub | CZB Cliahub Consortium |
| EPI_ISL_1185096 | CA DPH Viral and Rickettsial Disease Laboratory | Chan-Zuckerberg Biohub | CZB Cliahub Consortium |
| EPI_ISL_1185097 | Santa Clara County Public Health Laboratory | Chan-Zuckerberg Biohub | CZB Cliahub Consortium |
| EPI_ISL_1185098 | Orange County Public Health Lab | Chan-Zuckerberg Biohub | CZB Cliahub Consortium |
| EPI_ISL_1185099 | CA DPH Viral and Rickettsial Disease Laboratory | Chan-Zuckerberg Biohub | CZB Cliahub Consortium |
| EPI_ISL_1185100 | Alameda County Public Health Lab | Chan-Zuckerberg Biohub | CZB Cliahub Consortium |
| EPI_ISL_1185101 | Orange County Public Health Lab | Chan-Zuckerberg Biohub | CZB Cliahub Consortium |
| EPI_ISL_1185102 | Alameda County Public Health Lab | Chan-Zuckerberg Biohub | CZB Cliahub Consortium |
| EPI_ISL_1185103 | CA DPH Viral and Rickettsial Disease Laboratory | Chan-Zuckerberg Biohub | CZB Cliahub Consortium |
| EPI_ISL_1185104 | Orange County Public Health Lab | Chan-Zuckerberg Biohub | CZB Cliahub Consortium |
| EPI_ISL_1185105 | Alameda County Public Health Lab | Chan-Zuckerberg Biohub | CZB Cliahub Consortium |
| EPI_ISL_1185106, EPI_ISL_1185107, EPI_ISL_1185108 | Orange County Public Health Lab | Chan-Zuckerberg Biohub | CZB Cliahub Consortium |
| EPI_ISL_1185109 | Alameda County Public Health Lab | Chan-Zuckerberg Biohub | CZB Cliahub Consortium |
| EPI_ISL_1185110, EPI_ISL_1185111 | Santa Clara County Public Health Laboratory | Chan-Zuckerberg Biohub | CZB Cliahub Consortium |
| EPI_ISL_1185112, EPI_ISL_1185113, EPI_ISL_1185114, EPI_ISL_1185115 | Orange County Public Health Lab | Chan-Zuckerberg Biohub | CZB Cliahub Consortium |
| EPI_ISL_1185116 | Santa Clara County Public Health Laboratory | Chan-Zuckerberg Biohub | CZB Cliahub Consortium |
| EPI_ISL_1185117 | Alameda County Public Health Lab | Chan-Zuckerberg Biohub | CZB Cliahub Consortium |
| EPI_ISL_1185118 | Orange County Public Health Lab | Chan-Zuckerberg Biohub | CZB Cliahub Consortium |
| EPI_ISL_1185119 | CA DPH Viral and Rickettsial Disease Laboratory | Chan-Zuckerberg Biohub | CZB Cliahub Consortium |
| EPI_ISL_1185120 | Santa Clara County Public Health Laboratory | Chan-Zuckerberg Biohub | CZB Cliahub Consortium |
| EPI_ISL_1185121 | Alameda County Public Health Lab | Chan-Zuckerberg Biohub | CZB Cliahub Consortium |
| EPI_ISL_1185123 | Orange County Public Health Lab | Chan-Zuckerberg Biohub | CZB Cliahub Consortium |
| EPI_ISL_1185124 | Alameda County Public Health Lab | Chan-Zuckerberg Biohub | CZB Cliahub Consortium |
| EPI_ISL_1185125 | Orange County Public Health Lab | Chan-Zuckerberg Biohub | CZB Cliahub Consortium |
| EPI_ISL_1185126 | Alameda County Public Health Lab | Chan-Zuckerberg Biohub | CZB Cliahub Consortium |
| EPI_ISL_1185127 | Orange County Public Health Lab | Chan-Zuckerberg Biohub | CZB Cliahub Consortium |
| EPI_ISL_1185128 | Alameda County Public Health Lab | Chan-Zuckerberg Biohub | CZB Cliahub Consortium |

| | | | |
|---|---|---|---|
| EPI_ISL_1185129 | Santa Clara County Public Health Laboratory | Chan-Zuckerberg Biohub | CZB Cliahub Consortium |
| EPI_ISL_1185130 | Alameda County Public Health Lab | Chan-Zuckerberg Biohub | CZB Cliahub Consortium |
| EPI_ISL_1185131 | Orange County Public Health Lab | Chan-Zuckerberg Biohub | CZB Cliahub Consortium |
| EPI_ISL_1185133 | CA DPH Viral and Rickettsial Disease Laboratory | Chan-Zuckerberg Biohub | CZB Cliahub Consortium |
| EPI_ISL_1185134 | Orange County Public Health Lab | Chan-Zuckerberg Biohub | CZB Cliahub Consortium |
| EPI_ISL_1185135 | CA DPH Viral and Rickettsial Disease Laboratory | Chan-Zuckerberg Biohub | CZB Cliahub Consortium |
| EPI_ISL_1185136 | Alameda County Public Health Lab | Chan-Zuckerberg Biohub | CZB Cliahub Consortium |
| EPI_ISL_1185137 | Santa Clara County Public Health Laboratory | Chan-Zuckerberg Biohub | CZB Cliahub Consortium |
| EPI_ISL_1185138, EPI_ISL_1185139 | CA DPH Viral and Rickettsial Disease Laboratory | Chan-Zuckerberg Biohub | CZB Cliahub Consortium |
| EPI_ISL_1185140 | Orange County Public Health Lab | Chan-Zuckerberg Biohub | CZB Cliahub Consortium |
| EPI_ISL_1185142 | Alameda County Public Health Lab | Chan-Zuckerberg Biohub | CZB Cliahub Consortium |
| EPI_ISL_1185143 | Orange County Public Health Lab | Chan-Zuckerberg Biohub | CZB Cliahub Consortium |
| EPI_ISL_1185144 | CA DPH Viral and Rickettsial Disease Laboratory | Chan-Zuckerberg Biohub | CZB Cliahub Consortium |
| EPI_ISL_1185145, EPI_ISL_1185146 | Orange County Public Health Lab | Chan-Zuckerberg Biohub | CZB Cliahub Consortium |
| EPI_ISL_1185147 | CA DPH Viral and Rickettsial Disease Laboratory | Chan-Zuckerberg Biohub | CZB Cliahub Consortium |
| EPI_ISL_1185148, EPI_ISL_1185149 | Orange County Public Health Lab | Chan-Zuckerberg Biohub | CZB Cliahub Consortium |
| EPI_ISL_1185150 | CA DPH Viral and Rickettsial Disease Laboratory | Chan-Zuckerberg Biohub | CZB Cliahub Consortium |
| EPI_ISL_1185151 | Santa Clara County Public Health Laboratory | Chan-Zuckerberg Biohub | CZB Cliahub Consortium |
| EPI_ISL_1185152 | Orange County Public Health Lab | Chan-Zuckerberg Biohub | CZB Cliahub Consortium |
| EPI_ISL_1185153 | CA DPH Viral and Rickettsial Disease Laboratory | Chan-Zuckerberg Biohub | CZB Cliahub Consortium |
| EPI_ISL_1185154 | Orange County Public Health Lab | Chan-Zuckerberg Biohub | CZB Cliahub Consortium |
| EPI_ISL_1185155, EPI_ISL_1185156 | CA DPH Viral and Rickettsial Disease Laboratory | Chan-Zuckerberg Biohub | CZB Cliahub Consortium |
| EPI_ISL_1185157 | Alameda County Public Health Lab | Chan-Zuckerberg Biohub | CZB Cliahub Consortium |
| EPI_ISL_1185158 | CA DPH Viral and Rickettsial Disease Laboratory | Chan-Zuckerberg Biohub | CZB Cliahub Consortium |
| EPI_ISL_1185159, EPI_ISL_1185160 | Orange County Public Health Lab | Chan-Zuckerberg Biohub | CZB Cliahub Consortium |
| EPI_ISL_1185161, EPI_ISL_1185162 | Alameda County Public Health Lab | Chan-Zuckerberg Biohub | CZB Cliahub Consortium |
| EPI_ISL_1185163 | Orange County Public Health Lab | Chan-Zuckerberg Biohub | CZB Cliahub Consortium |
| EPI_ISL_1185164 | Alameda County Public Health Lab | Chan-Zuckerberg Biohub | CZB Cliahub Consortium |
| EPI_ISL_1185165 | Orange County Public Health Lab | Chan-Zuckerberg Biohub | CZB Cliahub Consortium |
| EPI_ISL_1185166 | CA DPH Viral and Rickettsial Disease Laboratory | Chan-Zuckerberg Biohub | CZB Cliahub Consortium |
| EPI_ISL_1185167 | Orange County Public Health Lab | Chan-Zuckerberg Biohub | CZB Cliahub Consortium |
| EPI_ISL_1185168, EPI_ISL_1185169 | CA DPH Viral and Rickettsial Disease Laboratory | Chan-Zuckerberg Biohub | CZB Cliahub Consortium |
| EPI_ISL_1185170 | Santa Clara County Public Health Laboratory | Chan-Zuckerberg Biohub | CZB Cliahub Consortium |
| EPI_ISL_1185171 | Orange County Public Health Lab | Chan-Zuckerberg Biohub | CZB Cliahub Consortium |
| EPI_ISL_1185172 | Alameda County Public Health Lab | Chan-Zuckerberg Biohub | CZB Cliahub Consortium |
| EPI_ISL_1185173 | CA DPH Viral and Rickettsial Disease Laboratory | Chan-Zuckerberg Biohub | CZB Cliahub Consortium |
| EPI_ISL_1185174, EPI_ISL_1185175 | Orange County Public Health Lab | Chan-Zuckerberg Biohub | CZB Cliahub Consortium |
| EPI_ISL_1185176 | Alameda County Public Health Lab | Chan-Zuckerberg Biohub | CZB Cliahub Consortium |
| EPI_ISL_1185177, EPI_ISL_1185178 | CA DPH Viral and Rickettsial Disease Laboratory | Chan-Zuckerberg Biohub | CZB Cliahub Consortium |
| EPI_ISL_1185179 | Alameda County Public Health Lab | Chan-Zuckerberg Biohub | CZB Cliahub Consortium |
| EPI_ISL_1185180 | Orange County Public Health Lab | Chan-Zuckerberg Biohub | CZB Cliahub Consortium |
| EPI_ISL_1185181 | Santa Clara County Public Health Laboratory | Chan-Zuckerberg Biohub | CZB Cliahub Consortium |
| EPI_ISL_1185182 | CA DPH Viral and Rickettsial Disease Laboratory | Chan-Zuckerberg Biohub | CZB Cliahub Consortium |
| EPI_ISL_1185183 | Santa Clara County Public Health Laboratory | Chan-Zuckerberg Biohub | CZB Cliahub Consortium |
| EPI_ISL_1185184 | CA DPH Viral and Rickettsial Disease Laboratory | Chan-Zuckerberg Biohub | CZB Cliahub Consortium |
| EPI_ISL_1185185 | Santa Clara County Public Health Laboratory | Chan-Zuckerberg Biohub | CZB Cliahub Consortium |
| EPI_ISL_1185186 | CA DPH Viral and Rickettsial Disease Laboratory | Chan-Zuckerberg Biohub | CZB Cliahub Consortium |
| EPI_ISL_1185187 | Alameda County Public Health Lab | Chan-Zuckerberg Biohub | CZB Cliahub Consortium |
| EPI_ISL_1185188 | CA DPH Viral and Rickettsial Disease Laboratory | Chan-Zuckerberg Biohub | CZB Cliahub Consortium |
| EPI_ISL_1185189 | Orange County Public Health Lab | Chan-Zuckerberg Biohub | CZB Cliahub Consortium |
| EPI_ISL_1185190 | CA DPH Viral and Rickettsial Disease Laboratory | Chan-Zuckerberg Biohub | CZB Cliahub Consortium |
| EPI_ISL_1185191 | Santa Clara County Public Health Laboratory | Chan-Zuckerberg Biohub | CZB Cliahub Consortium |
| EPI_ISL_1185192, EPI_ISL_1185193 | CA DPH Viral and Rickettsial Disease Laboratory | Chan-Zuckerberg Biohub | CZB Cliahub Consortium |
| EPI_ISL_1185194 | Orange County Public Health Lab | Chan-Zuckerberg Biohub | CZB Cliahub Consortium |

| | | | |
|---|---|---|---|
| EPI_ISL_1185195, EPI_ISL_1185196 | CA DPH Viral and Rickettsial Disease Laboratory | Chan-Zuckerberg Biohub | CZB Cliahub Consortium |
| EPI_ISL_1185197 | Santa Clara County Public Health Laboratory | Chan-Zuckerberg Biohub | CZB Cliahub Consortium |
| EPI_ISL_1185198, EPI_ISL_1185199 | Alameda County Public Health Lab | Chan-Zuckerberg Biohub | CZB Cliahub Consortium |
| EPI_ISL_1185200 | Santa Clara County Public Health Laboratory | Chan-Zuckerberg Biohub | CZB Cliahub Consortium |
| EPI_ISL_1185201 | Orange County Public Health Lab | Chan-Zuckerberg Biohub | CZB Cliahub Consortium |
| EPI_ISL_1185202, EPI_ISL_1185203 | CA DPH Viral and Rickettsial Disease Laboratory | Chan-Zuckerberg Biohub | CZB Cliahub Consortium |
| EPI_ISL_1185204 | Alameda County Public Health Lab | Chan-Zuckerberg Biohub | CZB Cliahub Consortium |
| EPI_ISL_1185205 | CA DPH Viral and Rickettsial Disease Laboratory | Chan-Zuckerberg Biohub | CZB Cliahub Consortium |
| EPI_ISL_1185206 | Orange County Public Health Lab | Chan-Zuckerberg Biohub | CZB Cliahub Consortium |
| EPI_ISL_1185207 | Alameda County Public Health Lab | Chan-Zuckerberg Biohub | CZB Cliahub Consortium |
| EPI_ISL_1185208 | CA DPH Viral and Rickettsial Disease Laboratory | Chan-Zuckerberg Biohub | CZB Cliahub Consortium |
| EPI_ISL_1185209 | Orange County Public Health Lab | Chan-Zuckerberg Biohub | CZB Cliahub Consortium |
| EPI_ISL_1185210, EPI_ISL_1185211 | CA DPH Viral and Rickettsial Disease Laboratory | Chan-Zuckerberg Biohub | CZB Cliahub Consortium |
| EPI_ISL_1185212 | Orange County Public Health Lab | Chan-Zuckerberg Biohub | CZB Cliahub Consortium |
| EPI_ISL_1185213 | CA DPH Viral and Rickettsial Disease Laboratory | Chan-Zuckerberg Biohub | CZB Cliahub Consortium |
| EPI_ISL_1185214 | Orange County Public Health Lab | Chan-Zuckerberg Biohub | CZB Cliahub Consortium |
| EPI_ISL_1185215 | Alameda County Public Health Lab | Chan-Zuckerberg Biohub | CZB Cliahub Consortium |
| EPI_ISL_1185216, EPI_ISL_1185217 | Orange County Public Health Lab | Chan-Zuckerberg Biohub | CZB Cliahub Consortium |
| EPI_ISL_1185218, EPI_ISL_1185219 | Santa Clara County Public Health Laboratory | Chan-Zuckerberg Biohub | CZB Cliahub Consortium |
| EPI_ISL_1185220 | CA DPH Viral and Rickettsial Disease Laboratory | Chan-Zuckerberg Biohub | CZB Cliahub Consortium |
| EPI_ISL_1185221 | Orange County Public Health Lab | Chan-Zuckerberg Biohub | CZB Cliahub Consortium |
| EPI_ISL_1185222 | Alameda County Public Health Lab | Chan-Zuckerberg Biohub | CZB Cliahub Consortium |
| EPI_ISL_1185223 | CA DPH Viral and Rickettsial Disease Laboratory | Chan-Zuckerberg Biohub | CZB Cliahub Consortium |
| EPI_ISL_1185224, EPI_ISL_1185225 | Orange County Public Health Lab | Chan-Zuckerberg Biohub | CZB Cliahub Consortium |
| EPI_ISL_1185226 | CA DPH Viral and Rickettsial Disease Laboratory | Chan-Zuckerberg Biohub | CZB Cliahub Consortium |
| EPI_ISL_1185227, EPI_ISL_1185228 | Orange County Public Health Lab | Chan-Zuckerberg Biohub | CZB Cliahub Consortium |
| EPI_ISL_1185229 | Alameda County Public Health Lab | Chan-Zuckerberg Biohub | CZB Cliahub Consortium |
| EPI_ISL_1185231 | Orange County Public Health Lab | Chan-Zuckerberg Biohub | CZB Cliahub Consortium |
| EPI_ISL_1185232 | Alameda County Public Health Lab | Chan-Zuckerberg Biohub | CZB Cliahub Consortium |
| EPI_ISL_1185233 | Orange County Public Health Lab | Chan-Zuckerberg Biohub | CZB Cliahub Consortium |
| EPI_ISL_1185234 | Santa Clara County Public Health Laboratory | Chan-Zuckerberg Biohub | CZB Cliahub Consortium |
| EPI_ISL_1185235, EPI_ISL_1185236, EPI_ISL_1185237, EPI_ISL_1185238 | Orange County Public Health Lab | Chan-Zuckerberg Biohub | CZB Cliahub Consortium |
| EPI_ISL_1185239 | Santa Clara County Public Health Laboratory | Chan-Zuckerberg Biohub | CZB Cliahub Consortium |
| EPI_ISL_1185240, EPI_ISL_1185241, EPI_ISL_1185242 | Orange County Public Health Lab | Chan-Zuckerberg Biohub | CZB Cliahub Consortium |
| EPI_ISL_1185243 | Alameda County Public Health Lab | Chan-Zuckerberg Biohub | CZB Cliahub Consortium |
| EPI_ISL_1185244, EPI_ISL_1185246, EPI_ISL_1185247, EPI_ISL_1185248, EPI_ISL_1185249 | CA DPH Viral and Rickettsial Disease Laboratory | Chan-Zuckerberg Biohub | CZB Cliahub Consortium |
| EPI_ISL_1185250 | Alameda County Public Health Lab | Chan-Zuckerberg Biohub | CZB Cliahub Consortium |
| EPI_ISL_1185251, EPI_ISL_1185252 | Santa Clara County Public Health Laboratory | Chan-Zuckerberg Biohub | CZB Cliahub Consortium |
| EPI_ISL_1185253, EPI_ISL_1185254 | CA DPH Viral and Rickettsial Disease Laboratory | Chan-Zuckerberg Biohub | CZB Cliahub Consortium |
| EPI_ISL_1185255 | Santa Clara County Public Health Laboratory | Chan-Zuckerberg Biohub | CZB Cliahub Consortium |
| EPI_ISL_1185256 | CA DPH Viral and Rickettsial Disease Laboratory | Chan-Zuckerberg Biohub | CZB Cliahub Consortium |
| EPI_ISL_1185257 | Orange County Public Health Lab | Chan-Zuckerberg Biohub | CZB Cliahub Consortium |
| EPI_ISL_1185258, EPI_ISL_1185259 | Alameda County Public Health Lab | Chan-Zuckerberg Biohub | CZB Cliahub Consortium |
| EPI_ISL_1185260 | CA DPH Viral and Rickettsial Disease Laboratory | Chan-Zuckerberg Biohub | CZB Cliahub Consortium |
| EPI_ISL_1185261 | Orange County Public Health Lab | Chan-Zuckerberg Biohub | CZB Cliahub Consortium |
| EPI_ISL_1185262 | Alameda County Public Health Lab | Chan-Zuckerberg Biohub | CZB Cliahub Consortium |
| EPI_ISL_1185263 | Orange County Public Health Lab | Chan-Zuckerberg Biohub | CZB Cliahub Consortium |
| EPI_ISL_1185264, EPI_ISL_1185265 | CA DPH Viral and Rickettsial Disease Laboratory | Chan-Zuckerberg Biohub | CZB Cliahub Consortium |
| EPI_ISL_1185266 | Orange County Public Health Lab | Chan-Zuckerberg Biohub | CZB Cliahub Consortium |
| EPI_ISL_1185267 | Santa Clara County Public Health Laboratory | Chan-Zuckerberg Biohub | CZB Cliahub Consortium |
| EPI_ISL_1185268 | CA DPH Viral and Rickettsial Disease Laboratory | Chan-Zuckerberg Biohub | CZB Cliahub Consortium |
| EPI_ISL_1185269 | Orange County Public Health Lab | Chan-Zuckerberg Biohub | CZB Cliahub Consortium |

| | | | |
|---|---|---|---|
| EPI_ISL_1185270 | CA DPH Viral and Rickettsial Disease Laboratory | Chan-Zuckerberg Biohub | CZB Cliahub Consortium |
| EPI_ISL_1185271 | Alameda County Public Health Lab | Chan-Zuckerberg Biohub | CZB Cliahub Consortium |
| EPI_ISL_1185272 | Orange County Public Health Lab | Chan-Zuckerberg Biohub | CZB Cliahub Consortium |
| EPI_ISL_1185273 | Santa Clara County Public Health Laboratory | Chan-Zuckerberg Biohub | CZB Cliahub Consortium |
| EPI_ISL_1185274 | Orange County Public Health Lab | Chan-Zuckerberg Biohub | CZB Cliahub Consortium |
| EPI_ISL_1185275, EPI_ISL_1185276 | CA DPH Viral and Rickettsial Disease Laboratory | Chan-Zuckerberg Biohub | CZB Cliahub Consortium |
| EPI_ISL_1185277 | Santa Clara County Public Health Laboratory | Chan-Zuckerberg Biohub | CZB Cliahub Consortium |
| EPI_ISL_1185278 | Orange County Public Health Lab | Chan-Zuckerberg Biohub | CZB Cliahub Consortium |
| EPI_ISL_1185279 | CA DPH Viral and Rickettsial Disease Laboratory | Chan-Zuckerberg Biohub | CZB Cliahub Consortium |
| EPI_ISL_1185280 | Orange County Public Health Lab | Chan-Zuckerberg Biohub | CZB Cliahub Consortium |
| EPI_ISL_1185281 | Alameda County Public Health Lab | Chan-Zuckerberg Biohub | CZB Cliahub Consortium |
| EPI_ISL_1185282 | CA DPH Viral and Rickettsial Disease Laboratory | Chan-Zuckerberg Biohub | CZB Cliahub Consortium |
| EPI_ISL_1185283 | Alameda County Public Health Lab | Chan-Zuckerberg Biohub | CZB Cliahub Consortium |
| EPI_ISL_1185284 | Santa Clara County Public Health Laboratory | Chan-Zuckerberg Biohub | CZB Cliahub Consortium |
| EPI_ISL_1185286 | Alameda County Public Health Lab | Chan-Zuckerberg Biohub | CZB Cliahub Consortium |
| EPI_ISL_1185287 | Humboldt County Public Health Laboratory | Chan-Zuckerberg Biohub | CZB Cliahub Consortium |
| EPI_ISL_1185288 | County of San Luis Obispo Public Health Laboratory | Chan-Zuckerberg Biohub | CZB Cliahub Consortium |
| EPI_ISL_1185289, EPI_ISL_1185290 | CA DPH Viral and Rickettsial Disease Laboratory | Chan-Zuckerberg Biohub | CZB Cliahub Consortium |
| EPI_ISL_1185291, EPI_ISL_1185292 | Humboldt County Public Health Laboratory | Chan-Zuckerberg Biohub | CZB Cliahub Consortium |
| EPI_ISL_1185293 | CA DPH Viral and Rickettsial Disease Laboratory | Chan-Zuckerberg Biohub | CZB Cliahub Consortium |
| EPI_ISL_1185294 | Humboldt County Public Health Laboratory | Chan-Zuckerberg Biohub | CZB Cliahub Consortium |
| EPI_ISL_1185295, EPI_ISL_1185296, EPI_ISL_1185297 | CA DPH Viral and Rickettsial Disease Laboratory | Chan-Zuckerberg Biohub | CZB Cliahub Consortium |
| EPI_ISL_1185298 | Humboldt County Public Health Laboratory | Chan-Zuckerberg Biohub | CZB Cliahub Consortium |
| EPI_ISL_1185299 | CA DPH Viral and Rickettsial Disease Laboratory | Chan-Zuckerberg Biohub | CZB Cliahub Consortium |
| EPI_ISL_1185300, EPI_ISL_1185301, EPI_ISL_1185302 | Humboldt County Public Health Laboratory | Chan-Zuckerberg Biohub | CZB Cliahub Consortium |
| EPI_ISL_1185303 | CA DPH Viral and Rickettsial Disease Laboratory | Chan-Zuckerberg Biohub | CZB Cliahub Consortium |
| EPI_ISL_1185304 | Humboldt County Public Health Laboratory | Chan-Zuckerberg Biohub | CZB Cliahub Consortium |
| EPI_ISL_1185305 | Alameda County Public Health Lab | Chan-Zuckerberg Biohub | CZB Cliahub Consortium |
| EPI_ISL_1185306 | Humboldt County Public Health Laboratory | Chan-Zuckerberg Biohub | CZB Cliahub Consortium |
| EPI_ISL_1185307 | CA DPH Viral and Rickettsial Disease Laboratory | Chan-Zuckerberg Biohub | CZB Cliahub Consortium |
| EPI_ISL_1185308, EPI_ISL_1185309 | Humboldt County Public Health Laboratory | Chan-Zuckerberg Biohub | CZB Cliahub Consortium |
| EPI_ISL_1185310, EPI_ISL_1185311 | CA DPH Viral and Rickettsial Disease Laboratory | Chan-Zuckerberg Biohub | CZB Cliahub Consortium |
| EPI_ISL_1185312 | Humboldt County Public Health Laboratory | Chan-Zuckerberg Biohub | CZB Cliahub Consortium |
| EPI_ISL_1185313 | Alameda County Public Health Lab | Chan-Zuckerberg Biohub | CZB Cliahub Consortium |
| EPI_ISL_1185314 | CA DPH Viral and Rickettsial Disease Laboratory | Chan-Zuckerberg Biohub | CZB Cliahub Consortium |
| EPI_ISL_1185315 | Alameda County Public Health Lab | Chan-Zuckerberg Biohub | CZB Cliahub Consortium |
| EPI_ISL_1185316, EPI_ISL_1185317 | CA DPH Viral and Rickettsial Disease Laboratory | Chan-Zuckerberg Biohub | CZB Cliahub Consortium |
| EPI_ISL_1185318, EPI_ISL_1185319 | Humboldt County Public Health Laboratory | Chan-Zuckerberg Biohub | CZB Cliahub Consortium |
| EPI_ISL_1185320, EPI_ISL_1185321 | CA DPH Viral and Rickettsial Disease Laboratory | Chan-Zuckerberg Biohub | CZB Cliahub Consortium |
| EPI_ISL_1185322 | Alameda County Public Health Lab | Chan-Zuckerberg Biohub | CZB Cliahub Consortium |
| EPI_ISL_1185323, EPI_ISL_1185325, EPI_ISL_1185326 | Humboldt County Public Health Laboratory | Chan-Zuckerberg Biohub | CZB Cliahub Consortium |
| EPI_ISL_1185327 | CA DPH Viral and Rickettsial Disease Laboratory | Chan-Zuckerberg Biohub | CZB Cliahub Consortium |
| EPI_ISL_1185328, EPI_ISL_1185329 | Humboldt County Public Health Laboratory | Chan-Zuckerberg Biohub | CZB Cliahub Consortium |
| EPI_ISL_1185330, EPI_ISL_1185331, EPI_ISL_1185332, EPI_ISL_1185333, EPI_ISL_1185334, EPI_ISL_1185336, EPI_ISL_1185337, EPI_ISL_1185338, EPI_ISL_1185339 | CA DPH Viral and Rickettsial Disease Laboratory | Chan-Zuckerberg Biohub | CZB Cliahub Consortium |
| EPI_ISL_1185340 | Humboldt County Public Health Laboratory | Chan-Zuckerberg Biohub | CZB Cliahub Consortium |
| EPI_ISL_1185341, | CA DPH Viral and Rickettsial Disease | Chan-Zuckerberg Biohub | CZB Cliahub Consortium |

| | | | |
|---|---|---|---|
| EPI_ISL_1185342, EPI_ISL_1185343, EPI_ISL_1185344 | Laboratory | | |
| EPI_ISL_1185345, EPI_ISL_1185346 | Humboldt County Public Health Laboratory | Chan-Zuckerberg Biohub | CZB Cliahub Consortium |
| EPI_ISL_1185347, EPI_ISL_1185348, EPI_ISL_1185349, EPI_ISL_1185350 | CA DPH Viral and Rickettsial Disease Laboratory | Chan-Zuckerberg Biohub | CZB Cliahub Consortium |
| EPI_ISL_1185351 | Alameda County Public Health Lab | Chan-Zuckerberg Biohub | CZB Cliahub Consortium |
| EPI_ISL_1185352, EPI_ISL_1185353 | Humboldt County Public Health Laboratory | Chan-Zuckerberg Biohub | CZB Cliahub Consortium |
| EPI_ISL_1185354 | CA DPH Viral and Rickettsial Disease Laboratory | Chan-Zuckerberg Biohub | CZB Cliahub Consortium |
| EPI_ISL_1185355 | Humboldt County Public Health Laboratory | Chan-Zuckerberg Biohub | CZB Cliahub Consortium |
| EPI_ISL_1185356, EPI_ISL_1185357, EPI_ISL_1185358 | Alameda County Public Health Lab | Chan-Zuckerberg Biohub | CZB Cliahub Consortium |
| EPI_ISL_1185359, EPI_ISL_1185360, EPI_ISL_1185361 | Humboldt County Public Health Laboratory | Chan-Zuckerberg Biohub | CZB Cliahub Consortium |
| EPI_ISL_1185362, EPI_ISL_1185363, EPI_ISL_1185364 | Alameda County Public Health Lab | Chan-Zuckerberg Biohub | CZB Cliahub Consortium |
| EPI_ISL_1185365 | Humboldt County Public Health Laboratory | Chan-Zuckerberg Biohub | CZB Cliahub Consortium |
| EPI_ISL_1185366 | CA DPH Viral and Rickettsial Disease Laboratory | Chan-Zuckerberg Biohub | CZB Cliahub Consortium |
| EPI_ISL_1185367, EPI_ISL_1185368 | Alameda County Public Health Lab | Chan-Zuckerberg Biohub | CZB Cliahub Consortium |
| EPI_ISL_1185369 | Humboldt County Public Health Laboratory | Chan-Zuckerberg Biohub | CZB Cliahub Consortium |
| EPI_ISL_1185371 | CA DPH Viral and Rickettsial Disease Laboratory | Chan-Zuckerberg Biohub | CZB Cliahub Consortium |
| EPI_ISL_1185372 | Alameda County Public Health Lab | Chan-Zuckerberg Biohub | CZB Cliahub Consortium |
| EPI_ISL_1185373, EPI_ISL_1185374, EPI_ISL_1185375, EPI_ISL_1185377, EPI_ISL_1185378, EPI_ISL_1185379, EPI_ISL_1185380, EPI_ISL_1185381 | CA DPH Viral and Rickettsial Disease Laboratory | Chan-Zuckerberg Biohub | CZB Cliahub Consortium |
| EPI_ISL_1185382 | Humboldt County Public Health Laboratory | Chan-Zuckerberg Biohub | CZB Cliahub Consortium |
| EPI_ISL_1185383 | Alameda County Public Health Lab | Chan-Zuckerberg Biohub | CZB Cliahub Consortium |
| EPI_ISL_1185384 | CA DPH Viral and Rickettsial Disease Laboratory | Chan-Zuckerberg Biohub | CZB Cliahub Consortium |
| EPI_ISL_1185385 | Humboldt County Public Health Laboratory | Chan-Zuckerberg Biohub | CZB Cliahub Consortium |
| EPI_ISL_1185386 | CA DPH Viral and Rickettsial Disease Laboratory | Chan-Zuckerberg Biohub | CZB Cliahub Consortium |
| EPI_ISL_1185387 | County of San Luis Obispo Public Health Laboratory | Chan-Zuckerberg Biohub | CZB Cliahub Consortium |
| EPI_ISL_1185388 | Alameda County Public Health Lab | Chan-Zuckerberg Biohub | CZB Cliahub Consortium |
| EPI_ISL_1185389, EPI_ISL_1185390, EPI_ISL_1185391, EPI_ISL_1185392 | CA DPH Viral and Rickettsial Disease Laboratory | Chan-Zuckerberg Biohub | CZB Cliahub Consortium |
| EPI_ISL_1185393 | Alameda County Public Health Lab | Chan-Zuckerberg Biohub | CZB Cliahub Consortium |
| EPI_ISL_1185395 | Humboldt County Public Health Laboratory | Chan-Zuckerberg Biohub | CZB Cliahub Consortium |
| EPI_ISL_1185396, EPI_ISL_1185397 | CA DPH Viral and Rickettsial Disease Laboratory | Chan-Zuckerberg Biohub | CZB Cliahub Consortium |
| EPI_ISL_1185399 | Humboldt County Public Health Laboratory | Chan-Zuckerberg Biohub | CZB Cliahub Consortium |
| EPI_ISL_1185401 | Alameda County Public Health Lab | Chan-Zuckerberg Biohub | CZB Cliahub Consortium |
| EPI_ISL_1185402, EPI_ISL_1185403, EPI_ISL_1185404, EPI_ISL_1185405, EPI_ISL_1185406 | CA DPH Viral and Rickettsial Disease Laboratory | Chan-Zuckerberg Biohub | CZB Cliahub Consortium |
| EPI_ISL_1185407, EPI_ISL_1185408, EPI_ISL_1185409, EPI_ISL_1185410 | Humboldt County Public Health Laboratory | Chan-Zuckerberg Biohub | CZB Cliahub Consortium |
| EPI_ISL_1185411 | CA DPH Viral and Rickettsial Disease Laboratory | Chan-Zuckerberg Biohub | CZB Cliahub Consortium |
| EPI_ISL_1185412, EPI_ISL_1185413 | Humboldt County Public Health Laboratory | Chan-Zuckerberg Biohub | CZB Cliahub Consortium |
| EPI_ISL_1185414 | Alameda County Public Health Lab | Chan-Zuckerberg Biohub | CZB Cliahub Consortium |
| EPI_ISL_1185415, EPI_ISL_1185416 | CA DPH Viral and Rickettsial Disease Laboratory | Chan-Zuckerberg Biohub | CZB Cliahub Consortium |
| EPI_ISL_1185417 | Humboldt County Public Health Laboratory | Chan-Zuckerberg Biohub | CZB Cliahub Consortium |
| EPI_ISL_1185418, EPI_ISL_1185419, EPI_ISL_1185420, EPI_ISL_1185421 | CA DPH Viral and Rickettsial Disease Laboratory | Chan-Zuckerberg Biohub | CZB Cliahub Consortium |
| EPI_ISL_1185422 | Alameda County Public Health Lab | Chan-Zuckerberg Biohub | CZB Cliahub Consortium |
| EPI_ISL_1185423 | CA DPH Viral and Rickettsial Disease | Chan-Zuckerberg Biohub | CZB Cliahub Consortium |

| | | | |
|---|---|---|---|
| | Laboratory | | |
| EPI_ISL_1185424, EPI_ISL_1185425 | Humboldt County Public Health Laboratory | Chan-Zuckerberg Biohub | CZB Cliahub Consortium |
| EPI_ISL_1185426 | CA DPH Viral and Rickettsial Disease Laboratory | Chan-Zuckerberg Biohub | CZB Cliahub Consortium |
| EPI_ISL_1185427 | Alameda County Public Health Lab | Chan-Zuckerberg Biohub | CZB Cliahub Consortium |
| EPI_ISL_1185428 | Humboldt County Public Health Laboratory | Chan-Zuckerberg Biohub | CZB Cliahub Consortium |
| EPI_ISL_1185429 | CA DPH Viral and Rickettsial Disease Laboratory | Chan-Zuckerberg Biohub | CZB Cliahub Consortium |
| EPI_ISL_1185430, EPI_ISL_1185431 | Alameda County Public Health Lab | Chan-Zuckerberg Biohub | CZB Cliahub Consortium |
| EPI_ISL_1185432 | Humboldt County Public Health Laboratory | Chan-Zuckerberg Biohub | CZB Cliahub Consortium |
| EPI_ISL_1185433, EPI_ISL_1185434, EPI_ISL_1185435 | CA DPH Viral and Rickettsial Disease Laboratory | Chan-Zuckerberg Biohub | CZB Cliahub Consortium |
| EPI_ISL_1185436, EPI_ISL_1185438, EPI_ISL_1185439, EPI_ISL_1185440 | Humboldt County Public Health Laboratory | Chan-Zuckerberg Biohub | CZB Cliahub Consortium |
| EPI_ISL_1185441, EPI_ISL_1185442 | CA DPH Viral and Rickettsial Disease Laboratory | Chan-Zuckerberg Biohub | CZB Cliahub Consortium |
| EPI_ISL_1185443 | County of San Luis Obispo Public Health Laboratory | Chan-Zuckerberg Biohub | CZB Cliahub Consortium |
| EPI_ISL_1185444, EPI_ISL_1185445, EPI_ISL_1185446 | CA DPH Viral and Rickettsial Disease Laboratory | Chan-Zuckerberg Biohub | CZB Cliahub Consortium |
| EPI_ISL_1185447 | Alameda County Public Health Lab | Chan-Zuckerberg Biohub | CZB Cliahub Consortium |
| EPI_ISL_1185448 | Humboldt County Public Health Laboratory | Chan-Zuckerberg Biohub | CZB Cliahub Consortium |
| EPI_ISL_1185449 | Alameda County Public Health Lab | Chan-Zuckerberg Biohub | CZB Cliahub Consortium |
| EPI_ISL_1185450, EPI_ISL_1185451 | CA DPH Viral and Rickettsial Disease Laboratory | Chan-Zuckerberg Biohub | CZB Cliahub Consortium |
| EPI_ISL_1185452 | Alameda County Public Health Lab | Chan-Zuckerberg Biohub | CZB Cliahub Consortium |
| EPI_ISL_1185453 | CA DPH Viral and Rickettsial Disease Laboratory | Chan-Zuckerberg Biohub | CZB Cliahub Consortium |
| EPI_ISL_1185454, EPI_ISL_1185455 | Humboldt County Public Health Laboratory | Chan-Zuckerberg Biohub | CZB Cliahub Consortium |
| EPI_ISL_1185456, EPI_ISL_1185457 | Alameda County Public Health Lab | Chan-Zuckerberg Biohub | CZB Cliahub Consortium |
| EPI_ISL_1185458 | CA DPH Viral and Rickettsial Disease Laboratory | Chan-Zuckerberg Biohub | CZB Cliahub Consortium |
| EPI_ISL_1185459 | Alameda County Public Health Lab | Chan-Zuckerberg Biohub | CZB Cliahub Consortium |
| EPI_ISL_1185460 | Humboldt County Public Health Laboratory | Chan-Zuckerberg Biohub | CZB Cliahub Consortium |
| EPI_ISL_1185461 | Alameda County Public Health Lab | Chan-Zuckerberg Biohub | CZB Cliahub Consortium |
| EPI_ISL_1185462 | CA DPH Viral and Rickettsial Disease Laboratory | Chan-Zuckerberg Biohub | CZB Cliahub Consortium |
| EPI_ISL_1185463 | Humboldt County Public Health Laboratory | Chan-Zuckerberg Biohub | CZB Cliahub Consortium |
| EPI_ISL_1185464, EPI_ISL_1185465 | CA DPH Viral and Rickettsial Disease Laboratory | Chan-Zuckerberg Biohub | CZB Cliahub Consortium |
| EPI_ISL_1185466 | Alameda County Public Health Lab | Chan-Zuckerberg Biohub | CZB Cliahub Consortium |
| EPI_ISL_1185467 | CA DPH Viral and Rickettsial Disease Laboratory | Chan-Zuckerberg Biohub | CZB Cliahub Consortium |
| EPI_ISL_1185468, EPI_ISL_1185469 | Humboldt County Public Health Laboratory | Chan-Zuckerberg Biohub | CZB Cliahub Consortium |
| EPI_ISL_1185470 | CA DPH Viral and Rickettsial Disease Laboratory | Chan-Zuckerberg Biohub | CZB Cliahub Consortium |
| EPI_ISL_1185471 | Humboldt County Public Health Laboratory | Chan-Zuckerberg Biohub | CZB Cliahub Consortium |
| EPI_ISL_1185472, EPI_ISL_1185473 | CA DPH Viral and Rickettsial Disease Laboratory | Chan-Zuckerberg Biohub | CZB Cliahub Consortium |
| EPI_ISL_1185474 | Humboldt County Public Health Laboratory | Chan-Zuckerberg Biohub | CZB Cliahub Consortium |
| EPI_ISL_1185475 | Alameda County Public Health Lab | Chan-Zuckerberg Biohub | CZB Cliahub Consortium |
| EPI_ISL_1185476 | Humboldt County Public Health Laboratory | Chan-Zuckerberg Biohub | CZB Cliahub Consortium |
| EPI_ISL_1185477, EPI_ISL_1185478 | CA DPH Viral and Rickettsial Disease Laboratory | Chan-Zuckerberg Biohub | CZB Cliahub Consortium |
| EPI_ISL_1185480 | Alameda County Public Health Lab | Chan-Zuckerberg Biohub | CZB Cliahub Consortium |
| EPI_ISL_1185481 | Humboldt County Public Health Laboratory | Chan-Zuckerberg Biohub | CZB Cliahub Consortium |
| EPI_ISL_1185482, EPI_ISL_1185483, EPI_ISL_1185484, EPI_ISL_1185485 | CA DPH Viral and Rickettsial Disease Laboratory | Chan-Zuckerberg Biohub | CZB Cliahub Consortium |
| EPI_ISL_1185486, EPI_ISL_1185487, EPI_ISL_1185488, EPI_ISL_1185489 | Humboldt County Public Health Laboratory | Chan-Zuckerberg Biohub | CZB Cliahub Consortium |
| EPI_ISL_1185490 | CA DPH Viral and Rickettsial Disease Laboratory | Chan-Zuckerberg Biohub | CZB Cliahub Consortium |
| EPI_ISL_1185491 | Alameda County Public Health Lab | Chan-Zuckerberg Biohub | CZB Cliahub Consortium |
| EPI_ISL_1185492 | CA DPH Viral and Rickettsial Disease Laboratory | Chan-Zuckerberg Biohub | CZB Cliahub Consortium |
| EPI_ISL_1185493 | Alameda County Public Health Lab | Chan-Zuckerberg Biohub | CZB Cliahub Consortium |
| EPI_ISL_1185494, EPI_ISL_1185495 | CA DPH Viral and Rickettsial Disease Laboratory | Chan-Zuckerberg Biohub | CZB Cliahub Consortium |

| | | | |
|---|---|---|---|
| EPI_ISL_1185496 | Alameda County Public Health Lab | Chan-Zuckerberg Biohub | CZB Cliahub Consortium |
| EPI_ISL_1185497, EPI_ISL_1185498 | Humboldt County Public Health Laboratory | Chan-Zuckerberg Biohub | CZB Cliahub Consortium |
| EPI_ISL_1185499 | Alameda County Public Health Lab | Chan-Zuckerberg Biohub | CZB Cliahub Consortium |
| EPI_ISL_1185500 | CA DPH Viral and Rickettsial Disease Laboratory | Chan-Zuckerberg Biohub | CZB Cliahub Consortium |
| EPI_ISL_1185501, EPI_ISL_1185502, EPI_ISL_1185503 | Alameda County Public Health Lab | Chan-Zuckerberg Biohub | CZB Cliahub Consortium |
| EPI_ISL_1185504, EPI_ISL_1185505 | Humboldt County Public Health Laboratory | Chan-Zuckerberg Biohub | CZB Cliahub Consortium |
| EPI_ISL_1185506 | CA DPH Viral and Rickettsial Disease Laboratory | Chan-Zuckerberg Biohub | CZB Cliahub Consortium |
| EPI_ISL_1185507 | Humboldt County Public Health Laboratory | Chan-Zuckerberg Biohub | CZB Cliahub Consortium |
| EPI_ISL_1185508 | CA DPH Viral and Rickettsial Disease Laboratory | Chan-Zuckerberg Biohub | CZB Cliahub Consortium |
| EPI_ISL_1185509 | Humboldt County Public Health Laboratory | Chan-Zuckerberg Biohub | CZB Cliahub Consortium |
| EPI_ISL_1185510, EPI_ISL_1185511, EPI_ISL_1185512, EPI_ISL_1185514, EPI_ISL_1185515 | CA DPH Viral and Rickettsial Disease Laboratory | Chan-Zuckerberg Biohub | CZB Cliahub Consortium |
| EPI_ISL_1185516 | Humboldt County Public Health Laboratory | Chan-Zuckerberg Biohub | CZB Cliahub Consortium |
| EPI_ISL_1185517 | Alameda County Public Health Lab | Chan-Zuckerberg Biohub | CZB Cliahub Consortium |
| EPI_ISL_1185518, EPI_ISL_1185519 | Humboldt County Public Health Laboratory | Chan-Zuckerberg Biohub | CZB Cliahub Consortium |
| EPI_ISL_1185520 | CA DPH Viral and Rickettsial Disease Laboratory | Chan-Zuckerberg Biohub | CZB Cliahub Consortium |
| EPI_ISL_1185521 | Humboldt County Public Health Laboratory | Chan-Zuckerberg Biohub | CZB Cliahub Consortium |
| EPI_ISL_1185522, EPI_ISL_1185523, EPI_ISL_1185524, EPI_ISL_1185525 | CA DPH Viral and Rickettsial Disease Laboratory | Chan-Zuckerberg Biohub | CZB Cliahub Consortium |
| EPI_ISL_1185526 | Humboldt County Public Health Laboratory | Chan-Zuckerberg Biohub | CZB Cliahub Consortium |
| EPI_ISL_1185527 | CA DPH Viral and Rickettsial Disease Laboratory | Chan-Zuckerberg Biohub | CZB Cliahub Consortium |
| EPI_ISL_1185528 | Humboldt County Public Health Laboratory | Chan-Zuckerberg Biohub | CZB Cliahub Consortium |
| EPI_ISL_1185529 | Alameda County Public Health Lab | Chan-Zuckerberg Biohub | CZB Cliahub Consortium |
| EPI_ISL_1185530, EPI_ISL_1185531, EPI_ISL_1185532 | CA DPH Viral and Rickettsial Disease Laboratory | Chan-Zuckerberg Biohub | CZB Cliahub Consortium |
| EPI_ISL_1185533, EPI_ISL_1185534 | Alameda County Public Health Lab | Chan-Zuckerberg Biohub | CZB Cliahub Consortium |
| EPI_ISL_1185854, EPI_ISL_1185855, EPI_ISL_1185856, EPI_ISL_1185857, EPI_ISL_1185858, EPI_ISL_1185859, EPI_ISL_1185860, EPI_ISL_1185861, EPI_ISL_1185862, EPI_ISL_1185863, EPI_ISL_1185864, EPI_ISL_1185865, EPI_ISL_1185866, EPI_ISL_1185867, EPI_ISL_1185868, EPI_ISL_1185869, EPI_ISL_1185870, EPI_ISL_1185871, EPI_ISL_1185872, EPI_ISL_1185873, EPI_ISL_1185874, EPI_ISL_1185875, EPI_ISL_1185876, EPI_ISL_1185877, EPI_ISL_1185878, EPI_ISL_1185879, EPI_ISL_1185880, EPI_ISL_1185881, EPI_ISL_1185882, EPI_ISL_1185883, EPI_ISL_1185884, EPI_ISL_1185885, EPI_ISL_1185886, EPI_ISL_1185887, EPI_ISL_1185888, EPI_ISL_1185889, EPI_ISL_1185890 | | | |
| see above | Scripps Medical Laboratory | Andersen lab at Scripps Research | SEARCH Alliance San Diego with Michael Quigley, Ellen Stefanski, Ian Mchardy |
| EPI_ISL_1185891, EPI_ISL_1185892, EPI_ISL_1185893, EPI_ISL_1185894, EPI_ISL_1185895, EPI_ISL_1185896, EPI_ISL_1185897, EPI_ISL_1185898, EPI_ISL_1185899, EPI_ISL_1185900, EPI_ISL_1185901, EPI_ISL_1185902, EPI_ISL_1185903, EPI_ISL_1185904, EPI_ISL_1185905, EPI_ISL_1185906, EPI_ISL_1185907, EPI_ISL_1185908, EPI_ISL_1185909, EPI_ISL_1185910, EPI_ISL_1185911, EPI_ISL_1185912, EPI_ISL_1185913, EPI_ISL_1185914, EPI_ISL_1185915, EPI_ISL_1185916, EPI_ISL_1185917, EPI_ISL_1185918, EPI_ISL_1185919, EPI_ISL_1185920, EPI_ISL_1185921, EPI_ISL_1185922, EPI_ISL_1185923, EPI_ISL_1185924, EPI_ISL_1185925, EPI_ISL_1185926, EPI_ISL_1185927, EPI_ISL_1185928, EPI_ISL_1185929, EPI_ISL_1185930, EPI_ISL_1185931, EPI_ISL_1185932, EPI_ISL_1185933, EPI_ISL_1185934, EPI_ISL_1185935, EPI_ISL_1185936, EPI_ISL_1185937, EPI_ISL_1185938, EPI_ISL_1185939, EPI_ISL_1185940, EPI_ISL_1185941, EPI_ISL_1185942, EPI_ISL_1185943, EPI_ISL_1185944, EPI_ISL_1185945, EPI_ISL_1185946, EPI_ISL_1185947, EPI_ISL_1185948, EPI_ISL_1185949, EPI_ISL_1185950, EPI_ISL_1185951, EPI_ISL_1185952, EPI_ISL_1185953, EPI_ISL_1185954, EPI_ISL_1185955, EPI_ISL_1185956, EPI_ISL_1185957, EPI_ISL_1185958, EPI_ISL_1185959 | | | |
| see above | Sharp HealthCare Laboratory | Andersen lab at Scripps Research | SEARCH Alliance San Diego with Aaron Harding, Jacquelyn Berumen, Cathy Woerle, Liam McGinnis, Art Mendoza, Omid Bakhtar |
| EPI_ISL_1185960, EPI_ISL_1185961, EPI_ISL_1185962, EPI_ISL_1185963, EPI_ISL_1185964, EPI_ISL_1185965, EPI_ISL_1185966, EPI_ISL_1185967, EPI_ISL_1185968, EPI_ISL_1185969, EPI_ISL_1185970, EPI_ISL_1185971, EPI_ISL_1185972, EPI_ISL_1185973, EPI_ISL_1185974, EPI_ISL_1185975, EPI_ISL_1185976, EPI_ISL_1185977, EPI_ISL_1185978, EPI_ISL_1185979, EPI_ISL_1185980, EPI_ISL_1185981, EPI_ISL_1185982, EPI_ISL_1185983, EPI_ISL_1185984, EPI_ISL_1185985, EPI_ISL_1185986, EPI_ISL_1185987, EPI_ISL_1185988, EPI_ISL_1185989, EPI_ISL_1185990, EPI_ISL_1185991, EPI_ISL_1185992, EPI_ISL_1185993, EPI_ISL_1185994, EPI_ISL_1185995, EPI_ISL_1185996, EPI_ISL_1185997, EPI_ISL_1185998, EPI_ISL_1185999, EPI_ISL_1186000, EPI_ISL_1186001, EPI_ISL_1186002, EPI_ISL_1186003, EPI_ISL_1186004, EPI_ISL_1186005, EPI_ISL_1186006, EPI_ISL_1186007, EPI_ISL_1186008, EPI_ISL_1186009, EPI_ISL_1186010, EPI_ISL_1186011, EPI_ISL_1186012, EPI_ISL_1186013, EPI_ISL_1186014, EPI_ISL_1186015, EPI_ISL_1186016, EPI_ISL_1186017, EPI_ISL_1186018, EPI_ISL_1186019, EPI_ISL_1186020, EPI_ISL_1186021, EPI_ISL_1186022 | | | |
| see above | San Diego County Public Health Laboratory | Andersen lab at Scripps Research | SEARCH Alliance San Diego with Tracy Basler, Jovan Shephard, Brett Austin |
| EPI_ISL_1186030, EPI_ISL_1186031, EPI_ISL_1186032, EPI_ISL_1186033, EPI_ISL_1186034, EPI_ISL_1186035, EPI_ISL_1186036, EPI_ISL_1186037, EPI_ISL_1186038, EPI_ISL_1186039, EPI_ISL_1186040, EPI_ISL_1186041, EPI_ISL_1186042, EPI_ISL_1186043, EPI_ISL_1186044, EPI_ISL_1186045, EPI_ISL_1186046, EPI_ISL_1186047, EPI_ISL_1186048, EPI_ISL_1186049, EPI_ISL_1186050, EPI_ISL_1186051, EPI_ISL_1186052, EPI_ISL_1186053, EPI_ISL_1186054, EPI_ISL_1186055, EPI_ISL_1186056, EPI_ISL_1186057, EPI_ISL_1186059, EPI_ISL_1186060, EPI_ISL_1186062, EPI_ISL_1186063, EPI_ISL_1186064, EPI_ISL_1186065, EPI_ISL_1186067 | | | |
| see above | New Mexico Department of Health Scientific Laboratory | New Mexico Department of Health Scientific Laboratory | Ellie Johnson, Anastacia Griego-Fisher, D'eldra Malone, Jennifer Benoit |
| EPI_ISL_1190729, EPI_ISL_1190730, EPI_ISL_1190731, EPI_ISL_1190732, EPI_ISL_1190733, EPI_ISL_1190734, EPI_ISL_1190735, EPI_ISL_1190736, EPI_ISL_1190737 | Idaho Bureau of Laboratories | Center for Global Health, University of New Mexico Health Sciences Center | Daryl Domman, Kurt Schwalm, Matthew Burns, Robert Voermans, Christopher Ball, Darrell Dinwiddie |
| EPI_ISL_1191094 | Baylor Esoteric + Molecular Lab | Baylor Esoteric + Molecular Lab | Elizabeth Forrester, Dawn Richards |
| EPI_ISL_1191109 | Los Angeles County Public Health Laboratories | Los Angeles County Public Health Laboratories | P. Hemarajata et al. |
| EPI_ISL_1191111, EPI_ISL_1191113, EPI_ISL_1191114, EPI_ISL_1191115, EPI_ISL_1191117, EPI_ISL_1191118, EPI_ISL_1191119, EPI_ISL_1191120, EPI_ISL_1191121, EPI_ISL_1191122, EPI_ISL_1191123, EPI_ISL_1191125, EPI_ISL_1191126, EPI_ISL_1191127, EPI_ISL_1191128 | | | |
| see above | UCLA Clinical Micro Lab | Los Angeles County Public Health Laboratories | P. Hemarajata et al. |
| EPI_ISL_1191462, EPI_ISL_1191463, EPI_ISL_1191464, EPI_ISL_1191465, EPI_ISL_1191466, EPI_ISL_1191467, EPI_ISL_1191468, EPI_ISL_1191469, EPI_ISL_1191470, EPI_ISL_1191471, EPI_ISL_1191472, EPI_ISL_1191473, EPI_ISL_1191474, EPI_ISL_1191475, EPI_ISL_1191476, EPI_ISL_1191477, EPI_ISL_1191478, EPI_ISL_1191479, EPI_ISL_1191480, EPI_ISL_1191481, EPI_ISL_1191482, EPI_ISL_1191483, EPI_ISL_1191484, EPI_ISL_1191485, EPI_ISL_1191486, EPI_ISL_1191487, EPI_ISL_1191488, EPI_ISL_1191489, EPI_ISL_1191490, EPI_ISL_1191491, EPI_ISL_1191492, EPI_ISL_1191493, EPI_ISL_1191494, EPI_ISL_1191495, EPI_ISL_1191496, EPI_ISL_1191497, EPI_ISL_1191498, EPI_ISL_1191499, EPI_ISL_1191500, EPI_ISL_1191501, EPI_ISL_1191502, EPI_ISL_1191503, EPI_ISL_1191504, EPI_ISL_1191506, EPI_ISL_1191507, EPI_ISL_1191508, EPI_ISL_1191509, EPI_ISL_1191510, EPI_ISL_1191511, EPI_ISL_1191513, EPI_ISL_1191514, EPI_ISL_1191516, EPI_ISL_1191518, EPI_ISL_1191519, EPI_ISL_1191521, EPI_ISL_1191523, EPI_ISL_1191524, EPI_ISL_1191525, EPI_ISL_1191526, EPI_ISL_1191527, EPI_ISL_1191528, EPI_ISL_1191529, EPI_ISL_1191530, EPI_ISL_1191531, EPI_ISL_1191532, EPI_ISL_1191533, EPI_ISL_1191534, EPI_ISL_1191536, EPI_ISL_1191537, EPI_ISL_1191538, EPI_ISL_1191539, EPI_ISL_1191541, EPI_ISL_1191542, EPI_ISL_1191543, EPI_ISL_1191545, EPI_ISL_1191546, EPI_ISL_1191547, EPI_ISL_1191548, EPI_ISL_1191549, EPI_ISL_1191550, EPI_ISL_1191551, EPI_ISL_1191552, EPI_ISL_1191553, EPI_ISL_1191554, EPI_ISL_1191555, EPI_ISL_1191556, EPI_ISL_1191557, EPI_ISL_1191558, EPI_ISL_1191560, EPI_ISL_1191562, EPI_ISL_1191563, EPI_ISL_1191564, EPI_ISL_1191565, EPI_ISL_1191567, EPI_ISL_1191568, EPI_ISL_1191569, EPI_ISL_1191570, EPI_ISL_1191571, EPI_ISL_1191573, EPI_ISL_1191574, EPI_ISL_1191575, EPI_ISL_1191576, EPI_ISL_1191579, EPI_ISL_1191582, EPI_ISL_1191583, EPI_ISL_1191584, EPI_ISL_1191586 | | | |
| see above | Florida Bureau of Public Health Laboratories | Florida Bureau of Public Health Laboratories | Sarah Schmedes, Jason Blanton |
| EPI_ISL_1191612, EPI_ISL_1191613, EPI_ISL_1191614, EPI_ISL_1191615, EPI_ISL_1191616, EPI_ISL_1191617, EPI_ISL_1191618, EPI_ISL_1191619, EPI_ISL_1191620, EPI_ISL_1191621, EPI_ISL_1191622, EPI_ISL_1191623 | | | |
| see above | DC Public Health Lab/ Dept. of Forensic Sciences | DC Public Health Lab/ Dept. of Forensic Sciences | Janis Doss, Scott Nguyen, Elizabeth Zelaya, Sarah Scott, Connie Maza, Monica Mann, Brittany Hamilton, David Payne, Jocelyn Hauser |
| EPI_ISL_1191727 | Coast Diagnostics | Synergy Laboratories, Inc. | Tina Louise Sutton-Young |

| | | | |
|---|---|---|---|
| EPI_ISL_1192336, EPI_ISL_1192337, EPI_ISL_1192338, EPI_ISL_1192339, EPI_ISL_1192340, EPI_ISL_1192341, EPI_ISL_1192342, EPI_ISL_1192343, EPI_ISL_1192344, EPI_ISL_1192345, EPI_ISL_1192346, EPI_ISL_1192347, EPI_ISL_1192348, EPI_ISL_1192349, EPI_ISL_1192350, EPI_ISL_1192351, EPI_ISL_1192352, EPI_ISL_1192353, EPI_ISL_1192354, EPI_ISL_1192355, EPI_ISL_1192356, EPI_ISL_1192357, EPI_ISL_1192358, EPI_ISL_1192359, EPI_ISL_1192360, EPI_ISL_1192361, EPI_ISL_1192362 | | | |
| see above | Clinical Molecular Microbiology Laboratory, UNC Hospitals | Jeremy Wang | Jeremy Wang, Alexander Rubinsteyn, Colleen Rice, Jason Smedberg, Shawn Hawken, Melissa Miller, Corbin Jones, Robert Hagan |
| EPI_ISL_1192363 | City of Hope Comprehensive Cancer Center | Los Angeles County Public Health Laboratories | P. Hemarajata et al. |
| EPI_ISL_1192364, EPI_ISL_1192365 | Huntington Memorial Hospital | Los Angeles County Public Health Laboratories | P. Hemarajata et al. |
| EPI_ISL_1192366 | Flow Diagnostics Lab | Los Angeles County Public Health Laboratories | P. Hemarajata et al. |
| EPI_ISL_1192367, EPI_ISL_1192368 | USC Clinical Lab | Los Angeles County Public Health Laboratories | P. Hemarajata et al. |
| EPI_ISL_1192369, EPI_ISL_1192370, EPI_ISL_1192371 | Flow Diagnostics Lab | Los Angeles County Public Health Laboratories | P. Hemarajata et al. |
| EPI_ISL_1193260, EPI_ISL_1193267, EPI_ISL_1193268, EPI_ISL_1193272, EPI_ISL_1193274, EPI_ISL_1193275, EPI_ISL_1193277, EPI_ISL_1193279, EPI_ISL_1193280, EPI_ISL_1193281, EPI_ISL_1193282, EPI_ISL_1193283, EPI_ISL_1193284, EPI_ISL_1193287, EPI_ISL_1193288, EPI_ISL_1193289, EPI_ISL_1193291, EPI_ISL_1193292, EPI_ISL_1193294, EPI_ISL_1193295, EPI_ISL_1193296, EPI_ISL_1193298, EPI_ISL_1193301, EPI_ISL_1193303, EPI_ISL_1193306, EPI_ISL_1193307, EPI_ISL_1193310, EPI_ISL_1193311, EPI_ISL_1193312, EPI_ISL_1193313, EPI_ISL_1193315, EPI_ISL_1193316, EPI_ISL_1193317, EPI_ISL_1193321, EPI_ISL_1193323, EPI_ISL_1193325, EPI_ISL_1193331, EPI_ISL_1193334, EPI_ISL_1193335, EPI_ISL_1193336, EPI_ISL_1193337, EPI_ISL_1193339, EPI_ISL_1193342, EPI_ISL_1193343, EPI_ISL_1193346, EPI_ISL_1193347, EPI_ISL_1193348, EPI_ISL_1193349, EPI_ISL_1193350, EPI_ISL_1193353, EPI_ISL_1193355, EPI_ISL_1193357, EPI_ISL_1193358, EPI_ISL_1193362, EPI_ISL_1193364, EPI_ISL_1193367 | | | |
| see above | Laboratory Corporation of America | Centers for Disease Control and Prevention Division of Viral Diseases, Pathogen Discovery | Peter W. Cook, Dakota Howard, Dhwani Batra, Ben L. Rambo-Martin, Minoo Agarwal, Eyad Almasri Debbie Boles, Ayla Burns, Nuthawin Charoensri, Oren Cohen, Susan Countryman, Mary Ann Cristobal, Bobbi Croy, Suzanne Dale, Hrushikesh Deshmukh, Amanda Douglas, Vincent Drouillon, Marcia Eisenberg, Howard Engler, Rama Ghatti, Prashant Gupta, Susan Hicks, Jake Humphrey, Lax Iyer, Manoj Jain, Mohan Kolli, Brian Krueger, Tim Kuphal, Stanley Letovsky, Michael Levandoski, Craig Lukasik, Jonathan Meltzer, Brian Norvell, Mindy Nye, Scott Parker, Christos Petropoulos, John Pruitt, Steven Ragan, Scott Ryan, Mike Sapeta, Jana Schroth, Suresh Babu Selvaraju, Goran Stevovic, Amanda Suchanek, Andrea Throop, Lyndon Tilson, Thomas Urban, Joe Voshell, Kimberly Wagner, Jonathan Williams, Mary Williamson, Qian Zeng, Tricia Zwiefelhofer, Clinton R. Paden, Suxiang Tong, Duncan MacCannell |
| EPI_ISL_1193374, EPI_ISL_1193375, EPI_ISL_1193376, EPI_ISL_1193377, EPI_ISL_1193378, EPI_ISL_1193379, EPI_ISL_1193380, EPI_ISL_1193381, EPI_ISL_1193382, EPI_ISL_1193383, EPI_ISL_1193384, EPI_ISL_1193385, EPI_ISL_1193386, EPI_ISL_1193387, EPI_ISL_1193388, EPI_ISL_1193389, EPI_ISL_1193390, EPI_ISL_1193391, EPI_ISL_1193392, EPI_ISL_1193393, EPI_ISL_1193394, EPI_ISL_1193395, EPI_ISL_1193396, EPI_ISL_1193397, EPI_ISL_1193400, EPI_ISL_1193403, EPI_ISL_1193405, EPI_ISL_1193408, EPI_ISL_1193411, EPI_ISL_1193413, EPI_ISL_1193414, EPI_ISL_1193415, EPI_ISL_1193416, EPI_ISL_1193417, EPI_ISL_1193418, EPI_ISL_1193419, EPI_ISL_1193420, EPI_ISL_1193421, EPI_ISL_1193422, EPI_ISL_1193423, EPI_ISL_1193424, EPI_ISL_1193425, EPI_ISL_1193426, EPI_ISL_1193427, EPI_ISL_1193428, EPI_ISL_1193429, EPI_ISL_1193430, EPI_ISL_1193431, EPI_ISL_1193432, EPI_ISL_1193433, EPI_ISL_1193434, EPI_ISL_1193435, EPI_ISL_1193436, EPI_ISL_1193437, EPI_ISL_1193438, EPI_ISL_1193439, EPI_ISL_1193440, EPI_ISL_1193441, EPI_ISL_1193442, EPI_ISL_1193443, EPI_ISL_1193444, EPI_ISL_1193446, EPI_ISL_1193449, EPI_ISL_1193452, EPI_ISL_1193454, EPI_ISL_1193457, EPI_ISL_1193460, EPI_ISL_1193463, EPI_ISL_1193464, EPI_ISL_1193465, EPI_ISL_1193466, EPI_ISL_1193467, EPI_ISL_1193468, EPI_ISL_1193469, EPI_ISL_1193470, EPI_ISL_1193471, EPI_ISL_1193472, EPI_ISL_1193473, EPI_ISL_1193474, EPI_ISL_1193475, EPI_ISL_1193476, EPI_ISL_1193477, EPI_ISL_1193478, EPI_ISL_1193479, EPI_ISL_1193480, EPI_ISL_1193481, EPI_ISL_1193482, EPI_ISL_1193483, EPI_ISL_1193484, EPI_ISL_1193485, EPI_ISL_1193486, EPI_ISL_1193487, EPI_ISL_1193488, EPI_ISL_1193489, EPI_ISL_1193490, EPI_ISL_1193491, EPI_ISL_1193492, EPI_ISL_1193493, EPI_ISL_1193494, EPI_ISL_1193495, EPI_ISL_1193496, EPI_ISL_1193497, EPI_ISL_1193498, EPI_ISL_1193499, EPI_ISL_1193500, EPI_ISL_1193501, EPI_ISL_1193502, EPI_ISL_1193503, EPI_ISL_1193504, EPI_ISL_1193505, EPI_ISL_1193506, EPI_ISL_1193507, EPI_ISL_1193508, EPI_ISL_1193509, EPI_ISL_1193510, EPI_ISL_1193511, EPI_ISL_1193512, EPI_ISL_1193513, EPI_ISL_1193514, EPI_ISL_1193515, EPI_ISL_1193516, EPI_ISL_1193517, EPI_ISL_1193518, EPI_ISL_1193519, EPI_ISL_1193520, EPI_ISL_1193521, EPI_ISL_1193522, EPI_ISL_1193523, EPI_ISL_1193524, EPI_ISL_1193525, EPI_ISL_1193526, EPI_ISL_1193527, EPI_ISL_1193528, EPI_ISL_1193529, EPI_ISL_1193530, EPI_ISL_1193531, EPI_ISL_1193532, EPI_ISL_1193533, EPI_ISL_1193534, EPI_ISL_1193535, EPI_ISL_1193536, EPI_ISL_1193537, EPI_ISL_1193538, EPI_ISL_1193539, EPI_ISL_1193540, EPI_ISL_1193541, EPI_ISL_1193542, EPI_ISL_1193543, EPI_ISL_1193544, EPI_ISL_1193545, EPI_ISL_1193546, EPI_ISL_1193547, EPI_ISL_1193548, EPI_ISL_1193549, EPI_ISL_1193550, EPI_ISL_1193551, EPI_ISL_1193552, EPI_ISL_1193553, EPI_ISL_1193554, EPI_ISL_1193555, EPI_ISL_1193556, EPI_ISL_1193557 | | | |
| see above | Quest Diagnostics Incorporated | Centers for Disease Control and Prevention Division of Viral Diseases, Pathogen Discovery | Peter W. Cook, Dakota Howard, Dhwani Batra, Ben L. Rambo-Martin, S. H. Rosenthal, A. Gerasimova, R. M. Kagan, B. Anderson, M. Hua, Y. Liu, L.E. Bernstein, K.E. Livingston, A. Perez, I. A. Shlyakhter, R. V. Rolando, R. Owen, P. Tanpaiboon, F. Lacbawan, Clinton R. Paden, Suxiang Tong, Duncan MacCannell |
| EPI_ISL_1193558 | Instituto Nacional de Medicina Genomica | Instituto Nacional de Medicina Genomica | Hidalgo-Miranda A, Mendoza-Vargas A, Reyes-Grajeda JP, Cedro-Tanda A, Alcaraz N, Gonzalez-Barrera D, Rangel-DeLeon D, Ramirez-Vega O, Sifuentes-Rojas C, Cisneros-Villanueva M, Herrera-Montalvo LA |
| EPI_ISL_1193559, EPI_ISL_1193560, EPI_ISL_1193561, EPI_ISL_1193562, EPI_ISL_1193563, EPI_ISL_1193564, EPI_ISL_1193565, EPI_ISL_1193566, EPI_ISL_1193567, EPI_ISL_1193568, EPI_ISL_1193569, EPI_ISL_1193570, EPI_ISL_1193571, EPI_ISL_1193572, EPI_ISL_1193573, EPI_ISL_1193574, EPI_ISL_1193575, EPI_ISL_1193576, EPI_ISL_1193577, EPI_ISL_1193578, EPI_ISL_1193579, EPI_ISL_1193580, EPI_ISL_1193581, EPI_ISL_1193582, EPI_ISL_1193583, EPI_ISL_1193584, EPI_ISL_1193585, EPI_ISL_1193586, EPI_ISL_1193587, EPI_ISL_1193588, EPI_ISL_1193589, EPI_ISL_1193590, EPI_ISL_1193591, EPI_ISL_1193592, EPI_ISL_1193593, EPI_ISL_1193594, EPI_ISL_1193595, EPI_ISL_1193596, EPI_ISL_1193597, EPI_ISL_1193598, EPI_ISL_1193599, EPI_ISL_1193600, EPI_ISL_1193601, EPI_ISL_1193602, EPI_ISL_1193603, EPI_ISL_1193604, EPI_ISL_1193605, EPI_ISL_1193606, EPI_ISL_1193607, EPI_ISL_1193608, EPI_ISL_1193609, EPI_ISL_1193610, EPI_ISL_1193611, EPI_ISL_1193612, EPI_ISL_1193613, EPI_ISL_1193614, EPI_ISL_1193615, EPI_ISL_1193616, EPI_ISL_1193617, EPI_ISL_1193618, EPI_ISL_1193619, EPI_ISL_1193620, EPI_ISL_1193621, EPI_ISL_1193622, EPI_ISL_1193623, EPI_ISL_1193624, EPI_ISL_1193625, EPI_ISL_1193626, EPI_ISL_1193627, EPI_ISL_1193628, EPI_ISL_1193629, EPI_ISL_1193630, EPI_ISL_1193631, EPI_ISL_1193632, EPI_ISL_1193633, EPI_ISL_1193634, EPI_ISL_1193635, EPI_ISL_1193636, EPI_ISL_1193637, EPI_ISL_1193638, EPI_ISL_1193639, EPI_ISL_1193640, EPI_ISL_1193641, EPI_ISL_1193642, EPI_ISL_1193643, EPI_ISL_1193644, EPI_ISL_1193645, EPI_ISL_1193646, EPI_ISL_1193647, EPI_ISL_1193648, EPI_ISL_1193649, EPI_ISL_1193650, EPI_ISL_1193651, EPI_ISL_1193652, EPI_ISL_1193653, EPI_ISL_1193654, EPI_ISL_1193655, EPI_ISL_1193656, EPI_ISL_1193657, EPI_ISL_1193658, EPI_ISL_1193659, EPI_ISL_1193660, EPI_ISL_1193661, EPI_ISL_1193662, EPI_ISL_1193663, EPI_ISL_1193664, EPI_ISL_1193665, EPI_ISL_1193666, EPI_ISL_1193667, EPI_ISL_1193668, EPI_ISL_1193669, EPI_ISL_1193670, EPI_ISL_1193671, EPI_ISL_1193672, EPI_ISL_1193673, EPI_ISL_1193674, EPI_ISL_1193675, EPI_ISL_1193676, EPI_ISL_1193677, EPI_ISL_1193678, EPI_ISL_1193679, EPI_ISL_1193680, EPI_ISL_1193681, EPI_ISL_1193682, EPI_ISL_1193683, EPI_ISL_1193684, EPI_ISL_1193685, EPI_ISL_1193686, EPI_ISL_1193687, EPI_ISL_1193688, EPI_ISL_1193689, EPI_ISL_1193690, EPI_ISL_1193691, EPI_ISL_1193692, EPI_ISL_1193693, EPI_ISL_1193694, EPI_ISL_1193695, EPI_ISL_1193696, EPI_ISL_1193697, EPI_ISL_1193698, EPI_ISL_1193699, EPI_ISL_1193700, EPI_ISL_1193701, EPI_ISL_1193702, EPI_ISL_1193703, EPI_ISL_1193704, EPI_ISL_1193705, EPI_ISL_1193706, EPI_ISL_1193707, EPI_ISL_1193708, EPI_ISL_1193709, EPI_ISL_1193710, EPI_ISL_1193711, EPI_ISL_1193712, EPI_ISL_1193713, EPI_ISL_1193714, EPI_ISL_1193715, EPI_ISL_1193716, EPI_ISL_1193717, EPI_ISL_1193718, EPI_ISL_1193719, EPI_ISL_1193720, EPI_ISL_1193721, EPI_ISL_1193722, EPI_ISL_1193723, EPI_ISL_1193724, EPI_ISL_1193725, EPI_ISL_1193726, EPI_ISL_1193727, EPI_ISL_1193728, EPI_ISL_1193729, EPI_ISL_1193730, EPI_ISL_1193731, EPI_ISL_1193732, EPI_ISL_1193733, EPI_ISL_1193734, EPI_ISL_1193735, EPI_ISL_1193736, EPI_ISL_1193737, EPI_ISL_1193738, EPI_ISL_1193739, EPI_ISL_1193740, EPI_ISL_1193741, EPI_ISL_1193742, EPI_ISL_1193743, EPI_ISL_1193744, EPI_ISL_1193745, EPI_ISL_1193746, EPI_ISL_1193747, EPI_ISL_1193748, EPI_ISL_1193749, EPI_ISL_1193750, EPI_ISL_1193751, EPI_ISL_1193752, EPI_ISL_1193753, EPI_ISL_1193754, EPI_ISL_1193755, EPI_ISL_1193756, EPI_ISL_1193757, EPI_ISL_1193758, EPI_ISL_1193759, EPI_ISL_1193760, EPI_ISL_1193761, EPI_ISL_1193762, EPI_ISL_1193763, EPI_ISL_1193764, EPI_ISL_1193765, EPI_ISL_1193766, EPI_ISL_1193767, EPI_ISL_1193768, EPI_ISL_1193769, EPI_ISL_1193770, EPI_ISL_1193771, EPI_ISL_1193772, EPI_ISL_1193773, EPI_ISL_1193774, EPI_ISL_1193775, EPI_ISL_1193776, EPI_ISL_1193777, EPI_ISL_1193778, EPI_ISL_1193779, EPI_ISL_1193780, EPI_ISL_1193781, EPI_ISL_1193782, EPI_ISL_1193783, EPI_ISL_1193784, EPI_ISL_1193785, EPI_ISL_1193786, EPI_ISL_1193787, EPI_ISL_1193788, EPI_ISL_1193789, EPI_ISL_1193790, EPI_ISL_1193791, EPI_ISL_1193792, EPI_ISL_1193793, EPI_ISL_1193794, EPI_ISL_1193795, EPI_ISL_1193796, EPI_ISL_1193797, EPI_ISL_1193798, EPI_ISL_1193799, EPI_ISL_1193800, EPI_ISL_1193801, EPI_ISL_1193802, EPI_ISL_1193803, EPI_ISL_1193804, EPI_ISL_1193805, EPI_ISL_1193806, EPI_ISL_1193807, EPI_ISL_1193808, EPI_ISL_1193809, EPI_ISL_1193810, EPI_ISL_1193811, EPI_ISL_1193812, EPI_ISL_1193813, EPI_ISL_1193814, EPI_ISL_1193815, EPI_ISL_1193816, EPI_ISL_1193817, EPI_ISL_1193818, EPI_ISL_1193819, EPI_ISL_1193820, EPI_ISL_1193821, EPI_ISL_1193822, EPI_ISL_1193823, EPI_ISL_1193824, EPI_ISL_1193825, EPI_ISL_1193826, EPI_ISL_1193827, EPI_ISL_1193828, EPI_ISL_1193829, EPI_ISL_1193830, EPI_ISL_1193831, EPI_ISL_1193832, EPI_ISL_1193833, EPI_ISL_1193834, EPI_ISL_1193835, EPI_ISL_1193836, EPI_ISL_1193837, EPI_ISL_1193838, EPI_ISL_1193839, EPI_ISL_1193840, EPI_ISL_1193841, EPI_ISL_1193842, EPI_ISL_1193843, EPI_ISL_1193844, EPI_ISL_1193845, EPI_ISL_1193846, EPI_ISL_1193847, EPI_ISL_1193848, EPI_ISL_1193849, EPI_ISL_1193850, EPI_ISL_1193851, EPI_ISL_1193852, EPI_ISL_1193853, EPI_ISL_1193854, EPI_ISL_1193855, EPI_ISL_1193856, EPI_ISL_1193857, EPI_ISL_1193858, EPI_ISL_1193859, EPI_ISL_1193860, EPI_ISL_1193861, EPI_ISL_1193862, EPI_ISL_1193863, EPI_ISL_1193864, EPI_ISL_1193865, EPI_ISL_1193866, EPI_ISL_1193867, EPI_ISL_1193868, EPI_ISL_1193869, EPI_ISL_1193870, EPI_ISL_1193871, EPI_ISL_1193872, EPI_ISL_1193873, EPI_ISL_1193874, EPI_ISL_1193875, EPI_ISL_1193876, EPI_ISL_1193877, EPI_ISL_1193878, EPI_ISL_1193879, EPI_ISL_1193880, EPI_ISL_1193881, EPI_ISL_1193882, EPI_ISL_1193883, EPI_ISL_1193884, EPI_ISL_1193885, EPI_ISL_1193886, EPI_ISL_1193887, EPI_ISL_1193888, EPI_ISL_1193889, EPI_ISL_1193890, EPI_ISL_1193891, EPI_ISL_1193892, EPI_ISL_1193893, EPI_ISL_1193894, EPI_ISL_1193895, EPI_ISL_1193896, EPI_ISL_1193897, EPI_ISL_1193898, EPI_ISL_1193899, EPI_ISL_1193900, EPI_ISL_1193901, EPI_ISL_1193902, EPI_ISL_1193903, EPI_ISL_1193904, EPI_ISL_1193905, EPI_ISL_1193906, EPI_ISL_1193907, EPI_ISL_1193908, EPI_ISL_1193909, EPI_ISL_1193910, EPI_ISL_1193911, EPI_ISL_1193912, EPI_ISL_1193913, EPI_ISL_1193914, EPI_ISL_1193915, EPI_ISL_1193916, EPI_ISL_1193917, EPI_ISL_1193918, EPI_ISL_1193919, EPI_ISL_1193920, EPI_ISL_1193921, EPI_ISL_1193922, EPI_ISL_1193923, EPI_ISL_1193924, EPI_ISL_1193925, EPI_ISL_1193926, EPI_ISL_1193927, EPI_ISL_1193928, EPI_ISL_1193929, EPI_ISL_1193930, EPI_ISL_1193931, EPI_ISL_1193932, EPI_ISL_1193933, EPI_ISL_1193934, EPI_ISL_1193935, EPI_ISL_1193936, EPI_ISL_1193937, EPI_ISL_1193938, EPI_ISL_1193939, EPI_ISL_1193940, EPI_ISL_1193941, EPI_ISL_1193942, EPI_ISL_1193943, EPI_ISL_1193944, EPI_ISL_1193945, EPI_ISL_1193946, EPI_ISL_1193947, EPI_ISL_1193948, EPI_ISL_1193949, EPI_ISL_1193950, EPI_ISL_1193951, EPI_ISL_1193952, EPI_ISL_1193953, EPI_ISL_1193954, EPI_ISL_1193955, EPI_ISL_1193956, EPI_ISL_1193957, EPI_ISL_1193958, EPI_ISL_1193959, EPI_ISL_1193960, EPI_ISL_1193961, EPI_ISL_1193962, EPI_ISL_1193963, EPI_ISL_1193964, EPI_ISL_1193965, EPI_ISL_1193966, EPI_ISL_1193967, EPI_ISL_1193968, EPI_ISL_1193969, EPI_ISL_1193970, EPI_ISL_1193971, EPI_ISL_1193972, EPI_ISL_1193973, EPI_ISL_1193974, EPI_ISL_1193975, EPI_ISL_1193976, EPI_ISL_1193977, EPI_ISL_1193978, EPI_ISL_1193979, EPI_ISL_1193980, EPI_ISL_1193981, EPI_ISL_1193982, EPI_ISL_1193983, EPI_ISL_1193984, EPI_ISL_1193985, EPI_ISL_1193986, EPI_ISL_1193987, EPI_ISL_1193988, EPI_ISL_1193989, EPI_ISL_1193990, EPI_ISL_1193991, EPI_ISL_1193992, EPI_ISL_1193993, EPI_ISL_1193994, EPI_ISL_1193995, EPI_ISL_1193996, EPI_ISL_1193997, EPI_ISL_1193998, EPI_ISL_1193999, EPI_ISL_1194000, EPI_ISL_1194001, EPI_ISL_1194002, EPI_ISL_1194003, EPI_ISL_1194004, EPI_ISL_1194005, EPI_ISL_1194006, EPI_ISL_1194007, EPI_ISL_1194008, EPI_ISL_1194009, EPI_ISL_1194010, EPI_ISL_1194011, EPI_ISL_1194012, EPI_ISL_1194013, EPI_ISL_1194014, EPI_ISL_1194015, EPI_ISL_1194016, EPI_ISL_1194017, EPI_ISL_1194018, EPI_ISL_1194019, EPI_ISL_1194020, EPI_ISL_1194021, EPI_ISL_1194022, EPI_ISL_1194023, EPI_ISL_1194024, EPI_ISL_1194025, EPI_ISL_1194026, EPI_ISL_1194027, EPI_ISL_1194028, EPI_ISL_1194029, EPI_ISL_1194030, EPI_ISL_1194031, EPI_ISL_1194032, EPI_ISL_1194033, EPI_ISL_1194034, EPI_ISL_1194035, EPI_ISL_1194036, EPI_ISL_1194037, EPI_ISL_1194038, EPI_ISL_1194039, EPI_ISL_1194040, EPI_ISL_1194041, EPI_ISL_1194042, EPI_ISL_1194043, EPI_ISL_1194044, EPI_ISL_1194045, EPI_ISL_1194048, EPI_ISL_1194051, EPI_ISL_1194054, EPI_ISL_1194056, EPI_ISL_1194059, EPI_ISL_1194062, EPI_ISL_1194065, EPI_ISL_1194068, EPI_ISL_1194071, EPI_ISL_1194074, EPI_ISL_1194077, EPI_ISL_1194080, EPI_ISL_1194083, EPI_ISL_1194086, EPI_ISL_1194089, EPI_ISL_1194092, EPI_ISL_1194094, EPI_ISL_1194097, EPI_ISL_1194100, EPI_ISL_1194103, EPI_ISL_1194107, EPI_ISL_1194110, EPI_ISL_1194113, EPI_ISL_1194116, EPI_ISL_1194119, EPI_ISL_1194122, EPI_ISL_1194125, EPI_ISL_1194128, EPI_ISL_1194131, EPI_ISL_1194134, EPI_ISL_1194137, EPI_ISL_1194140, EPI_ISL_1194143, EPI_ISL_1194145, EPI_ISL_1194148, EPI_ISL_1194151, EPI_ISL_1194154, EPI_ISL_1194157, EPI_ISL_1194160, EPI_ISL_1194162, EPI_ISL_1194165, EPI_ISL_1194167, EPI_ISL_1194168, EPI_ISL_1194169, EPI_ISL_1194170, EPI_ISL_1194171, EPI_ISL_1194172, EPI_ISL_1194173, EPI_ISL_1194174, EPI_ISL_1194175, EPI_ISL_1194176, EPI_ISL_1194177, EPI_ISL_1194178, EPI_ISL_1194179, EPI_ISL_1194180, EPI_ISL_1194181, EPI_ISL_1194182, EPI_ISL_1194183, EPI_ISL_1194184, EPI_ISL_1194185, EPI_ISL_1194186, EPI_ISL_1194187, EPI_ISL_1194188, EPI_ISL_1194189, EPI_ISL_1194190, EPI_ISL_1194191, EPI_ISL_1194192, EPI_ISL_1194193, EPI_ISL_1194194, EPI_ISL_1194195, EPI_ISL_1194196, EPI_ISL_1194197, EPI_ISL_1194198, EPI_ISL_1194199, EPI_ISL_1194200, EPI_ISL_1194201, EPI_ISL_1194202, EPI_ISL_1194203, EPI_ISL_1194204, EPI_ISL_1194205, EPI_ISL_1194206, EPI_ISL_1194207, EPI_ISL_1194208, EPI_ISL_1194209, EPI_ISL_1194210, EPI_ISL_1194211, EPI_ISL_1194212, EPI_ISL_1194213, EPI_ISL_1194214, EPI_ISL_1194215, EPI_ISL_1194216, EPI_ISL_1194217, EPI_ISL_1194218, EPI_ISL_1194219, EPI_ISL_1194220, EPI_ISL_1194221, EPI_ISL_1194222, EPI_ISL_1194223, EPI_ISL_1194224, EPI_ISL_1194225, EPI_ISL_1194226, EPI_ISL_1194227, EPI_ISL_1194228, EPI_ISL_1194229, EPI_ISL_1194230, EPI_ISL_1194231, EPI_ISL_1194232, EPI_ISL_1194233, EPI_ISL_1194234, EPI_ISL_1194235, EPI_ISL_1194236, EPI_ISL_1194237, EPI_ISL_1194238, EPI_ISL_1194239, EPI_ISL_1194240, EPI_ISL_1194241, EPI_ISL_1194242, EPI_ISL_1194243, EPI_ISL_1194244, EPI_ISL_1194245, EPI_ISL_1194246, EPI_ISL_1194247, EPI_ISL_1194248, EPI_ISL_1194249, EPI_ISL_1194250, EPI_ISL_1194251, EPI_ISL_1194252, EPI_ISL_1194253, EPI_ISL_1194254, EPI_ISL_1194255, EPI_ISL_1194256, EPI_ISL_1194257, EPI_ISL_1194258, EPI_ISL_1194259, EPI_ISL_1194260, EPI_ISL_1194261, EPI_ISL_1194262, EPI_ISL_1194263, EPI_ISL_1194264, EPI_ISL_1194265, EPI_ISL_1194266, EPI_ISL_1194267, EPI_ISL_1194268, EPI_ISL_1194269, EPI_ISL_1194270, EPI_ISL_1194271, EPI_ISL_1194272, EPI_ISL_1194273, EPI_ISL_1194274, EPI_ISL_1194275, EPI_ISL_1194276, EPI_ISL_1194277, EPI_ISL_1194278, EPI_ISL_1194279, EPI_ISL_1194280, EPI_ISL_1194281, EPI_ISL_1194282, EPI_ISL_1194283, EPI_ISL_1194284, EPI_ISL_1194285, EPI_ISL_1194286, EPI_ISL_1194287, EPI_ISL_1194288, EPI_ISL_1194289, EPI_ISL_1194290, EPI_ISL_1194291, EPI_ISL_1194292, EPI_ISL_1194293, EPI_ISL_1194294, EPI_ISL_1194295, EPI_ISL_1194296, EPI_ISL_1194297, EPI_ISL_1194298, EPI_ISL_1194299, EPI_ISL_1194300, EPI_ISL_1194301, EPI_ISL_1194302, EPI_ISL_1194303, EPI_ISL_1194304, EPI_ISL_1194305, EPI_ISL_1194306, EPI_ISL_1194307, EPI_ISL_1194308, EPI_ISL_1194309, EPI_ISL_1194310, EPI_ISL_1194311, EPI_ISL_1194312, EPI_ISL_1194313, EPI_ISL_1194314, EPI_ISL_1194315, EPI_ISL_1194316, EPI_ISL_1194317, EPI_ISL_1194318, EPI_ISL_1194319, EPI_ISL_1194320, EPI_ISL_1194321, EPI_ISL_1194322, EPI_ISL_1194323, EPI_ISL_1194324, EPI_ISL_1194325, EPI_ISL_1194326, EPI_ISL_1194327, EPI_ISL_1194328, EPI_ISL_1194329, EPI_ISL_1194330, EPI_ISL_1194331, EPI_ISL_1194332, EPI_ISL_1194333, EPI_ISL_1194334, EPI_ISL_1194335, EPI_ISL_1194336, EPI_ISL_1194337, EPI_ISL_1194338, EPI_ISL_1194339, EPI_ISL_1194340, EPI_ISL_1194341, EPI_ISL_1194342, EPI_ISL_1194343, EPI_ISL_1194344, EPI_ISL_1194345, EPI_ISL_1194346, EPI_ISL_1194347, EPI_ISL_1194348, EPI_ISL_1194349, EPI_ISL_1194350, EPI_ISL_1194351, EPI_ISL_1194352, EPI_ISL_1194353, EPI_ISL_1194354, EPI_ISL_1194355, EPI_ISL_1194356, EPI_ISL_1194357, EPI_ISL_1194358, EPI_ISL_1194359, EPI_ISL_1194360, EPI_ISL_1194361, EPI_ISL_1194362, EPI_ISL_1194363, EPI_ISL_1194364, EPI_ISL_1194365, EPI_ISL_1194366, EPI_ISL_1194367, EPI_ISL_1194368, EPI_ISL_1194369, EPI_ISL_1194370, EPI_ISL_1194371, EPI_ISL_1194372, EPI_ISL_1194373, EPI_ISL_1194374, EPI_ISL_1194375, EPI_ISL_1194376, EPI_ISL_1194377, EPI_ISL_1194378, EPI_ISL_1194379, EPI_ISL_1194380, EPI_ISL_1194381, EPI_ISL_1194382, EPI_ISL_1194383, EPI_ISL_1194384, EPI_ISL_1194385, EPI_ISL_1194386, EPI_ISL_1194387, EPI_ISL_1194388, EPI_ISL_1194389, EPI_ISL_1194390, EPI_ISL_1194391, EPI_ISL_1194392, EPI_ISL_1194393, EPI_ISL_1194394, EPI_ISL_1194395, EPI_ISL_1194396, EPI_ISL_1194397, EPI_ISL_1194398, EPI_ISL_1194399, EPI_ISL_1194400, EPI_ISL_1194401, EPI_ISL_1194402, EPI_ISL_1194403, EPI_ISL_1194404, EPI_ISL_1194405, EPI_ISL_1194406, EPI_ISL_1194407, EPI_ISL_1194408, EPI_ISL_1194409, EPI_ISL_1194410, EPI_ISL_1194411, EPI_ISL_1194412, EPI_ISL_1194413, EPI_ISL_1194414, EPI_ISL_1194415, EPI_ISL_1194416, EPI_ISL_1194417, EPI_ISL_1194418, EPI_ISL_1194419, EPI_ISL_1194420, EPI_ISL_1194421, EPI_ISL_1194422, EPI_ISL_1194423, EPI_ISL_1194424, EPI_ISL_1194425, EPI_ISL_1194426, EPI_ISL_1194427, EPI_ISL_1194428, EPI_ISL_1194429, EPI_ISL_1194430, EPI_ISL_1194431, EPI_ISL_1194432, EPI_ISL_1194433, EPI_ISL_1194434, EPI_ISL_1194435, EPI_ISL_1194436, EPI_ISL_1194437, EPI_ISL_1194438, EPI_ISL_1194439, EPI_ISL_1194440, EPI_ISL_1194441, EPI_ISL_1194442, EPI_ISL_1194443, EPI_ISL_1194444, EPI_ISL_1194445, EPI_ISL_1194446, EPI_ISL_1194447, EPI_ISL_1194448, EPI_ISL_1194449, EPI_ISL_1194450, EPI_ISL_1194451, EPI_ISL_1194452, EPI_ISL_1194453, EPI_ISL_1194454, EPI_ISL_1194455, EPI_ISL_1194456, EPI_ISL_1194457, EPI_ISL_1194458, EPI_ISL_1194459, EPI_ISL_1194460, EPI_ISL_1194461, EPI_ISL_1194462, EPI_ISL_1194463, EPI_ISL_1194464, EPI_ISL_1194465, EPI_ISL_1194466, EPI_ISL_1194467, EPI_ISL_1194468, EPI_ISL_1194469, EPI_ISL_1194470, EPI_ISL_1194471, EPI_ISL_1194472, EPI_ISL_1194473, EPI_ISL_1194474, EPI_ISL_1194475, EPI_ISL_1194476, EPI_ISL_1194477, EPI_ISL_1194478, EPI_ISL_1194479, EPI_ISL_1194480, EPI_ISL_1194481, EPI_ISL_1194482, EPI_ISL_1194483, EPI_ISL_1194484, EPI_ISL_1194485, EPI_ISL_1194486, EPI_ISL_1194487, EPI_ISL_1194488, EPI_ISL_1194489, EPI_ISL_1194490, EPI_ISL_1194491, EPI_ISL_1194492, EPI_ISL_1194493, EPI_ISL_1194494, EPI_ISL_1194495, EPI_ISL_1194496, EPI_ISL_1194498, EPI_ISL_1194501, EPI_ISL_1194504, EPI_ISL_1194506, EPI_ISL_1194509, EPI_ISL_1194512, EPI_ISL_1194515, EPI_ISL_1194518, EPI_ISL_1194521, EPI_ISL_1194524, EPI_ISL_1194527, EPI_ISL_1194538, EPI_ISL_1194540, EPI_ISL_1194543, EPI_ISL_1194546, EPI_ISL_1194548, EPI_ISL_1194559, EPI_ISL_1194561, EPI_ISL_1194564, EPI_ISL_1194567, EPI_ISL_1194570, EPI_ISL_1194573, EPI_ISL_1194575, EPI_ISL_1194578 | | | |
| see above | Quest Diagnostics Incorporated | Centers for Disease Control and Prevention Division of Viral Diseases, Pathogen Discovery | Peter W. Cook, Dakota Howard, Dhwani Batra, Ben L. Rambo-Martin, S. H. Rosenthal, A. Gerasimova, R. M. Kagan, B. Anderson, M. Hua, Y. Liu, L.E. Bernstein, K.E. Livingston, A. Perez, I. A. Shlyakhter, R. V. Rolando, R. Owen, P. Tanpaiboon, F. Lacbawan, Clinton R. Paden, Suxiang Tong, Duncan MacCannell |
| EPI_ISL_1194731, EPI_ISL_1194732, EPI_ISL_1194733, EPI_ISL_1194734, EPI_ISL_1194735, EPI_ISL_1194736, EPI_ISL_1194737, EPI_ISL_1194738, EPI_ISL_1194739, EPI_ISL_1194740, EPI_ISL_1194741, EPI_ISL_1194742, EPI_ISL_1194743, EPI_ISL_1194744, EPI_ISL_1194745, EPI_ISL_1194746, EPI_ISL_1194747, EPI_ISL_1194748, EPI_ISL_1194749, EPI_ISL_1194750, EPI_ISL_1194751, EPI_ISL_1194752, EPI_ISL_1194753, EPI_ISL_1194754, EPI_ISL_1194755, EPI_ISL_1194756, EPI_ISL_1194757, EPI_ISL_1194758, EPI_ISL_1194759, EPI_ISL_1194760, EPI_ISL_1194761, EPI_ISL_1194762, EPI_ISL_1194763, EPI_ISL_1194764, EPI_ISL_1194765, EPI_ISL_1194766, EPI_ISL_1194767, EPI_ISL_1194771, EPI_ISL_1194772, EPI_ISL_1194773, EPI_ISL_1194774, EPI_ISL_1194775, EPI_ISL_1194776, EPI_ISL_1194777, EPI_ISL_1194778, EPI_ISL_1194779, EPI_ISL_1194780, EPI_ISL_1194781, EPI_ISL_1194782, EPI_ISL_1194783, EPI_ISL_1194784, EPI_ISL_1194785, EPI_ISL_1194786, EPI_ISL_1194787, EPI_ISL_1194788, EPI_ISL_1194789, EPI_ISL_1194790, EPI_ISL_1194791, EPI_ISL_1194792, EPI_ISL_1194793, EPI_ISL_1194794, EPI_ISL_1194795, EPI_ISL_1194796, EPI_ISL_1194797, EPI_ISL_1194798, EPI_ISL_1194799, EPI_ISL_1194800, EPI_ISL_1194801, EPI_ISL_1194802, EPI_ISL_1194803, EPI_ISL_1194804, EPI_ISL_1194805, EPI_ISL_1194806, EPI_ISL_1194807, EPI_ISL_1194808, EPI_ISL_1194809, EPI_ISL_1194810, EPI_ISL_1194811, EPI_ISL_1194812, EPI_ISL_1194813, EPI_ISL_1194814, EPI_ISL_1194815, EPI_ISL_1194816, EPI_ISL_1194817, EPI_ISL_1194818, EPI_ISL_1194819, EPI_ISL_1194820, EPI_ISL_1194821, EPI_ISL_1194822, EPI_ISL_1194823, EPI_ISL_1194824, EPI_ISL_1194825, EPI_ISL_1194826, EPI_ISL_1194827, EPI_ISL_1194828, EPI_ISL_1194829, EPI_ISL_1194830, EPI_ISL_1194831, EPI_ISL_1194832, EPI_ISL_1194833, EPI_ISL_1194834, EPI_ISL_1194836, EPI_ISL_1194837, EPI_ISL_1194838, EPI_ISL_1194839, EPI_ISL_1194840, EPI_ISL_1194841, EPI_ISL_1194842, EPI_ISL_1194843, EPI_ISL_1194844, EPI_ISL_1194845, EPI_ISL_1194846, EPI_ISL_1194847, EPI_ISL_1194848, EPI_ISL_1194849, EPI_ISL_1194850, EPI_ISL_1194851, EPI_ISL_1194852, EPI_ISL_1194853, EPI_ISL_1194854, EPI_ISL_1194855, EPI_ISL_1194856, EPI_ISL_1194857, EPI_ISL_1194859, EPI_ISL_1194860, EPI_ISL_1194862, EPI_ISL_1194863, EPI_ISL_1194864, EPI_ISL_1194865 | | | |
| see above | Helix/Illumina | Centers for Disease Control and Prevention Division of Viral Diseases, Pathogen Discovery | Peter W. Cook, Dakota Howard, Dhwani Batra, Ben L. Rambo-Martin, Eileen de Feo, Jan Antico, Christine Tran, Matthew Tolentino, Shannon Wickline, Kim Gietzen, Brad Sickler, Jingtao Liu, Eric Allen, Phil Febbo, Summer Galloway, Nicole L. Washington, Simon White, Geraint Levan, Kelly Schiabor Barrett, Elizabeth Cirulli, Alexandre Bolze, Ary Ascencio, Charlotte Rivera-Garcia, Ryan Cho, Jason Nguyen, Sherry Wang, Jimmy Ramirez, Tyler Cassens, Efren Sandoval, Magnus Isaksson, William Lee, David Becker, Marc Laurent, James Lu, Clinton R. Paden, Suxiang Tong, Duncan MacCannell |
| EPI_ISL_1195294 | Baylor Esoteric + Molecular Lab | Baylor Esoteric + Molecular Lab | Elizabeth Forrester, Dawn Richards |
| EPI_ISL_1195478, EPI_ISL_1195479, EPI_ISL_1195480, EPI_ISL_1195481, EPI_ISL_1195482, EPI_ISL_1195483, EPI_ISL_1195484, EPI_ISL_1195485, EPI_ISL_1195486, EPI_ISL_1195487, EPI_ISL_1195488, EPI_ISL_1195489, EPI_ISL_1195490, EPI_ISL_1195491, EPI_ISL_1195492, EPI_ISL_1195493, EPI_ISL_1195494, EPI_ISL_1195495, EPI_ISL_1195496, EPI_ISL_1195497, EPI_ISL_1195498 | | | |
| see above | DC Public Health Lab/ Dept. of Forensic Sciences | DC Public Health Lab/ Dept. of Forensic Sciences | Janis Doss, Scott Nguyen, Elizabeth Zelaya, Sarah Scott, Connie Maza, Monica Mann, Brittany Hamilton, David Payne, Jocelyn Hauser |
| EPI_ISL_1195618, EPI_ISL_1195619, EPI_ISL_1195620, EPI_ISL_1195621, EPI_ISL_1195622, EPI_ISL_1195623, EPI_ISL_1195624, EPI_ISL_1195625, EPI_ISL_1195626, EPI_ISL_1195627, EPI_ISL_1195628, EPI_ISL_1195629, EPI_ISL_1195630, EPI_ISL_1195631, EPI_ISL_1195632, EPI_ISL_1195633, EPI_ISL_1195634, EPI_ISL_1195635, EPI_ISL_1195636, EPI_ISL_1195637, EPI_ISL_1195638, EPI_ISL_1195639, EPI_ISL_1195640, EPI_ISL_1195641, EPI_ISL_1195642, EPI_ISL_1195643, EPI_ISL_1195644, EPI_ISL_1195645, EPI_ISL_1195646, EPI_ISL_1195647, EPI_ISL_1195648, EPI_ISL_1195649, EPI_ISL_1195650, EPI_ISL_1195651, EPI_ISL_1195652, EPI_ISL_1195653, EPI_ISL_1195654, EPI_ISL_1195655, EPI_ISL_1195658, EPI_ISL_1195659, EPI_ISL_1195660 | | | |
| see above | North Dakota Department of Health, Public Health Laboratory | North Dakota Department of Health, Public Health Laboratory | Lisa Wingerter |
| EPI_ISL_1195744, EPI_ISL_1195745, EPI_ISL_1195746, EPI_ISL_1195747, EPI_ISL_1195748, EPI_ISL_1195749, EPI_ISL_1195751, EPI_ISL_1195752, EPI_ISL_1195753, EPI_ISL_1195754, EPI_ISL_1195755, EPI_ISL_1195757, EPI_ISL_1195758, EPI_ISL_1195759, EPI_ISL_1195760, EPI_ISL_1195761, EPI_ISL_1195762, EPI_ISL_1195763, EPI_ISL_1195764, EPI_ISL_1195765, EPI_ISL_1195766, EPI_ISL_1195767, EPI_ISL_1195768, EPI_ISL_1195769, EPI_ISL_1195770, EPI_ISL_1195771, EPI_ISL_1195772, EPI_ISL_1195773, EPI_ISL_1195774, EPI_ISL_1195775, EPI_ISL_1195776, EPI_ISL_1195777, EPI_ISL_1195778, EPI_ISL_1195779, EPI_ISL_1195780, EPI_ISL_1195781, EPI_ISL_1195782, EPI_ISL_1195783, EPI_ISL_1195785, EPI_ISL_1195786, EPI_ISL_1195787, EPI_ISL_1195788, EPI_ISL_1195789, EPI_ISL_1195790, EPI_ISL_1195791, EPI_ISL_1195792, EPI_ISL_1195793, EPI_ISL_1195794, EPI_ISL_1195795, EPI_ISL_1195796, EPI_ISL_1195797, EPI_ISL_1195798, EPI_ISL_1195799, EPI_ISL_1195800, EPI_ISL_1195801, EPI_ISL_1195802, EPI_ISL_1195806, EPI_ISL_1195807, EPI_ISL_1195808, EPI_ISL_1195809, EPI_ISL_1195810, EPI_ISL_1195811, EPI_ISL_1195812, EPI_ISL_1195813, EPI_ISL_1195815, EPI_ISL_1195816, EPI_ISL_1195817, EPI_ISL_1195818, EPI_ISL_1195819, EPI_ISL_1195820, EPI_ISL_1195821, EPI_ISL_1195822, EPI_ISL_1195823, EPI_ISL_1195824, EPI_ISL_1195825, EPI_ISL_1195826, EPI_ISL_1195827, EPI_ISL_1195828, EPI_ISL_1195829, EPI_ISL_1195830, EPI_ISL_1195831, EPI_ISL_1195832, EPI_ISL_1195833, EPI_ISL_1195835, EPI_ISL_1195836, EPI_ISL_1195837, EPI_ISL_1195838, EPI_ISL_1195839, EPI_ISL_1195840, EPI_ISL_1195841, EPI_ISL_1195842, EPI_ISL_1195843, EPI_ISL_1195844, EPI_ISL_1195845, EPI_ISL_1195846, EPI_ISL_1195847, EPI_ISL_1195848, EPI_ISL_1195849, EPI_ISL_1195850, EPI_ISL_1195851, EPI_ISL_1195852, EPI_ISL_1195853, EPI_ISL_1195854, EPI_ISL_1195855, EPI_ISL_1195856, EPI_ISL_1195857, EPI_ISL_1195858, EPI_ISL_1195859, EPI_ISL_1195861, EPI_ISL_1195862, | | | |

| | | | |
|---|---|---|---|
| EPI_ISL_1195863, EPI_ISL_1195864, EPI_ISL_1195865, EPI_ISL_1195866, EPI_ISL_1195867, EPI_ISL_1195868, EPI_ISL_1195869, EPI_ISL_1195871, EPI_ISL_1195872, EPI_ISL_1195873, EPI_ISL_1195875, EPI_ISL_1195876, EPI_ISL_1195877, EPI_ISL_1195878, EPI_ISL_1195880, EPI_ISL_1195882, EPI_ISL_1195883, EPI_ISL_1195884, EPI_ISL_1195885, EPI_ISL_1195887, EPI_ISL_1195888, EPI_ISL_1195889, EPI_ISL_1195890, EPI_ISL_1195891, EPI_ISL_1195892, EPI_ISL_1195893, EPI_ISL_1195894, EPI_ISL_1195895, EPI_ISL_1195897, EPI_ISL_1195899, EPI_ISL_1195901 | | | |
| see above | NYU Langone Health | Departments of Pathology and Medicine, New York University School of Medicine | Adriana Heguy, Dacia Dimartino, Emily Guzman, Christian Marier, Peter Meyn, Sitharam Ramaswami, Gael Westby, Paul Zappile, Yutong Zhang, Paolo Cotzia, Guiqing Wang |
| EPI_ISL_1195903, EPI_ISL_1195904, EPI_ISL_1195905, EPI_ISL_1195906, EPI_ISL_1195907, EPI_ISL_1195908, EPI_ISL_1195909 | Houston Health Dept. | Houston Health Dept. | Ryker Penn, Pamela Brown, Adolpho Lara |
| EPI_ISL_1195910, EPI_ISL_1195911, EPI_ISL_1195912, EPI_ISL_1195913, EPI_ISL_1195914, EPI_ISL_1195915, EPI_ISL_1195916, EPI_ISL_1195917, EPI_ISL_1195918, EPI_ISL_1195919, EPI_ISL_1195920, EPI_ISL_1195921, EPI_ISL_1195922, EPI_ISL_1195923, EPI_ISL_1195924, EPI_ISL_1195925, EPI_ISL_1195926, EPI_ISL_1195927, EPI_ISL_1195928, EPI_ISL_1195929, EPI_ISL_1195930, EPI_ISL_1195931, EPI_ISL_1195932, EPI_ISL_1195933, EPI_ISL_1195934, EPI_ISL_1195935, EPI_ISL_1195936, EPI_ISL_1195937, EPI_ISL_1195938, EPI_ISL_1195939, EPI_ISL_1195940, EPI_ISL_1195941, EPI_ISL_1195942, EPI_ISL_1195943, EPI_ISL_1195944, EPI_ISL_1195945 | | | |
| see above | Alaska State Virology Laboratory | Alaska State Virology Laboratory | Stephanie DeRonde, Elva House, Lisa Smith, Ph.D., Jack Chen, Ph.D. |
| EPI_ISL_1195946 | Curative Inc | Curative Inc | Katie Bond, Vladimir Slepnev |
| EPI_ISL_1195947, EPI_ISL_1195948, EPI_ISL_1195949 | Fulgent Genetics | Fulgent Genetics | Harry Gao, Mickey Li, John Gao, Joseph Fierro, Benafsh Sapra, Becky Tsai, Yan Meng, Doreen Ng, James Xie |
| EPI_ISL_1195964, EPI_ISL_1195965, EPI_ISL_1195966, EPI_ISL_1195967, EPI_ISL_1195968, EPI_ISL_1195969, EPI_ISL_1195970, EPI_ISL_1195971, EPI_ISL_1195972, EPI_ISL_1195973, EPI_ISL_1195974, EPI_ISL_1195975, EPI_ISL_1195976, EPI_ISL_1195977, EPI_ISL_1195978, EPI_ISL_1195979, EPI_ISL_1195980, EPI_ISL_1195981, EPI_ISL_1195982, EPI_ISL_1195983, EPI_ISL_1195984, EPI_ISL_1195985, EPI_ISL_1195986, EPI_ISL_1195987, EPI_ISL_1195988, EPI_ISL_1195989, EPI_ISL_1195990, EPI_ISL_1195991, EPI_ISL_1195992, EPI_ISL_1195993, EPI_ISL_1195994, EPI_ISL_1195995, EPI_ISL_1195996, EPI_ISL_1195997, EPI_ISL_1195998, EPI_ISL_1195999 | | | |
| see above | Alaska State Virology Laboratory | Alaska State Virology Laboratory | Stephanie DeRonde, Elva House, Lisa Smith, Ph.D., Jack Chen, Ph.D. |
| EPI_ISL_1196022, EPI_ISL_1196023, EPI_ISL_1196024, EPI_ISL_1196025, EPI_ISL_1196026, EPI_ISL_1196027, EPI_ISL_1196028, EPI_ISL_1196029, EPI_ISL_1196030, EPI_ISL_1196031, EPI_ISL_1196033, EPI_ISL_1196034, EPI_ISL_1196035, EPI_ISL_1196036, EPI_ISL_1196037, EPI_ISL_1196038, EPI_ISL_1196039, EPI_ISL_1196040, EPI_ISL_1196041, EPI_ISL_1196042, EPI_ISL_1196043, EPI_ISL_1196044, EPI_ISL_1196045, EPI_ISL_1196046, EPI_ISL_1196047, EPI_ISL_1196048, EPI_ISL_1196049, EPI_ISL_1196050, EPI_ISL_1196051, EPI_ISL_1196052, EPI_ISL_1196053, EPI_ISL_1196054, EPI_ISL_1196055, EPI_ISL_1196056, EPI_ISL_1196057, EPI_ISL_1196058, EPI_ISL_1196059, EPI_ISL_1196060, EPI_ISL_1196061, EPI_ISL_1196062, EPI_ISL_1196063, EPI_ISL_1196064, EPI_ISL_1196065, EPI_ISL_1196066, EPI_ISL_1196067, EPI_ISL_1196068, EPI_ISL_1196069, EPI_ISL_1196070, EPI_ISL_1196071, EPI_ISL_1196072, EPI_ISL_1196073, EPI_ISL_1196074, EPI_ISL_1196075, EPI_ISL_1196076, EPI_ISL_1196077, EPI_ISL_1196078, EPI_ISL_1196080, EPI_ISL_1196081, EPI_ISL_1196082, EPI_ISL_1196083, EPI_ISL_1196084, EPI_ISL_1196085, EPI_ISL_1196086, EPI_ISL_1196087, EPI_ISL_1196088, EPI_ISL_1196089, EPI_ISL_1196090, EPI_ISL_1196091, EPI_ISL_1196092, EPI_ISL_1196093, EPI_ISL_1196094, EPI_ISL_1196095, EPI_ISL_1196096, EPI_ISL_1196097, EPI_ISL_1196098, EPI_ISL_1196099, EPI_ISL_1196100, EPI_ISL_1196101, EPI_ISL_1196102 | | | |
| see above | Maryland Genomics, Institute for Genome Sciences, University of Maryland School of Medicine | Maryland Genomics, Institute for Genome Sciences, University of Maryland School of Medicine | Tallon, Luke J; Sadzewicz, Lisa D; Humphrys, Mike; Ott, Sandra; Roussey, Holly; Mehta, Aditya; Vavikolanu, Kranthi; Fraser, Claire M; Ravel, Jacques |
| EPI_ISL_1196198, EPI_ISL_1196199, EPI_ISL_1196200, EPI_ISL_1196201, EPI_ISL_1196202, EPI_ISL_1196203, EPI_ISL_1196204, EPI_ISL_1196205, EPI_ISL_1196207, EPI_ISL_1196208, EPI_ISL_1196209, EPI_ISL_1196210, EPI_ISL_1196211, EPI_ISL_1196212, EPI_ISL_1196213, EPI_ISL_1196214, EPI_ISL_1196215, EPI_ISL_1196216, EPI_ISL_1196217, EPI_ISL_1196218, EPI_ISL_1196219, EPI_ISL_1196220, EPI_ISL_1196221, EPI_ISL_1196222, EPI_ISL_1196223, EPI_ISL_1196224, EPI_ISL_1196225, EPI_ISL_1196226, EPI_ISL_1196227, EPI_ISL_1196228, EPI_ISL_1196229, EPI_ISL_1196230 | | | |
| see above | M Health Fairview | Minnesota Department of Health, Public Health Laboratory | Alexandra Lorentz, Jacob Garfin, Matt Plumb, and Xiong Wang |
| EPI_ISL_1196231, EPI_ISL_1196232, EPI_ISL_1196233, EPI_ISL_1196235, EPI_ISL_1196236, EPI_ISL_1196238, EPI_ISL_1196239, EPI_ISL_1196243, EPI_ISL_1196245, EPI_ISL_1196246, EPI_ISL_1196247, EPI_ISL_1196249, EPI_ISL_1196250, EPI_ISL_1196252, EPI_ISL_1196253, EPI_ISL_1196254, EPI_ISL_1196255, EPI_ISL_1196256, EPI_ISL_1196257, EPI_ISL_1196259, EPI_ISL_1196260, EPI_ISL_1196261, EPI_ISL_1196263, EPI_ISL_1196264, EPI_ISL_1196265, EPI_ISL_1196266, EPI_ISL_1196267, EPI_ISL_1196268, EPI_ISL_1196269, EPI_ISL_1196271, EPI_ISL_1196273, EPI_ISL_1196274, EPI_ISL_1196276, EPI_ISL_1196277, EPI_ISL_1196278 | | | |
| see above | Mayo Clinic & Mayo Clinic Laboratories | Minnesota Department of Health, Public Health Laboratory | Alexandra Lorentz, Jacob Garfin, Matt Plumb, and Xiong Wang |
| EPI_ISL_1196306, EPI_ISL_1196307, EPI_ISL_1196309, EPI_ISL_1196310, EPI_ISL_1196311, EPI_ISL_1196312, EPI_ISL_1196313, EPI_ISL_1196314, EPI_ISL_1196315, EPI_ISL_1196316, EPI_ISL_1196317, EPI_ISL_1196318, EPI_ISL_1196319, EPI_ISL_1196321, EPI_ISL_1196323, EPI_ISL_1196326, EPI_ISL_1196327, EPI_ISL_1196328, EPI_ISL_1196329, EPI_ISL_1196330, EPI_ISL_1196332, EPI_ISL_1196333, EPI_ISL_1196334, EPI_ISL_1196335, EPI_ISL_1196337, EPI_ISL_1196338, EPI_ISL_1196339, EPI_ISL_1196344, EPI_ISL_1196345, EPI_ISL_1196346, EPI_ISL_1196347, EPI_ISL_1196348, EPI_ISL_1196349, EPI_ISL_1196350, EPI_ISL_1196352, EPI_ISL_1196353, EPI_ISL_1196359, EPI_ISL_1196362, EPI_ISL_1196369, EPI_ISL_1196370, EPI_ISL_1196374, EPI_ISL_1196379, EPI_ISL_1196380, EPI_ISL_1196381, EPI_ISL_1196384 | | | |
| see above | Texas Department of State Health Services | Texas Department of State Health Services | Rashmi Tuladhar, Bonnie Oh, Jenny Zhang, Maliha Rahman, Mayela Pedrueza, Anita Pokharel, Myong Koag, Chun Wang, Rachel Lee, Grace Kubin |
| EPI_ISL_1196391, EPI_ISL_1196392, EPI_ISL_1196393, EPI_ISL_1196394, EPI_ISL_1196396, EPI_ISL_1196397, EPI_ISL_1196398, EPI_ISL_1196399, EPI_ISL_1196400, EPI_ISL_1196401, EPI_ISL_1196402, EPI_ISL_1196403, EPI_ISL_1196404, EPI_ISL_1196405, EPI_ISL_1196406, EPI_ISL_1196407, EPI_ISL_1196408, EPI_ISL_1196409, EPI_ISL_1196410, EPI_ISL_1196412 | | | |
| see above | Arizona State Public Health Laboratory | Arizona State Public Health Laboratory | Trung Huynh, Jessica Escobar, Katherine Fullerton, Nobuko Fukushima, Stacy White, Linda Getsinger, Victor Waddell |
| EPI_ISL_1196439, EPI_ISL_1196440, EPI_ISL_1196441, EPI_ISL_1196443, EPI_ISL_1196444, EPI_ISL_1196445, EPI_ISL_1196446, EPI_ISL_1196447, EPI_ISL_1196448, EPI_ISL_1196449, EPI_ISL_1196450, EPI_ISL_1196452, EPI_ISL_1196453, EPI_ISL_1196454, EPI_ISL_1196455, EPI_ISL_1196456, EPI_ISL_1196457, EPI_ISL_1196459, EPI_ISL_1196460, EPI_ISL_1196461, EPI_ISL_1196462, EPI_ISL_1196463, EPI_ISL_1196464, EPI_ISL_1196465, EPI_ISL_1196466, EPI_ISL_1196467, EPI_ISL_1196468, EPI_ISL_1196469, EPI_ISL_1196470, EPI_ISL_1196471, EPI_ISL_1196472, EPI_ISL_1196473, EPI_ISL_1196475, EPI_ISL_1196476, EPI_ISL_1196477, EPI_ISL_1196478, EPI_ISL_1196479, EPI_ISL_1196480, EPI_ISL_1196481, EPI_ISL_1196482, EPI_ISL_1196483 | | | |
| see above | New Mexico Department of Health Scientific Laboratory | New Mexico Department of Health Scientific Laboratory | Ellie Johnson, Anastacia Griego-Fisher, D'eldra Malone, Jennifer Benoit |
| EPI_ISL_1196751, EPI_ISL_1196752, EPI_ISL_1196753, EPI_ISL_1196754, EPI_ISL_1196755, EPI_ISL_1196756, EPI_ISL_1196757, EPI_ISL_1196758, EPI_ISL_1196759, EPI_ISL_1196760, EPI_ISL_1196761, EPI_ISL_1196762, EPI_ISL_1196763 | | | |
| see above | Ochsner Health | Bioinfoexperts, LLC | Rebecca Rose, Amy Feehan, David J. Nolan, Sissy Cross, David Moraga Amador, Tong Yang, Luke Caruso, Wayra Navia, Lydia Von Borstel, Xiao Hui Zhou, Julia-Garcia-Diaz, Susanna L. Lamers |
| EPI_ISL_1196775, EPI_ISL_1196776, EPI_ISL_1196777, EPI_ISL_1196778, EPI_ISL_1196779, EPI_ISL_1196780, EPI_ISL_1196781, EPI_ISL_1196782, EPI_ISL_1196783, EPI_ISL_1196784, EPI_ISL_1196785, EPI_ISL_1196786, EPI_ISL_1196787, EPI_ISL_1196788, EPI_ISL_1196789, EPI_ISL_1196790, EPI_ISL_1196791 | | | |
| see above | Alameda County Public Health Department | Alameda County Public Health Department | Kristna Hsieh, Maria Elma, Kevin Libuit |
| EPI_ISL_1196868 | IL Dept. of Public Health Springfield Laboratory | Pathogen Discovery, Respiratory Viruses Branch, Division of Viral Diseases, Centers for Disease Control and Prevention | Ying Tao, Jing Zhang, Yan Li, Brian Lynch, Anna Kelleher, Krista Queen, Anna Uehara, Peter Cook, Han Jia Justin Ng, Clinton R. Paden, Haibin Wang, Suxiang Tong |
| EPI_ISL_1196869 | OK Public Health Laboratory, Oklahoma State DOH | Pathogen Discovery, Respiratory Viruses Branch, Division of Viral Diseases, Centers for Disease Control and Prevention | Ying Tao, Jing Zhang, Yan Li, Brian Lynch, Anna Kelleher, Krista Queen, Anna Uehara, Peter Cook, Han Jia Justin Ng, Clinton R. Paden, Haibin Wang, Suxiang Tong |
| EPI_ISL_1196871 | NC State Laboratory of Public Health | Pathogen Discovery, Respiratory Viruses Branch, Division of Viral Diseases, Centers for Disease Control and Prevention | Ying Tao, Jing Zhang, Yan Li, Brian Lynch, Anna Kelleher, Krista Queen, Anna Uehara, Peter Cook, Han Jia Justin Ng, Clinton R. Paden, Haibin Wang, Suxiang Tong |
| EPI_ISL_1196872, EPI_ISL_1196873, EPI_ISL_1196874, EPI_ISL_1196876, EPI_ISL_1196877, EPI_ISL_1196878, EPI_ISL_1196879, EPI_ISL_1196880, EPI_ISL_1196881, EPI_ISL_1196882, EPI_ISL_1196883, EPI_ISL_1196884, EPI_ISL_1196885 | | | |
| see above | LA Office of Public Health Laboratories | Pathogen Discovery, Respiratory Viruses Branch, Division of Viral Diseases, Centers for Disease Control and Prevention | Ying Tao, Jing Zhang, Yan Li, Brian Lynch, Anna Kelleher, Krista Queen, Anna Uehara, Peter Cook, Han Jia Justin Ng, Clinton R. Paden, Haibin Wang, Suxiang Tong |
| EPI_ISL_1196886 | IL Department of Public Health Chicago Laboratory | Pathogen Discovery, Respiratory Viruses Branch, Division of Viral Diseases, Centers for Disease Control and Prevention | Ying Tao, Jing Zhang, Yan Li, Brian Lynch, Anna Kelleher, Krista Queen, Anna Uehara, Peter Cook, Han Jia Justin Ng, Clinton R. Paden, Haibin Wang, Suxiang Tong |
| EPI_ISL_1196887 | IL Dept. of Public Health Carbondale Laboratory | Pathogen Discovery, Respiratory Viruses Branch, Division of Viral Diseases, Centers for Disease Control and Prevention | Ying Tao, Jing Zhang, Yan Li, Brian Lynch, Anna Kelleher, Krista Queen, Anna Uehara, Peter Cook, Han Jia Justin Ng, Clinton R. Paden, Haibin Wang, Suxiang Tong |
| EPI_ISL_1196888, EPI_ISL_1196889, EPI_ISL_1196890 | RI State Health Laboratories | Pathogen Discovery, Respiratory Viruses Branch, Division of Viral Diseases, Centers for Disease Control and Prevention | Ying Tao, Jing Zhang, Yan Li, Brian Lynch, Anna Kelleher, Krista Queen, Anna Uehara, Peter Cook, Han Jia Justin Ng, Clinton R. Paden, Haibin Wang, Suxiang Tong |
| EPI_ISL_1196891, EPI_ISL_1196892 | TN Division of Laboratory Services | Pathogen Discovery, Respiratory Viruses Branch, Division of Viral Diseases, Centers for Disease Control and Prevention | Ying Tao, Jing Zhang, Yan Li, Brian Lynch, Anna Kelleher, Krista Queen, Anna Uehara, Peter Cook, Han Jia Justin Ng, Clinton R. Paden, Haibin Wang, Suxiang Tong |
| EPI_ISL_1196893, EPI_ISL_1196894 | MT Public Health Laboratory | Pathogen Discovery, Respiratory Viruses Branch, Division of Viral Diseases, Centers for Disease Control and Prevention | Ying Tao, Jing Zhang, Yan Li, Brian Lynch, Anna Kelleher, Krista Queen, Anna Uehara, Peter Cook, Han Jia Justin Ng, Clinton R. Paden, Haibin Wang, Suxiang Tong |
| EPI_ISL_1196895, EPI_ISL_1196896 | IL Dept. of Public Health Springfield Laboratory | Pathogen Discovery, Respiratory Viruses Branch, Division of Viral Diseases, Centers for Disease Control and Prevention | Ying Tao, Jing Zhang, Yan Li, Brian Lynch, Anna Kelleher, Krista Queen, Anna Uehara, Peter Cook, Han Jia Justin Ng, Clinton R. Paden, Haibin Wang, Suxiang Tong |
| EPI_ISL_1196897, EPI_ISL_1196898, EPI_ISL_1196899, EPI_ISL_1196900, EPI_ISL_1196901, EPI_ISL_1196902, EPI_ISL_1196903, EPI_ISL_1196904, EPI_ISL_1196905, EPI_ISL_1196906, EPI_ISL_1196907, EPI_ISL_1196908, EPI_ISL_1196909, EPI_ISL_1196910, EPI_ISL_1196911, EPI_ISL_1196912, EPI_ISL_1196913, EPI_ISL_1196914, EPI_ISL_1196915, EPI_ISL_1196916, EPI_ISL_1196917, EPI_ISL_1196918, EPI_ISL_1196919, EPI_ISL_1196920, EPI_ISL_1196921, EPI_ISL_1196922, EPI_ISL_1196923, EPI_ISL_1196924, EPI_ISL_1196925, EPI_ISL_1196926, EPI_ISL_1196927, EPI_ISL_1196928, EPI_ISL_1196929 | | | |
| see above | University of Michigan Clinical Microbiology Laboratory | Lauring Lab, University of Michigan, Department of Microbiology and Immunology | Valesano |
| EPI_ISL_1197042 | USC Clinical Lab | Los Angeles County Public Health Laboratories | P. Hemarajata et al. |
| EPI_ISL_1197043 | Martin Luther King Jr. Community Hospital | Los Angeles County Public Health Laboratories | P. Hemarajata et al. |
| EPI_ISL_1197044 | USC Clinical Lab | Los Angeles County Public Health | P. Hemarajata et al. |

| | | | |
|---|---|---|---|
| | | Laboratories | |
| EPI_ISL_1197045 | Martin Luther King Jr. Community Hospital | Los Angeles County Public Health Laboratories | P. Hemarajata et al. |
| EPI_ISL_1197046, EPI_ISL_1197047 | USC Clinical Lab | Los Angeles County Public Health Laboratories | P. Hemarajata et al. |
| EPI_ISL_1197048 | Martin Luther King Jr. Community Hospital | Los Angeles County Public Health Laboratories | P. Hemarajata et al. |
| EPI_ISL_1197049 | USC Clinical Lab | Los Angeles County Public Health Laboratories | P. Hemarajata et al. |
| EPI_ISL_1197050 | Martin Luther King Jr. Community Hospital | Los Angeles County Public Health Laboratories | P. Hemarajata et al. |
| EPI_ISL_1197051 | USC Clinical Lab | Los Angeles County Public Health Laboratories | P. Hemarajata et al. |
| EPI_ISL_1197052 | Martin Luther King Jr. Community Hospital | Los Angeles County Public Health Laboratories | P. Hemarajata et al. |
| EPI_ISL_1197053 | USC Clinical Lab | Los Angeles County Public Health Laboratories | P. Hemarajata et al. |
| EPI_ISL_1197054, EPI_ISL_1197055, EPI_ISL_1197056, EPI_ISL_1197057 | Martin Luther King Jr. Community Hospital | Los Angeles County Public Health Laboratories | P. Hemarajata et al. |
| EPI_ISL_1197058, EPI_ISL_1197059, EPI_ISL_1197060, EPI_ISL_1197061 | Flow Diagnostics Lab | Los Angeles County Public Health Laboratories | P. Hemarajata et al. |
| EPI_ISL_1197131, EPI_ISL_1197132, EPI_ISL_1197133, EPI_ISL_1197134, EPI_ISL_1197135, EPI_ISL_1197136, EPI_ISL_1197138, EPI_ISL_1197139, EPI_ISL_1197140, EPI_ISL_1197141, EPI_ISL_1197144, EPI_ISL_1197145, EPI_ISL_1197146, EPI_ISL_1197147, EPI_ISL_1197148 | | | |
| see above | USC Clinical Lab | Los Angeles County Public Health Laboratories | P. Hemarajata et al. |
| EPI_ISL_1197149 | UCLA Clinical Micro Lab | Los Angeles County Public Health Laboratories | P. Hemarajata et al. |
| EPI_ISL_1197236, EPI_ISL_1197691 | Ohio Department of Health Laboratory | Ohio Department of Health Laboratory | Holmes, Jennifer; Eric Brandt, Keoni Omura, Glen McGillivary, Caitlin McDonnell, Jade Mowery, Stephanie Mccracken, Tyler Payne, Kirtana Ramadugu, Erica Leasure, Brent Lee, Kelsey Florek, Heather Blankenship, Allison Black, Quanta Brown, and Tammy Bannerman |
| EPI_ISL_1197934, EPI_ISL_1197935, EPI_ISL_1197936, EPI_ISL_1197937, EPI_ISL_1197938, EPI_ISL_1197939, EPI_ISL_1197940, EPI_ISL_1197941, EPI_ISL_1197942, EPI_ISL_1197943, EPI_ISL_1197944, EPI_ISL_1197945, EPI_ISL_1197946, EPI_ISL_1197947, EPI_ISL_1197948, EPI_ISL_1197949, EPI_ISL_1197950, EPI_ISL_1197951, EPI_ISL_1197952, EPI_ISL_1197953, EPI_ISL_1197954, EPI_ISL_1197955, EPI_ISL_1197956, EPI_ISL_1197957, EPI_ISL_1197958, EPI_ISL_1197959, EPI_ISL_1197960, EPI_ISL_1197961, EPI_ISL_1197962, EPI_ISL_1197963, EPI_ISL_1197964, EPI_ISL_1197965, EPI_ISL_1197966, EPI_ISL_1197967, EPI_ISL_1197968, EPI_ISL_1197969, EPI_ISL_1197970, EPI_ISL_1197971, EPI_ISL_1197972, EPI_ISL_1197973, EPI_ISL_1197974, EPI_ISL_1197975, EPI_ISL_1197976, EPI_ISL_1197977, EPI_ISL_1197978, EPI_ISL_1197979, EPI_ISL_1197980, EPI_ISL_1197981, EPI_ISL_1197982, EPI_ISL_1197983, EPI_ISL_1197984, EPI_ISL_1197985, EPI_ISL_1197986, EPI_ISL_1197987, EPI_ISL_1197988, EPI_ISL_1197989, EPI_ISL_1197990, EPI_ISL_1197991, EPI_ISL_1197992, EPI_ISL_1197993, EPI_ISL_1197994, EPI_ISL_1197995, EPI_ISL_1197996, EPI_ISL_1197997, EPI_ISL_1197998, EPI_ISL_1197999, EPI_ISL_1198000, EPI_ISL_1198001, EPI_ISL_1198002, EPI_ISL_1198003 | | | |
| see above | New Mexico Department of Health Scientific Laboratory | Center for Global Health, University of New Mexico Health Sciences Center | Daryl Domman, Kurt Schwalm, Twila Kunde, Joseph Hicks, Anastacia Griego, Michael Edwards, Darrell Dinwiddie |
| EPI_ISL_1198004, EPI_ISL_1198008, EPI_ISL_1198793, EPI_ISL_1198797, EPI_ISL_1198798, EPI_ISL_1198799, EPI_ISL_1198804, EPI_ISL_1198805 | Ohio Department of Health Laboratory | Ohio Department of Health Laboratory | Holmes, Jennifer; Eric Brandt, Keoni Omura, Glen McGillivary, Caitlin McDonnell, Jade Mowery, Stephanie Mccracken, Tyler Payne, Kirtana Ramadugu, Erica Leasure, Brent Lee, Kelsey Florek, Heather Blankenship, Allison Black, Quanta Brown, and Tammy Bannerman |
| EPI_ISL_1200518 | DOHMH Chelsea | New York City Public Health Laboratory | Jade Wang, et al. |
| EPI_ISL_1200519 | HealthQuest Esoterics | New York City Public Health Laboratory | Jade Wang, et al. |
| EPI_ISL_1200520, EPI_ISL_1200521 | DOHMH Jamaica | New York City Public Health Laboratory | Jade Wang, et al. |
| EPI_ISL_1200522 | DOHMH Central Harlem | New York City Public Health Laboratory | Jade Wang, et al. |
| EPI_ISL_1200523 | DOHMH Fort Greene | New York City Public Health Laboratory | Jade Wang, et al. |
| EPI_ISL_1200524 | DOHMH Crown Heights | New York City Public Health Laboratory | Jade Wang, et al. |
| EPI_ISL_1200525 | DOHMH Riverside | New York City Public Health Laboratory | Jade Wang, et al. |
| EPI_ISL_1200526, EPI_ISL_1200527 | OCME Office Of Chief Medical Examiner | New York City Public Health Laboratory | Jade Wang, et al. |
| EPI_ISL_1200528, EPI_ISL_1200529, EPI_ISL_1200530 | DOHMH Chelsea | New York City Public Health Laboratory | Jade Wang, et al. |
| EPI_ISL_1200531, EPI_ISL_1200532 | OCME Office Of Chief Medical Examiner | New York City Public Health Laboratory | Jade Wang, et al. |
| EPI_ISL_1200533 | DOHMH Corona | New York City Public Health Laboratory | Jade Wang, et al. |
| EPI_ISL_1200534 | DOHMH Chelsea | New York City Public Health Laboratory | Jade Wang, et al. |
| EPI_ISL_1200535, EPI_ISL_1200536 | DOHMH Jamaica | New York City Public Health Laboratory | Jade Wang, et al. |
| EPI_ISL_1200537 | DOHMH Riverside | New York City Public Health Laboratory | Jade Wang, et al. |
| EPI_ISL_1200538 | DOHMH PHL | New York City Public Health Laboratory | Jade Wang, et al. |
| EPI_ISL_1200539 | OCME Office Of Chief Medical Examiner | New York City Public Health Laboratory | Jade Wang, et al. |
| EPI_ISL_1200540 | DOHMH Corona | New York City Public Health Laboratory | Jade Wang, et al. |
| EPI_ISL_1200541 | DOHMH Central Harlem | New York City Public Health Laboratory | Jade Wang, et al. |
| EPI_ISL_1200542 | DOHMH Jamaica | New York City Public Health Laboratory | Jade Wang, et al. |
| EPI_ISL_1200543 | DOHMH Crown Heights | New York City Public Health Laboratory | Jade Wang, et al. |
| EPI_ISL_1200544 | DOHMH Jamaica | New York City Public Health Laboratory | Jade Wang, et al. |
| EPI_ISL_1200545, EPI_ISL_1200546 | DOHMH Corona | New York City Public Health Laboratory | Jade Wang, et al. |
| EPI_ISL_1200547 | DOHMH Jamaica | New York City Public Health Laboratory | Jade Wang, et al. |
| EPI_ISL_1200548 | DOHMH PHL | New York City Public Health Laboratory | Jade Wang, et al. |
| EPI_ISL_1200549 | OCME Office Of Chief Medical Examiner | New York City Public Health Laboratory | Jade Wang, et al. |
| EPI_ISL_1200550 | DOHMH Morrisania | New York City Public Health Laboratory | Jade Wang, et al. |
| EPI_ISL_1200551 | DOHMH Jamaica | New York City Public Health Laboratory | Jade Wang, et al. |
| EPI_ISL_1200552 | DOHMH Central Harlem | New York City Public Health Laboratory | Jade Wang, et al. |
| EPI_ISL_1200553 | DOHMH Jamaica | New York City Public Health Laboratory | Jade Wang, et al. |
| EPI_ISL_1200554 | DOHMH Crown Heights | New York City Public Health Laboratory | Jade Wang, et al. |
| EPI_ISL_1200555 | OCME Office Of Chief Medical Examiner | New York City Public Health Laboratory | Jade Wang, et al. |
| EPI_ISL_1200556 | DOHMH Central Harlem | New York City Public Health Laboratory | Jade Wang, et al. |
| EPI_ISL_1200557, EPI_ISL_1200558, EPI_ISL_1200559 | DOHMH Morrisania | New York City Public Health Laboratory | Jade Wang, et al. |

| | | | |
|---|---|---|---|
| EPI_ISL_1200560 | DOHMH Chelsea | New York City Public Health Laboratory | Jade Wang, et al. |
| EPI_ISL_1200561 | Rockefeller University Hospital | New York City Public Health Laboratory | Jade Wang, et al. |
| EPI_ISL_1200562 | Department of Homeless Services | New York City Public Health Laboratory | Jade Wang, et al. |
| EPI_ISL_1200563 | DOHMH Jamaica | New York City Public Health Laboratory | Jade Wang, et al. |
| EPI_ISL_1200564 | DOHMH Chelsea | New York City Public Health Laboratory | Jade Wang, et al. |
| EPI_ISL_1200565 | DOHMH Corona | New York City Public Health Laboratory | Jade Wang, et al. |
| EPI_ISL_1200566 | DOHMH PHL | New York City Public Health Laboratory | Jade Wang, et al. |
| EPI_ISL_1200567 | DOHMH Morrisania | New York City Public Health Laboratory | Jade Wang, et al. |
| EPI_ISL_1200568, EPI_ISL_1200569, EPI_ISL_1200570 | DOHMH Corona | New York City Public Health Laboratory | Jade Wang, et al. |
| EPI_ISL_1200571 | DOHMH Crown Heights | New York City Public Health Laboratory | Jade Wang, et al. |
| EPI_ISL_1200572, EPI_ISL_1200573 | DOHMH Jamaica | New York City Public Health Laboratory | Jade Wang, et al. |
| EPI_ISL_1200574 | DOHMH Chelsea | New York City Public Health Laboratory | Jade Wang, et al. |
| EPI_ISL_1200575 | DOHMH Morrisania | New York City Public Health Laboratory | Jade Wang, et al. |
| EPI_ISL_1200576 | OCME Office Of Chief Medical Examiner | New York City Public Health Laboratory | Jade Wang, et al. |
| EPI_ISL_1200577 | DOHMH Central Harlem | New York City Public Health Laboratory | Jade Wang, et al. |
| EPI_ISL_1200578, EPI_ISL_1200579, EPI_ISL_1200580 | DOHMH Morrisania | New York City Public Health Laboratory | Jade Wang, et al. |
| EPI_ISL_1200581 | DOHMH Jamaica | New York City Public Health Laboratory | Jade Wang, et al. |
| EPI_ISL_1200582 | DOHMH Morrisania | New York City Public Health Laboratory | Jade Wang, et al. |
| EPI_ISL_1200583 | DOHMH Jamaica | New York City Public Health Laboratory | Jade Wang, et al. |
| EPI_ISL_1200584 | DOHMH Chelsea | New York City Public Health Laboratory | Jade Wang, et al. |
| EPI_ISL_1200585 | DOHMH Jamaica | New York City Public Health Laboratory | Jade Wang, et al. |
| EPI_ISL_1200586, EPI_ISL_1200587 | DOHMH PHL | New York City Public Health Laboratory | Jade Wang, et al. |
| EPI_ISL_1200588, EPI_ISL_1200589 | DOHMH Morrisania | New York City Public Health Laboratory | Jade Wang, et al. |
| EPI_ISL_1200590 | DOHMH Jamaica | New York City Public Health Laboratory | Jade Wang, et al. |
| EPI_ISL_1200591, EPI_ISL_1200592, EPI_ISL_1200593 | DOHMH Corona | New York City Public Health Laboratory | Jade Wang, et al. |
| EPI_ISL_1200594 | DOHMH Central Harlem | New York City Public Health Laboratory | Jade Wang, et al. |
| EPI_ISL_1200595, EPI_ISL_1200596 | DOHMH Jamaica | New York City Public Health Laboratory | Jade Wang, et al. |
| EPI_ISL_1200597, EPI_ISL_1200598 | DOHMH Corona | New York City Public Health Laboratory | Jade Wang, et al. |
| EPI_ISL_1200599 | Department of Homeless Services | New York City Public Health Laboratory | Jade Wang, et al. |
| EPI_ISL_1200600 | DOHMH Morrisania | New York City Public Health Laboratory | Jade Wang, et al. |
| EPI_ISL_1200601, EPI_ISL_1200602 | DOHMH PHL | New York City Public Health Laboratory | Jade Wang, et al. |
| EPI_ISL_1200603 | DOHMH Corona | New York City Public Health Laboratory | Jade Wang, et al. |
| EPI_ISL_1200604 | DOHMH Morrisania | New York City Public Health Laboratory | Jade Wang, et al. |
| EPI_ISL_1200605 | DOHMH Crown Heights | New York City Public Health Laboratory | Jade Wang, et al. |
| EPI_ISL_1200606 | DOHMH Riverside | New York City Public Health Laboratory | Jade Wang, et al. |
| EPI_ISL_1200607, EPI_ISL_1200608, EPI_ISL_1200609, EPI_ISL_1200610 | OCME Office Of Chief Medical Examiner | New York City Public Health Laboratory | Jade Wang, et al. |
| EPI_ISL_1200611 | DOHMH Riverside | New York City Public Health Laboratory | Jade Wang, et al. |
| EPI_ISL_1200612 | DOHMH Morrisania | New York City Public Health Laboratory | Jade Wang, et al. |
| EPI_ISL_1200613, EPI_ISL_1200614 | DOHMH Corona | New York City Public Health Laboratory | Jade Wang, et al. |
| EPI_ISL_1200615, EPI_ISL_1200616, EPI_ISL_1200617, EPI_ISL_1200618 | DOHMH Jamaica | New York City Public Health Laboratory | Jade Wang, et al. |
| EPI_ISL_1200619 | DOHMH PHL | New York City Public Health Laboratory | Jade Wang, et al. |
| EPI_ISL_1200620 | DOHMH Fort Greene | New York City Public Health Laboratory | Jade Wang, et al. |
| EPI_ISL_1200621, EPI_ISL_1200622 | DOHMH Central Harlem | New York City Public Health Laboratory | Jade Wang, et al. |
| EPI_ISL_1200623 | DOHMH PHL | New York City Public Health Laboratory | Jade Wang, et al. |
| EPI_ISL_1200624, EPI_ISL_1200625 | DOHMH Central Harlem | New York City Public Health Laboratory | Jade Wang, et al. |
| EPI_ISL_1200626 | DOHMH Corona | New York City Public Health Laboratory | Jade Wang, et al. |
| EPI_ISL_1200627 | OCME Office Of Chief Medical Examiner | New York City Public Health Laboratory | Jade Wang, et al. |
| EPI_ISL_1200655 | AIDS Vaccine Research Laboratories | AIDS Vaccine Research Laboratories | Gage Moreno, Katarina Braun, AVRL et al. |
| EPI_ISL_1200860, EPI_ISL_1200861, EPI_ISL_1200862, EPI_ISL_1200863, EPI_ISL_1200864, EPI_ISL_1200865, EPI_ISL_1200866, EPI_ISL_1200867, EPI_ISL_1200868, EPI_ISL_1200869, EPI_ISL_1200870, EPI_ISL_1200871, EPI_ISL_1200872, EPI_ISL_1200873, EPI_ISL_1200874, EPI_ISL_1200875, EPI_ISL_1200876, EPI_ISL_1200877, EPI_ISL_1200878, EPI_ISL_1200879, EPI_ISL_1200880, EPI_ISL_1200881, EPI_ISL_1200882, EPI_ISL_1200883, EPI_ISL_1200884, EPI_ISL_1200885, EPI_ISL_1200886, EPI_ISL_1200887, EPI_ISL_1200888, EPI_ISL_1200889, EPI_ISL_1200890, EPI_ISL_1200891, EPI_ISL_1200892, EPI_ISL_1200893, EPI_ISL_1200894, EPI_ISL_1200895, EPI_ISL_1200896, EPI_ISL_1200897, EPI_ISL_1200898, EPI_ISL_1200899, EPI_ISL_1200900, EPI_ISL_1200901, EPI_ISL_1200902, EPI_ISL_1200903, EPI_ISL_1200904, EPI_ISL_1200905, EPI_ISL_1200906, EPI_ISL_1200907, EPI_ISL_1200908, EPI_ISL_1200909, EPI_ISL_1200910, EPI_ISL_1200911, EPI_ISL_1200912, EPI_ISL_1200913, EPI_ISL_1200914, EPI_ISL_1200915, EPI_ISL_1200916, EPI_ISL_1200917, EPI_ISL_1200918, EPI_ISL_1200919, EPI_ISL_1200920, EPI_ISL_1200921, EPI_ISL_1200922, EPI_ISL_1200923, EPI_ISL_1200924, EPI_ISL_1200925, EPI_ISL_1200926, EPI_ISL_1200927, EPI_ISL_1200928, EPI_ISL_1200929, EPI_ISL_1200930, EPI_ISL_1200931, EPI_ISL_1200932, EPI_ISL_1200933, EPI_ISL_1200934, EPI_ISL_1200935, EPI_ISL_1200936, EPI_ISL_1200937, EPI_ISL_1200938, EPI_ISL_1200939, EPI_ISL_1200940, EPI_ISL_1200941, EPI_ISL_1200942, EPI_ISL_1200943, EPI_ISL_1200944, EPI_ISL_1200945, EPI_ISL_1200946, EPI_ISL_1200947, EPI_ISL_1200948, EPI_ISL_1200949, EPI_ISL_1200950, EPI_ISL_1200951, EPI_ISL_1200952, EPI_ISL_1200953, EPI_ISL_1200954, EPI_ISL_1200955, EPI_ISL_1200956, EPI_ISL_1200957, EPI_ISL_1200958, EPI_ISL_1200959, EPI_ISL_1200960, EPI_ISL_1200961, EPI_ISL_1200962, EPI_ISL_1200963, EPI_ISL_1200964, EPI_ISL_1200965, EPI_ISL_1200966, EPI_ISL_1200967, EPI_ISL_1200968, EPI_ISL_1200969, EPI_ISL_1200970, EPI_ISL_1200971, EPI_ISL_1200972, EPI_ISL_1200973, EPI_ISL_1200974, EPI_ISL_1200975, EPI_ISL_1200976, EPI_ISL_1200977, EPI_ISL_1200978, EPI_ISL_1200979, EPI_ISL_1200980, EPI_ISL_1200981, EPI_ISL_1200982, EPI_ISL_1200983, EPI_ISL_1200984, EPI_ISL_1200985, EPI_ISL_1200986, EPI_ISL_1200987, EPI_ISL_1200988, EPI_ISL_1200989, EPI_ISL_1200990, EPI_ISL_1200991, EPI_ISL_1200992, EPI_ISL_1200993, EPI_ISL_1200994, EPI_ISL_1200995, EPI_ISL_1200996, EPI_ISL_1200997, EPI_ISL_1200998, EPI_ISL_1200999, EPI_ISL_1201000, EPI_ISL_1201001 | | | |
| see above | University of Michigan Clinical Microbiology Laboratory | Lauring Lab, University of Michigan, Department of Microbiology and Immunology | Valesano |
| EPI_ISL_1201132, EPI_ISL_1201133, EPI_ISL_1201134, EPI_ISL_1201135, EPI_ISL_1201136, EPI_ISL_1201137, EPI_ISL_1201138, EPI_ISL_1201139, EPI_ISL_1201140, EPI_ISL_1201141, EPI_ISL_1201142, EPI_ISL_1201143, EPI_ISL_1201144, EPI_ISL_1201145, EPI_ISL_1201146, EPI_ISL_1201147, EPI_ISL_1201148, EPI_ISL_1201149, EPI_ISL_1201150, EPI_ISL_1201151, EPI_ISL_1201152, EPI_ISL_1201153, EPI_ISL_1201154, EPI_ISL_1201155, EPI_ISL_1201156, EPI_ISL_1201157, EPI_ISL_1201158, EPI_ISL_1201159, EPI_ISL_1201160, EPI_ISL_1201161, EPI_ISL_1201162, EPI_ISL_1201163, EPI_ISL_1201164, EPI_ISL_1201165, EPI_ISL_1201166, EPI_ISL_1201167, EPI_ISL_1201168, EPI_ISL_1201169, EPI_ISL_1201170, EPI_ISL_1201171, EPI_ISL_1201172, EPI_ISL_1201173, EPI_ISL_1201174, EPI_ISL_1201175, EPI_ISL_1201176, EPI_ISL_1201177, EPI_ISL_1201178, EPI_ISL_1201179, EPI_ISL_1201180, EPI_ISL_1201181, EPI_ISL_1201182, EPI_ISL_1201183, EPI_ISL_1201184, EPI_ISL_1201185, EPI_ISL_1201186, EPI_ISL_1201187, EPI_ISL_1201188, EPI_ISL_1201189, EPI_ISL_1201190, EPI_ISL_1201191, EPI_ISL_1201192, EPI_ISL_1201193, EPI_ISL_1201194, EPI_ISL_1201195, EPI_ISL_1201196, EPI_ISL_1201197, EPI_ISL_1201198, EPI_ISL_1201199, EPI_ISL_1201200, EPI_ISL_1201201, EPI_ISL_1201202 | | | |
| see above | UW Virology Lab | UW Virology Lab | Pavitra Roychoudhury, Hong Xie, Lasata Shrestha, Shah Mohamed Bakhash, Michelle Lin, Noah Baker, Sean Ellis, Saraswathi Sathees, Meei-Li Huang, Keith R Jerome, Alexander Greninger |
| EPI_ISL_1201287 | University of Michigan Clinical Microbiology Laboratory | Lauring Lab, University of Michigan, Department of Microbiology and Immunology | Valesano |
| EPI_ISL_1201288, EPI_ISL_1201289, | Nebraska Public Health Laboratory | NPHL COVID-19 Response Team | NPHL COVID-19 Response Team |

| | | | |
|---|---|---|---|
| EPI_ISL_1201290, EPI_ISL_1201291, EPI_ISL_1201292, EPI_ISL_1201293, EPI_ISL_1201294, EPI_ISL_1201295, EPI_ISL_1201296 | | | |
| EPI_ISL_1201297 | WVU Rapid Development Laboratory | WVU and Marshall University Combined Genomics Core Facilities | James Denvir, Peter Stoilov, Peter Perrotta, Wesley Kimble, Ryan Percifield |
| EPI_ISL_1201299, EPI_ISL_1201300, EPI_ISL_1201301, EPI_ISL_1201302, EPI_ISL_1201303, EPI_ISL_1201304, EPI_ISL_1201305, EPI_ISL_1201306, EPI_ISL_1201307, EPI_ISL_1201308, EPI_ISL_1201309, EPI_ISL_1201310, EPI_ISL_1201311, EPI_ISL_1201312, EPI_ISL_1201313, EPI_ISL_1201314, EPI_ISL_1201315, EPI_ISL_1201316, EPI_ISL_1201317, EPI_ISL_1201318, EPI_ISL_1201319, EPI_ISL_1201320 | | | |
| see above | Wyoming Public Health Laboratory | Wyoming Public Health Laboratory | Jim Mildenberger, Wanda Manley, Noah Hull, Taylor Fearing, Lynette Gumbleton, Channing Weber, Ashley Norberg, Chayse Rowley, Marley Goetz, Brian Dominguez, Elliot Thomasson, Cari Sloma, and Rob Christensen |
| EPI_ISL_1201321, EPI_ISL_1201322, EPI_ISL_1201323, EPI_ISL_1201324, EPI_ISL_1201325, EPI_ISL_1201326, EPI_ISL_1201327, EPI_ISL_1201328, EPI_ISL_1201329, EPI_ISL_1201330, EPI_ISL_1201331, EPI_ISL_1201332, EPI_ISL_1201335, EPI_ISL_1201336, EPI_ISL_1201337, EPI_ISL_1201338, EPI_ISL_1201339, EPI_ISL_1201340, EPI_ISL_1201341, EPI_ISL_1201342, EPI_ISL_1201343, EPI_ISL_1201344, EPI_ISL_1201345, EPI_ISL_1201346, EPI_ISL_1201347, EPI_ISL_1201348, EPI_ISL_1201349, EPI_ISL_1201350, EPI_ISL_1201351, EPI_ISL_1201352, EPI_ISL_1201353, EPI_ISL_1201354, EPI_ISL_1201355, EPI_ISL_1201356, EPI_ISL_1201357, EPI_ISL_1201358, EPI_ISL_1201359, EPI_ISL_1201360, EPI_ISL_1201361, EPI_ISL_1201362, EPI_ISL_1201363, EPI_ISL_1201364, EPI_ISL_1201365, EPI_ISL_1201366, EPI_ISL_1201367, EPI_ISL_1201368, EPI_ISL_1201369, EPI_ISL_1201370, EPI_ISL_1201371, EPI_ISL_1201372, EPI_ISL_1201373, EPI_ISL_1201374, EPI_ISL_1201375, EPI_ISL_1201376, EPI_ISL_1201377, EPI_ISL_1201378, EPI_ISL_1201379, EPI_ISL_1201380, EPI_ISL_1201381, EPI_ISL_1201382, EPI_ISL_1201383, EPI_ISL_1201384, EPI_ISL_1201385, EPI_ISL_1201386, EPI_ISL_1201387, EPI_ISL_1201388, EPI_ISL_1201389, EPI_ISL_1201390, EPI_ISL_1201391, EPI_ISL_1201392, EPI_ISL_1201393, EPI_ISL_1201394, EPI_ISL_1201395, EPI_ISL_1201396, EPI_ISL_1201397, EPI_ISL_1201398, EPI_ISL_1201399, EPI_ISL_1201400, EPI_ISL_1201401, EPI_ISL_1201402, EPI_ISL_1201403, EPI_ISL_1201404, EPI_ISL_1201405, EPI_ISL_1201406, EPI_ISL_1201407, EPI_ISL_1201408 | | | |
| see above | Texas Department of State Health Services | Texas Department of State Health Services | Rashmi Tuladhar, Bonnie Oh, Jenny Zhang, Maliha Rahman, Mayela Pedrueza, Anita Pokharel, Myong Koag, Chun Wang, Rachel Lee, Grace Kubin |
| EPI_ISL_1201409, EPI_ISL_1201410, EPI_ISL_1201411, EPI_ISL_1201412, EPI_ISL_1201413, EPI_ISL_1201414, EPI_ISL_1201415, EPI_ISL_1201416, EPI_ISL_1201417, EPI_ISL_1201418, EPI_ISL_1201419, EPI_ISL_1201420, EPI_ISL_1201421, EPI_ISL_1201422, EPI_ISL_1201423, EPI_ISL_1201424, EPI_ISL_1201425, EPI_ISL_1201426 | | | |
| see above | Wyoming Public Health Laboratory | Wyoming Public Health Laboratory | Jim Mildenberger, Wanda Manley, Noah Hull, Taylor Fearing, Lynette Gumbleton, Channing Weber, Ashley Norberg, Chayse Rowley, Marley Goetz, Brian Dominguez, Elliot Thomasson, Cari Sloma, and Rob Christensen |
| EPI_ISL_1201441, EPI_ISL_1201444, EPI_ISL_1201445, EPI_ISL_1201446, EPI_ISL_1201447, EPI_ISL_1201448, EPI_ISL_1201449, EPI_ISL_1201450, EPI_ISL_1201451, EPI_ISL_1201452, EPI_ISL_1201453, EPI_ISL_1201454, EPI_ISL_1201455, EPI_ISL_1201456, EPI_ISL_1201457, EPI_ISL_1201458, EPI_ISL_1201459, EPI_ISL_1201460 | | | |
| see above | Arizona State Public Health Laboratory | Arizona State Public Health Laboratory | Trung Huynh, Jessica Escobar, Katherine Fullerton, Nobuko Fukushima, Stacy White, Linda Getsinger, Victor Waddell |
| EPI_ISL_1201461, EPI_ISL_1201462, EPI_ISL_1201463, EPI_ISL_1201464, EPI_ISL_1201465, EPI_ISL_1201466, EPI_ISL_1201467, EPI_ISL_1201468, EPI_ISL_1201469, EPI_ISL_1201470, EPI_ISL_1201471, EPI_ISL_1201472, EPI_ISL_1201473, EPI_ISL_1201474, EPI_ISL_1201475, EPI_ISL_1201476, EPI_ISL_1201477, EPI_ISL_1201478, EPI_ISL_1201479 | | | |
| see above | Wyoming Public Health Laboratory | Wyoming Public Health Laboratory | Jim Mildenberger, Wanda Manley, Noah Hull, Taylor Fearing, Lynette Gumbleton, Channing Weber, Ashley Norberg, Chayse Rowley, Marley Goetz, Brian Dominguez, Elliot Thomasson, Cari Sloma, and Rob Christensen |
| EPI_ISL_1201486 | INNO Diagnostics Reference Laboratory | RCMI-Center for Research Resources, Ponce Research Institute | Vanessa Rivera-Amill, Andrea Arias-García, Raphael Sánchez-Torres |
| EPI_ISL_1201559, EPI_ISL_1201560 | USC Clinical Lab | Los Angeles County Public Health Laboratories | P. Hemarajata et al. |
| EPI_ISL_1201779, EPI_ISL_1201780 | New Mexico Department of Health Scientific Laboratory | New Mexico Department of Health Scientific Laboratory | Ellie Johnson, Anastacia Griego-Fisher, D'eldra Malone, Jennifer Benoit |
| EPI_ISL_1201898, EPI_ISL_1201899, EPI_ISL_1201900, EPI_ISL_1201901, EPI_ISL_1201902, EPI_ISL_1201903, EPI_ISL_1201904, EPI_ISL_1201905, EPI_ISL_1201906, EPI_ISL_1201907, EPI_ISL_1201908, EPI_ISL_1201909, EPI_ISL_1201910, EPI_ISL_1201911, EPI_ISL_1201912, EPI_ISL_1201913, EPI_ISL_1201914, EPI_ISL_1201915, EPI_ISL_1201916, EPI_ISL_1201917, EPI_ISL_1201918, EPI_ISL_1201919, EPI_ISL_1201920, EPI_ISL_1201921, EPI_ISL_1201922, EPI_ISL_1201923, EPI_ISL_1201924, EPI_ISL_1201925, EPI_ISL_1201926, EPI_ISL_1201927, EPI_ISL_1201928, EPI_ISL_1201929, EPI_ISL_1201930, EPI_ISL_1201931, EPI_ISL_1201932, EPI_ISL_1201933, EPI_ISL_1201934, EPI_ISL_1201935, EPI_ISL_1201936, EPI_ISL_1201937, EPI_ISL_1201938, EPI_ISL_1201939, EPI_ISL_1201940, EPI_ISL_1201941, EPI_ISL_1201942, EPI_ISL_1201943, EPI_ISL_1201944, EPI_ISL_1201945, EPI_ISL_1201946, EPI_ISL_1201947, EPI_ISL_1201948, EPI_ISL_1201949, EPI_ISL_1201950, EPI_ISL_1201951, EPI_ISL_1201952, EPI_ISL_1201953, EPI_ISL_1201954, EPI_ISL_1201955, EPI_ISL_1201956, EPI_ISL_1201957, EPI_ISL_1201958, EPI_ISL_1201959, EPI_ISL_1201960, EPI_ISL_1201961, EPI_ISL_1201962, EPI_ISL_1201963, EPI_ISL_1201964, EPI_ISL_1201965, EPI_ISL_1201966, EPI_ISL_1201967, EPI_ISL_1201968, EPI_ISL_1201969, EPI_ISL_1201970, EPI_ISL_1201971, EPI_ISL_1201972, EPI_ISL_1201973, EPI_ISL_1201974, EPI_ISL_1201975, EPI_ISL_1201976, EPI_ISL_1201977, EPI_ISL_1201978, EPI_ISL_1201979, EPI_ISL_1201980, EPI_ISL_1201981, EPI_ISL_1201982, EPI_ISL_1201983, EPI_ISL_1201984, EPI_ISL_1201985, EPI_ISL_1201986, EPI_ISL_1201987, EPI_ISL_1201988, EPI_ISL_1201989, EPI_ISL_1201990, EPI_ISL_1201991, EPI_ISL_1201992, EPI_ISL_1201993, EPI_ISL_1201994, EPI_ISL_1201995, EPI_ISL_1201996, EPI_ISL_1201997, EPI_ISL_1201999, EPI_ISL_1202000, EPI_ISL_1202001, EPI_ISL_1202002, EPI_ISL_1202003, EPI_ISL_1202004, EPI_ISL_1202005, EPI_ISL_1202006, EPI_ISL_1202007, EPI_ISL_1202008, EPI_ISL_1202009, EPI_ISL_1202010, EPI_ISL_1202011, EPI_ISL_1202012, EPI_ISL_1202013, EPI_ISL_1202014, EPI_ISL_1202015, EPI_ISL_1202016, EPI_ISL_1202017, EPI_ISL_1202018, EPI_ISL_1202019, EPI_ISL_1202020, EPI_ISL_1202021, EPI_ISL_1202022, EPI_ISL_1202023, EPI_ISL_1202024, EPI_ISL_1202025, EPI_ISL_1202026, EPI_ISL_1202027, EPI_ISL_1202028, EPI_ISL_1202029, EPI_ISL_1202030, EPI_ISL_1202031, EPI_ISL_1202032, EPI_ISL_1202033, EPI_ISL_1202034, EPI_ISL_1202035, EPI_ISL_1202036, EPI_ISL_1202037, EPI_ISL_1202038, EPI_ISL_1202039, EPI_ISL_1202040, EPI_ISL_1202041, EPI_ISL_1202042, EPI_ISL_1202043, EPI_ISL_1202044, EPI_ISL_1202045, EPI_ISL_1202046, EPI_ISL_1202047, EPI_ISL_1202048, EPI_ISL_1202049, EPI_ISL_1202050, EPI_ISL_1202051, EPI_ISL_1202052, EPI_ISL_1202053, EPI_ISL_1202054, EPI_ISL_1202055, EPI_ISL_1202056, EPI_ISL_1202057, EPI_ISL_1202058, EPI_ISL_1202059, EPI_ISL_1202060, EPI_ISL_1202061, EPI_ISL_1202062, EPI_ISL_1202063, EPI_ISL_1202064, EPI_ISL_1202065, EPI_ISL_1202066, EPI_ISL_1202067, EPI_ISL_1202068, EPI_ISL_1202069, EPI_ISL_1202070, EPI_ISL_1202071, EPI_ISL_1202072, EPI_ISL_1202073, EPI_ISL_1202074, EPI_ISL_1202075, EPI_ISL_1202076, EPI_ISL_1202077, EPI_ISL_1202078, EPI_ISL_1202079, EPI_ISL_1202080, EPI_ISL_1202081, EPI_ISL_1202082, EPI_ISL_1202083, EPI_ISL_1202084, EPI_ISL_1202085, EPI_ISL_1202086, EPI_ISL_1202087, EPI_ISL_1202088, EPI_ISL_1202089, EPI_ISL_1202090, EPI_ISL_1202091, EPI_ISL_1202092, EPI_ISL_1202093, EPI_ISL_1202094, EPI_ISL_1202095, EPI_ISL_1202096, EPI_ISL_1202097, EPI_ISL_1202098, EPI_ISL_1202099, EPI_ISL_1202100, EPI_ISL_1202101, EPI_ISL_1202102, EPI_ISL_1202103, EPI_ISL_1202104, EPI_ISL_1202105, EPI_ISL_1202106, EPI_ISL_1202107, EPI_ISL_1202108, EPI_ISL_1202109, EPI_ISL_1202110, EPI_ISL_1202111, EPI_ISL_1202112, EPI_ISL_1202113, EPI_ISL_1202114, EPI_ISL_1202115, EPI_ISL_1202116, EPI_ISL_1202117, EPI_ISL_1202118, EPI_ISL_1202119, EPI_ISL_1202120, EPI_ISL_1202121, EPI_ISL_1202122, EPI_ISL_1202123, EPI_ISL_1202124, EPI_ISL_1202125, EPI_ISL_1202126, EPI_ISL_1202127, EPI_ISL_1202128, EPI_ISL_1202129, EPI_ISL_1202130, EPI_ISL_1202131, EPI_ISL_1202132, EPI_ISL_1202133, EPI_ISL_1202134, EPI_ISL_1202135, EPI_ISL_1202136, EPI_ISL_1202137, EPI_ISL_1202138, EPI_ISL_1202139, EPI_ISL_1202140, EPI_ISL_1202141, EPI_ISL_1202142, EPI_ISL_1202143, EPI_ISL_1202144, EPI_ISL_1202145, EPI_ISL_1202146, EPI_ISL_1202147, EPI_ISL_1202148, EPI_ISL_1202149, EPI_ISL_1202150, EPI_ISL_1202151, EPI_ISL_1202152, EPI_ISL_1202153, EPI_ISL_1202154, EPI_ISL_1202155, EPI_ISL_1202156, EPI_ISL_1202157, EPI_ISL_1202158, EPI_ISL_1202159, EPI_ISL_1202160, EPI_ISL_1202161, EPI_ISL_1202162, EPI_ISL_1202163, EPI_ISL_1202164, EPI_ISL_1202165, EPI_ISL_1202166, EPI_ISL_1202167, EPI_ISL_1202168, EPI_ISL_1202169, EPI_ISL_1202170, EPI_ISL_1202171, EPI_ISL_1202172, EPI_ISL_1202173, EPI_ISL_1202174, EPI_ISL_1202175, EPI_ISL_1202176, EPI_ISL_1202177, EPI_ISL_1202178, EPI_ISL_1202179, EPI_ISL_1202180, EPI_ISL_1202181, EPI_ISL_1202182, EPI_ISL_1202183, EPI_ISL_1202184, EPI_ISL_1202185, EPI_ISL_1202186, EPI_ISL_1202187, EPI_ISL_1202188, EPI_ISL_1202189, EPI_ISL_1202190, EPI_ISL_1202191, EPI_ISL_1202192, EPI_ISL_1202193, EPI_ISL_1202194, EPI_ISL_1202195, EPI_ISL_1202196, EPI_ISL_1202197, EPI_ISL_1202198, EPI_ISL_1202199, EPI_ISL_1202200, EPI_ISL_1202201, EPI_ISL_1202202, EPI_ISL_1202203, EPI_ISL_1202204, EPI_ISL_1202205, EPI_ISL_1202206, EPI_ISL_1202207, EPI_ISL_1202208, EPI_ISL_1202209, EPI_ISL_1202210, EPI_ISL_1202211, EPI_ISL_1202212, EPI_ISL_1202213, EPI_ISL_1202214, EPI_ISL_1202215, EPI_ISL_1202216, EPI_ISL_1202217, EPI_ISL_1202218, EPI_ISL_1202219, EPI_ISL_1202220, EPI_ISL_1202221, EPI_ISL_1202222, EPI_ISL_1202223, EPI_ISL_1202224, EPI_ISL_1202225, EPI_ISL_1202226, EPI_ISL_1202227, EPI_ISL_1202228, EPI_ISL_1202229, EPI_ISL_1202230, EPI_ISL_1202231, EPI_ISL_1202232, EPI_ISL_1202233, EPI_ISL_1202234, EPI_ISL_1202235, EPI_ISL_1202236, EPI_ISL_1202237, EPI_ISL_1202238, EPI_ISL_1202239, EPI_ISL_1202240, EPI_ISL_1202241, EPI_ISL_1202242, EPI_ISL_1202243, EPI_ISL_1202244, EPI_ISL_1202245, EPI_ISL_1202246, EPI_ISL_1202247, EPI_ISL_1202248, EPI_ISL_1202249, EPI_ISL_1202250, EPI_ISL_1202251, EPI_ISL_1202252, EPI_ISL_1202253, EPI_ISL_1202254, EPI_ISL_1202255, EPI_ISL_1202256, EPI_ISL_1202257, EPI_ISL_1202258, EPI_ISL_1202259, EPI_ISL_1202260, EPI_ISL_1202261, EPI_ISL_1202262, EPI_ISL_1202263, EPI_ISL_1202264, EPI_ISL_1202265, EPI_ISL_1202266, EPI_ISL_1202267, EPI_ISL_1202268, EPI_ISL_1202269, EPI_ISL_1202270, EPI_ISL_1202271, EPI_ISL_1202272, EPI_ISL_1202273, EPI_ISL_1202274, EPI_ISL_1202275, EPI_ISL_1202276, EPI_ISL_1202277, EPI_ISL_1202278, EPI_ISL_1202279, EPI_ISL_1202280, EPI_ISL_1202281, EPI_ISL_1202282, EPI_ISL_1202283, EPI_ISL_1202284, EPI_ISL_1202285, EPI_ISL_1202286, EPI_ISL_1202287, EPI_ISL_1202288, EPI_ISL_1202289, EPI_ISL_1202290, EPI_ISL_1202291, EPI_ISL_1202292, EPI_ISL_1202293, EPI_ISL_1202294, EPI_ISL_1202295, EPI_ISL_1202296, EPI_ISL_1202297, EPI_ISL_1202298, EPI_ISL_1202299, EPI_ISL_1202300, EPI_ISL_1202301, EPI_ISL_1202302, EPI_ISL_1202303, EPI_ISL_1202304, EPI_ISL_1202305, EPI_ISL_1202306, EPI_ISL_1202307, EPI_ISL_1202308, EPI_ISL_1202309, EPI_ISL_1202310, EPI_ISL_1202311, EPI_ISL_1202312, EPI_ISL_1202313, EPI_ISL_1202314, EPI_ISL_1202315, EPI_ISL_1202316, EPI_ISL_1202317, EPI_ISL_1202318, EPI_ISL_1202319, EPI_ISL_1202320, EPI_ISL_1202321, EPI_ISL_1202322, EPI_ISL_1202323, EPI_ISL_1202324, EPI_ISL_1202325, EPI_ISL_1202326, EPI_ISL_1202327, EPI_ISL_1202328, EPI_ISL_1202329, EPI_ISL_1202330, EPI_ISL_1202331, EPI_ISL_1202332, EPI_ISL_1202333, EPI_ISL_1202334, EPI_ISL_1202335, EPI_ISL_1202336, EPI_ISL_1202337, EPI_ISL_1202338, EPI_ISL_1202339, EPI_ISL_1202340, EPI_ISL_1202341, EPI_ISL_1202342, EPI_ISL_1202343, EPI_ISL_1202344, EPI_ISL_1202345, EPI_ISL_1202346, EPI_ISL_1202347, EPI_ISL_1202348, EPI_ISL_1202349, EPI_ISL_1202350, EPI_ISL_1202351, EPI_ISL_1202352, EPI_ISL_1202353, EPI_ISL_1202354, EPI_ISL_1202355, EPI_ISL_1202356, EPI_ISL_1202357, EPI_ISL_1202358, EPI_ISL_1202359, EPI_ISL_1202360, EPI_ISL_1202361, EPI_ISL_1202362, EPI_ISL_1202363, EPI_ISL_1202364, EPI_ISL_1202365, EPI_ISL_1202366, EPI_ISL_1202367, EPI_ISL_1202368, EPI_ISL_1202369, EPI_ISL_1202370, EPI_ISL_1202371, EPI_ISL_1202372, EPI_ISL_1202373, EPI_ISL_1202374, EPI_ISL_1202375, EPI_ISL_1202376, EPI_ISL_1202377, EPI_ISL_1202378, EPI_ISL_1202379, EPI_ISL_1202380, EPI_ISL_1202381, EPI_ISL_1202382, EPI_ISL_1202383, EPI_ISL_1202384, EPI_ISL_1202385, EPI_ISL_1202386, EPI_ISL_1202387, EPI_ISL_1202388, EPI_ISL_1202389, EPI_ISL_1202390, EPI_ISL_1202391, EPI_ISL_1202392, EPI_ISL_1202393, EPI_ISL_1202394, EPI_ISL_1202395, EPI_ISL_1202396, EPI_ISL_1202397, EPI_ISL_1202398, EPI_ISL_1202399, EPI_ISL_1202400, EPI_ISL_1202401, EPI_ISL_1202402, EPI_ISL_1202403, EPI_ISL_1202404, EPI_ISL_1202405, EPI_ISL_1202406, EPI_ISL_1202407, EPI_ISL_1202408, EPI_ISL_1202409, EPI_ISL_1202410, EPI_ISL_1202411, EPI_ISL_1202412, EPI_ISL_1202413, EPI_ISL_1202414, EPI_ISL_1202415, EPI_ISL_1202416, EPI_ISL_1202417, EPI_ISL_1202418, EPI_ISL_1202419, EPI_ISL_1202420, EPI_ISL_1202421, EPI_ISL_1202422, EPI_ISL_1202423, EPI_ISL_1202424, EPI_ISL_1202425, EPI_ISL_1202426, EPI_ISL_1202427, EPI_ISL_1202428, EPI_ISL_1202429, EPI_ISL_1202430, EPI_ISL_1202431, EPI_ISL_1202432, EPI_ISL_1202433, EPI_ISL_1202434, EPI_ISL_1202435, EPI_ISL_1202436, EPI_ISL_1202437, EPI_ISL_1202438, EPI_ISL_1202439, EPI_ISL_1202440, EPI_ISL_1202441, EPI_ISL_1202442, EPI_ISL_1202443, EPI_ISL_1202444, EPI_ISL_1202445, EPI_ISL_1202446, EPI_ISL_1202447, EPI_ISL_1202448, EPI_ISL_1202449, EPI_ISL_1202450, EPI_ISL_1202451, EPI_ISL_1202452, EPI_ISL_1202453, EPI_ISL_1202454, EPI_ISL_1202455, EPI_ISL_1202456, EPI_ISL_1202457, EPI_ISL_1202458, EPI_ISL_1202459, EPI_ISL_1202460, EPI_ISL_1202461, EPI_ISL_1202462, EPI_ISL_1202463, EPI_ISL_1202464, EPI_ISL_1202465, EPI_ISL_1202466, EPI_ISL_1202467, EPI_ISL_1202468, EPI_ISL_1202469, EPI_ISL_1202470, EPI_ISL_1202471, EPI_ISL_1202472, EPI_ISL_1202473, EPI_ISL_1202474, EPI_ISL_1202475, EPI_ISL_1202476, EPI_ISL_1202477, EPI_ISL_1202478, EPI_ISL_1202479, EPI_ISL_1202480, EPI_ISL_1202481, EPI_ISL_1202482, EPI_ISL_1202483, EPI_ISL_1202484, EPI_ISL_1202485, EPI_ISL_1202486, EPI_ISL_1202487, EPI_ISL_1202488, EPI_ISL_1202489, EPI_ISL_1202490, EPI_ISL_1202491, EPI_ISL_1202492, EPI_ISL_1202493, EPI_ISL_1202494, EPI_ISL_1202495, EPI_ISL_1202496, EPI_ISL_1202497, EPI_ISL_1202498, EPI_ISL_1202499, EPI_ISL_1202500, EPI_ISL_1202501, EPI_ISL_1202502, EPI_ISL_1202503, EPI_ISL_1202504, EPI_ISL_1202505, EPI_ISL_1202506, EPI_ISL_1202507, EPI_ISL_1202508, EPI_ISL_1202509, EPI_ISL_1202510, EPI_ISL_1202511, EPI_ISL_1202512, EPI_ISL_1202513, EPI_ISL_1202514, EPI_ISL_1202515, EPI_ISL_1202516, EPI_ISL_1202517, EPI_ISL_1202518, EPI_ISL_1202519, EPI_ISL_1202520, EPI_ISL_1202521, EPI_ISL_1202522, EPI_ISL_1202523, EPI_ISL_1202524, EPI_ISL_1202525, EPI_ISL_1202526, EPI_ISL_1202527, EPI_ISL_1202528, EPI_ISL_1202529, EPI_ISL_1202530, EPI_ISL_1202531, EPI_ISL_1202532, EPI_ISL_1202533, EPI_ISL_1202534, EPI_ISL_1202535, EPI_ISL_1202536, EPI_ISL_1202537, EPI_ISL_1202538, EPI_ISL_1202539, EPI_ISL_1202540, EPI_ISL_1202541, EPI_ISL_1202542, EPI_ISL_1202543, EPI_ISL_1202544, EPI_ISL_1202545, EPI_ISL_1202546, EPI_ISL_1202547, EPI_ISL_1202548, EPI_ISL_1202549, EPI_ISL_1202550, EPI_ISL_1202551, EPI_ISL_1202552, EPI_ISL_1202553, EPI_ISL_1202554, EPI_ISL_1202555, EPI_ISL_1202556, EPI_ISL_1202557, EPI_ISL_1202558, EPI_ISL_1202559, EPI_ISL_1202560, EPI_ISL_1202561, EPI_ISL_1202562, EPI_ISL_1202563, EPI_ISL_1202564, EPI_ISL_1202565 | | | |
| see above | Houston Methodist Hospital | Houston Methodist Hospital | S. Wesley Long, Randall J. Olsen, Paul A. Christensen, Sishir Subedi, Robert Olson, James J. Davis, Matthew Ojeda Saavedra, Prasanti Yerramilli, Layne Pruitt, Kristina Reppond, Madison N. Shyer, Jessica Cambric, Ilya J. Finkelstein, Jimmy Gollihar, and James M. Musser |
| EPI_ISL_1202566 | Ohio State University Wexner Medical Center | James Molecular Laboratory | Jones D, Chang Y-S, Ru P, Chappell D, Snyder P, Koenig S, Corcoran S, Pancholi P |
| EPI_ISL_1202567, EPI_ISL_1202568, EPI_ISL_1202569, EPI_ISL_1202570, EPI_ISL_1202571, EPI_ISL_1202572, EPI_ISL_1202573, EPI_ISL_1202574, EPI_ISL_1202575, EPI_ISL_1202576, EPI_ISL_1202577, EPI_ISL_1202578, EPI_ISL_1202579, EPI_ISL_1202580, EPI_ISL_1202581, EPI_ISL_1202582, EPI_ISL_1202583, EPI_ISL_1202584, EPI_ISL_1202585, EPI_ISL_1202586, EPI_ISL_1202587, EPI_ISL_1202588, EPI_ISL_1202589, EPI_ISL_1202590, EPI_ISL_1202591, EPI_ISL_1202592, EPI_ISL_1202593, EPI_ISL_1202594, EPI_ISL_1202595, EPI_ISL_1202596, EPI_ISL_1202597, EPI_ISL_1202598, EPI_ISL_1202599, EPI_ISL_1202600, EPI_ISL_1202601, EPI_ISL_1202602, EPI_ISL_1202603, EPI_ISL_1202604, EPI_ISL_1202605, EPI_ISL_1202606, EPI_ISL_1202607, EPI_ISL_1202608, EPI_ISL_1202609, EPI_ISL_1202610, EPI_ISL_1202611, EPI_ISL_1202612, EPI_ISL_1202613, EPI_ISL_1202614, EPI_ISL_1202615, EPI_ISL_1202616, EPI_ISL_1202617, EPI_ISL_1202618, EPI_ISL_1202619, EPI_ISL_1202620, EPI_ISL_1202621, EPI_ISL_1202622, EPI_ISL_1202623, EPI_ISL_1202624, EPI_ISL_1202625, EPI_ISL_1202626, EPI_ISL_1202627, EPI_ISL_1202628, EPI_ISL_1202629, EPI_ISL_1202630, EPI_ISL_1202631, EPI_ISL_1202632, EPI_ISL_1202633, EPI_ISL_1202634, EPI_ISL_1202635, EPI_ISL_1202636, EPI_ISL_1202637, EPI_ISL_1202638, EPI_ISL_1202639, EPI_ISL_1202640, EPI_ISL_1202641, EPI_ISL_1202642, EPI_ISL_1202643, EPI_ISL_1202644, EPI_ISL_1202645, EPI_ISL_1202646, EPI_ISL_1202647, EPI_ISL_1202648, EPI_ISL_1202649, EPI_ISL_1202650, EPI_ISL_1202651, EPI_ISL_1202652, EPI_ISL_1202653, EPI_ISL_1202654, EPI_ISL_1202655, EPI_ISL_1202656, EPI_ISL_1202657, EPI_ISL_1202658, EPI_ISL_1202659, EPI_ISL_1202660, EPI_ISL_1202661, EPI_ISL_1202662, EPI_ISL_1202663, EPI_ISL_1202664, EPI_ISL_1202665, EPI_ISL_1202666, EPI_ISL_1202667, EPI_ISL_1202668, EPI_ISL_1202669, EPI_ISL_1202670, EPI_ISL_1202671, EPI_ISL_1202672, EPI_ISL_1202673, EPI_ISL_1202674, EPI_ISL_1202675, EPI_ISL_1202676, EPI_ISL_1202677, EPI_ISL_1202678, EPI_ISL_1202679, EPI_ISL_1202680, EPI_ISL_1202681, EPI_ISL_1202682, EPI_ISL_1202683, EPI_ISL_1202684, EPI_ISL_1202685, EPI_ISL_1202686, EPI_ISL_1202687, EPI_ISL_1202688, EPI_ISL_1202689, EPI_ISL_1202690, EPI_ISL_1202691, EPI_ISL_1202692, EPI_ISL_1202693, EPI_ISL_1202694, EPI_ISL_1202695, EPI_ISL_1202696, EPI_ISL_1202697, EPI_ISL_1202698, EPI_ISL_1202699, EPI_ISL_1202700, EPI_ISL_1202701, EPI_ISL_1202702, EPI_ISL_1202703, EPI_ISL_1202704, EPI_ISL_1202705, EPI_ISL_1202706, EPI_ISL_1202707, EPI_ISL_1202708, EPI_ISL_1202709, EPI_ISL_1202710, EPI_ISL_1202711, EPI_ISL_1202712, EPI_ISL_1202713, EPI_ISL_1202714, EPI_ISL_1202715, EPI_ISL_1202716, EPI_ISL_1202717, EPI_ISL_1202718, EPI_ISL_1202719, EPI_ISL_1202720, EPI_ISL_1202721, EPI_ISL_1202722, EPI_ISL_1202723, EPI_ISL_1202724, EPI_ISL_1202725, EPI_ISL_1202726, EPI_ISL_1202727, EPI_ISL_1202728, EPI_ISL_1202729, EPI_ISL_1202730, EPI_ISL_1202731, EPI_ISL_1202732, EPI_ISL_1202733, EPI_ISL_1202734, EPI_ISL_1202735, EPI_ISL_1202736, EPI_ISL_1202737, EPI_ISL_1202738, EPI_ISL_1202739, EPI_ISL_1202740, EPI_ISL_1202741, EPI_ISL_1202742, EPI_ISL_1202743, EPI_ISL_1202744, EPI_ISL_1202745, EPI_ISL_1202746, EPI_ISL_1202747, EPI_ISL_1202748, EPI_ISL_1202749, EPI_ISL_1202750, EPI_ISL_1202751, EPI_ISL_1202752, EPI_ISL_1202753, EPI_ISL_1202754, EPI_ISL_1202755, EPI_ISL_1202756, EPI_ISL_1202757, EPI_ISL_1202758, EPI_ISL_1202759, EPI_ISL_1202760, EPI_ISL_1202761, EPI_ISL_1202762, EPI_ISL_1202763, EPI_ISL_1202764, EPI_ISL_1202765, EPI_ISL_1202766, EPI_ISL_1202767, EPI_ISL_1202768, EPI_ISL_1202769, EPI_ISL_1202770, EPI_ISL_1202771, EPI_ISL_1202772, EPI_ISL_1202773, EPI_ISL_1202774, EPI_ISL_1202775, EPI_ISL_1202776, EPI_ISL_1202777, EPI_ISL_1202778, EPI_ISL_1202779, EPI_ISL_1202780, EPI_ISL_1202781, EPI_ISL_1202782, EPI_ISL_1202783, EPI_ISL_1202784, EPI_ISL_1202785, EPI_ISL_1202786, EPI_ISL_1202787, EPI_ISL_1202788, EPI_ISL_1202789, EPI_ISL_1202790, EPI_ISL_1202791, EPI_ISL_1202792, EPI_ISL_1202793, EPI_ISL_1202794, EPI_ISL_1202795, EPI_ISL_1202796, EPI_ISL_1202797, EPI_ISL_1202798, EPI_ISL_1202799, EPI_ISL_1202800, EPI_ISL_1202801, EPI_ISL_1202802, EPI_ISL_1202803, EPI_ISL_1202804, EPI_ISL_1202805, EPI_ISL_1202806, EPI_ISL_1202807, EPI_ISL_1202808, EPI_ISL_1202809, EPI_ISL_1202810, EPI_ISL_1202811, EPI_ISL_1202812, EPI_ISL_1202813, EPI_ISL_1202814, EPI_ISL_1202815, EPI_ISL_1202816, EPI_ISL_1202817, EPI_ISL_1202818, EPI_ISL_1202819, EPI_ISL_1202820, EPI_ISL_1202821, EPI_ISL_1202822, EPI_ISL_1202823, EPI_ISL_1202824, EPI_ISL_1202825, EPI_ISL_1202826, EPI_ISL_1202827, EPI_ISL_1202828, EPI_ISL_1202829, EPI_ISL_1202830, EPI_ISL_1202831, EPI_ISL_1202832, EPI_ISL_1202833, EPI_ISL_1202834, EPI_ISL_1202835, EPI_ISL_1202836, EPI_ISL_1202837, EPI_ISL_1202838, EPI_ISL_1202839, EPI_ISL_1202840, EPI_ISL_1202841, EPI_ISL_1202842, EPI_ISL_1202843, EPI_ISL_1202844, EPI_ISL_1202845, EPI_ISL_1202846, EPI_ISL_1202847, EPI_ISL_1202848, EPI_ISL_1202849, EPI_ISL_1202850, EPI_ISL_1202851, EPI_ISL_1202852, EPI_ISL_1202853, EPI_ISL_1202854, EPI_ISL_1202855, EPI_ISL_1202856, EPI_ISL_1202857, EPI_ISL_1202858, EPI_ISL_1202859, EPI_ISL_1202860, EPI_ISL_1202861, EPI_ISL_1202862, EPI_ISL_1202863, EPI_ISL_1202864, EPI_ISL_1202865, EPI_ISL_1202866, EPI_ISL_1202867, EPI_ISL_1202868, EPI_ISL_1202869, EPI_ISL_1202870, EPI_ISL_1202871, EPI_ISL_1202872, EPI_ISL_1202873, EPI_ISL_1202874, EPI_ISL_1202875, EPI_ISL_1202876, EPI_ISL_1202877, EPI_ISL_1202878, EPI_ISL_1202879, EPI_ISL_1202880, EPI_ISL_1202881, EPI_ISL_1202882, EPI_ISL_1202883, EPI_ISL_1202884, EPI_ISL_1202885, EPI_ISL_1202886, EPI_ISL_1202887, EPI_ISL_1202888, EPI_ISL_1202889, EPI_ISL_1202890, EPI_ISL_1202891, EPI_ISL_1202892, EPI_ISL_1202893, EPI_ISL_1202894, EPI_ISL_1202895, EPI_ISL_1202896, EPI_ISL_1202897, EPI_ISL_1202898, EPI_ISL_1202899, EPI_ISL_1202900, EPI_ISL_1202901, EPI_ISL_1202902, EPI_ISL_1202903, EPI_ISL_1202904, EPI_ISL_1202905, EPI_ISL_1202906, EPI_ISL_1202907, EPI_ISL_1202908, EPI_ISL_1202909, EPI_ISL_1202910, EPI_ISL_1202911, EPI_ISL_1202912, EPI_ISL_1202913, EPI_ISL_1202914, EPI_ISL_1202915, EPI_ISL_1202916, EPI_ISL_1202917, EPI_ISL_1202918, EPI_ISL_1202919, EPI_ISL_1202920, EPI_ISL_1202921, EPI_ISL_1202922, EPI_ISL_1202923, EPI_ISL_1202924, EPI_ISL_1202925, EPI_ISL_1202926, EPI_ISL_1202927, EPI_ISL_1202928, EPI_ISL_1202929, EPI_ISL_1202930, EPI_ISL_1202931, EPI_ISL_1202932, EPI_ISL_1202933, EPI_ISL_1202934, EPI_ISL_1202935, EPI_ISL_1202936, EPI_ISL_1202937, EPI_ISL_1202938, EPI_ISL_1202939, EPI_ISL_1202940, EPI_ISL_1202941, EPI_ISL_1202942, EPI_ISL_1202943, EPI_ISL_1202944, EPI_ISL_1202945, EPI_ISL_1202946, EPI_ISL_1202947, EPI_ISL_1202948, EPI_ISL_1202949, EPI_ISL_1202950, EPI_ISL_1202951, EPI_ISL_1202952, EPI_ISL_1202953, EPI_ISL_1202954, EPI_ISL_1202955, EPI_ISL_1202956, EPI_ISL_1202957, EPI_ISL_1202958, EPI_ISL_1202959, EPI_ISL_1202960, EPI_ISL_1202961, EPI_ISL_1202962, EPI_ISL_1202963, EPI_ISL_1202964, EPI_ISL_1202965, EPI_ISL_1202966, EPI_ISL_1202967, EPI_ISL_1202968, EPI_ISL_1202969, EPI_ISL_1202970, EPI_ISL_1202971, EPI_ISL_1202972, EPI_ISL_1202973, EPI_ISL_1202974, EPI_ISL_1202975, EPI_ISL_1202976, EPI_ISL_1202977, EPI_ISL_1202978, EPI_ISL_1202979, EPI_ISL_1202980, EPI_ISL_1202981, EPI_ISL_1202982, EPI_ISL_1202983, EPI_ISL_1202984, EPI_ISL_1202985, EPI_ISL_1202986, EPI_ISL_1202987, EPI_ISL_1202988, EPI_ISL_1202989, EPI_ISL_1202990, EPI_ISL_1202991, EPI_ISL_1202992, EPI_ISL_1202993, EPI_ISL_1202994, EPI_ISL_1202995, EPI_ISL_1202996, EPI_ISL_1202997, EPI_ISL_1202998, EPI_ISL_1202999, EPI_ISL_1203000, EPI_ISL_1203001, EPI_ISL_1203002, EPI_ISL_1203003, EPI_ISL_1203004, EPI_ISL_1203005, EPI_ISL_1203006, EPI_ISL_1203007, EPI_ISL_1203008, EPI_ISL_1203009, EPI_ISL_1203010, EPI_ISL_1203011, EPI_ISL_1203012, EPI_ISL_1203013, EPI_ISL_1203014, EPI_ISL_1203015, EPI_ISL_1203016, EPI_ISL_1203017, EPI_ISL_1203018, EPI_ISL_1203019, EPI_ISL_1203020, EPI_ISL_1203021, EPI_ISL_1203022, EPI_ISL_1203023, EPI_ISL_1203024, EPI_ISL_1203025, EPI_ISL_1203026, EPI_ISL_1203027, EPI_ISL_1203028, EPI_ISL_1203029, EPI_ISL_1203030, EPI_ISL_1203031, EPI_ISL_1203032, EPI_ISL_1203033, EPI_ISL_1203034, EPI_ISL_1203035, EPI_ISL_1203036, EPI_ISL_1203037, EPI_ISL_1203038, EPI_ISL_1203039, EPI_ISL_1203040, EPI_ISL_1203041, EPI_ISL_1203042, EPI_ISL_1203043, EPI_ISL_1203044, EPI_ISL_1203045, EPI_ISL_1203046, EPI_ISL_1203047, EPI_ISL_1203048, EPI_ISL_1203049, EPI_ISL_1203050, EPI_ISL_1203051, EPI_ISL_1203052, EPI_ISL_1203053, EPI_ISL_1203054, EPI_ISL_1203055, EPI_ISL_1203056, EPI_ISL_1203057, EPI_ISL_1203058, EPI_ISL_1203059, EPI_ISL_1203060, EPI_ISL_1203061, EPI_ISL_1203062, EPI_ISL_1203063, EPI_ISL_1203064, EPI_ISL_1203065, EPI_ISL_1203066, EPI_ISL_1203067, EPI_ISL_1203068, EPI_ISL_1203069, EPI_ISL_1203070, EPI_ISL_1203071, EPI_ISL_1203072, EPI_ISL_1203073, EPI_ISL_1203074, EPI_ISL_1203075, EPI_ISL_1203076, EPI_ISL_1203077, EPI_ISL_1203078, EPI_ISL_1203079, EPI_ISL_1203080, EPI_ISL_1203081, EPI_ISL_1203082, EPI_ISL_1203083, EPI_ISL_1203084, EPI_ISL_1203085, EPI_ISL_1203086, EPI_ISL_1203087, EPI_ISL_1203088, EPI_ISL_1203089, EPI_ISL_1203090, EPI_ISL_1203091, EPI_ISL_1203092, EPI_ISL_1203093, EPI_ISL_1203094, EPI_ISL_1203095, EPI_ISL_1203096, EPI_ISL_1203097, EPI_ISL_1203098, EPI_ISL_1203099, EPI_ISL_1203100, EPI_ISL_1203101, EPI_ISL_1203102, EPI_ISL_1203103, EPI_ISL_1203104, EPI_ISL_1203105, EPI_ISL_1203106, EPI_ISL_1203107, EPI_ISL_1203108, EPI_ISL_1203109, EPI_ISL_1203110, EPI_ISL_1203111, EPI_ISL_1203112, EPI_ISL_1203113, EPI_ISL_1203114, EPI_ISL_1203115, EPI_ISL_1203116, EPI_ISL_1203117, EPI_ISL_1203118, EPI_ISL_1203119, EPI_ISL_1203120, EPI_ISL_1203121, EPI_ISL_1203122, EPI_ISL_1203123, EPI_ISL_1203124, EPI_ISL_1203125, EPI_ISL_1203126, EPI_ISL_1203127, EPI_ISL_1203128, EPI_ISL_1203129, EPI_ISL_1203130, EPI_ISL_1203131, EPI_ISL_1203132, EPI_ISL_1203133, EPI_ISL_1203134, EPI_ISL_1203135, EPI_ISL_1203136, EPI_ISL_1203137, EPI_ISL_1203138, EPI_ISL_1203139, EPI_ISL_1203140, EPI_ISL_1203141, EPI_ISL_1203142, EPI_ISL_1203143, EPI_ISL_1203144, EPI_ISL_1203145, EPI_ISL_1203146, EPI_ISL_1203147, EPI_ISL_1203148, EPI_ISL_1203149, EPI_ISL_1203150, EPI_ISL_1203151, EPI_ISL_1203152, EPI_ISL_1203153, EPI_ISL_1203154, EPI_ISL_1203155, EPI_ISL_1203156, EPI_ISL_1203157, EPI_ISL_1203158, EPI_ISL_1203159, EPI_ISL_1203160, EPI_ISL_1203161, EPI_ISL_1203162, EPI_ISL_1203163, EPI_ISL_1203164, EPI_ISL_1203165, EPI_ISL_1203166, EPI_ISL_1203167, EPI_ISL_1203168, EPI_ISL_1203169, EPI_ISL_1203170, EPI_ISL_1203171, EPI_ISL_1203172, EPI_ISL_1203173, EPI_ISL_1203174, EPI_ISL_1203175, EPI_ISL_1203176, EPI_ISL_1203177, EPI_ISL_1203178, EPI_ISL_1203179, EPI_ISL_1203180, EPI_ISL_1203181, EPI_ISL_1203182, EPI_ISL_1203183, EPI_ISL_1203184, EPI_ISL_1203185, EPI_ISL_1203186, EPI_ISL_1203187, EPI_ISL_1203188, EPI_ISL_1203189, EPI_ISL_1203190, EPI_ISL_1203191, EPI_ISL_1203192, EPI_ISL_1203193, EPI_ISL_1203194, EPI_ISL_1203195, EPI_ISL_1203196, EPI_ISL_1203197, EPI_ISL_1203198, EPI_ISL_1203199, EPI_ISL_1203200, EPI_ISL_1203201, EPI_ISL_1203202, EPI_ISL_1203203, EPI_ISL_1203204, EPI_ISL_1203205, EPI_ISL_1203206, EPI_ISL_1203207, EPI_ISL_1203208, EPI_ISL_1203209, EPI_ISL_1203210, EPI_ISL_1203211, EPI_ISL_1203212, EPI_ISL_1203213, EPI_ISL_1203214, EPI_ISL_1203215, EPI_ISL_1203216, EPI_ISL_1203217, EPI_ISL_1203218, EPI_ISL_1203219, EPI_ISL_1203220, EPI_ISL_1203221, EPI_ISL_1203222, EPI_ISL_1203223, EPI_ISL_1203224, EPI_ISL_1203225, EPI_ISL_1203226, EPI_ISL_1203227, EPI_ISL_1203228, EPI_ISL_1203229, EPI_ISL_1203230, EPI_ISL_1203231, EPI_ISL_1203232, EPI_ISL_1203233, EPI_ISL_1203234, EPI_ISL_1203235, EPI_ISL_1203236, EPI_ISL_1203237, EPI_ISL_1203238, EPI_ISL_1203239, EPI_ISL_1203240, EPI_ISL_1203241, EPI_ISL_1203242, EPI_ISL_1203243, EPI_ISL_1203244, EPI_ISL_1203245, EPI_ISL_1203246, EPI_ISL_1203247, EPI_ISL_1203248, EPI_ISL_1203249, EPI_ISL_1203250, EPI_ISL_1203251, EPI_ISL_1203252, EPI_ISL_1203253, EPI_ISL_1203254, EPI_ISL_1203255, EPI_ISL_1203256, EPI_ISL_1203257, EPI_ISL_1203258, EPI_ISL_1203259, EPI_ISL_1203260, EPI_ISL_1203261, EPI_ISL_1203262, EPI_ISL_1203263, EPI_ISL_1203264, EPI_ISL_1203265, EPI_ISL_1203266, EPI_ISL_1203267, EPI_ISL_1203268, EPI_ISL_1203269, EPI_ISL_1203270, EPI_ISL_1203271, EPI_ISL_1203272, EPI_ISL_1203273, EPI_ISL_1203274, EPI_ISL_1203275, EPI_ISL_1203276, EPI_ISL_1203277, EPI_ISL_1203278, EPI_ISL_1203279, EPI_ISL_1203280, EPI_ISL_1203281, EPI_ISL_1203282, EPI_ISL_1203283, EPI_ISL_1203284, EPI_ISL_1203285, EPI_ISL_1203286, EPI_ISL_1203287, EPI_ISL_1203288, EPI_ISL_1203289, EPI_ISL_1203290, EPI_ISL_1203291, EPI_ISL_1203292, EPI_ISL_1203293, EPI_ISL_1203294, EPI_ISL_1203295, EPI_ISL_1203296, EPI_ISL_1203297, EPI_ISL_1203298, EPI_ISL_1203299, EPI_ISL_1203300, EPI_ISL_1203301, EPI_ISL_1203302, EPI_ISL_1203303, EPI_ISL_1203304, EPI_ISL_1203305, EPI_ISL_1203306, EPI_ISL_1203307, EPI_ISL_1203308, EPI_ISL_1203309, EPI_ISL_1203310, EPI_ISL_1203311, EPI_ISL_1203312, EPI_ISL_1203313, EPI_ISL_1203314, EPI_ISL_1203315, EPI_ISL_1203316, EPI_ISL_1203317, EPI_ISL_1203318, EPI_ISL_1203319, EPI_ISL_1203320, EPI_ISL_1203321, EPI_ISL_1203322, EPI_ISL_1203323, EPI_ISL_1203324, EPI_ISL_1203325, EPI_ISL_1203326, EPI_ISL_1203327, EPI_ISL_1203328, EPI_ISL_1203329, EPI_ISL_1203330, EPI_ISL_1203331, EPI_ISL_1203332, EPI_ISL_1203333, EPI_ISL_1203334, EPI_ISL_1203335, EPI_ISL_1203336, EPI_ISL_1203337, EPI_ISL_1203338, EPI_ISL_1203339, EPI_ISL_1203340, EPI_ISL_1203341, EPI_ISL_1203342, EPI_ISL_1203343, EPI_ISL_1203344, EPI_ISL_1203345, EPI_ISL_1203346, EPI_ISL_1203347, EPI_ISL_1203348, EPI_ISL_1203349, EPI_ISL_1203350, EPI_ISL_1203351, EPI_ISL_1203352, EPI_ISL_1203353, EPI_ISL_1203354, EPI_ISL_1203355, EPI_ISL_1203356, EPI_ISL_1203357, EPI_ISL_1203358, EPI_ISL_1203359, EPI_ISL_1203360, EPI_ISL_1203361, EPI_ISL_1203362, EPI_ISL_1203363, EPI_ISL_1203364, EPI_ISL_1203365, EPI_ISL_1203366, EPI_ISL_1203367, EPI_ISL_1203368, EPI_ISL_1203369, EPI_ISL_1203370, EPI_ISL_1203371, EPI_ISL_1203372, EPI_ISL_1203373, EPI_ISL_1203374, EPI_ISL_1203375, EPI_ISL_1203376, EPI_ISL_1203377, EPI_ISL_1203378, EPI_ISL_1203379, EPI_ISL_1203380, EPI_ISL_1203381, EPI_ISL_1203382, EPI_ISL_1203383, EPI_ISL_1203384, EPI_ISL_1203385, EPI_ISL_1203386, EPI_ISL_1203387, EPI_ISL_1203388, EPI_ISL_1203389, EPI_ISL_1203390, EPI_ISL_1203391, EPI_ISL_1203392, EPI_ISL_1203393, EPI_ISL_1203394, EPI_ISL_1203395, EPI_ISL_1203396, EPI_ISL_1203397, EPI_ISL_1203398, EPI_ISL_1203399, EPI_ISL_1203400, EPI_ISL_1203401, EPI_ISL_1203402, EPI_ISL_1203403, EPI_ISL_1203404, EPI_ISL_1203405, EPI_ISL_1203406, EPI_ISL_1203407, EPI_ISL_1203408, EPI_ISL_1203409, EPI_ISL_1203410, EPI_ISL_1203411, EPI_ISL_1203412, EPI_ISL_1203413, EPI_ISL_1203414, EPI_ISL_1203415, EPI_ISL_1203416, EPI_ISL_1203417, EPI_ISL_1203418, EPI_ISL_1203419, EPI_ISL_1203420, EPI_ISL_1203421, EPI_ISL_1203422, EPI_ISL_1203423, EPI_ISL_1203424, | | | |

| | | | |
|---|---|---|---|
| EPI_ISL_1203425, EPI_ISL_1203426, EPI_ISL_1203427, EPI_ISL_1203428, EPI_ISL_1203429, EPI_ISL_1203430, EPI_ISL_1203431, EPI_ISL_1203432, EPI_ISL_1203433, EPI_ISL_1203434, EPI_ISL_1203435, EPI_ISL_1203436, EPI_ISL_1203437, EPI_ISL_1203438, EPI_ISL_1203439, EPI_ISL_1203440, EPI_ISL_1203441, EPI_ISL_1203442, EPI_ISL_1203443, EPI_ISL_1203444, EPI_ISL_1203445, EPI_ISL_1203446, EPI_ISL_1203447, EPI_ISL_1203448, EPI_ISL_1203449, EPI_ISL_1203450, EPI_ISL_1203451, EPI_ISL_1203452, EPI_ISL_1203453, EPI_ISL_1203454, EPI_ISL_1203455, EPI_ISL_1203456, EPI_ISL_1203457, EPI_ISL_1203458, EPI_ISL_1203459, EPI_ISL_1203460, EPI_ISL_1203461, EPI_ISL_1203462, EPI_ISL_1203463, EPI_ISL_1203464, EPI_ISL_1203465, EPI_ISL_1203466, EPI_ISL_1203467, EPI_ISL_1203468, EPI_ISL_1203469, EPI_ISL_1203470, EPI_ISL_1203471, EPI_ISL_1203472, EPI_ISL_1203473, EPI_ISL_1203474, EPI_ISL_1203475, EPI_ISL_1203476, EPI_ISL_1203477, EPI_ISL_1203478, EPI_ISL_1203479, EPI_ISL_1203480, EPI_ISL_1203481, EPI_ISL_1203482, EPI_ISL_1203483, EPI_ISL_1203484, EPI_ISL_1203485, EPI_ISL_1203486, EPI_ISL_1203487, EPI_ISL_1203488, EPI_ISL_1203489, EPI_ISL_1203490, EPI_ISL_1203491, EPI_ISL_1203492, EPI_ISL_1203493, EPI_ISL_1203494, EPI_ISL_1203495, EPI_ISL_1203496, EPI_ISL_1203497, EPI_ISL_1203498, EPI_ISL_1203499, EPI_ISL_1203500, EPI_ISL_1203501, EPI_ISL_1203502, EPI_ISL_1203503, EPI_ISL_1203504, EPI_ISL_1203505, EPI_ISL_1203506, EPI_ISL_1203507, EPI_ISL_1203508, EPI_ISL_1203509, EPI_ISL_1203510, EPI_ISL_1203511, EPI_ISL_1203512, EPI_ISL_1203513, EPI_ISL_1203514, EPI_ISL_1203515, EPI_ISL_1203516, EPI_ISL_1203517, EPI_ISL_1203518, EPI_ISL_1203519, EPI_ISL_1203520, EPI_ISL_1203521, EPI_ISL_1203522, EPI_ISL_1203523, EPI_ISL_1203524, EPI_ISL_1203525, EPI_ISL_1203526, EPI_ISL_1203527, EPI_ISL_1203528, EPI_ISL_1203529, EPI_ISL_1203530, EPI_ISL_1203531, EPI_ISL_1203532, EPI_ISL_1203533, EPI_ISL_1203534, EPI_ISL_1203535, EPI_ISL_1203536, EPI_ISL_1203537, EPI_ISL_1203538, EPI_ISL_1203539, EPI_ISL_1203540, EPI_ISL_1203541, EPI_ISL_1203542, EPI_ISL_1203543, EPI_ISL_1203544, EPI_ISL_1203545, EPI_ISL_1203546, EPI_ISL_1203547, EPI_ISL_1203548, EPI_ISL_1203549, EPI_ISL_1203550, EPI_ISL_1203551, EPI_ISL_1203552, EPI_ISL_1203553, EPI_ISL_1203554, EPI_ISL_1203555, EPI_ISL_1203556, EPI_ISL_1203557, EPI_ISL_1203558, EPI_ISL_1203559, EPI_ISL_1203560, EPI_ISL_1203561, EPI_ISL_1203562, EPI_ISL_1203563, EPI_ISL_1203564, EPI_ISL_1203565, EPI_ISL_1203566, EPI_ISL_1203567, EPI_ISL_1203568, EPI_ISL_1203569, EPI_ISL_1203570, EPI_ISL_1203571, EPI_ISL_1203572, EPI_ISL_1203573, EPI_ISL_1203574, EPI_ISL_1203575, EPI_ISL_1203576, EPI_ISL_1203577, EPI_ISL_1203578, EPI_ISL_1203579, EPI_ISL_1203580, EPI_ISL_1203581, EPI_ISL_1203582, EPI_ISL_1203583, EPI_ISL_1203584, EPI_ISL_1203585, EPI_ISL_1203586, EPI_ISL_1203587, EPI_ISL_1203588, EPI_ISL_1203589, EPI_ISL_1203590, EPI_ISL_1203591, EPI_ISL_1203592, EPI_ISL_1203593, EPI_ISL_1203594, EPI_ISL_1203595, EPI_ISL_1203596, EPI_ISL_1203597, EPI_ISL_1203598, EPI_ISL_1203599, EPI_ISL_1203600, EPI_ISL_1203601, EPI_ISL_1203602, EPI_ISL_1203603, EPI_ISL_1203604, EPI_ISL_1203605, EPI_ISL_1203606, EPI_ISL_1203607, EPI_ISL_1203608, EPI_ISL_1203609, EPI_ISL_1203610, EPI_ISL_1203611, EPI_ISL_1203612, EPI_ISL_1203613, EPI_ISL_1203614, EPI_ISL_1203615, EPI_ISL_1203616, EPI_ISL_1203617, EPI_ISL_1203618, EPI_ISL_1203619, EPI_ISL_1203620, EPI_ISL_1203621, EPI_ISL_1203622, EPI_ISL_1203623, EPI_ISL_1203624, EPI_ISL_1203625, EPI_ISL_1203626, EPI_ISL_1203627, EPI_ISL_1203628, EPI_ISL_1203629, EPI_ISL_1203630, EPI_ISL_1203631, EPI_ISL_1203632, EPI_ISL_1203633, EPI_ISL_1203634, EPI_ISL_1203635, EPI_ISL_1203636, EPI_ISL_1203637, EPI_ISL_1203638, EPI_ISL_1203639, EPI_ISL_1203640, EPI_ISL_1203641, EPI_ISL_1203642, EPI_ISL_1203643, EPI_ISL_1203644, EPI_ISL_1203645, EPI_ISL_1203646, EPI_ISL_1203647, EPI_ISL_1203648, EPI_ISL_1203649, EPI_ISL_1203650, EPI_ISL_1203651, EPI_ISL_1203652, EPI_ISL_1203653, EPI_ISL_1203654, EPI_ISL_1203655, EPI_ISL_1203656, EPI_ISL_1203657, EPI_ISL_1203658, EPI_ISL_1203659, EPI_ISL_1203660, EPI_ISL_1203661, EPI_ISL_1203662, EPI_ISL_1203663, EPI_ISL_1203664, EPI_ISL_1203665, EPI_ISL_1203666, EPI_ISL_1203667, EPI_ISL_1203668, EPI_ISL_1203669, EPI_ISL_1203670, EPI_ISL_1203671, EPI_ISL_1203672, EPI_ISL_1203673, EPI_ISL_1203674, EPI_ISL_1203675, EPI_ISL_1203676, EPI_ISL_1203677, EPI_ISL_1203678, EPI_ISL_1203679, EPI_ISL_1203680, EPI_ISL_1203681, EPI_ISL_1203682, EPI_ISL_1203683, EPI_ISL_1203684, EPI_ISL_1203685, EPI_ISL_1203686, EPI_ISL_1203687, EPI_ISL_1203688, EPI_ISL_1203689, EPI_ISL_1203690, EPI_ISL_1203691, EPI_ISL_1203692, EPI_ISL_1203693, EPI_ISL_1203694, EPI_ISL_1203695, EPI_ISL_1203696, EPI_ISL_1203697, EPI_ISL_1203698, EPI_ISL_1203699, EPI_ISL_1203700, EPI_ISL_1203701, EPI_ISL_1203702, EPI_ISL_1203703, EPI_ISL_1203704, EPI_ISL_1203705, EPI_ISL_1203706, EPI_ISL_1203707, EPI_ISL_1203708, EPI_ISL_1203709, EPI_ISL_1203710, EPI_ISL_1203711, EPI_ISL_1203712, EPI_ISL_1203713, EPI_ISL_1203714, EPI_ISL_1203715, EPI_ISL_1203716, EPI_ISL_1203717, EPI_ISL_1203718, EPI_ISL_1203719, EPI_ISL_1203720, EPI_ISL_1203721, EPI_ISL_1203722, EPI_ISL_1203723, EPI_ISL_1203724, EPI_ISL_1203725, EPI_ISL_1203726, EPI_ISL_1203727, EPI_ISL_1203728, EPI_ISL_1203729, EPI_ISL_1203730, EPI_ISL_1203731, EPI_ISL_1203732, EPI_ISL_1203733, EPI_ISL_1203734, EPI_ISL_1203735, EPI_ISL_1203736, EPI_ISL_1203737, EPI_ISL_1203738, EPI_ISL_1203739, EPI_ISL_1203740, EPI_ISL_1203741, EPI_ISL_1203742, EPI_ISL_1203743, EPI_ISL_1203744, EPI_ISL_1203745, EPI_ISL_1203746, EPI_ISL_1203747, EPI_ISL_1203748, EPI_ISL_1203749, EPI_ISL_1203750, EPI_ISL_1203751, EPI_ISL_1203752, EPI_ISL_1203753, EPI_ISL_1203754, EPI_ISL_1203755, EPI_ISL_1203756, EPI_ISL_1203757, EPI_ISL_1203758, EPI_ISL_1203759, EPI_ISL_1203760, EPI_ISL_1203761, EPI_ISL_1203762, EPI_ISL_1203763, EPI_ISL_1203764, EPI_ISL_1203765, EPI_ISL_1203766, EPI_ISL_1203767, EPI_ISL_1203768, EPI_ISL_1203769, EPI_ISL_1203770, EPI_ISL_1203771, EPI_ISL_1203772, EPI_ISL_1203773, EPI_ISL_1203774, EPI_ISL_1203775, EPI_ISL_1203776, EPI_ISL_1203777, EPI_ISL_1203778, EPI_ISL_1203779, EPI_ISL_1203780, EPI_ISL_1203781, EPI_ISL_1203782, EPI_ISL_1203783, EPI_ISL_1203784, EPI_ISL_1203785, EPI_ISL_1203786, EPI_ISL_1203787, EPI_ISL_1203788, EPI_ISL_1203789, EPI_ISL_1203790, EPI_ISL_1203791, EPI_ISL_1203792, EPI_ISL_1203793, EPI_ISL_1203794, EPI_ISL_1203795, EPI_ISL_1203796, EPI_ISL_1203797, EPI_ISL_1203798, EPI_ISL_1203799, EPI_ISL_1203800, EPI_ISL_1203801, EPI_ISL_1203802, EPI_ISL_1203803, EPI_ISL_1203804, EPI_ISL_1203805, EPI_ISL_1203806, EPI_ISL_1203807, EPI_ISL_1203808, EPI_ISL_1203809, EPI_ISL_1203810, EPI_ISL_1203811, EPI_ISL_1203812, EPI_ISL_1203813, EPI_ISL_1203814, EPI_ISL_1203815 | | | |
| see above | Houston Methodist Hospital | Houston Methodist Hospital | S. Wesley Long, Randall J. Olsen, Paul A. Christensen, Sishir Subedi, Robert Olson, James J. Davis, Matthew Ojeda Saavedra, Prasanti Yerramilli, Layne Pruitt, Kristina Reppond, Madison N. Shyer, Jessica Cambric, Ilya J. Finkelstein, Jimmy Gollihar, and James M. Musser |
| EPI_ISL_1203817, EPI_ISL_1203818 | Santa Clara County Public Health Laboratory | Santa Clara County Public Health Laboratory | Santa Clara County Public Health Department |
| EPI_ISL_1203819, EPI_ISL_1203820, EPI_ISL_1203821, EPI_ISL_1203822, EPI_ISL_1203823, EPI_ISL_1203824, EPI_ISL_1203826, EPI_ISL_1203827, EPI_ISL_1203828, EPI_ISL_1203829, EPI_ISL_1203830, EPI_ISL_1203831, EPI_ISL_1203833, EPI_ISL_1203834, EPI_ISL_1203835, EPI_ISL_1203836, EPI_ISL_1203837, EPI_ISL_1203838, EPI_ISL_1203839, EPI_ISL_1203840, EPI_ISL_1203842 | | | |
| see above | The Jackson Laboratory | The Jackson Laboratory | Lloyd M, Sanderson B, Srivastava A, Maurya R, Renzette N, Omerza G, Kelly K, Li L, Wei C L, Adams M |
| EPI_ISL_1203843 | Clinical Microbiology Laboratory | James Molecular Laboratory | Huanyu Wang, Dan Jones, Pam Snyder, Zachary Rudy, Amy Leber |
| EPI_ISL_1205162, EPI_ISL_1205163, EPI_ISL_1205164, EPI_ISL_1205165, EPI_ISL_1205166, EPI_ISL_1205167, EPI_ISL_1205168, EPI_ISL_1205169, EPI_ISL_1205170, EPI_ISL_1205171, EPI_ISL_1205172, EPI_ISL_1205173, EPI_ISL_1205176, EPI_ISL_1205177, EPI_ISL_1205182, EPI_ISL_1205184, EPI_ISL_1205186, EPI_ISL_1205188, EPI_ISL_1205189, EPI_ISL_1205191, EPI_ISL_1205192, EPI_ISL_1205193, EPI_ISL_1205195, EPI_ISL_1205196, EPI_ISL_1205197, EPI_ISL_1205198, EPI_ISL_1205201, EPI_ISL_1205202 | | | |
| see above | Utah Public Health Laboratory | Utah Public Health Laboratory | Erin L. Young, Kelly F. Oakeson, Tara Gallagher |
| EPI_ISL_1205203, EPI_ISL_1205204, EPI_ISL_1205205, EPI_ISL_1205206, EPI_ISL_1205207, EPI_ISL_1205208, EPI_ISL_1205209, EPI_ISL_1205210, EPI_ISL_1205211, EPI_ISL_1205212, EPI_ISL_1205213, EPI_ISL_1205214, EPI_ISL_1205215, EPI_ISL_1205216, EPI_ISL_1205217, EPI_ISL_1205218 | | | |
| see above | Instituto Nacional de Medicina Genomica | Instituto Nacional de Medicina Genomica | Hidalgo-Miranda A, Mendoza-Vargas A, Reyes-Grajeda JP, Cedro-Tanda A, Alcaraz N, Gonzalez-Barrera D, Rangel-DeLeon D, Ramirez-Vega O, Sifuentes-Rojas C, Cisneros-Villanueva M, Herrera-Montalvo LA |
| EPI_ISL_1205220, EPI_ISL_1205221, EPI_ISL_1205222, EPI_ISL_1205223, EPI_ISL_1205224, EPI_ISL_1205225, EPI_ISL_1205226, EPI_ISL_1205227, EPI_ISL_1205228, EPI_ISL_1205229, EPI_ISL_1205230, EPI_ISL_1205231, EPI_ISL_1205232, EPI_ISL_1205233, EPI_ISL_1205234, EPI_ISL_1205235, EPI_ISL_1205236, EPI_ISL_1205237, EPI_ISL_1205238, EPI_ISL_1205239, EPI_ISL_1205240, EPI_ISL_1205241, EPI_ISL_1205242, EPI_ISL_1205243, EPI_ISL_1205244, EPI_ISL_1205245, EPI_ISL_1205246, EPI_ISL_1205247, EPI_ISL_1205248, EPI_ISL_1205249, EPI_ISL_1205250, EPI_ISL_1205251, EPI_ISL_1205252, EPI_ISL_1205253, EPI_ISL_1205254, EPI_ISL_1205255, EPI_ISL_1205256, EPI_ISL_1205257, EPI_ISL_1205258, EPI_ISL_1205259, EPI_ISL_1205260, EPI_ISL_1205261, EPI_ISL_1205262, EPI_ISL_1205263, EPI_ISL_1205264, EPI_ISL_1205265, EPI_ISL_1205266, EPI_ISL_1205267, EPI_ISL_1205268, EPI_ISL_1205269, EPI_ISL_1205270, EPI_ISL_1205271, EPI_ISL_1205272, EPI_ISL_1205273, EPI_ISL_1205274, EPI_ISL_1205275, EPI_ISL_1205276, EPI_ISL_1205277, EPI_ISL_1205278, EPI_ISL_1205279, EPI_ISL_1205280, EPI_ISL_1205281, EPI_ISL_1205282, EPI_ISL_1205283, EPI_ISL_1205285, EPI_ISL_1205286, EPI_ISL_1205287, EPI_ISL_1205288, EPI_ISL_1205289, EPI_ISL_1205290, EPI_ISL_1205291, EPI_ISL_1205292, EPI_ISL_1205293, EPI_ISL_1205294, EPI_ISL_1205295, EPI_ISL_1205296, EPI_ISL_1205297, EPI_ISL_1205298, EPI_ISL_1205299, EPI_ISL_1205300, EPI_ISL_1205301, EPI_ISL_1205302 | | | |
| see above | WVU Rapid Development Laboratory | WVU and Marshall University Combined Genomics Core Facilities | James Denvir, Peter Stoilov, Peter Perrotta, Wesley Kimble, Ryan Percifield |
| EPI_ISL_1205304, EPI_ISL_1205306, EPI_ISL_1205307, EPI_ISL_1205308, EPI_ISL_1205313, EPI_ISL_1205314, EPI_ISL_1205316, EPI_ISL_1205321, EPI_ISL_1205322, EPI_ISL_1205323, EPI_ISL_1205324, EPI_ISL_1205325, EPI_ISL_1205326, EPI_ISL_1205327, EPI_ISL_1205328, EPI_ISL_1205329, EPI_ISL_1205332, EPI_ISL_1205333, EPI_ISL_1205334, EPI_ISL_1205342, EPI_ISL_1205343, EPI_ISL_1205346, EPI_ISL_1205347, EPI_ISL_1205348, EPI_ISL_1205350, EPI_ISL_1205351, EPI_ISL_1205353, EPI_ISL_1205354 | | | |
| see above | Utah Public Health Laboratory | Utah Public Health Laboratory | Erin L. Young, Kelly F. Oakeson, Tara Gallagher |
| EPI_ISL_1205356, EPI_ISL_1205357, EPI_ISL_1205358, EPI_ISL_1205359, EPI_ISL_1205360, EPI_ISL_1205361, EPI_ISL_1205362, EPI_ISL_1205363, EPI_ISL_1205364, EPI_ISL_1205365, EPI_ISL_1205366, EPI_ISL_1205367, EPI_ISL_1205368, EPI_ISL_1205369, EPI_ISL_1205370, EPI_ISL_1205371, EPI_ISL_1205372, EPI_ISL_1205373, EPI_ISL_1205374, EPI_ISL_1205375, EPI_ISL_1205377, EPI_ISL_1205378, EPI_ISL_1205379, EPI_ISL_1205380 | | | |
| see above | Arizona State Public Health Laboratory | Arizona State Public Health Laboratory | Trung Huynh, Jessica Escobar, Katherine Fullerton, Nobuko Fukushima, Stacy White, Linda Getsinger, Victor Waddell |
| EPI_ISL_1205382, EPI_ISL_1205384, EPI_ISL_1205385, EPI_ISL_1205386, EPI_ISL_1205388, EPI_ISL_1205389, EPI_ISL_1205391, EPI_ISL_1205392, EPI_ISL_1205394, EPI_ISL_1205395, EPI_ISL_1205396, EPI_ISL_1205398, EPI_ISL_1205399, EPI_ISL_1205400, EPI_ISL_1205404, EPI_ISL_1205405, EPI_ISL_1205406, EPI_ISL_1205407, EPI_ISL_1205408, EPI_ISL_1205409, EPI_ISL_1205410, EPI_ISL_1205411, EPI_ISL_1205413, EPI_ISL_1205414, EPI_ISL_1205415, EPI_ISL_1205417, EPI_ISL_1205418, EPI_ISL_1205420, EPI_ISL_1205421, EPI_ISL_1205422, EPI_ISL_1205423, EPI_ISL_1205425, EPI_ISL_1205426, EPI_ISL_1205427, EPI_ISL_1205428, EPI_ISL_1205429, EPI_ISL_1205430, EPI_ISL_1205431, EPI_ISL_1205433, EPI_ISL_1205434, EPI_ISL_1205435, EPI_ISL_1205437, EPI_ISL_1205438, EPI_ISL_1205440, EPI_ISL_1205441, EPI_ISL_1205442, EPI_ISL_1205444, EPI_ISL_1205445, EPI_ISL_1205446, EPI_ISL_1205447, EPI_ISL_1205448, EPI_ISL_1205449, EPI_ISL_1205450, EPI_ISL_1205451, EPI_ISL_1205452, EPI_ISL_1205453, EPI_ISL_1205454, EPI_ISL_1205456 | | | |
| see above | Mayo Clinic & Mayo Clinic Laboratories | Minnesota Department of Health, Public Health Laboratory | Alexandra Lorentz, Jacob Garfin, Matt Plumb, and Xiong Wang |
| EPI_ISL_1205457, EPI_ISL_1205460 | University of Wisconsin-Madison AIDS Vaccine Research Laboratories | University of Wisconsin-Madison AIDS Vaccine Research Laboratories | Gage Moreno, Katarina Braun, et al. AIDS Vaccine Research Laboratories |
| EPI_ISL_1205472, EPI_ISL_1205474, EPI_ISL_1205476, EPI_ISL_1205477, EPI_ISL_1205478, EPI_ISL_1205480, EPI_ISL_1205481, EPI_ISL_1205482, EPI_ISL_1205483, EPI_ISL_1205484 | Alameda County Public Health Department | Alameda County Public Health Department | Kristna Hsieh, Maria Elma, Kevin Libuit |
| EPI_ISL_1209030, EPI_ISL_1209031, EPI_ISL_1209032, EPI_ISL_1209033, EPI_ISL_1209034, EPI_ISL_1209035, EPI_ISL_1209036, EPI_ISL_1209037, EPI_ISL_1209038, EPI_ISL_1209039, EPI_ISL_1209040, EPI_ISL_1209041, EPI_ISL_1209042, EPI_ISL_1209043, EPI_ISL_1209044, EPI_ISL_1209045, EPI_ISL_1209046, EPI_ISL_1209047, EPI_ISL_1209048, EPI_ISL_1209050, EPI_ISL_1209051, EPI_ISL_1209052, EPI_ISL_1209053, EPI_ISL_1209054, EPI_ISL_1209055, EPI_ISL_1209056, EPI_ISL_1209057, EPI_ISL_1209058, EPI_ISL_1209059, EPI_ISL_1209060, EPI_ISL_1209061, EPI_ISL_1209062, EPI_ISL_1209063, EPI_ISL_1209064, EPI_ISL_1209065, EPI_ISL_1209066, EPI_ISL_1209067, EPI_ISL_1209068, EPI_ISL_1209069, EPI_ISL_1209070, EPI_ISL_1209071, EPI_ISL_1209072, EPI_ISL_1209073, EPI_ISL_1209074, EPI_ISL_1209075, EPI_ISL_1209076, EPI_ISL_1209077, EPI_ISL_1209078, EPI_ISL_1209079, EPI_ISL_1209080, EPI_ISL_1209081, EPI_ISL_1209082, EPI_ISL_1209083, EPI_ISL_1209084, EPI_ISL_1209085, EPI_ISL_1209086, EPI_ISL_1209087, EPI_ISL_1209088, EPI_ISL_1209089, EPI_ISL_1209090, EPI_ISL_1209091, EPI_ISL_1209092, EPI_ISL_1209093, EPI_ISL_1209094, EPI_ISL_1209095, EPI_ISL_1209097, EPI_ISL_1209098, EPI_ISL_1209099, EPI_ISL_1209101, EPI_ISL_1209102, EPI_ISL_1209103, EPI_ISL_1209104, EPI_ISL_1209105, EPI_ISL_1209106, EPI_ISL_1209107, EPI_ISL_1209108, EPI_ISL_1209109, EPI_ISL_1209110, EPI_ISL_1209111, EPI_ISL_1209112, EPI_ISL_1209113, EPI_ISL_1209114, EPI_ISL_1209115, EPI_ISL_1209116, EPI_ISL_1209117, EPI_ISL_1209118, EPI_ISL_1209119, EPI_ISL_1209120, EPI_ISL_1209121, EPI_ISL_1209122, EPI_ISL_1209123, EPI_ISL_1209124, EPI_ISL_1209125, EPI_ISL_1209126, EPI_ISL_1209127, EPI_ISL_1209128, EPI_ISL_1209129, EPI_ISL_1209130, EPI_ISL_1209131, EPI_ISL_1209132, EPI_ISL_1209133, EPI_ISL_1209134, EPI_ISL_1209135, EPI_ISL_1209136, EPI_ISL_1209137, EPI_ISL_1209138, EPI_ISL_1209139, EPI_ISL_1209140, EPI_ISL_1209142, EPI_ISL_1209143, EPI_ISL_1209144, EPI_ISL_1209145, EPI_ISL_1209146, EPI_ISL_1209147, EPI_ISL_1209148, EPI_ISL_1209149, EPI_ISL_1209150, EPI_ISL_1209151, EPI_ISL_1209152, EPI_ISL_1209153, EPI_ISL_1209154, EPI_ISL_1209155, EPI_ISL_1209156, EPI_ISL_1209157, EPI_ISL_1209159, EPI_ISL_1209160, EPI_ISL_1209161, EPI_ISL_1209162, EPI_ISL_1209163, EPI_ISL_1209164, EPI_ISL_1209165, EPI_ISL_1209166, EPI_ISL_1209167, EPI_ISL_1209168, EPI_ISL_1209169, EPI_ISL_1209170, EPI_ISL_1209171, EPI_ISL_1209172, EPI_ISL_1209173, EPI_ISL_1209174, EPI_ISL_1209175, EPI_ISL_1209176, EPI_ISL_1209177, EPI_ISL_1209178, EPI_ISL_1209179, EPI_ISL_1209180, EPI_ISL_1209181, EPI_ISL_1209182, EPI_ISL_1209183, EPI_ISL_1209184, EPI_ISL_1209185, EPI_ISL_1209186, EPI_ISL_1209187, EPI_ISL_1209188, EPI_ISL_1209189, EPI_ISL_1209190, EPI_ISL_1209191, EPI_ISL_1209192, EPI_ISL_1209193, EPI_ISL_1209194, EPI_ISL_1209195, EPI_ISL_1209196, EPI_ISL_1209197, EPI_ISL_1209198, EPI_ISL_1209199, EPI_ISL_1209200, EPI_ISL_1209201, EPI_ISL_1209202, EPI_ISL_1209203, EPI_ISL_1209204, EPI_ISL_1209205, EPI_ISL_1209206, EPI_ISL_1209207, EPI_ISL_1209208, EPI_ISL_1209209, EPI_ISL_1209210, EPI_ISL_1209212, EPI_ISL_1209213, EPI_ISL_1209214, EPI_ISL_1209215, EPI_ISL_1209217, EPI_ISL_1209218, EPI_ISL_1209219, EPI_ISL_1209220, EPI_ISL_1209221, EPI_ISL_1209222, EPI_ISL_1209223, EPI_ISL_1209226, EPI_ISL_1209227, EPI_ISL_1209228, EPI_ISL_1209229, EPI_ISL_1209230, EPI_ISL_1209231, EPI_ISL_1209232, EPI_ISL_1209233, EPI_ISL_1209234, EPI_ISL_1209235, EPI_ISL_1209236, EPI_ISL_1209237, EPI_ISL_1209238, EPI_ISL_1209239, EPI_ISL_1209240, EPI_ISL_1209242 | | | |
| see above | UW Virology Lab | UW Virology Lab | Pavitra Roychoudhury, Hong Xie, Lasata Shrestha, Shah Mohamed Bakhash, Michelle Lin, Noah Baker, Sean Ellis, Saraswathi Sathees, Meei-Li Huang, Keith R Jerome, Alexander Greninger |
| EPI_ISL_1209454, EPI_ISL_1209455, EPI_ISL_1209456, EPI_ISL_1209457, EPI_ISL_1209458, EPI_ISL_1209459, EPI_ISL_1209460, EPI_ISL_1209462, EPI_ISL_1209464 | Univeristy of New Mexico Hospital | Center for Global Health, University of New Mexico Health Sciences Center | Daryl Domman, Kurt Schwalm, Justin Bacca, Jon Femling, Darrell Dinwiddie |
| EPI_ISL_1218653, EPI_ISL_1218654, EPI_ISL_1218655, EPI_ISL_1218656, EPI_ISL_1218658, EPI_ISL_1218659, EPI_ISL_1218662, EPI_ISL_1218663, EPI_ISL_1218664, EPI_ISL_1218665, EPI_ISL_1218666, EPI_ISL_1218668, EPI_ISL_1218669, EPI_ISL_1218670, EPI_ISL_1218671, EPI_ISL_1218672, EPI_ISL_1218673, EPI_ISL_1218674, EPI_ISL_1218676, EPI_ISL_1218677, EPI_ISL_1218678, EPI_ISL_1218680, EPI_ISL_1218681, EPI_ISL_1218683, EPI_ISL_1218685, EPI_ISL_1218687, EPI_ISL_1218688, EPI_ISL_1218689, EPI_ISL_1218690, EPI_ISL_1218691, EPI_ISL_1218692, EPI_ISL_1218694, EPI_ISL_1218695, EPI_ISL_1218696, EPI_ISL_1218697, EPI_ISL_1218698, EPI_ISL_1218699, EPI_ISL_1218700, EPI_ISL_1218701, EPI_ISL_1218702, EPI_ISL_1218704, EPI_ISL_1218705, EPI_ISL_1218706, EPI_ISL_1218707, EPI_ISL_1218708, EPI_ISL_1218709, EPI_ISL_1218711, EPI_ISL_1218713, EPI_ISL_1218714, EPI_ISL_1218715, EPI_ISL_1218716, EPI_ISL_1218719, EPI_ISL_1218720, EPI_ISL_1218721, EPI_ISL_1218722, EPI_ISL_1218723, EPI_ISL_1218724, EPI_ISL_1218725, EPI_ISL_1218726, EPI_ISL_1218728, EPI_ISL_1218731, EPI_ISL_1218732, EPI_ISL_1218733, EPI_ISL_1218734, EPI_ISL_1218735, EPI_ISL_1218736, EPI_ISL_1218737, EPI_ISL_1218738, EPI_ISL_1218739, EPI_ISL_1218740, EPI_ISL_1218741, EPI_ISL_1218742, EPI_ISL_1218743, EPI_ISL_1218744, EPI_ISL_1218745, EPI_ISL_1218746, EPI_ISL_1218747, EPI_ISL_1218749, EPI_ISL_1218750, EPI_ISL_1218751, EPI_ISL_1218752, EPI_ISL_1218753, EPI_ISL_1218754, EPI_ISL_1218755, EPI_ISL_1218757, EPI_ISL_1218758, EPI_ISL_1218760, EPI_ISL_1218761, EPI_ISL_1218762, EPI_ISL_1218763, EPI_ISL_1218764, EPI_ISL_1218765, EPI_ISL_1218766, EPI_ISL_1218768, EPI_ISL_1218769, EPI_ISL_1218770, EPI_ISL_1218771, EPI_ISL_1218772, EPI_ISL_1218773, EPI_ISL_1218774, EPI_ISL_1218775, EPI_ISL_1218776, EPI_ISL_1218777, EPI_ISL_1218778, EPI_ISL_1218779, EPI_ISL_1218780, EPI_ISL_1218781, EPI_ISL_1218782, EPI_ISL_1218783, EPI_ISL_1218784, EPI_ISL_1218785, EPI_ISL_1218786, EPI_ISL_1218787, EPI_ISL_1218788, EPI_ISL_1218789, EPI_ISL_1218790, EPI_ISL_1218791, EPI_ISL_1218792, EPI_ISL_1218793, EPI_ISL_1218794, EPI_ISL_1218795, EPI_ISL_1218796, EPI_ISL_1218797, EPI_ISL_1218798, EPI_ISL_1218800, EPI_ISL_1218801, EPI_ISL_1218802, EPI_ISL_1218803, EPI_ISL_1218804, EPI_ISL_1218805, EPI_ISL_1218806, EPI_ISL_1218807, EPI_ISL_1218809, EPI_ISL_1218810, EPI_ISL_1218811, EPI_ISL_1218812, EPI_ISL_1218813, EPI_ISL_1218814, EPI_ISL_1218816, EPI_ISL_1218817, EPI_ISL_1218818, EPI_ISL_1218819, EPI_ISL_1218820, EPI_ISL_1218821, EPI_ISL_1218822, EPI_ISL_1218823, EPI_ISL_1218824, EPI_ISL_1218825, EPI_ISL_1218826, EPI_ISL_1218827, EPI_ISL_1218828, EPI_ISL_1218832, EPI_ISL_1218833, EPI_ISL_1218834, EPI_ISL_1218835, EPI_ISL_1218836, EPI_ISL_1218837, EPI_ISL_1218839, EPI_ISL_1218840, EPI_ISL_1218841, EPI_ISL_1218842, EPI_ISL_1218843, EPI_ISL_1218844, EPI_ISL_1218846, EPI_ISL_1218847, EPI_ISL_1218848, EPI_ISL_1218849, EPI_ISL_1218850, EPI_ISL_1218851, EPI_ISL_1218852, EPI_ISL_1218853, EPI_ISL_1218857, EPI_ISL_1218858, EPI_ISL_1218860, EPI_ISL_1218861, EPI_ISL_1218862, EPI_ISL_1218864, EPI_ISL_1218866, EPI_ISL_1218868, EPI_ISL_1218869, EPI_ISL_1218870, EPI_ISL_1218871, EPI_ISL_1218872, EPI_ISL_1218873, EPI_ISL_1218875, EPI_ISL_1218876, EPI_ISL_1218877, EPI_ISL_1218878, EPI_ISL_1218879, EPI_ISL_1218881, EPI_ISL_1218885, EPI_ISL_1218886, EPI_ISL_1218888, EPI_ISL_1218889, EPI_ISL_1218890, EPI_ISL_1218891, EPI_ISL_1218892, EPI_ISL_1218893, EPI_ISL_1218894, EPI_ISL_1218895, EPI_ISL_1218896, EPI_ISL_1218897, EPI_ISL_1218898, EPI_ISL_1218899, EPI_ISL_1218900, EPI_ISL_1218901, EPI_ISL_1218902, EPI_ISL_1218903, EPI_ISL_1218904, EPI_ISL_1218905, EPI_ISL_1218906, EPI_ISL_1218907, EPI_ISL_1218908, EPI_ISL_1218909, EPI_ISL_1218910, EPI_ISL_1218911, EPI_ISL_1218912, EPI_ISL_1218913, EPI_ISL_1218914, EPI_ISL_1218915, EPI_ISL_1218916, EPI_ISL_1218917, EPI_ISL_1218918, EPI_ISL_1218921, EPI_ISL_1218922, EPI_ISL_1218923, EPI_ISL_1218924, EPI_ISL_1218925, EPI_ISL_1218926, EPI_ISL_1218927, EPI_ISL_1218928, EPI_ISL_1218929, EPI_ISL_1218930, EPI_ISL_1218931, EPI_ISL_1218932, EPI_ISL_1218933, EPI_ISL_1218934, EPI_ISL_1218935, EPI_ISL_1218936, EPI_ISL_1218937, EPI_ISL_1218939, EPI_ISL_1218940, EPI_ISL_1218941, EPI_ISL_1218942, EPI_ISL_1218943, EPI_ISL_1218944, EPI_ISL_1218945, EPI_ISL_1218946, EPI_ISL_1218947, EPI_ISL_1218948, EPI_ISL_1218950, EPI_ISL_1218951, EPI_ISL_1218953, EPI_ISL_1218955, EPI_ISL_1218956, EPI_ISL_1218958, EPI_ISL_1218959, EPI_ISL_1218960, EPI_ISL_1218961, EPI_ISL_1218962, EPI_ISL_1218963, EPI_ISL_1218964, EPI_ISL_1218965, EPI_ISL_1218966, EPI_ISL_1218967, EPI_ISL_1218970, EPI_ISL_1218971, EPI_ISL_1218972, EPI_ISL_1218973, EPI_ISL_1218974, EPI_ISL_1218975, EPI_ISL_1218976, EPI_ISL_1218977, EPI_ISL_1218978, EPI_ISL_1218979, EPI_ISL_1218980, EPI_ISL_1218981, EPI_ISL_1218982, EPI_ISL_1218983, EPI_ISL_1218985, EPI_ISL_1218986, EPI_ISL_1218987, EPI_ISL_1218988, EPI_ISL_1218989, EPI_ISL_1218990, EPI_ISL_1218991, EPI_ISL_1218992, EPI_ISL_1218993, EPI_ISL_1218994, EPI_ISL_1218995, EPI_ISL_1218996, EPI_ISL_1218997, EPI_ISL_1219000, EPI_ISL_1219001 | | | |
| see above | Florida Bureau of Public Health Laboratories | Florida Bureau of Public Health Laboratories | Sarah Schmedes, Jason Blanton |
| EPI_ISL_1219703, EPI_ISL_1219704, EPI_ISL_1219705, EPI_ISL_1219706 | School of Pharmacy, Shenandoah University | School of Pharmacy, Shenandoah University | Adams,S.M., Harralson,A.F., Kidd,R.S., Sawyer,G.W. |

| | | | |
|---|---|---|---|
| EPI_ISL_1219711, EPI_ISL_1219712, EPI_ISL_1219713 | Instituto de Diagnostico y Referencia Epidemiologicos INDRE_RNLSP | Instituto de Diagnostico y Referencia Epidemiologicos (INDRE) | Claudia Wong-Arambula, Abril Rodriguez-Maldonado, Vanessa Rivero-Arredondo, Ariadna Medina-Benitez, Joaquin Quiroz-Mercado,David Fragoso-Fonseca, Sergio Rangel-Guerrero, Natividad Cruz-Ortiz, Tatiana Nunez-Garcia, Gisela Barrera-Badillo, Lucia Hernandez-Rivas, Irma Lopez-Martinez, Ernesto Ramirez-Gonzalez. |
| EPI_ISL_1219714 | CICSaB/UASLP/LESP SLP | Instituto de Diagnostico y Referencia Epidemiologicos (INDRE) | Claudia Wong-Arambula, Abril Rodriguez-Maldonado, Vanessa Rivero-Arredondo, Ariadna Medina-Benitez, Joaquin Quiroz-Mercado,David Fragoso-Fonseca, Sergio Rangel-Guerrero, Andreu Comas-Garcia, Natividad Cruz-Ortiz, Tatiana Nunez-Garcia, Gisela Barrera-Badillo, Lucia Hernandez-Rivas, Irma Lopez-Martinez, Ernesto Ramirez-Gonzalez. |
| EPI_ISL_1219715, EPI_ISL_1219716, EPI_ISL_1219717, EPI_ISL_1219718 | Instituto de Diagnostico y Referencia Epidemiologicos INDRE_RNLSP | Instituto de Diagnostico y Referencia Epidemiologicos (INDRE) | Claudia Wong-Arambula, Abril Rodriguez-Maldonado, Vanessa Rivero-Arredondo, Ariadna Medina-Benitez, Joaquin Quiroz-Mercado,David Fragoso-Fonseca, Sergio Rangel-Guerrero, Natividad Cruz-Ortiz, Tatiana Nunez-Garcia, Gisela Barrera-Badillo, Lucia Hernandez-Rivas, Irma Lopez-Martinez, Ernesto Ramirez-Gonzalez. |
| EPI_ISL_1219802, EPI_ISL_1219803, EPI_ISL_1219804, EPI_ISL_1219805, EPI_ISL_1219806 | South Dakota Public Health Laboratory | South Dakota Public Health Laboratory | Jacob Garfin and Chris Carlson |
| EPI_ISL_1219807, EPI_ISL_1219808, EPI_ISL_1219809, EPI_ISL_1219810, EPI_ISL_1219811, EPI_ISL_1219812, EPI_ISL_1219813, EPI_ISL_1219814, EPI_ISL_1219815, EPI_ISL_1219816, EPI_ISL_1219817, EPI_ISL_1219818, EPI_ISL_1219819, EPI_ISL_1219820, EPI_ISL_1219821, EPI_ISL_1219822, EPI_ISL_1219823, EPI_ISL_1219824, EPI_ISL_1219825, EPI_ISL_1219826, EPI_ISL_1219834, EPI_ISL_1219835, EPI_ISL_1219836, EPI_ISL_1219837, EPI_ISL_1219838, EPI_ISL_1219839, EPI_ISL_1219840, EPI_ISL_1219841, EPI_ISL_1219842, EPI_ISL_1219843, EPI_ISL_1219844, EPI_ISL_1219845, EPI_ISL_1219846, EPI_ISL_1219847, EPI_ISL_1219848, EPI_ISL_1219849, EPI_ISL_1219850, EPI_ISL_1219851, EPI_ISL_1219852, EPI_ISL_1219853, EPI_ISL_1219854, EPI_ISL_1219855, EPI_ISL_1219856, EPI_ISL_1219857, EPI_ISL_1219858, EPI_ISL_1219859, EPI_ISL_1219860, EPI_ISL_1219861, EPI_ISL_1219863, EPI_ISL_1219864, EPI_ISL_1219865, EPI_ISL_1219866, EPI_ISL_1219867, EPI_ISL_1219868, EPI_ISL_1219869, EPI_ISL_1219870, EPI_ISL_1219871, EPI_ISL_1219872, EPI_ISL_1219873, EPI_ISL_1219874, EPI_ISL_1219875, EPI_ISL_1219876, EPI_ISL_1219877, EPI_ISL_1219878, EPI_ISL_1219879, EPI_ISL_1219880, EPI_ISL_1219881, EPI_ISL_1219882, EPI_ISL_1219883, EPI_ISL_1219884, EPI_ISL_1219885, EPI_ISL_1219886, EPI_ISL_1219887, EPI_ISL_1219888, EPI_ISL_1219889, EPI_ISL_1219890, EPI_ISL_1219891, EPI_ISL_1219892, EPI_ISL_1219893, EPI_ISL_1219894, EPI_ISL_1219895, EPI_ISL_1219896, EPI_ISL_1219897, EPI_ISL_1219898, EPI_ISL_1219899, EPI_ISL_1219900, EPI_ISL_1219901, EPI_ISL_1219902, EPI_ISL_1219903, EPI_ISL_1219904, EPI_ISL_1219905, EPI_ISL_1219906, EPI_ISL_1219907, EPI_ISL_1219908, EPI_ISL_1219909, EPI_ISL_1219910 | | | |
| see above | Nevada State Public Health Laboratory | Nevada State Public Health Laboratory | Andrew Gorzalski, Mark Pandori |
| EPI_ISL_1219911, EPI_ISL_1219912, EPI_ISL_1219913, EPI_ISL_1219914, EPI_ISL_1219915, EPI_ISL_1219916, EPI_ISL_1219917, EPI_ISL_1219918, EPI_ISL_1219919, EPI_ISL_1219920, EPI_ISL_1219921, EPI_ISL_1219922, EPI_ISL_1219923 | | | |
| see above | Idaho Bureau of Laboratories | Idaho Bureau of Laboratories | "R. Beukelman, Matthew Charles Burns, Aimee Ceniseros, Robert L. Voermans, Christopher Ball" |
| EPI_ISL_1219964, EPI_ISL_1219965, EPI_ISL_1219966, EPI_ISL_1219967, EPI_ISL_1219968, EPI_ISL_1219969, EPI_ISL_1219970, EPI_ISL_1219971, EPI_ISL_1219972, EPI_ISL_1219973, EPI_ISL_1219974, EPI_ISL_1219975, EPI_ISL_1219976, EPI_ISL_1219977, EPI_ISL_1219978, EPI_ISL_1219979, EPI_ISL_1219980, EPI_ISL_1219981, EPI_ISL_1219982, EPI_ISL_1219983, EPI_ISL_1219984, EPI_ISL_1219987, EPI_ISL_1219988, EPI_ISL_1219989, EPI_ISL_1219990, EPI_ISL_1219991, EPI_ISL_1219992, EPI_ISL_1219993, EPI_ISL_1219994, EPI_ISL_1219995, EPI_ISL_1219996, EPI_ISL_1219997, EPI_ISL_1219998, EPI_ISL_1219999, EPI_ISL_1220000, EPI_ISL_1220001, EPI_ISL_1220002, EPI_ISL_1220003, EPI_ISL_1220004, EPI_ISL_1220005, EPI_ISL_1220006, EPI_ISL_1220007, EPI_ISL_1220008, EPI_ISL_1220009, EPI_ISL_1220010, EPI_ISL_1220011, EPI_ISL_1220012, EPI_ISL_1220013, EPI_ISL_1220014, EPI_ISL_1220015, EPI_ISL_1220016, EPI_ISL_1220017, EPI_ISL_1220018, EPI_ISL_1220019, EPI_ISL_1220020, EPI_ISL_1220021, EPI_ISL_1220022, EPI_ISL_1220023, EPI_ISL_1220024, EPI_ISL_1220025, EPI_ISL_1220026, EPI_ISL_1220027, EPI_ISL_1220028, EPI_ISL_1220029, EPI_ISL_1220030, EPI_ISL_1220031, EPI_ISL_1220032, EPI_ISL_1220033, EPI_ISL_1220034, EPI_ISL_1220035, EPI_ISL_1220036, EPI_ISL_1220037, EPI_ISL_1220038, EPI_ISL_1220039, EPI_ISL_1220040, EPI_ISL_1220041, EPI_ISL_1220042, EPI_ISL_1220043, EPI_ISL_1220044 | | | |
| see above | Texas Department of State Health Services | Texas Department of State Health Services | Rashmi Tuladhar, Bonnie Oh, Jenny Zhang, Maliha Rahman, Mayela Pedrueza, Anita Pokharel, Myong Koag, Chun Wang, Rachel Lee, Grace Kubin |
| EPI_ISL_1220350, EPI_ISL_1220351, EPI_ISL_1220352, EPI_ISL_1220353, EPI_ISL_1220354, EPI_ISL_1220355, EPI_ISL_1220356, EPI_ISL_1220357, EPI_ISL_1220358, EPI_ISL_1220359, EPI_ISL_1220360, EPI_ISL_1220361, EPI_ISL_1220362, EPI_ISL_1220363, EPI_ISL_1220364, EPI_ISL_1220365, EPI_ISL_1220366, EPI_ISL_1220367, EPI_ISL_1220368, EPI_ISL_1220369, EPI_ISL_1220370, EPI_ISL_1220371, EPI_ISL_1220372, EPI_ISL_1220373, EPI_ISL_1220374, EPI_ISL_1220375, EPI_ISL_1220376, EPI_ISL_1220377, EPI_ISL_1220378, EPI_ISL_1220379, EPI_ISL_1220380, EPI_ISL_1220381, EPI_ISL_1220382, EPI_ISL_1220383, EPI_ISL_1220384, EPI_ISL_1220385, EPI_ISL_1220386, EPI_ISL_1220387, EPI_ISL_1220388, EPI_ISL_1220389, EPI_ISL_1220390, EPI_ISL_1220391, EPI_ISL_1220392, EPI_ISL_1220393, EPI_ISL_1220394, EPI_ISL_1220395, EPI_ISL_1220396, EPI_ISL_1220397, EPI_ISL_1220398, EPI_ISL_1220399, EPI_ISL_1220400, EPI_ISL_1220401, EPI_ISL_1220402, EPI_ISL_1220403, EPI_ISL_1220404, EPI_ISL_1220405, EPI_ISL_1220406, EPI_ISL_1220407, EPI_ISL_1220408, EPI_ISL_1220409, EPI_ISL_1220410, EPI_ISL_1220411, EPI_ISL_1220412, EPI_ISL_1220413, EPI_ISL_1220414, EPI_ISL_1220415, EPI_ISL_1220416, EPI_ISL_1220417, EPI_ISL_1220418, EPI_ISL_1220419, EPI_ISL_1220420, EPI_ISL_1220421, EPI_ISL_1220422, EPI_ISL_1220423, EPI_ISL_1220424, EPI_ISL_1220425, EPI_ISL_1220426, EPI_ISL_1220427, EPI_ISL_1220428, EPI_ISL_1220429, EPI_ISL_1220430, EPI_ISL_1220431, EPI_ISL_1220432, EPI_ISL_1220433, EPI_ISL_1220434, EPI_ISL_1220435, EPI_ISL_1220436, EPI_ISL_1220437, EPI_ISL_1220438, EPI_ISL_1220439, EPI_ISL_1220440, EPI_ISL_1220441, EPI_ISL_1220442, EPI_ISL_1220443, EPI_ISL_1220444, EPI_ISL_1220445, EPI_ISL_1220446, EPI_ISL_1220447, EPI_ISL_1220448, EPI_ISL_1220449, EPI_ISL_1220450, EPI_ISL_1220451, EPI_ISL_1220452, EPI_ISL_1220453, EPI_ISL_1220454, EPI_ISL_1220455, EPI_ISL_1220456, EPI_ISL_1220457, EPI_ISL_1220458, EPI_ISL_1220459, EPI_ISL_1220460, EPI_ISL_1220461, EPI_ISL_1220462, EPI_ISL_1220463, EPI_ISL_1220464, EPI_ISL_1220465, EPI_ISL_1220466, EPI_ISL_1220467, EPI_ISL_1220468, EPI_ISL_1220469, EPI_ISL_1220470, EPI_ISL_1220471, EPI_ISL_1220472, EPI_ISL_1220473, EPI_ISL_1220474, EPI_ISL_1220475, EPI_ISL_1220476, EPI_ISL_1220477, EPI_ISL_1220478, EPI_ISL_1220479, EPI_ISL_1220480, EPI_ISL_1220481, EPI_ISL_1220482, EPI_ISL_1220483, EPI_ISL_1220484, EPI_ISL_1220485, EPI_ISL_1220486, EPI_ISL_1220487, EPI_ISL_1220488, EPI_ISL_1220489, EPI_ISL_1220490, EPI_ISL_1220491, EPI_ISL_1220492, EPI_ISL_1220493, EPI_ISL_1220494, EPI_ISL_1220495, EPI_ISL_1220496, EPI_ISL_1220497, EPI_ISL_1220498, EPI_ISL_1220499, EPI_ISL_1220500, EPI_ISL_1220501, EPI_ISL_1220502, EPI_ISL_1220503, EPI_ISL_1220504, EPI_ISL_1220505, EPI_ISL_1220506, EPI_ISL_1220507, EPI_ISL_1220508, EPI_ISL_1220509, EPI_ISL_1220510, EPI_ISL_1220511, EPI_ISL_1220512, EPI_ISL_1220513, EPI_ISL_1220514, EPI_ISL_1220515, EPI_ISL_1220516, EPI_ISL_1220517, EPI_ISL_1220518, EPI_ISL_1220519, EPI_ISL_1220520, EPI_ISL_1220521, EPI_ISL_1220522, EPI_ISL_1220523, EPI_ISL_1220524, EPI_ISL_1220525, EPI_ISL_1220526, EPI_ISL_1220527, EPI_ISL_1220528, EPI_ISL_1220529, EPI_ISL_1220530, EPI_ISL_1220531, EPI_ISL_1220532, EPI_ISL_1220533, EPI_ISL_1220534, EPI_ISL_1220535, EPI_ISL_1220536, EPI_ISL_1220537, EPI_ISL_1220538, EPI_ISL_1220539, EPI_ISL_1220540, EPI_ISL_1220541, EPI_ISL_1220543, EPI_ISL_1220544, EPI_ISL_1220545, EPI_ISL_1220546, EPI_ISL_1220547, EPI_ISL_1220548, EPI_ISL_1220549, EPI_ISL_1220550, EPI_ISL_1220551, EPI_ISL_1220552, EPI_ISL_1220553, EPI_ISL_1220554, EPI_ISL_1220555, EPI_ISL_1220556, EPI_ISL_1220557, EPI_ISL_1220558, EPI_ISL_1220559, EPI_ISL_1220560, EPI_ISL_1220561, EPI_ISL_1220562, EPI_ISL_1220563, EPI_ISL_1220564, EPI_ISL_1220565, EPI_ISL_1220566, EPI_ISL_1220567, EPI_ISL_1220568, EPI_ISL_1220569, EPI_ISL_1220570, EPI_ISL_1220571, EPI_ISL_1220572, EPI_ISL_1220573, EPI_ISL_1220574, EPI_ISL_1220575, EPI_ISL_1220576, EPI_ISL_1220577, EPI_ISL_1220578, EPI_ISL_1220579, EPI_ISL_1220580, EPI_ISL_1220581, EPI_ISL_1220582, EPI_ISL_1220583, EPI_ISL_1220584, EPI_ISL_1220585, EPI_ISL_1220586, EPI_ISL_1220587, EPI_ISL_1220588, EPI_ISL_1220589, EPI_ISL_1220590, EPI_ISL_1220591, EPI_ISL_1220592, EPI_ISL_1220593, EPI_ISL_1220594, EPI_ISL_1220595, EPI_ISL_1220596, EPI_ISL_1220597, EPI_ISL_1220598, EPI_ISL_1220599, EPI_ISL_1220600, EPI_ISL_1220601, EPI_ISL_1220602, EPI_ISL_1220603, EPI_ISL_1220604, EPI_ISL_1220605, EPI_ISL_1220606, EPI_ISL_1220607, EPI_ISL_1220608, EPI_ISL_1220609, EPI_ISL_1220610, EPI_ISL_1220611, EPI_ISL_1220612, EPI_ISL_1220613, EPI_ISL_1220614, EPI_ISL_1220615, EPI_ISL_1220616, EPI_ISL_1220617, EPI_ISL_1220618, EPI_ISL_1220619, EPI_ISL_1220620, EPI_ISL_1220621, EPI_ISL_1220622, EPI_ISL_1220623, EPI_ISL_1220624, EPI_ISL_1220625, EPI_ISL_1220626, EPI_ISL_1220627, EPI_ISL_1220628, EPI_ISL_1220629, EPI_ISL_1220630, EPI_ISL_1220631, EPI_ISL_1220632, EPI_ISL_1220633, EPI_ISL_1220634, EPI_ISL_1220635, EPI_ISL_1220636, EPI_ISL_1220637, EPI_ISL_1220638, EPI_ISL_1220639, EPI_ISL_1220640, EPI_ISL_1220641, EPI_ISL_1220642, EPI_ISL_1220643, EPI_ISL_1220644, EPI_ISL_1220645, EPI_ISL_1220646, EPI_ISL_1220647, EPI_ISL_1220648, EPI_ISL_1220649, EPI_ISL_1220650, EPI_ISL_1220651, EPI_ISL_1220652, EPI_ISL_1220653, EPI_ISL_1220654, EPI_ISL_1220655, EPI_ISL_1220656, EPI_ISL_1220657, EPI_ISL_1220658, EPI_ISL_1220659, EPI_ISL_1220660, EPI_ISL_1220661, EPI_ISL_1220662, EPI_ISL_1220663, EPI_ISL_1220664, EPI_ISL_1220665, EPI_ISL_1220666, EPI_ISL_1220667, EPI_ISL_1220668, EPI_ISL_1220669, EPI_ISL_1220670, EPI_ISL_1220671, EPI_ISL_1220672, EPI_ISL_1220673, EPI_ISL_1220674, EPI_ISL_1220675, EPI_ISL_1220676, EPI_ISL_1220677, EPI_ISL_1220678, EPI_ISL_1220679, EPI_ISL_1220680, EPI_ISL_1220681, EPI_ISL_1220682, EPI_ISL_1220683, EPI_ISL_1220684, EPI_ISL_1220685, EPI_ISL_1220686, EPI_ISL_1220687, EPI_ISL_1220688, EPI_ISL_1220689, EPI_ISL_1220690, EPI_ISL_1220691, EPI_ISL_1220692, EPI_ISL_1220693, EPI_ISL_1220694, EPI_ISL_1220695, EPI_ISL_1220696, EPI_ISL_1220697, EPI_ISL_1220698, EPI_ISL_1220699, EPI_ISL_1220700, EPI_ISL_1220701, EPI_ISL_1220702, EPI_ISL_1220703, EPI_ISL_1220704, EPI_ISL_1220705, EPI_ISL_1220706, EPI_ISL_1220707, EPI_ISL_1220708, EPI_ISL_1220709, EPI_ISL_1220710, EPI_ISL_1220711, EPI_ISL_1220712, EPI_ISL_1220713, EPI_ISL_1220714, EPI_ISL_1220715, EPI_ISL_1220716, EPI_ISL_1220717, EPI_ISL_1220718, EPI_ISL_1220719, EPI_ISL_1220720, EPI_ISL_1220721, EPI_ISL_1220722, EPI_ISL_1220723, EPI_ISL_1220724, EPI_ISL_1220725, EPI_ISL_1220726, EPI_ISL_1220727, EPI_ISL_1220728, EPI_ISL_1220729, EPI_ISL_1220730, EPI_ISL_1220731, EPI_ISL_1220732, EPI_ISL_1220733, EPI_ISL_1220734, EPI_ISL_1220735, EPI_ISL_1220736, EPI_ISL_1220737, EPI_ISL_1220738, EPI_ISL_1220739, EPI_ISL_1220740, EPI_ISL_1220741, EPI_ISL_1220742, EPI_ISL_1220743, EPI_ISL_1220744, EPI_ISL_1220745, EPI_ISL_1220746, EPI_ISL_1220747, EPI_ISL_1220748, EPI_ISL_1220749, EPI_ISL_1220750, EPI_ISL_1220751, EPI_ISL_1220752, EPI_ISL_1220753, EPI_ISL_1220754, EPI_ISL_1220755, EPI_ISL_1220756, EPI_ISL_1220757, EPI_ISL_1220758, EPI_ISL_1220759, EPI_ISL_1220760, EPI_ISL_1220761, EPI_ISL_1220762, EPI_ISL_1220763, EPI_ISL_1220764, EPI_ISL_1220765, EPI_ISL_1220766, EPI_ISL_1220767, EPI_ISL_1220768, EPI_ISL_1220769, EPI_ISL_1220770, EPI_ISL_1220771, EPI_ISL_1220772, EPI_ISL_1220773, EPI_ISL_1220774, EPI_ISL_1220775, EPI_ISL_1220776, EPI_ISL_1220777, EPI_ISL_1220778, EPI_ISL_1220779, EPI_ISL_1220780, EPI_ISL_1220781, EPI_ISL_1220782, EPI_ISL_1220783, EPI_ISL_1220784, EPI_ISL_1220785, EPI_ISL_1220786, EPI_ISL_1220787, EPI_ISL_1220788, EPI_ISL_1220789, EPI_ISL_1220790, EPI_ISL_1220791, EPI_ISL_1220792, EPI_ISL_1220793, EPI_ISL_1220794, EPI_ISL_1220795, EPI_ISL_1220796, EPI_ISL_1220797, EPI_ISL_1220798, EPI_ISL_1220799, EPI_ISL_1220800, EPI_ISL_1220801, EPI_ISL_1220802, EPI_ISL_1220803, EPI_ISL_1220804, EPI_ISL_1220805, EPI_ISL_1220806, EPI_ISL_1220807, EPI_ISL_1220808, EPI_ISL_1220809, EPI_ISL_1220810, EPI_ISL_1220811, EPI_ISL_1220812, EPI_ISL_1220813, EPI_ISL_1220814, EPI_ISL_1220815, EPI_ISL_1220816, EPI_ISL_1220817, EPI_ISL_1220818, EPI_ISL_1220819, EPI_ISL_1220820, EPI_ISL_1220821, EPI_ISL_1220822, EPI_ISL_1220823, EPI_ISL_1220824, EPI_ISL_1220825, EPI_ISL_1220826, EPI_ISL_1220827, EPI_ISL_1220828, EPI_ISL_1220829, EPI_ISL_1220830, EPI_ISL_1220831, EPI_ISL_1220832, EPI_ISL_1220833, EPI_ISL_1220834, EPI_ISL_1220835, EPI_ISL_1220836, EPI_ISL_1220837, EPI_ISL_1220838, EPI_ISL_1220839, EPI_ISL_1220840, EPI_ISL_1220841, EPI_ISL_1220842, EPI_ISL_1220843, EPI_ISL_1220844, EPI_ISL_1220845, EPI_ISL_1220846, EPI_ISL_1220847, EPI_ISL_1220848, EPI_ISL_1220849, EPI_ISL_1220850, EPI_ISL_1220851, EPI_ISL_1220852, EPI_ISL_1220853, EPI_ISL_1220854, EPI_ISL_1220855, EPI_ISL_1220856, EPI_ISL_1220857, EPI_ISL_1220858, EPI_ISL_1220859, EPI_ISL_1220860, EPI_ISL_1220861 | | | |
| see above | Quest Diagnostics Incorporated | Centers for Disease Control and Prevention Division of Viral Diseases, Pathogen Discovery | Peter W. Cook, Dakota Howard, Dhwani Batra, Ben L. Rambo-Martin, S. H. Rosenthal, A. Gerasimova, R. M. Kagan, B. Anderson, M. Hua, Y. Liu, L.E. Bernstein, K.E. Livingston, A. Perez, I. A. Shlyakhter, R. V. Rolando, R. Owen, P. Tanpaiboon, F. Lacbawan, Clinton R. Paden, Suxiang Tong, Duncan MacCannell |
| EPI_ISL_1220863, EPI_ISL_1220864, EPI_ISL_1220865, EPI_ISL_1220866, EPI_ISL_1220867, EPI_ISL_1220868, EPI_ISL_1220869, EPI_ISL_1220870, EPI_ISL_1220871, EPI_ISL_1220872, EPI_ISL_1220873, EPI_ISL_1220874, EPI_ISL_1220875, EPI_ISL_1220876, EPI_ISL_1220877, EPI_ISL_1220878, EPI_ISL_1220879 | | | |
| see above | Nevada State Public Health Laboratory | Nevada State Public Health Laboratory | Andrew Gorzalski, Mark Pandori |
| EPI_ISL_1220902, EPI_ISL_1220905, EPI_ISL_1220916, EPI_ISL_1220919, EPI_ISL_1220921, EPI_ISL_1220923, EPI_ISL_1220931, EPI_ISL_1220934, EPI_ISL_1220935, EPI_ISL_1220936, EPI_ISL_1220937, EPI_ISL_1220949, EPI_ISL_1220954, EPI_ISL_1220957, EPI_ISL_1220958, EPI_ISL_1220965, EPI_ISL_1220967, EPI_ISL_1220968, EPI_ISL_1220969, EPI_ISL_1220970, EPI_ISL_1220974, EPI_ISL_1220975, EPI_ISL_1220976, EPI_ISL_1220978, EPI_ISL_1220979, EPI_ISL_1220981, EPI_ISL_1220983, EPI_ISL_1220990, EPI_ISL_1220992, EPI_ISL_1220993, EPI_ISL_1220996, EPI_ISL_1220999, EPI_ISL_1221000, EPI_ISL_1221014, EPI_ISL_1221017, EPI_ISL_1221018, EPI_ISL_1221025, EPI_ISL_1221026, EPI_ISL_1221028, EPI_ISL_1221033, EPI_ISL_1221035, EPI_ISL_1221036, EPI_ISL_1221038, EPI_ISL_1221042, EPI_ISL_1221043, EPI_ISL_1221047, EPI_ISL_1221050, EPI_ISL_1221051, EPI_ISL_1221056, EPI_ISL_1221064, EPI_ISL_1221068, EPI_ISL_1221069, EPI_ISL_1221076, EPI_ISL_1221080, EPI_ISL_1221081, EPI_ISL_1221082, EPI_ISL_1221083, EPI_ISL_1221085, EPI_ISL_1221086, EPI_ISL_1221088, EPI_ISL_1221089, EPI_ISL_1221091, EPI_ISL_1221097, EPI_ISL_1221106, EPI_ISL_1221110, EPI_ISL_1221113, EPI_ISL_1221117, EPI_ISL_1221118, EPI_ISL_1221129, EPI_ISL_1221135, EPI_ISL_1221140, EPI_ISL_1221142, EPI_ISL_1221143, EPI_ISL_1221153, EPI_ISL_1221154, EPI_ISL_1221159, EPI_ISL_1221161, EPI_ISL_1221164, EPI_ISL_1221172, EPI_ISL_1221174, EPI_ISL_1221187, EPI_ISL_1221188, EPI_ISL_1221189, EPI_ISL_1221197, EPI_ISL_1221199, EPI_ISL_1221204, EPI_ISL_1221206, EPI_ISL_1221207, EPI_ISL_1221211, EPI_ISL_1221212, EPI_ISL_1221214, EPI_ISL_1221215, EPI_ISL_1221217, EPI_ISL_1221222, EPI_ISL_1221224, EPI_ISL_1221231, EPI_ISL_1221234, EPI_ISL_1221236, EPI_ISL_1221239, EPI_ISL_1221240, EPI_ISL_1221244, EPI_ISL_1221245, EPI_ISL_1221249, EPI_ISL_1221250, EPI_ISL_1221251, EPI_ISL_1221252, EPI_ISL_1221253, EPI_ISL_1221257, EPI_ISL_1221264, EPI_ISL_1221271, EPI_ISL_1221272, EPI_ISL_1221274, EPI_ISL_1221275, EPI_ISL_1221286, EPI_ISL_1221290, EPI_ISL_1221292, EPI_ISL_1221297, EPI_ISL_1221302, EPI_ISL_1221303, EPI_ISL_1221309, EPI_ISL_1221310, EPI_ISL_1221311, EPI_ISL_1221315, EPI_ISL_1221319, EPI_ISL_1221320, EPI_ISL_1221321, EPI_ISL_1221322, EPI_ISL_1221324, EPI_ISL_1221326, EPI_ISL_1221329, EPI_ISL_1221331, EPI_ISL_1221334, EPI_ISL_1221337, EPI_ISL_1221342, EPI_ISL_1221344, EPI_ISL_1221345, EPI_ISL_1221346, EPI_ISL_1221347, EPI_ISL_1221349, EPI_ISL_1221350, EPI_ISL_1221357, EPI_ISL_1221358, EPI_ISL_1221362, EPI_ISL_1221363, EPI_ISL_1221364, EPI_ISL_1221365, EPI_ISL_1221366, EPI_ISL_1221372, EPI_ISL_1221373, EPI_ISL_1221374, EPI_ISL_1221375, EPI_ISL_1221376, EPI_ISL_1221377, EPI_ISL_1221379, EPI_ISL_1221381, EPI_ISL_1221387, EPI_ISL_1221390, EPI_ISL_1221392, EPI_ISL_1221393, EPI_ISL_1221396, EPI_ISL_1221398, EPI_ISL_1221401, EPI_ISL_1221402, EPI_ISL_1221405, EPI_ISL_1221406, EPI_ISL_1221407, EPI_ISL_1221409, EPI_ISL_1221410, EPI_ISL_1221411, EPI_ISL_1221412, EPI_ISL_1221413, EPI_ISL_1221415, EPI_ISL_1221416, EPI_ISL_1221417, EPI_ISL_1221418, EPI_ISL_1221419, EPI_ISL_1221420, EPI_ISL_1221425, EPI_ISL_1221430, EPI_ISL_1221431, EPI_ISL_1221441, EPI_ISL_1221442, EPI_ISL_1221443, EPI_ISL_1221444, EPI_ISL_1221446, EPI_ISL_1221448, EPI_ISL_1221451, EPI_ISL_1221461, EPI_ISL_1221463, EPI_ISL_1221464, EPI_ISL_1221465, EPI_ISL_1221466, EPI_ISL_1221468, EPI_ISL_1221471, EPI_ISL_1221477, EPI_ISL_1221482, EPI_ISL_1221484, EPI_ISL_1221486, EPI_ISL_1221488, EPI_ISL_1221489, EPI_ISL_1221491, EPI_ISL_1221492, EPI_ISL_1221493, EPI_ISL_1221494, EPI_ISL_1221498, EPI_ISL_1221500, EPI_ISL_1221501, EPI_ISL_1221513, EPI_ISL_1221515, EPI_ISL_1221520, EPI_ISL_1221522, EPI_ISL_1221523, EPI_ISL_1221527, EPI_ISL_1221528, EPI_ISL_1221530, EPI_ISL_1221531, EPI_ISL_1221534, EPI_ISL_1221535, EPI_ISL_1221536, EPI_ISL_1221537, EPI_ISL_1221538, EPI_ISL_1221539, EPI_ISL_1221548, EPI_ISL_1221551, EPI_ISL_1221552, EPI_ISL_1221560, EPI_ISL_1221561, EPI_ISL_1221562, EPI_ISL_1221564, EPI_ISL_1221565, EPI_ISL_1221569, EPI_ISL_1221571, EPI_ISL_1221573, EPI_ISL_1221574, EPI_ISL_1221575, EPI_ISL_1221576, EPI_ISL_1221579, EPI_ISL_1221586, EPI_ISL_1221592, EPI_ISL_1221593, EPI_ISL_1221596, EPI_ISL_1221598, EPI_ISL_1221599, EPI_ISL_1221605, EPI_ISL_1221606, EPI_ISL_1221609, EPI_ISL_1221613, EPI_ISL_1221619, EPI_ISL_1221621, EPI_ISL_1221626, EPI_ISL_1221627, EPI_ISL_1221630, EPI_ISL_1221635, EPI_ISL_1221636, EPI_ISL_1221637, EPI_ISL_1221638, EPI_ISL_1221639, EPI_ISL_1221650, EPI_ISL_1221651, EPI_ISL_1221652, EPI_ISL_1221655, EPI_ISL_1221658, EPI_ISL_1221661, EPI_ISL_1221662, EPI_ISL_1221665, EPI_ISL_1221668, EPI_ISL_1221670, EPI_ISL_1221672, EPI_ISL_1221674, EPI_ISL_1221675, EPI_ISL_1221680, EPI_ISL_1221681, EPI_ISL_1221684, EPI_ISL_1221685, EPI_ISL_1221690, EPI_ISL_1221695, EPI_ISL_1221696, EPI_ISL_1221698, EPI_ISL_1221700, EPI_ISL_1221701, EPI_ISL_1221702, EPI_ISL_1221704, EPI_ISL_1221706, EPI_ISL_1221710, EPI_ISL_1221712, EPI_ISL_1221715, EPI_ISL_1221719, EPI_ISL_1221722, EPI_ISL_1221735, EPI_ISL_1221737, EPI_ISL_1221739, EPI_ISL_1221740, EPI_ISL_1221745, EPI_ISL_1221746, EPI_ISL_1221755, EPI_ISL_1221757, EPI_ISL_1221760, EPI_ISL_1221763, EPI_ISL_1221773, EPI_ISL_1221775, EPI_ISL_1221779, EPI_ISL_1221784, EPI_ISL_1221789, EPI_ISL_1221790, EPI_ISL_1221791, EPI_ISL_1221794, EPI_ISL_1221796, EPI_ISL_1221798, EPI_ISL_1221805, EPI_ISL_1221806, EPI_ISL_1221808, EPI_ISL_1221810, EPI_ISL_1221812, EPI_ISL_1221814, EPI_ISL_1221815, EPI_ISL_1221816, EPI_ISL_1221820, EPI_ISL_1221822, EPI_ISL_1221825, EPI_ISL_1221827, EPI_ISL_1221828, EPI_ISL_1221831, EPI_ISL_1221835, EPI_ISL_1221843, EPI_ISL_1221844, EPI_ISL_1221852, EPI_ISL_1221855, EPI_ISL_1221856, EPI_ISL_1221858, EPI_ISL_1221859, EPI_ISL_1221862, EPI_ISL_1221864, EPI_ISL_1221868, EPI_ISL_1221870, EPI_ISL_1221871, EPI_ISL_1221874, EPI_ISL_1221878, EPI_ISL_1221880, EPI_ISL_1221883, EPI_ISL_1221887, EPI_ISL_1221891, EPI_ISL_1221896, EPI_ISL_1221897, EPI_ISL_1221901, EPI_ISL_1221905, EPI_ISL_1221908, EPI_ISL_1221909, EPI_ISL_1221914, EPI_ISL_1221915, EPI_ISL_1221919, EPI_ISL_1221923, EPI_ISL_1221925, EPI_ISL_1221927, EPI_ISL_1221930, EPI_ISL_1221932, EPI_ISL_1221935, EPI_ISL_1221936, EPI_ISL_1221939, EPI_ISL_1221941, EPI_ISL_1221947, EPI_ISL_1221950, EPI_ISL_1221951, EPI_ISL_1221953, EPI_ISL_1221955, EPI_ISL_1221956, EPI_ISL_1221959, EPI_ISL_1221961, EPI_ISL_1221962, EPI_ISL_1221963, EPI_ISL_1221966, EPI_ISL_1221969, EPI_ISL_1221974, EPI_ISL_1221975, EPI_ISL_1221978, EPI_ISL_1221986, EPI_ISL_1221988, EPI_ISL_1221991, EPI_ISL_1221993, EPI_ISL_1221994, EPI_ISL_1221997, EPI_ISL_1221999, EPI_ISL_1222004, EPI_ISL_1222007, EPI_ISL_1222009, EPI_ISL_1222011, EPI_ISL_1222013, EPI_ISL_1222014, EPI_ISL_1222017, EPI_ISL_1222020, EPI_ISL_1222024, EPI_ISL_1222028, EPI_ISL_1222031, EPI_ISL_1222035, EPI_ISL_1222037, EPI_ISL_1222038, EPI_ISL_1222039, EPI_ISL_1222040, EPI_ISL_1222043, EPI_ISL_1222044, EPI_ISL_1222053, EPI_ISL_1222056, EPI_ISL_1222057, EPI_ISL_1222061, EPI_ISL_1222063, EPI_ISL_1222066, EPI_ISL_1222067, EPI_ISL_1222069, EPI_ISL_1222072, EPI_ISL_1222073, EPI_ISL_1222087, EPI_ISL_1222089, EPI_ISL_1222094, EPI_ISL_1222097, EPI_ISL_1222098, EPI_ISL_1222101, EPI_ISL_1222106, EPI_ISL_1222108, EPI_ISL_1222109, EPI_ISL_1222110, EPI_ISL_1222112, EPI_ISL_1222114, EPI_ISL_1222117, EPI_ISL_1222119, EPI_ISL_1222122, EPI_ISL_1222125, EPI_ISL_1222126, EPI_ISL_1222130, EPI_ISL_1222137, EPI_ISL_1222141, EPI_ISL_1222145, EPI_ISL_1222150, EPI_ISL_1222154, EPI_ISL_1222155, EPI_ISL_1222161, EPI_ISL_1222162, EPI_ISL_1222165, EPI_ISL_1222168, EPI_ISL_1222170, EPI_ISL_1222174, EPI_ISL_1222175, EPI_ISL_1222177, EPI_ISL_1222180, EPI_ISL_1222181, EPI_ISL_1222184, EPI_ISL_1222187, EPI_ISL_1222188, EPI_ISL_1222192, EPI_ISL_1222196, EPI_ISL_1222197, EPI_ISL_1222200, EPI_ISL_1222201, EPI_ISL_1222204, EPI_ISL_1222206, EPI_ISL_1222208, EPI_ISL_1222215, EPI_ISL_1222216, EPI_ISL_1222218, EPI_ISL_1222219, EPI_ISL_1222222, EPI_ISL_1222223, EPI_ISL_1222225, EPI_ISL_1222228, EPI_ISL_1222231, EPI_ISL_1222232, EPI_ISL_1222233, EPI_ISL_1222235, EPI_ISL_1222237, EPI_ISL_1222238, EPI_ISL_1222242, EPI_ISL_1222247, EPI_ISL_1222250, EPI_ISL_1222252, EPI_ISL_1222253, EPI_ISL_1222255, EPI_ISL_1222256, EPI_ISL_1222260, EPI_ISL_1222265, EPI_ISL_1222266, EPI_ISL_1222267, EPI_ISL_1222269, EPI_ISL_1222280, EPI_ISL_1222281, EPI_ISL_1222285, EPI_ISL_1222290, EPI_ISL_1222293, EPI_ISL_1222294, EPI_ISL_1222295, EPI_ISL_1222297, EPI_ISL_1222301, EPI_ISL_1222303, EPI_ISL_1222308, EPI_ISL_1222309, EPI_ISL_1222310, EPI_ISL_1222314, EPI_ISL_1222315, EPI_ISL_1222319, EPI_ISL_1222320, EPI_ISL_1222324, EPI_ISL_1222329, EPI_ISL_1222331, EPI_ISL_1222337, EPI_ISL_1222338, EPI_ISL_1222345, EPI_ISL_1222348, EPI_ISL_1222349, EPI_ISL_1222352, EPI_ISL_1222355, EPI_ISL_1222358, EPI_ISL_1222361, EPI_ISL_1222363, EPI_ISL_1222367, EPI_ISL_1222371, EPI_ISL_1222373, EPI_ISL_1222374, EPI_ISL_1222377, EPI_ISL_1222379, EPI_ISL_1222388, EPI_ISL_1222389, EPI_ISL_1222392, EPI_ISL_1222393, EPI_ISL_1222394, EPI_ISL_1222397, EPI_ISL_1222400, EPI_ISL_1222401, EPI_ISL_1222403, EPI_ISL_1222404, EPI_ISL_1222410, EPI_ISL_1222414, EPI_ISL_1222419, EPI_ISL_1222423, EPI_ISL_1222424, EPI_ISL_1222427, EPI_ISL_1222432, EPI_ISL_1222440, EPI_ISL_1222441, EPI_ISL_1222442 | | | |
| see above | Laboratory Corporation of America | Centers for Disease Control and Prevention Division of Viral Diseases, Pathogen Discovery | Peter W. Cook, Dakota Howard, Dhwani Batra, Ben L. Rambo-Martin, Minoo Agarwal, Eyad Almasri, Debbie Boles, Ayla Burns, Nuthawin Charoensri, Oren Cohen, Susan Countryman, Mary Ann Cristobal, Bobbi Croy, Suzanne Dale, Hrushikesh Deshmukh, Amanda Douglas, Vincent Drouillon, Marcia Eisenberg, Howard Engler, Rama Ghatti, Prashant Gupta, Susan Hicks, Jake Humphrey, Lax Iyer, Manoj Jain, Mohan Kolli, Brian Krueger, Tim Kuphal, Stanley Letovsky, Michael Levandoski, Craig Lukasik, Jonathan Meltzer, Brian Norvell, Mindy Nye, Scott Parker, Christos Petropoulos, John Pruitt, Steven Ragan, Scott Ryan, Mike Sapeta, Jana Schroth, Suresh Babu Selvaraju, Goran Stevovic, Amanda Suchanek, Andrea Throop, Lyndon Tilson, Thomas Urban, Joe Voshell, Kimberly Wagner, Jonathan Williams, Mary Williamson, Qian Zeng, Tricia Zwiefelhofer, Clinton R. Paden, Suxiang Tong, Duncan MacCannell |
| EPI_ISL_1222447, EPI_ISL_1222448, EPI_ISL_1222450, EPI_ISL_1222452, EPI_ISL_1222453, EPI_ISL_1222454, EPI_ISL_1222455 | Seattle Flu Study | Seattle Flu Study | Deborah A. Nickerson, Chris D. Frazar, Jover Lee, Benjamin Pelle, Erica Ryke, Matthew Richardson, Amanda Adler, Elisabeth Brandstetter, Peter D. Han, Kairsten Fay, Misja Ilcisin, Kirsten Lacombe, Thomas R. Sibley, Melissa Truong, Caitlin R. Wolf, Karen Cowgill, Stephanie Schrag, Jeff Duchin, Michael Boeckh, Janet A. Englund, Michael Famulare, Barry R. Lutz, Mark J. Rieder, Lea M. Starita, Matthew Thompson, Helen Y. Chu, Trevor Bedford, Jay Shendure |
| EPI_ISL_1222456, EPI_ISL_1222457, EPI_ISL_1222458, EPI_ISL_1222459, EPI_ISL_1222460, EPI_ISL_1222461, EPI_ISL_1222462, EPI_ISL_1222463, EPI_ISL_1222464, EPI_ISL_1222465, EPI_ISL_1222466, EPI_ISL_1222467, EPI_ISL_1222468, EPI_ISL_1222470, EPI_ISL_1222472, EPI_ISL_1222473, EPI_ISL_1222474, EPI_ISL_1222475, EPI_ISL_1222477, EPI_ISL_1222478, EPI_ISL_1222479, EPI_ISL_1222480, EPI_ISL_1222482, EPI_ISL_1222483, EPI_ISL_1222484, EPI_ISL_1222485, EPI_ISL_1222486, EPI_ISL_1222487, EPI_ISL_1222488, EPI_ISL_1222489, EPI_ISL_1222490, EPI_ISL_1222491, EPI_ISL_1222492, EPI_ISL_1222493, EPI_ISL_1222495, EPI_ISL_1222496, EPI_ISL_1222499, EPI_ISL_1222501, EPI_ISL_1222502, EPI_ISL_1222503, EPI_ISL_1222505, EPI_ISL_1222506, EPI_ISL_1222507, EPI_ISL_1222508, EPI_ISL_1222509, EPI_ISL_1222510, EPI_ISL_1222511, EPI_ISL_1222512, EPI_ISL_1222513, EPI_ISL_1222514, EPI_ISL_1222515, EPI_ISL_1222516, EPI_ISL_1222517, EPI_ISL_1222518, EPI_ISL_1222519, EPI_ISL_1222520, EPI_ISL_1222521, EPI_ISL_1222522, EPI_ISL_1222523, EPI_ISL_1222524, EPI_ISL_1222525, EPI_ISL_1222526, EPI_ISL_1222527, EPI_ISL_1222528, EPI_ISL_1222529, EPI_ISL_1222530, EPI_ISL_1222531, EPI_ISL_1222532, EPI_ISL_1222533, EPI_ISL_1222534, EPI_ISL_1222535, EPI_ISL_1222537, EPI_ISL_1222540, EPI_ISL_1222541, EPI_ISL_1222542, EPI_ISL_1222543, EPI_ISL_1222544, EPI_ISL_1222545, EPI_ISL_1222546, EPI_ISL_1222547, EPI_ISL_1222548, EPI_ISL_1222549, EPI_ISL_1222550, EPI_ISL_1222551, EPI_ISL_1222553, EPI_ISL_1222554, EPI_ISL_1222555, EPI_ISL_1222557, EPI_ISL_1222558, EPI_ISL_1222559, EPI_ISL_1222560, EPI_ISL_1222561, EPI_ISL_1222563, EPI_ISL_1222564, EPI_ISL_1222565, EPI_ISL_1222566, EPI_ISL_1222567, EPI_ISL_1222568, EPI_ISL_1222569, EPI_ISL_1222570, EPI_ISL_1222571, EPI_ISL_1222573, EPI_ISL_1222574, EPI_ISL_1222575, EPI_ISL_1222577, EPI_ISL_1222578, EPI_ISL_1222580, EPI_ISL_1222581, EPI_ISL_1222582, EPI_ISL_1222584, EPI_ISL_1222585, EPI_ISL_1222587, EPI_ISL_1222588, EPI_ISL_1222590, EPI_ISL_1222591, EPI_ISL_1222593, EPI_ISL_1222595, EPI_ISL_1222596, EPI_ISL_1222598, EPI_ISL_1222599, EPI_ISL_1222600, EPI_ISL_1222602, EPI_ISL_1222603, EPI_ISL_1222604, EPI_ISL_1222605, EPI_ISL_1222606, EPI_ISL_1222607, EPI_ISL_1222608 | | | |
| see above | WA State PHL | Seattle Flu Study | Deborah A. Nickerson, Chris D. Frazar, Jover Lee, Benjamin Pelle, Erica Ryke, Matthew Richardson, Amanda Adler, Elisabeth Brandstetter, Peter D. Han, Kairsten Fay, Misja Ilcisin, Kirsten Lacombe, Thomas R. Sibley, Melissa Truong, Caitlin R. Wolf, Romesh Gautom, Geoff Melly, Brian Hiatt, Philip Dykema, Scott Lindquist, Michael Boeckh, Janet A. Englund, Michael Famulare, Barry R. Lutz, Mark J. Rieder, Lea M. Starita, Matthew Thompson, Helen Y. Chu, Jay Shendure, Trevor Bedford |

| | | | |
|---|---|---|---|
| EPI_ISL_1222610, EPI_ISL_1222611, EPI_ISL_1222612 | Seattle Flu Study | Seattle Flu Study | Deborah A. Nickerson, Chris D. Frazar, Jover Lee, Benjamin Pelle, Erica Ryke, Matthew Richardson, Amanda Adler, Elisabeth Brandstetter, Peter D. Han, Kairsten Fay, Misja Ilcisin, Kirsten Lacombe, Thomas R. Sibley, Melissa Truong, Caitlin R. Wolf, Michael Boeckh, Janet A. Englund, Michael Famulare, Barry R. Lutz, Mark J. Rieder, Lea M. Starita, Matthew Thompson, Jay Shendure, Trevor Bedford, Helen Y. Chu |
| EPI_ISL_1222613, EPI_ISL_1222614, EPI_ISL_1222615 | Evergreen Healthcare | Seattle Flu Study | Deborah A. Nickerson, Chris D. Frazar, Jover Lee, Benjamin Pelle, Erica Ryke, Matthew Richardson, Amanda Adler, Elisabeth Brandstetter, Peter D. Han, Kairsten Fay, Misja Ilcisin, Kirsten Lacombe, Thomas R. Sibley, Melissa Truong, Caitlin R. Wolf, Romesh Gautom, Geoff Melly, Brian Hiatt, Philip Dykema, Scott Lindquist, Michael Boeckh, Janet A. Englund, Michael Famulare, Barry R. Lutz, Mark J. Rieder, Lea M. Starita, Matthew Thompson, Helen Y. Chu, Jay Shendure, Trevor Bedford |
| EPI_ISL_1222617, EPI_ISL_1222618, EPI_ISL_1222620, EPI_ISL_1222622, EPI_ISL_1222624, EPI_ISL_1222626, EPI_ISL_1222632, EPI_ISL_1222633, EPI_ISL_1222635, EPI_ISL_1222637, EPI_ISL_1222644, EPI_ISL_1222645, EPI_ISL_1222649, EPI_ISL_1222650, EPI_ISL_1222651, EPI_ISL_1222652, EPI_ISL_1222654, EPI_ISL_1222656, EPI_ISL_1222657, EPI_ISL_1222658, EPI_ISL_1222659, EPI_ISL_1222660, EPI_ISL_1222661, EPI_ISL_1222664, EPI_ISL_1222665, EPI_ISL_1222666, EPI_ISL_1222667, EPI_ISL_1222669, EPI_ISL_1222670, EPI_ISL_1222671, EPI_ISL_1222672, EPI_ISL_1222674, EPI_ISL_1222675, EPI_ISL_1222677, EPI_ISL_1222679, EPI_ISL_1222681, EPI_ISL_1222682, EPI_ISL_1222684, EPI_ISL_1222685, EPI_ISL_1222686, EPI_ISL_1222687, EPI_ISL_1222688, EPI_ISL_1222689, EPI_ISL_1222690, EPI_ISL_1222692 | | | |
| see above | Altius Institute | Seattle Flu Study | Deborah A. Nickerson, Chris D. Frazar, Jover Lee, Benjamin Pelle, Erica Ryke, Matthew Richardson, Amanda Adler, Elisabeth Brandstetter, Peter D. Han, Kairsten Fay, Misja Ilcisin, Kirsten Lacombe, Thomas R. Sibley, Melissa Truong, Caitlin R. Wolf, Ryan Alexander, Daniel Bates, Rebecca Bruders, Stephanie DeBaun, Clem Green, Muhammad Halimun, Jessica Halow, Kneshay Harper, Matt Hartman, Andrew Meuser, Alex Nguyen, Truong Nguyen, Sofia Olsson, Sadie Patraw, Hannah Petersen, Tobias Ragoczy, Joshua Richards, Jacob Rodriguez, John Stamatoyannopoulos, Julia Wald, Olivia Waltner, Michael Boeckh, Janet A. Englund, Michael Famulare, Barry R. Lutz, Mark J. Rieder, Lea M. Starita, Matthew Thompson, Helen Y. Chu, Jay Shendure, Trevor Bedford |
| EPI_ISL_1222700, EPI_ISL_1222701, EPI_ISL_1222702, EPI_ISL_1222703 | Northwest Laboratories | Seattle Flu Study | Deborah A. Nickerson, Chris D. Frazar, Jover Lee, Benjamin Pelle, Erica Ryke, Matthew Richardson, Amanda Adler, Elisabeth Brandstetter, Peter D. Han, Kairsten Fay, Misja Ilcisin, Kirsten Lacombe, Thomas R. Sibley, Melissa Truong, Caitlin R. Wolf, Romesh Gautom, Geoff Melly, Brian Hiatt, Philip Dykema, Scott Lindquist, Michael Boeckh, Janet A. Englund, Michael Famulare, Barry R. Lutz, Mark J. Rieder, Lea M. Starita, Matthew Thompson, Helen Y. Chu, Jay Shendure, Trevor Bedford |
| EPI_ISL_1222704, EPI_ISL_1222705, EPI_ISL_1222706, EPI_ISL_1222707, EPI_ISL_1222708 | Evergreen Healthcare | Seattle Flu Study | Deborah A. Nickerson, Chris D. Frazar, Jover Lee, Benjamin Pelle, Erica Ryke, Matthew Richardson, Amanda Adler, Elisabeth Brandstetter, Peter D. Han, Kairsten Fay, Misja Ilcisin, Kirsten Lacombe, Thomas R. Sibley, Melissa Truong, Caitlin R. Wolf, Romesh Gautom, Geoff Melly, Brian Hiatt, Philip Dykema, Scott Lindquist, Michael Boeckh, Janet A. Englund, Michael Famulare, Barry R. Lutz, Mark J. Rieder, Lea M. Starita, Matthew Thompson, Helen Y. Chu, Jay Shendure, Trevor Bedford |
| EPI_ISL_1222757, EPI_ISL_1222758 | Santa Clara County Public Health Laboratory | Santa Clara County Public Health Laboratory | Santa Clara County Public Health Department |
| EPI_ISL_1224746, EPI_ISL_1224747, EPI_ISL_1224748, EPI_ISL_1224749, EPI_ISL_1224750, EPI_ISL_1224751, EPI_ISL_1224752, EPI_ISL_1224753, EPI_ISL_1224754, EPI_ISL_1224755, EPI_ISL_1224756, EPI_ISL_1224757, EPI_ISL_1224758, EPI_ISL_1224759, EPI_ISL_1224760, EPI_ISL_1224761, EPI_ISL_1224762, EPI_ISL_1224763, EPI_ISL_1224764, EPI_ISL_1224765, EPI_ISL_1224766, EPI_ISL_1224767 | | | |
| see above | Mayo Clinic & Mayo Clinic Laboratories | Minnesota Department of Health, Public Health Laboratory | Alexandra Lorentz, Jacob Garfin, Matt Plumb, and Xiong Wang |
| EPI_ISL_1224768 | M Health Fairview | Minnesota Department of Health, Public Health Laboratory | Alexandra Lorentz, Jacob Garfin, Matt Plumb, and Xiong Wang |
| EPI_ISL_1224769 | Mayo Clinic & Mayo Clinic Laboratories | Minnesota Department of Health, Public Health Laboratory | Alexandra Lorentz, Jacob Garfin, Matt Plumb, and Xiong Wang |
| EPI_ISL_1224770 | M Health Fairview | Minnesota Department of Health, Public Health Laboratory | Alexandra Lorentz, Jacob Garfin, Matt Plumb, and Xiong Wang |
| EPI_ISL_1224771 | Mayo Clinic & Mayo Clinic Laboratories | Minnesota Department of Health, Public Health Laboratory | Alexandra Lorentz, Jacob Garfin, Matt Plumb, and Xiong Wang |
| EPI_ISL_1224772 | M Health Fairview | Minnesota Department of Health, Public Health Laboratory | Alexandra Lorentz, Jacob Garfin, Matt Plumb, and Xiong Wang |
| EPI_ISL_1224773 | Mayo Clinic & Mayo Clinic Laboratories | Minnesota Department of Health, Public Health Laboratory | Alexandra Lorentz, Jacob Garfin, Matt Plumb, and Xiong Wang |
| EPI_ISL_1224774 | M Health Fairview | Minnesota Department of Health, Public Health Laboratory | Alexandra Lorentz, Jacob Garfin, Matt Plumb, and Xiong Wang |
| EPI_ISL_1224775, EPI_ISL_1224776 | Mayo Clinic & Mayo Clinic Laboratories | Minnesota Department of Health, Public Health Laboratory | Alexandra Lorentz, Jacob Garfin, Matt Plumb, and Xiong Wang |
| EPI_ISL_1224777 | M Health Fairview | Minnesota Department of Health, Public Health Laboratory | Alexandra Lorentz, Jacob Garfin, Matt Plumb, and Xiong Wang |
| EPI_ISL_1224778, EPI_ISL_1224779, EPI_ISL_1224780 | Mayo Clinic & Mayo Clinic Laboratories | Minnesota Department of Health, Public Health Laboratory | Alexandra Lorentz, Jacob Garfin, Matt Plumb, and Xiong Wang |
| EPI_ISL_1224781, EPI_ISL_1224782, EPI_ISL_1224783, EPI_ISL_1224784, EPI_ISL_1224785, EPI_ISL_1224786, EPI_ISL_1224787, EPI_ISL_1224788, EPI_ISL_1224789, EPI_ISL_1224790, EPI_ISL_1224791, EPI_ISL_1224792, EPI_ISL_1224793, EPI_ISL_1224794, EPI_ISL_1224795, EPI_ISL_1224796, EPI_ISL_1224797, EPI_ISL_1224798, EPI_ISL_1224799, EPI_ISL_1224800, EPI_ISL_1224801, EPI_ISL_1224802, EPI_ISL_1224803, EPI_ISL_1224804, EPI_ISL_1224805, EPI_ISL_1224806, EPI_ISL_1224807, EPI_ISL_1224808, EPI_ISL_1224809, EPI_ISL_1224810, EPI_ISL_1224811, EPI_ISL_1224812, EPI_ISL_1224813, EPI_ISL_1224814 | | | |
| see above | M Health Fairview | Minnesota Department of Health, Public Health Laboratory | Alexandra Lorentz, Jacob Garfin, Matt Plumb, and Xiong Wang |
| EPI_ISL_1224815, EPI_ISL_1224816, EPI_ISL_1224817, EPI_ISL_1224818, EPI_ISL_1224819, EPI_ISL_1224820, EPI_ISL_1224821 | Vault Health | Minnesota Department of Health, Public Health Laboratory | Alexandra Lorentz, Jacob Garfin, Matt Plumb, and Xiong Wang |
| EPI_ISL_1224822 | Minnesota Department of Health, Public Health Laboratory | Minnesota Department of Health, Public Health Laboratory | Alexandra Lorentz, Jacob Garfin, Matt Plumb, and Xiong Wang |
| EPI_ISL_1224824, EPI_ISL_1224825, EPI_ISL_1224826, EPI_ISL_1224827, EPI_ISL_1224828, EPI_ISL_1224829, EPI_ISL_1224830, EPI_ISL_1224831, EPI_ISL_1224832, EPI_ISL_1224833, EPI_ISL_1224834, EPI_ISL_1224835, EPI_ISL_1224836, EPI_ISL_1224837, EPI_ISL_1224838, EPI_ISL_1224840, EPI_ISL_1224841, EPI_ISL_1224842, EPI_ISL_1224843, EPI_ISL_1224844, EPI_ISL_1224845, EPI_ISL_1224846, EPI_ISL_1224847, EPI_ISL_1224848, EPI_ISL_1224849, EPI_ISL_1224850, EPI_ISL_1224851, EPI_ISL_1224852, EPI_ISL_1224853, EPI_ISL_1224854, EPI_ISL_1224855, EPI_ISL_1224856, EPI_ISL_1224857, EPI_ISL_1224858, EPI_ISL_1224859, EPI_ISL_1224860, EPI_ISL_1224861, EPI_ISL_1224862, EPI_ISL_1224863, EPI_ISL_1224864, EPI_ISL_1224865, EPI_ISL_1224866, EPI_ISL_1224867, EPI_ISL_1224868 | | | |
| see above | Vault Health | Minnesota Department of Health, Public Health Laboratory | Alexandra Lorentz, Jacob Garfin, Matt Plumb, and Xiong Wang |
| EPI_ISL_1224869, EPI_ISL_1224870, EPI_ISL_1224871, EPI_ISL_1224872, EPI_ISL_1224873, EPI_ISL_1224874, EPI_ISL_1224875, EPI_ISL_1224876, EPI_ISL_1224877, EPI_ISL_1224878, EPI_ISL_1224879, EPI_ISL_1224880, EPI_ISL_1224881, EPI_ISL_1224882, EPI_ISL_1224884, EPI_ISL_1224885, EPI_ISL_1224886, EPI_ISL_1224887, EPI_ISL_1224888, EPI_ISL_1224889, EPI_ISL_1224890, EPI_ISL_1224891, EPI_ISL_1224892, EPI_ISL_1224893, EPI_ISL_1224894, EPI_ISL_1224895, EPI_ISL_1224896, EPI_ISL_1224897, EPI_ISL_1224898, EPI_ISL_1224899, EPI_ISL_1224900, EPI_ISL_1224901, EPI_ISL_1224902, EPI_ISL_1224903, EPI_ISL_1224904, EPI_ISL_1224905, EPI_ISL_1224906, EPI_ISL_1224907, EPI_ISL_1224908 | | | |
| see above | Mayo Clinic & Mayo Clinic Laboratories | Minnesota Department of Health, Public Health Laboratory | Alexandra Lorentz, Jacob Garfin, Matt Plumb, and Xiong Wang |
| EPI_ISL_1224913, EPI_ISL_1224914, EPI_ISL_1224915, EPI_ISL_1224916, EPI_ISL_1224917, EPI_ISL_1224918, EPI_ISL_1224919, EPI_ISL_1224920, EPI_ISL_1224921, EPI_ISL_1224922, EPI_ISL_1224923, EPI_ISL_1224924, EPI_ISL_1224925, EPI_ISL_1224926, EPI_ISL_1224927, EPI_ISL_1224928, EPI_ISL_1224929, EPI_ISL_1224930, EPI_ISL_1224931, EPI_ISL_1224932, EPI_ISL_1224933, EPI_ISL_1224934, EPI_ISL_1224935, EPI_ISL_1224936, EPI_ISL_1224937, EPI_ISL_1224938, EPI_ISL_1224939, EPI_ISL_1224940 | | | |
| see above | Houston Health Dept. | Houston Health Dept. | Ryker Penn, Pamela Brown, Adolpho Lara |
| EPI_ISL_1224944, EPI_ISL_1224945, EPI_ISL_1224947, EPI_ISL_1224948 | Ohio Department of Health Laboratory | Ohio Department of Health Laboratory | Holmes, Jennifer; Eric Brandt, Keoni Omura, Glen McGillivary, Caitlin McDonnell, Jade Mowery, Stephanie Mccracken, Tyler Payne, Kirtana Ramadugu, Erica Leasure, Brent Lee, Kelsey Florek, Heather Blankenship, Allison Black, Quanta Brown, and Tammy Bannerman |
| EPI_ISL_1224991, EPI_ISL_1224992, EPI_ISL_1224993, EPI_ISL_1224994, EPI_ISL_1224995, EPI_ISL_1224996, EPI_ISL_1224997, EPI_ISL_1224998, EPI_ISL_1224999, EPI_ISL_1225000, EPI_ISL_1225001, EPI_ISL_1225002, EPI_ISL_1225003, EPI_ISL_1225004, EPI_ISL_1225005, EPI_ISL_1225006, EPI_ISL_1225007, EPI_ISL_1225008, EPI_ISL_1225009, EPI_ISL_1225010, EPI_ISL_1225011, EPI_ISL_1225012, EPI_ISL_1225013, EPI_ISL_1225014, EPI_ISL_1225015, EPI_ISL_1225016, EPI_ISL_1225017, EPI_ISL_1225018, EPI_ISL_1225019, EPI_ISL_1225020, EPI_ISL_1225024, EPI_ISL_1225025, EPI_ISL_1225026, EPI_ISL_1225027, EPI_ISL_1225028, EPI_ISL_1225029, EPI_ISL_1225030, EPI_ISL_1225031, EPI_ISL_1225032, EPI_ISL_1225033, EPI_ISL_1225034, EPI_ISL_1225035, EPI_ISL_1225036, EPI_ISL_1225037, EPI_ISL_1225038, EPI_ISL_1225039, EPI_ISL_1225040, EPI_ISL_1225041, EPI_ISL_1225042, EPI_ISL_1225043, EPI_ISL_1225044, EPI_ISL_1225045, EPI_ISL_1225046, EPI_ISL_1225047, EPI_ISL_1225048, EPI_ISL_1225049, EPI_ISL_1225050, EPI_ISL_1225051, EPI_ISL_1225052, EPI_ISL_1225053, EPI_ISL_1225054, EPI_ISL_1225055, EPI_ISL_1225056, EPI_ISL_1225057, EPI_ISL_1225058, EPI_ISL_1225059, EPI_ISL_1225060, EPI_ISL_1225061, EPI_ISL_1225062, EPI_ISL_1225063, EPI_ISL_1225064, EPI_ISL_1225065, EPI_ISL_1225066, EPI_ISL_1225067, EPI_ISL_1225068, EPI_ISL_1225069, EPI_ISL_1225070, EPI_ISL_1225071, EPI_ISL_1225072, EPI_ISL_1225073, EPI_ISL_1225074 | | | |
| see above | Texas Department of State Health Services | Texas Department of State Health Services | Rashmi Tuladhar, Bonnie Oh, Jenny Zhang, Maliha Rahman, Mayela Pedrueza, Anita Pokharel, Myong Koag, Chun Wang, Rachel Lee, Grace Kubin |
| EPI_ISL_1225075, EPI_ISL_1225076, EPI_ISL_1225078, EPI_ISL_1225155 | Ohio Department of Health Laboratory | Ohio Department of Health Laboratory | Holmes, Jennifer; Eric Brandt, Keoni Omura, Glen McGillivary, Caitlin McDonnell, Jade Mowery, Stephanie Mccracken, Tyler Payne, Kirtana Ramadugu, Erica Leasure, Brent Lee, Kelsey Florek, Heather Blankenship, Allison Black, Quanta Brown, and Tammy Bannerman |
| EPI_ISL_1225162, EPI_ISL_1225164, EPI_ISL_1225166, EPI_ISL_1225167, EPI_ISL_1225168, EPI_ISL_1225169, EPI_ISL_1225170, EPI_ISL_1225171, EPI_ISL_1225172, EPI_ISL_1225173, EPI_ISL_1225174, EPI_ISL_1225175, EPI_ISL_1225176, EPI_ISL_1225177, EPI_ISL_1225178, EPI_ISL_1225179, EPI_ISL_1225180, EPI_ISL_1225181, EPI_ISL_1225182, EPI_ISL_1225183, EPI_ISL_1225185, EPI_ISL_1225186, EPI_ISL_1225187, EPI_ISL_1225188, EPI_ISL_1225189, EPI_ISL_1225190, EPI_ISL_1225191, EPI_ISL_1225192, EPI_ISL_1225193, EPI_ISL_1225194, EPI_ISL_1225195, EPI_ISL_1225196, EPI_ISL_1225197, EPI_ISL_1225198, EPI_ISL_1225200, EPI_ISL_1225201, EPI_ISL_1225203 | | | |
| see above | Vault Health | Minnesota Department of Health, Public Health Laboratory | Alexandra Lorentz, Jacob Garfin, Matt Plumb, and Xiong Wang |
| EPI_ISL_1225204, EPI_ISL_1225205, EPI_ISL_1225206, EPI_ISL_1225208, EPI_ISL_1225209, EPI_ISL_1225210, EPI_ISL_1225211, EPI_ISL_1225212, EPI_ISL_1225213, EPI_ISL_1225214, EPI_ISL_1225216, EPI_ISL_1225217, EPI_ISL_1225219, EPI_ISL_1225220, EPI_ISL_1225221, EPI_ISL_1225222, EPI_ISL_1225223, EPI_ISL_1225225, EPI_ISL_1225226, EPI_ISL_1225228, EPI_ISL_1225229, EPI_ISL_1225230, EPI_ISL_1225231, EPI_ISL_1225232, EPI_ISL_1225233, EPI_ISL_1225234, EPI_ISL_1225235, EPI_ISL_1225236, EPI_ISL_1225237, EPI_ISL_1225239, EPI_ISL_1225240 | | | |
| see above | Mayo Clinic & Mayo Clinic Laboratories | Minnesota Department of Health, Public Health Laboratory | Alexandra Lorentz, Jacob Garfin, Matt Plumb, and Xiong Wang |
| EPI_ISL_1225241, EPI_ISL_1225242, EPI_ISL_1225243, EPI_ISL_1225244, EPI_ISL_1225245, EPI_ISL_1225246, EPI_ISL_1225247, EPI_ISL_1225248, EPI_ISL_1225249, EPI_ISL_1225250, EPI_ISL_1225251, EPI_ISL_1225252, EPI_ISL_1225253, EPI_ISL_1225254, EPI_ISL_1225259, EPI_ISL_1225260, EPI_ISL_1225261 | | | |

| | | | |
|---|---|---|---|
| see above | TriCore Reference Laboratories | Center for Global Health, University of New Mexico Health Sciences Center | Daryl Domman, Kurt Schwalm, Cecilia Thompson, Kendra Pesko, Karissa Culbreath, Darrell Dinwiddie |
| EPI_ISL_1225262 | TriCore Reference Laboratories | Center for Global Health, University of New Mexico Health Sciences Center | Daryl Domman, Kurt Schwalm, Cecilia Thompson, Kendra Pesko, Karissa Culbreath, Darrell Dinwiddie |
| EPI_ISL_1225263, EPI_ISL_1225264, EPI_ISL_1225265, EPI_ISL_1225266, EPI_ISL_1225267, EPI_ISL_1225268, EPI_ISL_1225269, EPI_ISL_1225270, EPI_ISL_1225271, EPI_ISL_1225272, EPI_ISL_1225273, EPI_ISL_1225274, EPI_ISL_1225275, EPI_ISL_1225276, EPI_ISL_1225277, EPI_ISL_1225278, EPI_ISL_1225279, EPI_ISL_1225280, EPI_ISL_1225281, EPI_ISL_1225282, EPI_ISL_1225283, EPI_ISL_1225284, EPI_ISL_1225285, EPI_ISL_1225286, EPI_ISL_1225287, EPI_ISL_1225288, EPI_ISL_1225289, EPI_ISL_1225290, EPI_ISL_1225291, EPI_ISL_1225292, EPI_ISL_1225293, EPI_ISL_1225294, EPI_ISL_1225295, EPI_ISL_1225296, EPI_ISL_1225297, EPI_ISL_1225298, EPI_ISL_1225299, EPI_ISL_1225300, EPI_ISL_1225301, EPI_ISL_1225302, EPI_ISL_1225303, EPI_ISL_1225304, EPI_ISL_1225305, EPI_ISL_1225306, EPI_ISL_1225307, EPI_ISL_1225308, EPI_ISL_1225309, EPI_ISL_1225310, EPI_ISL_1225311, EPI_ISL_1225312, EPI_ISL_1225313, EPI_ISL_1225314, EPI_ISL_1225315, EPI_ISL_1225316, EPI_ISL_1225317, EPI_ISL_1225318, EPI_ISL_1225319 | | | |
| see above | TriCore Reference Laboratories | Center for Global Health, University of New Mexico Health Sciences Center | Daryl Domman, Kurt Schwalm, Cecilia Thompson, Kendra Pesko, Karissa Culbreath, Darrell Dinwiddie |
| EPI_ISL_1225320, EPI_ISL_1225321, EPI_ISL_1225322, EPI_ISL_1225323, EPI_ISL_1225324, EPI_ISL_1225325, EPI_ISL_1225326, EPI_ISL_1225327, EPI_ISL_1225328, EPI_ISL_1225329 | GA Department of Public Health | GA Department of Public Health | Stacy Reeves, Jonathan Edwards, Cynthia Dixey, Tonia Parrott, Aliyah Fields, Taylor Smith |
| EPI_ISL_1225584, EPI_ISL_1225585, EPI_ISL_1225586, EPI_ISL_1225587, EPI_ISL_1225588, EPI_ISL_1225589, EPI_ISL_1225590, EPI_ISL_1225591, EPI_ISL_1225592 | Yale Clinical Virology Lab | Grubaugh Lab - Yale School of Public Health | Joseph Fauver, Mallery Breban, Isabell Ott, Tara Alpert, Mary Petrone, Anderson Brito, Chantal Vogels, Annie Watkins, Chaney Kalinich, Marie L. Landry, Nathan Grubaugh |
| EPI_ISL_1225593 | Tempus | Grubaugh Lab - Yale School of Public Health | Joseph Fauver, Mallery Breban, Isabell Ott, Tara Alpert, Mary Petrone, Anderson Brito, Chantal Vogels, Annie Watkins, Chaney Kalinich, Matthew J. MacKay, Gaurav Khullar, Jessica Metti, Joel T. Dudley, Megan Nash, Nike Beaubier, Christopher E. Mason, Nathan Grubaugh |
| EPI_ISL_1225594 | Yale Clinical Virology Lab | Grubaugh Lab - Yale School of Public Health | Joseph Fauver, Mallery Breban, Isabell Ott, Tara Alpert, Mary Petrone, Anderson Brito, Chantal Vogels, Annie Watkins, Chaney Kalinich, Marie L. Landry, Nathan Grubaugh |
| EPI_ISL_1225595 | Tempus | Grubaugh Lab - Yale School of Public Health | Joseph Fauver, Mallery Breban, Isabell Ott, Tara Alpert, Mary Petrone, Anderson Brito, Chantal Vogels, Annie Watkins, Chaney Kalinich, Matthew J. MacKay, Gaurav Khullar, Jessica Metti, Joel T. Dudley, Megan Nash, Nike Beaubier, Christopher E. Mason, Nathan Grubaugh |
| EPI_ISL_1225596, EPI_ISL_1225597, EPI_ISL_1225598 | Yale Clinical Virology Lab | Grubaugh Lab - Yale School of Public Health | Joseph Fauver, Mallery Breban, Isabell Ott, Tara Alpert, Mary Petrone, Anderson Brito, Chantal Vogels, Annie Watkins, Chaney Kalinich, Marie L. Landry, Nathan Grubaugh |
| EPI_ISL_1225599, EPI_ISL_1225600 | Tempus | Grubaugh Lab - Yale School of Public Health | Joseph Fauver, Mallery Breban, Isabell Ott, Tara Alpert, Mary Petrone, Anderson Brito, Chantal Vogels, Annie Watkins, Chaney Kalinich, Matthew J. MacKay, Gaurav Khullar, Jessica Metti, Joel T. Dudley, Megan Nash, Nike Beaubier, Christopher E. Mason, Nathan Grubaugh |
| EPI_ISL_1225601, EPI_ISL_1225602, EPI_ISL_1225603 | Yale Clinical Virology Lab | Grubaugh Lab - Yale School of Public Health | Joseph Fauver, Mallery Breban, Isabell Ott, Tara Alpert, Mary Petrone, Anderson Brito, Chantal Vogels, Annie Watkins, Chaney Kalinich, Marie L. Landry, Nathan Grubaugh |
| EPI_ISL_1225604, EPI_ISL_1225605, EPI_ISL_1225606 | Tempus | Grubaugh Lab - Yale School of Public Health | Joseph Fauver, Mallery Breban, Isabell Ott, Tara Alpert, Mary Petrone, Anderson Brito, Chantal Vogels, Annie Watkins, Chaney Kalinich, Matthew J. MacKay, Gaurav Khullar, Jessica Metti, Joel T. Dudley, Megan Nash, Nike Beaubier, Christopher E. Mason, Nathan Grubaugh |
| EPI_ISL_1225607, EPI_ISL_1225608, EPI_ISL_1225609, EPI_ISL_1225610 | Yale Clinical Virology Lab | Grubaugh Lab - Yale School of Public Health | Joseph Fauver, Mallery Breban, Isabell Ott, Tara Alpert, Mary Petrone, Anderson Brito, Chantal Vogels, Annie Watkins, Chaney Kalinich, Marie L. Landry, Nathan Grubaugh |
| EPI_ISL_1225611 | Tempus | Grubaugh Lab - Yale School of Public Health | Joseph Fauver, Mallery Breban, Isabell Ott, Tara Alpert, Mary Petrone, Anderson Brito, Chantal Vogels, Annie Watkins, Chaney Kalinich, Matthew J. MacKay, Gaurav Khullar, Jessica Metti, Joel T. Dudley, Megan Nash, Nike Beaubier, Christopher E. Mason, Nathan Grubaugh |
| EPI_ISL_1225612 | Yale Clinical Virology Lab | Grubaugh Lab - Yale School of Public Health | Joseph Fauver, Mallery Breban, Isabell Ott, Tara Alpert, Mary Petrone, Anderson Brito, Chantal Vogels, Annie Watkins, Chaney Kalinich, Marie L. Landry, Nathan Grubaugh |
| EPI_ISL_1225613 | Yale Pathology Lab | Grubaugh Lab - Yale School of Public Health | Joseph Fauver, Mallery Breban, Isabell Ott, Tara Alpert, Mary Petrone, Anderson Brito, Chantal Vogels, Annie Watkins, Chaney Kalinich, Jianhui Wang, Chen Liu, Pei Hui, Nathan Grubaugh |
| EPI_ISL_1225614 | Tempus | Grubaugh Lab - Yale School of Public Health | Joseph Fauver, Mallery Breban, Isabell Ott, Tara Alpert, Mary Petrone, Anderson Brito, Chantal Vogels, Annie Watkins, Chaney Kalinich, Matthew J. MacKay, Gaurav Khullar, Jessica Metti, Joel T. Dudley, Megan Nash, Nike Beaubier, Christopher E. Mason, Nathan Grubaugh |
| EPI_ISL_1225615, EPI_ISL_1225616, EPI_ISL_1225617 | Yale Clinical Virology Lab | Grubaugh Lab - Yale School of Public Health | Joseph Fauver, Mallery Breban, Isabell Ott, Tara Alpert, Mary Petrone, Anderson Brito, Chantal Vogels, Annie Watkins, Chaney Kalinich, Marie L. Landry, Nathan Grubaugh |
| EPI_ISL_1225618, EPI_ISL_1225619 | Tempus | Grubaugh Lab - Yale School of Public Health | Joseph Fauver, Mallery Breban, Isabell Ott, Tara Alpert, Mary Petrone, Anderson Brito, Chantal Vogels, Annie Watkins, Chaney Kalinich, Matthew J. MacKay, Gaurav Khullar, Jessica Metti, Joel T. Dudley, Megan Nash, Nike Beaubier, Christopher E. Mason, Nathan Grubaugh |
| EPI_ISL_1225620, EPI_ISL_1225621 | Yale Clinical Virology Lab | Grubaugh Lab - Yale School of Public Health | Joseph Fauver, Mallery Breban, Isabell Ott, Tara Alpert, Mary Petrone, Anderson Brito, Chantal Vogels, Annie Watkins, Chaney Kalinich, Marie L. Landry, Nathan Grubaugh |
| EPI_ISL_1225622 | Tempus | Grubaugh Lab - Yale School of Public Health | Joseph Fauver, Mallery Breban, Isabell Ott, Tara Alpert, Mary Petrone, Anderson Brito, Chantal Vogels, Annie Watkins, Chaney Kalinich, Matthew J. MacKay, Gaurav Khullar, Jessica Metti, Joel T. Dudley, Megan Nash, Nike Beaubier, Christopher E. Mason, Nathan Grubaugh |
| EPI_ISL_1225623, EPI_ISL_1225624 | Yale Clinical Virology Lab | Grubaugh Lab - Yale School of Public Health | Joseph Fauver, Mallery Breban, Isabell Ott, Tara Alpert, Mary Petrone, Anderson Brito, Chantal Vogels, Annie Watkins, Chaney Kalinich, Marie L. Landry, Nathan Grubaugh |
| EPI_ISL_1225625, EPI_ISL_1225626, EPI_ISL_1225627, EPI_ISL_1225628, EPI_ISL_1225629, EPI_ISL_1225630, EPI_ISL_1225631, EPI_ISL_1225632, EPI_ISL_1225633, EPI_ISL_1225634, EPI_ISL_1225635, EPI_ISL_1225636, EPI_ISL_1225637, EPI_ISL_1225638, EPI_ISL_1225639 | | | |
| see above | Tempus | Grubaugh Lab - Yale School of Public Health | Joseph Fauver, Mallery Breban, Isabell Ott, Tara Alpert, Mary Petrone, Anderson Brito, Chantal Vogels, Annie Watkins, Chaney Kalinich, Matthew J. MacKay, Gaurav Khullar, Jessica Metti, Joel T. Dudley, Megan Nash, Nike Beaubier, Christopher E. Mason, Nathan Grubaugh |
| EPI_ISL_1225640 | Connecticut DPH | Grubaugh Lab - Yale School of Public Health | Joseph Fauver, Mallery Breban, Isabell Ott, Tara Alpert, Mary Petrone, Anderson Brito, Chantal Vogels, Annie Watkins, Chaney Kalinich, Anthony Muyombwe, Randy Downing, Jafar Razeq, Stephen M. Bart, Nathan Grubaugh |
| EPI_ISL_1225641, EPI_ISL_1225642, EPI_ISL_1225643, EPI_ISL_1225644, EPI_ISL_1225645, EPI_ISL_1225646, EPI_ISL_1225647 | Tempus | Grubaugh Lab - Yale School of Public Health | Joseph Fauver, Mallery Breban, Isabell Ott, Tara Alpert, Mary Petrone, Anderson Brito, Chantal Vogels, Annie Watkins, Chaney Kalinich, Matthew J. MacKay, Gaurav Khullar, Jessica Metti, Joel T. Dudley, Megan Nash, Nike Beaubier, Christopher E. Mason, Nathan Grubaugh |
| EPI_ISL_1225648, EPI_ISL_1225649, EPI_ISL_1225650, EPI_ISL_1225651, EPI_ISL_1225652, EPI_ISL_1225653 | Yale Clinical Virology Lab | Grubaugh Lab - Yale School of Public Health | Joseph Fauver, Mallery Breban, Isabell Ott, Tara Alpert, Mary Petrone, Anderson Brito, Chantal Vogels, Annie Watkins, Chaney Kalinich, Marie L. Landry, Nathan Grubaugh |
| EPI_ISL_1225654 | Connecticut DPH | Grubaugh Lab - Yale School of Public Health | Joseph Fauver, Mallery Breban, Isabell Ott, Tara Alpert, Mary Petrone, Anderson Brito, Chantal Vogels, Annie Watkins, Chaney Kalinich, Anthony Muyombwe, Randy Downing, Jafar Razeq, Stephen M. Bart, Nathan Grubaugh |
| EPI_ISL_1225655, EPI_ISL_1225656, EPI_ISL_1225657 | Tempus | Grubaugh Lab - Yale School of Public Health | Joseph Fauver, Mallery Breban, Isabell Ott, Tara Alpert, Mary Petrone, Anderson Brito, Chantal Vogels, Annie Watkins, Chaney Kalinich, Matthew J. MacKay, Gaurav Khullar, Jessica Metti, Joel T. Dudley, Megan Nash, Nike Beaubier, Christopher E. Mason, Nathan Grubaugh |
| EPI_ISL_1225658, EPI_ISL_1225659 | Yale Clinical Virology Lab | Grubaugh Lab - Yale School of Public Health | Joseph Fauver, Mallery Breban, Isabell Ott, Tara Alpert, Mary Petrone, Anderson Brito, Chantal Vogels, Annie Watkins, Chaney Kalinich, Marie L. Landry, Nathan Grubaugh |
| EPI_ISL_1225727 | Yale Pathology Lab | Grubaugh Lab - Yale School of Public Health | Joseph Fauver, Mallery Breban, Isabell Ott, Tara Alpert, Mary Petrone, Anderson Brito, Chantal Vogels, Annie Watkins, Chaney Kalinich, Jianhui Wang, Chen Liu, Pei Hui, Nathan Grubaugh |
| EPI_ISL_1225728 | Connecticut DPH | Grubaugh Lab - Yale School of Public Health | Joseph Fauver, Mallery Breban, Isabell Ott, Tara Alpert, Mary Petrone, Anderson Brito, Chantal Vogels, Annie Watkins, Chaney Kalinich, Anthony Muyombwe, Randy Downing, Jafar Razeq, Stephen M. Bart, Nathan Grubaugh |
| EPI_ISL_1225729, EPI_ISL_1225730, EPI_ISL_1225731, EPI_ISL_1225732, EPI_ISL_1225733, EPI_ISL_1225734, EPI_ISL_1225735 | Yale Clinical Virology Lab | Grubaugh Lab - Yale School of Public Health | Joseph Fauver, Mallery Breban, Isabell Ott, Tara Alpert, Mary Petrone, Anderson Brito, Chantal Vogels, Annie Watkins, Chaney Kalinich, Marie L. Landry, Nathan Grubaugh |
| EPI_ISL_1225736, EPI_ISL_1225738, EPI_ISL_1225739, EPI_ISL_1225740, EPI_ISL_1225741, EPI_ISL_1225742, EPI_ISL_1225743, EPI_ISL_1225744, EPI_ISL_1225745, EPI_ISL_1225746, EPI_ISL_1225747, EPI_ISL_1225749, EPI_ISL_1225750, EPI_ISL_1225751, EPI_ISL_1225752, EPI_ISL_1225753, EPI_ISL_1225754, EPI_ISL_1225755, EPI_ISL_1225756, EPI_ISL_1225757, EPI_ISL_1225758, EPI_ISL_1225759, EPI_ISL_1225760, EPI_ISL_1225761, EPI_ISL_1225762, EPI_ISL_1225763, EPI_ISL_1225764, EPI_ISL_1225765, EPI_ISL_1225766, EPI_ISL_1225767, EPI_ISL_1225768, EPI_ISL_1225770, EPI_ISL_1225773, EPI_ISL_1225774, EPI_ISL_1225775, EPI_ISL_1225776, EPI_ISL_1225777, EPI_ISL_1225778, EPI_ISL_1225779 | | | |
| see above | Illinois Department of Public Health | Gagnon Lab, Southern Illinois University | Keith Gagnon |

| | | | |
|---|---|---|---|
| EPI_ISL_1225872 | IL Dept. of Public Health Springfield Laboratory | Genomics and Discovery, Respiratory Viruses Branch, Division of Viral Diseases, Centers for Disease Control and Prevention | Krista Queen, Yan Li, Ying Tao, Jing Zhang, Anna Uehara, Anna Montmayeur, Clinton R. Paden, Kristen Knipe, Matthew Schmerer, Shoshona Le, Katie Dillon, Peter W. Cook, Rachel Marine, Mili Sheth, Jasmine Padilla, Sarah Nobles, Mark Burroughs, Lori Rowe, Haibin Wang, Ben L. Rambo-Martin, Kristine Lacek, Sam Shepard, Dhwani Batra, Suxiang Tong, Justin Lee |
| EPI_ISL_1225873 | NC State Laboratory of Public Health | Genomics and Discovery, Respiratory Viruses Branch, Division of Viral Diseases, Centers for Disease Control and Prevention | Krista Queen, Yan Li, Ying Tao, Jing Zhang, Anna Uehara, Anna Montmayeur, Clinton R. Paden, Kristen Knipe, Matthew Schmerer, Shoshona Le, Katie Dillon, Peter W. Cook, Rachel Marine, Mili Sheth, Jasmine Padilla, Sarah Nobles, Mark Burroughs, Lori Rowe, Haibin Wang, Ben L. Rambo-Martin, Kristine Lacek, Sam Shepard, Dhwani Batra, Suxiang Tong, Justin Lee |
| EPI_ISL_1225874, EPI_ISL_1225875, EPI_ISL_1225876, EPI_ISL_1225877, EPI_ISL_1225878, EPI_ISL_1225879, EPI_ISL_1225880 | "AK State Public Health Lab, State Health Department" | Genomics and Discovery, Respiratory Viruses Branch, Division of Viral Diseases, Centers for Disease Control and Prevention | Krista Queen, Yan Li, Ying Tao, Jing Zhang, Anna Uehara, Anna Montmayeur, Clinton R. Paden, Kristen Knipe, Matthew Schmerer, Shoshona Le, Katie Dillon, Peter W. Cook, Rachel Marine, Mili Sheth, Jasmine Padilla, Sarah Nobles, Mark Burroughs, Lori Rowe, Haibin Wang, Ben L. Rambo-Martin, Kristine Lacek, Sam Shepard, Dhwani Batra, Suxiang Tong, Justin Lee |
| EPI_ISL_1225881 | SD Public Health Laboratory | Genomics and Discovery, Respiratory Viruses Branch, Division of Viral Diseases, Centers for Disease Control and Prevention | Krista Queen, Yan Li, Ying Tao, Jing Zhang, Anna Uehara, Anna Montmayeur, Clinton R. Paden, Kristen Knipe, Matthew Schmerer, Shoshona Le, Katie Dillon, Peter W. Cook, Rachel Marine, Mili Sheth, Jasmine Padilla, Sarah Nobles, Mark Burroughs, Lori Rowe, Haibin Wang, Ben L. Rambo-Martin, Kristine Lacek, Sam Shepard, Dhwani Batra, Suxiang Tong, Justin Lee |
| EPI_ISL_1225882, EPI_ISL_1225883, EPI_ISL_1225884 | ME Health and Environmental Testing Laboratory | Genomics and Discovery, Respiratory Viruses Branch, Division of Viral Diseases, Centers for Disease Control and Prevention | Krista Queen, Yan Li, Ying Tao, Jing Zhang, Anna Uehara, Anna Montmayeur, Clinton R. Paden, Kristen Knipe, Matthew Schmerer, Shoshona Le, Katie Dillon, Peter W. Cook, Rachel Marine, Mili Sheth, Jasmine Padilla, Sarah Nobles, Mark Burroughs, Lori Rowe, Haibin Wang, Ben L. Rambo-Martin, Kristine Lacek, Sam Shepard, Dhwani Batra, Suxiang Tong, Justin Lee |
| EPI_ISL_1225885, EPI_ISL_1225886, EPI_ISL_1225887, EPI_ISL_1225888 | MT Public Health Laboratory | Genomics and Discovery, Respiratory Viruses Branch, Division of Viral Diseases, Centers for Disease Control and Prevention | Krista Queen, Yan Li, Ying Tao, Jing Zhang, Anna Uehara, Anna Montmayeur, Clinton R. Paden, Kristen Knipe, Matthew Schmerer, Shoshona Le, Katie Dillon, Peter W. Cook, Rachel Marine, Mili Sheth, Jasmine Padilla, Sarah Nobles, Mark Burroughs, Lori Rowe, Haibin Wang, Ben L. Rambo-Martin, Kristine Lacek, Sam Shepard, Dhwani Batra, Suxiang Tong, Justin Lee |
| EPI_ISL_1225889, EPI_ISL_1225890, EPI_ISL_1225891, EPI_ISL_1225892, EPI_ISL_1225893, EPI_ISL_1225894, EPI_ISL_1225895, EPI_ISL_1225896, EPI_ISL_1225897, EPI_ISL_1225898, EPI_ISL_1225899, EPI_ISL_1225900, EPI_ISL_1225901, EPI_ISL_1225902, EPI_ISL_1225903, EPI_ISL_1225904, EPI_ISL_1225905, EPI_ISL_1225906 | | | |
| see above | NC State Laboratory of Public Health | Genomics and Discovery, Respiratory Viruses Branch, Division of Viral Diseases, Centers for Disease Control and Prevention | Krista Queen, Yan Li, Ying Tao, Jing Zhang, Anna Uehara, Anna Montmayeur, Clinton R. Paden, Kristen Knipe, Matthew Schmerer, Shoshona Le, Katie Dillon, Peter W. Cook, Rachel Marine, Mili Sheth, Jasmine Padilla, Sarah Nobles, Mark Burroughs, Lori Rowe, Haibin Wang, Ben L. Rambo-Martin, Kristine Lacek, Sam Shepard, Dhwani Batra, Suxiang Tong, Justin Lee |
| EPI_ISL_1225907 | RI State Health Laboratories | Genomics and Discovery, Respiratory Viruses Branch, Division of Viral Diseases, Centers for Disease Control and Prevention | Krista Queen, Yan Li, Ying Tao, Jing Zhang, Anna Uehara, Anna Montmayeur, Clinton R. Paden, Kristen Knipe, Matthew Schmerer, Shoshona Le, Katie Dillon, Peter W. Cook, Rachel Marine, Mili Sheth, Jasmine Padilla, Sarah Nobles, Mark Burroughs, Lori Rowe, Haibin Wang, Ben L. Rambo-Martin, Kristine Lacek, Sam Shepard, Dhwani Batra, Suxiang Tong, Justin Lee |
| EPI_ISL_1225908, EPI_ISL_1225909, EPI_ISL_1225910, EPI_ISL_1225911, EPI_ISL_1225912, EPI_ISL_1225913, EPI_ISL_1225914, EPI_ISL_1225915, EPI_ISL_1225916, EPI_ISL_1225917, EPI_ISL_1225918, EPI_ISL_1225919, EPI_ISL_1225920, EPI_ISL_1225921, EPI_ISL_1225922, EPI_ISL_1225923, EPI_ISL_1225924, EPI_ISL_1225925, EPI_ISL_1225926, EPI_ISL_1225927, EPI_ISL_1225928, EPI_ISL_1225929, EPI_ISL_1225930 | | | |
| see above | SC Dept of Health and Env. Control-Bureau of Laboratories | Genomics and Discovery, Respiratory Viruses Branch, Division of Viral Diseases, Centers for Disease Control and Prevention | Krista Queen, Yan Li, Ying Tao, Jing Zhang, Anna Uehara, Anna Montmayeur, Clinton R. Paden, Kristen Knipe, Matthew Schmerer, Shoshona Le, Katie Dillon, Peter W. Cook, Rachel Marine, Mili Sheth, Jasmine Padilla, Sarah Nobles, Mark Burroughs, Lori Rowe, Haibin Wang, Ben L. Rambo-Martin, Kristine Lacek, Sam Shepard, Dhwani Batra, Suxiang Tong, Justin Lee |
| EPI_ISL_1225931, EPI_ISL_1225932, EPI_ISL_1225933, EPI_ISL_1225934, EPI_ISL_1225935 | "TX DSHS, Lab Services Section MC 1947" | Genomics and Discovery, Respiratory Viruses Branch, Division of Viral Diseases, Centers for Disease Control and Prevention | Krista Queen, Yan Li, Ying Tao, Jing Zhang, Anna Uehara, Anna Montmayeur, Clinton R. Paden, Kristen Knipe, Matthew Schmerer, Shoshona Le, Katie Dillon, Peter W. Cook, Rachel Marine, Mili Sheth, Jasmine Padilla, Sarah Nobles, Mark Burroughs, Lori Rowe, Haibin Wang, Ben L. Rambo-Martin, Kristine Lacek, Sam Shepard, Dhwani Batra, Suxiang Tong, Justin Lee |
| EPI_ISL_1225936, EPI_ISL_1225937, EPI_ISL_1225938, EPI_ISL_1225939, EPI_ISL_1225940 | UT-Unified State Labs: Public Health Utah DOH | Genomics and Discovery, Respiratory Viruses Branch, Division of Viral Diseases, Centers for Disease Control and Prevention | Krista Queen, Yan Li, Ying Tao, Jing Zhang, Anna Uehara, Anna Montmayeur, Clinton R. Paden, Kristen Knipe, Matthew Schmerer, Shoshona Le, Katie Dillon, Peter W. Cook, Rachel Marine, Mili Sheth, Jasmine Padilla, Sarah Nobles, Mark Burroughs, Lori Rowe, Haibin Wang, Ben L. Rambo-Martin, Kristine Lacek, Sam Shepard, Dhwani Batra, Suxiang Tong, Justin Lee |
| EPI_ISL_1225941, EPI_ISL_1225942, EPI_ISL_1225943, EPI_ISL_1225944, EPI_ISL_1225945 | VT Dept. of Health Laboratory | Genomics and Discovery, Respiratory Viruses Branch, Division of Viral Diseases, Centers for Disease Control and Prevention | Krista Queen, Yan Li, Ying Tao, Jing Zhang, Anna Uehara, Anna Montmayeur, Clinton R. Paden, Kristen Knipe, Matthew Schmerer, Shoshona Le, Katie Dillon, Peter W. Cook, Rachel Marine, Mili Sheth, Jasmine Padilla, Sarah Nobles, Mark Burroughs, Lori Rowe, Haibin Wang, Ben L. Rambo-Martin, Kristine Lacek, Sam Shepard, Dhwani Batra, Suxiang Tong, Justin Lee |
| EPI_ISL_1225946, EPI_ISL_1225947, EPI_ISL_1225948 | WVDHHR - Office of Laboratory Services | Genomics and Discovery, Respiratory Viruses Branch, Division of Viral Diseases, Centers for Disease Control and Prevention | Krista Queen, Yan Li, Ying Tao, Jing Zhang, Anna Uehara, Anna Montmayeur, Clinton R. Paden, Kristen Knipe, Matthew Schmerer, Shoshona Le, Katie Dillon, Peter W. Cook, Rachel Marine, Mili Sheth, Jasmine Padilla, Sarah Nobles, Mark Burroughs, Lori Rowe, Haibin Wang, Ben L. Rambo-Martin, Kristine Lacek, Sam Shepard, Dhwani Batra, Suxiang Tong, Justin Lee |
| EPI_ISL_1225949, EPI_ISL_1225950, EPI_ISL_1225951, EPI_ISL_1225952, EPI_ISL_1225953, EPI_ISL_1225954, EPI_ISL_1225955, EPI_ISL_1225956, EPI_ISL_1225957 | CA-Los Angeles County Public Health Laboratory | Genomics and Discovery, Respiratory Viruses Branch, Division of Viral Diseases, Centers for Disease Control and Prevention | Krista Queen, Yan Li, Ying Tao, Jing Zhang, Anna Uehara, Anna Montmayeur, Clinton R. Paden, Kristen Knipe, Matthew Schmerer, Shoshona Le, Katie Dillon, Peter W. Cook, Rachel Marine, Mili Sheth, Jasmine Padilla, Sarah Nobles, Mark Burroughs, Lori Rowe, Haibin Wang, Ben L. Rambo-Martin, Kristine Lacek, Sam Shepard, Dhwani Batra, Suxiang Tong, Justin Lee |
| EPI_ISL_1225958, EPI_ISL_1225959, EPI_ISL_1225960, EPI_ISL_1225961, EPI_ISL_1225962, EPI_ISL_1225963, EPI_ISL_1225964, EPI_ISL_1225965, EPI_ISL_1225966, EPI_ISL_1225967, EPI_ISL_1225968, EPI_ISL_1225969, EPI_ISL_1225970, EPI_ISL_1225971, EPI_ISL_1225972, EPI_ISL_1225973, EPI_ISL_1225974 | | | |
| see above | "CDPH, Viral and Rickettsial Disease Laboratory" | Genomics and Discovery, Respiratory Viruses Branch, Division of Viral Diseases, Centers for Disease Control and Prevention | Krista Queen, Yan Li, Ying Tao, Jing Zhang, Anna Uehara, Anna Montmayeur, Clinton R. Paden, Kristen Knipe, Matthew Schmerer, Shoshona Le, Katie Dillon, Peter W. Cook, Rachel Marine, Mili Sheth, Jasmine Padilla, Sarah Nobles, Mark Burroughs, Lori Rowe, Haibin Wang, Ben L. Rambo-Martin, Kristine Lacek, Sam Shepard, Dhwani Batra, Suxiang Tong, Justin Lee |
| EPI_ISL_1225975, EPI_ISL_1225976, EPI_ISL_1225977, EPI_ISL_1225978, EPI_ISL_1225979, EPI_ISL_1225980, EPI_ISL_1225981, EPI_ISL_1225982, EPI_ISL_1225983, EPI_ISL_1225984, EPI_ISL_1225985, EPI_ISL_1225986 | | | |
| see above | "CO Dept. of Public Health and Environment, Lab Services Division" | Genomics and Discovery, Respiratory Viruses Branch, Division of Viral Diseases, Centers for Disease Control and Prevention | Krista Queen, Yan Li, Ying Tao, Jing Zhang, Anna Uehara, Anna Montmayeur, Clinton R. Paden, Kristen Knipe, Matthew Schmerer, Shoshona Le, Katie Dillon, Peter W. Cook, Rachel Marine, Mili Sheth, Jasmine Padilla, Sarah Nobles, Mark Burroughs, Lori Rowe, Haibin Wang, Ben L. Rambo-Martin, Kristine Lacek, Sam Shepard, Dhwani Batra, Suxiang Tong, Justin Lee |
| EPI_ISL_1225987, EPI_ISL_1225988, EPI_ISL_1225989, EPI_ISL_1225990, EPI_ISL_1225991 | IN State Department of Health Laboratory Services | Genomics and Discovery, Respiratory Viruses Branch, Division of Viral Diseases, Centers for Disease Control and Prevention | Krista Queen, Yan Li, Ying Tao, Jing Zhang, Anna Uehara, Anna Montmayeur, Clinton R. Paden, Kristen Knipe, Matthew Schmerer, Shoshona Le, Katie Dillon, Peter W. Cook, Rachel Marine, Mili Sheth, Jasmine Padilla, Sarah Nobles, Mark Burroughs, Lori Rowe, Haibin Wang, Ben L. Rambo-Martin, Kristine Lacek, Sam Shepard, Dhwani Batra, Suxiang Tong, Justin Lee |
| EPI_ISL_1225992, EPI_ISL_1225993, EPI_ISL_1225994, EPI_ISL_1225995, EPI_ISL_1225996, EPI_ISL_1225997, EPI_ISL_1225998, EPI_ISL_1225999, EPI_ISL_1226000, EPI_ISL_1226001, EPI_ISL_1226002 | | | |
| see above | KY State Public Health Lab | Genomics and Discovery, Respiratory Viruses Branch, Division of Viral Diseases, Centers for Disease Control and Prevention | Krista Queen, Yan Li, Ying Tao, Jing Zhang, Anna Uehara, Anna Montmayeur, Clinton R. Paden, Kristen Knipe, Matthew Schmerer, Shoshona Le, Katie Dillon, Peter W. Cook, Rachel Marine, Mili Sheth, Jasmine Padilla, Sarah Nobles, Mark Burroughs, Lori Rowe, Haibin Wang, Ben L. Rambo-Martin, Kristine Lacek, Sam Shepard, Dhwani Batra, Suxiang Tong, Justin Lee |
| EPI_ISL_1226003, EPI_ISL_1226004, EPI_ISL_1226005, EPI_ISL_1226006 | "NYSDOH Wadsworth Center, Virology Lab" | Genomics and Discovery, Respiratory Viruses Branch, Division of Viral Diseases, Centers for Disease Control and Prevention | Krista Queen, Yan Li, Ying Tao, Jing Zhang, Anna Uehara, Anna Montmayeur, Clinton R. Paden, Kristen Knipe, Matthew Schmerer, Shoshona Le, Katie Dillon, Peter W. Cook, Rachel Marine, Mili Sheth, Jasmine Padilla, Sarah Nobles, Mark Burroughs, Lori Rowe, Haibin Wang, Ben L. Rambo-Martin, Kristine Lacek, Sam Shepard, Dhwani Batra, Suxiang Tong, Justin Lee |
| EPI_ISL_1226007, EPI_ISL_1226008, EPI_ISL_1226009, EPI_ISL_1226010, EPI_ISL_1226011, EPI_ISL_1226012 | WA State Department of Health | Genomics and Discovery, Respiratory Viruses Branch, Division of Viral Diseases, Centers for Disease Control and Prevention | Krista Queen, Yan Li, Ying Tao, Jing Zhang, Anna Uehara, Anna Montmayeur, Clinton R. Paden, Kristen Knipe, Matthew Schmerer, Shoshona Le, Katie Dillon, Peter W. Cook, Rachel Marine, Mili Sheth, Jasmine Padilla, Sarah Nobles, Mark Burroughs, Lori Rowe, Haibin Wang, Ben L. Rambo-Martin, Kristine Lacek, Sam Shepard, Dhwani Batra, Suxiang Tong, Justin Lee |
| EPI_ISL_1226013, EPI_ISL_1226015 | NE Public Health Laboratory | Genomics and Discovery, Respiratory Viruses Branch, Division of Viral Diseases, Centers for Disease Control and Prevention | Krista Queen, Yan Li, Ying Tao, Jing Zhang, Anna Uehara, Anna Montmayeur, Clinton R. Paden, Kristen Knipe, Matthew Schmerer, Shoshona Le, Katie Dillon, Peter W. Cook, Rachel Marine, Mili Sheth, Jasmine Padilla, Sarah Nobles, Mark Burroughs, Lori Rowe, Haibin Wang, Ben L. Rambo-Martin, Kristine Lacek, Sam Shepard, Dhwani Batra, Suxiang Tong, Justin Lee |
| EPI_ISL_1226016, EPI_ISL_1226018, EPI_ISL_1226019, EPI_ISL_1226020, EPI_ISL_1226021, EPI_ISL_1226022, EPI_ISL_1226023, EPI_ISL_1226024, EPI_ISL_1226025, EPI_ISL_1226026, EPI_ISL_1226027, EPI_ISL_1226028, EPI_ISL_1226029, EPI_ISL_1226030, EPI_ISL_1226031, EPI_ISL_1226032, EPI_ISL_1226033, EPI_ISL_1226034 | | | |
| see above | "PA Department of Health, Bureau of Laboratories" | Genomics and Discovery, Respiratory Viruses Branch, Division of Viral Diseases, Centers for Disease Control and Prevention | Krista Queen, Yan Li, Ying Tao, Jing Zhang, Anna Uehara, Anna Montmayeur, Clinton R. Paden, Kristen Knipe, Matthew Schmerer, Shoshona Le, Katie Dillon, Peter W. Cook, Rachel Marine, Mili Sheth, Jasmine Padilla, Sarah Nobles, Mark Burroughs, Lori Rowe, Haibin Wang, Ben L. Rambo-Martin, Kristine Lacek, Sam Shepard, Dhwani Batra, Suxiang Tong, Justin Lee |
| EPI_ISL_1226035, EPI_ISL_1226036, | "MN PHL Division, Minnesota Department of Health" | Genomics and Discovery, Respiratory Viruses Branch, Division of Viral Diseases, | Krista Queen, Yan Li, Ying Tao, Jing Zhang, Anna Uehara, Anna Montmayeur, Clinton R. Paden, Kristen Knipe, Matthew Schmerer, Shoshona Le, Katie Dillon, Peter W. Cook, Rachel Marine, Mili Sheth, Jasmine Padilla, Sarah Nobles, Mark Burroughs, Lori Rowe, Haibin Wang, Ben L. Rambo-Martin, Kristine Lacek, Sam Shepard, Dhwani Batra, Suxiang Tong, Justin Lee |

| | | | |
|---|---|---|---|
| EPI_ISL_1226037, EPI_ISL_1226038 | | Centers for Disease Control and Prevention | |
| EPI_ISL_1226039, EPI_ISL_1226040, EPI_ISL_1226041, EPI_ISL_1226042, EPI_ISL_1226043, EPI_ISL_1226044, EPI_ISL_1226045, EPI_ISL_1226046, EPI_ISL_1226047 | SC Dept of Health and Env. Control-Bureau of Laboratories | Genomics and Discovery, Respiratory Viruses Branch, Division of Viral Diseases, Centers for Disease Control and Prevention | Krista Queen, Yan Li, Ying Tao, Jing Zhang, Anna Uehara, Anna Montmayeur, Clinton R. Paden, Kristen Knipe, Matthew Schmerer, Shoshona Le, Katie Dillon, Peter W. Cook, Rachel Marine, Mili Sheth, Jasmine Padilla, Sarah Nobles, Mark Burroughs, Lori Rowe, Haibin Wang, Ben L. Rambo-Martin, Kristine Lacek, Sam Shepard, Dhwani Batra, Suxiang Tong, Justin Lee |
| EPI_ISL_1226048, EPI_ISL_1226049, EPI_ISL_1226050, EPI_ISL_1226051, EPI_ISL_1226052, EPI_ISL_1226053, EPI_ISL_1226054, EPI_ISL_1226055, EPI_ISL_1226056, EPI_ISL_1226057, EPI_ISL_1226058, EPI_ISL_1226059, EPI_ISL_1226060, EPI_ISL_1226061, EPI_ISL_1226062, EPI_ISL_1226063, EPI_ISL_1226064, EPI_ISL_1226065, EPI_ISL_1226066, EPI_ISL_1226067, EPI_ISL_1226068, EPI_ISL_1226069, EPI_ISL_1226070, EPI_ISL_1226071, EPI_ISL_1226072, EPI_ISL_1226073, EPI_ISL_1226074, EPI_ISL_1226075, EPI_ISL_1226076, EPI_ISL_1226077, EPI_ISL_1226078, EPI_ISL_1226079, EPI_ISL_1226080, EPI_ISL_1226081 | | | |
| see above | "NYSDOH Wadsworth Center, Virology Lab" | Genomics and Discovery, Respiratory Viruses Branch, Division of Viral Diseases, Centers for Disease Control and Prevention | Krista Queen, Yan Li, Ying Tao, Jing Zhang, Anna Uehara, Anna Montmayeur, Clinton R. Paden, Kristen Knipe, Matthew Schmerer, Shoshona Le, Katie Dillon, Peter W. Cook, Rachel Marine, Mili Sheth, Jasmine Padilla, Sarah Nobles, Mark Burroughs, Lori Rowe, Haibin Wang, Ben L. Rambo-Martin, Kristine Lacek, Sam Shepard, Dhwani Batra, Suxiang Tong, Justin Lee |
| EPI_ISL_1226082, EPI_ISL_1226083, EPI_ISL_1226084, EPI_ISL_1226085, EPI_ISL_1226086, EPI_ISL_1226087, EPI_ISL_1226089, EPI_ISL_1226090, EPI_ISL_1226091 | GA Department of Public Health Laboratory | Genomics and Discovery, Respiratory Viruses Branch, Division of Viral Diseases, Centers for Disease Control and Prevention | Krista Queen, Yan Li, Ying Tao, Jing Zhang, Anna Uehara, Anna Montmayeur, Clinton R. Paden, Kristen Knipe, Matthew Schmerer, Shoshona Le, Katie Dillon, Peter W. Cook, Rachel Marine, Mili Sheth, Jasmine Padilla, Sarah Nobles, Mark Burroughs, Lori Rowe, Haibin Wang, Ben L. Rambo-Martin, Kristine Lacek, Sam Shepard, Dhwani Batra, Suxiang Tong, Justin Lee |
| EPI_ISL_1226092, EPI_ISL_1226093, EPI_ISL_1226094, EPI_ISL_1226095, EPI_ISL_1226096, EPI_ISL_1226097, EPI_ISL_1226098, EPI_ISL_1226099, EPI_ISL_1226100, EPI_ISL_1226101, EPI_ISL_1226102, EPI_ISL_1226103, EPI_ISL_1226104 | | | |
| see above | IL Dept. of Public Health Springfield Laboratory | Genomics and Discovery, Respiratory Viruses Branch, Division of Viral Diseases, Centers for Disease Control and Prevention | Krista Queen, Yan Li, Ying Tao, Jing Zhang, Anna Uehara, Anna Montmayeur, Clinton R. Paden, Kristen Knipe, Matthew Schmerer, Shoshona Le, Katie Dillon, Peter W. Cook, Rachel Marine, Mili Sheth, Jasmine Padilla, Sarah Nobles, Mark Burroughs, Lori Rowe, Haibin Wang, Ben L. Rambo-Martin, Kristine Lacek, Sam Shepard, Dhwani Batra, Suxiang Tong, Justin Lee |
| EPI_ISL_1226105 | GA Department of Public Health Laboratory | Genomics and Discovery, Respiratory Viruses Branch, Division of Viral Diseases, Centers for Disease Control and Prevention | Krista Queen, Yan Li, Ying Tao, Jing Zhang, Anna Uehara, Anna Montmayeur, Clinton R. Paden, Kristen Knipe, Matthew Schmerer, Shoshona Le, Katie Dillon, Peter W. Cook, Rachel Marine, Mili Sheth, Jasmine Padilla, Sarah Nobles, Mark Burroughs, Lori Rowe, Haibin Wang, Ben L. Rambo-Martin, Kristine Lacek, Sam Shepard, Dhwani Batra, Suxiang Tong, Justin Lee |
| EPI_ISL_1226106, EPI_ISL_1226107, EPI_ISL_1226108, EPI_ISL_1226109, EPI_ISL_1226110, EPI_ISL_1226111, EPI_ISL_1226112, EPI_ISL_1226113, EPI_ISL_1226114, EPI_ISL_1226115, EPI_ISL_1226116, EPI_ISL_1226117, EPI_ISL_1226118, EPI_ISL_1226119, EPI_ISL_1226120, EPI_ISL_1226121, EPI_ISL_1226122, EPI_ISL_1226123 | | | |
| see above | VA-Division of Consolidated Laboratory Services | Genomics and Discovery, Respiratory Viruses Branch, Division of Viral Diseases, Centers for Disease Control and Prevention | Krista Queen, Yan Li, Ying Tao, Jing Zhang, Anna Uehara, Anna Montmayeur, Clinton R. Paden, Kristen Knipe, Matthew Schmerer, Shoshona Le, Katie Dillon, Peter W. Cook, Rachel Marine, Mili Sheth, Jasmine Padilla, Sarah Nobles, Mark Burroughs, Lori Rowe, Haibin Wang, Ben L. Rambo-Martin, Kristine Lacek, Sam Shepard, Dhwani Batra, Suxiang Tong, Justin Lee |
| EPI_ISL_1226124, EPI_ISL_1226125, EPI_ISL_1226126, EPI_ISL_1226127, EPI_ISL_1226128, EPI_ISL_1226129, EPI_ISL_1226130, EPI_ISL_1226131, EPI_ISL_1226132, EPI_ISL_1226133, EPI_ISL_1226134, EPI_ISL_1226135, EPI_ISL_1226136, EPI_ISL_1226137, EPI_ISL_1226138, EPI_ISL_1226139, EPI_ISL_1226140, EPI_ISL_1226141, EPI_ISL_1226142, EPI_ISL_1226143 | | | |
| see above | MD DOH Laboratories Administration | Genomics and Discovery, Respiratory Viruses Branch, Division of Viral Diseases, Centers for Disease Control and Prevention | Krista Queen, Yan Li, Ying Tao, Jing Zhang, Anna Uehara, Anna Montmayeur, Clinton R. Paden, Kristen Knipe, Matthew Schmerer, Shoshona Le, Katie Dillon, Peter W. Cook, Rachel Marine, Mili Sheth, Jasmine Padilla, Sarah Nobles, Mark Burroughs, Lori Rowe, Haibin Wang, Ben L. Rambo-Martin, Kristine Lacek, Sam Shepard, Dhwani Batra, Suxiang Tong, Justin Lee |
| EPI_ISL_1226144, EPI_ISL_1226145, EPI_ISL_1226146, EPI_ISL_1226147, EPI_ISL_1226148, EPI_ISL_1226149, EPI_ISL_1226150, EPI_ISL_1226151, EPI_ISL_1226152, EPI_ISL_1226153, EPI_ISL_1226154, EPI_ISL_1226155, EPI_ISL_1226156, EPI_ISL_1226157, EPI_ISL_1226158, EPI_ISL_1226159, EPI_ISL_1226160, EPI_ISL_1226161, EPI_ISL_1226162, EPI_ISL_1226163, EPI_ISL_1226164, EPI_ISL_1226165, EPI_ISL_1226166, EPI_ISL_1226167, EPI_ISL_1226168 | | | |
| see above | OH Department of Health Laboratory | Genomics and Discovery, Respiratory Viruses Branch, Division of Viral Diseases, Centers for Disease Control and Prevention | Krista Queen, Yan Li, Ying Tao, Jing Zhang, Anna Uehara, Anna Montmayeur, Clinton R. Paden, Kristen Knipe, Matthew Schmerer, Shoshona Le, Katie Dillon, Peter W. Cook, Rachel Marine, Mili Sheth, Jasmine Padilla, Sarah Nobles, Mark Burroughs, Lori Rowe, Haibin Wang, Ben L. Rambo-Martin, Kristine Lacek, Sam Shepard, Dhwani Batra, Suxiang Tong, Justin Lee |
| EPI_ISL_1226169, EPI_ISL_1226170, EPI_ISL_1226171, EPI_ISL_1226172, EPI_ISL_1226173, EPI_ISL_1226174, EPI_ISL_1226175, EPI_ISL_1226176 | CA-Los Angeles County Public Health Laboratory | Genomics and Discovery, Respiratory Viruses Branch, Division of Viral Diseases, Centers for Disease Control and Prevention | Krista Queen, Yan Li, Ying Tao, Jing Zhang, Anna Uehara, Anna Montmayeur, Clinton R. Paden, Kristen Knipe, Matthew Schmerer, Shoshona Le, Katie Dillon, Peter W. Cook, Rachel Marine, Mili Sheth, Jasmine Padilla, Sarah Nobles, Mark Burroughs, Lori Rowe, Haibin Wang, Ben L. Rambo-Martin, Kristine Lacek, Sam Shepard, Dhwani Batra, Suxiang Tong, Justin Lee |
| EPI_ISL_1226177, EPI_ISL_1226178, EPI_ISL_1226179, EPI_ISL_1226180, EPI_ISL_1226181, EPI_ISL_1226183, EPI_ISL_1226184, EPI_ISL_1226185, EPI_ISL_1226186, EPI_ISL_1226187, EPI_ISL_1226188, EPI_ISL_1226189, EPI_ISL_1226190, EPI_ISL_1226191 | | | |
| see above | "AK State Public Health Lab, State Health Department" | Genomics and Discovery, Respiratory Viruses Branch, Division of Viral Diseases, Centers for Disease Control and Prevention | Krista Queen, Yan Li, Ying Tao, Jing Zhang, Anna Uehara, Anna Montmayeur, Clinton R. Paden, Kristen Knipe, Matthew Schmerer, Shoshona Le, Katie Dillon, Peter W. Cook, Rachel Marine, Mili Sheth, Jasmine Padilla, Sarah Nobles, Mark Burroughs, Lori Rowe, Haibin Wang, Ben L. Rambo-Martin, Kristine Lacek, Sam Shepard, Dhwani Batra, Suxiang Tong, Justin Lee |
| EPI_ISL_1226192, EPI_ISL_1226193, EPI_ISL_1226194, EPI_ISL_1226195, EPI_ISL_1226196, EPI_ISL_1226197, EPI_ISL_1226198, EPI_ISL_1226199, EPI_ISL_1226200, EPI_ISL_1226201, EPI_ISL_1226202, EPI_ISL_1226203, EPI_ISL_1226204, EPI_ISL_1226205, EPI_ISL_1226206, EPI_ISL_1226207, EPI_ISL_1226208, EPI_ISL_1226209, EPI_ISL_1226210, EPI_ISL_1226211 | | | |
| see above | "CO Dept. of Public Health and Environment, Lab Services Division" | Genomics and Discovery, Respiratory Viruses Branch, Division of Viral Diseases, Centers for Disease Control and Prevention | Krista Queen, Yan Li, Ying Tao, Jing Zhang, Anna Uehara, Anna Montmayeur, Clinton R. Paden, Kristen Knipe, Matthew Schmerer, Shoshona Le, Katie Dillon, Peter W. Cook, Rachel Marine, Mili Sheth, Jasmine Padilla, Sarah Nobles, Mark Burroughs, Lori Rowe, Haibin Wang, Ben L. Rambo-Martin, Kristine Lacek, Sam Shepard, Dhwani Batra, Suxiang Tong, Justin Lee |
| EPI_ISL_1226212, EPI_ISL_1226213 | CT-Dr. Katherine A. Kelley State Public Health Lab | Genomics and Discovery, Respiratory Viruses Branch, Division of Viral Diseases, Centers for Disease Control and Prevention | Krista Queen, Yan Li, Ying Tao, Jing Zhang, Anna Uehara, Anna Montmayeur, Clinton R. Paden, Kristen Knipe, Matthew Schmerer, Shoshona Le, Katie Dillon, Peter W. Cook, Rachel Marine, Mili Sheth, Jasmine Padilla, Sarah Nobles, Mark Burroughs, Lori Rowe, Haibin Wang, Ben L. Rambo-Martin, Kristine Lacek, Sam Shepard, Dhwani Batra, Suxiang Tong, Justin Lee |
| EPI_ISL_1226214, EPI_ISL_1226215, EPI_ISL_1226216, EPI_ISL_1226217, EPI_ISL_1226218, EPI_ISL_1226219, EPI_ISL_1226220, EPI_ISL_1226221, EPI_ISL_1226222, EPI_ISL_1226224, EPI_ISL_1226225, EPI_ISL_1226226 | | | |
| see above | WI State Laboratory of Hygiene | Genomics and Discovery, Respiratory Viruses Branch, Division of Viral Diseases, Centers for Disease Control and Prevention | Krista Queen, Yan Li, Ying Tao, Jing Zhang, Anna Uehara, Anna Montmayeur, Clinton R. Paden, Kristen Knipe, Matthew Schmerer, Shoshona Le, Katie Dillon, Peter W. Cook, Rachel Marine, Mili Sheth, Jasmine Padilla, Sarah Nobles, Mark Burroughs, Lori Rowe, Haibin Wang, Ben L. Rambo-Martin, Kristine Lacek, Sam Shepard, Dhwani Batra, Suxiang Tong, Justin Lee |
| EPI_ISL_1226227, EPI_ISL_1226228, EPI_ISL_1226229, EPI_ISL_1226230 | NJ Public Health and Environmental Laboratories | Genomics and Discovery, Respiratory Viruses Branch, Division of Viral Diseases, Centers for Disease Control and Prevention | Krista Queen, Yan Li, Ying Tao, Jing Zhang, Anna Uehara, Anna Montmayeur, Clinton R. Paden, Kristen Knipe, Matthew Schmerer, Shoshona Le, Katie Dillon, Peter W. Cook, Rachel Marine, Mili Sheth, Jasmine Padilla, Sarah Nobles, Mark Burroughs, Lori Rowe, Haibin Wang, Ben L. Rambo-Martin, Kristine Lacek, Sam Shepard, Dhwani Batra, Suxiang Tong, Justin Lee |
| EPI_ISL_1226231, EPI_ISL_1226232, EPI_ISL_1226233, EPI_ISL_1226234, EPI_ISL_1226235, EPI_ISL_1226236 | VT Dept. of Health Laboratory | Genomics and Discovery, Respiratory Viruses Branch, Division of Viral Diseases, Centers for Disease Control and Prevention | Krista Queen, Yan Li, Ying Tao, Jing Zhang, Anna Uehara, Anna Montmayeur, Clinton R. Paden, Kristen Knipe, Matthew Schmerer, Shoshona Le, Katie Dillon, Peter W. Cook, Rachel Marine, Mili Sheth, Jasmine Padilla, Sarah Nobles, Mark Burroughs, Lori Rowe, Haibin Wang, Ben L. Rambo-Martin, Kristine Lacek, Sam Shepard, Dhwani Batra, Suxiang Tong, Justin Lee |
| EPI_ISL_1226237, EPI_ISL_1226238, EPI_ISL_1226239, EPI_ISL_1226240, EPI_ISL_1226241, EPI_ISL_1226242, EPI_ISL_1226243, EPI_ISL_1226244, EPI_ISL_1226245, EPI_ISL_1226246, EPI_ISL_1226247, EPI_ISL_1226248, EPI_ISL_1226249, EPI_ISL_1226250, EPI_ISL_1226251, EPI_ISL_1226252, EPI_ISL_1226253, EPI_ISL_1226254, EPI_ISL_1226255, EPI_ISL_1226256, EPI_ISL_1226257, EPI_ISL_1226258, EPI_ISL_1226259, EPI_ISL_1226260, EPI_ISL_1226261, EPI_ISL_1226262, EPI_ISL_1226263, EPI_ISL_1226264, EPI_ISL_1226265, EPI_ISL_1226266, EPI_ISL_1226267, EPI_ISL_1226268, EPI_ISL_1226269, EPI_ISL_1226270, EPI_ISL_1226271, EPI_ISL_1226272, EPI_ISL_1226273, EPI_ISL_1226274, EPI_ISL_1226275 | | | |
| see above | "CDPH, Viral and Rickettsial Disease Laboratory" | Genomics and Discovery, Respiratory Viruses Branch, Division of Viral Diseases, Centers for Disease Control and Prevention | Krista Queen, Yan Li, Ying Tao, Jing Zhang, Anna Uehara, Anna Montmayeur, Clinton R. Paden, Kristen Knipe, Matthew Schmerer, Shoshona Le, Katie Dillon, Peter W. Cook, Rachel Marine, Mili Sheth, Jasmine Padilla, Sarah Nobles, Mark Burroughs, Lori Rowe, Haibin Wang, Ben L. Rambo-Martin, Kristine Lacek, Sam Shepard, Dhwani Batra, Suxiang Tong, Justin Lee |
| EPI_ISL_1226276, EPI_ISL_1226277 | MI - Michigan Department of Health and Human Services - Bureau of Laboratories | Genomics and Discovery, Respiratory Viruses Branch, Division of Viral Diseases, Centers for Disease Control and Prevention | Krista Queen, Yan Li, Ying Tao, Jing Zhang, Anna Uehara, Anna Montmayeur, Clinton R. Paden, Kristen Knipe, Matthew Schmerer, Shoshona Le, Katie Dillon, Peter W. Cook, Rachel Marine, Mili Sheth, Jasmine Padilla, Sarah Nobles, Mark Burroughs, Lori Rowe, Haibin Wang, Ben L. Rambo-Martin, Kristine Lacek, Sam Shepard, Dhwani Batra, Suxiang Tong, Justin Lee |
| EPI_ISL_1226278, EPI_ISL_1226279, EPI_ISL_1226280 | SD Public Health Laboratory | Genomics and Discovery, Respiratory Viruses Branch, Division of Viral Diseases, Centers for Disease Control and Prevention | Krista Queen, Yan Li, Ying Tao, Jing Zhang, Anna Uehara, Anna Montmayeur, Clinton R. Paden, Kristen Knipe, Matthew Schmerer, Shoshona Le, Katie Dillon, Peter W. Cook, Rachel Marine, Mili Sheth, Jasmine Padilla, Sarah Nobles, Mark Burroughs, Lori Rowe, Haibin Wang, Ben L. Rambo-Martin, Kristine Lacek, Sam Shepard, Dhwani Batra, Suxiang Tong, Justin Lee |
| EPI_ISL_1226281, EPI_ISL_1226282, EPI_ISL_1226283, EPI_ISL_1226284, EPI_ISL_1226285, EPI_ISL_1226286, EPI_ISL_1226287 | "NM Dept. Health, Scientific Laboratory Division " | Genomics and Discovery, Respiratory Viruses Branch, Division of Viral Diseases, Centers for Disease Control and Prevention | Krista Queen, Yan Li, Ying Tao, Jing Zhang, Anna Uehara, Anna Montmayeur, Clinton R. Paden, Kristen Knipe, Matthew Schmerer, Shoshona Le, Katie Dillon, Peter W. Cook, Rachel Marine, Mili Sheth, Jasmine Padilla, Sarah Nobles, Mark Burroughs, Lori Rowe, Haibin Wang, Ben L. Rambo-Martin, Kristine Lacek, Sam Shepard, Dhwani Batra, Suxiang Tong, Justin Lee |
| EPI_ISL_1226288, EPI_ISL_1226289, EPI_ISL_1226290, EPI_ISL_1226291, EPI_ISL_1226292, EPI_ISL_1226293, EPI_ISL_1226294, EPI_ISL_1226295, EPI_ISL_1226296, EPI_ISL_1226297, EPI_ISL_1226298, EPI_ISL_1226299, EPI_ISL_1226300, EPI_ISL_1226302 | | | |

| | | | |
|---|---|---|---|
| see above | MT Public Health Laboratory | Genomics and Discovery, Respiratory Viruses Branch, Division of Viral Diseases, Centers for Disease Control and Prevention | Krista Queen, Yan Li, Ying Tao, Jing Zhang, Anna Uehara, Anna Montmayeur, Clinton R. Paden, Kristen Knipe, Matthew Schmerer, Shoshona Le, Katie Dillon, Peter W. Cook, Rachel Marine, Mili Sheth, Jasmine Padilla, Sarah Nobles, Mark Burroughs, Lori Rowe, Haibin Wang, Ben L. Rambo-Martin, Kristine Lacek, Sam Shepard, Dhwani Batra, Suxiang Tong, Justin Lee |
| EPI_ISL_1226303, EPI_ISL_1226304, EPI_ISL_1226305, EPI_ISL_1226306 | MI - Michigan Department of Health and Human Services - Bureau of Laboratories | Genomics and Discovery, Respiratory Viruses Branch, Division of Viral Diseases, Centers for Disease Control and Prevention | Krista Queen, Yan Li, Ying Tao, Jing Zhang, Anna Uehara, Anna Montmayeur, Clinton R. Paden, Kristen Knipe, Matthew Schmerer, Shoshona Le, Katie Dillon, Peter W. Cook, Rachel Marine, Mili Sheth, Jasmine Padilla, Sarah Nobles, Mark Burroughs, Lori Rowe, Haibin Wang, Ben L. Rambo-Martin, Kristine Lacek, Sam Shepard, Dhwani Batra, Suxiang Tong, Justin Lee |
| EPI_ISL_1226307, EPI_ISL_1226308, EPI_ISL_1226309, EPI_ISL_1226310, EPI_ISL_1226311, EPI_ISL_1226312, EPI_ISL_1226313, EPI_ISL_1226314, EPI_ISL_1226315, EPI_ISL_1226316, EPI_ISL_1226317, EPI_ISL_1226318, EPI_ISL_1226319, EPI_ISL_1226320, EPI_ISL_1226321, EPI_ISL_1226322, EPI_ISL_1226323, EPI_ISL_1226324, EPI_ISL_1226325 | | | |
| see above | UT-Unified State Labs: Public Health Utah DOH | Genomics and Discovery, Respiratory Viruses Branch, Division of Viral Diseases, Centers for Disease Control and Prevention | Krista Queen, Yan Li, Ying Tao, Jing Zhang, Anna Uehara, Anna Montmayeur, Clinton R. Paden, Kristen Knipe, Matthew Schmerer, Shoshona Le, Katie Dillon, Peter W. Cook, Rachel Marine, Mili Sheth, Jasmine Padilla, Sarah Nobles, Mark Burroughs, Lori Rowe, Haibin Wang, Ben L. Rambo-Martin, Kristine Lacek, Sam Shepard, Dhwani Batra, Suxiang Tong, Justin Lee |
| EPI_ISL_1226326, EPI_ISL_1226327, EPI_ISL_1226328, EPI_ISL_1226329, EPI_ISL_1226330, EPI_ISL_1226331, EPI_ISL_1226332, EPI_ISL_1226333, EPI_ISL_1226334, EPI_ISL_1226335, EPI_ISL_1226336, EPI_ISL_1226337, EPI_ISL_1226338, EPI_ISL_1226339, EPI_ISL_1226340 | | | |
| see above | "OK Public Health Laboratory, Oklahoma State DOH" | Genomics and Discovery, Respiratory Viruses Branch, Division of Viral Diseases, Centers for Disease Control and Prevention | Krista Queen, Yan Li, Ying Tao, Jing Zhang, Anna Uehara, Anna Montmayeur, Clinton R. Paden, Kristen Knipe, Matthew Schmerer, Shoshona Le, Katie Dillon, Peter W. Cook, Rachel Marine, Mili Sheth, Jasmine Padilla, Sarah Nobles, Mark Burroughs, Lori Rowe, Haibin Wang, Ben L. Rambo-Martin, Kristine Lacek, Sam Shepard, Dhwani Batra, Suxiang Tong, Justin Lee |
| EPI_ISL_1226341, EPI_ISL_1226342, EPI_ISL_1226343 | "MN PHL Division, Minnesota Department of Health" | Genomics and Discovery, Respiratory Viruses Branch, Division of Viral Diseases, Centers for Disease Control and Prevention | Krista Queen, Yan Li, Ying Tao, Jing Zhang, Anna Uehara, Anna Montmayeur, Clinton R. Paden, Kristen Knipe, Matthew Schmerer, Shoshona Le, Katie Dillon, Peter W. Cook, Rachel Marine, Mili Sheth, Jasmine Padilla, Sarah Nobles, Mark Burroughs, Lori Rowe, Haibin Wang, Ben L. Rambo-Martin, Kristine Lacek, Sam Shepard, Dhwani Batra, Suxiang Tong, Justin Lee |
| EPI_ISL_1226344, EPI_ISL_1226345, EPI_ISL_1226346, EPI_ISL_1226347, EPI_ISL_1226348, EPI_ISL_1226349, EPI_ISL_1226350, EPI_ISL_1226351, EPI_ISL_1226352, EPI_ISL_1226353, EPI_ISL_1226354, EPI_ISL_1226355, EPI_ISL_1226356, EPI_ISL_1226357, EPI_ISL_1226358 | | | |
| see above | OR State PHL-Virology/Immunology Section | Genomics and Discovery, Respiratory Viruses Branch, Division of Viral Diseases, Centers for Disease Control and Prevention | Krista Queen, Yan Li, Ying Tao, Jing Zhang, Anna Uehara, Anna Montmayeur, Clinton R. Paden, Kristen Knipe, Matthew Schmerer, Shoshona Le, Katie Dillon, Peter W. Cook, Rachel Marine, Mili Sheth, Jasmine Padilla, Sarah Nobles, Mark Burroughs, Lori Rowe, Haibin Wang, Ben L. Rambo-Martin, Kristine Lacek, Sam Shepard, Dhwani Batra, Suxiang Tong, Justin Lee |
| EPI_ISL_1226359, EPI_ISL_1226360, EPI_ISL_1226361, EPI_ISL_1226362, EPI_ISL_1226363, EPI_ISL_1226364, EPI_ISL_1226365, EPI_ISL_1226366, EPI_ISL_1226367, EPI_ISL_1226368, EPI_ISL_1226369, EPI_ISL_1226370, EPI_ISL_1226371, EPI_ISL_1226372, EPI_ISL_1226373, EPI_ISL_1226374, EPI_ISL_1226375, EPI_ISL_1226376, EPI_ISL_1226377, EPI_ISL_1226378, EPI_ISL_1226379, EPI_ISL_1226380 | | | |
| see above | MO State Public Health Laboratory | Genomics and Discovery, Respiratory Viruses Branch, Division of Viral Diseases, Centers for Disease Control and Prevention | Krista Queen, Yan Li, Ying Tao, Jing Zhang, Anna Uehara, Anna Montmayeur, Clinton R. Paden, Kristen Knipe, Matthew Schmerer, Shoshona Le, Katie Dillon, Peter W. Cook, Rachel Marine, Mili Sheth, Jasmine Padilla, Sarah Nobles, Mark Burroughs, Lori Rowe, Haibin Wang, Ben L. Rambo-Martin, Kristine Lacek, Sam Shepard, Dhwani Batra, Suxiang Tong, Justin Lee |
| EPI_ISL_1226381 | "AZ SPHL, Arizona Department of Health Services" | Genomics and Discovery, Respiratory Viruses Branch, Division of Viral Diseases, Centers for Disease Control and Prevention | Krista Queen, Yan Li, Ying Tao, Jing Zhang, Anna Uehara, Anna Montmayeur, Clinton R. Paden, Kristen Knipe, Matthew Schmerer, Shoshona Le, Katie Dillon, Peter W. Cook, Rachel Marine, Mili Sheth, Jasmine Padilla, Sarah Nobles, Mark Burroughs, Lori Rowe, Haibin Wang, Ben L. Rambo-Martin, Kristine Lacek, Sam Shepard, Dhwani Batra, Suxiang Tong, Justin Lee |
| EPI_ISL_1226382, EPI_ISL_1226383, EPI_ISL_1226384 | "PA Department of Health, Bureau of Laboratories" | Genomics and Discovery, Respiratory Viruses Branch, Division of Viral Diseases, Centers for Disease Control and Prevention | Krista Queen, Yan Li, Ying Tao, Jing Zhang, Anna Uehara, Anna Montmayeur, Clinton R. Paden, Kristen Knipe, Matthew Schmerer, Shoshona Le, Katie Dillon, Peter W. Cook, Rachel Marine, Mili Sheth, Jasmine Padilla, Sarah Nobles, Mark Burroughs, Lori Rowe, Haibin Wang, Ben L. Rambo-Martin, Kristine Lacek, Sam Shepard, Dhwani Batra, Suxiang Tong, Justin Lee |
| EPI_ISL_1226385, EPI_ISL_1226386, EPI_ISL_1226387, EPI_ISL_1226388, EPI_ISL_1226389, EPI_ISL_1226390, EPI_ISL_1226391, EPI_ISL_1226392, EPI_ISL_1226393, EPI_ISL_1226394, EPI_ISL_1226395, EPI_ISL_1226396, EPI_ISL_1226397, EPI_ISL_1226398 | | | |
| see above | WA State Department of Health | Genomics and Discovery, Respiratory Viruses Branch, Division of Viral Diseases, Centers for Disease Control and Prevention | Krista Queen, Yan Li, Ying Tao, Jing Zhang, Anna Uehara, Anna Montmayeur, Clinton R. Paden, Kristen Knipe, Matthew Schmerer, Shoshona Le, Katie Dillon, Peter W. Cook, Rachel Marine, Mili Sheth, Jasmine Padilla, Sarah Nobles, Mark Burroughs, Lori Rowe, Haibin Wang, Ben L. Rambo-Martin, Kristine Lacek, Sam Shepard, Dhwani Batra, Suxiang Tong, Justin Lee |
| EPI_ISL_1226399, EPI_ISL_1226400, EPI_ISL_1226401, EPI_ISL_1226402, EPI_ISL_1226403, EPI_ISL_1226404, EPI_ISL_1226405, EPI_ISL_1226406 | NJ Public Health and Environmental Laboratories | Genomics and Discovery, Respiratory Viruses Branch, Division of Viral Diseases, Centers for Disease Control and Prevention | Krista Queen, Yan Li, Ying Tao, Jing Zhang, Anna Uehara, Anna Montmayeur, Clinton R. Paden, Kristen Knipe, Matthew Schmerer, Shoshona Le, Katie Dillon, Peter W. Cook, Rachel Marine, Mili Sheth, Jasmine Padilla, Sarah Nobles, Mark Burroughs, Lori Rowe, Haibin Wang, Ben L. Rambo-Martin, Kristine Lacek, Sam Shepard, Dhwani Batra, Suxiang Tong, Justin Lee |
| EPI_ISL_1226407 | "AZ SPHL, Arizona Department of Health Services" | Genomics and Discovery, Respiratory Viruses Branch, Division of Viral Diseases, Centers for Disease Control and Prevention | Krista Queen, Yan Li, Ying Tao, Jing Zhang, Anna Uehara, Anna Montmayeur, Clinton R. Paden, Kristen Knipe, Matthew Schmerer, Shoshona Le, Katie Dillon, Peter W. Cook, Rachel Marine, Mili Sheth, Jasmine Padilla, Sarah Nobles, Mark Burroughs, Lori Rowe, Haibin Wang, Ben L. Rambo-Martin, Kristine Lacek, Sam Shepard, Dhwani Batra, Suxiang Tong, Justin Lee |
| EPI_ISL_1226408, EPI_ISL_1226409, EPI_ISL_1226410, EPI_ISL_1226411, EPI_ISL_1226412, EPI_ISL_1226413, EPI_ISL_1226414, EPI_ISL_1226415, EPI_ISL_1226416 | MO State Public Health Laboratory | Genomics and Discovery, Respiratory Viruses Branch, Division of Viral Diseases, Centers for Disease Control and Prevention | Krista Queen, Yan Li, Ying Tao, Jing Zhang, Anna Uehara, Anna Montmayeur, Clinton R. Paden, Kristen Knipe, Matthew Schmerer, Shoshona Le, Katie Dillon, Peter W. Cook, Rachel Marine, Mili Sheth, Jasmine Padilla, Sarah Nobles, Mark Burroughs, Lori Rowe, Haibin Wang, Ben L. Rambo-Martin, Kristine Lacek, Sam Shepard, Dhwani Batra, Suxiang Tong, Justin Lee |
| EPI_ISL_1226417 | WESTCHESTER MEDICAL CENTER | Wadsworth Center, New York State Department of Health | Kirsten St. George, Daryl M. Lamson, Alexis Russel, Matthew Shudt, Melissa A Leisner, Jonathan Plitnick, Navjot Singh, John Kelly, Erasmus Schneider, Erica Lasek-Nesselquist |
| EPI_ISL_1226418 | GLENS FALLS HOSPITAL LABORATORY | Wadsworth Center, New York State Department of Health | Kirsten St. George, Daryl M. Lamson, Alexis Russel, Matthew Shudt, Melissa A Leisner, Jonathan Plitnick, Navjot Singh, John Kelly, Erasmus Schneider, Erica Lasek-Nesselquist |
| EPI_ISL_1226419 | NYC Pandemic Response Lab | Wadsworth Center, New York State Department of Health | Kirsten St. George, Daryl M. Lamson, Alexis Russel, Matthew Shudt, Melissa A Leisner, Jonathan Plitnick, Navjot Singh, John Kelly, Erasmus Schneider, Erica Lasek-Nesselquist |
| EPI_ISL_1226420 | NORTH SHORE UNIVERSITY HOSPITAL | Wadsworth Center, New York State Department of Health | Kirsten St. George, Daryl M. Lamson, Alexis Russel, Matthew Shudt, Melissa A Leisner, Jonathan Plitnick, Navjot Singh, John Kelly, Erasmus Schneider, Erica Lasek-Nesselquist |
| EPI_ISL_1226421 | URMC LABS | Wadsworth Center, New York State Department of Health | Kirsten St. George, Daryl M. Lamson, Alexis Russel, Matthew Shudt, Melissa A Leisner, Jonathan Plitnick, Navjot Singh, John Kelly, Erasmus Schneider, Erica Lasek-Nesselquist |
| EPI_ISL_1226422 | Masonic Care Community of NY | Wadsworth Center, New York State Department of Health | Kirsten St. George, Daryl M. Lamson, Alexis Russel, Matthew Shudt, Melissa A Leisner, Jonathan Plitnick, Navjot Singh, John Kelly, Erasmus Schneider, Erica Lasek-Nesselquist |
| EPI_ISL_1226423 | THE MARY IMOGENE BASSETT HOSPITAL | Wadsworth Center, New York State Department of Health | Kirsten St. George, Daryl M. Lamson, Alexis Russel, Matthew Shudt, Melissa A Leisner, Jonathan Plitnick, Navjot Singh, John Kelly, Erasmus Schneider, Erica Lasek-Nesselquist |
| EPI_ISL_1226424 | MONTEFIORE MEDICAL CENTER LABORATORIES | Wadsworth Center, New York State Department of Health | Kirsten St. George, Daryl M. Lamson, Alexis Russel, Matthew Shudt, Melissa A Leisner, Jonathan Plitnick, Navjot Singh, John Kelly, Erasmus Schneider, Erica Lasek-Nesselquist |
| EPI_ISL_1226425 | NORTHWELL HEALTH LABORATORIES | Wadsworth Center, New York State Department of Health | Kirsten St. George, Daryl M. Lamson, Alexis Russel, Matthew Shudt, Melissa A Leisner, Jonathan Plitnick, Navjot Singh, John Kelly, Erasmus Schneider, Erica Lasek-Nesselquist |
| EPI_ISL_1226426 | MONTEFIORE MEDICAL CENTER LABORATORIES | Wadsworth Center, New York State Department of Health | Kirsten St. George, Daryl M. Lamson, Alexis Russel, Matthew Shudt, Melissa A Leisner, Jonathan Plitnick, Navjot Singh, John Kelly, Erasmus Schneider, Erica Lasek-Nesselquist |
| EPI_ISL_1226427 | MEMORIAL SLOAN KETTERING CANCER CENTER | Wadsworth Center, New York State Department of Health | Kirsten St. George, Daryl M. Lamson, Alexis Russel, Matthew Shudt, Melissa A Leisner, Jonathan Plitnick, Navjot Singh, John Kelly, Erasmus Schneider, Erica Lasek-Nesselquist |
| EPI_ISL_1226428 | URMC LABS | Wadsworth Center, New York State Department of Health | Kirsten St. George, Daryl M. Lamson, Alexis Russel, Matthew Shudt, Melissa A Leisner, Jonathan Plitnick, Navjot Singh, John Kelly, Erasmus Schneider, Erica Lasek-Nesselquist |
| EPI_ISL_1226429 | Masonic Care Community of NY | Wadsworth Center, New York State Department of Health | Kirsten St. George, Daryl M. Lamson, Alexis Russel, Matthew Shudt, Melissa A Leisner, Jonathan Plitnick, Navjot Singh, John Kelly, Erasmus Schneider, Erica Lasek-Nesselquist |
| EPI_ISL_1226430 | NYC Pandemic Response Lab | Wadsworth Center, New York State Department of Health | Kirsten St. George, Daryl M. Lamson, Alexis Russel, Matthew Shudt, Melissa A Leisner, Jonathan Plitnick, Navjot Singh, John Kelly, Erasmus Schneider, Erica Lasek-Nesselquist |
| EPI_ISL_1226431 | MONTEFIORE MEDICAL CENTER LABORATORIES | Wadsworth Center, New York State Department of Health | Kirsten St. George, Daryl M. Lamson, Alexis Russel, Matthew Shudt, Melissa A Leisner, Jonathan Plitnick, Navjot Singh, John Kelly, Erasmus Schneider, Erica Lasek-Nesselquist |
| EPI_ISL_1226432 | URMC LABS | Wadsworth Center, New York State Department of Health | Kirsten St. George, Daryl M. Lamson, Alexis Russel, Matthew Shudt, Melissa A Leisner, Jonathan Plitnick, Navjot Singh, John Kelly, Erasmus Schneider, Erica Lasek-Nesselquist |
| EPI_ISL_1226433 | MONTEFIORE MEDICAL CENTER LABORATORIES | Wadsworth Center, New York State Department of Health | Kirsten St. George, Daryl M. Lamson, Alexis Russel, Matthew Shudt, Melissa A Leisner, Jonathan Plitnick, Navjot Singh, John Kelly, Erasmus Schneider, Erica Lasek-Nesselquist |
| EPI_ISL_1226434 | New York Presbyterian Hospital | Wadsworth Center, New York State Department of Health | Kirsten St. George, Daryl M. Lamson, Alexis Russel, Matthew Shudt, Melissa A Leisner, Jonathan Plitnick, Navjot Singh, John Kelly, Erasmus Schneider, Erica Lasek-Nesselquist |
| EPI_ISL_1226435 | URMC LABS | Wadsworth Center, New York State Department of Health | Kirsten St. George, Daryl M. Lamson, Alexis Russel, Matthew Shudt, Melissa A Leisner, Jonathan Plitnick, Navjot Singh, John Kelly, Erasmus Schneider, Erica Lasek-Nesselquist |

| | | | |
|---|---|---|---|
| EPI_ISL_1226436 | KALEIDA CENTER FOR LABORATORY MEDICINE | Wadsworth Center, New York State Department of Health | Kirsten St. George, Daryl M. Lamson, Alexis Russel, Matthew Shudt, Melissa A Leisner, Jonathan Plitnick, Navjot Singh, John Kelly, Erasmus Schneider, Erica Lasek-Nesselquist |
| EPI_ISL_1226437 | MEMORIAL SLOAN KETTERING CANCER CENTER | Wadsworth Center, New York State Department of Health | Kirsten St. George, Daryl M. Lamson, Alexis Russel, Matthew Shudt, Melissa A Leisner, Jonathan Plitnick, Navjot Singh, John Kelly, Erasmus Schneider, Erica Lasek-Nesselquist |
| EPI_ISL_1226438 | WESTCHESTER MEDICAL CENTER | Wadsworth Center, New York State Department of Health | Kirsten St. George, Daryl M. Lamson, Alexis Russel, Matthew Shudt, Melissa A Leisner, Jonathan Plitnick, Navjot Singh, John Kelly, Erasmus Schneider, Erica Lasek-Nesselquist |
| EPI_ISL_1226439, EPI_ISL_1226440, EPI_ISL_1226441 | NYC Pandemic Response Lab | Wadsworth Center, New York State Department of Health | Kirsten St. George, Daryl M. Lamson, Alexis Russel, Matthew Shudt, Melissa A Leisner, Jonathan Plitnick, Navjot Singh, John Kelly, Erasmus Schneider, Erica Lasek-Nesselquist |
| EPI_ISL_1226442 | MEMORIAL SLOAN KETTERING CANCER CENTER | Wadsworth Center, New York State Department of Health | Kirsten St. George, Daryl M. Lamson, Alexis Russel, Matthew Shudt, Melissa A Leisner, Jonathan Plitnick, Navjot Singh, John Kelly, Erasmus Schneider, Erica Lasek-Nesselquist |
| EPI_ISL_1226443 | NYC Pandemic Response Lab | Wadsworth Center, New York State Department of Health | Kirsten St. George, Daryl M. Lamson, Alexis Russel, Matthew Shudt, Melissa A Leisner, Jonathan Plitnick, Navjot Singh, John Kelly, Erasmus Schneider, Erica Lasek-Nesselquist |
| EPI_ISL_1226444 | NORTHWELL HEALTH LABORATORIES | Wadsworth Center, New York State Department of Health | Kirsten St. George, Daryl M. Lamson, Alexis Russel, Matthew Shudt, Melissa A Leisner, Jonathan Plitnick, Navjot Singh, John Kelly, Erasmus Schneider, Erica Lasek-Nesselquist |
| EPI_ISL_1226445 | URMC LABS | Wadsworth Center, New York State Department of Health | Kirsten St. George, Daryl M. Lamson, Alexis Russel, Matthew Shudt, Melissa A Leisner, Jonathan Plitnick, Navjot Singh, John Kelly, Erasmus Schneider, Erica Lasek-Nesselquist |
| EPI_ISL_1226446 | MONTEFIORE MEDICAL CENTER LABORATORIES | Wadsworth Center, New York State Department of Health | Kirsten St. George, Daryl M. Lamson, Alexis Russel, Matthew Shudt, Melissa A Leisner, Jonathan Plitnick, Navjot Singh, John Kelly, Erasmus Schneider, Erica Lasek-Nesselquist |
| EPI_ISL_1226447 | NORTH SHORE UNIVERSITY HOSPITAL | Wadsworth Center, New York State Department of Health | Kirsten St. George, Daryl M. Lamson, Alexis Russel, Matthew Shudt, Melissa A Leisner, Jonathan Plitnick, Navjot Singh, John Kelly, Erasmus Schneider, Erica Lasek-Nesselquist |
| EPI_ISL_1226448 | NYC Pandemic Response Lab | Wadsworth Center, New York State Department of Health | Kirsten St. George, Daryl M. Lamson, Alexis Russel, Matthew Shudt, Melissa A Leisner, Jonathan Plitnick, Navjot Singh, John Kelly, Erasmus Schneider, Erica Lasek-Nesselquist |
| EPI_ISL_1226449 | TEMPUS LABS INC | Wadsworth Center, New York State Department of Health | Kirsten St. George, Daryl M. Lamson, Alexis Russel, Matthew Shudt, Melissa A Leisner, Jonathan Plitnick, Navjot Singh, John Kelly, Erasmus Schneider, Erica Lasek-Nesselquist |
| EPI_ISL_1226450 | MONTEFIORE MEDICAL CENTER LABORATORIES | Wadsworth Center, New York State Department of Health | Kirsten St. George, Daryl M. Lamson, Alexis Russel, Matthew Shudt, Melissa A Leisner, Jonathan Plitnick, Navjot Singh, John Kelly, Erasmus Schneider, Erica Lasek-Nesselquist |
| EPI_ISL_1226451 | URMC LABS | Wadsworth Center, New York State Department of Health | Kirsten St. George, Daryl M. Lamson, Alexis Russel, Matthew Shudt, Melissa A Leisner, Jonathan Plitnick, Navjot Singh, John Kelly, Erasmus Schneider, Erica Lasek-Nesselquist |
| EPI_ISL_1226452 | GLENS FALLS HOSPITAL LABORATORY | Wadsworth Center, New York State Department of Health | Kirsten St. George, Daryl M. Lamson, Alexis Russel, Matthew Shudt, Melissa A Leisner, Jonathan Plitnick, Navjot Singh, John Kelly, Erasmus Schneider, Erica Lasek-Nesselquist |
| EPI_ISL_1226453 | WESTCHESTER MEDICAL CENTER | Wadsworth Center, New York State Department of Health | Kirsten St. George, Daryl M. Lamson, Alexis Russel, Matthew Shudt, Melissa A Leisner, Jonathan Plitnick, Navjot Singh, John Kelly, Erasmus Schneider, Erica Lasek-Nesselquist |
| EPI_ISL_1226454 | MONTEFIORE MEDICAL CENTER LABORATORIES | Wadsworth Center, New York State Department of Health | Kirsten St. George, Daryl M. Lamson, Alexis Russel, Matthew Shudt, Melissa A Leisner, Jonathan Plitnick, Navjot Singh, John Kelly, Erasmus Schneider, Erica Lasek-Nesselquist |
| EPI_ISL_1226455 | MEMORIAL SLOAN KETTERING CANCER CENTER | Wadsworth Center, New York State Department of Health | Kirsten St. George, Daryl M. Lamson, Alexis Russel, Matthew Shudt, Melissa A Leisner, Jonathan Plitnick, Navjot Singh, John Kelly, Erasmus Schneider, Erica Lasek-Nesselquist |
| EPI_ISL_1226456 | Columbia University Irving Medical Center | Wadsworth Center, New York State Department of Health | Kirsten St. George, Daryl M. Lamson, Alexis Russel, Matthew Shudt, Melissa A Leisner, Jonathan Plitnick, Navjot Singh, John Kelly, Erasmus Schneider, Erica Lasek-Nesselquist |
| EPI_ISL_1226457 | MONTEFIORE MEDICAL CENTER LABORATORIES | Wadsworth Center, New York State Department of Health | Kirsten St. George, Daryl M. Lamson, Alexis Russel, Matthew Shudt, Melissa A Leisner, Jonathan Plitnick, Navjot Singh, John Kelly, Erasmus Schneider, Erica Lasek-Nesselquist |
| EPI_ISL_1226458 | KALEIDA CENTER FOR LABORATORY MEDICINE | Wadsworth Center, New York State Department of Health | Kirsten St. George, Daryl M. Lamson, Alexis Russel, Matthew Shudt, Melissa A Leisner, Jonathan Plitnick, Navjot Singh, John Kelly, Erasmus Schneider, Erica Lasek-Nesselquist |
| EPI_ISL_1226459 | NYC Pandemic Response Lab | Wadsworth Center, New York State Department of Health | Kirsten St. George, Daryl M. Lamson, Alexis Russel, Matthew Shudt, Melissa A Leisner, Jonathan Plitnick, Navjot Singh, John Kelly, Erasmus Schneider, Erica Lasek-Nesselquist |
| EPI_ISL_1226460 | URMC LABS | Wadsworth Center, New York State Department of Health | Kirsten St. George, Daryl M. Lamson, Alexis Russel, Matthew Shudt, Melissa A Leisner, Jonathan Plitnick, Navjot Singh, John Kelly, Erasmus Schneider, Erica Lasek-Nesselquist |
| EPI_ISL_1226461 | Columbia University Irving Medical Center | Wadsworth Center, New York State Department of Health | Kirsten St. George, Daryl M. Lamson, Alexis Russel, Matthew Shudt, Melissa A Leisner, Jonathan Plitnick, Navjot Singh, John Kelly, Erasmus Schneider, Erica Lasek-Nesselquist |
| EPI_ISL_1226462 | WESTCHESTER MEDICAL CENTER | Wadsworth Center, New York State Department of Health | Kirsten St. George, Daryl M. Lamson, Alexis Russel, Matthew Shudt, Melissa A Leisner, Jonathan Plitnick, Navjot Singh, John Kelly, Erasmus Schneider, Erica Lasek-Nesselquist |
| EPI_ISL_1226463 | Columbia University Irving Medical Center | Wadsworth Center, New York State Department of Health | Kirsten St. George, Daryl M. Lamson, Alexis Russel, Matthew Shudt, Melissa A Leisner, Jonathan Plitnick, Navjot Singh, John Kelly, Erasmus Schneider, Erica Lasek-Nesselquist |
| EPI_ISL_1226464, EPI_ISL_1226465 | MONTEFIORE MEDICAL CENTER LABORATORIES | Wadsworth Center, New York State Department of Health | Kirsten St. George, Daryl M. Lamson, Alexis Russel, Matthew Shudt, Melissa A Leisner, Jonathan Plitnick, Navjot Singh, John Kelly, Erasmus Schneider, Erica Lasek-Nesselquist |
| EPI_ISL_1226466 | Columbia University Irving Medical Center | Wadsworth Center, New York State Department of Health | Kirsten St. George, Daryl M. Lamson, Alexis Russel, Matthew Shudt, Melissa A Leisner, Jonathan Plitnick, Navjot Singh, John Kelly, Erasmus Schneider, Erica Lasek-Nesselquist |
| EPI_ISL_1226467 | MEMORIAL SLOAN KETTERING CANCER CENTER | Wadsworth Center, New York State Department of Health | Kirsten St. George, Daryl M. Lamson, Alexis Russel, Matthew Shudt, Melissa A Leisner, Jonathan Plitnick, Navjot Singh, John Kelly, Erasmus Schneider, Erica Lasek-Nesselquist |
| EPI_ISL_1226468, EPI_ISL_1226469 | Wadsworth Center, New York State Department of Health | Wadsworth Center, New York State Department of Health | Kirsten St. George, Daryl M. Lamson, Alexis Russel, Matthew Shudt, Melissa A Leisner, Jonathan Plitnick, Navjot Singh, John Kelly, Erasmus Schneider, Erica Lasek-Nesselquist |
| EPI_ISL_1226470 | Masonic Care Community of NY | Wadsworth Center, New York State Department of Health | Kirsten St. George, Daryl M. Lamson, Alexis Russel, Matthew Shudt, Melissa A Leisner, Jonathan Plitnick, Navjot Singh, John Kelly, Erasmus Schneider, Erica Lasek-Nesselquist |
| EPI_ISL_1226471 | MEMORIAL SLOAN KETTERING CANCER CENTER | Wadsworth Center, New York State Department of Health | Kirsten St. George, Daryl M. Lamson, Alexis Russel, Matthew Shudt, Melissa A Leisner, Jonathan Plitnick, Navjot Singh, John Kelly, Erasmus Schneider, Erica Lasek-Nesselquist |
| EPI_ISL_1226472 | NORTHWELL HEALTH LABORATORIES | Wadsworth Center, New York State Department of Health | Kirsten St. George, Daryl M. Lamson, Alexis Russel, Matthew Shudt, Melissa A Leisner, Jonathan Plitnick, Navjot Singh, John Kelly, Erasmus Schneider, Erica Lasek-Nesselquist |
| EPI_ISL_1226473 | Masonic Care Community of NY | Wadsworth Center, New York State Department of Health | Kirsten St. George, Daryl M. Lamson, Alexis Russel, Matthew Shudt, Melissa A Leisner, Jonathan Plitnick, Navjot Singh, John Kelly, Erasmus Schneider, Erica Lasek-Nesselquist |
| EPI_ISL_1226474, EPI_ISL_1226475 | MEMORIAL SLOAN KETTERING CANCER CENTER | Wadsworth Center, New York State Department of Health | Kirsten St. George, Daryl M. Lamson, Alexis Russel, Matthew Shudt, Melissa A Leisner, Jonathan Plitnick, Navjot Singh, John Kelly, Erasmus Schneider, Erica Lasek-Nesselquist |
| EPI_ISL_1226476 | NYC Pandemic Response Lab | Wadsworth Center, New York State Department of Health | Kirsten St. George, Daryl M. Lamson, Alexis Russel, Matthew Shudt, Melissa A Leisner, Jonathan Plitnick, Navjot Singh, John Kelly, Erasmus Schneider, Erica Lasek-Nesselquist |
| EPI_ISL_1226477, EPI_ISL_1226478, EPI_ISL_1226479, EPI_ISL_1226480 | URMC LABS | Wadsworth Center, New York State Department of Health | Kirsten St. George, Daryl M. Lamson, Alexis Russel, Matthew Shudt, Melissa A Leisner, Jonathan Plitnick, Navjot Singh, John Kelly, Erasmus Schneider, Erica Lasek-Nesselquist |
| EPI_ISL_1226481 | MEMORIAL SLOAN KETTERING CANCER CENTER | Wadsworth Center, New York State Department of Health | Kirsten St. George, Daryl M. Lamson, Alexis Russel, Matthew Shudt, Melissa A Leisner, Jonathan Plitnick, Navjot Singh, John Kelly, Erasmus Schneider, Erica Lasek-Nesselquist |
| EPI_ISL_1226482, EPI_ISL_1226483, EPI_ISL_1226484 | Wadsworth Center, New York State Department of Health | Wadsworth Center, New York State Department of Health | Kirsten St. George, Daryl M. Lamson, Alexis Russel, Matthew Shudt, Melissa A Leisner, Jonathan Plitnick, Navjot Singh, John Kelly, Erasmus Schneider, Erica Lasek-Nesselquist |
| EPI_ISL_1226485 | ALBANY MEDICAL CENTER | Wadsworth Center, New York State Department of Health | Kirsten St. George, Daryl M. Lamson, Alexis Russel, Matthew Shudt, Melissa A Leisner, Jonathan Plitnick, Navjot Singh, John Kelly, Erasmus Schneider, Erica Lasek-Nesselquist |
| EPI_ISL_1226486 | URMC LABS | Wadsworth Center, New York State Department of Health | Kirsten St. George, Daryl M. Lamson, Alexis Russel, Matthew Shudt, Melissa A Leisner, Jonathan Plitnick, Navjot Singh, John Kelly, Erasmus Schneider, Erica Lasek-Nesselquist |
| EPI_ISL_1226487 | TEMPUS LABS INC | Wadsworth Center, New York State Department of Health | Kirsten St. George, Daryl M. Lamson, Alexis Russel, Matthew Shudt, Melissa A Leisner, Jonathan Plitnick, Navjot Singh, John Kelly, Erasmus Schneider, Erica Lasek-Nesselquist |
| EPI_ISL_1226488 | KALEIDA CENTER FOR LABORATORY MEDICINE | Wadsworth Center, New York State Department of Health | Kirsten St. George, Daryl M. Lamson, Alexis Russel, Matthew Shudt, Melissa A Leisner, Jonathan Plitnick, Navjot Singh, John Kelly, Erasmus Schneider, Erica Lasek-Nesselquist |
| EPI_ISL_1226489 | MONTEFIORE MEDICAL CENTER | Wadsworth Center, New York State | Kirsten St. George, Daryl M. Lamson, Alexis Russel, Matthew Shudt, Melissa A Leisner, Jonathan Plitnick, Navjot Singh, John Kelly, Erasmus Schneider, Erica Lasek-Nesselquist |

| | | | |
|---|---|---|---|
| | LABORATORIES | Department of Health | |
| EPI_ISL_1226490 | THE MARY IMOGENE BASSETT HOSPITAL | Wadsworth Center, New York State Department of Health | Kirsten St. George, Daryl M. Lamson, Alexis Russel, Matthew Shudt, Melissa A Leisner, Jonathan Plitnick, Navjot Singh, John Kelly, Erasmus Schneider, Erica Lasek-Nesselquist |
| EPI_ISL_1226491 | WESTCHESTER MEDICAL CENTER | Wadsworth Center, New York State Department of Health | Kirsten St. George, Daryl M. Lamson, Alexis Russel, Matthew Shudt, Melissa A Leisner, Jonathan Plitnick, Navjot Singh, John Kelly, Erasmus Schneider, Erica Lasek-Nesselquist |
| EPI_ISL_1226492, EPI_ISL_1226493 | MONTEFIORE MEDICAL CENTER LABORATORIES | Wadsworth Center, New York State Department of Health | Kirsten St. George, Daryl M. Lamson, Alexis Russel, Matthew Shudt, Melissa A Leisner, Jonathan Plitnick, Navjot Singh, John Kelly, Erasmus Schneider, Erica Lasek-Nesselquist |
| EPI_ISL_1226494 | MEMORIAL SLOAN KETTERING CANCER CENTER | Wadsworth Center, New York State Department of Health | Kirsten St. George, Daryl M. Lamson, Alexis Russel, Matthew Shudt, Melissa A Leisner, Jonathan Plitnick, Navjot Singh, John Kelly, Erasmus Schneider, Erica Lasek-Nesselquist |
| EPI_ISL_1226495 | TEMPUS LABS INC | Wadsworth Center, New York State Department of Health | Kirsten St. George, Daryl M. Lamson, Alexis Russel, Matthew Shudt, Melissa A Leisner, Jonathan Plitnick, Navjot Singh, John Kelly, Erasmus Schneider, Erica Lasek-Nesselquist |
| EPI_ISL_1226496, EPI_ISL_1226497 | WESTCHESTER MEDICAL CENTER | Wadsworth Center, New York State Department of Health | Kirsten St. George, Daryl M. Lamson, Alexis Russel, Matthew Shudt, Melissa A Leisner, Jonathan Plitnick, Navjot Singh, John Kelly, Erasmus Schneider, Erica Lasek-Nesselquist |
| EPI_ISL_1226498 | MONTEFIORE MEDICAL CENTER LABORATORIES | Wadsworth Center, New York State Department of Health | Kirsten St. George, Daryl M. Lamson, Alexis Russel, Matthew Shudt, Melissa A Leisner, Jonathan Plitnick, Navjot Singh, John Kelly, Erasmus Schneider, Erica Lasek-Nesselquist |
| EPI_ISL_1226499 | KALEIDA CENTER FOR LABORATORY MEDICINE | Wadsworth Center, New York State Department of Health | Kirsten St. George, Daryl M. Lamson, Alexis Russel, Matthew Shudt, Melissa A Leisner, Jonathan Plitnick, Navjot Singh, John Kelly, Erasmus Schneider, Erica Lasek-Nesselquist |
| EPI_ISL_1226500 | NYC Pandemic Response Lab | Wadsworth Center, New York State Department of Health | Kirsten St. George, Daryl M. Lamson, Alexis Russel, Matthew Shudt, Melissa A Leisner, Jonathan Plitnick, Navjot Singh, John Kelly, Erasmus Schneider, Erica Lasek-Nesselquist |
| EPI_ISL_1226501 | TEMPUS LABS INC | Wadsworth Center, New York State Department of Health | Kirsten St. George, Daryl M. Lamson, Alexis Russel, Matthew Shudt, Melissa A Leisner, Jonathan Plitnick, Navjot Singh, John Kelly, Erasmus Schneider, Erica Lasek-Nesselquist |
| EPI_ISL_1226502, EPI_ISL_1226503 | Columbia University Irving Medical Center | Wadsworth Center, New York State Department of Health | Kirsten St. George, Daryl M. Lamson, Alexis Russel, Matthew Shudt, Melissa A Leisner, Jonathan Plitnick, Navjot Singh, John Kelly, Erasmus Schneider, Erica Lasek-Nesselquist |
| EPI_ISL_1226504 | MEMORIAL SLOAN KETTERING CANCER CENTER | Wadsworth Center, New York State Department of Health | Kirsten St. George, Daryl M. Lamson, Alexis Russel, Matthew Shudt, Melissa A Leisner, Jonathan Plitnick, Navjot Singh, John Kelly, Erasmus Schneider, Erica Lasek-Nesselquist |
| EPI_ISL_1226505 | URMC LABS | Wadsworth Center, New York State Department of Health | Kirsten St. George, Daryl M. Lamson, Alexis Russel, Matthew Shudt, Melissa A Leisner, Jonathan Plitnick, Navjot Singh, John Kelly, Erasmus Schneider, Erica Lasek-Nesselquist |
| EPI_ISL_1226506 | NORTHWELL HEALTH LABORATORIES | Wadsworth Center, New York State Department of Health | Kirsten St. George, Daryl M. Lamson, Alexis Russel, Matthew Shudt, Melissa A Leisner, Jonathan Plitnick, Navjot Singh, John Kelly, Erasmus Schneider, Erica Lasek-Nesselquist |
| EPI_ISL_1226507, EPI_ISL_1226508 | NYC Pandemic Response Lab | Wadsworth Center, New York State Department of Health | Kirsten St. George, Daryl M. Lamson, Alexis Russel, Matthew Shudt, Melissa A Leisner, Jonathan Plitnick, Navjot Singh, John Kelly, Erasmus Schneider, Erica Lasek-Nesselquist |
| EPI_ISL_1226509 | MONTEFIORE MEDICAL CENTER LABORATORIES | Wadsworth Center, New York State Department of Health | Kirsten St. George, Daryl M. Lamson, Alexis Russel, Matthew Shudt, Melissa A Leisner, Jonathan Plitnick, Navjot Singh, John Kelly, Erasmus Schneider, Erica Lasek-Nesselquist |
| EPI_ISL_1226510 | NYC Pandemic Response Lab | Wadsworth Center, New York State Department of Health | Kirsten St. George, Daryl M. Lamson, Alexis Russel, Matthew Shudt, Melissa A Leisner, Jonathan Plitnick, Navjot Singh, John Kelly, Erasmus Schneider, Erica Lasek-Nesselquist |
| EPI_ISL_1226511 | ALBANY MEDICAL CENTER | Wadsworth Center, New York State Department of Health | Kirsten St. George, Daryl M. Lamson, Alexis Russel, Matthew Shudt, Melissa A Leisner, Jonathan Plitnick, Navjot Singh, John Kelly, Erasmus Schneider, Erica Lasek-Nesselquist |
| EPI_ISL_1226512 | Columbia University Irving Medical Center | Wadsworth Center, New York State Department of Health | Kirsten St. George, Daryl M. Lamson, Alexis Russel, Matthew Shudt, Melissa A Leisner, Jonathan Plitnick, Navjot Singh, John Kelly, Erasmus Schneider, Erica Lasek-Nesselquist |
| EPI_ISL_1226513 | NYC Pandemic Response Lab | Wadsworth Center, New York State Department of Health | Kirsten St. George, Daryl M. Lamson, Alexis Russel, Matthew Shudt, Melissa A Leisner, Jonathan Plitnick, Navjot Singh, John Kelly, Erasmus Schneider, Erica Lasek-Nesselquist |
| EPI_ISL_1226514 | URMC LABS | Wadsworth Center, New York State Department of Health | Kirsten St. George, Daryl M. Lamson, Alexis Russel, Matthew Shudt, Melissa A Leisner, Jonathan Plitnick, Navjot Singh, John Kelly, Erasmus Schneider, Erica Lasek-Nesselquist |
| EPI_ISL_1226515 | MONTEFIORE MEDICAL CENTER LABORATORIES | Wadsworth Center, New York State Department of Health | Kirsten St. George, Daryl M. Lamson, Alexis Russel, Matthew Shudt, Melissa A Leisner, Jonathan Plitnick, Navjot Singh, John Kelly, Erasmus Schneider, Erica Lasek-Nesselquist |
| EPI_ISL_1226516, EPI_ISL_1226517 | URMC LABS | Wadsworth Center, New York State Department of Health | Kirsten St. George, Daryl M. Lamson, Alexis Russel, Matthew Shudt, Melissa A Leisner, Jonathan Plitnick, Navjot Singh, John Kelly, Erasmus Schneider, Erica Lasek-Nesselquist |
| EPI_ISL_1226519 | Masonic Care Community of NY | Wadsworth Center, New York State Department of Health | Kirsten St. George, Daryl M. Lamson, Alexis Russel, Matthew Shudt, Melissa A Leisner, Jonathan Plitnick, Navjot Singh, John Kelly, Erasmus Schneider, Erica Lasek-Nesselquist |
| EPI_ISL_1226520 | URMC LABS | Wadsworth Center, New York State Department of Health | Kirsten St. George, Daryl M. Lamson, Alexis Russel, Matthew Shudt, Melissa A Leisner, Jonathan Plitnick, Navjot Singh, John Kelly, Erasmus Schneider, Erica Lasek-Nesselquist |
| EPI_ISL_1226521 | NYC Pandemic Response Lab | Wadsworth Center, New York State Department of Health | Kirsten St. George, Daryl M. Lamson, Alexis Russel, Matthew Shudt, Melissa A Leisner, Jonathan Plitnick, Navjot Singh, John Kelly, Erasmus Schneider, Erica Lasek-Nesselquist |
| EPI_ISL_1226522 | MEMORIAL SLOAN KETTERING CANCER CENTER | Wadsworth Center, New York State Department of Health | Kirsten St. George, Daryl M. Lamson, Alexis Russel, Matthew Shudt, Melissa A Leisner, Jonathan Plitnick, Navjot Singh, John Kelly, Erasmus Schneider, Erica Lasek-Nesselquist |
| EPI_ISL_1226523 | MONTEFIORE MEDICAL CENTER LABORATORIES | Wadsworth Center, New York State Department of Health | Kirsten St. George, Daryl M. Lamson, Alexis Russel, Matthew Shudt, Melissa A Leisner, Jonathan Plitnick, Navjot Singh, John Kelly, Erasmus Schneider, Erica Lasek-Nesselquist |
| EPI_ISL_1226524 | THE MARY IMOGENE BASSETT HOSPITAL | Wadsworth Center, New York State Department of Health | Kirsten St. George, Daryl M. Lamson, Alexis Russel, Matthew Shudt, Melissa A Leisner, Jonathan Plitnick, Navjot Singh, John Kelly, Erasmus Schneider, Erica Lasek-Nesselquist |
| EPI_ISL_1226525, EPI_ISL_1226526 | MEMORIAL SLOAN KETTERING CANCER CENTER | Wadsworth Center, New York State Department of Health | Kirsten St. George, Daryl M. Lamson, Alexis Russel, Matthew Shudt, Melissa A Leisner, Jonathan Plitnick, Navjot Singh, John Kelly, Erasmus Schneider, Erica Lasek-Nesselquist |
| EPI_ISL_1226527, EPI_ISL_1226528 | Wadsworth Center, New York State Department of Health | Wadsworth Center, New York State Department of Health | Kirsten St. George, Daryl M. Lamson, Alexis Russel, Matthew Shudt, Melissa A Leisner, Jonathan Plitnick, Navjot Singh, John Kelly, Erasmus Schneider, Erica Lasek-Nesselquist |
| EPI_ISL_1226529, EPI_ISL_1226530, EPI_ISL_1226531, EPI_ISL_1226532 | MONTEFIORE MEDICAL CENTER LABORATORIES | Wadsworth Center, New York State Department of Health | Kirsten St. George, Daryl M. Lamson, Alexis Russel, Matthew Shudt, Melissa A Leisner, Jonathan Plitnick, Navjot Singh, John Kelly, Erasmus Schneider, Erica Lasek-Nesselquist |
| EPI_ISL_1226533, EPI_ISL_1226534, EPI_ISL_1226535, EPI_ISL_1226536, EPI_ISL_1226537 | TEMPUS LABS INC | Wadsworth Center, New York State Department of Health | Kirsten St. George, Daryl M. Lamson, Alexis Russel, Matthew Shudt, Melissa A Leisner, Jonathan Plitnick, Navjot Singh, John Kelly, Erasmus Schneider, Erica Lasek-Nesselquist |
| EPI_ISL_1226538, EPI_ISL_1226539 | WESTCHESTER MEDICAL CENTER | Wadsworth Center, New York State Department of Health | Kirsten St. George, Daryl M. Lamson, Alexis Russel, Matthew Shudt, Melissa A Leisner, Jonathan Plitnick, Navjot Singh, John Kelly, Erasmus Schneider, Erica Lasek-Nesselquist |
| EPI_ISL_1226540 | BOSTON HEART DIAGNOSTICS CORP | Wadsworth Center, New York State Department of Health | Kirsten St. George, Daryl M. Lamson, Alexis Russel, Matthew Shudt, Melissa A Leisner, Jonathan Plitnick, Navjot Singh, John Kelly, Erasmus Schneider, Erica Lasek-Nesselquist |
| EPI_ISL_1226541, EPI_ISL_1226542 | KALEIDA CENTER FOR LABORATORY MEDICINE | Wadsworth Center, New York State Department of Health | Kirsten St. George, Daryl M. Lamson, Alexis Russel, Matthew Shudt, Melissa A Leisner, Jonathan Plitnick, Navjot Singh, John Kelly, Erasmus Schneider, Erica Lasek-Nesselquist |
| EPI_ISL_1226543 | MEMORIAL SLOAN KETTERING CANCER CENTER | Wadsworth Center, New York State Department of Health | Kirsten St. George, Daryl M. Lamson, Alexis Russel, Matthew Shudt, Melissa A Leisner, Jonathan Plitnick, Navjot Singh, John Kelly, Erasmus Schneider, Erica Lasek-Nesselquist |
| EPI_ISL_1226544 | New York Presbyterian Hospital | Wadsworth Center, New York State Department of Health | Kirsten St. George, Daryl M. Lamson, Alexis Russel, Matthew Shudt, Melissa A Leisner, Jonathan Plitnick, Navjot Singh, John Kelly, Erasmus Schneider, Erica Lasek-Nesselquist |
| EPI_ISL_1226545, EPI_ISL_1226546, EPI_ISL_1226547, EPI_ISL_1226548, EPI_ISL_1226549 | NYC Pandemic Response Lab | Wadsworth Center, New York State Department of Health | Kirsten St. George, Daryl M. Lamson, Alexis Russel, Matthew Shudt, Melissa A Leisner, Jonathan Plitnick, Navjot Singh, John Kelly, Erasmus Schneider, Erica Lasek-Nesselquist |
| EPI_ISL_1226550 | TEMPUS LABS INC | Wadsworth Center, New York State Department of Health | Kirsten St. George, Daryl M. Lamson, Alexis Russel, Matthew Shudt, Melissa A Leisner, Jonathan Plitnick, Navjot Singh, John Kelly, Erasmus Schneider, Erica Lasek-Nesselquist |
| EPI_ISL_1226551 | BIO-REFERENCE LABORATORIES | Wadsworth Center, New York State Department of Health | Kirsten St. George, Daryl M. Lamson, Alexis Russel, Matthew Shudt, Melissa A Leisner, Jonathan Plitnick, Navjot Singh, John Kelly, Erasmus Schneider, Erica Lasek-Nesselquist |
| EPI_ISL_1226552 | URMC LABS | Wadsworth Center, New York State | Kirsten St. George, Daryl M. Lamson, Alexis Russel, Matthew Shudt, Melissa A Leisner, Jonathan Plitnick, Navjot Singh, John Kelly, Erasmus Schneider, Erica Lasek-Nesselquist |

| | | | |
|---|---|---|---|
| | | Department of Health | |
| EPI_ISL_1226553 | NORTHWELL HEALTH LABORATORIES | Wadsworth Center, New York State Department of Health | Kirsten St. George, Daryl M. Lamson, Alexis Russel, Matthew Shudt, Melissa A Leisner, Jonathan Plitnick, Navjot Singh, John Kelly, Erasmus Schneider, Erica Lasek-Nesselquist |
| EPI_ISL_1226554 | NYC Pandemic Response Lab | Wadsworth Center, New York State Department of Health | Kirsten St. George, Daryl M. Lamson, Alexis Russel, Matthew Shudt, Melissa A Leisner, Jonathan Plitnick, Navjot Singh, John Kelly, Erasmus Schneider, Erica Lasek-Nesselquist |
| EPI_ISL_1226555 | TEMPUS LABS INC | Wadsworth Center, New York State Department of Health | Kirsten St. George, Daryl M. Lamson, Alexis Russel, Matthew Shudt, Melissa A Leisner, Jonathan Plitnick, Navjot Singh, John Kelly, Erasmus Schneider, Erica Lasek-Nesselquist |
| EPI_ISL_1226556 | BIO-REFERENCE LABORATORIES | Wadsworth Center, New York State Department of Health | Kirsten St. George, Daryl M. Lamson, Alexis Russel, Matthew Shudt, Melissa A Leisner, Jonathan Plitnick, Navjot Singh, John Kelly, Erasmus Schneider, Erica Lasek-Nesselquist |
| EPI_ISL_1226557 | TEMPUS LABS INC | Wadsworth Center, New York State Department of Health | Kirsten St. George, Daryl M. Lamson, Alexis Russel, Matthew Shudt, Melissa A Leisner, Jonathan Plitnick, Navjot Singh, John Kelly, Erasmus Schneider, Erica Lasek-Nesselquist |
| EPI_ISL_1226558 | WESTCHESTER MEDICAL CENTER | Wadsworth Center, New York State Department of Health | Kirsten St. George, Daryl M. Lamson, Alexis Russel, Matthew Shudt, Melissa A Leisner, Jonathan Plitnick, Navjot Singh, John Kelly, Erasmus Schneider, Erica Lasek-Nesselquist |
| EPI_ISL_1226559 | New York Presbyterian Hospital | Wadsworth Center, New York State Department of Health | Kirsten St. George, Daryl M. Lamson, Alexis Russel, Matthew Shudt, Melissa A Leisner, Jonathan Plitnick, Navjot Singh, John Kelly, Erasmus Schneider, Erica Lasek-Nesselquist |
| EPI_ISL_1226560, EPI_ISL_1226561, EPI_ISL_1226562 | NYC Pandemic Response Lab | Wadsworth Center, New York State Department of Health | Kirsten St. George, Daryl M. Lamson, Alexis Russel, Matthew Shudt, Melissa A Leisner, Jonathan Plitnick, Navjot Singh, John Kelly, Erasmus Schneider, Erica Lasek-Nesselquist |
| EPI_ISL_1226563 | ALBANY MEDICAL CENTER | Wadsworth Center, New York State Department of Health | Kirsten St. George, Daryl M. Lamson, Alexis Russel, Matthew Shudt, Melissa A Leisner, Jonathan Plitnick, Navjot Singh, John Kelly, Erasmus Schneider, Erica Lasek-Nesselquist |
| EPI_ISL_1226564, EPI_ISL_1226565, EPI_ISL_1226566 | NYC Pandemic Response Lab | Wadsworth Center, New York State Department of Health | Kirsten St. George, Daryl M. Lamson, Alexis Russel, Matthew Shudt, Melissa A Leisner, Jonathan Plitnick, Navjot Singh, John Kelly, Erasmus Schneider, Erica Lasek-Nesselquist |
| EPI_ISL_1226567 | TEMPUS LABS INC | Wadsworth Center, New York State Department of Health | Kirsten St. George, Daryl M. Lamson, Alexis Russel, Matthew Shudt, Melissa A Leisner, Jonathan Plitnick, Navjot Singh, John Kelly, Erasmus Schneider, Erica Lasek-Nesselquist |
| EPI_ISL_1226568 | WESTCHESTER MEDICAL CENTER | Wadsworth Center, New York State Department of Health | Kirsten St. George, Daryl M. Lamson, Alexis Russel, Matthew Shudt, Melissa A Leisner, Jonathan Plitnick, Navjot Singh, John Kelly, Erasmus Schneider, Erica Lasek-Nesselquist |
| EPI_ISL_1226569, EPI_ISL_1226570 | TEMPUS LABS INC | Wadsworth Center, New York State Department of Health | Kirsten St. George, Daryl M. Lamson, Alexis Russel, Matthew Shudt, Melissa A Leisner, Jonathan Plitnick, Navjot Singh, John Kelly, Erasmus Schneider, Erica Lasek-Nesselquist |
| EPI_ISL_1226571, EPI_ISL_1226572 | BIO-REFERENCE LABORATORIES | Wadsworth Center, New York State Department of Health | Kirsten St. George, Daryl M. Lamson, Alexis Russel, Matthew Shudt, Melissa A Leisner, Jonathan Plitnick, Navjot Singh, John Kelly, Erasmus Schneider, Erica Lasek-Nesselquist |
| EPI_ISL_1226573 | MONTEFIORE MEDICAL CENTER LABORATORIES | Wadsworth Center, New York State Department of Health | Kirsten St. George, Daryl M. Lamson, Alexis Russel, Matthew Shudt, Melissa A Leisner, Jonathan Plitnick, Navjot Singh, John Kelly, Erasmus Schneider, Erica Lasek-Nesselquist |
| EPI_ISL_1226574 | BIO-REFERENCE LABORATORIES | Wadsworth Center, New York State Department of Health | Kirsten St. George, Daryl M. Lamson, Alexis Russel, Matthew Shudt, Melissa A Leisner, Jonathan Plitnick, Navjot Singh, John Kelly, Erasmus Schneider, Erica Lasek-Nesselquist |
| EPI_ISL_1226575 | TEMPUS LABS INC | Wadsworth Center, New York State Department of Health | Kirsten St. George, Daryl M. Lamson, Alexis Russel, Matthew Shudt, Melissa A Leisner, Jonathan Plitnick, Navjot Singh, John Kelly, Erasmus Schneider, Erica Lasek-Nesselquist |
| EPI_ISL_1226576 | MONTEFIORE MEDICAL CENTER LABORATORIES | Wadsworth Center, New York State Department of Health | Kirsten St. George, Daryl M. Lamson, Alexis Russel, Matthew Shudt, Melissa A Leisner, Jonathan Plitnick, Navjot Singh, John Kelly, Erasmus Schneider, Erica Lasek-Nesselquist |
| EPI_ISL_1226577 | BIO-REFERENCE LABORATORIES | Wadsworth Center, New York State Department of Health | Kirsten St. George, Daryl M. Lamson, Alexis Russel, Matthew Shudt, Melissa A Leisner, Jonathan Plitnick, Navjot Singh, John Kelly, Erasmus Schneider, Erica Lasek-Nesselquist |
| EPI_ISL_1226578, EPI_ISL_1226579 | TEMPUS LABS INC | Wadsworth Center, New York State Department of Health | Kirsten St. George, Daryl M. Lamson, Alexis Russel, Matthew Shudt, Melissa A Leisner, Jonathan Plitnick, Navjot Singh, John Kelly, Erasmus Schneider, Erica Lasek-Nesselquist |
| EPI_ISL_1226580, EPI_ISL_1226581, EPI_ISL_1226582 | NYC Pandemic Response Lab | Wadsworth Center, New York State Department of Health | Kirsten St. George, Daryl M. Lamson, Alexis Russel, Matthew Shudt, Melissa A Leisner, Jonathan Plitnick, Navjot Singh, John Kelly, Erasmus Schneider, Erica Lasek-Nesselquist |
| EPI_ISL_1226583 | BIO-REFERENCE LABORATORIES | Wadsworth Center, New York State Department of Health | Kirsten St. George, Daryl M. Lamson, Alexis Russel, Matthew Shudt, Melissa A Leisner, Jonathan Plitnick, Navjot Singh, John Kelly, Erasmus Schneider, Erica Lasek-Nesselquist |
| EPI_ISL_1226584, EPI_ISL_1226585, EPI_ISL_1226586 | TEMPUS LABS INC | Wadsworth Center, New York State Department of Health | Kirsten St. George, Daryl M. Lamson, Alexis Russel, Matthew Shudt, Melissa A Leisner, Jonathan Plitnick, Navjot Singh, John Kelly, Erasmus Schneider, Erica Lasek-Nesselquist |
| EPI_ISL_1226587, EPI_ISL_1226588 | WESTCHESTER MEDICAL CENTER | Wadsworth Center, New York State Department of Health | Kirsten St. George, Daryl M. Lamson, Alexis Russel, Matthew Shudt, Melissa A Leisner, Jonathan Plitnick, Navjot Singh, John Kelly, Erasmus Schneider, Erica Lasek-Nesselquist |
| EPI_ISL_1226589 | TEMPUS LABS INC | Wadsworth Center, New York State Department of Health | Kirsten St. George, Daryl M. Lamson, Alexis Russel, Matthew Shudt, Melissa A Leisner, Jonathan Plitnick, Navjot Singh, John Kelly, Erasmus Schneider, Erica Lasek-Nesselquist |
| EPI_ISL_1226590, EPI_ISL_1226591 | WESTCHESTER MEDICAL CENTER | Wadsworth Center, New York State Department of Health | Kirsten St. George, Daryl M. Lamson, Alexis Russel, Matthew Shudt, Melissa A Leisner, Jonathan Plitnick, Navjot Singh, John Kelly, Erasmus Schneider, Erica Lasek-Nesselquist |
| EPI_ISL_1226592 | TEMPUS LABS INC | Wadsworth Center, New York State Department of Health | Kirsten St. George, Daryl M. Lamson, Alexis Russel, Matthew Shudt, Melissa A Leisner, Jonathan Plitnick, Navjot Singh, John Kelly, Erasmus Schneider, Erica Lasek-Nesselquist |
| EPI_ISL_1226593 | SUNY UPSTATE MEDICAL UNIVERSITY | Wadsworth Center, New York State Department of Health | Kirsten St. George, Daryl M. Lamson, Alexis Russel, Matthew Shudt, Melissa A Leisner, Jonathan Plitnick, Navjot Singh, John Kelly, Erasmus Schneider, Erica Lasek-Nesselquist |
| EPI_ISL_1226594 | MONTEFIORE MEDICAL CENTER LABORATORIES | Wadsworth Center, New York State Department of Health | Kirsten St. George, Daryl M. Lamson, Alexis Russel, Matthew Shudt, Melissa A Leisner, Jonathan Plitnick, Navjot Singh, John Kelly, Erasmus Schneider, Erica Lasek-Nesselquist |
| EPI_ISL_1226595 | NYC Pandemic Response Lab | Wadsworth Center, New York State Department of Health | Kirsten St. George, Daryl M. Lamson, Alexis Russel, Matthew Shudt, Melissa A Leisner, Jonathan Plitnick, Navjot Singh, John Kelly, Erasmus Schneider, Erica Lasek-Nesselquist |
| EPI_ISL_1226596 | Columbia University Irving Medical Center | Wadsworth Center, New York State Department of Health | Kirsten St. George, Daryl M. Lamson, Alexis Russel, Matthew Shudt, Melissa A Leisner, Jonathan Plitnick, Navjot Singh, John Kelly, Erasmus Schneider, Erica Lasek-Nesselquist |
| EPI_ISL_1226597 | MONTEFIORE MEDICAL CENTER LABORATORIES | Wadsworth Center, New York State Department of Health | Kirsten St. George, Daryl M. Lamson, Alexis Russel, Matthew Shudt, Melissa A Leisner, Jonathan Plitnick, Navjot Singh, John Kelly, Erasmus Schneider, Erica Lasek-Nesselquist |
| EPI_ISL_1226598 | URMC LABS | Wadsworth Center, New York State Department of Health | Kirsten St. George, Daryl M. Lamson, Alexis Russel, Matthew Shudt, Melissa A Leisner, Jonathan Plitnick, Navjot Singh, John Kelly, Erasmus Schneider, Erica Lasek-Nesselquist |
| EPI_ISL_1226599 | Columbia University Irving Medical Center | Wadsworth Center, New York State Department of Health | Kirsten St. George, Daryl M. Lamson, Alexis Russel, Matthew Shudt, Melissa A Leisner, Jonathan Plitnick, Navjot Singh, John Kelly, Erasmus Schneider, Erica Lasek-Nesselquist |
| EPI_ISL_1226600 | NYC Pandemic Response Lab | Wadsworth Center, New York State Department of Health | Kirsten St. George, Daryl M. Lamson, Alexis Russel, Matthew Shudt, Melissa A Leisner, Jonathan Plitnick, Navjot Singh, John Kelly, Erasmus Schneider, Erica Lasek-Nesselquist |
| EPI_ISL_1226601 | MONTEFIORE MEDICAL CENTER LABORATORIES | Wadsworth Center, New York State Department of Health | Kirsten St. George, Daryl M. Lamson, Alexis Russel, Matthew Shudt, Melissa A Leisner, Jonathan Plitnick, Navjot Singh, John Kelly, Erasmus Schneider, Erica Lasek-Nesselquist |
| EPI_ISL_1226602 | URMC LABS | Wadsworth Center, New York State Department of Health | Kirsten St. George, Daryl M. Lamson, Alexis Russel, Matthew Shudt, Melissa A Leisner, Jonathan Plitnick, Navjot Singh, John Kelly, Erasmus Schneider, Erica Lasek-Nesselquist |
| EPI_ISL_1226603 | TEMPUS LABS INC | Wadsworth Center, New York State Department of Health | Kirsten St. George, Daryl M. Lamson, Alexis Russel, Matthew Shudt, Melissa A Leisner, Jonathan Plitnick, Navjot Singh, John Kelly, Erasmus Schneider, Erica Lasek-Nesselquist |
| EPI_ISL_1226604, EPI_ISL_1226605, EPI_ISL_1226606, EPI_ISL_1226607, EPI_ISL_1226608, EPI_ISL_1226609, EPI_ISL_1226610, EPI_ISL_1226611, EPI_ISL_1226612, EPI_ISL_1226613, EPI_ISL_1226614 | | | |
| see above | MONTEFIORE MEDICAL CENTER LABORATORIES | Wadsworth Center, New York State Department of Health | Kirsten St. George, Daryl M. Lamson, Alexis Russel, Matthew Shudt, Melissa A Leisner, Jonathan Plitnick, Navjot Singh, John Kelly, Erasmus Schneider, Erica Lasek-Nesselquist |
| EPI_ISL_1226615 | URMC LABS | Wadsworth Center, New York State Department of Health | Kirsten St. George, Daryl M. Lamson, Alexis Russel, Matthew Shudt, Melissa A Leisner, Jonathan Plitnick, Navjot Singh, John Kelly, Erasmus Schneider, Erica Lasek-Nesselquist |
| EPI_ISL_1226616 | WESTCHESTER MEDICAL CENTER | Wadsworth Center, New York State Department of Health | Kirsten St. George, Daryl M. Lamson, Alexis Russel, Matthew Shudt, Melissa A Leisner, Jonathan Plitnick, Navjot Singh, John Kelly, Erasmus Schneider, Erica Lasek-Nesselquist |
| EPI_ISL_1226617 | KALEIDA CENTER FOR LABORATORY MEDICINE | Wadsworth Center, New York State Department of Health | Kirsten St. George, Daryl M. Lamson, Alexis Russel, Matthew Shudt, Melissa A Leisner, Jonathan Plitnick, Navjot Singh, John Kelly, Erasmus Schneider, Erica Lasek-Nesselquist |

| | | | |
|---|---|---|---|
| EPI_ISL_1226618, EPI_ISL_1226619 | TEMPUS LABS INC | Wadsworth Center, New York State Department of Health | Kirsten St. George, Daryl M. Lamson, Alexis Russel, Matthew Shudt, Melissa A Leisner, Jonathan Plitnick, Navjot Singh, John Kelly, Erasmus Schneider, Erica Lasek-Nesselquist |
| EPI_ISL_1226620, EPI_ISL_1226621 | NYC Pandemic Response Lab | Wadsworth Center, New York State Department of Health | Kirsten St. George, Daryl M. Lamson, Alexis Russel, Matthew Shudt, Melissa A Leisner, Jonathan Plitnick, Navjot Singh, John Kelly, Erasmus Schneider, Erica Lasek-Nesselquist |
| EPI_ISL_1226622 | BIO-REFERENCE LABORATORIES | Wadsworth Center, New York State Department of Health | Kirsten St. George, Daryl M. Lamson, Alexis Russel, Matthew Shudt, Melissa A Leisner, Jonathan Plitnick, Navjot Singh, John Kelly, Erasmus Schneider, Erica Lasek-Nesselquist |
| EPI_ISL_1226623 | URMC LABS | Wadsworth Center, New York State Department of Health | Kirsten St. George, Daryl M. Lamson, Alexis Russel, Matthew Shudt, Melissa A Leisner, Jonathan Plitnick, Navjot Singh, John Kelly, Erasmus Schneider, Erica Lasek-Nesselquist |
| EPI_ISL_1226624 | New York Presbyterian Hospital | Wadsworth Center, New York State Department of Health | Kirsten St. George, Daryl M. Lamson, Alexis Russel, Matthew Shudt, Melissa A Leisner, Jonathan Plitnick, Navjot Singh, John Kelly, Erasmus Schneider, Erica Lasek-Nesselquist |
| EPI_ISL_1226625 | NYC Pandemic Response Lab | Wadsworth Center, New York State Department of Health | Kirsten St. George, Daryl M. Lamson, Alexis Russel, Matthew Shudt, Melissa A Leisner, Jonathan Plitnick, Navjot Singh, John Kelly, Erasmus Schneider, Erica Lasek-Nesselquist |
| EPI_ISL_1226626, EPI_ISL_1226627, EPI_ISL_1226628, EPI_ISL_1226629, EPI_ISL_1226630, EPI_ISL_1226631, EPI_ISL_1226632, EPI_ISL_1226633, EPI_ISL_1226634, EPI_ISL_1226635 | TEMPUS LABS INC | Wadsworth Center, New York State Department of Health | Kirsten St. George, Daryl M. Lamson, Alexis Russel, Matthew Shudt, Melissa A Leisner, Jonathan Plitnick, Navjot Singh, John Kelly, Erasmus Schneider, Erica Lasek-Nesselquist |
| EPI_ISL_1226636 | NYC Pandemic Response Lab | Wadsworth Center, New York State Department of Health | Kirsten St. George, Daryl M. Lamson, Alexis Russel, Matthew Shudt, Melissa A Leisner, Jonathan Plitnick, Navjot Singh, John Kelly, Erasmus Schneider, Erica Lasek-Nesselquist |
| EPI_ISL_1226637, EPI_ISL_1226638 | Columbia University Irving Medical Center | Wadsworth Center, New York State Department of Health | Kirsten St. George, Daryl M. Lamson, Alexis Russel, Matthew Shudt, Melissa A Leisner, Jonathan Plitnick, Navjot Singh, John Kelly, Erasmus Schneider, Erica Lasek-Nesselquist |
| EPI_ISL_1226639, EPI_ISL_1226640, EPI_ISL_1226641, EPI_ISL_1226642, EPI_ISL_1226643, EPI_ISL_1226644 | NYC Pandemic Response Lab | Wadsworth Center, New York State Department of Health | Kirsten St. George, Daryl M. Lamson, Alexis Russel, Matthew Shudt, Melissa A Leisner, Jonathan Plitnick, Navjot Singh, John Kelly, Erasmus Schneider, Erica Lasek-Nesselquist |
| EPI_ISL_1226645, EPI_ISL_1226646 | BIO-REFERENCE LABORATORIES | Wadsworth Center, New York State Department of Health | Kirsten St. George, Daryl M. Lamson, Alexis Russel, Matthew Shudt, Melissa A Leisner, Jonathan Plitnick, Navjot Singh, John Kelly, Erasmus Schneider, Erica Lasek-Nesselquist |
| EPI_ISL_1226647, EPI_ISL_1226648 | NYC Pandemic Response Lab | Wadsworth Center, New York State Department of Health | Kirsten St. George, Daryl M. Lamson, Alexis Russel, Matthew Shudt, Melissa A Leisner, Jonathan Plitnick, Navjot Singh, John Kelly, Erasmus Schneider, Erica Lasek-Nesselquist |
| EPI_ISL_1226649, EPI_ISL_1226650 | TEMPUS LABS INC | Wadsworth Center, New York State Department of Health | Kirsten St. George, Daryl M. Lamson, Alexis Russel, Matthew Shudt, Melissa A Leisner, Jonathan Plitnick, Navjot Singh, John Kelly, Erasmus Schneider, Erica Lasek-Nesselquist |
| EPI_ISL_1226651, EPI_ISL_1226652 | NYC Pandemic Response Lab | Wadsworth Center, New York State Department of Health | Kirsten St. George, Daryl M. Lamson, Alexis Russel, Matthew Shudt, Melissa A Leisner, Jonathan Plitnick, Navjot Singh, John Kelly, Erasmus Schneider, Erica Lasek-Nesselquist |
| EPI_ISL_1226653 | TEMPUS LABS INC | Wadsworth Center, New York State Department of Health | Kirsten St. George, Daryl M. Lamson, Alexis Russel, Matthew Shudt, Melissa A Leisner, Jonathan Plitnick, Navjot Singh, John Kelly, Erasmus Schneider, Erica Lasek-Nesselquist |
| EPI_ISL_1226654 | MONTEFIORE MEDICAL CENTER LABORATORIES | Wadsworth Center, New York State Department of Health | Kirsten St. George, Daryl M. Lamson, Alexis Russel, Matthew Shudt, Melissa A Leisner, Jonathan Plitnick, Navjot Singh, John Kelly, Erasmus Schneider, Erica Lasek-Nesselquist |
| EPI_ISL_1226655, EPI_ISL_1226656, EPI_ISL_1226657 | WESTCHESTER MEDICAL CENTER | Wadsworth Center, New York State Department of Health | Kirsten St. George, Daryl M. Lamson, Alexis Russel, Matthew Shudt, Melissa A Leisner, Jonathan Plitnick, Navjot Singh, John Kelly, Erasmus Schneider, Erica Lasek-Nesselquist |
| EPI_ISL_1226658 | BIO-REFERENCE LABORATORIES | Wadsworth Center, New York State Department of Health | Kirsten St. George, Daryl M. Lamson, Alexis Russel, Matthew Shudt, Melissa A Leisner, Jonathan Plitnick, Navjot Singh, John Kelly, Erasmus Schneider, Erica Lasek-Nesselquist |
| EPI_ISL_1226659 | TEMPUS LABS INC | Wadsworth Center, New York State Department of Health | Kirsten St. George, Daryl M. Lamson, Alexis Russel, Matthew Shudt, Melissa A Leisner, Jonathan Plitnick, Navjot Singh, John Kelly, Erasmus Schneider, Erica Lasek-Nesselquist |
| EPI_ISL_1226660, EPI_ISL_1226661 | NYC Pandemic Response Lab | Wadsworth Center, New York State Department of Health | Kirsten St. George, Daryl M. Lamson, Alexis Russel, Matthew Shudt, Melissa A Leisner, Jonathan Plitnick, Navjot Singh, John Kelly, Erasmus Schneider, Erica Lasek-Nesselquist |
| EPI_ISL_1226662, EPI_ISL_1226663 | URMC LABS | Wadsworth Center, New York State Department of Health | Kirsten St. George, Daryl M. Lamson, Alexis Russel, Matthew Shudt, Melissa A Leisner, Jonathan Plitnick, Navjot Singh, John Kelly, Erasmus Schneider, Erica Lasek-Nesselquist |
| EPI_ISL_1226664 | MEMORIAL SLOAN KETTERING CANCER CENTER | Wadsworth Center, New York State Department of Health | Kirsten St. George, Daryl M. Lamson, Alexis Russel, Matthew Shudt, Melissa A Leisner, Jonathan Plitnick, Navjot Singh, John Kelly, Erasmus Schneider, Erica Lasek-Nesselquist |
| EPI_ISL_1226665 | NYC Pandemic Response Lab | Wadsworth Center, New York State Department of Health | Kirsten St. George, Daryl M. Lamson, Alexis Russel, Matthew Shudt, Melissa A Leisner, Jonathan Plitnick, Navjot Singh, John Kelly, Erasmus Schneider, Erica Lasek-Nesselquist |
| EPI_ISL_1226666, EPI_ISL_1226667 | TEMPUS LABS INC | Wadsworth Center, New York State Department of Health | Kirsten St. George, Daryl M. Lamson, Alexis Russel, Matthew Shudt, Melissa A Leisner, Jonathan Plitnick, Navjot Singh, John Kelly, Erasmus Schneider, Erica Lasek-Nesselquist |
| EPI_ISL_1226668 | WESTCHESTER MEDICAL CENTER | Wadsworth Center, New York State Department of Health | Kirsten St. George, Daryl M. Lamson, Alexis Russel, Matthew Shudt, Melissa A Leisner, Jonathan Plitnick, Navjot Singh, John Kelly, Erasmus Schneider, Erica Lasek-Nesselquist |
| EPI_ISL_1226669 | BIO-REFERENCE LABORATORIES | Wadsworth Center, New York State Department of Health | Kirsten St. George, Daryl M. Lamson, Alexis Russel, Matthew Shudt, Melissa A Leisner, Jonathan Plitnick, Navjot Singh, John Kelly, Erasmus Schneider, Erica Lasek-Nesselquist |
| EPI_ISL_1226670 | TEMPUS LABS INC | Wadsworth Center, New York State Department of Health | Kirsten St. George, Daryl M. Lamson, Alexis Russel, Matthew Shudt, Melissa A Leisner, Jonathan Plitnick, Navjot Singh, John Kelly, Erasmus Schneider, Erica Lasek-Nesselquist |
| EPI_ISL_1226671 | Columbia University Irving Medical Center | Wadsworth Center, New York State Department of Health | Kirsten St. George, Daryl M. Lamson, Alexis Russel, Matthew Shudt, Melissa A Leisner, Jonathan Plitnick, Navjot Singh, John Kelly, Erasmus Schneider, Erica Lasek-Nesselquist |
| EPI_ISL_1226672 | NORTHWELL HEALTH LABORATORIES | Wadsworth Center, New York State Department of Health | Kirsten St. George, Daryl M. Lamson, Alexis Russel, Matthew Shudt, Melissa A Leisner, Jonathan Plitnick, Navjot Singh, John Kelly, Erasmus Schneider, Erica Lasek-Nesselquist |
| EPI_ISL_1226673 | NYC Pandemic Response Lab | Wadsworth Center, New York State Department of Health | Kirsten St. George, Daryl M. Lamson, Alexis Russel, Matthew Shudt, Melissa A Leisner, Jonathan Plitnick, Navjot Singh, John Kelly, Erasmus Schneider, Erica Lasek-Nesselquist |
| EPI_ISL_1226674 | Columbia University Irving Medical Center | Wadsworth Center, New York State Department of Health | Kirsten St. George, Daryl M. Lamson, Alexis Russel, Matthew Shudt, Melissa A Leisner, Jonathan Plitnick, Navjot Singh, John Kelly, Erasmus Schneider, Erica Lasek-Nesselquist |
| EPI_ISL_1226675, EPI_ISL_1226676, EPI_ISL_1226677 | URMC LABS | Wadsworth Center, New York State Department of Health | Kirsten St. George, Daryl M. Lamson, Alexis Russel, Matthew Shudt, Melissa A Leisner, Jonathan Plitnick, Navjot Singh, John Kelly, Erasmus Schneider, Erica Lasek-Nesselquist |
| EPI_ISL_1226678, EPI_ISL_1226679 | WESTCHESTER MEDICAL CENTER | Wadsworth Center, New York State Department of Health | Kirsten St. George, Daryl M. Lamson, Alexis Russel, Matthew Shudt, Melissa A Leisner, Jonathan Plitnick, Navjot Singh, John Kelly, Erasmus Schneider, Erica Lasek-Nesselquist |
| EPI_ISL_1226680, EPI_ISL_1226681 | MONTEFIORE MEDICAL CENTER LABORATORIES | Wadsworth Center, New York State Department of Health | Kirsten St. George, Daryl M. Lamson, Alexis Russel, Matthew Shudt, Melissa A Leisner, Jonathan Plitnick, Navjot Singh, John Kelly, Erasmus Schneider, Erica Lasek-Nesselquist |
| EPI_ISL_1226682 | WESTCHESTER MEDICAL CENTER | Wadsworth Center, New York State Department of Health | Kirsten St. George, Daryl M. Lamson, Alexis Russel, Matthew Shudt, Melissa A Leisner, Jonathan Plitnick, Navjot Singh, John Kelly, Erasmus Schneider, Erica Lasek-Nesselquist |
| EPI_ISL_1226683 | SUNY UPSTATE MEDICAL UNIVERSITY | Wadsworth Center, New York State Department of Health | Kirsten St. George, Daryl M. Lamson, Alexis Russel, Matthew Shudt, Melissa A Leisner, Jonathan Plitnick, Navjot Singh, John Kelly, Erasmus Schneider, Erica Lasek-Nesselquist |
| EPI_ISL_1226684 | KALEIDA CENTER FOR LABORATORY MEDICINE | Wadsworth Center, New York State Department of Health | Kirsten St. George, Daryl M. Lamson, Alexis Russel, Matthew Shudt, Melissa A Leisner, Jonathan Plitnick, Navjot Singh, John Kelly, Erasmus Schneider, Erica Lasek-Nesselquist |
| EPI_ISL_1226685 | NYC Pandemic Response Lab | Wadsworth Center, New York State Department of Health | Kirsten St. George, Daryl M. Lamson, Alexis Russel, Matthew Shudt, Melissa A Leisner, Jonathan Plitnick, Navjot Singh, John Kelly, Erasmus Schneider, Erica Lasek-Nesselquist |
| EPI_ISL_1226686, EPI_ISL_1226687, EPI_ISL_1226688 | WESTCHESTER MEDICAL CENTER | Wadsworth Center, New York State Department of Health | Kirsten St. George, Daryl M. Lamson, Alexis Russel, Matthew Shudt, Melissa A Leisner, Jonathan Plitnick, Navjot Singh, John Kelly, Erasmus Schneider, Erica Lasek-Nesselquist |

| | | | |
|---|---|---|---|
| EPI_ISL_1226689, EPI_ISL_1226690, EPI_ISL_1226691 | MONTEFIORE MEDICAL CENTER LABORATORIES | Wadsworth Center, New York State Department of Health | Kirsten St. George, Daryl M. Lamson, Alexis Russel, Matthew Shudt, Melissa A Leisner, Jonathan Plitnick, Navjot Singh, John Kelly, Erasmus Schneider, Erica Lasek-Nesselquist |
| EPI_ISL_1226692 | NYC Pandemic Response Lab | Wadsworth Center, New York State Department of Health | Kirsten St. George, Daryl M. Lamson, Alexis Russel, Matthew Shudt, Melissa A Leisner, Jonathan Plitnick, Navjot Singh, John Kelly, Erasmus Schneider, Erica Lasek-Nesselquist |
| EPI_ISL_1226693, EPI_ISL_1226694, EPI_ISL_1226695 | WESTCHESTER MEDICAL CENTER | Wadsworth Center, New York State Department of Health | Kirsten St. George, Daryl M. Lamson, Alexis Russel, Matthew Shudt, Melissa A Leisner, Jonathan Plitnick, Navjot Singh, John Kelly, Erasmus Schneider, Erica Lasek-Nesselquist |
| EPI_ISL_1226696 | KALEIDA CENTER FOR LABORATORY MEDICINE | Wadsworth Center, New York State Department of Health | Kirsten St. George, Daryl M. Lamson, Alexis Russel, Matthew Shudt, Melissa A Leisner, Jonathan Plitnick, Navjot Singh, John Kelly, Erasmus Schneider, Erica Lasek-Nesselquist |
| EPI_ISL_1226697 | WESTCHESTER MEDICAL CENTER | Wadsworth Center, New York State Department of Health | Kirsten St. George, Daryl M. Lamson, Alexis Russel, Matthew Shudt, Melissa A Leisner, Jonathan Plitnick, Navjot Singh, John Kelly, Erasmus Schneider, Erica Lasek-Nesselquist |
| EPI_ISL_1226698 | Wadsworth Center, New York State Department of Health | Wadsworth Center, New York State Department of Health | Kirsten St. George, Daryl M. Lamson, Alexis Russel, Matthew Shudt, Melissa A Leisner, Jonathan Plitnick, Navjot Singh, John Kelly, Erasmus Schneider, Erica Lasek-Nesselquist |
| EPI_ISL_1226699 | NYC Pandemic Response Lab | Wadsworth Center, New York State Department of Health | Kirsten St. George, Daryl M. Lamson, Alexis Russel, Matthew Shudt, Melissa A Leisner, Jonathan Plitnick, Navjot Singh, John Kelly, Erasmus Schneider, Erica Lasek-Nesselquist |
| EPI_ISL_1226700, EPI_ISL_1226701 | MEMORIAL SLOAN KETTERING CANCER CENTER | Wadsworth Center, New York State Department of Health | Kirsten St. George, Daryl M. Lamson, Alexis Russel, Matthew Shudt, Melissa A Leisner, Jonathan Plitnick, Navjot Singh, John Kelly, Erasmus Schneider, Erica Lasek-Nesselquist |
| EPI_ISL_1226702 | WESTCHESTER MEDICAL CENTER | Wadsworth Center, New York State Department of Health | Kirsten St. George, Daryl M. Lamson, Alexis Russel, Matthew Shudt, Melissa A Leisner, Jonathan Plitnick, Navjot Singh, John Kelly, Erasmus Schneider, Erica Lasek-Nesselquist |
| EPI_ISL_1226703 | NYC Pandemic Response Lab | Wadsworth Center, New York State Department of Health | Kirsten St. George, Daryl M. Lamson, Alexis Russel, Matthew Shudt, Melissa A Leisner, Jonathan Plitnick, Navjot Singh, John Kelly, Erasmus Schneider, Erica Lasek-Nesselquist |
| EPI_ISL_1226704, EPI_ISL_1226705, EPI_ISL_1226706 | ALBANY MEDICAL CENTER | Wadsworth Center, New York State Department of Health | Kirsten St. George, Daryl M. Lamson, Alexis Russel, Matthew Shudt, Melissa A Leisner, Jonathan Plitnick, Navjot Singh, John Kelly, Erasmus Schneider, Erica Lasek-Nesselquist |
| EPI_ISL_1226707 | NORTHWELL HEALTH LABORATORIES | Wadsworth Center, New York State Department of Health | Kirsten St. George, Daryl M. Lamson, Alexis Russel, Matthew Shudt, Melissa A Leisner, Jonathan Plitnick, Navjot Singh, John Kelly, Erasmus Schneider, Erica Lasek-Nesselquist |
| EPI_ISL_1226708 | WESTCHESTER MEDICAL CENTER | Wadsworth Center, New York State Department of Health | Kirsten St. George, Daryl M. Lamson, Alexis Russel, Matthew Shudt, Melissa A Leisner, Jonathan Plitnick, Navjot Singh, John Kelly, Erasmus Schneider, Erica Lasek-Nesselquist |
| EPI_ISL_1226709 | TEMPUS LABS INC | Wadsworth Center, New York State Department of Health | Kirsten St. George, Daryl M. Lamson, Alexis Russel, Matthew Shudt, Melissa A Leisner, Jonathan Plitnick, Navjot Singh, John Kelly, Erasmus Schneider, Erica Lasek-Nesselquist |
| EPI_ISL_1226710 | MEMORIAL SLOAN KETTERING CANCER CENTER | Wadsworth Center, New York State Department of Health | Kirsten St. George, Daryl M. Lamson, Alexis Russel, Matthew Shudt, Melissa A Leisner, Jonathan Plitnick, Navjot Singh, John Kelly, Erasmus Schneider, Erica Lasek-Nesselquist |
| EPI_ISL_1226711 | NYC Pandemic Response Lab | Wadsworth Center, New York State Department of Health | Kirsten St. George, Daryl M. Lamson, Alexis Russel, Matthew Shudt, Melissa A Leisner, Jonathan Plitnick, Navjot Singh, John Kelly, Erasmus Schneider, Erica Lasek-Nesselquist |
| EPI_ISL_1226712 | TEMPUS LABS INC | Wadsworth Center, New York State Department of Health | Kirsten St. George, Daryl M. Lamson, Alexis Russel, Matthew Shudt, Melissa A Leisner, Jonathan Plitnick, Navjot Singh, John Kelly, Erasmus Schneider, Erica Lasek-Nesselquist |
| EPI_ISL_1226713 | URMC LABS | Wadsworth Center, New York State Department of Health | Kirsten St. George, Daryl M. Lamson, Alexis Russel, Matthew Shudt, Melissa A Leisner, Jonathan Plitnick, Navjot Singh, John Kelly, Erasmus Schneider, Erica Lasek-Nesselquist |
| EPI_ISL_1226714, EPI_ISL_1226715 | MONTEFIORE MEDICAL CENTER LABORATORIES | Wadsworth Center, New York State Department of Health | Kirsten St. George, Daryl M. Lamson, Alexis Russel, Matthew Shudt, Melissa A Leisner, Jonathan Plitnick, Navjot Singh, John Kelly, Erasmus Schneider, Erica Lasek-Nesselquist |
| EPI_ISL_1226716 | Wadsworth Center, New York State Department of Health | Wadsworth Center, New York State Department of Health | Kirsten St. George, Daryl M. Lamson, Alexis Russel, Matthew Shudt, Melissa A Leisner, Jonathan Plitnick, Navjot Singh, John Kelly, Erasmus Schneider, Erica Lasek-Nesselquist |
| EPI_ISL_1226717 | NORTHWELL HEALTH LABORATORIES | Wadsworth Center, New York State Department of Health | Kirsten St. George, Daryl M. Lamson, Alexis Russel, Matthew Shudt, Melissa A Leisner, Jonathan Plitnick, Navjot Singh, John Kelly, Erasmus Schneider, Erica Lasek-Nesselquist |
| EPI_ISL_1226718, EPI_ISL_1226719 | NYC Pandemic Response Lab | Wadsworth Center, New York State Department of Health | Kirsten St. George, Daryl M. Lamson, Alexis Russel, Matthew Shudt, Melissa A Leisner, Jonathan Plitnick, Navjot Singh, John Kelly, Erasmus Schneider, Erica Lasek-Nesselquist |
| EPI_ISL_1226720 | NORTHWELL HEALTH LABORATORIES | Wadsworth Center, New York State Department of Health | Kirsten St. George, Daryl M. Lamson, Alexis Russel, Matthew Shudt, Melissa A Leisner, Jonathan Plitnick, Navjot Singh, John Kelly, Erasmus Schneider, Erica Lasek-Nesselquist |
| EPI_ISL_1226721 | WESTCHESTER MEDICAL CENTER | Wadsworth Center, New York State Department of Health | Kirsten St. George, Daryl M. Lamson, Alexis Russel, Matthew Shudt, Melissa A Leisner, Jonathan Plitnick, Navjot Singh, John Kelly, Erasmus Schneider, Erica Lasek-Nesselquist |
| EPI_ISL_1226722 | MONTEFIORE MEDICAL CENTER LABORATORIES | Wadsworth Center, New York State Department of Health | Kirsten St. George, Daryl M. Lamson, Alexis Russel, Matthew Shudt, Melissa A Leisner, Jonathan Plitnick, Navjot Singh, John Kelly, Erasmus Schneider, Erica Lasek-Nesselquist |
| EPI_ISL_1226723 | WESTCHESTER MEDICAL CENTER | Wadsworth Center, New York State Department of Health | Kirsten St. George, Daryl M. Lamson, Alexis Russel, Matthew Shudt, Melissa A Leisner, Jonathan Plitnick, Navjot Singh, John Kelly, Erasmus Schneider, Erica Lasek-Nesselquist |
| EPI_ISL_1226724 | MONTEFIORE MEDICAL CENTER LABORATORIES | Wadsworth Center, New York State Department of Health | Kirsten St. George, Daryl M. Lamson, Alexis Russel, Matthew Shudt, Melissa A Leisner, Jonathan Plitnick, Navjot Singh, John Kelly, Erasmus Schneider, Erica Lasek-Nesselquist |
| EPI_ISL_1226725, EPI_ISL_1226726 | MEMORIAL SLOAN KETTERING CANCER CENTER | Wadsworth Center, New York State Department of Health | Kirsten St. George, Daryl M. Lamson, Alexis Russel, Matthew Shudt, Melissa A Leisner, Jonathan Plitnick, Navjot Singh, John Kelly, Erasmus Schneider, Erica Lasek-Nesselquist |
| EPI_ISL_1226727 | WESTCHESTER MEDICAL CENTER | Wadsworth Center, New York State Department of Health | Kirsten St. George, Daryl M. Lamson, Alexis Russel, Matthew Shudt, Melissa A Leisner, Jonathan Plitnick, Navjot Singh, John Kelly, Erasmus Schneider, Erica Lasek-Nesselquist |
| EPI_ISL_1226728 | Columbia University Irving Medical Center | Wadsworth Center, New York State Department of Health | Kirsten St. George, Daryl M. Lamson, Alexis Russel, Matthew Shudt, Melissa A Leisner, Jonathan Plitnick, Navjot Singh, John Kelly, Erasmus Schneider, Erica Lasek-Nesselquist |
| EPI_ISL_1226729 | NYC Pandemic Response Lab | Wadsworth Center, New York State Department of Health | Kirsten St. George, Daryl M. Lamson, Alexis Russel, Matthew Shudt, Melissa A Leisner, Jonathan Plitnick, Navjot Singh, John Kelly, Erasmus Schneider, Erica Lasek-Nesselquist |
| EPI_ISL_1226730 | WESTCHESTER MEDICAL CENTER | Wadsworth Center, New York State Department of Health | Kirsten St. George, Daryl M. Lamson, Alexis Russel, Matthew Shudt, Melissa A Leisner, Jonathan Plitnick, Navjot Singh, John Kelly, Erasmus Schneider, Erica Lasek-Nesselquist |
| EPI_ISL_1226731 | MEMORIAL SLOAN KETTERING CANCER CENTER | Wadsworth Center, New York State Department of Health | Kirsten St. George, Daryl M. Lamson, Alexis Russel, Matthew Shudt, Melissa A Leisner, Jonathan Plitnick, Navjot Singh, John Kelly, Erasmus Schneider, Erica Lasek-Nesselquist |
| EPI_ISL_1226732 | SUNY UPSTATE MEDICAL UNIVERSITY | Wadsworth Center, New York State Department of Health | Kirsten St. George, Daryl M. Lamson, Alexis Russel, Matthew Shudt, Melissa A Leisner, Jonathan Plitnick, Navjot Singh, John Kelly, Erasmus Schneider, Erica Lasek-Nesselquist |
| EPI_ISL_1226733 | MEMORIAL SLOAN KETTERING CANCER CENTER | Wadsworth Center, New York State Department of Health | Kirsten St. George, Daryl M. Lamson, Alexis Russel, Matthew Shudt, Melissa A Leisner, Jonathan Plitnick, Navjot Singh, John Kelly, Erasmus Schneider, Erica Lasek-Nesselquist |
| EPI_ISL_1226734, EPI_ISL_1226735 | MONTEFIORE MEDICAL CENTER LABORATORIES | Wadsworth Center, New York State Department of Health | Kirsten St. George, Daryl M. Lamson, Alexis Russel, Matthew Shudt, Melissa A Leisner, Jonathan Plitnick, Navjot Singh, John Kelly, Erasmus Schneider, Erica Lasek-Nesselquist |
| EPI_ISL_1226736 | MEMORIAL SLOAN KETTERING CANCER CENTER | Wadsworth Center, New York State Department of Health | Kirsten St. George, Daryl M. Lamson, Alexis Russel, Matthew Shudt, Melissa A Leisner, Jonathan Plitnick, Navjot Singh, John Kelly, Erasmus Schneider, Erica Lasek-Nesselquist |
| EPI_ISL_1226737, EPI_ISL_1226738 | MONTEFIORE MEDICAL CENTER LABORATORIES | Wadsworth Center, New York State Department of Health | Kirsten St. George, Daryl M. Lamson, Alexis Russel, Matthew Shudt, Melissa A Leisner, Jonathan Plitnick, Navjot Singh, John Kelly, Erasmus Schneider, Erica Lasek-Nesselquist |
| EPI_ISL_1226739 | NYC Pandemic Response Lab | Wadsworth Center, New York State Department of Health | Kirsten St. George, Daryl M. Lamson, Alexis Russel, Matthew Shudt, Melissa A Leisner, Jonathan Plitnick, Navjot Singh, John Kelly, Erasmus Schneider, Erica Lasek-Nesselquist |
| EPI_ISL_1226740, EPI_ISL_1226741, EPI_ISL_1226742 | MONTEFIORE MEDICAL CENTER LABORATORIES | Wadsworth Center, New York State Department of Health | Kirsten St. George, Daryl M. Lamson, Alexis Russel, Matthew Shudt, Melissa A Leisner, Jonathan Plitnick, Navjot Singh, John Kelly, Erasmus Schneider, Erica Lasek-Nesselquist |
| EPI_ISL_1226743 | Columbia University Irving Medical Center | Wadsworth Center, New York State Department of Health | Kirsten St. George, Daryl M. Lamson, Alexis Russel, Matthew Shudt, Melissa A Leisner, Jonathan Plitnick, Navjot Singh, John Kelly, Erasmus Schneider, Erica Lasek-Nesselquist |
| EPI_ISL_1226744, EPI_ISL_1226745, EPI_ISL_1226746, EPI_ISL_1226747, EPI_ISL_1226748, EPI_ISL_1226749, EPI_ISL_1226750, EPI_ISL_1226751, EPI_ISL_1226752, EPI_ISL_1226753, EPI_ISL_1226754, EPI_ISL_1226755 | | | |
| see above | MONTEFIORE MEDICAL CENTER LABORATORIES | Wadsworth Center, New York State Department of Health | Kirsten St. George, Daryl M. Lamson, Alexis Russel, Matthew Shudt, Melissa A Leisner, Jonathan Plitnick, Navjot Singh, John Kelly, Erasmus Schneider, Erica Lasek-Nesselquist |
| EPI_ISL_1226756 | WESTCHESTER MEDICAL CENTER | Wadsworth Center, New York State Department of Health | Kirsten St. George, Daryl M. Lamson, Alexis Russel, Matthew Shudt, Melissa A Leisner, Jonathan Plitnick, Navjot Singh, John Kelly, Erasmus Schneider, Erica Lasek-Nesselquist |

| | | | |
|---|---|---|---|
| EPI_ISL_1226757 | New York Presbyterian Hospital | Wadsworth Center, New York State Department of Health | Kirsten St. George, Daryl M. Lamson, Alexis Russel, Matthew Shudt, Melissa A Leisner, Jonathan Plitnick, Navjot Singh, John Kelly, Erasmus Schneider, Erica Lasek-Nesselquist |
| EPI_ISL_1226758 | MONTEFIORE MEDICAL CENTER LABORATORIES | Wadsworth Center, New York State Department of Health | Kirsten St. George, Daryl M. Lamson, Alexis Russel, Matthew Shudt, Melissa A Leisner, Jonathan Plitnick, Navjot Singh, John Kelly, Erasmus Schneider, Erica Lasek-Nesselquist |
| EPI_ISL_1226759 | Columbia University Irving Medical Center | Wadsworth Center, New York State Department of Health | Kirsten St. George, Daryl M. Lamson, Alexis Russel, Matthew Shudt, Melissa A Leisner, Jonathan Plitnick, Navjot Singh, John Kelly, Erasmus Schneider, Erica Lasek-Nesselquist |
| EPI_ISL_1226760, EPI_ISL_1226761, EPI_ISL_1226762, EPI_ISL_1226763, EPI_ISL_1226764 | MONTEFIORE MEDICAL CENTER LABORATORIES | Wadsworth Center, New York State Department of Health | Kirsten St. George, Daryl M. Lamson, Alexis Russel, Matthew Shudt, Melissa A Leisner, Jonathan Plitnick, Navjot Singh, John Kelly, Erasmus Schneider, Erica Lasek-Nesselquist |
| EPI_ISL_1226765 | Columbia University Irving Medical Center | Wadsworth Center, New York State Department of Health | Kirsten St. George, Daryl M. Lamson, Alexis Russel, Matthew Shudt, Melissa A Leisner, Jonathan Plitnick, Navjot Singh, John Kelly, Erasmus Schneider, Erica Lasek-Nesselquist |
| EPI_ISL_1226766, EPI_ISL_1226767 | MONTEFIORE MEDICAL CENTER LABORATORIES | Wadsworth Center, New York State Department of Health | Kirsten St. George, Daryl M. Lamson, Alexis Russel, Matthew Shudt, Melissa A Leisner, Jonathan Plitnick, Navjot Singh, John Kelly, Erasmus Schneider, Erica Lasek-Nesselquist |
| EPI_ISL_1226768, EPI_ISL_1226769 | NYC Pandemic Response Lab | Wadsworth Center, New York State Department of Health | Kirsten St. George, Daryl M. Lamson, Alexis Russel, Matthew Shudt, Melissa A Leisner, Jonathan Plitnick, Navjot Singh, John Kelly, Erasmus Schneider, Erica Lasek-Nesselquist |
| EPI_ISL_1226770, EPI_ISL_1226771 | MONTEFIORE MEDICAL CENTER LABORATORIES | Wadsworth Center, New York State Department of Health | Kirsten St. George, Daryl M. Lamson, Alexis Russel, Matthew Shudt, Melissa A Leisner, Jonathan Plitnick, Navjot Singh, John Kelly, Erasmus Schneider, Erica Lasek-Nesselquist |
| EPI_ISL_1226772 | WESTCHESTER MEDICAL CENTER | Wadsworth Center, New York State Department of Health | Kirsten St. George, Daryl M. Lamson, Alexis Russel, Matthew Shudt, Melissa A Leisner, Jonathan Plitnick, Navjot Singh, John Kelly, Erasmus Schneider, Erica Lasek-Nesselquist |
| EPI_ISL_1226773, EPI_ISL_1226774, EPI_ISL_1226775, EPI_ISL_1226776, EPI_ISL_1226777, EPI_ISL_1226778, EPI_ISL_1226779, EPI_ISL_1226780 | MONTEFIORE MEDICAL CENTER LABORATORIES | Wadsworth Center, New York State Department of Health | Kirsten St. George, Daryl M. Lamson, Alexis Russel, Matthew Shudt, Melissa A Leisner, Jonathan Plitnick, Navjot Singh, John Kelly, Erasmus Schneider, Erica Lasek-Nesselquist |
| EPI_ISL_1226781, EPI_ISL_1226782, EPI_ISL_1226783, EPI_ISL_1226784, EPI_ISL_1226785, EPI_ISL_1226786, EPI_ISL_1226787, EPI_ISL_1226788 | WESTCHESTER MEDICAL CENTER | Wadsworth Center, New York State Department of Health | Kirsten St. George, Daryl M. Lamson, Alexis Russel, Matthew Shudt, Melissa A Leisner, Jonathan Plitnick, Navjot Singh, John Kelly, Erasmus Schneider, Erica Lasek-Nesselquist |
| EPI_ISL_1226789, EPI_ISL_1226790, EPI_ISL_1226791, EPI_ISL_1226792, EPI_ISL_1226793, EPI_ISL_1226794, EPI_ISL_1226795, EPI_ISL_1226796, EPI_ISL_1226797 | NYC Pandemic Response Lab | Wadsworth Center, New York State Department of Health | Kirsten St. George, Daryl M. Lamson, Alexis Russel, Matthew Shudt, Melissa A Leisner, Jonathan Plitnick, Navjot Singh, John Kelly, Erasmus Schneider, Erica Lasek-Nesselquist |
| EPI_ISL_1226798 | NORTH SHORE UNIVERSITY HOSPITAL | Wadsworth Center, New York State Department of Health | Kirsten St. George, Daryl M. Lamson, Alexis Russel, Matthew Shudt, Melissa A Leisner, Jonathan Plitnick, Navjot Singh, John Kelly, Erasmus Schneider, Erica Lasek-Nesselquist |
| EPI_ISL_1226799 | MEMORIAL SLOAN KETTERING CANCER CENTER | Wadsworth Center, New York State Department of Health | Kirsten St. George, Daryl M. Lamson, Alexis Russel, Matthew Shudt, Melissa A Leisner, Jonathan Plitnick, Navjot Singh, John Kelly, Erasmus Schneider, Erica Lasek-Nesselquist |
| EPI_ISL_1226800, EPI_ISL_1226801, EPI_ISL_1226802, EPI_ISL_1226803, EPI_ISL_1226804, EPI_ISL_1226805, EPI_ISL_1226806, EPI_ISL_1226807 | Columbia University Irving Medical Center | Wadsworth Center, New York State Department of Health | Kirsten St. George, Daryl M. Lamson, Alexis Russel, Matthew Shudt, Melissa A Leisner, Jonathan Plitnick, Navjot Singh, John Kelly, Erasmus Schneider, Erica Lasek-Nesselquist |
| EPI_ISL_1226808, EPI_ISL_1226809, EPI_ISL_1226810 | NYC Pandemic Response Lab | Wadsworth Center, New York State Department of Health | Kirsten St. George, Daryl M. Lamson, Alexis Russel, Matthew Shudt, Melissa A Leisner, Jonathan Plitnick, Navjot Singh, John Kelly, Erasmus Schneider, Erica Lasek-Nesselquist |
| EPI_ISL_1226811, EPI_ISL_1226812 | New York Presbyterian Hospital | Wadsworth Center, New York State Department of Health | Kirsten St. George, Daryl M. Lamson, Alexis Russel, Matthew Shudt, Melissa A Leisner, Jonathan Plitnick, Navjot Singh, John Kelly, Erasmus Schneider, Erica Lasek-Nesselquist |
| EPI_ISL_1226813, EPI_ISL_1226814 | NYC Pandemic Response Lab | Wadsworth Center, New York State Department of Health | Kirsten St. George, Daryl M. Lamson, Alexis Russel, Matthew Shudt, Melissa A Leisner, Jonathan Plitnick, Navjot Singh, John Kelly, Erasmus Schneider, Erica Lasek-Nesselquist |
| EPI_ISL_1226815 | NORTH SHORE UNIVERSITY HOSPITAL | Wadsworth Center, New York State Department of Health | Kirsten St. George, Daryl M. Lamson, Alexis Russel, Matthew Shudt, Melissa A Leisner, Jonathan Plitnick, Navjot Singh, John Kelly, Erasmus Schneider, Erica Lasek-Nesselquist |
| EPI_ISL_1226816, EPI_ISL_1226817 | NYC Pandemic Response Lab | Wadsworth Center, New York State Department of Health | Kirsten St. George, Daryl M. Lamson, Alexis Russel, Matthew Shudt, Melissa A Leisner, Jonathan Plitnick, Navjot Singh, John Kelly, Erasmus Schneider, Erica Lasek-Nesselquist |
| EPI_ISL_1226818 | WESTCHESTER MEDICAL CENTER | Wadsworth Center, New York State Department of Health | Kirsten St. George, Daryl M. Lamson, Alexis Russel, Matthew Shudt, Melissa A Leisner, Jonathan Plitnick, Navjot Singh, John Kelly, Erasmus Schneider, Erica Lasek-Nesselquist |
| EPI_ISL_1226819 | ALBANY MEDICAL CENTER | Wadsworth Center, New York State Department of Health | Kirsten St. George, Daryl M. Lamson, Alexis Russel, Matthew Shudt, Melissa A Leisner, Jonathan Plitnick, Navjot Singh, John Kelly, Erasmus Schneider, Erica Lasek-Nesselquist |
| EPI_ISL_1226820 | MEMORIAL SLOAN KETTERING CANCER CENTER | Wadsworth Center, New York State Department of Health | Kirsten St. George, Daryl M. Lamson, Alexis Russel, Matthew Shudt, Melissa A Leisner, Jonathan Plitnick, Navjot Singh, John Kelly, Erasmus Schneider, Erica Lasek-Nesselquist |
| EPI_ISL_1226821, EPI_ISL_1226822 | WESTCHESTER MEDICAL CENTER | Wadsworth Center, New York State Department of Health | Kirsten St. George, Daryl M. Lamson, Alexis Russel, Matthew Shudt, Melissa A Leisner, Jonathan Plitnick, Navjot Singh, John Kelly, Erasmus Schneider, Erica Lasek-Nesselquist |
| EPI_ISL_1226823, EPI_ISL_1226824 | Columbia University Irving Medical Center | Wadsworth Center, New York State Department of Health | Kirsten St. George, Daryl M. Lamson, Alexis Russel, Matthew Shudt, Melissa A Leisner, Jonathan Plitnick, Navjot Singh, John Kelly, Erasmus Schneider, Erica Lasek-Nesselquist |
| EPI_ISL_1226825, EPI_ISL_1226826, EPI_ISL_1226827, EPI_ISL_1226828, EPI_ISL_1226829, EPI_ISL_1226830 | WESTCHESTER MEDICAL CENTER | Wadsworth Center, New York State Department of Health | Kirsten St. George, Daryl M. Lamson, Alexis Russel, Matthew Shudt, Melissa A Leisner, Jonathan Plitnick, Navjot Singh, John Kelly, Erasmus Schneider, Erica Lasek-Nesselquist |
| EPI_ISL_1226831 | MONTEFIORE MEDICAL CENTER LABORATORIES | Wadsworth Center, New York State Department of Health | Kirsten St. George, Daryl M. Lamson, Alexis Russel, Matthew Shudt, Melissa A Leisner, Jonathan Plitnick, Navjot Singh, John Kelly, Erasmus Schneider, Erica Lasek-Nesselquist |
| EPI_ISL_1226832, EPI_ISL_1226833 | NYC Pandemic Response Lab | Wadsworth Center, New York State Department of Health | Kirsten St. George, Daryl M. Lamson, Alexis Russel, Matthew Shudt, Melissa A Leisner, Jonathan Plitnick, Navjot Singh, John Kelly, Erasmus Schneider, Erica Lasek-Nesselquist |
| EPI_ISL_1226834 | WESTCHESTER MEDICAL CENTER | Wadsworth Center, New York State Department of Health | Kirsten St. George, Daryl M. Lamson, Alexis Russel, Matthew Shudt, Melissa A Leisner, Jonathan Plitnick, Navjot Singh, John Kelly, Erasmus Schneider, Erica Lasek-Nesselquist |
| EPI_ISL_1226835 | MONTEFIORE MEDICAL CENTER LABORATORIES | Wadsworth Center, New York State Department of Health | Kirsten St. George, Daryl M. Lamson, Alexis Russel, Matthew Shudt, Melissa A Leisner, Jonathan Plitnick, Navjot Singh, John Kelly, Erasmus Schneider, Erica Lasek-Nesselquist |
| EPI_ISL_1226836, EPI_ISL_1226837, EPI_ISL_1226838 | WESTCHESTER MEDICAL CENTER | Wadsworth Center, New York State Department of Health | Kirsten St. George, Daryl M. Lamson, Alexis Russel, Matthew Shudt, Melissa A Leisner, Jonathan Plitnick, Navjot Singh, John Kelly, Erasmus Schneider, Erica Lasek-Nesselquist |

| | | | |
|---|---|---|---|
| EPI_ISL_1226839 | MONTEFIORE MEDICAL CENTER LABORATORIES | Wadsworth Center, New York State Department of Health | Kirsten St. George, Daryl M. Lamson, Alexis Russel, Matthew Shudt, Melissa A Leisner, Jonathan Plitnick, Navjot Singh, John Kelly, Erasmus Schneider, Erica Lasek-Nesselquist |
| EPI_ISL_1226840, EPI_ISL_1226841 | WESTCHESTER MEDICAL CENTER | Wadsworth Center, New York State Department of Health | Kirsten St. George, Daryl M. Lamson, Alexis Russel, Matthew Shudt, Melissa A Leisner, Jonathan Plitnick, Navjot Singh, John Kelly, Erasmus Schneider, Erica Lasek-Nesselquist |
| EPI_ISL_1226842 | MONTEFIORE MEDICAL CENTER LABORATORIES | Wadsworth Center, New York State Department of Health | Kirsten St. George, Daryl M. Lamson, Alexis Russel, Matthew Shudt, Melissa A Leisner, Jonathan Plitnick, Navjot Singh, John Kelly, Erasmus Schneider, Erica Lasek-Nesselquist |
| EPI_ISL_1226843 | New York Presbyterian Hospital | Wadsworth Center, New York State Department of Health | Kirsten St. George, Daryl M. Lamson, Alexis Russel, Matthew Shudt, Melissa A Leisner, Jonathan Plitnick, Navjot Singh, John Kelly, Erasmus Schneider, Erica Lasek-Nesselquist |
| EPI_ISL_1226844 | MONTEFIORE MEDICAL CENTER LABORATORIES | Wadsworth Center, New York State Department of Health | Kirsten St. George, Daryl M. Lamson, Alexis Russel, Matthew Shudt, Melissa A Leisner, Jonathan Plitnick, Navjot Singh, John Kelly, Erasmus Schneider, Erica Lasek-Nesselquist |
| EPI_ISL_1226845 | WESTCHESTER MEDICAL CENTER | Wadsworth Center, New York State Department of Health | Kirsten St. George, Daryl M. Lamson, Alexis Russel, Matthew Shudt, Melissa A Leisner, Jonathan Plitnick, Navjot Singh, John Kelly, Erasmus Schneider, Erica Lasek-Nesselquist |
| EPI_ISL_1226846, EPI_ISL_1226847, EPI_ISL_1226848 | KALEIDA CENTER FOR LABORATORY MEDICINE | Wadsworth Center, New York State Department of Health | Kirsten St. George, Daryl M. Lamson, Alexis Russel, Matthew Shudt, Melissa A Leisner, Jonathan Plitnick, Navjot Singh, John Kelly, Erasmus Schneider, Erica Lasek-Nesselquist |
| EPI_ISL_1226849 | NORTHWELL HEALTH LABORATORIES | Wadsworth Center, New York State Department of Health | Kirsten St. George, Daryl M. Lamson, Alexis Russel, Matthew Shudt, Melissa A Leisner, Jonathan Plitnick, Navjot Singh, John Kelly, Erasmus Schneider, Erica Lasek-Nesselquist |
| EPI_ISL_1226850 | New York Presbyterian Hospital | Wadsworth Center, New York State Department of Health | Kirsten St. George, Daryl M. Lamson, Alexis Russel, Matthew Shudt, Melissa A Leisner, Jonathan Plitnick, Navjot Singh, John Kelly, Erasmus Schneider, Erica Lasek-Nesselquist |
| EPI_ISL_1226851, EPI_ISL_1226852 | URMC LABS | Wadsworth Center, New York State Department of Health | Kirsten St. George, Daryl M. Lamson, Alexis Russel, Matthew Shudt, Melissa A Leisner, Jonathan Plitnick, Navjot Singh, John Kelly, Erasmus Schneider, Erica Lasek-Nesselquist |
| EPI_ISL_1226853 | NORTHWELL HEALTH LABORATORIES | Wadsworth Center, New York State Department of Health | Kirsten St. George, Daryl M. Lamson, Alexis Russel, Matthew Shudt, Melissa A Leisner, Jonathan Plitnick, Navjot Singh, John Kelly, Erasmus Schneider, Erica Lasek-Nesselquist |
| EPI_ISL_1226854, EPI_ISL_1226855 | NYC Pandemic Response Lab | Wadsworth Center, New York State Department of Health | Kirsten St. George, Daryl M. Lamson, Alexis Russel, Matthew Shudt, Melissa A Leisner, Jonathan Plitnick, Navjot Singh, John Kelly, Erasmus Schneider, Erica Lasek-Nesselquist |
| EPI_ISL_1226856 | NORTHWELL HEALTH LABORATORIES | Wadsworth Center, New York State Department of Health | Kirsten St. George, Daryl M. Lamson, Alexis Russel, Matthew Shudt, Melissa A Leisner, Jonathan Plitnick, Navjot Singh, John Kelly, Erasmus Schneider, Erica Lasek-Nesselquist |
| EPI_ISL_1226857, EPI_ISL_1226858 | MEMORIAL SLOAN KETTERING CANCER CENTER | Wadsworth Center, New York State Department of Health | Kirsten St. George, Daryl M. Lamson, Alexis Russel, Matthew Shudt, Melissa A Leisner, Jonathan Plitnick, Navjot Singh, John Kelly, Erasmus Schneider, Erica Lasek-Nesselquist |
| EPI_ISL_1226859 | NORTHWELL HEALTH LABORATORIES | Wadsworth Center, New York State Department of Health | Kirsten St. George, Daryl M. Lamson, Alexis Russel, Matthew Shudt, Melissa A Leisner, Jonathan Plitnick, Navjot Singh, John Kelly, Erasmus Schneider, Erica Lasek-Nesselquist |
| EPI_ISL_1226860 | NYC Pandemic Response Lab | Wadsworth Center, New York State Department of Health | Kirsten St. George, Daryl M. Lamson, Alexis Russel, Matthew Shudt, Melissa A Leisner, Jonathan Plitnick, Navjot Singh, John Kelly, Erasmus Schneider, Erica Lasek-Nesselquist |
| EPI_ISL_1226861 | MEMORIAL SLOAN KETTERING CANCER CENTER | Wadsworth Center, New York State Department of Health | Kirsten St. George, Daryl M. Lamson, Alexis Russel, Matthew Shudt, Melissa A Leisner, Jonathan Plitnick, Navjot Singh, John Kelly, Erasmus Schneider, Erica Lasek-Nesselquist |
| EPI_ISL_1226862 | NYC Pandemic Response Lab | Wadsworth Center, New York State Department of Health | Kirsten St. George, Daryl M. Lamson, Alexis Russel, Matthew Shudt, Melissa A Leisner, Jonathan Plitnick, Navjot Singh, John Kelly, Erasmus Schneider, Erica Lasek-Nesselquist |
| EPI_ISL_1226863 | WESTCHESTER MEDICAL CENTER | Wadsworth Center, New York State Department of Health | Kirsten St. George, Daryl M. Lamson, Alexis Russel, Matthew Shudt, Melissa A Leisner, Jonathan Plitnick, Navjot Singh, John Kelly, Erasmus Schneider, Erica Lasek-Nesselquist |
| EPI_ISL_1226864 | MONTEFIORE MEDICAL CENTER LABORATORIES | Wadsworth Center, New York State Department of Health | Kirsten St. George, Daryl M. Lamson, Alexis Russel, Matthew Shudt, Melissa A Leisner, Jonathan Plitnick, Navjot Singh, John Kelly, Erasmus Schneider, Erica Lasek-Nesselquist |
| EPI_ISL_1226865 | NORTHWELL HEALTH LABORATORIES | Wadsworth Center, New York State Department of Health | Kirsten St. George, Daryl M. Lamson, Alexis Russel, Matthew Shudt, Melissa A Leisner, Jonathan Plitnick, Navjot Singh, John Kelly, Erasmus Schneider, Erica Lasek-Nesselquist |
| EPI_ISL_1226866 | URMC LABS | Wadsworth Center, New York State Department of Health | Kirsten St. George, Daryl M. Lamson, Alexis Russel, Matthew Shudt, Melissa A Leisner, Jonathan Plitnick, Navjot Singh, John Kelly, Erasmus Schneider, Erica Lasek-Nesselquist |
| EPI_ISL_1226867 | NYC Pandemic Response Lab | Wadsworth Center, New York State Department of Health | Kirsten St. George, Daryl M. Lamson, Alexis Russel, Matthew Shudt, Melissa A Leisner, Jonathan Plitnick, Navjot Singh, John Kelly, Erasmus Schneider, Erica Lasek-Nesselquist |
| EPI_ISL_1226868 | THE MARY IMOGENE BASSETT HOSPITAL | Wadsworth Center, New York State Department of Health | Kirsten St. George, Daryl M. Lamson, Alexis Russel, Matthew Shudt, Melissa A Leisner, Jonathan Plitnick, Navjot Singh, John Kelly, Erasmus Schneider, Erica Lasek-Nesselquist |
| EPI_ISL_1226869 | NYC Pandemic Response Lab | Wadsworth Center, New York State Department of Health | Kirsten St. George, Daryl M. Lamson, Alexis Russel, Matthew Shudt, Melissa A Leisner, Jonathan Plitnick, Navjot Singh, John Kelly, Erasmus Schneider, Erica Lasek-Nesselquist |
| EPI_ISL_1226870 | WESTCHESTER MEDICAL CENTER | Wadsworth Center, New York State Department of Health | Kirsten St. George, Daryl M. Lamson, Alexis Russel, Matthew Shudt, Melissa A Leisner, Jonathan Plitnick, Navjot Singh, John Kelly, Erasmus Schneider, Erica Lasek-Nesselquist |
| EPI_ISL_1226871 | MONTEFIORE MEDICAL CENTER LABORATORIES | Wadsworth Center, New York State Department of Health | Kirsten St. George, Daryl M. Lamson, Alexis Russel, Matthew Shudt, Melissa A Leisner, Jonathan Plitnick, Navjot Singh, John Kelly, Erasmus Schneider, Erica Lasek-Nesselquist |
| EPI_ISL_1226872, EPI_ISL_1226873 | SUNY UPSTATE MEDICAL UNIVERSITY | Wadsworth Center, New York State Department of Health | Kirsten St. George, Daryl M. Lamson, Alexis Russel, Matthew Shudt, Melissa A Leisner, Jonathan Plitnick, Navjot Singh, John Kelly, Erasmus Schneider, Erica Lasek-Nesselquist |
| EPI_ISL_1226874, EPI_ISL_1226875 | Columbia University Irving Medical Center | Wadsworth Center, New York State Department of Health | Kirsten St. George, Daryl M. Lamson, Alexis Russel, Matthew Shudt, Melissa A Leisner, Jonathan Plitnick, Navjot Singh, John Kelly, Erasmus Schneider, Erica Lasek-Nesselquist |
| EPI_ISL_1226876, EPI_ISL_1226877, EPI_ISL_1226878, EPI_ISL_1226879, EPI_ISL_1226880, EPI_ISL_1226881, EPI_ISL_1226882, EPI_ISL_1226883, EPI_ISL_1226884, EPI_ISL_1226885, EPI_ISL_1226886, EPI_ISL_1226887, EPI_ISL_1226888, EPI_ISL_1226889, EPI_ISL_1226890 | | | |
| see above | MONTEFIORE MEDICAL CENTER LABORATORIES | Wadsworth Center, New York State Department of Health | Kirsten St. George, Daryl M. Lamson, Alexis Russel, Matthew Shudt, Melissa A Leisner, Jonathan Plitnick, Navjot Singh, John Kelly, Erasmus Schneider, Erica Lasek-Nesselquist |
| EPI_ISL_1226891, EPI_ISL_1226892 | WESTCHESTER MEDICAL CENTER | Wadsworth Center, New York State Department of Health | Kirsten St. George, Daryl M. Lamson, Alexis Russel, Matthew Shudt, Melissa A Leisner, Jonathan Plitnick, Navjot Singh, John Kelly, Erasmus Schneider, Erica Lasek-Nesselquist |
| EPI_ISL_1226893, EPI_ISL_1226894, EPI_ISL_1226895, EPI_ISL_1226896 | NYC Pandemic Response Lab | Wadsworth Center, New York State Department of Health | Kirsten St. George, Daryl M. Lamson, Alexis Russel, Matthew Shudt, Melissa A Leisner, Jonathan Plitnick, Navjot Singh, John Kelly, Erasmus Schneider, Erica Lasek-Nesselquist |
| EPI_ISL_1226897 | URMC LABS | Wadsworth Center, New York State Department of Health | Kirsten St. George, Daryl M. Lamson, Alexis Russel, Matthew Shudt, Melissa A Leisner, Jonathan Plitnick, Navjot Singh, John Kelly, Erasmus Schneider, Erica Lasek-Nesselquist |
| EPI_ISL_1226898, EPI_ISL_1226899 | NORTHWELL HEALTH LABORATORIES | Wadsworth Center, New York State Department of Health | Kirsten St. George, Daryl M. Lamson, Alexis Russel, Matthew Shudt, Melissa A Leisner, Jonathan Plitnick, Navjot Singh, John Kelly, Erasmus Schneider, Erica Lasek-Nesselquist |
| EPI_ISL_1226900 | NYC Pandemic Response Lab | Wadsworth Center, New York State Department of Health | Kirsten St. George, Daryl M. Lamson, Alexis Russel, Matthew Shudt, Melissa A Leisner, Jonathan Plitnick, Navjot Singh, John Kelly, Erasmus Schneider, Erica Lasek-Nesselquist |
| EPI_ISL_1226901, EPI_ISL_1226902, EPI_ISL_1226903, EPI_ISL_1226904, EPI_ISL_1226905, EPI_ISL_1226906 | Columbia University Irving Medical Center | Wadsworth Center, New York State Department of Health | Kirsten St. George, Daryl M. Lamson, Alexis Russel, Matthew Shudt, Melissa A Leisner, Jonathan Plitnick, Navjot Singh, John Kelly, Erasmus Schneider, Erica Lasek-Nesselquist |
| EPI_ISL_1226907 | MEMORIAL SLOAN KETTERING CANCER CENTER | Wadsworth Center, New York State Department of Health | Kirsten St. George, Daryl M. Lamson, Alexis Russel, Matthew Shudt, Melissa A Leisner, Jonathan Plitnick, Navjot Singh, John Kelly, Erasmus Schneider, Erica Lasek-Nesselquist |
| EPI_ISL_1226908 | WESTCHESTER MEDICAL CENTER | Wadsworth Center, New York State Department of Health | Kirsten St. George, Daryl M. Lamson, Alexis Russel, Matthew Shudt, Melissa A Leisner, Jonathan Plitnick, Navjot Singh, John Kelly, Erasmus Schneider, Erica Lasek-Nesselquist |
| EPI_ISL_1226909 | MONTEFIORE MEDICAL CENTER LABORATORIES | Wadsworth Center, New York State Department of Health | Kirsten St. George, Daryl M. Lamson, Alexis Russel, Matthew Shudt, Melissa A Leisner, Jonathan Plitnick, Navjot Singh, John Kelly, Erasmus Schneider, Erica Lasek-Nesselquist |
| EPI_ISL_1226910 | MEMORIAL SLOAN KETTERING CANCER CENTER | Wadsworth Center, New York State Department of Health | Kirsten St. George, Daryl M. Lamson, Alexis Russel, Matthew Shudt, Melissa A Leisner, Jonathan Plitnick, Navjot Singh, John Kelly, Erasmus Schneider, Erica Lasek-Nesselquist |
| EPI_ISL_1226911, EPI_ISL_1226912 | NORTHWELL HEALTH LABORATORIES | Wadsworth Center, New York State Department of Health | Kirsten St. George, Daryl M. Lamson, Alexis Russel, Matthew Shudt, Melissa A Leisner, Jonathan Plitnick, Navjot Singh, John Kelly, Erasmus Schneider, Erica Lasek-Nesselquist |
| EPI_ISL_1226913, EPI_ISL_1226914 | NYC Pandemic Response Lab | Wadsworth Center, New York State Department of Health | Kirsten St. George, Daryl M. Lamson, Alexis Russel, Matthew Shudt, Melissa A Leisner, Jonathan Plitnick, Navjot Singh, John Kelly, Erasmus Schneider, Erica Lasek-Nesselquist |

| | | | |
|---|---|---|---|
| EPI_ISL_1226915 | New York Presbyterian Hospital | Wadsworth Center, New York State Department of Health | Kirsten St. George, Daryl M. Lamson, Alexis Russel, Matthew Shudt, Melissa A Leisner, Jonathan Plitnick, Navjot Singh, John Kelly, Erasmus Schneider, Erica Lasek-Nesselquist |
| EPI_ISL_1226916, EPI_ISL_1226917, EPI_ISL_1226918, EPI_ISL_1226919, EPI_ISL_1226920, EPI_ISL_1226921, EPI_ISL_1226922, EPI_ISL_1226923 | NYC Pandemic Response Lab | Wadsworth Center, New York State Department of Health | Kirsten St. George, Daryl M. Lamson, Alexis Russel, Matthew Shudt, Melissa A Leisner, Jonathan Plitnick, Navjot Singh, John Kelly, Erasmus Schneider, Erica Lasek-Nesselquist |
| EPI_ISL_1226924, EPI_ISL_1226925 | WESTCHESTER MEDICAL CENTER | Wadsworth Center, New York State Department of Health | Kirsten St. George, Daryl M. Lamson, Alexis Russel, Matthew Shudt, Melissa A Leisner, Jonathan Plitnick, Navjot Singh, John Kelly, Erasmus Schneider, Erica Lasek-Nesselquist |
| EPI_ISL_1226926 | Columbia University Irving Medical Center | Wadsworth Center, New York State Department of Health | Kirsten St. George, Daryl M. Lamson, Alexis Russel, Matthew Shudt, Melissa A Leisner, Jonathan Plitnick, Navjot Singh, John Kelly, Erasmus Schneider, Erica Lasek-Nesselquist |
| EPI_ISL_1226927, EPI_ISL_1226928, EPI_ISL_1226929 | WESTCHESTER MEDICAL CENTER | Wadsworth Center, New York State Department of Health | Kirsten St. George, Daryl M. Lamson, Alexis Russel, Matthew Shudt, Melissa A Leisner, Jonathan Plitnick, Navjot Singh, John Kelly, Erasmus Schneider, Erica Lasek-Nesselquist |
| EPI_ISL_1226930 | Columbia University Irving Medical Center | Wadsworth Center, New York State Department of Health | Kirsten St. George, Daryl M. Lamson, Alexis Russel, Matthew Shudt, Melissa A Leisner, Jonathan Plitnick, Navjot Singh, John Kelly, Erasmus Schneider, Erica Lasek-Nesselquist |
| EPI_ISL_1226931 | MONTEFIORE MEDICAL CENTER LABORATORIES | Wadsworth Center, New York State Department of Health | Kirsten St. George, Daryl M. Lamson, Alexis Russel, Matthew Shudt, Melissa A Leisner, Jonathan Plitnick, Navjot Singh, John Kelly, Erasmus Schneider, Erica Lasek-Nesselquist |
| EPI_ISL_1226932 | BIO-REFERENCE LABORATORIES | Wadsworth Center, New York State Department of Health | Kirsten St. George, Daryl M. Lamson, Alexis Russel, Matthew Shudt, Melissa A Leisner, Jonathan Plitnick, Navjot Singh, John Kelly, Erasmus Schneider, Erica Lasek-Nesselquist |
| EPI_ISL_1226933 | Wadsworth Center, New York State Department of Health | Wadsworth Center, New York State Department of Health | Kirsten St. George, Daryl M. Lamson, Alexis Russel, Matthew Shudt, Melissa A Leisner, Jonathan Plitnick, Navjot Singh, John Kelly, Erasmus Schneider, Erica Lasek-Nesselquist |
| EPI_ISL_1226934 | MONTEFIORE MEDICAL CENTER LABORATORIES | Wadsworth Center, New York State Department of Health | Kirsten St. George, Daryl M. Lamson, Alexis Russel, Matthew Shudt, Melissa A Leisner, Jonathan Plitnick, Navjot Singh, John Kelly, Erasmus Schneider, Erica Lasek-Nesselquist |
| EPI_ISL_1226935 | NYC Pandemic Response Lab | Wadsworth Center, New York State Department of Health | Kirsten St. George, Daryl M. Lamson, Alexis Russel, Matthew Shudt, Melissa A Leisner, Jonathan Plitnick, Navjot Singh, John Kelly, Erasmus Schneider, Erica Lasek-Nesselquist |
| EPI_ISL_1226936, EPI_ISL_1226937 | MONTEFIORE MEDICAL CENTER LABORATORIES | Wadsworth Center, New York State Department of Health | Kirsten St. George, Daryl M. Lamson, Alexis Russel, Matthew Shudt, Melissa A Leisner, Jonathan Plitnick, Navjot Singh, John Kelly, Erasmus Schneider, Erica Lasek-Nesselquist |
| EPI_ISL_1226938 | Wadsworth Center, New York State Department of Health | Wadsworth Center, New York State Department of Health | Kirsten St. George, Daryl M. Lamson, Alexis Russel, Matthew Shudt, Melissa A Leisner, Jonathan Plitnick, Navjot Singh, John Kelly, Erasmus Schneider, Erica Lasek-Nesselquist |
| EPI_ISL_1226939, EPI_ISL_1226940, EPI_ISL_1226941, EPI_ISL_1226942, EPI_ISL_1226943, EPI_ISL_1226944, EPI_ISL_1226945, EPI_ISL_1226946, EPI_ISL_1226947, EPI_ISL_1226948, EPI_ISL_1226949, EPI_ISL_1226950, EPI_ISL_1226951, EPI_ISL_1226952, EPI_ISL_1226953, EPI_ISL_1226954, EPI_ISL_1226955, EPI_ISL_1226956, EPI_ISL_1226957, EPI_ISL_1226958, EPI_ISL_1226959, EPI_ISL_1226960, EPI_ISL_1226961, EPI_ISL_1226962 | | | |
| see above | MONTEFIORE MEDICAL CENTER LABORATORIES | Wadsworth Center, New York State Department of Health | Kirsten St. George, Daryl M. Lamson, Alexis Russel, Matthew Shudt, Melissa A Leisner, Jonathan Plitnick, Navjot Singh, John Kelly, Erasmus Schneider, Erica Lasek-Nesselquist |
| EPI_ISL_1226963 | NYC Pandemic Response Lab | Wadsworth Center, New York State Department of Health | Kirsten St. George, Daryl M. Lamson, Alexis Russel, Matthew Shudt, Melissa A Leisner, Jonathan Plitnick, Navjot Singh, John Kelly, Erasmus Schneider, Erica Lasek-Nesselquist |
| EPI_ISL_1226964, EPI_ISL_1226965, EPI_ISL_1226966, EPI_ISL_1226967, EPI_ISL_1226968, EPI_ISL_1226969 | MONTEFIORE MEDICAL CENTER LABORATORIES | Wadsworth Center, New York State Department of Health | Kirsten St. George, Daryl M. Lamson, Alexis Russel, Matthew Shudt, Melissa A Leisner, Jonathan Plitnick, Navjot Singh, John Kelly, Erasmus Schneider, Erica Lasek-Nesselquist |
| EPI_ISL_1226970 | WESTCHESTER MEDICAL CENTER | Wadsworth Center, New York State Department of Health | Kirsten St. George, Daryl M. Lamson, Alexis Russel, Matthew Shudt, Melissa A Leisner, Jonathan Plitnick, Navjot Singh, John Kelly, Erasmus Schneider, Erica Lasek-Nesselquist |
| EPI_ISL_1226971, EPI_ISL_1226972 | MONTEFIORE MEDICAL CENTER LABORATORIES | Wadsworth Center, New York State Department of Health | Kirsten St. George, Daryl M. Lamson, Alexis Russel, Matthew Shudt, Melissa A Leisner, Jonathan Plitnick, Navjot Singh, John Kelly, Erasmus Schneider, Erica Lasek-Nesselquist |
| EPI_ISL_1226973 | WESTCHESTER MEDICAL CENTER | Wadsworth Center, New York State Department of Health | Kirsten St. George, Daryl M. Lamson, Alexis Russel, Matthew Shudt, Melissa A Leisner, Jonathan Plitnick, Navjot Singh, John Kelly, Erasmus Schneider, Erica Lasek-Nesselquist |
| EPI_ISL_1226974, EPI_ISL_1226975, EPI_ISL_1226976, EPI_ISL_1226977, EPI_ISL_1226978, EPI_ISL_1226979 | MONTEFIORE MEDICAL CENTER LABORATORIES | Wadsworth Center, New York State Department of Health | Kirsten St. George, Daryl M. Lamson, Alexis Russel, Matthew Shudt, Melissa A Leisner, Jonathan Plitnick, Navjot Singh, John Kelly, Erasmus Schneider, Erica Lasek-Nesselquist |
| EPI_ISL_1226980 | MEMORIAL SLOAN KETTERING CANCER CENTER | Wadsworth Center, New York State Department of Health | Kirsten St. George, Daryl M. Lamson, Alexis Russel, Matthew Shudt, Melissa A Leisner, Jonathan Plitnick, Navjot Singh, John Kelly, Erasmus Schneider, Erica Lasek-Nesselquist |
| EPI_ISL_1226981 | SUNY UPSTATE MEDICAL UNIVERSITY | Wadsworth Center, New York State Department of Health | Kirsten St. George, Daryl M. Lamson, Alexis Russel, Matthew Shudt, Melissa A Leisner, Jonathan Plitnick, Navjot Singh, John Kelly, Erasmus Schneider, Erica Lasek-Nesselquist |
| EPI_ISL_1226982, EPI_ISL_1226983, EPI_ISL_1226984 | NYC Pandemic Response Lab | Wadsworth Center, New York State Department of Health | Kirsten St. George, Daryl M. Lamson, Alexis Russel, Matthew Shudt, Melissa A Leisner, Jonathan Plitnick, Navjot Singh, John Kelly, Erasmus Schneider, Erica Lasek-Nesselquist |
| EPI_ISL_1226985 | BIO-REFERENCE LABORATORIES | Wadsworth Center, New York State Department of Health | Kirsten St. George, Daryl M. Lamson, Alexis Russel, Matthew Shudt, Melissa A Leisner, Jonathan Plitnick, Navjot Singh, John Kelly, Erasmus Schneider, Erica Lasek-Nesselquist |
| EPI_ISL_1226986 | MEMORIAL SLOAN KETTERING CANCER CENTER | Wadsworth Center, New York State Department of Health | Kirsten St. George, Daryl M. Lamson, Alexis Russel, Matthew Shudt, Melissa A Leisner, Jonathan Plitnick, Navjot Singh, John Kelly, Erasmus Schneider, Erica Lasek-Nesselquist |
| EPI_ISL_1226987 | NYC Pandemic Response Lab | Wadsworth Center, New York State Department of Health | Kirsten St. George, Daryl M. Lamson, Alexis Russel, Matthew Shudt, Melissa A Leisner, Jonathan Plitnick, Navjot Singh, John Kelly, Erasmus Schneider, Erica Lasek-Nesselquist |
| EPI_ISL_1226988, EPI_ISL_1226989, EPI_ISL_1226990, EPI_ISL_1226991, EPI_ISL_1226992, EPI_ISL_1226993, EPI_ISL_1226994, EPI_ISL_1226995, EPI_ISL_1226996, EPI_ISL_1226997 | Columbia University Irving Medical Center | Wadsworth Center, New York State Department of Health | Kirsten St. George, Daryl M. Lamson, Alexis Russel, Matthew Shudt, Melissa A Leisner, Jonathan Plitnick, Navjot Singh, John Kelly, Erasmus Schneider, Erica Lasek-Nesselquist |
| EPI_ISL_1226998, EPI_ISL_1226999 | NYC Pandemic Response Lab | Wadsworth Center, New York State Department of Health | Kirsten St. George, Daryl M. Lamson, Alexis Russel, Matthew Shudt, Melissa A Leisner, Jonathan Plitnick, Navjot Singh, John Kelly, Erasmus Schneider, Erica Lasek-Nesselquist |
| EPI_ISL_1227000 | New York Presbyterian Hospital | Wadsworth Center, New York State Department of Health | Kirsten St. George, Daryl M. Lamson, Alexis Russel, Matthew Shudt, Melissa A Leisner, Jonathan Plitnick, Navjot Singh, John Kelly, Erasmus Schneider, Erica Lasek-Nesselquist |
| EPI_ISL_1227001 | NYC Pandemic Response Lab | Wadsworth Center, New York State Department of Health | Kirsten St. George, Daryl M. Lamson, Alexis Russel, Matthew Shudt, Melissa A Leisner, Jonathan Plitnick, Navjot Singh, John Kelly, Erasmus Schneider, Erica Lasek-Nesselquist |
| EPI_ISL_1227002, EPI_ISL_1227003, EPI_ISL_1227004 | Columbia University Irving Medical Center | Wadsworth Center, New York State Department of Health | Kirsten St. George, Daryl M. Lamson, Alexis Russel, Matthew Shudt, Melissa A Leisner, Jonathan Plitnick, Navjot Singh, John Kelly, Erasmus Schneider, Erica Lasek-Nesselquist |
| EPI_ISL_1227005, EPI_ISL_1227006 | NYC Pandemic Response Lab | Wadsworth Center, New York State Department of Health | Kirsten St. George, Daryl M. Lamson, Alexis Russel, Matthew Shudt, Melissa A Leisner, Jonathan Plitnick, Navjot Singh, John Kelly, Erasmus Schneider, Erica Lasek-Nesselquist |
| EPI_ISL_1227007 | WESTCHESTER MEDICAL CENTER | Wadsworth Center, New York State Department of Health | Kirsten St. George, Daryl M. Lamson, Alexis Russel, Matthew Shudt, Melissa A Leisner, Jonathan Plitnick, Navjot Singh, John Kelly, Erasmus Schneider, Erica Lasek-Nesselquist |
| EPI_ISL_1227008 | NORTH SHORE UNIVERSITY HOSPITAL | Wadsworth Center, New York State | Kirsten St. George, Daryl M. Lamson, Alexis Russel, Matthew Shudt, Melissa A Leisner, Jonathan Plitnick, Navjot Singh, John Kelly, Erasmus Schneider, Erica Lasek-Nesselquist |

| | | | |
|---|---|---|---|
| | | Department of Health | |
| EPI_ISL_1227009, EPI_ISL_1227010, EPI_ISL_1227011, EPI_ISL_1227012, EPI_ISL_1227013 | NYC Pandemic Response Lab | Wadsworth Center, New York State Department of Health | Kirsten St. George, Daryl M. Lamson, Alexis Russel, Matthew Shudt, Melissa A Leisner, Jonathan Plitnick, Navjot Singh, John Kelly, Erasmus Schneider, Erica Lasek-Nesselquist |
| EPI_ISL_1227014 | NORTH SHORE UNIVERSITY HOSPITAL | Wadsworth Center, New York State Department of Health | Kirsten St. George, Daryl M. Lamson, Alexis Russel, Matthew Shudt, Melissa A Leisner, Jonathan Plitnick, Navjot Singh, John Kelly, Erasmus Schneider, Erica Lasek-Nesselquist |
| EPI_ISL_1227015 | NYC Pandemic Response Lab | Wadsworth Center, New York State Department of Health | Kirsten St. George, Daryl M. Lamson, Alexis Russel, Matthew Shudt, Melissa A Leisner, Jonathan Plitnick, Navjot Singh, John Kelly, Erasmus Schneider, Erica Lasek-Nesselquist |
| EPI_ISL_1227016, EPI_ISL_1227017, EPI_ISL_1227018 | MONTEFIORE MEDICAL CENTER LABORATORIES | Wadsworth Center, New York State Department of Health | Kirsten St. George, Daryl M. Lamson, Alexis Russel, Matthew Shudt, Melissa A Leisner, Jonathan Plitnick, Navjot Singh, John Kelly, Erasmus Schneider, Erica Lasek-Nesselquist |
| EPI_ISL_1227019, EPI_ISL_1227020 | WESTCHESTER MEDICAL CENTER | Wadsworth Center, New York State Department of Health | Kirsten St. George, Daryl M. Lamson, Alexis Russel, Matthew Shudt, Melissa A Leisner, Jonathan Plitnick, Navjot Singh, John Kelly, Erasmus Schneider, Erica Lasek-Nesselquist |
| EPI_ISL_1227021, EPI_ISL_1227022 | MONTEFIORE MEDICAL CENTER LABORATORIES | Wadsworth Center, New York State Department of Health | Kirsten St. George, Daryl M. Lamson, Alexis Russel, Matthew Shudt, Melissa A Leisner, Jonathan Plitnick, Navjot Singh, John Kelly, Erasmus Schneider, Erica Lasek-Nesselquist |
| EPI_ISL_1227023 | WESTCHESTER MEDICAL CENTER | Wadsworth Center, New York State Department of Health | Kirsten St. George, Daryl M. Lamson, Alexis Russel, Matthew Shudt, Melissa A Leisner, Jonathan Plitnick, Navjot Singh, John Kelly, Erasmus Schneider, Erica Lasek-Nesselquist |
| EPI_ISL_1227024, EPI_ISL_1227025 | MONTEFIORE MEDICAL CENTER LABORATORIES | Wadsworth Center, New York State Department of Health | Kirsten St. George, Daryl M. Lamson, Alexis Russel, Matthew Shudt, Melissa A Leisner, Jonathan Plitnick, Navjot Singh, John Kelly, Erasmus Schneider, Erica Lasek-Nesselquist |
| EPI_ISL_1227026 | NORTH SHORE UNIVERSITY HOSPITAL | Wadsworth Center, New York State Department of Health | Kirsten St. George, Daryl M. Lamson, Alexis Russel, Matthew Shudt, Melissa A Leisner, Jonathan Plitnick, Navjot Singh, John Kelly, Erasmus Schneider, Erica Lasek-Nesselquist |
| EPI_ISL_1227027, EPI_ISL_1227028 | NYC Pandemic Response Lab | Wadsworth Center, New York State Department of Health | Kirsten St. George, Daryl M. Lamson, Alexis Russel, Matthew Shudt, Melissa A Leisner, Jonathan Plitnick, Navjot Singh, John Kelly, Erasmus Schneider, Erica Lasek-Nesselquist |
| EPI_ISL_1227029 | ALBANY MEDICAL CENTER | Wadsworth Center, New York State Department of Health | Kirsten St. George, Daryl M. Lamson, Alexis Russel, Matthew Shudt, Melissa A Leisner, Jonathan Plitnick, Navjot Singh, John Kelly, Erasmus Schneider, Erica Lasek-Nesselquist |
| EPI_ISL_1227030 | Wadsworth Center, New York State Department of Health | Wadsworth Center, New York State Department of Health | Kirsten St. George, Daryl M. Lamson, Alexis Russel, Matthew Shudt, Melissa A Leisner, Jonathan Plitnick, Navjot Singh, John Kelly, Erasmus Schneider, Erica Lasek-Nesselquist |
| EPI_ISL_1227031, EPI_ISL_1227032 | ALBANY MEDICAL CENTER | Wadsworth Center, New York State Department of Health | Kirsten St. George, Daryl M. Lamson, Alexis Russel, Matthew Shudt, Melissa A Leisner, Jonathan Plitnick, Navjot Singh, John Kelly, Erasmus Schneider, Erica Lasek-Nesselquist |
| EPI_ISL_1227033, EPI_ISL_1227034 | MEMORIAL SLOAN KETTERING CANCER CENTER | Wadsworth Center, New York State Department of Health | Kirsten St. George, Daryl M. Lamson, Alexis Russel, Matthew Shudt, Melissa A Leisner, Jonathan Plitnick, Navjot Singh, John Kelly, Erasmus Schneider, Erica Lasek-Nesselquist |
| EPI_ISL_1227035, EPI_ISL_1227036, EPI_ISL_1227037, EPI_ISL_1227038, EPI_ISL_1227039 | MONTEFIORE MEDICAL CENTER LABORATORIES | Wadsworth Center, New York State Department of Health | Kirsten St. George, Daryl M. Lamson, Alexis Russel, Matthew Shudt, Melissa A Leisner, Jonathan Plitnick, Navjot Singh, John Kelly, Erasmus Schneider, Erica Lasek-Nesselquist |
| EPI_ISL_1227040 | WESTCHESTER MEDICAL CENTER | Wadsworth Center, New York State Department of Health | Kirsten St. George, Daryl M. Lamson, Alexis Russel, Matthew Shudt, Melissa A Leisner, Jonathan Plitnick, Navjot Singh, John Kelly, Erasmus Schneider, Erica Lasek-Nesselquist |
| EPI_ISL_1227041, EPI_ISL_1227042, EPI_ISL_1227043, EPI_ISL_1227044, EPI_ISL_1227045, EPI_ISL_1227046, EPI_ISL_1227047, EPI_ISL_1227048, EPI_ISL_1227049, EPI_ISL_1227050, EPI_ISL_1227051, EPI_ISL_1227052, EPI_ISL_1227053, EPI_ISL_1227054, EPI_ISL_1227055, EPI_ISL_1227056, EPI_ISL_1227057, EPI_ISL_1227058, EPI_ISL_1227059, EPI_ISL_1227060, EPI_ISL_1227061, EPI_ISL_1227062, EPI_ISL_1227063, EPI_ISL_1227064, EPI_ISL_1227065, EPI_ISL_1227066, EPI_ISL_1227067 | | | |
| see above | MONTEFIORE MEDICAL CENTER LABORATORIES | Wadsworth Center, New York State Department of Health | Kirsten St. George, Daryl M. Lamson, Alexis Russel, Matthew Shudt, Melissa A Leisner, Jonathan Plitnick, Navjot Singh, John Kelly, Erasmus Schneider, Erica Lasek-Nesselquist |
| EPI_ISL_1227068, EPI_ISL_1227069 | Columbia University Irving Medical Center | Wadsworth Center, New York State Department of Health | Kirsten St. George, Daryl M. Lamson, Alexis Russel, Matthew Shudt, Melissa A Leisner, Jonathan Plitnick, Navjot Singh, John Kelly, Erasmus Schneider, Erica Lasek-Nesselquist |
| EPI_ISL_1227070, EPI_ISL_1227071, EPI_ISL_1227072, EPI_ISL_1227073, EPI_ISL_1227074, EPI_ISL_1227075, EPI_ISL_1227076, EPI_ISL_1227077, EPI_ISL_1227078, EPI_ISL_1227079, EPI_ISL_1227080 | | | |
| see above | MONTEFIORE MEDICAL CENTER LABORATORIES | Wadsworth Center, New York State Department of Health | Kirsten St. George, Daryl M. Lamson, Alexis Russel, Matthew Shudt, Melissa A Leisner, Jonathan Plitnick, Navjot Singh, John Kelly, Erasmus Schneider, Erica Lasek-Nesselquist |
| EPI_ISL_1227081 | NYC Pandemic Response Lab | Wadsworth Center, New York State Department of Health | Kirsten St. George, Daryl M. Lamson, Alexis Russel, Matthew Shudt, Melissa A Leisner, Jonathan Plitnick, Navjot Singh, John Kelly, Erasmus Schneider, Erica Lasek-Nesselquist |
| EPI_ISL_1227082, EPI_ISL_1227083, EPI_ISL_1227084, EPI_ISL_1227085, EPI_ISL_1227086 | MONTEFIORE MEDICAL CENTER LABORATORIES | Wadsworth Center, New York State Department of Health | Kirsten St. George, Daryl M. Lamson, Alexis Russel, Matthew Shudt, Melissa A Leisner, Jonathan Plitnick, Navjot Singh, John Kelly, Erasmus Schneider, Erica Lasek-Nesselquist |
| EPI_ISL_1227087 | NYC Pandemic Response Lab | Wadsworth Center, New York State Department of Health | Kirsten St. George, Daryl M. Lamson, Alexis Russel, Matthew Shudt, Melissa A Leisner, Jonathan Plitnick, Navjot Singh, John Kelly, Erasmus Schneider, Erica Lasek-Nesselquist |
| EPI_ISL_1227088, EPI_ISL_1227089 | Columbia University Irving Medical Center | Wadsworth Center, New York State Department of Health | Kirsten St. George, Daryl M. Lamson, Alexis Russel, Matthew Shudt, Melissa A Leisner, Jonathan Plitnick, Navjot Singh, John Kelly, Erasmus Schneider, Erica Lasek-Nesselquist |
| EPI_ISL_1227090, EPI_ISL_1227091, EPI_ISL_1227092, EPI_ISL_1227093, EPI_ISL_1227094 | MONTEFIORE MEDICAL CENTER LABORATORIES | Wadsworth Center, New York State Department of Health | Kirsten St. George, Daryl M. Lamson, Alexis Russel, Matthew Shudt, Melissa A Leisner, Jonathan Plitnick, Navjot Singh, John Kelly, Erasmus Schneider, Erica Lasek-Nesselquist |
| EPI_ISL_1227095 | NYC Pandemic Response Lab | Wadsworth Center, New York State Department of Health | Kirsten St. George, Daryl M. Lamson, Alexis Russel, Matthew Shudt, Melissa A Leisner, Jonathan Plitnick, Navjot Singh, John Kelly, Erasmus Schneider, Erica Lasek-Nesselquist |
| EPI_ISL_1227096 | Columbia University Irving Medical Center | Wadsworth Center, New York State Department of Health | Kirsten St. George, Daryl M. Lamson, Alexis Russel, Matthew Shudt, Melissa A Leisner, Jonathan Plitnick, Navjot Singh, John Kelly, Erasmus Schneider, Erica Lasek-Nesselquist |
| EPI_ISL_1227097, EPI_ISL_1227098, EPI_ISL_1227099, EPI_ISL_1227100, EPI_ISL_1227101, EPI_ISL_1227102, EPI_ISL_1227103, EPI_ISL_1227104, EPI_ISL_1227105, EPI_ISL_1227106, EPI_ISL_1227107, EPI_ISL_1227108 | | | |
| see above | MONTEFIORE MEDICAL CENTER LABORATORIES | Wadsworth Center, New York State Department of Health | Kirsten St. George, Daryl M. Lamson, Alexis Russel, Matthew Shudt, Melissa A Leisner, Jonathan Plitnick, Navjot Singh, John Kelly, Erasmus Schneider, Erica Lasek-Nesselquist |
| EPI_ISL_1227109 | NYC Pandemic Response Lab | Wadsworth Center, New York State Department of Health | Kirsten St. George, Daryl M. Lamson, Alexis Russel, Matthew Shudt, Melissa A Leisner, Jonathan Plitnick, Navjot Singh, John Kelly, Erasmus Schneider, Erica Lasek-Nesselquist |
| EPI_ISL_1227110, EPI_ISL_1227111, EPI_ISL_1227112, EPI_ISL_1227113, EPI_ISL_1227114, EPI_ISL_1227115, EPI_ISL_1227116, EPI_ISL_1227117, EPI_ISL_1227118, EPI_ISL_1227119, EPI_ISL_1227120, EPI_ISL_1227121, EPI_ISL_1227122, EPI_ISL_1227123, EPI_ISL_1227124, EPI_ISL_1227125, EPI_ISL_1227126, EPI_ISL_1227127, EPI_ISL_1227128 | | | |
| see above | MONTEFIORE MEDICAL CENTER LABORATORIES | Wadsworth Center, New York State Department of Health | Kirsten St. George, Daryl M. Lamson, Alexis Russel, Matthew Shudt, Melissa A Leisner, Jonathan Plitnick, Navjot Singh, John Kelly, Erasmus Schneider, Erica Lasek-Nesselquist |
| EPI_ISL_1227129 | SUNY UPSTATE MEDICAL UNIVERSITY | Wadsworth Center, New York State Department of Health | Kirsten St. George, Daryl M. Lamson, Alexis Russel, Matthew Shudt, Melissa A Leisner, Jonathan Plitnick, Navjot Singh, John Kelly, Erasmus Schneider, Erica Lasek-Nesselquist |
| EPI_ISL_1227130, EPI_ISL_1227131, EPI_ISL_1227132, EPI_ISL_1227133 | WESTCHESTER MEDICAL CENTER | Wadsworth Center, New York State Department of Health | Kirsten St. George, Daryl M. Lamson, Alexis Russel, Matthew Shudt, Melissa A Leisner, Jonathan Plitnick, Navjot Singh, John Kelly, Erasmus Schneider, Erica Lasek-Nesselquist |
| EPI_ISL_1227134, EPI_ISL_1227135, EPI_ISL_1227136, EPI_ISL_1227137, EPI_ISL_1227138, EPI_ISL_1227139, EPI_ISL_1227140, EPI_ISL_1227141, EPI_ISL_1227142, EPI_ISL_1227143, EPI_ISL_1227144, EPI_ISL_1227145, EPI_ISL_1227146, EPI_ISL_1227147, EPI_ISL_1227148, EPI_ISL_1227149 | | | |
| see above | NYC Pandemic Response Lab | Wadsworth Center, New York State Department of Health | Kirsten St. George, Daryl M. Lamson, Alexis Russel, Matthew Shudt, Melissa A Leisner, Jonathan Plitnick, Navjot Singh, John Kelly, Erasmus Schneider, Erica Lasek-Nesselquist |
| EPI_ISL_1227150 | MONTEFIORE MEDICAL CENTER LABORATORIES | Wadsworth Center, New York State Department of Health | Kirsten St. George, Daryl M. Lamson, Alexis Russel, Matthew Shudt, Melissa A Leisner, Jonathan Plitnick, Navjot Singh, John Kelly, Erasmus Schneider, Erica Lasek-Nesselquist |
| EPI_ISL_1227151 | NORTH SHORE UNIVERSITY HOSPITAL | Wadsworth Center, New York State Department of Health | Kirsten St. George, Daryl M. Lamson, Alexis Russel, Matthew Shudt, Melissa A Leisner, Jonathan Plitnick, Navjot Singh, John Kelly, Erasmus Schneider, Erica Lasek-Nesselquist |
| EPI_ISL_1227152 | NYC Pandemic Response Lab | Wadsworth Center, New York State Department of Health | Kirsten St. George, Daryl M. Lamson, Alexis Russel, Matthew Shudt, Melissa A Leisner, Jonathan Plitnick, Navjot Singh, John Kelly, Erasmus Schneider, Erica Lasek-Nesselquist |

| | | | |
|---|---|---|---|
| EPI_ISL_1227153, EPI_ISL_1227154 | NORTHWELL HEALTH LABORATORIES | Wadsworth Center, New York State Department of Health | Kirsten St. George, Daryl M. Lamson, Alexis Russel, Matthew Shudt, Melissa A Leisner, Jonathan Plitnick, Navjot Singh, John Kelly, Erasmus Schneider, Erica Lasek-Nesselquist |
| EPI_ISL_1227155, EPI_ISL_1227156, EPI_ISL_1227157, EPI_ISL_1227158, EPI_ISL_1227159, EPI_ISL_1227160, EPI_ISL_1227161 | NYC Pandemic Response Lab | Wadsworth Center, New York State Department of Health | Kirsten St. George, Daryl M. Lamson, Alexis Russel, Matthew Shudt, Melissa A Leisner, Jonathan Plitnick, Navjot Singh, John Kelly, Erasmus Schneider, Erica Lasek-Nesselquist |
| EPI_ISL_1227162 | TEMPUS LABS INC | Wadsworth Center, New York State Department of Health | Kirsten St. George, Daryl M. Lamson, Alexis Russel, Matthew Shudt, Melissa A Leisner, Jonathan Plitnick, Navjot Singh, John Kelly, Erasmus Schneider, Erica Lasek-Nesselquist |
| EPI_ISL_1227163 | Columbia University Irving Medical Center | Wadsworth Center, New York State Department of Health | Kirsten St. George, Daryl M. Lamson, Alexis Russel, Matthew Shudt, Melissa A Leisner, Jonathan Plitnick, Navjot Singh, John Kelly, Erasmus Schneider, Erica Lasek-Nesselquist |
| EPI_ISL_1227164, EPI_ISL_1227165 | New York Presbyterian Hospital | Wadsworth Center, New York State Department of Health | Kirsten St. George, Daryl M. Lamson, Alexis Russel, Matthew Shudt, Melissa A Leisner, Jonathan Plitnick, Navjot Singh, John Kelly, Erasmus Schneider, Erica Lasek-Nesselquist |
| EPI_ISL_1227166 | Columbia University Irving Medical Center | Wadsworth Center, New York State Department of Health | Kirsten St. George, Daryl M. Lamson, Alexis Russel, Matthew Shudt, Melissa A Leisner, Jonathan Plitnick, Navjot Singh, John Kelly, Erasmus Schneider, Erica Lasek-Nesselquist |
| EPI_ISL_1227167, EPI_ISL_1227168, EPI_ISL_1227169, EPI_ISL_1227170, EPI_ISL_1227171, EPI_ISL_1227172 | NYC Pandemic Response Lab | Wadsworth Center, New York State Department of Health | Kirsten St. George, Daryl M. Lamson, Alexis Russel, Matthew Shudt, Melissa A Leisner, Jonathan Plitnick, Navjot Singh, John Kelly, Erasmus Schneider, Erica Lasek-Nesselquist |
| EPI_ISL_1227173 | SUNY UPSTATE MEDICAL UNIVERSITY | Wadsworth Center, New York State Department of Health | Kirsten St. George, Daryl M. Lamson, Alexis Russel, Matthew Shudt, Melissa A Leisner, Jonathan Plitnick, Navjot Singh, John Kelly, Erasmus Schneider, Erica Lasek-Nesselquist |
| EPI_ISL_1227174 | WESTCHESTER MEDICAL CENTER | Wadsworth Center, New York State Department of Health | Kirsten St. George, Daryl M. Lamson, Alexis Russel, Matthew Shudt, Melissa A Leisner, Jonathan Plitnick, Navjot Singh, John Kelly, Erasmus Schneider, Erica Lasek-Nesselquist |
| EPI_ISL_1227175 | MEMORIAL SLOAN KETTERING CANCER CENTER | Wadsworth Center, New York State Department of Health | Kirsten St. George, Daryl M. Lamson, Alexis Russel, Matthew Shudt, Melissa A Leisner, Jonathan Plitnick, Navjot Singh, John Kelly, Erasmus Schneider, Erica Lasek-Nesselquist |
| EPI_ISL_1227176, EPI_ISL_1227177 | WESTCHESTER MEDICAL CENTER | Wadsworth Center, New York State Department of Health | Kirsten St. George, Daryl M. Lamson, Alexis Russel, Matthew Shudt, Melissa A Leisner, Jonathan Plitnick, Navjot Singh, John Kelly, Erasmus Schneider, Erica Lasek-Nesselquist |
| EPI_ISL_1227178, EPI_ISL_1227179, EPI_ISL_1227180 | MEMORIAL SLOAN KETTERING CANCER CENTER | Wadsworth Center, New York State Department of Health | Kirsten St. George, Daryl M. Lamson, Alexis Russel, Matthew Shudt, Melissa A Leisner, Jonathan Plitnick, Navjot Singh, John Kelly, Erasmus Schneider, Erica Lasek-Nesselquist |
| EPI_ISL_1227181 | NORTHWELL HEALTH LABORATORIES | Wadsworth Center, New York State Department of Health | Kirsten St. George, Daryl M. Lamson, Alexis Russel, Matthew Shudt, Melissa A Leisner, Jonathan Plitnick, Navjot Singh, John Kelly, Erasmus Schneider, Erica Lasek-Nesselquist |
| EPI_ISL_1227182, EPI_ISL_1227183 | New York Presbyterian Hospital | Wadsworth Center, New York State Department of Health | Kirsten St. George, Daryl M. Lamson, Alexis Russel, Matthew Shudt, Melissa A Leisner, Jonathan Plitnick, Navjot Singh, John Kelly, Erasmus Schneider, Erica Lasek-Nesselquist |
| EPI_ISL_1227184 | NYC Pandemic Response Lab | Wadsworth Center, New York State Department of Health | Kirsten St. George, Daryl M. Lamson, Alexis Russel, Matthew Shudt, Melissa A Leisner, Jonathan Plitnick, Navjot Singh, John Kelly, Erasmus Schneider, Erica Lasek-Nesselquist |
| EPI_ISL_1227185 | New York Presbyterian Hospital | Wadsworth Center, New York State Department of Health | Kirsten St. George, Daryl M. Lamson, Alexis Russel, Matthew Shudt, Melissa A Leisner, Jonathan Plitnick, Navjot Singh, John Kelly, Erasmus Schneider, Erica Lasek-Nesselquist |
| EPI_ISL_1227186, EPI_ISL_1227187, EPI_ISL_1227188 | NYC Pandemic Response Lab | Wadsworth Center, New York State Department of Health | Kirsten St. George, Daryl M. Lamson, Alexis Russel, Matthew Shudt, Melissa A Leisner, Jonathan Plitnick, Navjot Singh, John Kelly, Erasmus Schneider, Erica Lasek-Nesselquist |
| EPI_ISL_1227189 | WESTCHESTER MEDICAL CENTER | Wadsworth Center, New York State Department of Health | Kirsten St. George, Daryl M. Lamson, Alexis Russel, Matthew Shudt, Melissa A Leisner, Jonathan Plitnick, Navjot Singh, John Kelly, Erasmus Schneider, Erica Lasek-Nesselquist |
| EPI_ISL_1227190, EPI_ISL_1227191, EPI_ISL_1227192, EPI_ISL_1227193, EPI_ISL_1227194, EPI_ISL_1227195, EPI_ISL_1227196, EPI_ISL_1227197, EPI_ISL_1227198, EPI_ISL_1227199, EPI_ISL_1227200, EPI_ISL_1227201, EPI_ISL_1227202, EPI_ISL_1227203, EPI_ISL_1227204, EPI_ISL_1227205 | | | |
| see above | NYC Pandemic Response Lab | Wadsworth Center, New York State Department of Health | Kirsten St. George, Daryl M. Lamson, Alexis Russel, Matthew Shudt, Melissa A Leisner, Jonathan Plitnick, Navjot Singh, John Kelly, Erasmus Schneider, Erica Lasek-Nesselquist |
| EPI_ISL_1227206, EPI_ISL_1227207 | WESTCHESTER MEDICAL CENTER | Wadsworth Center, New York State Department of Health | Kirsten St. George, Daryl M. Lamson, Alexis Russel, Matthew Shudt, Melissa A Leisner, Jonathan Plitnick, Navjot Singh, John Kelly, Erasmus Schneider, Erica Lasek-Nesselquist |
| EPI_ISL_1227208, EPI_ISL_1227209 | ALBANY MEDICAL CENTER | Wadsworth Center, New York State Department of Health | Kirsten St. George, Daryl M. Lamson, Alexis Russel, Matthew Shudt, Melissa A Leisner, Jonathan Plitnick, Navjot Singh, John Kelly, Erasmus Schneider, Erica Lasek-Nesselquist |
| EPI_ISL_1227210 | THE MARY IMOGENE BASSETT HOSPITAL | Wadsworth Center, New York State Department of Health | Kirsten St. George, Daryl M. Lamson, Alexis Russel, Matthew Shudt, Melissa A Leisner, Jonathan Plitnick, Navjot Singh, John Kelly, Erasmus Schneider, Erica Lasek-Nesselquist |
| EPI_ISL_1227211 | MEMORIAL SLOAN KETTERING CANCER CENTER | Wadsworth Center, New York State Department of Health | Kirsten St. George, Daryl M. Lamson, Alexis Russel, Matthew Shudt, Melissa A Leisner, Jonathan Plitnick, Navjot Singh, John Kelly, Erasmus Schneider, Erica Lasek-Nesselquist |
| EPI_ISL_1227212, EPI_ISL_1227213 | NYC Pandemic Response Lab | Wadsworth Center, New York State Department of Health | Kirsten St. George, Daryl M. Lamson, Alexis Russel, Matthew Shudt, Melissa A Leisner, Jonathan Plitnick, Navjot Singh, John Kelly, Erasmus Schneider, Erica Lasek-Nesselquist |
| EPI_ISL_1227214 | MEMORIAL SLOAN KETTERING CANCER CENTER | Wadsworth Center, New York State Department of Health | Kirsten St. George, Daryl M. Lamson, Alexis Russel, Matthew Shudt, Melissa A Leisner, Jonathan Plitnick, Navjot Singh, John Kelly, Erasmus Schneider, Erica Lasek-Nesselquist |
| EPI_ISL_1227215, EPI_ISL_1227216, EPI_ISL_1227217, EPI_ISL_1227218, EPI_ISL_1227219, EPI_ISL_1227220 | WESTCHESTER MEDICAL CENTER | Wadsworth Center, New York State Department of Health | Kirsten St. George, Daryl M. Lamson, Alexis Russel, Matthew Shudt, Melissa A Leisner, Jonathan Plitnick, Navjot Singh, John Kelly, Erasmus Schneider, Erica Lasek-Nesselquist |
| EPI_ISL_1227221 | NYC Pandemic Response Lab | Wadsworth Center, New York State Department of Health | Kirsten St. George, Daryl M. Lamson, Alexis Russel, Matthew Shudt, Melissa A Leisner, Jonathan Plitnick, Navjot Singh, John Kelly, Erasmus Schneider, Erica Lasek-Nesselquist |
| EPI_ISL_1227222 | Columbia University Irving Medical Center | Wadsworth Center, New York State Department of Health | Kirsten St. George, Daryl M. Lamson, Alexis Russel, Matthew Shudt, Melissa A Leisner, Jonathan Plitnick, Navjot Singh, John Kelly, Erasmus Schneider, Erica Lasek-Nesselquist |
| EPI_ISL_1227223, EPI_ISL_1227224, EPI_ISL_1227225 | WESTCHESTER MEDICAL CENTER | Wadsworth Center, New York State Department of Health | Kirsten St. George, Daryl M. Lamson, Alexis Russel, Matthew Shudt, Melissa A Leisner, Jonathan Plitnick, Navjot Singh, John Kelly, Erasmus Schneider, Erica Lasek-Nesselquist |
| EPI_ISL_1227226, EPI_ISL_1227227 | MONTEFIORE MEDICAL CENTER LABORATORIES | Wadsworth Center, New York State Department of Health | Kirsten St. George, Daryl M. Lamson, Alexis Russel, Matthew Shudt, Melissa A Leisner, Jonathan Plitnick, Navjot Singh, John Kelly, Erasmus Schneider, Erica Lasek-Nesselquist |
| EPI_ISL_1227228, EPI_ISL_1227229 | WESTCHESTER MEDICAL CENTER | Wadsworth Center, New York State Department of Health | Kirsten St. George, Daryl M. Lamson, Alexis Russel, Matthew Shudt, Melissa A Leisner, Jonathan Plitnick, Navjot Singh, John Kelly, Erasmus Schneider, Erica Lasek-Nesselquist |
| EPI_ISL_1227230 | MEMORIAL SLOAN KETTERING CANCER CENTER | Wadsworth Center, New York State Department of Health | Kirsten St. George, Daryl M. Lamson, Alexis Russel, Matthew Shudt, Melissa A Leisner, Jonathan Plitnick, Navjot Singh, John Kelly, Erasmus Schneider, Erica Lasek-Nesselquist |
| EPI_ISL_1227231 | URMC LABS | Wadsworth Center, New York State Department of Health | Kirsten St. George, Daryl M. Lamson, Alexis Russel, Matthew Shudt, Melissa A Leisner, Jonathan Plitnick, Navjot Singh, John Kelly, Erasmus Schneider, Erica Lasek-Nesselquist |
| EPI_ISL_1227232, EPI_ISL_1227233 | ALBANY MEDICAL CENTER | Wadsworth Center, New York State Department of Health | Kirsten St. George, Daryl M. Lamson, Alexis Russel, Matthew Shudt, Melissa A Leisner, Jonathan Plitnick, Navjot Singh, John Kelly, Erasmus Schneider, Erica Lasek-Nesselquist |
| EPI_ISL_1227234 | Wadsworth Center, New York State Department of Health | Wadsworth Center, New York State Department of Health | Kirsten St. George, Daryl M. Lamson, Alexis Russel, Matthew Shudt, Melissa A Leisner, Jonathan Plitnick, Navjot Singh, John Kelly, Erasmus Schneider, Erica Lasek-Nesselquist |
| EPI_ISL_1227235 | ALBANY MEDICAL CENTER | Wadsworth Center, New York State Department of Health | Kirsten St. George, Daryl M. Lamson, Alexis Russel, Matthew Shudt, Melissa A Leisner, Jonathan Plitnick, Navjot Singh, John Kelly, Erasmus Schneider, Erica Lasek-Nesselquist |
| EPI_ISL_1227236 | MEMORIAL SLOAN KETTERING CANCER CENTER | Wadsworth Center, New York State Department of Health | Kirsten St. George, Daryl M. Lamson, Alexis Russel, Matthew Shudt, Melissa A Leisner, Jonathan Plitnick, Navjot Singh, John Kelly, Erasmus Schneider, Erica Lasek-Nesselquist |
| EPI_ISL_1227237 | Columbia University Irving Medical Center | Wadsworth Center, New York State Department of Health | Kirsten St. George, Daryl M. Lamson, Alexis Russel, Matthew Shudt, Melissa A Leisner, Jonathan Plitnick, Navjot Singh, John Kelly, Erasmus Schneider, Erica Lasek-Nesselquist |

| | | | |
|---|---|---|---|
| EPI_ISL_1227238 | NYC Pandemic Response Lab | Wadsworth Center, New York State Department of Health | Kirsten St. George, Daryl M. Lamson, Alexis Russel, Matthew Shudt, Melissa A Leisner, Jonathan Plitnick, Navjot Singh, John Kelly, Erasmus Schneider, Erica Lasek-Nesselquist |
| EPI_ISL_1227239, EPI_ISL_1227240, EPI_ISL_1227241 | SUNY UPSTATE MEDICAL UNIVERSITY | Wadsworth Center, New York State Department of Health | Kirsten St. George, Daryl M. Lamson, Alexis Russel, Matthew Shudt, Melissa A Leisner, Jonathan Plitnick, Navjot Singh, John Kelly, Erasmus Schneider, Erica Lasek-Nesselquist |
| EPI_ISL_1227242, EPI_ISL_1227243 | MEMORIAL SLOAN KETTERING CANCER CENTER | Wadsworth Center, New York State Department of Health | Kirsten St. George, Daryl M. Lamson, Alexis Russel, Matthew Shudt, Melissa A Leisner, Jonathan Plitnick, Navjot Singh, John Kelly, Erasmus Schneider, Erica Lasek-Nesselquist |
| EPI_ISL_1227244 | NYC Pandemic Response Lab | Wadsworth Center, New York State Department of Health | Kirsten St. George, Daryl M. Lamson, Alexis Russel, Matthew Shudt, Melissa A Leisner, Jonathan Plitnick, Navjot Singh, John Kelly, Erasmus Schneider, Erica Lasek-Nesselquist |
| EPI_ISL_1227245 | SARATOGA HOSPITAL LABORATORY | Wadsworth Center, New York State Department of Health | Kirsten St. George, Daryl M. Lamson, Alexis Russel, Matthew Shudt, Melissa A Leisner, Jonathan Plitnick, Navjot Singh, John Kelly, Erasmus Schneider, Erica Lasek-Nesselquist |
| EPI_ISL_1227246 | Wadsworth Center, New York State Department of Health | Wadsworth Center, New York State Department of Health | Kirsten St. George, Daryl M. Lamson, Alexis Russel, Matthew Shudt, Melissa A Leisner, Jonathan Plitnick, Navjot Singh, John Kelly, Erasmus Schneider, Erica Lasek-Nesselquist |
| EPI_ISL_1227247, EPI_ISL_1227248 | WESTCHESTER MEDICAL CENTER | Wadsworth Center, New York State Department of Health | Kirsten St. George, Daryl M. Lamson, Alexis Russel, Matthew Shudt, Melissa A Leisner, Jonathan Plitnick, Navjot Singh, John Kelly, Erasmus Schneider, Erica Lasek-Nesselquist |
| EPI_ISL_1227249, EPI_ISL_1227250, EPI_ISL_1227251, EPI_ISL_1227252, EPI_ISL_1227253 | ALBANY MEDICAL CENTER | Wadsworth Center, New York State Department of Health | Kirsten St. George, Daryl M. Lamson, Alexis Russel, Matthew Shudt, Melissa A Leisner, Jonathan Plitnick, Navjot Singh, John Kelly, Erasmus Schneider, Erica Lasek-Nesselquist |
| EPI_ISL_1227254, EPI_ISL_1227255 | Wadsworth Center, New York State Department of Health | Wadsworth Center, New York State Department of Health | Kirsten St. George, Daryl M. Lamson, Alexis Russel, Matthew Shudt, Melissa A Leisner, Jonathan Plitnick, Navjot Singh, John Kelly, Erasmus Schneider, Erica Lasek-Nesselquist |
| EPI_ISL_1227256 | ALBANY MEDICAL CENTER | Wadsworth Center, New York State Department of Health | Kirsten St. George, Daryl M. Lamson, Alexis Russel, Matthew Shudt, Melissa A Leisner, Jonathan Plitnick, Navjot Singh, John Kelly, Erasmus Schneider, Erica Lasek-Nesselquist |
| EPI_ISL_1227257, EPI_ISL_1227258, EPI_ISL_1227259, EPI_ISL_1227260, EPI_ISL_1227261, EPI_ISL_1227262 | Wadsworth Center, New York State Department of Health | Wadsworth Center, New York State Department of Health | Kirsten St. George, Daryl M. Lamson, Alexis Russel, Matthew Shudt, Melissa A Leisner, Jonathan Plitnick, Navjot Singh, John Kelly, Erasmus Schneider, Erica Lasek-Nesselquist |
| EPI_ISL_1227263, EPI_ISL_1227264, EPI_ISL_1227265, EPI_ISL_1227266, EPI_ISL_1227267, EPI_ISL_1227268, EPI_ISL_1227269, EPI_ISL_1227270, EPI_ISL_1227271 | ALBANY MEDICAL CENTER | Wadsworth Center, New York State Department of Health | Kirsten St. George, Daryl M. Lamson, Alexis Russel, Matthew Shudt, Melissa A Leisner, Jonathan Plitnick, Navjot Singh, John Kelly, Erasmus Schneider, Erica Lasek-Nesselquist |
| EPI_ISL_1227272, EPI_ISL_1227273, EPI_ISL_1227274, EPI_ISL_1227275, EPI_ISL_1227276, EPI_ISL_1227277 | URMC LABS | Wadsworth Center, New York State Department of Health | Kirsten St. George, Daryl M. Lamson, Alexis Russel, Matthew Shudt, Melissa A Leisner, Jonathan Plitnick, Navjot Singh, John Kelly, Erasmus Schneider, Erica Lasek-Nesselquist |
| EPI_ISL_1227278, EPI_ISL_1227279, EPI_ISL_1227280, EPI_ISL_1227281, EPI_ISL_1227282 | MEMORIAL SLOAN KETTERING CANCER CENTER | Wadsworth Center, New York State Department of Health | Kirsten St. George, Daryl M. Lamson, Alexis Russel, Matthew Shudt, Melissa A Leisner, Jonathan Plitnick, Navjot Singh, John Kelly, Erasmus Schneider, Erica Lasek-Nesselquist |
| EPI_ISL_1227283 | Columbia University Irving Medical Center | Wadsworth Center, New York State Department of Health | Kirsten St. George, Daryl M. Lamson, Alexis Russel, Matthew Shudt, Melissa A Leisner, Jonathan Plitnick, Navjot Singh, John Kelly, Erasmus Schneider, Erica Lasek-Nesselquist |
| EPI_ISL_1227284, EPI_ISL_1227285, EPI_ISL_1227286, EPI_ISL_1227287, EPI_ISL_1227288 | MEMORIAL SLOAN KETTERING CANCER CENTER | Wadsworth Center, New York State Department of Health | Kirsten St. George, Daryl M. Lamson, Alexis Russel, Matthew Shudt, Melissa A Leisner, Jonathan Plitnick, Navjot Singh, John Kelly, Erasmus Schneider, Erica Lasek-Nesselquist |
| EPI_ISL_1227289, EPI_ISL_1227290 | Columbia University Irving Medical Center | Wadsworth Center, New York State Department of Health | Kirsten St. George, Daryl M. Lamson, Alexis Russel, Matthew Shudt, Melissa A Leisner, Jonathan Plitnick, Navjot Singh, John Kelly, Erasmus Schneider, Erica Lasek-Nesselquist |
| EPI_ISL_1227291, EPI_ISL_1227292 | MEMORIAL SLOAN KETTERING CANCER CENTER | Wadsworth Center, New York State Department of Health | Kirsten St. George, Daryl M. Lamson, Alexis Russel, Matthew Shudt, Melissa A Leisner, Jonathan Plitnick, Navjot Singh, John Kelly, Erasmus Schneider, Erica Lasek-Nesselquist |
| EPI_ISL_1227293 | Columbia University Irving Medical Center | Wadsworth Center, New York State Department of Health | Kirsten St. George, Daryl M. Lamson, Alexis Russel, Matthew Shudt, Melissa A Leisner, Jonathan Plitnick, Navjot Singh, John Kelly, Erasmus Schneider, Erica Lasek-Nesselquist |
| EPI_ISL_1227294 | WESTCHESTER MEDICAL CENTER | Wadsworth Center, New York State Department of Health | Kirsten St. George, Daryl M. Lamson, Alexis Russel, Matthew Shudt, Melissa A Leisner, Jonathan Plitnick, Navjot Singh, John Kelly, Erasmus Schneider, Erica Lasek-Nesselquist |
| EPI_ISL_1227295, EPI_ISL_1227296 | Columbia University Irving Medical Center | Wadsworth Center, New York State Department of Health | Kirsten St. George, Daryl M. Lamson, Alexis Russel, Matthew Shudt, Melissa A Leisner, Jonathan Plitnick, Navjot Singh, John Kelly, Erasmus Schneider, Erica Lasek-Nesselquist |
| EPI_ISL_1227297, EPI_ISL_1227298, EPI_ISL_1227299, EPI_ISL_1227300, EPI_ISL_1227301 | MONTEFIORE MEDICAL CENTER LABORATORIES | Wadsworth Center, New York State Department of Health | Kirsten St. George, Daryl M. Lamson, Alexis Russel, Matthew Shudt, Melissa A Leisner, Jonathan Plitnick, Navjot Singh, John Kelly, Erasmus Schneider, Erica Lasek-Nesselquist |
| EPI_ISL_1227302, EPI_ISL_1227303 | WESTCHESTER MEDICAL CENTER | Wadsworth Center, New York State Department of Health | Kirsten St. George, Daryl M. Lamson, Alexis Russel, Matthew Shudt, Melissa A Leisner, Jonathan Plitnick, Navjot Singh, John Kelly, Erasmus Schneider, Erica Lasek-Nesselquist |
| EPI_ISL_1227304, EPI_ISL_1227305 | Columbia University Irving Medical Center | Wadsworth Center, New York State Department of Health | Kirsten St. George, Daryl M. Lamson, Alexis Russel, Matthew Shudt, Melissa A Leisner, Jonathan Plitnick, Navjot Singh, John Kelly, Erasmus Schneider, Erica Lasek-Nesselquist |
| EPI_ISL_1227306, EPI_ISL_1227307, EPI_ISL_1227308, EPI_ISL_1227309, EPI_ISL_1227310, EPI_ISL_1227311, EPI_ISL_1227312, EPI_ISL_1227313 | MONTEFIORE MEDICAL CENTER LABORATORIES | Wadsworth Center, New York State Department of Health | Kirsten St. George, Daryl M. Lamson, Alexis Russel, Matthew Shudt, Melissa A Leisner, Jonathan Plitnick, Navjot Singh, John Kelly, Erasmus Schneider, Erica Lasek-Nesselquist |
| EPI_ISL_1227314 | Columbia University Irving Medical Center | Wadsworth Center, New York State Department of Health | Kirsten St. George, Daryl M. Lamson, Alexis Russel, Matthew Shudt, Melissa A Leisner, Jonathan Plitnick, Navjot Singh, John Kelly, Erasmus Schneider, Erica Lasek-Nesselquist |
| EPI_ISL_1227315, EPI_ISL_1227316, EPI_ISL_1227317, EPI_ISL_1227318, EPI_ISL_1227319 | MONTEFIORE MEDICAL CENTER LABORATORIES | Wadsworth Center, New York State Department of Health | Kirsten St. George, Daryl M. Lamson, Alexis Russel, Matthew Shudt, Melissa A Leisner, Jonathan Plitnick, Navjot Singh, John Kelly, Erasmus Schneider, Erica Lasek-Nesselquist |
| EPI_ISL_1227320 | Columbia University Irving Medical Center | Wadsworth Center, New York State Department of Health | Kirsten St. George, Daryl M. Lamson, Alexis Russel, Matthew Shudt, Melissa A Leisner, Jonathan Plitnick, Navjot Singh, John Kelly, Erasmus Schneider, Erica Lasek-Nesselquist |
| EPI_ISL_1227321, EPI_ISL_1227322, EPI_ISL_1227323, EPI_ISL_1227324, | MONTEFIORE MEDICAL CENTER LABORATORIES | Wadsworth Center, New York State Department of Health | Kirsten St. George, Daryl M. Lamson, Alexis Russel, Matthew Shudt, Melissa A Leisner, Jonathan Plitnick, Navjot Singh, John Kelly, Erasmus Schneider, Erica Lasek-Nesselquist |

| | | | |
|---|---|---|---|
| EPI_ISL_1227325 | | | |
| EPI_ISL_1227326 | Columbia University Irving Medical Center | Wadsworth Center, New York State Department of Health | Kirsten St. George, Daryl M. Lamson, Alexis Russel, Matthew Shudt, Melissa A Leisner, Jonathan Plitnick, Navjot Singh, John Kelly, Erasmus Schneider, Erica Lasek-Nesselquist |
| EPI_ISL_1227327, EPI_ISL_1227328, EPI_ISL_1227329, EPI_ISL_1227330, EPI_ISL_1227331, EPI_ISL_1227332, EPI_ISL_1227333, EPI_ISL_1227334, EPI_ISL_1227335, EPI_ISL_1227336 | MONTEFIORE MEDICAL CENTER LABORATORIES | Wadsworth Center, New York State Department of Health | Kirsten St. George, Daryl M. Lamson, Alexis Russel, Matthew Shudt, Melissa A Leisner, Jonathan Plitnick, Navjot Singh, John Kelly, Erasmus Schneider, Erica Lasek-Nesselquist |
| EPI_ISL_1227337 | WESTCHESTER MEDICAL CENTER | Wadsworth Center, New York State Department of Health | Kirsten St. George, Daryl M. Lamson, Alexis Russel, Matthew Shudt, Melissa A Leisner, Jonathan Plitnick, Navjot Singh, John Kelly, Erasmus Schneider, Erica Lasek-Nesselquist |
| EPI_ISL_1227338, EPI_ISL_1227339, EPI_ISL_1227340, EPI_ISL_1227341, EPI_ISL_1227342, EPI_ISL_1227343, EPI_ISL_1227344, EPI_ISL_1227345, EPI_ISL_1227346, EPI_ISL_1227347, EPI_ISL_1227348, EPI_ISL_1227349, EPI_ISL_1227350, EPI_ISL_1227351, EPI_ISL_1227352, EPI_ISL_1227353, EPI_ISL_1227354, EPI_ISL_1227355, EPI_ISL_1227356 | | | |
| see above | MONTEFIORE MEDICAL CENTER LABORATORIES | Wadsworth Center, New York State Department of Health | Kirsten St. George, Daryl M. Lamson, Alexis Russel, Matthew Shudt, Melissa A Leisner, Jonathan Plitnick, Navjot Singh, John Kelly, Erasmus Schneider, Erica Lasek-Nesselquist |
| EPI_ISL_1227357 | Columbia University Irving Medical Center | Wadsworth Center, New York State Department of Health | Kirsten St. George, Daryl M. Lamson, Alexis Russel, Matthew Shudt, Melissa A Leisner, Jonathan Plitnick, Navjot Singh, John Kelly, Erasmus Schneider, Erica Lasek-Nesselquist |
| EPI_ISL_1227358, EPI_ISL_1227359 | Wadsworth Center, New York State Department of Health | Wadsworth Center, New York State Department of Health | Kirsten St. George, Daryl M. Lamson, Alexis Russel, Matthew Shudt, Melissa A Leisner, Jonathan Plitnick, Navjot Singh, John Kelly, Erasmus Schneider, Erica Lasek-Nesselquist |
| EPI_ISL_1227360, EPI_ISL_1227361, EPI_ISL_1227362, EPI_ISL_1227363 | Columbia University Irving Medical Center | Wadsworth Center, New York State Department of Health | Kirsten St. George, Daryl M. Lamson, Alexis Russel, Matthew Shudt, Melissa A Leisner, Jonathan Plitnick, Navjot Singh, John Kelly, Erasmus Schneider, Erica Lasek-Nesselquist |
| EPI_ISL_1227364, EPI_ISL_1227365, EPI_ISL_1227366, EPI_ISL_1227367, EPI_ISL_1227368, EPI_ISL_1227369, EPI_ISL_1227370, EPI_ISL_1227371, EPI_ISL_1227372, EPI_ISL_1227373, EPI_ISL_1227374, EPI_ISL_1227375, EPI_ISL_1227376, EPI_ISL_1227377, EPI_ISL_1227378 | | | |
| see above | MONTEFIORE MEDICAL CENTER LABORATORIES | Wadsworth Center, New York State Department of Health | Kirsten St. George, Daryl M. Lamson, Alexis Russel, Matthew Shudt, Melissa A Leisner, Jonathan Plitnick, Navjot Singh, John Kelly, Erasmus Schneider, Erica Lasek-Nesselquist |
| EPI_ISL_1227379 | Wadsworth Center, New York State Department of Health | Wadsworth Center, New York State Department of Health | Kirsten St. George, Daryl M. Lamson, Alexis Russel, Matthew Shudt, Melissa A Leisner, Jonathan Plitnick, Navjot Singh, John Kelly, Erasmus Schneider, Erica Lasek-Nesselquist |
| EPI_ISL_1227380, EPI_ISL_1227381, EPI_ISL_1227382, EPI_ISL_1227383, EPI_ISL_1227384, EPI_ISL_1227385, EPI_ISL_1227386, EPI_ISL_1227387, EPI_ISL_1227388, EPI_ISL_1227389, EPI_ISL_1227390, EPI_ISL_1227391, EPI_ISL_1227392, EPI_ISL_1227393, EPI_ISL_1227394, EPI_ISL_1227395, EPI_ISL_1227396, EPI_ISL_1227397, EPI_ISL_1227398, EPI_ISL_1227399, EPI_ISL_1227400 | | | |
| see above | MONTEFIORE MEDICAL CENTER LABORATORIES | Wadsworth Center, New York State Department of Health | Kirsten St. George, Daryl M. Lamson, Alexis Russel, Matthew Shudt, Melissa A Leisner, Jonathan Plitnick, Navjot Singh, John Kelly, Erasmus Schneider, Erica Lasek-Nesselquist |
| EPI_ISL_1227401, EPI_ISL_1227402 | Columbia University Irving Medical Center | Wadsworth Center, New York State Department of Health | Kirsten St. George, Daryl M. Lamson, Alexis Russel, Matthew Shudt, Melissa A Leisner, Jonathan Plitnick, Navjot Singh, John Kelly, Erasmus Schneider, Erica Lasek-Nesselquist |
| EPI_ISL_1227403, EPI_ISL_1227404, EPI_ISL_1227405 | MONTEFIORE MEDICAL CENTER LABORATORIES | Wadsworth Center, New York State Department of Health | Kirsten St. George, Daryl M. Lamson, Alexis Russel, Matthew Shudt, Melissa A Leisner, Jonathan Plitnick, Navjot Singh, John Kelly, Erasmus Schneider, Erica Lasek-Nesselquist |
| EPI_ISL_1227406, EPI_ISL_1227407, EPI_ISL_1227408, EPI_ISL_1227409 | Columbia University Irving Medical Center | Wadsworth Center, New York State Department of Health | Kirsten St. George, Daryl M. Lamson, Alexis Russel, Matthew Shudt, Melissa A Leisner, Jonathan Plitnick, Navjot Singh, John Kelly, Erasmus Schneider, Erica Lasek-Nesselquist |
| EPI_ISL_1227410 | New York Presbyterian Hospital | Wadsworth Center, New York State Department of Health | Kirsten St. George, Daryl M. Lamson, Alexis Russel, Matthew Shudt, Melissa A Leisner, Jonathan Plitnick, Navjot Singh, John Kelly, Erasmus Schneider, Erica Lasek-Nesselquist |
| EPI_ISL_1227411, EPI_ISL_1227412, EPI_ISL_1227413, EPI_ISL_1227414, EPI_ISL_1227415 | MONTEFIORE MEDICAL CENTER LABORATORIES | Wadsworth Center, New York State Department of Health | Kirsten St. George, Daryl M. Lamson, Alexis Russel, Matthew Shudt, Melissa A Leisner, Jonathan Plitnick, Navjot Singh, John Kelly, Erasmus Schneider, Erica Lasek-Nesselquist |
| EPI_ISL_1227416 | Columbia University Irving Medical Center | Wadsworth Center, New York State Department of Health | Kirsten St. George, Daryl M. Lamson, Alexis Russel, Matthew Shudt, Melissa A Leisner, Jonathan Plitnick, Navjot Singh, John Kelly, Erasmus Schneider, Erica Lasek-Nesselquist |
| EPI_ISL_1227417 | New York Presbyterian Hospital | Wadsworth Center, New York State Department of Health | Kirsten St. George, Daryl M. Lamson, Alexis Russel, Matthew Shudt, Melissa A Leisner, Jonathan Plitnick, Navjot Singh, John Kelly, Erasmus Schneider, Erica Lasek-Nesselquist |
| EPI_ISL_1227418, EPI_ISL_1227419, EPI_ISL_1227420, EPI_ISL_1227421 | MONTEFIORE MEDICAL CENTER LABORATORIES | Wadsworth Center, New York State Department of Health | Kirsten St. George, Daryl M. Lamson, Alexis Russel, Matthew Shudt, Melissa A Leisner, Jonathan Plitnick, Navjot Singh, John Kelly, Erasmus Schneider, Erica Lasek-Nesselquist |
| EPI_ISL_1227422, EPI_ISL_1227423 | NYC Pandemic Response Lab | Wadsworth Center, New York State Department of Health | Kirsten St. George, Daryl M. Lamson, Alexis Russel, Matthew Shudt, Melissa A Leisner, Jonathan Plitnick, Navjot Singh, John Kelly, Erasmus Schneider, Erica Lasek-Nesselquist |
| EPI_ISL_1227424, EPI_ISL_1227425, EPI_ISL_1227426, EPI_ISL_1227427 | MONTEFIORE MEDICAL CENTER LABORATORIES | Wadsworth Center, New York State Department of Health | Kirsten St. George, Daryl M. Lamson, Alexis Russel, Matthew Shudt, Melissa A Leisner, Jonathan Plitnick, Navjot Singh, John Kelly, Erasmus Schneider, Erica Lasek-Nesselquist |
| EPI_ISL_1227428, EPI_ISL_1227429 | NYC Pandemic Response Lab | Wadsworth Center, New York State Department of Health | Kirsten St. George, Daryl M. Lamson, Alexis Russel, Matthew Shudt, Melissa A Leisner, Jonathan Plitnick, Navjot Singh, John Kelly, Erasmus Schneider, Erica Lasek-Nesselquist |
| EPI_ISL_1227430, EPI_ISL_1227431, EPI_ISL_1227432, EPI_ISL_1227433, EPI_ISL_1227434 | MONTEFIORE MEDICAL CENTER LABORATORIES | Wadsworth Center, New York State Department of Health | Kirsten St. George, Daryl M. Lamson, Alexis Russel, Matthew Shudt, Melissa A Leisner, Jonathan Plitnick, Navjot Singh, John Kelly, Erasmus Schneider, Erica Lasek-Nesselquist |
| EPI_ISL_1227435 | Columbia University Irving Medical Center | Wadsworth Center, New York State Department of Health | Kirsten St. George, Daryl M. Lamson, Alexis Russel, Matthew Shudt, Melissa A Leisner, Jonathan Plitnick, Navjot Singh, John Kelly, Erasmus Schneider, Erica Lasek-Nesselquist |
| EPI_ISL_1227436, EPI_ISL_1227437, EPI_ISL_1227438, EPI_ISL_1227439 | MONTEFIORE MEDICAL CENTER LABORATORIES | Wadsworth Center, New York State Department of Health | Kirsten St. George, Daryl M. Lamson, Alexis Russel, Matthew Shudt, Melissa A Leisner, Jonathan Plitnick, Navjot Singh, John Kelly, Erasmus Schneider, Erica Lasek-Nesselquist |
| EPI_ISL_1227440 | NYC Pandemic Response Lab | Wadsworth Center, New York State Department of Health | Kirsten St. George, Daryl M. Lamson, Alexis Russel, Matthew Shudt, Melissa A Leisner, Jonathan Plitnick, Navjot Singh, John Kelly, Erasmus Schneider, Erica Lasek-Nesselquist |
| EPI_ISL_1227441 | WESTCHESTER MEDICAL CENTER | Wadsworth Center, New York State Department of Health | Kirsten St. George, Daryl M. Lamson, Alexis Russel, Matthew Shudt, Melissa A Leisner, Jonathan Plitnick, Navjot Singh, John Kelly, Erasmus Schneider, Erica Lasek-Nesselquist |
| EPI_ISL_1227442, EPI_ISL_1227443, EPI_ISL_1227444, EPI_ISL_1227445, EPI_ISL_1227446 | MONTEFIORE MEDICAL CENTER LABORATORIES | Wadsworth Center, New York State Department of Health | Kirsten St. George, Daryl M. Lamson, Alexis Russel, Matthew Shudt, Melissa A Leisner, Jonathan Plitnick, Navjot Singh, John Kelly, Erasmus Schneider, Erica Lasek-Nesselquist |
| EPI_ISL_1227447 | SUNY UPSTATE MEDICAL UNIVERSITY | Wadsworth Center, New York State Department of Health | Kirsten St. George, Daryl M. Lamson, Alexis Russel, Matthew Shudt, Melissa A Leisner, Jonathan Plitnick, Navjot Singh, John Kelly, Erasmus Schneider, Erica Lasek-Nesselquist |
| EPI_ISL_1227448, EPI_ISL_1227449, EPI_ISL_1227450, EPI_ISL_1227451, EPI_ISL_1227452, EPI_ISL_1227453, EPI_ISL_1227454, EPI_ISL_1227455 | URMC LABS | Wadsworth Center, New York State Department of Health | Kirsten St. George, Daryl M. Lamson, Alexis Russel, Matthew Shudt, Melissa A Leisner, Jonathan Plitnick, Navjot Singh, John Kelly, Erasmus Schneider, Erica Lasek-Nesselquist |

| | | | |
|---|---|---|---|
| EPI_ISL_1227456, EPI_ISL_1227457 | THE MARY IMOGENE BASSETT HOSPITAL | Wadsworth Center, New York State Department of Health | Kirsten St. George, Daryl M. Lamson, Alexis Russel, Matthew Shudt, Melissa A Leisner, Jonathan Plitnick, Navjot Singh, John Kelly, Erasmus Schneider, Erica Lasek-Nesselquist |
| EPI_ISL_1227458, EPI_ISL_1227459, EPI_ISL_1227460, EPI_ISL_1227461, EPI_ISL_1227462 | ALBANY MEDICAL CENTER | Wadsworth Center, New York State Department of Health | Kirsten St. George, Daryl M. Lamson, Alexis Russel, Matthew Shudt, Melissa A Leisner, Jonathan Plitnick, Navjot Singh, John Kelly, Erasmus Schneider, Erica Lasek-Nesselquist |
| EPI_ISL_1227463, EPI_ISL_1227464, EPI_ISL_1227465 | SUNY UPSTATE MEDICAL UNIVERSITY | Wadsworth Center, New York State Department of Health | Kirsten St. George, Daryl M. Lamson, Alexis Russel, Matthew Shudt, Melissa A Leisner, Jonathan Plitnick, Navjot Singh, John Kelly, Erasmus Schneider, Erica Lasek-Nesselquist |
| EPI_ISL_1227466 | THE MARY IMOGENE BASSETT HOSPITAL | Wadsworth Center, New York State Department of Health | Kirsten St. George, Daryl M. Lamson, Alexis Russel, Matthew Shudt, Melissa A Leisner, Jonathan Plitnick, Navjot Singh, John Kelly, Erasmus Schneider, Erica Lasek-Nesselquist |
| EPI_ISL_1227467 | WESTCHESTER MEDICAL CENTER | Wadsworth Center, New York State Department of Health | Kirsten St. George, Daryl M. Lamson, Alexis Russel, Matthew Shudt, Melissa A Leisner, Jonathan Plitnick, Navjot Singh, John Kelly, Erasmus Schneider, Erica Lasek-Nesselquist |
| EPI_ISL_1227468, EPI_ISL_1227469 | MEMORIAL SLOAN KETTERING CANCER CENTER | Wadsworth Center, New York State Department of Health | Kirsten St. George, Daryl M. Lamson, Alexis Russel, Matthew Shudt, Melissa A Leisner, Jonathan Plitnick, Navjot Singh, John Kelly, Erasmus Schneider, Erica Lasek-Nesselquist |
| EPI_ISL_1227470 | WESTCHESTER MEDICAL CENTER | Wadsworth Center, New York State Department of Health | Kirsten St. George, Daryl M. Lamson, Alexis Russel, Matthew Shudt, Melissa A Leisner, Jonathan Plitnick, Navjot Singh, John Kelly, Erasmus Schneider, Erica Lasek-Nesselquist |
| EPI_ISL_1227471 | MEMORIAL SLOAN KETTERING CANCER CENTER | Wadsworth Center, New York State Department of Health | Kirsten St. George, Daryl M. Lamson, Alexis Russel, Matthew Shudt, Melissa A Leisner, Jonathan Plitnick, Navjot Singh, John Kelly, Erasmus Schneider, Erica Lasek-Nesselquist |
| EPI_ISL_1227472, EPI_ISL_1227473, EPI_ISL_1227474, EPI_ISL_1227475, EPI_ISL_1227476, EPI_ISL_1227477, EPI_ISL_1227478, EPI_ISL_1227479, EPI_ISL_1227480, EPI_ISL_1227481, EPI_ISL_1227482, EPI_ISL_1227483, EPI_ISL_1227484, EPI_ISL_1227485, EPI_ISL_1227486, EPI_ISL_1227487, EPI_ISL_1227488, EPI_ISL_1227489, EPI_ISL_1227490, EPI_ISL_1227491, EPI_ISL_1227492, EPI_ISL_1227493, EPI_ISL_1227494, EPI_ISL_1227495, EPI_ISL_1227496, EPI_ISL_1227497, EPI_ISL_1227498, EPI_ISL_1227499, EPI_ISL_1227500, EPI_ISL_1227501, EPI_ISL_1227502 | | | |
| see above | WESTCHESTER MEDICAL CENTER | Wadsworth Center, New York State Department of Health | Kirsten St. George, Daryl M. Lamson, Alexis Russel, Matthew Shudt, Melissa A Leisner, Jonathan Plitnick, Navjot Singh, John Kelly, Erasmus Schneider, Erica Lasek-Nesselquist |
| EPI_ISL_1227503 | MONTEFIORE MEDICAL CENTER LABORATORIES | Wadsworth Center, New York State Department of Health | Kirsten St. George, Daryl M. Lamson, Alexis Russel, Matthew Shudt, Melissa A Leisner, Jonathan Plitnick, Navjot Singh, John Kelly, Erasmus Schneider, Erica Lasek-Nesselquist |
| EPI_ISL_1227504, EPI_ISL_1227505, EPI_ISL_1227506, EPI_ISL_1227507, EPI_ISL_1227508, EPI_ISL_1227509, EPI_ISL_1227510, EPI_ISL_1227511, EPI_ISL_1227512, EPI_ISL_1227513, EPI_ISL_1227514, EPI_ISL_1227515, EPI_ISL_1227516, EPI_ISL_1227517, EPI_ISL_1227518, EPI_ISL_1227519, EPI_ISL_1227520, EPI_ISL_1227521, EPI_ISL_1227522, EPI_ISL_1227523, EPI_ISL_1227524, EPI_ISL_1227525, EPI_ISL_1227526, EPI_ISL_1227527, EPI_ISL_1227528, EPI_ISL_1227529, EPI_ISL_1227530, EPI_ISL_1227531, EPI_ISL_1227532, EPI_ISL_1227533, EPI_ISL_1227534, EPI_ISL_1227535 | | | |
| see above | WESTCHESTER MEDICAL CENTER | Wadsworth Center, New York State Department of Health | Kirsten St. George, Daryl M. Lamson, Alexis Russel, Matthew Shudt, Melissa A Leisner, Jonathan Plitnick, Navjot Singh, John Kelly, Erasmus Schneider, Erica Lasek-Nesselquist |
| EPI_ISL_1227536 | Columbia University Irving Medical Center | Wadsworth Center, New York State Department of Health | Kirsten St. George, Daryl M. Lamson, Alexis Russel, Matthew Shudt, Melissa A Leisner, Jonathan Plitnick, Navjot Singh, John Kelly, Erasmus Schneider, Erica Lasek-Nesselquist |
| EPI_ISL_1227537, EPI_ISL_1227538, EPI_ISL_1227539, EPI_ISL_1227540 | WESTCHESTER MEDICAL CENTER | Wadsworth Center, New York State Department of Health | Kirsten St. George, Daryl M. Lamson, Alexis Russel, Matthew Shudt, Melissa A Leisner, Jonathan Plitnick, Navjot Singh, John Kelly, Erasmus Schneider, Erica Lasek-Nesselquist |
| EPI_ISL_1227541, EPI_ISL_1227542, EPI_ISL_1227543, EPI_ISL_1227544, EPI_ISL_1227545 | THE MARY IMOGENE BASSETT HOSPITAL | Wadsworth Center, New York State Department of Health | Kirsten St. George, Daryl M. Lamson, Alexis Russel, Matthew Shudt, Melissa A Leisner, Jonathan Plitnick, Navjot Singh, John Kelly, Erasmus Schneider, Erica Lasek-Nesselquist |
| EPI_ISL_1227546, EPI_ISL_1227547, EPI_ISL_1227548, EPI_ISL_1227549, EPI_ISL_1227550, EPI_ISL_1227551, EPI_ISL_1227552, EPI_ISL_1227553, EPI_ISL_1227554 | KALEIDA CENTER FOR LABORATORY MEDICINE | Wadsworth Center, New York State Department of Health | Kirsten St. George, Daryl M. Lamson, Alexis Russel, Matthew Shudt, Melissa A Leisner, Jonathan Plitnick, Navjot Singh, John Kelly, Erasmus Schneider, Erica Lasek-Nesselquist |
| EPI_ISL_1227555 | ADIRONDACK MEDICAL CENTER | Wadsworth Center, New York State Department of Health | Kirsten St. George, Daryl M. Lamson, Alexis Russel, Matthew Shudt, Melissa A Leisner, Jonathan Plitnick, Navjot Singh, John Kelly, Erasmus Schneider, Erica Lasek-Nesselquist |
| EPI_ISL_1227556 | ALBANY MEDICAL CENTER | Wadsworth Center, New York State Department of Health | Kirsten St. George, Daryl M. Lamson, Alexis Russel, Matthew Shudt, Melissa A Leisner, Jonathan Plitnick, Navjot Singh, John Kelly, Erasmus Schneider, Erica Lasek-Nesselquist |
| EPI_ISL_1227557 | Wadsworth Center, New York State Department of Health | Wadsworth Center, New York State Department of Health | Kirsten St. George, Daryl M. Lamson, Alexis Russel, Matthew Shudt, Melissa A Leisner, Jonathan Plitnick, Navjot Singh, John Kelly, Erasmus Schneider, Erica Lasek-Nesselquist |
| EPI_ISL_1227558 | ALBANY MEDICAL CENTER | Wadsworth Center, New York State Department of Health | Kirsten St. George, Daryl M. Lamson, Alexis Russel, Matthew Shudt, Melissa A Leisner, Jonathan Plitnick, Navjot Singh, John Kelly, Erasmus Schneider, Erica Lasek-Nesselquist |
| EPI_ISL_1227559 | Wadsworth Center, New York State Department of Health | Wadsworth Center, New York State Department of Health | Kirsten St. George, Daryl M. Lamson, Alexis Russel, Matthew Shudt, Melissa A Leisner, Jonathan Plitnick, Navjot Singh, John Kelly, Erasmus Schneider, Erica Lasek-Nesselquist |
| EPI_ISL_1227560, EPI_ISL_1227561, EPI_ISL_1227562, EPI_ISL_1227563 | URMC LABS | Wadsworth Center, New York State Department of Health | Kirsten St. George, Daryl M. Lamson, Alexis Russel, Matthew Shudt, Melissa A Leisner, Jonathan Plitnick, Navjot Singh, John Kelly, Erasmus Schneider, Erica Lasek-Nesselquist |
| EPI_ISL_1227564, EPI_ISL_1227565 | ALBANY MEDICAL CENTER | Wadsworth Center, New York State Department of Health | Kirsten St. George, Daryl M. Lamson, Alexis Russel, Matthew Shudt, Melissa A Leisner, Jonathan Plitnick, Navjot Singh, John Kelly, Erasmus Schneider, Erica Lasek-Nesselquist |
| EPI_ISL_1227566, EPI_ISL_1227567, EPI_ISL_1227568 | Wadsworth Center, New York State Department of Health | Wadsworth Center, New York State Department of Health | Kirsten St. George, Daryl M. Lamson, Alexis Russel, Matthew Shudt, Melissa A Leisner, Jonathan Plitnick, Navjot Singh, John Kelly, Erasmus Schneider, Erica Lasek-Nesselquist |
| EPI_ISL_1227569 | WESTCHESTER MEDICAL CENTER | Wadsworth Center, New York State Department of Health | Kirsten St. George, Daryl M. Lamson, Alexis Russel, Matthew Shudt, Melissa A Leisner, Jonathan Plitnick, Navjot Singh, John Kelly, Erasmus Schneider, Erica Lasek-Nesselquist |
| EPI_ISL_1227570, EPI_ISL_1227571 | Wadsworth Center, New York State Department of Health | Wadsworth Center, New York State Department of Health | Kirsten St. George, Daryl M. Lamson, Alexis Russel, Matthew Shudt, Melissa A Leisner, Jonathan Plitnick, Navjot Singh, John Kelly, Erasmus Schneider, Erica Lasek-Nesselquist |
| EPI_ISL_1227572, EPI_ISL_1227573 | ALBANY MEDICAL CENTER | Wadsworth Center, New York State Department of Health | Kirsten St. George, Daryl M. Lamson, Alexis Russel, Matthew Shudt, Melissa A Leisner, Jonathan Plitnick, Navjot Singh, John Kelly, Erasmus Schneider, Erica Lasek-Nesselquist |
| EPI_ISL_1227574, EPI_ISL_1227575, EPI_ISL_1227576, EPI_ISL_1227577, EPI_ISL_1227578, EPI_ISL_1227579, EPI_ISL_1227580, EPI_ISL_1227581, EPI_ISL_1227582, EPI_ISL_1227583, EPI_ISL_1227584, EPI_ISL_1227585, EPI_ISL_1227586, EPI_ISL_1227587, EPI_ISL_1227588, EPI_ISL_1227589, EPI_ISL_1227590, EPI_ISL_1227591, EPI_ISL_1227592, EPI_ISL_1227593, EPI_ISL_1227594, EPI_ISL_1227595, EPI_ISL_1227596, EPI_ISL_1227597, EPI_ISL_1227598, EPI_ISL_1227599, EPI_ISL_1227600, EPI_ISL_1227601, EPI_ISL_1227602, EPI_ISL_1227603, EPI_ISL_1227604, EPI_ISL_1227605, EPI_ISL_1227606, EPI_ISL_1227607, EPI_ISL_1227608, EPI_ISL_1227609, EPI_ISL_1227610, EPI_ISL_1227611, EPI_ISL_1227612, EPI_ISL_1227613, EPI_ISL_1227614, EPI_ISL_1227615, EPI_ISL_1227616 | | | |
| see above | Wadsworth Center, New York State Department of Health | Wadsworth Center, New York State Department of Health | Kirsten St. George, Daryl M. Lamson, Alexis Russel, Matthew Shudt, Melissa A Leisner, Jonathan Plitnick, Navjot Singh, John Kelly, Erasmus Schneider, Erica Lasek-Nesselquist |
| EPI_ISL_1227617, EPI_ISL_1227618, EPI_ISL_1227619, EPI_ISL_1227620, EPI_ISL_1227621, EPI_ISL_1227622, EPI_ISL_1227623, EPI_ISL_1227624, EPI_ISL_1227625, EPI_ISL_1227626, EPI_ISL_1227627, EPI_ISL_1227628, EPI_ISL_1227629, EPI_ISL_1227630, EPI_ISL_1227631, EPI_ISL_1227632, EPI_ISL_1227633, EPI_ISL_1227634, EPI_ISL_1227635, EPI_ISL_1227636, EPI_ISL_1227637, EPI_ISL_1227638, EPI_ISL_1227639, EPI_ISL_1227640 | | | |
| see above | THE MARY IMOGENE BASSETT HOSPITAL | Wadsworth Center, New York State Department of Health | Kirsten St. George, Daryl M. Lamson, Alexis Russel, Matthew Shudt, Melissa A Leisner, Jonathan Plitnick, Navjot Singh, John Kelly, Erasmus Schneider, Erica Lasek-Nesselquist |
| EPI_ISL_1227641, EPI_ISL_1227642 | SUNY UPSTATE MEDICAL UNIVERSITY | Wadsworth Center, New York State Department of Health | Kirsten St. George, Daryl M. Lamson, Alexis Russel, Matthew Shudt, Melissa A Leisner, Jonathan Plitnick, Navjot Singh, John Kelly, Erasmus Schneider, Erica Lasek-Nesselquist |
| EPI_ISL_1227643, EPI_ISL_1227644, EPI_ISL_1227645, EPI_ISL_1227646, EPI_ISL_1227647, EPI_ISL_1227648, EPI_ISL_1227649, EPI_ISL_1227650, EPI_ISL_1227651, EPI_ISL_1227652, EPI_ISL_1227653, EPI_ISL_1227654, EPI_ISL_1227655, EPI_ISL_1227656, EPI_ISL_1227657, EPI_ISL_1227658, EPI_ISL_1227659, EPI_ISL_1227660, EPI_ISL_1227661, EPI_ISL_1227662 | | | |
| see above | MEMORIAL SLOAN KETTERING CANCER CENTER | Wadsworth Center, New York State Department of Health | Kirsten St. George, Daryl M. Lamson, Alexis Russel, Matthew Shudt, Melissa A Leisner, Jonathan Plitnick, Navjot Singh, John Kelly, Erasmus Schneider, Erica Lasek-Nesselquist |
| EPI_ISL_1227663 | NORTH SHORE UNIVERSITY HOSPITAL | Wadsworth Center, New York State Department of Health | Kirsten St. George, Daryl M. Lamson, Alexis Russel, Matthew Shudt, Melissa A Leisner, Jonathan Plitnick, Navjot Singh, John Kelly, Erasmus Schneider, Erica Lasek-Nesselquist |
| EPI_ISL_1227664 | NORTHWELL HEALTH LABORATORIES | Wadsworth Center, New York State Department of Health | Kirsten St. George, Daryl M. Lamson, Alexis Russel, Matthew Shudt, Melissa A Leisner, Jonathan Plitnick, Navjot Singh, John Kelly, Erasmus Schneider, Erica Lasek-Nesselquist |
| EPI_ISL_1227665 | MONTEFIORE MEDICAL CENTER | Wadsworth Center, New York State | Kirsten St. George, Daryl M. Lamson, Alexis Russel, Matthew Shudt, Melissa A Leisner, Jonathan Plitnick, Navjot Singh, John Kelly, Erasmus Schneider, Erica Lasek-Nesselquist |

| | | | |
|---|---|---|---|
| | LABORATORIES | Department of Health | |
| EPI_ISL_1227666, EPI_ISL_1227667, EPI_ISL_1227668, EPI_ISL_1227669, EPI_ISL_1227670, EPI_ISL_1227671, EPI_ISL_1227672, EPI_ISL_1227673, EPI_ISL_1227674 | NYC Pandemic Response Lab | Wadsworth Center, New York State Department of Health | Kirsten St. George, Daryl M. Lamson, Alexis Russel, Matthew Shudt, Melissa A Leisner, Jonathan Plitnick, Navjot Singh, John Kelly, Erasmus Schneider, Erica Lasek-Nesselquist |
| EPI_ISL_1227675 | Columbia University Irving Medical Center | Wadsworth Center, New York State Department of Health | Kirsten St. George, Daryl M. Lamson, Alexis Russel, Matthew Shudt, Melissa A Leisner, Jonathan Plitnick, Navjot Singh, John Kelly, Erasmus Schneider, Erica Lasek-Nesselquist |
| EPI_ISL_1227676 | WESTCHESTER MEDICAL CENTER | Wadsworth Center, New York State Department of Health | Kirsten St. George, Daryl M. Lamson, Alexis Russel, Matthew Shudt, Melissa A Leisner, Jonathan Plitnick, Navjot Singh, John Kelly, Erasmus Schneider, Erica Lasek-Nesselquist |
| EPI_ISL_1227677, EPI_ISL_1227678 | NORTHWELL HEALTH LABORATORIES | Wadsworth Center, New York State Department of Health | Kirsten St. George, Daryl M. Lamson, Alexis Russel, Matthew Shudt, Melissa A Leisner, Jonathan Plitnick, Navjot Singh, John Kelly, Erasmus Schneider, Erica Lasek-Nesselquist |
| EPI_ISL_1227679 | New York Presbyterian Hospital | Wadsworth Center, New York State Department of Health | Kirsten St. George, Daryl M. Lamson, Alexis Russel, Matthew Shudt, Melissa A Leisner, Jonathan Plitnick, Navjot Singh, John Kelly, Erasmus Schneider, Erica Lasek-Nesselquist |
| EPI_ISL_1227680, EPI_ISL_1227681, EPI_ISL_1227682, EPI_ISL_1227683 | NYC Pandemic Response Lab | Wadsworth Center, New York State Department of Health | Kirsten St. George, Daryl M. Lamson, Alexis Russel, Matthew Shudt, Melissa A Leisner, Jonathan Plitnick, Navjot Singh, John Kelly, Erasmus Schneider, Erica Lasek-Nesselquist |
| EPI_ISL_1227684, EPI_ISL_1227685 | New York Presbyterian Hospital | Wadsworth Center, New York State Department of Health | Kirsten St. George, Daryl M. Lamson, Alexis Russel, Matthew Shudt, Melissa A Leisner, Jonathan Plitnick, Navjot Singh, John Kelly, Erasmus Schneider, Erica Lasek-Nesselquist |
| EPI_ISL_1227686, EPI_ISL_1227687, EPI_ISL_1227688, EPI_ISL_1227689, EPI_ISL_1227690, EPI_ISL_1227691, EPI_ISL_1227692, EPI_ISL_1227693, EPI_ISL_1227694, EPI_ISL_1227695, EPI_ISL_1227696, EPI_ISL_1227697, EPI_ISL_1227698, EPI_ISL_1227699, EPI_ISL_1227700, EPI_ISL_1227701, EPI_ISL_1227702, EPI_ISL_1227703, EPI_ISL_1227704 | | | |
| see above | NYC Pandemic Response Lab | Wadsworth Center, New York State Department of Health | Kirsten St. George, Daryl M. Lamson, Alexis Russel, Matthew Shudt, Melissa A Leisner, Jonathan Plitnick, Navjot Singh, John Kelly, Erasmus Schneider, Erica Lasek-Nesselquist |
| EPI_ISL_1227705 | MONTEFIORE MEDICAL CENTER LABORATORIES | Wadsworth Center, New York State Department of Health | Kirsten St. George, Daryl M. Lamson, Alexis Russel, Matthew Shudt, Melissa A Leisner, Jonathan Plitnick, Navjot Singh, John Kelly, Erasmus Schneider, Erica Lasek-Nesselquist |
| EPI_ISL_1227706 | NYC Pandemic Response Lab | Wadsworth Center, New York State Department of Health | Kirsten St. George, Daryl M. Lamson, Alexis Russel, Matthew Shudt, Melissa A Leisner, Jonathan Plitnick, Navjot Singh, John Kelly, Erasmus Schneider, Erica Lasek-Nesselquist |
| EPI_ISL_1227707 | Masonic Care Community of NY | Wadsworth Center, New York State Department of Health | Kirsten St. George, Daryl M. Lamson, Alexis Russel, Matthew Shudt, Melissa A Leisner, Jonathan Plitnick, Navjot Singh, John Kelly, Erasmus Schneider, Erica Lasek-Nesselquist |
| EPI_ISL_1227708 | SUNY UPSTATE MEDICAL UNIVERSITY | Wadsworth Center, New York State Department of Health | Kirsten St. George, Daryl M. Lamson, Alexis Russel, Matthew Shudt, Melissa A Leisner, Jonathan Plitnick, Navjot Singh, John Kelly, Erasmus Schneider, Erica Lasek-Nesselquist |
| EPI_ISL_1227709, EPI_ISL_1227710, EPI_ISL_1227711, EPI_ISL_1227712, EPI_ISL_1227713, EPI_ISL_1227714, EPI_ISL_1227715, EPI_ISL_1227716, EPI_ISL_1227717, EPI_ISL_1227718, EPI_ISL_1227719 | | | |
| see above | URMC LABS | Wadsworth Center, New York State Department of Health | Kirsten St. George, Daryl M. Lamson, Alexis Russel, Matthew Shudt, Melissa A Leisner, Jonathan Plitnick, Navjot Singh, John Kelly, Erasmus Schneider, Erica Lasek-Nesselquist |
| EPI_ISL_1227720, EPI_ISL_1227721, EPI_ISL_1227722, EPI_ISL_1227723, EPI_ISL_1227724 | Masonic Care Community of NY | Wadsworth Center, New York State Department of Health | Kirsten St. George, Daryl M. Lamson, Alexis Russel, Matthew Shudt, Melissa A Leisner, Jonathan Plitnick, Navjot Singh, John Kelly, Erasmus Schneider, Erica Lasek-Nesselquist |
| EPI_ISL_1227725, EPI_ISL_1227726, EPI_ISL_1227727 | SUNY UPSTATE MEDICAL UNIVERSITY | Wadsworth Center, New York State Department of Health | Kirsten St. George, Daryl M. Lamson, Alexis Russel, Matthew Shudt, Melissa A Leisner, Jonathan Plitnick, Navjot Singh, John Kelly, Erasmus Schneider, Erica Lasek-Nesselquist |
| EPI_ISL_1227728, EPI_ISL_1227729, EPI_ISL_1227730, EPI_ISL_1227731, EPI_ISL_1227732, EPI_ISL_1227733, EPI_ISL_1227734, EPI_ISL_1227735, EPI_ISL_1227736, EPI_ISL_1227737, EPI_ISL_1227738, EPI_ISL_1227739, EPI_ISL_1227740, EPI_ISL_1227741, EPI_ISL_1227742, EPI_ISL_1227743, EPI_ISL_1227744, EPI_ISL_1227745, EPI_ISL_1227746, EPI_ISL_1227747, EPI_ISL_1227748, EPI_ISL_1227749, EPI_ISL_1227750, EPI_ISL_1227751, EPI_ISL_1227752, EPI_ISL_1227753, EPI_ISL_1227754, EPI_ISL_1227755, EPI_ISL_1227756, EPI_ISL_1227757, EPI_ISL_1227758, EPI_ISL_1227759, EPI_ISL_1227760, EPI_ISL_1227761, EPI_ISL_1227762, EPI_ISL_1227763, EPI_ISL_1227764, EPI_ISL_1227765, EPI_ISL_1227766, EPI_ISL_1227767, EPI_ISL_1227768, EPI_ISL_1227769, EPI_ISL_1227770, EPI_ISL_1227771, EPI_ISL_1227772, EPI_ISL_1227773, EPI_ISL_1227774, EPI_ISL_1227775, EPI_ISL_1227776, EPI_ISL_1227777, EPI_ISL_1227778, EPI_ISL_1227779, EPI_ISL_1227780, EPI_ISL_1227781, EPI_ISL_1227782, EPI_ISL_1227783, EPI_ISL_1227784, EPI_ISL_1227785, EPI_ISL_1227786, EPI_ISL_1227787, EPI_ISL_1227788, EPI_ISL_1227789 | | | |
| see above | URMC LABS | Wadsworth Center, New York State Department of Health | Kirsten St. George, Daryl M. Lamson, Alexis Russel, Matthew Shudt, Melissa A Leisner, Jonathan Plitnick, Navjot Singh, John Kelly, Erasmus Schneider, Erica Lasek-Nesselquist |
| EPI_ISL_1227790 | ALBANY MEDICAL CENTER | Wadsworth Center, New York State Department of Health | Kirsten St. George, Daryl M. Lamson, Alexis Russel, Matthew Shudt, Melissa A Leisner, Jonathan Plitnick, Navjot Singh, John Kelly, Erasmus Schneider, Erica Lasek-Nesselquist |
| EPI_ISL_1227791, EPI_ISL_1227792, EPI_ISL_1227793, EPI_ISL_1227794, EPI_ISL_1227795, EPI_ISL_1227796, EPI_ISL_1227797, EPI_ISL_1227798, EPI_ISL_1227799, EPI_ISL_1227800, EPI_ISL_1227801, EPI_ISL_1227802, EPI_ISL_1227803 | | | |
| see above | MEMORIAL SLOAN KETTERING CANCER CENTER | Wadsworth Center, New York State Department of Health | Kirsten St. George, Daryl M. Lamson, Alexis Russel, Matthew Shudt, Melissa A Leisner, Jonathan Plitnick, Navjot Singh, John Kelly, Erasmus Schneider, Erica Lasek-Nesselquist |
| EPI_ISL_1227804 | NORTH SHORE UNIVERSITY HOSPITAL | Wadsworth Center, New York State Department of Health | Kirsten St. George, Daryl M. Lamson, Alexis Russel, Matthew Shudt, Melissa A Leisner, Jonathan Plitnick, Navjot Singh, John Kelly, Erasmus Schneider, Erica Lasek-Nesselquist |
| EPI_ISL_1227805, EPI_ISL_1227806, EPI_ISL_1227807, EPI_ISL_1227808, EPI_ISL_1227809, EPI_ISL_1227810, EPI_ISL_1227811, EPI_ISL_1227812 | MEMORIAL SLOAN KETTERING CANCER CENTER | Wadsworth Center, New York State Department of Health | Kirsten St. George, Daryl M. Lamson, Alexis Russel, Matthew Shudt, Melissa A Leisner, Jonathan Plitnick, Navjot Singh, John Kelly, Erasmus Schneider, Erica Lasek-Nesselquist |
| EPI_ISL_1227813 | NORTH SHORE UNIVERSITY HOSPITAL | Wadsworth Center, New York State Department of Health | Kirsten St. George, Daryl M. Lamson, Alexis Russel, Matthew Shudt, Melissa A Leisner, Jonathan Plitnick, Navjot Singh, John Kelly, Erasmus Schneider, Erica Lasek-Nesselquist |
| EPI_ISL_1227814, EPI_ISL_1227815 | MEMORIAL SLOAN KETTERING CANCER CENTER | Wadsworth Center, New York State Department of Health | Kirsten St. George, Daryl M. Lamson, Alexis Russel, Matthew Shudt, Melissa A Leisner, Jonathan Plitnick, Navjot Singh, John Kelly, Erasmus Schneider, Erica Lasek-Nesselquist |
| EPI_ISL_1227816 | NORTH SHORE UNIVERSITY HOSPITAL | Wadsworth Center, New York State Department of Health | Kirsten St. George, Daryl M. Lamson, Alexis Russel, Matthew Shudt, Melissa A Leisner, Jonathan Plitnick, Navjot Singh, John Kelly, Erasmus Schneider, Erica Lasek-Nesselquist |
| EPI_ISL_1227817, EPI_ISL_1227818 | MEMORIAL SLOAN KETTERING CANCER CENTER | Wadsworth Center, New York State Department of Health | Kirsten St. George, Daryl M. Lamson, Alexis Russel, Matthew Shudt, Melissa A Leisner, Jonathan Plitnick, Navjot Singh, John Kelly, Erasmus Schneider, Erica Lasek-Nesselquist |
| EPI_ISL_1227819 | NORTHWELL HEALTH LABORATORIES | Wadsworth Center, New York State Department of Health | Kirsten St. George, Daryl M. Lamson, Alexis Russel, Matthew Shudt, Melissa A Leisner, Jonathan Plitnick, Navjot Singh, John Kelly, Erasmus Schneider, Erica Lasek-Nesselquist |
| EPI_ISL_1227820 | MEMORIAL SLOAN KETTERING CANCER CENTER | Wadsworth Center, New York State Department of Health | Kirsten St. George, Daryl M. Lamson, Alexis Russel, Matthew Shudt, Melissa A Leisner, Jonathan Plitnick, Navjot Singh, John Kelly, Erasmus Schneider, Erica Lasek-Nesselquist |
| EPI_ISL_1227821, EPI_ISL_1227822, EPI_ISL_1227823, EPI_ISL_1227824, EPI_ISL_1227825, EPI_ISL_1227826 | NORTHWELL HEALTH LABORATORIES | Wadsworth Center, New York State Department of Health | Kirsten St. George, Daryl M. Lamson, Alexis Russel, Matthew Shudt, Melissa A Leisner, Jonathan Plitnick, Navjot Singh, John Kelly, Erasmus Schneider, Erica Lasek-Nesselquist |
| EPI_ISL_1227827, EPI_ISL_1227828, EPI_ISL_1227829 | MEMORIAL SLOAN KETTERING CANCER CENTER | Wadsworth Center, New York State Department of Health | Kirsten St. George, Daryl M. Lamson, Alexis Russel, Matthew Shudt, Melissa A Leisner, Jonathan Plitnick, Navjot Singh, John Kelly, Erasmus Schneider, Erica Lasek-Nesselquist |
| EPI_ISL_1227830 | NORTH SHORE UNIVERSITY HOSPITAL | Wadsworth Center, New York State Department of Health | Kirsten St. George, Daryl M. Lamson, Alexis Russel, Matthew Shudt, Melissa A Leisner, Jonathan Plitnick, Navjot Singh, John Kelly, Erasmus Schneider, Erica Lasek-Nesselquist |
| EPI_ISL_1227831, EPI_ISL_1227832, EPI_ISL_1227833, EPI_ISL_1227834, EPI_ISL_1227835, | NORTHWELL HEALTH LABORATORIES | Wadsworth Center, New York State Department of Health | Kirsten St. George, Daryl M. Lamson, Alexis Russel, Matthew Shudt, Melissa A Leisner, Jonathan Plitnick, Navjot Singh, John Kelly, Erasmus Schneider, Erica Lasek-Nesselquist |

| | | | |
|---|---|---|---|
| EPI_ISL_1227836 | | | |
| EPI_ISL_1227837, EPI_ISL_1227838 | MEMORIAL SLOAN KETTERING CANCER CENTER | Wadsworth Center, New York State Department of Health | Kirsten St. George, Daryl M. Lamson, Alexis Russel, Matthew Shudt, Melissa A Leisner, Jonathan Plitnick, Navjot Singh, John Kelly, Erasmus Schneider, Erica Lasek-Nesselquist |
| EPI_ISL_1227839, EPI_ISL_1227840, EPI_ISL_1227841, EPI_ISL_1227842 | NORTHWELL HEALTH LABORATORIES | Wadsworth Center, New York State Department of Health | Kirsten St. George, Daryl M. Lamson, Alexis Russel, Matthew Shudt, Melissa A Leisner, Jonathan Plitnick, Navjot Singh, John Kelly, Erasmus Schneider, Erica Lasek-Nesselquist |
| EPI_ISL_1227843, EPI_ISL_1227844, EPI_ISL_1227845, EPI_ISL_1227846, EPI_ISL_1227847 | MEMORIAL SLOAN KETTERING CANCER CENTER | Wadsworth Center, New York State Department of Health | Kirsten St. George, Daryl M. Lamson, Alexis Russel, Matthew Shudt, Melissa A Leisner, Jonathan Plitnick, Navjot Singh, John Kelly, Erasmus Schneider, Erica Lasek-Nesselquist |
| EPI_ISL_1227848, EPI_ISL_1227849, EPI_ISL_1227850 | NORTHWELL HEALTH LABORATORIES | Wadsworth Center, New York State Department of Health | Kirsten St. George, Daryl M. Lamson, Alexis Russel, Matthew Shudt, Melissa A Leisner, Jonathan Plitnick, Navjot Singh, John Kelly, Erasmus Schneider, Erica Lasek-Nesselquist |
| EPI_ISL_1227851 | MEMORIAL SLOAN KETTERING CANCER CENTER | Wadsworth Center, New York State Department of Health | Kirsten St. George, Daryl M. Lamson, Alexis Russel, Matthew Shudt, Melissa A Leisner, Jonathan Plitnick, Navjot Singh, John Kelly, Erasmus Schneider, Erica Lasek-Nesselquist |
| EPI_ISL_1227852 | NORTH SHORE UNIVERSITY HOSPITAL | Wadsworth Center, New York State Department of Health | Kirsten St. George, Daryl M. Lamson, Alexis Russel, Matthew Shudt, Melissa A Leisner, Jonathan Plitnick, Navjot Singh, John Kelly, Erasmus Schneider, Erica Lasek-Nesselquist |
| EPI_ISL_1227853, EPI_ISL_1227854, EPI_ISL_1227855, EPI_ISL_1227856, EPI_ISL_1227857, EPI_ISL_1227858, EPI_ISL_1227859, EPI_ISL_1227860 | NORTHWELL HEALTH LABORATORIES | Wadsworth Center, New York State Department of Health | Kirsten St. George, Daryl M. Lamson, Alexis Russel, Matthew Shudt, Melissa A Leisner, Jonathan Plitnick, Navjot Singh, John Kelly, Erasmus Schneider, Erica Lasek-Nesselquist |
| EPI_ISL_1227861, EPI_ISL_1227862, EPI_ISL_1227863, EPI_ISL_1227864, EPI_ISL_1227865, EPI_ISL_1227866 | NYC Pandemic Response Lab | Wadsworth Center, New York State Department of Health | Kirsten St. George, Daryl M. Lamson, Alexis Russel, Matthew Shudt, Melissa A Leisner, Jonathan Plitnick, Navjot Singh, John Kelly, Erasmus Schneider, Erica Lasek-Nesselquist |
| EPI_ISL_1227867 | New York Presbyterian Hospital | Wadsworth Center, New York State Department of Health | Kirsten St. George, Daryl M. Lamson, Alexis Russel, Matthew Shudt, Melissa A Leisner, Jonathan Plitnick, Navjot Singh, John Kelly, Erasmus Schneider, Erica Lasek-Nesselquist |
| EPI_ISL_1227868, EPI_ISL_1227869 | NYC Pandemic Response Lab | Wadsworth Center, New York State Department of Health | Kirsten St. George, Daryl M. Lamson, Alexis Russel, Matthew Shudt, Melissa A Leisner, Jonathan Plitnick, Navjot Singh, John Kelly, Erasmus Schneider, Erica Lasek-Nesselquist |
| EPI_ISL_1227870 | NORTH SHORE UNIVERSITY HOSPITAL | Wadsworth Center, New York State Department of Health | Kirsten St. George, Daryl M. Lamson, Alexis Russel, Matthew Shudt, Melissa A Leisner, Jonathan Plitnick, Navjot Singh, John Kelly, Erasmus Schneider, Erica Lasek-Nesselquist |
| EPI_ISL_1227871, EPI_ISL_1227872, EPI_ISL_1227873, EPI_ISL_1227874, EPI_ISL_1227875, EPI_ISL_1227876, EPI_ISL_1227877, EPI_ISL_1227878, EPI_ISL_1227879 | NYC Pandemic Response Lab | Wadsworth Center, New York State Department of Health | Kirsten St. George, Daryl M. Lamson, Alexis Russel, Matthew Shudt, Melissa A Leisner, Jonathan Plitnick, Navjot Singh, John Kelly, Erasmus Schneider, Erica Lasek-Nesselquist |
| EPI_ISL_1227880, EPI_ISL_1227881, EPI_ISL_1227882, EPI_ISL_1227883, EPI_ISL_1227884, EPI_ISL_1227885, EPI_ISL_1227886, EPI_ISL_1227887, EPI_ISL_1227888, EPI_ISL_1227889, EPI_ISL_1227890, EPI_ISL_1227891, EPI_ISL_1227892, EPI_ISL_1227893, EPI_ISL_1227894 | | | |
| see above | MEMORIAL SLOAN KETTERING CANCER CENTER | Wadsworth Center, New York State Department of Health | Kirsten St. George, Daryl M. Lamson, Alexis Russel, Matthew Shudt, Melissa A Leisner, Jonathan Plitnick, Navjot Singh, John Kelly, Erasmus Schneider, Erica Lasek-Nesselquist |
| EPI_ISL_1227895, EPI_ISL_1227896, EPI_ISL_1227897, EPI_ISL_1227898, EPI_ISL_1227899, EPI_ISL_1227900, EPI_ISL_1227901, EPI_ISL_1227902 | Columbia University Irving Medical Center | Wadsworth Center, New York State Department of Health | Kirsten St. George, Daryl M. Lamson, Alexis Russel, Matthew Shudt, Melissa A Leisner, Jonathan Plitnick, Navjot Singh, John Kelly, Erasmus Schneider, Erica Lasek-Nesselquist |
| EPI_ISL_1227903 | NORTHWELL HEALTH LABORATORIES | Wadsworth Center, New York State Department of Health | Kirsten St. George, Daryl M. Lamson, Alexis Russel, Matthew Shudt, Melissa A Leisner, Jonathan Plitnick, Navjot Singh, John Kelly, Erasmus Schneider, Erica Lasek-Nesselquist |
| EPI_ISL_1227904, EPI_ISL_1227905, EPI_ISL_1227906, EPI_ISL_1227907, EPI_ISL_1227908 | Columbia University Irving Medical Center | Wadsworth Center, New York State Department of Health | Kirsten St. George, Daryl M. Lamson, Alexis Russel, Matthew Shudt, Melissa A Leisner, Jonathan Plitnick, Navjot Singh, John Kelly, Erasmus Schneider, Erica Lasek-Nesselquist |
| EPI_ISL_1227909 | MONTEFIORE MEDICAL CENTER LABORATORIES | Wadsworth Center, New York State Department of Health | Kirsten St. George, Daryl M. Lamson, Alexis Russel, Matthew Shudt, Melissa A Leisner, Jonathan Plitnick, Navjot Singh, John Kelly, Erasmus Schneider, Erica Lasek-Nesselquist |
| EPI_ISL_1227910 | Columbia University Irving Medical Center | Wadsworth Center, New York State Department of Health | Kirsten St. George, Daryl M. Lamson, Alexis Russel, Matthew Shudt, Melissa A Leisner, Jonathan Plitnick, Navjot Singh, John Kelly, Erasmus Schneider, Erica Lasek-Nesselquist |
| EPI_ISL_1227911, EPI_ISL_1227912 | MONTEFIORE MEDICAL CENTER LABORATORIES | Wadsworth Center, New York State Department of Health | Kirsten St. George, Daryl M. Lamson, Alexis Russel, Matthew Shudt, Melissa A Leisner, Jonathan Plitnick, Navjot Singh, John Kelly, Erasmus Schneider, Erica Lasek-Nesselquist |
| EPI_ISL_1227913, EPI_ISL_1227914, EPI_ISL_1227915, EPI_ISL_1227916, EPI_ISL_1227917, EPI_ISL_1227918, EPI_ISL_1227919 | Columbia University Irving Medical Center | Wadsworth Center, New York State Department of Health | Kirsten St. George, Daryl M. Lamson, Alexis Russel, Matthew Shudt, Melissa A Leisner, Jonathan Plitnick, Navjot Singh, John Kelly, Erasmus Schneider, Erica Lasek-Nesselquist |
| EPI_ISL_1227920 | NYC Pandemic Response Lab | Wadsworth Center, New York State Department of Health | Kirsten St. George, Daryl M. Lamson, Alexis Russel, Matthew Shudt, Melissa A Leisner, Jonathan Plitnick, Navjot Singh, John Kelly, Erasmus Schneider, Erica Lasek-Nesselquist |
| EPI_ISL_1227921, EPI_ISL_1227922, EPI_ISL_1227923, EPI_ISL_1227924, EPI_ISL_1227925, EPI_ISL_1227926, EPI_ISL_1227927, EPI_ISL_1227928, EPI_ISL_1227929, EPI_ISL_1227930, EPI_ISL_1227931 | | | |
| see above | Columbia University Irving Medical Center | Wadsworth Center, New York State Department of Health | Kirsten St. George, Daryl M. Lamson, Alexis Russel, Matthew Shudt, Melissa A Leisner, Jonathan Plitnick, Navjot Singh, John Kelly, Erasmus Schneider, Erica Lasek-Nesselquist |
| EPI_ISL_1227932, EPI_ISL_1227933 | MONTEFIORE MEDICAL CENTER LABORATORIES | Wadsworth Center, New York State Department of Health | Kirsten St. George, Daryl M. Lamson, Alexis Russel, Matthew Shudt, Melissa A Leisner, Jonathan Plitnick, Navjot Singh, John Kelly, Erasmus Schneider, Erica Lasek-Nesselquist |
| EPI_ISL_1227934 | Columbia University Irving Medical Center | Wadsworth Center, New York State Department of Health | Kirsten St. George, Daryl M. Lamson, Alexis Russel, Matthew Shudt, Melissa A Leisner, Jonathan Plitnick, Navjot Singh, John Kelly, Erasmus Schneider, Erica Lasek-Nesselquist |
| EPI_ISL_1227935, EPI_ISL_1227936, EPI_ISL_1227937 | New York Presbyterian Hospital | Wadsworth Center, New York State Department of Health | Kirsten St. George, Daryl M. Lamson, Alexis Russel, Matthew Shudt, Melissa A Leisner, Jonathan Plitnick, Navjot Singh, John Kelly, Erasmus Schneider, Erica Lasek-Nesselquist |
| EPI_ISL_1227938, EPI_ISL_1227939, EPI_ISL_1227940, EPI_ISL_1227941, EPI_ISL_1227942 | NYC Pandemic Response Lab | Wadsworth Center, New York State Department of Health | Kirsten St. George, Daryl M. Lamson, Alexis Russel, Matthew Shudt, Melissa A Leisner, Jonathan Plitnick, Navjot Singh, John Kelly, Erasmus Schneider, Erica Lasek-Nesselquist |
| EPI_ISL_1227943, | New York Presbyterian Hospital | Wadsworth Center, New York State | Kirsten St. George, Daryl M. Lamson, Alexis Russel, Matthew Shudt, Melissa A Leisner, Jonathan Plitnick, Navjot Singh, John Kelly, Erasmus Schneider, Erica Lasek-Nesselquist |

| | | | |
|---|---|---|---|
| EPI_ISL_1227944, EPI_ISL_1227945 | | Department of Health | |
| EPI_ISL_1227946, EPI_ISL_1227947, EPI_ISL_1227948, EPI_ISL_1227949, EPI_ISL_1227950, EPI_ISL_1227951, EPI_ISL_1227952, EPI_ISL_1227953, EPI_ISL_1227954, EPI_ISL_1227955, EPI_ISL_1227956, EPI_ISL_1227957, EPI_ISL_1227958 | | | |
| see above | NYC Pandemic Response Lab | Wadsworth Center, New York State Department of Health | Kirsten St. George, Daryl M. Lamson, Alexis Russel, Matthew Shudt, Melissa A Leisner, Jonathan Plitnick, Navjot Singh, John Kelly, Erasmus Schneider, Erica Lasek-Nesselquist |
| EPI_ISL_1227959 | MONTEFIORE MEDICAL CENTER LABORATORIES | Wadsworth Center, New York State Department of Health | Kirsten St. George, Daryl M. Lamson, Alexis Russel, Matthew Shudt, Melissa A Leisner, Jonathan Plitnick, Navjot Singh, John Kelly, Erasmus Schneider, Erica Lasek-Nesselquist |
| EPI_ISL_1227960 | Wadsworth Center, New York State Department of Health | Wadsworth Center, New York State Department of Health | Kirsten St. George, Daryl M. Lamson, Alexis Russel, Matthew Shudt, Melissa A Leisner, Jonathan Plitnick, Navjot Singh, John Kelly, Erasmus Schneider, Erica Lasek-Nesselquist |
| EPI_ISL_1227961 | Masonic Care Community of NY | Wadsworth Center, New York State Department of Health | Kirsten St. George, Daryl M. Lamson, Alexis Russel, Matthew Shudt, Melissa A Leisner, Jonathan Plitnick, Navjot Singh, John Kelly, Erasmus Schneider, Erica Lasek-Nesselquist |
| EPI_ISL_1227962, EPI_ISL_1227963, EPI_ISL_1227964 | THE MARY IMOGENE BASSETT HOSPITAL | Wadsworth Center, New York State Department of Health | Kirsten St. George, Daryl M. Lamson, Alexis Russel, Matthew Shudt, Melissa A Leisner, Jonathan Plitnick, Navjot Singh, John Kelly, Erasmus Schneider, Erica Lasek-Nesselquist |
| EPI_ISL_1227965, EPI_ISL_1227966, EPI_ISL_1227967, EPI_ISL_1227968, EPI_ISL_1227969, EPI_ISL_1227970, EPI_ISL_1227971, EPI_ISL_1227972 | KALEIDA CENTER FOR LABORATORY MEDICINE | Wadsworth Center, New York State Department of Health | Kirsten St. George, Daryl M. Lamson, Alexis Russel, Matthew Shudt, Melissa A Leisner, Jonathan Plitnick, Navjot Singh, John Kelly, Erasmus Schneider, Erica Lasek-Nesselquist |
| EPI_ISL_1227973 | URMC LABS | Wadsworth Center, New York State Department of Health | Kirsten St. George, Daryl M. Lamson, Alexis Russel, Matthew Shudt, Melissa A Leisner, Jonathan Plitnick, Navjot Singh, John Kelly, Erasmus Schneider, Erica Lasek-Nesselquist |
| EPI_ISL_1227974, EPI_ISL_1227975 | THE MARY IMOGENE BASSETT HOSPITAL | Wadsworth Center, New York State Department of Health | Kirsten St. George, Daryl M. Lamson, Alexis Russel, Matthew Shudt, Melissa A Leisner, Jonathan Plitnick, Navjot Singh, John Kelly, Erasmus Schneider, Erica Lasek-Nesselquist |
| EPI_ISL_1227976, EPI_ISL_1227977, EPI_ISL_1227978, EPI_ISL_1227979, EPI_ISL_1227980, EPI_ISL_1227981, EPI_ISL_1227982, EPI_ISL_1227983, EPI_ISL_1227984, EPI_ISL_1227985, EPI_ISL_1227986, EPI_ISL_1227987, EPI_ISL_1227988, EPI_ISL_1227989, EPI_ISL_1227990 | | | |
| see above | Masonic Care Community of NY | Wadsworth Center, New York State Department of Health | Kirsten St. George, Daryl M. Lamson, Alexis Russel, Matthew Shudt, Melissa A Leisner, Jonathan Plitnick, Navjot Singh, John Kelly, Erasmus Schneider, Erica Lasek-Nesselquist |
| EPI_ISL_1227991, EPI_ISL_1227992, EPI_ISL_1227993, EPI_ISL_1227994, EPI_ISL_1227995, EPI_ISL_1227996, EPI_ISL_1227997, EPI_ISL_1227998, EPI_ISL_1227999, EPI_ISL_1228000, EPI_ISL_1228001, EPI_ISL_1228002, EPI_ISL_1228003, EPI_ISL_1228004, EPI_ISL_1228005, EPI_ISL_1228006, EPI_ISL_1228007, EPI_ISL_1228008, EPI_ISL_1228009, EPI_ISL_1228010, EPI_ISL_1228011, EPI_ISL_1228012, EPI_ISL_1228013, EPI_ISL_1228014, EPI_ISL_1228015, EPI_ISL_1228016, EPI_ISL_1228017, EPI_ISL_1228018, EPI_ISL_1228019, EPI_ISL_1228020, EPI_ISL_1228021, EPI_ISL_1228022 | | | |
| see above | SUNY UPSTATE MEDICAL UNIVERSITY | Wadsworth Center, New York State Department of Health | Kirsten St. George, Daryl M. Lamson, Alexis Russel, Matthew Shudt, Melissa A Leisner, Jonathan Plitnick, Navjot Singh, John Kelly, Erasmus Schneider, Erica Lasek-Nesselquist |
| EPI_ISL_1228023, EPI_ISL_1228024, EPI_ISL_1228025, EPI_ISL_1228026, EPI_ISL_1228027, EPI_ISL_1228028, EPI_ISL_1228029, EPI_ISL_1228030, EPI_ISL_1228031 | URMC LABS | Wadsworth Center, New York State Department of Health | Kirsten St. George, Daryl M. Lamson, Alexis Russel, Matthew Shudt, Melissa A Leisner, Jonathan Plitnick, Navjot Singh, John Kelly, Erasmus Schneider, Erica Lasek-Nesselquist |
| EPI_ISL_1228032, EPI_ISL_1228033, EPI_ISL_1228034, EPI_ISL_1228035 | MEMORIAL SLOAN KETTERING CANCER CENTER | Wadsworth Center, New York State Department of Health | Kirsten St. George, Daryl M. Lamson, Alexis Russel, Matthew Shudt, Melissa A Leisner, Jonathan Plitnick, Navjot Singh, John Kelly, Erasmus Schneider, Erica Lasek-Nesselquist |
| EPI_ISL_1228036, EPI_ISL_1228037, EPI_ISL_1228038, EPI_ISL_1228039 | WESTCHESTER MEDICAL CENTER | Wadsworth Center, New York State Department of Health | Kirsten St. George, Daryl M. Lamson, Alexis Russel, Matthew Shudt, Melissa A Leisner, Jonathan Plitnick, Navjot Singh, John Kelly, Erasmus Schneider, Erica Lasek-Nesselquist |
| EPI_ISL_1228040, EPI_ISL_1228041 | URMC LABS | Wadsworth Center, New York State Department of Health | Kirsten St. George, Daryl M. Lamson, Alexis Russel, Matthew Shudt, Melissa A Leisner, Jonathan Plitnick, Navjot Singh, John Kelly, Erasmus Schneider, Erica Lasek-Nesselquist |
| EPI_ISL_1228042 | KALEIDA CENTER FOR LABORATORY MEDICINE | Wadsworth Center, New York State Department of Health | Kirsten St. George, Daryl M. Lamson, Alexis Russel, Matthew Shudt, Melissa A Leisner, Jonathan Plitnick, Navjot Singh, John Kelly, Erasmus Schneider, Erica Lasek-Nesselquist |
| EPI_ISL_1228043, EPI_ISL_1228044, EPI_ISL_1228045 | URMC LABS | Wadsworth Center, New York State Department of Health | Kirsten St. George, Daryl M. Lamson, Alexis Russel, Matthew Shudt, Melissa A Leisner, Jonathan Plitnick, Navjot Singh, John Kelly, Erasmus Schneider, Erica Lasek-Nesselquist |
| EPI_ISL_1228046 | Masonic Care Community of NY | Wadsworth Center, New York State Department of Health | Kirsten St. George, Daryl M. Lamson, Alexis Russel, Matthew Shudt, Melissa A Leisner, Jonathan Plitnick, Navjot Singh, John Kelly, Erasmus Schneider, Erica Lasek-Nesselquist |
| EPI_ISL_1228047, EPI_ISL_1228048 | SUNY UPSTATE MEDICAL UNIVERSITY | Wadsworth Center, New York State Department of Health | Kirsten St. George, Daryl M. Lamson, Alexis Russel, Matthew Shudt, Melissa A Leisner, Jonathan Plitnick, Navjot Singh, John Kelly, Erasmus Schneider, Erica Lasek-Nesselquist |
| EPI_ISL_1228049, EPI_ISL_1228050, EPI_ISL_1228051 | THE MARY IMOGENE BASSETT HOSPITAL | Wadsworth Center, New York State Department of Health | Kirsten St. George, Daryl M. Lamson, Alexis Russel, Matthew Shudt, Melissa A Leisner, Jonathan Plitnick, Navjot Singh, John Kelly, Erasmus Schneider, Erica Lasek-Nesselquist |
| EPI_ISL_1228052, EPI_ISL_1228053, EPI_ISL_1228054, EPI_ISL_1228055, EPI_ISL_1228056, EPI_ISL_1228057, EPI_ISL_1228058, EPI_ISL_1228059, EPI_ISL_1228060, EPI_ISL_1228061, EPI_ISL_1228062, EPI_ISL_1228063, EPI_ISL_1228064, EPI_ISL_1228065, EPI_ISL_1228066, EPI_ISL_1228067, EPI_ISL_1228068, EPI_ISL_1228069, EPI_ISL_1228070, EPI_ISL_1228071, EPI_ISL_1228072, EPI_ISL_1228073, EPI_ISL_1228074, EPI_ISL_1228075, EPI_ISL_1228076, EPI_ISL_1228077, EPI_ISL_1228078, EPI_ISL_1228079, EPI_ISL_1228080, EPI_ISL_1228081, EPI_ISL_1228082, EPI_ISL_1228083, EPI_ISL_1228084, EPI_ISL_1228085, EPI_ISL_1228086, EPI_ISL_1228087, EPI_ISL_1228088, EPI_ISL_1228089, EPI_ISL_1228090, EPI_ISL_1228091, EPI_ISL_1228092, EPI_ISL_1228093, EPI_ISL_1228094, EPI_ISL_1228095, EPI_ISL_1228096, EPI_ISL_1228097, EPI_ISL_1228098, EPI_ISL_1228099, EPI_ISL_1228100, EPI_ISL_1228101, EPI_ISL_1228102, EPI_ISL_1228103, EPI_ISL_1228104, EPI_ISL_1228105, EPI_ISL_1228106, EPI_ISL_1228107, EPI_ISL_1228108, EPI_ISL_1228109, EPI_ISL_1228110, EPI_ISL_1228111, EPI_ISL_1228112, EPI_ISL_1228113 | | | |
| see above | MEMORIAL SLOAN KETTERING CANCER CENTER | Wadsworth Center, New York State Department of Health | Kirsten St. George, Daryl M. Lamson, Alexis Russel, Matthew Shudt, Melissa A Leisner, Jonathan Plitnick, Navjot Singh, John Kelly, Erasmus Schneider, Erica Lasek-Nesselquist |
| EPI_ISL_1228114 | MONTEFIORE MEDICAL CENTER LABORATORIES | Wadsworth Center, New York State Department of Health | Kirsten St. George, Daryl M. Lamson, Alexis Russel, Matthew Shudt, Melissa A Leisner, Jonathan Plitnick, Navjot Singh, John Kelly, Erasmus Schneider, Erica Lasek-Nesselquist |
| EPI_ISL_1228115, EPI_ISL_1228116 | URMC LABS | Wadsworth Center, New York State Department of Health | Kirsten St. George, Daryl M. Lamson, Alexis Russel, Matthew Shudt, Melissa A Leisner, Jonathan Plitnick, Navjot Singh, John Kelly, Erasmus Schneider, Erica Lasek-Nesselquist |
| EPI_ISL_1228117, EPI_ISL_1228118 | MEMORIAL SLOAN KETTERING CANCER CENTER | Wadsworth Center, New York State Department of Health | Kirsten St. George, Daryl M. Lamson, Alexis Russel, Matthew Shudt, Melissa A Leisner, Jonathan Plitnick, Navjot Singh, John Kelly, Erasmus Schneider, Erica Lasek-Nesselquist |
| EPI_ISL_1228119, EPI_ISL_1228120, EPI_ISL_1228121 | WESTCHESTER MEDICAL CENTER | Wadsworth Center, New York State Department of Health | Kirsten St. George, Daryl M. Lamson, Alexis Russel, Matthew Shudt, Melissa A Leisner, Jonathan Plitnick, Navjot Singh, John Kelly, Erasmus Schneider, Erica Lasek-Nesselquist |
| EPI_ISL_1228122, EPI_ISL_1228123, EPI_ISL_1228124, EPI_ISL_1228125, EPI_ISL_1228126, EPI_ISL_1228127, EPI_ISL_1228128, EPI_ISL_1228129 | MEMORIAL SLOAN KETTERING CANCER CENTER | Wadsworth Center, New York State Department of Health | Kirsten St. George, Daryl M. Lamson, Alexis Russel, Matthew Shudt, Melissa A Leisner, Jonathan Plitnick, Navjot Singh, John Kelly, Erasmus Schneider, Erica Lasek-Nesselquist |
| EPI_ISL_1228130 | NORTH SHORE UNIVERSITY HOSPITAL | Wadsworth Center, New York State Department of Health | Kirsten St. George, Daryl M. Lamson, Alexis Russel, Matthew Shudt, Melissa A Leisner, Jonathan Plitnick, Navjot Singh, John Kelly, Erasmus Schneider, Erica Lasek-Nesselquist |
| EPI_ISL_1228131, EPI_ISL_1228132, EPI_ISL_1228133, EPI_ISL_1228134 | MEMORIAL SLOAN KETTERING CANCER CENTER | Wadsworth Center, New York State Department of Health | Kirsten St. George, Daryl M. Lamson, Alexis Russel, Matthew Shudt, Melissa A Leisner, Jonathan Plitnick, Navjot Singh, John Kelly, Erasmus Schneider, Erica Lasek-Nesselquist |
| EPI_ISL_1228135 | NORTH SHORE UNIVERSITY HOSPITAL | Wadsworth Center, New York State Department of Health | Kirsten St. George, Daryl M. Lamson, Alexis Russel, Matthew Shudt, Melissa A Leisner, Jonathan Plitnick, Navjot Singh, John Kelly, Erasmus Schneider, Erica Lasek-Nesselquist |
| EPI_ISL_1228136, | MEMORIAL SLOAN KETTERING CANCER | Wadsworth Center, New York State | Kirsten St. George, Daryl M. Lamson, Alexis Russel, Matthew Shudt, Melissa A Leisner, Jonathan Plitnick, Navjot Singh, John Kelly, Erasmus Schneider, Erica Lasek-Nesselquist |

| | | | |
|---|---|---|---|
| EPI_ISL_1228137, EPI_ISL_1228138 | CENTER | Department of Health | |
| EPI_ISL_1228139, EPI_ISL_1228140 | NORTH SHORE UNIVERSITY HOSPITAL | Wadsworth Center, New York State Department of Health | Kirsten St. George, Daryl M. Lamson, Alexis Russel, Matthew Shudt, Melissa A Leisner, Jonathan Plitnick, Navjot Singh, John Kelly, Erasmus Schneider, Erica Lasek-Nesselquist |
| EPI_ISL_1228141, EPI_ISL_1228142 | MEMORIAL SLOAN KETTERING CANCER CENTER | Wadsworth Center, New York State Department of Health | Kirsten St. George, Daryl M. Lamson, Alexis Russel, Matthew Shudt, Melissa A Leisner, Jonathan Plitnick, Navjot Singh, John Kelly, Erasmus Schneider, Erica Lasek-Nesselquist |
| EPI_ISL_1228143 | NYC Pandemic Response Lab | Wadsworth Center, New York State Department of Health | Kirsten St. George, Daryl M. Lamson, Alexis Russel, Matthew Shudt, Melissa A Leisner, Jonathan Plitnick, Navjot Singh, John Kelly, Erasmus Schneider, Erica Lasek-Nesselquist |
| EPI_ISL_1228144, EPI_ISL_1228145, EPI_ISL_1228146 | MEMORIAL SLOAN KETTERING CANCER CENTER | Wadsworth Center, New York State Department of Health | Kirsten St. George, Daryl M. Lamson, Alexis Russel, Matthew Shudt, Melissa A Leisner, Jonathan Plitnick, Navjot Singh, John Kelly, Erasmus Schneider, Erica Lasek-Nesselquist |
| EPI_ISL_1228147, EPI_ISL_1228148, EPI_ISL_1228149 | NORTH SHORE UNIVERSITY HOSPITAL | Wadsworth Center, New York State Department of Health | Kirsten St. George, Daryl M. Lamson, Alexis Russel, Matthew Shudt, Melissa A Leisner, Jonathan Plitnick, Navjot Singh, John Kelly, Erasmus Schneider, Erica Lasek-Nesselquist |
| EPI_ISL_1228150, EPI_ISL_1228151, EPI_ISL_1228152, EPI_ISL_1228153 | NORTHWELL HEALTH LABORATORIES | Wadsworth Center, New York State Department of Health | Kirsten St. George, Daryl M. Lamson, Alexis Russel, Matthew Shudt, Melissa A Leisner, Jonathan Plitnick, Navjot Singh, John Kelly, Erasmus Schneider, Erica Lasek-Nesselquist |
| EPI_ISL_1228154 | WESTCHESTER MEDICAL CENTER | Wadsworth Center, New York State Department of Health | Kirsten St. George, Daryl M. Lamson, Alexis Russel, Matthew Shudt, Melissa A Leisner, Jonathan Plitnick, Navjot Singh, John Kelly, Erasmus Schneider, Erica Lasek-Nesselquist |
| EPI_ISL_1228155, EPI_ISL_1228156, EPI_ISL_1228157 | NORTHWELL HEALTH LABORATORIES | Wadsworth Center, New York State Department of Health | Kirsten St. George, Daryl M. Lamson, Alexis Russel, Matthew Shudt, Melissa A Leisner, Jonathan Plitnick, Navjot Singh, John Kelly, Erasmus Schneider, Erica Lasek-Nesselquist |
| EPI_ISL_1228158 | Columbia University Irving Medical Center | Wadsworth Center, New York State Department of Health | Kirsten St. George, Daryl M. Lamson, Alexis Russel, Matthew Shudt, Melissa A Leisner, Jonathan Plitnick, Navjot Singh, John Kelly, Erasmus Schneider, Erica Lasek-Nesselquist |
| EPI_ISL_1228159, EPI_ISL_1228160, EPI_ISL_1228161 | NORTH SHORE UNIVERSITY HOSPITAL | Wadsworth Center, New York State Department of Health | Kirsten St. George, Daryl M. Lamson, Alexis Russel, Matthew Shudt, Melissa A Leisner, Jonathan Plitnick, Navjot Singh, John Kelly, Erasmus Schneider, Erica Lasek-Nesselquist |
| EPI_ISL_1228162, EPI_ISL_1228163, EPI_ISL_1228164, EPI_ISL_1228165, EPI_ISL_1228166, EPI_ISL_1228167, EPI_ISL_1228168, EPI_ISL_1228169 | NYC Pandemic Response Lab | Wadsworth Center, New York State Department of Health | Kirsten St. George, Daryl M. Lamson, Alexis Russel, Matthew Shudt, Melissa A Leisner, Jonathan Plitnick, Navjot Singh, John Kelly, Erasmus Schneider, Erica Lasek-Nesselquist |
| EPI_ISL_1228170 | NORTHWELL HEALTH LABORATORIES | Wadsworth Center, New York State Department of Health | Kirsten St. George, Daryl M. Lamson, Alexis Russel, Matthew Shudt, Melissa A Leisner, Jonathan Plitnick, Navjot Singh, John Kelly, Erasmus Schneider, Erica Lasek-Nesselquist |
| EPI_ISL_1228171, EPI_ISL_1228172 | NORTH SHORE UNIVERSITY HOSPITAL | Wadsworth Center, New York State Department of Health | Kirsten St. George, Daryl M. Lamson, Alexis Russel, Matthew Shudt, Melissa A Leisner, Jonathan Plitnick, Navjot Singh, John Kelly, Erasmus Schneider, Erica Lasek-Nesselquist |
| EPI_ISL_1228173 | MONTEFIORE MEDICAL CENTER LABORATORIES | Wadsworth Center, New York State Department of Health | Kirsten St. George, Daryl M. Lamson, Alexis Russel, Matthew Shudt, Melissa A Leisner, Jonathan Plitnick, Navjot Singh, John Kelly, Erasmus Schneider, Erica Lasek-Nesselquist |
| EPI_ISL_1228174, EPI_ISL_1228175 | NORTH SHORE UNIVERSITY HOSPITAL | Wadsworth Center, New York State Department of Health | Kirsten St. George, Daryl M. Lamson, Alexis Russel, Matthew Shudt, Melissa A Leisner, Jonathan Plitnick, Navjot Singh, John Kelly, Erasmus Schneider, Erica Lasek-Nesselquist |
| EPI_ISL_1228176 | MONTEFIORE MEDICAL CENTER LABORATORIES | Wadsworth Center, New York State Department of Health | Kirsten St. George, Daryl M. Lamson, Alexis Russel, Matthew Shudt, Melissa A Leisner, Jonathan Plitnick, Navjot Singh, John Kelly, Erasmus Schneider, Erica Lasek-Nesselquist |
| EPI_ISL_1228177, EPI_ISL_1228178, EPI_ISL_1228179, EPI_ISL_1228180 | NYC Pandemic Response Lab | Wadsworth Center, New York State Department of Health | Kirsten St. George, Daryl M. Lamson, Alexis Russel, Matthew Shudt, Melissa A Leisner, Jonathan Plitnick, Navjot Singh, John Kelly, Erasmus Schneider, Erica Lasek-Nesselquist |
| EPI_ISL_1228181 | New York Presbyterian Hospital | Wadsworth Center, New York State Department of Health | Kirsten St. George, Daryl M. Lamson, Alexis Russel, Matthew Shudt, Melissa A Leisner, Jonathan Plitnick, Navjot Singh, John Kelly, Erasmus Schneider, Erica Lasek-Nesselquist |
| EPI_ISL_1228182 | NYC Pandemic Response Lab | Wadsworth Center, New York State Department of Health | Kirsten St. George, Daryl M. Lamson, Alexis Russel, Matthew Shudt, Melissa A Leisner, Jonathan Plitnick, Navjot Singh, John Kelly, Erasmus Schneider, Erica Lasek-Nesselquist |
| EPI_ISL_1228183, EPI_ISL_1228184 | New York Presbyterian Hospital | Wadsworth Center, New York State Department of Health | Kirsten St. George, Daryl M. Lamson, Alexis Russel, Matthew Shudt, Melissa A Leisner, Jonathan Plitnick, Navjot Singh, John Kelly, Erasmus Schneider, Erica Lasek-Nesselquist |
| EPI_ISL_1228185, EPI_ISL_1228186, EPI_ISL_1228187 | NYC Pandemic Response Lab | Wadsworth Center, New York State Department of Health | Kirsten St. George, Daryl M. Lamson, Alexis Russel, Matthew Shudt, Melissa A Leisner, Jonathan Plitnick, Navjot Singh, John Kelly, Erasmus Schneider, Erica Lasek-Nesselquist |
| EPI_ISL_1228188 | MONTEFIORE MEDICAL CENTER LABORATORIES | Wadsworth Center, New York State Department of Health | Kirsten St. George, Daryl M. Lamson, Alexis Russel, Matthew Shudt, Melissa A Leisner, Jonathan Plitnick, Navjot Singh, John Kelly, Erasmus Schneider, Erica Lasek-Nesselquist |
| EPI_ISL_1228189, EPI_ISL_1228190, EPI_ISL_1228191, EPI_ISL_1228192, EPI_ISL_1228193, EPI_ISL_1228194, EPI_ISL_1228195, EPI_ISL_1228196, EPI_ISL_1228197, EPI_ISL_1228198, EPI_ISL_1228199, EPI_ISL_1228200, EPI_ISL_1228201, EPI_ISL_1228202, EPI_ISL_1228203, EPI_ISL_1228204, EPI_ISL_1228205, EPI_ISL_1228206, EPI_ISL_1228207, EPI_ISL_1228208, EPI_ISL_1228209, EPI_ISL_1228210, EPI_ISL_1228211, EPI_ISL_1228212, EPI_ISL_1228213, EPI_ISL_1228214, EPI_ISL_1228215, EPI_ISL_1228216, EPI_ISL_1228217 | | | |
| see above | NYC Pandemic Response Lab | Wadsworth Center, New York State Department of Health | Kirsten St. George, Daryl M. Lamson, Alexis Russel, Matthew Shudt, Melissa A Leisner, Jonathan Plitnick, Navjot Singh, John Kelly, Erasmus Schneider, Erica Lasek-Nesselquist |
| EPI_ISL_1228218, EPI_ISL_1228219, EPI_ISL_1228220, EPI_ISL_1228221 | ALBANY MEDICAL CENTER | Wadsworth Center, New York State Department of Health | Kirsten St. George, Daryl M. Lamson, Alexis Russel, Matthew Shudt, Melissa A Leisner, Jonathan Plitnick, Navjot Singh, John Kelly, Erasmus Schneider, Erica Lasek-Nesselquist |
| EPI_ISL_1228222, EPI_ISL_1228223 | Wadsworth Center, New York State Department of Health | Wadsworth Center, New York State Department of Health | Kirsten St. George, Daryl M. Lamson, Alexis Russel, Matthew Shudt, Melissa A Leisner, Jonathan Plitnick, Navjot Singh, John Kelly, Erasmus Schneider, Erica Lasek-Nesselquist |
| EPI_ISL_1228224, EPI_ISL_1228225, EPI_ISL_1228226, EPI_ISL_1228227 | MEMORIAL SLOAN KETTERING CANCER CENTER | Wadsworth Center, New York State Department of Health | Kirsten St. George, Daryl M. Lamson, Alexis Russel, Matthew Shudt, Melissa A Leisner, Jonathan Plitnick, Navjot Singh, John Kelly, Erasmus Schneider, Erica Lasek-Nesselquist |
| EPI_ISL_1228228 | NYC Pandemic Response Lab | Wadsworth Center, New York State Department of Health | Kirsten St. George, Daryl M. Lamson, Alexis Russel, Matthew Shudt, Melissa A Leisner, Jonathan Plitnick, Navjot Singh, John Kelly, Erasmus Schneider, Erica Lasek-Nesselquist |
| EPI_ISL_1228229 | MEMORIAL SLOAN KETTERING CANCER CENTER | Wadsworth Center, New York State Department of Health | Kirsten St. George, Daryl M. Lamson, Alexis Russel, Matthew Shudt, Melissa A Leisner, Jonathan Plitnick, Navjot Singh, John Kelly, Erasmus Schneider, Erica Lasek-Nesselquist |
| EPI_ISL_1228230, EPI_ISL_1228231, EPI_ISL_1228232, EPI_ISL_1228233, EPI_ISL_1228234 | NYC Pandemic Response Lab | Wadsworth Center, New York State Department of Health | Kirsten St. George, Daryl M. Lamson, Alexis Russel, Matthew Shudt, Melissa A Leisner, Jonathan Plitnick, Navjot Singh, John Kelly, Erasmus Schneider, Erica Lasek-Nesselquist |
| EPI_ISL_1228235 | NORTHWELL HEALTH LABORATORIES | Wadsworth Center, New York State Department of Health | Kirsten St. George, Daryl M. Lamson, Alexis Russel, Matthew Shudt, Melissa A Leisner, Jonathan Plitnick, Navjot Singh, John Kelly, Erasmus Schneider, Erica Lasek-Nesselquist |
| EPI_ISL_1228236 | MEMORIAL SLOAN KETTERING CANCER CENTER | Wadsworth Center, New York State Department of Health | Kirsten St. George, Daryl M. Lamson, Alexis Russel, Matthew Shudt, Melissa A Leisner, Jonathan Plitnick, Navjot Singh, John Kelly, Erasmus Schneider, Erica Lasek-Nesselquist |
| EPI_ISL_1228237, EPI_ISL_1228238, EPI_ISL_1228239 | NYC Pandemic Response Lab | Wadsworth Center, New York State Department of Health | Kirsten St. George, Daryl M. Lamson, Alexis Russel, Matthew Shudt, Melissa A Leisner, Jonathan Plitnick, Navjot Singh, John Kelly, Erasmus Schneider, Erica Lasek-Nesselquist |
| EPI_ISL_1228240, EPI_ISL_1228241, EPI_ISL_1228242, EPI_ISL_1228243, EPI_ISL_1228244, EPI_ISL_1228245, EPI_ISL_1228246, EPI_ISL_1228247, EPI_ISL_1228248, EPI_ISL_1228249, EPI_ISL_1228250, EPI_ISL_1228251 | | | |
| see above | MEMORIAL SLOAN KETTERING CANCER CENTER | Wadsworth Center, New York State Department of Health | Kirsten St. George, Daryl M. Lamson, Alexis Russel, Matthew Shudt, Melissa A Leisner, Jonathan Plitnick, Navjot Singh, John Kelly, Erasmus Schneider, Erica Lasek-Nesselquist |

| | | | |
|---|---|---|---|
| EPI_ISL_1228252 | WESTCHESTER MEDICAL CENTER | Wadsworth Center, New York State Department of Health | Kirsten St. George, Daryl M. Lamson, Alexis Russel, Matthew Shudt, Melissa A Leisner, Jonathan Plitnick, Navjot Singh, John Kelly, Erasmus Schneider, Erica Lasek-Nesselquist |
| EPI_ISL_1228253 | Columbia University Irving Medical Center | Wadsworth Center, New York State Department of Health | Kirsten St. George, Daryl M. Lamson, Alexis Russel, Matthew Shudt, Melissa A Leisner, Jonathan Plitnick, Navjot Singh, John Kelly, Erasmus Schneider, Erica Lasek-Nesselquist |
| EPI_ISL_1228254 | MEMORIAL SLOAN KETTERING CANCER CENTER | Wadsworth Center, New York State Department of Health | Kirsten St. George, Daryl M. Lamson, Alexis Russel, Matthew Shudt, Melissa A Leisner, Jonathan Plitnick, Navjot Singh, John Kelly, Erasmus Schneider, Erica Lasek-Nesselquist |
| EPI_ISL_1228255 | WESTCHESTER MEDICAL CENTER | Wadsworth Center, New York State Department of Health | Kirsten St. George, Daryl M. Lamson, Alexis Russel, Matthew Shudt, Melissa A Leisner, Jonathan Plitnick, Navjot Singh, John Kelly, Erasmus Schneider, Erica Lasek-Nesselquist |
| EPI_ISL_1228256 | MONTEFIORE MEDICAL CENTER LABORATORIES | Wadsworth Center, New York State Department of Health | Kirsten St. George, Daryl M. Lamson, Alexis Russel, Matthew Shudt, Melissa A Leisner, Jonathan Plitnick, Navjot Singh, John Kelly, Erasmus Schneider, Erica Lasek-Nesselquist |
| EPI_ISL_1228257, EPI_ISL_1228258, EPI_ISL_1228259 | WESTCHESTER MEDICAL CENTER | Wadsworth Center, New York State Department of Health | Kirsten St. George, Daryl M. Lamson, Alexis Russel, Matthew Shudt, Melissa A Leisner, Jonathan Plitnick, Navjot Singh, John Kelly, Erasmus Schneider, Erica Lasek-Nesselquist |
| EPI_ISL_1228260 | Wadsworth Center, New York State Department of Health | Wadsworth Center, New York State Department of Health | Kirsten St. George, Daryl M. Lamson, Alexis Russel, Matthew Shudt, Melissa A Leisner, Jonathan Plitnick, Navjot Singh, John Kelly, Erasmus Schneider, Erica Lasek-Nesselquist |
| EPI_ISL_1228261 | WESTCHESTER MEDICAL CENTER | Wadsworth Center, New York State Department of Health | Kirsten St. George, Daryl M. Lamson, Alexis Russel, Matthew Shudt, Melissa A Leisner, Jonathan Plitnick, Navjot Singh, John Kelly, Erasmus Schneider, Erica Lasek-Nesselquist |
| EPI_ISL_1228262 | MONTEFIORE MEDICAL CENTER LABORATORIES | Wadsworth Center, New York State Department of Health | Kirsten St. George, Daryl M. Lamson, Alexis Russel, Matthew Shudt, Melissa A Leisner, Jonathan Plitnick, Navjot Singh, John Kelly, Erasmus Schneider, Erica Lasek-Nesselquist |
| EPI_ISL_1228263, EPI_ISL_1228264 | WESTCHESTER MEDICAL CENTER | Wadsworth Center, New York State Department of Health | Kirsten St. George, Daryl M. Lamson, Alexis Russel, Matthew Shudt, Melissa A Leisner, Jonathan Plitnick, Navjot Singh, John Kelly, Erasmus Schneider, Erica Lasek-Nesselquist |
| EPI_ISL_1228265 | NYC Pandemic Response Lab | Wadsworth Center, New York State Department of Health | Kirsten St. George, Daryl M. Lamson, Alexis Russel, Matthew Shudt, Melissa A Leisner, Jonathan Plitnick, Navjot Singh, John Kelly, Erasmus Schneider, Erica Lasek-Nesselquist |
| EPI_ISL_1228266 | MONTEFIORE MEDICAL CENTER LABORATORIES | Wadsworth Center, New York State Department of Health | Kirsten St. George, Daryl M. Lamson, Alexis Russel, Matthew Shudt, Melissa A Leisner, Jonathan Plitnick, Navjot Singh, John Kelly, Erasmus Schneider, Erica Lasek-Nesselquist |
| EPI_ISL_1228267 | WESTCHESTER MEDICAL CENTER | Wadsworth Center, New York State Department of Health | Kirsten St. George, Daryl M. Lamson, Alexis Russel, Matthew Shudt, Melissa A Leisner, Jonathan Plitnick, Navjot Singh, John Kelly, Erasmus Schneider, Erica Lasek-Nesselquist |
| EPI_ISL_1228268, EPI_ISL_1228269 | GLENS FALLS HOSPITAL LABORATORY | Wadsworth Center, New York State Department of Health | Kirsten St. George, Daryl M. Lamson, Alexis Russel, Matthew Shudt, Melissa A Leisner, Jonathan Plitnick, Navjot Singh, John Kelly, Erasmus Schneider, Erica Lasek-Nesselquist |
| EPI_ISL_1228270 | ALBANY MEDICAL CENTER | Wadsworth Center, New York State Department of Health | Kirsten St. George, Daryl M. Lamson, Alexis Russel, Matthew Shudt, Melissa A Leisner, Jonathan Plitnick, Navjot Singh, John Kelly, Erasmus Schneider, Erica Lasek-Nesselquist |
| EPI_ISL_1228271 | Wadsworth Center, New York State Department of Health | Wadsworth Center, New York State Department of Health | Kirsten St. George, Daryl M. Lamson, Alexis Russel, Matthew Shudt, Melissa A Leisner, Jonathan Plitnick, Navjot Singh, John Kelly, Erasmus Schneider, Erica Lasek-Nesselquist |
| EPI_ISL_1228272, EPI_ISL_1228273, EPI_ISL_1228274 | GLENS FALLS HOSPITAL LABORATORY | Wadsworth Center, New York State Department of Health | Kirsten St. George, Daryl M. Lamson, Alexis Russel, Matthew Shudt, Melissa A Leisner, Jonathan Plitnick, Navjot Singh, John Kelly, Erasmus Schneider, Erica Lasek-Nesselquist |
| EPI_ISL_1228275, EPI_ISL_1228276, EPI_ISL_1228277 | ALBANY MEDICAL CENTER | Wadsworth Center, New York State Department of Health | Kirsten St. George, Daryl M. Lamson, Alexis Russel, Matthew Shudt, Melissa A Leisner, Jonathan Plitnick, Navjot Singh, John Kelly, Erasmus Schneider, Erica Lasek-Nesselquist |
| EPI_ISL_1228278 | SARATOGA HOSPITAL LABORATORY | Wadsworth Center, New York State Department of Health | Kirsten St. George, Daryl M. Lamson, Alexis Russel, Matthew Shudt, Melissa A Leisner, Jonathan Plitnick, Navjot Singh, John Kelly, Erasmus Schneider, Erica Lasek-Nesselquist |
| EPI_ISL_1228279 | ALBANY MEDICAL CENTER | Wadsworth Center, New York State Department of Health | Kirsten St. George, Daryl M. Lamson, Alexis Russel, Matthew Shudt, Melissa A Leisner, Jonathan Plitnick, Navjot Singh, John Kelly, Erasmus Schneider, Erica Lasek-Nesselquist |
| EPI_ISL_1228280 | Masonic Care Community of NY | Wadsworth Center, New York State Department of Health | Kirsten St. George, Daryl M. Lamson, Alexis Russel, Matthew Shudt, Melissa A Leisner, Jonathan Plitnick, Navjot Singh, John Kelly, Erasmus Schneider, Erica Lasek-Nesselquist |
| EPI_ISL_1228281 | Wadsworth Center, New York State Department of Health | Wadsworth Center, New York State Department of Health | Kirsten St. George, Daryl M. Lamson, Alexis Russel, Matthew Shudt, Melissa A Leisner, Jonathan Plitnick, Navjot Singh, John Kelly, Erasmus Schneider, Erica Lasek-Nesselquist |
| EPI_ISL_1228282, EPI_ISL_1228283 | ALBANY MEDICAL CENTER | Wadsworth Center, New York State Department of Health | Kirsten St. George, Daryl M. Lamson, Alexis Russel, Matthew Shudt, Melissa A Leisner, Jonathan Plitnick, Navjot Singh, John Kelly, Erasmus Schneider, Erica Lasek-Nesselquist |
| EPI_ISL_1228284, EPI_ISL_1228285, EPI_ISL_1228286, EPI_ISL_1228287, EPI_ISL_1228288, EPI_ISL_1228289, EPI_ISL_1228290, EPI_ISL_1228291 | THE MARY IMOGENE BASSETT HOSPITAL | Wadsworth Center, New York State Department of Health | Kirsten St. George, Daryl M. Lamson, Alexis Russel, Matthew Shudt, Melissa A Leisner, Jonathan Plitnick, Navjot Singh, John Kelly, Erasmus Schneider, Erica Lasek-Nesselquist |
| EPI_ISL_1228292 | ADIRONDACK MEDICAL CENTER | Wadsworth Center, New York State Department of Health | Kirsten St. George, Daryl M. Lamson, Alexis Russel, Matthew Shudt, Melissa A Leisner, Jonathan Plitnick, Navjot Singh, John Kelly, Erasmus Schneider, Erica Lasek-Nesselquist |
| EPI_ISL_1228293 | SUNY UPSTATE MEDICAL UNIVERSITY | Wadsworth Center, New York State Department of Health | Kirsten St. George, Daryl M. Lamson, Alexis Russel, Matthew Shudt, Melissa A Leisner, Jonathan Plitnick, Navjot Singh, John Kelly, Erasmus Schneider, Erica Lasek-Nesselquist |
| EPI_ISL_1228294, EPI_ISL_1228295 | Wadsworth Center, New York State Department of Health | Wadsworth Center, New York State Department of Health | Kirsten St. George, Daryl M. Lamson, Alexis Russel, Matthew Shudt, Melissa A Leisner, Jonathan Plitnick, Navjot Singh, John Kelly, Erasmus Schneider, Erica Lasek-Nesselquist |
| EPI_ISL_1228296, EPI_ISL_1228297, EPI_ISL_1228298, EPI_ISL_1228299, EPI_ISL_1228300, EPI_ISL_1228301, EPI_ISL_1228302, EPI_ISL_1228303, EPI_ISL_1228304, EPI_ISL_1228305, EPI_ISL_1228306 | | | |
| see above | URMC LABS | Wadsworth Center, New York State Department of Health | Kirsten St. George, Daryl M. Lamson, Alexis Russel, Matthew Shudt, Melissa A Leisner, Jonathan Plitnick, Navjot Singh, John Kelly, Erasmus Schneider, Erica Lasek-Nesselquist |
| EPI_ISL_1228307, EPI_ISL_1228308, EPI_ISL_1228309, EPI_ISL_1228310, EPI_ISL_1228311, EPI_ISL_1228312, EPI_ISL_1228313, EPI_ISL_1228314, EPI_ISL_1228315, EPI_ISL_1228316, EPI_ISL_1228317, EPI_ISL_1228318, EPI_ISL_1228319, EPI_ISL_1228320, EPI_ISL_1228321, EPI_ISL_1228322, EPI_ISL_1228323, EPI_ISL_1228324, EPI_ISL_1228325, EPI_ISL_1228326, EPI_ISL_1228327, EPI_ISL_1228328, EPI_ISL_1228329, EPI_ISL_1228330, EPI_ISL_1228331 | | | |
| see above | MEMORIAL SLOAN KETTERING CANCER CENTER | Wadsworth Center, New York State Department of Health | Kirsten St. George, Daryl M. Lamson, Alexis Russel, Matthew Shudt, Melissa A Leisner, Jonathan Plitnick, Navjot Singh, John Kelly, Erasmus Schneider, Erica Lasek-Nesselquist |
| EPI_ISL_1228332 | NORTH SHORE UNIVERSITY HOSPITAL | Wadsworth Center, New York State Department of Health | Kirsten St. George, Daryl M. Lamson, Alexis Russel, Matthew Shudt, Melissa A Leisner, Jonathan Plitnick, Navjot Singh, John Kelly, Erasmus Schneider, Erica Lasek-Nesselquist |
| EPI_ISL_1228333, EPI_ISL_1228334, EPI_ISL_1228335, EPI_ISL_1228336, EPI_ISL_1228337, EPI_ISL_1228338, EPI_ISL_1228339, EPI_ISL_1228340, EPI_ISL_1228341 | MEMORIAL SLOAN KETTERING CANCER CENTER | Wadsworth Center, New York State Department of Health | Kirsten St. George, Daryl M. Lamson, Alexis Russel, Matthew Shudt, Melissa A Leisner, Jonathan Plitnick, Navjot Singh, John Kelly, Erasmus Schneider, Erica Lasek-Nesselquist |
| EPI_ISL_1228342, EPI_ISL_1228343, EPI_ISL_1228344, EPI_ISL_1228345 | NORTHWELL HEALTH LABORATORIES | Wadsworth Center, New York State Department of Health | Kirsten St. George, Daryl M. Lamson, Alexis Russel, Matthew Shudt, Melissa A Leisner, Jonathan Plitnick, Navjot Singh, John Kelly, Erasmus Schneider, Erica Lasek-Nesselquist |
| EPI_ISL_1228346, EPI_ISL_1228347 | MEMORIAL SLOAN KETTERING CANCER CENTER | Wadsworth Center, New York State Department of Health | Kirsten St. George, Daryl M. Lamson, Alexis Russel, Matthew Shudt, Melissa A Leisner, Jonathan Plitnick, Navjot Singh, John Kelly, Erasmus Schneider, Erica Lasek-Nesselquist |
| EPI_ISL_1228348 | NORTHWELL HEALTH LABORATORIES | Wadsworth Center, New York State Department of Health | Kirsten St. George, Daryl M. Lamson, Alexis Russel, Matthew Shudt, Melissa A Leisner, Jonathan Plitnick, Navjot Singh, John Kelly, Erasmus Schneider, Erica Lasek-Nesselquist |
| EPI_ISL_1228349, EPI_ISL_1228350, EPI_ISL_1228351, EPI_ISL_1228352, EPI_ISL_1228353, | MEMORIAL SLOAN KETTERING CANCER CENTER | Wadsworth Center, New York State Department of Health | Kirsten St. George, Daryl M. Lamson, Alexis Russel, Matthew Shudt, Melissa A Leisner, Jonathan Plitnick, Navjot Singh, John Kelly, Erasmus Schneider, Erica Lasek-Nesselquist |

| | | | |
|---|---|---|---|
| EPI_ISL_1228354, EPI_ISL_1228355 | | | |
| EPI_ISL_1228356 | NORTH SHORE UNIVERSITY HOSPITAL | Wadsworth Center, New York State Department of Health | Kirsten St. George, Daryl M. Lamson, Alexis Russel, Matthew Shudt, Melissa A Leisner, Jonathan Plitnick, Navjot Singh, John Kelly, Erasmus Schneider, Erica Lasek-Nesselquist |
| EPI_ISL_1228357, EPI_ISL_1228358, EPI_ISL_1228359 | NORTHWELL HEALTH LABORATORIES | Wadsworth Center, New York State Department of Health | Kirsten St. George, Daryl M. Lamson, Alexis Russel, Matthew Shudt, Melissa A Leisner, Jonathan Plitnick, Navjot Singh, John Kelly, Erasmus Schneider, Erica Lasek-Nesselquist |
| EPI_ISL_1228360 | MEMORIAL SLOAN KETTERING CANCER CENTER | Wadsworth Center, New York State Department of Health | Kirsten St. George, Daryl M. Lamson, Alexis Russel, Matthew Shudt, Melissa A Leisner, Jonathan Plitnick, Navjot Singh, John Kelly, Erasmus Schneider, Erica Lasek-Nesselquist |
| EPI_ISL_1228361 | NORTHWELL HEALTH LABORATORIES | Wadsworth Center, New York State Department of Health | Kirsten St. George, Daryl M. Lamson, Alexis Russel, Matthew Shudt, Melissa A Leisner, Jonathan Plitnick, Navjot Singh, John Kelly, Erasmus Schneider, Erica Lasek-Nesselquist |
| EPI_ISL_1228362, EPI_ISL_1228363, EPI_ISL_1228364, EPI_ISL_1228365, EPI_ISL_1228366, EPI_ISL_1228367, EPI_ISL_1228368 | MEMORIAL SLOAN KETTERING CANCER CENTER | Wadsworth Center, New York State Department of Health | Kirsten St. George, Daryl M. Lamson, Alexis Russel, Matthew Shudt, Melissa A Leisner, Jonathan Plitnick, Navjot Singh, John Kelly, Erasmus Schneider, Erica Lasek-Nesselquist |
| EPI_ISL_1228369, EPI_ISL_1228370, EPI_ISL_1228371, EPI_ISL_1228372, EPI_ISL_1228373 | NYC Pandemic Response Lab | Wadsworth Center, New York State Department of Health | Kirsten St. George, Daryl M. Lamson, Alexis Russel, Matthew Shudt, Melissa A Leisner, Jonathan Plitnick, Navjot Singh, John Kelly, Erasmus Schneider, Erica Lasek-Nesselquist |
| EPI_ISL_1228374 | WESTCHESTER MEDICAL CENTER | Wadsworth Center, New York State Department of Health | Kirsten St. George, Daryl M. Lamson, Alexis Russel, Matthew Shudt, Melissa A Leisner, Jonathan Plitnick, Navjot Singh, John Kelly, Erasmus Schneider, Erica Lasek-Nesselquist |
| EPI_ISL_1228375, EPI_ISL_1228376, EPI_ISL_1228377 | DEPARTMENT OF VETERANS AFFAIR MEDICAL CENTER | Wadsworth Center, New York State Department of Health | Kirsten St. George, Daryl M. Lamson, Alexis Russel, Matthew Shudt, Melissa A Leisner, Jonathan Plitnick, Navjot Singh, John Kelly, Erasmus Schneider, Erica Lasek-Nesselquist |
| EPI_ISL_1228378, EPI_ISL_1228379 | WESTCHESTER MEDICAL CENTER | Wadsworth Center, New York State Department of Health | Kirsten St. George, Daryl M. Lamson, Alexis Russel, Matthew Shudt, Melissa A Leisner, Jonathan Plitnick, Navjot Singh, John Kelly, Erasmus Schneider, Erica Lasek-Nesselquist |
| EPI_ISL_1228380 | SUNY UPSTATE MEDICAL UNIVERSITY | Wadsworth Center, New York State Department of Health | Kirsten St. George, Daryl M. Lamson, Alexis Russel, Matthew Shudt, Melissa A Leisner, Jonathan Plitnick, Navjot Singh, John Kelly, Erasmus Schneider, Erica Lasek-Nesselquist |
| EPI_ISL_1228381, EPI_ISL_1228382, EPI_ISL_1228383, EPI_ISL_1228384, EPI_ISL_1228385, EPI_ISL_1228386, EPI_ISL_1228387, EPI_ISL_1228388, EPI_ISL_1228389, EPI_ISL_1228390, EPI_ISL_1228391, EPI_ISL_1228392, EPI_ISL_1228393, EPI_ISL_1228394, EPI_ISL_1228395, EPI_ISL_1228396, EPI_ISL_1228397, EPI_ISL_1228398, EPI_ISL_1228399, EPI_ISL_1228400, EPI_ISL_1228401, EPI_ISL_1228402, EPI_ISL_1228403, EPI_ISL_1228404, EPI_ISL_1228405, EPI_ISL_1228406, EPI_ISL_1228407, EPI_ISL_1228408, EPI_ISL_1228409, EPI_ISL_1228410, EPI_ISL_1228411, EPI_ISL_1228412, EPI_ISL_1228413, EPI_ISL_1228414, EPI_ISL_1228415, EPI_ISL_1228416, EPI_ISL_1228417, EPI_ISL_1228418, EPI_ISL_1228419, EPI_ISL_1228420, EPI_ISL_1228421, EPI_ISL_1228422, EPI_ISL_1228423, EPI_ISL_1228424, EPI_ISL_1228425, EPI_ISL_1228426, EPI_ISL_1228427, EPI_ISL_1228428, EPI_ISL_1228429 | | | |
| see above | WESTCHESTER MEDICAL CENTER | Wadsworth Center, New York State Department of Health | Kirsten St. George, Daryl M. Lamson, Alexis Russel, Matthew Shudt, Melissa A Leisner, Jonathan Plitnick, Navjot Singh, John Kelly, Erasmus Schneider, Erica Lasek-Nesselquist |
| EPI_ISL_1228430 | ALBANY MEDICAL CENTER | Wadsworth Center, New York State Department of Health | Kirsten St. George, Daryl M. Lamson, Alexis Russel, Matthew Shudt, Melissa A Leisner, Jonathan Plitnick, Navjot Singh, John Kelly, Erasmus Schneider, Erica Lasek-Nesselquist |
| EPI_ISL_1228431, EPI_ISL_1228432, EPI_ISL_1228433 | WESTCHESTER MEDICAL CENTER | Wadsworth Center, New York State Department of Health | Kirsten St. George, Daryl M. Lamson, Alexis Russel, Matthew Shudt, Melissa A Leisner, Jonathan Plitnick, Navjot Singh, John Kelly, Erasmus Schneider, Erica Lasek-Nesselquist |
| EPI_ISL_1228434, EPI_ISL_1228435, EPI_ISL_1228436, EPI_ISL_1228437, EPI_ISL_1228438, EPI_ISL_1228439, EPI_ISL_1228440, EPI_ISL_1228441, EPI_ISL_1228442, EPI_ISL_1228443 | GLENS FALLS HOSPITAL LABORATORY | Wadsworth Center, New York State Department of Health | Kirsten St. George, Daryl M. Lamson, Alexis Russel, Matthew Shudt, Melissa A Leisner, Jonathan Plitnick, Navjot Singh, John Kelly, Erasmus Schneider, Erica Lasek-Nesselquist |
| EPI_ISL_1228444, EPI_ISL_1228445, EPI_ISL_1228446 | Wadsworth Center, New York State Department of Health | Wadsworth Center, New York State Department of Health | Kirsten St. George, Daryl M. Lamson, Alexis Russel, Matthew Shudt, Melissa A Leisner, Jonathan Plitnick, Navjot Singh, John Kelly, Erasmus Schneider, Erica Lasek-Nesselquist |
| EPI_ISL_1228447, EPI_ISL_1228448, EPI_ISL_1228449 | GLENS FALLS HOSPITAL LABORATORY | Wadsworth Center, New York State Department of Health | Kirsten St. George, Daryl M. Lamson, Alexis Russel, Matthew Shudt, Melissa A Leisner, Jonathan Plitnick, Navjot Singh, John Kelly, Erasmus Schneider, Erica Lasek-Nesselquist |
| EPI_ISL_1228450 | Wadsworth Center, New York State Department of Health | Wadsworth Center, New York State Department of Health | Kirsten St. George, Daryl M. Lamson, Alexis Russel, Matthew Shudt, Melissa A Leisner, Jonathan Plitnick, Navjot Singh, John Kelly, Erasmus Schneider, Erica Lasek-Nesselquist |
| EPI_ISL_1228451 | GLENS FALLS HOSPITAL LABORATORY | Wadsworth Center, New York State Department of Health | Kirsten St. George, Daryl M. Lamson, Alexis Russel, Matthew Shudt, Melissa A Leisner, Jonathan Plitnick, Navjot Singh, John Kelly, Erasmus Schneider, Erica Lasek-Nesselquist |
| EPI_ISL_1228452 | Wadsworth Center, New York State Department of Health | Wadsworth Center, New York State Department of Health | Kirsten St. George, Daryl M. Lamson, Alexis Russel, Matthew Shudt, Melissa A Leisner, Jonathan Plitnick, Navjot Singh, John Kelly, Erasmus Schneider, Erica Lasek-Nesselquist |
| EPI_ISL_1228453 | THE MARY IMOGENE BASSETT HOSPITAL | Wadsworth Center, New York State Department of Health | Kirsten St. George, Daryl M. Lamson, Alexis Russel, Matthew Shudt, Melissa A Leisner, Jonathan Plitnick, Navjot Singh, John Kelly, Erasmus Schneider, Erica Lasek-Nesselquist |
| EPI_ISL_1228454, EPI_ISL_1228455, EPI_ISL_1228456, EPI_ISL_1228457, EPI_ISL_1228458, EPI_ISL_1228459 | GLENS FALLS HOSPITAL LABORATORY | Wadsworth Center, New York State Department of Health | Kirsten St. George, Daryl M. Lamson, Alexis Russel, Matthew Shudt, Melissa A Leisner, Jonathan Plitnick, Navjot Singh, John Kelly, Erasmus Schneider, Erica Lasek-Nesselquist |
| EPI_ISL_1228460, EPI_ISL_1228461, EPI_ISL_1228462 | Wadsworth Center, New York State Department of Health | Wadsworth Center, New York State Department of Health | Kirsten St. George, Daryl M. Lamson, Alexis Russel, Matthew Shudt, Melissa A Leisner, Jonathan Plitnick, Navjot Singh, John Kelly, Erasmus Schneider, Erica Lasek-Nesselquist |
| EPI_ISL_1228463, EPI_ISL_1228464 | GLENS FALLS HOSPITAL LABORATORY | Wadsworth Center, New York State Department of Health | Kirsten St. George, Daryl M. Lamson, Alexis Russel, Matthew Shudt, Melissa A Leisner, Jonathan Plitnick, Navjot Singh, John Kelly, Erasmus Schneider, Erica Lasek-Nesselquist |
| EPI_ISL_1228465, EPI_ISL_1228466 | Wadsworth Center, New York State Department of Health | Wadsworth Center, New York State Department of Health | Kirsten St. George, Daryl M. Lamson, Alexis Russel, Matthew Shudt, Melissa A Leisner, Jonathan Plitnick, Navjot Singh, John Kelly, Erasmus Schneider, Erica Lasek-Nesselquist |
| EPI_ISL_1228467, EPI_ISL_1228468 | GLENS FALLS HOSPITAL LABORATORY | Wadsworth Center, New York State Department of Health | Kirsten St. George, Daryl M. Lamson, Alexis Russel, Matthew Shudt, Melissa A Leisner, Jonathan Plitnick, Navjot Singh, John Kelly, Erasmus Schneider, Erica Lasek-Nesselquist |
| EPI_ISL_1228469 | Wadsworth Center, New York State Department of Health | Wadsworth Center, New York State Department of Health | Kirsten St. George, Daryl M. Lamson, Alexis Russel, Matthew Shudt, Melissa A Leisner, Jonathan Plitnick, Navjot Singh, John Kelly, Erasmus Schneider, Erica Lasek-Nesselquist |
| EPI_ISL_1228470, EPI_ISL_1228471, EPI_ISL_1228472, EPI_ISL_1228473 | GLENS FALLS HOSPITAL LABORATORY | Wadsworth Center, New York State Department of Health | Kirsten St. George, Daryl M. Lamson, Alexis Russel, Matthew Shudt, Melissa A Leisner, Jonathan Plitnick, Navjot Singh, John Kelly, Erasmus Schneider, Erica Lasek-Nesselquist |
| EPI_ISL_1228474, EPI_ISL_1228475 | Wadsworth Center, New York State Department of Health | Wadsworth Center, New York State Department of Health | Kirsten St. George, Daryl M. Lamson, Alexis Russel, Matthew Shudt, Melissa A Leisner, Jonathan Plitnick, Navjot Singh, John Kelly, Erasmus Schneider, Erica Lasek-Nesselquist |
| EPI_ISL_1228476 | GLENS FALLS HOSPITAL LABORATORY | Wadsworth Center, New York State Department of Health | Kirsten St. George, Daryl M. Lamson, Alexis Russel, Matthew Shudt, Melissa A Leisner, Jonathan Plitnick, Navjot Singh, John Kelly, Erasmus Schneider, Erica Lasek-Nesselquist |
| EPI_ISL_1228477, EPI_ISL_1228478, EPI_ISL_1228479 | ALBANY MEDICAL CENTER | Wadsworth Center, New York State Department of Health | Kirsten St. George, Daryl M. Lamson, Alexis Russel, Matthew Shudt, Melissa A Leisner, Jonathan Plitnick, Navjot Singh, John Kelly, Erasmus Schneider, Erica Lasek-Nesselquist |
| EPI_ISL_1228480, EPI_ISL_1228481, | URMC LABS | Wadsworth Center, New York State Department of Health | Kirsten St. George, Daryl M. Lamson, Alexis Russel, Matthew Shudt, Melissa A Leisner, Jonathan Plitnick, Navjot Singh, John Kelly, Erasmus Schneider, Erica Lasek-Nesselquist |

| | | | |
|---|---|---|---|
| EPI_ISL_1228482, EPI_ISL_1228483 | | | |
| EPI_ISL_1228484, EPI_ISL_1228485, EPI_ISL_1228486, EPI_ISL_1228487, EPI_ISL_1228488, EPI_ISL_1228489, EPI_ISL_1228490, EPI_ISL_1228491, EPI_ISL_1228492, EPI_ISL_1228493, EPI_ISL_1228494, EPI_ISL_1228495, EPI_ISL_1228496, EPI_ISL_1228497, EPI_ISL_1228498, EPI_ISL_1228499, EPI_ISL_1228500 | | | |
| see above | MEMORIAL SLOAN KETTERING CANCER CENTER | Wadsworth Center, New York State Department of Health | Kirsten St. George, Daryl M. Lamson, Alexis Russel, Matthew Shudt, Melissa A Leisner, Jonathan Plitnick, Navjot Singh, John Kelly, Erasmus Schneider, Erica Lasek-Nesselquist |
| EPI_ISL_1228501 | WESTCHESTER MEDICAL CENTER | Wadsworth Center, New York State Department of Health | Kirsten St. George, Daryl M. Lamson, Alexis Russel, Matthew Shudt, Melissa A Leisner, Jonathan Plitnick, Navjot Singh, John Kelly, Erasmus Schneider, Erica Lasek-Nesselquist |
| EPI_ISL_1228502, EPI_ISL_1228503 | MEMORIAL SLOAN KETTERING CANCER CENTER | Wadsworth Center, New York State Department of Health | Kirsten St. George, Daryl M. Lamson, Alexis Russel, Matthew Shudt, Melissa A Leisner, Jonathan Plitnick, Navjot Singh, John Kelly, Erasmus Schneider, Erica Lasek-Nesselquist |
| EPI_ISL_1228504, EPI_ISL_1228505, EPI_ISL_1228506, EPI_ISL_1228507, EPI_ISL_1228508 | WESTCHESTER MEDICAL CENTER | Wadsworth Center, New York State Department of Health | Kirsten St. George, Daryl M. Lamson, Alexis Russel, Matthew Shudt, Melissa A Leisner, Jonathan Plitnick, Navjot Singh, John Kelly, Erasmus Schneider, Erica Lasek-Nesselquist |
| EPI_ISL_1228509 | Columbia University Irving Medical Center | Wadsworth Center, New York State Department of Health | Kirsten St. George, Daryl M. Lamson, Alexis Russel, Matthew Shudt, Melissa A Leisner, Jonathan Plitnick, Navjot Singh, John Kelly, Erasmus Schneider, Erica Lasek-Nesselquist |
| EPI_ISL_1228510, EPI_ISL_1228511, EPI_ISL_1228512, EPI_ISL_1228513 | WESTCHESTER MEDICAL CENTER | Wadsworth Center, New York State Department of Health | Kirsten St. George, Daryl M. Lamson, Alexis Russel, Matthew Shudt, Melissa A Leisner, Jonathan Plitnick, Navjot Singh, John Kelly, Erasmus Schneider, Erica Lasek-Nesselquist |
| EPI_ISL_1228514 | Columbia University Irving Medical Center | Wadsworth Center, New York State Department of Health | Kirsten St. George, Daryl M. Lamson, Alexis Russel, Matthew Shudt, Melissa A Leisner, Jonathan Plitnick, Navjot Singh, John Kelly, Erasmus Schneider, Erica Lasek-Nesselquist |
| EPI_ISL_1228515, EPI_ISL_1228516, EPI_ISL_1228517, EPI_ISL_1228518, EPI_ISL_1228519 | WESTCHESTER MEDICAL CENTER | Wadsworth Center, New York State Department of Health | Kirsten St. George, Daryl M. Lamson, Alexis Russel, Matthew Shudt, Melissa A Leisner, Jonathan Plitnick, Navjot Singh, John Kelly, Erasmus Schneider, Erica Lasek-Nesselquist |
| EPI_ISL_1228520 | MONTEFIORE MEDICAL CENTER LABORATORIES | Wadsworth Center, New York State Department of Health | Kirsten St. George, Daryl M. Lamson, Alexis Russel, Matthew Shudt, Melissa A Leisner, Jonathan Plitnick, Navjot Singh, John Kelly, Erasmus Schneider, Erica Lasek-Nesselquist |
| EPI_ISL_1228521, EPI_ISL_1228522, EPI_ISL_1228523, EPI_ISL_1228524, EPI_ISL_1228525, EPI_ISL_1228526, EPI_ISL_1228527, EPI_ISL_1228528 | WESTCHESTER MEDICAL CENTER | Wadsworth Center, New York State Department of Health | Kirsten St. George, Daryl M. Lamson, Alexis Russel, Matthew Shudt, Melissa A Leisner, Jonathan Plitnick, Navjot Singh, John Kelly, Erasmus Schneider, Erica Lasek-Nesselquist |
| EPI_ISL_1228529 | Columbia University Irving Medical Center | Wadsworth Center, New York State Department of Health | Kirsten St. George, Daryl M. Lamson, Alexis Russel, Matthew Shudt, Melissa A Leisner, Jonathan Plitnick, Navjot Singh, John Kelly, Erasmus Schneider, Erica Lasek-Nesselquist |
| EPI_ISL_1228530, EPI_ISL_1228531 | MONTEFIORE MEDICAL CENTER LABORATORIES | Wadsworth Center, New York State Department of Health | Kirsten St. George, Daryl M. Lamson, Alexis Russel, Matthew Shudt, Melissa A Leisner, Jonathan Plitnick, Navjot Singh, John Kelly, Erasmus Schneider, Erica Lasek-Nesselquist |
| EPI_ISL_1228532, EPI_ISL_1228533, EPI_ISL_1228534, EPI_ISL_1228535 | WESTCHESTER MEDICAL CENTER | Wadsworth Center, New York State Department of Health | Kirsten St. George, Daryl M. Lamson, Alexis Russel, Matthew Shudt, Melissa A Leisner, Jonathan Plitnick, Navjot Singh, John Kelly, Erasmus Schneider, Erica Lasek-Nesselquist |
| EPI_ISL_1228536 | MONTEFIORE MEDICAL CENTER LABORATORIES | Wadsworth Center, New York State Department of Health | Kirsten St. George, Daryl M. Lamson, Alexis Russel, Matthew Shudt, Melissa A Leisner, Jonathan Plitnick, Navjot Singh, John Kelly, Erasmus Schneider, Erica Lasek-Nesselquist |
| EPI_ISL_1228537, EPI_ISL_1228538, EPI_ISL_1228539, EPI_ISL_1228540 | WESTCHESTER MEDICAL CENTER | Wadsworth Center, New York State Department of Health | Kirsten St. George, Daryl M. Lamson, Alexis Russel, Matthew Shudt, Melissa A Leisner, Jonathan Plitnick, Navjot Singh, John Kelly, Erasmus Schneider, Erica Lasek-Nesselquist |
| EPI_ISL_1228541, EPI_ISL_1228542 | MONTEFIORE MEDICAL CENTER LABORATORIES | Wadsworth Center, New York State Department of Health | Kirsten St. George, Daryl M. Lamson, Alexis Russel, Matthew Shudt, Melissa A Leisner, Jonathan Plitnick, Navjot Singh, John Kelly, Erasmus Schneider, Erica Lasek-Nesselquist |
| EPI_ISL_1228543, EPI_ISL_1228544 | Columbia University Irving Medical Center | Wadsworth Center, New York State Department of Health | Kirsten St. George, Daryl M. Lamson, Alexis Russel, Matthew Shudt, Melissa A Leisner, Jonathan Plitnick, Navjot Singh, John Kelly, Erasmus Schneider, Erica Lasek-Nesselquist |
| EPI_ISL_1228545, EPI_ISL_1228546, EPI_ISL_1228547, EPI_ISL_1228548, EPI_ISL_1228549 | WESTCHESTER MEDICAL CENTER | Wadsworth Center, New York State Department of Health | Kirsten St. George, Daryl M. Lamson, Alexis Russel, Matthew Shudt, Melissa A Leisner, Jonathan Plitnick, Navjot Singh, John Kelly, Erasmus Schneider, Erica Lasek-Nesselquist |
| EPI_ISL_1228550 | MONTEFIORE MEDICAL CENTER LABORATORIES | Wadsworth Center, New York State Department of Health | Kirsten St. George, Daryl M. Lamson, Alexis Russel, Matthew Shudt, Melissa A Leisner, Jonathan Plitnick, Navjot Singh, John Kelly, Erasmus Schneider, Erica Lasek-Nesselquist |
| EPI_ISL_1228551, EPI_ISL_1228552 | WESTCHESTER MEDICAL CENTER | Wadsworth Center, New York State Department of Health | Kirsten St. George, Daryl M. Lamson, Alexis Russel, Matthew Shudt, Melissa A Leisner, Jonathan Plitnick, Navjot Singh, John Kelly, Erasmus Schneider, Erica Lasek-Nesselquist |
| EPI_ISL_1228553 | MONTEFIORE MEDICAL CENTER LABORATORIES | Wadsworth Center, New York State Department of Health | Kirsten St. George, Daryl M. Lamson, Alexis Russel, Matthew Shudt, Melissa A Leisner, Jonathan Plitnick, Navjot Singh, John Kelly, Erasmus Schneider, Erica Lasek-Nesselquist |
| EPI_ISL_1228554, EPI_ISL_1228555, EPI_ISL_1228556, EPI_ISL_1228557 | WESTCHESTER MEDICAL CENTER | Wadsworth Center, New York State Department of Health | Kirsten St. George, Daryl M. Lamson, Alexis Russel, Matthew Shudt, Melissa A Leisner, Jonathan Plitnick, Navjot Singh, John Kelly, Erasmus Schneider, Erica Lasek-Nesselquist |
| EPI_ISL_1228558, EPI_ISL_1228559, EPI_ISL_1228560 | MONTEFIORE MEDICAL CENTER LABORATORIES | Wadsworth Center, New York State Department of Health | Kirsten St. George, Daryl M. Lamson, Alexis Russel, Matthew Shudt, Melissa A Leisner, Jonathan Plitnick, Navjot Singh, John Kelly, Erasmus Schneider, Erica Lasek-Nesselquist |
| EPI_ISL_1228561, EPI_ISL_1228562 | WESTCHESTER MEDICAL CENTER | Wadsworth Center, New York State Department of Health | Kirsten St. George, Daryl M. Lamson, Alexis Russel, Matthew Shudt, Melissa A Leisner, Jonathan Plitnick, Navjot Singh, John Kelly, Erasmus Schneider, Erica Lasek-Nesselquist |
| EPI_ISL_1228563 | NYC Pandemic Response Lab | Wadsworth Center, New York State Department of Health | Kirsten St. George, Daryl M. Lamson, Alexis Russel, Matthew Shudt, Melissa A Leisner, Jonathan Plitnick, Navjot Singh, John Kelly, Erasmus Schneider, Erica Lasek-Nesselquist |
| EPI_ISL_1228564 | New York Presbyterian Hospital | Wadsworth Center, New York State Department of Health | Kirsten St. George, Daryl M. Lamson, Alexis Russel, Matthew Shudt, Melissa A Leisner, Jonathan Plitnick, Navjot Singh, John Kelly, Erasmus Schneider, Erica Lasek-Nesselquist |
| EPI_ISL_1228565, EPI_ISL_1228566 | WESTCHESTER MEDICAL CENTER | Wadsworth Center, New York State Department of Health | Kirsten St. George, Daryl M. Lamson, Alexis Russel, Matthew Shudt, Melissa A Leisner, Jonathan Plitnick, Navjot Singh, John Kelly, Erasmus Schneider, Erica Lasek-Nesselquist |
| EPI_ISL_1228567 | MONTEFIORE MEDICAL CENTER LABORATORIES | Wadsworth Center, New York State Department of Health | Kirsten St. George, Daryl M. Lamson, Alexis Russel, Matthew Shudt, Melissa A Leisner, Jonathan Plitnick, Navjot Singh, John Kelly, Erasmus Schneider, Erica Lasek-Nesselquist |
| EPI_ISL_1228568 | New York Presbyterian Hospital | Wadsworth Center, New York State Department of Health | Kirsten St. George, Daryl M. Lamson, Alexis Russel, Matthew Shudt, Melissa A Leisner, Jonathan Plitnick, Navjot Singh, John Kelly, Erasmus Schneider, Erica Lasek-Nesselquist |
| EPI_ISL_1228569 | MONTEFIORE MEDICAL CENTER LABORATORIES | Wadsworth Center, New York State Department of Health | Kirsten St. George, Daryl M. Lamson, Alexis Russel, Matthew Shudt, Melissa A Leisner, Jonathan Plitnick, Navjot Singh, John Kelly, Erasmus Schneider, Erica Lasek-Nesselquist |
| EPI_ISL_1228570 | NYC Pandemic Response Lab | Wadsworth Center, New York State Department of Health | Kirsten St. George, Daryl M. Lamson, Alexis Russel, Matthew Shudt, Melissa A Leisner, Jonathan Plitnick, Navjot Singh, John Kelly, Erasmus Schneider, Erica Lasek-Nesselquist |
| EPI_ISL_1228571 | WESTCHESTER MEDICAL CENTER | Wadsworth Center, New York State Department of Health | Kirsten St. George, Daryl M. Lamson, Alexis Russel, Matthew Shudt, Melissa A Leisner, Jonathan Plitnick, Navjot Singh, John Kelly, Erasmus Schneider, Erica Lasek-Nesselquist |
| EPI_ISL_1228572 | MONTEFIORE MEDICAL CENTER LABORATORIES | Wadsworth Center, New York State Department of Health | Kirsten St. George, Daryl M. Lamson, Alexis Russel, Matthew Shudt, Melissa A Leisner, Jonathan Plitnick, Navjot Singh, John Kelly, Erasmus Schneider, Erica Lasek-Nesselquist |
| EPI_ISL_1228573 | WESTCHESTER MEDICAL CENTER | Wadsworth Center, New York State Department of Health | Kirsten St. George, Daryl M. Lamson, Alexis Russel, Matthew Shudt, Melissa A Leisner, Jonathan Plitnick, Navjot Singh, John Kelly, Erasmus Schneider, Erica Lasek-Nesselquist |

| | | | |
|---|---|---|---|
| EPI_ISL_1228574, EPI_ISL_1228575 | NYC Pandemic Response Lab | Wadsworth Center, New York State Department of Health | Kirsten St. George, Daryl M. Lamson, Alexis Russel, Matthew Shudt, Melissa A Leisner, Jonathan Plitnick, Navjot Singh, John Kelly, Erasmus Schneider, Erica Lasek-Nesselquist |
| EPI_ISL_1228576 | Columbia University Irving Medical Center | Wadsworth Center, New York State Department of Health | Kirsten St. George, Daryl M. Lamson, Alexis Russel, Matthew Shudt, Melissa A Leisner, Jonathan Plitnick, Navjot Singh, John Kelly, Erasmus Schneider, Erica Lasek-Nesselquist |
| EPI_ISL_1228577 | WESTCHESTER MEDICAL CENTER | Wadsworth Center, New York State Department of Health | Kirsten St. George, Daryl M. Lamson, Alexis Russel, Matthew Shudt, Melissa A Leisner, Jonathan Plitnick, Navjot Singh, John Kelly, Erasmus Schneider, Erica Lasek-Nesselquist |
| EPI_ISL_1228578 | MONTEFIORE MEDICAL CENTER LABORATORIES | Wadsworth Center, New York State Department of Health | Kirsten St. George, Daryl M. Lamson, Alexis Russel, Matthew Shudt, Melissa A Leisner, Jonathan Plitnick, Navjot Singh, John Kelly, Erasmus Schneider, Erica Lasek-Nesselquist |
| EPI_ISL_1228579, EPI_ISL_1228580, EPI_ISL_1228581, EPI_ISL_1228582, EPI_ISL_1228583 | WESTCHESTER MEDICAL CENTER | Wadsworth Center, New York State Department of Health | Kirsten St. George, Daryl M. Lamson, Alexis Russel, Matthew Shudt, Melissa A Leisner, Jonathan Plitnick, Navjot Singh, John Kelly, Erasmus Schneider, Erica Lasek-Nesselquist |
| EPI_ISL_1228584 | MONTEFIORE MEDICAL CENTER LABORATORIES | Wadsworth Center, New York State Department of Health | Kirsten St. George, Daryl M. Lamson, Alexis Russel, Matthew Shudt, Melissa A Leisner, Jonathan Plitnick, Navjot Singh, John Kelly, Erasmus Schneider, Erica Lasek-Nesselquist |
| EPI_ISL_1228585, EPI_ISL_1228586 | WESTCHESTER MEDICAL CENTER | Wadsworth Center, New York State Department of Health | Kirsten St. George, Daryl M. Lamson, Alexis Russel, Matthew Shudt, Melissa A Leisner, Jonathan Plitnick, Navjot Singh, John Kelly, Erasmus Schneider, Erica Lasek-Nesselquist |
| EPI_ISL_1229171, EPI_ISL_1229172, EPI_ISL_1229173 | WA State PHL | Seattle Flu Study | Deborah A. Nickerson, Chris D. Frazar, Jover Lee, Benjamin Pelle, Erica Ryke, Matthew Richardson, Amanda Adler, Elisabeth Brandstetter, Peter D. Han, Kairsten Fay, Misja Ilcisin, Kirsten Lacombe, Thomas R. Sibley, Melissa Truong, Caitlin R. Wolf, Romesh Gautom, Geoff Melly, Brian Hiatt, Philip Dykema, Scott Lindquist, Michael Boeckh, Janet A. Englund, Michael Famulare, Barry R. Lutz, Mark J. Rieder, Lea M. Starita, Matthew Thompson, Helen Y. Chu, Jay Shendure, Trevor Bedford |
| EPI_ISL_1229176 | Northwest Laboratories | Seattle Flu Study | Deborah A. Nickerson, Chris D. Frazar, Jover Lee, Benjamin Pelle, Erica Ryke, Matthew Richardson, Amanda Adler, Elisabeth Brandstetter, Peter D. Han, Kairsten Fay, Misja Ilcisin, Kirsten Lacombe, Thomas R. Sibley, Melissa Truong, Caitlin R. Wolf, Romesh Gautom, Geoff Melly, Brian Hiatt, Philip Dykema, Scott Lindquist, Michael Boeckh, Janet A. Englund, Michael Famulare, Barry R. Lutz, Mark J. Rieder, Lea M. Starita, Matthew Thompson, Helen Y. Chu, Jay Shendure, Trevor Bedford |
| EPI_ISL_1229369, EPI_ISL_1229370, EPI_ISL_1229371, EPI_ISL_1229372, EPI_ISL_1229373, EPI_ISL_1229374, EPI_ISL_1229375, EPI_ISL_1229376, EPI_ISL_1229377, EPI_ISL_1229378, EPI_ISL_1229379, EPI_ISL_1229380, EPI_ISL_1229381, EPI_ISL_1229382, EPI_ISL_1229383, EPI_ISL_1229384, EPI_ISL_1229385, EPI_ISL_1229386, EPI_ISL_1229387, EPI_ISL_1231192, EPI_ISL_1231193, EPI_ISL_1231194, EPI_ISL_1231195, EPI_ISL_1231196, EPI_ISL_1231197, EPI_ISL_1231198, EPI_ISL_1231199, EPI_ISL_1231200, EPI_ISL_1231201, EPI_ISL_1231202, EPI_ISL_1231203, EPI_ISL_1231204, EPI_ISL_1231205, EPI_ISL_1231206 | | | |
| see above | Kansas Health and Environmental Lab | Kansas Health and Environmental Lab | Mike Grose, Jonathan Barnell, Ben Olsen, and Phil Adam |
| EPI_ISL_1231436 | Baylor Esoteric + Molecular Lab | Baylor Esoteric + Molecular Lab | Elizabeth Forrester and Dawn Richards |
| EPI_ISL_1231437, EPI_ISL_1231438, EPI_ISL_1231439, EPI_ISL_1231440, EPI_ISL_1231441, EPI_ISL_1231442, EPI_ISL_1231443, EPI_ISL_1231444, EPI_ISL_1231445, EPI_ISL_1231446, EPI_ISL_1231447, EPI_ISL_1231448, EPI_ISL_1231449, EPI_ISL_1231450, EPI_ISL_1231451, EPI_ISL_1231452, EPI_ISL_1231453, EPI_ISL_1231454, EPI_ISL_1231455, EPI_ISL_1231456, EPI_ISL_1231457, EPI_ISL_1231458, EPI_ISL_1231459, EPI_ISL_1231460, EPI_ISL_1231461, EPI_ISL_1231462, EPI_ISL_1231463, EPI_ISL_1231464, EPI_ISL_1231465, EPI_ISL_1231466, EPI_ISL_1231467, EPI_ISL_1231468, EPI_ISL_1231469, EPI_ISL_1231470, EPI_ISL_1231471, EPI_ISL_1231472, EPI_ISL_1231473, EPI_ISL_1231474, EPI_ISL_1231475, EPI_ISL_1231476, EPI_ISL_1231477, EPI_ISL_1231478, EPI_ISL_1231479, EPI_ISL_1231480, EPI_ISL_1231481, EPI_ISL_1231482, EPI_ISL_1231483, EPI_ISL_1231484, EPI_ISL_1231485, EPI_ISL_1231486, EPI_ISL_1231487, EPI_ISL_1231488, EPI_ISL_1231489, EPI_ISL_1231490, EPI_ISL_1231491, EPI_ISL_1231492, EPI_ISL_1231493, EPI_ISL_1231494, EPI_ISL_1231495, EPI_ISL_1231496, EPI_ISL_1231497, EPI_ISL_1231498, EPI_ISL_1231499, EPI_ISL_1231500, EPI_ISL_1231501, EPI_ISL_1231502, EPI_ISL_1231503, EPI_ISL_1231504, EPI_ISL_1231505, EPI_ISL_1231506, EPI_ISL_1231507, EPI_ISL_1231508, EPI_ISL_1231509, EPI_ISL_1231510, EPI_ISL_1231511, EPI_ISL_1231512, EPI_ISL_1231513, EPI_ISL_1231514, EPI_ISL_1231515, EPI_ISL_1231516, EPI_ISL_1231517, EPI_ISL_1231518, EPI_ISL_1231519, EPI_ISL_1231520, EPI_ISL_1231521, EPI_ISL_1231522, EPI_ISL_1231523, EPI_ISL_1231524, EPI_ISL_1231525, EPI_ISL_1231526, EPI_ISL_1231527, EPI_ISL_1231528, EPI_ISL_1231529, EPI_ISL_1231530, EPI_ISL_1231533, EPI_ISL_1231537, EPI_ISL_1231541, EPI_ISL_1231545, EPI_ISL_1231549, EPI_ISL_1231553, EPI_ISL_1231557, EPI_ISL_1231561, EPI_ISL_1231565, EPI_ISL_1231569, EPI_ISL_1231573, EPI_ISL_1231577, EPI_ISL_1231581, EPI_ISL_1231585, EPI_ISL_1231590, EPI_ISL_1231594, EPI_ISL_1231598, EPI_ISL_1231602, EPI_ISL_1231606, EPI_ISL_1231610, EPI_ISL_1231615, EPI_ISL_1231618, EPI_ISL_1231623, EPI_ISL_1231626, EPI_ISL_1231630, EPI_ISL_1231634, EPI_ISL_1231638, EPI_ISL_1231642, EPI_ISL_1231646, EPI_ISL_1231651, EPI_ISL_1231655, EPI_ISL_1231659, EPI_ISL_1231663, EPI_ISL_1231666, EPI_ISL_1231670, EPI_ISL_1231675, EPI_ISL_1231678, EPI_ISL_1231682, EPI_ISL_1231686, EPI_ISL_1231690, EPI_ISL_1231694, EPI_ISL_1231698, EPI_ISL_1231702, EPI_ISL_1231706, EPI_ISL_1231710, EPI_ISL_1231714, EPI_ISL_1231719, EPI_ISL_1231723, EPI_ISL_1231727, EPI_ISL_1231730, EPI_ISL_1231735, EPI_ISL_1231739, EPI_ISL_1231743, EPI_ISL_1231747, EPI_ISL_1231751, EPI_ISL_1231755, EPI_ISL_1231758, EPI_ISL_1231762, EPI_ISL_1231767, EPI_ISL_1231771, EPI_ISL_1231774, EPI_ISL_1231775, EPI_ISL_1231776, EPI_ISL_1231777, EPI_ISL_1231778, EPI_ISL_1231779, EPI_ISL_1231780, EPI_ISL_1231781, EPI_ISL_1231782, EPI_ISL_1231783, EPI_ISL_1231784, EPI_ISL_1231785, EPI_ISL_1231786, EPI_ISL_1231787, EPI_ISL_1231788, EPI_ISL_1231789, EPI_ISL_1231790, EPI_ISL_1231791, EPI_ISL_1231793, EPI_ISL_1231794, EPI_ISL_1231795, EPI_ISL_1231796, EPI_ISL_1231797, EPI_ISL_1231798, EPI_ISL_1231799, EPI_ISL_1231800, EPI_ISL_1231801, EPI_ISL_1231802, EPI_ISL_1231803, EPI_ISL_1231804, EPI_ISL_1231805, EPI_ISL_1231806, EPI_ISL_1231807, EPI_ISL_1231808, EPI_ISL_1231809, EPI_ISL_1231810, EPI_ISL_1231811, EPI_ISL_1231813, EPI_ISL_1231814, EPI_ISL_1231815, EPI_ISL_1231816, EPI_ISL_1231818, EPI_ISL_1231819, EPI_ISL_1231820, EPI_ISL_1231821, EPI_ISL_1231822, EPI_ISL_1231823, EPI_ISL_1231824, EPI_ISL_1231825, EPI_ISL_1231826, EPI_ISL_1231827, EPI_ISL_1231828, EPI_ISL_1231829, EPI_ISL_1231830, EPI_ISL_1231831, EPI_ISL_1231832, EPI_ISL_1231833, EPI_ISL_1231834, EPI_ISL_1231835, EPI_ISL_1231836, EPI_ISL_1231837, EPI_ISL_1231838, EPI_ISL_1231839, EPI_ISL_1231840, EPI_ISL_1231841, EPI_ISL_1231842, EPI_ISL_1231843, EPI_ISL_1231844, EPI_ISL_1231845, EPI_ISL_1231846, EPI_ISL_1231847, EPI_ISL_1231848, EPI_ISL_1231849, EPI_ISL_1231850, EPI_ISL_1231851, EPI_ISL_1231852, EPI_ISL_1231853, EPI_ISL_1231854, EPI_ISL_1231855, EPI_ISL_1231856, EPI_ISL_1231857, EPI_ISL_1231858, EPI_ISL_1231859, EPI_ISL_1231860, EPI_ISL_1231861, EPI_ISL_1231862, EPI_ISL_1231863, EPI_ISL_1231864, EPI_ISL_1231865, EPI_ISL_1231866, EPI_ISL_1231867, EPI_ISL_1231868, EPI_ISL_1231869, EPI_ISL_1231870, EPI_ISL_1231871, EPI_ISL_1231872, EPI_ISL_1231873, EPI_ISL_1231874, EPI_ISL_1231875, EPI_ISL_1231876, EPI_ISL_1231877, EPI_ISL_1231878, EPI_ISL_1231879, EPI_ISL_1231880, EPI_ISL_1231881, EPI_ISL_1231882, EPI_ISL_1231883, EPI_ISL_1231884, EPI_ISL_1231885, EPI_ISL_1231886, EPI_ISL_1231887, EPI_ISL_1231888, EPI_ISL_1231889, EPI_ISL_1231890, EPI_ISL_1231891, EPI_ISL_1231892, EPI_ISL_1231893, EPI_ISL_1231894, EPI_ISL_1231895, EPI_ISL_1231896, EPI_ISL_1231897, EPI_ISL_1231898, EPI_ISL_1231899, EPI_ISL_1231900, EPI_ISL_1231901, EPI_ISL_1231902, EPI_ISL_1231903, EPI_ISL_1231904, EPI_ISL_1231905, EPI_ISL_1231906, EPI_ISL_1231907, EPI_ISL_1231908, EPI_ISL_1231909, EPI_ISL_1231910, EPI_ISL_1231911, EPI_ISL_1231912, EPI_ISL_1231913, EPI_ISL_1232029, EPI_ISL_1232030, EPI_ISL_1232031, EPI_ISL_1232032, EPI_ISL_1232033, EPI_ISL_1232034, EPI_ISL_1232035, EPI_ISL_1232036, EPI_ISL_1232037, EPI_ISL_1232038, EPI_ISL_1232039, EPI_ISL_1232040, EPI_ISL_1232041, EPI_ISL_1232042, EPI_ISL_1232043, EPI_ISL_1232044, EPI_ISL_1232045, EPI_ISL_1232046, EPI_ISL_1232047, EPI_ISL_1232048, EPI_ISL_1232049, EPI_ISL_1232050, EPI_ISL_1232051, EPI_ISL_1232052, EPI_ISL_1232053, EPI_ISL_1232054, EPI_ISL_1232055, EPI_ISL_1232056, EPI_ISL_1232057, EPI_ISL_1232058, EPI_ISL_1232059, EPI_ISL_1232060, EPI_ISL_1232061, EPI_ISL_1232062, EPI_ISL_1232063, EPI_ISL_1232064, EPI_ISL_1232065, EPI_ISL_1232066, EPI_ISL_1232067, EPI_ISL_1232068, EPI_ISL_1232069, EPI_ISL_1232070, EPI_ISL_1232071, EPI_ISL_1232072, EPI_ISL_1232073, EPI_ISL_1232074, EPI_ISL_1232075, EPI_ISL_1232076, EPI_ISL_1232077, EPI_ISL_1232078, EPI_ISL_1232079, EPI_ISL_1232080, EPI_ISL_1232081, EPI_ISL_1232082, EPI_ISL_1232083, EPI_ISL_1232084, EPI_ISL_1232085, EPI_ISL_1232086, EPI_ISL_1232087, EPI_ISL_1232088, EPI_ISL_1232089, EPI_ISL_1232090, EPI_ISL_1232091, EPI_ISL_1232092, EPI_ISL_1232093, EPI_ISL_1232094, EPI_ISL_1232095, EPI_ISL_1232096, EPI_ISL_1232097, EPI_ISL_1232098, EPI_ISL_1232099, EPI_ISL_1232100, EPI_ISL_1232101, EPI_ISL_1232102, EPI_ISL_1232103, EPI_ISL_1232104, EPI_ISL_1232105, EPI_ISL_1232106, EPI_ISL_1232107, EPI_ISL_1232108, EPI_ISL_1232109, EPI_ISL_1232110, EPI_ISL_1232111, EPI_ISL_1232112, EPI_ISL_1232113, EPI_ISL_1232114, EPI_ISL_1232115, EPI_ISL_1232116, EPI_ISL_1232117, EPI_ISL_1232118, EPI_ISL_1232119, EPI_ISL_1232120, EPI_ISL_1232121, EPI_ISL_1232122, EPI_ISL_1232123, EPI_ISL_1232124, EPI_ISL_1232125, EPI_ISL_1232126, EPI_ISL_1232127, EPI_ISL_1232128, EPI_ISL_1232129, EPI_ISL_1232130, EPI_ISL_1232131, EPI_ISL_1232132, EPI_ISL_1232133, EPI_ISL_1232134, EPI_ISL_1232135, EPI_ISL_1232136, EPI_ISL_1232137, EPI_ISL_1232138, EPI_ISL_1232139, EPI_ISL_1232140, EPI_ISL_1232141, EPI_ISL_1232142, EPI_ISL_1232143, EPI_ISL_1232144, EPI_ISL_1232145, EPI_ISL_1232146, EPI_ISL_1232147, EPI_ISL_1232148, EPI_ISL_1232149, EPI_ISL_1232150, EPI_ISL_1232151, EPI_ISL_1232152, EPI_ISL_1232153, EPI_ISL_1232154, EPI_ISL_1232155, EPI_ISL_1232156, EPI_ISL_1232157, EPI_ISL_1232158, EPI_ISL_1232159, EPI_ISL_1232160, EPI_ISL_1232161, EPI_ISL_1232162, EPI_ISL_1232163, EPI_ISL_1232164, EPI_ISL_1232165, EPI_ISL_1232166, EPI_ISL_1232167, EPI_ISL_1232168, EPI_ISL_1232169, EPI_ISL_1232170, EPI_ISL_1232171, EPI_ISL_1232172, EPI_ISL_1232173, EPI_ISL_1232174, EPI_ISL_1232175, EPI_ISL_1232176, EPI_ISL_1232177, EPI_ISL_1232178, EPI_ISL_1232179, EPI_ISL_1232180, EPI_ISL_1232181, EPI_ISL_1232182, EPI_ISL_1232183, EPI_ISL_1232184, EPI_ISL_1232185, EPI_ISL_1232186, EPI_ISL_1232187, EPI_ISL_1232188, EPI_ISL_1232189, EPI_ISL_1232190, EPI_ISL_1232191, EPI_ISL_1232192 | | | |
| see above | Fulgent Genetics | Fulgent Genetics | Harry Gao, Mickey Li, John Gao, Joseph Fierro, Benafsh Sapra, Becky Tsai, Yan Meng, Doreen Ng, James Xie |
| EPI_ISL_1232193, EPI_ISL_1232194, EPI_ISL_1232195, EPI_ISL_1232196, EPI_ISL_1232197, EPI_ISL_1232198, EPI_ISL_1232199, EPI_ISL_1232200, EPI_ISL_1232201, EPI_ISL_1232202, EPI_ISL_1232203, EPI_ISL_1232204, EPI_ISL_1232205, EPI_ISL_1232206, EPI_ISL_1232207, EPI_ISL_1232208, EPI_ISL_1232209, EPI_ISL_1232210, EPI_ISL_1232211, EPI_ISL_1232212, EPI_ISL_1232213, EPI_ISL_1232214, EPI_ISL_1232215, EPI_ISL_1232216, EPI_ISL_1232217, EPI_ISL_1232218, EPI_ISL_1232219, EPI_ISL_1232220, EPI_ISL_1232221, EPI_ISL_1232222, EPI_ISL_1232223, EPI_ISL_1232224, EPI_ISL_1232225, EPI_ISL_1232226, EPI_ISL_1232227, EPI_ISL_1232228, EPI_ISL_1232229, EPI_ISL_1232230, EPI_ISL_1232231, EPI_ISL_1232232, EPI_ISL_1232233, EPI_ISL_1232234, EPI_ISL_1232235, EPI_ISL_1232236, EPI_ISL_1232237, EPI_ISL_1232238, EPI_ISL_1232239 | | | |
| see above | Wyoming Public Health Laboratory | Wyoming Public Health Laboratory | Jim Mildenberger, Wanda Manley, Noah Hull, Taylor Fearing, Lynette Gumbleton, Channing Weber, Ashley Norberg, Chayse Rowley, Marley Goetz, Brian Dominguez, Elliot Thomasson, Cari Sloma, and Rob Christensen |
| EPI_ISL_1232240, EPI_ISL_1232241, EPI_ISL_1232242 | Oregon State University TRACE Project | OSU Center for Genome Research and Biocomputing | Oregon State University TRACE Project |
| EPI_ISL_1233008, EPI_ISL_1233009, EPI_ISL_1233010, EPI_ISL_1233011, EPI_ISL_1233012, EPI_ISL_1233013, EPI_ISL_1233014, EPI_ISL_1233015, EPI_ISL_1233016, EPI_ISL_1233017, EPI_ISL_1233018, EPI_ISL_1233019, EPI_ISL_1233020, EPI_ISL_1233021, EPI_ISL_1233022, EPI_ISL_1233023, EPI_ISL_1233024, EPI_ISL_1233025, EPI_ISL_1233026, EPI_ISL_1233027, EPI_ISL_1233028, EPI_ISL_1233030, EPI_ISL_1233031, EPI_ISL_1233032, EPI_ISL_1233033, EPI_ISL_1233034, EPI_ISL_1233035, EPI_ISL_1233036, EPI_ISL_1233037, EPI_ISL_1233038, EPI_ISL_1233039, EPI_ISL_1233040, EPI_ISL_1233041, EPI_ISL_1233042, EPI_ISL_1233043, EPI_ISL_1233044, EPI_ISL_1233045, EPI_ISL_1233046, EPI_ISL_1233047, EPI_ISL_1233048, EPI_ISL_1233049, EPI_ISL_1233050, EPI_ISL_1233051, EPI_ISL_1233052, EPI_ISL_1233053 | | | |
| see above | North Dakota Department of Health, Public Health Laboratory | North Dakota Department of Health, Public Health Laboratory | Lisa Wingerter |
| EPI_ISL_1233139, EPI_ISL_1233140, EPI_ISL_1233141, EPI_ISL_1233142, EPI_ISL_1233143, EPI_ISL_1233144, EPI_ISL_1233145, EPI_ISL_1233146, EPI_ISL_1233147, EPI_ISL_1233148, EPI_ISL_1233149, EPI_ISL_1233150, EPI_ISL_1233151, EPI_ISL_1233152, EPI_ISL_1233153, EPI_ISL_1233154, EPI_ISL_1233155, EPI_ISL_1233156, EPI_ISL_1233157, EPI_ISL_1233158, EPI_ISL_1233159, EPI_ISL_1233160, EPI_ISL_1233161, EPI_ISL_1233162, EPI_ISL_1233163, EPI_ISL_1233164, EPI_ISL_1233165, EPI_ISL_1233166, EPI_ISL_1233167, EPI_ISL_1233168, EPI_ISL_1233169, EPI_ISL_1233170, EPI_ISL_1233171, EPI_ISL_1233172, EPI_ISL_1233173, EPI_ISL_1233174, EPI_ISL_1233175, EPI_ISL_1233176, EPI_ISL_1233177, EPI_ISL_1233178, EPI_ISL_1233179, EPI_ISL_1233180, EPI_ISL_1233181, EPI_ISL_1233182, EPI_ISL_1233183, EPI_ISL_1233184, EPI_ISL_1233185, EPI_ISL_1233186 | | | |
| see above | UW Virology Lab | UW Virology Lab | Pavitra Roychoudhury, Hong Xie, Lasata Shrestha, Shah Mohamed Bakhash, Michelle Lin, Margaret Mills, Noah Baker, Sean Ellis, Saraswathi Sathees, Meei-Li Huang, Keith R Jerome, Alexander Greninger |
| EPI_ISL_1233187, EPI_ISL_1233222 | Swedish Medical Center, Seattle, WA, USA | Institute for Systems Biology | Jason D. Goldman, Chris Lausted, Kai Wang, Jared C. Roach, James R. Heath, Julie A. Wallick |
| EPI_ISL_1233255, EPI_ISL_1233256, EPI_ISL_1233257, EPI_ISL_1233258, EPI_ISL_1233259, EPI_ISL_1233261, EPI_ISL_1233263, EPI_ISL_1233264, EPI_ISL_1233266, EPI_ISL_1233267, EPI_ISL_1233270, EPI_ISL_1233271, EPI_ISL_1233272, EPI_ISL_1233273, EPI_ISL_1233276, EPI_ISL_1233277, EPI_ISL_1233278, EPI_ISL_1233279, EPI_ISL_1233280, EPI_ISL_1233281, EPI_ISL_1233282, EPI_ISL_1233285, EPI_ISL_1233286, EPI_ISL_1233287, EPI_ISL_1233288, EPI_ISL_1233289, EPI_ISL_1233290, EPI_ISL_1233291, EPI_ISL_1233292, EPI_ISL_1233293, EPI_ISL_1233294, EPI_ISL_1233295, EPI_ISL_1233298, EPI_ISL_1233299, EPI_ISL_1233300, EPI_ISL_1233301, EPI_ISL_1233302, EPI_ISL_1233303, EPI_ISL_1233304, EPI_ISL_1233306, EPI_ISL_1233307, EPI_ISL_1233311, EPI_ISL_1233313, EPI_ISL_1233318, EPI_ISL_1233319, EPI_ISL_1233320, EPI_ISL_1233324, EPI_ISL_1233325, EPI_ISL_1233326, EPI_ISL_1233328, EPI_ISL_1233329, EPI_ISL_1233331, EPI_ISL_1233332, EPI_ISL_1233333, EPI_ISL_1233334, EPI_ISL_1233335, EPI_ISL_1233336, EPI_ISL_1233338, EPI_ISL_1233339, EPI_ISL_1233341, EPI_ISL_1233342, EPI_ISL_1233343, EPI_ISL_1233344, EPI_ISL_1233345, EPI_ISL_1233347, EPI_ISL_1233349, EPI_ISL_1233350, EPI_ISL_1233351, EPI_ISL_1233352, EPI_ISL_1233353, EPI_ISL_1233354, EPI_ISL_1233355, EPI_ISL_1233358, EPI_ISL_1233359, EPI_ISL_1233360, EPI_ISL_1233361, EPI_ISL_1233362, EPI_ISL_1233363, EPI_ISL_1233364, EPI_ISL_1233365, EPI_ISL_1233366, EPI_ISL_1233368, EPI_ISL_1233370, EPI_ISL_1233373, EPI_ISL_1233374, EPI_ISL_1233375, EPI_ISL_1233376 | | | |
| see above | Massachusetts State Public Health Laboratory | Massachusetts State Public Health Laboratory | Andrew Lang, Timelia Fink, Glen Gallagher, Sandra Smole |
| EPI_ISL_1233424, EPI_ISL_1233425, EPI_ISL_1233436, EPI_ISL_1233438, EPI_ISL_1233483 | Kashi Clinical Laboratory | Kashi Clinical Laboratory | Zahra Kashi, Jenefer DeKoning |
| EPI_ISL_1233694, EPI_ISL_1233695, EPI_ISL_1233696, EPI_ISL_1233697, EPI_ISL_1233698, EPI_ISL_1233699, EPI_ISL_1233700, EPI_ISL_1233701, EPI_ISL_1233702, EPI_ISL_1233703, EPI_ISL_1233704, EPI_ISL_1233706, EPI_ISL_1233707, EPI_ISL_1233711, EPI_ISL_1233713, EPI_ISL_1233714, EPI_ISL_1233715, EPI_ISL_1233716, EPI_ISL_1233717, EPI_ISL_1233718, EPI_ISL_1233719, EPI_ISL_1233720, EPI_ISL_1233722, EPI_ISL_1233723, EPI_ISL_1233724, EPI_ISL_1233725, EPI_ISL_1233728, EPI_ISL_1233729 | | | |
| see above | North Dakota Department of Health, Public Health Laboratory | North Dakota Department of Health, Public Health Laboratory | Lisa Wingerter |
| EPI_ISL_1233730, EPI_ISL_1233731, EPI_ISL_1233732, EPI_ISL_1233733, EPI_ISL_1233734, EPI_ISL_1233735, EPI_ISL_1233736, EPI_ISL_1233737, EPI_ISL_1233738, EPI_ISL_1233739, EPI_ISL_1233740, EPI_ISL_1233741, EPI_ISL_1233742, EPI_ISL_1233743, EPI_ISL_1233744, EPI_ISL_1233746, EPI_ISL_1233747, EPI_ISL_1233749, EPI_ISL_1233750, EPI_ISL_1233751, EPI_ISL_1233752, EPI_ISL_1233753, EPI_ISL_1233754, EPI_ISL_1233756, EPI_ISL_1233757, EPI_ISL_1233758, EPI_ISL_1233759, EPI_ISL_1233760, EPI_ISL_1233761, EPI_ISL_1233762, EPI_ISL_1233763, EPI_ISL_1233764, EPI_ISL_1233765, EPI_ISL_1233766, EPI_ISL_1233767, EPI_ISL_1233768, EPI_ISL_1233769, EPI_ISL_1233770, EPI_ISL_1233771, EPI_ISL_1233773, EPI_ISL_1233774, EPI_ISL_1233775, EPI_ISL_1233776, EPI_ISL_1233777, EPI_ISL_1233778, EPI_ISL_1233779, EPI_ISL_1233780, EPI_ISL_1233781, EPI_ISL_1233782, EPI_ISL_1233783, EPI_ISL_1233784, EPI_ISL_1233785, EPI_ISL_1233786, EPI_ISL_1233787, EPI_ISL_1233788, EPI_ISL_1233789, EPI_ISL_1233790, EPI_ISL_1233791, EPI_ISL_1233792, EPI_ISL_1233793, EPI_ISL_1233794, EPI_ISL_1233795, EPI_ISL_1233796, EPI_ISL_1233797, EPI_ISL_1233798, EPI_ISL_1233799, EPI_ISL_1233800, EPI_ISL_1233801, EPI_ISL_1233803, EPI_ISL_1233804, EPI_ISL_1233805, EPI_ISL_1233806, EPI_ISL_1233807, EPI_ISL_1233808, EPI_ISL_1233809, EPI_ISL_1233810, EPI_ISL_1233811, EPI_ISL_1233818 | | | |
| see above | Michigan Department of Health and Human Services, Bureau of Laboratories | Michigan Department of Health and Human Services, Bureau of Laboratories | Blankenship HM, Riner D, Soehnlen MK |
| EPI_ISL_1233868, EPI_ISL_1233869, EPI_ISL_1233870, EPI_ISL_1233871, EPI_ISL_1233872 | Kashi Clinical Laboratory | Kashi Clinical Laboratory | Zahra Kashi, Jenefer DeKoning |
| EPI_ISL_1233874 | Department of Laboratory Medicine, Clinical Center, National Institutes of Health | Laboratory of Parasitic Diseases, Systems Genomics Section, National Institute of Allergy and Infectious Diseases, National Institutes of Health | Allison Roder, Stephanie Banakis, Matthew Chung, Jung-ho Youn, Rachel Mercado, Wei Wang, Tara Palmore, Michael Bell, Heike Bailin, Jessica McCormick-Ell, Sanchita Das, Jennifer Kwan, Elodie Ghedin |

| | | | |
|---|---|---|---|
| EPI_ISL_1233875, EPI_ISL_1233876 | Laboratory of Parasitic Diseases, Systems Genomics Section, National Institute of Allergy and Infectious Diseases, Nathional Institutes of Health | Laboratory of Parasitic Diseases, Systems Genomics Section, National Institute of Allergy and Infectious Diseases, Nathional Institutes of Health | Roder,A., Banakis,S., Chung,M., Youn,J.-h., Mercado,R., Wang,W., Palmore,T., Bell,M., Bailin,H., McCormick-Ell,J., Das,S., Kwan,J., Ghedin,E. |
| EPI_ISL_1233877, EPI_ISL_1233878 | Department of Laboratory Medicine, Clinical Center, National Institutes of Health | Laboratory of Parasitic Diseases, Systems Genomics Section, National Institute of Allergy and Infectious Diseases, National Institutes of Health | Allison Roder, Stephanie Banakis, Matthew Chung, Jung-ho Youn, Rachel Mercado, Wei Wang, Tara Palmore, Michael Bell, Heike Bailin, Jessica McCormick-Ell, Sanchita Das, Jennifer Kwan, Elodie Ghedin |
| EPI_ISL_1233880, EPI_ISL_1233881, EPI_ISL_1233882, EPI_ISL_1233883, EPI_ISL_1233884, EPI_ISL_1233885, EPI_ISL_1233886, EPI_ISL_1233887, EPI_ISL_1233889, EPI_ISL_1233890 | Respiratory Viruses Branch, Centers for Disease Control and Prevention | Respiratory Viruses Branch, Centers for Disease Control and Prevention | Cook,P.W., Howard,D.T., Batra,D., Rambo-Martin,B.L., de Feo,E., Antico,J., Tran,C., Tolentino,M., Wickline,S., Gietzen,K., Sickler,B., Liu,J., Allen,E., Febbo,P., Galloway,S., Washington,N.L., White,S., Levan,G., Barrett,K.S., Cirulli,E., Bolze,A., Ascencio,A., Rivera-Garcia,C., Cho,R., Nguyen,J., Wang,S., Ramirez,J., Cassens,T., Sandoval,E., Isaksson,M., Lee,W., Becker,D., Laurent,M., Lu,J., Paden,C.R., Tong,S., MacCannell,D. |
| EPI_ISL_1233893, EPI_ISL_1233894 | Infectious Diseases, Chan-Zuckerberg Biohub Covidtracker | Infectious Diseases, Chan-Zuckerberg Biohub Covidtracker | CZB Cliahub Consortium |
| EPI_ISL_1233895 | Pathology, University of Alabama at Birmingham | University of Alabama at Birmingham | Moates,D., Hendrickson,R.C., Lefkowitz,E.J., Leal,S.M. |
| EPI_ISL_1233896, EPI_ISL_1233897 | Public Health Laboratory - Infectious Disease Lab, Minnesota Department of Health Infectious Disease Laboratory Submission Group | Public Health Laboratory - Infectious Disease Lab, Minnesota Department of Health Infectious Disease Laboratory Submission Group | Lorentz,A., Garfin,J., Plumb,M., Wang,X. |
| EPI_ISL_1233898 | Clinical Division, Fred Hutchinson Cancer Research Center | Clinical Division, Fred Hutchinson Cancer Research Center | Pyo,C.-W., Nelson,W.C., Li,R., Wang,R., Ishitani,A., Mills,M.G., Randhawa,A.K., Roychoudhury,P., Greninger,A., Jerome,K., Zhao,L.P., Geraghty,D.E. |
| EPI_ISL_1233899 | Respiratory Viruses Branch, Centers for Disease Control and Prevention | Respiratory Viruses Branch, Centers for Disease Control and Prevention | Cook,P.W., Howard,D.T., Batra,D., Rambo-Martin,B.L., de Feo,E., Antico,J., Tran,C., Tolentino,M., Wickline,S., Gietzen,K., Sickler,B., Liu,J., Allen,E., Febbo,P., Galloway,S., Washington,N.L., White,S., Levan,G., Barrett,K.S., Cirulli,E., Bolze,A., Ascencio,A., Rivera-Garcia,C., Cho,R., Nguyen,J., Wang,S., Ramirez,J., Cassens,T., Sandoval,E., Isaksson,M., Lee,W., Becker,D., Laurent,M., Lu,J., Paden,C.R., Tong,S., MacCannell,D. |
| EPI_ISL_1233900 | Houston Health Department, Disease Prevention and Control | Houston Health Department | Penn,R., Brown,P., Lara,A. |
| EPI_ISL_1233901, EPI_ISL_1233902, EPI_ISL_1233903 | Infectious Diseases, Quest Diagnostics | Infectious Diseases, Quest Diagnostics | Rosenthal,S.H., Gerasimova,A., Kagan,R.M., Anderson,B., Hua,M., Liu,Y., Bernstein,L.E., Livingston,K.E., Perez,A., Shalhout,D.F., Shlyakhter,I.A., Owen,R., Tanpaiboon,P., Lacbawan,F. |
| EPI_ISL_1233904 | Public Health Laboratory - Infectious Disease Lab, Minnesota Department of Health Infectious Disease Laboratory Submission Group | Public Health Laboratory - Infectious Disease Lab, Minnesota Department of Health Infectious Disease Laboratory Submission Group | Lorentz,A., Garfin,J., Plumb,M., Wang,X. |
| EPI_ISL_1233919, EPI_ISL_1233920, EPI_ISL_1233921, EPI_ISL_1233924 | Infectious Diseases, Quest Diagnostics | Infectious Diseases, Quest Diagnostics | Rosenthal,S.H., Gerasimova,A., Kagan,R.M., Anderson,B., Bernstein,L.E., Livingston,K.E., Hua,M., Liu,Y., Shalhout,D.F., Owen,R., Lacbawan,F. |
| EPI_ISL_1233925, EPI_ISL_1233926, EPI_ISL_1233927, EPI_ISL_1233929, EPI_ISL_1233930, EPI_ISL_1233933, EPI_ISL_1233934, EPI_ISL_1233935 | Respiratory Viruses Branch, Centers for Disease Control and Prevention | Respiratory Viruses Branch, Centers for Disease Control and Prevention | Cook,P.W., Batra,D., Rambo-Martin,B.L., de Feo,E., Antico,J., Tran,C., Tolentino,M., Wickline,S., Gietzen,K., Sickler,B., Liu,J., Allen,E., Febbo,P., Galloway,S., Washington,N.L., White,S., Levan,G., Barret,K.S., Cirulli,E., Bolze,A., Ascencio,A., Rivera-Garcia,C., Cho,R., Nguyen,J., Wang,S., Ramirez,J., Cassens,T., Sandoval,E., Isaksson,M., Lee,W., Becker,D., Laurent,M., Lu,J., Paden,C.R., Tong,S., MacCannell,D. |
| EPI_ISL_1233936, EPI_ISL_1233937 | Division of Viral Diseases, CDC Pathogen Discovery Team | Division of Viral Diseases, CDC Pathogen Discovery Team | Queen,K., Li,Y., Tao,Y., Zhang,J., Uehara,A., Montmayeur,A., Paden,C.R., Cook,P.W., Marine,R., Sheth,M., Padilla,J., Nobles,S., Burroughs,M., Rowe,L., Wang,H., Rambo-Martin,B.L., Batra,D., Knipe,K., Lee,J.S., Tong,S. |
| EPI_ISL_1233938 | Respiratory Viruses Branch, Centers for Disease Control and Prevention | Respiratory Viruses Branch, Centers for Disease Control and Prevention | Cook,P.W., Batra,D., Rambo-Martin,B.L., de Feo,E., Antico,J., Tran,C., Tolentino,M., Wickline,S., Gietzen,K., Sickler,B., Liu,J., Allen,E., Febbo,P., Galloway,S., Washington,N.L., White,S., Levan,G., Barret,K.S., Cirulli,E., Bolze,A., Ascencio,A., Rivera-Garcia,C., Cho,R., Nguyen,J., Wang,S., Ramirez,J., Cassens,T., Sandoval,E., Isaksson,M., Lee,W., Becker,D., Laurent,M., Lu,J., Paden,C.R., Tong,S., MacCannell,D. |
| EPI_ISL_1233939, EPI_ISL_1233940 | Respiratory Viruses Branch, Centers for Disease Control and Prevention | Respiratory Viruses Branch, Centers for Disease Control and Prevention | Li,Y., Tao,Y., Montmayeur,A., Zhang,J., Lynch,B., Queen,K., Uehara,A., Marine,R., Cook,P., Paden,C.R., Wang,H., Tong,S. |
| EPI_ISL_1233941, EPI_ISL_1233942 | Respiratory Viruses Branch, Centers for Disease Control and Prevention | Respiratory Viruses Branch, Centers for Disease Control and Prevention | Queen,K., Li,Y., Tao,Y., Uehara,A., Montmayeur,A., Paden,C.R., Cook,P.W., Marine,R., Sheth,M., Wang,H., Lee,J., Tong,S. |
| EPI_ISL_1233943, EPI_ISL_1233944, EPI_ISL_1233945, EPI_ISL_1233946, EPI_ISL_1233947, EPI_ISL_1233948, EPI_ISL_1233949, EPI_ISL_1233950, EPI_ISL_1233951, EPI_ISL_1233952 | Maryland Public Health Laboratory | Maryland Public Health Laboratory | Keller,E., Loomis,J., Moore,T.L. |
| EPI_ISL_1233954, EPI_ISL_1233955, EPI_ISL_1233956, EPI_ISL_1233957, EPI_ISL_1233958, EPI_ISL_1233959, EPI_ISL_1233960 | Maryland Public Health Laboratory | Maryland Public Health Laboratory | Keller,E., Loomis,J. |
| EPI_ISL_1233961, EPI_ISL_1233962, EPI_ISL_1233963, EPI_ISL_1233964, EPI_ISL_1233965, EPI_ISL_1233966, EPI_ISL_1233967, EPI_ISL_1233968, EPI_ISL_1233969, EPI_ISL_1233970, EPI_ISL_1233971, EPI_ISL_1233972, EPI_ISL_1233973 | | | |
| see above | Maryland Public Health Laboratory | Maryland Public Health Laboratory | Keller,E., Loomis,J., Moore,T.L. |
| EPI_ISL_1233974 | Maryland Public Health Laboratory | Maryland Public Health Laboratory | Keller,E., Loomis,J. |
| EPI_ISL_1233975, EPI_ISL_1233976, EPI_ISL_1233977, EPI_ISL_1233978, EPI_ISL_1233979, EPI_ISL_1233980, EPI_ISL_1233981, EPI_ISL_1233982, EPI_ISL_1233983, EPI_ISL_1233984, EPI_ISL_1233985, EPI_ISL_1233986, EPI_ISL_1233987, EPI_ISL_1233988, EPI_ISL_1233989, EPI_ISL_1233990, EPI_ISL_1233991, EPI_ISL_1233992, EPI_ISL_1233994, EPI_ISL_1233995, EPI_ISL_1233996, EPI_ISL_1233998, EPI_ISL_1233999, EPI_ISL_1234000, EPI_ISL_1234001, EPI_ISL_1234002, EPI_ISL_1234003, EPI_ISL_1234004, EPI_ISL_1234005, EPI_ISL_1234006, EPI_ISL_1234007, EPI_ISL_1234008, EPI_ISL_1234009, EPI_ISL_1234010, EPI_ISL_1234011, EPI_ISL_1234012, EPI_ISL_1234013, EPI_ISL_1234014, EPI_ISL_1234015, EPI_ISL_1234016, EPI_ISL_1234017, EPI_ISL_1234018, EPI_ISL_1234019, EPI_ISL_1234020, EPI_ISL_1234021, EPI_ISL_1234022, EPI_ISL_1234023, EPI_ISL_1234024, EPI_ISL_1234025, EPI_ISL_1234026, EPI_ISL_1234027, EPI_ISL_1234028, EPI_ISL_1234029, EPI_ISL_1234030, EPI_ISL_1234032, EPI_ISL_1234033, EPI_ISL_1234034, EPI_ISL_1234035, EPI_ISL_1234036, EPI_ISL_1234037, EPI_ISL_1234038, EPI_ISL_1234039, EPI_ISL_1234040, EPI_ISL_1234041, EPI_ISL_1234042, EPI_ISL_1234043, EPI_ISL_1234046, EPI_ISL_1234047, EPI_ISL_1234048, EPI_ISL_1234049, EPI_ISL_1234052, EPI_ISL_1234053, EPI_ISL_1234054, EPI_ISL_1234057, EPI_ISL_1234058, EPI_ISL_1234059, EPI_ISL_1234060, EPI_ISL_1234061, EPI_ISL_1234062, EPI_ISL_1234063, EPI_ISL_1234064, EPI_ISL_1234065, EPI_ISL_1234066, EPI_ISL_1234067, EPI_ISL_1234068, EPI_ISL_1234069, EPI_ISL_1234070, EPI_ISL_1234071, EPI_ISL_1234072, EPI_ISL_1234073, EPI_ISL_1234074, EPI_ISL_1234075, EPI_ISL_1234076, EPI_ISL_1234077, EPI_ISL_1234078, EPI_ISL_1234079, EPI_ISL_1234080, EPI_ISL_1234081, EPI_ISL_1234082, EPI_ISL_1234083, EPI_ISL_1234085, EPI_ISL_1234086, EPI_ISL_1234087, EPI_ISL_1234088, EPI_ISL_1234089, EPI_ISL_1234090, EPI_ISL_1234091, EPI_ISL_1234092, EPI_ISL_1234093, EPI_ISL_1234094, EPI_ISL_1234095, EPI_ISL_1234096, EPI_ISL_1234097, EPI_ISL_1234099, EPI_ISL_1234100, EPI_ISL_1234101, EPI_ISL_1234102, EPI_ISL_1234103, EPI_ISL_1234104, EPI_ISL_1234105, EPI_ISL_1234106, EPI_ISL_1234107, EPI_ISL_1234108, EPI_ISL_1234109, EPI_ISL_1234110, EPI_ISL_1234111, EPI_ISL_1234112, EPI_ISL_1234113, EPI_ISL_1234114, EPI_ISL_1234115, EPI_ISL_1234116, EPI_ISL_1234117, EPI_ISL_1234118, EPI_ISL_1234119, EPI_ISL_1234120, EPI_ISL_1234121, EPI_ISL_1234122, EPI_ISL_1234123, EPI_ISL_1234124, EPI_ISL_1234125, EPI_ISL_1234126, EPI_ISL_1234127, EPI_ISL_1234128, EPI_ISL_1234129, EPI_ISL_1234130, EPI_ISL_1234131, EPI_ISL_1234132, EPI_ISL_1234133, EPI_ISL_1234135, EPI_ISL_1234136, EPI_ISL_1234137, EPI_ISL_1234138, EPI_ISL_1234139, EPI_ISL_1234140, EPI_ISL_1234141, EPI_ISL_1234143, EPI_ISL_1234144, EPI_ISL_1234145, EPI_ISL_1234146, EPI_ISL_1234147, EPI_ISL_1234148, EPI_ISL_1234149, EPI_ISL_1234150, EPI_ISL_1234151, EPI_ISL_1234152, EPI_ISL_1234153, EPI_ISL_1234154, EPI_ISL_1234155, EPI_ISL_1234156, EPI_ISL_1234157, EPI_ISL_1234158, EPI_ISL_1234159, EPI_ISL_1234160, EPI_ISL_1234161, EPI_ISL_1234162, EPI_ISL_1234164, EPI_ISL_1234165, EPI_ISL_1234166, EPI_ISL_1234167, EPI_ISL_1234168, EPI_ISL_1234169, EPI_ISL_1234171, EPI_ISL_1234172, EPI_ISL_1234173, EPI_ISL_1234174, EPI_ISL_1234175, EPI_ISL_1234176, EPI_ISL_1234177, EPI_ISL_1234178, EPI_ISL_1234179, EPI_ISL_1234180, EPI_ISL_1234181, EPI_ISL_1234182, EPI_ISL_1234183, EPI_ISL_1234184, EPI_ISL_1234185, EPI_ISL_1234186, EPI_ISL_1234187, EPI_ISL_1234188, EPI_ISL_1234189, EPI_ISL_1234190, EPI_ISL_1234191, EPI_ISL_1234192, EPI_ISL_1234193, EPI_ISL_1234194, EPI_ISL_1234195, EPI_ISL_1234196, EPI_ISL_1234197, EPI_ISL_1234198, EPI_ISL_1234199, EPI_ISL_1234200, EPI_ISL_1234201, EPI_ISL_1234202, EPI_ISL_1234203, EPI_ISL_1234204, EPI_ISL_1234205, EPI_ISL_1234206, EPI_ISL_1234207, EPI_ISL_1234208, EPI_ISL_1234209, EPI_ISL_1234210, EPI_ISL_1234211, EPI_ISL_1234212, EPI_ISL_1234213, EPI_ISL_1234214, EPI_ISL_1234215, EPI_ISL_1234216, EPI_ISL_1234217, EPI_ISL_1234220, EPI_ISL_1234221, EPI_ISL_1234222, EPI_ISL_1234223, EPI_ISL_1234224, EPI_ISL_1234225, EPI_ISL_1234226, EPI_ISL_1234227, EPI_ISL_1234228, EPI_ISL_1234229, EPI_ISL_1234230, EPI_ISL_1234231, EPI_ISL_1234233, EPI_ISL_1234234, EPI_ISL_1234235, EPI_ISL_1234236, EPI_ISL_1234237, EPI_ISL_1234238, EPI_ISL_1234239, EPI_ISL_1234240, EPI_ISL_1234241, EPI_ISL_1234242, EPI_ISL_1234243, EPI_ISL_1234244, EPI_ISL_1234245, EPI_ISL_1234247, EPI_ISL_1234248, EPI_ISL_1234249, EPI_ISL_1234250, EPI_ISL_1234251, EPI_ISL_1234252, EPI_ISL_1234253, EPI_ISL_1234254, EPI_ISL_1234256, EPI_ISL_1234257, EPI_ISL_1234258, EPI_ISL_1234259, EPI_ISL_1234260, EPI_ISL_1234261, EPI_ISL_1234262, EPI_ISL_1234263, EPI_ISL_1234264, EPI_ISL_1234266, EPI_ISL_1234267, EPI_ISL_1234268, EPI_ISL_1234269, EPI_ISL_1234270, EPI_ISL_1234271, EPI_ISL_1234272, EPI_ISL_1234273, EPI_ISL_1234274, EPI_ISL_1234275, EPI_ISL_1234276, EPI_ISL_1234277, EPI_ISL_1234278, EPI_ISL_1234279, EPI_ISL_1234281, EPI_ISL_1234282, EPI_ISL_1234283, EPI_ISL_1234284, EPI_ISL_1234285, EPI_ISL_1234286, EPI_ISL_1234287, EPI_ISL_1234288, EPI_ISL_1234289, EPI_ISL_1234290, EPI_ISL_1234291, EPI_ISL_1234292, EPI_ISL_1234293, EPI_ISL_1234294, EPI_ISL_1234295, EPI_ISL_1234296, EPI_ISL_1234297, EPI_ISL_1234298, EPI_ISL_1234299, EPI_ISL_1234300, EPI_ISL_1234301, EPI_ISL_1234302, EPI_ISL_1234303, EPI_ISL_1234304, EPI_ISL_1234305, EPI_ISL_1234306, EPI_ISL_1234307, EPI_ISL_1234308, EPI_ISL_1234309, EPI_ISL_1234310, EPI_ISL_1234312, EPI_ISL_1234313, EPI_ISL_1234314, EPI_ISL_1234315, EPI_ISL_1234316, EPI_ISL_1234317, EPI_ISL_1234318, EPI_ISL_1234319, EPI_ISL_1234320, EPI_ISL_1234321, EPI_ISL_1234322, EPI_ISL_1234323, EPI_ISL_1234324, EPI_ISL_1234325, EPI_ISL_1234326, EPI_ISL_1234327, EPI_ISL_1234328, EPI_ISL_1234329, EPI_ISL_1234330, EPI_ISL_1234331, EPI_ISL_1234332, EPI_ISL_1234333, EPI_ISL_1234334, EPI_ISL_1234335, EPI_ISL_1234336, EPI_ISL_1234337, EPI_ISL_1234338, EPI_ISL_1234339, EPI_ISL_1234340, EPI_ISL_1234341, EPI_ISL_1234343, EPI_ISL_1234344, EPI_ISL_1234345, EPI_ISL_1234346, EPI_ISL_1234347, EPI_ISL_1234348, EPI_ISL_1234349, EPI_ISL_1234350, EPI_ISL_1234351, EPI_ISL_1234353, EPI_ISL_1234354, EPI_ISL_1234355, EPI_ISL_1234356, EPI_ISL_1234357, EPI_ISL_1234358, EPI_ISL_1234359, EPI_ISL_1234361, EPI_ISL_1234362, EPI_ISL_1234363, EPI_ISL_1234364, EPI_ISL_1234365, EPI_ISL_1234366, EPI_ISL_1234367, EPI_ISL_1234369, EPI_ISL_1234370, EPI_ISL_1234371, EPI_ISL_1234372, EPI_ISL_1234373, EPI_ISL_1234374, EPI_ISL_1234375, EPI_ISL_1234376, EPI_ISL_1234377, EPI_ISL_1234378, EPI_ISL_1234379, EPI_ISL_1234380 | | | |
| see above | Colorado Department of Public Health and Environment | Colorado Department of Public Health and Environment | Laura Bankers, Molly C. Hetherington-Rauth, Diana Ir, Shannon Ely, Shannon R. Matzinger, Sarah Elizabeth Totten, Emily A. Travanty |

EPI_ISL_1234474, EPI_ISL_1234475, EPI_ISL_1234476, EPI_ISL_1234477, EPI_ISL_1234478, EPI_ISL_1234479, EPI_ISL_1234480, EPI_ISL_1234481, EPI_ISL_1234482, EPI_ISL_1234483, EPI_ISL_1234484, EPI_ISL_1234485, EPI_ISL_1234486, EPI_ISL_1234487, EPI_ISL_1234488, EPI_ISL_1234489, EPI_ISL_1234490, EPI_ISL_1234491, EPI_ISL_1234492, EPI_ISL_1234493, EPI_ISL_1234494, EPI_ISL_1234495, EPI_ISL_1234496, EPI_ISL_1234497

| | | | |
|---|---|---|---|
| see above | Nebraska Public Health Laboratory | UNMC COVID-19 Response Team | UNMC COVID-19 Response Team |
| EPI_ISL_1234537 | Santa Clara County Public Health Laboratory | Chan-Zuckerberg Biohub | CZB Cliahub Consortium |
| EPI_ISL_1234538 | Contra Costa County Public Health Lab | Chan-Zuckerberg Biohub | CZB Cliahub Consortium |
| EPI_ISL_1234539 | Santa Clara County Public Health Laboratory | Chan-Zuckerberg Biohub | CZB Cliahub Consortium |
| EPI_ISL_1234540 | Contra Costa County Public Health Lab | Chan-Zuckerberg Biohub | CZB Cliahub Consortium |
| EPI_ISL_1234542, EPI_ISL_1234543 | Alameda County Public Health Lab | Chan-Zuckerberg Biohub | CZB Cliahub Consortium |
| EPI_ISL_1234544, EPI_ISL_1234545 | CA DPH Viral and Rickettsial Disease Laboratory | Chan-Zuckerberg Biohub | CZB Cliahub Consortium |
| EPI_ISL_1234546 | Contra Costa County Public Health Lab | Chan-Zuckerberg Biohub | CZB Cliahub Consortium |
| EPI_ISL_1234547, EPI_ISL_1234548, EPI_ISL_1234549 | Santa Clara County Public Health Laboratory | Chan-Zuckerberg Biohub | CZB Cliahub Consortium |
| EPI_ISL_1234550, EPI_ISL_1234551 | CA DPH Viral and Rickettsial Disease Laboratory | Chan-Zuckerberg Biohub | CZB Cliahub Consortium |
| EPI_ISL_1234552, EPI_ISL_1234553 | Contra Costa County Public Health Lab | Chan-Zuckerberg Biohub | CZB Cliahub Consortium |
| EPI_ISL_1234554 | CA DPH Viral and Rickettsial Disease Laboratory | Chan-Zuckerberg Biohub | CZB Cliahub Consortium |
| EPI_ISL_1234555, EPI_ISL_1234556 | Contra Costa County Public Health Lab | Chan-Zuckerberg Biohub | CZB Cliahub Consortium |
| EPI_ISL_1234557 | CA DPH Viral and Rickettsial Disease Laboratory | Chan-Zuckerberg Biohub | CZB Cliahub Consortium |
| EPI_ISL_1234558, EPI_ISL_1234559 | Contra Costa County Public Health Lab | Chan-Zuckerberg Biohub | CZB Cliahub Consortium |
| EPI_ISL_1234560, EPI_ISL_1234561 | Santa Clara County Public Health Laboratory | Chan-Zuckerberg Biohub | CZB Cliahub Consortium |
| EPI_ISL_1234562 | Alameda County Public Health Lab | Chan-Zuckerberg Biohub | CZB Cliahub Consortium |
| EPI_ISL_1234563 | Contra Costa County Public Health Lab | Chan-Zuckerberg Biohub | CZB Cliahub Consortium |
| EPI_ISL_1234564, EPI_ISL_1234565, EPI_ISL_1234566, EPI_ISL_1234567, EPI_ISL_1234568 | Alameda County Public Health Lab | Chan-Zuckerberg Biohub | CZB Cliahub Consortium |
| EPI_ISL_1234569, EPI_ISL_1234570 | Contra Costa County Public Health Lab | Chan-Zuckerberg Biohub | CZB Cliahub Consortium |
| EPI_ISL_1234571 | CA DPH Viral and Rickettsial Disease Laboratory | Chan-Zuckerberg Biohub | CZB Cliahub Consortium |
| EPI_ISL_1234572, EPI_ISL_1234573 | Contra Costa County Public Health Lab | Chan-Zuckerberg Biohub | CZB Cliahub Consortium |
| EPI_ISL_1234574 | Alameda County Public Health Lab | Chan-Zuckerberg Biohub | CZB Cliahub Consortium |
| EPI_ISL_1234575, EPI_ISL_1234576 | CA DPH Viral and Rickettsial Disease Laboratory | Chan-Zuckerberg Biohub | CZB Cliahub Consortium |
| EPI_ISL_1234577 | Contra Costa County Public Health Lab | Chan-Zuckerberg Biohub | CZB Cliahub Consortium |
| EPI_ISL_1234578 | Santa Clara County Public Health Laboratory | Chan-Zuckerberg Biohub | CZB Cliahub Consortium |
| EPI_ISL_1234579, EPI_ISL_1234580 | Contra Costa County Public Health Lab | Chan-Zuckerberg Biohub | CZB Cliahub Consortium |
| EPI_ISL_1234581, EPI_ISL_1234582 | CA DPH Viral and Rickettsial Disease Laboratory | Chan-Zuckerberg Biohub | CZB Cliahub Consortium |
| EPI_ISL_1234583 | Contra Costa County Public Health Lab | Chan-Zuckerberg Biohub | CZB Cliahub Consortium |
| EPI_ISL_1234584 | Santa Clara County Public Health Laboratory | Chan-Zuckerberg Biohub | CZB Cliahub Consortium |
| EPI_ISL_1234585, EPI_ISL_1234587 | Contra Costa County Public Health Lab | Chan-Zuckerberg Biohub | CZB Cliahub Consortium |
| EPI_ISL_1234588 | Santa Clara County Public Health Laboratory | Chan-Zuckerberg Biohub | CZB Cliahub Consortium |
| EPI_ISL_1234589 | CA DPH Viral and Rickettsial Disease Laboratory | Chan-Zuckerberg Biohub | CZB Cliahub Consortium |
| EPI_ISL_1234590 | Contra Costa County Public Health Lab | Chan-Zuckerberg Biohub | CZB Cliahub Consortium |
| EPI_ISL_1234591, EPI_ISL_1234592, EPI_ISL_1234593, EPI_ISL_1234594, EPI_ISL_1234595 | CA DPH Viral and Rickettsial Disease Laboratory | Chan-Zuckerberg Biohub | CZB Cliahub Consortium |
| EPI_ISL_1234596 | Contra Costa County Public Health Lab | Chan-Zuckerberg Biohub | CZB Cliahub Consortium |
| EPI_ISL_1234597 | Alameda County Public Health Lab | Chan-Zuckerberg Biohub | CZB Cliahub Consortium |
| EPI_ISL_1234598 | Contra Costa County Public Health Lab | Chan-Zuckerberg Biohub | CZB Cliahub Consortium |
| EPI_ISL_1234599 | CA DPH Viral and Rickettsial Disease Laboratory | Chan-Zuckerberg Biohub | CZB Cliahub Consortium |
| EPI_ISL_1234600 | Alameda County Public Health Lab | Chan-Zuckerberg Biohub | CZB Cliahub Consortium |
| EPI_ISL_1234601 | CA DPH Viral and Rickettsial Disease Laboratory | Chan-Zuckerberg Biohub | CZB Cliahub Consortium |
| EPI_ISL_1234602 | Contra Costa County Public Health Lab | Chan-Zuckerberg Biohub | CZB Cliahub Consortium |
| EPI_ISL_1234603, EPI_ISL_1234604 | Alameda County Public Health Lab | Chan-Zuckerberg Biohub | CZB Cliahub Consortium |
| EPI_ISL_1234605 | CA DPH Viral and Rickettsial Disease Laboratory | Chan-Zuckerberg Biohub | CZB Cliahub Consortium |
| EPI_ISL_1234606 | Santa Clara County Public Health Laboratory | Chan-Zuckerberg Biohub | CZB Cliahub Consortium |
| EPI_ISL_1234607 | Contra Costa County Public Health Lab | Chan-Zuckerberg Biohub | CZB Cliahub Consortium |
| EPI_ISL_1234608 | CA DPH Viral and Rickettsial Disease Laboratory | Chan-Zuckerberg Biohub | CZB Cliahub Consortium |
| EPI_ISL_1234609 | Santa Clara County Public Health Laboratory | Chan-Zuckerberg Biohub | CZB Cliahub Consortium |

| | | | |
|---|---|---|---|
| EPI_ISL_1234610 | Contra Costa County Public Health Lab | Chan-Zuckerberg Biohub | CZB Cliahub Consortium |
| EPI_ISL_1234611, EPI_ISL_1234612 | Alameda County Public Health Lab | Chan-Zuckerberg Biohub | CZB Cliahub Consortium |
| EPI_ISL_1234613 | Santa Clara County Public Health Laboratory | Chan-Zuckerberg Biohub | CZB Cliahub Consortium |
| EPI_ISL_1234614, EPI_ISL_1234615, EPI_ISL_1234616 | Contra Costa County Public Health Lab | Chan-Zuckerberg Biohub | CZB Cliahub Consortium |
| EPI_ISL_1234617 | Alameda County Public Health Lab | Chan-Zuckerberg Biohub | CZB Cliahub Consortium |
| EPI_ISL_1234618 | Contra Costa County Public Health Lab | Chan-Zuckerberg Biohub | CZB Cliahub Consortium |
| EPI_ISL_1234619, EPI_ISL_1234620 | CA DPH Viral and Rickettsial Disease Laboratory | Chan-Zuckerberg Biohub | CZB Cliahub Consortium |
| EPI_ISL_1234621 | Alameda County Public Health Lab | Chan-Zuckerberg Biohub | CZB Cliahub Consortium |
| EPI_ISL_1234622 | Contra Costa County Public Health Lab | Chan-Zuckerberg Biohub | CZB Cliahub Consortium |
| EPI_ISL_1234623, EPI_ISL_1234624, EPI_ISL_1234625 | CA DPH Viral and Rickettsial Disease Laboratory | Chan-Zuckerberg Biohub | CZB Cliahub Consortium |
| EPI_ISL_1234626 | Contra Costa County Public Health Lab | Chan-Zuckerberg Biohub | CZB Cliahub Consortium |
| EPI_ISL_1234627 | CA DPH Viral and Rickettsial Disease Laboratory | Chan-Zuckerberg Biohub | CZB Cliahub Consortium |
| EPI_ISL_1234628, EPI_ISL_1234629, EPI_ISL_1234630, EPI_ISL_1234631, EPI_ISL_1234632 | Contra Costa County Public Health Lab | Chan-Zuckerberg Biohub | CZB Cliahub Consortium |
| EPI_ISL_1234633, EPI_ISL_1234634, EPI_ISL_1234635 | CA DPH Viral and Rickettsial Disease Laboratory | Chan-Zuckerberg Biohub | CZB Cliahub Consortium |
| EPI_ISL_1234636 | Contra Costa County Public Health Lab | Chan-Zuckerberg Biohub | CZB Cliahub Consortium |
| EPI_ISL_1234637 | Alameda County Public Health Lab | Chan-Zuckerberg Biohub | CZB Cliahub Consortium |
| EPI_ISL_1234639 | CA DPH Viral and Rickettsial Disease Laboratory | Chan-Zuckerberg Biohub | CZB Cliahub Consortium |
| EPI_ISL_1234640 | Alameda County Public Health Lab | Chan-Zuckerberg Biohub | CZB Cliahub Consortium |
| EPI_ISL_1234641 | Santa Clara County Public Health Laboratory | Chan-Zuckerberg Biohub | CZB Cliahub Consortium |
| EPI_ISL_1234642, EPI_ISL_1234643 | Contra Costa County Public Health Lab | Chan-Zuckerberg Biohub | CZB Cliahub Consortium |
| EPI_ISL_1234644 | CA DPH Viral and Rickettsial Disease Laboratory | Chan-Zuckerberg Biohub | CZB Cliahub Consortium |
| EPI_ISL_1234645 | Santa Clara County Public Health Laboratory | Chan-Zuckerberg Biohub | CZB Cliahub Consortium |
| EPI_ISL_1234646 | Alameda County Public Health Lab | Chan-Zuckerberg Biohub | CZB Cliahub Consortium |
| EPI_ISL_1234647 | Contra Costa County Public Health Lab | Chan-Zuckerberg Biohub | CZB Cliahub Consortium |
| EPI_ISL_1234648 | Alameda County Public Health Lab | Chan-Zuckerberg Biohub | CZB Cliahub Consortium |
| EPI_ISL_1234649, EPI_ISL_1234650, EPI_ISL_1234651, EPI_ISL_1234652 | CA DPH Viral and Rickettsial Disease Laboratory | Chan-Zuckerberg Biohub | CZB Cliahub Consortium |
| EPI_ISL_1234653 | Santa Clara County Public Health Laboratory | Chan-Zuckerberg Biohub | CZB Cliahub Consortium |
| EPI_ISL_1234654, EPI_ISL_1234655 | Contra Costa County Public Health Lab | Chan-Zuckerberg Biohub | CZB Cliahub Consortium |
| EPI_ISL_1234656 | CA DPH Viral and Rickettsial Disease Laboratory | Chan-Zuckerberg Biohub | CZB Cliahub Consortium |
| EPI_ISL_1234657 | Santa Clara County Public Health Laboratory | Chan-Zuckerberg Biohub | CZB Cliahub Consortium |
| EPI_ISL_1234658 | CA DPH Viral and Rickettsial Disease Laboratory | Chan-Zuckerberg Biohub | CZB Cliahub Consortium |
| EPI_ISL_1234659 | Contra Costa County Public Health Lab | Chan-Zuckerberg Biohub | CZB Cliahub Consortium |
| EPI_ISL_1234660 | Santa Clara County Public Health Laboratory | Chan-Zuckerberg Biohub | CZB Cliahub Consortium |
| EPI_ISL_1234661 | Alameda County Public Health Lab | Chan-Zuckerberg Biohub | CZB Cliahub Consortium |
| EPI_ISL_1234662 | Contra Costa County Public Health Lab | Chan-Zuckerberg Biohub | CZB Cliahub Consortium |
| EPI_ISL_1234663 | CA DPH Viral and Rickettsial Disease Laboratory | Chan-Zuckerberg Biohub | CZB Cliahub Consortium |
| EPI_ISL_1234664 | Contra Costa County Public Health Lab | Chan-Zuckerberg Biohub | CZB Cliahub Consortium |
| EPI_ISL_1234665, EPI_ISL_1234666 | Alameda County Public Health Lab | Chan-Zuckerberg Biohub | CZB Cliahub Consortium |
| EPI_ISL_1234667 | Santa Clara County Public Health Laboratory | Chan-Zuckerberg Biohub | CZB Cliahub Consortium |
| EPI_ISL_1234668, EPI_ISL_1234669 | CA DPH Viral and Rickettsial Disease Laboratory | Chan-Zuckerberg Biohub | CZB Cliahub Consortium |
| EPI_ISL_1234670 | Alameda County Public Health Lab | Chan-Zuckerberg Biohub | CZB Cliahub Consortium |
| EPI_ISL_1234671 | Santa Clara County Public Health Laboratory | Chan-Zuckerberg Biohub | CZB Cliahub Consortium |
| EPI_ISL_1234672 | Alameda County Public Health Lab | Chan-Zuckerberg Biohub | CZB Cliahub Consortium |
| EPI_ISL_1234673 | CA DPH Viral and Rickettsial Disease Laboratory | Chan-Zuckerberg Biohub | CZB Cliahub Consortium |
| EPI_ISL_1234674, EPI_ISL_1234675 | Contra Costa County Public Health Lab | Chan-Zuckerberg Biohub | CZB Cliahub Consortium |
| EPI_ISL_1234676, EPI_ISL_1234677 | Alameda County Public Health Lab | Chan-Zuckerberg Biohub | CZB Cliahub Consortium |
| EPI_ISL_1234678 | Santa Clara County Public Health Laboratory | Chan-Zuckerberg Biohub | CZB Cliahub Consortium |
| EPI_ISL_1234679 | Alameda County Public Health Lab | Chan-Zuckerberg Biohub | CZB Cliahub Consortium |
| EPI_ISL_1234680 | Contra Costa County Public Health Lab | Chan-Zuckerberg Biohub | CZB Cliahub Consortium |
| EPI_ISL_1234681 | CA DPH Viral and Rickettsial Disease Laboratory | Chan-Zuckerberg Biohub | CZB Cliahub Consortium |

| | | | |
|---|---|---|---|
| EPI_ISL_1234682 | Contra Costa County Public Health Lab | Chan-Zuckerberg Biohub | CZB Cliahub Consortium |
| EPI_ISL_1234683 | Alameda County Public Health Lab | Chan-Zuckerberg Biohub | CZB Cliahub Consortium |
| EPI_ISL_1234684, EPI_ISL_1234685, EPI_ISL_1234686 | Contra Costa County Public Health Lab | Chan-Zuckerberg Biohub | CZB Cliahub Consortium |
| EPI_ISL_1234687 | Santa Clara County Public Health Laboratory | Chan-Zuckerberg Biohub | CZB Cliahub Consortium |
| EPI_ISL_1234688 | Contra Costa County Public Health Lab | Chan-Zuckerberg Biohub | CZB Cliahub Consortium |
| EPI_ISL_1234689 | Santa Clara County Public Health Laboratory | Chan-Zuckerberg Biohub | CZB Cliahub Consortium |
| EPI_ISL_1234690, EPI_ISL_1234691, EPI_ISL_1234692, EPI_ISL_1234693 | Contra Costa County Public Health Lab | Chan-Zuckerberg Biohub | CZB Cliahub Consortium |
| EPI_ISL_1234694 | Santa Clara County Public Health Laboratory | Chan-Zuckerberg Biohub | CZB Cliahub Consortium |
| EPI_ISL_1234695 | CA DPH Viral and Rickettsial Disease Laboratory | Chan-Zuckerberg Biohub | CZB Cliahub Consortium |
| EPI_ISL_1234696, EPI_ISL_1234697 | Contra Costa County Public Health Lab | Chan-Zuckerberg Biohub | CZB Cliahub Consortium |
| EPI_ISL_1234698 | Alameda County Public Health Lab | Chan-Zuckerberg Biohub | CZB Cliahub Consortium |
| EPI_ISL_1234699, EPI_ISL_1234700 | CA DPH Viral and Rickettsial Disease Laboratory | Chan-Zuckerberg Biohub | CZB Cliahub Consortium |
| EPI_ISL_1234701 | Alameda County Public Health Lab | Chan-Zuckerberg Biohub | CZB Cliahub Consortium |
| EPI_ISL_1234702 | Contra Costa County Public Health Lab | Chan-Zuckerberg Biohub | CZB Cliahub Consortium |
| EPI_ISL_1234703, EPI_ISL_1234704 | CA DPH Viral and Rickettsial Disease Laboratory | Chan-Zuckerberg Biohub | CZB Cliahub Consortium |
| EPI_ISL_1234705 | Contra Costa County Public Health Lab | Chan-Zuckerberg Biohub | CZB Cliahub Consortium |
| EPI_ISL_1234706 | Santa Clara County Public Health Laboratory | Chan-Zuckerberg Biohub | CZB Cliahub Consortium |
| EPI_ISL_1234707, EPI_ISL_1234708, EPI_ISL_1234709, EPI_ISL_1234710 | Contra Costa County Public Health Lab | Chan-Zuckerberg Biohub | CZB Cliahub Consortium |
| EPI_ISL_1234711 | Santa Clara County Public Health Laboratory | Chan-Zuckerberg Biohub | CZB Cliahub Consortium |
| EPI_ISL_1234712, EPI_ISL_1234713, EPI_ISL_1234714, EPI_ISL_1234715 | CA DPH Viral and Rickettsial Disease Laboratory | Chan-Zuckerberg Biohub | CZB Cliahub Consortium |
| EPI_ISL_1234716 | Alameda County Public Health Lab | Chan-Zuckerberg Biohub | CZB Cliahub Consortium |
| EPI_ISL_1234717 | CA DPH Viral and Rickettsial Disease Laboratory | Chan-Zuckerberg Biohub | CZB Cliahub Consortium |
| EPI_ISL_1234718 | Alameda County Public Health Lab | Chan-Zuckerberg Biohub | CZB Cliahub Consortium |
| EPI_ISL_1234719 | CA DPH Viral and Rickettsial Disease Laboratory | Chan-Zuckerberg Biohub | CZB Cliahub Consortium |
| EPI_ISL_1234721 | Santa Clara County Public Health Laboratory | Chan-Zuckerberg Biohub | CZB Cliahub Consortium |
| EPI_ISL_1234722 | CA DPH Viral and Rickettsial Disease Laboratory | Chan-Zuckerberg Biohub | CZB Cliahub Consortium |
| EPI_ISL_1234723 | Alameda County Public Health Lab | Chan-Zuckerberg Biohub | CZB Cliahub Consortium |
| EPI_ISL_1234724 | Santa Clara County Public Health Laboratory | Chan-Zuckerberg Biohub | CZB Cliahub Consortium |
| EPI_ISL_1234726, EPI_ISL_1234727 | Contra Costa County Public Health Lab | Chan-Zuckerberg Biohub | CZB Cliahub Consortium |
| EPI_ISL_1234728, EPI_ISL_1234729 | Santa Clara County Public Health Laboratory | Chan-Zuckerberg Biohub | CZB Cliahub Consortium |
| EPI_ISL_1234730, EPI_ISL_1234731 | CA DPH Viral and Rickettsial Disease Laboratory | Chan-Zuckerberg Biohub | CZB Cliahub Consortium |
| EPI_ISL_1234732 | Alameda County Public Health Lab | Chan-Zuckerberg Biohub | CZB Cliahub Consortium |
| EPI_ISL_1234733, EPI_ISL_1234734, EPI_ISL_1234735 | CA DPH Viral and Rickettsial Disease Laboratory | Chan-Zuckerberg Biohub | CZB Cliahub Consortium |
| EPI_ISL_1234736 | Contra Costa County Public Health Lab | Chan-Zuckerberg Biohub | CZB Cliahub Consortium |
| EPI_ISL_1234737 | Santa Clara County Public Health Laboratory | Chan-Zuckerberg Biohub | CZB Cliahub Consortium |
| EPI_ISL_1234738 | Contra Costa County Public Health Lab | Chan-Zuckerberg Biohub | CZB Cliahub Consortium |
| EPI_ISL_1234739 | Alameda County Public Health Lab | Chan-Zuckerberg Biohub | CZB Cliahub Consortium |
| EPI_ISL_1234740 | CA DPH Viral and Rickettsial Disease Laboratory | Chan-Zuckerberg Biohub | CZB Cliahub Consortium |
| EPI_ISL_1234741 | Contra Costa County Public Health Lab | Chan-Zuckerberg Biohub | CZB Cliahub Consortium |
| EPI_ISL_1234742, EPI_ISL_1234743 | Alameda County Public Health Lab | Chan-Zuckerberg Biohub | CZB Cliahub Consortium |
| EPI_ISL_1234745, EPI_ISL_1234746 | Contra Costa County Public Health Lab | Chan-Zuckerberg Biohub | CZB Cliahub Consortium |
| EPI_ISL_1234747, EPI_ISL_1234748 | CA DPH Viral and Rickettsial Disease Laboratory | Chan-Zuckerberg Biohub | CZB Cliahub Consortium |
| EPI_ISL_1234749 | Contra Costa County Public Health Lab | Chan-Zuckerberg Biohub | CZB Cliahub Consortium |
| EPI_ISL_1234750 | CA DPH Viral and Rickettsial Disease Laboratory | Chan-Zuckerberg Biohub | CZB Cliahub Consortium |
| EPI_ISL_1234751 | Alameda County Public Health Lab | Chan-Zuckerberg Biohub | CZB Cliahub Consortium |
| EPI_ISL_1234752, EPI_ISL_1234753, EPI_ISL_1234754 | CA DPH Viral and Rickettsial Disease Laboratory | Chan-Zuckerberg Biohub | CZB Cliahub Consortium |
| EPI_ISL_1234755 | Alameda County Public Health Lab | Chan-Zuckerberg Biohub | CZB Cliahub Consortium |
| EPI_ISL_1234756, EPI_ISL_1234757 | Contra Costa County Public Health Lab | Chan-Zuckerberg Biohub | CZB Cliahub Consortium |
| EPI_ISL_1234758 | Alameda County Public Health Lab | Chan-Zuckerberg Biohub | CZB Cliahub Consortium |

| | | | |
|---|---|---|---|
| EPI_ISL_1234759, EPI_ISL_1234760 | Santa Clara County Public Health Laboratory | Chan-Zuckerberg Biohub | CZB Cliahub Consortium |
| EPI_ISL_1234761 | CA DPH Viral and Rickettsial Disease Laboratory | Chan-Zuckerberg Biohub | CZB Cliahub Consortium |
| EPI_ISL_1234762 | Alameda County Public Health Lab | Chan-Zuckerberg Biohub | CZB Cliahub Consortium |
| EPI_ISL_1234763, EPI_ISL_1234764 | Contra Costa County Public Health Lab | Chan-Zuckerberg Biohub | CZB Cliahub Consortium |
| EPI_ISL_1234765 | Alameda County Public Health Lab | Chan-Zuckerberg Biohub | CZB Cliahub Consortium |
| EPI_ISL_1234766 | CA DPH Viral and Rickettsial Disease Laboratory | Chan-Zuckerberg Biohub | CZB Cliahub Consortium |
| EPI_ISL_1234767 | Alameda County Public Health Lab | Chan-Zuckerberg Biohub | CZB Cliahub Consortium |
| EPI_ISL_1234768 | Contra Costa County Public Health Lab | Chan-Zuckerberg Biohub | CZB Cliahub Consortium |
| EPI_ISL_1234769 | Santa Clara County Public Health Laboratory | Chan-Zuckerberg Biohub | CZB Cliahub Consortium |
| EPI_ISL_1234770 | Contra Costa County Public Health Lab | Chan-Zuckerberg Biohub | CZB Cliahub Consortium |
| EPI_ISL_1234771 | CA DPH Viral and Rickettsial Disease Laboratory | Chan-Zuckerberg Biohub | CZB Cliahub Consortium |
| EPI_ISL_1234772 | Contra Costa County Public Health Lab | Chan-Zuckerberg Biohub | CZB Cliahub Consortium |
| EPI_ISL_1234773 | Alameda County Public Health Lab | Chan-Zuckerberg Biohub | CZB Cliahub Consortium |
| EPI_ISL_1234774 | CA DPH Viral and Rickettsial Disease Laboratory | Chan-Zuckerberg Biohub | CZB Cliahub Consortium |
| EPI_ISL_1234775 | Contra Costa County Public Health Lab | Chan-Zuckerberg Biohub | CZB Cliahub Consortium |
| EPI_ISL_1234776 | Alameda County Public Health Lab | Chan-Zuckerberg Biohub | CZB Cliahub Consortium |
| EPI_ISL_1234777 | Contra Costa County Public Health Lab | Chan-Zuckerberg Biohub | CZB Cliahub Consortium |
| EPI_ISL_1234778 | CA DPH Viral and Rickettsial Disease Laboratory | Chan-Zuckerberg Biohub | CZB Cliahub Consortium |
| EPI_ISL_1234779 | Alameda County Public Health Lab | Chan-Zuckerberg Biohub | CZB Cliahub Consortium |
| EPI_ISL_1234780, EPI_ISL_1234781 | CA DPH Viral and Rickettsial Disease Laboratory | Chan-Zuckerberg Biohub | CZB Cliahub Consortium |
| EPI_ISL_1234782 | Alameda County Public Health Lab | Chan-Zuckerberg Biohub | CZB Cliahub Consortium |
| EPI_ISL_1234783 | Contra Costa County Public Health Lab | Chan-Zuckerberg Biohub | CZB Cliahub Consortium |
| EPI_ISL_1234784 | CA DPH Viral and Rickettsial Disease Laboratory | Chan-Zuckerberg Biohub | CZB Cliahub Consortium |
| EPI_ISL_1234785 | Alameda County Public Health Lab | Chan-Zuckerberg Biohub | CZB Cliahub Consortium |
| EPI_ISL_1234786 | Humboldt County Public Health Laboratory | Chan-Zuckerberg Biohub | CZB Cliahub Consortium |
| EPI_ISL_1234787 | Santa Clara County Public Health Laboratory | Chan-Zuckerberg Biohub | CZB Cliahub Consortium |
| EPI_ISL_1234788, EPI_ISL_1234789 | Humboldt County Public Health Laboratory | Chan-Zuckerberg Biohub | CZB Cliahub Consortium |
| EPI_ISL_1234790 | Alameda County Public Health Lab | Chan-Zuckerberg Biohub | CZB Cliahub Consortium |
| EPI_ISL_1234791 | Orange County Public Health Lab | Chan-Zuckerberg Biohub | CZB Cliahub Consortium |
| EPI_ISL_1234792, EPI_ISL_1234793, EPI_ISL_1234794 | Humboldt County Public Health Laboratory | Chan-Zuckerberg Biohub | CZB Cliahub Consortium |
| EPI_ISL_1234795 | Alameda County Public Health Lab | Chan-Zuckerberg Biohub | CZB Cliahub Consortium |
| EPI_ISL_1234796, EPI_ISL_1234797 | Orange County Public Health Lab | Chan-Zuckerberg Biohub | CZB Cliahub Consortium |
| EPI_ISL_1234798 | Santa Clara County Public Health Laboratory | Chan-Zuckerberg Biohub | CZB Cliahub Consortium |
| EPI_ISL_1234799 | Humboldt County Public Health Laboratory | Chan-Zuckerberg Biohub | CZB Cliahub Consortium |
| EPI_ISL_1234800 | Santa Clara County Public Health Laboratory | Chan-Zuckerberg Biohub | CZB Cliahub Consortium |
| EPI_ISL_1234801 | Humboldt County Public Health Laboratory | Chan-Zuckerberg Biohub | CZB Cliahub Consortium |
| EPI_ISL_1234802 | Alameda County Public Health Lab | Chan-Zuckerberg Biohub | CZB Cliahub Consortium |
| EPI_ISL_1234803 | Humboldt County Public Health Laboratory | Chan-Zuckerberg Biohub | CZB Cliahub Consortium |
| EPI_ISL_1234804 | Orange County Public Health Lab | Chan-Zuckerberg Biohub | CZB Cliahub Consortium |
| EPI_ISL_1234805 | Humboldt County Public Health Laboratory | Chan-Zuckerberg Biohub | CZB Cliahub Consortium |
| EPI_ISL_1234806 | County of San Luis Obispo Public Health Laboratory | Chan-Zuckerberg Biohub | CZB Cliahub Consortium |
| EPI_ISL_1234807 | Humboldt County Public Health Laboratory | Chan-Zuckerberg Biohub | CZB Cliahub Consortium |
| EPI_ISL_1234808, EPI_ISL_1234809 | Alameda County Public Health Lab | Chan-Zuckerberg Biohub | CZB Cliahub Consortium |
| EPI_ISL_1234810 | Santa Clara County Public Health Laboratory | Chan-Zuckerberg Biohub | CZB Cliahub Consortium |
| EPI_ISL_1234811 | Humboldt County Public Health Laboratory | Chan-Zuckerberg Biohub | CZB Cliahub Consortium |
| EPI_ISL_1234812 | Santa Clara County Public Health Laboratory | Chan-Zuckerberg Biohub | CZB Cliahub Consortium |
| EPI_ISL_1234813 | California Institute of Technology | Chan-Zuckerberg Biohub | CZB Cliahub Consortium |
| EPI_ISL_1234814 | Humboldt County Public Health Laboratory | Chan-Zuckerberg Biohub | CZB Cliahub Consortium |
| EPI_ISL_1234815 | Alameda County Public Health Lab | Chan-Zuckerberg Biohub | CZB Cliahub Consortium |
| EPI_ISL_1234816, EPI_ISL_1234817 | Humboldt County Public Health Laboratory | Chan-Zuckerberg Biohub | CZB Cliahub Consortium |
| EPI_ISL_1234818 | Santa Clara County Public Health Laboratory | Chan-Zuckerberg Biohub | CZB Cliahub Consortium |
| EPI_ISL_1234819, EPI_ISL_1234820 | Alameda County Public Health Lab | Chan-Zuckerberg Biohub | CZB Cliahub Consortium |
| EPI_ISL_1234821, EPI_ISL_1234822, EPI_ISL_1234823, | Humboldt County Public Health Laboratory | Chan-Zuckerberg Biohub | CZB Cliahub Consortium |

| | | | |
|---|---|---|---|
| EPI_ISL_1234824 | | | |
| EPI_ISL_1234825 | Orange County Public Health Lab | Chan-Zuckerberg Biohub | CZB Cliahub Consortium |
| EPI_ISL_1234826 | Humboldt County Public Health Laboratory | Chan-Zuckerberg Biohub | CZB Cliahub Consortium |
| EPI_ISL_1234827 | Alameda County Public Health Lab | Chan-Zuckerberg Biohub | CZB Cliahub Consortium |
| EPI_ISL_1234828 | Orange County Public Health Lab | Chan-Zuckerberg Biohub | CZB Cliahub Consortium |
| EPI_ISL_1234829, EPI_ISL_1234830 | Alameda County Public Health Lab | Chan-Zuckerberg Biohub | CZB Cliahub Consortium |
| EPI_ISL_1234831 | Humboldt County Public Health Laboratory | Chan-Zuckerberg Biohub | CZB Cliahub Consortium |
| EPI_ISL_1234832 | Alameda County Public Health Lab | Chan-Zuckerberg Biohub | CZB Cliahub Consortium |
| EPI_ISL_1234833 | Humboldt County Public Health Laboratory | Chan-Zuckerberg Biohub | CZB Cliahub Consortium |
| EPI_ISL_1234834, EPI_ISL_1234835 | Alameda County Public Health Lab | Chan-Zuckerberg Biohub | CZB Cliahub Consortium |
| EPI_ISL_1234836 | Humboldt County Public Health Laboratory | Chan-Zuckerberg Biohub | CZB Cliahub Consortium |
| EPI_ISL_1234837 | Santa Clara County Public Health Laboratory | Chan-Zuckerberg Biohub | CZB Cliahub Consortium |
| EPI_ISL_1234838 | Orange County Public Health Lab | Chan-Zuckerberg Biohub | CZB Cliahub Consortium |
| EPI_ISL_1234839 | Alameda County Public Health Lab | Chan-Zuckerberg Biohub | CZB Cliahub Consortium |
| EPI_ISL_1234840, EPI_ISL_1234841 | Santa Clara County Public Health Laboratory | Chan-Zuckerberg Biohub | CZB Cliahub Consortium |
| EPI_ISL_1234842 | Orange County Public Health Lab | Chan-Zuckerberg Biohub | CZB Cliahub Consortium |
| EPI_ISL_1234843 | Santa Clara County Public Health Laboratory | Chan-Zuckerberg Biohub | CZB Cliahub Consortium |
| EPI_ISL_1234844 | Alameda County Public Health Lab | Chan-Zuckerberg Biohub | CZB Cliahub Consortium |
| EPI_ISL_1234845 | California Institute of Technology | Chan-Zuckerberg Biohub | CZB Cliahub Consortium |
| EPI_ISL_1234846 | Santa Clara County Public Health Laboratory | Chan-Zuckerberg Biohub | CZB Cliahub Consortium |
| EPI_ISL_1234847 | Orange County Public Health Lab | Chan-Zuckerberg Biohub | CZB Cliahub Consortium |
| EPI_ISL_1234848 | Alameda County Public Health Lab | Chan-Zuckerberg Biohub | CZB Cliahub Consortium |
| EPI_ISL_1234849, EPI_ISL_1234850, EPI_ISL_1234851 | Santa Clara County Public Health Laboratory | Chan-Zuckerberg Biohub | CZB Cliahub Consortium |
| EPI_ISL_1234852, EPI_ISL_1234853 | Humboldt County Public Health Laboratory | Chan-Zuckerberg Biohub | CZB Cliahub Consortium |
| EPI_ISL_1234854 | Santa Clara County Public Health Laboratory | Chan-Zuckerberg Biohub | CZB Cliahub Consortium |
| EPI_ISL_1234855 | Orange County Public Health Lab | Chan-Zuckerberg Biohub | CZB Cliahub Consortium |
| EPI_ISL_1234856 | Humboldt County Public Health Laboratory | Chan-Zuckerberg Biohub | CZB Cliahub Consortium |
| EPI_ISL_1234857 | Santa Clara County Public Health Laboratory | Chan-Zuckerberg Biohub | CZB Cliahub Consortium |
| EPI_ISL_1234858 | Alameda County Public Health Lab | Chan-Zuckerberg Biohub | CZB Cliahub Consortium |
| EPI_ISL_1234859 | Humboldt County Public Health Laboratory | Chan-Zuckerberg Biohub | CZB Cliahub Consortium |
| EPI_ISL_1234860 | Alameda County Public Health Lab | Chan-Zuckerberg Biohub | CZB Cliahub Consortium |
| EPI_ISL_1234861 | Humboldt County Public Health Laboratory | Chan-Zuckerberg Biohub | CZB Cliahub Consortium |
| EPI_ISL_1234862 | Alameda County Public Health Lab | Chan-Zuckerberg Biohub | CZB Cliahub Consortium |
| EPI_ISL_1234863 | Humboldt County Public Health Laboratory | Chan-Zuckerberg Biohub | CZB Cliahub Consortium |
| EPI_ISL_1234864 | Orange County Public Health Lab | Chan-Zuckerberg Biohub | CZB Cliahub Consortium |
| EPI_ISL_1234865, EPI_ISL_1234866 | Santa Clara County Public Health Laboratory | Chan-Zuckerberg Biohub | CZB Cliahub Consortium |
| EPI_ISL_1234867 | California Institute of Technology | Chan-Zuckerberg Biohub | CZB Cliahub Consortium |
| EPI_ISL_1234868 | Humboldt County Public Health Laboratory | Chan-Zuckerberg Biohub | CZB Cliahub Consortium |
| EPI_ISL_1234869 | Orange County Public Health Lab | Chan-Zuckerberg Biohub | CZB Cliahub Consortium |
| EPI_ISL_1234870 | Alameda County Public Health Lab | Chan-Zuckerberg Biohub | CZB Cliahub Consortium |
| EPI_ISL_1234871 | Humboldt County Public Health Laboratory | Chan-Zuckerberg Biohub | CZB Cliahub Consortium |
| EPI_ISL_1234872, EPI_ISL_1234873 | Alameda County Public Health Lab | Chan-Zuckerberg Biohub | CZB Cliahub Consortium |
| EPI_ISL_1234874 | Humboldt County Public Health Laboratory | Chan-Zuckerberg Biohub | CZB Cliahub Consortium |
| EPI_ISL_1234875, EPI_ISL_1234876 | Santa Clara County Public Health Laboratory | Chan-Zuckerberg Biohub | CZB Cliahub Consortium |
| EPI_ISL_1234877 | Alameda County Public Health Lab | Chan-Zuckerberg Biohub | CZB Cliahub Consortium |
| EPI_ISL_1234878, EPI_ISL_1234879, EPI_ISL_1234880, EPI_ISL_1234881 | Santa Clara County Public Health Laboratory | Chan-Zuckerberg Biohub | CZB Cliahub Consortium |
| EPI_ISL_1234882 | Humboldt County Public Health Laboratory | Chan-Zuckerberg Biohub | CZB Cliahub Consortium |
| EPI_ISL_1234884 | Santa Clara County Public Health Laboratory | Chan-Zuckerberg Biohub | CZB Cliahub Consortium |
| EPI_ISL_1234885 | California Institute of Technology | Chan-Zuckerberg Biohub | CZB Cliahub Consortium |
| EPI_ISL_1234886 | Santa Clara County Public Health Laboratory | Chan-Zuckerberg Biohub | CZB Cliahub Consortium |
| EPI_ISL_1234887 | Orange County Public Health Lab | Chan-Zuckerberg Biohub | CZB Cliahub Consortium |
| EPI_ISL_1234888, EPI_ISL_1234889, EPI_ISL_1234890 | Humboldt County Public Health Laboratory | Chan-Zuckerberg Biohub | CZB Cliahub Consortium |
| EPI_ISL_1234891 | California Institute of Technology | Chan-Zuckerberg Biohub | CZB Cliahub Consortium |

| | | | |
|---|---|---|---|
| EPI_ISL_1234892 | Alameda County Public Health Lab | Chan-Zuckerberg Biohub | CZB Cliahub Consortium |
| EPI_ISL_1234893 | Humboldt County Public Health Laboratory | Chan-Zuckerberg Biohub | CZB Cliahub Consortium |
| EPI_ISL_1234894, EPI_ISL_1234895 | Alameda County Public Health Lab | Chan-Zuckerberg Biohub | CZB Cliahub Consortium |
| EPI_ISL_1234896 | Humboldt County Public Health Laboratory | Chan-Zuckerberg Biohub | CZB Cliahub Consortium |
| EPI_ISL_1234897 | Santa Clara County Public Health Laboratory | Chan-Zuckerberg Biohub | CZB Cliahub Consortium |
| EPI_ISL_1234898 | Humboldt County Public Health Laboratory | Chan-Zuckerberg Biohub | CZB Cliahub Consortium |
| EPI_ISL_1234899 | Santa Clara County Public Health Laboratory | Chan-Zuckerberg Biohub | CZB Cliahub Consortium |
| EPI_ISL_1234900, EPI_ISL_1234901 | Alameda County Public Health Lab | Chan-Zuckerberg Biohub | CZB Cliahub Consortium |
| EPI_ISL_1234902 | Santa Clara County Public Health Laboratory | Chan-Zuckerberg Biohub | CZB Cliahub Consortium |
| EPI_ISL_1234903 | Humboldt County Public Health Laboratory | Chan-Zuckerberg Biohub | CZB Cliahub Consortium |
| EPI_ISL_1234904 | Alameda County Public Health Lab | Chan-Zuckerberg Biohub | CZB Cliahub Consortium |
| EPI_ISL_1234905, EPI_ISL_1234906 | Humboldt County Public Health Laboratory | Chan-Zuckerberg Biohub | CZB Cliahub Consortium |
| EPI_ISL_1234907 | Alameda County Public Health Lab | Chan-Zuckerberg Biohub | CZB Cliahub Consortium |
| EPI_ISL_1234908 | Humboldt County Public Health Laboratory | Chan-Zuckerberg Biohub | CZB Cliahub Consortium |
| EPI_ISL_1234909 | Orange County Public Health Lab | Chan-Zuckerberg Biohub | CZB Cliahub Consortium |
| EPI_ISL_1234910, EPI_ISL_1234911, EPI_ISL_1234912 | Humboldt County Public Health Laboratory | Chan-Zuckerberg Biohub | CZB Cliahub Consortium |
| EPI_ISL_1234913 | Santa Clara County Public Health Laboratory | Chan-Zuckerberg Biohub | CZB Cliahub Consortium |
| EPI_ISL_1234914 | Humboldt County Public Health Laboratory | Chan-Zuckerberg Biohub | CZB Cliahub Consortium |
| EPI_ISL_1234915 | California Institute of Technology | Chan-Zuckerberg Biohub | CZB Cliahub Consortium |
| EPI_ISL_1234916 | Humboldt County Public Health Laboratory | Chan-Zuckerberg Biohub | CZB Cliahub Consortium |
| EPI_ISL_1234917 | Santa Clara County Public Health Laboratory | Chan-Zuckerberg Biohub | CZB Cliahub Consortium |
| EPI_ISL_1234918 | Orange County Public Health Lab | Chan-Zuckerberg Biohub | CZB Cliahub Consortium |
| EPI_ISL_1234919 | Santa Clara County Public Health Laboratory | Chan-Zuckerberg Biohub | CZB Cliahub Consortium |
| EPI_ISL_1234920, EPI_ISL_1234921 | Alameda County Public Health Lab | Chan-Zuckerberg Biohub | CZB Cliahub Consortium |
| EPI_ISL_1234922 | Humboldt County Public Health Laboratory | Chan-Zuckerberg Biohub | CZB Cliahub Consortium |
| EPI_ISL_1234923 | Orange County Public Health Lab | Chan-Zuckerberg Biohub | CZB Cliahub Consortium |
| EPI_ISL_1234924 | Alameda County Public Health Lab | Chan-Zuckerberg Biohub | CZB Cliahub Consortium |
| EPI_ISL_1234926 | Orange County Public Health Lab | Chan-Zuckerberg Biohub | CZB Cliahub Consortium |
| EPI_ISL_1234927, EPI_ISL_1234928 | Humboldt County Public Health Laboratory | Chan-Zuckerberg Biohub | CZB Cliahub Consortium |
| EPI_ISL_1234929 | Orange County Public Health Lab | Chan-Zuckerberg Biohub | CZB Cliahub Consortium |
| EPI_ISL_1234930, EPI_ISL_1234931 | Humboldt County Public Health Laboratory | Chan-Zuckerberg Biohub | CZB Cliahub Consortium |
| EPI_ISL_1234932, EPI_ISL_1234933 | Alameda County Public Health Lab | Chan-Zuckerberg Biohub | CZB Cliahub Consortium |
| EPI_ISL_1234934 | Humboldt County Public Health Laboratory | Chan-Zuckerberg Biohub | CZB Cliahub Consortium |
| EPI_ISL_1234935 | Santa Clara County Public Health Laboratory | Chan-Zuckerberg Biohub | CZB Cliahub Consortium |
| EPI_ISL_1234936 | Humboldt County Public Health Laboratory | Chan-Zuckerberg Biohub | CZB Cliahub Consortium |
| EPI_ISL_1234937 | Orange County Public Health Lab | Chan-Zuckerberg Biohub | CZB Cliahub Consortium |
| EPI_ISL_1234938 | County of San Luis Obispo Public Health Laboratory | Chan-Zuckerberg Biohub | CZB Cliahub Consortium |
| EPI_ISL_1234939 | Alameda County Public Health Lab | Chan-Zuckerberg Biohub | CZB Cliahub Consortium |
| EPI_ISL_1234940 | Orange County Public Health Lab | Chan-Zuckerberg Biohub | CZB Cliahub Consortium |
| EPI_ISL_1234941 | Alameda County Public Health Lab | Chan-Zuckerberg Biohub | CZB Cliahub Consortium |
| EPI_ISL_1234942, EPI_ISL_1234943, EPI_ISL_1234944, EPI_ISL_1234945 | Orange County Public Health Lab | Chan-Zuckerberg Biohub | CZB Cliahub Consortium |
| EPI_ISL_1234946 | Santa Clara County Public Health Laboratory | Chan-Zuckerberg Biohub | CZB Cliahub Consortium |
| EPI_ISL_1234947 | California Institute of Technology | Chan-Zuckerberg Biohub | CZB Cliahub Consortium |
| EPI_ISL_1234948, EPI_ISL_1234949 | Humboldt County Public Health Laboratory | Chan-Zuckerberg Biohub | CZB Cliahub Consortium |
| EPI_ISL_1234950 | Alameda County Public Health Lab | Chan-Zuckerberg Biohub | CZB Cliahub Consortium |
| EPI_ISL_1234951 | California Institute of Technology | Chan-Zuckerberg Biohub | CZB Cliahub Consortium |
| EPI_ISL_1234952, EPI_ISL_1234953 | Alameda County Public Health Lab | Chan-Zuckerberg Biohub | CZB Cliahub Consortium |
| EPI_ISL_1234954 | Orange County Public Health Lab | Chan-Zuckerberg Biohub | CZB Cliahub Consortium |
| EPI_ISL_1234955 | Humboldt County Public Health Laboratory | Chan-Zuckerberg Biohub | CZB Cliahub Consortium |
| EPI_ISL_1234956 | Alameda County Public Health Lab | Chan-Zuckerberg Biohub | CZB Cliahub Consortium |
| EPI_ISL_1234957 | Santa Clara County Public Health Laboratory | Chan-Zuckerberg Biohub | CZB Cliahub Consortium |
| EPI_ISL_1234958 | Orange County Public Health Lab | Chan-Zuckerberg Biohub | CZB Cliahub Consortium |

| | | | |
|---|---|---|---|
| EPI_ISL_1234959 | Alameda County Public Health Lab | Chan-Zuckerberg Biohub | CZB Cliahub Consortium |
| EPI_ISL_1234960 | Humboldt County Public Health Laboratory | Chan-Zuckerberg Biohub | CZB Cliahub Consortium |
| EPI_ISL_1234961, EPI_ISL_1234962 | Santa Clara County Public Health Laboratory | Chan-Zuckerberg Biohub | CZB Cliahub Consortium |
| EPI_ISL_1234963 | Humboldt County Public Health Laboratory | Chan-Zuckerberg Biohub | CZB Cliahub Consortium |
| EPI_ISL_1234964 | California Institute of Technology | Chan-Zuckerberg Biohub | CZB Cliahub Consortium |
| EPI_ISL_1234965 | Orange County Public Health Lab | Chan-Zuckerberg Biohub | CZB Cliahub Consortium |
| EPI_ISL_1234966 | Humboldt County Public Health Laboratory | Chan-Zuckerberg Biohub | CZB Cliahub Consortium |
| EPI_ISL_1234967 | California Institute of Technology | Chan-Zuckerberg Biohub | CZB Cliahub Consortium |
| EPI_ISL_1234968, EPI_ISL_1234969 | Alameda County Public Health Lab | Chan-Zuckerberg Biohub | CZB Cliahub Consortium |
| EPI_ISL_1234970 | Orange County Public Health Lab | Chan-Zuckerberg Biohub | CZB Cliahub Consortium |
| EPI_ISL_1234971 | Humboldt County Public Health Laboratory | Chan-Zuckerberg Biohub | CZB Cliahub Consortium |
| EPI_ISL_1234972, EPI_ISL_1234973 | Santa Clara County Public Health Laboratory | Chan-Zuckerberg Biohub | CZB Cliahub Consortium |
| EPI_ISL_1234974 | Humboldt County Public Health Laboratory | Chan-Zuckerberg Biohub | CZB Cliahub Consortium |
| EPI_ISL_1234975, EPI_ISL_1234976 | Santa Clara County Public Health Laboratory | Chan-Zuckerberg Biohub | CZB Cliahub Consortium |
| EPI_ISL_1234977 | Orange County Public Health Lab | Chan-Zuckerberg Biohub | CZB Cliahub Consortium |
| EPI_ISL_1234978 | California Institute of Technology | Chan-Zuckerberg Biohub | CZB Cliahub Consortium |
| EPI_ISL_1234979 | Alameda County Public Health Lab | Chan-Zuckerberg Biohub | CZB Cliahub Consortium |
| EPI_ISL_1234981 | Humboldt County Public Health Laboratory | Chan-Zuckerberg Biohub | CZB Cliahub Consortium |
| EPI_ISL_1234982 | Alameda County Public Health Lab | Chan-Zuckerberg Biohub | CZB Cliahub Consortium |
| EPI_ISL_1234983 | Santa Clara County Public Health Laboratory | Chan-Zuckerberg Biohub | CZB Cliahub Consortium |
| EPI_ISL_1234984 | Humboldt County Public Health Laboratory | Chan-Zuckerberg Biohub | CZB Cliahub Consortium |
| EPI_ISL_1234985 | Santa Clara County Public Health Laboratory | Chan-Zuckerberg Biohub | CZB Cliahub Consortium |
| EPI_ISL_1234986, EPI_ISL_1234987 | Alameda County Public Health Lab | Chan-Zuckerberg Biohub | CZB Cliahub Consortium |
| EPI_ISL_1234988 | Santa Clara County Public Health Laboratory | Chan-Zuckerberg Biohub | CZB Cliahub Consortium |
| EPI_ISL_1234989 | Humboldt County Public Health Laboratory | Chan-Zuckerberg Biohub | CZB Cliahub Consortium |
| EPI_ISL_1234990 | Alameda County Public Health Lab | Chan-Zuckerberg Biohub | CZB Cliahub Consortium |
| EPI_ISL_1234991 | Santa Clara County Public Health Laboratory | Chan-Zuckerberg Biohub | CZB Cliahub Consortium |
| EPI_ISL_1234992 | California Institute of Technology | Chan-Zuckerberg Biohub | CZB Cliahub Consortium |
| EPI_ISL_1234993 | Santa Clara County Public Health Laboratory | Chan-Zuckerberg Biohub | CZB Cliahub Consortium |
| EPI_ISL_1234994 | California Institute of Technology | Chan-Zuckerberg Biohub | CZB Cliahub Consortium |
| EPI_ISL_1234995 | Alameda County Public Health Lab | Chan-Zuckerberg Biohub | CZB Cliahub Consortium |
| EPI_ISL_1234996 | Santa Clara County Public Health Laboratory | Chan-Zuckerberg Biohub | CZB Cliahub Consortium |
| EPI_ISL_1234997 | Orange County Public Health Lab | Chan-Zuckerberg Biohub | CZB Cliahub Consortium |
| EPI_ISL_1234998 | Humboldt County Public Health Laboratory | Chan-Zuckerberg Biohub | CZB Cliahub Consortium |
| EPI_ISL_1234999, EPI_ISL_1235000 | Santa Clara County Public Health Laboratory | Chan-Zuckerberg Biohub | CZB Cliahub Consortium |
| EPI_ISL_1235001, EPI_ISL_1235002, EPI_ISL_1235003, EPI_ISL_1235004, EPI_ISL_1235005 | Humboldt County Public Health Laboratory | Chan-Zuckerberg Biohub | CZB Cliahub Consortium |
| EPI_ISL_1235006 | Alameda County Public Health Lab | Chan-Zuckerberg Biohub | CZB Cliahub Consortium |
| EPI_ISL_1235007, EPI_ISL_1235008, EPI_ISL_1235009 | Orange County Public Health Lab | Chan-Zuckerberg Biohub | CZB Cliahub Consortium |
| EPI_ISL_1235011, EPI_ISL_1235012 | Humboldt County Public Health Laboratory | Chan-Zuckerberg Biohub | CZB Cliahub Consortium |
| EPI_ISL_1235013 | Alameda County Public Health Lab | Chan-Zuckerberg Biohub | CZB Cliahub Consortium |
| EPI_ISL_1235014 | California Institute of Technology | Chan-Zuckerberg Biohub | CZB Cliahub Consortium |
| EPI_ISL_1235015 | Santa Clara County Public Health Laboratory | Chan-Zuckerberg Biohub | CZB Cliahub Consortium |
| EPI_ISL_1235016 | Orange County Public Health Lab | Chan-Zuckerberg Biohub | CZB Cliahub Consortium |
| EPI_ISL_1235017, EPI_ISL_1235018, EPI_ISL_1235019, EPI_ISL_1235020 | Santa Clara County Public Health Laboratory | Chan-Zuckerberg Biohub | CZB Cliahub Consortium |
| EPI_ISL_1235021, EPI_ISL_1235022 | Alameda County Public Health Lab | Chan-Zuckerberg Biohub | CZB Cliahub Consortium |
| EPI_ISL_1235023 | Humboldt County Public Health Laboratory | Chan-Zuckerberg Biohub | CZB Cliahub Consortium |
| EPI_ISL_1235024 | Santa Clara County Public Health Laboratory | Chan-Zuckerberg Biohub | CZB Cliahub Consortium |
| EPI_ISL_1235025 | Alameda County Public Health Lab | Chan-Zuckerberg Biohub | CZB Cliahub Consortium |
| EPI_ISL_1235026, EPI_ISL_1235027 | California Institute of Technology | Chan-Zuckerberg Biohub | CZB Cliahub Consortium |
| EPI_ISL_1235028 | Alameda County Public Health Lab | Chan-Zuckerberg Biohub | CZB Cliahub Consortium |
| EPI_ISL_1235029 | County of San Luis Obispo Public Health Laboratory | Chan-Zuckerberg Biohub | CZB Cliahub Consortium |

| | | | |
|---|---|---|---|
| EPI_ISL_1235030 | Humboldt County Public Health Laboratory | Chan-Zuckerberg Biohub | CZB Cliahub Consortium |
| EPI_ISL_1235031 | Santa Clara County Public Health Laboratory | Chan-Zuckerberg Biohub | CZB Cliahub Consortium |
| EPI_ISL_1235032 | Alameda County Public Health Lab | Chan-Zuckerberg Biohub | CZB Cliahub Consortium |
| EPI_ISL_1235033 | Humboldt County Public Health Laboratory | Chan-Zuckerberg Biohub | CZB Cliahub Consortium |
| EPI_ISL_1235034 | Santa Clara County Public Health Laboratory | Chan-Zuckerberg Biohub | CZB Cliahub Consortium |
| EPI_ISL_1235035, EPI_ISL_1235036 | California Institute of Technology | Chan-Zuckerberg Biohub | CZB Cliahub Consortium |
| EPI_ISL_1235037 | Humboldt County Public Health Laboratory | Chan-Zuckerberg Biohub | CZB Cliahub Consortium |
| EPI_ISL_1235038 | Santa Clara County Public Health Laboratory | Chan-Zuckerberg Biohub | CZB Cliahub Consortium |
| EPI_ISL_1235039, EPI_ISL_1235040 | Humboldt County Public Health Laboratory | Chan-Zuckerberg Biohub | CZB Cliahub Consortium |
| EPI_ISL_1235041 | Orange County Public Health Lab | Chan-Zuckerberg Biohub | CZB Cliahub Consortium |
| EPI_ISL_1235042 | Alameda County Public Health Lab | Chan-Zuckerberg Biohub | CZB Cliahub Consortium |
| EPI_ISL_1235043 | Santa Clara County Public Health Laboratory | Chan-Zuckerberg Biohub | CZB Cliahub Consortium |
| EPI_ISL_1235044 | Orange County Public Health Lab | Chan-Zuckerberg Biohub | CZB Cliahub Consortium |
| EPI_ISL_1235045, EPI_ISL_1235046 | Humboldt County Public Health Laboratory | Chan-Zuckerberg Biohub | CZB Cliahub Consortium |
| EPI_ISL_1235047 | Santa Clara County Public Health Laboratory | Chan-Zuckerberg Biohub | CZB Cliahub Consortium |
| EPI_ISL_1235048 | Alameda County Public Health Lab | Chan-Zuckerberg Biohub | CZB Cliahub Consortium |
| EPI_ISL_1235049, EPI_ISL_1235050, EPI_ISL_1235051 | Santa Clara County Public Health Laboratory | Chan-Zuckerberg Biohub | CZB Cliahub Consortium |
| EPI_ISL_1235052 | Orange County Public Health Lab | Chan-Zuckerberg Biohub | CZB Cliahub Consortium |
| EPI_ISL_1235053 | Humboldt County Public Health Laboratory | Chan-Zuckerberg Biohub | CZB Cliahub Consortium |
| EPI_ISL_1235054 | Alameda County Public Health Lab | Chan-Zuckerberg Biohub | CZB Cliahub Consortium |
| EPI_ISL_1235055, EPI_ISL_1235056 | California Institute of Technology | Chan-Zuckerberg Biohub | CZB Cliahub Consortium |
| EPI_ISL_1235057, EPI_ISL_1235058 | Orange County Public Health Lab | Chan-Zuckerberg Biohub | CZB Cliahub Consortium |
| EPI_ISL_1235059 | Alameda County Public Health Lab | Chan-Zuckerberg Biohub | CZB Cliahub Consortium |
| EPI_ISL_1235060 | CA DPH Viral and Rickettsial Disease Laboratory | Chan-Zuckerberg Biohub | CZB Cliahub Consortium |
| EPI_ISL_1235061, EPI_ISL_1235062 | Contra Costa County Public Health Lab | Chan-Zuckerberg Biohub | CZB Cliahub Consortium |
| EPI_ISL_1235063 | CA DPH Viral and Rickettsial Disease Laboratory | Chan-Zuckerberg Biohub | CZB Cliahub Consortium |
| EPI_ISL_1235064, EPI_ISL_1235065, EPI_ISL_1235066, EPI_ISL_1235067 | Contra Costa County Public Health Lab | Chan-Zuckerberg Biohub | CZB Cliahub Consortium |
| EPI_ISL_1235068 | CA DPH Viral and Rickettsial Disease Laboratory | Chan-Zuckerberg Biohub | CZB Cliahub Consortium |
| EPI_ISL_1235069 | Contra Costa County Public Health Lab | Chan-Zuckerberg Biohub | CZB Cliahub Consortium |
| EPI_ISL_1235070 | CA DPH Viral and Rickettsial Disease Laboratory | Chan-Zuckerberg Biohub | CZB Cliahub Consortium |
| EPI_ISL_1235071, EPI_ISL_1235072, EPI_ISL_1235073 | Contra Costa County Public Health Lab | Chan-Zuckerberg Biohub | CZB Cliahub Consortium |
| EPI_ISL_1235074 | CA DPH Viral and Rickettsial Disease Laboratory | Chan-Zuckerberg Biohub | CZB Cliahub Consortium |
| EPI_ISL_1235075 | Alameda County Public Health Lab | Chan-Zuckerberg Biohub | CZB Cliahub Consortium |
| EPI_ISL_1235076 | Contra Costa County Public Health Lab | Chan-Zuckerberg Biohub | CZB Cliahub Consortium |
| EPI_ISL_1235077 | Alameda County Public Health Lab | Chan-Zuckerberg Biohub | CZB Cliahub Consortium |
| EPI_ISL_1235078 | Contra Costa County Public Health Lab | Chan-Zuckerberg Biohub | CZB Cliahub Consortium |
| EPI_ISL_1235079 | CA DPH Viral and Rickettsial Disease Laboratory | Chan-Zuckerberg Biohub | CZB Cliahub Consortium |
| EPI_ISL_1235080, EPI_ISL_1235081, EPI_ISL_1235082 | Contra Costa County Public Health Lab | Chan-Zuckerberg Biohub | CZB Cliahub Consortium |
| EPI_ISL_1235083 | Alameda County Public Health Lab | Chan-Zuckerberg Biohub | CZB Cliahub Consortium |
| EPI_ISL_1235084 | CA DPH Viral and Rickettsial Disease Laboratory | Chan-Zuckerberg Biohub | CZB Cliahub Consortium |
| EPI_ISL_1235085, EPI_ISL_1235086, EPI_ISL_1235087, EPI_ISL_1235088, EPI_ISL_1235089, EPI_ISL_1235090 | Contra Costa County Public Health Lab | Chan-Zuckerberg Biohub | CZB Cliahub Consortium |
| EPI_ISL_1235091 | Alameda County Public Health Lab | Chan-Zuckerberg Biohub | CZB Cliahub Consortium |
| EPI_ISL_1235092 | CA DPH Viral and Rickettsial Disease Laboratory | Chan-Zuckerberg Biohub | CZB Cliahub Consortium |
| EPI_ISL_1235093, EPI_ISL_1235094 | Alameda County Public Health Lab | Chan-Zuckerberg Biohub | CZB Cliahub Consortium |
| EPI_ISL_1235095 | CA DPH Viral and Rickettsial Disease Laboratory | Chan-Zuckerberg Biohub | CZB Cliahub Consortium |
| EPI_ISL_1235096 | Contra Costa County Public Health Lab | Chan-Zuckerberg Biohub | CZB Cliahub Consortium |
| EPI_ISL_1235097 | CA DPH Viral and Rickettsial Disease Laboratory | Chan-Zuckerberg Biohub | CZB Cliahub Consortium |
| EPI_ISL_1235098, EPI_ISL_1235099 | Alameda County Public Health Lab | Chan-Zuckerberg Biohub | CZB Cliahub Consortium |

| | | | |
|---|---|---|---|
| EPI_ISL_1235100, EPI_ISL_1235101 | Contra Costa County Public Health Lab | Chan-Zuckerberg Biohub | CZB Cliahub Consortium |
| EPI_ISL_1235102 | Alameda County Public Health Lab | Chan-Zuckerberg Biohub | CZB Cliahub Consortium |
| EPI_ISL_1235103 | Contra Costa County Public Health Lab | Chan-Zuckerberg Biohub | CZB Cliahub Consortium |
| EPI_ISL_1235104 | CA DPH Viral and Rickettsial Disease Laboratory | Chan-Zuckerberg Biohub | CZB Cliahub Consortium |
| EPI_ISL_1235105, EPI_ISL_1235106 | Contra Costa County Public Health Lab | Chan-Zuckerberg Biohub | CZB Cliahub Consortium |
| EPI_ISL_1235107 | CA DPH Viral and Rickettsial Disease Laboratory | Chan-Zuckerberg Biohub | CZB Cliahub Consortium |
| EPI_ISL_1235108, EPI_ISL_1235109, EPI_ISL_1235110 | Contra Costa County Public Health Lab | Chan-Zuckerberg Biohub | CZB Cliahub Consortium |
| EPI_ISL_1235111 | Alameda County Public Health Lab | Chan-Zuckerberg Biohub | CZB Cliahub Consortium |
| EPI_ISL_1235112 | CA DPH Viral and Rickettsial Disease Laboratory | Chan-Zuckerberg Biohub | CZB Cliahub Consortium |
| EPI_ISL_1235113, EPI_ISL_1235114 | Contra Costa County Public Health Lab | Chan-Zuckerberg Biohub | CZB Cliahub Consortium |
| EPI_ISL_1235115, EPI_ISL_1235116 | Alameda County Public Health Lab | Chan-Zuckerberg Biohub | CZB Cliahub Consortium |
| EPI_ISL_1235117 | Contra Costa County Public Health Lab | Chan-Zuckerberg Biohub | CZB Cliahub Consortium |
| EPI_ISL_1235118, EPI_ISL_1235119 | CA DPH Viral and Rickettsial Disease Laboratory | Chan-Zuckerberg Biohub | CZB Cliahub Consortium |
| EPI_ISL_1235120 | Contra Costa County Public Health Lab | Chan-Zuckerberg Biohub | CZB Cliahub Consortium |
| EPI_ISL_1235121 | CA DPH Viral and Rickettsial Disease Laboratory | Chan-Zuckerberg Biohub | CZB Cliahub Consortium |
| EPI_ISL_1235122 | Alameda County Public Health Lab | Chan-Zuckerberg Biohub | CZB Cliahub Consortium |
| EPI_ISL_1235123 | CA DPH Viral and Rickettsial Disease Laboratory | Chan-Zuckerberg Biohub | CZB Cliahub Consortium |
| EPI_ISL_1235124 | Alameda County Public Health Lab | Chan-Zuckerberg Biohub | CZB Cliahub Consortium |
| EPI_ISL_1235126, EPI_ISL_1235127, EPI_ISL_1235128 | CA DPH Viral and Rickettsial Disease Laboratory | Chan-Zuckerberg Biohub | CZB Cliahub Consortium |
| EPI_ISL_1235129, EPI_ISL_1235130, EPI_ISL_1235131 | Contra Costa County Public Health Lab | Chan-Zuckerberg Biohub | CZB Cliahub Consortium |
| EPI_ISL_1235132, EPI_ISL_1235133 | CA DPH Viral and Rickettsial Disease Laboratory | Chan-Zuckerberg Biohub | CZB Cliahub Consortium |
| EPI_ISL_1235134 | Contra Costa County Public Health Lab | Chan-Zuckerberg Biohub | CZB Cliahub Consortium |
| EPI_ISL_1235135, EPI_ISL_1235136 | CA DPH Viral and Rickettsial Disease Laboratory | Chan-Zuckerberg Biohub | CZB Cliahub Consortium |
| EPI_ISL_1235137 | Contra Costa County Public Health Lab | Chan-Zuckerberg Biohub | CZB Cliahub Consortium |
| EPI_ISL_1235138 | CA DPH Viral and Rickettsial Disease Laboratory | Chan-Zuckerberg Biohub | CZB Cliahub Consortium |
| EPI_ISL_1235139 | Contra Costa County Public Health Lab | Chan-Zuckerberg Biohub | CZB Cliahub Consortium |
| EPI_ISL_1235140 | CA DPH Viral and Rickettsial Disease Laboratory | Chan-Zuckerberg Biohub | CZB Cliahub Consortium |
| EPI_ISL_1235141, EPI_ISL_1235142 | Alameda County Public Health Lab | Chan-Zuckerberg Biohub | CZB Cliahub Consortium |
| EPI_ISL_1235143 | Contra Costa County Public Health Lab | Chan-Zuckerberg Biohub | CZB Cliahub Consortium |
| EPI_ISL_1235144 | CA DPH Viral and Rickettsial Disease Laboratory | Chan-Zuckerberg Biohub | CZB Cliahub Consortium |
| EPI_ISL_1235145, EPI_ISL_1235146 | Contra Costa County Public Health Lab | Chan-Zuckerberg Biohub | CZB Cliahub Consortium |
| EPI_ISL_1235147 | Alameda County Public Health Lab | Chan-Zuckerberg Biohub | CZB Cliahub Consortium |
| EPI_ISL_1235148 | CA DPH Viral and Rickettsial Disease Laboratory | Chan-Zuckerberg Biohub | CZB Cliahub Consortium |
| EPI_ISL_1235149 | Alameda County Public Health Lab | Chan-Zuckerberg Biohub | CZB Cliahub Consortium |
| EPI_ISL_1235150 | CA DPH Viral and Rickettsial Disease Laboratory | Chan-Zuckerberg Biohub | CZB Cliahub Consortium |
| EPI_ISL_1235151 | Contra Costa County Public Health Lab | Chan-Zuckerberg Biohub | CZB Cliahub Consortium |
| EPI_ISL_1235152 | CA DPH Viral and Rickettsial Disease Laboratory | Chan-Zuckerberg Biohub | CZB Cliahub Consortium |
| EPI_ISL_1235153, EPI_ISL_1235154, EPI_ISL_1235155, EPI_ISL_1235156, EPI_ISL_1235157 | Contra Costa County Public Health Lab | Chan-Zuckerberg Biohub | CZB Cliahub Consortium |
| EPI_ISL_1235158 | Alameda County Public Health Lab | Chan-Zuckerberg Biohub | CZB Cliahub Consortium |
| EPI_ISL_1235159, EPI_ISL_1235160, EPI_ISL_1235161 | Contra Costa County Public Health Lab | Chan-Zuckerberg Biohub | CZB Cliahub Consortium |
| EPI_ISL_1235162 | CA DPH Viral and Rickettsial Disease Laboratory | Chan-Zuckerberg Biohub | CZB Cliahub Consortium |
| EPI_ISL_1235163, EPI_ISL_1235164 | Contra Costa County Public Health Lab | Chan-Zuckerberg Biohub | CZB Cliahub Consortium |
| EPI_ISL_1235165 | Alameda County Public Health Lab | Chan-Zuckerberg Biohub | CZB Cliahub Consortium |
| EPI_ISL_1235166, EPI_ISL_1235167, EPI_ISL_1235168, EPI_ISL_1235169, EPI_ISL_1235170 | Contra Costa County Public Health Lab | Chan-Zuckerberg Biohub | CZB Cliahub Consortium |
| EPI_ISL_1235171 | Alameda County Public Health Lab | Chan-Zuckerberg Biohub | CZB Cliahub Consortium |
| EPI_ISL_1235172, EPI_ISL_1235173, EPI_ISL_1235174 | Contra Costa County Public Health Lab | Chan-Zuckerberg Biohub | CZB Cliahub Consortium |
| EPI_ISL_1235175 | Alameda County Public Health Lab | Chan-Zuckerberg Biohub | CZB Cliahub Consortium |
| EPI_ISL_1235176 | CA DPH Viral and Rickettsial Disease Laboratory | Chan-Zuckerberg Biohub | CZB Cliahub Consortium |

| | | | |
|---|---|---|---|
| EPI_ISL_1235177, EPI_ISL_1235179, EPI_ISL_1235180 | Contra Costa County Public Health Lab | Chan-Zuckerberg Biohub | CZB Cliahub Consortium |
| EPI_ISL_1235181 | Alameda County Public Health Lab | Chan-Zuckerberg Biohub | CZB Cliahub Consortium |
| EPI_ISL_1235182 | CA DPH Viral and Rickettsial Disease Laboratory | Chan-Zuckerberg Biohub | CZB Cliahub Consortium |
| EPI_ISL_1235183 | Alameda County Public Health Lab | Chan-Zuckerberg Biohub | CZB Cliahub Consortium |
| EPI_ISL_1235184 | Contra Costa County Public Health Lab | Chan-Zuckerberg Biohub | CZB Cliahub Consortium |
| EPI_ISL_1235185 | Alameda County Public Health Lab | Chan-Zuckerberg Biohub | CZB Cliahub Consortium |
| EPI_ISL_1235186 | CA DPH Viral and Rickettsial Disease Laboratory | Chan-Zuckerberg Biohub | CZB Cliahub Consortium |
| EPI_ISL_1235187 | Contra Costa County Public Health Lab | Chan-Zuckerberg Biohub | CZB Cliahub Consortium |
| EPI_ISL_1235188 | Alameda County Public Health Lab | Chan-Zuckerberg Biohub | CZB Cliahub Consortium |
| EPI_ISL_1235189 | Contra Costa County Public Health Lab | Chan-Zuckerberg Biohub | CZB Cliahub Consortium |
| EPI_ISL_1235190 | CA DPH Viral and Rickettsial Disease Laboratory | Chan-Zuckerberg Biohub | CZB Cliahub Consortium |
| EPI_ISL_1235191 | Contra Costa County Public Health Lab | Chan-Zuckerberg Biohub | CZB Cliahub Consortium |
| EPI_ISL_1235192, EPI_ISL_1235193 | CA DPH Viral and Rickettsial Disease Laboratory | Chan-Zuckerberg Biohub | CZB Cliahub Consortium |
| EPI_ISL_1235194 | Alameda County Public Health Lab | Chan-Zuckerberg Biohub | CZB Cliahub Consortium |
| EPI_ISL_1235195 | CA DPH Viral and Rickettsial Disease Laboratory | Chan-Zuckerberg Biohub | CZB Cliahub Consortium |
| EPI_ISL_1235196 | Contra Costa County Public Health Lab | Chan-Zuckerberg Biohub | CZB Cliahub Consortium |
| EPI_ISL_1235197 | CA DPH Viral and Rickettsial Disease Laboratory | Chan-Zuckerberg Biohub | CZB Cliahub Consortium |
| EPI_ISL_1235198 | Alameda County Public Health Lab | Chan-Zuckerberg Biohub | CZB Cliahub Consortium |
| EPI_ISL_1235199 | CA DPH Viral and Rickettsial Disease Laboratory | Chan-Zuckerberg Biohub | CZB Cliahub Consortium |
| EPI_ISL_1235200, EPI_ISL_1235201, EPI_ISL_1235202 | Contra Costa County Public Health Lab | Chan-Zuckerberg Biohub | CZB Cliahub Consortium |
| EPI_ISL_1235203 | CA DPH Viral and Rickettsial Disease Laboratory | Chan-Zuckerberg Biohub | CZB Cliahub Consortium |
| EPI_ISL_1235204, EPI_ISL_1235205 | Contra Costa County Public Health Lab | Chan-Zuckerberg Biohub | CZB Cliahub Consortium |
| EPI_ISL_1235206 | CA DPH Viral and Rickettsial Disease Laboratory | Chan-Zuckerberg Biohub | CZB Cliahub Consortium |
| EPI_ISL_1235207, EPI_ISL_1235208, EPI_ISL_1235209 | Contra Costa County Public Health Lab | Chan-Zuckerberg Biohub | CZB Cliahub Consortium |
| EPI_ISL_1235210 | Alameda County Public Health Lab | Chan-Zuckerberg Biohub | CZB Cliahub Consortium |
| EPI_ISL_1235211 | Contra Costa County Public Health Lab | Chan-Zuckerberg Biohub | CZB Cliahub Consortium |
| EPI_ISL_1235212 | CA DPH Viral and Rickettsial Disease Laboratory | Chan-Zuckerberg Biohub | CZB Cliahub Consortium |
| EPI_ISL_1235213 | Alameda County Public Health Lab | Chan-Zuckerberg Biohub | CZB Cliahub Consortium |
| EPI_ISL_1235214, EPI_ISL_1235215, EPI_ISL_1235216 | Contra Costa County Public Health Lab | Chan-Zuckerberg Biohub | CZB Cliahub Consortium |
| EPI_ISL_1235217 | Alameda County Public Health Lab | Chan-Zuckerberg Biohub | CZB Cliahub Consortium |
| EPI_ISL_1235218 | Contra Costa County Public Health Lab | Chan-Zuckerberg Biohub | CZB Cliahub Consortium |
| EPI_ISL_1235219 | Alameda County Public Health Lab | Chan-Zuckerberg Biohub | CZB Cliahub Consortium |
| EPI_ISL_1235220 | CA DPH Viral and Rickettsial Disease Laboratory | Chan-Zuckerberg Biohub | CZB Cliahub Consortium |
| EPI_ISL_1235221 | Contra Costa County Public Health Lab | Chan-Zuckerberg Biohub | CZB Cliahub Consortium |
| EPI_ISL_1235222 | CA DPH Viral and Rickettsial Disease Laboratory | Chan-Zuckerberg Biohub | CZB Cliahub Consortium |
| EPI_ISL_1235223 | Alameda County Public Health Lab | Chan-Zuckerberg Biohub | CZB Cliahub Consortium |
| EPI_ISL_1235224, EPI_ISL_1235225 | Contra Costa County Public Health Lab | Chan-Zuckerberg Biohub | CZB Cliahub Consortium |
| EPI_ISL_1235226 | CA DPH Viral and Rickettsial Disease Laboratory | Chan-Zuckerberg Biohub | CZB Cliahub Consortium |
| EPI_ISL_1235227, EPI_ISL_1235228 | Contra Costa County Public Health Lab | Chan-Zuckerberg Biohub | CZB Cliahub Consortium |
| EPI_ISL_1235229 | CA DPH Viral and Rickettsial Disease Laboratory | Chan-Zuckerberg Biohub | CZB Cliahub Consortium |
| EPI_ISL_1235230 | Alameda County Public Health Lab | Chan-Zuckerberg Biohub | CZB Cliahub Consortium |
| EPI_ISL_1235231 | Contra Costa County Public Health Lab | Chan-Zuckerberg Biohub | CZB Cliahub Consortium |
| EPI_ISL_1235232 | CA DPH Viral and Rickettsial Disease Laboratory | Chan-Zuckerberg Biohub | CZB Cliahub Consortium |
| EPI_ISL_1235233 | Contra Costa County Public Health Lab | Chan-Zuckerberg Biohub | CZB Cliahub Consortium |
| EPI_ISL_1235234 | CA DPH Viral and Rickettsial Disease Laboratory | Chan-Zuckerberg Biohub | CZB Cliahub Consortium |
| EPI_ISL_1235235 | Alameda County Public Health Lab | Chan-Zuckerberg Biohub | CZB Cliahub Consortium |
| EPI_ISL_1235236 | CA DPH Viral and Rickettsial Disease Laboratory | Chan-Zuckerberg Biohub | CZB Cliahub Consortium |
| EPI_ISL_1235237 | Contra Costa County Public Health Lab | Chan-Zuckerberg Biohub | CZB Cliahub Consortium |
| EPI_ISL_1235238 | CA DPH Viral and Rickettsial Disease Laboratory | Chan-Zuckerberg Biohub | CZB Cliahub Consortium |
| EPI_ISL_1235239, EPI_ISL_1235240 | Contra Costa County Public Health Lab | Chan-Zuckerberg Biohub | CZB Cliahub Consortium |
| EPI_ISL_1235241 | Alameda County Public Health Lab | Chan-Zuckerberg Biohub | CZB Cliahub Consortium |
| EPI_ISL_1235242 | CA DPH Viral and Rickettsial Disease Laboratory | Chan-Zuckerberg Biohub | CZB Cliahub Consortium |
| EPI_ISL_1235243, EPI_ISL_1235244 | Alameda County Public Health Lab | Chan-Zuckerberg Biohub | CZB Cliahub Consortium |
| EPI_ISL_1235245, EPI_ISL_1235246, | Contra Costa County Public Health Lab | Chan-Zuckerberg Biohub | CZB Cliahub Consortium |

| | | | |
|---|---|---|---|
| EPI_ISL_1235247 | | | |
| EPI_ISL_1235248 | CA DPH Viral and Rickettsial Disease Laboratory | Chan-Zuckerberg Biohub | CZB Cliahub Consortium |
| EPI_ISL_1235249 | Alameda County Public Health Lab | Chan-Zuckerberg Biohub | CZB Cliahub Consortium |
| EPI_ISL_1235250 | Contra Costa County Public Health Lab | Chan-Zuckerberg Biohub | CZB Cliahub Consortium |
| EPI_ISL_1235251, EPI_ISL_1235252 | CA DPH Viral and Rickettsial Disease Laboratory | Chan-Zuckerberg Biohub | CZB Cliahub Consortium |
| EPI_ISL_1235253 | Contra Costa County Public Health Lab | Chan-Zuckerberg Biohub | CZB Cliahub Consortium |
| EPI_ISL_1235254 | CA DPH Viral and Rickettsial Disease Laboratory | Chan-Zuckerberg Biohub | CZB Cliahub Consortium |
| EPI_ISL_1235255, EPI_ISL_1235256 | Alameda County Public Health Lab | Chan-Zuckerberg Biohub | CZB Cliahub Consortium |
| EPI_ISL_1235257, EPI_ISL_1235258, EPI_ISL_1235259, EPI_ISL_1235260, EPI_ISL_1235261, EPI_ISL_1235262 | Contra Costa County Public Health Lab | Chan-Zuckerberg Biohub | CZB Cliahub Consortium |
| EPI_ISL_1235263 | Alameda County Public Health Lab | Chan-Zuckerberg Biohub | CZB Cliahub Consortium |
| EPI_ISL_1235264, EPI_ISL_1235265, EPI_ISL_1235266, EPI_ISL_1235267, EPI_ISL_1235268 | Contra Costa County Public Health Lab | Chan-Zuckerberg Biohub | CZB Cliahub Consortium |
| EPI_ISL_1235269 | CA DPH Viral and Rickettsial Disease Laboratory | Chan-Zuckerberg Biohub | CZB Cliahub Consortium |
| EPI_ISL_1235270 | Alameda County Public Health Lab | Chan-Zuckerberg Biohub | CZB Cliahub Consortium |
| EPI_ISL_1235271 | Contra Costa County Public Health Lab | Chan-Zuckerberg Biohub | CZB Cliahub Consortium |
| EPI_ISL_1235272, EPI_ISL_1235273, EPI_ISL_1235274 | CA DPH Viral and Rickettsial Disease Laboratory | Chan-Zuckerberg Biohub | CZB Cliahub Consortium |
| EPI_ISL_1235275 | Contra Costa County Public Health Lab | Chan-Zuckerberg Biohub | CZB Cliahub Consortium |
| EPI_ISL_1235276 | Alameda County Public Health Lab | Chan-Zuckerberg Biohub | CZB Cliahub Consortium |
| EPI_ISL_1235277, EPI_ISL_1235278 | Contra Costa County Public Health Lab | Chan-Zuckerberg Biohub | CZB Cliahub Consortium |
| EPI_ISL_1235279 | Alameda County Public Health Lab | Chan-Zuckerberg Biohub | CZB Cliahub Consortium |
| EPI_ISL_1235280 | Contra Costa County Public Health Lab | Chan-Zuckerberg Biohub | CZB Cliahub Consortium |
| EPI_ISL_1235281 | Alameda County Public Health Lab | Chan-Zuckerberg Biohub | CZB Cliahub Consortium |
| EPI_ISL_1235282 | CA DPH Viral and Rickettsial Disease Laboratory | Chan-Zuckerberg Biohub | CZB Cliahub Consortium |
| EPI_ISL_1235283, EPI_ISL_1235284 | Contra Costa County Public Health Lab | Chan-Zuckerberg Biohub | CZB Cliahub Consortium |
| EPI_ISL_1235285 | CA DPH Viral and Rickettsial Disease Laboratory | Chan-Zuckerberg Biohub | CZB Cliahub Consortium |
| EPI_ISL_1235286 | Contra Costa County Public Health Lab | Chan-Zuckerberg Biohub | CZB Cliahub Consortium |
| EPI_ISL_1235287 | CA DPH Viral and Rickettsial Disease Laboratory | Chan-Zuckerberg Biohub | CZB Cliahub Consortium |
| EPI_ISL_1235288 | Contra Costa County Public Health Lab | Chan-Zuckerberg Biohub | CZB Cliahub Consortium |
| EPI_ISL_1235289 | Alameda County Public Health Lab | Chan-Zuckerberg Biohub | CZB Cliahub Consortium |
| EPI_ISL_1235290, EPI_ISL_1235291 | CA DPH Viral and Rickettsial Disease Laboratory | Chan-Zuckerberg Biohub | CZB Cliahub Consortium |
| EPI_ISL_1235292 | Alameda County Public Health Lab | Chan-Zuckerberg Biohub | CZB Cliahub Consortium |
| EPI_ISL_1235293 | Contra Costa County Public Health Lab | Chan-Zuckerberg Biohub | CZB Cliahub Consortium |
| EPI_ISL_1235295 | CA DPH Viral and Rickettsial Disease Laboratory | Chan-Zuckerberg Biohub | CZB Cliahub Consortium |
| EPI_ISL_1235296, EPI_ISL_1235297 | Contra Costa County Public Health Lab | Chan-Zuckerberg Biohub | CZB Cliahub Consortium |
| EPI_ISL_1235298 | CA DPH Viral and Rickettsial Disease Laboratory | Chan-Zuckerberg Biohub | CZB Cliahub Consortium |
| EPI_ISL_1235300, EPI_ISL_1235301, EPI_ISL_1235302 | Contra Costa County Public Health Lab | Chan-Zuckerberg Biohub | CZB Cliahub Consortium |
| EPI_ISL_1235303 | Alameda County Public Health Lab | Chan-Zuckerberg Biohub | CZB Cliahub Consortium |
| EPI_ISL_1235304, EPI_ISL_1235305, EPI_ISL_1235306 | Contra Costa County Public Health Lab | Chan-Zuckerberg Biohub | CZB Cliahub Consortium |
| EPI_ISL_1235307 | CA DPH Viral and Rickettsial Disease Laboratory | Chan-Zuckerberg Biohub | CZB Cliahub Consortium |
| EPI_ISL_1235308, EPI_ISL_1235309 | Alameda County Public Health Lab | Chan-Zuckerberg Biohub | CZB Cliahub Consortium |
| EPI_ISL_1235310 | CA DPH Viral and Rickettsial Disease Laboratory | Chan-Zuckerberg Biohub | CZB Cliahub Consortium |
| EPI_ISL_1235311 | Contra Costa County Public Health Lab | Chan-Zuckerberg Biohub | CZB Cliahub Consortium |
| EPI_ISL_1235312, EPI_ISL_1235314 | Alameda County Public Health Lab | Chan-Zuckerberg Biohub | CZB Cliahub Consortium |
| EPI_ISL_1235315, EPI_ISL_1235316 | Contra Costa County Public Health Lab | Chan-Zuckerberg Biohub | CZB Cliahub Consortium |
| EPI_ISL_1235317 | CA DPH Viral and Rickettsial Disease Laboratory | Chan-Zuckerberg Biohub | CZB Cliahub Consortium |
| EPI_ISL_1235318 | Alameda County Public Health Lab | Chan-Zuckerberg Biohub | CZB Cliahub Consortium |
| EPI_ISL_1235319, EPI_ISL_1235320, EPI_ISL_1235321, EPI_ISL_1235323, EPI_ISL_1235324, EPI_ISL_1235325, EPI_ISL_1235326, EPI_ISL_1235327, EPI_ISL_1235328, EPI_ISL_1235329, EPI_ISL_1235330, EPI_ISL_1235331, EPI_ISL_1235332, EPI_ISL_1235333, EPI_ISL_1235334 | | | |
| see above | Contra Costa County Public Health Lab | Chan-Zuckerberg Biohub | CZB Cliahub Consortium |
| EPI_ISL_1235335 | Alameda County Public Health Lab | Chan-Zuckerberg Biohub | CZB Cliahub Consortium |
| EPI_ISL_1235336, EPI_ISL_1235337 | Contra Costa County Public Health Lab | Chan-Zuckerberg Biohub | CZB Cliahub Consortium |
| EPI_ISL_1235338, EPI_ISL_1235339 | Alameda County Public Health Lab | Chan-Zuckerberg Biohub | CZB Cliahub Consortium |
| EPI_ISL_1235340, | Contra Costa County Public Health Lab | Chan-Zuckerberg Biohub | CZB Cliahub Consortium |

| | | | |
|---|---|---|---|
| EPI_ISL_1235341, EPI_ISL_1235342, EPI_ISL_1235343, EPI_ISL_1235344, EPI_ISL_1235345 | | | |
| EPI_ISL_1235346 | CA DPH Viral and Rickettsial Disease Laboratory | Chan-Zuckerberg Biohub | CZB Cliahub Consortium |
| EPI_ISL_1235347, EPI_ISL_1235348, EPI_ISL_1235349, EPI_ISL_1235350 | Contra Costa County Public Health Lab | Chan-Zuckerberg Biohub | CZB Cliahub Consortium |
| EPI_ISL_1235351 | CA DPH Viral and Rickettsial Disease Laboratory | Chan-Zuckerberg Biohub | CZB Cliahub Consortium |
| EPI_ISL_1235352, EPI_ISL_1235353 | Contra Costa County Public Health Lab | Chan-Zuckerberg Biohub | CZB Cliahub Consortium |
| EPI_ISL_1235354 | CA DPH Viral and Rickettsial Disease Laboratory | Chan-Zuckerberg Biohub | CZB Cliahub Consortium |
| EPI_ISL_1235355, EPI_ISL_1235356, EPI_ISL_1235357, EPI_ISL_1235358 | Contra Costa County Public Health Lab | Chan-Zuckerberg Biohub | CZB Cliahub Consortium |
| EPI_ISL_1235359 | Alameda County Public Health Lab | Chan-Zuckerberg Biohub | CZB Cliahub Consortium |
| EPI_ISL_1235360 | CA DPH Viral and Rickettsial Disease Laboratory | Chan-Zuckerberg Biohub | CZB Cliahub Consortium |
| EPI_ISL_1235361 | Alameda County Public Health Lab | Chan-Zuckerberg Biohub | CZB Cliahub Consortium |
| EPI_ISL_1235362 | Santa Clara County Public Health Laboratory | Chan-Zuckerberg Biohub | CZB Cliahub Consortium |
| EPI_ISL_1235363, EPI_ISL_1235364, EPI_ISL_1235365, EPI_ISL_1235366 | Alameda County Public Health Lab | Chan-Zuckerberg Biohub | CZB Cliahub Consortium |
| EPI_ISL_1235367 | CA DPH Viral and Rickettsial Disease Laboratory | Chan-Zuckerberg Biohub | CZB Cliahub Consortium |
| EPI_ISL_1235368 | Alameda County Public Health Lab | Chan-Zuckerberg Biohub | CZB Cliahub Consortium |
| EPI_ISL_1235370 | Santa Clara County Public Health Laboratory | Chan-Zuckerberg Biohub | CZB Cliahub Consortium |
| EPI_ISL_1235371 | Fresno County Public Health Laboratory | Chan-Zuckerberg Biohub | CZB Cliahub Consortium |
| EPI_ISL_1235372, EPI_ISL_1235373 | CA DPH Viral and Rickettsial Disease Laboratory | Chan-Zuckerberg Biohub | CZB Cliahub Consortium |
| EPI_ISL_1235374 | Alameda County Public Health Lab | Chan-Zuckerberg Biohub | CZB Cliahub Consortium |
| EPI_ISL_1235375 | CA DPH Viral and Rickettsial Disease Laboratory | Chan-Zuckerberg Biohub | CZB Cliahub Consortium |
| EPI_ISL_1235376 | Alameda County Public Health Lab | Chan-Zuckerberg Biohub | CZB Cliahub Consortium |
| EPI_ISL_1235377, EPI_ISL_1235378 | CA DPH Viral and Rickettsial Disease Laboratory | Chan-Zuckerberg Biohub | CZB Cliahub Consortium |
| EPI_ISL_1235379 | Santa Clara County Public Health Laboratory | Chan-Zuckerberg Biohub | CZB Cliahub Consortium |
| EPI_ISL_1235380 | Alameda County Public Health Lab | Chan-Zuckerberg Biohub | CZB Cliahub Consortium |
| EPI_ISL_1235381 | CA DPH Viral and Rickettsial Disease Laboratory | Chan-Zuckerberg Biohub | CZB Cliahub Consortium |
| EPI_ISL_1235382 | Santa Clara County Public Health Laboratory | Chan-Zuckerberg Biohub | CZB Cliahub Consortium |
| EPI_ISL_1235383, EPI_ISL_1235384 | CA DPH Viral and Rickettsial Disease Laboratory | Chan-Zuckerberg Biohub | CZB Cliahub Consortium |
| EPI_ISL_1235385, EPI_ISL_1235386, EPI_ISL_1235387, EPI_ISL_1235388, EPI_ISL_1235389, EPI_ISL_1235390, EPI_ISL_1235391, EPI_ISL_1235392, EPI_ISL_1235393 | Alameda County Public Health Lab | Chan-Zuckerberg Biohub | CZB Cliahub Consortium |
| EPI_ISL_1235394, EPI_ISL_1235395, EPI_ISL_1235396 | Santa Clara County Public Health Laboratory | Chan-Zuckerberg Biohub | CZB Cliahub Consortium |
| EPI_ISL_1235397, EPI_ISL_1235398, EPI_ISL_1235399 | Alameda County Public Health Lab | Chan-Zuckerberg Biohub | CZB Cliahub Consortium |
| EPI_ISL_1235400, EPI_ISL_1235401 | CA DPH Viral and Rickettsial Disease Laboratory | Chan-Zuckerberg Biohub | CZB Cliahub Consortium |
| EPI_ISL_1235402 | Alameda County Public Health Lab | Chan-Zuckerberg Biohub | CZB Cliahub Consortium |
| EPI_ISL_1235403 | CA DPH Viral and Rickettsial Disease Laboratory | Chan-Zuckerberg Biohub | CZB Cliahub Consortium |
| EPI_ISL_1235404, EPI_ISL_1235405, EPI_ISL_1235406, EPI_ISL_1235407 | Alameda County Public Health Lab | Chan-Zuckerberg Biohub | CZB Cliahub Consortium |
| EPI_ISL_1235408 | Fresno County Public Health Laboratory | Chan-Zuckerberg Biohub | CZB Cliahub Consortium |
| EPI_ISL_1235409 | Santa Clara County Public Health Laboratory | Chan-Zuckerberg Biohub | CZB Cliahub Consortium |
| EPI_ISL_1235410 | CA DPH Viral and Rickettsial Disease Laboratory | Chan-Zuckerberg Biohub | CZB Cliahub Consortium |
| EPI_ISL_1235411, EPI_ISL_1235412 | Santa Clara County Public Health Laboratory | Chan-Zuckerberg Biohub | CZB Cliahub Consortium |
| EPI_ISL_1235413 | Fresno County Public Health Laboratory | Chan-Zuckerberg Biohub | CZB Cliahub Consortium |
| EPI_ISL_1235414 | Alameda County Public Health Lab | Chan-Zuckerberg Biohub | CZB Cliahub Consortium |
| EPI_ISL_1235415 | Santa Clara County Public Health Laboratory | Chan-Zuckerberg Biohub | CZB Cliahub Consortium |
| EPI_ISL_1235416, EPI_ISL_1235417, EPI_ISL_1235418, | CA DPH Viral and Rickettsial Disease Laboratory | Chan-Zuckerberg Biohub | CZB Cliahub Consortium |

| | | | |
|---|---|---|---|
| EPI_ISL_1235419 | | | |
| EPI_ISL_1235420 | Santa Clara County Public Health Laboratory | Chan-Zuckerberg Biohub | CZB Cliahub Consortium |
| EPI_ISL_1235421 | Alameda County Public Health Lab | Chan-Zuckerberg Biohub | CZB Cliahub Consortium |
| EPI_ISL_1235422 | Fresno County Public Health Laboratory | Chan-Zuckerberg Biohub | CZB Cliahub Consortium |
| EPI_ISL_1235424 | CA DPH Viral and Rickettsial Disease Laboratory | Chan-Zuckerberg Biohub | CZB Cliahub Consortium |
| EPI_ISL_1235425, EPI_ISL_1235426 | Alameda County Public Health Lab | Chan-Zuckerberg Biohub | CZB Cliahub Consortium |
| EPI_ISL_1235427 | CA DPH Viral and Rickettsial Disease Laboratory | Chan-Zuckerberg Biohub | CZB Cliahub Consortium |
| EPI_ISL_1235428, EPI_ISL_1235429, EPI_ISL_1235430, EPI_ISL_1235431 | Santa Clara County Public Health Laboratory | Chan-Zuckerberg Biohub | CZB Cliahub Consortium |
| EPI_ISL_1235432, EPI_ISL_1235433 | Alameda County Public Health Lab | Chan-Zuckerberg Biohub | CZB Cliahub Consortium |
| EPI_ISL_1235434, EPI_ISL_1235435 | CA DPH Viral and Rickettsial Disease Laboratory | Chan-Zuckerberg Biohub | CZB Cliahub Consortium |
| EPI_ISL_1235436 | Santa Clara County Public Health Laboratory | Chan-Zuckerberg Biohub | CZB Cliahub Consortium |
| EPI_ISL_1235437 | Alameda County Public Health Lab | Chan-Zuckerberg Biohub | CZB Cliahub Consortium |
| EPI_ISL_1235438 | CA DPH Viral and Rickettsial Disease Laboratory | Chan-Zuckerberg Biohub | CZB Cliahub Consortium |
| EPI_ISL_1235439 | Santa Clara County Public Health Laboratory | Chan-Zuckerberg Biohub | CZB Cliahub Consortium |
| EPI_ISL_1235440 | Alameda County Public Health Lab | Chan-Zuckerberg Biohub | CZB Cliahub Consortium |
| EPI_ISL_1235441, EPI_ISL_1235442 | CA DPH Viral and Rickettsial Disease Laboratory | Chan-Zuckerberg Biohub | CZB Cliahub Consortium |
| EPI_ISL_1235443 | Alameda County Public Health Lab | Chan-Zuckerberg Biohub | CZB Cliahub Consortium |
| EPI_ISL_1235444, EPI_ISL_1235445 | Santa Clara County Public Health Laboratory | Chan-Zuckerberg Biohub | CZB Cliahub Consortium |
| EPI_ISL_1235446, EPI_ISL_1235447, EPI_ISL_1235448 | Alameda County Public Health Lab | Chan-Zuckerberg Biohub | CZB Cliahub Consortium |
| EPI_ISL_1235449 | Santa Clara County Public Health Laboratory | Chan-Zuckerberg Biohub | CZB Cliahub Consortium |
| EPI_ISL_1235450, EPI_ISL_1235451 | CA DPH Viral and Rickettsial Disease Laboratory | Chan-Zuckerberg Biohub | CZB Cliahub Consortium |
| EPI_ISL_1235452, EPI_ISL_1235453, EPI_ISL_1235454 | Santa Clara County Public Health Laboratory | Chan-Zuckerberg Biohub | CZB Cliahub Consortium |
| EPI_ISL_1235455 | CA DPH Viral and Rickettsial Disease Laboratory | Chan-Zuckerberg Biohub | CZB Cliahub Consortium |
| EPI_ISL_1235456, EPI_ISL_1235457, EPI_ISL_1235458, EPI_ISL_1235459, EPI_ISL_1235460 | Alameda County Public Health Lab | Chan-Zuckerberg Biohub | CZB Cliahub Consortium |
| EPI_ISL_1235461 | Santa Clara County Public Health Laboratory | Chan-Zuckerberg Biohub | CZB Cliahub Consortium |
| EPI_ISL_1235462, EPI_ISL_1235463, EPI_ISL_1235464, EPI_ISL_1235465 | Alameda County Public Health Lab | Chan-Zuckerberg Biohub | CZB Cliahub Consortium |
| EPI_ISL_1235466, EPI_ISL_1235467 | Santa Clara County Public Health Laboratory | Chan-Zuckerberg Biohub | CZB Cliahub Consortium |
| EPI_ISL_1235468, EPI_ISL_1235469 | CA DPH Viral and Rickettsial Disease Laboratory | Chan-Zuckerberg Biohub | CZB Cliahub Consortium |
| EPI_ISL_1235470 | Santa Clara County Public Health Laboratory | Chan-Zuckerberg Biohub | CZB Cliahub Consortium |
| EPI_ISL_1235471 | Alameda County Public Health Lab | Chan-Zuckerberg Biohub | CZB Cliahub Consortium |
| EPI_ISL_1235472, EPI_ISL_1235473 | CA DPH Viral and Rickettsial Disease Laboratory | Chan-Zuckerberg Biohub | CZB Cliahub Consortium |
| EPI_ISL_1235474 | Santa Clara County Public Health Laboratory | Chan-Zuckerberg Biohub | CZB Cliahub Consortium |
| EPI_ISL_1235475, EPI_ISL_1235476, EPI_ISL_1235477 | Alameda County Public Health Lab | Chan-Zuckerberg Biohub | CZB Cliahub Consortium |
| EPI_ISL_1235478 | CA DPH Viral and Rickettsial Disease Laboratory | Chan-Zuckerberg Biohub | CZB Cliahub Consortium |
| EPI_ISL_1235479 | Santa Clara County Public Health Laboratory | Chan-Zuckerberg Biohub | CZB Cliahub Consortium |
| EPI_ISL_1235480, EPI_ISL_1235481, EPI_ISL_1235482 | Alameda County Public Health Lab | Chan-Zuckerberg Biohub | CZB Cliahub Consortium |
| EPI_ISL_1235483, EPI_ISL_1235484 | Santa Clara County Public Health Laboratory | Chan-Zuckerberg Biohub | CZB Cliahub Consortium |
| EPI_ISL_1235485, EPI_ISL_1235486 | Alameda County Public Health Lab | Chan-Zuckerberg Biohub | CZB Cliahub Consortium |
| EPI_ISL_1235487 | Santa Clara County Public Health Laboratory | Chan-Zuckerberg Biohub | CZB Cliahub Consortium |
| EPI_ISL_1235488 | CA DPH Viral and Rickettsial Disease Laboratory | Chan-Zuckerberg Biohub | CZB Cliahub Consortium |
| EPI_ISL_1235489 | Alameda County Public Health Lab | Chan-Zuckerberg Biohub | CZB Cliahub Consortium |
| EPI_ISL_1235490 | CA DPH Viral and Rickettsial Disease Laboratory | Chan-Zuckerberg Biohub | CZB Cliahub Consortium |
| EPI_ISL_1235491 | Santa Clara County Public Health Laboratory | Chan-Zuckerberg Biohub | CZB Cliahub Consortium |
| EPI_ISL_1235492, EPI_ISL_1235493, | Alameda County Public Health Lab | Chan-Zuckerberg Biohub | CZB Cliahub Consortium |

| | | | |
|---|---|---|---|
| EPI_ISL_1235494, EPI_ISL_1235495 | | | |
| EPI_ISL_1235496 | CA DPH Viral and Rickettsial Disease Laboratory | Chan-Zuckerberg Biohub | CZB Cliahub Consortium |
| EPI_ISL_1235497 | Alameda County Public Health Lab | Chan-Zuckerberg Biohub | CZB Cliahub Consortium |
| EPI_ISL_1235498 | CA DPH Viral and Rickettsial Disease Laboratory | Chan-Zuckerberg Biohub | CZB Cliahub Consortium |
| EPI_ISL_1235499, EPI_ISL_1235500 | Alameda County Public Health Lab | Chan-Zuckerberg Biohub | CZB Cliahub Consortium |
| EPI_ISL_1235501, EPI_ISL_1235502 | CA DPH Viral and Rickettsial Disease Laboratory | Chan-Zuckerberg Biohub | CZB Cliahub Consortium |
| EPI_ISL_1235503, EPI_ISL_1235504, EPI_ISL_1235505 | Alameda County Public Health Lab | Chan-Zuckerberg Biohub | CZB Cliahub Consortium |
| EPI_ISL_1235506, EPI_ISL_1235507 | CA DPH Viral and Rickettsial Disease Laboratory | Chan-Zuckerberg Biohub | CZB Cliahub Consortium |
| EPI_ISL_1235508 | Santa Clara County Public Health Laboratory | Chan-Zuckerberg Biohub | CZB Cliahub Consortium |
| EPI_ISL_1235509 | CA DPH Viral and Rickettsial Disease Laboratory | Chan-Zuckerberg Biohub | CZB Cliahub Consortium |
| EPI_ISL_1235510 | Alameda County Public Health Lab | Chan-Zuckerberg Biohub | CZB Cliahub Consortium |
| EPI_ISL_1235511 | CA DPH Viral and Rickettsial Disease Laboratory | Chan-Zuckerberg Biohub | CZB Cliahub Consortium |
| EPI_ISL_1235512, EPI_ISL_1235513, EPI_ISL_1235514 | Alameda County Public Health Lab | Chan-Zuckerberg Biohub | CZB Cliahub Consortium |
| EPI_ISL_1235515 | CA DPH Viral and Rickettsial Disease Laboratory | Chan-Zuckerberg Biohub | CZB Cliahub Consortium |
| EPI_ISL_1235516, EPI_ISL_1235517 | Alameda County Public Health Lab | Chan-Zuckerberg Biohub | CZB Cliahub Consortium |
| EPI_ISL_1235518, EPI_ISL_1235519 | Santa Clara County Public Health Laboratory | Chan-Zuckerberg Biohub | CZB Cliahub Consortium |
| EPI_ISL_1235520 | Alameda County Public Health Lab | Chan-Zuckerberg Biohub | CZB Cliahub Consortium |
| EPI_ISL_1235521, EPI_ISL_1235522 | CA DPH Viral and Rickettsial Disease Laboratory | Chan-Zuckerberg Biohub | CZB Cliahub Consortium |
| EPI_ISL_1235523, EPI_ISL_1235524 | Alameda County Public Health Lab | Chan-Zuckerberg Biohub | CZB Cliahub Consortium |
| EPI_ISL_1235525, EPI_ISL_1235526 | CA DPH Viral and Rickettsial Disease Laboratory | Chan-Zuckerberg Biohub | CZB Cliahub Consortium |
| EPI_ISL_1235527 | Alameda County Public Health Lab | Chan-Zuckerberg Biohub | CZB Cliahub Consortium |
| EPI_ISL_1235528 | CA DPH Viral and Rickettsial Disease Laboratory | Chan-Zuckerberg Biohub | CZB Cliahub Consortium |
| EPI_ISL_1235529 | Alameda County Public Health Lab | Chan-Zuckerberg Biohub | CZB Cliahub Consortium |
| EPI_ISL_1235530, EPI_ISL_1235531 | Santa Clara County Public Health Laboratory | Chan-Zuckerberg Biohub | CZB Cliahub Consortium |
| EPI_ISL_1235532 | Alameda County Public Health Lab | Chan-Zuckerberg Biohub | CZB Cliahub Consortium |
| EPI_ISL_1235534, EPI_ISL_1235535, EPI_ISL_1235536 | Santa Clara County Public Health Laboratory | Chan-Zuckerberg Biohub | CZB Cliahub Consortium |
| EPI_ISL_1235537, EPI_ISL_1235538 | Alameda County Public Health Lab | Chan-Zuckerberg Biohub | CZB Cliahub Consortium |
| EPI_ISL_1235539, EPI_ISL_1235540 | CA DPH Viral and Rickettsial Disease Laboratory | Chan-Zuckerberg Biohub | CZB Cliahub Consortium |
| EPI_ISL_1235541, EPI_ISL_1235542 | Santa Clara County Public Health Laboratory | Chan-Zuckerberg Biohub | CZB Cliahub Consortium |
| EPI_ISL_1235543 | Alameda County Public Health Lab | Chan-Zuckerberg Biohub | CZB Cliahub Consortium |
| EPI_ISL_1235544, EPI_ISL_1235545, EPI_ISL_1235546, EPI_ISL_1235547 | CA DPH Viral and Rickettsial Disease Laboratory | Chan-Zuckerberg Biohub | CZB Cliahub Consortium |
| EPI_ISL_1235548 | Alameda County Public Health Lab | Chan-Zuckerberg Biohub | CZB Cliahub Consortium |
| EPI_ISL_1235549 | CA DPH Viral and Rickettsial Disease Laboratory | Chan-Zuckerberg Biohub | CZB Cliahub Consortium |
| EPI_ISL_1235550 | Alameda County Public Health Lab | Chan-Zuckerberg Biohub | CZB Cliahub Consortium |
| EPI_ISL_1235551 | CA DPH Viral and Rickettsial Disease Laboratory | Chan-Zuckerberg Biohub | CZB Cliahub Consortium |
| EPI_ISL_1235552, EPI_ISL_1235553, EPI_ISL_1235554, EPI_ISL_1235555 | Alameda County Public Health Lab | Chan-Zuckerberg Biohub | CZB Cliahub Consortium |
| EPI_ISL_1235556 | Santa Clara County Public Health Laboratory | Chan-Zuckerberg Biohub | CZB Cliahub Consortium |
| EPI_ISL_1235557 | Alameda County Public Health Lab | Chan-Zuckerberg Biohub | CZB Cliahub Consortium |
| EPI_ISL_1235558 | CA DPH Viral and Rickettsial Disease Laboratory | Chan-Zuckerberg Biohub | CZB Cliahub Consortium |
| EPI_ISL_1235559 | Alameda County Public Health Lab | Chan-Zuckerberg Biohub | CZB Cliahub Consortium |
| EPI_ISL_1235560 | Fresno County Public Health Laboratory | Chan-Zuckerberg Biohub | CZB Cliahub Consortium |
| EPI_ISL_1235561, EPI_ISL_1235562 | Alameda County Public Health Lab | Chan-Zuckerberg Biohub | CZB Cliahub Consortium |
| EPI_ISL_1235563 | Santa Clara County Public Health Laboratory | Chan-Zuckerberg Biohub | CZB Cliahub Consortium |
| EPI_ISL_1235564, EPI_ISL_1235565, EPI_ISL_1235566, EPI_ISL_1235567, EPI_ISL_1235568, EPI_ISL_1235569 | Alameda County Public Health Lab | Chan-Zuckerberg Biohub | CZB Cliahub Consortium |
| EPI_ISL_1235570 | CA DPH Viral and Rickettsial Disease Laboratory | Chan-Zuckerberg Biohub | CZB Cliahub Consortium |
| EPI_ISL_1235571 | Alameda County Public Health Lab | Chan-Zuckerberg Biohub | CZB Cliahub Consortium |

| | | | |
|---|---|---|---|
| EPI_ISL_1235572 | CA DPH Viral and Rickettsial Disease Laboratory | Chan-Zuckerberg Biohub | CZB Cliahub Consortium |
| EPI_ISL_1235573, EPI_ISL_1235574, EPI_ISL_1235575, EPI_ISL_1235576, EPI_ISL_1235577 | Alameda County Public Health Lab | Chan-Zuckerberg Biohub | CZB Cliahub Consortium |
| EPI_ISL_1235578 | CA DPH Viral and Rickettsial Disease Laboratory | Chan-Zuckerberg Biohub | CZB Cliahub Consortium |
| EPI_ISL_1235579, EPI_ISL_1235580 | Alameda County Public Health Lab | Chan-Zuckerberg Biohub | CZB Cliahub Consortium |
| EPI_ISL_1235581 | Santa Clara County Public Health Laboratory | Chan-Zuckerberg Biohub | CZB Cliahub Consortium |
| EPI_ISL_1235582 | CA DPH Viral and Rickettsial Disease Laboratory | Chan-Zuckerberg Biohub | CZB Cliahub Consortium |
| EPI_ISL_1235583 | Santa Clara County Public Health Laboratory | Chan-Zuckerberg Biohub | CZB Cliahub Consortium |
| EPI_ISL_1235585, EPI_ISL_1235586, EPI_ISL_1235587 | Alameda County Public Health Lab | Chan-Zuckerberg Biohub | CZB Cliahub Consortium |
| EPI_ISL_1235588 | CA DPH Viral and Rickettsial Disease Laboratory | Chan-Zuckerberg Biohub | CZB Cliahub Consortium |
| EPI_ISL_1235589, EPI_ISL_1235590 | Alameda County Public Health Lab | Chan-Zuckerberg Biohub | CZB Cliahub Consortium |
| EPI_ISL_1235591 | CA DPH Viral and Rickettsial Disease Laboratory | Chan-Zuckerberg Biohub | CZB Cliahub Consortium |
| EPI_ISL_1235592 | Alameda County Public Health Lab | Chan-Zuckerberg Biohub | CZB Cliahub Consortium |
| EPI_ISL_1235593 | Santa Clara County Public Health Laboratory | Chan-Zuckerberg Biohub | CZB Cliahub Consortium |
| EPI_ISL_1235594, EPI_ISL_1235595 | Alameda County Public Health Lab | Chan-Zuckerberg Biohub | CZB Cliahub Consortium |
| EPI_ISL_1235596 | CA DPH Viral and Rickettsial Disease Laboratory | Chan-Zuckerberg Biohub | CZB Cliahub Consortium |
| EPI_ISL_1235597, EPI_ISL_1235598, EPI_ISL_1235599 | Alameda County Public Health Lab | Chan-Zuckerberg Biohub | CZB Cliahub Consortium |
| EPI_ISL_1235600 | CA DPH Viral and Rickettsial Disease Laboratory | Chan-Zuckerberg Biohub | CZB Cliahub Consortium |
| EPI_ISL_1235601 | Santa Clara County Public Health Laboratory | Chan-Zuckerberg Biohub | CZB Cliahub Consortium |
| EPI_ISL_1235602 | CA DPH Viral and Rickettsial Disease Laboratory | Chan-Zuckerberg Biohub | CZB Cliahub Consortium |
| EPI_ISL_1235603 | Alameda County Public Health Lab | Chan-Zuckerberg Biohub | CZB Cliahub Consortium |
| EPI_ISL_1235604 | CA DPH Viral and Rickettsial Disease Laboratory | Chan-Zuckerberg Biohub | CZB Cliahub Consortium |
| EPI_ISL_1235605 | Alameda County Public Health Lab | Chan-Zuckerberg Biohub | CZB Cliahub Consortium |
| EPI_ISL_1235606 | CA DPH Viral and Rickettsial Disease Laboratory | Chan-Zuckerberg Biohub | CZB Cliahub Consortium |
| EPI_ISL_1235607 | Fresno County Public Health Laboratory | Chan-Zuckerberg Biohub | CZB Cliahub Consortium |
| EPI_ISL_1235608 | CA DPH Viral and Rickettsial Disease Laboratory | Chan-Zuckerberg Biohub | CZB Cliahub Consortium |
| EPI_ISL_1235609, EPI_ISL_1235610, EPI_ISL_1235611 | Alameda County Public Health Lab | Chan-Zuckerberg Biohub | CZB Cliahub Consortium |
| EPI_ISL_1235612 | Santa Clara County Public Health Laboratory | Chan-Zuckerberg Biohub | CZB Cliahub Consortium |
| EPI_ISL_1235613, EPI_ISL_1235614, EPI_ISL_1235615, EPI_ISL_1235616, EPI_ISL_1235617, EPI_ISL_1235618, EPI_ISL_1235619, EPI_ISL_1235621, EPI_ISL_1235622, EPI_ISL_1235623, EPI_ISL_1235624, EPI_ISL_1235625, EPI_ISL_1235626, EPI_ISL_1235627, EPI_ISL_1235629, EPI_ISL_1235630, EPI_ISL_1235632 | | | |
| see above | University of Wisconsin-Madison AIDS Vaccine Research Laboratories | University of Wisconsin-Madison AIDS Vaccine Research Laboratories | Gage Moreno, Katarina Braun, et al. AIDS Vaccine Research Laboratories |
| EPI_ISL_1235633, EPI_ISL_1235634, EPI_ISL_1235635, EPI_ISL_1235636, EPI_ISL_1235638, EPI_ISL_1235639, EPI_ISL_1235640, EPI_ISL_1235641 | Santa Clara County Public Health Laboratory | Santa Clara County Public Health Laboratory | Santa Clara County Public Health Department |
| EPI_ISL_1235668, EPI_ISL_1235669, EPI_ISL_1235670, EPI_ISL_1235671, EPI_ISL_1235672, EPI_ISL_1235673, EPI_ISL_1235674, EPI_ISL_1235675, EPI_ISL_1235676, EPI_ISL_1235677 | INNO Diagnostics Reference Laboratory | RCMI-Center for Research Resources, Ponce Research Institute | Vanessa Rivera-Amill, Andrea Arias-García, Raphael Sánchez-Torres |
| EPI_ISL_1235678, EPI_ISL_1235679 | Baylor Scott & White-Temple | Baylor Scott & White-Temple | Ari Rao, Linden Morales, Kimberly Walker, Marcus Volz, Shelby Hendrickson, Caitlin Maloney |
| EPI_ISL_1235680 | Baylor Scott & White-Irving | Baylor Scott & White-Temple | Ari Rao, Linden Morales, Kimberly Walker, Marcus Volz, Shelby Hendrickson, Caitlin Maloney |
| EPI_ISL_1235681, EPI_ISL_1235682, EPI_ISL_1235683, EPI_ISL_1235684, EPI_ISL_1235685 | Baylor Scott & White-Temple | Baylor Scott & White-Temple | Ari Rao, Linden Morales, Kimberly Walker, Marcus Volz, Shelby Hendrickson, Caitlin Maloney |
| EPI_ISL_1235691, EPI_ISL_1235692, EPI_ISL_1235693, EPI_ISL_1235694, EPI_ISL_1235695, EPI_ISL_1235696, EPI_ISL_1235697, EPI_ISL_1235698, EPI_ISL_1235699, EPI_ISL_1235700, EPI_ISL_1235701, EPI_ISL_1235702, EPI_ISL_1235703, EPI_ISL_1235704, EPI_ISL_1235705, EPI_ISL_1235706, EPI_ISL_1235707, EPI_ISL_1235708, EPI_ISL_1235709, EPI_ISL_1235710, EPI_ISL_1235711, EPI_ISL_1235712, EPI_ISL_1235713, EPI_ISL_1235714, EPI_ISL_1235715, EPI_ISL_1235716, EPI_ISL_1235717, EPI_ISL_1235718, EPI_ISL_1235719, EPI_ISL_1235720, EPI_ISL_1235721, EPI_ISL_1235722, EPI_ISL_1235723, EPI_ISL_1235724, EPI_ISL_1235725, EPI_ISL_1235726, EPI_ISL_1235727, EPI_ISL_1235728, EPI_ISL_1235729, EPI_ISL_1235730, EPI_ISL_1235731, EPI_ISL_1235732, EPI_ISL_1235733, EPI_ISL_1235734, EPI_ISL_1235735, EPI_ISL_1235736, EPI_ISL_1235737, EPI_ISL_1235738, EPI_ISL_1235739, EPI_ISL_1235740, EPI_ISL_1235741, EPI_ISL_1235742, EPI_ISL_1235743, EPI_ISL_1235744, EPI_ISL_1235745, EPI_ISL_1235746, EPI_ISL_1235747, EPI_ISL_1235748, EPI_ISL_1235749, EPI_ISL_1235750, EPI_ISL_1235751, EPI_ISL_1235752, EPI_ISL_1235753, EPI_ISL_1235754, EPI_ISL_1235755, EPI_ISL_1235756, EPI_ISL_1235757, EPI_ISL_1235758, EPI_ISL_1235759, EPI_ISL_1235760, EPI_ISL_1235761, EPI_ISL_1235762, EPI_ISL_1235763, EPI_ISL_1235764, EPI_ISL_1235765, EPI_ISL_1235766, EPI_ISL_1235767, EPI_ISL_1235768, EPI_ISL_1235769, EPI_ISL_1235770, EPI_ISL_1235771, EPI_ISL_1235772, EPI_ISL_1235773, EPI_ISL_1235774, EPI_ISL_1235775, EPI_ISL_1235776, EPI_ISL_1235777, EPI_ISL_1235778, EPI_ISL_1235779, EPI_ISL_1235780, EPI_ISL_1235781, EPI_ISL_1235782, EPI_ISL_1235783, EPI_ISL_1235784, EPI_ISL_1235785, EPI_ISL_1235786, EPI_ISL_1235787, EPI_ISL_1235788, EPI_ISL_1235789, EPI_ISL_1235790, EPI_ISL_1235791, EPI_ISL_1235792, EPI_ISL_1235793, EPI_ISL_1235794, EPI_ISL_1235795, EPI_ISL_1235796, EPI_ISL_1235797, EPI_ISL_1235798, EPI_ISL_1235799, EPI_ISL_1235800, EPI_ISL_1235801, EPI_ISL_1235802, EPI_ISL_1235803, EPI_ISL_1235804, EPI_ISL_1235805, EPI_ISL_1235806, EPI_ISL_1235807, EPI_ISL_1235808, EPI_ISL_1235809, EPI_ISL_1235810, EPI_ISL_1235811, EPI_ISL_1235812, EPI_ISL_1235813, EPI_ISL_1235814, EPI_ISL_1235815, EPI_ISL_1235816, EPI_ISL_1235817, EPI_ISL_1235818, EPI_ISL_1235819, EPI_ISL_1235820, EPI_ISL_1235821, EPI_ISL_1235822, EPI_ISL_1235823, EPI_ISL_1235824, EPI_ISL_1235825, EPI_ISL_1235826, EPI_ISL_1235827, EPI_ISL_1235828, EPI_ISL_1235829, EPI_ISL_1235830, EPI_ISL_1235831, EPI_ISL_1235832, EPI_ISL_1235833, EPI_ISL_1235834, EPI_ISL_1235835, EPI_ISL_1235836, EPI_ISL_1235837, EPI_ISL_1235838, EPI_ISL_1235839, EPI_ISL_1235840, EPI_ISL_1235841, EPI_ISL_1235842, EPI_ISL_1235843, EPI_ISL_1235844, EPI_ISL_1235845, EPI_ISL_1235846, EPI_ISL_1235847, EPI_ISL_1235848, EPI_ISL_1235849, EPI_ISL_1235850, EPI_ISL_1235851, EPI_ISL_1235852, EPI_ISL_1235853, EPI_ISL_1235854, EPI_ISL_1235855, EPI_ISL_1235856, EPI_ISL_1235857, EPI_ISL_1235858, EPI_ISL_1235859, EPI_ISL_1235860, EPI_ISL_1235861, EPI_ISL_1235862, EPI_ISL_1235863, EPI_ISL_1235864, EPI_ISL_1235865, EPI_ISL_1235866, EPI_ISL_1235867, EPI_ISL_1235868, EPI_ISL_1235869, EPI_ISL_1235870, EPI_ISL_1235871, EPI_ISL_1235872, EPI_ISL_1235873, EPI_ISL_1235874, EPI_ISL_1235875, EPI_ISL_1235876, EPI_ISL_1235877, EPI_ISL_1235878, EPI_ISL_1235879, EPI_ISL_1235880, EPI_ISL_1235881, EPI_ISL_1235882, EPI_ISL_1235883, EPI_ISL_1235884, EPI_ISL_1235885, EPI_ISL_1235886, EPI_ISL_1235887, EPI_ISL_1235888, | | | |

EPI_ISL_1235889, EPI_ISL_1235890, EPI_ISL_1235891, EPI_ISL_1235892, EPI_ISL_1235893, EPI_ISL_1235894, EPI_ISL_1235895, EPI_ISL_1235896, EPI_ISL_1235897, EPI_ISL_1235898, EPI_ISL_1235899, EPI_ISL_1235900, EPI_ISL_1235901, EPI_ISL_1235902, EPI_ISL_1235903, EPI_ISL_1235904, EPI_ISL_1235905, EPI_ISL_1235906, EPI_ISL_1235907, EPI_ISL_1235908, EPI_ISL_1235909, EPI_ISL_1235910,
EPI_ISL_1235911, EPI_ISL_1235912, EPI_ISL_1235913, EPI_ISL_1235914, EPI_ISL_1235915, EPI_ISL_1235916, EPI_ISL_1235917, EPI_ISL_1235918, EPI_ISL_1235919, EPI_ISL_1235920, EPI_ISL_1235921, EPI_ISL_1235922, EPI_ISL_1235923, EPI_ISL_1235924, EPI_ISL_1235925, EPI_ISL_1235926, EPI_ISL_1235927, EPI_ISL_1235928, EPI_ISL_1235929, EPI_ISL_1235930, EPI_ISL_1235931, EPI_ISL_1235932,
EPI_ISL_1235933, EPI_ISL_1235934, EPI_ISL_1235935, EPI_ISL_1235936, EPI_ISL_1235937, EPI_ISL_1235938, EPI_ISL_1235939, EPI_ISL_1235940, EPI_ISL_1235941, EPI_ISL_1235942, EPI_ISL_1235943, EPI_ISL_1235944, EPI_ISL_1235945, EPI_ISL_1235946, EPI_ISL_1235947, EPI_ISL_1235948, EPI_ISL_1235949, EPI_ISL_1235950, EPI_ISL_1235951, EPI_ISL_1235952, EPI_ISL_1235953, EPI_ISL_1235954,
EPI_ISL_1235955, EPI_ISL_1235956, EPI_ISL_1235957, EPI_ISL_1235958, EPI_ISL_1235959, EPI_ISL_1235960, EPI_ISL_1235961, EPI_ISL_1235962, EPI_ISL_1235963, EPI_ISL_1235964, EPI_ISL_1235965, EPI_ISL_1235966, EPI_ISL_1235967, EPI_ISL_1235968, EPI_ISL_1235969, EPI_ISL_1235970, EPI_ISL_1235971, EPI_ISL_1235972, EPI_ISL_1235973, EPI_ISL_1235974, EPI_ISL_1235975, EPI_ISL_1235976,
EPI_ISL_1235977, EPI_ISL_1235978, EPI_ISL_1235979, EPI_ISL_1235980, EPI_ISL_1235981, EPI_ISL_1235982, EPI_ISL_1235983, EPI_ISL_1235984, EPI_ISL_1235985, EPI_ISL_1235986, EPI_ISL_1235987, EPI_ISL_1235988, EPI_ISL_1235989, EPI_ISL_1235990, EPI_ISL_1235991, EPI_ISL_1235992, EPI_ISL_1235993, EPI_ISL_1235994, EPI_ISL_1235995, EPI_ISL_1235996, EPI_ISL_1235997, EPI_ISL_1235998,
EPI_ISL_1235999, EPI_ISL_1236000, EPI_ISL_1236001, EPI_ISL_1236002, EPI_ISL_1236003, EPI_ISL_1236004, EPI_ISL_1236005, EPI_ISL_1236006, EPI_ISL_1236007, EPI_ISL_1236008, EPI_ISL_1236009, EPI_ISL_1236010, EPI_ISL_1236011, EPI_ISL_1236012, EPI_ISL_1236013, EPI_ISL_1236014, EPI_ISL_1236015, EPI_ISL_1236016, EPI_ISL_1236017, EPI_ISL_1236018, EPI_ISL_1236019, EPI_ISL_1236020,
EPI_ISL_1236021, EPI_ISL_1236022, EPI_ISL_1236023, EPI_ISL_1236024, EPI_ISL_1236025, EPI_ISL_1236026, EPI_ISL_1236027, EPI_ISL_1236028, EPI_ISL_1236029, EPI_ISL_1236030, EPI_ISL_1236031, EPI_ISL_1236032, EPI_ISL_1236033, EPI_ISL_1236034, EPI_ISL_1236035, EPI_ISL_1236036, EPI_ISL_1236037, EPI_ISL_1236038, EPI_ISL_1236039, EPI_ISL_1236040, EPI_ISL_1236041, EPI_ISL_1236042,
EPI_ISL_1236043, EPI_ISL_1236044, EPI_ISL_1236045, EPI_ISL_1236046, EPI_ISL_1236047, EPI_ISL_1236048, EPI_ISL_1236049, EPI_ISL_1236050, EPI_ISL_1236051, EPI_ISL_1236052, EPI_ISL_1236053, EPI_ISL_1236054, EPI_ISL_1236055, EPI_ISL_1236056, EPI_ISL_1236057, EPI_ISL_1236058, EPI_ISL_1236059, EPI_ISL_1236060, EPI_ISL_1236061, EPI_ISL_1236062, EPI_ISL_1236063, EPI_ISL_1236064,
EPI_ISL_1236065, EPI_ISL_1236066, EPI_ISL_1236067, EPI_ISL_1236068, EPI_ISL_1236069, EPI_ISL_1236070, EPI_ISL_1236071, EPI_ISL_1236072, EPI_ISL_1236073, EPI_ISL_1236074, EPI_ISL_1236075, EPI_ISL_1236076, EPI_ISL_1236077, EPI_ISL_1236078, EPI_ISL_1236079, EPI_ISL_1236080, EPI_ISL_1236081, EPI_ISL_1236082, EPI_ISL_1236083, EPI_ISL_1236084, EPI_ISL_1236085, EPI_ISL_1236086,
EPI_ISL_1236087, EPI_ISL_1236088, EPI_ISL_1236089, EPI_ISL_1236090, EPI_ISL_1236091, EPI_ISL_1236092, EPI_ISL_1236093, EPI_ISL_1236094, EPI_ISL_1236095, EPI_ISL_1236096, EPI_ISL_1236097, EPI_ISL_1236098, EPI_ISL_1236099, EPI_ISL_1236100, EPI_ISL_1236101, EPI_ISL_1236102, EPI_ISL_1236103, EPI_ISL_1236104, EPI_ISL_1236105, EPI_ISL_1236106, EPI_ISL_1236107, EPI_ISL_1236108,
EPI_ISL_1236109, EPI_ISL_1236110, EPI_ISL_1236111, EPI_ISL_1236112, EPI_ISL_1236113, EPI_ISL_1236114, EPI_ISL_1236115, EPI_ISL_1236116, EPI_ISL_1236117, EPI_ISL_1236118, EPI_ISL_1236119, EPI_ISL_1236120, EPI_ISL_1236121, EPI_ISL_1236122, EPI_ISL_1236123, EPI_ISL_1236124, EPI_ISL_1236125, EPI_ISL_1236126, EPI_ISL_1236127, EPI_ISL_1236128, EPI_ISL_1236129, EPI_ISL_1236130,
EPI_ISL_1236131, EPI_ISL_1236132, EPI_ISL_1236133, EPI_ISL_1236134, EPI_ISL_1236135, EPI_ISL_1236136, EPI_ISL_1236137, EPI_ISL_1236138, EPI_ISL_1236139, EPI_ISL_1236140, EPI_ISL_1236141, EPI_ISL_1236142, EPI_ISL_1236143, EPI_ISL_1236144, EPI_ISL_1236145, EPI_ISL_1236146, EPI_ISL_1236147, EPI_ISL_1236148, EPI_ISL_1236149, EPI_ISL_1236150, EPI_ISL_1236151, EPI_ISL_1236152,
EPI_ISL_1236153, EPI_ISL_1236154, EPI_ISL_1236155, EPI_ISL_1236156, EPI_ISL_1236157, EPI_ISL_1236158, EPI_ISL_1236159, EPI_ISL_1236160, EPI_ISL_1236161, EPI_ISL_1236162, EPI_ISL_1236163, EPI_ISL_1236164, EPI_ISL_1236165, EPI_ISL_1236166, EPI_ISL_1236167, EPI_ISL_1236168, EPI_ISL_1236169, EPI_ISL_1236170, EPI_ISL_1236171, EPI_ISL_1236172, EPI_ISL_1236173, EPI_ISL_1236174,
EPI_ISL_1236175, EPI_ISL_1236176, EPI_ISL_1236177, EPI_ISL_1236178, EPI_ISL_1236179, EPI_ISL_1236180, EPI_ISL_1236181, EPI_ISL_1236182, EPI_ISL_1236183, EPI_ISL_1236184, EPI_ISL_1236185, EPI_ISL_1236186, EPI_ISL_1236187, EPI_ISL_1236188, EPI_ISL_1236189, EPI_ISL_1236190, EPI_ISL_1236191, EPI_ISL_1236192, EPI_ISL_1236193, EPI_ISL_1236194, EPI_ISL_1236195, EPI_ISL_1236196,
EPI_ISL_1236197, EPI_ISL_1236198, EPI_ISL_1236199, EPI_ISL_1236200, EPI_ISL_1236201, EPI_ISL_1236202, EPI_ISL_1236203, EPI_ISL_1236204, EPI_ISL_1236205, EPI_ISL_1236206, EPI_ISL_1236207, EPI_ISL_1236208, EPI_ISL_1236209, EPI_ISL_1236210, EPI_ISL_1236211, EPI_ISL_1236212, EPI_ISL_1236213, EPI_ISL_1236214, EPI_ISL_1236215, EPI_ISL_1236216, EPI_ISL_1236217, EPI_ISL_1236218,
EPI_ISL_1236219, EPI_ISL_1236220, EPI_ISL_1236221, EPI_ISL_1236222, EPI_ISL_1236223, EPI_ISL_1236224, EPI_ISL_1236225, EPI_ISL_1236226, EPI_ISL_1236227, EPI_ISL_1236228, EPI_ISL_1236229, EPI_ISL_1236230, EPI_ISL_1236231, EPI_ISL_1236232, EPI_ISL_1236233, EPI_ISL_1236234, EPI_ISL_1236235, EPI_ISL_1236236, EPI_ISL_1236237, EPI_ISL_1236238, EPI_ISL_1236239, EPI_ISL_1236240,
EPI_ISL_1236241, EPI_ISL_1236242, EPI_ISL_1236243, EPI_ISL_1236244, EPI_ISL_1236245, EPI_ISL_1236246, EPI_ISL_1236247, EPI_ISL_1236248, EPI_ISL_1236249, EPI_ISL_1236250, EPI_ISL_1236251, EPI_ISL_1236252, EPI_ISL_1236253, EPI_ISL_1236254, EPI_ISL_1236255, EPI_ISL_1236256, EPI_ISL_1236257, EPI_ISL_1236258, EPI_ISL_1236259, EPI_ISL_1236260, EPI_ISL_1236261, EPI_ISL_1236262,
EPI_ISL_1236263, EPI_ISL_1236264, EPI_ISL_1236265, EPI_ISL_1236266, EPI_ISL_1236267, EPI_ISL_1236268, EPI_ISL_1236269, EPI_ISL_1236270, EPI_ISL_1236271, EPI_ISL_1236272, EPI_ISL_1236273, EPI_ISL_1236274, EPI_ISL_1236275, EPI_ISL_1236276, EPI_ISL_1236277, EPI_ISL_1236278, EPI_ISL_1236279, EPI_ISL_1236280, EPI_ISL_1236281, EPI_ISL_1236282, EPI_ISL_1236283, EPI_ISL_1236284,
EPI_ISL_1236285, EPI_ISL_1236286, EPI_ISL_1236287, EPI_ISL_1236288, EPI_ISL_1236289, EPI_ISL_1236290, EPI_ISL_1236291, EPI_ISL_1236292, EPI_ISL_1236293, EPI_ISL_1236294, EPI_ISL_1236295, EPI_ISL_1236296, EPI_ISL_1236297, EPI_ISL_1236298, EPI_ISL_1236299, EPI_ISL_1236300, EPI_ISL_1236301, EPI_ISL_1236302, EPI_ISL_1236303, EPI_ISL_1236304, EPI_ISL_1236305, EPI_ISL_1236306,
EPI_ISL_1236307, EPI_ISL_1236308, EPI_ISL_1236309, EPI_ISL_1236310, EPI_ISL_1236311, EPI_ISL_1236312, EPI_ISL_1236313, EPI_ISL_1236314, EPI_ISL_1236315, EPI_ISL_1236316, EPI_ISL_1236317, EPI_ISL_1236318, EPI_ISL_1236319, EPI_ISL_1236320, EPI_ISL_1236321, EPI_ISL_1236322, EPI_ISL_1236323, EPI_ISL_1236324, EPI_ISL_1236325, EPI_ISL_1236326, EPI_ISL_1236327, EPI_ISL_1236328,
EPI_ISL_1236329, EPI_ISL_1236330, EPI_ISL_1236331, EPI_ISL_1236332, EPI_ISL_1236333, EPI_ISL_1236334, EPI_ISL_1236335, EPI_ISL_1236336, EPI_ISL_1236337, EPI_ISL_1236338, EPI_ISL_1236339, EPI_ISL_1236340, EPI_ISL_1236341, EPI_ISL_1236342, EPI_ISL_1236343, EPI_ISL_1236344, EPI_ISL_1236345, EPI_ISL_1236346, EPI_ISL_1236347, EPI_ISL_1236348, EPI_ISL_1236349, EPI_ISL_1236350,
EPI_ISL_1236351, EPI_ISL_1236352, EPI_ISL_1236353, EPI_ISL_1236354, EPI_ISL_1236355, EPI_ISL_1236356, EPI_ISL_1236357, EPI_ISL_1236358, EPI_ISL_1236359, EPI_ISL_1236360, EPI_ISL_1236361, EPI_ISL_1236362, EPI_ISL_1236363, EPI_ISL_1236364, EPI_ISL_1236365, EPI_ISL_1236366, EPI_ISL_1236367, EPI_ISL_1236368, EPI_ISL_1236369, EPI_ISL_1236370, EPI_ISL_1236371, EPI_ISL_1236372,
EPI_ISL_1236373, EPI_ISL_1236374, EPI_ISL_1236375, EPI_ISL_1236376, EPI_ISL_1236377, EPI_ISL_1236378, EPI_ISL_1236379, EPI_ISL_1236380, EPI_ISL_1236381, EPI_ISL_1236382, EPI_ISL_1236383, EPI_ISL_1236384, EPI_ISL_1236385, EPI_ISL_1236386, EPI_ISL_1236387, EPI_ISL_1236388, EPI_ISL_1236389, EPI_ISL_1236390, EPI_ISL_1236391, EPI_ISL_1236392, EPI_ISL_1236393, EPI_ISL_1236394,
EPI_ISL_1236395, EPI_ISL_1236396, EPI_ISL_1236397, EPI_ISL_1236398, EPI_ISL_1236399, EPI_ISL_1236400, EPI_ISL_1236401, EPI_ISL_1236402, EPI_ISL_1236403, EPI_ISL_1236404, EPI_ISL_1236405, EPI_ISL_1236406, EPI_ISL_1236407, EPI_ISL_1236408, EPI_ISL_1236409, EPI_ISL_1236410, EPI_ISL_1236411, EPI_ISL_1236412, EPI_ISL_1236413, EPI_ISL_1236414, EPI_ISL_1236415, EPI_ISL_1236416,
EPI_ISL_1236417, EPI_ISL_1236418, EPI_ISL_1236419, EPI_ISL_1236420, EPI_ISL_1236421, EPI_ISL_1236422, EPI_ISL_1236423, EPI_ISL_1236424, EPI_ISL_1236425, EPI_ISL_1236426, EPI_ISL_1236427, EPI_ISL_1236428, EPI_ISL_1236429, EPI_ISL_1236430, EPI_ISL_1236431, EPI_ISL_1236432, EPI_ISL_1236433, EPI_ISL_1236434, EPI_ISL_1236435, EPI_ISL_1236436, EPI_ISL_1236437, EPI_ISL_1236438,
EPI_ISL_1236439, EPI_ISL_1236440, EPI_ISL_1236441, EPI_ISL_1236442, EPI_ISL_1236443, EPI_ISL_1236444, EPI_ISL_1236446, EPI_ISL_1236447, EPI_ISL_1236448, EPI_ISL_1236449, EPI_ISL_1236450, EPI_ISL_1236451, EPI_ISL_1236452, EPI_ISL_1236453, EPI_ISL_1236454, EPI_ISL_1236455, EPI_ISL_1236456, EPI_ISL_1236457, EPI_ISL_1236458, EPI_ISL_1236459, EPI_ISL_1236460, EPI_ISL_1236461,
EPI_ISL_1236462, EPI_ISL_1236463, EPI_ISL_1236464, EPI_ISL_1236465, EPI_ISL_1236466, EPI_ISL_1236467, EPI_ISL_1236468, EPI_ISL_1236469, EPI_ISL_1236470, EPI_ISL_1236471, EPI_ISL_1236472, EPI_ISL_1236473, EPI_ISL_1236474, EPI_ISL_1236475, EPI_ISL_1236476, EPI_ISL_1236477, EPI_ISL_1236478, EPI_ISL_1236479, EPI_ISL_1236480, EPI_ISL_1236481, EPI_ISL_1236482, EPI_ISL_1236483,
EPI_ISL_1236484, EPI_ISL_1236485, EPI_ISL_1236486, EPI_ISL_1236487, EPI_ISL_1236488, EPI_ISL_1236489, EPI_ISL_1236490, EPI_ISL_1236491, EPI_ISL_1236492, EPI_ISL_1236493, EPI_ISL_1236494, EPI_ISL_1236495, EPI_ISL_1236496, EPI_ISL_1236497, EPI_ISL_1236498, EPI_ISL_1236499, EPI_ISL_1236500, EPI_ISL_1236501, EPI_ISL_1236502, EPI_ISL_1236503, EPI_ISL_1236504, EPI_ISL_1236505,
EPI_ISL_1236506, EPI_ISL_1236507, EPI_ISL_1236508, EPI_ISL_1236509, EPI_ISL_1236510, EPI_ISL_1236511, EPI_ISL_1236512, EPI_ISL_1236513, EPI_ISL_1236514, EPI_ISL_1236515, EPI_ISL_1236516, EPI_ISL_1236517, EPI_ISL_1236518, EPI_ISL_1236519, EPI_ISL_1236520, EPI_ISL_1236521, EPI_ISL_1236522, EPI_ISL_1236523, EPI_ISL_1236524, EPI_ISL_1236525, EPI_ISL_1236526, EPI_ISL_1236527,
EPI_ISL_1236528, EPI_ISL_1236529, EPI_ISL_1236530, EPI_ISL_1236531, EPI_ISL_1236532, EPI_ISL_1236533, EPI_ISL_1236534, EPI_ISL_1236535, EPI_ISL_1236536, EPI_ISL_1236537, EPI_ISL_1236538, EPI_ISL_1236539, EPI_ISL_1236540, EPI_ISL_1236541, EPI_ISL_1236542, EPI_ISL_1236543, EPI_ISL_1236544, EPI_ISL_1236545, EPI_ISL_1236546, EPI_ISL_1236547, EPI_ISL_1236548, EPI_ISL_1236549,
EPI_ISL_1236550, EPI_ISL_1236551, EPI_ISL_1236552, EPI_ISL_1236553, EPI_ISL_1236554, EPI_ISL_1236555, EPI_ISL_1236556, EPI_ISL_1236557, EPI_ISL_1236558, EPI_ISL_1236559, EPI_ISL_1236560, EPI_ISL_1236561, EPI_ISL_1236562, EPI_ISL_1236563, EPI_ISL_1236564, EPI_ISL_1236565, EPI_ISL_1236566, EPI_ISL_1236567, EPI_ISL_1236568, EPI_ISL_1236569, EPI_ISL_1236570, EPI_ISL_1236571,
EPI_ISL_1236572, EPI_ISL_1236573, EPI_ISL_1236574, EPI_ISL_1236575, EPI_ISL_1236576, EPI_ISL_1236577, EPI_ISL_1236578, EPI_ISL_1236579, EPI_ISL_1236580, EPI_ISL_1236581, EPI_ISL_1236582, EPI_ISL_1236583, EPI_ISL_1236584, EPI_ISL_1236585, EPI_ISL_1236586, EPI_ISL_1236587, EPI_ISL_1236588, EPI_ISL_1236589, EPI_ISL_1236590, EPI_ISL_1236591, EPI_ISL_1236592, EPI_ISL_1236593,
EPI_ISL_1236594, EPI_ISL_1236595, EPI_ISL_1236596, EPI_ISL_1236597, EPI_ISL_1236598, EPI_ISL_1236599, EPI_ISL_1236600, EPI_ISL_1236601, EPI_ISL_1236602, EPI_ISL_1236603, EPI_ISL_1236604, EPI_ISL_1236605, EPI_ISL_1236606, EPI_ISL_1236607, EPI_ISL_1236608, EPI_ISL_1236609, EPI_ISL_1236610, EPI_ISL_1236611, EPI_ISL_1236612, EPI_ISL_1236613, EPI_ISL_1236614, EPI_ISL_1236615,
EPI_ISL_1236616, EPI_ISL_1236617, EPI_ISL_1236618, EPI_ISL_1236619, EPI_ISL_1236620, EPI_ISL_1236621, EPI_ISL_1236622, EPI_ISL_1236623, EPI_ISL_1236624, EPI_ISL_1236625, EPI_ISL_1236626, EPI_ISL_1236627, EPI_ISL_1236628, EPI_ISL_1236629, EPI_ISL_1236630, EPI_ISL_1236631, EPI_ISL_1236632, EPI_ISL_1236633, EPI_ISL_1236634, EPI_ISL_1236635, EPI_ISL_1236636, EPI_ISL_1236637,
EPI_ISL_1236638, EPI_ISL_1236639, EPI_ISL_1236640, EPI_ISL_1236641, EPI_ISL_1236642, EPI_ISL_1236643, EPI_ISL_1236644, EPI_ISL_1236645, EPI_ISL_1236646, EPI_ISL_1236647, EPI_ISL_1236648, EPI_ISL_1236649, EPI_ISL_1236650, EPI_ISL_1236651, EPI_ISL_1236652, EPI_ISL_1236653, EPI_ISL_1236654, EPI_ISL_1236655, EPI_ISL_1236656, EPI_ISL_1236657, EPI_ISL_1236658, EPI_ISL_1236659,
EPI_ISL_1236660, EPI_ISL_1236661, EPI_ISL_1236662, EPI_ISL_1236663, EPI_ISL_1236664, EPI_ISL_1236665, EPI_ISL_1236666, EPI_ISL_1236667, EPI_ISL_1236668, EPI_ISL_1236669, EPI_ISL_1236670, EPI_ISL_1236671, EPI_ISL_1236672, EPI_ISL_1236673, EPI_ISL_1236674, EPI_ISL_1236675, EPI_ISL_1236676, EPI_ISL_1236677, EPI_ISL_1236678, EPI_ISL_1236679, EPI_ISL_1236680, EPI_ISL_1236681,
EPI_ISL_1236682, EPI_ISL_1236683, EPI_ISL_1236684, EPI_ISL_1236685, EPI_ISL_1236686, EPI_ISL_1236687, EPI_ISL_1236688, EPI_ISL_1236689, EPI_ISL_1236690, EPI_ISL_1236691, EPI_ISL_1236692, EPI_ISL_1236693, EPI_ISL_1236694, EPI_ISL_1236695, EPI_ISL_1236696, EPI_ISL_1236697, EPI_ISL_1236698, EPI_ISL_1236699, EPI_ISL_1236700, EPI_ISL_1236701, EPI_ISL_1236702, EPI_ISL_1236703,
EPI_ISL_1236704, EPI_ISL_1236705, EPI_ISL_1236706, EPI_ISL_1236707, EPI_ISL_1236708, EPI_ISL_1236709, EPI_ISL_1236710, EPI_ISL_1236711, EPI_ISL_1236712, EPI_ISL_1236713, EPI_ISL_1236714, EPI_ISL_1236715, EPI_ISL_1236716, EPI_ISL_1236717, EPI_ISL_1236718, EPI_ISL_1236719, EPI_ISL_1236720, EPI_ISL_1236721, EPI_ISL_1236722, EPI_ISL_1236723, EPI_ISL_1236724, EPI_ISL_1236725,
EPI_ISL_1236726, EPI_ISL_1236727, EPI_ISL_1236728, EPI_ISL_1236729, EPI_ISL_1236730, EPI_ISL_1236731, EPI_ISL_1236732, EPI_ISL_1236733, EPI_ISL_1236734, EPI_ISL_1236735, EPI_ISL_1236736, EPI_ISL_1236737, EPI_ISL_1236738, EPI_ISL_1236739, EPI_ISL_1236740, EPI_ISL_1236741, EPI_ISL_1236742, EPI_ISL_1236743, EPI_ISL_1236744, EPI_ISL_1236745, EPI_ISL_1236746, EPI_ISL_1236747,
EPI_ISL_1236748, EPI_ISL_1236749, EPI_ISL_1236750, EPI_ISL_1236751, EPI_ISL_1236752, EPI_ISL_1236753, EPI_ISL_1236754, EPI_ISL_1236755, EPI_ISL_1236756, EPI_ISL_1236757, EPI_ISL_1236758, EPI_ISL_1236759, EPI_ISL_1236760, EPI_ISL_1236761, EPI_ISL_1236762, EPI_ISL_1236763, EPI_ISL_1236764, EPI_ISL_1236765, EPI_ISL_1236766, EPI_ISL_1236767, EPI_ISL_1236768, EPI_ISL_1236769,
EPI_ISL_1236770, EPI_ISL_1236771, EPI_ISL_1236772, EPI_ISL_1236773, EPI_ISL_1236774, EPI_ISL_1236775, EPI_ISL_1236776, EPI_ISL_1236777, EPI_ISL_1236778, EPI_ISL_1236779, EPI_ISL_1236780, EPI_ISL_1236781, EPI_ISL_1236782, EPI_ISL_1236783, EPI_ISL_1236784, EPI_ISL_1236785, EPI_ISL_1236786, EPI_ISL_1236787, EPI_ISL_1236788, EPI_ISL_1236789, EPI_ISL_1236790, EPI_ISL_1236791,
EPI_ISL_1236792, EPI_ISL_1236793, EPI_ISL_1236794, EPI_ISL_1236795, EPI_ISL_1236796, EPI_ISL_1236797, EPI_ISL_1236798, EPI_ISL_1236799, EPI_ISL_1236800, EPI_ISL_1236801, EPI_ISL_1236802, EPI_ISL_1236803, EPI_ISL_1236804, EPI_ISL_1236805, EPI_ISL_1236806, EPI_ISL_1236807, EPI_ISL_1236808, EPI_ISL_1236809, EPI_ISL_1236810, EPI_ISL_1236811, EPI_ISL_1236812, EPI_ISL_1236813,
EPI_ISL_1236814, EPI_ISL_1236815, EPI_ISL_1236816, EPI_ISL_1236817, EPI_ISL_1236818, EPI_ISL_1236819, EPI_ISL_1236820, EPI_ISL_1236821, EPI_ISL_1236822, EPI_ISL_1236823, EPI_ISL_1236824, EPI_ISL_1236825, EPI_ISL_1236826, EPI_ISL_1236827, EPI_ISL_1236828, EPI_ISL_1236829, EPI_ISL_1236830, EPI_ISL_1236831, EPI_ISL_1236832, EPI_ISL_1236833, EPI_ISL_1236834, EPI_ISL_1236835,
EPI_ISL_1236836, EPI_ISL_1236837, EPI_ISL_1236838, EPI_ISL_1236839, EPI_ISL_1236840, EPI_ISL_1236841, EPI_ISL_1236842, EPI_ISL_1236843, EPI_ISL_1236844, EPI_ISL_1236845, EPI_ISL_1236846, EPI_ISL_1236847, EPI_ISL_1236848, EPI_ISL_1236849, EPI_ISL_1236850, EPI_ISL_1236851, EPI_ISL_1236852, EPI_ISL_1236853, EPI_ISL_1236854, EPI_ISL_1236855, EPI_ISL_1236856, EPI_ISL_1236857,
EPI_ISL_1236858, EPI_ISL_1236859, EPI_ISL_1236860, EPI_ISL_1236861, EPI_ISL_1236862, EPI_ISL_1236863, EPI_ISL_1236864, EPI_ISL_1236865, EPI_ISL_1236866, EPI_ISL_1236867, EPI_ISL_1236868, EPI_ISL_1236869, EPI_ISL_1236870, EPI_ISL_1236871, EPI_ISL_1236872, EPI_ISL_1236873, EPI_ISL_1236874, EPI_ISL_1236875, EPI_ISL_1236876, EPI_ISL_1236877, EPI_ISL_1236878, EPI_ISL_1236879,
EPI_ISL_1236880, EPI_ISL_1236881, EPI_ISL_1236882, EPI_ISL_1236883, EPI_ISL_1236884, EPI_ISL_1236885, EPI_ISL_1236886, EPI_ISL_1236887, EPI_ISL_1236888, EPI_ISL_1236889, EPI_ISL_1236890, EPI_ISL_1236891, EPI_ISL_1236892, EPI_ISL_1236893, EPI_ISL_1236894, EPI_ISL_1236895, EPI_ISL_1236896, EPI_ISL_1236897, EPI_ISL_1236898, EPI_ISL_1236899, EPI_ISL_1236900, EPI_ISL_1236902,
EPI_ISL_1236903, EPI_ISL_1236904, EPI_ISL_1236905, EPI_ISL_1236906, EPI_ISL_1236907, EPI_ISL_1236908, EPI_ISL_1236909, EPI_ISL_1236910, EPI_ISL_1236911, EPI_ISL_1236912, EPI_ISL_1236913, EPI_ISL_1236914, EPI_ISL_1236915, EPI_ISL_1236916, EPI_ISL_1236917, EPI_ISL_1236918, EPI_ISL_1236919, EPI_ISL_1236920, EPI_ISL_1236921, EPI_ISL_1236922, EPI_ISL_1236923, EPI_ISL_1236924,
EPI_ISL_1236925, EPI_ISL_1236926, EPI_ISL_1236927, EPI_ISL_1236928, EPI_ISL_1236929, EPI_ISL_1236930, EPI_ISL_1236931, EPI_ISL_1236932, EPI_ISL_1236933, EPI_ISL_1236934, EPI_ISL_1236935, EPI_ISL_1236936, EPI_ISL_1236937, EPI_ISL_1236938, EPI_ISL_1236939, EPI_ISL_1236940, EPI_ISL_1236941, EPI_ISL_1236942, EPI_ISL_1236943, EPI_ISL_1236944, EPI_ISL_1236945, EPI_ISL_1236946,
EPI_ISL_1236947, EPI_ISL_1236948, EPI_ISL_1236949, EPI_ISL_1236950, EPI_ISL_1236951, EPI_ISL_1236952, EPI_ISL_1236953, EPI_ISL_1236954, EPI_ISL_1236955, EPI_ISL_1236956, EPI_ISL_1236957, EPI_ISL_1236958, EPI_ISL_1236959, EPI_ISL_1236960, EPI_ISL_1236961, EPI_ISL_1236962, EPI_ISL_1236963, EPI_ISL_1236964, EPI_ISL_1236965, EPI_ISL_1236966, EPI_ISL_1236967, EPI_ISL_1236968,
EPI_ISL_1236969, EPI_ISL_1236970, EPI_ISL_1236971, EPI_ISL_1236972, EPI_ISL_1236973, EPI_ISL_1236974, EPI_ISL_1236975, EPI_ISL_1236976, EPI_ISL_1236977, EPI_ISL_1236978, EPI_ISL_1236979, EPI_ISL_1236980, EPI_ISL_1236981, EPI_ISL_1236982, EPI_ISL_1236983, EPI_ISL_1236984, EPI_ISL_1236985, EPI_ISL_1236986, EPI_ISL_1236987, EPI_ISL_1236988, EPI_ISL_1236989, EPI_ISL_1236990,
EPI_ISL_1236991, EPI_ISL_1236992, EPI_ISL_1236993, EPI_ISL_1236994, EPI_ISL_1236995, EPI_ISL_1236996, EPI_ISL_1236997, EPI_ISL_1236998, EPI_ISL_1236999, EPI_ISL_1237000, EPI_ISL_1237001, EPI_ISL_1237002, EPI_ISL_1237003, EPI_ISL_1237004, EPI_ISL_1237005, EPI_ISL_1237006, EPI_ISL_1237007, EPI_ISL_1237008, EPI_ISL_1237009, EPI_ISL_1237010, EPI_ISL_1237011, EPI_ISL_1237012,
EPI_ISL_1237013, EPI_ISL_1237014, EPI_ISL_1237015, EPI_ISL_1237016, EPI_ISL_1237017, EPI_ISL_1237018, EPI_ISL_1237019, EPI_ISL_1237020, EPI_ISL_1237021, EPI_ISL_1237022, EPI_ISL_1237023, EPI_ISL_1237024, EPI_ISL_1237025, EPI_ISL_1237026, EPI_ISL_1237027, EPI_ISL_1237028, EPI_ISL_1237029, EPI_ISL_1237030, EPI_ISL_1237031, EPI_ISL_1237032, EPI_ISL_1237033, EPI_ISL_1237034,
EPI_ISL_1237035, EPI_ISL_1237036, EPI_ISL_1237037, EPI_ISL_1237038, EPI_ISL_1237039, EPI_ISL_1237040, EPI_ISL_1237041, EPI_ISL_1237042, EPI_ISL_1237043, EPI_ISL_1237044, EPI_ISL_1237045, EPI_ISL_1237046, EPI_ISL_1237047, EPI_ISL_1237048, EPI_ISL_1237049, EPI_ISL_1237050, EPI_ISL_1237051, EPI_ISL_1237052, EPI_ISL_1237053, EPI_ISL_1237054, EPI_ISL_1237055, EPI_ISL_1237056,
EPI_ISL_1237057, EPI_ISL_1237058, EPI_ISL_1237059, EPI_ISL_1237060, EPI_ISL_1237061, EPI_ISL_1237062, EPI_ISL_1237063, EPI_ISL_1237064, EPI_ISL_1237065, EPI_ISL_1237066, EPI_ISL_1237067, EPI_ISL_1237068, EPI_ISL_1237069, EPI_ISL_1237070, EPI_ISL_1237071, EPI_ISL_1237072, EPI_ISL_1237073, EPI_ISL_1237074, EPI_ISL_1237075, EPI_ISL_1237076, EPI_ISL_1237077, EPI_ISL_1237078,
EPI_ISL_1237079, EPI_ISL_1237080, EPI_ISL_1237081, EPI_ISL_1237082, EPI_ISL_1237083, EPI_ISL_1237084, EPI_ISL_1237085, EPI_ISL_1237086, EPI_ISL_1237087, EPI_ISL_1237088, EPI_ISL_1237089, EPI_ISL_1237090, EPI_ISL_1237091, EPI_ISL_1237092, EPI_ISL_1237093, EPI_ISL_1237094, EPI_ISL_1237095, EPI_ISL_1237096, EPI_ISL_1237097, EPI_ISL_1237098, EPI_ISL_1237099, EPI_ISL_1237100,
EPI_ISL_1237101, EPI_ISL_1237102, EPI_ISL_1237103, EPI_ISL_1237104, EPI_ISL_1237105, EPI_ISL_1237106, EPI_ISL_1237107, EPI_ISL_1237108, EPI_ISL_1237109, EPI_ISL_1237110, EPI_ISL_1237111, EPI_ISL_1237112, EPI_ISL_1237113, EPI_ISL_1237114, EPI_ISL_1237115, EPI_ISL_1237116, EPI_ISL_1237117, EPI_ISL_1237118, EPI_ISL_1237119, EPI_ISL_1237120, EPI_ISL_1237121, EPI_ISL_1237122,
EPI_ISL_1237123, EPI_ISL_1237124, EPI_ISL_1237125, EPI_ISL_1237126, EPI_ISL_1237127, EPI_ISL_1237128, EPI_ISL_1237129, EPI_ISL_1237130, EPI_ISL_1237131, EPI_ISL_1237132, EPI_ISL_1237133, EPI_ISL_1237134, EPI_ISL_1237135, EPI_ISL_1237136, EPI_ISL_1237137, EPI_ISL_1237138, EPI_ISL_1237139, EPI_ISL_1237140, EPI_ISL_1237141, EPI_ISL_1237142, EPI_ISL_1237143, EPI_ISL_1237144,
EPI_ISL_1237145, EPI_ISL_1237146, EPI_ISL_1237147, EPI_ISL_1237148, EPI_ISL_1237149, EPI_ISL_1237150, EPI_ISL_1237151, EPI_ISL_1237152, EPI_ISL_1237153, EPI_ISL_1237154, EPI_ISL_1237155, EPI_ISL_1237156, EPI_ISL_1237157, EPI_ISL_1237158, EPI_ISL_1237159, EPI_ISL_1237160, EPI_ISL_1237161, EPI_ISL_1237162, EPI_ISL_1237163, EPI_ISL_1237164, EPI_ISL_1237165, EPI_ISL_1237166,
EPI_ISL_1237167, EPI_ISL_1237168, EPI_ISL_1237169, EPI_ISL_1237170, EPI_ISL_1237171, EPI_ISL_1237172, EPI_ISL_1237173, EPI_ISL_1237174, EPI_ISL_1237175, EPI_ISL_1237176, EPI_ISL_1237177, EPI_ISL_1237178, EPI_ISL_1237179, EPI_ISL_1237180, EPI_ISL_1237181, EPI_ISL_1237182, EPI_ISL_1237183, EPI_ISL_1237184, EPI_ISL_1237185, EPI_ISL_1237186, EPI_ISL_1237187, EPI_ISL_1237188,
EPI_ISL_1237189, EPI_ISL_1237190, EPI_ISL_1237191, EPI_ISL_1237192, EPI_ISL_1237193, EPI_ISL_1237194, EPI_ISL_1237195, EPI_ISL_1237196, EPI_ISL_1237197, EPI_ISL_1237198, EPI_ISL_1237199, EPI_ISL_1237200, EPI_ISL_1237201, EPI_ISL_1237202, EPI_ISL_1237203, EPI_ISL_1237204, EPI_ISL_1237205, EPI_ISL_1237206, EPI_ISL_1237207, EPI_ISL_1237208, EPI_ISL_1237209, EPI_ISL_1237210,
EPI_ISL_1237211, EPI_ISL_1237212, EPI_ISL_1237213, EPI_ISL_1237214, EPI_ISL_1237215, EPI_ISL_1237216, EPI_ISL_1237217, EPI_ISL_1237218, EPI_ISL_1237219, EPI_ISL_1237220, EPI_ISL_1237221, EPI_ISL_1237222, EPI_ISL_1237223, EPI_ISL_1237224, EPI_ISL_1237225, EPI_ISL_1237226, EPI_ISL_1237227, EPI_ISL_1237228, EPI_ISL_1237229, EPI_ISL_1237230, EPI_ISL_1237231, EPI_ISL_1237232,
EPI_ISL_1237233, EPI_ISL_1237234, EPI_ISL_1237235, EPI_ISL_1237236, EPI_ISL_1237237, EPI_ISL_1237238, EPI_ISL_1237239, EPI_ISL_1237240, EPI_ISL_1237241, EPI_ISL_1237242, EPI_ISL_1237243, EPI_ISL_1237244, EPI_ISL_1237245, EPI_ISL_1237246, EPI_ISL_1237247, EPI_ISL_1237248, EPI_ISL_1237249, EPI_ISL_1237250, EPI_ISL_1237251, EPI_ISL_1237252, EPI_ISL_1237253, EPI_ISL_1237254,
EPI_ISL_1237255, EPI_ISL_1237256, EPI_ISL_1237257, EPI_ISL_1237258, EPI_ISL_1237259, EPI_ISL_1237260, EPI_ISL_1237261, EPI_ISL_1237262, EPI_ISL_1237263, EPI_ISL_1237264, EPI_ISL_1237265, EPI_ISL_1237266, EPI_ISL_1237267, EPI_ISL_1237268, EPI_ISL_1237269, EPI_ISL_1237270, EPI_ISL_1237271, EPI_ISL_1237272, EPI_ISL_1237273, EPI_ISL_1237274, EPI_ISL_1237275, EPI_ISL_1237276,
EPI_ISL_1237277, EPI_ISL_1237278, EPI_ISL_1237279, EPI_ISL_1237280, EPI_ISL_1237281, EPI_ISL_1237282, EPI_ISL_1237283, EPI_ISL_1237284, EPI_ISL_1237285, EPI_ISL_1237286, EPI_ISL_1237287, EPI_ISL_1237288, EPI_ISL_1237289, EPI_ISL_1237290, EPI_ISL_1237291, EPI_ISL_1237292, EPI_ISL_1237293, EPI_ISL_1237294, EPI_ISL_1237295, EPI_ISL_1237296, EPI_ISL_1237297, EPI_ISL_1237298,
EPI_ISL_1237299, EPI_ISL_1237300, EPI_ISL_1237301, EPI_ISL_1237302, EPI_ISL_1237303, EPI_ISL_1237304, EPI_ISL_1237305, EPI_ISL_1237306, EPI_ISL_1237307, EPI_ISL_1237308, EPI_ISL_1237309, EPI_ISL_1237310, EPI_ISL_1237311, EPI_ISL_1237312, EPI_ISL_1237313, EPI_ISL_1237314, EPI_ISL_1237315, EPI_ISL_1237316, EPI_ISL_1237317, EPI_ISL_1237318, EPI_ISL_1237319, EPI_ISL_1237320,
EPI_ISL_1237321, EPI_ISL_1237322, EPI_ISL_1237323, EPI_ISL_1237324, EPI_ISL_1237325, EPI_ISL_1237326, EPI_ISL_1237327, EPI_ISL_1237328, EPI_ISL_1237329, EPI_ISL_1237330, EPI_ISL_1237331, EPI_ISL_1237332, EPI_ISL_1237333, EPI_ISL_1237334, EPI_ISL_1237335, EPI_ISL_1237336, EPI_ISL_1237337, EPI_ISL_1237338, EPI_ISL_1237339, EPI_ISL_1237340, EPI_ISL_1237341, EPI_ISL_1237342,
EPI_ISL_1237343, EPI_ISL_1237344, EPI_ISL_1237345, EPI_ISL_1237346, EPI_ISL_1237347, EPI_ISL_1237348, EPI_ISL_1237349, EPI_ISL_1237350, EPI_ISL_1237351, EPI_ISL_1237352, EPI_ISL_1237353, EPI_ISL_1237354, EPI_ISL_1237355, EPI_ISL_1237356, EPI_ISL_1237357, EPI_ISL_1237358, EPI_ISL_1237359, EPI_ISL_1237360, EPI_ISL_1237361, EPI_ISL_1237362, EPI_ISL_1237363, EPI_ISL_1237364,
EPI_ISL_1237365, EPI_ISL_1237366, EPI_ISL_1237367, EPI_ISL_1237368, EPI_ISL_1237369, EPI_ISL_1237370, EPI_ISL_1237371, EPI_ISL_1237372, EPI_ISL_1237373, EPI_ISL_1237374, EPI_ISL_1237375, EPI_ISL_1237376, EPI_ISL_1237377, EPI_ISL_1237378, EPI_ISL_1237379, EPI_ISL_1237380, EPI_ISL_1237381, EPI_ISL_1237382, EPI_ISL_1237383, EPI_ISL_1237384, EPI_ISL_1237385, EPI_ISL_1237386,
EPI_ISL_1237387, EPI_ISL_1237388, EPI_ISL_1237389, EPI_ISL_1237390, EPI_ISL_1237391, EPI_ISL_1237392, EPI_ISL_1237393, EPI_ISL_1237394, EPI_ISL_1237395, EPI_ISL_1237396, EPI_ISL_1237397, EPI_ISL_1237398, EPI_ISL_1237399, EPI_ISL_1237400, EPI_ISL_1237401, EPI_ISL_1237402, EPI_ISL_1237403, EPI_ISL_1237404, EPI_ISL_1237405, EPI_ISL_1237406, EPI_ISL_1237407, EPI_ISL_1237408,
EPI_ISL_1237409, EPI_ISL_1237410, EPI_ISL_1237411, EPI_ISL_1237412, EPI_ISL_1237413, EPI_ISL_1237414, EPI_ISL_1237415, EPI_ISL_1237416, EPI_ISL_1237417, EPI_ISL_1237418, EPI_ISL_1237419, EPI_ISL_1237420, EPI_ISL_1237421, EPI_ISL_1237422, EPI_ISL_1237423, EPI_ISL_1237424, EPI_ISL_1237425, EPI_ISL_1237426, EPI_ISL_1237427, EPI_ISL_1237428, EPI_ISL_1237429, EPI_ISL_1237430,
EPI_ISL_1237431, EPI_ISL_1237432, EPI_ISL_1237433, EPI_ISL_1237434, EPI_ISL_1237435, EPI_ISL_1237436, EPI_ISL_1237437, EPI_ISL_1237438, EPI_ISL_1237439, EPI_ISL_1237440, EPI_ISL_1237441, EPI_ISL_1237442, EPI_ISL_1237443, EPI_ISL_1237444, EPI_ISL_1237445, EPI_ISL_1237446, EPI_ISL_1237447, EPI_ISL_1237448, EPI_ISL_1237449, EPI_ISL_1237450, EPI_ISL_1237451, EPI_ISL_1237452,
EPI_ISL_1237453, EPI_ISL_1237454, EPI_ISL_1237455, EPI_ISL_1237456, EPI_ISL_1237457, EPI_ISL_1237458, EPI_ISL_1237459, EPI_ISL_1237460, EPI_ISL_1237461, EPI_ISL_1237462, EPI_ISL_1237463, EPI_ISL_1237464, EPI_ISL_1237465, EPI_ISL_1237466, EPI_ISL_1237467, EPI_ISL_1237468, EPI_ISL_1237469, EPI_ISL_1237470, EPI_ISL_1237471, EPI_ISL_1237472, EPI_ISL_1237473, EPI_ISL_1237474,
EPI_ISL_1237475, EPI_ISL_1237476, EPI_ISL_1237477, EPI_ISL_1237478, EPI_ISL_1237479, EPI_ISL_1237480, EPI_ISL_1237481, EPI_ISL_1237482, EPI_ISL_1237483, EPI_ISL_1237484, EPI_ISL_1237485, EPI_ISL_1237486, EPI_ISL_1237487, EPI_ISL_1237488, EPI_ISL_1237489, EPI_ISL_1237490, EPI_ISL_1237491, EPI_ISL_1237492, EPI_ISL_1237493, EPI_ISL_1237494, EPI_ISL_1237495, EPI_ISL_1237496,
EPI_ISL_1237497, EPI_ISL_1237498, EPI_ISL_1237499, EPI_ISL_1237500, EPI_ISL_1237501, EPI_ISL_1237502, EPI_ISL_1237503, EPI_ISL_1237504, EPI_ISL_1237505, EPI_ISL_1237506, EPI_ISL_1237507, EPI_ISL_1237508, EPI_ISL_1237509, EPI_ISL_1237510, EPI_ISL_1237511, EPI_ISL_1237512, EPI_ISL_1237513, EPI_ISL_1237514, EPI_ISL_1237515, EPI_ISL_1237516, EPI_ISL_1237517, EPI_ISL_1237518,
EPI_ISL_1237519, EPI_ISL_1237520, EPI_ISL_1237521, EPI_ISL_1237522, EPI_ISL_1237523, EPI_ISL_1237524, EPI_ISL_1237525, EPI_ISL_1237526, EPI_ISL_1237527, EPI_ISL_1237528, EPI_ISL_1237529, EPI_ISL_1237530, EPI_ISL_1237531, EPI_ISL_1237532, EPI_ISL_1237533, EPI_ISL_1237534, EPI_ISL_1237535, EPI_ISL_1237536, EPI_ISL_1237537, EPI_ISL_1237538, EPI_ISL_1237539, EPI_ISL_1237540,
EPI_ISL_1237541, EPI_ISL_1237542, EPI_ISL_1237543, EPI_ISL_1237544, EPI_ISL_1237545, EPI_ISL_1237546, EPI_ISL_1237547, EPI_ISL_1237548, EPI_ISL_1237549, EPI_ISL_1237550, EPI_ISL_1237551, EPI_ISL_1237552, EPI_ISL_1237553, EPI_ISL_1237554, EPI_ISL_1237555, EPI_ISL_1237556, EPI_ISL_1237557, EPI_ISL_1237558, EPI_ISL_1237559, EPI_ISL_1237560, EPI_ISL_1237561, EPI_ISL_1237562,
EPI_ISL_1237563, EPI_ISL_1237564, EPI_ISL_1237565, EPI_ISL_1237566, EPI_ISL_1237567, EPI_ISL_1237568, EPI_ISL_1237569, EPI_ISL_1237570, EPI_ISL_1237571, EPI_ISL_1237572, EPI_ISL_1237573, EPI_ISL_1237574, EPI_ISL_1237575, EPI_ISL_1237576, EPI_ISL_1237577, EPI_ISL_1237578, EPI_ISL_1237579, EPI_ISL_1237580, EPI_ISL_1237581, EPI_ISL_1237582, EPI_ISL_1237583, EPI_ISL_1237584,
EPI_ISL_1237585, EPI_ISL_1237586, EPI_ISL_1237587, EPI_ISL_1237588, EPI_ISL_1237589, EPI_ISL_1237590, EPI_ISL_1237591, EPI_ISL_1237592, EPI_ISL_1237593, EPI_ISL_1237594, EPI_ISL_1237595, EPI_ISL_1237596, EPI_ISL_1237597, EPI_ISL_1237598, EPI_ISL_1237599, EPI_ISL_1237600, EPI_ISL_1237601, EPI_ISL_1237602, EPI_ISL_1237603, EPI_ISL_1237604, EPI_ISL_1237605, EPI_ISL_1237606,
EPI_ISL_1237607, EPI_ISL_1237608, EPI_ISL_1237609, EPI_ISL_1237610, EPI_ISL_1237611, EPI_ISL_1237612, EPI_ISL_1237613, EPI_ISL_1237614, EPI_ISL_1237615, EPI_ISL_1237616, EPI_ISL_1237617, EPI_ISL_1237618, EPI_ISL_1237619, EPI_ISL_1237620, EPI_ISL_1237621, EPI_ISL_1237622, EPI_ISL_1237623, EPI_ISL_1237624, EPI_ISL_1237625, EPI_ISL_1237626, EPI_ISL_1237627, EPI_ISL_1237628,
EPI_ISL_1237629, EPI_ISL_1237630, EPI_ISL_1237631, EPI_ISL_1237632, EPI_ISL_1237633, EPI_ISL_1237634, EPI_ISL_1237635, EPI_ISL_1237636, EPI_ISL_1237637, EPI_ISL_1237638, EPI_ISL_1237639, EPI_ISL_1237640, EPI_ISL_1237641, EPI_ISL_1237642, EPI_ISL_1237643, EPI_ISL_1237644, EPI_ISL_1237645, EPI_ISL_1237646, EPI_ISL_1237647, EPI_ISL_1237648, EPI_ISL_1237649, EPI_ISL_1237650,
EPI_ISL_1237651, EPI_ISL_1237652, EPI_ISL_1237653, EPI_ISL_1237654, EPI_ISL_1237655, EPI_ISL_1237656, EPI_ISL_1237657, EPI_ISL_1237658, EPI_ISL_1237659, EPI_ISL_1237660, EPI_ISL_1237661, EPI_ISL_1237662, EPI_ISL_1237663, EPI_ISL_1237664, EPI_ISL_1237665, EPI_ISL_1237666, EPI_ISL_1237667, EPI_ISL_1237668, EPI_ISL_1237669, EPI_ISL_1237670, EPI_ISL_1237671, EPI_ISL_1237672,
EPI_ISL_1237673, EPI_ISL_1237674, EPI_ISL_1237675, EPI_ISL_1237676, EPI_ISL_1237677, EPI_ISL_1237678, EPI_ISL_1237679, EPI_ISL_1237680, EPI_ISL_1237681, EPI_ISL_1237682, EPI_ISL_1237683, EPI_ISL_1237684, EPI_ISL_1237685, EPI_ISL_1237686, EPI_ISL_1237687, EPI_ISL_1237688, EPI_ISL_1237689, EPI_ISL_1237690, EPI_ISL_1237691, EPI_ISL_1237692, EPI_ISL_1237693, EPI_ISL_1237694,
EPI_ISL_1237695, EPI_ISL_1237696, EPI_ISL_1237697, EPI_ISL_1237698, EPI_ISL_1237699, EPI_ISL_1237700, EPI_ISL_1237701, EPI_ISL_1237702, EPI_ISL_1237703, EPI_ISL_1237704, EPI_ISL_1237705, EPI_ISL_1237706, EPI_ISL_1237707, EPI_ISL_1237708, EPI_ISL_1237709, EPI_ISL_1237710, EPI_ISL_1237711, EPI_ISL_1237712, EPI_ISL_1237713, EPI_ISL_1237714, EPI_ISL_1237715, EPI_ISL_1237716,
EPI_ISL_1237717, EPI_ISL_1237718, EPI_ISL_1237719, EPI_ISL_1237720, EPI_ISL_1237721, EPI_ISL_1237722, EPI_ISL_1237723, EPI_ISL_1237724, EPI_ISL_1237725, EPI_ISL_1237726, EPI_ISL_1237727, EPI_ISL_1237728, EPI_ISL_1237729, EPI_ISL_1237730, EPI_ISL_1237731, EPI_ISL_1237732, EPI_ISL_1237733, EPI_ISL_1237734, EPI_ISL_1237735, EPI_ISL_1237736, EPI_ISL_1237737, EPI_ISL_1237738,
EPI_ISL_1237739, EPI_ISL_1237740, EPI_ISL_1237741, EPI_ISL_1237742, EPI_ISL_1237743, EPI_ISL_1237744, EPI_ISL_1237745, EPI_ISL_1237746, EPI_ISL_1237747, EPI_ISL_1237748, EPI_ISL_1237749, EPI_ISL_1237750, EPI_ISL_1237751, EPI_ISL_1237752, EPI_ISL_1237753, EPI_ISL_1237754, EPI_ISL_1237755, EPI_ISL_1237756, EPI_ISL_1237757, EPI_ISL_1237758, EPI_ISL_1237759, EPI_ISL_1237760,
EPI_ISL_1237761, EPI_ISL_1237762, EPI_ISL_1237763, EPI_ISL_1237764, EPI_ISL_1237765, EPI_ISL_1237766, EPI_ISL_1237767, EPI_ISL_1237768, EPI_ISL_1237769, EPI_ISL_1237770, EPI_ISL_1237771, EPI_ISL_1237772, EPI_ISL_1237773, EPI_ISL_1237774, EPI_ISL_1237775, EPI_ISL_1237776, EPI_ISL_1237777, EPI_ISL_1237778, EPI_ISL_1237779, EPI_ISL_1237780, EPI_ISL_1237781, EPI_ISL_1237782,
EPI_ISL_1237783, EPI_ISL_1237784, EPI_ISL_1237785, EPI_ISL_1237786, EPI_ISL_1237787, EPI_ISL_1237788, EPI_ISL_1237789, EPI_ISL_1237790, EPI_ISL_1237791, EPI_ISL_1237792, EPI_ISL_1237793, EPI_ISL_1237794, EPI_ISL_1237795, EPI_ISL_1237796, EPI_ISL_1237797, EPI_ISL_1237798, EPI_ISL_1237799, EPI_ISL_1237800, EPI_ISL_1237801, EPI_ISL_1237802, EPI_ISL_1237803, EPI_ISL_1237804,
EPI_ISL_1237805, EPI_ISL_1237806, EPI_ISL_1237807, EPI_ISL_1237808, EPI_ISL_1237809, EPI_ISL_1237810, EPI_ISL_1237811, EPI_ISL_1237812, EPI_ISL_1237813, EPI_ISL_1237814, EPI_ISL_1237815, EPI_ISL_1237816, EPI_ISL_1237817, EPI_ISL_1237818, EPI_ISL_1237819, EPI_ISL_1237820, EPI_ISL_1237821, EPI_ISL_1237822, EPI_ISL_1237823, EPI_ISL_1237824, EPI_ISL_1237825, EPI_ISL_1237826,
EPI_ISL_1237827, EPI_ISL_1237828, EPI_ISL_1237829, EPI_ISL_1237830, EPI_ISL_1237831, EPI_ISL_1237832, EPI_ISL_1237833, EPI_ISL_1237834, EPI_ISL_1237835, EPI_ISL_1237836, EPI_ISL_1237837, EPI_ISL_1237838, EPI_ISL_1237839, EPI_ISL_1237840, EPI_ISL_1237841, EPI_ISL_1237842, EPI_ISL_1237843, EPI_ISL_1237844, EPI_ISL_1237845, EPI_ISL_1237846, EPI_ISL_1237847, EPI_ISL_1237848,
EPI_ISL_1237849, EPI_ISL_1237850, EPI_ISL_1237851, EPI_ISL_1237852, EPI_ISL_1237853, EPI_ISL_1237854, EPI_ISL_1237855, EPI_ISL_1237856, EPI_ISL_1237857, EPI_ISL_1237858, EPI_ISL_1237859, EPI_ISL_1237860, EPI_ISL_1237861, EPI_ISL_1237862, EPI_ISL_1237863, EPI_ISL_1237864, EPI_ISL_1237865, EPI_ISL_1237866, EPI_ISL_1237867, EPI_ISL_1237868, EPI_ISL_1237869, EPI_ISL_1237870,
EPI_ISL_1237871, EPI_ISL_1237872, EPI_ISL_1237873, EPI_ISL_1237874, EPI_ISL_1237875, EPI_ISL_1237876, EPI_ISL_1237877, EPI_ISL_1237878, EPI_ISL_1237879, EPI_ISL_1237880, EPI_ISL_1237881, EPI_ISL_1237882, EPI_ISL_1237883, EPI_ISL_1237884, EPI_ISL_1237885, EPI_ISL_1237886, EPI_ISL_1237887, EPI_ISL_1237888, EPI_ISL_1237889, EPI_ISL_1237890, EPI_ISL_1237891, EPI_ISL_1237892,
EPI_ISL_1237893, EPI_ISL_1237894, EPI_ISL_1237895, EPI_ISL_1237896, EPI_ISL_1237897, EPI_ISL_1237898, EPI_ISL_1237899, EPI_ISL_1237900, EPI_ISL_1237901, EPI_ISL_1237902, EPI_ISL_1237903, EPI_ISL_1237904, EPI_ISL_1237905, EPI_ISL_1237906, EPI_ISL_1237907, EPI_ISL_1237908, EPI_ISL_1237909, EPI_ISL_1237910, EPI_ISL_1237911, EPI_ISL_1237912, EPI_ISL_1237913, EPI_ISL_1237914,
EPI_ISL_1237915, EPI_ISL_1237916, EPI_ISL_1237917, EPI_ISL_1237918, EPI_ISL_1237919, EPI_ISL_1237920, EPI_ISL_1237921, EPI_ISL_1237922, EPI_ISL_1237923, EPI_ISL_1237924, EPI_ISL_1237925, EPI_ISL_1237926, EPI_ISL_1237927, EPI_ISL_1237928, EPI_ISL_1237929, EPI_ISL_1237930, EPI_ISL_1237931, EPI_ISL_1237932, EPI_ISL_1237933, EPI_ISL_1237934, EPI_ISL_1237935, EPI_ISL_1237936,
EPI_ISL_1237937, EPI_ISL_1237938, EPI_ISL_1237939, EPI_ISL_1237940, EPI_ISL_1237941, EPI_ISL_1237942, EPI_ISL_1237943, EPI_ISL_1237944, EPI_ISL_1237945, EPI_ISL_1237946, EPI_ISL_1237947, EPI_ISL_1237948, EPI_ISL_1237949, EPI_ISL_1237950, EPI_ISL_1237951, EPI_ISL_1237952, EPI_ISL_1237953, EPI_ISL_1237954, EPI_ISL_1237955, EPI_ISL_1237956, EPI_ISL_1237957, EPI_ISL_1237958,
EPI_ISL_1237959, EPI_ISL_1237960, EPI_ISL_1237961, EPI_ISL_1237962, EPI_ISL_1237963, EPI_ISL_1237964, EPI_ISL_1237965, EPI_ISL_1237966, EPI_ISL_1237967, EPI_ISL_1237968, EPI_ISL_1237969, EPI_ISL_1237970, EPI_ISL_1237971, EPI_ISL_1237972, EPI_ISL_1237973, EPI_ISL_1237974, EPI_ISL_1237975, EPI_ISL_1237976, EPI_ISL_1237977, EPI_ISL_1237978, EPI_ISL_1237979, EPI_ISL_1237980,
EPI_ISL_1237981, EPI_ISL_1237982, EPI_ISL_1237983, EPI_ISL_1237984, EPI_ISL_1237985, EPI_ISL_1237986, EPI_ISL_1237987, EPI_ISL_1237988, EPI_ISL_1237989, EPI_ISL_1237990, EPI_ISL_1237991, EPI_ISL_1237992, EPI_ISL_1237993, EPI_ISL_1237994, EPI_ISL_1237995, EPI_ISL_1237996, EPI_ISL_1237997, EPI_ISL_1237998, EPI_ISL_1237999, EPI_ISL_1238000, EPI_ISL_1238001, EPI_ISL_1238002,
EPI_ISL_1238003, EPI_ISL_1238004, EPI_ISL_1238005, EPI_ISL_1238006, EPI_ISL_1238007, EPI_ISL_1238008, EPI_ISL_1238009, EPI_ISL_1238010, EPI_ISL_1238011, EPI_ISL_1238012, EPI_ISL_1238013, EPI_ISL_1238014, EPI_ISL_1238015, EPI_ISL_1238016, EPI_ISL_1238017, EPI_ISL_1238018, EPI_ISL_1238019, EPI_ISL_1238020, EPI_ISL_1238021, EPI_ISL_1238022, EPI_ISL_1238023, EPI_ISL_1238024,
EPI_ISL_1238025, EPI_ISL_1238026, EPI_ISL_1238027, EPI_ISL_1238028, EPI_ISL_1238029, EPI_ISL_1238030, EPI_ISL_1238031, EPI_ISL_1238032, EPI_ISL_1238033, EPI_ISL_1238034, EPI_ISL_1238035, EPI_ISL_1238036, EPI_ISL_1238037, EPI_ISL_1238038, EPI_ISL_1238039, EPI_ISL_1238040, EPI_ISL_1238041, EPI_ISL_1238042, EPI_ISL_1238043, EPI_ISL_1238044, EPI_ISL_1238045, EPI_ISL_1238046,
EPI_ISL_1238047, EPI_ISL_1238048, EPI_ISL_1238049, EPI_ISL_1238050, EPI_ISL_1238051, EPI_ISL_1238052, EPI_ISL_1238053, EPI_ISL_1238054, EPI_ISL_1238055, EPI_ISL_1238056, EPI_ISL_1238057, EPI_ISL_1238058, EPI_ISL_1238059, EPI_ISL_1238060, EPI_ISL_1238061, EPI_ISL_1238062, EPI_ISL_1238063, EPI_ISL_1238064, EPI_ISL_1238065, EPI_ISL_1238066, EPI_ISL_1238067, EPI_ISL_1238068,
EPI_ISL_1238069, EPI_ISL_1238070, EPI_ISL_1238071, EPI_ISL_1238072, EPI_ISL_1238073, EPI_ISL_1238074, EPI_ISL_1238075, EPI_ISL_1238076, EPI_ISL_1238077, EPI_ISL_1238078, EPI_ISL_1238079, EPI_ISL_1238080, EPI_ISL_1238081, EPI_ISL_1238082, EPI_ISL_1238083, EPI_ISL_1238084, EPI_ISL_1238085, EPI_ISL_1238086, EPI_ISL_1238087, EPI_ISL_1238088, EPI_ISL_1238089, EPI_ISL_1238090,
EPI_ISL_1238091, EPI_ISL_1238092, EPI_ISL_1238093, EPI_ISL_1238094, EPI_ISL_1238095, EPI_ISL_1238096, EPI_ISL_1238097, EPI_ISL_1238098, EPI_ISL_1238099, EPI_ISL_1238100, EPI_ISL_1238101, EPI_ISL_1238102, EPI_ISL_1238103, EPI_ISL_1238104, EPI_ISL_1238105, EPI_ISL_1238106, EPI_ISL_1238107, EPI_ISL_1238108, EPI_ISL_1238109, EPI_ISL_1238110, EPI_ISL_1238111, EPI_ISL_1238112,
EPI_ISL_1238113, EPI_ISL_1238114, EPI_ISL_1238115, EPI_ISL_1238116, EPI_ISL_1238117, EPI_ISL_1238118, EPI_ISL_1238119, EPI_ISL_1238120, EPI_ISL_1238121, EPI_ISL_1238122, EPI_ISL_1238123, EPI_ISL_1238124, EPI_ISL_1238125, EPI_ISL_1238126, EPI_ISL_1238127, EPI_ISL_1238128, EPI_ISL_1238129, EPI_ISL_1238130, EPI_ISL_1238131, EPI_ISL_1238132, EPI_ISL_1238133, EPI_ISL_1238134,
EPI_ISL_1238135, EPI_ISL_1238136, EPI_ISL_1238137, EPI_ISL_1238138, EPI_ISL_1238139, EPI_ISL_1238140, EPI_ISL_1238141, EPI_ISL_1238142, EPI_ISL_1238143, EPI_ISL_1238144, EPI_ISL_1238145, EPI_ISL_1238146, EPI_ISL_1238147, EPI_ISL_1238148, EPI_ISL_1238149, EPI_ISL_1238150, EPI_ISL_1238151, EPI_ISL_1238152, EPI_ISL_1238153, EPI_ISL_1238154, EPI_ISL_1238155, EPI_ISL_1238156,
EPI_ISL_1238157, EPI_ISL_1238158, EPI_ISL_1238159, EPI_ISL_1238160, EPI_ISL_1238161, EPI_ISL_1238162, EPI_ISL_1238163, EPI_ISL_1238164, EPI_ISL_1238165, EPI_ISL_1238166, EPI_ISL_1238167, EPI_ISL_1238168, EPI_ISL_1238169, EPI_ISL_1238170, EPI_ISL_1238171, EPI_ISL_1238172, EPI_ISL_1238173, EPI_ISL_1238174, EPI_ISL_1238175, EPI_ISL_1238176, EPI_ISL_1238177, EPI_ISL_1238178,
EPI_ISL_1238179, EPI_ISL_1238180, EPI_ISL_1238181, EPI_ISL_1238182, EPI_ISL_1238183, EPI_ISL_1238184, EPI_ISL_1238185, EPI_ISL_1238186, EPI_ISL_1238187, EPI_ISL_1238188, EPI_ISL_1238189, EPI_ISL_1238190, EPI_ISL_1238191, EPI_ISL_1238192, EPI_ISL_1238193, EPI_ISL_1238194, EPI_ISL_1238196, EPI_ISL_1238197, EPI_ISL_1238198, EPI_ISL_1238199, EPI_ISL_1238200, EPI_ISL_1238201,
EPI_ISL_1238202, EPI_ISL_1238203, EPI_ISL_1238204, EPI_ISL_1238205, EPI_ISL_1238206, EPI_ISL_1238207, EPI_ISL_1238208, EPI_ISL_1238209, EPI_ISL_1238210, EPI_ISL_1238211, EPI_ISL_1238212, EPI_ISL_1238213, EPI_ISL_1238214, EPI_ISL_1238215, EPI_ISL_1238216, EPI_ISL_1238217, EPI_ISL_1238218, EPI_ISL_1238219, EPI_ISL_1238220, EPI_ISL_1238221, EPI_ISL_1238222, EPI_ISL_1238223,
EPI_ISL_1238224, EPI_ISL_1238225, EPI_ISL_1238226, EPI_ISL_1238227, EPI_ISL_1238228, EPI_ISL_1238229, EPI_ISL_1238230, EPI_ISL_1238231, EPI_ISL_1238232, EPI_ISL_1238233, EPI_ISL_1238234, EPI_ISL_1238235, EPI_ISL_1238236, EPI_ISL_1238237, EPI_ISL_1238238, EPI_ISL_1238239, EPI_ISL_1238240, EPI_ISL_1238241, EPI_ISL_1238242, EPI_ISL_1238243, EPI_ISL_1238244, EPI_ISL_1238245,
EPI_ISL_1238246, EPI_ISL_1238247, EPI_ISL_1238248, EPI_ISL_1238249, EPI_ISL_1238250, EPI_ISL_1238251, EPI_ISL_1238252, EPI_ISL_1238253, EPI_ISL_1238254, EPI_ISL_1238255, EPI_ISL_1238256, EPI_ISL_1238257, EPI_ISL_1238258, EPI_ISL_1238259, EPI_ISL_1238260, EPI_ISL_1238261, EPI_ISL_1238262, EPI_ISL_1238263, EPI_ISL_1238264, EPI_ISL_1238265, EPI_ISL_1238266, EPI_ISL_1238267,
EPI_ISL_1238268, EPI_ISL_1238269, EPI_ISL_1238270, EPI_ISL_1238271, EPI_ISL_1238272, EPI_ISL_1238273, EPI_ISL_1238274, EPI_ISL_1238275, EPI_ISL_1238276, EPI_ISL_1238277, EPI_ISL_1238278, EPI_ISL_1238279, EPI_ISL_1238280, EPI_ISL_1238281, EPI_ISL_1238282, EPI_ISL_1238283, EPI_ISL_1238284, EPI_ISL_1238285, EPI_ISL_1238286, EPI_ISL_1238287, EPI_ISL_1238288, EPI_ISL_1238289,
EPI_ISL_1238290, EPI_ISL_1238291, EPI_ISL_1238292, EPI_ISL_1238293, EPI_ISL_1238294, EPI_ISL_1238295, EPI_ISL_1238296, EPI_ISL_1238297, EPI_ISL_1238298, EPI_ISL_1238299, EPI_ISL_1238300, EPI_ISL_1238301, EPI_ISL_1238302, EPI_ISL_1238303, EPI_ISL_1238305, EPI_ISL_1238306, EPI_ISL_1238307, EPI_ISL_1238308, EPI_ISL_1238309, EPI_ISL_1238310, EPI_ISL_1238311, EPI_ISL_1238312,

| | | | |
|---|---|---|---|
| EPI_ISL_1238313, EPI_ISL_1238314, EPI_ISL_1238315, EPI_ISL_1238316, EPI_ISL_1238317, EPI_ISL_1238318, EPI_ISL_1238319, EPI_ISL_1238320, EPI_ISL_1238321, EPI_ISL_1238322, EPI_ISL_1238323, EPI_ISL_1238324, EPI_ISL_1238325, EPI_ISL_1238326, EPI_ISL_1238327, EPI_ISL_1238328, EPI_ISL_1238329, EPI_ISL_1238330, EPI_ISL_1238331, EPI_ISL_1238332, EPI_ISL_1238333, EPI_ISL_1238334, EPI_ISL_1238335, EPI_ISL_1238336, EPI_ISL_1238337, EPI_ISL_1238338, EPI_ISL_1238339, EPI_ISL_1238340, EPI_ISL_1238341, EPI_ISL_1238342, EPI_ISL_1238343, EPI_ISL_1238344, EPI_ISL_1238345, EPI_ISL_1238346, EPI_ISL_1238347, EPI_ISL_1238348, EPI_ISL_1238349, EPI_ISL_1238350, EPI_ISL_1238351, EPI_ISL_1238352, EPI_ISL_1238353, EPI_ISL_1238354, EPI_ISL_1238355, EPI_ISL_1238356, EPI_ISL_1238357, EPI_ISL_1238358, EPI_ISL_1238359, EPI_ISL_1238360, EPI_ISL_1238361, EPI_ISL_1238362, EPI_ISL_1238363, EPI_ISL_1238364, EPI_ISL_1238365, EPI_ISL_1238366, EPI_ISL_1238367, EPI_ISL_1238368, EPI_ISL_1238369, EPI_ISL_1238370, EPI_ISL_1238371, EPI_ISL_1238372, EPI_ISL_1238373, EPI_ISL_1238374, EPI_ISL_1238375, EPI_ISL_1238376, EPI_ISL_1238377, EPI_ISL_1238378, EPI_ISL_1238379, EPI_ISL_1238380, EPI_ISL_1238381, EPI_ISL_1238382, EPI_ISL_1238383, EPI_ISL_1238384, EPI_ISL_1238385, EPI_ISL_1238386, EPI_ISL_1238387, EPI_ISL_1238388, EPI_ISL_1238389, EPI_ISL_1238390, EPI_ISL_1238391, EPI_ISL_1238392, EPI_ISL_1238393, EPI_ISL_1238394, EPI_ISL_1238395, EPI_ISL_1238396, EPI_ISL_1238397, EPI_ISL_1238398, EPI_ISL_1238399, EPI_ISL_1238400, EPI_ISL_1238401, EPI_ISL_1238402, EPI_ISL_1238403, EPI_ISL_1238404, EPI_ISL_1238405, EPI_ISL_1238406, EPI_ISL_1238407, EPI_ISL_1238408, EPI_ISL_1238409, EPI_ISL_1238410, EPI_ISL_1238411, EPI_ISL_1238412, EPI_ISL_1238413, EPI_ISL_1238414, EPI_ISL_1238415, EPI_ISL_1238416, EPI_ISL_1238417, EPI_ISL_1238418, EPI_ISL_1238419, EPI_ISL_1238420, EPI_ISL_1238421, EPI_ISL_1238422, EPI_ISL_1238423, EPI_ISL_1238424, EPI_ISL_1238425, EPI_ISL_1238426, EPI_ISL_1238427, EPI_ISL_1238428, EPI_ISL_1238429, EPI_ISL_1238430, EPI_ISL_1238431, EPI_ISL_1238432, EPI_ISL_1238433, EPI_ISL_1238434, EPI_ISL_1238435, EPI_ISL_1238436, EPI_ISL_1238437, EPI_ISL_1238438, EPI_ISL_1238439, EPI_ISL_1238440, EPI_ISL_1238441, EPI_ISL_1238442, EPI_ISL_1238443, EPI_ISL_1238444, EPI_ISL_1238445, EPI_ISL_1238446, EPI_ISL_1238447, EPI_ISL_1238448, EPI_ISL_1238449, EPI_ISL_1238450, EPI_ISL_1238451, EPI_ISL_1238452, EPI_ISL_1238453, EPI_ISL_1238454, EPI_ISL_1238455, EPI_ISL_1238456, EPI_ISL_1238457, EPI_ISL_1238458, EPI_ISL_1238459, EPI_ISL_1238460, EPI_ISL_1238461, EPI_ISL_1238462, EPI_ISL_1238463, EPI_ISL_1238464, EPI_ISL_1238465, EPI_ISL_1238466, EPI_ISL_1238467, EPI_ISL_1238468, EPI_ISL_1238469, EPI_ISL_1238470, EPI_ISL_1238471, EPI_ISL_1238472, EPI_ISL_1238473, EPI_ISL_1238474, EPI_ISL_1238475, EPI_ISL_1238476, EPI_ISL_1238477, EPI_ISL_1238478, EPI_ISL_1238479, EPI_ISL_1238480, EPI_ISL_1238481, EPI_ISL_1238482, EPI_ISL_1238483, EPI_ISL_1238484, EPI_ISL_1238485, EPI_ISL_1238486, EPI_ISL_1238487, EPI_ISL_1238488, EPI_ISL_1238489, EPI_ISL_1238490, EPI_ISL_1238491, EPI_ISL_1238492, EPI_ISL_1238493, EPI_ISL_1238494, EPI_ISL_1238495, EPI_ISL_1238496, EPI_ISL_1238497, EPI_ISL_1238498, EPI_ISL_1238499, EPI_ISL_1238500, EPI_ISL_1238501, EPI_ISL_1238502, EPI_ISL_1238503, EPI_ISL_1238504, EPI_ISL_1238505, EPI_ISL_1238506, EPI_ISL_1238507, EPI_ISL_1238508, EPI_ISL_1238509, EPI_ISL_1238510, EPI_ISL_1238511, EPI_ISL_1238512, EPI_ISL_1238513, EPI_ISL_1238514, EPI_ISL_1238515, EPI_ISL_1238516, EPI_ISL_1238517, EPI_ISL_1238518, EPI_ISL_1238519, EPI_ISL_1238520, EPI_ISL_1238521, EPI_ISL_1238522, EPI_ISL_1238523, EPI_ISL_1238524, EPI_ISL_1238525, EPI_ISL_1238526, EPI_ISL_1238527, EPI_ISL_1238528, EPI_ISL_1238529, EPI_ISL_1238530, EPI_ISL_1238531, EPI_ISL_1238532, EPI_ISL_1238533, EPI_ISL_1238534, EPI_ISL_1238535, EPI_ISL_1238536, EPI_ISL_1238537, EPI_ISL_1238538, EPI_ISL_1238539, EPI_ISL_1238540, EPI_ISL_1238541, EPI_ISL_1238542, EPI_ISL_1238543, EPI_ISL_1238544, EPI_ISL_1238545, EPI_ISL_1238546, EPI_ISL_1238547, EPI_ISL_1238548, EPI_ISL_1238549, EPI_ISL_1238550, EPI_ISL_1238551, EPI_ISL_1238552, EPI_ISL_1238553, EPI_ISL_1238554, EPI_ISL_1238555, EPI_ISL_1238556, EPI_ISL_1238557, EPI_ISL_1238558, EPI_ISL_1238559, EPI_ISL_1238560, EPI_ISL_1238561, EPI_ISL_1238562, EPI_ISL_1238563, EPI_ISL_1238564, EPI_ISL_1238565, EPI_ISL_1238566, EPI_ISL_1238567, EPI_ISL_1238568, EPI_ISL_1238569, EPI_ISL_1238570, EPI_ISL_1238571, EPI_ISL_1238572, EPI_ISL_1238573, EPI_ISL_1238574, EPI_ISL_1238575, EPI_ISL_1238576, EPI_ISL_1238577, EPI_ISL_1238578, EPI_ISL_1238579, EPI_ISL_1238580, EPI_ISL_1238581, EPI_ISL_1238582, EPI_ISL_1238583, EPI_ISL_1238584, EPI_ISL_1238585, EPI_ISL_1238586, EPI_ISL_1238587, EPI_ISL_1238588, EPI_ISL_1238589, EPI_ISL_1238590, EPI_ISL_1238591, EPI_ISL_1238592, EPI_ISL_1238593, EPI_ISL_1238594, EPI_ISL_1238595, EPI_ISL_1238596, EPI_ISL_1238597, EPI_ISL_1238598, EPI_ISL_1238599, EPI_ISL_1238600, EPI_ISL_1238601, EPI_ISL_1238602, EPI_ISL_1238603, EPI_ISL_1238604, EPI_ISL_1238605, EPI_ISL_1238606, EPI_ISL_1238607, EPI_ISL_1238608, EPI_ISL_1238609, EPI_ISL_1238610, EPI_ISL_1238611, EPI_ISL_1238612, EPI_ISL_1238613, EPI_ISL_1238614, EPI_ISL_1238615, EPI_ISL_1238616, EPI_ISL_1238617, EPI_ISL_1238618, EPI_ISL_1238619, EPI_ISL_1238620, EPI_ISL_1238621, EPI_ISL_1238622, EPI_ISL_1238623, EPI_ISL_1238624, EPI_ISL_1238625, EPI_ISL_1238626, EPI_ISL_1238627, EPI_ISL_1238628, EPI_ISL_1238629, EPI_ISL_1238630, EPI_ISL_1238631, EPI_ISL_1238632, EPI_ISL_1238633, EPI_ISL_1238634, EPI_ISL_1238635, EPI_ISL_1238636, EPI_ISL_1238637, EPI_ISL_1238638, EPI_ISL_1238639, EPI_ISL_1238640, EPI_ISL_1238641, EPI_ISL_1238642, EPI_ISL_1238643, EPI_ISL_1238644, EPI_ISL_1238645, EPI_ISL_1238646, EPI_ISL_1238647, EPI_ISL_1238648, EPI_ISL_1238649, EPI_ISL_1238650, EPI_ISL_1238651, EPI_ISL_1238652, EPI_ISL_1238653, EPI_ISL_1238654, EPI_ISL_1238655, EPI_ISL_1238656, EPI_ISL_1238657, EPI_ISL_1238658, EPI_ISL_1238659, EPI_ISL_1238660, EPI_ISL_1238661, EPI_ISL_1238662, EPI_ISL_1238663, EPI_ISL_1238664, EPI_ISL_1238665, EPI_ISL_1238666, EPI_ISL_1238667, EPI_ISL_1238668, EPI_ISL_1238669, EPI_ISL_1238670, EPI_ISL_1238671, EPI_ISL_1238672, EPI_ISL_1238673, EPI_ISL_1238674, EPI_ISL_1238675, EPI_ISL_1238676, EPI_ISL_1238677, EPI_ISL_1238678, EPI_ISL_1238679, EPI_ISL_1238680, EPI_ISL_1238681, EPI_ISL_1238682, EPI_ISL_1238683, EPI_ISL_1238684, EPI_ISL_1238685, EPI_ISL_1238686, EPI_ISL_1238687, EPI_ISL_1238688, EPI_ISL_1238689, EPI_ISL_1238690, EPI_ISL_1238691, EPI_ISL_1238692, EPI_ISL_1238693, EPI_ISL_1238694, EPI_ISL_1238695, EPI_ISL_1238696, EPI_ISL_1238697, EPI_ISL_1238698, EPI_ISL_1238699, EPI_ISL_1238700, EPI_ISL_1238701, EPI_ISL_1238702, EPI_ISL_1238703, EPI_ISL_1238704, EPI_ISL_1238705, EPI_ISL_1238706, EPI_ISL_1238707, EPI_ISL_1238708, EPI_ISL_1238709, EPI_ISL_1238710, EPI_ISL_1238711, EPI_ISL_1238712, EPI_ISL_1238713, EPI_ISL_1238714, EPI_ISL_1238715, EPI_ISL_1238716, EPI_ISL_1238717, EPI_ISL_1238718, EPI_ISL_1238719, EPI_ISL_1238720, EPI_ISL_1238721, EPI_ISL_1238722, EPI_ISL_1238723, EPI_ISL_1238724, EPI_ISL_1238725, EPI_ISL_1238726, EPI_ISL_1238727, EPI_ISL_1238728, EPI_ISL_1238729, EPI_ISL_1238730, EPI_ISL_1238731, EPI_ISL_1238732, EPI_ISL_1238733, EPI_ISL_1238734, EPI_ISL_1238735, EPI_ISL_1238736, EPI_ISL_1238737, EPI_ISL_1238738, EPI_ISL_1238739, EPI_ISL_1238740, EPI_ISL_1238741, EPI_ISL_1238742, EPI_ISL_1238743, EPI_ISL_1238744, EPI_ISL_1238745, EPI_ISL_1238746, EPI_ISL_1238747, EPI_ISL_1238748, EPI_ISL_1238749, EPI_ISL_1238750, EPI_ISL_1238751, EPI_ISL_1238752, EPI_ISL_1238753, EPI_ISL_1238754, EPI_ISL_1238755, EPI_ISL_1238756, EPI_ISL_1238757, EPI_ISL_1238758, EPI_ISL_1238759, EPI_ISL_1238760, EPI_ISL_1238761, EPI_ISL_1238762, EPI_ISL_1238763, EPI_ISL_1238764, EPI_ISL_1238765 | | | |
| see above | Houston Methodist Hospital | Houston Methodist Hospital | S. Wesley Long, Randall J. Olsen, Paul A. Christensen, Sishir Subedi, Robert Olson, James J. Davis, Matthew Ojeda Saavedra, Prasanti Yerramilli, Layne Pruitt, Kristina Reppond, Madison N. Shyer, Jessica Cambric, Ilya J. Finkelstein, Jimmy Gollihar, and James M. Musser |
| EPI_ISL_1238766 | Florida Bureau of Public Health Laboratories | Florida Bureau of Public Health Laboratories | Sarah Schmedes, Jason Blanton |
| EPI_ISL_1238767, EPI_ISL_1238768, EPI_ISL_1238769, EPI_ISL_1238770, EPI_ISL_1238771, EPI_ISL_1238772, EPI_ISL_1238773, EPI_ISL_1238774, EPI_ISL_1238775, EPI_ISL_1238776, EPI_ISL_1238777, EPI_ISL_1238778, EPI_ISL_1238779, EPI_ISL_1238780, EPI_ISL_1238781, EPI_ISL_1238782, EPI_ISL_1238784, EPI_ISL_1238785, EPI_ISL_1238786 | | | |
| see above | North Dakota Department of Health, Public Health Laboratory | North Dakota Department of Health, Public Health Laboratory | Lisa Wingerter |
| EPI_ISL_1238787, EPI_ISL_1238788, EPI_ISL_1238789, EPI_ISL_1238790, EPI_ISL_1238791, EPI_ISL_1238792, EPI_ISL_1238793 | Ecological and Evolutionary Genomics. UGA-CINVESTAV | Unidad Universitaria de Secuenciación Masiva y Bioinformática (UUSMB). IBT-UNAM | Angelica Cibrian |
| EPI_ISL_1238794, EPI_ISL_1238795, EPI_ISL_1238796, EPI_ISL_1238797, EPI_ISL_1238798, EPI_ISL_1238799, EPI_ISL_1238800, EPI_ISL_1238801, EPI_ISL_1238802, EPI_ISL_1238803, EPI_ISL_1238804, EPI_ISL_1238805, EPI_ISL_1238810, EPI_ISL_1238811, EPI_ISL_1238812, EPI_ISL_1238814, EPI_ISL_1238815, EPI_ISL_1238816, EPI_ISL_1238817, EPI_ISL_1238818, EPI_ISL_1238819, EPI_ISL_1238820, EPI_ISL_1238821, EPI_ISL_1238822, EPI_ISL_1238823, EPI_ISL_1238824, EPI_ISL_1238825, EPI_ISL_1238826, EPI_ISL_1238827, EPI_ISL_1238828, EPI_ISL_1238829, EPI_ISL_1238830, EPI_ISL_1238831, EPI_ISL_1238832, EPI_ISL_1238833, EPI_ISL_1238834, EPI_ISL_1238835, EPI_ISL_1238836, EPI_ISL_1238837, EPI_ISL_1238838, EPI_ISL_1238839 | | | |
| see above | MD PHL | MD PHL | Maryland Department of Health Laboratories Administration |
| EPI_ISL_1238840, EPI_ISL_1238842, EPI_ISL_1238843, EPI_ISL_1238845, EPI_ISL_1238846, EPI_ISL_1238847, EPI_ISL_1238848, EPI_ISL_1238849, EPI_ISL_1238850, EPI_ISL_1238851, EPI_ISL_1238852, EPI_ISL_1238853, EPI_ISL_1238855, EPI_ISL_1238856, EPI_ISL_1238857, EPI_ISL_1238858, EPI_ISL_1238859, EPI_ISL_1238864, EPI_ISL_1238866, EPI_ISL_1238868, EPI_ISL_1238869, EPI_ISL_1238870, EPI_ISL_1238871, EPI_ISL_1238872, EPI_ISL_1238873, EPI_ISL_1238874, EPI_ISL_1238875, EPI_ISL_1238876, EPI_ISL_1238877, EPI_ISL_1238878, EPI_ISL_1238879, EPI_ISL_1238880, EPI_ISL_1238881, EPI_ISL_1238882, EPI_ISL_1238886, EPI_ISL_1238887, EPI_ISL_1238888, EPI_ISL_1238890, EPI_ISL_1238892, EPI_ISL_1238894, EPI_ISL_1238895, EPI_ISL_1238896, EPI_ISL_1238897, EPI_ISL_1238898, EPI_ISL_1238899 | | | |
| see above | Utah Public Health Laboratory | Utah Public Health Laboratory | Erin L. Young, Kelly F. Oakeson, Tara Gallagher |
| EPI_ISL_1238904, EPI_ISL_1238906, EPI_ISL_1238907, EPI_ISL_1238909, EPI_ISL_1238910, EPI_ISL_1238911, EPI_ISL_1238913, EPI_ISL_1238914, EPI_ISL_1238915, EPI_ISL_1238916, EPI_ISL_1238917, EPI_ISL_1238918, EPI_ISL_1238919, EPI_ISL_1238920, EPI_ISL_1238922, EPI_ISL_1238923, EPI_ISL_1238924, EPI_ISL_1238925, EPI_ISL_1238926, EPI_ISL_1238927, EPI_ISL_1238928, EPI_ISL_1238929, EPI_ISL_1238930, EPI_ISL_1238931, EPI_ISL_1238932, EPI_ISL_1238933, EPI_ISL_1238934, EPI_ISL_1238935, EPI_ISL_1238936, EPI_ISL_1238937, EPI_ISL_1238938, EPI_ISL_1238939, EPI_ISL_1238941, EPI_ISL_1238942, EPI_ISL_1238943, EPI_ISL_1238944, EPI_ISL_1238945, EPI_ISL_1238946, EPI_ISL_1238947, EPI_ISL_1238948, EPI_ISL_1238949, EPI_ISL_1238950, EPI_ISL_1238951, EPI_ISL_1238952, EPI_ISL_1238953, EPI_ISL_1238954, EPI_ISL_1238955, EPI_ISL_1238956, EPI_ISL_1238957, EPI_ISL_1238958, EPI_ISL_1238959, EPI_ISL_1238960, EPI_ISL_1238961, EPI_ISL_1238962, EPI_ISL_1238963, EPI_ISL_1238964, EPI_ISL_1238966, EPI_ISL_1238967, EPI_ISL_1238968, EPI_ISL_1238969, EPI_ISL_1238970, EPI_ISL_1238971, EPI_ISL_1238973, EPI_ISL_1238974, EPI_ISL_1238975, EPI_ISL_1238976, EPI_ISL_1238977, EPI_ISL_1238978, EPI_ISL_1238979, EPI_ISL_1238980, EPI_ISL_1238981, EPI_ISL_1238982, EPI_ISL_1238983, EPI_ISL_1238984, EPI_ISL_1238985, EPI_ISL_1238986, EPI_ISL_1238987, EPI_ISL_1238988, EPI_ISL_1238989, EPI_ISL_1238990, EPI_ISL_1238991, EPI_ISL_1238992, EPI_ISL_1238993 | | | |
| see above | Johns Hopkins Hospital Department of Pathology | Johns Hopkins Hospital Department of Pathology | C. Paul Morris, Chun Huai Luo, Adannaya Amadi, Matthew Schwartz, Nicholas Gallagher, Heba H. Mostafa |
| EPI_ISL_1238994, EPI_ISL_1238995, EPI_ISL_1238996, EPI_ISL_1238997, EPI_ISL_1238998, EPI_ISL_1238999, EPI_ISL_1239000, EPI_ISL_1239001, EPI_ISL_1239002, EPI_ISL_1239003, EPI_ISL_1239004, EPI_ISL_1239005, EPI_ISL_1239006, EPI_ISL_1239007, EPI_ISL_1239008, EPI_ISL_1239009, EPI_ISL_1239010, EPI_ISL_1239011 | | | |
| see above | GA Department of Public Health | GA Department of Public Health | Stacy Reeves, Jonathan Edwards, Cynthia Dixey, Tonia Parrott, Aliyah Fields, Taylor Smith |
| EPI_ISL_1239017, EPI_ISL_1239018, EPI_ISL_1239019, EPI_ISL_1239020, EPI_ISL_1239022, EPI_ISL_1239024, EPI_ISL_1239025, EPI_ISL_1239028, EPI_ISL_1239030, EPI_ISL_1239031 | Wyoming Public Health Laboratory | Wyoming Public Health Laboratory | Jim Mildenberger, Wanda Manley, Noah Hull, Taylor Fearing, Lynette Gumbleton, Channing Weber, Ashley Norberg, Chayse Rowley, Marley Goetz, Brian Dominguez, Elliot Thomasson, Cari Sloma, and Rob Christensen |
| EPI_ISL_1239048, EPI_ISL_1239049, EPI_ISL_1239050, EPI_ISL_1239051, EPI_ISL_1239052, EPI_ISL_1239053, EPI_ISL_1239054, EPI_ISL_1239056, EPI_ISL_1239058, EPI_ISL_1239060, EPI_ISL_1239061, EPI_ISL_1239062, EPI_ISL_1239063, EPI_ISL_1239064, EPI_ISL_1239065, EPI_ISL_1239066, EPI_ISL_1239067, EPI_ISL_1239068, EPI_ISL_1239069, EPI_ISL_1239070, EPI_ISL_1239071, EPI_ISL_1239072, EPI_ISL_1239073, EPI_ISL_1239074, EPI_ISL_1239075, EPI_ISL_1239076, EPI_ISL_1239077, EPI_ISL_1239078, EPI_ISL_1239079, EPI_ISL_1239080, EPI_ISL_1239081, EPI_ISL_1239082, EPI_ISL_1239083, EPI_ISL_1239084, EPI_ISL_1239085, EPI_ISL_1239086, EPI_ISL_1239087, EPI_ISL_1239088, EPI_ISL_1239089, EPI_ISL_1239090, EPI_ISL_1239091, EPI_ISL_1239092, EPI_ISL_1239093, EPI_ISL_1239094, EPI_ISL_1239095, EPI_ISL_1239096, EPI_ISL_1239097, EPI_ISL_1239098, EPI_ISL_1239099, EPI_ISL_1239100, EPI_ISL_1239101, EPI_ISL_1239102, EPI_ISL_1239103, EPI_ISL_1239104, EPI_ISL_1239105, EPI_ISL_1239106, EPI_ISL_1239107, EPI_ISL_1239108 | | | |
| see above | Arizona State Public Health Laboratory | Arizona State Public Health Laboratory | Trung Huynh, Jessica Escobar, Katherine Fullerton, Nobuko Fukushima, Stacy White, Linda Getsinger, Victor Waddell |
| EPI_ISL_1239140 | Ohio State University Wexner Medical Center | Polaris Molecular Laboratory | Ru P, Chappell D, Chang Y-S, Tu H, Snyder P, Pancholi P, Koenig S, Corcoran S, Jones D |
| EPI_ISL_1239141 | Ohio State University Wexner Medical Center | Polaris Molecular Laboratory | Chappell D, Ru P, Chang Y-S, Tu H, Snyder P, Pancholi P, Koenig S, Corcoran S, Jones D |
| EPI_ISL_1239142 | Ohio State University Wexner Medical Cente | Polaris Molecular Laboratory | Chang Y-S, Ru P, Chappell D, Tu H, Snyder P, Pancholi P, Koenig S, Corcoran S, Jones D |
| EPI_ISL_1239453, EPI_ISL_1239454, EPI_ISL_1239455, EPI_ISL_1239456, EPI_ISL_1239457 | Gundersen Molecular Diagnostics Laboratory | Kabara Cancer Research Institute | Craig S. Richmond, Paraic A. Kenny |
| EPI_ISL_1239769, EPI_ISL_1239770, EPI_ISL_1239771, EPI_ISL_1239772, EPI_ISL_1239773, EPI_ISL_1239774, EPI_ISL_1239775, EPI_ISL_1239776, EPI_ISL_1239777, EPI_ISL_1239778, EPI_ISL_1239779, EPI_ISL_1239780, EPI_ISL_1239781, EPI_ISL_1239782, EPI_ISL_1239783, EPI_ISL_1239784, EPI_ISL_1239785, EPI_ISL_1239786, EPI_ISL_1239787, EPI_ISL_1239788, EPI_ISL_1239789, EPI_ISL_1239790, EPI_ISL_1239791, EPI_ISL_1239792, EPI_ISL_1239793, EPI_ISL_1239794, EPI_ISL_1239795, EPI_ISL_1239796, EPI_ISL_1239797, EPI_ISL_1239798, EPI_ISL_1239799, EPI_ISL_1239800, EPI_ISL_1239801, EPI_ISL_1239802, EPI_ISL_1239803, EPI_ISL_1239804, EPI_ISL_1239805, EPI_ISL_1239806, EPI_ISL_1239807, EPI_ISL_1239808, EPI_ISL_1239809, EPI_ISL_1239810, EPI_ISL_1239811, EPI_ISL_1239812, EPI_ISL_1239813, EPI_ISL_1239814, EPI_ISL_1239815, EPI_ISL_1239816, EPI_ISL_1239817, EPI_ISL_1239818, EPI_ISL_1239819, EPI_ISL_1239820, EPI_ISL_1239821, EPI_ISL_1239822, EPI_ISL_1239823, EPI_ISL_1239824, EPI_ISL_1239825, EPI_ISL_1239826, EPI_ISL_1239827, EPI_ISL_1239828, EPI_ISL_1239829, EPI_ISL_1239830, EPI_ISL_1239831, EPI_ISL_1239832, EPI_ISL_1239833, EPI_ISL_1239834, EPI_ISL_1239835, EPI_ISL_1239836, EPI_ISL_1239837, EPI_ISL_1239838, EPI_ISL_1239839, EPI_ISL_1239840, EPI_ISL_1239841, EPI_ISL_1239842, EPI_ISL_1239843, EPI_ISL_1239844, EPI_ISL_1239845, EPI_ISL_1239846 | | | |
| see above | TriCore Reference Laboratories | Center for Global Health, University of New Mexico Health Sciences Center | Daryl Domman, Kurt Schwalm, Cecilia Thompson, Kendra Pesko, Karissa Culbreath, Darrell Dinwiddie |
| EPI_ISL_1239847 | Connecticut Veterans' Affairs Hospital | Grubaugh Lab - Yale School of Public Health | Mary Petrone, Joseph Fauver, Tara Alpert, Chantal Vogels, Isabel Ott, Shaili Gupta, Danielle Plank, Nathan Grubaugh |
| EPI_ISL_1239848 | New York Presbetyrian / Weill Cornell Medicine | Grubaugh Lab - Yale School of Public Health | Mary Petrone, Joseph Fauver, Tara Alpert, Anderson Brito, Mallery Breban, Anne Wyllie, Chantal Vogels, Annie Watkins, Chaney Kalinich, Isabel Ott, Nathan Grubaugh |
| EPI_ISL_1239883, EPI_ISL_1239884, EPI_ISL_1239885, EPI_ISL_1239886, EPI_ISL_1239887, EPI_ISL_1239888, EPI_ISL_1239889, EPI_ISL_1239890, EPI_ISL_1239891, EPI_ISL_1239892, EPI_ISL_1239893, EPI_ISL_1239894, EPI_ISL_1239895, EPI_ISL_1239896, EPI_ISL_1239897, EPI_ISL_1239898, EPI_ISL_1239899, EPI_ISL_1239900, EPI_ISL_1239901, EPI_ISL_1239902, EPI_ISL_1239903, EPI_ISL_1239904, EPI_ISL_1239905, EPI_ISL_1239906, EPI_ISL_1239907, EPI_ISL_1239908, EPI_ISL_1239909, EPI_ISL_1239910, EPI_ISL_1239911, EPI_ISL_1239912, EPI_ISL_1239913, EPI_ISL_1239914, EPI_ISL_1239915, EPI_ISL_1239916, EPI_ISL_1239917, EPI_ISL_1239918, EPI_ISL_1239919, EPI_ISL_1239920, EPI_ISL_1239921, EPI_ISL_1239922, EPI_ISL_1239923, EPI_ISL_1239924, EPI_ISL_1239925, EPI_ISL_1239926, EPI_ISL_1239927, EPI_ISL_1239928, EPI_ISL_1239929, EPI_ISL_1239930, EPI_ISL_1239931, EPI_ISL_1239932, EPI_ISL_1239933, EPI_ISL_1239934, EPI_ISL_1239935, EPI_ISL_1239936, EPI_ISL_1239937, EPI_ISL_1239938, EPI_ISL_1239939, EPI_ISL_1239940, EPI_ISL_1239941, EPI_ISL_1239942, EPI_ISL_1239943, EPI_ISL_1239944, EPI_ISL_1239972, EPI_ISL_1239973, EPI_ISL_1239974, EPI_ISL_1239975, EPI_ISL_1239976, EPI_ISL_1239977, EPI_ISL_1239978, EPI_ISL_1239979, EPI_ISL_1239980, EPI_ISL_1239981, EPI_ISL_1239982, EPI_ISL_1239983, EPI_ISL_1239984, EPI_ISL_1239985, EPI_ISL_1239986, EPI_ISL_1239987, EPI_ISL_1239988, EPI_ISL_1239989, EPI_ISL_1239990, EPI_ISL_1239991, EPI_ISL_1239992, EPI_ISL_1239993, EPI_ISL_1239994, EPI_ISL_1239995, EPI_ISL_1239996, EPI_ISL_1239997, EPI_ISL_1239998, EPI_ISL_1239999, EPI_ISL_1240000, EPI_ISL_1240001, EPI_ISL_1240002, EPI_ISL_1240003, EPI_ISL_1240004, EPI_ISL_1240005, EPI_ISL_1240006, EPI_ISL_1240007, EPI_ISL_1240008, EPI_ISL_1240009, EPI_ISL_1240010, EPI_ISL_1240011, EPI_ISL_1240012, EPI_ISL_1240013, EPI_ISL_1240014, EPI_ISL_1240015, EPI_ISL_1240016, EPI_ISL_1240017, EPI_ISL_1240018, EPI_ISL_1240019, EPI_ISL_1240020, EPI_ISL_1240021, EPI_ISL_1240022, EPI_ISL_1240023, EPI_ISL_1240024, EPI_ISL_1240025, EPI_ISL_1240026, EPI_ISL_1240027, EPI_ISL_1240028, EPI_ISL_1240029, EPI_ISL_1240030, EPI_ISL_1240031, EPI_ISL_1240032, EPI_ISL_1240033, EPI_ISL_1240034, EPI_ISL_1240035, EPI_ISL_1240036, EPI_ISL_1240037, EPI_ISL_1240038, EPI_ISL_1240039, EPI_ISL_1240040, EPI_ISL_1240041, EPI_ISL_1240042, EPI_ISL_1240043, EPI_ISL_1240044, EPI_ISL_1240045, EPI_ISL_1240046, EPI_ISL_1240047, EPI_ISL_1240048, EPI_ISL_1240049, EPI_ISL_1240050, EPI_ISL_1240051, EPI_ISL_1240052, EPI_ISL_1240053, EPI_ISL_1240054, EPI_ISL_1240055, EPI_ISL_1240056, EPI_ISL_1240057, EPI_ISL_1240058, EPI_ISL_1240059, EPI_ISL_1240060, EPI_ISL_1240061, EPI_ISL_1240062, EPI_ISL_1240063, EPI_ISL_1240064, EPI_ISL_1240065, EPI_ISL_1240066, EPI_ISL_1240067, EPI_ISL_1240068, EPI_ISL_1240069, EPI_ISL_1240070 | | | |
| see above | Florida Bureau of Public Health Laboratories | Florida Bureau of Public Health Laboratories | Sarah Schmedes, Jason Blanton |
| EPI_ISL_1240726, EPI_ISL_1240727, EPI_ISL_1240728, EPI_ISL_1240729, EPI_ISL_1240730, EPI_ISL_1240731, EPI_ISL_1240732, EPI_ISL_1240733, EPI_ISL_1240734, EPI_ISL_1240735, EPI_ISL_1240736, EPI_ISL_1240737, EPI_ISL_1240738, EPI_ISL_1240739, EPI_ISL_1240740, EPI_ISL_1240741, EPI_ISL_1240742, EPI_ISL_1240743, EPI_ISL_1240744, EPI_ISL_1240745, EPI_ISL_1240746, EPI_ISL_1240747, EPI_ISL_1240748, EPI_ISL_1240749, EPI_ISL_1240750, EPI_ISL_1240751, EPI_ISL_1240752, EPI_ISL_1240753, EPI_ISL_1240754, EPI_ISL_1240755, EPI_ISL_1240756, EPI_ISL_1240757, EPI_ISL_1240758, EPI_ISL_1240759, EPI_ISL_1240760, EPI_ISL_1240761, EPI_ISL_1240762, EPI_ISL_1240763, EPI_ISL_1240764, EPI_ISL_1240765, EPI_ISL_1240766, EPI_ISL_1240767, EPI_ISL_1240768, EPI_ISL_1240769, EPI_ISL_1240770, EPI_ISL_1240771, EPI_ISL_1240772, EPI_ISL_1240773, EPI_ISL_1240774, EPI_ISL_1240775, EPI_ISL_1240776, EPI_ISL_1240777, EPI_ISL_1240778, EPI_ISL_1240779, EPI_ISL_1240780, EPI_ISL_1240781, EPI_ISL_1240782, EPI_ISL_1240783, EPI_ISL_1240784, EPI_ISL_1240785, EPI_ISL_1240786, EPI_ISL_1240787, EPI_ISL_1240788, EPI_ISL_1240789, EPI_ISL_1240790, EPI_ISL_1240791, EPI_ISL_1240792, EPI_ISL_1240793, EPI_ISL_1240794 | | | |
| see above | University Hospitals Translational | University Hospitals Translational Laboratory | Sadri,N., Alouani,D., Song,X. |

| | | | |
|---|---|---|---|
| | Laboratory (UHTL), University Hospitals | (UHTL), University Hospitals | |
| EPI_ISL_1240795, EPI_ISL_1240796, EPI_ISL_1240797, EPI_ISL_1240798, EPI_ISL_1240799, EPI_ISL_1240800, EPI_ISL_1240801, EPI_ISL_1240802, EPI_ISL_1240803, EPI_ISL_1240804, EPI_ISL_1240805, EPI_ISL_1240806, EPI_ISL_1240807, EPI_ISL_1240808, EPI_ISL_1240809, EPI_ISL_1240810, EPI_ISL_1240811, EPI_ISL_1240813, EPI_ISL_1240814, EPI_ISL_1240815, EPI_ISL_1240816, EPI_ISL_1240817, EPI_ISL_1240818, EPI_ISL_1240819, EPI_ISL_1240820, EPI_ISL_1240821, EPI_ISL_1240822, EPI_ISL_1240823, EPI_ISL_1240824, EPI_ISL_1240825, EPI_ISL_1240826, EPI_ISL_1240827, EPI_ISL_1240828, EPI_ISL_1240829, EPI_ISL_1240830, EPI_ISL_1240832, EPI_ISL_1240833, EPI_ISL_1240834, EPI_ISL_1240835, EPI_ISL_1240836, EPI_ISL_1240837, EPI_ISL_1240839, EPI_ISL_1240840, EPI_ISL_1240841, EPI_ISL_1240842, EPI_ISL_1240843, EPI_ISL_1240844, EPI_ISL_1240845, EPI_ISL_1240846, EPI_ISL_1240847, EPI_ISL_1240848, EPI_ISL_1240849, EPI_ISL_1240850, EPI_ISL_1240851, EPI_ISL_1240852, EPI_ISL_1240853, EPI_ISL_1240854, EPI_ISL_1240855, EPI_ISL_1240856, EPI_ISL_1240857, EPI_ISL_1240858, EPI_ISL_1240859, EPI_ISL_1240860, EPI_ISL_1240861, EPI_ISL_1240862, EPI_ISL_1240863, EPI_ISL_1240864, EPI_ISL_1240865, EPI_ISL_1240866, EPI_ISL_1240868, EPI_ISL_1240869, EPI_ISL_1240870, EPI_ISL_1240871, EPI_ISL_1240872, EPI_ISL_1240874, EPI_ISL_1240875, EPI_ISL_1240876, EPI_ISL_1240878, EPI_ISL_1240879, EPI_ISL_1240880, EPI_ISL_1240881, EPI_ISL_1240882, EPI_ISL_1240883, EPI_ISL_1240884, EPI_ISL_1240886, EPI_ISL_1240887, EPI_ISL_1240888, EPI_ISL_1240889, EPI_ISL_1240890, EPI_ISL_1240891, EPI_ISL_1240892, EPI_ISL_1240893, EPI_ISL_1240894, EPI_ISL_1240895, EPI_ISL_1240896, EPI_ISL_1240897, EPI_ISL_1240900, EPI_ISL_1240901, EPI_ISL_1240902, EPI_ISL_1240903, EPI_ISL_1240904, EPI_ISL_1240908, EPI_ISL_1240909, EPI_ISL_1240910, EPI_ISL_1240911, EPI_ISL_1240913, EPI_ISL_1240914, EPI_ISL_1240915, EPI_ISL_1240916, EPI_ISL_1240917, EPI_ISL_1240918, EPI_ISL_1240919, EPI_ISL_1240920, EPI_ISL_1240923, EPI_ISL_1240924, EPI_ISL_1240925, EPI_ISL_1240926, EPI_ISL_1240928, EPI_ISL_1240930, EPI_ISL_1240931, EPI_ISL_1240932, EPI_ISL_1240933, EPI_ISL_1240934 | | | |
| see above | Clinical Molecular Microbiology Laboratory, UNC Hospitals | Jeremy Wang | Jeremy Wang, Alexander Rubinsteyn, Colleen Rice, Jason Smedberg, Shawn Hawken, Melissa Miller, Corbin Jones, Robert Hagan |
| EPI_ISL_1247017, EPI_ISL_1247018, EPI_ISL_1247359, EPI_ISL_1247746, EPI_ISL_1248090, EPI_ISL_1248476 | Ohio Department of Health Laboratory | Ohio Department of Health Laboratory | Holmes, Jennifer; Eric Brandt, Keoni Omura, Glen McGillivary, Caitlin McDonnell, Jade Mowery, Stephanie Mccracken, Tyler Payne, Kirtana Ramadugu, Erica Leasure, Brent Lee, Kelsey Florek, Heather Blankenship, Allison Black, Quanta Brown, and Tammy Bannerman |
| EPI_ISL_1250320, EPI_ISL_1250321, EPI_ISL_1250322, EPI_ISL_1250323, EPI_ISL_1250324, EPI_ISL_1250325, EPI_ISL_1250326, EPI_ISL_1250327, EPI_ISL_1250328, EPI_ISL_1250329, EPI_ISL_1250330, EPI_ISL_1250331, EPI_ISL_1250332, EPI_ISL_1250333, EPI_ISL_1250334, EPI_ISL_1250336, EPI_ISL_1250337, EPI_ISL_1250338, EPI_ISL_1250339, EPI_ISL_1250340, EPI_ISL_1250341, EPI_ISL_1250342, EPI_ISL_1250343, EPI_ISL_1250344, EPI_ISL_1250345, EPI_ISL_1250347, EPI_ISL_1250348, EPI_ISL_1250349, EPI_ISL_1250350, EPI_ISL_1250351, EPI_ISL_1250352, EPI_ISL_1250353, EPI_ISL_1250354, EPI_ISL_1250355, EPI_ISL_1250357, EPI_ISL_1250360, EPI_ISL_1250363, EPI_ISL_1250365, EPI_ISL_1250368, EPI_ISL_1250372, EPI_ISL_1250374, EPI_ISL_1250377, EPI_ISL_1250380, EPI_ISL_1250382, EPI_ISL_1250384, EPI_ISL_1250387, EPI_ISL_1250389, EPI_ISL_1250391, EPI_ISL_1250395, EPI_ISL_1250398, EPI_ISL_1250400, EPI_ISL_1250403, EPI_ISL_1250406, EPI_ISL_1250408, EPI_ISL_1250415, EPI_ISL_1250421, EPI_ISL_1250423, EPI_ISL_1250426, EPI_ISL_1250429, EPI_ISL_1250433, EPI_ISL_1250435, EPI_ISL_1250438, EPI_ISL_1250440, EPI_ISL_1250443, EPI_ISL_1250445, EPI_ISL_1250451, EPI_ISL_1250454, EPI_ISL_1250457, EPI_ISL_1250470, EPI_ISL_1250473, EPI_ISL_1250478, EPI_ISL_1250481, EPI_ISL_1250484, EPI_ISL_1250487, EPI_ISL_1250489, EPI_ISL_1250492, EPI_ISL_1250495, EPI_ISL_1250498, EPI_ISL_1250500, EPI_ISL_1250506, EPI_ISL_1250508, EPI_ISL_1250510, EPI_ISL_1250511, EPI_ISL_1250513, EPI_ISL_1250516, EPI_ISL_1250519, EPI_ISL_1250521, EPI_ISL_1250524, EPI_ISL_1250527, EPI_ISL_1250532, EPI_ISL_1250534, EPI_ISL_1250536, EPI_ISL_1250539, EPI_ISL_1250541, EPI_ISL_1250544, EPI_ISL_1250546, EPI_ISL_1250549, EPI_ISL_1250554, EPI_ISL_1250555, EPI_ISL_1250556, EPI_ISL_1250557, EPI_ISL_1250558, EPI_ISL_1250559, EPI_ISL_1250560, EPI_ISL_1250561, EPI_ISL_1250562, EPI_ISL_1250563, EPI_ISL_1250564, EPI_ISL_1250565, EPI_ISL_1250566, EPI_ISL_1250567, EPI_ISL_1250568, EPI_ISL_1250569, EPI_ISL_1250570, EPI_ISL_1250571, EPI_ISL_1250572, EPI_ISL_1250573, EPI_ISL_1250575, EPI_ISL_1250576, EPI_ISL_1250577, EPI_ISL_1250578 | | | |
| see above | Michigan Department of Health and Human Services, Bureau of Laboratories | Michigan Department of Health and Human Services, Bureau of Laboratories | Blankenship HM, Riner D, Soehnlen MK |
| EPI_ISL_1250843, EPI_ISL_1250847, EPI_ISL_1250848, EPI_ISL_1250849, EPI_ISL_1250850 | NCSLPH | NCSLPH | Chase K, Miller MC, Greene S, Glover W |
| EPI_ISL_1250852 | Florida Bureau of Public Health Laboratories | Florida Bureau of Public Health Laboratories | Sarah Schmedes, Jason Blanton |
| EPI_ISL_1251226 | Microbiology Division, South Carolina Department of Health and Environmental Control Public Health Laboratory (SC DHEC PHL) | Microbiology Division, South Carolina Department of Health and Environmental Control Public Health Laboratory (SC DHEC PHL) | Haley Flores, Jessica Freeman |
| EPI_ISL_1251260, EPI_ISL_1251273, EPI_ISL_1252050 | Fulgent Genetics | Fulgent Genetics | Harry Gao, Mickey Li, John Gao, Joseph Fierro, Benafsh Sapra, Becky Tsai, Yan Meng, Doreen Ng, James Xie |
| EPI_ISL_1252345, EPI_ISL_1252350, EPI_ISL_1252351, EPI_ISL_1252352, EPI_ISL_1252353, EPI_ISL_1252354, EPI_ISL_1252355, EPI_ISL_1252356, EPI_ISL_1252357, EPI_ISL_1252358, EPI_ISL_1252359, EPI_ISL_1252360, EPI_ISL_1252361, EPI_ISL_1252362, EPI_ISL_1252363, EPI_ISL_1252456, EPI_ISL_1252457, EPI_ISL_1252458, EPI_ISL_1252459, EPI_ISL_1252460, EPI_ISL_1252461, EPI_ISL_1252462, EPI_ISL_1252463, EPI_ISL_1252464, EPI_ISL_1252465, EPI_ISL_1252468, EPI_ISL_1252469, EPI_ISL_1252470, EPI_ISL_1252471, EPI_ISL_1252472, EPI_ISL_1252473, EPI_ISL_1252556, EPI_ISL_1252557, EPI_ISL_1252558, EPI_ISL_1252559, EPI_ISL_1252560, EPI_ISL_1252561, EPI_ISL_1252562, EPI_ISL_1252563, EPI_ISL_1252564, EPI_ISL_1252565, EPI_ISL_1252566, EPI_ISL_1252567, EPI_ISL_1252568, EPI_ISL_1252569, EPI_ISL_1252570, EPI_ISL_1252571, EPI_ISL_1252572, EPI_ISL_1252573, EPI_ISL_1252574, EPI_ISL_1252575, EPI_ISL_1252576, EPI_ISL_1252577, EPI_ISL_1252578, EPI_ISL_1252579, EPI_ISL_1252580, EPI_ISL_1252581, EPI_ISL_1252582, EPI_ISL_1252583, EPI_ISL_1252584, EPI_ISL_1252585, EPI_ISL_1252586, EPI_ISL_1252587 | | | |
| see above | University of Wisconsin-Madison AIDS Vaccine Research Laboratories | University of Wisconsin-Madison AIDS Vaccine Research Laboratories | Gage Moreno, Katarina Braun, et al. AIDS Vaccine Research Laboratories |
| EPI_ISL_1252588, EPI_ISL_1252589, EPI_ISL_1252590, EPI_ISL_1252591, EPI_ISL_1252592, EPI_ISL_1252593, EPI_ISL_1252594, EPI_ISL_1252595, EPI_ISL_1252596, EPI_ISL_1252597, EPI_ISL_1252598, EPI_ISL_1252599, EPI_ISL_1252600, EPI_ISL_1252601, EPI_ISL_1252602, EPI_ISL_1252603, EPI_ISL_1252604, EPI_ISL_1252605, EPI_ISL_1252606, EPI_ISL_1252607, EPI_ISL_1252608, EPI_ISL_1252609, EPI_ISL_1252610, EPI_ISL_1252611, EPI_ISL_1252612, EPI_ISL_1252613, EPI_ISL_1252614, EPI_ISL_1252615, EPI_ISL_1252616, EPI_ISL_1252617, EPI_ISL_1252618, EPI_ISL_1252619, EPI_ISL_1252620, EPI_ISL_1252621, EPI_ISL_1252622, EPI_ISL_1252623, EPI_ISL_1252624, EPI_ISL_1252625, EPI_ISL_1252626, EPI_ISL_1252627, EPI_ISL_1252628, EPI_ISL_1252629, EPI_ISL_1252630, EPI_ISL_1252631, EPI_ISL_1252632, EPI_ISL_1252633, EPI_ISL_1252634, EPI_ISL_1252636, EPI_ISL_1252637, EPI_ISL_1252638, EPI_ISL_1252639, EPI_ISL_1252640, EPI_ISL_1252641, EPI_ISL_1252642, EPI_ISL_1252643, EPI_ISL_1252644, EPI_ISL_1252645, EPI_ISL_1252646, EPI_ISL_1252647, EPI_ISL_1252648, EPI_ISL_1252649, EPI_ISL_1252650, EPI_ISL_1252651, EPI_ISL_1252652, EPI_ISL_1252653, EPI_ISL_1252654, EPI_ISL_1252655, EPI_ISL_1252656, EPI_ISL_1252657, EPI_ISL_1252658, EPI_ISL_1252659, EPI_ISL_1252661, EPI_ISL_1252662, EPI_ISL_1252663, EPI_ISL_1252664, EPI_ISL_1252665, EPI_ISL_1252666, EPI_ISL_1252667, EPI_ISL_1252668, EPI_ISL_1252669, EPI_ISL_1252670 | | | |
| see above | TXDSHS | TXDSHS | Rashmi Tuladhar, Bonnie Oh, Jenny Zhang, Maliha Rahman, Mayela Pedrueza, Anita Pokharel, Myong Koag, Chun Wang, Rachel Lee, Grace Kubin |
| EPI_ISL_1252844, EPI_ISL_1252845, EPI_ISL_1252846 | INNO Diagnostics Reference Laboratory | RCMI-Center for Research Resources, Ponce Research Institute | Vanessa Rivera-Amill, Andrea Arias-García, Raphael Sánchez-Torres |
| EPI_ISL_1252847, EPI_ISL_1252848, EPI_ISL_1252849, EPI_ISL_1252850, EPI_ISL_1252851, EPI_ISL_1252852, EPI_ISL_1252853, EPI_ISL_1252854, EPI_ISL_1252855, EPI_ISL_1252856, EPI_ISL_1252857, EPI_ISL_1252858, EPI_ISL_1252859, EPI_ISL_1252860, EPI_ISL_1252861, EPI_ISL_1252862, EPI_ISL_1252863, EPI_ISL_1252864, EPI_ISL_1252865, EPI_ISL_1252866, EPI_ISL_1252867, EPI_ISL_1252868, EPI_ISL_1252869, EPI_ISL_1252870, EPI_ISL_1252871, EPI_ISL_1252872, EPI_ISL_1252873, EPI_ISL_1252874, EPI_ISL_1252875, EPI_ISL_1252876, EPI_ISL_1252877, EPI_ISL_1252878, EPI_ISL_1252879, EPI_ISL_1252880, EPI_ISL_1252881, EPI_ISL_1252882, EPI_ISL_1252883, EPI_ISL_1252884, EPI_ISL_1252885, EPI_ISL_1252886, EPI_ISL_1252887, EPI_ISL_1252888, EPI_ISL_1252889, EPI_ISL_1252890, EPI_ISL_1252891, EPI_ISL_1252905, EPI_ISL_1252907, EPI_ISL_1252911 | | | |
| see above | University of Wisconsin-Madison AIDS Vaccine Research Laboratories | University of Wisconsin-Madison AIDS Vaccine Research Laboratories | Gage Moreno, Katarina Braun, et al. AIDS Vaccine Research Laboratories |
| EPI_ISL_1252917, EPI_ISL_1252918, EPI_ISL_1252919, EPI_ISL_1252920, EPI_ISL_1252921, EPI_ISL_1252922, EPI_ISL_1252923, EPI_ISL_1252924, EPI_ISL_1252925, EPI_ISL_1252926, EPI_ISL_1252927, EPI_ISL_1252928, EPI_ISL_1252929, EPI_ISL_1252930, EPI_ISL_1252931, EPI_ISL_1252932, EPI_ISL_1252933, EPI_ISL_1252934, EPI_ISL_1252935, EPI_ISL_1252936, EPI_ISL_1252937, EPI_ISL_1252938, EPI_ISL_1252939, EPI_ISL_1252940, EPI_ISL_1252941, EPI_ISL_1252942, EPI_ISL_1252943, EPI_ISL_1252944, EPI_ISL_1252945, EPI_ISL_1252946, EPI_ISL_1252947, EPI_ISL_1252948, EPI_ISL_1252949, EPI_ISL_1252950, EPI_ISL_1252951, EPI_ISL_1252952, EPI_ISL_1252953, EPI_ISL_1252954, EPI_ISL_1252955, EPI_ISL_1252956, EPI_ISL_1252957, EPI_ISL_1252958, EPI_ISL_1252959, EPI_ISL_1252960, EPI_ISL_1252961, EPI_ISL_1252962, EPI_ISL_1252963, EPI_ISL_1252964, EPI_ISL_1252965, EPI_ISL_1252966, EPI_ISL_1252967, EPI_ISL_1252968, EPI_ISL_1252969, EPI_ISL_1252970, EPI_ISL_1252971, EPI_ISL_1252972, EPI_ISL_1252973, EPI_ISL_1252974, EPI_ISL_1252975, EPI_ISL_1252976, EPI_ISL_1252977, EPI_ISL_1252978, EPI_ISL_1252979, EPI_ISL_1252980, EPI_ISL_1252981, EPI_ISL_1252982, EPI_ISL_1252983, EPI_ISL_1252984, EPI_ISL_1252985, EPI_ISL_1252986, EPI_ISL_1252987, EPI_ISL_1252988, EPI_ISL_1252989, EPI_ISL_1252990, EPI_ISL_1252991, EPI_ISL_1252992, EPI_ISL_1252993, EPI_ISL_1252994, EPI_ISL_1252995, EPI_ISL_1252996, EPI_ISL_1252997, EPI_ISL_1252998, EPI_ISL_1252999, EPI_ISL_1253000, EPI_ISL_1253001, EPI_ISL_1253002, EPI_ISL_1253003, EPI_ISL_1253004, EPI_ISL_1253005, EPI_ISL_1253006, EPI_ISL_1253007, EPI_ISL_1253008, EPI_ISL_1253009, EPI_ISL_1253010, EPI_ISL_1253011, EPI_ISL_1253012, EPI_ISL_1253013, EPI_ISL_1253014, EPI_ISL_1253015, EPI_ISL_1253016, EPI_ISL_1253017, EPI_ISL_1253018, EPI_ISL_1253019, EPI_ISL_1253020, EPI_ISL_1253021, EPI_ISL_1253022, EPI_ISL_1253023, EPI_ISL_1253024, EPI_ISL_1253025, EPI_ISL_1253026, EPI_ISL_1253027, EPI_ISL_1253028, EPI_ISL_1253029, EPI_ISL_1253030, EPI_ISL_1253031, EPI_ISL_1253032, EPI_ISL_1253033, EPI_ISL_1253034, EPI_ISL_1253035, EPI_ISL_1253036, EPI_ISL_1253037, EPI_ISL_1253038, EPI_ISL_1253039, EPI_ISL_1253040, EPI_ISL_1253041, EPI_ISL_1253042, EPI_ISL_1253043, EPI_ISL_1253044, EPI_ISL_1253045, EPI_ISL_1253046, EPI_ISL_1253047, EPI_ISL_1253048, EPI_ISL_1253049, EPI_ISL_1253050, EPI_ISL_1253051, EPI_ISL_1253052, EPI_ISL_1253053, EPI_ISL_1253054, EPI_ISL_1253055, EPI_ISL_1253056, EPI_ISL_1253057, EPI_ISL_1253058, EPI_ISL_1253059, EPI_ISL_1253060, EPI_ISL_1253061, EPI_ISL_1253062, EPI_ISL_1253063, EPI_ISL_1253064, EPI_ISL_1253065, EPI_ISL_1253066, EPI_ISL_1253067, EPI_ISL_1253069, EPI_ISL_1253070, EPI_ISL_1253071, EPI_ISL_1253072, EPI_ISL_1253073, EPI_ISL_1253074, EPI_ISL_1253075, EPI_ISL_1253076, EPI_ISL_1253077, EPI_ISL_1253078, EPI_ISL_1253079, EPI_ISL_1253080, EPI_ISL_1253081, EPI_ISL_1253082, EPI_ISL_1253083, EPI_ISL_1253084, EPI_ISL_1253085, EPI_ISL_1253086, EPI_ISL_1253087, EPI_ISL_1253088, EPI_ISL_1253089, EPI_ISL_1253090, EPI_ISL_1253091, EPI_ISL_1253092, EPI_ISL_1253093, EPI_ISL_1253094, EPI_ISL_1253095, EPI_ISL_1253096, EPI_ISL_1253097, EPI_ISL_1253098, EPI_ISL_1253099, EPI_ISL_1253100, EPI_ISL_1253101, EPI_ISL_1253102, EPI_ISL_1253103, EPI_ISL_1253104, EPI_ISL_1253105, EPI_ISL_1253106, EPI_ISL_1253107, EPI_ISL_1253108, EPI_ISL_1253109, EPI_ISL_1253110, EPI_ISL_1253111, EPI_ISL_1253112, EPI_ISL_1253113, EPI_ISL_1253114, EPI_ISL_1253115, EPI_ISL_1253116, EPI_ISL_1253117, EPI_ISL_1253118, EPI_ISL_1253119, EPI_ISL_1253120, EPI_ISL_1253121, EPI_ISL_1253122, EPI_ISL_1253123, EPI_ISL_1253124, EPI_ISL_1253125, EPI_ISL_1253126, EPI_ISL_1253127, EPI_ISL_1253128, EPI_ISL_1253129, EPI_ISL_1253130, EPI_ISL_1253131, EPI_ISL_1253132, EPI_ISL_1253133, EPI_ISL_1253134, EPI_ISL_1253135, EPI_ISL_1253136, EPI_ISL_1253137, EPI_ISL_1253138, EPI_ISL_1253139, EPI_ISL_1253140, EPI_ISL_1253141, EPI_ISL_1253142, EPI_ISL_1253143, EPI_ISL_1253144, EPI_ISL_1253145, EPI_ISL_1253146, EPI_ISL_1253147, EPI_ISL_1253148, EPI_ISL_1253149, EPI_ISL_1253150, EPI_ISL_1253151, EPI_ISL_1253152, EPI_ISL_1253153, EPI_ISL_1253154, EPI_ISL_1253155, EPI_ISL_1253156, EPI_ISL_1253157, EPI_ISL_1253158, EPI_ISL_1253159, EPI_ISL_1253160, EPI_ISL_1253161, EPI_ISL_1253162, EPI_ISL_1253163, EPI_ISL_1253164, EPI_ISL_1253165, EPI_ISL_1253166, EPI_ISL_1253167, EPI_ISL_1253168, EPI_ISL_1253169, EPI_ISL_1253170, EPI_ISL_1253171, EPI_ISL_1253172, EPI_ISL_1253173, EPI_ISL_1253174, EPI_ISL_1253175, EPI_ISL_1253176, EPI_ISL_1253177, EPI_ISL_1253178, EPI_ISL_1253179, EPI_ISL_1253180, EPI_ISL_1253181, EPI_ISL_1253182, EPI_ISL_1253183, EPI_ISL_1253184, EPI_ISL_1253185, EPI_ISL_1253186, EPI_ISL_1253187, EPI_ISL_1253188, EPI_ISL_1253189, EPI_ISL_1253190, EPI_ISL_1253191, EPI_ISL_1253192, EPI_ISL_1253193, EPI_ISL_1253194, EPI_ISL_1253195, EPI_ISL_1253196, EPI_ISL_1253197, EPI_ISL_1253198, EPI_ISL_1253199, EPI_ISL_1253200, EPI_ISL_1253201, EPI_ISL_1253202, EPI_ISL_1253203, EPI_ISL_1253204, EPI_ISL_1253205, EPI_ISL_1253206, EPI_ISL_1253207, EPI_ISL_1253208, EPI_ISL_1253209, EPI_ISL_1253210, EPI_ISL_1253211, EPI_ISL_1253212, EPI_ISL_1253213, EPI_ISL_1253214, EPI_ISL_1253215, EPI_ISL_1253216, EPI_ISL_1253217, EPI_ISL_1253218, EPI_ISL_1253219, EPI_ISL_1253220, EPI_ISL_1253221, EPI_ISL_1253222, EPI_ISL_1253223, EPI_ISL_1253224, EPI_ISL_1253225, EPI_ISL_1253226, EPI_ISL_1253227, EPI_ISL_1253228, EPI_ISL_1253229, EPI_ISL_1253230, EPI_ISL_1253231, EPI_ISL_1253232, EPI_ISL_1253233, EPI_ISL_1253234, EPI_ISL_1253235, EPI_ISL_1253236, EPI_ISL_1253237, EPI_ISL_1253238, EPI_ISL_1253239, EPI_ISL_1253240, EPI_ISL_1253241, EPI_ISL_1253242, EPI_ISL_1253243, EPI_ISL_1253244, EPI_ISL_1253245, EPI_ISL_1253246, EPI_ISL_1253247, EPI_ISL_1253248, EPI_ISL_1253249, EPI_ISL_1253250, EPI_ISL_1253251, EPI_ISL_1253252, EPI_ISL_1253253, EPI_ISL_1253254, EPI_ISL_1253255, EPI_ISL_1253256, EPI_ISL_1253257, EPI_ISL_1253258, EPI_ISL_1253259, EPI_ISL_1253260, EPI_ISL_1253261, EPI_ISL_1253262, EPI_ISL_1253263, EPI_ISL_1253264, EPI_ISL_1253265, EPI_ISL_1253266, EPI_ISL_1253267, EPI_ISL_1253268, EPI_ISL_1253269, EPI_ISL_1253270, EPI_ISL_1253271, EPI_ISL_1253272, EPI_ISL_1253273, EPI_ISL_1253274, EPI_ISL_1253275, EPI_ISL_1253276, EPI_ISL_1253277, EPI_ISL_1253278, EPI_ISL_1253279, EPI_ISL_1253280, EPI_ISL_1253281, EPI_ISL_1253282, EPI_ISL_1253283, EPI_ISL_1253284, EPI_ISL_1253285, EPI_ISL_1253286, EPI_ISL_1253287, EPI_ISL_1253288, EPI_ISL_1253289, EPI_ISL_1253290 | | | |
| see above | Quest Diagnostics Incorporated | Centers for Disease Control and Prevention Division of Viral Diseases, Pathogen Discovery | Peter W. Cook, Dakota Howard, Dhwani Batra, Ben L. Rambo-Martin, S. H. Rosenthal, A. Gerasimova, R. M. Kagan, B. Anderson, M. Hua, Y. Liu, L.E. Bernstein, K.E. Livingston, A. Perez, I. A. Shlyakhter, R. V. Rolando, R. Owen, P. Tanpaiboon, F. Lacbawan, Clinton R. Paden, Suxiang Tong, Duncan MacCannell |
| EPI_ISL_1253291 | University of Wisconsin-Madison AIDS Vaccine Research Laboratories | University of Wisconsin-Madison AIDS Vaccine Research Laboratories | Gage Moreno, Katarina Braun, et al. AIDS Vaccine Research Laboratories |
| EPI_ISL_1253292 | Quest Diagnostics Incorporated | Centers for Disease Control and Prevention Division of Viral Diseases, Pathogen Discovery | Peter W. Cook, Dakota Howard, Dhwani Batra, Ben L. Rambo-Martin, S. H. Rosenthal, A. Gerasimova, R. M. Kagan, B. Anderson, M. Hua, Y. Liu, L.E. Bernstein, K.E. Livingston, A. Perez, I. A. Shlyakhter, R. V. Rolando, R. Owen, P. Tanpaiboon, F. Lacbawan, Clinton R. Paden, Suxiang Tong, Duncan MacCannell |
| EPI_ISL_1253293 | University of Wisconsin-Madison AIDS Vaccine Research Laboratories | University of Wisconsin-Madison AIDS Vaccine Research Laboratories | Gage Moreno, Katarina Braun, et al. AIDS Vaccine Research Laboratories |
| EPI_ISL_1253294 | Quest Diagnostics Incorporated | Centers for Disease Control and Prevention Division of Viral Diseases, Pathogen Discovery | Peter W. Cook, Dakota Howard, Dhwani Batra, Ben L. Rambo-Martin, S. H. Rosenthal, A. Gerasimova, R. M. Kagan, B. Anderson, M. Hua, Y. Liu, L.E. Bernstein, K.E. Livingston, A. Perez, I. A. Shlyakhter, R. V. Rolando, R. Owen, P. Tanpaiboon, F. Lacbawan, Clinton R. Paden, Suxiang Tong, Duncan MacCannell |
| EPI_ISL_1253295, EPI_ISL_1253296 | University of Wisconsin-Madison AIDS Vaccine Research Laboratories | University of Wisconsin-Madison AIDS Vaccine Research Laboratories | Gage Moreno, Katarina Braun, et al. AIDS Vaccine Research Laboratories |
| EPI_ISL_1253297 | Quest Diagnostics Incorporated | Centers for Disease Control and Prevention Division of Viral Diseases, Pathogen Discovery | Peter W. Cook, Dakota Howard, Dhwani Batra, Ben L. Rambo-Martin, S. H. Rosenthal, A. Gerasimova, R. M. Kagan, B. Anderson, M. Hua, Y. Liu, L.E. Bernstein, K.E. Livingston, A. Perez, I. A. Shlyakhter, R. V. Rolando, R. Owen, P. Tanpaiboon, F. Lacbawan, Clinton R. Paden, Suxiang Tong, Duncan MacCannell |
| EPI_ISL_1253298, EPI_ISL_1253299 | University of Wisconsin-Madison AIDS Vaccine Research Laboratories | University of Wisconsin-Madison AIDS Vaccine Research Laboratories | Gage Moreno, Katarina Braun, et al. AIDS Vaccine Research Laboratories |
| EPI_ISL_1253300 | Quest Diagnostics Incorporated | Centers for Disease Control and Prevention Division of Viral Diseases, Pathogen Discovery | Peter W. Cook, Dakota Howard, Dhwani Batra, Ben L. Rambo-Martin, S. H. Rosenthal, A. Gerasimova, R. M. Kagan, B. Anderson, M. Hua, Y. Liu, L.E. Bernstein, K.E. Livingston, A. Perez, I. A. Shlyakhter, R. V. Rolando, R. Owen, P. Tanpaiboon, F. Lacbawan, Clinton R. Paden, Suxiang Tong, Duncan MacCannell |
| EPI_ISL_1253301, EPI_ISL_1253302 | University of Wisconsin-Madison AIDS Vaccine Research Laboratories | University of Wisconsin-Madison AIDS Vaccine Research Laboratories | Gage Moreno, Katarina Braun, et al. AIDS Vaccine Research Laboratories |

| | | | |
|---|---|---|---|
| EPI_ISL_1253303 | Quest Diagnostics Incorporated | Centers for Disease Control and Prevention Division of Viral Diseases, Pathogen Discovery | Peter W. Cook, Dakota Howard, Dhwani Batra, Ben L. Rambo-Martin, S. H. Rosenthal, A. Gerasimova, R. M. Kagan, B. Anderson, M. Hua, Y. Liu, L.E. Bernstein, K.E. Livingston, A. Perez, I. A. Shlyakhter, R. V. Rolando, R. Owen, P. Tanpaiboon, F. Lacbawan, Clinton R. Paden, Suxiang Tong, Duncan MacCannell |
| EPI_ISL_1253304, EPI_ISL_1253305 | University of Wisconsin-Madison AIDS Vaccine Research Laboratories | University of Wisconsin-Madison AIDS Vaccine Research Laboratories | Gage Moreno, Katarina Braun, et al. AIDS Vaccine Research Laboratories |
| EPI_ISL_1253306, EPI_ISL_1253307, EPI_ISL_1253308, EPI_ISL_1253309, EPI_ISL_1253310, EPI_ISL_1253311, EPI_ISL_1253312, EPI_ISL_1253313, EPI_ISL_1253314, EPI_ISL_1253315, EPI_ISL_1253316, EPI_ISL_1253317, EPI_ISL_1253318, EPI_ISL_1253319, EPI_ISL_1253320, EPI_ISL_1253321, EPI_ISL_1253322, EPI_ISL_1253323, EPI_ISL_1253324, EPI_ISL_1253325, EPI_ISL_1253326, EPI_ISL_1253327, EPI_ISL_1253328, EPI_ISL_1253329, EPI_ISL_1253330, EPI_ISL_1253331, EPI_ISL_1253332, EPI_ISL_1253333, EPI_ISL_1253334, EPI_ISL_1253335, EPI_ISL_1253336, EPI_ISL_1253337, EPI_ISL_1253338, EPI_ISL_1253339, EPI_ISL_1253340, EPI_ISL_1253341, EPI_ISL_1253342, EPI_ISL_1253343, EPI_ISL_1253344, EPI_ISL_1253345, EPI_ISL_1253346, EPI_ISL_1253347, EPI_ISL_1253348, EPI_ISL_1253349, EPI_ISL_1253350, EPI_ISL_1253351, EPI_ISL_1253352, EPI_ISL_1253353, EPI_ISL_1253354, EPI_ISL_1253355, EPI_ISL_1253356, EPI_ISL_1253357, EPI_ISL_1253358, EPI_ISL_1253359, EPI_ISL_1253360, EPI_ISL_1253361, EPI_ISL_1253362, EPI_ISL_1253363, EPI_ISL_1253364, EPI_ISL_1253365, EPI_ISL_1253366, EPI_ISL_1253367, EPI_ISL_1253368, EPI_ISL_1253369, EPI_ISL_1253370, EPI_ISL_1253371, EPI_ISL_1253372, EPI_ISL_1253373, EPI_ISL_1253374, EPI_ISL_1253375, EPI_ISL_1253376, EPI_ISL_1253377, EPI_ISL_1253378, EPI_ISL_1253379, EPI_ISL_1253380, EPI_ISL_1253381, EPI_ISL_1253382, EPI_ISL_1253383, EPI_ISL_1253384, EPI_ISL_1253385, EPI_ISL_1253386, EPI_ISL_1253387, EPI_ISL_1253388, EPI_ISL_1253389, EPI_ISL_1253390, EPI_ISL_1253391, EPI_ISL_1253392, EPI_ISL_1253393, EPI_ISL_1253394, EPI_ISL_1253395, EPI_ISL_1253396, EPI_ISL_1253397, EPI_ISL_1253398, EPI_ISL_1253400, EPI_ISL_1253401, EPI_ISL_1253402, EPI_ISL_1253403, EPI_ISL_1253404, EPI_ISL_1253405, EPI_ISL_1253406, EPI_ISL_1253407, EPI_ISL_1253408, EPI_ISL_1253409, EPI_ISL_1253410, EPI_ISL_1253411, EPI_ISL_1253412, EPI_ISL_1253413, EPI_ISL_1253414, EPI_ISL_1253415, EPI_ISL_1253416, EPI_ISL_1253417, EPI_ISL_1253418, EPI_ISL_1253419, EPI_ISL_1253420, EPI_ISL_1253421, EPI_ISL_1253422, EPI_ISL_1253423, EPI_ISL_1253424, EPI_ISL_1253425, EPI_ISL_1253426, EPI_ISL_1253427, EPI_ISL_1253428, EPI_ISL_1253429, EPI_ISL_1253430, EPI_ISL_1253431, EPI_ISL_1253432, EPI_ISL_1253433, EPI_ISL_1253434, EPI_ISL_1253435, EPI_ISL_1253436, EPI_ISL_1253437, EPI_ISL_1253438, EPI_ISL_1253439, EPI_ISL_1253440, EPI_ISL_1253441, EPI_ISL_1253442, EPI_ISL_1253443, EPI_ISL_1253444, EPI_ISL_1253445, EPI_ISL_1253446, EPI_ISL_1253447, EPI_ISL_1253448, EPI_ISL_1253449, EPI_ISL_1253450, EPI_ISL_1253451, EPI_ISL_1253452, EPI_ISL_1253453, EPI_ISL_1253454, EPI_ISL_1253455, EPI_ISL_1253456, EPI_ISL_1253457, EPI_ISL_1253458, EPI_ISL_1253459, EPI_ISL_1253460, EPI_ISL_1253461, EPI_ISL_1253462, EPI_ISL_1253463, EPI_ISL_1253464, EPI_ISL_1253465, EPI_ISL_1253466, EPI_ISL_1253467, EPI_ISL_1253468, EPI_ISL_1253469, EPI_ISL_1253470, EPI_ISL_1253471, EPI_ISL_1253472, EPI_ISL_1253473, EPI_ISL_1253474, EPI_ISL_1253475, EPI_ISL_1253476, EPI_ISL_1253477, EPI_ISL_1253478, EPI_ISL_1253479, EPI_ISL_1253480, EPI_ISL_1253481, EPI_ISL_1253482, EPI_ISL_1253483, EPI_ISL_1253484, EPI_ISL_1253485, EPI_ISL_1253486, EPI_ISL_1253487, EPI_ISL_1253488, EPI_ISL_1253489, EPI_ISL_1253490, EPI_ISL_1253491, EPI_ISL_1253492, EPI_ISL_1253493, EPI_ISL_1253494, EPI_ISL_1253495, EPI_ISL_1253496, EPI_ISL_1253497, EPI_ISL_1253498, EPI_ISL_1253499, EPI_ISL_1253500, EPI_ISL_1253501, EPI_ISL_1253502, EPI_ISL_1253503, EPI_ISL_1253504, EPI_ISL_1253505, EPI_ISL_1253506, EPI_ISL_1253507, EPI_ISL_1253508, EPI_ISL_1253509, EPI_ISL_1253510, EPI_ISL_1253511, EPI_ISL_1253512, EPI_ISL_1253513, EPI_ISL_1253514, EPI_ISL_1253515, EPI_ISL_1253516, EPI_ISL_1253517, EPI_ISL_1253518, EPI_ISL_1253519, EPI_ISL_1253520, EPI_ISL_1253521, EPI_ISL_1253522, EPI_ISL_1253523, EPI_ISL_1253524, EPI_ISL_1253525, EPI_ISL_1253526, EPI_ISL_1253527, EPI_ISL_1253528, EPI_ISL_1253529, EPI_ISL_1253530, EPI_ISL_1253531, EPI_ISL_1253532, EPI_ISL_1253533, EPI_ISL_1253534, EPI_ISL_1253535, EPI_ISL_1253536, EPI_ISL_1253537 | | | |
| see above | Quest Diagnostics Incorporated | Centers for Disease Control and Prevention Division of Viral Diseases, Pathogen Discovery | Peter W. Cook, Dakota Howard, Dhwani Batra, Ben L. Rambo-Martin, S. H. Rosenthal, A. Gerasimova, R. M. Kagan, B. Anderson, M. Hua, Y. Liu, L.E. Bernstein, K.E. Livingston, A. Perez, I. A. Shlyakhter, R. V. Rolando, R. Owen, P. Tanpaiboon, F. Lacbawan, Clinton R. Paden, Suxiang Tong, Duncan MacCannell |
| EPI_ISL_1253580, EPI_ISL_1253581, EPI_ISL_1253582, EPI_ISL_1253583, EPI_ISL_1253584, EPI_ISL_1253585, EPI_ISL_1253586, EPI_ISL_1253587, EPI_ISL_1253588, EPI_ISL_1253589, EPI_ISL_1253590, EPI_ISL_1253591, EPI_ISL_1253592, EPI_ISL_1253593, EPI_ISL_1253594, EPI_ISL_1253595, EPI_ISL_1253596, EPI_ISL_1253597, EPI_ISL_1253599, EPI_ISL_1253600, EPI_ISL_1253601, EPI_ISL_1253602, EPI_ISL_1253603, EPI_ISL_1253604, EPI_ISL_1253605, EPI_ISL_1253606, EPI_ISL_1253607, EPI_ISL_1253608 | | | |
| see above | University of Wisconsin-Madison AIDS Vaccine Research Laboratories | University of Wisconsin-Madison AIDS Vaccine Research Laboratories | Gage Moreno, Katarina Braun, et al. AIDS Vaccine Research Laboratories |
| EPI_ISL_1253663, EPI_ISL_1253664, EPI_ISL_1253665, EPI_ISL_1253666, EPI_ISL_1253667, EPI_ISL_1253668, EPI_ISL_1253669, EPI_ISL_1253670, EPI_ISL_1253671, EPI_ISL_1253672, EPI_ISL_1253673, EPI_ISL_1253674, EPI_ISL_1253675, EPI_ISL_1253676, EPI_ISL_1253677, EPI_ISL_1253678, EPI_ISL_1253679, EPI_ISL_1253680, EPI_ISL_1253681, EPI_ISL_1253682, EPI_ISL_1253684, EPI_ISL_1253685, EPI_ISL_1253686, EPI_ISL_1253687, EPI_ISL_1253688, EPI_ISL_1253689, EPI_ISL_1253690, EPI_ISL_1253691, EPI_ISL_1253692, EPI_ISL_1253693, EPI_ISL_1253694, EPI_ISL_1253695, EPI_ISL_1253696, EPI_ISL_1253697, EPI_ISL_1253698, EPI_ISL_1253699, EPI_ISL_1253700, EPI_ISL_1253701, EPI_ISL_1253702, EPI_ISL_1253703, EPI_ISL_1253704 | | | |
| see above | South Dakota Public Health Laboratory | South Dakota Public Health Laboratory | Jacob Garfin and Chris Carlson |
| EPI_ISL_1253706 | Microbiology Division, South Carolina Department of Health and Environmental Control Public Health Laboratory (SC DHEC PHL) | Microbiology Division, South Carolina Department of Health and Environmental Control Public Health Laboratory (SC DHEC PHL) | Haley Flores, Jessica Freeman |
| EPI_ISL_1253707, EPI_ISL_1253708, EPI_ISL_1253709 | Massachusetts General Hospital | Infectious Disease Program, Broad Institute of Harvard and MIT | Lemieux,J.E., Siddle,K.J., Shaw,B., Adams,G., Pierce,V., Turbett,S., Anahtar,M., Branda,J., Slater,D., Harris,J., Lin,A.E., Gladden-Young,A., Lagerborg,K., Rudy,M., DeRuff,K., Carter,A., Normandin,E., Bauer,M., Reilly,S., Tomkins-Tinch,C., Loreth,C., Chaluvadi,S., Neumann,A., Cusick,C., Chapman,S.B., Gnirke,A., Flowers,K., Cerrato,F., Birren,B.W., Gallagher,G., Smole,S., Park,D.J., MacInnis,B.L., Ryan,E., LaRocque,R., Rosenberg,E. and Sabeti,P.C. |
| EPI_ISL_1253714, EPI_ISL_1253715, EPI_ISL_1253716 | Flow Health | Infectious Disease Program, Broad Institute of Harvard and MIT | Lemieux,J.E., Siddle,K.J., Adams,G., Gladden-Young,A., Lagerborg,K., Rudy,M., DeRuff,K., Carter,A., Normandin,E., Bauer,M., Reilly,S., Tomkins-Tinch,C., Loreth,C., Chaluvadi,S., Birren,B.W., Gallagher,G., Smole,S., Park,D.J., MacInnis,B.L., and Sabeti,P.C. |
| EPI_ISL_1253719, EPI_ISL_1253720, EPI_ISL_1253721, EPI_ISL_1253722, EPI_ISL_1253723, EPI_ISL_1253724, EPI_ISL_1253725, EPI_ISL_1253726, EPI_ISL_1253727, EPI_ISL_1253728, EPI_ISL_1253729, EPI_ISL_1253730, EPI_ISL_1253731, EPI_ISL_1253732, EPI_ISL_1253733, EPI_ISL_1253734, EPI_ISL_1253735, EPI_ISL_1253736, EPI_ISL_1253737, EPI_ISL_1253738, EPI_ISL_1253739, EPI_ISL_1253740, EPI_ISL_1253741, EPI_ISL_1253742, EPI_ISL_1253743, EPI_ISL_1253744, EPI_ISL_1253745, EPI_ISL_1253746, EPI_ISL_1253747, EPI_ISL_1253749, EPI_ISL_1253750, EPI_ISL_1253751, EPI_ISL_1253752, EPI_ISL_1253753, EPI_ISL_1253754, EPI_ISL_1253756, EPI_ISL_1253757, EPI_ISL_1253758, EPI_ISL_1253759, EPI_ISL_1253760, EPI_ISL_1253761, EPI_ISL_1253762, EPI_ISL_1253763, EPI_ISL_1253764, EPI_ISL_1253765, EPI_ISL_1253766, EPI_ISL_1253767, EPI_ISL_1253768, EPI_ISL_1253769, EPI_ISL_1253770, EPI_ISL_1253771, EPI_ISL_1253772, EPI_ISL_1253773, EPI_ISL_1253774, EPI_ISL_1253775, EPI_ISL_1253776, EPI_ISL_1253777, EPI_ISL_1253778, EPI_ISL_1253779, EPI_ISL_1253780, EPI_ISL_1253781, EPI_ISL_1253782, EPI_ISL_1253783, EPI_ISL_1253784, EPI_ISL_1253785, EPI_ISL_1253786, EPI_ISL_1253787, EPI_ISL_1253788, EPI_ISL_1253789, EPI_ISL_1253790, EPI_ISL_1253791, EPI_ISL_1253792, EPI_ISL_1253793, EPI_ISL_1253794, EPI_ISL_1253795, EPI_ISL_1253796, EPI_ISL_1253797, EPI_ISL_1253798, EPI_ISL_1253799, EPI_ISL_1253800, EPI_ISL_1253801, EPI_ISL_1253802, EPI_ISL_1253803, EPI_ISL_1253804, EPI_ISL_1253805, EPI_ISL_1253806, EPI_ISL_1253807, EPI_ISL_1253808, EPI_ISL_1253809, EPI_ISL_1253810, EPI_ISL_1253811, EPI_ISL_1253812, EPI_ISL_1253813, EPI_ISL_1253814, EPI_ISL_1253815, EPI_ISL_1253816, EPI_ISL_1253817, EPI_ISL_1253818, EPI_ISL_1253819, EPI_ISL_1253820, EPI_ISL_1253821, EPI_ISL_1253822, EPI_ISL_1253823, EPI_ISL_1253824, EPI_ISL_1253825, EPI_ISL_1253826, EPI_ISL_1253827, EPI_ISL_1253828, EPI_ISL_1253829, EPI_ISL_1253830, EPI_ISL_1253831, EPI_ISL_1253832, EPI_ISL_1253833, EPI_ISL_1253834, EPI_ISL_1253835, EPI_ISL_1253836, EPI_ISL_1253837, EPI_ISL_1253838, EPI_ISL_1253839, EPI_ISL_1253840, EPI_ISL_1253841, EPI_ISL_1253842, EPI_ISL_1253843, EPI_ISL_1253844, EPI_ISL_1253845, EPI_ISL_1253846, EPI_ISL_1253847, EPI_ISL_1253848, EPI_ISL_1253849, EPI_ISL_1253850, EPI_ISL_1253851, EPI_ISL_1253852, EPI_ISL_1253853, EPI_ISL_1253854, EPI_ISL_1253855, EPI_ISL_1253856, EPI_ISL_1253857, EPI_ISL_1253858, EPI_ISL_1253859, EPI_ISL_1253860, EPI_ISL_1253861, EPI_ISL_1253862, EPI_ISL_1253863, EPI_ISL_1253864, EPI_ISL_1253865, EPI_ISL_1253866, EPI_ISL_1253867, EPI_ISL_1253868, EPI_ISL_1253869, EPI_ISL_1253870, EPI_ISL_1253871, EPI_ISL_1253872, EPI_ISL_1253873, EPI_ISL_1253874, EPI_ISL_1253875, EPI_ISL_1253876, EPI_ISL_1253877, EPI_ISL_1253878, EPI_ISL_1253879, EPI_ISL_1253880, EPI_ISL_1253881, EPI_ISL_1253882, EPI_ISL_1253883, EPI_ISL_1253884 | | | |
| see above | Broad Institute Clinical Research Sequencing Platform | Infectious Disease Program, Broad Institute of Harvard and MIT | Lemieux,J.E., Siddle,K.J., Adams,G., Gladden-Young,A., Lagerborg,K., Rudy,M., DeRuff,K., Carter,A., Normandin,E., Bauer,M., Reilly,S., Tomkins-Tinch,C., Loreth,C., Chaluvadi,S., Birren,B.W., Gallagher,G., Smole,S., Park,D.J., MacInnis,B.L., and Sabeti,P.C. |
| EPI_ISL_1253885, EPI_ISL_1253886, EPI_ISL_1253887, EPI_ISL_1253888, EPI_ISL_1253889, EPI_ISL_1253890, EPI_ISL_1253892, EPI_ISL_1253893, EPI_ISL_1253894, EPI_ISL_1253895, EPI_ISL_1253896, EPI_ISL_1253897, EPI_ISL_1253898, EPI_ISL_1253899, EPI_ISL_1253900, EPI_ISL_1253901, EPI_ISL_1253902, EPI_ISL_1253903, EPI_ISL_1253904, EPI_ISL_1253905, EPI_ISL_1253906, EPI_ISL_1253907, EPI_ISL_1253908, EPI_ISL_1253909, EPI_ISL_1253910, EPI_ISL_1253911, EPI_ISL_1253912, EPI_ISL_1253913, EPI_ISL_1253914, EPI_ISL_1253915, EPI_ISL_1253916, EPI_ISL_1253917, EPI_ISL_1253918, EPI_ISL_1253919, EPI_ISL_1253920, EPI_ISL_1253921, EPI_ISL_1253923, EPI_ISL_1253924, EPI_ISL_1253925, EPI_ISL_1253926, EPI_ISL_1253928, EPI_ISL_1253930, EPI_ISL_1253931, EPI_ISL_1253933, EPI_ISL_1253934, EPI_ISL_1253935, EPI_ISL_1253936, EPI_ISL_1253937, EPI_ISL_1253938, EPI_ISL_1253939, EPI_ISL_1253940 | | | |
| see above | Warrior Diagnostics Laboratory | Infectious Disease Program, Broad Institute of Harvard and MIT | Lemieux,J.E., Siddle,K.J., Marshall,J., ONeill,M., Bronson,A., Adams,G., Gladden-Young,A., Lagerborg,K., Rudy,M., DeRuff,K., Carter,A., Normandin,E., Bauer,M., Reilly,S., Tomkins-Tinch,C., Loreth,C., Chaluvadi,S., Birren,B.W., Gallagher,G., Smole,S., Park,D.J., MacInnis,B.L., and Sabeti,P.C. |
| EPI_ISL_1253942, EPI_ISL_1253943, EPI_ISL_1253944, EPI_ISL_1253945, EPI_ISL_1253946, EPI_ISL_1253947, EPI_ISL_1253948, EPI_ISL_1253949, EPI_ISL_1253950, EPI_ISL_1253951, EPI_ISL_1253952, EPI_ISL_1253953, EPI_ISL_1253954, EPI_ISL_1253955, EPI_ISL_1253956, EPI_ISL_1253957, EPI_ISL_1253958, EPI_ISL_1253959, EPI_ISL_1253960, EPI_ISL_1253961, EPI_ISL_1253962, EPI_ISL_1253963, EPI_ISL_1253965, EPI_ISL_1253966, EPI_ISL_1253967, EPI_ISL_1253968, EPI_ISL_1253969, EPI_ISL_1253970, EPI_ISL_1253971, EPI_ISL_1253972, EPI_ISL_1253973, EPI_ISL_1253974, EPI_ISL_1253975, EPI_ISL_1253976, EPI_ISL_1253977, EPI_ISL_1253978, EPI_ISL_1253979, EPI_ISL_1253980, EPI_ISL_1253981, EPI_ISL_1253982, EPI_ISL_1253983, EPI_ISL_1253984, EPI_ISL_1253985, EPI_ISL_1253986, EPI_ISL_1253987, EPI_ISL_1253988, EPI_ISL_1253989, EPI_ISL_1253990, EPI_ISL_1253991, EPI_ISL_1253992, EPI_ISL_1253993, EPI_ISL_1253994, EPI_ISL_1253995, EPI_ISL_1253996, EPI_ISL_1253998, EPI_ISL_1253999, EPI_ISL_1254000, EPI_ISL_1254001, EPI_ISL_1254002, EPI_ISL_1254003, EPI_ISL_1254004, EPI_ISL_1254005, EPI_ISL_1254006, EPI_ISL_1254007, EPI_ISL_1254008 | | | |
| see above | Rhode Island Department of Health | Infectious Disease Program, Broad Institute of Harvard and MIT | Lemieux,J.E., Siddle,K.J., Huard,R., King,E., Azevedo,K., Miller,A., Adams,G., Gladden-Young,A., Lagerborg,K., Rudy,M., DeRuff,K., Carter,A., Normandin,E., Bauer,M., Reilly,S., Tomkins-Tinch,C., Loreth,C., Chaluvadi,S., Birren,B.W., Gallagher,G., Smole,S., Park,D.J., MacInnis,B.L., and Sabeti,P.C. |
| EPI_ISL_1254009, EPI_ISL_1254011 | Flow Health | Infectious Disease Program, Broad Institute of Harvard and MIT | Lemieux,J.E., Siddle,K.J., Adams,G., Gladden-Young,A., Lagerborg,K., Rudy,M., DeRuff,K., Carter,A., Normandin,E., Bauer,M., Reilly,S., Tomkins-Tinch,C., Loreth,C., Chaluvadi,S., Birren,B.W., Gallagher,G., Smole,S., Park,D.J., MacInnis,B.L., and Sabeti,P.C. |
| EPI_ISL_1254013, EPI_ISL_1254014, EPI_ISL_1254015, EPI_ISL_1254016, EPI_ISL_1254017, EPI_ISL_1254018, EPI_ISL_1254019, EPI_ISL_1254020, EPI_ISL_1254021, EPI_ISL_1254022, EPI_ISL_1254023, EPI_ISL_1254024, EPI_ISL_1254025, EPI_ISL_1254026, EPI_ISL_1254027, EPI_ISL_1254028, EPI_ISL_1254029, EPI_ISL_1254030, EPI_ISL_1254031, EPI_ISL_1254032, EPI_ISL_1254033, EPI_ISL_1254034, EPI_ISL_1254035, EPI_ISL_1254036, EPI_ISL_1254037, EPI_ISL_1254038, EPI_ISL_1254039, EPI_ISL_1254040, EPI_ISL_1254041, EPI_ISL_1254042, EPI_ISL_1254043, EPI_ISL_1254044, EPI_ISL_1254045, EPI_ISL_1254046, EPI_ISL_1254047, EPI_ISL_1254048, EPI_ISL_1254049, EPI_ISL_1254050, EPI_ISL_1254051, EPI_ISL_1254052, EPI_ISL_1254053, EPI_ISL_1254054, EPI_ISL_1254055, EPI_ISL_1254056, EPI_ISL_1254057, EPI_ISL_1254058, EPI_ISL_1254059, EPI_ISL_1254060, EPI_ISL_1254061, EPI_ISL_1254062, EPI_ISL_1254063, EPI_ISL_1254064, EPI_ISL_1254065, EPI_ISL_1254066, EPI_ISL_1254067, EPI_ISL_1254068, EPI_ISL_1254069, EPI_ISL_1254070, EPI_ISL_1254071, EPI_ISL_1254072, EPI_ISL_1254073, EPI_ISL_1254074, EPI_ISL_1254075, EPI_ISL_1254076, EPI_ISL_1254077, EPI_ISL_1254078, EPI_ISL_1254079, EPI_ISL_1254080, EPI_ISL_1254081, EPI_ISL_1254082, EPI_ISL_1254083, EPI_ISL_1254084, EPI_ISL_1254085, EPI_ISL_1254086, EPI_ISL_1254087, EPI_ISL_1254088, EPI_ISL_1254089, EPI_ISL_1254090, EPI_ISL_1254091, EPI_ISL_1254092, EPI_ISL_1254093, EPI_ISL_1254094, EPI_ISL_1254095, EPI_ISL_1254096, EPI_ISL_1254097, EPI_ISL_1254098, EPI_ISL_1254099, EPI_ISL_1254100, EPI_ISL_1254101, EPI_ISL_1254102, EPI_ISL_1254103, EPI_ISL_1254104, EPI_ISL_1254105, EPI_ISL_1254106, EPI_ISL_1254107, EPI_ISL_1254108, EPI_ISL_1254109, EPI_ISL_1254110, EPI_ISL_1254111, EPI_ISL_1254112, EPI_ISL_1254113, EPI_ISL_1254114, EPI_ISL_1254115, EPI_ISL_1254116, EPI_ISL_1254117, EPI_ISL_1254118, EPI_ISL_1254119, EPI_ISL_1254120, EPI_ISL_1254121, EPI_ISL_1254122, EPI_ISL_1254123, EPI_ISL_1254124, EPI_ISL_1254125, EPI_ISL_1254126, EPI_ISL_1254127, EPI_ISL_1254128, EPI_ISL_1254129, EPI_ISL_1254130, EPI_ISL_1254131, EPI_ISL_1254132, EPI_ISL_1254133, EPI_ISL_1254134, EPI_ISL_1254135, EPI_ISL_1254136, EPI_ISL_1254137, EPI_ISL_1254138, EPI_ISL_1254139, EPI_ISL_1254140, EPI_ISL_1254141, EPI_ISL_1254142, EPI_ISL_1254143, EPI_ISL_1254144, EPI_ISL_1254145, EPI_ISL_1254146, EPI_ISL_1254147, EPI_ISL_1254148, EPI_ISL_1254149, EPI_ISL_1254150, EPI_ISL_1254151, EPI_ISL_1254152, EPI_ISL_1254153, EPI_ISL_1254154, EPI_ISL_1254155, EPI_ISL_1254156, EPI_ISL_1254157, EPI_ISL_1254158, EPI_ISL_1254159, EPI_ISL_1254160, EPI_ISL_1254161, EPI_ISL_1254162, EPI_ISL_1254163, EPI_ISL_1254164, EPI_ISL_1254165, EPI_ISL_1254166, EPI_ISL_1254167, EPI_ISL_1254168, EPI_ISL_1254169, EPI_ISL_1254170, EPI_ISL_1254171, EPI_ISL_1254172, EPI_ISL_1254173, EPI_ISL_1254174, EPI_ISL_1254175, EPI_ISL_1254176, EPI_ISL_1254177, EPI_ISL_1254178, EPI_ISL_1254179, EPI_ISL_1254180, EPI_ISL_1254181, EPI_ISL_1254182, EPI_ISL_1254183, EPI_ISL_1254184, EPI_ISL_1254185, EPI_ISL_1254186, EPI_ISL_1254187, EPI_ISL_1254188, EPI_ISL_1254189, EPI_ISL_1254190, EPI_ISL_1254191, EPI_ISL_1254192, EPI_ISL_1254193, EPI_ISL_1254194, EPI_ISL_1254195, EPI_ISL_1254196, EPI_ISL_1254197, EPI_ISL_1254198, EPI_ISL_1254199, EPI_ISL_1254200, EPI_ISL_1254201, EPI_ISL_1254202, EPI_ISL_1254203, EPI_ISL_1254204, EPI_ISL_1254205, EPI_ISL_1254206, EPI_ISL_1254207, EPI_ISL_1254208, EPI_ISL_1254209, EPI_ISL_1254210, EPI_ISL_1254211, EPI_ISL_1254212, EPI_ISL_1254213, EPI_ISL_1254214, EPI_ISL_1254215, EPI_ISL_1254216, EPI_ISL_1254217, EPI_ISL_1254218, EPI_ISL_1254219, EPI_ISL_1254220, EPI_ISL_1254221, EPI_ISL_1254222, EPI_ISL_1254223, EPI_ISL_1254224 | | | |
| see above | Quest Diagnostics Incorporated | Centers for Disease Control and Prevention Division of Viral Diseases, Pathogen Discovery | Peter W. Cook, Dakota Howard, Dhwani Batra, Ben L. Rambo-Martin, S. H. Rosenthal, A. Gerasimova, R. M. Kagan, B. Anderson, M. Hua, Y. Liu, L.E. Bernstein, K.E. Livingston, A. Perez, I. A. Shlyakhter, R. V. Rolando, R. Owen, P. Tanpaiboon, F. Lacbawan, Clinton R. Paden, Suxiang Tong, Duncan MacCannell |
| EPI_ISL_1254226 | NJDOH, Public Health and Environmental Laboratories | NJ_PHEL | Lindsey Bodnar, Shiv K. Verma, Dana Woell, Byeong Jeong |
| EPI_ISL_1254227 | Quest Diagnostics Incorporated | Centers for Disease Control and Prevention Division of Viral Diseases, Pathogen Discovery | Peter W. Cook, Dakota Howard, Dhwani Batra, Ben L. Rambo-Martin, S. H. Rosenthal, A. Gerasimova, R. M. Kagan, B. Anderson, M. Hua, Y. Liu, L.E. Bernstein, K.E. Livingston, A. Perez, I. A. Shlyakhter, R. V. Rolando, R. Owen, P. Tanpaiboon, F. Lacbawan, Clinton R. Paden, Suxiang Tong, Duncan MacCannell |
| EPI_ISL_1254228 | NJDOH, Public Health and Environmental Laboratories | NJ_PHEL | Lindsey Bodnar, Shiv K. Verma, Dana Woell, Byeong Jeong |
| EPI_ISL_1254229 | Quest Diagnostics Incorporated | Centers for Disease Control and Prevention Division of Viral Diseases, Pathogen Discovery | Peter W. Cook, Dakota Howard, Dhwani Batra, Ben L. Rambo-Martin, S. H. Rosenthal, A. Gerasimova, R. M. Kagan, B. Anderson, M. Hua, Y. Liu, L.E. Bernstein, K.E. Livingston, A. Perez, I. A. Shlyakhter, R. V. Rolando, R. Owen, P. Tanpaiboon, F. Lacbawan, Clinton R. Paden, Suxiang Tong, Duncan MacCannell |
| EPI_ISL_1254231 | NJDOH, Public Health and Environmental Laboratories | NJ_PHEL | Lindsey Bodnar, Shiv K. Verma, Dana Woell, Byeong Jeong |
| EPI_ISL_1254232 | Quest Diagnostics Incorporated | Centers for Disease Control and Prevention Division of Viral Diseases, Pathogen Discovery | Peter W. Cook, Dakota Howard, Dhwani Batra, Ben L. Rambo-Martin, S. H. Rosenthal, A. Gerasimova, R. M. Kagan, B. Anderson, M. Hua, Y. Liu, L.E. Bernstein, K.E. Livingston, A. Perez, I. A. Shlyakhter, R. V. Rolando, R. Owen, P. Tanpaiboon, F. Lacbawan, Clinton R. Paden, Suxiang Tong, Duncan MacCannell |
| EPI_ISL_1254233, EPI_ISL_1254234 | NJDOH, Public Health and Environmental Laboratories | NJ_PHEL | Lindsey Bodnar, Shiv K. Verma, Dana Woell, Byeong Jeong |
| EPI_ISL_1254235 | Quest Diagnostics Incorporated | Centers for Disease Control and Prevention Division of Viral Diseases, Pathogen Discovery | Peter W. Cook, Dakota Howard, Dhwani Batra, Ben L. Rambo-Martin, S. H. Rosenthal, A. Gerasimova, R. M. Kagan, B. Anderson, M. Hua, Y. Liu, L.E. Bernstein, K.E. Livingston, A. Perez, I. A. Shlyakhter, R. V. Rolando, R. Owen, P. Tanpaiboon, F. Lacbawan, Clinton R. Paden, Suxiang Tong, Duncan MacCannell |
| EPI_ISL_1254236, EPI_ISL_1254237 | NJDOH, Public Health and Environmental Laboratories | NJ_PHEL | Lindsey Bodnar, Shiv K. Verma, Dana Woell, Byeong Jeong |
| EPI_ISL_1254238 | Quest Diagnostics Incorporated | Centers for Disease Control and Prevention Division of Viral Diseases, Pathogen Discovery | Peter W. Cook, Dakota Howard, Dhwani Batra, Ben L. Rambo-Martin, S. H. Rosenthal, A. Gerasimova, R. M. Kagan, B. Anderson, M. Hua, Y. Liu, L.E. Bernstein, K.E. Livingston, A. Perez, I. A. Shlyakhter, R. V. Rolando, R. Owen, P. Tanpaiboon, F. Lacbawan, Clinton R. Paden, Suxiang Tong, Duncan MacCannell |
| EPI_ISL_1254239, | NJDOH, Public Health and | NJ_PHEL | Lindsey Bodnar, Shiv K. Verma, Dana Woell, Byeong Jeong |

| | | | |
|---|---|---|---|
| EPI_ISL_1254240 | Environmental Laboratories | | |
| EPI_ISL_1254241 | Quest Diagnostics Incorporated | Centers for Disease Control and Prevention Division of Viral Diseases, Pathogen Discovery | Peter W. Cook, Dakota Howard, Dhwani Batra, Ben L. Rambo-Martin, S. H. Rosenthal, A. Gerasimova, R. M. Kagan, B. Anderson, M. Hua, Y. Liu, L.E. Bernstein, K.E. Livingston, A. Perez, I. A. Shlyakhter, R. V. Rolando, R. Owen, P. Tanpaiboon, F. Lacbawan, Clinton R. Paden, Suxiang Tong, Duncan MacCannell |
| EPI_ISL_1254242, EPI_ISL_1254243 | NJDOH, Public Health and Environmental Laboratories | NJ_PHEL | Lindsey Bodnar, Shiv K. Verma, Dana Woell, Byeong Jeong |
| EPI_ISL_1254244, EPI_ISL_1254246, EPI_ISL_1254249, EPI_ISL_1254251 | Quest Diagnostics Incorporated | Centers for Disease Control and Prevention Division of Viral Diseases, Pathogen Discovery | Peter W. Cook, Dakota Howard, Dhwani Batra, Ben L. Rambo-Martin, S. H. Rosenthal, A. Gerasimova, R. M. Kagan, B. Anderson, M. Hua, Y. Liu, L.E. Bernstein, K.E. Livingston, A. Perez, I. A. Shlyakhter, R. V. Rolando, R. Owen, P. Tanpaiboon, F. Lacbawan, Clinton R. Paden, Suxiang Tong, Duncan MacCannell |
| EPI_ISL_1254253 | NJDOH, Public Health and Environmental Laboratories | NJ_PHEL | Lindsey Bodnar, Shiv K. Verma, Dana Woell, Byeong Jeong |
| EPI_ISL_1254254 | Quest Diagnostics Incorporated | Centers for Disease Control and Prevention Division of Viral Diseases, Pathogen Discovery | Peter W. Cook, Dakota Howard, Dhwani Batra, Ben L. Rambo-Martin, S. H. Rosenthal, A. Gerasimova, R. M. Kagan, B. Anderson, M. Hua, Y. Liu, L.E. Bernstein, K.E. Livingston, A. Perez, I. A. Shlyakhter, R. V. Rolando, R. Owen, P. Tanpaiboon, F. Lacbawan, Clinton R. Paden, Suxiang Tong, Duncan MacCannell |
| EPI_ISL_1254255 | NJDOH, Public Health and Environmental Laboratories | NJ_PHEL | Lindsey Bodnar, Shiv K. Verma, Dana Woell, Byeong Jeong |
| EPI_ISL_1254257 | Quest Diagnostics Incorporated | Centers for Disease Control and Prevention Division of Viral Diseases, Pathogen Discovery | Peter W. Cook, Dakota Howard, Dhwani Batra, Ben L. Rambo-Martin, S. H. Rosenthal, A. Gerasimova, R. M. Kagan, B. Anderson, M. Hua, Y. Liu, L.E. Bernstein, K.E. Livingston, A. Perez, I. A. Shlyakhter, R. V. Rolando, R. Owen, P. Tanpaiboon, F. Lacbawan, Clinton R. Paden, Suxiang Tong, Duncan MacCannell |
| EPI_ISL_1254258, EPI_ISL_1254259 | NJDOH, Public Health and Environmental Laboratories | NJ_PHEL | Lindsey Bodnar, Shiv K. Verma, Dana Woell, Byeong Jeong |
| EPI_ISL_1254260, EPI_ISL_1254262 | Quest Diagnostics Incorporated | Centers for Disease Control and Prevention Division of Viral Diseases, Pathogen Discovery | Peter W. Cook, Dakota Howard, Dhwani Batra, Ben L. Rambo-Martin, S. H. Rosenthal, A. Gerasimova, R. M. Kagan, B. Anderson, M. Hua, Y. Liu, L.E. Bernstein, K.E. Livingston, A. Perez, I. A. Shlyakhter, R. V. Rolando, R. Owen, P. Tanpaiboon, F. Lacbawan, Clinton R. Paden, Suxiang Tong, Duncan MacCannell |
| EPI_ISL_1254263, EPI_ISL_1254264 | NJDOH, Public Health and Environmental Laboratories | NJ_PHEL | Lindsey Bodnar, Shiv K. Verma, Dana Woell, Byeong Jeong |
| EPI_ISL_1254265 | Quest Diagnostics Incorporated | Centers for Disease Control and Prevention Division of Viral Diseases, Pathogen Discovery | Peter W. Cook, Dakota Howard, Dhwani Batra, Ben L. Rambo-Martin, S. H. Rosenthal, A. Gerasimova, R. M. Kagan, B. Anderson, M. Hua, Y. Liu, L.E. Bernstein, K.E. Livingston, A. Perez, I. A. Shlyakhter, R. V. Rolando, R. Owen, P. Tanpaiboon, F. Lacbawan, Clinton R. Paden, Suxiang Tong, Duncan MacCannell |
| EPI_ISL_1254266 | NJDOH, Public Health and Environmental Laboratories | NJ_PHEL | Lindsey Bodnar, Shiv K. Verma, Dana Woell, Byeong Jeong |
| EPI_ISL_1254267, EPI_ISL_1254270 | Quest Diagnostics Incorporated | Centers for Disease Control and Prevention Division of Viral Diseases, Pathogen Discovery | Peter W. Cook, Dakota Howard, Dhwani Batra, Ben L. Rambo-Martin, S. H. Rosenthal, A. Gerasimova, R. M. Kagan, B. Anderson, M. Hua, Y. Liu, L.E. Bernstein, K.E. Livingston, A. Perez, I. A. Shlyakhter, R. V. Rolando, R. Owen, P. Tanpaiboon, F. Lacbawan, Clinton R. Paden, Suxiang Tong, Duncan MacCannell |
| EPI_ISL_1254271, EPI_ISL_1254272 | NJDOH, Public Health and Environmental Laboratories | NJ_PHEL | Lindsey Bodnar, Shiv K. Verma, Dana Woell, Byeong Jeong |
| EPI_ISL_1254273 | Quest Diagnostics Incorporated | Centers for Disease Control and Prevention Division of Viral Diseases, Pathogen Discovery | Peter W. Cook, Dakota Howard, Dhwani Batra, Ben L. Rambo-Martin, S. H. Rosenthal, A. Gerasimova, R. M. Kagan, B. Anderson, M. Hua, Y. Liu, L.E. Bernstein, K.E. Livingston, A. Perez, I. A. Shlyakhter, R. V. Rolando, R. Owen, P. Tanpaiboon, F. Lacbawan, Clinton R. Paden, Suxiang Tong, Duncan MacCannell |
| EPI_ISL_1254274 | NJDOH, Public Health and Environmental Laboratories | NJ_PHEL | Lindsey Bodnar, Shiv K. Verma, Dana Woell, Byeong Jeong |
| EPI_ISL_1254275 | Quest Diagnostics Incorporated | Centers for Disease Control and Prevention Division of Viral Diseases, Pathogen Discovery | Peter W. Cook, Dakota Howard, Dhwani Batra, Ben L. Rambo-Martin, S. H. Rosenthal, A. Gerasimova, R. M. Kagan, B. Anderson, M. Hua, Y. Liu, L.E. Bernstein, K.E. Livingston, A. Perez, I. A. Shlyakhter, R. V. Rolando, R. Owen, P. Tanpaiboon, F. Lacbawan, Clinton R. Paden, Suxiang Tong, Duncan MacCannell |
| EPI_ISL_1254276, EPI_ISL_1254277 | NJDOH, Public Health and Environmental Laboratories | NJ_PHEL | Lindsey Bodnar, Shiv K. Verma, Dana Woell, Byeong Jeong |
| EPI_ISL_1254278 | Quest Diagnostics Incorporated | Centers for Disease Control and Prevention Division of Viral Diseases, Pathogen Discovery | Peter W. Cook, Dakota Howard, Dhwani Batra, Ben L. Rambo-Martin, S. H. Rosenthal, A. Gerasimova, R. M. Kagan, B. Anderson, M. Hua, Y. Liu, L.E. Bernstein, K.E. Livingston, A. Perez, I. A. Shlyakhter, R. V. Rolando, R. Owen, P. Tanpaiboon, F. Lacbawan, Clinton R. Paden, Suxiang Tong, Duncan MacCannell |
| EPI_ISL_1254279, EPI_ISL_1254280 | NJDOH, Public Health and Environmental Laboratories | NJ_PHEL | Lindsey Bodnar, Shiv K. Verma, Dana Woell, Byeong Jeong |
| EPI_ISL_1254281 | Quest Diagnostics Incorporated | Centers for Disease Control and Prevention Division of Viral Diseases, Pathogen Discovery | Peter W. Cook, Dakota Howard, Dhwani Batra, Ben L. Rambo-Martin, S. H. Rosenthal, A. Gerasimova, R. M. Kagan, B. Anderson, M. Hua, Y. Liu, L.E. Bernstein, K.E. Livingston, A. Perez, I. A. Shlyakhter, R. V. Rolando, R. Owen, P. Tanpaiboon, F. Lacbawan, Clinton R. Paden, Suxiang Tong, Duncan MacCannell |
| EPI_ISL_1254282 | NJDOH, Public Health and Environmental Laboratories | NJ_PHEL | Lindsey Bodnar, Shiv K. Verma, Dana Woell, Byeong Jeong |
| EPI_ISL_1254284, EPI_ISL_1254286 | Quest Diagnostics Incorporated | Centers for Disease Control and Prevention Division of Viral Diseases, Pathogen Discovery | Peter W. Cook, Dakota Howard, Dhwani Batra, Ben L. Rambo-Martin, S. H. Rosenthal, A. Gerasimova, R. M. Kagan, B. Anderson, M. Hua, Y. Liu, L.E. Bernstein, K.E. Livingston, A. Perez, I. A. Shlyakhter, R. V. Rolando, R. Owen, P. Tanpaiboon, F. Lacbawan, Clinton R. Paden, Suxiang Tong, Duncan MacCannell |
| EPI_ISL_1254288, EPI_ISL_1254289 | NJDOH, Public Health and Environmental Laboratories | NJ_PHEL | Lindsey Bodnar, Shiv K. Verma, Dana Woell, Byeong Jeong |
| EPI_ISL_1254290 | Quest Diagnostics Incorporated | Centers for Disease Control and Prevention Division of Viral Diseases, Pathogen Discovery | Peter W. Cook, Dakota Howard, Dhwani Batra, Ben L. Rambo-Martin, S. H. Rosenthal, A. Gerasimova, R. M. Kagan, B. Anderson, M. Hua, Y. Liu, L.E. Bernstein, K.E. Livingston, A. Perez, I. A. Shlyakhter, R. V. Rolando, R. Owen, P. Tanpaiboon, F. Lacbawan, Clinton R. Paden, Suxiang Tong, Duncan MacCannell |
| EPI_ISL_1254291 | NJDOH, Public Health and Environmental Laboratories | NJ_PHEL | Lindsey Bodnar, Shiv K. Verma, Dana Woell, Byeong Jeong |
| EPI_ISL_1254292 | Quest Diagnostics Incorporated | Centers for Disease Control and Prevention Division of Viral Diseases, Pathogen Discovery | Peter W. Cook, Dakota Howard, Dhwani Batra, Ben L. Rambo-Martin, S. H. Rosenthal, A. Gerasimova, R. M. Kagan, B. Anderson, M. Hua, Y. Liu, L.E. Bernstein, K.E. Livingston, A. Perez, I. A. Shlyakhter, R. V. Rolando, R. Owen, P. Tanpaiboon, F. Lacbawan, Clinton R. Paden, Suxiang Tong, Duncan MacCannell |
| EPI_ISL_1254293 | NJDOH, Public Health and Environmental Laboratories | NJ_PHEL | Lindsey Bodnar, Shiv K. Verma, Dana Woell, Byeong Jeong |
| EPI_ISL_1254294, EPI_ISL_1254295, EPI_ISL_1254296, EPI_ISL_1254297, EPI_ISL_1254298, EPI_ISL_1254299, EPI_ISL_1254300, EPI_ISL_1254301, EPI_ISL_1254302, EPI_ISL_1254303, EPI_ISL_1254304, EPI_ISL_1254305, EPI_ISL_1254306, EPI_ISL_1254307, EPI_ISL_1254308, EPI_ISL_1254309, EPI_ISL_1254310, EPI_ISL_1254311, EPI_ISL_1254312, EPI_ISL_1254313, EPI_ISL_1254314, EPI_ISL_1254315, EPI_ISL_1254316, EPI_ISL_1254317, EPI_ISL_1254318, EPI_ISL_1254319, EPI_ISL_1254320, EPI_ISL_1254321, EPI_ISL_1254322, EPI_ISL_1254323, EPI_ISL_1254324, EPI_ISL_1254325, EPI_ISL_1254326, EPI_ISL_1254327, EPI_ISL_1254328, EPI_ISL_1254329, EPI_ISL_1254330, EPI_ISL_1254331, EPI_ISL_1254332, EPI_ISL_1254333, EPI_ISL_1254334, EPI_ISL_1254335, EPI_ISL_1254336, EPI_ISL_1254337, EPI_ISL_1254338, EPI_ISL_1254339, EPI_ISL_1254340, EPI_ISL_1254341, EPI_ISL_1254342, EPI_ISL_1254343, EPI_ISL_1254344, EPI_ISL_1254345, EPI_ISL_1254346, EPI_ISL_1254347, EPI_ISL_1254348, EPI_ISL_1254349, EPI_ISL_1254350, EPI_ISL_1254351, EPI_ISL_1254352, EPI_ISL_1254353, EPI_ISL_1254354, EPI_ISL_1254355, EPI_ISL_1254356, EPI_ISL_1254357, EPI_ISL_1254358, EPI_ISL_1254359, EPI_ISL_1254360, EPI_ISL_1254361, EPI_ISL_1254362, EPI_ISL_1254363, EPI_ISL_1254364, EPI_ISL_1254365, EPI_ISL_1254366, EPI_ISL_1254367, EPI_ISL_1254368, EPI_ISL_1254369, EPI_ISL_1254370, EPI_ISL_1254371, EPI_ISL_1254372, EPI_ISL_1254373, EPI_ISL_1254374, EPI_ISL_1254375, EPI_ISL_1254376, EPI_ISL_1254377, EPI_ISL_1254378, EPI_ISL_1254379, EPI_ISL_1254380, EPI_ISL_1254381, EPI_ISL_1254382, EPI_ISL_1254383, EPI_ISL_1254384, EPI_ISL_1254385, EPI_ISL_1254386, EPI_ISL_1254387, EPI_ISL_1254388, EPI_ISL_1254389, EPI_ISL_1254390, EPI_ISL_1254391, EPI_ISL_1254392, EPI_ISL_1254393, EPI_ISL_1254394, EPI_ISL_1254395, EPI_ISL_1254396, EPI_ISL_1254397, EPI_ISL_1254398, EPI_ISL_1254399, EPI_ISL_1254400, EPI_ISL_1254401, EPI_ISL_1254402, EPI_ISL_1254403, EPI_ISL_1254404, EPI_ISL_1254405, EPI_ISL_1254406, EPI_ISL_1254407, EPI_ISL_1254408, EPI_ISL_1254409, EPI_ISL_1254410, EPI_ISL_1254411, EPI_ISL_1254412, EPI_ISL_1254413, EPI_ISL_1254414, EPI_ISL_1254415, EPI_ISL_1254416, EPI_ISL_1254417, EPI_ISL_1254418, EPI_ISL_1254419, EPI_ISL_1254420, EPI_ISL_1254421, EPI_ISL_1254422, EPI_ISL_1254423, EPI_ISL_1254424, EPI_ISL_1254425, EPI_ISL_1254426, EPI_ISL_1254427, EPI_ISL_1254428, EPI_ISL_1254429, EPI_ISL_1254430, EPI_ISL_1254431, EPI_ISL_1254432, EPI_ISL_1254433, EPI_ISL_1254434, EPI_ISL_1254435, EPI_ISL_1254436, EPI_ISL_1254437, EPI_ISL_1254438, EPI_ISL_1254439, EPI_ISL_1254440, EPI_ISL_1254441, EPI_ISL_1254442, EPI_ISL_1254443, EPI_ISL_1254444, EPI_ISL_1254445, EPI_ISL_1254446, EPI_ISL_1254447, EPI_ISL_1254448, EPI_ISL_1254449, EPI_ISL_1254450, EPI_ISL_1254451, EPI_ISL_1254452, EPI_ISL_1254453, EPI_ISL_1254454, EPI_ISL_1254455, EPI_ISL_1254456, EPI_ISL_1254457, EPI_ISL_1254458, EPI_ISL_1254459, EPI_ISL_1254460, EPI_ISL_1254461, EPI_ISL_1254462, EPI_ISL_1254463, EPI_ISL_1254464, EPI_ISL_1254465, EPI_ISL_1254466, EPI_ISL_1254467, EPI_ISL_1254468, EPI_ISL_1254469, EPI_ISL_1254470, EPI_ISL_1254471, EPI_ISL_1254472, EPI_ISL_1254473, EPI_ISL_1254474, EPI_ISL_1254475, EPI_ISL_1254476, EPI_ISL_1254477, EPI_ISL_1254478, EPI_ISL_1254479, EPI_ISL_1254480, EPI_ISL_1254481, EPI_ISL_1254482, EPI_ISL_1254483, EPI_ISL_1254484, EPI_ISL_1254485, EPI_ISL_1254486, EPI_ISL_1254487, EPI_ISL_1254488, EPI_ISL_1254489, EPI_ISL_1254490, EPI_ISL_1254491, EPI_ISL_1254492, EPI_ISL_1254493, EPI_ISL_1254494, EPI_ISL_1254495, EPI_ISL_1254496, EPI_ISL_1254497, EPI_ISL_1254498, EPI_ISL_1254499, EPI_ISL_1254500, EPI_ISL_1254501, EPI_ISL_1254502, EPI_ISL_1254503, EPI_ISL_1254504, EPI_ISL_1254505, EPI_ISL_1254506, EPI_ISL_1254507, EPI_ISL_1254508, EPI_ISL_1254509, EPI_ISL_1254510, EPI_ISL_1254511, EPI_ISL_1254512, EPI_ISL_1254513, EPI_ISL_1254514, EPI_ISL_1254515, EPI_ISL_1254516, EPI_ISL_1254517, EPI_ISL_1254518, EPI_ISL_1254519, EPI_ISL_1254520, EPI_ISL_1254521, EPI_ISL_1254522, EPI_ISL_1254523, EPI_ISL_1254524, EPI_ISL_1254525, EPI_ISL_1254526, EPI_ISL_1254527, EPI_ISL_1254528, EPI_ISL_1254529, EPI_ISL_1254530 | | | |
| see above | Quest Diagnostics Incorporated | Centers for Disease Control and Prevention Division of Viral Diseases, Pathogen Discovery | Peter W. Cook, Dakota Howard, Dhwani Batra, Ben L. Rambo-Martin, S. H. Rosenthal, A. Gerasimova, R. M. Kagan, B. Anderson, M. Hua, Y. Liu, L.E. Bernstein, K.E. Livingston, A. Perez, I. A. Shlyakhter, R. V. Rolando, R. Owen, P. Tanpaiboon, F. Lacbawan, Clinton R. Paden, Suxiang Tong, Duncan MacCannell |
| EPI_ISL_1254640, EPI_ISL_1254641, EPI_ISL_1254642 | TGen North | TGen North | "Jolene Bowers, Heather Centner, Chris French, Hayley Yaglom, Ashlyn Pfeiffer, Darrin Lemmer, Dave Engelthaler, The Arizona COVID Genomics Union (ACGU)" |
| EPI_ISL_1254645 | Sonora Quest Laboratories | TGen North | "Jolene Bowers, Heather Centner, Chris French, Hayley Yaglom, Ashlyn Pfeiffer, Darrin Lemmer, Dave Engelthaler, The Arizona COVID Genomics Union (ACGU)" |
| EPI_ISL_1254646 | Diagnostic Genomics Lab and Functional Genomics Core University of South Carolina | Diagnostic Genomics Lab and Functional Genomics Core, University of South Carolina | Carolyn Banister, Hao Ji, Diego Altomare, Herr Doktor Professor Genomiklaborsdirektor Universitätkovidenpandemieretter Phillip Buckhaults, Michael Shtutman, Helmut Albrecht |
| EPI_ISL_1254704, EPI_ISL_1254705, EPI_ISL_1254706, EPI_ISL_1254707, EPI_ISL_1254708, EPI_ISL_1254709, EPI_ISL_1254710, EPI_ISL_1254711, EPI_ISL_1254712, EPI_ISL_1254713, EPI_ISL_1254714 | | | |
| see above | Oregon State University TRACE Project | OSU Center for Genome Research and | Oregon State University TRACE Project |

| | | | |
|---|---|---|---|
| | | Biocomputing | |
| EPI_ISL_1255052, EPI_ISL_1255053, EPI_ISL_1255054, EPI_ISL_1255055, EPI_ISL_1255056, EPI_ISL_1255057, EPI_ISL_1255058, EPI_ISL_1255059 | Oregon State Public Health Laboratory | Oregon State Public Health Laboratory | LaDonna Grenz, Karim Morey, John Fontana, Shane Sevey |
| EPI_ISL_1255087, EPI_ISL_1255089, EPI_ISL_1255090, EPI_ISL_1255091, EPI_ISL_1255092, EPI_ISL_1255098, EPI_ISL_1255100, EPI_ISL_1255101 | Laboratory Corporation of America | Centers for Disease Control and Prevention Division of Viral Diseases, Pathogen Discovery | Peter W. Cook, Dakota Howard, Dhwani Batra, Ben L. Rambo-Martin, Minoo Agarwal, Eyad Almasri Debbie Boles, Ayla Burns, Nuthawin Charoensri, Oren Cohen, Susan Countryman, Mary Ann Cristobal, Bobbi Croy, Suzanne Dale, Hrushikesh Deshmukh, Amanda Douglas, Vincent Drouillon, Marcia Eisenberg, Howard Engler, Rama Ghatti, Prashant Gupta, Susan Hicks, Jake Humphrey, Lax Iyer, Manoj Jain, Mohan Kolli, Brian Krueger, Tim Kuphal, Stanley Letovsky, Michael Levandoski, Craig Lukasik, Jonathan Meltzer, Brian Norvell, Mindy Nye, Scott Parker, Christos Petropoulos, John Pruitt, Steven Ragan, Scott Ryan, Mike Sapeta, Jana Schroth, Suresh Babu Selvaraju, Goran Stevovic, Amanda Suchanek, Andrea Throop, Lyndon Tilson, Thomas Urban, Joe Voshell, Kimberly Wagner, Jonathan Williams, Mary Williamson, Qian Zeng, Tricia Zwiefelhofer, Clinton R. Paden, Suxiang Tong, Duncan MacCannell |
| EPI_ISL_1255102 | Curative Inc | Curative Inc | Katie Bond |
| EPI_ISL_1255103 | Sonora Quest Laboratories | TGen North | Jolene Bowers, Heather Centner, Chris French, Hayley Yaglom, Ashlyn Pfeiffer, Darrin Lemmer, Dave Engelthaler, The Arizona COVID Genomics Union (ACGU) |
| EPI_ISL_1255279, EPI_ISL_1255280, EPI_ISL_1255281, EPI_ISL_1255282, EPI_ISL_1255283, EPI_ISL_1255284, EPI_ISL_1255285, EPI_ISL_1255286, EPI_ISL_1255287, EPI_ISL_1255288, EPI_ISL_1255289, EPI_ISL_1255290, EPI_ISL_1255291, EPI_ISL_1255292, EPI_ISL_1255293, EPI_ISL_1255294, EPI_ISL_1255295 | | | |
| see above | Indiana Animal Disease Diagnostic Laboratory | Carpi Laboratory - Purdue University | Jack Dorman, Ilinca I Ciubotariu, Nicole M Perry, Jobin J Kattoor, Lev Gorenstein, Abebe A Fola, G Kenitra Hendrix, Rebecca P Wilkes, Giovanna Carpi |
| EPI_ISL_1255296, EPI_ISL_1255298, EPI_ISL_1255299, EPI_ISL_1255300, EPI_ISL_1255301, EPI_ISL_1255302, EPI_ISL_1255303, EPI_ISL_1255304, EPI_ISL_1255306, EPI_ISL_1255392, EPI_ISL_1255394, EPI_ISL_1255397 | | | |
| see above | Laboratory Corporation of America | Respiratory Viruses Branch, Division of Viral Diseases, Centers for Disease Control and Prevention | Peter W. Cook, Dakota Howard, Dhwani Batra, Ben L. Rambo-Martin, Minoo Agarwal, Eyad Almasri Debbie Boles, Ayla Burns, Nuthawin Charoensri, Oren Cohen, Susan Countryman, Mary Ann Cristobal, Bobbi Croy, Suzanne Dale, Hrushikesh Deshmukh, Amanda Douglas, Vincent Drouillon, Marcia Eisenberg, Howard Engler, Rama Ghatti, Prashant Gupta, Susan Hicks, Jake Humphrey, Lax Iyer, Manoj Jain, Mohan Kolli, Brian Krueger, Tim Kuphal, Stanley Letovsky, Michael Levandoski, Craig Lukasik, Jonathan Meltzer, Brian Norvell, Mindy Nye, Scott Parker, Christos Petropoulos, John Pruitt, Steven Ragan, Scott Ryan, Mike Sapeta, Jana Schroth, Suresh Babu Selvaraju, Goran Stevovic, Amanda Suchanek, Andrea Throop, Lyndon Tilson, Thomas Urban, Joe Voshell, Kimberly Wagner, Jonathan Williams, Mary Williamson, Qian Zeng, Tricia Zwiefelhofer, Clinton R. Paden, Suxiang Tong, Duncan MacCannell |
| EPI_ISL_1255398, EPI_ISL_1255399, EPI_ISL_1255400, EPI_ISL_1255401, EPI_ISL_1255402, EPI_ISL_1255403, EPI_ISL_1255404, EPI_ISL_1255405, EPI_ISL_1255406, EPI_ISL_1255407, EPI_ISL_1255408, EPI_ISL_1255409, EPI_ISL_1255410, EPI_ISL_1255411, EPI_ISL_1255412, EPI_ISL_1255413 | | | |
| see above | Helix / Illumina | Respiratory Viruses Branch, Division of Viral Diseases, Centers for Disease Control and Prevention | Peter W. Cook, Dakota Howard, Dhwani Batra, Ben L. Rambo-Martin, Eileen de Feo, Jan Antico, Christine Tran, Matthew Tolentino, Shannon Wickline, Kim Gietzen, Brad Sickler, Jingtao Liu, Eric Allen, Phil Febbo, Summer Galloway, Nicole L. Washington, Simon White, Geraint Levan, Kelly Schiabor Barrett, Elizabeth Cirulli, Alexandre Bolze, Ary Ascencio, Charlotte Rivera-Garcia, Ryan Cho, Jason Nguyen, Sherry Wang, Jimmy Ramirez, Tyler Cassens, Efren Sandoval, Magnus Isaksson, William Lee, David Becker, Marc Laurent, James Lu, Clinton R. Paden, Suxiang Tong, Duncan MacCannell |
| EPI_ISL_1255415 | Helix/Illumina | Respiratory Viruses Branch, Division of Viral Diseases, Centers for Disease Control and Prevention | Peter W. Cook, Dakota Howard, Dhwani Batra, Ben L. Rambo-Martin, Eileen de Feo, Jan Antico, Christine Tran, Matthew Tolentino, Shannon Wickline, Kim Gietzen, Brad Sickler, Jingtao Liu, Eric Allen, Phil Febbo, Summer Galloway, Nicole L. Washington, Simon White, Geraint Levan, Kelly Schiabor Barrett, Elizabeth Cirulli, Alexandre Bolze, Ary Ascencio, Charlotte Rivera-Garcia, Ryan Cho, Jason Nguyen, Sherry Wang, Jimmy Ramirez, Tyler Cassens, Efren Sandoval, Magnus Isaksson, William Lee, David Becker, Marc Laurent, James Lu, Clinton R. Paden, Suxiang Tong, Duncan MacCannell |
| EPI_ISL_1255416, EPI_ISL_1255417, EPI_ISL_1255418, EPI_ISL_1255419, EPI_ISL_1255420, EPI_ISL_1255421, EPI_ISL_1255422, EPI_ISL_1255423 | Atlas Genomics | Atlas Genomics | Michael Kalnoski, MD |
| EPI_ISL_1255932, EPI_ISL_1255935, EPI_ISL_1255937, EPI_ISL_1255940, EPI_ISL_1255943, EPI_ISL_1255946, EPI_ISL_1255949, EPI_ISL_1255951, EPI_ISL_1255954, EPI_ISL_1255957, EPI_ISL_1255960, EPI_ISL_1255963, EPI_ISL_1255966, EPI_ISL_1255969, EPI_ISL_1255972, EPI_ISL_1255975, EPI_ISL_1255977, EPI_ISL_1255979, EPI_ISL_1255985, EPI_ISL_1255988, EPI_ISL_1255991, EPI_ISL_1255997, EPI_ISL_1255999, EPI_ISL_1256002, EPI_ISL_1256005, EPI_ISL_1256008, EPI_ISL_1256011, EPI_ISL_1256014, EPI_ISL_1256016, EPI_ISL_1256019, EPI_ISL_1256021, EPI_ISL_1256024, EPI_ISL_1256027, EPI_ISL_1256029, EPI_ISL_1256032, EPI_ISL_1256035, EPI_ISL_1256039, EPI_ISL_1256042, EPI_ISL_1256045, EPI_ISL_1256048, EPI_ISL_1256051, EPI_ISL_1256053 | | | |
| see above | New Mexico Department of Health Scientific Laboratory | New Mexico Department of Health Scientific Laboratory | Ellie Johnson, Anastacia Griego-Fisher, D'eldra Malone, Jennifer Benoit |
| EPI_ISL_1258466, EPI_ISL_1258467, EPI_ISL_1258468, EPI_ISL_1258469, EPI_ISL_1258470, EPI_ISL_1258471, EPI_ISL_1258472, EPI_ISL_1258473, EPI_ISL_1258474, EPI_ISL_1258475, EPI_ISL_1258476, EPI_ISL_1258477, EPI_ISL_1258478, EPI_ISL_1258479, EPI_ISL_1258480, EPI_ISL_1258481, EPI_ISL_1258482, EPI_ISL_1258483, EPI_ISL_1258484, EPI_ISL_1258485, EPI_ISL_1258486, EPI_ISL_1258487, EPI_ISL_1258488, EPI_ISL_1258489, EPI_ISL_1258490, EPI_ISL_1258491, EPI_ISL_1258492, EPI_ISL_1258493, EPI_ISL_1258494, EPI_ISL_1258495, EPI_ISL_1258496, EPI_ISL_1258497, EPI_ISL_1258498, EPI_ISL_1258499, EPI_ISL_1258500, EPI_ISL_1258501, EPI_ISL_1258502, EPI_ISL_1258503, EPI_ISL_1258504, EPI_ISL_1258505, EPI_ISL_1258506, EPI_ISL_1258507, EPI_ISL_1258508, EPI_ISL_1258509, EPI_ISL_1258510, EPI_ISL_1258511, EPI_ISL_1258512, EPI_ISL_1258513, EPI_ISL_1258514, EPI_ISL_1258515, EPI_ISL_1258516, EPI_ISL_1258517, EPI_ISL_1258518, EPI_ISL_1258519, EPI_ISL_1258520, EPI_ISL_1258521, EPI_ISL_1258522, EPI_ISL_1258523, EPI_ISL_1258524, EPI_ISL_1258525, EPI_ISL_1258526, EPI_ISL_1258527, EPI_ISL_1258528, EPI_ISL_1258529, EPI_ISL_1258530, EPI_ISL_1258531, EPI_ISL_1258532, EPI_ISL_1258533, EPI_ISL_1258534, EPI_ISL_1258535, EPI_ISL_1258536, EPI_ISL_1258537, EPI_ISL_1258538, EPI_ISL_1258539, EPI_ISL_1258540, EPI_ISL_1258541, EPI_ISL_1258542, EPI_ISL_1258543, EPI_ISL_1258544, EPI_ISL_1258545, EPI_ISL_1258546, EPI_ISL_1258547, EPI_ISL_1258548, EPI_ISL_1258549, EPI_ISL_1258550, EPI_ISL_1258551, EPI_ISL_1258552, EPI_ISL_1258553, EPI_ISL_1258554, EPI_ISL_1258555, EPI_ISL_1258556, EPI_ISL_1258557, EPI_ISL_1258558, EPI_ISL_1258559, EPI_ISL_1258560, EPI_ISL_1258561, EPI_ISL_1258562, EPI_ISL_1258563, EPI_ISL_1258564, EPI_ISL_1258565, EPI_ISL_1258566, EPI_ISL_1258567, EPI_ISL_1258568, EPI_ISL_1258569, EPI_ISL_1258570, EPI_ISL_1258571, EPI_ISL_1258572, EPI_ISL_1258573, EPI_ISL_1258574, EPI_ISL_1258575, EPI_ISL_1258576, EPI_ISL_1258577, EPI_ISL_1258578, EPI_ISL_1258579, EPI_ISL_1258580, EPI_ISL_1258581, EPI_ISL_1258582, EPI_ISL_1258583, EPI_ISL_1258584, EPI_ISL_1258585, EPI_ISL_1258586, EPI_ISL_1258587, EPI_ISL_1258588, EPI_ISL_1258589, EPI_ISL_1258590, EPI_ISL_1258591, EPI_ISL_1258592, EPI_ISL_1258593, EPI_ISL_1258594, EPI_ISL_1258595, EPI_ISL_1258596, EPI_ISL_1258597, EPI_ISL_1258598, EPI_ISL_1258599, EPI_ISL_1258600, EPI_ISL_1258601, EPI_ISL_1258602, EPI_ISL_1258603, EPI_ISL_1258604, EPI_ISL_1258605, EPI_ISL_1258606, EPI_ISL_1258607, EPI_ISL_1258608, EPI_ISL_1258609, EPI_ISL_1258610, EPI_ISL_1258611, EPI_ISL_1258612, EPI_ISL_1258613, EPI_ISL_1258614, EPI_ISL_1258615, EPI_ISL_1258616, EPI_ISL_1258617, EPI_ISL_1258618, EPI_ISL_1258619, EPI_ISL_1258620, EPI_ISL_1258621, EPI_ISL_1258622, EPI_ISL_1258623, EPI_ISL_1258624, EPI_ISL_1258625, EPI_ISL_1258626, EPI_ISL_1258627, EPI_ISL_1258628, EPI_ISL_1258629, EPI_ISL_1258630, EPI_ISL_1258631, EPI_ISL_1258632, EPI_ISL_1258633, EPI_ISL_1258634, EPI_ISL_1258689, EPI_ISL_1258690, EPI_ISL_1258691, EPI_ISL_1258692, EPI_ISL_1258693, EPI_ISL_1258694, EPI_ISL_1258695, EPI_ISL_1258696, EPI_ISL_1258697, EPI_ISL_1258698, EPI_ISL_1258699, EPI_ISL_1258700, EPI_ISL_1258701, EPI_ISL_1258702, EPI_ISL_1258703, EPI_ISL_1258704, EPI_ISL_1258705, EPI_ISL_1258706, EPI_ISL_1258707, EPI_ISL_1258708, EPI_ISL_1258709, EPI_ISL_1258710, EPI_ISL_1258711, EPI_ISL_1258712, EPI_ISL_1258713, EPI_ISL_1258714, EPI_ISL_1258715, EPI_ISL_1258716, EPI_ISL_1258717, EPI_ISL_1258718, EPI_ISL_1258719, EPI_ISL_1258720, EPI_ISL_1258721, EPI_ISL_1258722, EPI_ISL_1258723, EPI_ISL_1258724, EPI_ISL_1258725, EPI_ISL_1258726, EPI_ISL_1258727, EPI_ISL_1258728, EPI_ISL_1258729, EPI_ISL_1258730, EPI_ISL_1258731, EPI_ISL_1258732, EPI_ISL_1258733, EPI_ISL_1258734, EPI_ISL_1258735, EPI_ISL_1258736, EPI_ISL_1258737, EPI_ISL_1258738, EPI_ISL_1258739, EPI_ISL_1258740, EPI_ISL_1258741, EPI_ISL_1258742, EPI_ISL_1258743, EPI_ISL_1258744, EPI_ISL_1258745, EPI_ISL_1258746, EPI_ISL_1258747, EPI_ISL_1258748, EPI_ISL_1258749, EPI_ISL_1258750, EPI_ISL_1258751, EPI_ISL_1258752, EPI_ISL_1258753, EPI_ISL_1258754, EPI_ISL_1258755, EPI_ISL_1258756, EPI_ISL_1258757, EPI_ISL_1258758, EPI_ISL_1258759, EPI_ISL_1258760, EPI_ISL_1258761, EPI_ISL_1258762, EPI_ISL_1258763, EPI_ISL_1258764, EPI_ISL_1258765, EPI_ISL_1258766, EPI_ISL_1258767, EPI_ISL_1258768, EPI_ISL_1258769, EPI_ISL_1258770, EPI_ISL_1258771, EPI_ISL_1258772, EPI_ISL_1258773, EPI_ISL_1258774, EPI_ISL_1258775, EPI_ISL_1258776, EPI_ISL_1258777, EPI_ISL_1258778, EPI_ISL_1258779, EPI_ISL_1258780, EPI_ISL_1258781, EPI_ISL_1258782, EPI_ISL_1258783, EPI_ISL_1258784, EPI_ISL_1258785, EPI_ISL_1258786, EPI_ISL_1258787, EPI_ISL_1258788, EPI_ISL_1258789, EPI_ISL_1258790, EPI_ISL_1258791, EPI_ISL_1258792, EPI_ISL_1258793, EPI_ISL_1258794, EPI_ISL_1258795, EPI_ISL_1258796, EPI_ISL_1258797, EPI_ISL_1258798, EPI_ISL_1258799, EPI_ISL_1258800, EPI_ISL_1258801, EPI_ISL_1258802, EPI_ISL_1258803, EPI_ISL_1258804, EPI_ISL_1258805, EPI_ISL_1258806, EPI_ISL_1258807, EPI_ISL_1258808, EPI_ISL_1258809, EPI_ISL_1258810, EPI_ISL_1258811, EPI_ISL_1258812, EPI_ISL_1258813, EPI_ISL_1258814, EPI_ISL_1258815, EPI_ISL_1258816, EPI_ISL_1258817, EPI_ISL_1258818, EPI_ISL_1258819, EPI_ISL_1258820, EPI_ISL_1258821, EPI_ISL_1258822, EPI_ISL_1258823, EPI_ISL_1258824, EPI_ISL_1258825, EPI_ISL_1258826, EPI_ISL_1258827, EPI_ISL_1258828, EPI_ISL_1258829, EPI_ISL_1258830, EPI_ISL_1258831, EPI_ISL_1258832, EPI_ISL_1258833, EPI_ISL_1258834, EPI_ISL_1258835, EPI_ISL_1258836, EPI_ISL_1258837, EPI_ISL_1258838, EPI_ISL_1258839, EPI_ISL_1258840, EPI_ISL_1258841, EPI_ISL_1258842, EPI_ISL_1258843, EPI_ISL_1258844, EPI_ISL_1258845, EPI_ISL_1258846, EPI_ISL_1258847, EPI_ISL_1258848, EPI_ISL_1258849, EPI_ISL_1258850, EPI_ISL_1258851, EPI_ISL_1258852, EPI_ISL_1258853, EPI_ISL_1258854, EPI_ISL_1258855, EPI_ISL_1258856, EPI_ISL_1258857, EPI_ISL_1258858, EPI_ISL_1258859, EPI_ISL_1258860, EPI_ISL_1258861, EPI_ISL_1258862, EPI_ISL_1258863, EPI_ISL_1258864, EPI_ISL_1258865, EPI_ISL_1258866, EPI_ISL_1258867, EPI_ISL_1258868, EPI_ISL_1258869, EPI_ISL_1258870, EPI_ISL_1258871, EPI_ISL_1258872, EPI_ISL_1258873, EPI_ISL_1258874, EPI_ISL_1258875, EPI_ISL_1258876, EPI_ISL_1258877, EPI_ISL_1258878, EPI_ISL_1258879, EPI_ISL_1258880, EPI_ISL_1258881, EPI_ISL_1258882, EPI_ISL_1258883, EPI_ISL_1258884, EPI_ISL_1258885, EPI_ISL_1258886, EPI_ISL_1258887, EPI_ISL_1258888, EPI_ISL_1258889, EPI_ISL_1258890, EPI_ISL_1258891, EPI_ISL_1258892, EPI_ISL_1258893, EPI_ISL_1258894, EPI_ISL_1258895, EPI_ISL_1258896, EPI_ISL_1258897, EPI_ISL_1258898, EPI_ISL_1258899, EPI_ISL_1258900, EPI_ISL_1258901, EPI_ISL_1258902, EPI_ISL_1258903, EPI_ISL_1258904, EPI_ISL_1258905, EPI_ISL_1258906, EPI_ISL_1258907, EPI_ISL_1258908, EPI_ISL_1258909, EPI_ISL_1258910, EPI_ISL_1258911, EPI_ISL_1258912, EPI_ISL_1258913, EPI_ISL_1258914, EPI_ISL_1258915, EPI_ISL_1258916, EPI_ISL_1258917, EPI_ISL_1258918, EPI_ISL_1258919, EPI_ISL_1258920, EPI_ISL_1258921, EPI_ISL_1258922, EPI_ISL_1258923, EPI_ISL_1258924, EPI_ISL_1258925, EPI_ISL_1258926, EPI_ISL_1258927, EPI_ISL_1258928, EPI_ISL_1258929, EPI_ISL_1258930, EPI_ISL_1258931, EPI_ISL_1258932, EPI_ISL_1258933, EPI_ISL_1258934, EPI_ISL_1258935, EPI_ISL_1258936, EPI_ISL_1258937, EPI_ISL_1258938, EPI_ISL_1258939, EPI_ISL_1258940, EPI_ISL_1258941, EPI_ISL_1258942, EPI_ISL_1258943, EPI_ISL_1258944, EPI_ISL_1258945, EPI_ISL_1258946, EPI_ISL_1258947, EPI_ISL_1258948, EPI_ISL_1258949, EPI_ISL_1258950, EPI_ISL_1258951, EPI_ISL_1258952, EPI_ISL_1258953, EPI_ISL_1258954, EPI_ISL_1258955, EPI_ISL_1258956, EPI_ISL_1258957, EPI_ISL_1258958, EPI_ISL_1258959, EPI_ISL_1258960, EPI_ISL_1258961, EPI_ISL_1258962, EPI_ISL_1258963, EPI_ISL_1258964, EPI_ISL_1258965, EPI_ISL_1258966, EPI_ISL_1258967, EPI_ISL_1258968, EPI_ISL_1258969, EPI_ISL_1258970, EPI_ISL_1258971, EPI_ISL_1258972, EPI_ISL_1258973, EPI_ISL_1258974, EPI_ISL_1258975, EPI_ISL_1258976, EPI_ISL_1258977, EPI_ISL_1258978, EPI_ISL_1258979, EPI_ISL_1258980, EPI_ISL_1258981, EPI_ISL_1258982, EPI_ISL_1258983, EPI_ISL_1258984, EPI_ISL_1258985, EPI_ISL_1258986, EPI_ISL_1258987 | | | |
| see above | Pandemic Response Lab - NYC | Pandemic Response Lab, R&D | Henry Lee, Michael Hammerling, Melissa Hopkins, Cybill del Castillo, Shinyoung Clair Kang, William Ward, Pradeep Bugga, Haiping Hao, Jon Laurent |
| EPI_ISL_1259311, EPI_ISL_1259312, EPI_ISL_1259314, EPI_ISL_1259372, EPI_ISL_1259396, EPI_ISL_1259398 | Ohio Department of Health Laboratory | Ohio Department of Health Laboratory | Holmes, Jennifer; Eric Brandt, Keoni Omura, Glen McGillivary, Caitlin McDonnell, Jade Mowery, Stephanie Mccracken, Tyler Payne, Kirtana Ramadugu, Erica Leasure, Brent Lee, Kelsey Florek, Heather Blankenship, Allison Black, Quanta Brown, and Tammy Bannerman |
| EPI_ISL_1259401, EPI_ISL_1259402, EPI_ISL_1259403, EPI_ISL_1259404 | Florida Bureau of Public Health Laboratories | Florida Bureau of Public Health Laboratories | Sarah Schmedes, Jason Blanton |
| EPI_ISL_1260610, EPI_ISL_1260611, EPI_ISL_1260612, EPI_ISL_1260613, EPI_ISL_1260614, EPI_ISL_1260615, EPI_ISL_1260616, EPI_ISL_1260617, EPI_ISL_1260618, EPI_ISL_1260619, EPI_ISL_1260620, EPI_ISL_1260621, EPI_ISL_1260622, EPI_ISL_1260623, EPI_ISL_1260624, EPI_ISL_1260625, EPI_ISL_1260626, EPI_ISL_1260627, EPI_ISL_1260628, EPI_ISL_1260629, EPI_ISL_1260630, EPI_ISL_1260631, EPI_ISL_1260632 | | | |
| see above | Houston Health Dept. | Houston Health Dept. | Ryker Penn, Pamela Brown, Adolpho Lara |
| EPI_ISL_1260633 | Ohio Department of Health Laboratory | Ohio Department of Health Laboratory | Holmes, Jennifer; Eric Brandt, Keoni Omura, Glen McGillivary, Caitlin McDonnell, Jade Mowery, Stephanie Mccracken, Tyler Payne, Kirtana Ramadugu, Erica Leasure, Brent Lee, Kelsey Florek, Heather Blankenship, Allison Black, Quanta Brown, and Tammy Bannerman |
| EPI_ISL_1260634, EPI_ISL_1260635, EPI_ISL_1260636, EPI_ISL_1260637, EPI_ISL_1260638, EPI_ISL_1260639, EPI_ISL_1260640, EPI_ISL_1260641, EPI_ISL_1260642, EPI_ISL_1260643, EPI_ISL_1260644, EPI_ISL_1260645, EPI_ISL_1260646, EPI_ISL_1260647, EPI_ISL_1260648, EPI_ISL_1260649, EPI_ISL_1260650, EPI_ISL_1260651, EPI_ISL_1260652, EPI_ISL_1260653, EPI_ISL_1260654, EPI_ISL_1260655, EPI_ISL_1260656, EPI_ISL_1260657, EPI_ISL_1260658, EPI_ISL_1260659, EPI_ISL_1260660 | | | |
| see above | Houston Health Dept. | Houston Health Dept. | Ryker Penn, Pamela Brown, Adolpho Lara |
| EPI_ISL_1260661, EPI_ISL_1260695, EPI_ISL_1260696 | Ohio Department of Health Laboratory | Ohio Department of Health Laboratory | Holmes, Jennifer; Eric Brandt, Keoni Omura, Glen McGillivary, Caitlin McDonnell, Jade Mowery, Stephanie Mccracken, Tyler Payne, Kirtana Ramadugu, Erica Leasure, Brent Lee, Kelsey Florek, Heather Blankenship, Allison Black, Quanta Brown, and Tammy Bannerman |
| EPI_ISL_1260701, EPI_ISL_1260702, EPI_ISL_1260704, EPI_ISL_1260705, EPI_ISL_1260706, EPI_ISL_1260707, EPI_ISL_1260708, EPI_ISL_1260710, EPI_ISL_1260711, EPI_ISL_1260712, EPI_ISL_1260714, EPI_ISL_1260716, EPI_ISL_1260717, EPI_ISL_1260718 | | | |
| see above | US Air Force School of Aerospace Medicine | US Air Force School of Aerospace Medicine | Anthony Fries, Jennifer Meyer, William Gruner, William Buggele, Amanda Javorina, Sarah Purves, Clarise Starr, Elizabeth Macias |
| EPI_ISL_1260719, EPI_ISL_1260720, EPI_ISL_1260721, EPI_ISL_1260722, EPI_ISL_1260723, EPI_ISL_1260724, EPI_ISL_1260725, EPI_ISL_1260726, EPI_ISL_1260727, EPI_ISL_1260728, EPI_ISL_1260729, EPI_ISL_1260730, EPI_ISL_1260731, EPI_ISL_1260732, EPI_ISL_1260733, EPI_ISL_1260734, EPI_ISL_1260735, EPI_ISL_1260736, EPI_ISL_1260737, EPI_ISL_1260738, EPI_ISL_1260739, EPI_ISL_1260740, EPI_ISL_1260741, EPI_ISL_1260742, EPI_ISL_1260743 | | | |
| see above | United States Air Force Academy | US Air Force School of Aerospace Medicine | Anthony Fries, Jennifer Meyer, William Gruner, William Buggele, Amanda Javorina, Sarah Purves, Clarise Starr, Elizabeth Macias |
| EPI_ISL_1260966, EPI_ISL_1260967, EPI_ISL_1260968, EPI_ISL_1260969, EPI_ISL_1260970, EPI_ISL_1260971, EPI_ISL_1260972, EPI_ISL_1260973, EPI_ISL_1260974, EPI_ISL_1260975, EPI_ISL_1260976, EPI_ISL_1260977, EPI_ISL_1260978, EPI_ISL_1260979, EPI_ISL_1260980, EPI_ISL_1260981, EPI_ISL_1260982, EPI_ISL_1260983, EPI_ISL_1260984, EPI_ISL_1260985, EPI_ISL_1260986, EPI_ISL_1260987, EPI_ISL_1260988, EPI_ISL_1260989, EPI_ISL_1260990, EPI_ISL_1260991, EPI_ISL_1260992, EPI_ISL_1260993, EPI_ISL_1260994, EPI_ISL_1260995, EPI_ISL_1260996, EPI_ISL_1260997, EPI_ISL_1260998, EPI_ISL_1260999, EPI_ISL_1261000, EPI_ISL_1261001, EPI_ISL_1261002, EPI_ISL_1261003, EPI_ISL_1261004, EPI_ISL_1261005, EPI_ISL_1261006, EPI_ISL_1261007, EPI_ISL_1261008, EPI_ISL_1261009, | | | |

| | | | |
|---|---|---|---|
| EPI_ISL_1261010, EPI_ISL_1261011, EPI_ISL_1261012, EPI_ISL_1261013, EPI_ISL_1261014, EPI_ISL_1261015, EPI_ISL_1261016, EPI_ISL_1261017, EPI_ISL_1261018, EPI_ISL_1261019, EPI_ISL_1261020, EPI_ISL_1261021, EPI_ISL_1261022, EPI_ISL_1261023, EPI_ISL_1261024, EPI_ISL_1261025, EPI_ISL_1261026, EPI_ISL_1261027, EPI_ISL_1261028, EPI_ISL_1261029, EPI_ISL_1261030, EPI_ISL_1261031, EPI_ISL_1261032, EPI_ISL_1261033, EPI_ISL_1261034, EPI_ISL_1261035, EPI_ISL_1261036, EPI_ISL_1261037, EPI_ISL_1261038, EPI_ISL_1261039, EPI_ISL_1261040, EPI_ISL_1261041, EPI_ISL_1261042, EPI_ISL_1261043, EPI_ISL_1261044, EPI_ISL_1261045, EPI_ISL_1261046, EPI_ISL_1261047, EPI_ISL_1261048, EPI_ISL_1261049, EPI_ISL_1261050, EPI_ISL_1261051, EPI_ISL_1261052, EPI_ISL_1261053, EPI_ISL_1261054, EPI_ISL_1261055, EPI_ISL_1261056, EPI_ISL_1261057, EPI_ISL_1261058, EPI_ISL_1261059, EPI_ISL_1261060, EPI_ISL_1261061, EPI_ISL_1261062, EPI_ISL_1261063, EPI_ISL_1261064, EPI_ISL_1261065, EPI_ISL_1261066, EPI_ISL_1261067, EPI_ISL_1261068, EPI_ISL_1261069, EPI_ISL_1261070, EPI_ISL_1261071, EPI_ISL_1261072, EPI_ISL_1261073, EPI_ISL_1261074, EPI_ISL_1261075, EPI_ISL_1261076, EPI_ISL_1261077, EPI_ISL_1261078, EPI_ISL_1261079, EPI_ISL_1261080, EPI_ISL_1261081, EPI_ISL_1261082, EPI_ISL_1261083, EPI_ISL_1261084, EPI_ISL_1261085, EPI_ISL_1261086, EPI_ISL_1261087, EPI_ISL_1261088, EPI_ISL_1261089, EPI_ISL_1261090, EPI_ISL_1261091, EPI_ISL_1261092, EPI_ISL_1261093, EPI_ISL_1261094, EPI_ISL_1261095, EPI_ISL_1261096, EPI_ISL_1261097, EPI_ISL_1261098, EPI_ISL_1261099, EPI_ISL_1261100, EPI_ISL_1261101, EPI_ISL_1261102, EPI_ISL_1261103, EPI_ISL_1261104, EPI_ISL_1261105, EPI_ISL_1261106, EPI_ISL_1261107, EPI_ISL_1261108, EPI_ISL_1261109, EPI_ISL_1261110, EPI_ISL_1261111, EPI_ISL_1261112, EPI_ISL_1261113, EPI_ISL_1261114, EPI_ISL_1261115, EPI_ISL_1261116, EPI_ISL_1261117, EPI_ISL_1261118, EPI_ISL_1261119, EPI_ISL_1261120, EPI_ISL_1261121 | | | |
| see above | University of Michigan Clinical Microbiology Laboratory | Lauring Lab, University of Michigan, Department of Microbiology and Immunology | Valesano |
| EPI_ISL_1261124, EPI_ISL_1261125, EPI_ISL_1261126, EPI_ISL_1261127, EPI_ISL_1261128, EPI_ISL_1261129, EPI_ISL_1261130, EPI_ISL_1261131, EPI_ISL_1261132, EPI_ISL_1261133, EPI_ISL_1261134 | | | |
| see above | Kansas Health and Environmental Lab | Kansas Health and Environmental Lab | Mike Grose, Jonathan Barnell, Ben Olsen, and Phil Adam |
| EPI_ISL_1261137, EPI_ISL_1261138, EPI_ISL_1261139, EPI_ISL_1261140, EPI_ISL_1261141, EPI_ISL_1261142, EPI_ISL_1261143, EPI_ISL_1261144, EPI_ISL_1261145, EPI_ISL_1261146, EPI_ISL_1261147, EPI_ISL_1261148, EPI_ISL_1261149, EPI_ISL_1261150, EPI_ISL_1261151, EPI_ISL_1261152, EPI_ISL_1261153, EPI_ISL_1261154, EPI_ISL_1261156, EPI_ISL_1261157, EPI_ISL_1261158, EPI_ISL_1261159, EPI_ISL_1261160, EPI_ISL_1261161 | | | |
| see above | Molecular & Genomic Pathology Laboratory, Thomas Jefferson University Hospital | Molecular & Genomic Pathology Laboratory, Thomas Jefferson University Hospital | Nitika Badjatia, Run Jin, Zi-xuan Wang |
| EPI_ISL_1261425, EPI_ISL_1261426, EPI_ISL_1261428, EPI_ISL_1261429, EPI_ISL_1261430, EPI_ISL_1261433, EPI_ISL_1261434, EPI_ISL_1261435, EPI_ISL_1261438, EPI_ISL_1261440, EPI_ISL_1261441, EPI_ISL_1261444, EPI_ISL_1261446, EPI_ISL_1261448, EPI_ISL_1261449, EPI_ISL_1261451, EPI_ISL_1261452, EPI_ISL_1261453, EPI_ISL_1261456 | | | |
| see above | US Air Force School of Aerospace Medicine | US Air Force School of Aerospace Medicine | Anthony Fries, Jennifer Meyer, William Gruner, William Buggele, Amanda Javorina, Sarah Purves, Clarise Starr, Elizabeth Macias |
| EPI_ISL_1261541, EPI_ISL_1261542 | Microbiology Division, South Carolina Department of Health and Environmental Control Public Health Laboratory (SC DHEC PHL) | Microbiology Division, South Carolina Department of Health and Environmental Control Public Health Laboratory (SC DHEC PHL) | Haley Flores, Jessica Freeman |
| EPI_ISL_1261543, EPI_ISL_1261544, EPI_ISL_1261545, EPI_ISL_1261546, EPI_ISL_1261547, EPI_ISL_1261548, EPI_ISL_1261549, EPI_ISL_1261550, EPI_ISL_1261551, EPI_ISL_1261552, EPI_ISL_1261558, EPI_ISL_1261559, EPI_ISL_1261560, EPI_ISL_1261561, EPI_ISL_1261562, EPI_ISL_1261563, EPI_ISL_1261564, EPI_ISL_1261565, EPI_ISL_1261566, EPI_ISL_1261567, EPI_ISL_1261568, EPI_ISL_1261569, EPI_ISL_1261570, EPI_ISL_1261571, EPI_ISL_1261572, EPI_ISL_1261573, EPI_ISL_1261574, EPI_ISL_1261575, EPI_ISL_1261576, EPI_ISL_1261577, EPI_ISL_1261578, EPI_ISL_1261579, EPI_ISL_1261580, EPI_ISL_1261581, EPI_ISL_1261582, EPI_ISL_1261583, EPI_ISL_1261584, EPI_ISL_1261585 | | | |
| see above | New Mexico Department of Health Scientific Laboratory | New Mexico Department of Health Scientific Laboratory | Ellie Johnson, Anastacia Griego-Fisher, D'eldra Malone, Jennifer Benoit |
| EPI_ISL_1261586, EPI_ISL_1261587, EPI_ISL_1261588, EPI_ISL_1261589, EPI_ISL_1261590, EPI_ISL_1261591, EPI_ISL_1261592, EPI_ISL_1261593, EPI_ISL_1261594, EPI_ISL_1261595, EPI_ISL_1261596, EPI_ISL_1261597, EPI_ISL_1261598, EPI_ISL_1261602, EPI_ISL_1261603, EPI_ISL_1261604, EPI_ISL_1261605, EPI_ISL_1261606, EPI_ISL_1261607, EPI_ISL_1261608, EPI_ISL_1261609, EPI_ISL_1261610, EPI_ISL_1261611, EPI_ISL_1261612, EPI_ISL_1261613, EPI_ISL_1261614, EPI_ISL_1261615, EPI_ISL_1261616, EPI_ISL_1261617, EPI_ISL_1261618, EPI_ISL_1261619, EPI_ISL_1261620, EPI_ISL_1261621, EPI_ISL_1261622, EPI_ISL_1261623, EPI_ISL_1261624, EPI_ISL_1261625, EPI_ISL_1261626, EPI_ISL_1261627, EPI_ISL_1261628, EPI_ISL_1261629, EPI_ISL_1261630, EPI_ISL_1261631, EPI_ISL_1261632, EPI_ISL_1261633, EPI_ISL_1261634, EPI_ISL_1261635, EPI_ISL_1261636, EPI_ISL_1261637, EPI_ISL_1261638, EPI_ISL_1261639, EPI_ISL_1261640, EPI_ISL_1261641, EPI_ISL_1261642, EPI_ISL_1261643, EPI_ISL_1261644, EPI_ISL_1261645, EPI_ISL_1261646, EPI_ISL_1261647, EPI_ISL_1261648, EPI_ISL_1261649, EPI_ISL_1261650, EPI_ISL_1261651, EPI_ISL_1261652, EPI_ISL_1261653, EPI_ISL_1261654, EPI_ISL_1261655, EPI_ISL_1261656, EPI_ISL_1261657, EPI_ISL_1261658, EPI_ISL_1261659, EPI_ISL_1261660, EPI_ISL_1261661, EPI_ISL_1261662, EPI_ISL_1261663, EPI_ISL_1261664, EPI_ISL_1261665, EPI_ISL_1261666, EPI_ISL_1261667, EPI_ISL_1261668, EPI_ISL_1261669, EPI_ISL_1261670, EPI_ISL_1261671, EPI_ISL_1261672 | | | |
| see above | TXDSHS | TXDSHS | Rashmi Tuladhar, Bonnie Oh, Jenny Zhang, Maliha Rahman, Mayela Pedrueza, Anita Pokharel, Myong Koag, Chun Wang, Rachel Lee, Grace Kubin |
| EPI_ISL_1261775, EPI_ISL_1261776 | State Hygienic Laboratory at the University of Iowa | State Hygienic Laboratory at the University of Iowa | Valerie Reeb, Wes Hottel, Alankar Kampoowale |
| EPI_ISL_1261777 | Test Iowa | State Hygienic Laboratory at the University of Iowa | Valerie Reeb, Wes Hottel, Alankar Kampoowale |
| EPI_ISL_1261778, EPI_ISL_1261779 | State Hygienic Laboratory at the University of Iowa | State Hygienic Laboratory at the University of Iowa | Valerie Reeb, Wes Hottel, Alankar Kampoowale |
| EPI_ISL_1261780 | Test Iowa | State Hygienic Laboratory at the University of Iowa | Valerie Reeb, Wes Hottel, Alankar Kampoowale |
| EPI_ISL_1261781, EPI_ISL_1261782, EPI_ISL_1261783 | State Hygienic Laboratory at the University of Iowa | State Hygienic Laboratory at the University of Iowa | Valerie Reeb, Wes Hottel, Alankar Kampoowale |
| EPI_ISL_1261784, EPI_ISL_1261785, EPI_ISL_1261786, EPI_ISL_1261787, EPI_ISL_1261788, EPI_ISL_1261789 | State Hygienic Laboratory at the University of Iowa | State Hygienic Laboratory at the University of Iowa | Valerie Reeb, Erik Twait, Wes Hottel, Alankar Kampoowale |
| EPI_ISL_1261790, EPI_ISL_1261791, EPI_ISL_1261792, EPI_ISL_1261793, EPI_ISL_1261794 | DOHMH Jamaica | New York City Public Health Laboratory | Jade Wang, et al. |
| EPI_ISL_1261795 | DOHMH Riverside | New York City Public Health Laboratory | Jade Wang, et al. |
| EPI_ISL_1261796, EPI_ISL_1261797 | OCME Office Of Chief Medical Examiner | New York City Public Health Laboratory | Jade Wang, et al. |
| EPI_ISL_1261798 | DOHMH PHL | New York City Public Health Laboratory | Jade Wang, et al. |
| EPI_ISL_1261799 | DOHMH Riverside | New York City Public Health Laboratory | Jade Wang, et al. |
| EPI_ISL_1261800 | Department of Homeless Services | New York City Public Health Laboratory | Jade Wang, et al. |
| EPI_ISL_1261801 | DOHMH Central Harlem | New York City Public Health Laboratory | Jade Wang, et al. |
| EPI_ISL_1261802 | DOHMH Jamaica | New York City Public Health Laboratory | Jade Wang, et al. |
| EPI_ISL_1261803 | DOHMH Crown Heights | New York City Public Health Laboratory | Jade Wang, et al. |
| EPI_ISL_1261804 | DOHMH Central Harlem | New York City Public Health Laboratory | Jade Wang, et al. |
| EPI_ISL_1261805 | DOHMH PHL | New York City Public Health Laboratory | Jade Wang, et al. |
| EPI_ISL_1261806 | DOHMH Corona | New York City Public Health Laboratory | Jade Wang, et al. |
| EPI_ISL_1261807, EPI_ISL_1261808 | DOHMH Central Harlem | New York City Public Health Laboratory | Jade Wang, et al. |
| EPI_ISL_1261809 | DOHMH Fort Greene | New York City Public Health Laboratory | Jade Wang, et al. |
| EPI_ISL_1261810 | DOHMH Corona | New York City Public Health Laboratory | Jade Wang, et al. |
| EPI_ISL_1261811 | OCME Office Of Chief Medical Examiner | New York City Public Health Laboratory | Jade Wang, et al. |
| EPI_ISL_1261812, EPI_ISL_1261813 | DOHMH Corona | New York City Public Health Laboratory | Jade Wang, et al. |
| EPI_ISL_1261814 | DOHMH Morrisania | New York City Public Health Laboratory | Jade Wang, et al. |
| EPI_ISL_1261815 | DOHMH Jamaica | New York City Public Health Laboratory | Jade Wang, et al. |
| EPI_ISL_1261816 | DOHMH Riverside | New York City Public Health Laboratory | Jade Wang, et al. |
| EPI_ISL_1261817 | DOHMH Chelsea | New York City Public Health Laboratory | Jade Wang, et al. |
| EPI_ISL_1261818 | DOHMH Fort Greene | New York City Public Health Laboratory | Jade Wang, et al. |
| EPI_ISL_1261819 | OCME Office Of Chief Medical Examiner | New York City Public Health Laboratory | Jade Wang, et al. |
| EPI_ISL_1261820 | DOHMH Jamaica | New York City Public Health Laboratory | Jade Wang, et al. |
| EPI_ISL_1261821 | DOHMH Crown Heights | New York City Public Health Laboratory | Jade Wang, et al. |
| EPI_ISL_1261822 | DOHMH Jamaica | New York City Public Health Laboratory | Jade Wang, et al. |
| EPI_ISL_1261823 | DOHMH PHL | New York City Public Health Laboratory | Jade Wang, et al. |
| EPI_ISL_1261824 | DOHMH Central Harlem | New York City Public Health Laboratory | Jade Wang, et al. |
| EPI_ISL_1261825 | DOHMH Crown Heights | New York City Public Health Laboratory | Jade Wang, et al. |
| EPI_ISL_1261826 | DOHMH Morrisania | New York City Public Health Laboratory | Jade Wang, et al. |
| EPI_ISL_1261827, EPI_ISL_1261828, EPI_ISL_1261829 | OCME Office Of Chief Medical Examiner | New York City Public Health Laboratory | Jade Wang, et al. |

| | | | |
|---|---|---|---|
| EPI_ISL_1261830 | Department of Homeless Services | New York City Public Health Laboratory | Jade Wang, et al. |
| EPI_ISL_1261831 | DOHMH Jamaica | New York City Public Health Laboratory | Jade Wang, et al. |
| EPI_ISL_1261832, EPI_ISL_1261833 | DOHMH Corona | New York City Public Health Laboratory | Jade Wang, et al. |
| EPI_ISL_1261834 | DOHMH Crown Heights | New York City Public Health Laboratory | Jade Wang, et al. |
| EPI_ISL_1261835, EPI_ISL_1261836, EPI_ISL_1261837 | DOHMH Corona | New York City Public Health Laboratory | Jade Wang, et al. |
| EPI_ISL_1261838, EPI_ISL_1261839 | DOHMH Morrisania | New York City Public Health Laboratory | Jade Wang, et al. |
| EPI_ISL_1261840, EPI_ISL_1261841, EPI_ISL_1261842 | DOHMH Riverside | New York City Public Health Laboratory | Jade Wang, et al. |
| EPI_ISL_1261843 | DOHMH Central Harlem | New York City Public Health Laboratory | Jade Wang, et al. |
| EPI_ISL_1261844 | DOHMH Corona | New York City Public Health Laboratory | Jade Wang, et al. |
| EPI_ISL_1261845 | DOHMH Riverside | New York City Public Health Laboratory | Jade Wang, et al. |
| EPI_ISL_1261846 | OCME Office Of Chief Medical Examiner | New York City Public Health Laboratory | Jade Wang, et al. |
| EPI_ISL_1261847 | DOHMH Fort Greene | New York City Public Health Laboratory | Jade Wang, et al. |
| EPI_ISL_1261848 | DOHMH Morrisania | New York City Public Health Laboratory | Jade Wang, et al. |
| EPI_ISL_1261849 | DOHMH Riverside | New York City Public Health Laboratory | Jade Wang, et al. |
| EPI_ISL_1261850 | DOHMH Fort Greene | New York City Public Health Laboratory | Jade Wang, et al. |
| EPI_ISL_1261851 | DOHMH Morrisania | New York City Public Health Laboratory | Jade Wang, et al. |
| EPI_ISL_1261852 | DOHMH Chelsea | New York City Public Health Laboratory | Jade Wang, et al. |
| EPI_ISL_1261853 | Department of Homeless Services | New York City Public Health Laboratory | Jade Wang, et al. |
| EPI_ISL_1261854 | OCME Office Of Chief Medical Examiner | New York City Public Health Laboratory | Jade Wang, et al. |
| EPI_ISL_1261855 | DOHMH Chelsea | New York City Public Health Laboratory | Jade Wang, et al. |
| EPI_ISL_1261856 | DOHMH Central Harlem | New York City Public Health Laboratory | Jade Wang, et al. |
| EPI_ISL_1261857 | DOHMH Jamaica | New York City Public Health Laboratory | Jade Wang, et al. |
| EPI_ISL_1261858 | DOHMH Central Harlem | New York City Public Health Laboratory | Jade Wang, et al. |
| EPI_ISL_1261859 | DOHMH Corona | New York City Public Health Laboratory | Jade Wang, et al. |
| EPI_ISL_1261860 | DOHMH Jamaica | New York City Public Health Laboratory | Jade Wang, et al. |
| EPI_ISL_1261861 | DOHMH Crown Heights | New York City Public Health Laboratory | Jade Wang, et al. |
| EPI_ISL_1261862, EPI_ISL_1261863 | DOHMH Chelsea | New York City Public Health Laboratory | Jade Wang, et al. |
| EPI_ISL_1261864 | DOHMH Central Harlem | New York City Public Health Laboratory | Jade Wang, et al. |
| EPI_ISL_1261865 | OCME Office Of Chief Medical Examiner | New York City Public Health Laboratory | Jade Wang, et al. |
| EPI_ISL_1261866 | DOHMH Jamaica | New York City Public Health Laboratory | Jade Wang, et al. |
| EPI_ISL_1261867 | Department of Homeless Services | New York City Public Health Laboratory | Jade Wang, et al. |
| EPI_ISL_1261871, EPI_ISL_1261872, EPI_ISL_1261873, EPI_ISL_1261874, EPI_ISL_1261875, EPI_ISL_1261876, EPI_ISL_1261877, EPI_ISL_1261878, EPI_ISL_1261879, EPI_ISL_1261880, EPI_ISL_1261881, EPI_ISL_1261882, EPI_ISL_1261883, EPI_ISL_1261884, EPI_ISL_1261886, EPI_ISL_1261887, EPI_ISL_1261888, EPI_ISL_1261889, EPI_ISL_1261890, EPI_ISL_1261891, EPI_ISL_1261892, EPI_ISL_1261893, EPI_ISL_1261894 | | | |
| see above | UNC-CH COVID Surveillance Lab | Jeremy Wang | Jeremy Wang, Alexander Rubinsteyn, Melissa Miller, Corbin Jones, Amy James Loftis, Amir Barzin, Susan Fiscus |
| EPI_ISL_1261895, EPI_ISL_1261896, EPI_ISL_1261897, EPI_ISL_1261898, EPI_ISL_1261899, EPI_ISL_1261900, EPI_ISL_1261901, EPI_ISL_1261902, EPI_ISL_1261903, EPI_ISL_1261904, EPI_ISL_1261905, EPI_ISL_1261906, EPI_ISL_1261907, EPI_ISL_1261908, EPI_ISL_1261909, EPI_ISL_1261910, EPI_ISL_1261911, EPI_ISL_1261912, EPI_ISL_1261913, EPI_ISL_1261914, EPI_ISL_1261915, EPI_ISL_1261916, EPI_ISL_1261917, EPI_ISL_1261918, EPI_ISL_1261919, EPI_ISL_1261920, EPI_ISL_1261921, EPI_ISL_1261922, EPI_ISL_1261923, EPI_ISL_1261924, EPI_ISL_1261925, EPI_ISL_1261926, EPI_ISL_1261927, EPI_ISL_1261928, EPI_ISL_1261929, EPI_ISL_1261930, EPI_ISL_1261931, EPI_ISL_1261932, EPI_ISL_1261933, EPI_ISL_1261934, EPI_ISL_1261935, EPI_ISL_1261936, EPI_ISL_1261937, EPI_ISL_1261938, EPI_ISL_1261939, EPI_ISL_1261940, EPI_ISL_1261941, EPI_ISL_1261942, EPI_ISL_1261943, EPI_ISL_1261944, EPI_ISL_1261945, EPI_ISL_1261946, EPI_ISL_1261947, EPI_ISL_1261948, EPI_ISL_1261949, EPI_ISL_1261950, EPI_ISL_1261951, EPI_ISL_1261952, EPI_ISL_1261953, EPI_ISL_1261954, EPI_ISL_1261955, EPI_ISL_1261956, EPI_ISL_1261957, EPI_ISL_1261958, EPI_ISL_1261959, EPI_ISL_1261960, EPI_ISL_1261961, EPI_ISL_1261962, EPI_ISL_1261963, EPI_ISL_1261964, EPI_ISL_1261965, EPI_ISL_1261966, EPI_ISL_1261967, EPI_ISL_1261968, EPI_ISL_1261969, EPI_ISL_1261970, EPI_ISL_1261971, EPI_ISL_1261972, EPI_ISL_1261973, EPI_ISL_1261974, EPI_ISL_1261975, EPI_ISL_1261976, EPI_ISL_1261977, EPI_ISL_1261978, EPI_ISL_1261979, EPI_ISL_1261980, EPI_ISL_1261981, EPI_ISL_1261982, EPI_ISL_1261983, EPI_ISL_1261984, EPI_ISL_1261985, EPI_ISL_1261986, EPI_ISL_1261987, EPI_ISL_1261988, EPI_ISL_1261989, EPI_ISL_1261990, EPI_ISL_1261991, EPI_ISL_1261992, EPI_ISL_1261993, EPI_ISL_1261994, EPI_ISL_1261995, EPI_ISL_1261996, EPI_ISL_1261997, EPI_ISL_1261998, EPI_ISL_1261999, EPI_ISL_1262000, EPI_ISL_1262001, EPI_ISL_1262002, EPI_ISL_1262003, EPI_ISL_1262004, EPI_ISL_1262005, EPI_ISL_1262006, EPI_ISL_1262007, EPI_ISL_1262008, EPI_ISL_1262009, EPI_ISL_1262010, EPI_ISL_1262011, EPI_ISL_1262012, EPI_ISL_1262013, EPI_ISL_1262014, EPI_ISL_1262015, EPI_ISL_1262016, EPI_ISL_1262017, EPI_ISL_1262018, EPI_ISL_1262019, EPI_ISL_1262020, EPI_ISL_1262021, EPI_ISL_1262022, EPI_ISL_1262023, EPI_ISL_1262025, EPI_ISL_1262026, EPI_ISL_1262027, EPI_ISL_1262028, EPI_ISL_1262029, EPI_ISL_1262030, EPI_ISL_1262031, EPI_ISL_1262032, EPI_ISL_1262033, EPI_ISL_1262034, EPI_ISL_1262035, EPI_ISL_1262036, EPI_ISL_1262037, EPI_ISL_1262038, EPI_ISL_1262041, EPI_ISL_1262045, EPI_ISL_1262048, EPI_ISL_1262051, EPI_ISL_1262054, EPI_ISL_1262057, EPI_ISL_1262061, EPI_ISL_1262067, EPI_ISL_1262070, EPI_ISL_1262073, EPI_ISL_1262076, EPI_ISL_1262080, EPI_ISL_1262083, EPI_ISL_1262086, EPI_ISL_1262089, EPI_ISL_1262092, EPI_ISL_1262095, EPI_ISL_1262098, EPI_ISL_1262101, EPI_ISL_1262104, EPI_ISL_1262107, EPI_ISL_1262110, EPI_ISL_1262113, EPI_ISL_1262116, EPI_ISL_1262119, EPI_ISL_1262122, EPI_ISL_1262125, EPI_ISL_1262127, EPI_ISL_1262128, EPI_ISL_1262129, EPI_ISL_1262130, EPI_ISL_1262131, EPI_ISL_1262132, EPI_ISL_1262133, EPI_ISL_1262134, EPI_ISL_1262135, EPI_ISL_1262136, EPI_ISL_1262137, EPI_ISL_1262138, EPI_ISL_1262139, EPI_ISL_1262140, EPI_ISL_1262141, EPI_ISL_1262142, EPI_ISL_1262143, EPI_ISL_1262144, EPI_ISL_1262145, EPI_ISL_1262146, EPI_ISL_1262147, EPI_ISL_1262148, EPI_ISL_1262149, EPI_ISL_1262150, EPI_ISL_1262151, EPI_ISL_1262152, EPI_ISL_1262153, EPI_ISL_1262154, EPI_ISL_1262155, EPI_ISL_1262156, EPI_ISL_1262157, EPI_ISL_1262158, EPI_ISL_1262159, EPI_ISL_1262160, EPI_ISL_1262161, EPI_ISL_1262162, EPI_ISL_1262163, EPI_ISL_1262164, EPI_ISL_1262165, EPI_ISL_1262166, EPI_ISL_1262167, EPI_ISL_1262168, EPI_ISL_1262169, EPI_ISL_1262170, EPI_ISL_1262171, EPI_ISL_1262172, EPI_ISL_1262173, EPI_ISL_1262174, EPI_ISL_1262175, EPI_ISL_1262176, EPI_ISL_1262177, EPI_ISL_1262178, EPI_ISL_1262179, EPI_ISL_1262180, EPI_ISL_1262181, EPI_ISL_1262182, EPI_ISL_1262183, EPI_ISL_1262184, EPI_ISL_1262185, EPI_ISL_1262187, EPI_ISL_1262189, EPI_ISL_1262190, EPI_ISL_1262191, EPI_ISL_1262192, EPI_ISL_1262193, EPI_ISL_1262194, EPI_ISL_1262195, EPI_ISL_1262196, EPI_ISL_1262197, EPI_ISL_1262198, EPI_ISL_1262199, EPI_ISL_1262200, EPI_ISL_1262201, EPI_ISL_1262202, EPI_ISL_1262203, EPI_ISL_1262204, EPI_ISL_1262205, EPI_ISL_1262206, EPI_ISL_1262207, EPI_ISL_1262208, EPI_ISL_1262209, EPI_ISL_1262210, EPI_ISL_1262211, EPI_ISL_1262212, EPI_ISL_1262213, EPI_ISL_1262214, EPI_ISL_1262215, EPI_ISL_1262216, EPI_ISL_1262217, EPI_ISL_1262218, EPI_ISL_1262219, EPI_ISL_1262220, EPI_ISL_1262221, EPI_ISL_1262222, EPI_ISL_1262223, EPI_ISL_1262224, EPI_ISL_1262225, EPI_ISL_1262226, EPI_ISL_1262227, EPI_ISL_1262228, EPI_ISL_1262229, EPI_ISL_1262230, EPI_ISL_1262231, EPI_ISL_1262232, EPI_ISL_1262233, EPI_ISL_1262234, EPI_ISL_1262235, EPI_ISL_1262236, EPI_ISL_1262237, EPI_ISL_1262238, EPI_ISL_1262239, EPI_ISL_1262240, EPI_ISL_1262241, EPI_ISL_1262242, EPI_ISL_1262243, EPI_ISL_1262244, EPI_ISL_1262245, EPI_ISL_1262246, EPI_ISL_1262247, EPI_ISL_1262248, EPI_ISL_1262249, EPI_ISL_1262250, EPI_ISL_1262251, EPI_ISL_1262252, EPI_ISL_1262253, EPI_ISL_1262254, EPI_ISL_1262255, EPI_ISL_1262256, EPI_ISL_1262257, EPI_ISL_1262258, EPI_ISL_1262259, EPI_ISL_1262260, EPI_ISL_1262261, EPI_ISL_1262262, EPI_ISL_1262263, EPI_ISL_1262264, EPI_ISL_1262265, EPI_ISL_1262266, EPI_ISL_1262267, EPI_ISL_1262268, EPI_ISL_1262269, EPI_ISL_1262270, EPI_ISL_1262271, EPI_ISL_1262272, EPI_ISL_1262273, EPI_ISL_1262274, EPI_ISL_1262275, EPI_ISL_1262276, EPI_ISL_1262277, EPI_ISL_1262278, EPI_ISL_1262279, EPI_ISL_1262280, EPI_ISL_1262281, EPI_ISL_1262282, EPI_ISL_1262283, EPI_ISL_1262284, EPI_ISL_1262285, EPI_ISL_1262286, EPI_ISL_1262287, EPI_ISL_1262288, EPI_ISL_1262289, EPI_ISL_1262290, EPI_ISL_1262291, EPI_ISL_1262292, EPI_ISL_1262293, EPI_ISL_1262294, EPI_ISL_1262295, EPI_ISL_1262296, EPI_ISL_1262297, EPI_ISL_1262298, EPI_ISL_1262299, EPI_ISL_1262300, EPI_ISL_1262301, EPI_ISL_1262302, EPI_ISL_1262303, EPI_ISL_1262304, EPI_ISL_1262305, EPI_ISL_1262306, EPI_ISL_1262307, EPI_ISL_1262308, EPI_ISL_1262309, EPI_ISL_1262310, EPI_ISL_1262311, EPI_ISL_1262312, EPI_ISL_1262313, EPI_ISL_1262314, EPI_ISL_1262315, EPI_ISL_1262316, EPI_ISL_1262317, EPI_ISL_1262319, EPI_ISL_1262320, EPI_ISL_1262322, EPI_ISL_1262323, EPI_ISL_1262324, EPI_ISL_1262325, EPI_ISL_1262327, EPI_ISL_1262328, EPI_ISL_1262329, EPI_ISL_1262330, EPI_ISL_1262331, EPI_ISL_1262332, EPI_ISL_1262333, EPI_ISL_1262334, EPI_ISL_1262335, EPI_ISL_1262336, EPI_ISL_1262337, EPI_ISL_1262338, EPI_ISL_1262339, EPI_ISL_1262340, EPI_ISL_1262341, EPI_ISL_1262342, EPI_ISL_1262343, EPI_ISL_1262344, EPI_ISL_1262345, EPI_ISL_1262346, EPI_ISL_1262347, EPI_ISL_1262348, EPI_ISL_1262349, EPI_ISL_1262350, EPI_ISL_1262351, EPI_ISL_1262352, EPI_ISL_1262353, EPI_ISL_1262354, EPI_ISL_1262355, EPI_ISL_1262356, EPI_ISL_1262357, EPI_ISL_1262358, EPI_ISL_1262359, EPI_ISL_1262360, EPI_ISL_1262361, EPI_ISL_1262362, EPI_ISL_1262363, EPI_ISL_1262364, EPI_ISL_1262365, EPI_ISL_1262366, EPI_ISL_1262367, EPI_ISL_1262369, EPI_ISL_1262370, EPI_ISL_1262372, EPI_ISL_1262373, EPI_ISL_1262374, EPI_ISL_1262375, EPI_ISL_1262376, EPI_ISL_1262377, EPI_ISL_1262378, EPI_ISL_1262379, EPI_ISL_1262380, EPI_ISL_1262381, EPI_ISL_1262383, EPI_ISL_1262384, EPI_ISL_1262385, EPI_ISL_1262386, EPI_ISL_1262387, EPI_ISL_1262388, EPI_ISL_1262389, EPI_ISL_1262390, EPI_ISL_1262391, EPI_ISL_1262392, EPI_ISL_1262393, EPI_ISL_1262394, EPI_ISL_1262395, EPI_ISL_1262396, EPI_ISL_1262397, EPI_ISL_1262398, EPI_ISL_1262399, EPI_ISL_1262400, EPI_ISL_1262401, EPI_ISL_1262403, EPI_ISL_1262404, EPI_ISL_1262405, EPI_ISL_1262406, EPI_ISL_1262407, EPI_ISL_1262408, EPI_ISL_1262409, EPI_ISL_1262410, EPI_ISL_1262411, EPI_ISL_1262412, EPI_ISL_1262413, EPI_ISL_1262414, EPI_ISL_1262415, EPI_ISL_1262416, EPI_ISL_1262417, EPI_ISL_1262418, EPI_ISL_1262419, EPI_ISL_1262420, EPI_ISL_1262421, EPI_ISL_1262422, EPI_ISL_1262423, EPI_ISL_1262424, EPI_ISL_1262425, EPI_ISL_1262426, EPI_ISL_1262427, EPI_ISL_1262428, EPI_ISL_1262429, EPI_ISL_1262430, EPI_ISL_1262431, EPI_ISL_1262432, EPI_ISL_1262433, EPI_ISL_1262434, EPI_ISL_1262435, EPI_ISL_1262436, EPI_ISL_1262437, EPI_ISL_1262438, EPI_ISL_1262439, EPI_ISL_1262440, EPI_ISL_1262441, EPI_ISL_1262442, EPI_ISL_1262443, EPI_ISL_1262444, EPI_ISL_1262445, EPI_ISL_1262446, EPI_ISL_1262447, EPI_ISL_1262448, EPI_ISL_1262449, EPI_ISL_1262450, EPI_ISL_1262451, EPI_ISL_1262452, EPI_ISL_1262453, EPI_ISL_1262454 | | | |
| see above | Helix/Illumina | Centers for Disease Control and Prevention Division of Viral Diseases, Pathogen Discovery | Peter W. Cook, Dakota Howard, Dhwani Batra, Ben L. Rambo-Martin, Eileen de Feo, Jan Antico, Christine Tran, Matthew Tolentino, Shannon Wickline, Kim Gietzen, Brad Sickler, Jingtao Liu, Eric Allen, Phil Febbo, Summer Galloway, Nicole L. Washington, Simon White, Geraint Levan, Kelly Schiabor Barrett, Elizabeth Cirulli, Alexandre Bolze, Ary Ascencio, Charlotte Rivera-Garcia, Ryan Cho, Jason Nguyen, Sherry Wang, Jimmy Ramirez, Tyler Cassens, Efren Sandoval, Magnus Isaksson, William Lee, David Becker, Marc Laurent, James Lu, Clinton R. Paden, Suxiang Tong, Duncan MacCannell |
| EPI_ISL_1262455, EPI_ISL_1262456, EPI_ISL_1262457, EPI_ISL_1262458 | University of Michigan Clinical Microbiology Laboratory | Lauring Lab, University of Michigan, Department of Microbiology and Immunology | Valesano |
| EPI_ISL_1262459, EPI_ISL_1262460, EPI_ISL_1262461, EPI_ISL_1262462, EPI_ISL_1262463, EPI_ISL_1262464, EPI_ISL_1262465, EPI_ISL_1262466, EPI_ISL_1262467, EPI_ISL_1262468, EPI_ISL_1262469, EPI_ISL_1262470, EPI_ISL_1262471, EPI_ISL_1262472, EPI_ISL_1262473, EPI_ISL_1262474, EPI_ISL_1262475, EPI_ISL_1262476, EPI_ISL_1262477, EPI_ISL_1262478, EPI_ISL_1262479, EPI_ISL_1262480, EPI_ISL_1262481, EPI_ISL_1262482, EPI_ISL_1262483, EPI_ISL_1262484, EPI_ISL_1262485, EPI_ISL_1262486, EPI_ISL_1262487, EPI_ISL_1262488, EPI_ISL_1262489, EPI_ISL_1262490, EPI_ISL_1262491, EPI_ISL_1262492, EPI_ISL_1262493, EPI_ISL_1262494, EPI_ISL_1262495, EPI_ISL_1262496, EPI_ISL_1262497, EPI_ISL_1262498, EPI_ISL_1262499, EPI_ISL_1262500 | | | |
| see above | GA Department of Public Health | GA Department of Public Health | Stacy Reeves, Jonathan Edwards, Cynthia Dixey, Tonia Parrott, Aliyah Fields, Taylor Smith |
| EPI_ISL_1262501, EPI_ISL_1262502, EPI_ISL_1262503, EPI_ISL_1262504, EPI_ISL_1262505, EPI_ISL_1262506, EPI_ISL_1262507, EPI_ISL_1262508, EPI_ISL_1262509, EPI_ISL_1262510, EPI_ISL_1262511, EPI_ISL_1262512, EPI_ISL_1262513, EPI_ISL_1262514, EPI_ISL_1262515, EPI_ISL_1262516, EPI_ISL_1262517, EPI_ISL_1262518, EPI_ISL_1262519, EPI_ISL_1262520, EPI_ISL_1262521, EPI_ISL_1262522, EPI_ISL_1262523, EPI_ISL_1262524, EPI_ISL_1262525, EPI_ISL_1262526, EPI_ISL_1262527, EPI_ISL_1262528, EPI_ISL_1262529, EPI_ISL_1262530, EPI_ISL_1262531, EPI_ISL_1262532, EPI_ISL_1262533, EPI_ISL_1262534, EPI_ISL_1262543, EPI_ISL_1262544, EPI_ISL_1262545, EPI_ISL_1262546, EPI_ISL_1262547, EPI_ISL_1262548, EPI_ISL_1262549, EPI_ISL_1262550, EPI_ISL_1262551, EPI_ISL_1262552, EPI_ISL_1262553, EPI_ISL_1262554, EPI_ISL_1262555, EPI_ISL_1262556, EPI_ISL_1262557, EPI_ISL_1262558, EPI_ISL_1262559, EPI_ISL_1262560, EPI_ISL_1262561, EPI_ISL_1262562, EPI_ISL_1262563, EPI_ISL_1262564, EPI_ISL_1262565, EPI_ISL_1262566, EPI_ISL_1262567, EPI_ISL_1262568, EPI_ISL_1262569, EPI_ISL_1262570, EPI_ISL_1262571, EPI_ISL_1262572, EPI_ISL_1262573, EPI_ISL_1262574, EPI_ISL_1262575, EPI_ISL_1262576, EPI_ISL_1262577, EPI_ISL_1262578, EPI_ISL_1262579, EPI_ISL_1262580, EPI_ISL_1262581, EPI_ISL_1262582, EPI_ISL_1262583, EPI_ISL_1262584, EPI_ISL_1262585 | | | |
| see above | NYU Langone Health | Departments of Pathology and Medicine, New York University School of Medicine | Adriana Heguy, Dacia Dimartino, Emily Guzman, Christian Marier, Peter Meyn, Sitharam Ramaswami, Gael Westby, Paul Zappile, Yutong Zhang, Paolo Cotzia, Guiqing Wang |
| EPI_ISL_1262586, EPI_ISL_1262587, EPI_ISL_1262588, EPI_ISL_1262591, EPI_ISL_1262593, EPI_ISL_1262594, EPI_ISL_1262595, EPI_ISL_1262596, EPI_ISL_1262597, EPI_ISL_1262598, EPI_ISL_1262599, EPI_ISL_1262600, EPI_ISL_1262601, EPI_ISL_1262604, EPI_ISL_1262605, EPI_ISL_1262606, EPI_ISL_1262607 | | | |
| see above | City of Milwaukee Health Department Laboratory | City of Milwaukee Health Department Laboratory | Sanjib Bhattacharyya |
| EPI_ISL_1262630, EPI_ISL_1262631, EPI_ISL_1262632, EPI_ISL_1262633, EPI_ISL_1262634, EPI_ISL_1262635, EPI_ISL_1262636, EPI_ISL_1262637, EPI_ISL_1262638, EPI_ISL_1262640, EPI_ISL_1262641, EPI_ISL_1262642, EPI_ISL_1262643, EPI_ISL_1262644, EPI_ISL_1262645, EPI_ISL_1262646, EPI_ISL_1262647, EPI_ISL_1262648, EPI_ISL_1262649, EPI_ISL_1262650, EPI_ISL_1262651, EPI_ISL_1262652, EPI_ISL_1262653, EPI_ISL_1262654, EPI_ISL_1262655, EPI_ISL_1262656, EPI_ISL_1262657, EPI_ISL_1262658, EPI_ISL_1262659, EPI_ISL_1262660, EPI_ISL_1262661, EPI_ISL_1262662, EPI_ISL_1262663, EPI_ISL_1262664, EPI_ISL_1262665, EPI_ISL_1262666, EPI_ISL_1262667, EPI_ISL_1262668, EPI_ISL_1262669, EPI_ISL_1262670, EPI_ISL_1262671, EPI_ISL_1262672, EPI_ISL_1262673, EPI_ISL_1262674, EPI_ISL_1262675, EPI_ISL_1262676, EPI_ISL_1262677, EPI_ISL_1262678, EPI_ISL_1262679, EPI_ISL_1262680, EPI_ISL_1262681, EPI_ISL_1262682, EPI_ISL_1262683, EPI_ISL_1262684, EPI_ISL_1262685, EPI_ISL_1262686, EPI_ISL_1262687, EPI_ISL_1262688, EPI_ISL_1262689, EPI_ISL_1262690, EPI_ISL_1262691, EPI_ISL_1262692, EPI_ISL_1262693, EPI_ISL_1262694, EPI_ISL_1262695, EPI_ISL_1262696, EPI_ISL_1262697, EPI_ISL_1262698, EPI_ISL_1262699, EPI_ISL_1262700, EPI_ISL_1262701, EPI_ISL_1262702, EPI_ISL_1262703, EPI_ISL_1262704, EPI_ISL_1262705, EPI_ISL_1262706, EPI_ISL_1262707, EPI_ISL_1262708, EPI_ISL_1262709, EPI_ISL_1262710, EPI_ISL_1262711, EPI_ISL_1262712, EPI_ISL_1262713, EPI_ISL_1262714, EPI_ISL_1262715, EPI_ISL_1262716, EPI_ISL_1262717 | | | |
| see above | Instituto Nacional de Medicina Genomica | Instituto Nacional de Medicina Genomica | Hidalgo-Miranda A, Mendoza-Vargas A, Reyes-Grajeda JP, Cedro-Tanda A, Alcaraz N, Gonzalez-Barrera D, Rangel-DeLeon D, Ramirez-Vega O, Herrera-Montalvo LA |
| EPI_ISL_1263103, EPI_ISL_1263104, EPI_ISL_1263105, EPI_ISL_1263106, EPI_ISL_1263107, EPI_ISL_1263108, EPI_ISL_1263109, EPI_ISL_1263110, EPI_ISL_1263111, EPI_ISL_1263112, EPI_ISL_1263113, EPI_ISL_1263114, EPI_ISL_1263115, EPI_ISL_1263116, EPI_ISL_1263117, EPI_ISL_1263118, EPI_ISL_1263119, EPI_ISL_1263120, EPI_ISL_1263121, EPI_ISL_1263123, EPI_ISL_1263124, EPI_ISL_1263125, EPI_ISL_1263126, EPI_ISL_1263127, EPI_ISL_1263128, EPI_ISL_1263129, EPI_ISL_1263130, EPI_ISL_1263131, EPI_ISL_1263132, EPI_ISL_1263133, EPI_ISL_1263134, EPI_ISL_1263135, EPI_ISL_1263136, EPI_ISL_1263137, EPI_ISL_1263138, EPI_ISL_1263139, EPI_ISL_1263140, EPI_ISL_1263141, EPI_ISL_1263142, EPI_ISL_1263143, EPI_ISL_1263144, EPI_ISL_1263145, EPI_ISL_1263146, EPI_ISL_1263147, | | | |

| | | | |
|---|---|---|---|
| EPI_ISL_1263148, EPI_ISL_1263149, EPI_ISL_1263150, EPI_ISL_1263151, EPI_ISL_1263152, EPI_ISL_1263153, EPI_ISL_1263154, EPI_ISL_1263156, EPI_ISL_1263157, EPI_ISL_1263158, EPI_ISL_1263159, EPI_ISL_1263160, EPI_ISL_1263161, EPI_ISL_1263162, EPI_ISL_1263163, EPI_ISL_1263164, EPI_ISL_1263165, EPI_ISL_1263166, EPI_ISL_1263167, EPI_ISL_1263168, EPI_ISL_1263169, EPI_ISL_1263170, EPI_ISL_1263171, EPI_ISL_1263172, EPI_ISL_1263173, EPI_ISL_1263174, EPI_ISL_1263175, EPI_ISL_1263176, EPI_ISL_1263177, EPI_ISL_1263178, EPI_ISL_1263179, EPI_ISL_1263180, EPI_ISL_1263181, EPI_ISL_1263182, EPI_ISL_1263183, EPI_ISL_1263184, EPI_ISL_1263185, EPI_ISL_1263186, EPI_ISL_1263187, EPI_ISL_1263188, EPI_ISL_1263189, EPI_ISL_1263190, EPI_ISL_1263191 | | | |
| see above | Nevada State Public Health Laboratory | Nevada State Public Health Laboratory | Andrew Gorzalski, Mark Pandori |
| EPI_ISL_1263207, EPI_ISL_1263208, EPI_ISL_1263209, EPI_ISL_1263210, EPI_ISL_1263211, EPI_ISL_1263212, EPI_ISL_1263213, EPI_ISL_1263214, EPI_ISL_1263215, EPI_ISL_1263216, EPI_ISL_1263217, EPI_ISL_1263218, EPI_ISL_1263219, EPI_ISL_1263220, EPI_ISL_1263221, EPI_ISL_1263222, EPI_ISL_1263223, EPI_ISL_1263224, EPI_ISL_1263225, EPI_ISL_1263226, EPI_ISL_1263227, EPI_ISL_1263228, EPI_ISL_1263229, EPI_ISL_1263230, EPI_ISL_1263231, EPI_ISL_1263232, EPI_ISL_1263233, EPI_ISL_1263234, EPI_ISL_1263235 | | | |
| see above | Monterey County Public Health Laboratory | Monterey County Public Health Laboratory | Monterey County Public Health Laboratory |
| EPI_ISL_1263324, EPI_ISL_1263325, EPI_ISL_1263326, EPI_ISL_1263327, EPI_ISL_1263328, EPI_ISL_1263329 | Kansas Health and Environmental Lab | Kansas Health and Environmental Lab | Mike Grose, Jonathan Barnell, Ben Olsen, and Phil Adam |
| EPI_ISL_1263334, EPI_ISL_1263335, EPI_ISL_1263336, EPI_ISL_1263337, EPI_ISL_1263338, EPI_ISL_1263339, EPI_ISL_1263340, EPI_ISL_1263341, EPI_ISL_1263342, EPI_ISL_1263343, EPI_ISL_1263344, EPI_ISL_1263345, EPI_ISL_1263346, EPI_ISL_1263347, EPI_ISL_1263348, EPI_ISL_1263349, EPI_ISL_1263350, EPI_ISL_1263351, EPI_ISL_1263352, EPI_ISL_1263353, EPI_ISL_1263354, EPI_ISL_1263355, EPI_ISL_1263356, EPI_ISL_1263357, EPI_ISL_1263358, EPI_ISL_1263359, EPI_ISL_1263360, EPI_ISL_1263361, EPI_ISL_1263362, EPI_ISL_1263363 | | | |
| see above | Washington State Department of Health Public Health Laboratories | Washington State Department of Health Public Health Laboratories | Drew MacKellar, Philip Dykema, Denny Russell, Joenice Gonzalez, Geoff Melly, Vanessa De Los Santos, Darren Lucas, JohnAric Peterson, Avi Singh, Rebecca Cao |
| EPI_ISL_1263461, EPI_ISL_1263462, EPI_ISL_1263463, EPI_ISL_1263464, EPI_ISL_1263465, EPI_ISL_1263466, EPI_ISL_1263467, EPI_ISL_1263468, EPI_ISL_1263469, EPI_ISL_1263470, EPI_ISL_1263471, EPI_ISL_1263472, EPI_ISL_1263473, EPI_ISL_1263474, EPI_ISL_1263475, EPI_ISL_1263476, EPI_ISL_1263477, EPI_ISL_1263478, EPI_ISL_1263479, EPI_ISL_1263480, EPI_ISL_1263481, EPI_ISL_1263482, EPI_ISL_1263483, EPI_ISL_1263484, EPI_ISL_1263485, EPI_ISL_1263486, EPI_ISL_1263487, EPI_ISL_1263488, EPI_ISL_1263489, EPI_ISL_1263490, EPI_ISL_1263491, EPI_ISL_1263492, EPI_ISL_1263493, EPI_ISL_1263494, EPI_ISL_1263495, EPI_ISL_1263496, EPI_ISL_1263497, EPI_ISL_1263498, EPI_ISL_1263499, EPI_ISL_1263500, EPI_ISL_1263501, EPI_ISL_1263502, EPI_ISL_1263503, EPI_ISL_1263504, EPI_ISL_1263505, EPI_ISL_1263506, EPI_ISL_1263507, EPI_ISL_1263508, EPI_ISL_1263509, EPI_ISL_1263510, EPI_ISL_1263511, EPI_ISL_1263512, EPI_ISL_1263513, EPI_ISL_1263514, EPI_ISL_1263515, EPI_ISL_1263516, EPI_ISL_1263517, EPI_ISL_1263518, EPI_ISL_1263519, EPI_ISL_1263520, EPI_ISL_1263521, EPI_ISL_1263522, EPI_ISL_1263523, EPI_ISL_1263524, EPI_ISL_1263525, EPI_ISL_1263526, EPI_ISL_1263527, EPI_ISL_1263528, EPI_ISL_1263529, EPI_ISL_1263530, EPI_ISL_1263531, EPI_ISL_1263532, EPI_ISL_1263533, EPI_ISL_1263534, EPI_ISL_1263535, EPI_ISL_1263536, EPI_ISL_1263537, EPI_ISL_1263538, EPI_ISL_1263539 | | | |
| see above | UW Virology Lab | UW Virology Lab | Pavitra Roychoudhury, Hong Xie, Lasata Shrestha, Shah Mohamed Bakhash, Michelle Lin, Noah Baker, Sean Ellis, Saraswathi Sathees, Meei-Li Huang, Keith R Jerome, Alexander Greninger |
| EPI_ISL_1264247, EPI_ISL_1265066, EPI_ISL_1265142, EPI_ISL_1265162 | Ohio Department of Health Laboratory | Ohio Department of Health Laboratory | Holmes, Jennifer; Eric Brandt, Keoni Omura, Glen McGillivary, Caitlin McDonnell, Jade Mowery, Stephanie Mccracken, Tyler Payne, Kirtana Ramadugu, Erica Leasure, Brent Lee, Kelsey Florek, Heather Blankenship, Allison Black, Quanta Brown, and Tammy Bannerman |
| EPI_ISL_1265374, EPI_ISL_1265375, EPI_ISL_1265376, EPI_ISL_1265377, EPI_ISL_1265378, EPI_ISL_1265379, EPI_ISL_1265380, EPI_ISL_1265381, EPI_ISL_1265382, EPI_ISL_1265383, EPI_ISL_1265384, EPI_ISL_1265385, EPI_ISL_1265386, EPI_ISL_1265387, EPI_ISL_1265388, EPI_ISL_1265389, EPI_ISL_1265390, EPI_ISL_1265391, EPI_ISL_1265392 | | | |
| see above | Nebraska Public Health Laboratory | NPHL COVID-19 Response Team | NPHL COVID-19 Response Team |
| EPI_ISL_1265670, EPI_ISL_1265671, EPI_ISL_1265739, EPI_ISL_1265751, EPI_ISL_1265900 | Ohio Department of Health Laboratory | Ohio Department of Health Laboratory | Holmes, Jennifer; Eric Brandt, Keoni Omura, Glen McGillivary, Caitlin McDonnell, Jade Mowery, Stephanie Mccracken, Tyler Payne, Kirtana Ramadugu, Erica Leasure, Brent Lee, Kelsey Florek, Heather Blankenship, Allison Black, Quanta Brown, and Tammy Bannerman |
| EPI_ISL_1265902, EPI_ISL_1265903, EPI_ISL_1265904, EPI_ISL_1265905, EPI_ISL_1265906, EPI_ISL_1265907, EPI_ISL_1265908 | Instituto Nacional de Medicina Genomica | Instituto Nacional de Medicina Genomica | Hidalgo-Miranda A, Mendoza-Vargas A, Reyes-Grajeda JP, Cedro-Tanda A, Alcaraz N, Gonzalez-Barrera D, Rangel-DeLeon D, Ramirez-Vega O, Herrera-Montalvo LA |
| EPI_ISL_1265909 | Ohio Department of Health Laboratory | Ohio Department of Health Laboratory | Holmes, Jennifer; Eric Brandt, Keoni Omura, Glen McGillivary, Caitlin McDonnell, Jade Mowery, Stephanie Mccracken, Tyler Payne, Kirtana Ramadugu, Erica Leasure, Brent Lee, Kelsey Florek, Heather Blankenship, Allison Black, Quanta Brown, and Tammy Bannerman |
| EPI_ISL_1265910, EPI_ISL_1265911, EPI_ISL_1265912, EPI_ISL_1265913, EPI_ISL_1265914, EPI_ISL_1265915, EPI_ISL_1265916, EPI_ISL_1265917, EPI_ISL_1265918, EPI_ISL_1265919, EPI_ISL_1265920, EPI_ISL_1265921, EPI_ISL_1265922, EPI_ISL_1265923, EPI_ISL_1265924, EPI_ISL_1265925, EPI_ISL_1265926, EPI_ISL_1265927, EPI_ISL_1265928, EPI_ISL_1265929, EPI_ISL_1265930, EPI_ISL_1265931, EPI_ISL_1265932, EPI_ISL_1265933, EPI_ISL_1265934, EPI_ISL_1265935, EPI_ISL_1265936, EPI_ISL_1265937, EPI_ISL_1265938, EPI_ISL_1265939, EPI_ISL_1265940, EPI_ISL_1265941, EPI_ISL_1265942, EPI_ISL_1265943, EPI_ISL_1265944, EPI_ISL_1265945, EPI_ISL_1265946, EPI_ISL_1265947, EPI_ISL_1265948, EPI_ISL_1265949, EPI_ISL_1265950, EPI_ISL_1265951, EPI_ISL_1265952, EPI_ISL_1265953, EPI_ISL_1265954, EPI_ISL_1265955, EPI_ISL_1265956, EPI_ISL_1265957, EPI_ISL_1265958, EPI_ISL_1265959, EPI_ISL_1265960, EPI_ISL_1265961, EPI_ISL_1265962, EPI_ISL_1265963, EPI_ISL_1265964, EPI_ISL_1265965, EPI_ISL_1265966, EPI_ISL_1265967, EPI_ISL_1265968, EPI_ISL_1265969, EPI_ISL_1265970, EPI_ISL_1265971, EPI_ISL_1265972, EPI_ISL_1265973, EPI_ISL_1265974, EPI_ISL_1265975, EPI_ISL_1265976, EPI_ISL_1265977, EPI_ISL_1265978, EPI_ISL_1265979, EPI_ISL_1265980, EPI_ISL_1265981, EPI_ISL_1265982, EPI_ISL_1265983, EPI_ISL_1265984, EPI_ISL_1265985, EPI_ISL_1265986, EPI_ISL_1265987, EPI_ISL_1265988, EPI_ISL_1265989, EPI_ISL_1265990, EPI_ISL_1265991, EPI_ISL_1265992, EPI_ISL_1265993, EPI_ISL_1265994, EPI_ISL_1265995, EPI_ISL_1265996, EPI_ISL_1265997, EPI_ISL_1265998, EPI_ISL_1265999, EPI_ISL_1266000, EPI_ISL_1266001, EPI_ISL_1266002, EPI_ISL_1266003, EPI_ISL_1266004, EPI_ISL_1266005, EPI_ISL_1266006, EPI_ISL_1266007, EPI_ISL_1266008, EPI_ISL_1266009, EPI_ISL_1266010, EPI_ISL_1266011, EPI_ISL_1266012, EPI_ISL_1266013, EPI_ISL_1266014, EPI_ISL_1266015, EPI_ISL_1266016, EPI_ISL_1266017, EPI_ISL_1266018, EPI_ISL_1266019, EPI_ISL_1266020, EPI_ISL_1266021, EPI_ISL_1266022, EPI_ISL_1266023, EPI_ISL_1266024, EPI_ISL_1266025, EPI_ISL_1266026, EPI_ISL_1266027, EPI_ISL_1266028, EPI_ISL_1266029, EPI_ISL_1266030, EPI_ISL_1266031, EPI_ISL_1266032, EPI_ISL_1266033, EPI_ISL_1266034, EPI_ISL_1266035, EPI_ISL_1266036, EPI_ISL_1266037, EPI_ISL_1266038, EPI_ISL_1266039, EPI_ISL_1266040, EPI_ISL_1266041, EPI_ISL_1266042, EPI_ISL_1266043, EPI_ISL_1266044, EPI_ISL_1266045, EPI_ISL_1266046, EPI_ISL_1266047, EPI_ISL_1266048, EPI_ISL_1266049, EPI_ISL_1266050, EPI_ISL_1266051, EPI_ISL_1266052, EPI_ISL_1266053, EPI_ISL_1266054, EPI_ISL_1266055, EPI_ISL_1266056, EPI_ISL_1266057, EPI_ISL_1266058, EPI_ISL_1266059, EPI_ISL_1266060, EPI_ISL_1266061, EPI_ISL_1266062, EPI_ISL_1266063, EPI_ISL_1266064, EPI_ISL_1266065, EPI_ISL_1266066, EPI_ISL_1266067, EPI_ISL_1266068, EPI_ISL_1266069, EPI_ISL_1266070, EPI_ISL_1266071, EPI_ISL_1266072, EPI_ISL_1266073, EPI_ISL_1266074, EPI_ISL_1266075, EPI_ISL_1266076, EPI_ISL_1266077, EPI_ISL_1266078, EPI_ISL_1266079, EPI_ISL_1266080, EPI_ISL_1266081, EPI_ISL_1266082, EPI_ISL_1266083, EPI_ISL_1266084, EPI_ISL_1266085, EPI_ISL_1266086, EPI_ISL_1266087, EPI_ISL_1266088, EPI_ISL_1266089, EPI_ISL_1266090, EPI_ISL_1266091, EPI_ISL_1266092, EPI_ISL_1266093, EPI_ISL_1266094, EPI_ISL_1266095, EPI_ISL_1266096, EPI_ISL_1266097, EPI_ISL_1266098, EPI_ISL_1266099, EPI_ISL_1266100, EPI_ISL_1266101, EPI_ISL_1266102, EPI_ISL_1266103, EPI_ISL_1266104, EPI_ISL_1266105, EPI_ISL_1266106, EPI_ISL_1266107, EPI_ISL_1266108, EPI_ISL_1266109, EPI_ISL_1266110, EPI_ISL_1266111, EPI_ISL_1266112, EPI_ISL_1266113 | | | |
| see above | Quest Diagnostics Incorporated | Centers for Disease Control and Prevention Division of Viral Diseases, Pathogen Discovery | Peter W. Cook, Dakota Howard, Dhwani Batra, Ben L. Rambo-Martin, S. H. Rosenthal, A. Gerasimova, R. M. Kagan, B. Anderson, M. Hua, Y. Liu, L.E. Bernstein, K.E. Livingston, A. Perez, I. A. Shlyakhter, R. V. Rolando, R. Owen, P. Tanpaiboon, F. Lacbawan, Clinton R. Paden, Suxiang Tong, Duncan MacCannell |
| EPI_ISL_1266114 | Nebraska Public Health Laboratory | NPHL COVID-19 Response Team | NPHL COVID-19 Response Team |
| EPI_ISL_1266115 | Quest Diagnostics Incorporated | Centers for Disease Control and Prevention Division of Viral Diseases, Pathogen Discovery | Peter W. Cook, Dakota Howard, Dhwani Batra, Ben L. Rambo-Martin, S. H. Rosenthal, A. Gerasimova, R. M. Kagan, B. Anderson, M. Hua, Y. Liu, L.E. Bernstein, K.E. Livingston, A. Perez, I. A. Shlyakhter, R. V. Rolando, R. Owen, P. Tanpaiboon, F. Lacbawan, Clinton R. Paden, Suxiang Tong, Duncan MacCannell |
| EPI_ISL_1266116, EPI_ISL_1266117 | Nebraska Public Health Laboratory | NPHL COVID-19 Response Team | NPHL COVID-19 Response Team |
| EPI_ISL_1266118 | Quest Diagnostics Incorporated | Centers for Disease Control and Prevention Division of Viral Diseases, Pathogen Discovery | Peter W. Cook, Dakota Howard, Dhwani Batra, Ben L. Rambo-Martin, S. H. Rosenthal, A. Gerasimova, R. M. Kagan, B. Anderson, M. Hua, Y. Liu, L.E. Bernstein, K.E. Livingston, A. Perez, I. A. Shlyakhter, R. V. Rolando, R. Owen, P. Tanpaiboon, F. Lacbawan, Clinton R. Paden, Suxiang Tong, Duncan MacCannell |
| EPI_ISL_1266119, EPI_ISL_1266120 | Nebraska Public Health Laboratory | NPHL COVID-19 Response Team | NPHL COVID-19 Response Team |
| EPI_ISL_1266121 | Quest Diagnostics Incorporated | Centers for Disease Control and Prevention Division of Viral Diseases, Pathogen Discovery | Peter W. Cook, Dakota Howard, Dhwani Batra, Ben L. Rambo-Martin, S. H. Rosenthal, A. Gerasimova, R. M. Kagan, B. Anderson, M. Hua, Y. Liu, L.E. Bernstein, K.E. Livingston, A. Perez, I. A. Shlyakhter, R. V. Rolando, R. Owen, P. Tanpaiboon, F. Lacbawan, Clinton R. Paden, Suxiang Tong, Duncan MacCannell |
| EPI_ISL_1266122, EPI_ISL_1266123 | Nebraska Public Health Laboratory | NPHL COVID-19 Response Team | NPHL COVID-19 Response Team |
| EPI_ISL_1266124 | Quest Diagnostics Incorporated | Centers for Disease Control and Prevention Division of Viral Diseases, Pathogen Discovery | Peter W. Cook, Dakota Howard, Dhwani Batra, Ben L. Rambo-Martin, S. H. Rosenthal, A. Gerasimova, R. M. Kagan, B. Anderson, M. Hua, Y. Liu, L.E. Bernstein, K.E. Livingston, A. Perez, I. A. Shlyakhter, R. V. Rolando, R. Owen, P. Tanpaiboon, F. Lacbawan, Clinton R. Paden, Suxiang Tong, Duncan MacCannell |
| EPI_ISL_1266125, EPI_ISL_1266126 | Nebraska Public Health Laboratory | NPHL COVID-19 Response Team | NPHL COVID-19 Response Team |
| EPI_ISL_1266127 | Quest Diagnostics Incorporated | Centers for Disease Control and Prevention Division of Viral Diseases, Pathogen Discovery | Peter W. Cook, Dakota Howard, Dhwani Batra, Ben L. Rambo-Martin, S. H. Rosenthal, A. Gerasimova, R. M. Kagan, B. Anderson, M. Hua, Y. Liu, L.E. Bernstein, K.E. Livingston, A. Perez, I. A. Shlyakhter, R. V. Rolando, R. Owen, P. Tanpaiboon, F. Lacbawan, Clinton R. Paden, Suxiang Tong, Duncan MacCannell |
| EPI_ISL_1266128, EPI_ISL_1266129 | Nebraska Public Health Laboratory | NPHL COVID-19 Response Team | NPHL COVID-19 Response Team |
| EPI_ISL_1266130 | Quest Diagnostics Incorporated | Centers for Disease Control and Prevention Division of Viral Diseases, Pathogen Discovery | Peter W. Cook, Dakota Howard, Dhwani Batra, Ben L. Rambo-Martin, S. H. Rosenthal, A. Gerasimova, R. M. Kagan, B. Anderson, M. Hua, Y. Liu, L.E. Bernstein, K.E. Livingston, A. Perez, I. A. Shlyakhter, R. V. Rolando, R. Owen, P. Tanpaiboon, F. Lacbawan, Clinton R. Paden, Suxiang Tong, Duncan MacCannell |
| EPI_ISL_1266131 | Nebraska Public Health Laboratory | NPHL COVID-19 Response Team | NPHL COVID-19 Response Team |
| EPI_ISL_1266132, EPI_ISL_1266133, EPI_ISL_1266134, EPI_ISL_1266135, EPI_ISL_1266136, EPI_ISL_1266137, EPI_ISL_1266138, EPI_ISL_1266139, EPI_ISL_1266140, EPI_ISL_1266141, EPI_ISL_1266142, EPI_ISL_1266143, EPI_ISL_1266144, EPI_ISL_1266145, EPI_ISL_1266146, EPI_ISL_1266147, EPI_ISL_1266148, EPI_ISL_1266149, EPI_ISL_1266150, EPI_ISL_1266151, EPI_ISL_1266152, EPI_ISL_1266153, EPI_ISL_1266154, EPI_ISL_1266155, EPI_ISL_1266156, EPI_ISL_1266157, EPI_ISL_1266158, EPI_ISL_1266159, EPI_ISL_1266160, EPI_ISL_1266161, EPI_ISL_1266162, EPI_ISL_1266163, EPI_ISL_1266164, EPI_ISL_1266165, EPI_ISL_1266166, EPI_ISL_1266167, EPI_ISL_1266168, EPI_ISL_1266169, EPI_ISL_1266170, EPI_ISL_1266171, EPI_ISL_1266172, EPI_ISL_1266173, EPI_ISL_1266174, EPI_ISL_1266175, EPI_ISL_1266176, EPI_ISL_1266177, EPI_ISL_1266178, EPI_ISL_1266179, EPI_ISL_1266180, EPI_ISL_1266181, EPI_ISL_1266182, EPI_ISL_1266183, EPI_ISL_1266184, EPI_ISL_1266185 | | | |
| see above | Quest Diagnostics Incorporated | Centers for Disease Control and Prevention Division of Viral Diseases, Pathogen Discovery | Peter W. Cook, Dakota Howard, Dhwani Batra, Ben L. Rambo-Martin, S. H. Rosenthal, A. Gerasimova, R. M. Kagan, B. Anderson, M. Hua, Y. Liu, L.E. Bernstein, K.E. Livingston, A. Perez, I. A. Shlyakhter, R. V. Rolando, R. Owen, P. Tanpaiboon, F. Lacbawan, Clinton R. Paden, Suxiang Tong, Duncan MacCannell |
| EPI_ISL_1266186 | Ohio Department of Health Laboratory | Ohio Department of Health Laboratory | Holmes, Jennifer; Eric Brandt, Keoni Omura, Glen McGillivary, Caitlin McDonnell, Jade Mowery, Stephanie Mccracken, Tyler Payne, Kirtana Ramadugu, Erica Leasure, Brent Lee, Kelsey Florek, Heather Blankenship, Allison Black, Quanta Brown, and Tammy Bannerman |
| EPI_ISL_1266187, EPI_ISL_1266188, EPI_ISL_1266189, EPI_ISL_1266190, EPI_ISL_1266191, EPI_ISL_1266192, EPI_ISL_1266193, EPI_ISL_1266194, EPI_ISL_1266195, EPI_ISL_1266196, EPI_ISL_1266197, EPI_ISL_1266198, EPI_ISL_1266199, EPI_ISL_1266200, EPI_ISL_1266201, EPI_ISL_1266202, EPI_ISL_1266203, EPI_ISL_1266204, EPI_ISL_1266205, EPI_ISL_1266206, EPI_ISL_1266207, EPI_ISL_1266208, EPI_ISL_1266209, EPI_ISL_1266210, EPI_ISL_1266211, EPI_ISL_1266212, EPI_ISL_1266213, EPI_ISL_1266214, EPI_ISL_1266215, EPI_ISL_1266216, EPI_ISL_1266217, EPI_ISL_1266218, EPI_ISL_1266219, EPI_ISL_1266220, EPI_ISL_1266221, EPI_ISL_1266222, EPI_ISL_1266223, EPI_ISL_1266224, EPI_ISL_1266225, EPI_ISL_1266226, EPI_ISL_1266227, EPI_ISL_1266446, EPI_ISL_1266449, EPI_ISL_1266452, EPI_ISL_1266455, EPI_ISL_1266458, EPI_ISL_1266461, EPI_ISL_1266465, EPI_ISL_1266468, EPI_ISL_1266471, EPI_ISL_1266474, EPI_ISL_1266476, EPI_ISL_1266479, EPI_ISL_1266482, EPI_ISL_1266485, EPI_ISL_1266488, EPI_ISL_1266491, EPI_ISL_1266494, EPI_ISL_1266497, EPI_ISL_1266500, EPI_ISL_1266503, EPI_ISL_1266506, EPI_ISL_1266509, EPI_ISL_1266512, EPI_ISL_1266515, EPI_ISL_1266518, EPI_ISL_1266521, EPI_ISL_1266524, EPI_ISL_1266527, EPI_ISL_1266530, EPI_ISL_1266533, EPI_ISL_1266536, EPI_ISL_1266539, EPI_ISL_1266541, EPI_ISL_1266544, EPI_ISL_1266547, EPI_ISL_1266550, EPI_ISL_1266553, EPI_ISL_1266556, EPI_ISL_1266559, EPI_ISL_1266562, EPI_ISL_1266565, EPI_ISL_1266568, EPI_ISL_1266571, EPI_ISL_1266574, EPI_ISL_1266577, EPI_ISL_1266580, EPI_ISL_1266584, EPI_ISL_1266587, EPI_ISL_1266590, EPI_ISL_1266593, EPI_ISL_1266596, EPI_ISL_1266599, EPI_ISL_1266602, EPI_ISL_1266605, EPI_ISL_1266608, EPI_ISL_1266611, EPI_ISL_1266613, EPI_ISL_1266616, EPI_ISL_1266619, EPI_ISL_1266622, EPI_ISL_1266625, EPI_ISL_1266628, EPI_ISL_1266631, EPI_ISL_1266634, EPI_ISL_1266637, EPI_ISL_1266640, EPI_ISL_1266643, EPI_ISL_1266646, EPI_ISL_1266649, | | | |

EPI_ISL_1266651, EPI_ISL_1266654, EPI_ISL_1266657, EPI_ISL_1266660, EPI_ISL_1266663, EPI_ISL_1266667, EPI_ISL_1266670, EPI_ISL_1266673, EPI_ISL_1266676, EPI_ISL_1266679, EPI_ISL_1266682, EPI_ISL_1266685, EPI_ISL_1266688, EPI_ISL_1266691, EPI_ISL_1266694, EPI_ISL_1266697, EPI_ISL_1266700, EPI_ISL_1266703, EPI_ISL_1266706, EPI_ISL_1266709, EPI_ISL_1266712, EPI_ISL_1266715, EPI_ISL_1266718, EPI_ISL_1266721, EPI_ISL_1266724, EPI_ISL_1266728, EPI_ISL_1266731, EPI_ISL_1266734, EPI_ISL_1266737, EPI_ISL_1266739, EPI_ISL_1266742, EPI_ISL_1266745, EPI_ISL_1266748, EPI_ISL_1266751, EPI_ISL_1266754, EPI_ISL_1266757, EPI_ISL_1266760, EPI_ISL_1266763, EPI_ISL_1266766, EPI_ISL_1266769, EPI_ISL_1266772, EPI_ISL_1266775, EPI_ISL_1266778, EPI_ISL_1266781, EPI_ISL_1266784, EPI_ISL_1266787, EPI_ISL_1266790, EPI_ISL_1266793, EPI_ISL_1266796, EPI_ISL_1266799, EPI_ISL_1266802, EPI_ISL_1266804, EPI_ISL_1266808, EPI_ISL_1266810, EPI_ISL_1266813, EPI_ISL_1266816, EPI_ISL_1266819, EPI_ISL_1266823, EPI_ISL_1266826, EPI_ISL_1266829, EPI_ISL_1266832, EPI_ISL_1266835, EPI_ISL_1266839, EPI_ISL_1266842, EPI_ISL_1266844, EPI_ISL_1266847, EPI_ISL_1266851, EPI_ISL_1266853, EPI_ISL_1266856, EPI_ISL_1266859, EPI_ISL_1266862, EPI_ISL_1266866, EPI_ISL_1266869, EPI_ISL_1266872, EPI_ISL_1266875, EPI_ISL_1266878, EPI_ISL_1266881, EPI_ISL_1266884, EPI_ISL_1266887, EPI_ISL_1266890, EPI_ISL_1266893, EPI_ISL_1266896, EPI_ISL_1266899, EPI_ISL_1266902, EPI_ISL_1266906, EPI_ISL_1266909, EPI_ISL_1266912, EPI_ISL_1266915, EPI_ISL_1266918, EPI_ISL_1266921, EPI_ISL_1266923, EPI_ISL_1266927, EPI_ISL_1266930, EPI_ISL_1266933, EPI_ISL_1266936, EPI_ISL_1266939, EPI_ISL_1266941, EPI_ISL_1266945, EPI_ISL_1266947, EPI_ISL_1266950, EPI_ISL_1266953, EPI_ISL_1266957, EPI_ISL_1266960, EPI_ISL_1266962, EPI_ISL_1266966, EPI_ISL_1266969, EPI_ISL_1266972, EPI_ISL_1266975, EPI_ISL_1266976, EPI_ISL_1266978, EPI_ISL_1266981, EPI_ISL_1266984, EPI_ISL_1266987, EPI_ISL_1266991, EPI_ISL_1266993, EPI_ISL_1266997, EPI_ISL_1267000, EPI_ISL_1267003, EPI_ISL_1267006, EPI_ISL_1267009, EPI_ISL_1267012, EPI_ISL_1267015, EPI_ISL_1267018, EPI_ISL_1267021, EPI_ISL_1267024, EPI_ISL_1267027, EPI_ISL_1267030, EPI_ISL_1267033, EPI_ISL_1267036, EPI_ISL_1267039, EPI_ISL_1267042, EPI_ISL_1267045, EPI_ISL_1267048, EPI_ISL_1267051, EPI_ISL_1267054, EPI_ISL_1267058, EPI_ISL_1267061, EPI_ISL_1267064, EPI_ISL_1267067, EPI_ISL_1267070, EPI_ISL_1267073, EPI_ISL_1267075, EPI_ISL_1267078, EPI_ISL_1267081, EPI_ISL_1267084, EPI_ISL_1267087, EPI_ISL_1267090, EPI_ISL_1267093, EPI_ISL_1267096, EPI_ISL_1267099, EPI_ISL_1267102, EPI_ISL_1267105, EPI_ISL_1267108, EPI_ISL_1267111, EPI_ISL_1267114, EPI_ISL_1267117, EPI_ISL_1267119, EPI_ISL_1267120, EPI_ISL_1267121, EPI_ISL_1267122, EPI_ISL_1267123, EPI_ISL_1267124, EPI_ISL_1267125, EPI_ISL_1267126, EPI_ISL_1267127, EPI_ISL_1267128, EPI_ISL_1267129, EPI_ISL_1267130, EPI_ISL_1267131, EPI_ISL_1267132, EPI_ISL_1267133, EPI_ISL_1267134, EPI_ISL_1267135, EPI_ISL_1267136, EPI_ISL_1267137, EPI_ISL_1267138, EPI_ISL_1267139, EPI_ISL_1267140, EPI_ISL_1267141, EPI_ISL_1267142, EPI_ISL_1267143, EPI_ISL_1267144, EPI_ISL_1267145, EPI_ISL_1267146, EPI_ISL_1267147, EPI_ISL_1267148, EPI_ISL_1267149, EPI_ISL_1267150, EPI_ISL_1267151, EPI_ISL_1267152, EPI_ISL_1267153, EPI_ISL_1267154, EPI_ISL_1267155, EPI_ISL_1267156, EPI_ISL_1267157, EPI_ISL_1267158, EPI_ISL_1267159, EPI_ISL_1267160, EPI_ISL_1267161, EPI_ISL_1267162, EPI_ISL_1267163, EPI_ISL_1267164, EPI_ISL_1267165, EPI_ISL_1267166, EPI_ISL_1267167, EPI_ISL_1267168, EPI_ISL_1267169, EPI_ISL_1267170, EPI_ISL_1267171, EPI_ISL_1267172, EPI_ISL_1267173, EPI_ISL_1267174, EPI_ISL_1267175, EPI_ISL_1267176, EPI_ISL_1267177, EPI_ISL_1267178, EPI_ISL_1267179, EPI_ISL_1267180, EPI_ISL_1267181, EPI_ISL_1267182, EPI_ISL_1267183, EPI_ISL_1267184, EPI_ISL_1267185, EPI_ISL_1267186, EPI_ISL_1267187, EPI_ISL_1267188, EPI_ISL_1267189, EPI_ISL_1267190, EPI_ISL_1267191, EPI_ISL_1267192, EPI_ISL_1267193, EPI_ISL_1267194, EPI_ISL_1267195, EPI_ISL_1267196, EPI_ISL_1267197, EPI_ISL_1267198, EPI_ISL_1267199, EPI_ISL_1267200, EPI_ISL_1267201, EPI_ISL_1267202, EPI_ISL_1267203, EPI_ISL_1267204, EPI_ISL_1267205, EPI_ISL_1267206, EPI_ISL_1267207, EPI_ISL_1267208, EPI_ISL_1267209, EPI_ISL_1267210, EPI_ISL_1267211, EPI_ISL_1267212, EPI_ISL_1267213, EPI_ISL_1267214, EPI_ISL_1267215, EPI_ISL_1267216, EPI_ISL_1267217

| | | | |
|---|---|---|---|
| see above | Quest Diagnostics Incorporated | Centers for Disease Control and Prevention Division of Viral Diseases, Pathogen Discovery | Peter W. Cook, Dakota Howard, Dhwani Batra, Ben L. Rambo-Martin, S. H. Rosenthal, A. Gerasimova, R. M. Kagan, B. Anderson, M. Hua, Y. Liu, L.E. Bernstein, K.E. Livingston, A. Perez, I. A. Shlyakhter, R. V. Rolando, R. Owen, P. Tanpaiboon, F. Lacbawan, Clinton R. Paden, Suxiang Tong, Duncan MacCannell |
| EPI_ISL_1267218 | Vault Health | Minnesota Department of Health, Public Health Laboratory | Alexandra Lorentz, Jacob Garfin, Matt Plumb, and Xiong Wang |
| EPI_ISL_1267219 | Quest Diagnostics Incorporated | Centers for Disease Control and Prevention Division of Viral Diseases, Pathogen Discovery | Peter W. Cook, Dakota Howard, Dhwani Batra, Ben L. Rambo-Martin, S. H. Rosenthal, A. Gerasimova, R. M. Kagan, B. Anderson, M. Hua, Y. Liu, L.E. Bernstein, K.E. Livingston, A. Perez, I. A. Shlyakhter, R. V. Rolando, R. Owen, P. Tanpaiboon, F. Lacbawan, Clinton R. Paden, Suxiang Tong, Duncan MacCannell |
| EPI_ISL_1267221 | Vault Health | Minnesota Department of Health, Public Health Laboratory | Alexandra Lorentz, Jacob Garfin, Matt Plumb, and Xiong Wang |
| EPI_ISL_1267222 | Quest Diagnostics Incorporated | Centers for Disease Control and Prevention Division of Viral Diseases, Pathogen Discovery | Peter W. Cook, Dakota Howard, Dhwani Batra, Ben L. Rambo-Martin, S. H. Rosenthal, A. Gerasimova, R. M. Kagan, B. Anderson, M. Hua, Y. Liu, L.E. Bernstein, K.E. Livingston, A. Perez, I. A. Shlyakhter, R. V. Rolando, R. Owen, P. Tanpaiboon, F. Lacbawan, Clinton R. Paden, Suxiang Tong, Duncan MacCannell |
| EPI_ISL_1267223, EPI_ISL_1267224, EPI_ISL_1267225 | Vault Health | Minnesota Department of Health, Public Health Laboratory | Alexandra Lorentz, Jacob Garfin, Matt Plumb, and Xiong Wang |
| EPI_ISL_1267226 | Quest Diagnostics Incorporated | Centers for Disease Control and Prevention Division of Viral Diseases, Pathogen Discovery | Peter W. Cook, Dakota Howard, Dhwani Batra, Ben L. Rambo-Martin, S. H. Rosenthal, A. Gerasimova, R. M. Kagan, B. Anderson, M. Hua, Y. Liu, L.E. Bernstein, K.E. Livingston, A. Perez, I. A. Shlyakhter, R. V. Rolando, R. Owen, P. Tanpaiboon, F. Lacbawan, Clinton R. Paden, Suxiang Tong, Duncan MacCannell |
| EPI_ISL_1267227, EPI_ISL_1267228 | Vault Health | Minnesota Department of Health, Public Health Laboratory | Alexandra Lorentz, Jacob Garfin, Matt Plumb, and Xiong Wang |
| EPI_ISL_1267229 | Quest Diagnostics Incorporated | Centers for Disease Control and Prevention Division of Viral Diseases, Pathogen Discovery | Peter W. Cook, Dakota Howard, Dhwani Batra, Ben L. Rambo-Martin, S. H. Rosenthal, A. Gerasimova, R. M. Kagan, B. Anderson, M. Hua, Y. Liu, L.E. Bernstein, K.E. Livingston, A. Perez, I. A. Shlyakhter, R. V. Rolando, R. Owen, P. Tanpaiboon, F. Lacbawan, Clinton R. Paden, Suxiang Tong, Duncan MacCannell |
| EPI_ISL_1267230, EPI_ISL_1267231 | Vault Health | Minnesota Department of Health, Public Health Laboratory | Alexandra Lorentz, Jacob Garfin, Matt Plumb, and Xiong Wang |
| EPI_ISL_1267232 | Quest Diagnostics Incorporated | Centers for Disease Control and Prevention Division of Viral Diseases, Pathogen Discovery | Peter W. Cook, Dakota Howard, Dhwani Batra, Ben L. Rambo-Martin, S. H. Rosenthal, A. Gerasimova, R. M. Kagan, B. Anderson, M. Hua, Y. Liu, L.E. Bernstein, K.E. Livingston, A. Perez, I. A. Shlyakhter, R. V. Rolando, R. Owen, P. Tanpaiboon, F. Lacbawan, Clinton R. Paden, Suxiang Tong, Duncan MacCannell |
| EPI_ISL_1267233, EPI_ISL_1267234 | Vault Health | Minnesota Department of Health, Public Health Laboratory | Alexandra Lorentz, Jacob Garfin, Matt Plumb, and Xiong Wang |
| EPI_ISL_1267235 | Quest Diagnostics Incorporated | Centers for Disease Control and Prevention Division of Viral Diseases, Pathogen Discovery | Peter W. Cook, Dakota Howard, Dhwani Batra, Ben L. Rambo-Martin, S. H. Rosenthal, A. Gerasimova, R. M. Kagan, B. Anderson, M. Hua, Y. Liu, L.E. Bernstein, K.E. Livingston, A. Perez, I. A. Shlyakhter, R. V. Rolando, R. Owen, P. Tanpaiboon, F. Lacbawan, Clinton R. Paden, Suxiang Tong, Duncan MacCannell |
| EPI_ISL_1267236, EPI_ISL_1267237 | Vault Health | Minnesota Department of Health, Public Health Laboratory | Alexandra Lorentz, Jacob Garfin, Matt Plumb, and Xiong Wang |
| EPI_ISL_1267238 | Quest Diagnostics Incorporated | Centers for Disease Control and Prevention Division of Viral Diseases, Pathogen Discovery | Peter W. Cook, Dakota Howard, Dhwani Batra, Ben L. Rambo-Martin, S. H. Rosenthal, A. Gerasimova, R. M. Kagan, B. Anderson, M. Hua, Y. Liu, L.E. Bernstein, K.E. Livingston, A. Perez, I. A. Shlyakhter, R. V. Rolando, R. Owen, P. Tanpaiboon, F. Lacbawan, Clinton R. Paden, Suxiang Tong, Duncan MacCannell |
| EPI_ISL_1267239, EPI_ISL_1267240 | Vault Health | Minnesota Department of Health, Public Health Laboratory | Alexandra Lorentz, Jacob Garfin, Matt Plumb, and Xiong Wang |
| EPI_ISL_1267241 | Quest Diagnostics Incorporated | Centers for Disease Control and Prevention Division of Viral Diseases, Pathogen Discovery | Peter W. Cook, Dakota Howard, Dhwani Batra, Ben L. Rambo-Martin, S. H. Rosenthal, A. Gerasimova, R. M. Kagan, B. Anderson, M. Hua, Y. Liu, L.E. Bernstein, K.E. Livingston, A. Perez, I. A. Shlyakhter, R. V. Rolando, R. Owen, P. Tanpaiboon, F. Lacbawan, Clinton R. Paden, Suxiang Tong, Duncan MacCannell |
| EPI_ISL_1267242 | Vault Health | Minnesota Department of Health, Public Health Laboratory | Alexandra Lorentz, Jacob Garfin, Matt Plumb, and Xiong Wang |
| EPI_ISL_1267244 | Quest Diagnostics Incorporated | Centers for Disease Control and Prevention Division of Viral Diseases, Pathogen Discovery | Peter W. Cook, Dakota Howard, Dhwani Batra, Ben L. Rambo-Martin, S. H. Rosenthal, A. Gerasimova, R. M. Kagan, B. Anderson, M. Hua, Y. Liu, L.E. Bernstein, K.E. Livingston, A. Perez, I. A. Shlyakhter, R. V. Rolando, R. Owen, P. Tanpaiboon, F. Lacbawan, Clinton R. Paden, Suxiang Tong, Duncan MacCannell |
| EPI_ISL_1267245, EPI_ISL_1267246 | Vault Health | Minnesota Department of Health, Public Health Laboratory | Alexandra Lorentz, Jacob Garfin, Matt Plumb, and Xiong Wang |
| EPI_ISL_1267247 | Quest Diagnostics Incorporated | Centers for Disease Control and Prevention Division of Viral Diseases, Pathogen Discovery | Peter W. Cook, Dakota Howard, Dhwani Batra, Ben L. Rambo-Martin, S. H. Rosenthal, A. Gerasimova, R. M. Kagan, B. Anderson, M. Hua, Y. Liu, L.E. Bernstein, K.E. Livingston, A. Perez, I. A. Shlyakhter, R. V. Rolando, R. Owen, P. Tanpaiboon, F. Lacbawan, Clinton R. Paden, Suxiang Tong, Duncan MacCannell |
| EPI_ISL_1267248, EPI_ISL_1267249 | Vault Health | Minnesota Department of Health, Public Health Laboratory | Alexandra Lorentz, Jacob Garfin, Matt Plumb, and Xiong Wang |
| EPI_ISL_1267250 | Quest Diagnostics Incorporated | Centers for Disease Control and Prevention Division of Viral Diseases, Pathogen Discovery | Peter W. Cook, Dakota Howard, Dhwani Batra, Ben L. Rambo-Martin, S. H. Rosenthal, A. Gerasimova, R. M. Kagan, B. Anderson, M. Hua, Y. Liu, L.E. Bernstein, K.E. Livingston, A. Perez, I. A. Shlyakhter, R. V. Rolando, R. Owen, P. Tanpaiboon, F. Lacbawan, Clinton R. Paden, Suxiang Tong, Duncan MacCannell |
| EPI_ISL_1267251, EPI_ISL_1267252 | Vault Health | Minnesota Department of Health, Public Health Laboratory | Alexandra Lorentz, Jacob Garfin, Matt Plumb, and Xiong Wang |
| EPI_ISL_1267253 | Quest Diagnostics Incorporated | Centers for Disease Control and Prevention Division of Viral Diseases, Pathogen Discovery | Peter W. Cook, Dakota Howard, Dhwani Batra, Ben L. Rambo-Martin, S. H. Rosenthal, A. Gerasimova, R. M. Kagan, B. Anderson, M. Hua, Y. Liu, L.E. Bernstein, K.E. Livingston, A. Perez, I. A. Shlyakhter, R. V. Rolando, R. Owen, P. Tanpaiboon, F. Lacbawan, Clinton R. Paden, Suxiang Tong, Duncan MacCannell |
| EPI_ISL_1267254, EPI_ISL_1267255 | Vault Health | Minnesota Department of Health, Public Health Laboratory | Alexandra Lorentz, Jacob Garfin, Matt Plumb, and Xiong Wang |
| EPI_ISL_1267256 | Quest Diagnostics Incorporated | Centers for Disease Control and Prevention Division of Viral Diseases, Pathogen Discovery | Peter W. Cook, Dakota Howard, Dhwani Batra, Ben L. Rambo-Martin, S. H. Rosenthal, A. Gerasimova, R. M. Kagan, B. Anderson, M. Hua, Y. Liu, L.E. Bernstein, K.E. Livingston, A. Perez, I. A. Shlyakhter, R. V. Rolando, R. Owen, P. Tanpaiboon, F. Lacbawan, Clinton R. Paden, Suxiang Tong, Duncan MacCannell |
| EPI_ISL_1267257, EPI_ISL_1267258 | Vault Health | Minnesota Department of Health, Public Health Laboratory | Alexandra Lorentz, Jacob Garfin, Matt Plumb, and Xiong Wang |
| EPI_ISL_1267259 | Quest Diagnostics Incorporated | Centers for Disease Control and Prevention Division of Viral Diseases, Pathogen Discovery | Peter W. Cook, Dakota Howard, Dhwani Batra, Ben L. Rambo-Martin, S. H. Rosenthal, A. Gerasimova, R. M. Kagan, B. Anderson, M. Hua, Y. Liu, L.E. Bernstein, K.E. Livingston, A. Perez, I. A. Shlyakhter, R. V. Rolando, R. Owen, P. Tanpaiboon, F. Lacbawan, Clinton R. Paden, Suxiang Tong, Duncan MacCannell |
| EPI_ISL_1267260, EPI_ISL_1267261 | Vault Health | Minnesota Department of Health, Public Health Laboratory | Alexandra Lorentz, Jacob Garfin, Matt Plumb, and Xiong Wang |
| EPI_ISL_1267262 | Quest Diagnostics Incorporated | Centers for Disease Control and Prevention Division of Viral Diseases, Pathogen Discovery | Peter W. Cook, Dakota Howard, Dhwani Batra, Ben L. Rambo-Martin, S. H. Rosenthal, A. Gerasimova, R. M. Kagan, B. Anderson, M. Hua, Y. Liu, L.E. Bernstein, K.E. Livingston, A. Perez, I. A. Shlyakhter, R. V. Rolando, R. Owen, P. Tanpaiboon, F. Lacbawan, Clinton R. Paden, Suxiang Tong, Duncan MacCannell |
| EPI_ISL_1267263, EPI_ISL_1267264 | Vault Health | Minnesota Department of Health, Public Health Laboratory | Alexandra Lorentz, Jacob Garfin, Matt Plumb, and Xiong Wang |
| EPI_ISL_1267265 | Quest Diagnostics Incorporated | Centers for Disease Control and Prevention Division of Viral Diseases, Pathogen | Peter W. Cook, Dakota Howard, Dhwani Batra, Ben L. Rambo-Martin, S. H. Rosenthal, A. Gerasimova, R. M. Kagan, B. Anderson, M. Hua, Y. Liu, L.E. Bernstein, K.E. Livingston, A. Perez, I. A. Shlyakhter, R. V. Rolando, R. Owen, P. Tanpaiboon, F. Lacbawan, Clinton R. Paden, Suxiang Tong, Duncan MacCannell |

| | | | |
|---|---|---|---|
| | | Discovery | |
| EPI_ISL_1267266, EPI_ISL_1267267 | Vault Health | Minnesota Department of Health, Public Health Laboratory | Alexandra Lorentz, Jacob Garfin, Matt Plumb, and Xiong Wang |
| EPI_ISL_1267268 | Quest Diagnostics Incorporated | Centers for Disease Control and Prevention Division of Viral Diseases, Pathogen Discovery | Peter W. Cook, Dakota Howard, Dhwani Batra, Ben L. Rambo-Martin, S. H. Rosenthal, A. Gerasimova, R. M. Kagan, B. Anderson, M. Hua, Y. Liu, L.E. Bernstein, K.E. Livingston, A. Perez, I. A. Shlyakhter, R. V. Rolando, R. Owen, P. Tanpaiboon, F. Lacbawan, Clinton R. Paden, Suxiang Tong, Duncan MacCannell |
| EPI_ISL_1267269, EPI_ISL_1267270 | Vault Health | Minnesota Department of Health, Public Health Laboratory | Alexandra Lorentz, Jacob Garfin, Matt Plumb, and Xiong Wang |
| EPI_ISL_1267271 | Quest Diagnostics Incorporated | Centers for Disease Control and Prevention Division of Viral Diseases, Pathogen Discovery | Peter W. Cook, Dakota Howard, Dhwani Batra, Ben L. Rambo-Martin, S. H. Rosenthal, A. Gerasimova, R. M. Kagan, B. Anderson, M. Hua, Y. Liu, L.E. Bernstein, K.E. Livingston, A. Perez, I. A. Shlyakhter, R. V. Rolando, R. Owen, P. Tanpaiboon, F. Lacbawan, Clinton R. Paden, Suxiang Tong, Duncan MacCannell |
| EPI_ISL_1267272, EPI_ISL_1267273 | Vault Health | Minnesota Department of Health, Public Health Laboratory | Alexandra Lorentz, Jacob Garfin, Matt Plumb, and Xiong Wang |
| EPI_ISL_1267274 | Quest Diagnostics Incorporated | Centers for Disease Control and Prevention Division of Viral Diseases, Pathogen Discovery | Peter W. Cook, Dakota Howard, Dhwani Batra, Ben L. Rambo-Martin, S. H. Rosenthal, A. Gerasimova, R. M. Kagan, B. Anderson, M. Hua, Y. Liu, L.E. Bernstein, K.E. Livingston, A. Perez, I. A. Shlyakhter, R. V. Rolando, R. Owen, P. Tanpaiboon, F. Lacbawan, Clinton R. Paden, Suxiang Tong, Duncan MacCannell |
| EPI_ISL_1267275, EPI_ISL_1267276 | Vault Health | Minnesota Department of Health, Public Health Laboratory | Alexandra Lorentz, Jacob Garfin, Matt Plumb, and Xiong Wang |
| EPI_ISL_1267277 | Quest Diagnostics Incorporated | Centers for Disease Control and Prevention Division of Viral Diseases, Pathogen Discovery | Peter W. Cook, Dakota Howard, Dhwani Batra, Ben L. Rambo-Martin, S. H. Rosenthal, A. Gerasimova, R. M. Kagan, B. Anderson, M. Hua, Y. Liu, L.E. Bernstein, K.E. Livingston, A. Perez, I. A. Shlyakhter, R. V. Rolando, R. Owen, P. Tanpaiboon, F. Lacbawan, Clinton R. Paden, Suxiang Tong, Duncan MacCannell |
| EPI_ISL_1267278, EPI_ISL_1267279 | Vault Health | Minnesota Department of Health, Public Health Laboratory | Alexandra Lorentz, Jacob Garfin, Matt Plumb, and Xiong Wang |
| EPI_ISL_1267280 | Quest Diagnostics Incorporated | Centers for Disease Control and Prevention Division of Viral Diseases, Pathogen Discovery | Peter W. Cook, Dakota Howard, Dhwani Batra, Ben L. Rambo-Martin, S. H. Rosenthal, A. Gerasimova, R. M. Kagan, B. Anderson, M. Hua, Y. Liu, L.E. Bernstein, K.E. Livingston, A. Perez, I. A. Shlyakhter, R. V. Rolando, R. Owen, P. Tanpaiboon, F. Lacbawan, Clinton R. Paden, Suxiang Tong, Duncan MacCannell |
| EPI_ISL_1267281, EPI_ISL_1267282 | Vault Health | Minnesota Department of Health, Public Health Laboratory | Alexandra Lorentz, Jacob Garfin, Matt Plumb, and Xiong Wang |
| EPI_ISL_1267283 | Quest Diagnostics Incorporated | Centers for Disease Control and Prevention Division of Viral Diseases, Pathogen Discovery | Peter W. Cook, Dakota Howard, Dhwani Batra, Ben L. Rambo-Martin, S. H. Rosenthal, A. Gerasimova, R. M. Kagan, B. Anderson, M. Hua, Y. Liu, L.E. Bernstein, K.E. Livingston, A. Perez, I. A. Shlyakhter, R. V. Rolando, R. Owen, P. Tanpaiboon, F. Lacbawan, Clinton R. Paden, Suxiang Tong, Duncan MacCannell |
| EPI_ISL_1267284, EPI_ISL_1267285 | Vault Health | Minnesota Department of Health, Public Health Laboratory | Alexandra Lorentz, Jacob Garfin, Matt Plumb, and Xiong Wang |
| EPI_ISL_1267286 | Quest Diagnostics Incorporated | Centers for Disease Control and Prevention Division of Viral Diseases, Pathogen Discovery | Peter W. Cook, Dakota Howard, Dhwani Batra, Ben L. Rambo-Martin, S. H. Rosenthal, A. Gerasimova, R. M. Kagan, B. Anderson, M. Hua, Y. Liu, L.E. Bernstein, K.E. Livingston, A. Perez, I. A. Shlyakhter, R. V. Rolando, R. Owen, P. Tanpaiboon, F. Lacbawan, Clinton R. Paden, Suxiang Tong, Duncan MacCannell |
| EPI_ISL_1267287, EPI_ISL_1267288 | Vault Health | Minnesota Department of Health, Public Health Laboratory | Alexandra Lorentz, Jacob Garfin, Matt Plumb, and Xiong Wang |
| EPI_ISL_1267289 | Quest Diagnostics Incorporated | Centers for Disease Control and Prevention Division of Viral Diseases, Pathogen Discovery | Peter W. Cook, Dakota Howard, Dhwani Batra, Ben L. Rambo-Martin, S. H. Rosenthal, A. Gerasimova, R. M. Kagan, B. Anderson, M. Hua, Y. Liu, L.E. Bernstein, K.E. Livingston, A. Perez, I. A. Shlyakhter, R. V. Rolando, R. Owen, P. Tanpaiboon, F. Lacbawan, Clinton R. Paden, Suxiang Tong, Duncan MacCannell |
| EPI_ISL_1267290, EPI_ISL_1267291 | Vault Health | Minnesota Department of Health, Public Health Laboratory | Alexandra Lorentz, Jacob Garfin, Matt Plumb, and Xiong Wang |
| EPI_ISL_1267292 | Quest Diagnostics Incorporated | Centers for Disease Control and Prevention Division of Viral Diseases, Pathogen Discovery | Peter W. Cook, Dakota Howard, Dhwani Batra, Ben L. Rambo-Martin, S. H. Rosenthal, A. Gerasimova, R. M. Kagan, B. Anderson, M. Hua, Y. Liu, L.E. Bernstein, K.E. Livingston, A. Perez, I. A. Shlyakhter, R. V. Rolando, R. Owen, P. Tanpaiboon, F. Lacbawan, Clinton R. Paden, Suxiang Tong, Duncan MacCannell |
| EPI_ISL_1267293, EPI_ISL_1267294 | Vault Health | Minnesota Department of Health, Public Health Laboratory | Alexandra Lorentz, Jacob Garfin, Matt Plumb, and Xiong Wang |
| EPI_ISL_1267295 | Quest Diagnostics Incorporated | Centers for Disease Control and Prevention Division of Viral Diseases, Pathogen Discovery | Peter W. Cook, Dakota Howard, Dhwani Batra, Ben L. Rambo-Martin, S. H. Rosenthal, A. Gerasimova, R. M. Kagan, B. Anderson, M. Hua, Y. Liu, L.E. Bernstein, K.E. Livingston, A. Perez, I. A. Shlyakhter, R. V. Rolando, R. Owen, P. Tanpaiboon, F. Lacbawan, Clinton R. Paden, Suxiang Tong, Duncan MacCannell |
| EPI_ISL_1267296, EPI_ISL_1267297 | Vault Health | Minnesota Department of Health, Public Health Laboratory | Alexandra Lorentz, Jacob Garfin, Matt Plumb, and Xiong Wang |
| EPI_ISL_1267298 | Quest Diagnostics Incorporated | Centers for Disease Control and Prevention Division of Viral Diseases, Pathogen Discovery | Peter W. Cook, Dakota Howard, Dhwani Batra, Ben L. Rambo-Martin, S. H. Rosenthal, A. Gerasimova, R. M. Kagan, B. Anderson, M. Hua, Y. Liu, L.E. Bernstein, K.E. Livingston, A. Perez, I. A. Shlyakhter, R. V. Rolando, R. Owen, P. Tanpaiboon, F. Lacbawan, Clinton R. Paden, Suxiang Tong, Duncan MacCannell |
| EPI_ISL_1267300 | Mayo Clinic & Mayo Clinic Laboratories | Minnesota Department of Health, Public Health Laboratory | Alexandra Lorentz, Jacob Garfin, Matt Plumb, and Xiong Wang |
| EPI_ISL_1267301 | Quest Diagnostics Incorporated | Centers for Disease Control and Prevention Division of Viral Diseases, Pathogen Discovery | Peter W. Cook, Dakota Howard, Dhwani Batra, Ben L. Rambo-Martin, S. H. Rosenthal, A. Gerasimova, R. M. Kagan, B. Anderson, M. Hua, Y. Liu, L.E. Bernstein, K.E. Livingston, A. Perez, I. A. Shlyakhter, R. V. Rolando, R. Owen, P. Tanpaiboon, F. Lacbawan, Clinton R. Paden, Suxiang Tong, Duncan MacCannell |
| EPI_ISL_1267302 | Mayo Clinic & Mayo Clinic Laboratories | Minnesota Department of Health, Public Health Laboratory | Alexandra Lorentz, Jacob Garfin, Matt Plumb, and Xiong Wang |
| EPI_ISL_1267303 | Quest Diagnostics Incorporated | Centers for Disease Control and Prevention Division of Viral Diseases, Pathogen Discovery | Peter W. Cook, Dakota Howard, Dhwani Batra, Ben L. Rambo-Martin, S. H. Rosenthal, A. Gerasimova, R. M. Kagan, B. Anderson, M. Hua, Y. Liu, L.E. Bernstein, K.E. Livingston, A. Perez, I. A. Shlyakhter, R. V. Rolando, R. Owen, P. Tanpaiboon, F. Lacbawan, Clinton R. Paden, Suxiang Tong, Duncan MacCannell |
| EPI_ISL_1267304, EPI_ISL_1267305 | Mayo Clinic & Mayo Clinic Laboratories | Minnesota Department of Health, Public Health Laboratory | Alexandra Lorentz, Jacob Garfin, Matt Plumb, and Xiong Wang |
| EPI_ISL_1267306 | Quest Diagnostics Incorporated | Centers for Disease Control and Prevention Division of Viral Diseases, Pathogen Discovery | Peter W. Cook, Dakota Howard, Dhwani Batra, Ben L. Rambo-Martin, S. H. Rosenthal, A. Gerasimova, R. M. Kagan, B. Anderson, M. Hua, Y. Liu, L.E. Bernstein, K.E. Livingston, A. Perez, I. A. Shlyakhter, R. V. Rolando, R. Owen, P. Tanpaiboon, F. Lacbawan, Clinton R. Paden, Suxiang Tong, Duncan MacCannell |
| EPI_ISL_1267307, EPI_ISL_1267308 | Mayo Clinic & Mayo Clinic Laboratories | Minnesota Department of Health, Public Health Laboratory | Alexandra Lorentz, Jacob Garfin, Matt Plumb, and Xiong Wang |
| EPI_ISL_1267309 | Quest Diagnostics Incorporated | Centers for Disease Control and Prevention Division of Viral Diseases, Pathogen Discovery | Peter W. Cook, Dakota Howard, Dhwani Batra, Ben L. Rambo-Martin, S. H. Rosenthal, A. Gerasimova, R. M. Kagan, B. Anderson, M. Hua, Y. Liu, L.E. Bernstein, K.E. Livingston, A. Perez, I. A. Shlyakhter, R. V. Rolando, R. Owen, P. Tanpaiboon, F. Lacbawan, Clinton R. Paden, Suxiang Tong, Duncan MacCannell |
| EPI_ISL_1267310, EPI_ISL_1267311, EPI_ISL_1267312 | Mayo Clinic & Mayo Clinic Laboratories | Minnesota Department of Health, Public Health Laboratory | Alexandra Lorentz, Jacob Garfin, Matt Plumb, and Xiong Wang |
| EPI_ISL_1267313 | Quest Diagnostics Incorporated | Centers for Disease Control and Prevention Division of Viral Diseases, Pathogen Discovery | Peter W. Cook, Dakota Howard, Dhwani Batra, Ben L. Rambo-Martin, S. H. Rosenthal, A. Gerasimova, R. M. Kagan, B. Anderson, M. Hua, Y. Liu, L.E. Bernstein, K.E. Livingston, A. Perez, I. A. Shlyakhter, R. V. Rolando, R. Owen, P. Tanpaiboon, F. Lacbawan, Clinton R. Paden, Suxiang Tong, Duncan MacCannell |
| EPI_ISL_1267314, EPI_ISL_1267315 | Mayo Clinic & Mayo Clinic Laboratories | Minnesota Department of Health, Public Health Laboratory | Alexandra Lorentz, Jacob Garfin, Matt Plumb, and Xiong Wang |
| EPI_ISL_1267316 | Quest Diagnostics Incorporated | Centers for Disease Control and Prevention Division of Viral Diseases, Pathogen Discovery | Peter W. Cook, Dakota Howard, Dhwani Batra, Ben L. Rambo-Martin, S. H. Rosenthal, A. Gerasimova, R. M. Kagan, B. Anderson, M. Hua, Y. Liu, L.E. Bernstein, K.E. Livingston, A. Perez, I. A. Shlyakhter, R. V. Rolando, R. Owen, P. Tanpaiboon, F. Lacbawan, Clinton R. Paden, Suxiang Tong, Duncan MacCannell |
| EPI_ISL_1267317, EPI_ISL_1267318 | Mayo Clinic & Mayo Clinic Laboratories | Minnesota Department of Health, Public Health Laboratory | Alexandra Lorentz, Jacob Garfin, Matt Plumb, and Xiong Wang |
| EPI_ISL_1267319 | Quest Diagnostics Incorporated | Centers for Disease Control and Prevention Division of Viral Diseases, Pathogen Discovery | Peter W. Cook, Dakota Howard, Dhwani Batra, Ben L. Rambo-Martin, S. H. Rosenthal, A. Gerasimova, R. M. Kagan, B. Anderson, M. Hua, Y. Liu, L.E. Bernstein, K.E. Livingston, A. Perez, I. A. Shlyakhter, R. V. Rolando, R. Owen, P. Tanpaiboon, F. Lacbawan, Clinton R. Paden, Suxiang Tong, Duncan MacCannell |
| EPI_ISL_1267320 | Mayo Clinic & Mayo Clinic Laboratories | Minnesota Department of Health, Public | Alexandra Lorentz, Jacob Garfin, Matt Plumb, and Xiong Wang |

| | | | |
|---|---|---|---|
| | | Health Laboratory | |
| EPI_ISL_1267321 | Quest Diagnostics Incorporated | Centers for Disease Control and Prevention Division of Viral Diseases, Pathogen Discovery | Peter W. Cook, Dakota Howard, Dhwani Batra, Ben L. Rambo-Martin, S. H. Rosenthal, A. Gerasimova, R. M. Kagan, B. Anderson, M. Hua, Y. Liu, L.E. Bernstein, K.E. Livingston, A. Perez, I. A. Shlyakhter, R. V. Rolando, R. Owen, P. Tanpaiboon, F. Lacbawan, Clinton R. Paden, Suxiang Tong, Duncan MacCannell |
| EPI_ISL_1267322, EPI_ISL_1267323 | Mayo Clinic & Mayo Clinic Laboratories | Minnesota Department of Health, Public Health Laboratory | Alexandra Lorentz, Jacob Garfin, Matt Plumb, and Xiong Wang |
| EPI_ISL_1267324 | Quest Diagnostics Incorporated | Centers for Disease Control and Prevention Division of Viral Diseases, Pathogen Discovery | Peter W. Cook, Dakota Howard, Dhwani Batra, Ben L. Rambo-Martin, S. H. Rosenthal, A. Gerasimova, R. M. Kagan, B. Anderson, M. Hua, Y. Liu, L.E. Bernstein, K.E. Livingston, A. Perez, I. A. Shlyakhter, R. V. Rolando, R. Owen, P. Tanpaiboon, F. Lacbawan, Clinton R. Paden, Suxiang Tong, Duncan MacCannell |
| EPI_ISL_1267325, EPI_ISL_1267326 | Mayo Clinic & Mayo Clinic Laboratories | Minnesota Department of Health, Public Health Laboratory | Alexandra Lorentz, Jacob Garfin, Matt Plumb, and Xiong Wang |
| EPI_ISL_1267327, EPI_ISL_1267330 | Quest Diagnostics Incorporated | Centers for Disease Control and Prevention Division of Viral Diseases, Pathogen Discovery | Peter W. Cook, Dakota Howard, Dhwani Batra, Ben L. Rambo-Martin, S. H. Rosenthal, A. Gerasimova, R. M. Kagan, B. Anderson, M. Hua, Y. Liu, L.E. Bernstein, K.E. Livingston, A. Perez, I. A. Shlyakhter, R. V. Rolando, R. Owen, P. Tanpaiboon, F. Lacbawan, Clinton R. Paden, Suxiang Tong, Duncan MacCannell |
| EPI_ISL_1267331 | Vault Health | Minnesota Department of Health, Public Health Laboratory | Alexandra Lorentz, Jacob Garfin, Matt Plumb, and Xiong Wang |
| EPI_ISL_1267333 | Quest Diagnostics Incorporated | Centers for Disease Control and Prevention Division of Viral Diseases, Pathogen Discovery | Peter W. Cook, Dakota Howard, Dhwani Batra, Ben L. Rambo-Martin, S. H. Rosenthal, A. Gerasimova, R. M. Kagan, B. Anderson, M. Hua, Y. Liu, L.E. Bernstein, K.E. Livingston, A. Perez, I. A. Shlyakhter, R. V. Rolando, R. Owen, P. Tanpaiboon, F. Lacbawan, Clinton R. Paden, Suxiang Tong, Duncan MacCannell |
| EPI_ISL_1267334, EPI_ISL_1267335 | Vault Health | Minnesota Department of Health, Public Health Laboratory | Alexandra Lorentz, Jacob Garfin, Matt Plumb, and Xiong Wang |
| EPI_ISL_1267336 | Quest Diagnostics Incorporated | Centers for Disease Control and Prevention Division of Viral Diseases, Pathogen Discovery | Peter W. Cook, Dakota Howard, Dhwani Batra, Ben L. Rambo-Martin, S. H. Rosenthal, A. Gerasimova, R. M. Kagan, B. Anderson, M. Hua, Y. Liu, L.E. Bernstein, K.E. Livingston, A. Perez, I. A. Shlyakhter, R. V. Rolando, R. Owen, P. Tanpaiboon, F. Lacbawan, Clinton R. Paden, Suxiang Tong, Duncan MacCannell |
| EPI_ISL_1267337 | Vault Health | Minnesota Department of Health, Public Health Laboratory | Alexandra Lorentz, Jacob Garfin, Matt Plumb, and Xiong Wang |
| EPI_ISL_1267338, EPI_ISL_1267339, EPI_ISL_1267340, EPI_ISL_1267341, EPI_ISL_1267342, EPI_ISL_1267343, EPI_ISL_1267344, EPI_ISL_1267345, EPI_ISL_1267346, EPI_ISL_1267347, EPI_ISL_1267348, EPI_ISL_1267349, EPI_ISL_1267350, EPI_ISL_1267351, EPI_ISL_1267352, EPI_ISL_1267353, EPI_ISL_1267354, EPI_ISL_1267355, EPI_ISL_1267356, EPI_ISL_1267357, EPI_ISL_1267358, EPI_ISL_1267359, EPI_ISL_1267360, EPI_ISL_1267361, EPI_ISL_1267362, EPI_ISL_1267363, EPI_ISL_1267364, EPI_ISL_1267365, EPI_ISL_1267366, EPI_ISL_1267367, EPI_ISL_1267368, EPI_ISL_1267369, EPI_ISL_1267370, EPI_ISL_1267371, EPI_ISL_1267372, EPI_ISL_1267373, EPI_ISL_1267374, EPI_ISL_1267375, EPI_ISL_1267376, EPI_ISL_1267377, EPI_ISL_1267378, EPI_ISL_1267379, EPI_ISL_1267380, EPI_ISL_1267381, EPI_ISL_1267382, EPI_ISL_1267383, EPI_ISL_1267384, EPI_ISL_1267385, EPI_ISL_1267386, EPI_ISL_1267387, EPI_ISL_1267388, EPI_ISL_1267389, EPI_ISL_1267390, EPI_ISL_1267391, EPI_ISL_1267392, EPI_ISL_1267393, EPI_ISL_1267394, EPI_ISL_1267395, EPI_ISL_1267396, EPI_ISL_1267397, EPI_ISL_1267398, EPI_ISL_1267399, EPI_ISL_1267400, EPI_ISL_1267401, EPI_ISL_1267402, EPI_ISL_1267403, EPI_ISL_1267404, EPI_ISL_1267405, EPI_ISL_1267406, EPI_ISL_1267407, EPI_ISL_1267408, EPI_ISL_1267409, EPI_ISL_1267410, EPI_ISL_1267411, EPI_ISL_1267412, EPI_ISL_1267413, EPI_ISL_1267414, EPI_ISL_1267415, EPI_ISL_1267416, EPI_ISL_1267417, EPI_ISL_1267418, EPI_ISL_1267419, EPI_ISL_1267420, EPI_ISL_1267421, EPI_ISL_1267422, EPI_ISL_1267423, EPI_ISL_1267424, EPI_ISL_1267425, EPI_ISL_1267426, EPI_ISL_1267427, EPI_ISL_1267428, EPI_ISL_1267429, EPI_ISL_1267430, EPI_ISL_1267431, EPI_ISL_1267432, EPI_ISL_1267433, EPI_ISL_1267434, EPI_ISL_1267435, EPI_ISL_1267436, EPI_ISL_1267437, EPI_ISL_1267438, EPI_ISL_1267439, EPI_ISL_1267440, EPI_ISL_1267441, EPI_ISL_1267442, EPI_ISL_1267443, EPI_ISL_1267444, EPI_ISL_1267445, EPI_ISL_1267446, EPI_ISL_1267447, EPI_ISL_1267448, EPI_ISL_1267449, EPI_ISL_1267450, EPI_ISL_1267451, EPI_ISL_1267452, EPI_ISL_1267453, EPI_ISL_1267454, EPI_ISL_1267455, EPI_ISL_1267456, EPI_ISL_1267457, EPI_ISL_1267458, EPI_ISL_1267459, EPI_ISL_1267460, EPI_ISL_1267461, EPI_ISL_1267462, EPI_ISL_1267463, EPI_ISL_1267464, EPI_ISL_1267465, EPI_ISL_1267466, EPI_ISL_1267467, EPI_ISL_1267468, EPI_ISL_1267469, EPI_ISL_1267470, EPI_ISL_1267471, EPI_ISL_1267472, EPI_ISL_1267473, EPI_ISL_1267474, EPI_ISL_1267475, EPI_ISL_1267476, EPI_ISL_1267477, EPI_ISL_1267478, EPI_ISL_1267479, EPI_ISL_1267480, EPI_ISL_1267481, EPI_ISL_1267482, EPI_ISL_1267483, EPI_ISL_1267484, EPI_ISL_1267485, EPI_ISL_1267486, EPI_ISL_1267487, EPI_ISL_1267488, EPI_ISL_1267489, EPI_ISL_1267490, EPI_ISL_1267491, EPI_ISL_1267492, EPI_ISL_1267493, EPI_ISL_1267494, EPI_ISL_1267495, EPI_ISL_1267496, EPI_ISL_1267497, EPI_ISL_1267498, EPI_ISL_1267499, EPI_ISL_1267500, EPI_ISL_1267501 | | | |
| see above | Quest Diagnostics Incorporated | Centers for Disease Control and Prevention Division of Viral Diseases, Pathogen Discovery | Peter W. Cook, Dakota Howard, Dhwani Batra, Ben L. Rambo-Martin, S. H. Rosenthal, A. Gerasimova, R. M. Kagan, B. Anderson, M. Hua, Y. Liu, L.E. Bernstein, K.E. Livingston, A. Perez, I. A. Shlyakhter, R. V. Rolando, R. Owen, P. Tanpaiboon, F. Lacbawan, Clinton R. Paden, Suxiang Tong, Duncan MacCannell |
| EPI_ISL_1267502, EPI_ISL_1267503, EPI_ISL_1267504, EPI_ISL_1267505, EPI_ISL_1267506, EPI_ISL_1267507, EPI_ISL_1267508, EPI_ISL_1267509, EPI_ISL_1267510, EPI_ISL_1267511, EPI_ISL_1267512, EPI_ISL_1267513, EPI_ISL_1267514, EPI_ISL_1267515, EPI_ISL_1267516, EPI_ISL_1267517, EPI_ISL_1267519, EPI_ISL_1267520, EPI_ISL_1267521, EPI_ISL_1267522, EPI_ISL_1267523, EPI_ISL_1267524, EPI_ISL_1267525, EPI_ISL_1267526, EPI_ISL_1267527, EPI_ISL_1267528, EPI_ISL_1267529, EPI_ISL_1267530, EPI_ISL_1267531, EPI_ISL_1267532, EPI_ISL_1267533, EPI_ISL_1267534, EPI_ISL_1267535, EPI_ISL_1267536, EPI_ISL_1267537, EPI_ISL_1267538, EPI_ISL_1267539, EPI_ISL_1267540, EPI_ISL_1267541, EPI_ISL_1267542, EPI_ISL_1267543, EPI_ISL_1267544, EPI_ISL_1267545, EPI_ISL_1267546, EPI_ISL_1267547, EPI_ISL_1267548, EPI_ISL_1267549, EPI_ISL_1267550, EPI_ISL_1267551, EPI_ISL_1267552, EPI_ISL_1267553, EPI_ISL_1267554, EPI_ISL_1267555, EPI_ISL_1267556, EPI_ISL_1267557, EPI_ISL_1267558, EPI_ISL_1267559, EPI_ISL_1267560, EPI_ISL_1267561, EPI_ISL_1267562, EPI_ISL_1267563, EPI_ISL_1267564, EPI_ISL_1267565, EPI_ISL_1267566, EPI_ISL_1267567, EPI_ISL_1267568, EPI_ISL_1267569, EPI_ISL_1267570, EPI_ISL_1267571, EPI_ISL_1267572, EPI_ISL_1267573, EPI_ISL_1267574, EPI_ISL_1267575, EPI_ISL_1267576, EPI_ISL_1267577, EPI_ISL_1267578, EPI_ISL_1267579, EPI_ISL_1267580, EPI_ISL_1267581, EPI_ISL_1267582, EPI_ISL_1267583, EPI_ISL_1267584, EPI_ISL_1267585, EPI_ISL_1267586, EPI_ISL_1267587, EPI_ISL_1267588, EPI_ISL_1267589 | | | |
| see above | Helix/Illumina | Centers for Disease Control and Prevention Division of Viral Diseases, Pathogen Discovery | Peter W. Cook, Dakota Howard, Dhwani Batra, Ben L. Rambo-Martin, Eileen de Feo, Jan Antico, Christine Tran, Matthew Tolentino, Shannon Wickline, Kim Gietzen, Brad Sickler, Jingtao Liu, Eric Allen, Phil Febbo, Summer Galloway, Nicole L. Washington, Simon White, Geraint Levan, Kelly Schiabor Barrett, Elizabeth Cirulli, Alexandre Bolze, Ary Ascencio, Charlotte Rivera-Garcia, Ryan Cho, Jason Nguyen, Sherry Wang, Jimmy Ramirez, Tyler Cassens, Efren Sandoval, Magnus Isaksson, William Lee, David Becker, Marc Laurent, James Lu, Clinton R. Paden, Suxiang Tong, Duncan MacCannell |
| EPI_ISL_1267590, EPI_ISL_1267591 | Microbiology Division, South Carolina Department of Health and Environmental Control Public Health Laboratory (SC DHEC PHL) | Microbiology Division, South Carolina Department of Health and Environmental Control Public Health Laboratory (SC DHEC PHL) | Haley Flores, Jessica Freeman |
| EPI_ISL_1267592 | Helix/Illumina | Centers for Disease Control and Prevention Division of Viral Diseases, Pathogen Discovery | Peter W. Cook, Dakota Howard, Dhwani Batra, Ben L. Rambo-Martin, Eileen de Feo, Jan Antico, Christine Tran, Matthew Tolentino, Shannon Wickline, Kim Gietzen, Brad Sickler, Jingtao Liu, Eric Allen, Phil Febbo, Summer Galloway, Nicole L. Washington, Simon White, Geraint Levan, Kelly Schiabor Barrett, Elizabeth Cirulli, Alexandre Bolze, Ary Ascencio, Charlotte Rivera-Garcia, Ryan Cho, Jason Nguyen, Sherry Wang, Jimmy Ramirez, Tyler Cassens, Efren Sandoval, Magnus Isaksson, William Lee, David Becker, Marc Laurent, James Lu, Clinton R. Paden, Suxiang Tong, Duncan MacCannell |
| EPI_ISL_1267593, EPI_ISL_1267594 | Microbiology Division, South Carolina Department of Health and Environmental Control Public Health Laboratory (SC DHEC PHL) | Microbiology Division, South Carolina Department of Health and Environmental Control Public Health Laboratory (SC DHEC PHL) | Haley Flores, Jessica Freeman |
| EPI_ISL_1267595 | Helix/Illumina | Centers for Disease Control and Prevention Division of Viral Diseases, Pathogen Discovery | Peter W. Cook, Dakota Howard, Dhwani Batra, Ben L. Rambo-Martin, Eileen de Feo, Jan Antico, Christine Tran, Matthew Tolentino, Shannon Wickline, Kim Gietzen, Brad Sickler, Jingtao Liu, Eric Allen, Phil Febbo, Summer Galloway, Nicole L. Washington, Simon White, Geraint Levan, Kelly Schiabor Barrett, Elizabeth Cirulli, Alexandre Bolze, Ary Ascencio, Charlotte Rivera-Garcia, Ryan Cho, Jason Nguyen, Sherry Wang, Jimmy Ramirez, Tyler Cassens, Efren Sandoval, Magnus Isaksson, William Lee, David Becker, Marc Laurent, James Lu, Clinton R. Paden, Suxiang Tong, Duncan MacCannell |
| EPI_ISL_1267596, EPI_ISL_1267597 | Microbiology Division, South Carolina Department of Health and Environmental Control Public Health Laboratory (SC DHEC PHL) | Microbiology Division, South Carolina Department of Health and Environmental Control Public Health Laboratory (SC DHEC PHL) | Haley Flores, Jessica Freeman |
| EPI_ISL_1267598 | Helix/Illumina | Centers for Disease Control and Prevention Division of Viral Diseases, Pathogen Discovery | Peter W. Cook, Dakota Howard, Dhwani Batra, Ben L. Rambo-Martin, Eileen de Feo, Jan Antico, Christine Tran, Matthew Tolentino, Shannon Wickline, Kim Gietzen, Brad Sickler, Jingtao Liu, Eric Allen, Phil Febbo, Summer Galloway, Nicole L. Washington, Simon White, Geraint Levan, Kelly Schiabor Barrett, Elizabeth Cirulli, Alexandre Bolze, Ary Ascencio, Charlotte Rivera-Garcia, Ryan Cho, Jason Nguyen, Sherry Wang, Jimmy Ramirez, Tyler Cassens, Efren Sandoval, Magnus Isaksson, William Lee, David Becker, Marc Laurent, James Lu, Clinton R. Paden, Suxiang Tong, Duncan MacCannell |
| EPI_ISL_1267599, EPI_ISL_1267600 | Microbiology Division, South Carolina Department of Health and Environmental Control Public Health Laboratory (SC DHEC PHL) | Microbiology Division, South Carolina Department of Health and Environmental Control Public Health Laboratory (SC DHEC PHL) | Haley Flores, Jessica Freeman |
| EPI_ISL_1267601 | Helix/Illumina | Centers for Disease Control and Prevention Division of Viral Diseases, Pathogen Discovery | Peter W. Cook, Dakota Howard, Dhwani Batra, Ben L. Rambo-Martin, Eileen de Feo, Jan Antico, Christine Tran, Matthew Tolentino, Shannon Wickline, Kim Gietzen, Brad Sickler, Jingtao Liu, Eric Allen, Phil Febbo, Summer Galloway, Nicole L. Washington, Simon White, Geraint Levan, Kelly Schiabor Barrett, Elizabeth Cirulli, Alexandre Bolze, Ary Ascencio, Charlotte Rivera-Garcia, Ryan Cho, Jason Nguyen, Sherry Wang, Jimmy Ramirez, Tyler Cassens, Efren Sandoval, Magnus Isaksson, William Lee, David Becker, Marc Laurent, James Lu, Clinton R. Paden, Suxiang Tong, Duncan MacCannell |
| EPI_ISL_1267602 | Microbiology Division, South Carolina Department of Health and Environmental Control Public Health Laboratory (SC DHEC PHL) | Microbiology Division, South Carolina Department of Health and Environmental Control Public Health Laboratory (SC DHEC PHL) | Haley Flores, Jessica Freeman |
| EPI_ISL_1267603, EPI_ISL_1267604, EPI_ISL_1267605, EPI_ISL_1267606, EPI_ISL_1267607, EPI_ISL_1267608, EPI_ISL_1267609, EPI_ISL_1267610, EPI_ISL_1267611, EPI_ISL_1267612, EPI_ISL_1267613, EPI_ISL_1267614, EPI_ISL_1267615, EPI_ISL_1267616, EPI_ISL_1267617, EPI_ISL_1267618, EPI_ISL_1267619, EPI_ISL_1267620, EPI_ISL_1267621, EPI_ISL_1267622, EPI_ISL_1267623, EPI_ISL_1267624, EPI_ISL_1267625, EPI_ISL_1267626, EPI_ISL_1267627, EPI_ISL_1267628, EPI_ISL_1267629, EPI_ISL_1267631, EPI_ISL_1267632, EPI_ISL_1267633, EPI_ISL_1267634, EPI_ISL_1267635, EPI_ISL_1267636, EPI_ISL_1267637, EPI_ISL_1267638, EPI_ISL_1267639, EPI_ISL_1267640, EPI_ISL_1267641, EPI_ISL_1267642, EPI_ISL_1267643, EPI_ISL_1267644, EPI_ISL_1267645, EPI_ISL_1267646, EPI_ISL_1267647, EPI_ISL_1267648, EPI_ISL_1267649, EPI_ISL_1267650, EPI_ISL_1267651, EPI_ISL_1267652, EPI_ISL_1267653, EPI_ISL_1267654, EPI_ISL_1267655, EPI_ISL_1267657, EPI_ISL_1267658, EPI_ISL_1267659, EPI_ISL_1267660, EPI_ISL_1267661, EPI_ISL_1267662, EPI_ISL_1267663, EPI_ISL_1267664, EPI_ISL_1267665, EPI_ISL_1267666, EPI_ISL_1267667, EPI_ISL_1267668, EPI_ISL_1267669, EPI_ISL_1267670, EPI_ISL_1267671, EPI_ISL_1267672, EPI_ISL_1267673, EPI_ISL_1267674, EPI_ISL_1267675, EPI_ISL_1267676, EPI_ISL_1267677, EPI_ISL_1267678, EPI_ISL_1267679, EPI_ISL_1267680, EPI_ISL_1267681, EPI_ISL_1267682, EPI_ISL_1267683, EPI_ISL_1267684, EPI_ISL_1267685, EPI_ISL_1267687, EPI_ISL_1267688, EPI_ISL_1267689, EPI_ISL_1267690, EPI_ISL_1267691, EPI_ISL_1267692, EPI_ISL_1267693, EPI_ISL_1267694, EPI_ISL_1267695, EPI_ISL_1267696, EPI_ISL_1267697, EPI_ISL_1267698, EPI_ISL_1267699, EPI_ISL_1267700, EPI_ISL_1267701, EPI_ISL_1267702, EPI_ISL_1267703, EPI_ISL_1267704, EPI_ISL_1267705, EPI_ISL_1267706, EPI_ISL_1267707, EPI_ISL_1267708, EPI_ISL_1267709, EPI_ISL_1267710, EPI_ISL_1267711, EPI_ISL_1267712, EPI_ISL_1267713, EPI_ISL_1267714, EPI_ISL_1267715, EPI_ISL_1267716, EPI_ISL_1267717, EPI_ISL_1267718, EPI_ISL_1267720, EPI_ISL_1267721, EPI_ISL_1267722, EPI_ISL_1267723, EPI_ISL_1267724, EPI_ISL_1267725, EPI_ISL_1267727, EPI_ISL_1267728, EPI_ISL_1267729, EPI_ISL_1267730, EPI_ISL_1267731, EPI_ISL_1267732, EPI_ISL_1267733, EPI_ISL_1267734, EPI_ISL_1267735, EPI_ISL_1267736, EPI_ISL_1267737, EPI_ISL_1267738, EPI_ISL_1267739, EPI_ISL_1267740, EPI_ISL_1267741, EPI_ISL_1267742, EPI_ISL_1267743, EPI_ISL_1267744, EPI_ISL_1267745, EPI_ISL_1267746, EPI_ISL_1267747, EPI_ISL_1267748, EPI_ISL_1267749, EPI_ISL_1267750, EPI_ISL_1267751, EPI_ISL_1267752, EPI_ISL_1267753, EPI_ISL_1267754, EPI_ISL_1267755, EPI_ISL_1267756, EPI_ISL_1267757, EPI_ISL_1267758, EPI_ISL_1267759, EPI_ISL_1267760, EPI_ISL_1267761, EPI_ISL_1267762, EPI_ISL_1267763, EPI_ISL_1267764, EPI_ISL_1267765, EPI_ISL_1267766, EPI_ISL_1267767, EPI_ISL_1267768, EPI_ISL_1267769, EPI_ISL_1267770, EPI_ISL_1267771, EPI_ISL_1267772, EPI_ISL_1267773, EPI_ISL_1267774, EPI_ISL_1267775, EPI_ISL_1267776, EPI_ISL_1267777, EPI_ISL_1267778, EPI_ISL_1267779, EPI_ISL_1267780, EPI_ISL_1267781, EPI_ISL_1267782, EPI_ISL_1267783, EPI_ISL_1267784, EPI_ISL_1267785, EPI_ISL_1267786, EPI_ISL_1267787, EPI_ISL_1267788, EPI_ISL_1267789, EPI_ISL_1267790, EPI_ISL_1267791, EPI_ISL_1267792, EPI_ISL_1267793, EPI_ISL_1267794, EPI_ISL_1267796, EPI_ISL_1267797, EPI_ISL_1267798, EPI_ISL_1267799, EPI_ISL_1267800, EPI_ISL_1267801, EPI_ISL_1267803, EPI_ISL_1267804, EPI_ISL_1267805, EPI_ISL_1267806, EPI_ISL_1267807, EPI_ISL_1267808, EPI_ISL_1267809, EPI_ISL_1267810, EPI_ISL_1267811, EPI_ISL_1267812, EPI_ISL_1267813, EPI_ISL_1267814, EPI_ISL_1267815, EPI_ISL_1267816, EPI_ISL_1267817, EPI_ISL_1267818, EPI_ISL_1267819, EPI_ISL_1267820, EPI_ISL_1267821, EPI_ISL_1267822, EPI_ISL_1267823, EPI_ISL_1267824, EPI_ISL_1267825, EPI_ISL_1267826, EPI_ISL_1267827, EPI_ISL_1267828, EPI_ISL_1267829, EPI_ISL_1267830, EPI_ISL_1267831, EPI_ISL_1267832, EPI_ISL_1267833, EPI_ISL_1267834, EPI_ISL_1267835, EPI_ISL_1267836, EPI_ISL_1267837, EPI_ISL_1267838, EPI_ISL_1267839, EPI_ISL_1267840, EPI_ISL_1267841, EPI_ISL_1267842, EPI_ISL_1267843, EPI_ISL_1267844, EPI_ISL_1267845, EPI_ISL_1267846, EPI_ISL_1267847, EPI_ISL_1267848, EPI_ISL_1267849, EPI_ISL_1267850, EPI_ISL_1267851, EPI_ISL_1267852, EPI_ISL_1267853, EPI_ISL_1267854, EPI_ISL_1267855, EPI_ISL_1267856, EPI_ISL_1267857, EPI_ISL_1267858, EPI_ISL_1267859, EPI_ISL_1267860, EPI_ISL_1267861, EPI_ISL_1267862, EPI_ISL_1267863, EPI_ISL_1267864, EPI_ISL_1267865, EPI_ISL_1267866, EPI_ISL_1267867, EPI_ISL_1267868, EPI_ISL_1267869, EPI_ISL_1267870, EPI_ISL_1267871, EPI_ISL_1267872, EPI_ISL_1267873, EPI_ISL_1267874, EPI_ISL_1267875, EPI_ISL_1267876, EPI_ISL_1267877, EPI_ISL_1267878, EPI_ISL_1267879, EPI_ISL_1267880, EPI_ISL_1267881, EPI_ISL_1267882, EPI_ISL_1267883, EPI_ISL_1267884, EPI_ISL_1267885, EPI_ISL_1267886, EPI_ISL_1267887, EPI_ISL_1267888, EPI_ISL_1267889, EPI_ISL_1267890, EPI_ISL_1267891, EPI_ISL_1267892, EPI_ISL_1267893, EPI_ISL_1267894, EPI_ISL_1267895, EPI_ISL_1267896, EPI_ISL_1267897, EPI_ISL_1267898, EPI_ISL_1267899, EPI_ISL_1267900, EPI_ISL_1267901, EPI_ISL_1267902, EPI_ISL_1267903, EPI_ISL_1267904, EPI_ISL_1267905, EPI_ISL_1267906, EPI_ISL_1267907, EPI_ISL_1267908, EPI_ISL_1267909, EPI_ISL_1267910, EPI_ISL_1267911, EPI_ISL_1267912, EPI_ISL_1267913, EPI_ISL_1267914, EPI_ISL_1267915, EPI_ISL_1267916, EPI_ISL_1267917, EPI_ISL_1267918, EPI_ISL_1267919, EPI_ISL_1267920, EPI_ISL_1267921, EPI_ISL_1267922, EPI_ISL_1267923, EPI_ISL_1267924, EPI_ISL_1267925, EPI_ISL_1267926, EPI_ISL_1267927, EPI_ISL_1267928, EPI_ISL_1267929, EPI_ISL_1267930, EPI_ISL_1267931, EPI_ISL_1267932, EPI_ISL_1267933, EPI_ISL_1267934, EPI_ISL_1267935, EPI_ISL_1267936, EPI_ISL_1267937, EPI_ISL_1267938, EPI_ISL_1267939, EPI_ISL_1267940, EPI_ISL_1267941, EPI_ISL_1267942, EPI_ISL_1267943, EPI_ISL_1267944, EPI_ISL_1267945, EPI_ISL_1267946, EPI_ISL_1267947, EPI_ISL_1267948, EPI_ISL_1267949, EPI_ISL_1267950, EPI_ISL_1267951, EPI_ISL_1267952, EPI_ISL_1267953, EPI_ISL_1267954, EPI_ISL_1267955, EPI_ISL_1267956, EPI_ISL_1267957, EPI_ISL_1267958, EPI_ISL_1267959, EPI_ISL_1267960, EPI_ISL_1267961, EPI_ISL_1267962, EPI_ISL_1267963, EPI_ISL_1267964, EPI_ISL_1267965, EPI_ISL_1267966, EPI_ISL_1267967, EPI_ISL_1267968, EPI_ISL_1267969, EPI_ISL_1267970, EPI_ISL_1267971, EPI_ISL_1267972, EPI_ISL_1267973, EPI_ISL_1267974, EPI_ISL_1267975, EPI_ISL_1267976, EPI_ISL_1267977, EPI_ISL_1267978, EPI_ISL_1267979, EPI_ISL_1267980, EPI_ISL_1267981, EPI_ISL_1267982, EPI_ISL_1267983, EPI_ISL_1267984, EPI_ISL_1267985, EPI_ISL_1267986, EPI_ISL_1267987, EPI_ISL_1267988, EPI_ISL_1267989, EPI_ISL_1267990, EPI_ISL_1267991, EPI_ISL_1267992, EPI_ISL_1267993, EPI_ISL_1267994, EPI_ISL_1267995, EPI_ISL_1267996, EPI_ISL_1267997, EPI_ISL_1267998, EPI_ISL_1267999, EPI_ISL_1268000, EPI_ISL_1268001, EPI_ISL_1268002, EPI_ISL_1268003, EPI_ISL_1268004, EPI_ISL_1268005, EPI_ISL_1268006, EPI_ISL_1268007, EPI_ISL_1268008, EPI_ISL_1268009, EPI_ISL_1268010, EPI_ISL_1268011, EPI_ISL_1268012, EPI_ISL_1268013, EPI_ISL_1268014, EPI_ISL_1268015, EPI_ISL_1268016, EPI_ISL_1268017, EPI_ISL_1268018, EPI_ISL_1268019, EPI_ISL_1268020, EPI_ISL_1268021, EPI_ISL_1268022, EPI_ISL_1268023, EPI_ISL_1268024, EPI_ISL_1268025, EPI_ISL_1268026, EPI_ISL_1268027, EPI_ISL_1268028, EPI_ISL_1268029, EPI_ISL_1268030, EPI_ISL_1268031, EPI_ISL_1268032, EPI_ISL_1268033, EPI_ISL_1268034, EPI_ISL_1268035, EPI_ISL_1268036, EPI_ISL_1268037, EPI_ISL_1268038, EPI_ISL_1268039, EPI_ISL_1268040, EPI_ISL_1268041, EPI_ISL_1268042, EPI_ISL_1268043, EPI_ISL_1268044, EPI_ISL_1268045, EPI_ISL_1268046, EPI_ISL_1268047, EPI_ISL_1268048, EPI_ISL_1268049, EPI_ISL_1268050, EPI_ISL_1268051, EPI_ISL_1268052, EPI_ISL_1268053, EPI_ISL_1268054, EPI_ISL_1268055, EPI_ISL_1268056, EPI_ISL_1268057, EPI_ISL_1268058, EPI_ISL_1268059, EPI_ISL_1268060, EPI_ISL_1268061, EPI_ISL_1268063, EPI_ISL_1268064, EPI_ISL_1268065, EPI_ISL_1268066, EPI_ISL_1268067, EPI_ISL_1268068, EPI_ISL_1268069, EPI_ISL_1268070, EPI_ISL_1268071, EPI_ISL_1268072, EPI_ISL_1268073, EPI_ISL_1268074, EPI_ISL_1268075, EPI_ISL_1268076, EPI_ISL_1268077, EPI_ISL_1268078, EPI_ISL_1268079, EPI_ISL_1268080, EPI_ISL_1268081, EPI_ISL_1268082, EPI_ISL_1268083, EPI_ISL_1268084, EPI_ISL_1268086, EPI_ISL_1268087, EPI_ISL_1268088, EPI_ISL_1268089, EPI_ISL_1268090, EPI_ISL_1268091, EPI_ISL_1268092, EPI_ISL_1268093, EPI_ISL_1268094, EPI_ISL_1268095, EPI_ISL_1268096, EPI_ISL_1268097, EPI_ISL_1268098, EPI_ISL_1268099, EPI_ISL_1268100, EPI_ISL_1268101, EPI_ISL_1268102, EPI_ISL_1268103, EPI_ISL_1268104, EPI_ISL_1268105, EPI_ISL_1268106, EPI_ISL_1268107, EPI_ISL_1268108, EPI_ISL_1268109, EPI_ISL_1268110, EPI_ISL_1268111, EPI_ISL_1268112, EPI_ISL_1268113, EPI_ISL_1268114, EPI_ISL_1268115, EPI_ISL_1268116, EPI_ISL_1268117, | | | |

EPI_ISL_1268118, EPI_ISL_1268119, EPI_ISL_1268120, EPI_ISL_1268121, EPI_ISL_1268122, EPI_ISL_1268123, EPI_ISL_1268124, EPI_ISL_1268125, EPI_ISL_1268126, EPI_ISL_1268127, EPI_ISL_1268128, EPI_ISL_1268129, EPI_ISL_1268130, EPI_ISL_1268131, EPI_ISL_1268132, EPI_ISL_1268133, EPI_ISL_1268134, EPI_ISL_1268135, EPI_ISL_1268136, EPI_ISL_1268137, EPI_ISL_1268138, EPI_ISL_1268139, EPI_ISL_1268140, EPI_ISL_1268141, EPI_ISL_1268142, EPI_ISL_1268143, EPI_ISL_1268144, EPI_ISL_1268145, EPI_ISL_1268146, EPI_ISL_1268147, EPI_ISL_1268148, EPI_ISL_1268149, EPI_ISL_1268150, EPI_ISL_1268151, EPI_ISL_1268152, EPI_ISL_1268153, EPI_ISL_1268154, EPI_ISL_1268155, EPI_ISL_1268156, EPI_ISL_1268157, EPI_ISL_1268158, EPI_ISL_1268159, EPI_ISL_1268160, EPI_ISL_1268161, EPI_ISL_1268162, EPI_ISL_1268163, EPI_ISL_1268164, EPI_ISL_1268165, EPI_ISL_1268166, EPI_ISL_1268167, EPI_ISL_1268168, EPI_ISL_1268169, EPI_ISL_1268170, EPI_ISL_1268171, EPI_ISL_1268172, EPI_ISL_1268173, EPI_ISL_1268174, EPI_ISL_1268175, EPI_ISL_1268176, EPI_ISL_1268177, EPI_ISL_1268178, EPI_ISL_1268179, EPI_ISL_1268180, EPI_ISL_1268181, EPI_ISL_1268182, EPI_ISL_1268183, EPI_ISL_1268184, EPI_ISL_1268185, EPI_ISL_1268186, EPI_ISL_1268187, EPI_ISL_1268188, EPI_ISL_1268189, EPI_ISL_1268190, EPI_ISL_1268191, EPI_ISL_1268192, EPI_ISL_1268193

| | | | |
|---|---|---|---|
| see above | Helix/Illumina | Centers for Disease Control and Prevention Division of Viral Diseases, Pathogen Discovery | Peter W. Cook, Dakota Howard, Dhwani Batra, Ben L. Rambo-Martin, Eileen de Feo, Jan Antico, Christine Tran, Matthew Tolentino, Shannon Wickline, Kim Gietzen, Brad Sickler, Jingtao Liu, Eric Allen, Phil Febbo, Summer Galloway, Nicole L. Washington, Simon White, Geraint Levan, Kelly Schiabor Barrett, Elizabeth Cirulli, Alexandre Bolze, Ary Ascencio, Charlotte Rivera-Garcia, Ryan Cho, Jason Nguyen, Sherry Wang, Jimmy Ramirez, Tyler Cassens, Efren Sandoval, Magnus Isaksson, William Lee, David Becker, Marc Laurent, James Lu, Clinton R. Paden, Suxiang Tong, Duncan MacCannell |
| EPI_ISL_1268194, EPI_ISL_1268195 | DPHL | Delaware Public Health Lab | Rebecca Savage |
| EPI_ISL_1268196, EPI_ISL_1268197 | Helix/Illumina | Centers for Disease Control and Prevention Division of Viral Diseases, Pathogen Discovery | Peter W. Cook, Dakota Howard, Dhwani Batra, Ben L. Rambo-Martin, Eileen de Feo, Jan Antico, Christine Tran, Matthew Tolentino, Shannon Wickline, Kim Gietzen, Brad Sickler, Jingtao Liu, Eric Allen, Phil Febbo, Summer Galloway, Nicole L. Washington, Simon White, Geraint Levan, Kelly Schiabor Barrett, Elizabeth Cirulli, Alexandre Bolze, Ary Ascencio, Charlotte Rivera-Garcia, Ryan Cho, Jason Nguyen, Sherry Wang, Jimmy Ramirez, Tyler Cassens, Efren Sandoval, Magnus Isaksson, William Lee, David Becker, Marc Laurent, James Lu, Clinton R. Paden, Suxiang Tong, Duncan MacCannell |
| EPI_ISL_1268198 | DPHL | Delaware Public Health Lab | Rebecca Savage |
| EPI_ISL_1268199 | Helix/Illumina | Centers for Disease Control and Prevention Division of Viral Diseases, Pathogen Discovery | Peter W. Cook, Dakota Howard, Dhwani Batra, Ben L. Rambo-Martin, Eileen de Feo, Jan Antico, Christine Tran, Matthew Tolentino, Shannon Wickline, Kim Gietzen, Brad Sickler, Jingtao Liu, Eric Allen, Phil Febbo, Summer Galloway, Nicole L. Washington, Simon White, Geraint Levan, Kelly Schiabor Barrett, Elizabeth Cirulli, Alexandre Bolze, Ary Ascencio, Charlotte Rivera-Garcia, Ryan Cho, Jason Nguyen, Sherry Wang, Jimmy Ramirez, Tyler Cassens, Efren Sandoval, Magnus Isaksson, William Lee, David Becker, Marc Laurent, James Lu, Clinton R. Paden, Suxiang Tong, Duncan MacCannell |
| EPI_ISL_1268200 | DPHL | Delaware Public Health Lab | Rebecca Savage |
| EPI_ISL_1268201 | Helix/Illumina | Centers for Disease Control and Prevention Division of Viral Diseases, Pathogen Discovery | Peter W. Cook, Dakota Howard, Dhwani Batra, Ben L. Rambo-Martin, Eileen de Feo, Jan Antico, Christine Tran, Matthew Tolentino, Shannon Wickline, Kim Gietzen, Brad Sickler, Jingtao Liu, Eric Allen, Phil Febbo, Summer Galloway, Nicole L. Washington, Simon White, Geraint Levan, Kelly Schiabor Barrett, Elizabeth Cirulli, Alexandre Bolze, Ary Ascencio, Charlotte Rivera-Garcia, Ryan Cho, Jason Nguyen, Sherry Wang, Jimmy Ramirez, Tyler Cassens, Efren Sandoval, Magnus Isaksson, William Lee, David Becker, Marc Laurent, James Lu, Clinton R. Paden, Suxiang Tong, Duncan MacCannell |
| EPI_ISL_1268202 | DPHL | Delaware Public Health Lab | Rebecca Savage |
| EPI_ISL_1268203 | Helix/Illumina | Centers for Disease Control and Prevention Division of Viral Diseases, Pathogen Discovery | Peter W. Cook, Dakota Howard, Dhwani Batra, Ben L. Rambo-Martin, Eileen de Feo, Jan Antico, Christine Tran, Matthew Tolentino, Shannon Wickline, Kim Gietzen, Brad Sickler, Jingtao Liu, Eric Allen, Phil Febbo, Summer Galloway, Nicole L. Washington, Simon White, Geraint Levan, Kelly Schiabor Barrett, Elizabeth Cirulli, Alexandre Bolze, Ary Ascencio, Charlotte Rivera-Garcia, Ryan Cho, Jason Nguyen, Sherry Wang, Jimmy Ramirez, Tyler Cassens, Efren Sandoval, Magnus Isaksson, William Lee, David Becker, Marc Laurent, James Lu, Clinton R. Paden, Suxiang Tong, Duncan MacCannell |
| EPI_ISL_1268204, EPI_ISL_1268205 | DPHL | Delaware Public Health Lab | Rebecca Savage |
| EPI_ISL_1268206 | Helix/Illumina | Centers for Disease Control and Prevention Division of Viral Diseases, Pathogen Discovery | Peter W. Cook, Dakota Howard, Dhwani Batra, Ben L. Rambo-Martin, Eileen de Feo, Jan Antico, Christine Tran, Matthew Tolentino, Shannon Wickline, Kim Gietzen, Brad Sickler, Jingtao Liu, Eric Allen, Phil Febbo, Summer Galloway, Nicole L. Washington, Simon White, Geraint Levan, Kelly Schiabor Barrett, Elizabeth Cirulli, Alexandre Bolze, Ary Ascencio, Charlotte Rivera-Garcia, Ryan Cho, Jason Nguyen, Sherry Wang, Jimmy Ramirez, Tyler Cassens, Efren Sandoval, Magnus Isaksson, William Lee, David Becker, Marc Laurent, James Lu, Clinton R. Paden, Suxiang Tong, Duncan MacCannell |
| EPI_ISL_1268207 | DPHL | Delaware Public Health Lab | Rebecca Savage |
| EPI_ISL_1268208 | Helix/Illumina | Centers for Disease Control and Prevention Division of Viral Diseases, Pathogen Discovery | Peter W. Cook, Dakota Howard, Dhwani Batra, Ben L. Rambo-Martin, Eileen de Feo, Jan Antico, Christine Tran, Matthew Tolentino, Shannon Wickline, Kim Gietzen, Brad Sickler, Jingtao Liu, Eric Allen, Phil Febbo, Summer Galloway, Nicole L. Washington, Simon White, Geraint Levan, Kelly Schiabor Barrett, Elizabeth Cirulli, Alexandre Bolze, Ary Ascencio, Charlotte Rivera-Garcia, Ryan Cho, Jason Nguyen, Sherry Wang, Jimmy Ramirez, Tyler Cassens, Efren Sandoval, Magnus Isaksson, William Lee, David Becker, Marc Laurent, James Lu, Clinton R. Paden, Suxiang Tong, Duncan MacCannell |
| EPI_ISL_1268209 | DPHL | Delaware Public Health Lab | Rebecca Savage |
| EPI_ISL_1268210 | Helix/Illumina | Centers for Disease Control and Prevention Division of Viral Diseases, Pathogen Discovery | Peter W. Cook, Dakota Howard, Dhwani Batra, Ben L. Rambo-Martin, Eileen de Feo, Jan Antico, Christine Tran, Matthew Tolentino, Shannon Wickline, Kim Gietzen, Brad Sickler, Jingtao Liu, Eric Allen, Phil Febbo, Summer Galloway, Nicole L. Washington, Simon White, Geraint Levan, Kelly Schiabor Barrett, Elizabeth Cirulli, Alexandre Bolze, Ary Ascencio, Charlotte Rivera-Garcia, Ryan Cho, Jason Nguyen, Sherry Wang, Jimmy Ramirez, Tyler Cassens, Efren Sandoval, Magnus Isaksson, William Lee, David Becker, Marc Laurent, James Lu, Clinton R. Paden, Suxiang Tong, Duncan MacCannell |
| EPI_ISL_1268211 | DPHL | Delaware Public Health Lab | Rebecca Savage |
| EPI_ISL_1268212 | Helix/Illumina | Centers for Disease Control and Prevention Division of Viral Diseases, Pathogen Discovery | Peter W. Cook, Dakota Howard, Dhwani Batra, Ben L. Rambo-Martin, Eileen de Feo, Jan Antico, Christine Tran, Matthew Tolentino, Shannon Wickline, Kim Gietzen, Brad Sickler, Jingtao Liu, Eric Allen, Phil Febbo, Summer Galloway, Nicole L. Washington, Simon White, Geraint Levan, Kelly Schiabor Barrett, Elizabeth Cirulli, Alexandre Bolze, Ary Ascencio, Charlotte Rivera-Garcia, Ryan Cho, Jason Nguyen, Sherry Wang, Jimmy Ramirez, Tyler Cassens, Efren Sandoval, Magnus Isaksson, William Lee, David Becker, Marc Laurent, James Lu, Clinton R. Paden, Suxiang Tong, Duncan MacCannell |
| EPI_ISL_1268213 | DPHL | Delaware Public Health Lab | Rebecca Savage |
| EPI_ISL_1268214, EPI_ISL_1268216 | Helix/Illumina | Centers for Disease Control and Prevention Division of Viral Diseases, Pathogen Discovery | Peter W. Cook, Dakota Howard, Dhwani Batra, Ben L. Rambo-Martin, Eileen de Feo, Jan Antico, Christine Tran, Matthew Tolentino, Shannon Wickline, Kim Gietzen, Brad Sickler, Jingtao Liu, Eric Allen, Phil Febbo, Summer Galloway, Nicole L. Washington, Simon White, Geraint Levan, Kelly Schiabor Barrett, Elizabeth Cirulli, Alexandre Bolze, Ary Ascencio, Charlotte Rivera-Garcia, Ryan Cho, Jason Nguyen, Sherry Wang, Jimmy Ramirez, Tyler Cassens, Efren Sandoval, Magnus Isaksson, William Lee, David Becker, Marc Laurent, James Lu, Clinton R. Paden, Suxiang Tong, Duncan MacCannell |
| EPI_ISL_1268217 | DPHL | Delaware Public Health Lab | Rebecca Savage |
| EPI_ISL_1268218 | Helix/Illumina | Centers for Disease Control and Prevention Division of Viral Diseases, Pathogen Discovery | Peter W. Cook, Dakota Howard, Dhwani Batra, Ben L. Rambo-Martin, Eileen de Feo, Jan Antico, Christine Tran, Matthew Tolentino, Shannon Wickline, Kim Gietzen, Brad Sickler, Jingtao Liu, Eric Allen, Phil Febbo, Summer Galloway, Nicole L. Washington, Simon White, Geraint Levan, Kelly Schiabor Barrett, Elizabeth Cirulli, Alexandre Bolze, Ary Ascencio, Charlotte Rivera-Garcia, Ryan Cho, Jason Nguyen, Sherry Wang, Jimmy Ramirez, Tyler Cassens, Efren Sandoval, Magnus Isaksson, William Lee, David Becker, Marc Laurent, James Lu, Clinton R. Paden, Suxiang Tong, Duncan MacCannell |
| EPI_ISL_1268219 | DPHL | Delaware Public Health Lab | Rebecca Savage |
| EPI_ISL_1268220 | Helix/Illumina | Centers for Disease Control and Prevention Division of Viral Diseases, Pathogen Discovery | Peter W. Cook, Dakota Howard, Dhwani Batra, Ben L. Rambo-Martin, Eileen de Feo, Jan Antico, Christine Tran, Matthew Tolentino, Shannon Wickline, Kim Gietzen, Brad Sickler, Jingtao Liu, Eric Allen, Phil Febbo, Summer Galloway, Nicole L. Washington, Simon White, Geraint Levan, Kelly Schiabor Barrett, Elizabeth Cirulli, Alexandre Bolze, Ary Ascencio, Charlotte Rivera-Garcia, Ryan Cho, Jason Nguyen, Sherry Wang, Jimmy Ramirez, Tyler Cassens, Efren Sandoval, Magnus Isaksson, William Lee, David Becker, Marc Laurent, James Lu, Clinton R. Paden, Suxiang Tong, Duncan MacCannell |
| EPI_ISL_1268221 | DPHL | Delaware Public Health Lab | Rebecca Savage |
| EPI_ISL_1268222 | Helix/Illumina | Centers for Disease Control and Prevention Division of Viral Diseases, Pathogen Discovery | Peter W. Cook, Dakota Howard, Dhwani Batra, Ben L. Rambo-Martin, Eileen de Feo, Jan Antico, Christine Tran, Matthew Tolentino, Shannon Wickline, Kim Gietzen, Brad Sickler, Jingtao Liu, Eric Allen, Phil Febbo, Summer Galloway, Nicole L. Washington, Simon White, Geraint Levan, Kelly Schiabor Barrett, Elizabeth Cirulli, Alexandre Bolze, Ary Ascencio, Charlotte Rivera-Garcia, Ryan Cho, Jason Nguyen, Sherry Wang, Jimmy Ramirez, Tyler Cassens, Efren Sandoval, Magnus Isaksson, William Lee, David Becker, Marc Laurent, James Lu, Clinton R. Paden, Suxiang Tong, Duncan MacCannell |
| EPI_ISL_1268223, EPI_ISL_1268224 | DPHL | Delaware Public Health Lab | Rebecca Savage |
| EPI_ISL_1268225 | Helix/Illumina | Centers for Disease Control and Prevention Division of Viral Diseases, Pathogen Discovery | Peter W. Cook, Dakota Howard, Dhwani Batra, Ben L. Rambo-Martin, Eileen de Feo, Jan Antico, Christine Tran, Matthew Tolentino, Shannon Wickline, Kim Gietzen, Brad Sickler, Jingtao Liu, Eric Allen, Phil Febbo, Summer Galloway, Nicole L. Washington, Simon White, Geraint Levan, Kelly Schiabor Barrett, Elizabeth Cirulli, Alexandre Bolze, Ary Ascencio, Charlotte Rivera-Garcia, Ryan Cho, Jason Nguyen, Sherry Wang, Jimmy Ramirez, Tyler Cassens, Efren Sandoval, Magnus Isaksson, William Lee, David Becker, Marc Laurent, James Lu, Clinton R. Paden, Suxiang Tong, Duncan MacCannell |
| EPI_ISL_1268226 | DPHL | Delaware Public Health Lab | Rebecca Savage |
| EPI_ISL_1268227 | Helix/Illumina | Centers for Disease Control and Prevention Division of Viral Diseases, Pathogen Discovery | Peter W. Cook, Dakota Howard, Dhwani Batra, Ben L. Rambo-Martin, Eileen de Feo, Jan Antico, Christine Tran, Matthew Tolentino, Shannon Wickline, Kim Gietzen, Brad Sickler, Jingtao Liu, Eric Allen, Phil Febbo, Summer Galloway, Nicole L. Washington, Simon White, Geraint Levan, Kelly Schiabor Barrett, Elizabeth Cirulli, Alexandre Bolze, Ary Ascencio, Charlotte Rivera-Garcia, Ryan Cho, Jason Nguyen, Sherry Wang, Jimmy Ramirez, Tyler Cassens, Efren Sandoval, Magnus Isaksson, William Lee, David Becker, Marc Laurent, James Lu, Clinton R. Paden, Suxiang Tong, Duncan MacCannell |
| EPI_ISL_1268228 | DPHL | Delaware Public Health Lab | Rebecca Savage |
| EPI_ISL_1268229 | Helix/Illumina | Centers for Disease Control and Prevention Division of Viral Diseases, Pathogen Discovery | Peter W. Cook, Dakota Howard, Dhwani Batra, Ben L. Rambo-Martin, Eileen de Feo, Jan Antico, Christine Tran, Matthew Tolentino, Shannon Wickline, Kim Gietzen, Brad Sickler, Jingtao Liu, Eric Allen, Phil Febbo, Summer Galloway, Nicole L. Washington, Simon White, Geraint Levan, Kelly Schiabor Barrett, Elizabeth Cirulli, Alexandre Bolze, Ary Ascencio, Charlotte Rivera-Garcia, Ryan Cho, Jason Nguyen, Sherry Wang, Jimmy Ramirez, Tyler Cassens, Efren Sandoval, Magnus Isaksson, William Lee, David Becker, Marc Laurent, James Lu, Clinton R. Paden, Suxiang Tong, Duncan MacCannell |
| EPI_ISL_1268230 | DPHL | Delaware Public Health Lab | Rebecca Savage |
| EPI_ISL_1268231 | Helix/Illumina | Centers for Disease Control and Prevention Division of Viral Diseases, Pathogen Discovery | Peter W. Cook, Dakota Howard, Dhwani Batra, Ben L. Rambo-Martin, Eileen de Feo, Jan Antico, Christine Tran, Matthew Tolentino, Shannon Wickline, Kim Gietzen, Brad Sickler, Jingtao Liu, Eric Allen, Phil Febbo, Summer Galloway, Nicole L. Washington, Simon White, Geraint Levan, Kelly Schiabor Barrett, Elizabeth Cirulli, Alexandre Bolze, Ary Ascencio, Charlotte Rivera-Garcia, Ryan Cho, Jason Nguyen, Sherry Wang, Jimmy Ramirez, Tyler Cassens, Efren Sandoval, Magnus Isaksson, William Lee, David Becker, Marc Laurent, James Lu, Clinton R. Paden, Suxiang Tong, Duncan MacCannell |
| EPI_ISL_1268232 | DPHL | Delaware Public Health Lab | Rebecca Savage |
| EPI_ISL_1268233 | Helix/Illumina | Centers for Disease Control and Prevention Division of Viral Diseases, Pathogen Discovery | Peter W. Cook, Dakota Howard, Dhwani Batra, Ben L. Rambo-Martin, Eileen de Feo, Jan Antico, Christine Tran, Matthew Tolentino, Shannon Wickline, Kim Gietzen, Brad Sickler, Jingtao Liu, Eric Allen, Phil Febbo, Summer Galloway, Nicole L. Washington, Simon White, Geraint Levan, Kelly Schiabor Barrett, Elizabeth Cirulli, Alexandre Bolze, Ary Ascencio, Charlotte Rivera-Garcia, Ryan Cho, Jason Nguyen, Sherry Wang, Jimmy Ramirez, Tyler Cassens, Efren Sandoval, Magnus Isaksson, William Lee, David Becker, Marc Laurent, James Lu, Clinton R. Paden, Suxiang Tong, Duncan MacCannell |
| EPI_ISL_1268234 | DPHL | Delaware Public Health Lab | Rebecca Savage |
| EPI_ISL_1268235 | Helix/Illumina | Centers for Disease Control and Prevention Division of Viral Diseases, Pathogen Discovery | Peter W. Cook, Dakota Howard, Dhwani Batra, Ben L. Rambo-Martin, Eileen de Feo, Jan Antico, Christine Tran, Matthew Tolentino, Shannon Wickline, Kim Gietzen, Brad Sickler, Jingtao Liu, Eric Allen, Phil Febbo, Summer Galloway, Nicole L. Washington, Simon White, Geraint Levan, Kelly Schiabor Barrett, Elizabeth Cirulli, Alexandre Bolze, Ary Ascencio, Charlotte Rivera-Garcia, Ryan Cho, Jason Nguyen, Sherry Wang, Jimmy Ramirez, Tyler Cassens, Efren Sandoval, Magnus Isaksson, William Lee, David Becker, Marc Laurent, James Lu, Clinton R. Paden, Suxiang Tong, Duncan MacCannell |
| EPI_ISL_1268236 | DPHL | Delaware Public Health Lab | Rebecca Savage |
| EPI_ISL_1268237 | Helix/Illumina | Centers for Disease Control and Prevention Division of Viral Diseases, Pathogen Discovery | Peter W. Cook, Dakota Howard, Dhwani Batra, Ben L. Rambo-Martin, Eileen de Feo, Jan Antico, Christine Tran, Matthew Tolentino, Shannon Wickline, Kim Gietzen, Brad Sickler, Jingtao Liu, Eric Allen, Phil Febbo, Summer Galloway, Nicole L. Washington, Simon White, Geraint Levan, Kelly Schiabor Barrett, Elizabeth Cirulli, Alexandre Bolze, Ary Ascencio, Charlotte Rivera-Garcia, Ryan Cho, Jason Nguyen, Sherry Wang, Jimmy Ramirez, Tyler Cassens, Efren Sandoval, Magnus Isaksson, William Lee, David Becker, Marc Laurent, James Lu, Clinton R. Paden, Suxiang Tong, Duncan MacCannell |
| EPI_ISL_1268238 | DPHL | Delaware Public Health Lab | Rebecca Savage |
| EPI_ISL_1268239 | Helix/Illumina | Centers for Disease Control and Prevention Division of Viral Diseases, Pathogen Discovery | Peter W. Cook, Dakota Howard, Dhwani Batra, Ben L. Rambo-Martin, Eileen de Feo, Jan Antico, Christine Tran, Matthew Tolentino, Shannon Wickline, Kim Gietzen, Brad Sickler, Jingtao Liu, Eric Allen, Phil Febbo, Summer Galloway, Nicole L. Washington, Simon White, Geraint Levan, Kelly Schiabor Barrett, Elizabeth Cirulli, Alexandre Bolze, Ary Ascencio, Charlotte Rivera-Garcia, Ryan Cho, Jason Nguyen, Sherry Wang, Jimmy Ramirez, Tyler Cassens, Efren Sandoval, Magnus Isaksson, William Lee, David Becker, Marc Laurent, James Lu, Clinton R. Paden, Suxiang Tong, Duncan MacCannell |
| EPI_ISL_1268240 | DPHL | Delaware Public Health Lab | Rebecca Savage |

| | | | |
|---|---|---|---|
| EPI_ISL_1268241, EPI_ISL_1268242 | Helix/Illumina | Centers for Disease Control and Prevention Division of Viral Diseases, Pathogen Discovery | Peter W. Cook, Dakota Howard, Dhwani Batra, Ben L. Rambo-Martin, Eileen de Feo, Jan Antico, Christine Tran, Matthew Tolentino, Shannon Wickline, Kim Gietzen, Brad Sickler, Jingtao Liu, Eric Allen, Phil Febbo, Summer Galloway, Nicole L. Washington, Simon White, Geraint Levan, Kelly Schiabor Barrett, Elizabeth Cirulli, Alexandre Bolze, Ary Ascencio, Charlotte Rivera-Garcia, Ryan Cho, Jason Nguyen, Sherry Wang, Jimmy Ramirez, Tyler Cassens, Efren Sandoval, Magnus Isaksson, William Lee, David Becker, Marc Laurent, James Lu, Clinton R. Paden, Suxiang Tong, Duncan MacCannell |
| EPI_ISL_1268243, EPI_ISL_1268244, EPI_ISL_1268246 | DPHL | Delaware Public Health Lab | Rebecca Savage |
| EPI_ISL_1268247 | Helix/Illumina | Centers for Disease Control and Prevention Division of Viral Diseases, Pathogen Discovery | Peter W. Cook, Dakota Howard, Dhwani Batra, Ben L. Rambo-Martin, Eileen de Feo, Jan Antico, Christine Tran, Matthew Tolentino, Shannon Wickline, Kim Gietzen, Brad Sickler, Jingtao Liu, Eric Allen, Phil Febbo, Summer Galloway, Nicole L. Washington, Simon White, Geraint Levan, Kelly Schiabor Barrett, Elizabeth Cirulli, Alexandre Bolze, Ary Ascencio, Charlotte Rivera-Garcia, Ryan Cho, Jason Nguyen, Sherry Wang, Jimmy Ramirez, Tyler Cassens, Efren Sandoval, Magnus Isaksson, William Lee, David Becker, Marc Laurent, James Lu, Clinton R. Paden, Suxiang Tong, Duncan MacCannell |
| EPI_ISL_1268248 | DPHL | Delaware Public Health Lab | Rebecca Savage |
| EPI_ISL_1268249 | Helix/Illumina | Centers for Disease Control and Prevention Division of Viral Diseases, Pathogen Discovery | Peter W. Cook, Dakota Howard, Dhwani Batra, Ben L. Rambo-Martin, Eileen de Feo, Jan Antico, Christine Tran, Matthew Tolentino, Shannon Wickline, Kim Gietzen, Brad Sickler, Jingtao Liu, Eric Allen, Phil Febbo, Summer Galloway, Nicole L. Washington, Simon White, Geraint Levan, Kelly Schiabor Barrett, Elizabeth Cirulli, Alexandre Bolze, Ary Ascencio, Charlotte Rivera-Garcia, Ryan Cho, Jason Nguyen, Sherry Wang, Jimmy Ramirez, Tyler Cassens, Efren Sandoval, Magnus Isaksson, William Lee, David Becker, Marc Laurent, James Lu, Clinton R. Paden, Suxiang Tong, Duncan MacCannell |
| EPI_ISL_1268250 | DPHL | Delaware Public Health Lab | Rebecca Savage |
| EPI_ISL_1268251, EPI_ISL_1268252 | Helix/Illumina | Centers for Disease Control and Prevention Division of Viral Diseases, Pathogen Discovery | Peter W. Cook, Dakota Howard, Dhwani Batra, Ben L. Rambo-Martin, Eileen de Feo, Jan Antico, Christine Tran, Matthew Tolentino, Shannon Wickline, Kim Gietzen, Brad Sickler, Jingtao Liu, Eric Allen, Phil Febbo, Summer Galloway, Nicole L. Washington, Simon White, Geraint Levan, Kelly Schiabor Barrett, Elizabeth Cirulli, Alexandre Bolze, Ary Ascencio, Charlotte Rivera-Garcia, Ryan Cho, Jason Nguyen, Sherry Wang, Jimmy Ramirez, Tyler Cassens, Efren Sandoval, Magnus Isaksson, William Lee, David Becker, Marc Laurent, James Lu, Clinton R. Paden, Suxiang Tong, Duncan MacCannell |
| EPI_ISL_1268253 | DPHL | Delaware Public Health Lab | Rebecca Savage |
| EPI_ISL_1268254 | Helix/Illumina | Centers for Disease Control and Prevention Division of Viral Diseases, Pathogen Discovery | Peter W. Cook, Dakota Howard, Dhwani Batra, Ben L. Rambo-Martin, Eileen de Feo, Jan Antico, Christine Tran, Matthew Tolentino, Shannon Wickline, Kim Gietzen, Brad Sickler, Jingtao Liu, Eric Allen, Phil Febbo, Summer Galloway, Nicole L. Washington, Simon White, Geraint Levan, Kelly Schiabor Barrett, Elizabeth Cirulli, Alexandre Bolze, Ary Ascencio, Charlotte Rivera-Garcia, Ryan Cho, Jason Nguyen, Sherry Wang, Jimmy Ramirez, Tyler Cassens, Efren Sandoval, Magnus Isaksson, William Lee, David Becker, Marc Laurent, James Lu, Clinton R. Paden, Suxiang Tong, Duncan MacCannell |
| EPI_ISL_1268255 | DPHL | Delaware Public Health Lab | Rebecca Savage |
| EPI_ISL_1268256 | Helix/Illumina | Centers for Disease Control and Prevention Division of Viral Diseases, Pathogen Discovery | Peter W. Cook, Dakota Howard, Dhwani Batra, Ben L. Rambo-Martin, Eileen de Feo, Jan Antico, Christine Tran, Matthew Tolentino, Shannon Wickline, Kim Gietzen, Brad Sickler, Jingtao Liu, Eric Allen, Phil Febbo, Summer Galloway, Nicole L. Washington, Simon White, Geraint Levan, Kelly Schiabor Barrett, Elizabeth Cirulli, Alexandre Bolze, Ary Ascencio, Charlotte Rivera-Garcia, Ryan Cho, Jason Nguyen, Sherry Wang, Jimmy Ramirez, Tyler Cassens, Efren Sandoval, Magnus Isaksson, William Lee, David Becker, Marc Laurent, James Lu, Clinton R. Paden, Suxiang Tong, Duncan MacCannell |
| EPI_ISL_1268257 | DPHL | Delaware Public Health Lab | Rebecca Savage |
| EPI_ISL_1268258 | Helix/Illumina | Centers for Disease Control and Prevention Division of Viral Diseases, Pathogen Discovery | Peter W. Cook, Dakota Howard, Dhwani Batra, Ben L. Rambo-Martin, Eileen de Feo, Jan Antico, Christine Tran, Matthew Tolentino, Shannon Wickline, Kim Gietzen, Brad Sickler, Jingtao Liu, Eric Allen, Phil Febbo, Summer Galloway, Nicole L. Washington, Simon White, Geraint Levan, Kelly Schiabor Barrett, Elizabeth Cirulli, Alexandre Bolze, Ary Ascencio, Charlotte Rivera-Garcia, Ryan Cho, Jason Nguyen, Sherry Wang, Jimmy Ramirez, Tyler Cassens, Efren Sandoval, Magnus Isaksson, William Lee, David Becker, Marc Laurent, James Lu, Clinton R. Paden, Suxiang Tong, Duncan MacCannell |
| EPI_ISL_1268259 | DPHL | Delaware Public Health Lab | Rebecca Savage |
| EPI_ISL_1268260 | Helix/Illumina | Centers for Disease Control and Prevention Division of Viral Diseases, Pathogen Discovery | Peter W. Cook, Dakota Howard, Dhwani Batra, Ben L. Rambo-Martin, Eileen de Feo, Jan Antico, Christine Tran, Matthew Tolentino, Shannon Wickline, Kim Gietzen, Brad Sickler, Jingtao Liu, Eric Allen, Phil Febbo, Summer Galloway, Nicole L. Washington, Simon White, Geraint Levan, Kelly Schiabor Barrett, Elizabeth Cirulli, Alexandre Bolze, Ary Ascencio, Charlotte Rivera-Garcia, Ryan Cho, Jason Nguyen, Sherry Wang, Jimmy Ramirez, Tyler Cassens, Efren Sandoval, Magnus Isaksson, William Lee, David Becker, Marc Laurent, James Lu, Clinton R. Paden, Suxiang Tong, Duncan MacCannell |
| EPI_ISL_1268261 | DPHL | Delaware Public Health Lab | Rebecca Savage |
| EPI_ISL_1268262 | Helix/Illumina | Centers for Disease Control and Prevention Division of Viral Diseases, Pathogen Discovery | Peter W. Cook, Dakota Howard, Dhwani Batra, Ben L. Rambo-Martin, Eileen de Feo, Jan Antico, Christine Tran, Matthew Tolentino, Shannon Wickline, Kim Gietzen, Brad Sickler, Jingtao Liu, Eric Allen, Phil Febbo, Summer Galloway, Nicole L. Washington, Simon White, Geraint Levan, Kelly Schiabor Barrett, Elizabeth Cirulli, Alexandre Bolze, Ary Ascencio, Charlotte Rivera-Garcia, Ryan Cho, Jason Nguyen, Sherry Wang, Jimmy Ramirez, Tyler Cassens, Efren Sandoval, Magnus Isaksson, William Lee, David Becker, Marc Laurent, James Lu, Clinton R. Paden, Suxiang Tong, Duncan MacCannell |
| EPI_ISL_1268263 | DPHL | Delaware Public Health Lab | Rebecca Savage |
| EPI_ISL_1268264, EPI_ISL_1268265, EPI_ISL_1268266, EPI_ISL_1268267, EPI_ISL_1268268, EPI_ISL_1268269, EPI_ISL_1268270, EPI_ISL_1268271, EPI_ISL_1268272, EPI_ISL_1268273, EPI_ISL_1268274, EPI_ISL_1268275, EPI_ISL_1268276, EPI_ISL_1268277, EPI_ISL_1268278, EPI_ISL_1268279, EPI_ISL_1268280, EPI_ISL_1268281, EPI_ISL_1268282, EPI_ISL_1268283, EPI_ISL_1268284, EPI_ISL_1268285, EPI_ISL_1268286, EPI_ISL_1268287, EPI_ISL_1268288, EPI_ISL_1268289, EPI_ISL_1268290, EPI_ISL_1268291, EPI_ISL_1268292, EPI_ISL_1268293, EPI_ISL_1268294, EPI_ISL_1268295, EPI_ISL_1268296, EPI_ISL_1268297, EPI_ISL_1268298, EPI_ISL_1268299, EPI_ISL_1268300, EPI_ISL_1268301, EPI_ISL_1268302, EPI_ISL_1268303, EPI_ISL_1268304, EPI_ISL_1268305, EPI_ISL_1268306, EPI_ISL_1268307, EPI_ISL_1268308, EPI_ISL_1268309, EPI_ISL_1268310, EPI_ISL_1268311, EPI_ISL_1268312, EPI_ISL_1268313, EPI_ISL_1268314, EPI_ISL_1268315, EPI_ISL_1268316, EPI_ISL_1268317, EPI_ISL_1268318, EPI_ISL_1268319, EPI_ISL_1268320, EPI_ISL_1268321, EPI_ISL_1268322, EPI_ISL_1268323, EPI_ISL_1268324, EPI_ISL_1268325, EPI_ISL_1268326, EPI_ISL_1268327, EPI_ISL_1268328, EPI_ISL_1268329, EPI_ISL_1268330, EPI_ISL_1268331, EPI_ISL_1268332, EPI_ISL_1268333, EPI_ISL_1268334, EPI_ISL_1268335, EPI_ISL_1268336, EPI_ISL_1268337, EPI_ISL_1268338, EPI_ISL_1268339, EPI_ISL_1268340, EPI_ISL_1268341, EPI_ISL_1268342, EPI_ISL_1268343, EPI_ISL_1268344, EPI_ISL_1268345, EPI_ISL_1268346, EPI_ISL_1268347, EPI_ISL_1268348, EPI_ISL_1268349, EPI_ISL_1268350, EPI_ISL_1268351, EPI_ISL_1268352, EPI_ISL_1268353, EPI_ISL_1268354, EPI_ISL_1268355, EPI_ISL_1268356, EPI_ISL_1268357, EPI_ISL_1268358, EPI_ISL_1268359, EPI_ISL_1268360, EPI_ISL_1268361, EPI_ISL_1268362, EPI_ISL_1268363, EPI_ISL_1268364, EPI_ISL_1268365, EPI_ISL_1268366, EPI_ISL_1268367, EPI_ISL_1268368, EPI_ISL_1268369, EPI_ISL_1268370, EPI_ISL_1268371, EPI_ISL_1268372, EPI_ISL_1268373, EPI_ISL_1268374, EPI_ISL_1268375, EPI_ISL_1268376, EPI_ISL_1268377, EPI_ISL_1268378, EPI_ISL_1268379, EPI_ISL_1268380, EPI_ISL_1268381, EPI_ISL_1268382, EPI_ISL_1268383, EPI_ISL_1268384, EPI_ISL_1268385, EPI_ISL_1268386, EPI_ISL_1268387, EPI_ISL_1268388, EPI_ISL_1268389, EPI_ISL_1268390, EPI_ISL_1268391, EPI_ISL_1268392, EPI_ISL_1268393, EPI_ISL_1268394, EPI_ISL_1268395, EPI_ISL_1268396, EPI_ISL_1268397, EPI_ISL_1268398, EPI_ISL_1268399, EPI_ISL_1268400, EPI_ISL_1268401, EPI_ISL_1268402, EPI_ISL_1268403, EPI_ISL_1268404, EPI_ISL_1268405, EPI_ISL_1268406, EPI_ISL_1268407, EPI_ISL_1268408, EPI_ISL_1268409, EPI_ISL_1268410, EPI_ISL_1268411, EPI_ISL_1268412, EPI_ISL_1268413, EPI_ISL_1268414, EPI_ISL_1268415, EPI_ISL_1268416, EPI_ISL_1268417, EPI_ISL_1268418, EPI_ISL_1268419, EPI_ISL_1268420, EPI_ISL_1268421, EPI_ISL_1268422, EPI_ISL_1268423, EPI_ISL_1268424, EPI_ISL_1268425, EPI_ISL_1268426, EPI_ISL_1268427, EPI_ISL_1268428, EPI_ISL_1268429, EPI_ISL_1268430, EPI_ISL_1268431, EPI_ISL_1268432, EPI_ISL_1268433, EPI_ISL_1268434, EPI_ISL_1268435, EPI_ISL_1268436, EPI_ISL_1268437, EPI_ISL_1268438, EPI_ISL_1268439, EPI_ISL_1268440, EPI_ISL_1268441, EPI_ISL_1268442, EPI_ISL_1268443, EPI_ISL_1268444, EPI_ISL_1268445, EPI_ISL_1268446, EPI_ISL_1268447, EPI_ISL_1268448, EPI_ISL_1268449, EPI_ISL_1268450, EPI_ISL_1268451, EPI_ISL_1268452, EPI_ISL_1268453, EPI_ISL_1268454, EPI_ISL_1268455, EPI_ISL_1268456, EPI_ISL_1268457, EPI_ISL_1268458, EPI_ISL_1268459, EPI_ISL_1268461, EPI_ISL_1268462, EPI_ISL_1268463, EPI_ISL_1268464, EPI_ISL_1268465, EPI_ISL_1268466, EPI_ISL_1268467, EPI_ISL_1268468, EPI_ISL_1268469, EPI_ISL_1268470, EPI_ISL_1268471, EPI_ISL_1268472, EPI_ISL_1268473, EPI_ISL_1268474, EPI_ISL_1268475, EPI_ISL_1268476, EPI_ISL_1268477, EPI_ISL_1268478, EPI_ISL_1268479, EPI_ISL_1268480, EPI_ISL_1268481, EPI_ISL_1268482, EPI_ISL_1268483, EPI_ISL_1268484, EPI_ISL_1268485, EPI_ISL_1268486, EPI_ISL_1268487, EPI_ISL_1268488, EPI_ISL_1268489, EPI_ISL_1268490, EPI_ISL_1268491, EPI_ISL_1268492, EPI_ISL_1268493, EPI_ISL_1268494, EPI_ISL_1268495, EPI_ISL_1268496, EPI_ISL_1268497, EPI_ISL_1268498, EPI_ISL_1268499, EPI_ISL_1268500, EPI_ISL_1268501, EPI_ISL_1268502, EPI_ISL_1268503, EPI_ISL_1268504, EPI_ISL_1268505, EPI_ISL_1268506, EPI_ISL_1268507, EPI_ISL_1268508, EPI_ISL_1268509, EPI_ISL_1268510, EPI_ISL_1268511, EPI_ISL_1268512, EPI_ISL_1268513, EPI_ISL_1268514, EPI_ISL_1268515, EPI_ISL_1268516, EPI_ISL_1268517, EPI_ISL_1268518, EPI_ISL_1268519, EPI_ISL_1268520, EPI_ISL_1268521, EPI_ISL_1268522, EPI_ISL_1268523, EPI_ISL_1268524, EPI_ISL_1268525, EPI_ISL_1268526, EPI_ISL_1268527, EPI_ISL_1268528, EPI_ISL_1268529, EPI_ISL_1268530, EPI_ISL_1268531, EPI_ISL_1268532, EPI_ISL_1268533, EPI_ISL_1268534, EPI_ISL_1268535, EPI_ISL_1268536, EPI_ISL_1268537, EPI_ISL_1268538, EPI_ISL_1268539, EPI_ISL_1268540, EPI_ISL_1268541, EPI_ISL_1268542, EPI_ISL_1268543, EPI_ISL_1268544, EPI_ISL_1268545, EPI_ISL_1268546, EPI_ISL_1268547, EPI_ISL_1268548, EPI_ISL_1268549, EPI_ISL_1268550, EPI_ISL_1268551, EPI_ISL_1268552, EPI_ISL_1268553, EPI_ISL_1268554, EPI_ISL_1268555, EPI_ISL_1268556, EPI_ISL_1268557, EPI_ISL_1268558, EPI_ISL_1268559, EPI_ISL_1268560, EPI_ISL_1268561, EPI_ISL_1268562, EPI_ISL_1268563, EPI_ISL_1268564, EPI_ISL_1268565, EPI_ISL_1268566, EPI_ISL_1268567, EPI_ISL_1268568, EPI_ISL_1268569, EPI_ISL_1268570, EPI_ISL_1268571, EPI_ISL_1268572, EPI_ISL_1268573, EPI_ISL_1268574, EPI_ISL_1268575, EPI_ISL_1268576, EPI_ISL_1268577, EPI_ISL_1268578, EPI_ISL_1268579, EPI_ISL_1268580, EPI_ISL_1268581, EPI_ISL_1268582, EPI_ISL_1268583, EPI_ISL_1268584, EPI_ISL_1268585, EPI_ISL_1268586, EPI_ISL_1268587, EPI_ISL_1268588, EPI_ISL_1268589, EPI_ISL_1268590, EPI_ISL_1268591, EPI_ISL_1268592, EPI_ISL_1268593, EPI_ISL_1268594, EPI_ISL_1268595, EPI_ISL_1268596, EPI_ISL_1268597, EPI_ISL_1268598, EPI_ISL_1268599, EPI_ISL_1268600, EPI_ISL_1268601, EPI_ISL_1268602, EPI_ISL_1268603, EPI_ISL_1268604, EPI_ISL_1268605, EPI_ISL_1268606, EPI_ISL_1268607, EPI_ISL_1268608, EPI_ISL_1268609, EPI_ISL_1268610, EPI_ISL_1268611, EPI_ISL_1268612, EPI_ISL_1268613, EPI_ISL_1268614, EPI_ISL_1268615, EPI_ISL_1268616, EPI_ISL_1268617, EPI_ISL_1268618, EPI_ISL_1268619, EPI_ISL_1268620, EPI_ISL_1268621, EPI_ISL_1268622, EPI_ISL_1268623, EPI_ISL_1268624, EPI_ISL_1268625, EPI_ISL_1268626, EPI_ISL_1268627, EPI_ISL_1268628, EPI_ISL_1268629, EPI_ISL_1268630, EPI_ISL_1268631, EPI_ISL_1268632, EPI_ISL_1268633, EPI_ISL_1268634, EPI_ISL_1268635, EPI_ISL_1268636, EPI_ISL_1268637, EPI_ISL_1268638, EPI_ISL_1268639, EPI_ISL_1268640, EPI_ISL_1268641, EPI_ISL_1268642, EPI_ISL_1268643, EPI_ISL_1268644, EPI_ISL_1268645, EPI_ISL_1268646, EPI_ISL_1268647, EPI_ISL_1268654, EPI_ISL_1268657, EPI_ISL_1268660, EPI_ISL_1268663, EPI_ISL_1268666, EPI_ISL_1268669, EPI_ISL_1268672, EPI_ISL_1268675, EPI_ISL_1268678, EPI_ISL_1268684, EPI_ISL_1268687, EPI_ISL_1268690, EPI_ISL_1268693, EPI_ISL_1268696, EPI_ISL_1268699, EPI_ISL_1268701, EPI_ISL_1268704, EPI_ISL_1268706, EPI_ISL_1268708, EPI_ISL_1268710, EPI_ISL_1268712, EPI_ISL_1268714, EPI_ISL_1268716, EPI_ISL_1268718, EPI_ISL_1268720, EPI_ISL_1268722, EPI_ISL_1268724, EPI_ISL_1268726, EPI_ISL_1268730, EPI_ISL_1268732, EPI_ISL_1268734, EPI_ISL_1268736, EPI_ISL_1268738, EPI_ISL_1268740, EPI_ISL_1268742, EPI_ISL_1268744, EPI_ISL_1268746, EPI_ISL_1268748, EPI_ISL_1268751, EPI_ISL_1268752, EPI_ISL_1268754, EPI_ISL_1268756, EPI_ISL_1268758, EPI_ISL_1268760, EPI_ISL_1268762, EPI_ISL_1268764, EPI_ISL_1268766, EPI_ISL_1268768, EPI_ISL_1268770, EPI_ISL_1268772, EPI_ISL_1268774, EPI_ISL_1268777, EPI_ISL_1268779, EPI_ISL_1268781, EPI_ISL_1268783, EPI_ISL_1268785, EPI_ISL_1268787, EPI_ISL_1268789, EPI_ISL_1268791, EPI_ISL_1268793, EPI_ISL_1268795, EPI_ISL_1268798, EPI_ISL_1268800, EPI_ISL_1268802, EPI_ISL_1268804, EPI_ISL_1268806, EPI_ISL_1268808, EPI_ISL_1268810, EPI_ISL_1268812, EPI_ISL_1268814, EPI_ISL_1268816, EPI_ISL_1268818, EPI_ISL_1268820, EPI_ISL_1268822, EPI_ISL_1268824, EPI_ISL_1268826, EPI_ISL_1268827, EPI_ISL_1268829, EPI_ISL_1268833, EPI_ISL_1268835, EPI_ISL_1268839, EPI_ISL_1268840, EPI_ISL_1268842, EPI_ISL_1268844, EPI_ISL_1268847, EPI_ISL_1268848, EPI_ISL_1268851, EPI_ISL_1268854, EPI_ISL_1268857, EPI_ISL_1268860, EPI_ISL_1268863, EPI_ISL_1268866, EPI_ISL_1268870, EPI_ISL_1268873, EPI_ISL_1268876, EPI_ISL_1268879, EPI_ISL_1268882, EPI_ISL_1268885, EPI_ISL_1268888, EPI_ISL_1268891, EPI_ISL_1268894, EPI_ISL_1268895, EPI_ISL_1268896, EPI_ISL_1268897, EPI_ISL_1268898, EPI_ISL_1268899, EPI_ISL_1268900, EPI_ISL_1268901, EPI_ISL_1268902, EPI_ISL_1268903, EPI_ISL_1268904, EPI_ISL_1268905, EPI_ISL_1268906, EPI_ISL_1268907, EPI_ISL_1268908, EPI_ISL_1268909, EPI_ISL_1268910, EPI_ISL_1268911, EPI_ISL_1268912, EPI_ISL_1268913, EPI_ISL_1268914, EPI_ISL_1268915, EPI_ISL_1268916, EPI_ISL_1268917, EPI_ISL_1268918, EPI_ISL_1268919, EPI_ISL_1268920, EPI_ISL_1268921, EPI_ISL_1268922, EPI_ISL_1268923, EPI_ISL_1268924, EPI_ISL_1268925, EPI_ISL_1268926, EPI_ISL_1268927, EPI_ISL_1268928, EPI_ISL_1268929, EPI_ISL_1268930, EPI_ISL_1268931, EPI_ISL_1268932, EPI_ISL_1268933, EPI_ISL_1268934, EPI_ISL_1268935, EPI_ISL_1268936, EPI_ISL_1268937, EPI_ISL_1268938, EPI_ISL_1268939, EPI_ISL_1268940, EPI_ISL_1268941, EPI_ISL_1268942, EPI_ISL_1268943, EPI_ISL_1268944, EPI_ISL_1268945, EPI_ISL_1268946, EPI_ISL_1268947, EPI_ISL_1268948, EPI_ISL_1268949, EPI_ISL_1268950, EPI_ISL_1268951, EPI_ISL_1268952, EPI_ISL_1268953, EPI_ISL_1268954, EPI_ISL_1268955, EPI_ISL_1268956, EPI_ISL_1268957, EPI_ISL_1268958, EPI_ISL_1268959, EPI_ISL_1268960, EPI_ISL_1268961, EPI_ISL_1268962, EPI_ISL_1268963, EPI_ISL_1268964, EPI_ISL_1268965, EPI_ISL_1268966, EPI_ISL_1268967, EPI_ISL_1268968, EPI_ISL_1268969, EPI_ISL_1268970, EPI_ISL_1268971, EPI_ISL_1268972, EPI_ISL_1268973, EPI_ISL_1268974, EPI_ISL_1268975, EPI_ISL_1268976, EPI_ISL_1268977, EPI_ISL_1268978, EPI_ISL_1268979, EPI_ISL_1268980, EPI_ISL_1268981, EPI_ISL_1268982, EPI_ISL_1268983, EPI_ISL_1268984, EPI_ISL_1268985, EPI_ISL_1268986, EPI_ISL_1268987, EPI_ISL_1268988, EPI_ISL_1268989, EPI_ISL_1268990, EPI_ISL_1268991, EPI_ISL_1268992, EPI_ISL_1268993, EPI_ISL_1268994, EPI_ISL_1268995, EPI_ISL_1268996, EPI_ISL_1268997, EPI_ISL_1268998, EPI_ISL_1268999, EPI_ISL_1269000, EPI_ISL_1269001, EPI_ISL_1269002, EPI_ISL_1269003, EPI_ISL_1269004, EPI_ISL_1269005, EPI_ISL_1269006, EPI_ISL_1269007, EPI_ISL_1269008, EPI_ISL_1269009, EPI_ISL_1269010, EPI_ISL_1269011, EPI_ISL_1269012, EPI_ISL_1269013, EPI_ISL_1269014, EPI_ISL_1269015, EPI_ISL_1269016, EPI_ISL_1269017 | | | |
| see above | Helix/Illumina | Centers for Disease Control and Prevention Division of Viral Diseases, Pathogen Discovery | Peter W. Cook, Dakota Howard, Dhwani Batra, Ben L. Rambo-Martin, Eileen de Feo, Jan Antico, Christine Tran, Matthew Tolentino, Shannon Wickline, Kim Gietzen, Brad Sickler, Jingtao Liu, Eric Allen, Phil Febbo, Summer Galloway, Nicole L. Washington, Simon White, Geraint Levan, Kelly Schiabor Barrett, Elizabeth Cirulli, Alexandre Bolze, Ary Ascencio, Charlotte Rivera-Garcia, Ryan Cho, Jason Nguyen, Sherry Wang, Jimmy Ramirez, Tyler Cassens, Efren Sandoval, Magnus Isaksson, William Lee, David Becker, Marc Laurent, James Lu, Clinton R. Paden, Suxiang Tong, Duncan MacCannell |
| EPI_ISL_1269018 | Microbiology Division, South Carolina Department of Health and Environmental Control Public Health Laboratory (SC DHEC PHL) | Microbiology Division, South Carolina Department of Health and Environmental Control Public Health Laboratory (SC DHEC PHL) | Haley Flores, Jessica Freeman |
| EPI_ISL_1269019, EPI_ISL_1269020, EPI_ISL_1269021, EPI_ISL_1269022, EPI_ISL_1269023, EPI_ISL_1269024, EPI_ISL_1269025, EPI_ISL_1269026, EPI_ISL_1269027, EPI_ISL_1269028, EPI_ISL_1269029, EPI_ISL_1269030, EPI_ISL_1269031, EPI_ISL_1269032, EPI_ISL_1269033, EPI_ISL_1269034, EPI_ISL_1269035, EPI_ISL_1269036, EPI_ISL_1269037, EPI_ISL_1269038, EPI_ISL_1269039, EPI_ISL_1269040, EPI_ISL_1269041, EPI_ISL_1269042, EPI_ISL_1269043, EPI_ISL_1269044, EPI_ISL_1269045, EPI_ISL_1269046, EPI_ISL_1269047, EPI_ISL_1269048, EPI_ISL_1269049, EPI_ISL_1269050, EPI_ISL_1269051, EPI_ISL_1269052, EPI_ISL_1269053, EPI_ISL_1269054, EPI_ISL_1269055, EPI_ISL_1269056, EPI_ISL_1269057, EPI_ISL_1269058, EPI_ISL_1269059, EPI_ISL_1269060, EPI_ISL_1269061, EPI_ISL_1269062, EPI_ISL_1269063, EPI_ISL_1269064, EPI_ISL_1269065, EPI_ISL_1269066, EPI_ISL_1269067, EPI_ISL_1269068, EPI_ISL_1269069, EPI_ISL_1269070, EPI_ISL_1269071, EPI_ISL_1269072, EPI_ISL_1269073, EPI_ISL_1269074, EPI_ISL_1269075, EPI_ISL_1269076, EPI_ISL_1269078, EPI_ISL_1269079, EPI_ISL_1269080, EPI_ISL_1269081, EPI_ISL_1269082, EPI_ISL_1269083, EPI_ISL_1269084, EPI_ISL_1269085, EPI_ISL_1269086, EPI_ISL_1269087, EPI_ISL_1269088, EPI_ISL_1269089, EPI_ISL_1269090, EPI_ISL_1269091, EPI_ISL_1269092, EPI_ISL_1269093, EPI_ISL_1269094, EPI_ISL_1269095, EPI_ISL_1269096, EPI_ISL_1269097, EPI_ISL_1269098, EPI_ISL_1269099, EPI_ISL_1269100, EPI_ISL_1269101, EPI_ISL_1269102, EPI_ISL_1269103, EPI_ISL_1269104, EPI_ISL_1269105, EPI_ISL_1269106, EPI_ISL_1269107, EPI_ISL_1269108, EPI_ISL_1269109, EPI_ISL_1269110, EPI_ISL_1269111, EPI_ISL_1269112, EPI_ISL_1269113, EPI_ISL_1269114, EPI_ISL_1269115, EPI_ISL_1269116, EPI_ISL_1269117, EPI_ISL_1269118, EPI_ISL_1269119, EPI_ISL_1269120, EPI_ISL_1269121, EPI_ISL_1269122, EPI_ISL_1269123, EPI_ISL_1269124, EPI_ISL_1269125, EPI_ISL_1269126, EPI_ISL_1269127, EPI_ISL_1269128, EPI_ISL_1269130, EPI_ISL_1269131, EPI_ISL_1269132, EPI_ISL_1269133, EPI_ISL_1269134, EPI_ISL_1269135, EPI_ISL_1269136, EPI_ISL_1269137, EPI_ISL_1269138, EPI_ISL_1269139, EPI_ISL_1269140, EPI_ISL_1269141, EPI_ISL_1269142, EPI_ISL_1269143, EPI_ISL_1269144, EPI_ISL_1269145, EPI_ISL_1269146, EPI_ISL_1269147, EPI_ISL_1269148, EPI_ISL_1269149, EPI_ISL_1269151, EPI_ISL_1269152, EPI_ISL_1269153, EPI_ISL_1269154, EPI_ISL_1269155, EPI_ISL_1269156, EPI_ISL_1269157, EPI_ISL_1269158, EPI_ISL_1269159, EPI_ISL_1269160, EPI_ISL_1269161, EPI_ISL_1269162, EPI_ISL_1269163, EPI_ISL_1269164, EPI_ISL_1269165, EPI_ISL_1269166, EPI_ISL_1269167, EPI_ISL_1269168, EPI_ISL_1269169, EPI_ISL_1269170, EPI_ISL_1269171, EPI_ISL_1269172, EPI_ISL_1269173, EPI_ISL_1269174, EPI_ISL_1269175, EPI_ISL_1269176, EPI_ISL_1269177, EPI_ISL_1269178, EPI_ISL_1269179, EPI_ISL_1269180, EPI_ISL_1269181, EPI_ISL_1269182, EPI_ISL_1269183, EPI_ISL_1269184, EPI_ISL_1269185, EPI_ISL_1269186, EPI_ISL_1269187, EPI_ISL_1269188, EPI_ISL_1269189, EPI_ISL_1269190, EPI_ISL_1269191, EPI_ISL_1269192, EPI_ISL_1269193, EPI_ISL_1269194, EPI_ISL_1269195, EPI_ISL_1269196, EPI_ISL_1269197, EPI_ISL_1269198, EPI_ISL_1269199, EPI_ISL_1269200, EPI_ISL_1269201, EPI_ISL_1269202, EPI_ISL_1269203, EPI_ISL_1269204, EPI_ISL_1269205, EPI_ISL_1269206, EPI_ISL_1269207, EPI_ISL_1269208, EPI_ISL_1269209, EPI_ISL_1269210, EPI_ISL_1269211, EPI_ISL_1269212, EPI_ISL_1269213, EPI_ISL_1269214, EPI_ISL_1269215, EPI_ISL_1269216, EPI_ISL_1269217, EPI_ISL_1269218, EPI_ISL_1269219, EPI_ISL_1269220, EPI_ISL_1269221, EPI_ISL_1269222, EPI_ISL_1269223, EPI_ISL_1269224, EPI_ISL_1269225, EPI_ISL_1269226, EPI_ISL_1269227, EPI_ISL_1269228, EPI_ISL_1269229, EPI_ISL_1269230, EPI_ISL_1269231, EPI_ISL_1269232, EPI_ISL_1269233, EPI_ISL_1269234, EPI_ISL_1269235, EPI_ISL_1269236, EPI_ISL_1269237, EPI_ISL_1269238, EPI_ISL_1269239, EPI_ISL_1269240, EPI_ISL_1269241, EPI_ISL_1269242, EPI_ISL_1269243, EPI_ISL_1269244, EPI_ISL_1269245, EPI_ISL_1269246, EPI_ISL_1269247, EPI_ISL_1269248, EPI_ISL_1269249, EPI_ISL_1269250, EPI_ISL_1269251, EPI_ISL_1269252, EPI_ISL_1269253, EPI_ISL_1269254, EPI_ISL_1269255, EPI_ISL_1269256, EPI_ISL_1269257, EPI_ISL_1269258, EPI_ISL_1269259, EPI_ISL_1269261, EPI_ISL_1269262, EPI_ISL_1269263, EPI_ISL_1269264, EPI_ISL_1269265, EPI_ISL_1269266, EPI_ISL_1269267, EPI_ISL_1269268, EPI_ISL_1269269, EPI_ISL_1269270, EPI_ISL_1269271, EPI_ISL_1269272, EPI_ISL_1269273, EPI_ISL_1269275, EPI_ISL_1269276, EPI_ISL_1269277, EPI_ISL_1269278, EPI_ISL_1269279, EPI_ISL_1269280, EPI_ISL_1269281, EPI_ISL_1269282, EPI_ISL_1269283, EPI_ISL_1269284, EPI_ISL_1269285, EPI_ISL_1269286, EPI_ISL_1269287, EPI_ISL_1269288, EPI_ISL_1269289, EPI_ISL_1269290, EPI_ISL_1269291, EPI_ISL_1269292, EPI_ISL_1269293, EPI_ISL_1269294, EPI_ISL_1269295, EPI_ISL_1269296, EPI_ISL_1269297, EPI_ISL_1269298, EPI_ISL_1269299, EPI_ISL_1269300, EPI_ISL_1269301, EPI_ISL_1269302, EPI_ISL_1269303, EPI_ISL_1269304, EPI_ISL_1269305, EPI_ISL_1269306, EPI_ISL_1269307, EPI_ISL_1269308, EPI_ISL_1269309, EPI_ISL_1269310, EPI_ISL_1269311, EPI_ISL_1269312, EPI_ISL_1269313 | | | |
| see above | Helix/Illumina | Centers for Disease Control and Prevention Division of Viral Diseases, Pathogen Discovery | Peter W. Cook, Dakota Howard, Dhwani Batra, Ben L. Rambo-Martin, Eileen de Feo, Jan Antico, Christine Tran, Matthew Tolentino, Shannon Wickline, Kim Gietzen, Brad Sickler, Jingtao Liu, Eric Allen, Phil Febbo, Summer Galloway, Nicole L. Washington, Simon White, Geraint Levan, Kelly Schiabor Barrett, Elizabeth Cirulli, Alexandre Bolze, Ary Ascencio, Charlotte Rivera-Garcia, Ryan Cho, Jason Nguyen, Sherry Wang, Jimmy Ramirez, Tyler Cassens, Efren Sandoval, Magnus Isaksson, William Lee, David Becker, Marc Laurent, James Lu, Clinton R. Paden, Suxiang Tong, Duncan MacCannell |
| EPI_ISL_1269314, EPI_ISL_1269315 | Wisconsin State Laboratory of Hygiene Communicable Disease Division | Ginkgo Bioworks Clinical Laboratory | Malaika Mckenzie-Bennett, James McGann, Jim Griffin, Keith Robison, Alex Plocik, Becky Schilling, Martha Pierson, Rebecca Littlefield, Michelle Spencer, Amanda Riccardi, Birgitte Simen, Kelsey R. Florek, Abigail C. Shockey |
| EPI_ISL_1269316 | Helix/Illumina | Centers for Disease Control and Prevention Division of Viral Diseases, Pathogen Discovery | Peter W. Cook, Dakota Howard, Dhwani Batra, Ben L. Rambo-Martin, Eileen de Feo, Jan Antico, Christine Tran, Matthew Tolentino, Shannon Wickline, Kim Gietzen, Brad Sickler, Jingtao Liu, Eric Allen, Phil Febbo, Summer Galloway, Nicole L. Washington, Simon White, Geraint Levan, Kelly Schiabor Barrett, Elizabeth Cirulli, Alexandre Bolze, Ary Ascencio, Charlotte Rivera-Garcia, Ryan Cho, Jason Nguyen, Sherry Wang, Jimmy Ramirez, Tyler Cassens, Efren Sandoval, Magnus Isaksson, William Lee, David Becker, Marc Laurent, James Lu, Clinton R. Paden, Suxiang Tong, Duncan MacCannell |
| EPI_ISL_1269317, EPI_ISL_1269318, EPI_ISL_1269319 | Wisconsin State Laboratory of Hygiene Communicable Disease Division | Ginkgo Bioworks Clinical Laboratory | Malaika Mckenzie-Bennett, James McGann, Jim Griffin, Keith Robison, Alex Plocik, Becky Schilling, Martha Pierson, Rebecca Littlefield, Michelle Spencer, Amanda Riccardi, Birgitte Simen, Kelsey R. Florek, Abigail C. Shockey |

| | | | |
|---|---|---|---|
| EPI_ISL_1269320 | Helix/Illumina | Centers for Disease Control and Prevention Division of Viral Diseases, Pathogen Discovery | Peter W. Cook, Dakota Howard, Dhwani Batra, Ben L. Rambo-Martin, Eileen de Feo, Jan Antico, Christine Tran, Matthew Tolentino, Shannon Wickline, Kim Gietzen, Brad Sickler, Jingtao Liu, Eric Allen, Phil Febbo, Summer Galloway, Nicole L. Washington, Simon White, Geraint Levan, Kelly Schiabor Barrett, Elizabeth Cirulli, Alexandre Bolze, Ary Ascencio, Charlotte Rivera-Garcia, Ryan Cho, Jason Nguyen, Sherry Wang, Jimmy Ramirez, Tyler Cassens, Efren Sandoval, Magnus Isaksson, William Lee, David Becker, Marc Laurent, James Lu, Clinton R. Paden, Suxiang Tong, Duncan MacCannell |
| EPI_ISL_1269321, EPI_ISL_1269322 | Wisconsin State Laboratory of Hygiene Communicable Disease Division | Ginkgo Bioworks Clinical Laboratory | Malaika Mckenzie-Bennett, James McGann, Jim Griffin, Keith Robison, Alex Plocik, Becky Schilling, Martha Pierson, Rebecca Littlefield, Michelle Spencer, Amanda Riccardi, Birgitte Simen, Kelsey R. Florek, Abigail C. Shockey |
| EPI_ISL_1269323 | Helix/Illumina | Centers for Disease Control and Prevention Division of Viral Diseases, Pathogen Discovery | Peter W. Cook, Dakota Howard, Dhwani Batra, Ben L. Rambo-Martin, Eileen de Feo, Jan Antico, Christine Tran, Matthew Tolentino, Shannon Wickline, Kim Gietzen, Brad Sickler, Jingtao Liu, Eric Allen, Phil Febbo, Summer Galloway, Nicole L. Washington, Simon White, Geraint Levan, Kelly Schiabor Barrett, Elizabeth Cirulli, Alexandre Bolze, Ary Ascencio, Charlotte Rivera-Garcia, Ryan Cho, Jason Nguyen, Sherry Wang, Jimmy Ramirez, Tyler Cassens, Efren Sandoval, Magnus Isaksson, William Lee, David Becker, Marc Laurent, James Lu, Clinton R. Paden, Suxiang Tong, Duncan MacCannell |
| EPI_ISL_1269324, EPI_ISL_1269325 | Wisconsin State Laboratory of Hygiene Communicable Disease Division | Ginkgo Bioworks Clinical Laboratory | Malaika Mckenzie-Bennett, James McGann, Jim Griffin, Keith Robison, Alex Plocik, Becky Schilling, Martha Pierson, Rebecca Littlefield, Michelle Spencer, Amanda Riccardi, Birgitte Simen, Kelsey R. Florek, Abigail C. Shockey |
| EPI_ISL_1269326 | Helix/Illumina | Centers for Disease Control and Prevention Division of Viral Diseases, Pathogen Discovery | Peter W. Cook, Dakota Howard, Dhwani Batra, Ben L. Rambo-Martin, Eileen de Feo, Jan Antico, Christine Tran, Matthew Tolentino, Shannon Wickline, Kim Gietzen, Brad Sickler, Jingtao Liu, Eric Allen, Phil Febbo, Summer Galloway, Nicole L. Washington, Simon White, Geraint Levan, Kelly Schiabor Barrett, Elizabeth Cirulli, Alexandre Bolze, Ary Ascencio, Charlotte Rivera-Garcia, Ryan Cho, Jason Nguyen, Sherry Wang, Jimmy Ramirez, Tyler Cassens, Efren Sandoval, Magnus Isaksson, William Lee, David Becker, Marc Laurent, James Lu, Clinton R. Paden, Suxiang Tong, Duncan MacCannell |
| EPI_ISL_1269327, EPI_ISL_1269328 | Wisconsin State Laboratory of Hygiene Communicable Disease Division | Ginkgo Bioworks Clinical Laboratory | Malaika Mckenzie-Bennett, James McGann, Jim Griffin, Keith Robison, Alex Plocik, Becky Schilling, Martha Pierson, Rebecca Littlefield, Michelle Spencer, Amanda Riccardi, Birgitte Simen, Kelsey R. Florek, Abigail C. Shockey |
| EPI_ISL_1269329 | Helix/Illumina | Centers for Disease Control and Prevention Division of Viral Diseases, Pathogen Discovery | Peter W. Cook, Dakota Howard, Dhwani Batra, Ben L. Rambo-Martin, Eileen de Feo, Jan Antico, Christine Tran, Matthew Tolentino, Shannon Wickline, Kim Gietzen, Brad Sickler, Jingtao Liu, Eric Allen, Phil Febbo, Summer Galloway, Nicole L. Washington, Simon White, Geraint Levan, Kelly Schiabor Barrett, Elizabeth Cirulli, Alexandre Bolze, Ary Ascencio, Charlotte Rivera-Garcia, Ryan Cho, Jason Nguyen, Sherry Wang, Jimmy Ramirez, Tyler Cassens, Efren Sandoval, Magnus Isaksson, William Lee, David Becker, Marc Laurent, James Lu, Clinton R. Paden, Suxiang Tong, Duncan MacCannell |
| EPI_ISL_1269330, EPI_ISL_1269331 | Wisconsin State Laboratory of Hygiene Communicable Disease Division | Ginkgo Bioworks Clinical Laboratory | Malaika Mckenzie-Bennett, James McGann, Jim Griffin, Keith Robison, Alex Plocik, Becky Schilling, Martha Pierson, Rebecca Littlefield, Michelle Spencer, Amanda Riccardi, Birgitte Simen, Kelsey R. Florek, Abigail C. Shockey |
| EPI_ISL_1269332 | Helix/Illumina | Centers for Disease Control and Prevention Division of Viral Diseases, Pathogen Discovery | Peter W. Cook, Dakota Howard, Dhwani Batra, Ben L. Rambo-Martin, Eileen de Feo, Jan Antico, Christine Tran, Matthew Tolentino, Shannon Wickline, Kim Gietzen, Brad Sickler, Jingtao Liu, Eric Allen, Phil Febbo, Summer Galloway, Nicole L. Washington, Simon White, Geraint Levan, Kelly Schiabor Barrett, Elizabeth Cirulli, Alexandre Bolze, Ary Ascencio, Charlotte Rivera-Garcia, Ryan Cho, Jason Nguyen, Sherry Wang, Jimmy Ramirez, Tyler Cassens, Efren Sandoval, Magnus Isaksson, William Lee, David Becker, Marc Laurent, James Lu, Clinton R. Paden, Suxiang Tong, Duncan MacCannell |
| EPI_ISL_1269333, EPI_ISL_1269334 | Wisconsin State Laboratory of Hygiene Communicable Disease Division | Ginkgo Bioworks Clinical Laboratory | Malaika Mckenzie-Bennett, James McGann, Jim Griffin, Keith Robison, Alex Plocik, Becky Schilling, Martha Pierson, Rebecca Littlefield, Michelle Spencer, Amanda Riccardi, Birgitte Simen, Kelsey R. Florek, Abigail C. Shockey |
| EPI_ISL_1269335 | Helix/Illumina | Centers for Disease Control and Prevention Division of Viral Diseases, Pathogen Discovery | Peter W. Cook, Dakota Howard, Dhwani Batra, Ben L. Rambo-Martin, Eileen de Feo, Jan Antico, Christine Tran, Matthew Tolentino, Shannon Wickline, Kim Gietzen, Brad Sickler, Jingtao Liu, Eric Allen, Phil Febbo, Summer Galloway, Nicole L. Washington, Simon White, Geraint Levan, Kelly Schiabor Barrett, Elizabeth Cirulli, Alexandre Bolze, Ary Ascencio, Charlotte Rivera-Garcia, Ryan Cho, Jason Nguyen, Sherry Wang, Jimmy Ramirez, Tyler Cassens, Efren Sandoval, Magnus Isaksson, William Lee, David Becker, Marc Laurent, James Lu, Clinton R. Paden, Suxiang Tong, Duncan MacCannell |
| EPI_ISL_1269336, EPI_ISL_1269337, EPI_ISL_1269338 | Wisconsin State Laboratory of Hygiene Communicable Disease Division | Ginkgo Bioworks Clinical Laboratory | Malaika Mckenzie-Bennett, James McGann, Jim Griffin, Keith Robison, Alex Plocik, Becky Schilling, Martha Pierson, Rebecca Littlefield, Michelle Spencer, Amanda Riccardi, Birgitte Simen, Kelsey R. Florek, Abigail C. Shockey |
| EPI_ISL_1269339 | Helix/Illumina | Centers for Disease Control and Prevention Division of Viral Diseases, Pathogen Discovery | Peter W. Cook, Dakota Howard, Dhwani Batra, Ben L. Rambo-Martin, Eileen de Feo, Jan Antico, Christine Tran, Matthew Tolentino, Shannon Wickline, Kim Gietzen, Brad Sickler, Jingtao Liu, Eric Allen, Phil Febbo, Summer Galloway, Nicole L. Washington, Simon White, Geraint Levan, Kelly Schiabor Barrett, Elizabeth Cirulli, Alexandre Bolze, Ary Ascencio, Charlotte Rivera-Garcia, Ryan Cho, Jason Nguyen, Sherry Wang, Jimmy Ramirez, Tyler Cassens, Efren Sandoval, Magnus Isaksson, William Lee, David Becker, Marc Laurent, James Lu, Clinton R. Paden, Suxiang Tong, Duncan MacCannell |
| EPI_ISL_1269340, EPI_ISL_1269341 | Wisconsin State Laboratory of Hygiene Communicable Disease Division | Ginkgo Bioworks Clinical Laboratory | Malaika Mckenzie-Bennett, James McGann, Jim Griffin, Keith Robison, Alex Plocik, Becky Schilling, Martha Pierson, Rebecca Littlefield, Michelle Spencer, Amanda Riccardi, Birgitte Simen, Kelsey R. Florek, Abigail C. Shockey |
| EPI_ISL_1269342 | Helix/Illumina | Centers for Disease Control and Prevention Division of Viral Diseases, Pathogen Discovery | Peter W. Cook, Dakota Howard, Dhwani Batra, Ben L. Rambo-Martin, Eileen de Feo, Jan Antico, Christine Tran, Matthew Tolentino, Shannon Wickline, Kim Gietzen, Brad Sickler, Jingtao Liu, Eric Allen, Phil Febbo, Summer Galloway, Nicole L. Washington, Simon White, Geraint Levan, Kelly Schiabor Barrett, Elizabeth Cirulli, Alexandre Bolze, Ary Ascencio, Charlotte Rivera-Garcia, Ryan Cho, Jason Nguyen, Sherry Wang, Jimmy Ramirez, Tyler Cassens, Efren Sandoval, Magnus Isaksson, William Lee, David Becker, Marc Laurent, James Lu, Clinton R. Paden, Suxiang Tong, Duncan MacCannell |
| EPI_ISL_1269343, EPI_ISL_1269344 | Wisconsin State Laboratory of Hygiene Communicable Disease Division | Ginkgo Bioworks Clinical Laboratory | Malaika Mckenzie-Bennett, James McGann, Jim Griffin, Keith Robison, Alex Plocik, Becky Schilling, Martha Pierson, Rebecca Littlefield, Michelle Spencer, Amanda Riccardi, Birgitte Simen, Kelsey R. Florek, Abigail C. Shockey |
| EPI_ISL_1269345 | Helix/Illumina | Centers for Disease Control and Prevention Division of Viral Diseases, Pathogen Discovery | Peter W. Cook, Dakota Howard, Dhwani Batra, Ben L. Rambo-Martin, Eileen de Feo, Jan Antico, Christine Tran, Matthew Tolentino, Shannon Wickline, Kim Gietzen, Brad Sickler, Jingtao Liu, Eric Allen, Phil Febbo, Summer Galloway, Nicole L. Washington, Simon White, Geraint Levan, Kelly Schiabor Barrett, Elizabeth Cirulli, Alexandre Bolze, Ary Ascencio, Charlotte Rivera-Garcia, Ryan Cho, Jason Nguyen, Sherry Wang, Jimmy Ramirez, Tyler Cassens, Efren Sandoval, Magnus Isaksson, William Lee, David Becker, Marc Laurent, James Lu, Clinton R. Paden, Suxiang Tong, Duncan MacCannell |
| EPI_ISL_1269346, EPI_ISL_1269347, EPI_ISL_1269348 | Wisconsin State Laboratory of Hygiene Communicable Disease Division | Ginkgo Bioworks Clinical Laboratory | Malaika Mckenzie-Bennett, James McGann, Jim Griffin, Keith Robison, Alex Plocik, Becky Schilling, Martha Pierson, Rebecca Littlefield, Michelle Spencer, Amanda Riccardi, Birgitte Simen, Kelsey R. Florek, Abigail C. Shockey |
| EPI_ISL_1269349 | Helix/Illumina | Centers for Disease Control and Prevention Division of Viral Diseases, Pathogen Discovery | Peter W. Cook, Dakota Howard, Dhwani Batra, Ben L. Rambo-Martin, Eileen de Feo, Jan Antico, Christine Tran, Matthew Tolentino, Shannon Wickline, Kim Gietzen, Brad Sickler, Jingtao Liu, Eric Allen, Phil Febbo, Summer Galloway, Nicole L. Washington, Simon White, Geraint Levan, Kelly Schiabor Barrett, Elizabeth Cirulli, Alexandre Bolze, Ary Ascencio, Charlotte Rivera-Garcia, Ryan Cho, Jason Nguyen, Sherry Wang, Jimmy Ramirez, Tyler Cassens, Efren Sandoval, Magnus Isaksson, William Lee, David Becker, Marc Laurent, James Lu, Clinton R. Paden, Suxiang Tong, Duncan MacCannell |
| EPI_ISL_1269350, EPI_ISL_1269351 | Wisconsin State Laboratory of Hygiene Communicable Disease Division | Ginkgo Bioworks Clinical Laboratory | Malaika Mckenzie-Bennett, James McGann, Jim Griffin, Keith Robison, Alex Plocik, Becky Schilling, Martha Pierson, Rebecca Littlefield, Michelle Spencer, Amanda Riccardi, Birgitte Simen, Kelsey R. Florek, Abigail C. Shockey |
| EPI_ISL_1269352 | Helix/Illumina | Centers for Disease Control and Prevention Division of Viral Diseases, Pathogen Discovery | Peter W. Cook, Dakota Howard, Dhwani Batra, Ben L. Rambo-Martin, Eileen de Feo, Jan Antico, Christine Tran, Matthew Tolentino, Shannon Wickline, Kim Gietzen, Brad Sickler, Jingtao Liu, Eric Allen, Phil Febbo, Summer Galloway, Nicole L. Washington, Simon White, Geraint Levan, Kelly Schiabor Barrett, Elizabeth Cirulli, Alexandre Bolze, Ary Ascencio, Charlotte Rivera-Garcia, Ryan Cho, Jason Nguyen, Sherry Wang, Jimmy Ramirez, Tyler Cassens, Efren Sandoval, Magnus Isaksson, William Lee, David Becker, Marc Laurent, James Lu, Clinton R. Paden, Suxiang Tong, Duncan MacCannell |
| EPI_ISL_1269353, EPI_ISL_1269354 | Wisconsin State Laboratory of Hygiene Communicable Disease Division | Ginkgo Bioworks Clinical Laboratory | Malaika Mckenzie-Bennett, James McGann, Jim Griffin, Keith Robison, Alex Plocik, Becky Schilling, Martha Pierson, Rebecca Littlefield, Michelle Spencer, Amanda Riccardi, Birgitte Simen, Kelsey R. Florek, Abigail C. Shockey |
| EPI_ISL_1269355 | Helix/Illumina | Centers for Disease Control and Prevention Division of Viral Diseases, Pathogen Discovery | Peter W. Cook, Dakota Howard, Dhwani Batra, Ben L. Rambo-Martin, Eileen de Feo, Jan Antico, Christine Tran, Matthew Tolentino, Shannon Wickline, Kim Gietzen, Brad Sickler, Jingtao Liu, Eric Allen, Phil Febbo, Summer Galloway, Nicole L. Washington, Simon White, Geraint Levan, Kelly Schiabor Barrett, Elizabeth Cirulli, Alexandre Bolze, Ary Ascencio, Charlotte Rivera-Garcia, Ryan Cho, Jason Nguyen, Sherry Wang, Jimmy Ramirez, Tyler Cassens, Efren Sandoval, Magnus Isaksson, William Lee, David Becker, Marc Laurent, James Lu, Clinton R. Paden, Suxiang Tong, Duncan MacCannell |
| EPI_ISL_1269356, EPI_ISL_1269357 | Wisconsin State Laboratory of Hygiene Communicable Disease Division | Ginkgo Bioworks Clinical Laboratory | Malaika Mckenzie-Bennett, James McGann, Jim Griffin, Keith Robison, Alex Plocik, Becky Schilling, Martha Pierson, Rebecca Littlefield, Michelle Spencer, Amanda Riccardi, Birgitte Simen, Kelsey R. Florek, Abigail C. Shockey |
| EPI_ISL_1269358 | Helix/Illumina | Centers for Disease Control and Prevention Division of Viral Diseases, Pathogen Discovery | Peter W. Cook, Dakota Howard, Dhwani Batra, Ben L. Rambo-Martin, Eileen de Feo, Jan Antico, Christine Tran, Matthew Tolentino, Shannon Wickline, Kim Gietzen, Brad Sickler, Jingtao Liu, Eric Allen, Phil Febbo, Summer Galloway, Nicole L. Washington, Simon White, Geraint Levan, Kelly Schiabor Barrett, Elizabeth Cirulli, Alexandre Bolze, Ary Ascencio, Charlotte Rivera-Garcia, Ryan Cho, Jason Nguyen, Sherry Wang, Jimmy Ramirez, Tyler Cassens, Efren Sandoval, Magnus Isaksson, William Lee, David Becker, Marc Laurent, James Lu, Clinton R. Paden, Suxiang Tong, Duncan MacCannell |
| EPI_ISL_1269359, EPI_ISL_1269360 | Wisconsin State Laboratory of Hygiene Communicable Disease Division | Ginkgo Bioworks Clinical Laboratory | Malaika Mckenzie-Bennett, James McGann, Jim Griffin, Keith Robison, Alex Plocik, Becky Schilling, Martha Pierson, Rebecca Littlefield, Michelle Spencer, Amanda Riccardi, Birgitte Simen, Kelsey R. Florek, Abigail C. Shockey |
| EPI_ISL_1269361 | Helix/Illumina | Centers for Disease Control and Prevention Division of Viral Diseases, Pathogen Discovery | Peter W. Cook, Dakota Howard, Dhwani Batra, Ben L. Rambo-Martin, Eileen de Feo, Jan Antico, Christine Tran, Matthew Tolentino, Shannon Wickline, Kim Gietzen, Brad Sickler, Jingtao Liu, Eric Allen, Phil Febbo, Summer Galloway, Nicole L. Washington, Simon White, Geraint Levan, Kelly Schiabor Barrett, Elizabeth Cirulli, Alexandre Bolze, Ary Ascencio, Charlotte Rivera-Garcia, Ryan Cho, Jason Nguyen, Sherry Wang, Jimmy Ramirez, Tyler Cassens, Efren Sandoval, Magnus Isaksson, William Lee, David Becker, Marc Laurent, James Lu, Clinton R. Paden, Suxiang Tong, Duncan MacCannell |
| EPI_ISL_1269362, EPI_ISL_1269363 | Wisconsin State Laboratory of Hygiene Communicable Disease Division | Ginkgo Bioworks Clinical Laboratory | Malaika Mckenzie-Bennett, James McGann, Jim Griffin, Keith Robison, Alex Plocik, Becky Schilling, Martha Pierson, Rebecca Littlefield, Michelle Spencer, Amanda Riccardi, Birgitte Simen, Kelsey R. Florek, Abigail C. Shockey |
| EPI_ISL_1269364 | Helix/Illumina | Centers for Disease Control and Prevention Division of Viral Diseases, Pathogen Discovery | Peter W. Cook, Dakota Howard, Dhwani Batra, Ben L. Rambo-Martin, Eileen de Feo, Jan Antico, Christine Tran, Matthew Tolentino, Shannon Wickline, Kim Gietzen, Brad Sickler, Jingtao Liu, Eric Allen, Phil Febbo, Summer Galloway, Nicole L. Washington, Simon White, Geraint Levan, Kelly Schiabor Barrett, Elizabeth Cirulli, Alexandre Bolze, Ary Ascencio, Charlotte Rivera-Garcia, Ryan Cho, Jason Nguyen, Sherry Wang, Jimmy Ramirez, Tyler Cassens, Efren Sandoval, Magnus Isaksson, William Lee, David Becker, Marc Laurent, James Lu, Clinton R. Paden, Suxiang Tong, Duncan MacCannell |
| EPI_ISL_1269365, EPI_ISL_1269366, EPI_ISL_1269367 | Wisconsin State Laboratory of Hygiene Communicable Disease Division | Ginkgo Bioworks Clinical Laboratory | Malaika Mckenzie-Bennett, James McGann, Jim Griffin, Keith Robison, Alex Plocik, Becky Schilling, Martha Pierson, Rebecca Littlefield, Michelle Spencer, Amanda Riccardi, Birgitte Simen, Kelsey R. Florek, Abigail C. Shockey |
| EPI_ISL_1269368 | Helix/Illumina | Centers for Disease Control and Prevention Division of Viral Diseases, Pathogen Discovery | Peter W. Cook, Dakota Howard, Dhwani Batra, Ben L. Rambo-Martin, Eileen de Feo, Jan Antico, Christine Tran, Matthew Tolentino, Shannon Wickline, Kim Gietzen, Brad Sickler, Jingtao Liu, Eric Allen, Phil Febbo, Summer Galloway, Nicole L. Washington, Simon White, Geraint Levan, Kelly Schiabor Barrett, Elizabeth Cirulli, Alexandre Bolze, Ary Ascencio, Charlotte Rivera-Garcia, Ryan Cho, Jason Nguyen, Sherry Wang, Jimmy Ramirez, Tyler Cassens, Efren Sandoval, Magnus Isaksson, William Lee, David Becker, Marc Laurent, James Lu, Clinton R. Paden, Suxiang Tong, Duncan MacCannell |
| EPI_ISL_1269369, EPI_ISL_1269370 | Wisconsin State Laboratory of Hygiene Communicable Disease Division | Ginkgo Bioworks Clinical Laboratory | Malaika Mckenzie-Bennett, James McGann, Jim Griffin, Keith Robison, Alex Plocik, Becky Schilling, Martha Pierson, Rebecca Littlefield, Michelle Spencer, Amanda Riccardi, Birgitte Simen, Kelsey R. Florek, Abigail C. Shockey |
| EPI_ISL_1269371 | Helix/Illumina | Centers for Disease Control and Prevention Division of Viral Diseases, Pathogen Discovery | Peter W. Cook, Dakota Howard, Dhwani Batra, Ben L. Rambo-Martin, Eileen de Feo, Jan Antico, Christine Tran, Matthew Tolentino, Shannon Wickline, Kim Gietzen, Brad Sickler, Jingtao Liu, Eric Allen, Phil Febbo, Summer Galloway, Nicole L. Washington, Simon White, Geraint Levan, Kelly Schiabor Barrett, Elizabeth Cirulli, Alexandre Bolze, Ary Ascencio, Charlotte Rivera-Garcia, Ryan Cho, Jason Nguyen, Sherry Wang, Jimmy Ramirez, Tyler Cassens, Efren Sandoval, Magnus Isaksson, William Lee, David Becker, Marc Laurent, James Lu, Clinton R. Paden, Suxiang Tong, Duncan MacCannell |
| EPI_ISL_1269372, EPI_ISL_1269373 | Wisconsin State Laboratory of Hygiene Communicable Disease Division | Ginkgo Bioworks Clinical Laboratory | Malaika Mckenzie-Bennett, James McGann, Jim Griffin, Keith Robison, Alex Plocik, Becky Schilling, Martha Pierson, Rebecca Littlefield, Michelle Spencer, Amanda Riccardi, Birgitte Simen, Kelsey R. Florek, Abigail C. Shockey |
| EPI_ISL_1269374 | Helix/Illumina | Centers for Disease Control and Prevention Division of Viral Diseases, Pathogen Discovery | Peter W. Cook, Dakota Howard, Dhwani Batra, Ben L. Rambo-Martin, Eileen de Feo, Jan Antico, Christine Tran, Matthew Tolentino, Shannon Wickline, Kim Gietzen, Brad Sickler, Jingtao Liu, Eric Allen, Phil Febbo, Summer Galloway, Nicole L. Washington, Simon White, Geraint Levan, Kelly Schiabor Barrett, Elizabeth Cirulli, Alexandre Bolze, Ary Ascencio, Charlotte Rivera-Garcia, Ryan Cho, Jason Nguyen, Sherry Wang, Jimmy Ramirez, Tyler Cassens, Efren Sandoval, Magnus Isaksson, William Lee, David Becker, Marc Laurent, James Lu, Clinton R. Paden, Suxiang Tong, Duncan MacCannell |
| EPI_ISL_1269375, EPI_ISL_1269376 | Wisconsin State Laboratory of Hygiene Communicable Disease Division | Ginkgo Bioworks Clinical Laboratory | Malaika Mckenzie-Bennett, James McGann, Jim Griffin, Keith Robison, Alex Plocik, Becky Schilling, Martha Pierson, Rebecca Littlefield, Michelle Spencer, Amanda Riccardi, Birgitte Simen, Kelsey R. Florek, Abigail C. Shockey |
| EPI_ISL_1269377 | Helix/Illumina | Centers for Disease Control and Prevention | Peter W. Cook, Dakota Howard, Dhwani Batra, Ben L. Rambo-Martin, Eileen de Feo, Jan Antico, Christine Tran, Matthew Tolentino, Shannon Wickline, Kim Gietzen, Brad Sickler, Jingtao Liu, Eric Allen, Phil Febbo, Summer Galloway, Nicole L. Washington, Simon White, Geraint Levan, Kelly |

| | | | |
|---|---|---|---|
| | | Division of Viral Diseases, Pathogen Discovery | Schiabor Barrett, Elizabeth Cirulli, Alexandre Bolze, Ary Ascencio, Charlotte Rivera-Garcia, Ryan Cho, Jason Nguyen, Sherry Wang, Jimmy Ramirez, Tyler Cassens, Efren Sandoval, Magnus Isaksson, William Lee, David Becker, Marc Laurent, James Lu, Clinton R. Paden, Suxiang Tong, Duncan MacCannell |
| EPI_ISL_1269378, EPI_ISL_1269379 | Wisconsin State Laboratory of Hygiene Communicable Disease Division | Ginkgo Bioworks Clinical Laboratory | Malaika Mckenzie-Bennett, James McGann, Jim Griffin, Keith Robison, Alex Plocik, Becky Schilling, Martha Pierson, Rebecca Littlefield, Michelle Spencer, Amanda Riccardi, Birgitte Simen, Kelsey R. Florek, Abigail C. Shockey |
| EPI_ISL_1269380 | Helix/Illumina | Centers for Disease Control and Prevention Division of Viral Diseases, Pathogen Discovery | Peter W. Cook, Dakota Howard, Dhwani Batra, Ben L. Rambo-Martin, Eileen de Feo, Jan Antico, Christine Tran, Matthew Tolentino, Shannon Wickline, Kim Gietzen, Brad Sickler, Jingtao Liu, Eric Allen, Phil Febbo, Summer Galloway, Nicole L. Washington, Simon White, Geraint Levan, Kelly Schiabor Barrett, Elizabeth Cirulli, Alexandre Bolze, Ary Ascencio, Charlotte Rivera-Garcia, Ryan Cho, Jason Nguyen, Sherry Wang, Jimmy Ramirez, Tyler Cassens, Efren Sandoval, Magnus Isaksson, William Lee, David Becker, Marc Laurent, James Lu, Clinton R. Paden, Suxiang Tong, Duncan MacCannell |
| EPI_ISL_1269381, EPI_ISL_1269382 | Wisconsin State Laboratory of Hygiene Communicable Disease Division | Ginkgo Bioworks Clinical Laboratory | Malaika Mckenzie-Bennett, James McGann, Jim Griffin, Keith Robison, Alex Plocik, Becky Schilling, Martha Pierson, Rebecca Littlefield, Michelle Spencer, Amanda Riccardi, Birgitte Simen, Kelsey R. Florek, Abigail C. Shockey |
| EPI_ISL_1269383 | Helix/Illumina | Centers for Disease Control and Prevention Division of Viral Diseases, Pathogen Discovery | Peter W. Cook, Dakota Howard, Dhwani Batra, Ben L. Rambo-Martin, Eileen de Feo, Jan Antico, Christine Tran, Matthew Tolentino, Shannon Wickline, Kim Gietzen, Brad Sickler, Jingtao Liu, Eric Allen, Phil Febbo, Summer Galloway, Nicole L. Washington, Simon White, Geraint Levan, Kelly Schiabor Barrett, Elizabeth Cirulli, Alexandre Bolze, Ary Ascencio, Charlotte Rivera-Garcia, Ryan Cho, Jason Nguyen, Sherry Wang, Jimmy Ramirez, Tyler Cassens, Efren Sandoval, Magnus Isaksson, William Lee, David Becker, Marc Laurent, James Lu, Clinton R. Paden, Suxiang Tong, Duncan MacCannell |
| EPI_ISL_1269384, EPI_ISL_1269385 | Wisconsin State Laboratory of Hygiene Communicable Disease Division | Ginkgo Bioworks Clinical Laboratory | Malaika Mckenzie-Bennett, James McGann, Jim Griffin, Keith Robison, Alex Plocik, Becky Schilling, Martha Pierson, Rebecca Littlefield, Michelle Spencer, Amanda Riccardi, Birgitte Simen, Kelsey R. Florek, Abigail C. Shockey |
| EPI_ISL_1269386 | Helix/Illumina | Centers for Disease Control and Prevention Division of Viral Diseases, Pathogen Discovery | Peter W. Cook, Dakota Howard, Dhwani Batra, Ben L. Rambo-Martin, Eileen de Feo, Jan Antico, Christine Tran, Matthew Tolentino, Shannon Wickline, Kim Gietzen, Brad Sickler, Jingtao Liu, Eric Allen, Phil Febbo, Summer Galloway, Nicole L. Washington, Simon White, Geraint Levan, Kelly Schiabor Barrett, Elizabeth Cirulli, Alexandre Bolze, Ary Ascencio, Charlotte Rivera-Garcia, Ryan Cho, Jason Nguyen, Sherry Wang, Jimmy Ramirez, Tyler Cassens, Efren Sandoval, Magnus Isaksson, William Lee, David Becker, Marc Laurent, James Lu, Clinton R. Paden, Suxiang Tong, Duncan MacCannell |
| EPI_ISL_1269387, EPI_ISL_1269388 | Wisconsin State Laboratory of Hygiene Communicable Disease Division | Ginkgo Bioworks Clinical Laboratory | Malaika Mckenzie-Bennett, James McGann, Jim Griffin, Keith Robison, Alex Plocik, Becky Schilling, Martha Pierson, Rebecca Littlefield, Michelle Spencer, Amanda Riccardi, Birgitte Simen, Kelsey R. Florek, Abigail C. Shockey |
| EPI_ISL_1269389 | Helix/Illumina | Centers for Disease Control and Prevention Division of Viral Diseases, Pathogen Discovery | Peter W. Cook, Dakota Howard, Dhwani Batra, Ben L. Rambo-Martin, Eileen de Feo, Jan Antico, Christine Tran, Matthew Tolentino, Shannon Wickline, Kim Gietzen, Brad Sickler, Jingtao Liu, Eric Allen, Phil Febbo, Summer Galloway, Nicole L. Washington, Simon White, Geraint Levan, Kelly Schiabor Barrett, Elizabeth Cirulli, Alexandre Bolze, Ary Ascencio, Charlotte Rivera-Garcia, Ryan Cho, Jason Nguyen, Sherry Wang, Jimmy Ramirez, Tyler Cassens, Efren Sandoval, Magnus Isaksson, William Lee, David Becker, Marc Laurent, James Lu, Clinton R. Paden, Suxiang Tong, Duncan MacCannell |
| EPI_ISL_1269390, EPI_ISL_1269391 | Wisconsin State Laboratory of Hygiene Communicable Disease Division | Ginkgo Bioworks Clinical Laboratory | Malaika Mckenzie-Bennett, James McGann, Jim Griffin, Keith Robison, Alex Plocik, Becky Schilling, Martha Pierson, Rebecca Littlefield, Michelle Spencer, Amanda Riccardi, Birgitte Simen, Kelsey R. Florek, Abigail C. Shockey |
| EPI_ISL_1269392 | Helix/Illumina | Centers for Disease Control and Prevention Division of Viral Diseases, Pathogen Discovery | Peter W. Cook, Dakota Howard, Dhwani Batra, Ben L. Rambo-Martin, Eileen de Feo, Jan Antico, Christine Tran, Matthew Tolentino, Shannon Wickline, Kim Gietzen, Brad Sickler, Jingtao Liu, Eric Allen, Phil Febbo, Summer Galloway, Nicole L. Washington, Simon White, Geraint Levan, Kelly Schiabor Barrett, Elizabeth Cirulli, Alexandre Bolze, Ary Ascencio, Charlotte Rivera-Garcia, Ryan Cho, Jason Nguyen, Sherry Wang, Jimmy Ramirez, Tyler Cassens, Efren Sandoval, Magnus Isaksson, William Lee, David Becker, Marc Laurent, James Lu, Clinton R. Paden, Suxiang Tong, Duncan MacCannell |
| EPI_ISL_1269393, EPI_ISL_1269394 | Wisconsin State Laboratory of Hygiene Communicable Disease Division | Ginkgo Bioworks Clinical Laboratory | Malaika Mckenzie-Bennett, James McGann, Jim Griffin, Keith Robison, Alex Plocik, Becky Schilling, Martha Pierson, Rebecca Littlefield, Michelle Spencer, Amanda Riccardi, Birgitte Simen, Kelsey R. Florek, Abigail C. Shockey |
| EPI_ISL_1269395 | Helix/Illumina | Centers for Disease Control and Prevention Division of Viral Diseases, Pathogen Discovery | Peter W. Cook, Dakota Howard, Dhwani Batra, Ben L. Rambo-Martin, Eileen de Feo, Jan Antico, Christine Tran, Matthew Tolentino, Shannon Wickline, Kim Gietzen, Brad Sickler, Jingtao Liu, Eric Allen, Phil Febbo, Summer Galloway, Nicole L. Washington, Simon White, Geraint Levan, Kelly Schiabor Barrett, Elizabeth Cirulli, Alexandre Bolze, Ary Ascencio, Charlotte Rivera-Garcia, Ryan Cho, Jason Nguyen, Sherry Wang, Jimmy Ramirez, Tyler Cassens, Efren Sandoval, Magnus Isaksson, William Lee, David Becker, Marc Laurent, James Lu, Clinton R. Paden, Suxiang Tong, Duncan MacCannell |
| EPI_ISL_1269396, EPI_ISL_1269397, EPI_ISL_1269398 | Wisconsin State Laboratory of Hygiene Communicable Disease Division | Ginkgo Bioworks Clinical Laboratory | Malaika Mckenzie-Bennett, James McGann, Jim Griffin, Keith Robison, Alex Plocik, Becky Schilling, Martha Pierson, Rebecca Littlefield, Michelle Spencer, Amanda Riccardi, Birgitte Simen, Kelsey R. Florek, Abigail C. Shockey |
| EPI_ISL_1269399 | Helix/Illumina | Centers for Disease Control and Prevention Division of Viral Diseases, Pathogen Discovery | Peter W. Cook, Dakota Howard, Dhwani Batra, Ben L. Rambo-Martin, Eileen de Feo, Jan Antico, Christine Tran, Matthew Tolentino, Shannon Wickline, Kim Gietzen, Brad Sickler, Jingtao Liu, Eric Allen, Phil Febbo, Summer Galloway, Nicole L. Washington, Simon White, Geraint Levan, Kelly Schiabor Barrett, Elizabeth Cirulli, Alexandre Bolze, Ary Ascencio, Charlotte Rivera-Garcia, Ryan Cho, Jason Nguyen, Sherry Wang, Jimmy Ramirez, Tyler Cassens, Efren Sandoval, Magnus Isaksson, William Lee, David Becker, Marc Laurent, James Lu, Clinton R. Paden, Suxiang Tong, Duncan MacCannell |
| EPI_ISL_1269400, EPI_ISL_1269401 | Wisconsin State Laboratory of Hygiene Communicable Disease Division | Ginkgo Bioworks Clinical Laboratory | Malaika Mckenzie-Bennett, James McGann, Jim Griffin, Keith Robison, Alex Plocik, Becky Schilling, Martha Pierson, Rebecca Littlefield, Michelle Spencer, Amanda Riccardi, Birgitte Simen, Kelsey R. Florek, Abigail C. Shockey |
| EPI_ISL_1269402 | Helix/Illumina | Centers for Disease Control and Prevention Division of Viral Diseases, Pathogen Discovery | Peter W. Cook, Dakota Howard, Dhwani Batra, Ben L. Rambo-Martin, Eileen de Feo, Jan Antico, Christine Tran, Matthew Tolentino, Shannon Wickline, Kim Gietzen, Brad Sickler, Jingtao Liu, Eric Allen, Phil Febbo, Summer Galloway, Nicole L. Washington, Simon White, Geraint Levan, Kelly Schiabor Barrett, Elizabeth Cirulli, Alexandre Bolze, Ary Ascencio, Charlotte Rivera-Garcia, Ryan Cho, Jason Nguyen, Sherry Wang, Jimmy Ramirez, Tyler Cassens, Efren Sandoval, Magnus Isaksson, William Lee, David Becker, Marc Laurent, James Lu, Clinton R. Paden, Suxiang Tong, Duncan MacCannell |
| EPI_ISL_1269403, EPI_ISL_1269404 | Wisconsin State Laboratory of Hygiene Communicable Disease Division | Ginkgo Bioworks Clinical Laboratory | Malaika Mckenzie-Bennett, James McGann, Jim Griffin, Keith Robison, Alex Plocik, Becky Schilling, Martha Pierson, Rebecca Littlefield, Michelle Spencer, Amanda Riccardi, Birgitte Simen, Kelsey R. Florek, Abigail C. Shockey |
| EPI_ISL_1269405 | Helix/Illumina | Centers for Disease Control and Prevention Division of Viral Diseases, Pathogen Discovery | Peter W. Cook, Dakota Howard, Dhwani Batra, Ben L. Rambo-Martin, Eileen de Feo, Jan Antico, Christine Tran, Matthew Tolentino, Shannon Wickline, Kim Gietzen, Brad Sickler, Jingtao Liu, Eric Allen, Phil Febbo, Summer Galloway, Nicole L. Washington, Simon White, Geraint Levan, Kelly Schiabor Barrett, Elizabeth Cirulli, Alexandre Bolze, Ary Ascencio, Charlotte Rivera-Garcia, Ryan Cho, Jason Nguyen, Sherry Wang, Jimmy Ramirez, Tyler Cassens, Efren Sandoval, Magnus Isaksson, William Lee, David Becker, Marc Laurent, James Lu, Clinton R. Paden, Suxiang Tong, Duncan MacCannell |
| EPI_ISL_1269406, EPI_ISL_1269407 | Wisconsin State Laboratory of Hygiene Communicable Disease Division | Ginkgo Bioworks Clinical Laboratory | Malaika Mckenzie-Bennett, James McGann, Jim Griffin, Keith Robison, Alex Plocik, Becky Schilling, Martha Pierson, Rebecca Littlefield, Michelle Spencer, Amanda Riccardi, Birgitte Simen, Kelsey R. Florek, Abigail C. Shockey |
| EPI_ISL_1269408 | Helix/Illumina | Centers for Disease Control and Prevention Division of Viral Diseases, Pathogen Discovery | Peter W. Cook, Dakota Howard, Dhwani Batra, Ben L. Rambo-Martin, Eileen de Feo, Jan Antico, Christine Tran, Matthew Tolentino, Shannon Wickline, Kim Gietzen, Brad Sickler, Jingtao Liu, Eric Allen, Phil Febbo, Summer Galloway, Nicole L. Washington, Simon White, Geraint Levan, Kelly Schiabor Barrett, Elizabeth Cirulli, Alexandre Bolze, Ary Ascencio, Charlotte Rivera-Garcia, Ryan Cho, Jason Nguyen, Sherry Wang, Jimmy Ramirez, Tyler Cassens, Efren Sandoval, Magnus Isaksson, William Lee, David Becker, Marc Laurent, James Lu, Clinton R. Paden, Suxiang Tong, Duncan MacCannell |
| EPI_ISL_1269409, EPI_ISL_1269410, EPI_ISL_1269411 | Wisconsin State Laboratory of Hygiene Communicable Disease Division | Ginkgo Bioworks Clinical Laboratory | Malaika Mckenzie-Bennett, James McGann, Jim Griffin, Keith Robison, Alex Plocik, Becky Schilling, Martha Pierson, Rebecca Littlefield, Michelle Spencer, Amanda Riccardi, Birgitte Simen, Kelsey R. Florek, Abigail C. Shockey |
| EPI_ISL_1269412 | Helix/Illumina | Centers for Disease Control and Prevention Division of Viral Diseases, Pathogen Discovery | Peter W. Cook, Dakota Howard, Dhwani Batra, Ben L. Rambo-Martin, Eileen de Feo, Jan Antico, Christine Tran, Matthew Tolentino, Shannon Wickline, Kim Gietzen, Brad Sickler, Jingtao Liu, Eric Allen, Phil Febbo, Summer Galloway, Nicole L. Washington, Simon White, Geraint Levan, Kelly Schiabor Barrett, Elizabeth Cirulli, Alexandre Bolze, Ary Ascencio, Charlotte Rivera-Garcia, Ryan Cho, Jason Nguyen, Sherry Wang, Jimmy Ramirez, Tyler Cassens, Efren Sandoval, Magnus Isaksson, William Lee, David Becker, Marc Laurent, James Lu, Clinton R. Paden, Suxiang Tong, Duncan MacCannell |
| EPI_ISL_1269413, EPI_ISL_1269414 | Wisconsin State Laboratory of Hygiene Communicable Disease Division | Ginkgo Bioworks Clinical Laboratory | Malaika Mckenzie-Bennett, James McGann, Jim Griffin, Keith Robison, Alex Plocik, Becky Schilling, Martha Pierson, Rebecca Littlefield, Michelle Spencer, Amanda Riccardi, Birgitte Simen, Kelsey R. Florek, Abigail C. Shockey |
| EPI_ISL_1269415 | Helix/Illumina | Centers for Disease Control and Prevention Division of Viral Diseases, Pathogen Discovery | Peter W. Cook, Dakota Howard, Dhwani Batra, Ben L. Rambo-Martin, Eileen de Feo, Jan Antico, Christine Tran, Matthew Tolentino, Shannon Wickline, Kim Gietzen, Brad Sickler, Jingtao Liu, Eric Allen, Phil Febbo, Summer Galloway, Nicole L. Washington, Simon White, Geraint Levan, Kelly Schiabor Barrett, Elizabeth Cirulli, Alexandre Bolze, Ary Ascencio, Charlotte Rivera-Garcia, Ryan Cho, Jason Nguyen, Sherry Wang, Jimmy Ramirez, Tyler Cassens, Efren Sandoval, Magnus Isaksson, William Lee, David Becker, Marc Laurent, James Lu, Clinton R. Paden, Suxiang Tong, Duncan MacCannell |
| EPI_ISL_1269416, EPI_ISL_1269417 | Wisconsin State Laboratory of Hygiene Communicable Disease Division | Ginkgo Bioworks Clinical Laboratory | Malaika Mckenzie-Bennett, James McGann, Jim Griffin, Keith Robison, Alex Plocik, Becky Schilling, Martha Pierson, Rebecca Littlefield, Michelle Spencer, Amanda Riccardi, Birgitte Simen, Kelsey R. Florek, Abigail C. Shockey |
| EPI_ISL_1269418 | Helix/Illumina | Centers for Disease Control and Prevention Division of Viral Diseases, Pathogen Discovery | Peter W. Cook, Dakota Howard, Dhwani Batra, Ben L. Rambo-Martin, Eileen de Feo, Jan Antico, Christine Tran, Matthew Tolentino, Shannon Wickline, Kim Gietzen, Brad Sickler, Jingtao Liu, Eric Allen, Phil Febbo, Summer Galloway, Nicole L. Washington, Simon White, Geraint Levan, Kelly Schiabor Barrett, Elizabeth Cirulli, Alexandre Bolze, Ary Ascencio, Charlotte Rivera-Garcia, Ryan Cho, Jason Nguyen, Sherry Wang, Jimmy Ramirez, Tyler Cassens, Efren Sandoval, Magnus Isaksson, William Lee, David Becker, Marc Laurent, James Lu, Clinton R. Paden, Suxiang Tong, Duncan MacCannell |
| EPI_ISL_1269419, EPI_ISL_1269420, EPI_ISL_1269421 | Wisconsin State Laboratory of Hygiene Communicable Disease Division | Ginkgo Bioworks Clinical Laboratory | Malaika Mckenzie-Bennett, James McGann, Jim Griffin, Keith Robison, Alex Plocik, Becky Schilling, Martha Pierson, Rebecca Littlefield, Michelle Spencer, Amanda Riccardi, Birgitte Simen, Kelsey R. Florek, Abigail C. Shockey |
| EPI_ISL_1269422 | Helix/Illumina | Centers for Disease Control and Prevention Division of Viral Diseases, Pathogen Discovery | Peter W. Cook, Dakota Howard, Dhwani Batra, Ben L. Rambo-Martin, Eileen de Feo, Jan Antico, Christine Tran, Matthew Tolentino, Shannon Wickline, Kim Gietzen, Brad Sickler, Jingtao Liu, Eric Allen, Phil Febbo, Summer Galloway, Nicole L. Washington, Simon White, Geraint Levan, Kelly Schiabor Barrett, Elizabeth Cirulli, Alexandre Bolze, Ary Ascencio, Charlotte Rivera-Garcia, Ryan Cho, Jason Nguyen, Sherry Wang, Jimmy Ramirez, Tyler Cassens, Efren Sandoval, Magnus Isaksson, William Lee, David Becker, Marc Laurent, James Lu, Clinton R. Paden, Suxiang Tong, Duncan MacCannell |
| EPI_ISL_1269423, EPI_ISL_1269424 | Wisconsin State Laboratory of Hygiene Communicable Disease Division | Ginkgo Bioworks Clinical Laboratory | Malaika Mckenzie-Bennett, James McGann, Jim Griffin, Keith Robison, Alex Plocik, Becky Schilling, Martha Pierson, Rebecca Littlefield, Michelle Spencer, Amanda Riccardi, Birgitte Simen, Kelsey R. Florek, Abigail C. Shockey |
| EPI_ISL_1269425 | Helix/Illumina | Centers for Disease Control and Prevention Division of Viral Diseases, Pathogen Discovery | Peter W. Cook, Dakota Howard, Dhwani Batra, Ben L. Rambo-Martin, Eileen de Feo, Jan Antico, Christine Tran, Matthew Tolentino, Shannon Wickline, Kim Gietzen, Brad Sickler, Jingtao Liu, Eric Allen, Phil Febbo, Summer Galloway, Nicole L. Washington, Simon White, Geraint Levan, Kelly Schiabor Barrett, Elizabeth Cirulli, Alexandre Bolze, Ary Ascencio, Charlotte Rivera-Garcia, Ryan Cho, Jason Nguyen, Sherry Wang, Jimmy Ramirez, Tyler Cassens, Efren Sandoval, Magnus Isaksson, William Lee, David Becker, Marc Laurent, James Lu, Clinton R. Paden, Suxiang Tong, Duncan MacCannell |
| EPI_ISL_1269426, EPI_ISL_1269427 | Wisconsin State Laboratory of Hygiene Communicable Disease Division | Ginkgo Bioworks Clinical Laboratory | Malaika Mckenzie-Bennett, James McGann, Jim Griffin, Keith Robison, Alex Plocik, Becky Schilling, Martha Pierson, Rebecca Littlefield, Michelle Spencer, Amanda Riccardi, Birgitte Simen, Kelsey R. Florek, Abigail C. Shockey |
| EPI_ISL_1269428 | Helix/Illumina | Centers for Disease Control and Prevention Division of Viral Diseases, Pathogen Discovery | Peter W. Cook, Dakota Howard, Dhwani Batra, Ben L. Rambo-Martin, Eileen de Feo, Jan Antico, Christine Tran, Matthew Tolentino, Shannon Wickline, Kim Gietzen, Brad Sickler, Jingtao Liu, Eric Allen, Phil Febbo, Summer Galloway, Nicole L. Washington, Simon White, Geraint Levan, Kelly Schiabor Barrett, Elizabeth Cirulli, Alexandre Bolze, Ary Ascencio, Charlotte Rivera-Garcia, Ryan Cho, Jason Nguyen, Sherry Wang, Jimmy Ramirez, Tyler Cassens, Efren Sandoval, Magnus Isaksson, William Lee, David Becker, Marc Laurent, James Lu, Clinton R. Paden, Suxiang Tong, Duncan MacCannell |
| EPI_ISL_1269429, EPI_ISL_1269430 | Wisconsin State Laboratory of Hygiene Communicable Disease Division | Ginkgo Bioworks Clinical Laboratory | Malaika Mckenzie-Bennett, James McGann, Jim Griffin, Keith Robison, Alex Plocik, Becky Schilling, Martha Pierson, Rebecca Littlefield, Michelle Spencer, Amanda Riccardi, Birgitte Simen, Kelsey R. Florek, Abigail C. Shockey |
| EPI_ISL_1269431 | Helix/Illumina | Centers for Disease Control and Prevention Division of Viral Diseases, Pathogen Discovery | Peter W. Cook, Dakota Howard, Dhwani Batra, Ben L. Rambo-Martin, Eileen de Feo, Jan Antico, Christine Tran, Matthew Tolentino, Shannon Wickline, Kim Gietzen, Brad Sickler, Jingtao Liu, Eric Allen, Phil Febbo, Summer Galloway, Nicole L. Washington, Simon White, Geraint Levan, Kelly Schiabor Barrett, Elizabeth Cirulli, Alexandre Bolze, Ary Ascencio, Charlotte Rivera-Garcia, Ryan Cho, Jason Nguyen, Sherry Wang, Jimmy Ramirez, Tyler Cassens, Efren Sandoval, Magnus Isaksson, William Lee, David Becker, Marc Laurent, James Lu, Clinton R. Paden, Suxiang Tong, Duncan MacCannell |
| EPI_ISL_1269432, EPI_ISL_1269433 | Wisconsin State Laboratory of Hygiene Communicable Disease Division | Ginkgo Bioworks Clinical Laboratory | Malaika Mckenzie-Bennett, James McGann, Jim Griffin, Keith Robison, Alex Plocik, Becky Schilling, Martha Pierson, Rebecca Littlefield, Michelle Spencer, Amanda Riccardi, Birgitte Simen, Kelsey R. Florek, Abigail C. Shockey |
| EPI_ISL_1269434 | Helix/Illumina | Centers for Disease Control and Prevention Division of Viral Diseases, Pathogen | Peter W. Cook, Dakota Howard, Dhwani Batra, Ben L. Rambo-Martin, Eileen de Feo, Jan Antico, Christine Tran, Matthew Tolentino, Shannon Wickline, Kim Gietzen, Brad Sickler, Jingtao Liu, Eric Allen, Phil Febbo, Summer Galloway, Nicole L. Washington, Simon White, Geraint Levan, Kelly Schiabor Barrett, Elizabeth Cirulli, Alexandre Bolze, Ary Ascencio, Charlotte Rivera-Garcia, Ryan Cho, Jason Nguyen, Sherry Wang, Jimmy Ramirez, Tyler Cassens, Efren Sandoval, Magnus Isaksson, William Lee, David Becker, Marc Laurent, James Lu, Clinton R. Paden, Suxiang Tong, |

| | | | |
|---|---|---|---|
| | | Discovery | Duncan MacCannell |
| EPI_ISL_1269435, EPI_ISL_1269436 | Wisconsin State Laboratory of Hygiene Communicable Disease Division | Ginkgo Bioworks Clinical Laboratory | Malaika Mckenzie-Bennett, James McGann, Jim Griffin, Keith Robison, Alex Plocik, Becky Schilling, Martha Pierson, Rebecca Littlefield, Michelle Spencer, Amanda Riccardi, Birgitte Simen, Kelsey R. Florek, Abigail C. Shockey |
| EPI_ISL_1269437 | Helix/Illumina | Centers for Disease Control and Prevention Division of Viral Diseases, Pathogen Discovery | Peter W. Cook, Dakota Howard, Dhwani Batra, Ben L. Rambo-Martin, Eileen de Feo, Jan Antico, Christine Tran, Matthew Tolentino, Shannon Wickline, Kim Gietzen, Brad Sickler, Jingtao Liu, Eric Allen, Phil Febbo, Summer Galloway, Nicole L. Washington, Simon White, Geraint Levan, Kelly Schiabor Barrett, Elizabeth Cirulli, Alexandre Bolze, Ary Ascencio, Charlotte Rivera-Garcia, Ryan Cho, Jason Nguyen, Sherry Wang, Jimmy Ramirez, Tyler Cassens, Efren Sandoval, Magnus Isaksson, William Lee, David Becker, Marc Laurent, James Lu, Clinton R. Paden, Suxiang Tong, Duncan MacCannell |
| EPI_ISL_1269438, EPI_ISL_1269439, EPI_ISL_1269440 | Wisconsin State Laboratory of Hygiene Communicable Disease Division | Ginkgo Bioworks Clinical Laboratory | Malaika Mckenzie-Bennett, James McGann, Jim Griffin, Keith Robison, Alex Plocik, Becky Schilling, Martha Pierson, Rebecca Littlefield, Michelle Spencer, Amanda Riccardi, Birgitte Simen, Kelsey R. Florek, Abigail C. Shockey |
| EPI_ISL_1269441 | Helix/Illumina | Centers for Disease Control and Prevention Division of Viral Diseases, Pathogen Discovery | Peter W. Cook, Dakota Howard, Dhwani Batra, Ben L. Rambo-Martin, Eileen de Feo, Jan Antico, Christine Tran, Matthew Tolentino, Shannon Wickline, Kim Gietzen, Brad Sickler, Jingtao Liu, Eric Allen, Phil Febbo, Summer Galloway, Nicole L. Washington, Simon White, Geraint Levan, Kelly Schiabor Barrett, Elizabeth Cirulli, Alexandre Bolze, Ary Ascencio, Charlotte Rivera-Garcia, Ryan Cho, Jason Nguyen, Sherry Wang, Jimmy Ramirez, Tyler Cassens, Efren Sandoval, Magnus Isaksson, William Lee, David Becker, Marc Laurent, James Lu, Clinton R. Paden, Suxiang Tong, Duncan MacCannell |
| EPI_ISL_1269442, EPI_ISL_1269443 | Wisconsin State Laboratory of Hygiene Communicable Disease Division | Ginkgo Bioworks Clinical Laboratory | Malaika Mckenzie-Bennett, James McGann, Jim Griffin, Keith Robison, Alex Plocik, Becky Schilling, Martha Pierson, Rebecca Littlefield, Michelle Spencer, Amanda Riccardi, Birgitte Simen, Kelsey R. Florek, Abigail C. Shockey |
| EPI_ISL_1269444 | Helix/Illumina | Centers for Disease Control and Prevention Division of Viral Diseases, Pathogen Discovery | Peter W. Cook, Dakota Howard, Dhwani Batra, Ben L. Rambo-Martin, Eileen de Feo, Jan Antico, Christine Tran, Matthew Tolentino, Shannon Wickline, Kim Gietzen, Brad Sickler, Jingtao Liu, Eric Allen, Phil Febbo, Summer Galloway, Nicole L. Washington, Simon White, Geraint Levan, Kelly Schiabor Barrett, Elizabeth Cirulli, Alexandre Bolze, Ary Ascencio, Charlotte Rivera-Garcia, Ryan Cho, Jason Nguyen, Sherry Wang, Jimmy Ramirez, Tyler Cassens, Efren Sandoval, Magnus Isaksson, William Lee, David Becker, Marc Laurent, James Lu, Clinton R. Paden, Suxiang Tong, Duncan MacCannell |
| EPI_ISL_1269445, EPI_ISL_1269446 | Wisconsin State Laboratory of Hygiene Communicable Disease Division | Ginkgo Bioworks Clinical Laboratory | Malaika Mckenzie-Bennett, James McGann, Jim Griffin, Keith Robison, Alex Plocik, Becky Schilling, Martha Pierson, Rebecca Littlefield, Michelle Spencer, Amanda Riccardi, Birgitte Simen, Kelsey R. Florek, Abigail C. Shockey |
| EPI_ISL_1269447 | Helix/Illumina | Centers for Disease Control and Prevention Division of Viral Diseases, Pathogen Discovery | Peter W. Cook, Dakota Howard, Dhwani Batra, Ben L. Rambo-Martin, Eileen de Feo, Jan Antico, Christine Tran, Matthew Tolentino, Shannon Wickline, Kim Gietzen, Brad Sickler, Jingtao Liu, Eric Allen, Phil Febbo, Summer Galloway, Nicole L. Washington, Simon White, Geraint Levan, Kelly Schiabor Barrett, Elizabeth Cirulli, Alexandre Bolze, Ary Ascencio, Charlotte Rivera-Garcia, Ryan Cho, Jason Nguyen, Sherry Wang, Jimmy Ramirez, Tyler Cassens, Efren Sandoval, Magnus Isaksson, William Lee, David Becker, Marc Laurent, James Lu, Clinton R. Paden, Suxiang Tong, Duncan MacCannell |
| EPI_ISL_1269448, EPI_ISL_1269449 | Wisconsin State Laboratory of Hygiene Communicable Disease Division | Ginkgo Bioworks Clinical Laboratory | Malaika Mckenzie-Bennett, James McGann, Jim Griffin, Keith Robison, Alex Plocik, Becky Schilling, Martha Pierson, Rebecca Littlefield, Michelle Spencer, Amanda Riccardi, Birgitte Simen, Kelsey R. Florek, Abigail C. Shockey |
| EPI_ISL_1269450 | Helix/Illumina | Centers for Disease Control and Prevention Division of Viral Diseases, Pathogen Discovery | Peter W. Cook, Dakota Howard, Dhwani Batra, Ben L. Rambo-Martin, Eileen de Feo, Jan Antico, Christine Tran, Matthew Tolentino, Shannon Wickline, Kim Gietzen, Brad Sickler, Jingtao Liu, Eric Allen, Phil Febbo, Summer Galloway, Nicole L. Washington, Simon White, Geraint Levan, Kelly Schiabor Barrett, Elizabeth Cirulli, Alexandre Bolze, Ary Ascencio, Charlotte Rivera-Garcia, Ryan Cho, Jason Nguyen, Sherry Wang, Jimmy Ramirez, Tyler Cassens, Efren Sandoval, Magnus Isaksson, William Lee, David Becker, Marc Laurent, James Lu, Clinton R. Paden, Suxiang Tong, Duncan MacCannell |
| EPI_ISL_1269451, EPI_ISL_1269452 | Wisconsin State Laboratory of Hygiene Communicable Disease Division | Ginkgo Bioworks Clinical Laboratory | Malaika Mckenzie-Bennett, James McGann, Jim Griffin, Keith Robison, Alex Plocik, Becky Schilling, Martha Pierson, Rebecca Littlefield, Michelle Spencer, Amanda Riccardi, Birgitte Simen, Kelsey R. Florek, Abigail C. Shockey |
| EPI_ISL_1269453 | Helix/Illumina | Centers for Disease Control and Prevention Division of Viral Diseases, Pathogen Discovery | Peter W. Cook, Dakota Howard, Dhwani Batra, Ben L. Rambo-Martin, Eileen de Feo, Jan Antico, Christine Tran, Matthew Tolentino, Shannon Wickline, Kim Gietzen, Brad Sickler, Jingtao Liu, Eric Allen, Phil Febbo, Summer Galloway, Nicole L. Washington, Simon White, Geraint Levan, Kelly Schiabor Barrett, Elizabeth Cirulli, Alexandre Bolze, Ary Ascencio, Charlotte Rivera-Garcia, Ryan Cho, Jason Nguyen, Sherry Wang, Jimmy Ramirez, Tyler Cassens, Efren Sandoval, Magnus Isaksson, William Lee, David Becker, Marc Laurent, James Lu, Clinton R. Paden, Suxiang Tong, Duncan MacCannell |
| EPI_ISL_1269454, EPI_ISL_1269455, EPI_ISL_1269456 | Wisconsin State Laboratory of Hygiene Communicable Disease Division | Ginkgo Bioworks Clinical Laboratory | Malaika Mckenzie-Bennett, James McGann, Jim Griffin, Keith Robison, Alex Plocik, Becky Schilling, Martha Pierson, Rebecca Littlefield, Michelle Spencer, Amanda Riccardi, Birgitte Simen, Kelsey R. Florek, Abigail C. Shockey |
| EPI_ISL_1269457 | Helix/Illumina | Centers for Disease Control and Prevention Division of Viral Diseases, Pathogen Discovery | Peter W. Cook, Dakota Howard, Dhwani Batra, Ben L. Rambo-Martin, Eileen de Feo, Jan Antico, Christine Tran, Matthew Tolentino, Shannon Wickline, Kim Gietzen, Brad Sickler, Jingtao Liu, Eric Allen, Phil Febbo, Summer Galloway, Nicole L. Washington, Simon White, Geraint Levan, Kelly Schiabor Barrett, Elizabeth Cirulli, Alexandre Bolze, Ary Ascencio, Charlotte Rivera-Garcia, Ryan Cho, Jason Nguyen, Sherry Wang, Jimmy Ramirez, Tyler Cassens, Efren Sandoval, Magnus Isaksson, William Lee, David Becker, Marc Laurent, James Lu, Clinton R. Paden, Suxiang Tong, Duncan MacCannell |
| EPI_ISL_1269458, EPI_ISL_1269459 | Wisconsin State Laboratory of Hygiene Communicable Disease Division | Ginkgo Bioworks Clinical Laboratory | Malaika Mckenzie-Bennett, James McGann, Jim Griffin, Keith Robison, Alex Plocik, Becky Schilling, Martha Pierson, Rebecca Littlefield, Michelle Spencer, Amanda Riccardi, Birgitte Simen, Kelsey R. Florek, Abigail C. Shockey |
| EPI_ISL_1269460 | Helix/Illumina | Centers for Disease Control and Prevention Division of Viral Diseases, Pathogen Discovery | Peter W. Cook, Dakota Howard, Dhwani Batra, Ben L. Rambo-Martin, Eileen de Feo, Jan Antico, Christine Tran, Matthew Tolentino, Shannon Wickline, Kim Gietzen, Brad Sickler, Jingtao Liu, Eric Allen, Phil Febbo, Summer Galloway, Nicole L. Washington, Simon White, Geraint Levan, Kelly Schiabor Barrett, Elizabeth Cirulli, Alexandre Bolze, Ary Ascencio, Charlotte Rivera-Garcia, Ryan Cho, Jason Nguyen, Sherry Wang, Jimmy Ramirez, Tyler Cassens, Efren Sandoval, Magnus Isaksson, William Lee, David Becker, Marc Laurent, James Lu, Clinton R. Paden, Suxiang Tong, Duncan MacCannell |
| EPI_ISL_1269461, EPI_ISL_1269462 | Wisconsin State Laboratory of Hygiene Communicable Disease Division | Ginkgo Bioworks Clinical Laboratory | Malaika Mckenzie-Bennett, James McGann, Jim Griffin, Keith Robison, Alex Plocik, Becky Schilling, Martha Pierson, Rebecca Littlefield, Michelle Spencer, Amanda Riccardi, Birgitte Simen, Kelsey R. Florek, Abigail C. Shockey |
| EPI_ISL_1269463 | Helix/Illumina | Centers for Disease Control and Prevention Division of Viral Diseases, Pathogen Discovery | Peter W. Cook, Dakota Howard, Dhwani Batra, Ben L. Rambo-Martin, Eileen de Feo, Jan Antico, Christine Tran, Matthew Tolentino, Shannon Wickline, Kim Gietzen, Brad Sickler, Jingtao Liu, Eric Allen, Phil Febbo, Summer Galloway, Nicole L. Washington, Simon White, Geraint Levan, Kelly Schiabor Barrett, Elizabeth Cirulli, Alexandre Bolze, Ary Ascencio, Charlotte Rivera-Garcia, Ryan Cho, Jason Nguyen, Sherry Wang, Jimmy Ramirez, Tyler Cassens, Efren Sandoval, Magnus Isaksson, William Lee, David Becker, Marc Laurent, James Lu, Clinton R. Paden, Suxiang Tong, Duncan MacCannell |
| EPI_ISL_1269464, EPI_ISL_1269465 | Wisconsin State Laboratory of Hygiene Communicable Disease Division | Ginkgo Bioworks Clinical Laboratory | Malaika Mckenzie-Bennett, James McGann, Jim Griffin, Keith Robison, Alex Plocik, Becky Schilling, Martha Pierson, Rebecca Littlefield, Michelle Spencer, Amanda Riccardi, Birgitte Simen, Kelsey R. Florek, Abigail C. Shockey |
| EPI_ISL_1269466 | Helix/Illumina | Centers for Disease Control and Prevention Division of Viral Diseases, Pathogen Discovery | Peter W. Cook, Dakota Howard, Dhwani Batra, Ben L. Rambo-Martin, Eileen de Feo, Jan Antico, Christine Tran, Matthew Tolentino, Shannon Wickline, Kim Gietzen, Brad Sickler, Jingtao Liu, Eric Allen, Phil Febbo, Summer Galloway, Nicole L. Washington, Simon White, Geraint Levan, Kelly Schiabor Barrett, Elizabeth Cirulli, Alexandre Bolze, Ary Ascencio, Charlotte Rivera-Garcia, Ryan Cho, Jason Nguyen, Sherry Wang, Jimmy Ramirez, Tyler Cassens, Efren Sandoval, Magnus Isaksson, William Lee, David Becker, Marc Laurent, James Lu, Clinton R. Paden, Suxiang Tong, Duncan MacCannell |
| EPI_ISL_1269467 | Wisconsin State Laboratory of Hygiene Communicable Disease Division | Ginkgo Bioworks Clinical Laboratory | Malaika Mckenzie-Bennett, James McGann, Jim Griffin, Keith Robison, Alex Plocik, Becky Schilling, Martha Pierson, Rebecca Littlefield, Michelle Spencer, Amanda Riccardi, Birgitte Simen, Kelsey R. Florek, Abigail C. Shockey |
| EPI_ISL_1269470, EPI_ISL_1269473, EPI_ISL_1269476, EPI_ISL_1269479, EPI_ISL_1269482, EPI_ISL_1269485, EPI_ISL_1269488, EPI_ISL_1269491, EPI_ISL_1269495 | Helix/Illumina | Centers for Disease Control and Prevention Division of Viral Diseases, Pathogen Discovery | Peter W. Cook, Dakota Howard, Dhwani Batra, Ben L. Rambo-Martin, Eileen de Feo, Jan Antico, Christine Tran, Matthew Tolentino, Shannon Wickline, Kim Gietzen, Brad Sickler, Jingtao Liu, Eric Allen, Phil Febbo, Summer Galloway, Nicole L. Washington, Simon White, Geraint Levan, Kelly Schiabor Barrett, Elizabeth Cirulli, Alexandre Bolze, Ary Ascencio, Charlotte Rivera-Garcia, Ryan Cho, Jason Nguyen, Sherry Wang, Jimmy Ramirez, Tyler Cassens, Efren Sandoval, Magnus Isaksson, William Lee, David Becker, Marc Laurent, James Lu, Clinton R. Paden, Suxiang Tong, Duncan MacCannell |
| EPI_ISL_1269497 | Wisconsin State Laboratory of Hygiene Communicable Disease Division | Ginkgo Bioworks Clinical Laboratory | Malaika Mckenzie-Bennett, James McGann, Jim Griffin, Keith Robison, Alex Plocik, Becky Schilling, Martha Pierson, Rebecca Littlefield, Michelle Spencer, Amanda Riccardi, Birgitte Simen, Kelsey R. Florek, Abigail C. Shockey |
| EPI_ISL_1269498 | Helix/Illumina | Centers for Disease Control and Prevention Division of Viral Diseases, Pathogen Discovery | Peter W. Cook, Dakota Howard, Dhwani Batra, Ben L. Rambo-Martin, Eileen de Feo, Jan Antico, Christine Tran, Matthew Tolentino, Shannon Wickline, Kim Gietzen, Brad Sickler, Jingtao Liu, Eric Allen, Phil Febbo, Summer Galloway, Nicole L. Washington, Simon White, Geraint Levan, Kelly Schiabor Barrett, Elizabeth Cirulli, Alexandre Bolze, Ary Ascencio, Charlotte Rivera-Garcia, Ryan Cho, Jason Nguyen, Sherry Wang, Jimmy Ramirez, Tyler Cassens, Efren Sandoval, Magnus Isaksson, William Lee, David Becker, Marc Laurent, James Lu, Clinton R. Paden, Suxiang Tong, Duncan MacCannell |
| EPI_ISL_1269499, EPI_ISL_1269500, EPI_ISL_1269501, EPI_ISL_1269502 | Wisconsin State Laboratory of Hygiene Communicable Disease Division | Ginkgo Bioworks Clinical Laboratory | Malaika Mckenzie-Bennett, James McGann, Jim Griffin, Keith Robison, Alex Plocik, Becky Schilling, Martha Pierson, Rebecca Littlefield, Michelle Spencer, Amanda Riccardi, Birgitte Simen, Kelsey R. Florek, Abigail C. Shockey |
| EPI_ISL_1269503 | Helix/Illumina | Centers for Disease Control and Prevention Division of Viral Diseases, Pathogen Discovery | Peter W. Cook, Dakota Howard, Dhwani Batra, Ben L. Rambo-Martin, Eileen de Feo, Jan Antico, Christine Tran, Matthew Tolentino, Shannon Wickline, Kim Gietzen, Brad Sickler, Jingtao Liu, Eric Allen, Phil Febbo, Summer Galloway, Nicole L. Washington, Simon White, Geraint Levan, Kelly Schiabor Barrett, Elizabeth Cirulli, Alexandre Bolze, Ary Ascencio, Charlotte Rivera-Garcia, Ryan Cho, Jason Nguyen, Sherry Wang, Jimmy Ramirez, Tyler Cassens, Efren Sandoval, Magnus Isaksson, William Lee, David Becker, Marc Laurent, James Lu, Clinton R. Paden, Suxiang Tong, Duncan MacCannell |
| EPI_ISL_1269504, EPI_ISL_1269505, EPI_ISL_1269506 | Wisconsin State Laboratory of Hygiene Communicable Disease Division | Ginkgo Bioworks Clinical Laboratory | Malaika Mckenzie-Bennett, James McGann, Jim Griffin, Keith Robison, Alex Plocik, Becky Schilling, Martha Pierson, Rebecca Littlefield, Michelle Spencer, Amanda Riccardi, Birgitte Simen, Kelsey R. Florek, Abigail C. Shockey |
| EPI_ISL_1269507, EPI_ISL_1269509 | Helix/Illumina | Centers for Disease Control and Prevention Division of Viral Diseases, Pathogen Discovery | Peter W. Cook, Dakota Howard, Dhwani Batra, Ben L. Rambo-Martin, Eileen de Feo, Jan Antico, Christine Tran, Matthew Tolentino, Shannon Wickline, Kim Gietzen, Brad Sickler, Jingtao Liu, Eric Allen, Phil Febbo, Summer Galloway, Nicole L. Washington, Simon White, Geraint Levan, Kelly Schiabor Barrett, Elizabeth Cirulli, Alexandre Bolze, Ary Ascencio, Charlotte Rivera-Garcia, Ryan Cho, Jason Nguyen, Sherry Wang, Jimmy Ramirez, Tyler Cassens, Efren Sandoval, Magnus Isaksson, William Lee, David Becker, Marc Laurent, James Lu, Clinton R. Paden, Suxiang Tong, Duncan MacCannell |
| EPI_ISL_1269510, EPI_ISL_1269511, EPI_ISL_1269512 | Wisconsin State Laboratory of Hygiene Communicable Disease Division | Ginkgo Bioworks Clinical Laboratory | Malaika Mckenzie-Bennett, James McGann, Jim Griffin, Keith Robison, Alex Plocik, Becky Schilling, Martha Pierson, Rebecca Littlefield, Michelle Spencer, Amanda Riccardi, Birgitte Simen, Kelsey R. Florek, Abigail C. Shockey |
| EPI_ISL_1269513 | Helix/Illumina | Centers for Disease Control and Prevention Division of Viral Diseases, Pathogen Discovery | Peter W. Cook, Dakota Howard, Dhwani Batra, Ben L. Rambo-Martin, Eileen de Feo, Jan Antico, Christine Tran, Matthew Tolentino, Shannon Wickline, Kim Gietzen, Brad Sickler, Jingtao Liu, Eric Allen, Phil Febbo, Summer Galloway, Nicole L. Washington, Simon White, Geraint Levan, Kelly Schiabor Barrett, Elizabeth Cirulli, Alexandre Bolze, Ary Ascencio, Charlotte Rivera-Garcia, Ryan Cho, Jason Nguyen, Sherry Wang, Jimmy Ramirez, Tyler Cassens, Efren Sandoval, Magnus Isaksson, William Lee, David Becker, Marc Laurent, James Lu, Clinton R. Paden, Suxiang Tong, Duncan MacCannell |
| EPI_ISL_1269514, EPI_ISL_1269515 | Wisconsin State Laboratory of Hygiene Communicable Disease Division | Ginkgo Bioworks Clinical Laboratory | Malaika Mckenzie-Bennett, James McGann, Jim Griffin, Keith Robison, Alex Plocik, Becky Schilling, Martha Pierson, Rebecca Littlefield, Michelle Spencer, Amanda Riccardi, Birgitte Simen, Kelsey R. Florek, Abigail C. Shockey |
| EPI_ISL_1269516 | Helix/Illumina | Centers for Disease Control and Prevention Division of Viral Diseases, Pathogen Discovery | Peter W. Cook, Dakota Howard, Dhwani Batra, Ben L. Rambo-Martin, Eileen de Feo, Jan Antico, Christine Tran, Matthew Tolentino, Shannon Wickline, Kim Gietzen, Brad Sickler, Jingtao Liu, Eric Allen, Phil Febbo, Summer Galloway, Nicole L. Washington, Simon White, Geraint Levan, Kelly Schiabor Barrett, Elizabeth Cirulli, Alexandre Bolze, Ary Ascencio, Charlotte Rivera-Garcia, Ryan Cho, Jason Nguyen, Sherry Wang, Jimmy Ramirez, Tyler Cassens, Efren Sandoval, Magnus Isaksson, William Lee, David Becker, Marc Laurent, James Lu, Clinton R. Paden, Suxiang Tong, Duncan MacCannell |
| EPI_ISL_1269517, EPI_ISL_1269518 | Wisconsin State Laboratory of Hygiene Communicable Disease Division | Ginkgo Bioworks Clinical Laboratory | Malaika Mckenzie-Bennett, James McGann, Jim Griffin, Keith Robison, Alex Plocik, Becky Schilling, Martha Pierson, Rebecca Littlefield, Michelle Spencer, Amanda Riccardi, Birgitte Simen, Kelsey R. Florek, Abigail C. Shockey |

| | | | |
|---|---|---|---|
| EPI_ISL_1269519 | Helix/Illumina | Centers for Disease Control and Prevention Division of Viral Diseases, Pathogen Discovery | Peter W. Cook, Dakota Howard, Dhwani Batra, Ben L. Rambo-Martin, Eileen de Feo, Jan Antico, Christine Tran, Matthew Tolentino, Shannon Wickline, Kim Gietzen, Brad Sickler, Jingtao Liu, Eric Allen, Phil Febbo, Summer Galloway, Nicole L. Washington, Simon White, Geraint Levan, Kelly Schiabor Barrett, Elizabeth Cirulli, Alexandre Bolze, Ary Ascencio, Charlotte Rivera-Garcia, Ryan Cho, Jason Nguyen, Sherry Wang, Jimmy Ramirez, Tyler Cassens, Efren Sandoval, Magnus Isaksson, William Lee, David Becker, Marc Laurent, James Lu, Clinton R. Paden, Suxiang Tong, Duncan MacCannell |
| EPI_ISL_1269520, EPI_ISL_1269521 | Wisconsin State Laboratory of Hygiene Communicable Disease Division | Ginkgo Bioworks Clinical Laboratory | Malaika Mckenzie-Bennett, James McGann, Jim Griffin, Keith Robison, Alex Plocik, Becky Schilling, Martha Pierson, Rebecca Littlefield, Michelle Spencer, Amanda Riccardi, Birgitte Simen, Kelsey R. Florek, Abigail C. Shockey |
| EPI_ISL_1269522 | Helix/Illumina | Centers for Disease Control and Prevention Division of Viral Diseases, Pathogen Discovery | Peter W. Cook, Dakota Howard, Dhwani Batra, Ben L. Rambo-Martin, Eileen de Feo, Jan Antico, Christine Tran, Matthew Tolentino, Shannon Wickline, Kim Gietzen, Brad Sickler, Jingtao Liu, Eric Allen, Phil Febbo, Summer Galloway, Nicole L. Washington, Simon White, Geraint Levan, Kelly Schiabor Barrett, Elizabeth Cirulli, Alexandre Bolze, Ary Ascencio, Charlotte Rivera-Garcia, Ryan Cho, Jason Nguyen, Sherry Wang, Jimmy Ramirez, Tyler Cassens, Efren Sandoval, Magnus Isaksson, William Lee, David Becker, Marc Laurent, James Lu, Clinton R. Paden, Suxiang Tong, Duncan MacCannell |
| EPI_ISL_1269523, EPI_ISL_1269524 | Wisconsin State Laboratory of Hygiene Communicable Disease Division | Ginkgo Bioworks Clinical Laboratory | Malaika Mckenzie-Bennett, James McGann, Jim Griffin, Keith Robison, Alex Plocik, Becky Schilling, Martha Pierson, Rebecca Littlefield, Michelle Spencer, Amanda Riccardi, Birgitte Simen, Kelsey R. Florek, Abigail C. Shockey |
| EPI_ISL_1269525 | Helix/Illumina | Centers for Disease Control and Prevention Division of Viral Diseases, Pathogen Discovery | Peter W. Cook, Dakota Howard, Dhwani Batra, Ben L. Rambo-Martin, Eileen de Feo, Jan Antico, Christine Tran, Matthew Tolentino, Shannon Wickline, Kim Gietzen, Brad Sickler, Jingtao Liu, Eric Allen, Phil Febbo, Summer Galloway, Nicole L. Washington, Simon White, Geraint Levan, Kelly Schiabor Barrett, Elizabeth Cirulli, Alexandre Bolze, Ary Ascencio, Charlotte Rivera-Garcia, Ryan Cho, Jason Nguyen, Sherry Wang, Jimmy Ramirez, Tyler Cassens, Efren Sandoval, Magnus Isaksson, William Lee, David Becker, Marc Laurent, James Lu, Clinton R. Paden, Suxiang Tong, Duncan MacCannell |
| EPI_ISL_1269526, EPI_ISL_1269527 | Wisconsin State Laboratory of Hygiene Communicable Disease Division | Ginkgo Bioworks Clinical Laboratory | Malaika Mckenzie-Bennett, James McGann, Jim Griffin, Keith Robison, Alex Plocik, Becky Schilling, Martha Pierson, Rebecca Littlefield, Michelle Spencer, Amanda Riccardi, Birgitte Simen, Kelsey R. Florek, Abigail C. Shockey |
| EPI_ISL_1269528 | Helix/Illumina | Centers for Disease Control and Prevention Division of Viral Diseases, Pathogen Discovery | Peter W. Cook, Dakota Howard, Dhwani Batra, Ben L. Rambo-Martin, Eileen de Feo, Jan Antico, Christine Tran, Matthew Tolentino, Shannon Wickline, Kim Gietzen, Brad Sickler, Jingtao Liu, Eric Allen, Phil Febbo, Summer Galloway, Nicole L. Washington, Simon White, Geraint Levan, Kelly Schiabor Barrett, Elizabeth Cirulli, Alexandre Bolze, Ary Ascencio, Charlotte Rivera-Garcia, Ryan Cho, Jason Nguyen, Sherry Wang, Jimmy Ramirez, Tyler Cassens, Efren Sandoval, Magnus Isaksson, William Lee, David Becker, Marc Laurent, James Lu, Clinton R. Paden, Suxiang Tong, Duncan MacCannell |
| EPI_ISL_1269529, EPI_ISL_1269530, EPI_ISL_1269531 | Wisconsin State Laboratory of Hygiene Communicable Disease Division | Ginkgo Bioworks Clinical Laboratory | Malaika Mckenzie-Bennett, James McGann, Jim Griffin, Keith Robison, Alex Plocik, Becky Schilling, Martha Pierson, Rebecca Littlefield, Michelle Spencer, Amanda Riccardi, Birgitte Simen, Kelsey R. Florek, Abigail C. Shockey |
| EPI_ISL_1269532 | Helix/Illumina | Centers for Disease Control and Prevention Division of Viral Diseases, Pathogen Discovery | Peter W. Cook, Dakota Howard, Dhwani Batra, Ben L. Rambo-Martin, Eileen de Feo, Jan Antico, Christine Tran, Matthew Tolentino, Shannon Wickline, Kim Gietzen, Brad Sickler, Jingtao Liu, Eric Allen, Phil Febbo, Summer Galloway, Nicole L. Washington, Simon White, Geraint Levan, Kelly Schiabor Barrett, Elizabeth Cirulli, Alexandre Bolze, Ary Ascencio, Charlotte Rivera-Garcia, Ryan Cho, Jason Nguyen, Sherry Wang, Jimmy Ramirez, Tyler Cassens, Efren Sandoval, Magnus Isaksson, William Lee, David Becker, Marc Laurent, James Lu, Clinton R. Paden, Suxiang Tong, Duncan MacCannell |
| EPI_ISL_1269533 | Wisconsin State Laboratory of Hygiene Communicable Disease Division | Ginkgo Bioworks Clinical Laboratory | Malaika Mckenzie-Bennett, James McGann, Jim Griffin, Keith Robison, Alex Plocik, Becky Schilling, Martha Pierson, Rebecca Littlefield, Michelle Spencer, Amanda Riccardi, Birgitte Simen, Kelsey R. Florek, Abigail C. Shockey |
| EPI_ISL_1269534 | Helix/Illumina | Centers for Disease Control and Prevention Division of Viral Diseases, Pathogen Discovery | Peter W. Cook, Dakota Howard, Dhwani Batra, Ben L. Rambo-Martin, Eileen de Feo, Jan Antico, Christine Tran, Matthew Tolentino, Shannon Wickline, Kim Gietzen, Brad Sickler, Jingtao Liu, Eric Allen, Phil Febbo, Summer Galloway, Nicole L. Washington, Simon White, Geraint Levan, Kelly Schiabor Barrett, Elizabeth Cirulli, Alexandre Bolze, Ary Ascencio, Charlotte Rivera-Garcia, Ryan Cho, Jason Nguyen, Sherry Wang, Jimmy Ramirez, Tyler Cassens, Efren Sandoval, Magnus Isaksson, William Lee, David Becker, Marc Laurent, James Lu, Clinton R. Paden, Suxiang Tong, Duncan MacCannell |
| EPI_ISL_1269535, EPI_ISL_1269536, EPI_ISL_1269537 | Wisconsin State Laboratory of Hygiene Communicable Disease Division | Ginkgo Bioworks Clinical Laboratory | Malaika Mckenzie-Bennett, James McGann, Jim Griffin, Keith Robison, Alex Plocik, Becky Schilling, Martha Pierson, Rebecca Littlefield, Michelle Spencer, Amanda Riccardi, Birgitte Simen, Kelsey R. Florek, Abigail C. Shockey |
| EPI_ISL_1269538 | Helix/Illumina | Centers for Disease Control and Prevention Division of Viral Diseases, Pathogen Discovery | Peter W. Cook, Dakota Howard, Dhwani Batra, Ben L. Rambo-Martin, Eileen de Feo, Jan Antico, Christine Tran, Matthew Tolentino, Shannon Wickline, Kim Gietzen, Brad Sickler, Jingtao Liu, Eric Allen, Phil Febbo, Summer Galloway, Nicole L. Washington, Simon White, Geraint Levan, Kelly Schiabor Barrett, Elizabeth Cirulli, Alexandre Bolze, Ary Ascencio, Charlotte Rivera-Garcia, Ryan Cho, Jason Nguyen, Sherry Wang, Jimmy Ramirez, Tyler Cassens, Efren Sandoval, Magnus Isaksson, William Lee, David Becker, Marc Laurent, James Lu, Clinton R. Paden, Suxiang Tong, Duncan MacCannell |
| EPI_ISL_1269539, EPI_ISL_1269540 | Wisconsin State Laboratory of Hygiene Communicable Disease Division | Ginkgo Bioworks Clinical Laboratory | Malaika Mckenzie-Bennett, James McGann, Jim Griffin, Keith Robison, Alex Plocik, Becky Schilling, Martha Pierson, Rebecca Littlefield, Michelle Spencer, Amanda Riccardi, Birgitte Simen, Kelsey R. Florek, Abigail C. Shockey |
| EPI_ISL_1269541 | Helix/Illumina | Centers for Disease Control and Prevention Division of Viral Diseases, Pathogen Discovery | Peter W. Cook, Dakota Howard, Dhwani Batra, Ben L. Rambo-Martin, Eileen de Feo, Jan Antico, Christine Tran, Matthew Tolentino, Shannon Wickline, Kim Gietzen, Brad Sickler, Jingtao Liu, Eric Allen, Phil Febbo, Summer Galloway, Nicole L. Washington, Simon White, Geraint Levan, Kelly Schiabor Barrett, Elizabeth Cirulli, Alexandre Bolze, Ary Ascencio, Charlotte Rivera-Garcia, Ryan Cho, Jason Nguyen, Sherry Wang, Jimmy Ramirez, Tyler Cassens, Efren Sandoval, Magnus Isaksson, William Lee, David Becker, Marc Laurent, James Lu, Clinton R. Paden, Suxiang Tong, Duncan MacCannell |
| EPI_ISL_1269542, EPI_ISL_1269543 | Wisconsin State Laboratory of Hygiene Communicable Disease Division | Ginkgo Bioworks Clinical Laboratory | Malaika Mckenzie-Bennett, James McGann, Jim Griffin, Keith Robison, Alex Plocik, Becky Schilling, Martha Pierson, Rebecca Littlefield, Michelle Spencer, Amanda Riccardi, Birgitte Simen, Kelsey R. Florek, Abigail C. Shockey |
| EPI_ISL_1269544 | Helix/Illumina | Centers for Disease Control and Prevention Division of Viral Diseases, Pathogen Discovery | Peter W. Cook, Dakota Howard, Dhwani Batra, Ben L. Rambo-Martin, Eileen de Feo, Jan Antico, Christine Tran, Matthew Tolentino, Shannon Wickline, Kim Gietzen, Brad Sickler, Jingtao Liu, Eric Allen, Phil Febbo, Summer Galloway, Nicole L. Washington, Simon White, Geraint Levan, Kelly Schiabor Barrett, Elizabeth Cirulli, Alexandre Bolze, Ary Ascencio, Charlotte Rivera-Garcia, Ryan Cho, Jason Nguyen, Sherry Wang, Jimmy Ramirez, Tyler Cassens, Efren Sandoval, Magnus Isaksson, William Lee, David Becker, Marc Laurent, James Lu, Clinton R. Paden, Suxiang Tong, Duncan MacCannell |
| EPI_ISL_1269545, EPI_ISL_1269546 | Wisconsin State Laboratory of Hygiene Communicable Disease Division | Ginkgo Bioworks Clinical Laboratory | Malaika Mckenzie-Bennett, James McGann, Jim Griffin, Keith Robison, Alex Plocik, Becky Schilling, Martha Pierson, Rebecca Littlefield, Michelle Spencer, Amanda Riccardi, Birgitte Simen, Kelsey R. Florek, Abigail C. Shockey |
| EPI_ISL_1269547 | Helix/Illumina | Centers for Disease Control and Prevention Division of Viral Diseases, Pathogen Discovery | Peter W. Cook, Dakota Howard, Dhwani Batra, Ben L. Rambo-Martin, Eileen de Feo, Jan Antico, Christine Tran, Matthew Tolentino, Shannon Wickline, Kim Gietzen, Brad Sickler, Jingtao Liu, Eric Allen, Phil Febbo, Summer Galloway, Nicole L. Washington, Simon White, Geraint Levan, Kelly Schiabor Barrett, Elizabeth Cirulli, Alexandre Bolze, Ary Ascencio, Charlotte Rivera-Garcia, Ryan Cho, Jason Nguyen, Sherry Wang, Jimmy Ramirez, Tyler Cassens, Efren Sandoval, Magnus Isaksson, William Lee, David Becker, Marc Laurent, James Lu, Clinton R. Paden, Suxiang Tong, Duncan MacCannell |
| EPI_ISL_1269548, EPI_ISL_1269549, EPI_ISL_1269550 | Wisconsin State Laboratory of Hygiene Communicable Disease Division | Ginkgo Bioworks Clinical Laboratory | Malaika Mckenzie-Bennett, James McGann, Jim Griffin, Keith Robison, Alex Plocik, Becky Schilling, Martha Pierson, Rebecca Littlefield, Michelle Spencer, Amanda Riccardi, Birgitte Simen, Kelsey R. Florek, Abigail C. Shockey |
| EPI_ISL_1269551 | Helix/Illumina | Centers for Disease Control and Prevention Division of Viral Diseases, Pathogen Discovery | Peter W. Cook, Dakota Howard, Dhwani Batra, Ben L. Rambo-Martin, Eileen de Feo, Jan Antico, Christine Tran, Matthew Tolentino, Shannon Wickline, Kim Gietzen, Brad Sickler, Jingtao Liu, Eric Allen, Phil Febbo, Summer Galloway, Nicole L. Washington, Simon White, Geraint Levan, Kelly Schiabor Barrett, Elizabeth Cirulli, Alexandre Bolze, Ary Ascencio, Charlotte Rivera-Garcia, Ryan Cho, Jason Nguyen, Sherry Wang, Jimmy Ramirez, Tyler Cassens, Efren Sandoval, Magnus Isaksson, William Lee, David Becker, Marc Laurent, James Lu, Clinton R. Paden, Suxiang Tong, Duncan MacCannell |
| EPI_ISL_1269552, EPI_ISL_1269553 | Wisconsin State Laboratory of Hygiene Communicable Disease Division | Ginkgo Bioworks Clinical Laboratory | Malaika Mckenzie-Bennett, James McGann, Jim Griffin, Keith Robison, Alex Plocik, Becky Schilling, Martha Pierson, Rebecca Littlefield, Michelle Spencer, Amanda Riccardi, Birgitte Simen, Kelsey R. Florek, Abigail C. Shockey |
| EPI_ISL_1269554 | Helix/Illumina | Centers for Disease Control and Prevention Division of Viral Diseases, Pathogen Discovery | Peter W. Cook, Dakota Howard, Dhwani Batra, Ben L. Rambo-Martin, Eileen de Feo, Jan Antico, Christine Tran, Matthew Tolentino, Shannon Wickline, Kim Gietzen, Brad Sickler, Jingtao Liu, Eric Allen, Phil Febbo, Summer Galloway, Nicole L. Washington, Simon White, Geraint Levan, Kelly Schiabor Barrett, Elizabeth Cirulli, Alexandre Bolze, Ary Ascencio, Charlotte Rivera-Garcia, Ryan Cho, Jason Nguyen, Sherry Wang, Jimmy Ramirez, Tyler Cassens, Efren Sandoval, Magnus Isaksson, William Lee, David Becker, Marc Laurent, James Lu, Clinton R. Paden, Suxiang Tong, Duncan MacCannell |
| EPI_ISL_1269555, EPI_ISL_1269556 | Wisconsin State Laboratory of Hygiene Communicable Disease Division | Ginkgo Bioworks Clinical Laboratory | Malaika Mckenzie-Bennett, James McGann, Jim Griffin, Keith Robison, Alex Plocik, Becky Schilling, Martha Pierson, Rebecca Littlefield, Michelle Spencer, Amanda Riccardi, Birgitte Simen, Kelsey R. Florek, Abigail C. Shockey |
| EPI_ISL_1269557 | Helix/Illumina | Centers for Disease Control and Prevention Division of Viral Diseases, Pathogen Discovery | Peter W. Cook, Dakota Howard, Dhwani Batra, Ben L. Rambo-Martin, Eileen de Feo, Jan Antico, Christine Tran, Matthew Tolentino, Shannon Wickline, Kim Gietzen, Brad Sickler, Jingtao Liu, Eric Allen, Phil Febbo, Summer Galloway, Nicole L. Washington, Simon White, Geraint Levan, Kelly Schiabor Barrett, Elizabeth Cirulli, Alexandre Bolze, Ary Ascencio, Charlotte Rivera-Garcia, Ryan Cho, Jason Nguyen, Sherry Wang, Jimmy Ramirez, Tyler Cassens, Efren Sandoval, Magnus Isaksson, William Lee, David Becker, Marc Laurent, James Lu, Clinton R. Paden, Suxiang Tong, Duncan MacCannell |
| EPI_ISL_1269558, EPI_ISL_1269559 | Wisconsin State Laboratory of Hygiene Communicable Disease Division | Ginkgo Bioworks Clinical Laboratory | Malaika Mckenzie-Bennett, James McGann, Jim Griffin, Keith Robison, Alex Plocik, Becky Schilling, Martha Pierson, Rebecca Littlefield, Michelle Spencer, Amanda Riccardi, Birgitte Simen, Kelsey R. Florek, Abigail C. Shockey |
| EPI_ISL_1269560 | Helix/Illumina | Centers for Disease Control and Prevention Division of Viral Diseases, Pathogen Discovery | Peter W. Cook, Dakota Howard, Dhwani Batra, Ben L. Rambo-Martin, Eileen de Feo, Jan Antico, Christine Tran, Matthew Tolentino, Shannon Wickline, Kim Gietzen, Brad Sickler, Jingtao Liu, Eric Allen, Phil Febbo, Summer Galloway, Nicole L. Washington, Simon White, Geraint Levan, Kelly Schiabor Barrett, Elizabeth Cirulli, Alexandre Bolze, Ary Ascencio, Charlotte Rivera-Garcia, Ryan Cho, Jason Nguyen, Sherry Wang, Jimmy Ramirez, Tyler Cassens, Efren Sandoval, Magnus Isaksson, William Lee, David Becker, Marc Laurent, James Lu, Clinton R. Paden, Suxiang Tong, Duncan MacCannell |
| EPI_ISL_1269561, EPI_ISL_1269562, EPI_ISL_1269563 | Wisconsin State Laboratory of Hygiene Communicable Disease Division | Ginkgo Bioworks Clinical Laboratory | Malaika Mckenzie-Bennett, James McGann, Jim Griffin, Keith Robison, Alex Plocik, Becky Schilling, Martha Pierson, Rebecca Littlefield, Michelle Spencer, Amanda Riccardi, Birgitte Simen, Kelsey R. Florek, Abigail C. Shockey |
| EPI_ISL_1269564 | Helix/Illumina | Centers for Disease Control and Prevention Division of Viral Diseases, Pathogen Discovery | Peter W. Cook, Dakota Howard, Dhwani Batra, Ben L. Rambo-Martin, Eileen de Feo, Jan Antico, Christine Tran, Matthew Tolentino, Shannon Wickline, Kim Gietzen, Brad Sickler, Jingtao Liu, Eric Allen, Phil Febbo, Summer Galloway, Nicole L. Washington, Simon White, Geraint Levan, Kelly Schiabor Barrett, Elizabeth Cirulli, Alexandre Bolze, Ary Ascencio, Charlotte Rivera-Garcia, Ryan Cho, Jason Nguyen, Sherry Wang, Jimmy Ramirez, Tyler Cassens, Efren Sandoval, Magnus Isaksson, William Lee, David Becker, Marc Laurent, James Lu, Clinton R. Paden, Suxiang Tong, Duncan MacCannell |
| EPI_ISL_1269565, EPI_ISL_1269566 | Wisconsin State Laboratory of Hygiene Communicable Disease Division | Ginkgo Bioworks Clinical Laboratory | Malaika Mckenzie-Bennett, James McGann, Jim Griffin, Keith Robison, Alex Plocik, Becky Schilling, Martha Pierson, Rebecca Littlefield, Michelle Spencer, Amanda Riccardi, Birgitte Simen, Kelsey R. Florek, Abigail C. Shockey |
| EPI_ISL_1269567 | Helix/Illumina | Centers for Disease Control and Prevention Division of Viral Diseases, Pathogen Discovery | Peter W. Cook, Dakota Howard, Dhwani Batra, Ben L. Rambo-Martin, Eileen de Feo, Jan Antico, Christine Tran, Matthew Tolentino, Shannon Wickline, Kim Gietzen, Brad Sickler, Jingtao Liu, Eric Allen, Phil Febbo, Summer Galloway, Nicole L. Washington, Simon White, Geraint Levan, Kelly Schiabor Barrett, Elizabeth Cirulli, Alexandre Bolze, Ary Ascencio, Charlotte Rivera-Garcia, Ryan Cho, Jason Nguyen, Sherry Wang, Jimmy Ramirez, Tyler Cassens, Efren Sandoval, Magnus Isaksson, William Lee, David Becker, Marc Laurent, James Lu, Clinton R. Paden, Suxiang Tong, Duncan MacCannell |
| EPI_ISL_1269568, EPI_ISL_1269569 | Wisconsin State Laboratory of Hygiene Communicable Disease Division | Ginkgo Bioworks Clinical Laboratory | Malaika Mckenzie-Bennett, James McGann, Jim Griffin, Keith Robison, Alex Plocik, Becky Schilling, Martha Pierson, Rebecca Littlefield, Michelle Spencer, Amanda Riccardi, Birgitte Simen, Kelsey R. Florek, Abigail C. Shockey |
| EPI_ISL_1269570 | Helix/Illumina | Centers for Disease Control and Prevention Division of Viral Diseases, Pathogen Discovery | Peter W. Cook, Dakota Howard, Dhwani Batra, Ben L. Rambo-Martin, Eileen de Feo, Jan Antico, Christine Tran, Matthew Tolentino, Shannon Wickline, Kim Gietzen, Brad Sickler, Jingtao Liu, Eric Allen, Phil Febbo, Summer Galloway, Nicole L. Washington, Simon White, Geraint Levan, Kelly Schiabor Barrett, Elizabeth Cirulli, Alexandre Bolze, Ary Ascencio, Charlotte Rivera-Garcia, Ryan Cho, Jason Nguyen, Sherry Wang, Jimmy Ramirez, Tyler Cassens, Efren Sandoval, Magnus Isaksson, William Lee, David Becker, Marc Laurent, James Lu, Clinton R. Paden, Suxiang Tong, Duncan MacCannell |
| EPI_ISL_1269571, EPI_ISL_1269572 | Wisconsin State Laboratory of Hygiene Communicable Disease Division | Ginkgo Bioworks Clinical Laboratory | Malaika Mckenzie-Bennett, James McGann, Jim Griffin, Keith Robison, Alex Plocik, Becky Schilling, Martha Pierson, Rebecca Littlefield, Michelle Spencer, Amanda Riccardi, Birgitte Simen, Kelsey R. Florek, Abigail C. Shockey |
| EPI_ISL_1269573 | Helix/Illumina | Centers for Disease Control and Prevention Division of Viral Diseases, Pathogen Discovery | Peter W. Cook, Dakota Howard, Dhwani Batra, Ben L. Rambo-Martin, Eileen de Feo, Jan Antico, Christine Tran, Matthew Tolentino, Shannon Wickline, Kim Gietzen, Brad Sickler, Jingtao Liu, Eric Allen, Phil Febbo, Summer Galloway, Nicole L. Washington, Simon White, Geraint Levan, Kelly Schiabor Barrett, Elizabeth Cirulli, Alexandre Bolze, Ary Ascencio, Charlotte Rivera-Garcia, Ryan Cho, Jason Nguyen, Sherry Wang, Jimmy Ramirez, Tyler Cassens, Efren Sandoval, Magnus Isaksson, William Lee, David Becker, Marc Laurent, James Lu, Clinton R. Paden, Suxiang Tong, Duncan MacCannell |
| EPI_ISL_1269574, EPI_ISL_1269575 | Wisconsin State Laboratory of Hygiene Communicable Disease Division | Ginkgo Bioworks Clinical Laboratory | Malaika Mckenzie-Bennett, James McGann, Jim Griffin, Keith Robison, Alex Plocik, Becky Schilling, Martha Pierson, Rebecca Littlefield, Michelle Spencer, Amanda Riccardi, Birgitte Simen, Kelsey R. Florek, Abigail C. Shockey |

| | | | |
|---|---|---|---|
| EPI_ISL_1269576 | Helix/Illumina | Centers for Disease Control and Prevention Division of Viral Diseases, Pathogen Discovery | Peter W. Cook, Dakota Howard, Dhwani Batra, Ben L. Rambo-Martin, Eileen de Feo, Jan Antico, Christine Tran, Matthew Tolentino, Shannon Wickline, Kim Gietzen, Brad Sickler, Jingtao Liu, Eric Allen, Phil Febbo, Summer Galloway, Nicole L. Washington, Simon White, Geraint Levan, Kelly Schiabor Barrett, Elizabeth Cirulli, Alexandre Bolze, Ary Ascencio, Charlotte Rivera-Garcia, Ryan Cho, Jason Nguyen, Sherry Wang, Jimmy Ramirez, Tyler Cassens, Efren Sandoval, Magnus Isaksson, William Lee, David Becker, Marc Laurent, James Lu, Clinton R. Paden, Suxiang Tong, Duncan MacCannell |
| EPI_ISL_1269577, EPI_ISL_1269578 | Wisconsin State Laboratory of Hygiene Communicable Disease Division | Ginkgo Bioworks Clinical Laboratory | Malaika Mckenzie-Bennett, James McGann, Jim Griffin, Keith Robison, Alex Plocik, Becky Schilling, Martha Pierson, Rebecca Littlefield, Michelle Spencer, Amanda Riccardi, Birgitte Simen, Kelsey R. Florek, Abigail C. Shockey |
| EPI_ISL_1269579 | Helix/Illumina | Centers for Disease Control and Prevention Division of Viral Diseases, Pathogen Discovery | Peter W. Cook, Dakota Howard, Dhwani Batra, Ben L. Rambo-Martin, Eileen de Feo, Jan Antico, Christine Tran, Matthew Tolentino, Shannon Wickline, Kim Gietzen, Brad Sickler, Jingtao Liu, Eric Allen, Phil Febbo, Summer Galloway, Nicole L. Washington, Simon White, Geraint Levan, Kelly Schiabor Barrett, Elizabeth Cirulli, Alexandre Bolze, Ary Ascencio, Charlotte Rivera-Garcia, Ryan Cho, Jason Nguyen, Sherry Wang, Jimmy Ramirez, Tyler Cassens, Efren Sandoval, Magnus Isaksson, William Lee, David Becker, Marc Laurent, James Lu, Clinton R. Paden, Suxiang Tong, Duncan MacCannell |
| EPI_ISL_1269580, EPI_ISL_1269581 | Wisconsin State Laboratory of Hygiene Communicable Disease Division | Ginkgo Bioworks Clinical Laboratory | Malaika Mckenzie-Bennett, James McGann, Jim Griffin, Keith Robison, Alex Plocik, Becky Schilling, Martha Pierson, Rebecca Littlefield, Michelle Spencer, Amanda Riccardi, Birgitte Simen, Kelsey R. Florek, Abigail C. Shockey |
| EPI_ISL_1269582 | Helix/Illumina | Centers for Disease Control and Prevention Division of Viral Diseases, Pathogen Discovery | Peter W. Cook, Dakota Howard, Dhwani Batra, Ben L. Rambo-Martin, Eileen de Feo, Jan Antico, Christine Tran, Matthew Tolentino, Shannon Wickline, Kim Gietzen, Brad Sickler, Jingtao Liu, Eric Allen, Phil Febbo, Summer Galloway, Nicole L. Washington, Simon White, Geraint Levan, Kelly Schiabor Barrett, Elizabeth Cirulli, Alexandre Bolze, Ary Ascencio, Charlotte Rivera-Garcia, Ryan Cho, Jason Nguyen, Sherry Wang, Jimmy Ramirez, Tyler Cassens, Efren Sandoval, Magnus Isaksson, William Lee, David Becker, Marc Laurent, James Lu, Clinton R. Paden, Suxiang Tong, Duncan MacCannell |
| EPI_ISL_1269583, EPI_ISL_1269584 | Wisconsin State Laboratory of Hygiene Communicable Disease Division | Ginkgo Bioworks Clinical Laboratory | Malaika Mckenzie-Bennett, James McGann, Jim Griffin, Keith Robison, Alex Plocik, Becky Schilling, Martha Pierson, Rebecca Littlefield, Michelle Spencer, Amanda Riccardi, Birgitte Simen, Kelsey R. Florek, Abigail C. Shockey |
| EPI_ISL_1269585 | Helix/Illumina | Centers for Disease Control and Prevention Division of Viral Diseases, Pathogen Discovery | Peter W. Cook, Dakota Howard, Dhwani Batra, Ben L. Rambo-Martin, Eileen de Feo, Jan Antico, Christine Tran, Matthew Tolentino, Shannon Wickline, Kim Gietzen, Brad Sickler, Jingtao Liu, Eric Allen, Phil Febbo, Summer Galloway, Nicole L. Washington, Simon White, Geraint Levan, Kelly Schiabor Barrett, Elizabeth Cirulli, Alexandre Bolze, Ary Ascencio, Charlotte Rivera-Garcia, Ryan Cho, Jason Nguyen, Sherry Wang, Jimmy Ramirez, Tyler Cassens, Efren Sandoval, Magnus Isaksson, William Lee, David Becker, Marc Laurent, James Lu, Clinton R. Paden, Suxiang Tong, Duncan MacCannell |
| EPI_ISL_1269586, EPI_ISL_1269587 | Wisconsin State Laboratory of Hygiene Communicable Disease Division | Ginkgo Bioworks Clinical Laboratory | Malaika Mckenzie-Bennett, James McGann, Jim Griffin, Keith Robison, Alex Plocik, Becky Schilling, Martha Pierson, Rebecca Littlefield, Michelle Spencer, Amanda Riccardi, Birgitte Simen, Kelsey R. Florek, Abigail C. Shockey |
| EPI_ISL_1269588 | Helix/Illumina | Centers for Disease Control and Prevention Division of Viral Diseases, Pathogen Discovery | Peter W. Cook, Dakota Howard, Dhwani Batra, Ben L. Rambo-Martin, Eileen de Feo, Jan Antico, Christine Tran, Matthew Tolentino, Shannon Wickline, Kim Gietzen, Brad Sickler, Jingtao Liu, Eric Allen, Phil Febbo, Summer Galloway, Nicole L. Washington, Simon White, Geraint Levan, Kelly Schiabor Barrett, Elizabeth Cirulli, Alexandre Bolze, Ary Ascencio, Charlotte Rivera-Garcia, Ryan Cho, Jason Nguyen, Sherry Wang, Jimmy Ramirez, Tyler Cassens, Efren Sandoval, Magnus Isaksson, William Lee, David Becker, Marc Laurent, James Lu, Clinton R. Paden, Suxiang Tong, Duncan MacCannell |
| EPI_ISL_1269589, EPI_ISL_1269590 | Wisconsin State Laboratory of Hygiene Communicable Disease Division | Ginkgo Bioworks Clinical Laboratory | Malaika Mckenzie-Bennett, James McGann, Jim Griffin, Keith Robison, Alex Plocik, Becky Schilling, Martha Pierson, Rebecca Littlefield, Michelle Spencer, Amanda Riccardi, Birgitte Simen, Kelsey R. Florek, Abigail C. Shockey |
| EPI_ISL_1269591 | Helix/Illumina | Centers for Disease Control and Prevention Division of Viral Diseases, Pathogen Discovery | Peter W. Cook, Dakota Howard, Dhwani Batra, Ben L. Rambo-Martin, Eileen de Feo, Jan Antico, Christine Tran, Matthew Tolentino, Shannon Wickline, Kim Gietzen, Brad Sickler, Jingtao Liu, Eric Allen, Phil Febbo, Summer Galloway, Nicole L. Washington, Simon White, Geraint Levan, Kelly Schiabor Barrett, Elizabeth Cirulli, Alexandre Bolze, Ary Ascencio, Charlotte Rivera-Garcia, Ryan Cho, Jason Nguyen, Sherry Wang, Jimmy Ramirez, Tyler Cassens, Efren Sandoval, Magnus Isaksson, William Lee, David Becker, Marc Laurent, James Lu, Clinton R. Paden, Suxiang Tong, Duncan MacCannell |
| EPI_ISL_1269592 | Wisconsin State Laboratory of Hygiene Communicable Disease Division | Ginkgo Bioworks Clinical Laboratory | Malaika Mckenzie-Bennett, James McGann, Jim Griffin, Keith Robison, Alex Plocik, Becky Schilling, Martha Pierson, Rebecca Littlefield, Michelle Spencer, Amanda Riccardi, Birgitte Simen, Kelsey R. Florek, Abigail C. Shockey |
| EPI_ISL_1269593 | Helix/Illumina | Centers for Disease Control and Prevention Division of Viral Diseases, Pathogen Discovery | Peter W. Cook, Dakota Howard, Dhwani Batra, Ben L. Rambo-Martin, Eileen de Feo, Jan Antico, Christine Tran, Matthew Tolentino, Shannon Wickline, Kim Gietzen, Brad Sickler, Jingtao Liu, Eric Allen, Phil Febbo, Summer Galloway, Nicole L. Washington, Simon White, Geraint Levan, Kelly Schiabor Barrett, Elizabeth Cirulli, Alexandre Bolze, Ary Ascencio, Charlotte Rivera-Garcia, Ryan Cho, Jason Nguyen, Sherry Wang, Jimmy Ramirez, Tyler Cassens, Efren Sandoval, Magnus Isaksson, William Lee, David Becker, Marc Laurent, James Lu, Clinton R. Paden, Suxiang Tong, Duncan MacCannell |
| EPI_ISL_1269594, EPI_ISL_1269595 | Wisconsin State Laboratory of Hygiene Communicable Disease Division | Ginkgo Bioworks Clinical Laboratory | Malaika Mckenzie-Bennett, James McGann, Jim Griffin, Keith Robison, Alex Plocik, Becky Schilling, Martha Pierson, Rebecca Littlefield, Michelle Spencer, Amanda Riccardi, Birgitte Simen, Kelsey R. Florek, Abigail C. Shockey |
| EPI_ISL_1269596 | Helix/Illumina | Centers for Disease Control and Prevention Division of Viral Diseases, Pathogen Discovery | Peter W. Cook, Dakota Howard, Dhwani Batra, Ben L. Rambo-Martin, Eileen de Feo, Jan Antico, Christine Tran, Matthew Tolentino, Shannon Wickline, Kim Gietzen, Brad Sickler, Jingtao Liu, Eric Allen, Phil Febbo, Summer Galloway, Nicole L. Washington, Simon White, Geraint Levan, Kelly Schiabor Barrett, Elizabeth Cirulli, Alexandre Bolze, Ary Ascencio, Charlotte Rivera-Garcia, Ryan Cho, Jason Nguyen, Sherry Wang, Jimmy Ramirez, Tyler Cassens, Efren Sandoval, Magnus Isaksson, William Lee, David Becker, Marc Laurent, James Lu, Clinton R. Paden, Suxiang Tong, Duncan MacCannell |
| EPI_ISL_1269597, EPI_ISL_1269598 | Wisconsin State Laboratory of Hygiene Communicable Disease Division | Ginkgo Bioworks Clinical Laboratory | Malaika Mckenzie-Bennett, James McGann, Jim Griffin, Keith Robison, Alex Plocik, Becky Schilling, Martha Pierson, Rebecca Littlefield, Michelle Spencer, Amanda Riccardi, Birgitte Simen, Kelsey R. Florek, Abigail C. Shockey |
| EPI_ISL_1269599 | Helix/Illumina | Centers for Disease Control and Prevention Division of Viral Diseases, Pathogen Discovery | Peter W. Cook, Dakota Howard, Dhwani Batra, Ben L. Rambo-Martin, Eileen de Feo, Jan Antico, Christine Tran, Matthew Tolentino, Shannon Wickline, Kim Gietzen, Brad Sickler, Jingtao Liu, Eric Allen, Phil Febbo, Summer Galloway, Nicole L. Washington, Simon White, Geraint Levan, Kelly Schiabor Barrett, Elizabeth Cirulli, Alexandre Bolze, Ary Ascencio, Charlotte Rivera-Garcia, Ryan Cho, Jason Nguyen, Sherry Wang, Jimmy Ramirez, Tyler Cassens, Efren Sandoval, Magnus Isaksson, William Lee, David Becker, Marc Laurent, James Lu, Clinton R. Paden, Suxiang Tong, Duncan MacCannell |
| EPI_ISL_1269600, EPI_ISL_1269601, EPI_ISL_1269602 | Wisconsin State Laboratory of Hygiene Communicable Disease Division | Ginkgo Bioworks Clinical Laboratory | Malaika Mckenzie-Bennett, James McGann, Jim Griffin, Keith Robison, Alex Plocik, Becky Schilling, Martha Pierson, Rebecca Littlefield, Michelle Spencer, Amanda Riccardi, Birgitte Simen, Kelsey R. Florek, Abigail C. Shockey |
| EPI_ISL_1269603 | Helix/Illumina | Centers for Disease Control and Prevention Division of Viral Diseases, Pathogen Discovery | Peter W. Cook, Dakota Howard, Dhwani Batra, Ben L. Rambo-Martin, Eileen de Feo, Jan Antico, Christine Tran, Matthew Tolentino, Shannon Wickline, Kim Gietzen, Brad Sickler, Jingtao Liu, Eric Allen, Phil Febbo, Summer Galloway, Nicole L. Washington, Simon White, Geraint Levan, Kelly Schiabor Barrett, Elizabeth Cirulli, Alexandre Bolze, Ary Ascencio, Charlotte Rivera-Garcia, Ryan Cho, Jason Nguyen, Sherry Wang, Jimmy Ramirez, Tyler Cassens, Efren Sandoval, Magnus Isaksson, William Lee, David Becker, Marc Laurent, James Lu, Clinton R. Paden, Suxiang Tong, Duncan MacCannell |
| EPI_ISL_1269604, EPI_ISL_1269605 | Wisconsin State Laboratory of Hygiene Communicable Disease Division | Ginkgo Bioworks Clinical Laboratory | Malaika Mckenzie-Bennett, James McGann, Jim Griffin, Keith Robison, Alex Plocik, Becky Schilling, Martha Pierson, Rebecca Littlefield, Michelle Spencer, Amanda Riccardi, Birgitte Simen, Kelsey R. Florek, Abigail C. Shockey |
| EPI_ISL_1269606 | Helix/Illumina | Centers for Disease Control and Prevention Division of Viral Diseases, Pathogen Discovery | Peter W. Cook, Dakota Howard, Dhwani Batra, Ben L. Rambo-Martin, Eileen de Feo, Jan Antico, Christine Tran, Matthew Tolentino, Shannon Wickline, Kim Gietzen, Brad Sickler, Jingtao Liu, Eric Allen, Phil Febbo, Summer Galloway, Nicole L. Washington, Simon White, Geraint Levan, Kelly Schiabor Barrett, Elizabeth Cirulli, Alexandre Bolze, Ary Ascencio, Charlotte Rivera-Garcia, Ryan Cho, Jason Nguyen, Sherry Wang, Jimmy Ramirez, Tyler Cassens, Efren Sandoval, Magnus Isaksson, William Lee, David Becker, Marc Laurent, James Lu, Clinton R. Paden, Suxiang Tong, Duncan MacCannell |
| EPI_ISL_1269607, EPI_ISL_1269608 | Wisconsin State Laboratory of Hygiene Communicable Disease Division | Ginkgo Bioworks Clinical Laboratory | Malaika Mckenzie-Bennett, James McGann, Jim Griffin, Keith Robison, Alex Plocik, Becky Schilling, Martha Pierson, Rebecca Littlefield, Michelle Spencer, Amanda Riccardi, Birgitte Simen, Kelsey R. Florek, Abigail C. Shockey |
| EPI_ISL_1269609 | Helix/Illumina | Centers for Disease Control and Prevention Division of Viral Diseases, Pathogen Discovery | Peter W. Cook, Dakota Howard, Dhwani Batra, Ben L. Rambo-Martin, Eileen de Feo, Jan Antico, Christine Tran, Matthew Tolentino, Shannon Wickline, Kim Gietzen, Brad Sickler, Jingtao Liu, Eric Allen, Phil Febbo, Summer Galloway, Nicole L. Washington, Simon White, Geraint Levan, Kelly Schiabor Barrett, Elizabeth Cirulli, Alexandre Bolze, Ary Ascencio, Charlotte Rivera-Garcia, Ryan Cho, Jason Nguyen, Sherry Wang, Jimmy Ramirez, Tyler Cassens, Efren Sandoval, Magnus Isaksson, William Lee, David Becker, Marc Laurent, James Lu, Clinton R. Paden, Suxiang Tong, Duncan MacCannell |
| EPI_ISL_1269610 | Wisconsin State Laboratory of Hygiene Communicable Disease Division | Ginkgo Bioworks Clinical Laboratory | Malaika Mckenzie-Bennett, James McGann, Jim Griffin, Keith Robison, Alex Plocik, Becky Schilling, Martha Pierson, Rebecca Littlefield, Michelle Spencer, Amanda Riccardi, Birgitte Simen, Kelsey R. Florek, Abigail C. Shockey |
| EPI_ISL_1269611 | Helix/Illumina | Centers for Disease Control and Prevention Division of Viral Diseases, Pathogen Discovery | Peter W. Cook, Dakota Howard, Dhwani Batra, Ben L. Rambo-Martin, Eileen de Feo, Jan Antico, Christine Tran, Matthew Tolentino, Shannon Wickline, Kim Gietzen, Brad Sickler, Jingtao Liu, Eric Allen, Phil Febbo, Summer Galloway, Nicole L. Washington, Simon White, Geraint Levan, Kelly Schiabor Barrett, Elizabeth Cirulli, Alexandre Bolze, Ary Ascencio, Charlotte Rivera-Garcia, Ryan Cho, Jason Nguyen, Sherry Wang, Jimmy Ramirez, Tyler Cassens, Efren Sandoval, Magnus Isaksson, William Lee, David Becker, Marc Laurent, James Lu, Clinton R. Paden, Suxiang Tong, Duncan MacCannell |
| EPI_ISL_1269612, EPI_ISL_1269613, EPI_ISL_1269614 | Wisconsin State Laboratory of Hygiene Communicable Disease Division | Ginkgo Bioworks Clinical Laboratory | Malaika Mckenzie-Bennett, James McGann, Jim Griffin, Keith Robison, Alex Plocik, Becky Schilling, Martha Pierson, Rebecca Littlefield, Michelle Spencer, Amanda Riccardi, Birgitte Simen, Kelsey R. Florek, Abigail C. Shockey |
| EPI_ISL_1269615 | Helix/Illumina | Centers for Disease Control and Prevention Division of Viral Diseases, Pathogen Discovery | Peter W. Cook, Dakota Howard, Dhwani Batra, Ben L. Rambo-Martin, Eileen de Feo, Jan Antico, Christine Tran, Matthew Tolentino, Shannon Wickline, Kim Gietzen, Brad Sickler, Jingtao Liu, Eric Allen, Phil Febbo, Summer Galloway, Nicole L. Washington, Simon White, Geraint Levan, Kelly Schiabor Barrett, Elizabeth Cirulli, Alexandre Bolze, Ary Ascencio, Charlotte Rivera-Garcia, Ryan Cho, Jason Nguyen, Sherry Wang, Jimmy Ramirez, Tyler Cassens, Efren Sandoval, Magnus Isaksson, William Lee, David Becker, Marc Laurent, James Lu, Clinton R. Paden, Suxiang Tong, Duncan MacCannell |
| EPI_ISL_1269616, EPI_ISL_1269617 | Wisconsin State Laboratory of Hygiene Communicable Disease Division | Ginkgo Bioworks Clinical Laboratory | Malaika Mckenzie-Bennett, James McGann, Jim Griffin, Keith Robison, Alex Plocik, Becky Schilling, Martha Pierson, Rebecca Littlefield, Michelle Spencer, Amanda Riccardi, Birgitte Simen, Kelsey R. Florek, Abigail C. Shockey |
| EPI_ISL_1269618 | Helix/Illumina | Centers for Disease Control and Prevention Division of Viral Diseases, Pathogen Discovery | Peter W. Cook, Dakota Howard, Dhwani Batra, Ben L. Rambo-Martin, Eileen de Feo, Jan Antico, Christine Tran, Matthew Tolentino, Shannon Wickline, Kim Gietzen, Brad Sickler, Jingtao Liu, Eric Allen, Phil Febbo, Summer Galloway, Nicole L. Washington, Simon White, Geraint Levan, Kelly Schiabor Barrett, Elizabeth Cirulli, Alexandre Bolze, Ary Ascencio, Charlotte Rivera-Garcia, Ryan Cho, Jason Nguyen, Sherry Wang, Jimmy Ramirez, Tyler Cassens, Efren Sandoval, Magnus Isaksson, William Lee, David Becker, Marc Laurent, James Lu, Clinton R. Paden, Suxiang Tong, Duncan MacCannell |
| EPI_ISL_1269619, EPI_ISL_1269620 | Wisconsin State Laboratory of Hygiene Communicable Disease Division | Ginkgo Bioworks Clinical Laboratory | Malaika Mckenzie-Bennett, James McGann, Jim Griffin, Keith Robison, Alex Plocik, Becky Schilling, Martha Pierson, Rebecca Littlefield, Michelle Spencer, Amanda Riccardi, Birgitte Simen, Kelsey R. Florek, Abigail C. Shockey |
| EPI_ISL_1269621 | Helix/Illumina | Centers for Disease Control and Prevention Division of Viral Diseases, Pathogen Discovery | Peter W. Cook, Dakota Howard, Dhwani Batra, Ben L. Rambo-Martin, Eileen de Feo, Jan Antico, Christine Tran, Matthew Tolentino, Shannon Wickline, Kim Gietzen, Brad Sickler, Jingtao Liu, Eric Allen, Phil Febbo, Summer Galloway, Nicole L. Washington, Simon White, Geraint Levan, Kelly Schiabor Barrett, Elizabeth Cirulli, Alexandre Bolze, Ary Ascencio, Charlotte Rivera-Garcia, Ryan Cho, Jason Nguyen, Sherry Wang, Jimmy Ramirez, Tyler Cassens, Efren Sandoval, Magnus Isaksson, William Lee, David Becker, Marc Laurent, James Lu, Clinton R. Paden, Suxiang Tong, Duncan MacCannell |
| EPI_ISL_1269622, EPI_ISL_1269623 | Wisconsin State Laboratory of Hygiene Communicable Disease Division | Ginkgo Bioworks Clinical Laboratory | Malaika Mckenzie-Bennett, James McGann, Jim Griffin, Keith Robison, Alex Plocik, Becky Schilling, Martha Pierson, Rebecca Littlefield, Michelle Spencer, Amanda Riccardi, Birgitte Simen, Kelsey R. Florek, Abigail C. Shockey |
| EPI_ISL_1269624 | Helix/Illumina | Centers for Disease Control and Prevention Division of Viral Diseases, Pathogen Discovery | Peter W. Cook, Dakota Howard, Dhwani Batra, Ben L. Rambo-Martin, Eileen de Feo, Jan Antico, Christine Tran, Matthew Tolentino, Shannon Wickline, Kim Gietzen, Brad Sickler, Jingtao Liu, Eric Allen, Phil Febbo, Summer Galloway, Nicole L. Washington, Simon White, Geraint Levan, Kelly Schiabor Barrett, Elizabeth Cirulli, Alexandre Bolze, Ary Ascencio, Charlotte Rivera-Garcia, Ryan Cho, Jason Nguyen, Sherry Wang, Jimmy Ramirez, Tyler Cassens, Efren Sandoval, Magnus Isaksson, William Lee, David Becker, Marc Laurent, James Lu, Clinton R. Paden, Suxiang Tong, Duncan MacCannell |
| EPI_ISL_1269625, EPI_ISL_1269626 | Wisconsin State Laboratory of Hygiene Communicable Disease Division | Ginkgo Bioworks Clinical Laboratory | Malaika Mckenzie-Bennett, James McGann, Jim Griffin, Keith Robison, Alex Plocik, Becky Schilling, Martha Pierson, Rebecca Littlefield, Michelle Spencer, Amanda Riccardi, Birgitte Simen, Kelsey R. Florek, Abigail C. Shockey |
| EPI_ISL_1269627 | Helix/Illumina | Centers for Disease Control and Prevention Division of Viral Diseases, Pathogen Discovery | Peter W. Cook, Dakota Howard, Dhwani Batra, Ben L. Rambo-Martin, Eileen de Feo, Jan Antico, Christine Tran, Matthew Tolentino, Shannon Wickline, Kim Gietzen, Brad Sickler, Jingtao Liu, Eric Allen, Phil Febbo, Summer Galloway, Nicole L. Washington, Simon White, Geraint Levan, Kelly Schiabor Barrett, Elizabeth Cirulli, Alexandre Bolze, Ary Ascencio, Charlotte Rivera-Garcia, Ryan Cho, Jason Nguyen, Sherry Wang, Jimmy Ramirez, Tyler Cassens, Efren Sandoval, Magnus Isaksson, William Lee, David Becker, Marc Laurent, James Lu, Clinton R. Paden, Suxiang Tong, Duncan MacCannell |
| EPI_ISL_1269628, EPI_ISL_1269629 | Wisconsin State Laboratory of Hygiene Communicable Disease Division | Ginkgo Bioworks Clinical Laboratory | Malaika Mckenzie-Bennett, James McGann, Jim Griffin, Keith Robison, Alex Plocik, Becky Schilling, Martha Pierson, Rebecca Littlefield, Michelle Spencer, Amanda Riccardi, Birgitte Simen, Kelsey R. Florek, Abigail C. Shockey |
| EPI_ISL_1269630 | Helix/Illumina | Centers for Disease Control and Prevention Division of Viral Diseases, Pathogen | Peter W. Cook, Dakota Howard, Dhwani Batra, Ben L. Rambo-Martin, Eileen de Feo, Jan Antico, Christine Tran, Matthew Tolentino, Shannon Wickline, Kim Gietzen, Brad Sickler, Jingtao Liu, Eric Allen, Phil Febbo, Summer Galloway, Nicole L. Washington, Simon White, Geraint Levan, Kelly Schiabor Barrett, Elizabeth Cirulli, Alexandre Bolze, Ary Ascencio, Charlotte Rivera-Garcia, Ryan Cho, Jason Nguyen, Sherry Wang, Jimmy Ramirez, Tyler Cassens, Efren Sandoval, Magnus Isaksson, William Lee, David Becker, Marc Laurent, James Lu, Clinton R. Paden, Suxiang Tong, |

| | | | |
|---|---|---|---|
| | | Discovery | Duncan MacCannell |
| EPI_ISL_1269631, EPI_ISL_1269632 | Wisconsin State Laboratory of Hygiene Communicable Disease Division | Ginkgo Bioworks Clinical Laboratory | Malaika Mckenzie-Bennett, James McGann, Jim Griffin, Keith Robison, Alex Plocik, Becky Schilling, Martha Pierson, Rebecca Littlefield, Michelle Spencer, Amanda Riccardi, Birgitte Simen, Kelsey R. Florek, Abigail C. Shockey |
| EPI_ISL_1269633 | Helix/Illumina | Centers for Disease Control and Prevention Division of Viral Diseases, Pathogen Discovery | Peter W. Cook, Dakota Howard, Dhwani Batra, Ben L. Rambo-Martin, Eileen de Feo, Jan Antico, Christine Tran, Matthew Tolentino, Shannon Wickline, Kim Gietzen, Brad Sickler, Jingtao Liu, Eric Allen, Phil Febbo, Summer Galloway, Nicole L. Washington, Simon White, Geraint Levan, Kelly Schiabor Barrett, Elizabeth Cirulli, Alexandre Bolze, Ary Ascencio, Charlotte Rivera-Garcia, Ryan Cho, Jason Nguyen, Sherry Wang, Jimmy Ramirez, Tyler Cassens, Efren Sandoval, Magnus Isaksson, William Lee, David Becker, Marc Laurent, James Lu, Clinton R. Paden, Suxiang Tong, Duncan MacCannell |
| EPI_ISL_1269634, EPI_ISL_1269635 | Wisconsin State Laboratory of Hygiene Communicable Disease Division | Ginkgo Bioworks Clinical Laboratory | Malaika Mckenzie-Bennett, James McGann, Jim Griffin, Keith Robison, Alex Plocik, Becky Schilling, Martha Pierson, Rebecca Littlefield, Michelle Spencer, Amanda Riccardi, Birgitte Simen, Kelsey R. Florek, Abigail C. Shockey |
| EPI_ISL_1269636 | Helix/Illumina | Centers for Disease Control and Prevention Division of Viral Diseases, Pathogen Discovery | Peter W. Cook, Dakota Howard, Dhwani Batra, Ben L. Rambo-Martin, Eileen de Feo, Jan Antico, Christine Tran, Matthew Tolentino, Shannon Wickline, Kim Gietzen, Brad Sickler, Jingtao Liu, Eric Allen, Phil Febbo, Summer Galloway, Nicole L. Washington, Simon White, Geraint Levan, Kelly Schiabor Barrett, Elizabeth Cirulli, Alexandre Bolze, Ary Ascencio, Charlotte Rivera-Garcia, Ryan Cho, Jason Nguyen, Sherry Wang, Jimmy Ramirez, Tyler Cassens, Efren Sandoval, Magnus Isaksson, William Lee, David Becker, Marc Laurent, James Lu, Clinton R. Paden, Suxiang Tong, Duncan MacCannell |
| EPI_ISL_1269637, EPI_ISL_1269638 | Wisconsin State Laboratory of Hygiene Communicable Disease Division | Ginkgo Bioworks Clinical Laboratory | Malaika Mckenzie-Bennett, James McGann, Jim Griffin, Keith Robison, Alex Plocik, Becky Schilling, Martha Pierson, Rebecca Littlefield, Michelle Spencer, Amanda Riccardi, Birgitte Simen, Kelsey R. Florek, Abigail C. Shockey |
| EPI_ISL_1269639 | Helix/Illumina | Centers for Disease Control and Prevention Division of Viral Diseases, Pathogen Discovery | Peter W. Cook, Dakota Howard, Dhwani Batra, Ben L. Rambo-Martin, Eileen de Feo, Jan Antico, Christine Tran, Matthew Tolentino, Shannon Wickline, Kim Gietzen, Brad Sickler, Jingtao Liu, Eric Allen, Phil Febbo, Summer Galloway, Nicole L. Washington, Simon White, Geraint Levan, Kelly Schiabor Barrett, Elizabeth Cirulli, Alexandre Bolze, Ary Ascencio, Charlotte Rivera-Garcia, Ryan Cho, Jason Nguyen, Sherry Wang, Jimmy Ramirez, Tyler Cassens, Efren Sandoval, Magnus Isaksson, William Lee, David Becker, Marc Laurent, James Lu, Clinton R. Paden, Suxiang Tong, Duncan MacCannell |
| EPI_ISL_1269640, EPI_ISL_1269641 | Wisconsin State Laboratory of Hygiene Communicable Disease Division | Ginkgo Bioworks Clinical Laboratory | Malaika Mckenzie-Bennett, James McGann, Jim Griffin, Keith Robison, Alex Plocik, Becky Schilling, Martha Pierson, Rebecca Littlefield, Michelle Spencer, Amanda Riccardi, Birgitte Simen, Kelsey R. Florek, Abigail C. Shockey |
| EPI_ISL_1269642 | Helix/Illumina | Centers for Disease Control and Prevention Division of Viral Diseases, Pathogen Discovery | Peter W. Cook, Dakota Howard, Dhwani Batra, Ben L. Rambo-Martin, Eileen de Feo, Jan Antico, Christine Tran, Matthew Tolentino, Shannon Wickline, Kim Gietzen, Brad Sickler, Jingtao Liu, Eric Allen, Phil Febbo, Summer Galloway, Nicole L. Washington, Simon White, Geraint Levan, Kelly Schiabor Barrett, Elizabeth Cirulli, Alexandre Bolze, Ary Ascencio, Charlotte Rivera-Garcia, Ryan Cho, Jason Nguyen, Sherry Wang, Jimmy Ramirez, Tyler Cassens, Efren Sandoval, Magnus Isaksson, William Lee, David Becker, Marc Laurent, James Lu, Clinton R. Paden, Suxiang Tong, Duncan MacCannell |
| EPI_ISL_1269643, EPI_ISL_1269644, EPI_ISL_1269645 | Wisconsin State Laboratory of Hygiene Communicable Disease Division | Ginkgo Bioworks Clinical Laboratory | Malaika Mckenzie-Bennett, James McGann, Jim Griffin, Keith Robison, Alex Plocik, Becky Schilling, Martha Pierson, Rebecca Littlefield, Michelle Spencer, Amanda Riccardi, Birgitte Simen, Kelsey R. Florek, Abigail C. Shockey |
| EPI_ISL_1269646 | Helix/Illumina | Centers for Disease Control and Prevention Division of Viral Diseases, Pathogen Discovery | Peter W. Cook, Dakota Howard, Dhwani Batra, Ben L. Rambo-Martin, Eileen de Feo, Jan Antico, Christine Tran, Matthew Tolentino, Shannon Wickline, Kim Gietzen, Brad Sickler, Jingtao Liu, Eric Allen, Phil Febbo, Summer Galloway, Nicole L. Washington, Simon White, Geraint Levan, Kelly Schiabor Barrett, Elizabeth Cirulli, Alexandre Bolze, Ary Ascencio, Charlotte Rivera-Garcia, Ryan Cho, Jason Nguyen, Sherry Wang, Jimmy Ramirez, Tyler Cassens, Efren Sandoval, Magnus Isaksson, William Lee, David Becker, Marc Laurent, James Lu, Clinton R. Paden, Suxiang Tong, Duncan MacCannell |
| EPI_ISL_1269647 | Wisconsin State Laboratory of Hygiene Communicable Disease Division | Ginkgo Bioworks Clinical Laboratory | Malaika Mckenzie-Bennett, James McGann, Jim Griffin, Keith Robison, Alex Plocik, Becky Schilling, Martha Pierson, Rebecca Littlefield, Michelle Spencer, Amanda Riccardi, Birgitte Simen, Kelsey R. Florek, Abigail C. Shockey |
| EPI_ISL_1269648 | Helix/Illumina | Centers for Disease Control and Prevention Division of Viral Diseases, Pathogen Discovery | Peter W. Cook, Dakota Howard, Dhwani Batra, Ben L. Rambo-Martin, Eileen de Feo, Jan Antico, Christine Tran, Matthew Tolentino, Shannon Wickline, Kim Gietzen, Brad Sickler, Jingtao Liu, Eric Allen, Phil Febbo, Summer Galloway, Nicole L. Washington, Simon White, Geraint Levan, Kelly Schiabor Barrett, Elizabeth Cirulli, Alexandre Bolze, Ary Ascencio, Charlotte Rivera-Garcia, Ryan Cho, Jason Nguyen, Sherry Wang, Jimmy Ramirez, Tyler Cassens, Efren Sandoval, Magnus Isaksson, William Lee, David Becker, Marc Laurent, James Lu, Clinton R. Paden, Suxiang Tong, Duncan MacCannell |
| EPI_ISL_1269649, EPI_ISL_1269650 | Wisconsin State Laboratory of Hygiene Communicable Disease Division | Ginkgo Bioworks Clinical Laboratory | Malaika Mckenzie-Bennett, James McGann, Jim Griffin, Keith Robison, Alex Plocik, Becky Schilling, Martha Pierson, Rebecca Littlefield, Michelle Spencer, Amanda Riccardi, Birgitte Simen, Kelsey R. Florek, Abigail C. Shockey |
| EPI_ISL_1269651 | Helix/Illumina | Centers for Disease Control and Prevention Division of Viral Diseases, Pathogen Discovery | Peter W. Cook, Dakota Howard, Dhwani Batra, Ben L. Rambo-Martin, Eileen de Feo, Jan Antico, Christine Tran, Matthew Tolentino, Shannon Wickline, Kim Gietzen, Brad Sickler, Jingtao Liu, Eric Allen, Phil Febbo, Summer Galloway, Nicole L. Washington, Simon White, Geraint Levan, Kelly Schiabor Barrett, Elizabeth Cirulli, Alexandre Bolze, Ary Ascencio, Charlotte Rivera-Garcia, Ryan Cho, Jason Nguyen, Sherry Wang, Jimmy Ramirez, Tyler Cassens, Efren Sandoval, Magnus Isaksson, William Lee, David Becker, Marc Laurent, James Lu, Clinton R. Paden, Suxiang Tong, Duncan MacCannell |
| EPI_ISL_1269652, EPI_ISL_1269653 | Wisconsin State Laboratory of Hygiene Communicable Disease Division | Ginkgo Bioworks Clinical Laboratory | Malaika Mckenzie-Bennett, James McGann, Jim Griffin, Keith Robison, Alex Plocik, Becky Schilling, Martha Pierson, Rebecca Littlefield, Michelle Spencer, Amanda Riccardi, Birgitte Simen, Kelsey R. Florek, Abigail C. Shockey |
| EPI_ISL_1269654 | Helix/Illumina | Centers for Disease Control and Prevention Division of Viral Diseases, Pathogen Discovery | Peter W. Cook, Dakota Howard, Dhwani Batra, Ben L. Rambo-Martin, Eileen de Feo, Jan Antico, Christine Tran, Matthew Tolentino, Shannon Wickline, Kim Gietzen, Brad Sickler, Jingtao Liu, Eric Allen, Phil Febbo, Summer Galloway, Nicole L. Washington, Simon White, Geraint Levan, Kelly Schiabor Barrett, Elizabeth Cirulli, Alexandre Bolze, Ary Ascencio, Charlotte Rivera-Garcia, Ryan Cho, Jason Nguyen, Sherry Wang, Jimmy Ramirez, Tyler Cassens, Efren Sandoval, Magnus Isaksson, William Lee, David Becker, Marc Laurent, James Lu, Clinton R. Paden, Suxiang Tong, Duncan MacCannell |
| EPI_ISL_1269655, EPI_ISL_1269656 | Wisconsin State Laboratory of Hygiene Communicable Disease Division | Ginkgo Bioworks Clinical Laboratory | Malaika Mckenzie-Bennett, James McGann, Jim Griffin, Keith Robison, Alex Plocik, Becky Schilling, Martha Pierson, Rebecca Littlefield, Michelle Spencer, Amanda Riccardi, Birgitte Simen, Kelsey R. Florek, Abigail C. Shockey |
| EPI_ISL_1269657 | Helix/Illumina | Centers for Disease Control and Prevention Division of Viral Diseases, Pathogen Discovery | Peter W. Cook, Dakota Howard, Dhwani Batra, Ben L. Rambo-Martin, Eileen de Feo, Jan Antico, Christine Tran, Matthew Tolentino, Shannon Wickline, Kim Gietzen, Brad Sickler, Jingtao Liu, Eric Allen, Phil Febbo, Summer Galloway, Nicole L. Washington, Simon White, Geraint Levan, Kelly Schiabor Barrett, Elizabeth Cirulli, Alexandre Bolze, Ary Ascencio, Charlotte Rivera-Garcia, Ryan Cho, Jason Nguyen, Sherry Wang, Jimmy Ramirez, Tyler Cassens, Efren Sandoval, Magnus Isaksson, William Lee, David Becker, Marc Laurent, James Lu, Clinton R. Paden, Suxiang Tong, Duncan MacCannell |
| EPI_ISL_1269658, EPI_ISL_1269659 | Wisconsin State Laboratory of Hygiene Communicable Disease Division | Ginkgo Bioworks Clinical Laboratory | Malaika Mckenzie-Bennett, James McGann, Jim Griffin, Keith Robison, Alex Plocik, Becky Schilling, Martha Pierson, Rebecca Littlefield, Michelle Spencer, Amanda Riccardi, Birgitte Simen, Kelsey R. Florek, Abigail C. Shockey |
| EPI_ISL_1269660 | Helix/Illumina | Centers for Disease Control and Prevention Division of Viral Diseases, Pathogen Discovery | Peter W. Cook, Dakota Howard, Dhwani Batra, Ben L. Rambo-Martin, Eileen de Feo, Jan Antico, Christine Tran, Matthew Tolentino, Shannon Wickline, Kim Gietzen, Brad Sickler, Jingtao Liu, Eric Allen, Phil Febbo, Summer Galloway, Nicole L. Washington, Simon White, Geraint Levan, Kelly Schiabor Barrett, Elizabeth Cirulli, Alexandre Bolze, Ary Ascencio, Charlotte Rivera-Garcia, Ryan Cho, Jason Nguyen, Sherry Wang, Jimmy Ramirez, Tyler Cassens, Efren Sandoval, Magnus Isaksson, William Lee, David Becker, Marc Laurent, James Lu, Clinton R. Paden, Suxiang Tong, Duncan MacCannell |
| EPI_ISL_1269661, EPI_ISL_1269662 | Wisconsin State Laboratory of Hygiene Communicable Disease Division | Ginkgo Bioworks Clinical Laboratory | Malaika Mckenzie-Bennett, James McGann, Jim Griffin, Keith Robison, Alex Plocik, Becky Schilling, Martha Pierson, Rebecca Littlefield, Michelle Spencer, Amanda Riccardi, Birgitte Simen, Kelsey R. Florek, Abigail C. Shockey |
| EPI_ISL_1269663 | Helix/Illumina | Centers for Disease Control and Prevention Division of Viral Diseases, Pathogen Discovery | Peter W. Cook, Dakota Howard, Dhwani Batra, Ben L. Rambo-Martin, Eileen de Feo, Jan Antico, Christine Tran, Matthew Tolentino, Shannon Wickline, Kim Gietzen, Brad Sickler, Jingtao Liu, Eric Allen, Phil Febbo, Summer Galloway, Nicole L. Washington, Simon White, Geraint Levan, Kelly Schiabor Barrett, Elizabeth Cirulli, Alexandre Bolze, Ary Ascencio, Charlotte Rivera-Garcia, Ryan Cho, Jason Nguyen, Sherry Wang, Jimmy Ramirez, Tyler Cassens, Efren Sandoval, Magnus Isaksson, William Lee, David Becker, Marc Laurent, James Lu, Clinton R. Paden, Suxiang Tong, Duncan MacCannell |
| EPI_ISL_1269664, EPI_ISL_1269665 | Wisconsin State Laboratory of Hygiene Communicable Disease Division | Ginkgo Bioworks Clinical Laboratory | Malaika Mckenzie-Bennett, James McGann, Jim Griffin, Keith Robison, Alex Plocik, Becky Schilling, Martha Pierson, Rebecca Littlefield, Michelle Spencer, Amanda Riccardi, Birgitte Simen, Kelsey R. Florek, Abigail C. Shockey |
| EPI_ISL_1269666 | Helix/Illumina | Centers for Disease Control and Prevention Division of Viral Diseases, Pathogen Discovery | Peter W. Cook, Dakota Howard, Dhwani Batra, Ben L. Rambo-Martin, Eileen de Feo, Jan Antico, Christine Tran, Matthew Tolentino, Shannon Wickline, Kim Gietzen, Brad Sickler, Jingtao Liu, Eric Allen, Phil Febbo, Summer Galloway, Nicole L. Washington, Simon White, Geraint Levan, Kelly Schiabor Barrett, Elizabeth Cirulli, Alexandre Bolze, Ary Ascencio, Charlotte Rivera-Garcia, Ryan Cho, Jason Nguyen, Sherry Wang, Jimmy Ramirez, Tyler Cassens, Efren Sandoval, Magnus Isaksson, William Lee, David Becker, Marc Laurent, James Lu, Clinton R. Paden, Suxiang Tong, Duncan MacCannell |
| EPI_ISL_1269667, EPI_ISL_1269668, EPI_ISL_1269669 | Wisconsin State Laboratory of Hygiene Communicable Disease Division | Ginkgo Bioworks Clinical Laboratory | Malaika Mckenzie-Bennett, James McGann, Jim Griffin, Keith Robison, Alex Plocik, Becky Schilling, Martha Pierson, Rebecca Littlefield, Michelle Spencer, Amanda Riccardi, Birgitte Simen, Kelsey R. Florek, Abigail C. Shockey |
| EPI_ISL_1269670 | Helix/Illumina | Centers for Disease Control and Prevention Division of Viral Diseases, Pathogen Discovery | Peter W. Cook, Dakota Howard, Dhwani Batra, Ben L. Rambo-Martin, Eileen de Feo, Jan Antico, Christine Tran, Matthew Tolentino, Shannon Wickline, Kim Gietzen, Brad Sickler, Jingtao Liu, Eric Allen, Phil Febbo, Summer Galloway, Nicole L. Washington, Simon White, Geraint Levan, Kelly Schiabor Barrett, Elizabeth Cirulli, Alexandre Bolze, Ary Ascencio, Charlotte Rivera-Garcia, Ryan Cho, Jason Nguyen, Sherry Wang, Jimmy Ramirez, Tyler Cassens, Efren Sandoval, Magnus Isaksson, William Lee, David Becker, Marc Laurent, James Lu, Clinton R. Paden, Suxiang Tong, Duncan MacCannell |
| EPI_ISL_1269671, EPI_ISL_1269672 | Wisconsin State Laboratory of Hygiene Communicable Disease Division | Ginkgo Bioworks Clinical Laboratory | Malaika Mckenzie-Bennett, James McGann, Jim Griffin, Keith Robison, Alex Plocik, Becky Schilling, Martha Pierson, Rebecca Littlefield, Michelle Spencer, Amanda Riccardi, Birgitte Simen, Kelsey R. Florek, Abigail C. Shockey |
| EPI_ISL_1269673 | Helix/Illumina | Centers for Disease Control and Prevention Division of Viral Diseases, Pathogen Discovery | Peter W. Cook, Dakota Howard, Dhwani Batra, Ben L. Rambo-Martin, Eileen de Feo, Jan Antico, Christine Tran, Matthew Tolentino, Shannon Wickline, Kim Gietzen, Brad Sickler, Jingtao Liu, Eric Allen, Phil Febbo, Summer Galloway, Nicole L. Washington, Simon White, Geraint Levan, Kelly Schiabor Barrett, Elizabeth Cirulli, Alexandre Bolze, Ary Ascencio, Charlotte Rivera-Garcia, Ryan Cho, Jason Nguyen, Sherry Wang, Jimmy Ramirez, Tyler Cassens, Efren Sandoval, Magnus Isaksson, William Lee, David Becker, Marc Laurent, James Lu, Clinton R. Paden, Suxiang Tong, Duncan MacCannell |
| EPI_ISL_1269674, EPI_ISL_1269675 | Wisconsin State Laboratory of Hygiene Communicable Disease Division | Ginkgo Bioworks Clinical Laboratory | Malaika Mckenzie-Bennett, James McGann, Jim Griffin, Keith Robison, Alex Plocik, Becky Schilling, Martha Pierson, Rebecca Littlefield, Michelle Spencer, Amanda Riccardi, Birgitte Simen, Kelsey R. Florek, Abigail C. Shockey |
| EPI_ISL_1269676 | Helix/Illumina | Centers for Disease Control and Prevention Division of Viral Diseases, Pathogen Discovery | Peter W. Cook, Dakota Howard, Dhwani Batra, Ben L. Rambo-Martin, Eileen de Feo, Jan Antico, Christine Tran, Matthew Tolentino, Shannon Wickline, Kim Gietzen, Brad Sickler, Jingtao Liu, Eric Allen, Phil Febbo, Summer Galloway, Nicole L. Washington, Simon White, Geraint Levan, Kelly Schiabor Barrett, Elizabeth Cirulli, Alexandre Bolze, Ary Ascencio, Charlotte Rivera-Garcia, Ryan Cho, Jason Nguyen, Sherry Wang, Jimmy Ramirez, Tyler Cassens, Efren Sandoval, Magnus Isaksson, William Lee, David Becker, Marc Laurent, James Lu, Clinton R. Paden, Suxiang Tong, Duncan MacCannell |
| EPI_ISL_1269677, EPI_ISL_1269678 | Wisconsin State Laboratory of Hygiene Communicable Disease Division | Ginkgo Bioworks Clinical Laboratory | Malaika Mckenzie-Bennett, James McGann, Jim Griffin, Keith Robison, Alex Plocik, Becky Schilling, Martha Pierson, Rebecca Littlefield, Michelle Spencer, Amanda Riccardi, Birgitte Simen, Kelsey R. Florek, Abigail C. Shockey |
| EPI_ISL_1269679 | Helix/Illumina | Centers for Disease Control and Prevention Division of Viral Diseases, Pathogen Discovery | Peter W. Cook, Dakota Howard, Dhwani Batra, Ben L. Rambo-Martin, Eileen de Feo, Jan Antico, Christine Tran, Matthew Tolentino, Shannon Wickline, Kim Gietzen, Brad Sickler, Jingtao Liu, Eric Allen, Phil Febbo, Summer Galloway, Nicole L. Washington, Simon White, Geraint Levan, Kelly Schiabor Barrett, Elizabeth Cirulli, Alexandre Bolze, Ary Ascencio, Charlotte Rivera-Garcia, Ryan Cho, Jason Nguyen, Sherry Wang, Jimmy Ramirez, Tyler Cassens, Efren Sandoval, Magnus Isaksson, William Lee, David Becker, Marc Laurent, James Lu, Clinton R. Paden, Suxiang Tong, Duncan MacCannell |
| EPI_ISL_1269680, EPI_ISL_1269681, EPI_ISL_1269683, EPI_ISL_1269684 | Wisconsin State Laboratory of Hygiene Communicable Disease Division | Ginkgo Bioworks Clinical Laboratory | Malaika Mckenzie-Bennett, James McGann, Jim Griffin, Keith Robison, Alex Plocik, Becky Schilling, Martha Pierson, Rebecca Littlefield, Michelle Spencer, Amanda Riccardi, Birgitte Simen, Kelsey R. Florek, Abigail C. Shockey |
| EPI_ISL_1269685 | Helix/Illumina | Centers for Disease Control and Prevention Division of Viral Diseases, Pathogen Discovery | Peter W. Cook, Dakota Howard, Dhwani Batra, Ben L. Rambo-Martin, Eileen de Feo, Jan Antico, Christine Tran, Matthew Tolentino, Shannon Wickline, Kim Gietzen, Brad Sickler, Jingtao Liu, Eric Allen, Phil Febbo, Summer Galloway, Nicole L. Washington, Simon White, Geraint Levan, Kelly Schiabor Barrett, Elizabeth Cirulli, Alexandre Bolze, Ary Ascencio, Charlotte Rivera-Garcia, Ryan Cho, Jason Nguyen, Sherry Wang, Jimmy Ramirez, Tyler Cassens, Efren Sandoval, Magnus Isaksson, William Lee, David Becker, Marc Laurent, James Lu, Clinton R. Paden, Suxiang Tong, Duncan MacCannell |
| EPI_ISL_1269686, EPI_ISL_1269687 | Wisconsin State Laboratory of Hygiene Communicable Disease Division | Ginkgo Bioworks Clinical Laboratory | Malaika Mckenzie-Bennett, James McGann, Jim Griffin, Keith Robison, Alex Plocik, Becky Schilling, Martha Pierson, Rebecca Littlefield, Michelle Spencer, Amanda Riccardi, Birgitte Simen, Kelsey R. Florek, Abigail C. Shockey |
| EPI_ISL_1269688 | Helix/Illumina | Centers for Disease Control and Prevention Division of Viral Diseases, Pathogen | Peter W. Cook, Dakota Howard, Dhwani Batra, Ben L. Rambo-Martin, Eileen de Feo, Jan Antico, Christine Tran, Matthew Tolentino, Shannon Wickline, Kim Gietzen, Brad Sickler, Jingtao Liu, Eric Allen, Phil Febbo, Summer Galloway, Nicole L. Washington, Simon White, Geraint Levan, Kelly Schiabor Barrett, Elizabeth Cirulli, Alexandre Bolze, Ary Ascencio, Charlotte Rivera-Garcia, Ryan Cho, Jason Nguyen, Sherry Wang, Jimmy Ramirez, Tyler Cassens, Efren Sandoval, Magnus Isaksson, William Lee, David Becker, Marc Laurent, James Lu, Clinton R. Paden, Suxiang Tong, |

| | | | |
|---|---|---|---|
| | | Discovery | Duncan MacCannell |
| EPI_ISL_1269689, EPI_ISL_1269690 | Wisconsin State Laboratory of Hygiene Communicable Disease Division | Ginkgo Bioworks Clinical Laboratory | Malaika Mckenzie-Bennett, James McGann, Jim Griffin, Keith Robison, Alex Plocik, Becky Schilling, Martha Pierson, Rebecca Littlefield, Michelle Spencer, Amanda Riccardi, Birgitte Simen, Kelsey R. Florek, Abigail C. Shockey |
| EPI_ISL_1269691 | Helix/Illumina | Centers for Disease Control and Prevention Division of Viral Diseases, Pathogen Discovery | Peter W. Cook, Dakota Howard, Dhwani Batra, Ben L. Rambo-Martin, Eileen de Feo, Jan Antico, Christine Tran, Matthew Tolentino, Shannon Wickline, Kim Gietzen, Brad Sickler, Jingtao Liu, Eric Allen, Phil Febbo, Summer Galloway, Nicole L. Washington, Simon White, Geraint Levan, Kelly Schiabor Barrett, Elizabeth Cirulli, Alexandre Bolze, Ary Ascencio, Charlotte Rivera-Garcia, Ryan Cho, Jason Nguyen, Sherry Wang, Jimmy Ramirez, Tyler Cassens, Efren Sandoval, Magnus Isaksson, William Lee, David Becker, Marc Laurent, James Lu, Clinton R. Paden, Suxiang Tong, Duncan MacCannell |
| EPI_ISL_1269692, EPI_ISL_1269693 | Wisconsin State Laboratory of Hygiene Communicable Disease Division | Ginkgo Bioworks Clinical Laboratory | Malaika Mckenzie-Bennett, James McGann, Jim Griffin, Keith Robison, Alex Plocik, Becky Schilling, Martha Pierson, Rebecca Littlefield, Michelle Spencer, Amanda Riccardi, Birgitte Simen, Kelsey R. Florek, Abigail C. Shockey |
| EPI_ISL_1269694 | Helix/Illumina | Centers for Disease Control and Prevention Division of Viral Diseases, Pathogen Discovery | Peter W. Cook, Dakota Howard, Dhwani Batra, Ben L. Rambo-Martin, Eileen de Feo, Jan Antico, Christine Tran, Matthew Tolentino, Shannon Wickline, Kim Gietzen, Brad Sickler, Jingtao Liu, Eric Allen, Phil Febbo, Summer Galloway, Nicole L. Washington, Simon White, Geraint Levan, Kelly Schiabor Barrett, Elizabeth Cirulli, Alexandre Bolze, Ary Ascencio, Charlotte Rivera-Garcia, Ryan Cho, Jason Nguyen, Sherry Wang, Jimmy Ramirez, Tyler Cassens, Efren Sandoval, Magnus Isaksson, William Lee, David Becker, Marc Laurent, James Lu, Clinton R. Paden, Suxiang Tong, Duncan MacCannell |
| EPI_ISL_1269695, EPI_ISL_1269696 | Wisconsin State Laboratory of Hygiene Communicable Disease Division | Ginkgo Bioworks Clinical Laboratory | Malaika Mckenzie-Bennett, James McGann, Jim Griffin, Keith Robison, Alex Plocik, Becky Schilling, Martha Pierson, Rebecca Littlefield, Michelle Spencer, Amanda Riccardi, Birgitte Simen, Kelsey R. Florek, Abigail C. Shockey |
| EPI_ISL_1269697 | Helix/Illumina | Centers for Disease Control and Prevention Division of Viral Diseases, Pathogen Discovery | Peter W. Cook, Dakota Howard, Dhwani Batra, Ben L. Rambo-Martin, Eileen de Feo, Jan Antico, Christine Tran, Matthew Tolentino, Shannon Wickline, Kim Gietzen, Brad Sickler, Jingtao Liu, Eric Allen, Phil Febbo, Summer Galloway, Nicole L. Washington, Simon White, Geraint Levan, Kelly Schiabor Barrett, Elizabeth Cirulli, Alexandre Bolze, Ary Ascencio, Charlotte Rivera-Garcia, Ryan Cho, Jason Nguyen, Sherry Wang, Jimmy Ramirez, Tyler Cassens, Efren Sandoval, Magnus Isaksson, William Lee, David Becker, Marc Laurent, James Lu, Clinton R. Paden, Suxiang Tong, Duncan MacCannell |
| EPI_ISL_1269698, EPI_ISL_1269699 | Wisconsin State Laboratory of Hygiene Communicable Disease Division | Ginkgo Bioworks Clinical Laboratory | Malaika Mckenzie-Bennett, James McGann, Jim Griffin, Keith Robison, Alex Plocik, Becky Schilling, Martha Pierson, Rebecca Littlefield, Michelle Spencer, Amanda Riccardi, Birgitte Simen, Kelsey R. Florek, Abigail C. Shockey |
| EPI_ISL_1269700 | Helix/Illumina | Centers for Disease Control and Prevention Division of Viral Diseases, Pathogen Discovery | Peter W. Cook, Dakota Howard, Dhwani Batra, Ben L. Rambo-Martin, Eileen de Feo, Jan Antico, Christine Tran, Matthew Tolentino, Shannon Wickline, Kim Gietzen, Brad Sickler, Jingtao Liu, Eric Allen, Phil Febbo, Summer Galloway, Nicole L. Washington, Simon White, Geraint Levan, Kelly Schiabor Barrett, Elizabeth Cirulli, Alexandre Bolze, Ary Ascencio, Charlotte Rivera-Garcia, Ryan Cho, Jason Nguyen, Sherry Wang, Jimmy Ramirez, Tyler Cassens, Efren Sandoval, Magnus Isaksson, William Lee, David Becker, Marc Laurent, James Lu, Clinton R. Paden, Suxiang Tong, Duncan MacCannell |
| EPI_ISL_1269701, EPI_ISL_1269702, EPI_ISL_1269703 | Wisconsin State Laboratory of Hygiene Communicable Disease Division | Ginkgo Bioworks Clinical Laboratory | Malaika Mckenzie-Bennett, James McGann, Jim Griffin, Keith Robison, Alex Plocik, Becky Schilling, Martha Pierson, Rebecca Littlefield, Michelle Spencer, Amanda Riccardi, Birgitte Simen, Kelsey R. Florek, Abigail C. Shockey |
| EPI_ISL_1269704 | Helix/Illumina | Centers for Disease Control and Prevention Division of Viral Diseases, Pathogen Discovery | Peter W. Cook, Dakota Howard, Dhwani Batra, Ben L. Rambo-Martin, Eileen de Feo, Jan Antico, Christine Tran, Matthew Tolentino, Shannon Wickline, Kim Gietzen, Brad Sickler, Jingtao Liu, Eric Allen, Phil Febbo, Summer Galloway, Nicole L. Washington, Simon White, Geraint Levan, Kelly Schiabor Barrett, Elizabeth Cirulli, Alexandre Bolze, Ary Ascencio, Charlotte Rivera-Garcia, Ryan Cho, Jason Nguyen, Sherry Wang, Jimmy Ramirez, Tyler Cassens, Efren Sandoval, Magnus Isaksson, William Lee, David Becker, Marc Laurent, James Lu, Clinton R. Paden, Suxiang Tong, Duncan MacCannell |
| EPI_ISL_1269705, EPI_ISL_1269706 | Wisconsin State Laboratory of Hygiene Communicable Disease Division | Ginkgo Bioworks Clinical Laboratory | Malaika Mckenzie-Bennett, James McGann, Jim Griffin, Keith Robison, Alex Plocik, Becky Schilling, Martha Pierson, Rebecca Littlefield, Michelle Spencer, Amanda Riccardi, Birgitte Simen, Kelsey R. Florek, Abigail C. Shockey |
| EPI_ISL_1269707 | Helix/Illumina | Centers for Disease Control and Prevention Division of Viral Diseases, Pathogen Discovery | Peter W. Cook, Dakota Howard, Dhwani Batra, Ben L. Rambo-Martin, Eileen de Feo, Jan Antico, Christine Tran, Matthew Tolentino, Shannon Wickline, Kim Gietzen, Brad Sickler, Jingtao Liu, Eric Allen, Phil Febbo, Summer Galloway, Nicole L. Washington, Simon White, Geraint Levan, Kelly Schiabor Barrett, Elizabeth Cirulli, Alexandre Bolze, Ary Ascencio, Charlotte Rivera-Garcia, Ryan Cho, Jason Nguyen, Sherry Wang, Jimmy Ramirez, Tyler Cassens, Efren Sandoval, Magnus Isaksson, William Lee, David Becker, Marc Laurent, James Lu, Clinton R. Paden, Suxiang Tong, Duncan MacCannell |
| EPI_ISL_1269708, EPI_ISL_1269709 | Wisconsin State Laboratory of Hygiene Communicable Disease Division | Ginkgo Bioworks Clinical Laboratory | Malaika Mckenzie-Bennett, James McGann, Jim Griffin, Keith Robison, Alex Plocik, Becky Schilling, Martha Pierson, Rebecca Littlefield, Michelle Spencer, Amanda Riccardi, Birgitte Simen, Kelsey R. Florek, Abigail C. Shockey |
| EPI_ISL_1269710 | Helix/Illumina | Centers for Disease Control and Prevention Division of Viral Diseases, Pathogen Discovery | Peter W. Cook, Dakota Howard, Dhwani Batra, Ben L. Rambo-Martin, Eileen de Feo, Jan Antico, Christine Tran, Matthew Tolentino, Shannon Wickline, Kim Gietzen, Brad Sickler, Jingtao Liu, Eric Allen, Phil Febbo, Summer Galloway, Nicole L. Washington, Simon White, Geraint Levan, Kelly Schiabor Barrett, Elizabeth Cirulli, Alexandre Bolze, Ary Ascencio, Charlotte Rivera-Garcia, Ryan Cho, Jason Nguyen, Sherry Wang, Jimmy Ramirez, Tyler Cassens, Efren Sandoval, Magnus Isaksson, William Lee, David Becker, Marc Laurent, James Lu, Clinton R. Paden, Suxiang Tong, Duncan MacCannell |
| EPI_ISL_1269711, EPI_ISL_1269712 | Wisconsin State Laboratory of Hygiene Communicable Disease Division | Ginkgo Bioworks Clinical Laboratory | Malaika Mckenzie-Bennett, James McGann, Jim Griffin, Keith Robison, Alex Plocik, Becky Schilling, Martha Pierson, Rebecca Littlefield, Michelle Spencer, Amanda Riccardi, Birgitte Simen, Kelsey R. Florek, Abigail C. Shockey |
| EPI_ISL_1269713 | Helix/Illumina | Centers for Disease Control and Prevention Division of Viral Diseases, Pathogen Discovery | Peter W. Cook, Dakota Howard, Dhwani Batra, Ben L. Rambo-Martin, Eileen de Feo, Jan Antico, Christine Tran, Matthew Tolentino, Shannon Wickline, Kim Gietzen, Brad Sickler, Jingtao Liu, Eric Allen, Phil Febbo, Summer Galloway, Nicole L. Washington, Simon White, Geraint Levan, Kelly Schiabor Barrett, Elizabeth Cirulli, Alexandre Bolze, Ary Ascencio, Charlotte Rivera-Garcia, Ryan Cho, Jason Nguyen, Sherry Wang, Jimmy Ramirez, Tyler Cassens, Efren Sandoval, Magnus Isaksson, William Lee, David Becker, Marc Laurent, James Lu, Clinton R. Paden, Suxiang Tong, Duncan MacCannell |
| EPI_ISL_1269714, EPI_ISL_1269715 | Wisconsin State Laboratory of Hygiene Communicable Disease Division | Ginkgo Bioworks Clinical Laboratory | Malaika Mckenzie-Bennett, James McGann, Jim Griffin, Keith Robison, Alex Plocik, Becky Schilling, Martha Pierson, Rebecca Littlefield, Michelle Spencer, Amanda Riccardi, Birgitte Simen, Kelsey R. Florek, Abigail C. Shockey |
| EPI_ISL_1269716 | Helix/Illumina | Centers for Disease Control and Prevention Division of Viral Diseases, Pathogen Discovery | Peter W. Cook, Dakota Howard, Dhwani Batra, Ben L. Rambo-Martin, Eileen de Feo, Jan Antico, Christine Tran, Matthew Tolentino, Shannon Wickline, Kim Gietzen, Brad Sickler, Jingtao Liu, Eric Allen, Phil Febbo, Summer Galloway, Nicole L. Washington, Simon White, Geraint Levan, Kelly Schiabor Barrett, Elizabeth Cirulli, Alexandre Bolze, Ary Ascencio, Charlotte Rivera-Garcia, Ryan Cho, Jason Nguyen, Sherry Wang, Jimmy Ramirez, Tyler Cassens, Efren Sandoval, Magnus Isaksson, William Lee, David Becker, Marc Laurent, James Lu, Clinton R. Paden, Suxiang Tong, Duncan MacCannell |
| EPI_ISL_1269717 | Wisconsin State Laboratory of Hygiene Communicable Disease Division | Ginkgo Bioworks Clinical Laboratory | Malaika Mckenzie-Bennett, James McGann, Jim Griffin, Keith Robison, Alex Plocik, Becky Schilling, Martha Pierson, Rebecca Littlefield, Michelle Spencer, Amanda Riccardi, Birgitte Simen, Kelsey R. Florek, Abigail C. Shockey |
| EPI_ISL_1269718 | Helix/Illumina | Centers for Disease Control and Prevention Division of Viral Diseases, Pathogen Discovery | Peter W. Cook, Dakota Howard, Dhwani Batra, Ben L. Rambo-Martin, Eileen de Feo, Jan Antico, Christine Tran, Matthew Tolentino, Shannon Wickline, Kim Gietzen, Brad Sickler, Jingtao Liu, Eric Allen, Phil Febbo, Summer Galloway, Nicole L. Washington, Simon White, Geraint Levan, Kelly Schiabor Barrett, Elizabeth Cirulli, Alexandre Bolze, Ary Ascencio, Charlotte Rivera-Garcia, Ryan Cho, Jason Nguyen, Sherry Wang, Jimmy Ramirez, Tyler Cassens, Efren Sandoval, Magnus Isaksson, William Lee, David Becker, Marc Laurent, James Lu, Clinton R. Paden, Suxiang Tong, Duncan MacCannell |
| EPI_ISL_1269719, EPI_ISL_1269720, EPI_ISL_1269721 | Wisconsin State Laboratory of Hygiene Communicable Disease Division | Ginkgo Bioworks Clinical Laboratory | Malaika Mckenzie-Bennett, James McGann, Jim Griffin, Keith Robison, Alex Plocik, Becky Schilling, Martha Pierson, Rebecca Littlefield, Michelle Spencer, Amanda Riccardi, Birgitte Simen, Kelsey R. Florek, Abigail C. Shockey |
| EPI_ISL_1269722 | Helix/Illumina | Centers for Disease Control and Prevention Division of Viral Diseases, Pathogen Discovery | Peter W. Cook, Dakota Howard, Dhwani Batra, Ben L. Rambo-Martin, Eileen de Feo, Jan Antico, Christine Tran, Matthew Tolentino, Shannon Wickline, Kim Gietzen, Brad Sickler, Jingtao Liu, Eric Allen, Phil Febbo, Summer Galloway, Nicole L. Washington, Simon White, Geraint Levan, Kelly Schiabor Barrett, Elizabeth Cirulli, Alexandre Bolze, Ary Ascencio, Charlotte Rivera-Garcia, Ryan Cho, Jason Nguyen, Sherry Wang, Jimmy Ramirez, Tyler Cassens, Efren Sandoval, Magnus Isaksson, William Lee, David Becker, Marc Laurent, James Lu, Clinton R. Paden, Suxiang Tong, Duncan MacCannell |
| EPI_ISL_1269723, EPI_ISL_1269725, EPI_ISL_1269726 | Wisconsin State Laboratory of Hygiene Communicable Disease Division | Ginkgo Bioworks Clinical Laboratory | Malaika Mckenzie-Bennett, James McGann, Jim Griffin, Keith Robison, Alex Plocik, Becky Schilling, Martha Pierson, Rebecca Littlefield, Michelle Spencer, Amanda Riccardi, Birgitte Simen, Kelsey R. Florek, Abigail C. Shockey |
| EPI_ISL_1269727 | Helix/Illumina | Centers for Disease Control and Prevention Division of Viral Diseases, Pathogen Discovery | Peter W. Cook, Dakota Howard, Dhwani Batra, Ben L. Rambo-Martin, Eileen de Feo, Jan Antico, Christine Tran, Matthew Tolentino, Shannon Wickline, Kim Gietzen, Brad Sickler, Jingtao Liu, Eric Allen, Phil Febbo, Summer Galloway, Nicole L. Washington, Simon White, Geraint Levan, Kelly Schiabor Barrett, Elizabeth Cirulli, Alexandre Bolze, Ary Ascencio, Charlotte Rivera-Garcia, Ryan Cho, Jason Nguyen, Sherry Wang, Jimmy Ramirez, Tyler Cassens, Efren Sandoval, Magnus Isaksson, William Lee, David Becker, Marc Laurent, James Lu, Clinton R. Paden, Suxiang Tong, Duncan MacCannell |
| EPI_ISL_1269728, EPI_ISL_1269729 | Wisconsin State Laboratory of Hygiene Communicable Disease Division | Ginkgo Bioworks Clinical Laboratory | Malaika Mckenzie-Bennett, James McGann, Jim Griffin, Keith Robison, Alex Plocik, Becky Schilling, Martha Pierson, Rebecca Littlefield, Michelle Spencer, Amanda Riccardi, Birgitte Simen, Kelsey R. Florek, Abigail C. Shockey |
| EPI_ISL_1269730 | Helix/Illumina | Centers for Disease Control and Prevention Division of Viral Diseases, Pathogen Discovery | Peter W. Cook, Dakota Howard, Dhwani Batra, Ben L. Rambo-Martin, Eileen de Feo, Jan Antico, Christine Tran, Matthew Tolentino, Shannon Wickline, Kim Gietzen, Brad Sickler, Jingtao Liu, Eric Allen, Phil Febbo, Summer Galloway, Nicole L. Washington, Simon White, Geraint Levan, Kelly Schiabor Barrett, Elizabeth Cirulli, Alexandre Bolze, Ary Ascencio, Charlotte Rivera-Garcia, Ryan Cho, Jason Nguyen, Sherry Wang, Jimmy Ramirez, Tyler Cassens, Efren Sandoval, Magnus Isaksson, William Lee, David Becker, Marc Laurent, James Lu, Clinton R. Paden, Suxiang Tong, Duncan MacCannell |
| EPI_ISL_1269731, EPI_ISL_1269732 | Wisconsin State Laboratory of Hygiene Communicable Disease Division | Ginkgo Bioworks Clinical Laboratory | Malaika Mckenzie-Bennett, James McGann, Jim Griffin, Keith Robison, Alex Plocik, Becky Schilling, Martha Pierson, Rebecca Littlefield, Michelle Spencer, Amanda Riccardi, Birgitte Simen, Kelsey R. Florek, Abigail C. Shockey |
| EPI_ISL_1269733 | Helix/Illumina | Centers for Disease Control and Prevention Division of Viral Diseases, Pathogen Discovery | Peter W. Cook, Dakota Howard, Dhwani Batra, Ben L. Rambo-Martin, Eileen de Feo, Jan Antico, Christine Tran, Matthew Tolentino, Shannon Wickline, Kim Gietzen, Brad Sickler, Jingtao Liu, Eric Allen, Phil Febbo, Summer Galloway, Nicole L. Washington, Simon White, Geraint Levan, Kelly Schiabor Barrett, Elizabeth Cirulli, Alexandre Bolze, Ary Ascencio, Charlotte Rivera-Garcia, Ryan Cho, Jason Nguyen, Sherry Wang, Jimmy Ramirez, Tyler Cassens, Efren Sandoval, Magnus Isaksson, William Lee, David Becker, Marc Laurent, James Lu, Clinton R. Paden, Suxiang Tong, Duncan MacCannell |
| EPI_ISL_1269734, EPI_ISL_1269735 | Wisconsin State Laboratory of Hygiene Communicable Disease Division | Ginkgo Bioworks Clinical Laboratory | Malaika Mckenzie-Bennett, James McGann, Jim Griffin, Keith Robison, Alex Plocik, Becky Schilling, Martha Pierson, Rebecca Littlefield, Michelle Spencer, Amanda Riccardi, Birgitte Simen, Kelsey R. Florek, Abigail C. Shockey |
| EPI_ISL_1269736 | Helix/Illumina | Centers for Disease Control and Prevention Division of Viral Diseases, Pathogen Discovery | Peter W. Cook, Dakota Howard, Dhwani Batra, Ben L. Rambo-Martin, Eileen de Feo, Jan Antico, Christine Tran, Matthew Tolentino, Shannon Wickline, Kim Gietzen, Brad Sickler, Jingtao Liu, Eric Allen, Phil Febbo, Summer Galloway, Nicole L. Washington, Simon White, Geraint Levan, Kelly Schiabor Barrett, Elizabeth Cirulli, Alexandre Bolze, Ary Ascencio, Charlotte Rivera-Garcia, Ryan Cho, Jason Nguyen, Sherry Wang, Jimmy Ramirez, Tyler Cassens, Efren Sandoval, Magnus Isaksson, William Lee, David Becker, Marc Laurent, James Lu, Clinton R. Paden, Suxiang Tong, Duncan MacCannell |
| EPI_ISL_1269737, EPI_ISL_1269738 | Wisconsin State Laboratory of Hygiene Communicable Disease Division | Ginkgo Bioworks Clinical Laboratory | Malaika Mckenzie-Bennett, James McGann, Jim Griffin, Keith Robison, Alex Plocik, Becky Schilling, Martha Pierson, Rebecca Littlefield, Michelle Spencer, Amanda Riccardi, Birgitte Simen, Kelsey R. Florek, Abigail C. Shockey |
| EPI_ISL_1269739 | Helix/Illumina | Centers for Disease Control and Prevention Division of Viral Diseases, Pathogen Discovery | Peter W. Cook, Dakota Howard, Dhwani Batra, Ben L. Rambo-Martin, Eileen de Feo, Jan Antico, Christine Tran, Matthew Tolentino, Shannon Wickline, Kim Gietzen, Brad Sickler, Jingtao Liu, Eric Allen, Phil Febbo, Summer Galloway, Nicole L. Washington, Simon White, Geraint Levan, Kelly Schiabor Barrett, Elizabeth Cirulli, Alexandre Bolze, Ary Ascencio, Charlotte Rivera-Garcia, Ryan Cho, Jason Nguyen, Sherry Wang, Jimmy Ramirez, Tyler Cassens, Efren Sandoval, Magnus Isaksson, William Lee, David Becker, Marc Laurent, James Lu, Clinton R. Paden, Suxiang Tong, Duncan MacCannell |
| EPI_ISL_1269740, EPI_ISL_1269741 | Wisconsin State Laboratory of Hygiene Communicable Disease Division | Ginkgo Bioworks Clinical Laboratory | Malaika Mckenzie-Bennett, James McGann, Jim Griffin, Keith Robison, Alex Plocik, Becky Schilling, Martha Pierson, Rebecca Littlefield, Michelle Spencer, Amanda Riccardi, Birgitte Simen, Kelsey R. Florek, Abigail C. Shockey |
| EPI_ISL_1269742 | Helix/Illumina | Centers for Disease Control and Prevention Division of Viral Diseases, Pathogen Discovery | Peter W. Cook, Dakota Howard, Dhwani Batra, Ben L. Rambo-Martin, Eileen de Feo, Jan Antico, Christine Tran, Matthew Tolentino, Shannon Wickline, Kim Gietzen, Brad Sickler, Jingtao Liu, Eric Allen, Phil Febbo, Summer Galloway, Nicole L. Washington, Simon White, Geraint Levan, Kelly Schiabor Barrett, Elizabeth Cirulli, Alexandre Bolze, Ary Ascencio, Charlotte Rivera-Garcia, Ryan Cho, Jason Nguyen, Sherry Wang, Jimmy Ramirez, Tyler Cassens, Efren Sandoval, Magnus Isaksson, William Lee, David Becker, Marc Laurent, James Lu, Clinton R. Paden, Suxiang Tong, Duncan MacCannell |
| EPI_ISL_1269743, EPI_ISL_1269744 | Wisconsin State Laboratory of Hygiene Communicable Disease Division | Ginkgo Bioworks Clinical Laboratory | Malaika Mckenzie-Bennett, James McGann, Jim Griffin, Keith Robison, Alex Plocik, Becky Schilling, Martha Pierson, Rebecca Littlefield, Michelle Spencer, Amanda Riccardi, Birgitte Simen, Kelsey R. Florek, Abigail C. Shockey |
| EPI_ISL_1269745 | Helix/Illumina | Centers for Disease Control and Prevention Division of Viral Diseases, Pathogen Discovery | Peter W. Cook, Dakota Howard, Dhwani Batra, Ben L. Rambo-Martin, Eileen de Feo, Jan Antico, Christine Tran, Matthew Tolentino, Shannon Wickline, Kim Gietzen, Brad Sickler, Jingtao Liu, Eric Allen, Phil Febbo, Summer Galloway, Nicole L. Washington, Simon White, Geraint Levan, Kelly Schiabor Barrett, Elizabeth Cirulli, Alexandre Bolze, Ary Ascencio, Charlotte Rivera-Garcia, Ryan Cho, Jason Nguyen, Sherry Wang, Jimmy Ramirez, Tyler Cassens, Efren Sandoval, Magnus Isaksson, William Lee, David Becker, Marc Laurent, James Lu, Clinton R. Paden, Suxiang Tong, Duncan MacCannell |

| | | | |
|---|---|---|---|
| EPI_ISL_1269746, EPI_ISL_1269747, EPI_ISL_1269748 | Wisconsin State Laboratory of Hygiene Communicable Disease Division | Ginkgo Bioworks Clinical Laboratory | Malaika Mckenzie-Bennett, James McGann, Jim Griffin, Keith Robison, Alex Plocik, Becky Schilling, Martha Pierson, Rebecca Littlefield, Michelle Spencer, Amanda Riccardi, Birgitte Simen, Kelsey R. Florek, Abigail C. Shockey |
| EPI_ISL_1269749 | Helix/Illumina | Centers for Disease Control and Prevention Division of Viral Diseases, Pathogen Discovery | Peter W. Cook, Dakota Howard, Dhwani Batra, Ben L. Rambo-Martin, Eileen de Feo, Jan Antico, Christine Tran, Matthew Tolentino, Shannon Wickline, Kim Gietzen, Brad Sickler, Jingtao Liu, Eric Allen, Phil Febbo, Summer Galloway, Nicole L. Washington, Simon White, Geraint Levan, Kelly Schiabor Barrett, Elizabeth Cirulli, Alexandre Bolze, Ary Ascencio, Charlotte Rivera-Garcia, Ryan Cho, Jason Nguyen, Sherry Wang, Jimmy Ramirez, Tyler Cassens, Efren Sandoval, Magnus Isaksson, William Lee, David Becker, Marc Laurent, James Lu, Clinton R. Paden, Suxiang Tong, Duncan MacCannell |
| EPI_ISL_1269750, EPI_ISL_1269751 | Wisconsin State Laboratory of Hygiene Communicable Disease Division | Ginkgo Bioworks Clinical Laboratory | Malaika Mckenzie-Bennett, James McGann, Jim Griffin, Keith Robison, Alex Plocik, Becky Schilling, Martha Pierson, Rebecca Littlefield, Michelle Spencer, Amanda Riccardi, Birgitte Simen, Kelsey R. Florek, Abigail C. Shockey |
| EPI_ISL_1269752 | Helix/Illumina | Centers for Disease Control and Prevention Division of Viral Diseases, Pathogen Discovery | Peter W. Cook, Dakota Howard, Dhwani Batra, Ben L. Rambo-Martin, Eileen de Feo, Jan Antico, Christine Tran, Matthew Tolentino, Shannon Wickline, Kim Gietzen, Brad Sickler, Jingtao Liu, Eric Allen, Phil Febbo, Summer Galloway, Nicole L. Washington, Simon White, Geraint Levan, Kelly Schiabor Barrett, Elizabeth Cirulli, Alexandre Bolze, Ary Ascencio, Charlotte Rivera-Garcia, Ryan Cho, Jason Nguyen, Sherry Wang, Jimmy Ramirez, Tyler Cassens, Efren Sandoval, Magnus Isaksson, William Lee, David Becker, Marc Laurent, James Lu, Clinton R. Paden, Suxiang Tong, Duncan MacCannell |
| EPI_ISL_1269753, EPI_ISL_1269754 | Wisconsin State Laboratory of Hygiene Communicable Disease Division | Ginkgo Bioworks Clinical Laboratory | Malaika Mckenzie-Bennett, James McGann, Jim Griffin, Keith Robison, Alex Plocik, Becky Schilling, Martha Pierson, Rebecca Littlefield, Michelle Spencer, Amanda Riccardi, Birgitte Simen, Kelsey R. Florek, Abigail C. Shockey |
| EPI_ISL_1269755 | Helix/Illumina | Centers for Disease Control and Prevention Division of Viral Diseases, Pathogen Discovery | Peter W. Cook, Dakota Howard, Dhwani Batra, Ben L. Rambo-Martin, Eileen de Feo, Jan Antico, Christine Tran, Matthew Tolentino, Shannon Wickline, Kim Gietzen, Brad Sickler, Jingtao Liu, Eric Allen, Phil Febbo, Summer Galloway, Nicole L. Washington, Simon White, Geraint Levan, Kelly Schiabor Barrett, Elizabeth Cirulli, Alexandre Bolze, Ary Ascencio, Charlotte Rivera-Garcia, Ryan Cho, Jason Nguyen, Sherry Wang, Jimmy Ramirez, Tyler Cassens, Efren Sandoval, Magnus Isaksson, William Lee, David Becker, Marc Laurent, James Lu, Clinton R. Paden, Suxiang Tong, Duncan MacCannell |
| EPI_ISL_1269756, EPI_ISL_1269757, EPI_ISL_1269758 | Wisconsin State Laboratory of Hygiene Communicable Disease Division | Ginkgo Bioworks Clinical Laboratory | Malaika Mckenzie-Bennett, James McGann, Jim Griffin, Keith Robison, Alex Plocik, Becky Schilling, Martha Pierson, Rebecca Littlefield, Michelle Spencer, Amanda Riccardi, Birgitte Simen, Kelsey R. Florek, Abigail C. Shockey |
| EPI_ISL_1269759 | Helix/Illumina | Centers for Disease Control and Prevention Division of Viral Diseases, Pathogen Discovery | Peter W. Cook, Dakota Howard, Dhwani Batra, Ben L. Rambo-Martin, Eileen de Feo, Jan Antico, Christine Tran, Matthew Tolentino, Shannon Wickline, Kim Gietzen, Brad Sickler, Jingtao Liu, Eric Allen, Phil Febbo, Summer Galloway, Nicole L. Washington, Simon White, Geraint Levan, Kelly Schiabor Barrett, Elizabeth Cirulli, Alexandre Bolze, Ary Ascencio, Charlotte Rivera-Garcia, Ryan Cho, Jason Nguyen, Sherry Wang, Jimmy Ramirez, Tyler Cassens, Efren Sandoval, Magnus Isaksson, William Lee, David Becker, Marc Laurent, James Lu, Clinton R. Paden, Suxiang Tong, Duncan MacCannell |
| EPI_ISL_1269760, EPI_ISL_1269761 | Wisconsin State Laboratory of Hygiene Communicable Disease Division | Ginkgo Bioworks Clinical Laboratory | Malaika Mckenzie-Bennett, James McGann, Jim Griffin, Keith Robison, Alex Plocik, Becky Schilling, Martha Pierson, Rebecca Littlefield, Michelle Spencer, Amanda Riccardi, Birgitte Simen, Kelsey R. Florek, Abigail C. Shockey |
| EPI_ISL_1269762 | Helix/Illumina | Centers for Disease Control and Prevention Division of Viral Diseases, Pathogen Discovery | Peter W. Cook, Dakota Howard, Dhwani Batra, Ben L. Rambo-Martin, Eileen de Feo, Jan Antico, Christine Tran, Matthew Tolentino, Shannon Wickline, Kim Gietzen, Brad Sickler, Jingtao Liu, Eric Allen, Phil Febbo, Summer Galloway, Nicole L. Washington, Simon White, Geraint Levan, Kelly Schiabor Barrett, Elizabeth Cirulli, Alexandre Bolze, Ary Ascencio, Charlotte Rivera-Garcia, Ryan Cho, Jason Nguyen, Sherry Wang, Jimmy Ramirez, Tyler Cassens, Efren Sandoval, Magnus Isaksson, William Lee, David Becker, Marc Laurent, James Lu, Clinton R. Paden, Suxiang Tong, Duncan MacCannell |
| EPI_ISL_1269763, EPI_ISL_1269765, EPI_ISL_1269766 | Wisconsin State Laboratory of Hygiene Communicable Disease Division | Ginkgo Bioworks Clinical Laboratory | Malaika Mckenzie-Bennett, James McGann, Jim Griffin, Keith Robison, Alex Plocik, Becky Schilling, Martha Pierson, Rebecca Littlefield, Michelle Spencer, Amanda Riccardi, Birgitte Simen, Kelsey R. Florek, Abigail C. Shockey |
| EPI_ISL_1269767 | Helix/Illumina | Centers for Disease Control and Prevention Division of Viral Diseases, Pathogen Discovery | Peter W. Cook, Dakota Howard, Dhwani Batra, Ben L. Rambo-Martin, Eileen de Feo, Jan Antico, Christine Tran, Matthew Tolentino, Shannon Wickline, Kim Gietzen, Brad Sickler, Jingtao Liu, Eric Allen, Phil Febbo, Summer Galloway, Nicole L. Washington, Simon White, Geraint Levan, Kelly Schiabor Barrett, Elizabeth Cirulli, Alexandre Bolze, Ary Ascencio, Charlotte Rivera-Garcia, Ryan Cho, Jason Nguyen, Sherry Wang, Jimmy Ramirez, Tyler Cassens, Efren Sandoval, Magnus Isaksson, William Lee, David Becker, Marc Laurent, James Lu, Clinton R. Paden, Suxiang Tong, Duncan MacCannell |
| EPI_ISL_1269768, EPI_ISL_1269769 | Wisconsin State Laboratory of Hygiene Communicable Disease Division | Ginkgo Bioworks Clinical Laboratory | Malaika Mckenzie-Bennett, James McGann, Jim Griffin, Keith Robison, Alex Plocik, Becky Schilling, Martha Pierson, Rebecca Littlefield, Michelle Spencer, Amanda Riccardi, Birgitte Simen, Kelsey R. Florek, Abigail C. Shockey |
| EPI_ISL_1269770 | Helix/Illumina | Centers for Disease Control and Prevention Division of Viral Diseases, Pathogen Discovery | Peter W. Cook, Dakota Howard, Dhwani Batra, Ben L. Rambo-Martin, Eileen de Feo, Jan Antico, Christine Tran, Matthew Tolentino, Shannon Wickline, Kim Gietzen, Brad Sickler, Jingtao Liu, Eric Allen, Phil Febbo, Summer Galloway, Nicole L. Washington, Simon White, Geraint Levan, Kelly Schiabor Barrett, Elizabeth Cirulli, Alexandre Bolze, Ary Ascencio, Charlotte Rivera-Garcia, Ryan Cho, Jason Nguyen, Sherry Wang, Jimmy Ramirez, Tyler Cassens, Efren Sandoval, Magnus Isaksson, William Lee, David Becker, Marc Laurent, James Lu, Clinton R. Paden, Suxiang Tong, Duncan MacCannell |
| EPI_ISL_1269771, EPI_ISL_1269772, EPI_ISL_1269773 | Wisconsin State Laboratory of Hygiene Communicable Disease Division | Ginkgo Bioworks Clinical Laboratory | Malaika Mckenzie-Bennett, James McGann, Jim Griffin, Keith Robison, Alex Plocik, Becky Schilling, Martha Pierson, Rebecca Littlefield, Michelle Spencer, Amanda Riccardi, Birgitte Simen, Kelsey R. Florek, Abigail C. Shockey |
| EPI_ISL_1269774 | Helix/Illumina | Centers for Disease Control and Prevention Division of Viral Diseases, Pathogen Discovery | Peter W. Cook, Dakota Howard, Dhwani Batra, Ben L. Rambo-Martin, Eileen de Feo, Jan Antico, Christine Tran, Matthew Tolentino, Shannon Wickline, Kim Gietzen, Brad Sickler, Jingtao Liu, Eric Allen, Phil Febbo, Summer Galloway, Nicole L. Washington, Simon White, Geraint Levan, Kelly Schiabor Barrett, Elizabeth Cirulli, Alexandre Bolze, Ary Ascencio, Charlotte Rivera-Garcia, Ryan Cho, Jason Nguyen, Sherry Wang, Jimmy Ramirez, Tyler Cassens, Efren Sandoval, Magnus Isaksson, William Lee, David Becker, Marc Laurent, James Lu, Clinton R. Paden, Suxiang Tong, Duncan MacCannell |
| EPI_ISL_1269775, EPI_ISL_1269776 | Wisconsin State Laboratory of Hygiene Communicable Disease Division | Ginkgo Bioworks Clinical Laboratory | Malaika Mckenzie-Bennett, James McGann, Jim Griffin, Keith Robison, Alex Plocik, Becky Schilling, Martha Pierson, Rebecca Littlefield, Michelle Spencer, Amanda Riccardi, Birgitte Simen, Kelsey R. Florek, Abigail C. Shockey |
| EPI_ISL_1269777 | Helix/Illumina | Centers for Disease Control and Prevention Division of Viral Diseases, Pathogen Discovery | Peter W. Cook, Dakota Howard, Dhwani Batra, Ben L. Rambo-Martin, Eileen de Feo, Jan Antico, Christine Tran, Matthew Tolentino, Shannon Wickline, Kim Gietzen, Brad Sickler, Jingtao Liu, Eric Allen, Phil Febbo, Summer Galloway, Nicole L. Washington, Simon White, Geraint Levan, Kelly Schiabor Barrett, Elizabeth Cirulli, Alexandre Bolze, Ary Ascencio, Charlotte Rivera-Garcia, Ryan Cho, Jason Nguyen, Sherry Wang, Jimmy Ramirez, Tyler Cassens, Efren Sandoval, Magnus Isaksson, William Lee, David Becker, Marc Laurent, James Lu, Clinton R. Paden, Suxiang Tong, Duncan MacCannell |
| EPI_ISL_1269778 | Wisconsin State Laboratory of Hygiene Communicable Disease Division | Ginkgo Bioworks Clinical Laboratory | Malaika Mckenzie-Bennett, James McGann, Jim Griffin, Keith Robison, Alex Plocik, Becky Schilling, Martha Pierson, Rebecca Littlefield, Michelle Spencer, Amanda Riccardi, Birgitte Simen, Kelsey R. Florek, Abigail C. Shockey |
| EPI_ISL_1269779 | Helix/Illumina | Centers for Disease Control and Prevention Division of Viral Diseases, Pathogen Discovery | Peter W. Cook, Dakota Howard, Dhwani Batra, Ben L. Rambo-Martin, Eileen de Feo, Jan Antico, Christine Tran, Matthew Tolentino, Shannon Wickline, Kim Gietzen, Brad Sickler, Jingtao Liu, Eric Allen, Phil Febbo, Summer Galloway, Nicole L. Washington, Simon White, Geraint Levan, Kelly Schiabor Barrett, Elizabeth Cirulli, Alexandre Bolze, Ary Ascencio, Charlotte Rivera-Garcia, Ryan Cho, Jason Nguyen, Sherry Wang, Jimmy Ramirez, Tyler Cassens, Efren Sandoval, Magnus Isaksson, William Lee, David Becker, Marc Laurent, James Lu, Clinton R. Paden, Suxiang Tong, Duncan MacCannell |
| EPI_ISL_1269780, EPI_ISL_1269781 | Wisconsin State Laboratory of Hygiene Communicable Disease Division | Ginkgo Bioworks Clinical Laboratory | Malaika Mckenzie-Bennett, James McGann, Jim Griffin, Keith Robison, Alex Plocik, Becky Schilling, Martha Pierson, Rebecca Littlefield, Michelle Spencer, Amanda Riccardi, Birgitte Simen, Kelsey R. Florek, Abigail C. Shockey |
| EPI_ISL_1269782 | Helix/Illumina | Centers for Disease Control and Prevention Division of Viral Diseases, Pathogen Discovery | Peter W. Cook, Dakota Howard, Dhwani Batra, Ben L. Rambo-Martin, Eileen de Feo, Jan Antico, Christine Tran, Matthew Tolentino, Shannon Wickline, Kim Gietzen, Brad Sickler, Jingtao Liu, Eric Allen, Phil Febbo, Summer Galloway, Nicole L. Washington, Simon White, Geraint Levan, Kelly Schiabor Barrett, Elizabeth Cirulli, Alexandre Bolze, Ary Ascencio, Charlotte Rivera-Garcia, Ryan Cho, Jason Nguyen, Sherry Wang, Jimmy Ramirez, Tyler Cassens, Efren Sandoval, Magnus Isaksson, William Lee, David Becker, Marc Laurent, James Lu, Clinton R. Paden, Suxiang Tong, Duncan MacCannell |
| EPI_ISL_1269783, EPI_ISL_1269784, EPI_ISL_1269785 | Wisconsin State Laboratory of Hygiene Communicable Disease Division | Ginkgo Bioworks Clinical Laboratory | Malaika Mckenzie-Bennett, James McGann, Jim Griffin, Keith Robison, Alex Plocik, Becky Schilling, Martha Pierson, Rebecca Littlefield, Michelle Spencer, Amanda Riccardi, Birgitte Simen, Kelsey R. Florek, Abigail C. Shockey |
| EPI_ISL_1269786 | Helix/Illumina | Centers for Disease Control and Prevention Division of Viral Diseases, Pathogen Discovery | Peter W. Cook, Dakota Howard, Dhwani Batra, Ben L. Rambo-Martin, Eileen de Feo, Jan Antico, Christine Tran, Matthew Tolentino, Shannon Wickline, Kim Gietzen, Brad Sickler, Jingtao Liu, Eric Allen, Phil Febbo, Summer Galloway, Nicole L. Washington, Simon White, Geraint Levan, Kelly Schiabor Barrett, Elizabeth Cirulli, Alexandre Bolze, Ary Ascencio, Charlotte Rivera-Garcia, Ryan Cho, Jason Nguyen, Sherry Wang, Jimmy Ramirez, Tyler Cassens, Efren Sandoval, Magnus Isaksson, William Lee, David Becker, Marc Laurent, James Lu, Clinton R. Paden, Suxiang Tong, Duncan MacCannell |
| EPI_ISL_1269787, EPI_ISL_1269788 | Wisconsin State Laboratory of Hygiene Communicable Disease Division | Ginkgo Bioworks Clinical Laboratory | Malaika Mckenzie-Bennett, James McGann, Jim Griffin, Keith Robison, Alex Plocik, Becky Schilling, Martha Pierson, Rebecca Littlefield, Michelle Spencer, Amanda Riccardi, Birgitte Simen, Kelsey R. Florek, Abigail C. Shockey |
| EPI_ISL_1269789 | Helix/Illumina | Centers for Disease Control and Prevention Division of Viral Diseases, Pathogen Discovery | Peter W. Cook, Dakota Howard, Dhwani Batra, Ben L. Rambo-Martin, Eileen de Feo, Jan Antico, Christine Tran, Matthew Tolentino, Shannon Wickline, Kim Gietzen, Brad Sickler, Jingtao Liu, Eric Allen, Phil Febbo, Summer Galloway, Nicole L. Washington, Simon White, Geraint Levan, Kelly Schiabor Barrett, Elizabeth Cirulli, Alexandre Bolze, Ary Ascencio, Charlotte Rivera-Garcia, Ryan Cho, Jason Nguyen, Sherry Wang, Jimmy Ramirez, Tyler Cassens, Efren Sandoval, Magnus Isaksson, William Lee, David Becker, Marc Laurent, James Lu, Clinton R. Paden, Suxiang Tong, Duncan MacCannell |
| EPI_ISL_1269790, EPI_ISL_1269791 | Wisconsin State Laboratory of Hygiene Communicable Disease Division | Ginkgo Bioworks Clinical Laboratory | Malaika Mckenzie-Bennett, James McGann, Jim Griffin, Keith Robison, Alex Plocik, Becky Schilling, Martha Pierson, Rebecca Littlefield, Michelle Spencer, Amanda Riccardi, Birgitte Simen, Kelsey R. Florek, Abigail C. Shockey |
| EPI_ISL_1269792 | Helix/Illumina | Centers for Disease Control and Prevention Division of Viral Diseases, Pathogen Discovery | Peter W. Cook, Dakota Howard, Dhwani Batra, Ben L. Rambo-Martin, Eileen de Feo, Jan Antico, Christine Tran, Matthew Tolentino, Shannon Wickline, Kim Gietzen, Brad Sickler, Jingtao Liu, Eric Allen, Phil Febbo, Summer Galloway, Nicole L. Washington, Simon White, Geraint Levan, Kelly Schiabor Barrett, Elizabeth Cirulli, Alexandre Bolze, Ary Ascencio, Charlotte Rivera-Garcia, Ryan Cho, Jason Nguyen, Sherry Wang, Jimmy Ramirez, Tyler Cassens, Efren Sandoval, Magnus Isaksson, William Lee, David Becker, Marc Laurent, James Lu, Clinton R. Paden, Suxiang Tong, Duncan MacCannell |
| EPI_ISL_1269793, EPI_ISL_1269794 | Wisconsin State Laboratory of Hygiene Communicable Disease Division | Ginkgo Bioworks Clinical Laboratory | Malaika Mckenzie-Bennett, James McGann, Jim Griffin, Keith Robison, Alex Plocik, Becky Schilling, Martha Pierson, Rebecca Littlefield, Michelle Spencer, Amanda Riccardi, Birgitte Simen, Kelsey R. Florek, Abigail C. Shockey |
| EPI_ISL_1269795 | Helix/Illumina | Centers for Disease Control and Prevention Division of Viral Diseases, Pathogen Discovery | Peter W. Cook, Dakota Howard, Dhwani Batra, Ben L. Rambo-Martin, Eileen de Feo, Jan Antico, Christine Tran, Matthew Tolentino, Shannon Wickline, Kim Gietzen, Brad Sickler, Jingtao Liu, Eric Allen, Phil Febbo, Summer Galloway, Nicole L. Washington, Simon White, Geraint Levan, Kelly Schiabor Barrett, Elizabeth Cirulli, Alexandre Bolze, Ary Ascencio, Charlotte Rivera-Garcia, Ryan Cho, Jason Nguyen, Sherry Wang, Jimmy Ramirez, Tyler Cassens, Efren Sandoval, Magnus Isaksson, William Lee, David Becker, Marc Laurent, James Lu, Clinton R. Paden, Suxiang Tong, Duncan MacCannell |
| EPI_ISL_1269796, EPI_ISL_1269797 | Wisconsin State Laboratory of Hygiene Communicable Disease Division | Ginkgo Bioworks Clinical Laboratory | Malaika Mckenzie-Bennett, James McGann, Jim Griffin, Keith Robison, Alex Plocik, Becky Schilling, Martha Pierson, Rebecca Littlefield, Michelle Spencer, Amanda Riccardi, Birgitte Simen, Kelsey R. Florek, Abigail C. Shockey |
| EPI_ISL_1269798 | Helix/Illumina | Centers for Disease Control and Prevention Division of Viral Diseases, Pathogen Discovery | Peter W. Cook, Dakota Howard, Dhwani Batra, Ben L. Rambo-Martin, Eileen de Feo, Jan Antico, Christine Tran, Matthew Tolentino, Shannon Wickline, Kim Gietzen, Brad Sickler, Jingtao Liu, Eric Allen, Phil Febbo, Summer Galloway, Nicole L. Washington, Simon White, Geraint Levan, Kelly Schiabor Barrett, Elizabeth Cirulli, Alexandre Bolze, Ary Ascencio, Charlotte Rivera-Garcia, Ryan Cho, Jason Nguyen, Sherry Wang, Jimmy Ramirez, Tyler Cassens, Efren Sandoval, Magnus Isaksson, William Lee, David Becker, Marc Laurent, James Lu, Clinton R. Paden, Suxiang Tong, Duncan MacCannell |
| EPI_ISL_1269799, EPI_ISL_1269800 | Wisconsin State Laboratory of Hygiene Communicable Disease Division | Ginkgo Bioworks Clinical Laboratory | Malaika Mckenzie-Bennett, James McGann, Jim Griffin, Keith Robison, Alex Plocik, Becky Schilling, Martha Pierson, Rebecca Littlefield, Michelle Spencer, Amanda Riccardi, Birgitte Simen, Kelsey R. Florek, Abigail C. Shockey |
| EPI_ISL_1269801 | Helix/Illumina | Centers for Disease Control and Prevention Division of Viral Diseases, Pathogen Discovery | Peter W. Cook, Dakota Howard, Dhwani Batra, Ben L. Rambo-Martin, Eileen de Feo, Jan Antico, Christine Tran, Matthew Tolentino, Shannon Wickline, Kim Gietzen, Brad Sickler, Jingtao Liu, Eric Allen, Phil Febbo, Summer Galloway, Nicole L. Washington, Simon White, Geraint Levan, Kelly Schiabor Barrett, Elizabeth Cirulli, Alexandre Bolze, Ary Ascencio, Charlotte Rivera-Garcia, Ryan Cho, Jason Nguyen, Sherry Wang, Jimmy Ramirez, Tyler Cassens, Efren Sandoval, Magnus Isaksson, William Lee, David Becker, Marc Laurent, James Lu, Clinton R. Paden, Suxiang Tong, Duncan MacCannell |
| EPI_ISL_1269802, EPI_ISL_1269803 | Wisconsin State Laboratory of Hygiene Communicable Disease Division | Ginkgo Bioworks Clinical Laboratory | Malaika Mckenzie-Bennett, James McGann, Jim Griffin, Keith Robison, Alex Plocik, Becky Schilling, Martha Pierson, Rebecca Littlefield, Michelle Spencer, Amanda Riccardi, Birgitte Simen, Kelsey R. Florek, Abigail C. Shockey |
| EPI_ISL_1269804 | Helix/Illumina | Centers for Disease Control and Prevention Division of Viral Diseases, Pathogen | Peter W. Cook, Dakota Howard, Dhwani Batra, Ben L. Rambo-Martin, Eileen de Feo, Jan Antico, Christine Tran, Matthew Tolentino, Shannon Wickline, Kim Gietzen, Brad Sickler, Jingtao Liu, Eric Allen, Phil Febbo, Summer Galloway, Nicole L. Washington, Simon White, Geraint Levan, Kelly Schiabor Barrett, Elizabeth Cirulli, Alexandre Bolze, Ary Ascencio, Charlotte Rivera-Garcia, Ryan Cho, Jason Nguyen, Sherry Wang, Jimmy Ramirez, Tyler Cassens, Efren Sandoval, Magnus Isaksson, William Lee, David Becker, Marc Laurent, James Lu, Clinton R. Paden, Suxiang Tong, |

| | | | |
|---|---|---|---|
| | | Discovery | Duncan MacCannell |
| EPI_ISL_1269805, EPI_ISL_1269806, EPI_ISL_1269807 | Wisconsin State Laboratory of Hygiene Communicable Disease Division | Ginkgo Bioworks Clinical Laboratory | Malaika Mckenzie-Bennett, James McGann, Jim Griffin, Keith Robison, Alex Plocik, Becky Schilling, Martha Pierson, Rebecca Littlefield, Michelle Spencer, Amanda Riccardi, Birgitte Simen, Kelsey R. Florek, Abigail C. Shockey |
| EPI_ISL_1269808 | Helix/Illumina | Centers for Disease Control and Prevention Division of Viral Diseases, Pathogen Discovery | Peter W. Cook, Dakota Howard, Dhwani Batra, Ben L. Rambo-Martin, Eileen de Feo, Jan Antico, Christine Tran, Matthew Tolentino, Shannon Wickline, Kim Gietzen, Brad Sickler, Jingtao Liu, Eric Allen, Phil Febbo, Summer Galloway, Nicole L. Washington, Simon White, Geraint Levan, Kelly Schiabor Barrett, Elizabeth Cirulli, Alexandre Bolze, Ary Ascencio, Charlotte Rivera-Garcia, Ryan Cho, Jason Nguyen, Sherry Wang, Jimmy Ramirez, Tyler Cassens, Efren Sandoval, Magnus Isaksson, William Lee, David Becker, Marc Laurent, James Lu, Clinton R. Paden, Suxiang Tong, Duncan MacCannell |
| EPI_ISL_1269809, EPI_ISL_1269810 | Wisconsin State Laboratory of Hygiene Communicable Disease Division | Ginkgo Bioworks Clinical Laboratory | Malaika Mckenzie-Bennett, James McGann, Jim Griffin, Keith Robison, Alex Plocik, Becky Schilling, Martha Pierson, Rebecca Littlefield, Michelle Spencer, Amanda Riccardi, Birgitte Simen, Kelsey R. Florek, Abigail C. Shockey |
| EPI_ISL_1269811 | Helix/Illumina | Centers for Disease Control and Prevention Division of Viral Diseases, Pathogen Discovery | Peter W. Cook, Dakota Howard, Dhwani Batra, Ben L. Rambo-Martin, Eileen de Feo, Jan Antico, Christine Tran, Matthew Tolentino, Shannon Wickline, Kim Gietzen, Brad Sickler, Jingtao Liu, Eric Allen, Phil Febbo, Summer Galloway, Nicole L. Washington, Simon White, Geraint Levan, Kelly Schiabor Barrett, Elizabeth Cirulli, Alexandre Bolze, Ary Ascencio, Charlotte Rivera-Garcia, Ryan Cho, Jason Nguyen, Sherry Wang, Jimmy Ramirez, Tyler Cassens, Efren Sandoval, Magnus Isaksson, William Lee, David Becker, Marc Laurent, James Lu, Clinton R. Paden, Suxiang Tong, Duncan MacCannell |
| EPI_ISL_1269812, EPI_ISL_1269813 | Wisconsin State Laboratory of Hygiene Communicable Disease Division | Ginkgo Bioworks Clinical Laboratory | Malaika Mckenzie-Bennett, James McGann, Jim Griffin, Keith Robison, Alex Plocik, Becky Schilling, Martha Pierson, Rebecca Littlefield, Michelle Spencer, Amanda Riccardi, Birgitte Simen, Kelsey R. Florek, Abigail C. Shockey |
| EPI_ISL_1269814 | Helix/Illumina | Centers for Disease Control and Prevention Division of Viral Diseases, Pathogen Discovery | Peter W. Cook, Dakota Howard, Dhwani Batra, Ben L. Rambo-Martin, Eileen de Feo, Jan Antico, Christine Tran, Matthew Tolentino, Shannon Wickline, Kim Gietzen, Brad Sickler, Jingtao Liu, Eric Allen, Phil Febbo, Summer Galloway, Nicole L. Washington, Simon White, Geraint Levan, Kelly Schiabor Barrett, Elizabeth Cirulli, Alexandre Bolze, Ary Ascencio, Charlotte Rivera-Garcia, Ryan Cho, Jason Nguyen, Sherry Wang, Jimmy Ramirez, Tyler Cassens, Efren Sandoval, Magnus Isaksson, William Lee, David Becker, Marc Laurent, James Lu, Clinton R. Paden, Suxiang Tong, Duncan MacCannell |
| EPI_ISL_1269815, EPI_ISL_1269816 | Wisconsin State Laboratory of Hygiene Communicable Disease Division | Ginkgo Bioworks Clinical Laboratory | Malaika Mckenzie-Bennett, James McGann, Jim Griffin, Keith Robison, Alex Plocik, Becky Schilling, Martha Pierson, Rebecca Littlefield, Michelle Spencer, Amanda Riccardi, Birgitte Simen, Kelsey R. Florek, Abigail C. Shockey |
| EPI_ISL_1269817 | Helix/Illumina | Centers for Disease Control and Prevention Division of Viral Diseases, Pathogen Discovery | Peter W. Cook, Dakota Howard, Dhwani Batra, Ben L. Rambo-Martin, Eileen de Feo, Jan Antico, Christine Tran, Matthew Tolentino, Shannon Wickline, Kim Gietzen, Brad Sickler, Jingtao Liu, Eric Allen, Phil Febbo, Summer Galloway, Nicole L. Washington, Simon White, Geraint Levan, Kelly Schiabor Barrett, Elizabeth Cirulli, Alexandre Bolze, Ary Ascencio, Charlotte Rivera-Garcia, Ryan Cho, Jason Nguyen, Sherry Wang, Jimmy Ramirez, Tyler Cassens, Efren Sandoval, Magnus Isaksson, William Lee, David Becker, Marc Laurent, James Lu, Clinton R. Paden, Suxiang Tong, Duncan MacCannell |
| EPI_ISL_1269818, EPI_ISL_1269819 | Wisconsin State Laboratory of Hygiene Communicable Disease Division | Ginkgo Bioworks Clinical Laboratory | Malaika Mckenzie-Bennett, James McGann, Jim Griffin, Keith Robison, Alex Plocik, Becky Schilling, Martha Pierson, Rebecca Littlefield, Michelle Spencer, Amanda Riccardi, Birgitte Simen, Kelsey R. Florek, Abigail C. Shockey |
| EPI_ISL_1269820 | Helix/Illumina | Centers for Disease Control and Prevention Division of Viral Diseases, Pathogen Discovery | Peter W. Cook, Dakota Howard, Dhwani Batra, Ben L. Rambo-Martin, Eileen de Feo, Jan Antico, Christine Tran, Matthew Tolentino, Shannon Wickline, Kim Gietzen, Brad Sickler, Jingtao Liu, Eric Allen, Phil Febbo, Summer Galloway, Nicole L. Washington, Simon White, Geraint Levan, Kelly Schiabor Barrett, Elizabeth Cirulli, Alexandre Bolze, Ary Ascencio, Charlotte Rivera-Garcia, Ryan Cho, Jason Nguyen, Sherry Wang, Jimmy Ramirez, Tyler Cassens, Efren Sandoval, Magnus Isaksson, William Lee, David Becker, Marc Laurent, James Lu, Clinton R. Paden, Suxiang Tong, Duncan MacCannell |
| EPI_ISL_1269821, EPI_ISL_1269822 | Wisconsin State Laboratory of Hygiene Communicable Disease Division | Ginkgo Bioworks Clinical Laboratory | Malaika Mckenzie-Bennett, James McGann, Jim Griffin, Keith Robison, Alex Plocik, Becky Schilling, Martha Pierson, Rebecca Littlefield, Michelle Spencer, Amanda Riccardi, Birgitte Simen, Kelsey R. Florek, Abigail C. Shockey |
| EPI_ISL_1269823 | Helix/Illumina | Centers for Disease Control and Prevention Division of Viral Diseases, Pathogen Discovery | Peter W. Cook, Dakota Howard, Dhwani Batra, Ben L. Rambo-Martin, Eileen de Feo, Jan Antico, Christine Tran, Matthew Tolentino, Shannon Wickline, Kim Gietzen, Brad Sickler, Jingtao Liu, Eric Allen, Phil Febbo, Summer Galloway, Nicole L. Washington, Simon White, Geraint Levan, Kelly Schiabor Barrett, Elizabeth Cirulli, Alexandre Bolze, Ary Ascencio, Charlotte Rivera-Garcia, Ryan Cho, Jason Nguyen, Sherry Wang, Jimmy Ramirez, Tyler Cassens, Efren Sandoval, Magnus Isaksson, William Lee, David Becker, Marc Laurent, James Lu, Clinton R. Paden, Suxiang Tong, Duncan MacCannell |
| EPI_ISL_1269824, EPI_ISL_1269825 | Wisconsin State Laboratory of Hygiene Communicable Disease Division | Ginkgo Bioworks Clinical Laboratory | Malaika Mckenzie-Bennett, James McGann, Jim Griffin, Keith Robison, Alex Plocik, Becky Schilling, Martha Pierson, Rebecca Littlefield, Michelle Spencer, Amanda Riccardi, Birgitte Simen, Kelsey R. Florek, Abigail C. Shockey |
| EPI_ISL_1269826 | Helix/Illumina | Centers for Disease Control and Prevention Division of Viral Diseases, Pathogen Discovery | Peter W. Cook, Dakota Howard, Dhwani Batra, Ben L. Rambo-Martin, Eileen de Feo, Jan Antico, Christine Tran, Matthew Tolentino, Shannon Wickline, Kim Gietzen, Brad Sickler, Jingtao Liu, Eric Allen, Phil Febbo, Summer Galloway, Nicole L. Washington, Simon White, Geraint Levan, Kelly Schiabor Barrett, Elizabeth Cirulli, Alexandre Bolze, Ary Ascencio, Charlotte Rivera-Garcia, Ryan Cho, Jason Nguyen, Sherry Wang, Jimmy Ramirez, Tyler Cassens, Efren Sandoval, Magnus Isaksson, William Lee, David Becker, Marc Laurent, James Lu, Clinton R. Paden, Suxiang Tong, Duncan MacCannell |
| EPI_ISL_1269827, EPI_ISL_1269828 | Wisconsin State Laboratory of Hygiene Communicable Disease Division | Ginkgo Bioworks Clinical Laboratory | Malaika Mckenzie-Bennett, James McGann, Jim Griffin, Keith Robison, Alex Plocik, Becky Schilling, Martha Pierson, Rebecca Littlefield, Michelle Spencer, Amanda Riccardi, Birgitte Simen, Kelsey R. Florek, Abigail C. Shockey |
| EPI_ISL_1269829 | Helix/Illumina | Centers for Disease Control and Prevention Division of Viral Diseases, Pathogen Discovery | Peter W. Cook, Dakota Howard, Dhwani Batra, Ben L. Rambo-Martin, Eileen de Feo, Jan Antico, Christine Tran, Matthew Tolentino, Shannon Wickline, Kim Gietzen, Brad Sickler, Jingtao Liu, Eric Allen, Phil Febbo, Summer Galloway, Nicole L. Washington, Simon White, Geraint Levan, Kelly Schiabor Barrett, Elizabeth Cirulli, Alexandre Bolze, Ary Ascencio, Charlotte Rivera-Garcia, Ryan Cho, Jason Nguyen, Sherry Wang, Jimmy Ramirez, Tyler Cassens, Efren Sandoval, Magnus Isaksson, William Lee, David Becker, Marc Laurent, James Lu, Clinton R. Paden, Suxiang Tong, Duncan MacCannell |
| EPI_ISL_1269830, EPI_ISL_1269831 | Wisconsin State Laboratory of Hygiene Communicable Disease Division | Ginkgo Bioworks Clinical Laboratory | Malaika Mckenzie-Bennett, James McGann, Jim Griffin, Keith Robison, Alex Plocik, Becky Schilling, Martha Pierson, Rebecca Littlefield, Michelle Spencer, Amanda Riccardi, Birgitte Simen, Kelsey R. Florek, Abigail C. Shockey |
| EPI_ISL_1269832 | Helix/Illumina | Centers for Disease Control and Prevention Division of Viral Diseases, Pathogen Discovery | Peter W. Cook, Dakota Howard, Dhwani Batra, Ben L. Rambo-Martin, Eileen de Feo, Jan Antico, Christine Tran, Matthew Tolentino, Shannon Wickline, Kim Gietzen, Brad Sickler, Jingtao Liu, Eric Allen, Phil Febbo, Summer Galloway, Nicole L. Washington, Simon White, Geraint Levan, Kelly Schiabor Barrett, Elizabeth Cirulli, Alexandre Bolze, Ary Ascencio, Charlotte Rivera-Garcia, Ryan Cho, Jason Nguyen, Sherry Wang, Jimmy Ramirez, Tyler Cassens, Efren Sandoval, Magnus Isaksson, William Lee, David Becker, Marc Laurent, James Lu, Clinton R. Paden, Suxiang Tong, Duncan MacCannell |
| EPI_ISL_1269833, EPI_ISL_1269834, EPI_ISL_1269835 | Wisconsin State Laboratory of Hygiene Communicable Disease Division | Ginkgo Bioworks Clinical Laboratory | Malaika Mckenzie-Bennett, James McGann, Jim Griffin, Keith Robison, Alex Plocik, Becky Schilling, Martha Pierson, Rebecca Littlefield, Michelle Spencer, Amanda Riccardi, Birgitte Simen, Kelsey R. Florek, Abigail C. Shockey |
| EPI_ISL_1269836 | Helix/Illumina | Centers for Disease Control and Prevention Division of Viral Diseases, Pathogen Discovery | Peter W. Cook, Dakota Howard, Dhwani Batra, Ben L. Rambo-Martin, Eileen de Feo, Jan Antico, Christine Tran, Matthew Tolentino, Shannon Wickline, Kim Gietzen, Brad Sickler, Jingtao Liu, Eric Allen, Phil Febbo, Summer Galloway, Nicole L. Washington, Simon White, Geraint Levan, Kelly Schiabor Barrett, Elizabeth Cirulli, Alexandre Bolze, Ary Ascencio, Charlotte Rivera-Garcia, Ryan Cho, Jason Nguyen, Sherry Wang, Jimmy Ramirez, Tyler Cassens, Efren Sandoval, Magnus Isaksson, William Lee, David Becker, Marc Laurent, James Lu, Clinton R. Paden, Suxiang Tong, Duncan MacCannell |
| EPI_ISL_1269837, EPI_ISL_1269838 | Wisconsin State Laboratory of Hygiene Communicable Disease Division | Ginkgo Bioworks Clinical Laboratory | Malaika Mckenzie-Bennett, James McGann, Jim Griffin, Keith Robison, Alex Plocik, Becky Schilling, Martha Pierson, Rebecca Littlefield, Michelle Spencer, Amanda Riccardi, Birgitte Simen, Kelsey R. Florek, Abigail C. Shockey |
| EPI_ISL_1269839 | Helix/Illumina | Centers for Disease Control and Prevention Division of Viral Diseases, Pathogen Discovery | Peter W. Cook, Dakota Howard, Dhwani Batra, Ben L. Rambo-Martin, Eileen de Feo, Jan Antico, Christine Tran, Matthew Tolentino, Shannon Wickline, Kim Gietzen, Brad Sickler, Jingtao Liu, Eric Allen, Phil Febbo, Summer Galloway, Nicole L. Washington, Simon White, Geraint Levan, Kelly Schiabor Barrett, Elizabeth Cirulli, Alexandre Bolze, Ary Ascencio, Charlotte Rivera-Garcia, Ryan Cho, Jason Nguyen, Sherry Wang, Jimmy Ramirez, Tyler Cassens, Efren Sandoval, Magnus Isaksson, William Lee, David Becker, Marc Laurent, James Lu, Clinton R. Paden, Suxiang Tong, Duncan MacCannell |
| EPI_ISL_1269840, EPI_ISL_1269841 | Wisconsin State Laboratory of Hygiene Communicable Disease Division | Ginkgo Bioworks Clinical Laboratory | Malaika Mckenzie-Bennett, James McGann, Jim Griffin, Keith Robison, Alex Plocik, Becky Schilling, Martha Pierson, Rebecca Littlefield, Michelle Spencer, Amanda Riccardi, Birgitte Simen, Kelsey R. Florek, Abigail C. Shockey |
| EPI_ISL_1269842 | Helix/Illumina | Centers for Disease Control and Prevention Division of Viral Diseases, Pathogen Discovery | Peter W. Cook, Dakota Howard, Dhwani Batra, Ben L. Rambo-Martin, Eileen de Feo, Jan Antico, Christine Tran, Matthew Tolentino, Shannon Wickline, Kim Gietzen, Brad Sickler, Jingtao Liu, Eric Allen, Phil Febbo, Summer Galloway, Nicole L. Washington, Simon White, Geraint Levan, Kelly Schiabor Barrett, Elizabeth Cirulli, Alexandre Bolze, Ary Ascencio, Charlotte Rivera-Garcia, Ryan Cho, Jason Nguyen, Sherry Wang, Jimmy Ramirez, Tyler Cassens, Efren Sandoval, Magnus Isaksson, William Lee, David Becker, Marc Laurent, James Lu, Clinton R. Paden, Suxiang Tong, Duncan MacCannell |
| EPI_ISL_1269843, EPI_ISL_1269844, EPI_ISL_1269845 | Wisconsin State Laboratory of Hygiene Communicable Disease Division | Ginkgo Bioworks Clinical Laboratory | Malaika Mckenzie-Bennett, James McGann, Jim Griffin, Keith Robison, Alex Plocik, Becky Schilling, Martha Pierson, Rebecca Littlefield, Michelle Spencer, Amanda Riccardi, Birgitte Simen, Kelsey R. Florek, Abigail C. Shockey |
| EPI_ISL_1269846 | Helix/Illumina | Centers for Disease Control and Prevention Division of Viral Diseases, Pathogen Discovery | Peter W. Cook, Dakota Howard, Dhwani Batra, Ben L. Rambo-Martin, Eileen de Feo, Jan Antico, Christine Tran, Matthew Tolentino, Shannon Wickline, Kim Gietzen, Brad Sickler, Jingtao Liu, Eric Allen, Phil Febbo, Summer Galloway, Nicole L. Washington, Simon White, Geraint Levan, Kelly Schiabor Barrett, Elizabeth Cirulli, Alexandre Bolze, Ary Ascencio, Charlotte Rivera-Garcia, Ryan Cho, Jason Nguyen, Sherry Wang, Jimmy Ramirez, Tyler Cassens, Efren Sandoval, Magnus Isaksson, William Lee, David Becker, Marc Laurent, James Lu, Clinton R. Paden, Suxiang Tong, Duncan MacCannell |
| EPI_ISL_1269847, EPI_ISL_1269848 | Wisconsin State Laboratory of Hygiene Communicable Disease Division | Ginkgo Bioworks Clinical Laboratory | Malaika Mckenzie-Bennett, James McGann, Jim Griffin, Keith Robison, Alex Plocik, Becky Schilling, Martha Pierson, Rebecca Littlefield, Michelle Spencer, Amanda Riccardi, Birgitte Simen, Kelsey R. Florek, Abigail C. Shockey |
| EPI_ISL_1269849 | Helix/Illumina | Centers for Disease Control and Prevention Division of Viral Diseases, Pathogen Discovery | Peter W. Cook, Dakota Howard, Dhwani Batra, Ben L. Rambo-Martin, Eileen de Feo, Jan Antico, Christine Tran, Matthew Tolentino, Shannon Wickline, Kim Gietzen, Brad Sickler, Jingtao Liu, Eric Allen, Phil Febbo, Summer Galloway, Nicole L. Washington, Simon White, Geraint Levan, Kelly Schiabor Barrett, Elizabeth Cirulli, Alexandre Bolze, Ary Ascencio, Charlotte Rivera-Garcia, Ryan Cho, Jason Nguyen, Sherry Wang, Jimmy Ramirez, Tyler Cassens, Efren Sandoval, Magnus Isaksson, William Lee, David Becker, Marc Laurent, James Lu, Clinton R. Paden, Suxiang Tong, Duncan MacCannell |
| EPI_ISL_1269850, EPI_ISL_1269851 | Wisconsin State Laboratory of Hygiene Communicable Disease Division | Ginkgo Bioworks Clinical Laboratory | Malaika Mckenzie-Bennett, James McGann, Jim Griffin, Keith Robison, Alex Plocik, Becky Schilling, Martha Pierson, Rebecca Littlefield, Michelle Spencer, Amanda Riccardi, Birgitte Simen, Kelsey R. Florek, Abigail C. Shockey |
| EPI_ISL_1269852 | Helix/Illumina | Centers for Disease Control and Prevention Division of Viral Diseases, Pathogen Discovery | Peter W. Cook, Dakota Howard, Dhwani Batra, Ben L. Rambo-Martin, Eileen de Feo, Jan Antico, Christine Tran, Matthew Tolentino, Shannon Wickline, Kim Gietzen, Brad Sickler, Jingtao Liu, Eric Allen, Phil Febbo, Summer Galloway, Nicole L. Washington, Simon White, Geraint Levan, Kelly Schiabor Barrett, Elizabeth Cirulli, Alexandre Bolze, Ary Ascencio, Charlotte Rivera-Garcia, Ryan Cho, Jason Nguyen, Sherry Wang, Jimmy Ramirez, Tyler Cassens, Efren Sandoval, Magnus Isaksson, William Lee, David Becker, Marc Laurent, James Lu, Clinton R. Paden, Suxiang Tong, Duncan MacCannell |
| EPI_ISL_1269853, EPI_ISL_1269854 | Wisconsin State Laboratory of Hygiene Communicable Disease Division | Ginkgo Bioworks Clinical Laboratory | Malaika Mckenzie-Bennett, James McGann, Jim Griffin, Keith Robison, Alex Plocik, Becky Schilling, Martha Pierson, Rebecca Littlefield, Michelle Spencer, Amanda Riccardi, Birgitte Simen, Kelsey R. Florek, Abigail C. Shockey |
| EPI_ISL_1269855 | Helix/Illumina | Centers for Disease Control and Prevention Division of Viral Diseases, Pathogen Discovery | Peter W. Cook, Dakota Howard, Dhwani Batra, Ben L. Rambo-Martin, Eileen de Feo, Jan Antico, Christine Tran, Matthew Tolentino, Shannon Wickline, Kim Gietzen, Brad Sickler, Jingtao Liu, Eric Allen, Phil Febbo, Summer Galloway, Nicole L. Washington, Simon White, Geraint Levan, Kelly Schiabor Barrett, Elizabeth Cirulli, Alexandre Bolze, Ary Ascencio, Charlotte Rivera-Garcia, Ryan Cho, Jason Nguyen, Sherry Wang, Jimmy Ramirez, Tyler Cassens, Efren Sandoval, Magnus Isaksson, William Lee, David Becker, Marc Laurent, James Lu, Clinton R. Paden, Suxiang Tong, Duncan MacCannell |
| EPI_ISL_1269856, EPI_ISL_1269857 | Wisconsin State Laboratory of Hygiene Communicable Disease Division | Ginkgo Bioworks Clinical Laboratory | Malaika Mckenzie-Bennett, James McGann, Jim Griffin, Keith Robison, Alex Plocik, Becky Schilling, Martha Pierson, Rebecca Littlefield, Michelle Spencer, Amanda Riccardi, Birgitte Simen, Kelsey R. Florek, Abigail C. Shockey |
| EPI_ISL_1269858 | Helix/Illumina | Centers for Disease Control and Prevention Division of Viral Diseases, Pathogen Discovery | Peter W. Cook, Dakota Howard, Dhwani Batra, Ben L. Rambo-Martin, Eileen de Feo, Jan Antico, Christine Tran, Matthew Tolentino, Shannon Wickline, Kim Gietzen, Brad Sickler, Jingtao Liu, Eric Allen, Phil Febbo, Summer Galloway, Nicole L. Washington, Simon White, Geraint Levan, Kelly Schiabor Barrett, Elizabeth Cirulli, Alexandre Bolze, Ary Ascencio, Charlotte Rivera-Garcia, Ryan Cho, Jason Nguyen, Sherry Wang, Jimmy Ramirez, Tyler Cassens, Efren Sandoval, Magnus Isaksson, William Lee, David Becker, Marc Laurent, James Lu, Clinton R. Paden, Suxiang Tong, Duncan MacCannell |
| EPI_ISL_1269859, EPI_ISL_1269860 | Wisconsin State Laboratory of Hygiene Communicable Disease Division | Ginkgo Bioworks Clinical Laboratory | Malaika Mckenzie-Bennett, James McGann, Jim Griffin, Keith Robison, Alex Plocik, Becky Schilling, Martha Pierson, Rebecca Littlefield, Michelle Spencer, Amanda Riccardi, Birgitte Simen, Kelsey R. Florek, Abigail C. Shockey |
| EPI_ISL_1269861 | Helix/Illumina | Centers for Disease Control and Prevention Division of Viral Diseases, Pathogen Discovery | Peter W. Cook, Dakota Howard, Dhwani Batra, Ben L. Rambo-Martin, Eileen de Feo, Jan Antico, Christine Tran, Matthew Tolentino, Shannon Wickline, Kim Gietzen, Brad Sickler, Jingtao Liu, Eric Allen, Phil Febbo, Summer Galloway, Nicole L. Washington, Simon White, Geraint Levan, Kelly Schiabor Barrett, Elizabeth Cirulli, Alexandre Bolze, Ary Ascencio, Charlotte Rivera-Garcia, Ryan Cho, Jason Nguyen, Sherry Wang, Jimmy Ramirez, Tyler Cassens, Efren Sandoval, Magnus Isaksson, William Lee, David Becker, Marc Laurent, James Lu, Clinton R. Paden, Suxiang Tong, Duncan MacCannell |

| | | | |
|---|---|---|---|
| EPI_ISL_1269862, EPI_ISL_1269863 | Wisconsin State Laboratory of Hygiene Communicable Disease Division | Ginkgo Bioworks Clinical Laboratory | Malaika Mckenzie-Bennett, James McGann, Jim Griffin, Keith Robison, Alex Plocik, Becky Schilling, Martha Pierson, Rebecca Littlefield, Michelle Spencer, Amanda Riccardi, Birgitte Simen, Kelsey R. Florek, Abigail C. Shockey |
| EPI_ISL_1269864 | Helix/Illumina | Centers for Disease Control and Prevention Division of Viral Diseases, Pathogen Discovery | Peter W. Cook, Dakota Howard, Dhwani Batra, Ben L. Rambo-Martin, Eileen de Feo, Jan Antico, Christine Tran, Matthew Tolentino, Shannon Wickline, Kim Gietzen, Brad Sickler, Jingtao Liu, Eric Allen, Phil Febbo, Summer Galloway, Nicole L. Washington, Simon White, Geraint Levan, Kelly Schiabor Barrett, Elizabeth Cirulli, Alexandre Bolze, Ary Ascencio, Charlotte Rivera-Garcia, Ryan Cho, Jason Nguyen, Sherry Wang, Jimmy Ramirez, Tyler Cassens, Efren Sandoval, Magnus Isaksson, William Lee, David Becker, Marc Laurent, James Lu, Clinton R. Paden, Suxiang Tong, Duncan MacCannell |
| EPI_ISL_1269865, EPI_ISL_1269866 | Wisconsin State Laboratory of Hygiene Communicable Disease Division | Ginkgo Bioworks Clinical Laboratory | Malaika Mckenzie-Bennett, James McGann, Jim Griffin, Keith Robison, Alex Plocik, Becky Schilling, Martha Pierson, Rebecca Littlefield, Michelle Spencer, Amanda Riccardi, Birgitte Simen, Kelsey R. Florek, Abigail C. Shockey |
| EPI_ISL_1269867 | Helix/Illumina | Centers for Disease Control and Prevention Division of Viral Diseases, Pathogen Discovery | Peter W. Cook, Dakota Howard, Dhwani Batra, Ben L. Rambo-Martin, Eileen de Feo, Jan Antico, Christine Tran, Matthew Tolentino, Shannon Wickline, Kim Gietzen, Brad Sickler, Jingtao Liu, Eric Allen, Phil Febbo, Summer Galloway, Nicole L. Washington, Simon White, Geraint Levan, Kelly Schiabor Barrett, Elizabeth Cirulli, Alexandre Bolze, Ary Ascencio, Charlotte Rivera-Garcia, Ryan Cho, Jason Nguyen, Sherry Wang, Jimmy Ramirez, Tyler Cassens, Efren Sandoval, Magnus Isaksson, William Lee, David Becker, Marc Laurent, James Lu, Clinton R. Paden, Suxiang Tong, Duncan MacCannell |
| EPI_ISL_1269868, EPI_ISL_1269869 | Wisconsin State Laboratory of Hygiene Communicable Disease Division | Ginkgo Bioworks Clinical Laboratory | Malaika Mckenzie-Bennett, James McGann, Jim Griffin, Keith Robison, Alex Plocik, Becky Schilling, Martha Pierson, Rebecca Littlefield, Michelle Spencer, Amanda Riccardi, Birgitte Simen, Kelsey R. Florek, Abigail C. Shockey |
| EPI_ISL_1269870 | Helix/Illumina | Centers for Disease Control and Prevention Division of Viral Diseases, Pathogen Discovery | Peter W. Cook, Dakota Howard, Dhwani Batra, Ben L. Rambo-Martin, Eileen de Feo, Jan Antico, Christine Tran, Matthew Tolentino, Shannon Wickline, Kim Gietzen, Brad Sickler, Jingtao Liu, Eric Allen, Phil Febbo, Summer Galloway, Nicole L. Washington, Simon White, Geraint Levan, Kelly Schiabor Barrett, Elizabeth Cirulli, Alexandre Bolze, Ary Ascencio, Charlotte Rivera-Garcia, Ryan Cho, Jason Nguyen, Sherry Wang, Jimmy Ramirez, Tyler Cassens, Efren Sandoval, Magnus Isaksson, William Lee, David Becker, Marc Laurent, James Lu, Clinton R. Paden, Suxiang Tong, Duncan MacCannell |
| EPI_ISL_1269871, EPI_ISL_1269872 | Wisconsin State Laboratory of Hygiene Communicable Disease Division | Ginkgo Bioworks Clinical Laboratory | Malaika Mckenzie-Bennett, James McGann, Jim Griffin, Keith Robison, Alex Plocik, Becky Schilling, Martha Pierson, Rebecca Littlefield, Michelle Spencer, Amanda Riccardi, Birgitte Simen, Kelsey R. Florek, Abigail C. Shockey |
| EPI_ISL_1269873 | Helix/Illumina | Centers for Disease Control and Prevention Division of Viral Diseases, Pathogen Discovery | Peter W. Cook, Dakota Howard, Dhwani Batra, Ben L. Rambo-Martin, Eileen de Feo, Jan Antico, Christine Tran, Matthew Tolentino, Shannon Wickline, Kim Gietzen, Brad Sickler, Jingtao Liu, Eric Allen, Phil Febbo, Summer Galloway, Nicole L. Washington, Simon White, Geraint Levan, Kelly Schiabor Barrett, Elizabeth Cirulli, Alexandre Bolze, Ary Ascencio, Charlotte Rivera-Garcia, Ryan Cho, Jason Nguyen, Sherry Wang, Jimmy Ramirez, Tyler Cassens, Efren Sandoval, Magnus Isaksson, William Lee, David Becker, Marc Laurent, James Lu, Clinton R. Paden, Suxiang Tong, Duncan MacCannell |
| EPI_ISL_1269874, EPI_ISL_1269875 | Wisconsin State Laboratory of Hygiene Communicable Disease Division | Ginkgo Bioworks Clinical Laboratory | Malaika Mckenzie-Bennett, James McGann, Jim Griffin, Keith Robison, Alex Plocik, Becky Schilling, Martha Pierson, Rebecca Littlefield, Michelle Spencer, Amanda Riccardi, Birgitte Simen, Kelsey R. Florek, Abigail C. Shockey |
| EPI_ISL_1269876 | Helix/Illumina | Centers for Disease Control and Prevention Division of Viral Diseases, Pathogen Discovery | Peter W. Cook, Dakota Howard, Dhwani Batra, Ben L. Rambo-Martin, Eileen de Feo, Jan Antico, Christine Tran, Matthew Tolentino, Shannon Wickline, Kim Gietzen, Brad Sickler, Jingtao Liu, Eric Allen, Phil Febbo, Summer Galloway, Nicole L. Washington, Simon White, Geraint Levan, Kelly Schiabor Barrett, Elizabeth Cirulli, Alexandre Bolze, Ary Ascencio, Charlotte Rivera-Garcia, Ryan Cho, Jason Nguyen, Sherry Wang, Jimmy Ramirez, Tyler Cassens, Efren Sandoval, Magnus Isaksson, William Lee, David Becker, Marc Laurent, James Lu, Clinton R. Paden, Suxiang Tong, Duncan MacCannell |
| EPI_ISL_1269877, EPI_ISL_1269878 | Wisconsin State Laboratory of Hygiene Communicable Disease Division | Ginkgo Bioworks Clinical Laboratory | Malaika Mckenzie-Bennett, James McGann, Jim Griffin, Keith Robison, Alex Plocik, Becky Schilling, Martha Pierson, Rebecca Littlefield, Michelle Spencer, Amanda Riccardi, Birgitte Simen, Kelsey R. Florek, Abigail C. Shockey |
| EPI_ISL_1269879 | Helix/Illumina | Centers for Disease Control and Prevention Division of Viral Diseases, Pathogen Discovery | Peter W. Cook, Dakota Howard, Dhwani Batra, Ben L. Rambo-Martin, Eileen de Feo, Jan Antico, Christine Tran, Matthew Tolentino, Shannon Wickline, Kim Gietzen, Brad Sickler, Jingtao Liu, Eric Allen, Phil Febbo, Summer Galloway, Nicole L. Washington, Simon White, Geraint Levan, Kelly Schiabor Barrett, Elizabeth Cirulli, Alexandre Bolze, Ary Ascencio, Charlotte Rivera-Garcia, Ryan Cho, Jason Nguyen, Sherry Wang, Jimmy Ramirez, Tyler Cassens, Efren Sandoval, Magnus Isaksson, William Lee, David Becker, Marc Laurent, James Lu, Clinton R. Paden, Suxiang Tong, Duncan MacCannell |
| EPI_ISL_1269880, EPI_ISL_1269881 | Wisconsin State Laboratory of Hygiene Communicable Disease Division | Ginkgo Bioworks Clinical Laboratory | Malaika Mckenzie-Bennett, James McGann, Jim Griffin, Keith Robison, Alex Plocik, Becky Schilling, Martha Pierson, Rebecca Littlefield, Michelle Spencer, Amanda Riccardi, Birgitte Simen, Kelsey R. Florek, Abigail C. Shockey |
| EPI_ISL_1269882 | Helix/Illumina | Centers for Disease Control and Prevention Division of Viral Diseases, Pathogen Discovery | Peter W. Cook, Dakota Howard, Dhwani Batra, Ben L. Rambo-Martin, Eileen de Feo, Jan Antico, Christine Tran, Matthew Tolentino, Shannon Wickline, Kim Gietzen, Brad Sickler, Jingtao Liu, Eric Allen, Phil Febbo, Summer Galloway, Nicole L. Washington, Simon White, Geraint Levan, Kelly Schiabor Barrett, Elizabeth Cirulli, Alexandre Bolze, Ary Ascencio, Charlotte Rivera-Garcia, Ryan Cho, Jason Nguyen, Sherry Wang, Jimmy Ramirez, Tyler Cassens, Efren Sandoval, Magnus Isaksson, William Lee, David Becker, Marc Laurent, James Lu, Clinton R. Paden, Suxiang Tong, Duncan MacCannell |
| EPI_ISL_1269883, EPI_ISL_1269884 | Wisconsin State Laboratory of Hygiene Communicable Disease Division | Ginkgo Bioworks Clinical Laboratory | Malaika Mckenzie-Bennett, James McGann, Jim Griffin, Keith Robison, Alex Plocik, Becky Schilling, Martha Pierson, Rebecca Littlefield, Michelle Spencer, Amanda Riccardi, Birgitte Simen, Kelsey R. Florek, Abigail C. Shockey |
| EPI_ISL_1269885 | Helix/Illumina | Centers for Disease Control and Prevention Division of Viral Diseases, Pathogen Discovery | Peter W. Cook, Dakota Howard, Dhwani Batra, Ben L. Rambo-Martin, Eileen de Feo, Jan Antico, Christine Tran, Matthew Tolentino, Shannon Wickline, Kim Gietzen, Brad Sickler, Jingtao Liu, Eric Allen, Phil Febbo, Summer Galloway, Nicole L. Washington, Simon White, Geraint Levan, Kelly Schiabor Barrett, Elizabeth Cirulli, Alexandre Bolze, Ary Ascencio, Charlotte Rivera-Garcia, Ryan Cho, Jason Nguyen, Sherry Wang, Jimmy Ramirez, Tyler Cassens, Efren Sandoval, Magnus Isaksson, William Lee, David Becker, Marc Laurent, James Lu, Clinton R. Paden, Suxiang Tong, Duncan MacCannell |
| EPI_ISL_1269886, EPI_ISL_1269887, EPI_ISL_1269888 | Wisconsin State Laboratory of Hygiene Communicable Disease Division | Ginkgo Bioworks Clinical Laboratory | Malaika Mckenzie-Bennett, James McGann, Jim Griffin, Keith Robison, Alex Plocik, Becky Schilling, Martha Pierson, Rebecca Littlefield, Michelle Spencer, Amanda Riccardi, Birgitte Simen, Kelsey R. Florek, Abigail C. Shockey |
| EPI_ISL_1269889 | Helix/Illumina | Centers for Disease Control and Prevention Division of Viral Diseases, Pathogen Discovery | Peter W. Cook, Dakota Howard, Dhwani Batra, Ben L. Rambo-Martin, Eileen de Feo, Jan Antico, Christine Tran, Matthew Tolentino, Shannon Wickline, Kim Gietzen, Brad Sickler, Jingtao Liu, Eric Allen, Phil Febbo, Summer Galloway, Nicole L. Washington, Simon White, Geraint Levan, Kelly Schiabor Barrett, Elizabeth Cirulli, Alexandre Bolze, Ary Ascencio, Charlotte Rivera-Garcia, Ryan Cho, Jason Nguyen, Sherry Wang, Jimmy Ramirez, Tyler Cassens, Efren Sandoval, Magnus Isaksson, William Lee, David Becker, Marc Laurent, James Lu, Clinton R. Paden, Suxiang Tong, Duncan MacCannell |
| EPI_ISL_1269890, EPI_ISL_1269891 | Wisconsin State Laboratory of Hygiene Communicable Disease Division | Ginkgo Bioworks Clinical Laboratory | Malaika Mckenzie-Bennett, James McGann, Jim Griffin, Keith Robison, Alex Plocik, Becky Schilling, Martha Pierson, Rebecca Littlefield, Michelle Spencer, Amanda Riccardi, Birgitte Simen, Kelsey R. Florek, Abigail C. Shockey |
| EPI_ISL_1269892 | Helix/Illumina | Centers for Disease Control and Prevention Division of Viral Diseases, Pathogen Discovery | Peter W. Cook, Dakota Howard, Dhwani Batra, Ben L. Rambo-Martin, Eileen de Feo, Jan Antico, Christine Tran, Matthew Tolentino, Shannon Wickline, Kim Gietzen, Brad Sickler, Jingtao Liu, Eric Allen, Phil Febbo, Summer Galloway, Nicole L. Washington, Simon White, Geraint Levan, Kelly Schiabor Barrett, Elizabeth Cirulli, Alexandre Bolze, Ary Ascencio, Charlotte Rivera-Garcia, Ryan Cho, Jason Nguyen, Sherry Wang, Jimmy Ramirez, Tyler Cassens, Efren Sandoval, Magnus Isaksson, William Lee, David Becker, Marc Laurent, James Lu, Clinton R. Paden, Suxiang Tong, Duncan MacCannell |
| EPI_ISL_1269893 | Wisconsin State Laboratory of Hygiene Communicable Disease Division | Ginkgo Bioworks Clinical Laboratory | Malaika Mckenzie-Bennett, James McGann, Jim Griffin, Keith Robison, Alex Plocik, Becky Schilling, Martha Pierson, Rebecca Littlefield, Michelle Spencer, Amanda Riccardi, Birgitte Simen, Kelsey R. Florek, Abigail C. Shockey |
| EPI_ISL_1269894 | Helix/Illumina | Centers for Disease Control and Prevention Division of Viral Diseases, Pathogen Discovery | Peter W. Cook, Dakota Howard, Dhwani Batra, Ben L. Rambo-Martin, Eileen de Feo, Jan Antico, Christine Tran, Matthew Tolentino, Shannon Wickline, Kim Gietzen, Brad Sickler, Jingtao Liu, Eric Allen, Phil Febbo, Summer Galloway, Nicole L. Washington, Simon White, Geraint Levan, Kelly Schiabor Barrett, Elizabeth Cirulli, Alexandre Bolze, Ary Ascencio, Charlotte Rivera-Garcia, Ryan Cho, Jason Nguyen, Sherry Wang, Jimmy Ramirez, Tyler Cassens, Efren Sandoval, Magnus Isaksson, William Lee, David Becker, Marc Laurent, James Lu, Clinton R. Paden, Suxiang Tong, Duncan MacCannell |
| EPI_ISL_1269895, EPI_ISL_1269896 | Wisconsin State Laboratory of Hygiene Communicable Disease Division | Ginkgo Bioworks Clinical Laboratory | Malaika Mckenzie-Bennett, James McGann, Jim Griffin, Keith Robison, Alex Plocik, Becky Schilling, Martha Pierson, Rebecca Littlefield, Michelle Spencer, Amanda Riccardi, Birgitte Simen, Kelsey R. Florek, Abigail C. Shockey |
| EPI_ISL_1269897 | Helix/Illumina | Centers for Disease Control and Prevention Division of Viral Diseases, Pathogen Discovery | Peter W. Cook, Dakota Howard, Dhwani Batra, Ben L. Rambo-Martin, Eileen de Feo, Jan Antico, Christine Tran, Matthew Tolentino, Shannon Wickline, Kim Gietzen, Brad Sickler, Jingtao Liu, Eric Allen, Phil Febbo, Summer Galloway, Nicole L. Washington, Simon White, Geraint Levan, Kelly Schiabor Barrett, Elizabeth Cirulli, Alexandre Bolze, Ary Ascencio, Charlotte Rivera-Garcia, Ryan Cho, Jason Nguyen, Sherry Wang, Jimmy Ramirez, Tyler Cassens, Efren Sandoval, Magnus Isaksson, William Lee, David Becker, Marc Laurent, James Lu, Clinton R. Paden, Suxiang Tong, Duncan MacCannell |
| EPI_ISL_1269898, EPI_ISL_1269899 | Wisconsin State Laboratory of Hygiene Communicable Disease Division | Ginkgo Bioworks Clinical Laboratory | Malaika Mckenzie-Bennett, James McGann, Jim Griffin, Keith Robison, Alex Plocik, Becky Schilling, Martha Pierson, Rebecca Littlefield, Michelle Spencer, Amanda Riccardi, Birgitte Simen, Kelsey R. Florek, Abigail C. Shockey |
| EPI_ISL_1269900 | Helix/Illumina | Centers for Disease Control and Prevention Division of Viral Diseases, Pathogen Discovery | Peter W. Cook, Dakota Howard, Dhwani Batra, Ben L. Rambo-Martin, Eileen de Feo, Jan Antico, Christine Tran, Matthew Tolentino, Shannon Wickline, Kim Gietzen, Brad Sickler, Jingtao Liu, Eric Allen, Phil Febbo, Summer Galloway, Nicole L. Washington, Simon White, Geraint Levan, Kelly Schiabor Barrett, Elizabeth Cirulli, Alexandre Bolze, Ary Ascencio, Charlotte Rivera-Garcia, Ryan Cho, Jason Nguyen, Sherry Wang, Jimmy Ramirez, Tyler Cassens, Efren Sandoval, Magnus Isaksson, William Lee, David Becker, Marc Laurent, James Lu, Clinton R. Paden, Suxiang Tong, Duncan MacCannell |
| EPI_ISL_1269901, EPI_ISL_1269902 | Wisconsin State Laboratory of Hygiene Communicable Disease Division | Ginkgo Bioworks Clinical Laboratory | Malaika Mckenzie-Bennett, James McGann, Jim Griffin, Keith Robison, Alex Plocik, Becky Schilling, Martha Pierson, Rebecca Littlefield, Michelle Spencer, Amanda Riccardi, Birgitte Simen, Kelsey R. Florek, Abigail C. Shockey |
| EPI_ISL_1269903 | Helix/Illumina | Centers for Disease Control and Prevention Division of Viral Diseases, Pathogen Discovery | Peter W. Cook, Dakota Howard, Dhwani Batra, Ben L. Rambo-Martin, Eileen de Feo, Jan Antico, Christine Tran, Matthew Tolentino, Shannon Wickline, Kim Gietzen, Brad Sickler, Jingtao Liu, Eric Allen, Phil Febbo, Summer Galloway, Nicole L. Washington, Simon White, Geraint Levan, Kelly Schiabor Barrett, Elizabeth Cirulli, Alexandre Bolze, Ary Ascencio, Charlotte Rivera-Garcia, Ryan Cho, Jason Nguyen, Sherry Wang, Jimmy Ramirez, Tyler Cassens, Efren Sandoval, Magnus Isaksson, William Lee, David Becker, Marc Laurent, James Lu, Clinton R. Paden, Suxiang Tong, Duncan MacCannell |
| EPI_ISL_1269904, EPI_ISL_1269905, EPI_ISL_1269906 | Wisconsin State Laboratory of Hygiene Communicable Disease Division | Ginkgo Bioworks Clinical Laboratory | Malaika Mckenzie-Bennett, James McGann, Jim Griffin, Keith Robison, Alex Plocik, Becky Schilling, Martha Pierson, Rebecca Littlefield, Michelle Spencer, Amanda Riccardi, Birgitte Simen, Kelsey R. Florek, Abigail C. Shockey |
| EPI_ISL_1269907 | Helix/Illumina | Centers for Disease Control and Prevention Division of Viral Diseases, Pathogen Discovery | Peter W. Cook, Dakota Howard, Dhwani Batra, Ben L. Rambo-Martin, Eileen de Feo, Jan Antico, Christine Tran, Matthew Tolentino, Shannon Wickline, Kim Gietzen, Brad Sickler, Jingtao Liu, Eric Allen, Phil Febbo, Summer Galloway, Nicole L. Washington, Simon White, Geraint Levan, Kelly Schiabor Barrett, Elizabeth Cirulli, Alexandre Bolze, Ary Ascencio, Charlotte Rivera-Garcia, Ryan Cho, Jason Nguyen, Sherry Wang, Jimmy Ramirez, Tyler Cassens, Efren Sandoval, Magnus Isaksson, William Lee, David Becker, Marc Laurent, James Lu, Clinton R. Paden, Suxiang Tong, Duncan MacCannell |
| EPI_ISL_1269908, EPI_ISL_1269909 | Wisconsin State Laboratory of Hygiene Communicable Disease Division | Ginkgo Bioworks Clinical Laboratory | Malaika Mckenzie-Bennett, James McGann, Jim Griffin, Keith Robison, Alex Plocik, Becky Schilling, Martha Pierson, Rebecca Littlefield, Michelle Spencer, Amanda Riccardi, Birgitte Simen, Kelsey R. Florek, Abigail C. Shockey |
| EPI_ISL_1269910 | Helix/Illumina | Centers for Disease Control and Prevention Division of Viral Diseases, Pathogen Discovery | Peter W. Cook, Dakota Howard, Dhwani Batra, Ben L. Rambo-Martin, Eileen de Feo, Jan Antico, Christine Tran, Matthew Tolentino, Shannon Wickline, Kim Gietzen, Brad Sickler, Jingtao Liu, Eric Allen, Phil Febbo, Summer Galloway, Nicole L. Washington, Simon White, Geraint Levan, Kelly Schiabor Barrett, Elizabeth Cirulli, Alexandre Bolze, Ary Ascencio, Charlotte Rivera-Garcia, Ryan Cho, Jason Nguyen, Sherry Wang, Jimmy Ramirez, Tyler Cassens, Efren Sandoval, Magnus Isaksson, William Lee, David Becker, Marc Laurent, James Lu, Clinton R. Paden, Suxiang Tong, Duncan MacCannell |
| EPI_ISL_1269911, EPI_ISL_1269912 | Wisconsin State Laboratory of Hygiene Communicable Disease Division | Ginkgo Bioworks Clinical Laboratory | Malaika Mckenzie-Bennett, James McGann, Jim Griffin, Keith Robison, Alex Plocik, Becky Schilling, Martha Pierson, Rebecca Littlefield, Michelle Spencer, Amanda Riccardi, Birgitte Simen, Kelsey R. Florek, Abigail C. Shockey |
| EPI_ISL_1269913 | Helix/Illumina | Centers for Disease Control and Prevention Division of Viral Diseases, Pathogen Discovery | Peter W. Cook, Dakota Howard, Dhwani Batra, Ben L. Rambo-Martin, Eileen de Feo, Jan Antico, Christine Tran, Matthew Tolentino, Shannon Wickline, Kim Gietzen, Brad Sickler, Jingtao Liu, Eric Allen, Phil Febbo, Summer Galloway, Nicole L. Washington, Simon White, Geraint Levan, Kelly Schiabor Barrett, Elizabeth Cirulli, Alexandre Bolze, Ary Ascencio, Charlotte Rivera-Garcia, Ryan Cho, Jason Nguyen, Sherry Wang, Jimmy Ramirez, Tyler Cassens, Efren Sandoval, Magnus Isaksson, William Lee, David Becker, Marc Laurent, James Lu, Clinton R. Paden, Suxiang Tong, Duncan MacCannell |
| EPI_ISL_1269914, EPI_ISL_1269915 | Wisconsin State Laboratory of Hygiene Communicable Disease Division | Ginkgo Bioworks Clinical Laboratory | Malaika Mckenzie-Bennett, James McGann, Jim Griffin, Keith Robison, Alex Plocik, Becky Schilling, Martha Pierson, Rebecca Littlefield, Michelle Spencer, Amanda Riccardi, Birgitte Simen, Kelsey R. Florek, Abigail C. Shockey |
| EPI_ISL_1269916 | Helix/Illumina | Centers for Disease Control and Prevention Division of Viral Diseases, Pathogen Discovery | Peter W. Cook, Dakota Howard, Dhwani Batra, Ben L. Rambo-Martin, Eileen de Feo, Jan Antico, Christine Tran, Matthew Tolentino, Shannon Wickline, Kim Gietzen, Brad Sickler, Jingtao Liu, Eric Allen, Phil Febbo, Summer Galloway, Nicole L. Washington, Simon White, Geraint Levan, Kelly Schiabor Barrett, Elizabeth Cirulli, Alexandre Bolze, Ary Ascencio, Charlotte Rivera-Garcia, Ryan Cho, Jason Nguyen, Sherry Wang, Jimmy Ramirez, Tyler Cassens, Efren Sandoval, Magnus Isaksson, William Lee, David Becker, Marc Laurent, James Lu, Clinton R. Paden, Suxiang Tong, Duncan MacCannell |
| EPI_ISL_1269917, EPI_ISL_1269918 | Wisconsin State Laboratory of Hygiene Communicable Disease Division | Ginkgo Bioworks Clinical Laboratory | Malaika Mckenzie-Bennett, James McGann, Jim Griffin, Keith Robison, Alex Plocik, Becky Schilling, Martha Pierson, Rebecca Littlefield, Michelle Spencer, Amanda Riccardi, Birgitte Simen, Kelsey R. Florek, Abigail C. Shockey |

| | | | |
|---|---|---|---|
| EPI_ISL_1269919 | Helix/Illumina | Centers for Disease Control and Prevention Division of Viral Diseases, Pathogen Discovery | Peter W. Cook, Dakota Howard, Dhwani Batra, Ben L. Rambo-Martin, Eileen de Feo, Jan Antico, Christine Tran, Matthew Tolentino, Shannon Wickline, Kim Gietzen, Brad Sickler, Jingtao Liu, Eric Allen, Phil Febbo, Summer Galloway, Nicole L. Washington, Simon White, Geraint Levan, Kelly Schiabor Barrett, Elizabeth Cirulli, Alexandre Bolze, Ary Ascencio, Charlotte Rivera-Garcia, Ryan Cho, Jason Nguyen, Sherry Wang, Jimmy Ramirez, Tyler Cassens, Efren Sandoval, Magnus Isaksson, William Lee, David Becker, Marc Laurent, James Lu, Clinton R. Paden, Suxiang Tong, Duncan MacCannell |
| EPI_ISL_1269920, EPI_ISL_1269921 | Wisconsin State Laboratory of Hygiene Communicable Disease Division | Ginkgo Bioworks Clinical Laboratory | Malaika Mckenzie-Bennett, James McGann, Jim Griffin, Keith Robison, Alex Plocik, Becky Schilling, Martha Pierson, Rebecca Littlefield, Michelle Spencer, Amanda Riccardi, Birgitte Simen, Kelsey R. Florek, Abigail C. Shockey |
| EPI_ISL_1269922 | Helix/Illumina | Centers for Disease Control and Prevention Division of Viral Diseases, Pathogen Discovery | Peter W. Cook, Dakota Howard, Dhwani Batra, Ben L. Rambo-Martin, Eileen de Feo, Jan Antico, Christine Tran, Matthew Tolentino, Shannon Wickline, Kim Gietzen, Brad Sickler, Jingtao Liu, Eric Allen, Phil Febbo, Summer Galloway, Nicole L. Washington, Simon White, Geraint Levan, Kelly Schiabor Barrett, Elizabeth Cirulli, Alexandre Bolze, Ary Ascencio, Charlotte Rivera-Garcia, Ryan Cho, Jason Nguyen, Sherry Wang, Jimmy Ramirez, Tyler Cassens, Efren Sandoval, Magnus Isaksson, William Lee, David Becker, Marc Laurent, James Lu, Clinton R. Paden, Suxiang Tong, Duncan MacCannell |
| EPI_ISL_1269923, EPI_ISL_1269924 | Wisconsin State Laboratory of Hygiene Communicable Disease Division | Ginkgo Bioworks Clinical Laboratory | Malaika Mckenzie-Bennett, James McGann, Jim Griffin, Keith Robison, Alex Plocik, Becky Schilling, Martha Pierson, Rebecca Littlefield, Michelle Spencer, Amanda Riccardi, Birgitte Simen, Kelsey R. Florek, Abigail C. Shockey |
| EPI_ISL_1269925 | Helix/Illumina | Centers for Disease Control and Prevention Division of Viral Diseases, Pathogen Discovery | Peter W. Cook, Dakota Howard, Dhwani Batra, Ben L. Rambo-Martin, Eileen de Feo, Jan Antico, Christine Tran, Matthew Tolentino, Shannon Wickline, Kim Gietzen, Brad Sickler, Jingtao Liu, Eric Allen, Phil Febbo, Summer Galloway, Nicole L. Washington, Simon White, Geraint Levan, Kelly Schiabor Barrett, Elizabeth Cirulli, Alexandre Bolze, Ary Ascencio, Charlotte Rivera-Garcia, Ryan Cho, Jason Nguyen, Sherry Wang, Jimmy Ramirez, Tyler Cassens, Efren Sandoval, Magnus Isaksson, William Lee, David Becker, Marc Laurent, James Lu, Clinton R. Paden, Suxiang Tong, Duncan MacCannell |
| EPI_ISL_1269926, EPI_ISL_1269927 | Wisconsin State Laboratory of Hygiene Communicable Disease Division | Ginkgo Bioworks Clinical Laboratory | Malaika Mckenzie-Bennett, James McGann, Jim Griffin, Keith Robison, Alex Plocik, Becky Schilling, Martha Pierson, Rebecca Littlefield, Michelle Spencer, Amanda Riccardi, Birgitte Simen, Kelsey R. Florek, Abigail C. Shockey |
| EPI_ISL_1269928 | Helix/Illumina | Centers for Disease Control and Prevention Division of Viral Diseases, Pathogen Discovery | Peter W. Cook, Dakota Howard, Dhwani Batra, Ben L. Rambo-Martin, Eileen de Feo, Jan Antico, Christine Tran, Matthew Tolentino, Shannon Wickline, Kim Gietzen, Brad Sickler, Jingtao Liu, Eric Allen, Phil Febbo, Summer Galloway, Nicole L. Washington, Simon White, Geraint Levan, Kelly Schiabor Barrett, Elizabeth Cirulli, Alexandre Bolze, Ary Ascencio, Charlotte Rivera-Garcia, Ryan Cho, Jason Nguyen, Sherry Wang, Jimmy Ramirez, Tyler Cassens, Efren Sandoval, Magnus Isaksson, William Lee, David Becker, Marc Laurent, James Lu, Clinton R. Paden, Suxiang Tong, Duncan MacCannell |
| EPI_ISL_1269929, EPI_ISL_1269930 | Wisconsin State Laboratory of Hygiene Communicable Disease Division | Ginkgo Bioworks Clinical Laboratory | Malaika Mckenzie-Bennett, James McGann, Jim Griffin, Keith Robison, Alex Plocik, Becky Schilling, Martha Pierson, Rebecca Littlefield, Michelle Spencer, Amanda Riccardi, Birgitte Simen, Kelsey R. Florek, Abigail C. Shockey |
| EPI_ISL_1269931 | Helix/Illumina | Centers for Disease Control and Prevention Division of Viral Diseases, Pathogen Discovery | Peter W. Cook, Dakota Howard, Dhwani Batra, Ben L. Rambo-Martin, Eileen de Feo, Jan Antico, Christine Tran, Matthew Tolentino, Shannon Wickline, Kim Gietzen, Brad Sickler, Jingtao Liu, Eric Allen, Phil Febbo, Summer Galloway, Nicole L. Washington, Simon White, Geraint Levan, Kelly Schiabor Barrett, Elizabeth Cirulli, Alexandre Bolze, Ary Ascencio, Charlotte Rivera-Garcia, Ryan Cho, Jason Nguyen, Sherry Wang, Jimmy Ramirez, Tyler Cassens, Efren Sandoval, Magnus Isaksson, William Lee, David Becker, Marc Laurent, James Lu, Clinton R. Paden, Suxiang Tong, Duncan MacCannell |
| EPI_ISL_1269932, EPI_ISL_1269933 | Wisconsin State Laboratory of Hygiene Communicable Disease Division | Ginkgo Bioworks Clinical Laboratory | Malaika Mckenzie-Bennett, James McGann, Jim Griffin, Keith Robison, Alex Plocik, Becky Schilling, Martha Pierson, Rebecca Littlefield, Michelle Spencer, Amanda Riccardi, Birgitte Simen, Kelsey R. Florek, Abigail C. Shockey |
| EPI_ISL_1269934 | Helix/Illumina | Centers for Disease Control and Prevention Division of Viral Diseases, Pathogen Discovery | Peter W. Cook, Dakota Howard, Dhwani Batra, Ben L. Rambo-Martin, Eileen de Feo, Jan Antico, Christine Tran, Matthew Tolentino, Shannon Wickline, Kim Gietzen, Brad Sickler, Jingtao Liu, Eric Allen, Phil Febbo, Summer Galloway, Nicole L. Washington, Simon White, Geraint Levan, Kelly Schiabor Barrett, Elizabeth Cirulli, Alexandre Bolze, Ary Ascencio, Charlotte Rivera-Garcia, Ryan Cho, Jason Nguyen, Sherry Wang, Jimmy Ramirez, Tyler Cassens, Efren Sandoval, Magnus Isaksson, William Lee, David Becker, Marc Laurent, James Lu, Clinton R. Paden, Suxiang Tong, Duncan MacCannell |
| EPI_ISL_1269935 | Wisconsin State Laboratory of Hygiene Communicable Disease Division | Ginkgo Bioworks Clinical Laboratory | Malaika Mckenzie-Bennett, James McGann, Jim Griffin, Keith Robison, Alex Plocik, Becky Schilling, Martha Pierson, Rebecca Littlefield, Michelle Spencer, Amanda Riccardi, Birgitte Simen, Kelsey R. Florek, Abigail C. Shockey |
| EPI_ISL_1269936 | Helix/Illumina | Centers for Disease Control and Prevention Division of Viral Diseases, Pathogen Discovery | Peter W. Cook, Dakota Howard, Dhwani Batra, Ben L. Rambo-Martin, Eileen de Feo, Jan Antico, Christine Tran, Matthew Tolentino, Shannon Wickline, Kim Gietzen, Brad Sickler, Jingtao Liu, Eric Allen, Phil Febbo, Summer Galloway, Nicole L. Washington, Simon White, Geraint Levan, Kelly Schiabor Barrett, Elizabeth Cirulli, Alexandre Bolze, Ary Ascencio, Charlotte Rivera-Garcia, Ryan Cho, Jason Nguyen, Sherry Wang, Jimmy Ramirez, Tyler Cassens, Efren Sandoval, Magnus Isaksson, William Lee, David Becker, Marc Laurent, James Lu, Clinton R. Paden, Suxiang Tong, Duncan MacCannell |
| EPI_ISL_1269937, EPI_ISL_1269938, EPI_ISL_1269939 | Wisconsin State Laboratory of Hygiene Communicable Disease Division | Ginkgo Bioworks Clinical Laboratory | Malaika Mckenzie-Bennett, James McGann, Jim Griffin, Keith Robison, Alex Plocik, Becky Schilling, Martha Pierson, Rebecca Littlefield, Michelle Spencer, Amanda Riccardi, Birgitte Simen, Kelsey R. Florek, Abigail C. Shockey |
| EPI_ISL_1269940 | Helix/Illumina | Centers for Disease Control and Prevention Division of Viral Diseases, Pathogen Discovery | Peter W. Cook, Dakota Howard, Dhwani Batra, Ben L. Rambo-Martin, Eileen de Feo, Jan Antico, Christine Tran, Matthew Tolentino, Shannon Wickline, Kim Gietzen, Brad Sickler, Jingtao Liu, Eric Allen, Phil Febbo, Summer Galloway, Nicole L. Washington, Simon White, Geraint Levan, Kelly Schiabor Barrett, Elizabeth Cirulli, Alexandre Bolze, Ary Ascencio, Charlotte Rivera-Garcia, Ryan Cho, Jason Nguyen, Sherry Wang, Jimmy Ramirez, Tyler Cassens, Efren Sandoval, Magnus Isaksson, William Lee, David Becker, Marc Laurent, James Lu, Clinton R. Paden, Suxiang Tong, Duncan MacCannell |
| EPI_ISL_1269941, EPI_ISL_1269942 | Wisconsin State Laboratory of Hygiene Communicable Disease Division | Ginkgo Bioworks Clinical Laboratory | Malaika Mckenzie-Bennett, James McGann, Jim Griffin, Keith Robison, Alex Plocik, Becky Schilling, Martha Pierson, Rebecca Littlefield, Michelle Spencer, Amanda Riccardi, Birgitte Simen, Kelsey R. Florek, Abigail C. Shockey |
| EPI_ISL_1269943 | Helix/Illumina | Centers for Disease Control and Prevention Division of Viral Diseases, Pathogen Discovery | Peter W. Cook, Dakota Howard, Dhwani Batra, Ben L. Rambo-Martin, Eileen de Feo, Jan Antico, Christine Tran, Matthew Tolentino, Shannon Wickline, Kim Gietzen, Brad Sickler, Jingtao Liu, Eric Allen, Phil Febbo, Summer Galloway, Nicole L. Washington, Simon White, Geraint Levan, Kelly Schiabor Barrett, Elizabeth Cirulli, Alexandre Bolze, Ary Ascencio, Charlotte Rivera-Garcia, Ryan Cho, Jason Nguyen, Sherry Wang, Jimmy Ramirez, Tyler Cassens, Efren Sandoval, Magnus Isaksson, William Lee, David Becker, Marc Laurent, James Lu, Clinton R. Paden, Suxiang Tong, Duncan MacCannell |
| EPI_ISL_1269944, EPI_ISL_1269945 | Wisconsin State Laboratory of Hygiene Communicable Disease Division | Ginkgo Bioworks Clinical Laboratory | Malaika Mckenzie-Bennett, James McGann, Jim Griffin, Keith Robison, Alex Plocik, Becky Schilling, Martha Pierson, Rebecca Littlefield, Michelle Spencer, Amanda Riccardi, Birgitte Simen, Kelsey R. Florek, Abigail C. Shockey |
| EPI_ISL_1269946 | Helix/Illumina | Centers for Disease Control and Prevention Division of Viral Diseases, Pathogen Discovery | Peter W. Cook, Dakota Howard, Dhwani Batra, Ben L. Rambo-Martin, Eileen de Feo, Jan Antico, Christine Tran, Matthew Tolentino, Shannon Wickline, Kim Gietzen, Brad Sickler, Jingtao Liu, Eric Allen, Phil Febbo, Summer Galloway, Nicole L. Washington, Simon White, Geraint Levan, Kelly Schiabor Barrett, Elizabeth Cirulli, Alexandre Bolze, Ary Ascencio, Charlotte Rivera-Garcia, Ryan Cho, Jason Nguyen, Sherry Wang, Jimmy Ramirez, Tyler Cassens, Efren Sandoval, Magnus Isaksson, William Lee, David Becker, Marc Laurent, James Lu, Clinton R. Paden, Suxiang Tong, Duncan MacCannell |
| EPI_ISL_1269947 | Wisconsin State Laboratory of Hygiene Communicable Disease Division | Ginkgo Bioworks Clinical Laboratory | Malaika Mckenzie-Bennett, James McGann, Jim Griffin, Keith Robison, Alex Plocik, Becky Schilling, Martha Pierson, Rebecca Littlefield, Michelle Spencer, Amanda Riccardi, Birgitte Simen, Kelsey R. Florek, Abigail C. Shockey |
| EPI_ISL_1269948, EPI_ISL_1269949, EPI_ISL_1269950, EPI_ISL_1269951, EPI_ISL_1269952, EPI_ISL_1269953, EPI_ISL_1269954, EPI_ISL_1269955, EPI_ISL_1269956, EPI_ISL_1269957, EPI_ISL_1269958, EPI_ISL_1269959, EPI_ISL_1269960, EPI_ISL_1269961, EPI_ISL_1269962, EPI_ISL_1269963, EPI_ISL_1269964, EPI_ISL_1269965, EPI_ISL_1269966, EPI_ISL_1269967, EPI_ISL_1269968, EPI_ISL_1269969, EPI_ISL_1269970, EPI_ISL_1269971, EPI_ISL_1269972, EPI_ISL_1269973, EPI_ISL_1269974, EPI_ISL_1269975, EPI_ISL_1269976, EPI_ISL_1269977, EPI_ISL_1269978, EPI_ISL_1269979, EPI_ISL_1269980, EPI_ISL_1269981, EPI_ISL_1269982, EPI_ISL_1269983, EPI_ISL_1269984, EPI_ISL_1269985, EPI_ISL_1269986, EPI_ISL_1269987, EPI_ISL_1269988, EPI_ISL_1269989, EPI_ISL_1269990, EPI_ISL_1269991, EPI_ISL_1269992, EPI_ISL_1269993, EPI_ISL_1269994, EPI_ISL_1269995, EPI_ISL_1269996, EPI_ISL_1269998, EPI_ISL_1269999, EPI_ISL_1270000, EPI_ISL_1270001, EPI_ISL_1270002, EPI_ISL_1270003, EPI_ISL_1270004, EPI_ISL_1270005, EPI_ISL_1270006, EPI_ISL_1270007, EPI_ISL_1270008, EPI_ISL_1270009, EPI_ISL_1270010, EPI_ISL_1270011, EPI_ISL_1270012, EPI_ISL_1270013, EPI_ISL_1270014, EPI_ISL_1270015, EPI_ISL_1270016, EPI_ISL_1270017, EPI_ISL_1270018, EPI_ISL_1270019, EPI_ISL_1270020, EPI_ISL_1270021, EPI_ISL_1270022, EPI_ISL_1270023, EPI_ISL_1270024, EPI_ISL_1270025, EPI_ISL_1270026, EPI_ISL_1270027, EPI_ISL_1270028, EPI_ISL_1270029, EPI_ISL_1270030, EPI_ISL_1270031, EPI_ISL_1270032, EPI_ISL_1270033, EPI_ISL_1270034, EPI_ISL_1270035, EPI_ISL_1270036, EPI_ISL_1270037, EPI_ISL_1270038, EPI_ISL_1270039, EPI_ISL_1270040, EPI_ISL_1270041, EPI_ISL_1270042, EPI_ISL_1270043, EPI_ISL_1270044, EPI_ISL_1270045, EPI_ISL_1270046, EPI_ISL_1270047, EPI_ISL_1270048, EPI_ISL_1270049, EPI_ISL_1270050, EPI_ISL_1270051, EPI_ISL_1270052, EPI_ISL_1270053, EPI_ISL_1270054, EPI_ISL_1270055, EPI_ISL_1270056, EPI_ISL_1270057, EPI_ISL_1270058, EPI_ISL_1270059, EPI_ISL_1270060, EPI_ISL_1270061, EPI_ISL_1270062, EPI_ISL_1270063, EPI_ISL_1270064, EPI_ISL_1270065, EPI_ISL_1270066, EPI_ISL_1270067, EPI_ISL_1270068, EPI_ISL_1270069, EPI_ISL_1270070, EPI_ISL_1270071, EPI_ISL_1270072, EPI_ISL_1270073, EPI_ISL_1270074, EPI_ISL_1270075, EPI_ISL_1270076, EPI_ISL_1270077, EPI_ISL_1270078, EPI_ISL_1270079, EPI_ISL_1270080, EPI_ISL_1270081, EPI_ISL_1270082, EPI_ISL_1270083, EPI_ISL_1270084, EPI_ISL_1270085, EPI_ISL_1270086, EPI_ISL_1270087, EPI_ISL_1270088, EPI_ISL_1270089, EPI_ISL_1270090, EPI_ISL_1270091, EPI_ISL_1270092, EPI_ISL_1270093, EPI_ISL_1270094, EPI_ISL_1270095, EPI_ISL_1270096, EPI_ISL_1270097, EPI_ISL_1270098, EPI_ISL_1270099, EPI_ISL_1270100, EPI_ISL_1270101, EPI_ISL_1270102, EPI_ISL_1270103, EPI_ISL_1270104, EPI_ISL_1270105, EPI_ISL_1270106, EPI_ISL_1270107, EPI_ISL_1270108, EPI_ISL_1270109, EPI_ISL_1270110, EPI_ISL_1270112, EPI_ISL_1270113, EPI_ISL_1270114, EPI_ISL_1270115, EPI_ISL_1270116, EPI_ISL_1270117, EPI_ISL_1270118, EPI_ISL_1270119, EPI_ISL_1270120, EPI_ISL_1270121, EPI_ISL_1270122, EPI_ISL_1270123, EPI_ISL_1270124, EPI_ISL_1270125, EPI_ISL_1270126, EPI_ISL_1270127, EPI_ISL_1270128, EPI_ISL_1270129, EPI_ISL_1270130, EPI_ISL_1270131, EPI_ISL_1270132, EPI_ISL_1270133, EPI_ISL_1270134, EPI_ISL_1270135, EPI_ISL_1270136, EPI_ISL_1270137, EPI_ISL_1270138, EPI_ISL_1270139, EPI_ISL_1270140, EPI_ISL_1270141, EPI_ISL_1270142, EPI_ISL_1270143, EPI_ISL_1270144, EPI_ISL_1270145, EPI_ISL_1270146, EPI_ISL_1270147, EPI_ISL_1270148, EPI_ISL_1270149, EPI_ISL_1270150, EPI_ISL_1270151, EPI_ISL_1270152, EPI_ISL_1270153, EPI_ISL_1270154, EPI_ISL_1270155, EPI_ISL_1270156, EPI_ISL_1270157, EPI_ISL_1270158, EPI_ISL_1270159, EPI_ISL_1270160, EPI_ISL_1270161, EPI_ISL_1270162, EPI_ISL_1270163, EPI_ISL_1270165, EPI_ISL_1270166, EPI_ISL_1270167, EPI_ISL_1270168, EPI_ISL_1270169, EPI_ISL_1270170, EPI_ISL_1270171, EPI_ISL_1270172, EPI_ISL_1270173, EPI_ISL_1270174, EPI_ISL_1270175, EPI_ISL_1270176, EPI_ISL_1270177, EPI_ISL_1270178, EPI_ISL_1270179, EPI_ISL_1270180, EPI_ISL_1270181, EPI_ISL_1270182, EPI_ISL_1270183, EPI_ISL_1270184, EPI_ISL_1270186, EPI_ISL_1270187, EPI_ISL_1270188, EPI_ISL_1270189, EPI_ISL_1270190, EPI_ISL_1270192, EPI_ISL_1270193, EPI_ISL_1270194, EPI_ISL_1270195, EPI_ISL_1270197, EPI_ISL_1270198, EPI_ISL_1270199, EPI_ISL_1270200, EPI_ISL_1270201 | | | |
| see above | Helix/Illumina | Centers for Disease Control and Prevention Division of Viral Diseases, Pathogen Discovery | Peter W. Cook, Dakota Howard, Dhwani Batra, Ben L. Rambo-Martin, Eileen de Feo, Jan Antico, Christine Tran, Matthew Tolentino, Shannon Wickline, Kim Gietzen, Brad Sickler, Jingtao Liu, Eric Allen, Phil Febbo, Summer Galloway, Nicole L. Washington, Simon White, Geraint Levan, Kelly Schiabor Barrett, Elizabeth Cirulli, Alexandre Bolze, Ary Ascencio, Charlotte Rivera-Garcia, Ryan Cho, Jason Nguyen, Sherry Wang, Jimmy Ramirez, Tyler Cassens, Efren Sandoval, Magnus Isaksson, William Lee, David Becker, Marc Laurent, James Lu, Clinton R. Paden, Suxiang Tong, Duncan MacCannell |
| EPI_ISL_1270202 | Arizona State Public Health Laboratory | Arizona State Public Health Laboratory | Trung Huynh, Jessica Escobar, Katherine Fullerton, Nobuko Fukushima, Stacy White, Linda Getsinger, Victor Waddell |
| EPI_ISL_1270203 | Helix/Illumina | Centers for Disease Control and Prevention Division of Viral Diseases, Pathogen Discovery | Peter W. Cook, Dakota Howard, Dhwani Batra, Ben L. Rambo-Martin, Eileen de Feo, Jan Antico, Christine Tran, Matthew Tolentino, Shannon Wickline, Kim Gietzen, Brad Sickler, Jingtao Liu, Eric Allen, Phil Febbo, Summer Galloway, Nicole L. Washington, Simon White, Geraint Levan, Kelly Schiabor Barrett, Elizabeth Cirulli, Alexandre Bolze, Ary Ascencio, Charlotte Rivera-Garcia, Ryan Cho, Jason Nguyen, Sherry Wang, Jimmy Ramirez, Tyler Cassens, Efren Sandoval, Magnus Isaksson, William Lee, David Becker, Marc Laurent, James Lu, Clinton R. Paden, Suxiang Tong, Duncan MacCannell |
| EPI_ISL_1270204 | Arizona State Public Health Laboratory | Arizona State Public Health Laboratory | Trung Huynh, Jessica Escobar, Katherine Fullerton, Nobuko Fukushima, Stacy White, Linda Getsinger, Victor Waddell |
| EPI_ISL_1270205 | Helix/Illumina | Centers for Disease Control and Prevention Division of Viral Diseases, Pathogen Discovery | Peter W. Cook, Dakota Howard, Dhwani Batra, Ben L. Rambo-Martin, Eileen de Feo, Jan Antico, Christine Tran, Matthew Tolentino, Shannon Wickline, Kim Gietzen, Brad Sickler, Jingtao Liu, Eric Allen, Phil Febbo, Summer Galloway, Nicole L. Washington, Simon White, Geraint Levan, Kelly Schiabor Barrett, Elizabeth Cirulli, Alexandre Bolze, Ary Ascencio, Charlotte Rivera-Garcia, Ryan Cho, Jason Nguyen, Sherry Wang, Jimmy Ramirez, Tyler Cassens, Efren Sandoval, Magnus Isaksson, William Lee, David Becker, Marc Laurent, James Lu, Clinton R. Paden, Suxiang Tong, Duncan MacCannell |
| EPI_ISL_1270206 | Arizona State Public Health Laboratory | Arizona State Public Health Laboratory | Trung Huynh, Jessica Escobar, Katherine Fullerton, Nobuko Fukushima, Stacy White, Linda Getsinger, Victor Waddell |
| EPI_ISL_1270207 | Helix/Illumina | Centers for Disease Control and Prevention Division of Viral Diseases, Pathogen Discovery | Peter W. Cook, Dakota Howard, Dhwani Batra, Ben L. Rambo-Martin, Eileen de Feo, Jan Antico, Christine Tran, Matthew Tolentino, Shannon Wickline, Kim Gietzen, Brad Sickler, Jingtao Liu, Eric Allen, Phil Febbo, Summer Galloway, Nicole L. Washington, Simon White, Geraint Levan, Kelly Schiabor Barrett, Elizabeth Cirulli, Alexandre Bolze, Ary Ascencio, Charlotte Rivera-Garcia, Ryan Cho, Jason Nguyen, Sherry Wang, Jimmy Ramirez, Tyler Cassens, Efren Sandoval, Magnus Isaksson, William Lee, David Becker, Marc Laurent, James Lu, Clinton R. Paden, Suxiang Tong, Duncan MacCannell |
| EPI_ISL_1270208 | Arizona State Public Health Laboratory | Arizona State Public Health Laboratory | Trung Huynh, Jessica Escobar, Katherine Fullerton, Nobuko Fukushima, Stacy White, Linda Getsinger, Victor Waddell |
| EPI_ISL_1270209 | Helix/Illumina | Centers for Disease Control and Prevention Division of Viral Diseases, Pathogen Discovery | Peter W. Cook, Dakota Howard, Dhwani Batra, Ben L. Rambo-Martin, Eileen de Feo, Jan Antico, Christine Tran, Matthew Tolentino, Shannon Wickline, Kim Gietzen, Brad Sickler, Jingtao Liu, Eric Allen, Phil Febbo, Summer Galloway, Nicole L. Washington, Simon White, Geraint Levan, Kelly Schiabor Barrett, Elizabeth Cirulli, Alexandre Bolze, Ary Ascencio, Charlotte Rivera-Garcia, Ryan Cho, Jason Nguyen, Sherry Wang, Jimmy Ramirez, Tyler Cassens, Efren Sandoval, Magnus Isaksson, William Lee, David Becker, Marc Laurent, James Lu, Clinton R. Paden, Suxiang Tong, Duncan MacCannell |
| EPI_ISL_1270210 | Arizona State Public Health Laboratory | Arizona State Public Health Laboratory | Trung Huynh, Jessica Escobar, Katherine Fullerton, Nobuko Fukushima, Stacy White, Linda Getsinger, Victor Waddell |
| EPI_ISL_1270211 | Helix/Illumina | Centers for Disease Control and Prevention Division of Viral Diseases, Pathogen Discovery | Peter W. Cook, Dakota Howard, Dhwani Batra, Ben L. Rambo-Martin, Eileen de Feo, Jan Antico, Christine Tran, Matthew Tolentino, Shannon Wickline, Kim Gietzen, Brad Sickler, Jingtao Liu, Eric Allen, Phil Febbo, Summer Galloway, Nicole L. Washington, Simon White, Geraint Levan, Kelly Schiabor Barrett, Elizabeth Cirulli, Alexandre Bolze, Ary Ascencio, Charlotte Rivera-Garcia, Ryan Cho, Jason Nguyen, Sherry Wang, Jimmy Ramirez, Tyler Cassens, Efren Sandoval, Magnus Isaksson, William Lee, David Becker, Marc Laurent, James Lu, Clinton R. Paden, Suxiang Tong, Duncan MacCannell |
| EPI_ISL_1270212 | Arizona State Public Health Laboratory | Arizona State Public Health Laboratory | Trung Huynh, Jessica Escobar, Katherine Fullerton, Nobuko Fukushima, Stacy White, Linda Getsinger, Victor Waddell |
| EPI_ISL_1270213 | Helix/Illumina | Centers for Disease Control and Prevention Division of Viral Diseases, Pathogen Discovery | Peter W. Cook, Dakota Howard, Dhwani Batra, Ben L. Rambo-Martin, Eileen de Feo, Jan Antico, Christine Tran, Matthew Tolentino, Shannon Wickline, Kim Gietzen, Brad Sickler, Jingtao Liu, Eric Allen, Phil Febbo, Summer Galloway, Nicole L. Washington, Simon White, Geraint Levan, Kelly Schiabor Barrett, Elizabeth Cirulli, Alexandre Bolze, Ary Ascencio, Charlotte Rivera-Garcia, Ryan Cho, Jason Nguyen, Sherry Wang, Jimmy Ramirez, Tyler Cassens, Efren Sandoval, Magnus Isaksson, William Lee, David Becker, Marc Laurent, James Lu, Clinton R. Paden, Suxiang Tong, Duncan MacCannell |
| EPI_ISL_1270214 | Arizona State Public Health Laboratory | Arizona State Public Health Laboratory | Trung Huynh, Jessica Escobar, Katherine Fullerton, Nobuko Fukushima, Stacy White, Linda Getsinger, Victor Waddell |
| EPI_ISL_1270215 | Helix/Illumina | Centers for Disease Control and Prevention Division of Viral Diseases, Pathogen Discovery | Peter W. Cook, Dakota Howard, Dhwani Batra, Ben L. Rambo-Martin, Eileen de Feo, Jan Antico, Christine Tran, Matthew Tolentino, Shannon Wickline, Kim Gietzen, Brad Sickler, Jingtao Liu, Eric Allen, Phil Febbo, Summer Galloway, Nicole L. Washington, Simon White, Geraint Levan, Kelly Schiabor Barrett, Elizabeth Cirulli, Alexandre Bolze, Ary Ascencio, Charlotte Rivera-Garcia, Ryan Cho, Jason Nguyen, Sherry Wang, Jimmy Ramirez, Tyler Cassens, Efren Sandoval, Magnus Isaksson, William Lee, David Becker, Marc Laurent, James Lu, Clinton R. Paden, Suxiang Tong, Duncan MacCannell |

| | | | |
|---|---|---|---|
| EPI_ISL_1270216 | Arizona State Public Health Laboratory | Arizona State Public Health Laboratory | Trung Huynh, Jessica Escobar, Katherine Fullerton, Nobuko Fukushima, Stacy White, Linda Getsinger, Victor Waddell |
| EPI_ISL_1270217 | Helix/Illumina | Centers for Disease Control and Prevention Division of Viral Diseases, Pathogen Discovery | Peter W. Cook, Dakota Howard, Dhwani Batra, Ben L. Rambo-Martin, Eileen de Feo, Jan Antico, Christine Tran, Matthew Tolentino, Shannon Wickline, Kim Gietzen, Brad Sickler, Jingtao Liu, Eric Allen, Phil Febbo, Summer Galloway, Nicole L. Washington, Simon White, Geraint Levan, Kelly Schiabor Barrett, Elizabeth Cirulli, Alexandre Bolze, Ary Ascencio, Charlotte Rivera-Garcia, Ryan Cho, Jason Nguyen, Sherry Wang, Jimmy Ramirez, Tyler Cassens, Efren Sandoval, Magnus Isaksson, William Lee, David Becker, Marc Laurent, James Lu, Clinton R. Paden, Suxiang Tong, Duncan MacCannell |
| EPI_ISL_1270218 | Arizona State Public Health Laboratory | Arizona State Public Health Laboratory | Trung Huynh, Jessica Escobar, Katherine Fullerton, Nobuko Fukushima, Stacy White, Linda Getsinger, Victor Waddell |
| EPI_ISL_1270219 | Helix/Illumina | Centers for Disease Control and Prevention Division of Viral Diseases, Pathogen Discovery | Peter W. Cook, Dakota Howard, Dhwani Batra, Ben L. Rambo-Martin, Eileen de Feo, Jan Antico, Christine Tran, Matthew Tolentino, Shannon Wickline, Kim Gietzen, Brad Sickler, Jingtao Liu, Eric Allen, Phil Febbo, Summer Galloway, Nicole L. Washington, Simon White, Geraint Levan, Kelly Schiabor Barrett, Elizabeth Cirulli, Alexandre Bolze, Ary Ascencio, Charlotte Rivera-Garcia, Ryan Cho, Jason Nguyen, Sherry Wang, Jimmy Ramirez, Tyler Cassens, Efren Sandoval, Magnus Isaksson, William Lee, David Becker, Marc Laurent, James Lu, Clinton R. Paden, Suxiang Tong, Duncan MacCannell |
| EPI_ISL_1270220 | Arizona State Public Health Laboratory | Arizona State Public Health Laboratory | Trung Huynh, Jessica Escobar, Katherine Fullerton, Nobuko Fukushima, Stacy White, Linda Getsinger, Victor Waddell |
| EPI_ISL_1270221 | Helix/Illumina | Centers for Disease Control and Prevention Division of Viral Diseases, Pathogen Discovery | Peter W. Cook, Dakota Howard, Dhwani Batra, Ben L. Rambo-Martin, Eileen de Feo, Jan Antico, Christine Tran, Matthew Tolentino, Shannon Wickline, Kim Gietzen, Brad Sickler, Jingtao Liu, Eric Allen, Phil Febbo, Summer Galloway, Nicole L. Washington, Simon White, Geraint Levan, Kelly Schiabor Barrett, Elizabeth Cirulli, Alexandre Bolze, Ary Ascencio, Charlotte Rivera-Garcia, Ryan Cho, Jason Nguyen, Sherry Wang, Jimmy Ramirez, Tyler Cassens, Efren Sandoval, Magnus Isaksson, William Lee, David Becker, Marc Laurent, James Lu, Clinton R. Paden, Suxiang Tong, Duncan MacCannell |
| EPI_ISL_1270222 | Arizona State Public Health Laboratory | Arizona State Public Health Laboratory | Trung Huynh, Jessica Escobar, Katherine Fullerton, Nobuko Fukushima, Stacy White, Linda Getsinger, Victor Waddell |
| EPI_ISL_1270223 | Helix/Illumina | Centers for Disease Control and Prevention Division of Viral Diseases, Pathogen Discovery | Peter W. Cook, Dakota Howard, Dhwani Batra, Ben L. Rambo-Martin, Eileen de Feo, Jan Antico, Christine Tran, Matthew Tolentino, Shannon Wickline, Kim Gietzen, Brad Sickler, Jingtao Liu, Eric Allen, Phil Febbo, Summer Galloway, Nicole L. Washington, Simon White, Geraint Levan, Kelly Schiabor Barrett, Elizabeth Cirulli, Alexandre Bolze, Ary Ascencio, Charlotte Rivera-Garcia, Ryan Cho, Jason Nguyen, Sherry Wang, Jimmy Ramirez, Tyler Cassens, Efren Sandoval, Magnus Isaksson, William Lee, David Becker, Marc Laurent, James Lu, Clinton R. Paden, Suxiang Tong, Duncan MacCannell |
| EPI_ISL_1270224 | Arizona State Public Health Laboratory | Arizona State Public Health Laboratory | Trung Huynh, Jessica Escobar, Katherine Fullerton, Nobuko Fukushima, Stacy White, Linda Getsinger, Victor Waddell |
| EPI_ISL_1270225, EPI_ISL_1270226 | Helix/Illumina | Centers for Disease Control and Prevention Division of Viral Diseases, Pathogen Discovery | Peter W. Cook, Dakota Howard, Dhwani Batra, Ben L. Rambo-Martin, Eileen de Feo, Jan Antico, Christine Tran, Matthew Tolentino, Shannon Wickline, Kim Gietzen, Brad Sickler, Jingtao Liu, Eric Allen, Phil Febbo, Summer Galloway, Nicole L. Washington, Simon White, Geraint Levan, Kelly Schiabor Barrett, Elizabeth Cirulli, Alexandre Bolze, Ary Ascencio, Charlotte Rivera-Garcia, Ryan Cho, Jason Nguyen, Sherry Wang, Jimmy Ramirez, Tyler Cassens, Efren Sandoval, Magnus Isaksson, William Lee, David Becker, Marc Laurent, James Lu, Clinton R. Paden, Suxiang Tong, Duncan MacCannell |
| EPI_ISL_1270227 | Arizona State Public Health Laboratory | Arizona State Public Health Laboratory | Trung Huynh, Jessica Escobar, Katherine Fullerton, Nobuko Fukushima, Stacy White, Linda Getsinger, Victor Waddell |
| EPI_ISL_1270228 | Helix/Illumina | Centers for Disease Control and Prevention Division of Viral Diseases, Pathogen Discovery | Peter W. Cook, Dakota Howard, Dhwani Batra, Ben L. Rambo-Martin, Eileen de Feo, Jan Antico, Christine Tran, Matthew Tolentino, Shannon Wickline, Kim Gietzen, Brad Sickler, Jingtao Liu, Eric Allen, Phil Febbo, Summer Galloway, Nicole L. Washington, Simon White, Geraint Levan, Kelly Schiabor Barrett, Elizabeth Cirulli, Alexandre Bolze, Ary Ascencio, Charlotte Rivera-Garcia, Ryan Cho, Jason Nguyen, Sherry Wang, Jimmy Ramirez, Tyler Cassens, Efren Sandoval, Magnus Isaksson, William Lee, David Becker, Marc Laurent, James Lu, Clinton R. Paden, Suxiang Tong, Duncan MacCannell |
| EPI_ISL_1270229 | Arizona State Public Health Laboratory | Arizona State Public Health Laboratory | Trung Huynh, Jessica Escobar, Katherine Fullerton, Nobuko Fukushima, Stacy White, Linda Getsinger, Victor Waddell |
| EPI_ISL_1270230 | Helix/Illumina | Centers for Disease Control and Prevention Division of Viral Diseases, Pathogen Discovery | Peter W. Cook, Dakota Howard, Dhwani Batra, Ben L. Rambo-Martin, Eileen de Feo, Jan Antico, Christine Tran, Matthew Tolentino, Shannon Wickline, Kim Gietzen, Brad Sickler, Jingtao Liu, Eric Allen, Phil Febbo, Summer Galloway, Nicole L. Washington, Simon White, Geraint Levan, Kelly Schiabor Barrett, Elizabeth Cirulli, Alexandre Bolze, Ary Ascencio, Charlotte Rivera-Garcia, Ryan Cho, Jason Nguyen, Sherry Wang, Jimmy Ramirez, Tyler Cassens, Efren Sandoval, Magnus Isaksson, William Lee, David Becker, Marc Laurent, James Lu, Clinton R. Paden, Suxiang Tong, Duncan MacCannell |
| EPI_ISL_1270231 | Arizona State Public Health Laboratory | Arizona State Public Health Laboratory | Trung Huynh, Jessica Escobar, Katherine Fullerton, Nobuko Fukushima, Stacy White, Linda Getsinger, Victor Waddell |
| EPI_ISL_1270232 | Helix/Illumina | Centers for Disease Control and Prevention Division of Viral Diseases, Pathogen Discovery | Peter W. Cook, Dakota Howard, Dhwani Batra, Ben L. Rambo-Martin, Eileen de Feo, Jan Antico, Christine Tran, Matthew Tolentino, Shannon Wickline, Kim Gietzen, Brad Sickler, Jingtao Liu, Eric Allen, Phil Febbo, Summer Galloway, Nicole L. Washington, Simon White, Geraint Levan, Kelly Schiabor Barrett, Elizabeth Cirulli, Alexandre Bolze, Ary Ascencio, Charlotte Rivera-Garcia, Ryan Cho, Jason Nguyen, Sherry Wang, Jimmy Ramirez, Tyler Cassens, Efren Sandoval, Magnus Isaksson, William Lee, David Becker, Marc Laurent, James Lu, Clinton R. Paden, Suxiang Tong, Duncan MacCannell |
| EPI_ISL_1270233 | Arizona State Public Health Laboratory | Arizona State Public Health Laboratory | Trung Huynh, Jessica Escobar, Katherine Fullerton, Nobuko Fukushima, Stacy White, Linda Getsinger, Victor Waddell |
| EPI_ISL_1270234 | Helix/Illumina | Centers for Disease Control and Prevention Division of Viral Diseases, Pathogen Discovery | Peter W. Cook, Dakota Howard, Dhwani Batra, Ben L. Rambo-Martin, Eileen de Feo, Jan Antico, Christine Tran, Matthew Tolentino, Shannon Wickline, Kim Gietzen, Brad Sickler, Jingtao Liu, Eric Allen, Phil Febbo, Summer Galloway, Nicole L. Washington, Simon White, Geraint Levan, Kelly Schiabor Barrett, Elizabeth Cirulli, Alexandre Bolze, Ary Ascencio, Charlotte Rivera-Garcia, Ryan Cho, Jason Nguyen, Sherry Wang, Jimmy Ramirez, Tyler Cassens, Efren Sandoval, Magnus Isaksson, William Lee, David Becker, Marc Laurent, James Lu, Clinton R. Paden, Suxiang Tong, Duncan MacCannell |
| EPI_ISL_1270235 | Arizona State Public Health Laboratory | Arizona State Public Health Laboratory | Trung Huynh, Jessica Escobar, Katherine Fullerton, Nobuko Fukushima, Stacy White, Linda Getsinger, Victor Waddell |
| EPI_ISL_1270236 | Helix/Illumina | Centers for Disease Control and Prevention Division of Viral Diseases, Pathogen Discovery | Peter W. Cook, Dakota Howard, Dhwani Batra, Ben L. Rambo-Martin, Eileen de Feo, Jan Antico, Christine Tran, Matthew Tolentino, Shannon Wickline, Kim Gietzen, Brad Sickler, Jingtao Liu, Eric Allen, Phil Febbo, Summer Galloway, Nicole L. Washington, Simon White, Geraint Levan, Kelly Schiabor Barrett, Elizabeth Cirulli, Alexandre Bolze, Ary Ascencio, Charlotte Rivera-Garcia, Ryan Cho, Jason Nguyen, Sherry Wang, Jimmy Ramirez, Tyler Cassens, Efren Sandoval, Magnus Isaksson, William Lee, David Becker, Marc Laurent, James Lu, Clinton R. Paden, Suxiang Tong, Duncan MacCannell |
| EPI_ISL_1270237 | Arizona State Public Health Laboratory | Arizona State Public Health Laboratory | Trung Huynh, Jessica Escobar, Katherine Fullerton, Nobuko Fukushima, Stacy White, Linda Getsinger, Victor Waddell |
| EPI_ISL_1270238 | Helix/Illumina | Centers for Disease Control and Prevention Division of Viral Diseases, Pathogen Discovery | Peter W. Cook, Dakota Howard, Dhwani Batra, Ben L. Rambo-Martin, Eileen de Feo, Jan Antico, Christine Tran, Matthew Tolentino, Shannon Wickline, Kim Gietzen, Brad Sickler, Jingtao Liu, Eric Allen, Phil Febbo, Summer Galloway, Nicole L. Washington, Simon White, Geraint Levan, Kelly Schiabor Barrett, Elizabeth Cirulli, Alexandre Bolze, Ary Ascencio, Charlotte Rivera-Garcia, Ryan Cho, Jason Nguyen, Sherry Wang, Jimmy Ramirez, Tyler Cassens, Efren Sandoval, Magnus Isaksson, William Lee, David Becker, Marc Laurent, James Lu, Clinton R. Paden, Suxiang Tong, Duncan MacCannell |
| EPI_ISL_1270239 | Arizona State Public Health Laboratory | Arizona State Public Health Laboratory | Trung Huynh, Jessica Escobar, Katherine Fullerton, Nobuko Fukushima, Stacy White, Linda Getsinger, Victor Waddell |
| EPI_ISL_1270240 | Helix/Illumina | Centers for Disease Control and Prevention Division of Viral Diseases, Pathogen Discovery | Peter W. Cook, Dakota Howard, Dhwani Batra, Ben L. Rambo-Martin, Eileen de Feo, Jan Antico, Christine Tran, Matthew Tolentino, Shannon Wickline, Kim Gietzen, Brad Sickler, Jingtao Liu, Eric Allen, Phil Febbo, Summer Galloway, Nicole L. Washington, Simon White, Geraint Levan, Kelly Schiabor Barrett, Elizabeth Cirulli, Alexandre Bolze, Ary Ascencio, Charlotte Rivera-Garcia, Ryan Cho, Jason Nguyen, Sherry Wang, Jimmy Ramirez, Tyler Cassens, Efren Sandoval, Magnus Isaksson, William Lee, David Becker, Marc Laurent, James Lu, Clinton R. Paden, Suxiang Tong, Duncan MacCannell |
| EPI_ISL_1270241 | Arizona State Public Health Laboratory | Arizona State Public Health Laboratory | Trung Huynh, Jessica Escobar, Katherine Fullerton, Nobuko Fukushima, Stacy White, Linda Getsinger, Victor Waddell |
| EPI_ISL_1270242 | Helix/Illumina | Centers for Disease Control and Prevention Division of Viral Diseases, Pathogen Discovery | Peter W. Cook, Dakota Howard, Dhwani Batra, Ben L. Rambo-Martin, Eileen de Feo, Jan Antico, Christine Tran, Matthew Tolentino, Shannon Wickline, Kim Gietzen, Brad Sickler, Jingtao Liu, Eric Allen, Phil Febbo, Summer Galloway, Nicole L. Washington, Simon White, Geraint Levan, Kelly Schiabor Barrett, Elizabeth Cirulli, Alexandre Bolze, Ary Ascencio, Charlotte Rivera-Garcia, Ryan Cho, Jason Nguyen, Sherry Wang, Jimmy Ramirez, Tyler Cassens, Efren Sandoval, Magnus Isaksson, William Lee, David Becker, Marc Laurent, James Lu, Clinton R. Paden, Suxiang Tong, Duncan MacCannell |
| EPI_ISL_1270243 | Arizona State Public Health Laboratory | Arizona State Public Health Laboratory | Trung Huynh, Jessica Escobar, Katherine Fullerton, Nobuko Fukushima, Stacy White, Linda Getsinger, Victor Waddell |
| EPI_ISL_1270244 | Helix/Illumina | Centers for Disease Control and Prevention Division of Viral Diseases, Pathogen Discovery | Peter W. Cook, Dakota Howard, Dhwani Batra, Ben L. Rambo-Martin, Eileen de Feo, Jan Antico, Christine Tran, Matthew Tolentino, Shannon Wickline, Kim Gietzen, Brad Sickler, Jingtao Liu, Eric Allen, Phil Febbo, Summer Galloway, Nicole L. Washington, Simon White, Geraint Levan, Kelly Schiabor Barrett, Elizabeth Cirulli, Alexandre Bolze, Ary Ascencio, Charlotte Rivera-Garcia, Ryan Cho, Jason Nguyen, Sherry Wang, Jimmy Ramirez, Tyler Cassens, Efren Sandoval, Magnus Isaksson, William Lee, David Becker, Marc Laurent, James Lu, Clinton R. Paden, Suxiang Tong, Duncan MacCannell |
| EPI_ISL_1270245 | Arizona State Public Health Laboratory | Arizona State Public Health Laboratory | Trung Huynh, Jessica Escobar, Katherine Fullerton, Nobuko Fukushima, Stacy White, Linda Getsinger, Victor Waddell |
| EPI_ISL_1270246 | Helix/Illumina | Centers for Disease Control and Prevention Division of Viral Diseases, Pathogen Discovery | Peter W. Cook, Dakota Howard, Dhwani Batra, Ben L. Rambo-Martin, Eileen de Feo, Jan Antico, Christine Tran, Matthew Tolentino, Shannon Wickline, Kim Gietzen, Brad Sickler, Jingtao Liu, Eric Allen, Phil Febbo, Summer Galloway, Nicole L. Washington, Simon White, Geraint Levan, Kelly Schiabor Barrett, Elizabeth Cirulli, Alexandre Bolze, Ary Ascencio, Charlotte Rivera-Garcia, Ryan Cho, Jason Nguyen, Sherry Wang, Jimmy Ramirez, Tyler Cassens, Efren Sandoval, Magnus Isaksson, William Lee, David Becker, Marc Laurent, James Lu, Clinton R. Paden, Suxiang Tong, Duncan MacCannell |
| EPI_ISL_1270247 | Arizona State Public Health Laboratory | Arizona State Public Health Laboratory | Trung Huynh, Jessica Escobar, Katherine Fullerton, Nobuko Fukushima, Stacy White, Linda Getsinger, Victor Waddell |
| EPI_ISL_1270248, EPI_ISL_1270249 | Helix/Illumina | Centers for Disease Control and Prevention Division of Viral Diseases, Pathogen Discovery | Peter W. Cook, Dakota Howard, Dhwani Batra, Ben L. Rambo-Martin, Eileen de Feo, Jan Antico, Christine Tran, Matthew Tolentino, Shannon Wickline, Kim Gietzen, Brad Sickler, Jingtao Liu, Eric Allen, Phil Febbo, Summer Galloway, Nicole L. Washington, Simon White, Geraint Levan, Kelly Schiabor Barrett, Elizabeth Cirulli, Alexandre Bolze, Ary Ascencio, Charlotte Rivera-Garcia, Ryan Cho, Jason Nguyen, Sherry Wang, Jimmy Ramirez, Tyler Cassens, Efren Sandoval, Magnus Isaksson, William Lee, David Becker, Marc Laurent, James Lu, Clinton R. Paden, Suxiang Tong, Duncan MacCannell |
| EPI_ISL_1270250 | Arizona State Public Health Laboratory | Arizona State Public Health Laboratory | Trung Huynh, Jessica Escobar, Katherine Fullerton, Nobuko Fukushima, Stacy White, Linda Getsinger, Victor Waddell |
| EPI_ISL_1270251, EPI_ISL_1270253 | Helix/Illumina | Centers for Disease Control and Prevention Division of Viral Diseases, Pathogen Discovery | Peter W. Cook, Dakota Howard, Dhwani Batra, Ben L. Rambo-Martin, Eileen de Feo, Jan Antico, Christine Tran, Matthew Tolentino, Shannon Wickline, Kim Gietzen, Brad Sickler, Jingtao Liu, Eric Allen, Phil Febbo, Summer Galloway, Nicole L. Washington, Simon White, Geraint Levan, Kelly Schiabor Barrett, Elizabeth Cirulli, Alexandre Bolze, Ary Ascencio, Charlotte Rivera-Garcia, Ryan Cho, Jason Nguyen, Sherry Wang, Jimmy Ramirez, Tyler Cassens, Efren Sandoval, Magnus Isaksson, William Lee, David Becker, Marc Laurent, James Lu, Clinton R. Paden, Suxiang Tong, Duncan MacCannell |
| EPI_ISL_1270254 | Arizona State Public Health Laboratory | Arizona State Public Health Laboratory | Trung Huynh, Jessica Escobar, Katherine Fullerton, Nobuko Fukushima, Stacy White, Linda Getsinger, Victor Waddell |

EPI_ISL_1270255, EPI_ISL_1270256, EPI_ISL_1270257, EPI_ISL_1270258, EPI_ISL_1270259, EPI_ISL_1270260, EPI_ISL_1270261, EPI_ISL_1270262, EPI_ISL_1270263, EPI_ISL_1270264, EPI_ISL_1270265, EPI_ISL_1270266, EPI_ISL_1270267, EPI_ISL_1270268, EPI_ISL_1270269, EPI_ISL_1270270, EPI_ISL_1270271, EPI_ISL_1270272, EPI_ISL_1270273, EPI_ISL_1270274, EPI_ISL_1270275, EPI_ISL_1270276, EPI_ISL_1270277, EPI_ISL_1270278, EPI_ISL_1270279, EPI_ISL_1270280, EPI_ISL_1270281, EPI_ISL_1270282, EPI_ISL_1270283, EPI_ISL_1270284, EPI_ISL_1270285, EPI_ISL_1270286, EPI_ISL_1270287, EPI_ISL_1270288, EPI_ISL_1270289, EPI_ISL_1270290, EPI_ISL_1270291, EPI_ISL_1270292, EPI_ISL_1270293, EPI_ISL_1270294, EPI_ISL_1270295, EPI_ISL_1270296, EPI_ISL_1270297, EPI_ISL_1270298, EPI_ISL_1270299, EPI_ISL_1270300, EPI_ISL_1270301, EPI_ISL_1270302, EPI_ISL_1270303, EPI_ISL_1270304, EPI_ISL_1270305, EPI_ISL_1270306, EPI_ISL_1270307, EPI_ISL_1270308, EPI_ISL_1270309, EPI_ISL_1270310, EPI_ISL_1270311, EPI_ISL_1270312, EPI_ISL_1270313, EPI_ISL_1270314, EPI_ISL_1270315, EPI_ISL_1270316, EPI_ISL_1270317, EPI_ISL_1270318, EPI_ISL_1270319, EPI_ISL_1270320, EPI_ISL_1270321, EPI_ISL_1270322, EPI_ISL_1270323, EPI_ISL_1270324, EPI_ISL_1270325, EPI_ISL_1270326, EPI_ISL_1270327, EPI_ISL_1270328, EPI_ISL_1270329, EPI_ISL_1270330, EPI_ISL_1270331, EPI_ISL_1270332, EPI_ISL_1270333, EPI_ISL_1270334, EPI_ISL_1270335, EPI_ISL_1270336, EPI_ISL_1270337, EPI_ISL_1270338, EPI_ISL_1270339, EPI_ISL_1270340, EPI_ISL_1270341, EPI_ISL_1270342, EPI_ISL_1270343, EPI_ISL_1270344, EPI_ISL_1270345, EPI_ISL_1270346, EPI_ISL_1270347, EPI_ISL_1270348, EPI_ISL_1270349, EPI_ISL_1270350, EPI_ISL_1270351, EPI_ISL_1270352, EPI_ISL_1270353, EPI_ISL_1270354, EPI_ISL_1270355, EPI_ISL_1270356, EPI_ISL_1270357, EPI_ISL_1270358, EPI_ISL_1270359, EPI_ISL_1270360, EPI_ISL_1270361, EPI_ISL_1270362, EPI_ISL_1270363, EPI_ISL_1270364, EPI_ISL_1270365, EPI_ISL_1270366, EPI_ISL_1270367, EPI_ISL_1270368, EPI_ISL_1270369, EPI_ISL_1270370, EPI_ISL_1270371, EPI_ISL_1270372, EPI_ISL_1270373, EPI_ISL_1270374, EPI_ISL_1270375, EPI_ISL_1270376, EPI_ISL_1270377, EPI_ISL_1270378, EPI_ISL_1270379, EPI_ISL_1270380, EPI_ISL_1270381, EPI_ISL_1270382, EPI_ISL_1270383, EPI_ISL_1270384, EPI_ISL_1270385, EPI_ISL_1270386, EPI_ISL_1270387, EPI_ISL_1270388, EPI_ISL_1270389, EPI_ISL_1270390, EPI_ISL_1270391, EPI_ISL_1270392, EPI_ISL_1270393, EPI_ISL_1270394, EPI_ISL_1270395, EPI_ISL_1270396, EPI_ISL_1270397, EPI_ISL_1270398, EPI_ISL_1270399, EPI_ISL_1270400, EPI_ISL_1270401, EPI_ISL_1270402, EPI_ISL_1270403, EPI_ISL_1270404, EPI_ISL_1270406, EPI_ISL_1270407, EPI_ISL_1270408, EPI_ISL_1270409, EPI_ISL_1270410, EPI_ISL_1270411, EPI_ISL_1270412, EPI_ISL_1270413, EPI_ISL_1270414, EPI_ISL_1270415, EPI_ISL_1270416, EPI_ISL_1270417, EPI_ISL_1270418, EPI_ISL_1270419, EPI_ISL_1270420, EPI_ISL_1270421, EPI_ISL_1270422, EPI_ISL_1270423, EPI_ISL_1270424, EPI_ISL_1270425, EPI_ISL_1270426, EPI_ISL_1270427, EPI_ISL_1270428, EPI_ISL_1270429, EPI_ISL_1270430, EPI_ISL_1270431, EPI_ISL_1270432, EPI_ISL_1270433, EPI_ISL_1270434, EPI_ISL_1270435, EPI_ISL_1270436, EPI_ISL_1270437, EPI_ISL_1270438, EPI_ISL_1270439, EPI_ISL_1270440, EPI_ISL_1270441, EPI_ISL_1270442, EPI_ISL_1270443, EPI_ISL_1270444, EPI_ISL_1270445, EPI_ISL_1270446, EPI_ISL_1270447, EPI_ISL_1270448, EPI_ISL_1270449, EPI_ISL_1270450, EPI_ISL_1270451, EPI_ISL_1270452, EPI_ISL_1270453, EPI_ISL_1270454, EPI_ISL_1270455, EPI_ISL_1270456, EPI_ISL_1270457, EPI_ISL_1270458, EPI_ISL_1270459, EPI_ISL_1270460, EPI_ISL_1270461, EPI_ISL_1270462, EPI_ISL_1270463, EPI_ISL_1270464, EPI_ISL_1270465, EPI_ISL_1270466, EPI_ISL_1270467, EPI_ISL_1270468, EPI_ISL_1270469, EPI_ISL_1270470, EPI_ISL_1270471, EPI_ISL_1270472, EPI_ISL_1270473, EPI_ISL_1270474, EPI_ISL_1270475, EPI_ISL_1270476, EPI_ISL_1270477, EPI_ISL_1270478, EPI_ISL_1270479, EPI_ISL_1270480, EPI_ISL_1270481, EPI_ISL_1270482, EPI_ISL_1270483, EPI_ISL_1270484, EPI_ISL_1270485, EPI_ISL_1270486, EPI_ISL_1270487, EPI_ISL_1270488, EPI_ISL_1270489, EPI_ISL_1270490, EPI_ISL_1270491, EPI_ISL_1270492, EPI_ISL_1270493, EPI_ISL_1270494, EPI_ISL_1270496, EPI_ISL_1270497, EPI_ISL_1270498, EPI_ISL_1270499, EPI_ISL_1270500, EPI_ISL_1270502, EPI_ISL_1270503, EPI_ISL_1270504, EPI_ISL_1270505, EPI_ISL_1270506, EPI_ISL_1270507, EPI_ISL_1270508, EPI_ISL_1270509, EPI_ISL_1270510, EPI_ISL_1270511, EPI_ISL_1270512, EPI_ISL_1270513, EPI_ISL_1270514, EPI_ISL_1270515, EPI_ISL_1270516, EPI_ISL_1270517, EPI_ISL_1270518, EPI_ISL_1270519, EPI_ISL_1270520, EPI_ISL_1270521, EPI_ISL_1270522, EPI_ISL_1270523, EPI_ISL_1270524, EPI_ISL_1270525, EPI_ISL_1270526, EPI_ISL_1270527, EPI_ISL_1270528, EPI_ISL_1270529, EPI_ISL_1270530, EPI_ISL_1270531, EPI_ISL_1270532, EPI_ISL_1270533, EPI_ISL_1270534, EPI_ISL_1270535, EPI_ISL_1270536, EPI_ISL_1270537, EPI_ISL_1270538, EPI_ISL_1270539, EPI_ISL_1270540, EPI_ISL_1270541, EPI_ISL_1270542, EPI_ISL_1270543, EPI_ISL_1270544, EPI_ISL_1270545, EPI_ISL_1270546, EPI_ISL_1270547, EPI_ISL_1270548, EPI_ISL_1270549, EPI_ISL_1270551, EPI_ISL_1270552, EPI_ISL_1270553, EPI_ISL_1270554, EPI_ISL_1270555, EPI_ISL_1270556, EPI_ISL_1270557, EPI_ISL_1270558, EPI_ISL_1270559, EPI_ISL_1270560, EPI_ISL_1270561, EPI_ISL_1270562, EPI_ISL_1270563, EPI_ISL_1270564, EPI_ISL_1270565, EPI_ISL_1270566, EPI_ISL_1270567, EPI_ISL_1270568, EPI_ISL_1270569, EPI_ISL_1270570, EPI_ISL_1270571, EPI_ISL_1270572, EPI_ISL_1270573, EPI_ISL_1270574, EPI_ISL_1270575, EPI_ISL_1270576, EPI_ISL_1270577, EPI_ISL_1270578, EPI_ISL_1270579, EPI_ISL_1270580, EPI_ISL_1270581, EPI_ISL_1270582, EPI_ISL_1270583, EPI_ISL_1270584, EPI_ISL_1270585, EPI_ISL_1270586, EPI_ISL_1270588, EPI_ISL_1270589, EPI_ISL_1270590, EPI_ISL_1270591, EPI_ISL_1270592, EPI_ISL_1270593, EPI_ISL_1270594, EPI_ISL_1270595, EPI_ISL_1270596, EPI_ISL_1270597, EPI_ISL_1270598, EPI_ISL_1270599, EPI_ISL_1270600, EPI_ISL_1270601, EPI_ISL_1270602, EPI_ISL_1270603, EPI_ISL_1270604, EPI_ISL_1270606, EPI_ISL_1270607, EPI_ISL_1270608, EPI_ISL_1270609, EPI_ISL_1270610, EPI_ISL_1270611, EPI_ISL_1270612, EPI_ISL_1270613, EPI_ISL_1270614, EPI_ISL_1270615, EPI_ISL_1270616, EPI_ISL_1270617, EPI_ISL_1270618, EPI_ISL_1270619, EPI_ISL_1270620, EPI_ISL_1270621, EPI_ISL_1270622, EPI_ISL_1270623, EPI_ISL_1270624, EPI_ISL_1270625, EPI_ISL_1270626, EPI_ISL_1270627, EPI_ISL_1270628, EPI_ISL_1270629, EPI_ISL_1270630, EPI_ISL_1270631, EPI_ISL_1270632, EPI_ISL_1270633, EPI_ISL_1270634, EPI_ISL_1270635, EPI_ISL_1270636, EPI_ISL_1270637, EPI_ISL_1270638, EPI_ISL_1270639, EPI_ISL_1270640, EPI_ISL_1270641, EPI_ISL_1270642, EPI_ISL_1270643, EPI_ISL_1270644, EPI_ISL_1270645, EPI_ISL_1270646, EPI_ISL_1270647, EPI_ISL_1270648, EPI_ISL_1270649, EPI_ISL_1270650, EPI_ISL_1270651, EPI_ISL_1270652, EPI_ISL_1270653, EPI_ISL_1270654, EPI_ISL_1270655, EPI_ISL_1270656, EPI_ISL_1270657, EPI_ISL_1270658, EPI_ISL_1270659, EPI_ISL_1270660, EPI_ISL_1270661, EPI_ISL_1270662, EPI_ISL_1270663, EPI_ISL_1270664, EPI_ISL_1270665, EPI_ISL_1270666, EPI_ISL_1270667, EPI_ISL_1270668, EPI_ISL_1270669, EPI_ISL_1270670, EPI_ISL_1270671, EPI_ISL_1270672, EPI_ISL_1270673, EPI_ISL_1270674, EPI_ISL_1270675, EPI_ISL_1270676, EPI_ISL_1270677, EPI_ISL_1270678, EPI_ISL_1270679, EPI_ISL_1270680, EPI_ISL_1270681, EPI_ISL_1270682, EPI_ISL_1270683, EPI_ISL_1270684, EPI_ISL_1270685, EPI_ISL_1270688, EPI_ISL_1270689, EPI_ISL_1270690, EPI_ISL_1270691, EPI_ISL_1270692, EPI_ISL_1270693, EPI_ISL_1270694, EPI_ISL_1270695, EPI_ISL_1270696, EPI_ISL_1270697, EPI_ISL_1270699, EPI_ISL_1270700, EPI_ISL_1270701, EPI_ISL_1270702, EPI_ISL_1270703, EPI_ISL_1270704, EPI_ISL_1270705, EPI_ISL_1270706, EPI_ISL_1270708, EPI_ISL_1270709, EPI_ISL_1270710, EPI_ISL_1270711, EPI_ISL_1270712, EPI_ISL_1270713, EPI_ISL_1270714, EPI_ISL_1270716, EPI_ISL_1270717, EPI_ISL_1270718, EPI_ISL_1270719, EPI_ISL_1270720, EPI_ISL_1270722, EPI_ISL_1270723, EPI_ISL_1270724, EPI_ISL_1270728, EPI_ISL_1270729, EPI_ISL_1270730, EPI_ISL_1270731, EPI_ISL_1270732, EPI_ISL_1270734, EPI_ISL_1270735, EPI_ISL_1270739, EPI_ISL_1270740, EPI_ISL_1270741, EPI_ISL_1270742, EPI_ISL_1270743, EPI_ISL_1270744, EPI_ISL_1270745, EPI_ISL_1270746, EPI_ISL_1270747, EPI_ISL_1270748, EPI_ISL_1270749, EPI_ISL_1270750, EPI_ISL_1270751, EPI_ISL_1270752, EPI_ISL_1270753, EPI_ISL_1270754, EPI_ISL_1270755, EPI_ISL_1270756, EPI_ISL_1270757, EPI_ISL_1270758, EPI_ISL_1270759, EPI_ISL_1270760, EPI_ISL_1270761, EPI_ISL_1270763, EPI_ISL_1270764, EPI_ISL_1270765, EPI_ISL_1270766, EPI_ISL_1270767, EPI_ISL_1270768, EPI_ISL_1270769, EPI_ISL_1270770, EPI_ISL_1270771, EPI_ISL_1270772, EPI_ISL_1270773, EPI_ISL_1270774, EPI_ISL_1270775, EPI_ISL_1270776, EPI_ISL_1270777, EPI_ISL_1270778, EPI_ISL_1270779, EPI_ISL_1270780, EPI_ISL_1270781, EPI_ISL_1270782, EPI_ISL_1270783, EPI_ISL_1270784, EPI_ISL_1270785, EPI_ISL_1270786

| | | | |
|---|---|---|---|
| see above | Helix/Illumina | Centers for Disease Control and Prevention Division of Viral Diseases, Pathogen Discovery | Peter W. Cook, Dakota Howard, Dhwani Batra, Ben L. Rambo-Martin, Eileen de Feo, Jan Antico, Christine Tran, Matthew Tolentino, Shannon Wickline, Kim Gietzen, Brad Sickler, Jingtao Liu, Eric Allen, Phil Febbo, Summer Galloway, Nicole L. Washington, Simon White, Geraint Levan, Kelly Schiabor Barrett, Elizabeth Cirulli, Alexandre Bolze, Ary Ascencio, Charlotte Rivera-Garcia, Ryan Cho, Jason Nguyen, Sherry Wang, Jimmy Ramirez, Tyler Cassens, Efren Sandoval, Magnus Isaksson, William Lee, David Becker, Marc Laurent, James Lu, Clinton R. Paden, Suxiang Tong, Duncan MacCannell |
| EPI_ISL_1270787, EPI_ISL_1270788, EPI_ISL_1270790, EPI_ISL_1270791, EPI_ISL_1270792, EPI_ISL_1270793, EPI_ISL_1270794, EPI_ISL_1270795, EPI_ISL_1270796 | Nevada State Public Health Laboratory | Nevada State Public Health Laboratory | Andrew Gorzalski, Mark Pandori |
| EPI_ISL_1270804, EPI_ISL_1270805, EPI_ISL_1270806, EPI_ISL_1270807, EPI_ISL_1270808, EPI_ISL_1270809, EPI_ISL_1270810, EPI_ISL_1270811, EPI_ISL_1270812, EPI_ISL_1270813, EPI_ISL_1270814, EPI_ISL_1270815, EPI_ISL_1270816, EPI_ISL_1270817, EPI_ISL_1270818, EPI_ISL_1270819, EPI_ISL_1270820, EPI_ISL_1270821, EPI_ISL_1271968, EPI_ISL_1271970, EPI_ISL_1271971, EPI_ISL_1271972, EPI_ISL_1271973, EPI_ISL_1271974, EPI_ISL_1271975, EPI_ISL_1271976, EPI_ISL_1271977, EPI_ISL_1271978, EPI_ISL_1271979, EPI_ISL_1271980, EPI_ISL_1271981, EPI_ISL_1271982, EPI_ISL_1271983, EPI_ISL_1271984, EPI_ISL_1271985, EPI_ISL_1271986, EPI_ISL_1272077, EPI_ISL_1272078, EPI_ISL_1272079, EPI_ISL_1272080, EPI_ISL_1272081, EPI_ISL_1272086, EPI_ISL_1272087, EPI_ISL_1272088, EPI_ISL_1272089, EPI_ISL_1272094 | | | |
| see above | Arizona State Public Health Laboratory | Arizona State Public Health Laboratory | Trung Huynh, Jessica Escobar, Katherine Fullerton, Nobuko Fukushima, Stacy White, Linda Getsinger, Victor Waddell |
| EPI_ISL_1272189, EPI_ISL_1272190, EPI_ISL_1272191, EPI_ISL_1272192, EPI_ISL_1272193, EPI_ISL_1272194, EPI_ISL_1272195, EPI_ISL_1272196, EPI_ISL_1272197, EPI_ISL_1272198, EPI_ISL_1272199, EPI_ISL_1272200, EPI_ISL_1272201, EPI_ISL_1272202, EPI_ISL_1272203, EPI_ISL_1272204, EPI_ISL_1272205, EPI_ISL_1272206, EPI_ISL_1272207, EPI_ISL_1272208, EPI_ISL_1272209, EPI_ISL_1272210, EPI_ISL_1272211, EPI_ISL_1272212, EPI_ISL_1272213, EPI_ISL_1272214, EPI_ISL_1272215, EPI_ISL_1272216, EPI_ISL_1272217, EPI_ISL_1272218, EPI_ISL_1272219, EPI_ISL_1272220, EPI_ISL_1272221, EPI_ISL_1272222, EPI_ISL_1272223, EPI_ISL_1272224, EPI_ISL_1272226, EPI_ISL_1272227, EPI_ISL_1272228, EPI_ISL_1272229, EPI_ISL_1272230, EPI_ISL_1272231, EPI_ISL_1272232, EPI_ISL_1272233, EPI_ISL_1272234, EPI_ISL_1272235 | | | |
| see above | MD PHL | MD PHL | Maryland Department of Health Laboratories Administration |
| EPI_ISL_1272276, EPI_ISL_1272277, EPI_ISL_1272278, EPI_ISL_1272279, EPI_ISL_1272280, EPI_ISL_1272281, EPI_ISL_1272282, EPI_ISL_1272283, EPI_ISL_1272284, EPI_ISL_1272285, EPI_ISL_1272286, EPI_ISL_1272287, EPI_ISL_1272288, EPI_ISL_1272289, EPI_ISL_1272290, EPI_ISL_1272291, EPI_ISL_1272292, EPI_ISL_1272293, EPI_ISL_1272294, EPI_ISL_1272295, EPI_ISL_1272296, EPI_ISL_1272297, EPI_ISL_1272298, EPI_ISL_1272299 | | | |
| see above | Arizona State Public Health Laboratory | Arizona State Public Health Laboratory | Trung Huynh, Jessica Escobar, Katherine Fullerton, Nobuko Fukushima, Stacy White, Linda Getsinger, Victor Waddell |
| EPI_ISL_1272594, EPI_ISL_1272595, EPI_ISL_1272596, EPI_ISL_1272597, EPI_ISL_1272598 | NJ Public Health and Environmental Laboratories | Centers for Disease Control and Prevention Division of Viral Diseases, Pathogen Discovery | Krista Queen, Yan Li, Ying Tao, Jing Zhang, Anna Uehara, Anna Montmayeur, Clinton R. Paden, Kristen Knipe, Matthew Schmerer, Shoshona Le, Katie Dillon, Peter W. Cook, Rachel Marine, Mili Sheth, Jasmine Padilla, Sarah Nobles, Mark Burroughs, Lori Rowe, Haibin Wang, Ben L. Rambo-Martin, Kristine Lacek, Sam Shepard, Dhwani Batra, Suxiang Tong, Justin Lee |
| EPI_ISL_1272599, EPI_ISL_1272600, EPI_ISL_1272601, EPI_ISL_1272602, EPI_ISL_1272603, EPI_ISL_1272604, EPI_ISL_1272605, EPI_ISL_1272606, EPI_ISL_1272607, EPI_ISL_1272608, EPI_ISL_1272609, EPI_ISL_1272610, EPI_ISL_1272611, EPI_ISL_1272612, EPI_ISL_1272613 | | | |
| see above | NM Dept. Health, Scientific Laboratory Division | Centers for Disease Control and Prevention Division of Viral Diseases, Pathogen Discovery | Krista Queen, Yan Li, Ying Tao, Jing Zhang, Anna Uehara, Anna Montmayeur, Clinton R. Paden, Kristen Knipe, Matthew Schmerer, Shoshona Le, Katie Dillon, Peter W. Cook, Rachel Marine, Mili Sheth, Jasmine Padilla, Sarah Nobles, Mark Burroughs, Lori Rowe, Haibin Wang, Ben L. Rambo-Martin, Kristine Lacek, Sam Shepard, Dhwani Batra, Suxiang Tong, Justin Lee |
| EPI_ISL_1272614, EPI_ISL_1272615, EPI_ISL_1272616, EPI_ISL_1272617, EPI_ISL_1272618, EPI_ISL_1272619, EPI_ISL_1272620, EPI_ISL_1272621, EPI_ISL_1272622, EPI_ISL_1272623, EPI_ISL_1272624, EPI_ISL_1272625, EPI_ISL_1272626, EPI_ISL_1272627, EPI_ISL_1272628, EPI_ISL_1272629, EPI_ISL_1272630, EPI_ISL_1272631, EPI_ISL_1272632, EPI_ISL_1272633, EPI_ISL_1272634, EPI_ISL_1272635, EPI_ISL_1272636, EPI_ISL_1272637 | | | |
| see above | MS Public Health Laboratory | Centers for Disease Control and Prevention Division of Viral Diseases, Pathogen Discovery | Krista Queen, Yan Li, Ying Tao, Jing Zhang, Anna Uehara, Anna Montmayeur, Clinton R. Paden, Kristen Knipe, Matthew Schmerer, Shoshona Le, Katie Dillon, Peter W. Cook, Rachel Marine, Mili Sheth, Jasmine Padilla, Sarah Nobles, Mark Burroughs, Lori Rowe, Haibin Wang, Ben L. Rambo-Martin, Kristine Lacek, Sam Shepard, Dhwani Batra, Suxiang Tong, Justin Lee |
| EPI_ISL_1272638, EPI_ISL_1272639, EPI_ISL_1272640, EPI_ISL_1272641, EPI_ISL_1272642, EPI_ISL_1272643, EPI_ISL_1272644, EPI_ISL_1272645, EPI_ISL_1272646, EPI_ISL_1272647, EPI_ISL_1272648, EPI_ISL_1272649, EPI_ISL_1272650, EPI_ISL_1272651, EPI_ISL_1272652 | | | |
| see above | HI Dept. of Health, State Laboratories Division | Centers for Disease Control and Prevention Division of Viral Diseases, Pathogen Discovery | Krista Queen, Yan Li, Ying Tao, Jing Zhang, Anna Uehara, Anna Montmayeur, Clinton R. Paden, Kristen Knipe, Matthew Schmerer, Shoshona Le, Katie Dillon, Peter W. Cook, Rachel Marine, Mili Sheth, Jasmine Padilla, Sarah Nobles, Mark Burroughs, Lori Rowe, Haibin Wang, Ben L. Rambo-Martin, Kristine Lacek, Sam Shepard, Dhwani Batra, Suxiang Tong, Justin Lee |
| EPI_ISL_1272653, EPI_ISL_1272654, EPI_ISL_1272655, EPI_ISL_1272656, EPI_ISL_1272657, EPI_ISL_1272658, EPI_ISL_1272659 | IN State Department of Health Laboratory Services | Centers for Disease Control and Prevention Division of Viral Diseases, Pathogen Discovery | Krista Queen, Yan Li, Ying Tao, Jing Zhang, Anna Uehara, Anna Montmayeur, Clinton R. Paden, Kristen Knipe, Matthew Schmerer, Shoshona Le, Katie Dillon, Peter W. Cook, Rachel Marine, Mili Sheth, Jasmine Padilla, Sarah Nobles, Mark Burroughs, Lori Rowe, Haibin Wang, Ben L. Rambo-Martin, Kristine Lacek, Sam Shepard, Dhwani Batra, Suxiang Tong, Justin Lee |
| EPI_ISL_1272660, EPI_ISL_1272661, EPI_ISL_1272662, EPI_ISL_1272663, EPI_ISL_1272664, EPI_ISL_1272665, EPI_ISL_1272666, EPI_ISL_1272667, EPI_ISL_1272668, EPI_ISL_1272669 | OR State PHL-Virology/Immunology Section | Centers for Disease Control and Prevention Division of Viral Diseases, Pathogen Discovery | Krista Queen, Yan Li, Ying Tao, Jing Zhang, Anna Uehara, Anna Montmayeur, Clinton R. Paden, Kristen Knipe, Matthew Schmerer, Shoshona Le, Katie Dillon, Peter W. Cook, Rachel Marine, Mili Sheth, Jasmine Padilla, Sarah Nobles, Mark Burroughs, Lori Rowe, Haibin Wang, Ben L. Rambo-Martin, Kristine Lacek, Sam Shepard, Dhwani Batra, Suxiang Tong, Justin Lee |
| EPI_ISL_1272670, EPI_ISL_1272671, EPI_ISL_1272672, EPI_ISL_1272673 | ME Health and Environmental Testing Laboratory | Centers for Disease Control and Prevention Division of Viral Diseases, Pathogen Discovery | Krista Queen, Yan Li, Ying Tao, Jing Zhang, Anna Uehara, Anna Montmayeur, Clinton R. Paden, Kristen Knipe, Matthew Schmerer, Shoshona Le, Katie Dillon, Peter W. Cook, Rachel Marine, Mili Sheth, Jasmine Padilla, Sarah Nobles, Mark Burroughs, Lori Rowe, Haibin Wang, Ben L. Rambo-Martin, Kristine Lacek, Sam Shepard, Dhwani Batra, Suxiang Tong, Justin Lee |
| EPI_ISL_1272674, EPI_ISL_1272675, EPI_ISL_1272676, EPI_ISL_1272677, EPI_ISL_1272678, EPI_ISL_1272679, EPI_ISL_1272680 | RI State Health Laboratories | Centers for Disease Control and Prevention Division of Viral Diseases, Pathogen Discovery | Krista Queen, Yan Li, Ying Tao, Jing Zhang, Anna Uehara, Anna Montmayeur, Clinton R. Paden, Kristen Knipe, Matthew Schmerer, Shoshona Le, Katie Dillon, Peter W. Cook, Rachel Marine, Mili Sheth, Jasmine Padilla, Sarah Nobles, Mark Burroughs, Lori Rowe, Haibin Wang, Ben L. Rambo-Martin, Kristine Lacek, Sam Shepard, Dhwani Batra, Suxiang Tong, Justin Lee |
| EPI_ISL_1272681, EPI_ISL_1272682, EPI_ISL_1272683, EPI_ISL_1272684, EPI_ISL_1272685, EPI_ISL_1272686, EPI_ISL_1272687, EPI_ISL_1272688 | AK State Public Health Lab, State Health Department | Centers for Disease Control and Prevention Division of Viral Diseases, Pathogen Discovery | Krista Queen, Yan Li, Ying Tao, Jing Zhang, Anna Uehara, Anna Montmayeur, Clinton R. Paden, Kristen Knipe, Matthew Schmerer, Shoshona Le, Katie Dillon, Peter W. Cook, Rachel Marine, Mili Sheth, Jasmine Padilla, Sarah Nobles, Mark Burroughs, Lori Rowe, Haibin Wang, Ben L. Rambo-Martin, Kristine Lacek, Sam Shepard, Dhwani Batra, Suxiang Tong, Justin Lee |
| EPI_ISL_1272689, EPI_ISL_1272690, EPI_ISL_1272691, EPI_ISL_1272692, EPI_ISL_1272693, EPI_ISL_1272694, EPI_ISL_1272695, EPI_ISL_1272696 | TX DSHS, Lab Services Section MC 1947 | Centers for Disease Control and Prevention Division of Viral Diseases, Pathogen Discovery | Krista Queen, Yan Li, Ying Tao, Jing Zhang, Anna Uehara, Anna Montmayeur, Clinton R. Paden, Kristen Knipe, Matthew Schmerer, Shoshona Le, Katie Dillon, Peter W. Cook, Rachel Marine, Mili Sheth, Jasmine Padilla, Sarah Nobles, Mark Burroughs, Lori Rowe, Haibin Wang, Ben L. Rambo-Martin, Kristine Lacek, Sam Shepard, Dhwani Batra, Suxiang Tong, Justin Lee |
| EPI_ISL_1272697, EPI_ISL_1272698, EPI_ISL_1272699, EPI_ISL_1272700, EPI_ISL_1272701, EPI_ISL_1272702, EPI_ISL_1272703, EPI_ISL_1272704, EPI_ISL_1272705, EPI_ISL_1272706, EPI_ISL_1272707, EPI_ISL_1272708, EPI_ISL_1272709, EPI_ISL_1272710, EPI_ISL_1272711, EPI_ISL_1272712 | | | |
| see above | IL Dept. of Public Health Springfield Laboratory | Centers for Disease Control and Prevention Division of Viral Diseases, Pathogen Discovery | Krista Queen, Yan Li, Ying Tao, Jing Zhang, Anna Uehara, Anna Montmayeur, Clinton R. Paden, Kristen Knipe, Matthew Schmerer, Shoshona Le, Katie Dillon, Peter W. Cook, Rachel Marine, Mili Sheth, Jasmine Padilla, Sarah Nobles, Mark Burroughs, Lori Rowe, Haibin Wang, Ben L. Rambo-Martin, Kristine Lacek, Sam Shepard, Dhwani Batra, Suxiang Tong, Justin Lee |
| EPI_ISL_1272713, EPI_ISL_1272714, EPI_ISL_1272715, EPI_ISL_1272716, EPI_ISL_1272717, EPI_ISL_1272718, EPI_ISL_1272719 | FL Bureau of Public Health Laboratories-Tampa | Centers for Disease Control and Prevention Division of Viral Diseases, Pathogen Discovery | Krista Queen, Yan Li, Ying Tao, Jing Zhang, Anna Uehara, Anna Montmayeur, Clinton R. Paden, Kristen Knipe, Matthew Schmerer, Shoshona Le, Katie Dillon, Peter W. Cook, Rachel Marine, Mili Sheth, Jasmine Padilla, Sarah Nobles, Mark Burroughs, Lori Rowe, Haibin Wang, Ben L. Rambo-Martin, Kristine Lacek, Sam Shepard, Dhwani Batra, Suxiang Tong, Justin Lee |
| EPI_ISL_1272720, EPI_ISL_1272721, EPI_ISL_1272722, | CT-Dr. Katherine A. Kelley State Public Health Lab | Centers for Disease Control and Prevention Division of Viral Diseases, Pathogen Discovery | Krista Queen, Yan Li, Ying Tao, Jing Zhang, Anna Uehara, Anna Montmayeur, Clinton R. Paden, Kristen Knipe, Matthew Schmerer, Shoshona Le, Katie Dillon, Peter W. Cook, Rachel Marine, Mili Sheth, Jasmine Padilla, Sarah Nobles, Mark Burroughs, Lori Rowe, Haibin Wang, Ben L. Rambo-Martin, Kristine Lacek, Sam Shepard, Dhwani Batra, Suxiang Tong, Justin Lee |

| | | | |
|---|---|---|---|
| EPI_ISL_1272723, EPI_ISL_1272724, EPI_ISL_1272725, EPI_ISL_1272726 | | | |
| EPI_ISL_1272727 | MI - Michigan Department of Health and Human Services - Bureau of Laboratories | Centers for Disease Control and Prevention Division of Viral Diseases, Pathogen Discovery | Krista Queen, Yan Li, Ying Tao, Jing Zhang, Anna Uehara, Anna Montmayeur, Clinton R. Paden, Kristen Knipe, Matthew Schmerer, Shoshona Le, Katie Dillon, Peter W. Cook, Rachel Marine, Mili Sheth, Jasmine Padilla, Sarah Nobles, Mark Burroughs, Lori Rowe, Haibin Wang, Ben L. Rambo-Martin, Kristine Lacek, Sam Shepard, Dhwani Batra, Suxiang Tong, Justin Lee |
| EPI_ISL_1272728, EPI_ISL_1272729, EPI_ISL_1272730, EPI_ISL_1272731, EPI_ISL_1272732 | VA-Division of Consolidated Laboratory Services | Centers for Disease Control and Prevention Division of Viral Diseases, Pathogen Discovery | Krista Queen, Yan Li, Ying Tao, Jing Zhang, Anna Uehara, Anna Montmayeur, Clinton R. Paden, Kristen Knipe, Matthew Schmerer, Shoshona Le, Katie Dillon, Peter W. Cook, Rachel Marine, Mili Sheth, Jasmine Padilla, Sarah Nobles, Mark Burroughs, Lori Rowe, Haibin Wang, Ben L. Rambo-Martin, Kristine Lacek, Sam Shepard, Dhwani Batra, Suxiang Tong, Justin Lee |
| EPI_ISL_1272733, EPI_ISL_1272734, EPI_ISL_1272735, EPI_ISL_1272736, EPI_ISL_1272737, EPI_ISL_1272738, EPI_ISL_1272739, EPI_ISL_1272740, EPI_ISL_1272741, EPI_ISL_1272742, EPI_ISL_1272743, EPI_ISL_1272744, EPI_ISL_1272745, EPI_ISL_1272746, EPI_ISL_1272747, EPI_ISL_1272748, EPI_ISL_1272749, EPI_ISL_1272750, EPI_ISL_1272751, EPI_ISL_1272752, EPI_ISL_1272753, EPI_ISL_1272754, EPI_ISL_1272755 | | | |
| see above | WA State Department of Health | Centers for Disease Control and Prevention Division of Viral Diseases, Pathogen Discovery | Krista Queen, Yan Li, Ying Tao, Jing Zhang, Anna Uehara, Anna Montmayeur, Clinton R. Paden, Kristen Knipe, Matthew Schmerer, Shoshona Le, Katie Dillon, Peter W. Cook, Rachel Marine, Mili Sheth, Jasmine Padilla, Sarah Nobles, Mark Burroughs, Lori Rowe, Haibin Wang, Ben L. Rambo-Martin, Kristine Lacek, Sam Shepard, Dhwani Batra, Suxiang Tong, Justin Lee |
| EPI_ISL_1272756, EPI_ISL_1272757, EPI_ISL_1272758, EPI_ISL_1272759, EPI_ISL_1272760, EPI_ISL_1272761, EPI_ISL_1272762, EPI_ISL_1272763, EPI_ISL_1272764, EPI_ISL_1272765, EPI_ISL_1272766, EPI_ISL_1272767, EPI_ISL_1272768, EPI_ISL_1272769, EPI_ISL_1272770, EPI_ISL_1272771, EPI_ISL_1272772, EPI_ISL_1272773, EPI_ISL_1272774 | | | |
| see above | SC Dept of Health and Env. Control-Bureau of Laboratories | Centers for Disease Control and Prevention Division of Viral Diseases, Pathogen Discovery | Krista Queen, Yan Li, Ying Tao, Jing Zhang, Anna Uehara, Anna Montmayeur, Clinton R. Paden, Kristen Knipe, Matthew Schmerer, Shoshona Le, Katie Dillon, Peter W. Cook, Rachel Marine, Mili Sheth, Jasmine Padilla, Sarah Nobles, Mark Burroughs, Lori Rowe, Haibin Wang, Ben L. Rambo-Martin, Kristine Lacek, Sam Shepard, Dhwani Batra, Suxiang Tong, Justin Lee |
| EPI_ISL_1272775, EPI_ISL_1272776, EPI_ISL_1272777, EPI_ISL_1272778, EPI_ISL_1272779, EPI_ISL_1272780, EPI_ISL_1272781 | TX-Houston Health Department Laboratory | Centers for Disease Control and Prevention Division of Viral Diseases, Pathogen Discovery | Krista Queen, Yan Li, Ying Tao, Jing Zhang, Anna Uehara, Anna Montmayeur, Clinton R. Paden, Kristen Knipe, Matthew Schmerer, Shoshona Le, Katie Dillon, Peter W. Cook, Rachel Marine, Mili Sheth, Jasmine Padilla, Sarah Nobles, Mark Burroughs, Lori Rowe, Haibin Wang, Ben L. Rambo-Martin, Kristine Lacek, Sam Shepard, Dhwani Batra, Suxiang Tong, Justin Lee |
| EPI_ISL_1272782, EPI_ISL_1272783, EPI_ISL_1272784, EPI_ISL_1272785, EPI_ISL_1272786, EPI_ISL_1272787, EPI_ISL_1272788, EPI_ISL_1272789, EPI_ISL_1272790, EPI_ISL_1272791, EPI_ISL_1272792, EPI_ISL_1272793, EPI_ISL_1272794, EPI_ISL_1272795, EPI_ISL_1272796, EPI_ISL_1272797 | | | |
| see above | LA Office of Public Health Laboratories | Centers for Disease Control and Prevention Division of Viral Diseases, Pathogen Discovery | Krista Queen, Yan Li, Ying Tao, Jing Zhang, Anna Uehara, Anna Montmayeur, Clinton R. Paden, Kristen Knipe, Matthew Schmerer, Shoshona Le, Katie Dillon, Peter W. Cook, Rachel Marine, Mili Sheth, Jasmine Padilla, Sarah Nobles, Mark Burroughs, Lori Rowe, Haibin Wang, Ben L. Rambo-Martin, Kristine Lacek, Sam Shepard, Dhwani Batra, Suxiang Tong, Justin Lee |
| EPI_ISL_1272798, EPI_ISL_1272799, EPI_ISL_1272800, EPI_ISL_1272801, EPI_ISL_1272802, EPI_ISL_1272803, EPI_ISL_1272804, EPI_ISL_1272805, EPI_ISL_1272806, EPI_ISL_1272807 | NV State Public Health Laboratory | Centers for Disease Control and Prevention Division of Viral Diseases, Pathogen Discovery | Krista Queen, Yan Li, Ying Tao, Jing Zhang, Anna Uehara, Anna Montmayeur, Clinton R. Paden, Kristen Knipe, Matthew Schmerer, Shoshona Le, Katie Dillon, Peter W. Cook, Rachel Marine, Mili Sheth, Jasmine Padilla, Sarah Nobles, Mark Burroughs, Lori Rowe, Haibin Wang, Ben L. Rambo-Martin, Kristine Lacek, Sam Shepard, Dhwani Batra, Suxiang Tong, Justin Lee |
| EPI_ISL_1272808, EPI_ISL_1272809, EPI_ISL_1272810, EPI_ISL_1272811, EPI_ISL_1272812, EPI_ISL_1272813 | OK Public Health Laboratory, Oklahoma State DOH | Centers for Disease Control and Prevention Division of Viral Diseases, Pathogen Discovery | Krista Queen, Yan Li, Ying Tao, Jing Zhang, Anna Uehara, Anna Montmayeur, Clinton R. Paden, Kristen Knipe, Matthew Schmerer, Shoshona Le, Katie Dillon, Peter W. Cook, Rachel Marine, Mili Sheth, Jasmine Padilla, Sarah Nobles, Mark Burroughs, Lori Rowe, Haibin Wang, Ben L. Rambo-Martin, Kristine Lacek, Sam Shepard, Dhwani Batra, Suxiang Tong, Justin Lee |
| EPI_ISL_1272814, EPI_ISL_1272815, EPI_ISL_1272816, EPI_ISL_1272817, EPI_ISL_1272818, EPI_ISL_1272819, EPI_ISL_1272820, EPI_ISL_1272821, EPI_ISL_1272822 | FL Bur. of Public Health Laboratories-Jacksonville | Centers for Disease Control and Prevention Division of Viral Diseases, Pathogen Discovery | Krista Queen, Yan Li, Ying Tao, Jing Zhang, Anna Uehara, Anna Montmayeur, Clinton R. Paden, Kristen Knipe, Matthew Schmerer, Shoshona Le, Katie Dillon, Peter W. Cook, Rachel Marine, Mili Sheth, Jasmine Padilla, Sarah Nobles, Mark Burroughs, Lori Rowe, Haibin Wang, Ben L. Rambo-Martin, Kristine Lacek, Sam Shepard, Dhwani Batra, Suxiang Tong, Justin Lee |
| EPI_ISL_1272823, EPI_ISL_1272824, EPI_ISL_1272825, EPI_ISL_1272826, EPI_ISL_1272827, EPI_ISL_1272828, EPI_ISL_1272829, EPI_ISL_1272830, EPI_ISL_1272831, EPI_ISL_1272832, EPI_ISL_1272833, EPI_ISL_1272834, EPI_ISL_1272835, EPI_ISL_1272836, EPI_ISL_1272837, EPI_ISL_1272838, EPI_ISL_1272839, EPI_ISL_1272840, EPI_ISL_1272841 | | | |
| see above | UT-Unified State Labs: Public Health Utah DOH | Centers for Disease Control and Prevention Division of Viral Diseases, Pathogen Discovery | Krista Queen, Yan Li, Ying Tao, Jing Zhang, Anna Uehara, Anna Montmayeur, Clinton R. Paden, Kristen Knipe, Matthew Schmerer, Shoshona Le, Katie Dillon, Peter W. Cook, Rachel Marine, Mili Sheth, Jasmine Padilla, Sarah Nobles, Mark Burroughs, Lori Rowe, Haibin Wang, Ben L. Rambo-Martin, Kristine Lacek, Sam Shepard, Dhwani Batra, Suxiang Tong, Justin Lee |
| EPI_ISL_1272842, EPI_ISL_1272843, EPI_ISL_1272844, EPI_ISL_1272845, EPI_ISL_1272846, EPI_ISL_1272847, EPI_ISL_1272848, EPI_ISL_1272849, EPI_ISL_1272850, EPI_ISL_1272851, EPI_ISL_1272852, EPI_ISL_1272853, EPI_ISL_1272854, EPI_ISL_1272855 | | | |
| see above | DC Public Health Lab/ Dept. of Forensic Sciences | Centers for Disease Control and Prevention Division of Viral Diseases, Pathogen Discovery | Krista Queen, Yan Li, Ying Tao, Jing Zhang, Anna Uehara, Anna Montmayeur, Clinton R. Paden, Kristen Knipe, Matthew Schmerer, Shoshona Le, Katie Dillon, Peter W. Cook, Rachel Marine, Mili Sheth, Jasmine Padilla, Sarah Nobles, Mark Burroughs, Lori Rowe, Haibin Wang, Ben L. Rambo-Martin, Kristine Lacek, Sam Shepard, Dhwani Batra, Suxiang Tong, Justin Lee |
| EPI_ISL_1272856, EPI_ISL_1272857, EPI_ISL_1272858, EPI_ISL_1272859, EPI_ISL_1272860, EPI_ISL_1272861, EPI_ISL_1272862, EPI_ISL_1272863, EPI_ISL_1272864, EPI_ISL_1272865, EPI_ISL_1272866, EPI_ISL_1272867, EPI_ISL_1272868 | | | |
| see above | AR Dept. of Health-PHL, Molecular Diagnostics | Centers for Disease Control and Prevention Division of Viral Diseases, Pathogen Discovery | Krista Queen, Yan Li, Ying Tao, Jing Zhang, Anna Uehara, Anna Montmayeur, Clinton R. Paden, Kristen Knipe, Matthew Schmerer, Shoshona Le, Katie Dillon, Peter W. Cook, Rachel Marine, Mili Sheth, Jasmine Padilla, Sarah Nobles, Mark Burroughs, Lori Rowe, Haibin Wang, Ben L. Rambo-Martin, Kristine Lacek, Sam Shepard, Dhwani Batra, Suxiang Tong, Justin Lee |
| EPI_ISL_1272869, EPI_ISL_1272870, EPI_ISL_1272871, EPI_ISL_1272872 | VT Dept. of Health Laboratory | Centers for Disease Control and Prevention Division of Viral Diseases, Pathogen Discovery | Krista Queen, Yan Li, Ying Tao, Jing Zhang, Anna Uehara, Anna Montmayeur, Clinton R. Paden, Kristen Knipe, Matthew Schmerer, Shoshona Le, Katie Dillon, Peter W. Cook, Rachel Marine, Mili Sheth, Jasmine Padilla, Sarah Nobles, Mark Burroughs, Lori Rowe, Haibin Wang, Ben L. Rambo-Martin, Kristine Lacek, Sam Shepard, Dhwani Batra, Suxiang Tong, Justin Lee |
| EPI_ISL_1272873, EPI_ISL_1272874, EPI_ISL_1272875, EPI_ISL_1272876, EPI_ISL_1272877, EPI_ISL_1272878, EPI_ISL_1272879, EPI_ISL_1272880, EPI_ISL_1272881, EPI_ISL_1272882, EPI_ISL_1272883, EPI_ISL_1272884, EPI_ISL_1272885, EPI_ISL_1272886, EPI_ISL_1272887, EPI_ISL_1272888, EPI_ISL_1272889, EPI_ISL_1272890, EPI_ISL_1272891, EPI_ISL_1272892, EPI_ISL_1272893, EPI_ISL_1272894, EPI_ISL_1272895 | | | |
| see above | CA-Los Angeles County Public Health Laboratory | Centers for Disease Control and Prevention Division of Viral Diseases, Pathogen Discovery | Krista Queen, Yan Li, Ying Tao, Jing Zhang, Anna Uehara, Anna Montmayeur, Clinton R. Paden, Kristen Knipe, Matthew Schmerer, Shoshona Le, Katie Dillon, Peter W. Cook, Rachel Marine, Mili Sheth, Jasmine Padilla, Sarah Nobles, Mark Burroughs, Lori Rowe, Haibin Wang, Ben L. Rambo-Martin, Kristine Lacek, Sam Shepard, Dhwani Batra, Suxiang Tong, Justin Lee |
| EPI_ISL_1272896, EPI_ISL_1272897, EPI_ISL_1272898, EPI_ISL_1272899, EPI_ISL_1272900, EPI_ISL_1272901, EPI_ISL_1272902, EPI_ISL_1272903, EPI_ISL_1272904, EPI_ISL_1272905, EPI_ISL_1272906, EPI_ISL_1272907, EPI_ISL_1272908, EPI_ISL_1272909, EPI_ISL_1272910, EPI_ISL_1272911, EPI_ISL_1272912, EPI_ISL_1272913, EPI_ISL_1272914, EPI_ISL_1272915 | | | |
| see above | MD DOH Laboratories Administration | Centers for Disease Control and Prevention Division of Viral Diseases, Pathogen Discovery | Krista Queen, Yan Li, Ying Tao, Jing Zhang, Anna Uehara, Anna Montmayeur, Clinton R. Paden, Kristen Knipe, Matthew Schmerer, Shoshona Le, Katie Dillon, Peter W. Cook, Rachel Marine, Mili Sheth, Jasmine Padilla, Sarah Nobles, Mark Burroughs, Lori Rowe, Haibin Wang, Ben L. Rambo-Martin, Kristine Lacek, Sam Shepard, Dhwani Batra, Suxiang Tong, Justin Lee |
| EPI_ISL_1272916, EPI_ISL_1272917, EPI_ISL_1272918, EPI_ISL_1272919 | PR Public Health Lab | Centers for Disease Control and Prevention Division of Viral Diseases, Pathogen Discovery | Krista Queen, Yan Li, Ying Tao, Jing Zhang, Anna Uehara, Anna Montmayeur, Clinton R. Paden, Kristen Knipe, Matthew Schmerer, Shoshona Le, Katie Dillon, Peter W. Cook, Rachel Marine, Mili Sheth, Jasmine Padilla, Sarah Nobles, Mark Burroughs, Lori Rowe, Haibin Wang, Ben L. Rambo-Martin, Kristine Lacek, Sam Shepard, Dhwani Batra, Suxiang Tong, Justin Lee |
| EPI_ISL_1272920, EPI_ISL_1272921 | WVDHHR - Office of Laboratory Services | Centers for Disease Control and Prevention Division of Viral Diseases, Pathogen Discovery | Krista Queen, Yan Li, Ying Tao, Jing Zhang, Anna Uehara, Anna Montmayeur, Clinton R. Paden, Kristen Knipe, Matthew Schmerer, Shoshona Le, Katie Dillon, Peter W. Cook, Rachel Marine, Mili Sheth, Jasmine Padilla, Sarah Nobles, Mark Burroughs, Lori Rowe, Haibin Wang, Ben L. Rambo-Martin, Kristine Lacek, Sam Shepard, Dhwani Batra, Suxiang Tong, Justin Lee |
| EPI_ISL_1272922, EPI_ISL_1272923, EPI_ISL_1272924, EPI_ISL_1272925, EPI_ISL_1272926, EPI_ISL_1272927 | NYSDOH Wadsworth Center, Virology Lab | Centers for Disease Control and Prevention Division of Viral Diseases, Pathogen Discovery | Krista Queen, Yan Li, Ying Tao, Jing Zhang, Anna Uehara, Anna Montmayeur, Clinton R. Paden, Kristen Knipe, Matthew Schmerer, Shoshona Le, Katie Dillon, Peter W. Cook, Rachel Marine, Mili Sheth, Jasmine Padilla, Sarah Nobles, Mark Burroughs, Lori Rowe, Haibin Wang, Ben L. Rambo-Martin, Kristine Lacek, Sam Shepard, Dhwani Batra, Suxiang Tong, Justin Lee |
| EPI_ISL_1272928, EPI_ISL_1272929, EPI_ISL_1272930, EPI_ISL_1272931, EPI_ISL_1272932, EPI_ISL_1272933, EPI_ISL_1272934, EPI_ISL_1272935, EPI_ISL_1272936, EPI_ISL_1272937, EPI_ISL_1272938, EPI_ISL_1272939, EPI_ISL_1272940, EPI_ISL_1272941 | | | |
| see above | IA State Hygienic Laboratory | Centers for Disease Control and Prevention Division of Viral Diseases, Pathogen | Krista Queen, Yan Li, Ying Tao, Jing Zhang, Anna Uehara, Anna Montmayeur, Clinton R. Paden, Kristen Knipe, Matthew Schmerer, Shoshona Le, Katie Dillon, Peter W. Cook, Rachel Marine, Mili Sheth, Jasmine Padilla, Sarah Nobles, Mark Burroughs, Lori Rowe, Haibin Wang, Ben L. Rambo-Martin, Kristine Lacek, Sam Shepard, Dhwani Batra, Suxiang Tong, Justin Lee |

| | | | |
|---|---|---|---|
| | | Discovery | |
| EPI_ISL_1272942, EPI_ISL_1272943, EPI_ISL_1272944, EPI_ISL_1272945, EPI_ISL_1272946, EPI_ISL_1272947, EPI_ISL_1272948, EPI_ISL_1272949, EPI_ISL_1272950, EPI_ISL_1272951, EPI_ISL_1272952, EPI_ISL_1272953, EPI_ISL_1272954, EPI_ISL_1272955, EPI_ISL_1272956, EPI_ISL_1272957, EPI_ISL_1272958, EPI_ISL_1272959, EPI_ISL_1272960, EPI_ISL_1272961, EPI_ISL_1272962, EPI_ISL_1272963, EPI_ISL_1272964, EPI_ISL_1272965, EPI_ISL_1272966, EPI_ISL_1272967, EPI_ISL_1272968, EPI_ISL_1272969, EPI_ISL_1272970 | | | |
| see above | WY Public Health Laboratory | Centers for Disease Control and Prevention Division of Viral Diseases, Pathogen Discovery | Krista Queen, Yan Li, Ying Tao, Jing Zhang, Anna Uehara, Anna Montmayeur, Clinton R. Paden, Kristen Knipe, Matthew Schmerer, Shoshona Le, Katie Dillon, Peter W. Cook, Rachel Marine, Mili Sheth, Jasmine Padilla, Sarah Nobles, Mark Burroughs, Lori Rowe, Haibin Wang, Ben L. Rambo-Martin, Kristine Lacek, Sam Shepard, Dhwani Batra, Suxiang Tong, Justin Lee |
| EPI_ISL_1272971 | FL Bureau of Public Health Laboratories-Miami | Centers for Disease Control and Prevention Division of Viral Diseases, Pathogen Discovery | Krista Queen, Yan Li, Ying Tao, Jing Zhang, Anna Uehara, Anna Montmayeur, Clinton R. Paden, Kristen Knipe, Matthew Schmerer, Shoshona Le, Katie Dillon, Peter W. Cook, Rachel Marine, Mili Sheth, Jasmine Padilla, Sarah Nobles, Mark Burroughs, Lori Rowe, Haibin Wang, Ben L. Rambo-Martin, Kristine Lacek, Sam Shepard, Dhwani Batra, Suxiang Tong, Justin Lee |
| EPI_ISL_1272972, EPI_ISL_1272973, EPI_ISL_1272974, EPI_ISL_1272975, EPI_ISL_1272976, EPI_ISL_1272977, EPI_ISL_1272978, EPI_ISL_1272979, EPI_ISL_1272980, EPI_ISL_1272981, EPI_ISL_1272982, EPI_ISL_1272983, EPI_ISL_1272984, EPI_ISL_1272985, EPI_ISL_1272986, EPI_ISL_1272987, EPI_ISL_1272988 | | | |
| see above | NV-Southern Nevada Public Health Laboratory | Centers for Disease Control and Prevention Division of Viral Diseases, Pathogen Discovery | Krista Queen, Yan Li, Ying Tao, Jing Zhang, Anna Uehara, Anna Montmayeur, Clinton R. Paden, Kristen Knipe, Matthew Schmerer, Shoshona Le, Katie Dillon, Peter W. Cook, Rachel Marine, Mili Sheth, Jasmine Padilla, Sarah Nobles, Mark Burroughs, Lori Rowe, Haibin Wang, Ben L. Rambo-Martin, Kristine Lacek, Sam Shepard, Dhwani Batra, Suxiang Tong, Justin Lee |
| EPI_ISL_1272989, EPI_ISL_1272990, EPI_ISL_1272991, EPI_ISL_1272992, EPI_ISL_1272993, EPI_ISL_1272994, EPI_ISL_1272995, EPI_ISL_1272996, EPI_ISL_1272997 | MT Public Health Laboratory | Centers for Disease Control and Prevention Division of Viral Diseases, Pathogen Discovery | Krista Queen, Yan Li, Ying Tao, Jing Zhang, Anna Uehara, Anna Montmayeur, Clinton R. Paden, Kristen Knipe, Matthew Schmerer, Shoshona Le, Katie Dillon, Peter W. Cook, Rachel Marine, Mili Sheth, Jasmine Padilla, Sarah Nobles, Mark Burroughs, Lori Rowe, Haibin Wang, Ben L. Rambo-Martin, Kristine Lacek, Sam Shepard, Dhwani Batra, Suxiang Tong, Justin Lee |
| EPI_ISL_1272998, EPI_ISL_1272999, EPI_ISL_1273000, EPI_ISL_1273001, EPI_ISL_1273002, EPI_ISL_1273003, EPI_ISL_1273004, EPI_ISL_1273005, EPI_ISL_1273006, EPI_ISL_1273007, EPI_ISL_1273008, EPI_ISL_1273009, EPI_ISL_1273010, EPI_ISL_1273011, EPI_ISL_1273012, EPI_ISL_1273013, EPI_ISL_1273014, EPI_ISL_1273015, EPI_ISL_1273016, EPI_ISL_1273017, EPI_ISL_1273018, EPI_ISL_1273019, EPI_ISL_1273020, EPI_ISL_1273021, EPI_ISL_1273022, EPI_ISL_1273023, EPI_ISL_1273024, EPI_ISL_1273025, EPI_ISL_1273026, EPI_ISL_1273027, EPI_ISL_1273028, EPI_ISL_1273029, EPI_ISL_1273030, EPI_ISL_1273031, EPI_ISL_1273032, EPI_ISL_1273033, EPI_ISL_1273034, EPI_ISL_1273035, EPI_ISL_1273036 | | | |
| see above | GA Department of Public Health Laboratory | Centers for Disease Control and Prevention Division of Viral Diseases, Pathogen Discovery | Krista Queen, Yan Li, Ying Tao, Jing Zhang, Anna Uehara, Anna Montmayeur, Clinton R. Paden, Kristen Knipe, Matthew Schmerer, Shoshona Le, Katie Dillon, Peter W. Cook, Rachel Marine, Mili Sheth, Jasmine Padilla, Sarah Nobles, Mark Burroughs, Lori Rowe, Haibin Wang, Ben L. Rambo-Martin, Kristine Lacek, Sam Shepard, Dhwani Batra, Suxiang Tong, Justin Lee |
| EPI_ISL_1273037, EPI_ISL_1273038, EPI_ISL_1273039, EPI_ISL_1273040, EPI_ISL_1273041, EPI_ISL_1273042 | NH Dept. of Health and Human Services Public Health Labs | Centers for Disease Control and Prevention Division of Viral Diseases, Pathogen Discovery | Krista Queen, Yan Li, Ying Tao, Jing Zhang, Anna Uehara, Anna Montmayeur, Clinton R. Paden, Kristen Knipe, Matthew Schmerer, Shoshona Le, Katie Dillon, Peter W. Cook, Rachel Marine, Mili Sheth, Jasmine Padilla, Sarah Nobles, Mark Burroughs, Lori Rowe, Haibin Wang, Ben L. Rambo-Martin, Kristine Lacek, Sam Shepard, Dhwani Batra, Suxiang Tong, Justin Lee |
| EPI_ISL_1273043, EPI_ISL_1273044, EPI_ISL_1273045, EPI_ISL_1273046, EPI_ISL_1273047 | NYC Department of Health and Mental Hygiene | Centers for Disease Control and Prevention Division of Viral Diseases, Pathogen Discovery | Krista Queen, Yan Li, Ying Tao, Jing Zhang, Anna Uehara, Anna Montmayeur, Clinton R. Paden, Kristen Knipe, Matthew Schmerer, Shoshona Le, Katie Dillon, Peter W. Cook, Rachel Marine, Mili Sheth, Jasmine Padilla, Sarah Nobles, Mark Burroughs, Lori Rowe, Haibin Wang, Ben L. Rambo-Martin, Kristine Lacek, Sam Shepard, Dhwani Batra, Suxiang Tong, Justin Lee |
| EPI_ISL_1273226 | Texas Department of State Health Services (Texas Department of State Health Services (TXDSHS)) | Texas Department of State Health Services (TXDSHS) | Rashmi Tuladhar, Bonnie Oh, Jenny Zhang, Maliha Rahman, Mayela Pedrueza, Anita Pokharel, Myong Koag, Chun Wang, Rachel Lee, Grace Kubin |
| EPI_ISL_1273227, EPI_ISL_1273228, EPI_ISL_1273229, EPI_ISL_1273231, EPI_ISL_1273232, EPI_ISL_1273233, EPI_ISL_1273234, EPI_ISL_1273235, EPI_ISL_1273236, EPI_ISL_1273237, EPI_ISL_1273238, EPI_ISL_1273239, EPI_ISL_1273240, EPI_ISL_1273241, EPI_ISL_1273242, EPI_ISL_1273243, EPI_ISL_1273246, EPI_ISL_1273247, EPI_ISL_1273248, EPI_ISL_1273249, EPI_ISL_1273250, EPI_ISL_1273251, EPI_ISL_1273252, EPI_ISL_1273253, EPI_ISL_1273254, EPI_ISL_1273255, EPI_ISL_1273256, EPI_ISL_1273257, EPI_ISL_1273258, EPI_ISL_1273259, EPI_ISL_1273260, EPI_ISL_1273261, EPI_ISL_1273262, EPI_ISL_1273263, EPI_ISL_1273264, EPI_ISL_1273265, EPI_ISL_1273266, EPI_ISL_1273269, EPI_ISL_1273270, EPI_ISL_1273271, EPI_ISL_1273272, EPI_ISL_1273273, EPI_ISL_1273274, EPI_ISL_1273275, EPI_ISL_1273276, EPI_ISL_1273277, EPI_ISL_1273278, EPI_ISL_1273279, EPI_ISL_1273280, EPI_ISL_1273281, EPI_ISL_1273282, EPI_ISL_1273283, EPI_ISL_1273284, EPI_ISL_1273285, EPI_ISL_1273286, EPI_ISL_1273287, EPI_ISL_1273288, EPI_ISL_1273289, EPI_ISL_1273290, EPI_ISL_1273291, EPI_ISL_1273292, EPI_ISL_1273293, EPI_ISL_1273294, EPI_ISL_1273295, EPI_ISL_1273296, EPI_ISL_1273297, EPI_ISL_1273298, EPI_ISL_1273299, EPI_ISL_1273300, EPI_ISL_1273301, EPI_ISL_1273302, EPI_ISL_1273303, EPI_ISL_1273304, EPI_ISL_1273305 | | | |
| see above | Texas Department of State Health Services (TXDSHS) | Texas Department of State Health Services (TXDSHS) | Rashmi Tuladhar, Bonnie Oh, Jenny Zhang, Maliha Rahman, Mayela Pedrueza, Anita Pokharel, Myong Koag, Chun Wang, Rachel Lee, Grace Kubin |
| EPI_ISL_1273307, EPI_ISL_1273308, EPI_ISL_1273309, EPI_ISL_1273310, EPI_ISL_1273311, EPI_ISL_1273312, EPI_ISL_1273313, EPI_ISL_1273314, EPI_ISL_1273315, EPI_ISL_1273316, EPI_ISL_1273317, EPI_ISL_1273318, EPI_ISL_1273319, EPI_ISL_1273320, EPI_ISL_1273321, EPI_ISL_1273322, EPI_ISL_1273323, EPI_ISL_1273324, EPI_ISL_1273325, EPI_ISL_1273326, EPI_ISL_1273327, EPI_ISL_1273328, EPI_ISL_1273329, EPI_ISL_1273330, EPI_ISL_1273331, EPI_ISL_1273332, EPI_ISL_1273333, EPI_ISL_1273334, EPI_ISL_1273335, EPI_ISL_1273336, EPI_ISL_1273337, EPI_ISL_1273338, EPI_ISL_1273339, EPI_ISL_1273340, EPI_ISL_1273341, EPI_ISL_1273342, EPI_ISL_1273343, EPI_ISL_1273344, EPI_ISL_1273345, EPI_ISL_1273346, EPI_ISL_1273347, EPI_ISL_1273348, EPI_ISL_1273349, EPI_ISL_1273350, EPI_ISL_1273351, EPI_ISL_1273352, EPI_ISL_1273353, EPI_ISL_1273354, EPI_ISL_1273355, EPI_ISL_1273356, EPI_ISL_1273357, EPI_ISL_1273358, EPI_ISL_1273359, EPI_ISL_1273360, EPI_ISL_1273361, EPI_ISL_1273362, EPI_ISL_1273363, EPI_ISL_1273364, EPI_ISL_1273365, EPI_ISL_1273366, EPI_ISL_1273367, EPI_ISL_1273368, EPI_ISL_1273369, EPI_ISL_1273370, EPI_ISL_1273371, EPI_ISL_1273372, EPI_ISL_1273373, EPI_ISL_1273374, EPI_ISL_1273375, EPI_ISL_1273376, EPI_ISL_1273377, EPI_ISL_1273378 | | | |
| see above | The Jackson Laboratory | The Jackson Laboratory | Bergeron D, Renzette N, Adams M, Omerza G, Kelly K, Li L |
| EPI_ISL_1273379, EPI_ISL_1273380, EPI_ISL_1273381 | Atlas Genomics | Atlas Genomics | Michael Kalnoski, MD |
| EPI_ISL_1278007, EPI_ISL_1278008, EPI_ISL_1278009, EPI_ISL_1278010, EPI_ISL_1278011, EPI_ISL_1278012, EPI_ISL_1278013, EPI_ISL_1278014, EPI_ISL_1278015, EPI_ISL_1278016, EPI_ISL_1278017, EPI_ISL_1278018, EPI_ISL_1278019, EPI_ISL_1278020, EPI_ISL_1278021, EPI_ISL_1278022, EPI_ISL_1278023, EPI_ISL_1278024, EPI_ISL_1278025, EPI_ISL_1278026, EPI_ISL_1278027, EPI_ISL_1278028 | | | |
| see above | GA Department of Public Health | GA Department of Public Health | Stacy Reeves, Jonathan Edwards, Cynthia Dixey, Tonia Parrott, Aliyah Fields, Taylor Smith |
| EPI_ISL_1278055, EPI_ISL_1278056, EPI_ISL_1278057, EPI_ISL_1278058, EPI_ISL_1278059, EPI_ISL_1278061, EPI_ISL_1278062, EPI_ISL_1278063, EPI_ISL_1278064, EPI_ISL_1278065, EPI_ISL_1278066, EPI_ISL_1278068, EPI_ISL_1278069, EPI_ISL_1278070, EPI_ISL_1278071, EPI_ISL_1278072, EPI_ISL_1278073, EPI_ISL_1278074, EPI_ISL_1278075, EPI_ISL_1278076, EPI_ISL_1278077, EPI_ISL_1278078, EPI_ISL_1278079, EPI_ISL_1278080, EPI_ISL_1278081, EPI_ISL_1278082, EPI_ISL_1278083, EPI_ISL_1278084, EPI_ISL_1278085, EPI_ISL_1278086, EPI_ISL_1278087, EPI_ISL_1278088, EPI_ISL_1278089, EPI_ISL_1278090, EPI_ISL_1278091, EPI_ISL_1278092, EPI_ISL_1278093, EPI_ISL_1278094, EPI_ISL_1278095, EPI_ISL_1278096, EPI_ISL_1278097, EPI_ISL_1278098, EPI_ISL_1278099, EPI_ISL_1278100 | | | |
| see above | Emory Molecular Diagnostics Laboratory, Emory Healthcare | Piantadosi Lab, Emory Department of Pathology | Ahmed Babiker, Anne Piantadosi |
| EPI_ISL_1279263 | Laboratorio Central de Epidemiología (LCE) | Instituto Nacional de Enfermedades Respiratorias (INER): Centro de Investigación en Enfermedades Infecciosas (CIENI) | Consorcio Mexicano de Vigilancia Genómica (CoViGen-Mex). Authors (in alphabetical order): Julio Elias Alvarado-Yaah, Carlos F. Arias, Santiago Ávila-Ríos, Víctor Hugo Borja-Aburto, Celia Boukadida, Juan Bautista Chale-Dzul , José Antonio Enciso-Moreno, Gloria Elena Espinoza-Ayala, Fernando Fontove-Herrera, Concepción Grajales-Muñiz, Ricardo Grande, Alfredo Herrera-Estrella, Carla Ivón Herrera-Najera, Pavel Isa, Brenda Irasema Maldonado-Meza, Bernardo Martínez-Miguel, Margarita Matías-Florentino, María Guadalupe de Jesús Mireles-Rivera, Gloria María Molina-Salinas, Hector Montoya-Fuentes, José Esteban Muñoz-Medina, José de Jesús Nuñez-Contreras, Alicia Ocaña-Mondragón, Luis Alberto Ochoa-Carrera, Hector Esteban Paz-Juárez, Francisco Pulido, Helen Haydee Fernanda Ramírez-Plascencia, Angel Gustavo Salas-Lais, Jorge Ivan Salinal-Nevarez, Alejandro Sanchez-Flores, Clara Esperanza Santacruz-Tinoco, María Guadalupe Santiago-Mauricio , Nelly Sélem-Mojica, Blanca Taboada , Gloria Vazquez |
| EPI_ISL_1279264 | Centro de Investigación Biomédica del Noreste (CIBIN) | Instituto Nacional de Enfermedades Respiratorias (INER): Centro de Investigación en Enfermedades Infecciosas (CIENI) | Consorcio Mexicano de Vigilancia Genómica (CoViGen-Mex). Authors (in alphabetical order): Julio Elias Alvarado-Yaah, Carlos F. Arias, Santiago Ávila-Ríos, Víctor Hugo Borja-Aburto, Celia Boukadida, Juan Bautista Chale-Dzul , José Antonio Enciso-Moreno, Gloria Elena Espinoza-Ayala, Fernando Fontove-Herrera, Concepción Grajales-Muñiz, Ricardo Grande, Alfredo Herrera-Estrella, Carla Ivón Herrera-Najera, Pavel Isa, Brenda Irasema Maldonado-Meza, Bernardo Martínez-Miguel, Margarita Matías-Florentino, María Guadalupe de Jesús Mireles-Rivera, Gloria María Molina-Salinas, Hector Montoya-Fuentes, José Esteban Muñoz-Medina, José de Jesús Nuñez-Contreras, Alicia Ocaña-Mondragón, Luis Alberto Ochoa-Carrera, Hector Esteban Paz-Juárez, Francisco Pulido, Helen Haydee Fernanda Ramírez-Plascencia, Angel Gustavo Salas-Lais, Jorge Ivan Salinal-Nevarez, Alejandro Sanchez-Flores, Clara Esperanza Santacruz-Tinoco, María Guadalupe Santiago-Mauricio , Nelly Sélem-Mojica, Blanca Taboada , Gloria Vazquez |
| EPI_ISL_1279265 | Unidad de Investigación Biomédica de Zacatecas (UIBZ) | Instituto Nacional de Enfermedades Respiratorias (INER): Centro de Investigación en Enfermedades Infecciosas (CIENI) | Consorcio Mexicano de Vigilancia Genómica (CoViGen-Mex). Authors (in alphabetical order): Julio Elias Alvarado-Yaah, Carlos F. Arias, Santiago Ávila-Ríos, Víctor Hugo Borja-Aburto, Celia Boukadida, Juan Bautista Chale-Dzul , José Antonio Enciso-Moreno, Gloria Elena Espinoza-Ayala, Fernando Fontove-Herrera, Concepción Grajales-Muñiz, Ricardo Grande, Alfredo Herrera-Estrella, Carla Ivón Herrera-Najera, Pavel Isa, Brenda Irasema Maldonado-Meza, Bernardo Martínez-Miguel, Margarita Matías-Florentino, María Guadalupe de Jesús Mireles-Rivera, Gloria María Molina-Salinas, Hector Montoya-Fuentes, José Esteban Muñoz-Medina, José de Jesús Nuñez-Contreras, Alicia Ocaña-Mondragón, Luis Alberto Ochoa-Carrera, Hector Esteban Paz-Juárez, Francisco Pulido, Helen Haydee Fernanda Ramírez-Plascencia, Angel Gustavo Salas-Lais, Jorge Ivan Salinal-Nevarez, Alejandro Sanchez-Flores, Clara Esperanza Santacruz-Tinoco, María Guadalupe Santiago-Mauricio , Nelly Sélem-Mojica, Blanca Taboada , Gloria Vazquez |
| EPI_ISL_1279266, EPI_ISL_1279267 | Centro de Investigación Biomédica del Noreste (CIBIN) | Instituto Nacional de Enfermedades Respiratorias (INER): Centro de Investigación en Enfermedades Infecciosas (CIENI) | Consorcio Mexicano de Vigilancia Genómica (CoViGen-Mex). Authors (in alphabetical order): Julio Elias Alvarado-Yaah, Carlos F. Arias, Santiago Ávila-Ríos, Víctor Hugo Borja-Aburto, Celia Boukadida, Juan Bautista Chale-Dzul , José Antonio Enciso-Moreno, Gloria Elena Espinoza-Ayala, Fernando Fontove-Herrera, Concepción Grajales-Muñiz, Ricardo Grande, Alfredo Herrera-Estrella, Carla Ivón Herrera-Najera, Pavel Isa, Brenda Irasema Maldonado-Meza, Bernardo Martínez-Miguel, Margarita Matías-Florentino, María Guadalupe de Jesús Mireles-Rivera, Gloria María Molina-Salinas, Hector Montoya-Fuentes, José Esteban Muñoz-Medina, José de Jesús Nuñez-Contreras, Alicia Ocaña-Mondragón, Luis Alberto Ochoa-Carrera, Hector Esteban Paz-Juárez, Francisco Pulido, Helen Haydee Fernanda Ramírez-Plascencia, Angel Gustavo Salas-Lais, Jorge Ivan Salinal-Nevarez, Alejandro Sanchez-Flores, Clara Esperanza Santacruz-Tinoco, María Guadalupe Santiago-Mauricio , Nelly Sélem-Mojica, Blanca Taboada , Gloria Vazquez |
| EPI_ISL_1279268, EPI_ISL_1279269 | Laboratorio Central de Epidemiología (LCE) | Instituto Nacional de Enfermedades Respiratorias (INER): Centro de Investigación en Enfermedades Infecciosas (CIENI) | Consorcio Mexicano de Vigilancia Genómica (CoViGen-Mex). Authors (in alphabetical order): Julio Elias Alvarado-Yaah, Carlos F. Arias, Santiago Ávila-Ríos, Víctor Hugo Borja-Aburto, Celia Boukadida, Juan Bautista Chale-Dzul , José Antonio Enciso-Moreno, Gloria Elena Espinoza-Ayala, Fernando Fontove-Herrera, Concepción Grajales-Muñiz, Ricardo Grande, Alfredo Herrera-Estrella, Carla Ivón Herrera-Najera, Pavel Isa, Brenda Irasema Maldonado-Meza, Bernardo Martínez-Miguel, Margarita Matías-Florentino, María Guadalupe de Jesús Mireles-Rivera, Gloria María Molina-Salinas, Hector Montoya-Fuentes, José Esteban Muñoz-Medina, José de Jesús Nuñez-Contreras, Alicia Ocaña-Mondragón, Luis Alberto Ochoa-Carrera, Hector Esteban Paz-Juárez, Francisco Pulido, Helen Haydee Fernanda Ramírez-Plascencia, Angel Gustavo Salas-Lais, Jorge Ivan Salinal-Nevarez, Alejandro Sanchez-Flores, Clara Esperanza Santacruz-Tinoco, María Guadalupe Santiago-Mauricio , Nelly Sélem-Mojica, Blanca Taboada , Gloria Vazquez |
| EPI_ISL_1279270, EPI_ISL_1279271, EPI_ISL_1279272, EPI_ISL_1279273 | Unidad de Investigación Biomédica de Zacatecas (UIBZ) | Instituto Nacional de Enfermedades Respiratorias (INER): Centro de Investigación en Enfermedades Infecciosas (CIENI) | Consorcio Mexicano de Vigilancia Genómica (CoViGen-Mex). Authors (in alphabetical order): Julio Elias Alvarado-Yaah, Carlos F. Arias, Santiago Ávila-Ríos, Víctor Hugo Borja-Aburto, Celia Boukadida, Juan Bautista Chale-Dzul , José Antonio Enciso-Moreno, Gloria Elena Espinoza-Ayala, Fernando Fontove-Herrera, Concepción Grajales-Muñiz, Ricardo Grande, Alfredo Herrera-Estrella, Carla Ivón Herrera-Najera, Pavel Isa, Brenda Irasema Maldonado-Meza, Bernardo Martínez-Miguel, Margarita Matías-Florentino, María Guadalupe de Jesús Mireles-Rivera, Gloria María Molina-Salinas, Hector Montoya-Fuentes, José Esteban Muñoz-Medina, José de Jesús Nuñez-Contreras, Alicia Ocaña-Mondragón, Luis Alberto Ochoa-Carrera, Hector Esteban Paz-Juárez, Francisco Pulido, Helen Haydee Fernanda Ramírez-Plascencia, Angel Gustavo Salas-Lais, Jorge Ivan Salinal-Nevarez, Alejandro Sanchez-Flores, Clara Esperanza Santacruz-Tinoco, María Guadalupe Santiago-Mauricio , Nelly Sélem-Mojica, Blanca Taboada , Gloria Vazquez |
| EPI_ISL_1279274 | Laboratorio Central de Epidemiología (LCE) | Instituto Nacional de Enfermedades Respiratorias (INER): Centro de Investigación en Enfermedades Infecciosas (CIENI) | Consorcio Mexicano de Vigilancia Genómica (CoViGen-Mex). Authors (in alphabetical order): Julio Elias Alvarado-Yaah, Carlos F. Arias, Santiago Ávila-Ríos, Víctor Hugo Borja-Aburto, Celia Boukadida, Juan Bautista Chale-Dzul , José Antonio Enciso-Moreno, Gloria Elena Espinoza-Ayala, Fernando Fontove-Herrera, Concepción Grajales-Muñiz, Ricardo Grande, Alfredo Herrera-Estrella, Carla Ivón Herrera-Najera, Pavel Isa, Brenda Irasema Maldonado-Meza, Bernardo Martínez-Miguel, Margarita Matías-Florentino, María Guadalupe de Jesús Mireles-Rivera, Gloria María Molina-Salinas, Hector Montoya-Fuentes, José Esteban Muñoz-Medina, José de Jesús Nuñez-Contreras, Alicia Ocaña-Mondragón, Luis Alberto Ochoa-Carrera, Hector Esteban Paz-Juárez, Francisco Pulido, Helen Haydee Fernanda Ramírez-Plascencia, Angel Gustavo Salas-Lais, Jorge Ivan Salinal-Nevarez, Alejandro Sanchez-Flores, Clara Esperanza Santacruz-Tinoco, María Guadalupe Santiago-Mauricio , Nelly Sélem-Mojica, Blanca Taboada , Gloria Vazquez |
| EPI_ISL_1279275, EPI_ISL_1279276 | Centro de Investigación Biomédica del Noreste (CIBIN) | Instituto Nacional de Enfermedades Respiratorias (INER): Centro de Investigación en Enfermedades Infecciosas (CIENI) | Consorcio Mexicano de Vigilancia Genómica (CoViGen-Mex). Authors (in alphabetical order): Julio Elias Alvarado-Yaah, Carlos F. Arias, Santiago Ávila-Ríos, Víctor Hugo Borja-Aburto, Celia Boukadida, Juan Bautista Chale-Dzul , José Antonio Enciso-Moreno, Gloria Elena Espinoza-Ayala, Fernando Fontove-Herrera, Concepción Grajales-Muñiz, Ricardo Grande, Alfredo Herrera-Estrella, Carla Ivón Herrera-Najera, Pavel Isa, Brenda Irasema Maldonado-Meza, Bernardo Martínez-Miguel, Margarita Matías-Florentino, María Guadalupe de Jesús Mireles-Rivera, Gloria María Molina-Salinas, Hector Montoya-Fuentes, José Esteban Muñoz-Medina, José de Jesús Nuñez-Contreras, Alicia Ocaña-Mondragón, Luis Alberto Ochoa-Carrera, Hector Esteban Paz-Juárez, Francisco Pulido, Helen Haydee Fernanda Ramírez-Plascencia, Angel Gustavo Salas-Lais, Jorge Ivan Salinal-Nevarez, Alejandro Sanchez-Flores, Clara Esperanza Santacruz-Tinoco, María Guadalupe Santiago-Mauricio , Nelly Sélem-Mojica, Blanca Taboada , Gloria Vazquez |
| EPI_ISL_1279277, EPI_ISL_1279278 | Laboratorio Central de Epidemiología (LCE) | Instituto Nacional de Enfermedades Respiratorias (INER): Centro de Investigación en Enfermedades Infecciosas (CIENI) | Consorcio Mexicano de Vigilancia Genómica (CoViGen-Mex). Authors (in alphabetical order): Julio Elias Alvarado-Yaah, Carlos F. Arias, Santiago Ávila-Ríos, Víctor Hugo Borja-Aburto, Celia Boukadida, Juan Bautista Chale-Dzul , José Antonio Enciso-Moreno, Gloria Elena Espinoza-Ayala, Fernando Fontove-Herrera, Concepción Grajales-Muñiz, Ricardo Grande, Alfredo Herrera-Estrella, Carla Ivón Herrera-Najera, Pavel Isa, Brenda Irasema Maldonado-Meza, Bernardo Martínez-Miguel, Margarita Matías-Florentino, María Guadalupe de Jesús Mireles-Rivera, Gloria María Molina-Salinas, Hector Montoya-Fuentes, José Esteban Muñoz-Medina, José de Jesús Nuñez-Contreras, Alicia Ocaña-Mondragón, Luis Alberto Ochoa-Carrera, Hector Esteban Paz-Juárez, Francisco Pulido, Helen Haydee Fernanda Ramírez-Plascencia, Angel Gustavo Salas-Lais, Jorge Ivan Salinal-Nevarez, Alejandro Sanchez-Flores, Clara Esperanza Santacruz-Tinoco, María Guadalupe Santiago-Mauricio , Nelly Sélem-Mojica, Blanca Taboada , Gloria Vazquez |

| | | | |
|---|---|---|---|
| EPI_ISL_1279279, EPI_ISL_1279280, EPI_ISL_1279281 | Unidad de Investigación Biomédica de Zacatecas (UIBZ) | Instituto Nacional de Enfermedades Respiratorias (INER): Centro de Investigación en Enfermedades Infecciosas (CIENI) | Consorcio Mexicano de Vigilancia Genómica (CoViGen-Mex). Authors (in alphabetical order): Julio Elias Alvarado-Yaah, Carlos F. Arias, Santiago Ávila-Ríos, Víctor Hugo Borja-Aburto, Celia Boukadida, Juan Bautista Chale-Dzul , José Antonio Enciso-Moreno, Gloria Elena Espinoza-Ayala, Fernando Fontove-Herrera, Concepción Grajales-Muñiz, Ricardo Grande, Alfredo Herrera-Estrella, Carla Ivón Herrera-Najera, Pavel Isa, Brenda Irasema Maldonado-Meza, Bernardo Martínez-Miguel, Margarita Matías-Florentino, María Guadalupe de Jesús Mireles-Rivera, Gloria María Molina-Salinas, Hector Montoya-Fuentes, José Esteban Muñoz-Medina, José de Jesús Nuñez-Contreras, Alicia Ocaña-Mondragón, Luis Alberto Ochoa-Carrera, Hector Esteban Paz-Juárez, Francisco Pulido, Helen Haydee Fernanda Ramírez-Plascencia, Angel Gustavo Salas-Lais, Jorge Ivan Salinal-Nevarez, Alejandro Sanchez-Flores, Clara Esperanza Santacruz-Tinoco, María Guadalupe Santiago-Mauricio , Nelly Sélem-Mojica, Blanca Taboada , Gloria Vazquez |
| EPI_ISL_1279282 | Unidad de Investigación Médica de Yucatán (UIMY) | Instituto Nacional de Enfermedades Respiratorias (INER): Centro de Investigación en Enfermedades Infecciosas (CIENI) | Consorcio Mexicano de Vigilancia Genómica (CoViGen-Mex). Authors (in alphabetical order): Julio Elias Alvarado-Yaah, Carlos F. Arias, Santiago Ávila-Ríos, Víctor Hugo Borja-Aburto, Celia Boukadida, Juan Bautista Chale-Dzul , José Antonio Enciso-Moreno, Gloria Elena Espinoza-Ayala, Fernando Fontove-Herrera, Concepción Grajales-Muñiz, Ricardo Grande, Alfredo Herrera-Estrella, Carla Ivón Herrera-Najera, Pavel Isa, Brenda Irasema Maldonado-Meza, Bernardo Martínez-Miguel, Margarita Matías-Florentino, María Guadalupe de Jesús Mireles-Rivera, Gloria María Molina-Salinas, Hector Montoya-Fuentes, José Esteban Muñoz-Medina, José de Jesús Nuñez-Contreras, Alicia Ocaña-Mondragón, Luis Alberto Ochoa-Carrera, Hector Esteban Paz-Juárez, Francisco Pulido, Helen Haydee Fernanda Ramírez-Plascencia, Angel Gustavo Salas-Lais, Jorge Ivan Salinal-Nevarez, Alejandro Sanchez-Flores, Clara Esperanza Santacruz-Tinoco, María Guadalupe Santiago-Mauricio , Nelly Sélem-Mojica, Blanca Taboada , Gloria Vazquez |
| EPI_ISL_1279283 | Laboratorio Central de Epidemiología (LCE) | Instituto Nacional de Enfermedades Respiratorias (INER): Centro de Investigación en Enfermedades Infecciosas (CIENI) | Consorcio Mexicano de Vigilancia Genómica (CoViGen-Mex). Authors (in alphabetical order): Julio Elias Alvarado-Yaah, Carlos F. Arias, Santiago Ávila-Ríos, Víctor Hugo Borja-Aburto, Celia Boukadida, Juan Bautista Chale-Dzul , José Antonio Enciso-Moreno, Gloria Elena Espinoza-Ayala, Fernando Fontove-Herrera, Concepción Grajales-Muñiz, Ricardo Grande, Alfredo Herrera-Estrella, Carla Ivón Herrera-Najera, Pavel Isa, Brenda Irasema Maldonado-Meza, Bernardo Martínez-Miguel, Margarita Matías-Florentino, María Guadalupe de Jesús Mireles-Rivera, Gloria María Molina-Salinas, Hector Montoya-Fuentes, José Esteban Muñoz-Medina, José de Jesús Nuñez-Contreras, Alicia Ocaña-Mondragón, Luis Alberto Ochoa-Carrera, Hector Esteban Paz-Juárez, Francisco Pulido, Helen Haydee Fernanda Ramírez-Plascencia, Angel Gustavo Salas-Lais, Jorge Ivan Salinal-Nevarez, Alejandro Sanchez-Flores, Clara Esperanza Santacruz-Tinoco, María Guadalupe Santiago-Mauricio , Nelly Sélem-Mojica, Blanca Taboada , Gloria Vazquez |
| EPI_ISL_1279284, EPI_ISL_1279285 | Centro de Investigación Biomédica del Noreste (CIBIN) | Instituto Nacional de Enfermedades Respiratorias (INER): Centro de Investigación en Enfermedades Infecciosas (CIENI) | Consorcio Mexicano de Vigilancia Genómica (CoViGen-Mex). Authors (in alphabetical order): Julio Elias Alvarado-Yaah, Carlos F. Arias, Santiago Ávila-Ríos, Víctor Hugo Borja-Aburto, Celia Boukadida, Juan Bautista Chale-Dzul , José Antonio Enciso-Moreno, Gloria Elena Espinoza-Ayala, Fernando Fontove-Herrera, Concepción Grajales-Muñiz, Ricardo Grande, Alfredo Herrera-Estrella, Carla Ivón Herrera-Najera, Pavel Isa, Brenda Irasema Maldonado-Meza, Bernardo Martínez-Miguel, Margarita Matías-Florentino, María Guadalupe de Jesús Mireles-Rivera, Gloria María Molina-Salinas, Hector Montoya-Fuentes, José Esteban Muñoz-Medina, José de Jesús Nuñez-Contreras, Alicia Ocaña-Mondragón, Luis Alberto Ochoa-Carrera, Hector Esteban Paz-Juárez, Francisco Pulido, Helen Haydee Fernanda Ramírez-Plascencia, Angel Gustavo Salas-Lais, Jorge Ivan Salinal-Nevarez, Alejandro Sanchez-Flores, Clara Esperanza Santacruz-Tinoco, María Guadalupe Santiago-Mauricio , Nelly Sélem-Mojica, Blanca Taboada , Gloria Vazquez |
| EPI_ISL_1279286 | Laboratorio Central de Epidemiología (LCE) | Instituto Nacional de Enfermedades Respiratorias (INER): Centro de Investigación en Enfermedades Infecciosas (CIENI) | Consorcio Mexicano de Vigilancia Genómica (CoViGen-Mex). Authors (in alphabetical order): Julio Elias Alvarado-Yaah, Carlos F. Arias, Santiago Ávila-Ríos, Víctor Hugo Borja-Aburto, Celia Boukadida, Juan Bautista Chale-Dzul , José Antonio Enciso-Moreno, Gloria Elena Espinoza-Ayala, Fernando Fontove-Herrera, Concepción Grajales-Muñiz, Ricardo Grande, Alfredo Herrera-Estrella, Carla Ivón Herrera-Najera, Pavel Isa, Brenda Irasema Maldonado-Meza, Bernardo Martínez-Miguel, Margarita Matías-Florentino, María Guadalupe de Jesús Mireles-Rivera, Gloria María Molina-Salinas, Hector Montoya-Fuentes, José Esteban Muñoz-Medina, José de Jesús Nuñez-Contreras, Alicia Ocaña-Mondragón, Luis Alberto Ochoa-Carrera, Hector Esteban Paz-Juárez, Francisco Pulido, Helen Haydee Fernanda Ramírez-Plascencia, Angel Gustavo Salas-Lais, Jorge Ivan Salinal-Nevarez, Alejandro Sanchez-Flores, Clara Esperanza Santacruz-Tinoco, María Guadalupe Santiago-Mauricio , Nelly Sélem-Mojica, Blanca Taboada , Gloria Vazquez |
| EPI_ISL_1279287 | Centro de Investigación Biomédica del Noreste (CIBIN) | Instituto Nacional de Enfermedades Respiratorias (INER): Centro de Investigación en Enfermedades Infecciosas (CIENI) | Consorcio Mexicano de Vigilancia Genómica (CoViGen-Mex). Authors (in alphabetical order): Julio Elias Alvarado-Yaah, Carlos F. Arias, Santiago Ávila-Ríos, Víctor Hugo Borja-Aburto, Celia Boukadida, Juan Bautista Chale-Dzul , José Antonio Enciso-Moreno, Gloria Elena Espinoza-Ayala, Fernando Fontove-Herrera, Concepción Grajales-Muñiz, Ricardo Grande, Alfredo Herrera-Estrella, Carla Ivón Herrera-Najera, Pavel Isa, Brenda Irasema Maldonado-Meza, Bernardo Martínez-Miguel, Margarita Matías-Florentino, María Guadalupe de Jesús Mireles-Rivera, Gloria María Molina-Salinas, Hector Montoya-Fuentes, José Esteban Muñoz-Medina, José de Jesús Nuñez-Contreras, Alicia Ocaña-Mondragón, Luis Alberto Ochoa-Carrera, Hector Esteban Paz-Juárez, Francisco Pulido, Helen Haydee Fernanda Ramírez-Plascencia, Angel Gustavo Salas-Lais, Jorge Ivan Salinal-Nevarez, Alejandro Sanchez-Flores, Clara Esperanza Santacruz-Tinoco, María Guadalupe Santiago-Mauricio , Nelly Sélem-Mojica, Blanca Taboada , Gloria Vazquez |
| EPI_ISL_1279288, EPI_ISL_1279289, EPI_ISL_1279290, EPI_ISL_1279291, EPI_ISL_1279292, EPI_ISL_1279293 | Laboratorio Central de Epidemiología (LCE) | Instituto Nacional de Enfermedades Respiratorias (INER): Centro de Investigación en Enfermedades Infecciosas (CIENI) | Consorcio Mexicano de Vigilancia Genómica (CoViGen-Mex). Authors (in alphabetical order): Julio Elias Alvarado-Yaah, Carlos F. Arias, Santiago Ávila-Ríos, Víctor Hugo Borja-Aburto, Celia Boukadida, Juan Bautista Chale-Dzul , José Antonio Enciso-Moreno, Gloria Elena Espinoza-Ayala, Fernando Fontove-Herrera, Concepción Grajales-Muñiz, Ricardo Grande, Alfredo Herrera-Estrella, Carla Ivón Herrera-Najera, Pavel Isa, Brenda Irasema Maldonado-Meza, Bernardo Martínez-Miguel, Margarita Matías-Florentino, María Guadalupe de Jesús Mireles-Rivera, Gloria María Molina-Salinas, Hector Montoya-Fuentes, José Esteban Muñoz-Medina, José de Jesús Nuñez-Contreras, Alicia Ocaña-Mondragón, Luis Alberto Ochoa-Carrera, Hector Esteban Paz-Juárez, Francisco Pulido, Helen Haydee Fernanda Ramírez-Plascencia, Angel Gustavo Salas-Lais, Jorge Ivan Salinal-Nevarez, Alejandro Sanchez-Flores, Clara Esperanza Santacruz-Tinoco, María Guadalupe Santiago-Mauricio , Nelly Sélem-Mojica, Blanca Taboada , Gloria Vazquez |
| EPI_ISL_1279294 | Unidad de Investigación Médica de Yucatán (UIMY) | Instituto Nacional de Enfermedades Respiratorias (INER): Centro de Investigación en Enfermedades Infecciosas (CIENI) | Consorcio Mexicano de Vigilancia Genómica (CoViGen-Mex). Authors (in alphabetical order): Julio Elias Alvarado-Yaah, Carlos F. Arias, Santiago Ávila-Ríos, Víctor Hugo Borja-Aburto, Celia Boukadida, Juan Bautista Chale-Dzul , José Antonio Enciso-Moreno, Gloria Elena Espinoza-Ayala, Fernando Fontove-Herrera, Concepción Grajales-Muñiz, Ricardo Grande, Alfredo Herrera-Estrella, Carla Ivón Herrera-Najera, Pavel Isa, Brenda Irasema Maldonado-Meza, Bernardo Martínez-Miguel, Margarita Matías-Florentino, María Guadalupe de Jesús Mireles-Rivera, Gloria María Molina-Salinas, Hector Montoya-Fuentes, José Esteban Muñoz-Medina, José de Jesús Nuñez-Contreras, Alicia Ocaña-Mondragón, Luis Alberto Ochoa-Carrera, Hector Esteban Paz-Juárez, Francisco Pulido, Helen Haydee Fernanda Ramírez-Plascencia, Angel Gustavo Salas-Lais, Jorge Ivan Salinal-Nevarez, Alejandro Sanchez-Flores, Clara Esperanza Santacruz-Tinoco, María Guadalupe Santiago-Mauricio , Nelly Sélem-Mojica, Blanca Taboada , Gloria Vazquez |
| EPI_ISL_1279295 | Unidad de Investigación Biomédica de Zacatecas (UIBZ) | Instituto Nacional de Enfermedades Respiratorias (INER): Centro de Investigación en Enfermedades Infecciosas (CIENI) | Consorcio Mexicano de Vigilancia Genómica (CoViGen-Mex). Authors (in alphabetical order): Julio Elias Alvarado-Yaah, Carlos F. Arias, Santiago Ávila-Ríos, Víctor Hugo Borja-Aburto, Celia Boukadida, Juan Bautista Chale-Dzul , José Antonio Enciso-Moreno, Gloria Elena Espinoza-Ayala, Fernando Fontove-Herrera, Concepción Grajales-Muñiz, Ricardo Grande, Alfredo Herrera-Estrella, Carla Ivón Herrera-Najera, Pavel Isa, Brenda Irasema Maldonado-Meza, Bernardo Martínez-Miguel, Margarita Matías-Florentino, María Guadalupe de Jesús Mireles-Rivera, Gloria María Molina-Salinas, Hector Montoya-Fuentes, José Esteban Muñoz-Medina, José de Jesús Nuñez-Contreras, Alicia Ocaña-Mondragón, Luis Alberto Ochoa-Carrera, Hector Esteban Paz-Juárez, Francisco Pulido, Helen Haydee Fernanda Ramírez-Plascencia, Angel Gustavo Salas-Lais, Jorge Ivan Salinal-Nevarez, Alejandro Sanchez-Flores, Clara Esperanza Santacruz-Tinoco, María Guadalupe Santiago-Mauricio , Nelly Sélem-Mojica, Blanca Taboada , Gloria Vazquez |
| EPI_ISL_1279296, EPI_ISL_1279297 | Laboratorio Central de Epidemiología (LCE) | Instituto Nacional de Enfermedades Respiratorias (INER): Centro de Investigación en Enfermedades Infecciosas (CIENI) | Consorcio Mexicano de Vigilancia Genómica (CoViGen-Mex). Authors (in alphabetical order): Julio Elias Alvarado-Yaah, Carlos F. Arias, Santiago Ávila-Ríos, Víctor Hugo Borja-Aburto, Celia Boukadida, Juan Bautista Chale-Dzul , José Antonio Enciso-Moreno, Gloria Elena Espinoza-Ayala, Fernando Fontove-Herrera, Concepción Grajales-Muñiz, Ricardo Grande, Alfredo Herrera-Estrella, Carla Ivón Herrera-Najera, Pavel Isa, Brenda Irasema Maldonado-Meza, Bernardo Martínez-Miguel, Margarita Matías-Florentino, María Guadalupe de Jesús Mireles-Rivera, Gloria María Molina-Salinas, Hector Montoya-Fuentes, José Esteban Muñoz-Medina, José de Jesús Nuñez-Contreras, Alicia Ocaña-Mondragón, Luis Alberto Ochoa-Carrera, Hector Esteban Paz-Juárez, Francisco Pulido, Helen Haydee Fernanda Ramírez-Plascencia, Angel Gustavo Salas-Lais, Jorge Ivan Salinal-Nevarez, Alejandro Sanchez-Flores, Clara Esperanza Santacruz-Tinoco, María Guadalupe Santiago-Mauricio , Nelly Sélem-Mojica, Blanca Taboada , Gloria Vazquez |
| EPI_ISL_1279298 | Centro de Investigación Biomédica del Noreste (CIBIN) | Instituto Nacional de Enfermedades Respiratorias (INER): Centro de Investigación en Enfermedades Infecciosas (CIENI) | Consorcio Mexicano de Vigilancia Genómica (CoViGen-Mex). Authors (in alphabetical order): Julio Elias Alvarado-Yaah, Carlos F. Arias, Santiago Ávila-Ríos, Víctor Hugo Borja-Aburto, Celia Boukadida, Juan Bautista Chale-Dzul , José Antonio Enciso-Moreno, Gloria Elena Espinoza-Ayala, Fernando Fontove-Herrera, Concepción Grajales-Muñiz, Ricardo Grande, Alfredo Herrera-Estrella, Carla Ivón Herrera-Najera, Pavel Isa, Brenda Irasema Maldonado-Meza, Bernardo Martínez-Miguel, Margarita Matías-Florentino, María Guadalupe de Jesús Mireles-Rivera, Gloria María Molina-Salinas, Hector Montoya-Fuentes, José Esteban Muñoz-Medina, José de Jesús Nuñez-Contreras, Alicia Ocaña-Mondragón, Luis Alberto Ochoa-Carrera, Hector Esteban Paz-Juárez, Francisco Pulido, Helen Haydee Fernanda Ramírez-Plascencia, Angel Gustavo Salas-Lais, Jorge Ivan Salinal-Nevarez, Alejandro Sanchez-Flores, Clara Esperanza Santacruz-Tinoco, María Guadalupe Santiago-Mauricio , Nelly Sélem-Mojica, Blanca Taboada , Gloria Vazquez |
| EPI_ISL_1279299 | Laboratorio Central de Epidemiología (LCE) | Instituto Nacional de Enfermedades Respiratorias (INER): Centro de Investigación en Enfermedades Infecciosas (CIENI) | Consorcio Mexicano de Vigilancia Genómica (CoViGen-Mex). Authors (in alphabetical order): Julio Elias Alvarado-Yaah, Carlos F. Arias, Santiago Ávila-Ríos, Víctor Hugo Borja-Aburto, Celia Boukadida, Juan Bautista Chale-Dzul , José Antonio Enciso-Moreno, Gloria Elena Espinoza-Ayala, Fernando Fontove-Herrera, Concepción Grajales-Muñiz, Ricardo Grande, Alfredo Herrera-Estrella, Carla Ivón Herrera-Najera, Pavel Isa, Brenda Irasema Maldonado-Meza, Bernardo Martínez-Miguel, Margarita Matías-Florentino, María Guadalupe de Jesús Mireles-Rivera, Gloria María Molina-Salinas, Hector Montoya-Fuentes, José Esteban Muñoz-Medina, José de Jesús Nuñez-Contreras, Alicia Ocaña-Mondragón, Luis Alberto Ochoa-Carrera, Hector Esteban Paz-Juárez, Francisco Pulido, Helen Haydee Fernanda Ramírez-Plascencia, Angel Gustavo Salas-Lais, Jorge Ivan Salinal-Nevarez, Alejandro Sanchez-Flores, Clara Esperanza Santacruz-Tinoco, María Guadalupe Santiago-Mauricio , Nelly Sélem-Mojica, Blanca Taboada , Gloria Vazquez |
| EPI_ISL_1279300, EPI_ISL_1279301 | Unidad de Investigación Biomédica de Zacatecas (UIBZ) | Instituto Nacional de Enfermedades Respiratorias (INER): Centro de Investigación en Enfermedades Infecciosas (CIENI) | Consorcio Mexicano de Vigilancia Genómica (CoViGen-Mex). Authors (in alphabetical order): Julio Elias Alvarado-Yaah, Carlos F. Arias, Santiago Ávila-Ríos, Víctor Hugo Borja-Aburto, Celia Boukadida, Juan Bautista Chale-Dzul , José Antonio Enciso-Moreno, Gloria Elena Espinoza-Ayala, Fernando Fontove-Herrera, Concepción Grajales-Muñiz, Ricardo Grande, Alfredo Herrera-Estrella, Carla Ivón Herrera-Najera, Pavel Isa, Brenda Irasema Maldonado-Meza, Bernardo Martínez-Miguel, Margarita Matías-Florentino, María Guadalupe de Jesús Mireles-Rivera, Gloria María Molina-Salinas, Hector Montoya-Fuentes, José Esteban Muñoz-Medina, José de Jesús Nuñez-Contreras, Alicia Ocaña-Mondragón, Luis Alberto Ochoa-Carrera, Hector Esteban Paz-Juárez, Francisco Pulido, Helen Haydee Fernanda Ramírez-Plascencia, Angel Gustavo Salas-Lais, Jorge Ivan Salinal-Nevarez, Alejandro Sanchez-Flores, Clara Esperanza Santacruz-Tinoco, María Guadalupe Santiago-Mauricio , Nelly Sélem-Mojica, Blanca Taboada , Gloria Vazquez |
| EPI_ISL_1279302 | Centro de Investigación Biomédica del Noreste (CIBIN) | Instituto Nacional de Enfermedades Respiratorias (INER): Centro de Investigación en Enfermedades Infecciosas (CIENI) | Consorcio Mexicano de Vigilancia Genómica (CoViGen-Mex). Authors (in alphabetical order): Julio Elias Alvarado-Yaah, Carlos F. Arias, Santiago Ávila-Ríos, Víctor Hugo Borja-Aburto, Celia Boukadida, Juan Bautista Chale-Dzul , José Antonio Enciso-Moreno, Gloria Elena Espinoza-Ayala, Fernando Fontove-Herrera, Concepción Grajales-Muñiz, Ricardo Grande, Alfredo Herrera-Estrella, Carla Ivón Herrera-Najera, Pavel Isa, Brenda Irasema Maldonado-Meza, Bernardo Martínez-Miguel, Margarita Matías-Florentino, María Guadalupe de Jesús Mireles-Rivera, Gloria María Molina-Salinas, Hector Montoya-Fuentes, José Esteban Muñoz-Medina, José de Jesús Nuñez-Contreras, Alicia Ocaña-Mondragón, Luis Alberto Ochoa-Carrera, Hector Esteban Paz-Juárez, Francisco Pulido, Helen Haydee Fernanda Ramírez-Plascencia, Angel Gustavo Salas-Lais, Jorge Ivan Salinal-Nevarez, Alejandro Sanchez-Flores, Clara Esperanza Santacruz-Tinoco, María Guadalupe Santiago-Mauricio , Nelly Sélem-Mojica, Blanca Taboada , Gloria Vazquez |
| EPI_ISL_1279303 | Laboratorio Central de Epidemiología (LCE) | Instituto Nacional de Enfermedades Respiratorias (INER): Centro de Investigación en Enfermedades Infecciosas (CIENI) | Consorcio Mexicano de Vigilancia Genómica (CoViGen-Mex). Authors (in alphabetical order): Julio Elias Alvarado-Yaah, Carlos F. Arias, Santiago Ávila-Ríos, Víctor Hugo Borja-Aburto, Celia Boukadida, Juan Bautista Chale-Dzul , José Antonio Enciso-Moreno, Gloria Elena Espinoza-Ayala, Fernando Fontove-Herrera, Concepción Grajales-Muñiz, Ricardo Grande, Alfredo Herrera-Estrella, Carla Ivón Herrera-Najera, Pavel Isa, Brenda Irasema Maldonado-Meza, Bernardo Martínez-Miguel, Margarita Matías-Florentino, María Guadalupe de Jesús Mireles-Rivera, Gloria María Molina-Salinas, Hector Montoya-Fuentes, José Esteban Muñoz-Medina, José de Jesús Nuñez-Contreras, Alicia Ocaña-Mondragón, Luis Alberto Ochoa-Carrera, Hector Esteban Paz-Juárez, Francisco Pulido, Helen Haydee Fernanda Ramírez-Plascencia, Angel Gustavo Salas-Lais, Jorge Ivan Salinal-Nevarez, Alejandro Sanchez-Flores, Clara Esperanza Santacruz-Tinoco, María Guadalupe Santiago-Mauricio , Nelly Sélem-Mojica, Blanca Taboada , Gloria Vazquez |
| EPI_ISL_1279304, EPI_ISL_1279305, EPI_ISL_1279306, EPI_ISL_1279307, EPI_ISL_1279308, EPI_ISL_1279309, EPI_ISL_1279310, EPI_ISL_1279311, EPI_ISL_1279312, EPI_ISL_1279313, EPI_ISL_1279314, EPI_ISL_1279315, EPI_ISL_1279316, EPI_ISL_1279317, EPI_ISL_1279318, EPI_ISL_1279319, EPI_ISL_1279320, EPI_ISL_1279321, EPI_ISL_1279322, EPI_ISL_1279323, EPI_ISL_1279324, EPI_ISL_1279325, EPI_ISL_1279326, EPI_ISL_1279327, EPI_ISL_1279328, EPI_ISL_1279329, EPI_ISL_1279330, EPI_ISL_1279331, EPI_ISL_1279332, EPI_ISL_1279333, EPI_ISL_1279334, EPI_ISL_1279335, EPI_ISL_1279336, EPI_ISL_1279337, EPI_ISL_1279338, EPI_ISL_1279339, EPI_ISL_1279340, EPI_ISL_1279341, EPI_ISL_1279342, EPI_ISL_1279343, EPI_ISL_1279344, EPI_ISL_1279345, EPI_ISL_1279346, EPI_ISL_1279347, EPI_ISL_1279348, EPI_ISL_1279349, EPI_ISL_1279350, EPI_ISL_1279351, EPI_ISL_1279352, EPI_ISL_1279353, EPI_ISL_1279354, EPI_ISL_1279355, EPI_ISL_1279356, EPI_ISL_1279357, EPI_ISL_1279358, EPI_ISL_1279359, EPI_ISL_1279360, EPI_ISL_1279361, EPI_ISL_1279362, EPI_ISL_1279363, EPI_ISL_1279364, EPI_ISL_1279365, EPI_ISL_1279366, EPI_ISL_1279367, EPI_ISL_1279368, EPI_ISL_1279369, EPI_ISL_1279370, EPI_ISL_1279371, EPI_ISL_1279372, EPI_ISL_1279373, EPI_ISL_1279374, EPI_ISL_1279375, EPI_ISL_1279376, EPI_ISL_1279377, EPI_ISL_1279378, EPI_ISL_1279379, EPI_ISL_1279380, EPI_ISL_1279381, EPI_ISL_1279382 | | | |
| see above | Unidad de Investigación Biomédica de Zacatecas (UIBZ) | Instituto Nacional de Enfermedades Respiratorias (INER): Centro de Investigación en Enfermedades Infecciosas (CIENI) | Consorcio Mexicano de Vigilancia Genómica (CoViGen-Mex). Authors (in alphabetical order): Julio Elias Alvarado-Yaah, Carlos F. Arias, Santiago Ávila-Ríos, Víctor Hugo Borja-Aburto, Celia Boukadida, Juan Bautista Chale-Dzul , José Antonio Enciso-Moreno, Gloria Elena Espinoza-Ayala, Fernando Fontove-Herrera, Concepción Grajales-Muñiz, Ricardo Grande, Alfredo Herrera-Estrella, Carla Ivón Herrera-Najera, Pavel Isa, Brenda Irasema Maldonado-Meza, Bernardo Martínez-Miguel, Margarita Matías-Florentino, María Guadalupe de Jesús Mireles-Rivera, Gloria María Molina-Salinas, Hector Montoya-Fuentes, José Esteban Muñoz-Medina, José de Jesús Nuñez-Contreras, Alicia Ocaña-Mondragón, Luis Alberto Ochoa-Carrera, Hector Esteban Paz-Juárez, Francisco Pulido, Helen Haydee Fernanda Ramírez-Plascencia, Angel Gustavo Salas-Lais, Jorge Ivan Salinal-Nevarez, Alejandro Sanchez-Flores, Clara Esperanza Santacruz-Tinoco, María Guadalupe Santiago-Mauricio , Nelly Sélem-Mojica, Blanca Taboada , Gloria Vazquez |
| EPI_ISL_1279383, EPI_ISL_1279384, EPI_ISL_1279385, EPI_ISL_1279386, EPI_ISL_1279387, EPI_ISL_1279388, EPI_ISL_1279389, EPI_ISL_1279390, EPI_ISL_1279391, EPI_ISL_1279392, EPI_ISL_1279393, EPI_ISL_1279394, EPI_ISL_1279395, EPI_ISL_1279396, EPI_ISL_1279397, EPI_ISL_1279398, EPI_ISL_1279399, EPI_ISL_1279400, EPI_ISL_1279401, EPI_ISL_1279402, EPI_ISL_1279403, EPI_ISL_1279404, EPI_ISL_1279405, EPI_ISL_1279406, EPI_ISL_1279407, EPI_ISL_1279408, EPI_ISL_1279409, EPI_ISL_1279410, EPI_ISL_1279411, EPI_ISL_1279412, EPI_ISL_1279413, EPI_ISL_1279414, EPI_ISL_1279415, EPI_ISL_1279416, EPI_ISL_1279417, EPI_ISL_1279418, EPI_ISL_1279419, EPI_ISL_1279420, EPI_ISL_1279421, EPI_ISL_1279422, EPI_ISL_1279423, EPI_ISL_1279424, EPI_ISL_1279425, EPI_ISL_1279426, EPI_ISL_1279427, EPI_ISL_1279428, EPI_ISL_1279429, EPI_ISL_1279430, EPI_ISL_1279431, EPI_ISL_1279432, EPI_ISL_1279433, EPI_ISL_1279434, EPI_ISL_1279435, EPI_ISL_1279436, EPI_ISL_1279437, EPI_ISL_1279438, EPI_ISL_1279439 | | | |
| see above | Laboratorio Central de Epidemiología (LCE) | Instituto Nacional de Enfermedades Respiratorias (INER): Centro de Investigación en Enfermedades Infecciosas (CIENI) | Consorcio Mexicano de Vigilancia Genómica (CoViGen-Mex). Authors (in alphabetical order): Julio Elias Alvarado-Yaah, Carlos F. Arias, Santiago Ávila-Ríos, Víctor Hugo Borja-Aburto, Celia Boukadida, Juan Bautista Chale-Dzul , José Antonio Enciso-Moreno, Gloria Elena Espinoza-Ayala, Fernando Fontove-Herrera, Concepción Grajales-Muñiz, Ricardo Grande, Alfredo Herrera-Estrella, Carla Ivón Herrera-Najera, Pavel Isa, Brenda Irasema Maldonado-Meza, Bernardo Martínez-Miguel, Margarita Matías-Florentino, María Guadalupe de Jesús Mireles-Rivera, Gloria María Molina-Salinas, Hector Montoya-Fuentes, José Esteban Muñoz-Medina, José de Jesús Nuñez-Contreras, Alicia Ocaña-Mondragón, Luis Alberto Ochoa-Carrera, Hector Esteban Paz-Juárez, Francisco Pulido, Helen Haydee Fernanda Ramírez-Plascencia, Angel Gustavo Salas-Lais, Jorge Ivan Salinal-Nevarez, Alejandro Sanchez-Flores, Clara Esperanza Santacruz-Tinoco, María Guadalupe Santiago-Mauricio , Nelly Sélem-Mojica, Blanca Taboada , Gloria Vazquez |
| EPI_ISL_1279440 | Unidad de Investigación Médica de Yucatán (UIMY) | Instituto Nacional de Enfermedades Respiratorias (INER): Centro de Investigación en Enfermedades Infecciosas (CIENI) | Consorcio Mexicano de Vigilancia Genómica (CoViGen-Mex). Authors (in alphabetical order): Julio Elias Alvarado-Yaah, Carlos F. Arias, Santiago Ávila-Ríos, Víctor Hugo Borja-Aburto, Celia Boukadida, Juan Bautista Chale-Dzul , José Antonio Enciso-Moreno, Gloria Elena Espinoza-Ayala, Fernando Fontove-Herrera, Concepción Grajales-Muñiz, Ricardo Grande, Alfredo Herrera-Estrella, Carla Ivón Herrera-Najera, Pavel Isa, Brenda Irasema Maldonado-Meza, Bernardo Martínez-Miguel, Margarita Matías-Florentino, María Guadalupe de Jesús Mireles-Rivera, Gloria María Molina-Salinas, Hector Montoya-Fuentes, José Esteban Muñoz-Medina, José de Jesús Nuñez-Contreras, Alicia Ocaña-Mondragón, Luis Alberto Ochoa-Carrera, Hector Esteban Paz-Juárez, Francisco Pulido, Helen Haydee Fernanda Ramírez-Plascencia, Angel Gustavo Salas-Lais, Jorge Ivan Salinal-Nevarez, Alejandro Sanchez-Flores, Clara Esperanza Santacruz-Tinoco, María Guadalupe Santiago-Mauricio , Nelly Sélem-Mojica, Blanca Taboada , Gloria Vazquez |
| EPI_ISL_1279441, EPI_ISL_1279442, EPI_ISL_1279443, EPI_ISL_1279444, EPI_ISL_1279445, EPI_ISL_1279446, EPI_ISL_1279447, EPI_ISL_1279448, EPI_ISL_1279449, EPI_ISL_1279450, EPI_ISL_1279451, EPI_ISL_1279452, EPI_ISL_1279453, EPI_ISL_1279454, EPI_ISL_1279455, EPI_ISL_1279456, EPI_ISL_1279457, EPI_ISL_1279458, EPI_ISL_1279459, EPI_ISL_1279460, EPI_ISL_1279461 | | | |
| see above | Laboratorio Central de Epidemiología (LCE) | Instituto Nacional de Enfermedades Respiratorias (INER): Centro de Investigación en Enfermedades Infecciosas (CIENI) | Consorcio Mexicano de Vigilancia Genómica (CoViGen-Mex). Authors (in alphabetical order): Julio Elias Alvarado-Yaah, Carlos F. Arias, Santiago Ávila-Ríos, Víctor Hugo Borja-Aburto, Celia Boukadida, Juan Bautista Chale-Dzul , José Antonio Enciso-Moreno, Gloria Elena Espinoza-Ayala, Fernando Fontove-Herrera, Concepción Grajales-Muñiz, Ricardo Grande, Alfredo Herrera-Estrella, Carla Ivón Herrera-Najera, Pavel Isa, Brenda Irasema Maldonado-Meza, Bernardo Martínez-Miguel, Margarita Matías-Florentino, María Guadalupe de Jesús Mireles-Rivera, Gloria María Molina-Salinas, Hector Montoya-Fuentes, José Esteban Muñoz-Medina, José de Jesús Nuñez-Contreras, Alicia Ocaña-Mondragón, Luis Alberto Ochoa-Carrera, Hector Esteban Paz-Juárez, Francisco Pulido, Helen Haydee Fernanda Ramírez-Plascencia, Angel Gustavo Salas-Lais, Jorge Ivan Salinal-Nevarez, Alejandro Sanchez-Flores, Clara Esperanza Santacruz-Tinoco, María Guadalupe Santiago-Mauricio , Nelly Sélem-Mojica, Blanca Taboada , Gloria Vazquez |
| EPI_ISL_1279462, EPI_ISL_1279463, EPI_ISL_1279464, EPI_ISL_1279465, EPI_ISL_1279466, EPI_ISL_1279467, EPI_ISL_1279468, EPI_ISL_1279469, EPI_ISL_1279470, EPI_ISL_1279471, EPI_ISL_1279472, EPI_ISL_1279473, EPI_ISL_1279474, EPI_ISL_1279475, EPI_ISL_1279476, EPI_ISL_1279477, EPI_ISL_1279478, EPI_ISL_1279479, EPI_ISL_1279480, EPI_ISL_1279481, EPI_ISL_1279482, EPI_ISL_1279483, EPI_ISL_1279484, EPI_ISL_1279485, EPI_ISL_1279486, EPI_ISL_1279487, EPI_ISL_1279488, EPI_ISL_1279489, EPI_ISL_1279490, EPI_ISL_1279491, EPI_ISL_1279492, EPI_ISL_1279493, EPI_ISL_1279494, EPI_ISL_1279495, EPI_ISL_1279496, EPI_ISL_1279497, EPI_ISL_1279498, EPI_ISL_1279499, EPI_ISL_1279500, EPI_ISL_1279501, EPI_ISL_1279502, EPI_ISL_1279503, EPI_ISL_1279504, EPI_ISL_1279505, EPI_ISL_1279506, EPI_ISL_1279507, EPI_ISL_1279508, EPI_ISL_1279509, EPI_ISL_1279510, EPI_ISL_1279511, EPI_ISL_1279512, EPI_ISL_1279513, EPI_ISL_1279514, EPI_ISL_1279515, EPI_ISL_1279516, EPI_ISL_1279517, EPI_ISL_1279518, EPI_ISL_1279519, EPI_ISL_1279520, EPI_ISL_1279521, EPI_ISL_1279522, EPI_ISL_1279523, EPI_ISL_1279524, EPI_ISL_1279525, EPI_ISL_1279526, EPI_ISL_1279527, EPI_ISL_1279528, EPI_ISL_1279529, EPI_ISL_1279530, EPI_ISL_1279531, EPI_ISL_1279532, EPI_ISL_1279533, EPI_ISL_1279534, EPI_ISL_1279535, EPI_ISL_1279536 | | | |
| see above | Centro de Investigación Biomédica del Noreste (CIBIN) | Instituto Nacional de Enfermedades Respiratorias (INER): Centro de Investigación en Enfermedades Infecciosas (CIENI) | Consorcio Mexicano de Vigilancia Genómica (CoViGen-Mex). Authors (in alphabetical order): Julio Elias Alvarado-Yaah, Carlos F. Arias, Santiago Ávila-Ríos, Víctor Hugo Borja-Aburto, Celia Boukadida, Juan Bautista Chale-Dzul , José Antonio Enciso-Moreno, Gloria Elena Espinoza-Ayala, Fernando Fontove-Herrera, Concepción Grajales-Muñiz, Ricardo Grande, Alfredo Herrera-Estrella, Carla Ivón Herrera-Najera, Pavel Isa, Brenda Irasema Maldonado-Meza, Bernardo Martínez-Miguel, Margarita Matías-Florentino, María Guadalupe de Jesús Mireles-Rivera, Gloria María Molina-Salinas, Hector Montoya-Fuentes, José Esteban Muñoz-Medina, José de Jesús Nuñez-Contreras, Alicia Ocaña-Mondragón, Luis Alberto Ochoa-Carrera, Hector Esteban Paz-Juárez, Francisco Pulido, Helen Haydee Fernanda Ramírez-Plascencia, Angel Gustavo Salas-Lais, Jorge Ivan Salinal-Nevarez, Alejandro Sanchez-Flores, Clara Esperanza Santacruz-Tinoco, María Guadalupe Santiago-Mauricio , Nelly Sélem-Mojica, Blanca Taboada , Gloria Vazquez |
| EPI_ISL_1279537, EPI_ISL_1279538, EPI_ISL_1279539, EPI_ISL_1279540, EPI_ISL_1279541, EPI_ISL_1279542, EPI_ISL_1279543, EPI_ISL_1279544, EPI_ISL_1279545, EPI_ISL_1279546, EPI_ISL_1279547, EPI_ISL_1279548, EPI_ISL_1279549, EPI_ISL_1279550, EPI_ISL_1279551, EPI_ISL_1279552, EPI_ISL_1279553, EPI_ISL_1279554, EPI_ISL_1279555, EPI_ISL_1279556, EPI_ISL_1279557, EPI_ISL_1279558, EPI_ISL_1279559, EPI_ISL_1279560, EPI_ISL_1279561, EPI_ISL_1279562, EPI_ISL_1279563, EPI_ISL_1279564, EPI_ISL_1279565, EPI_ISL_1279566, EPI_ISL_1279567, EPI_ISL_1279568, EPI_ISL_1279569, EPI_ISL_1279570, EPI_ISL_1279571, EPI_ISL_1279572, EPI_ISL_1279573, EPI_ISL_1279574, EPI_ISL_1279575, EPI_ISL_1279576, EPI_ISL_1279577, EPI_ISL_1279578, EPI_ISL_1279579, EPI_ISL_1279580, EPI_ISL_1279581, EPI_ISL_1279582, EPI_ISL_1279583, EPI_ISL_1279584, EPI_ISL_1279585, EPI_ISL_1279586, EPI_ISL_1279587, EPI_ISL_1279588, EPI_ISL_1279589, EPI_ISL_1279590, EPI_ISL_1279591, EPI_ISL_1279592, EPI_ISL_1279593, EPI_ISL_1279594, EPI_ISL_1279595, EPI_ISL_1279596, EPI_ISL_1279597, EPI_ISL_1279598, EPI_ISL_1279599, EPI_ISL_1279600, EPI_ISL_1279601, EPI_ISL_1279602, EPI_ISL_1279603 | | | |
| see above | Laboratorio Central de Epidemiología | Instituto Nacional de Enfermedades | Consorcio Mexicano de Vigilancia Genómica (CoViGen-Mex). Authors (in alphabetical order): Julio Elias Alvarado-Yaah, Carlos F. Arias, Santiago Ávila-Ríos, Víctor Hugo Borja-Aburto, Celia Boukadida, Juan Bautista Chale-Dzul , José Antonio Enciso-Moreno, Gloria Elena Espinoza-Ayala, |

| | | | |
|---|---|---|---|
| | (LCE) | Respiratorias (INER): Centro de Investigación en Enfermedades Infecciosas (CIENI) | Fernando Fontove-Herrera, Concepción Grajales-Muñiz, Ricardo Grande, Alfredo Herrera-Estrella, Carla Ivón Herrera-Najera, Pavel Isa, Brenda Irasema Maldonado-Meza, Bernardo Martínez-Miguel, Margarita Matías-Florentino, María Guadalupe de Jesús Mireles-Rivera, Gloria María Molina-Salinas, Hector Montoya-Fuentes, José Esteban Muñoz-Medina, José de Jesús Nuñez-Contreras, Alicia Ocaña-Mondragón, Luis Alberto Ochoa-Carrera, Hector Esteban Paz-Juárez, Francisco Pulido, Helen Haydee Fernanda Ramírez-Plascencia, Angel Gustavo Salas-Lais, Jorge Ivan Salinal-Nevarez, Alejandro Sanchez-Flores, Clara Esperanza Santacruz-Tinoco, María Guadalupe Santiago-Mauricio , Nelly Sélem-Mojica, Blanca Taboada , Gloria Vazquez |
| EPI_ISL_1279604, EPI_ISL_1279605, EPI_ISL_1279606, EPI_ISL_1279607, EPI_ISL_1279608, EPI_ISL_1279609, EPI_ISL_1279610, EPI_ISL_1279611, EPI_ISL_1279612, EPI_ISL_1279613, EPI_ISL_1279614 | | | |
| see above | Unidad de Investigación Médica de Yucatán (UIMY) | Instituto Nacional de Enfermedades Respiratorias (INER): Centro de Investigación en Enfermedades Infecciosas (CIENI) | Consorcio Mexicano de Vigilancia Genómica (CoViGen-Mex). Authors (in alphabetical order): Julio Elias Alvarado-Yaah, Carlos F. Arias, Santiago Ávila-Ríos, Víctor Hugo Borja-Aburto, Celia Boukadida, Juan Bautista Chale-Dzul , José Antonio Enciso-Moreno, Gloria Elena Espinoza-Ayala, Fernando Fontove-Herrera, Concepción Grajales-Muñiz, Ricardo Grande, Alfredo Herrera-Estrella, Carla Ivón Herrera-Najera, Pavel Isa, Brenda Irasema Maldonado-Meza, Bernardo Martínez-Miguel, Margarita Matías-Florentino, María Guadalupe de Jesús Mireles-Rivera, Gloria María Molina-Salinas, Hector Montoya-Fuentes, José Esteban Muñoz-Medina, José de Jesús Nuñez-Contreras, Alicia Ocaña-Mondragón, Luis Alberto Ochoa-Carrera, Hector Esteban Paz-Juárez, Francisco Pulido, Helen Haydee Fernanda Ramírez-Plascencia, Angel Gustavo Salas-Lais, Jorge Ivan Salinal-Nevarez, Alejandro Sanchez-Flores, Clara Esperanza Santacruz-Tinoco, María Guadalupe Santiago-Mauricio , Nelly Sélem-Mojica, Blanca Taboada , Gloria Vazquez |
| EPI_ISL_1279891, EPI_ISL_1279892, EPI_ISL_1279893, EPI_ISL_1279894, EPI_ISL_1279895, EPI_ISL_1279896, EPI_ISL_1279897, EPI_ISL_1279898, EPI_ISL_1279899, EPI_ISL_1279900, EPI_ISL_1279901, EPI_ISL_1279902 | | | |
| see above | Nebraska Public Health Laboratory | NPHL COVID-19 Response Team | NPHL COVID-19 Response Team |
| EPI_ISL_1283377, EPI_ISL_1283379, EPI_ISL_1283382, EPI_ISL_1283384, EPI_ISL_1283387, EPI_ISL_1283389, EPI_ISL_1283392, EPI_ISL_1283394, EPI_ISL_1283397, EPI_ISL_1283399, EPI_ISL_1283402, EPI_ISL_1283404, EPI_ISL_1283407, EPI_ISL_1283409, EPI_ISL_1283412, EPI_ISL_1283414, EPI_ISL_1283417, EPI_ISL_1283419, EPI_ISL_1283422, EPI_ISL_1283425, EPI_ISL_1283427, EPI_ISL_1283430, EPI_ISL_1283433, EPI_ISL_1283435, EPI_ISL_1283438, EPI_ISL_1283440, EPI_ISL_1283443, EPI_ISL_1283445, EPI_ISL_1283448, EPI_ISL_1283450, EPI_ISL_1283453, EPI_ISL_1283456, EPI_ISL_1283458, EPI_ISL_1283460, EPI_ISL_1283463, EPI_ISL_1283465, EPI_ISL_1283468, EPI_ISL_1283470, EPI_ISL_1283473, EPI_ISL_1283476, EPI_ISL_1283478, EPI_ISL_1283481, EPI_ISL_1283484, EPI_ISL_1283486, EPI_ISL_1283489, EPI_ISL_1283492, EPI_ISL_1283494, EPI_ISL_1283497, EPI_ISL_1283499, EPI_ISL_1283502, EPI_ISL_1283504, EPI_ISL_1283507, EPI_ISL_1283509, EPI_ISL_1283512, EPI_ISL_1283515, EPI_ISL_1283517, EPI_ISL_1283520, EPI_ISL_1283522, EPI_ISL_1283524, EPI_ISL_1283527, EPI_ISL_1283529, EPI_ISL_1283531, EPI_ISL_1283534, EPI_ISL_1283537, EPI_ISL_1283540, EPI_ISL_1283542, EPI_ISL_1283544, EPI_ISL_1283547, EPI_ISL_1283549, EPI_ISL_1283552, EPI_ISL_1283554, EPI_ISL_1283557, EPI_ISL_1283559, EPI_ISL_1283562, EPI_ISL_1283564, EPI_ISL_1283567, EPI_ISL_1283570, EPI_ISL_1283572, EPI_ISL_1283575, EPI_ISL_1283577, EPI_ISL_1283580, EPI_ISL_1283582, EPI_ISL_1283585, EPI_ISL_1283587, EPI_ISL_1283590, EPI_ISL_1283592, EPI_ISL_1283595, EPI_ISL_1283598, EPI_ISL_1283600, EPI_ISL_1283602, EPI_ISL_1283605, EPI_ISL_1283607, EPI_ISL_1283610, EPI_ISL_1283612, EPI_ISL_1283615, EPI_ISL_1283617, EPI_ISL_1283620, EPI_ISL_1283622, EPI_ISL_1283625, EPI_ISL_1283627, EPI_ISL_1283629, EPI_ISL_1283631, EPI_ISL_1283634, EPI_ISL_1283636, EPI_ISL_1283639, EPI_ISL_1283642, EPI_ISL_1283644, EPI_ISL_1283647, EPI_ISL_1283650, EPI_ISL_1283652, EPI_ISL_1283655, EPI_ISL_1283658, EPI_ISL_1283660, EPI_ISL_1283662, EPI_ISL_1283664, EPI_ISL_1283667, EPI_ISL_1283670, EPI_ISL_1283672, EPI_ISL_1283674, EPI_ISL_1283677, EPI_ISL_1283679, EPI_ISL_1283682, EPI_ISL_1283685, EPI_ISL_1283687, EPI_ISL_1283689, EPI_ISL_1283692, EPI_ISL_1283695, EPI_ISL_1283697, EPI_ISL_1283699, EPI_ISL_1283702, EPI_ISL_1283704, EPI_ISL_1283707, EPI_ISL_1283709, EPI_ISL_1283712, EPI_ISL_1283714, EPI_ISL_1283717, EPI_ISL_1283719, EPI_ISL_1283722, EPI_ISL_1283724, EPI_ISL_1283727, EPI_ISL_1283730, EPI_ISL_1283732, EPI_ISL_1283735, EPI_ISL_1283737, EPI_ISL_1283740, EPI_ISL_1283742, EPI_ISL_1283745, EPI_ISL_1283747, EPI_ISL_1283750, EPI_ISL_1283752, EPI_ISL_1283754, EPI_ISL_1283756, EPI_ISL_1283759, EPI_ISL_1283762, EPI_ISL_1283764, EPI_ISL_1283767, EPI_ISL_1283771, EPI_ISL_1283773, EPI_ISL_1283776, EPI_ISL_1283778, EPI_ISL_1283781, EPI_ISL_1283783, EPI_ISL_1283786, EPI_ISL_1283788, EPI_ISL_1283791, EPI_ISL_1283794, EPI_ISL_1283796, EPI_ISL_1283799, EPI_ISL_1283804, EPI_ISL_1283806, EPI_ISL_1283808, EPI_ISL_1283813, EPI_ISL_1283816, EPI_ISL_1283819, EPI_ISL_1283821, EPI_ISL_1283823, EPI_ISL_1283826, EPI_ISL_1283828, EPI_ISL_1283831, EPI_ISL_1283833, EPI_ISL_1283835, EPI_ISL_1283838, EPI_ISL_1283840, EPI_ISL_1283843, EPI_ISL_1283845, EPI_ISL_1283848, EPI_ISL_1283851, EPI_ISL_1283853, EPI_ISL_1283856, EPI_ISL_1283859, EPI_ISL_1283862, EPI_ISL_1283864, EPI_ISL_1283867, EPI_ISL_1283869, EPI_ISL_1283872, EPI_ISL_1283875, EPI_ISL_1283877, EPI_ISL_1283878, EPI_ISL_1283879, EPI_ISL_1283882, EPI_ISL_1283884, EPI_ISL_1283886, EPI_ISL_1283889, EPI_ISL_1283891, EPI_ISL_1283893, EPI_ISL_1283896, EPI_ISL_1283898, EPI_ISL_1283901, EPI_ISL_1283904, EPI_ISL_1283906, EPI_ISL_1283909, EPI_ISL_1283911, EPI_ISL_1283914, EPI_ISL_1283916, EPI_ISL_1283919, EPI_ISL_1283921, EPI_ISL_1283926, EPI_ISL_1283930, EPI_ISL_1283935, EPI_ISL_1283940, EPI_ISL_1283943, EPI_ISL_1283946, EPI_ISL_1283952, EPI_ISL_1283956, EPI_ISL_1283959, EPI_ISL_1283962, EPI_ISL_1283965, EPI_ISL_1283968, EPI_ISL_1283971, EPI_ISL_1283975, EPI_ISL_1283977, EPI_ISL_1283985, EPI_ISL_1283990, EPI_ISL_1283993, EPI_ISL_1283997, EPI_ISL_1284001, EPI_ISL_1284006, EPI_ISL_1284010, EPI_ISL_1284013, EPI_ISL_1284016, EPI_ISL_1284019, EPI_ISL_1284023, EPI_ISL_1284028, EPI_ISL_1284032, EPI_ISL_1284036, EPI_ISL_1284039, EPI_ISL_1284043, EPI_ISL_1284047, EPI_ISL_1284051, EPI_ISL_1284056, EPI_ISL_1284060, EPI_ISL_1284065, EPI_ISL_1284070, EPI_ISL_1284075, EPI_ISL_1284079, EPI_ISL_1284082, EPI_ISL_1284085, EPI_ISL_1284088, EPI_ISL_1284094, EPI_ISL_1284097, EPI_ISL_1284100, EPI_ISL_1284103, EPI_ISL_1284107, EPI_ISL_1284110, EPI_ISL_1284113, EPI_ISL_1284116, EPI_ISL_1284120, EPI_ISL_1284123, EPI_ISL_1284127, EPI_ISL_1284131, EPI_ISL_1284134, EPI_ISL_1284138, EPI_ISL_1284141, EPI_ISL_1284144, EPI_ISL_1284148, EPI_ISL_1284151, EPI_ISL_1284154, EPI_ISL_1284158, EPI_ISL_1284161, EPI_ISL_1284166, EPI_ISL_1284171, EPI_ISL_1284175, EPI_ISL_1284179, EPI_ISL_1284185, EPI_ISL_1284189, EPI_ISL_1284192, EPI_ISL_1284195, EPI_ISL_1284199, EPI_ISL_1284202, EPI_ISL_1284205, EPI_ISL_1284209, EPI_ISL_1284213, EPI_ISL_1284217, EPI_ISL_1284220, EPI_ISL_1284224, EPI_ISL_1284227, EPI_ISL_1284231, EPI_ISL_1284233, EPI_ISL_1284236, EPI_ISL_1284239, EPI_ISL_1284243, EPI_ISL_1284246, EPI_ISL_1284249, EPI_ISL_1284253, EPI_ISL_1284256, EPI_ISL_1284260, EPI_ISL_1284264, EPI_ISL_1284267, EPI_ISL_1284271, EPI_ISL_1284275, EPI_ISL_1284278, EPI_ISL_1284282, EPI_ISL_1284285, EPI_ISL_1284289, EPI_ISL_1284291, EPI_ISL_1284295, EPI_ISL_1284298, EPI_ISL_1284302, EPI_ISL_1284306, EPI_ISL_1284309, EPI_ISL_1284312, EPI_ISL_1284316, EPI_ISL_1284320, EPI_ISL_1284323, EPI_ISL_1284326, EPI_ISL_1284329, EPI_ISL_1284333, EPI_ISL_1284336, EPI_ISL_1284339, EPI_ISL_1284343, EPI_ISL_1284346, EPI_ISL_1284349, EPI_ISL_1284351, EPI_ISL_1284354, EPI_ISL_1284356, EPI_ISL_1284359, EPI_ISL_1284361, EPI_ISL_1284363, EPI_ISL_1284366, EPI_ISL_1284368, EPI_ISL_1284370, EPI_ISL_1284372, EPI_ISL_1284375, EPI_ISL_1284377, EPI_ISL_1284380, EPI_ISL_1284382, EPI_ISL_1284385, EPI_ISL_1284387, EPI_ISL_1284389, EPI_ISL_1284392, EPI_ISL_1284394, EPI_ISL_1284397, EPI_ISL_1284400, EPI_ISL_1284402, EPI_ISL_1284405, EPI_ISL_1284407, EPI_ISL_1284409, EPI_ISL_1284410, EPI_ISL_1284412, EPI_ISL_1284419, EPI_ISL_1284422, EPI_ISL_1284424, EPI_ISL_1284426, EPI_ISL_1284429, EPI_ISL_1284432, EPI_ISL_1284434, EPI_ISL_1284437, EPI_ISL_1284439, EPI_ISL_1284442, EPI_ISL_1284444, EPI_ISL_1284447, EPI_ISL_1284449, EPI_ISL_1284451, EPI_ISL_1284454, EPI_ISL_1284456 | | | |
| see above | Helix/Illumina | Centers for Disease Control and Prevention Division of Viral Diseases, Pathogen Discovery | Peter W. Cook, Dakota Howard, Dhwani Batra, Ben L. Rambo-Martin, Eileen de Feo, Jan Antico, Christine Tran, Matthew Tolentino, Shannon Wickline, Kim Gietzen, Brad Sickler, Jingtao Liu, Eric Allen, Phil Febbo, Summer Galloway, Nicole L. Washington, Simon White, Geraint Levan, Kelly Schiabor Barrett, Elizabeth Cirulli, Alexandre Bolze, Ary Ascencio, Charlotte Rivera-Garcia, Ryan Cho, Jason Nguyen, Sherry Wang, Jimmy Ramirez, Tyler Cassens, Efren Sandoval, Magnus Isaksson, William Lee, David Becker, Marc Laurent, James Lu, Clinton R. Paden, Suxiang Tong, Duncan MacCannell |
| EPI_ISL_1287761, EPI_ISL_1287762, EPI_ISL_1287763, EPI_ISL_1287764, EPI_ISL_1287765, EPI_ISL_1287766, EPI_ISL_1287767 | Unidad de Investigación Biomédica de Zacatecas (UIBZ) | Instituto Nacional de Enfermedades Respiratorias (INER): Centro de Investigación en Enfermedades Infecciosas (CIENI) | Consorcio Mexicano de Vigilancia Genómica (CoViGen-Mex). Authors (in alphabetical order): Julio Elias Alvarado-Yaah, Carlos F. Arias, Santiago Ávila-Ríos, Víctor Hugo Borja-Aburto, Celia Boukadida, Juan Bautista Chale-Dzul , José Antonio Enciso-Moreno, Gloria Elena Espinoza-Ayala, Fernando Fontove-Herrera, Concepción Grajales-Muñiz, Ricardo Grande, Alfredo Herrera-Estrella, Carla Ivón Herrera-Najera, Pavel Isa, Brenda Irasema Maldonado-Meza, Bernardo Martínez-Miguel, Margarita Matías-Florentino, María Guadalupe de Jesús Mireles-Rivera, Gloria María Molina-Salinas, Hector Montoya-Fuentes, José Esteban Muñoz-Medina, José de Jesús Nuñez-Contreras, Alicia Ocaña-Mondragón, Luis Alberto Ochoa-Carrera, Hector Esteban Paz-Juárez, Francisco Pulido, Helen Haydee Fernanda Ramírez-Plascencia, Angel Gustavo Salas-Lais, Jorge Ivan Salinal-Nevarez, Alejandro Sanchez-Flores, Clara Esperanza Santacruz-Tinoco, María Guadalupe Santiago-Mauricio , Nelly Sélem-Mojica, Blanca Taboada , Gloria Vazquez |
| EPI_ISL_1287768, EPI_ISL_1287769, EPI_ISL_1287770 | Laboratorio Central de Epidemiología (LCE) | Instituto Nacional de Enfermedades Respiratorias (INER): Centro de Investigación en Enfermedades Infecciosas (CIENI) | Consorcio Mexicano de Vigilancia Genómica (CoViGen-Mex). Authors (in alphabetical order): Julio Elias Alvarado-Yaah, Carlos F. Arias, Santiago Ávila-Ríos, Víctor Hugo Borja-Aburto, Celia Boukadida, Juan Bautista Chale-Dzul , José Antonio Enciso-Moreno, Gloria Elena Espinoza-Ayala, Fernando Fontove-Herrera, Concepción Grajales-Muñiz, Ricardo Grande, Alfredo Herrera-Estrella, Carla Ivón Herrera-Najera, Pavel Isa, Brenda Irasema Maldonado-Meza, Bernardo Martínez-Miguel, Margarita Matías-Florentino, María Guadalupe de Jesús Mireles-Rivera, Gloria María Molina-Salinas, Hector Montoya-Fuentes, José Esteban Muñoz-Medina, José de Jesús Nuñez-Contreras, Alicia Ocaña-Mondragón, Luis Alberto Ochoa-Carrera, Hector Esteban Paz-Juárez, Francisco Pulido, Helen Haydee Fernanda Ramírez-Plascencia, Angel Gustavo Salas-Lais, Jorge Ivan Salinal-Nevarez, Alejandro Sanchez-Flores, Clara Esperanza Santacruz-Tinoco, María Guadalupe Santiago-Mauricio , Nelly Sélem-Mojica, Blanca Taboada , Gloria Vazquez |
| EPI_ISL_1287771, EPI_ISL_1287772, EPI_ISL_1287773 | Centro de Investigación Biomédica del Noreste (CIBIN) | Instituto Nacional de Enfermedades Respiratorias (INER): Centro de Investigación en Enfermedades Infecciosas (CIENI) | Consorcio Mexicano de Vigilancia Genómica (CoViGen-Mex). Authors (in alphabetical order): Julio Elias Alvarado-Yaah, Carlos F. Arias, Santiago Ávila-Ríos, Víctor Hugo Borja-Aburto, Celia Boukadida, Juan Bautista Chale-Dzul , José Antonio Enciso-Moreno, Gloria Elena Espinoza-Ayala, Fernando Fontove-Herrera, Concepción Grajales-Muñiz, Ricardo Grande, Alfredo Herrera-Estrella, Carla Ivón Herrera-Najera, Pavel Isa, Brenda Irasema Maldonado-Meza, Bernardo Martínez-Miguel, Margarita Matías-Florentino, María Guadalupe de Jesús Mireles-Rivera, Gloria María Molina-Salinas, Hector Montoya-Fuentes, José Esteban Muñoz-Medina, José de Jesús Nuñez-Contreras, Alicia Ocaña-Mondragón, Luis Alberto Ochoa-Carrera, Hector Esteban Paz-Juárez, Francisco Pulido, Helen Haydee Fernanda Ramírez-Plascencia, Angel Gustavo Salas-Lais, Jorge Ivan Salinal-Nevarez, Alejandro Sanchez-Flores, Clara Esperanza Santacruz-Tinoco, María Guadalupe Santiago-Mauricio , Nelly Sélem-Mojica, Blanca Taboada , Gloria Vazquez |
| EPI_ISL_1287774, EPI_ISL_1287775, EPI_ISL_1287776, EPI_ISL_1287777 | Laboratorio Central de Epidemiología (LCE) | Instituto Nacional de Enfermedades Respiratorias (INER): Centro de Investigación en Enfermedades Infecciosas (CIENI) | Consorcio Mexicano de Vigilancia Genómica (CoViGen-Mex). Authors (in alphabetical order): Julio Elias Alvarado-Yaah, Carlos F. Arias, Santiago Ávila-Ríos, Víctor Hugo Borja-Aburto, Celia Boukadida, Juan Bautista Chale-Dzul , José Antonio Enciso-Moreno, Gloria Elena Espinoza-Ayala, Fernando Fontove-Herrera, Concepción Grajales-Muñiz, Ricardo Grande, Alfredo Herrera-Estrella, Carla Ivón Herrera-Najera, Pavel Isa, Brenda Irasema Maldonado-Meza, Bernardo Martínez-Miguel, Margarita Matías-Florentino, María Guadalupe de Jesús Mireles-Rivera, Gloria María Molina-Salinas, Hector Montoya-Fuentes, José Esteban Muñoz-Medina, José de Jesús Nuñez-Contreras, Alicia Ocaña-Mondragón, Luis Alberto Ochoa-Carrera, Hector Esteban Paz-Juárez, Francisco Pulido, Helen Haydee Fernanda Ramírez-Plascencia, Angel Gustavo Salas-Lais, Jorge Ivan Salinal-Nevarez, Alejandro Sanchez-Flores, Clara Esperanza Santacruz-Tinoco, María Guadalupe Santiago-Mauricio , Nelly Sélem-Mojica, Blanca Taboada , Gloria Vazquez |
| EPI_ISL_1287778, EPI_ISL_1287779, EPI_ISL_1287780, EPI_ISL_1287781, EPI_ISL_1287782 | Kansas Health and Environmental Lab | Cytocheck Laboratory | Mike Grose, Jonathan Barnell, Ben Olsen, and Phil Adam |
| EPI_ISL_1288153, EPI_ISL_1288154, EPI_ISL_1288155, EPI_ISL_1288157, EPI_ISL_1288158, EPI_ISL_1288159, EPI_ISL_1288160, EPI_ISL_1288161, EPI_ISL_1288162, EPI_ISL_1288163, EPI_ISL_1288164, EPI_ISL_1288165, EPI_ISL_1288166, EPI_ISL_1288167, EPI_ISL_1288168, EPI_ISL_1288169, EPI_ISL_1288170, EPI_ISL_1288171, EPI_ISL_1288172, EPI_ISL_1288173, EPI_ISL_1288174, EPI_ISL_1288175, EPI_ISL_1288176, EPI_ISL_1288177, EPI_ISL_1288178, EPI_ISL_1288179, EPI_ISL_1288180, EPI_ISL_1288181, EPI_ISL_1288182, EPI_ISL_1288183, EPI_ISL_1288184, EPI_ISL_1288185, EPI_ISL_1288186, EPI_ISL_1288187, EPI_ISL_1288188, EPI_ISL_1288189, EPI_ISL_1288190, EPI_ISL_1288191, EPI_ISL_1288192, EPI_ISL_1288193, EPI_ISL_1288194, EPI_ISL_1288195, EPI_ISL_1288196, EPI_ISL_1288197, EPI_ISL_1288198, EPI_ISL_1288199, EPI_ISL_1288200, EPI_ISL_1288201, EPI_ISL_1288202, EPI_ISL_1288203, EPI_ISL_1288204, EPI_ISL_1288205, EPI_ISL_1288206, EPI_ISL_1288207, EPI_ISL_1288208, EPI_ISL_1288209, EPI_ISL_1288210, EPI_ISL_1288211, EPI_ISL_1288212, EPI_ISL_1288213, EPI_ISL_1288214, EPI_ISL_1288215, EPI_ISL_1288216, EPI_ISL_1288217, EPI_ISL_1288218, EPI_ISL_1288219, EPI_ISL_1288220, EPI_ISL_1288221, EPI_ISL_1288222, EPI_ISL_1288223, EPI_ISL_1288224, EPI_ISL_1288225, EPI_ISL_1288226, EPI_ISL_1288227, EPI_ISL_1288228, EPI_ISL_1288229, EPI_ISL_1288230, EPI_ISL_1288231, EPI_ISL_1288232, EPI_ISL_1288233, EPI_ISL_1288234, EPI_ISL_1288235, EPI_ISL_1288236, EPI_ISL_1288237, EPI_ISL_1288238, EPI_ISL_1288239, EPI_ISL_1288240, EPI_ISL_1288241, EPI_ISL_1288242, EPI_ISL_1288243, EPI_ISL_1288244, EPI_ISL_1288245, EPI_ISL_1288246, EPI_ISL_1288247, EPI_ISL_1288248, EPI_ISL_1288249, EPI_ISL_1288250, EPI_ISL_1288251, EPI_ISL_1288252, EPI_ISL_1288253, EPI_ISL_1288254, EPI_ISL_1288255, EPI_ISL_1288256, EPI_ISL_1288257, EPI_ISL_1288258, EPI_ISL_1288259, EPI_ISL_1288260, EPI_ISL_1288261, EPI_ISL_1288262, EPI_ISL_1288263, EPI_ISL_1288264, EPI_ISL_1288265, EPI_ISL_1288266, EPI_ISL_1288267, EPI_ISL_1288268, EPI_ISL_1288269, EPI_ISL_1288270, EPI_ISL_1288271, EPI_ISL_1288272, EPI_ISL_1288273, EPI_ISL_1288274, EPI_ISL_1288275, EPI_ISL_1288276, EPI_ISL_1288277, EPI_ISL_1288278, EPI_ISL_1288279, EPI_ISL_1288280, EPI_ISL_1288281, EPI_ISL_1288282, EPI_ISL_1288283, EPI_ISL_1288284, EPI_ISL_1288285, EPI_ISL_1288286, EPI_ISL_1288287, EPI_ISL_1288288, EPI_ISL_1288289, EPI_ISL_1288290, EPI_ISL_1288291, EPI_ISL_1288292, EPI_ISL_1288293, EPI_ISL_1288294, EPI_ISL_1288295, EPI_ISL_1288296, EPI_ISL_1288297, EPI_ISL_1288298, EPI_ISL_1288299, EPI_ISL_1288300, EPI_ISL_1288301, EPI_ISL_1288302, EPI_ISL_1288303, EPI_ISL_1288304, EPI_ISL_1288306, EPI_ISL_1288307, EPI_ISL_1288308, EPI_ISL_1288309, EPI_ISL_1288310, EPI_ISL_1288311, EPI_ISL_1288312, EPI_ISL_1288313, EPI_ISL_1288314, EPI_ISL_1288315, EPI_ISL_1288316, EPI_ISL_1288317, EPI_ISL_1288318, EPI_ISL_1288319, EPI_ISL_1288320, EPI_ISL_1288321, EPI_ISL_1288322, EPI_ISL_1288323, EPI_ISL_1288324, EPI_ISL_1288325, EPI_ISL_1288326, EPI_ISL_1288327, EPI_ISL_1288328, EPI_ISL_1288329, EPI_ISL_1288330, EPI_ISL_1288331, EPI_ISL_1288332, EPI_ISL_1288333, EPI_ISL_1288334, EPI_ISL_1288335, EPI_ISL_1288336, EPI_ISL_1288337, EPI_ISL_1288338, EPI_ISL_1288339, EPI_ISL_1288340, EPI_ISL_1288341, EPI_ISL_1288342, EPI_ISL_1288343, EPI_ISL_1288344, EPI_ISL_1288345, EPI_ISL_1288346, EPI_ISL_1288348, EPI_ISL_1288349, EPI_ISL_1288350, EPI_ISL_1288351, EPI_ISL_1288352, EPI_ISL_1288353, EPI_ISL_1288354, EPI_ISL_1288355, EPI_ISL_1288356, EPI_ISL_1288357, EPI_ISL_1288358, EPI_ISL_1288359, EPI_ISL_1288360, EPI_ISL_1288361, EPI_ISL_1288362, EPI_ISL_1288363, EPI_ISL_1288364, EPI_ISL_1288365, EPI_ISL_1288366, EPI_ISL_1288367, EPI_ISL_1288368, EPI_ISL_1288369, EPI_ISL_1288370, EPI_ISL_1288371, EPI_ISL_1288372, EPI_ISL_1288373, EPI_ISL_1288374, EPI_ISL_1288375, EPI_ISL_1288376, EPI_ISL_1288377, EPI_ISL_1288378, EPI_ISL_1288379, EPI_ISL_1288380, EPI_ISL_1288381, EPI_ISL_1288382, EPI_ISL_1288383, EPI_ISL_1288384, EPI_ISL_1288385, EPI_ISL_1288386, EPI_ISL_1288387, EPI_ISL_1288388, EPI_ISL_1288389, EPI_ISL_1288390, EPI_ISL_1288391, EPI_ISL_1288392, EPI_ISL_1288393, EPI_ISL_1288394, EPI_ISL_1288395, EPI_ISL_1288396, EPI_ISL_1288397, EPI_ISL_1288398, EPI_ISL_1288399, EPI_ISL_1288400, EPI_ISL_1288401, EPI_ISL_1288402, EPI_ISL_1288403, EPI_ISL_1288404, EPI_ISL_1288405, EPI_ISL_1288406, EPI_ISL_1288407, EPI_ISL_1288408, EPI_ISL_1288409, EPI_ISL_1288410, EPI_ISL_1288411, EPI_ISL_1288412, EPI_ISL_1288413, EPI_ISL_1288414, EPI_ISL_1288415, EPI_ISL_1288416, EPI_ISL_1288417, EPI_ISL_1288418, EPI_ISL_1288419, EPI_ISL_1288420, EPI_ISL_1288421, EPI_ISL_1288422, EPI_ISL_1288423, EPI_ISL_1288424, EPI_ISL_1288425, EPI_ISL_1288426, EPI_ISL_1288427, EPI_ISL_1288428, EPI_ISL_1288429, EPI_ISL_1288430, EPI_ISL_1288431, EPI_ISL_1288432, EPI_ISL_1288433, EPI_ISL_1288434, EPI_ISL_1288435, EPI_ISL_1288436, EPI_ISL_1288437, EPI_ISL_1288438, EPI_ISL_1288439, EPI_ISL_1288440, EPI_ISL_1288441, EPI_ISL_1288442, EPI_ISL_1288443, EPI_ISL_1288444, EPI_ISL_1288445, EPI_ISL_1288446, EPI_ISL_1288447, EPI_ISL_1288448, EPI_ISL_1288449, EPI_ISL_1288450, EPI_ISL_1288451, EPI_ISL_1288452, EPI_ISL_1288453, EPI_ISL_1288454, EPI_ISL_1288455, EPI_ISL_1288456, EPI_ISL_1288457, EPI_ISL_1288458, EPI_ISL_1288459, EPI_ISL_1288460, EPI_ISL_1288461, EPI_ISL_1288462, EPI_ISL_1288463, EPI_ISL_1288464, EPI_ISL_1288465, EPI_ISL_1288466, EPI_ISL_1288467, EPI_ISL_1288468, EPI_ISL_1288469, EPI_ISL_1288470, EPI_ISL_1288471, EPI_ISL_1288472, EPI_ISL_1288473, EPI_ISL_1288474, EPI_ISL_1288475, EPI_ISL_1288476, EPI_ISL_1288477, EPI_ISL_1288478, EPI_ISL_1288479, EPI_ISL_1288480, EPI_ISL_1288481, EPI_ISL_1288482, EPI_ISL_1288483, EPI_ISL_1288484, EPI_ISL_1288485, EPI_ISL_1288486, EPI_ISL_1288487, EPI_ISL_1288488, EPI_ISL_1288489, EPI_ISL_1288490, EPI_ISL_1288491, EPI_ISL_1288492, EPI_ISL_1288493, EPI_ISL_1288494, EPI_ISL_1288495, EPI_ISL_1288496, EPI_ISL_1288497, EPI_ISL_1288498, EPI_ISL_1288499, EPI_ISL_1288500, EPI_ISL_1288501, EPI_ISL_1288502, EPI_ISL_1288503, EPI_ISL_1288504, EPI_ISL_1288505, EPI_ISL_1288506, EPI_ISL_1288507, EPI_ISL_1288508, EPI_ISL_1288509, EPI_ISL_1288510, EPI_ISL_1288511, EPI_ISL_1288512, EPI_ISL_1288513, EPI_ISL_1288514, EPI_ISL_1288515, EPI_ISL_1288516, EPI_ISL_1288517, EPI_ISL_1288518, EPI_ISL_1288519, EPI_ISL_1288520, EPI_ISL_1288521, EPI_ISL_1288522, EPI_ISL_1288523, EPI_ISL_1288524, EPI_ISL_1288525, EPI_ISL_1288526, EPI_ISL_1288527, EPI_ISL_1288528, EPI_ISL_1288529, EPI_ISL_1288530, EPI_ISL_1288531, EPI_ISL_1288533 | | | |
| see above | Laboratorio Central de Epidemiología (LCE) | Instituto de Biotecnología de la UNAM | Consorcio Mexicano de Vigilancia Genómica (CoViGen-Mex). Authors (in alphabetical order): Julio Elias Alvarado-Yaah, Carlos F. Arias, Santiago Ávila-Ríos, Víctor Hugo Borja-Aburto, Celia Boukadida, Juan Bautista Chale-Dzul , José Antonio Enciso-Moreno, Gloria Elena Espinoza-Ayala, Fernando Fontove-Herrera, Concepción Grajales-Muñiz, Ricardo Grande, Alfredo Herrera-Estrella, Carla Ivón Herrera-Najera, Pavel Isa, Brenda Irasema Maldonado-Meza, Bernardo Martínez-Miguel, Margarita Matías-Florentino, María Guadalupe de Jesús Mireles-Rivera, Gloria María Molina-Salinas, Hector Montoya-Fuentes, José Esteban Muñoz-Medina, José de Jesús Nuñez-Contreras, Alicia Ocaña-Mondragón, Luis Alberto Ochoa-Carrera, Hector Esteban Paz-Juárez, Francisco Pulido, Helen Haydee Fernanda Ramírez-Plascencia, Angel Gustavo Salas-Lais, Jorge Ivan Salinal-Nevarez, Alejandro Sanchez-Flores, Clara Esperanza Santacruz-Tinoco, María Guadalupe Santiago-Mauricio , Nelly Sélem-Mojica, Blanca Taboada , Gloria Vazquez |
| EPI_ISL_1289447, EPI_ISL_1289450, EPI_ISL_1289451, EPI_ISL_1289452, EPI_ISL_1289453, EPI_ISL_1289454, EPI_ISL_1289455, EPI_ISL_1289456, EPI_ISL_1289457, EPI_ISL_1289458, EPI_ISL_1289459, EPI_ISL_1289460, EPI_ISL_1289461, EPI_ISL_1289477, EPI_ISL_1289478, EPI_ISL_1289479, EPI_ISL_1289480, EPI_ISL_1289481, EPI_ISL_1289482, EPI_ISL_1289483, EPI_ISL_1289485, EPI_ISL_1289486, EPI_ISL_1289487, EPI_ISL_1289488, EPI_ISL_1289489, EPI_ISL_1289490, EPI_ISL_1289491, EPI_ISL_1289492, EPI_ISL_1289493, EPI_ISL_1289494, EPI_ISL_1289496, EPI_ISL_1289497, EPI_ISL_1289498, EPI_ISL_1289499, EPI_ISL_1289500, EPI_ISL_1289501, EPI_ISL_1289503, EPI_ISL_1289504, EPI_ISL_1289505, EPI_ISL_1289507, EPI_ISL_1289508, EPI_ISL_1289509, EPI_ISL_1289510, EPI_ISL_1289511, EPI_ISL_1289512, EPI_ISL_1289513, EPI_ISL_1289514, EPI_ISL_1289515, EPI_ISL_1289516, EPI_ISL_1289517, EPI_ISL_1289518, EPI_ISL_1289519, EPI_ISL_1289520, EPI_ISL_1289523, EPI_ISL_1289524, EPI_ISL_1289525, EPI_ISL_1289526, EPI_ISL_1289527, EPI_ISL_1289528, EPI_ISL_1289529, EPI_ISL_1289530, EPI_ISL_1289531, EPI_ISL_1289532, EPI_ISL_1289533, EPI_ISL_1289534, EPI_ISL_1289535, EPI_ISL_1289536, EPI_ISL_1289537, EPI_ISL_1289538, EPI_ISL_1289539, EPI_ISL_1289540, EPI_ISL_1289541, EPI_ISL_1289542, EPI_ISL_1289543, EPI_ISL_1289544, EPI_ISL_1289545, EPI_ISL_1289546, EPI_ISL_1289547, EPI_ISL_1289548, EPI_ISL_1289549, EPI_ISL_1289550, EPI_ISL_1289551, EPI_ISL_1289552, EPI_ISL_1289553, EPI_ISL_1289554, EPI_ISL_1289555, EPI_ISL_1289556, EPI_ISL_1289557, EPI_ISL_1289558, EPI_ISL_1289559, EPI_ISL_1289560, EPI_ISL_1289561, EPI_ISL_1289562, EPI_ISL_1289563, EPI_ISL_1289568, EPI_ISL_1289569, EPI_ISL_1289570, EPI_ISL_1289571, EPI_ISL_1289572, EPI_ISL_1289573, EPI_ISL_1289574, EPI_ISL_1289575, EPI_ISL_1289577, EPI_ISL_1289578, EPI_ISL_1289579, EPI_ISL_1289580, EPI_ISL_1289581, EPI_ISL_1289582, EPI_ISL_1289583, EPI_ISL_1289584, EPI_ISL_1289585, EPI_ISL_1289586, EPI_ISL_1289587, EPI_ISL_1289588, EPI_ISL_1289589, EPI_ISL_1289590, EPI_ISL_1289591, EPI_ISL_1289592, EPI_ISL_1289593, EPI_ISL_1289594, EPI_ISL_1289597, EPI_ISL_1289598, EPI_ISL_1289599, EPI_ISL_1289600, EPI_ISL_1289601, EPI_ISL_1289602, EPI_ISL_1289603, EPI_ISL_1289604, EPI_ISL_1289605, EPI_ISL_1289606, EPI_ISL_1289607, EPI_ISL_1289608, EPI_ISL_1289609, EPI_ISL_1289610, EPI_ISL_1289611, EPI_ISL_1289613, EPI_ISL_1289614, EPI_ISL_1289615, EPI_ISL_1289616, EPI_ISL_1289617, EPI_ISL_1289618, EPI_ISL_1289619, EPI_ISL_1289620, EPI_ISL_1289621, EPI_ISL_1289622, EPI_ISL_1289623, EPI_ISL_1289624, EPI_ISL_1289625, EPI_ISL_1289626, EPI_ISL_1289627, EPI_ISL_1289628, EPI_ISL_1289630, EPI_ISL_1289633, EPI_ISL_1289634, EPI_ISL_1289636, EPI_ISL_1289637, EPI_ISL_1289638, EPI_ISL_1289639, EPI_ISL_1289640, EPI_ISL_1289641, EPI_ISL_1289642, EPI_ISL_1289643, EPI_ISL_1289644, EPI_ISL_1289645, EPI_ISL_1289646, EPI_ISL_1289649, EPI_ISL_1289650, EPI_ISL_1289651, EPI_ISL_1289652, EPI_ISL_1289653, EPI_ISL_1289654, EPI_ISL_1289655, EPI_ISL_1289656, EPI_ISL_1289657, EPI_ISL_1289658, EPI_ISL_1289659, EPI_ISL_1289660, EPI_ISL_1289662, EPI_ISL_1289663, EPI_ISL_1289664, EPI_ISL_1289665, EPI_ISL_1289666, EPI_ISL_1289667, EPI_ISL_1289668, EPI_ISL_1289669, EPI_ISL_1289670, EPI_ISL_1289671, EPI_ISL_1289672, EPI_ISL_1289673, EPI_ISL_1289674, EPI_ISL_1289675, EPI_ISL_1289676, EPI_ISL_1289678, EPI_ISL_1289680, EPI_ISL_1289681, EPI_ISL_1289682, EPI_ISL_1289683, EPI_ISL_1289684, EPI_ISL_1289685, EPI_ISL_1289686, EPI_ISL_1289687, EPI_ISL_1289688, EPI_ISL_1289689, EPI_ISL_1289692, EPI_ISL_1289693, EPI_ISL_1289694, EPI_ISL_1289695, EPI_ISL_1289701, EPI_ISL_1289705, EPI_ISL_1289706, EPI_ISL_1289707, EPI_ISL_1289708, EPI_ISL_1289709, EPI_ISL_1289710, EPI_ISL_1289711, EPI_ISL_1289712, EPI_ISL_1289713, EPI_ISL_1289714, EPI_ISL_1289715, EPI_ISL_1289716, EPI_ISL_1289717, EPI_ISL_1289718, EPI_ISL_1289719, EPI_ISL_1289720, EPI_ISL_1289721, EPI_ISL_1289722, EPI_ISL_1289723, EPI_ISL_1289724, EPI_ISL_1289725, EPI_ISL_1289727, EPI_ISL_1289728, EPI_ISL_1289729, EPI_ISL_1289730, EPI_ISL_1289731, EPI_ISL_1289732, EPI_ISL_1289733, EPI_ISL_1289734, EPI_ISL_1289735, EPI_ISL_1289736, EPI_ISL_1289737, EPI_ISL_1289738, EPI_ISL_1289739, EPI_ISL_1289740, EPI_ISL_1289741, EPI_ISL_1289742, EPI_ISL_1289743, EPI_ISL_1289744, EPI_ISL_1289745, EPI_ISL_1289746, EPI_ISL_1289748, EPI_ISL_1289749, EPI_ISL_1289750, EPI_ISL_1289751, EPI_ISL_1289752, EPI_ISL_1289753, EPI_ISL_1289754, EPI_ISL_1289755, EPI_ISL_1289757, EPI_ISL_1289758, EPI_ISL_1289759, EPI_ISL_1289760, EPI_ISL_1289761, EPI_ISL_1289762, EPI_ISL_1289763, EPI_ISL_1289764, EPI_ISL_1289765, EPI_ISL_1289766, EPI_ISL_1289767, EPI_ISL_1289768, EPI_ISL_1289769, EPI_ISL_1289770, EPI_ISL_1289771, EPI_ISL_1289772, EPI_ISL_1289773, EPI_ISL_1289774, EPI_ISL_1289775, EPI_ISL_1289776, EPI_ISL_1289777, EPI_ISL_1289778, EPI_ISL_1289779, EPI_ISL_1289780, EPI_ISL_1289781, EPI_ISL_1289782, EPI_ISL_1289783, EPI_ISL_1289784 | | | |
| see above | Michigan Department of Health and Human Services, Bureau of Laboratories | Michigan Department of Health and Human Services, Bureau of Laboratories | Blankenship HM, Riner D, Soehnlen MK |
| EPI_ISL_1289786, EPI_ISL_1289787, EPI_ISL_1289788 | The Jackson Laboratory | The Jackson Laboratory | Bergeron D, Renzette N, Adams M, Omerza G, Kelly K, Li L |
| EPI_ISL_1290809, EPI_ISL_1290810, EPI_ISL_1290811, EPI_ISL_1290812, EPI_ISL_1290813, EPI_ISL_1290814, EPI_ISL_1290815, EPI_ISL_1290816, EPI_ISL_1290817, EPI_ISL_1290818, EPI_ISL_1290819, EPI_ISL_1290820, EPI_ISL_1290821, EPI_ISL_1290822, EPI_ISL_1290823, EPI_ISL_1290824, EPI_ISL_1290825, EPI_ISL_1290826, EPI_ISL_1290827, EPI_ISL_1290828, EPI_ISL_1290829, EPI_ISL_1290830, EPI_ISL_1290831, EPI_ISL_1290832, EPI_ISL_1290833, EPI_ISL_1290834, EPI_ISL_1290835 | | | |
| see above | Oregon State University TRACE Project | OSU Center for Genome Research and Biocomputing | Oregon State University TRACE Project |
| EPI_ISL_1290855, EPI_ISL_1290856 | Microbiology Division, South Carolina Department of Health and Environmental Control Public Health | Microbiology Division, South Carolina Department of Health and Environmental Control Public Health Laboratory (SC DHEC | Haley Flores, Jessica Freeman |

| | | | |
|---|---|---|---|
| | Laboratory (SC DHEC PHL) | PHL) | |
| EPI_ISL_1290857, EPI_ISL_1290858, EPI_ISL_1290859, EPI_ISL_1290860 | Quest Diagnostics Incorporated | Respiratory Viruses Branch, Division of Viral Diseases, Centers for Disease Control and Prevention | Peter W. Cook, Dakota Howard, Dhwani Batra, Ben L. Rambo-Martin, S. H. Rosenthal, A. Gerasimova, R. M. Kagan, B. Anderson, M. Hua, Y. Liu, L.E. Bernstein, K.E. Livingston, A. Perez, I. A. Shlyakhter, R. V. Rolando, R. Owen, P. Tanpaiboon, F. Lacbawan, Clinton R. Paden, Suxiang Tong, Duncan MacCannell |
| EPI_ISL_1290896, EPI_ISL_1290922, EPI_ISL_1290956 | Montana Public Health Laboratory | Montana Public Health Laboratory | Joy Ritter, Michelle Mozer, Carrie Biskupiak, Deborah Gibson |
| EPI_ISL_1290976, EPI_ISL_1290977, EPI_ISL_1290978, EPI_ISL_1290979, EPI_ISL_1290980, EPI_ISL_1290981, EPI_ISL_1290982, EPI_ISL_1290983, EPI_ISL_1290984, EPI_ISL_1290985, EPI_ISL_1290986, EPI_ISL_1290987, EPI_ISL_1290988, EPI_ISL_1290989, EPI_ISL_1290990, EPI_ISL_1290991, EPI_ISL_1290992, EPI_ISL_1290993, EPI_ISL_1290994, EPI_ISL_1290995, EPI_ISL_1290996, EPI_ISL_1290997, EPI_ISL_1290998, EPI_ISL_1290999, EPI_ISL_1291000, EPI_ISL_1291001, EPI_ISL_1291002, EPI_ISL_1291003, EPI_ISL_1291004, EPI_ISL_1291005, EPI_ISL_1291006, EPI_ISL_1291007, EPI_ISL_1291008, EPI_ISL_1291009, EPI_ISL_1291010, EPI_ISL_1291011, EPI_ISL_1291012, EPI_ISL_1291013, EPI_ISL_1291014, EPI_ISL_1291015, EPI_ISL_1291016, EPI_ISL_1291017, EPI_ISL_1291018, EPI_ISL_1291019, EPI_ISL_1291020, EPI_ISL_1291021, EPI_ISL_1291022, EPI_ISL_1291023, EPI_ISL_1291024, EPI_ISL_1291025, EPI_ISL_1291026, EPI_ISL_1291027, EPI_ISL_1291028, EPI_ISL_1291029, EPI_ISL_1291030, EPI_ISL_1291031, EPI_ISL_1291032, EPI_ISL_1291033, EPI_ISL_1291034, EPI_ISL_1291035, EPI_ISL_1291036, EPI_ISL_1291037, EPI_ISL_1291038, EPI_ISL_1291039, EPI_ISL_1291040, EPI_ISL_1291041, EPI_ISL_1291042, EPI_ISL_1291043, EPI_ISL_1291044, EPI_ISL_1291045, EPI_ISL_1291046, EPI_ISL_1291047, EPI_ISL_1291048, EPI_ISL_1291049, EPI_ISL_1291050, EPI_ISL_1291051, EPI_ISL_1291052, EPI_ISL_1291053, EPI_ISL_1291054, EPI_ISL_1291055, EPI_ISL_1291056, EPI_ISL_1291057, EPI_ISL_1291058, EPI_ISL_1291059, EPI_ISL_1291060, EPI_ISL_1291061, EPI_ISL_1291062, EPI_ISL_1291063, EPI_ISL_1291064, EPI_ISL_1291065, EPI_ISL_1291066, EPI_ISL_1291067, EPI_ISL_1291068, EPI_ISL_1291069, EPI_ISL_1291070, EPI_ISL_1291071, EPI_ISL_1291072, EPI_ISL_1291073, EPI_ISL_1291074, EPI_ISL_1291075, EPI_ISL_1291076, EPI_ISL_1291077, EPI_ISL_1291078, EPI_ISL_1291079, EPI_ISL_1291080, EPI_ISL_1291081, EPI_ISL_1291082, EPI_ISL_1291083, EPI_ISL_1291084, EPI_ISL_1291085, EPI_ISL_1291086, EPI_ISL_1291087, EPI_ISL_1291088, EPI_ISL_1291089, EPI_ISL_1291090, EPI_ISL_1291091, EPI_ISL_1291092, EPI_ISL_1291093, EPI_ISL_1291094, EPI_ISL_1291095, EPI_ISL_1291096, EPI_ISL_1291097, EPI_ISL_1291098, EPI_ISL_1291099, EPI_ISL_1291100, EPI_ISL_1291101, EPI_ISL_1291102, EPI_ISL_1291103, EPI_ISL_1291104, EPI_ISL_1291105, EPI_ISL_1291106, EPI_ISL_1291107, EPI_ISL_1291108, EPI_ISL_1291109, EPI_ISL_1291110, EPI_ISL_1291111, EPI_ISL_1291112, EPI_ISL_1291113, EPI_ISL_1291114, EPI_ISL_1291115, EPI_ISL_1291116, EPI_ISL_1291117, EPI_ISL_1291118, EPI_ISL_1291119, EPI_ISL_1291120, EPI_ISL_1291121, EPI_ISL_1291122, EPI_ISL_1291123, EPI_ISL_1291124, EPI_ISL_1291125, EPI_ISL_1291126, EPI_ISL_1291127, EPI_ISL_1291128, EPI_ISL_1291129, EPI_ISL_1291130, EPI_ISL_1291131, EPI_ISL_1291132 | | | |
| see above | Quest Diagnostics Incorporated | Centers for Disease Control and Prevention Division of Viral Diseases, Pathogen Discovery | Peter W. Cook, Dakota Howard, Dhwani Batra, Ben L. Rambo-Martin, S. H. Rosenthal, A. Gerasimova, R. M. Kagan, B. Anderson, M. Hua, Y. Liu, L.E. Bernstein, K.E. Livingston, A. Perez, I. A. Shlyakhter, R. V. Rolando, R. Owen, P. Tanpaiboon, F. Lacbawan, Clinton R. Paden, Suxiang Tong, Duncan MacCannell |
| EPI_ISL_1291133 | Montana Public Health Laboratory | Montana Public Health Laboratory | Joy Ritter, Michelle Mozer, Carrie Biskupiak, Deborah Gibson |
| EPI_ISL_1291134, EPI_ISL_1291135, EPI_ISL_1291136, EPI_ISL_1291137, EPI_ISL_1291138, EPI_ISL_1291139, EPI_ISL_1291140, EPI_ISL_1291141, EPI_ISL_1291142, EPI_ISL_1291143, EPI_ISL_1291144, EPI_ISL_1291145, EPI_ISL_1291146, EPI_ISL_1291147, EPI_ISL_1291148, EPI_ISL_1291149, EPI_ISL_1291150, EPI_ISL_1291151, EPI_ISL_1291152, EPI_ISL_1291153, EPI_ISL_1291154, EPI_ISL_1291155, EPI_ISL_1291156, EPI_ISL_1291157, EPI_ISL_1291158, EPI_ISL_1291159, EPI_ISL_1291160, EPI_ISL_1291161, EPI_ISL_1291162, EPI_ISL_1291163, EPI_ISL_1291164, EPI_ISL_1291165, EPI_ISL_1291166, EPI_ISL_1291167, EPI_ISL_1291168, EPI_ISL_1291169, EPI_ISL_1291170, EPI_ISL_1291171, EPI_ISL_1291172, EPI_ISL_1291173, EPI_ISL_1291174, EPI_ISL_1291175, EPI_ISL_1291176, EPI_ISL_1291177, EPI_ISL_1291178, EPI_ISL_1291179, EPI_ISL_1291180, EPI_ISL_1291181, EPI_ISL_1291182, EPI_ISL_1291183, EPI_ISL_1291184, EPI_ISL_1291185, EPI_ISL_1291186, EPI_ISL_1291187, EPI_ISL_1291188 | | | |
| see above | Quest Diagnostics Incorporated | Centers for Disease Control and Prevention Division of Viral Diseases, Pathogen Discovery | Peter W. Cook, Dakota Howard, Dhwani Batra, Ben L. Rambo-Martin, S. H. Rosenthal, A. Gerasimova, R. M. Kagan, B. Anderson, M. Hua, Y. Liu, L.E. Bernstein, K.E. Livingston, A. Perez, I. A. Shlyakhter, R. V. Rolando, R. Owen, P. Tanpaiboon, F. Lacbawan, Clinton R. Paden, Suxiang Tong, Duncan MacCannell |
| EPI_ISL_1291189, EPI_ISL_1291190, EPI_ISL_1291191 | Montana Public Health Laboratory | Montana Public Health Laboratory | Joy Ritter, Michelle Mozer, Carrie Biskupiak, Deborah Gibson |
| EPI_ISL_1291199, EPI_ISL_1291201, EPI_ISL_1291203, EPI_ISL_1291206, EPI_ISL_1291207, EPI_ISL_1291208, EPI_ISL_1291209, EPI_ISL_1291214, EPI_ISL_1291215, EPI_ISL_1291216, EPI_ISL_1291219, EPI_ISL_1291220, EPI_ISL_1291221, EPI_ISL_1291224, EPI_ISL_1291226, EPI_ISL_1291227, EPI_ISL_1291231, EPI_ISL_1291233, EPI_ISL_1291234, EPI_ISL_1291239, EPI_ISL_1291240, EPI_ISL_1291242, EPI_ISL_1291243, EPI_ISL_1291244, EPI_ISL_1291245, EPI_ISL_1291246, EPI_ISL_1291247, EPI_ISL_1291248, EPI_ISL_1291249, EPI_ISL_1291251, EPI_ISL_1291252, EPI_ISL_1291253, EPI_ISL_1291254, EPI_ISL_1291255, EPI_ISL_1291256, EPI_ISL_1291257, EPI_ISL_1291258, EPI_ISL_1291261, EPI_ISL_1291263, EPI_ISL_1291265, EPI_ISL_1291266, EPI_ISL_1291268, EPI_ISL_1291270, EPI_ISL_1291274, EPI_ISL_1291277, EPI_ISL_1291279, EPI_ISL_1291281, EPI_ISL_1291284, EPI_ISL_1291291, EPI_ISL_1291294, EPI_ISL_1291295, EPI_ISL_1291296, EPI_ISL_1291297, EPI_ISL_1291298, EPI_ISL_1291299, EPI_ISL_1291300, EPI_ISL_1291301, EPI_ISL_1291302, EPI_ISL_1291303, EPI_ISL_1291304, EPI_ISL_1291305, EPI_ISL_1291306 | | | |
| see above | Laboratory Corporation of America | Centers for Disease Control and Prevention Division of Viral Diseases, Pathogen Discovery | Peter W. Cook, Dakota Howard, Dhwani Batra, Ben L. Rambo-Martin, Minoo Agarwal, Eyad Almasri Debbie Boles, Ayla Burns, Nuthawin Charoensri, Oren Cohen, Susan Countryman, Mary Ann Cristobal, Bobbi Croy, Suzanne Dale, Hrushikesh Deshmukh, Amanda Douglas, Vincent Drouillon, Marcia Eisenberg, Howard Engler, Rama Ghatti, Prashant Gupta, Susan Hicks, Jake Humphrey, Lax Iyer, Manoj Jain, Mohan Kolli, Brian Krueger, Tim Kuphal, Stanley Letovsky, Michael Levandoski, Craig Lukasik, Jonathan Meltzer, Brian Norvell, Mindy Nye, Scott Parker, Christos Petropoulos, John Pruitt, Steven Ragan, Scott Ryan, Mike Sapeta, Jana Schroth, Suresh Babu Selvaraju, Goran Stevovic, Amanda Suchanek, Andrea Throop, Lyndon Tilson, Thomas Urban, Joe Voshell, Kimberly Wagner, Jonathan Williams, Mary Williamson, Qian Zeng, Tricia Zwiefelhofer, Clinton R. Paden, Suxiang Tong, Duncan MacCannell |
| EPI_ISL_1291307, EPI_ISL_1291308, EPI_ISL_1291309, EPI_ISL_1291310, EPI_ISL_1291311, EPI_ISL_1291312, EPI_ISL_1291313, EPI_ISL_1291314, EPI_ISL_1291315, EPI_ISL_1291316, EPI_ISL_1291317, EPI_ISL_1291318, EPI_ISL_1291319, EPI_ISL_1291320, EPI_ISL_1291321, EPI_ISL_1291322, EPI_ISL_1291323, EPI_ISL_1291324, EPI_ISL_1291325, EPI_ISL_1291326, EPI_ISL_1291327, EPI_ISL_1291328, EPI_ISL_1291329, EPI_ISL_1291330, EPI_ISL_1291331, EPI_ISL_1291333, EPI_ISL_1291334, EPI_ISL_1291335, EPI_ISL_1291336, EPI_ISL_1291337, EPI_ISL_1291338, EPI_ISL_1291339, EPI_ISL_1291340, EPI_ISL_1291341, EPI_ISL_1291342, EPI_ISL_1291343, EPI_ISL_1291344, EPI_ISL_1291346, EPI_ISL_1291347, EPI_ISL_1291348, EPI_ISL_1291349, EPI_ISL_1291350, EPI_ISL_1291351, EPI_ISL_1291352, EPI_ISL_1291353, EPI_ISL_1291354, EPI_ISL_1291355, EPI_ISL_1291356, EPI_ISL_1291357, EPI_ISL_1291358, EPI_ISL_1291359, EPI_ISL_1291360, EPI_ISL_1291361, EPI_ISL_1291362, EPI_ISL_1291363, EPI_ISL_1291364, EPI_ISL_1291365, EPI_ISL_1291366, EPI_ISL_1291367, EPI_ISL_1291368, EPI_ISL_1291369, EPI_ISL_1291370, EPI_ISL_1291371, EPI_ISL_1291372, EPI_ISL_1291373, EPI_ISL_1291374, EPI_ISL_1291375, EPI_ISL_1291376 | | | |
| see above | TXDSHS | TXDSHS | Rashmi Tuladhar, Bonnie Oh, Jenny Zhang, Maliha Rahman, Mayela Pedrueza, Anita Pokharel, Myong Koag, Chun Wang, Rachel Lee, Grace Kubin |
| EPI_ISL_1291378, EPI_ISL_1291379, EPI_ISL_1291380, EPI_ISL_1291381, EPI_ISL_1291382, EPI_ISL_1291383, EPI_ISL_1291384, EPI_ISL_1291385, EPI_ISL_1291386, EPI_ISL_1291387, EPI_ISL_1291388, EPI_ISL_1291389, EPI_ISL_1291390, EPI_ISL_1291391, EPI_ISL_1291392, EPI_ISL_1291393, EPI_ISL_1291394, EPI_ISL_1291395, EPI_ISL_1291396, EPI_ISL_1291397, EPI_ISL_1291398, EPI_ISL_1291399, EPI_ISL_1291400, EPI_ISL_1291401, EPI_ISL_1291402, EPI_ISL_1291403, EPI_ISL_1291404, EPI_ISL_1291405, EPI_ISL_1291406, EPI_ISL_1291407, EPI_ISL_1291408, EPI_ISL_1291409, EPI_ISL_1291410, EPI_ISL_1291411, EPI_ISL_1291412, EPI_ISL_1291413, EPI_ISL_1291414, EPI_ISL_1291415, EPI_ISL_1291416, EPI_ISL_1291417, EPI_ISL_1291418, EPI_ISL_1291419, EPI_ISL_1291420, EPI_ISL_1291421, EPI_ISL_1291422, EPI_ISL_1291423, EPI_ISL_1291424, EPI_ISL_1291425, EPI_ISL_1291426, EPI_ISL_1291427, EPI_ISL_1291428, EPI_ISL_1291429, EPI_ISL_1291430, EPI_ISL_1291431, EPI_ISL_1291432, EPI_ISL_1291433, EPI_ISL_1291434, EPI_ISL_1291435, EPI_ISL_1291436, EPI_ISL_1291437, EPI_ISL_1291438, EPI_ISL_1291439, EPI_ISL_1291440, EPI_ISL_1291441, EPI_ISL_1291442, EPI_ISL_1291443, EPI_ISL_1291444, EPI_ISL_1291445, EPI_ISL_1291446, EPI_ISL_1291447, EPI_ISL_1291448, EPI_ISL_1291449, EPI_ISL_1291450, EPI_ISL_1291451, EPI_ISL_1291452, EPI_ISL_1291453, EPI_ISL_1291454, EPI_ISL_1291455, EPI_ISL_1291456, EPI_ISL_1291457, EPI_ISL_1291458, EPI_ISL_1291459, EPI_ISL_1291460, EPI_ISL_1291461, EPI_ISL_1291462, EPI_ISL_1291463, EPI_ISL_1291464, EPI_ISL_1291465, EPI_ISL_1291466, EPI_ISL_1291467, EPI_ISL_1291468, EPI_ISL_1291469, EPI_ISL_1291470, EPI_ISL_1291471, EPI_ISL_1291472, EPI_ISL_1291473, EPI_ISL_1291474, EPI_ISL_1291475, EPI_ISL_1291476, EPI_ISL_1291477, EPI_ISL_1291478, EPI_ISL_1291479, EPI_ISL_1291480, EPI_ISL_1291481, EPI_ISL_1291482, EPI_ISL_1291483, EPI_ISL_1291484, EPI_ISL_1291485, EPI_ISL_1291486, EPI_ISL_1291487, EPI_ISL_1291488, EPI_ISL_1291489, EPI_ISL_1291490, EPI_ISL_1291491, EPI_ISL_1291492, EPI_ISL_1291493, EPI_ISL_1291494, EPI_ISL_1291495, EPI_ISL_1291496, EPI_ISL_1291497, EPI_ISL_1291498, EPI_ISL_1291499, EPI_ISL_1291500, EPI_ISL_1291501, EPI_ISL_1291502, EPI_ISL_1291503, EPI_ISL_1291504, EPI_ISL_1291505, EPI_ISL_1291506, EPI_ISL_1291507, EPI_ISL_1291508, EPI_ISL_1291509, EPI_ISL_1291510, EPI_ISL_1291511, EPI_ISL_1291512, EPI_ISL_1291513, EPI_ISL_1291514, EPI_ISL_1291515, EPI_ISL_1291516, EPI_ISL_1291517, EPI_ISL_1291518, EPI_ISL_1291519, EPI_ISL_1291520, EPI_ISL_1291521, EPI_ISL_1291522, EPI_ISL_1291523, EPI_ISL_1291524, EPI_ISL_1291525, EPI_ISL_1291526, EPI_ISL_1291527, EPI_ISL_1291528, EPI_ISL_1291529, EPI_ISL_1291530, EPI_ISL_1291531, EPI_ISL_1291532, EPI_ISL_1291533, EPI_ISL_1291534, EPI_ISL_1291535, EPI_ISL_1291536, EPI_ISL_1291537, EPI_ISL_1291538, EPI_ISL_1291539, EPI_ISL_1291540, EPI_ISL_1291541 | | | |
| see above | Quest Diagnostics Incorporated | Centers for Disease Control and Prevention Division of Viral Diseases, Pathogen Discovery | Peter W. Cook, Dakota Howard, Dhwani Batra, Ben L. Rambo-Martin, S. H. Rosenthal, A. Gerasimova, R. M. Kagan, B. Anderson, M. Hua, Y. Liu, L.E. Bernstein, K.E. Livingston, A. Perez, I. A. Shlyakhter, R. V. Rolando, R. Owen, P. Tanpaiboon, F. Lacbawan, Clinton R. Paden, Suxiang Tong, Duncan MacCannell |
| EPI_ISL_1291542, EPI_ISL_1291544, EPI_ISL_1291545, EPI_ISL_1291546, EPI_ISL_1291547, EPI_ISL_1291548 | Nebraska Public Health Laboratory | NPHL COVID-19 Response Team | NPHL COVID-19 Response Team |
| EPI_ISL_1291558, EPI_ISL_1291559, EPI_ISL_1291560, EPI_ISL_1291561, EPI_ISL_1291562, EPI_ISL_1291563, EPI_ISL_1291564, EPI_ISL_1291565, EPI_ISL_1291566, EPI_ISL_1291567, EPI_ISL_1291568, EPI_ISL_1291569, EPI_ISL_1291570, EPI_ISL_1291571, EPI_ISL_1291572, EPI_ISL_1291573, EPI_ISL_1291574, EPI_ISL_1291575, EPI_ISL_1291576, EPI_ISL_1291577, EPI_ISL_1291578, EPI_ISL_1291579, EPI_ISL_1291580, EPI_ISL_1291581, EPI_ISL_1291582 | | | |
| see above | Quest Diagnostics Incorporated | Centers for Disease Control and Prevention Division of Viral Diseases, Pathogen Discovery | Peter W. Cook, Dakota Howard, Dhwani Batra, Ben L. Rambo-Martin, S. H. Rosenthal, A. Gerasimova, R. M. Kagan, B. Anderson, M. Hua, Y. Liu, L.E. Bernstein, K.E. Livingston, A. Perez, I. A. Shlyakhter, R. V. Rolando, R. Owen, P. Tanpaiboon, F. Lacbawan, Clinton R. Paden, Suxiang Tong, Duncan MacCannell |
| EPI_ISL_1291583, EPI_ISL_1291584, EPI_ISL_1291585, EPI_ISL_1291586, EPI_ISL_1291587, EPI_ISL_1291588, EPI_ISL_1291589, EPI_ISL_1291590, EPI_ISL_1291591, EPI_ISL_1291592, EPI_ISL_1291593, EPI_ISL_1291594, EPI_ISL_1291595, EPI_ISL_1291596, EPI_ISL_1291597, EPI_ISL_1291598, EPI_ISL_1291599, EPI_ISL_1291600, EPI_ISL_1291601, EPI_ISL_1291602, EPI_ISL_1291603, EPI_ISL_1291604, EPI_ISL_1291605, EPI_ISL_1291606 | | | |
| see above | University of Michigan Clinical Microbiology Laboratory | Lauring Lab, University of Michigan, Department of Microbiology and Immunology | Valesano |
| EPI_ISL_1291607, EPI_ISL_1291608, EPI_ISL_1291609, EPI_ISL_1291610, EPI_ISL_1291611, EPI_ISL_1291612, EPI_ISL_1291613, EPI_ISL_1291614, EPI_ISL_1291615, EPI_ISL_1291616, EPI_ISL_1291617, EPI_ISL_1291618, EPI_ISL_1291619, EPI_ISL_1291620, EPI_ISL_1291621, EPI_ISL_1291622, EPI_ISL_1291623, EPI_ISL_1291625, EPI_ISL_1291626, EPI_ISL_1291627, EPI_ISL_1291628, EPI_ISL_1291629, EPI_ISL_1291630, EPI_ISL_1291631, EPI_ISL_1291632, EPI_ISL_1291633, EPI_ISL_1291634, EPI_ISL_1291635, EPI_ISL_1291636, EPI_ISL_1291637, EPI_ISL_1291638, EPI_ISL_1291639, EPI_ISL_1291640, EPI_ISL_1291641, EPI_ISL_1291642, EPI_ISL_1291643, EPI_ISL_1291644, EPI_ISL_1291645, EPI_ISL_1291646, EPI_ISL_1291647, EPI_ISL_1291648, EPI_ISL_1291649, EPI_ISL_1291650, EPI_ISL_1291651 | | | |
| see above | Univeristy of New Mexico Hospital | Center for Global Health, University of New Mexico Health Sciences Center | Daryl Domman, Kurt Schwalm, Justin Bacca, Jon Femling, Darrell Dinwiddie |
| EPI_ISL_1291652 | Microbiology Division, South Carolina Department of Health and Environmental Control Public Health Laboratory (SC DHEC PHL) | Microbiology Division, South Carolina Department of Health and Environmental Control Public Health Laboratory (SC DHEC PHL) | Haley Flores, Jessica Freeman |
| EPI_ISL_1291653, EPI_ISL_1291654, EPI_ISL_1291655, EPI_ISL_1291656 | Maryland Genomics, Institute for Genome Sciences, University of Maryland School of Medicine | Maryland Genomics, Institute for Genome Sciences, University of Maryland School of Medicine | Tallon, Luke J; Sadzewicz, Lisa D; Humphrys, Mike; Ott, Sandra; Roussey, Holly; Mehta, Aditya; Vavikolanu, Kranthi; Fraser, Claire M; Ravel, Jacques |
| EPI_ISL_1291657, EPI_ISL_1291658, EPI_ISL_1291659, EPI_ISL_1291660, EPI_ISL_1291661, EPI_ISL_1291662, EPI_ISL_1291663, EPI_ISL_1291664, EPI_ISL_1291665, EPI_ISL_1291666, EPI_ISL_1291667, EPI_ISL_1291668, EPI_ISL_1291669, EPI_ISL_1291670, EPI_ISL_1291671, EPI_ISL_1291672, EPI_ISL_1291673, EPI_ISL_1291674, EPI_ISL_1291675, EPI_ISL_1291676, EPI_ISL_1291677, EPI_ISL_1291678, EPI_ISL_1291679, EPI_ISL_1291680, EPI_ISL_1291681, EPI_ISL_1291682, EPI_ISL_1291683, EPI_ISL_1291684, EPI_ISL_1291685, EPI_ISL_1291686, EPI_ISL_1291687, EPI_ISL_1291688, EPI_ISL_1291689, EPI_ISL_1291690, EPI_ISL_1291691, EPI_ISL_1291692, EPI_ISL_1291693, EPI_ISL_1291694, EPI_ISL_1291695, EPI_ISL_1291696, EPI_ISL_1291697, EPI_ISL_1291698, EPI_ISL_1291699, EPI_ISL_1291700, EPI_ISL_1291701, EPI_ISL_1291702, EPI_ISL_1291703, EPI_ISL_1291704, EPI_ISL_1291705, EPI_ISL_1291706, EPI_ISL_1291707, EPI_ISL_1291708, EPI_ISL_1291709, EPI_ISL_1291710, EPI_ISL_1291711, EPI_ISL_1291712, EPI_ISL_1291713, EPI_ISL_1291715, EPI_ISL_1291716, EPI_ISL_1291717, EPI_ISL_1291718, EPI_ISL_1291719, EPI_ISL_1291721, EPI_ISL_1291722, EPI_ISL_1291723, EPI_ISL_1291724, EPI_ISL_1291725, EPI_ISL_1291726, EPI_ISL_1291727, EPI_ISL_1291728, EPI_ISL_1291729, EPI_ISL_1291730, EPI_ISL_1291731, EPI_ISL_1291732, EPI_ISL_1291733, EPI_ISL_1291734, EPI_ISL_1291735, EPI_ISL_1291736, EPI_ISL_1291737, EPI_ISL_1291738, EPI_ISL_1291739, EPI_ISL_1291740 | | | |
| see above | Arizona State University | Arizona State University | Peter T. Skidmore, LaRinda A. Holland, Rabia Maqsood, Emily A. Kaelin, Nicholas J. Mellor, Kristina Buss, Joy M. Blain, Neal W Woodbury, Valerie Harris, Joshua LaBaer, Vel Murugan, Efrem S. Lim |
| EPI_ISL_1291741 | Montana Public Health Laboratory | Montana Public Health Laboratory | Joy Ritter, Michelle Mozer, Carrie Biskupiak, Deborah Gibson. |
| EPI_ISL_1291742 | Montana Public Health Laboratory | Montana Public Health Laboratory | Joy Ritter, Michelle Mozer, Carrie Biskupiak, Deborah Gibson |
| EPI_ISL_1291743 | Santa Clara County Public Health Laboratory | Santa Clara County Public Health Laboratory | Santa Clara County Public Health Department |
| EPI_ISL_1291744, EPI_ISL_1291746, EPI_ISL_1291748, EPI_ISL_1291749, EPI_ISL_1291750, EPI_ISL_1291751, EPI_ISL_1291752, EPI_ISL_1291753, EPI_ISL_1291754 | Montana Public Health Laboratory | Montana Public Health Laboratory | Joy Ritter, Michelle Mozer, Carrie Biskupiak, Deborah Gibson |

| | | | |
|---|---|---|---|
| EPI_ISL_1292089 | Baylor Scott & White-Irving | Baylor Scott & White-Temple | Ari Rao, Linden Morales, Kimberly Walker, Marcus Volz, Shelby Hendrickson, Caitlin Maloney |
| EPI_ISL_1292090, EPI_ISL_1292091, EPI_ISL_1292096, EPI_ISL_1292097, EPI_ISL_1292099, EPI_ISL_1292100, EPI_ISL_1292101, EPI_ISL_1292102, EPI_ISL_1292103, EPI_ISL_1292104, EPI_ISL_1292109, EPI_ISL_1292110, EPI_ISL_1292112, EPI_ISL_1292114, EPI_ISL_1292117, EPI_ISL_1292119, EPI_ISL_1292122, EPI_ISL_1292123, EPI_ISL_1292124, EPI_ISL_1292125, EPI_ISL_1292126, EPI_ISL_1292131, EPI_ISL_1292132, EPI_ISL_1292134, EPI_ISL_1292136, EPI_ISL_1292137, EPI_ISL_1292138, EPI_ISL_1292139, EPI_ISL_1292140, EPI_ISL_1292143, EPI_ISL_1292144, EPI_ISL_1292147, EPI_ISL_1292151, EPI_ISL_1292153, EPI_ISL_1292155, EPI_ISL_1292156, EPI_ISL_1292157, EPI_ISL_1292158, EPI_ISL_1292159, EPI_ISL_1292160, EPI_ISL_1292161, EPI_ISL_1292163, EPI_ISL_1292166, EPI_ISL_1292168, EPI_ISL_1292171, EPI_ISL_1292172, EPI_ISL_1292173, EPI_ISL_1292175, EPI_ISL_1292176, EPI_ISL_1292177, EPI_ISL_1292179, EPI_ISL_1292181, EPI_ISL_1292184, EPI_ISL_1292185, EPI_ISL_1292186, EPI_ISL_1292187, EPI_ISL_1292188, EPI_ISL_1292189, EPI_ISL_1292190, EPI_ISL_1292191, EPI_ISL_1292192, EPI_ISL_1292193, EPI_ISL_1292194, EPI_ISL_1292195, EPI_ISL_1292196, EPI_ISL_1292197, EPI_ISL_1292198, EPI_ISL_1292199, EPI_ISL_1292200, EPI_ISL_1292201, EPI_ISL_1292202, EPI_ISL_1292203, EPI_ISL_1292204, EPI_ISL_1292205, EPI_ISL_1292206, EPI_ISL_1292207, EPI_ISL_1292208, EPI_ISL_1292209, EPI_ISL_1292210, EPI_ISL_1292211, EPI_ISL_1292212, EPI_ISL_1292213, EPI_ISL_1292214, EPI_ISL_1292215, EPI_ISL_1292216, EPI_ISL_1292218, EPI_ISL_1292219, EPI_ISL_1292220, EPI_ISL_1292222, EPI_ISL_1292223, EPI_ISL_1292226, EPI_ISL_1292228, EPI_ISL_1292231, EPI_ISL_1292234, EPI_ISL_1292235, EPI_ISL_1292238, EPI_ISL_1292242, EPI_ISL_1292243, EPI_ISL_1292244, EPI_ISL_1292245, EPI_ISL_1292248, EPI_ISL_1292250, EPI_ISL_1292251, EPI_ISL_1292252, EPI_ISL_1292253, EPI_ISL_1292254, EPI_ISL_1292255, EPI_ISL_1292256, EPI_ISL_1292257, EPI_ISL_1292258, EPI_ISL_1292261, EPI_ISL_1292262, EPI_ISL_1292263, EPI_ISL_1292265, EPI_ISL_1292268, EPI_ISL_1292270, EPI_ISL_1292274, EPI_ISL_1292275, EPI_ISL_1292277, EPI_ISL_1292279, EPI_ISL_1292281, EPI_ISL_1292282, EPI_ISL_1292283, EPI_ISL_1292285, EPI_ISL_1292286, EPI_ISL_1292287, EPI_ISL_1292288, EPI_ISL_1292289, EPI_ISL_1292290, EPI_ISL_1292291, EPI_ISL_1292292, EPI_ISL_1292293, EPI_ISL_1292295, EPI_ISL_1292296, EPI_ISL_1292298, EPI_ISL_1292299, EPI_ISL_1292301, EPI_ISL_1292303, EPI_ISL_1292305, EPI_ISL_1292306, EPI_ISL_1292307, EPI_ISL_1292312, EPI_ISL_1292314, EPI_ISL_1292317, EPI_ISL_1292318, EPI_ISL_1292320, EPI_ISL_1292321, EPI_ISL_1292323, EPI_ISL_1292324, EPI_ISL_1292325, EPI_ISL_1292326, EPI_ISL_1292327, EPI_ISL_1292329, EPI_ISL_1292330, EPI_ISL_1292331, EPI_ISL_1292333, EPI_ISL_1292334, EPI_ISL_1292339, EPI_ISL_1292341, EPI_ISL_1292342, EPI_ISL_1292343, EPI_ISL_1292344, EPI_ISL_1292347, EPI_ISL_1292349, EPI_ISL_1292350, EPI_ISL_1292351, EPI_ISL_1292352, EPI_ISL_1292354, EPI_ISL_1292355, EPI_ISL_1292356, EPI_ISL_1292357, EPI_ISL_1292358, EPI_ISL_1292359 | | | |
| see above | Sonora Quest Laboratories | TGen North | "Jolene Bowers, Heather Centner, Chris French, Hayley Yaglom, Ashlyn Pfeiffer, Darrin Lemmer, Dave Engelthaler, The Arizona COVID Genomics Union (ACGU)" |
| EPI_ISL_1292360 | TGen North | TGen North | "Jolene Bowers, Heather Centner, Chris French, Hayley Yaglom, Ashlyn Pfeiffer, Darrin Lemmer, Dave Engelthaler, The Arizona COVID Genomics Union (ACGU)" |
| EPI_ISL_1292362, EPI_ISL_1292363, EPI_ISL_1292364, EPI_ISL_1292365, EPI_ISL_1292366 | Sonora Quest Laboratories | TGen North | "Jolene Bowers, Heather Centner, Chris French, Hayley Yaglom, Ashlyn Pfeiffer, Darrin Lemmer, Dave Engelthaler, The Arizona COVID Genomics Union (ACGU)" |
| EPI_ISL_1292367, EPI_ISL_1292370 | TGen North | TGen North | "Jolene Bowers, Heather Centner, Chris French, Hayley Yaglom, Ashlyn Pfeiffer, Darrin Lemmer, Dave Engelthaler, The Arizona COVID Genomics Union (ACGU)" |
| EPI_ISL_1292374, EPI_ISL_1292376, EPI_ISL_1292377, EPI_ISL_1292380, EPI_ISL_1292381, EPI_ISL_1292382, EPI_ISL_1292383 | Sonora Quest Laboratories | TGen North | "Jolene Bowers, Heather Centner, Chris French, Hayley Yaglom, Ashlyn Pfeiffer, Darrin Lemmer, Dave Engelthaler, The Arizona COVID Genomics Union (ACGU)" |
| EPI_ISL_1292384, EPI_ISL_1292386, EPI_ISL_1292387 | TGen North | TGen North | "Jolene Bowers, Heather Centner, Chris French, Hayley Yaglom, Ashlyn Pfeiffer, Darrin Lemmer, Dave Engelthaler, The Arizona COVID Genomics Union (ACGU)" |
| EPI_ISL_1292390, EPI_ISL_1292392, EPI_ISL_1292394 | Sonora Quest Laboratories | TGen North | "Jolene Bowers, Heather Centner, Chris French, Hayley Yaglom, Ashlyn Pfeiffer, Darrin Lemmer, Dave Engelthaler, The Arizona COVID Genomics Union (ACGU)" |
| EPI_ISL_1292397, EPI_ISL_1292398 | TGen North | TGen North | "Jolene Bowers, Heather Centner, Chris French, Hayley Yaglom, Ashlyn Pfeiffer, Darrin Lemmer, Dave Engelthaler, The Arizona COVID Genomics Union (ACGU)" |
| EPI_ISL_1292399, EPI_ISL_1292400, EPI_ISL_1292402 | Sonora Quest Laboratories | TGen North | "Jolene Bowers, Heather Centner, Chris French, Hayley Yaglom, Ashlyn Pfeiffer, Darrin Lemmer, Dave Engelthaler, The Arizona COVID Genomics Union (ACGU)" |
| EPI_ISL_1292403, EPI_ISL_1292404 | TGen North | TGen North | "Jolene Bowers, Heather Centner, Chris French, Hayley Yaglom, Ashlyn Pfeiffer, Darrin Lemmer, Dave Engelthaler, The Arizona COVID Genomics Union (ACGU)" |
| EPI_ISL_1292405, EPI_ISL_1292408, EPI_ISL_1292409, EPI_ISL_1292410 | Sonora Quest Laboratories | TGen North | "Jolene Bowers, Heather Centner, Chris French, Hayley Yaglom, Ashlyn Pfeiffer, Darrin Lemmer, Dave Engelthaler, The Arizona COVID Genomics Union (ACGU)" |
| EPI_ISL_1292411 | TGen North | TGen North | "Jolene Bowers, Heather Centner, Chris French, Hayley Yaglom, Ashlyn Pfeiffer, Darrin Lemmer, Dave Engelthaler, The Arizona COVID Genomics Union (ACGU)" |
| EPI_ISL_1292412, EPI_ISL_1292414, EPI_ISL_1292416, EPI_ISL_1292417, EPI_ISL_1292418, EPI_ISL_1292419 | Sonora Quest Laboratories | TGen North | "Jolene Bowers, Heather Centner, Chris French, Hayley Yaglom, Ashlyn Pfeiffer, Darrin Lemmer, Dave Engelthaler, The Arizona COVID Genomics Union (ACGU)" |
| EPI_ISL_1292420 | TGen North | TGen North | "Jolene Bowers, Heather Centner, Chris French, Hayley Yaglom, Ashlyn Pfeiffer, Darrin Lemmer, Dave Engelthaler, The Arizona COVID Genomics Union (ACGU)" |
| EPI_ISL_1292421, EPI_ISL_1292424 | Sonora Quest Laboratories | TGen North | "Jolene Bowers, Heather Centner, Chris French, Hayley Yaglom, Ashlyn Pfeiffer, Darrin Lemmer, Dave Engelthaler, The Arizona COVID Genomics Union (ACGU)" |
| EPI_ISL_1292425, EPI_ISL_1292426, EPI_ISL_1292427, EPI_ISL_1292428, EPI_ISL_1292429, EPI_ISL_1292430, EPI_ISL_1292431, EPI_ISL_1292432, EPI_ISL_1292433, EPI_ISL_1292434, EPI_ISL_1292435, EPI_ISL_1292436, EPI_ISL_1292437, EPI_ISL_1292438, EPI_ISL_1292439, EPI_ISL_1292440, EPI_ISL_1292441, EPI_ISL_1292442, EPI_ISL_1292443, EPI_ISL_1292444, EPI_ISL_1292445, EPI_ISL_1292448, EPI_ISL_1292449, EPI_ISL_1292450, EPI_ISL_1292451, EPI_ISL_1292452, EPI_ISL_1292454, EPI_ISL_1292455, EPI_ISL_1292456, EPI_ISL_1292457, EPI_ISL_1292458, EPI_ISL_1292459, EPI_ISL_1292460, EPI_ISL_1292461, EPI_ISL_1292462, EPI_ISL_1292463, EPI_ISL_1292464, EPI_ISL_1292465, EPI_ISL_1292467, EPI_ISL_1292468, EPI_ISL_1292469, EPI_ISL_1292471, EPI_ISL_1292472, EPI_ISL_1292473, EPI_ISL_1292474, EPI_ISL_1292475, EPI_ISL_1292476, EPI_ISL_1292477, EPI_ISL_1292478, EPI_ISL_1292479, EPI_ISL_1292480, EPI_ISL_1292481, EPI_ISL_1292482, EPI_ISL_1292483, EPI_ISL_1292484, EPI_ISL_1292485, EPI_ISL_1292486, EPI_ISL_1292487, EPI_ISL_1292488, EPI_ISL_1292489, EPI_ISL_1292490, EPI_ISL_1292491, EPI_ISL_1292492, EPI_ISL_1292494, EPI_ISL_1292496, EPI_ISL_1292497, EPI_ISL_1292499, EPI_ISL_1292500, EPI_ISL_1292501, EPI_ISL_1292502, EPI_ISL_1292503, EPI_ISL_1292504, EPI_ISL_1292506, EPI_ISL_1292507, EPI_ISL_1292508, EPI_ISL_1292509, EPI_ISL_1292511, EPI_ISL_1292512, EPI_ISL_1292513, EPI_ISL_1292514, EPI_ISL_1292515, EPI_ISL_1292516, EPI_ISL_1292517, EPI_ISL_1292518, EPI_ISL_1292520, EPI_ISL_1292522, EPI_ISL_1292523, EPI_ISL_1292524, EPI_ISL_1292525, EPI_ISL_1292526, EPI_ISL_1292527, EPI_ISL_1292528, EPI_ISL_1292529, EPI_ISL_1292530, EPI_ISL_1292531, EPI_ISL_1292532, EPI_ISL_1292533, EPI_ISL_1292534, EPI_ISL_1292536, EPI_ISL_1292537, EPI_ISL_1292538, EPI_ISL_1292539, EPI_ISL_1292540, EPI_ISL_1292541, EPI_ISL_1292542, EPI_ISL_1292543, EPI_ISL_1292544, EPI_ISL_1292545, EPI_ISL_1292548, EPI_ISL_1292549, EPI_ISL_1292551, EPI_ISL_1292554, EPI_ISL_1292555, EPI_ISL_1292556, EPI_ISL_1292558, EPI_ISL_1292559, EPI_ISL_1292560, EPI_ISL_1292562, EPI_ISL_1292563, EPI_ISL_1292564, EPI_ISL_1292566, EPI_ISL_1292567, EPI_ISL_1292568, EPI_ISL_1292569, EPI_ISL_1292570, EPI_ISL_1292571, EPI_ISL_1292572, EPI_ISL_1292573, EPI_ISL_1292574, EPI_ISL_1292575, EPI_ISL_1292576, EPI_ISL_1292577, EPI_ISL_1292578, EPI_ISL_1292579, EPI_ISL_1292580, EPI_ISL_1292581, EPI_ISL_1292582, EPI_ISL_1292583, EPI_ISL_1292584, EPI_ISL_1292585, EPI_ISL_1292586, EPI_ISL_1292587, EPI_ISL_1292588, EPI_ISL_1292589, EPI_ISL_1292590, EPI_ISL_1292591, EPI_ISL_1292592, EPI_ISL_1292593, EPI_ISL_1292594, EPI_ISL_1292595, EPI_ISL_1292596, EPI_ISL_1292597, EPI_ISL_1292598, EPI_ISL_1292599, EPI_ISL_1292600, EPI_ISL_1292601, EPI_ISL_1292602, EPI_ISL_1292603, EPI_ISL_1292604, EPI_ISL_1292605, EPI_ISL_1292606, EPI_ISL_1292607, EPI_ISL_1292608, EPI_ISL_1292609, EPI_ISL_1292611, EPI_ISL_1292612, EPI_ISL_1292613, EPI_ISL_1292614, EPI_ISL_1292615, EPI_ISL_1292616, EPI_ISL_1292617, EPI_ISL_1292618, EPI_ISL_1292619, EPI_ISL_1292620, EPI_ISL_1292621, EPI_ISL_1292622, EPI_ISL_1292623, EPI_ISL_1292624, EPI_ISL_1292625, EPI_ISL_1292626, EPI_ISL_1292627, EPI_ISL_1292628, EPI_ISL_1292629, EPI_ISL_1292630, EPI_ISL_1292631, EPI_ISL_1292632, EPI_ISL_1292633, EPI_ISL_1292634, EPI_ISL_1292635, EPI_ISL_1292636, EPI_ISL_1292637, EPI_ISL_1292638, EPI_ISL_1292639, EPI_ISL_1292640, EPI_ISL_1292641, EPI_ISL_1292642, EPI_ISL_1292643, EPI_ISL_1292644, EPI_ISL_1292645, EPI_ISL_1292646, EPI_ISL_1292647, EPI_ISL_1292648, EPI_ISL_1292649, EPI_ISL_1292650, EPI_ISL_1292652, EPI_ISL_1292653, EPI_ISL_1292654, EPI_ISL_1292655, EPI_ISL_1292656, EPI_ISL_1292657, EPI_ISL_1292658, EPI_ISL_1292659, EPI_ISL_1292661, EPI_ISL_1292662, EPI_ISL_1292663, EPI_ISL_1292664, EPI_ISL_1292665, EPI_ISL_1292666, EPI_ISL_1292667, EPI_ISL_1292668, EPI_ISL_1292669, EPI_ISL_1292670, EPI_ISL_1292671, EPI_ISL_1292672, EPI_ISL_1292673, EPI_ISL_1292674, EPI_ISL_1292675, EPI_ISL_1292676, EPI_ISL_1292677, EPI_ISL_1292678, EPI_ISL_1292679, EPI_ISL_1292680, EPI_ISL_1292681, EPI_ISL_1292682, EPI_ISL_1292683, EPI_ISL_1292684, EPI_ISL_1292685, EPI_ISL_1292686, EPI_ISL_1292687, EPI_ISL_1292688, EPI_ISL_1292689, EPI_ISL_1292690, EPI_ISL_1292691, EPI_ISL_1292692, EPI_ISL_1292693, EPI_ISL_1292694, EPI_ISL_1292695, EPI_ISL_1292696, EPI_ISL_1292697, EPI_ISL_1292698, EPI_ISL_1292699, EPI_ISL_1292700, EPI_ISL_1292701, EPI_ISL_1292702, EPI_ISL_1292703, EPI_ISL_1292704, EPI_ISL_1292705, EPI_ISL_1292706, EPI_ISL_1292707, EPI_ISL_1292708, EPI_ISL_1292709, EPI_ISL_1292710, EPI_ISL_1292711, EPI_ISL_1292713, EPI_ISL_1292714, EPI_ISL_1292715, EPI_ISL_1292716, EPI_ISL_1292717, EPI_ISL_1292720, EPI_ISL_1292721, EPI_ISL_1292722, EPI_ISL_1292723, EPI_ISL_1292724, EPI_ISL_1292725, EPI_ISL_1292726, EPI_ISL_1292727, EPI_ISL_1292728, EPI_ISL_1292729, EPI_ISL_1292730, EPI_ISL_1292731, EPI_ISL_1292732, EPI_ISL_1292733, EPI_ISL_1292734, EPI_ISL_1292735, EPI_ISL_1292736, EPI_ISL_1292737, EPI_ISL_1292738, EPI_ISL_1292739, EPI_ISL_1292740, EPI_ISL_1292741, EPI_ISL_1292742, EPI_ISL_1292744, EPI_ISL_1292745, EPI_ISL_1292746, EPI_ISL_1292747, EPI_ISL_1292748, EPI_ISL_1292749, EPI_ISL_1292750, EPI_ISL_1292751, EPI_ISL_1292752, EPI_ISL_1292754, EPI_ISL_1292755, EPI_ISL_1292756, EPI_ISL_1292757, EPI_ISL_1292758, EPI_ISL_1292759, EPI_ISL_1292760, EPI_ISL_1292761, EPI_ISL_1292762, EPI_ISL_1292763, EPI_ISL_1292764, EPI_ISL_1292765, EPI_ISL_1292766, EPI_ISL_1292767, EPI_ISL_1292768, EPI_ISL_1292769, EPI_ISL_1292770, EPI_ISL_1292771, EPI_ISL_1292772, EPI_ISL_1292773, EPI_ISL_1292774, EPI_ISL_1292775, EPI_ISL_1292776, EPI_ISL_1292777, EPI_ISL_1292778, EPI_ISL_1292779, EPI_ISL_1292780, EPI_ISL_1292782, EPI_ISL_1292783, EPI_ISL_1292785, EPI_ISL_1292786, EPI_ISL_1292787, EPI_ISL_1292788, EPI_ISL_1292789, EPI_ISL_1292790, EPI_ISL_1292791, EPI_ISL_1292792, EPI_ISL_1292793, EPI_ISL_1292794, EPI_ISL_1292795, EPI_ISL_1292796, EPI_ISL_1292797, EPI_ISL_1292798, EPI_ISL_1292799, EPI_ISL_1292800, EPI_ISL_1292801, EPI_ISL_1292802, EPI_ISL_1292803, EPI_ISL_1292804, EPI_ISL_1292805, EPI_ISL_1292806 | | | |
| see above | State Laboratories Division, Hawaii State Department of Health | State Laboratories Division, Hawaii State Department of Health | Pamela O'Brien, Drew Kuwazaki, Ayana Garnet, Razvan Sultana, Edward Desmond |
| EPI_ISL_1292810, EPI_ISL_1292811 | Arizona State University | Arizona State University | Rabia Maqsood, LaRinda A. Holland, Peter T. Skidmore, Emily A. Kaelin, Nicholas J. Mellor, Kristina Buss, Joy M. Blain, Neal W Woodbury, Valerie Harris, Joshua LaBaer, Vel Murugan, Efrem S. Lim |
| EPI_ISL_1292815, EPI_ISL_1292820, EPI_ISL_1292821, EPI_ISL_1292822, EPI_ISL_1292823, EPI_ISL_1292824, EPI_ISL_1292825, EPI_ISL_1292826, EPI_ISL_1292827, EPI_ISL_1292828, EPI_ISL_1292829, EPI_ISL_1292830, EPI_ISL_1292831, EPI_ISL_1292832, EPI_ISL_1292833, EPI_ISL_1292834, EPI_ISL_1292835, EPI_ISL_1292836, EPI_ISL_1292837, EPI_ISL_1292838, EPI_ISL_1292840, EPI_ISL_1292841, EPI_ISL_1292842, EPI_ISL_1292843, EPI_ISL_1292844, EPI_ISL_1292845, EPI_ISL_1292846, EPI_ISL_1292847, EPI_ISL_1292848, EPI_ISL_1292849, EPI_ISL_1292851, EPI_ISL_1292852, EPI_ISL_1292853, EPI_ISL_1292854, EPI_ISL_1292855, EPI_ISL_1292856, EPI_ISL_1292857, EPI_ISL_1292858, EPI_ISL_1292859, EPI_ISL_1292861, EPI_ISL_1292862, EPI_ISL_1292863, EPI_ISL_1292864, EPI_ISL_1292865, EPI_ISL_1292866, EPI_ISL_1292867, EPI_ISL_1292868, EPI_ISL_1292869, EPI_ISL_1292870, EPI_ISL_1292871, EPI_ISL_1292872, EPI_ISL_1292874, EPI_ISL_1292875, EPI_ISL_1292876, EPI_ISL_1292877, EPI_ISL_1292878, EPI_ISL_1292879, EPI_ISL_1292880, EPI_ISL_1292881, EPI_ISL_1292882, EPI_ISL_1292883, EPI_ISL_1292884, EPI_ISL_1292886, EPI_ISL_1292887, EPI_ISL_1292888, EPI_ISL_1292889, EPI_ISL_1292891, EPI_ISL_1292892, EPI_ISL_1292893, EPI_ISL_1292894, EPI_ISL_1292895, EPI_ISL_1292896, EPI_ISL_1292897, EPI_ISL_1292898, EPI_ISL_1292899, EPI_ISL_1292900, EPI_ISL_1292901, EPI_ISL_1292902, EPI_ISL_1292904, EPI_ISL_1292905, EPI_ISL_1292906, EPI_ISL_1292907, EPI_ISL_1292908, EPI_ISL_1292909, EPI_ISL_1292911, EPI_ISL_1292912, EPI_ISL_1292913, EPI_ISL_1292915, EPI_ISL_1292917, EPI_ISL_1292918, EPI_ISL_1292919, EPI_ISL_1292920, EPI_ISL_1292921, EPI_ISL_1292922, EPI_ISL_1292924, EPI_ISL_1292926, EPI_ISL_1292927, EPI_ISL_1292928, EPI_ISL_1292929, EPI_ISL_1292930, EPI_ISL_1292931, EPI_ISL_1292932, EPI_ISL_1292934, EPI_ISL_1292935, EPI_ISL_1292936, EPI_ISL_1292937, EPI_ISL_1292938, EPI_ISL_1292939, EPI_ISL_1292940, EPI_ISL_1292941, EPI_ISL_1292942, EPI_ISL_1292943, EPI_ISL_1292944, EPI_ISL_1292945, EPI_ISL_1292946, EPI_ISL_1292947, EPI_ISL_1292948, EPI_ISL_1292949, EPI_ISL_1292950, EPI_ISL_1292951, EPI_ISL_1292954, EPI_ISL_1292955, EPI_ISL_1292956, EPI_ISL_1292957, EPI_ISL_1292958, EPI_ISL_1292959, EPI_ISL_1292961, EPI_ISL_1292962, EPI_ISL_1292963, EPI_ISL_1292965, EPI_ISL_1292966, EPI_ISL_1292967, EPI_ISL_1292968, EPI_ISL_1292970 | | | |
| see above | Maryland Genomics, Institute for Genome Sciences, University of Maryland School of Medicine | Maryland Genomics, Institute for Genome Sciences, University of Maryland School of Medicine | Tallon, Luke J; Sadzewicz, Lisa D; Humphrys, Mike; Ott, Sandra; Roussey, Holly; Mehta, Aditya; Vavikolanu, Kranthi; Fraser, Claire M; Ravel, Jacques |
| EPI_ISL_1292974, EPI_ISL_1292975, EPI_ISL_1292976, EPI_ISL_1292977, EPI_ISL_1292978 | Connecticut DPH | Grubaugh Lab - Yale School of Public Health | Joseph Fauver, Mallery Breban, Isabell Ott, Tara Alpert, Mary Petrone, Anderson Brito, Chantal Vogels, Annie Watkins, Chaney Kalinich, Jessica Rothman, Anthony Muyombwe, Randy Downing, Jafar Razeq, Stephen M. Bart, Nathan Grubaugh |
| EPI_ISL_1292979, EPI_ISL_1292982, EPI_ISL_1292983, EPI_ISL_1292984 | Yale Clinical Virology Lab | Grubaugh Lab - Yale School of Public Health | Joseph Fauver, Mallery Breban, Isabell Ott, Tara Alpert, Mary Petrone, Anderson Brito, Chantal Vogels, Annie Watkins, Chaney Kalinich, Jessica Rothman, Marie L. Landry, Nathan Grubaugh |
| EPI_ISL_1292985 | Murphy Medical Association | Grubaugh Lab - Yale School of Public Health | Joseph Fauver, Mallery Breban, Isabell Ott, Tara Alpert, Mary Petrone, Anderson Brito, Chantal Vogels, Annie Watkins, Chaney Kalinich, Jessica Rothman, Caleb Neal, Eva Laszlo, Steven Murphy, Nathan Grubaugh |
| EPI_ISL_1292986, EPI_ISL_1292987, EPI_ISL_1292988, EPI_ISL_1292989, EPI_ISL_1292990, EPI_ISL_1292991, EPI_ISL_1292992, | Yale Clinical Virology Lab | Grubaugh Lab - Yale School of Public Health | Joseph Fauver, Mallery Breban, Isabell Ott, Tara Alpert, Mary Petrone, Anderson Brito, Chantal Vogels, Annie Watkins, Chaney Kalinich, Jessica Rothman, Marie L. Landry, Nathan Grubaugh |

| | | | |
|---|---|---|---|
| EPI_ISL_1292993, EPI_ISL_1292994, EPI_ISL_1292995 | | | |
| EPI_ISL_1292998, EPI_ISL_1292999, EPI_ISL_1293000, EPI_ISL_1293001, EPI_ISL_1293002, EPI_ISL_1293003, EPI_ISL_1293004, EPI_ISL_1293005, EPI_ISL_1293006, EPI_ISL_1293007, EPI_ISL_1293008, EPI_ISL_1293009, EPI_ISL_1293010, EPI_ISL_1293011, EPI_ISL_1293012, EPI_ISL_1293013, EPI_ISL_1293014, EPI_ISL_1293015, EPI_ISL_1293016, EPI_ISL_1293017, EPI_ISL_1293018, EPI_ISL_1293019, EPI_ISL_1293020, EPI_ISL_1293021, EPI_ISL_1293022, EPI_ISL_1293023, EPI_ISL_1293024, EPI_ISL_1293025, EPI_ISL_1293026, EPI_ISL_1293027, EPI_ISL_1293028, EPI_ISL_1293029, EPI_ISL_1293030, EPI_ISL_1293031, EPI_ISL_1293032, EPI_ISL_1293033, EPI_ISL_1293034, EPI_ISL_1293035, EPI_ISL_1293036, EPI_ISL_1293037, EPI_ISL_1293038, EPI_ISL_1293039, EPI_ISL_1293040, EPI_ISL_1293041, EPI_ISL_1293042, EPI_ISL_1293043, EPI_ISL_1293044 | | | |
| see above | MD Public Health Laboratory | MD Public Health Laboratory | Maryland Department of Health Laboratories Administration |
| EPI_ISL_1293082, EPI_ISL_1293083, EPI_ISL_1293084, EPI_ISL_1293086, EPI_ISL_1293087, EPI_ISL_1293088 | Tempus | Grubaugh Lab - Yale School of Public Health | Joseph Fauver, Mallery Breban, Isabell Ott, Tara Alpert, Mary Petrone, Anderson Brito, Chantal Vogels, Annie Watkins, Chaney Kalinich, Jessica Rothman, Matthew J. MacKay, Gaurav Khullar, Jessica Metti, Joel T. Dudley, Megan Nash, Nike Beaubier, Christopher E. Mason, Nathan Grubaugh |
| EPI_ISL_1293089, EPI_ISL_1293090, EPI_ISL_1293091, EPI_ISL_1293092 | Yale Pathology Lab | Grubaugh Lab - Yale School of Public Health | Joseph Fauver, Mallery Breban, Isabell Ott, Tara Alpert, Mary Petrone, Anderson Brito, Chantal Vogels, Annie Watkins, Chaney Kalinich, Jessica Rothman, Jianhui Wang, Chen Liu, Pei Hui, Nathan Grubaugh |
| EPI_ISL_1293093 | Oak Ridge | Grubaugh Lab - Yale School of Public Health | Joseph Fauver, Mallery Breban, Isabell Ott, Tara Alpert, Mary Petrone, Anderson Brito, Chantal Vogels, Annie Watkins, Chaney Kalinich, Jessica Rothman, Daniel Jones, Nathan Grubaugh |
| EPI_ISL_1293095, EPI_ISL_1293096, EPI_ISL_1293097, EPI_ISL_1293098, EPI_ISL_1293099, EPI_ISL_1293101, EPI_ISL_1293104, EPI_ISL_1293105, EPI_ISL_1293106, EPI_ISL_1293107, EPI_ISL_1293108, EPI_ISL_1293109, EPI_ISL_1293110, EPI_ISL_1293111, EPI_ISL_1293112, EPI_ISL_1293113, EPI_ISL_1293114, EPI_ISL_1293115, EPI_ISL_1293116, EPI_ISL_1293118, EPI_ISL_1293119, EPI_ISL_1293120, EPI_ISL_1293121, EPI_ISL_1293123, EPI_ISL_1293125, EPI_ISL_1293126, EPI_ISL_1293127, EPI_ISL_1293128 | | | |
| see above | Tempus | Grubaugh Lab - Yale School of Public Health | Joseph Fauver, Mallery Breban, Isabell Ott, Tara Alpert, Mary Petrone, Anderson Brito, Chantal Vogels, Annie Watkins, Chaney Kalinich, Jessica Rothman, Matthew J. MacKay, Gaurav Khullar, Jessica Metti, Joel T. Dudley, Megan Nash, Nike Beaubier, Christopher E. Mason, Nathan Grubaugh |
| EPI_ISL_1293129, EPI_ISL_1293134, EPI_ISL_1293135, EPI_ISL_1293137 | Murphy Medical Association | Grubaugh Lab - Yale School of Public Health | Joseph Fauver, Mallery Breban, Isabell Ott, Tara Alpert, Mary Petrone, Anderson Brito, Chantal Vogels, Annie Watkins, Chaney Kalinich, Jessica Rothman, Caleb Neal, Eva Laszlo, Steven Murphy, Nathan Grubaugh |
| EPI_ISL_1293144, EPI_ISL_1293145, EPI_ISL_1293146, EPI_ISL_1293147, EPI_ISL_1293148 | Yale Pathology Lab | Grubaugh Lab - Yale School of Public Health | Joseph Fauver, Mallery Breban, Isabell Ott, Tara Alpert, Mary Petrone, Anderson Brito, Chantal Vogels, Annie Watkins, Chaney Kalinich, Jessica Rothman, Jianhui Wang, Chen Liu, Pei Hui, Nathan Grubaugh |
| EPI_ISL_1293149, EPI_ISL_1293150, EPI_ISL_1293151, EPI_ISL_1293152, EPI_ISL_1293154, EPI_ISL_1293155, EPI_ISL_1293157, EPI_ISL_1293158, EPI_ISL_1293159, EPI_ISL_1293161, EPI_ISL_1293162, EPI_ISL_1293163, EPI_ISL_1293166, EPI_ISL_1293168, EPI_ISL_1293171, EPI_ISL_1293172, EPI_ISL_1293173, EPI_ISL_1293175, EPI_ISL_1293176, EPI_ISL_1293178, EPI_ISL_1293179, EPI_ISL_1293180, EPI_ISL_1293181, EPI_ISL_1293182, EPI_ISL_1293184, EPI_ISL_1293185, EPI_ISL_1293186, EPI_ISL_1293187, EPI_ISL_1293188, EPI_ISL_1293189, EPI_ISL_1293190, EPI_ISL_1293191, EPI_ISL_1293192, EPI_ISL_1293193, EPI_ISL_1293194, EPI_ISL_1293195, EPI_ISL_1293196, EPI_ISL_1293197, EPI_ISL_1293198, EPI_ISL_1293199, EPI_ISL_1293200, EPI_ISL_1293201, EPI_ISL_1293202, EPI_ISL_1293203, EPI_ISL_1293205, EPI_ISL_1293207, EPI_ISL_1293208, EPI_ISL_1293210, EPI_ISL_1293211, EPI_ISL_1293212, EPI_ISL_1293213, EPI_ISL_1293214, EPI_ISL_1293215, EPI_ISL_1293216 | | | |
| see above | Yale Clinical Virology Lab | Grubaugh Lab - Yale School of Public Health | Joseph Fauver, Mallery Breban, Isabell Ott, Tara Alpert, Mary Petrone, Anderson Brito, Chantal Vogels, Annie Watkins, Chaney Kalinich, Jessica Rothman, Marie L. Landry, Nathan Grubaugh |
| EPI_ISL_1293218 | Oak Ridge | Grubaugh Lab - Yale School of Public Health | Joseph Fauver, Mallery Breban, Isabell Ott, Tara Alpert, Mary Petrone, Anderson Brito, Chantal Vogels, Annie Watkins, Chaney Kalinich, Jessica Rothman, Daniel Jones, Nathan Grubaugh |
| EPI_ISL_1293327, EPI_ISL_1293328, EPI_ISL_1293329, EPI_ISL_1293330 | University of Michigan Clinical Microbiology Laboratory | Lauring Lab, University of Michigan, Department of Microbiology and Immunology | Valesano |
| EPI_ISL_1293332, EPI_ISL_1293333, EPI_ISL_1293334, EPI_ISL_1293335, EPI_ISL_1293336, EPI_ISL_1293337, EPI_ISL_1293340, EPI_ISL_1293341, EPI_ISL_1293342 | Quest Diagnostics Incorporated | Respiratory Viruses Branch, Division of Viral Diseases, Centers for Disease Control and Prevention | Peter W. Cook, Dakota Howard, Dhwani Batra, Ben L. Rambo-Martin, S. H. Rosenthal, A. Gerasimova, R. M. Kagan, B. Anderson, M. Hua, Y. Liu, L.E. Bernstein, K.E. Livingston, A. Perez, I. A. Shlyakhter, R. V. Rolando, R. Owen, P. Tanpaiboon, F. Lacbawan, Clinton R. Paden, Suxiang Tong, Duncan MacCannell |
| EPI_ISL_1295182, EPI_ISL_1295183, EPI_ISL_1295184, EPI_ISL_1295185, EPI_ISL_1295187, EPI_ISL_1295188, EPI_ISL_1295189, EPI_ISL_1295190, EPI_ISL_1295191, EPI_ISL_1295192, EPI_ISL_1295193, EPI_ISL_1295194, EPI_ISL_1295195, EPI_ISL_1295196, EPI_ISL_1295197, EPI_ISL_1295198, EPI_ISL_1295199, EPI_ISL_1295200, EPI_ISL_1295201, EPI_ISL_1295202, EPI_ISL_1295203, EPI_ISL_1295204, EPI_ISL_1295205, EPI_ISL_1295206, EPI_ISL_1295207, EPI_ISL_1295208, EPI_ISL_1295209, EPI_ISL_1295210, EPI_ISL_1295211, EPI_ISL_1295213, EPI_ISL_1295214, EPI_ISL_1295215, EPI_ISL_1295216, EPI_ISL_1295217, EPI_ISL_1295219, EPI_ISL_1295220, EPI_ISL_1295222, EPI_ISL_1295223, EPI_ISL_1295225, EPI_ISL_1295226, EPI_ISL_1295228, EPI_ISL_1295229, EPI_ISL_1295230, EPI_ISL_1295231, EPI_ISL_1295232, EPI_ISL_1295233, EPI_ISL_1295234, EPI_ISL_1295235, EPI_ISL_1295236, EPI_ISL_1295237, EPI_ISL_1295238, EPI_ISL_1295239, EPI_ISL_1295240, EPI_ISL_1295241, EPI_ISL_1295242, EPI_ISL_1295243, EPI_ISL_1295244, EPI_ISL_1295245, EPI_ISL_1295246, EPI_ISL_1295247, EPI_ISL_1295248, EPI_ISL_1295249, EPI_ISL_1295250, EPI_ISL_1295251, EPI_ISL_1295252, EPI_ISL_1295253, EPI_ISL_1295254, EPI_ISL_1295255, EPI_ISL_1295256, EPI_ISL_1295257, EPI_ISL_1295258, EPI_ISL_1295259, EPI_ISL_1295260, EPI_ISL_1295261, EPI_ISL_1295262, EPI_ISL_1295263, EPI_ISL_1295264, EPI_ISL_1295265, EPI_ISL_1295266, EPI_ISL_1295267, EPI_ISL_1295268, EPI_ISL_1295269, EPI_ISL_1295270, EPI_ISL_1295271, EPI_ISL_1295272, EPI_ISL_1295273, EPI_ISL_1295274, EPI_ISL_1295275, EPI_ISL_1295276, EPI_ISL_1295277, EPI_ISL_1295278, EPI_ISL_1295279, EPI_ISL_1295280, EPI_ISL_1295281, EPI_ISL_1295282, EPI_ISL_1295283, EPI_ISL_1295284, EPI_ISL_1295285, EPI_ISL_1295286, EPI_ISL_1295287, EPI_ISL_1295288, EPI_ISL_1295289, EPI_ISL_1295290, EPI_ISL_1295291, EPI_ISL_1295292, EPI_ISL_1295293, EPI_ISL_1295294, EPI_ISL_1295295, EPI_ISL_1295296, EPI_ISL_1295297, EPI_ISL_1295298, EPI_ISL_1295300, EPI_ISL_1295301, EPI_ISL_1295302, EPI_ISL_1295303, EPI_ISL_1295304, EPI_ISL_1295305, EPI_ISL_1295306, EPI_ISL_1295307, EPI_ISL_1295308, EPI_ISL_1295309, EPI_ISL_1295310, EPI_ISL_1295311, EPI_ISL_1295312, EPI_ISL_1295313, EPI_ISL_1295314, EPI_ISL_1295315, EPI_ISL_1295316, EPI_ISL_1295317, EPI_ISL_1295318, EPI_ISL_1295319, EPI_ISL_1295320, EPI_ISL_1295321, EPI_ISL_1295322, EPI_ISL_1295323, EPI_ISL_1295324, EPI_ISL_1295325, EPI_ISL_1295326, EPI_ISL_1295327, EPI_ISL_1295328, EPI_ISL_1295329, EPI_ISL_1295330, EPI_ISL_1295331, EPI_ISL_1295332, EPI_ISL_1295333, EPI_ISL_1295334, EPI_ISL_1295335, EPI_ISL_1295336, EPI_ISL_1295337, EPI_ISL_1295338, EPI_ISL_1295339, EPI_ISL_1295340, EPI_ISL_1295341, EPI_ISL_1295342, EPI_ISL_1295343, EPI_ISL_1295344, EPI_ISL_1295345, EPI_ISL_1295346, EPI_ISL_1295347, EPI_ISL_1295348, EPI_ISL_1295349, EPI_ISL_1295351, EPI_ISL_1295352, EPI_ISL_1295353, EPI_ISL_1295354, EPI_ISL_1295355, EPI_ISL_1295356, EPI_ISL_1295357, EPI_ISL_1295358, EPI_ISL_1295359, EPI_ISL_1295360, EPI_ISL_1295361, EPI_ISL_1295362, EPI_ISL_1295363, EPI_ISL_1295364, EPI_ISL_1295365, EPI_ISL_1295366, EPI_ISL_1295367, EPI_ISL_1295368, EPI_ISL_1295369, EPI_ISL_1295370, EPI_ISL_1295371, EPI_ISL_1295372, EPI_ISL_1295373, EPI_ISL_1295374, EPI_ISL_1295375, EPI_ISL_1295376, EPI_ISL_1295377, EPI_ISL_1295378, EPI_ISL_1295379, EPI_ISL_1295380, EPI_ISL_1295381, EPI_ISL_1295382, EPI_ISL_1295383, EPI_ISL_1295384, EPI_ISL_1295385, EPI_ISL_1295386, EPI_ISL_1295387, EPI_ISL_1295388, EPI_ISL_1295389, EPI_ISL_1295390, EPI_ISL_1295391, EPI_ISL_1295392, EPI_ISL_1295393, EPI_ISL_1295394, EPI_ISL_1295395, EPI_ISL_1295396, EPI_ISL_1295397, EPI_ISL_1295398, EPI_ISL_1295399, EPI_ISL_1295400, EPI_ISL_1295401, EPI_ISL_1295402, EPI_ISL_1295403, EPI_ISL_1295404, EPI_ISL_1295405, EPI_ISL_1295406, EPI_ISL_1295407, EPI_ISL_1295408, EPI_ISL_1295409, EPI_ISL_1295410, EPI_ISL_1295411, EPI_ISL_1295412, EPI_ISL_1295413, EPI_ISL_1295414, EPI_ISL_1295415, EPI_ISL_1295416, EPI_ISL_1295417, EPI_ISL_1295418, EPI_ISL_1295419, EPI_ISL_1295420, EPI_ISL_1295421, EPI_ISL_1295422, EPI_ISL_1295424, EPI_ISL_1295425, EPI_ISL_1295426, EPI_ISL_1295427, EPI_ISL_1295428, EPI_ISL_1295430, EPI_ISL_1295431, EPI_ISL_1295432, EPI_ISL_1295433, EPI_ISL_1295434, EPI_ISL_1295435, EPI_ISL_1295436, EPI_ISL_1295437, EPI_ISL_1295438, EPI_ISL_1295439, EPI_ISL_1295440, EPI_ISL_1295442, EPI_ISL_1295443, EPI_ISL_1295445, EPI_ISL_1295446, EPI_ISL_1295447, EPI_ISL_1295448, EPI_ISL_1295449, EPI_ISL_1295450, EPI_ISL_1295451, EPI_ISL_1295452, EPI_ISL_1295453, EPI_ISL_1295454, EPI_ISL_1295455, EPI_ISL_1295457, EPI_ISL_1295458, EPI_ISL_1295459, EPI_ISL_1295460, EPI_ISL_1295461, EPI_ISL_1295462, EPI_ISL_1295464, EPI_ISL_1295465, EPI_ISL_1295466, EPI_ISL_1295467, EPI_ISL_1295468, EPI_ISL_1295469, EPI_ISL_1295470, EPI_ISL_1295471, EPI_ISL_1295472, EPI_ISL_1295474, EPI_ISL_1295475, EPI_ISL_1295476, EPI_ISL_1295477, EPI_ISL_1295478, EPI_ISL_1295479, EPI_ISL_1295480, EPI_ISL_1295481, EPI_ISL_1295482, EPI_ISL_1295483, EPI_ISL_1295484, EPI_ISL_1295485, EPI_ISL_1295486, EPI_ISL_1295487, EPI_ISL_1295488, EPI_ISL_1295489, EPI_ISL_1295491, EPI_ISL_1295492, EPI_ISL_1295493, EPI_ISL_1295494, EPI_ISL_1295495, EPI_ISL_1295496, EPI_ISL_1295497, EPI_ISL_1295498, EPI_ISL_1295499, EPI_ISL_1295500, EPI_ISL_1295501, EPI_ISL_1295502, EPI_ISL_1295503, EPI_ISL_1295504, EPI_ISL_1295505, EPI_ISL_1295506, EPI_ISL_1295507, EPI_ISL_1295508, EPI_ISL_1295509, EPI_ISL_1295510, EPI_ISL_1295511, EPI_ISL_1295512, EPI_ISL_1295513, EPI_ISL_1295514, EPI_ISL_1295515, EPI_ISL_1295517, EPI_ISL_1295518, EPI_ISL_1295520, EPI_ISL_1295521, EPI_ISL_1295522, EPI_ISL_1295523, EPI_ISL_1295524, EPI_ISL_1295525, EPI_ISL_1295526, EPI_ISL_1295527, EPI_ISL_1295528, EPI_ISL_1295529, EPI_ISL_1295530, EPI_ISL_1295531, EPI_ISL_1295533, EPI_ISL_1295535, EPI_ISL_1295536, EPI_ISL_1295537, EPI_ISL_1295538, EPI_ISL_1295541, EPI_ISL_1295542, EPI_ISL_1295543, EPI_ISL_1295544, EPI_ISL_1295545, EPI_ISL_1295546, EPI_ISL_1295547, EPI_ISL_1295549, EPI_ISL_1295550, EPI_ISL_1295551, EPI_ISL_1295552, EPI_ISL_1295553, EPI_ISL_1295554, EPI_ISL_1295555, EPI_ISL_1295556, EPI_ISL_1295557, EPI_ISL_1295558, EPI_ISL_1295559, EPI_ISL_1295561, EPI_ISL_1295562, EPI_ISL_1295563 | | | |
| see above | Wisconsin State Laboratory of Hygiene Communicable Disease Division | Wisconsin State Laboratory of Hygiene Communicable Disease Division | Kelsey R. Florek, Abigail C. Shockey |
| EPI_ISL_1295564, EPI_ISL_1295565, EPI_ISL_1295566, EPI_ISL_1295567, EPI_ISL_1295568, EPI_ISL_1295569, EPI_ISL_1295570, EPI_ISL_1295571, EPI_ISL_1295572, EPI_ISL_1295573, EPI_ISL_1295574, EPI_ISL_1295575, EPI_ISL_1295576, EPI_ISL_1295577, EPI_ISL_1295578, EPI_ISL_1295579, EPI_ISL_1295580, EPI_ISL_1295581, EPI_ISL_1295582, EPI_ISL_1295583, EPI_ISL_1295584 | | | |
| see above | Clinical virology Laboratory, Children's Hospital Los Angeles | Center for Personalized Medicine, Children's Hospital Los Angeles | Alex Ryutov, Alexander R. Judkins, Ananthanarayanan Govindarajan, David Ruble, Dejerianne Ostrow, Dennis Maglinte, Jaclyn A. Biegel, Jennifer Dien Bard, Jennifer Han, Lishuang Shen, Moiz Bootwalla, Timothy J. Triche, Utsav Pandey, Xiaowu Gai |
| EPI_ISL_1295619, EPI_ISL_1295620, EPI_ISL_1295621, EPI_ISL_1295622, EPI_ISL_1295623, EPI_ISL_1295624, EPI_ISL_1295625, EPI_ISL_1295626, EPI_ISL_1295627, EPI_ISL_1295628, EPI_ISL_1295629, EPI_ISL_1295630, EPI_ISL_1295631, EPI_ISL_1295632, EPI_ISL_1295633, EPI_ISL_1295637 | | | |
| see above | DC Public Health Lab/ Dept. of Forensic Sciences | DC Public Health Lab/ Dept. of Forensic Sciences | Scott Nguyen, Janis Doss, Elizabeth Zelaya, Sarah Scott, Connie Maza, Monica Mann, Brittany Hamilton, David Payne, Jocelyn Hauser |
| EPI_ISL_1295640 | Ohio Department of Health Laboratory | Ohio Department of Health Laboratory | Holmes, Jennifer; Eric Brandt, Keoni Omura, Glen McGillivary, Caitlin McDonnell, Jade Mowery, Stephanie Mccracken, Tyler Payne, Kirtana Ramadugu, Erica Leasure, Brent Lee, Kelsey Florek, Heather Blankenship, Allison Black, Quanta Brown, and Tammy Bannerman |
| EPI_ISL_1295646, EPI_ISL_1295647, EPI_ISL_1295648, EPI_ISL_1295649, EPI_ISL_1295650, EPI_ISL_1295651, EPI_ISL_1295652, EPI_ISL_1295653, EPI_ISL_1295654, EPI_ISL_1295655, EPI_ISL_1295656, EPI_ISL_1295657, EPI_ISL_1295658, EPI_ISL_1295659, EPI_ISL_1295660, EPI_ISL_1295661, EPI_ISL_1295662, EPI_ISL_1295663, EPI_ISL_1295664, EPI_ISL_1295665, EPI_ISL_1295666, EPI_ISL_1295667, EPI_ISL_1295668, EPI_ISL_1295669, EPI_ISL_1295670, EPI_ISL_1295671, EPI_ISL_1295672, EPI_ISL_1295673, EPI_ISL_1295674, EPI_ISL_1295675, EPI_ISL_1295676, EPI_ISL_1295677, EPI_ISL_1295678, EPI_ISL_1295679, EPI_ISL_1295680, EPI_ISL_1295681, EPI_ISL_1295682, EPI_ISL_1295683, EPI_ISL_1295684, EPI_ISL_1295685, EPI_ISL_1295686, EPI_ISL_1295687, EPI_ISL_1295688, EPI_ISL_1295689, EPI_ISL_1295690, EPI_ISL_1295691, EPI_ISL_1295692, EPI_ISL_1295693, EPI_ISL_1295694, EPI_ISL_1295695, EPI_ISL_1295696, EPI_ISL_1295697, EPI_ISL_1295698, EPI_ISL_1295699, EPI_ISL_1295700, EPI_ISL_1295701, EPI_ISL_1295702, EPI_ISL_1295703, EPI_ISL_1295704, EPI_ISL_1295705, EPI_ISL_1295706, EPI_ISL_1295707, EPI_ISL_1295708, EPI_ISL_1295709, EPI_ISL_1295710, EPI_ISL_1295711, EPI_ISL_1295712, EPI_ISL_1295713, EPI_ISL_1295714, EPI_ISL_1295715, EPI_ISL_1295716, EPI_ISL_1295717, EPI_ISL_1295718, EPI_ISL_1295719, EPI_ISL_1295720, EPI_ISL_1295721, EPI_ISL_1295722, EPI_ISL_1295723, EPI_ISL_1295724, EPI_ISL_1295725, EPI_ISL_1295726, EPI_ISL_1295727, EPI_ISL_1295728, EPI_ISL_1295729, EPI_ISL_1295730, EPI_ISL_1295731, EPI_ISL_1295732, EPI_ISL_1295733, EPI_ISL_1295734, EPI_ISL_1295735, EPI_ISL_1295736, EPI_ISL_1295737, EPI_ISL_1295738, EPI_ISL_1295739, EPI_ISL_1295740, EPI_ISL_1295741, EPI_ISL_1295742, EPI_ISL_1295743, EPI_ISL_1295744, EPI_ISL_1295745, EPI_ISL_1295746, EPI_ISL_1295747, EPI_ISL_1295748, EPI_ISL_1295749, EPI_ISL_1295750, EPI_ISL_1295751, EPI_ISL_1295752, EPI_ISL_1295753, EPI_ISL_1295754, EPI_ISL_1295755, EPI_ISL_1295756, EPI_ISL_1295757, EPI_ISL_1295758, EPI_ISL_1295759, EPI_ISL_1295760, EPI_ISL_1295761, EPI_ISL_1295762, EPI_ISL_1295763, EPI_ISL_1295764, EPI_ISL_1295765, EPI_ISL_1295766, EPI_ISL_1295767, EPI_ISL_1295768, EPI_ISL_1295769, EPI_ISL_1295770, EPI_ISL_1295771, EPI_ISL_1295772, EPI_ISL_1295773, EPI_ISL_1295774, EPI_ISL_1295775, EPI_ISL_1295776, EPI_ISL_1295777, EPI_ISL_1295778, EPI_ISL_1295779, EPI_ISL_1295780, EPI_ISL_1295781, EPI_ISL_1295782, EPI_ISL_1295783, EPI_ISL_1295784, EPI_ISL_1295785, EPI_ISL_1295786, EPI_ISL_1295787, EPI_ISL_1295788, EPI_ISL_1295789, EPI_ISL_1295790, EPI_ISL_1295791, EPI_ISL_1295792, EPI_ISL_1295793, EPI_ISL_1295794, EPI_ISL_1295795 | | | |
| see above | Sharp HealthCare Laboratory | Andersen lab at Scripps Research | SEARCH Alliance San Diego with Aaron Harding, Jacquelyn Berumen, Cathy Woerle, Liam McGinnis, Art Mendoza, Omid Bakhtar |
| EPI_ISL_1295796, EPI_ISL_1295797, EPI_ISL_1295798, EPI_ISL_1295799, EPI_ISL_1295800, EPI_ISL_1295801, EPI_ISL_1295802, EPI_ISL_1295803, EPI_ISL_1295804, EPI_ISL_1295805, EPI_ISL_1295806, EPI_ISL_1295807, EPI_ISL_1295808, EPI_ISL_1295809, EPI_ISL_1295810, EPI_ISL_1295811, EPI_ISL_1295812, EPI_ISL_1295813, EPI_ISL_1295814 | | | |
| see above | Rady Children's Hospital - San Diego | Andersen lab at Scripps Research | SEARCH Alliance San Diego with Nanda Radamchar, David Dimmock, Linda Luo, Christina Clarke, Kathryn Bouic, Teresa Mueller, Denise Malicki |
| EPI_ISL_1295815, EPI_ISL_1295816, EPI_ISL_1295817 | San Diego County Public Health Laboratory | Andersen lab at Scripps Research | SEARCH Alliance San Diego with Tracy Basler, Jovan Shephard, Brett Austin |
| EPI_ISL_1295818, EPI_ISL_1295819, EPI_ISL_1295820, EPI_ISL_1295821, EPI_ISL_1295822, EPI_ISL_1295823, EPI_ISL_1295824, EPI_ISL_1295825, EPI_ISL_1295826, EPI_ISL_1295827, EPI_ISL_1295828, EPI_ISL_1295829, EPI_ISL_1295830, EPI_ISL_1295831, EPI_ISL_1295832, EPI_ISL_1295833, EPI_ISL_1295834, EPI_ISL_1295835, EPI_ISL_1295836, EPI_ISL_1295837, EPI_ISL_1295838, EPI_ISL_1295839, EPI_ISL_1295840, EPI_ISL_1295841, EPI_ISL_1295842, EPI_ISL_1295843, EPI_ISL_1295844, EPI_ISL_1295845, EPI_ISL_1295846, EPI_ISL_1295847, EPI_ISL_1295848, EPI_ISL_1295849, EPI_ISL_1295850, EPI_ISL_1295851, EPI_ISL_1295852, EPI_ISL_1295853, EPI_ISL_1295854, EPI_ISL_1295855, EPI_ISL_1295856, EPI_ISL_1295857, EPI_ISL_1295858, EPI_ISL_1295859, EPI_ISL_1295860, EPI_ISL_1295861, EPI_ISL_1295862, EPI_ISL_1295863, EPI_ISL_1295864, EPI_ISL_1295865, EPI_ISL_1295866, EPI_ISL_1295867, EPI_ISL_1295868, EPI_ISL_1295869, EPI_ISL_1295870, EPI_ISL_1295871, EPI_ISL_1295872, EPI_ISL_1295873, EPI_ISL_1295874, EPI_ISL_1295875, EPI_ISL_1295876, EPI_ISL_1295877, EPI_ISL_1295878, EPI_ISL_1295879, EPI_ISL_1295880, EPI_ISL_1295881, EPI_ISL_1295882, EPI_ISL_1295883, EPI_ISL_1295884, EPI_ISL_1295885, EPI_ISL_1295886, EPI_ISL_1295887, EPI_ISL_1295888, EPI_ISL_1295889, EPI_ISL_1295890, EPI_ISL_1295891, EPI_ISL_1295892, EPI_ISL_1295893, EPI_ISL_1295894, EPI_ISL_1295895, EPI_ISL_1295896, EPI_ISL_1295897, EPI_ISL_1295898, EPI_ISL_1295899, EPI_ISL_1295900 | | | |
| see above | Sharp HealthCare Laboratory | Andersen lab at Scripps Research | SEARCH Alliance San Diego with Aaron Harding, Jacquelyn Berumen, Cathy Woerle, Liam McGinnis, Art Mendoza, Omid Bakhtar |
| EPI_ISL_1295902, EPI_ISL_1295904, EPI_ISL_1295905, EPI_ISL_1295906, EPI_ISL_1295907, EPI_ISL_1295908, EPI_ISL_1295910, EPI_ISL_1295911, EPI_ISL_1295913, EPI_ISL_1295914, EPI_ISL_1295915, EPI_ISL_1295916, EPI_ISL_1295917, EPI_ISL_1295918, EPI_ISL_1295919, EPI_ISL_1295920, EPI_ISL_1295922, EPI_ISL_1295923, EPI_ISL_1295924, EPI_ISL_1295925, EPI_ISL_1295926, EPI_ISL_1295927, EPI_ISL_1295928 | | | |
| see above | Nevada State Public Health Laboratory | Nevada State Public Health Laboratory | Andrew Gorzalski, Mark Pandori |

| | | | |
|---|---|---|---|
| EPI_ISL_1296234, EPI_ISL_1296235 | Ohio Department of Health Laboratory | Ohio Department of Health Laboratory | Holmes, Jennifer; Eric Brandt, Keoni Omura, Glen McGillivary, Caitlin McDonnell, Jade Mowery, Stephanie Mccracken, Tyler Payne, Kirtana Ramadugu, Erica Leasure, Brent Lee, Kelsey Florek, Heather Blankenship, Allison Black, Quanta Brown, and Tammy Bannerman |
| EPI_ISL_1296294, EPI_ISL_1296296, EPI_ISL_1296480, EPI_ISL_1296653 | Montana Public Health Laboratory | Montana Public Health Laboratory | Joy Ritter, Michelle Mozer, Carrie Biskupiak, Deborah Gibson |
| EPI_ISL_1296686, EPI_ISL_1296687, EPI_ISL_1296688, EPI_ISL_1296690, EPI_ISL_1296691, EPI_ISL_1296692, EPI_ISL_1296693, EPI_ISL_1296694, EPI_ISL_1296695, EPI_ISL_1296696, EPI_ISL_1296697, EPI_ISL_1296698, EPI_ISL_1296699, EPI_ISL_1296700, EPI_ISL_1296701, EPI_ISL_1296702 | | | |
| see above | Sonora Quest Laboratories | TGen North | "Jolene Bowers, Heather Centner, Chris French, Hayley Yaglom, Ashlyn Pfeiffer, Darrin Lemmer, Dave Engelthaler, The Arizona COVID Genomics Union (ACGU)" |
| EPI_ISL_1296703 | TGen North | TGen North | "Jolene Bowers, Heather Centner, Chris French, Hayley Yaglom, Ashlyn Pfeiffer, Darrin Lemmer, Dave Engelthaler, The Arizona COVID Genomics Union (ACGU)" |
| EPI_ISL_1296704, EPI_ISL_1296705, EPI_ISL_1296706, EPI_ISL_1296707, EPI_ISL_1296708, EPI_ISL_1296709, EPI_ISL_1296710, EPI_ISL_1296711, EPI_ISL_1296712, EPI_ISL_1296713, EPI_ISL_1296715, EPI_ISL_1296716, EPI_ISL_1296717, EPI_ISL_1296718, EPI_ISL_1296719, EPI_ISL_1296720, EPI_ISL_1296722 | | | |
| see above | Sonora Quest Laboratories | TGen North | "Jolene Bowers, Heather Centner, Chris French, Hayley Yaglom, Ashlyn Pfeiffer, Darrin Lemmer, Dave Engelthaler, The Arizona COVID Genomics Union (ACGU)" |
| EPI_ISL_1296724 | TGen North | TGen North | "Jolene Bowers, Heather Centner, Chris French, Hayley Yaglom, Ashlyn Pfeiffer, Darrin Lemmer, Dave Engelthaler, The Arizona COVID Genomics Union (ACGU)" |
| EPI_ISL_1296725, EPI_ISL_1296726 | Sonora Quest Laboratories | TGen North | "Jolene Bowers, Heather Centner, Chris French, Hayley Yaglom, Ashlyn Pfeiffer, Darrin Lemmer, Dave Engelthaler, The Arizona COVID Genomics Union (ACGU)" |
| EPI_ISL_1296727 | TGen North | TGen North | "Jolene Bowers, Heather Centner, Chris French, Hayley Yaglom, Ashlyn Pfeiffer, Darrin Lemmer, Dave Engelthaler, The Arizona COVID Genomics Union (ACGU)" |
| EPI_ISL_1296729, EPI_ISL_1296730, EPI_ISL_1296731, EPI_ISL_1296732, EPI_ISL_1296733, EPI_ISL_1296734, EPI_ISL_1296735, EPI_ISL_1296736, EPI_ISL_1296737 | Sonora Quest Laboratories | TGen North | "Jolene Bowers, Heather Centner, Chris French, Hayley Yaglom, Ashlyn Pfeiffer, Darrin Lemmer, Dave Engelthaler, The Arizona COVID Genomics Union (ACGU)" |
| EPI_ISL_1296738 | TGen North | TGen North | "Jolene Bowers, Heather Centner, Chris French, Hayley Yaglom, Ashlyn Pfeiffer, Darrin Lemmer, Dave Engelthaler, The Arizona COVID Genomics Union (ACGU)" |
| EPI_ISL_1296739 | Sonora Quest Laboratories | TGen North | "Jolene Bowers, Heather Centner, Chris French, Hayley Yaglom, Ashlyn Pfeiffer, Darrin Lemmer, Dave Engelthaler, The Arizona COVID Genomics Union (ACGU)" |
| EPI_ISL_1296742 | TGen North | TGen North | "Jolene Bowers, Heather Centner, Chris French, Hayley Yaglom, Ashlyn Pfeiffer, Darrin Lemmer, Dave Engelthaler, The Arizona COVID Genomics Union (ACGU)" |
| EPI_ISL_1296743, EPI_ISL_1296744, EPI_ISL_1296745, EPI_ISL_1296746, EPI_ISL_1296747, EPI_ISL_1296750, EPI_ISL_1296751, EPI_ISL_1296752, EPI_ISL_1296753, EPI_ISL_1296754, EPI_ISL_1296755, EPI_ISL_1296756, EPI_ISL_1296757, EPI_ISL_1296758, EPI_ISL_1296759, EPI_ISL_1296760, EPI_ISL_1296761, EPI_ISL_1296762, EPI_ISL_1296763, EPI_ISL_1296764, EPI_ISL_1296766, EPI_ISL_1296767, EPI_ISL_1296768, EPI_ISL_1296769, EPI_ISL_1296770, EPI_ISL_1296771, EPI_ISL_1296772, EPI_ISL_1296773 | | | |
| see above | Sonora Quest Laboratories | TGen North | "Jolene Bowers, Heather Centner, Chris French, Hayley Yaglom, Ashlyn Pfeiffer, Darrin Lemmer, Dave Engelthaler, The Arizona COVID Genomics Union (ACGU)" |
| EPI_ISL_1296774 | TGen North | TGen North | "Jolene Bowers, Heather Centner, Chris French, Hayley Yaglom, Ashlyn Pfeiffer, Darrin Lemmer, Dave Engelthaler, The Arizona COVID Genomics Union (ACGU)" |
| EPI_ISL_1296775, EPI_ISL_1296776, EPI_ISL_1296777, EPI_ISL_1296778, EPI_ISL_1296779, EPI_ISL_1296780, EPI_ISL_1296781, EPI_ISL_1296782, EPI_ISL_1296784, EPI_ISL_1296785, EPI_ISL_1296787, EPI_ISL_1296788, EPI_ISL_1296789, EPI_ISL_1296790 | | | |
| see above | Sonora Quest Laboratories | TGen North | "Jolene Bowers, Heather Centner, Chris French, Hayley Yaglom, Ashlyn Pfeiffer, Darrin Lemmer, Dave Engelthaler, The Arizona COVID Genomics Union (ACGU)" |
| EPI_ISL_1296791 | TGen North | TGen North | "Jolene Bowers, Heather Centner, Chris French, Hayley Yaglom, Ashlyn Pfeiffer, Darrin Lemmer, Dave Engelthaler, The Arizona COVID Genomics Union (ACGU)" |
| EPI_ISL_1296792, EPI_ISL_1296793, EPI_ISL_1296794, EPI_ISL_1296795, EPI_ISL_1296796, EPI_ISL_1296798 | Sonora Quest Laboratories | TGen North | "Jolene Bowers, Heather Centner, Chris French, Hayley Yaglom, Ashlyn Pfeiffer, Darrin Lemmer, Dave Engelthaler, The Arizona COVID Genomics Union (ACGU)" |
| EPI_ISL_1296799 | TGen North | TGen North | "Jolene Bowers, Heather Centner, Chris French, Hayley Yaglom, Ashlyn Pfeiffer, Darrin Lemmer, Dave Engelthaler, The Arizona COVID Genomics Union (ACGU)" |
| EPI_ISL_1296800, EPI_ISL_1296801, EPI_ISL_1296803, EPI_ISL_1296804, EPI_ISL_1296805, EPI_ISL_1296807, EPI_ISL_1296808, EPI_ISL_1296809, EPI_ISL_1296810, EPI_ISL_1296812, EPI_ISL_1296813, EPI_ISL_1296817, EPI_ISL_1296818, EPI_ISL_1296820 | | | |
| see above | Sonora Quest Laboratories | TGen North | "Jolene Bowers, Heather Centner, Chris French, Hayley Yaglom, Ashlyn Pfeiffer, Darrin Lemmer, Dave Engelthaler, The Arizona COVID Genomics Union (ACGU)" |
| EPI_ISL_1296823 | TGen North | TGen North | "Jolene Bowers, Heather Centner, Chris French, Hayley Yaglom, Ashlyn Pfeiffer, Darrin Lemmer, Dave Engelthaler, The Arizona COVID Genomics Union (ACGU)" |
| EPI_ISL_1296831, EPI_ISL_1296835, EPI_ISL_1296839, EPI_ISL_1296842, EPI_ISL_1296844, EPI_ISL_1296845, EPI_ISL_1296846, EPI_ISL_1296847, EPI_ISL_1296848, EPI_ISL_1296850, EPI_ISL_1296851, EPI_ISL_1296853, EPI_ISL_1296854, EPI_ISL_1296855, EPI_ISL_1296857, EPI_ISL_1296858, EPI_ISL_1296859, EPI_ISL_1296860, EPI_ISL_1296861, EPI_ISL_1296862, EPI_ISL_1296864, EPI_ISL_1296865, EPI_ISL_1296866, EPI_ISL_1296867, EPI_ISL_1296870, EPI_ISL_1296873, EPI_ISL_1296874, EPI_ISL_1296875, EPI_ISL_1296876, EPI_ISL_1296879, EPI_ISL_1296881, EPI_ISL_1296882, EPI_ISL_1296884, EPI_ISL_1296886, EPI_ISL_1296887, EPI_ISL_1296889, EPI_ISL_1296890, EPI_ISL_1296891, EPI_ISL_1296892, EPI_ISL_1296893, EPI_ISL_1296894, EPI_ISL_1296895, EPI_ISL_1296896 | | | |
| see above | Sonora Quest Laboratories | TGen North | "Jolene Bowers, Heather Centner, Chris French, Hayley Yaglom, Ashlyn Pfeiffer, Darrin Lemmer, Dave Engelthaler, The Arizona COVID Genomics Union (ACGU)" |
| EPI_ISL_1296898 | TGen North | TGen North | "Jolene Bowers, Heather Centner, Chris French, Hayley Yaglom, Ashlyn Pfeiffer, Darrin Lemmer, Dave Engelthaler, The Arizona COVID Genomics Union (ACGU)" |
| EPI_ISL_1296900, EPI_ISL_1296901, EPI_ISL_1296902, EPI_ISL_1296903, EPI_ISL_1296904, EPI_ISL_1296905, EPI_ISL_1296907, EPI_ISL_1296908, EPI_ISL_1296910, EPI_ISL_1296911, EPI_ISL_1296913, EPI_ISL_1296914, EPI_ISL_1296915, EPI_ISL_1296916, EPI_ISL_1296917, EPI_ISL_1296918, EPI_ISL_1296919, EPI_ISL_1296920, EPI_ISL_1296921, EPI_ISL_1296922, EPI_ISL_1296923, EPI_ISL_1296924, EPI_ISL_1296925, EPI_ISL_1296928, EPI_ISL_1296930, EPI_ISL_1296931, EPI_ISL_1296932, EPI_ISL_1296933, EPI_ISL_1296934, EPI_ISL_1296935, EPI_ISL_1296938, EPI_ISL_1296941, EPI_ISL_1296942, EPI_ISL_1296944, EPI_ISL_1296945, EPI_ISL_1296946, EPI_ISL_1296947, EPI_ISL_1296948, EPI_ISL_1296951, EPI_ISL_1296952, EPI_ISL_1296953 | | | |
| see above | Sonora Quest Laboratories | TGen North | "Jolene Bowers, Heather Centner, Chris French, Hayley Yaglom, Ashlyn Pfeiffer, Darrin Lemmer, Dave Engelthaler, The Arizona COVID Genomics Union (ACGU)" |
| EPI_ISL_1296955 | TGen North | TGen North | "Jolene Bowers, Heather Centner, Chris French, Hayley Yaglom, Ashlyn Pfeiffer, Darrin Lemmer, Dave Engelthaler, The Arizona COVID Genomics Union (ACGU)" |
| EPI_ISL_1296956, EPI_ISL_1296957 | Sonora Quest Laboratories | TGen North | "Jolene Bowers, Heather Centner, Chris French, Hayley Yaglom, Ashlyn Pfeiffer, Darrin Lemmer, Dave Engelthaler, The Arizona COVID Genomics Union (ACGU)" |
| EPI_ISL_1296959 | TGen North | TGen North | "Jolene Bowers, Heather Centner, Chris French, Hayley Yaglom, Ashlyn Pfeiffer, Darrin Lemmer, Dave Engelthaler, The Arizona COVID Genomics Union (ACGU)" |
| EPI_ISL_1296960, EPI_ISL_1296961, EPI_ISL_1296964, EPI_ISL_1296965, EPI_ISL_1296966 | Sonora Quest Laboratories | TGen North | "Jolene Bowers, Heather Centner, Chris French, Hayley Yaglom, Ashlyn Pfeiffer, Darrin Lemmer, Dave Engelthaler, The Arizona COVID Genomics Union (ACGU)" |
| EPI_ISL_1296968 | TGen North | TGen North | "Jolene Bowers, Heather Centner, Chris French, Hayley Yaglom, Ashlyn Pfeiffer, Darrin Lemmer, Dave Engelthaler, The Arizona COVID Genomics Union (ACGU)" |
| EPI_ISL_1296970, EPI_ISL_1296971, EPI_ISL_1296972, EPI_ISL_1296973 | Sonora Quest Laboratories | TGen North | "Jolene Bowers, Heather Centner, Chris French, Hayley Yaglom, Ashlyn Pfeiffer, Darrin Lemmer, Dave Engelthaler, The Arizona COVID Genomics Union (ACGU)" |
| EPI_ISL_1296974 | TGen North | TGen North | "Jolene Bowers, Heather Centner, Chris French, Hayley Yaglom, Ashlyn Pfeiffer, Darrin Lemmer, Dave Engelthaler, The Arizona COVID Genomics Union (ACGU)" |
| EPI_ISL_1296975, EPI_ISL_1296976, EPI_ISL_1296978, EPI_ISL_1296979, EPI_ISL_1296980, EPI_ISL_1296981, EPI_ISL_1296982, EPI_ISL_1296983, EPI_ISL_1296984, EPI_ISL_1296985, EPI_ISL_1296986, EPI_ISL_1296988, EPI_ISL_1296989, EPI_ISL_1296990, EPI_ISL_1296992 | | | |
| see above | Sonora Quest Laboratories | TGen North | "Jolene Bowers, Heather Centner, Chris French, Hayley Yaglom, Ashlyn Pfeiffer, Darrin Lemmer, Dave Engelthaler, The Arizona COVID Genomics Union (ACGU)" |
| EPI_ISL_1296993 | TGen North | TGen North | "Jolene Bowers, Heather Centner, Chris French, Hayley Yaglom, Ashlyn Pfeiffer, Darrin Lemmer, Dave Engelthaler, The Arizona COVID Genomics Union (ACGU)" |
| EPI_ISL_1296994, EPI_ISL_1296995, EPI_ISL_1296996, EPI_ISL_1296997, EPI_ISL_1296998, EPI_ISL_1296999, EPI_ISL_1297000, EPI_ISL_1297001, EPI_ISL_1297002, EPI_ISL_1297003, EPI_ISL_1297004, EPI_ISL_1297005, EPI_ISL_1297006, EPI_ISL_1297007, EPI_ISL_1297008, EPI_ISL_1297009, EPI_ISL_1297010, EPI_ISL_1297011, EPI_ISL_1297013, EPI_ISL_1297014, EPI_ISL_1297015, EPI_ISL_1297016, EPI_ISL_1297017, EPI_ISL_1297018, EPI_ISL_1297019, EPI_ISL_1297020, EPI_ISL_1297021, EPI_ISL_1297022, EPI_ISL_1297023, EPI_ISL_1297024, EPI_ISL_1297025, EPI_ISL_1297028, EPI_ISL_1297030, EPI_ISL_1297031, EPI_ISL_1297032, EPI_ISL_1297033, EPI_ISL_1297034, EPI_ISL_1297035, EPI_ISL_1297036, EPI_ISL_1297037, EPI_ISL_1297038, EPI_ISL_1297039, EPI_ISL_1297040, EPI_ISL_1297042, EPI_ISL_1297043, EPI_ISL_1297044, EPI_ISL_1297045, EPI_ISL_1297046 | | | |
| see above | Sonora Quest Laboratories | TGen North | "Jolene Bowers, Heather Centner, Chris French, Hayley Yaglom, Ashlyn Pfeiffer, Darrin Lemmer, Dave Engelthaler, The Arizona COVID Genomics Union (ACGU)" |
| EPI_ISL_1297047 | TGen North | TGen North | "Jolene Bowers, Heather Centner, Chris French, Hayley Yaglom, Ashlyn Pfeiffer, Darrin Lemmer, Dave Engelthaler, The Arizona COVID Genomics Union (ACGU)" |
| EPI_ISL_1297048, EPI_ISL_1297051, EPI_ISL_1297052, EPI_ISL_1297053, EPI_ISL_1297054 | Sonora Quest Laboratories | TGen North | "Jolene Bowers, Heather Centner, Chris French, Hayley Yaglom, Ashlyn Pfeiffer, Darrin Lemmer, Dave Engelthaler, The Arizona COVID Genomics Union (ACGU)" |
| EPI_ISL_1297056 | TGen North | TGen North | "Jolene Bowers, Heather Centner, Chris French, Hayley Yaglom, Ashlyn Pfeiffer, Darrin Lemmer, Dave Engelthaler, The Arizona COVID Genomics Union (ACGU)" |
| EPI_ISL_1297057, EPI_ISL_1297058, EPI_ISL_1297059, EPI_ISL_1297060, EPI_ISL_1297061, EPI_ISL_1297063, EPI_ISL_1297064, EPI_ISL_1297065, EPI_ISL_1297066, EPI_ISL_1297068, EPI_ISL_1297070, EPI_ISL_1297071, EPI_ISL_1297072, EPI_ISL_1297073, EPI_ISL_1297075, EPI_ISL_1297076 | | | |
| see above | Sonora Quest Laboratories | TGen North | "Jolene Bowers, Heather Centner, Chris French, Hayley Yaglom, Ashlyn Pfeiffer, Darrin Lemmer, Dave Engelthaler, The Arizona COVID Genomics Union (ACGU)" |
| EPI_ISL_1297077 | TGen North | TGen North | "Jolene Bowers, Heather Centner, Chris French, Hayley Yaglom, Ashlyn Pfeiffer, Darrin Lemmer, Dave Engelthaler, The Arizona COVID Genomics Union (ACGU)" |
| EPI_ISL_1297078 | Sonora Quest Laboratories | TGen North | "Jolene Bowers, Heather Centner, Chris French, Hayley Yaglom, Ashlyn Pfeiffer, Darrin Lemmer, Dave Engelthaler, The Arizona COVID Genomics Union (ACGU)" |
| EPI_ISL_1297082 | TGen North | TGen North | "Jolene Bowers, Heather Centner, Chris French, Hayley Yaglom, Ashlyn Pfeiffer, Darrin Lemmer, Dave Engelthaler, The Arizona COVID Genomics Union (ACGU)" |
| EPI_ISL_1297083, EPI_ISL_1297084, EPI_ISL_1297085, EPI_ISL_1297086, EPI_ISL_1297088, EPI_ISL_1297089, EPI_ISL_1297090, EPI_ISL_1297091, EPI_ISL_1297093, EPI_ISL_1297094, EPI_ISL_1297095 | | | |
| see above | Sonora Quest Laboratories | TGen North | "Jolene Bowers, Heather Centner, Chris French, Hayley Yaglom, Ashlyn Pfeiffer, Darrin Lemmer, Dave Engelthaler, The Arizona COVID Genomics Union (ACGU)" |

| | | | |
|---|---|---|---|
| EPI_ISL_1297096 | TGen North | TGen North | "Jolene Bowers, Heather Centner, Chris French, Hayley Yaglom, Ashlyn Pfeiffer, Darrin Lemmer, Dave Engelthaler, The Arizona COVID Genomics Union (ACGU)" |
| EPI_ISL_1297097, EPI_ISL_1297098, EPI_ISL_1297100, EPI_ISL_1297101, EPI_ISL_1297102, EPI_ISL_1297103, EPI_ISL_1297104, EPI_ISL_1297105, EPI_ISL_1297106, EPI_ISL_1297107, EPI_ISL_1297108, EPI_ISL_1297109, EPI_ISL_1297110, EPI_ISL_1297111, EPI_ISL_1297112, EPI_ISL_1297114, EPI_ISL_1297115, EPI_ISL_1297116, EPI_ISL_1297117 | | | |
| see above | Sonora Quest Laboratories | TGen North | "Jolene Bowers, Heather Centner, Chris French, Hayley Yaglom, Ashlyn Pfeiffer, Darrin Lemmer, Dave Engelthaler, The Arizona COVID Genomics Union (ACGU)" |
| EPI_ISL_1297118 | TGen North | TGen North | "Jolene Bowers, Heather Centner, Chris French, Hayley Yaglom, Ashlyn Pfeiffer, Darrin Lemmer, Dave Engelthaler, The Arizona COVID Genomics Union (ACGU)" |
| EPI_ISL_1297119, EPI_ISL_1297120, EPI_ISL_1297121, EPI_ISL_1297122, EPI_ISL_1297125, EPI_ISL_1297126, EPI_ISL_1297128, EPI_ISL_1297129, EPI_ISL_1297131, EPI_ISL_1297132, EPI_ISL_1297133, EPI_ISL_1297134, EPI_ISL_1297135, EPI_ISL_1297136, EPI_ISL_1297137, EPI_ISL_1297138, EPI_ISL_1297139, EPI_ISL_1297140, EPI_ISL_1297143, EPI_ISL_1297145, EPI_ISL_1297146, EPI_ISL_1297148, EPI_ISL_1297149, EPI_ISL_1297150, EPI_ISL_1297151, EPI_ISL_1297152, EPI_ISL_1297153, EPI_ISL_1297155, EPI_ISL_1297156, EPI_ISL_1297157, EPI_ISL_1297158, EPI_ISL_1297159, EPI_ISL_1297160, EPI_ISL_1297161, EPI_ISL_1297162, EPI_ISL_1297163 | | | |
| see above | Sonora Quest Laboratories | TGen North | "Jolene Bowers, Heather Centner, Chris French, Hayley Yaglom, Ashlyn Pfeiffer, Darrin Lemmer, Dave Engelthaler, The Arizona COVID Genomics Union (ACGU)" |
| EPI_ISL_1297197 | Microbiology Division, South Carolina Department of Health and Environmental Control Public Health Laboratory (SC DHEC PHL) | Microbiology Division, South Carolina Department of Health and Environmental Control Public Health Laboratory (SC DHEC PHL) | Haley Flores, Jessica Freeman |
| EPI_ISL_1297198, EPI_ISL_1297199, EPI_ISL_1297200, EPI_ISL_1297201 | DC Public Health Lab/ Dept. of Forensic Sciences | DC Public Health Lab/ Dept. of Forensic Sciences | Scott Nguyen, Janis Doss, Elizabeth Zelaya, Sarah Scott, Connie Maza, Monica Mann, Brittany Hamilton, David Payne, Jocelyn Hauser |
| EPI_ISL_1297492, EPI_ISL_1297493, EPI_ISL_1297494 | Quest Diagnostics Incorporated | Centers for Disease Control and Prevention Division of Viral Diseases, Pathogen Discovery | Peter W. Cook, Dakota Howard, Dhwani Batra, Ben L. Rambo-Martin, S. H. Rosenthal, A. Gerasimova, R. M. Kagan, B. Anderson, M. Hua, Y. Liu, L.E. Bernstein, K.E. Livingston, A. Perez, I. A. Shlyakhter, R. V. Rolando, R. Owen, P. Tanpaiboon, F. Lacbawan, Clinton R. Paden, Suxiang Tong, Duncan MacCannell |
| EPI_ISL_1297495, EPI_ISL_1297496 | UW Virology Lab | UW Virology Lab | Pavitra Roychoudhury, Hong Xie, Lasata Shrestha, Shah Mohamed Bakhash, Michelle Lin, Noah Baker, Sean Ellis, Saraswathi Sathees, Meei-Li Huang, Keith R Jerome, Alexander Greninger |
| EPI_ISL_1297497, EPI_ISL_1297498, EPI_ISL_1297499, EPI_ISL_1297500, EPI_ISL_1297501, EPI_ISL_1297502, EPI_ISL_1297503, EPI_ISL_1297504, EPI_ISL_1297505, EPI_ISL_1297506, EPI_ISL_1297507, EPI_ISL_1297508, EPI_ISL_1297509, EPI_ISL_1297510, EPI_ISL_1297511, EPI_ISL_1297512, EPI_ISL_1297513, EPI_ISL_1297514, EPI_ISL_1297515, EPI_ISL_1297516, EPI_ISL_1297517, EPI_ISL_1297518, EPI_ISL_1297519, EPI_ISL_1297520, EPI_ISL_1297521, EPI_ISL_1297522, EPI_ISL_1297523, EPI_ISL_1297524, EPI_ISL_1297525, EPI_ISL_1297526, EPI_ISL_1297527, EPI_ISL_1297528, EPI_ISL_1297529, EPI_ISL_1297530, EPI_ISL_1297531, EPI_ISL_1297532, EPI_ISL_1297533, EPI_ISL_1297534, EPI_ISL_1297535, EPI_ISL_1297536, EPI_ISL_1297537, EPI_ISL_1297538, EPI_ISL_1297539, EPI_ISL_1297540, EPI_ISL_1297541, EPI_ISL_1297542, EPI_ISL_1297543, EPI_ISL_1297544, EPI_ISL_1297545, EPI_ISL_1297546, EPI_ISL_1297547, EPI_ISL_1297548, EPI_ISL_1297549, EPI_ISL_1297550, EPI_ISL_1297551, EPI_ISL_1297552, EPI_ISL_1297553, EPI_ISL_1297554, EPI_ISL_1297555, EPI_ISL_1297556, EPI_ISL_1297557, EPI_ISL_1297558, EPI_ISL_1297559, EPI_ISL_1297560, EPI_ISL_1297561, EPI_ISL_1297562, EPI_ISL_1297563, EPI_ISL_1297564, EPI_ISL_1297565, EPI_ISL_1297566, EPI_ISL_1297567, EPI_ISL_1297568, EPI_ISL_1297569, EPI_ISL_1297570, EPI_ISL_1297571, EPI_ISL_1297572, EPI_ISL_1297573, EPI_ISL_1297574, EPI_ISL_1297575, EPI_ISL_1297576, EPI_ISL_1297577, EPI_ISL_1297578, EPI_ISL_1297579, EPI_ISL_1297580, EPI_ISL_1297581, EPI_ISL_1297582, EPI_ISL_1297583, EPI_ISL_1297584, EPI_ISL_1297585, EPI_ISL_1297586, EPI_ISL_1297587, EPI_ISL_1297588, EPI_ISL_1297589, EPI_ISL_1297590, EPI_ISL_1297591, EPI_ISL_1297592, EPI_ISL_1297593, EPI_ISL_1297594, EPI_ISL_1297595, EPI_ISL_1297596, EPI_ISL_1297597, EPI_ISL_1297598, EPI_ISL_1297599, EPI_ISL_1297600, EPI_ISL_1297601, EPI_ISL_1297602, EPI_ISL_1297603, EPI_ISL_1297604, EPI_ISL_1297605, EPI_ISL_1297606, EPI_ISL_1297607, EPI_ISL_1297608, EPI_ISL_1297609, EPI_ISL_1297610, EPI_ISL_1297611, EPI_ISL_1297612, EPI_ISL_1297613, EPI_ISL_1297614, EPI_ISL_1297615, EPI_ISL_1297616, EPI_ISL_1297617, EPI_ISL_1297618, EPI_ISL_1297619, EPI_ISL_1297620, EPI_ISL_1297621, EPI_ISL_1297622, EPI_ISL_1297623, EPI_ISL_1297624, EPI_ISL_1297625, EPI_ISL_1297626, EPI_ISL_1297627, EPI_ISL_1297628, EPI_ISL_1297629, EPI_ISL_1297630, EPI_ISL_1297631, EPI_ISL_1297632, EPI_ISL_1297633, EPI_ISL_1297634, EPI_ISL_1297635, EPI_ISL_1297636, EPI_ISL_1297637, EPI_ISL_1297638, EPI_ISL_1297639, EPI_ISL_1297640, EPI_ISL_1297641, EPI_ISL_1297642, EPI_ISL_1297643, EPI_ISL_1297644, EPI_ISL_1297645, EPI_ISL_1297646, EPI_ISL_1297647, EPI_ISL_1297648, EPI_ISL_1297649, EPI_ISL_1297650, EPI_ISL_1297651, EPI_ISL_1297652, EPI_ISL_1297653, EPI_ISL_1297654, EPI_ISL_1297655, EPI_ISL_1297656, EPI_ISL_1297657, EPI_ISL_1297658, EPI_ISL_1297659, EPI_ISL_1297660, EPI_ISL_1297661, EPI_ISL_1297662, EPI_ISL_1297663, EPI_ISL_1297664, EPI_ISL_1297665, EPI_ISL_1297666, EPI_ISL_1297667, EPI_ISL_1297668, EPI_ISL_1297669, EPI_ISL_1297670, EPI_ISL_1297671, EPI_ISL_1297672, EPI_ISL_1297673, EPI_ISL_1297674, EPI_ISL_1297675, EPI_ISL_1297676, EPI_ISL_1297677, EPI_ISL_1297678, EPI_ISL_1297679, EPI_ISL_1297680, EPI_ISL_1297681, EPI_ISL_1297682, EPI_ISL_1297683, EPI_ISL_1297684, EPI_ISL_1297685, EPI_ISL_1297686, EPI_ISL_1297687, EPI_ISL_1297688, EPI_ISL_1297689, EPI_ISL_1297690, EPI_ISL_1297691, EPI_ISL_1297692, EPI_ISL_1297693, EPI_ISL_1297694, EPI_ISL_1297695, EPI_ISL_1297696, EPI_ISL_1297697, EPI_ISL_1297698, EPI_ISL_1297699, EPI_ISL_1297700, EPI_ISL_1297701, EPI_ISL_1297702, EPI_ISL_1297703, EPI_ISL_1297704, EPI_ISL_1297705, EPI_ISL_1297706, EPI_ISL_1297707, EPI_ISL_1297708, EPI_ISL_1297709, EPI_ISL_1297710, EPI_ISL_1297711, EPI_ISL_1297712, EPI_ISL_1297713, EPI_ISL_1297714, EPI_ISL_1297715, EPI_ISL_1297716, EPI_ISL_1297717, EPI_ISL_1297718, EPI_ISL_1297719, EPI_ISL_1297720, EPI_ISL_1297721, EPI_ISL_1297722, EPI_ISL_1297723, EPI_ISL_1297724, EPI_ISL_1297725, EPI_ISL_1297726, EPI_ISL_1297727, EPI_ISL_1297728, EPI_ISL_1297729, EPI_ISL_1297730, EPI_ISL_1297731, EPI_ISL_1297732, EPI_ISL_1297733, EPI_ISL_1297734, EPI_ISL_1297735, EPI_ISL_1297736, EPI_ISL_1297737, EPI_ISL_1297738, EPI_ISL_1297739, EPI_ISL_1297740, EPI_ISL_1297741, EPI_ISL_1297742, EPI_ISL_1297743, EPI_ISL_1297744, EPI_ISL_1297745, EPI_ISL_1297746, EPI_ISL_1297747, EPI_ISL_1297748, EPI_ISL_1297749, EPI_ISL_1297750, EPI_ISL_1297751, EPI_ISL_1297752, EPI_ISL_1297753, EPI_ISL_1297754, EPI_ISL_1297755, EPI_ISL_1297756, EPI_ISL_1297757, EPI_ISL_1297758, EPI_ISL_1297759, EPI_ISL_1297760, EPI_ISL_1297761, EPI_ISL_1297762, EPI_ISL_1297763, EPI_ISL_1297764, EPI_ISL_1297765, EPI_ISL_1297766, EPI_ISL_1297767, EPI_ISL_1297768, EPI_ISL_1297769, EPI_ISL_1297770, EPI_ISL_1297771, EPI_ISL_1297772, EPI_ISL_1297773, EPI_ISL_1297774, EPI_ISL_1297775, EPI_ISL_1297776, EPI_ISL_1297777, EPI_ISL_1297778, EPI_ISL_1297779, EPI_ISL_1297780, EPI_ISL_1297781, EPI_ISL_1297782, EPI_ISL_1297783, EPI_ISL_1297784, EPI_ISL_1297785, EPI_ISL_1297786, EPI_ISL_1297787, EPI_ISL_1297788, EPI_ISL_1297789, EPI_ISL_1297790, EPI_ISL_1297791, EPI_ISL_1297792, EPI_ISL_1297793, EPI_ISL_1297794, EPI_ISL_1297795, EPI_ISL_1297796, EPI_ISL_1297797, EPI_ISL_1297798, EPI_ISL_1297799, EPI_ISL_1297800, EPI_ISL_1297801, EPI_ISL_1297802, EPI_ISL_1297803, EPI_ISL_1297804, EPI_ISL_1297805, EPI_ISL_1297806, EPI_ISL_1297807, EPI_ISL_1297808, EPI_ISL_1297809, EPI_ISL_1297810, EPI_ISL_1297811, EPI_ISL_1297812, EPI_ISL_1297813, EPI_ISL_1297814, EPI_ISL_1297815, EPI_ISL_1297816, EPI_ISL_1297817, EPI_ISL_1297818, EPI_ISL_1297819, EPI_ISL_1297820, EPI_ISL_1297821, EPI_ISL_1297822, EPI_ISL_1297823, EPI_ISL_1297824, EPI_ISL_1297825, EPI_ISL_1297826 | | | |
| see above | Quest Diagnostics Incorporated | Centers for Disease Control and Prevention Division of Viral Diseases, Pathogen Discovery | Peter W. Cook, Dakota Howard, Dhwani Batra, Ben L. Rambo-Martin, S. H. Rosenthal, A. Gerasimova, R. M. Kagan, B. Anderson, M. Hua, Y. Liu, L.E. Bernstein, K.E. Livingston, A. Perez, I. A. Shlyakhter, R. V. Rolando, R. Owen, P. Tanpaiboon, F. Lacbawan, Clinton R. Paden, Suxiang Tong, Duncan MacCannell |
| EPI_ISL_1297851, EPI_ISL_1297852, EPI_ISL_1297853, EPI_ISL_1297854, EPI_ISL_1297855, EPI_ISL_1297856, EPI_ISL_1297857, EPI_ISL_1297858, EPI_ISL_1297859, EPI_ISL_1297860, EPI_ISL_1297861, EPI_ISL_1297862, EPI_ISL_1297863, EPI_ISL_1297864, EPI_ISL_1297865, EPI_ISL_1297866, EPI_ISL_1297867, EPI_ISL_1297868, EPI_ISL_1297869, EPI_ISL_1297870, EPI_ISL_1297871, EPI_ISL_1297872, EPI_ISL_1297873, EPI_ISL_1297874, EPI_ISL_1297875, EPI_ISL_1297876, EPI_ISL_1297877, EPI_ISL_1297878, EPI_ISL_1297879, EPI_ISL_1297880, EPI_ISL_1297881, EPI_ISL_1297882, EPI_ISL_1297883, EPI_ISL_1297884, EPI_ISL_1297885, EPI_ISL_1297886, EPI_ISL_1297887, EPI_ISL_1297888, EPI_ISL_1297889, EPI_ISL_1297890, EPI_ISL_1297892, EPI_ISL_1297893, EPI_ISL_1297894, EPI_ISL_1297895, EPI_ISL_1297896, EPI_ISL_1297897, EPI_ISL_1297898, EPI_ISL_1297899, EPI_ISL_1297900, EPI_ISL_1297901, EPI_ISL_1297902, EPI_ISL_1297903, EPI_ISL_1297904, EPI_ISL_1297905, EPI_ISL_1297906, EPI_ISL_1297907, EPI_ISL_1297908, EPI_ISL_1297909, EPI_ISL_1297910, EPI_ISL_1297912, EPI_ISL_1297913, EPI_ISL_1297914, EPI_ISL_1297915, EPI_ISL_1297916, EPI_ISL_1297917, EPI_ISL_1297918, EPI_ISL_1297919, EPI_ISL_1297920, EPI_ISL_1297921, EPI_ISL_1297922, EPI_ISL_1297923, EPI_ISL_1297924, EPI_ISL_1297925, EPI_ISL_1297926, EPI_ISL_1297927, EPI_ISL_1297928, EPI_ISL_1297929, EPI_ISL_1297930, EPI_ISL_1297931, EPI_ISL_1297932, EPI_ISL_1297933, EPI_ISL_1297934, EPI_ISL_1297935, EPI_ISL_1297936, EPI_ISL_1297937, EPI_ISL_1297938, EPI_ISL_1297939, EPI_ISL_1297940, EPI_ISL_1297941, EPI_ISL_1297942, EPI_ISL_1297943, EPI_ISL_1297944, EPI_ISL_1297945, EPI_ISL_1297946, EPI_ISL_1297947, EPI_ISL_1297948, EPI_ISL_1297949, EPI_ISL_1297950, EPI_ISL_1297951, EPI_ISL_1297952, EPI_ISL_1297953, EPI_ISL_1297954, EPI_ISL_1297955, EPI_ISL_1297956, EPI_ISL_1297957, EPI_ISL_1297958, EPI_ISL_1297959, EPI_ISL_1297960, EPI_ISL_1297961, EPI_ISL_1297962, EPI_ISL_1297963, EPI_ISL_1297964, EPI_ISL_1297965, EPI_ISL_1297966, EPI_ISL_1297967, EPI_ISL_1297968, EPI_ISL_1297969, EPI_ISL_1297970, EPI_ISL_1297971, EPI_ISL_1297972, EPI_ISL_1297973, EPI_ISL_1297974, EPI_ISL_1297975, EPI_ISL_1297976, EPI_ISL_1297977, EPI_ISL_1297978, EPI_ISL_1297979, EPI_ISL_1297980, EPI_ISL_1297981, EPI_ISL_1297982, EPI_ISL_1297983, EPI_ISL_1297984, EPI_ISL_1297985, EPI_ISL_1297986, EPI_ISL_1297987, EPI_ISL_1297988, EPI_ISL_1297989, EPI_ISL_1297990, EPI_ISL_1297991, EPI_ISL_1297992, EPI_ISL_1297993, EPI_ISL_1297994, EPI_ISL_1297995, EPI_ISL_1297996, EPI_ISL_1297997, EPI_ISL_1297998, EPI_ISL_1297999, EPI_ISL_1298000, EPI_ISL_1298001, EPI_ISL_1298002, EPI_ISL_1298003, EPI_ISL_1298004, EPI_ISL_1298005, EPI_ISL_1298006, EPI_ISL_1298007, EPI_ISL_1298008, EPI_ISL_1298009, EPI_ISL_1298010, EPI_ISL_1298011, EPI_ISL_1298012, EPI_ISL_1298013, EPI_ISL_1298014, EPI_ISL_1298015, EPI_ISL_1298016, EPI_ISL_1298017, EPI_ISL_1298018, EPI_ISL_1298019, EPI_ISL_1298020, EPI_ISL_1298021, EPI_ISL_1298022, EPI_ISL_1298023, EPI_ISL_1298024, EPI_ISL_1298025, EPI_ISL_1298026, EPI_ISL_1298027, EPI_ISL_1298028, EPI_ISL_1298029, EPI_ISL_1298030, EPI_ISL_1298031, EPI_ISL_1298032, EPI_ISL_1298033, EPI_ISL_1298034, EPI_ISL_1298035, EPI_ISL_1298036, EPI_ISL_1298037, EPI_ISL_1298038, EPI_ISL_1298039, EPI_ISL_1298040, EPI_ISL_1298041, EPI_ISL_1298042, EPI_ISL_1298043, EPI_ISL_1298044, EPI_ISL_1298045, EPI_ISL_1298046, EPI_ISL_1298047, EPI_ISL_1298048, EPI_ISL_1298049, EPI_ISL_1298050, EPI_ISL_1298051, EPI_ISL_1298052, EPI_ISL_1298053, EPI_ISL_1298054, EPI_ISL_1298055, EPI_ISL_1298056, EPI_ISL_1298057, EPI_ISL_1298058, EPI_ISL_1298059, EPI_ISL_1298060, EPI_ISL_1298061, EPI_ISL_1298062, EPI_ISL_1298063, EPI_ISL_1298065, EPI_ISL_1298066, EPI_ISL_1298067, EPI_ISL_1298068, EPI_ISL_1298069, EPI_ISL_1298070, EPI_ISL_1298071, EPI_ISL_1298072, EPI_ISL_1298073, EPI_ISL_1298074, EPI_ISL_1298075, EPI_ISL_1298076, EPI_ISL_1298077, EPI_ISL_1298078, EPI_ISL_1298079, EPI_ISL_1298080, EPI_ISL_1298081, EPI_ISL_1298082, EPI_ISL_1298083, EPI_ISL_1298084, EPI_ISL_1298085, EPI_ISL_1298086, EPI_ISL_1298087, EPI_ISL_1298088, EPI_ISL_1298089, EPI_ISL_1298090, EPI_ISL_1298091, EPI_ISL_1298092, EPI_ISL_1298093, EPI_ISL_1298094, EPI_ISL_1298095, EPI_ISL_1298096, EPI_ISL_1298097, EPI_ISL_1298098, EPI_ISL_1298099, EPI_ISL_1298100, EPI_ISL_1298101, EPI_ISL_1298102, EPI_ISL_1298103, EPI_ISL_1298104, EPI_ISL_1298105, EPI_ISL_1298106, EPI_ISL_1298107, EPI_ISL_1298108, EPI_ISL_1298109, EPI_ISL_1298110, EPI_ISL_1298111, EPI_ISL_1298112, EPI_ISL_1298113, EPI_ISL_1298114, EPI_ISL_1298115, EPI_ISL_1298116, EPI_ISL_1298117, EPI_ISL_1298118, EPI_ISL_1298119, EPI_ISL_1298120, EPI_ISL_1298121, EPI_ISL_1298122, EPI_ISL_1298123, EPI_ISL_1298124, EPI_ISL_1298125, EPI_ISL_1298126, EPI_ISL_1298127, EPI_ISL_1298128, EPI_ISL_1298129, EPI_ISL_1298130, EPI_ISL_1298131, EPI_ISL_1298132, EPI_ISL_1298133, EPI_ISL_1298134, EPI_ISL_1298135, EPI_ISL_1298136, EPI_ISL_1298137, EPI_ISL_1298138, EPI_ISL_1298139, EPI_ISL_1298140, EPI_ISL_1298141, EPI_ISL_1298142, EPI_ISL_1298143, EPI_ISL_1298144, EPI_ISL_1298145, EPI_ISL_1298146, EPI_ISL_1298147, EPI_ISL_1298148, EPI_ISL_1298149, EPI_ISL_1298150, EPI_ISL_1298151, EPI_ISL_1298152, EPI_ISL_1298153, EPI_ISL_1298154, EPI_ISL_1298155, EPI_ISL_1298156, EPI_ISL_1298157, EPI_ISL_1298158, EPI_ISL_1298159, EPI_ISL_1298160, EPI_ISL_1298161, EPI_ISL_1298162, EPI_ISL_1298163, EPI_ISL_1298164, EPI_ISL_1298165, EPI_ISL_1298166, EPI_ISL_1298167, EPI_ISL_1298168, EPI_ISL_1298169, EPI_ISL_1298171, EPI_ISL_1298172, EPI_ISL_1298173, EPI_ISL_1298174, EPI_ISL_1298175, EPI_ISL_1298177, EPI_ISL_1298179, EPI_ISL_1298182, EPI_ISL_1298185, EPI_ISL_1298187, EPI_ISL_1298190, EPI_ISL_1298193, EPI_ISL_1298195, EPI_ISL_1298196, EPI_ISL_1298197, EPI_ISL_1298198, EPI_ISL_1298199, EPI_ISL_1298200, EPI_ISL_1298201, EPI_ISL_1298202, EPI_ISL_1298203, EPI_ISL_1298204, EPI_ISL_1298205, EPI_ISL_1298206, EPI_ISL_1298207, EPI_ISL_1298208, EPI_ISL_1298209, EPI_ISL_1298210, EPI_ISL_1298211, EPI_ISL_1298212, EPI_ISL_1298213, EPI_ISL_1298214, EPI_ISL_1298215, EPI_ISL_1298216, EPI_ISL_1298218, EPI_ISL_1298219, EPI_ISL_1298220, EPI_ISL_1298221, EPI_ISL_1298222, EPI_ISL_1298223, EPI_ISL_1298224, EPI_ISL_1298225 | | | |
| see above | Helix/Illumina | Centers for Disease Control and Prevention Division of Viral Diseases, Pathogen Discovery | Peter W. Cook, Dakota Howard, Dhwani Batra, Ben L. Rambo-Martin, Eileen de Feo, Jan Antico, Christine Tran, Matthew Tolentino, Shannon Wickline, Kim Gietzen, Brad Sickler, Jingtao Liu, Eric Allen, Phil Febbo, Summer Galloway, Nicole L. Washington, Simon White, Geraint Levan, Kelly Schiabor Barrett, Elizabeth Cirulli, Alexandre Bolze, Ary Ascencio, Charlotte Rivera-Garcia, Ryan Cho, Jason Nguyen, Sherry Wang, Jimmy Ramirez, Tyler Cassens, Efren Sandoval, Magnus Isaksson, William Lee, David Becker, Marc Laurent, James Lu, Clinton R. Paden, Suxiang Tong, Duncan MacCannell |
| EPI_ISL_1298226, EPI_ISL_1298227, EPI_ISL_1298228, EPI_ISL_1298229, EPI_ISL_1298230, EPI_ISL_1298231, EPI_ISL_1298232, EPI_ISL_1298233, EPI_ISL_1298234, EPI_ISL_1298235, EPI_ISL_1298236, EPI_ISL_1298237, EPI_ISL_1298238, EPI_ISL_1298239, EPI_ISL_1298240, EPI_ISL_1298241, EPI_ISL_1298242, EPI_ISL_1298243, EPI_ISL_1298244, EPI_ISL_1298245, EPI_ISL_1298246 | | | |
| see above | Quest Diagnostics Incorporated | Centers for Disease Control and Prevention Division of Viral Diseases, Pathogen Discovery | Peter W. Cook, Dakota Howard, Dhwani Batra, Ben L. Rambo-Martin, S. H. Rosenthal, A. Gerasimova, R. M. Kagan, B. Anderson, M. Hua, Y. Liu, L.E. Bernstein, K.E. Livingston, A. Perez, I. A. Shlyakhter, R. V. Rolando, R. Owen, P. Tanpaiboon, F. Lacbawan, Clinton R. Paden, Suxiang Tong, Duncan MacCannell |
| EPI_ISL_1298251, EPI_ISL_1298252, EPI_ISL_1298253, EPI_ISL_1298254, EPI_ISL_1298255, EPI_ISL_1298256, EPI_ISL_1298258, EPI_ISL_1298259, EPI_ISL_1298264, EPI_ISL_1298267, EPI_ISL_1298269, EPI_ISL_1298271, EPI_ISL_1298272, EPI_ISL_1298276, EPI_ISL_1298279, EPI_ISL_1298282, EPI_ISL_1298285, EPI_ISL_1298286, EPI_ISL_1298287, EPI_ISL_1298288, EPI_ISL_1298289, EPI_ISL_1298290, EPI_ISL_1298291, EPI_ISL_1298292, EPI_ISL_1298296, EPI_ISL_1298297, EPI_ISL_1298298, EPI_ISL_1298303, EPI_ISL_1298304, EPI_ISL_1298307, EPI_ISL_1298315, EPI_ISL_1298317, EPI_ISL_1298318, EPI_ISL_1298319, EPI_ISL_1298320, EPI_ISL_1298321, EPI_ISL_1298323, EPI_ISL_1298325, EPI_ISL_1298327, EPI_ISL_1298329, EPI_ISL_1298331, EPI_ISL_1298332, EPI_ISL_1298334, EPI_ISL_1298339, EPI_ISL_1298340, EPI_ISL_1298341, EPI_ISL_1298342, EPI_ISL_1298343, EPI_ISL_1298345, EPI_ISL_1298346, EPI_ISL_1298347, EPI_ISL_1298348, EPI_ISL_1298351, EPI_ISL_1298354, EPI_ISL_1298358, EPI_ISL_1298359, EPI_ISL_1298362, EPI_ISL_1298366, EPI_ISL_1298368, EPI_ISL_1298369, EPI_ISL_1298370, EPI_ISL_1298371, EPI_ISL_1298372, EPI_ISL_1298374, EPI_ISL_1298375, EPI_ISL_1298383, EPI_ISL_1298384, EPI_ISL_1298385, EPI_ISL_1298386, EPI_ISL_1298387, EPI_ISL_1298391, EPI_ISL_1298394, EPI_ISL_1298395, EPI_ISL_1298396, EPI_ISL_1298398, EPI_ISL_1298399, EPI_ISL_1298401, EPI_ISL_1298402, EPI_ISL_1298403, EPI_ISL_1298405, EPI_ISL_1298406, EPI_ISL_1298407, EPI_ISL_1298408, EPI_ISL_1298409, EPI_ISL_1298410, EPI_ISL_1298412, EPI_ISL_1298413, EPI_ISL_1298414, EPI_ISL_1298415, EPI_ISL_1298416, EPI_ISL_1298417, EPI_ISL_1298418, EPI_ISL_1298424, EPI_ISL_1298425, EPI_ISL_1298427, EPI_ISL_1298428, EPI_ISL_1298429 | | | |
| see above | Laboratory Corporation of America | Centers for Disease Control and Prevention Division of Viral Diseases, Pathogen Discovery | Peter W. Cook, Dakota Howard, Dhwani Batra, Ben L. Rambo-Martin, Minoo Agarwal, Eyad Almasri, Debbie Boles, Ayla Burns, Nuthawin Charoensri, Oren Cohen, Susan Countryman, Mary Ann Cristobal, Bobbi Croy, Suzanne Dale, Hrushikesh Deshmukh, Amanda Douglas, Vincent Drouillon, Marcia Eisenberg, Howard Engler, Rama Ghatti, Prashant Gupta, Susan Hicks, Jake Humphrey, Lax Iyer, Manoj Jain, Mohan Kolli, Brian Krueger, Tim Kuphal, Stanley Letovsky, Michael Levandoski, Craig Lukasik, Jonathan Meltzer, Brian Norvell, Mindy Nye, Scott Parker, Christos Petropoulos, John Pruitt, Steven Ragan, Scott Ryan, Mike Sapeta, Jana Schroth, Suresh Babu Selvaraju, Goran Stevovic, Amanda Suchanek, Andrea Throop, Lyndon Tilson, Thomas Urban, Joe Voshell, Kimberly Wagner, Jonathan Williams, Mary Williamson, Qian Zeng, Tricia Zwiefelhofer, Clinton R. Paden, Suxiang Tong, Duncan MacCannell |
| EPI_ISL_1298451, EPI_ISL_1298452, EPI_ISL_1298453, EPI_ISL_1298454, EPI_ISL_1298455, EPI_ISL_1298456, EPI_ISL_1298457, EPI_ISL_1298458, EPI_ISL_1298459, EPI_ISL_1298460, EPI_ISL_1298461, EPI_ISL_1298462, EPI_ISL_1298463, EPI_ISL_1298464, EPI_ISL_1298465, EPI_ISL_1298466, EPI_ISL_1298467, EPI_ISL_1298468, EPI_ISL_1298469, EPI_ISL_1298470, EPI_ISL_1298471, EPI_ISL_1298472, EPI_ISL_1298473 | | | |
| see above | Instituto Nacional de Medicina Genomica | Instituto Nacional de Medicina Genomica | Hidalgo-Miranda A, Mendoza-Vargas A, Reyes-Grajeda JP, Cedro-Tanda A, Alcaraz N, Gonzalez-Barrera D, Rangel-DeLeon D, Ramirez-Vega O, Herrera-Montalvo LA |
| EPI_ISL_1298489 | Kansas Health and Environmental Lab | Kansas Health and Environmental Lab | Mike Grose, Jonathan Barnell, Ben Olsen, and Phil Adam |
| EPI_ISL_1299220, EPI_ISL_1299221 | Cytocheck Laboratory | Kansas Health and Environmental Lab | Mike Grose, Jonathan Barnell, Ben Olsen, and Phil Adam |
| EPI_ISL_1299222, EPI_ISL_1299223, EPI_ISL_1299224, EPI_ISL_1299225, EPI_ISL_1299226 | Kansas Health and Environmental Lab | Kansas Health and Environmental Lab | Mike Grose, Jonathan Barnell, Ben Olsen, and Phil Adam |
| EPI_ISL_1299257, EPI_ISL_1299259, EPI_ISL_1299260, EPI_ISL_1299261, EPI_ISL_1299262, EPI_ISL_1299266, EPI_ISL_1299267, EPI_ISL_1299268, EPI_ISL_1299269, EPI_ISL_1299270, EPI_ISL_1299271, EPI_ISL_1299272 | | | |
| see above | Wyoming Public Health Laboratory | Wyoming Public Health Laboratory | Jim Mildenberger, Wanda Manley, Noah Hull, Taylor Fearing, Lynette Gumbleton, Channing Weber, Ashley Norberg, Chayse Rowley, Marley Goetz, Brian Dominguez, Elliot Thomasson, Cari Sloma, and Rob Christensen |
| EPI_ISL_1299316, EPI_ISL_1299317, EPI_ISL_1299318, EPI_ISL_1299319, EPI_ISL_1299320, EPI_ISL_1299321, EPI_ISL_1299322, EPI_ISL_1299323, EPI_ISL_1299324 | Arizona State University | Arizona State University | Peter T. Skidmore, LaRinda A. Holland, Rabia Maqsood, Emily A. Kaelin, Nicholas J. Mellor, Kristina Buss, Joy M. Blain, Neal W Woodbury, Valerie Harris, Joshua LaBaer, Vel Murugan, Efrem S. Lim |
| EPI_ISL_1300031, EPI_ISL_1300032, EPI_ISL_1300033, EPI_ISL_1300034, EPI_ISL_1300035, EPI_ISL_1300036, EPI_ISL_1300037, EPI_ISL_1300038, EPI_ISL_1300039, EPI_ISL_1300040, EPI_ISL_1300041, EPI_ISL_1300042, EPI_ISL_1300043, EPI_ISL_1300044, EPI_ISL_1300045, EPI_ISL_1300046, EPI_ISL_1300047, EPI_ISL_1300048, EPI_ISL_1300049, EPI_ISL_1300050, EPI_ISL_1300051, EPI_ISL_1300052 | | | |
| see above | Wyoming Public Health Laboratory | Wyoming Public Health Laboratory | Jim Mildenberger, Wanda Manley, Noah Hull, Taylor Fearing, Lynette Gumbleton, Channing Weber, Ashley Norberg, Chayse Rowley, Marley Goetz, Brian Dominguez, Elliot Thomasson, Cari Sloma, and Rob Christensen |
| EPI_ISL_1300053, EPI_ISL_1300054, EPI_ISL_1300055, EPI_ISL_1300056, EPI_ISL_1300057, EPI_ISL_1300058, EPI_ISL_1300059, EPI_ISL_1300060, EPI_ISL_1300061, EPI_ISL_1300062, EPI_ISL_1300063, EPI_ISL_1300064, EPI_ISL_1300065, EPI_ISL_1300066, EPI_ISL_1300067, EPI_ISL_1300068, EPI_ISL_1300069, EPI_ISL_1300070, EPI_ISL_1300071, EPI_ISL_1300073, EPI_ISL_1300074, EPI_ISL_1300075, | | | |

| | | | |
|---|---|---|---|
| EPI_ISL_1300077, EPI_ISL_1300078, EPI_ISL_1300079, EPI_ISL_1300080, EPI_ISL_1300081, EPI_ISL_1300082, EPI_ISL_1300085, EPI_ISL_1300086, EPI_ISL_1300087, EPI_ISL_1300088, EPI_ISL_1300089, EPI_ISL_1300091, EPI_ISL_1300093, EPI_ISL_1300094, EPI_ISL_1300095, EPI_ISL_1300096, EPI_ISL_1300097, EPI_ISL_1300098, EPI_ISL_1300099, EPI_ISL_1300100, EPI_ISL_1300103, EPI_ISL_1300104, EPI_ISL_1300105, EPI_ISL_1300107, EPI_ISL_1300108, EPI_ISL_1300110, EPI_ISL_1300111, EPI_ISL_1300112, EPI_ISL_1300113, EPI_ISL_1300114, EPI_ISL_1300115, EPI_ISL_1300116, EPI_ISL_1300117, EPI_ISL_1300118, EPI_ISL_1300119, EPI_ISL_1300120, EPI_ISL_1300121, EPI_ISL_1300122, EPI_ISL_1300123, EPI_ISL_1300124, EPI_ISL_1300125, EPI_ISL_1300126, EPI_ISL_1300127, EPI_ISL_1300128, EPI_ISL_1300129, EPI_ISL_1300130, EPI_ISL_1300131, EPI_ISL_1300132, EPI_ISL_1300133, EPI_ISL_1300134, EPI_ISL_1300135, EPI_ISL_1300136, EPI_ISL_1300137, EPI_ISL_1300138, EPI_ISL_1300139, EPI_ISL_1300140, EPI_ISL_1300141, EPI_ISL_1300142, EPI_ISL_1300143, EPI_ISL_1300145, EPI_ISL_1300146, EPI_ISL_1300147, EPI_ISL_1300148, EPI_ISL_1300149, EPI_ISL_1300150, EPI_ISL_1300151, EPI_ISL_1300152, EPI_ISL_1300153, EPI_ISL_1300154, EPI_ISL_1300155, EPI_ISL_1300156, EPI_ISL_1300157, EPI_ISL_1300158, EPI_ISL_1300159, EPI_ISL_1300160, EPI_ISL_1300161, EPI_ISL_1300162, EPI_ISL_1300163, EPI_ISL_1300165, EPI_ISL_1300166, EPI_ISL_1300167, EPI_ISL_1300168, EPI_ISL_1300169, EPI_ISL_1300170, EPI_ISL_1300171, EPI_ISL_1300172, EPI_ISL_1300173, EPI_ISL_1300174 | | | |
| see above | Colorado Department of Public Health and Environment | Colorado Department of Public Health and Environment | Laura Bankers, Molly C. Hetherington-Rauth, Diana Ir, Shannon Ely, Shannon R. Matzinger, Sarah Elizabeth Totten, Emily A. Travanty |
| EPI_ISL_1300175, EPI_ISL_1300176, EPI_ISL_1300177, EPI_ISL_1300178, EPI_ISL_1300179, EPI_ISL_1300180, EPI_ISL_1300181, EPI_ISL_1300182, EPI_ISL_1300183, EPI_ISL_1300184, EPI_ISL_1300185, EPI_ISL_1300186, EPI_ISL_1300187, EPI_ISL_1300188, EPI_ISL_1300189, EPI_ISL_1300190, EPI_ISL_1300191, EPI_ISL_1300192, EPI_ISL_1300193, EPI_ISL_1300194, EPI_ISL_1300195, EPI_ISL_1300196, EPI_ISL_1300197, EPI_ISL_1300198, EPI_ISL_1300199, EPI_ISL_1300200, EPI_ISL_1300201, EPI_ISL_1300202, EPI_ISL_1300203, EPI_ISL_1300204, EPI_ISL_1300205, EPI_ISL_1300206, EPI_ISL_1300207, EPI_ISL_1300208, EPI_ISL_1300209, EPI_ISL_1300210, EPI_ISL_1300211, EPI_ISL_1300213, EPI_ISL_1300214, EPI_ISL_1300215, EPI_ISL_1300216, EPI_ISL_1300217, EPI_ISL_1300218, EPI_ISL_1300219, EPI_ISL_1300220, EPI_ISL_1300221, EPI_ISL_1300222, EPI_ISL_1300223, EPI_ISL_1300224, EPI_ISL_1300225, EPI_ISL_1300226, EPI_ISL_1300227, EPI_ISL_1300228, EPI_ISL_1300229, EPI_ISL_1300230, EPI_ISL_1300231, EPI_ISL_1300232, EPI_ISL_1300233, EPI_ISL_1300234, EPI_ISL_1300235, EPI_ISL_1300236, EPI_ISL_1300237, EPI_ISL_1300238, EPI_ISL_1300239, EPI_ISL_1300240, EPI_ISL_1300241, EPI_ISL_1300242, EPI_ISL_1300243, EPI_ISL_1300244, EPI_ISL_1300245, EPI_ISL_1300246, EPI_ISL_1300247, EPI_ISL_1300248, EPI_ISL_1300249, EPI_ISL_1300250, EPI_ISL_1300251, EPI_ISL_1300252, EPI_ISL_1300253, EPI_ISL_1300254, EPI_ISL_1300255, EPI_ISL_1300256, EPI_ISL_1300257, EPI_ISL_1300258, EPI_ISL_1300259, EPI_ISL_1300260, EPI_ISL_1300262, EPI_ISL_1300263, EPI_ISL_1300264, EPI_ISL_1300265, EPI_ISL_1300266, EPI_ISL_1300267, EPI_ISL_1300268, EPI_ISL_1300269, EPI_ISL_1300270, EPI_ISL_1300271, EPI_ISL_1300272, EPI_ISL_1300273, EPI_ISL_1300274, EPI_ISL_1300275, EPI_ISL_1300276 | | | |
| see above | NYU Langone Health | Departments of Pathology and Medicine, New York University School of Medicine | Adriana Heguy, Dacia Dimartino, Emily Guzman, Christian Marier, Peter Meyn, Sitharam Ramaswami, Gael Westby, Paul Zappile, Yutong Zhang, Paolo Cotzia, Guiqing Wang |
| EPI_ISL_1300335, EPI_ISL_1300336, EPI_ISL_1300337, EPI_ISL_1300338, EPI_ISL_1300339, EPI_ISL_1300340, EPI_ISL_1300341, EPI_ISL_1300342, EPI_ISL_1300343, EPI_ISL_1300344, EPI_ISL_1300345, EPI_ISL_1300346, EPI_ISL_1300347, EPI_ISL_1300348, EPI_ISL_1300349, EPI_ISL_1300350, EPI_ISL_1300351, EPI_ISL_1300352, EPI_ISL_1300353, EPI_ISL_1300354, EPI_ISL_1300355, EPI_ISL_1300356, EPI_ISL_1300357, EPI_ISL_1300358, EPI_ISL_1300359, EPI_ISL_1300360, EPI_ISL_1300361, EPI_ISL_1300362, EPI_ISL_1300363, EPI_ISL_1300364, EPI_ISL_1300365, EPI_ISL_1300366, EPI_ISL_1300367, EPI_ISL_1300368, EPI_ISL_1300369, EPI_ISL_1300370, EPI_ISL_1300371, EPI_ISL_1300372, EPI_ISL_1300373, EPI_ISL_1300374, EPI_ISL_1300375, EPI_ISL_1300376, EPI_ISL_1300377, EPI_ISL_1300378, EPI_ISL_1300379, EPI_ISL_1300380, EPI_ISL_1300381, EPI_ISL_1300382, EPI_ISL_1300383, EPI_ISL_1300384, EPI_ISL_1300385, EPI_ISL_1300386, EPI_ISL_1300387, EPI_ISL_1300388, EPI_ISL_1300389, EPI_ISL_1300390, EPI_ISL_1300391, EPI_ISL_1300392, EPI_ISL_1300393, EPI_ISL_1300394, EPI_ISL_1300395, EPI_ISL_1300396, EPI_ISL_1300397, EPI_ISL_1300398, EPI_ISL_1300399, EPI_ISL_1300400, EPI_ISL_1300401, EPI_ISL_1300402, EPI_ISL_1300403, EPI_ISL_1300404, EPI_ISL_1300405, EPI_ISL_1300406, EPI_ISL_1300407, EPI_ISL_1300408, EPI_ISL_1300409, EPI_ISL_1300410, EPI_ISL_1300411, EPI_ISL_1300415, EPI_ISL_1300416, EPI_ISL_1300418, EPI_ISL_1300419, EPI_ISL_1300420, EPI_ISL_1300421, EPI_ISL_1300422, EPI_ISL_1300423, EPI_ISL_1300424 | | | |
| see above | Texas Department of State Health Services - TXDSHS | Texas Department of State Health Services - TXDSHS | Rashmi Tuladhar, Bonnie Oh, Jenny Zhang, Maliha Rahman, Mayela Pedrueza, Anita Pokharel, Myong Koag, Chun Wang, Rachel Lee, Grace Kubin |
| EPI_ISL_1300425 | Instituto Nacional de Medicina Genomica | Instituto Nacional de Medicina Genomica | Hidalgo-Miranda A, Mendoza-Vargas A, Reyes-Grajeda JP, Cedro-Tanda A, Alcaraz N, Gonzalez-Barrera D, Rangel-DeLeon D, Ramirez-Vega O, Herrera-Montalvo LA |
| EPI_ISL_1300427, EPI_ISL_1300428, EPI_ISL_1300429, EPI_ISL_1300430, EPI_ISL_1300431, EPI_ISL_1300433, EPI_ISL_1300434, EPI_ISL_1300435, EPI_ISL_1300436, EPI_ISL_1300437, EPI_ISL_1300438, EPI_ISL_1300439, EPI_ISL_1300440, EPI_ISL_1300441, EPI_ISL_1300442, EPI_ISL_1300443, EPI_ISL_1300444, EPI_ISL_1300445, EPI_ISL_1300446, EPI_ISL_1300447 | | | |
| see above | South Dakota Public Health Laboratory | South Dakota Public Health Laboratory | Jacob Garfin and Chris Carlson |
| EPI_ISL_1300448 | Epidemiology of Microbial Diseases, Yale School of Public Health | Yale School of Public Health | Fauver,J.R. |
| EPI_ISL_1300537, EPI_ISL_1300538, EPI_ISL_1300539 | Arizona State University | Arizona State University | Rabia Maqsood, LaRinda A. Holland, Peter T. Skidmore, Emily A. Kaelin, Nicholas J. Mellor, Kristina Buss, Joy M. Blain, Neal W Woodbury, Valerie Harris, Joshua LaBaer, Vel Murugan, Efrem S. Lim |
| EPI_ISL_1300541, EPI_ISL_1300542, EPI_ISL_1300543, EPI_ISL_1300544, EPI_ISL_1300545, EPI_ISL_1300546, EPI_ISL_1300547, EPI_ISL_1300548, EPI_ISL_1300549, EPI_ISL_1300550, EPI_ISL_1300551, EPI_ISL_1300552, EPI_ISL_1300553, EPI_ISL_1300554, EPI_ISL_1300555, EPI_ISL_1300556, EPI_ISL_1300557, EPI_ISL_1300558, EPI_ISL_1300559, EPI_ISL_1300560, EPI_ISL_1300561, EPI_ISL_1300562, EPI_ISL_1300563, EPI_ISL_1300564, EPI_ISL_1300565, EPI_ISL_1300566, EPI_ISL_1300567, EPI_ISL_1300568, EPI_ISL_1300569, EPI_ISL_1300570, EPI_ISL_1300571, EPI_ISL_1300572, EPI_ISL_1300573, EPI_ISL_1300574, EPI_ISL_1300575, EPI_ISL_1300576, EPI_ISL_1300577, EPI_ISL_1300578, EPI_ISL_1300579, EPI_ISL_1300580, EPI_ISL_1300581, EPI_ISL_1300582, EPI_ISL_1300583, EPI_ISL_1300584, EPI_ISL_1300585, EPI_ISL_1300586, EPI_ISL_1300587, EPI_ISL_1300588, EPI_ISL_1300589, EPI_ISL_1300590, EPI_ISL_1300591, EPI_ISL_1300592, EPI_ISL_1300593, EPI_ISL_1300594, EPI_ISL_1300595, EPI_ISL_1300596, EPI_ISL_1300597, EPI_ISL_1300598, EPI_ISL_1300599, EPI_ISL_1300600, EPI_ISL_1300601, EPI_ISL_1300602, EPI_ISL_1300603, EPI_ISL_1300604, EPI_ISL_1300605, EPI_ISL_1300606, EPI_ISL_1300607, EPI_ISL_1300608, EPI_ISL_1300609, EPI_ISL_1300610, EPI_ISL_1300611, EPI_ISL_1300612, EPI_ISL_1300613, EPI_ISL_1300614, EPI_ISL_1300615, EPI_ISL_1300616, EPI_ISL_1300617, EPI_ISL_1300618, EPI_ISL_1300619, EPI_ISL_1300620, EPI_ISL_1300621, EPI_ISL_1300622, EPI_ISL_1300623, EPI_ISL_1300624, EPI_ISL_1300625, EPI_ISL_1300626, EPI_ISL_1300627, EPI_ISL_1300628, EPI_ISL_1300629, EPI_ISL_1300630, EPI_ISL_1300631, EPI_ISL_1300632, EPI_ISL_1300633, EPI_ISL_1300634, EPI_ISL_1300635, EPI_ISL_1300636, EPI_ISL_1300637, EPI_ISL_1300638, EPI_ISL_1300639, EPI_ISL_1300640, EPI_ISL_1300641, EPI_ISL_1300642, EPI_ISL_1300643 | | | |
| see above | Utah Public Health Laboratory | Utah Public Health Laboratory | Erin L. Young, Kelly F. Oakeson, Tara Gallagher |
| EPI_ISL_1300756, EPI_ISL_1300757, EPI_ISL_1300758, EPI_ISL_1300759, EPI_ISL_1300760, EPI_ISL_1300761, EPI_ISL_1300762, EPI_ISL_1300763, EPI_ISL_1300764, EPI_ISL_1300765, EPI_ISL_1300766, EPI_ISL_1300767, EPI_ISL_1300768, EPI_ISL_1300769, EPI_ISL_1300770, EPI_ISL_1300771, EPI_ISL_1300772, EPI_ISL_1300773, EPI_ISL_1300774, EPI_ISL_1300775, EPI_ISL_1300776, EPI_ISL_1300777, EPI_ISL_1300778, EPI_ISL_1300779, EPI_ISL_1300780, EPI_ISL_1300781, EPI_ISL_1300782, EPI_ISL_1300783, EPI_ISL_1300784, EPI_ISL_1300785, EPI_ISL_1300786, EPI_ISL_1300787, EPI_ISL_1300788, EPI_ISL_1300789, EPI_ISL_1300790, EPI_ISL_1300791, EPI_ISL_1300792, EPI_ISL_1300793, EPI_ISL_1300794, EPI_ISL_1300795, EPI_ISL_1300796, EPI_ISL_1300797, EPI_ISL_1300798, EPI_ISL_1300799, EPI_ISL_1300800, EPI_ISL_1300801, EPI_ISL_1300802, EPI_ISL_1300803, EPI_ISL_1300804, EPI_ISL_1300805, EPI_ISL_1300806, EPI_ISL_1300807, EPI_ISL_1300808, EPI_ISL_1300809, EPI_ISL_1300810, EPI_ISL_1300811, EPI_ISL_1300812, EPI_ISL_1300813, EPI_ISL_1300814, EPI_ISL_1300815, EPI_ISL_1300816, EPI_ISL_1300817, EPI_ISL_1300818, EPI_ISL_1300821, EPI_ISL_1300822, EPI_ISL_1300823, EPI_ISL_1300824, EPI_ISL_1300825, EPI_ISL_1300826, EPI_ISL_1300827, EPI_ISL_1300828, EPI_ISL_1300829, EPI_ISL_1300830, EPI_ISL_1300831, EPI_ISL_1300832, EPI_ISL_1300833, EPI_ISL_1300834, EPI_ISL_1300835, EPI_ISL_1300836, EPI_ISL_1300837, EPI_ISL_1300838, EPI_ISL_1300839, EPI_ISL_1300840, EPI_ISL_1300841, EPI_ISL_1300842, EPI_ISL_1300843, EPI_ISL_1300844, EPI_ISL_1300845, EPI_ISL_1300846, EPI_ISL_1300847, EPI_ISL_1300848, EPI_ISL_1300850, EPI_ISL_1300851, EPI_ISL_1300852, EPI_ISL_1300853, EPI_ISL_1300854, EPI_ISL_1300855, EPI_ISL_1300856, EPI_ISL_1300857, EPI_ISL_1300858, EPI_ISL_1300859, EPI_ISL_1300860, EPI_ISL_1300861, EPI_ISL_1300862, EPI_ISL_1300863, EPI_ISL_1300864, EPI_ISL_1300865, EPI_ISL_1300866, EPI_ISL_1300867, EPI_ISL_1300868, EPI_ISL_1300869, EPI_ISL_1300870, EPI_ISL_1300871, EPI_ISL_1300872, EPI_ISL_1300873, EPI_ISL_1300874, EPI_ISL_1300875, EPI_ISL_1300876, EPI_ISL_1300877, EPI_ISL_1300878, EPI_ISL_1300879, EPI_ISL_1300880, EPI_ISL_1300881, EPI_ISL_1300882, EPI_ISL_1300883, EPI_ISL_1300884, EPI_ISL_1300885, EPI_ISL_1300886, EPI_ISL_1300887, EPI_ISL_1300888, EPI_ISL_1300889, EPI_ISL_1300890, EPI_ISL_1300891, EPI_ISL_1300892, EPI_ISL_1300893, EPI_ISL_1300894, EPI_ISL_1300895, EPI_ISL_1300896, EPI_ISL_1300897, EPI_ISL_1300898, EPI_ISL_1300899, EPI_ISL_1300900, EPI_ISL_1300901, EPI_ISL_1300902, EPI_ISL_1300903, EPI_ISL_1300904, EPI_ISL_1300905, EPI_ISL_1300906, EPI_ISL_1300907, EPI_ISL_1300908, EPI_ISL_1300909, EPI_ISL_1300910, EPI_ISL_1300911, EPI_ISL_1300912, EPI_ISL_1300913, EPI_ISL_1300914, EPI_ISL_1300915, EPI_ISL_1300916, EPI_ISL_1300917, EPI_ISL_1300918, EPI_ISL_1300919, EPI_ISL_1300920, EPI_ISL_1300921, EPI_ISL_1300922, EPI_ISL_1300923, EPI_ISL_1300924, EPI_ISL_1300925, EPI_ISL_1300926, EPI_ISL_1300927, EPI_ISL_1300928, EPI_ISL_1300929, EPI_ISL_1300930, EPI_ISL_1300931, EPI_ISL_1300932, EPI_ISL_1300933, EPI_ISL_1300934, EPI_ISL_1300935, EPI_ISL_1300936, EPI_ISL_1300937, EPI_ISL_1300938, EPI_ISL_1300939, EPI_ISL_1300940, EPI_ISL_1300941, EPI_ISL_1300942, EPI_ISL_1300943, EPI_ISL_1300944, EPI_ISL_1300945, EPI_ISL_1300946, EPI_ISL_1300947, EPI_ISL_1300948, EPI_ISL_1300949, EPI_ISL_1300950, EPI_ISL_1300951, EPI_ISL_1300952, EPI_ISL_1300953, EPI_ISL_1300954, EPI_ISL_1300955, EPI_ISL_1300956, EPI_ISL_1300957, EPI_ISL_1300958, EPI_ISL_1300959, EPI_ISL_1300960, EPI_ISL_1300961, EPI_ISL_1300962, EPI_ISL_1300963, EPI_ISL_1300964, EPI_ISL_1300965, EPI_ISL_1300966, EPI_ISL_1300967, EPI_ISL_1300968, EPI_ISL_1300969, EPI_ISL_1300970, EPI_ISL_1300971, EPI_ISL_1300972, EPI_ISL_1300973, EPI_ISL_1300974, EPI_ISL_1300975, EPI_ISL_1300976, EPI_ISL_1300977, EPI_ISL_1300978, EPI_ISL_1300979, EPI_ISL_1300980, EPI_ISL_1300981, EPI_ISL_1300982, EPI_ISL_1300983, EPI_ISL_1300984, EPI_ISL_1300985, EPI_ISL_1300986, EPI_ISL_1300987, EPI_ISL_1300988, EPI_ISL_1300989, EPI_ISL_1300990, EPI_ISL_1300991, EPI_ISL_1300992, EPI_ISL_1300993, EPI_ISL_1300994, EPI_ISL_1300995, EPI_ISL_1300996, EPI_ISL_1300997, EPI_ISL_1300998, EPI_ISL_1300999, EPI_ISL_1301000, EPI_ISL_1301001, EPI_ISL_1301002, EPI_ISL_1301003, EPI_ISL_1301004, EPI_ISL_1301005, EPI_ISL_1301006, EPI_ISL_1301007, EPI_ISL_1301008, EPI_ISL_1301009, EPI_ISL_1301010, EPI_ISL_1301011, EPI_ISL_1301012, EPI_ISL_1301013, EPI_ISL_1301014, EPI_ISL_1301015, EPI_ISL_1301016, EPI_ISL_1301017, EPI_ISL_1301018, EPI_ISL_1301019, EPI_ISL_1301020, EPI_ISL_1301021, EPI_ISL_1301022, EPI_ISL_1301023, EPI_ISL_1301024, EPI_ISL_1301025, EPI_ISL_1301026, EPI_ISL_1301027, EPI_ISL_1301028, EPI_ISL_1301029, EPI_ISL_1301030, EPI_ISL_1301031, EPI_ISL_1301032, EPI_ISL_1301033, EPI_ISL_1301034, EPI_ISL_1301035, EPI_ISL_1301036, EPI_ISL_1301037, EPI_ISL_1301038, EPI_ISL_1301039, EPI_ISL_1301040, EPI_ISL_1301041, EPI_ISL_1301042, EPI_ISL_1301043, EPI_ISL_1301044, EPI_ISL_1301045, EPI_ISL_1301046, EPI_ISL_1301047, EPI_ISL_1301048, EPI_ISL_1301049, EPI_ISL_1301050, EPI_ISL_1301051, EPI_ISL_1301052, EPI_ISL_1301053, EPI_ISL_1301054, EPI_ISL_1301055, EPI_ISL_1301056, EPI_ISL_1301057, EPI_ISL_1301058, EPI_ISL_1301059, EPI_ISL_1301060, EPI_ISL_1301061, EPI_ISL_1301062, EPI_ISL_1301063, EPI_ISL_1301064, EPI_ISL_1301065, EPI_ISL_1301066, EPI_ISL_1301067, EPI_ISL_1301068, EPI_ISL_1301069, EPI_ISL_1301070, EPI_ISL_1301071, EPI_ISL_1301072, EPI_ISL_1301073, EPI_ISL_1301074, EPI_ISL_1301075, EPI_ISL_1301076, EPI_ISL_1301077, EPI_ISL_1301078, EPI_ISL_1301079, EPI_ISL_1301080, EPI_ISL_1301081, EPI_ISL_1301082, EPI_ISL_1301083, EPI_ISL_1301084, EPI_ISL_1301085, EPI_ISL_1301086, EPI_ISL_1301087, EPI_ISL_1301088, EPI_ISL_1301089, EPI_ISL_1301090, EPI_ISL_1301091, EPI_ISL_1301092, EPI_ISL_1301093, EPI_ISL_1301094, EPI_ISL_1301095, EPI_ISL_1301096, EPI_ISL_1301097, EPI_ISL_1301098, EPI_ISL_1301099, EPI_ISL_1301100, EPI_ISL_1301101, EPI_ISL_1301102, EPI_ISL_1301103, EPI_ISL_1301104, EPI_ISL_1301105, EPI_ISL_1301106, EPI_ISL_1301107, EPI_ISL_1301108, EPI_ISL_1301109, EPI_ISL_1301110, EPI_ISL_1301111, EPI_ISL_1301112, EPI_ISL_1301113, EPI_ISL_1301114, EPI_ISL_1301115, EPI_ISL_1301116, EPI_ISL_1301117, EPI_ISL_1301118, EPI_ISL_1301119, EPI_ISL_1301120, EPI_ISL_1301121, EPI_ISL_1301122, EPI_ISL_1301123, EPI_ISL_1301124, EPI_ISL_1301125, EPI_ISL_1301126, EPI_ISL_1301127, EPI_ISL_1301128, EPI_ISL_1301129, EPI_ISL_1301130, EPI_ISL_1301131, EPI_ISL_1301132, EPI_ISL_1301133, EPI_ISL_1301134, EPI_ISL_1301135, EPI_ISL_1301136, EPI_ISL_1301137, EPI_ISL_1301138, EPI_ISL_1301139, EPI_ISL_1301140, EPI_ISL_1301141, EPI_ISL_1301142, EPI_ISL_1301143, EPI_ISL_1301144, EPI_ISL_1301145, EPI_ISL_1301146, EPI_ISL_1301147, EPI_ISL_1301148, EPI_ISL_1301149, EPI_ISL_1301150, EPI_ISL_1301151, EPI_ISL_1301152, EPI_ISL_1301153, EPI_ISL_1301154, EPI_ISL_1301155, EPI_ISL_1301156, EPI_ISL_1301157, EPI_ISL_1301158, EPI_ISL_1301159, EPI_ISL_1301160, EPI_ISL_1301161, EPI_ISL_1301162, EPI_ISL_1301163, EPI_ISL_1301164, EPI_ISL_1301165, EPI_ISL_1301166, EPI_ISL_1301167, EPI_ISL_1301168, EPI_ISL_1301169, EPI_ISL_1301170, EPI_ISL_1301171, EPI_ISL_1301172, EPI_ISL_1301173, EPI_ISL_1301174, EPI_ISL_1301175, EPI_ISL_1301176, EPI_ISL_1301177, EPI_ISL_1301178, EPI_ISL_1301179, EPI_ISL_1301180, EPI_ISL_1301181, EPI_ISL_1301182, EPI_ISL_1301183, EPI_ISL_1301184, EPI_ISL_1301185, EPI_ISL_1301186, EPI_ISL_1301187, EPI_ISL_1301188, EPI_ISL_1301189, EPI_ISL_1301190, EPI_ISL_1301191, EPI_ISL_1301192, EPI_ISL_1301193, EPI_ISL_1301194, EPI_ISL_1301195, EPI_ISL_1301196, EPI_ISL_1301197, EPI_ISL_1301198, EPI_ISL_1301199, EPI_ISL_1301200, EPI_ISL_1301201, EPI_ISL_1301202, EPI_ISL_1301203, EPI_ISL_1301204, EPI_ISL_1301205, EPI_ISL_1301206, EPI_ISL_1301207, EPI_ISL_1301208, EPI_ISL_1301209, EPI_ISL_1301210, EPI_ISL_1301211, EPI_ISL_1301212, EPI_ISL_1301213, EPI_ISL_1301214, EPI_ISL_1301215, EPI_ISL_1301216, EPI_ISL_1301217, EPI_ISL_1301218, EPI_ISL_1301219, EPI_ISL_1301220, EPI_ISL_1301221, EPI_ISL_1301222, EPI_ISL_1301223, EPI_ISL_1301224, EPI_ISL_1301225, EPI_ISL_1301226, EPI_ISL_1301227, EPI_ISL_1301228, EPI_ISL_1301229, EPI_ISL_1301230, EPI_ISL_1301231, EPI_ISL_1301232, EPI_ISL_1301233, EPI_ISL_1301234, EPI_ISL_1301235, EPI_ISL_1301236, EPI_ISL_1301237, EPI_ISL_1301238, EPI_ISL_1301239, EPI_ISL_1301240, EPI_ISL_1301241, EPI_ISL_1301242, EPI_ISL_1301243, EPI_ISL_1301244, EPI_ISL_1301245, EPI_ISL_1301246, EPI_ISL_1301247, EPI_ISL_1301248, EPI_ISL_1301249, EPI_ISL_1301250, EPI_ISL_1301251, EPI_ISL_1301252, EPI_ISL_1301253, EPI_ISL_1301254, EPI_ISL_1301255, EPI_ISL_1301256, EPI_ISL_1301257, EPI_ISL_1301258, EPI_ISL_1301259, EPI_ISL_1301260, EPI_ISL_1301261, EPI_ISL_1301262, EPI_ISL_1301263, EPI_ISL_1301264, EPI_ISL_1301265, EPI_ISL_1301266, EPI_ISL_1301267, EPI_ISL_1301268, EPI_ISL_1301269, EPI_ISL_1301270, EPI_ISL_1301271, EPI_ISL_1301272, EPI_ISL_1301273, EPI_ISL_1301274, EPI_ISL_1301275, EPI_ISL_1301276, EPI_ISL_1301277, EPI_ISL_1301278, EPI_ISL_1301279, EPI_ISL_1301280, EPI_ISL_1301281, EPI_ISL_1301282, EPI_ISL_1301283, EPI_ISL_1301284, EPI_ISL_1301285, EPI_ISL_1301286, EPI_ISL_1301287, EPI_ISL_1301288, EPI_ISL_1301289, EPI_ISL_1301290, EPI_ISL_1301291, EPI_ISL_1301292, EPI_ISL_1301293, EPI_ISL_1301294, EPI_ISL_1301295, EPI_ISL_1301296, EPI_ISL_1301297, EPI_ISL_1301298, EPI_ISL_1301299, EPI_ISL_1301300, EPI_ISL_1301301, EPI_ISL_1301302, EPI_ISL_1301303, EPI_ISL_1301304, EPI_ISL_1301305, EPI_ISL_1301306, EPI_ISL_1301307, EPI_ISL_1301308, EPI_ISL_1301309, EPI_ISL_1301310, EPI_ISL_1301311, EPI_ISL_1301312, EPI_ISL_1301313, EPI_ISL_1301314, EPI_ISL_1301315, EPI_ISL_1301316, EPI_ISL_1301317, EPI_ISL_1301318, EPI_ISL_1301319, EPI_ISL_1301320, EPI_ISL_1301321, EPI_ISL_1301322, EPI_ISL_1301323, EPI_ISL_1301324, EPI_ISL_1301325, EPI_ISL_1301326, EPI_ISL_1301327, EPI_ISL_1301328, EPI_ISL_1301329, EPI_ISL_1301330, EPI_ISL_1301331, EPI_ISL_1301332, EPI_ISL_1301333, EPI_ISL_1301334, EPI_ISL_1301335, EPI_ISL_1301336, EPI_ISL_1301337, EPI_ISL_1301338, EPI_ISL_1301339, EPI_ISL_1301340, EPI_ISL_1301341, EPI_ISL_1301342, EPI_ISL_1301343, EPI_ISL_1301344, EPI_ISL_1301345, EPI_ISL_1301346, EPI_ISL_1301347, EPI_ISL_1301348, EPI_ISL_1301349, EPI_ISL_1301350, EPI_ISL_1301351, EPI_ISL_1301352, EPI_ISL_1301353, EPI_ISL_1301354, EPI_ISL_1301355, EPI_ISL_1301356, EPI_ISL_1301357, EPI_ISL_1301358, EPI_ISL_1301359, EPI_ISL_1301360, EPI_ISL_1301361, EPI_ISL_1301362, EPI_ISL_1301363, EPI_ISL_1301364, EPI_ISL_1301365, EPI_ISL_1301366, EPI_ISL_1301367, EPI_ISL_1301368, EPI_ISL_1301369, EPI_ISL_1301370, EPI_ISL_1301371, EPI_ISL_1301372, EPI_ISL_1301373, EPI_ISL_1301374, EPI_ISL_1301375, EPI_ISL_1301376, EPI_ISL_1301377, EPI_ISL_1301378, EPI_ISL_1301379, EPI_ISL_1301380, EPI_ISL_1301381, EPI_ISL_1301382, EPI_ISL_1301383, EPI_ISL_1301384, EPI_ISL_1301385, EPI_ISL_1301386, EPI_ISL_1301387, EPI_ISL_1301388, EPI_ISL_1301389, EPI_ISL_1301390, EPI_ISL_1301391, EPI_ISL_1301392, EPI_ISL_1301393, EPI_ISL_1301394, EPI_ISL_1301395, EPI_ISL_1301396, EPI_ISL_1301397, EPI_ISL_1301398, EPI_ISL_1301399, EPI_ISL_1301400, EPI_ISL_1301401, EPI_ISL_1301402, EPI_ISL_1301403, EPI_ISL_1301404, EPI_ISL_1301405, EPI_ISL_1301406, EPI_ISL_1301407, EPI_ISL_1301408, EPI_ISL_1301409, EPI_ISL_1301410, EPI_ISL_1301411, EPI_ISL_1301412, EPI_ISL_1301413, EPI_ISL_1301414, EPI_ISL_1301415, EPI_ISL_1301416, EPI_ISL_1301417, EPI_ISL_1301418, EPI_ISL_1301419, EPI_ISL_1301420, EPI_ISL_1301421, EPI_ISL_1301422, EPI_ISL_1301423, EPI_ISL_1301424, EPI_ISL_1301425, EPI_ISL_1301426, EPI_ISL_1301427, EPI_ISL_1301428, EPI_ISL_1301429, EPI_ISL_1301430, EPI_ISL_1301431, EPI_ISL_1301432, EPI_ISL_1301433, EPI_ISL_1301434, EPI_ISL_1301435, EPI_ISL_1301436, EPI_ISL_1301437, EPI_ISL_1301438, EPI_ISL_1301439 | | | |
| see above | MSHS Clinical Microbiology Laboratories | MSHS Pathogen Surveillance Program | Ana S. Gonzalez-Reiche, Hala Alshammary, Mitchell J. Sullivan, Brianne Ciferri, Ajay Obla, Angela Amoako, Mahmoud Awawda, Daniel Floda, Julia Matthews, Ashley Salimbangon, Levy Sominsky, Katherine Beach, Kayla Russo, Charles Gleason, Shelcie Fabre, Giulio Kleiner, Zenab Khan, Bremy Alburquerque, Adriana van de Guchte, Komal Srivastava, Matthew M. Hernandez, Jayeeta Dutta, Denise Jurczyszak, Nancy Francoeur, Betsaida Salom Melo, Irina Oussenko, Gintaras Deikus, Juan Soto, Shwetha Hara Sridhar, Ying-Chih Wang, Kathryn Twyman, Deena R. Altman, Robert Sebra, Adolfo García-Sastre, Marta Luksza, Gopi Patel, Sarah Schaefer, Melissa Gitman, Michael D. Nowak, Alberto Paniz-Mondolfi, Emilia Mia Sordillo, Viviana Simon, Harm van Bakel |
| EPI_ISL_1301440 | MSHS Clinical Microbiology Laboratories | NBACC | Michael Schuit, Jennifer Biryukov, Katie Beck, Jason Yolitz, Jordan Bohannon, Wade Weaver, David Miller, Brian Holland, Melissa Krause, Denise Freeburger, Gregory Williams, Stewart Wood, Amanda Graham, M.J. Rosovitz, Adam Bazinet, Aaron Phillips, Sean Lovett, Karla Garcia, Elyse Abbott, Victoria Wahl, Shanna Ratnesar-Shumate, Paul Dabisch |
| EPI_ISL_1301442, EPI_ISL_1301443 | UCSD EXCITE | NBACC | Michael Schuit, Jennifer Biryukov, Katie Beck, Jason Yolitz, Jordan Bohannon, Wade Weaver, David Miller, Brian Holland, Melissa Krause, Denise Freeburger, Gregory Williams, Stewart Wood, Amanda Graham, M.J. Rosovitz, Adam Bazinet, Aaron Phillips, Sean Lovett, Karla Garcia, Elyse Abbott, Victoria Wahl, Shanna Ratnesar-Shumate, Paul Dabisch |
| EPI_ISL_1301444 | Instituto de Diagnostico y Referencia Epidemiologicos InDRE_RNLSP | Instituto de Biotecnología de la UNAM | Authors from IBT, IMSS, InDRE and INER (in alphabetical order): Carlos F. Arias, Santiago Ávila-Ríos, Gisela Barrera-Badillo, Eduardo Becerril-Vargas, Celia Boukadida, Natividad Cruz-Ortiz, Larissa Fernandes-Matano, Ricardo Grande, Lucia Hernandez-Rivas, Alejandra Hernández-Terán, Pavel Isa, Irma Lopez-Martinez, José Arturo Martínez-Orozco, Margarita Matías-Florentino, Fidencio Mejía-Nepomuceno, Edgar Mendieta-Condado, Mario Mújica-Sánchez, José Esteban Muñoz-Medina, Tatiana Nunez-Garcia, Luis Alberto Ochoa-Carrera, Hector Esteban Paz-Juárez, Francisco Pulido, José Ernesto Ramírez-González, Alma Rincón-Rubio, Teresita Rojas-Mendoza, Jorge Salas-Hernández, Alejandro Sanchez-Flores, Clara Esperanza Santacruz-Tinoco, Andrea Santos Coy-Arechavaleta, Blanca Taboada, Gloria Vazquez, Joel Armando Vázquez-Pérez, Jerome Jean Verleyen |
| EPI_ISL_1301445 | Instituto Nacional de Enfermedades Respiratorias (INER) | Instituto de Biotecnología de la UNAM | Authors from IBT, IMSS, InDRE and INER (in alphabetical order): Carlos F. Arias, Santiago Ávila-Ríos, Gisela Barrera-Badillo, Eduardo Becerril-Vargas, Celia Boukadida, Natividad Cruz-Ortiz, Larissa Fernandes-Matano, Ricardo Grande, Lucia Hernandez-Rivas, Alejandra Hernández-Terán, Pavel Isa, Irma Lopez-Martinez, José Arturo Martínez-Orozco, Margarita Matías-Florentino, Fidencio Mejía-Nepomuceno, Edgar Mendieta-Condado, Mario Mújica-Sánchez, José Esteban Muñoz-Medina, Tatiana Nunez-Garcia, Luis Alberto Ochoa-Carrera, Hector Esteban Paz-Juárez, Francisco Pulido, José Ernesto Ramírez-González, Alma Rincón-Rubio, Teresita Rojas-Mendoza, Jorge Salas-Hernández, Alejandro Sanchez-Flores, Clara Esperanza Santacruz-Tinoco, Andrea Santos Coy-Arechavaleta, Blanca Taboada, Gloria Vazquez, Joel Armando Vázquez-Pérez, Jerome Jean Verleyen |
| EPI_ISL_1301446 | Laboratorio Central de Epidemiología IMSS | Instituto de Biotecnología de la UNAM | Authors from IBT, IMSS, InDRE and INER (in alphabetical order): Carlos F. Arias, Santiago Ávila-Ríos, Gisela Barrera-Badillo, Eduardo Becerril-Vargas, Celia Boukadida, Natividad Cruz-Ortiz, Larissa Fernandes-Matano, Ricardo Grande, Lucia Hernandez-Rivas, Alejandra Hernández-Terán, Pavel Isa, Irma Lopez-Martinez, José Arturo Martínez-Orozco, Margarita Matías-Florentino, Fidencio Mejía-Nepomuceno, Edgar Mendieta-Condado, Mario Mújica-Sánchez, José Esteban Muñoz-Medina, Tatiana Nunez-Garcia, Luis Alberto Ochoa-Carrera, Hector Esteban Paz-Juárez, Francisco Pulido, José Ernesto Ramírez-González, Alma Rincón-Rubio, Teresita Rojas-Mendoza, Jorge Salas-Hernández, Alejandro Sanchez-Flores, Clara Esperanza Santacruz-Tinoco, Andrea Santos Coy-Arechavaleta, Blanca Taboada, Gloria Vazquez, Joel Armando Vázquez-Pérez, Jerome Jean Verleyen |
| EPI_ISL_1301447, EPI_ISL_1301448, EPI_ISL_1301449 | Instituto de Diagnostico y Referencia Epidemiologicos InDRE_RNLSP | Instituto de Biotecnología de la UNAM | Authors from IBT, IMSS, InDRE and INER (in alphabetical order): Carlos F. Arias, Santiago Ávila-Ríos, Gisela Barrera-Badillo, Eduardo Becerril-Vargas, Celia Boukadida, Natividad Cruz-Ortiz, Larissa Fernandes-Matano, Ricardo Grande, Lucia Hernandez-Rivas, Alejandra Hernández-Terán, Pavel Isa, Irma Lopez-Martinez, José Arturo Martínez-Orozco, Margarita Matías-Florentino, Fidencio Mejía-Nepomuceno, Edgar Mendieta-Condado, Mario Mújica-Sánchez, José Esteban Muñoz-Medina, Tatiana Nunez-Garcia, Luis Alberto Ochoa-Carrera, Hector Esteban Paz-Juárez, Francisco Pulido, José Ernesto Ramírez-González, Alma Rincón-Rubio, Teresita Rojas-Mendoza, Jorge Salas-Hernández, Alejandro Sanchez-Flores, Clara Esperanza Santacruz-Tinoco, Andrea Santos Coy-Arechavaleta, Blanca Taboada, Gloria Vazquez, Joel Armando Vázquez-Pérez, Jerome Jean Verleyen |
| EPI_ISL_1301450 | Laboratorio Central de Epidemiología IMSS | Instituto de Biotecnología de la UNAM | Authors from IBT, IMSS, InDRE and INER (in alphabetical order): Carlos F. Arias, Santiago Ávila-Ríos, Gisela Barrera-Badillo, Eduardo Becerril-Vargas, Celia Boukadida, Natividad Cruz-Ortiz, Larissa Fernandes-Matano, Ricardo Grande, Lucia Hernandez-Rivas, Alejandra Hernández-Terán, Pavel Isa, Irma Lopez-Martinez, José Arturo Martínez-Orozco, Margarita Matías-Florentino, Fidencio Mejía-Nepomuceno, Edgar Mendieta-Condado, Mario Mújica-Sánchez, José Esteban Muñoz-Medina, Tatiana Nunez-Garcia, Luis Alberto Ochoa-Carrera, Hector Esteban Paz-Juárez, Francisco Pulido, José Ernesto Ramírez-González, Alma Rincón-Rubio, Teresita Rojas-Mendoza, Jorge Salas-Hernández, Alejandro Sanchez-Flores, Clara Esperanza Santacruz-Tinoco, Andrea Santos Coy-Arechavaleta, Blanca Taboada, Gloria Vazquez, Joel Armando Vázquez-Pérez, Jerome Jean Verleyen |
| EPI_ISL_1301451 | Instituto de Diagnostico y Referencia Epidemiologicos InDRE_RNLSP | Instituto de Biotecnología de la UNAM | Authors from IBT, IMSS, InDRE and INER (in alphabetical order): Carlos F. Arias, Santiago Ávila-Ríos, Gisela Barrera-Badillo, Eduardo Becerril-Vargas, Celia Boukadida, Natividad Cruz-Ortiz, Larissa Fernandes-Matano, Ricardo Grande, Lucia Hernandez-Rivas, Alejandra Hernández-Terán, Pavel Isa, Irma Lopez-Martinez, José Arturo Martínez-Orozco, Margarita Matías-Florentino, Fidencio Mejía-Nepomuceno, Edgar Mendieta-Condado, Mario Mújica-Sánchez, José Esteban Muñoz-Medina, Tatiana Nunez-Garcia, Luis Alberto Ochoa-Carrera, Hector Esteban Paz-Juárez, Francisco Pulido, José Ernesto Ramírez-González, Alma Rincón-Rubio, Teresita Rojas-Mendoza, Jorge Salas-Hernández, Alejandro Sanchez-Flores, Clara Esperanza Santacruz-Tinoco, Andrea Santos Coy-Arechavaleta, Blanca Taboada, Gloria Vazquez, Joel Armando Vázquez-Pérez, Jerome Jean Verleyen |
| EPI_ISL_1301452, EPI_ISL_1301453 | Instituto Nacional de Enfermedades Respiratorias (INER) | Instituto de Biotecnología de la UNAM | Authors from IBT, IMSS, InDRE and INER (in alphabetical order): Carlos F. Arias, Santiago Ávila-Ríos, Gisela Barrera-Badillo, Eduardo Becerril-Vargas, Celia Boukadida, Natividad Cruz-Ortiz, Larissa Fernandes-Matano, Ricardo Grande, Lucia Hernandez-Rivas, Alejandra Hernández-Terán, Pavel Isa, Irma Lopez-Martinez, José Arturo Martínez-Orozco, Margarita Matías-Florentino, Fidencio Mejía-Nepomuceno, Edgar Mendieta-Condado, Mario Mújica-Sánchez, José Esteban Muñoz-Medina, Tatiana Nunez-Garcia, Luis Alberto Ochoa-Carrera, Hector Esteban Paz-Juárez, Francisco Pulido, José Ernesto Ramírez-González, Alma Rincón-Rubio, Teresita Rojas-Mendoza, Jorge Salas-Hernández, Alejandro Sanchez-Flores, Clara Esperanza Santacruz-Tinoco, Andrea Santos Coy-Arechavaleta, Blanca Taboada, Gloria Vazquez, Joel Armando Vázquez-Pérez, Jerome Jean Verleyen |

| | | | |
|---|---|---|---|
| EPI_ISL_1301454, EPI_ISL_1301455, EPI_ISL_1301456, EPI_ISL_1301457, EPI_ISL_1301458, EPI_ISL_1301459, EPI_ISL_1301460, EPI_ISL_1301461 | Instituto de Diagnostico y Referencia Epidemiologicos InDRE_RNLSP | Instituto de Biotecnología de la UNAM | Authors from IBT, IMSS, InDRE and INER (in alphabetical order): Carlos F. Arias, Santiago Ávila-Ríos, Gisela Barrera-Badillo, Eduardo Becerril-Vargas, Celia Boukadida, Natividad Cruz-Ortiz, Larissa Fernandes-Matano, Ricardo Grande, Lucia Hernandez-Rivas, Alejandra Hernández-Terán, Pavel Isa, Irma Lopez-Martinez, José Arturo Martínez-Orozco, Margarita Matías-Florentino, Fidencio Mejía-Nepomuceno, Edgar Mendieta-Condado, Mario Mújica-Sánchez, José Esteban Muñoz-Medina, Tatiana Nunez-Garcia, Luis Alberto Ochoa-Carrera, Hector Esteban Paz-Juárez, Francisco Pulido, José Ernesto Ramírez-González, Alma Rincón-Rubio, Teresita Rojas-Mendoza, Jorge Salas-Hernández, Alejandro Sanchez-Flores, Clara Esperanza Santacruz-Tinoco, Andrea Santos Coy-Arechavaleta, Blanca Taboada, Gloria Vazquez, Joel Armando Vázquez-Pérez, Jerome Jean Verleyen |
| EPI_ISL_1301462, EPI_ISL_1301463 | Laboratorio Central de Epidemiología IMSS | Instituto de Biotecnología de la UNAM | Authors from IBT, IMSS, InDRE and INER (in alphabetical order): Carlos F. Arias, Santiago Ávila-Ríos, Gisela Barrera-Badillo, Eduardo Becerril-Vargas, Celia Boukadida, Natividad Cruz-Ortiz, Larissa Fernandes-Matano, Ricardo Grande, Lucia Hernandez-Rivas, Alejandra Hernández-Terán, Pavel Isa, Irma Lopez-Martinez, José Arturo Martínez-Orozco, Margarita Matías-Florentino, Fidencio Mejía-Nepomuceno, Edgar Mendieta-Condado, Mario Mújica-Sánchez, José Esteban Muñoz-Medina, Tatiana Nunez-Garcia, Luis Alberto Ochoa-Carrera, Hector Esteban Paz-Juárez, Francisco Pulido, José Ernesto Ramírez-González, Alma Rincón-Rubio, Teresita Rojas-Mendoza, Jorge Salas-Hernández, Alejandro Sanchez-Flores, Clara Esperanza Santacruz-Tinoco, Andrea Santos Coy-Arechavaleta, Blanca Taboada, Gloria Vazquez, Joel Armando Vázquez-Pérez, Jerome Jean Verleyen |
| EPI_ISL_1301464 | Instituto de Diagnostico y Referencia Epidemiologicos InDRE_RNLSP | Instituto de Biotecnología de la UNAM | Authors from IBT, IMSS, InDRE and INER (in alphabetical order): Carlos F. Arias, Santiago Ávila-Ríos, Gisela Barrera-Badillo, Eduardo Becerril-Vargas, Celia Boukadida, Natividad Cruz-Ortiz, Larissa Fernandes-Matano, Ricardo Grande, Lucia Hernandez-Rivas, Alejandra Hernández-Terán, Pavel Isa, Irma Lopez-Martinez, José Arturo Martínez-Orozco, Margarita Matías-Florentino, Fidencio Mejía-Nepomuceno, Edgar Mendieta-Condado, Mario Mújica-Sánchez, José Esteban Muñoz-Medina, Tatiana Nunez-Garcia, Luis Alberto Ochoa-Carrera, Hector Esteban Paz-Juárez, Francisco Pulido, José Ernesto Ramírez-González, Alma Rincón-Rubio, Teresita Rojas-Mendoza, Jorge Salas-Hernández, Alejandro Sanchez-Flores, Clara Esperanza Santacruz-Tinoco, Andrea Santos Coy-Arechavaleta, Blanca Taboada, Gloria Vazquez, Joel Armando Vázquez-Pérez, Jerome Jean Verleyen |
| EPI_ISL_1301465 | Instituto Nacional de Enfermedades Respiratorias (INER) | Instituto de Biotecnología de la UNAM | Authors from IBT, IMSS, InDRE and INER (in alphabetical order): Carlos F. Arias, Santiago Ávila-Ríos, Gisela Barrera-Badillo, Eduardo Becerril-Vargas, Celia Boukadida, Natividad Cruz-Ortiz, Larissa Fernandes-Matano, Ricardo Grande, Lucia Hernandez-Rivas, Alejandra Hernández-Terán, Pavel Isa, Irma Lopez-Martinez, José Arturo Martínez-Orozco, Margarita Matías-Florentino, Fidencio Mejía-Nepomuceno, Edgar Mendieta-Condado, Mario Mújica-Sánchez, José Esteban Muñoz-Medina, Tatiana Nunez-Garcia, Luis Alberto Ochoa-Carrera, Hector Esteban Paz-Juárez, Francisco Pulido, José Ernesto Ramírez-González, Alma Rincón-Rubio, Teresita Rojas-Mendoza, Jorge Salas-Hernández, Alejandro Sanchez-Flores, Clara Esperanza Santacruz-Tinoco, Andrea Santos Coy-Arechavaleta, Blanca Taboada, Gloria Vazquez, Joel Armando Vázquez-Pérez, Jerome Jean Verleyen |
| EPI_ISL_1301466, EPI_ISL_1301467, EPI_ISL_1301468 | Laboratorio Central de Epidemiología IMSS | Instituto de Biotecnología de la UNAM | Authors from IBT, IMSS, InDRE and INER (in alphabetical order): Carlos F. Arias, Santiago Ávila-Ríos, Gisela Barrera-Badillo, Eduardo Becerril-Vargas, Celia Boukadida, Natividad Cruz-Ortiz, Larissa Fernandes-Matano, Ricardo Grande, Lucia Hernandez-Rivas, Alejandra Hernández-Terán, Pavel Isa, Irma Lopez-Martinez, José Arturo Martínez-Orozco, Margarita Matías-Florentino, Fidencio Mejía-Nepomuceno, Edgar Mendieta-Condado, Mario Mújica-Sánchez, José Esteban Muñoz-Medina, Tatiana Nunez-Garcia, Luis Alberto Ochoa-Carrera, Hector Esteban Paz-Juárez, Francisco Pulido, José Ernesto Ramírez-González, Alma Rincón-Rubio, Teresita Rojas-Mendoza, Jorge Salas-Hernández, Alejandro Sanchez-Flores, Clara Esperanza Santacruz-Tinoco, Andrea Santos Coy-Arechavaleta, Blanca Taboada, Gloria Vazquez, Joel Armando Vázquez-Pérez, Jerome Jean Verleyen |
| EPI_ISL_1301469, EPI_ISL_1301470 | Instituto de Diagnostico y Referencia Epidemiologicos InDRE_RNLSP | Instituto de Biotecnología de la UNAM | Authors from IBT, IMSS, InDRE and INER (in alphabetical order): Carlos F. Arias, Santiago Ávila-Ríos, Gisela Barrera-Badillo, Eduardo Becerril-Vargas, Celia Boukadida, Natividad Cruz-Ortiz, Larissa Fernandes-Matano, Ricardo Grande, Lucia Hernandez-Rivas, Alejandra Hernández-Terán, Pavel Isa, Irma Lopez-Martinez, José Arturo Martínez-Orozco, Margarita Matías-Florentino, Fidencio Mejía-Nepomuceno, Edgar Mendieta-Condado, Mario Mújica-Sánchez, José Esteban Muñoz-Medina, Tatiana Nunez-Garcia, Luis Alberto Ochoa-Carrera, Hector Esteban Paz-Juárez, Francisco Pulido, José Ernesto Ramírez-González, Alma Rincón-Rubio, Teresita Rojas-Mendoza, Jorge Salas-Hernández, Alejandro Sanchez-Flores, Clara Esperanza Santacruz-Tinoco, Andrea Santos Coy-Arechavaleta, Blanca Taboada, Gloria Vazquez, Joel Armando Vázquez-Pérez, Jerome Jean Verleyen |
| EPI_ISL_1301471, EPI_ISL_1301472 | Instituto Nacional de Enfermedades Respiratorias (INER) | Instituto de Biotecnología de la UNAM | Authors from IBT, IMSS, InDRE and INER (in alphabetical order): Carlos F. Arias, Santiago Ávila-Ríos, Gisela Barrera-Badillo, Eduardo Becerril-Vargas, Celia Boukadida, Natividad Cruz-Ortiz, Larissa Fernandes-Matano, Ricardo Grande, Lucia Hernandez-Rivas, Alejandra Hernández-Terán, Pavel Isa, Irma Lopez-Martinez, José Arturo Martínez-Orozco, Margarita Matías-Florentino, Fidencio Mejía-Nepomuceno, Edgar Mendieta-Condado, Mario Mújica-Sánchez, José Esteban Muñoz-Medina, Tatiana Nunez-Garcia, Luis Alberto Ochoa-Carrera, Hector Esteban Paz-Juárez, Francisco Pulido, José Ernesto Ramírez-González, Alma Rincón-Rubio, Teresita Rojas-Mendoza, Jorge Salas-Hernández, Alejandro Sanchez-Flores, Clara Esperanza Santacruz-Tinoco, Andrea Santos Coy-Arechavaleta, Blanca Taboada, Gloria Vazquez, Joel Armando Vázquez-Pérez, Jerome Jean Verleyen |
| EPI_ISL_1301473 | Laboratorio Central de Epidemiología IMSS | Instituto de Biotecnología de la UNAM | Authors from IBT, IMSS, InDRE and INER (in alphabetical order): Carlos F. Arias, Santiago Ávila-Ríos, Gisela Barrera-Badillo, Eduardo Becerril-Vargas, Celia Boukadida, Natividad Cruz-Ortiz, Larissa Fernandes-Matano, Ricardo Grande, Lucia Hernandez-Rivas, Alejandra Hernández-Terán, Pavel Isa, Irma Lopez-Martinez, José Arturo Martínez-Orozco, Margarita Matías-Florentino, Fidencio Mejía-Nepomuceno, Edgar Mendieta-Condado, Mario Mújica-Sánchez, José Esteban Muñoz-Medina, Tatiana Nunez-Garcia, Luis Alberto Ochoa-Carrera, Hector Esteban Paz-Juárez, Francisco Pulido, José Ernesto Ramírez-González, Alma Rincón-Rubio, Teresita Rojas-Mendoza, Jorge Salas-Hernández, Alejandro Sanchez-Flores, Clara Esperanza Santacruz-Tinoco, Andrea Santos Coy-Arechavaleta, Blanca Taboada, Gloria Vazquez, Joel Armando Vázquez-Pérez, Jerome Jean Verleyen |
| EPI_ISL_1301474, EPI_ISL_1301475, EPI_ISL_1301476, EPI_ISL_1301477, EPI_ISL_1301478, EPI_ISL_1301479, EPI_ISL_1301480, EPI_ISL_1301481, EPI_ISL_1301482, EPI_ISL_1301483 | Instituto de Diagnostico y Referencia Epidemiologicos InDRE_RNLSP | Instituto de Biotecnología de la UNAM | Authors from IBT, IMSS, InDRE and INER (in alphabetical order): Carlos F. Arias, Santiago Ávila-Ríos, Gisela Barrera-Badillo, Eduardo Becerril-Vargas, Celia Boukadida, Natividad Cruz-Ortiz, Larissa Fernandes-Matano, Ricardo Grande, Lucia Hernandez-Rivas, Alejandra Hernández-Terán, Pavel Isa, Irma Lopez-Martinez, José Arturo Martínez-Orozco, Margarita Matías-Florentino, Fidencio Mejía-Nepomuceno, Edgar Mendieta-Condado, Mario Mújica-Sánchez, José Esteban Muñoz-Medina, Tatiana Nunez-Garcia, Luis Alberto Ochoa-Carrera, Hector Esteban Paz-Juárez, Francisco Pulido, José Ernesto Ramírez-González, Alma Rincón-Rubio, Teresita Rojas-Mendoza, Jorge Salas-Hernández, Alejandro Sanchez-Flores, Clara Esperanza Santacruz-Tinoco, Andrea Santos Coy-Arechavaleta, Blanca Taboada, Gloria Vazquez, Joel Armando Vázquez-Pérez, Jerome Jean Verleyen |
| EPI_ISL_1301484 | Laboratorio Central de Epidemiología IMSS | Instituto de Biotecnología de la UNAM | Authors from IBT, IMSS, InDRE and INER (in alphabetical order): Carlos F. Arias, Santiago Ávila-Ríos, Gisela Barrera-Badillo, Eduardo Becerril-Vargas, Celia Boukadida, Natividad Cruz-Ortiz, Larissa Fernandes-Matano, Ricardo Grande, Lucia Hernandez-Rivas, Alejandra Hernández-Terán, Pavel Isa, Irma Lopez-Martinez, José Arturo Martínez-Orozco, Margarita Matías-Florentino, Fidencio Mejía-Nepomuceno, Edgar Mendieta-Condado, Mario Mújica-Sánchez, José Esteban Muñoz-Medina, Tatiana Nunez-Garcia, Luis Alberto Ochoa-Carrera, Hector Esteban Paz-Juárez, Francisco Pulido, José Ernesto Ramírez-González, Alma Rincón-Rubio, Teresita Rojas-Mendoza, Jorge Salas-Hernández, Alejandro Sanchez-Flores, Clara Esperanza Santacruz-Tinoco, Andrea Santos Coy-Arechavaleta, Blanca Taboada, Gloria Vazquez, Joel Armando Vázquez-Pérez, Jerome Jean Verleyen |
| EPI_ISL_1301485, EPI_ISL_1301486, EPI_ISL_1301487 | Instituto de Diagnostico y Referencia Epidemiologicos InDRE_RNLSP | Instituto de Biotecnología de la UNAM | Authors from IBT, IMSS, InDRE and INER (in alphabetical order): Carlos F. Arias, Santiago Ávila-Ríos, Gisela Barrera-Badillo, Eduardo Becerril-Vargas, Celia Boukadida, Natividad Cruz-Ortiz, Larissa Fernandes-Matano, Ricardo Grande, Lucia Hernandez-Rivas, Alejandra Hernández-Terán, Pavel Isa, Irma Lopez-Martinez, José Arturo Martínez-Orozco, Margarita Matías-Florentino, Fidencio Mejía-Nepomuceno, Edgar Mendieta-Condado, Mario Mújica-Sánchez, José Esteban Muñoz-Medina, Tatiana Nunez-Garcia, Luis Alberto Ochoa-Carrera, Hector Esteban Paz-Juárez, Francisco Pulido, José Ernesto Ramírez-González, Alma Rincón-Rubio, Teresita Rojas-Mendoza, Jorge Salas-Hernández, Alejandro Sanchez-Flores, Clara Esperanza Santacruz-Tinoco, Andrea Santos Coy-Arechavaleta, Blanca Taboada, Gloria Vazquez, Joel Armando Vázquez-Pérez, Jerome Jean Verleyen |
| EPI_ISL_1301488, EPI_ISL_1301489, EPI_ISL_1301490 | Instituto Nacional de Enfermedades Respiratorias (INER) | Instituto de Biotecnología de la UNAM | Authors from IBT, IMSS, InDRE and INER (in alphabetical order): Carlos F. Arias, Santiago Ávila-Ríos, Gisela Barrera-Badillo, Eduardo Becerril-Vargas, Celia Boukadida, Natividad Cruz-Ortiz, Larissa Fernandes-Matano, Ricardo Grande, Lucia Hernandez-Rivas, Alejandra Hernández-Terán, Pavel Isa, Irma Lopez-Martinez, José Arturo Martínez-Orozco, Margarita Matías-Florentino, Fidencio Mejía-Nepomuceno, Edgar Mendieta-Condado, Mario Mújica-Sánchez, José Esteban Muñoz-Medina, Tatiana Nunez-Garcia, Luis Alberto Ochoa-Carrera, Hector Esteban Paz-Juárez, Francisco Pulido, José Ernesto Ramírez-González, Alma Rincón-Rubio, Teresita Rojas-Mendoza, Jorge Salas-Hernández, Alejandro Sanchez-Flores, Clara Esperanza Santacruz-Tinoco, Andrea Santos Coy-Arechavaleta, Blanca Taboada, Gloria Vazquez, Joel Armando Vázquez-Pérez, Jerome Jean Verleyen |
| EPI_ISL_1301491, EPI_ISL_1301492, EPI_ISL_1301493, EPI_ISL_1301494, EPI_ISL_1301495, EPI_ISL_1301497 | Instituto de Diagnostico y Referencia Epidemiologicos InDRE_RNLSP | Instituto de Biotecnología de la UNAM | Authors from IBT, IMSS, InDRE and INER (in alphabetical order): Carlos F. Arias, Santiago Ávila-Ríos, Gisela Barrera-Badillo, Eduardo Becerril-Vargas, Celia Boukadida, Natividad Cruz-Ortiz, Larissa Fernandes-Matano, Ricardo Grande, Lucia Hernandez-Rivas, Alejandra Hernández-Terán, Pavel Isa, Irma Lopez-Martinez, José Arturo Martínez-Orozco, Margarita Matías-Florentino, Fidencio Mejía-Nepomuceno, Edgar Mendieta-Condado, Mario Mújica-Sánchez, José Esteban Muñoz-Medina, Tatiana Nunez-Garcia, Luis Alberto Ochoa-Carrera, Hector Esteban Paz-Juárez, Francisco Pulido, José Ernesto Ramírez-González, Alma Rincón-Rubio, Teresita Rojas-Mendoza, Jorge Salas-Hernández, Alejandro Sanchez-Flores, Clara Esperanza Santacruz-Tinoco, Andrea Santos Coy-Arechavaleta, Blanca Taboada, Gloria Vazquez, Joel Armando Vázquez-Pérez, Jerome Jean Verleyen |
| EPI_ISL_1301498 | Laboratorio Central de Epidemiología IMSS | Instituto de Biotecnología de la UNAM | Authors from IBT, IMSS, InDRE and INER (in alphabetical order): Carlos F. Arias, Santiago Ávila-Ríos, Gisela Barrera-Badillo, Eduardo Becerril-Vargas, Celia Boukadida, Natividad Cruz-Ortiz, Larissa Fernandes-Matano, Ricardo Grande, Lucia Hernandez-Rivas, Alejandra Hernández-Terán, Pavel Isa, Irma Lopez-Martinez, José Arturo Martínez-Orozco, Margarita Matías-Florentino, Fidencio Mejía-Nepomuceno, Edgar Mendieta-Condado, Mario Mújica-Sánchez, José Esteban Muñoz-Medina, Tatiana Nunez-Garcia, Luis Alberto Ochoa-Carrera, Hector Esteban Paz-Juárez, Francisco Pulido, José Ernesto Ramírez-González, Alma Rincón-Rubio, Teresita Rojas-Mendoza, Jorge Salas-Hernández, Alejandro Sanchez-Flores, Clara Esperanza Santacruz-Tinoco, Andrea Santos Coy-Arechavaleta, Blanca Taboada, Gloria Vazquez, Joel Armando Vázquez-Pérez, Jerome Jean Verleyen |
| EPI_ISL_1301499 | Instituto Nacional de Enfermedades Respiratorias (INER) | Instituto de Biotecnología de la UNAM | Authors from IBT, IMSS, InDRE and INER (in alphabetical order): Carlos F. Arias, Santiago Ávila-Ríos, Gisela Barrera-Badillo, Eduardo Becerril-Vargas, Celia Boukadida, Natividad Cruz-Ortiz, Larissa Fernandes-Matano, Ricardo Grande, Lucia Hernandez-Rivas, Alejandra Hernández-Terán, Pavel Isa, Irma Lopez-Martinez, José Arturo Martínez-Orozco, Margarita Matías-Florentino, Fidencio Mejía-Nepomuceno, Edgar Mendieta-Condado, Mario Mújica-Sánchez, José Esteban Muñoz-Medina, Tatiana Nunez-Garcia, Luis Alberto Ochoa-Carrera, Hector Esteban Paz-Juárez, Francisco Pulido, José Ernesto Ramírez-González, Alma Rincón-Rubio, Teresita Rojas-Mendoza, Jorge Salas-Hernández, Alejandro Sanchez-Flores, Clara Esperanza Santacruz-Tinoco, Andrea Santos Coy-Arechavaleta, Blanca Taboada, Gloria Vazquez, Joel Armando Vázquez-Pérez, Jerome Jean Verleyen |
| EPI_ISL_1301500, EPI_ISL_1301501, EPI_ISL_1301502 | Instituto de Diagnostico y Referencia Epidemiologicos InDRE_RNLSP | Instituto de Biotecnología de la UNAM | Authors from IBT, IMSS, InDRE and INER (in alphabetical order): Carlos F. Arias, Santiago Ávila-Ríos, Gisela Barrera-Badillo, Eduardo Becerril-Vargas, Celia Boukadida, Natividad Cruz-Ortiz, Larissa Fernandes-Matano, Ricardo Grande, Lucia Hernandez-Rivas, Alejandra Hernández-Terán, Pavel Isa, Irma Lopez-Martinez, José Arturo Martínez-Orozco, Margarita Matías-Florentino, Fidencio Mejía-Nepomuceno, Edgar Mendieta-Condado, Mario Mújica-Sánchez, José Esteban Muñoz-Medina, Tatiana Nunez-Garcia, Luis Alberto Ochoa-Carrera, Hector Esteban Paz-Juárez, Francisco Pulido, José Ernesto Ramírez-González, Alma Rincón-Rubio, Teresita Rojas-Mendoza, Jorge Salas-Hernández, Alejandro Sanchez-Flores, Clara Esperanza Santacruz-Tinoco, Andrea Santos Coy-Arechavaleta, Blanca Taboada, Gloria Vazquez, Joel Armando Vázquez-Pérez, Jerome Jean Verleyen |
| EPI_ISL_1301503, EPI_ISL_1301504 | Instituto Nacional de Enfermedades Respiratorias (INER) | Instituto de Biotecnología de la UNAM | Authors from IBT, IMSS, InDRE and INER (in alphabetical order): Carlos F. Arias, Santiago Ávila-Ríos, Gisela Barrera-Badillo, Eduardo Becerril-Vargas, Celia Boukadida, Natividad Cruz-Ortiz, Larissa Fernandes-Matano, Ricardo Grande, Lucia Hernandez-Rivas, Alejandra Hernández-Terán, Pavel Isa, Irma Lopez-Martinez, José Arturo Martínez-Orozco, Margarita Matías-Florentino, Fidencio Mejía-Nepomuceno, Edgar Mendieta-Condado, Mario Mújica-Sánchez, José Esteban Muñoz-Medina, Tatiana Nunez-Garcia, Luis Alberto Ochoa-Carrera, Hector Esteban Paz-Juárez, Francisco Pulido, José Ernesto Ramírez-González, Alma Rincón-Rubio, Teresita Rojas-Mendoza, Jorge Salas-Hernández, Alejandro Sanchez-Flores, Clara Esperanza Santacruz-Tinoco, Andrea Santos Coy-Arechavaleta, Blanca Taboada, Gloria Vazquez, Joel Armando Vázquez-Pérez, Jerome Jean Verleyen |
| EPI_ISL_1301505 | Instituto de Diagnostico y Referencia Epidemiologicos InDRE_RNLSP | Instituto de Biotecnología de la UNAM | Authors from IBT, IMSS, InDRE and INER (in alphabetical order): Carlos F. Arias, Santiago Ávila-Ríos, Gisela Barrera-Badillo, Eduardo Becerril-Vargas, Celia Boukadida, Natividad Cruz-Ortiz, Larissa Fernandes-Matano, Ricardo Grande, Lucia Hernandez-Rivas, Alejandra Hernández-Terán, Pavel Isa, Irma Lopez-Martinez, José Arturo Martínez-Orozco, Margarita Matías-Florentino, Fidencio Mejía-Nepomuceno, Edgar Mendieta-Condado, Mario Mújica-Sánchez, José Esteban Muñoz-Medina, Tatiana Nunez-Garcia, Luis Alberto Ochoa-Carrera, Hector Esteban Paz-Juárez, Francisco Pulido, José Ernesto Ramírez-González, Alma Rincón-Rubio, Teresita Rojas-Mendoza, Jorge Salas-Hernández, Alejandro Sanchez-Flores, Clara Esperanza Santacruz-Tinoco, Andrea Santos Coy-Arechavaleta, Blanca Taboada, Gloria Vazquez, Joel Armando Vázquez-Pérez, Jerome Jean Verleyen |
| EPI_ISL_1301506, EPI_ISL_1301507 | Instituto Nacional de Enfermedades Respiratorias (INER) | Instituto de Biotecnología de la UNAM | Authors from IBT, IMSS, InDRE and INER (in alphabetical order): Carlos F. Arias, Santiago Ávila-Ríos, Gisela Barrera-Badillo, Eduardo Becerril-Vargas, Celia Boukadida, Natividad Cruz-Ortiz, Larissa Fernandes-Matano, Ricardo Grande, Lucia Hernandez-Rivas, Alejandra Hernández-Terán, Pavel Isa, Irma Lopez-Martinez, José Arturo Martínez-Orozco, Margarita Matías-Florentino, Fidencio Mejía-Nepomuceno, Edgar Mendieta-Condado, Mario Mújica-Sánchez, José Esteban Muñoz-Medina, Tatiana Nunez-Garcia, Luis Alberto Ochoa-Carrera, Hector Esteban Paz-Juárez, Francisco Pulido, José Ernesto Ramírez-González, Alma Rincón-Rubio, Teresita Rojas-Mendoza, Jorge Salas-Hernández, Alejandro Sanchez-Flores, Clara Esperanza Santacruz-Tinoco, Andrea Santos Coy-Arechavaleta, Blanca Taboada, Gloria Vazquez, Joel Armando Vázquez-Pérez, Jerome Jean Verleyen |
| EPI_ISL_1301508 | Instituto de Diagnostico y Referencia Epidemiologicos InDRE_RNLSP | Instituto de Biotecnología de la UNAM | Authors from IBT, IMSS, InDRE and INER (in alphabetical order): Carlos F. Arias, Santiago Ávila-Ríos, Gisela Barrera-Badillo, Eduardo Becerril-Vargas, Celia Boukadida, Natividad Cruz-Ortiz, Larissa Fernandes-Matano, Ricardo Grande, Lucia Hernandez-Rivas, Alejandra Hernández-Terán, Pavel Isa, Irma Lopez-Martinez, José Arturo Martínez-Orozco, Margarita Matías-Florentino, Fidencio Mejía-Nepomuceno, Edgar Mendieta-Condado, Mario Mújica-Sánchez, José Esteban Muñoz-Medina, Tatiana Nunez-Garcia, Luis Alberto Ochoa-Carrera, Hector Esteban Paz-Juárez, Francisco Pulido, José Ernesto Ramírez-González, Alma Rincón-Rubio, Teresita Rojas-Mendoza, Jorge Salas-Hernández, Alejandro Sanchez-Flores, Clara Esperanza Santacruz-Tinoco, Andrea Santos Coy-Arechavaleta, Blanca Taboada, Gloria Vazquez, Joel Armando Vázquez-Pérez, Jerome Jean Verleyen |
| EPI_ISL_1301509 | Laboratorio Central de Epidemiología IMSS | Instituto de Biotecnología de la UNAM | Authors from IBT, IMSS, InDRE and INER (in alphabetical order): Carlos F. Arias, Santiago Ávila-Ríos, Gisela Barrera-Badillo, Eduardo Becerril-Vargas, Celia Boukadida, Natividad Cruz-Ortiz, Larissa Fernandes-Matano, Ricardo Grande, Lucia Hernandez-Rivas, Alejandra Hernández-Terán, Pavel Isa, Irma Lopez-Martinez, José Arturo Martínez-Orozco, Margarita Matías-Florentino, Fidencio Mejía-Nepomuceno, Edgar Mendieta-Condado, Mario Mújica-Sánchez, José Esteban Muñoz-Medina, Tatiana Nunez-Garcia, Luis Alberto Ochoa-Carrera, Hector Esteban Paz-Juárez, Francisco Pulido, José Ernesto Ramírez-González, Alma Rincón-Rubio, Teresita Rojas-Mendoza, Jorge Salas-Hernández, Alejandro Sanchez-Flores, Clara Esperanza Santacruz-Tinoco, Andrea Santos Coy-Arechavaleta, Blanca Taboada, Gloria Vazquez, Joel Armando Vázquez-Pérez, Jerome Jean Verleyen |
| EPI_ISL_1301510 | Instituto Nacional de Enfermedades Respiratorias (INER) | Instituto de Biotecnología de la UNAM | Authors from IBT, IMSS, InDRE and INER (in alphabetical order): Carlos F. Arias, Santiago Ávila-Ríos, Gisela Barrera-Badillo, Eduardo Becerril-Vargas, Celia Boukadida, Natividad Cruz-Ortiz, Larissa Fernandes-Matano, Ricardo Grande, Lucia Hernandez-Rivas, Alejandra Hernández-Terán, Pavel Isa, Irma Lopez-Martinez, José Arturo Martínez-Orozco, Margarita Matías-Florentino, Fidencio Mejía-Nepomuceno, Edgar Mendieta-Condado, Mario Mújica-Sánchez, José Esteban Muñoz-Medina, Tatiana Nunez-Garcia, Luis Alberto Ochoa-Carrera, Hector Esteban Paz-Juárez, Francisco Pulido, José Ernesto Ramírez-González, Alma Rincón-Rubio, Teresita Rojas-Mendoza, Jorge Salas-Hernández, Alejandro Sanchez-Flores, Clara Esperanza Santacruz-Tinoco, Andrea Santos Coy-Arechavaleta, Blanca Taboada, Gloria Vazquez, Joel Armando Vázquez-Pérez, Jerome Jean Verleyen |

| | | | |
|---|---|---|---|
| EPI_ISL_1301511, EPI_ISL_1301512, EPI_ISL_1301513, EPI_ISL_1301514 | Instituto de Diagnostico y Referencia Epidemiologicos InDRE_RNLSP | Instituto de Biotecnología de la UNAM | Authors from IBT, IMSS, InDRE and INER (in alphabetical order): Carlos F. Arias, Santiago Ávila-Ríos, Gisela Barrera-Badillo, Eduardo Becerril-Vargas, Celia Boukadida, Natividad Cruz-Ortiz, Larissa Fernandes-Matano, Ricardo Grande, Lucia Hernandez-Rivas, Alejandra Hernández-Terán, Pavel Isa, Irma Lopez-Martinez, José Arturo Martínez-Orozco, Margarita Matías-Florentino, Fidencio Mejía-Nepomuceno, Edgar Mendieta-Condado, Mario Mújica-Sánchez, José Esteban Muñoz-Medina, Tatiana Nunez-Garcia, Luis Alberto Ochoa-Carrera, Hector Esteban Paz-Juárez, Francisco Pulido, José Ernesto Ramírez-González, Alma Rincón-Rubio, Teresita Rojas-Mendoza, Jorge Salas-Hernández, Alejandro Sanchez-Flores, Clara Esperanza Santacruz-Tinoco, Andrea Santos Coy-Arechavaleta, Blanca Taboada, Gloria Vazquez, Joel Armando Vázquez-Pérez, Jerome Jean Verleyen |
| EPI_ISL_1301515 | Instituto Nacional de Enfermedades Respiratorias (INER) | Instituto de Biotecnología de la UNAM | Authors from IBT, IMSS, InDRE and INER (in alphabetical order): Carlos F. Arias, Santiago Ávila-Ríos, Gisela Barrera-Badillo, Eduardo Becerril-Vargas, Celia Boukadida, Natividad Cruz-Ortiz, Larissa Fernandes-Matano, Ricardo Grande, Lucia Hernandez-Rivas, Alejandra Hernández-Terán, Pavel Isa, Irma Lopez-Martinez, José Arturo Martínez-Orozco, Margarita Matías-Florentino, Fidencio Mejía-Nepomuceno, Edgar Mendieta-Condado, Mario Mújica-Sánchez, José Esteban Muñoz-Medina, Tatiana Nunez-Garcia, Luis Alberto Ochoa-Carrera, Hector Esteban Paz-Juárez, Francisco Pulido, José Ernesto Ramírez-González, Alma Rincón-Rubio, Teresita Rojas-Mendoza, Jorge Salas-Hernández, Alejandro Sanchez-Flores, Clara Esperanza Santacruz-Tinoco, Andrea Santos Coy-Arechavaleta, Blanca Taboada, Gloria Vazquez, Joel Armando Vázquez-Pérez, Jerome Jean Verleyen |
| EPI_ISL_1301516 | Laboratorio Central de Epidemiología IMSS | Instituto de Biotecnología de la UNAM | Authors from IBT, IMSS, InDRE and INER (in alphabetical order): Carlos F. Arias, Santiago Ávila-Ríos, Gisela Barrera-Badillo, Eduardo Becerril-Vargas, Celia Boukadida, Natividad Cruz-Ortiz, Larissa Fernandes-Matano, Ricardo Grande, Lucia Hernandez-Rivas, Alejandra Hernández-Terán, Pavel Isa, Irma Lopez-Martinez, José Arturo Martínez-Orozco, Margarita Matías-Florentino, Fidencio Mejía-Nepomuceno, Edgar Mendieta-Condado, Mario Mújica-Sánchez, José Esteban Muñoz-Medina, Tatiana Nunez-Garcia, Luis Alberto Ochoa-Carrera, Hector Esteban Paz-Juárez, Francisco Pulido, José Ernesto Ramírez-González, Alma Rincón-Rubio, Teresita Rojas-Mendoza, Jorge Salas-Hernández, Alejandro Sanchez-Flores, Clara Esperanza Santacruz-Tinoco, Andrea Santos Coy-Arechavaleta, Blanca Taboada, Gloria Vazquez, Joel Armando Vázquez-Pérez, Jerome Jean Verleyen |
| EPI_ISL_1301517 | Instituto de Diagnostico y Referencia Epidemiologicos InDRE_RNLSP | Instituto de Biotecnología de la UNAM | Authors from IBT, IMSS, InDRE and INER (in alphabetical order): Carlos F. Arias, Santiago Ávila-Ríos, Gisela Barrera-Badillo, Eduardo Becerril-Vargas, Celia Boukadida, Natividad Cruz-Ortiz, Larissa Fernandes-Matano, Ricardo Grande, Lucia Hernandez-Rivas, Alejandra Hernández-Terán, Pavel Isa, Irma Lopez-Martinez, José Arturo Martínez-Orozco, Margarita Matías-Florentino, Fidencio Mejía-Nepomuceno, Edgar Mendieta-Condado, Mario Mújica-Sánchez, José Esteban Muñoz-Medina, Tatiana Nunez-Garcia, Luis Alberto Ochoa-Carrera, Hector Esteban Paz-Juárez, Francisco Pulido, José Ernesto Ramírez-González, Alma Rincón-Rubio, Teresita Rojas-Mendoza, Jorge Salas-Hernández, Alejandro Sanchez-Flores, Clara Esperanza Santacruz-Tinoco, Andrea Santos Coy-Arechavaleta, Blanca Taboada, Gloria Vazquez, Joel Armando Vázquez-Pérez, Jerome Jean Verleyen |
| EPI_ISL_1301518 | Laboratorio Central de Epidemiología IMSS | Instituto de Biotecnología de la UNAM | Authors from IBT, IMSS, InDRE and INER (in alphabetical order): Carlos F. Arias, Santiago Ávila-Ríos, Gisela Barrera-Badillo, Eduardo Becerril-Vargas, Celia Boukadida, Natividad Cruz-Ortiz, Larissa Fernandes-Matano, Ricardo Grande, Lucia Hernandez-Rivas, Alejandra Hernández-Terán, Pavel Isa, Irma Lopez-Martinez, José Arturo Martínez-Orozco, Margarita Matías-Florentino, Fidencio Mejía-Nepomuceno, Edgar Mendieta-Condado, Mario Mújica-Sánchez, José Esteban Muñoz-Medina, Tatiana Nunez-Garcia, Luis Alberto Ochoa-Carrera, Hector Esteban Paz-Juárez, Francisco Pulido, José Ernesto Ramírez-González, Alma Rincón-Rubio, Teresita Rojas-Mendoza, Jorge Salas-Hernández, Alejandro Sanchez-Flores, Clara Esperanza Santacruz-Tinoco, Andrea Santos Coy-Arechavaleta, Blanca Taboada, Gloria Vazquez, Joel Armando Vázquez-Pérez, Jerome Jean Verleyen |
| EPI_ISL_1301519 | Instituto de Diagnostico y Referencia Epidemiologicos InDRE_RNLSP | Instituto de Biotecnología de la UNAM | Authors from IBT, IMSS, InDRE and INER (in alphabetical order): Carlos F. Arias, Santiago Ávila-Ríos, Gisela Barrera-Badillo, Eduardo Becerril-Vargas, Celia Boukadida, Natividad Cruz-Ortiz, Larissa Fernandes-Matano, Ricardo Grande, Lucia Hernandez-Rivas, Alejandra Hernández-Terán, Pavel Isa, Irma Lopez-Martinez, José Arturo Martínez-Orozco, Margarita Matías-Florentino, Fidencio Mejía-Nepomuceno, Edgar Mendieta-Condado, Mario Mújica-Sánchez, José Esteban Muñoz-Medina, Tatiana Nunez-Garcia, Luis Alberto Ochoa-Carrera, Hector Esteban Paz-Juárez, Francisco Pulido, José Ernesto Ramírez-González, Alma Rincón-Rubio, Teresita Rojas-Mendoza, Jorge Salas-Hernández, Alejandro Sanchez-Flores, Clara Esperanza Santacruz-Tinoco, Andrea Santos Coy-Arechavaleta, Blanca Taboada, Gloria Vazquez, Joel Armando Vázquez-Pérez, Jerome Jean Verleyen |
| EPI_ISL_1301520 | Instituto Nacional de Enfermedades Respiratorias (INER) | Instituto de Biotecnología de la UNAM | Authors from IBT, IMSS, InDRE and INER (in alphabetical order): Carlos F. Arias, Santiago Ávila-Ríos, Gisela Barrera-Badillo, Eduardo Becerril-Vargas, Celia Boukadida, Natividad Cruz-Ortiz, Larissa Fernandes-Matano, Ricardo Grande, Lucia Hernandez-Rivas, Alejandra Hernández-Terán, Pavel Isa, Irma Lopez-Martinez, José Arturo Martínez-Orozco, Margarita Matías-Florentino, Fidencio Mejía-Nepomuceno, Edgar Mendieta-Condado, Mario Mújica-Sánchez, José Esteban Muñoz-Medina, Tatiana Nunez-Garcia, Luis Alberto Ochoa-Carrera, Hector Esteban Paz-Juárez, Francisco Pulido, José Ernesto Ramírez-González, Alma Rincón-Rubio, Teresita Rojas-Mendoza, Jorge Salas-Hernández, Alejandro Sanchez-Flores, Clara Esperanza Santacruz-Tinoco, Andrea Santos Coy-Arechavaleta, Blanca Taboada, Gloria Vazquez, Joel Armando Vázquez-Pérez, Jerome Jean Verleyen |
| EPI_ISL_1301521 | Instituto de Diagnostico y Referencia Epidemiologicos InDRE_RNLSP | Instituto de Biotecnología de la UNAM | Authors from IBT, IMSS, InDRE and INER (in alphabetical order): Carlos F. Arias, Santiago Ávila-Ríos, Gisela Barrera-Badillo, Eduardo Becerril-Vargas, Celia Boukadida, Natividad Cruz-Ortiz, Larissa Fernandes-Matano, Ricardo Grande, Lucia Hernandez-Rivas, Alejandra Hernández-Terán, Pavel Isa, Irma Lopez-Martinez, José Arturo Martínez-Orozco, Margarita Matías-Florentino, Fidencio Mejía-Nepomuceno, Edgar Mendieta-Condado, Mario Mújica-Sánchez, José Esteban Muñoz-Medina, Tatiana Nunez-Garcia, Luis Alberto Ochoa-Carrera, Hector Esteban Paz-Juárez, Francisco Pulido, José Ernesto Ramírez-González, Alma Rincón-Rubio, Teresita Rojas-Mendoza, Jorge Salas-Hernández, Alejandro Sanchez-Flores, Clara Esperanza Santacruz-Tinoco, Andrea Santos Coy-Arechavaleta, Blanca Taboada, Gloria Vazquez, Joel Armando Vázquez-Pérez, Jerome Jean Verleyen |
| EPI_ISL_1301522 | Instituto Nacional de Enfermedades Respiratorias (INER) | Instituto de Biotecnología de la UNAM | Authors from IBT, IMSS, InDRE and INER (in alphabetical order): Carlos F. Arias, Santiago Ávila-Ríos, Gisela Barrera-Badillo, Eduardo Becerril-Vargas, Celia Boukadida, Natividad Cruz-Ortiz, Larissa Fernandes-Matano, Ricardo Grande, Lucia Hernandez-Rivas, Alejandra Hernández-Terán, Pavel Isa, Irma Lopez-Martinez, José Arturo Martínez-Orozco, Margarita Matías-Florentino, Fidencio Mejía-Nepomuceno, Edgar Mendieta-Condado, Mario Mújica-Sánchez, José Esteban Muñoz-Medina, Tatiana Nunez-Garcia, Luis Alberto Ochoa-Carrera, Hector Esteban Paz-Juárez, Francisco Pulido, José Ernesto Ramírez-González, Alma Rincón-Rubio, Teresita Rojas-Mendoza, Jorge Salas-Hernández, Alejandro Sanchez-Flores, Clara Esperanza Santacruz-Tinoco, Andrea Santos Coy-Arechavaleta, Blanca Taboada, Gloria Vazquez, Joel Armando Vázquez-Pérez, Jerome Jean Verleyen |
| EPI_ISL_1301523 | Instituto de Diagnostico y Referencia Epidemiologicos InDRE_RNLSP | Instituto de Biotecnología de la UNAM | Authors from IBT, IMSS, InDRE and INER (in alphabetical order): Carlos F. Arias, Santiago Ávila-Ríos, Gisela Barrera-Badillo, Eduardo Becerril-Vargas, Celia Boukadida, Natividad Cruz-Ortiz, Larissa Fernandes-Matano, Ricardo Grande, Lucia Hernandez-Rivas, Alejandra Hernández-Terán, Pavel Isa, Irma Lopez-Martinez, José Arturo Martínez-Orozco, Margarita Matías-Florentino, Fidencio Mejía-Nepomuceno, Edgar Mendieta-Condado, Mario Mújica-Sánchez, José Esteban Muñoz-Medina, Tatiana Nunez-Garcia, Luis Alberto Ochoa-Carrera, Hector Esteban Paz-Juárez, Francisco Pulido, José Ernesto Ramírez-González, Alma Rincón-Rubio, Teresita Rojas-Mendoza, Jorge Salas-Hernández, Alejandro Sanchez-Flores, Clara Esperanza Santacruz-Tinoco, Andrea Santos Coy-Arechavaleta, Blanca Taboada, Gloria Vazquez, Joel Armando Vázquez-Pérez, Jerome Jean Verleyen |
| EPI_ISL_1301524 | Instituto Nacional de Enfermedades Respiratorias (INER) | Instituto de Biotecnología de la UNAM | Authors from IBT, IMSS, InDRE and INER (in alphabetical order): Carlos F. Arias, Santiago Ávila-Ríos, Gisela Barrera-Badillo, Eduardo Becerril-Vargas, Celia Boukadida, Natividad Cruz-Ortiz, Larissa Fernandes-Matano, Ricardo Grande, Lucia Hernandez-Rivas, Alejandra Hernández-Terán, Pavel Isa, Irma Lopez-Martinez, José Arturo Martínez-Orozco, Margarita Matías-Florentino, Fidencio Mejía-Nepomuceno, Edgar Mendieta-Condado, Mario Mújica-Sánchez, José Esteban Muñoz-Medina, Tatiana Nunez-Garcia, Luis Alberto Ochoa-Carrera, Hector Esteban Paz-Juárez, Francisco Pulido, José Ernesto Ramírez-González, Alma Rincón-Rubio, Teresita Rojas-Mendoza, Jorge Salas-Hernández, Alejandro Sanchez-Flores, Clara Esperanza Santacruz-Tinoco, Andrea Santos Coy-Arechavaleta, Blanca Taboada, Gloria Vazquez, Joel Armando Vázquez-Pérez, Jerome Jean Verleyen |
| EPI_ISL_1301525 | Laboratorio Central de Epidemiología IMSS | Instituto de Biotecnología de la UNAM | Authors from IBT, IMSS, InDRE and INER (in alphabetical order): Carlos F. Arias, Santiago Ávila-Ríos, Gisela Barrera-Badillo, Eduardo Becerril-Vargas, Celia Boukadida, Natividad Cruz-Ortiz, Larissa Fernandes-Matano, Ricardo Grande, Lucia Hernandez-Rivas, Alejandra Hernández-Terán, Pavel Isa, Irma Lopez-Martinez, José Arturo Martínez-Orozco, Margarita Matías-Florentino, Fidencio Mejía-Nepomuceno, Edgar Mendieta-Condado, Mario Mújica-Sánchez, José Esteban Muñoz-Medina, Tatiana Nunez-Garcia, Luis Alberto Ochoa-Carrera, Hector Esteban Paz-Juárez, Francisco Pulido, José Ernesto Ramírez-González, Alma Rincón-Rubio, Teresita Rojas-Mendoza, Jorge Salas-Hernández, Alejandro Sanchez-Flores, Clara Esperanza Santacruz-Tinoco, Andrea Santos Coy-Arechavaleta, Blanca Taboada, Gloria Vazquez, Joel Armando Vázquez-Pérez, Jerome Jean Verleyen |
| EPI_ISL_1301526, EPI_ISL_1301527, EPI_ISL_1301528, EPI_ISL_1301529 | Instituto de Diagnostico y Referencia Epidemiologicos InDRE_RNLSP | Instituto de Biotecnología de la UNAM | Authors from IBT, IMSS, InDRE and INER (in alphabetical order): Carlos F. Arias, Santiago Ávila-Ríos, Gisela Barrera-Badillo, Eduardo Becerril-Vargas, Celia Boukadida, Natividad Cruz-Ortiz, Larissa Fernandes-Matano, Ricardo Grande, Lucia Hernandez-Rivas, Alejandra Hernández-Terán, Pavel Isa, Irma Lopez-Martinez, José Arturo Martínez-Orozco, Margarita Matías-Florentino, Fidencio Mejía-Nepomuceno, Edgar Mendieta-Condado, Mario Mújica-Sánchez, José Esteban Muñoz-Medina, Tatiana Nunez-Garcia, Luis Alberto Ochoa-Carrera, Hector Esteban Paz-Juárez, Francisco Pulido, José Ernesto Ramírez-González, Alma Rincón-Rubio, Teresita Rojas-Mendoza, Jorge Salas-Hernández, Alejandro Sanchez-Flores, Clara Esperanza Santacruz-Tinoco, Andrea Santos Coy-Arechavaleta, Blanca Taboada, Gloria Vazquez, Joel Armando Vázquez-Pérez, Jerome Jean Verleyen |
| EPI_ISL_1301530 | Laboratorio Central de Epidemiología IMSS | Instituto de Biotecnología de la UNAM | Authors from IBT, IMSS, InDRE and INER (in alphabetical order): Carlos F. Arias, Santiago Ávila-Ríos, Gisela Barrera-Badillo, Eduardo Becerril-Vargas, Celia Boukadida, Natividad Cruz-Ortiz, Larissa Fernandes-Matano, Ricardo Grande, Lucia Hernandez-Rivas, Alejandra Hernández-Terán, Pavel Isa, Irma Lopez-Martinez, José Arturo Martínez-Orozco, Margarita Matías-Florentino, Fidencio Mejía-Nepomuceno, Edgar Mendieta-Condado, Mario Mújica-Sánchez, José Esteban Muñoz-Medina, Tatiana Nunez-Garcia, Luis Alberto Ochoa-Carrera, Hector Esteban Paz-Juárez, Francisco Pulido, José Ernesto Ramírez-González, Alma Rincón-Rubio, Teresita Rojas-Mendoza, Jorge Salas-Hernández, Alejandro Sanchez-Flores, Clara Esperanza Santacruz-Tinoco, Andrea Santos Coy-Arechavaleta, Blanca Taboada, Gloria Vazquez, Joel Armando Vázquez-Pérez, Jerome Jean Verleyen |
| EPI_ISL_1301531, EPI_ISL_1301532 | Instituto de Diagnostico y Referencia Epidemiologicos InDRE_RNLSP | Instituto de Biotecnología de la UNAM | Authors from IBT, IMSS, InDRE and INER (in alphabetical order): Carlos F. Arias, Santiago Ávila-Ríos, Gisela Barrera-Badillo, Eduardo Becerril-Vargas, Celia Boukadida, Natividad Cruz-Ortiz, Larissa Fernandes-Matano, Ricardo Grande, Lucia Hernandez-Rivas, Alejandra Hernández-Terán, Pavel Isa, Irma Lopez-Martinez, José Arturo Martínez-Orozco, Margarita Matías-Florentino, Fidencio Mejía-Nepomuceno, Edgar Mendieta-Condado, Mario Mújica-Sánchez, José Esteban Muñoz-Medina, Tatiana Nunez-Garcia, Luis Alberto Ochoa-Carrera, Hector Esteban Paz-Juárez, Francisco Pulido, José Ernesto Ramírez-González, Alma Rincón-Rubio, Teresita Rojas-Mendoza, Jorge Salas-Hernández, Alejandro Sanchez-Flores, Clara Esperanza Santacruz-Tinoco, Andrea Santos Coy-Arechavaleta, Blanca Taboada, Gloria Vazquez, Joel Armando Vázquez-Pérez, Jerome Jean Verleyen |
| EPI_ISL_1301533 | Instituto Nacional de Enfermedades Respiratorias (INER) | Instituto de Biotecnología de la UNAM | Authors from IBT, IMSS, InDRE and INER (in alphabetical order): Carlos F. Arias, Santiago Ávila-Ríos, Gisela Barrera-Badillo, Eduardo Becerril-Vargas, Celia Boukadida, Natividad Cruz-Ortiz, Larissa Fernandes-Matano, Ricardo Grande, Lucia Hernandez-Rivas, Alejandra Hernández-Terán, Pavel Isa, Irma Lopez-Martinez, José Arturo Martínez-Orozco, Margarita Matías-Florentino, Fidencio Mejía-Nepomuceno, Edgar Mendieta-Condado, Mario Mújica-Sánchez, José Esteban Muñoz-Medina, Tatiana Nunez-Garcia, Luis Alberto Ochoa-Carrera, Hector Esteban Paz-Juárez, Francisco Pulido, José Ernesto Ramírez-González, Alma Rincón-Rubio, Teresita Rojas-Mendoza, Jorge Salas-Hernández, Alejandro Sanchez-Flores, Clara Esperanza Santacruz-Tinoco, Andrea Santos Coy-Arechavaleta, Blanca Taboada, Gloria Vazquez, Joel Armando Vázquez-Pérez, Jerome Jean Verleyen |
| EPI_ISL_1301534 | Instituto de Diagnostico y Referencia Epidemiologicos InDRE_RNLSP | Instituto de Biotecnología de la UNAM | Authors from IBT, IMSS, InDRE and INER (in alphabetical order): Carlos F. Arias, Santiago Ávila-Ríos, Gisela Barrera-Badillo, Eduardo Becerril-Vargas, Celia Boukadida, Natividad Cruz-Ortiz, Larissa Fernandes-Matano, Ricardo Grande, Lucia Hernandez-Rivas, Alejandra Hernández-Terán, Pavel Isa, Irma Lopez-Martinez, José Arturo Martínez-Orozco, Margarita Matías-Florentino, Fidencio Mejía-Nepomuceno, Edgar Mendieta-Condado, Mario Mújica-Sánchez, José Esteban Muñoz-Medina, Tatiana Nunez-Garcia, Luis Alberto Ochoa-Carrera, Hector Esteban Paz-Juárez, Francisco Pulido, José Ernesto Ramírez-González, Alma Rincón-Rubio, Teresita Rojas-Mendoza, Jorge Salas-Hernández, Alejandro Sanchez-Flores, Clara Esperanza Santacruz-Tinoco, Andrea Santos Coy-Arechavaleta, Blanca Taboada, Gloria Vazquez, Joel Armando Vázquez-Pérez, Jerome Jean Verleyen |
| EPI_ISL_1301535, EPI_ISL_1301536 | Laboratorio Central de Epidemiología IMSS | Instituto de Biotecnología de la UNAM | Authors from IBT, IMSS, InDRE and INER (in alphabetical order): Carlos F. Arias, Santiago Ávila-Ríos, Gisela Barrera-Badillo, Eduardo Becerril-Vargas, Celia Boukadida, Natividad Cruz-Ortiz, Larissa Fernandes-Matano, Ricardo Grande, Lucia Hernandez-Rivas, Alejandra Hernández-Terán, Pavel Isa, Irma Lopez-Martinez, José Arturo Martínez-Orozco, Margarita Matías-Florentino, Fidencio Mejía-Nepomuceno, Edgar Mendieta-Condado, Mario Mújica-Sánchez, José Esteban Muñoz-Medina, Tatiana Nunez-Garcia, Luis Alberto Ochoa-Carrera, Hector Esteban Paz-Juárez, Francisco Pulido, José Ernesto Ramírez-González, Alma Rincón-Rubio, Teresita Rojas-Mendoza, Jorge Salas-Hernández, Alejandro Sanchez-Flores, Clara Esperanza Santacruz-Tinoco, Andrea Santos Coy-Arechavaleta, Blanca Taboada, Gloria Vazquez, Joel Armando Vázquez-Pérez, Jerome Jean Verleyen |
| EPI_ISL_1301537 | Instituto de Diagnostico y Referencia Epidemiologicos InDRE_RNLSP | Instituto de Biotecnología de la UNAM | Authors from IBT, IMSS, InDRE and INER (in alphabetical order): Carlos F. Arias, Santiago Ávila-Ríos, Gisela Barrera-Badillo, Eduardo Becerril-Vargas, Celia Boukadida, Natividad Cruz-Ortiz, Larissa Fernandes-Matano, Ricardo Grande, Lucia Hernandez-Rivas, Alejandra Hernández-Terán, Pavel Isa, Irma Lopez-Martinez, José Arturo Martínez-Orozco, Margarita Matías-Florentino, Fidencio Mejía-Nepomuceno, Edgar Mendieta-Condado, Mario Mújica-Sánchez, José Esteban Muñoz-Medina, Tatiana Nunez-Garcia, Luis Alberto Ochoa-Carrera, Hector Esteban Paz-Juárez, Francisco Pulido, José Ernesto Ramírez-González, Alma Rincón-Rubio, Teresita Rojas-Mendoza, Jorge Salas-Hernández, Alejandro Sanchez-Flores, Clara Esperanza Santacruz-Tinoco, Andrea Santos Coy-Arechavaleta, Blanca Taboada, Gloria Vazquez, Joel Armando Vázquez-Pérez, Jerome Jean Verleyen |
| EPI_ISL_1301538 | Laboratorio Central de Epidemiología IMSS | Instituto de Biotecnología de la UNAM | Authors from IBT, IMSS, InDRE and INER (in alphabetical order): Carlos F. Arias, Santiago Ávila-Ríos, Gisela Barrera-Badillo, Eduardo Becerril-Vargas, Celia Boukadida, Natividad Cruz-Ortiz, Larissa Fernandes-Matano, Ricardo Grande, Lucia Hernandez-Rivas, Alejandra Hernández-Terán, Pavel Isa, Irma Lopez-Martinez, José Arturo Martínez-Orozco, Margarita Matías-Florentino, Fidencio Mejía-Nepomuceno, Edgar Mendieta-Condado, Mario Mújica-Sánchez, José Esteban Muñoz-Medina, Tatiana Nunez-Garcia, Luis Alberto Ochoa-Carrera, Hector Esteban Paz-Juárez, Francisco Pulido, José Ernesto Ramírez-González, Alma Rincón-Rubio, Teresita Rojas-Mendoza, Jorge Salas-Hernández, Alejandro Sanchez-Flores, Clara Esperanza Santacruz-Tinoco, Andrea Santos Coy-Arechavaleta, Blanca Taboada, Gloria Vazquez, Joel Armando Vázquez-Pérez, Jerome Jean Verleyen |
| EPI_ISL_1301539 | Instituto Nacional de Enfermedades Respiratorias (INER) | Instituto de Biotecnología de la UNAM | Authors from IBT, IMSS, InDRE and INER (in alphabetical order): Carlos F. Arias, Santiago Ávila-Ríos, Gisela Barrera-Badillo, Eduardo Becerril-Vargas, Celia Boukadida, Natividad Cruz-Ortiz, Larissa Fernandes-Matano, Ricardo Grande, Lucia Hernandez-Rivas, Alejandra Hernández-Terán, Pavel Isa, Irma Lopez-Martinez, José Arturo Martínez-Orozco, Margarita Matías-Florentino, Fidencio Mejía-Nepomuceno, Edgar Mendieta-Condado, Mario Mújica-Sánchez, José Esteban Muñoz-Medina, Tatiana Nunez-Garcia, Luis Alberto Ochoa-Carrera, Hector Esteban Paz-Juárez, Francisco Pulido, José Ernesto Ramírez-González, Alma Rincón-Rubio, Teresita Rojas-Mendoza, Jorge Salas-Hernández, Alejandro Sanchez-Flores, Clara Esperanza Santacruz-Tinoco, Andrea Santos Coy-Arechavaleta, Blanca Taboada, Gloria Vazquez, Joel Armando Vázquez-Pérez, Jerome Jean Verleyen |
| EPI_ISL_1301540, EPI_ISL_1301541, EPI_ISL_1301542, EPI_ISL_1301543, EPI_ISL_1301544 | Instituto de Diagnostico y Referencia Epidemiologicos InDRE_RNLSP | Instituto de Biotecnología de la UNAM | Authors from IBT, IMSS, InDRE and INER (in alphabetical order): Carlos F. Arias, Santiago Ávila-Ríos, Gisela Barrera-Badillo, Eduardo Becerril-Vargas, Celia Boukadida, Natividad Cruz-Ortiz, Larissa Fernandes-Matano, Ricardo Grande, Lucia Hernandez-Rivas, Alejandra Hernández-Terán, Pavel Isa, Irma Lopez-Martinez, José Arturo Martínez-Orozco, Margarita Matías-Florentino, Fidencio Mejía-Nepomuceno, Edgar Mendieta-Condado, Mario Mújica-Sánchez, José Esteban Muñoz-Medina, Tatiana Nunez-Garcia, Luis Alberto Ochoa-Carrera, Hector Esteban Paz-Juárez, Francisco Pulido, José Ernesto Ramírez-González, Alma Rincón-Rubio, Teresita Rojas-Mendoza, Jorge Salas-Hernández, Alejandro Sanchez-Flores, Clara Esperanza Santacruz-Tinoco, Andrea Santos Coy-Arechavaleta, Blanca Taboada, Gloria Vazquez, Joel Armando Vázquez-Pérez, Jerome Jean Verleyen |
| EPI_ISL_1301545 | Instituto Nacional de Enfermedades Respiratorias (INER) | Instituto de Biotecnología de la UNAM | Authors from IBT, IMSS, InDRE and INER (in alphabetical order): Carlos F. Arias, Santiago Ávila-Ríos, Gisela Barrera-Badillo, Eduardo Becerril-Vargas, Celia Boukadida, Natividad Cruz-Ortiz, Larissa Fernandes-Matano, Ricardo Grande, Lucia Hernandez-Rivas, Alejandra Hernández-Terán, Pavel Isa, Irma Lopez-Martinez, José Arturo Martínez-Orozco, Margarita Matías-Florentino, Fidencio Mejía-Nepomuceno, Edgar Mendieta-Condado, Mario Mújica-Sánchez, José Esteban Muñoz-Medina, Tatiana Nunez-Garcia, Luis Alberto Ochoa-Carrera, Hector Esteban Paz-Juárez, Francisco Pulido, José Ernesto Ramírez-González, Alma Rincón-Rubio, Teresita Rojas-Mendoza, Jorge Salas-Hernández, Alejandro Sanchez-Flores, Clara Esperanza Santacruz-Tinoco, Andrea Santos Coy-Arechavaleta, Blanca Taboada, Gloria Vazquez, Joel Armando Vázquez-Pérez, Jerome Jean Verleyen |
| EPI_ISL_1301546, EPI_ISL_1301547, EPI_ISL_1301548, EPI_ISL_1301549 | Instituto de Diagnostico y Referencia Epidemiologicos InDRE_RNLSP | Instituto de Biotecnología de la UNAM | Authors from IBT, IMSS, InDRE and INER (in alphabetical order): Carlos F. Arias, Santiago Ávila-Ríos, Gisela Barrera-Badillo, Eduardo Becerril-Vargas, Celia Boukadida, Natividad Cruz-Ortiz, Larissa Fernandes-Matano, Ricardo Grande, Lucia Hernandez-Rivas, Alejandra Hernández-Terán, Pavel Isa, Irma Lopez-Martinez, José Arturo Martínez-Orozco, Margarita Matías-Florentino, Fidencio Mejía-Nepomuceno, Edgar Mendieta-Condado, Mario Mújica-Sánchez, José Esteban Muñoz-Medina, Tatiana Nunez-Garcia, Luis Alberto Ochoa-Carrera, Hector Esteban Paz-Juárez, Francisco Pulido, José Ernesto Ramírez-González, Alma Rincón-Rubio, Teresita Rojas-Mendoza, Jorge Salas-Hernández, Alejandro Sanchez-Flores, Clara Esperanza Santacruz-Tinoco, Andrea Santos Coy-Arechavaleta, Blanca Taboada, Gloria Vazquez, Joel Armando Vázquez-Pérez, Jerome Jean Verleyen |
| EPI_ISL_1301550, EPI_ISL_1301551 | Instituto Nacional de Enfermedades Respiratorias (INER) | Instituto de Biotecnología de la UNAM | Authors from IBT, IMSS, InDRE and INER (in alphabetical order): Carlos F. Arias, Santiago Ávila-Ríos, Gisela Barrera-Badillo, Eduardo Becerril-Vargas, Celia Boukadida, Natividad Cruz-Ortiz, Larissa Fernandes-Matano, Ricardo Grande, Lucia Hernandez-Rivas, Alejandra Hernández-Terán, Pavel Isa, Irma Lopez-Martinez, José Arturo Martínez-Orozco, Margarita Matías-Florentino, Fidencio Mejía-Nepomuceno, Edgar Mendieta-Condado, Mario Mújica-Sánchez, José Esteban Muñoz-Medina, Tatiana Nunez-Garcia, Luis Alberto Ochoa-Carrera, Hector Esteban Paz-Juárez, Francisco |

| | | | |
|---|---|---|---|
| | | | Pulido, José Ernesto Ramírez-González, Alma Rincón-Rubio, Teresita Rojas-Mendoza, Jorge Salas-Hernández, Alejandro Sanchez-Flores, Clara Esperanza Santacruz-Tinoco, Andrea Santos Coy-Arechavaleta, Blanca Taboada, Gloria Vazquez, Joel Armando Vázquez-Pérez, Jerome Jean Verleyen |
| EPI_ISL_1301552, EPI_ISL_1301553 | Laboratorio Central de Epidemiología IMSS | Instituto de Biotecnología de la UNAM | Authors from IBT, IMSS, InDRE and INER (in alphabetical order): Carlos F. Arias, Santiago Ávila-Ríos, Gisela Barrera-Badillo, Eduardo Becerril-Vargas, Celia Boukadida, Natividad Cruz-Ortiz, Larissa Fernandes-Matano, Ricardo Grande, Lucia Hernandez-Rivas, Alejandra Hernández-Terán, Pavel Isa, Irma Lopez-Martinez, José Arturo Martínez-Orozco, Margarita Matías-Florentino, Fidencio Mejía-Nepomuceno, Edgar Mendieta-Condado, Mario Mújica-Sánchez, José Esteban Muñoz-Medina, Tatiana Nunez-Garcia, Luis Alberto Ochoa-Carrera, Hector Esteban Paz-Juárez, Francisco Pulido, José Ernesto Ramírez-González, Alma Rincón-Rubio, Teresita Rojas-Mendoza, Jorge Salas-Hernández, Alejandro Sanchez-Flores, Clara Esperanza Santacruz-Tinoco, Andrea Santos Coy-Arechavaleta, Blanca Taboada, Gloria Vazquez, Joel Armando Vázquez-Pérez, Jerome Jean Verleyen |
| EPI_ISL_1301554 | Instituto Nacional de Enfermedades Respiratorias (INER) | Instituto de Biotecnología de la UNAM | Authors from IBT, IMSS, InDRE and INER (in alphabetical order): Carlos F. Arias, Santiago Ávila-Ríos, Gisela Barrera-Badillo, Eduardo Becerril-Vargas, Celia Boukadida, Natividad Cruz-Ortiz, Larissa Fernandes-Matano, Ricardo Grande, Lucia Hernandez-Rivas, Alejandra Hernández-Terán, Pavel Isa, Irma Lopez-Martinez, José Arturo Martínez-Orozco, Margarita Matías-Florentino, Fidencio Mejía-Nepomuceno, Edgar Mendieta-Condado, Mario Mújica-Sánchez, José Esteban Muñoz-Medina, Tatiana Nunez-Garcia, Luis Alberto Ochoa-Carrera, Hector Esteban Paz-Juárez, Francisco Pulido, José Ernesto Ramírez-González, Alma Rincón-Rubio, Teresita Rojas-Mendoza, Jorge Salas-Hernández, Alejandro Sanchez-Flores, Clara Esperanza Santacruz-Tinoco, Andrea Santos Coy-Arechavaleta, Blanca Taboada, Gloria Vazquez, Joel Armando Vázquez-Pérez, Jerome Jean Verleyen |
| EPI_ISL_1301555, EPI_ISL_1301556, EPI_ISL_1301557, EPI_ISL_1301558, EPI_ISL_1301559, EPI_ISL_1301560, EPI_ISL_1301561, EPI_ISL_1301562, EPI_ISL_1301563, EPI_ISL_1301564, EPI_ISL_1301565, EPI_ISL_1301566, EPI_ISL_1301567, EPI_ISL_1301568, EPI_ISL_1301569, EPI_ISL_1301570, EPI_ISL_1301571, EPI_ISL_1301572, EPI_ISL_1301573, EPI_ISL_1301574, EPI_ISL_1301575, EPI_ISL_1301576, EPI_ISL_1301577, EPI_ISL_1301578, EPI_ISL_1301579, EPI_ISL_1301580, EPI_ISL_1301581, EPI_ISL_1301582, EPI_ISL_1301583, EPI_ISL_1301584, EPI_ISL_1301585, EPI_ISL_1301586, EPI_ISL_1301587, EPI_ISL_1301588, EPI_ISL_1301589, EPI_ISL_1301590, EPI_ISL_1301591, EPI_ISL_1301592 | | | |
| see above | Instituto de Diagnostico y Referencia Epidemiologicos InDRE_RNLSP | Instituto de Biotecnología de la UNAM | Authors from IBT, IMSS, InDRE and INER (in alphabetical order): Carlos F. Arias, Santiago Ávila-Ríos, Gisela Barrera-Badillo, Eduardo Becerril-Vargas, Celia Boukadida, Natividad Cruz-Ortiz, Larissa Fernandes-Matano, Ricardo Grande, Lucia Hernandez-Rivas, Alejandra Hernández-Terán, Pavel Isa, Irma Lopez-Martinez, José Arturo Martínez-Orozco, Margarita Matías-Florentino, Fidencio Mejía-Nepomuceno, Edgar Mendieta-Condado, Mario Mújica-Sánchez, José Esteban Muñoz-Medina, Tatiana Nunez-Garcia, Luis Alberto Ochoa-Carrera, Hector Esteban Paz-Juárez, Francisco Pulido, José Ernesto Ramírez-González, Alma Rincón-Rubio, Teresita Rojas-Mendoza, Jorge Salas-Hernández, Alejandro Sanchez-Flores, Clara Esperanza Santacruz-Tinoco, Andrea Santos Coy-Arechavaleta, Blanca Taboada, Gloria Vazquez, Joel Armando Vázquez-Pérez, Jerome Jean Verleyen |
| EPI_ISL_1301593, EPI_ISL_1301594, EPI_ISL_1301595, EPI_ISL_1301596, EPI_ISL_1301597 | Laboratorio Central de Epidemiología IMSS | Instituto de Biotecnología de la UNAM | Authors from IBT, IMSS, InDRE and INER (in alphabetical order): Carlos F. Arias, Santiago Ávila-Ríos, Gisela Barrera-Badillo, Eduardo Becerril-Vargas, Celia Boukadida, Natividad Cruz-Ortiz, Larissa Fernandes-Matano, Ricardo Grande, Lucia Hernandez-Rivas, Alejandra Hernández-Terán, Pavel Isa, Irma Lopez-Martinez, José Arturo Martínez-Orozco, Margarita Matías-Florentino, Fidencio Mejía-Nepomuceno, Edgar Mendieta-Condado, Mario Mújica-Sánchez, José Esteban Muñoz-Medina, Tatiana Nunez-Garcia, Luis Alberto Ochoa-Carrera, Hector Esteban Paz-Juárez, Francisco Pulido, José Ernesto Ramírez-González, Alma Rincón-Rubio, Teresita Rojas-Mendoza, Jorge Salas-Hernández, Alejandro Sanchez-Flores, Clara Esperanza Santacruz-Tinoco, Andrea Santos Coy-Arechavaleta, Blanca Taboada, Gloria Vazquez, Joel Armando Vázquez-Pérez, Jerome Jean Verleyen |
| EPI_ISL_1301598, EPI_ISL_1301600, EPI_ISL_1301602, EPI_ISL_1301603, EPI_ISL_1301604, EPI_ISL_1301605, EPI_ISL_1301606, EPI_ISL_1301607, EPI_ISL_1301608, EPI_ISL_1301609, EPI_ISL_1301610, EPI_ISL_1301611, EPI_ISL_1301612, EPI_ISL_1301613, EPI_ISL_1301614, EPI_ISL_1301615, EPI_ISL_1301616, EPI_ISL_1301617, EPI_ISL_1301618, EPI_ISL_1301619 | | | |
| see above | Instituto Nacional de Enfermedades Respiratorias (INER) | Instituto de Biotecnología de la UNAM | Authors from IBT, IMSS, InDRE and INER (in alphabetical order): Carlos F. Arias, Santiago Ávila-Ríos, Gisela Barrera-Badillo, Eduardo Becerril-Vargas, Celia Boukadida, Natividad Cruz-Ortiz, Larissa Fernandes-Matano, Ricardo Grande, Lucia Hernandez-Rivas, Alejandra Hernández-Terán, Pavel Isa, Irma Lopez-Martinez, José Arturo Martínez-Orozco, Margarita Matías-Florentino, Fidencio Mejía-Nepomuceno, Edgar Mendieta-Condado, Mario Mújica-Sánchez, José Esteban Muñoz-Medina, Tatiana Nunez-Garcia, Luis Alberto Ochoa-Carrera, Hector Esteban Paz-Juárez, Francisco Pulido, José Ernesto Ramírez-González, Alma Rincón-Rubio, Teresita Rojas-Mendoza, Jorge Salas-Hernández, Alejandro Sanchez-Flores, Clara Esperanza Santacruz-Tinoco, Andrea Santos Coy-Arechavaleta, Blanca Taboada, Gloria Vazquez, Joel Armando Vázquez-Pérez, Jerome Jean Verleyen |
| EPI_ISL_1301620, EPI_ISL_1301621, EPI_ISL_1301622, EPI_ISL_1301623, EPI_ISL_1301624, EPI_ISL_1301625, EPI_ISL_1301626, EPI_ISL_1301627, EPI_ISL_1301628, EPI_ISL_1301629, EPI_ISL_1301630, EPI_ISL_1301631, EPI_ISL_1301632, EPI_ISL_1301633, EPI_ISL_1301634, EPI_ISL_1301635, EPI_ISL_1301636, EPI_ISL_1301637, EPI_ISL_1301638, EPI_ISL_1301639, EPI_ISL_1301640, EPI_ISL_1301641, EPI_ISL_1301642, EPI_ISL_1301643, EPI_ISL_1301644, EPI_ISL_1301645, EPI_ISL_1301646, EPI_ISL_1301647, EPI_ISL_1301648, EPI_ISL_1301649, EPI_ISL_1301650, EPI_ISL_1301651, EPI_ISL_1301652, EPI_ISL_1301653, EPI_ISL_1301654, EPI_ISL_1301655, EPI_ISL_1301656, EPI_ISL_1301657, EPI_ISL_1301658, EPI_ISL_1301659, EPI_ISL_1301660, EPI_ISL_1301661, EPI_ISL_1301662, EPI_ISL_1301663, EPI_ISL_1301664, EPI_ISL_1301665, EPI_ISL_1301666, EPI_ISL_1301667, EPI_ISL_1301668, EPI_ISL_1301669, EPI_ISL_1301670, EPI_ISL_1301671, EPI_ISL_1301672, EPI_ISL_1301673, EPI_ISL_1301674, EPI_ISL_1301675, EPI_ISL_1301676, EPI_ISL_1301677, EPI_ISL_1301678 | | | |
| see above | Laboratorio Central de Epidemiología IMSS | Instituto de Biotecnología de la UNAM | Authors from IBT, IMSS, InDRE and INER (in alphabetical order): Carlos F. Arias, Santiago Ávila-Ríos, Gisela Barrera-Badillo, Eduardo Becerril-Vargas, Celia Boukadida, Natividad Cruz-Ortiz, Larissa Fernandes-Matano, Ricardo Grande, Lucia Hernandez-Rivas, Alejandra Hernández-Terán, Pavel Isa, Irma Lopez-Martinez, José Arturo Martínez-Orozco, Margarita Matías-Florentino, Fidencio Mejía-Nepomuceno, Edgar Mendieta-Condado, Mario Mújica-Sánchez, José Esteban Muñoz-Medina, Tatiana Nunez-Garcia, Luis Alberto Ochoa-Carrera, Hector Esteban Paz-Juárez, Francisco Pulido, José Ernesto Ramírez-González, Alma Rincón-Rubio, Teresita Rojas-Mendoza, Jorge Salas-Hernández, Alejandro Sanchez-Flores, Clara Esperanza Santacruz-Tinoco, Andrea Santos Coy-Arechavaleta, Blanca Taboada, Gloria Vazquez, Joel Armando Vázquez-Pérez, Jerome Jean Verleyen |
| EPI_ISL_1301679, EPI_ISL_1301680, EPI_ISL_1301681, EPI_ISL_1301682, EPI_ISL_1301683, EPI_ISL_1301684, EPI_ISL_1301685, EPI_ISL_1301686, EPI_ISL_1301687, EPI_ISL_1301688, EPI_ISL_1301689, EPI_ISL_1301690, EPI_ISL_1301691, EPI_ISL_1301692, EPI_ISL_1301693, EPI_ISL_1301694, EPI_ISL_1301695, EPI_ISL_1301696, EPI_ISL_1301697, EPI_ISL_1301698, EPI_ISL_1301699, EPI_ISL_1301700, EPI_ISL_1301701, EPI_ISL_1301702, EPI_ISL_1301703, EPI_ISL_1301704, EPI_ISL_1301705, EPI_ISL_1301706, EPI_ISL_1301707, EPI_ISL_1301708, EPI_ISL_1301709, EPI_ISL_1301710, EPI_ISL_1301711, EPI_ISL_1301712, EPI_ISL_1301713 | | | |
| see above | Instituto de Diagnostico y Referencia Epidemiologicos InDRE_RNLSP | Instituto de Biotecnología de la UNAM | Authors from IBT, IMSS, InDRE and INER (in alphabetical order): Carlos F. Arias, Santiago Ávila-Ríos, Gisela Barrera-Badillo, Eduardo Becerril-Vargas, Celia Boukadida, Natividad Cruz-Ortiz, Larissa Fernandes-Matano, Ricardo Grande, Lucia Hernandez-Rivas, Alejandra Hernández-Terán, Pavel Isa, Irma Lopez-Martinez, José Arturo Martínez-Orozco, Margarita Matías-Florentino, Fidencio Mejía-Nepomuceno, Edgar Mendieta-Condado, Mario Mújica-Sánchez, José Esteban Muñoz-Medina, Tatiana Nunez-Garcia, Luis Alberto Ochoa-Carrera, Hector Esteban Paz-Juárez, Francisco Pulido, José Ernesto Ramírez-González, Alma Rincón-Rubio, Teresita Rojas-Mendoza, Jorge Salas-Hernández, Alejandro Sanchez-Flores, Clara Esperanza Santacruz-Tinoco, Andrea Santos Coy-Arechavaleta, Blanca Taboada, Gloria Vazquez, Joel Armando Vázquez-Pérez, Jerome Jean Verleyen |
| EPI_ISL_1301714, EPI_ISL_1301715, EPI_ISL_1301716 | Laboratorio Central de Epidemiología IMSS | Instituto de Biotecnología de la UNAM | Authors from IBT, IMSS, InDRE and INER (in alphabetical order): Carlos F. Arias, Santiago Ávila-Ríos, Gisela Barrera-Badillo, Eduardo Becerril-Vargas, Celia Boukadida, Natividad Cruz-Ortiz, Larissa Fernandes-Matano, Ricardo Grande, Lucia Hernandez-Rivas, Alejandra Hernández-Terán, Pavel Isa, Irma Lopez-Martinez, José Arturo Martínez-Orozco, Margarita Matías-Florentino, Fidencio Mejía-Nepomuceno, Edgar Mendieta-Condado, Mario Mújica-Sánchez, José Esteban Muñoz-Medina, Tatiana Nunez-Garcia, Luis Alberto Ochoa-Carrera, Hector Esteban Paz-Juárez, Francisco Pulido, José Ernesto Ramírez-González, Alma Rincón-Rubio, Teresita Rojas-Mendoza, Jorge Salas-Hernández, Alejandro Sanchez-Flores, Clara Esperanza Santacruz-Tinoco, Andrea Santos Coy-Arechavaleta, Blanca Taboada, Gloria Vazquez, Joel Armando Vázquez-Pérez, Jerome Jean Verleyen |
| EPI_ISL_1301717, EPI_ISL_1301718, EPI_ISL_1301719, EPI_ISL_1301720, EPI_ISL_1301721, EPI_ISL_1301722, EPI_ISL_1301723, EPI_ISL_1301724, EPI_ISL_1301725, EPI_ISL_1301726, EPI_ISL_1301727, EPI_ISL_1301728, EPI_ISL_1301729 | | | |
| see above | Instituto Nacional de Enfermedades Respiratorias (INER) | Instituto de Biotecnología de la UNAM | Authors from IBT, IMSS, InDRE and INER (in alphabetical order): Carlos F. Arias, Santiago Ávila-Ríos, Gisela Barrera-Badillo, Eduardo Becerril-Vargas, Celia Boukadida, Natividad Cruz-Ortiz, Larissa Fernandes-Matano, Ricardo Grande, Lucia Hernandez-Rivas, Alejandra Hernández-Terán, Pavel Isa, Irma Lopez-Martinez, José Arturo Martínez-Orozco, Margarita Matías-Florentino, Fidencio Mejía-Nepomuceno, Edgar Mendieta-Condado, Mario Mújica-Sánchez, José Esteban Muñoz-Medina, Tatiana Nunez-Garcia, Luis Alberto Ochoa-Carrera, Hector Esteban Paz-Juárez, Francisco Pulido, José Ernesto Ramírez-González, Alma Rincón-Rubio, Teresita Rojas-Mendoza, Jorge Salas-Hernández, Alejandro Sanchez-Flores, Clara Esperanza Santacruz-Tinoco, Andrea Santos Coy-Arechavaleta, Blanca Taboada, Gloria Vazquez, Joel Armando Vázquez-Pérez, Jerome Jean Verleyen |
| EPI_ISL_1301730 | Instituto de Diagnostico y Referencia Epidemiologicos InDRE_RNLSP | Instituto de Biotecnología de la UNAM | Authors from IBT, IMSS, InDRE and INER (in alphabetical order): Carlos F. Arias, Santiago Ávila-Ríos, Gisela Barrera-Badillo, Eduardo Becerril-Vargas, Celia Boukadida, Natividad Cruz-Ortiz, Larissa Fernandes-Matano, Ricardo Grande, Lucia Hernandez-Rivas, Alejandra Hernández-Terán, Pavel Isa, Irma Lopez-Martinez, José Arturo Martínez-Orozco, Margarita Matías-Florentino, Fidencio Mejía-Nepomuceno, Edgar Mendieta-Condado, Mario Mújica-Sánchez, José Esteban Muñoz-Medina, Tatiana Nunez-Garcia, Luis Alberto Ochoa-Carrera, Hector Esteban Paz-Juárez, Francisco Pulido, José Ernesto Ramírez-González, Alma Rincón-Rubio, Teresita Rojas-Mendoza, Jorge Salas-Hernández, Alejandro Sanchez-Flores, Clara Esperanza Santacruz-Tinoco, Andrea Santos Coy-Arechavaleta, Blanca Taboada, Gloria Vazquez, Joel Armando Vázquez-Pérez, Jerome Jean Verleyen |
| EPI_ISL_1301731, EPI_ISL_1301732, EPI_ISL_1301733, EPI_ISL_1302152 | Instituto Nacional de Enfermedades Respiratorias (INER) | Instituto de Biotecnología de la UNAM | Authors from IBT, IMSS, InDRE and INER (in alphabetical order): Carlos F. Arias, Santiago Ávila-Ríos, Gisela Barrera-Badillo, Eduardo Becerril-Vargas, Celia Boukadida, Natividad Cruz-Ortiz, Larissa Fernandes-Matano, Ricardo Grande, Lucia Hernandez-Rivas, Alejandra Hernández-Terán, Pavel Isa, Irma Lopez-Martinez, José Arturo Martínez-Orozco, Margarita Matías-Florentino, Fidencio Mejía-Nepomuceno, Edgar Mendieta-Condado, Mario Mújica-Sánchez, José Esteban Muñoz-Medina, Tatiana Nunez-Garcia, Luis Alberto Ochoa-Carrera, Hector Esteban Paz-Juárez, Francisco Pulido, José Ernesto Ramírez-González, Alma Rincón-Rubio, Teresita Rojas-Mendoza, Jorge Salas-Hernández, Alejandro Sanchez-Flores, Clara Esperanza Santacruz-Tinoco, Andrea Santos Coy-Arechavaleta, Blanca Taboada, Gloria Vazquez, Joel Armando Vázquez-Pérez, Jerome Jean Verleyen |
| EPI_ISL_1302153 | Laboratorio Central de Epidemiología IMSS | Instituto de Biotecnología de la UNAM | Authors from IBT, IMSS, InDRE and INER (in alphabetical order): Carlos F. Arias, Santiago Ávila-Ríos, Gisela Barrera-Badillo, Eduardo Becerril-Vargas, Celia Boukadida, Natividad Cruz-Ortiz, Larissa Fernandes-Matano, Ricardo Grande, Lucia Hernandez-Rivas, Alejandra Hernández-Terán, Pavel Isa, Irma Lopez-Martinez, José Arturo Martínez-Orozco, Margarita Matías-Florentino, Fidencio Mejía-Nepomuceno, Edgar Mendieta-Condado, Mario Mújica-Sánchez, José Esteban Muñoz-Medina, Tatiana Nunez-Garcia, Luis Alberto Ochoa-Carrera, Hector Esteban Paz-Juárez, Francisco Pulido, José Ernesto Ramírez-González, Alma Rincón-Rubio, Teresita Rojas-Mendoza, Jorge Salas-Hernández, Alejandro Sanchez-Flores, Clara Esperanza Santacruz-Tinoco, Andrea Santos Coy-Arechavaleta, Blanca Taboada, Gloria Vazquez, Joel Armando Vázquez-Pérez, Jerome Jean Verleyen |
| EPI_ISL_1302154 | Instituto de Diagnostico y Referencia Epidemiologicos InDRE_RNLSP | Instituto de Biotecnología de la UNAM | Authors from IBT, IMSS, InDRE and INER (in alphabetical order): Carlos F. Arias, Santiago Ávila-Ríos, Gisela Barrera-Badillo, Eduardo Becerril-Vargas, Celia Boukadida, Natividad Cruz-Ortiz, Larissa Fernandes-Matano, Ricardo Grande, Lucia Hernandez-Rivas, Alejandra Hernández-Terán, Pavel Isa, Irma Lopez-Martinez, José Arturo Martínez-Orozco, Margarita Matías-Florentino, Fidencio Mejía-Nepomuceno, Edgar Mendieta-Condado, Mario Mújica-Sánchez, José Esteban Muñoz-Medina, Tatiana Nunez-Garcia, Luis Alberto Ochoa-Carrera, Hector Esteban Paz-Juárez, Francisco Pulido, José Ernesto Ramírez-González, Alma Rincón-Rubio, Teresita Rojas-Mendoza, Jorge Salas-Hernández, Alejandro Sanchez-Flores, Clara Esperanza Santacruz-Tinoco, Andrea Santos Coy-Arechavaleta, Blanca Taboada, Gloria Vazquez, Joel Armando Vázquez-Pérez, Jerome Jean Verleyen |
| EPI_ISL_1302155 | Instituto Nacional de Enfermedades Respiratorias (INER) | Instituto de Biotecnología de la UNAM | Authors from IBT, IMSS, InDRE and INER (in alphabetical order): Carlos F. Arias, Santiago Ávila-Ríos, Gisela Barrera-Badillo, Eduardo Becerril-Vargas, Celia Boukadida, Natividad Cruz-Ortiz, Larissa Fernandes-Matano, Ricardo Grande, Lucia Hernandez-Rivas, Alejandra Hernández-Terán, Pavel Isa, Irma Lopez-Martinez, José Arturo Martínez-Orozco, Margarita Matías-Florentino, Fidencio Mejía-Nepomuceno, Edgar Mendieta-Condado, Mario Mújica-Sánchez, José Esteban Muñoz-Medina, Tatiana Nunez-Garcia, Luis Alberto Ochoa-Carrera, Hector Esteban Paz-Juárez, Francisco Pulido, José Ernesto Ramírez-González, Alma Rincón-Rubio, Teresita Rojas-Mendoza, Jorge Salas-Hernández, Alejandro Sanchez-Flores, Clara Esperanza Santacruz-Tinoco, Andrea Santos Coy-Arechavaleta, Blanca Taboada, Gloria Vazquez, Joel Armando Vázquez-Pérez, Jerome Jean Verleyen |
| EPI_ISL_1302156, EPI_ISL_1302157, EPI_ISL_1302158 | Laboratorio Central de Epidemiología IMSS | Instituto de Biotecnología de la UNAM | Authors from IBT, IMSS, InDRE and INER (in alphabetical order): Carlos F. Arias, Santiago Ávila-Ríos, Gisela Barrera-Badillo, Eduardo Becerril-Vargas, Celia Boukadida, Natividad Cruz-Ortiz, Larissa Fernandes-Matano, Ricardo Grande, Lucia Hernandez-Rivas, Alejandra Hernández-Terán, Pavel Isa, Irma Lopez-Martinez, José Arturo Martínez-Orozco, Margarita Matías-Florentino, Fidencio Mejía-Nepomuceno, Edgar Mendieta-Condado, Mario Mújica-Sánchez, José Esteban Muñoz-Medina, Tatiana Nunez-Garcia, Luis Alberto Ochoa-Carrera, Hector Esteban Paz-Juárez, Francisco Pulido, José Ernesto Ramírez-González, Alma Rincón-Rubio, Teresita Rojas-Mendoza, Jorge Salas-Hernández, Alejandro Sanchez-Flores, Clara Esperanza Santacruz-Tinoco, Andrea Santos Coy-Arechavaleta, Blanca Taboada, Gloria Vazquez, Joel Armando Vázquez-Pérez, Jerome Jean Verleyen |
| EPI_ISL_1302159, EPI_ISL_1302160 | Instituto Nacional de Enfermedades Respiratorias (INER) | Instituto de Biotecnología de la UNAM | Authors from IBT, IMSS, InDRE and INER (in alphabetical order): Carlos F. Arias, Santiago Ávila-Ríos, Gisela Barrera-Badillo, Eduardo Becerril-Vargas, Celia Boukadida, Natividad Cruz-Ortiz, Larissa Fernandes-Matano, Ricardo Grande, Lucia Hernandez-Rivas, Alejandra Hernández-Terán, Pavel Isa, Irma Lopez-Martinez, José Arturo Martínez-Orozco, Margarita Matías-Florentino, Fidencio Mejía-Nepomuceno, Edgar Mendieta-Condado, Mario Mújica-Sánchez, José Esteban Muñoz-Medina, Tatiana Nunez-Garcia, Luis Alberto Ochoa-Carrera, Hector Esteban Paz-Juárez, Francisco Pulido, José Ernesto Ramírez-González, Alma Rincón-Rubio, Teresita Rojas-Mendoza, Jorge Salas-Hernández, Alejandro Sanchez-Flores, Clara Esperanza Santacruz-Tinoco, Andrea Santos Coy-Arechavaleta, Blanca Taboada, Gloria Vazquez, Joel Armando Vázquez-Pérez, Jerome Jean Verleyen |
| EPI_ISL_1302161 | Laboratorio Central de Epidemiología IMSS | Instituto de Biotecnología de la UNAM | Authors from IBT, IMSS, InDRE and INER (in alphabetical order): Carlos F. Arias, Santiago Ávila-Ríos, Gisela Barrera-Badillo, Eduardo Becerril-Vargas, Celia Boukadida, Natividad Cruz-Ortiz, Larissa Fernandes-Matano, Ricardo Grande, Lucia Hernandez-Rivas, Alejandra Hernández-Terán, Pavel Isa, Irma Lopez-Martinez, José Arturo Martínez-Orozco, Margarita Matías-Florentino, Fidencio Mejía-Nepomuceno, Edgar Mendieta-Condado, Mario Mújica-Sánchez, José Esteban Muñoz-Medina, Tatiana Nunez-Garcia, Luis Alberto Ochoa-Carrera, Hector Esteban Paz-Juárez, Francisco Pulido, José Ernesto Ramírez-González, Alma Rincón-Rubio, Teresita Rojas-Mendoza, Jorge Salas-Hernández, Alejandro Sanchez-Flores, Clara Esperanza Santacruz-Tinoco, Andrea Santos Coy-Arechavaleta, Blanca Taboada, Gloria Vazquez, Joel Armando Vázquez-Pérez, Jerome Jean Verleyen |
| EPI_ISL_1302162 | Instituto de Diagnostico y Referencia Epidemiologicos InDRE_RNLSP | Instituto de Biotecnología de la UNAM | Authors from IBT, IMSS, InDRE and INER (in alphabetical order): Carlos F. Arias, Santiago Ávila-Ríos, Gisela Barrera-Badillo, Eduardo Becerril-Vargas, Celia Boukadida, Natividad Cruz-Ortiz, Larissa Fernandes-Matano, Ricardo Grande, Lucia Hernandez-Rivas, Alejandra Hernández-Terán, Pavel Isa, Irma Lopez-Martinez, José Arturo Martínez-Orozco, Margarita Matías-Florentino, Fidencio Mejía-Nepomuceno, Edgar Mendieta-Condado, Mario Mújica-Sánchez, José Esteban Muñoz-Medina, Tatiana Nunez-Garcia, Luis Alberto Ochoa-Carrera, Hector Esteban Paz-Juárez, Francisco Pulido, José Ernesto Ramírez-González, Alma Rincón-Rubio, Teresita Rojas-Mendoza, Jorge Salas-Hernández, Alejandro Sanchez-Flores, Clara Esperanza Santacruz-Tinoco, Andrea Santos Coy-Arechavaleta, Blanca Taboada, Gloria Vazquez, Joel Armando Vázquez-Pérez, Jerome Jean Verleyen |
| EPI_ISL_1302163 | Instituto Nacional de Enfermedades Respiratorias (INER) | Instituto de Biotecnología de la UNAM | Authors from IBT, IMSS, InDRE and INER (in alphabetical order): Carlos F. Arias, Santiago Ávila-Ríos, Gisela Barrera-Badillo, Eduardo Becerril-Vargas, Celia Boukadida, Natividad Cruz-Ortiz, Larissa Fernandes-Matano, Ricardo Grande, Lucia Hernandez-Rivas, Alejandra Hernández-Terán, Pavel Isa, Irma Lopez-Martinez, José Arturo Martínez-Orozco, Margarita Matías-Florentino, Fidencio Mejía-Nepomuceno, Edgar Mendieta-Condado, Mario Mújica-Sánchez, José Esteban Muñoz-Medina, Tatiana Nunez-Garcia, Luis Alberto Ochoa-Carrera, Hector Esteban Paz-Juárez, Francisco Pulido, José Ernesto Ramírez-González, Alma Rincón-Rubio, Teresita Rojas-Mendoza, Jorge Salas-Hernández, Alejandro Sanchez-Flores, Clara Esperanza Santacruz-Tinoco, Andrea Santos Coy-Arechavaleta, Blanca Taboada, Gloria Vazquez, Joel Armando Vázquez-Pérez, Jerome Jean Verleyen |
| EPI_ISL_1302165 | Laboratorio Central de Epidemiología IMSS | Instituto de Biotecnología de la UNAM | Authors from IBT, IMSS, InDRE and INER (in alphabetical order): Carlos F. Arias, Santiago Ávila-Ríos, Gisela Barrera-Badillo, Eduardo Becerril-Vargas, Celia Boukadida, Natividad Cruz-Ortiz, Larissa Fernandes-Matano, Ricardo Grande, Lucia Hernandez-Rivas, Alejandra Hernández-Terán, Pavel Isa, Irma Lopez-Martinez, José Arturo Martínez-Orozco, Margarita Matías-Florentino, Fidencio Mejía-Nepomuceno, Edgar Mendieta-Condado, Mario Mújica-Sánchez, José Esteban Muñoz-Medina, Tatiana Nunez-Garcia, Luis Alberto Ochoa-Carrera, Hector Esteban Paz-Juárez, Francisco Pulido, José Ernesto Ramírez-González, Alma Rincón-Rubio, Teresita Rojas-Mendoza, Jorge Salas-Hernández, Alejandro Sanchez-Flores, Clara Esperanza Santacruz-Tinoco, Andrea Santos Coy-Arechavaleta, Blanca Taboada, Gloria Vazquez, Joel Armando Vázquez-Pérez, Jerome Jean Verleyen |
| EPI_ISL_1302166, EPI_ISL_1302167, EPI_ISL_1302168 | Instituto de Diagnostico y Referencia Epidemiologicos InDRE_RNLSP | Instituto de Biotecnología de la UNAM | Authors from IBT, IMSS, InDRE and INER (in alphabetical order): Carlos F. Arias, Santiago Ávila-Ríos, Gisela Barrera-Badillo, Eduardo Becerril-Vargas, Celia Boukadida, Natividad Cruz-Ortiz, Larissa Fernandes-Matano, Ricardo Grande, Lucia Hernandez-Rivas, Alejandra Hernández-Terán, Pavel Isa, Irma Lopez-Martinez, José Arturo Martínez-Orozco, Margarita Matías-Florentino, Fidencio Mejía-Nepomuceno, Edgar Mendieta-Condado, Mario Mújica-Sánchez, José Esteban Muñoz-Medina, Tatiana Nunez-Garcia, Luis Alberto Ochoa-Carrera, Hector Esteban Paz-Juárez, Francisco Pulido, José Ernesto Ramírez-González, Alma Rincón-Rubio, Teresita Rojas-Mendoza, Jorge Salas-Hernández, Alejandro Sanchez-Flores, Clara Esperanza Santacruz-Tinoco, Andrea Santos Coy-Arechavaleta, Blanca Taboada, Gloria Vazquez, Joel Armando Vázquez-Pérez, Jerome Jean Verleyen |
| EPI_ISL_1302169 | Laboratorio Central de Epidemiología IMSS | Instituto de Biotecnología de la UNAM | Authors from IBT, IMSS, InDRE and INER (in alphabetical order): Carlos F. Arias, Santiago Ávila-Ríos, Gisela Barrera-Badillo, Eduardo Becerril-Vargas, Celia Boukadida, Natividad Cruz-Ortiz, Larissa Fernandes-Matano, Ricardo Grande, Lucia Hernandez-Rivas, Alejandra Hernández-Terán, Pavel Isa, Irma Lopez-Martinez, José Arturo Martínez-Orozco, Margarita Matías-Florentino, Fidencio Mejía-Nepomuceno, Edgar Mendieta-Condado, Mario Mújica-Sánchez, José Esteban Muñoz-Medina, Tatiana Nunez-Garcia, Luis Alberto Ochoa-Carrera, Hector Esteban Paz-Juárez, Francisco |

| | | | |
|---|---|---|---|
| | | | Pulido, José Ernesto Ramírez-González, Alma Rincón-Rubio, Teresita Rojas-Mendoza, Jorge Salas-Hernández, Alejandro Sanchez-Flores, Clara Esperanza Santacruz-Tinoco, Andrea Santos Coy-Arechavaleta, Blanca Taboada, Gloria Vazquez, Joel Armando Vázquez-Pérez, Jerome Jean Verleyen |
| EPI_ISL_1302170, EPI_ISL_1302171 | Instituto Nacional de Enfermedades Respiratorias (INER) | Instituto de Biotecnología de la UNAM | Authors from IBT, IMSS, InDRE and INER (in alphabetical order): Carlos F. Arias, Santiago Ávila-Ríos, Gisela Barrera-Badillo, Eduardo Becerril-Vargas, Celia Boukadida, Natividad Cruz-Ortiz, Larissa Fernandes-Matano, Ricardo Grande, Lucia Hernandez-Rivas, Alejandra Hernández-Terán, Pavel Isa, Irma Lopez-Martinez, José Arturo Martínez-Orozco, Margarita Matías-Florentino, Fidencio Mejía-Nepomuceno, Edgar Mendieta-Condado, Mario Mújica-Sánchez, José Esteban Muñoz-Medina, Tatiana Nunez-Garcia, Luis Alberto Ochoa-Carrera, Hector Esteban Paz-Juárez, Francisco Pulido, José Ernesto Ramírez-González, Alma Rincón-Rubio, Teresita Rojas-Mendoza, Jorge Salas-Hernández, Alejandro Sanchez-Flores, Clara Esperanza Santacruz-Tinoco, Andrea Santos Coy-Arechavaleta, Blanca Taboada, Gloria Vazquez, Joel Armando Vázquez-Pérez, Jerome Jean Verleyen |
| EPI_ISL_1302172, EPI_ISL_1302173 | Laboratorio Central de Epidemiología IMSS | Instituto de Biotecnología de la UNAM | Authors from IBT, IMSS, InDRE and INER (in alphabetical order): Carlos F. Arias, Santiago Ávila-Ríos, Gisela Barrera-Badillo, Eduardo Becerril-Vargas, Celia Boukadida, Natividad Cruz-Ortiz, Larissa Fernandes-Matano, Ricardo Grande, Lucia Hernandez-Rivas, Alejandra Hernández-Terán, Pavel Isa, Irma Lopez-Martinez, José Arturo Martínez-Orozco, Margarita Matías-Florentino, Fidencio Mejía-Nepomuceno, Edgar Mendieta-Condado, Mario Mújica-Sánchez, José Esteban Muñoz-Medina, Tatiana Nunez-Garcia, Luis Alberto Ochoa-Carrera, Hector Esteban Paz-Juárez, Francisco Pulido, José Ernesto Ramírez-González, Alma Rincón-Rubio, Teresita Rojas-Mendoza, Jorge Salas-Hernández, Alejandro Sanchez-Flores, Clara Esperanza Santacruz-Tinoco, Andrea Santos Coy-Arechavaleta, Blanca Taboada, Gloria Vazquez, Joel Armando Vázquez-Pérez, Jerome Jean Verleyen |
| EPI_ISL_1302174, EPI_ISL_1302175, EPI_ISL_1302176, EPI_ISL_1302177 | Instituto de Diagnostico y Referencia Epidemiologicos InDRE_RNLSP | Instituto de Biotecnología de la UNAM | Authors from IBT, IMSS, InDRE and INER (in alphabetical order): Carlos F. Arias, Santiago Ávila-Ríos, Gisela Barrera-Badillo, Eduardo Becerril-Vargas, Celia Boukadida, Natividad Cruz-Ortiz, Larissa Fernandes-Matano, Ricardo Grande, Lucia Hernandez-Rivas, Alejandra Hernández-Terán, Pavel Isa, Irma Lopez-Martinez, José Arturo Martínez-Orozco, Margarita Matías-Florentino, Fidencio Mejía-Nepomuceno, Edgar Mendieta-Condado, Mario Mújica-Sánchez, José Esteban Muñoz-Medina, Tatiana Nunez-Garcia, Luis Alberto Ochoa-Carrera, Hector Esteban Paz-Juárez, Francisco Pulido, José Ernesto Ramírez-González, Alma Rincón-Rubio, Teresita Rojas-Mendoza, Jorge Salas-Hernández, Alejandro Sanchez-Flores, Clara Esperanza Santacruz-Tinoco, Andrea Santos Coy-Arechavaleta, Blanca Taboada, Gloria Vazquez, Joel Armando Vázquez-Pérez, Jerome Jean Verleyen |
| EPI_ISL_1302178 | Laboratorio Central de Epidemiología IMSS | Instituto de Biotecnología de la UNAM | Authors from IBT, IMSS, InDRE and INER (in alphabetical order): Carlos F. Arias, Santiago Ávila-Ríos, Gisela Barrera-Badillo, Eduardo Becerril-Vargas, Celia Boukadida, Natividad Cruz-Ortiz, Larissa Fernandes-Matano, Ricardo Grande, Lucia Hernandez-Rivas, Alejandra Hernández-Terán, Pavel Isa, Irma Lopez-Martinez, José Arturo Martínez-Orozco, Margarita Matías-Florentino, Fidencio Mejía-Nepomuceno, Edgar Mendieta-Condado, Mario Mújica-Sánchez, José Esteban Muñoz-Medina, Tatiana Nunez-Garcia, Luis Alberto Ochoa-Carrera, Hector Esteban Paz-Juárez, Francisco Pulido, José Ernesto Ramírez-González, Alma Rincón-Rubio, Teresita Rojas-Mendoza, Jorge Salas-Hernández, Alejandro Sanchez-Flores, Clara Esperanza Santacruz-Tinoco, Andrea Santos Coy-Arechavaleta, Blanca Taboada, Gloria Vazquez, Joel Armando Vázquez-Pérez, Jerome Jean Verleyen |
| EPI_ISL_1302179, EPI_ISL_1302180 | Instituto Nacional de Enfermedades Respiratorias (INER) | Instituto de Biotecnología de la UNAM | Authors from IBT, IMSS, InDRE and INER (in alphabetical order): Carlos F. Arias, Santiago Ávila-Ríos, Gisela Barrera-Badillo, Eduardo Becerril-Vargas, Celia Boukadida, Natividad Cruz-Ortiz, Larissa Fernandes-Matano, Ricardo Grande, Lucia Hernandez-Rivas, Alejandra Hernández-Terán, Pavel Isa, Irma Lopez-Martinez, José Arturo Martínez-Orozco, Margarita Matías-Florentino, Fidencio Mejía-Nepomuceno, Edgar Mendieta-Condado, Mario Mújica-Sánchez, José Esteban Muñoz-Medina, Tatiana Nunez-Garcia, Luis Alberto Ochoa-Carrera, Hector Esteban Paz-Juárez, Francisco Pulido, José Ernesto Ramírez-González, Alma Rincón-Rubio, Teresita Rojas-Mendoza, Jorge Salas-Hernández, Alejandro Sanchez-Flores, Clara Esperanza Santacruz-Tinoco, Andrea Santos Coy-Arechavaleta, Blanca Taboada, Gloria Vazquez, Joel Armando Vázquez-Pérez, Jerome Jean Verleyen |
| EPI_ISL_1302181, EPI_ISL_1302182, EPI_ISL_1302183, EPI_ISL_1302184 | Instituto de Diagnostico y Referencia Epidemiologicos InDRE_RNLSP | Instituto de Biotecnología de la UNAM | Authors from IBT, IMSS, InDRE and INER (in alphabetical order): Carlos F. Arias, Santiago Ávila-Ríos, Gisela Barrera-Badillo, Eduardo Becerril-Vargas, Celia Boukadida, Natividad Cruz-Ortiz, Larissa Fernandes-Matano, Ricardo Grande, Lucia Hernandez-Rivas, Alejandra Hernández-Terán, Pavel Isa, Irma Lopez-Martinez, José Arturo Martínez-Orozco, Margarita Matías-Florentino, Fidencio Mejía-Nepomuceno, Edgar Mendieta-Condado, Mario Mújica-Sánchez, José Esteban Muñoz-Medina, Tatiana Nunez-Garcia, Luis Alberto Ochoa-Carrera, Hector Esteban Paz-Juárez, Francisco Pulido, José Ernesto Ramírez-González, Alma Rincón-Rubio, Teresita Rojas-Mendoza, Jorge Salas-Hernández, Alejandro Sanchez-Flores, Clara Esperanza Santacruz-Tinoco, Andrea Santos Coy-Arechavaleta, Blanca Taboada, Gloria Vazquez, Joel Armando Vázquez-Pérez, Jerome Jean Verleyen |
| EPI_ISL_1302185, EPI_ISL_1302186 | Instituto Nacional de Enfermedades Respiratorias (INER) | Instituto de Biotecnología de la UNAM | Authors from IBT, IMSS, InDRE and INER (in alphabetical order): Carlos F. Arias, Santiago Ávila-Ríos, Gisela Barrera-Badillo, Eduardo Becerril-Vargas, Celia Boukadida, Natividad Cruz-Ortiz, Larissa Fernandes-Matano, Ricardo Grande, Lucia Hernandez-Rivas, Alejandra Hernández-Terán, Pavel Isa, Irma Lopez-Martinez, José Arturo Martínez-Orozco, Margarita Matías-Florentino, Fidencio Mejía-Nepomuceno, Edgar Mendieta-Condado, Mario Mújica-Sánchez, José Esteban Muñoz-Medina, Tatiana Nunez-Garcia, Luis Alberto Ochoa-Carrera, Hector Esteban Paz-Juárez, Francisco Pulido, José Ernesto Ramírez-González, Alma Rincón-Rubio, Teresita Rojas-Mendoza, Jorge Salas-Hernández, Alejandro Sanchez-Flores, Clara Esperanza Santacruz-Tinoco, Andrea Santos Coy-Arechavaleta, Blanca Taboada, Gloria Vazquez, Joel Armando Vázquez-Pérez, Jerome Jean Verleyen |
| EPI_ISL_1302187, EPI_ISL_1302188, EPI_ISL_1302189 | Instituto de Diagnostico y Referencia Epidemiologicos InDRE_RNLSP | Instituto de Biotecnología de la UNAM | Authors from IBT, IMSS, InDRE and INER (in alphabetical order): Carlos F. Arias, Santiago Ávila-Ríos, Gisela Barrera-Badillo, Eduardo Becerril-Vargas, Celia Boukadida, Natividad Cruz-Ortiz, Larissa Fernandes-Matano, Ricardo Grande, Lucia Hernandez-Rivas, Alejandra Hernández-Terán, Pavel Isa, Irma Lopez-Martinez, José Arturo Martínez-Orozco, Margarita Matías-Florentino, Fidencio Mejía-Nepomuceno, Edgar Mendieta-Condado, Mario Mújica-Sánchez, José Esteban Muñoz-Medina, Tatiana Nunez-Garcia, Luis Alberto Ochoa-Carrera, Hector Esteban Paz-Juárez, Francisco Pulido, José Ernesto Ramírez-González, Alma Rincón-Rubio, Teresita Rojas-Mendoza, Jorge Salas-Hernández, Alejandro Sanchez-Flores, Clara Esperanza Santacruz-Tinoco, Andrea Santos Coy-Arechavaleta, Blanca Taboada, Gloria Vazquez, Joel Armando Vázquez-Pérez, Jerome Jean Verleyen |
| EPI_ISL_1302190 | Laboratorio Central de Epidemiología IMSS | Instituto de Biotecnología de la UNAM | Authors from IBT, IMSS, InDRE and INER (in alphabetical order): Carlos F. Arias, Santiago Ávila-Ríos, Gisela Barrera-Badillo, Eduardo Becerril-Vargas, Celia Boukadida, Natividad Cruz-Ortiz, Larissa Fernandes-Matano, Ricardo Grande, Lucia Hernandez-Rivas, Alejandra Hernández-Terán, Pavel Isa, Irma Lopez-Martinez, José Arturo Martínez-Orozco, Margarita Matías-Florentino, Fidencio Mejía-Nepomuceno, Edgar Mendieta-Condado, Mario Mújica-Sánchez, José Esteban Muñoz-Medina, Tatiana Nunez-Garcia, Luis Alberto Ochoa-Carrera, Hector Esteban Paz-Juárez, Francisco Pulido, José Ernesto Ramírez-González, Alma Rincón-Rubio, Teresita Rojas-Mendoza, Jorge Salas-Hernández, Alejandro Sanchez-Flores, Clara Esperanza Santacruz-Tinoco, Andrea Santos Coy-Arechavaleta, Blanca Taboada, Gloria Vazquez, Joel Armando Vázquez-Pérez, Jerome Jean Verleyen |
| EPI_ISL_1302191, EPI_ISL_1302192, EPI_ISL_1302193, EPI_ISL_1302194 | Instituto Nacional de Enfermedades Respiratorias (INER) | Instituto de Biotecnología de la UNAM | Authors from IBT, IMSS, InDRE and INER (in alphabetical order): Carlos F. Arias, Santiago Ávila-Ríos, Gisela Barrera-Badillo, Eduardo Becerril-Vargas, Celia Boukadida, Natividad Cruz-Ortiz, Larissa Fernandes-Matano, Ricardo Grande, Lucia Hernandez-Rivas, Alejandra Hernández-Terán, Pavel Isa, Irma Lopez-Martinez, José Arturo Martínez-Orozco, Margarita Matías-Florentino, Fidencio Mejía-Nepomuceno, Edgar Mendieta-Condado, Mario Mújica-Sánchez, José Esteban Muñoz-Medina, Tatiana Nunez-Garcia, Luis Alberto Ochoa-Carrera, Hector Esteban Paz-Juárez, Francisco Pulido, José Ernesto Ramírez-González, Alma Rincón-Rubio, Teresita Rojas-Mendoza, Jorge Salas-Hernández, Alejandro Sanchez-Flores, Clara Esperanza Santacruz-Tinoco, Andrea Santos Coy-Arechavaleta, Blanca Taboada, Gloria Vazquez, Joel Armando Vázquez-Pérez, Jerome Jean Verleyen |
| EPI_ISL_1302195, EPI_ISL_1302196 | Laboratorio Central de Epidemiología IMSS | Instituto de Biotecnología de la UNAM | Authors from IBT, IMSS, InDRE and INER (in alphabetical order): Carlos F. Arias, Santiago Ávila-Ríos, Gisela Barrera-Badillo, Eduardo Becerril-Vargas, Celia Boukadida, Natividad Cruz-Ortiz, Larissa Fernandes-Matano, Ricardo Grande, Lucia Hernandez-Rivas, Alejandra Hernández-Terán, Pavel Isa, Irma Lopez-Martinez, José Arturo Martínez-Orozco, Margarita Matías-Florentino, Fidencio Mejía-Nepomuceno, Edgar Mendieta-Condado, Mario Mújica-Sánchez, José Esteban Muñoz-Medina, Tatiana Nunez-Garcia, Luis Alberto Ochoa-Carrera, Hector Esteban Paz-Juárez, Francisco Pulido, José Ernesto Ramírez-González, Alma Rincón-Rubio, Teresita Rojas-Mendoza, Jorge Salas-Hernández, Alejandro Sanchez-Flores, Clara Esperanza Santacruz-Tinoco, Andrea Santos Coy-Arechavaleta, Blanca Taboada, Gloria Vazquez, Joel Armando Vázquez-Pérez, Jerome Jean Verleyen |
| EPI_ISL_1302197 | Instituto de Diagnostico y Referencia Epidemiologicos InDRE_RNLSP | Instituto de Biotecnología de la UNAM | Authors from IBT, IMSS, InDRE and INER (in alphabetical order): Carlos F. Arias, Santiago Ávila-Ríos, Gisela Barrera-Badillo, Eduardo Becerril-Vargas, Celia Boukadida, Natividad Cruz-Ortiz, Larissa Fernandes-Matano, Ricardo Grande, Lucia Hernandez-Rivas, Alejandra Hernández-Terán, Pavel Isa, Irma Lopez-Martinez, José Arturo Martínez-Orozco, Margarita Matías-Florentino, Fidencio Mejía-Nepomuceno, Edgar Mendieta-Condado, Mario Mújica-Sánchez, José Esteban Muñoz-Medina, Tatiana Nunez-Garcia, Luis Alberto Ochoa-Carrera, Hector Esteban Paz-Juárez, Francisco Pulido, José Ernesto Ramírez-González, Alma Rincón-Rubio, Teresita Rojas-Mendoza, Jorge Salas-Hernández, Alejandro Sanchez-Flores, Clara Esperanza Santacruz-Tinoco, Andrea Santos Coy-Arechavaleta, Blanca Taboada, Gloria Vazquez, Joel Armando Vázquez-Pérez, Jerome Jean Verleyen |
| EPI_ISL_1302198 | Instituto Nacional de Enfermedades Respiratorias (INER) | Instituto de Biotecnología de la UNAM | Authors from IBT, IMSS, InDRE and INER (in alphabetical order): Carlos F. Arias, Santiago Ávila-Ríos, Gisela Barrera-Badillo, Eduardo Becerril-Vargas, Celia Boukadida, Natividad Cruz-Ortiz, Larissa Fernandes-Matano, Ricardo Grande, Lucia Hernandez-Rivas, Alejandra Hernández-Terán, Pavel Isa, Irma Lopez-Martinez, José Arturo Martínez-Orozco, Margarita Matías-Florentino, Fidencio Mejía-Nepomuceno, Edgar Mendieta-Condado, Mario Mújica-Sánchez, José Esteban Muñoz-Medina, Tatiana Nunez-Garcia, Luis Alberto Ochoa-Carrera, Hector Esteban Paz-Juárez, Francisco Pulido, José Ernesto Ramírez-González, Alma Rincón-Rubio, Teresita Rojas-Mendoza, Jorge Salas-Hernández, Alejandro Sanchez-Flores, Clara Esperanza Santacruz-Tinoco, Andrea Santos Coy-Arechavaleta, Blanca Taboada, Gloria Vazquez, Joel Armando Vázquez-Pérez, Jerome Jean Verleyen |
| EPI_ISL_1302199, EPI_ISL_1302200, EPI_ISL_1302201, EPI_ISL_1302202 | Instituto de Diagnostico y Referencia Epidemiologicos InDRE_RNLSP | Instituto de Biotecnología de la UNAM | Authors from IBT, IMSS, InDRE and INER (in alphabetical order): Carlos F. Arias, Santiago Ávila-Ríos, Gisela Barrera-Badillo, Eduardo Becerril-Vargas, Celia Boukadida, Natividad Cruz-Ortiz, Larissa Fernandes-Matano, Ricardo Grande, Lucia Hernandez-Rivas, Alejandra Hernández-Terán, Pavel Isa, Irma Lopez-Martinez, José Arturo Martínez-Orozco, Margarita Matías-Florentino, Fidencio Mejía-Nepomuceno, Edgar Mendieta-Condado, Mario Mújica-Sánchez, José Esteban Muñoz-Medina, Tatiana Nunez-Garcia, Luis Alberto Ochoa-Carrera, Hector Esteban Paz-Juárez, Francisco Pulido, José Ernesto Ramírez-González, Alma Rincón-Rubio, Teresita Rojas-Mendoza, Jorge Salas-Hernández, Alejandro Sanchez-Flores, Clara Esperanza Santacruz-Tinoco, Andrea Santos Coy-Arechavaleta, Blanca Taboada, Gloria Vazquez, Joel Armando Vázquez-Pérez, Jerome Jean Verleyen |
| EPI_ISL_1302203, EPI_ISL_1302204 | Laboratorio Central de Epidemiología IMSS | Instituto de Biotecnología de la UNAM | Authors from IBT, IMSS, InDRE and INER (in alphabetical order): Carlos F. Arias, Santiago Ávila-Ríos, Gisela Barrera-Badillo, Eduardo Becerril-Vargas, Celia Boukadida, Natividad Cruz-Ortiz, Larissa Fernandes-Matano, Ricardo Grande, Lucia Hernandez-Rivas, Alejandra Hernández-Terán, Pavel Isa, Irma Lopez-Martinez, José Arturo Martínez-Orozco, Margarita Matías-Florentino, Fidencio Mejía-Nepomuceno, Edgar Mendieta-Condado, Mario Mújica-Sánchez, José Esteban Muñoz-Medina, Tatiana Nunez-Garcia, Luis Alberto Ochoa-Carrera, Hector Esteban Paz-Juárez, Francisco Pulido, José Ernesto Ramírez-González, Alma Rincón-Rubio, Teresita Rojas-Mendoza, Jorge Salas-Hernández, Alejandro Sanchez-Flores, Clara Esperanza Santacruz-Tinoco, Andrea Santos Coy-Arechavaleta, Blanca Taboada, Gloria Vazquez, Joel Armando Vázquez-Pérez, Jerome Jean Verleyen |
| EPI_ISL_1302205, EPI_ISL_1302206, EPI_ISL_1302207, EPI_ISL_1302208 | Instituto Nacional de Enfermedades Respiratorias (INER) | Instituto de Biotecnología de la UNAM | Authors from IBT, IMSS, InDRE and INER (in alphabetical order): Carlos F. Arias, Santiago Ávila-Ríos, Gisela Barrera-Badillo, Eduardo Becerril-Vargas, Celia Boukadida, Natividad Cruz-Ortiz, Larissa Fernandes-Matano, Ricardo Grande, Lucia Hernandez-Rivas, Alejandra Hernández-Terán, Pavel Isa, Irma Lopez-Martinez, José Arturo Martínez-Orozco, Margarita Matías-Florentino, Fidencio Mejía-Nepomuceno, Edgar Mendieta-Condado, Mario Mújica-Sánchez, José Esteban Muñoz-Medina, Tatiana Nunez-Garcia, Luis Alberto Ochoa-Carrera, Hector Esteban Paz-Juárez, Francisco Pulido, José Ernesto Ramírez-González, Alma Rincón-Rubio, Teresita Rojas-Mendoza, Jorge Salas-Hernández, Alejandro Sanchez-Flores, Clara Esperanza Santacruz-Tinoco, Andrea Santos Coy-Arechavaleta, Blanca Taboada, Gloria Vazquez, Joel Armando Vázquez-Pérez, Jerome Jean Verleyen |
| EPI_ISL_1302209, EPI_ISL_1302210, EPI_ISL_1302211, EPI_ISL_1302212, EPI_ISL_1302213, EPI_ISL_1302214, EPI_ISL_1302215, EPI_ISL_1302216, EPI_ISL_1302217, EPI_ISL_1302218, EPI_ISL_1302219 | | | |
| see above | Instituto de Diagnostico y Referencia Epidemiologicos InDRE_RNLSP | Instituto de Biotecnología de la UNAM | Authors from IBT, IMSS, InDRE and INER (in alphabetical order): Carlos F. Arias, Santiago Ávila-Ríos, Gisela Barrera-Badillo, Eduardo Becerril-Vargas, Celia Boukadida, Natividad Cruz-Ortiz, Larissa Fernandes-Matano, Ricardo Grande, Lucia Hernandez-Rivas, Alejandra Hernández-Terán, Pavel Isa, Irma Lopez-Martinez, José Arturo Martínez-Orozco, Margarita Matías-Florentino, Fidencio Mejía-Nepomuceno, Edgar Mendieta-Condado, Mario Mújica-Sánchez, José Esteban Muñoz-Medina, Tatiana Nunez-Garcia, Luis Alberto Ochoa-Carrera, Hector Esteban Paz-Juárez, Francisco Pulido, José Ernesto Ramírez-González, Alma Rincón-Rubio, Teresita Rojas-Mendoza, Jorge Salas-Hernández, Alejandro Sanchez-Flores, Clara Esperanza Santacruz-Tinoco, Andrea Santos Coy-Arechavaleta, Blanca Taboada, Gloria Vazquez, Joel Armando Vázquez-Pérez, Jerome Jean Verleyen |
| EPI_ISL_1302220, EPI_ISL_1302221, EPI_ISL_1302222, EPI_ISL_1302223, EPI_ISL_1302224, EPI_ISL_1302225, EPI_ISL_1302226 | Laboratorio Central de Epidemiología IMSS | Instituto de Biotecnología de la UNAM | Authors from IBT, IMSS, InDRE and INER (in alphabetical order): Carlos F. Arias, Santiago Ávila-Ríos, Gisela Barrera-Badillo, Eduardo Becerril-Vargas, Celia Boukadida, Natividad Cruz-Ortiz, Larissa Fernandes-Matano, Ricardo Grande, Lucia Hernandez-Rivas, Alejandra Hernández-Terán, Pavel Isa, Irma Lopez-Martinez, José Arturo Martínez-Orozco, Margarita Matías-Florentino, Fidencio Mejía-Nepomuceno, Edgar Mendieta-Condado, Mario Mújica-Sánchez, José Esteban Muñoz-Medina, Tatiana Nunez-Garcia, Luis Alberto Ochoa-Carrera, Hector Esteban Paz-Juárez, Francisco Pulido, José Ernesto Ramírez-González, Alma Rincón-Rubio, Teresita Rojas-Mendoza, Jorge Salas-Hernández, Alejandro Sanchez-Flores, Clara Esperanza Santacruz-Tinoco, Andrea Santos Coy-Arechavaleta, Blanca Taboada, Gloria Vazquez, Joel Armando Vázquez-Pérez, Jerome Jean Verleyen |
| EPI_ISL_1302227, EPI_ISL_1302228, EPI_ISL_1302229, EPI_ISL_1302230, EPI_ISL_1302231, EPI_ISL_1302232, EPI_ISL_1302233, EPI_ISL_1302234 | Instituto Nacional de Enfermedades Respiratorias (INER) | Instituto de Biotecnología de la UNAM | Authors from IBT, IMSS, InDRE and INER (in alphabetical order): Carlos F. Arias, Santiago Ávila-Ríos, Gisela Barrera-Badillo, Eduardo Becerril-Vargas, Celia Boukadida, Natividad Cruz-Ortiz, Larissa Fernandes-Matano, Ricardo Grande, Lucia Hernandez-Rivas, Alejandra Hernández-Terán, Pavel Isa, Irma Lopez-Martinez, José Arturo Martínez-Orozco, Margarita Matías-Florentino, Fidencio Mejía-Nepomuceno, Edgar Mendieta-Condado, Mario Mújica-Sánchez, José Esteban Muñoz-Medina, Tatiana Nunez-Garcia, Luis Alberto Ochoa-Carrera, Hector Esteban Paz-Juárez, Francisco Pulido, José Ernesto Ramírez-González, Alma Rincón-Rubio, Teresita Rojas-Mendoza, Jorge Salas-Hernández, Alejandro Sanchez-Flores, Clara Esperanza Santacruz-Tinoco, Andrea Santos Coy-Arechavaleta, Blanca Taboada, Gloria Vazquez, Joel Armando Vázquez-Pérez, Jerome Jean Verleyen |
| EPI_ISL_1302235, EPI_ISL_1302236, EPI_ISL_1302237, EPI_ISL_1302238, EPI_ISL_1302239, EPI_ISL_1302240 | Laboratorio Central de Epidemiología IMSS | Instituto de Biotecnología de la UNAM | Authors from IBT, IMSS, InDRE and INER (in alphabetical order): Carlos F. Arias, Santiago Ávila-Ríos, Gisela Barrera-Badillo, Eduardo Becerril-Vargas, Celia Boukadida, Natividad Cruz-Ortiz, Larissa Fernandes-Matano, Ricardo Grande, Lucia Hernandez-Rivas, Alejandra Hernández-Terán, Pavel Isa, Irma Lopez-Martinez, José Arturo Martínez-Orozco, Margarita Matías-Florentino, Fidencio Mejía-Nepomuceno, Edgar Mendieta-Condado, Mario Mújica-Sánchez, José Esteban Muñoz-Medina, Tatiana Nunez-Garcia, Luis Alberto Ochoa-Carrera, Hector Esteban Paz-Juárez, Francisco Pulido, José Ernesto Ramírez-González, Alma Rincón-Rubio, Teresita Rojas-Mendoza, Jorge Salas-Hernández, Alejandro Sanchez-Flores, Clara Esperanza Santacruz-Tinoco, Andrea Santos Coy-Arechavaleta, Blanca Taboada, Gloria Vazquez, Joel Armando Vázquez-Pérez, Jerome Jean Verleyen |
| EPI_ISL_1302241, EPI_ISL_1302242 | Instituto de Diagnostico y Referencia Epidemiologicos InDRE_RNLSP | Instituto de Biotecnología de la UNAM | Authors from IBT, IMSS, InDRE and INER (in alphabetical order): Carlos F. Arias, Santiago Ávila-Ríos, Gisela Barrera-Badillo, Eduardo Becerril-Vargas, Celia Boukadida, Natividad Cruz-Ortiz, Larissa Fernandes-Matano, Ricardo Grande, Lucia Hernandez-Rivas, Alejandra Hernández-Terán, Pavel Isa, Irma Lopez-Martinez, José Arturo Martínez-Orozco, Margarita Matías-Florentino, Fidencio Mejía-Nepomuceno, Edgar Mendieta-Condado, Mario Mújica-Sánchez, José Esteban Muñoz-Medina, Tatiana Nunez-Garcia, Luis Alberto Ochoa-Carrera, Hector Esteban Paz-Juárez, Francisco Pulido, José Ernesto Ramírez-González, Alma Rincón-Rubio, Teresita Rojas-Mendoza, Jorge Salas-Hernández, Alejandro Sanchez-Flores, Clara Esperanza Santacruz-Tinoco, Andrea Santos Coy-Arechavaleta, Blanca Taboada, Gloria Vazquez, Joel Armando Vázquez-Pérez, Jerome Jean Verleyen |
| EPI_ISL_1302243, EPI_ISL_1302244 | Laboratorio Central de Epidemiología IMSS | Instituto de Biotecnología de la UNAM | Authors from IBT, IMSS, InDRE and INER (in alphabetical order): Carlos F. Arias, Santiago Ávila-Ríos, Gisela Barrera-Badillo, Eduardo Becerril-Vargas, Celia Boukadida, Natividad Cruz-Ortiz, Larissa Fernandes-Matano, Ricardo Grande, Lucia Hernandez-Rivas, Alejandra Hernández-Terán, Pavel Isa, Irma Lopez-Martinez, José Arturo Martínez-Orozco, Margarita Matías-Florentino, Fidencio Mejía-Nepomuceno, Edgar Mendieta-Condado, Mario Mújica-Sánchez, José Esteban Muñoz-Medina, Tatiana Nunez-Garcia, Luis Alberto Ochoa-Carrera, Hector Esteban Paz-Juárez, Francisco Pulido, José Ernesto Ramírez-González, Alma Rincón-Rubio, Teresita Rojas-Mendoza, Jorge Salas-Hernández, Alejandro Sanchez-Flores, Clara Esperanza Santacruz-Tinoco, Andrea Santos Coy-Arechavaleta, Blanca Taboada, Gloria Vazquez, Joel Armando Vázquez-Pérez, Jerome Jean Verleyen |

| | | | |
|---|---|---|---|
| EPI_ISL_1302245, EPI_ISL_1302246, EPI_ISL_1302247 | Instituto Nacional de Enfermedades Respiratorias (INER) | Instituto de Biotecnología de la UNAM | Authors from IBT, IMSS, InDRE and INER (in alphabetical order): Carlos F. Arias, Santiago Ávila-Ríos, Gisela Barrera-Badillo, Eduardo Becerril-Vargas, Celia Boukadida, Natividad Cruz-Ortiz, Larissa Fernandes-Matano, Ricardo Grande, Lucia Hernandez-Rivas, Alejandra Hernández-Terán, Pavel Isa, Irma Lopez-Martinez, José Arturo Martínez-Orozco, Margarita Matías-Florentino, Fidencio Mejía-Nepomuceno, Edgar Mendieta-Condado, Mario Mújica-Sánchez, José Esteban Muñoz-Medina, Tatiana Nunez-Garcia, Luis Alberto Ochoa-Carrera, Hector Esteban Paz-Juárez, Francisco Pulido, José Ernesto Ramírez-González, Alma Rincón-Rubio, Teresita Rojas-Mendoza, Jorge Salas-Hernández, Alejandro Sanchez-Flores, Clara Esperanza Santacruz-Tinoco, Andrea Santos Coy-Arechavaleta, Blanca Taboada, Gloria Vazquez, Joel Armando Vázquez-Pérez, Jerome Jean Verleyen |
| EPI_ISL_1302248 | Laboratorio Central de Epidemiología IMSS | Instituto de Biotecnología de la UNAM | Authors from IBT, IMSS, InDRE and INER (in alphabetical order): Carlos F. Arias, Santiago Ávila-Ríos, Gisela Barrera-Badillo, Eduardo Becerril-Vargas, Celia Boukadida, Natividad Cruz-Ortiz, Larissa Fernandes-Matano, Ricardo Grande, Lucia Hernandez-Rivas, Alejandra Hernández-Terán, Pavel Isa, Irma Lopez-Martinez, José Arturo Martínez-Orozco, Margarita Matías-Florentino, Fidencio Mejía-Nepomuceno, Edgar Mendieta-Condado, Mario Mújica-Sánchez, José Esteban Muñoz-Medina, Tatiana Nunez-Garcia, Luis Alberto Ochoa-Carrera, Hector Esteban Paz-Juárez, Francisco Pulido, José Ernesto Ramírez-González, Alma Rincón-Rubio, Teresita Rojas-Mendoza, Jorge Salas-Hernández, Alejandro Sanchez-Flores, Clara Esperanza Santacruz-Tinoco, Andrea Santos Coy-Arechavaleta, Blanca Taboada, Gloria Vazquez, Joel Armando Vázquez-Pérez, Jerome Jean Verleyen |
| EPI_ISL_1302249, EPI_ISL_1302250 | Instituto Nacional de Enfermedades Respiratorias (INER) | Instituto de Biotecnología de la UNAM | Authors from IBT, IMSS, InDRE and INER (in alphabetical order): Carlos F. Arias, Santiago Ávila-Ríos, Gisela Barrera-Badillo, Eduardo Becerril-Vargas, Celia Boukadida, Natividad Cruz-Ortiz, Larissa Fernandes-Matano, Ricardo Grande, Lucia Hernandez-Rivas, Alejandra Hernández-Terán, Pavel Isa, Irma Lopez-Martinez, José Arturo Martínez-Orozco, Margarita Matías-Florentino, Fidencio Mejía-Nepomuceno, Edgar Mendieta-Condado, Mario Mújica-Sánchez, José Esteban Muñoz-Medina, Tatiana Nunez-Garcia, Luis Alberto Ochoa-Carrera, Hector Esteban Paz-Juárez, Francisco Pulido, José Ernesto Ramírez-González, Alma Rincón-Rubio, Teresita Rojas-Mendoza, Jorge Salas-Hernández, Alejandro Sanchez-Flores, Clara Esperanza Santacruz-Tinoco, Andrea Santos Coy-Arechavaleta, Blanca Taboada, Gloria Vazquez, Joel Armando Vázquez-Pérez, Jerome Jean Verleyen |
| EPI_ISL_1302251, EPI_ISL_1302252 | Laboratorio Central de Epidemiología IMSS | Instituto de Biotecnología de la UNAM | Authors from IBT, IMSS, InDRE and INER (in alphabetical order): Carlos F. Arias, Santiago Ávila-Ríos, Gisela Barrera-Badillo, Eduardo Becerril-Vargas, Celia Boukadida, Natividad Cruz-Ortiz, Larissa Fernandes-Matano, Ricardo Grande, Lucia Hernandez-Rivas, Alejandra Hernández-Terán, Pavel Isa, Irma Lopez-Martinez, José Arturo Martínez-Orozco, Margarita Matías-Florentino, Fidencio Mejía-Nepomuceno, Edgar Mendieta-Condado, Mario Mújica-Sánchez, José Esteban Muñoz-Medina, Tatiana Nunez-Garcia, Luis Alberto Ochoa-Carrera, Hector Esteban Paz-Juárez, Francisco Pulido, José Ernesto Ramírez-González, Alma Rincón-Rubio, Teresita Rojas-Mendoza, Jorge Salas-Hernández, Alejandro Sanchez-Flores, Clara Esperanza Santacruz-Tinoco, Andrea Santos Coy-Arechavaleta, Blanca Taboada, Gloria Vazquez, Joel Armando Vázquez-Pérez, Jerome Jean Verleyen |
| EPI_ISL_1302253 | Instituto Nacional de Enfermedades Respiratorias (INER) | Instituto de Biotecnología de la UNAM | Authors from IBT, IMSS, InDRE and INER (in alphabetical order): Carlos F. Arias, Santiago Ávila-Ríos, Gisela Barrera-Badillo, Eduardo Becerril-Vargas, Celia Boukadida, Natividad Cruz-Ortiz, Larissa Fernandes-Matano, Ricardo Grande, Lucia Hernandez-Rivas, Alejandra Hernández-Terán, Pavel Isa, Irma Lopez-Martinez, José Arturo Martínez-Orozco, Margarita Matías-Florentino, Fidencio Mejía-Nepomuceno, Edgar Mendieta-Condado, Mario Mújica-Sánchez, José Esteban Muñoz-Medina, Tatiana Nunez-Garcia, Luis Alberto Ochoa-Carrera, Hector Esteban Paz-Juárez, Francisco Pulido, José Ernesto Ramírez-González, Alma Rincón-Rubio, Teresita Rojas-Mendoza, Jorge Salas-Hernández, Alejandro Sanchez-Flores, Clara Esperanza Santacruz-Tinoco, Andrea Santos Coy-Arechavaleta, Blanca Taboada, Gloria Vazquez, Joel Armando Vázquez-Pérez, Jerome Jean Verleyen |
| EPI_ISL_1302254 | Instituto de Diagnostico y Referencia Epidemiologicos InDRE_RNLSP | Instituto de Biotecnología de la UNAM | Authors from IBT, IMSS, InDRE and INER (in alphabetical order): Carlos F. Arias, Santiago Ávila-Ríos, Gisela Barrera-Badillo, Eduardo Becerril-Vargas, Celia Boukadida, Natividad Cruz-Ortiz, Larissa Fernandes-Matano, Ricardo Grande, Lucia Hernandez-Rivas, Alejandra Hernández-Terán, Pavel Isa, Irma Lopez-Martinez, José Arturo Martínez-Orozco, Margarita Matías-Florentino, Fidencio Mejía-Nepomuceno, Edgar Mendieta-Condado, Mario Mújica-Sánchez, José Esteban Muñoz-Medina, Tatiana Nunez-Garcia, Luis Alberto Ochoa-Carrera, Hector Esteban Paz-Juárez, Francisco Pulido, José Ernesto Ramírez-González, Alma Rincón-Rubio, Teresita Rojas-Mendoza, Jorge Salas-Hernández, Alejandro Sanchez-Flores, Clara Esperanza Santacruz-Tinoco, Andrea Santos Coy-Arechavaleta, Blanca Taboada, Gloria Vazquez, Joel Armando Vázquez-Pérez, Jerome Jean Verleyen |
| EPI_ISL_1302255, EPI_ISL_1302256, EPI_ISL_1302257, EPI_ISL_1302258 | Instituto Nacional de Enfermedades Respiratorias (INER) | Instituto de Biotecnología de la UNAM | Authors from IBT, IMSS, InDRE and INER (in alphabetical order): Carlos F. Arias, Santiago Ávila-Ríos, Gisela Barrera-Badillo, Eduardo Becerril-Vargas, Celia Boukadida, Natividad Cruz-Ortiz, Larissa Fernandes-Matano, Ricardo Grande, Lucia Hernandez-Rivas, Alejandra Hernández-Terán, Pavel Isa, Irma Lopez-Martinez, José Arturo Martínez-Orozco, Margarita Matías-Florentino, Fidencio Mejía-Nepomuceno, Edgar Mendieta-Condado, Mario Mújica-Sánchez, José Esteban Muñoz-Medina, Tatiana Nunez-Garcia, Luis Alberto Ochoa-Carrera, Hector Esteban Paz-Juárez, Francisco Pulido, José Ernesto Ramírez-González, Alma Rincón-Rubio, Teresita Rojas-Mendoza, Jorge Salas-Hernández, Alejandro Sanchez-Flores, Clara Esperanza Santacruz-Tinoco, Andrea Santos Coy-Arechavaleta, Blanca Taboada, Gloria Vazquez, Joel Armando Vázquez-Pérez, Jerome Jean Verleyen |
| EPI_ISL_1302259 | Instituto de Diagnostico y Referencia Epidemiologicos InDRE_RNLSP | Instituto de Biotecnología de la UNAM | Authors from IBT, IMSS, InDRE and INER (in alphabetical order): Carlos F. Arias, Santiago Ávila-Ríos, Gisela Barrera-Badillo, Eduardo Becerril-Vargas, Celia Boukadida, Natividad Cruz-Ortiz, Larissa Fernandes-Matano, Ricardo Grande, Lucia Hernandez-Rivas, Alejandra Hernández-Terán, Pavel Isa, Irma Lopez-Martinez, José Arturo Martínez-Orozco, Margarita Matías-Florentino, Fidencio Mejía-Nepomuceno, Edgar Mendieta-Condado, Mario Mújica-Sánchez, José Esteban Muñoz-Medina, Tatiana Nunez-Garcia, Luis Alberto Ochoa-Carrera, Hector Esteban Paz-Juárez, Francisco Pulido, José Ernesto Ramírez-González, Alma Rincón-Rubio, Teresita Rojas-Mendoza, Jorge Salas-Hernández, Alejandro Sanchez-Flores, Clara Esperanza Santacruz-Tinoco, Andrea Santos Coy-Arechavaleta, Blanca Taboada, Gloria Vazquez, Joel Armando Vázquez-Pérez, Jerome Jean Verleyen |
| EPI_ISL_1302260, EPI_ISL_1302261 | Laboratorio Central de Epidemiología IMSS | Instituto de Biotecnología de la UNAM | Authors from IBT, IMSS, InDRE and INER (in alphabetical order): Carlos F. Arias, Santiago Ávila-Ríos, Gisela Barrera-Badillo, Eduardo Becerril-Vargas, Celia Boukadida, Natividad Cruz-Ortiz, Larissa Fernandes-Matano, Ricardo Grande, Lucia Hernandez-Rivas, Alejandra Hernández-Terán, Pavel Isa, Irma Lopez-Martinez, José Arturo Martínez-Orozco, Margarita Matías-Florentino, Fidencio Mejía-Nepomuceno, Edgar Mendieta-Condado, Mario Mújica-Sánchez, José Esteban Muñoz-Medina, Tatiana Nunez-Garcia, Luis Alberto Ochoa-Carrera, Hector Esteban Paz-Juárez, Francisco Pulido, José Ernesto Ramírez-González, Alma Rincón-Rubio, Teresita Rojas-Mendoza, Jorge Salas-Hernández, Alejandro Sanchez-Flores, Clara Esperanza Santacruz-Tinoco, Andrea Santos Coy-Arechavaleta, Blanca Taboada, Gloria Vazquez, Joel Armando Vázquez-Pérez, Jerome Jean Verleyen |
| EPI_ISL_1302262 | Instituto de Diagnostico y Referencia Epidemiologicos InDRE_RNLSP | Instituto de Biotecnología de la UNAM | Authors from IBT, IMSS, InDRE and INER (in alphabetical order): Carlos F. Arias, Santiago Ávila-Ríos, Gisela Barrera-Badillo, Eduardo Becerril-Vargas, Celia Boukadida, Natividad Cruz-Ortiz, Larissa Fernandes-Matano, Ricardo Grande, Lucia Hernandez-Rivas, Alejandra Hernández-Terán, Pavel Isa, Irma Lopez-Martinez, José Arturo Martínez-Orozco, Margarita Matías-Florentino, Fidencio Mejía-Nepomuceno, Edgar Mendieta-Condado, Mario Mújica-Sánchez, José Esteban Muñoz-Medina, Tatiana Nunez-Garcia, Luis Alberto Ochoa-Carrera, Hector Esteban Paz-Juárez, Francisco Pulido, José Ernesto Ramírez-González, Alma Rincón-Rubio, Teresita Rojas-Mendoza, Jorge Salas-Hernández, Alejandro Sanchez-Flores, Clara Esperanza Santacruz-Tinoco, Andrea Santos Coy-Arechavaleta, Blanca Taboada, Gloria Vazquez, Joel Armando Vázquez-Pérez, Jerome Jean Verleyen |
| EPI_ISL_1302263 | Instituto Nacional de Enfermedades Respiratorias (INER) | Instituto de Biotecnología de la UNAM | Authors from IBT, IMSS, InDRE and INER (in alphabetical order): Carlos F. Arias, Santiago Ávila-Ríos, Gisela Barrera-Badillo, Eduardo Becerril-Vargas, Celia Boukadida, Natividad Cruz-Ortiz, Larissa Fernandes-Matano, Ricardo Grande, Lucia Hernandez-Rivas, Alejandra Hernández-Terán, Pavel Isa, Irma Lopez-Martinez, José Arturo Martínez-Orozco, Margarita Matías-Florentino, Fidencio Mejía-Nepomuceno, Edgar Mendieta-Condado, Mario Mújica-Sánchez, José Esteban Muñoz-Medina, Tatiana Nunez-Garcia, Luis Alberto Ochoa-Carrera, Hector Esteban Paz-Juárez, Francisco Pulido, José Ernesto Ramírez-González, Alma Rincón-Rubio, Teresita Rojas-Mendoza, Jorge Salas-Hernández, Alejandro Sanchez-Flores, Clara Esperanza Santacruz-Tinoco, Andrea Santos Coy-Arechavaleta, Blanca Taboada, Gloria Vazquez, Joel Armando Vázquez-Pérez, Jerome Jean Verleyen |
| EPI_ISL_1302264 | Laboratorio Central de Epidemiología IMSS | Instituto de Biotecnología de la UNAM | Authors from IBT, IMSS, InDRE and INER (in alphabetical order): Carlos F. Arias, Santiago Ávila-Ríos, Gisela Barrera-Badillo, Eduardo Becerril-Vargas, Celia Boukadida, Natividad Cruz-Ortiz, Larissa Fernandes-Matano, Ricardo Grande, Lucia Hernandez-Rivas, Alejandra Hernández-Terán, Pavel Isa, Irma Lopez-Martinez, José Arturo Martínez-Orozco, Margarita Matías-Florentino, Fidencio Mejía-Nepomuceno, Edgar Mendieta-Condado, Mario Mújica-Sánchez, José Esteban Muñoz-Medina, Tatiana Nunez-Garcia, Luis Alberto Ochoa-Carrera, Hector Esteban Paz-Juárez, Francisco Pulido, José Ernesto Ramírez-González, Alma Rincón-Rubio, Teresita Rojas-Mendoza, Jorge Salas-Hernández, Alejandro Sanchez-Flores, Clara Esperanza Santacruz-Tinoco, Andrea Santos Coy-Arechavaleta, Blanca Taboada, Gloria Vazquez, Joel Armando Vázquez-Pérez, Jerome Jean Verleyen |
| EPI_ISL_1302265 | Instituto de Diagnostico y Referencia Epidemiologicos InDRE_RNLSP | Instituto de Biotecnología de la UNAM | Authors from IBT, IMSS, InDRE and INER (in alphabetical order): Carlos F. Arias, Santiago Ávila-Ríos, Gisela Barrera-Badillo, Eduardo Becerril-Vargas, Celia Boukadida, Natividad Cruz-Ortiz, Larissa Fernandes-Matano, Ricardo Grande, Lucia Hernandez-Rivas, Alejandra Hernández-Terán, Pavel Isa, Irma Lopez-Martinez, José Arturo Martínez-Orozco, Margarita Matías-Florentino, Fidencio Mejía-Nepomuceno, Edgar Mendieta-Condado, Mario Mújica-Sánchez, José Esteban Muñoz-Medina, Tatiana Nunez-Garcia, Luis Alberto Ochoa-Carrera, Hector Esteban Paz-Juárez, Francisco Pulido, José Ernesto Ramírez-González, Alma Rincón-Rubio, Teresita Rojas-Mendoza, Jorge Salas-Hernández, Alejandro Sanchez-Flores, Clara Esperanza Santacruz-Tinoco, Andrea Santos Coy-Arechavaleta, Blanca Taboada, Gloria Vazquez, Joel Armando Vázquez-Pérez, Jerome Jean Verleyen |
| EPI_ISL_1302266, EPI_ISL_1302267 | Instituto Nacional de Enfermedades Respiratorias (INER) | Instituto de Biotecnología de la UNAM | Authors from IBT, IMSS, InDRE and INER (in alphabetical order): Carlos F. Arias, Santiago Ávila-Ríos, Gisela Barrera-Badillo, Eduardo Becerril-Vargas, Celia Boukadida, Natividad Cruz-Ortiz, Larissa Fernandes-Matano, Ricardo Grande, Lucia Hernandez-Rivas, Alejandra Hernández-Terán, Pavel Isa, Irma Lopez-Martinez, José Arturo Martínez-Orozco, Margarita Matías-Florentino, Fidencio Mejía-Nepomuceno, Edgar Mendieta-Condado, Mario Mújica-Sánchez, José Esteban Muñoz-Medina, Tatiana Nunez-Garcia, Luis Alberto Ochoa-Carrera, Hector Esteban Paz-Juárez, Francisco Pulido, José Ernesto Ramírez-González, Alma Rincón-Rubio, Teresita Rojas-Mendoza, Jorge Salas-Hernández, Alejandro Sanchez-Flores, Clara Esperanza Santacruz-Tinoco, Andrea Santos Coy-Arechavaleta, Blanca Taboada, Gloria Vazquez, Joel Armando Vázquez-Pérez, Jerome Jean Verleyen |
| EPI_ISL_1302268, EPI_ISL_1302270 | Laboratorio Central de Epidemiología IMSS | Instituto de Biotecnología de la UNAM | Authors from IBT, IMSS, InDRE and INER (in alphabetical order): Carlos F. Arias, Santiago Ávila-Ríos, Gisela Barrera-Badillo, Eduardo Becerril-Vargas, Celia Boukadida, Natividad Cruz-Ortiz, Larissa Fernandes-Matano, Ricardo Grande, Lucia Hernandez-Rivas, Alejandra Hernández-Terán, Pavel Isa, Irma Lopez-Martinez, José Arturo Martínez-Orozco, Margarita Matías-Florentino, Fidencio Mejía-Nepomuceno, Edgar Mendieta-Condado, Mario Mújica-Sánchez, José Esteban Muñoz-Medina, Tatiana Nunez-Garcia, Luis Alberto Ochoa-Carrera, Hector Esteban Paz-Juárez, Francisco Pulido, José Ernesto Ramírez-González, Alma Rincón-Rubio, Teresita Rojas-Mendoza, Jorge Salas-Hernández, Alejandro Sanchez-Flores, Clara Esperanza Santacruz-Tinoco, Andrea Santos Coy-Arechavaleta, Blanca Taboada, Gloria Vazquez, Joel Armando Vázquez-Pérez, Jerome Jean Verleyen |
| EPI_ISL_1302271 | Instituto Nacional de Enfermedades Respiratorias (INER) | Instituto de Biotecnología de la UNAM | Authors from IBT, IMSS, InDRE and INER (in alphabetical order): Carlos F. Arias, Santiago Ávila-Ríos, Gisela Barrera-Badillo, Eduardo Becerril-Vargas, Celia Boukadida, Natividad Cruz-Ortiz, Larissa Fernandes-Matano, Ricardo Grande, Lucia Hernandez-Rivas, Alejandra Hernández-Terán, Pavel Isa, Irma Lopez-Martinez, José Arturo Martínez-Orozco, Margarita Matías-Florentino, Fidencio Mejía-Nepomuceno, Edgar Mendieta-Condado, Mario Mújica-Sánchez, José Esteban Muñoz-Medina, Tatiana Nunez-Garcia, Luis Alberto Ochoa-Carrera, Hector Esteban Paz-Juárez, Francisco Pulido, José Ernesto Ramírez-González, Alma Rincón-Rubio, Teresita Rojas-Mendoza, Jorge Salas-Hernández, Alejandro Sanchez-Flores, Clara Esperanza Santacruz-Tinoco, Andrea Santos Coy-Arechavaleta, Blanca Taboada, Gloria Vazquez, Joel Armando Vázquez-Pérez, Jerome Jean Verleyen |
| EPI_ISL_1302272 | Laboratorio Central de Epidemiología IMSS | Instituto de Biotecnología de la UNAM | Authors from IBT, IMSS, InDRE and INER (in alphabetical order): Carlos F. Arias, Santiago Ávila-Ríos, Gisela Barrera-Badillo, Eduardo Becerril-Vargas, Celia Boukadida, Natividad Cruz-Ortiz, Larissa Fernandes-Matano, Ricardo Grande, Lucia Hernandez-Rivas, Alejandra Hernández-Terán, Pavel Isa, Irma Lopez-Martinez, José Arturo Martínez-Orozco, Margarita Matías-Florentino, Fidencio Mejía-Nepomuceno, Edgar Mendieta-Condado, Mario Mújica-Sánchez, José Esteban Muñoz-Medina, Tatiana Nunez-Garcia, Luis Alberto Ochoa-Carrera, Hector Esteban Paz-Juárez, Francisco Pulido, José Ernesto Ramírez-González, Alma Rincón-Rubio, Teresita Rojas-Mendoza, Jorge Salas-Hernández, Alejandro Sanchez-Flores, Clara Esperanza Santacruz-Tinoco, Andrea Santos Coy-Arechavaleta, Blanca Taboada, Gloria Vazquez, Joel Armando Vázquez-Pérez, Jerome Jean Verleyen |
| EPI_ISL_1302273 | Instituto Nacional de Enfermedades Respiratorias (INER) | Instituto de Biotecnología de la UNAM | Authors from IBT, IMSS, InDRE and INER (in alphabetical order): Carlos F. Arias, Santiago Ávila-Ríos, Gisela Barrera-Badillo, Eduardo Becerril-Vargas, Celia Boukadida, Natividad Cruz-Ortiz, Larissa Fernandes-Matano, Ricardo Grande, Lucia Hernandez-Rivas, Alejandra Hernández-Terán, Pavel Isa, Irma Lopez-Martinez, José Arturo Martínez-Orozco, Margarita Matías-Florentino, Fidencio Mejía-Nepomuceno, Edgar Mendieta-Condado, Mario Mújica-Sánchez, José Esteban Muñoz-Medina, Tatiana Nunez-Garcia, Luis Alberto Ochoa-Carrera, Hector Esteban Paz-Juárez, Francisco Pulido, José Ernesto Ramírez-González, Alma Rincón-Rubio, Teresita Rojas-Mendoza, Jorge Salas-Hernández, Alejandro Sanchez-Flores, Clara Esperanza Santacruz-Tinoco, Andrea Santos Coy-Arechavaleta, Blanca Taboada, Gloria Vazquez, Joel Armando Vázquez-Pérez, Jerome Jean Verleyen |
| EPI_ISL_1302274 | Instituto de Diagnostico y Referencia Epidemiologicos InDRE_RNLSP | Instituto de Biotecnología de la UNAM | Authors from IBT, IMSS, InDRE and INER (in alphabetical order): Carlos F. Arias, Santiago Ávila-Ríos, Gisela Barrera-Badillo, Eduardo Becerril-Vargas, Celia Boukadida, Natividad Cruz-Ortiz, Larissa Fernandes-Matano, Ricardo Grande, Lucia Hernandez-Rivas, Alejandra Hernández-Terán, Pavel Isa, Irma Lopez-Martinez, José Arturo Martínez-Orozco, Margarita Matías-Florentino, Fidencio Mejía-Nepomuceno, Edgar Mendieta-Condado, Mario Mújica-Sánchez, José Esteban Muñoz-Medina, Tatiana Nunez-Garcia, Luis Alberto Ochoa-Carrera, Hector Esteban Paz-Juárez, Francisco Pulido, José Ernesto Ramírez-González, Alma Rincón-Rubio, Teresita Rojas-Mendoza, Jorge Salas-Hernández, Alejandro Sanchez-Flores, Clara Esperanza Santacruz-Tinoco, Andrea Santos Coy-Arechavaleta, Blanca Taboada, Gloria Vazquez, Joel Armando Vázquez-Pérez, Jerome Jean Verleyen |
| EPI_ISL_1302275 | Laboratorio Central de Epidemiología IMSS | Instituto de Biotecnología de la UNAM | Authors from IBT, IMSS, InDRE and INER (in alphabetical order): Carlos F. Arias, Santiago Ávila-Ríos, Gisela Barrera-Badillo, Eduardo Becerril-Vargas, Celia Boukadida, Natividad Cruz-Ortiz, Larissa Fernandes-Matano, Ricardo Grande, Lucia Hernandez-Rivas, Alejandra Hernández-Terán, Pavel Isa, Irma Lopez-Martinez, José Arturo Martínez-Orozco, Margarita Matías-Florentino, Fidencio Mejía-Nepomuceno, Edgar Mendieta-Condado, Mario Mújica-Sánchez, José Esteban Muñoz-Medina, Tatiana Nunez-Garcia, Luis Alberto Ochoa-Carrera, Hector Esteban Paz-Juárez, Francisco Pulido, José Ernesto Ramírez-González, Alma Rincón-Rubio, Teresita Rojas-Mendoza, Jorge Salas-Hernández, Alejandro Sanchez-Flores, Clara Esperanza Santacruz-Tinoco, Andrea Santos Coy-Arechavaleta, Blanca Taboada, Gloria Vazquez, Joel Armando Vázquez-Pérez, Jerome Jean Verleyen |
| EPI_ISL_1302276 | Instituto de Diagnostico y Referencia Epidemiologicos InDRE_RNLSP | Instituto de Biotecnología de la UNAM | Authors from IBT, IMSS, InDRE and INER (in alphabetical order): Carlos F. Arias, Santiago Ávila-Ríos, Gisela Barrera-Badillo, Eduardo Becerril-Vargas, Celia Boukadida, Natividad Cruz-Ortiz, Larissa Fernandes-Matano, Ricardo Grande, Lucia Hernandez-Rivas, Alejandra Hernández-Terán, Pavel Isa, Irma Lopez-Martinez, José Arturo Martínez-Orozco, Margarita Matías-Florentino, Fidencio Mejía-Nepomuceno, Edgar Mendieta-Condado, Mario Mújica-Sánchez, José Esteban Muñoz-Medina, Tatiana Nunez-Garcia, Luis Alberto Ochoa-Carrera, Hector Esteban Paz-Juárez, Francisco Pulido, José Ernesto Ramírez-González, Alma Rincón-Rubio, Teresita Rojas-Mendoza, Jorge Salas-Hernández, Alejandro Sanchez-Flores, Clara Esperanza Santacruz-Tinoco, Andrea Santos Coy-Arechavaleta, Blanca Taboada, Gloria Vazquez, Joel Armando Vázquez-Pérez, Jerome Jean Verleyen |
| EPI_ISL_1302277, EPI_ISL_1302278 | Instituto Nacional de Enfermedades Respiratorias (INER) | Instituto de Biotecnología de la UNAM | Authors from IBT, IMSS, InDRE and INER (in alphabetical order): Carlos F. Arias, Santiago Ávila-Ríos, Gisela Barrera-Badillo, Eduardo Becerril-Vargas, Celia Boukadida, Natividad Cruz-Ortiz, Larissa Fernandes-Matano, Ricardo Grande, Lucia Hernandez-Rivas, Alejandra Hernández-Terán, Pavel Isa, Irma Lopez-Martinez, José Arturo Martínez-Orozco, Margarita Matías-Florentino, Fidencio Mejía-Nepomuceno, Edgar Mendieta-Condado, Mario Mújica-Sánchez, José Esteban Muñoz-Medina, Tatiana Nunez-Garcia, Luis Alberto Ochoa-Carrera, Hector Esteban Paz-Juárez, Francisco Pulido, José Ernesto Ramírez-González, Alma Rincón-Rubio, Teresita Rojas-Mendoza, Jorge Salas-Hernández, Alejandro Sanchez-Flores, Clara Esperanza Santacruz-Tinoco, Andrea Santos Coy-Arechavaleta, Blanca Taboada, Gloria Vazquez, Joel Armando Vázquez-Pérez, Jerome Jean Verleyen |
| EPI_ISL_1302279 | Instituto de Diagnostico y Referencia Epidemiologicos InDRE_RNLSP | Instituto de Biotecnología de la UNAM | Authors from IBT, IMSS, InDRE and INER (in alphabetical order): Carlos F. Arias, Santiago Ávila-Ríos, Gisela Barrera-Badillo, Eduardo Becerril-Vargas, Celia Boukadida, Natividad Cruz-Ortiz, Larissa Fernandes-Matano, Ricardo Grande, Lucia Hernandez-Rivas, Alejandra Hernández-Terán, Pavel Isa, Irma Lopez-Martinez, José Arturo Martínez-Orozco, Margarita Matías-Florentino, Fidencio Mejía-Nepomuceno, Edgar Mendieta-Condado, Mario Mújica-Sánchez, José Esteban Muñoz-Medina, Tatiana Nunez-Garcia, Luis Alberto Ochoa-Carrera, Hector Esteban Paz-Juárez, Francisco Pulido, José Ernesto Ramírez-González, Alma Rincón-Rubio, Teresita Rojas-Mendoza, Jorge Salas-Hernández, Alejandro Sanchez-Flores, Clara Esperanza Santacruz-Tinoco, Andrea Santos Coy-Arechavaleta, Blanca Taboada, Gloria Vazquez, Joel Armando Vázquez-Pérez, Jerome Jean Verleyen |
| EPI_ISL_1302280 | Instituto Nacional de Enfermedades Respiratorias (INER) | Instituto de Biotecnología de la UNAM | Authors from IBT, IMSS, InDRE and INER (in alphabetical order): Carlos F. Arias, Santiago Ávila-Ríos, Gisela Barrera-Badillo, Eduardo Becerril-Vargas, Celia Boukadida, Natividad Cruz-Ortiz, Larissa Fernandes-Matano, Ricardo Grande, Lucia Hernandez-Rivas, Alejandra Hernández-Terán, Pavel Isa, Irma Lopez-Martinez, José Arturo Martínez-Orozco, Margarita Matías-Florentino, Fidencio Mejía-Nepomuceno, Edgar Mendieta-Condado, Mario Mújica-Sánchez, José Esteban Muñoz-Medina, Tatiana Nunez-Garcia, Luis Alberto Ochoa-Carrera, Hector Esteban Paz-Juárez, Francisco Pulido, José Ernesto Ramírez-González, Alma Rincón-Rubio, Teresita Rojas-Mendoza, Jorge Salas-Hernández, Alejandro Sanchez-Flores, Clara Esperanza Santacruz-Tinoco, Andrea Santos Coy-Arechavaleta, Blanca Taboada, Gloria Vazquez, Joel Armando Vázquez-Pérez, Jerome Jean Verleyen |
| EPI_ISL_1302281, EPI_ISL_1302282 | Laboratorio Central de Epidemiología IMSS | Instituto de Biotecnología de la UNAM | Authors from IBT, IMSS, InDRE and INER (in alphabetical order): Carlos F. Arias, Santiago Ávila-Ríos, Gisela Barrera-Badillo, Eduardo Becerril-Vargas, Celia Boukadida, Natividad Cruz-Ortiz, Larissa Fernandes-Matano, Ricardo Grande, Lucia Hernandez-Rivas, Alejandra Hernández-Terán, Pavel Isa, Irma Lopez-Martinez, José Arturo Martínez-Orozco, Margarita Matías-Florentino, Fidencio Mejía-Nepomuceno, Edgar Mendieta-Condado, Mario Mújica-Sánchez, José Esteban Muñoz-Medina, Tatiana Nunez-Garcia, Luis Alberto Ochoa-Carrera, Hector Esteban Paz-Juárez, Francisco Pulido, José Ernesto Ramírez-González, Alma Rincón-Rubio, Teresita Rojas-Mendoza, Jorge Salas-Hernández, Alejandro Sanchez-Flores, Clara Esperanza Santacruz-Tinoco, Andrea Santos Coy-Arechavaleta, Blanca Taboada, Gloria Vazquez, Joel Armando Vázquez-Pérez, Jerome Jean |

| | | | |
|---|---|---|---|
| | | | Verleyen |
| EPI_ISL_1302283 | Instituto de Diagnostico y Referencia Epidemiologicos InDRE_RNLSP | Instituto de Biotecnología de la UNAM | Authors from IBT, IMSS, InDRE and INER (in alphabetical order): Carlos F. Arias, Santiago Ávila-Ríos, Gisela Barrera-Badillo, Eduardo Becerril-Vargas, Celia Boukadida, Natividad Cruz-Ortiz, Larissa Fernandes-Matano, Ricardo Grande, Lucia Hernandez-Rivas, Alejandra Hernández-Terán, Pavel Isa, Irma Lopez-Martinez, José Arturo Martínez-Orozco, Margarita Matías-Florentino, Fidencio Mejía-Nepomuceno, Edgar Mendieta-Condado, Mario Mújica-Sánchez, José Esteban Muñoz-Medina, Tatiana Nunez-Garcia, Luis Alberto Ochoa-Carrera, Hector Esteban Paz-Juárez, Francisco Pulido, José Ernesto Ramírez-González, Alma Rincón-Rubio, Teresita Rojas-Mendoza, Jorge Salas-Hernández, Alejandro Sanchez-Flores, Clara Esperanza Santacruz-Tinoco, Andrea Santos Coy-Arechavaleta, Blanca Taboada, Gloria Vazquez, Joel Armando Vázquez-Pérez, Jerome Jean Verleyen |
| EPI_ISL_1302284, EPI_ISL_1302285 | Laboratorio Central de Epidemiología IMSS | Instituto de Biotecnología de la UNAM | Authors from IBT, IMSS, InDRE and INER (in alphabetical order): Carlos F. Arias, Santiago Ávila-Ríos, Gisela Barrera-Badillo, Eduardo Becerril-Vargas, Celia Boukadida, Natividad Cruz-Ortiz, Larissa Fernandes-Matano, Ricardo Grande, Lucia Hernandez-Rivas, Alejandra Hernández-Terán, Pavel Isa, Irma Lopez-Martinez, José Arturo Martínez-Orozco, Margarita Matías-Florentino, Fidencio Mejía-Nepomuceno, Edgar Mendieta-Condado, Mario Mújica-Sánchez, José Esteban Muñoz-Medina, Tatiana Nunez-Garcia, Luis Alberto Ochoa-Carrera, Hector Esteban Paz-Juárez, Francisco Pulido, José Ernesto Ramírez-González, Alma Rincón-Rubio, Teresita Rojas-Mendoza, Jorge Salas-Hernández, Alejandro Sanchez-Flores, Clara Esperanza Santacruz-Tinoco, Andrea Santos Coy-Arechavaleta, Blanca Taboada, Gloria Vazquez, Joel Armando Vázquez-Pérez, Jerome Jean Verleyen |
| EPI_ISL_1302286, EPI_ISL_1302287 | Instituto de Diagnostico y Referencia Epidemiologicos InDRE_RNLSP | Instituto de Biotecnología de la UNAM | Authors from IBT, IMSS, InDRE and INER (in alphabetical order): Carlos F. Arias, Santiago Ávila-Ríos, Gisela Barrera-Badillo, Eduardo Becerril-Vargas, Celia Boukadida, Natividad Cruz-Ortiz, Larissa Fernandes-Matano, Ricardo Grande, Lucia Hernandez-Rivas, Alejandra Hernández-Terán, Pavel Isa, Irma Lopez-Martinez, José Arturo Martínez-Orozco, Margarita Matías-Florentino, Fidencio Mejía-Nepomuceno, Edgar Mendieta-Condado, Mario Mújica-Sánchez, José Esteban Muñoz-Medina, Tatiana Nunez-Garcia, Luis Alberto Ochoa-Carrera, Hector Esteban Paz-Juárez, Francisco Pulido, José Ernesto Ramírez-González, Alma Rincón-Rubio, Teresita Rojas-Mendoza, Jorge Salas-Hernández, Alejandro Sanchez-Flores, Clara Esperanza Santacruz-Tinoco, Andrea Santos Coy-Arechavaleta, Blanca Taboada, Gloria Vazquez, Joel Armando Vázquez-Pérez, Jerome Jean Verleyen |
| EPI_ISL_1302288 | Laboratorio Central de Epidemiología IMSS | Instituto de Biotecnología de la UNAM | Authors from IBT, IMSS, InDRE and INER (in alphabetical order): Carlos F. Arias, Santiago Ávila-Ríos, Gisela Barrera-Badillo, Eduardo Becerril-Vargas, Celia Boukadida, Natividad Cruz-Ortiz, Larissa Fernandes-Matano, Ricardo Grande, Lucia Hernandez-Rivas, Alejandra Hernández-Terán, Pavel Isa, Irma Lopez-Martinez, José Arturo Martínez-Orozco, Margarita Matías-Florentino, Fidencio Mejía-Nepomuceno, Edgar Mendieta-Condado, Mario Mújica-Sánchez, José Esteban Muñoz-Medina, Tatiana Nunez-Garcia, Luis Alberto Ochoa-Carrera, Hector Esteban Paz-Juárez, Francisco Pulido, José Ernesto Ramírez-González, Alma Rincón-Rubio, Teresita Rojas-Mendoza, Jorge Salas-Hernández, Alejandro Sanchez-Flores, Clara Esperanza Santacruz-Tinoco, Andrea Santos Coy-Arechavaleta, Blanca Taboada, Gloria Vazquez, Joel Armando Vázquez-Pérez, Jerome Jean Verleyen |
| EPI_ISL_1302289 | Instituto de Diagnostico y Referencia Epidemiologicos InDRE_RNLSP | Instituto de Biotecnología de la UNAM | Authors from IBT, IMSS, InDRE and INER (in alphabetical order): Carlos F. Arias, Santiago Ávila-Ríos, Gisela Barrera-Badillo, Eduardo Becerril-Vargas, Celia Boukadida, Natividad Cruz-Ortiz, Larissa Fernandes-Matano, Ricardo Grande, Lucia Hernandez-Rivas, Alejandra Hernández-Terán, Pavel Isa, Irma Lopez-Martinez, José Arturo Martínez-Orozco, Margarita Matías-Florentino, Fidencio Mejía-Nepomuceno, Edgar Mendieta-Condado, Mario Mújica-Sánchez, José Esteban Muñoz-Medina, Tatiana Nunez-Garcia, Luis Alberto Ochoa-Carrera, Hector Esteban Paz-Juárez, Francisco Pulido, José Ernesto Ramírez-González, Alma Rincón-Rubio, Teresita Rojas-Mendoza, Jorge Salas-Hernández, Alejandro Sanchez-Flores, Clara Esperanza Santacruz-Tinoco, Andrea Santos Coy-Arechavaleta, Blanca Taboada, Gloria Vazquez, Joel Armando Vázquez-Pérez, Jerome Jean Verleyen |
| EPI_ISL_1302290, EPI_ISL_1302291, EPI_ISL_1302292 | Instituto Nacional de Enfermedades Respiratorias (INER) | Instituto de Biotecnología de la UNAM | Authors from IBT, IMSS, InDRE and INER (in alphabetical order): Carlos F. Arias, Santiago Ávila-Ríos, Gisela Barrera-Badillo, Eduardo Becerril-Vargas, Celia Boukadida, Natividad Cruz-Ortiz, Larissa Fernandes-Matano, Ricardo Grande, Lucia Hernandez-Rivas, Alejandra Hernández-Terán, Pavel Isa, Irma Lopez-Martinez, José Arturo Martínez-Orozco, Margarita Matías-Florentino, Fidencio Mejía-Nepomuceno, Edgar Mendieta-Condado, Mario Mújica-Sánchez, José Esteban Muñoz-Medina, Tatiana Nunez-Garcia, Luis Alberto Ochoa-Carrera, Hector Esteban Paz-Juárez, Francisco Pulido, José Ernesto Ramírez-González, Alma Rincón-Rubio, Teresita Rojas-Mendoza, Jorge Salas-Hernández, Alejandro Sanchez-Flores, Clara Esperanza Santacruz-Tinoco, Andrea Santos Coy-Arechavaleta, Blanca Taboada, Gloria Vazquez, Joel Armando Vázquez-Pérez, Jerome Jean Verleyen |
| EPI_ISL_1302293 | Instituto de Diagnostico y Referencia Epidemiologicos InDRE_RNLSP | Instituto de Biotecnología de la UNAM | Authors from IBT, IMSS, InDRE and INER (in alphabetical order): Carlos F. Arias, Santiago Ávila-Ríos, Gisela Barrera-Badillo, Eduardo Becerril-Vargas, Celia Boukadida, Natividad Cruz-Ortiz, Larissa Fernandes-Matano, Ricardo Grande, Lucia Hernandez-Rivas, Alejandra Hernández-Terán, Pavel Isa, Irma Lopez-Martinez, José Arturo Martínez-Orozco, Margarita Matías-Florentino, Fidencio Mejía-Nepomuceno, Edgar Mendieta-Condado, Mario Mújica-Sánchez, José Esteban Muñoz-Medina, Tatiana Nunez-Garcia, Luis Alberto Ochoa-Carrera, Hector Esteban Paz-Juárez, Francisco Pulido, José Ernesto Ramírez-González, Alma Rincón-Rubio, Teresita Rojas-Mendoza, Jorge Salas-Hernández, Alejandro Sanchez-Flores, Clara Esperanza Santacruz-Tinoco, Andrea Santos Coy-Arechavaleta, Blanca Taboada, Gloria Vazquez, Joel Armando Vázquez-Pérez, Jerome Jean Verleyen |
| EPI_ISL_1302294 | Laboratorio Central de Epidemiología IMSS | Instituto de Biotecnología de la UNAM | Authors from IBT, IMSS, InDRE and INER (in alphabetical order): Carlos F. Arias, Santiago Ávila-Ríos, Gisela Barrera-Badillo, Eduardo Becerril-Vargas, Celia Boukadida, Natividad Cruz-Ortiz, Larissa Fernandes-Matano, Ricardo Grande, Lucia Hernandez-Rivas, Alejandra Hernández-Terán, Pavel Isa, Irma Lopez-Martinez, José Arturo Martínez-Orozco, Margarita Matías-Florentino, Fidencio Mejía-Nepomuceno, Edgar Mendieta-Condado, Mario Mújica-Sánchez, José Esteban Muñoz-Medina, Tatiana Nunez-Garcia, Luis Alberto Ochoa-Carrera, Hector Esteban Paz-Juárez, Francisco Pulido, José Ernesto Ramírez-González, Alma Rincón-Rubio, Teresita Rojas-Mendoza, Jorge Salas-Hernández, Alejandro Sanchez-Flores, Clara Esperanza Santacruz-Tinoco, Andrea Santos Coy-Arechavaleta, Blanca Taboada, Gloria Vazquez, Joel Armando Vázquez-Pérez, Jerome Jean Verleyen |
| EPI_ISL_1302295, EPI_ISL_1302296 | Instituto de Diagnostico y Referencia Epidemiologicos InDRE_RNLSP | Instituto de Biotecnología de la UNAM | Authors from IBT, IMSS, InDRE and INER (in alphabetical order): Carlos F. Arias, Santiago Ávila-Ríos, Gisela Barrera-Badillo, Eduardo Becerril-Vargas, Celia Boukadida, Natividad Cruz-Ortiz, Larissa Fernandes-Matano, Ricardo Grande, Lucia Hernandez-Rivas, Alejandra Hernández-Terán, Pavel Isa, Irma Lopez-Martinez, José Arturo Martínez-Orozco, Margarita Matías-Florentino, Fidencio Mejía-Nepomuceno, Edgar Mendieta-Condado, Mario Mújica-Sánchez, José Esteban Muñoz-Medina, Tatiana Nunez-Garcia, Luis Alberto Ochoa-Carrera, Hector Esteban Paz-Juárez, Francisco Pulido, José Ernesto Ramírez-González, Alma Rincón-Rubio, Teresita Rojas-Mendoza, Jorge Salas-Hernández, Alejandro Sanchez-Flores, Clara Esperanza Santacruz-Tinoco, Andrea Santos Coy-Arechavaleta, Blanca Taboada, Gloria Vazquez, Joel Armando Vázquez-Pérez, Jerome Jean Verleyen |
| EPI_ISL_1302297, EPI_ISL_1302298, EPI_ISL_1302299 | Instituto Nacional de Enfermedades Respiratorias (INER) | Instituto de Biotecnología de la UNAM | Authors from IBT, IMSS, InDRE and INER (in alphabetical order): Carlos F. Arias, Santiago Ávila-Ríos, Gisela Barrera-Badillo, Eduardo Becerril-Vargas, Celia Boukadida, Natividad Cruz-Ortiz, Larissa Fernandes-Matano, Ricardo Grande, Lucia Hernandez-Rivas, Alejandra Hernández-Terán, Pavel Isa, Irma Lopez-Martinez, José Arturo Martínez-Orozco, Margarita Matías-Florentino, Fidencio Mejía-Nepomuceno, Edgar Mendieta-Condado, Mario Mújica-Sánchez, José Esteban Muñoz-Medina, Tatiana Nunez-Garcia, Luis Alberto Ochoa-Carrera, Hector Esteban Paz-Juárez, Francisco Pulido, José Ernesto Ramírez-González, Alma Rincón-Rubio, Teresita Rojas-Mendoza, Jorge Salas-Hernández, Alejandro Sanchez-Flores, Clara Esperanza Santacruz-Tinoco, Andrea Santos Coy-Arechavaleta, Blanca Taboada, Gloria Vazquez, Joel Armando Vázquez-Pérez, Jerome Jean Verleyen |
| EPI_ISL_1302300 | Instituto de Diagnostico y Referencia Epidemiologicos InDRE_RNLSP | Instituto de Biotecnología de la UNAM | Authors from IBT, IMSS, InDRE and INER (in alphabetical order): Carlos F. Arias, Santiago Ávila-Ríos, Gisela Barrera-Badillo, Eduardo Becerril-Vargas, Celia Boukadida, Natividad Cruz-Ortiz, Larissa Fernandes-Matano, Ricardo Grande, Lucia Hernandez-Rivas, Alejandra Hernández-Terán, Pavel Isa, Irma Lopez-Martinez, José Arturo Martínez-Orozco, Margarita Matías-Florentino, Fidencio Mejía-Nepomuceno, Edgar Mendieta-Condado, Mario Mújica-Sánchez, José Esteban Muñoz-Medina, Tatiana Nunez-Garcia, Luis Alberto Ochoa-Carrera, Hector Esteban Paz-Juárez, Francisco Pulido, José Ernesto Ramírez-González, Alma Rincón-Rubio, Teresita Rojas-Mendoza, Jorge Salas-Hernández, Alejandro Sanchez-Flores, Clara Esperanza Santacruz-Tinoco, Andrea Santos Coy-Arechavaleta, Blanca Taboada, Gloria Vazquez, Joel Armando Vázquez-Pérez, Jerome Jean Verleyen |
| EPI_ISL_1302301, EPI_ISL_1302302, EPI_ISL_1302303, EPI_ISL_1302304 | Instituto Nacional de Enfermedades Respiratorias (INER) | Instituto de Biotecnología de la UNAM | Authors from IBT, IMSS, InDRE and INER (in alphabetical order): Carlos F. Arias, Santiago Ávila-Ríos, Gisela Barrera-Badillo, Eduardo Becerril-Vargas, Celia Boukadida, Natividad Cruz-Ortiz, Larissa Fernandes-Matano, Ricardo Grande, Lucia Hernandez-Rivas, Alejandra Hernández-Terán, Pavel Isa, Irma Lopez-Martinez, José Arturo Martínez-Orozco, Margarita Matías-Florentino, Fidencio Mejía-Nepomuceno, Edgar Mendieta-Condado, Mario Mújica-Sánchez, José Esteban Muñoz-Medina, Tatiana Nunez-Garcia, Luis Alberto Ochoa-Carrera, Hector Esteban Paz-Juárez, Francisco Pulido, José Ernesto Ramírez-González, Alma Rincón-Rubio, Teresita Rojas-Mendoza, Jorge Salas-Hernández, Alejandro Sanchez-Flores, Clara Esperanza Santacruz-Tinoco, Andrea Santos Coy-Arechavaleta, Blanca Taboada, Gloria Vazquez, Joel Armando Vázquez-Pérez, Jerome Jean Verleyen |
| EPI_ISL_1302305 | Laboratorio Central de Epidemiología IMSS | Instituto de Biotecnología de la UNAM | Authors from IBT, IMSS, InDRE and INER (in alphabetical order): Carlos F. Arias, Santiago Ávila-Ríos, Gisela Barrera-Badillo, Eduardo Becerril-Vargas, Celia Boukadida, Natividad Cruz-Ortiz, Larissa Fernandes-Matano, Ricardo Grande, Lucia Hernandez-Rivas, Alejandra Hernández-Terán, Pavel Isa, Irma Lopez-Martinez, José Arturo Martínez-Orozco, Margarita Matías-Florentino, Fidencio Mejía-Nepomuceno, Edgar Mendieta-Condado, Mario Mújica-Sánchez, José Esteban Muñoz-Medina, Tatiana Nunez-Garcia, Luis Alberto Ochoa-Carrera, Hector Esteban Paz-Juárez, Francisco Pulido, José Ernesto Ramírez-González, Alma Rincón-Rubio, Teresita Rojas-Mendoza, Jorge Salas-Hernández, Alejandro Sanchez-Flores, Clara Esperanza Santacruz-Tinoco, Andrea Santos Coy-Arechavaleta, Blanca Taboada, Gloria Vazquez, Joel Armando Vázquez-Pérez, Jerome Jean Verleyen |
| EPI_ISL_1302306, EPI_ISL_1302307, EPI_ISL_1302308 | Instituto Nacional de Enfermedades Respiratorias (INER) | Instituto de Biotecnología de la UNAM | Authors from IBT, IMSS, InDRE and INER (in alphabetical order): Carlos F. Arias, Santiago Ávila-Ríos, Gisela Barrera-Badillo, Eduardo Becerril-Vargas, Celia Boukadida, Natividad Cruz-Ortiz, Larissa Fernandes-Matano, Ricardo Grande, Lucia Hernandez-Rivas, Alejandra Hernández-Terán, Pavel Isa, Irma Lopez-Martinez, José Arturo Martínez-Orozco, Margarita Matías-Florentino, Fidencio Mejía-Nepomuceno, Edgar Mendieta-Condado, Mario Mújica-Sánchez, José Esteban Muñoz-Medina, Tatiana Nunez-Garcia, Luis Alberto Ochoa-Carrera, Hector Esteban Paz-Juárez, Francisco Pulido, José Ernesto Ramírez-González, Alma Rincón-Rubio, Teresita Rojas-Mendoza, Jorge Salas-Hernández, Alejandro Sanchez-Flores, Clara Esperanza Santacruz-Tinoco, Andrea Santos Coy-Arechavaleta, Blanca Taboada, Gloria Vazquez, Joel Armando Vázquez-Pérez, Jerome Jean Verleyen |
| EPI_ISL_1302309, EPI_ISL_1302310 | Instituto de Diagnostico y Referencia Epidemiologicos InDRE_RNLSP | Instituto de Biotecnología de la UNAM | Authors from IBT, IMSS, InDRE and INER (in alphabetical order): Carlos F. Arias, Santiago Ávila-Ríos, Gisela Barrera-Badillo, Eduardo Becerril-Vargas, Celia Boukadida, Natividad Cruz-Ortiz, Larissa Fernandes-Matano, Ricardo Grande, Lucia Hernandez-Rivas, Alejandra Hernández-Terán, Pavel Isa, Irma Lopez-Martinez, José Arturo Martínez-Orozco, Margarita Matías-Florentino, Fidencio Mejía-Nepomuceno, Edgar Mendieta-Condado, Mario Mújica-Sánchez, José Esteban Muñoz-Medina, Tatiana Nunez-Garcia, Luis Alberto Ochoa-Carrera, Hector Esteban Paz-Juárez, Francisco Pulido, José Ernesto Ramírez-González, Alma Rincón-Rubio, Teresita Rojas-Mendoza, Jorge Salas-Hernández, Alejandro Sanchez-Flores, Clara Esperanza Santacruz-Tinoco, Andrea Santos Coy-Arechavaleta, Blanca Taboada, Gloria Vazquez, Joel Armando Vázquez-Pérez, Jerome Jean Verleyen |
| EPI_ISL_1302311 | Laboratorio Central de Epidemiología IMSS | Instituto de Biotecnología de la UNAM | Authors from IBT, IMSS, InDRE and INER (in alphabetical order): Carlos F. Arias, Santiago Ávila-Ríos, Gisela Barrera-Badillo, Eduardo Becerril-Vargas, Celia Boukadida, Natividad Cruz-Ortiz, Larissa Fernandes-Matano, Ricardo Grande, Lucia Hernandez-Rivas, Alejandra Hernández-Terán, Pavel Isa, Irma Lopez-Martinez, José Arturo Martínez-Orozco, Margarita Matías-Florentino, Fidencio Mejía-Nepomuceno, Edgar Mendieta-Condado, Mario Mújica-Sánchez, José Esteban Muñoz-Medina, Tatiana Nunez-Garcia, Luis Alberto Ochoa-Carrera, Hector Esteban Paz-Juárez, Francisco Pulido, José Ernesto Ramírez-González, Alma Rincón-Rubio, Teresita Rojas-Mendoza, Jorge Salas-Hernández, Alejandro Sanchez-Flores, Clara Esperanza Santacruz-Tinoco, Andrea Santos Coy-Arechavaleta, Blanca Taboada, Gloria Vazquez, Joel Armando Vázquez-Pérez, Jerome Jean Verleyen |
| EPI_ISL_1302312 | Instituto Nacional de Enfermedades Respiratorias (INER) | Instituto de Biotecnología de la UNAM | Authors from IBT, IMSS, InDRE and INER (in alphabetical order): Carlos F. Arias, Santiago Ávila-Ríos, Gisela Barrera-Badillo, Eduardo Becerril-Vargas, Celia Boukadida, Natividad Cruz-Ortiz, Larissa Fernandes-Matano, Ricardo Grande, Lucia Hernandez-Rivas, Alejandra Hernández-Terán, Pavel Isa, Irma Lopez-Martinez, José Arturo Martínez-Orozco, Margarita Matías-Florentino, Fidencio Mejía-Nepomuceno, Edgar Mendieta-Condado, Mario Mújica-Sánchez, José Esteban Muñoz-Medina, Tatiana Nunez-Garcia, Luis Alberto Ochoa-Carrera, Hector Esteban Paz-Juárez, Francisco Pulido, José Ernesto Ramírez-González, Alma Rincón-Rubio, Teresita Rojas-Mendoza, Jorge Salas-Hernández, Alejandro Sanchez-Flores, Clara Esperanza Santacruz-Tinoco, Andrea Santos Coy-Arechavaleta, Blanca Taboada, Gloria Vazquez, Joel Armando Vázquez-Pérez, Jerome Jean Verleyen |
| EPI_ISL_1302313, EPI_ISL_1302314 | Instituto de Diagnostico y Referencia Epidemiologicos InDRE_RNLSP | Instituto de Biotecnología de la UNAM | Authors from IBT, IMSS, InDRE and INER (in alphabetical order): Carlos F. Arias, Santiago Ávila-Ríos, Gisela Barrera-Badillo, Eduardo Becerril-Vargas, Celia Boukadida, Natividad Cruz-Ortiz, Larissa Fernandes-Matano, Ricardo Grande, Lucia Hernandez-Rivas, Alejandra Hernández-Terán, Pavel Isa, Irma Lopez-Martinez, José Arturo Martínez-Orozco, Margarita Matías-Florentino, Fidencio Mejía-Nepomuceno, Edgar Mendieta-Condado, Mario Mújica-Sánchez, José Esteban Muñoz-Medina, Tatiana Nunez-Garcia, Luis Alberto Ochoa-Carrera, Hector Esteban Paz-Juárez, Francisco Pulido, José Ernesto Ramírez-González, Alma Rincón-Rubio, Teresita Rojas-Mendoza, Jorge Salas-Hernández, Alejandro Sanchez-Flores, Clara Esperanza Santacruz-Tinoco, Andrea Santos Coy-Arechavaleta, Blanca Taboada, Gloria Vazquez, Joel Armando Vázquez-Pérez, Jerome Jean Verleyen |
| EPI_ISL_1302315 | Laboratorio Central de Epidemiología IMSS | Instituto de Biotecnología de la UNAM | Authors from IBT, IMSS, InDRE and INER (in alphabetical order): Carlos F. Arias, Santiago Ávila-Ríos, Gisela Barrera-Badillo, Eduardo Becerril-Vargas, Celia Boukadida, Natividad Cruz-Ortiz, Larissa Fernandes-Matano, Ricardo Grande, Lucia Hernandez-Rivas, Alejandra Hernández-Terán, Pavel Isa, Irma Lopez-Martinez, José Arturo Martínez-Orozco, Margarita Matías-Florentino, Fidencio Mejía-Nepomuceno, Edgar Mendieta-Condado, Mario Mújica-Sánchez, José Esteban Muñoz-Medina, Tatiana Nunez-Garcia, Luis Alberto Ochoa-Carrera, Hector Esteban Paz-Juárez, Francisco Pulido, José Ernesto Ramírez-González, Alma Rincón-Rubio, Teresita Rojas-Mendoza, Jorge Salas-Hernández, Alejandro Sanchez-Flores, Clara Esperanza Santacruz-Tinoco, Andrea Santos Coy-Arechavaleta, Blanca Taboada, Gloria Vazquez, Joel Armando Vázquez-Pérez, Jerome Jean Verleyen |
| EPI_ISL_1302316, EPI_ISL_1302317 | Instituto Nacional de Enfermedades Respiratorias (INER) | Instituto de Biotecnología de la UNAM | Authors from IBT, IMSS, InDRE and INER (in alphabetical order): Carlos F. Arias, Santiago Ávila-Ríos, Gisela Barrera-Badillo, Eduardo Becerril-Vargas, Celia Boukadida, Natividad Cruz-Ortiz, Larissa Fernandes-Matano, Ricardo Grande, Lucia Hernandez-Rivas, Alejandra Hernández-Terán, Pavel Isa, Irma Lopez-Martinez, José Arturo Martínez-Orozco, Margarita Matías-Florentino, Fidencio Mejía-Nepomuceno, Edgar Mendieta-Condado, Mario Mújica-Sánchez, José Esteban Muñoz-Medina, Tatiana Nunez-Garcia, Luis Alberto Ochoa-Carrera, Hector Esteban Paz-Juárez, Francisco Pulido, José Ernesto Ramírez-González, Alma Rincón-Rubio, Teresita Rojas-Mendoza, Jorge Salas-Hernández, Alejandro Sanchez-Flores, Clara Esperanza Santacruz-Tinoco, Andrea Santos Coy-Arechavaleta, Blanca Taboada, Gloria Vazquez, Joel Armando Vázquez-Pérez, Jerome Jean Verleyen |
| EPI_ISL_1302318 | Laboratorio Central de Epidemiología IMSS | Instituto de Biotecnología de la UNAM | Authors from IBT, IMSS, InDRE and INER (in alphabetical order): Carlos F. Arias, Santiago Ávila-Ríos, Gisela Barrera-Badillo, Eduardo Becerril-Vargas, Celia Boukadida, Natividad Cruz-Ortiz, Larissa Fernandes-Matano, Ricardo Grande, Lucia Hernandez-Rivas, Alejandra Hernández-Terán, Pavel Isa, Irma Lopez-Martinez, José Arturo Martínez-Orozco, Margarita Matías-Florentino, Fidencio Mejía-Nepomuceno, Edgar Mendieta-Condado, Mario Mújica-Sánchez, José Esteban Muñoz-Medina, Tatiana Nunez-Garcia, Luis Alberto Ochoa-Carrera, Hector Esteban Paz-Juárez, Francisco Pulido, José Ernesto Ramírez-González, Alma Rincón-Rubio, Teresita Rojas-Mendoza, Jorge Salas-Hernández, Alejandro Sanchez-Flores, Clara Esperanza Santacruz-Tinoco, Andrea Santos Coy-Arechavaleta, Blanca Taboada, Gloria Vazquez, Joel Armando Vázquez-Pérez, Jerome Jean Verleyen |
| EPI_ISL_1302319, EPI_ISL_1302320 | Instituto Nacional de Enfermedades Respiratorias (INER) | Instituto de Biotecnología de la UNAM | Authors from IBT, IMSS, InDRE and INER (in alphabetical order): Carlos F. Arias, Santiago Ávila-Ríos, Gisela Barrera-Badillo, Eduardo Becerril-Vargas, Celia Boukadida, Natividad Cruz-Ortiz, Larissa Fernandes-Matano, Ricardo Grande, Lucia Hernandez-Rivas, Alejandra Hernández-Terán, Pavel Isa, Irma Lopez-Martinez, José Arturo Martínez-Orozco, Margarita Matías-Florentino, Fidencio Mejía-Nepomuceno, Edgar Mendieta-Condado, Mario Mújica-Sánchez, José Esteban Muñoz-Medina, Tatiana Nunez-Garcia, Luis Alberto Ochoa-Carrera, Hector Esteban Paz-Juárez, Francisco Pulido, José Ernesto Ramírez-González, Alma Rincón-Rubio, Teresita Rojas-Mendoza, Jorge Salas-Hernández, Alejandro Sanchez-Flores, Clara Esperanza Santacruz-Tinoco, Andrea Santos Coy-Arechavaleta, Blanca Taboada, Gloria Vazquez, Joel Armando Vázquez-Pérez, Jerome Jean Verleyen |
| EPI_ISL_1302321, EPI_ISL_1302322, EPI_ISL_1302323 | Instituto de Diagnostico y Referencia Epidemiologicos InDRE_RNLSP | Instituto de Biotecnología de la UNAM | Authors from IBT, IMSS, InDRE and INER (in alphabetical order): Carlos F. Arias, Santiago Ávila-Ríos, Gisela Barrera-Badillo, Eduardo Becerril-Vargas, Celia Boukadida, Natividad Cruz-Ortiz, Larissa Fernandes-Matano, Ricardo Grande, Lucia Hernandez-Rivas, Alejandra Hernández-Terán, Pavel Isa, Irma Lopez-Martinez, José Arturo Martínez-Orozco, Margarita Matías-Florentino, Fidencio Mejía-Nepomuceno, Edgar Mendieta-Condado, Mario Mújica-Sánchez, José Esteban Muñoz-Medina, Tatiana Nunez-Garcia, Luis Alberto Ochoa-Carrera, Hector Esteban Paz-Juárez, Francisco Pulido, José Ernesto Ramírez-González, Alma Rincón-Rubio, Teresita Rojas-Mendoza, Jorge Salas-Hernández, Alejandro Sanchez-Flores, Clara Esperanza Santacruz-Tinoco, Andrea Santos Coy-Arechavaleta, Blanca Taboada, Gloria Vazquez, Joel Armando Vázquez-Pérez, Jerome Jean Verleyen |
| EPI_ISL_1302324 | Laboratorio Central de Epidemiología IMSS | Instituto de Biotecnología de la UNAM | Authors from IBT, IMSS, InDRE and INER (in alphabetical order): Carlos F. Arias, Santiago Ávila-Ríos, Gisela Barrera-Badillo, Eduardo Becerril-Vargas, Celia Boukadida, Natividad Cruz-Ortiz, Larissa Fernandes-Matano, Ricardo Grande, Lucia Hernandez-Rivas, Alejandra Hernández-Terán, Pavel Isa, Irma Lopez-Martinez, José Arturo Martínez-Orozco, Margarita Matías-Florentino, Fidencio Mejía-Nepomuceno, Edgar Mendieta-Condado, Mario Mújica-Sánchez, José Esteban Muñoz-Medina, Tatiana Nunez-Garcia, Luis Alberto Ochoa-Carrera, Hector Esteban Paz-Juárez, Francisco Pulido, José Ernesto Ramírez-González, Alma Rincón-Rubio, Teresita Rojas-Mendoza, Jorge Salas-Hernández, Alejandro Sanchez-Flores, Clara Esperanza Santacruz-Tinoco, Andrea Santos Coy-Arechavaleta, Blanca Taboada, Gloria Vazquez, Joel Armando Vázquez-Pérez, Jerome Jean Verleyen |
| EPI_ISL_1302325, EPI_ISL_1302326, | Instituto Nacional de Enfermedades Respiratorias (INER) | Instituto de Biotecnología de la UNAM | Authors from IBT, IMSS, InDRE and INER (in alphabetical order): Carlos F. Arias, Santiago Ávila-Ríos, Gisela Barrera-Badillo, Eduardo Becerril-Vargas, Celia Boukadida, Natividad Cruz-Ortiz, Larissa Fernandes-Matano, Ricardo Grande, Lucia Hernandez-Rivas, Alejandra Hernández-Terán, Pavel Isa, Irma Lopez-Martinez, José Arturo Martínez-Orozco, Margarita Matías-Florentino, Fidencio Mejía-Nepomuceno, Edgar Mendieta-Condado, Mario Mújica-Sánchez, José Esteban Muñoz-Medina, Tatiana Nunez-Garcia, Luis Alberto Ochoa-Carrera, Hector Esteban Paz-Juárez, Francisco |

| | | | |
|---|---|---|---|
| EPI_ISL_1302327, EPI_ISL_1302328, EPI_ISL_1302329, EPI_ISL_1302330 | | | Pulido, José Ernesto Ramírez-González, Alma Rincón-Rubio, Teresita Rojas-Mendoza, Jorge Salas-Hernández, Alejandro Sanchez-Flores, Clara Esperanza Santacruz-Tinoco, Andrea Santos Coy-Arechavaleta, Blanca Taboada, Gloria Vazquez, Joel Armando Vázquez-Pérez, Jerome Jean Verleyen |
| EPI_ISL_1302331 | Instituto de Diagnostico y Referencia Epidemiologicos InDRE_RNLSP | Instituto de Biotecnología de la UNAM | Authors from IBT, IMSS, InDRE and INER (in alphabetical order): Carlos F. Arias, Santiago Ávila-Ríos, Gisela Barrera-Badillo, Eduardo Becerril-Vargas, Celia Boukadida, Natividad Cruz-Ortiz, Larissa Fernandes-Matano, Ricardo Grande, Lucia Hernandez-Rivas, Alejandra Hernández-Terán, Pavel Isa, Irma Lopez-Martinez, José Arturo Martínez-Orozco, Margarita Matías-Florentino, Fidencio Mejía-Nepomuceno, Edgar Mendieta-Condado, Mario Mújica-Sánchez, José Esteban Muñoz-Medina, Tatiana Nunez-Garcia, Luis Alberto Ochoa-Carrera, Hector Esteban Paz-Juárez, Francisco Pulido, José Ernesto Ramírez-González, Alma Rincón-Rubio, Teresita Rojas-Mendoza, Jorge Salas-Hernández, Alejandro Sanchez-Flores, Clara Esperanza Santacruz-Tinoco, Andrea Santos Coy-Arechavaleta, Blanca Taboada, Gloria Vazquez, Joel Armando Vázquez-Pérez, Jerome Jean Verleyen |
| EPI_ISL_1302332 | Laboratorio Central de Epidemiología IMSS | Instituto de Biotecnología de la UNAM | Authors from IBT, IMSS, InDRE and INER (in alphabetical order): Carlos F. Arias, Santiago Ávila-Ríos, Gisela Barrera-Badillo, Eduardo Becerril-Vargas, Celia Boukadida, Natividad Cruz-Ortiz, Larissa Fernandes-Matano, Ricardo Grande, Lucia Hernandez-Rivas, Alejandra Hernández-Terán, Pavel Isa, Irma Lopez-Martinez, José Arturo Martínez-Orozco, Margarita Matías-Florentino, Fidencio Mejía-Nepomuceno, Edgar Mendieta-Condado, Mario Mújica-Sánchez, José Esteban Muñoz-Medina, Tatiana Nunez-Garcia, Luis Alberto Ochoa-Carrera, Hector Esteban Paz-Juárez, Francisco Pulido, José Ernesto Ramírez-González, Alma Rincón-Rubio, Teresita Rojas-Mendoza, Jorge Salas-Hernández, Alejandro Sanchez-Flores, Clara Esperanza Santacruz-Tinoco, Andrea Santos Coy-Arechavaleta, Blanca Taboada, Gloria Vazquez, Joel Armando Vázquez-Pérez, Jerome Jean Verleyen |
| EPI_ISL_1302333 | Instituto de Diagnostico y Referencia Epidemiologicos InDRE_RNLSP | Instituto de Biotecnología de la UNAM | Authors from IBT, IMSS, InDRE and INER (in alphabetical order): Carlos F. Arias, Santiago Ávila-Ríos, Gisela Barrera-Badillo, Eduardo Becerril-Vargas, Celia Boukadida, Natividad Cruz-Ortiz, Larissa Fernandes-Matano, Ricardo Grande, Lucia Hernandez-Rivas, Alejandra Hernández-Terán, Pavel Isa, Irma Lopez-Martinez, José Arturo Martínez-Orozco, Margarita Matías-Florentino, Fidencio Mejía-Nepomuceno, Edgar Mendieta-Condado, Mario Mújica-Sánchez, José Esteban Muñoz-Medina, Tatiana Nunez-Garcia, Luis Alberto Ochoa-Carrera, Hector Esteban Paz-Juárez, Francisco Pulido, José Ernesto Ramírez-González, Alma Rincón-Rubio, Teresita Rojas-Mendoza, Jorge Salas-Hernández, Alejandro Sanchez-Flores, Clara Esperanza Santacruz-Tinoco, Andrea Santos Coy-Arechavaleta, Blanca Taboada, Gloria Vazquez, Joel Armando Vázquez-Pérez, Jerome Jean Verleyen |
| EPI_ISL_1302334 | Instituto Nacional de Enfermedades Respiratorias (INER) | Instituto de Biotecnología de la UNAM | Authors from IBT, IMSS, InDRE and INER (in alphabetical order): Carlos F. Arias, Santiago Ávila-Ríos, Gisela Barrera-Badillo, Eduardo Becerril-Vargas, Celia Boukadida, Natividad Cruz-Ortiz, Larissa Fernandes-Matano, Ricardo Grande, Lucia Hernandez-Rivas, Alejandra Hernández-Terán, Pavel Isa, Irma Lopez-Martinez, José Arturo Martínez-Orozco, Margarita Matías-Florentino, Fidencio Mejía-Nepomuceno, Edgar Mendieta-Condado, Mario Mújica-Sánchez, José Esteban Muñoz-Medina, Tatiana Nunez-Garcia, Luis Alberto Ochoa-Carrera, Hector Esteban Paz-Juárez, Francisco Pulido, José Ernesto Ramírez-González, Alma Rincón-Rubio, Teresita Rojas-Mendoza, Jorge Salas-Hernández, Alejandro Sanchez-Flores, Clara Esperanza Santacruz-Tinoco, Andrea Santos Coy-Arechavaleta, Blanca Taboada, Gloria Vazquez, Joel Armando Vázquez-Pérez, Jerome Jean Verleyen |
| EPI_ISL_1302335 | Instituto de Diagnostico y Referencia Epidemiologicos InDRE_RNLSP | Instituto de Biotecnología de la UNAM | Authors from IBT, IMSS, InDRE and INER (in alphabetical order): Carlos F. Arias, Santiago Ávila-Ríos, Gisela Barrera-Badillo, Eduardo Becerril-Vargas, Celia Boukadida, Natividad Cruz-Ortiz, Larissa Fernandes-Matano, Ricardo Grande, Lucia Hernandez-Rivas, Alejandra Hernández-Terán, Pavel Isa, Irma Lopez-Martinez, José Arturo Martínez-Orozco, Margarita Matías-Florentino, Fidencio Mejía-Nepomuceno, Edgar Mendieta-Condado, Mario Mújica-Sánchez, José Esteban Muñoz-Medina, Tatiana Nunez-Garcia, Luis Alberto Ochoa-Carrera, Hector Esteban Paz-Juárez, Francisco Pulido, José Ernesto Ramírez-González, Alma Rincón-Rubio, Teresita Rojas-Mendoza, Jorge Salas-Hernández, Alejandro Sanchez-Flores, Clara Esperanza Santacruz-Tinoco, Andrea Santos Coy-Arechavaleta, Blanca Taboada, Gloria Vazquez, Joel Armando Vázquez-Pérez, Jerome Jean Verleyen |
| EPI_ISL_1302336 | Instituto Nacional de Enfermedades Respiratorias (INER) | Instituto de Biotecnología de la UNAM | Authors from IBT, IMSS, InDRE and INER (in alphabetical order): Carlos F. Arias, Santiago Ávila-Ríos, Gisela Barrera-Badillo, Eduardo Becerril-Vargas, Celia Boukadida, Natividad Cruz-Ortiz, Larissa Fernandes-Matano, Ricardo Grande, Lucia Hernandez-Rivas, Alejandra Hernández-Terán, Pavel Isa, Irma Lopez-Martinez, José Arturo Martínez-Orozco, Margarita Matías-Florentino, Fidencio Mejía-Nepomuceno, Edgar Mendieta-Condado, Mario Mújica-Sánchez, José Esteban Muñoz-Medina, Tatiana Nunez-Garcia, Luis Alberto Ochoa-Carrera, Hector Esteban Paz-Juárez, Francisco Pulido, José Ernesto Ramírez-González, Alma Rincón-Rubio, Teresita Rojas-Mendoza, Jorge Salas-Hernández, Alejandro Sanchez-Flores, Clara Esperanza Santacruz-Tinoco, Andrea Santos Coy-Arechavaleta, Blanca Taboada, Gloria Vazquez, Joel Armando Vázquez-Pérez, Jerome Jean Verleyen |
| EPI_ISL_1302337 | Instituto de Diagnostico y Referencia Epidemiologicos InDRE_RNLSP | Instituto de Biotecnología de la UNAM | Authors from IBT, IMSS, InDRE and INER (in alphabetical order): Carlos F. Arias, Santiago Ávila-Ríos, Gisela Barrera-Badillo, Eduardo Becerril-Vargas, Celia Boukadida, Natividad Cruz-Ortiz, Larissa Fernandes-Matano, Ricardo Grande, Lucia Hernandez-Rivas, Alejandra Hernández-Terán, Pavel Isa, Irma Lopez-Martinez, José Arturo Martínez-Orozco, Margarita Matías-Florentino, Fidencio Mejía-Nepomuceno, Edgar Mendieta-Condado, Mario Mújica-Sánchez, José Esteban Muñoz-Medina, Tatiana Nunez-Garcia, Luis Alberto Ochoa-Carrera, Hector Esteban Paz-Juárez, Francisco Pulido, José Ernesto Ramírez-González, Alma Rincón-Rubio, Teresita Rojas-Mendoza, Jorge Salas-Hernández, Alejandro Sanchez-Flores, Clara Esperanza Santacruz-Tinoco, Andrea Santos Coy-Arechavaleta, Blanca Taboada, Gloria Vazquez, Joel Armando Vázquez-Pérez, Jerome Jean Verleyen |
| EPI_ISL_1302338, EPI_ISL_1302339 | Instituto Nacional de Enfermedades Respiratorias (INER) | Instituto de Biotecnología de la UNAM | Authors from IBT, IMSS, InDRE and INER (in alphabetical order): Carlos F. Arias, Santiago Ávila-Ríos, Gisela Barrera-Badillo, Eduardo Becerril-Vargas, Celia Boukadida, Natividad Cruz-Ortiz, Larissa Fernandes-Matano, Ricardo Grande, Lucia Hernandez-Rivas, Alejandra Hernández-Terán, Pavel Isa, Irma Lopez-Martinez, José Arturo Martínez-Orozco, Margarita Matías-Florentino, Fidencio Mejía-Nepomuceno, Edgar Mendieta-Condado, Mario Mújica-Sánchez, José Esteban Muñoz-Medina, Tatiana Nunez-Garcia, Luis Alberto Ochoa-Carrera, Hector Esteban Paz-Juárez, Francisco Pulido, José Ernesto Ramírez-González, Alma Rincón-Rubio, Teresita Rojas-Mendoza, Jorge Salas-Hernández, Alejandro Sanchez-Flores, Clara Esperanza Santacruz-Tinoco, Andrea Santos Coy-Arechavaleta, Blanca Taboada, Gloria Vazquez, Joel Armando Vázquez-Pérez, Jerome Jean Verleyen |
| EPI_ISL_1302340 | Instituto de Diagnostico y Referencia Epidemiologicos InDRE_RNLSP | Instituto de Biotecnología de la UNAM | Authors from IBT, IMSS, InDRE and INER (in alphabetical order): Carlos F. Arias, Santiago Ávila-Ríos, Gisela Barrera-Badillo, Eduardo Becerril-Vargas, Celia Boukadida, Natividad Cruz-Ortiz, Larissa Fernandes-Matano, Ricardo Grande, Lucia Hernandez-Rivas, Alejandra Hernández-Terán, Pavel Isa, Irma Lopez-Martinez, José Arturo Martínez-Orozco, Margarita Matías-Florentino, Fidencio Mejía-Nepomuceno, Edgar Mendieta-Condado, Mario Mújica-Sánchez, José Esteban Muñoz-Medina, Tatiana Nunez-Garcia, Luis Alberto Ochoa-Carrera, Hector Esteban Paz-Juárez, Francisco Pulido, José Ernesto Ramírez-González, Alma Rincón-Rubio, Teresita Rojas-Mendoza, Jorge Salas-Hernández, Alejandro Sanchez-Flores, Clara Esperanza Santacruz-Tinoco, Andrea Santos Coy-Arechavaleta, Blanca Taboada, Gloria Vazquez, Joel Armando Vázquez-Pérez, Jerome Jean Verleyen |
| EPI_ISL_1302341 | Laboratorio Central de Epidemiología IMSS | Instituto de Biotecnología de la UNAM | Authors from IBT, IMSS, InDRE and INER (in alphabetical order): Carlos F. Arias, Santiago Ávila-Ríos, Gisela Barrera-Badillo, Eduardo Becerril-Vargas, Celia Boukadida, Natividad Cruz-Ortiz, Larissa Fernandes-Matano, Ricardo Grande, Lucia Hernandez-Rivas, Alejandra Hernández-Terán, Pavel Isa, Irma Lopez-Martinez, José Arturo Martínez-Orozco, Margarita Matías-Florentino, Fidencio Mejía-Nepomuceno, Edgar Mendieta-Condado, Mario Mújica-Sánchez, José Esteban Muñoz-Medina, Tatiana Nunez-Garcia, Luis Alberto Ochoa-Carrera, Hector Esteban Paz-Juárez, Francisco Pulido, José Ernesto Ramírez-González, Alma Rincón-Rubio, Teresita Rojas-Mendoza, Jorge Salas-Hernández, Alejandro Sanchez-Flores, Clara Esperanza Santacruz-Tinoco, Andrea Santos Coy-Arechavaleta, Blanca Taboada, Gloria Vazquez, Joel Armando Vázquez-Pérez, Jerome Jean Verleyen |
| EPI_ISL_1302342, EPI_ISL_1302343, EPI_ISL_1302344 | Instituto de Diagnostico y Referencia Epidemiologicos InDRE_RNLSP | Instituto de Biotecnología de la UNAM | Authors from IBT, IMSS, InDRE and INER (in alphabetical order): Carlos F. Arias, Santiago Ávila-Ríos, Gisela Barrera-Badillo, Eduardo Becerril-Vargas, Celia Boukadida, Natividad Cruz-Ortiz, Larissa Fernandes-Matano, Ricardo Grande, Lucia Hernandez-Rivas, Alejandra Hernández-Terán, Pavel Isa, Irma Lopez-Martinez, José Arturo Martínez-Orozco, Margarita Matías-Florentino, Fidencio Mejía-Nepomuceno, Edgar Mendieta-Condado, Mario Mújica-Sánchez, José Esteban Muñoz-Medina, Tatiana Nunez-Garcia, Luis Alberto Ochoa-Carrera, Hector Esteban Paz-Juárez, Francisco Pulido, José Ernesto Ramírez-González, Alma Rincón-Rubio, Teresita Rojas-Mendoza, Jorge Salas-Hernández, Alejandro Sanchez-Flores, Clara Esperanza Santacruz-Tinoco, Andrea Santos Coy-Arechavaleta, Blanca Taboada, Gloria Vazquez, Joel Armando Vázquez-Pérez, Jerome Jean Verleyen |
| EPI_ISL_1302345, EPI_ISL_1302346 | Instituto Nacional de Enfermedades Respiratorias (INER) | Instituto de Biotecnología de la UNAM | Authors from IBT, IMSS, InDRE and INER (in alphabetical order): Carlos F. Arias, Santiago Ávila-Ríos, Gisela Barrera-Badillo, Eduardo Becerril-Vargas, Celia Boukadida, Natividad Cruz-Ortiz, Larissa Fernandes-Matano, Ricardo Grande, Lucia Hernandez-Rivas, Alejandra Hernández-Terán, Pavel Isa, Irma Lopez-Martinez, José Arturo Martínez-Orozco, Margarita Matías-Florentino, Fidencio Mejía-Nepomuceno, Edgar Mendieta-Condado, Mario Mújica-Sánchez, José Esteban Muñoz-Medina, Tatiana Nunez-Garcia, Luis Alberto Ochoa-Carrera, Hector Esteban Paz-Juárez, Francisco Pulido, José Ernesto Ramírez-González, Alma Rincón-Rubio, Teresita Rojas-Mendoza, Jorge Salas-Hernández, Alejandro Sanchez-Flores, Clara Esperanza Santacruz-Tinoco, Andrea Santos Coy-Arechavaleta, Blanca Taboada, Gloria Vazquez, Joel Armando Vázquez-Pérez, Jerome Jean Verleyen |
| EPI_ISL_1302347, EPI_ISL_1302348 | Laboratorio Central de Epidemiología IMSS | Instituto de Biotecnología de la UNAM | Authors from IBT, IMSS, InDRE and INER (in alphabetical order): Carlos F. Arias, Santiago Ávila-Ríos, Gisela Barrera-Badillo, Eduardo Becerril-Vargas, Celia Boukadida, Natividad Cruz-Ortiz, Larissa Fernandes-Matano, Ricardo Grande, Lucia Hernandez-Rivas, Alejandra Hernández-Terán, Pavel Isa, Irma Lopez-Martinez, José Arturo Martínez-Orozco, Margarita Matías-Florentino, Fidencio Mejía-Nepomuceno, Edgar Mendieta-Condado, Mario Mújica-Sánchez, José Esteban Muñoz-Medina, Tatiana Nunez-Garcia, Luis Alberto Ochoa-Carrera, Hector Esteban Paz-Juárez, Francisco Pulido, José Ernesto Ramírez-González, Alma Rincón-Rubio, Teresita Rojas-Mendoza, Jorge Salas-Hernández, Alejandro Sanchez-Flores, Clara Esperanza Santacruz-Tinoco, Andrea Santos Coy-Arechavaleta, Blanca Taboada, Gloria Vazquez, Joel Armando Vázquez-Pérez, Jerome Jean Verleyen |
| EPI_ISL_1302349, EPI_ISL_1302350 | Instituto Nacional de Enfermedades Respiratorias (INER) | Instituto de Biotecnología de la UNAM | Authors from IBT, IMSS, InDRE and INER (in alphabetical order): Carlos F. Arias, Santiago Ávila-Ríos, Gisela Barrera-Badillo, Eduardo Becerril-Vargas, Celia Boukadida, Natividad Cruz-Ortiz, Larissa Fernandes-Matano, Ricardo Grande, Lucia Hernandez-Rivas, Alejandra Hernández-Terán, Pavel Isa, Irma Lopez-Martinez, José Arturo Martínez-Orozco, Margarita Matías-Florentino, Fidencio Mejía-Nepomuceno, Edgar Mendieta-Condado, Mario Mújica-Sánchez, José Esteban Muñoz-Medina, Tatiana Nunez-Garcia, Luis Alberto Ochoa-Carrera, Hector Esteban Paz-Juárez, Francisco Pulido, José Ernesto Ramírez-González, Alma Rincón-Rubio, Teresita Rojas-Mendoza, Jorge Salas-Hernández, Alejandro Sanchez-Flores, Clara Esperanza Santacruz-Tinoco, Andrea Santos Coy-Arechavaleta, Blanca Taboada, Gloria Vazquez, Joel Armando Vázquez-Pérez, Jerome Jean Verleyen |
| EPI_ISL_1302351, EPI_ISL_1302352, EPI_ISL_1302353, EPI_ISL_1302354, EPI_ISL_1302355 | Laboratorio Central de Epidemiología IMSS | Instituto de Biotecnología de la UNAM | Authors from IBT, IMSS, InDRE and INER (in alphabetical order): Carlos F. Arias, Santiago Ávila-Ríos, Gisela Barrera-Badillo, Eduardo Becerril-Vargas, Celia Boukadida, Natividad Cruz-Ortiz, Larissa Fernandes-Matano, Ricardo Grande, Lucia Hernandez-Rivas, Alejandra Hernández-Terán, Pavel Isa, Irma Lopez-Martinez, José Arturo Martínez-Orozco, Margarita Matías-Florentino, Fidencio Mejía-Nepomuceno, Edgar Mendieta-Condado, Mario Mújica-Sánchez, José Esteban Muñoz-Medina, Tatiana Nunez-Garcia, Luis Alberto Ochoa-Carrera, Hector Esteban Paz-Juárez, Francisco Pulido, José Ernesto Ramírez-González, Alma Rincón-Rubio, Teresita Rojas-Mendoza, Jorge Salas-Hernández, Alejandro Sanchez-Flores, Clara Esperanza Santacruz-Tinoco, Andrea Santos Coy-Arechavaleta, Blanca Taboada, Gloria Vazquez, Joel Armando Vázquez-Pérez, Jerome Jean Verleyen |
| EPI_ISL_1302356, EPI_ISL_1302357 | Instituto Nacional de Enfermedades Respiratorias (INER) | Instituto de Biotecnología de la UNAM | Authors from IBT, IMSS, InDRE and INER (in alphabetical order): Carlos F. Arias, Santiago Ávila-Ríos, Gisela Barrera-Badillo, Eduardo Becerril-Vargas, Celia Boukadida, Natividad Cruz-Ortiz, Larissa Fernandes-Matano, Ricardo Grande, Lucia Hernandez-Rivas, Alejandra Hernández-Terán, Pavel Isa, Irma Lopez-Martinez, José Arturo Martínez-Orozco, Margarita Matías-Florentino, Fidencio Mejía-Nepomuceno, Edgar Mendieta-Condado, Mario Mújica-Sánchez, José Esteban Muñoz-Medina, Tatiana Nunez-Garcia, Luis Alberto Ochoa-Carrera, Hector Esteban Paz-Juárez, Francisco Pulido, José Ernesto Ramírez-González, Alma Rincón-Rubio, Teresita Rojas-Mendoza, Jorge Salas-Hernández, Alejandro Sanchez-Flores, Clara Esperanza Santacruz-Tinoco, Andrea Santos Coy-Arechavaleta, Blanca Taboada, Gloria Vazquez, Joel Armando Vázquez-Pérez, Jerome Jean Verleyen |
| EPI_ISL_1302358 | Instituto de Diagnostico y Referencia Epidemiologicos InDRE_RNLSP | Instituto de Biotecnología de la UNAM | Authors from IBT, IMSS, InDRE and INER (in alphabetical order): Carlos F. Arias, Santiago Ávila-Ríos, Gisela Barrera-Badillo, Eduardo Becerril-Vargas, Celia Boukadida, Natividad Cruz-Ortiz, Larissa Fernandes-Matano, Ricardo Grande, Lucia Hernandez-Rivas, Alejandra Hernández-Terán, Pavel Isa, Irma Lopez-Martinez, José Arturo Martínez-Orozco, Margarita Matías-Florentino, Fidencio Mejía-Nepomuceno, Edgar Mendieta-Condado, Mario Mújica-Sánchez, José Esteban Muñoz-Medina, Tatiana Nunez-Garcia, Luis Alberto Ochoa-Carrera, Hector Esteban Paz-Juárez, Francisco Pulido, José Ernesto Ramírez-González, Alma Rincón-Rubio, Teresita Rojas-Mendoza, Jorge Salas-Hernández, Alejandro Sanchez-Flores, Clara Esperanza Santacruz-Tinoco, Andrea Santos Coy-Arechavaleta, Blanca Taboada, Gloria Vazquez, Joel Armando Vázquez-Pérez, Jerome Jean Verleyen |
| EPI_ISL_1302359 | Laboratorio Central de Epidemiología IMSS | Instituto de Biotecnología de la UNAM | Authors from IBT, IMSS, InDRE and INER (in alphabetical order): Carlos F. Arias, Santiago Ávila-Ríos, Gisela Barrera-Badillo, Eduardo Becerril-Vargas, Celia Boukadida, Natividad Cruz-Ortiz, Larissa Fernandes-Matano, Ricardo Grande, Lucia Hernandez-Rivas, Alejandra Hernández-Terán, Pavel Isa, Irma Lopez-Martinez, José Arturo Martínez-Orozco, Margarita Matías-Florentino, Fidencio Mejía-Nepomuceno, Edgar Mendieta-Condado, Mario Mújica-Sánchez, José Esteban Muñoz-Medina, Tatiana Nunez-Garcia, Luis Alberto Ochoa-Carrera, Hector Esteban Paz-Juárez, Francisco Pulido, José Ernesto Ramírez-González, Alma Rincón-Rubio, Teresita Rojas-Mendoza, Jorge Salas-Hernández, Alejandro Sanchez-Flores, Clara Esperanza Santacruz-Tinoco, Andrea Santos Coy-Arechavaleta, Blanca Taboada, Gloria Vazquez, Joel Armando Vázquez-Pérez, Jerome Jean Verleyen |
| EPI_ISL_1302360, EPI_ISL_1302361 | Instituto Nacional de Enfermedades Respiratorias (INER) | Instituto de Biotecnología de la UNAM | Authors from IBT, IMSS, InDRE and INER (in alphabetical order): Carlos F. Arias, Santiago Ávila-Ríos, Gisela Barrera-Badillo, Eduardo Becerril-Vargas, Celia Boukadida, Natividad Cruz-Ortiz, Larissa Fernandes-Matano, Ricardo Grande, Lucia Hernandez-Rivas, Alejandra Hernández-Terán, Pavel Isa, Irma Lopez-Martinez, José Arturo Martínez-Orozco, Margarita Matías-Florentino, Fidencio Mejía-Nepomuceno, Edgar Mendieta-Condado, Mario Mújica-Sánchez, José Esteban Muñoz-Medina, Tatiana Nunez-Garcia, Luis Alberto Ochoa-Carrera, Hector Esteban Paz-Juárez, Francisco Pulido, José Ernesto Ramírez-González, Alma Rincón-Rubio, Teresita Rojas-Mendoza, Jorge Salas-Hernández, Alejandro Sanchez-Flores, Clara Esperanza Santacruz-Tinoco, Andrea Santos Coy-Arechavaleta, Blanca Taboada, Gloria Vazquez, Joel Armando Vázquez-Pérez, Jerome Jean Verleyen |
| EPI_ISL_1302362 | Laboratorio Central de Epidemiología IMSS | Instituto de Biotecnología de la UNAM | Authors from IBT, IMSS, InDRE and INER (in alphabetical order): Carlos F. Arias, Santiago Ávila-Ríos, Gisela Barrera-Badillo, Eduardo Becerril-Vargas, Celia Boukadida, Natividad Cruz-Ortiz, Larissa Fernandes-Matano, Ricardo Grande, Lucia Hernandez-Rivas, Alejandra Hernández-Terán, Pavel Isa, Irma Lopez-Martinez, José Arturo Martínez-Orozco, Margarita Matías-Florentino, Fidencio Mejía-Nepomuceno, Edgar Mendieta-Condado, Mario Mújica-Sánchez, José Esteban Muñoz-Medina, Tatiana Nunez-Garcia, Luis Alberto Ochoa-Carrera, Hector Esteban Paz-Juárez, Francisco Pulido, José Ernesto Ramírez-González, Alma Rincón-Rubio, Teresita Rojas-Mendoza, Jorge Salas-Hernández, Alejandro Sanchez-Flores, Clara Esperanza Santacruz-Tinoco, Andrea Santos Coy-Arechavaleta, Blanca Taboada, Gloria Vazquez, Joel Armando Vázquez-Pérez, Jerome Jean Verleyen |
| EPI_ISL_1302363 | Instituto de Diagnostico y Referencia Epidemiologicos InDRE_RNLSP | Instituto de Biotecnología de la UNAM | Authors from IBT, IMSS, InDRE and INER (in alphabetical order): Carlos F. Arias, Santiago Ávila-Ríos, Gisela Barrera-Badillo, Eduardo Becerril-Vargas, Celia Boukadida, Natividad Cruz-Ortiz, Larissa Fernandes-Matano, Ricardo Grande, Lucia Hernandez-Rivas, Alejandra Hernández-Terán, Pavel Isa, Irma Lopez-Martinez, José Arturo Martínez-Orozco, Margarita Matías-Florentino, Fidencio Mejía-Nepomuceno, Edgar Mendieta-Condado, Mario Mújica-Sánchez, José Esteban Muñoz-Medina, Tatiana Nunez-Garcia, Luis Alberto Ochoa-Carrera, Hector Esteban Paz-Juárez, Francisco Pulido, José Ernesto Ramírez-González, Alma Rincón-Rubio, Teresita Rojas-Mendoza, Jorge Salas-Hernández, Alejandro Sanchez-Flores, Clara Esperanza Santacruz-Tinoco, Andrea Santos Coy-Arechavaleta, Blanca Taboada, Gloria Vazquez, Joel Armando Vázquez-Pérez, Jerome Jean Verleyen |
| EPI_ISL_1302364, EPI_ISL_1302365, EPI_ISL_1302366 | Instituto Nacional de Enfermedades Respiratorias (INER) | Instituto de Biotecnología de la UNAM | Authors from IBT, IMSS, InDRE and INER (in alphabetical order): Carlos F. Arias, Santiago Ávila-Ríos, Gisela Barrera-Badillo, Eduardo Becerril-Vargas, Celia Boukadida, Natividad Cruz-Ortiz, Larissa Fernandes-Matano, Ricardo Grande, Lucia Hernandez-Rivas, Alejandra Hernández-Terán, Pavel Isa, Irma Lopez-Martinez, José Arturo Martínez-Orozco, Margarita Matías-Florentino, Fidencio Mejía-Nepomuceno, Edgar Mendieta-Condado, Mario Mújica-Sánchez, José Esteban Muñoz-Medina, Tatiana Nunez-Garcia, Luis Alberto Ochoa-Carrera, Hector Esteban Paz-Juárez, Francisco Pulido, José Ernesto Ramírez-González, Alma Rincón-Rubio, Teresita Rojas-Mendoza, Jorge Salas-Hernández, Alejandro Sanchez-Flores, Clara Esperanza Santacruz-Tinoco, Andrea Santos Coy-Arechavaleta, Blanca Taboada, Gloria Vazquez, Joel Armando Vázquez-Pérez, Jerome Jean Verleyen |
| EPI_ISL_1302367 | Laboratorio Central de Epidemiología IMSS | Instituto de Biotecnología de la UNAM | Authors from IBT, IMSS, InDRE and INER (in alphabetical order): Carlos F. Arias, Santiago Ávila-Ríos, Gisela Barrera-Badillo, Eduardo Becerril-Vargas, Celia Boukadida, Natividad Cruz-Ortiz, Larissa Fernandes-Matano, Ricardo Grande, Lucia Hernandez-Rivas, Alejandra Hernández-Terán, Pavel Isa, Irma Lopez-Martinez, José Arturo Martínez-Orozco, Margarita Matías-Florentino, Fidencio Mejía-Nepomuceno, Edgar Mendieta-Condado, Mario Mújica-Sánchez, José Esteban Muñoz-Medina, Tatiana Nunez-Garcia, Luis Alberto Ochoa-Carrera, Hector Esteban Paz-Juárez, Francisco Pulido, José Ernesto Ramírez-González, Alma Rincón-Rubio, Teresita Rojas-Mendoza, Jorge Salas-Hernández, Alejandro Sanchez-Flores, Clara Esperanza Santacruz-Tinoco, Andrea Santos Coy-Arechavaleta, Blanca Taboada, Gloria Vazquez, Joel Armando Vázquez-Pérez, Jerome Jean Verleyen |
| EPI_ISL_1302368, EPI_ISL_1302369 | Instituto Nacional de Enfermedades Respiratorias (INER) | Instituto de Biotecnología de la UNAM | Authors from IBT, IMSS, InDRE and INER (in alphabetical order): Carlos F. Arias, Santiago Ávila-Ríos, Gisela Barrera-Badillo, Eduardo Becerril-Vargas, Celia Boukadida, Natividad Cruz-Ortiz, Larissa Fernandes-Matano, Ricardo Grande, Lucia Hernandez-Rivas, Alejandra Hernández-Terán, Pavel Isa, Irma Lopez-Martinez, José Arturo Martínez-Orozco, Margarita Matías-Florentino, Fidencio Mejía-Nepomuceno, Edgar Mendieta-Condado, Mario Mújica-Sánchez, José Esteban Muñoz-Medina, Tatiana Nunez-Garcia, Luis Alberto Ochoa-Carrera, Hector Esteban Paz-Juárez, Francisco |

| | | | |
|---|---|---|---|
| | | | Pulido, José Ernesto Ramírez-González, Alma Rincón-Rubio, Teresita Rojas-Mendoza, Jorge Salas-Hernández, Alejandro Sanchez-Flores, Clara Esperanza Santacruz-Tinoco, Andrea Santos Coy-Arechavaleta, Blanca Taboada, Gloria Vazquez, Joel Armando Vázquez-Pérez, Jerome Jean Verleyen |
| EPI_ISL_1302370 | Laboratorio Central de Epidemiología IMSS | Instituto de Biotecnología de la UNAM | Authors from IBT, IMSS, InDRE and INER (in alphabetical order): Carlos F. Arias, Santiago Ávila-Ríos, Gisela Barrera-Badillo, Eduardo Becerril-Vargas, Celia Boukadida, Natividad Cruz-Ortiz, Larissa Fernandes-Matano, Ricardo Grande, Lucia Hernandez-Rivas, Alejandra Hernández-Terán, Pavel Isa, Irma Lopez-Martinez, José Arturo Martínez-Orozco, Margarita Matías-Florentino, Fidencio Mejía-Nepomuceno, Edgar Mendieta-Condado, Mario Mújica-Sánchez, José Esteban Muñoz-Medina, Tatiana Nunez-Garcia, Luis Alberto Ochoa-Carrera, Hector Esteban Paz-Juárez, Francisco Pulido, José Ernesto Ramírez-González, Alma Rincón-Rubio, Teresita Rojas-Mendoza, Jorge Salas-Hernández, Alejandro Sanchez-Flores, Clara Esperanza Santacruz-Tinoco, Andrea Santos Coy-Arechavaleta, Blanca Taboada, Gloria Vazquez, Joel Armando Vázquez-Pérez, Jerome Jean Verleyen |
| EPI_ISL_1302371 | Instituto de Diagnostico y Referencia Epidemiologicos InDRE_RNLSP | Instituto de Biotecnología de la UNAM | Authors from IBT, IMSS, InDRE and INER (in alphabetical order): Carlos F. Arias, Santiago Ávila-Ríos, Gisela Barrera-Badillo, Eduardo Becerril-Vargas, Celia Boukadida, Natividad Cruz-Ortiz, Larissa Fernandes-Matano, Ricardo Grande, Lucia Hernandez-Rivas, Alejandra Hernández-Terán, Pavel Isa, Irma Lopez-Martinez, José Arturo Martínez-Orozco, Margarita Matías-Florentino, Fidencio Mejía-Nepomuceno, Edgar Mendieta-Condado, Mario Mújica-Sánchez, José Esteban Muñoz-Medina, Tatiana Nunez-Garcia, Luis Alberto Ochoa-Carrera, Hector Esteban Paz-Juárez, Francisco Pulido, José Ernesto Ramírez-González, Alma Rincón-Rubio, Teresita Rojas-Mendoza, Jorge Salas-Hernández, Alejandro Sanchez-Flores, Clara Esperanza Santacruz-Tinoco, Andrea Santos Coy-Arechavaleta, Blanca Taboada, Gloria Vazquez, Joel Armando Vázquez-Pérez, Jerome Jean Verleyen |
| EPI_ISL_1302372 | Laboratorio Central de Epidemiología IMSS | Instituto de Biotecnología de la UNAM | Authors from IBT, IMSS, InDRE and INER (in alphabetical order): Carlos F. Arias, Santiago Ávila-Ríos, Gisela Barrera-Badillo, Eduardo Becerril-Vargas, Celia Boukadida, Natividad Cruz-Ortiz, Larissa Fernandes-Matano, Ricardo Grande, Lucia Hernandez-Rivas, Alejandra Hernández-Terán, Pavel Isa, Irma Lopez-Martinez, José Arturo Martínez-Orozco, Margarita Matías-Florentino, Fidencio Mejía-Nepomuceno, Edgar Mendieta-Condado, Mario Mújica-Sánchez, José Esteban Muñoz-Medina, Tatiana Nunez-Garcia, Luis Alberto Ochoa-Carrera, Hector Esteban Paz-Juárez, Francisco Pulido, José Ernesto Ramírez-González, Alma Rincón-Rubio, Teresita Rojas-Mendoza, Jorge Salas-Hernández, Alejandro Sanchez-Flores, Clara Esperanza Santacruz-Tinoco, Andrea Santos Coy-Arechavaleta, Blanca Taboada, Gloria Vazquez, Joel Armando Vázquez-Pérez, Jerome Jean Verleyen |
| EPI_ISL_1302373, EPI_ISL_1302374, EPI_ISL_1302375, EPI_ISL_1302376, EPI_ISL_1302377 | Instituto Nacional de Enfermedades Respiratorias (INER) | Instituto de Biotecnología de la UNAM | Authors from IBT, IMSS, InDRE and INER (in alphabetical order): Carlos F. Arias, Santiago Ávila-Ríos, Gisela Barrera-Badillo, Eduardo Becerril-Vargas, Celia Boukadida, Natividad Cruz-Ortiz, Larissa Fernandes-Matano, Ricardo Grande, Lucia Hernandez-Rivas, Alejandra Hernández-Terán, Pavel Isa, Irma Lopez-Martinez, José Arturo Martínez-Orozco, Margarita Matías-Florentino, Fidencio Mejía-Nepomuceno, Edgar Mendieta-Condado, Mario Mújica-Sánchez, José Esteban Muñoz-Medina, Tatiana Nunez-Garcia, Luis Alberto Ochoa-Carrera, Hector Esteban Paz-Juárez, Francisco Pulido, José Ernesto Ramírez-González, Alma Rincón-Rubio, Teresita Rojas-Mendoza, Jorge Salas-Hernández, Alejandro Sanchez-Flores, Clara Esperanza Santacruz-Tinoco, Andrea Santos Coy-Arechavaleta, Blanca Taboada, Gloria Vazquez, Joel Armando Vázquez-Pérez, Jerome Jean Verleyen |
| EPI_ISL_1302378, EPI_ISL_1302379 | Laboratorio Central de Epidemiología IMSS | Instituto de Biotecnología de la UNAM | Authors from IBT, IMSS, InDRE and INER (in alphabetical order): Carlos F. Arias, Santiago Ávila-Ríos, Gisela Barrera-Badillo, Eduardo Becerril-Vargas, Celia Boukadida, Natividad Cruz-Ortiz, Larissa Fernandes-Matano, Ricardo Grande, Lucia Hernandez-Rivas, Alejandra Hernández-Terán, Pavel Isa, Irma Lopez-Martinez, José Arturo Martínez-Orozco, Margarita Matías-Florentino, Fidencio Mejía-Nepomuceno, Edgar Mendieta-Condado, Mario Mújica-Sánchez, José Esteban Muñoz-Medina, Tatiana Nunez-Garcia, Luis Alberto Ochoa-Carrera, Hector Esteban Paz-Juárez, Francisco Pulido, José Ernesto Ramírez-González, Alma Rincón-Rubio, Teresita Rojas-Mendoza, Jorge Salas-Hernández, Alejandro Sanchez-Flores, Clara Esperanza Santacruz-Tinoco, Andrea Santos Coy-Arechavaleta, Blanca Taboada, Gloria Vazquez, Joel Armando Vázquez-Pérez, Jerome Jean Verleyen |
| EPI_ISL_1302380 | Instituto de Diagnostico y Referencia Epidemiologicos InDRE_RNLSP | Instituto de Biotecnología de la UNAM | Authors from IBT, IMSS, InDRE and INER (in alphabetical order): Carlos F. Arias, Santiago Ávila-Ríos, Gisela Barrera-Badillo, Eduardo Becerril-Vargas, Celia Boukadida, Natividad Cruz-Ortiz, Larissa Fernandes-Matano, Ricardo Grande, Lucia Hernandez-Rivas, Alejandra Hernández-Terán, Pavel Isa, Irma Lopez-Martinez, José Arturo Martínez-Orozco, Margarita Matías-Florentino, Fidencio Mejía-Nepomuceno, Edgar Mendieta-Condado, Mario Mújica-Sánchez, José Esteban Muñoz-Medina, Tatiana Nunez-Garcia, Luis Alberto Ochoa-Carrera, Hector Esteban Paz-Juárez, Francisco Pulido, José Ernesto Ramírez-González, Alma Rincón-Rubio, Teresita Rojas-Mendoza, Jorge Salas-Hernández, Alejandro Sanchez-Flores, Clara Esperanza Santacruz-Tinoco, Andrea Santos Coy-Arechavaleta, Blanca Taboada, Gloria Vazquez, Joel Armando Vázquez-Pérez, Jerome Jean Verleyen |
| EPI_ISL_1302381 | Instituto Nacional de Enfermedades Respiratorias (INER) | Instituto de Biotecnología de la UNAM | Authors from IBT, IMSS, InDRE and INER (in alphabetical order): Carlos F. Arias, Santiago Ávila-Ríos, Gisela Barrera-Badillo, Eduardo Becerril-Vargas, Celia Boukadida, Natividad Cruz-Ortiz, Larissa Fernandes-Matano, Ricardo Grande, Lucia Hernandez-Rivas, Alejandra Hernández-Terán, Pavel Isa, Irma Lopez-Martinez, José Arturo Martínez-Orozco, Margarita Matías-Florentino, Fidencio Mejía-Nepomuceno, Edgar Mendieta-Condado, Mario Mújica-Sánchez, José Esteban Muñoz-Medina, Tatiana Nunez-Garcia, Luis Alberto Ochoa-Carrera, Hector Esteban Paz-Juárez, Francisco Pulido, José Ernesto Ramírez-González, Alma Rincón-Rubio, Teresita Rojas-Mendoza, Jorge Salas-Hernández, Alejandro Sanchez-Flores, Clara Esperanza Santacruz-Tinoco, Andrea Santos Coy-Arechavaleta, Blanca Taboada, Gloria Vazquez, Joel Armando Vázquez-Pérez, Jerome Jean Verleyen |
| EPI_ISL_1302382 | Instituto de Diagnostico y Referencia Epidemiologicos InDRE_RNLSP | Instituto de Biotecnología de la UNAM | Authors from IBT, IMSS, InDRE and INER (in alphabetical order): Carlos F. Arias, Santiago Ávila-Ríos, Gisela Barrera-Badillo, Eduardo Becerril-Vargas, Celia Boukadida, Natividad Cruz-Ortiz, Larissa Fernandes-Matano, Ricardo Grande, Lucia Hernandez-Rivas, Alejandra Hernández-Terán, Pavel Isa, Irma Lopez-Martinez, José Arturo Martínez-Orozco, Margarita Matías-Florentino, Fidencio Mejía-Nepomuceno, Edgar Mendieta-Condado, Mario Mújica-Sánchez, José Esteban Muñoz-Medina, Tatiana Nunez-Garcia, Luis Alberto Ochoa-Carrera, Hector Esteban Paz-Juárez, Francisco Pulido, José Ernesto Ramírez-González, Alma Rincón-Rubio, Teresita Rojas-Mendoza, Jorge Salas-Hernández, Alejandro Sanchez-Flores, Clara Esperanza Santacruz-Tinoco, Andrea Santos Coy-Arechavaleta, Blanca Taboada, Gloria Vazquez, Joel Armando Vázquez-Pérez, Jerome Jean Verleyen |
| EPI_ISL_1302383, EPI_ISL_1302384, EPI_ISL_1302385, EPI_ISL_1302386, EPI_ISL_1302387 | Instituto Nacional de Enfermedades Respiratorias (INER) | Instituto de Biotecnología de la UNAM | Authors from IBT, IMSS, InDRE and INER (in alphabetical order): Carlos F. Arias, Santiago Ávila-Ríos, Gisela Barrera-Badillo, Eduardo Becerril-Vargas, Celia Boukadida, Natividad Cruz-Ortiz, Larissa Fernandes-Matano, Ricardo Grande, Lucia Hernandez-Rivas, Alejandra Hernández-Terán, Pavel Isa, Irma Lopez-Martinez, José Arturo Martínez-Orozco, Margarita Matías-Florentino, Fidencio Mejía-Nepomuceno, Edgar Mendieta-Condado, Mario Mújica-Sánchez, José Esteban Muñoz-Medina, Tatiana Nunez-Garcia, Luis Alberto Ochoa-Carrera, Hector Esteban Paz-Juárez, Francisco Pulido, José Ernesto Ramírez-González, Alma Rincón-Rubio, Teresita Rojas-Mendoza, Jorge Salas-Hernández, Alejandro Sanchez-Flores, Clara Esperanza Santacruz-Tinoco, Andrea Santos Coy-Arechavaleta, Blanca Taboada, Gloria Vazquez, Joel Armando Vázquez-Pérez, Jerome Jean Verleyen |
| EPI_ISL_1302388 | Instituto de Diagnostico y Referencia Epidemiologicos InDRE_RNLSP | Instituto de Biotecnología de la UNAM | Authors from IBT, IMSS, InDRE and INER (in alphabetical order): Carlos F. Arias, Santiago Ávila-Ríos, Gisela Barrera-Badillo, Eduardo Becerril-Vargas, Celia Boukadida, Natividad Cruz-Ortiz, Larissa Fernandes-Matano, Ricardo Grande, Lucia Hernandez-Rivas, Alejandra Hernández-Terán, Pavel Isa, Irma Lopez-Martinez, José Arturo Martínez-Orozco, Margarita Matías-Florentino, Fidencio Mejía-Nepomuceno, Edgar Mendieta-Condado, Mario Mújica-Sánchez, José Esteban Muñoz-Medina, Tatiana Nunez-Garcia, Luis Alberto Ochoa-Carrera, Hector Esteban Paz-Juárez, Francisco Pulido, José Ernesto Ramírez-González, Alma Rincón-Rubio, Teresita Rojas-Mendoza, Jorge Salas-Hernández, Alejandro Sanchez-Flores, Clara Esperanza Santacruz-Tinoco, Andrea Santos Coy-Arechavaleta, Blanca Taboada, Gloria Vazquez, Joel Armando Vázquez-Pérez, Jerome Jean Verleyen |
| EPI_ISL_1302389 | Instituto Nacional de Enfermedades Respiratorias (INER) | Instituto de Biotecnología de la UNAM | Authors from IBT, IMSS, InDRE and INER (in alphabetical order): Carlos F. Arias, Santiago Ávila-Ríos, Gisela Barrera-Badillo, Eduardo Becerril-Vargas, Celia Boukadida, Natividad Cruz-Ortiz, Larissa Fernandes-Matano, Ricardo Grande, Lucia Hernandez-Rivas, Alejandra Hernández-Terán, Pavel Isa, Irma Lopez-Martinez, José Arturo Martínez-Orozco, Margarita Matías-Florentino, Fidencio Mejía-Nepomuceno, Edgar Mendieta-Condado, Mario Mújica-Sánchez, José Esteban Muñoz-Medina, Tatiana Nunez-Garcia, Luis Alberto Ochoa-Carrera, Hector Esteban Paz-Juárez, Francisco Pulido, José Ernesto Ramírez-González, Alma Rincón-Rubio, Teresita Rojas-Mendoza, Jorge Salas-Hernández, Alejandro Sanchez-Flores, Clara Esperanza Santacruz-Tinoco, Andrea Santos Coy-Arechavaleta, Blanca Taboada, Gloria Vazquez, Joel Armando Vázquez-Pérez, Jerome Jean Verleyen |
| EPI_ISL_1302390 | Laboratorio Central de Epidemiología IMSS | Instituto de Biotecnología de la UNAM | Authors from IBT, IMSS, InDRE and INER (in alphabetical order): Carlos F. Arias, Santiago Ávila-Ríos, Gisela Barrera-Badillo, Eduardo Becerril-Vargas, Celia Boukadida, Natividad Cruz-Ortiz, Larissa Fernandes-Matano, Ricardo Grande, Lucia Hernandez-Rivas, Alejandra Hernández-Terán, Pavel Isa, Irma Lopez-Martinez, José Arturo Martínez-Orozco, Margarita Matías-Florentino, Fidencio Mejía-Nepomuceno, Edgar Mendieta-Condado, Mario Mújica-Sánchez, José Esteban Muñoz-Medina, Tatiana Nunez-Garcia, Luis Alberto Ochoa-Carrera, Hector Esteban Paz-Juárez, Francisco Pulido, José Ernesto Ramírez-González, Alma Rincón-Rubio, Teresita Rojas-Mendoza, Jorge Salas-Hernández, Alejandro Sanchez-Flores, Clara Esperanza Santacruz-Tinoco, Andrea Santos Coy-Arechavaleta, Blanca Taboada, Gloria Vazquez, Joel Armando Vázquez-Pérez, Jerome Jean Verleyen |
| EPI_ISL_1302391 | Instituto Nacional de Enfermedades Respiratorias (INER) | Instituto de Biotecnología de la UNAM | Authors from IBT, IMSS, InDRE and INER (in alphabetical order): Carlos F. Arias, Santiago Ávila-Ríos, Gisela Barrera-Badillo, Eduardo Becerril-Vargas, Celia Boukadida, Natividad Cruz-Ortiz, Larissa Fernandes-Matano, Ricardo Grande, Lucia Hernandez-Rivas, Alejandra Hernández-Terán, Pavel Isa, Irma Lopez-Martinez, José Arturo Martínez-Orozco, Margarita Matías-Florentino, Fidencio Mejía-Nepomuceno, Edgar Mendieta-Condado, Mario Mújica-Sánchez, José Esteban Muñoz-Medina, Tatiana Nunez-Garcia, Luis Alberto Ochoa-Carrera, Hector Esteban Paz-Juárez, Francisco Pulido, José Ernesto Ramírez-González, Alma Rincón-Rubio, Teresita Rojas-Mendoza, Jorge Salas-Hernández, Alejandro Sanchez-Flores, Clara Esperanza Santacruz-Tinoco, Andrea Santos Coy-Arechavaleta, Blanca Taboada, Gloria Vazquez, Joel Armando Vázquez-Pérez, Jerome Jean Verleyen |
| EPI_ISL_1302392, EPI_ISL_1302393 | Laboratorio Central de Epidemiología IMSS | Instituto de Biotecnología de la UNAM | Authors from IBT, IMSS, InDRE and INER (in alphabetical order): Carlos F. Arias, Santiago Ávila-Ríos, Gisela Barrera-Badillo, Eduardo Becerril-Vargas, Celia Boukadida, Natividad Cruz-Ortiz, Larissa Fernandes-Matano, Ricardo Grande, Lucia Hernandez-Rivas, Alejandra Hernández-Terán, Pavel Isa, Irma Lopez-Martinez, José Arturo Martínez-Orozco, Margarita Matías-Florentino, Fidencio Mejía-Nepomuceno, Edgar Mendieta-Condado, Mario Mújica-Sánchez, José Esteban Muñoz-Medina, Tatiana Nunez-Garcia, Luis Alberto Ochoa-Carrera, Hector Esteban Paz-Juárez, Francisco Pulido, José Ernesto Ramírez-González, Alma Rincón-Rubio, Teresita Rojas-Mendoza, Jorge Salas-Hernández, Alejandro Sanchez-Flores, Clara Esperanza Santacruz-Tinoco, Andrea Santos Coy-Arechavaleta, Blanca Taboada, Gloria Vazquez, Joel Armando Vázquez-Pérez, Jerome Jean Verleyen |
| EPI_ISL_1302394, EPI_ISL_1302395 | Instituto Nacional de Enfermedades Respiratorias (INER) | Instituto de Biotecnología de la UNAM | Authors from IBT, IMSS, InDRE and INER (in alphabetical order): Carlos F. Arias, Santiago Ávila-Ríos, Gisela Barrera-Badillo, Eduardo Becerril-Vargas, Celia Boukadida, Natividad Cruz-Ortiz, Larissa Fernandes-Matano, Ricardo Grande, Lucia Hernandez-Rivas, Alejandra Hernández-Terán, Pavel Isa, Irma Lopez-Martinez, José Arturo Martínez-Orozco, Margarita Matías-Florentino, Fidencio Mejía-Nepomuceno, Edgar Mendieta-Condado, Mario Mújica-Sánchez, José Esteban Muñoz-Medina, Tatiana Nunez-Garcia, Luis Alberto Ochoa-Carrera, Hector Esteban Paz-Juárez, Francisco Pulido, José Ernesto Ramírez-González, Alma Rincón-Rubio, Teresita Rojas-Mendoza, Jorge Salas-Hernández, Alejandro Sanchez-Flores, Clara Esperanza Santacruz-Tinoco, Andrea Santos Coy-Arechavaleta, Blanca Taboada, Gloria Vazquez, Joel Armando Vázquez-Pérez, Jerome Jean Verleyen |
| EPI_ISL_1302396, EPI_ISL_1302397, EPI_ISL_1302398 | Instituto de Diagnostico y Referencia Epidemiologicos InDRE_RNLSP | Instituto de Biotecnología de la UNAM | Authors from IBT, IMSS, InDRE and INER (in alphabetical order): Carlos F. Arias, Santiago Ávila-Ríos, Gisela Barrera-Badillo, Eduardo Becerril-Vargas, Celia Boukadida, Natividad Cruz-Ortiz, Larissa Fernandes-Matano, Ricardo Grande, Lucia Hernandez-Rivas, Alejandra Hernández-Terán, Pavel Isa, Irma Lopez-Martinez, José Arturo Martínez-Orozco, Margarita Matías-Florentino, Fidencio Mejía-Nepomuceno, Edgar Mendieta-Condado, Mario Mújica-Sánchez, José Esteban Muñoz-Medina, Tatiana Nunez-Garcia, Luis Alberto Ochoa-Carrera, Hector Esteban Paz-Juárez, Francisco Pulido, José Ernesto Ramírez-González, Alma Rincón-Rubio, Teresita Rojas-Mendoza, Jorge Salas-Hernández, Alejandro Sanchez-Flores, Clara Esperanza Santacruz-Tinoco, Andrea Santos Coy-Arechavaleta, Blanca Taboada, Gloria Vazquez, Joel Armando Vázquez-Pérez, Jerome Jean Verleyen |
| EPI_ISL_1302399, EPI_ISL_1302400 | Instituto Nacional de Enfermedades Respiratorias (INER) | Instituto de Biotecnología de la UNAM | Authors from IBT, IMSS, InDRE and INER (in alphabetical order): Carlos F. Arias, Santiago Ávila-Ríos, Gisela Barrera-Badillo, Eduardo Becerril-Vargas, Celia Boukadida, Natividad Cruz-Ortiz, Larissa Fernandes-Matano, Ricardo Grande, Lucia Hernandez-Rivas, Alejandra Hernández-Terán, Pavel Isa, Irma Lopez-Martinez, José Arturo Martínez-Orozco, Margarita Matías-Florentino, Fidencio Mejía-Nepomuceno, Edgar Mendieta-Condado, Mario Mújica-Sánchez, José Esteban Muñoz-Medina, Tatiana Nunez-Garcia, Luis Alberto Ochoa-Carrera, Hector Esteban Paz-Juárez, Francisco Pulido, José Ernesto Ramírez-González, Alma Rincón-Rubio, Teresita Rojas-Mendoza, Jorge Salas-Hernández, Alejandro Sanchez-Flores, Clara Esperanza Santacruz-Tinoco, Andrea Santos Coy-Arechavaleta, Blanca Taboada, Gloria Vazquez, Joel Armando Vázquez-Pérez, Jerome Jean Verleyen |
| EPI_ISL_1302401 | Laboratorio Central de Epidemiología IMSS | Instituto de Biotecnología de la UNAM | Authors from IBT, IMSS, InDRE and INER (in alphabetical order): Carlos F. Arias, Santiago Ávila-Ríos, Gisela Barrera-Badillo, Eduardo Becerril-Vargas, Celia Boukadida, Natividad Cruz-Ortiz, Larissa Fernandes-Matano, Ricardo Grande, Lucia Hernandez-Rivas, Alejandra Hernández-Terán, Pavel Isa, Irma Lopez-Martinez, José Arturo Martínez-Orozco, Margarita Matías-Florentino, Fidencio Mejía-Nepomuceno, Edgar Mendieta-Condado, Mario Mújica-Sánchez, José Esteban Muñoz-Medina, Tatiana Nunez-Garcia, Luis Alberto Ochoa-Carrera, Hector Esteban Paz-Juárez, Francisco Pulido, José Ernesto Ramírez-González, Alma Rincón-Rubio, Teresita Rojas-Mendoza, Jorge Salas-Hernández, Alejandro Sanchez-Flores, Clara Esperanza Santacruz-Tinoco, Andrea Santos Coy-Arechavaleta, Blanca Taboada, Gloria Vazquez, Joel Armando Vázquez-Pérez, Jerome Jean Verleyen |
| EPI_ISL_1302682, EPI_ISL_1302683, EPI_ISL_1302684 | Kentucky State Public Health Lab | Kentucky State Public Health Lab | Stephanie Lunn, Karim George, Joshua Tobias, William Grooms, Vaneet Arora, Matthew Johnson, Rachel Zinner, Rhonda Lucas |
| EPI_ISL_1302685, EPI_ISL_1302686 | Gravity Diagnostics | Kentucky State Public Health Lab | Stephanie Lunn, Karim George, Joshua Tobias, William Grooms, Vaneet Arora, Matthew Johnson, Rachel Zinner, Rhonda Lucas |
| EPI_ISL_1302687, EPI_ISL_1302688, EPI_ISL_1302689, EPI_ISL_1302690, EPI_ISL_1302691, EPI_ISL_1302692, EPI_ISL_1302693 | Kentucky State Public Health Lab | Kentucky State Public Health Lab | Stephanie Lunn, Karim George, Joshua Tobias, William Grooms, Vaneet Arora, Matthew Johnson, Rachel Zinner, Rhonda Lucas |
| EPI_ISL_1302694, EPI_ISL_1302695, EPI_ISL_1302696, EPI_ISL_1302697 | Gravity Diagnostics | Kentucky State Public Health Lab | Stephanie Lunn, Karim George, Joshua Tobias, William Grooms, Vaneet Arora, Matthew Johnson, Rachel Zinner, Rhonda Lucas |
| EPI_ISL_1302698 | Kentucky State Public Health Lab | Kentucky State Public Health Lab | Stephanie Lunn, Karim George, Joshua Tobias, William Grooms, Vaneet Arora, Matthew Johnson, Rachel Zinner, Rhonda Lucas |
| EPI_ISL_1302699, EPI_ISL_1302700 | Gravity Diagnostics | Kentucky State Public Health Lab | Stephanie Lunn, Karim George, Joshua Tobias, William Grooms, Vaneet Arora, Matthew Johnson, Rachel Zinner, Rhonda Lucas |
| EPI_ISL_1302701, EPI_ISL_1302702, EPI_ISL_1302703, EPI_ISL_1302704, | Kentucky State Public Health Lab | Kentucky State Public Health Lab | Stephanie Lunn, Karim George, Joshua Tobias, William Grooms, Vaneet Arora, Matthew Johnson, Rachel Zinner, Rhonda Lucas |

| | | | |
|---|---|---|---|
| EPI_ISL_1302705, EPI_ISL_1302706, EPI_ISL_1302707 | | | |
| EPI_ISL_1302708, EPI_ISL_1302709, EPI_ISL_1302710, EPI_ISL_1302711, EPI_ISL_1302712, EPI_ISL_1302713, EPI_ISL_1302714, EPI_ISL_1302715, EPI_ISL_1302716 | Gravity Diagnostics | Kentucky State Public Health Lab | Stephanie Lunn, Karim George, Joshua Tobias, William Grooms, Vaneet Arora, Matthew Johnson, Rachel Zinner, Rhonda Lucas |
| EPI_ISL_1303033, EPI_ISL_1303034, EPI_ISL_1303038, EPI_ISL_1303040, EPI_ISL_1303041, EPI_ISL_1303042, EPI_ISL_1303043, EPI_ISL_1303044, EPI_ISL_1303045, EPI_ISL_1303046, EPI_ISL_1303047, EPI_ISL_1303048, EPI_ISL_1303049, EPI_ISL_1303050, EPI_ISL_1303051, EPI_ISL_1303052, EPI_ISL_1303053, EPI_ISL_1303054, EPI_ISL_1303055, EPI_ISL_1303056, EPI_ISL_1303057, EPI_ISL_1303058, EPI_ISL_1303059, EPI_ISL_1303060, EPI_ISL_1303061, EPI_ISL_1303062, EPI_ISL_1303063, EPI_ISL_1303064, EPI_ISL_1303065, EPI_ISL_1303066, EPI_ISL_1303067, EPI_ISL_1303069, EPI_ISL_1303070, EPI_ISL_1303071, EPI_ISL_1303072, EPI_ISL_1303073, EPI_ISL_1303074, EPI_ISL_1303075, EPI_ISL_1303076, EPI_ISL_1303077, EPI_ISL_1303078, EPI_ISL_1303079, EPI_ISL_1303080, EPI_ISL_1303081, EPI_ISL_1303082, EPI_ISL_1303083, EPI_ISL_1303084, EPI_ISL_1303086, EPI_ISL_1303087, EPI_ISL_1303088, EPI_ISL_1303089, EPI_ISL_1303090, EPI_ISL_1303092, EPI_ISL_1303093, EPI_ISL_1303094, EPI_ISL_1303095, EPI_ISL_1303096, EPI_ISL_1303097, EPI_ISL_1303098, EPI_ISL_1303099, EPI_ISL_1303100, EPI_ISL_1303101, EPI_ISL_1303102, EPI_ISL_1303103, EPI_ISL_1303104, EPI_ISL_1303105, EPI_ISL_1303106, EPI_ISL_1303107, EPI_ISL_1303109, EPI_ISL_1303110, EPI_ISL_1303111, EPI_ISL_1303112, EPI_ISL_1303114, EPI_ISL_1303115, EPI_ISL_1303116, EPI_ISL_1303118, EPI_ISL_1303119, EPI_ISL_1303120, EPI_ISL_1303121, EPI_ISL_1303122, EPI_ISL_1303123, EPI_ISL_1303124, EPI_ISL_1303125, EPI_ISL_1303126, EPI_ISL_1303127, EPI_ISL_1303128, EPI_ISL_1303129, EPI_ISL_1303130, EPI_ISL_1303131, EPI_ISL_1303132, EPI_ISL_1303133, EPI_ISL_1303134, EPI_ISL_1303135, EPI_ISL_1303136, EPI_ISL_1303137, EPI_ISL_1303138, EPI_ISL_1303139, EPI_ISL_1303140, EPI_ISL_1303141, EPI_ISL_1303142, EPI_ISL_1303143, EPI_ISL_1303144, EPI_ISL_1303145, EPI_ISL_1303146, EPI_ISL_1303147, EPI_ISL_1303148, EPI_ISL_1303149, EPI_ISL_1303150, EPI_ISL_1303151, EPI_ISL_1303152, EPI_ISL_1303153, EPI_ISL_1303154, EPI_ISL_1303155, EPI_ISL_1303156, EPI_ISL_1303157, EPI_ISL_1303158, EPI_ISL_1303159, EPI_ISL_1303160, EPI_ISL_1303161, EPI_ISL_1303162, EPI_ISL_1303163, EPI_ISL_1303164, EPI_ISL_1303165, EPI_ISL_1303168, EPI_ISL_1303169, EPI_ISL_1303170, EPI_ISL_1303171, EPI_ISL_1303172, EPI_ISL_1303173, EPI_ISL_1303174, EPI_ISL_1303175, EPI_ISL_1303176, EPI_ISL_1303177, EPI_ISL_1303178, EPI_ISL_1303180, EPI_ISL_1303181, EPI_ISL_1303182, EPI_ISL_1303183, EPI_ISL_1303184, EPI_ISL_1303185, EPI_ISL_1303186, EPI_ISL_1303187, EPI_ISL_1303188, EPI_ISL_1303190, EPI_ISL_1303191, EPI_ISL_1303192, EPI_ISL_1303193, EPI_ISL_1303194, EPI_ISL_1303195, EPI_ISL_1303196, EPI_ISL_1303197, EPI_ISL_1303198, EPI_ISL_1303199, EPI_ISL_1303200, EPI_ISL_1303201, EPI_ISL_1303202, EPI_ISL_1303203, EPI_ISL_1303204, EPI_ISL_1303205, EPI_ISL_1303206, EPI_ISL_1303207, EPI_ISL_1303208, EPI_ISL_1303209, EPI_ISL_1303210, EPI_ISL_1303211, EPI_ISL_1303212, EPI_ISL_1303213, EPI_ISL_1303214, EPI_ISL_1303215, EPI_ISL_1303216, EPI_ISL_1303217, EPI_ISL_1303218, EPI_ISL_1303219, EPI_ISL_1303220, EPI_ISL_1303221, EPI_ISL_1303222, EPI_ISL_1303223, EPI_ISL_1303224, EPI_ISL_1303225, EPI_ISL_1303226, EPI_ISL_1303227, EPI_ISL_1303228, EPI_ISL_1303229, EPI_ISL_1303230, EPI_ISL_1303231, EPI_ISL_1303232, EPI_ISL_1303233, EPI_ISL_1303234, EPI_ISL_1303235, EPI_ISL_1303236, EPI_ISL_1303237, EPI_ISL_1303239, EPI_ISL_1303240, EPI_ISL_1303241, EPI_ISL_1303242, EPI_ISL_1303243, EPI_ISL_1303245, EPI_ISL_1303246, EPI_ISL_1303247, EPI_ISL_1303248, EPI_ISL_1303249, EPI_ISL_1303251, EPI_ISL_1303252, EPI_ISL_1303253, EPI_ISL_1303254, EPI_ISL_1303255, EPI_ISL_1303256, EPI_ISL_1303257, EPI_ISL_1303258, EPI_ISL_1303259, EPI_ISL_1303260, EPI_ISL_1303261, EPI_ISL_1303262, EPI_ISL_1303263, EPI_ISL_1303264, EPI_ISL_1303265, EPI_ISL_1303266, EPI_ISL_1303267, EPI_ISL_1303268, EPI_ISL_1303269, EPI_ISL_1303271, EPI_ISL_1303272, EPI_ISL_1303273, EPI_ISL_1303274, EPI_ISL_1303275, EPI_ISL_1303276, EPI_ISL_1303277, EPI_ISL_1303278, EPI_ISL_1303279, EPI_ISL_1303280, EPI_ISL_1303281, EPI_ISL_1303282, EPI_ISL_1303283, EPI_ISL_1303285, EPI_ISL_1303286, EPI_ISL_1303287, EPI_ISL_1303288, EPI_ISL_1303289, EPI_ISL_1303290, EPI_ISL_1303292, EPI_ISL_1303293, EPI_ISL_1303294, EPI_ISL_1303295, EPI_ISL_1303296, EPI_ISL_1303297, EPI_ISL_1303298, EPI_ISL_1303299, EPI_ISL_1303300, EPI_ISL_1303301, EPI_ISL_1303302, EPI_ISL_1303303, EPI_ISL_1303304, EPI_ISL_1303305, EPI_ISL_1303306, EPI_ISL_1303307, EPI_ISL_1303308, EPI_ISL_1303309, EPI_ISL_1303310, EPI_ISL_1303312, EPI_ISL_1303313, EPI_ISL_1303314, EPI_ISL_1303315, EPI_ISL_1303316, EPI_ISL_1303317, EPI_ISL_1303319, EPI_ISL_1303320, EPI_ISL_1303321, EPI_ISL_1303322, EPI_ISL_1303323, EPI_ISL_1303324, EPI_ISL_1303325, EPI_ISL_1303327, EPI_ISL_1303328, EPI_ISL_1303329, EPI_ISL_1303330, EPI_ISL_1303331, EPI_ISL_1303332, EPI_ISL_1303334, EPI_ISL_1303335, EPI_ISL_1303336, EPI_ISL_1303338, EPI_ISL_1303339, EPI_ISL_1303340, EPI_ISL_1303341, EPI_ISL_1303342, EPI_ISL_1303343, EPI_ISL_1303344, EPI_ISL_1303345, EPI_ISL_1303346, EPI_ISL_1303347, EPI_ISL_1303348, EPI_ISL_1303349, EPI_ISL_1303350, EPI_ISL_1303351, EPI_ISL_1303352, EPI_ISL_1303353, EPI_ISL_1303354, EPI_ISL_1303355, EPI_ISL_1303356, EPI_ISL_1303357, EPI_ISL_1303358, EPI_ISL_1303359, EPI_ISL_1303360, EPI_ISL_1303361, EPI_ISL_1303362, EPI_ISL_1303363, EPI_ISL_1303364 | | | |
| see above | Microbiology Division, South Carolina Department of Health and Environmental Control | Microbiology Division, South Carolina Department of Health and Environmental Control | Flores,H., Freeman,J. |
| EPI_ISL_1303404, EPI_ISL_1303405, EPI_ISL_1303406 | Pomona Valley Hospital Medical Center | Los Angeles County Public Health Laboratories | P. Hemarajata et al. |
| EPI_ISL_1303407, EPI_ISL_1303408, EPI_ISL_1303409, EPI_ISL_1303410, EPI_ISL_1303411, EPI_ISL_1303412, EPI_ISL_1303413 | USC Clinical Lab | Los Angeles County Public Health Laboratories | P. Hemarajata et al. |
| EPI_ISL_1303414 | Martin Luther King Jr. Community Hospital | Los Angeles County Public Health Laboratories | P. Hemarajata et al. |
| EPI_ISL_1303416, EPI_ISL_1303417, EPI_ISL_1303418, EPI_ISL_1303419, EPI_ISL_1303420, EPI_ISL_1303421, EPI_ISL_1303422 | MD Laboratories | Los Angeles County Public Health Laboratories | P. Hemarajata et al. |
| EPI_ISL_1303424, EPI_ISL_1303425 | USC Clinical Lab | Los Angeles County Public Health Laboratories | P. Hemarajata et al. |
| EPI_ISL_1303426 | Flow Diagnostics Lab | Los Angeles County Public Health Laboratories | P. Hemarajata et al. |
| EPI_ISL_1303432, EPI_ISL_1303433, EPI_ISL_1303434, EPI_ISL_1303435, EPI_ISL_1303436, EPI_ISL_1303437 | UCLA Clinical Micro Lab | Los Angeles County Public Health Laboratories | P. Hemarajata et al. |
| EPI_ISL_1303438 | USC Clinical Lab | Los Angeles County Public Health Laboratories | P. Hemarajata et al. |
| EPI_ISL_1303439 | Martin Luther King Jr. Community Hospital | Los Angeles County Public Health Laboratories | P. Hemarajata et al. |
| EPI_ISL_1303441, EPI_ISL_1303442, EPI_ISL_1303443, EPI_ISL_1303445, EPI_ISL_1303446 | USC Clinical Lab | Los Angeles County Public Health Laboratories | P. Hemarajata et al. |
| EPI_ISL_1303447 | Martin Luther King Jr. Community Hospital | Los Angeles County Public Health Laboratories | P. Hemarajata et al. |
| EPI_ISL_1303449, EPI_ISL_1303450, EPI_ISL_1303451, EPI_ISL_1303452, EPI_ISL_1303453, EPI_ISL_1303454, EPI_ISL_1303455, EPI_ISL_1303456, EPI_ISL_1303457, EPI_ISL_1303458, EPI_ISL_1303459, EPI_ISL_1303460, EPI_ISL_1303461 | | | |
| see above | UCLA Clinical Micro Lab | Los Angeles County Public Health Laboratories | P. Hemarajata et al. |
| EPI_ISL_1303464, EPI_ISL_1303465, EPI_ISL_1303466, EPI_ISL_1303467, EPI_ISL_1303468, EPI_ISL_1303469, EPI_ISL_1303470 | USC Clinical Lab | Los Angeles County Public Health Laboratories | P. Hemarajata et al. |
| EPI_ISL_1303471 | Henry Mayo Newhall Hospital | Los Angeles County Public Health Laboratories | P. Hemarajata et al. |
| EPI_ISL_1303472, EPI_ISL_1303473 | Pomona Valley Hospital Medical Center | Los Angeles County Public Health Laboratories | P. Hemarajata et al. |
| EPI_ISL_1303474, EPI_ISL_1303475, EPI_ISL_1303476, EPI_ISL_1303477, EPI_ISL_1303478, EPI_ISL_1303479 | Flow Diagnostics Lab | Los Angeles County Public Health Laboratories | P. Hemarajata et al. |
| EPI_ISL_1303480, EPI_ISL_1303481 | USC Clinical Lab | Los Angeles County Public Health Laboratories | P. Hemarajata et al. |
| EPI_ISL_1303482 | Martin Luther King Jr. Community Hospital | Los Angeles County Public Health Laboratories | P. Hemarajata et al. |
| EPI_ISL_1303483 | MD Laboratories | Los Angeles County Public Health Laboratories | P. Hemarajata et al. |
| EPI_ISL_1303484, EPI_ISL_1303485, EPI_ISL_1303486, EPI_ISL_1303487, | Flow Diagnostics Lab | Los Angeles County Public Health Laboratories | P. Hemarajata et al. |

| | | | |
|---|---|---|---|
| EPI_ISL_1303488 | | | |
| EPI_ISL_1303489, EPI_ISL_1303490, EPI_ISL_1303491, EPI_ISL_1303492, EPI_ISL_1303493, EPI_ISL_1303494, EPI_ISL_1303495, EPI_ISL_1303496, EPI_ISL_1303497, EPI_ISL_1303498 | USC Clinical Lab | Los Angeles County Public Health Laboratories | P. Hemarajata et al. |

EPI_ISL_1303551, EPI_ISL_1303552, EPI_ISL_1303553, EPI_ISL_1303554, EPI_ISL_1303555, EPI_ISL_1303556, EPI_ISL_1303557, EPI_ISL_1303558, EPI_ISL_1303559, EPI_ISL_1303560, EPI_ISL_1303561, EPI_ISL_1303562, EPI_ISL_1303563, EPI_ISL_1303564, EPI_ISL_1303565, EPI_ISL_1303566, EPI_ISL_1303567, EPI_ISL_1303568, EPI_ISL_1303569, EPI_ISL_1303570, EPI_ISL_1303571, EPI_ISL_1303572,
EPI_ISL_1303573, EPI_ISL_1303574, EPI_ISL_1303575, EPI_ISL_1303576, EPI_ISL_1303577, EPI_ISL_1303578, EPI_ISL_1303579, EPI_ISL_1303580, EPI_ISL_1303581, EPI_ISL_1303582, EPI_ISL_1303583, EPI_ISL_1303584, EPI_ISL_1303585, EPI_ISL_1303586, EPI_ISL_1303587, EPI_ISL_1303588, EPI_ISL_1303589, EPI_ISL_1303590, EPI_ISL_1303591, EPI_ISL_1303592, EPI_ISL_1303593, EPI_ISL_1303594,
EPI_ISL_1303595, EPI_ISL_1303596, EPI_ISL_1303597, EPI_ISL_1303598, EPI_ISL_1303599, EPI_ISL_1303600, EPI_ISL_1303601, EPI_ISL_1303602, EPI_ISL_1303603, EPI_ISL_1303604, EPI_ISL_1303605, EPI_ISL_1303606, EPI_ISL_1303607, EPI_ISL_1303608, EPI_ISL_1303609, EPI_ISL_1303610, EPI_ISL_1303611, EPI_ISL_1303612, EPI_ISL_1303613, EPI_ISL_1303614, EPI_ISL_1303615, EPI_ISL_1303616,
EPI_ISL_1303617, EPI_ISL_1303618, EPI_ISL_1303619, EPI_ISL_1303620, EPI_ISL_1303621, EPI_ISL_1303622, EPI_ISL_1303623, EPI_ISL_1303624, EPI_ISL_1303625, EPI_ISL_1303626, EPI_ISL_1303627, EPI_ISL_1303628, EPI_ISL_1303629, EPI_ISL_1303630, EPI_ISL_1303631, EPI_ISL_1303632, EPI_ISL_1303633, EPI_ISL_1303634, EPI_ISL_1303635, EPI_ISL_1303636, EPI_ISL_1303637, EPI_ISL_1303638,
EPI_ISL_1303639, EPI_ISL_1303640, EPI_ISL_1303641, EPI_ISL_1303642, EPI_ISL_1303643, EPI_ISL_1303644, EPI_ISL_1303645, EPI_ISL_1303646, EPI_ISL_1303647, EPI_ISL_1303648, EPI_ISL_1303649, EPI_ISL_1303650, EPI_ISL_1303651, EPI_ISL_1303652, EPI_ISL_1303653, EPI_ISL_1303654, EPI_ISL_1303655, EPI_ISL_1303656, EPI_ISL_1303657, EPI_ISL_1303658, EPI_ISL_1303659, EPI_ISL_1303660,
EPI_ISL_1303661, EPI_ISL_1303662, EPI_ISL_1303663, EPI_ISL_1303664, EPI_ISL_1303665, EPI_ISL_1303666, EPI_ISL_1303667, EPI_ISL_1303668, EPI_ISL_1303669, EPI_ISL_1303670, EPI_ISL_1303671, EPI_ISL_1303672, EPI_ISL_1303673, EPI_ISL_1303674, EPI_ISL_1303675, EPI_ISL_1303676, EPI_ISL_1303677, EPI_ISL_1303678, EPI_ISL_1303679, EPI_ISL_1303680, EPI_ISL_1303681, EPI_ISL_1303682,
EPI_ISL_1303683, EPI_ISL_1303684, EPI_ISL_1303685, EPI_ISL_1303686, EPI_ISL_1303687, EPI_ISL_1303688, EPI_ISL_1303689, EPI_ISL_1303690, EPI_ISL_1303691, EPI_ISL_1303692, EPI_ISL_1303693, EPI_ISL_1303694, EPI_ISL_1303695, EPI_ISL_1303696, EPI_ISL_1303697, EPI_ISL_1303698, EPI_ISL_1303699, EPI_ISL_1303701, EPI_ISL_1303702, EPI_ISL_1303703, EPI_ISL_1303704, EPI_ISL_1303705,
EPI_ISL_1303706, EPI_ISL_1303707, EPI_ISL_1303708, EPI_ISL_1303709, EPI_ISL_1303710, EPI_ISL_1303711, EPI_ISL_1303712, EPI_ISL_1303713, EPI_ISL_1303714, EPI_ISL_1303715, EPI_ISL_1303716, EPI_ISL_1303717, EPI_ISL_1303718, EPI_ISL_1303719, EPI_ISL_1303720, EPI_ISL_1303721, EPI_ISL_1303722, EPI_ISL_1303723, EPI_ISL_1303724, EPI_ISL_1303725, EPI_ISL_1303726, EPI_ISL_1303727,
EPI_ISL_1303728, EPI_ISL_1303729, EPI_ISL_1303730, EPI_ISL_1303731, EPI_ISL_1303732, EPI_ISL_1303733, EPI_ISL_1303734, EPI_ISL_1303735, EPI_ISL_1303736, EPI_ISL_1303737, EPI_ISL_1303738, EPI_ISL_1303739, EPI_ISL_1303740, EPI_ISL_1303741, EPI_ISL_1303742, EPI_ISL_1303743, EPI_ISL_1303744, EPI_ISL_1303745, EPI_ISL_1303746, EPI_ISL_1303747, EPI_ISL_1303748, EPI_ISL_1303749,
EPI_ISL_1303750, EPI_ISL_1303751, EPI_ISL_1303752, EPI_ISL_1303753, EPI_ISL_1303754, EPI_ISL_1303755, EPI_ISL_1303756, EPI_ISL_1303757, EPI_ISL_1303758, EPI_ISL_1303759, EPI_ISL_1303760, EPI_ISL_1303761, EPI_ISL_1303762, EPI_ISL_1303763, EPI_ISL_1303764, EPI_ISL_1303765, EPI_ISL_1303766, EPI_ISL_1303767, EPI_ISL_1303768, EPI_ISL_1303769, EPI_ISL_1303770, EPI_ISL_1303771,
EPI_ISL_1303772, EPI_ISL_1303773, EPI_ISL_1303774, EPI_ISL_1303775, EPI_ISL_1303777, EPI_ISL_1303778, EPI_ISL_1303779, EPI_ISL_1303780, EPI_ISL_1303781, EPI_ISL_1303782, EPI_ISL_1303783, EPI_ISL_1303784, EPI_ISL_1303785, EPI_ISL_1303786, EPI_ISL_1303787, EPI_ISL_1303788, EPI_ISL_1303789, EPI_ISL_1303790, EPI_ISL_1303791, EPI_ISL_1303792, EPI_ISL_1303793, EPI_ISL_1303794,
EPI_ISL_1303795, EPI_ISL_1303796, EPI_ISL_1303797, EPI_ISL_1303798, EPI_ISL_1303799, EPI_ISL_1303800, EPI_ISL_1303801, EPI_ISL_1303802, EPI_ISL_1303803, EPI_ISL_1303804, EPI_ISL_1303805, EPI_ISL_1303806, EPI_ISL_1303807, EPI_ISL_1303808, EPI_ISL_1303809, EPI_ISL_1303810, EPI_ISL_1303811, EPI_ISL_1303812, EPI_ISL_1303813, EPI_ISL_1303814, EPI_ISL_1303815, EPI_ISL_1303816,
EPI_ISL_1303817, EPI_ISL_1303818, EPI_ISL_1303819, EPI_ISL_1303820, EPI_ISL_1303821, EPI_ISL_1303822, EPI_ISL_1303823, EPI_ISL_1303824, EPI_ISL_1303825, EPI_ISL_1303826, EPI_ISL_1303827, EPI_ISL_1303828, EPI_ISL_1303829, EPI_ISL_1303830, EPI_ISL_1303831, EPI_ISL_1303832, EPI_ISL_1303833, EPI_ISL_1303834, EPI_ISL_1303835, EPI_ISL_1303836, EPI_ISL_1303837, EPI_ISL_1303838,
EPI_ISL_1303839, EPI_ISL_1303840, EPI_ISL_1303841, EPI_ISL_1303842, EPI_ISL_1303843, EPI_ISL_1303844, EPI_ISL_1303845, EPI_ISL_1303846, EPI_ISL_1303847, EPI_ISL_1303848, EPI_ISL_1303849, EPI_ISL_1303850, EPI_ISL_1303851, EPI_ISL_1303852, EPI_ISL_1303853, EPI_ISL_1303854, EPI_ISL_1303855, EPI_ISL_1303856, EPI_ISL_1303857, EPI_ISL_1303858, EPI_ISL_1303859, EPI_ISL_1303860,
EPI_ISL_1303861, EPI_ISL_1303862, EPI_ISL_1303863, EPI_ISL_1303864, EPI_ISL_1303865, EPI_ISL_1303866, EPI_ISL_1303867, EPI_ISL_1303868, EPI_ISL_1303869, EPI_ISL_1303870, EPI_ISL_1303871, EPI_ISL_1303872, EPI_ISL_1303873, EPI_ISL_1303874, EPI_ISL_1303875, EPI_ISL_1303876, EPI_ISL_1303877, EPI_ISL_1303878, EPI_ISL_1303879, EPI_ISL_1303880, EPI_ISL_1303881, EPI_ISL_1303882,
EPI_ISL_1303883, EPI_ISL_1303884, EPI_ISL_1303885, EPI_ISL_1303886, EPI_ISL_1303887, EPI_ISL_1303888, EPI_ISL_1303889, EPI_ISL_1303890, EPI_ISL_1303891, EPI_ISL_1303892, EPI_ISL_1303893, EPI_ISL_1303894, EPI_ISL_1303895, EPI_ISL_1303896, EPI_ISL_1303897, EPI_ISL_1303898, EPI_ISL_1303899, EPI_ISL_1303900, EPI_ISL_1303901, EPI_ISL_1303902, EPI_ISL_1303903, EPI_ISL_1303904,
EPI_ISL_1303905, EPI_ISL_1303906, EPI_ISL_1303907, EPI_ISL_1303908, EPI_ISL_1303909, EPI_ISL_1303910, EPI_ISL_1303911, EPI_ISL_1303912, EPI_ISL_1303913, EPI_ISL_1303914, EPI_ISL_1303915, EPI_ISL_1303916, EPI_ISL_1303917, EPI_ISL_1303918, EPI_ISL_1303919, EPI_ISL_1303920, EPI_ISL_1303921, EPI_ISL_1303922, EPI_ISL_1303923, EPI_ISL_1303924, EPI_ISL_1303925, EPI_ISL_1303926,
EPI_ISL_1303927, EPI_ISL_1303928, EPI_ISL_1303929, EPI_ISL_1303930, EPI_ISL_1303931, EPI_ISL_1303932, EPI_ISL_1303933, EPI_ISL_1303934, EPI_ISL_1303935, EPI_ISL_1303936, EPI_ISL_1303937, EPI_ISL_1303938, EPI_ISL_1303939, EPI_ISL_1303940, EPI_ISL_1303941, EPI_ISL_1303942, EPI_ISL_1303943, EPI_ISL_1303944, EPI_ISL_1303945, EPI_ISL_1303946, EPI_ISL_1303947, EPI_ISL_1303948,
EPI_ISL_1303949, EPI_ISL_1303950, EPI_ISL_1303951, EPI_ISL_1303952, EPI_ISL_1303953, EPI_ISL_1303954, EPI_ISL_1303955, EPI_ISL_1303956, EPI_ISL_1303957, EPI_ISL_1303958, EPI_ISL_1303959, EPI_ISL_1303960, EPI_ISL_1303961, EPI_ISL_1303962, EPI_ISL_1303963, EPI_ISL_1303964, EPI_ISL_1303965, EPI_ISL_1303966, EPI_ISL_1303967, EPI_ISL_1303968, EPI_ISL_1303969, EPI_ISL_1303970,
EPI_ISL_1303971, EPI_ISL_1303972, EPI_ISL_1303973, EPI_ISL_1303974, EPI_ISL_1303975, EPI_ISL_1303976, EPI_ISL_1303977, EPI_ISL_1303978, EPI_ISL_1303979, EPI_ISL_1303980, EPI_ISL_1303981, EPI_ISL_1303982, EPI_ISL_1303983, EPI_ISL_1303984, EPI_ISL_1303985, EPI_ISL_1303986, EPI_ISL_1303987, EPI_ISL_1303988, EPI_ISL_1303989, EPI_ISL_1303990, EPI_ISL_1303991, EPI_ISL_1303992,
EPI_ISL_1303993, EPI_ISL_1303994, EPI_ISL_1303995, EPI_ISL_1303996, EPI_ISL_1303997, EPI_ISL_1303998, EPI_ISL_1303999, EPI_ISL_1304000, EPI_ISL_1304001, EPI_ISL_1304002, EPI_ISL_1304003, EPI_ISL_1304004, EPI_ISL_1304005, EPI_ISL_1304006, EPI_ISL_1304007, EPI_ISL_1304008, EPI_ISL_1304009, EPI_ISL_1304010, EPI_ISL_1304011, EPI_ISL_1304012, EPI_ISL_1304013, EPI_ISL_1304014,
EPI_ISL_1304015, EPI_ISL_1304016, EPI_ISL_1304017, EPI_ISL_1304018, EPI_ISL_1304019, EPI_ISL_1304020, EPI_ISL_1304021, EPI_ISL_1304022, EPI_ISL_1304023, EPI_ISL_1304024, EPI_ISL_1304025, EPI_ISL_1304026, EPI_ISL_1304027, EPI_ISL_1304028, EPI_ISL_1304029, EPI_ISL_1304030, EPI_ISL_1304031, EPI_ISL_1304032, EPI_ISL_1304033, EPI_ISL_1304034, EPI_ISL_1304035, EPI_ISL_1304036,
EPI_ISL_1304037, EPI_ISL_1304038, EPI_ISL_1304039, EPI_ISL_1304040, EPI_ISL_1304041, EPI_ISL_1304042, EPI_ISL_1304043, EPI_ISL_1304044, EPI_ISL_1304045, EPI_ISL_1304046, EPI_ISL_1304047, EPI_ISL_1304048, EPI_ISL_1304049, EPI_ISL_1304050, EPI_ISL_1304051, EPI_ISL_1304052, EPI_ISL_1304053, EPI_ISL_1304054, EPI_ISL_1304055, EPI_ISL_1304056, EPI_ISL_1304057, EPI_ISL_1304058,
EPI_ISL_1304059, EPI_ISL_1304060, EPI_ISL_1304061, EPI_ISL_1304062, EPI_ISL_1304063, EPI_ISL_1304064, EPI_ISL_1304065, EPI_ISL_1304066, EPI_ISL_1304067, EPI_ISL_1304068, EPI_ISL_1304069, EPI_ISL_1304070, EPI_ISL_1304071, EPI_ISL_1304072, EPI_ISL_1304073, EPI_ISL_1304074, EPI_ISL_1304075, EPI_ISL_1304076, EPI_ISL_1304077, EPI_ISL_1304078, EPI_ISL_1304079, EPI_ISL_1304080,
EPI_ISL_1304081, EPI_ISL_1304082, EPI_ISL_1304083, EPI_ISL_1304084, EPI_ISL_1304085, EPI_ISL_1304086, EPI_ISL_1304087, EPI_ISL_1304088, EPI_ISL_1304089, EPI_ISL_1304090, EPI_ISL_1304091, EPI_ISL_1304092, EPI_ISL_1304093, EPI_ISL_1304094, EPI_ISL_1304095, EPI_ISL_1304096, EPI_ISL_1304097, EPI_ISL_1304098, EPI_ISL_1304099, EPI_ISL_1304100, EPI_ISL_1304101, EPI_ISL_1304102,
EPI_ISL_1304103, EPI_ISL_1304104, EPI_ISL_1304105, EPI_ISL_1304106, EPI_ISL_1304107, EPI_ISL_1304108, EPI_ISL_1304109, EPI_ISL_1304110, EPI_ISL_1304111, EPI_ISL_1304112, EPI_ISL_1304113, EPI_ISL_1304114, EPI_ISL_1304115, EPI_ISL_1304116, EPI_ISL_1304117, EPI_ISL_1304118, EPI_ISL_1304119, EPI_ISL_1304120, EPI_ISL_1304121, EPI_ISL_1304122, EPI_ISL_1304123, EPI_ISL_1304124,
EPI_ISL_1304125, EPI_ISL_1304126, EPI_ISL_1304127, EPI_ISL_1304128, EPI_ISL_1304129, EPI_ISL_1304130, EPI_ISL_1304131, EPI_ISL_1304132, EPI_ISL_1304133, EPI_ISL_1304134, EPI_ISL_1304135, EPI_ISL_1304136, EPI_ISL_1304137, EPI_ISL_1304138, EPI_ISL_1304139, EPI_ISL_1304140, EPI_ISL_1304141, EPI_ISL_1304142, EPI_ISL_1304143, EPI_ISL_1304144, EPI_ISL_1304145, EPI_ISL_1304146,
EPI_ISL_1304147, EPI_ISL_1304148, EPI_ISL_1304149, EPI_ISL_1304150, EPI_ISL_1304151, EPI_ISL_1304152, EPI_ISL_1304153, EPI_ISL_1304154, EPI_ISL_1304155, EPI_ISL_1304156, EPI_ISL_1304157, EPI_ISL_1304158, EPI_ISL_1304159, EPI_ISL_1304160, EPI_ISL_1304161, EPI_ISL_1304162, EPI_ISL_1304163, EPI_ISL_1304164, EPI_ISL_1304165, EPI_ISL_1304166, EPI_ISL_1304167, EPI_ISL_1304168,
EPI_ISL_1304169, EPI_ISL_1304170, EPI_ISL_1304171, EPI_ISL_1304172, EPI_ISL_1304173, EPI_ISL_1304174, EPI_ISL_1304175, EPI_ISL_1304176, EPI_ISL_1304177, EPI_ISL_1304178, EPI_ISL_1304179, EPI_ISL_1304180, EPI_ISL_1304181, EPI_ISL_1304182, EPI_ISL_1304183, EPI_ISL_1304184, EPI_ISL_1304185, EPI_ISL_1304186, EPI_ISL_1304187, EPI_ISL_1304188, EPI_ISL_1304189, EPI_ISL_1304190,
EPI_ISL_1304191, EPI_ISL_1304192, EPI_ISL_1304193, EPI_ISL_1304194, EPI_ISL_1304195, EPI_ISL_1304196, EPI_ISL_1304197, EPI_ISL_1304198, EPI_ISL_1304199, EPI_ISL_1304200, EPI_ISL_1304201, EPI_ISL_1304202, EPI_ISL_1304203, EPI_ISL_1304204, EPI_ISL_1304205, EPI_ISL_1304206, EPI_ISL_1304207, EPI_ISL_1304208, EPI_ISL_1304209, EPI_ISL_1304210, EPI_ISL_1304211, EPI_ISL_1304212,
EPI_ISL_1304213, EPI_ISL_1304214, EPI_ISL_1304215, EPI_ISL_1304216, EPI_ISL_1304217, EPI_ISL_1304218, EPI_ISL_1304219, EPI_ISL_1304220, EPI_ISL_1304221, EPI_ISL_1304222, EPI_ISL_1304223, EPI_ISL_1304224, EPI_ISL_1304225, EPI_ISL_1304226, EPI_ISL_1304227, EPI_ISL_1304228, EPI_ISL_1304229, EPI_ISL_1304230, EPI_ISL_1304231, EPI_ISL_1304232, EPI_ISL_1304233, EPI_ISL_1304234,
EPI_ISL_1304235, EPI_ISL_1304236, EPI_ISL_1304237, EPI_ISL_1304238, EPI_ISL_1304239, EPI_ISL_1304240, EPI_ISL_1304241, EPI_ISL_1304242, EPI_ISL_1304243, EPI_ISL_1304244, EPI_ISL_1304245, EPI_ISL_1304246, EPI_ISL_1304247, EPI_ISL_1304248, EPI_ISL_1304249, EPI_ISL_1304250, EPI_ISL_1304251, EPI_ISL_1304252, EPI_ISL_1304253, EPI_ISL_1304254, EPI_ISL_1304255, EPI_ISL_1304256,
EPI_ISL_1304257, EPI_ISL_1304258, EPI_ISL_1304259, EPI_ISL_1304260, EPI_ISL_1304261, EPI_ISL_1304262, EPI_ISL_1304263, EPI_ISL_1304264, EPI_ISL_1304265, EPI_ISL_1304266, EPI_ISL_1304267, EPI_ISL_1304268, EPI_ISL_1304269, EPI_ISL_1304270, EPI_ISL_1304271, EPI_ISL_1304272, EPI_ISL_1304273, EPI_ISL_1304274, EPI_ISL_1304275, EPI_ISL_1304276, EPI_ISL_1304277, EPI_ISL_1304278,
EPI_ISL_1304279, EPI_ISL_1304280, EPI_ISL_1304281, EPI_ISL_1304282, EPI_ISL_1304283, EPI_ISL_1304284, EPI_ISL_1304285, EPI_ISL_1304286, EPI_ISL_1304287, EPI_ISL_1304288, EPI_ISL_1304289, EPI_ISL_1304290, EPI_ISL_1304291, EPI_ISL_1304292, EPI_ISL_1304293, EPI_ISL_1304294, EPI_ISL_1304295, EPI_ISL_1304296, EPI_ISL_1304297, EPI_ISL_1304298, EPI_ISL_1304299, EPI_ISL_1304300,
EPI_ISL_1304301, EPI_ISL_1304302, EPI_ISL_1304303, EPI_ISL_1304304, EPI_ISL_1304305, EPI_ISL_1304306, EPI_ISL_1304307, EPI_ISL_1304308, EPI_ISL_1304309, EPI_ISL_1304310, EPI_ISL_1304311, EPI_ISL_1304312, EPI_ISL_1304313, EPI_ISL_1304314, EPI_ISL_1304315, EPI_ISL_1304316, EPI_ISL_1304317, EPI_ISL_1304318, EPI_ISL_1304319, EPI_ISL_1304320, EPI_ISL_1304321, EPI_ISL_1304322,
EPI_ISL_1304323, EPI_ISL_1304324, EPI_ISL_1304325, EPI_ISL_1304326, EPI_ISL_1304327, EPI_ISL_1304328, EPI_ISL_1304329, EPI_ISL_1304330, EPI_ISL_1304331, EPI_ISL_1304332, EPI_ISL_1304333, EPI_ISL_1304334, EPI_ISL_1304335, EPI_ISL_1304336, EPI_ISL_1304337, EPI_ISL_1304338, EPI_ISL_1304339, EPI_ISL_1304340, EPI_ISL_1304341, EPI_ISL_1304342, EPI_ISL_1304343, EPI_ISL_1304344,
EPI_ISL_1304345, EPI_ISL_1304346, EPI_ISL_1304347, EPI_ISL_1304348, EPI_ISL_1304349, EPI_ISL_1304350, EPI_ISL_1304351, EPI_ISL_1304352, EPI_ISL_1304353, EPI_ISL_1304354, EPI_ISL_1304355, EPI_ISL_1304356, EPI_ISL_1304357, EPI_ISL_1304358, EPI_ISL_1304359, EPI_ISL_1304360, EPI_ISL_1304361, EPI_ISL_1304362, EPI_ISL_1304363, EPI_ISL_1304364, EPI_ISL_1304365, EPI_ISL_1304366,
EPI_ISL_1304367, EPI_ISL_1304368, EPI_ISL_1304369, EPI_ISL_1304370, EPI_ISL_1304371, EPI_ISL_1304372, EPI_ISL_1304373, EPI_ISL_1304374, EPI_ISL_1304375, EPI_ISL_1304376, EPI_ISL_1304377, EPI_ISL_1304378, EPI_ISL_1304379, EPI_ISL_1304380, EPI_ISL_1304381, EPI_ISL_1304382, EPI_ISL_1304383, EPI_ISL_1304384, EPI_ISL_1304385, EPI_ISL_1304386, EPI_ISL_1304387, EPI_ISL_1304388,
EPI_ISL_1304389, EPI_ISL_1304390, EPI_ISL_1304391, EPI_ISL_1304392, EPI_ISL_1304393, EPI_ISL_1304394, EPI_ISL_1304395, EPI_ISL_1304396, EPI_ISL_1304397, EPI_ISL_1304398, EPI_ISL_1304399, EPI_ISL_1304400, EPI_ISL_1304401, EPI_ISL_1304402, EPI_ISL_1304403, EPI_ISL_1304404, EPI_ISL_1304405, EPI_ISL_1304406, EPI_ISL_1304407, EPI_ISL_1304408, EPI_ISL_1304409, EPI_ISL_1304410,
EPI_ISL_1304411, EPI_ISL_1304412, EPI_ISL_1304413, EPI_ISL_1304414, EPI_ISL_1304415, EPI_ISL_1304416, EPI_ISL_1304417, EPI_ISL_1304418, EPI_ISL_1304419, EPI_ISL_1304420, EPI_ISL_1304421, EPI_ISL_1304422, EPI_ISL_1304423, EPI_ISL_1304424, EPI_ISL_1304425, EPI_ISL_1304426, EPI_ISL_1304427, EPI_ISL_1304428, EPI_ISL_1304429, EPI_ISL_1304430, EPI_ISL_1304431, EPI_ISL_1304432,
EPI_ISL_1304433, EPI_ISL_1304434, EPI_ISL_1304435, EPI_ISL_1304436, EPI_ISL_1304437, EPI_ISL_1304438, EPI_ISL_1304439, EPI_ISL_1304440, EPI_ISL_1304441, EPI_ISL_1304442, EPI_ISL_1304443, EPI_ISL_1304444, EPI_ISL_1304445, EPI_ISL_1304446, EPI_ISL_1304447, EPI_ISL_1304448, EPI_ISL_1304449, EPI_ISL_1304450, EPI_ISL_1304451, EPI_ISL_1304452, EPI_ISL_1304453, EPI_ISL_1304454,
EPI_ISL_1304455, EPI_ISL_1304456, EPI_ISL_1304457, EPI_ISL_1304458, EPI_ISL_1304459, EPI_ISL_1304460, EPI_ISL_1304461, EPI_ISL_1304462, EPI_ISL_1304463, EPI_ISL_1304464, EPI_ISL_1304465, EPI_ISL_1304466, EPI_ISL_1304467, EPI_ISL_1304468, EPI_ISL_1304469, EPI_ISL_1304470, EPI_ISL_1304471, EPI_ISL_1304472, EPI_ISL_1304473, EPI_ISL_1304474, EPI_ISL_1304475, EPI_ISL_1304476,
EPI_ISL_1304477, EPI_ISL_1304478, EPI_ISL_1304479, EPI_ISL_1304480, EPI_ISL_1304481, EPI_ISL_1304482, EPI_ISL_1304483, EPI_ISL_1304484, EPI_ISL_1304485, EPI_ISL_1304486, EPI_ISL_1304487, EPI_ISL_1304488, EPI_ISL_1304489, EPI_ISL_1304490, EPI_ISL_1304491, EPI_ISL_1304492, EPI_ISL_1304493, EPI_ISL_1304494, EPI_ISL_1304495, EPI_ISL_1304496, EPI_ISL_1304497, EPI_ISL_1304498,
EPI_ISL_1304499, EPI_ISL_1304500, EPI_ISL_1304501, EPI_ISL_1304502, EPI_ISL_1304503, EPI_ISL_1304504, EPI_ISL_1304505, EPI_ISL_1304506, EPI_ISL_1304507, EPI_ISL_1304508, EPI_ISL_1304509, EPI_ISL_1304510, EPI_ISL_1304511, EPI_ISL_1304512, EPI_ISL_1304513, EPI_ISL_1304514, EPI_ISL_1304515, EPI_ISL_1304516, EPI_ISL_1304517, EPI_ISL_1304518, EPI_ISL_1304519, EPI_ISL_1304520,
EPI_ISL_1304521, EPI_ISL_1304522, EPI_ISL_1304523, EPI_ISL_1304524, EPI_ISL_1304525, EPI_ISL_1304526, EPI_ISL_1304527, EPI_ISL_1304528, EPI_ISL_1304529, EPI_ISL_1304530, EPI_ISL_1304531, EPI_ISL_1304532, EPI_ISL_1304533, EPI_ISL_1304534, EPI_ISL_1304535, EPI_ISL_1304536, EPI_ISL_1304537, EPI_ISL_1304538, EPI_ISL_1304539, EPI_ISL_1304540, EPI_ISL_1304541, EPI_ISL_1304542,
EPI_ISL_1304543, EPI_ISL_1304544, EPI_ISL_1304545, EPI_ISL_1304546, EPI_ISL_1304547, EPI_ISL_1304548, EPI_ISL_1304549, EPI_ISL_1304550, EPI_ISL_1304551, EPI_ISL_1304552, EPI_ISL_1304553, EPI_ISL_1304554, EPI_ISL_1304555, EPI_ISL_1304556, EPI_ISL_1304557, EPI_ISL_1304558, EPI_ISL_1304559, EPI_ISL_1304560, EPI_ISL_1304561, EPI_ISL_1304562, EPI_ISL_1304563, EPI_ISL_1304564,
EPI_ISL_1304565, EPI_ISL_1304566, EPI_ISL_1304567, EPI_ISL_1304568, EPI_ISL_1304569, EPI_ISL_1304570, EPI_ISL_1304571, EPI_ISL_1304572, EPI_ISL_1304573, EPI_ISL_1304574, EPI_ISL_1304575, EPI_ISL_1304576, EPI_ISL_1304577, EPI_ISL_1304578, EPI_ISL_1304579, EPI_ISL_1304580, EPI_ISL_1304581, EPI_ISL_1304582, EPI_ISL_1304583, EPI_ISL_1304584, EPI_ISL_1304585, EPI_ISL_1304586,
EPI_ISL_1304587, EPI_ISL_1304588, EPI_ISL_1304589, EPI_ISL_1304590, EPI_ISL_1304591, EPI_ISL_1304592, EPI_ISL_1304593, EPI_ISL_1304594, EPI_ISL_1304595, EPI_ISL_1304596, EPI_ISL_1304597, EPI_ISL_1304598, EPI_ISL_1304599, EPI_ISL_1304600, EPI_ISL_1304601, EPI_ISL_1304602, EPI_ISL_1304603, EPI_ISL_1304604, EPI_ISL_1304605, EPI_ISL_1304606, EPI_ISL_1304607, EPI_ISL_1304608,
EPI_ISL_1304609, EPI_ISL_1304610, EPI_ISL_1304611, EPI_ISL_1304612, EPI_ISL_1304613, EPI_ISL_1304614, EPI_ISL_1304615, EPI_ISL_1304616, EPI_ISL_1304617, EPI_ISL_1304618, EPI_ISL_1304619, EPI_ISL_1304620, EPI_ISL_1304621, EPI_ISL_1304622, EPI_ISL_1304623, EPI_ISL_1304624, EPI_ISL_1304625, EPI_ISL_1304626, EPI_ISL_1304627, EPI_ISL_1304628, EPI_ISL_1304629, EPI_ISL_1304630,
EPI_ISL_1304631, EPI_ISL_1304632, EPI_ISL_1304633, EPI_ISL_1304634, EPI_ISL_1304635, EPI_ISL_1304636, EPI_ISL_1304637, EPI_ISL_1304638, EPI_ISL_1304639, EPI_ISL_1304640, EPI_ISL_1304641, EPI_ISL_1304642, EPI_ISL_1304643, EPI_ISL_1304644, EPI_ISL_1304645, EPI_ISL_1304646, EPI_ISL_1304647, EPI_ISL_1304648, EPI_ISL_1304649, EPI_ISL_1304650, EPI_ISL_1304651, EPI_ISL_1304652,
EPI_ISL_1304653, EPI_ISL_1304654, EPI_ISL_1304655, EPI_ISL_1304656, EPI_ISL_1304657, EPI_ISL_1304658, EPI_ISL_1304659, EPI_ISL_1304660, EPI_ISL_1304661, EPI_ISL_1304662, EPI_ISL_1304663, EPI_ISL_1304664, EPI_ISL_1304665, EPI_ISL_1304666, EPI_ISL_1304667, EPI_ISL_1304668, EPI_ISL_1304669, EPI_ISL_1304670, EPI_ISL_1304671, EPI_ISL_1304672, EPI_ISL_1304673, EPI_ISL_1304674,
EPI_ISL_1304675, EPI_ISL_1304676, EPI_ISL_1304677, EPI_ISL_1304678, EPI_ISL_1304679, EPI_ISL_1304680, EPI_ISL_1304681, EPI_ISL_1304682, EPI_ISL_1304683, EPI_ISL_1304684, EPI_ISL_1304685, EPI_ISL_1304686, EPI_ISL_1304687, EPI_ISL_1304688, EPI_ISL_1304689, EPI_ISL_1304690, EPI_ISL_1304691, EPI_ISL_1304692, EPI_ISL_1304693, EPI_ISL_1304694, EPI_ISL_1304695, EPI_ISL_1304696,
EPI_ISL_1304697, EPI_ISL_1304698, EPI_ISL_1304699, EPI_ISL_1304700, EPI_ISL_1304701, EPI_ISL_1304702, EPI_ISL_1304703, EPI_ISL_1304704, EPI_ISL_1304705, EPI_ISL_1304706, EPI_ISL_1304707, EPI_ISL_1304708, EPI_ISL_1304709, EPI_ISL_1304710, EPI_ISL_1304711, EPI_ISL_1304712, EPI_ISL_1304713, EPI_ISL_1304714, EPI_ISL_1304715, EPI_ISL_1304716, EPI_ISL_1304717, EPI_ISL_1304718,
EPI_ISL_1304719, EPI_ISL_1304720, EPI_ISL_1304721, EPI_ISL_1304722, EPI_ISL_1304723, EPI_ISL_1304724, EPI_ISL_1304725, EPI_ISL_1304726, EPI_ISL_1304727, EPI_ISL_1304728, EPI_ISL_1304729, EPI_ISL_1304730, EPI_ISL_1304731, EPI_ISL_1304732, EPI_ISL_1304733, EPI_ISL_1304734, EPI_ISL_1304735, EPI_ISL_1304736, EPI_ISL_1304737, EPI_ISL_1304738, EPI_ISL_1304739, EPI_ISL_1304740,
EPI_ISL_1304741, EPI_ISL_1304742, EPI_ISL_1304743, EPI_ISL_1304744, EPI_ISL_1304745, EPI_ISL_1304746, EPI_ISL_1304747, EPI_ISL_1304748, EPI_ISL_1304749, EPI_ISL_1304750, EPI_ISL_1304751, EPI_ISL_1304752, EPI_ISL_1304753, EPI_ISL_1304754, EPI_ISL_1304755, EPI_ISL_1304756, EPI_ISL_1304757, EPI_ISL_1304758, EPI_ISL_1304759, EPI_ISL_1304760, EPI_ISL_1304761, EPI_ISL_1304762,
EPI_ISL_1304763, EPI_ISL_1304764, EPI_ISL_1304765, EPI_ISL_1304766, EPI_ISL_1304767, EPI_ISL_1304768, EPI_ISL_1304769, EPI_ISL_1304770, EPI_ISL_1304771, EPI_ISL_1304772, EPI_ISL_1304773, EPI_ISL_1304774, EPI_ISL_1304775, EPI_ISL_1304776, EPI_ISL_1304777, EPI_ISL_1304778, EPI_ISL_1304779, EPI_ISL_1304780, EPI_ISL_1304781, EPI_ISL_1304782, EPI_ISL_1304783, EPI_ISL_1304784,
EPI_ISL_1304785, EPI_ISL_1304786, EPI_ISL_1304787, EPI_ISL_1304788, EPI_ISL_1304789, EPI_ISL_1304790, EPI_ISL_1304791, EPI_ISL_1304792, EPI_ISL_1304793, EPI_ISL_1304794, EPI_ISL_1304795, EPI_ISL_1304796, EPI_ISL_1304797, EPI_ISL_1304798, EPI_ISL_1304799, EPI_ISL_1304800, EPI_ISL_1304801, EPI_ISL_1304802, EPI_ISL_1304803, EPI_ISL_1304804, EPI_ISL_1304805, EPI_ISL_1304806,
EPI_ISL_1304807, EPI_ISL_1304808, EPI_ISL_1304809, EPI_ISL_1304810, EPI_ISL_1304811, EPI_ISL_1304812, EPI_ISL_1304813, EPI_ISL_1304814, EPI_ISL_1304815, EPI_ISL_1304816, EPI_ISL_1304817, EPI_ISL_1304818, EPI_ISL_1304819, EPI_ISL_1304820, EPI_ISL_1304821, EPI_ISL_1304822, EPI_ISL_1304823, EPI_ISL_1304824, EPI_ISL_1304825, EPI_ISL_1304826, EPI_ISL_1304827, EPI_ISL_1304828,
EPI_ISL_1304829, EPI_ISL_1304830, EPI_ISL_1304831, EPI_ISL_1304832, EPI_ISL_1304833, EPI_ISL_1304834, EPI_ISL_1304835, EPI_ISL_1304836, EPI_ISL_1304837, EPI_ISL_1304838, EPI_ISL_1304839, EPI_ISL_1304840, EPI_ISL_1304841, EPI_ISL_1304842, EPI_ISL_1304843, EPI_ISL_1304844, EPI_ISL_1304845, EPI_ISL_1304846, EPI_ISL_1304847, EPI_ISL_1304848, EPI_ISL_1304849, EPI_ISL_1304850,
EPI_ISL_1304851, EPI_ISL_1304852, EPI_ISL_1304853, EPI_ISL_1304854, EPI_ISL_1304855, EPI_ISL_1304856, EPI_ISL_1304857, EPI_ISL_1304858, EPI_ISL_1304859, EPI_ISL_1304860, EPI_ISL_1304861, EPI_ISL_1304862, EPI_ISL_1304863, EPI_ISL_1304864, EPI_ISL_1304865, EPI_ISL_1304866, EPI_ISL_1304867, EPI_ISL_1304868, EPI_ISL_1304869, EPI_ISL_1304870, EPI_ISL_1304871, EPI_ISL_1304872,
EPI_ISL_1304873, EPI_ISL_1304874, EPI_ISL_1304875, EPI_ISL_1304876, EPI_ISL_1304877, EPI_ISL_1304878, EPI_ISL_1304879, EPI_ISL_1304880, EPI_ISL_1304881, EPI_ISL_1304882, EPI_ISL_1304883, EPI_ISL_1304884, EPI_ISL_1304885, EPI_ISL_1304886, EPI_ISL_1304887, EPI_ISL_1304888, EPI_ISL_1304889, EPI_ISL_1304890, EPI_ISL_1304891, EPI_ISL_1304892, EPI_ISL_1304893, EPI_ISL_1304894,
EPI_ISL_1304895, EPI_ISL_1304896, EPI_ISL_1304897, EPI_ISL_1304898, EPI_ISL_1304899, EPI_ISL_1304900, EPI_ISL_1304901, EPI_ISL_1304902, EPI_ISL_1304903, EPI_ISL_1304904, EPI_ISL_1304905, EPI_ISL_1304906, EPI_ISL_1304907, EPI_ISL_1304908, EPI_ISL_1304909, EPI_ISL_1304910, EPI_ISL_1304911, EPI_ISL_1304912, EPI_ISL_1304913, EPI_ISL_1304914, EPI_ISL_1304915, EPI_ISL_1304916,
EPI_ISL_1304917, EPI_ISL_1304918, EPI_ISL_1304919, EPI_ISL_1304920, EPI_ISL_1304921, EPI_ISL_1304922, EPI_ISL_1304923, EPI_ISL_1304924, EPI_ISL_1304925, EPI_ISL_1304926, EPI_ISL_1304927, EPI_ISL_1304928, EPI_ISL_1304929, EPI_ISL_1304930, EPI_ISL_1304931, EPI_ISL_1304932, EPI_ISL_1304933, EPI_ISL_1304934, EPI_ISL_1304935, EPI_ISL_1304936, EPI_ISL_1304937, EPI_ISL_1304938,
EPI_ISL_1304939, EPI_ISL_1304940, EPI_ISL_1304941, EPI_ISL_1304942, EPI_ISL_1304943, EPI_ISL_1304944, EPI_ISL_1304945, EPI_ISL_1304946, EPI_ISL_1304947, EPI_ISL_1304948, EPI_ISL_1304949, EPI_ISL_1304950, EPI_ISL_1304951, EPI_ISL_1304952, EPI_ISL_1304953, EPI_ISL_1304954, EPI_ISL_1304955, EPI_ISL_1304956, EPI_ISL_1304957, EPI_ISL_1304958, EPI_ISL_1304959, EPI_ISL_1304960,
EPI_ISL_1304961, EPI_ISL_1304962, EPI_ISL_1304963, EPI_ISL_1304964, EPI_ISL_1304965, EPI_ISL_1304966, EPI_ISL_1304967, EPI_ISL_1304968, EPI_ISL_1304969, EPI_ISL_1304970, EPI_ISL_1304971, EPI_ISL_1304972, EPI_ISL_1304973, EPI_ISL_1304974, EPI_ISL_1304975, EPI_ISL_1304976, EPI_ISL_1304977, EPI_ISL_1304978, EPI_ISL_1304979, EPI_ISL_1304980, EPI_ISL_1304981, EPI_ISL_1304982,
EPI_ISL_1304983, EPI_ISL_1304984, EPI_ISL_1304985, EPI_ISL_1304986, EPI_ISL_1304987, EPI_ISL_1304988, EPI_ISL_1304989, EPI_ISL_1304990, EPI_ISL_1304991, EPI_ISL_1304992, EPI_ISL_1304993, EPI_ISL_1304994, EPI_ISL_1304995, EPI_ISL_1304996, EPI_ISL_1304997, EPI_ISL_1304998, EPI_ISL_1304999, EPI_ISL_1305000, EPI_ISL_1305001, EPI_ISL_1305002, EPI_ISL_1305003, EPI_ISL_1305004,
EPI_ISL_1305005, EPI_ISL_1305006, EPI_ISL_1305007, EPI_ISL_1305008, EPI_ISL_1305009, EPI_ISL_1305010, EPI_ISL_1305011, EPI_ISL_1305012, EPI_ISL_1305013, EPI_ISL_1305014, EPI_ISL_1305015, EPI_ISL_1305016, EPI_ISL_1305017, EPI_ISL_1305018, EPI_ISL_1305019, EPI_ISL_1305020, EPI_ISL_1305021, EPI_ISL_1305022, EPI_ISL_1305023, EPI_ISL_1305024, EPI_ISL_1305025, EPI_ISL_1305026,
EPI_ISL_1305027, EPI_ISL_1305028, EPI_ISL_1305029, EPI_ISL_1305030, EPI_ISL_1305031, EPI_ISL_1305032, EPI_ISL_1305033, EPI_ISL_1305034, EPI_ISL_1305035, EPI_ISL_1305036, EPI_ISL_1305037, EPI_ISL_1305038, EPI_ISL_1305039, EPI_ISL_1305040, EPI_ISL_1305041, EPI_ISL_1305042, EPI_ISL_1305043, EPI_ISL_1305044, EPI_ISL_1305045, EPI_ISL_1305046, EPI_ISL_1305047, EPI_ISL_1305048,
EPI_ISL_1305049, EPI_ISL_1305050, EPI_ISL_1305051, EPI_ISL_1305052, EPI_ISL_1305053, EPI_ISL_1305054, EPI_ISL_1305055, EPI_ISL_1305056, EPI_ISL_1305057, EPI_ISL_1305058, EPI_ISL_1305059, EPI_ISL_1305060, EPI_ISL_1305061, EPI_ISL_1305062, EPI_ISL_1305063, EPI_ISL_1305064, EPI_ISL_1305065, EPI_ISL_1305066, EPI_ISL_1305067, EPI_ISL_1305068, EPI_ISL_1305069, EPI_ISL_1305070,
EPI_ISL_1305071, EPI_ISL_1305072, EPI_ISL_1305073, EPI_ISL_1305074, EPI_ISL_1305075, EPI_ISL_1305076, EPI_ISL_1305077, EPI_ISL_1305078, EPI_ISL_1305079, EPI_ISL_1305080, EPI_ISL_1305081, EPI_ISL_1305082, EPI_ISL_1305083, EPI_ISL_1305084, EPI_ISL_1305085, EPI_ISL_1305086, EPI_ISL_1305087, EPI_ISL_1305088, EPI_ISL_1305089, EPI_ISL_1305090, EPI_ISL_1305091, EPI_ISL_1305092,
EPI_ISL_1305093, EPI_ISL_1305094, EPI_ISL_1305095, EPI_ISL_1305096, EPI_ISL_1305097, EPI_ISL_1305098, EPI_ISL_1305099, EPI_ISL_1305100, EPI_ISL_1305101, EPI_ISL_1305102, EPI_ISL_1305103, EPI_ISL_1305104, EPI_ISL_1305105, EPI_ISL_1305106, EPI_ISL_1305107, EPI_ISL_1305108, EPI_ISL_1305109, EPI_ISL_1305110, EPI_ISL_1305111, EPI_ISL_1305112, EPI_ISL_1305113, EPI_ISL_1305114,
EPI_ISL_1305115, EPI_ISL_1305116, EPI_ISL_1305117, EPI_ISL_1305118, EPI_ISL_1305119, EPI_ISL_1305120, EPI_ISL_1305121, EPI_ISL_1305122, EPI_ISL_1305123, EPI_ISL_1305124, EPI_ISL_1305125, EPI_ISL_1305126, EPI_ISL_1305127, EPI_ISL_1305128, EPI_ISL_1305129, EPI_ISL_1305130, EPI_ISL_1305131, EPI_ISL_1305132, EPI_ISL_1305133, EPI_ISL_1305134, EPI_ISL_1305135, EPI_ISL_1305136,
EPI_ISL_1305137, EPI_ISL_1305138, EPI_ISL_1305139, EPI_ISL_1305140, EPI_ISL_1305141, EPI_ISL_1305142, EPI_ISL_1305143, EPI_ISL_1305144, EPI_ISL_1305145, EPI_ISL_1305146, EPI_ISL_1305147, EPI_ISL_1305148, EPI_ISL_1305149, EPI_ISL_1305150, EPI_ISL_1305151, EPI_ISL_1305152, EPI_ISL_1305153, EPI_ISL_1305154, EPI_ISL_1305155, EPI_ISL_1305156, EPI_ISL_1305157, EPI_ISL_1305158,
EPI_ISL_1305159, EPI_ISL_1305160, EPI_ISL_1305161, EPI_ISL_1305162, EPI_ISL_1305163, EPI_ISL_1305164, EPI_ISL_1305165, EPI_ISL_1305166, EPI_ISL_1305167, EPI_ISL_1305168, EPI_ISL_1305169, EPI_ISL_1305170, EPI_ISL_1305171, EPI_ISL_1305172, EPI_ISL_1305173, EPI_ISL_1305174, EPI_ISL_1305175, EPI_ISL_1305176, EPI_ISL_1305177, EPI_ISL_1305178, EPI_ISL_1305179, EPI_ISL_1305180,
EPI_ISL_1305181, EPI_ISL_1305182, EPI_ISL_1305183, EPI_ISL_1305184, EPI_ISL_1305185, EPI_ISL_1305186, EPI_ISL_1305187, EPI_ISL_1305188, EPI_ISL_1305189, EPI_ISL_1305190, EPI_ISL_1305191, EPI_ISL_1305192, EPI_ISL_1305193, EPI_ISL_1305194, EPI_ISL_1305195, EPI_ISL_1305196, EPI_ISL_1305197, EPI_ISL_1305198, EPI_ISL_1305199, EPI_ISL_1305200, EPI_ISL_1305201, EPI_ISL_1305202,
EPI_ISL_1305203, EPI_ISL_1305204, EPI_ISL_1305205, EPI_ISL_1305206, EPI_ISL_1305207, EPI_ISL_1305208, EPI_ISL_1305209, EPI_ISL_1305210, EPI_ISL_1305211, EPI_ISL_1305212, EPI_ISL_1305213, EPI_ISL_1305214, EPI_ISL_1305215, EPI_ISL_1305216, EPI_ISL_1305217, EPI_ISL_1305218, EPI_ISL_1305219, EPI_ISL_1305220, EPI_ISL_1305221, EPI_ISL_1305222, EPI_ISL_1305223, EPI_ISL_1305224,
EPI_ISL_1305225, EPI_ISL_1305226, EPI_ISL_1305227, EPI_ISL_1305228, EPI_ISL_1305229, EPI_ISL_1305230, EPI_ISL_1305231, EPI_ISL_1305232, EPI_ISL_1305233, EPI_ISL_1305234, EPI_ISL_1305235, EPI_ISL_1305236, EPI_ISL_1305237, EPI_ISL_1305238, EPI_ISL_1305239, EPI_ISL_1305240, EPI_ISL_1305241, EPI_ISL_1305242, EPI_ISL_1305243, EPI_ISL_1305244, EPI_ISL_1305245, EPI_ISL_1305246,
EPI_ISL_1305247, EPI_ISL_1305248, EPI_ISL_1305249, EPI_ISL_1305250, EPI_ISL_1305251, EPI_ISL_1305252, EPI_ISL_1305253, EPI_ISL_1305254, EPI_ISL_1305255, EPI_ISL_1305256, EPI_ISL_1305257, EPI_ISL_1305258, EPI_ISL_1305259, EPI_ISL_1305260, EPI_ISL_1305261, EPI_ISL_1305262, EPI_ISL_1305263, EPI_ISL_1305264, EPI_ISL_1305265, EPI_ISL_1305266, EPI_ISL_1305267, EPI_ISL_1305268,
EPI_ISL_1305269, EPI_ISL_1305270, EPI_ISL_1305271, EPI_ISL_1305272, EPI_ISL_1305273, EPI_ISL_1305274, EPI_ISL_1305275, EPI_ISL_1305276, EPI_ISL_1305277, EPI_ISL_1305278, EPI_ISL_1305279, EPI_ISL_1305280, EPI_ISL_1305281, EPI_ISL_1305282, EPI_ISL_1305283, EPI_ISL_1305284, EPI_ISL_1305285, EPI_ISL_1305286, EPI_ISL_1305287, EPI_ISL_1305288, EPI_ISL_1305289, EPI_ISL_1305290,
EPI_ISL_1305291, EPI_ISL_1305292, EPI_ISL_1305293, EPI_ISL_1305294, EPI_ISL_1305295, EPI_ISL_1305296, EPI_ISL_1305297, EPI_ISL_1305298, EPI_ISL_1305299, EPI_ISL_1305300, EPI_ISL_1305301, EPI_ISL_1305302, EPI_ISL_1305303, EPI_ISL_1305304, EPI_ISL_1305305, EPI_ISL_1305306, EPI_ISL_1305307, EPI_ISL_1305308, EPI_ISL_1305309, EPI_ISL_1305310, EPI_ISL_1305311, EPI_ISL_1305312,
EPI_ISL_1305313, EPI_ISL_1305314, EPI_ISL_1305315, EPI_ISL_1305316, EPI_ISL_1305317, EPI_ISL_1305318, EPI_ISL_1305319, EPI_ISL_1305320, EPI_ISL_1305321, EPI_ISL_1305322, EPI_ISL_1305323, EPI_ISL_1305324, EPI_ISL_1305325, EPI_ISL_1305326, EPI_ISL_1305327, EPI_ISL_1305328, EPI_ISL_1305329, EPI_ISL_1305330, EPI_ISL_1305331, EPI_ISL_1305332, EPI_ISL_1305333, EPI_ISL_1305334,
EPI_ISL_1305335, EPI_ISL_1305336, EPI_ISL_1305337, EPI_ISL_1305338, EPI_ISL_1305339, EPI_ISL_1305340, EPI_ISL_1305341, EPI_ISL_1305342, EPI_ISL_1305343, EPI_ISL_1305344, EPI_ISL_1305345, EPI_ISL_1305346, EPI_ISL_1305347, EPI_ISL_1305348, EPI_ISL_1305349, EPI_ISL_1305350, EPI_ISL_1305351, EPI_ISL_1305352, EPI_ISL_1305353, EPI_ISL_1305354, EPI_ISL_1305355, EPI_ISL_1305356,
EPI_ISL_1305357, EPI_ISL_1305358, EPI_ISL_1305359, EPI_ISL_1305360, EPI_ISL_1305361, EPI_ISL_1305362, EPI_ISL_1305363, EPI_ISL_1305364, EPI_ISL_1305365, EPI_ISL_1305366, EPI_ISL_1305367, EPI_ISL_1305368, EPI_ISL_1305369, EPI_ISL_1305370, EPI_ISL_1305371, EPI_ISL_1305372, EPI_ISL_1305373, EPI_ISL_1305374, EPI_ISL_1305375, EPI_ISL_1305376, EPI_ISL_1305377, EPI_ISL_1305378,
EPI_ISL_1305379, EPI_ISL_1305380, EPI_ISL_1305381, EPI_ISL_1305382, EPI_ISL_1305383, EPI_ISL_1305384, EPI_ISL_1305385, EPI_ISL_1305386, EPI_ISL_1305387, EPI_ISL_1305388, EPI_ISL_1305389, EPI_ISL_1305390, EPI_ISL_1305391, EPI_ISL_1305392, EPI_ISL_1305393, EPI_ISL_1305394, EPI_ISL_1305395, EPI_ISL_1305396, EPI_ISL_1305397, EPI_ISL_1305398, EPI_ISL_1305399, EPI_ISL_1305400,
EPI_ISL_1305401, EPI_ISL_1305402, EPI_ISL_1305403, EPI_ISL_1305404, EPI_ISL_1305405, EPI_ISL_1305406, EPI_ISL_1305407, EPI_ISL_1305408, EPI_ISL_1305409, EPI_ISL_1305410, EPI_ISL_1305411, EPI_ISL_1305412, EPI_ISL_1305413, EPI_ISL_1305414, EPI_ISL_1305415, EPI_ISL_1305416, EPI_ISL_1305417, EPI_ISL_1305418, EPI_ISL_1305419, EPI_ISL_1305420, EPI_ISL_1305421, EPI_ISL_1305422,
EPI_ISL_1305423, EPI_ISL_1305424, EPI_ISL_1305425, EPI_ISL_1305426, EPI_ISL_1305427, EPI_ISL_1305428, EPI_ISL_1305429, EPI_ISL_1305430, EPI_ISL_1305431, EPI_ISL_1305432, EPI_ISL_1305433, EPI_ISL_1305434, EPI_ISL_1305435, EPI_ISL_1305436, EPI_ISL_1305437, EPI_ISL_1305438, EPI_ISL_1305439, EPI_ISL_1305440, EPI_ISL_1305441, EPI_ISL_1305442, EPI_ISL_1305443, EPI_ISL_1305444,
EPI_ISL_1305445, EPI_ISL_1305446, EPI_ISL_1305447, EPI_ISL_1305448, EPI_ISL_1305449, EPI_ISL_1305450, EPI_ISL_1305451, EPI_ISL_1305452, EPI_ISL_1305453, EPI_ISL_1305454, EPI_ISL_1305455, EPI_ISL_1305456, EPI_ISL_1305457, EPI_ISL_1305458, EPI_ISL_1305459, EPI_ISL_1305460, EPI_ISL_1305461, EPI_ISL_1305462, EPI_ISL_1305463, EPI_ISL_1305464, EPI_ISL_1305465, EPI_ISL_1305466,
EPI_ISL_1305467, EPI_ISL_1305468, EPI_ISL_1305469, EPI_ISL_1305470, EPI_ISL_1305471, EPI_ISL_1305472, EPI_ISL_1305473, EPI_ISL_1305474, EPI_ISL_1305475, EPI_ISL_1305476, EPI_ISL_1305477, EPI_ISL_1305478, EPI_ISL_1305479, EPI_ISL_1305480, EPI_ISL_1305481, EPI_ISL_1305482, EPI_ISL_1305483, EPI_ISL_1305484, EPI_ISL_1305485, EPI_ISL_1305486, EPI_ISL_1305487, EPI_ISL_1305488,
EPI_ISL_1305489, EPI_ISL_1305490, EPI_ISL_1305491, EPI_ISL_1305492, EPI_ISL_1305493, EPI_ISL_1305494, EPI_ISL_1305495, EPI_ISL_1305496, EPI_ISL_1305497, EPI_ISL_1305498, EPI_ISL_1305499, EPI_ISL_1305500, EPI_ISL_1305501, EPI_ISL_1305502, EPI_ISL_1305503, EPI_ISL_1305504, EPI_ISL_1305505, EPI_ISL_1305506, EPI_ISL_1305507, EPI_ISL_1305508, EPI_ISL_1305509, EPI_ISL_1305510,
EPI_ISL_1305511, EPI_ISL_1305512, EPI_ISL_1305513, EPI_ISL_1305514, EPI_ISL_1305515, EPI_ISL_1305516, EPI_ISL_1305517, EPI_ISL_1305518, EPI_ISL_1305519, EPI_ISL_1305520, EPI_ISL_1305521, EPI_ISL_1305522, EPI_ISL_1305523, EPI_ISL_1305524, EPI_ISL_1305525, EPI_ISL_1305526, EPI_ISL_1305527, EPI_ISL_1305528, EPI_ISL_1305529, EPI_ISL_1305530, EPI_ISL_1305531, EPI_ISL_1305532,
EPI_ISL_1305533, EPI_ISL_1305534, EPI_ISL_1305535, EPI_ISL_1305536, EPI_ISL_1305537, EPI_ISL_1305538, EPI_ISL_1305539, EPI_ISL_1305540, EPI_ISL_1305541, EPI_ISL_1305542, EPI_ISL_1305543, EPI_ISL_1305544, EPI_ISL_1305545, EPI_ISL_1305546, EPI_ISL_1305547, EPI_ISL_1305548, EPI_ISL_1305549, EPI_ISL_1305550, EPI_ISL_1305551, EPI_ISL_1305552, EPI_ISL_1305553, EPI_ISL_1305554,
EPI_ISL_1305555, EPI_ISL_1305556, EPI_ISL_1305557, EPI_ISL_1305558, EPI_ISL_1305559, EPI_ISL_1305560, EPI_ISL_1305561, EPI_ISL_1305562, EPI_ISL_1305563, EPI_ISL_1305564, EPI_ISL_1305565, EPI_ISL_1305566, EPI_ISL_1305567, EPI_ISL_1305568, EPI_ISL_1305569, EPI_ISL_1305570, EPI_ISL_1305571, EPI_ISL_1305572, EPI_ISL_1305573, EPI_ISL_1305574, EPI_ISL_1305575, EPI_ISL_1305576,
EPI_ISL_1305577, EPI_ISL_1305578, EPI_ISL_1305579, EPI_ISL_1305580, EPI_ISL_1305581, EPI_ISL_1305582, EPI_ISL_1305583, EPI_ISL_1305584, EPI_ISL_1305585, EPI_ISL_1305586, EPI_ISL_1305587, EPI_ISL_1305588, EPI_ISL_1305589, EPI_ISL_1305590, EPI_ISL_1305591, EPI_ISL_1305592, EPI_ISL_1305593, EPI_ISL_1305594, EPI_ISL_1305595, EPI_ISL_1305596, EPI_ISL_1305597, EPI_ISL_1305598,
EPI_ISL_1305599, EPI_ISL_1305600, EPI_ISL_1305601, EPI_ISL_1305602, EPI_ISL_1305603, EPI_ISL_1305604, EPI_ISL_1305605, EPI_ISL_1305606, EPI_ISL_1305607, EPI_ISL_1305608, EPI_ISL_1305609, EPI_ISL_1305610, EPI_ISL_1305611, EPI_ISL_1305612, EPI_ISL_1305613, EPI_ISL_1305614, EPI_ISL_1305615, EPI_ISL_1305616, EPI_ISL_1305617, EPI_ISL_1305618, EPI_ISL_1305619, EPI_ISL_1305620,
EPI_ISL_1305621, EPI_ISL_1305622, EPI_ISL_1305623, EPI_ISL_1305624, EPI_ISL_1305625, EPI_ISL_1305626, EPI_ISL_1305627, EPI_ISL_1305628, EPI_ISL_1305629, EPI_ISL_1305630, EPI_ISL_1305631, EPI_ISL_1305632, EPI_ISL_1305633, EPI_ISL_1305634, EPI_ISL_1305635, EPI_ISL_1305636, EPI_ISL_1305637, EPI_ISL_1305638, EPI_ISL_1305639, EPI_ISL_1305640, EPI_ISL_1305641, EPI_ISL_1305642,
EPI_ISL_1305643, EPI_ISL_1305644, EPI_ISL_1305645, EPI_ISL_1305646, EPI_ISL_1305647, EPI_ISL_1305648, EPI_ISL_1305649, EPI_ISL_1305650, EPI_ISL_1305651, EPI_ISL_1305652, EPI_ISL_1305653, EPI_ISL_1305654, EPI_ISL_1305655, EPI_ISL_1305656, EPI_ISL_1305657, EPI_ISL_1305658, EPI_ISL_1305659, EPI_ISL_1305660, EPI_ISL_1305661, EPI_ISL_1305662, EPI_ISL_1305663, EPI_ISL_1305664,
EPI_ISL_1305665, EPI_ISL_1305666, EPI_ISL_1305667, EPI_ISL_1305668, EPI_ISL_1305669, EPI_ISL_1305670, EPI_ISL_1305671, EPI_ISL_1305672, EPI_ISL_1305673, EPI_ISL_1305674, EPI_ISL_1305675, EPI_ISL_1305676, EPI_ISL_1305677, EPI_ISL_1305678, EPI_ISL_1305679, EPI_ISL_1305680, EPI_ISL_1305681, EPI_ISL_1305682, EPI_ISL_1305683, EPI_ISL_1305684, EPI_ISL_1305685, EPI_ISL_1305686,
EPI_ISL_1305687, EPI_ISL_1305688, EPI_ISL_1305689, EPI_ISL_1305690, EPI_ISL_1305691, EPI_ISL_1305692, EPI_ISL_1305693, EPI_ISL_1305694, EPI_ISL_1305695, EPI_ISL_1305696, EPI_ISL_1305697, EPI_ISL_1305698, EPI_ISL_1305699, EPI_ISL_1305700, EPI_ISL_1305701, EPI_ISL_1305702, EPI_ISL_1305703, EPI_ISL_1305704, EPI_ISL_1305705, EPI_ISL_1305706, EPI_ISL_1305707, EPI_ISL_1305708,

| | | | |
|---|---|---|---|
| EPI_ISL_1305709, EPI_ISL_1305710, EPI_ISL_1305711, EPI_ISL_1305712, EPI_ISL_1305713, EPI_ISL_1305714, EPI_ISL_1305715, EPI_ISL_1305716, EPI_ISL_1305717, EPI_ISL_1305718, EPI_ISL_1305719, EPI_ISL_1305720, EPI_ISL_1305721, EPI_ISL_1305722, EPI_ISL_1305723, EPI_ISL_1305724, EPI_ISL_1305725, EPI_ISL_1305726, EPI_ISL_1305727, EPI_ISL_1305728, EPI_ISL_1305729, EPI_ISL_1305730, EPI_ISL_1305731, EPI_ISL_1305732, EPI_ISL_1305733, EPI_ISL_1305734, EPI_ISL_1305735, EPI_ISL_1305736, EPI_ISL_1305737, EPI_ISL_1305738, EPI_ISL_1305739, EPI_ISL_1305740, EPI_ISL_1305741, EPI_ISL_1305742, EPI_ISL_1305743, EPI_ISL_1305744, EPI_ISL_1305745, EPI_ISL_1305746, EPI_ISL_1305747, EPI_ISL_1305748, EPI_ISL_1305749, EPI_ISL_1305750, EPI_ISL_1305751, EPI_ISL_1305752, EPI_ISL_1305753, EPI_ISL_1305754, EPI_ISL_1305755, EPI_ISL_1305756, EPI_ISL_1305757, EPI_ISL_1305758, EPI_ISL_1305759, EPI_ISL_1305760, EPI_ISL_1305761, EPI_ISL_1305762, EPI_ISL_1305763, EPI_ISL_1305764, EPI_ISL_1305765, EPI_ISL_1305766, EPI_ISL_1305767, EPI_ISL_1305768, EPI_ISL_1305769, EPI_ISL_1305770, EPI_ISL_1305771, EPI_ISL_1305772, EPI_ISL_1305773 | | | |
| see above | Houston Methodist Hospital | Houston Methodist Hospital | S. Wesley Long, Randall J. Olsen, Paul A. Christensen, Sishir Subedi, Robert Olson, James J. Davis, Matthew Ojeda Saavedra, Prasanti Yerramilli, Layne Pruitt, Kristina Reppond, Madison N. Shyer, Jessica Cambric, Ilya J. Finkelstein, Jimmy Gollihar, and James M. Musser |
| EPI_ISL_1305819, EPI_ISL_1305820, EPI_ISL_1305821, EPI_ISL_1305822, EPI_ISL_1305823, EPI_ISL_1305824, EPI_ISL_1305825, EPI_ISL_1305826, EPI_ISL_1305827, EPI_ISL_1305828, EPI_ISL_1305829, EPI_ISL_1305830, EPI_ISL_1305831, EPI_ISL_1305832, EPI_ISL_1305833, EPI_ISL_1305834, EPI_ISL_1305835, EPI_ISL_1305836, EPI_ISL_1305837, EPI_ISL_1305838, EPI_ISL_1305839, EPI_ISL_1305840, EPI_ISL_1305841, EPI_ISL_1305842, EPI_ISL_1305843, EPI_ISL_1305844, EPI_ISL_1305845, EPI_ISL_1305846, EPI_ISL_1305847, EPI_ISL_1305848, EPI_ISL_1305849, EPI_ISL_1305850, EPI_ISL_1305851, EPI_ISL_1305852, EPI_ISL_1305853, EPI_ISL_1305854, EPI_ISL_1305855, EPI_ISL_1305856, EPI_ISL_1305857, EPI_ISL_1305858, EPI_ISL_1305859, EPI_ISL_1305860, EPI_ISL_1305861, EPI_ISL_1305862, EPI_ISL_1305863, EPI_ISL_1305864, EPI_ISL_1305865, EPI_ISL_1305866, EPI_ISL_1305867, EPI_ISL_1305868, EPI_ISL_1305869, EPI_ISL_1305870, EPI_ISL_1305871, EPI_ISL_1305872, EPI_ISL_1305873, EPI_ISL_1305874, EPI_ISL_1305875, EPI_ISL_1305876, EPI_ISL_1305877, EPI_ISL_1305878, EPI_ISL_1305879, EPI_ISL_1305880, EPI_ISL_1305881, EPI_ISL_1305882, EPI_ISL_1305883, EPI_ISL_1305884, EPI_ISL_1305885, EPI_ISL_1305886, EPI_ISL_1305887, EPI_ISL_1305888, EPI_ISL_1305889, EPI_ISL_1305890, EPI_ISL_1305891, EPI_ISL_1305892, EPI_ISL_1305893, EPI_ISL_1305894, EPI_ISL_1305895, EPI_ISL_1305896, EPI_ISL_1305897, EPI_ISL_1305898, EPI_ISL_1305899, EPI_ISL_1305900, EPI_ISL_1305901, EPI_ISL_1305902, EPI_ISL_1305903, EPI_ISL_1305908, EPI_ISL_1305909, EPI_ISL_1305910, EPI_ISL_1305911, EPI_ISL_1305912, EPI_ISL_1305913, EPI_ISL_1305914, EPI_ISL_1305915, EPI_ISL_1305916, EPI_ISL_1305917, EPI_ISL_1305918, EPI_ISL_1305919, EPI_ISL_1305920, EPI_ISL_1305921, EPI_ISL_1305922, EPI_ISL_1305923, EPI_ISL_1305924, EPI_ISL_1305925, EPI_ISL_1305926, EPI_ISL_1305927, EPI_ISL_1305928, EPI_ISL_1305929, EPI_ISL_1305930, EPI_ISL_1305931, EPI_ISL_1305932, EPI_ISL_1305933, EPI_ISL_1305934, EPI_ISL_1305935, EPI_ISL_1305936, EPI_ISL_1305937, EPI_ISL_1305938, EPI_ISL_1305939, EPI_ISL_1305940, EPI_ISL_1305941, EPI_ISL_1305942, EPI_ISL_1305943, EPI_ISL_1305944, EPI_ISL_1305945, EPI_ISL_1305946, EPI_ISL_1305947, EPI_ISL_1305948, EPI_ISL_1305949, EPI_ISL_1305950, EPI_ISL_1305951, EPI_ISL_1305952, EPI_ISL_1305953, EPI_ISL_1305954, EPI_ISL_1305955, EPI_ISL_1305956, EPI_ISL_1305957, EPI_ISL_1305958, EPI_ISL_1305959, EPI_ISL_1305960, EPI_ISL_1305961, EPI_ISL_1305962, EPI_ISL_1305963, EPI_ISL_1305964, EPI_ISL_1305965, EPI_ISL_1305966, EPI_ISL_1305967, EPI_ISL_1305968, EPI_ISL_1305969, EPI_ISL_1305970, EPI_ISL_1305971, EPI_ISL_1305972, EPI_ISL_1305973 | | | |
| see above | Johns Hopkins Hospital Department of Pathology | Johns Hopkins Hospital Department of Pathology | C. Paul Morris, Chun Huai Luo, Adannaya Amadi, Matthew Schwartz, Heba H. Mostafa |
| EPI_ISL_1306092 | UW Virology Lab | UW Virology Lab | Pavitra Roychoudhury, Hong Xie, Lasata Shrestha, Shah Mohamed Bakhash, Michelle Lin, Margaret Mills, Noah Baker, Sean Ellis, Saraswathi Sathees, Meei-Li Huang, Keith R Jerome, Alexander Greninger |
| EPI_ISL_1306093, EPI_ISL_1306094, EPI_ISL_1306095, EPI_ISL_1306096, EPI_ISL_1306097, EPI_ISL_1306098, EPI_ISL_1306099, EPI_ISL_1306101, EPI_ISL_1306102, EPI_ISL_1306103, EPI_ISL_1306105, EPI_ISL_1306106, EPI_ISL_1306107, EPI_ISL_1306108, EPI_ISL_1306109, EPI_ISL_1306113, EPI_ISL_1306114, EPI_ISL_1306115, EPI_ISL_1306116, EPI_ISL_1306117, EPI_ISL_1306118, EPI_ISL_1306119 | | | |
| see above | Arizona State Public Health Laboratory | Arizona State Public Health Laboratory | Trung Huynh, Jessica Escobar, Katherine Fullerton, Nobuko Fukushima, Stacy White, Linda Getsinger, Victor Waddell |
| EPI_ISL_1306139, EPI_ISL_1306140, EPI_ISL_1306141, EPI_ISL_1306142, EPI_ISL_1306143, EPI_ISL_1306144, EPI_ISL_1306145, EPI_ISL_1306146, EPI_ISL_1306147, EPI_ISL_1306148, EPI_ISL_1306149, EPI_ISL_1306150, EPI_ISL_1306151, EPI_ISL_1306152, EPI_ISL_1306153, EPI_ISL_1306154, EPI_ISL_1306155, EPI_ISL_1306156, EPI_ISL_1306157, EPI_ISL_1306158, EPI_ISL_1306159, EPI_ISL_1306160, EPI_ISL_1306161, EPI_ISL_1306162, EPI_ISL_1306163, EPI_ISL_1306164, EPI_ISL_1306165, EPI_ISL_1306166, EPI_ISL_1306167, EPI_ISL_1306168, EPI_ISL_1306169, EPI_ISL_1306170, EPI_ISL_1306171, EPI_ISL_1306172, EPI_ISL_1306173, EPI_ISL_1306174, EPI_ISL_1306175, EPI_ISL_1306176, EPI_ISL_1306177, EPI_ISL_1306178, EPI_ISL_1306179, EPI_ISL_1306180, EPI_ISL_1306181, EPI_ISL_1306182, EPI_ISL_1306183, EPI_ISL_1306184, EPI_ISL_1306185, EPI_ISL_1306186, EPI_ISL_1306187, EPI_ISL_1306188, EPI_ISL_1306189, EPI_ISL_1306190, EPI_ISL_1306191, EPI_ISL_1306192, EPI_ISL_1306193, EPI_ISL_1306194, EPI_ISL_1306195, EPI_ISL_1306196, EPI_ISL_1306197, EPI_ISL_1306198, EPI_ISL_1306199, EPI_ISL_1306200, EPI_ISL_1306201, EPI_ISL_1306202, EPI_ISL_1306203, EPI_ISL_1306204, EPI_ISL_1306205, EPI_ISL_1306206, EPI_ISL_1306207, EPI_ISL_1306208, EPI_ISL_1306209, EPI_ISL_1306210, EPI_ISL_1306211, EPI_ISL_1306212, EPI_ISL_1306213, EPI_ISL_1306214, EPI_ISL_1306215, EPI_ISL_1306216, EPI_ISL_1306217, EPI_ISL_1306218, EPI_ISL_1306219, EPI_ISL_1306220, EPI_ISL_1306221, EPI_ISL_1306222, EPI_ISL_1306223, EPI_ISL_1306224, EPI_ISL_1306225, EPI_ISL_1306226, EPI_ISL_1306227, EPI_ISL_1306228, EPI_ISL_1306229, EPI_ISL_1306230, EPI_ISL_1306231, EPI_ISL_1306232, EPI_ISL_1306233, EPI_ISL_1306234, EPI_ISL_1306235, EPI_ISL_1306236, EPI_ISL_1306237, EPI_ISL_1306238, EPI_ISL_1306239, EPI_ISL_1306240, EPI_ISL_1306241, EPI_ISL_1306242, EPI_ISL_1306243, EPI_ISL_1306244, EPI_ISL_1306245, EPI_ISL_1306246, EPI_ISL_1306247, EPI_ISL_1306248, EPI_ISL_1306249, EPI_ISL_1306250, EPI_ISL_1306251, EPI_ISL_1306252 | | | |
| see above | Department of Laboratory Medicine, Clinical Center, National Institutes of Health | Laboratory of Parasitic Diseases, Systems Genomics Section, National Institute of Allergy and Infectious Diseases, National Institutes of Health | Allison Roder, Stephanie Banakis, Matthew Chung, Jung-ho Youn, Rachel Mercado, Wei Wang, Tara Palmore, Michael Bell, Heike Bailin, Jessica McCormick-Ell, Sanchita Das, Jennifer Kwan, Elodie Ghedin |
| EPI_ISL_1306253, EPI_ISL_1306254, EPI_ISL_1306255, EPI_ISL_1306256, EPI_ISL_1306257, EPI_ISL_1306258, EPI_ISL_1306259, EPI_ISL_1306260, EPI_ISL_1306261, EPI_ISL_1306262, EPI_ISL_1306264, EPI_ISL_1306265, EPI_ISL_1306266, EPI_ISL_1306267, EPI_ISL_1306268, EPI_ISL_1306269, EPI_ISL_1306271, EPI_ISL_1306272, EPI_ISL_1306273, EPI_ISL_1306274, EPI_ISL_1306275, EPI_ISL_1306276, EPI_ISL_1306277, EPI_ISL_1306278, EPI_ISL_1306279, EPI_ISL_1306280, EPI_ISL_1306281, EPI_ISL_1306282, EPI_ISL_1306283, EPI_ISL_1306284, EPI_ISL_1306286, EPI_ISL_1306287, EPI_ISL_1306288, EPI_ISL_1306289, EPI_ISL_1306291, EPI_ISL_1306293, EPI_ISL_1306294, EPI_ISL_1306295, EPI_ISL_1306296, EPI_ISL_1306298, EPI_ISL_1306299, EPI_ISL_1306300, EPI_ISL_1306301, EPI_ISL_1306302, EPI_ISL_1306303, EPI_ISL_1306305, EPI_ISL_1306306, EPI_ISL_1306307, EPI_ISL_1306308, EPI_ISL_1306309, EPI_ISL_1306310, EPI_ISL_1306311, EPI_ISL_1306312, EPI_ISL_1306313, EPI_ISL_1306314, EPI_ISL_1306315, EPI_ISL_1306316, EPI_ISL_1306317, EPI_ISL_1306318, EPI_ISL_1306319, EPI_ISL_1306320, EPI_ISL_1306321, EPI_ISL_1306322, EPI_ISL_1306323, EPI_ISL_1306324, EPI_ISL_1306325, EPI_ISL_1306326, EPI_ISL_1306328, EPI_ISL_1306329, EPI_ISL_1306330, EPI_ISL_1306331, EPI_ISL_1306332, EPI_ISL_1306333, EPI_ISL_1306334, EPI_ISL_1306335, EPI_ISL_1306336, EPI_ISL_1306337, EPI_ISL_1306338, EPI_ISL_1306339, EPI_ISL_1306340, EPI_ISL_1306341, EPI_ISL_1306342, EPI_ISL_1306343, EPI_ISL_1306344, EPI_ISL_1306345, EPI_ISL_1306347, EPI_ISL_1306348, EPI_ISL_1306349, EPI_ISL_1306350, EPI_ISL_1306351, EPI_ISL_1306352, EPI_ISL_1306353, EPI_ISL_1306354, EPI_ISL_1306355, EPI_ISL_1306356, EPI_ISL_1306357, EPI_ISL_1306358, EPI_ISL_1306359, EPI_ISL_1306360, EPI_ISL_1306361, EPI_ISL_1306362, EPI_ISL_1306363, EPI_ISL_1306364, EPI_ISL_1306365, EPI_ISL_1306366, EPI_ISL_1306367, EPI_ISL_1306368, EPI_ISL_1306369, EPI_ISL_1306370, EPI_ISL_1306371, EPI_ISL_1306372, EPI_ISL_1306373, EPI_ISL_1306374, EPI_ISL_1306375, EPI_ISL_1306376, EPI_ISL_1306377, EPI_ISL_1306378, EPI_ISL_1306379, EPI_ISL_1306380, EPI_ISL_1306381, EPI_ISL_1306382, EPI_ISL_1306384, EPI_ISL_1306385, EPI_ISL_1306386, EPI_ISL_1306387, EPI_ISL_1306388, EPI_ISL_1306389, EPI_ISL_1306391, EPI_ISL_1306392, EPI_ISL_1306394, EPI_ISL_1306396, EPI_ISL_1306397, EPI_ISL_1306398, EPI_ISL_1306399, EPI_ISL_1306400, EPI_ISL_1306401, EPI_ISL_1306402, EPI_ISL_1306403, EPI_ISL_1306404, EPI_ISL_1306405, EPI_ISL_1306406, EPI_ISL_1306407, EPI_ISL_1306408, EPI_ISL_1306409, EPI_ISL_1306410, EPI_ISL_1306411, EPI_ISL_1306412, EPI_ISL_1306413, EPI_ISL_1306414, EPI_ISL_1306415, EPI_ISL_1306416, EPI_ISL_1306417, EPI_ISL_1306418, EPI_ISL_1306419, EPI_ISL_1306420, EPI_ISL_1306421, EPI_ISL_1306424, EPI_ISL_1306425, EPI_ISL_1306426, EPI_ISL_1306427, EPI_ISL_1306428, EPI_ISL_1306429, EPI_ISL_1306430, EPI_ISL_1306431, EPI_ISL_1306432, EPI_ISL_1306433, EPI_ISL_1306434, EPI_ISL_1306435, EPI_ISL_1306436, EPI_ISL_1306437, EPI_ISL_1306438, EPI_ISL_1306439, EPI_ISL_1306440, EPI_ISL_1306441, EPI_ISL_1306442, EPI_ISL_1306443, EPI_ISL_1306445, EPI_ISL_1306446, EPI_ISL_1306447, EPI_ISL_1306448, EPI_ISL_1306449, EPI_ISL_1306450, EPI_ISL_1306454, EPI_ISL_1306455, EPI_ISL_1306456, EPI_ISL_1306457, EPI_ISL_1306460, EPI_ISL_1306462, EPI_ISL_1306463, EPI_ISL_1306464, EPI_ISL_1306465, EPI_ISL_1306466, EPI_ISL_1306467, EPI_ISL_1306468, EPI_ISL_1306469, EPI_ISL_1306470, EPI_ISL_1306471, EPI_ISL_1306472, EPI_ISL_1306473, EPI_ISL_1306474, EPI_ISL_1306475, EPI_ISL_1306477, EPI_ISL_1306478, EPI_ISL_1306480, EPI_ISL_1306481, EPI_ISL_1306482, EPI_ISL_1306483, EPI_ISL_1306484, EPI_ISL_1306485, EPI_ISL_1306486, EPI_ISL_1306487, EPI_ISL_1306488, EPI_ISL_1306489, EPI_ISL_1306490, EPI_ISL_1306491, EPI_ISL_1306492, EPI_ISL_1306493, EPI_ISL_1306495, EPI_ISL_1306496, EPI_ISL_1306498, EPI_ISL_1306499, EPI_ISL_1306500, EPI_ISL_1306501, EPI_ISL_1306502, EPI_ISL_1306503, EPI_ISL_1306505, EPI_ISL_1306506, EPI_ISL_1306507, EPI_ISL_1306508, EPI_ISL_1306509, EPI_ISL_1306510, EPI_ISL_1306511, EPI_ISL_1306512, EPI_ISL_1306513, EPI_ISL_1306514, EPI_ISL_1306519, EPI_ISL_1306520, EPI_ISL_1306521, EPI_ISL_1306522, EPI_ISL_1306524, EPI_ISL_1306526, EPI_ISL_1306527, EPI_ISL_1306528, EPI_ISL_1306529, EPI_ISL_1306530, EPI_ISL_1306531, EPI_ISL_1306532, EPI_ISL_1306533, EPI_ISL_1306536, EPI_ISL_1306537, EPI_ISL_1306538, EPI_ISL_1306539, EPI_ISL_1306540, EPI_ISL_1306542, EPI_ISL_1306547, EPI_ISL_1306548, EPI_ISL_1306549, EPI_ISL_1306550, EPI_ISL_1306551, EPI_ISL_1306553, EPI_ISL_1306554, EPI_ISL_1306555, EPI_ISL_1306556, EPI_ISL_1306557, EPI_ISL_1306558, EPI_ISL_1306560, EPI_ISL_1306561, EPI_ISL_1306562, EPI_ISL_1306563, EPI_ISL_1306564, EPI_ISL_1306565, EPI_ISL_1306566, EPI_ISL_1306567, EPI_ISL_1306568, EPI_ISL_1306569, EPI_ISL_1306570, EPI_ISL_1306571, EPI_ISL_1306572, EPI_ISL_1306573, EPI_ISL_1306574, EPI_ISL_1306575, EPI_ISL_1306576, EPI_ISL_1306577, EPI_ISL_1306578, EPI_ISL_1306579, EPI_ISL_1306580, EPI_ISL_1306582, EPI_ISL_1306583, EPI_ISL_1306584, EPI_ISL_1306585, EPI_ISL_1306587, EPI_ISL_1306588, EPI_ISL_1306589, EPI_ISL_1306590, EPI_ISL_1306591, EPI_ISL_1306592, EPI_ISL_1306593, EPI_ISL_1306594, EPI_ISL_1306595, EPI_ISL_1306596, EPI_ISL_1306597, EPI_ISL_1306598, EPI_ISL_1306599, EPI_ISL_1306600, EPI_ISL_1306601, EPI_ISL_1306602, EPI_ISL_1306603, EPI_ISL_1306606, EPI_ISL_1306607, EPI_ISL_1306609, EPI_ISL_1306611, EPI_ISL_1306612, EPI_ISL_1306613, EPI_ISL_1306614, EPI_ISL_1306615, EPI_ISL_1306616, EPI_ISL_1306617, EPI_ISL_1306618, EPI_ISL_1306619, EPI_ISL_1306620, EPI_ISL_1306621, EPI_ISL_1306622, EPI_ISL_1306623, EPI_ISL_1306624, EPI_ISL_1306626, EPI_ISL_1306627, EPI_ISL_1306628, EPI_ISL_1306629, EPI_ISL_1306630, EPI_ISL_1306631, EPI_ISL_1306632, EPI_ISL_1306633, EPI_ISL_1306634, EPI_ISL_1306636, EPI_ISL_1306637, EPI_ISL_1306638, EPI_ISL_1306639, EPI_ISL_1306642, EPI_ISL_1306643, EPI_ISL_1306644, EPI_ISL_1306645, EPI_ISL_1306646, EPI_ISL_1306647, EPI_ISL_1306648, EPI_ISL_1306649, EPI_ISL_1306650, EPI_ISL_1306651, EPI_ISL_1306652, EPI_ISL_1306653, EPI_ISL_1306654, EPI_ISL_1306655, EPI_ISL_1306656, EPI_ISL_1306657, EPI_ISL_1306658, EPI_ISL_1306659, EPI_ISL_1306661, EPI_ISL_1306662, EPI_ISL_1306664, EPI_ISL_1306665, EPI_ISL_1306666, EPI_ISL_1306667, EPI_ISL_1306668, EPI_ISL_1306669, EPI_ISL_1306670, EPI_ISL_1306671, EPI_ISL_1306672, EPI_ISL_1306673, EPI_ISL_1306674, EPI_ISL_1306676, EPI_ISL_1306677, EPI_ISL_1306678, EPI_ISL_1306681, EPI_ISL_1306683, EPI_ISL_1306684, EPI_ISL_1306685, EPI_ISL_1306686, EPI_ISL_1306687, EPI_ISL_1306688, EPI_ISL_1306689, EPI_ISL_1306690, EPI_ISL_1306691, EPI_ISL_1306692, EPI_ISL_1306693, EPI_ISL_1306694, EPI_ISL_1306695, EPI_ISL_1306696, EPI_ISL_1306698, EPI_ISL_1306699, EPI_ISL_1306701, EPI_ISL_1306703, EPI_ISL_1306704, EPI_ISL_1306705, EPI_ISL_1306706, EPI_ISL_1306707, EPI_ISL_1306708, EPI_ISL_1306709, EPI_ISL_1306710, EPI_ISL_1306711, EPI_ISL_1306712, EPI_ISL_1306713, EPI_ISL_1306714, EPI_ISL_1306715, EPI_ISL_1306716, EPI_ISL_1306718, EPI_ISL_1306720, EPI_ISL_1306722, EPI_ISL_1306727, EPI_ISL_1306728, EPI_ISL_1306729, EPI_ISL_1306730, EPI_ISL_1306731, EPI_ISL_1306732, EPI_ISL_1306733, EPI_ISL_1306734, EPI_ISL_1306735, EPI_ISL_1306736, EPI_ISL_1306737, EPI_ISL_1306738, EPI_ISL_1306739, EPI_ISL_1306741, EPI_ISL_1306743, EPI_ISL_1306744, EPI_ISL_1306745, EPI_ISL_1306746, EPI_ISL_1306747, EPI_ISL_1306748, EPI_ISL_1306749, EPI_ISL_1306750, EPI_ISL_1306751, EPI_ISL_1306752, EPI_ISL_1306755, EPI_ISL_1306756, EPI_ISL_1306757, EPI_ISL_1306758, EPI_ISL_1306759, EPI_ISL_1306760, EPI_ISL_1306762, EPI_ISL_1306763, EPI_ISL_1306764, EPI_ISL_1306765, EPI_ISL_1306766, EPI_ISL_1306767, EPI_ISL_1306768, EPI_ISL_1306769, EPI_ISL_1306770, EPI_ISL_1306771, EPI_ISL_1306773, EPI_ISL_1306774, EPI_ISL_1306775, EPI_ISL_1306776, EPI_ISL_1306777, EPI_ISL_1306778, EPI_ISL_1306780, EPI_ISL_1306781, EPI_ISL_1306783, EPI_ISL_1306784, EPI_ISL_1306785, EPI_ISL_1306786, EPI_ISL_1306787, EPI_ISL_1306788, EPI_ISL_1306789, EPI_ISL_1306790, EPI_ISL_1306791, EPI_ISL_1306792, EPI_ISL_1306793, EPI_ISL_1306794, EPI_ISL_1306796, EPI_ISL_1306797, EPI_ISL_1306798, EPI_ISL_1306800, EPI_ISL_1306803, EPI_ISL_1306804, EPI_ISL_1306805, EPI_ISL_1306806, EPI_ISL_1306807, EPI_ISL_1306808, EPI_ISL_1306809, EPI_ISL_1306811, EPI_ISL_1306812, EPI_ISL_1306813, EPI_ISL_1306814, EPI_ISL_1306815, EPI_ISL_1306816, EPI_ISL_1306817, EPI_ISL_1306818, EPI_ISL_1306819, EPI_ISL_1306820, EPI_ISL_1306821, EPI_ISL_1306822, EPI_ISL_1306823, EPI_ISL_1306824, EPI_ISL_1306825, EPI_ISL_1306826, EPI_ISL_1306827, EPI_ISL_1306828, EPI_ISL_1306829, EPI_ISL_1306830, EPI_ISL_1306831, EPI_ISL_1306832, EPI_ISL_1306833, EPI_ISL_1306834, EPI_ISL_1306835, EPI_ISL_1306836, EPI_ISL_1306837, EPI_ISL_1306838, EPI_ISL_1306839, EPI_ISL_1306840, EPI_ISL_1306841, EPI_ISL_1306842, EPI_ISL_1306844, EPI_ISL_1306845, EPI_ISL_1306846, EPI_ISL_1306847, EPI_ISL_1306848, EPI_ISL_1306849, EPI_ISL_1306850, EPI_ISL_1306851, EPI_ISL_1306852, EPI_ISL_1306853, EPI_ISL_1306854, EPI_ISL_1306855, EPI_ISL_1306857, EPI_ISL_1306858, EPI_ISL_1306859, EPI_ISL_1306861, EPI_ISL_1306862, EPI_ISL_1306863, EPI_ISL_1306864, EPI_ISL_1306865, EPI_ISL_1306866, EPI_ISL_1306867, EPI_ISL_1306868, EPI_ISL_1306869, EPI_ISL_1306870, EPI_ISL_1306871, EPI_ISL_1306872, EPI_ISL_1306873, EPI_ISL_1306874, EPI_ISL_1306875, EPI_ISL_1306876, EPI_ISL_1306877, EPI_ISL_1306878, EPI_ISL_1306879, EPI_ISL_1306880, EPI_ISL_1306881, EPI_ISL_1306882, EPI_ISL_1306883, EPI_ISL_1306884, EPI_ISL_1306885, EPI_ISL_1306886, EPI_ISL_1306887, EPI_ISL_1306888, EPI_ISL_1306889, EPI_ISL_1306890, EPI_ISL_1306891, EPI_ISL_1306892, EPI_ISL_1306893, EPI_ISL_1306894, EPI_ISL_1306895, EPI_ISL_1306896, EPI_ISL_1306898, EPI_ISL_1306899, EPI_ISL_1306900, EPI_ISL_1306901, EPI_ISL_1306902, EPI_ISL_1306903, EPI_ISL_1306904, EPI_ISL_1306905, EPI_ISL_1306906, EPI_ISL_1306907, EPI_ISL_1306908, EPI_ISL_1306909, EPI_ISL_1306910, EPI_ISL_1306911, EPI_ISL_1306912, EPI_ISL_1306914, EPI_ISL_1306915, EPI_ISL_1306916, EPI_ISL_1306917, EPI_ISL_1306918, EPI_ISL_1306920, EPI_ISL_1306921, EPI_ISL_1306922, EPI_ISL_1306923, EPI_ISL_1306924, EPI_ISL_1306925, EPI_ISL_1306926, EPI_ISL_1306928, EPI_ISL_1306929, EPI_ISL_1306930, EPI_ISL_1306931, EPI_ISL_1306932, EPI_ISL_1306935, EPI_ISL_1306936, EPI_ISL_1306937, EPI_ISL_1306938, EPI_ISL_1306939, EPI_ISL_1306940, EPI_ISL_1306941, EPI_ISL_1306942, EPI_ISL_1306943, EPI_ISL_1306944, EPI_ISL_1306945, EPI_ISL_1306946, EPI_ISL_1306947, EPI_ISL_1306948, EPI_ISL_1306949, EPI_ISL_1306950, EPI_ISL_1306952, EPI_ISL_1306953, EPI_ISL_1306955, EPI_ISL_1306956, EPI_ISL_1306957, EPI_ISL_1306958, EPI_ISL_1306959, EPI_ISL_1306960, EPI_ISL_1306961, EPI_ISL_1306962, EPI_ISL_1306963, EPI_ISL_1306964, EPI_ISL_1306965, EPI_ISL_1306966, EPI_ISL_1306967, EPI_ISL_1306968, EPI_ISL_1306969, EPI_ISL_1306970, EPI_ISL_1306972, EPI_ISL_1306973, EPI_ISL_1306974, EPI_ISL_1306975, EPI_ISL_1306976, EPI_ISL_1306977, EPI_ISL_1306978, EPI_ISL_1306979, EPI_ISL_1306980, EPI_ISL_1306981, EPI_ISL_1306982, EPI_ISL_1306983, EPI_ISL_1306984, EPI_ISL_1306986, EPI_ISL_1306987, EPI_ISL_1306988, EPI_ISL_1306989, EPI_ISL_1306990, EPI_ISL_1306991, EPI_ISL_1306992, EPI_ISL_1306993, EPI_ISL_1306994, EPI_ISL_1306995, EPI_ISL_1306996, EPI_ISL_1306997, EPI_ISL_1306998, EPI_ISL_1306999, EPI_ISL_1307000, EPI_ISL_1307001, EPI_ISL_1307002, EPI_ISL_1307003, EPI_ISL_1307005, EPI_ISL_1307006, EPI_ISL_1307007, EPI_ISL_1307008, EPI_ISL_1307009, EPI_ISL_1307010, EPI_ISL_1307011, EPI_ISL_1307012, EPI_ISL_1307014, EPI_ISL_1307015, EPI_ISL_1307016, EPI_ISL_1307017, EPI_ISL_1307018, EPI_ISL_1307019, EPI_ISL_1307020, EPI_ISL_1307021, EPI_ISL_1307023, EPI_ISL_1307027, EPI_ISL_1307028, EPI_ISL_1307029, EPI_ISL_1307030, EPI_ISL_1307031, EPI_ISL_1307032, EPI_ISL_1307033, EPI_ISL_1307034, EPI_ISL_1307036, EPI_ISL_1307038, EPI_ISL_1307039, EPI_ISL_1307040, EPI_ISL_1307041, EPI_ISL_1307042, EPI_ISL_1307043, EPI_ISL_1307044, EPI_ISL_1307045, EPI_ISL_1307046, EPI_ISL_1307047, EPI_ISL_1307048, EPI_ISL_1307049, EPI_ISL_1307052, EPI_ISL_1307053, EPI_ISL_1307054, EPI_ISL_1307055, EPI_ISL_1307057, EPI_ISL_1307058, EPI_ISL_1307059, EPI_ISL_1307060, EPI_ISL_1307061, EPI_ISL_1307062, EPI_ISL_1307063, EPI_ISL_1307064, EPI_ISL_1307065, EPI_ISL_1307066, EPI_ISL_1307067, EPI_ISL_1307068, EPI_ISL_1307069, EPI_ISL_1307070, EPI_ISL_1307071, EPI_ISL_1307072, EPI_ISL_1307073, EPI_ISL_1307074, EPI_ISL_1307076, EPI_ISL_1307077, EPI_ISL_1307078, EPI_ISL_1307079, EPI_ISL_1307080, EPI_ISL_1307081, EPI_ISL_1307082, EPI_ISL_1307083, EPI_ISL_1307084, EPI_ISL_1307085, EPI_ISL_1307086, EPI_ISL_1307087, EPI_ISL_1307088, EPI_ISL_1307089, EPI_ISL_1307090, EPI_ISL_1307091, EPI_ISL_1307093, EPI_ISL_1307094, EPI_ISL_1307095, EPI_ISL_1307096, EPI_ISL_1307097, EPI_ISL_1307098, EPI_ISL_1307099, EPI_ISL_1307100, EPI_ISL_1307101, EPI_ISL_1307102, EPI_ISL_1307103, EPI_ISL_1307104, EPI_ISL_1307105, EPI_ISL_1307106, EPI_ISL_1307108, EPI_ISL_1307109, EPI_ISL_1307110, EPI_ISL_1307112, EPI_ISL_1307113, EPI_ISL_1307114, EPI_ISL_1307115, EPI_ISL_1307116, EPI_ISL_1307117, EPI_ISL_1307118, EPI_ISL_1307119, EPI_ISL_1307121, EPI_ISL_1307122, EPI_ISL_1307124, EPI_ISL_1307125, EPI_ISL_1307126, EPI_ISL_1307127, EPI_ISL_1307128, EPI_ISL_1307129, EPI_ISL_1307131, EPI_ISL_1307132, EPI_ISL_1307133, EPI_ISL_1307134, EPI_ISL_1307135, EPI_ISL_1307136, EPI_ISL_1307137, EPI_ISL_1307138, EPI_ISL_1307139, EPI_ISL_1307140, EPI_ISL_1307141, EPI_ISL_1307142, EPI_ISL_1307143, EPI_ISL_1307145, EPI_ISL_1307146, EPI_ISL_1307147, EPI_ISL_1307150, EPI_ISL_1307151, EPI_ISL_1307152, EPI_ISL_1307153, EPI_ISL_1307154, EPI_ISL_1307155, EPI_ISL_1307156, EPI_ISL_1307157, EPI_ISL_1307158, EPI_ISL_1307160, EPI_ISL_1307161, EPI_ISL_1307163, EPI_ISL_1307165, EPI_ISL_1307166, EPI_ISL_1307167, EPI_ISL_1307168, EPI_ISL_1307169, EPI_ISL_1307170, EPI_ISL_1307171, EPI_ISL_1307172, EPI_ISL_1307174, EPI_ISL_1307175, EPI_ISL_1307176, EPI_ISL_1307178, EPI_ISL_1307179, EPI_ISL_1307180, EPI_ISL_1307182, EPI_ISL_1307183, EPI_ISL_1307184, EPI_ISL_1307185, EPI_ISL_1307186, EPI_ISL_1307187, EPI_ISL_1307191, EPI_ISL_1307192, EPI_ISL_1307193, EPI_ISL_1307194, EPI_ISL_1307195, EPI_ISL_1307196, EPI_ISL_1307197, EPI_ISL_1307198, EPI_ISL_1307199, EPI_ISL_1307200, EPI_ISL_1307201, EPI_ISL_1307202, EPI_ISL_1307204, EPI_ISL_1307205, EPI_ISL_1307206, EPI_ISL_1307207, EPI_ISL_1307208, EPI_ISL_1307209, EPI_ISL_1307210, EPI_ISL_1307211, EPI_ISL_1307212, EPI_ISL_1307213, EPI_ISL_1307214, EPI_ISL_1307216, EPI_ISL_1307217, EPI_ISL_1307218, EPI_ISL_1307219, EPI_ISL_1307220, EPI_ISL_1307221, EPI_ISL_1307222, EPI_ISL_1307225, EPI_ISL_1307226, EPI_ISL_1307227, EPI_ISL_1307228, EPI_ISL_1307229, EPI_ISL_1307230, EPI_ISL_1307231, EPI_ISL_1307232, EPI_ISL_1307233, EPI_ISL_1307234, EPI_ISL_1307235, EPI_ISL_1307240, EPI_ISL_1307241, EPI_ISL_1307242, EPI_ISL_1307243, EPI_ISL_1307244, EPI_ISL_1307245, EPI_ISL_1307246, EPI_ISL_1307247, EPI_ISL_1307248, EPI_ISL_1307249, EPI_ISL_1307251, EPI_ISL_1307253, EPI_ISL_1307254, EPI_ISL_1307255, EPI_ISL_1307256, EPI_ISL_1307259, EPI_ISL_1307260, EPI_ISL_1307261, EPI_ISL_1307262, EPI_ISL_1307263, EPI_ISL_1307264, EPI_ISL_1307265, EPI_ISL_1307266, EPI_ISL_1307267, EPI_ISL_1307268, EPI_ISL_1307270, EPI_ISL_1307271, EPI_ISL_1307272, EPI_ISL_1307273, EPI_ISL_1307274, EPI_ISL_1307275, EPI_ISL_1307276, EPI_ISL_1307278, EPI_ISL_1307282, EPI_ISL_1307283, EPI_ISL_1307284, EPI_ISL_1307285, EPI_ISL_1307286, EPI_ISL_1307288, EPI_ISL_1307289, EPI_ISL_1307290, EPI_ISL_1307291, EPI_ISL_1307292, EPI_ISL_1307293, EPI_ISL_1307294, EPI_ISL_1307295, EPI_ISL_1307296, EPI_ISL_1307297, EPI_ISL_1307298, EPI_ISL_1307299, EPI_ISL_1307300, EPI_ISL_1307301, EPI_ISL_1307302, EPI_ISL_1307303, EPI_ISL_1307304, EPI_ISL_1307305, EPI_ISL_1307306, EPI_ISL_1307307, EPI_ISL_1307308, EPI_ISL_1307309, EPI_ISL_1307310, EPI_ISL_1307311, EPI_ISL_1307312, EPI_ISL_1307313, EPI_ISL_1307314, EPI_ISL_1307315, EPI_ISL_1307316, EPI_ISL_1307317, EPI_ISL_1307318, EPI_ISL_1307319, EPI_ISL_1307320, EPI_ISL_1307321, EPI_ISL_1307322, EPI_ISL_1307323, EPI_ISL_1307324, EPI_ISL_1307325, EPI_ISL_1307326, EPI_ISL_1307327, EPI_ISL_1307328, EPI_ISL_1307329, EPI_ISL_1307330, EPI_ISL_1307331, EPI_ISL_1307332, EPI_ISL_1307333, EPI_ISL_1307334, EPI_ISL_1307335, EPI_ISL_1307336, EPI_ISL_1307337, EPI_ISL_1307338, EPI_ISL_1307339, EPI_ISL_1307340, EPI_ISL_1307341, EPI_ISL_1307342, EPI_ISL_1307343, EPI_ISL_1307344, EPI_ISL_1307345, EPI_ISL_1307346, EPI_ISL_1307347, EPI_ISL_1307348, EPI_ISL_1307349, EPI_ISL_1307350, EPI_ISL_1307351, EPI_ISL_1307352, EPI_ISL_1307354, EPI_ISL_1307355, EPI_ISL_1307356, EPI_ISL_1307357, EPI_ISL_1307358, EPI_ISL_1307359, EPI_ISL_1307360, EPI_ISL_1307361, EPI_ISL_1307362, EPI_ISL_1307363, EPI_ISL_1307364, EPI_ISL_1307365, EPI_ISL_1307366, EPI_ISL_1307367, EPI_ISL_1307368, EPI_ISL_1307369, EPI_ISL_1307370, EPI_ISL_1307371, EPI_ISL_1307372, EPI_ISL_1307373, EPI_ISL_1307374, EPI_ISL_1307375, EPI_ISL_1307376, EPI_ISL_1307377, EPI_ISL_1307378, EPI_ISL_1307379, EPI_ISL_1307380, EPI_ISL_1307381, EPI_ISL_1307382, EPI_ISL_1307383, EPI_ISL_1307384, EPI_ISL_1307385, EPI_ISL_1307386, EPI_ISL_1307387, EPI_ISL_1307388, EPI_ISL_1307389, EPI_ISL_1307390, EPI_ISL_1307391, EPI_ISL_1307392, EPI_ISL_1307393, EPI_ISL_1307394, EPI_ISL_1307395, EPI_ISL_1307396, EPI_ISL_1307397, EPI_ISL_1307399, EPI_ISL_1307400, EPI_ISL_1307401, EPI_ISL_1307402, EPI_ISL_1307403, EPI_ISL_1307404, EPI_ISL_1307405, EPI_ISL_1307406, EPI_ISL_1307407, EPI_ISL_1307408, EPI_ISL_1307409, EPI_ISL_1307410, EPI_ISL_1307411, EPI_ISL_1307412, EPI_ISL_1307413, EPI_ISL_1307414, EPI_ISL_1307415, EPI_ISL_1307416, EPI_ISL_1307417, EPI_ISL_1307418, EPI_ISL_1307420, EPI_ISL_1307421, EPI_ISL_1307422, EPI_ISL_1307423, EPI_ISL_1307424, EPI_ISL_1307425, EPI_ISL_1307426, EPI_ISL_1307427, EPI_ISL_1307428, EPI_ISL_1307429, EPI_ISL_1307430, EPI_ISL_1307431, EPI_ISL_1307432, EPI_ISL_1307433, EPI_ISL_1307434, EPI_ISL_1307435, EPI_ISL_1307436, EPI_ISL_1307437, EPI_ISL_1307438, EPI_ISL_1307439, EPI_ISL_1307441, EPI_ISL_1307442, EPI_ISL_1307443, EPI_ISL_1307444, EPI_ISL_1307445, EPI_ISL_1307446, EPI_ISL_1307447, EPI_ISL_1307448, EPI_ISL_1307449, EPI_ISL_1307450, EPI_ISL_1307451, EPI_ISL_1307452, EPI_ISL_1307453, EPI_ISL_1307454, EPI_ISL_1307455, EPI_ISL_1307456, EPI_ISL_1307457, EPI_ISL_1307458, EPI_ISL_1307459, EPI_ISL_1307460, EPI_ISL_1307461, EPI_ISL_1307463, EPI_ISL_1307464, EPI_ISL_1307465, EPI_ISL_1307466, EPI_ISL_1307467, EPI_ISL_1307468, EPI_ISL_1307469, EPI_ISL_1307470, EPI_ISL_1307471, EPI_ISL_1307472, EPI_ISL_1307473, EPI_ISL_1307474, EPI_ISL_1307475, EPI_ISL_1307476, EPI_ISL_1307477, EPI_ISL_1307478, EPI_ISL_1307479, EPI_ISL_1307480, EPI_ISL_1307481, EPI_ISL_1307482, EPI_ISL_1307484, EPI_ISL_1307485, EPI_ISL_1307486, EPI_ISL_1307487, EPI_ISL_1307488, EPI_ISL_1307489, EPI_ISL_1307490, EPI_ISL_1307491, EPI_ISL_1307492, EPI_ISL_1307493, EPI_ISL_1307495, EPI_ISL_1307496, EPI_ISL_1307497, EPI_ISL_1307498, EPI_ISL_1307499, EPI_ISL_1307500, EPI_ISL_1307501, EPI_ISL_1307502, EPI_ISL_1307503, EPI_ISL_1307504, EPI_ISL_1307505, EPI_ISL_1307506, EPI_ISL_1307507, EPI_ISL_1307508, EPI_ISL_1307509, EPI_ISL_1307510, EPI_ISL_1307511, EPI_ISL_1307512, EPI_ISL_1307513, EPI_ISL_1307514, EPI_ISL_1307515, EPI_ISL_1307516, EPI_ISL_1307517, EPI_ISL_1307518, EPI_ISL_1307519, EPI_ISL_1307520, EPI_ISL_1307521, EPI_ISL_1307522, EPI_ISL_1307523, EPI_ISL_1307524, EPI_ISL_1307525, EPI_ISL_1307526, EPI_ISL_1307527, EPI_ISL_1307528, EPI_ISL_1307529, EPI_ISL_1307530, EPI_ISL_1307531, EPI_ISL_1307532, EPI_ISL_1307533, EPI_ISL_1307534, EPI_ISL_1307535, EPI_ISL_1307536, EPI_ISL_1307537, EPI_ISL_1307538, EPI_ISL_1307539, EPI_ISL_1307540, EPI_ISL_1307541, EPI_ISL_1307542, EPI_ISL_1307543, EPI_ISL_1307544, EPI_ISL_1307545, EPI_ISL_1307546, EPI_ISL_1307547, EPI_ISL_1307548, EPI_ISL_1307550, EPI_ISL_1307551, EPI_ISL_1307552, EPI_ISL_1307553, EPI_ISL_1307554, EPI_ISL_1307555, EPI_ISL_1307556, EPI_ISL_1307557, EPI_ISL_1307558, EPI_ISL_1307559, EPI_ISL_1307560, EPI_ISL_1307561, EPI_ISL_1307562, EPI_ISL_1307563, EPI_ISL_1307564, EPI_ISL_1307565, EPI_ISL_1307566, EPI_ISL_1307567, EPI_ISL_1307568, EPI_ISL_1307570, EPI_ISL_1307571, EPI_ISL_1307572, EPI_ISL_1307573, EPI_ISL_1307574, EPI_ISL_1307575, EPI_ISL_1307576, EPI_ISL_1307577, EPI_ISL_1307578, EPI_ISL_1307579, EPI_ISL_1307580, EPI_ISL_1307581, EPI_ISL_1307582, EPI_ISL_1307583, EPI_ISL_1307584, EPI_ISL_1307585, EPI_ISL_1307586, EPI_ISL_1307587, EPI_ISL_1307588, EPI_ISL_1307589, EPI_ISL_1307590, EPI_ISL_1307593, EPI_ISL_1307594, EPI_ISL_1307595, EPI_ISL_1307596, EPI_ISL_1307597, EPI_ISL_1307598, EPI_ISL_1307599, EPI_ISL_1307600, EPI_ISL_1307601, EPI_ISL_1307602, EPI_ISL_1307603, EPI_ISL_1307604, EPI_ISL_1307605, EPI_ISL_1307606, EPI_ISL_1307607, EPI_ISL_1307608, EPI_ISL_1307609, EPI_ISL_1307610, EPI_ISL_1307611, EPI_ISL_1307612, EPI_ISL_1307613, EPI_ISL_1307614, EPI_ISL_1307615, EPI_ISL_1307616, EPI_ISL_1307617, EPI_ISL_1307618, EPI_ISL_1307619, EPI_ISL_1307620, EPI_ISL_1307621, EPI_ISL_1307622, EPI_ISL_1307623, EPI_ISL_1307624, EPI_ISL_1307625, EPI_ISL_1307626, EPI_ISL_1307627, EPI_ISL_1307628, EPI_ISL_1307629, EPI_ISL_1307630, EPI_ISL_1307631, EPI_ISL_1307632, EPI_ISL_1307633, EPI_ISL_1307634, EPI_ISL_1307635 | | | |
| see above | Pandemic Response Lab - NYC | Pandemic Response Lab, R&D | Henry Lee, Michael Hammerling, Melissa Hopkins, Cybill del Castillo, Shinyoung Clair Kang, William Ward, Pradeep Bugga, Sol Rey, Dylan Law, Haiping Hao, Jon Laurent |
| EPI_ISL_1307638 | VA Medical Center | Los Angeles County Public Health Laboratories | P. Hemarajata et al. |
| EPI_ISL_1307682 | South Dakota Public Health Laboratory, South Dakota Department of Health | South Dakota Public Health Laboratory, South Dakota Department of Health | Carlson,C. and Garfin,J. |
| EPI_ISL_1307717 | Baylor Esoteric + Molecular Lab | Baylor Esoteric + Molecular Lab | Elizabeth Forrester, Dawn Richards |
| EPI_ISL_1310787 | Ohio State University Wexner Medical Center | James Polaris Molecular Laboratory | Garee J, Ru P, Chappell D, Chang Y-S, Tu H, Snyder P, Pancholi P, Koenig S, Corcoran S, Jones D |
| EPI_ISL_1310788 | Ohio State University Wexner Medical Center | James Polaris Molecular Laboratory | Garee J, Tu H, Snyder P, Pancholi P, Koenig S, Corcoran S, Jones D |
| EPI_ISL_1310797, EPI_ISL_1310798, EPI_ISL_1310800, EPI_ISL_1310801, EPI_ISL_1310803, EPI_ISL_1310804, EPI_ISL_1310805, EPI_ISL_1310807 | Ohio Department of Health Laboratory | Ohio Department of Health Laboratory | Holmes, Jennifer; Eric Brandt, Keoni Omura, Glen McGillivary, Caitlin McDonnell, Jade Mowery, Stephanie Mccracken, Tyler Payne, Kirtana Ramadugu, Erica Leasure, Brent Lee, Kelsey Florek, Heather Blankenship, Allison Black, Quanta Brown, and Tammy Bannerman |
| EPI_ISL_1310808 | Instituto Nacional de Medicina Genomica | Instituto Nacional de Medicina Genomica | Hidalgo-Miranda A, Mendoza-Vargas A, Reyes-Grajeda JP, Cedro-Tanda A, Alcaraz N, Gonzalez-Barrera D, Rangel-DeLeon D, Ramirez-Vega O, Herrera-Montalvo LA |

| | | | |
|---|---|---|---|
| EPI_ISL_1310809, EPI_ISL_1310810 | Ohio Department of Health Laboratory | Ohio Department of Health Laboratory | Holmes, Jennifer; Eric Brandt, Keoni Omura, Glen McGillivary, Caitlin McDonnell, Jade Mowery, Stephanie Mccracken, Tyler Payne, Kirtana Ramadugu, Erica Leasure, Brent Lee, Kelsey Florek, Heather Blankenship, Allison Black, Quanta Brown, and Tammy Bannerman |
| EPI_ISL_1310811 | Instituto Nacional de Medicina Genomica | Instituto Nacional de Medicina Genomica | Hidalgo-Miranda A, Mendoza-Vargas A, Reyes-Grajeda JP, Cedro-Tanda A, Alcaraz N, Gonzalez-Barrera D, Rangel-DeLeon D, Ramirez-Vega O, Herrera-Montalvo LA |
| EPI_ISL_1310832 | UHTL | UHTL | Xiao Song |
| EPI_ISL_1311848 | Baylor Esoteric + Molecular Lab | Baylor Esoteric + Molecular Lab | Elizabeth Forrester, Dawn Richards |
| EPI_ISL_1311849, EPI_ISL_1311864, EPI_ISL_1311865, EPI_ISL_1311866 | Ohio Department of Health Laboratory | Ohio Department of Health Laboratory | Holmes, Jennifer; Eric Brandt, Keoni Omura, Glen McGillivary, Caitlin McDonnell, Jade Mowery, Stephanie Mccracken, Tyler Payne, Kirtana Ramadugu, Erica Leasure, Brent Lee, Kelsey Florek, Heather Blankenship, Allison Black, Quanta Brown, and Tammy Bannerman |
| EPI_ISL_1311965 | The Ohio State University Applied Microbiology Services Laboratory | The Ohio State University Applied Microbiology Services Laboratory | Seth A. Faith PhD |
| EPI_ISL_1312344, EPI_ISL_1312345 | Gravity Diagnostics | Kentucky State Public Health Lab | Stephanie Lunn, Karim George, Joshua Tobias, William Grooms, Vaneet Arora, Matthew Johnson, Rachel Zinner, Rhonda Lucas |
| EPI_ISL_1312346, EPI_ISL_1312347, EPI_ISL_1312348, EPI_ISL_1312350, EPI_ISL_1312351 | Kentucky State Public Health Lab | Kentucky State Public Health Lab | Stephanie Lunn, Karim George, Joshua Tobias, William Grooms, Vaneet Arora, Matthew Johnson, Rachel Zinner, Rhonda Lucas |
| EPI_ISL_1312354, EPI_ISL_1312357, EPI_ISL_1312358, EPI_ISL_1312359, EPI_ISL_1312360, EPI_ISL_1312361, EPI_ISL_1312362 | Gravity Diagnostics | Kentucky State Public Health Lab | Stephanie Lunn, Karim George, Joshua Tobias, William Grooms, Vaneet Arora, Matthew Johnson, Rachel Zinner, Rhonda Lucas |
| EPI_ISL_1312364 | Cytocheck Laboratory | Kansas Health and Environmental Lab | Mike Grose, Jonathan Barnell, Ben Olsen, and Phil Adam |
| EPI_ISL_1312773, EPI_ISL_1312774, EPI_ISL_1312775, EPI_ISL_1312776, EPI_ISL_1312777, EPI_ISL_1312778, EPI_ISL_1312779, EPI_ISL_1312780, EPI_ISL_1312781, EPI_ISL_1312782, EPI_ISL_1312783, EPI_ISL_1312784, EPI_ISL_1312785, EPI_ISL_1312786, EPI_ISL_1312787, EPI_ISL_1312788, EPI_ISL_1312789, EPI_ISL_1312790, EPI_ISL_1312791, EPI_ISL_1312792, EPI_ISL_1312793, EPI_ISL_1312794, EPI_ISL_1312795, EPI_ISL_1312796 | | | |
| see above | Molecular & Genomic Pathology Laboratory, Thomas Jefferson University Hospital | Molecular & Genomic Pathology Laboratory, Thomas Jefferson University Hospital | Run Jin, Nitika Badjatia, Zi-xuan Wang |
| EPI_ISL_1313542, EPI_ISL_1313543, EPI_ISL_1313547, EPI_ISL_1313549, EPI_ISL_1313551, EPI_ISL_1313553, EPI_ISL_1313554, EPI_ISL_1313555, EPI_ISL_1313556, EPI_ISL_1313557 | NJDOH, Public Health and Environmental Laboratories | NJ_PHEL | Lindsey Bodnar, Shiv K. Verma, Dana Woell, Byeong Jeong |
| EPI_ISL_1313714, EPI_ISL_1313715, EPI_ISL_1313716, EPI_ISL_1313717, EPI_ISL_1313718, EPI_ISL_1313719, EPI_ISL_1313720, EPI_ISL_1313721, EPI_ISL_1313722, EPI_ISL_1313723, EPI_ISL_1313724, EPI_ISL_1313725, EPI_ISL_1313726, EPI_ISL_1313727, EPI_ISL_1313728, EPI_ISL_1313729, EPI_ISL_1313730, EPI_ISL_1313731, EPI_ISL_1313732, EPI_ISL_1313733, EPI_ISL_1313734, EPI_ISL_1313735, EPI_ISL_1313736, EPI_ISL_1313737, EPI_ISL_1313738, EPI_ISL_1313739, EPI_ISL_1313740, EPI_ISL_1313741, EPI_ISL_1313742, EPI_ISL_1313743, EPI_ISL_1313744, EPI_ISL_1313745, EPI_ISL_1313746, EPI_ISL_1313747, EPI_ISL_1313748, EPI_ISL_1313749, EPI_ISL_1313750, EPI_ISL_1313751, EPI_ISL_1313752, EPI_ISL_1313753, EPI_ISL_1313754, EPI_ISL_1313755, EPI_ISL_1313756, EPI_ISL_1313757, EPI_ISL_1313758, EPI_ISL_1313759, EPI_ISL_1313760, EPI_ISL_1313761, EPI_ISL_1313762, EPI_ISL_1313763, EPI_ISL_1313764, EPI_ISL_1313765, EPI_ISL_1313766, EPI_ISL_1313767, EPI_ISL_1313768, EPI_ISL_1313769, EPI_ISL_1313770, EPI_ISL_1313771, EPI_ISL_1313772, EPI_ISL_1313773, EPI_ISL_1313774, EPI_ISL_1313775, EPI_ISL_1313776, EPI_ISL_1313777, EPI_ISL_1313778, EPI_ISL_1313779, EPI_ISL_1313780, EPI_ISL_1313781, EPI_ISL_1313782, EPI_ISL_1313783, EPI_ISL_1313784, EPI_ISL_1313785, EPI_ISL_1313786, EPI_ISL_1313787, EPI_ISL_1313788, EPI_ISL_1313789, EPI_ISL_1313790, EPI_ISL_1313791, EPI_ISL_1313792, EPI_ISL_1313793, EPI_ISL_1313794, EPI_ISL_1313795, EPI_ISL_1313796, EPI_ISL_1313797, EPI_ISL_1313798, EPI_ISL_1313799, EPI_ISL_1313800, EPI_ISL_1313801, EPI_ISL_1313802, EPI_ISL_1313803, EPI_ISL_1313804, EPI_ISL_1313805, EPI_ISL_1313806, EPI_ISL_1313807, EPI_ISL_1313808, EPI_ISL_1313809, EPI_ISL_1313810, EPI_ISL_1313811, EPI_ISL_1313812, EPI_ISL_1313813, EPI_ISL_1313814, EPI_ISL_1313815, EPI_ISL_1313816, EPI_ISL_1313817, EPI_ISL_1313818, EPI_ISL_1313819, EPI_ISL_1313820, EPI_ISL_1313821, EPI_ISL_1313822, EPI_ISL_1313823, EPI_ISL_1313824, EPI_ISL_1313825, EPI_ISL_1313826, EPI_ISL_1313827, EPI_ISL_1313828, EPI_ISL_1313829, EPI_ISL_1313830, EPI_ISL_1313831, EPI_ISL_1313832, EPI_ISL_1313833, EPI_ISL_1313834, EPI_ISL_1313835, EPI_ISL_1313836, EPI_ISL_1313837, EPI_ISL_1313838, EPI_ISL_1313839, EPI_ISL_1313840, EPI_ISL_1313841, EPI_ISL_1313842, EPI_ISL_1313843, EPI_ISL_1313844, EPI_ISL_1313845, EPI_ISL_1313846, EPI_ISL_1313847, EPI_ISL_1313848, EPI_ISL_1313849, EPI_ISL_1313850, EPI_ISL_1313851, EPI_ISL_1313852, EPI_ISL_1313853, EPI_ISL_1313854, EPI_ISL_1313855, EPI_ISL_1313856, EPI_ISL_1313857, EPI_ISL_1313858, EPI_ISL_1313859, EPI_ISL_1313860, EPI_ISL_1313861, EPI_ISL_1313862, EPI_ISL_1313863, EPI_ISL_1313864, EPI_ISL_1313865, EPI_ISL_1313866, EPI_ISL_1313867, EPI_ISL_1313868, EPI_ISL_1313869, EPI_ISL_1313870, EPI_ISL_1313871, EPI_ISL_1313872, EPI_ISL_1313873, EPI_ISL_1313874, EPI_ISL_1313875, EPI_ISL_1313876, EPI_ISL_1313877, EPI_ISL_1313878, EPI_ISL_1313879, EPI_ISL_1313880, EPI_ISL_1313881, EPI_ISL_1313882, EPI_ISL_1313883 | | | |
| see above | Utah Public Health Laboratory | Utah Public Health Laboratory | Erin L. Young, Kelly F. Oakeson, Tara Gallagher |
| EPI_ISL_1314057, EPI_ISL_1314058, EPI_ISL_1314059, EPI_ISL_1314060, EPI_ISL_1314061, EPI_ISL_1314062, EPI_ISL_1314067, EPI_ISL_1314068, EPI_ISL_1314069 | Wyoming Public Health Laboratory | Wyoming Public Health Laboratory | Jim Mildenberger, Wanda Manley, Noah Hull, Taylor Fearing, Lynette Gumbleton, Channing Weber, Ashley Norberg, Chayse Rowley, Marley Goetz, Brian Dominguez, Elliot Thomasson, Cari Sloma, and Rob Christensen |
| EPI_ISL_1314070, EPI_ISL_1314071, EPI_ISL_1314072, EPI_ISL_1314073, EPI_ISL_1314074, EPI_ISL_1314075, EPI_ISL_1314076, EPI_ISL_1314077, EPI_ISL_1314078, EPI_ISL_1314079, EPI_ISL_1314080, EPI_ISL_1314081, EPI_ISL_1314082, EPI_ISL_1314083, EPI_ISL_1314084, EPI_ISL_1314085, EPI_ISL_1314086, EPI_ISL_1314087, EPI_ISL_1314088 | | | |
| see above | University of Wisconsin-Madison AIDS Vaccine Research Laboratories | University of Wisconsin-Madison AIDS Vaccine Research Laboratories | Gage Moreno, Katarina Braun, et al. AIDS Vaccine Research Laboratories |
| EPI_ISL_1314091, EPI_ISL_1314092, EPI_ISL_1314094, EPI_ISL_1314095, EPI_ISL_1314096, EPI_ISL_1314097, EPI_ISL_1314098, EPI_ISL_1314099, EPI_ISL_1314104, EPI_ISL_1314108, EPI_ISL_1314110, EPI_ISL_1314112, EPI_ISL_1314115, EPI_ISL_1314119, EPI_ISL_1314120, EPI_ISL_1314122, EPI_ISL_1314123, EPI_ISL_1314124, EPI_ISL_1314125, EPI_ISL_1314126, EPI_ISL_1314127, EPI_ISL_1314128, EPI_ISL_1314129, EPI_ISL_1314130, EPI_ISL_1314131, EPI_ISL_1314132, EPI_ISL_1314133, EPI_ISL_1314134, EPI_ISL_1314135, EPI_ISL_1314136, EPI_ISL_1314137, EPI_ISL_1314138, EPI_ISL_1314139, EPI_ISL_1314140, EPI_ISL_1314141, EPI_ISL_1314142, EPI_ISL_1314143 | | | |
| see above | Wyoming Public Health Laboratory | Wyoming Public Health Laboratory | Jim Mildenberger, Wanda Manley, Noah Hull, Taylor Fearing, Lynette Gumbleton, Channing Weber, Ashley Norberg, Chayse Rowley, Marley Goetz, Brian Dominguez, Elliot Thomasson, Cari Sloma, and Rob Christensen |
| EPI_ISL_1314144, EPI_ISL_1314145, EPI_ISL_1314146, EPI_ISL_1314147, EPI_ISL_1314148, EPI_ISL_1314149, EPI_ISL_1314150, EPI_ISL_1314151, EPI_ISL_1314152, EPI_ISL_1314153, EPI_ISL_1314154, EPI_ISL_1314155, EPI_ISL_1314156, EPI_ISL_1314157, EPI_ISL_1314158, EPI_ISL_1314159, EPI_ISL_1314160, EPI_ISL_1314161, EPI_ISL_1314162, EPI_ISL_1314163, EPI_ISL_1314164, EPI_ISL_1314165, EPI_ISL_1314166, EPI_ISL_1314167 | | | |
| see above | University of Wisconsin-Madison AIDS Vaccine Research Laboratories | University of Wisconsin-Madison AIDS Vaccine Research Laboratories | Gage Moreno, Katarina Braun, et al. AIDS Vaccine Research Laboratories |
| EPI_ISL_1314168, EPI_ISL_1314169 | Gravity Diagnostics | Kentucky State Public Health Lab | Stephanie Lunn, Karim George, Joshua Tobias, William Grooms, Vaneet Arora, Matthew Johnson, Rachel Zinner, Rhonda Lucas |
| EPI_ISL_1314170, EPI_ISL_1314171, EPI_ISL_1314172, EPI_ISL_1314173, EPI_ISL_1314174, EPI_ISL_1314175, EPI_ISL_1314176, EPI_ISL_1314177 | Kentucky State Public Health Lab | Kentucky State Public Health Lab | Stephanie Lunn, Karim George, Joshua Tobias, William Grooms, Vaneet Arora, Matthew Johnson, Rachel Zinner, Rhonda Lucas |
| EPI_ISL_1314178, EPI_ISL_1314179, EPI_ISL_1314180, EPI_ISL_1314181, EPI_ISL_1314182, EPI_ISL_1314183, EPI_ISL_1314184, EPI_ISL_1314185, EPI_ISL_1314186, EPI_ISL_1314187, EPI_ISL_1314188, EPI_ISL_1314189 | | | |
| see above | Gravity Diagnostics | Kentucky State Public Health Lab | Stephanie Lunn, Karim George, Joshua Tobias, William Grooms, Vaneet Arora, Matthew Johnson, Rachel Zinner, Rhonda Lucas |
| EPI_ISL_1314192, EPI_ISL_1314193, EPI_ISL_1314196, EPI_ISL_1314197 | Kentucky State Public Health Lab | Kentucky State Public Health Lab | Stephanie Lunn, Karim George, Joshua Tobias, William Grooms, Vaneet Arora, Matthew Johnson, Rachel Zinner, Rhonda Lucas |
| EPI_ISL_1314198, EPI_ISL_1314199, EPI_ISL_1314200, EPI_ISL_1314201, EPI_ISL_1314202, EPI_ISL_1314203, EPI_ISL_1314204, EPI_ISL_1314205, EPI_ISL_1314206, EPI_ISL_1314207 | Gravity Diagnostics | Kentucky State Public Health Lab | Stephanie Lunn, Karim George, Joshua Tobias, William Grooms, Vaneet Arora, Matthew Johnson, Rachel Zinner, Rhonda Lucas |
| EPI_ISL_1314233, EPI_ISL_1314234, EPI_ISL_1314235, EPI_ISL_1314236, EPI_ISL_1314237, EPI_ISL_1314238, EPI_ISL_1314239, EPI_ISL_1314240, EPI_ISL_1314241, EPI_ISL_1314242, EPI_ISL_1314243, EPI_ISL_1314244, EPI_ISL_1314245, EPI_ISL_1314246, EPI_ISL_1314247, EPI_ISL_1314248, EPI_ISL_1314249, EPI_ISL_1314250, EPI_ISL_1314251, EPI_ISL_1314252, EPI_ISL_1314253, EPI_ISL_1314254, | | | |

| | | | |
|---|---|---|---|
| EPI_ISL_1314255, EPI_ISL_1314256, EPI_ISL_1314257 | | | |
| see above | University of Wisconsin-Madison AIDS Vaccine Research Laboratories | University of Wisconsin-Madison AIDS Vaccine Research Laboratories | Gage Moreno, Katarina Braun, et al. AIDS Vaccine Research Laboratories |
| EPI_ISL_1314258, EPI_ISL_1314259, EPI_ISL_1314260, EPI_ISL_1314261 | Kansas Health and Environmental Lab | Kansas Health and Environmental Lab | Mike Grose, Jonathan Barnell, Ben Olsen, and Phil Adam |
| EPI_ISL_1314262, EPI_ISL_1314263 | Cytocheck Laboratory | Kansas Health and Environmental Lab | Mike Grose, Jonathan Barnell, Ben Olsen, and Phil Adam |
| EPI_ISL_1314264, EPI_ISL_1314265, EPI_ISL_1314266, EPI_ISL_1314267, EPI_ISL_1314268, EPI_ISL_1314269, EPI_ISL_1314270, EPI_ISL_1314271, EPI_ISL_1314272, EPI_ISL_1314273, EPI_ISL_1314274, EPI_ISL_1314275, EPI_ISL_1314276, EPI_ISL_1314277, EPI_ISL_1314278, EPI_ISL_1314279, EPI_ISL_1314280, EPI_ISL_1314281, EPI_ISL_1314282, EPI_ISL_1314283, EPI_ISL_1314284, EPI_ISL_1314285, EPI_ISL_1314286, EPI_ISL_1314287, EPI_ISL_1314288, EPI_ISL_1314289, EPI_ISL_1314290, EPI_ISL_1314291, EPI_ISL_1314292, EPI_ISL_1314293, EPI_ISL_1314294, EPI_ISL_1314295, EPI_ISL_1314296 | | | |
| see above | University of Michigan Clinical Microbiology Laboratory | Lauring Lab, University of Michigan, Department of Microbiology and Immunology | Valesano |
| EPI_ISL_1314297 | Vault Health | Minnesota Department of Health, Public Health Laboratory | Alexandra Lorentz, Jacob Garfin, Matt Plumb, and Xiong Wang |
| EPI_ISL_1314299 | Regions Hospital | Minnesota Department of Health, Public Health Laboratory | Alexandra Lorentz, Jacob Garfin, Matt Plumb, and Xiong Wang |
| EPI_ISL_1314300 | Children's Hospitals and Clinics of Minnesota | Minnesota Department of Health, Public Health Laboratory | Alexandra Lorentz, Jacob Garfin, Matt Plumb, and Xiong Wang |
| EPI_ISL_1314301, EPI_ISL_1314302, EPI_ISL_1314303, EPI_ISL_1314304, EPI_ISL_1314305, EPI_ISL_1314306, EPI_ISL_1314307, EPI_ISL_1314308, EPI_ISL_1314310, EPI_ISL_1314311, EPI_ISL_1314312 | | | |
| see above | M Health Fairview | Minnesota Department of Health, Public Health Laboratory | Alexandra Lorentz, Jacob Garfin, Matt Plumb, and Xiong Wang |
| EPI_ISL_1314314, EPI_ISL_1314315, EPI_ISL_1314316, EPI_ISL_1314317, EPI_ISL_1314319, EPI_ISL_1314320, EPI_ISL_1314321 | Mayo Clinic & Mayo Clinic Laboratories | Minnesota Department of Health, Public Health Laboratory | Alexandra Lorentz, Jacob Garfin, Matt Plumb, and Xiong Wang |
| EPI_ISL_1314322, EPI_ISL_1314323, EPI_ISL_1314324, EPI_ISL_1314326, EPI_ISL_1314327, EPI_ISL_1314328, EPI_ISL_1314329, EPI_ISL_1314330, EPI_ISL_1314332, EPI_ISL_1314333, EPI_ISL_1314334, EPI_ISL_1314335, EPI_ISL_1314336, EPI_ISL_1314337, EPI_ISL_1314338, EPI_ISL_1314339, EPI_ISL_1314340, EPI_ISL_1314341, EPI_ISL_1314342, EPI_ISL_1314343, EPI_ISL_1314344, EPI_ISL_1314345, EPI_ISL_1314348, EPI_ISL_1314349, EPI_ISL_1314351, EPI_ISL_1314352, EPI_ISL_1314353, EPI_ISL_1314354, EPI_ISL_1314355, EPI_ISL_1314356, EPI_ISL_1314357, EPI_ISL_1314358, EPI_ISL_1314359, EPI_ISL_1314360, EPI_ISL_1314361, EPI_ISL_1314362, EPI_ISL_1314363 | | | |
| see above | Vault Health | Minnesota Department of Health, Public Health Laboratory | Alexandra Lorentz, Jacob Garfin, Matt Plumb, and Xiong Wang |
| EPI_ISL_1314542, EPI_ISL_1314543, EPI_ISL_1314544, EPI_ISL_1314545, EPI_ISL_1314546, EPI_ISL_1314547, EPI_ISL_1314548, EPI_ISL_1314549, EPI_ISL_1314550, EPI_ISL_1314551, EPI_ISL_1314552, EPI_ISL_1314553, EPI_ISL_1314554, EPI_ISL_1314555, EPI_ISL_1314556, EPI_ISL_1314557, EPI_ISL_1314558, EPI_ISL_1314559, EPI_ISL_1314560, EPI_ISL_1314561, EPI_ISL_1314562, EPI_ISL_1314563, EPI_ISL_1314564, EPI_ISL_1314565, EPI_ISL_1314566, EPI_ISL_1314567, EPI_ISL_1314568, EPI_ISL_1314569, EPI_ISL_1314570, EPI_ISL_1314571, EPI_ISL_1314572, EPI_ISL_1314573, EPI_ISL_1314574, EPI_ISL_1314575, EPI_ISL_1314576, EPI_ISL_1314577, EPI_ISL_1314578, EPI_ISL_1314579, EPI_ISL_1314580, EPI_ISL_1314581, EPI_ISL_1314582, EPI_ISL_1314583, EPI_ISL_1314584, EPI_ISL_1314585, EPI_ISL_1314586, EPI_ISL_1314587, EPI_ISL_1314588, EPI_ISL_1314589, EPI_ISL_1314590, EPI_ISL_1314591, EPI_ISL_1314592, EPI_ISL_1314593, EPI_ISL_1314594, EPI_ISL_1314595, EPI_ISL_1314596, EPI_ISL_1314597, EPI_ISL_1314598, EPI_ISL_1314599, EPI_ISL_1314600, EPI_ISL_1314601, EPI_ISL_1314602, EPI_ISL_1314603, EPI_ISL_1314604, EPI_ISL_1314605, EPI_ISL_1314606, EPI_ISL_1314607, EPI_ISL_1314608, EPI_ISL_1314609, EPI_ISL_1314610, EPI_ISL_1314611, EPI_ISL_1314612, EPI_ISL_1314613, EPI_ISL_1314614, EPI_ISL_1314615, EPI_ISL_1314616, EPI_ISL_1314617, EPI_ISL_1314618, EPI_ISL_1314619, EPI_ISL_1314620, EPI_ISL_1314621, EPI_ISL_1314622, EPI_ISL_1314623, EPI_ISL_1314624, EPI_ISL_1314625, EPI_ISL_1314626, EPI_ISL_1314627, EPI_ISL_1314628, EPI_ISL_1314629, EPI_ISL_1314630, EPI_ISL_1314631, EPI_ISL_1314632, EPI_ISL_1314633, EPI_ISL_1314634, EPI_ISL_1314635, EPI_ISL_1314636, EPI_ISL_1314637, EPI_ISL_1314638, EPI_ISL_1314639, EPI_ISL_1314640, EPI_ISL_1314641, EPI_ISL_1314642, EPI_ISL_1314643, EPI_ISL_1314644, EPI_ISL_1314645, EPI_ISL_1314646, EPI_ISL_1314647, EPI_ISL_1314648, EPI_ISL_1314649, EPI_ISL_1314650, EPI_ISL_1314651, EPI_ISL_1314652, EPI_ISL_1314653, EPI_ISL_1314654, EPI_ISL_1314655, EPI_ISL_1314656, EPI_ISL_1314657, EPI_ISL_1314658, EPI_ISL_1314659, EPI_ISL_1314660, EPI_ISL_1314661, EPI_ISL_1314662, EPI_ISL_1314663, EPI_ISL_1314664, EPI_ISL_1314665, EPI_ISL_1314666, EPI_ISL_1314667, EPI_ISL_1314668, EPI_ISL_1314669, EPI_ISL_1314670, EPI_ISL_1314671, EPI_ISL_1314672, EPI_ISL_1314673, EPI_ISL_1314674, EPI_ISL_1314675, EPI_ISL_1314676, EPI_ISL_1314677, EPI_ISL_1314678, EPI_ISL_1314679, EPI_ISL_1314680, EPI_ISL_1314681, EPI_ISL_1314682, EPI_ISL_1314683, EPI_ISL_1314684, EPI_ISL_1314685, EPI_ISL_1314686, EPI_ISL_1314687, EPI_ISL_1314688, EPI_ISL_1314689, EPI_ISL_1314690, EPI_ISL_1314691, EPI_ISL_1314692, EPI_ISL_1314693, EPI_ISL_1314694, EPI_ISL_1314695, EPI_ISL_1314696, EPI_ISL_1314697, EPI_ISL_1314698, EPI_ISL_1314699, EPI_ISL_1314700, EPI_ISL_1314701, EPI_ISL_1314702, EPI_ISL_1314703, EPI_ISL_1314704, EPI_ISL_1314705, EPI_ISL_1314706, EPI_ISL_1314707, EPI_ISL_1314708, EPI_ISL_1314709, EPI_ISL_1314710, EPI_ISL_1314711, EPI_ISL_1314712, EPI_ISL_1314713, EPI_ISL_1314714, EPI_ISL_1314715, EPI_ISL_1314716, EPI_ISL_1314717, EPI_ISL_1314718, EPI_ISL_1314719, EPI_ISL_1314720, EPI_ISL_1314721, EPI_ISL_1314722, EPI_ISL_1314723, EPI_ISL_1314724, EPI_ISL_1314725, EPI_ISL_1314726, EPI_ISL_1314727, EPI_ISL_1314728, EPI_ISL_1314729, EPI_ISL_1314730, EPI_ISL_1314731, EPI_ISL_1314732, EPI_ISL_1314733, EPI_ISL_1314734, EPI_ISL_1314735, EPI_ISL_1314736, EPI_ISL_1314737, EPI_ISL_1314738, EPI_ISL_1314739, EPI_ISL_1314740, EPI_ISL_1314741, EPI_ISL_1314742, EPI_ISL_1314743, EPI_ISL_1314744, EPI_ISL_1314745, EPI_ISL_1314746, EPI_ISL_1314747, EPI_ISL_1314748, EPI_ISL_1314749, EPI_ISL_1314750, EPI_ISL_1314751, EPI_ISL_1314752, EPI_ISL_1314753, EPI_ISL_1314754, EPI_ISL_1314755, EPI_ISL_1314756, EPI_ISL_1314757, EPI_ISL_1314758, EPI_ISL_1314759, EPI_ISL_1314760, EPI_ISL_1314761, EPI_ISL_1314762, EPI_ISL_1314763, EPI_ISL_1314764, EPI_ISL_1314765, EPI_ISL_1314766, EPI_ISL_1314767, EPI_ISL_1314768, EPI_ISL_1314769, EPI_ISL_1314770, EPI_ISL_1314771, EPI_ISL_1314772, EPI_ISL_1314773, EPI_ISL_1314774, EPI_ISL_1314775, EPI_ISL_1314776, EPI_ISL_1314777, EPI_ISL_1314778, EPI_ISL_1314779, EPI_ISL_1314780, EPI_ISL_1314781, EPI_ISL_1314782, EPI_ISL_1314783, EPI_ISL_1314784, EPI_ISL_1314785, EPI_ISL_1314786, EPI_ISL_1314787, EPI_ISL_1314788, EPI_ISL_1314789, EPI_ISL_1314790, EPI_ISL_1314791, EPI_ISL_1314792, EPI_ISL_1314793, EPI_ISL_1314794, EPI_ISL_1314795, EPI_ISL_1314796, EPI_ISL_1314797, EPI_ISL_1314798, EPI_ISL_1314799, EPI_ISL_1314800, EPI_ISL_1314801, EPI_ISL_1314802, EPI_ISL_1314803, EPI_ISL_1314804, EPI_ISL_1314805, EPI_ISL_1314806, EPI_ISL_1314807, EPI_ISL_1314808, EPI_ISL_1314809, EPI_ISL_1314810, EPI_ISL_1314811, EPI_ISL_1314812, EPI_ISL_1314813, EPI_ISL_1314814, EPI_ISL_1314815, EPI_ISL_1314816, EPI_ISL_1314817, EPI_ISL_1314818, EPI_ISL_1314819, EPI_ISL_1314820, EPI_ISL_1314821, EPI_ISL_1314822, EPI_ISL_1314823, EPI_ISL_1314824, EPI_ISL_1314825, EPI_ISL_1314826, EPI_ISL_1314827, EPI_ISL_1314828, EPI_ISL_1314829, EPI_ISL_1314830, EPI_ISL_1314831, EPI_ISL_1314832, EPI_ISL_1314833, EPI_ISL_1314834, EPI_ISL_1314835, EPI_ISL_1314836, EPI_ISL_1314837, EPI_ISL_1314838, EPI_ISL_1314839, EPI_ISL_1314840, EPI_ISL_1314841, EPI_ISL_1314842, EPI_ISL_1314843, EPI_ISL_1314844, EPI_ISL_1314845, EPI_ISL_1314846, EPI_ISL_1314847, EPI_ISL_1314848, EPI_ISL_1314849, EPI_ISL_1314850, EPI_ISL_1314851, EPI_ISL_1314852, EPI_ISL_1314853, EPI_ISL_1314854 | | | |
| see above | Quest Diagnostics Incorporated | Centers for Disease Control and Prevention Division of Viral Diseases, Pathogen Discovery | Peter W. Cook, Dakota Howard, Dhwani Batra, Ben L. Rambo-Martin, S. H. Rosenthal, A. Gerasimova, R. M. Kagan, B. Anderson, M. Hua, Y. Liu, L.E. Bernstein, K.E. Livingston, A. Perez, I. A. Shlyakhter, R. V. Rolando, R. Owen, P. Tanpaiboon, F. Lacbawan, Clinton R. Paden, Suxiang Tong, Duncan MacCannell |
| EPI_ISL_1314855 | South Dakota Public Health Laboratory | South Dakota Public Health Laboratory | Jacob Garfin and Chris Carlson |
| EPI_ISL_1314856 | Quest Diagnostics Incorporated | Centers for Disease Control and Prevention Division of Viral Diseases, Pathogen Discovery | Peter W. Cook, Dakota Howard, Dhwani Batra, Ben L. Rambo-Martin, S. H. Rosenthal, A. Gerasimova, R. M. Kagan, B. Anderson, M. Hua, Y. Liu, L.E. Bernstein, K.E. Livingston, A. Perez, I. A. Shlyakhter, R. V. Rolando, R. Owen, P. Tanpaiboon, F. Lacbawan, Clinton R. Paden, Suxiang Tong, Duncan MacCannell |
| EPI_ISL_1314857, EPI_ISL_1314858 | South Dakota Public Health Laboratory | South Dakota Public Health Laboratory | Jacob Garfin and Chris Carlson |
| EPI_ISL_1314859 | Quest Diagnostics Incorporated | Centers for Disease Control and Prevention Division of Viral Diseases, Pathogen Discovery | Peter W. Cook, Dakota Howard, Dhwani Batra, Ben L. Rambo-Martin, S. H. Rosenthal, A. Gerasimova, R. M. Kagan, B. Anderson, M. Hua, Y. Liu, L.E. Bernstein, K.E. Livingston, A. Perez, I. A. Shlyakhter, R. V. Rolando, R. Owen, P. Tanpaiboon, F. Lacbawan, Clinton R. Paden, Suxiang Tong, Duncan MacCannell |
| EPI_ISL_1314860, EPI_ISL_1314861, EPI_ISL_1314862, EPI_ISL_1314863, EPI_ISL_1314864, EPI_ISL_1314865, EPI_ISL_1314866, EPI_ISL_1314867, EPI_ISL_1314868, EPI_ISL_1314869, EPI_ISL_1314870, EPI_ISL_1314871, EPI_ISL_1314872, EPI_ISL_1314873, EPI_ISL_1314874, EPI_ISL_1314875 | | | |
| see above | South Dakota Public Health Laboratory | South Dakota Public Health Laboratory | Jacob Garfin and Chris Carlson |
| EPI_ISL_1314876, EPI_ISL_1314877, EPI_ISL_1314878, EPI_ISL_1314879, EPI_ISL_1314880, EPI_ISL_1314881, EPI_ISL_1314882, EPI_ISL_1314883, EPI_ISL_1314884, EPI_ISL_1314885, EPI_ISL_1314886, EPI_ISL_1314887, EPI_ISL_1314888, EPI_ISL_1314889, EPI_ISL_1314890, EPI_ISL_1314891, EPI_ISL_1314892, EPI_ISL_1314893, EPI_ISL_1314894, EPI_ISL_1314895, EPI_ISL_1314896, EPI_ISL_1314897, EPI_ISL_1314898, EPI_ISL_1314899, EPI_ISL_1314900, EPI_ISL_1314901, EPI_ISL_1314902, EPI_ISL_1314903, EPI_ISL_1314904, EPI_ISL_1314905, EPI_ISL_1314906, EPI_ISL_1314907, EPI_ISL_1314908, EPI_ISL_1314909, EPI_ISL_1314910, EPI_ISL_1314911, EPI_ISL_1314912, EPI_ISL_1314913, EPI_ISL_1314914, EPI_ISL_1314915, EPI_ISL_1314916, EPI_ISL_1314917, EPI_ISL_1314918, EPI_ISL_1314919, EPI_ISL_1314920, EPI_ISL_1314921, EPI_ISL_1314922, EPI_ISL_1314923, EPI_ISL_1314924, EPI_ISL_1314925, EPI_ISL_1314926, EPI_ISL_1314927, EPI_ISL_1314928, EPI_ISL_1314929, EPI_ISL_1314930, EPI_ISL_1314931, EPI_ISL_1314932, EPI_ISL_1314933, EPI_ISL_1314934, EPI_ISL_1314935, EPI_ISL_1314936, EPI_ISL_1314937, EPI_ISL_1314938, EPI_ISL_1314939, EPI_ISL_1314940, EPI_ISL_1314941, EPI_ISL_1314942, EPI_ISL_1314943, EPI_ISL_1314944, EPI_ISL_1314945, EPI_ISL_1314946, EPI_ISL_1314947, EPI_ISL_1314948, EPI_ISL_1314949, EPI_ISL_1314950, EPI_ISL_1314951, EPI_ISL_1314952, EPI_ISL_1314953, EPI_ISL_1314954, EPI_ISL_1314955, EPI_ISL_1314956, EPI_ISL_1314957, EPI_ISL_1314958, EPI_ISL_1314959, EPI_ISL_1314960, EPI_ISL_1314961, EPI_ISL_1314962, EPI_ISL_1314963, EPI_ISL_1314964, EPI_ISL_1314965, EPI_ISL_1314966, EPI_ISL_1314967 | | | |
| see above | Quest Diagnostics Incorporated | Centers for Disease Control and Prevention Division of Viral Diseases, Pathogen Discovery | Peter W. Cook, Dakota Howard, Dhwani Batra, Ben L. Rambo-Martin, S. H. Rosenthal, A. Gerasimova, R. M. Kagan, B. Anderson, M. Hua, Y. Liu, L.E. Bernstein, K.E. Livingston, A. Perez, I. A. Shlyakhter, R. V. Rolando, R. Owen, P. Tanpaiboon, F. Lacbawan, Clinton R. Paden, Suxiang Tong, Duncan MacCannell |
| EPI_ISL_1315073 | San Diego County Public Health Laboratory | Andersen lab at Scripps Research | SEARCH Alliance San Diego with Tracy Basler, Jovan Shephard, Brett Austin |
| EPI_ISL_1315076, EPI_ISL_1315077, EPI_ISL_1315078, EPI_ISL_1315079, EPI_ISL_1315080, EPI_ISL_1315081, EPI_ISL_1315082, EPI_ISL_1315083, EPI_ISL_1315084, EPI_ISL_1315085, EPI_ISL_1315086, EPI_ISL_1315087, EPI_ISL_1315088, EPI_ISL_1315089, EPI_ISL_1315090, EPI_ISL_1315091 | | | |
| see above | Nebraska Public Health Laboratory | NPHL COVID-19 Response Team | NPHL COVID-19 Response Team |
| EPI_ISL_1315092, EPI_ISL_1315093, EPI_ISL_1315094, EPI_ISL_1315095, EPI_ISL_1315096, EPI_ISL_1315097, EPI_ISL_1315098, EPI_ISL_1315099, EPI_ISL_1315100, EPI_ISL_1315101, EPI_ISL_1315102, EPI_ISL_1315103, EPI_ISL_1315104, EPI_ISL_1315105, EPI_ISL_1315106, EPI_ISL_1315107, EPI_ISL_1315108, EPI_ISL_1315109, EPI_ISL_1315110, EPI_ISL_1315111, EPI_ISL_1315112, EPI_ISL_1315113, EPI_ISL_1315114, EPI_ISL_1315115, EPI_ISL_1315116, EPI_ISL_1315117, EPI_ISL_1315118, EPI_ISL_1315119, EPI_ISL_1315120, EPI_ISL_1315121, EPI_ISL_1315122, EPI_ISL_1315123, EPI_ISL_1315124, EPI_ISL_1315125 | | | |
| see above | Nevada State Public Health Laboratory | Nevada State Public Health Laboratory | Andrew Gorzalski, Mark Pandori |
| EPI_ISL_1315126, EPI_ISL_1315127, EPI_ISL_1315128, EPI_ISL_1315129, EPI_ISL_1315130, EPI_ISL_1315131, EPI_ISL_1315132, EPI_ISL_1315133, EPI_ISL_1315134, EPI_ISL_1315135, EPI_ISL_1315136, EPI_ISL_1315137, EPI_ISL_1315138, EPI_ISL_1315139, EPI_ISL_1315140, EPI_ISL_1315141, EPI_ISL_1315142, EPI_ISL_1315143, EPI_ISL_1315144, EPI_ISL_1315145, EPI_ISL_1315146, EPI_ISL_1315147, EPI_ISL_1315148, EPI_ISL_1315149, EPI_ISL_1315150, EPI_ISL_1315151, EPI_ISL_1315152, EPI_ISL_1315153, EPI_ISL_1315154, EPI_ISL_1315155, EPI_ISL_1315156, EPI_ISL_1315157, EPI_ISL_1315158, EPI_ISL_1315159, EPI_ISL_1315160, EPI_ISL_1315161, EPI_ISL_1315162, EPI_ISL_1315163, EPI_ISL_1315164, EPI_ISL_1315165, EPI_ISL_1315166, EPI_ISL_1315167, EPI_ISL_1315168, EPI_ISL_1315169, EPI_ISL_1315170, EPI_ISL_1315171, EPI_ISL_1315172, EPI_ISL_1315173, EPI_ISL_1315174, EPI_ISL_1315175, EPI_ISL_1315176, EPI_ISL_1315177, EPI_ISL_1315178, EPI_ISL_1315179, EPI_ISL_1315180, EPI_ISL_1315181, EPI_ISL_1315182, EPI_ISL_1315183, EPI_ISL_1315184, EPI_ISL_1315185, EPI_ISL_1315186, EPI_ISL_1315187, EPI_ISL_1315188, EPI_ISL_1315189, EPI_ISL_1315190, EPI_ISL_1315191, EPI_ISL_1315192, EPI_ISL_1315193, EPI_ISL_1315194, EPI_ISL_1315195, EPI_ISL_1315196, EPI_ISL_1315197, EPI_ISL_1315198, EPI_ISL_1315199, EPI_ISL_1315200, EPI_ISL_1315201, EPI_ISL_1315202, EPI_ISL_1315203, EPI_ISL_1315204, EPI_ISL_1315205, EPI_ISL_1315206, EPI_ISL_1315207, EPI_ISL_1315208, EPI_ISL_1315209, EPI_ISL_1315210, EPI_ISL_1315211, EPI_ISL_1315212, EPI_ISL_1315213, EPI_ISL_1315214, EPI_ISL_1315215, EPI_ISL_1315216, EPI_ISL_1315217, EPI_ISL_1315218, EPI_ISL_1315219, EPI_ISL_1315220, EPI_ISL_1315221, EPI_ISL_1315222, EPI_ISL_1315223 | | | |
| see above | The Jackson Laboratory | The Jackson Laboratory | Bergeron D, Renzette N, Adams M, Omerza G, Kelly K, Li L |
| EPI_ISL_1315479, EPI_ISL_1315480, EPI_ISL_1315481, EPI_ISL_1315482, EPI_ISL_1315483, EPI_ISL_1315484, EPI_ISL_1315485, EPI_ISL_1315486, EPI_ISL_1315487, EPI_ISL_1315488, EPI_ISL_1315489, EPI_ISL_1315490, EPI_ISL_1315491, EPI_ISL_1315492, EPI_ISL_1315493, EPI_ISL_1315494, EPI_ISL_1315495, EPI_ISL_1315496, EPI_ISL_1315497, EPI_ISL_1315498, EPI_ISL_1315499, EPI_ISL_1315500, EPI_ISL_1315501, EPI_ISL_1315502, EPI_ISL_1315504, EPI_ISL_1315505, EPI_ISL_1315506, EPI_ISL_1315507, EPI_ISL_1315508, EPI_ISL_1315509, EPI_ISL_1315510, EPI_ISL_1315511, EPI_ISL_1315512, EPI_ISL_1315513, EPI_ISL_1315514, EPI_ISL_1315515, EPI_ISL_1315516, EPI_ISL_1315517, EPI_ISL_1315518, EPI_ISL_1315519, EPI_ISL_1315520, EPI_ISL_1315521, EPI_ISL_1315522, EPI_ISL_1315523, EPI_ISL_1315524, EPI_ISL_1315525, EPI_ISL_1315526, EPI_ISL_1315527, EPI_ISL_1315528, EPI_ISL_1315529, EPI_ISL_1315530, EPI_ISL_1315531, EPI_ISL_1315532, EPI_ISL_1315533, EPI_ISL_1315534, EPI_ISL_1315535, EPI_ISL_1315536, EPI_ISL_1315537, EPI_ISL_1315539, EPI_ISL_1315540, EPI_ISL_1315541, EPI_ISL_1315542, EPI_ISL_1315543, EPI_ISL_1315544, EPI_ISL_1315545, EPI_ISL_1315546, EPI_ISL_1315547, EPI_ISL_1315548, EPI_ISL_1315549, EPI_ISL_1315550, EPI_ISL_1315551, EPI_ISL_1315552, EPI_ISL_1315553, EPI_ISL_1315554, EPI_ISL_1315555, EPI_ISL_1315556, EPI_ISL_1315557, EPI_ISL_1315558, EPI_ISL_1315559, EPI_ISL_1315560, EPI_ISL_1315561, EPI_ISL_1315562, EPI_ISL_1315563, EPI_ISL_1315564, EPI_ISL_1315566, EPI_ISL_1315567, EPI_ISL_1315568, EPI_ISL_1315569, EPI_ISL_1315570, EPI_ISL_1315571, EPI_ISL_1315572 | | | |
| see above | Instituto Nacional de Medicina Genomica | Instituto Nacional de Medicina Genomica | Hidalgo-Miranda A, Mendoza-Vargas A, Reyes-Grajeda JP, Cedro-Tanda A, Alcaraz N, Arriaga-Canon C, Frías-Jimenez E, Gonzalez-Barrera D, Rangel-DeLeon D, Ramirez-Vega O, Munguia-Garza, Garcia-Cárdenas FJ, González-Woge MA, Herrera-Montalvo LA |
| EPI_ISL_1315583, EPI_ISL_1315584, EPI_ISL_1315585, EPI_ISL_1315586, EPI_ISL_1315587, EPI_ISL_1315588, EPI_ISL_1315589, EPI_ISL_1315590, EPI_ISL_1315591, EPI_ISL_1315592, EPI_ISL_1315593, EPI_ISL_1315594, EPI_ISL_1315596, EPI_ISL_1315597, EPI_ISL_1315598, EPI_ISL_1315599, EPI_ISL_1315600, EPI_ISL_1315601, EPI_ISL_1315602, EPI_ISL_1315603, EPI_ISL_1315604 | | | |
| see above | Arizona State Public Health Laboratory | Arizona State Public Health Laboratory | Trung Huynh, Jessica Escobar, Katherine Fullerton, Nobuko Fukushima, Stacy White, Linda Getsinger, Victor Waddell |
| EPI_ISL_1318220 | Ohio Department of Health Laboratory | Ohio Department of Health Laboratory | Holmes, Jennifer; Eric Brandt, Keoni Omura, Glen McGillivary, Caitlin McDonnell, Jade Mowery, Stephanie Mccracken, Tyler Payne, Kirtana Ramadugu, Erica Leasure, Brent Lee, Kelsey Florek, Heather Blankenship, Allison Black, Quanta Brown, and Tammy Bannerman |
| EPI_ISL_1318221 | University of Illinois Veterinary | Roy J. Carver Biotechnology Center | Alvaro Hernandez, Chris Wright, Christopher Fields, Mireille Farjo, Christopher Brooke |

| | | | |
|---|---|---|---|
| | Diagnostic Laboratory | | |
| EPI_ISL_1318222, EPI_ISL_1318223 | LA Office of Public Health Laboratories | Genomics and Discovery, Respiratory Viruses Branch, Division of Viral Diseases, Centers for Disease Control and Prevention | Ying Tao, Jing Zhang, Yan Li, Brian Lynch, Anna Kelleher, Krista Queen, Anna Uehara, Peter Cook, Han Jia Justin Ng, Clinton R. Paden, Haibin Wang, Suxiang Tong |
| EPI_ISL_1318224 | IL Department of Public Health Chicago Laboratory | Genomics and Discovery, Respiratory Viruses Branch, Division of Viral Diseases, Centers for Disease Control and Prevention | Ying Tao, Jing Zhang, Yan Li, Brian Lynch, Anna Kelleher, Krista Queen, Anna Uehara, Peter Cook, Han Jia Justin Ng, Clinton R. Paden, Haibin Wang, Suxiang Tong |
| EPI_ISL_1318225 | LA Office of Public Health Laboratories | Genomics and Discovery, Respiratory Viruses Branch, Division of Viral Diseases, Centers for Disease Control and Prevention | Ying Tao, Jing Zhang, Yan Li, Brian Lynch, Anna Kelleher, Krista Queen, Anna Uehara, Peter Cook, Han Jia Justin Ng, Clinton R. Paden, Haibin Wang, Suxiang Tong |
| EPI_ISL_1318226 | IN State Department of Health Laboratory Services | Genomics and Discovery, Respiratory Viruses Branch, Division of Viral Diseases, Centers for Disease Control and Prevention | Ying Tao, Jing Zhang, Yan Li, Brian Lynch, Anna Kelleher, Krista Queen, Anna Uehara, Peter Cook, Han Jia Justin Ng, Clinton R. Paden, Haibin Wang, Suxiang Tong |
| EPI_ISL_1318227, EPI_ISL_1318228, EPI_ISL_1318229 | LA Office of Public Health Laboratories | Genomics and Discovery, Respiratory Viruses Branch, Division of Viral Diseases, Centers for Disease Control and Prevention | Ying Tao, Jing Zhang, Yan Li, Brian Lynch, Anna Kelleher, Krista Queen, Anna Uehara, Peter Cook, Han Jia Justin Ng, Clinton R. Paden, Haibin Wang, Suxiang Tong |
| EPI_ISL_1318230 | "AR Dept. of Health-PHL, Molecular Diagnostics" | Genomics and Discovery, Respiratory Viruses Branch, Division of Viral Diseases, Centers for Disease Control and Prevention | Ying Tao, Jing Zhang, Yan Li, Brian Lynch, Anna Kelleher, Krista Queen, Anna Uehara, Peter Cook, Han Jia Justin Ng, Clinton R. Paden, Haibin Wang, Suxiang Tong |
| EPI_ISL_1318231 | NC State Laboratory of Public Health | Genomics and Discovery, Respiratory Viruses Branch, Division of Viral Diseases, Centers for Disease Control and Prevention | Ying Tao, Jing Zhang, Yan Li, Brian Lynch, Anna Kelleher, Krista Queen, Anna Uehara, Peter Cook, Han Jia Justin Ng, Clinton R. Paden, Haibin Wang, Suxiang Tong |
| EPI_ISL_1318232 | LA Office of Public Health Laboratories | Genomics and Discovery, Respiratory Viruses Branch, Division of Viral Diseases, Centers for Disease Control and Prevention | Ying Tao, Jing Zhang, Yan Li, Brian Lynch, Anna Kelleher, Krista Queen, Anna Uehara, Peter Cook, Han Jia Justin Ng, Clinton R. Paden, Haibin Wang, Suxiang Tong |
| EPI_ISL_1318233 | TN Division of Laboratory Services | Genomics and Discovery, Respiratory Viruses Branch, Division of Viral Diseases, Centers for Disease Control and Prevention | Ying Tao, Jing Zhang, Yan Li, Brian Lynch, Anna Kelleher, Krista Queen, Anna Uehara, Peter Cook, Han Jia Justin Ng, Clinton R. Paden, Haibin Wang, Suxiang Tong |
| EPI_ISL_1318234 | IL Department of Public Health Chicago Laboratory | Genomics and Discovery, Respiratory Viruses Branch, Division of Viral Diseases, Centers for Disease Control and Prevention | Ying Tao, Jing Zhang, Yan Li, Brian Lynch, Anna Kelleher, Krista Queen, Anna Uehara, Peter Cook, Han Jia Justin Ng, Clinton R. Paden, Haibin Wang, Suxiang Tong |
| EPI_ISL_1318235 | LA Office of Public Health Laboratories | Genomics and Discovery, Respiratory Viruses Branch, Division of Viral Diseases, Centers for Disease Control and Prevention | Ying Tao, Jing Zhang, Yan Li, Brian Lynch, Anna Kelleher, Krista Queen, Anna Uehara, Peter Cook, Han Jia Justin Ng, Clinton R. Paden, Haibin Wang, Suxiang Tong |
| EPI_ISL_1318236 | IN State Department of Health Laboratory Services | Genomics and Discovery, Respiratory Viruses Branch, Division of Viral Diseases, Centers for Disease Control and Prevention | Ying Tao, Jing Zhang, Yan Li, Brian Lynch, Anna Kelleher, Krista Queen, Anna Uehara, Peter Cook, Han Jia Justin Ng, Clinton R. Paden, Haibin Wang, Suxiang Tong |
| EPI_ISL_1318237 | LA Office of Public Health Laboratories | Genomics and Discovery, Respiratory Viruses Branch, Division of Viral Diseases, Centers for Disease Control and Prevention | Ying Tao, Jing Zhang, Yan Li, Brian Lynch, Anna Kelleher, Krista Queen, Anna Uehara, Peter Cook, Han Jia Justin Ng, Clinton R. Paden, Haibin Wang, Suxiang Tong |
| EPI_ISL_1318238 | "OK Public Health Laboratory, Oklahoma State DOH" | Genomics and Discovery, Respiratory Viruses Branch, Division of Viral Diseases, Centers for Disease Control and Prevention | Ying Tao, Jing Zhang, Yan Li, Brian Lynch, Anna Kelleher, Krista Queen, Anna Uehara, Peter Cook, Han Jia Justin Ng, Clinton R. Paden, Haibin Wang, Suxiang Tong |
| EPI_ISL_1318239 | "AR Dept. of Health-PHL, Molecular Diagnostics" | Genomics and Discovery, Respiratory Viruses Branch, Division of Viral Diseases, Centers for Disease Control and Prevention | Ying Tao, Jing Zhang, Yan Li, Brian Lynch, Anna Kelleher, Krista Queen, Anna Uehara, Peter Cook, Han Jia Justin Ng, Clinton R. Paden, Haibin Wang, Suxiang Tong |
| EPI_ISL_1318240, EPI_ISL_1318241, EPI_ISL_1318242, EPI_ISL_1318243, EPI_ISL_1318244 | LA Office of Public Health Laboratories | Genomics and Discovery, Respiratory Viruses Branch, Division of Viral Diseases, Centers for Disease Control and Prevention | Ying Tao, Jing Zhang, Yan Li, Brian Lynch, Anna Kelleher, Krista Queen, Anna Uehara, Peter Cook, Han Jia Justin Ng, Clinton R. Paden, Haibin Wang, Suxiang Tong |
| EPI_ISL_1318245 | NC State Laboratory of Public Health | Genomics and Discovery, Respiratory Viruses Branch, Division of Viral Diseases, Centers for Disease Control and Prevention | Ying Tao, Jing Zhang, Yan Li, Brian Lynch, Anna Kelleher, Krista Queen, Anna Uehara, Peter Cook, Han Jia Justin Ng, Clinton R. Paden, Haibin Wang, Suxiang Tong |
| EPI_ISL_1318246 | LA Office of Public Health Laboratories | Genomics and Discovery, Respiratory Viruses Branch, Division of Viral Diseases, Centers for Disease Control and Prevention | Ying Tao, Jing Zhang, Yan Li, Brian Lynch, Anna Kelleher, Krista Queen, Anna Uehara, Peter Cook, Han Jia Justin Ng, Clinton R. Paden, Haibin Wang, Suxiang Tong |
| EPI_ISL_1318247 | TN Division of Laboratory Services | Genomics and Discovery, Respiratory Viruses Branch, Division of Viral Diseases, Centers for Disease Control and Prevention | Ying Tao, Jing Zhang, Yan Li, Brian Lynch, Anna Kelleher, Krista Queen, Anna Uehara, Peter Cook, Han Jia Justin Ng, Clinton R. Paden, Haibin Wang, Suxiang Tong |
| EPI_ISL_1318248 | SD Public Health Laboratory | Genomics and Discovery, Respiratory Viruses Branch, Division of Viral Diseases, Centers for Disease Control and Prevention | Ying Tao, Jing Zhang, Yan Li, Brian Lynch, Anna Kelleher, Krista Queen, Anna Uehara, Peter Cook, Han Jia Justin Ng, Clinton R. Paden, Haibin Wang, Suxiang Tong |
| EPI_ISL_1318249 | IL Dept. of Public Health Springfield Laboratory | Genomics and Discovery, Respiratory Viruses Branch, Division of Viral Diseases, Centers for Disease Control and Prevention | Ying Tao, Jing Zhang, Yan Li, Brian Lynch, Anna Kelleher, Krista Queen, Anna Uehara, Peter Cook, Han Jia Justin Ng, Clinton R. Paden, Haibin Wang, Suxiang Tong |
| EPI_ISL_1318250 | LA Office of Public Health Laboratories | Genomics and Discovery, Respiratory Viruses Branch, Division of Viral Diseases, Centers for Disease Control and Prevention | Ying Tao, Jing Zhang, Yan Li, Brian Lynch, Anna Kelleher, Krista Queen, Anna Uehara, Peter Cook, Han Jia Justin Ng, Clinton R. Paden, Haibin Wang, Suxiang Tong |
| EPI_ISL_1318251 | IN State Department of Health Laboratory Services | Genomics and Discovery, Respiratory Viruses Branch, Division of Viral Diseases, Centers for Disease Control and Prevention | Ying Tao, Jing Zhang, Yan Li, Brian Lynch, Anna Kelleher, Krista Queen, Anna Uehara, Peter Cook, Han Jia Justin Ng, Clinton R. Paden, Haibin Wang, Suxiang Tong |
| EPI_ISL_1318252 | TN Division of Laboratory Services | Genomics and Discovery, Respiratory Viruses Branch, Division of Viral Diseases, Centers for Disease Control and Prevention | Ying Tao, Jing Zhang, Yan Li, Brian Lynch, Anna Kelleher, Krista Queen, Anna Uehara, Peter Cook, Han Jia Justin Ng, Clinton R. Paden, Haibin Wang, Suxiang Tong |
| EPI_ISL_1318253 | MT Public Health Laboratory | Genomics and Discovery, Respiratory Viruses Branch, Division of Viral Diseases, Centers for Disease Control and Prevention | Ying Tao, Jing Zhang, Yan Li, Brian Lynch, Anna Kelleher, Krista Queen, Anna Uehara, Peter Cook, Han Jia Justin Ng, Clinton R. Paden, Haibin Wang, Suxiang Tong |
| EPI_ISL_1318254 | LA Office of Public Health Laboratories | Genomics and Discovery, Respiratory Viruses Branch, Division of Viral Diseases, Centers for Disease Control and Prevention | Ying Tao, Jing Zhang, Yan Li, Brian Lynch, Anna Kelleher, Krista Queen, Anna Uehara, Peter Cook, Han Jia Justin Ng, Clinton R. Paden, Haibin Wang, Suxiang Tong |
| EPI_ISL_1318255 | "OK Public Health Laboratory, Oklahoma State DOH" | Genomics and Discovery, Respiratory Viruses Branch, Division of Viral Diseases, Centers for Disease Control and Prevention | Ying Tao, Jing Zhang, Yan Li, Brian Lynch, Anna Kelleher, Krista Queen, Anna Uehara, Peter Cook, Han Jia Justin Ng, Clinton R. Paden, Haibin Wang, Suxiang Tong |
| EPI_ISL_1318256 | IN State Department of Health Laboratory Services | Genomics and Discovery, Respiratory Viruses Branch, Division of Viral Diseases, Centers for Disease Control and Prevention | Ying Tao, Jing Zhang, Yan Li, Brian Lynch, Anna Kelleher, Krista Queen, Anna Uehara, Peter Cook, Han Jia Justin Ng, Clinton R. Paden, Haibin Wang, Suxiang Tong |
| EPI_ISL_1318257 | TN Division of Laboratory Services | Genomics and Discovery, Respiratory Viruses Branch, Division of Viral Diseases, Centers for Disease Control and Prevention | Ying Tao, Jing Zhang, Yan Li, Brian Lynch, Anna Kelleher, Krista Queen, Anna Uehara, Peter Cook, Han Jia Justin Ng, Clinton R. Paden, Haibin Wang, Suxiang Tong |
| EPI_ISL_1318258 | IL Dept. of Public Health Springfield Laboratory | Genomics and Discovery, Respiratory Viruses Branch, Division of Viral Diseases, Centers for Disease Control and Prevention | Ying Tao, Jing Zhang, Yan Li, Brian Lynch, Anna Kelleher, Krista Queen, Anna Uehara, Peter Cook, Han Jia Justin Ng, Clinton R. Paden, Haibin Wang, Suxiang Tong |
| EPI_ISL_1318259 | IL Department of Public Health Chicago Laboratory | Genomics and Discovery, Respiratory Viruses Branch, Division of Viral Diseases, Centers for Disease Control and Prevention | Ying Tao, Jing Zhang, Yan Li, Brian Lynch, Anna Kelleher, Krista Queen, Anna Uehara, Peter Cook, Han Jia Justin Ng, Clinton R. Paden, Haibin Wang, Suxiang Tong |

| | | | |
|---|---|---|---|
| EPI_ISL_1318260, EPI_ISL_1318261 | LA Office of Public Health Laboratories | Genomics and Discovery, Respiratory Viruses Branch, Division of Viral Diseases, Centers for Disease Control and Prevention | Ying Tao, Jing Zhang, Yan Li, Brian Lynch, Anna Kelleher, Krista Queen, Anna Uehara, Peter Cook, Han Jia Justin Ng, Clinton R. Paden, Haibin Wang, Suxiang Tong |
| EPI_ISL_1318262 | "OK Public Health Laboratory, Oklahoma State DOH" | Genomics and Discovery, Respiratory Viruses Branch, Division of Viral Diseases, Centers for Disease Control and Prevention | Ying Tao, Jing Zhang, Yan Li, Brian Lynch, Anna Kelleher, Krista Queen, Anna Uehara, Peter Cook, Han Jia Justin Ng, Clinton R. Paden, Haibin Wang, Suxiang Tong |
| EPI_ISL_1318263, EPI_ISL_1318264 | LA Office of Public Health Laboratories | Genomics and Discovery, Respiratory Viruses Branch, Division of Viral Diseases, Centers for Disease Control and Prevention | Ying Tao, Jing Zhang, Yan Li, Brian Lynch, Anna Kelleher, Krista Queen, Anna Uehara, Peter Cook, Han Jia Justin Ng, Clinton R. Paden, Haibin Wang, Suxiang Tong |
| EPI_ISL_1318265, EPI_ISL_1318267, EPI_ISL_1318268, EPI_ISL_1318269, EPI_ISL_1318270, EPI_ISL_1318271, EPI_ISL_1318272, EPI_ISL_1318273, EPI_ISL_1318275, EPI_ISL_1318276, EPI_ISL_1318277, EPI_ISL_1318278, EPI_ISL_1318279, EPI_ISL_1318280, EPI_ISL_1318283, EPI_ISL_1318284, EPI_ISL_1318285, EPI_ISL_1318288 | | | |
| see above | Wyoming Public Health Laboratory | Wyoming Public Health Laboratory | Jim Mildenberger, Wanda Manley, Noah Hull, Taylor Fearing, Lynette Gumbleton, Channing Weber, Ashley Norberg, Chayse Rowley, Marley Goetz, Brian Dominguez, Elliot Thomasson, Cari Sloma, and Rob Christensen |
| EPI_ISL_1318290, EPI_ISL_1318291, EPI_ISL_1318292 | Ohio Department of Health Laboratory | Ohio Department of Health Laboratory | Holmes, Jennifer; Eric Brandt, Keoni Omura, Glen McGillivary, Caitlin McDonnell, Jade Mowery, Stephanie Mccracken, Tyler Payne, Kirtana Ramadugu, Erica Leasure, Brent Lee, Kelsey Florek, Heather Blankenship, Allison Black, Quanta Brown, and Tammy Bannerman |
| EPI_ISL_1318293, EPI_ISL_1318294, EPI_ISL_1318295, EPI_ISL_1318296, EPI_ISL_1318297, EPI_ISL_1318298, EPI_ISL_1318299, EPI_ISL_1318300, EPI_ISL_1318301, EPI_ISL_1318302, EPI_ISL_1318303, EPI_ISL_1318304, EPI_ISL_1318305, EPI_ISL_1318306, EPI_ISL_1318307, EPI_ISL_1318308, EPI_ISL_1318309, EPI_ISL_1318310, EPI_ISL_1318311, EPI_ISL_1318312, EPI_ISL_1318313, EPI_ISL_1318314, EPI_ISL_1318315, EPI_ISL_1318316, EPI_ISL_1318317 | | | |
| see above | TN Division of Laboratory Services | Centers for Disease Control and Prevention Division of Viral Diseases, Pathogen Discovery | Krista Queen, Yan Li, Ying Tao, Jing Zhang, Anna Uehara, Anna Montmayeur, Clinton R. Paden, Kristen Knipe, Matthew Schmerer, Shoshona Le, Katie Dillon, Peter W. Cook, Rachel Marine, Mili Sheth, Jasmine Padilla, Sarah Nobles, Mark Burroughs, Lori Rowe, Haibin Wang, Ben L. Rambo-Martin, Kristine Lacek, Sam Shepard, Dhwani Batra, Suxiang Tong, Justin Lee |
| EPI_ISL_1318318, EPI_ISL_1318319, EPI_ISL_1318320, EPI_ISL_1318321, EPI_ISL_1318322 | MI - Michigan Department of Health and Human Services - Bureau of Laboratories | Centers for Disease Control and Prevention Division of Viral Diseases, Pathogen Discovery | Krista Queen, Yan Li, Ying Tao, Jing Zhang, Anna Uehara, Anna Montmayeur, Clinton R. Paden, Kristen Knipe, Matthew Schmerer, Shoshona Le, Katie Dillon, Peter W. Cook, Rachel Marine, Mili Sheth, Jasmine Padilla, Sarah Nobles, Mark Burroughs, Lori Rowe, Haibin Wang, Ben L. Rambo-Martin, Kristine Lacek, Sam Shepard, Dhwani Batra, Suxiang Tong, Justin Lee |
| EPI_ISL_1318323, EPI_ISL_1318324, EPI_ISL_1318325, EPI_ISL_1318326, EPI_ISL_1318327 | UT-Unified State Labs: Public Health Utah DOH | Centers for Disease Control and Prevention Division of Viral Diseases, Pathogen Discovery | Krista Queen, Yan Li, Ying Tao, Jing Zhang, Anna Uehara, Anna Montmayeur, Clinton R. Paden, Kristen Knipe, Matthew Schmerer, Shoshona Le, Katie Dillon, Peter W. Cook, Rachel Marine, Mili Sheth, Jasmine Padilla, Sarah Nobles, Mark Burroughs, Lori Rowe, Haibin Wang, Ben L. Rambo-Martin, Kristine Lacek, Sam Shepard, Dhwani Batra, Suxiang Tong, Justin Lee |
| EPI_ISL_1318328, EPI_ISL_1318329, EPI_ISL_1318330, EPI_ISL_1318331, EPI_ISL_1318332, EPI_ISL_1318333, EPI_ISL_1318334, EPI_ISL_1318335, EPI_ISL_1318336, EPI_ISL_1318337, EPI_ISL_1318338, EPI_ISL_1318339, EPI_ISL_1318340, EPI_ISL_1318341, EPI_ISL_1318342, EPI_ISL_1318343, EPI_ISL_1318344, EPI_ISL_1318345, EPI_ISL_1318346 | | | |
| see above | DC Public Health Lab/ Dept. of Forensic Sciences | Centers for Disease Control and Prevention Division of Viral Diseases, Pathogen Discovery | Krista Queen, Yan Li, Ying Tao, Jing Zhang, Anna Uehara, Anna Montmayeur, Clinton R. Paden, Kristen Knipe, Matthew Schmerer, Shoshona Le, Katie Dillon, Peter W. Cook, Rachel Marine, Mili Sheth, Jasmine Padilla, Sarah Nobles, Mark Burroughs, Lori Rowe, Haibin Wang, Ben L. Rambo-Martin, Kristine Lacek, Sam Shepard, Dhwani Batra, Suxiang Tong, Justin Lee |
| EPI_ISL_1318347, EPI_ISL_1318348, EPI_ISL_1318349 | NYC Department of Health and Mental Hygiene | Centers for Disease Control and Prevention Division of Viral Diseases, Pathogen Discovery | Krista Queen, Yan Li, Ying Tao, Jing Zhang, Anna Uehara, Anna Montmayeur, Clinton R. Paden, Kristen Knipe, Matthew Schmerer, Shoshona Le, Katie Dillon, Peter W. Cook, Rachel Marine, Mili Sheth, Jasmine Padilla, Sarah Nobles, Mark Burroughs, Lori Rowe, Haibin Wang, Ben L. Rambo-Martin, Kristine Lacek, Sam Shepard, Dhwani Batra, Suxiang Tong, Justin Lee |
| EPI_ISL_1318350, EPI_ISL_1318351, EPI_ISL_1318352, EPI_ISL_1318353, EPI_ISL_1318354, EPI_ISL_1318355 | KS Health and Environmental Laboratories | Centers for Disease Control and Prevention Division of Viral Diseases, Pathogen Discovery | Krista Queen, Yan Li, Ying Tao, Jing Zhang, Anna Uehara, Anna Montmayeur, Clinton R. Paden, Kristen Knipe, Matthew Schmerer, Shoshona Le, Katie Dillon, Peter W. Cook, Rachel Marine, Mili Sheth, Jasmine Padilla, Sarah Nobles, Mark Burroughs, Lori Rowe, Haibin Wang, Ben L. Rambo-Martin, Kristine Lacek, Sam Shepard, Dhwani Batra, Suxiang Tong, Justin Lee |
| EPI_ISL_1318356, EPI_ISL_1318357, EPI_ISL_1318358, EPI_ISL_1318359, EPI_ISL_1318360, EPI_ISL_1318361, EPI_ISL_1318362, EPI_ISL_1318363, EPI_ISL_1318364, EPI_ISL_1318365, EPI_ISL_1318366, EPI_ISL_1318367, EPI_ISL_1318368 | | | |
| see above | VA-Division of Consolidated Laboratory Services | Centers for Disease Control and Prevention Division of Viral Diseases, Pathogen Discovery | Krista Queen, Yan Li, Ying Tao, Jing Zhang, Anna Uehara, Anna Montmayeur, Clinton R. Paden, Kristen Knipe, Matthew Schmerer, Shoshona Le, Katie Dillon, Peter W. Cook, Rachel Marine, Mili Sheth, Jasmine Padilla, Sarah Nobles, Mark Burroughs, Lori Rowe, Haibin Wang, Ben L. Rambo-Martin, Kristine Lacek, Sam Shepard, Dhwani Batra, Suxiang Tong, Justin Lee |
| EPI_ISL_1318369, EPI_ISL_1318370, EPI_ISL_1318371, EPI_ISL_1318372, EPI_ISL_1318373, EPI_ISL_1318374, EPI_ISL_1318375, EPI_ISL_1318376 | SC Dept of Health and Env. Control-Bureau of Laboratories | Centers for Disease Control and Prevention Division of Viral Diseases, Pathogen Discovery | Krista Queen, Yan Li, Ying Tao, Jing Zhang, Anna Uehara, Anna Montmayeur, Clinton R. Paden, Kristen Knipe, Matthew Schmerer, Shoshona Le, Katie Dillon, Peter W. Cook, Rachel Marine, Mili Sheth, Jasmine Padilla, Sarah Nobles, Mark Burroughs, Lori Rowe, Haibin Wang, Ben L. Rambo-Martin, Kristine Lacek, Sam Shepard, Dhwani Batra, Suxiang Tong, Justin Lee |
| EPI_ISL_1318377, EPI_ISL_1318378, EPI_ISL_1318379, EPI_ISL_1318380 | CA-Los Angeles County Public Health Laboratory | Centers for Disease Control and Prevention Division of Viral Diseases, Pathogen Discovery | Krista Queen, Yan Li, Ying Tao, Jing Zhang, Anna Uehara, Anna Montmayeur, Clinton R. Paden, Kristen Knipe, Matthew Schmerer, Shoshona Le, Katie Dillon, Peter W. Cook, Rachel Marine, Mili Sheth, Jasmine Padilla, Sarah Nobles, Mark Burroughs, Lori Rowe, Haibin Wang, Ben L. Rambo-Martin, Kristine Lacek, Sam Shepard, Dhwani Batra, Suxiang Tong, Justin Lee |
| EPI_ISL_1318381, EPI_ISL_1318382, EPI_ISL_1318383, EPI_ISL_1318384, EPI_ISL_1318385, EPI_ISL_1318386, EPI_ISL_1318387, EPI_ISL_1318388, EPI_ISL_1318389, EPI_ISL_1318390, EPI_ISL_1318391, EPI_ISL_1318392, EPI_ISL_1318393, EPI_ISL_1318394, EPI_ISL_1318395, EPI_ISL_1318396, EPI_ISL_1318397, EPI_ISL_1318398, EPI_ISL_1318399, EPI_ISL_1318400, EPI_ISL_1318401 | | | |
| see above | "CO Dept. of Public Health and Environment, Lab Services Division" | Centers for Disease Control and Prevention Division of Viral Diseases, Pathogen Discovery | Krista Queen, Yan Li, Ying Tao, Jing Zhang, Anna Uehara, Anna Montmayeur, Clinton R. Paden, Kristen Knipe, Matthew Schmerer, Shoshona Le, Katie Dillon, Peter W. Cook, Rachel Marine, Mili Sheth, Jasmine Padilla, Sarah Nobles, Mark Burroughs, Lori Rowe, Haibin Wang, Ben L. Rambo-Martin, Kristine Lacek, Sam Shepard, Dhwani Batra, Suxiang Tong, Justin Lee |
| EPI_ISL_1318402, EPI_ISL_1318403, EPI_ISL_1318404, EPI_ISL_1318405, EPI_ISL_1318406, EPI_ISL_1318407 | ID Bureau of Laboratories | Centers for Disease Control and Prevention Division of Viral Diseases, Pathogen Discovery | Krista Queen, Yan Li, Ying Tao, Jing Zhang, Anna Uehara, Anna Montmayeur, Clinton R. Paden, Kristen Knipe, Matthew Schmerer, Shoshona Le, Katie Dillon, Peter W. Cook, Rachel Marine, Mili Sheth, Jasmine Padilla, Sarah Nobles, Mark Burroughs, Lori Rowe, Haibin Wang, Ben L. Rambo-Martin, Kristine Lacek, Sam Shepard, Dhwani Batra, Suxiang Tong, Justin Lee |
| EPI_ISL_1318408 | Ohio Department of Health Laboratory | Ohio Department of Health Laboratory | Holmes, Jennifer; Eric Brandt, Keoni Omura, Glen McGillivary, Caitlin McDonnell, Jade Mowery, Stephanie Mccracken, Tyler Payne, Kirtana Ramadugu, Erica Leasure, Brent Lee, Kelsey Florek, Heather Blankenship, Allison Black, Quanta Brown, and Tammy Bannerman |
| EPI_ISL_1318409, EPI_ISL_1318410, EPI_ISL_1318411, EPI_ISL_1318412, EPI_ISL_1318413, EPI_ISL_1318414, EPI_ISL_1318415, EPI_ISL_1318416, EPI_ISL_1318417, EPI_ISL_1318418, EPI_ISL_1318419 | | | |
| see above | ID Bureau of Laboratories | Centers for Disease Control and Prevention Division of Viral Diseases, Pathogen Discovery | Krista Queen, Yan Li, Ying Tao, Jing Zhang, Anna Uehara, Anna Montmayeur, Clinton R. Paden, Kristen Knipe, Matthew Schmerer, Shoshona Le, Katie Dillon, Peter W. Cook, Rachel Marine, Mili Sheth, Jasmine Padilla, Sarah Nobles, Mark Burroughs, Lori Rowe, Haibin Wang, Ben L. Rambo-Martin, Kristine Lacek, Sam Shepard, Dhwani Batra, Suxiang Tong, Justin Lee |
| EPI_ISL_1318420, EPI_ISL_1318421, EPI_ISL_1318422, EPI_ISL_1318423 | NH Dept. of Health and Human Services Public Health Labs | Centers for Disease Control and Prevention Division of Viral Diseases, Pathogen Discovery | Krista Queen, Yan Li, Ying Tao, Jing Zhang, Anna Uehara, Anna Montmayeur, Clinton R. Paden, Kristen Knipe, Matthew Schmerer, Shoshona Le, Katie Dillon, Peter W. Cook, Rachel Marine, Mili Sheth, Jasmine Padilla, Sarah Nobles, Mark Burroughs, Lori Rowe, Haibin Wang, Ben L. Rambo-Martin, Kristine Lacek, Sam Shepard, Dhwani Batra, Suxiang Tong, Justin Lee |
| EPI_ISL_1318424 | IL Department of Public Health Chicago Laboratory | Centers for Disease Control and Prevention Division of Viral Diseases, Pathogen Discovery | Krista Queen, Yan Li, Ying Tao, Jing Zhang, Anna Uehara, Anna Montmayeur, Clinton R. Paden, Kristen Knipe, Matthew Schmerer, Shoshona Le, Katie Dillon, Peter W. Cook, Rachel Marine, Mili Sheth, Jasmine Padilla, Sarah Nobles, Mark Burroughs, Lori Rowe, Haibin Wang, Ben L. Rambo-Martin, Kristine Lacek, Sam Shepard, Dhwani Batra, Suxiang Tong, Justin Lee |
| EPI_ISL_1318425, EPI_ISL_1318426, EPI_ISL_1318427 | TX-Houston Health Department Laboratory | Centers for Disease Control and Prevention Division of Viral Diseases, Pathogen Discovery | Krista Queen, Yan Li, Ying Tao, Jing Zhang, Anna Uehara, Anna Montmayeur, Clinton R. Paden, Kristen Knipe, Matthew Schmerer, Shoshona Le, Katie Dillon, Peter W. Cook, Rachel Marine, Mili Sheth, Jasmine Padilla, Sarah Nobles, Mark Burroughs, Lori Rowe, Haibin Wang, Ben L. Rambo-Martin, Kristine Lacek, Sam Shepard, Dhwani Batra, Suxiang Tong, Justin Lee |
| EPI_ISL_1318428, EPI_ISL_1318429, EPI_ISL_1318430, EPI_ISL_1318431, EPI_ISL_1318432, EPI_ISL_1318433, EPI_ISL_1318434, EPI_ISL_1318435, EPI_ISL_1318436, EPI_ISL_1318437, EPI_ISL_1318438, EPI_ISL_1318439, EPI_ISL_1318440, EPI_ISL_1318441, EPI_ISL_1318442, EPI_ISL_1318443, EPI_ISL_1318444, EPI_ISL_1318445 | | | |
| see above | ND Dept. of Health Laboratory Services-Microbiology | Centers for Disease Control and Prevention Division of Viral Diseases, Pathogen Discovery | Krista Queen, Yan Li, Ying Tao, Jing Zhang, Anna Uehara, Anna Montmayeur, Clinton R. Paden, Kristen Knipe, Matthew Schmerer, Shoshona Le, Katie Dillon, Peter W. Cook, Rachel Marine, Mili Sheth, Jasmine Padilla, Sarah Nobles, Mark Burroughs, Lori Rowe, Haibin Wang, Ben L. Rambo-Martin, Kristine Lacek, Sam Shepard, Dhwani Batra, Suxiang Tong, Justin Lee |
| EPI_ISL_1318446, EPI_ISL_1318447, EPI_ISL_1318448 | "TX DSHS, Lab Services Section MC 1947" | Centers for Disease Control and Prevention Division of Viral Diseases, Pathogen Discovery | Krista Queen, Yan Li, Ying Tao, Jing Zhang, Anna Uehara, Anna Montmayeur, Clinton R. Paden, Kristen Knipe, Matthew Schmerer, Shoshona Le, Katie Dillon, Peter W. Cook, Rachel Marine, Mili Sheth, Jasmine Padilla, Sarah Nobles, Mark Burroughs, Lori Rowe, Haibin Wang, Ben L. Rambo-Martin, Kristine Lacek, Sam Shepard, Dhwani Batra, Suxiang Tong, Justin Lee |
| EPI_ISL_1318449, EPI_ISL_1318450, EPI_ISL_1318451, EPI_ISL_1318452, EPI_ISL_1318453, EPI_ISL_1318454, EPI_ISL_1318455, EPI_ISL_1318456, EPI_ISL_1318457, EPI_ISL_1318458, EPI_ISL_1318459, EPI_ISL_1318460, EPI_ISL_1318461, EPI_ISL_1318462, EPI_ISL_1318463, EPI_ISL_1318464 | | | |
| see above | RI State Health Laboratories | Centers for Disease Control and Prevention Division of Viral Diseases, Pathogen Discovery | Krista Queen, Yan Li, Ying Tao, Jing Zhang, Anna Uehara, Anna Montmayeur, Clinton R. Paden, Kristen Knipe, Matthew Schmerer, Shoshona Le, Katie Dillon, Peter W. Cook, Rachel Marine, Mili Sheth, Jasmine Padilla, Sarah Nobles, Mark Burroughs, Lori Rowe, Haibin Wang, Ben L. Rambo-Martin, Kristine Lacek, Sam Shepard, Dhwani Batra, Suxiang Tong, Justin Lee |
| EPI_ISL_1318465, EPI_ISL_1318466, | CT-Dr. Katherine A. Kelley State Public Health Lab | Centers for Disease Control and Prevention Division of Viral Diseases, Pathogen | Krista Queen, Yan Li, Ying Tao, Jing Zhang, Anna Uehara, Anna Montmayeur, Clinton R. Paden, Kristen Knipe, Matthew Schmerer, Shoshona Le, Katie Dillon, Peter W. Cook, Rachel Marine, Mili Sheth, Jasmine Padilla, Sarah Nobles, Mark Burroughs, Lori Rowe, Haibin Wang, Ben L. Rambo-Martin, Kristine Lacek, Sam Shepard, Dhwani Batra, Suxiang Tong, Justin Lee |

| | | | |
|---|---|---|---|
| EPI_ISL_1318467 | | Discovery | |
| EPI_ISL_1318468, EPI_ISL_1318469, EPI_ISL_1318470, EPI_ISL_1318471, EPI_ISL_1318472, EPI_ISL_1318473, EPI_ISL_1318474, EPI_ISL_1318475, EPI_ISL_1318476, EPI_ISL_1318477 | MT Public Health Laboratory | Centers for Disease Control and Prevention Division of Viral Diseases, Pathogen Discovery | Krista Queen, Yan Li, Ying Tao, Jing Zhang, Anna Uehara, Anna Montmayeur, Clinton R. Paden, Kristen Knipe, Matthew Schmerer, Shoshona Le, Katie Dillon, Peter W. Cook, Rachel Marine, Mili Sheth, Jasmine Padilla, Sarah Nobles, Mark Burroughs, Lori Rowe, Haibin Wang, Ben L. Rambo-Martin, Kristine Lacek, Sam Shepard, Dhwani Batra, Suxiang Tong, Justin Lee |
| EPI_ISL_1318478 | WVDHHR - Office of Laboratory Services | Centers for Disease Control and Prevention Division of Viral Diseases, Pathogen Discovery | Krista Queen, Yan Li, Ying Tao, Jing Zhang, Anna Uehara, Anna Montmayeur, Clinton R. Paden, Kristen Knipe, Matthew Schmerer, Shoshona Le, Katie Dillon, Peter W. Cook, Rachel Marine, Mili Sheth, Jasmine Padilla, Sarah Nobles, Mark Burroughs, Lori Rowe, Haibin Wang, Ben L. Rambo-Martin, Kristine Lacek, Sam Shepard, Dhwani Batra, Suxiang Tong, Justin Lee |
| EPI_ISL_1318479, EPI_ISL_1318480, EPI_ISL_1318481, EPI_ISL_1318482, EPI_ISL_1318483, EPI_ISL_1318484, EPI_ISL_1318485, EPI_ISL_1318486, EPI_ISL_1318487 | FL Bureau of Public Health Laboratories-Tampa | Centers for Disease Control and Prevention Division of Viral Diseases, Pathogen Discovery | Krista Queen, Yan Li, Ying Tao, Jing Zhang, Anna Uehara, Anna Montmayeur, Clinton R. Paden, Kristen Knipe, Matthew Schmerer, Shoshona Le, Katie Dillon, Peter W. Cook, Rachel Marine, Mili Sheth, Jasmine Padilla, Sarah Nobles, Mark Burroughs, Lori Rowe, Haibin Wang, Ben L. Rambo-Martin, Kristine Lacek, Sam Shepard, Dhwani Batra, Suxiang Tong, Justin Lee |
| EPI_ISL_1318488, EPI_ISL_1318489, EPI_ISL_1318490, EPI_ISL_1318491, EPI_ISL_1318492, EPI_ISL_1318493, EPI_ISL_1318494, EPI_ISL_1318495, EPI_ISL_1318496, EPI_ISL_1318497, EPI_ISL_1318498, EPI_ISL_1318499, EPI_ISL_1318500 | | | |
| see above | "HI Dept. of Health, State Laboratories Division" | Centers for Disease Control and Prevention Division of Viral Diseases, Pathogen Discovery | Krista Queen, Yan Li, Ying Tao, Jing Zhang, Anna Uehara, Anna Montmayeur, Clinton R. Paden, Kristen Knipe, Matthew Schmerer, Shoshona Le, Katie Dillon, Peter W. Cook, Rachel Marine, Mili Sheth, Jasmine Padilla, Sarah Nobles, Mark Burroughs, Lori Rowe, Haibin Wang, Ben L. Rambo-Martin, Kristine Lacek, Sam Shepard, Dhwani Batra, Suxiang Tong, Justin Lee |
| EPI_ISL_1318501, EPI_ISL_1318502, EPI_ISL_1318503, EPI_ISL_1318504 | SD Public Health Laboratory | Centers for Disease Control and Prevention Division of Viral Diseases, Pathogen Discovery | Krista Queen, Yan Li, Ying Tao, Jing Zhang, Anna Uehara, Anna Montmayeur, Clinton R. Paden, Kristen Knipe, Matthew Schmerer, Shoshona Le, Katie Dillon, Peter W. Cook, Rachel Marine, Mili Sheth, Jasmine Padilla, Sarah Nobles, Mark Burroughs, Lori Rowe, Haibin Wang, Ben L. Rambo-Martin, Kristine Lacek, Sam Shepard, Dhwani Batra, Suxiang Tong, Justin Lee |
| EPI_ISL_1318505, EPI_ISL_1318506, EPI_ISL_1318507, EPI_ISL_1318508, EPI_ISL_1318509 | MA State Public Health Laboratory | Centers for Disease Control and Prevention Division of Viral Diseases, Pathogen Discovery | Krista Queen, Yan Li, Ying Tao, Jing Zhang, Anna Uehara, Anna Montmayeur, Clinton R. Paden, Kristen Knipe, Matthew Schmerer, Shoshona Le, Katie Dillon, Peter W. Cook, Rachel Marine, Mili Sheth, Jasmine Padilla, Sarah Nobles, Mark Burroughs, Lori Rowe, Haibin Wang, Ben L. Rambo-Martin, Kristine Lacek, Sam Shepard, Dhwani Batra, Suxiang Tong, Justin Lee |
| EPI_ISL_1318510, EPI_ISL_1318511, EPI_ISL_1318512, EPI_ISL_1318513, EPI_ISL_1318514, EPI_ISL_1318515, EPI_ISL_1318516, EPI_ISL_1318517, EPI_ISL_1318518, EPI_ISL_1318519, EPI_ISL_1318520, EPI_ISL_1318521, EPI_ISL_1318522, EPI_ISL_1318523, EPI_ISL_1318524 | | | |
| see above | "AR Dept. of Health-PHL, Molecular Diagnostics" | Centers for Disease Control and Prevention Division of Viral Diseases, Pathogen Discovery | Krista Queen, Yan Li, Ying Tao, Jing Zhang, Anna Uehara, Anna Montmayeur, Clinton R. Paden, Kristen Knipe, Matthew Schmerer, Shoshona Le, Katie Dillon, Peter W. Cook, Rachel Marine, Mili Sheth, Jasmine Padilla, Sarah Nobles, Mark Burroughs, Lori Rowe, Haibin Wang, Ben L. Rambo-Martin, Kristine Lacek, Sam Shepard, Dhwani Batra, Suxiang Tong, Justin Lee |
| EPI_ISL_1318525, EPI_ISL_1318527, EPI_ISL_1318528, EPI_ISL_1318529, EPI_ISL_1318530, EPI_ISL_1318531 | FL Bur. of Public Health Laboratories-Jacksonville | Centers for Disease Control and Prevention Division of Viral Diseases, Pathogen Discovery | Krista Queen, Yan Li, Ying Tao, Jing Zhang, Anna Uehara, Anna Montmayeur, Clinton R. Paden, Kristen Knipe, Matthew Schmerer, Shoshona Le, Katie Dillon, Peter W. Cook, Rachel Marine, Mili Sheth, Jasmine Padilla, Sarah Nobles, Mark Burroughs, Lori Rowe, Haibin Wang, Ben L. Rambo-Martin, Kristine Lacek, Sam Shepard, Dhwani Batra, Suxiang Tong, Justin Lee |
| EPI_ISL_1318532, EPI_ISL_1318533, EPI_ISL_1318534, EPI_ISL_1318535, EPI_ISL_1318536, EPI_ISL_1318537 | WI State Laboratory of Hygiene | Centers for Disease Control and Prevention Division of Viral Diseases, Pathogen Discovery | Krista Queen, Yan Li, Ying Tao, Jing Zhang, Anna Uehara, Anna Montmayeur, Clinton R. Paden, Kristen Knipe, Matthew Schmerer, Shoshona Le, Katie Dillon, Peter W. Cook, Rachel Marine, Mili Sheth, Jasmine Padilla, Sarah Nobles, Mark Burroughs, Lori Rowe, Haibin Wang, Ben L. Rambo-Martin, Kristine Lacek, Sam Shepard, Dhwani Batra, Suxiang Tong, Justin Lee |
| EPI_ISL_1318538, EPI_ISL_1318539, EPI_ISL_1318540, EPI_ISL_1318541, EPI_ISL_1318542, EPI_ISL_1318543, EPI_ISL_1318544, EPI_ISL_1318545, EPI_ISL_1318546, EPI_ISL_1318547 | AL Dept. of Public Health Bureau of Clinical Laboratories | Centers for Disease Control and Prevention Division of Viral Diseases, Pathogen Discovery | Krista Queen, Yan Li, Ying Tao, Jing Zhang, Anna Uehara, Anna Montmayeur, Clinton R. Paden, Kristen Knipe, Matthew Schmerer, Shoshona Le, Katie Dillon, Peter W. Cook, Rachel Marine, Mili Sheth, Jasmine Padilla, Sarah Nobles, Mark Burroughs, Lori Rowe, Haibin Wang, Ben L. Rambo-Martin, Kristine Lacek, Sam Shepard, Dhwani Batra, Suxiang Tong, Justin Lee |
| EPI_ISL_1318548, EPI_ISL_1318549, EPI_ISL_1318550, EPI_ISL_1318551, EPI_ISL_1318552, EPI_ISL_1318553, EPI_ISL_1318554, EPI_ISL_1318555, EPI_ISL_1318556, EPI_ISL_1318557, EPI_ISL_1318558, EPI_ISL_1318559, EPI_ISL_1318560, EPI_ISL_1318561, EPI_ISL_1318562, EPI_ISL_1318563, EPI_ISL_1318564, EPI_ISL_1318565, EPI_ISL_1318566, EPI_ISL_1318567, EPI_ISL_1318568, EPI_ISL_1318569, EPI_ISL_1318570, EPI_ISL_1318571, EPI_ISL_1318572, EPI_ISL_1318573, EPI_ISL_1318574, EPI_ISL_1318575, EPI_ISL_1318576, EPI_ISL_1318577, EPI_ISL_1318578, EPI_ISL_1318579, EPI_ISL_1318580, EPI_ISL_1318581, EPI_ISL_1318582, EPI_ISL_1318583, EPI_ISL_1318584, EPI_ISL_1318585, EPI_ISL_1318586, EPI_ISL_1318587, EPI_ISL_1318588, EPI_ISL_1318589, EPI_ISL_1318590, EPI_ISL_1318591, EPI_ISL_1318592, EPI_ISL_1318593, EPI_ISL_1318594 | | | |
| see above | GA Department of Public Health Laboratory | Centers for Disease Control and Prevention Division of Viral Diseases, Pathogen Discovery | Krista Queen, Yan Li, Ying Tao, Jing Zhang, Anna Uehara, Anna Montmayeur, Clinton R. Paden, Kristen Knipe, Matthew Schmerer, Shoshona Le, Katie Dillon, Peter W. Cook, Rachel Marine, Mili Sheth, Jasmine Padilla, Sarah Nobles, Mark Burroughs, Lori Rowe, Haibin Wang, Ben L. Rambo-Martin, Kristine Lacek, Sam Shepard, Dhwani Batra, Suxiang Tong, Justin Lee |
| EPI_ISL_1318595, EPI_ISL_1318596, EPI_ISL_1318597, EPI_ISL_1318598, EPI_ISL_1318599, EPI_ISL_1318600, EPI_ISL_1318601, EPI_ISL_1318602, EPI_ISL_1318603, EPI_ISL_1318604 | NC State Laboratory of Public Health | Centers for Disease Control and Prevention Division of Viral Diseases, Pathogen Discovery | Krista Queen, Yan Li, Ying Tao, Jing Zhang, Anna Uehara, Anna Montmayeur, Clinton R. Paden, Kristen Knipe, Matthew Schmerer, Shoshona Le, Katie Dillon, Peter W. Cook, Rachel Marine, Mili Sheth, Jasmine Padilla, Sarah Nobles, Mark Burroughs, Lori Rowe, Haibin Wang, Ben L. Rambo-Martin, Kristine Lacek, Sam Shepard, Dhwani Batra, Suxiang Tong, Justin Lee |
| EPI_ISL_1318605 | Ohio Department of Health Laboratory | Ohio Department of Health Laboratory | Holmes, Jennifer; Eric Brandt, Keoni Omura, Glen McGillivary, Caitlin McDonnell, Jade Mowery, Stephanie Mccracken, Tyler Payne, Kirtana Ramadugu, Erica Leasure, Brent Lee, Kelsey Florek, Heather Blankenship, Allison Black, Quanta Brown, and Tammy Bannerman |
| EPI_ISL_1318606, EPI_ISL_1318607, EPI_ISL_1318608, EPI_ISL_1318609, EPI_ISL_1318610, EPI_ISL_1318611, EPI_ISL_1318612, EPI_ISL_1318613 | NC State Laboratory of Public Health | Centers for Disease Control and Prevention Division of Viral Diseases, Pathogen Discovery | Krista Queen, Yan Li, Ying Tao, Jing Zhang, Anna Uehara, Anna Montmayeur, Clinton R. Paden, Kristen Knipe, Matthew Schmerer, Shoshona Le, Katie Dillon, Peter W. Cook, Rachel Marine, Mili Sheth, Jasmine Padilla, Sarah Nobles, Mark Burroughs, Lori Rowe, Haibin Wang, Ben L. Rambo-Martin, Kristine Lacek, Sam Shepard, Dhwani Batra, Suxiang Tong, Justin Lee |
| EPI_ISL_1318614, EPI_ISL_1318615, EPI_ISL_1318616, EPI_ISL_1318617, EPI_ISL_1318618, EPI_ISL_1318619, EPI_ISL_1318620, EPI_ISL_1318621, EPI_ISL_1318622 | IA State Hygienic Laboratory | Centers for Disease Control and Prevention Division of Viral Diseases, Pathogen Discovery | Krista Queen, Yan Li, Ying Tao, Jing Zhang, Anna Uehara, Anna Montmayeur, Clinton R. Paden, Kristen Knipe, Matthew Schmerer, Shoshona Le, Katie Dillon, Peter W. Cook, Rachel Marine, Mili Sheth, Jasmine Padilla, Sarah Nobles, Mark Burroughs, Lori Rowe, Haibin Wang, Ben L. Rambo-Martin, Kristine Lacek, Sam Shepard, Dhwani Batra, Suxiang Tong, Justin Lee |
| EPI_ISL_1318623, EPI_ISL_1318624, EPI_ISL_1318625, EPI_ISL_1318626, EPI_ISL_1318627, EPI_ISL_1318628, EPI_ISL_1318629, EPI_ISL_1318630, EPI_ISL_1318631, EPI_ISL_1318632, EPI_ISL_1318633 | | | |
| see above | LA Office of Public Health Laboratories | Centers for Disease Control and Prevention Division of Viral Diseases, Pathogen Discovery | Krista Queen, Yan Li, Ying Tao, Jing Zhang, Anna Uehara, Anna Montmayeur, Clinton R. Paden, Kristen Knipe, Matthew Schmerer, Shoshona Le, Katie Dillon, Peter W. Cook, Rachel Marine, Mili Sheth, Jasmine Padilla, Sarah Nobles, Mark Burroughs, Lori Rowe, Haibin Wang, Ben L. Rambo-Martin, Kristine Lacek, Sam Shepard, Dhwani Batra, Suxiang Tong, Justin Lee |
| EPI_ISL_1318634, EPI_ISL_1318635, EPI_ISL_1318636, EPI_ISL_1318637, EPI_ISL_1318638, EPI_ISL_1318639, EPI_ISL_1318640, EPI_ISL_1318641, EPI_ISL_1318642, EPI_ISL_1318643, EPI_ISL_1318644, EPI_ISL_1318645, EPI_ISL_1318646, EPI_ISL_1318647, EPI_ISL_1318648, EPI_ISL_1318649, EPI_ISL_1318650, EPI_ISL_1318651, EPI_ISL_1318652, EPI_ISL_1318653, EPI_ISL_1318654, EPI_ISL_1318655, | | | |

| | | | |
|---|---|---|---|
| EPI_ISL_1318656, EPI_ISL_1318657 | | | |
| see above | "CDPH, Viral and Rickettsial Disease Laboratory" | Centers for Disease Control and Prevention Division of Viral Diseases, Pathogen Discovery | Krista Queen, Yan Li, Ying Tao, Jing Zhang, Anna Uehara, Anna Montmayeur, Clinton R. Paden, Kristen Knipe, Matthew Schmerer, Shoshona Le, Katie Dillon, Peter W. Cook, Rachel Marine, Mili Sheth, Jasmine Padilla, Sarah Nobles, Mark Burroughs, Lori Rowe, Haibin Wang, Ben L. Rambo-Martin, Kristine Lacek, Sam Shepard, Dhwani Batra, Suxiang Tong, Justin Lee |
| EPI_ISL_1318658 | Ohio Department of Health Laboratory | Ohio Department of Health Laboratory | Holmes, Jennifer; Eric Brandt, Keoni Omura, Glen McGillivary, Caitlin McDonnell, Jade Mowery, Stephanie Mccracken, Tyler Payne, Kirtana Ramadugu, Erica Leasure, Brent Lee, Kelsey Florek, Heather Blankenship, Allison Black, Quanta Brown, and Tammy Bannerman |
| EPI_ISL_1318659, EPI_ISL_1318660, EPI_ISL_1318661, EPI_ISL_1318662, EPI_ISL_1318663, EPI_ISL_1318664, EPI_ISL_1318665, EPI_ISL_1318666, EPI_ISL_1318667 | "CDPH, Viral and Rickettsial Disease Laboratory" | Centers for Disease Control and Prevention Division of Viral Diseases, Pathogen Discovery | Krista Queen, Yan Li, Ying Tao, Jing Zhang, Anna Uehara, Anna Montmayeur, Clinton R. Paden, Kristen Knipe, Matthew Schmerer, Shoshona Le, Katie Dillon, Peter W. Cook, Rachel Marine, Mili Sheth, Jasmine Padilla, Sarah Nobles, Mark Burroughs, Lori Rowe, Haibin Wang, Ben L. Rambo-Martin, Kristine Lacek, Sam Shepard, Dhwani Batra, Suxiang Tong, Justin Lee |
| EPI_ISL_1318668, EPI_ISL_1318669, EPI_ISL_1318670, EPI_ISL_1318671, EPI_ISL_1318672, EPI_ISL_1318673, EPI_ISL_1318674, EPI_ISL_1318675, EPI_ISL_1318676, EPI_ISL_1318677, EPI_ISL_1318678, EPI_ISL_1318679, EPI_ISL_1318680 | | | |
| see above | "PA Department of Health, Bureau of Laboratories" | Centers for Disease Control and Prevention Division of Viral Diseases, Pathogen Discovery | Krista Queen, Yan Li, Ying Tao, Jing Zhang, Anna Uehara, Anna Montmayeur, Clinton R. Paden, Kristen Knipe, Matthew Schmerer, Shoshona Le, Katie Dillon, Peter W. Cook, Rachel Marine, Mili Sheth, Jasmine Padilla, Sarah Nobles, Mark Burroughs, Lori Rowe, Haibin Wang, Ben L. Rambo-Martin, Kristine Lacek, Sam Shepard, Dhwani Batra, Suxiang Tong, Justin Lee |
| EPI_ISL_1318681, EPI_ISL_1318682, EPI_ISL_1318683, EPI_ISL_1318684, EPI_ISL_1318685, EPI_ISL_1318686, EPI_ISL_1318687, EPI_ISL_1318688, EPI_ISL_1318689, EPI_ISL_1318690, EPI_ISL_1318691 | | | |
| see above | NE Public Health Laboratory | Centers for Disease Control and Prevention Division of Viral Diseases, Pathogen Discovery | Krista Queen, Yan Li, Ying Tao, Jing Zhang, Anna Uehara, Anna Montmayeur, Clinton R. Paden, Kristen Knipe, Matthew Schmerer, Shoshona Le, Katie Dillon, Peter W. Cook, Rachel Marine, Mili Sheth, Jasmine Padilla, Sarah Nobles, Mark Burroughs, Lori Rowe, Haibin Wang, Ben L. Rambo-Martin, Kristine Lacek, Sam Shepard, Dhwani Batra, Suxiang Tong, Justin Lee |
| EPI_ISL_1318692, EPI_ISL_1318693, EPI_ISL_1318695 | Ohio Department of Health Laboratory | Ohio Department of Health Laboratory | Holmes, Jennifer; Eric Brandt, Keoni Omura, Glen McGillivary, Caitlin McDonnell, Jade Mowery, Stephanie Mccracken, Tyler Payne, Kirtana Ramadugu, Erica Leasure, Brent Lee, Kelsey Florek, Heather Blankenship, Allison Black, Quanta Brown, and Tammy Bannerman |
| EPI_ISL_1318696 | University of Illinois Veterinary Diagnostic Laboratory | Roy J. Carver Biotechnology Center | Alvaro Hernandez, Chris Wright, Christopher Fields, Mireille Farjo, Christopher Brooke |
| EPI_ISL_1318697, EPI_ISL_1318698, EPI_ISL_1318699, EPI_ISL_1318700, EPI_ISL_1318701, EPI_ISL_1318702, EPI_ISL_1318703, EPI_ISL_1318704, EPI_ISL_1318705, EPI_ISL_1318706, EPI_ISL_1318707, EPI_ISL_1318708, EPI_ISL_1318709, EPI_ISL_1318710, EPI_ISL_1318711, EPI_ISL_1318712, EPI_ISL_1318713, EPI_ISL_1318714, EPI_ISL_1318715, EPI_ISL_1318716, EPI_ISL_1318717, EPI_ISL_1318718, EPI_ISL_1318719, EPI_ISL_1318720, EPI_ISL_1318721, EPI_ISL_1318722, EPI_ISL_1318723, EPI_ISL_1318724, EPI_ISL_1318725, EPI_ISL_1318726, EPI_ISL_1318727, EPI_ISL_1318728, EPI_ISL_1318729, EPI_ISL_1318730, EPI_ISL_1318731, EPI_ISL_1318732, EPI_ISL_1318733, EPI_ISL_1318734, EPI_ISL_1318735, EPI_ISL_1318736, EPI_ISL_1318737, EPI_ISL_1318738, EPI_ISL_1318739, EPI_ISL_1318740, EPI_ISL_1318741, EPI_ISL_1318742, EPI_ISL_1318743, EPI_ISL_1318744, EPI_ISL_1318745, EPI_ISL_1318746, EPI_ISL_1318747, EPI_ISL_1318748, EPI_ISL_1318749, EPI_ISL_1318750, EPI_ISL_1318751, EPI_ISL_1318752, EPI_ISL_1318753, EPI_ISL_1318754, EPI_ISL_1318755, EPI_ISL_1318756, EPI_ISL_1318757, EPI_ISL_1318758, EPI_ISL_1318759, EPI_ISL_1318760, EPI_ISL_1318761, EPI_ISL_1318762, EPI_ISL_1318763, EPI_ISL_1318764, EPI_ISL_1318765, EPI_ISL_1318766, EPI_ISL_1318767, EPI_ISL_1318768, EPI_ISL_1318769, EPI_ISL_1318770, EPI_ISL_1318771, EPI_ISL_1318772, EPI_ISL_1318773, EPI_ISL_1318774, EPI_ISL_1318775, EPI_ISL_1318776, EPI_ISL_1318777, EPI_ISL_1318778, EPI_ISL_1318779, EPI_ISL_1318780, EPI_ISL_1318781, EPI_ISL_1318782, EPI_ISL_1318783, EPI_ISL_1318784, EPI_ISL_1318785, EPI_ISL_1318786, EPI_ISL_1318787, EPI_ISL_1318788, EPI_ISL_1318789, EPI_ISL_1318790, EPI_ISL_1318791, EPI_ISL_1318792, EPI_ISL_1318793, EPI_ISL_1318794, EPI_ISL_1318795, EPI_ISL_1318796, EPI_ISL_1318797, EPI_ISL_1318798, EPI_ISL_1318799, EPI_ISL_1318800, EPI_ISL_1318801, EPI_ISL_1318802, EPI_ISL_1318803, EPI_ISL_1318804, EPI_ISL_1318805, EPI_ISL_1318806, EPI_ISL_1318807, EPI_ISL_1318808, EPI_ISL_1318809, EPI_ISL_1318810, EPI_ISL_1318811, EPI_ISL_1318812, EPI_ISL_1318813, EPI_ISL_1318814, EPI_ISL_1318815, EPI_ISL_1318816, EPI_ISL_1318817, EPI_ISL_1318818, EPI_ISL_1318819, EPI_ISL_1318820, EPI_ISL_1318821, EPI_ISL_1318822, EPI_ISL_1318823, EPI_ISL_1318824, EPI_ISL_1318825, EPI_ISL_1318826, EPI_ISL_1318827, EPI_ISL_1318828, EPI_ISL_1318829, EPI_ISL_1318830, EPI_ISL_1318831, EPI_ISL_1318832, EPI_ISL_1318833, EPI_ISL_1318834, EPI_ISL_1318835, EPI_ISL_1318836, EPI_ISL_1318837, EPI_ISL_1318838, EPI_ISL_1318839, EPI_ISL_1318840, EPI_ISL_1318841, EPI_ISL_1318842, EPI_ISL_1318843, EPI_ISL_1318844, EPI_ISL_1318845, EPI_ISL_1318846, EPI_ISL_1318847, EPI_ISL_1318848, EPI_ISL_1318849, EPI_ISL_1318850, EPI_ISL_1318851, EPI_ISL_1318852, EPI_ISL_1318853, EPI_ISL_1318854, EPI_ISL_1318855, EPI_ISL_1318856, EPI_ISL_1318857, EPI_ISL_1318858, EPI_ISL_1318859, EPI_ISL_1318860, EPI_ISL_1318861, EPI_ISL_1318862, EPI_ISL_1318863, EPI_ISL_1318864, EPI_ISL_1318865, EPI_ISL_1318866, EPI_ISL_1318867, EPI_ISL_1318868, EPI_ISL_1318869, EPI_ISL_1318870, EPI_ISL_1318871, EPI_ISL_1318872, EPI_ISL_1318873, EPI_ISL_1318874, EPI_ISL_1318875, EPI_ISL_1318876, EPI_ISL_1318877, EPI_ISL_1318878, EPI_ISL_1318879, EPI_ISL_1318880, EPI_ISL_1318881, EPI_ISL_1318882, EPI_ISL_1318883, EPI_ISL_1318884, EPI_ISL_1318885, EPI_ISL_1318886, EPI_ISL_1318887, EPI_ISL_1318888, EPI_ISL_1318889, EPI_ISL_1318890, EPI_ISL_1318891, EPI_ISL_1318892, EPI_ISL_1318893, EPI_ISL_1318894, EPI_ISL_1318895, EPI_ISL_1318896, EPI_ISL_1318897, EPI_ISL_1318898, EPI_ISL_1318899, EPI_ISL_1318900, EPI_ISL_1318901, EPI_ISL_1318902, EPI_ISL_1318903, EPI_ISL_1318904, EPI_ISL_1318905, EPI_ISL_1318906, EPI_ISL_1318907, EPI_ISL_1318908, EPI_ISL_1318909, EPI_ISL_1318910, EPI_ISL_1318911, EPI_ISL_1318912, EPI_ISL_1318913, EPI_ISL_1318914, EPI_ISL_1318915, EPI_ISL_1318916, EPI_ISL_1318917, EPI_ISL_1318918, EPI_ISL_1318919, EPI_ISL_1318920, EPI_ISL_1318921, EPI_ISL_1318922, EPI_ISL_1318923, EPI_ISL_1318924, EPI_ISL_1318925, EPI_ISL_1318926, EPI_ISL_1318927, EPI_ISL_1318928, EPI_ISL_1318929, EPI_ISL_1318930, EPI_ISL_1318931, EPI_ISL_1318932, EPI_ISL_1318933, EPI_ISL_1318934, EPI_ISL_1318935, EPI_ISL_1318936, EPI_ISL_1318937, EPI_ISL_1318938, EPI_ISL_1318939, EPI_ISL_1318940, EPI_ISL_1318941, EPI_ISL_1318942, EPI_ISL_1318943, EPI_ISL_1318944, EPI_ISL_1318945, EPI_ISL_1318946, EPI_ISL_1318947, EPI_ISL_1318948, EPI_ISL_1318949, EPI_ISL_1318950, EPI_ISL_1318951, EPI_ISL_1318952, EPI_ISL_1318953, EPI_ISL_1318954, EPI_ISL_1318955, EPI_ISL_1318956, EPI_ISL_1318957, EPI_ISL_1318958, EPI_ISL_1318959, EPI_ISL_1318960, EPI_ISL_1318961, EPI_ISL_1318962, EPI_ISL_1318963, EPI_ISL_1318964, EPI_ISL_1318965, EPI_ISL_1318966, EPI_ISL_1318967, EPI_ISL_1318968, EPI_ISL_1318969, EPI_ISL_1318970, EPI_ISL_1318971, EPI_ISL_1318972, EPI_ISL_1318973, EPI_ISL_1318974, EPI_ISL_1318975, EPI_ISL_1318976, EPI_ISL_1318977, EPI_ISL_1318978, EPI_ISL_1318979, EPI_ISL_1318980, EPI_ISL_1318981, EPI_ISL_1318982, EPI_ISL_1318983, EPI_ISL_1318984, EPI_ISL_1318985, EPI_ISL_1318986, EPI_ISL_1318988 | | | |
| see above | DeRisi Lab, University of California, San Francisco | DeRisi Lab, University of California, San Francisco | Sabrina Mann, Anthea Mitchell, Jamin Liu, Sara Sunshine, Matthew Laurie, Genay Pilarowski, Amy Kistler, Manu Vanaerschot, Patrick Ayscue, Lucy Li, Eric Chow, IDseq Team, James Peng, Carina Marquez, Luis Rubio, Gabriel Chamie, Diane Jones, Jon Jacobo, Susana Rojas, Susy Rojas, Valerie Tulier-Laiwa, Douglas Black, Jackie Martinez, Maya Petersen, Diane Havlir, and Joseph DeRisi |
| EPI_ISL_1319044, EPI_ISL_1319047, EPI_ISL_1319051, EPI_ISL_1319052, EPI_ISL_1319056, EPI_ISL_1319057, EPI_ISL_1319059, EPI_ISL_1319060, EPI_ISL_1319062, EPI_ISL_1319065, EPI_ISL_1319068, EPI_ISL_1319069, EPI_ISL_1319071, EPI_ISL_1319073, EPI_ISL_1319074, EPI_ISL_1319075, EPI_ISL_1319076, EPI_ISL_1319077, EPI_ISL_1319079, EPI_ISL_1319080, EPI_ISL_1319083, EPI_ISL_1319084, EPI_ISL_1319085, EPI_ISL_1319086, EPI_ISL_1319090, EPI_ISL_1319091, EPI_ISL_1319093, EPI_ISL_1319111, EPI_ISL_1319115, EPI_ISL_1319117, EPI_ISL_1319122, EPI_ISL_1319124, EPI_ISL_1319130, EPI_ISL_1319135, EPI_ISL_1319137, EPI_ISL_1319139, EPI_ISL_1319140, EPI_ISL_1319141, EPI_ISL_1319144, EPI_ISL_1319147, EPI_ISL_1319152, EPI_ISL_1319153, EPI_ISL_1319154, EPI_ISL_1319158, EPI_ISL_1319175, EPI_ISL_1319176, EPI_ISL_1319177, EPI_ISL_1319178, EPI_ISL_1319179, EPI_ISL_1319180, EPI_ISL_1319182, EPI_ISL_1319183, EPI_ISL_1319189, EPI_ISL_1319190, EPI_ISL_1319194, EPI_ISL_1319207, EPI_ISL_1319211, EPI_ISL_1319215, EPI_ISL_1319216, EPI_ISL_1319222, EPI_ISL_1319224, EPI_ISL_1319234, EPI_ISL_1319237, EPI_ISL_1319238, EPI_ISL_1319240, EPI_ISL_1319241, EPI_ISL_1319246, EPI_ISL_1319248, EPI_ISL_1319253, EPI_ISL_1319257, EPI_ISL_1319278, EPI_ISL_1319282, EPI_ISL_1319326, EPI_ISL_1319349, EPI_ISL_1319368, EPI_ISL_1319389, EPI_ISL_1319391, EPI_ISL_1319396, EPI_ISL_1319398, EPI_ISL_1319405, EPI_ISL_1319408, EPI_ISL_1319410, EPI_ISL_1319414, EPI_ISL_1319442, EPI_ISL_1319444, EPI_ISL_1319448, EPI_ISL_1319458, EPI_ISL_1319459, EPI_ISL_1319460, EPI_ISL_1319461, EPI_ISL_1319464, EPI_ISL_1319466, EPI_ISL_1319468, EPI_ISL_1319471, EPI_ISL_1319472, EPI_ISL_1319474, EPI_ISL_1319475, EPI_ISL_1319476, EPI_ISL_1319480, EPI_ISL_1319488, EPI_ISL_1319489, EPI_ISL_1319491, EPI_ISL_1319497, EPI_ISL_1319503, EPI_ISL_1319506, EPI_ISL_1319514, EPI_ISL_1319516, EPI_ISL_1319517, EPI_ISL_1319523, EPI_ISL_1319524, EPI_ISL_1319525, EPI_ISL_1319527, EPI_ISL_1319528, EPI_ISL_1319530, EPI_ISL_1319532, EPI_ISL_1319539, EPI_ISL_1319540, EPI_ISL_1319542, EPI_ISL_1319543, EPI_ISL_1319547, EPI_ISL_1319549, EPI_ISL_1319551, EPI_ISL_1319558, EPI_ISL_1319559, EPI_ISL_1319561, EPI_ISL_1319564, EPI_ISL_1319570, EPI_ISL_1319573, EPI_ISL_1319576, EPI_ISL_1319577, EPI_ISL_1319579, EPI_ISL_1319580, EPI_ISL_1319581, EPI_ISL_1319584, EPI_ISL_1319585, EPI_ISL_1319591, EPI_ISL_1319592, EPI_ISL_1319594, EPI_ISL_1319601, EPI_ISL_1319602, EPI_ISL_1319603, EPI_ISL_1319606, EPI_ISL_1319607, EPI_ISL_1319608, EPI_ISL_1319612, EPI_ISL_1319619, EPI_ISL_1319634, EPI_ISL_1319635, EPI_ISL_1319639, EPI_ISL_1319646, EPI_ISL_1319647, EPI_ISL_1319658, EPI_ISL_1319667, EPI_ISL_1319669, EPI_ISL_1319678, EPI_ISL_1319680, EPI_ISL_1319686, EPI_ISL_1319688, EPI_ISL_1319692, EPI_ISL_1319693, EPI_ISL_1319713, EPI_ISL_1319715, EPI_ISL_1319717, EPI_ISL_1319719, EPI_ISL_1319720, EPI_ISL_1319734, EPI_ISL_1319744, EPI_ISL_1319747, EPI_ISL_1319748, EPI_ISL_1319749, EPI_ISL_1319750, EPI_ISL_1319763, EPI_ISL_1319772, EPI_ISL_1319778, EPI_ISL_1319781, EPI_ISL_1319783, EPI_ISL_1319784, EPI_ISL_1319787, EPI_ISL_1319788, EPI_ISL_1319790, EPI_ISL_1319791, EPI_ISL_1319799, EPI_ISL_1319801, EPI_ISL_1319804, EPI_ISL_1319805, EPI_ISL_1319806, EPI_ISL_1319807, EPI_ISL_1319811, EPI_ISL_1319816, EPI_ISL_1319819, EPI_ISL_1319831, EPI_ISL_1319837, EPI_ISL_1319841, EPI_ISL_1319842, EPI_ISL_1319844, EPI_ISL_1319846, EPI_ISL_1319848, EPI_ISL_1319850, EPI_ISL_1319852, EPI_ISL_1319855, EPI_ISL_1319858, EPI_ISL_1319865, EPI_ISL_1319867, EPI_ISL_1319871, EPI_ISL_1319872, EPI_ISL_1319877, EPI_ISL_1319878, EPI_ISL_1319880, EPI_ISL_1319887, EPI_ISL_1319889, EPI_ISL_1319890, EPI_ISL_1319897, EPI_ISL_1319898, EPI_ISL_1319901, EPI_ISL_1319902, EPI_ISL_1319903, EPI_ISL_1319904, EPI_ISL_1319905, EPI_ISL_1319906, EPI_ISL_1319909, EPI_ISL_1319913, EPI_ISL_1319914, EPI_ISL_1319916, EPI_ISL_1319918, EPI_ISL_1319919, EPI_ISL_1319920, EPI_ISL_1319922, EPI_ISL_1319932, EPI_ISL_1319933, EPI_ISL_1319935, EPI_ISL_1319940, EPI_ISL_1319943, EPI_ISL_1319944, EPI_ISL_1319945, EPI_ISL_1319949, EPI_ISL_1319950, EPI_ISL_1319954, EPI_ISL_1319956, EPI_ISL_1319957, EPI_ISL_1319960, EPI_ISL_1319964, EPI_ISL_1319966, EPI_ISL_1319973, EPI_ISL_1319978, EPI_ISL_1319979, EPI_ISL_1319980, EPI_ISL_1319981, EPI_ISL_1319986, EPI_ISL_1319987, EPI_ISL_1319988, EPI_ISL_1319990, EPI_ISL_1319991, EPI_ISL_1319993, EPI_ISL_1319994, EPI_ISL_1319995, EPI_ISL_1320000, EPI_ISL_1320004, EPI_ISL_1320008, EPI_ISL_1320013, EPI_ISL_1320017, EPI_ISL_1320018, EPI_ISL_1320019, EPI_ISL_1320020, EPI_ISL_1320023, EPI_ISL_1320032, EPI_ISL_1320036, EPI_ISL_1320037, EPI_ISL_1320043, EPI_ISL_1320046, EPI_ISL_1320047, EPI_ISL_1320048, EPI_ISL_1320049, EPI_ISL_1320053, EPI_ISL_1320054, EPI_ISL_1320055, EPI_ISL_1320056, EPI_ISL_1320058, EPI_ISL_1320059, EPI_ISL_1320060, EPI_ISL_1320065, EPI_ISL_1320067, EPI_ISL_1320072, EPI_ISL_1320073, EPI_ISL_1320081, EPI_ISL_1320082, EPI_ISL_1320084, EPI_ISL_1320089, EPI_ISL_1320090, EPI_ISL_1320092, EPI_ISL_1320095, EPI_ISL_1320097, EPI_ISL_1320102, EPI_ISL_1320103, EPI_ISL_1320105, EPI_ISL_1320109, EPI_ISL_1320110, EPI_ISL_1320111, EPI_ISL_1320112, EPI_ISL_1320116, EPI_ISL_1320119, EPI_ISL_1320120, EPI_ISL_1320121, EPI_ISL_1320123, EPI_ISL_1320134, EPI_ISL_1320139, EPI_ISL_1320145, EPI_ISL_1320146, EPI_ISL_1320149, EPI_ISL_1320151, EPI_ISL_1320152, EPI_ISL_1320153, EPI_ISL_1320154, EPI_ISL_1320156, EPI_ISL_1320158, EPI_ISL_1320159, EPI_ISL_1320161, EPI_ISL_1320165, EPI_ISL_1320166, EPI_ISL_1320167, EPI_ISL_1320186, EPI_ISL_1320196, EPI_ISL_1320201, EPI_ISL_1320202, EPI_ISL_1320213, EPI_ISL_1320214, EPI_ISL_1320216, EPI_ISL_1320218, EPI_ISL_1320221, EPI_ISL_1320222, EPI_ISL_1320223, EPI_ISL_1320224, EPI_ISL_1320226, EPI_ISL_1320229, EPI_ISL_1320230, EPI_ISL_1320238, EPI_ISL_1320241, EPI_ISL_1320242, EPI_ISL_1320246, EPI_ISL_1320248, EPI_ISL_1320251, EPI_ISL_1320252, EPI_ISL_1320254, EPI_ISL_1320256, EPI_ISL_1320257, EPI_ISL_1320261, EPI_ISL_1320264, EPI_ISL_1320266, EPI_ISL_1320267, EPI_ISL_1320271, EPI_ISL_1320277, EPI_ISL_1320279, EPI_ISL_1320282, EPI_ISL_1320283, EPI_ISL_1320289, EPI_ISL_1320290, EPI_ISL_1320293, EPI_ISL_1320298, EPI_ISL_1320301, EPI_ISL_1320302, EPI_ISL_1320304, EPI_ISL_1320305, EPI_ISL_1320307, EPI_ISL_1320310, EPI_ISL_1320311, EPI_ISL_1320312, EPI_ISL_1320313, EPI_ISL_1320316, EPI_ISL_1320317, EPI_ISL_1320318, EPI_ISL_1320319, EPI_ISL_1320323, EPI_ISL_1320325, EPI_ISL_1320328, EPI_ISL_1320329, EPI_ISL_1320335, EPI_ISL_1320339, EPI_ISL_1320340, EPI_ISL_1320347, EPI_ISL_1320348, EPI_ISL_1320349, EPI_ISL_1320353, EPI_ISL_1320359, EPI_ISL_1320361, EPI_ISL_1320362, EPI_ISL_1320363, EPI_ISL_1320364, EPI_ISL_1320365, EPI_ISL_1320366, EPI_ISL_1320367, EPI_ISL_1320368, EPI_ISL_1320371, EPI_ISL_1320375, EPI_ISL_1320377, EPI_ISL_1320379, EPI_ISL_1320380, EPI_ISL_1320382, EPI_ISL_1320383, EPI_ISL_1320385, EPI_ISL_1320395, EPI_ISL_1320398, EPI_ISL_1320404, EPI_ISL_1320409, EPI_ISL_1320412, EPI_ISL_1320414, EPI_ISL_1320416, EPI_ISL_1320419, EPI_ISL_1320420, EPI_ISL_1320426, EPI_ISL_1320427, EPI_ISL_1320429, EPI_ISL_1320431, EPI_ISL_1320432, EPI_ISL_1320440, EPI_ISL_1320443, EPI_ISL_1320446, EPI_ISL_1320449, EPI_ISL_1320454, EPI_ISL_1320455, EPI_ISL_1320460, EPI_ISL_1320463, EPI_ISL_1320466, EPI_ISL_1320467, EPI_ISL_1320468, EPI_ISL_1320469, EPI_ISL_1320470, EPI_ISL_1320471, EPI_ISL_1320472, EPI_ISL_1320473, EPI_ISL_1320477, EPI_ISL_1320478, EPI_ISL_1320485, EPI_ISL_1320492, EPI_ISL_1320496, EPI_ISL_1320498, EPI_ISL_1320499, EPI_ISL_1320500, EPI_ISL_1320503, EPI_ISL_1320504, EPI_ISL_1320507, EPI_ISL_1320511, EPI_ISL_1320513, EPI_ISL_1320516, EPI_ISL_1320517, EPI_ISL_1320518, EPI_ISL_1320523, EPI_ISL_1320524, EPI_ISL_1320525, EPI_ISL_1320526, EPI_ISL_1320527, EPI_ISL_1320528, EPI_ISL_1320531, EPI_ISL_1320537, EPI_ISL_1320538, EPI_ISL_1320539, EPI_ISL_1320542, EPI_ISL_1320543, EPI_ISL_1320546, EPI_ISL_1320550, EPI_ISL_1320551, EPI_ISL_1320552, EPI_ISL_1320553, EPI_ISL_1320555, EPI_ISL_1320558, EPI_ISL_1320559, EPI_ISL_1320563, EPI_ISL_1320564, EPI_ISL_1320565, EPI_ISL_1320566, EPI_ISL_1320567, EPI_ISL_1320568, EPI_ISL_1320570, EPI_ISL_1320571, EPI_ISL_1320572, EPI_ISL_1320574, EPI_ISL_1320578, EPI_ISL_1320579, EPI_ISL_1320580, EPI_ISL_1320585, EPI_ISL_1320587, EPI_ISL_1320588, EPI_ISL_1320591, EPI_ISL_1320593, EPI_ISL_1320594, EPI_ISL_1320595, EPI_ISL_1320596, EPI_ISL_1320599, EPI_ISL_1320601, EPI_ISL_1320602, EPI_ISL_1320604, EPI_ISL_1320606, EPI_ISL_1320607, EPI_ISL_1320610, EPI_ISL_1320612, EPI_ISL_1320621, EPI_ISL_1320623, EPI_ISL_1320624, EPI_ISL_1320625, EPI_ISL_1320633, EPI_ISL_1320635, EPI_ISL_1320636, EPI_ISL_1320638, EPI_ISL_1320639, EPI_ISL_1320640, EPI_ISL_1320641, EPI_ISL_1320646, EPI_ISL_1320647, EPI_ISL_1320649, EPI_ISL_1320653, EPI_ISL_1320654, EPI_ISL_1320655, EPI_ISL_1320658, EPI_ISL_1320659, EPI_ISL_1320661, EPI_ISL_1320663, EPI_ISL_1320667, EPI_ISL_1320668, EPI_ISL_1320669, EPI_ISL_1320670, EPI_ISL_1320672, EPI_ISL_1320674, EPI_ISL_1320675, EPI_ISL_1320680, EPI_ISL_1320683, EPI_ISL_1320692, EPI_ISL_1320693, EPI_ISL_1320696, EPI_ISL_1320697, EPI_ISL_1320699, EPI_ISL_1320705, EPI_ISL_1320708, EPI_ISL_1320713, EPI_ISL_1320714, EPI_ISL_1320715, EPI_ISL_1320717, EPI_ISL_1320718, EPI_ISL_1320719, EPI_ISL_1320724, EPI_ISL_1320728, EPI_ISL_1320732, EPI_ISL_1320735, EPI_ISL_1320738, EPI_ISL_1320742, EPI_ISL_1320745, EPI_ISL_1320746, EPI_ISL_1320747, EPI_ISL_1320754, EPI_ISL_1320756, EPI_ISL_1320763, EPI_ISL_1320764, EPI_ISL_1320769, EPI_ISL_1320772, EPI_ISL_1320775, EPI_ISL_1320779, EPI_ISL_1320780, EPI_ISL_1320781, EPI_ISL_1320783, EPI_ISL_1320790, EPI_ISL_1320796, EPI_ISL_1320797, EPI_ISL_1320799, EPI_ISL_1320800, EPI_ISL_1320801, EPI_ISL_1320802, EPI_ISL_1320803, EPI_ISL_1320804, EPI_ISL_1320806, EPI_ISL_1320808, EPI_ISL_1320809, EPI_ISL_1320813, EPI_ISL_1320815, EPI_ISL_1320817, EPI_ISL_1320818, EPI_ISL_1320820, EPI_ISL_1320824, EPI_ISL_1320825, EPI_ISL_1320831, EPI_ISL_1320834, EPI_ISL_1320835, EPI_ISL_1320839, EPI_ISL_1320845, EPI_ISL_1320846, EPI_ISL_1320848, EPI_ISL_1320849, EPI_ISL_1320850, EPI_ISL_1320852, EPI_ISL_1320858, EPI_ISL_1320860, EPI_ISL_1320861, EPI_ISL_1320864, EPI_ISL_1320866, EPI_ISL_1320871, EPI_ISL_1320875, EPI_ISL_1320878, EPI_ISL_1320879, EPI_ISL_1320881, EPI_ISL_1320882, EPI_ISL_1320890, EPI_ISL_1320891, EPI_ISL_1320894, EPI_ISL_1320896, EPI_ISL_1320897, EPI_ISL_1320902, EPI_ISL_1320903, EPI_ISL_1320909, EPI_ISL_1320911, EPI_ISL_1320912, EPI_ISL_1320918, EPI_ISL_1320919, EPI_ISL_1320922, EPI_ISL_1320926, EPI_ISL_1320930, EPI_ISL_1320931, EPI_ISL_1320940, EPI_ISL_1320941, EPI_ISL_1320942, EPI_ISL_1320943, EPI_ISL_1320945, EPI_ISL_1320948, EPI_ISL_1320950, EPI_ISL_1320951, EPI_ISL_1320952, EPI_ISL_1320953, EPI_ISL_1320962, EPI_ISL_1320964, EPI_ISL_1320967, EPI_ISL_1320969, EPI_ISL_1320970, EPI_ISL_1320971, EPI_ISL_1320973, EPI_ISL_1320975, EPI_ISL_1320980, EPI_ISL_1320986, EPI_ISL_1320993, EPI_ISL_1320995, EPI_ISL_1320999, EPI_ISL_1321000, EPI_ISL_1321001, EPI_ISL_1321004, EPI_ISL_1321009, EPI_ISL_1321010, EPI_ISL_1321011, EPI_ISL_1321013, EPI_ISL_1321014, EPI_ISL_1321016, EPI_ISL_1321017, EPI_ISL_1321023, EPI_ISL_1321024, EPI_ISL_1321026, EPI_ISL_1321030, EPI_ISL_1321031, EPI_ISL_1321033, EPI_ISL_1321034, EPI_ISL_1321035, EPI_ISL_1321036, EPI_ISL_1321040, EPI_ISL_1321044, EPI_ISL_1321048, EPI_ISL_1321054, EPI_ISL_1321055, EPI_ISL_1321057, EPI_ISL_1321058, EPI_ISL_1321060, EPI_ISL_1321061, EPI_ISL_1321062, EPI_ISL_1321064, EPI_ISL_1321066, EPI_ISL_1321067, EPI_ISL_1321070, EPI_ISL_1321072, EPI_ISL_1321073, EPI_ISL_1321075, EPI_ISL_1321079, EPI_ISL_1321086, EPI_ISL_1321090, EPI_ISL_1321091, EPI_ISL_1321092, EPI_ISL_1321094, EPI_ISL_1321095, EPI_ISL_1321098, EPI_ISL_1321099, EPI_ISL_1321100, EPI_ISL_1321101, EPI_ISL_1321102, EPI_ISL_1321103, EPI_ISL_1321104, EPI_ISL_1321106, EPI_ISL_1321110, EPI_ISL_1321111, EPI_ISL_1321112, EPI_ISL_1321114, EPI_ISL_1321117, EPI_ISL_1321119, EPI_ISL_1321120, EPI_ISL_1321122, EPI_ISL_1321125, EPI_ISL_1321128, EPI_ISL_1321130, EPI_ISL_1321132, EPI_ISL_1321133, EPI_ISL_1321134, EPI_ISL_1321135, EPI_ISL_1321141, EPI_ISL_1321144, EPI_ISL_1321145, EPI_ISL_1321148, EPI_ISL_1321152, EPI_ISL_1321153, EPI_ISL_1321154, EPI_ISL_1321155 | | | |
| see above | Laboratory Corporation of America | Centers for Disease Control and Prevention Division of Viral Diseases, Pathogen Discovery | Peter W. Cook, Dakota Howard, Dhwani Batra, Ben L. Rambo-Martin, Minoo Agarwal, Eyad Almasri, Debbie Boles, Ayla Burns, Nuthawin Charoensri, Oren Cohen, Susan Countryman, Mary Ann Cristobal, Bobbi Croy, Suzanne Dale, Hrushikesh Deshmukh, Amanda Douglas, Vincent Drouillon, Marcia Eisenberg, Howard Engler, Rama Ghatti, Prashant Gupta, Susan Hicks, Jake Humphrey, Lax Iyer, Manoj Jain, Mohan Kolli, Brian Krueger, Tim Kuphal, Stanley Letovsky, Michael Levandoski, Craig Lukasik, Jonathan Meltzer, Brian Norvell, Mindy Nye, Scott Parker, Christos Petropoulos, John Pruitt, Steven Ragan, Scott Ryan, Mike Sapeta, Jana Schroth, Suresh Babu Selvaraju, Goran Stevovic, Amanda Suchanek, Andrea Throop, Lyndon Tilson, Thomas Urban, Joe Voshell, Kimberly Wagner, Jonathan Williams, Mary Williamson, Qian Zeng, Tricia Zwiefelhofer, Clinton R. Paden, Suxiang Tong, Duncan MacCannell |
| EPI_ISL_1321156 | Ohio Department of Health Laboratory | Ohio Department of Health Laboratory | Holmes, Jennifer; Eric Brandt, Keoni Omura, Glen McGillivary, Caitlin McDonnell, Jade Mowery, Stephanie Mccracken, Tyler Payne, Kirtana Ramadugu, Erica Leasure, Brent Lee, Kelsey Florek, Heather Blankenship, Allison Black, Quanta Brown, and Tammy Bannerman |
| EPI_ISL_1321158, EPI_ISL_1321159, EPI_ISL_1321160, EPI_ISL_1321162, EPI_ISL_1321164, EPI_ISL_1321167, EPI_ISL_1321170, EPI_ISL_1321171, EPI_ISL_1321172, EPI_ISL_1321175, EPI_ISL_1321178, EPI_ISL_1321181, EPI_ISL_1321182, EPI_ISL_1321187, EPI_ISL_1321191, EPI_ISL_1321198, EPI_ISL_1321202, EPI_ISL_1321203, EPI_ISL_1321207, EPI_ISL_1321209, EPI_ISL_1321210, EPI_ISL_1321211, EPI_ISL_1321214, EPI_ISL_1321215, EPI_ISL_1321216, EPI_ISL_1321222, EPI_ISL_1321225, EPI_ISL_1321226, EPI_ISL_1321229, EPI_ISL_1321230, EPI_ISL_1321236, EPI_ISL_1321237, EPI_ISL_1321240, EPI_ISL_1321242, EPI_ISL_1321243, EPI_ISL_1321247, EPI_ISL_1321248, EPI_ISL_1321249, EPI_ISL_1321250, EPI_ISL_1321255, EPI_ISL_1321258, EPI_ISL_1321259, EPI_ISL_1321263, EPI_ISL_1321264, EPI_ISL_1321265, EPI_ISL_1321267, EPI_ISL_1321269, EPI_ISL_1321273, EPI_ISL_1321277, EPI_ISL_1321279, EPI_ISL_1321289, EPI_ISL_1321290, EPI_ISL_1321293, EPI_ISL_1321296, EPI_ISL_1321297, EPI_ISL_1321300, EPI_ISL_1321302, EPI_ISL_1321304, EPI_ISL_1321306, EPI_ISL_1321307, EPI_ISL_1321308 | | | |
| see above | Laboratory Corporation of America | Centers for Disease Control and Prevention Division of Viral Diseases, Pathogen Discovery | Peter W. Cook, Dakota Howard, Dhwani Batra, Ben L. Rambo-Martin, Minoo Agarwal, Eyad Almasri, Debbie Boles, Ayla Burns, Nuthawin Charoensri, Oren Cohen, Susan Countryman, Mary Ann Cristobal, Bobbi Croy, Suzanne Dale, Hrushikesh Deshmukh, Amanda Douglas, Vincent Drouillon, Marcia Eisenberg, Howard Engler, Rama Ghatti, Prashant Gupta, Susan Hicks, Jake Humphrey, Lax Iyer, Manoj Jain, Mohan Kolli, Brian Krueger, Tim Kuphal, Stanley Letovsky, Michael Levandoski, Craig Lukasik, Jonathan Meltzer, Brian Norvell, Mindy Nye, Scott Parker, Christos Petropoulos, John Pruitt, Steven Ragan, Scott Ryan, Mike Sapeta, Jana Schroth, Suresh Babu Selvaraju, Goran Stevovic, Amanda Suchanek, Andrea Throop, Lyndon Tilson, Thomas Urban, Joe Voshell, Kimberly Wagner, Jonathan Williams, Mary Williamson, Qian Zeng, Tricia Zwiefelhofer, Clinton R. Paden, Suxiang Tong, Duncan MacCannell |
| EPI_ISL_1321309 | Ohio Department of Health Laboratory | Ohio Department of Health Laboratory | Holmes, Jennifer; Eric Brandt, Keoni Omura, Glen McGillivary, Caitlin McDonnell, Jade Mowery, Stephanie Mccracken, Tyler Payne, Kirtana Ramadugu, Erica Leasure, Brent Lee, Kelsey Florek, Heather Blankenship, Allison Black, Quanta Brown, and Tammy Bannerman |
| EPI_ISL_1321310, EPI_ISL_1321316, EPI_ISL_1321322, EPI_ISL_1321324, EPI_ISL_1321325, EPI_ISL_1321327, EPI_ISL_1321328, EPI_ISL_1321329, EPI_ISL_1321331, EPI_ISL_1321332, EPI_ISL_1321334, EPI_ISL_1321336, EPI_ISL_1321337, EPI_ISL_1321338, EPI_ISL_1321340, EPI_ISL_1321342, EPI_ISL_1321346, EPI_ISL_1321347, EPI_ISL_1321349, EPI_ISL_1321350, EPI_ISL_1321351, EPI_ISL_1321352, EPI_ISL_1321355 | | | |
| see above | Laboratory Corporation of America | Centers for Disease Control and Prevention Division of Viral Diseases, Pathogen Discovery | Peter W. Cook, Dakota Howard, Dhwani Batra, Ben L. Rambo-Martin, Minoo Agarwal, Eyad Almasri, Debbie Boles, Ayla Burns, Nuthawin Charoensri, Oren Cohen, Susan Countryman, Mary Ann Cristobal, Bobbi Croy, Suzanne Dale, Hrushikesh Deshmukh, Amanda Douglas, Vincent Drouillon, Marcia Eisenberg, Howard Engler, Rama Ghatti, Prashant Gupta, Susan Hicks, Jake Humphrey, Lax Iyer, Manoj Jain, Mohan Kolli, Brian Krueger, Tim Kuphal, Stanley Letovsky, Michael Levandoski, Craig Lukasik, Jonathan Meltzer, Brian Norvell, Mindy Nye, Scott Parker, Christos Petropoulos, John Pruitt, Steven Ragan, Scott Ryan, Mike Sapeta, Jana Schroth, Suresh Babu Selvaraju, Goran Stevovic, Amanda Suchanek, Andrea Throop, Lyndon Tilson, Thomas Urban, Joe Voshell, Kimberly Wagner, Jonathan Williams, Mary Williamson, Qian Zeng, Tricia Zwiefelhofer, Clinton R. Paden, Suxiang Tong, Duncan MacCannell |
| EPI_ISL_1321446 | Ohio Department of Health Laboratories | Ohio Department of Health Laboratories | Holmes, Jennifer; Eric Brandt, Keoni Omura, Glen McGillivary, Caitlin McDonnell, Jade Mowery, Stephanie Mccracken, Tyler Payne, Kirtana Ramadugu, Erica Leasure, Brent Lee, Kelsey Florek, Heather Blankenship, Allison Black, Quanta Brown, and Tammy Bannerman |
| EPI_ISL_1321601, EPI_ISL_1321602, EPI_ISL_1321603 | Ohio Department of Health Laboratory | Ohio Department of Health Laboratory | Holmes, Jennifer; Eric Brandt, Keoni Omura, Glen McGillivary, Caitlin McDonnell, Jade Mowery, Stephanie Mccracken, Tyler Payne, Kirtana Ramadugu, Erica Leasure, Brent Lee, Kelsey Florek, Heather Blankenship, Allison Black, Quanta Brown, and Tammy Bannerman |
| EPI_ISL_1321646, EPI_ISL_1321647, EPI_ISL_1321648, EPI_ISL_1321649, EPI_ISL_1321651, EPI_ISL_1321652, EPI_ISL_1321653, EPI_ISL_1321655, EPI_ISL_1321656, EPI_ISL_1321657, EPI_ISL_1321658, EPI_ISL_1321659, EPI_ISL_1321660, EPI_ISL_1321661, EPI_ISL_1321662, EPI_ISL_1321663, EPI_ISL_1321664, EPI_ISL_1321665, EPI_ISL_1321666, EPI_ISL_1321667, EPI_ISL_1321668, EPI_ISL_1321669, EPI_ISL_1321670, EPI_ISL_1321671, EPI_ISL_1321672, EPI_ISL_1321673, EPI_ISL_1321674, EPI_ISL_1321675, EPI_ISL_1321676, EPI_ISL_1321677, EPI_ISL_1321678, EPI_ISL_1321679, EPI_ISL_1321680, EPI_ISL_1321681, EPI_ISL_1321682, EPI_ISL_1321683, EPI_ISL_1321684, EPI_ISL_1321685, EPI_ISL_1321686, EPI_ISL_1321687, EPI_ISL_1321688, EPI_ISL_1321689, EPI_ISL_1321690, EPI_ISL_1321691, | | | |

| | | | |
|---|---|---|---|
| EPI_ISL_1321693, EPI_ISL_1321694, EPI_ISL_1321695, EPI_ISL_1321696, EPI_ISL_1321697, EPI_ISL_1321698, EPI_ISL_1321699, EPI_ISL_1321700, EPI_ISL_1321701, EPI_ISL_1321703, EPI_ISL_1321704, EPI_ISL_1321705, EPI_ISL_1321706, EPI_ISL_1321707, EPI_ISL_1321708, EPI_ISL_1321709, EPI_ISL_1321710, EPI_ISL_1321711, EPI_ISL_1321712, EPI_ISL_1321714, EPI_ISL_1321715, EPI_ISL_1321717, EPI_ISL_1321718, EPI_ISL_1321720, EPI_ISL_1321721, EPI_ISL_1321722, EPI_ISL_1321723, EPI_ISL_1321724, EPI_ISL_1321728, EPI_ISL_1321731, EPI_ISL_1321732, EPI_ISL_1321733 | | | |
| see above | Clinical Molecular Microbiology Laboratory, UNC Hospitals | Jeremy Wang | Jeremy Wang, Alexander Rubinsteyn, Colleen Rice, Jason Smedberg, Shawn Hawken, Melissa Miller, Corbin Jones, Robert Hagan |
| EPI_ISL_1321807, EPI_ISL_1321808 | Ohio Department of Health Laboratory | Ohio Department of Health Laboratory | Holmes, Jennifer; Eric Brandt, Keoni Omura, Glen McGillivary, Caitlin McDonnell, Jade Mowery, Stephanie Mccracken, Tyler Payne, Kirtana Ramadugu, Erica Leasure, Brent Lee, Kelsey Florek, Heather Blankenship, Allison Black, Quanta Brown, and Tammy Bannerman |
| EPI_ISL_1321810, EPI_ISL_1321811, EPI_ISL_1321812, EPI_ISL_1321813, EPI_ISL_1321815, EPI_ISL_1321816, EPI_ISL_1321817, EPI_ISL_1321818, EPI_ISL_1321819, EPI_ISL_1321820, EPI_ISL_1321821, EPI_ISL_1321822, EPI_ISL_1321823, EPI_ISL_1321824, EPI_ISL_1321825, EPI_ISL_1321826, EPI_ISL_1321827, EPI_ISL_1321828, EPI_ISL_1321829, EPI_ISL_1321830, EPI_ISL_1321831, EPI_ISL_1321832, EPI_ISL_1321833, EPI_ISL_1321834, EPI_ISL_1321835, EPI_ISL_1321836, EPI_ISL_1321837, EPI_ISL_1321838, EPI_ISL_1321839, EPI_ISL_1321840, EPI_ISL_1321841, EPI_ISL_1321842, EPI_ISL_1321843, EPI_ISL_1321844, EPI_ISL_1321845, EPI_ISL_1321846, EPI_ISL_1321847, EPI_ISL_1321848, EPI_ISL_1321849, EPI_ISL_1321850, EPI_ISL_1321851, EPI_ISL_1321852, EPI_ISL_1321853, EPI_ISL_1321854, EPI_ISL_1321855, EPI_ISL_1321856, EPI_ISL_1321857, EPI_ISL_1321858, EPI_ISL_1321859, EPI_ISL_1321860, EPI_ISL_1321861, EPI_ISL_1321862, EPI_ISL_1321863, EPI_ISL_1321864, EPI_ISL_1321865, EPI_ISL_1321866, EPI_ISL_1321867, EPI_ISL_1321868, EPI_ISL_1321869, EPI_ISL_1321870, EPI_ISL_1321871, EPI_ISL_1321872, EPI_ISL_1321873, EPI_ISL_1321874, EPI_ISL_1321875, EPI_ISL_1321876, EPI_ISL_1321877, EPI_ISL_1321878, EPI_ISL_1321879, EPI_ISL_1321880, EPI_ISL_1321881, EPI_ISL_1321882, EPI_ISL_1321883, EPI_ISL_1321884, EPI_ISL_1321885, EPI_ISL_1321886, EPI_ISL_1321887, EPI_ISL_1321888, EPI_ISL_1321889, EPI_ISL_1321890, EPI_ISL_1321891, EPI_ISL_1321892, EPI_ISL_1321893, EPI_ISL_1321894, EPI_ISL_1321895, EPI_ISL_1321896, EPI_ISL_1321897 | | | |
| see above | TXDSHS | TXDSHS | Rashmi Tuladhar, Bonnie Oh, Jenny Zhang, Maliha Rahman, Mayela Pedrueza, Anita Pokharel, Myong Koag, Chun Wang, Rachel Lee, Grace Kubin |
| EPI_ISL_1322432, EPI_ISL_1322433, EPI_ISL_1322434, EPI_ISL_1322435, EPI_ISL_1322436, EPI_ISL_1322437, EPI_ISL_1322438, EPI_ISL_1322439, EPI_ISL_1322440, EPI_ISL_1322441, EPI_ISL_1322442, EPI_ISL_1322443, EPI_ISL_1322444, EPI_ISL_1322445, EPI_ISL_1322446, EPI_ISL_1322447, EPI_ISL_1322448, EPI_ISL_1322449, EPI_ISL_1322450, EPI_ISL_1322451, EPI_ISL_1322452, EPI_ISL_1322453, EPI_ISL_1322454, EPI_ISL_1322455, EPI_ISL_1322456, EPI_ISL_1322457, EPI_ISL_1322458, EPI_ISL_1322459, EPI_ISL_1322460, EPI_ISL_1322461, EPI_ISL_1322462, EPI_ISL_1322463, EPI_ISL_1322464 | | | |
| see above | Kansas Health and Environmental Lab | Kansas Health and Environmental Lab | Mike Grose, Jonathan Barnell, Ben Olsen, and Phil Adam |
| EPI_ISL_1322465 | Springhill Medical Center Laboratories | Synergy Laboratories | Megan Cornwell |
| EPI_ISL_1322744 | Springhill Medical Center Clinical Laboratory | Synergy Laboratories | Megan Cornwell |
| EPI_ISL_1322745, EPI_ISL_1322746, EPI_ISL_1322747, EPI_ISL_1322748, EPI_ISL_1322749, EPI_ISL_1322750, EPI_ISL_1322751, EPI_ISL_1322752, EPI_ISL_1322753, EPI_ISL_1322755, EPI_ISL_1322756, EPI_ISL_1322757, EPI_ISL_1322758, EPI_ISL_1322759, EPI_ISL_1322760, EPI_ISL_1322762, EPI_ISL_1322763, EPI_ISL_1322764, EPI_ISL_1322765, EPI_ISL_1322766, EPI_ISL_1322767, EPI_ISL_1322768, EPI_ISL_1322770, EPI_ISL_1322771, EPI_ISL_1322772, EPI_ISL_1322773, EPI_ISL_1322774, EPI_ISL_1322777, EPI_ISL_1322778, EPI_ISL_1322779, EPI_ISL_1322780, EPI_ISL_1322781, EPI_ISL_1322782, EPI_ISL_1322783, EPI_ISL_1322785, EPI_ISL_1322786, EPI_ISL_1322787, EPI_ISL_1322788, EPI_ISL_1322789, EPI_ISL_1322790, EPI_ISL_1322791, EPI_ISL_1322792, EPI_ISL_1322793, EPI_ISL_1322794, EPI_ISL_1322795, EPI_ISL_1322796, EPI_ISL_1322797, EPI_ISL_1322798, EPI_ISL_1322799, EPI_ISL_1322800, EPI_ISL_1322802, EPI_ISL_1322804, EPI_ISL_1322805, EPI_ISL_1322807, EPI_ISL_1322808, EPI_ISL_1322809, EPI_ISL_1322810, EPI_ISL_1322811, EPI_ISL_1322812, EPI_ISL_1322813, EPI_ISL_1322814, EPI_ISL_1322815, EPI_ISL_1322816, EPI_ISL_1322817, EPI_ISL_1322818, EPI_ISL_1322820, EPI_ISL_1322821, EPI_ISL_1322822, EPI_ISL_1322824, EPI_ISL_1322826, EPI_ISL_1322827, EPI_ISL_1322829, EPI_ISL_1322830, EPI_ISL_1322831, EPI_ISL_1322832, EPI_ISL_1322833 | | | |
| see above | NYU Langone Health | Departments of Pathology and Medicine, New York University School of Medicine | Adriana Heguy, Dacia Dimartino, Emily Guzman, Christian Marier, Peter Meyn, Sitharam Ramaswami, Gael Westby, Paul Zappile, Yutong Zhang, Paolo Cotzia, Guiqing Wang |
| EPI_ISL_1322835, EPI_ISL_1322836, EPI_ISL_1322837, EPI_ISL_1322838, EPI_ISL_1322839, EPI_ISL_1322840, EPI_ISL_1322841, EPI_ISL_1322842, EPI_ISL_1322843, EPI_ISL_1322844, EPI_ISL_1322845, EPI_ISL_1322846, EPI_ISL_1322847, EPI_ISL_1322848, EPI_ISL_1322849, EPI_ISL_1322850, EPI_ISL_1322851, EPI_ISL_1322852, EPI_ISL_1322853, EPI_ISL_1322854, EPI_ISL_1322855, EPI_ISL_1322856, EPI_ISL_1322857, EPI_ISL_1322858, EPI_ISL_1322859, EPI_ISL_1322860, EPI_ISL_1322861, EPI_ISL_1322862, EPI_ISL_1322863, EPI_ISL_1322864, EPI_ISL_1322865, EPI_ISL_1322866, EPI_ISL_1322867, EPI_ISL_1322868, EPI_ISL_1322869, EPI_ISL_1322870, EPI_ISL_1322871, EPI_ISL_1322872, EPI_ISL_1322873, EPI_ISL_1322874, EPI_ISL_1322875, EPI_ISL_1322876, EPI_ISL_1322877, EPI_ISL_1322878, EPI_ISL_1322879, EPI_ISL_1322880, EPI_ISL_1322881, EPI_ISL_1322882, EPI_ISL_1322883, EPI_ISL_1322884, EPI_ISL_1322885, EPI_ISL_1322886, EPI_ISL_1322887, EPI_ISL_1322888, EPI_ISL_1322889, EPI_ISL_1322890, EPI_ISL_1322891, EPI_ISL_1322892, EPI_ISL_1322893, EPI_ISL_1322894, EPI_ISL_1322895, EPI_ISL_1322896, EPI_ISL_1322897, EPI_ISL_1322898, EPI_ISL_1322899, EPI_ISL_1322900, EPI_ISL_1322901, EPI_ISL_1322902, EPI_ISL_1322903, EPI_ISL_1322904, EPI_ISL_1322905, EPI_ISL_1322906, EPI_ISL_1322907, EPI_ISL_1322908, EPI_ISL_1322909, EPI_ISL_1322910, EPI_ISL_1322911, EPI_ISL_1322912, EPI_ISL_1322913, EPI_ISL_1322914, EPI_ISL_1322915, EPI_ISL_1322916, EPI_ISL_1322917, EPI_ISL_1322918, EPI_ISL_1322919, EPI_ISL_1322920, EPI_ISL_1322921, EPI_ISL_1322922, EPI_ISL_1322923, EPI_ISL_1322924, EPI_ISL_1322925 | | | |
| see above | TXDSHS | TXDSHS | Rashmi Tuladhar, Bonnie Oh, Jenny Zhang, Maliha Rahman, Mayela Pedrueza, Anita Pokharel, Myong Koag, Chun Wang, Rachel Lee, Grace Kubin |
| EPI_ISL_1323116 | IL Department of Public Health Chicago Laboratory | Genomics and Discovery, Respiratory Viruses Branch, Division of Viral Diseases, Centers for Disease Control and Prevention | Ying Tao, Jing Zhang, Yan Li, Brian Lynch, Anna Kelleher, Krista Queen, Anna Uehara, Peter Cook, Han Jia Justin Ng, Clinton R. Paden, Haibin Wang, Suxiang Tong |
| EPI_ISL_1323117 | "OK Public Health Laboratory, Oklahoma State DOH" | Genomics and Discovery, Respiratory Viruses Branch, Division of Viral Diseases, Centers for Disease Control and Prevention | Ying Tao, Jing Zhang, Yan Li, Brian Lynch, Anna Kelleher, Krista Queen, Anna Uehara, Peter Cook, Han Jia Justin Ng, Clinton R. Paden, Haibin Wang, Suxiang Tong |
| EPI_ISL_1323118 | LA Office of Public Health Laboratories | Genomics and Discovery, Respiratory Viruses Branch, Division of Viral Diseases, Centers for Disease Control and Prevention | Ying Tao, Jing Zhang, Yan Li, Brian Lynch, Anna Kelleher, Krista Queen, Anna Uehara, Peter Cook, Han Jia Justin Ng, Clinton R. Paden, Haibin Wang, Suxiang Tong |
| EPI_ISL_1323192, EPI_ISL_1323196, EPI_ISL_1323199, EPI_ISL_1323200, EPI_ISL_1323201, EPI_ISL_1323202, EPI_ISL_1323204 | Wyoming Public Health Laboratory | Wyoming Public Health Laboratory | Jim Mildenberger, Wanda Manley, Noah Hull, Taylor Fearing, Lynette Gumbleton, Channing Weber, Ashley Norberg, Chayse Rowley, Marley Goetz, Brian Dominguez, Elliot Thomasson, Cari Sloma, and Rob Christensen |
| EPI_ISL_1323205, EPI_ISL_1323206, EPI_ISL_1323208, EPI_ISL_1323209 | Illinois Department of Public Health | Gagnon Lab, Southern Illinois University | Keith Gagnon |
| EPI_ISL_1323210 | Montana Public Health Laboratory | Montana Public Health Laboratory | Joy Ritter, Michelle Mozer, Carrie Biskupiak, Deborah Gibson |
| EPI_ISL_1323211, EPI_ISL_1323212, EPI_ISL_1323213, EPI_ISL_1323214, EPI_ISL_1323215, EPI_ISL_1323216, EPI_ISL_1323217, EPI_ISL_1323218, EPI_ISL_1323219, EPI_ISL_1323220, EPI_ISL_1323221, EPI_ISL_1323222, EPI_ISL_1323223, EPI_ISL_1323224, EPI_ISL_1323225, EPI_ISL_1323226, EPI_ISL_1323227, EPI_ISL_1323228, EPI_ISL_1323229, EPI_ISL_1323230, EPI_ISL_1323231, EPI_ISL_1323232, EPI_ISL_1323233, EPI_ISL_1323234, EPI_ISL_1323235, EPI_ISL_1323236, EPI_ISL_1323237, EPI_ISL_1323238, EPI_ISL_1323239, EPI_ISL_1323241, EPI_ISL_1323242, EPI_ISL_1323245, EPI_ISL_1323246, EPI_ISL_1323247, EPI_ISL_1323248, EPI_ISL_1323249, EPI_ISL_1323250, EPI_ISL_1323251, EPI_ISL_1323252, EPI_ISL_1323253, EPI_ISL_1323254, EPI_ISL_1323255, EPI_ISL_1323256, EPI_ISL_1323257, EPI_ISL_1323258, EPI_ISL_1323259, EPI_ISL_1323260, EPI_ISL_1323263, EPI_ISL_1323264, EPI_ISL_1323265, EPI_ISL_1323266, EPI_ISL_1323267, EPI_ISL_1323268, EPI_ISL_1323269, EPI_ISL_1323270, EPI_ISL_1323272, EPI_ISL_1323273, EPI_ISL_1323274, EPI_ISL_1323275, EPI_ISL_1323277, EPI_ISL_1323278, EPI_ISL_1323279, EPI_ISL_1323280, EPI_ISL_1323281, EPI_ISL_1323282, EPI_ISL_1323283, EPI_ISL_1323284, EPI_ISL_1323287, EPI_ISL_1323288, EPI_ISL_1323289, EPI_ISL_1323290, EPI_ISL_1323291, EPI_ISL_1323292, EPI_ISL_1323293, EPI_ISL_1323294, EPI_ISL_1323295, EPI_ISL_1323296, EPI_ISL_1323297, EPI_ISL_1323298, EPI_ISL_1323299, EPI_ISL_1323302, EPI_ISL_1323303, EPI_ISL_1323305, EPI_ISL_1323306, EPI_ISL_1323307, EPI_ISL_1323308, EPI_ISL_1323309, EPI_ISL_1323311, EPI_ISL_1323312, EPI_ISL_1323313, EPI_ISL_1323314, EPI_ISL_1323315, EPI_ISL_1323317, EPI_ISL_1323318, EPI_ISL_1323319, EPI_ISL_1323320, EPI_ISL_1323323, EPI_ISL_1323324, EPI_ISL_1323325, EPI_ISL_1323326, EPI_ISL_1323328, EPI_ISL_1323329, EPI_ISL_1323330, EPI_ISL_1323331, EPI_ISL_1323332, EPI_ISL_1323333, EPI_ISL_1323334, EPI_ISL_1323335, EPI_ISL_1323336, EPI_ISL_1323337, EPI_ISL_1323338, EPI_ISL_1323339, EPI_ISL_1323340, EPI_ISL_1323341, EPI_ISL_1323342, EPI_ISL_1323343, EPI_ISL_1323344, EPI_ISL_1323345, EPI_ISL_1323346, EPI_ISL_1323347, EPI_ISL_1323348, EPI_ISL_1323349, EPI_ISL_1323351, EPI_ISL_1323352, EPI_ISL_1323353, EPI_ISL_1323354, EPI_ISL_1323355, EPI_ISL_1323356, EPI_ISL_1323358, EPI_ISL_1323359, EPI_ISL_1323360, EPI_ISL_1323361, EPI_ISL_1323362, EPI_ISL_1323363, EPI_ISL_1323364, EPI_ISL_1323365, EPI_ISL_1323366, EPI_ISL_1323367, EPI_ISL_1323368, EPI_ISL_1323369, EPI_ISL_1323370, EPI_ISL_1323371, EPI_ISL_1323372, EPI_ISL_1323373, EPI_ISL_1323374, EPI_ISL_1323375, EPI_ISL_1323376, EPI_ISL_1323378, EPI_ISL_1323379, EPI_ISL_1323380, EPI_ISL_1323381, EPI_ISL_1323383, EPI_ISL_1323384, EPI_ISL_1323385, EPI_ISL_1323386, EPI_ISL_1323388, EPI_ISL_1323389, EPI_ISL_1323390, EPI_ISL_1323391, EPI_ISL_1323392, EPI_ISL_1323393, EPI_ISL_1323395, EPI_ISL_1323396, EPI_ISL_1323398, EPI_ISL_1323399, EPI_ISL_1323400, EPI_ISL_1323401, EPI_ISL_1323403, EPI_ISL_1323404, EPI_ISL_1323405, EPI_ISL_1323406, EPI_ISL_1323407, EPI_ISL_1323408, EPI_ISL_1323409, EPI_ISL_1323410, EPI_ISL_1323411, EPI_ISL_1323412, EPI_ISL_1323413, EPI_ISL_1323414, EPI_ISL_1323415, EPI_ISL_1323416, EPI_ISL_1323417, EPI_ISL_1323418, EPI_ISL_1323419, EPI_ISL_1323420, EPI_ISL_1323421, EPI_ISL_1323422, EPI_ISL_1323424, EPI_ISL_1323425, EPI_ISL_1323426, EPI_ISL_1323427, EPI_ISL_1323428, EPI_ISL_1323429, EPI_ISL_1323430, EPI_ISL_1323432, EPI_ISL_1323433, EPI_ISL_1323434, EPI_ISL_1323435, EPI_ISL_1323436, EPI_ISL_1323437, EPI_ISL_1323439, EPI_ISL_1323440, EPI_ISL_1323441, EPI_ISL_1323442, EPI_ISL_1323443, EPI_ISL_1323447, EPI_ISL_1323448, EPI_ISL_1323449, EPI_ISL_1323450, EPI_ISL_1323451, EPI_ISL_1323452, EPI_ISL_1323453, EPI_ISL_1323454, EPI_ISL_1323455, EPI_ISL_1323456, EPI_ISL_1323457, EPI_ISL_1323459, EPI_ISL_1323460, EPI_ISL_1323461, EPI_ISL_1323462, EPI_ISL_1323464, EPI_ISL_1323465, EPI_ISL_1323467, EPI_ISL_1323471, EPI_ISL_1323472, EPI_ISL_1323473, EPI_ISL_1323474, EPI_ISL_1323476, EPI_ISL_1323477, EPI_ISL_1323478, EPI_ISL_1323479, EPI_ISL_1323481, EPI_ISL_1323483, EPI_ISL_1323484, EPI_ISL_1323485, EPI_ISL_1323486, EPI_ISL_1323487, EPI_ISL_1323488, EPI_ISL_1323489, EPI_ISL_1323491, EPI_ISL_1323492, EPI_ISL_1323493, EPI_ISL_1323494, EPI_ISL_1323496, EPI_ISL_1323497, EPI_ISL_1323498, EPI_ISL_1323499, EPI_ISL_1323502, EPI_ISL_1323503, EPI_ISL_1323505, EPI_ISL_1323506, EPI_ISL_1323507, EPI_ISL_1323509, EPI_ISL_1323510, EPI_ISL_1323511, EPI_ISL_1323512, EPI_ISL_1323513, EPI_ISL_1323514, EPI_ISL_1323515, EPI_ISL_1323516, EPI_ISL_1323517, EPI_ISL_1323518, EPI_ISL_1323519, EPI_ISL_1323520, EPI_ISL_1323521, EPI_ISL_1323522, EPI_ISL_1323523, EPI_ISL_1323524, EPI_ISL_1323525, EPI_ISL_1323526, EPI_ISL_1323527, EPI_ISL_1323528, EPI_ISL_1323529, EPI_ISL_1323530, EPI_ISL_1323531, EPI_ISL_1323532, EPI_ISL_1323534, EPI_ISL_1323535, EPI_ISL_1323536, EPI_ISL_1323537, EPI_ISL_1323538, EPI_ISL_1323539, EPI_ISL_1323540, EPI_ISL_1323541, EPI_ISL_1323542, EPI_ISL_1323543, EPI_ISL_1323544, EPI_ISL_1323545, EPI_ISL_1323546 | | | |
| see above | Illinois Department of Public Health | Gagnon Lab, Southern Illinois University | Keith Gagnon |
| EPI_ISL_1323620, EPI_ISL_1323621, EPI_ISL_1323622, EPI_ISL_1323623, EPI_ISL_1323624, EPI_ISL_1323625, EPI_ISL_1323626, EPI_ISL_1323627, EPI_ISL_1323628, EPI_ISL_1323629, EPI_ISL_1323631, EPI_ISL_1323632, EPI_ISL_1323633, EPI_ISL_1323634, EPI_ISL_1323635 | | | |
| see above | Montana Public Health Laboratory | Montana Public Health Laboratory | Joy Ritter, Michelle Mozer, Carrie Biskupiak, Deborah Gibson |
| EPI_ISL_1323636, EPI_ISL_1323637, EPI_ISL_1323638, EPI_ISL_1323639, EPI_ISL_1323640, EPI_ISL_1323641, EPI_ISL_1323642, EPI_ISL_1323643, EPI_ISL_1323644, EPI_ISL_1323645, EPI_ISL_1323647, EPI_ISL_1323648, EPI_ISL_1323649, EPI_ISL_1323650, EPI_ISL_1323651 | | | |
| see above | Nevada State Public Health Laboratory | Nevada State Public Health Laboratory | Andrew Gorzalski, Mark Pandori |
| EPI_ISL_1323653, EPI_ISL_1323654, EPI_ISL_1323655, EPI_ISL_1323656, EPI_ISL_1323657 | Montana Public Health Laboratory | Montana Public Health Laboratory | Joy Ritter, Michelle Mozer, Carrie Biskupiak, Deborah Gibson |
| EPI_ISL_1323846, EPI_ISL_1323847, EPI_ISL_1323848, EPI_ISL_1323849, EPI_ISL_1323850, EPI_ISL_1323851, EPI_ISL_1323852, EPI_ISL_1323853, EPI_ISL_1323854, EPI_ISL_1323855, EPI_ISL_1323856, EPI_ISL_1323857, EPI_ISL_1323858, EPI_ISL_1323859, EPI_ISL_1323860, EPI_ISL_1323861 | | | |
| see above | Kansas Health and Environmental Lab | Kansas Health and Environmental Lab | Mike Grose, Jonathan Barnell, Ben Olsen, and Phil Adam |
| EPI_ISL_1324132, EPI_ISL_1324133, EPI_ISL_1324146, EPI_ISL_1324150, EPI_ISL_1324151, EPI_ISL_1324152 | UW Virology Lab | UW Virology Lab | Pavitra Roychoudhury, Hong Xie, Lasata Shrestha, Shah Mohamed Bakhash, Michelle Lin, Margaret Mills, Noah Baker, Sean Ellis, Saraswathi Sathees, Meei-Li Huang, Keith R Jerome, Alexander Greninger |
| EPI_ISL_1324153 | LSUHS Emerging Viral Threat Laboratory | Microbial Genome Sequencing Center | Jeremy P. Kamil, Jennifer L. Carroll, Rona S. Scott, Maarten Van Diest, Andrew D. Yurochko, Christopher G. Kevil, Martin J. Sapp, Daniel J. Snyder, Vaughn S. Cooper, John A. Vanchiere |
| EPI_ISL_1324154, EPI_ISL_1324155, EPI_ISL_1324156, EPI_ISL_1324157, EPI_ISL_1324158, EPI_ISL_1324159, EPI_ISL_1324160, EPI_ISL_1324161, EPI_ISL_1324162, EPI_ISL_1324163, EPI_ISL_1324164, EPI_ISL_1324165, EPI_ISL_1324166, EPI_ISL_1324167, EPI_ISL_1324168 | | | |
| see above | LSUHS Emerging Viral Threat Laboratory | Microbial Genome Sequencing Center | Maarten Van Diest, Jennifer L. Carroll, Rona S. Scott, Andrew D. Yurochko, Christopher G. Kevil, Martin J. Sapp, Daniel J. Snyder, Vaughn S. Cooper, John A. Vanchiere, Jeremy P. Kamil |
| EPI_ISL_1324169, EPI_ISL_1324170, EPI_ISL_1324171, EPI_ISL_1324172, EPI_ISL_1324173, EPI_ISL_1324174, EPI_ISL_1324175, EPI_ISL_1324176, EPI_ISL_1324177, EPI_ISL_1324178, EPI_ISL_1324179, EPI_ISL_1324180, EPI_ISL_1324181, EPI_ISL_1324182, EPI_ISL_1324183 | | | |
| see above | LSUHS Emerging Viral Threat Laboratory | Microbial Genome Sequencing Center | Rona S. Scott, Jennifer L. Carroll, Maarten Van Diest, Andrew D. Yurochko, Christopher G. Kevil, Martin J. Sapp, Daniel J. Snyder, Vaughn S. Cooper, John A. Vanchiere, Jeremy P. Kamil |
| EPI_ISL_1324184, EPI_ISL_1324185, EPI_ISL_1324186, EPI_ISL_1324187, EPI_ISL_1324188, | LSUHS Emerging Viral Threat Laboratory | Microbial Genome Sequencing Center | Daniel J. Snyder, Jennifer L. Carroll, Rona S. Scott, Maarten Van Diest, Andrew D. Yurochko, Christopher G. Kevil, Martin J. Sapp, Vaughn S. Cooper, John A. Vanchiere, Jeremy P. Kamil |

| | | | |
|---|---|---|---|
| EPI_ISL_1324189, EPI_ISL_1324190, EPI_ISL_1324191, EPI_ISL_1324192, EPI_ISL_1324193 | | | |
| EPI_ISL_1324194, EPI_ISL_1324195, EPI_ISL_1324196, EPI_ISL_1324197, EPI_ISL_1324198, EPI_ISL_1324199, EPI_ISL_1324200, EPI_ISL_1324201, EPI_ISL_1324202, EPI_ISL_1324203 | LSUHS Emerging Viral Threat Laboratory | Microbial Genome Sequencing Center | Jennifer L. Carroll, Rona S. Scott, Maarten Van Diest, Andrew D. Yurochko, Christopher G. Kevil, Martin J. Sapp, Daniel J. Snyder, Vaughn S. Cooper, John A. Vanchiere, Jeremy P. Kamil |
| EPI_ISL_1324204, EPI_ISL_1324205, EPI_ISL_1324206, EPI_ISL_1324207, EPI_ISL_1324208, EPI_ISL_1324209, EPI_ISL_1324210, EPI_ISL_1324211, EPI_ISL_1324212, EPI_ISL_1324213, EPI_ISL_1324214, EPI_ISL_1324215, EPI_ISL_1324216, EPI_ISL_1324217, EPI_ISL_1324218, EPI_ISL_1324219, EPI_ISL_1324220, EPI_ISL_1324221, EPI_ISL_1324222, EPI_ISL_1324223, EPI_ISL_1324224, EPI_ISL_1324225, EPI_ISL_1324226, EPI_ISL_1324227, EPI_ISL_1324228, EPI_ISL_1324229, EPI_ISL_1324230, EPI_ISL_1324231, EPI_ISL_1324232, EPI_ISL_1324233, EPI_ISL_1324234, EPI_ISL_1324235, EPI_ISL_1324236, EPI_ISL_1324237, EPI_ISL_1324238, EPI_ISL_1324239, EPI_ISL_1324240, EPI_ISL_1324241, EPI_ISL_1324242, EPI_ISL_1324243, EPI_ISL_1324244, EPI_ISL_1324245, EPI_ISL_1324246, EPI_ISL_1324247, EPI_ISL_1324248, EPI_ISL_1324249, EPI_ISL_1324250, EPI_ISL_1324251, EPI_ISL_1324252, EPI_ISL_1324253, EPI_ISL_1324254, EPI_ISL_1324255, EPI_ISL_1324256, EPI_ISL_1324257, EPI_ISL_1324258, EPI_ISL_1324259, EPI_ISL_1324260, EPI_ISL_1324261, EPI_ISL_1324262, EPI_ISL_1324263, EPI_ISL_1324264, EPI_ISL_1324265, EPI_ISL_1324266, EPI_ISL_1324267, EPI_ISL_1324268, EPI_ISL_1324269, EPI_ISL_1324270, EPI_ISL_1324271, EPI_ISL_1324272, EPI_ISL_1324273, EPI_ISL_1324274, EPI_ISL_1324275, EPI_ISL_1324276, EPI_ISL_1324277, EPI_ISL_1324278, EPI_ISL_1324279, EPI_ISL_1324280, EPI_ISL_1324281, EPI_ISL_1324282, EPI_ISL_1324283, EPI_ISL_1324284, EPI_ISL_1324285, EPI_ISL_1324286, EPI_ISL_1324287, EPI_ISL_1324288, EPI_ISL_1324289, EPI_ISL_1324290, EPI_ISL_1324291, EPI_ISL_1324292, EPI_ISL_1324293, EPI_ISL_1324294, EPI_ISL_1324295, EPI_ISL_1324296, EPI_ISL_1324297, EPI_ISL_1324298, EPI_ISL_1324299, EPI_ISL_1324300, EPI_ISL_1324301, EPI_ISL_1324302, EPI_ISL_1324303, EPI_ISL_1324304, EPI_ISL_1324305, EPI_ISL_1324306, EPI_ISL_1324307, EPI_ISL_1324308, EPI_ISL_1324309, EPI_ISL_1324310, EPI_ISL_1324311, EPI_ISL_1324312, EPI_ISL_1324313, EPI_ISL_1324314, EPI_ISL_1324315, EPI_ISL_1324316, EPI_ISL_1324317, EPI_ISL_1324318, EPI_ISL_1324319, EPI_ISL_1324320, EPI_ISL_1324321, EPI_ISL_1324322, EPI_ISL_1324323, EPI_ISL_1324324, EPI_ISL_1324325, EPI_ISL_1324326, EPI_ISL_1324327, EPI_ISL_1324328, EPI_ISL_1324329, EPI_ISL_1324330, EPI_ISL_1324331, EPI_ISL_1324332, EPI_ISL_1324333, EPI_ISL_1324334, EPI_ISL_1324335, EPI_ISL_1324336, EPI_ISL_1324337, EPI_ISL_1324338, EPI_ISL_1324339, EPI_ISL_1324340, EPI_ISL_1324341, EPI_ISL_1324342, EPI_ISL_1324343, EPI_ISL_1324344, EPI_ISL_1324345, EPI_ISL_1324346, EPI_ISL_1324347, EPI_ISL_1324348, EPI_ISL_1324349, EPI_ISL_1324350, EPI_ISL_1324351, EPI_ISL_1324352, EPI_ISL_1324353, EPI_ISL_1324354, EPI_ISL_1324355, EPI_ISL_1324356, EPI_ISL_1324357, EPI_ISL_1324358, EPI_ISL_1324359, EPI_ISL_1324360, EPI_ISL_1324361, EPI_ISL_1324362, EPI_ISL_1324363, EPI_ISL_1324364, EPI_ISL_1324365, EPI_ISL_1324366, EPI_ISL_1324367, EPI_ISL_1324368, EPI_ISL_1324369, EPI_ISL_1324370, EPI_ISL_1324371, EPI_ISL_1324372, EPI_ISL_1324373, EPI_ISL_1324374, EPI_ISL_1324375, EPI_ISL_1324376, EPI_ISL_1324377, EPI_ISL_1324378, EPI_ISL_1324379, EPI_ISL_1324380, EPI_ISL_1324381, EPI_ISL_1324382, EPI_ISL_1324383 | | | |
| see above | LSUHS Emerging Viral Threat Laboratory | Microbial Genome Sequencing Center | Jeremy P. Kamil, Jennifer L. Carroll, Rona S. Scott, Maarten Van Diest, Andrew D. Yurochko, Christopher G. Kevil, Martin J. Sapp, Daniel J. Snyder, Vaughn S. Cooper, John A. Vanchiere |
| EPI_ISL_1324758, EPI_ISL_1324759, EPI_ISL_1324760, EPI_ISL_1324761, EPI_ISL_1324762, EPI_ISL_1324763, EPI_ISL_1324764, EPI_ISL_1324765, EPI_ISL_1324766 | Laboratorio de Diagnostico de Enfermedades Emergentes y Reemergentes | Instituto de Diagnostico y Referencia Epidemiologicos (INDRE) | Claudia Wong-Arambula, Abril Rodriguez-Maldonado, Vanessa Rivero-Arredondo, Ariadna Medina-Benitez, Joaquin Quiroz-Mercado, Sergio Rangel-Guerrero,Natali Vega-Magana, Natividad Cruz-Ortiz, Tatiana Nunez-Garcia, Gisela Barrera-Badillo, Lucia Hernandez-Rivas, Irma Lopez-Martinez, Ernesto Ramirez-Gonzalez. |
| EPI_ISL_1324767 | LESP Quintana Roo | Instituto de Diagnostico y Referencia Epidemiologicos (INDRE) | Claudia Wong-Arambula, Abril Rodriguez-Maldonado, Vanessa Rivero-Arredondo, Ariadna Medina-Benitez, Joaquin Quiroz-Mercado, Sergio Rangel-Guerrero,Natali Vega-Magana, Natividad Cruz-Ortiz, Tatiana Nunez-Garcia, Gisela Barrera-Badillo, Lucia Hernandez-Rivas, Irma Lopez-Martinez, Ernesto Ramirez-Gonzalez. |
| EPI_ISL_1324774, EPI_ISL_1324776, EPI_ISL_1324777, EPI_ISL_1324778 | Wyoming Public Health Laboratory | Wyoming Public Health Laboratory | Jim Mildenberger, Wanda Manley, Noah Hull, Taylor Fearing, Lynette Gumbleton, Channing Weber, Ashley Norberg, Chayse Rowley, Marley Goetz, Brian Dominguez, Elliot Thomasson, Cari Sloma, and Rob Christensen |
| EPI_ISL_1324784, EPI_ISL_1324785, EPI_ISL_1324786, EPI_ISL_1324787, EPI_ISL_1324788, EPI_ISL_1324789, EPI_ISL_1324790, EPI_ISL_1324791, EPI_ISL_1324793, EPI_ISL_1324794, EPI_ISL_1324795, EPI_ISL_1324796, EPI_ISL_1324797, EPI_ISL_1324798 | | | |
| see above | Nevada State Public Health Laboratory | Nevada State Public Health Laboratory | Andrew Gorzalski, Mark Pandori |
| EPI_ISL_1334287, EPI_ISL_1334288, EPI_ISL_1334289, EPI_ISL_1334290, EPI_ISL_1334291, EPI_ISL_1334292, EPI_ISL_1334293, EPI_ISL_1334294, EPI_ISL_1334295, EPI_ISL_1334296, EPI_ISL_1334297, EPI_ISL_1334298, EPI_ISL_1334299, EPI_ISL_1334300, EPI_ISL_1334301, EPI_ISL_1334302, EPI_ISL_1334303, EPI_ISL_1334304, EPI_ISL_1334305, EPI_ISL_1334306, EPI_ISL_1334307, EPI_ISL_1334308, EPI_ISL_1334309, EPI_ISL_1334310, EPI_ISL_1334311, EPI_ISL_1334312, EPI_ISL_1334313, EPI_ISL_1334314, EPI_ISL_1334315, EPI_ISL_1334316 | | | |
| see above | Wyoming Public Health Laboratory | Wyoming Public Health Laboratory | Jim Mildenberger, Wanda Manley, Noah Hull, Taylor Fearing, Lynette Gumbleton, Channing Weber, Ashley Norberg, Chayse Rowley, Marley Goetz, Brian Dominguez, Elliot Thomasson, Cari Sloma, and Rob Christensen |
| EPI_ISL_1334366, EPI_ISL_1334367, EPI_ISL_1334368, EPI_ISL_1334369, EPI_ISL_1334371, EPI_ISL_1334372, EPI_ISL_1334373, EPI_ISL_1334374, EPI_ISL_1334375, EPI_ISL_1334376, EPI_ISL_1334377, EPI_ISL_1334378, EPI_ISL_1334379 | | | |
| see above | Washington State Department of Health Public Health Laboratories | Washington State Department of Health Public Health Laboratories | Drew MacKellar, Philip Dykema, Denny Russell, Joenice Gonzalez, Geoff Melly, Vanessa De Los Santos, Darren Lucas, JohnAric Peterson, Avi Singh, Rebecca Cao |
| EPI_ISL_1334380 | Laboratorios Lister | Instituto de Diagnostico y Referencia Epidemiologicos (INDRE) | Claudia Wong-Arambula, Abril Rodriguez-Maldonado, Vanessa Rivero-Arredondo, Ariadna Medina-Benitez, Joaquin Quiroz-Mercado, Sergio Rangel-Guerrero, Natividad Cruz-Ortiz, Tatiana Nunez-Garcia, Gisela Barrera-Badillo, Lucia Hernandez-Rivas, Irma Lopez-Martinez, Ernesto Ramirez-Gonzalez. |
| EPI_ISL_1334381 | LESP Guerrero | Instituto de Diagnostico y Referencia Epidemiologicos (INDRE) | Claudia Wong-Arambula, Abril Rodriguez-Maldonado, Vanessa Rivero-Arredondo, Ariadna Medina-Benitez, Joaquin Quiroz-Mercado, Sergio Rangel-Guerrero, Natividad Cruz-Ortiz, Tatiana Nunez-Garcia, Gisela Barrera-Badillo, Lucia Hernandez-Rivas, Irma Lopez-Martinez, Ernesto Ramirez-Gonzalez. |
| EPI_ISL_1334382, EPI_ISL_1334383, EPI_ISL_1334384, EPI_ISL_1334385 | Laboratorios Lister | Instituto de Diagnostico y Referencia Epidemiologicos (INDRE) | Claudia Wong-Arambula, Abril Rodriguez-Maldonado, Vanessa Rivero-Arredondo, Ariadna Medina-Benitez, Joaquin Quiroz-Mercado, Sergio Rangel-Guerrero, Natividad Cruz-Ortiz, Tatiana Nunez-Garcia, Gisela Barrera-Badillo, Lucia Hernandez-Rivas, Irma Lopez-Martinez, Ernesto Ramirez-Gonzalez. |
| EPI_ISL_1334386 | LESP Guanajuato | Instituto de Diagnostico y Referencia Epidemiologicos (INDRE) | Claudia Wong-Arambula, Abril Rodriguez-Maldonado, Vanessa Rivero-Arredondo, Ariadna Medina-Benitez, Joaquin Quiroz-Mercado, Sergio Rangel-Guerrero, Natividad Cruz-Ortiz, Tatiana Nunez-Garcia, Gisela Barrera-Badillo, Lucia Hernandez-Rivas, Irma Lopez-Martinez, Ernesto Ramirez-Gonzalez. |
| EPI_ISL_1334387, EPI_ISL_1334388, EPI_ISL_1334389, EPI_ISL_1334390, EPI_ISL_1334391, EPI_ISL_1334393, EPI_ISL_1334394, EPI_ISL_1334395, EPI_ISL_1334397, EPI_ISL_1334398, EPI_ISL_1334399, EPI_ISL_1334400, EPI_ISL_1334401, EPI_ISL_1334402, EPI_ISL_1334403, EPI_ISL_1334404, EPI_ISL_1334405, EPI_ISL_1334406, EPI_ISL_1334407, EPI_ISL_1334408, EPI_ISL_1334409, EPI_ISL_1334410, EPI_ISL_1334411, EPI_ISL_1334412 | | | |
| see above | Wyoming Public Health Laboratory | Wyoming Public Health Laboratory | Jim Mildenberger, Wanda Manley, Noah Hull, Taylor Fearing, Lynette Gumbleton, Channing Weber, Ashley Norberg, Chayse Rowley, Marley Goetz, Brian Dominguez, Elliot Thomasson, Cari Sloma, and Rob Christensen |
| EPI_ISL_1334518, EPI_ISL_1334519, EPI_ISL_1334520, EPI_ISL_1334521, EPI_ISL_1334522, EPI_ISL_1334523, EPI_ISL_1334524, EPI_ISL_1334525, EPI_ISL_1334526, EPI_ISL_1334527, EPI_ISL_1334528, EPI_ISL_1334529, EPI_ISL_1334530, EPI_ISL_1334531, EPI_ISL_1334532, EPI_ISL_1334533, EPI_ISL_1334534, EPI_ISL_1334535, EPI_ISL_1334536, EPI_ISL_1334537, EPI_ISL_1334538, EPI_ISL_1334539, EPI_ISL_1334540, EPI_ISL_1334541, EPI_ISL_1334542, EPI_ISL_1334543, EPI_ISL_1334544, EPI_ISL_1334545, EPI_ISL_1334546, EPI_ISL_1334547, EPI_ISL_1334548, EPI_ISL_1334549, EPI_ISL_1334550, EPI_ISL_1334551, EPI_ISL_1334552, EPI_ISL_1334553, EPI_ISL_1334554, EPI_ISL_1334555, EPI_ISL_1334556, EPI_ISL_1334557, EPI_ISL_1334558, EPI_ISL_1334559, EPI_ISL_1334560, EPI_ISL_1334561, EPI_ISL_1334562, EPI_ISL_1334563, EPI_ISL_1334564, EPI_ISL_1334565, EPI_ISL_1334566, EPI_ISL_1334567, EPI_ISL_1334568 | | | |
| see above | Clinical Molecular Microbiology Laboratory, UNC Hospital | Dirk Dittmer | Justin T. Landis, Razia Moorad, Brent A. Eason, Melissa B. Miller, Linda Pluta, Dirk Dittmer, Angelica Juarez, Cameroon Grant, Evelyn Hoffman, Patricio Cano, Jason Wong, Carolina Caro-Vegas, Ryan McNamara, Danielle Chappell, Cecilia Thompson, Shawn Hawken, Jason Smedberg, Andrea McDaniel-Harper, Billy Fisher, Ashwini Roy-Chaudhury, Blossom Damania. |
| EPI_ISL_1334571 | Kansas Health and Environmental Lab | Kansas Health and Environmental Lab | Mike Grose, Jonathan Barnell, Ben Olsen, and Phil Adam |
| EPI_ISL_1334581, EPI_ISL_1334582, EPI_ISL_1334583, EPI_ISL_1334584, EPI_ISL_1334585, EPI_ISL_1334586, EPI_ISL_1334588, EPI_ISL_1334589, EPI_ISL_1334591, EPI_ISL_1334592, EPI_ISL_1334593, EPI_ISL_1334594, EPI_ISL_1334595, EPI_ISL_1334596, EPI_ISL_1334597, EPI_ISL_1334598, EPI_ISL_1334599 | | | |
| see above | Arizona State Public Health Laboratory | Arizona State Public Health Laboratory | Trung Huynh, Jessica Escobar, Katherine Fullerton, Nobuko Fukushima, Stacy White, Linda Getsinger, Victor Waddell |
| EPI_ISL_1335757 | LESP Zacatecas | Instituto de diagnóstico y Referencia Epidemiologicos (INDRE) Departamento de Virología | Claudia Wong-Arambula, Abril Rodriguez-Maldonado, Vanessa Rivero-Arredondo, Ariadna Medina-Benitez, Joaquin Quiroz-Mercado, Sergio Rangel-Guerrero, Natividad Cruz-Ortiz, Tatiana Nunez-Garcia, Gisela Barrera-Badillo, Lucia Hernandez-Rivas, Irma Lopez-Martinez, Ernesto Ramirez-Gonzalez. |
| EPI_ISL_1335761, EPI_ISL_1335762, EPI_ISL_1335764, EPI_ISL_1335765, EPI_ISL_1335766, EPI_ISL_1335767, EPI_ISL_1335768, EPI_ISL_1335769, EPI_ISL_1335770, EPI_ISL_1335772, EPI_ISL_1335773, EPI_ISL_1335774, EPI_ISL_1335777, EPI_ISL_1335778, EPI_ISL_1335780, EPI_ISL_1335781, EPI_ISL_1335782, EPI_ISL_1335783, EPI_ISL_1335785, EPI_ISL_1335786, EPI_ISL_1335787, EPI_ISL_1335789, EPI_ISL_1335790, EPI_ISL_1335791, EPI_ISL_1335792, EPI_ISL_1335793, EPI_ISL_1335794, EPI_ISL_1335795, EPI_ISL_1335797, EPI_ISL_1335798, EPI_ISL_1335799, EPI_ISL_1335800, EPI_ISL_1335802 | | | |
| see above | Wyoming Public Health Laboratory | Wyoming Public Health Laboratory | Jim Mildenberger, Wanda Manley, Noah Hull, Taylor Fearing, Lynette Gumbleton, Channing Weber, Ashley Norberg, Chayse Rowley, Marley Goetz, Brian Dominguez, Elliot Thomasson, Cari Sloma, and Rob Christensen |
| EPI_ISL_1335804, EPI_ISL_1335805, EPI_ISL_1335806, EPI_ISL_1335807, EPI_ISL_1335808, EPI_ISL_1335809, EPI_ISL_1335810, EPI_ISL_1335811, EPI_ISL_1335812, EPI_ISL_1335813, EPI_ISL_1335814, EPI_ISL_1335815, EPI_ISL_1335817, EPI_ISL_1335818, EPI_ISL_1335819, EPI_ISL_1335820, EPI_ISL_1335822, EPI_ISL_1335823, EPI_ISL_1335824, EPI_ISL_1335825, EPI_ISL_1335826, EPI_ISL_1335827, EPI_ISL_1335828, EPI_ISL_1335829, EPI_ISL_1335830, EPI_ISL_1335831, EPI_ISL_1335832, EPI_ISL_1335833, EPI_ISL_1335834, EPI_ISL_1335835, EPI_ISL_1335836, EPI_ISL_1335837, EPI_ISL_1335838, EPI_ISL_1335839, EPI_ISL_1335840, EPI_ISL_1335841, EPI_ISL_1335842, EPI_ISL_1335843, EPI_ISL_1335844, EPI_ISL_1335845, EPI_ISL_1335848, EPI_ISL_1335849, EPI_ISL_1335850, EPI_ISL_1335851, EPI_ISL_1335852, EPI_ISL_1335854, EPI_ISL_1335855, EPI_ISL_1335856, EPI_ISL_1335857, EPI_ISL_1335858, EPI_ISL_1335859, EPI_ISL_1335860, EPI_ISL_1335861, EPI_ISL_1335862, EPI_ISL_1335864, EPI_ISL_1335866, EPI_ISL_1335867, EPI_ISL_1335868, EPI_ISL_1335869, EPI_ISL_1335870 | | | |
| see above | Santa Clara County Public Health Laboratory | Santa Clara County Public Health Laboratory | Santa Clara County Public Health Department |
| EPI_ISL_1335871, EPI_ISL_1335872 | Stanford Health Care | Clinical Virology Laboratory, Stanford University School or Medicine | Malaya K. Sahoo, Fumiko Yamamoto, Mamdouh Sibai, Daniel Solis, Michelle Vergese, ChunHong Huang, James Zehnder, and Benjamin A. Pinsky |
| EPI_ISL_1335900, EPI_ISL_1335901, EPI_ISL_1335902, EPI_ISL_1335903, EPI_ISL_1335904, EPI_ISL_1335905, EPI_ISL_1335906, EPI_ISL_1335907, EPI_ISL_1335908, EPI_ISL_1335909, EPI_ISL_1335910, EPI_ISL_1335911, EPI_ISL_1335912, EPI_ISL_1335913, EPI_ISL_1335914, EPI_ISL_1335915, EPI_ISL_1335916, EPI_ISL_1335917, EPI_ISL_1335918, EPI_ISL_1335919, EPI_ISL_1335920, EPI_ISL_1335921, EPI_ISL_1335922, EPI_ISL_1335923, EPI_ISL_1335924, EPI_ISL_1335925, EPI_ISL_1335926, EPI_ISL_1335927, EPI_ISL_1335928, EPI_ISL_1335929, EPI_ISL_1335930, EPI_ISL_1335931, EPI_ISL_1335932, EPI_ISL_1335933, EPI_ISL_1335934, EPI_ISL_1335935, EPI_ISL_1335936, EPI_ISL_1335937, EPI_ISL_1335938, EPI_ISL_1335939, EPI_ISL_1335940, EPI_ISL_1335941, EPI_ISL_1335942, EPI_ISL_1335943, EPI_ISL_1335944, EPI_ISL_1335945, EPI_ISL_1335946, EPI_ISL_1335947, EPI_ISL_1335948, EPI_ISL_1335949, EPI_ISL_1335950, EPI_ISL_1335951, EPI_ISL_1335952, EPI_ISL_1335953, EPI_ISL_1335954, EPI_ISL_1335955, EPI_ISL_1335956, EPI_ISL_1335957, EPI_ISL_1335958, EPI_ISL_1335959, EPI_ISL_1335960, EPI_ISL_1335961, EPI_ISL_1335962, EPI_ISL_1335963, EPI_ISL_1335964, EPI_ISL_1335965, EPI_ISL_1335966, EPI_ISL_1335967, EPI_ISL_1335968, EPI_ISL_1335969, EPI_ISL_1335970, EPI_ISL_1335971, EPI_ISL_1335972, EPI_ISL_1335973, EPI_ISL_1335974, EPI_ISL_1335975, EPI_ISL_1335976, EPI_ISL_1335977, EPI_ISL_1335978, EPI_ISL_1335979, EPI_ISL_1335980, EPI_ISL_1335981, EPI_ISL_1335982, EPI_ISL_1335983, EPI_ISL_1335984, EPI_ISL_1335985, EPI_ISL_1335986, EPI_ISL_1335987, EPI_ISL_1335988, EPI_ISL_1335989, EPI_ISL_1335990, EPI_ISL_1335991, EPI_ISL_1335992, EPI_ISL_1335993, EPI_ISL_1335994, EPI_ISL_1335995, EPI_ISL_1335996, EPI_ISL_1335997, EPI_ISL_1335998, EPI_ISL_1335999, EPI_ISL_1336000, EPI_ISL_1336001, EPI_ISL_1336002, EPI_ISL_1336003, EPI_ISL_1336004, EPI_ISL_1336005, EPI_ISL_1336006, EPI_ISL_1336007, EPI_ISL_1336008, EPI_ISL_1336009, EPI_ISL_1336010, EPI_ISL_1336011, EPI_ISL_1336012, EPI_ISL_1336013, EPI_ISL_1336014, EPI_ISL_1336015, EPI_ISL_1336016, EPI_ISL_1336017, EPI_ISL_1336018, EPI_ISL_1336019, EPI_ISL_1336020, EPI_ISL_1336021, EPI_ISL_1336022, EPI_ISL_1336023, EPI_ISL_1336024, EPI_ISL_1336025, EPI_ISL_1336026, EPI_ISL_1336027, EPI_ISL_1336028, EPI_ISL_1336029, EPI_ISL_1336030, EPI_ISL_1336031, EPI_ISL_1336032, EPI_ISL_1336033, EPI_ISL_1336034, EPI_ISL_1336035, EPI_ISL_1336036, EPI_ISL_1336037, EPI_ISL_1336038, EPI_ISL_1336039, EPI_ISL_1336040, EPI_ISL_1336041, EPI_ISL_1336042, EPI_ISL_1336043, EPI_ISL_1336044, EPI_ISL_1336045, EPI_ISL_1336046, EPI_ISL_1336047, EPI_ISL_1336048, EPI_ISL_1336049, EPI_ISL_1336050, EPI_ISL_1336051, EPI_ISL_1336052, EPI_ISL_1336053, EPI_ISL_1336054, EPI_ISL_1336055, EPI_ISL_1336056, EPI_ISL_1336057, EPI_ISL_1336058, EPI_ISL_1336059, EPI_ISL_1336060, EPI_ISL_1336061, EPI_ISL_1336062, EPI_ISL_1336063, EPI_ISL_1336064, EPI_ISL_1336065, EPI_ISL_1336066, EPI_ISL_1336067, EPI_ISL_1336068, EPI_ISL_1336069, EPI_ISL_1336070, EPI_ISL_1336071, EPI_ISL_1336072, EPI_ISL_1336073, EPI_ISL_1336074, EPI_ISL_1336075, EPI_ISL_1336076, EPI_ISL_1336077, EPI_ISL_1336078, EPI_ISL_1336079, EPI_ISL_1336080, EPI_ISL_1336081, EPI_ISL_1336082, EPI_ISL_1336085, EPI_ISL_1336086, EPI_ISL_1336087, EPI_ISL_1336088, EPI_ISL_1336089, EPI_ISL_1336090, EPI_ISL_1336091, EPI_ISL_1336092, EPI_ISL_1336093, EPI_ISL_1336094, EPI_ISL_1336095, EPI_ISL_1336096, EPI_ISL_1336097, EPI_ISL_1336098, EPI_ISL_1336099, EPI_ISL_1336100, EPI_ISL_1336101, EPI_ISL_1336102, EPI_ISL_1336103, EPI_ISL_1336104, EPI_ISL_1336105, EPI_ISL_1336106, EPI_ISL_1336107, EPI_ISL_1336108, EPI_ISL_1336109, EPI_ISL_1336110, EPI_ISL_1336111, EPI_ISL_1336112, EPI_ISL_1336113, EPI_ISL_1336114, EPI_ISL_1336115, EPI_ISL_1336116, EPI_ISL_1336117, EPI_ISL_1336118, EPI_ISL_1336119, EPI_ISL_1336120, EPI_ISL_1336121, EPI_ISL_1336122, EPI_ISL_1336123, EPI_ISL_1336124, EPI_ISL_1336125, EPI_ISL_1336126, EPI_ISL_1336127, EPI_ISL_1336128, EPI_ISL_1336129, EPI_ISL_1336130, EPI_ISL_1336131, EPI_ISL_1336132, EPI_ISL_1336133, EPI_ISL_1336134, EPI_ISL_1336135, EPI_ISL_1336136, EPI_ISL_1336137, EPI_ISL_1336138, EPI_ISL_1336139, EPI_ISL_1336140, EPI_ISL_1336141, EPI_ISL_1336142, EPI_ISL_1336143, EPI_ISL_1336144, EPI_ISL_1336145, EPI_ISL_1336146, EPI_ISL_1336147, EPI_ISL_1336148, EPI_ISL_1336149, EPI_ISL_1336150, EPI_ISL_1336151, EPI_ISL_1336152, EPI_ISL_1336153, EPI_ISL_1336154, EPI_ISL_1336155, EPI_ISL_1336156, EPI_ISL_1336157, EPI_ISL_1336158, EPI_ISL_1336159, EPI_ISL_1336160, EPI_ISL_1336161, EPI_ISL_1336162, EPI_ISL_1336163, EPI_ISL_1336164, EPI_ISL_1336165, EPI_ISL_1336166, EPI_ISL_1336167, EPI_ISL_1336168, EPI_ISL_1336169, EPI_ISL_1336170, EPI_ISL_1336171, EPI_ISL_1336172, EPI_ISL_1336173, EPI_ISL_1336174, EPI_ISL_1336175, EPI_ISL_1336176, EPI_ISL_1336177 | | | |
| see above | Quest Diagnostics Incorporated | Centers for Disease Control and Prevention | Peter W. Cook, Dakota Howard, Dhwani Batra, Ben L. Rambo-Martin, S. H. Rosenthal, A. Gerasimova, R. M. Kagan, B. Anderson, M. Hua, Y. Liu, L.E. Bernstein, K.E. Livingston, A. Perez, I. A. Shlyakhter, R. V. Rolando, R. Owen, P. Tanpaiboon, F. Lacbawan, Clinton R. Paden, Suxiang Tong, |

| | | | |
|---|---|---|---|
| | | Division of Viral Diseases, Pathogen Discovery | Duncan MacCannell |
| EPI_ISL_1336238, EPI_ISL_1336240, EPI_ISL_1336246, EPI_ISL_1336248, EPI_ISL_1336250, EPI_ISL_1336252, EPI_ISL_1336257 | Wyoming Public Health Laboratory | Wyoming Public Health Laboratory | Jim Mildenberger, Wanda Manley, Noah Hull, Taylor Fearing, Lynette Gumbleton, Channing Weber, Ashley Norberg, Chayse Rowley, Marley Goetz, Brian Dominguez, Elliot Thomasson, Cari Sloma, and Rob Christensen |
| EPI_ISL_1336282, EPI_ISL_1336283, EPI_ISL_1336284, EPI_ISL_1336285, EPI_ISL_1336286, EPI_ISL_1336287, EPI_ISL_1336288, EPI_ISL_1336289, EPI_ISL_1336290, EPI_ISL_1336291, EPI_ISL_1336292, EPI_ISL_1336293, EPI_ISL_1336294, EPI_ISL_1336295, EPI_ISL_1336296, EPI_ISL_1336297, EPI_ISL_1336298, EPI_ISL_1336299, EPI_ISL_1336300, EPI_ISL_1336301, EPI_ISL_1336302, EPI_ISL_1336303, EPI_ISL_1336304, EPI_ISL_1336305, EPI_ISL_1336306, EPI_ISL_1336307, EPI_ISL_1336308, EPI_ISL_1336309, EPI_ISL_1336310, EPI_ISL_1336311, EPI_ISL_1336312, EPI_ISL_1336313, EPI_ISL_1336314, EPI_ISL_1336315, EPI_ISL_1336316, EPI_ISL_1336317, EPI_ISL_1336318, EPI_ISL_1336319, EPI_ISL_1336320, EPI_ISL_1336321, EPI_ISL_1336322, EPI_ISL_1336323, EPI_ISL_1336324, EPI_ISL_1336325, EPI_ISL_1336326, EPI_ISL_1336327, EPI_ISL_1336328 | | | |
| see above | MD PHL | MD PHL | Maryland Department of Health Laboratories Administration |
| EPI_ISL_1336352, EPI_ISL_1336353 | Kansas Health and Environmental Lab | Kansas Health and Environmental Lab | Mike Grose, Jonathan Barnell, Ben Olsen, and Phil Adam |
| EPI_ISL_1336964, EPI_ISL_1336965, EPI_ISL_1336966, EPI_ISL_1336967, EPI_ISL_1336968, EPI_ISL_1336969, EPI_ISL_1336970, EPI_ISL_1336971, EPI_ISL_1336972, EPI_ISL_1336973, EPI_ISL_1336974, EPI_ISL_1336975, EPI_ISL_1336976, EPI_ISL_1336977, EPI_ISL_1336978, EPI_ISL_1336980, EPI_ISL_1336981, EPI_ISL_1336982, EPI_ISL_1336983, EPI_ISL_1336984, EPI_ISL_1336985, EPI_ISL_1336986, EPI_ISL_1336987, EPI_ISL_1336988, EPI_ISL_1336989, EPI_ISL_1336990, EPI_ISL_1336991, EPI_ISL_1336992, EPI_ISL_1336993, EPI_ISL_1336994, EPI_ISL_1336995, EPI_ISL_1336996, EPI_ISL_1336997, EPI_ISL_1336998, EPI_ISL_1336999, EPI_ISL_1337000, EPI_ISL_1337001, EPI_ISL_1337002, EPI_ISL_1337003, EPI_ISL_1337004, EPI_ISL_1337005, EPI_ISL_1337006, EPI_ISL_1337007, EPI_ISL_1337009, EPI_ISL_1337010, EPI_ISL_1337011, EPI_ISL_1337012, EPI_ISL_1337013, EPI_ISL_1337014, EPI_ISL_1337015, EPI_ISL_1337016, EPI_ISL_1337017, EPI_ISL_1337018, EPI_ISL_1337019, EPI_ISL_1337020, EPI_ISL_1337021, EPI_ISL_1337022, EPI_ISL_1337023, EPI_ISL_1337024, EPI_ISL_1337025, EPI_ISL_1337026, EPI_ISL_1337027, EPI_ISL_1337028, EPI_ISL_1337029, EPI_ISL_1337030, EPI_ISL_1337031, EPI_ISL_1337032, EPI_ISL_1337033, EPI_ISL_1337034, EPI_ISL_1337035, EPI_ISL_1337036, EPI_ISL_1337037, EPI_ISL_1337038, EPI_ISL_1337039, EPI_ISL_1337040, EPI_ISL_1337041, EPI_ISL_1337042, EPI_ISL_1337043, EPI_ISL_1337044, EPI_ISL_1337045, EPI_ISL_1337046, EPI_ISL_1337047, EPI_ISL_1337048, EPI_ISL_1337049, EPI_ISL_1337050, EPI_ISL_1337051, EPI_ISL_1337052, EPI_ISL_1337053, EPI_ISL_1337054, EPI_ISL_1337055, EPI_ISL_1337056, EPI_ISL_1337057, EPI_ISL_1337058, EPI_ISL_1337059, EPI_ISL_1337060, EPI_ISL_1337061, EPI_ISL_1337062, EPI_ISL_1337063, EPI_ISL_1337064, EPI_ISL_1337065, EPI_ISL_1337066, EPI_ISL_1337067, EPI_ISL_1337068, EPI_ISL_1337069, EPI_ISL_1337070, EPI_ISL_1337071, EPI_ISL_1337073, EPI_ISL_1337074, EPI_ISL_1337075, EPI_ISL_1337076, EPI_ISL_1337077, EPI_ISL_1337078, EPI_ISL_1337079, EPI_ISL_1337080, EPI_ISL_1337081, EPI_ISL_1337082, EPI_ISL_1337083, EPI_ISL_1337084, EPI_ISL_1337085, EPI_ISL_1337086, EPI_ISL_1337087, EPI_ISL_1337089, EPI_ISL_1337090, EPI_ISL_1337091, EPI_ISL_1337092, EPI_ISL_1337093, EPI_ISL_1337094, EPI_ISL_1337095, EPI_ISL_1337096, EPI_ISL_1337097, EPI_ISL_1337098, EPI_ISL_1337099, EPI_ISL_1337100, EPI_ISL_1337101, EPI_ISL_1337102, EPI_ISL_1337103, EPI_ISL_1337104, EPI_ISL_1337105, EPI_ISL_1337106, EPI_ISL_1337107, EPI_ISL_1337108, EPI_ISL_1337109, EPI_ISL_1337110, EPI_ISL_1337111, EPI_ISL_1337112, EPI_ISL_1337113, EPI_ISL_1337114, EPI_ISL_1337115, EPI_ISL_1337116, EPI_ISL_1337117, EPI_ISL_1337118, EPI_ISL_1337119, EPI_ISL_1337120, EPI_ISL_1337121, EPI_ISL_1337122, EPI_ISL_1337123, EPI_ISL_1337124, EPI_ISL_1337125, EPI_ISL_1337126, EPI_ISL_1337127, EPI_ISL_1337128, EPI_ISL_1337129, EPI_ISL_1337130, EPI_ISL_1337131, EPI_ISL_1337132, EPI_ISL_1337133, EPI_ISL_1337134, EPI_ISL_1337135, EPI_ISL_1337136, EPI_ISL_1337137, EPI_ISL_1337138, EPI_ISL_1337139, EPI_ISL_1337140, EPI_ISL_1337141, EPI_ISL_1337142, EPI_ISL_1337143, EPI_ISL_1337144, EPI_ISL_1337145, EPI_ISL_1337146, EPI_ISL_1337147, EPI_ISL_1337148, EPI_ISL_1337149, EPI_ISL_1337150, EPI_ISL_1337151, EPI_ISL_1337152, EPI_ISL_1337153, EPI_ISL_1337154, EPI_ISL_1337155, EPI_ISL_1337156, EPI_ISL_1337157, EPI_ISL_1337158, EPI_ISL_1337159, EPI_ISL_1337160, EPI_ISL_1337161, EPI_ISL_1337162, EPI_ISL_1337163, EPI_ISL_1337164, EPI_ISL_1337165, EPI_ISL_1337166, EPI_ISL_1337167, EPI_ISL_1337168, EPI_ISL_1337169, EPI_ISL_1337170, EPI_ISL_1337171, EPI_ISL_1337172, EPI_ISL_1337173, EPI_ISL_1337174, EPI_ISL_1337175, EPI_ISL_1337176, EPI_ISL_1337177, EPI_ISL_1337178, EPI_ISL_1337179, EPI_ISL_1337180, EPI_ISL_1337181, EPI_ISL_1337182, EPI_ISL_1337183, EPI_ISL_1337184, EPI_ISL_1337185, EPI_ISL_1337186, EPI_ISL_1337187, EPI_ISL_1337188, EPI_ISL_1337189, EPI_ISL_1337190, EPI_ISL_1337191, EPI_ISL_1337192, EPI_ISL_1337193, EPI_ISL_1337194, EPI_ISL_1337195, EPI_ISL_1337196, EPI_ISL_1337197, EPI_ISL_1337198, EPI_ISL_1337199, EPI_ISL_1337200, EPI_ISL_1337201, EPI_ISL_1337202, EPI_ISL_1337203, EPI_ISL_1337204, EPI_ISL_1337205, EPI_ISL_1337206, EPI_ISL_1337207, EPI_ISL_1337208, EPI_ISL_1337209, EPI_ISL_1337210, EPI_ISL_1337211, EPI_ISL_1337212, EPI_ISL_1337213, EPI_ISL_1337214, EPI_ISL_1337215, EPI_ISL_1337216, EPI_ISL_1337217, EPI_ISL_1337218, EPI_ISL_1337219, EPI_ISL_1337220, EPI_ISL_1337221, EPI_ISL_1337222, EPI_ISL_1337223 | | | |
| see above | Helix/Illumina | Centers for Disease Control and Prevention Division of Viral Diseases, Pathogen Discovery | Peter W. Cook, Dakota Howard, Dhwani Batra, Ben L. Rambo-Martin, Eileen de Feo, Jan Antico, Christine Tran, Matthew Tolentino, Shannon Wickline, Kim Gietzen, Brad Sickler, Jingtao Liu, Eric Allen, Phil Febbo, Summer Galloway, Nicole L. Washington, Simon White, Geraint Levan, Kelly Schiabor Barrett, Elizabeth Cirulli, Alexandre Bolze, Ary Ascencio, Charlotte Rivera-Garcia, Ryan Cho, Jason Nguyen, Sherry Wang, Jimmy Ramirez, Tyler Cassens, Efren Sandoval, Magnus Isaksson, William Lee, David Becker, Marc Laurent, James Lu, Clinton R. Paden, Suxiang Tong, Duncan MacCannell |
| EPI_ISL_1337224 | Kansas Health and Environmental Lab | Kansas Health and Environmental Lab | Mike Grose, Jonathan Barnell, Ben Olsen, and Phil Adam |
| EPI_ISL_1337225 | Helix/Illumina | Centers for Disease Control and Prevention Division of Viral Diseases, Pathogen Discovery | Peter W. Cook, Dakota Howard, Dhwani Batra, Ben L. Rambo-Martin, Eileen de Feo, Jan Antico, Christine Tran, Matthew Tolentino, Shannon Wickline, Kim Gietzen, Brad Sickler, Jingtao Liu, Eric Allen, Phil Febbo, Summer Galloway, Nicole L. Washington, Simon White, Geraint Levan, Kelly Schiabor Barrett, Elizabeth Cirulli, Alexandre Bolze, Ary Ascencio, Charlotte Rivera-Garcia, Ryan Cho, Jason Nguyen, Sherry Wang, Jimmy Ramirez, Tyler Cassens, Efren Sandoval, Magnus Isaksson, William Lee, David Becker, Marc Laurent, James Lu, Clinton R. Paden, Suxiang Tong, Duncan MacCannell |
| EPI_ISL_1337226, EPI_ISL_1337227 | Kansas Health and Environmental Lab | Kansas Health and Environmental Lab | Mike Grose, Jonathan Barnell, Ben Olsen, and Phil Adam |
| EPI_ISL_1337228 | Helix/Illumina | Centers for Disease Control and Prevention Division of Viral Diseases, Pathogen Discovery | Peter W. Cook, Dakota Howard, Dhwani Batra, Ben L. Rambo-Martin, Eileen de Feo, Jan Antico, Christine Tran, Matthew Tolentino, Shannon Wickline, Kim Gietzen, Brad Sickler, Jingtao Liu, Eric Allen, Phil Febbo, Summer Galloway, Nicole L. Washington, Simon White, Geraint Levan, Kelly Schiabor Barrett, Elizabeth Cirulli, Alexandre Bolze, Ary Ascencio, Charlotte Rivera-Garcia, Ryan Cho, Jason Nguyen, Sherry Wang, Jimmy Ramirez, Tyler Cassens, Efren Sandoval, Magnus Isaksson, William Lee, David Becker, Marc Laurent, James Lu, Clinton R. Paden, Suxiang Tong, Duncan MacCannell |
| EPI_ISL_1337229, EPI_ISL_1337230 | Kansas Health and Environmental Lab | Kansas Health and Environmental Lab | Mike Grose, Jonathan Barnell, Ben Olsen, and Phil Adam |
| EPI_ISL_1337231 | Helix/Illumina | Centers for Disease Control and Prevention Division of Viral Diseases, Pathogen Discovery | Peter W. Cook, Dakota Howard, Dhwani Batra, Ben L. Rambo-Martin, Eileen de Feo, Jan Antico, Christine Tran, Matthew Tolentino, Shannon Wickline, Kim Gietzen, Brad Sickler, Jingtao Liu, Eric Allen, Phil Febbo, Summer Galloway, Nicole L. Washington, Simon White, Geraint Levan, Kelly Schiabor Barrett, Elizabeth Cirulli, Alexandre Bolze, Ary Ascencio, Charlotte Rivera-Garcia, Ryan Cho, Jason Nguyen, Sherry Wang, Jimmy Ramirez, Tyler Cassens, Efren Sandoval, Magnus Isaksson, William Lee, David Becker, Marc Laurent, James Lu, Clinton R. Paden, Suxiang Tong, Duncan MacCannell |
| EPI_ISL_1337232 | Kansas Health and Environmental Lab | Kansas Health and Environmental Lab | Mike Grose, Jonathan Barnell, Ben Olsen, and Phil Adam |
| EPI_ISL_1337233, EPI_ISL_1337234 | Helix/Illumina | Centers for Disease Control and Prevention Division of Viral Diseases, Pathogen Discovery | Peter W. Cook, Dakota Howard, Dhwani Batra, Ben L. Rambo-Martin, Eileen de Feo, Jan Antico, Christine Tran, Matthew Tolentino, Shannon Wickline, Kim Gietzen, Brad Sickler, Jingtao Liu, Eric Allen, Phil Febbo, Summer Galloway, Nicole L. Washington, Simon White, Geraint Levan, Kelly Schiabor Barrett, Elizabeth Cirulli, Alexandre Bolze, Ary Ascencio, Charlotte Rivera-Garcia, Ryan Cho, Jason Nguyen, Sherry Wang, Jimmy Ramirez, Tyler Cassens, Efren Sandoval, Magnus Isaksson, William Lee, David Becker, Marc Laurent, James Lu, Clinton R. Paden, Suxiang Tong, Duncan MacCannell |
| EPI_ISL_1337235 | Kansas Health and Environmental Lab | Kansas Health and Environmental Lab | Mike Grose, Jonathan Barnell, Ben Olsen, and Phil Adam |
| EPI_ISL_1337236 | Helix/Illumina | Centers for Disease Control and Prevention Division of Viral Diseases, Pathogen Discovery | Peter W. Cook, Dakota Howard, Dhwani Batra, Ben L. Rambo-Martin, Eileen de Feo, Jan Antico, Christine Tran, Matthew Tolentino, Shannon Wickline, Kim Gietzen, Brad Sickler, Jingtao Liu, Eric Allen, Phil Febbo, Summer Galloway, Nicole L. Washington, Simon White, Geraint Levan, Kelly Schiabor Barrett, Elizabeth Cirulli, Alexandre Bolze, Ary Ascencio, Charlotte Rivera-Garcia, Ryan Cho, Jason Nguyen, Sherry Wang, Jimmy Ramirez, Tyler Cassens, Efren Sandoval, Magnus Isaksson, William Lee, David Becker, Marc Laurent, James Lu, Clinton R. Paden, Suxiang Tong, Duncan MacCannell |
| EPI_ISL_1337237, EPI_ISL_1337238 | Kansas Health and Environmental Lab | Kansas Health and Environmental Lab | Mike Grose, Jonathan Barnell, Ben Olsen, and Phil Adam |
| EPI_ISL_1337239 | Helix/Illumina | Centers for Disease Control and Prevention Division of Viral Diseases, Pathogen Discovery | Peter W. Cook, Dakota Howard, Dhwani Batra, Ben L. Rambo-Martin, Eileen de Feo, Jan Antico, Christine Tran, Matthew Tolentino, Shannon Wickline, Kim Gietzen, Brad Sickler, Jingtao Liu, Eric Allen, Phil Febbo, Summer Galloway, Nicole L. Washington, Simon White, Geraint Levan, Kelly Schiabor Barrett, Elizabeth Cirulli, Alexandre Bolze, Ary Ascencio, Charlotte Rivera-Garcia, Ryan Cho, Jason Nguyen, Sherry Wang, Jimmy Ramirez, Tyler Cassens, Efren Sandoval, Magnus Isaksson, William Lee, David Becker, Marc Laurent, James Lu, Clinton R. Paden, Suxiang Tong, Duncan MacCannell |
| EPI_ISL_1337240, EPI_ISL_1337241, EPI_ISL_1337242 | Kansas Health and Environmental Lab | Kansas Health and Environmental Lab | Mike Grose, Jonathan Barnell, Ben Olsen, and Phil Adam |
| EPI_ISL_1337243, EPI_ISL_1337244, EPI_ISL_1337245, EPI_ISL_1337246, EPI_ISL_1337247, EPI_ISL_1337248, EPI_ISL_1337249, EPI_ISL_1337250, EPI_ISL_1337251, EPI_ISL_1337252, EPI_ISL_1337253, EPI_ISL_1337254, EPI_ISL_1337255, EPI_ISL_1337256, EPI_ISL_1337257, EPI_ISL_1337258, EPI_ISL_1337259, EPI_ISL_1337260, EPI_ISL_1337261, EPI_ISL_1337262, EPI_ISL_1337263, EPI_ISL_1337264, EPI_ISL_1337265, EPI_ISL_1337266, EPI_ISL_1337267, EPI_ISL_1337268, EPI_ISL_1337269, EPI_ISL_1337270, EPI_ISL_1337271, EPI_ISL_1337272, EPI_ISL_1337273, EPI_ISL_1337274, EPI_ISL_1337275, EPI_ISL_1337276, EPI_ISL_1337277, EPI_ISL_1337278, EPI_ISL_1337279, EPI_ISL_1337280, EPI_ISL_1337281, EPI_ISL_1337282, EPI_ISL_1337283, EPI_ISL_1337284, EPI_ISL_1337285, EPI_ISL_1337286, EPI_ISL_1337287, EPI_ISL_1337288, EPI_ISL_1337289, EPI_ISL_1337290, EPI_ISL_1337291, EPI_ISL_1337292, EPI_ISL_1337293, EPI_ISL_1337294, EPI_ISL_1337295, EPI_ISL_1337296, EPI_ISL_1337297, EPI_ISL_1337298, EPI_ISL_1337299, EPI_ISL_1337300, EPI_ISL_1337301, EPI_ISL_1337302, EPI_ISL_1337303, EPI_ISL_1337304, EPI_ISL_1337305, EPI_ISL_1337306, EPI_ISL_1337307, EPI_ISL_1337308, EPI_ISL_1337309, EPI_ISL_1337310, EPI_ISL_1337311, EPI_ISL_1337312, EPI_ISL_1337313, EPI_ISL_1337314, EPI_ISL_1337315, EPI_ISL_1337316, EPI_ISL_1337317, EPI_ISL_1337318, EPI_ISL_1337319, EPI_ISL_1337320, EPI_ISL_1337321, EPI_ISL_1337322, EPI_ISL_1337323, EPI_ISL_1337324, EPI_ISL_1337325, EPI_ISL_1337326, EPI_ISL_1337327, EPI_ISL_1337328, EPI_ISL_1337329, EPI_ISL_1337330, EPI_ISL_1337331, EPI_ISL_1337332, EPI_ISL_1337333, EPI_ISL_1337334, EPI_ISL_1337335, EPI_ISL_1337336, EPI_ISL_1337337, EPI_ISL_1337338, EPI_ISL_1337339, EPI_ISL_1337340, EPI_ISL_1337341, EPI_ISL_1337342, EPI_ISL_1337343, EPI_ISL_1337344, EPI_ISL_1337345, EPI_ISL_1337346, EPI_ISL_1337347, EPI_ISL_1337348, EPI_ISL_1337349, EPI_ISL_1337350, EPI_ISL_1337351, EPI_ISL_1337352, EPI_ISL_1337353, EPI_ISL_1337354, EPI_ISL_1337355, EPI_ISL_1337356, EPI_ISL_1337357, EPI_ISL_1337358, EPI_ISL_1337359 | | | |
| see above | Helix/Illumina | Centers for Disease Control and Prevention Division of Viral Diseases, Pathogen Discovery | Peter W. Cook, Dakota Howard, Dhwani Batra, Ben L. Rambo-Martin, Eileen de Feo, Jan Antico, Christine Tran, Matthew Tolentino, Shannon Wickline, Kim Gietzen, Brad Sickler, Jingtao Liu, Eric Allen, Phil Febbo, Summer Galloway, Nicole L. Washington, Simon White, Geraint Levan, Kelly Schiabor Barrett, Elizabeth Cirulli, Alexandre Bolze, Ary Ascencio, Charlotte Rivera-Garcia, Ryan Cho, Jason Nguyen, Sherry Wang, Jimmy Ramirez, Tyler Cassens, Efren Sandoval, Magnus Isaksson, William Lee, David Becker, Marc Laurent, James Lu, Clinton R. Paden, Suxiang Tong, Duncan MacCannell |
| EPI_ISL_1337360 | LESP Queretaro | Instituto de Diagnostico y Referencia Epidemiologicos (INDRE) | Claudia Wong-Arambula, Abril Rodriguez-Maldonado, Vanessa Rivero-Arredondo, Ariadna Medina-Benitez, Joaquin Quiroz-Mercado, Sergio Rangel-Guerrero, Natividad Cruz-Ortiz, Tatiana Nunez-Garcia, Gisela Barrera-Badillo, Lucia Hernandez-Rivas, Irma Lopez-Martinez, Ernesto Ramirez-Gonzalez. |
| EPI_ISL_1337361 | Helix/Illumina | Centers for Disease Control and Prevention Division of Viral Diseases, Pathogen Discovery | Peter W. Cook, Dakota Howard, Dhwani Batra, Ben L. Rambo-Martin, Eileen de Feo, Jan Antico, Christine Tran, Matthew Tolentino, Shannon Wickline, Kim Gietzen, Brad Sickler, Jingtao Liu, Eric Allen, Phil Febbo, Summer Galloway, Nicole L. Washington, Simon White, Geraint Levan, Kelly Schiabor Barrett, Elizabeth Cirulli, Alexandre Bolze, Ary Ascencio, Charlotte Rivera-Garcia, Ryan Cho, Jason Nguyen, Sherry Wang, Jimmy Ramirez, Tyler Cassens, Efren Sandoval, Magnus Isaksson, William Lee, David Becker, Marc Laurent, James Lu, Clinton R. Paden, Suxiang Tong, Duncan MacCannell |
| EPI_ISL_1337362 | LESP Queretaro | Instituto de Diagnostico y Referencia Epidemiologicos (INDRE) | Claudia Wong-Arambula, Abril Rodriguez-Maldonado, Vanessa Rivero-Arredondo, Ariadna Medina-Benitez, Joaquin Quiroz-Mercado, Sergio Rangel-Guerrero, Natividad Cruz-Ortiz, Tatiana Nunez-Garcia, Gisela Barrera-Badillo, Lucia Hernandez-Rivas, Irma Lopez-Martinez, Ernesto Ramirez-Gonzalez. |
| EPI_ISL_1337363 | LESP Guanajuato | Instituto de Diagnostico y Referencia Epidemiologicos (INDRE) | Claudia Wong-Arambula, Abril Rodriguez-Maldonado, Vanessa Rivero-Arredondo, Ariadna Medina-Benitez, Joaquin Quiroz-Mercado, Sergio Rangel-Guerrero, Natividad Cruz-Ortiz, Tatiana Nunez-Garcia, Gisela Barrera-Badillo, Lucia Hernandez-Rivas, Irma Lopez-Martinez, Ernesto Ramirez-Gonzalez. |
| EPI_ISL_1337364 | Helix/Illumina | Centers for Disease Control and Prevention Division of Viral Diseases, Pathogen Discovery | Peter W. Cook, Dakota Howard, Dhwani Batra, Ben L. Rambo-Martin, Eileen de Feo, Jan Antico, Christine Tran, Matthew Tolentino, Shannon Wickline, Kim Gietzen, Brad Sickler, Jingtao Liu, Eric Allen, Phil Febbo, Summer Galloway, Nicole L. Washington, Simon White, Geraint Levan, Kelly Schiabor Barrett, Elizabeth Cirulli, Alexandre Bolze, Ary Ascencio, Charlotte Rivera-Garcia, Ryan Cho, Jason Nguyen, Sherry Wang, Jimmy Ramirez, Tyler Cassens, Efren Sandoval, Magnus Isaksson, William Lee, David Becker, Marc Laurent, James Lu, Clinton R. Paden, Suxiang Tong, Duncan MacCannell |
| EPI_ISL_1337365 | LESP Guanajuato | Instituto de Diagnostico y Referencia Epidemiologicos (INDRE) | Claudia Wong-Arambula, Abril Rodriguez-Maldonado, Vanessa Rivero-Arredondo, Ariadna Medina-Benitez, Joaquin Quiroz-Mercado, Sergio Rangel-Guerrero, Natividad Cruz-Ortiz, Tatiana Nunez-Garcia, Gisela Barrera-Badillo, Lucia Hernandez-Rivas, Irma Lopez-Martinez, Ernesto Ramirez-Gonzalez. |
| EPI_ISL_1337366 | Helix/Illumina | Centers for Disease Control and Prevention Division of Viral Diseases, Pathogen Discovery | Peter W. Cook, Dakota Howard, Dhwani Batra, Ben L. Rambo-Martin, Eileen de Feo, Jan Antico, Christine Tran, Matthew Tolentino, Shannon Wickline, Kim Gietzen, Brad Sickler, Jingtao Liu, Eric Allen, Phil Febbo, Summer Galloway, Nicole L. Washington, Simon White, Geraint Levan, Kelly Schiabor Barrett, Elizabeth Cirulli, Alexandre Bolze, Ary Ascencio, Charlotte Rivera-Garcia, Ryan Cho, Jason Nguyen, Sherry Wang, Jimmy Ramirez, Tyler Cassens, Efren Sandoval, Magnus Isaksson, William Lee, David Becker, Marc Laurent, James Lu, Clinton R. Paden, Suxiang Tong, Duncan MacCannell |
| EPI_ISL_1337367 | LESP Guanajuato | Instituto de Diagnostico y Referencia Epidemiologicos (INDRE) | Claudia Wong-Arambula, Abril Rodriguez-Maldonado, Vanessa Rivero-Arredondo, Ariadna Medina-Benitez, Joaquin Quiroz-Mercado, Sergio Rangel-Guerrero, Natividad Cruz-Ortiz, Tatiana Nunez-Garcia, Gisela Barrera-Badillo, Lucia Hernandez-Rivas, Irma Lopez-Martinez, Ernesto Ramirez-Gonzalez. |
| EPI_ISL_1337368 | LESP Nuevo Leon | Instituto de Diagnostico y Referencia Epidemiologicos (INDRE) | Claudia Wong-Arambula, Abril Rodriguez-Maldonado, Vanessa Rivero-Arredondo, Ariadna Medina-Benitez, Joaquin Quiroz-Mercado, Sergio Rangel-Guerrero, Natividad Cruz-Ortiz, Tatiana Nunez-Garcia, Gisela Barrera-Badillo, Lucia Hernandez-Rivas, Irma Lopez-Martinez, Ernesto Ramirez-Gonzalez. |
| EPI_ISL_1337369 | Helix/Illumina | Centers for Disease Control and Prevention Division of Viral Diseases, Pathogen Discovery | Peter W. Cook, Dakota Howard, Dhwani Batra, Ben L. Rambo-Martin, Eileen de Feo, Jan Antico, Christine Tran, Matthew Tolentino, Shannon Wickline, Kim Gietzen, Brad Sickler, Jingtao Liu, Eric Allen, Phil Febbo, Summer Galloway, Nicole L. Washington, Simon White, Geraint Levan, Kelly Schiabor Barrett, Elizabeth Cirulli, Alexandre Bolze, Ary Ascencio, Charlotte Rivera-Garcia, Ryan Cho, Jason Nguyen, Sherry Wang, Jimmy Ramirez, Tyler Cassens, Efren Sandoval, Magnus Isaksson, William Lee, David Becker, Marc Laurent, James Lu, Clinton R. Paden, Suxiang Tong, Duncan MacCannell |
| EPI_ISL_1337370, EPI_ISL_1337371 | LESP Baja California Sur | Instituto de Diagnostico y Referencia Epidemiologicos (INDRE) | Claudia Wong-Arambula, Abril Rodriguez-Maldonado, Vanessa Rivero-Arredondo, Ariadna Medina-Benitez, Joaquin Quiroz-Mercado, Sergio Rangel-Guerrero, Natividad Cruz-Ortiz, Tatiana Nunez-Garcia, Gisela Barrera-Badillo, Lucia Hernandez-Rivas, Irma Lopez-Martinez, Ernesto Ramirez-Gonzalez. |
| EPI_ISL_1337372 | Helix/Illumina | Centers for Disease Control and Prevention | Peter W. Cook, Dakota Howard, Dhwani Batra, Ben L. Rambo-Martin, Eileen de Feo, Jan Antico, Christine Tran, Matthew Tolentino, Shannon Wickline, Kim Gietzen, Brad Sickler, Jingtao Liu, Eric Allen, Phil Febbo, Summer Galloway, Nicole L. Washington, Simon White, Geraint Levan, Kelly |

| | | | |
|---|---|---|---|
| | | Division of Viral Diseases, Pathogen Discovery | Schiabor Barrett, Elizabeth Cirulli, Alexandre Bolze, Ary Ascencio, Charlotte Rivera-Garcia, Ryan Cho, Jason Nguyen, Sherry Wang, Jimmy Ramirez, Tyler Cassens, Efren Sandoval, Magnus Isaksson, William Lee, David Becker, Marc Laurent, James Lu, Clinton R. Paden, Suxiang Tong, Duncan MacCannell |
| EPI_ISL_1337373, EPI_ISL_1337374 | LESP Baja California Sur | Instituto de Diagnostico y Referencia Epidemiologicos (INDRE) | Claudia Wong-Arambula, Abril Rodriguez-Maldonado, Vanessa Rivero-Arredondo, Ariadna Medina-Benitez, Joaquin Quiroz-Mercado, Sergio Rangel-Guerrero, Natividad Cruz-Ortiz, Tatiana Nunez-Garcia, Gisela Barrera-Badillo, Lucia Hernandez-Rivas, Irma Lopez-Martinez, Ernesto Ramirez-Gonzalez. |
| EPI_ISL_1337375 | Helix/Illumina | Centers for Disease Control and Prevention Division of Viral Diseases, Pathogen Discovery | Peter W. Cook, Dakota Howard, Dhwani Batra, Ben L. Rambo-Martin, Eileen de Feo, Jan Antico, Christine Tran, Matthew Tolentino, Shannon Wickline, Kim Gietzen, Brad Sickler, Jingtao Liu, Eric Allen, Phil Febbo, Summer Galloway, Nicole L. Washington, Simon White, Geraint Levan, Kelly Schiabor Barrett, Elizabeth Cirulli, Alexandre Bolze, Ary Ascencio, Charlotte Rivera-Garcia, Ryan Cho, Jason Nguyen, Sherry Wang, Jimmy Ramirez, Tyler Cassens, Efren Sandoval, Magnus Isaksson, William Lee, David Becker, Marc Laurent, James Lu, Clinton R. Paden, Suxiang Tong, Duncan MacCannell |
| EPI_ISL_1337376, EPI_ISL_1337377 | LESP Guanajuato | Instituto de Diagnostico y Referencia Epidemiologicos (INDRE) | Claudia Wong-Arambula, Abril Rodriguez-Maldonado, Vanessa Rivero-Arredondo, Ariadna Medina-Benitez, Joaquin Quiroz-Mercado, Sergio Rangel-Guerrero, Natividad Cruz-Ortiz, Tatiana Nunez-Garcia, Gisela Barrera-Badillo, Lucia Hernandez-Rivas, Irma Lopez-Martinez, Ernesto Ramirez-Gonzalez. |
| EPI_ISL_1337378 | Helix/Illumina | Centers for Disease Control and Prevention Division of Viral Diseases, Pathogen Discovery | Peter W. Cook, Dakota Howard, Dhwani Batra, Ben L. Rambo-Martin, Eileen de Feo, Jan Antico, Christine Tran, Matthew Tolentino, Shannon Wickline, Kim Gietzen, Brad Sickler, Jingtao Liu, Eric Allen, Phil Febbo, Summer Galloway, Nicole L. Washington, Simon White, Geraint Levan, Kelly Schiabor Barrett, Elizabeth Cirulli, Alexandre Bolze, Ary Ascencio, Charlotte Rivera-Garcia, Ryan Cho, Jason Nguyen, Sherry Wang, Jimmy Ramirez, Tyler Cassens, Efren Sandoval, Magnus Isaksson, William Lee, David Becker, Marc Laurent, James Lu, Clinton R. Paden, Suxiang Tong, Duncan MacCannell |
| EPI_ISL_1337379, EPI_ISL_1337380 | LESP Guanajuato | Instituto de Diagnostico y Referencia Epidemiologicos (INDRE) | Claudia Wong-Arambula, Abril Rodriguez-Maldonado, Vanessa Rivero-Arredondo, Ariadna Medina-Benitez, Joaquin Quiroz-Mercado, Sergio Rangel-Guerrero, Natividad Cruz-Ortiz, Tatiana Nunez-Garcia, Gisela Barrera-Badillo, Lucia Hernandez-Rivas, Irma Lopez-Martinez, Ernesto Ramirez-Gonzalez. |
| EPI_ISL_1337381 | Helix/Illumina | Centers for Disease Control and Prevention Division of Viral Diseases, Pathogen Discovery | Peter W. Cook, Dakota Howard, Dhwani Batra, Ben L. Rambo-Martin, Eileen de Feo, Jan Antico, Christine Tran, Matthew Tolentino, Shannon Wickline, Kim Gietzen, Brad Sickler, Jingtao Liu, Eric Allen, Phil Febbo, Summer Galloway, Nicole L. Washington, Simon White, Geraint Levan, Kelly Schiabor Barrett, Elizabeth Cirulli, Alexandre Bolze, Ary Ascencio, Charlotte Rivera-Garcia, Ryan Cho, Jason Nguyen, Sherry Wang, Jimmy Ramirez, Tyler Cassens, Efren Sandoval, Magnus Isaksson, William Lee, David Becker, Marc Laurent, James Lu, Clinton R. Paden, Suxiang Tong, Duncan MacCannell |
| EPI_ISL_1337382, EPI_ISL_1337383, EPI_ISL_1337384 | LESP Guanajuato | Instituto de Diagnostico y Referencia Epidemiologicos (INDRE) | Claudia Wong-Arambula, Abril Rodriguez-Maldonado, Vanessa Rivero-Arredondo, Ariadna Medina-Benitez, Joaquin Quiroz-Mercado, Sergio Rangel-Guerrero, Natividad Cruz-Ortiz, Tatiana Nunez-Garcia, Gisela Barrera-Badillo, Lucia Hernandez-Rivas, Irma Lopez-Martinez, Ernesto Ramirez-Gonzalez. |
| EPI_ISL_1337385 | Helix/Illumina | Centers for Disease Control and Prevention Division of Viral Diseases, Pathogen Discovery | Peter W. Cook, Dakota Howard, Dhwani Batra, Ben L. Rambo-Martin, Eileen de Feo, Jan Antico, Christine Tran, Matthew Tolentino, Shannon Wickline, Kim Gietzen, Brad Sickler, Jingtao Liu, Eric Allen, Phil Febbo, Summer Galloway, Nicole L. Washington, Simon White, Geraint Levan, Kelly Schiabor Barrett, Elizabeth Cirulli, Alexandre Bolze, Ary Ascencio, Charlotte Rivera-Garcia, Ryan Cho, Jason Nguyen, Sherry Wang, Jimmy Ramirez, Tyler Cassens, Efren Sandoval, Magnus Isaksson, William Lee, David Becker, Marc Laurent, James Lu, Clinton R. Paden, Suxiang Tong, Duncan MacCannell |
| EPI_ISL_1337386, EPI_ISL_1337387 | LESP Guanajuato | Instituto de Diagnostico y Referencia Epidemiologicos (INDRE) | Claudia Wong-Arambula, Abril Rodriguez-Maldonado, Vanessa Rivero-Arredondo, Ariadna Medina-Benitez, Joaquin Quiroz-Mercado, Sergio Rangel-Guerrero, Natividad Cruz-Ortiz, Tatiana Nunez-Garcia, Gisela Barrera-Badillo, Lucia Hernandez-Rivas, Irma Lopez-Martinez, Ernesto Ramirez-Gonzalez. |
| EPI_ISL_1337388 | Helix/Illumina | Centers for Disease Control and Prevention Division of Viral Diseases, Pathogen Discovery | Peter W. Cook, Dakota Howard, Dhwani Batra, Ben L. Rambo-Martin, Eileen de Feo, Jan Antico, Christine Tran, Matthew Tolentino, Shannon Wickline, Kim Gietzen, Brad Sickler, Jingtao Liu, Eric Allen, Phil Febbo, Summer Galloway, Nicole L. Washington, Simon White, Geraint Levan, Kelly Schiabor Barrett, Elizabeth Cirulli, Alexandre Bolze, Ary Ascencio, Charlotte Rivera-Garcia, Ryan Cho, Jason Nguyen, Sherry Wang, Jimmy Ramirez, Tyler Cassens, Efren Sandoval, Magnus Isaksson, William Lee, David Becker, Marc Laurent, James Lu, Clinton R. Paden, Suxiang Tong, Duncan MacCannell |
| EPI_ISL_1337389 | LESP Sinaloa | Instituto de Diagnostico y Referencia Epidemiologicos (INDRE) | Claudia Wong-Arambula, Abril Rodriguez-Maldonado, Vanessa Rivero-Arredondo, Ariadna Medina-Benitez, Joaquin Quiroz-Mercado, Sergio Rangel-Guerrero, Natividad Cruz-Ortiz, Tatiana Nunez-Garcia, Gisela Barrera-Badillo, Lucia Hernandez-Rivas, Irma Lopez-Martinez, Ernesto Ramirez-Gonzalez. |
| EPI_ISL_1337390 | LESP Baja California | Instituto de Diagnostico y Referencia Epidemiologicos (INDRE) | Claudia Wong-Arambula, Abril Rodriguez-Maldonado, Vanessa Rivero-Arredondo, Ariadna Medina-Benitez, Joaquin Quiroz-Mercado, Sergio Rangel-Guerrero, Natividad Cruz-Ortiz, Tatiana Nunez-Garcia, Gisela Barrera-Badillo, Lucia Hernandez-Rivas, Irma Lopez-Martinez, Ernesto Ramirez-Gonzalez. |
| EPI_ISL_1337391 | Helix/Illumina | Centers for Disease Control and Prevention Division of Viral Diseases, Pathogen Discovery | Peter W. Cook, Dakota Howard, Dhwani Batra, Ben L. Rambo-Martin, Eileen de Feo, Jan Antico, Christine Tran, Matthew Tolentino, Shannon Wickline, Kim Gietzen, Brad Sickler, Jingtao Liu, Eric Allen, Phil Febbo, Summer Galloway, Nicole L. Washington, Simon White, Geraint Levan, Kelly Schiabor Barrett, Elizabeth Cirulli, Alexandre Bolze, Ary Ascencio, Charlotte Rivera-Garcia, Ryan Cho, Jason Nguyen, Sherry Wang, Jimmy Ramirez, Tyler Cassens, Efren Sandoval, Magnus Isaksson, William Lee, David Becker, Marc Laurent, James Lu, Clinton R. Paden, Suxiang Tong, Duncan MacCannell |
| EPI_ISL_1337392, EPI_ISL_1337393 | LESP Baja California Sur | Instituto de Diagnostico y Referencia Epidemiologicos (INDRE) | Claudia Wong-Arambula, Abril Rodriguez-Maldonado, Vanessa Rivero-Arredondo, Ariadna Medina-Benitez, Joaquin Quiroz-Mercado, Sergio Rangel-Guerrero, Natividad Cruz-Ortiz, Tatiana Nunez-Garcia, Gisela Barrera-Badillo, Lucia Hernandez-Rivas, Irma Lopez-Martinez, Ernesto Ramirez-Gonzalez. |
| EPI_ISL_1337394 | Helix/Illumina | Centers for Disease Control and Prevention Division of Viral Diseases, Pathogen Discovery | Peter W. Cook, Dakota Howard, Dhwani Batra, Ben L. Rambo-Martin, Eileen de Feo, Jan Antico, Christine Tran, Matthew Tolentino, Shannon Wickline, Kim Gietzen, Brad Sickler, Jingtao Liu, Eric Allen, Phil Febbo, Summer Galloway, Nicole L. Washington, Simon White, Geraint Levan, Kelly Schiabor Barrett, Elizabeth Cirulli, Alexandre Bolze, Ary Ascencio, Charlotte Rivera-Garcia, Ryan Cho, Jason Nguyen, Sherry Wang, Jimmy Ramirez, Tyler Cassens, Efren Sandoval, Magnus Isaksson, William Lee, David Becker, Marc Laurent, James Lu, Clinton R. Paden, Suxiang Tong, Duncan MacCannell |
| EPI_ISL_1337395 | LESP Colima | Instituto de Diagnostico y Referencia Epidemiologicos (INDRE) | Claudia Wong-Arambula, Abril Rodriguez-Maldonado, Vanessa Rivero-Arredondo, Ariadna Medina-Benitez, Joaquin Quiroz-Mercado, Sergio Rangel-Guerrero, Natividad Cruz-Ortiz, Tatiana Nunez-Garcia, Gisela Barrera-Badillo, Lucia Hernandez-Rivas, Irma Lopez-Martinez, Ernesto Ramirez-Gonzalez. |
| EPI_ISL_1337396, EPI_ISL_1337397, EPI_ISL_1337398, EPI_ISL_1337399, EPI_ISL_1337400, EPI_ISL_1337401, EPI_ISL_1337402, EPI_ISL_1337403, EPI_ISL_1337404, EPI_ISL_1337405, EPI_ISL_1337406, EPI_ISL_1337407, EPI_ISL_1337408, EPI_ISL_1337409, EPI_ISL_1337410, EPI_ISL_1337411, EPI_ISL_1337412, EPI_ISL_1337413, EPI_ISL_1337414, EPI_ISL_1337415, EPI_ISL_1337416, EPI_ISL_1337417, EPI_ISL_1337418, EPI_ISL_1337419, EPI_ISL_1337420, EPI_ISL_1337421, EPI_ISL_1337422, EPI_ISL_1337423, EPI_ISL_1337424, EPI_ISL_1337425, EPI_ISL_1337426, EPI_ISL_1337427, EPI_ISL_1337428, EPI_ISL_1337429, EPI_ISL_1337430, EPI_ISL_1337431, EPI_ISL_1337432, EPI_ISL_1337433, EPI_ISL_1337434, EPI_ISL_1337435, EPI_ISL_1337436, EPI_ISL_1337437, EPI_ISL_1337438, EPI_ISL_1337440, EPI_ISL_1337441 | | | |
| see above | Helix/Illumina | Centers for Disease Control and Prevention Division of Viral Diseases, Pathogen Discovery | Peter W. Cook, Dakota Howard, Dhwani Batra, Ben L. Rambo-Martin, Eileen de Feo, Jan Antico, Christine Tran, Matthew Tolentino, Shannon Wickline, Kim Gietzen, Brad Sickler, Jingtao Liu, Eric Allen, Phil Febbo, Summer Galloway, Nicole L. Washington, Simon White, Geraint Levan, Kelly Schiabor Barrett, Elizabeth Cirulli, Alexandre Bolze, Ary Ascencio, Charlotte Rivera-Garcia, Ryan Cho, Jason Nguyen, Sherry Wang, Jimmy Ramirez, Tyler Cassens, Efren Sandoval, Magnus Isaksson, William Lee, David Becker, Marc Laurent, James Lu, Clinton R. Paden, Suxiang Tong, Duncan MacCannell |
| EPI_ISL_1337463 | DOHMH Morrisania | New York City Public Health Laboratory | Jade Wang, et al. |
| EPI_ISL_1337464 | DOHMH Chelsea | New York City Public Health Laboratory | Jade Wang, et al. |
| EPI_ISL_1337465 | DOHMH Central Harlem | New York City Public Health Laboratory | Jade Wang, et al. |
| EPI_ISL_1337466 | Department of Homeless Services | New York City Public Health Laboratory | Jade Wang, et al. |
| EPI_ISL_1337467 | DOHMH Corona | New York City Public Health Laboratory | Jade Wang, et al. |
| EPI_ISL_1337468 | DOHMH Fort Greene | New York City Public Health Laboratory | Jade Wang, et al. |
| EPI_ISL_1337469 | DOHMH Jamaica | New York City Public Health Laboratory | Jade Wang, et al. |
| EPI_ISL_1337470 | DOHMH Morrisania | New York City Public Health Laboratory | Jade Wang, et al. |
| EPI_ISL_1337471 | DOHMH Fort Greene | New York City Public Health Laboratory | Jade Wang, et al. |
| EPI_ISL_1337472 | Department of Homeless Services | New York City Public Health Laboratory | Jade Wang, et al. |
| EPI_ISL_1337473 | DOHMH Morrisania | New York City Public Health Laboratory | Jade Wang, et al. |
| EPI_ISL_1337474 | DOHMH Crown Heights | New York City Public Health Laboratory | Jade Wang, et al. |
| EPI_ISL_1337475 | DOHMH Corona | New York City Public Health Laboratory | Jade Wang, et al. |
| EPI_ISL_1337476 | DOHMH Central Harlem | New York City Public Health Laboratory | Jade Wang, et al. |
| EPI_ISL_1337477 | DOHMH Corona | New York City Public Health Laboratory | Jade Wang, et al. |
| EPI_ISL_1337478 | DOHMH Morrisania | New York City Public Health Laboratory | Jade Wang, et al. |
| EPI_ISL_1337479 | DOHMH Chelsea | New York City Public Health Laboratory | Jade Wang, et al. |
| EPI_ISL_1337480 | DOHMH Riverside | New York City Public Health Laboratory | Jade Wang, et al. |
| EPI_ISL_1337481 | DOHMH Morrisania | New York City Public Health Laboratory | Jade Wang, et al. |
| EPI_ISL_1337482 | OCME Office Of Chief Medical Examiner | New York City Public Health Laboratory | Jade Wang, et al. |
| EPI_ISL_1337483 | DOHMH Riverside | New York City Public Health Laboratory | Jade Wang, et al. |
| EPI_ISL_1337484 | DOHMH Corona | New York City Public Health Laboratory | Jade Wang, et al. |
| EPI_ISL_1337485 | DOHMH Morrisania | New York City Public Health Laboratory | Jade Wang, et al. |
| EPI_ISL_1337486, EPI_ISL_1337487 | DOHMH Jamaica | New York City Public Health Laboratory | Jade Wang, et al. |
| EPI_ISL_1337488 | DOHMH Crown Heights | New York City Public Health Laboratory | Jade Wang, et al. |
| EPI_ISL_1337489 | DOHMH Central Harlem | New York City Public Health Laboratory | Jade Wang, et al. |
| EPI_ISL_1337490 | DOHMH Crown Heights | New York City Public Health Laboratory | Jade Wang, et al. |
| EPI_ISL_1337491 | DOHMH Corona | New York City Public Health Laboratory | Jade Wang, et al. |
| EPI_ISL_1337492, EPI_ISL_1337493, EPI_ISL_1337494 | DOHMH Crown Heights | New York City Public Health Laboratory | Jade Wang, et al. |
| EPI_ISL_1337495, EPI_ISL_1337496, EPI_ISL_1337497, EPI_ISL_1337498 | DOHMH Jamaica | New York City Public Health Laboratory | Jade Wang, et al. |
| EPI_ISL_1337499 | DOHMH Crown Heights | New York City Public Health Laboratory | Jade Wang, et al. |
| EPI_ISL_1337500 | DOHMH Chelsea | New York City Public Health Laboratory | Jade Wang, et al. |
| EPI_ISL_1337501 | DOHMH Jamaica | New York City Public Health Laboratory | Jade Wang, et al. |

| | | | |
|---|---|---|---|
| EPI_ISL_1337502 | DOHMH Morrisania | New York City Public Health Laboratory | Jade Wang, et al. |
| EPI_ISL_1337503 | DOHMH Riverside | New York City Public Health Laboratory | Jade Wang, et al. |
| EPI_ISL_1337504, EPI_ISL_1337505 | DOHMH Jamaica | New York City Public Health Laboratory | Jade Wang, et al. |
| EPI_ISL_1337506 | DOHMH Morrisania | New York City Public Health Laboratory | Jade Wang, et al. |
| EPI_ISL_1337507 | DOHMH Corona | New York City Public Health Laboratory | Jade Wang, et al. |
| EPI_ISL_1337508, EPI_ISL_1337509 | DOHMH PHL | New York City Public Health Laboratory | Jade Wang, et al. |
| EPI_ISL_1337510 | DOHMH Fort Greene | New York City Public Health Laboratory | Jade Wang, et al. |
| EPI_ISL_1337511, EPI_ISL_1337512 | DOHMH Corona | New York City Public Health Laboratory | Jade Wang, et al. |
| EPI_ISL_1337513 | DOHMH Jamaica | New York City Public Health Laboratory | Jade Wang, et al. |
| EPI_ISL_1337514, EPI_ISL_1337515 | DOHMH Corona | New York City Public Health Laboratory | Jade Wang, et al. |
| EPI_ISL_1337516 | DOHMH PHL | New York City Public Health Laboratory | Jade Wang, et al. |
| EPI_ISL_1337517 | DOHMH Central Harlem | New York City Public Health Laboratory | Jade Wang, et al. |
| EPI_ISL_1337518, EPI_ISL_1337519 | OCME Office Of Chief Medical Examiner | New York City Public Health Laboratory | Jade Wang, et al. |
| EPI_ISL_1337520 | DOHMH Central Harlem | New York City Public Health Laboratory | Jade Wang, et al. |
| EPI_ISL_1337521, EPI_ISL_1337522 | DOHMH Jamaica | New York City Public Health Laboratory | Jade Wang, et al. |
| EPI_ISL_1337523 | DOHMH Morrisania | New York City Public Health Laboratory | Jade Wang, et al. |
| EPI_ISL_1337524 | OCME Office Of Chief Medical Examiner | New York City Public Health Laboratory | Jade Wang, et al. |
| EPI_ISL_1337525 | DOHMH Jamaica | New York City Public Health Laboratory | Jade Wang, et al. |
| EPI_ISL_1337526 | DOHMH Riverside | New York City Public Health Laboratory | Jade Wang, et al. |
| EPI_ISL_1337527 | OCME Office Of Chief Medical Examiner | New York City Public Health Laboratory | Jade Wang, et al. |
| EPI_ISL_1337528 | DOHMH Crown Heights | New York City Public Health Laboratory | Jade Wang, et al. |
| EPI_ISL_1337529 | OCME Office Of Chief Medical Examiner | New York City Public Health Laboratory | Jade Wang, et al. |
| EPI_ISL_1337530 | DOHMH PHL | New York City Public Health Laboratory | Jade Wang, et al. |
| EPI_ISL_1337531 | DOHMH Corona | New York City Public Health Laboratory | Jade Wang, et al. |
| EPI_ISL_1337532, EPI_ISL_1337533 | DOHMH Jamaica | New York City Public Health Laboratory | Jade Wang, et al. |
| EPI_ISL_1337534, EPI_ISL_1337535 | DOHMH Crown Heights | New York City Public Health Laboratory | Jade Wang, et al. |
| EPI_ISL_1337536 | DOHMH Central Harlem | New York City Public Health Laboratory | Jade Wang, et al. |
| EPI_ISL_1337537, EPI_ISL_1337538 | DOHMH Jamaica | New York City Public Health Laboratory | Jade Wang, et al. |
| EPI_ISL_1337539 | OCME Office Of Chief Medical Examiner | New York City Public Health Laboratory | Jade Wang, et al. |
| EPI_ISL_1337540 | DOHMH Morrisania | New York City Public Health Laboratory | Jade Wang, et al. |
| EPI_ISL_1337541 | OCME Office Of Chief Medical Examiner | New York City Public Health Laboratory | Jade Wang, et al. |
| EPI_ISL_1337542 | DOHMH Chelsea | New York City Public Health Laboratory | Jade Wang, et al. |
| EPI_ISL_1337543 | DOHMH Riverside | New York City Public Health Laboratory | Jade Wang, et al. |
| EPI_ISL_1337544 | DOHMH Crown Heights | New York City Public Health Laboratory | Jade Wang, et al. |
| EPI_ISL_1337545 | DOHMH Morrisania | New York City Public Health Laboratory | Jade Wang, et al. |
| EPI_ISL_1337546 | OCME Office Of Chief Medical Examiner | New York City Public Health Laboratory | Jade Wang, et al. |
| EPI_ISL_1337547 | DOHMH Riverside | New York City Public Health Laboratory | Jade Wang, et al. |
| EPI_ISL_1337548, EPI_ISL_1337549 | DOHMH Central Harlem | New York City Public Health Laboratory | Jade Wang, et al. |
| EPI_ISL_1337550 | DOHMH Jamaica | New York City Public Health Laboratory | Jade Wang, et al. |
| EPI_ISL_1337551 | DOHMH Corona | New York City Public Health Laboratory | Jade Wang, et al. |
| EPI_ISL_1337556, EPI_ISL_1337572, EPI_ISL_1337573, EPI_ISL_1337576, EPI_ISL_1337578, EPI_ISL_1337585, EPI_ISL_1337586, EPI_ISL_1337588, EPI_ISL_1337590, EPI_ISL_1337595, EPI_ISL_1337600, EPI_ISL_1337601, EPI_ISL_1337602, EPI_ISL_1337609, EPI_ISL_1337610, EPI_ISL_1337611, EPI_ISL_1337612, EPI_ISL_1337614, EPI_ISL_1337616, EPI_ISL_1337617, EPI_ISL_1337618, EPI_ISL_1337619, EPI_ISL_1337620, EPI_ISL_1337621, EPI_ISL_1337622, EPI_ISL_1337623, EPI_ISL_1337624, EPI_ISL_1337625, EPI_ISL_1337626, EPI_ISL_1337627, EPI_ISL_1337628, EPI_ISL_1337629, EPI_ISL_1337631, EPI_ISL_1337632, EPI_ISL_1337633, EPI_ISL_1337634, EPI_ISL_1337635, EPI_ISL_1337636, EPI_ISL_1337637, EPI_ISL_1337638, EPI_ISL_1337639, EPI_ISL_1337640, EPI_ISL_1337641, EPI_ISL_1337642, EPI_ISL_1337643, EPI_ISL_1337644, EPI_ISL_1337645, EPI_ISL_1337646, EPI_ISL_1337647, EPI_ISL_1337648, EPI_ISL_1337650, EPI_ISL_1337652, EPI_ISL_1337653, EPI_ISL_1337654, EPI_ISL_1337655, EPI_ISL_1337656, EPI_ISL_1337657, EPI_ISL_1337658, EPI_ISL_1337659, EPI_ISL_1337660, EPI_ISL_1337662, EPI_ISL_1337663, EPI_ISL_1337664, EPI_ISL_1337665 | | | |
| see above | Wyoming Public Health Laboratory | Wyoming Public Health Laboratory | Jim Mildenberger, Wanda Manley, Noah Hull, Taylor Fearing, Lynette Gumbleton, Channing Weber, Ashley Norberg, Chayse Rowley, Marley Goetz, Brian Dominguez, Elliot Thomasson, Cari Sloma, and Rob Christensen |
| EPI_ISL_1337669, EPI_ISL_1337670, EPI_ISL_1337671, EPI_ISL_1337672, EPI_ISL_1337673, EPI_ISL_1337674, EPI_ISL_1337675, EPI_ISL_1337676, EPI_ISL_1337677, EPI_ISL_1337678, EPI_ISL_1337679, EPI_ISL_1337680, EPI_ISL_1337681, EPI_ISL_1337682, EPI_ISL_1337683, EPI_ISL_1337684, EPI_ISL_1337685, EPI_ISL_1337686, EPI_ISL_1337687, EPI_ISL_1337688, EPI_ISL_1337689, EPI_ISL_1337690, EPI_ISL_1337691, EPI_ISL_1337692, EPI_ISL_1337693, EPI_ISL_1337694, EPI_ISL_1337695, EPI_ISL_1337696, EPI_ISL_1337697, EPI_ISL_1337698, EPI_ISL_1337699, EPI_ISL_1337700, EPI_ISL_1337701, EPI_ISL_1337702, EPI_ISL_1337703, EPI_ISL_1337704, EPI_ISL_1337705, EPI_ISL_1337706, EPI_ISL_1337707, EPI_ISL_1337708, EPI_ISL_1337709, EPI_ISL_1337710, EPI_ISL_1337711, EPI_ISL_1337712, EPI_ISL_1337713, EPI_ISL_1337714, EPI_ISL_1337715, EPI_ISL_1337716, EPI_ISL_1337717, EPI_ISL_1337718, EPI_ISL_1337719, EPI_ISL_1337720, EPI_ISL_1337721, EPI_ISL_1337722, EPI_ISL_1337723, EPI_ISL_1337724, EPI_ISL_1337725, EPI_ISL_1337726, EPI_ISL_1337727, EPI_ISL_1337728, EPI_ISL_1337729, EPI_ISL_1337730, EPI_ISL_1337731, EPI_ISL_1337732, EPI_ISL_1337733, EPI_ISL_1337734, EPI_ISL_1337735, EPI_ISL_1337736, EPI_ISL_1337737, EPI_ISL_1337738, EPI_ISL_1337739, EPI_ISL_1337740, EPI_ISL_1337741, EPI_ISL_1337742, EPI_ISL_1337743, EPI_ISL_1337744, EPI_ISL_1337745, EPI_ISL_1337746, EPI_ISL_1337747, EPI_ISL_1337748, EPI_ISL_1337749, EPI_ISL_1337750, EPI_ISL_1337751, EPI_ISL_1337752, EPI_ISL_1337753, EPI_ISL_1337754, EPI_ISL_1337755, EPI_ISL_1337756, EPI_ISL_1337757, EPI_ISL_1337758, EPI_ISL_1337759, EPI_ISL_1337760, EPI_ISL_1337761, EPI_ISL_1337762 | | | |
| see above | TXDSHS | TXDSHS | Rashmi Tuladhar, Bonnie Oh, Jenny Zhang, Maliha Rahman, Mayela Pedrueza, Anita Pokharel, Loarraine Rodriguez, Myong Koag, Chun Wang, Rachel Lee, Grace Kubin |
| EPI_ISL_1337989, EPI_ISL_1337990, EPI_ISL_1337991, EPI_ISL_1337992, EPI_ISL_1337993, EPI_ISL_1337994, EPI_ISL_1337995, EPI_ISL_1337996, EPI_ISL_1337997, EPI_ISL_1337998, EPI_ISL_1337999, EPI_ISL_1338000, EPI_ISL_1338001, EPI_ISL_1338002 | | | |
| see above | DC Public Health Lab/ Dept. of Forensic Sciences | DC Public Health Lab/ Dept. of Forensic Sciences | Janis Doss, Scott Nguyen, Elizabeth Zelaya, Sarah Scott, Connie Maza, Monica Mann, Brittany Hamilton, David Payne, Jocelyn Hauser |
| EPI_ISL_1338003 | Kansas Health and Environmental Lab | Kansas Health and Environmental Lab | Mike Grose, Jonathan Barnell, Ben Olsen, and Phil Adam |
| EPI_ISL_1338004, EPI_ISL_1338005, EPI_ISL_1338006, EPI_ISL_1338007, EPI_ISL_1338008, EPI_ISL_1338009, EPI_ISL_1338010, EPI_ISL_1338011, EPI_ISL_1338012, EPI_ISL_1338013, EPI_ISL_1338014, EPI_ISL_1338015 | | | |
| see above | Cytocheck Laboratory | Kansas Health and Environmental Lab | Mike Grose, Jonathan Barnell, Ben Olsen, and Phil Adam |
| EPI_ISL_1338016, EPI_ISL_1338017, EPI_ISL_1338018, EPI_ISL_1338019, EPI_ISL_1338020, EPI_ISL_1338021, EPI_ISL_1338022, EPI_ISL_1338023, EPI_ISL_1338024, EPI_ISL_1338025, EPI_ISL_1338026, EPI_ISL_1338027, EPI_ISL_1338028, EPI_ISL_1338029, EPI_ISL_1338030, EPI_ISL_1338031, EPI_ISL_1338032, EPI_ISL_1338033, EPI_ISL_1338034, EPI_ISL_1338035, EPI_ISL_1338036, EPI_ISL_1338037, EPI_ISL_1338038, EPI_ISL_1338039, EPI_ISL_1338040, EPI_ISL_1338041, EPI_ISL_1338042, EPI_ISL_1338043, EPI_ISL_1338044, EPI_ISL_1338045, EPI_ISL_1338046, EPI_ISL_1338047, EPI_ISL_1338048, EPI_ISL_1338049, EPI_ISL_1338050, EPI_ISL_1338051, EPI_ISL_1338052, EPI_ISL_1338053, EPI_ISL_1338054, EPI_ISL_1338055, EPI_ISL_1338056, EPI_ISL_1338057, EPI_ISL_1338058, EPI_ISL_1338059, EPI_ISL_1338060, EPI_ISL_1338061, EPI_ISL_1338062, EPI_ISL_1338063, EPI_ISL_1338064, EPI_ISL_1338065, EPI_ISL_1338066, EPI_ISL_1338068, EPI_ISL_1338069, EPI_ISL_1338070, EPI_ISL_1338071, EPI_ISL_1338072, EPI_ISL_1338073, EPI_ISL_1338074, EPI_ISL_1338075, EPI_ISL_1338076, EPI_ISL_1338079, EPI_ISL_1338080, EPI_ISL_1338081, EPI_ISL_1338082, EPI_ISL_1338083, EPI_ISL_1338084, EPI_ISL_1338085, EPI_ISL_1338086, EPI_ISL_1338087, EPI_ISL_1338088, EPI_ISL_1338089, EPI_ISL_1338090, EPI_ISL_1338091, EPI_ISL_1338092, EPI_ISL_1338093, EPI_ISL_1338094, EPI_ISL_1338095, EPI_ISL_1338096, EPI_ISL_1338097, EPI_ISL_1338098, EPI_ISL_1338099, EPI_ISL_1338100, EPI_ISL_1338101, EPI_ISL_1338102, EPI_ISL_1338103, EPI_ISL_1338104, EPI_ISL_1338105, EPI_ISL_1338106, EPI_ISL_1338107, EPI_ISL_1338108, EPI_ISL_1338109, EPI_ISL_1338110, EPI_ISL_1338111, EPI_ISL_1338112, EPI_ISL_1338113, EPI_ISL_1338114, EPI_ISL_1338115, EPI_ISL_1338116, EPI_ISL_1338117, EPI_ISL_1338118, EPI_ISL_1338119, EPI_ISL_1338120, EPI_ISL_1338121, EPI_ISL_1338122, EPI_ISL_1338123, EPI_ISL_1338124, EPI_ISL_1338125, EPI_ISL_1338126, EPI_ISL_1338127, EPI_ISL_1338128, EPI_ISL_1338129, EPI_ISL_1338130, EPI_ISL_1338131, EPI_ISL_1338132, EPI_ISL_1338133, EPI_ISL_1338134, EPI_ISL_1338135, EPI_ISL_1338136, EPI_ISL_1338137, EPI_ISL_1338138, EPI_ISL_1338139, EPI_ISL_1338140, EPI_ISL_1338141, EPI_ISL_1338142, EPI_ISL_1338143, EPI_ISL_1338144, EPI_ISL_1338145, EPI_ISL_1338146, EPI_ISL_1338147, EPI_ISL_1338148, EPI_ISL_1338149, EPI_ISL_1338150, EPI_ISL_1338151, EPI_ISL_1338152, EPI_ISL_1338153, EPI_ISL_1338154, EPI_ISL_1338155, EPI_ISL_1338156, EPI_ISL_1338157, EPI_ISL_1338159, EPI_ISL_1338160, EPI_ISL_1338161, EPI_ISL_1338163, EPI_ISL_1338164, EPI_ISL_1338165, EPI_ISL_1338166, EPI_ISL_1338167, EPI_ISL_1338168, EPI_ISL_1338169, EPI_ISL_1338170, EPI_ISL_1338171, EPI_ISL_1338172, EPI_ISL_1338173, EPI_ISL_1338174, EPI_ISL_1338175, EPI_ISL_1338176, EPI_ISL_1338177, EPI_ISL_1338178, EPI_ISL_1338179, EPI_ISL_1338180, EPI_ISL_1338181, EPI_ISL_1338182, EPI_ISL_1338183, EPI_ISL_1338184, EPI_ISL_1338185, EPI_ISL_1338186, EPI_ISL_1338187, EPI_ISL_1338188, EPI_ISL_1338189, EPI_ISL_1338190, EPI_ISL_1338191, EPI_ISL_1338192, EPI_ISL_1338193, EPI_ISL_1338194, EPI_ISL_1338195, EPI_ISL_1338196, EPI_ISL_1338197, EPI_ISL_1338198, EPI_ISL_1338199, EPI_ISL_1338200, EPI_ISL_1338201, EPI_ISL_1338202, EPI_ISL_1338203, EPI_ISL_1338204, EPI_ISL_1338205, EPI_ISL_1338206, EPI_ISL_1338207, EPI_ISL_1338208, EPI_ISL_1338209, EPI_ISL_1338210, EPI_ISL_1338211, EPI_ISL_1338212, EPI_ISL_1338213, EPI_ISL_1338214, EPI_ISL_1338215, EPI_ISL_1338216, EPI_ISL_1338217, EPI_ISL_1338218, EPI_ISL_1338219, EPI_ISL_1338220, EPI_ISL_1338221, EPI_ISL_1338222, EPI_ISL_1338223, EPI_ISL_1338224, EPI_ISL_1338225, EPI_ISL_1338226, EPI_ISL_1338227, EPI_ISL_1338228, EPI_ISL_1338229, EPI_ISL_1338230, EPI_ISL_1338231, EPI_ISL_1338232, EPI_ISL_1338233, EPI_ISL_1338234, EPI_ISL_1338235, EPI_ISL_1338236, EPI_ISL_1338237, EPI_ISL_1338238, EPI_ISL_1338239, EPI_ISL_1338240, EPI_ISL_1338241, EPI_ISL_1338242, EPI_ISL_1338243, EPI_ISL_1338244, EPI_ISL_1338245, EPI_ISL_1338246, EPI_ISL_1338247, EPI_ISL_1338248, EPI_ISL_1338249, EPI_ISL_1338250, EPI_ISL_1338251, EPI_ISL_1338252, EPI_ISL_1338253, EPI_ISL_1338254, EPI_ISL_1338255, EPI_ISL_1338256, EPI_ISL_1338257, EPI_ISL_1338258, EPI_ISL_1338259, EPI_ISL_1338260, EPI_ISL_1338261, EPI_ISL_1338262, EPI_ISL_1338263, EPI_ISL_1338264, EPI_ISL_1338265, EPI_ISL_1338266, EPI_ISL_1338267, EPI_ISL_1338268, EPI_ISL_1338269, EPI_ISL_1338270, EPI_ISL_1338271, EPI_ISL_1338272, EPI_ISL_1338273, EPI_ISL_1338274, EPI_ISL_1338275, EPI_ISL_1338276, EPI_ISL_1338277, EPI_ISL_1338278, EPI_ISL_1338279, EPI_ISL_1338280, EPI_ISL_1338281, EPI_ISL_1338282, EPI_ISL_1338283, EPI_ISL_1338284, EPI_ISL_1338285, EPI_ISL_1338286, EPI_ISL_1338287, EPI_ISL_1338288, EPI_ISL_1338289, EPI_ISL_1338290, EPI_ISL_1338291, EPI_ISL_1338292, EPI_ISL_1338294, EPI_ISL_1338295, EPI_ISL_1338296, EPI_ISL_1338297, EPI_ISL_1338298, EPI_ISL_1338299, EPI_ISL_1338300, EPI_ISL_1338301, EPI_ISL_1338302, EPI_ISL_1338303, EPI_ISL_1338304, EPI_ISL_1338305, EPI_ISL_1338306, EPI_ISL_1338307, EPI_ISL_1338308, EPI_ISL_1338309, EPI_ISL_1338310, EPI_ISL_1338311, EPI_ISL_1338312, EPI_ISL_1338313, EPI_ISL_1338314, EPI_ISL_1338315, EPI_ISL_1338316, EPI_ISL_1338317, EPI_ISL_1338318, EPI_ISL_1338319, EPI_ISL_1338320, EPI_ISL_1338321, EPI_ISL_1338322, EPI_ISL_1338323, EPI_ISL_1338324, EPI_ISL_1338325, EPI_ISL_1338326, EPI_ISL_1338327, EPI_ISL_1338328, EPI_ISL_1338329, EPI_ISL_1338330, EPI_ISL_1338331, EPI_ISL_1338332, EPI_ISL_1338333, EPI_ISL_1338334, EPI_ISL_1338335, EPI_ISL_1338336, EPI_ISL_1338337, EPI_ISL_1338338, EPI_ISL_1338339, EPI_ISL_1338340, EPI_ISL_1338341, EPI_ISL_1338342, EPI_ISL_1338343, EPI_ISL_1338344, EPI_ISL_1338345, EPI_ISL_1338346, EPI_ISL_1338347, EPI_ISL_1338348, EPI_ISL_1338349, EPI_ISL_1338350, EPI_ISL_1338351, EPI_ISL_1338352, EPI_ISL_1338353, EPI_ISL_1338354, EPI_ISL_1338355, EPI_ISL_1338356, EPI_ISL_1338357, EPI_ISL_1338358, EPI_ISL_1338359, EPI_ISL_1338360, EPI_ISL_1338361, EPI_ISL_1338362, EPI_ISL_1338363, EPI_ISL_1338364, EPI_ISL_1338365, EPI_ISL_1338366, EPI_ISL_1338367, EPI_ISL_1338368, EPI_ISL_1338369, EPI_ISL_1338370, EPI_ISL_1338371, EPI_ISL_1338372, EPI_ISL_1338373, EPI_ISL_1338374, EPI_ISL_1338375, EPI_ISL_1338376, EPI_ISL_1338377, EPI_ISL_1338378, EPI_ISL_1338379, EPI_ISL_1338380, EPI_ISL_1338381, EPI_ISL_1338382, EPI_ISL_1338383, EPI_ISL_1338384, EPI_ISL_1338385, EPI_ISL_1338386, EPI_ISL_1338387, EPI_ISL_1338388, EPI_ISL_1338389, EPI_ISL_1338390, EPI_ISL_1338391, EPI_ISL_1338392, EPI_ISL_1338393, EPI_ISL_1338394, EPI_ISL_1338395, EPI_ISL_1338396, EPI_ISL_1338397, EPI_ISL_1338398 | | | |
| see above | Helix/Illumina | Centers for Disease Control and Prevention Division of Viral Diseases, Pathogen Discovery | Peter W. Cook, Dakota Howard, Dhwani Batra, Ben L. Rambo-Martin, Eileen de Feo, Jan Antico, Christine Tran, Matthew Tolentino, Shannon Wickline, Kim Gietzen, Brad Sickler, Jingtao Liu, Eric Allen, Phil Febbo, Summer Galloway, Nicole L. Washington, Simon White, Geraint Levan, Kelly Schiabor Barrett, Elizabeth Cirulli, Alexandre Bolze, Ary Ascencio, Charlotte Rivera-Garcia, Ryan Cho, Jason Nguyen, Sherry Wang, Jimmy Ramirez, Tyler Cassens, Efren Sandoval, Magnus Isaksson, William Lee, David Becker, Marc Laurent, James Lu, Clinton R. Paden, Suxiang Tong, Duncan MacCannell |
| EPI_ISL_1338407, EPI_ISL_1338409, EPI_ISL_1338410, EPI_ISL_1338411, EPI_ISL_1338414, EPI_ISL_1338415, EPI_ISL_1338416, EPI_ISL_1338417, EPI_ISL_1338418, EPI_ISL_1338420, EPI_ISL_1338422, EPI_ISL_1338426, EPI_ISL_1338427, EPI_ISL_1338430, EPI_ISL_1338432, EPI_ISL_1338436, EPI_ISL_1338442, EPI_ISL_1338444, EPI_ISL_1338445, EPI_ISL_1338446, EPI_ISL_1338451, EPI_ISL_1338452, | | | |

| | | | |
|---|---|---|---|
| EPI_ISL_1338453, EPI_ISL_1338455, EPI_ISL_1338457, EPI_ISL_1338458, EPI_ISL_1338461, EPI_ISL_1338463, EPI_ISL_1338464, EPI_ISL_1338466, EPI_ISL_1338469, EPI_ISL_1338474, EPI_ISL_1338475, EPI_ISL_1338476, EPI_ISL_1338477, EPI_ISL_1338480, EPI_ISL_1338482, EPI_ISL_1338487, EPI_ISL_1338488, EPI_ISL_1338491, EPI_ISL_1338492, EPI_ISL_1338494, EPI_ISL_1338495, EPI_ISL_1338498, EPI_ISL_1338503, EPI_ISL_1338505, EPI_ISL_1338512, EPI_ISL_1338515, EPI_ISL_1338516, EPI_ISL_1338520, EPI_ISL_1338521, EPI_ISL_1338522, EPI_ISL_1338523, EPI_ISL_1338525, EPI_ISL_1338526, EPI_ISL_1338530, EPI_ISL_1338532, EPI_ISL_1338537, EPI_ISL_1338543, EPI_ISL_1338544, EPI_ISL_1338547, EPI_ISL_1338550, EPI_ISL_1338551, EPI_ISL_1338553, EPI_ISL_1338555, EPI_ISL_1338559, EPI_ISL_1338561, EPI_ISL_1338562, EPI_ISL_1338567, EPI_ISL_1338569, EPI_ISL_1338570, EPI_ISL_1338573, EPI_ISL_1338577, EPI_ISL_1338579, EPI_ISL_1338580, EPI_ISL_1338583, EPI_ISL_1338586, EPI_ISL_1338589, EPI_ISL_1338592, EPI_ISL_1338595, EPI_ISL_1338597, EPI_ISL_1338603, EPI_ISL_1338609, EPI_ISL_1338611 | | | |
| see above | Laboratory Corporation of America | Centers for Disease Control and Prevention Division of Viral Diseases, Pathogen Discovery | Peter W. Cook, Dakota Howard, Dhwani Batra, Ben L. Rambo-Martin, Minoo Agarwal, Eyad Almasri, Debbie Boles, Ayla Burns, Nuthawin Charoensri, Oren Cohen, Susan Countryman, Mary Ann Cristobal, Bobbi Croy, Suzanne Dale, Hrushikesh Deshmukh, Amanda Douglas, Vincent Drouillon, Marcia Eisenberg, Howard Engler, Rama Ghatti, Prashant Gupta, Susan Hicks, Jake Humphrey, Lax Iyer, Manoj Jain, Mohan Kolli, Brian Krueger, Tim Kuphal, Stanley Letovsky, Michael Levandoski, Craig Lukasik, Jonathan Meltzer, Brian Norvell, Mindy Nye, Scott Parker, Christos Petropoulos, John Pruitt, Steven Ragan, Scott Ryan, Mike Sapeta, Jana Schroth, Suresh Babu Selvaraju, Goran Stevovic, Amanda Suchanek, Andrea Throop, Lyndon Tilson, Thomas Urban, Joe Voshell, Kimberly Wagner, Jonathan Williams, Mary Williamson, Qian Zeng, Tricia Zwiefelhofer, Clinton R. Paden, Suxiang Tong, Duncan MacCannell |
| EPI_ISL_1338612, EPI_ISL_1338613, EPI_ISL_1338615, EPI_ISL_1338616 | WSU Molecular Diagnostics Laboratory | Kansas Health and Environmental Lab | Mike Grose, Jonathan Barnell, Ben Olsen, and Phil Adam |
| EPI_ISL_1338618, EPI_ISL_1338620, EPI_ISL_1338622, EPI_ISL_1338626, EPI_ISL_1338628, EPI_ISL_1338629, EPI_ISL_1338630, EPI_ISL_1338632, EPI_ISL_1338633, EPI_ISL_1338634, EPI_ISL_1338637, EPI_ISL_1338639, EPI_ISL_1338640, EPI_ISL_1338641, EPI_ISL_1338642, EPI_ISL_1338645, EPI_ISL_1338646, EPI_ISL_1338648, EPI_ISL_1338653, EPI_ISL_1338657, EPI_ISL_1338662, EPI_ISL_1338664, EPI_ISL_1338665, EPI_ISL_1338666, EPI_ISL_1338668, EPI_ISL_1338669, EPI_ISL_1338670, EPI_ISL_1338671, EPI_ISL_1338676, EPI_ISL_1338682, EPI_ISL_1338688, EPI_ISL_1338696, EPI_ISL_1338700, EPI_ISL_1338703, EPI_ISL_1338711, EPI_ISL_1338713, EPI_ISL_1338714, EPI_ISL_1338716, EPI_ISL_1338717, EPI_ISL_1338718, EPI_ISL_1338719, EPI_ISL_1338725, EPI_ISL_1338729, EPI_ISL_1338730, EPI_ISL_1338731, EPI_ISL_1338732, EPI_ISL_1338733, EPI_ISL_1338734, EPI_ISL_1338738, EPI_ISL_1338741, EPI_ISL_1338745, EPI_ISL_1338746, EPI_ISL_1338748, EPI_ISL_1338752, EPI_ISL_1338753, EPI_ISL_1338755, EPI_ISL_1338757, EPI_ISL_1338764, EPI_ISL_1338765, EPI_ISL_1338768, EPI_ISL_1338774, EPI_ISL_1338776, EPI_ISL_1338779, EPI_ISL_1338782, EPI_ISL_1338788, EPI_ISL_1338789, EPI_ISL_1338790, EPI_ISL_1338793, EPI_ISL_1338801, EPI_ISL_1338803, EPI_ISL_1338813, EPI_ISL_1338815, EPI_ISL_1338817, EPI_ISL_1338818, EPI_ISL_1338821, EPI_ISL_1338827, EPI_ISL_1338833, EPI_ISL_1338839, EPI_ISL_1338842, EPI_ISL_1338844, EPI_ISL_1338846, EPI_ISL_1338848, EPI_ISL_1338852, EPI_ISL_1338856, EPI_ISL_1338861, EPI_ISL_1338864, EPI_ISL_1338866, EPI_ISL_1338867, EPI_ISL_1338868, EPI_ISL_1338871, EPI_ISL_1338873, EPI_ISL_1338878, EPI_ISL_1338886, EPI_ISL_1338887, EPI_ISL_1338891, EPI_ISL_1338897, EPI_ISL_1338898, EPI_ISL_1338900, EPI_ISL_1338901, EPI_ISL_1338902, EPI_ISL_1338904, EPI_ISL_1338906, EPI_ISL_1338907, EPI_ISL_1338908, EPI_ISL_1338909, EPI_ISL_1338912, EPI_ISL_1338919, EPI_ISL_1338920, EPI_ISL_1338923, EPI_ISL_1338924, EPI_ISL_1338925, EPI_ISL_1338926, EPI_ISL_1338927, EPI_ISL_1338928, EPI_ISL_1338930, EPI_ISL_1338932, EPI_ISL_1338933, EPI_ISL_1338934, EPI_ISL_1338935, EPI_ISL_1338936, EPI_ISL_1338938, EPI_ISL_1338939, EPI_ISL_1338940, EPI_ISL_1338948, EPI_ISL_1338953, EPI_ISL_1338954, EPI_ISL_1338960, EPI_ISL_1338961, EPI_ISL_1338962, EPI_ISL_1338965, EPI_ISL_1338968, EPI_ISL_1338970, EPI_ISL_1338971, EPI_ISL_1338974, EPI_ISL_1338976, EPI_ISL_1338977, EPI_ISL_1338982, EPI_ISL_1338984, EPI_ISL_1338988, EPI_ISL_1338990, EPI_ISL_1338993, EPI_ISL_1338996, EPI_ISL_1338998, EPI_ISL_1339002, EPI_ISL_1339003, EPI_ISL_1339004, EPI_ISL_1339005, EPI_ISL_1339006, EPI_ISL_1339012, EPI_ISL_1339014, EPI_ISL_1339021, EPI_ISL_1339029, EPI_ISL_1339030, EPI_ISL_1339040, EPI_ISL_1339043, EPI_ISL_1339048, EPI_ISL_1339050, EPI_ISL_1339051, EPI_ISL_1339054, EPI_ISL_1339055, EPI_ISL_1339057, EPI_ISL_1339059, EPI_ISL_1339060, EPI_ISL_1339061, EPI_ISL_1339062, EPI_ISL_1339063, EPI_ISL_1339068, EPI_ISL_1339070, EPI_ISL_1339073, EPI_ISL_1339078, EPI_ISL_1339080, EPI_ISL_1339085, EPI_ISL_1339086, EPI_ISL_1339091, EPI_ISL_1339094, EPI_ISL_1339095, EPI_ISL_1339096, EPI_ISL_1339098, EPI_ISL_1339101, EPI_ISL_1339105, EPI_ISL_1339106, EPI_ISL_1339107, EPI_ISL_1339112, EPI_ISL_1339114, EPI_ISL_1339115, EPI_ISL_1339120, EPI_ISL_1339122, EPI_ISL_1339124, EPI_ISL_1339125, EPI_ISL_1339126, EPI_ISL_1339127, EPI_ISL_1339128, EPI_ISL_1339131, EPI_ISL_1339135, EPI_ISL_1339136, EPI_ISL_1339137, EPI_ISL_1339138, EPI_ISL_1339143, EPI_ISL_1339144, EPI_ISL_1339145, EPI_ISL_1339146, EPI_ISL_1339151, EPI_ISL_1339152, EPI_ISL_1339158, EPI_ISL_1339159, EPI_ISL_1339160, EPI_ISL_1339161, EPI_ISL_1339162, EPI_ISL_1339166, EPI_ISL_1339167, EPI_ISL_1339168, EPI_ISL_1339170, EPI_ISL_1339173, EPI_ISL_1339182, EPI_ISL_1339184, EPI_ISL_1339186, EPI_ISL_1339187, EPI_ISL_1339188, EPI_ISL_1339191, EPI_ISL_1339194, EPI_ISL_1339196, EPI_ISL_1339200, EPI_ISL_1339201, EPI_ISL_1339202, EPI_ISL_1339203, EPI_ISL_1339204, EPI_ISL_1339205, EPI_ISL_1339207, EPI_ISL_1339228, EPI_ISL_1339229, EPI_ISL_1339234, EPI_ISL_1339235, EPI_ISL_1339236, EPI_ISL_1339239, EPI_ISL_1339244, EPI_ISL_1339245, EPI_ISL_1339250, EPI_ISL_1339251, EPI_ISL_1339255, EPI_ISL_1339258, EPI_ISL_1339262, EPI_ISL_1339263, EPI_ISL_1339264, EPI_ISL_1339273, EPI_ISL_1339274, EPI_ISL_1339276, EPI_ISL_1339278, EPI_ISL_1339279, EPI_ISL_1339289, EPI_ISL_1339290, EPI_ISL_1339291, EPI_ISL_1339293, EPI_ISL_1339300, EPI_ISL_1339301, EPI_ISL_1339302 | | | |
| see above | Laboratory Corporation of America | Centers for Disease Control and Prevention Division of Viral Diseases, Pathogen Discovery | Peter W. Cook, Dakota Howard, Dhwani Batra, Ben L. Rambo-Martin, Minoo Agarwal, Eyad Almasri, Debbie Boles, Ayla Burns, Nuthawin Charoensri, Oren Cohen, Susan Countryman, Mary Ann Cristobal, Bobbi Croy, Suzanne Dale, Hrushikesh Deshmukh, Amanda Douglas, Vincent Drouillon, Marcia Eisenberg, Howard Engler, Rama Ghatti, Prashant Gupta, Susan Hicks, Jake Humphrey, Lax Iyer, Manoj Jain, Mohan Kolli, Brian Krueger, Tim Kuphal, Stanley Letovsky, Michael Levandoski, Craig Lukasik, Jonathan Meltzer, Brian Norvell, Mindy Nye, Scott Parker, Christos Petropoulos, John Pruitt, Steven Ragan, Scott Ryan, Mike Sapeta, Jana Schroth, Suresh Babu Selvaraju, Goran Stevovic, Amanda Suchanek, Andrea Throop, Lyndon Tilson, Thomas Urban, Joe Voshell, Kimberly Wagner, Jonathan Williams, Mary Williamson, Qian Zeng, Tricia Zwiefelhofer, Clinton R. Paden, Suxiang Tong, Duncan MacCannell |
| EPI_ISL_1339303, EPI_ISL_1339304, EPI_ISL_1339306, EPI_ISL_1339307, EPI_ISL_1339309, EPI_ISL_1339310, EPI_ISL_1339312, EPI_ISL_1339313 | Minnesota Department of Health, Public Health Laboratory | Minnesota Department of Health, Public Health Laboratory | Alexandra Lorentz, Jacob Garfin, Matt Plumb, and Xiong Wang |
| EPI_ISL_1339314 | Laboratory Corporation of America | Centers for Disease Control and Prevention Division of Viral Diseases, Pathogen Discovery | Peter W. Cook, Dakota Howard, Dhwani Batra, Ben L. Rambo-Martin, Minoo Agarwal, Eyad Almasri, Debbie Boles, Ayla Burns, Nuthawin Charoensri, Oren Cohen, Susan Countryman, Mary Ann Cristobal, Bobbi Croy, Suzanne Dale, Hrushikesh Deshmukh, Amanda Douglas, Vincent Drouillon, Marcia Eisenberg, Howard Engler, Rama Ghatti, Prashant Gupta, Susan Hicks, Jake Humphrey, Lax Iyer, Manoj Jain, Mohan Kolli, Brian Krueger, Tim Kuphal, Stanley Letovsky, Michael Levandoski, Craig Lukasik, Jonathan Meltzer, Brian Norvell, Mindy Nye, Scott Parker, Christos Petropoulos, John Pruitt, Steven Ragan, Scott Ryan, Mike Sapeta, Jana Schroth, Suresh Babu Selvaraju, Goran Stevovic, Amanda Suchanek, Andrea Throop, Lyndon Tilson, Thomas Urban, Joe Voshell, Kimberly Wagner, Jonathan Williams, Mary Williamson, Qian Zeng, Tricia Zwiefelhofer, Clinton R. Paden, Suxiang Tong, Duncan MacCannell |
| EPI_ISL_1339318, EPI_ISL_1339319, EPI_ISL_1339320, EPI_ISL_1339322, EPI_ISL_1339323 | Minnesota Department of Health, Public Health Laboratory | Minnesota Department of Health, Public Health Laboratory | Alexandra Lorentz, Jacob Garfin, Matt Plumb, and Xiong Wang |
| EPI_ISL_1339324 | Laboratory Corporation of America | Centers for Disease Control and Prevention Division of Viral Diseases, Pathogen Discovery | Peter W. Cook, Dakota Howard, Dhwani Batra, Ben L. Rambo-Martin, Minoo Agarwal, Eyad Almasri, Debbie Boles, Ayla Burns, Nuthawin Charoensri, Oren Cohen, Susan Countryman, Mary Ann Cristobal, Bobbi Croy, Suzanne Dale, Hrushikesh Deshmukh, Amanda Douglas, Vincent Drouillon, Marcia Eisenberg, Howard Engler, Rama Ghatti, Prashant Gupta, Susan Hicks, Jake Humphrey, Lax Iyer, Manoj Jain, Mohan Kolli, Brian Krueger, Tim Kuphal, Stanley Letovsky, Michael Levandoski, Craig Lukasik, Jonathan Meltzer, Brian Norvell, Mindy Nye, Scott Parker, Christos Petropoulos, John Pruitt, Steven Ragan, Scott Ryan, Mike Sapeta, Jana Schroth, Suresh Babu Selvaraju, Goran Stevovic, Amanda Suchanek, Andrea Throop, Lyndon Tilson, Thomas Urban, Joe Voshell, Kimberly Wagner, Jonathan Williams, Mary Williamson, Qian Zeng, Tricia Zwiefelhofer, Clinton R. Paden, Suxiang Tong, Duncan MacCannell |
| EPI_ISL_1339325, EPI_ISL_1339326 | Minnesota Department of Health, Public Health Laboratory | Minnesota Department of Health, Public Health Laboratory | Alexandra Lorentz, Jacob Garfin, Matt Plumb, and Xiong Wang |
| EPI_ISL_1339327 | Laboratory Corporation of America | Centers for Disease Control and Prevention Division of Viral Diseases, Pathogen Discovery | Peter W. Cook, Dakota Howard, Dhwani Batra, Ben L. Rambo-Martin, Minoo Agarwal, Eyad Almasri, Debbie Boles, Ayla Burns, Nuthawin Charoensri, Oren Cohen, Susan Countryman, Mary Ann Cristobal, Bobbi Croy, Suzanne Dale, Hrushikesh Deshmukh, Amanda Douglas, Vincent Drouillon, Marcia Eisenberg, Howard Engler, Rama Ghatti, Prashant Gupta, Susan Hicks, Jake Humphrey, Lax Iyer, Manoj Jain, Mohan Kolli, Brian Krueger, Tim Kuphal, Stanley Letovsky, Michael Levandoski, Craig Lukasik, Jonathan Meltzer, Brian Norvell, Mindy Nye, Scott Parker, Christos Petropoulos, John Pruitt, Steven Ragan, Scott Ryan, Mike Sapeta, Jana Schroth, Suresh Babu Selvaraju, Goran Stevovic, Amanda Suchanek, Andrea Throop, Lyndon Tilson, Thomas Urban, Joe Voshell, Kimberly Wagner, Jonathan Williams, Mary Williamson, Qian Zeng, Tricia Zwiefelhofer, Clinton R. Paden, Suxiang Tong, Duncan MacCannell |
| EPI_ISL_1339328, EPI_ISL_1339329, EPI_ISL_1339330 | Minnesota Department of Health, Public Health Laboratory | Minnesota Department of Health, Public Health Laboratory | Alexandra Lorentz, Jacob Garfin, Matt Plumb, and Xiong Wang |
| EPI_ISL_1339331 | Laboratory Corporation of America | Centers for Disease Control and Prevention Division of Viral Diseases, Pathogen Discovery | Peter W. Cook, Dakota Howard, Dhwani Batra, Ben L. Rambo-Martin, Minoo Agarwal, Eyad Almasri, Debbie Boles, Ayla Burns, Nuthawin Charoensri, Oren Cohen, Susan Countryman, Mary Ann Cristobal, Bobbi Croy, Suzanne Dale, Hrushikesh Deshmukh, Amanda Douglas, Vincent Drouillon, Marcia Eisenberg, Howard Engler, Rama Ghatti, Prashant Gupta, Susan Hicks, Jake Humphrey, Lax Iyer, Manoj Jain, Mohan Kolli, Brian Krueger, Tim Kuphal, Stanley Letovsky, Michael Levandoski, Craig Lukasik, Jonathan Meltzer, Brian Norvell, Mindy Nye, Scott Parker, Christos Petropoulos, John Pruitt, Steven Ragan, Scott Ryan, Mike Sapeta, Jana Schroth, Suresh Babu Selvaraju, Goran Stevovic, Amanda Suchanek, Andrea Throop, Lyndon Tilson, Thomas Urban, Joe Voshell, Kimberly Wagner, Jonathan Williams, Mary Williamson, Qian Zeng, Tricia Zwiefelhofer, Clinton R. Paden, Suxiang Tong, Duncan MacCannell |
| EPI_ISL_1339332, EPI_ISL_1339333, EPI_ISL_1339335, EPI_ISL_1339336, EPI_ISL_1339338, EPI_ISL_1339339, EPI_ISL_1339341, EPI_ISL_1339342, EPI_ISL_1339344, EPI_ISL_1339345, EPI_ISL_1339346, EPI_ISL_1339348 | | | |
| see above | Minnesota Department of Health, Public Health Laboratory | Minnesota Department of Health, Public Health Laboratory | Alexandra Lorentz, Jacob Garfin, Matt Plumb, and Xiong Wang |
| EPI_ISL_1339349 | Minneapolis VA Health Care System | Minnesota Department of Health, Public Health Laboratory | Alexandra Lorentz, Jacob Garfin, Matt Plumb, and Xiong Wang |
| EPI_ISL_1339350 | Laboratory Corporation of America | Centers for Disease Control and Prevention Division of Viral Diseases, Pathogen Discovery | Peter W. Cook, Dakota Howard, Dhwani Batra, Ben L. Rambo-Martin, Minoo Agarwal, Eyad Almasri, Debbie Boles, Ayla Burns, Nuthawin Charoensri, Oren Cohen, Susan Countryman, Mary Ann Cristobal, Bobbi Croy, Suzanne Dale, Hrushikesh Deshmukh, Amanda Douglas, Vincent Drouillon, Marcia Eisenberg, Howard Engler, Rama Ghatti, Prashant Gupta, Susan Hicks, Jake Humphrey, Lax Iyer, Manoj Jain, Mohan Kolli, Brian Krueger, Tim Kuphal, Stanley Letovsky, Michael Levandoski, Craig Lukasik, Jonathan Meltzer, Brian Norvell, Mindy Nye, Scott Parker, Christos Petropoulos, John Pruitt, Steven Ragan, Scott Ryan, Mike Sapeta, Jana Schroth, Suresh Babu Selvaraju, Goran Stevovic, Amanda Suchanek, Andrea Throop, Lyndon Tilson, Thomas Urban, Joe Voshell, Kimberly Wagner, Jonathan Williams, Mary Williamson, Qian Zeng, Tricia Zwiefelhofer, Clinton R. Paden, Suxiang Tong, Duncan MacCannell |
| EPI_ISL_1339351, EPI_ISL_1339352, EPI_ISL_1339354 | Minnesota Department of Health, Public Health Laboratory | Minnesota Department of Health, Public Health Laboratory | Alexandra Lorentz, Jacob Garfin, Matt Plumb, and Xiong Wang |
| EPI_ISL_1339355, EPI_ISL_1339358, EPI_ISL_1339359, EPI_ISL_1339361, EPI_ISL_1339362 | Vault Health | Minnesota Department of Health, Public Health Laboratory | Alexandra Lorentz, Jacob Garfin, Matt Plumb, and Xiong Wang |
| EPI_ISL_1339363 | Laboratory Corporation of America | Centers for Disease Control and Prevention Division of Viral Diseases, Pathogen Discovery | Peter W. Cook, Dakota Howard, Dhwani Batra, Ben L. Rambo-Martin, Minoo Agarwal, Eyad Almasri, Debbie Boles, Ayla Burns, Nuthawin Charoensri, Oren Cohen, Susan Countryman, Mary Ann Cristobal, Bobbi Croy, Suzanne Dale, Hrushikesh Deshmukh, Amanda Douglas, Vincent Drouillon, Marcia Eisenberg, Howard Engler, Rama Ghatti, Prashant Gupta, Susan Hicks, Jake Humphrey, Lax Iyer, Manoj Jain, Mohan Kolli, Brian Krueger, Tim Kuphal, Stanley Letovsky, Michael Levandoski, Craig Lukasik, Jonathan Meltzer, Brian Norvell, Mindy Nye, Scott Parker, Christos Petropoulos, John Pruitt, Steven Ragan, Scott Ryan, Mike Sapeta, Jana Schroth, Suresh Babu Selvaraju, Goran Stevovic, Amanda Suchanek, Andrea Throop, Lyndon Tilson, Thomas Urban, Joe Voshell, Kimberly Wagner, Jonathan Williams, Mary Williamson, Qian Zeng, Tricia Zwiefelhofer, Clinton R. Paden, Suxiang Tong, Duncan MacCannell |
| EPI_ISL_1339364, EPI_ISL_1339365, EPI_ISL_1339367, EPI_ISL_1339368, EPI_ISL_1339371, EPI_ISL_1339372, EPI_ISL_1339374, EPI_ISL_1339375, EPI_ISL_1339377, EPI_ISL_1339378 | Vault Health | Minnesota Department of Health, Public Health Laboratory | Alexandra Lorentz, Jacob Garfin, Matt Plumb, and Xiong Wang |
| EPI_ISL_1339379 | Laboratory Corporation of America | Centers for Disease Control and Prevention Division of Viral Diseases, Pathogen Discovery | Peter W. Cook, Dakota Howard, Dhwani Batra, Ben L. Rambo-Martin, Minoo Agarwal, Eyad Almasri, Debbie Boles, Ayla Burns, Nuthawin Charoensri, Oren Cohen, Susan Countryman, Mary Ann Cristobal, Bobbi Croy, Suzanne Dale, Hrushikesh Deshmukh, Amanda Douglas, Vincent Drouillon, Marcia Eisenberg, Howard Engler, Rama Ghatti, Prashant Gupta, Susan Hicks, Jake Humphrey, Lax Iyer, Manoj Jain, Mohan Kolli, Brian Krueger, Tim Kuphal, Stanley Letovsky, Michael Levandoski, Craig Lukasik, Jonathan Meltzer, Brian Norvell, Mindy Nye, Scott Parker, Christos Petropoulos, John Pruitt, Steven Ragan, Scott Ryan, Mike Sapeta, Jana Schroth, Suresh Babu Selvaraju, Goran Stevovic, Amanda Suchanek, Andrea Throop, Lyndon Tilson, Thomas Urban, Joe Voshell, Kimberly Wagner, Jonathan Williams, Mary Williamson, Qian Zeng, Tricia Zwiefelhofer, Clinton R. Paden, Suxiang Tong, Duncan MacCannell |
| EPI_ISL_1339380, EPI_ISL_1339381, EPI_ISL_1339383, EPI_ISL_1339385, EPI_ISL_1339386 | Vault Health | Minnesota Department of Health, Public Health Laboratory | Alexandra Lorentz, Jacob Garfin, Matt Plumb, and Xiong Wang |

| | | | |
|---|---|---|---|
| EPI_ISL_1339387 | Laboratory Corporation of America | Centers for Disease Control and Prevention Division of Viral Diseases, Pathogen Discovery | Peter W. Cook, Dakota Howard, Dhwani Batra, Ben L. Rambo-Martin, Minoo Agarwal, Eyad Almasri, Debbie Boles, Ayla Burns, Nuthawin Charoensri, Oren Cohen, Susan Countryman, Mary Ann Cristobal, Bobbi Croy, Suzanne Dale, Hrushikesh Deshmukh, Amanda Douglas, Vincent Drouillon, Marcia Eisenberg, Howard Engler, Rama Ghatti, Prashant Gupta, Susan Hicks, Jake Humphrey, Lax Iyer, Manoj Jain, Mohan Kolli, Brian Krueger, Tim Kuphal, Stanley Letovsky, Michael Levandoski, Craig Lukasik, Jonathan Meltzer, Brian Norvell, Mindy Nye, Scott Parker, Christos Petropoulos, John Pruitt, Steven Ragan, Scott Ryan, Mike Sapeta, Jana Schroth, Suresh Babu Selvaraju, Goran Stevovic, Amanda Suchanek, Andrea Throop, Lyndon Tilson, Thomas Urban, Joe Voshell, Kimberly Wagner, Jonathan Williams, Mary Williamson, Qian Zeng, Tricia Zwiefelhofer, Clinton R. Paden, Suxiang Tong, Duncan MacCannell |
| EPI_ISL_1339388, EPI_ISL_1339389, EPI_ISL_1339391, EPI_ISL_1339392, EPI_ISL_1339394, EPI_ISL_1339395, EPI_ISL_1339396 | Vault Health | Minnesota Department of Health, Public Health Laboratory | Alexandra Lorentz, Jacob Garfin, Matt Plumb, and Xiong Wang |
| EPI_ISL_1339397 | Laboratory Corporation of America | Centers for Disease Control and Prevention Division of Viral Diseases, Pathogen Discovery | Peter W. Cook, Dakota Howard, Dhwani Batra, Ben L. Rambo-Martin, Minoo Agarwal, Eyad Almasri, Debbie Boles, Ayla Burns, Nuthawin Charoensri, Oren Cohen, Susan Countryman, Mary Ann Cristobal, Bobbi Croy, Suzanne Dale, Hrushikesh Deshmukh, Amanda Douglas, Vincent Drouillon, Marcia Eisenberg, Howard Engler, Rama Ghatti, Prashant Gupta, Susan Hicks, Jake Humphrey, Lax Iyer, Manoj Jain, Mohan Kolli, Brian Krueger, Tim Kuphal, Stanley Letovsky, Michael Levandoski, Craig Lukasik, Jonathan Meltzer, Brian Norvell, Mindy Nye, Scott Parker, Christos Petropoulos, John Pruitt, Steven Ragan, Scott Ryan, Mike Sapeta, Jana Schroth, Suresh Babu Selvaraju, Goran Stevovic, Amanda Suchanek, Andrea Throop, Lyndon Tilson, Thomas Urban, Joe Voshell, Kimberly Wagner, Jonathan Williams, Mary Williamson, Qian Zeng, Tricia Zwiefelhofer, Clinton R. Paden, Suxiang Tong, Duncan MacCannell |
| EPI_ISL_1339398, EPI_ISL_1339399 | Vault Health | Minnesota Department of Health, Public Health Laboratory | Alexandra Lorentz, Jacob Garfin, Matt Plumb, and Xiong Wang |
| EPI_ISL_1339400 | Laboratory Corporation of America | Centers for Disease Control and Prevention Division of Viral Diseases, Pathogen Discovery | Peter W. Cook, Dakota Howard, Dhwani Batra, Ben L. Rambo-Martin, Minoo Agarwal, Eyad Almasri, Debbie Boles, Ayla Burns, Nuthawin Charoensri, Oren Cohen, Susan Countryman, Mary Ann Cristobal, Bobbi Croy, Suzanne Dale, Hrushikesh Deshmukh, Amanda Douglas, Vincent Drouillon, Marcia Eisenberg, Howard Engler, Rama Ghatti, Prashant Gupta, Susan Hicks, Jake Humphrey, Lax Iyer, Manoj Jain, Mohan Kolli, Brian Krueger, Tim Kuphal, Stanley Letovsky, Michael Levandoski, Craig Lukasik, Jonathan Meltzer, Brian Norvell, Mindy Nye, Scott Parker, Christos Petropoulos, John Pruitt, Steven Ragan, Scott Ryan, Mike Sapeta, Jana Schroth, Suresh Babu Selvaraju, Goran Stevovic, Amanda Suchanek, Andrea Throop, Lyndon Tilson, Thomas Urban, Joe Voshell, Kimberly Wagner, Jonathan Williams, Mary Williamson, Qian Zeng, Tricia Zwiefelhofer, Clinton R. Paden, Suxiang Tong, Duncan MacCannell |
| EPI_ISL_1339401, EPI_ISL_1339402, EPI_ISL_1339404, EPI_ISL_1339405 | Vault Health | Minnesota Department of Health, Public Health Laboratory | Alexandra Lorentz, Jacob Garfin, Matt Plumb, and Xiong Wang |
| EPI_ISL_1339406 | Laboratory Corporation of America | Centers for Disease Control and Prevention Division of Viral Diseases, Pathogen Discovery | Peter W. Cook, Dakota Howard, Dhwani Batra, Ben L. Rambo-Martin, Minoo Agarwal, Eyad Almasri, Debbie Boles, Ayla Burns, Nuthawin Charoensri, Oren Cohen, Susan Countryman, Mary Ann Cristobal, Bobbi Croy, Suzanne Dale, Hrushikesh Deshmukh, Amanda Douglas, Vincent Drouillon, Marcia Eisenberg, Howard Engler, Rama Ghatti, Prashant Gupta, Susan Hicks, Jake Humphrey, Lax Iyer, Manoj Jain, Mohan Kolli, Brian Krueger, Tim Kuphal, Stanley Letovsky, Michael Levandoski, Craig Lukasik, Jonathan Meltzer, Brian Norvell, Mindy Nye, Scott Parker, Christos Petropoulos, John Pruitt, Steven Ragan, Scott Ryan, Mike Sapeta, Jana Schroth, Suresh Babu Selvaraju, Goran Stevovic, Amanda Suchanek, Andrea Throop, Lyndon Tilson, Thomas Urban, Joe Voshell, Kimberly Wagner, Jonathan Williams, Mary Williamson, Qian Zeng, Tricia Zwiefelhofer, Clinton R. Paden, Suxiang Tong, Duncan MacCannell |
| EPI_ISL_1339407, EPI_ISL_1339408, EPI_ISL_1339411, EPI_ISL_1339413, EPI_ISL_1339414 | Vault Health | Minnesota Department of Health, Public Health Laboratory | Alexandra Lorentz, Jacob Garfin, Matt Plumb, and Xiong Wang |
| EPI_ISL_1339415, EPI_ISL_1339418 | Laboratory Corporation of America | Centers for Disease Control and Prevention Division of Viral Diseases, Pathogen Discovery | Peter W. Cook, Dakota Howard, Dhwani Batra, Ben L. Rambo-Martin, Minoo Agarwal, Eyad Almasri, Debbie Boles, Ayla Burns, Nuthawin Charoensri, Oren Cohen, Susan Countryman, Mary Ann Cristobal, Bobbi Croy, Suzanne Dale, Hrushikesh Deshmukh, Amanda Douglas, Vincent Drouillon, Marcia Eisenberg, Howard Engler, Rama Ghatti, Prashant Gupta, Susan Hicks, Jake Humphrey, Lax Iyer, Manoj Jain, Mohan Kolli, Brian Krueger, Tim Kuphal, Stanley Letovsky, Michael Levandoski, Craig Lukasik, Jonathan Meltzer, Brian Norvell, Mindy Nye, Scott Parker, Christos Petropoulos, John Pruitt, Steven Ragan, Scott Ryan, Mike Sapeta, Jana Schroth, Suresh Babu Selvaraju, Goran Stevovic, Amanda Suchanek, Andrea Throop, Lyndon Tilson, Thomas Urban, Joe Voshell, Kimberly Wagner, Jonathan Williams, Mary Williamson, Qian Zeng, Tricia Zwiefelhofer, Clinton R. Paden, Suxiang Tong, Duncan MacCannell |
| EPI_ISL_1339419, EPI_ISL_1339420 | Vault Health | Minnesota Department of Health, Public Health Laboratory | Alexandra Lorentz, Jacob Garfin, Matt Plumb, and Xiong Wang |
| EPI_ISL_1339426, EPI_ISL_1339431, EPI_ISL_1339433, EPI_ISL_1339434, EPI_ISL_1339435, EPI_ISL_1339436, EPI_ISL_1339438, EPI_ISL_1339439, EPI_ISL_1339442, EPI_ISL_1339443, EPI_ISL_1339447, EPI_ISL_1339449, EPI_ISL_1339450, EPI_ISL_1339451, EPI_ISL_1339452, EPI_ISL_1339453, EPI_ISL_1339454, EPI_ISL_1339456, EPI_ISL_1339457, EPI_ISL_1339458, EPI_ISL_1339459, EPI_ISL_1339461, EPI_ISL_1339462, EPI_ISL_1339463, EPI_ISL_1339465, EPI_ISL_1339466, EPI_ISL_1339468, EPI_ISL_1339472, EPI_ISL_1339474, EPI_ISL_1339475, EPI_ISL_1339476, EPI_ISL_1339477, EPI_ISL_1339479, EPI_ISL_1339480, EPI_ISL_1339482, EPI_ISL_1339483, EPI_ISL_1339484, EPI_ISL_1339485, EPI_ISL_1339487, EPI_ISL_1339490, EPI_ISL_1339491, EPI_ISL_1339493, EPI_ISL_1339495, EPI_ISL_1339496, EPI_ISL_1339499, EPI_ISL_1339500, EPI_ISL_1339501, EPI_ISL_1339502, EPI_ISL_1339506, EPI_ISL_1339508, EPI_ISL_1339509, EPI_ISL_1339515, EPI_ISL_1339528, EPI_ISL_1339530, EPI_ISL_1339531, EPI_ISL_1339534, EPI_ISL_1339537, EPI_ISL_1339540, EPI_ISL_1339541, EPI_ISL_1339542, EPI_ISL_1339545, EPI_ISL_1339548, EPI_ISL_1339549, EPI_ISL_1339550, EPI_ISL_1339551, EPI_ISL_1339552, EPI_ISL_1339553, EPI_ISL_1339554, EPI_ISL_1339556, EPI_ISL_1339557, EPI_ISL_1339558, EPI_ISL_1339560, EPI_ISL_1339565, EPI_ISL_1339570, EPI_ISL_1339572, EPI_ISL_1339573, EPI_ISL_1339582, EPI_ISL_1339583, EPI_ISL_1339592, EPI_ISL_1339597, EPI_ISL_1339598, EPI_ISL_1339600, EPI_ISL_1339601, EPI_ISL_1339602, EPI_ISL_1339603, EPI_ISL_1339604, EPI_ISL_1339606, EPI_ISL_1339607, EPI_ISL_1339609, EPI_ISL_1339611, EPI_ISL_1339612, EPI_ISL_1339615, EPI_ISL_1339618, EPI_ISL_1339621, EPI_ISL_1339622, EPI_ISL_1339624, EPI_ISL_1339625, EPI_ISL_1339626, EPI_ISL_1339634, EPI_ISL_1339636, EPI_ISL_1339637, EPI_ISL_1339638, EPI_ISL_1339640, EPI_ISL_1339645, EPI_ISL_1339649, EPI_ISL_1339652, EPI_ISL_1339656, EPI_ISL_1339658, EPI_ISL_1339663, EPI_ISL_1339666, EPI_ISL_1339675, EPI_ISL_1339679, EPI_ISL_1339683, EPI_ISL_1339684, EPI_ISL_1339688, EPI_ISL_1339690, EPI_ISL_1339691, EPI_ISL_1339696, EPI_ISL_1339698, EPI_ISL_1339699, EPI_ISL_1339700, EPI_ISL_1339707, EPI_ISL_1339708, EPI_ISL_1339712, EPI_ISL_1339719, EPI_ISL_1339724, EPI_ISL_1339737, EPI_ISL_1339738, EPI_ISL_1339739, EPI_ISL_1339740, EPI_ISL_1339741, EPI_ISL_1339742, EPI_ISL_1339747, EPI_ISL_1339750, EPI_ISL_1339751, EPI_ISL_1339757, EPI_ISL_1339758, EPI_ISL_1339761, EPI_ISL_1339766, EPI_ISL_1339768, EPI_ISL_1339769, EPI_ISL_1339775, EPI_ISL_1339778, EPI_ISL_1339779, EPI_ISL_1339781, EPI_ISL_1339782, EPI_ISL_1339789, EPI_ISL_1339790, EPI_ISL_1339792, EPI_ISL_1339795, EPI_ISL_1339796, EPI_ISL_1339797, EPI_ISL_1339798, EPI_ISL_1339799, EPI_ISL_1339801, EPI_ISL_1339802, EPI_ISL_1339803, EPI_ISL_1339806, EPI_ISL_1339807, EPI_ISL_1339812, EPI_ISL_1339817, EPI_ISL_1339818, EPI_ISL_1339820 | | | |
| see above | Laboratory Corporation of America | Centers for Disease Control and Prevention Division of Viral Diseases, Pathogen Discovery | Peter W. Cook, Dakota Howard, Dhwani Batra, Ben L. Rambo-Martin, Minoo Agarwal, Eyad Almasri, Debbie Boles, Ayla Burns, Nuthawin Charoensri, Oren Cohen, Susan Countryman, Mary Ann Cristobal, Bobbi Croy, Suzanne Dale, Hrushikesh Deshmukh, Amanda Douglas, Vincent Drouillon, Marcia Eisenberg, Howard Engler, Rama Ghatti, Prashant Gupta, Susan Hicks, Jake Humphrey, Lax Iyer, Manoj Jain, Mohan Kolli, Brian Krueger, Tim Kuphal, Stanley Letovsky, Michael Levandoski, Craig Lukasik, Jonathan Meltzer, Brian Norvell, Mindy Nye, Scott Parker, Christos Petropoulos, John Pruitt, Steven Ragan, Scott Ryan, Mike Sapeta, Jana Schroth, Suresh Babu Selvaraju, Goran Stevovic, Amanda Suchanek, Andrea Throop, Lyndon Tilson, Thomas Urban, Joe Voshell, Kimberly Wagner, Jonathan Williams, Mary Williamson, Qian Zeng, Tricia Zwiefelhofer, Clinton R. Paden, Suxiang Tong, Duncan MacCannell |
| EPI_ISL_1339823, EPI_ISL_1339824, EPI_ISL_1339825, EPI_ISL_1339826, EPI_ISL_1339827, EPI_ISL_1339828, EPI_ISL_1339829, EPI_ISL_1339830, EPI_ISL_1339831, EPI_ISL_1339832, EPI_ISL_1339833, EPI_ISL_1339834, EPI_ISL_1339835, EPI_ISL_1339836, EPI_ISL_1339837, EPI_ISL_1339838, EPI_ISL_1339839, EPI_ISL_1339840, EPI_ISL_1339841, EPI_ISL_1339842, EPI_ISL_1339843, EPI_ISL_1339844, EPI_ISL_1339845, EPI_ISL_1339846, EPI_ISL_1339847, EPI_ISL_1339848, EPI_ISL_1339849, EPI_ISL_1339850, EPI_ISL_1339851, EPI_ISL_1339852, EPI_ISL_1339853, EPI_ISL_1339854, EPI_ISL_1339855, EPI_ISL_1339856, EPI_ISL_1339857, EPI_ISL_1339858, EPI_ISL_1339859, EPI_ISL_1339860, EPI_ISL_1339861, EPI_ISL_1339862, EPI_ISL_1339863, EPI_ISL_1339864, EPI_ISL_1339865, EPI_ISL_1339866, EPI_ISL_1339867, EPI_ISL_1339868, EPI_ISL_1339869, EPI_ISL_1339870, EPI_ISL_1339871, EPI_ISL_1339872, EPI_ISL_1339873, EPI_ISL_1339874, EPI_ISL_1339875, EPI_ISL_1339876, EPI_ISL_1339877, EPI_ISL_1339878, EPI_ISL_1339879, EPI_ISL_1339880, EPI_ISL_1339881, EPI_ISL_1339882, EPI_ISL_1339883, EPI_ISL_1339884, EPI_ISL_1339885, EPI_ISL_1339886, EPI_ISL_1339887, EPI_ISL_1339888, EPI_ISL_1339889, EPI_ISL_1339890, EPI_ISL_1339891, EPI_ISL_1339892, EPI_ISL_1339893, EPI_ISL_1339894, EPI_ISL_1339895, EPI_ISL_1339896, EPI_ISL_1339897, EPI_ISL_1339898, EPI_ISL_1339899, EPI_ISL_1339900, EPI_ISL_1339901, EPI_ISL_1339902, EPI_ISL_1339903, EPI_ISL_1339904, EPI_ISL_1339905, EPI_ISL_1339906, EPI_ISL_1339907, EPI_ISL_1339908, EPI_ISL_1339909, EPI_ISL_1339910, EPI_ISL_1339911, EPI_ISL_1339912, EPI_ISL_1339913, EPI_ISL_1339914, EPI_ISL_1339915, EPI_ISL_1339916, EPI_ISL_1339917, EPI_ISL_1339918, EPI_ISL_1339919, EPI_ISL_1339920, EPI_ISL_1339921, EPI_ISL_1339922, EPI_ISL_1339923, EPI_ISL_1339924, EPI_ISL_1339925, EPI_ISL_1339926, EPI_ISL_1339927, EPI_ISL_1339928, EPI_ISL_1339929, EPI_ISL_1339930, EPI_ISL_1339931, EPI_ISL_1339932, EPI_ISL_1339933, EPI_ISL_1339934, EPI_ISL_1339935, EPI_ISL_1339936, EPI_ISL_1339937, EPI_ISL_1339938, EPI_ISL_1339939, EPI_ISL_1339940, EPI_ISL_1339941, EPI_ISL_1339942, EPI_ISL_1339943, EPI_ISL_1339944, EPI_ISL_1339945, EPI_ISL_1339946, EPI_ISL_1339947, EPI_ISL_1339948, EPI_ISL_1339949, EPI_ISL_1339950, EPI_ISL_1339951, EPI_ISL_1339952, EPI_ISL_1339953, EPI_ISL_1339954, EPI_ISL_1339955, EPI_ISL_1339956, EPI_ISL_1339957, EPI_ISL_1339958, EPI_ISL_1339959, EPI_ISL_1339960, EPI_ISL_1339961, EPI_ISL_1339962, EPI_ISL_1339963, EPI_ISL_1339964, EPI_ISL_1339965, EPI_ISL_1339966, EPI_ISL_1339967 | | | |
| see above | Quest Diagnostics Incorporated | Centers for Disease Control and Prevention Division of Viral Diseases, Pathogen Discovery | Peter W. Cook, Dakota Howard, Dhwani Batra, Ben L. Rambo-Martin, S. H. Rosenthal, A. Gerasimova, R. M. Kagan, B. Anderson, M. Hua, Y. Liu, L.E. Bernstein, K.E. Livingston, A. Perez, I. A. Shlyakhter, R. V. Rolando, R. Owen, P. Tanpaiboon, F. Lacbawan, Clinton R. Paden, Suxiang Tong, Duncan MacCannell |
| EPI_ISL_1339968, EPI_ISL_1339969, EPI_ISL_1339970, EPI_ISL_1339971, EPI_ISL_1339972, EPI_ISL_1339973, EPI_ISL_1339975, EPI_ISL_1339976, EPI_ISL_1339977, EPI_ISL_1339978, EPI_ISL_1339979, EPI_ISL_1339980, EPI_ISL_1339982, EPI_ISL_1339983, EPI_ISL_1339985, EPI_ISL_1339986, EPI_ISL_1339987, EPI_ISL_1339988, EPI_ISL_1339989, EPI_ISL_1339990, EPI_ISL_1339991, EPI_ISL_1339992, EPI_ISL_1339993, EPI_ISL_1339994, EPI_ISL_1339995, EPI_ISL_1339996, EPI_ISL_1339997, EPI_ISL_1339998, EPI_ISL_1339999, EPI_ISL_1340000, EPI_ISL_1340001, EPI_ISL_1340002, EPI_ISL_1340003, EPI_ISL_1340004, EPI_ISL_1340005, EPI_ISL_1340006, EPI_ISL_1340007, EPI_ISL_1340008, EPI_ISL_1340009, EPI_ISL_1340010, EPI_ISL_1340011, EPI_ISL_1340012, EPI_ISL_1340013, EPI_ISL_1340014, EPI_ISL_1340015, EPI_ISL_1340016, EPI_ISL_1340017, EPI_ISL_1340018, EPI_ISL_1340019, EPI_ISL_1340020, EPI_ISL_1340021 | | | |
| see above | DPHL | Delaware Public Health Lab | Rebecca Savage |
| EPI_ISL_1340022, EPI_ISL_1340023, EPI_ISL_1340024, EPI_ISL_1340025, EPI_ISL_1340026, EPI_ISL_1340027, EPI_ISL_1340028, EPI_ISL_1340029, EPI_ISL_1340030, EPI_ISL_1340031, EPI_ISL_1340032, EPI_ISL_1340033, EPI_ISL_1340034, EPI_ISL_1340035, EPI_ISL_1340036, EPI_ISL_1340037, EPI_ISL_1340038, EPI_ISL_1340039 | | | |
| see above | Houston Health Dept. | Houston Health Dept. | Ryker Penn, Pamela Brown, Adolpho Lara, Yanlai Lai |
| EPI_ISL_1340040, EPI_ISL_1340041, EPI_ISL_1340042, EPI_ISL_1340043, EPI_ISL_1340044, EPI_ISL_1340045, EPI_ISL_1340046, EPI_ISL_1340047, EPI_ISL_1340048, EPI_ISL_1340049, EPI_ISL_1340050, EPI_ISL_1340051, EPI_ISL_1340052, EPI_ISL_1340053, EPI_ISL_1340054 | | | |
| see above | Nebraska Public Health Laboratory | NPHL COVID-19 Response Team | NPHL COVID-19 Response Team |
| EPI_ISL_1340055, EPI_ISL_1340056, EPI_ISL_1340057, EPI_ISL_1340058, EPI_ISL_1340059, EPI_ISL_1340060, EPI_ISL_1340061, EPI_ISL_1340062, EPI_ISL_1340063, EPI_ISL_1340064, EPI_ISL_1340065, EPI_ISL_1340066, EPI_ISL_1340067, EPI_ISL_1340068, EPI_ISL_1340069, EPI_ISL_1340070, EPI_ISL_1340071, EPI_ISL_1340072, EPI_ISL_1340073, EPI_ISL_1340074, EPI_ISL_1340075, EPI_ISL_1340076, EPI_ISL_1340077, EPI_ISL_1340078, EPI_ISL_1340079, EPI_ISL_1340080, EPI_ISL_1340081, EPI_ISL_1340082, EPI_ISL_1340083, EPI_ISL_1340084, EPI_ISL_1340085, EPI_ISL_1340086, EPI_ISL_1340087, EPI_ISL_1340088, EPI_ISL_1340089, EPI_ISL_1340090, EPI_ISL_1340091, EPI_ISL_1340092, EPI_ISL_1340093, EPI_ISL_1340094, EPI_ISL_1340095, EPI_ISL_1340096, EPI_ISL_1340097, EPI_ISL_1340098, EPI_ISL_1340099, EPI_ISL_1340100, EPI_ISL_1340101, EPI_ISL_1340102, EPI_ISL_1340103, EPI_ISL_1340104, EPI_ISL_1340105, EPI_ISL_1340106, EPI_ISL_1340107, EPI_ISL_1340108, EPI_ISL_1340109, EPI_ISL_1340110, EPI_ISL_1340111, EPI_ISL_1340112, EPI_ISL_1340113, EPI_ISL_1340114, EPI_ISL_1340115, EPI_ISL_1340116, EPI_ISL_1340117, EPI_ISL_1340118, EPI_ISL_1340119, EPI_ISL_1340121, EPI_ISL_1340122, EPI_ISL_1340123, EPI_ISL_1340124, EPI_ISL_1340125, EPI_ISL_1340126, EPI_ISL_1340127, EPI_ISL_1340128, EPI_ISL_1340129, EPI_ISL_1340130, EPI_ISL_1340131, EPI_ISL_1340132, EPI_ISL_1340133, EPI_ISL_1340134, EPI_ISL_1340135, EPI_ISL_1340136, EPI_ISL_1340137, EPI_ISL_1340139, EPI_ISL_1340140, EPI_ISL_1340141, EPI_ISL_1340142, EPI_ISL_1340143, EPI_ISL_1340144, EPI_ISL_1340145, EPI_ISL_1340146, EPI_ISL_1340147, EPI_ISL_1340148, EPI_ISL_1340149, EPI_ISL_1340150, EPI_ISL_1340151, EPI_ISL_1340152, EPI_ISL_1340153, EPI_ISL_1340154, EPI_ISL_1340155, EPI_ISL_1340156, EPI_ISL_1340157, EPI_ISL_1340158, EPI_ISL_1340159, EPI_ISL_1340160, EPI_ISL_1340161, EPI_ISL_1340162, EPI_ISL_1340163, EPI_ISL_1340164, EPI_ISL_1340165, EPI_ISL_1340166, EPI_ISL_1340167, EPI_ISL_1340168, EPI_ISL_1340169, EPI_ISL_1340170, EPI_ISL_1340171, EPI_ISL_1340172 | | | |
| see above | Helix/Illumina | Centers for Disease Control and Prevention Division of Viral Diseases, Pathogen Discovery | Peter W. Cook, Dakota Howard, Dhwani Batra, Ben L. Rambo-Martin, Eileen de Feo, Jan Antico, Christine Tran, Matthew Tolentino, Shannon Wickline, Kim Gietzen, Brad Sickler, Jingtao Liu, Eric Allen, Phil Febbo, Summer Galloway, Nicole L. Washington, Simon White, Geraint Levan, Kelly Schiabor Barrett, Elizabeth Cirulli, Alexandre Bolze, Ary Ascencio, Charlotte Rivera-Garcia, Ryan Cho, Jason Nguyen, Sherry Wang, Jimmy Ramirez, Tyler Cassens, Efren Sandoval, Magnus Isaksson, William Lee, David Becker, Marc Laurent, James Lu, Clinton R. Paden, Suxiang Tong, Duncan MacCannell |
| EPI_ISL_1340173 | NJDOH, Public Health and Environmental Laboratories | New Jersey Public Health and Environmental Laboratories (PHEL) | Lindsey Bodnar, Shiv K. Verma, Dana Woell, Byeong Jeong |
| EPI_ISL_1340174 | Helix/Illumina | Centers for Disease Control and Prevention Division of Viral Diseases, Pathogen Discovery | Peter W. Cook, Dakota Howard, Dhwani Batra, Ben L. Rambo-Martin, Eileen de Feo, Jan Antico, Christine Tran, Matthew Tolentino, Shannon Wickline, Kim Gietzen, Brad Sickler, Jingtao Liu, Eric Allen, Phil Febbo, Summer Galloway, Nicole L. Washington, Simon White, Geraint Levan, Kelly Schiabor Barrett, Elizabeth Cirulli, Alexandre Bolze, Ary Ascencio, Charlotte Rivera-Garcia, Ryan Cho, Jason Nguyen, Sherry Wang, Jimmy Ramirez, Tyler Cassens, Efren Sandoval, Magnus Isaksson, William Lee, David Becker, Marc Laurent, James Lu, Clinton R. Paden, Suxiang Tong, Duncan MacCannell |
| EPI_ISL_1340175, EPI_ISL_1340176 | NJDOH, Public Health and Environmental Laboratories | New Jersey Public Health and Environmental Laboratories (PHEL) | Lindsey Bodnar, Shiv K. Verma, Dana Woell, Byeong Jeong |
| EPI_ISL_1340177 | Helix/Illumina | Centers for Disease Control and Prevention Division of Viral Diseases, Pathogen Discovery | Peter W. Cook, Dakota Howard, Dhwani Batra, Ben L. Rambo-Martin, Eileen de Feo, Jan Antico, Christine Tran, Matthew Tolentino, Shannon Wickline, Kim Gietzen, Brad Sickler, Jingtao Liu, Eric Allen, Phil Febbo, Summer Galloway, Nicole L. Washington, Simon White, Geraint Levan, Kelly Schiabor Barrett, Elizabeth Cirulli, Alexandre Bolze, Ary Ascencio, Charlotte Rivera-Garcia, Ryan Cho, Jason Nguyen, Sherry Wang, Jimmy Ramirez, Tyler Cassens, Efren Sandoval, Magnus Isaksson, William Lee, David Becker, Marc Laurent, James Lu, Clinton R. Paden, Suxiang Tong, Duncan MacCannell |
| EPI_ISL_1340178, EPI_ISL_1340179 | NJDOH, Public Health and Environmental Laboratories | New Jersey Public Health and Environmental Laboratories (PHEL) | Lindsey Bodnar, Shiv K. Verma, Dana Woell, Byeong Jeong |
| EPI_ISL_1340180, EPI_ISL_1340182, EPI_ISL_1340186, EPI_ISL_1340188, EPI_ISL_1340192 | Helix/Illumina | Centers for Disease Control and Prevention Division of Viral Diseases, Pathogen Discovery | Peter W. Cook, Dakota Howard, Dhwani Batra, Ben L. Rambo-Martin, Eileen de Feo, Jan Antico, Christine Tran, Matthew Tolentino, Shannon Wickline, Kim Gietzen, Brad Sickler, Jingtao Liu, Eric Allen, Phil Febbo, Summer Galloway, Nicole L. Washington, Simon White, Geraint Levan, Kelly Schiabor Barrett, Elizabeth Cirulli, Alexandre Bolze, Ary Ascencio, Charlotte Rivera-Garcia, Ryan Cho, Jason Nguyen, Sherry Wang, Jimmy Ramirez, Tyler Cassens, Efren Sandoval, Magnus Isaksson, William Lee, David Becker, Marc Laurent, James Lu, Clinton R. Paden, Suxiang Tong, Duncan MacCannell |
| EPI_ISL_1340194 | NJDOH, Public Health and | New Jersey Public Health and Environmental | Lindsey Bodnar, Shiv K. Verma, Dana Woell, Byeong Jeong |

| | | | |
|---|---|---|---|
| | Environmental Laboratories | Laboratories (PHEL) | |
| EPI_ISL_1340195 | Helix/Illumina | Centers for Disease Control and Prevention Division of Viral Diseases, Pathogen Discovery | Peter W. Cook, Dakota Howard, Dhwani Batra, Ben L. Rambo-Martin, Eileen de Feo, Jan Antico, Christine Tran, Matthew Tolentino, Shannon Wickline, Kim Gietzen, Brad Sickler, Jingtao Liu, Eric Allen, Phil Febbo, Summer Galloway, Nicole L. Washington, Simon White, Geraint Levan, Kelly Schiabor Barrett, Elizabeth Cirulli, Alexandre Bolze, Ary Ascencio, Charlotte Rivera-Garcia, Ryan Cho, Jason Nguyen, Sherry Wang, Jimmy Ramirez, Tyler Cassens, Efren Sandoval, Magnus Isaksson, William Lee, David Becker, Marc Laurent, James Lu, Clinton R. Paden, Suxiang Tong, Duncan MacCannell |
| EPI_ISL_1340196 | NJDOH, Public Health and Environmental Laboratories | New Jersey Public Health and Environmental Laboratories (PHEL) | Lindsey Bodnar, Shiv K. Verma, Dana Woell, Byeong Jeong |
| EPI_ISL_1340197 | Helix/Illumina | Centers for Disease Control and Prevention Division of Viral Diseases, Pathogen Discovery | Peter W. Cook, Dakota Howard, Dhwani Batra, Ben L. Rambo-Martin, Eileen de Feo, Jan Antico, Christine Tran, Matthew Tolentino, Shannon Wickline, Kim Gietzen, Brad Sickler, Jingtao Liu, Eric Allen, Phil Febbo, Summer Galloway, Nicole L. Washington, Simon White, Geraint Levan, Kelly Schiabor Barrett, Elizabeth Cirulli, Alexandre Bolze, Ary Ascencio, Charlotte Rivera-Garcia, Ryan Cho, Jason Nguyen, Sherry Wang, Jimmy Ramirez, Tyler Cassens, Efren Sandoval, Magnus Isaksson, William Lee, David Becker, Marc Laurent, James Lu, Clinton R. Paden, Suxiang Tong, Duncan MacCannell |
| EPI_ISL_1340198, EPI_ISL_1340199 | NJDOH, Public Health and Environmental Laboratories | New Jersey Public Health and Environmental Laboratories (PHEL) | Lindsey Bodnar, Shiv K. Verma, Dana Woell, Byeong Jeong |
| EPI_ISL_1340200 | Helix/Illumina | Centers for Disease Control and Prevention Division of Viral Diseases, Pathogen Discovery | Peter W. Cook, Dakota Howard, Dhwani Batra, Ben L. Rambo-Martin, Eileen de Feo, Jan Antico, Christine Tran, Matthew Tolentino, Shannon Wickline, Kim Gietzen, Brad Sickler, Jingtao Liu, Eric Allen, Phil Febbo, Summer Galloway, Nicole L. Washington, Simon White, Geraint Levan, Kelly Schiabor Barrett, Elizabeth Cirulli, Alexandre Bolze, Ary Ascencio, Charlotte Rivera-Garcia, Ryan Cho, Jason Nguyen, Sherry Wang, Jimmy Ramirez, Tyler Cassens, Efren Sandoval, Magnus Isaksson, William Lee, David Becker, Marc Laurent, James Lu, Clinton R. Paden, Suxiang Tong, Duncan MacCannell |
| EPI_ISL_1340201, EPI_ISL_1340202 | NJDOH, Public Health and Environmental Laboratories | New Jersey Public Health and Environmental Laboratories (PHEL) | Lindsey Bodnar, Shiv K. Verma, Dana Woell, Byeong Jeong |
| EPI_ISL_1340203 | Helix/Illumina | Centers for Disease Control and Prevention Division of Viral Diseases, Pathogen Discovery | Peter W. Cook, Dakota Howard, Dhwani Batra, Ben L. Rambo-Martin, Eileen de Feo, Jan Antico, Christine Tran, Matthew Tolentino, Shannon Wickline, Kim Gietzen, Brad Sickler, Jingtao Liu, Eric Allen, Phil Febbo, Summer Galloway, Nicole L. Washington, Simon White, Geraint Levan, Kelly Schiabor Barrett, Elizabeth Cirulli, Alexandre Bolze, Ary Ascencio, Charlotte Rivera-Garcia, Ryan Cho, Jason Nguyen, Sherry Wang, Jimmy Ramirez, Tyler Cassens, Efren Sandoval, Magnus Isaksson, William Lee, David Becker, Marc Laurent, James Lu, Clinton R. Paden, Suxiang Tong, Duncan MacCannell |
| EPI_ISL_1340204, EPI_ISL_1340205 | NJDOH, Public Health and Environmental Laboratories | New Jersey Public Health and Environmental Laboratories (PHEL) | Lindsey Bodnar, Shiv K. Verma, Dana Woell, Byeong Jeong |
| EPI_ISL_1340206 | Helix/Illumina | Centers for Disease Control and Prevention Division of Viral Diseases, Pathogen Discovery | Peter W. Cook, Dakota Howard, Dhwani Batra, Ben L. Rambo-Martin, Eileen de Feo, Jan Antico, Christine Tran, Matthew Tolentino, Shannon Wickline, Kim Gietzen, Brad Sickler, Jingtao Liu, Eric Allen, Phil Febbo, Summer Galloway, Nicole L. Washington, Simon White, Geraint Levan, Kelly Schiabor Barrett, Elizabeth Cirulli, Alexandre Bolze, Ary Ascencio, Charlotte Rivera-Garcia, Ryan Cho, Jason Nguyen, Sherry Wang, Jimmy Ramirez, Tyler Cassens, Efren Sandoval, Magnus Isaksson, William Lee, David Becker, Marc Laurent, James Lu, Clinton R. Paden, Suxiang Tong, Duncan MacCannell |
| EPI_ISL_1340207, EPI_ISL_1340208, EPI_ISL_1340209 | NJDOH, Public Health and Environmental Laboratories | New Jersey Public Health and Environmental Laboratories (PHEL) | Lindsey Bodnar, Shiv K. Verma, Dana Woell, Byeong Jeong |
| EPI_ISL_1340210 | Helix/Illumina | Centers for Disease Control and Prevention Division of Viral Diseases, Pathogen Discovery | Peter W. Cook, Dakota Howard, Dhwani Batra, Ben L. Rambo-Martin, Eileen de Feo, Jan Antico, Christine Tran, Matthew Tolentino, Shannon Wickline, Kim Gietzen, Brad Sickler, Jingtao Liu, Eric Allen, Phil Febbo, Summer Galloway, Nicole L. Washington, Simon White, Geraint Levan, Kelly Schiabor Barrett, Elizabeth Cirulli, Alexandre Bolze, Ary Ascencio, Charlotte Rivera-Garcia, Ryan Cho, Jason Nguyen, Sherry Wang, Jimmy Ramirez, Tyler Cassens, Efren Sandoval, Magnus Isaksson, William Lee, David Becker, Marc Laurent, James Lu, Clinton R. Paden, Suxiang Tong, Duncan MacCannell |
| EPI_ISL_1340211, EPI_ISL_1340212 | NJDOH, Public Health and Environmental Laboratories | New Jersey Public Health and Environmental Laboratories (PHEL) | Lindsey Bodnar, Shiv K. Verma, Dana Woell, Byeong Jeong |
| EPI_ISL_1340213 | Helix/Illumina | Centers for Disease Control and Prevention Division of Viral Diseases, Pathogen Discovery | Peter W. Cook, Dakota Howard, Dhwani Batra, Ben L. Rambo-Martin, Eileen de Feo, Jan Antico, Christine Tran, Matthew Tolentino, Shannon Wickline, Kim Gietzen, Brad Sickler, Jingtao Liu, Eric Allen, Phil Febbo, Summer Galloway, Nicole L. Washington, Simon White, Geraint Levan, Kelly Schiabor Barrett, Elizabeth Cirulli, Alexandre Bolze, Ary Ascencio, Charlotte Rivera-Garcia, Ryan Cho, Jason Nguyen, Sherry Wang, Jimmy Ramirez, Tyler Cassens, Efren Sandoval, Magnus Isaksson, William Lee, David Becker, Marc Laurent, James Lu, Clinton R. Paden, Suxiang Tong, Duncan MacCannell |
| EPI_ISL_1340214, EPI_ISL_1340215 | NJDOH, Public Health and Environmental Laboratories | New Jersey Public Health and Environmental Laboratories (PHEL) | Lindsey Bodnar, Shiv K. Verma, Dana Woell, Byeong Jeong |
| EPI_ISL_1340216 | Helix/Illumina | Centers for Disease Control and Prevention Division of Viral Diseases, Pathogen Discovery | Peter W. Cook, Dakota Howard, Dhwani Batra, Ben L. Rambo-Martin, Eileen de Feo, Jan Antico, Christine Tran, Matthew Tolentino, Shannon Wickline, Kim Gietzen, Brad Sickler, Jingtao Liu, Eric Allen, Phil Febbo, Summer Galloway, Nicole L. Washington, Simon White, Geraint Levan, Kelly Schiabor Barrett, Elizabeth Cirulli, Alexandre Bolze, Ary Ascencio, Charlotte Rivera-Garcia, Ryan Cho, Jason Nguyen, Sherry Wang, Jimmy Ramirez, Tyler Cassens, Efren Sandoval, Magnus Isaksson, William Lee, David Becker, Marc Laurent, James Lu, Clinton R. Paden, Suxiang Tong, Duncan MacCannell |
| EPI_ISL_1340217 | NJDOH, Public Health and Environmental Laboratories | New Jersey Public Health and Environmental Laboratories (PHEL) | Lindsey Bodnar, Shiv K. Verma, Dana Woell, Byeong Jeong |
| EPI_ISL_1340218 | Helix/Illumina | Centers for Disease Control and Prevention Division of Viral Diseases, Pathogen Discovery | Peter W. Cook, Dakota Howard, Dhwani Batra, Ben L. Rambo-Martin, Eileen de Feo, Jan Antico, Christine Tran, Matthew Tolentino, Shannon Wickline, Kim Gietzen, Brad Sickler, Jingtao Liu, Eric Allen, Phil Febbo, Summer Galloway, Nicole L. Washington, Simon White, Geraint Levan, Kelly Schiabor Barrett, Elizabeth Cirulli, Alexandre Bolze, Ary Ascencio, Charlotte Rivera-Garcia, Ryan Cho, Jason Nguyen, Sherry Wang, Jimmy Ramirez, Tyler Cassens, Efren Sandoval, Magnus Isaksson, William Lee, David Becker, Marc Laurent, James Lu, Clinton R. Paden, Suxiang Tong, Duncan MacCannell |
| EPI_ISL_1340219, EPI_ISL_1340220 | NJDOH, Public Health and Environmental Laboratories | New Jersey Public Health and Environmental Laboratories (PHEL) | Lindsey Bodnar, Shiv K. Verma, Dana Woell, Byeong Jeong |
| EPI_ISL_1340221 | Helix/Illumina | Centers for Disease Control and Prevention Division of Viral Diseases, Pathogen Discovery | Peter W. Cook, Dakota Howard, Dhwani Batra, Ben L. Rambo-Martin, Eileen de Feo, Jan Antico, Christine Tran, Matthew Tolentino, Shannon Wickline, Kim Gietzen, Brad Sickler, Jingtao Liu, Eric Allen, Phil Febbo, Summer Galloway, Nicole L. Washington, Simon White, Geraint Levan, Kelly Schiabor Barrett, Elizabeth Cirulli, Alexandre Bolze, Ary Ascencio, Charlotte Rivera-Garcia, Ryan Cho, Jason Nguyen, Sherry Wang, Jimmy Ramirez, Tyler Cassens, Efren Sandoval, Magnus Isaksson, William Lee, David Becker, Marc Laurent, James Lu, Clinton R. Paden, Suxiang Tong, Duncan MacCannell |
| EPI_ISL_1340222, EPI_ISL_1340223, EPI_ISL_1340224 | NJDOH, Public Health and Environmental Laboratories | New Jersey Public Health and Environmental Laboratories (PHEL) | Lindsey Bodnar, Shiv K. Verma, Dana Woell, Byeong Jeong |
| EPI_ISL_1340225 | Helix/Illumina | Centers for Disease Control and Prevention Division of Viral Diseases, Pathogen Discovery | Peter W. Cook, Dakota Howard, Dhwani Batra, Ben L. Rambo-Martin, Eileen de Feo, Jan Antico, Christine Tran, Matthew Tolentino, Shannon Wickline, Kim Gietzen, Brad Sickler, Jingtao Liu, Eric Allen, Phil Febbo, Summer Galloway, Nicole L. Washington, Simon White, Geraint Levan, Kelly Schiabor Barrett, Elizabeth Cirulli, Alexandre Bolze, Ary Ascencio, Charlotte Rivera-Garcia, Ryan Cho, Jason Nguyen, Sherry Wang, Jimmy Ramirez, Tyler Cassens, Efren Sandoval, Magnus Isaksson, William Lee, David Becker, Marc Laurent, James Lu, Clinton R. Paden, Suxiang Tong, Duncan MacCannell |
| EPI_ISL_1340226, EPI_ISL_1340227, EPI_ISL_1340229, EPI_ISL_1340230 | NJDOH, Public Health and Environmental Laboratories | New Jersey Public Health and Environmental Laboratories (PHEL) | Lindsey Bodnar, Shiv K. Verma, Dana Woell, Byeong Jeong |
| EPI_ISL_1340231 | Helix/Illumina | Centers for Disease Control and Prevention Division of Viral Diseases, Pathogen Discovery | Peter W. Cook, Dakota Howard, Dhwani Batra, Ben L. Rambo-Martin, Eileen de Feo, Jan Antico, Christine Tran, Matthew Tolentino, Shannon Wickline, Kim Gietzen, Brad Sickler, Jingtao Liu, Eric Allen, Phil Febbo, Summer Galloway, Nicole L. Washington, Simon White, Geraint Levan, Kelly Schiabor Barrett, Elizabeth Cirulli, Alexandre Bolze, Ary Ascencio, Charlotte Rivera-Garcia, Ryan Cho, Jason Nguyen, Sherry Wang, Jimmy Ramirez, Tyler Cassens, Efren Sandoval, Magnus Isaksson, William Lee, David Becker, Marc Laurent, James Lu, Clinton R. Paden, Suxiang Tong, Duncan MacCannell |
| EPI_ISL_1340232, EPI_ISL_1340233 | NJDOH, Public Health and Environmental Laboratories | New Jersey Public Health and Environmental Laboratories (PHEL) | Lindsey Bodnar, Shiv K. Verma, Dana Woell, Byeong Jeong |
| EPI_ISL_1340234, EPI_ISL_1340235, EPI_ISL_1340236, EPI_ISL_1340237, EPI_ISL_1340238, EPI_ISL_1340239, EPI_ISL_1340240, EPI_ISL_1340241, EPI_ISL_1340242, EPI_ISL_1340243, EPI_ISL_1340244, EPI_ISL_1340245, EPI_ISL_1340246, EPI_ISL_1340247, EPI_ISL_1340248, EPI_ISL_1340249, EPI_ISL_1340250, EPI_ISL_1340251, EPI_ISL_1340252, EPI_ISL_1340253, EPI_ISL_1340254, EPI_ISL_1340255, EPI_ISL_1340256, EPI_ISL_1340257, EPI_ISL_1340258, EPI_ISL_1340259, EPI_ISL_1340260, EPI_ISL_1340261, EPI_ISL_1340262, EPI_ISL_1340263, EPI_ISL_1340264, EPI_ISL_1340265, EPI_ISL_1340266, EPI_ISL_1340267, EPI_ISL_1340268, EPI_ISL_1340269, EPI_ISL_1340270, EPI_ISL_1340271, EPI_ISL_1340272, EPI_ISL_1340273, EPI_ISL_1340274, EPI_ISL_1340275, EPI_ISL_1340276, EPI_ISL_1340277, EPI_ISL_1340278, EPI_ISL_1340279, EPI_ISL_1340280, EPI_ISL_1340281, EPI_ISL_1340282, EPI_ISL_1340283, EPI_ISL_1340284, EPI_ISL_1340285, EPI_ISL_1340287, EPI_ISL_1340288, EPI_ISL_1340289, EPI_ISL_1340290, EPI_ISL_1340291, EPI_ISL_1340292, EPI_ISL_1340293, EPI_ISL_1340294, EPI_ISL_1340295, EPI_ISL_1340296, EPI_ISL_1340297, EPI_ISL_1340298, EPI_ISL_1340299, EPI_ISL_1340300, EPI_ISL_1340301, EPI_ISL_1340302, EPI_ISL_1340303, EPI_ISL_1340304, EPI_ISL_1340305, EPI_ISL_1340306, EPI_ISL_1340307, EPI_ISL_1340308, EPI_ISL_1340309, EPI_ISL_1340310, EPI_ISL_1340311, EPI_ISL_1340312, EPI_ISL_1340313, EPI_ISL_1340314, EPI_ISL_1340315, EPI_ISL_1340316, EPI_ISL_1340317, EPI_ISL_1340318, EPI_ISL_1340319, EPI_ISL_1340320, EPI_ISL_1340321, EPI_ISL_1340322, EPI_ISL_1340323, EPI_ISL_1340324, EPI_ISL_1340325, EPI_ISL_1340326, EPI_ISL_1340327, EPI_ISL_1340328, EPI_ISL_1340329, EPI_ISL_1340330, EPI_ISL_1340331, EPI_ISL_1340332, EPI_ISL_1340333, EPI_ISL_1340334, EPI_ISL_1340335, EPI_ISL_1340336, EPI_ISL_1340337, EPI_ISL_1340338, EPI_ISL_1340339, EPI_ISL_1340340, EPI_ISL_1340341, EPI_ISL_1340342, EPI_ISL_1340343, EPI_ISL_1340344, EPI_ISL_1340345, EPI_ISL_1340346, EPI_ISL_1340347, EPI_ISL_1340348, EPI_ISL_1340349, EPI_ISL_1340350, EPI_ISL_1340351, EPI_ISL_1340352, EPI_ISL_1340353, EPI_ISL_1340354, EPI_ISL_1340355, EPI_ISL_1340356, EPI_ISL_1340357, EPI_ISL_1340358, EPI_ISL_1340359, EPI_ISL_1340360, EPI_ISL_1340361, EPI_ISL_1340362, EPI_ISL_1340363, EPI_ISL_1340364, EPI_ISL_1340365, EPI_ISL_1340366, EPI_ISL_1340367, EPI_ISL_1340368, EPI_ISL_1340369, EPI_ISL_1340370, EPI_ISL_1340371, EPI_ISL_1340372, EPI_ISL_1340373, EPI_ISL_1340375, EPI_ISL_1340376, EPI_ISL_1340377, EPI_ISL_1340378, EPI_ISL_1340379, EPI_ISL_1340380, EPI_ISL_1340381, EPI_ISL_1340382, EPI_ISL_1340383, EPI_ISL_1340384, EPI_ISL_1340385, EPI_ISL_1340386, EPI_ISL_1340387, EPI_ISL_1340388, EPI_ISL_1340389, EPI_ISL_1340390, EPI_ISL_1340391, EPI_ISL_1340392, EPI_ISL_1340393, EPI_ISL_1340394, EPI_ISL_1340395, EPI_ISL_1340396, EPI_ISL_1340397, EPI_ISL_1340398, EPI_ISL_1340399, EPI_ISL_1340400, EPI_ISL_1340401, EPI_ISL_1340402, EPI_ISL_1340403, EPI_ISL_1340404, EPI_ISL_1340405, EPI_ISL_1340406, EPI_ISL_1340407, EPI_ISL_1340408, EPI_ISL_1340409, EPI_ISL_1340410, EPI_ISL_1340411, EPI_ISL_1340412, EPI_ISL_1340413, EPI_ISL_1340414, EPI_ISL_1340416, EPI_ISL_1340417, EPI_ISL_1340418, EPI_ISL_1340419, EPI_ISL_1340420, EPI_ISL_1340421, EPI_ISL_1340422, EPI_ISL_1340423, EPI_ISL_1340424, EPI_ISL_1340425, EPI_ISL_1340426, EPI_ISL_1340427, EPI_ISL_1340428, EPI_ISL_1340429, EPI_ISL_1340430, EPI_ISL_1340431, EPI_ISL_1340432, EPI_ISL_1340433, EPI_ISL_1340434, EPI_ISL_1340435, EPI_ISL_1340436, EPI_ISL_1340437, EPI_ISL_1340438, EPI_ISL_1340439, EPI_ISL_1340440, EPI_ISL_1340441, EPI_ISL_1340442, EPI_ISL_1340443, EPI_ISL_1340444, EPI_ISL_1340445, EPI_ISL_1340446, EPI_ISL_1340447, EPI_ISL_1340448, EPI_ISL_1340449 | | | |
| see above | Helix/Illumina | Centers for Disease Control and Prevention Division of Viral Diseases, Pathogen Discovery | Peter W. Cook, Dakota Howard, Dhwani Batra, Ben L. Rambo-Martin, Eileen de Feo, Jan Antico, Christine Tran, Matthew Tolentino, Shannon Wickline, Kim Gietzen, Brad Sickler, Jingtao Liu, Eric Allen, Phil Febbo, Summer Galloway, Nicole L. Washington, Simon White, Geraint Levan, Kelly Schiabor Barrett, Elizabeth Cirulli, Alexandre Bolze, Ary Ascencio, Charlotte Rivera-Garcia, Ryan Cho, Jason Nguyen, Sherry Wang, Jimmy Ramirez, Tyler Cassens, Efren Sandoval, Magnus Isaksson, William Lee, David Becker, Marc Laurent, James Lu, Clinton R. Paden, Suxiang Tong, Duncan MacCannell |
| EPI_ISL_1340450, EPI_ISL_1340451, EPI_ISL_1340452, EPI_ISL_1340453, EPI_ISL_1340454, EPI_ISL_1340455, EPI_ISL_1340456, EPI_ISL_1340457, EPI_ISL_1340458, EPI_ISL_1340459, EPI_ISL_1340460, EPI_ISL_1340461, EPI_ISL_1340462, EPI_ISL_1340463, EPI_ISL_1340464, EPI_ISL_1340465, EPI_ISL_1340466, EPI_ISL_1340467, EPI_ISL_1340468, EPI_ISL_1340469, EPI_ISL_1340470, EPI_ISL_1340471, EPI_ISL_1340472, EPI_ISL_1340473, EPI_ISL_1340474, EPI_ISL_1340475, EPI_ISL_1340476, EPI_ISL_1340477, EPI_ISL_1340478, EPI_ISL_1340479, EPI_ISL_1340480, EPI_ISL_1340481, EPI_ISL_1340482, EPI_ISL_1340483, EPI_ISL_1340484, EPI_ISL_1340485, EPI_ISL_1340486, EPI_ISL_1340487, EPI_ISL_1340488, EPI_ISL_1340489, EPI_ISL_1340490, EPI_ISL_1340491, EPI_ISL_1340492, EPI_ISL_1340493, EPI_ISL_1340494, EPI_ISL_1340495, EPI_ISL_1340496, EPI_ISL_1340497, EPI_ISL_1340498, EPI_ISL_1340499, EPI_ISL_1340500, EPI_ISL_1340501, EPI_ISL_1340502, EPI_ISL_1340503, EPI_ISL_1340504, EPI_ISL_1340505, EPI_ISL_1340506, EPI_ISL_1340507, EPI_ISL_1340508, EPI_ISL_1340509, EPI_ISL_1340510, EPI_ISL_1340511, EPI_ISL_1340512, EPI_ISL_1340513, EPI_ISL_1340514, EPI_ISL_1340515, EPI_ISL_1340516, EPI_ISL_1340517, EPI_ISL_1340518, EPI_ISL_1340519, EPI_ISL_1340520, EPI_ISL_1340521, EPI_ISL_1340522, EPI_ISL_1340523, EPI_ISL_1340524, EPI_ISL_1340525, EPI_ISL_1340526, EPI_ISL_1340527, EPI_ISL_1340528, EPI_ISL_1340529, EPI_ISL_1340530, EPI_ISL_1340531, EPI_ISL_1340532, EPI_ISL_1340533, EPI_ISL_1340534, EPI_ISL_1340535, EPI_ISL_1340536, EPI_ISL_1340537, EPI_ISL_1340538, EPI_ISL_1340539, EPI_ISL_1340540, EPI_ISL_1340541, EPI_ISL_1340542, EPI_ISL_1340543, EPI_ISL_1340544, EPI_ISL_1340545, EPI_ISL_1340546, EPI_ISL_1340547, EPI_ISL_1340548, EPI_ISL_1340549, EPI_ISL_1340550, EPI_ISL_1340551, EPI_ISL_1340552, EPI_ISL_1340553, EPI_ISL_1340554, EPI_ISL_1340555, EPI_ISL_1340556, EPI_ISL_1340557, EPI_ISL_1340558, EPI_ISL_1340559, EPI_ISL_1340560, EPI_ISL_1340561, EPI_ISL_1340562, EPI_ISL_1340563, EPI_ISL_1340564, EPI_ISL_1340565, EPI_ISL_1340566, EPI_ISL_1340567, EPI_ISL_1340568, EPI_ISL_1340569, EPI_ISL_1340570, EPI_ISL_1340571, EPI_ISL_1340572, EPI_ISL_1340573, EPI_ISL_1340574, EPI_ISL_1340575, EPI_ISL_1340576, EPI_ISL_1340577, EPI_ISL_1340578, EPI_ISL_1340579, EPI_ISL_1340580, EPI_ISL_1340581, EPI_ISL_1340582, EPI_ISL_1340583, EPI_ISL_1340584, EPI_ISL_1340585, EPI_ISL_1340586, EPI_ISL_1340587, EPI_ISL_1340588, EPI_ISL_1340589, EPI_ISL_1340590, EPI_ISL_1340591, EPI_ISL_1340592, EPI_ISL_1340593, EPI_ISL_1340594, EPI_ISL_1340595, EPI_ISL_1340596, EPI_ISL_1340597, EPI_ISL_1340598, EPI_ISL_1340599, EPI_ISL_1340600, EPI_ISL_1340601, EPI_ISL_1340602, EPI_ISL_1340603, EPI_ISL_1340604, EPI_ISL_1340605, EPI_ISL_1340606, EPI_ISL_1340607 | | | |
| see above | Quest Diagnostics Incorporated | Centers for Disease Control and Prevention Division of Viral Diseases, Pathogen Discovery | Peter W. Cook, Dakota Howard, Dhwani Batra, Ben L. Rambo-Martin, S. H. Rosenthal, A. Gerasimova, R. M. Kagan, B. Anderson, M. Hua, Y. Liu, L.E. Bernstein, K.E. Livingston, A. Perez, I. A. Shlyakhter, R. V. Rolando, R. Owen, P. Tanpaiboon, F. Lacbawan, Clinton R. Paden, Suxiang Tong, Duncan MacCannell |
| EPI_ISL_1340608 | LESP Guanajuato | Instituto de Diagnostico y Referencia Epidemiologicos (INDRE) | Claudia Wong-Arambula, Abril Rodriguez-Maldonado, Vanessa Rivero-Arredondo, Ariadna Medina-Benitez, Joaquin Quiroz-Mercado, Sergio Rangel-Guerrero, Natividad Cruz-Ortiz, Tatiana Nunez-Garcia, Gisela Barrera-Badillo, Lucia Hernandez-Rivas, Irma Lopez-Martinez, Ernesto Ramirez-Gonzalez. |
| EPI_ISL_1340609 | Quest Diagnostics Incorporated | Centers for Disease Control and Prevention Division of Viral Diseases, Pathogen Discovery | Peter W. Cook, Dakota Howard, Dhwani Batra, Ben L. Rambo-Martin, S. H. Rosenthal, A. Gerasimova, R. M. Kagan, B. Anderson, M. Hua, Y. Liu, L.E. Bernstein, K.E. Livingston, A. Perez, I. A. Shlyakhter, R. V. Rolando, R. Owen, P. Tanpaiboon, F. Lacbawan, Clinton R. Paden, Suxiang Tong, Duncan MacCannell |
| EPI_ISL_1340610, EPI_ISL_1340611 | LESP Guanajuato | Instituto de Diagnostico y Referencia Epidemiologicos (INDRE) | Claudia Wong-Arambula, Abril Rodriguez-Maldonado, Vanessa Rivero-Arredondo, Ariadna Medina-Benitez, Joaquin Quiroz-Mercado, Sergio Rangel-Guerrero, Natividad Cruz-Ortiz, Tatiana Nunez-Garcia, Gisela Barrera-Badillo, Lucia Hernandez-Rivas, Irma Lopez-Martinez, Ernesto Ramirez-Gonzalez. |

| | | | |
|---|---|---|---|
| EPI_ISL_1340612 | Quest Diagnostics Incorporated | Centers for Disease Control and Prevention Division of Viral Diseases, Pathogen Discovery | Peter W. Cook, Dakota Howard, Dhwani Batra, Ben L. Rambo-Martin, S. H. Rosenthal, A. Gerasimova, R. M. Kagan, B. Anderson, M. Hua, Y. Liu, L.E. Bernstein, K.E. Livingston, A. Perez, I. A. Shlyakhter, R. V. Rolando, R. Owen, P. Tanpaiboon, F. Lacbawan, Clinton R. Paden, Suxiang Tong, Duncan MacCannell |
| EPI_ISL_1340613 | Laboratorios DIAGNOMOL | Instituto de Diagnostico y Referencia Epidemiologicos (INDRE) | Claudia Wong-Arambula, Abril Rodriguez-Maldonado, Vanessa Rivero-Arredondo, Ariadna Medina-Benitez, Joaquin Quiroz-Mercado, Sergio Rangel-Guerrero, Natividad Cruz-Ortiz, Tatiana Nunez-Garcia, Gisela Barrera-Badillo, Lucia Hernandez-Rivas, Irma Lopez-Martinez, Ernesto Ramirez-Gonzalez. |
| EPI_ISL_1340614 | LESP Guanajuato | Instituto de Diagnostico y Referencia Epidemiologicos (INDRE) | Claudia Wong-Arambula, Abril Rodriguez-Maldonado, Vanessa Rivero-Arredondo, Ariadna Medina-Benitez, Joaquin Quiroz-Mercado, Sergio Rangel-Guerrero, Natividad Cruz-Ortiz, Tatiana Nunez-Garcia, Gisela Barrera-Badillo, Lucia Hernandez-Rivas, Irma Lopez-Martinez, Ernesto Ramirez-Gonzalez. |
| EPI_ISL_1340615 | Quest Diagnostics Incorporated | Centers for Disease Control and Prevention Division of Viral Diseases, Pathogen Discovery | Peter W. Cook, Dakota Howard, Dhwani Batra, Ben L. Rambo-Martin, S. H. Rosenthal, A. Gerasimova, R. M. Kagan, B. Anderson, M. Hua, Y. Liu, L.E. Bernstein, K.E. Livingston, A. Perez, I. A. Shlyakhter, R. V. Rolando, R. Owen, P. Tanpaiboon, F. Lacbawan, Clinton R. Paden, Suxiang Tong, Duncan MacCannell |
| EPI_ISL_1340616, EPI_ISL_1340617 | LESP Guanajuato | Instituto de Diagnostico y Referencia Epidemiologicos (INDRE) | Claudia Wong-Arambula, Abril Rodriguez-Maldonado, Vanessa Rivero-Arredondo, Ariadna Medina-Benitez, Joaquin Quiroz-Mercado, Sergio Rangel-Guerrero, Natividad Cruz-Ortiz, Tatiana Nunez-Garcia, Gisela Barrera-Badillo, Lucia Hernandez-Rivas, Irma Lopez-Martinez, Ernesto Ramirez-Gonzalez. |
| EPI_ISL_1340618 | Quest Diagnostics Incorporated | Centers for Disease Control and Prevention Division of Viral Diseases, Pathogen Discovery | Peter W. Cook, Dakota Howard, Dhwani Batra, Ben L. Rambo-Martin, S. H. Rosenthal, A. Gerasimova, R. M. Kagan, B. Anderson, M. Hua, Y. Liu, L.E. Bernstein, K.E. Livingston, A. Perez, I. A. Shlyakhter, R. V. Rolando, R. Owen, P. Tanpaiboon, F. Lacbawan, Clinton R. Paden, Suxiang Tong, Duncan MacCannell |
| EPI_ISL_1340619 | Laboratorios BIMODI | Instituto de Diagnostico y Referencia Epidemiologicos (INDRE) | Claudia Wong-Arambula, Abril Rodriguez-Maldonado, Vanessa Rivero-Arredondo, Ariadna Medina-Benitez, Joaquin Quiroz-Mercado, Sergio Rangel-Guerrero, Natividad Cruz-Ortiz, Tatiana Nunez-Garcia, Gisela Barrera-Badillo, Lucia Hernandez-Rivas, Irma Lopez-Martinez, Ernesto Ramirez-Gonzalez. |
| EPI_ISL_1340620, EPI_ISL_1340621 | Laboratorio BIOMEDICA | Instituto de Diagnostico y Referencia Epidemiologicos (INDRE) | Claudia Wong-Arambula, Abril Rodriguez-Maldonado, Vanessa Rivero-Arredondo, Ariadna Medina-Benitez, Joaquin Quiroz-Mercado, Sergio Rangel-Guerrero, Natividad Cruz-Ortiz, Tatiana Nunez-Garcia, Gisela Barrera-Badillo, Lucia Hernandez-Rivas, Irma Lopez-Martinez, Ernesto Ramirez-Gonzalez. |
| EPI_ISL_1340622 | Quest Diagnostics Incorporated | Centers for Disease Control and Prevention Division of Viral Diseases, Pathogen Discovery | Peter W. Cook, Dakota Howard, Dhwani Batra, Ben L. Rambo-Martin, S. H. Rosenthal, A. Gerasimova, R. M. Kagan, B. Anderson, M. Hua, Y. Liu, L.E. Bernstein, K.E. Livingston, A. Perez, I. A. Shlyakhter, R. V. Rolando, R. Owen, P. Tanpaiboon, F. Lacbawan, Clinton R. Paden, Suxiang Tong, Duncan MacCannell |
| EPI_ISL_1340623 | Laboratorio BIOMEDICA | Instituto de Diagnostico y Referencia Epidemiologicos (INDRE) | Claudia Wong-Arambula, Abril Rodriguez-Maldonado, Vanessa Rivero-Arredondo, Ariadna Medina-Benitez, Joaquin Quiroz-Mercado, Sergio Rangel-Guerrero, Natividad Cruz-Ortiz, Tatiana Nunez-Garcia, Gisela Barrera-Badillo, Lucia Hernandez-Rivas, Irma Lopez-Martinez, Ernesto Ramirez-Gonzalez. |
| EPI_ISL_1340624 | LESP Guanajuato | Instituto de Diagnostico y Referencia Epidemiologicos (INDRE) | Claudia Wong-Arambula, Abril Rodriguez-Maldonado, Vanessa Rivero-Arredondo, Ariadna Medina-Benitez, Joaquin Quiroz-Mercado, Sergio Rangel-Guerrero, Natividad Cruz-Ortiz, Tatiana Nunez-Garcia, Gisela Barrera-Badillo, Lucia Hernandez-Rivas, Irma Lopez-Martinez, Ernesto Ramirez-Gonzalez. |
| EPI_ISL_1340625 | Quest Diagnostics Incorporated | Centers for Disease Control and Prevention Division of Viral Diseases, Pathogen Discovery | Peter W. Cook, Dakota Howard, Dhwani Batra, Ben L. Rambo-Martin, S. H. Rosenthal, A. Gerasimova, R. M. Kagan, B. Anderson, M. Hua, Y. Liu, L.E. Bernstein, K.E. Livingston, A. Perez, I. A. Shlyakhter, R. V. Rolando, R. Owen, P. Tanpaiboon, F. Lacbawan, Clinton R. Paden, Suxiang Tong, Duncan MacCannell |
| EPI_ISL_1340626, EPI_ISL_1340627 | LESP Guanajuato | Instituto de Diagnostico y Referencia Epidemiologicos (INDRE) | Claudia Wong-Arambula, Abril Rodriguez-Maldonado, Vanessa Rivero-Arredondo, Ariadna Medina-Benitez, Joaquin Quiroz-Mercado, Sergio Rangel-Guerrero, Natividad Cruz-Ortiz, Tatiana Nunez-Garcia, Gisela Barrera-Badillo, Lucia Hernandez-Rivas, Irma Lopez-Martinez, Ernesto Ramirez-Gonzalez. |
| EPI_ISL_1340628 | Quest Diagnostics Incorporated | Centers for Disease Control and Prevention Division of Viral Diseases, Pathogen Discovery | Peter W. Cook, Dakota Howard, Dhwani Batra, Ben L. Rambo-Martin, S. H. Rosenthal, A. Gerasimova, R. M. Kagan, B. Anderson, M. Hua, Y. Liu, L.E. Bernstein, K.E. Livingston, A. Perez, I. A. Shlyakhter, R. V. Rolando, R. Owen, P. Tanpaiboon, F. Lacbawan, Clinton R. Paden, Suxiang Tong, Duncan MacCannell |
| EPI_ISL_1340629 | INER | Instituto de Diagnostico y Referencia Epidemiologicos (INDRE) | Claudia Wong-Arambula, Abril Rodriguez-Maldonado, Vanessa Rivero-Arredondo, Ariadna Medina-Benitez, Joaquin Quiroz-Mercado, Sergio Rangel-Guerrero, Natividad Cruz-Ortiz, Tatiana Nunez-Garcia, Gisela Barrera-Badillo, Lucia Hernandez-Rivas, Irma Lopez-Martinez, Ernesto Ramirez-Gonzalez. |
| EPI_ISL_1340630, EPI_ISL_1340631 | LESP Guanajuato | Instituto de Diagnostico y Referencia Epidemiologicos (INDRE) | Claudia Wong-Arambula, Abril Rodriguez-Maldonado, Vanessa Rivero-Arredondo, Ariadna Medina-Benitez, Joaquin Quiroz-Mercado, Sergio Rangel-Guerrero, Natividad Cruz-Ortiz, Tatiana Nunez-Garcia, Gisela Barrera-Badillo, Lucia Hernandez-Rivas, Irma Lopez-Martinez, Ernesto Ramirez-Gonzalez. |
| EPI_ISL_1340632 | Quest Diagnostics Incorporated | Centers for Disease Control and Prevention Division of Viral Diseases, Pathogen Discovery | Peter W. Cook, Dakota Howard, Dhwani Batra, Ben L. Rambo-Martin, S. H. Rosenthal, A. Gerasimova, R. M. Kagan, B. Anderson, M. Hua, Y. Liu, L.E. Bernstein, K.E. Livingston, A. Perez, I. A. Shlyakhter, R. V. Rolando, R. Owen, P. Tanpaiboon, F. Lacbawan, Clinton R. Paden, Suxiang Tong, Duncan MacCannell |
| EPI_ISL_1340633 | LESP Guanajuato | Instituto de Diagnostico y Referencia Epidemiologicos (INDRE) | Claudia Wong-Arambula, Abril Rodriguez-Maldonado, Vanessa Rivero-Arredondo, Ariadna Medina-Benitez, Joaquin Quiroz-Mercado, Sergio Rangel-Guerrero, Natividad Cruz-Ortiz, Tatiana Nunez-Garcia, Gisela Barrera-Badillo, Lucia Hernandez-Rivas, Irma Lopez-Martinez, Ernesto Ramirez-Gonzalez. |
| EPI_ISL_1340634 | Laboratorios BIMODI | Instituto de Diagnostico y Referencia Epidemiologicos (INDRE) | Claudia Wong-Arambula, Abril Rodriguez-Maldonado, Vanessa Rivero-Arredondo, Ariadna Medina-Benitez, Joaquin Quiroz-Mercado, Sergio Rangel-Guerrero, Natividad Cruz-Ortiz, Tatiana Nunez-Garcia, Gisela Barrera-Badillo, Lucia Hernandez-Rivas, Irma Lopez-Martinez, Ernesto Ramirez-Gonzalez. |
| EPI_ISL_1340635 | Quest Diagnostics Incorporated | Centers for Disease Control and Prevention Division of Viral Diseases, Pathogen Discovery | Peter W. Cook, Dakota Howard, Dhwani Batra, Ben L. Rambo-Martin, S. H. Rosenthal, A. Gerasimova, R. M. Kagan, B. Anderson, M. Hua, Y. Liu, L.E. Bernstein, K.E. Livingston, A. Perez, I. A. Shlyakhter, R. V. Rolando, R. Owen, P. Tanpaiboon, F. Lacbawan, Clinton R. Paden, Suxiang Tong, Duncan MacCannell |
| EPI_ISL_1340636, EPI_ISL_1340637 | LESP Queretaro | Instituto de Diagnostico y Referencia Epidemiologicos (INDRE) | Claudia Wong-Arambula, Abril Rodriguez-Maldonado, Vanessa Rivero-Arredondo, Ariadna Medina-Benitez, Joaquin Quiroz-Mercado, Sergio Rangel-Guerrero, Natividad Cruz-Ortiz, Tatiana Nunez-Garcia, Gisela Barrera-Badillo, Lucia Hernandez-Rivas, Irma Lopez-Martinez, Ernesto Ramirez-Gonzalez. |
| EPI_ISL_1340638 | Quest Diagnostics Incorporated | Centers for Disease Control and Prevention Division of Viral Diseases, Pathogen Discovery | Peter W. Cook, Dakota Howard, Dhwani Batra, Ben L. Rambo-Martin, S. H. Rosenthal, A. Gerasimova, R. M. Kagan, B. Anderson, M. Hua, Y. Liu, L.E. Bernstein, K.E. Livingston, A. Perez, I. A. Shlyakhter, R. V. Rolando, R. Owen, P. Tanpaiboon, F. Lacbawan, Clinton R. Paden, Suxiang Tong, Duncan MacCannell |
| EPI_ISL_1340639 | LESP Queretaro | Instituto de Diagnostico y Referencia Epidemiologicos (INDRE) | Claudia Wong-Arambula, Abril Rodriguez-Maldonado, Vanessa Rivero-Arredondo, Ariadna Medina-Benitez, Joaquin Quiroz-Mercado, Sergio Rangel-Guerrero, Natividad Cruz-Ortiz, Tatiana Nunez-Garcia, Gisela Barrera-Badillo, Lucia Hernandez-Rivas, Irma Lopez-Martinez, Ernesto Ramirez-Gonzalez. |
| EPI_ISL_1340640 | LESP Hidalgo | Instituto de Diagnostico y Referencia Epidemiologicos (INDRE) | Claudia Wong-Arambula, Abril Rodriguez-Maldonado, Vanessa Rivero-Arredondo, Ariadna Medina-Benitez, Joaquin Quiroz-Mercado, Sergio Rangel-Guerrero, Natividad Cruz-Ortiz, Tatiana Nunez-Garcia, Gisela Barrera-Badillo, Lucia Hernandez-Rivas, Irma Lopez-Martinez, Ernesto Ramirez-Gonzalez. |
| EPI_ISL_1340641 | Quest Diagnostics Incorporated | Centers for Disease Control and Prevention Division of Viral Diseases, Pathogen Discovery | Peter W. Cook, Dakota Howard, Dhwani Batra, Ben L. Rambo-Martin, S. H. Rosenthal, A. Gerasimova, R. M. Kagan, B. Anderson, M. Hua, Y. Liu, L.E. Bernstein, K.E. Livingston, A. Perez, I. A. Shlyakhter, R. V. Rolando, R. Owen, P. Tanpaiboon, F. Lacbawan, Clinton R. Paden, Suxiang Tong, Duncan MacCannell |
| EPI_ISL_1340642 | LESP Hidalgo | Instituto de Diagnostico y Referencia Epidemiologicos (INDRE) | Claudia Wong-Arambula, Abril Rodriguez-Maldonado, Vanessa Rivero-Arredondo, Ariadna Medina-Benitez, Joaquin Quiroz-Mercado, Sergio Rangel-Guerrero, Natividad Cruz-Ortiz, Tatiana Nunez-Garcia, Gisela Barrera-Badillo, Lucia Hernandez-Rivas, Irma Lopez-Martinez, Ernesto Ramirez-Gonzalez. |
| EPI_ISL_1340643, EPI_ISL_1340644 | LESP Guanajuato | Instituto de Diagnostico y Referencia Epidemiologicos (INDRE) | Claudia Wong-Arambula, Abril Rodriguez-Maldonado, Vanessa Rivero-Arredondo, Ariadna Medina-Benitez, Joaquin Quiroz-Mercado, Sergio Rangel-Guerrero, Natividad Cruz-Ortiz, Tatiana Nunez-Garcia, Gisela Barrera-Badillo, Lucia Hernandez-Rivas, Irma Lopez-Martinez, Ernesto Ramirez-Gonzalez. |
| EPI_ISL_1340645 | Quest Diagnostics Incorporated | Centers for Disease Control and Prevention Division of Viral Diseases, Pathogen Discovery | Peter W. Cook, Dakota Howard, Dhwani Batra, Ben L. Rambo-Martin, S. H. Rosenthal, A. Gerasimova, R. M. Kagan, B. Anderson, M. Hua, Y. Liu, L.E. Bernstein, K.E. Livingston, A. Perez, I. A. Shlyakhter, R. V. Rolando, R. Owen, P. Tanpaiboon, F. Lacbawan, Clinton R. Paden, Suxiang Tong, Duncan MacCannell |
| EPI_ISL_1340646, EPI_ISL_1340647, EPI_ISL_1340648 | LESP Guanajuato | Instituto de Diagnostico y Referencia Epidemiologicos (INDRE) | Claudia Wong-Arambula, Abril Rodriguez-Maldonado, Vanessa Rivero-Arredondo, Ariadna Medina-Benitez, Joaquin Quiroz-Mercado, Sergio Rangel-Guerrero, Natividad Cruz-Ortiz, Tatiana Nunez-Garcia, Gisela Barrera-Badillo, Lucia Hernandez-Rivas, Irma Lopez-Martinez, Ernesto Ramirez-Gonzalez. |
| EPI_ISL_1340649 | Quest Diagnostics Incorporated | Centers for Disease Control and Prevention Division of Viral Diseases, Pathogen Discovery | Peter W. Cook, Dakota Howard, Dhwani Batra, Ben L. Rambo-Martin, S. H. Rosenthal, A. Gerasimova, R. M. Kagan, B. Anderson, M. Hua, Y. Liu, L.E. Bernstein, K.E. Livingston, A. Perez, I. A. Shlyakhter, R. V. Rolando, R. Owen, P. Tanpaiboon, F. Lacbawan, Clinton R. Paden, Suxiang Tong, Duncan MacCannell |
| EPI_ISL_1340650 | LESP Guanajuato | Instituto de Diagnostico y Referencia Epidemiologicos (INDRE) | Claudia Wong-Arambula, Abril Rodriguez-Maldonado, Vanessa Rivero-Arredondo, Ariadna Medina-Benitez, Joaquin Quiroz-Mercado, Sergio Rangel-Guerrero, Natividad Cruz-Ortiz, Tatiana Nunez-Garcia, Gisela Barrera-Badillo, Lucia Hernandez-Rivas, Irma Lopez-Martinez, Ernesto Ramirez-Gonzalez. |
| EPI_ISL_1340651, EPI_ISL_1340652 | LESP Puebla | Instituto de Diagnostico y Referencia Epidemiologicos (INDRE) | Claudia Wong-Arambula, Abril Rodriguez-Maldonado, Vanessa Rivero-Arredondo, Ariadna Medina-Benitez, Joaquin Quiroz-Mercado, Sergio Rangel-Guerrero, Natividad Cruz-Ortiz, Tatiana Nunez-Garcia, Gisela Barrera-Badillo, Lucia Hernandez-Rivas, Irma Lopez-Martinez, Ernesto Ramirez-Gonzalez. |
| EPI_ISL_1340653 | Quest Diagnostics Incorporated | Centers for Disease Control and Prevention Division of Viral Diseases, Pathogen Discovery | Peter W. Cook, Dakota Howard, Dhwani Batra, Ben L. Rambo-Martin, S. H. Rosenthal, A. Gerasimova, R. M. Kagan, B. Anderson, M. Hua, Y. Liu, L.E. Bernstein, K.E. Livingston, A. Perez, I. A. Shlyakhter, R. V. Rolando, R. Owen, P. Tanpaiboon, F. Lacbawan, Clinton R. Paden, Suxiang Tong, Duncan MacCannell |
| EPI_ISL_1340654 | Laboratorios BIMODI | Instituto de Diagnostico y Referencia Epidemiologicos (INDRE) | Claudia Wong-Arambula, Abril Rodriguez-Maldonado, Vanessa Rivero-Arredondo, Ariadna Medina-Benitez, Joaquin Quiroz-Mercado, Sergio Rangel-Guerrero, Natividad Cruz-Ortiz, Tatiana Nunez-Garcia, Gisela Barrera-Badillo, Lucia Hernandez-Rivas, Irma Lopez-Martinez, Ernesto Ramirez-Gonzalez. |
| EPI_ISL_1340655 | LESP Queretaro | Instituto de Diagnostico y Referencia Epidemiologicos (INDRE) | Claudia Wong-Arambula, Abril Rodriguez-Maldonado, Vanessa Rivero-Arredondo, Ariadna Medina-Benitez, Joaquin Quiroz-Mercado, Sergio Rangel-Guerrero, Natividad Cruz-Ortiz, Tatiana Nunez-Garcia, Gisela Barrera-Badillo, Lucia Hernandez-Rivas, Irma Lopez-Martinez, Ernesto Ramirez-Gonzalez. |
| EPI_ISL_1340656 | Quest Diagnostics Incorporated | Centers for Disease Control and Prevention Division of Viral Diseases, Pathogen Discovery | Peter W. Cook, Dakota Howard, Dhwani Batra, Ben L. Rambo-Martin, S. H. Rosenthal, A. Gerasimova, R. M. Kagan, B. Anderson, M. Hua, Y. Liu, L.E. Bernstein, K.E. Livingston, A. Perez, I. A. Shlyakhter, R. V. Rolando, R. Owen, P. Tanpaiboon, F. Lacbawan, Clinton R. Paden, Suxiang Tong, Duncan MacCannell |
| EPI_ISL_1340657 | LESP Queretaro | Instituto de Diagnostico y Referencia Epidemiologicos (INDRE) | Claudia Wong-Arambula, Abril Rodriguez-Maldonado, Vanessa Rivero-Arredondo, Ariadna Medina-Benitez, Joaquin Quiroz-Mercado, Sergio Rangel-Guerrero, Natividad Cruz-Ortiz, Tatiana Nunez-Garcia, Gisela Barrera-Badillo, Lucia Hernandez-Rivas, Irma Lopez-Martinez, Ernesto Ramirez-Gonzalez. |
| EPI_ISL_1340658 | LESP Yucatan | Instituto de Diagnostico y Referencia Epidemiologicos (INDRE) | Claudia Wong-Arambula, Abril Rodriguez-Maldonado, Vanessa Rivero-Arredondo, Ariadna Medina-Benitez, Joaquin Quiroz-Mercado, Sergio Rangel-Guerrero, Natividad Cruz-Ortiz, Tatiana Nunez-Garcia, Gisela Barrera-Badillo, Lucia Hernandez-Rivas, Irma Lopez-Martinez, Ernesto Ramirez-Gonzalez. |
| EPI_ISL_1340659 | Quest Diagnostics Incorporated | Centers for Disease Control and Prevention Division of Viral Diseases, Pathogen | Peter W. Cook, Dakota Howard, Dhwani Batra, Ben L. Rambo-Martin, S. H. Rosenthal, A. Gerasimova, R. M. Kagan, B. Anderson, M. Hua, Y. Liu, L.E. Bernstein, K.E. Livingston, A. Perez, I. A. Shlyakhter, R. V. Rolando, R. Owen, P. Tanpaiboon, F. Lacbawan, Clinton R. Paden, Suxiang Tong, Duncan MacCannell |

| | | | |
|---|---|---|---|
| | | Discovery | |
| EPI_ISL_1340660 | LESP Zacatecas | Instituto de Diagnostico y Referencia Epidemiologicos (INDRE) | Claudia Wong-Arambula, Abril Rodriguez-Maldonado, Vanessa Rivero-Arredondo, Ariadna Medina-Benitez, Joaquin Quiroz-Mercado, Sergio Rangel-Guerrero, Natividad Cruz-Ortiz, Tatiana Nunez-Garcia, Gisela Barrera-Badillo, Lucia Hernandez-Rivas, Irma Lopez-Martinez, Ernesto Ramirez-Gonzalez. |
| EPI_ISL_1340661 | LESP Yucatan | Instituto de Diagnostico y Referencia Epidemiologicos (INDRE) | Claudia Wong-Arambula, Abril Rodriguez-Maldonado, Vanessa Rivero-Arredondo, Ariadna Medina-Benitez, Joaquin Quiroz-Mercado, Sergio Rangel-Guerrero, Natividad Cruz-Ortiz, Tatiana Nunez-Garcia, Gisela Barrera-Badillo, Lucia Hernandez-Rivas, Irma Lopez-Martinez, Ernesto Ramirez-Gonzalez. |
| EPI_ISL_1340662, EPI_ISL_1340663, EPI_ISL_1340664, EPI_ISL_1340665, EPI_ISL_1340666, EPI_ISL_1340667, EPI_ISL_1340668, EPI_ISL_1340669, EPI_ISL_1340670, EPI_ISL_1340671, EPI_ISL_1340672, EPI_ISL_1340673, EPI_ISL_1340674, EPI_ISL_1340675, EPI_ISL_1340676, EPI_ISL_1340677, EPI_ISL_1340678, EPI_ISL_1340679, EPI_ISL_1340680, EPI_ISL_1340681, EPI_ISL_1340682, EPI_ISL_1340683, EPI_ISL_1340684, EPI_ISL_1340685, EPI_ISL_1340686, EPI_ISL_1340687, EPI_ISL_1340688, EPI_ISL_1340689, EPI_ISL_1340690, EPI_ISL_1340691, EPI_ISL_1340692, EPI_ISL_1340693, EPI_ISL_1340694, EPI_ISL_1340695, EPI_ISL_1340696, EPI_ISL_1340697, EPI_ISL_1340698, EPI_ISL_1340699, EPI_ISL_1340700, EPI_ISL_1340701, EPI_ISL_1340702, EPI_ISL_1340703, EPI_ISL_1340704, EPI_ISL_1340705, EPI_ISL_1340706, EPI_ISL_1340707, EPI_ISL_1340708, EPI_ISL_1340709, EPI_ISL_1340710, EPI_ISL_1340711, EPI_ISL_1340712, EPI_ISL_1340713, EPI_ISL_1340714, EPI_ISL_1340715, EPI_ISL_1340716, EPI_ISL_1340717, EPI_ISL_1340718, EPI_ISL_1340719, EPI_ISL_1340720, EPI_ISL_1340721, EPI_ISL_1340722, EPI_ISL_1340723, EPI_ISL_1340724, EPI_ISL_1340725, EPI_ISL_1340726, EPI_ISL_1340727, EPI_ISL_1340728, EPI_ISL_1340729, EPI_ISL_1340730, EPI_ISL_1340731, EPI_ISL_1340732, EPI_ISL_1340733, EPI_ISL_1340734, EPI_ISL_1340735, EPI_ISL_1340736, EPI_ISL_1340737, EPI_ISL_1340738, EPI_ISL_1340739, EPI_ISL_1340740, EPI_ISL_1340741, EPI_ISL_1340742, EPI_ISL_1340743, EPI_ISL_1340744, EPI_ISL_1340745, EPI_ISL_1340746, EPI_ISL_1340747, EPI_ISL_1340748, EPI_ISL_1340749 | | | |
| see above | Quest Diagnostics Incorporated | Centers for Disease Control and Prevention Division of Viral Diseases, Pathogen Discovery | Peter W. Cook, Dakota Howard, Dhwani Batra, Ben L. Rambo-Martin, S. H. Rosenthal, A. Gerasimova, R. M. Kagan, B. Anderson, M. Hua, Y. Liu, L.E. Bernstein, K.E. Livingston, A. Perez, I. A. Shlyakhter, R. V. Rolando, R. Owen, P. Tanpaiboon, F. Lacbawan, Clinton R. Paden, Suxiang Tong, Duncan MacCannell |
| EPI_ISL_1340765, EPI_ISL_1340766, EPI_ISL_1340767, EPI_ISL_1340768, EPI_ISL_1340769, EPI_ISL_1340771, EPI_ISL_1340772, EPI_ISL_1340773, EPI_ISL_1340774, EPI_ISL_1340775, EPI_ISL_1340776, EPI_ISL_1340777, EPI_ISL_1340780 | | | |
| see above | Wyoming Public Health Laboratory | Wyoming Public Health Laboratory | Jim Mildenberger, Wanda Manley, Noah Hull, Taylor Fearing, Lynette Gumbleton, Channing Weber, Ashley Norberg, Chayse Rowley, Marley Goetz, Brian Dominguez, Elliot Thomasson, Cari Sloma, and Rob Christensen |
| EPI_ISL_1340782, EPI_ISL_1340783, EPI_ISL_1340784, EPI_ISL_1340785, EPI_ISL_1340786, EPI_ISL_1340787, EPI_ISL_1340788, EPI_ISL_1340789, EPI_ISL_1340790, EPI_ISL_1340791, EPI_ISL_1340792, EPI_ISL_1340793, EPI_ISL_1340794, EPI_ISL_1340795, EPI_ISL_1340796, EPI_ISL_1340798, EPI_ISL_1340799, EPI_ISL_1340800, EPI_ISL_1340801, EPI_ISL_1340802, EPI_ISL_1340803, EPI_ISL_1340804, EPI_ISL_1340805, EPI_ISL_1340806, EPI_ISL_1340807, EPI_ISL_1340808, EPI_ISL_1340809, EPI_ISL_1340810, EPI_ISL_1340811, EPI_ISL_1340812, EPI_ISL_1340813, EPI_ISL_1340814, EPI_ISL_1340815, EPI_ISL_1340816, EPI_ISL_1340817, EPI_ISL_1340818, EPI_ISL_1340819, EPI_ISL_1340820, EPI_ISL_1340821, EPI_ISL_1340822, EPI_ISL_1340823, EPI_ISL_1340824, EPI_ISL_1340825, EPI_ISL_1340826, EPI_ISL_1340827, EPI_ISL_1340828, EPI_ISL_1340829, EPI_ISL_1340830, EPI_ISL_1340831, EPI_ISL_1340832, EPI_ISL_1340833, EPI_ISL_1340835, EPI_ISL_1340836, EPI_ISL_1340837, EPI_ISL_1340838, EPI_ISL_1340839, EPI_ISL_1340840, EPI_ISL_1340841, EPI_ISL_1340842, EPI_ISL_1340843, EPI_ISL_1340844, EPI_ISL_1340845, EPI_ISL_1340846, EPI_ISL_1340847, EPI_ISL_1340848, EPI_ISL_1340849, EPI_ISL_1340850, EPI_ISL_1340851, EPI_ISL_1340852, EPI_ISL_1340853, EPI_ISL_1340854, EPI_ISL_1340855, EPI_ISL_1340856, EPI_ISL_1340857, EPI_ISL_1340858, EPI_ISL_1340860, EPI_ISL_1340861 | | | |
| see above | Northwestern Memorial Hospital | Northwestern University - Ozer Lab | Ramon Lorenzo-Redondo, Lacy M. Simons, Taylor J. Dean, Chad J. Achenbach, Lawrence J. Jennings, Chao Qi, Michael G. Ison, Judd F. Hultquist, Egon A. Ozer |
| EPI_ISL_1340880, EPI_ISL_1340881, EPI_ISL_1340882, EPI_ISL_1340883, EPI_ISL_1340884, EPI_ISL_1340885, EPI_ISL_1340886, EPI_ISL_1340887, EPI_ISL_1340888, EPI_ISL_1340889, EPI_ISL_1340890, EPI_ISL_1340891, EPI_ISL_1340892, EPI_ISL_1340893 | | | |
| see above | New Mexico Department of Health Scientific Laboratory | New Mexico Department of Health Scientific Laboratory | Ellie Johnson, Anastacia Griego-Fisher, D'eldra Malone, Jennifer Benoit |
| EPI_ISL_1340894, EPI_ISL_1340895, EPI_ISL_1340896, EPI_ISL_1340897 | TriCore Reference Laboratory Inc. | New Mexico Department of Health Scientific Laboratory | Ellie Johnson, Anastacia Griego-Fisher, D'eldra Malone, Jennifer Benoit |
| EPI_ISL_1340898, EPI_ISL_1340899 | New Mexico Department of Health Scientific Laboratory | New Mexico Department of Health Scientific Laboratory | Ellie Johnson, Anastacia Griego-Fisher, D'eldra Malone, Jennifer Benoit |
| EPI_ISL_1340900 | VA Hospital Path & Lab | New Mexico Department of Health Scientific Laboratory | Ellie Johnson, Anastacia Griego-Fisher, D'eldra Malone, Jennifer Benoit |
| EPI_ISL_1340901, EPI_ISL_1340903, EPI_ISL_1340904, EPI_ISL_1340905, EPI_ISL_1340906, EPI_ISL_1340907, EPI_ISL_1340908, EPI_ISL_1340909, EPI_ISL_1340910, EPI_ISL_1340911 | New Mexico Department of Health Scientific Laboratory | New Mexico Department of Health Scientific Laboratory | Ellie Johnson, Anastacia Griego-Fisher, D'eldra Malone, Jennifer Benoit |
| EPI_ISL_1340912, EPI_ISL_1340913 | San Juan Regional Medical Center | New Mexico Department of Health Scientific Laboratory | Ellie Johnson, Anastacia Griego-Fisher, D'eldra Malone, Jennifer Benoit |
| EPI_ISL_1340914, EPI_ISL_1340915, EPI_ISL_1340916 | PHS/IHS Hospital-Jicarilla Service Unit | New Mexico Department of Health Scientific Laboratory | Ellie Johnson, Anastacia Griego-Fisher, D'eldra Malone, Jennifer Benoit |
| EPI_ISL_1340917, EPI_ISL_1340918, EPI_ISL_1340919, EPI_ISL_1340920, EPI_ISL_1340921 | New Mexico Department of Health Scientific Laboratory | New Mexico Department of Health Scientific Laboratory | Ellie Johnson, Anastacia Griego-Fisher, D'eldra Malone, Jennifer Benoit |
| EPI_ISL_1340922 | Gallup Indian Medical Center | New Mexico Department of Health Scientific Laboratory | Ellie Johnson, Anastacia Griego-Fisher, D'eldra Malone, Jennifer Benoit |
| EPI_ISL_1340923, EPI_ISL_1340924, EPI_ISL_1340925, EPI_ISL_1340926 | New Mexico Department of Health Scientific Laboratory | New Mexico Department of Health Scientific Laboratory | Ellie Johnson, Anastacia Griego-Fisher, D'eldra Malone, Jennifer Benoit |
| EPI_ISL_1341151, EPI_ISL_1341152, EPI_ISL_1341153, EPI_ISL_1341154, EPI_ISL_1341155, EPI_ISL_1341156, EPI_ISL_1341157, EPI_ISL_1341158, EPI_ISL_1341159, EPI_ISL_1341160, EPI_ISL_1341161, EPI_ISL_1341162, EPI_ISL_1341163, EPI_ISL_1341164, EPI_ISL_1341165, EPI_ISL_1341166, EPI_ISL_1341167, EPI_ISL_1341168, EPI_ISL_1341169, EPI_ISL_1341170, EPI_ISL_1341171, EPI_ISL_1341172, EPI_ISL_1341173, EPI_ISL_1341174, EPI_ISL_1341175, EPI_ISL_1341176, EPI_ISL_1341177, EPI_ISL_1341178, EPI_ISL_1341179, EPI_ISL_1341180, EPI_ISL_1341181, EPI_ISL_1341182, EPI_ISL_1341183, EPI_ISL_1341184, EPI_ISL_1341185, EPI_ISL_1341186, EPI_ISL_1341187, EPI_ISL_1341188, EPI_ISL_1341189, EPI_ISL_1341190, EPI_ISL_1341191, EPI_ISL_1341192, EPI_ISL_1341193, EPI_ISL_1341194, EPI_ISL_1341195, EPI_ISL_1341196, EPI_ISL_1341197, EPI_ISL_1341198, EPI_ISL_1341199, EPI_ISL_1341200, EPI_ISL_1341201, EPI_ISL_1341202, EPI_ISL_1341203, EPI_ISL_1341204, EPI_ISL_1341205, EPI_ISL_1341206, EPI_ISL_1341207, EPI_ISL_1341208, EPI_ISL_1341209, EPI_ISL_1341210, EPI_ISL_1341211, EPI_ISL_1341212, EPI_ISL_1341213, EPI_ISL_1341214, EPI_ISL_1341215, EPI_ISL_1341216, EPI_ISL_1341217, EPI_ISL_1341218, EPI_ISL_1341219, EPI_ISL_1341220, EPI_ISL_1341221, EPI_ISL_1341222, EPI_ISL_1341223, EPI_ISL_1341224, EPI_ISL_1341225, EPI_ISL_1341226, EPI_ISL_1341227, EPI_ISL_1341228, EPI_ISL_1341229, EPI_ISL_1341230, EPI_ISL_1341231, EPI_ISL_1341232, EPI_ISL_1341233, EPI_ISL_1341234, EPI_ISL_1341235, EPI_ISL_1341236, EPI_ISL_1341237, EPI_ISL_1341238, EPI_ISL_1341239, EPI_ISL_1341240, EPI_ISL_1341241, EPI_ISL_1341242, EPI_ISL_1341243, EPI_ISL_1341244, EPI_ISL_1341245, EPI_ISL_1341246, EPI_ISL_1341247, EPI_ISL_1341248, EPI_ISL_1341249, EPI_ISL_1341250, EPI_ISL_1341251, EPI_ISL_1341252, EPI_ISL_1341253, EPI_ISL_1341254, EPI_ISL_1341255, EPI_ISL_1341256, EPI_ISL_1341257, EPI_ISL_1341258, EPI_ISL_1341259, EPI_ISL_1341260, EPI_ISL_1341261, EPI_ISL_1341262, EPI_ISL_1341263, EPI_ISL_1341264, EPI_ISL_1341265, EPI_ISL_1341266, EPI_ISL_1341267, EPI_ISL_1341268, EPI_ISL_1341269, EPI_ISL_1341270, EPI_ISL_1341271, EPI_ISL_1341272, EPI_ISL_1341273, EPI_ISL_1341274, EPI_ISL_1341275, EPI_ISL_1341276, EPI_ISL_1341277, EPI_ISL_1341278, EPI_ISL_1341279, EPI_ISL_1341280, EPI_ISL_1341281, EPI_ISL_1341282, EPI_ISL_1341283, EPI_ISL_1341284, EPI_ISL_1341285, EPI_ISL_1341286, EPI_ISL_1341287, EPI_ISL_1341288, EPI_ISL_1341289, EPI_ISL_1341290, EPI_ISL_1341291, EPI_ISL_1341292, EPI_ISL_1341293, EPI_ISL_1341294, EPI_ISL_1341295, EPI_ISL_1341296, EPI_ISL_1341297, EPI_ISL_1341298, EPI_ISL_1341299, EPI_ISL_1341300, EPI_ISL_1341301, EPI_ISL_1341302, EPI_ISL_1341303, EPI_ISL_1341304, EPI_ISL_1341305, EPI_ISL_1341306, EPI_ISL_1341307, EPI_ISL_1341308, EPI_ISL_1341309, EPI_ISL_1341310, EPI_ISL_1341311, EPI_ISL_1341314, EPI_ISL_1341315, EPI_ISL_1341317, EPI_ISL_1341318, EPI_ISL_1341319, EPI_ISL_1341320, EPI_ISL_1341321, EPI_ISL_1341322, EPI_ISL_1341323, EPI_ISL_1341324, EPI_ISL_1341325, EPI_ISL_1341326, EPI_ISL_1341327, EPI_ISL_1341328, EPI_ISL_1341329, EPI_ISL_1341330, EPI_ISL_1341331, EPI_ISL_1341332, EPI_ISL_1341333, EPI_ISL_1341334, EPI_ISL_1341335, EPI_ISL_1341336, EPI_ISL_1341337, EPI_ISL_1341338, EPI_ISL_1341339, EPI_ISL_1341340, EPI_ISL_1341341, EPI_ISL_1341342, EPI_ISL_1341343, EPI_ISL_1341344, EPI_ISL_1341345, EPI_ISL_1341346, EPI_ISL_1341347, EPI_ISL_1341348, EPI_ISL_1341349, EPI_ISL_1341350, EPI_ISL_1341351, EPI_ISL_1341352, EPI_ISL_1341353, EPI_ISL_1341355, EPI_ISL_1341356, EPI_ISL_1341357, EPI_ISL_1341358, EPI_ISL_1341359, EPI_ISL_1341360, EPI_ISL_1341361, EPI_ISL_1341362, EPI_ISL_1341363, EPI_ISL_1341364, EPI_ISL_1341365, EPI_ISL_1341366, EPI_ISL_1341367, EPI_ISL_1341368, EPI_ISL_1341369, EPI_ISL_1341370, EPI_ISL_1341371, EPI_ISL_1341372, EPI_ISL_1341373, EPI_ISL_1341374, EPI_ISL_1341375, EPI_ISL_1341376, EPI_ISL_1341382, EPI_ISL_1341383, EPI_ISL_1341384, EPI_ISL_1341385, EPI_ISL_1341386, EPI_ISL_1341387, EPI_ISL_1341388, EPI_ISL_1341389, EPI_ISL_1341390, EPI_ISL_1341391, EPI_ISL_1341392, EPI_ISL_1341393, EPI_ISL_1341394, EPI_ISL_1341395, EPI_ISL_1341396, EPI_ISL_1341397, EPI_ISL_1341398, EPI_ISL_1341399, EPI_ISL_1341400, EPI_ISL_1341401, EPI_ISL_1341402, EPI_ISL_1341404, EPI_ISL_1341405, EPI_ISL_1341406, EPI_ISL_1341407, EPI_ISL_1341408, EPI_ISL_1341409, EPI_ISL_1341411, EPI_ISL_1341412, EPI_ISL_1341413, EPI_ISL_1341414, EPI_ISL_1341415, EPI_ISL_1341416, EPI_ISL_1341417, EPI_ISL_1341418, EPI_ISL_1341419, EPI_ISL_1341420, EPI_ISL_1341421, EPI_ISL_1341422, EPI_ISL_1341423, EPI_ISL_1341425, EPI_ISL_1341426, EPI_ISL_1341427, EPI_ISL_1341428, EPI_ISL_1341429, EPI_ISL_1341430, EPI_ISL_1341431, EPI_ISL_1341432, EPI_ISL_1341433, EPI_ISL_1341434, EPI_ISL_1341435, EPI_ISL_1341436, EPI_ISL_1341437, EPI_ISL_1341438, EPI_ISL_1341439, EPI_ISL_1341440, EPI_ISL_1341441, EPI_ISL_1341442, EPI_ISL_1341443, EPI_ISL_1341444, EPI_ISL_1341445, EPI_ISL_1341446, EPI_ISL_1341447, EPI_ISL_1341448, EPI_ISL_1341449, EPI_ISL_1341450, EPI_ISL_1341451, EPI_ISL_1341452, EPI_ISL_1341453, EPI_ISL_1341454, EPI_ISL_1341455, EPI_ISL_1341456, EPI_ISL_1341457, EPI_ISL_1341458, EPI_ISL_1341459, EPI_ISL_1341460, EPI_ISL_1341462, EPI_ISL_1341463, EPI_ISL_1341464, EPI_ISL_1341465, EPI_ISL_1341466, EPI_ISL_1341467, EPI_ISL_1341468, EPI_ISL_1341469, EPI_ISL_1341470, EPI_ISL_1341471, EPI_ISL_1341472, EPI_ISL_1341473, EPI_ISL_1341474, EPI_ISL_1341475, EPI_ISL_1341476, EPI_ISL_1341477, EPI_ISL_1341478, EPI_ISL_1341479, EPI_ISL_1341480, EPI_ISL_1341481, EPI_ISL_1341482, EPI_ISL_1341483, EPI_ISL_1341484, EPI_ISL_1341485, EPI_ISL_1341486, EPI_ISL_1341487, EPI_ISL_1341488, EPI_ISL_1341489, EPI_ISL_1341490, EPI_ISL_1341491, EPI_ISL_1341492, EPI_ISL_1341493, EPI_ISL_1341494, EPI_ISL_1341495, EPI_ISL_1341496, EPI_ISL_1341497, EPI_ISL_1341498, EPI_ISL_1341499, EPI_ISL_1341500, EPI_ISL_1341509, EPI_ISL_1341510, EPI_ISL_1341511, EPI_ISL_1341512, EPI_ISL_1341513, EPI_ISL_1341514, EPI_ISL_1341515, EPI_ISL_1341516, EPI_ISL_1341517, EPI_ISL_1341518, EPI_ISL_1341519, EPI_ISL_1341520, EPI_ISL_1341521, EPI_ISL_1341522, EPI_ISL_1341523, EPI_ISL_1341524, EPI_ISL_1341525, EPI_ISL_1341526, EPI_ISL_1341527, EPI_ISL_1341528, EPI_ISL_1341529, EPI_ISL_1341530, EPI_ISL_1341531, EPI_ISL_1341532, EPI_ISL_1341533, EPI_ISL_1341534, EPI_ISL_1341535, EPI_ISL_1341536, EPI_ISL_1341537, EPI_ISL_1341538, EPI_ISL_1341539, EPI_ISL_1341540, EPI_ISL_1341541, EPI_ISL_1341542, EPI_ISL_1341543, EPI_ISL_1341544, EPI_ISL_1341545, EPI_ISL_1341546, EPI_ISL_1341547, EPI_ISL_1341548, EPI_ISL_1341549, EPI_ISL_1341550, EPI_ISL_1341551, EPI_ISL_1341552, EPI_ISL_1341553, EPI_ISL_1341554, EPI_ISL_1341555, EPI_ISL_1341556, EPI_ISL_1341557, EPI_ISL_1341558, EPI_ISL_1341559, EPI_ISL_1341560, EPI_ISL_1341561, EPI_ISL_1341562, EPI_ISL_1341563, EPI_ISL_1341564, EPI_ISL_1341565, EPI_ISL_1341566, EPI_ISL_1341567, EPI_ISL_1341568, EPI_ISL_1341569, EPI_ISL_1341570, EPI_ISL_1341571, EPI_ISL_1341572, EPI_ISL_1341573, EPI_ISL_1341574, EPI_ISL_1341575, EPI_ISL_1341577, EPI_ISL_1341579, EPI_ISL_1341580, EPI_ISL_1341581, EPI_ISL_1341582, EPI_ISL_1341583, EPI_ISL_1341584, EPI_ISL_1341585, EPI_ISL_1341586, EPI_ISL_1341587, EPI_ISL_1341588, EPI_ISL_1341589, EPI_ISL_1341591, EPI_ISL_1341592, EPI_ISL_1341593, EPI_ISL_1341594, EPI_ISL_1341595, EPI_ISL_1341596, EPI_ISL_1341597, EPI_ISL_1341598, EPI_ISL_1341599, EPI_ISL_1341600, EPI_ISL_1341601, EPI_ISL_1341602, EPI_ISL_1341603, EPI_ISL_1341604, EPI_ISL_1341605, EPI_ISL_1341606, EPI_ISL_1341607, EPI_ISL_1341608, EPI_ISL_1341609, EPI_ISL_1341610, EPI_ISL_1341611, EPI_ISL_1341612, EPI_ISL_1341613, EPI_ISL_1341614, EPI_ISL_1341616, EPI_ISL_1341617, EPI_ISL_1341618, EPI_ISL_1341619, EPI_ISL_1341620, EPI_ISL_1341621, EPI_ISL_1341622, EPI_ISL_1341623, EPI_ISL_1341624, EPI_ISL_1341625, EPI_ISL_1341626, EPI_ISL_1341627, EPI_ISL_1341629, EPI_ISL_1341630, EPI_ISL_1341631, EPI_ISL_1341632, EPI_ISL_1341633, EPI_ISL_1341634, EPI_ISL_1341635, EPI_ISL_1341636, EPI_ISL_1341638, EPI_ISL_1341639, EPI_ISL_1341640 | | | |
| see above | UW Virology Lab | UW Virology Lab | Pavitra Roychoudhury, Hong Xie, Lasata Shrestha, Shah Mohamed Bakhash, Michelle Lin, Noah Baker, Sean Ellis, Saraswathi Sathees, Meei-Li Huang, Keith R Jerome, Alexander Greninger |
| EPI_ISL_1341644, EPI_ISL_1341645, EPI_ISL_1341646, EPI_ISL_1341647, EPI_ISL_1341648, EPI_ISL_1341649, EPI_ISL_1341650, EPI_ISL_1341651, EPI_ISL_1341652, EPI_ISL_1341653, EPI_ISL_1341655, EPI_ISL_1341656, EPI_ISL_1341657, EPI_ISL_1341658, EPI_ISL_1341659, EPI_ISL_1341660, EPI_ISL_1341661, EPI_ISL_1341662, EPI_ISL_1341663, EPI_ISL_1341664, EPI_ISL_1341665, EPI_ISL_1341666, EPI_ISL_1341667, EPI_ISL_1341668, EPI_ISL_1341669, EPI_ISL_1341670, EPI_ISL_1341671, EPI_ISL_1341672, EPI_ISL_1341673, EPI_ISL_1341674, EPI_ISL_1341675, EPI_ISL_1341676, EPI_ISL_1341677, EPI_ISL_1341678, EPI_ISL_1341679, EPI_ISL_1341681, EPI_ISL_1341682, EPI_ISL_1341683, EPI_ISL_1341684, EPI_ISL_1341685, EPI_ISL_1341686, EPI_ISL_1341687, EPI_ISL_1341688, EPI_ISL_1341689, EPI_ISL_1341690, EPI_ISL_1341691, EPI_ISL_1341692, EPI_ISL_1341693, EPI_ISL_1341694, EPI_ISL_1341695, EPI_ISL_1341696, EPI_ISL_1341697, EPI_ISL_1341698, EPI_ISL_1341699, EPI_ISL_1341700, EPI_ISL_1341701, EPI_ISL_1341702, EPI_ISL_1341703, EPI_ISL_1341704, EPI_ISL_1341705, EPI_ISL_1341706, EPI_ISL_1341707, EPI_ISL_1341708, EPI_ISL_1341709, EPI_ISL_1341710, EPI_ISL_1341711, EPI_ISL_1341712, EPI_ISL_1341713, EPI_ISL_1341714, EPI_ISL_1341715, EPI_ISL_1341716, EPI_ISL_1341717, EPI_ISL_1341718, EPI_ISL_1341719, EPI_ISL_1341720, EPI_ISL_1341721, EPI_ISL_1341722, EPI_ISL_1341723, EPI_ISL_1341724, EPI_ISL_1341725, EPI_ISL_1341726, EPI_ISL_1341728, EPI_ISL_1341729, EPI_ISL_1341730, EPI_ISL_1341731, EPI_ISL_1341732, EPI_ISL_1341733, EPI_ISL_1341734, EPI_ISL_1341735, EPI_ISL_1341736, EPI_ISL_1341737, EPI_ISL_1341738, EPI_ISL_1341739, EPI_ISL_1341740, EPI_ISL_1341741, EPI_ISL_1341743, EPI_ISL_1341744, EPI_ISL_1341745, EPI_ISL_1341746, EPI_ISL_1341747, EPI_ISL_1341748, EPI_ISL_1341749, EPI_ISL_1341750, EPI_ISL_1341751, EPI_ISL_1341752, EPI_ISL_1341753, EPI_ISL_1341754, EPI_ISL_1341755, EPI_ISL_1341756, EPI_ISL_1341757, EPI_ISL_1341758, EPI_ISL_1341759, EPI_ISL_1341760, EPI_ISL_1341761, EPI_ISL_1341762, EPI_ISL_1341763, EPI_ISL_1341764, EPI_ISL_1341765, EPI_ISL_1341766, EPI_ISL_1341767, EPI_ISL_1341768, EPI_ISL_1341769, EPI_ISL_1341770, EPI_ISL_1341771, EPI_ISL_1341772, EPI_ISL_1341773, EPI_ISL_1341774, EPI_ISL_1341775, EPI_ISL_1341776, EPI_ISL_1341777, EPI_ISL_1341778, EPI_ISL_1341779, EPI_ISL_1341780, EPI_ISL_1341781, EPI_ISL_1341782 | | | |
| see above | UW Virology Lab | UW Virology Lab | Pavitra Roychoudhury, Hong Xie, Lasata Shrestha, Shah Mohamed Bakhash, Michelle Lin, Noah R. Baker, Sean Ellis, Saraswathi Sathees, Meei-Li Huang, Keith R Jerome, Alexander Greninger |
| EPI_ISL_1341783, EPI_ISL_1341784, EPI_ISL_1341785, EPI_ISL_1341786, EPI_ISL_1341787, EPI_ISL_1341788, EPI_ISL_1341789, EPI_ISL_1341790, EPI_ISL_1341791 | LSUHS Emerging Viral Threat Laboratory | Microbial Genome Sequencing Center | Jeremy P. Kamil, Jennifer L. Carroll, Rona S. Scott, Maarten Van Diest, Andrew D. Yurochko, Christopher G. Kevil, Martin J. Sapp, Daniel J. Snyder, Vaughn S. Cooper, John A. Vanchiere |
| EPI_ISL_1346490, EPI_ISL_1346491, EPI_ISL_1346492, EPI_ISL_1346494, EPI_ISL_1346495, EPI_ISL_1346496, EPI_ISL_1346497, EPI_ISL_1346498, EPI_ISL_1346499, EPI_ISL_1346500, EPI_ISL_1346501, EPI_ISL_1346502, EPI_ISL_1346503, EPI_ISL_1346504, EPI_ISL_1346505, EPI_ISL_1346507, EPI_ISL_1346508, EPI_ISL_1346509, EPI_ISL_1346510, EPI_ISL_1346511, EPI_ISL_1346512, EPI_ISL_1346513, EPI_ISL_1346514, EPI_ISL_1346515, EPI_ISL_1346516, EPI_ISL_1346517, EPI_ISL_1346518, EPI_ISL_1346519, EPI_ISL_1346520, EPI_ISL_1346521, EPI_ISL_1346522, EPI_ISL_1346523, EPI_ISL_1346524, EPI_ISL_1346525, EPI_ISL_1346526, EPI_ISL_1346527, EPI_ISL_1346528, EPI_ISL_1346529, EPI_ISL_1346530, EPI_ISL_1346531, EPI_ISL_1346532, EPI_ISL_1346534, EPI_ISL_1346535, EPI_ISL_1346536, | | | |

| | | | |
|---|---|---|---|
| EPI_ISL_1346537, EPI_ISL_1346538, EPI_ISL_1346539, EPI_ISL_1346540, EPI_ISL_1346541, EPI_ISL_1346542, EPI_ISL_1346543, EPI_ISL_1346544, EPI_ISL_1346545, EPI_ISL_1346546, EPI_ISL_1346547, EPI_ISL_1346548, EPI_ISL_1346549, EPI_ISL_1346550, EPI_ISL_1346551, EPI_ISL_1346552, EPI_ISL_1346553, EPI_ISL_1346555, EPI_ISL_1346556, EPI_ISL_1346557, EPI_ISL_1346558, EPI_ISL_1346559, EPI_ISL_1346560, EPI_ISL_1346561, EPI_ISL_1346562, EPI_ISL_1346563, EPI_ISL_1346564, EPI_ISL_1346565, EPI_ISL_1346566, EPI_ISL_1346567, EPI_ISL_1346568, EPI_ISL_1346569, EPI_ISL_1346570, EPI_ISL_1346571, EPI_ISL_1346572, EPI_ISL_1346574, EPI_ISL_1346575, EPI_ISL_1346576, EPI_ISL_1346577, EPI_ISL_1346578, EPI_ISL_1346579, EPI_ISL_1346580, EPI_ISL_1346581, EPI_ISL_1346582, EPI_ISL_1346583, EPI_ISL_1346584, EPI_ISL_1346585, EPI_ISL_1346586, EPI_ISL_1346587, EPI_ISL_1346588, EPI_ISL_1346589, EPI_ISL_1346590, EPI_ISL_1346591, EPI_ISL_1346592, EPI_ISL_1346593, EPI_ISL_1346594, EPI_ISL_1346595, EPI_ISL_1346596, EPI_ISL_1346597, EPI_ISL_1346598, EPI_ISL_1346599, EPI_ISL_1346600, EPI_ISL_1346601, EPI_ISL_1346602, EPI_ISL_1346603, EPI_ISL_1346604, EPI_ISL_1346605, EPI_ISL_1346606, EPI_ISL_1346607, EPI_ISL_1346609, EPI_ISL_1346610, EPI_ISL_1346611, EPI_ISL_1346612, EPI_ISL_1346613, EPI_ISL_1346614, EPI_ISL_1346615, EPI_ISL_1346616, EPI_ISL_1346617, EPI_ISL_1346618, EPI_ISL_1346620, EPI_ISL_1346621, EPI_ISL_1346622 | | | |
| see above | UW Virology Lab | UW Virology Lab | Pavitra Roychoudhury, Hong Xie, Lasata Shrestha, Shah Mohamed Bakhash, Michelle Lin, Noah R. Baker, Sean Ellis, Saraswathi Sathees, Meei-Li Huang, Keith R Jerome, Alexander Greninger |
| EPI_ISL_1347778, EPI_ISL_1347779, EPI_ISL_1347780, EPI_ISL_1347781, EPI_ISL_1347782, EPI_ISL_1347783, EPI_ISL_1347784, EPI_ISL_1347785, EPI_ISL_1347786, EPI_ISL_1347787, EPI_ISL_1347788, EPI_ISL_1347789 | | | |
| see above | DeRisi Lab, University of California, San Francisco | DeRisi Lab, University of California, San Francisco | Sabrina Mann, Anthea Mitchell, Jamin Liu, Sara Sunshine, Matthew Laurie, Genay Pilarowski, Amy Kistler, Manu Vanaerschot, Patrick Ayscue, Lucy Li, Eric Chow, IDseq Team, James Peng, Carina Marquez, Luis Rubio, Gabriel Chamie, Diane Jones, Jon Jacobo, Susana Rojas, Susy Rojas, Valerie Tulier-Laiwa, Douglas Black, Jackie Martinez, Maya Petersen, Diane Havlir, and Joseph DeRisi |
| EPI_ISL_1348107, EPI_ISL_1348109, EPI_ISL_1348110, EPI_ISL_1348112, EPI_ISL_1348114, EPI_ISL_1348116, EPI_ISL_1348118, EPI_ISL_1348120, EPI_ISL_1348121, EPI_ISL_1348123, EPI_ISL_1348125, EPI_ISL_1348127, EPI_ISL_1348129, EPI_ISL_1348130, EPI_ISL_1348132, EPI_ISL_1348134, EPI_ISL_1348138, EPI_ISL_1348140, EPI_ISL_1348142, EPI_ISL_1348144, EPI_ISL_1348146, EPI_ISL_1348148, EPI_ISL_1348149, EPI_ISL_1348151, EPI_ISL_1348153, EPI_ISL_1348155, EPI_ISL_1348157, EPI_ISL_1348158, EPI_ISL_1348160, EPI_ISL_1348164, EPI_ISL_1348166, EPI_ISL_1348168, EPI_ISL_1348170, EPI_ISL_1348171, EPI_ISL_1348173, EPI_ISL_1348175, EPI_ISL_1348177, EPI_ISL_1348179, EPI_ISL_1348180, EPI_ISL_1348182, EPI_ISL_1348184, EPI_ISL_1348186, EPI_ISL_1348188, EPI_ISL_1348189, EPI_ISL_1348191, EPI_ISL_1348193, EPI_ISL_1348197, EPI_ISL_1348198, EPI_ISL_1348201, EPI_ISL_1348202, EPI_ISL_1348204, EPI_ISL_1348206, EPI_ISL_1348210, EPI_ISL_1348211, EPI_ISL_1348213, EPI_ISL_1348215, EPI_ISL_1348217, EPI_ISL_1348218, EPI_ISL_1348220, EPI_ISL_1348222, EPI_ISL_1348224, EPI_ISL_1348226, EPI_ISL_1348228, EPI_ISL_1348229, EPI_ISL_1348231, EPI_ISL_1348233, EPI_ISL_1348235, EPI_ISL_1348236, EPI_ISL_1348238, EPI_ISL_1348240, EPI_ISL_1348242, EPI_ISL_1348244, EPI_ISL_1348245, EPI_ISL_1348247, EPI_ISL_1348249, EPI_ISL_1348250, EPI_ISL_1348254, EPI_ISL_1348256, EPI_ISL_1348258, EPI_ISL_1348259, EPI_ISL_1348261, EPI_ISL_1348263, EPI_ISL_1348265, EPI_ISL_1348267, EPI_ISL_1348268, EPI_ISL_1348270, EPI_ISL_1348272, EPI_ISL_1348274, EPI_ISL_1348276, EPI_ISL_1348278, EPI_ISL_1348279, EPI_ISL_1348281, EPI_ISL_1348283, EPI_ISL_1348284, EPI_ISL_1348286, EPI_ISL_1348288, EPI_ISL_1348292, EPI_ISL_1348294, EPI_ISL_1348295, EPI_ISL_1348297, EPI_ISL_1348299, EPI_ISL_1348301, EPI_ISL_1348303, EPI_ISL_1348304, EPI_ISL_1348306, EPI_ISL_1348308, EPI_ISL_1348310, EPI_ISL_1348311, EPI_ISL_1348313, EPI_ISL_1348315, EPI_ISL_1348317, EPI_ISL_1348319, EPI_ISL_1348320, EPI_ISL_1348322, EPI_ISL_1348324, EPI_ISL_1348326, EPI_ISL_1348328, EPI_ISL_1348329, EPI_ISL_1348331, EPI_ISL_1348333, EPI_ISL_1348335, EPI_ISL_1348336, EPI_ISL_1348338, EPI_ISL_1348340 | | | |
| see above | Maryland Genomics, Institute for Genome Sciences, University of Maryland School of Medicine | Maryland Genomics, Institute for Genome Sciences, University of Maryland School of Medicine | Tallon, Luke J; Sadzewicz, Lisa D; Humphrys, Mike; Ott, Sandra; Roussey, Holly; Mehta, Aditya; Vavikolanu, Kranthi; Fraser, Claire M; Ravel, Jacques |
| EPI_ISL_1349075, EPI_ISL_1349077, EPI_ISL_1349078, EPI_ISL_1349080, EPI_ISL_1349082, EPI_ISL_1349084, EPI_ISL_1349085, EPI_ISL_1349087 | DeRisi Lab, University of California, San Francisco | DeRisi Lab, University of California, San Francisco | Sabrina Mann, Anthea Mitchell, Jamin Liu, Sara Sunshine, Matthew Laurie, Genay Pilarowski, Amy Kistler, Manu Vanaerschot, Patrick Ayscue, Lucy Li, Eric Chow, IDseq Team, James Peng, Carina Marquez, Luis Rubio, Gabriel Chamie, Diane Jones, Jon Jacobo, Susana Rojas, Susy Rojas, Valerie Tulier-Laiwa, Douglas Black, Jackie Martinez, Maya Petersen, Diane Havlir, and Joseph DeRisi |
| EPI_ISL_1351416, EPI_ISL_1351418, EPI_ISL_1351420, EPI_ISL_1351422, EPI_ISL_1351424, EPI_ISL_1351426, EPI_ISL_1351428, EPI_ISL_1351430, EPI_ISL_1351433, EPI_ISL_1351435, EPI_ISL_1351437, EPI_ISL_1351439, EPI_ISL_1351441, EPI_ISL_1351443, EPI_ISL_1351445, EPI_ISL_1351448, EPI_ISL_1351451, EPI_ISL_1351454, EPI_ISL_1351457, EPI_ISL_1351460, EPI_ISL_1351462 | | | |
| see above | Centro de InvestigaciÃ³n BiomÃ©dica del Noreste (CIBIN) | Unidad de Genomica Avanzada | Consorcio Mexicano de Vigilancia Genomica (CoViGen-Mex). Authors (in alphabetical order): Julio Elias Alvarado-Yaah, Carlos F. Arias, Santiago Avila-Rios, Victor Hugo Borja-Aburto, Celia Boukadida, Juan Bautista Chale-Dzul , Jose Antonio Enciso-Moreno, Gloria Elena Espinoza-Ayala, Fernando Fontove-Herrera, Concepcion Grajales-Muniz, Ricardo Grande, Alfredo Herrera-Estrella, Carla Ivon Herrera-Najera, Pavel Isa, Brenda Irasema Maldonado-Meza, Bernardo Martinez-Miguel, Margarita Matias-Florentino, Maria Guadalupe de Jesus Mireles-Rivera, Gloria Maria Molina-Salinas, Hector Montoya-Fuentes, Jose Esteban Munoz-Medina, Jose de Jesus Nunez-Contreras, Alicia Ocana-Mondragon, Luis Alberto Ochoa-Carrera, Hector Esteban Paz-Juarez, Francisco Pulido, Helen Haydee Fernanda Ramirez-Plascencia, Angel Gustavo Salas-Lais, Jorge Ivan Salinal-Nevarez, Alejandro Sanchez-Flores, Clara Esperanza Santacruz-Tinoco, Maria Guadalupe Santiago-Mauricio, Nelly Selem-Mojica, Blanca Taboada, Gloria Vazquez |
| EPI_ISL_1351465, EPI_ISL_1351468, EPI_ISL_1351471, EPI_ISL_1351474, EPI_ISL_1351477, EPI_ISL_1351480, EPI_ISL_1351483, EPI_ISL_1351486, EPI_ISL_1351489, EPI_ISL_1351492, EPI_ISL_1351494, EPI_ISL_1351498, EPI_ISL_1351500, EPI_ISL_1351503, EPI_ISL_1351506, EPI_ISL_1351509 | | | |
| see above | Unidad de InvestigaciÃ³n MÃ©dica de YucatÃ¡n (UIMY) | Unidad de Genomica Avanzada | Consorcio Mexicano de Vigilancia Genomica (CoViGen-Mex). Authors (in alphabetical order): Julio Elias Alvarado-Yaah, Carlos F. Arias, Santiago Avila-Rios, Victor Hugo Borja-Aburto, Celia Boukadida, Juan Bautista Chale-Dzul , Jose Antonio Enciso-Moreno, Gloria Elena Espinoza-Ayala, Fernando Fontove-Herrera, Concepcion Grajales-Muniz, Ricardo Grande, Alfredo Herrera-Estrella, Carla Ivon Herrera-Najera, Pavel Isa, Brenda Irasema Maldonado-Meza, Bernardo Martinez-Miguel, Margarita Matias-Florentino, Maria Guadalupe de Jesus Mireles-Rivera, Gloria Maria Molina-Salinas, Hector Montoya-Fuentes, Jose Esteban Munoz-Medina, Jose de Jesus Nunez-Contreras, Alicia Ocana-Mondragon, Luis Alberto Ochoa-Carrera, Hector Esteban Paz-Juarez, Francisco Pulido, Helen Haydee Fernanda Ramirez-Plascencia, Angel Gustavo Salas-Lais, Jorge Ivan Salinal-Nevarez, Alejandro Sanchez-Flores, Clara Esperanza Santacruz-Tinoco, Maria Guadalupe Santiago-Mauricio, Nelly Selem-Mojica, Blanca Taboada, Gloria Vazquez |
| EPI_ISL_1351511, EPI_ISL_1351514, EPI_ISL_1351517, EPI_ISL_1351520, EPI_ISL_1351523, EPI_ISL_1351526, EPI_ISL_1351529, EPI_ISL_1351532, EPI_ISL_1351535, EPI_ISL_1351538, EPI_ISL_1351540, EPI_ISL_1351543, EPI_ISL_1351547, EPI_ISL_1351549, EPI_ISL_1351552, EPI_ISL_1351555, EPI_ISL_1351558, EPI_ISL_1351561, EPI_ISL_1351564, EPI_ISL_1351567, EPI_ISL_1351571, EPI_ISL_1351574, EPI_ISL_1351577, EPI_ISL_1351580, EPI_ISL_1351583, EPI_ISL_1351585, EPI_ISL_1351589, EPI_ISL_1351592, EPI_ISL_1351595, EPI_ISL_1351598, EPI_ISL_1351601, EPI_ISL_1351605, EPI_ISL_1351608, EPI_ISL_1351611, EPI_ISL_1351614, EPI_ISL_1351617, EPI_ISL_1351620, EPI_ISL_1351623, EPI_ISL_1351626, EPI_ISL_1351629, EPI_ISL_1351632, EPI_ISL_1351635, EPI_ISL_1351638, EPI_ISL_1351642, EPI_ISL_1351645, EPI_ISL_1351648, EPI_ISL_1351651, EPI_ISL_1351654, EPI_ISL_1351656, EPI_ISL_1351659, EPI_ISL_1351662, EPI_ISL_1351665, EPI_ISL_1351668, EPI_ISL_1351670, EPI_ISL_1351672, EPI_ISL_1351675, EPI_ISL_1351679, EPI_ISL_1351682, EPI_ISL_1351685, EPI_ISL_1351688, EPI_ISL_1351691, EPI_ISL_1351694, EPI_ISL_1351697, EPI_ISL_1351700, EPI_ISL_1351703, EPI_ISL_1351706, EPI_ISL_1351709 | | | |
| see above | Centro de InvestigaciÃ³n BiomÃ©dica de Occidente (CIBO) | Unidad de Genomica Avanzada | Consorcio Mexicano de Vigilancia Genomica (CoViGen-Mex). Authors (in alphabetical order): Julio Elias Alvarado-Yaah, Carlos F. Arias, Santiago Avila-Rios, Victor Hugo Borja-Aburto, Celia Boukadida, Juan Bautista Chale-Dzul , Jose Antonio Enciso-Moreno, Gloria Elena Espinoza-Ayala, Fernando Fontove-Herrera, Concepcion Grajales-Muniz, Ricardo Grande, Alfredo Herrera-Estrella, Carla Ivon Herrera-Najera, Pavel Isa, Brenda Irasema Maldonado-Meza, Bernardo Martinez-Miguel, Margarita Matias-Florentino, Maria Guadalupe de Jesus Mireles-Rivera, Gloria Maria Molina-Salinas, Hector Montoya-Fuentes, Jose Esteban Munoz-Medina, Jose de Jesus Nunez-Contreras, Alicia Ocana-Mondragon, Luis Alberto Ochoa-Carrera, Hector Esteban Paz-Juarez, Francisco Pulido, Helen Haydee Fernanda Ramirez-Plascencia, Angel Gustavo Salas-Lais, Jorge Ivan Salinal-Nevarez, Alejandro Sanchez-Flores, Clara Esperanza Santacruz-Tinoco, Maria Guadalupe Santiago-Mauricio, Nelly Selem-Mojica, Blanca Taboada, Gloria Vazquez |
| EPI_ISL_1351713, EPI_ISL_1351716, EPI_ISL_1351719, EPI_ISL_1351725, EPI_ISL_1351728, EPI_ISL_1351731, EPI_ISL_1351734, EPI_ISL_1351737, EPI_ISL_1351739, EPI_ISL_1351743, EPI_ISL_1351746, EPI_ISL_1351749, EPI_ISL_1351752, EPI_ISL_1351754, EPI_ISL_1351757, EPI_ISL_1351760, EPI_ISL_1351764, EPI_ISL_1351767, EPI_ISL_1351770, EPI_ISL_1351774, EPI_ISL_1351777, EPI_ISL_1351780, EPI_ISL_1351783, EPI_ISL_1351787, EPI_ISL_1351790, EPI_ISL_1351793, EPI_ISL_1351796, EPI_ISL_1351799, EPI_ISL_1351802, EPI_ISL_1351805, EPI_ISL_1351809, EPI_ISL_1351810, EPI_ISL_1351811, EPI_ISL_1351815, EPI_ISL_1351818, EPI_ISL_1351821, EPI_ISL_1351824, EPI_ISL_1351827, EPI_ISL_1351830, EPI_ISL_1351833, EPI_ISL_1351836, EPI_ISL_1351839, EPI_ISL_1351842, EPI_ISL_1351845, EPI_ISL_1351848, EPI_ISL_1351851, EPI_ISL_1351854, EPI_ISL_1351857, EPI_ISL_1351860, EPI_ISL_1351863, EPI_ISL_1351866, EPI_ISL_1351869, EPI_ISL_1351872, EPI_ISL_1351875, EPI_ISL_1351878, EPI_ISL_1351880, EPI_ISL_1351883, EPI_ISL_1351886, EPI_ISL_1351890, EPI_ISL_1351893, EPI_ISL_1351896, EPI_ISL_1351899, EPI_ISL_1351902, EPI_ISL_1351905, EPI_ISL_1351908, EPI_ISL_1351911, EPI_ISL_1351914, EPI_ISL_1351917, EPI_ISL_1351921, EPI_ISL_1351924, EPI_ISL_1351927, EPI_ISL_1351930, EPI_ISL_1351931, EPI_ISL_1351933, EPI_ISL_1351935, EPI_ISL_1351937, EPI_ISL_1351941, EPI_ISL_1351944, EPI_ISL_1351947, EPI_ISL_1351950, EPI_ISL_1351952, EPI_ISL_1351954, EPI_ISL_1351956, EPI_ISL_1351958, EPI_ISL_1351960, EPI_ISL_1351963, EPI_ISL_1351966, EPI_ISL_1351969, EPI_ISL_1351972, EPI_ISL_1351975, EPI_ISL_1351978, EPI_ISL_1351981, EPI_ISL_1351984, EPI_ISL_1351987, EPI_ISL_1351990, EPI_ISL_1351993, EPI_ISL_1351997, EPI_ISL_1352000, EPI_ISL_1352002, EPI_ISL_1352006, EPI_ISL_1352009, EPI_ISL_1352012, EPI_ISL_1352015, EPI_ISL_1352018, EPI_ISL_1352020, EPI_ISL_1352023, EPI_ISL_1352026, EPI_ISL_1352029, EPI_ISL_1352032, EPI_ISL_1352035, EPI_ISL_1352038, EPI_ISL_1352041, EPI_ISL_1352043, EPI_ISL_1352047, EPI_ISL_1352048, EPI_ISL_1352050, EPI_ISL_1352051, EPI_ISL_1352053, EPI_ISL_1352055, EPI_ISL_1352057, EPI_ISL_1352059, EPI_ISL_1352060, EPI_ISL_1352062, EPI_ISL_1352066, EPI_ISL_1352068, EPI_ISL_1352070, EPI_ISL_1352072 | | | |
| see above | Laboratorio Central de EpidemiologÃa (LCE) | Unidad de Genomica Avanzada | Consorcio Mexicano de Vigilancia Genomica (CoViGen-Mex). Authors (in alphabetical order): Julio Elias Alvarado-Yaah, Carlos F. Arias, Santiago Avila-Rios, Victor Hugo Borja-Aburto, Celia Boukadida, Juan Bautista Chale-Dzul , Jose Antonio Enciso-Moreno, Gloria Elena Espinoza-Ayala, Fernando Fontove-Herrera, Concepcion Grajales-Muniz, Ricardo Grande, Alfredo Herrera-Estrella, Carla Ivon Herrera-Najera, Pavel Isa, Brenda Irasema Maldonado-Meza, Bernardo Martinez-Miguel, Margarita Matias-Florentino, Maria Guadalupe de Jesus Mireles-Rivera, Gloria Maria Molina-Salinas, Hector Montoya-Fuentes, Jose Esteban Munoz-Medina, Jose de Jesus Nunez-Contreras, Alicia Ocana-Mondragon, Luis Alberto Ochoa-Carrera, Hector Esteban Paz-Juarez, Francisco Pulido, Helen Haydee Fernanda Ramirez-Plascencia, Angel Gustavo Salas-Lais, Jorge Ivan Salinal-Nevarez, Alejandro Sanchez-Flores, Clara Esperanza Santacruz-Tinoco, Maria Guadalupe Santiago-Mauricio, Nelly Selem-Mojica, Blanca Taboada, Gloria Vazquez |
| EPI_ISL_1352076, EPI_ISL_1352079, EPI_ISL_1352083, EPI_ISL_1352085 | Unidad de InvestigaciÃ³n BiomÃ©dica de Zacatecas (UIBZ) | Unidad de Genomica Avanzada | Consorcio Mexicano de Vigilancia Genomica (CoViGen-Mex). Authors (in alphabetical order): Julio Elias Alvarado-Yaah, Carlos F. Arias, Santiago Avila-Rios, Victor Hugo Borja-Aburto, Celia Boukadida, Juan Bautista Chale-Dzul , Jose Antonio Enciso-Moreno, Gloria Elena Espinoza-Ayala, Fernando Fontove-Herrera, Concepcion Grajales-Muniz, Ricardo Grande, Alfredo Herrera-Estrella, Carla Ivon Herrera-Najera, Pavel Isa, Brenda Irasema Maldonado-Meza, Bernardo Martinez-Miguel, Margarita Matias-Florentino, Maria Guadalupe de Jesus Mireles-Rivera, Gloria Maria Molina-Salinas, Hector Montoya-Fuentes, Jose Esteban Munoz-Medina, Jose de Jesus Nunez-Contreras, Alicia Ocana-Mondragon, Luis Alberto Ochoa-Carrera, Hector Esteban Paz-Juarez, Francisco Pulido, Helen Haydee Fernanda Ramirez-Plascencia, Angel Gustavo Salas-Lais, Jorge Ivan Salinal-Nevarez, Alejandro Sanchez-Flores, Clara Esperanza Santacruz-Tinoco, Maria Guadalupe Santiago-Mauricio, Nelly Selem-Mojica, Blanca Taboada, Gloria Vazquez |
| EPI_ISL_1357605 | Dr. Ann Chahroudi's Lab | Dr. Thomas Vanderford's Lab | Thomas Vanderford, Maud Mavigner, Nils Schoof, Ann Chahroudi |
| EPI_ISL_1357842, EPI_ISL_1357843, EPI_ISL_1357844 | DC Public Health Lab/ Dept. of Forensic Sciences | DC Public Health Lab/ Dept. of Forensic Sciences | Janis Doss, Scott Nguyen, Elizabeth Zelaya, Sarah Scott, Connie Maza, Monica Mann, Brittany Hamilton, David Payne, Jocelyn Hauser |
| EPI_ISL_1357846, EPI_ISL_1357847, EPI_ISL_1357848, EPI_ISL_1357849, EPI_ISL_1357850, EPI_ISL_1357851, EPI_ISL_1357852, EPI_ISL_1357853, EPI_ISL_1357854, EPI_ISL_1357855, EPI_ISL_1357856, EPI_ISL_1357857, EPI_ISL_1357858, EPI_ISL_1357859 | | | |
| see above | Kansas Health and Environmental Lab | Kansas Health and Environmental Lab | Mike Grose, Jonathan Barnell, Ben Olsen, and Phil Adam |
| EPI_ISL_1358408, EPI_ISL_1358409 | Wyoming Public Health Laboratory | Wyoming Public Health Laboratory | Jim Mildenberger, Wanda Manley, Noah Hull, Taylor Fearing, Lynette Gumbleton, Channing Weber, Ashley Norberg, Chayse Rowley, Marley Goetz, Brian Dominguez, Elliot Thomasson, Cari Sloma, and Rob Christensen |
| EPI_ISL_1358904, EPI_ISL_1358905, EPI_ISL_1358906, EPI_ISL_1358907, EPI_ISL_1358908, EPI_ISL_1358909, EPI_ISL_1358910, EPI_ISL_1358911, EPI_ISL_1358912, EPI_ISL_1358914, EPI_ISL_1358915, EPI_ISL_1358916, EPI_ISL_1358917 | | | |
| see above | IN State Department of Health Laboratory Services | IN State Department of Health Laboratory Services | Cassandra Campion, Jamie Yeadon, Brian Pope, Lixia Liu, Kyle Brownlee, Melissa Hindenlang, Ankita Kashikar, Mark Glazier |
| EPI_ISL_1358942, EPI_ISL_1358943, EPI_ISL_1358944, EPI_ISL_1358945, EPI_ISL_1358947, EPI_ISL_1358948, EPI_ISL_1358949, EPI_ISL_1358950, EPI_ISL_1358951, EPI_ISL_1358952, EPI_ISL_1358953 | | | |
| see above | Minnesota Department of Health, Public Health Laboratory | Minnesota Department of Health, Public Health Laboratory | Alexandra Lorentz, Jacob Garfin, Matt Plumb, and Xiong Wang |
| EPI_ISL_1358954, EPI_ISL_1358955, EPI_ISL_1358956, EPI_ISL_1358957, EPI_ISL_1358958 | Hennepin County Medical Center | Minnesota Department of Health, Public Health Laboratory | Alexandra Lorentz, Jacob Garfin, Matt Plumb, and Xiong Wang |
| EPI_ISL_1358960, EPI_ISL_1358961, EPI_ISL_1358962, EPI_ISL_1358963, EPI_ISL_1358964, EPI_ISL_1358965, EPI_ISL_1358966, EPI_ISL_1358967, EPI_ISL_1358968, EPI_ISL_1358969, EPI_ISL_1358970, EPI_ISL_1358971, EPI_ISL_1358972, EPI_ISL_1358973, EPI_ISL_1358974, EPI_ISL_1358975, EPI_ISL_1358976, EPI_ISL_1358977, EPI_ISL_1358978, EPI_ISL_1358979, EPI_ISL_1358980, EPI_ISL_1358981, EPI_ISL_1358982, EPI_ISL_1358983, EPI_ISL_1358984, EPI_ISL_1358985, EPI_ISL_1358986, EPI_ISL_1358987, EPI_ISL_1358988, EPI_ISL_1358989, EPI_ISL_1358990, EPI_ISL_1358991, EPI_ISL_1358992, EPI_ISL_1358993, EPI_ISL_1358994, EPI_ISL_1358995, EPI_ISL_1358996, EPI_ISL_1358997, EPI_ISL_1358998, EPI_ISL_1358999, EPI_ISL_1359000, EPI_ISL_1359001, EPI_ISL_1359002, EPI_ISL_1359003, EPI_ISL_1359004, EPI_ISL_1359005, EPI_ISL_1359006, EPI_ISL_1359007, EPI_ISL_1359008, EPI_ISL_1359009, EPI_ISL_1359010, EPI_ISL_1359011, EPI_ISL_1359012, EPI_ISL_1359013, EPI_ISL_1359014, EPI_ISL_1359015, EPI_ISL_1359016, EPI_ISL_1359017, EPI_ISL_1359018, EPI_ISL_1359019 | | | |
| see above | Vault Health | Minnesota Department of Health, Public Health Laboratory | Alexandra Lorentz, Jacob Garfin, Matt Plumb, and Xiong Wang |
| EPI_ISL_1359020, EPI_ISL_1359021, EPI_ISL_1359022, EPI_ISL_1359023, EPI_ISL_1359024, EPI_ISL_1359025, EPI_ISL_1359026, EPI_ISL_1359027, EPI_ISL_1359028, EPI_ISL_1359029, EPI_ISL_1359030, EPI_ISL_1359031, EPI_ISL_1359032, EPI_ISL_1359033, EPI_ISL_1359034, EPI_ISL_1359035, EPI_ISL_1359036, EPI_ISL_1359037, EPI_ISL_1359038, EPI_ISL_1359039, EPI_ISL_1359040, EPI_ISL_1359041, EPI_ISL_1359042, EPI_ISL_1359043, EPI_ISL_1359044, EPI_ISL_1359055, EPI_ISL_1359056, EPI_ISL_1359057, EPI_ISL_1359058, EPI_ISL_1359059, EPI_ISL_1359060, EPI_ISL_1359061 | | | |
| see above | Wyoming Public Health Laboratory | Wyoming Public Health Laboratory | Jim Mildenberger, Wanda Manley, Noah Hull, Taylor Fearing, Lynette Gumbleton, Channing Weber, Ashley Norberg, Chayse Rowley, Marley Goetz, Brian Dominguez, Elliot Thomasson, Cari Sloma, and Rob Christensen |
| EPI_ISL_1359062 | Laboratorios LISTER | Instituto de Diagnostico y Referencia Epidemiologicos (INDRE) | Claudia Wong-Arambula, Abril Rodriguez-Maldonado, Vanessa Rivero-Arredondo, Ariadna Medina-Benitez, Joaquin Quiroz-Mercado, Sergio Rangel-Guerrero, Natividad Cruz-Ortiz, Tatiana Nunez-Garcia, Gisela Barrera-Badillo, Lucia Hernandez-Rivas, Irma Lopez-Martinez, Ernesto Ramirez-Gonzalez. |
| EPI_ISL_1359063 | Laboratorio BIOMEDICA | Instituto de Diagnostico y Referencia Epidemiologicos (INDRE) | Claudia Wong-Arambula, Abril Rodriguez-Maldonado, Vanessa Rivero-Arredondo, Ariadna Medina-Benitez, Joaquin Quiroz-Mercado, Sergio Rangel-Guerrero, Natividad Cruz-Ortiz, Tatiana Nunez-Garcia, Gisela Barrera-Badillo, Lucia Hernandez-Rivas, Irma Lopez-Martinez, Ernesto Ramirez-Gonzalez. |
| EPI_ISL_1359064, EPI_ISL_1359065 | LESP Queretaro | Instituto de Diagnostico y Referencia Epidemiologicos (INDRE) | Claudia Wong-Arambula, Abril Rodriguez-Maldonado, Vanessa Rivero-Arredondo, Ariadna Medina-Benitez, Joaquin Quiroz-Mercado, Sergio Rangel-Guerrero, Natividad Cruz-Ortiz, Tatiana Nunez-Garcia, Gisela Barrera-Badillo, Lucia Hernandez-Rivas, Irma Lopez-Martinez, Ernesto Ramirez-Gonzalez. |
| EPI_ISL_1359066, EPI_ISL_1359067 | LESP Yucatan | Instituto de Diagnostico y Referencia Epidemiologicos (INDRE) | Claudia Wong-Arambula, Abril Rodriguez-Maldonado, Vanessa Rivero-Arredondo, Ariadna Medina-Benitez, Joaquin Quiroz-Mercado, Sergio Rangel-Guerrero, Natividad Cruz-Ortiz, Tatiana Nunez-Garcia, Gisela Barrera-Badillo, Lucia Hernandez-Rivas, Irma Lopez-Martinez, Ernesto Ramirez-Gonzalez. |
| EPI_ISL_1359068, EPI_ISL_1359069 | LESP Zacatecas | Instituto de Diagnostico y Referencia Epidemiologicos (INDRE) | Claudia Wong-Arambula, Abril Rodriguez-Maldonado, Vanessa Rivero-Arredondo, Ariadna Medina-Benitez, Joaquin Quiroz-Mercado, Sergio Rangel-Guerrero, Natividad Cruz-Ortiz, Tatiana Nunez-Garcia, Gisela Barrera-Badillo, Lucia Hernandez-Rivas, Irma Lopez-Martinez, Ernesto Ramirez-Gonzalez. |

| | | | |
|---|---|---|---|
| EPI_ISL_1359070, EPI_ISL_1359071, EPI_ISL_1359072, EPI_ISL_1359073, EPI_ISL_1359074, EPI_ISL_1359075, EPI_ISL_1359076, EPI_ISL_1359077, EPI_ISL_1359078, EPI_ISL_1359079, EPI_ISL_1359080, EPI_ISL_1359081, EPI_ISL_1359082, EPI_ISL_1359083, EPI_ISL_1359084, EPI_ISL_1359085, EPI_ISL_1359086, EPI_ISL_1359087 | | | |
| see above | LESP Guanajuato | Instituto de Diagnostico y Referencia Epidemiologicos (INDRE) | Claudia Wong-Arambula, Abril Rodriguez-Maldonado, Vanessa Rivero-Arredondo, Ariadna Medina-Benitez, Joaquin Quiroz-Mercado, Sergio Rangel-Guerrero, Natividad Cruz-Ortiz, Tatiana Nunez-Garcia, Gisela Barrera-Badillo, Lucia Hernandez-Rivas, Irma Lopez-Martinez, Ernesto Ramirez-Gonzalez. |
| EPI_ISL_1360407, EPI_ISL_1360408, EPI_ISL_1360409, EPI_ISL_1360410, EPI_ISL_1360411, EPI_ISL_1360412 | Laboratorio de Enfermedades Emergentes y Reemergentes | Laboratorio de Enfermedades Emergentes y Reemergentes | Vega-Magaña Natali, Peña-Rodriguez Marcela, Viera-Segura Oliver, García-Chagollan Mariel, Hernández-Bello Jorge, Muñoz-Valle José Francisco |
| EPI_ISL_1362552, EPI_ISL_1362553, EPI_ISL_1362554, EPI_ISL_1362555, EPI_ISL_1362556, EPI_ISL_1362557, EPI_ISL_1362558, EPI_ISL_1362559, EPI_ISL_1362560, EPI_ISL_1362561, EPI_ISL_1362562, EPI_ISL_1362563, EPI_ISL_1362564, EPI_ISL_1362565, EPI_ISL_1362566, EPI_ISL_1362567, EPI_ISL_1362568, EPI_ISL_1362569, EPI_ISL_1362570, EPI_ISL_1362571, EPI_ISL_1362572, EPI_ISL_1362573, EPI_ISL_1362574, EPI_ISL_1362575, EPI_ISL_1362576, EPI_ISL_1362577, EPI_ISL_1362578, EPI_ISL_1362579, EPI_ISL_1362580, EPI_ISL_1362581, EPI_ISL_1362582, EPI_ISL_1362583, EPI_ISL_1362584, EPI_ISL_1362585, EPI_ISL_1362586, EPI_ISL_1362587, EPI_ISL_1362588, EPI_ISL_1362589, EPI_ISL_1362591, EPI_ISL_1362592, EPI_ISL_1362593, EPI_ISL_1362594, EPI_ISL_1362595, EPI_ISL_1362596, EPI_ISL_1362597, EPI_ISL_1362598, EPI_ISL_1362599, EPI_ISL_1362600, EPI_ISL_1362601, EPI_ISL_1362602, EPI_ISL_1362603, EPI_ISL_1362604, EPI_ISL_1362605, EPI_ISL_1362606, EPI_ISL_1362607, EPI_ISL_1362609, EPI_ISL_1362610, EPI_ISL_1362611, EPI_ISL_1362612, EPI_ISL_1362613, EPI_ISL_1362614, EPI_ISL_1362615, EPI_ISL_1362616, EPI_ISL_1362617, EPI_ISL_1362618, EPI_ISL_1362619, EPI_ISL_1362620, EPI_ISL_1362621, EPI_ISL_1362622, EPI_ISL_1362623, EPI_ISL_1362624, EPI_ISL_1362626, EPI_ISL_1362627, EPI_ISL_1362628, EPI_ISL_1362629, EPI_ISL_1362630, EPI_ISL_1362631, EPI_ISL_1362632, EPI_ISL_1362633, EPI_ISL_1362634, EPI_ISL_1362635, EPI_ISL_1362636, EPI_ISL_1362637, EPI_ISL_1362639, EPI_ISL_1362640, EPI_ISL_1362641, EPI_ISL_1362642, EPI_ISL_1362643, EPI_ISL_1362644, EPI_ISL_1362645, EPI_ISL_1362646, EPI_ISL_1362647, EPI_ISL_1362648, EPI_ISL_1362649, EPI_ISL_1362650, EPI_ISL_1362651, EPI_ISL_1362652, EPI_ISL_1362653, EPI_ISL_1362654, EPI_ISL_1362655, EPI_ISL_1362656, EPI_ISL_1362657, EPI_ISL_1362658, EPI_ISL_1362659, EPI_ISL_1362660, EPI_ISL_1362661, EPI_ISL_1362662, EPI_ISL_1362663, EPI_ISL_1362664, EPI_ISL_1362665, EPI_ISL_1362666, EPI_ISL_1362667, EPI_ISL_1362668, EPI_ISL_1362669, EPI_ISL_1362670, EPI_ISL_1362671, EPI_ISL_1362672, EPI_ISL_1362673, EPI_ISL_1362674, EPI_ISL_1362675, EPI_ISL_1362676, EPI_ISL_1362677, EPI_ISL_1362678, EPI_ISL_1362679, EPI_ISL_1362680, EPI_ISL_1362681, EPI_ISL_1362682, EPI_ISL_1362683, EPI_ISL_1362684, EPI_ISL_1362685, EPI_ISL_1362686, EPI_ISL_1362687, EPI_ISL_1362688, EPI_ISL_1362689, EPI_ISL_1362690, EPI_ISL_1362691, EPI_ISL_1362692, EPI_ISL_1362693, EPI_ISL_1362694, EPI_ISL_1362695, EPI_ISL_1362696, EPI_ISL_1362697, EPI_ISL_1362698, EPI_ISL_1362699, EPI_ISL_1362700, EPI_ISL_1362701, EPI_ISL_1362702, EPI_ISL_1362703, EPI_ISL_1362704, EPI_ISL_1362705, EPI_ISL_1362706, EPI_ISL_1362707, EPI_ISL_1362708, EPI_ISL_1362709, EPI_ISL_1362710, EPI_ISL_1362711, EPI_ISL_1362712, EPI_ISL_1362713, EPI_ISL_1362714, EPI_ISL_1362715, EPI_ISL_1362716, EPI_ISL_1362717, EPI_ISL_1362718, EPI_ISL_1362719, EPI_ISL_1362720, EPI_ISL_1362722, EPI_ISL_1362723, EPI_ISL_1362724, EPI_ISL_1362725, EPI_ISL_1362726, EPI_ISL_1362727, EPI_ISL_1362729, EPI_ISL_1362730, EPI_ISL_1362731, EPI_ISL_1362732, EPI_ISL_1362733, EPI_ISL_1362734, EPI_ISL_1362735, EPI_ISL_1362736, EPI_ISL_1362737, EPI_ISL_1362738, EPI_ISL_1362739, EPI_ISL_1362740, EPI_ISL_1362741, EPI_ISL_1362742, EPI_ISL_1362743, EPI_ISL_1362744, EPI_ISL_1362745, EPI_ISL_1362746, EPI_ISL_1362747, EPI_ISL_1362748, EPI_ISL_1362749, EPI_ISL_1362750, EPI_ISL_1362751, EPI_ISL_1362752, EPI_ISL_1362753, EPI_ISL_1362754, EPI_ISL_1362756, EPI_ISL_1362757, EPI_ISL_1362758, EPI_ISL_1362759, EPI_ISL_1362760, EPI_ISL_1362761, EPI_ISL_1362762, EPI_ISL_1362763, EPI_ISL_1362764, EPI_ISL_1362765, EPI_ISL_1362766, EPI_ISL_1362767, EPI_ISL_1362768, EPI_ISL_1362770, EPI_ISL_1362771, EPI_ISL_1362772, EPI_ISL_1362773, EPI_ISL_1362774, EPI_ISL_1362775, EPI_ISL_1362776, EPI_ISL_1362777, EPI_ISL_1362778, EPI_ISL_1362779, EPI_ISL_1362780, EPI_ISL_1362781, EPI_ISL_1362782, EPI_ISL_1362783, EPI_ISL_1362784, EPI_ISL_1362785, EPI_ISL_1362786, EPI_ISL_1362787, EPI_ISL_1362788, EPI_ISL_1362789, EPI_ISL_1362790, EPI_ISL_1362791, EPI_ISL_1362792, EPI_ISL_1362793, EPI_ISL_1362794, EPI_ISL_1362795, EPI_ISL_1362796, EPI_ISL_1362797, EPI_ISL_1362798, EPI_ISL_1362799, EPI_ISL_1362800, EPI_ISL_1362801, EPI_ISL_1362802, EPI_ISL_1362803, EPI_ISL_1362804, EPI_ISL_1362805, EPI_ISL_1362806, EPI_ISL_1362807, EPI_ISL_1362808, EPI_ISL_1362809, EPI_ISL_1362810, EPI_ISL_1362811, EPI_ISL_1362812, EPI_ISL_1362813, EPI_ISL_1362814, EPI_ISL_1362815, EPI_ISL_1362816, EPI_ISL_1362817, EPI_ISL_1362818, EPI_ISL_1362819, EPI_ISL_1362820, EPI_ISL_1362821, EPI_ISL_1362822, EPI_ISL_1362823, EPI_ISL_1362824, EPI_ISL_1362825, EPI_ISL_1362826, EPI_ISL_1362827, EPI_ISL_1362828, EPI_ISL_1362829, EPI_ISL_1362830, EPI_ISL_1362831, EPI_ISL_1362832, EPI_ISL_1362833, EPI_ISL_1362834, EPI_ISL_1362835, EPI_ISL_1362836, EPI_ISL_1362837, EPI_ISL_1362838, EPI_ISL_1362839, EPI_ISL_1362840, EPI_ISL_1362841, EPI_ISL_1362842, EPI_ISL_1362843, EPI_ISL_1362844, EPI_ISL_1362845, EPI_ISL_1362846, EPI_ISL_1362847, EPI_ISL_1362848, EPI_ISL_1362849, EPI_ISL_1362850, EPI_ISL_1362852, EPI_ISL_1362853, EPI_ISL_1362854, EPI_ISL_1362855, EPI_ISL_1362856, EPI_ISL_1362857, EPI_ISL_1362858, EPI_ISL_1362859, EPI_ISL_1362860, EPI_ISL_1362861, EPI_ISL_1362862, EPI_ISL_1362863, EPI_ISL_1362864, EPI_ISL_1362865, EPI_ISL_1362866, EPI_ISL_1362867, EPI_ISL_1362869, EPI_ISL_1362870, EPI_ISL_1362871, EPI_ISL_1362872, EPI_ISL_1362873, EPI_ISL_1362874, EPI_ISL_1362875, EPI_ISL_1362876, EPI_ISL_1362877, EPI_ISL_1362878, EPI_ISL_1362879, EPI_ISL_1362880, EPI_ISL_1362881, EPI_ISL_1362882, EPI_ISL_1362883, EPI_ISL_1362884, EPI_ISL_1362885, EPI_ISL_1362886, EPI_ISL_1362887, EPI_ISL_1362888, EPI_ISL_1362889, EPI_ISL_1362890, EPI_ISL_1362891, EPI_ISL_1362892, EPI_ISL_1362893, EPI_ISL_1362894, EPI_ISL_1362895, EPI_ISL_1362896, EPI_ISL_1362897, EPI_ISL_1362898, EPI_ISL_1362899, EPI_ISL_1362900, EPI_ISL_1362901, EPI_ISL_1362902, EPI_ISL_1362903, EPI_ISL_1362904, EPI_ISL_1362905, EPI_ISL_1362906, EPI_ISL_1362908, EPI_ISL_1362909, EPI_ISL_1362910, EPI_ISL_1362911, EPI_ISL_1362912, EPI_ISL_1362913, EPI_ISL_1362914, EPI_ISL_1362915, EPI_ISL_1362916, EPI_ISL_1362917, EPI_ISL_1362918, EPI_ISL_1362919, EPI_ISL_1362920, EPI_ISL_1362921, EPI_ISL_1362922, EPI_ISL_1362923, EPI_ISL_1362924, EPI_ISL_1362925, EPI_ISL_1362926, EPI_ISL_1362927, EPI_ISL_1362928, EPI_ISL_1362929, EPI_ISL_1362930, EPI_ISL_1362931, EPI_ISL_1362932, EPI_ISL_1362933, EPI_ISL_1362934, EPI_ISL_1362935, EPI_ISL_1362936, EPI_ISL_1362937, EPI_ISL_1362938, EPI_ISL_1362939, EPI_ISL_1362940, EPI_ISL_1362941, EPI_ISL_1362942, EPI_ISL_1362943, EPI_ISL_1362944, EPI_ISL_1362945, EPI_ISL_1362946, EPI_ISL_1362947, EPI_ISL_1362948, EPI_ISL_1362949, EPI_ISL_1362950, EPI_ISL_1362951, EPI_ISL_1362952, EPI_ISL_1362953, EPI_ISL_1362954, EPI_ISL_1362956, EPI_ISL_1362957, EPI_ISL_1362958, EPI_ISL_1362960, EPI_ISL_1362961, EPI_ISL_1362962, EPI_ISL_1362963, EPI_ISL_1362964, EPI_ISL_1362965, EPI_ISL_1362966, EPI_ISL_1362967, EPI_ISL_1362968, EPI_ISL_1362969, EPI_ISL_1362970, EPI_ISL_1362971, EPI_ISL_1362972, EPI_ISL_1362973, EPI_ISL_1362974, EPI_ISL_1362975, EPI_ISL_1362976, EPI_ISL_1362977, EPI_ISL_1362978, EPI_ISL_1362979, EPI_ISL_1362980, EPI_ISL_1362981 | | | |
| see above | Missouri State Public Health Laboratory | Missouri State Public Health Laboratory | Matthew Sinn, Joshua Barry, Ashley New |
| EPI_ISL_1362982, EPI_ISL_1362983, EPI_ISL_1362984, EPI_ISL_1362985 | Kansas Health and Environmental Lab | Kansas Health and Environmental Lab | Mike Grose, Jonathan Barnell, Ben Olsen, and Phil Adam |
| EPI_ISL_1362994, EPI_ISL_1362995, EPI_ISL_1362996, EPI_ISL_1362997, EPI_ISL_1362998, EPI_ISL_1362999, EPI_ISL_1363000, EPI_ISL_1363001, EPI_ISL_1363002, EPI_ISL_1363003, EPI_ISL_1363004, EPI_ISL_1363005, EPI_ISL_1363006, EPI_ISL_1363007, EPI_ISL_1363008, EPI_ISL_1363009, EPI_ISL_1363010, EPI_ISL_1363011, EPI_ISL_1363012, EPI_ISL_1363013, EPI_ISL_1363014, EPI_ISL_1363015, EPI_ISL_1363016, EPI_ISL_1363017, EPI_ISL_1363018, EPI_ISL_1363019, EPI_ISL_1363020, EPI_ISL_1363021, EPI_ISL_1363022, EPI_ISL_1363023, EPI_ISL_1363024, EPI_ISL_1363025, EPI_ISL_1363026, EPI_ISL_1363027, EPI_ISL_1363029, EPI_ISL_1363030, EPI_ISL_1363032, EPI_ISL_1363033, EPI_ISL_1363034, EPI_ISL_1363035, EPI_ISL_1363036, EPI_ISL_1363037, EPI_ISL_1363039, EPI_ISL_1363040, EPI_ISL_1363041, EPI_ISL_1363042, EPI_ISL_1363043, EPI_ISL_1363044, EPI_ISL_1363045, EPI_ISL_1363046, EPI_ISL_1363047, EPI_ISL_1363048, EPI_ISL_1363049, EPI_ISL_1363050, EPI_ISL_1363051, EPI_ISL_1363052, EPI_ISL_1363053, EPI_ISL_1363054, EPI_ISL_1363055, EPI_ISL_1363056, EPI_ISL_1363057, EPI_ISL_1363058, EPI_ISL_1363059, EPI_ISL_1363060, EPI_ISL_1363061, EPI_ISL_1363062, EPI_ISL_1363063, EPI_ISL_1363064, EPI_ISL_1363065, EPI_ISL_1363066, EPI_ISL_1363067, EPI_ISL_1363069, EPI_ISL_1363070, EPI_ISL_1363071, EPI_ISL_1363072, EPI_ISL_1363073, EPI_ISL_1363074, EPI_ISL_1363075, EPI_ISL_1363076, EPI_ISL_1363077, EPI_ISL_1363078, EPI_ISL_1363079, EPI_ISL_1363080, EPI_ISL_1363081 | | | |
| see above | TXDSHS | TXDSHS | Rashmi Tuladhar, Bonnie Oh, Jenny Zhang, Maliha Rahman, Mayela Pedrueza, Anita Pokharel, Loarraine Rodriguez, Myong Koag, Chun Wang, Rachel Lee, Grace Kubin |
| EPI_ISL_1363151, EPI_ISL_1363152, EPI_ISL_1363153, EPI_ISL_1363156, EPI_ISL_1363157, EPI_ISL_1363158, EPI_ISL_1363159, EPI_ISL_1363160, EPI_ISL_1363161, EPI_ISL_1363162, EPI_ISL_1363163, EPI_ISL_1363164, EPI_ISL_1363165, EPI_ISL_1363166 | | | |
| see above | IN State Department of Health Laboratory Services | IN State Department of Health Laboratory Services | Cassandra Campion, Jamie Yeadon, Brian Pope, Lixia Liu, Kyle Brownlee, Melissa Hindenlang, Ankita Kashikar, Mark Glazier |
| EPI_ISL_1363168, EPI_ISL_1363169, EPI_ISL_1363170, EPI_ISL_1363171, EPI_ISL_1363175, EPI_ISL_1363176, EPI_ISL_1363177, EPI_ISL_1363178, EPI_ISL_1363179, EPI_ISL_1363181, EPI_ISL_1363182, EPI_ISL_1363183, EPI_ISL_1363186, EPI_ISL_1363187, EPI_ISL_1363188, EPI_ISL_1363189, EPI_ISL_1363190, EPI_ISL_1363191, EPI_ISL_1363192, EPI_ISL_1363194 | | | |
| see above | Idaho Bureau of Laboratories | Idaho Bureau of Laboratories | "R. Beukelman, Matthew Charles Burns, Aimee Ceniseros, Robert L. Voermans, Christopher Ball" |
| EPI_ISL_1363219, EPI_ISL_1363220, EPI_ISL_1363221, EPI_ISL_1363222 | Sonora Quest Laboratories | TGen North | Jolene Bowers, Heather Centner, Chris French, Hayley Yaglom, Ashlyn Pfeiffer, Darrin Lemmer, Dave Engelthaler, The Arizona COVID Genomics Union (ACGU) |
| EPI_ISL_1363223 | TGen North | TGen North | Jolene Bowers, Heather Centner, Chris French, Hayley Yaglom, Ashlyn Pfeiffer, Darrin Lemmer, Dave Engelthaler, The Arizona COVID Genomics Union (ACGU) |
| EPI_ISL_1363224, EPI_ISL_1363227, EPI_ISL_1363229, EPI_ISL_1363230, EPI_ISL_1363231, EPI_ISL_1363232, EPI_ISL_1363233, EPI_ISL_1363234, EPI_ISL_1363235, EPI_ISL_1363236, EPI_ISL_1363237, EPI_ISL_1363243, EPI_ISL_1363244, EPI_ISL_1363245, EPI_ISL_1363246, EPI_ISL_1363247, EPI_ISL_1363250, EPI_ISL_1363252, EPI_ISL_1363253, EPI_ISL_1363254, EPI_ISL_1363255, EPI_ISL_1363256, EPI_ISL_1363257, EPI_ISL_1363259, EPI_ISL_1363260, EPI_ISL_1363261, EPI_ISL_1363262, EPI_ISL_1363263, EPI_ISL_1363264, EPI_ISL_1363265, EPI_ISL_1363267, EPI_ISL_1363268, EPI_ISL_1363269, EPI_ISL_1363271 | | | |
| see above | Sonora Quest Laboratories | TGen North | Jolene Bowers, Heather Centner, Chris French, Hayley Yaglom, Ashlyn Pfeiffer, Darrin Lemmer, Dave Engelthaler, The Arizona COVID Genomics Union (ACGU) |
| EPI_ISL_1363274 | TGen North | TGen North | Jolene Bowers, Heather Centner, Chris French, Hayley Yaglom, Ashlyn Pfeiffer, Darrin Lemmer, Dave Engelthaler, The Arizona COVID Genomics Union (ACGU) |
| EPI_ISL_1363275, EPI_ISL_1363277, EPI_ISL_1363279, EPI_ISL_1363280, EPI_ISL_1363281, EPI_ISL_1363282, EPI_ISL_1363284, EPI_ISL_1363285, EPI_ISL_1363286, EPI_ISL_1363288, EPI_ISL_1363289, EPI_ISL_1363291, EPI_ISL_1363292, EPI_ISL_1363293, EPI_ISL_1363295, EPI_ISL_1363296, EPI_ISL_1363298, EPI_ISL_1363299, EPI_ISL_1363300, EPI_ISL_1363302, EPI_ISL_1363303, EPI_ISL_1363304, EPI_ISL_1363305, EPI_ISL_1363306, EPI_ISL_1363307, EPI_ISL_1363308, EPI_ISL_1363310, EPI_ISL_1363311, EPI_ISL_1363313 | | | |
| see above | Sonora Quest Laboratories | TGen North | Jolene Bowers, Heather Centner, Chris French, Hayley Yaglom, Ashlyn Pfeiffer, Darrin Lemmer, Dave Engelthaler, The Arizona COVID Genomics Union (ACGU) |
| EPI_ISL_1363314 | TGen North | TGen North | Jolene Bowers, Heather Centner, Chris French, Hayley Yaglom, Ashlyn Pfeiffer, Darrin Lemmer, Dave Engelthaler, The Arizona COVID Genomics Union (ACGU) |
| EPI_ISL_1363316, EPI_ISL_1363317, EPI_ISL_1363318 | Sonora Quest Laboratories | TGen North | Jolene Bowers, Heather Centner, Chris French, Hayley Yaglom, Ashlyn Pfeiffer, Darrin Lemmer, Dave Engelthaler, The Arizona COVID Genomics Union (ACGU) |
| EPI_ISL_1363319 | TGen North | TGen North | Jolene Bowers, Heather Centner, Chris French, Hayley Yaglom, Ashlyn Pfeiffer, Darrin Lemmer, Dave Engelthaler, The Arizona COVID Genomics Union (ACGU) |
| EPI_ISL_1363320, EPI_ISL_1363321, EPI_ISL_1363322, EPI_ISL_1363323, EPI_ISL_1363325, EPI_ISL_1363326, EPI_ISL_1363328, EPI_ISL_1363329, EPI_ISL_1363331, EPI_ISL_1363332, EPI_ISL_1363333, EPI_ISL_1363334, EPI_ISL_1363335, EPI_ISL_1363337, EPI_ISL_1363338, EPI_ISL_1363339, EPI_ISL_1363343, EPI_ISL_1363344, EPI_ISL_1363345, EPI_ISL_1363346, EPI_ISL_1363349, EPI_ISL_1363350, EPI_ISL_1363352, EPI_ISL_1363353, EPI_ISL_1363354, EPI_ISL_1363355, EPI_ISL_1363356, EPI_ISL_1363357, EPI_ISL_1363358, EPI_ISL_1363359, EPI_ISL_1363360, EPI_ISL_1363361, EPI_ISL_1363362, EPI_ISL_1363363, EPI_ISL_1363366, EPI_ISL_1363367 | | | |
| see above | Sonora Quest Laboratories | TGen North | Jolene Bowers, Heather Centner, Chris French, Hayley Yaglom, Ashlyn Pfeiffer, Darrin Lemmer, Dave Engelthaler, The Arizona COVID Genomics Union (ACGU) |
| EPI_ISL_1363368 | TGen North | TGen North | Jolene Bowers, Heather Centner, Chris French, Hayley Yaglom, Ashlyn Pfeiffer, Darrin Lemmer, Dave Engelthaler, The Arizona COVID Genomics Union (ACGU) |
| EPI_ISL_1363370, EPI_ISL_1363371, EPI_ISL_1363372, EPI_ISL_1363373, EPI_ISL_1363374, EPI_ISL_1363375, EPI_ISL_1363376, EPI_ISL_1363377, EPI_ISL_1363378, EPI_ISL_1363379, EPI_ISL_1363380, EPI_ISL_1363382, EPI_ISL_1363383, EPI_ISL_1363386, EPI_ISL_1363387, EPI_ISL_1363389, EPI_ISL_1363390, EPI_ISL_1363391, EPI_ISL_1363392, EPI_ISL_1363393, EPI_ISL_1363395, EPI_ISL_1363397, EPI_ISL_1363398, EPI_ISL_1363399, EPI_ISL_1363400, EPI_ISL_1363401 | | | |
| see above | Sonora Quest Laboratories | TGen North | Jolene Bowers, Heather Centner, Chris French, Hayley Yaglom, Ashlyn Pfeiffer, Darrin Lemmer, Dave Engelthaler, The Arizona COVID Genomics Union (ACGU) |
| EPI_ISL_1363403 | TGen North | TGen North | Jolene Bowers, Heather Centner, Chris French, Hayley Yaglom, Ashlyn Pfeiffer, Darrin Lemmer, Dave Engelthaler, The Arizona COVID Genomics Union (ACGU) |
| EPI_ISL_1363404, EPI_ISL_1363405, EPI_ISL_1363408, EPI_ISL_1363410, EPI_ISL_1363411, EPI_ISL_1363412, EPI_ISL_1363413, EPI_ISL_1363414, EPI_ISL_1363415, EPI_ISL_1363416, EPI_ISL_1363417, EPI_ISL_1363419, EPI_ISL_1363421, EPI_ISL_1363422, EPI_ISL_1363423, EPI_ISL_1363424, EPI_ISL_1363425, EPI_ISL_1363426, EPI_ISL_1363427, EPI_ISL_1363428, EPI_ISL_1363429, EPI_ISL_1363430, EPI_ISL_1363431, EPI_ISL_1363432, EPI_ISL_1363435 | | | |
| see above | Sonora Quest Laboratories | TGen North | Jolene Bowers, Heather Centner, Chris French, Hayley Yaglom, Ashlyn Pfeiffer, Darrin Lemmer, Dave Engelthaler, The Arizona COVID Genomics Union (ACGU) |
| EPI_ISL_1363436 | TGen North | TGen North | Jolene Bowers, Heather Centner, Chris French, Hayley Yaglom, Ashlyn Pfeiffer, Darrin Lemmer, Dave Engelthaler, The Arizona COVID Genomics Union (ACGU) |
| EPI_ISL_1363437, EPI_ISL_1363438, EPI_ISL_1363440, EPI_ISL_1363442, EPI_ISL_1363444, EPI_ISL_1363445 | Sonora Quest Laboratories | TGen North | Jolene Bowers, Heather Centner, Chris French, Hayley Yaglom, Ashlyn Pfeiffer, Darrin Lemmer, Dave Engelthaler, The Arizona COVID Genomics Union (ACGU) |
| EPI_ISL_1363447 | TGen North | TGen North | Jolene Bowers, Heather Centner, Chris French, Hayley Yaglom, Ashlyn Pfeiffer, Darrin Lemmer, Dave Engelthaler, The Arizona COVID Genomics Union (ACGU) |
| EPI_ISL_1363448, EPI_ISL_1363449, EPI_ISL_1363450, EPI_ISL_1363453 | Sonora Quest Laboratories | TGen North | Jolene Bowers, Heather Centner, Chris French, Hayley Yaglom, Ashlyn Pfeiffer, Darrin Lemmer, Dave Engelthaler, The Arizona COVID Genomics Union (ACGU) |
| EPI_ISL_1363454 | TGen North | TGen North | Jolene Bowers, Heather Centner, Chris French, Hayley Yaglom, Ashlyn Pfeiffer, Darrin Lemmer, Dave Engelthaler, The Arizona COVID Genomics Union (ACGU) |
| EPI_ISL_1363456, EPI_ISL_1363457, EPI_ISL_1363458, EPI_ISL_1363459, EPI_ISL_1363460, EPI_ISL_1363461, EPI_ISL_1363463, EPI_ISL_1363465, EPI_ISL_1363466, EPI_ISL_1363467, EPI_ISL_1363468, EPI_ISL_1363469, EPI_ISL_1363470, EPI_ISL_1363471, EPI_ISL_1363472, EPI_ISL_1363474, EPI_ISL_1363475, EPI_ISL_1363478, EPI_ISL_1363479, EPI_ISL_1363480, EPI_ISL_1363481, EPI_ISL_1363482, EPI_ISL_1363483, EPI_ISL_1363484, EPI_ISL_1363485, EPI_ISL_1363486, EPI_ISL_1363487, EPI_ISL_1363488, EPI_ISL_1363489, EPI_ISL_1363490, EPI_ISL_1363492, EPI_ISL_1363493, EPI_ISL_1363494, EPI_ISL_1363495, EPI_ISL_1363496, EPI_ISL_1363497, EPI_ISL_1363498, EPI_ISL_1363499 | | | |
| see above | Sonora Quest Laboratories | TGen North | Jolene Bowers, Heather Centner, Chris French, Hayley Yaglom, Ashlyn Pfeiffer, Darrin Lemmer, Dave Engelthaler, The Arizona COVID Genomics Union (ACGU) |
| EPI_ISL_1363502, EPI_ISL_1363503 | TGen North | TGen North | Jolene Bowers, Heather Centner, Chris French, Hayley Yaglom, Ashlyn Pfeiffer, Darrin Lemmer, Dave Engelthaler, The Arizona COVID Genomics Union (ACGU) |
| EPI_ISL_1363504, EPI_ISL_1363506, EPI_ISL_1363508, EPI_ISL_1363509, EPI_ISL_1363510, EPI_ISL_1363511, EPI_ISL_1363512, EPI_ISL_1363513, EPI_ISL_1363514, EPI_ISL_1363515, EPI_ISL_1363516, EPI_ISL_1363519, EPI_ISL_1363520, EPI_ISL_1363521, EPI_ISL_1363523, EPI_ISL_1363525, EPI_ISL_1363526, EPI_ISL_1363527, EPI_ISL_1363528, EPI_ISL_1363529, EPI_ISL_1363530, EPI_ISL_1363531, EPI_ISL_1363532, EPI_ISL_1363534, EPI_ISL_1363535, EPI_ISL_1363536, EPI_ISL_1363538, EPI_ISL_1363539, EPI_ISL_1363540, EPI_ISL_1363541, EPI_ISL_1363542 | | | |
| see above | Sonora Quest Laboratories | TGen North | Jolene Bowers, Heather Centner, Chris French, Hayley Yaglom, Ashlyn Pfeiffer, Darrin Lemmer, Dave Engelthaler, The Arizona COVID Genomics Union (ACGU) |
| EPI_ISL_1363543 | TGen North | TGen North | Jolene Bowers, Heather Centner, Chris French, Hayley Yaglom, Ashlyn Pfeiffer, Darrin Lemmer, Dave Engelthaler, The Arizona COVID Genomics Union (ACGU) |

| | | | |
|---|---|---|---|
| EPI_ISL_1363545, EPI_ISL_1363547, EPI_ISL_1363549 | Sonora Quest Laboratories | TGen North | Jolene Bowers, Heather Centner, Chris French, Hayley Yaglom, Ashlyn Pfeiffer, Darrin Lemmer, Dave Engelthaler, The Arizona COVID Genomics Union (ACGU) |
| EPI_ISL_1363550 | TGen North | TGen North | Jolene Bowers, Heather Centner, Chris French, Hayley Yaglom, Ashlyn Pfeiffer, Darrin Lemmer, Dave Engelthaler, The Arizona COVID Genomics Union (ACGU) |
| EPI_ISL_1363551, EPI_ISL_1363552, EPI_ISL_1363553, EPI_ISL_1363555, EPI_ISL_1363556, EPI_ISL_1363557, EPI_ISL_1363559, EPI_ISL_1363560, EPI_ISL_1363561, EPI_ISL_1363563, EPI_ISL_1363564, EPI_ISL_1363565, EPI_ISL_1363566, EPI_ISL_1363567, EPI_ISL_1363568, EPI_ISL_1363569, EPI_ISL_1363570, EPI_ISL_1363571, EPI_ISL_1363573, EPI_ISL_1363574, EPI_ISL_1363575 | | | |
| see above | Sonora Quest Laboratories | TGen North | Jolene Bowers, Heather Centner, Chris French, Hayley Yaglom, Ashlyn Pfeiffer, Darrin Lemmer, Dave Engelthaler, The Arizona COVID Genomics Union (ACGU) |
| EPI_ISL_1363576 | TGen North | TGen North | Jolene Bowers, Heather Centner, Chris French, Hayley Yaglom, Ashlyn Pfeiffer, Darrin Lemmer, Dave Engelthaler, The Arizona COVID Genomics Union (ACGU) |
| EPI_ISL_1363577, EPI_ISL_1363578 | Sonora Quest Laboratories | TGen North | Jolene Bowers, Heather Centner, Chris French, Hayley Yaglom, Ashlyn Pfeiffer, Darrin Lemmer, Dave Engelthaler, The Arizona COVID Genomics Union (ACGU) |
| EPI_ISL_1363579 | TGen North | TGen North | Jolene Bowers, Heather Centner, Chris French, Hayley Yaglom, Ashlyn Pfeiffer, Darrin Lemmer, Dave Engelthaler, The Arizona COVID Genomics Union (ACGU) |
| EPI_ISL_1363581, EPI_ISL_1363582, EPI_ISL_1363583, EPI_ISL_1363584, EPI_ISL_1363587, EPI_ISL_1363589, EPI_ISL_1363590, EPI_ISL_1363591, EPI_ISL_1363592, EPI_ISL_1363593 | Sonora Quest Laboratories | TGen North | Jolene Bowers, Heather Centner, Chris French, Hayley Yaglom, Ashlyn Pfeiffer, Darrin Lemmer, Dave Engelthaler, The Arizona COVID Genomics Union (ACGU) |
| EPI_ISL_1363594 | TGen North | TGen North | Jolene Bowers, Heather Centner, Chris French, Hayley Yaglom, Ashlyn Pfeiffer, Darrin Lemmer, Dave Engelthaler, The Arizona COVID Genomics Union (ACGU) |
| EPI_ISL_1363596, EPI_ISL_1363597, EPI_ISL_1363598, EPI_ISL_1363599, EPI_ISL_1363601, EPI_ISL_1363602, EPI_ISL_1363606 | Sonora Quest Laboratories | TGen North | Jolene Bowers, Heather Centner, Chris French, Hayley Yaglom, Ashlyn Pfeiffer, Darrin Lemmer, Dave Engelthaler, The Arizona COVID Genomics Union (ACGU) |
| EPI_ISL_1363608 | TGen North | TGen North | Jolene Bowers, Heather Centner, Chris French, Hayley Yaglom, Ashlyn Pfeiffer, Darrin Lemmer, Dave Engelthaler, The Arizona COVID Genomics Union (ACGU) |
| EPI_ISL_1363609, EPI_ISL_1363610, EPI_ISL_1363611, EPI_ISL_1363612, EPI_ISL_1363613, EPI_ISL_1363615, EPI_ISL_1363616, EPI_ISL_1363617, EPI_ISL_1363618, EPI_ISL_1363619, EPI_ISL_1363620, EPI_ISL_1363622, EPI_ISL_1363623, EPI_ISL_1363624, EPI_ISL_1363625, EPI_ISL_1363626, EPI_ISL_1363629, EPI_ISL_1363632, EPI_ISL_1363633, EPI_ISL_1363634, EPI_ISL_1363635, EPI_ISL_1363636, EPI_ISL_1363637, EPI_ISL_1363638, EPI_ISL_1363639, EPI_ISL_1363640, EPI_ISL_1363641, EPI_ISL_1363642, EPI_ISL_1363643, EPI_ISL_1363644, EPI_ISL_1363646, EPI_ISL_1363647, EPI_ISL_1363648, EPI_ISL_1363649, EPI_ISL_1363650, EPI_ISL_1363651, EPI_ISL_1363652, EPI_ISL_1363653, EPI_ISL_1363654, EPI_ISL_1363655, EPI_ISL_1363657 | | | |
| see above | Sonora Quest Laboratories | TGen North | Jolene Bowers, Heather Centner, Chris French, Hayley Yaglom, Ashlyn Pfeiffer, Darrin Lemmer, Dave Engelthaler, The Arizona COVID Genomics Union (ACGU) |
| EPI_ISL_1363659 | TGen North | TGen North | Jolene Bowers, Heather Centner, Chris French, Hayley Yaglom, Ashlyn Pfeiffer, Darrin Lemmer, Dave Engelthaler, The Arizona COVID Genomics Union (ACGU) |
| EPI_ISL_1363662, EPI_ISL_1363664, EPI_ISL_1363665, EPI_ISL_1363666, EPI_ISL_1363667, EPI_ISL_1363668, EPI_ISL_1363669, EPI_ISL_1363670, EPI_ISL_1363672, EPI_ISL_1363673 | Sonora Quest Laboratories | TGen North | Jolene Bowers, Heather Centner, Chris French, Hayley Yaglom, Ashlyn Pfeiffer, Darrin Lemmer, Dave Engelthaler, The Arizona COVID Genomics Union (ACGU) |
| EPI_ISL_1363674 | TGen North | TGen North | Jolene Bowers, Heather Centner, Chris French, Hayley Yaglom, Ashlyn Pfeiffer, Darrin Lemmer, Dave Engelthaler, The Arizona COVID Genomics Union (ACGU) |
| EPI_ISL_1363675, EPI_ISL_1363676, EPI_ISL_1363677, EPI_ISL_1363678, EPI_ISL_1363680, EPI_ISL_1363681, EPI_ISL_1363682, EPI_ISL_1363683, EPI_ISL_1363685, EPI_ISL_1363686, EPI_ISL_1363687, EPI_ISL_1363688, EPI_ISL_1363690, EPI_ISL_1363691, EPI_ISL_1363692, EPI_ISL_1363693, EPI_ISL_1363694, EPI_ISL_1363696, EPI_ISL_1363697, EPI_ISL_1363698, EPI_ISL_1363699, EPI_ISL_1363700, EPI_ISL_1363701 | | | |
| see above | Sonora Quest Laboratories | TGen North | Jolene Bowers, Heather Centner, Chris French, Hayley Yaglom, Ashlyn Pfeiffer, Darrin Lemmer, Dave Engelthaler, The Arizona COVID Genomics Union (ACGU) |
| EPI_ISL_1363702 | TGen North | TGen North | Jolene Bowers, Heather Centner, Chris French, Hayley Yaglom, Ashlyn Pfeiffer, Darrin Lemmer, Dave Engelthaler, The Arizona COVID Genomics Union (ACGU) |
| EPI_ISL_1363704, EPI_ISL_1363705 | Sonora Quest Laboratories | TGen North | Jolene Bowers, Heather Centner, Chris French, Hayley Yaglom, Ashlyn Pfeiffer, Darrin Lemmer, Dave Engelthaler, The Arizona COVID Genomics Union (ACGU) |
| EPI_ISL_1363707 | TGen North | TGen North | Jolene Bowers, Heather Centner, Chris French, Hayley Yaglom, Ashlyn Pfeiffer, Darrin Lemmer, Dave Engelthaler, The Arizona COVID Genomics Union (ACGU) |
| EPI_ISL_1363708, EPI_ISL_1363709, EPI_ISL_1363712 | Sonora Quest Laboratories | TGen North | Jolene Bowers, Heather Centner, Chris French, Hayley Yaglom, Ashlyn Pfeiffer, Darrin Lemmer, Dave Engelthaler, The Arizona COVID Genomics Union (ACGU) |
| EPI_ISL_1363714 | TGen North | TGen North | Jolene Bowers, Heather Centner, Chris French, Hayley Yaglom, Ashlyn Pfeiffer, Darrin Lemmer, Dave Engelthaler, The Arizona COVID Genomics Union (ACGU) |
| EPI_ISL_1363715, EPI_ISL_1363716, EPI_ISL_1363717, EPI_ISL_1363718, EPI_ISL_1363719, EPI_ISL_1363720, EPI_ISL_1363722, EPI_ISL_1363723, EPI_ISL_1363724, EPI_ISL_1363725, EPI_ISL_1363726, EPI_ISL_1363727, EPI_ISL_1363728, EPI_ISL_1363729, EPI_ISL_1363730, EPI_ISL_1363731, EPI_ISL_1363732, EPI_ISL_1363733, EPI_ISL_1363734, EPI_ISL_1363736, EPI_ISL_1363738, EPI_ISL_1363740, EPI_ISL_1363741, EPI_ISL_1363742, EPI_ISL_1363743, EPI_ISL_1363744, EPI_ISL_1363745, EPI_ISL_1363746, EPI_ISL_1363747, EPI_ISL_1363748, EPI_ISL_1363749 | | | |
| see above | Sonora Quest Laboratories | TGen North | Jolene Bowers, Heather Centner, Chris French, Hayley Yaglom, Ashlyn Pfeiffer, Darrin Lemmer, Dave Engelthaler, The Arizona COVID Genomics Union (ACGU) |
| EPI_ISL_1363750 | TGen North | TGen North | Jolene Bowers, Heather Centner, Chris French, Hayley Yaglom, Ashlyn Pfeiffer, Darrin Lemmer, Dave Engelthaler, The Arizona COVID Genomics Union (ACGU) |
| EPI_ISL_1363751, EPI_ISL_1363752, EPI_ISL_1363753, EPI_ISL_1363755 | Sonora Quest Laboratories | TGen North | Jolene Bowers, Heather Centner, Chris French, Hayley Yaglom, Ashlyn Pfeiffer, Darrin Lemmer, Dave Engelthaler, The Arizona COVID Genomics Union (ACGU) |
| EPI_ISL_1363756 | TGen North | TGen North | Jolene Bowers, Heather Centner, Chris French, Hayley Yaglom, Ashlyn Pfeiffer, Darrin Lemmer, Dave Engelthaler, The Arizona COVID Genomics Union (ACGU) |
| EPI_ISL_1363763, EPI_ISL_1363765, EPI_ISL_1363768, EPI_ISL_1363771, EPI_ISL_1363772, EPI_ISL_1363774, EPI_ISL_1363775, EPI_ISL_1363776, EPI_ISL_1363777, EPI_ISL_1363778, EPI_ISL_1363779, EPI_ISL_1363782 | | | |
| see above | Maryland Genomics, Institute for Genome Sciences, University of Maryland School of Medicine | Maryland Genomics, Institute for Genome Sciences, University of Maryland School of Medicine | Tallon, Luke J; Sadzewicz, Lisa D; Humphrys, Mike; Ott, Sandra; Roussey, Holly; Mehta, Aditya; Vavikolanu, Kranthi; Fraser, Claire M; Ravel, Jacques |
| EPI_ISL_1363788, EPI_ISL_1363789, EPI_ISL_1363790, EPI_ISL_1363791, EPI_ISL_1363792, EPI_ISL_1363793, EPI_ISL_1363794, EPI_ISL_1363795, EPI_ISL_1363796, EPI_ISL_1363797, EPI_ISL_1363798, EPI_ISL_1363799, EPI_ISL_1363800, EPI_ISL_1363801, EPI_ISL_1363802 | | | |
| see above | IN State Department of Health Laboratory Services | IN State Department of Health Laboratory Services | Cassandra Campion, Jamie Yeadon, Brian Pope, Lixia Liu, Kyle Brownlee, Melissa Hindenlang, Ankita Kashikar, Mark Glazier |
| EPI_ISL_1364430, EPI_ISL_1364431, EPI_ISL_1364432, EPI_ISL_1364433, EPI_ISL_1364434, EPI_ISL_1364435, EPI_ISL_1364436, EPI_ISL_1364437, EPI_ISL_1364438, EPI_ISL_1364439, EPI_ISL_1364441, EPI_ISL_1364442, EPI_ISL_1364443, EPI_ISL_1364444, EPI_ISL_1364445, EPI_ISL_1364446, EPI_ISL_1364447, EPI_ISL_1364449 | | | |
| see above | Nevada State Public Health Laboratory | Nevada State Public Health Laboratory | Andrew Gorzalski, Mark Pandori |
| EPI_ISL_1364450, EPI_ISL_1364451, EPI_ISL_1364452, EPI_ISL_1364453, EPI_ISL_1364454, EPI_ISL_1364455, EPI_ISL_1364456, EPI_ISL_1364457, EPI_ISL_1364458, EPI_ISL_1364459, EPI_ISL_1364460, EPI_ISL_1364461, EPI_ISL_1364462, EPI_ISL_1364463, EPI_ISL_1364464, EPI_ISL_1364465 | | | |
| see above | Arizona State University | Arizona State University | Rabia Maqsood, LaRinda A. Holland, Peter T. Skidmore, Emily A. Kaelin, Nicholas J. Mellor, Kristina Buss, Joy M. Blain, Neal W Woodbury, Valerie Harris, Joshua LaBaer, Vel Murugan, Efrem S. Lim |
| EPI_ISL_1364469, EPI_ISL_1364470, EPI_ISL_1364471, EPI_ISL_1364472, EPI_ISL_1364473, EPI_ISL_1364474, EPI_ISL_1364475 | Oregon State University TRACE Project | OSU Center for Genome Research and Biocomputing | Oregon State University TRACE Project |
| EPI_ISL_1364488, EPI_ISL_1364489, EPI_ISL_1364490, EPI_ISL_1364491, EPI_ISL_1364492, EPI_ISL_1364493, EPI_ISL_1364494, EPI_ISL_1364495, EPI_ISL_1364496, EPI_ISL_1364497, EPI_ISL_1364498, EPI_ISL_1364499, EPI_ISL_1364500, EPI_ISL_1364501, EPI_ISL_1364502, EPI_ISL_1364503, EPI_ISL_1364504, EPI_ISL_1364505, EPI_ISL_1364506, EPI_ISL_1364507, EPI_ISL_1364508, EPI_ISL_1364509, EPI_ISL_1364510, EPI_ISL_1364511, EPI_ISL_1364512, EPI_ISL_1364513, EPI_ISL_1364514, EPI_ISL_1364515, EPI_ISL_1364516, EPI_ISL_1364517, EPI_ISL_1364518, EPI_ISL_1364519, EPI_ISL_1364520, EPI_ISL_1364521, EPI_ISL_1364522, EPI_ISL_1364523, EPI_ISL_1364524, EPI_ISL_1364525 | | | |
| see above | Stanford Health Care | Stanford University School of Medicine, Clinical Virology Laboratory | Daniel Solis, Mamdouh Sibai, Fumiko Yamamoto, Malaya K. Sahoo, Michelle Verghese, ChunHong Huang, James Zehnder, and Benjamin A. Pinsky |
| EPI_ISL_1364526 | Stanford Health Care | Stanford University School of Medicine, Clinical Virology Laboratory | Daniel Solis, Mamdouh Sibai, Fumiko Yamamoto, Becky Jiang, Malaya K. Sahoo, Michelle Verghese, ChunHong Huang, James Zehnder, and Benjamin A. Pinsky |
| EPI_ISL_1364536 | Wyoming Public Health Laboratory | Wyoming Public Health Laboratory | Jim Mildenberger, Wanda Manley, Noah Hull, Taylor Fearing, Lynette Gumbleton, Channing Weber, Ashley Norberg, Chayse Rowley, Marley Goetz, Brian Dominguez, Elliot Thomasson, Cari Sloma, and Rob Christensen |
| EPI_ISL_1364542, EPI_ISL_1364543, EPI_ISL_1364544, EPI_ISL_1364545, EPI_ISL_1364546, EPI_ISL_1364547, EPI_ISL_1364549, EPI_ISL_1364550, EPI_ISL_1364551, EPI_ISL_1364552, EPI_ISL_1364553, EPI_ISL_1364554, EPI_ISL_1364555, EPI_ISL_1364556, EPI_ISL_1364557, EPI_ISL_1364558, EPI_ISL_1364559, EPI_ISL_1364560, EPI_ISL_1364561, EPI_ISL_1364562, EPI_ISL_1364563, EPI_ISL_1364564, EPI_ISL_1364565, EPI_ISL_1364566, EPI_ISL_1364567, EPI_ISL_1364568, EPI_ISL_1364569, EPI_ISL_1364570, EPI_ISL_1364571, EPI_ISL_1364572, EPI_ISL_1364573, EPI_ISL_1364574, EPI_ISL_1364575, EPI_ISL_1364576, EPI_ISL_1364577, EPI_ISL_1364578, EPI_ISL_1364579, EPI_ISL_1364580, EPI_ISL_1364581, EPI_ISL_1364582, EPI_ISL_1364583, EPI_ISL_1364584, EPI_ISL_1364585, EPI_ISL_1364586, | | | |

| | | | |
|---|---|---|---|
| EPI_ISL_1364587, EPI_ISL_1364588, EPI_ISL_1364589, EPI_ISL_1364590, EPI_ISL_1364591, EPI_ISL_1364592, EPI_ISL_1364593, EPI_ISL_1364594, EPI_ISL_1364595, EPI_ISL_1364596, EPI_ISL_1364597, EPI_ISL_1364599, EPI_ISL_1364600, EPI_ISL_1364601, EPI_ISL_1364602, EPI_ISL_1364603, EPI_ISL_1364604, EPI_ISL_1364605, EPI_ISL_1364606, EPI_ISL_1364607, EPI_ISL_1364608, EPI_ISL_1364609, EPI_ISL_1364610, EPI_ISL_1364611 | | | |
| see above | Virginia Division of Consolidated Laboratory Services | Virginia Division of Consolidated Laboratory Services | Virginia DCLS |
| EPI_ISL_1364622, EPI_ISL_1364623, EPI_ISL_1364624, EPI_ISL_1364625, EPI_ISL_1364626, EPI_ISL_1364627, EPI_ISL_1364628, EPI_ISL_1364629, EPI_ISL_1364630, EPI_ISL_1364631, EPI_ISL_1364632, EPI_ISL_1364633, EPI_ISL_1364634, EPI_ISL_1364636, EPI_ISL_1364637, EPI_ISL_1364638, EPI_ISL_1364639, EPI_ISL_1364640, EPI_ISL_1364641, EPI_ISL_1364643, EPI_ISL_1364644, EPI_ISL_1364646, EPI_ISL_1364647, EPI_ISL_1364648, EPI_ISL_1364649, EPI_ISL_1364650, EPI_ISL_1364652, EPI_ISL_1364653, EPI_ISL_1364654, EPI_ISL_1364656, EPI_ISL_1364657, EPI_ISL_1364658, EPI_ISL_1364659, EPI_ISL_1364660, EPI_ISL_1364661, EPI_ISL_1364662, EPI_ISL_1364663, EPI_ISL_1364664, EPI_ISL_1364665, EPI_ISL_1364667, EPI_ISL_1364668, EPI_ISL_1364669, EPI_ISL_1364670, EPI_ISL_1364671, EPI_ISL_1364672, EPI_ISL_1364673, EPI_ISL_1364675, EPI_ISL_1364677, EPI_ISL_1364678, EPI_ISL_1364679, EPI_ISL_1364680, EPI_ISL_1364681, EPI_ISL_1364682, EPI_ISL_1364683, EPI_ISL_1364684, EPI_ISL_1364685, EPI_ISL_1364686, EPI_ISL_1364689, EPI_ISL_1364690, EPI_ISL_1364691, EPI_ISL_1364692, EPI_ISL_1364693, EPI_ISL_1364694, EPI_ISL_1364695, EPI_ISL_1364696, EPI_ISL_1364697, EPI_ISL_1364698, EPI_ISL_1364700, EPI_ISL_1364702, EPI_ISL_1364703, EPI_ISL_1364704, EPI_ISL_1364706, EPI_ISL_1364707, EPI_ISL_1364709, EPI_ISL_1364711, EPI_ISL_1364712, EPI_ISL_1364713, EPI_ISL_1364714, EPI_ISL_1364715, EPI_ISL_1364716, EPI_ISL_1364717, EPI_ISL_1364718, EPI_ISL_1364719, EPI_ISL_1364720, EPI_ISL_1364721, EPI_ISL_1364722, EPI_ISL_1364723, EPI_ISL_1364724, EPI_ISL_1364726, EPI_ISL_1364727, EPI_ISL_1364728, EPI_ISL_1364729, EPI_ISL_1364730, EPI_ISL_1364731, EPI_ISL_1364732, EPI_ISL_1364733, EPI_ISL_1364736, EPI_ISL_1364737, EPI_ISL_1364740, EPI_ISL_1364741, EPI_ISL_1364742, EPI_ISL_1364743, EPI_ISL_1364744, EPI_ISL_1364745, EPI_ISL_1364746, EPI_ISL_1364747, EPI_ISL_1364748, EPI_ISL_1364749, EPI_ISL_1364751, EPI_ISL_1364752, EPI_ISL_1364753, EPI_ISL_1364755, EPI_ISL_1364756, EPI_ISL_1364757, EPI_ISL_1364758, EPI_ISL_1364759, EPI_ISL_1364760, EPI_ISL_1364761, EPI_ISL_1364762, EPI_ISL_1364763, EPI_ISL_1364764, EPI_ISL_1364765, EPI_ISL_1364767, EPI_ISL_1364769, EPI_ISL_1364771, EPI_ISL_1364772, EPI_ISL_1364773, EPI_ISL_1364774, EPI_ISL_1364775, EPI_ISL_1364776, EPI_ISL_1364777, EPI_ISL_1364778, EPI_ISL_1364779, EPI_ISL_1364780, EPI_ISL_1364781, EPI_ISL_1364782, EPI_ISL_1364783, EPI_ISL_1364784, EPI_ISL_1364785, EPI_ISL_1364786, EPI_ISL_1364787, EPI_ISL_1364788, EPI_ISL_1364789, EPI_ISL_1364790, EPI_ISL_1364791, EPI_ISL_1364792, EPI_ISL_1364793, EPI_ISL_1364794, EPI_ISL_1364795, EPI_ISL_1364796, EPI_ISL_1364797, EPI_ISL_1364798, EPI_ISL_1364800, EPI_ISL_1364805, EPI_ISL_1364806, EPI_ISL_1364807, EPI_ISL_1364808, EPI_ISL_1364809, EPI_ISL_1364810, EPI_ISL_1364811, EPI_ISL_1364812, EPI_ISL_1364813, EPI_ISL_1364814, EPI_ISL_1364815, EPI_ISL_1364816, EPI_ISL_1364819, EPI_ISL_1364820, EPI_ISL_1364821, EPI_ISL_1364822, EPI_ISL_1364823, EPI_ISL_1364824, EPI_ISL_1364825, EPI_ISL_1364828, EPI_ISL_1364830, EPI_ISL_1364831, EPI_ISL_1364832, EPI_ISL_1364833, EPI_ISL_1364834, EPI_ISL_1364835, EPI_ISL_1364836, EPI_ISL_1364837, EPI_ISL_1364838, EPI_ISL_1364839, EPI_ISL_1364840, EPI_ISL_1364841, EPI_ISL_1364843, EPI_ISL_1364844, EPI_ISL_1364846, EPI_ISL_1364847, EPI_ISL_1364848, EPI_ISL_1364849, EPI_ISL_1364850, EPI_ISL_1364852, EPI_ISL_1364853, EPI_ISL_1364855, EPI_ISL_1364856, EPI_ISL_1364857, EPI_ISL_1364858, EPI_ISL_1364859, EPI_ISL_1364861, EPI_ISL_1364862 | | | |
| see above | Arizona State University | Arizona State University | Peter T. Skidmore, LaRinda A. Holland, Rabia Maqsood, Emily A. Kaelin, Nicholas J. Mellor, Kristina Buss, Joy M. Blain, Neal W Woodbury, Valerie Harris, Joshua LaBaer, Vel Murugan, Efrem S. Lim |
| EPI_ISL_1364866, EPI_ISL_1364867, EPI_ISL_1364868, EPI_ISL_1364869, EPI_ISL_1364870, EPI_ISL_1364871, EPI_ISL_1364872, EPI_ISL_1364873, EPI_ISL_1364874, EPI_ISL_1364875, EPI_ISL_1364876, EPI_ISL_1364877, EPI_ISL_1364878 | | | |
| see above | Kansas Health and Environmental Lab | Kansas Health and Environmental Lab | Mike Grose, Jonathan Barnell, Ben Olsen, and Phil Adam |
| EPI_ISL_1365366, EPI_ISL_1365368, EPI_ISL_1365370, EPI_ISL_1365371, EPI_ISL_1365372, EPI_ISL_1365373, EPI_ISL_1365374, EPI_ISL_1365375, EPI_ISL_1365376, EPI_ISL_1365377, EPI_ISL_1365378, EPI_ISL_1365379, EPI_ISL_1365380, EPI_ISL_1365381, EPI_ISL_1365382, EPI_ISL_1365383, EPI_ISL_1365384, EPI_ISL_1365385, EPI_ISL_1365386, EPI_ISL_1365387, EPI_ISL_1365388, EPI_ISL_1365389, EPI_ISL_1365390, EPI_ISL_1365391, EPI_ISL_1365392, EPI_ISL_1365393, EPI_ISL_1365395, EPI_ISL_1365396, EPI_ISL_1365398, EPI_ISL_1365399, EPI_ISL_1365400, EPI_ISL_1365401, EPI_ISL_1365402, EPI_ISL_1365403, EPI_ISL_1365404, EPI_ISL_1365405, EPI_ISL_1365406, EPI_ISL_1365408 | | | |
| see above | M Health Fairview | Minnesota Department of Health, Public Health Laboratory | Alexandra Lorentz, Jacob Garfin, Matt Plumb, and Xiong Wang |
| EPI_ISL_1365409, EPI_ISL_1365410, EPI_ISL_1365411, EPI_ISL_1365412, EPI_ISL_1365413, EPI_ISL_1365414, EPI_ISL_1365415, EPI_ISL_1365416, EPI_ISL_1365417, EPI_ISL_1365418, EPI_ISL_1365419, EPI_ISL_1365420, EPI_ISL_1365421, EPI_ISL_1365422, EPI_ISL_1365423, EPI_ISL_1365424, EPI_ISL_1365425, EPI_ISL_1365426, EPI_ISL_1365427, EPI_ISL_1365428, EPI_ISL_1365429, EPI_ISL_1365430, EPI_ISL_1365431, EPI_ISL_1365432, EPI_ISL_1365433, EPI_ISL_1365434, EPI_ISL_1365435, EPI_ISL_1365436, EPI_ISL_1365437, EPI_ISL_1365438, EPI_ISL_1365439, EPI_ISL_1365440, EPI_ISL_1365441, EPI_ISL_1365442, EPI_ISL_1365443, EPI_ISL_1365444, EPI_ISL_1365445, EPI_ISL_1365446, EPI_ISL_1365447, EPI_ISL_1365448 | | | |
| see above | Mayo Clinic & Mayo Clinic Laboratories | Minnesota Department of Health, Public Health Laboratory | Alexandra Lorentz, Jacob Garfin, Matt Plumb, and Xiong Wang |
| EPI_ISL_1365452, EPI_ISL_1365453, EPI_ISL_1365454, EPI_ISL_1365455, EPI_ISL_1365456, EPI_ISL_1365457, EPI_ISL_1365458, EPI_ISL_1365459, EPI_ISL_1365460, EPI_ISL_1365461, EPI_ISL_1365462, EPI_ISL_1365463, EPI_ISL_1365464, EPI_ISL_1365465, EPI_ISL_1365466, EPI_ISL_1365467, EPI_ISL_1365468, EPI_ISL_1365469, EPI_ISL_1365470, EPI_ISL_1365471, EPI_ISL_1365472, EPI_ISL_1365474, EPI_ISL_1365475, EPI_ISL_1365476, EPI_ISL_1365477, EPI_ISL_1365478, EPI_ISL_1365479, EPI_ISL_1365480, EPI_ISL_1365481, EPI_ISL_1365482, EPI_ISL_1365483, EPI_ISL_1365484, EPI_ISL_1365485, EPI_ISL_1365486, EPI_ISL_1365487, EPI_ISL_1365488, EPI_ISL_1365489, EPI_ISL_1365490, EPI_ISL_1365491, EPI_ISL_1365492, EPI_ISL_1365493, EPI_ISL_1365494, EPI_ISL_1365495, EPI_ISL_1365496, EPI_ISL_1365497, EPI_ISL_1365498, EPI_ISL_1365499, EPI_ISL_1365500, EPI_ISL_1365501, EPI_ISL_1365502, EPI_ISL_1365503, EPI_ISL_1365504, EPI_ISL_1365505, EPI_ISL_1365506, EPI_ISL_1365507, EPI_ISL_1365509, EPI_ISL_1365511, EPI_ISL_1365512, EPI_ISL_1365513, EPI_ISL_1365514, EPI_ISL_1365515, EPI_ISL_1365516, EPI_ISL_1365517, EPI_ISL_1365518, EPI_ISL_1365519, EPI_ISL_1365520, EPI_ISL_1365521, EPI_ISL_1365522, EPI_ISL_1365523, EPI_ISL_1365524, EPI_ISL_1365525, EPI_ISL_1365526, EPI_ISL_1365527, EPI_ISL_1365528, EPI_ISL_1365529, EPI_ISL_1365530, EPI_ISL_1365531, EPI_ISL_1365532, EPI_ISL_1365533, EPI_ISL_1365534, EPI_ISL_1365535, EPI_ISL_1365536, EPI_ISL_1365537, EPI_ISL_1365538, EPI_ISL_1365539, EPI_ISL_1365540, EPI_ISL_1365541, EPI_ISL_1365542, EPI_ISL_1365543, EPI_ISL_1365544, EPI_ISL_1365545, EPI_ISL_1365547, EPI_ISL_1365548, EPI_ISL_1365549, EPI_ISL_1365550, EPI_ISL_1365551, EPI_ISL_1365552, EPI_ISL_1365553, EPI_ISL_1365554, EPI_ISL_1365555, EPI_ISL_1365556, EPI_ISL_1365557, EPI_ISL_1365558, EPI_ISL_1365559, EPI_ISL_1365561, EPI_ISL_1365562, EPI_ISL_1365563, EPI_ISL_1365564, EPI_ISL_1365565, EPI_ISL_1365566, EPI_ISL_1365567, EPI_ISL_1365568, EPI_ISL_1365569, EPI_ISL_1365570, EPI_ISL_1365571, EPI_ISL_1365572, EPI_ISL_1365573, EPI_ISL_1365574, EPI_ISL_1365575, EPI_ISL_1365576, EPI_ISL_1365577, EPI_ISL_1365578, EPI_ISL_1365579, EPI_ISL_1365580, EPI_ISL_1365581, EPI_ISL_1365582, EPI_ISL_1365583, EPI_ISL_1365584, EPI_ISL_1365585, EPI_ISL_1365586, EPI_ISL_1365587, EPI_ISL_1365588, EPI_ISL_1365589, EPI_ISL_1365590, EPI_ISL_1365591, EPI_ISL_1365592, EPI_ISL_1365593, EPI_ISL_1365594, EPI_ISL_1365595, EPI_ISL_1365596, EPI_ISL_1365597, EPI_ISL_1365598, EPI_ISL_1365599, EPI_ISL_1365600, EPI_ISL_1365601, EPI_ISL_1365602, EPI_ISL_1365603, EPI_ISL_1365604, EPI_ISL_1365605, EPI_ISL_1365606, EPI_ISL_1365607, EPI_ISL_1365608, EPI_ISL_1365609, EPI_ISL_1365610, EPI_ISL_1365611, EPI_ISL_1365612, EPI_ISL_1365613, EPI_ISL_1365614, EPI_ISL_1365615, EPI_ISL_1365616, EPI_ISL_1365617, EPI_ISL_1365618, EPI_ISL_1365619, EPI_ISL_1365620, EPI_ISL_1365621, EPI_ISL_1365623, EPI_ISL_1365624, EPI_ISL_1365626, EPI_ISL_1365627, EPI_ISL_1365628, EPI_ISL_1365629, EPI_ISL_1365630, EPI_ISL_1365631, EPI_ISL_1365632, EPI_ISL_1365633, EPI_ISL_1365634, EPI_ISL_1365635, EPI_ISL_1365636, EPI_ISL_1365637 | | | |
| see above | Arizona State University | Arizona State University | Peter T. Skidmore, LaRinda A. Holland, Rabia Maqsood, Emily A. Kaelin, Nicholas J. Mellor, Kristina Buss, Joy M. Blain, Neal W Woodbury, Valerie Harris, Joshua LaBaer, Vel Murugan, Efrem S. Lim |
| EPI_ISL_1365639, EPI_ISL_1365640, EPI_ISL_1365641, EPI_ISL_1365642, EPI_ISL_1365643, EPI_ISL_1365644, EPI_ISL_1365645, EPI_ISL_1365646, EPI_ISL_1365647, EPI_ISL_1365648 | LESP Nuevo Leon | Instituto de Diagnostico y Referencia Epidemiologicos (INDRE) | Claudia Wong-Arambula, Abril Rodriguez-Maldonado, Vanessa Rivero-Arredondo, Ariadna Medina-Benitez, Joaquin Quiroz-Mercado, Sergio Rangel-Guerrero, Natividad Cruz-Ortiz, Tatiana Nunez-Garcia, Gisela Barrera-Badillo, Lucia Hernandez-Rivas, Irma Lopez-Martinez, Ernesto Ramirez-Gonzalez. |
| EPI_ISL_1365649 | LESP Tamaulipas | Instituto de Diagnostico y Referencia Epidemiologicos (INDRE) | Claudia Wong-Arambula, Abril Rodriguez-Maldonado, Vanessa Rivero-Arredondo, Ariadna Medina-Benitez, Joaquin Quiroz-Mercado, Sergio Rangel-Guerrero, Natividad Cruz-Ortiz, Tatiana Nunez-Garcia, Gisela Barrera-Badillo, Lucia Hernandez-Rivas, Irma Lopez-Martinez, Ernesto Ramirez-Gonzalez. |
| EPI_ISL_1365650, EPI_ISL_1365651, EPI_ISL_1365652, EPI_ISL_1365653, EPI_ISL_1365654, EPI_ISL_1365655 | LESP Morelos | Instituto de Diagnostico y Referencia Epidemiologicos (INDRE) | Claudia Wong-Arambula, Abril Rodriguez-Maldonado, Vanessa Rivero-Arredondo, Ariadna Medina-Benitez, Joaquin Quiroz-Mercado, Sergio Rangel-Guerrero, Natividad Cruz-Ortiz, Tatiana Nunez-Garcia, Gisela Barrera-Badillo, Lucia Hernandez-Rivas, Irma Lopez-Martinez, Ernesto Ramirez-Gonzalez. |
| EPI_ISL_1365656 | LESP Veracruz | Instituto de Diagnostico y Referencia Epidemiologicos (INDRE) | Claudia Wong-Arambula, Abril Rodriguez-Maldonado, Vanessa Rivero-Arredondo, Ariadna Medina-Benitez, Joaquin Quiroz-Mercado, Sergio Rangel-Guerrero, Natividad Cruz-Ortiz, Tatiana Nunez-Garcia, Gisela Barrera-Badillo, Lucia Hernandez-Rivas, Irma Lopez-Martinez, Ernesto Ramirez-Gonzalez. |
| EPI_ISL_1365657 | LESP Ciudad de Mexico | Instituto de Diagnostico y Referencia Epidemiologicos (INDRE) | Claudia Wong-Arambula, Abril Rodriguez-Maldonado, Vanessa Rivero-Arredondo, Ariadna Medina-Benitez, Joaquin Quiroz-Mercado, Sergio Rangel-Guerrero, Natividad Cruz-Ortiz, Tatiana Nunez-Garcia, Gisela Barrera-Badillo, Lucia Hernandez-Rivas, Irma Lopez-Martinez, Ernesto Ramirez-Gonzalez. |
| EPI_ISL_1365658, EPI_ISL_1365659, EPI_ISL_1365660 | LESP Baja California | Instituto de Diagnostico y Referencia Epidemiologicos (INDRE) | Claudia Wong-Arambula, Abril Rodriguez-Maldonado, Vanessa Rivero-Arredondo, Ariadna Medina-Benitez, Joaquin Quiroz-Mercado, Sergio Rangel-Guerrero, Natividad Cruz-Ortiz, Tatiana Nunez-Garcia, Gisela Barrera-Badillo, Lucia Hernandez-Rivas, Irma Lopez-Martinez, Ernesto Ramirez-Gonzalez. |
| EPI_ISL_1365661 | LESP Quintana Roo | Instituto de Diagnostico y Referencia Epidemiologicos (INDRE) | Claudia Wong-Arambula, Abril Rodriguez-Maldonado, Vanessa Rivero-Arredondo, Ariadna Medina-Benitez, Joaquin Quiroz-Mercado, Sergio Rangel-Guerrero, Natividad Cruz-Ortiz, Tatiana Nunez-Garcia, Gisela Barrera-Badillo, Lucia Hernandez-Rivas, Irma Lopez-Martinez, Ernesto Ramirez-Gonzalez. |
| EPI_ISL_1365662, EPI_ISL_1365663, EPI_ISL_1365664, EPI_ISL_1365665, EPI_ISL_1365666, EPI_ISL_1365667 | LESP Coahuila | Instituto de Diagnostico y Referencia Epidemiologicos (INDRE) | Claudia Wong-Arambula, Abril Rodriguez-Maldonado, Vanessa Rivero-Arredondo, Ariadna Medina-Benitez, Joaquin Quiroz-Mercado, Sergio Rangel-Guerrero, Natividad Cruz-Ortiz, Tatiana Nunez-Garcia, Gisela Barrera-Badillo, Lucia Hernandez-Rivas, Irma Lopez-Martinez, Ernesto Ramirez-Gonzalez. |
| EPI_ISL_1365668, EPI_ISL_1365669, EPI_ISL_1365670, EPI_ISL_1365671, EPI_ISL_1365672, EPI_ISL_1365673, EPI_ISL_1365674 | LESP Nuevo Leon | Instituto de Diagnostico y Referencia Epidemiologicos (INDRE) | Claudia Wong-Arambula, Abril Rodriguez-Maldonado, Vanessa Rivero-Arredondo, Ariadna Medina-Benitez, Joaquin Quiroz-Mercado, Sergio Rangel-Guerrero, Natividad Cruz-Ortiz, Tatiana Nunez-Garcia, Gisela Barrera-Badillo, Lucia Hernandez-Rivas, Irma Lopez-Martinez, Ernesto Ramirez-Gonzalez. |
| EPI_ISL_1365675 | LESP Guanajuato | Instituto de Diagnostico y Referencia Epidemiologicos (INDRE) | Claudia Wong-Arambula, Abril Rodriguez-Maldonado, Vanessa Rivero-Arredondo, Ariadna Medina-Benitez, Joaquin Quiroz-Mercado, Sergio Rangel-Guerrero, Natividad Cruz-Ortiz, Tatiana Nunez-Garcia, Gisela Barrera-Badillo, Lucia Hernandez-Rivas, Irma Lopez-Martinez, Ernesto Ramirez-Gonzalez. |
| EPI_ISL_1365699, EPI_ISL_1365700, EPI_ISL_1365701, EPI_ISL_1365702, EPI_ISL_1365703, EPI_ISL_1365704, EPI_ISL_1365705, EPI_ISL_1365706, EPI_ISL_1365707, EPI_ISL_1365708, EPI_ISL_1365709, EPI_ISL_1365710, EPI_ISL_1365711, EPI_ISL_1365712, EPI_ISL_1365713, EPI_ISL_1365714, EPI_ISL_1365715, EPI_ISL_1365716, EPI_ISL_1365717, EPI_ISL_1365718, EPI_ISL_1365719, EPI_ISL_1365720, EPI_ISL_1365721, EPI_ISL_1365722, EPI_ISL_1365723, EPI_ISL_1365725, EPI_ISL_1365726, EPI_ISL_1365727, EPI_ISL_1365728, EPI_ISL_1365729, EPI_ISL_1365730, EPI_ISL_1365731, EPI_ISL_1365732 | | | |
| see above | Virginia Division of Consolidated Laboratory Services | Virginia Division of Consolidated Laboratory Services | Virginia DCLS |
| EPI_ISL_1366071, EPI_ISL_1366072, EPI_ISL_1366073, EPI_ISL_1366074, EPI_ISL_1366075, EPI_ISL_1366076, EPI_ISL_1366077, EPI_ISL_1366078, EPI_ISL_1366079, EPI_ISL_1366080, EPI_ISL_1366081, EPI_ISL_1366082, EPI_ISL_1366084, EPI_ISL_1366085, EPI_ISL_1366086, EPI_ISL_1366087, EPI_ISL_1366088, EPI_ISL_1366089, EPI_ISL_1366090, EPI_ISL_1366091, EPI_ISL_1366092, EPI_ISL_1366093, EPI_ISL_1366094, EPI_ISL_1366095, EPI_ISL_1366096, EPI_ISL_1366097, EPI_ISL_1366098, EPI_ISL_1366102, EPI_ISL_1366103, EPI_ISL_1366104, EPI_ISL_1366105, EPI_ISL_1366106, EPI_ISL_1366107, EPI_ISL_1366108, EPI_ISL_1366109, EPI_ISL_1366110, EPI_ISL_1366111, EPI_ISL_1366112, EPI_ISL_1366113, EPI_ISL_1366114, EPI_ISL_1366115, EPI_ISL_1366116, EPI_ISL_1366117, EPI_ISL_1366118, EPI_ISL_1366119, EPI_ISL_1366120, EPI_ISL_1366121, EPI_ISL_1366122, EPI_ISL_1366123, EPI_ISL_1366124, EPI_ISL_1366125, EPI_ISL_1366126, EPI_ISL_1366127, EPI_ISL_1366128, EPI_ISL_1366129, EPI_ISL_1366130, EPI_ISL_1366131, EPI_ISL_1366132, EPI_ISL_1366133, EPI_ISL_1366134, EPI_ISL_1366135, EPI_ISL_1366136, EPI_ISL_1366137, EPI_ISL_1366138, EPI_ISL_1366139, EPI_ISL_1366140, EPI_ISL_1366141, EPI_ISL_1366142, EPI_ISL_1366143, EPI_ISL_1366144, EPI_ISL_1366145, EPI_ISL_1366146, EPI_ISL_1366147, EPI_ISL_1366148, EPI_ISL_1366149, EPI_ISL_1366150, EPI_ISL_1366151, EPI_ISL_1366152, EPI_ISL_1366153, EPI_ISL_1366154, EPI_ISL_1366155, EPI_ISL_1366156 | | | |
| see above | TXDSHS | TXDSHS | Rashmi Tuladhar, Bonnie Oh, Jenny Zhang, Maliha Rahman, Mayela Pedrueza, Anita Pokharel, Loarraine Rodriguez, Myong Koag, Chun Wang, Rachel Lee, Grace Kubin |
| EPI_ISL_1366157, EPI_ISL_1366158, EPI_ISL_1366159, EPI_ISL_1366160, EPI_ISL_1366161, EPI_ISL_1366162, EPI_ISL_1366163, EPI_ISL_1366164, EPI_ISL_1366165, EPI_ISL_1366166, EPI_ISL_1366167, EPI_ISL_1366168, EPI_ISL_1366169, EPI_ISL_1366170, EPI_ISL_1366171, EPI_ISL_1366172, EPI_ISL_1366173, EPI_ISL_1366174, EPI_ISL_1366175, EPI_ISL_1366176, EPI_ISL_1366177, EPI_ISL_1366178, EPI_ISL_1366179, EPI_ISL_1366180, EPI_ISL_1366181, EPI_ISL_1366182, EPI_ISL_1366183, EPI_ISL_1366184, EPI_ISL_1366185, EPI_ISL_1366186, EPI_ISL_1366187, EPI_ISL_1366188, EPI_ISL_1366189, EPI_ISL_1366190, EPI_ISL_1366191, EPI_ISL_1366192, EPI_ISL_1366193, EPI_ISL_1366194, EPI_ISL_1366195, EPI_ISL_1366196, EPI_ISL_1366197, EPI_ISL_1366198, EPI_ISL_1366199, EPI_ISL_1366200, EPI_ISL_1366201, EPI_ISL_1366202, EPI_ISL_1366203, EPI_ISL_1366204, EPI_ISL_1366205, EPI_ISL_1366206, EPI_ISL_1366207, EPI_ISL_1366208, EPI_ISL_1366209, EPI_ISL_1366210, EPI_ISL_1366211, EPI_ISL_1366212, EPI_ISL_1366213, EPI_ISL_1366214, EPI_ISL_1366215, EPI_ISL_1366216, EPI_ISL_1366217, EPI_ISL_1366218, EPI_ISL_1366219, EPI_ISL_1366220, EPI_ISL_1366221, EPI_ISL_1366222, EPI_ISL_1366223, EPI_ISL_1366224, EPI_ISL_1366225, EPI_ISL_1366226, EPI_ISL_1366227, EPI_ISL_1366228, EPI_ISL_1366229, EPI_ISL_1366230, EPI_ISL_1366231, EPI_ISL_1366232, EPI_ISL_1366233, EPI_ISL_1366234, EPI_ISL_1366235, EPI_ISL_1366236, EPI_ISL_1366237 | | | |
| see above | UW Virology Lab | UW Virology Lab | Pavitra Roychoudhury, Hong Xie, Lasata Shrestha, Shah Mohamed Bakhash, Michelle Lin, Noah R. Baker, Sean Ellis, Saraswathi Sathees, Meei-Li Huang, Keith R Jerome, Alexander Greninger |
| EPI_ISL_1366240, EPI_ISL_1366241, EPI_ISL_1366242, EPI_ISL_1366243, EPI_ISL_1366244, EPI_ISL_1366245, EPI_ISL_1366246, EPI_ISL_1366247, EPI_ISL_1366248, EPI_ISL_1366249, EPI_ISL_1366250, EPI_ISL_1366251, EPI_ISL_1366252, EPI_ISL_1366253, EPI_ISL_1366254, EPI_ISL_1366255, EPI_ISL_1366256, EPI_ISL_1366257, EPI_ISL_1366258, EPI_ISL_1366259, EPI_ISL_1366260, EPI_ISL_1366261, EPI_ISL_1366262, EPI_ISL_1366263, EPI_ISL_1366264, EPI_ISL_1366265, EPI_ISL_1366266, EPI_ISL_1366267, EPI_ISL_1366268 | | | |
| see above | Wyoming Public Health Laboratory | Wyoming Public Health Laboratory | Jim Mildenberger, Wanda Manley, Noah Hull, Taylor Fearing, Lynette Gumbleton, Channing Weber, Ashley Norberg, Chayse Rowley, Marley Goetz, Brian Dominguez, Elliot Thomasson, Cari Sloma, and Rob Christensen |
| EPI_ISL_1366275 | Scripps Medical Laboratory | Andersen lab at Scripps Research | SEARCH Alliance San Diego with Michael Quigley, Ellen Stefanski, Ian Mchardy |

| | | | |
|---|---|---|---|
| EPI_ISL_1366276, EPI_ISL_1366277 | Sharp HealthCare Laboratory | Andersen lab at Scripps Research | SEARCH Alliance San Diego with Aaron Harding, Jacquelyn Berumen, Cathy Woerle, Liam McGinnis, Art Mendoza, Omid Bakhtar |
| EPI_ISL_1366278 | Scripps Medical Laboratory | Andersen lab at Scripps Research | SEARCH Alliance San Diego with Michael Quigley, Ellen Stefanski, Ian Mchardy |
| EPI_ISL_1366279, EPI_ISL_1366280, EPI_ISL_1366281 | Sharp HealthCare Laboratory | Andersen lab at Scripps Research | SEARCH Alliance San Diego with Aaron Harding, Jacquelyn Berumen, Cathy Woerle, Liam McGinnis, Art Mendoza, Omid Bakhtar |
| EPI_ISL_1366282 | Scripps Medical Laboratory | Andersen lab at Scripps Research | SEARCH Alliance San Diego with Michael Quigley, Ellen Stefanski, Ian Mchardy |
| EPI_ISL_1366283, EPI_ISL_1366284 | Sharp HealthCare Laboratory | Andersen lab at Scripps Research | SEARCH Alliance San Diego with Aaron Harding, Jacquelyn Berumen, Cathy Woerle, Liam McGinnis, Art Mendoza, Omid Bakhtar |
| EPI_ISL_1366285 | Scripps Medical Laboratory | Andersen lab at Scripps Research | SEARCH Alliance San Diego with Michael Quigley, Ellen Stefanski, Ian Mchardy |
| EPI_ISL_1366286, EPI_ISL_1366287, EPI_ISL_1366288, EPI_ISL_1366289, EPI_ISL_1366290, EPI_ISL_1366291, EPI_ISL_1366292, EPI_ISL_1366293, EPI_ISL_1366294, EPI_ISL_1366295, EPI_ISL_1366296 | | | |
| see above | Sharp HealthCare Laboratory | Andersen lab at Scripps Research | SEARCH Alliance San Diego with Aaron Harding, Jacquelyn Berumen, Cathy Woerle, Liam McGinnis, Art Mendoza, Omid Bakhtar |
| EPI_ISL_1366297 | Scripps Medical Laboratory | Andersen lab at Scripps Research | SEARCH Alliance San Diego with Michael Quigley, Ellen Stefanski, Ian Mchardy |
| EPI_ISL_1366298, EPI_ISL_1366299, EPI_ISL_1366300, EPI_ISL_1366301, EPI_ISL_1366302, EPI_ISL_1366303, EPI_ISL_1366304, EPI_ISL_1366305, EPI_ISL_1366306, EPI_ISL_1366307, EPI_ISL_1366308, EPI_ISL_1366309, EPI_ISL_1366310, EPI_ISL_1366311, EPI_ISL_1366312, EPI_ISL_1366313, EPI_ISL_1366314, EPI_ISL_1366315, EPI_ISL_1366316, EPI_ISL_1366317 | | | |
| see above | Sharp HealthCare Laboratory | Andersen lab at Scripps Research | SEARCH Alliance San Diego with Aaron Harding, Jacquelyn Berumen, Cathy Woerle, Liam McGinnis, Art Mendoza, Omid Bakhtar |
| EPI_ISL_1366318 | Scripps Medical Laboratory | Andersen lab at Scripps Research | SEARCH Alliance San Diego with Michael Quigley, Ellen Stefanski, Ian Mchardy |
| EPI_ISL_1366319 | Sharp HealthCare Laboratory | Andersen lab at Scripps Research | SEARCH Alliance San Diego with Aaron Harding, Jacquelyn Berumen, Cathy Woerle, Liam McGinnis, Art Mendoza, Omid Bakhtar |
| EPI_ISL_1366320 | Scripps Medical Laboratory | Andersen lab at Scripps Research | SEARCH Alliance San Diego with Michael Quigley, Ellen Stefanski, Ian Mchardy |
| EPI_ISL_1366321 | Sharp HealthCare Laboratory | Andersen lab at Scripps Research | SEARCH Alliance San Diego with Aaron Harding, Jacquelyn Berumen, Cathy Woerle, Liam McGinnis, Art Mendoza, Omid Bakhtar |
| EPI_ISL_1366322 | Scripps Medical Laboratory | Andersen lab at Scripps Research | SEARCH Alliance San Diego with Michael Quigley, Ellen Stefanski, Ian Mchardy |
| EPI_ISL_1366323, EPI_ISL_1366324, EPI_ISL_1366325 | Sharp HealthCare Laboratory | Andersen lab at Scripps Research | SEARCH Alliance San Diego with Aaron Harding, Jacquelyn Berumen, Cathy Woerle, Liam McGinnis, Art Mendoza, Omid Bakhtar |
| EPI_ISL_1366326, EPI_ISL_1366327, EPI_ISL_1366328, EPI_ISL_1366329, EPI_ISL_1366330, EPI_ISL_1366331, EPI_ISL_1366332, EPI_ISL_1366333, EPI_ISL_1366334 | Genomica Lab Molecular, Mexico | Andersen lab at Scripps Research | SEARCH Alliance San Diego with Jonathan Gonzalez Garcia, Jose Roman Chavez Mendez, Jose Horacio Reyna Verdugo, Martin Gonzalez Ibarra, Luis Alberto Rangel Gonzalez |
| EPI_ISL_1366335, EPI_ISL_1366336, EPI_ISL_1366337, EPI_ISL_1366338, EPI_ISL_1366339, EPI_ISL_1366340, EPI_ISL_1366341, EPI_ISL_1366342, EPI_ISL_1366343, EPI_ISL_1366344, EPI_ISL_1366345, EPI_ISL_1366346, EPI_ISL_1366347, EPI_ISL_1366348, EPI_ISL_1366349, EPI_ISL_1366350, EPI_ISL_1366351, EPI_ISL_1366352, EPI_ISL_1366353, EPI_ISL_1366354, EPI_ISL_1366355, EPI_ISL_1366356, EPI_ISL_1366357, EPI_ISL_1366358, EPI_ISL_1366359, EPI_ISL_1366360, EPI_ISL_1366361, EPI_ISL_1366362, EPI_ISL_1366363, EPI_ISL_1366364, EPI_ISL_1366365, EPI_ISL_1366366, EPI_ISL_1366367, EPI_ISL_1366368, EPI_ISL_1366369, EPI_ISL_1366370, EPI_ISL_1366371, EPI_ISL_1366372, EPI_ISL_1366373, EPI_ISL_1366374, EPI_ISL_1366375, EPI_ISL_1366376 | | | |
| see above | Sharp HealthCare Laboratory | Andersen lab at Scripps Research | SEARCH Alliance San Diego with Aaron Harding, Jacquelyn Berumen, Cathy Woerle, Liam McGinnis, Art Mendoza, Omid Bakhtar |
| EPI_ISL_1366377, EPI_ISL_1366378 | San Diego County Public Health Laboratory | Andersen lab at Scripps Research | SEARCH Alliance San Diego with Tracy Basler, Jovan Shephard, Brett Austin |
| EPI_ISL_1366379, EPI_ISL_1366380, EPI_ISL_1366381, EPI_ISL_1366382, EPI_ISL_1366383, EPI_ISL_1366384, EPI_ISL_1366385, EPI_ISL_1366386, EPI_ISL_1366387, EPI_ISL_1366388, EPI_ISL_1366389, EPI_ISL_1366390, EPI_ISL_1366391, EPI_ISL_1366392, EPI_ISL_1366393, EPI_ISL_1366394, EPI_ISL_1366395, EPI_ISL_1366396, EPI_ISL_1366397, EPI_ISL_1366398, EPI_ISL_1366399, EPI_ISL_1366400, EPI_ISL_1366401, EPI_ISL_1366402, EPI_ISL_1366403, EPI_ISL_1366404, EPI_ISL_1366405, EPI_ISL_1366406, EPI_ISL_1366407, EPI_ISL_1366408, EPI_ISL_1366409, EPI_ISL_1366410, EPI_ISL_1366411, EPI_ISL_1366412, EPI_ISL_1366413, EPI_ISL_1366414, EPI_ISL_1366415, EPI_ISL_1366416, EPI_ISL_1366417, EPI_ISL_1366418, EPI_ISL_1366419, EPI_ISL_1366420, EPI_ISL_1366421, EPI_ISL_1366422, EPI_ISL_1366423 | | | |
| see above | Scripps Medical Laboratory | Andersen lab at Scripps Research | SEARCH Alliance San Diego with Michael Quigley, Ellen Stefanski, Ian Mchardy |
| EPI_ISL_1366424, EPI_ISL_1366425, EPI_ISL_1366426, EPI_ISL_1366427, EPI_ISL_1366428, EPI_ISL_1366429, EPI_ISL_1366430, EPI_ISL_1366431, EPI_ISL_1366432, EPI_ISL_1366433, EPI_ISL_1366434, EPI_ISL_1366435, EPI_ISL_1366436, EPI_ISL_1366437, EPI_ISL_1366438, EPI_ISL_1366439, EPI_ISL_1366440, EPI_ISL_1366441, EPI_ISL_1366442, EPI_ISL_1366443, EPI_ISL_1366444, EPI_ISL_1366445, EPI_ISL_1366446, EPI_ISL_1366447, EPI_ISL_1366448, EPI_ISL_1366449, EPI_ISL_1366450, EPI_ISL_1366451, EPI_ISL_1366452, EPI_ISL_1366453, EPI_ISL_1366454, EPI_ISL_1366455, EPI_ISL_1366456, EPI_ISL_1366457, EPI_ISL_1366458, EPI_ISL_1366459, EPI_ISL_1366460, EPI_ISL_1366461, EPI_ISL_1366462, EPI_ISL_1366463, EPI_ISL_1366464, EPI_ISL_1366465, EPI_ISL_1366466, EPI_ISL_1366467, EPI_ISL_1366468, EPI_ISL_1366469, EPI_ISL_1366470, EPI_ISL_1366471, EPI_ISL_1366472, EPI_ISL_1366473, EPI_ISL_1366474, EPI_ISL_1366475, EPI_ISL_1366476, EPI_ISL_1366477, EPI_ISL_1366478, EPI_ISL_1366479, EPI_ISL_1366480, EPI_ISL_1366481, EPI_ISL_1366482, EPI_ISL_1366483, EPI_ISL_1366484, EPI_ISL_1366485, EPI_ISL_1366486, EPI_ISL_1366487, EPI_ISL_1366488, EPI_ISL_1366489, EPI_ISL_1366490, EPI_ISL_1366491, EPI_ISL_1366492, EPI_ISL_1366493, EPI_ISL_1366494, EPI_ISL_1366495, EPI_ISL_1366496 | | | |
| see above | Sharp HealthCare Laboratory | Andersen lab at Scripps Research | SEARCH Alliance San Diego with Aaron Harding, Jacquelyn Berumen, Cathy Woerle, Liam McGinnis, Art Mendoza, Omid Bakhtar |
| EPI_ISL_1366614, EPI_ISL_1366615, EPI_ISL_1366616, EPI_ISL_1366617, EPI_ISL_1366618, EPI_ISL_1366619, EPI_ISL_1366620, EPI_ISL_1366621, EPI_ISL_1366622, EPI_ISL_1366623, EPI_ISL_1366624, EPI_ISL_1366625, EPI_ISL_1366626, EPI_ISL_1366627, EPI_ISL_1366628, EPI_ISL_1366629, EPI_ISL_1366630, EPI_ISL_1366631, EPI_ISL_1366632, EPI_ISL_1366633, EPI_ISL_1366634, EPI_ISL_1366635, EPI_ISL_1366636, EPI_ISL_1366637, EPI_ISL_1366638, EPI_ISL_1366639, EPI_ISL_1366640, EPI_ISL_1366641, EPI_ISL_1366642, EPI_ISL_1366643, EPI_ISL_1366644, EPI_ISL_1366645, EPI_ISL_1366646, EPI_ISL_1366647, EPI_ISL_1366648, EPI_ISL_1366649, EPI_ISL_1366650, EPI_ISL_1366651, EPI_ISL_1366652, EPI_ISL_1366653 | | | |
| see above | Gundersen Clinical Microbiology Laboratory | Kabara Cancer Research Institute | Craig S. Richmond, Paraic A. Kenny |
| EPI_ISL_1366660 | LESP Puebla | Instituto de Diagnostico y Referencia Epidemiologicos (INDRE) | Claudia Wong-Arambula, Abril Rodriguez-Maldonado, Vanessa Rivero-Arredondo, Ariadna Medina-Benitez, Joaquin Quiroz-Mercado, Sergio Rangel-Guerrero, Natividad Cruz-Ortiz, Tatiana Nunez-Garcia, Gisela Barrera-Badillo, Lucia Hernandez-Rivas, Irma Lopez-Martinez, Ernesto Ramirez-Gonzalez. |
| EPI_ISL_1366661, EPI_ISL_1366662 | LESP Jalisco | Instituto de Diagnostico y Referencia Epidemiologicos (INDRE) | Claudia Wong-Arambula, Abril Rodriguez-Maldonado, Vanessa Rivero-Arredondo, Ariadna Medina-Benitez, Joaquin Quiroz-Mercado, Sergio Rangel-Guerrero, Natividad Cruz-Ortiz, Tatiana Nunez-Garcia, Gisela Barrera-Badillo, Lucia Hernandez-Rivas, Irma Lopez-Martinez, Ernesto Ramirez-Gonzalez. |
| EPI_ISL_1366663, EPI_ISL_1366664, EPI_ISL_1366665 | LESP Nuevo Leon | Instituto de Diagnostico y Referencia Epidemiologicos (INDRE) | Claudia Wong-Arambula, Abril Rodriguez-Maldonado, Vanessa Rivero-Arredondo, Ariadna Medina-Benitez, Joaquin Quiroz-Mercado, Sergio Rangel-Guerrero, Natividad Cruz-Ortiz, Tatiana Nunez-Garcia, Gisela Barrera-Badillo, Lucia Hernandez-Rivas, Irma Lopez-Martinez, Ernesto Ramirez-Gonzalez. |
| EPI_ISL_1366666 | LESP Yucatan | Instituto de Diagnostico y Referencia Epidemiologicos (INDRE) | Claudia Wong-Arambula, Abril Rodriguez-Maldonado, Vanessa Rivero-Arredondo, Ariadna Medina-Benitez, Joaquin Quiroz-Mercado, Sergio Rangel-Guerrero, Natividad Cruz-Ortiz, Tatiana Nunez-Garcia, Gisela Barrera-Badillo, Lucia Hernandez-Rivas, Irma Lopez-Martinez, Ernesto Ramirez-Gonzalez. |
| EPI_ISL_1366667, EPI_ISL_1366668, EPI_ISL_1366669, EPI_ISL_1366670, EPI_ISL_1366671, EPI_ISL_1366672, EPI_ISL_1366673, EPI_ISL_1366674, EPI_ISL_1366675, EPI_ISL_1366676 | LESP Queretaro | Instituto de Diagnostico y Referencia Epidemiologicos (INDRE) | Claudia Wong-Arambula, Abril Rodriguez-Maldonado, Vanessa Rivero-Arredondo, Ariadna Medina-Benitez, Joaquin Quiroz-Mercado, Sergio Rangel-Guerrero, Natividad Cruz-Ortiz, Tatiana Nunez-Garcia, Gisela Barrera-Badillo, Lucia Hernandez-Rivas, Irma Lopez-Martinez, Ernesto Ramirez-Gonzalez. |
| EPI_ISL_1366677, EPI_ISL_1366678, EPI_ISL_1366679, EPI_ISL_1366680, EPI_ISL_1366681 | LESP Morelos | Instituto de Diagnostico y Referencia Epidemiologicos (INDRE) | Claudia Wong-Arambula, Abril Rodriguez-Maldonado, Vanessa Rivero-Arredondo, Ariadna Medina-Benitez, Joaquin Quiroz-Mercado, Sergio Rangel-Guerrero, Natividad Cruz-Ortiz, Tatiana Nunez-Garcia, Gisela Barrera-Badillo, Lucia Hernandez-Rivas, Irma Lopez-Martinez, Ernesto Ramirez-Gonzalez. |
| EPI_ISL_1366682, EPI_ISL_1366683, EPI_ISL_1366684, EPI_ISL_1366685, EPI_ISL_1366686 | LESP Veracruz | Instituto de Diagnostico y Referencia Epidemiologicos (INDRE) | Claudia Wong-Arambula, Abril Rodriguez-Maldonado, Vanessa Rivero-Arredondo, Ariadna Medina-Benitez, Joaquin Quiroz-Mercado, Sergio Rangel-Guerrero, Natividad Cruz-Ortiz, Tatiana Nunez-Garcia, Gisela Barrera-Badillo, Lucia Hernandez-Rivas, Irma Lopez-Martinez, Ernesto Ramirez-Gonzalez. |
| EPI_ISL_1366687, EPI_ISL_1366688, EPI_ISL_1366689 | LESP Ciudad de Mexico | Instituto de Diagnostico y Referencia Epidemiologicos (INDRE) | Claudia Wong-Arambula, Abril Rodriguez-Maldonado, Vanessa Rivero-Arredondo, Ariadna Medina-Benitez, Joaquin Quiroz-Mercado, Sergio Rangel-Guerrero, Natividad Cruz-Ortiz, Tatiana Nunez-Garcia, Gisela Barrera-Badillo, Lucia Hernandez-Rivas, Irma Lopez-Martinez, Ernesto Ramirez-Gonzalez. |
| EPI_ISL_1366690, EPI_ISL_1366691, EPI_ISL_1366692 | LESP Hidalgo | Instituto de Diagnostico y Referencia Epidemiologicos (INDRE) | Claudia Wong-Arambula, Abril Rodriguez-Maldonado, Vanessa Rivero-Arredondo, Ariadna Medina-Benitez, Joaquin Quiroz-Mercado, Sergio Rangel-Guerrero, Natividad Cruz-Ortiz, Tatiana Nunez-Garcia, Gisela Barrera-Badillo, Lucia Hernandez-Rivas, Irma Lopez-Martinez, Ernesto Ramirez-Gonzalez. |
| EPI_ISL_1366693, EPI_ISL_1366694, EPI_ISL_1366695, EPI_ISL_1366696, EPI_ISL_1366697, EPI_ISL_1366698, EPI_ISL_1366699 | LESP Nuevo Leon | Instituto de Diagnostico y Referencia Epidemiologicos (INDRE) | Claudia Wong-Arambula, Abril Rodriguez-Maldonado, Vanessa Rivero-Arredondo, Ariadna Medina-Benitez, Joaquin Quiroz-Mercado, Sergio Rangel-Guerrero, Natividad Cruz-Ortiz, Tatiana Nunez-Garcia, Gisela Barrera-Badillo, Lucia Hernandez-Rivas, Irma Lopez-Martinez, Ernesto Ramirez-Gonzalez. |
| EPI_ISL_1366700 | LESP Guanajuato | Instituto de Diagnostico y Referencia Epidemiologicos (INDRE) | Claudia Wong-Arambula, Abril Rodriguez-Maldonado, Vanessa Rivero-Arredondo, Ariadna Medina-Benitez, Joaquin Quiroz-Mercado, Sergio Rangel-Guerrero, Natividad Cruz-Ortiz, Tatiana Nunez-Garcia, Gisela Barrera-Badillo, Lucia Hernandez-Rivas, Irma Lopez-Martinez, Ernesto Ramirez-Gonzalez. |

| | | | |
|---|---|---|---|
| EPI_ISL_1366702, EPI_ISL_1366703, EPI_ISL_1366705, EPI_ISL_1366706, EPI_ISL_1366707, EPI_ISL_1366709, EPI_ISL_1366710, EPI_ISL_1366711, EPI_ISL_1366712, EPI_ISL_1366713 | Idaho Bureau of Laboratories | Idaho Bureau of Laboratories | "R. Beukelman, Matthew Charles Burns, Aimee Ceniseros, Robert L. Voermans, Christopher Ball" |
| EPI_ISL_1366747, EPI_ISL_1366748, EPI_ISL_1366749, EPI_ISL_1366750, EPI_ISL_1366751 | San Diego County Public Health Laboratory | Andersen lab at Scripps Research | SEARCH Alliance San Diego with Tracy Basler, Jovan Shephard, Brett Austin |
| EPI_ISL_1367163, EPI_ISL_1367164, EPI_ISL_1367165, EPI_ISL_1367166, EPI_ISL_1367167, EPI_ISL_1367168, EPI_ISL_1367169, EPI_ISL_1367170, EPI_ISL_1367171, EPI_ISL_1367172, EPI_ISL_1367173, EPI_ISL_1367174, EPI_ISL_1367175, EPI_ISL_1367176, EPI_ISL_1367177, EPI_ISL_1367178, EPI_ISL_1367179, EPI_ISL_1367180, EPI_ISL_1367181, EPI_ISL_1367182, EPI_ISL_1367183, EPI_ISL_1367184, EPI_ISL_1367185, EPI_ISL_1367186, EPI_ISL_1367187, EPI_ISL_1367188, EPI_ISL_1367189, EPI_ISL_1367190, EPI_ISL_1367191, EPI_ISL_1367192, EPI_ISL_1367193, EPI_ISL_1367194, EPI_ISL_1367195, EPI_ISL_1367196, EPI_ISL_1367197, EPI_ISL_1367198, EPI_ISL_1367199, EPI_ISL_1367200, EPI_ISL_1367201, EPI_ISL_1367202, EPI_ISL_1367203, EPI_ISL_1367204, EPI_ISL_1367205, EPI_ISL_1367206, EPI_ISL_1367207, EPI_ISL_1367208, EPI_ISL_1367209, EPI_ISL_1367210, EPI_ISL_1367211, EPI_ISL_1367212, EPI_ISL_1367213, EPI_ISL_1367214, EPI_ISL_1367215, EPI_ISL_1367216, EPI_ISL_1367217, EPI_ISL_1367218, EPI_ISL_1367219, EPI_ISL_1367220, EPI_ISL_1367221, EPI_ISL_1367222, EPI_ISL_1367223, EPI_ISL_1367224, EPI_ISL_1367225, EPI_ISL_1367226, EPI_ISL_1367227, EPI_ISL_1367228, EPI_ISL_1367229, EPI_ISL_1367230, EPI_ISL_1367231, EPI_ISL_1367232, EPI_ISL_1367233, EPI_ISL_1367234, EPI_ISL_1367235, EPI_ISL_1367236, EPI_ISL_1367237, EPI_ISL_1367238, EPI_ISL_1367239, EPI_ISL_1367240, EPI_ISL_1367241, EPI_ISL_1367242, EPI_ISL_1367243, EPI_ISL_1367244, EPI_ISL_1367245, EPI_ISL_1367246, EPI_ISL_1367247, EPI_ISL_1367248, EPI_ISL_1367249, EPI_ISL_1367250, EPI_ISL_1367251, EPI_ISL_1367252, EPI_ISL_1367253, EPI_ISL_1367254, EPI_ISL_1367255, EPI_ISL_1367256, EPI_ISL_1367257, EPI_ISL_1367258, EPI_ISL_1367259, EPI_ISL_1367260, EPI_ISL_1367261, EPI_ISL_1367262, EPI_ISL_1367263, EPI_ISL_1367264, EPI_ISL_1367265, EPI_ISL_1367266, EPI_ISL_1367267, EPI_ISL_1367268, EPI_ISL_1367269, EPI_ISL_1367270, EPI_ISL_1367271, EPI_ISL_1367272, EPI_ISL_1367273, EPI_ISL_1367274, EPI_ISL_1367275, EPI_ISL_1367276, EPI_ISL_1367277, EPI_ISL_1367278, EPI_ISL_1367279, EPI_ISL_1367280, EPI_ISL_1367281, EPI_ISL_1367282, EPI_ISL_1367283, EPI_ISL_1367284, EPI_ISL_1367285, EPI_ISL_1367286, EPI_ISL_1367287, EPI_ISL_1367288, EPI_ISL_1367289, EPI_ISL_1367290, EPI_ISL_1367291, EPI_ISL_1367292, EPI_ISL_1367293, EPI_ISL_1367294, EPI_ISL_1367295, EPI_ISL_1367296, EPI_ISL_1367297, EPI_ISL_1367298, EPI_ISL_1367299, EPI_ISL_1367300, EPI_ISL_1367301, EPI_ISL_1367302, EPI_ISL_1367303, EPI_ISL_1367304, EPI_ISL_1367305, EPI_ISL_1367306, EPI_ISL_1367307, EPI_ISL_1367308, EPI_ISL_1367309, EPI_ISL_1367310, EPI_ISL_1367311, EPI_ISL_1367312, EPI_ISL_1367313, EPI_ISL_1367314, EPI_ISL_1367315, EPI_ISL_1367316, EPI_ISL_1367317, EPI_ISL_1367318, EPI_ISL_1367319, EPI_ISL_1367320, EPI_ISL_1367321, EPI_ISL_1367322, EPI_ISL_1367323, EPI_ISL_1367324, EPI_ISL_1367325, EPI_ISL_1367326, EPI_ISL_1367327, EPI_ISL_1367328, EPI_ISL_1367329, EPI_ISL_1367330, EPI_ISL_1367331, EPI_ISL_1367332, EPI_ISL_1367333, EPI_ISL_1367334, EPI_ISL_1367335, EPI_ISL_1367336, EPI_ISL_1367337, EPI_ISL_1367338, EPI_ISL_1367339, EPI_ISL_1367340, EPI_ISL_1367341, EPI_ISL_1367342, EPI_ISL_1367343, EPI_ISL_1367344, EPI_ISL_1367345, EPI_ISL_1367346, EPI_ISL_1367347, EPI_ISL_1367348, EPI_ISL_1367349, EPI_ISL_1367350, EPI_ISL_1367351, EPI_ISL_1367352, EPI_ISL_1367353, EPI_ISL_1367354, EPI_ISL_1367355, EPI_ISL_1367356, EPI_ISL_1367357, EPI_ISL_1367358, EPI_ISL_1367359, EPI_ISL_1367360, EPI_ISL_1367361, EPI_ISL_1367362, EPI_ISL_1367363, EPI_ISL_1367364, EPI_ISL_1367365, EPI_ISL_1367366, EPI_ISL_1367367, EPI_ISL_1367368, EPI_ISL_1367369, EPI_ISL_1367370, EPI_ISL_1367371, EPI_ISL_1367372, EPI_ISL_1367373, EPI_ISL_1367374, EPI_ISL_1367375, EPI_ISL_1367376, EPI_ISL_1367377, EPI_ISL_1367378, EPI_ISL_1367379, EPI_ISL_1367380, EPI_ISL_1367381, EPI_ISL_1367382, EPI_ISL_1367383, EPI_ISL_1367384, EPI_ISL_1367385, EPI_ISL_1367386, EPI_ISL_1367387, EPI_ISL_1367388, EPI_ISL_1367389, EPI_ISL_1367390, EPI_ISL_1367391, EPI_ISL_1367392, EPI_ISL_1367393, EPI_ISL_1367394, EPI_ISL_1367395, EPI_ISL_1367396, EPI_ISL_1367397, EPI_ISL_1367398, EPI_ISL_1367399, EPI_ISL_1367400, EPI_ISL_1367401, EPI_ISL_1367402, EPI_ISL_1367403, EPI_ISL_1367404, EPI_ISL_1367405, EPI_ISL_1367406, EPI_ISL_1367407, EPI_ISL_1367408, EPI_ISL_1367409, EPI_ISL_1367410, EPI_ISL_1367411, EPI_ISL_1367412, EPI_ISL_1367413, EPI_ISL_1367414, EPI_ISL_1367415, EPI_ISL_1367416, EPI_ISL_1367417, EPI_ISL_1367418, EPI_ISL_1367419, EPI_ISL_1367420, EPI_ISL_1367421, EPI_ISL_1367422, EPI_ISL_1367423, EPI_ISL_1367424, EPI_ISL_1367425, EPI_ISL_1367426, EPI_ISL_1367427, EPI_ISL_1367428, EPI_ISL_1367429, EPI_ISL_1367430, EPI_ISL_1367431, EPI_ISL_1367432, EPI_ISL_1367433, EPI_ISL_1367434, EPI_ISL_1367435, EPI_ISL_1367436, EPI_ISL_1367437, EPI_ISL_1367438, EPI_ISL_1367439, EPI_ISL_1367440, EPI_ISL_1367441, EPI_ISL_1367442, EPI_ISL_1367443, EPI_ISL_1367444, EPI_ISL_1367445, EPI_ISL_1367446, EPI_ISL_1367447, EPI_ISL_1367448, EPI_ISL_1367449, EPI_ISL_1367450, EPI_ISL_1367451, EPI_ISL_1367452, EPI_ISL_1367453, EPI_ISL_1367454, EPI_ISL_1367455, EPI_ISL_1367456, EPI_ISL_1367457, EPI_ISL_1367458, EPI_ISL_1367459, EPI_ISL_1367460, EPI_ISL_1367461, EPI_ISL_1367462, EPI_ISL_1367463, EPI_ISL_1367464, EPI_ISL_1367465, EPI_ISL_1367466, EPI_ISL_1367467, EPI_ISL_1367468, EPI_ISL_1367469, EPI_ISL_1367470, EPI_ISL_1367471, EPI_ISL_1367472, EPI_ISL_1367473, EPI_ISL_1367474, EPI_ISL_1367475, EPI_ISL_1367476, EPI_ISL_1367477, EPI_ISL_1367478, EPI_ISL_1367479, EPI_ISL_1367480, EPI_ISL_1367481, EPI_ISL_1367482 | | | |
| see above | Quest Diagnostics Incorporated | Centers for Disease Control and Prevention Division of Viral Diseases, Pathogen Discovery | Peter W. Cook, Dakota Howard, Dhwani Batra, Ben L. Rambo-Martin, S. H. Rosenthal, A. Gerasimova, R. M. Kagan, B. Anderson, M. Hua, Y. Liu, L.E. Bernstein, K.E. Livingston, A. Perez, I. A. Shlyakhter, R. V. Rolando, R. Owen, P. Tanpaiboon, F. Lacbawan, Clinton R. Paden, Suxiang Tong, Duncan MacCannell |
| EPI_ISL_1367595, EPI_ISL_1367597, EPI_ISL_1367604, EPI_ISL_1367606, EPI_ISL_1367608, EPI_ISL_1367613, EPI_ISL_1367614, EPI_ISL_1367615, EPI_ISL_1367621, EPI_ISL_1367622, EPI_ISL_1367626, EPI_ISL_1367637, EPI_ISL_1367641, EPI_ISL_1367642, EPI_ISL_1367645, EPI_ISL_1367646, EPI_ISL_1367647, EPI_ISL_1367648, EPI_ISL_1367649, EPI_ISL_1367651, EPI_ISL_1367652, EPI_ISL_1367653, EPI_ISL_1367654, EPI_ISL_1367656, EPI_ISL_1367657, EPI_ISL_1367660, EPI_ISL_1367662, EPI_ISL_1367663, EPI_ISL_1367664, EPI_ISL_1367665, EPI_ISL_1367667, EPI_ISL_1367669, EPI_ISL_1367670, EPI_ISL_1367672, EPI_ISL_1367674, EPI_ISL_1367675, EPI_ISL_1367679, EPI_ISL_1367684, EPI_ISL_1367687, EPI_ISL_1367693, EPI_ISL_1367696, EPI_ISL_1367699, EPI_ISL_1367703, EPI_ISL_1367706, EPI_ISL_1367707, EPI_ISL_1367708, EPI_ISL_1367709, EPI_ISL_1367710, EPI_ISL_1367711, EPI_ISL_1367712, EPI_ISL_1367713, EPI_ISL_1367714, EPI_ISL_1367715, EPI_ISL_1367716, EPI_ISL_1367717, EPI_ISL_1367718, EPI_ISL_1367719, EPI_ISL_1367720, EPI_ISL_1367722, EPI_ISL_1367723, EPI_ISL_1367724, EPI_ISL_1367725, EPI_ISL_1367727, EPI_ISL_1367728, EPI_ISL_1367729, EPI_ISL_1367730, EPI_ISL_1367731, EPI_ISL_1367732, EPI_ISL_1367734, EPI_ISL_1367736, EPI_ISL_1367737, EPI_ISL_1367738, EPI_ISL_1367739, EPI_ISL_1367740, EPI_ISL_1367741, EPI_ISL_1367742, EPI_ISL_1367743, EPI_ISL_1367745, EPI_ISL_1367746, EPI_ISL_1367748, EPI_ISL_1367753, EPI_ISL_1367760, EPI_ISL_1367763, EPI_ISL_1367770, EPI_ISL_1367771, EPI_ISL_1367774, EPI_ISL_1367775, EPI_ISL_1367776, EPI_ISL_1367777, EPI_ISL_1367778, EPI_ISL_1367779, EPI_ISL_1367780, EPI_ISL_1367782, EPI_ISL_1367783, EPI_ISL_1367784, EPI_ISL_1367785, EPI_ISL_1367786, EPI_ISL_1367788, EPI_ISL_1367789, EPI_ISL_1367790, EPI_ISL_1367791, EPI_ISL_1367792, EPI_ISL_1367793, EPI_ISL_1367794, EPI_ISL_1367795, EPI_ISL_1367796, EPI_ISL_1367797, EPI_ISL_1367798, EPI_ISL_1367800, EPI_ISL_1367801, EPI_ISL_1367802, EPI_ISL_1367803, EPI_ISL_1367804, EPI_ISL_1367805, EPI_ISL_1367806, EPI_ISL_1367808, EPI_ISL_1367809, EPI_ISL_1367811, EPI_ISL_1367812, EPI_ISL_1367813, EPI_ISL_1367815, EPI_ISL_1367816, EPI_ISL_1367817, EPI_ISL_1367818, EPI_ISL_1367819, EPI_ISL_1367821, EPI_ISL_1367822, EPI_ISL_1367823, EPI_ISL_1367824, EPI_ISL_1367825, EPI_ISL_1367827, EPI_ISL_1367828, EPI_ISL_1367829, EPI_ISL_1367830, EPI_ISL_1367832, EPI_ISL_1367833, EPI_ISL_1367834, EPI_ISL_1367835, EPI_ISL_1367836, EPI_ISL_1367837, EPI_ISL_1367838, EPI_ISL_1367839, EPI_ISL_1367840, EPI_ISL_1367841, EPI_ISL_1367842, EPI_ISL_1367843, EPI_ISL_1367845, EPI_ISL_1367846, EPI_ISL_1367847, EPI_ISL_1367848, EPI_ISL_1367849, EPI_ISL_1367850, EPI_ISL_1367851, EPI_ISL_1367852, EPI_ISL_1367854, EPI_ISL_1367855, EPI_ISL_1367857, EPI_ISL_1367858, EPI_ISL_1367859, EPI_ISL_1367860, EPI_ISL_1367861, EPI_ISL_1367862, EPI_ISL_1367863, EPI_ISL_1367864, EPI_ISL_1367865, EPI_ISL_1367866, EPI_ISL_1367867, EPI_ISL_1367868, EPI_ISL_1367869, EPI_ISL_1367870, EPI_ISL_1367871, EPI_ISL_1367872, EPI_ISL_1367873, EPI_ISL_1367874, EPI_ISL_1367875, EPI_ISL_1367877, EPI_ISL_1367878, EPI_ISL_1367879, EPI_ISL_1367880, EPI_ISL_1367881, EPI_ISL_1367882, EPI_ISL_1367885, EPI_ISL_1367886, EPI_ISL_1367887, EPI_ISL_1367888, EPI_ISL_1367889, EPI_ISL_1367890, EPI_ISL_1367891, EPI_ISL_1367892, EPI_ISL_1367893, EPI_ISL_1367894, EPI_ISL_1367895, EPI_ISL_1367896, EPI_ISL_1367897, EPI_ISL_1367898, EPI_ISL_1367899, EPI_ISL_1367900, EPI_ISL_1367901, EPI_ISL_1367902, EPI_ISL_1367903, EPI_ISL_1367904, EPI_ISL_1367905, EPI_ISL_1367906, EPI_ISL_1367907, EPI_ISL_1367908, EPI_ISL_1367909, EPI_ISL_1367910, EPI_ISL_1367911, EPI_ISL_1367912, EPI_ISL_1367913, EPI_ISL_1367914, EPI_ISL_1367916, EPI_ISL_1367917 | | | |
| see above | Michigan Department of Health and Human Services, Bureau of Laboratories | Michigan Department of Health and Human Services, Bureau of Laboratories | Blankenship HM, Riner D, Soehnlen MK |
| EPI_ISL_1371707, EPI_ISL_1371708, EPI_ISL_1371709, EPI_ISL_1371710, EPI_ISL_1371711, EPI_ISL_1371712, EPI_ISL_1371713, EPI_ISL_1371714, EPI_ISL_1371715, EPI_ISL_1371716, EPI_ISL_1371717, EPI_ISL_1371718, EPI_ISL_1371719, EPI_ISL_1371720, EPI_ISL_1371721, EPI_ISL_1371722, EPI_ISL_1371723, EPI_ISL_1371724, EPI_ISL_1371725, EPI_ISL_1371726, EPI_ISL_1371727, EPI_ISL_1371728, EPI_ISL_1371729, EPI_ISL_1371733, EPI_ISL_1371734, EPI_ISL_1371735, EPI_ISL_1371736, EPI_ISL_1371737, EPI_ISL_1371738, EPI_ISL_1371739, EPI_ISL_1371740, EPI_ISL_1371741, EPI_ISL_1371742, EPI_ISL_1371743, EPI_ISL_1371744, EPI_ISL_1371745, EPI_ISL_1371746, EPI_ISL_1371747, EPI_ISL_1371748, EPI_ISL_1371749, EPI_ISL_1371750, EPI_ISL_1371751, EPI_ISL_1371752, EPI_ISL_1371753, EPI_ISL_1371754, EPI_ISL_1371755, EPI_ISL_1371756, EPI_ISL_1371757, EPI_ISL_1371758, EPI_ISL_1371759, EPI_ISL_1371760 | | | |
| see above | Virginia Division of Consolidated Laboratory Services | Virginia Division of Consolidated Laboratory Services | Virginia DCLS |
| EPI_ISL_1372097 | Minnesota Department of Health, Public Health Laboratory | Minnesota Department of Health, Public Health Laboratory | Alexandra Lorentz, Jacob Garfin, Matt Plumb, and Xiong Wang |
| EPI_ISL_1372098 | Pathnostics | Minnesota Department of Health, Public Health Laboratory | Alexandra Lorentz, Jacob Garfin, Matt Plumb, and Xiong Wang |
| EPI_ISL_1372099 | Minnesota Department of Health, Public Health Laboratory | Minnesota Department of Health, Public Health Laboratory | Alexandra Lorentz, Jacob Garfin, Matt Plumb, and Xiong Wang |
| EPI_ISL_1372100 | M Health Fairview | Minnesota Department of Health, Public Health Laboratory | Alexandra Lorentz, Jacob Garfin, Matt Plumb, and Xiong Wang |
| EPI_ISL_1372101, EPI_ISL_1372102, EPI_ISL_1372103 | Minnesota Department of Health, Public Health Laboratory | Minnesota Department of Health, Public Health Laboratory | Alexandra Lorentz, Jacob Garfin, Matt Plumb, and Xiong Wang |
| EPI_ISL_1372104, EPI_ISL_1372105, EPI_ISL_1372106 | M Health Fairview | Minnesota Department of Health, Public Health Laboratory | Alexandra Lorentz, Jacob Garfin, Matt Plumb, and Xiong Wang |
| EPI_ISL_1372107 | Minnesota Department of Health, Public Health Laboratory | Minnesota Department of Health, Public Health Laboratory | Alexandra Lorentz, Jacob Garfin, Matt Plumb, and Xiong Wang |
| EPI_ISL_1372108, EPI_ISL_1372109, EPI_ISL_1372110 | M Health Fairview | Minnesota Department of Health, Public Health Laboratory | Alexandra Lorentz, Jacob Garfin, Matt Plumb, and Xiong Wang |
| EPI_ISL_1372111 | LabCorp Lakewood | Minnesota Department of Health, Public Health Laboratory | Alexandra Lorentz, Jacob Garfin, Matt Plumb, and Xiong Wang |
| EPI_ISL_1372112, EPI_ISL_1372113, EPI_ISL_1372114 | Minnesota Department of Health, Public Health Laboratory | Minnesota Department of Health, Public Health Laboratory | Alexandra Lorentz, Jacob Garfin, Matt Plumb, and Xiong Wang |
| EPI_ISL_1372115 | M Health Fairview | Minnesota Department of Health, Public Health Laboratory | Alexandra Lorentz, Jacob Garfin, Matt Plumb, and Xiong Wang |
| EPI_ISL_1372116, EPI_ISL_1372117 | Minnesota Department of Health, Public Health Laboratory | Minnesota Department of Health, Public Health Laboratory | Alexandra Lorentz, Jacob Garfin, Matt Plumb, and Xiong Wang |
| EPI_ISL_1372118, EPI_ISL_1372119, EPI_ISL_1372120, EPI_ISL_1372121, EPI_ISL_1372122, EPI_ISL_1372123, EPI_ISL_1372124, EPI_ISL_1372125, EPI_ISL_1372126, EPI_ISL_1372131, EPI_ISL_1372132, EPI_ISL_1372133 | | | |
| see above | M Health Fairview | Minnesota Department of Health, Public Health Laboratory | Alexandra Lorentz, Jacob Garfin, Matt Plumb, and Xiong Wang |
| EPI_ISL_1372134, EPI_ISL_1372135, EPI_ISL_1372136, EPI_ISL_1372137, EPI_ISL_1372138, EPI_ISL_1372139, EPI_ISL_1372140, EPI_ISL_1372141, EPI_ISL_1372142, EPI_ISL_1372143, EPI_ISL_1372144, EPI_ISL_1372145, EPI_ISL_1372146, EPI_ISL_1372147, EPI_ISL_1372148, EPI_ISL_1372149, EPI_ISL_1372150, EPI_ISL_1372151, EPI_ISL_1372152, EPI_ISL_1372153, EPI_ISL_1372154, EPI_ISL_1372155, EPI_ISL_1372156 | | | |
| see above | Minnesota Department of Health, Public Health Laboratory | Minnesota Department of Health, Public Health Laboratory | Alexandra Lorentz, Jacob Garfin, Matt Plumb, and Xiong Wang |
| EPI_ISL_1372157 | M Health Fairview | Minnesota Department of Health, Public Health Laboratory | Alexandra Lorentz, Jacob Garfin, Matt Plumb, and Xiong Wang |
| EPI_ISL_1372158 | Minnesota Department of Health, Public Health Laboratory | Minnesota Department of Health, Public Health Laboratory | Alexandra Lorentz, Jacob Garfin, Matt Plumb, and Xiong Wang |
| EPI_ISL_1372159, EPI_ISL_1372160, EPI_ISL_1372161, EPI_ISL_1372162, EPI_ISL_1372163, EPI_ISL_1372164 | M Health Fairview | Minnesota Department of Health, Public Health Laboratory | Alexandra Lorentz, Jacob Garfin, Matt Plumb, and Xiong Wang |
| EPI_ISL_1372165, EPI_ISL_1372166, EPI_ISL_1372167 | Minnesota Department of Health, Public Health Laboratory | Minnesota Department of Health, Public Health Laboratory | Alexandra Lorentz, Jacob Garfin, Matt Plumb, and Xiong Wang |

| | | | |
|---|---|---|---|
| EPI_ISL_1372168 | M Health Fairview | Minnesota Department of Health, Public Health Laboratory | Alexandra Lorentz, Jacob Garfin, Matt Plumb, and Xiong Wang |
| EPI_ISL_1372169 | Minnesota Department of Health, Public Health Laboratory | Minnesota Department of Health, Public Health Laboratory | Alexandra Lorentz, Jacob Garfin, Matt Plumb, and Xiong Wang |
| EPI_ISL_1372170, EPI_ISL_1372171, EPI_ISL_1372172, EPI_ISL_1372173, EPI_ISL_1372174, EPI_ISL_1372175, EPI_ISL_1372176 | M Health Fairview | Minnesota Department of Health, Public Health Laboratory | Alexandra Lorentz, Jacob Garfin, Matt Plumb, and Xiong Wang |
| EPI_ISL_1372177 | Minnesota Department of Health, Public Health Laboratory | Minnesota Department of Health, Public Health Laboratory | Alexandra Lorentz, Jacob Garfin, Matt Plumb, and Xiong Wang |
| EPI_ISL_1372178, EPI_ISL_1372179, EPI_ISL_1372180 | M Health Fairview | Minnesota Department of Health, Public Health Laboratory | Alexandra Lorentz, Jacob Garfin, Matt Plumb, and Xiong Wang |
| EPI_ISL_1372188 | Minnesota Department of Health, Public Health Laboratory | Minnesota Department of Health, Public Health Laboratory | Alexandra Lorentz, Jacob Garfin, Matt Plumb, and Xiong Wang |
| EPI_ISL_1372189, EPI_ISL_1372190 | Vault Health | Minnesota Department of Health, Public Health Laboratory | Alexandra Lorentz, Jacob Garfin, Matt Plumb, and Xiong Wang |
| EPI_ISL_1372191, EPI_ISL_1372192 | Mayo Clinic & Mayo Clinic Laboratories | Minnesota Department of Health, Public Health Laboratory | Alexandra Lorentz, Jacob Garfin, Matt Plumb, and Xiong Wang |
| EPI_ISL_1372193 | Vault Health | Minnesota Department of Health, Public Health Laboratory | Alexandra Lorentz, Jacob Garfin, Matt Plumb, and Xiong Wang |
| EPI_ISL_1372194, EPI_ISL_1372195, EPI_ISL_1372196 | Mayo Clinic & Mayo Clinic Laboratories | Minnesota Department of Health, Public Health Laboratory | Alexandra Lorentz, Jacob Garfin, Matt Plumb, and Xiong Wang |
| EPI_ISL_1372197 | Vault Health | Minnesota Department of Health, Public Health Laboratory | Alexandra Lorentz, Jacob Garfin, Matt Plumb, and Xiong Wang |
| EPI_ISL_1372198 | Mayo Clinic & Mayo Clinic Laboratories | Minnesota Department of Health, Public Health Laboratory | Alexandra Lorentz, Jacob Garfin, Matt Plumb, and Xiong Wang |
| EPI_ISL_1372199 | Carris Health Rice Memorial Hospital | Minnesota Department of Health, Public Health Laboratory | Alexandra Lorentz, Jacob Garfin, Matt Plumb, and Xiong Wang |
| EPI_ISL_1372200 | Vault Health | Minnesota Department of Health, Public Health Laboratory | Alexandra Lorentz, Jacob Garfin, Matt Plumb, and Xiong Wang |
| EPI_ISL_1372201, EPI_ISL_1372202, EPI_ISL_1372203, EPI_ISL_1372204, EPI_ISL_1372205, EPI_ISL_1372206 | Mayo Clinic & Mayo Clinic Laboratories | Minnesota Department of Health, Public Health Laboratory | Alexandra Lorentz, Jacob Garfin, Matt Plumb, and Xiong Wang |
| EPI_ISL_1372207, EPI_ISL_1372208 | M Health Fairview | Minnesota Department of Health, Public Health Laboratory | Alexandra Lorentz, Jacob Garfin, Matt Plumb, and Xiong Wang |
| EPI_ISL_1372209, EPI_ISL_1372210, EPI_ISL_1372214, EPI_ISL_1372215, EPI_ISL_1372216, EPI_ISL_1372217, EPI_ISL_1372218 | Mayo Clinic & Mayo Clinic Laboratories | Minnesota Department of Health, Public Health Laboratory | Alexandra Lorentz, Jacob Garfin, Matt Plumb, and Xiong Wang |
| EPI_ISL_1372219, EPI_ISL_1372220, EPI_ISL_1372221, EPI_ISL_1372222, EPI_ISL_1372223, EPI_ISL_1372224, EPI_ISL_1372225 | Minnesota Department of Health, Public Health Laboratory | Minnesota Department of Health, Public Health Laboratory | Alexandra Lorentz, Jacob Garfin, Matt Plumb, and Xiong Wang |
| EPI_ISL_1372226, EPI_ISL_1372227, EPI_ISL_1372228, EPI_ISL_1372229 | M Health Fairview | Minnesota Department of Health, Public Health Laboratory | Alexandra Lorentz, Jacob Garfin, Matt Plumb, and Xiong Wang |
| EPI_ISL_1372230 | Mayo Clinic & Mayo Clinic Laboratories | Minnesota Department of Health, Public Health Laboratory | Alexandra Lorentz, Jacob Garfin, Matt Plumb, and Xiong Wang |
| EPI_ISL_1372231 | Vault Health | Minnesota Department of Health, Public Health Laboratory | Alexandra Lorentz, Jacob Garfin, Matt Plumb, and Xiong Wang |
| EPI_ISL_1372232, EPI_ISL_1372233, EPI_ISL_1372234, EPI_ISL_1372235, EPI_ISL_1372236, EPI_ISL_1372237, EPI_ISL_1372238, EPI_ISL_1372239, EPI_ISL_1372240, EPI_ISL_1372241, EPI_ISL_1372242, EPI_ISL_1372243, EPI_ISL_1372244, EPI_ISL_1372245, EPI_ISL_1372246, EPI_ISL_1372247, EPI_ISL_1372248, EPI_ISL_1372249, EPI_ISL_1372250, EPI_ISL_1372251, EPI_ISL_1372252, EPI_ISL_1372253, EPI_ISL_1372254, EPI_ISL_1372255, EPI_ISL_1372256, EPI_ISL_1372257, EPI_ISL_1372258, EPI_ISL_1372259, EPI_ISL_1372260, EPI_ISL_1372261, EPI_ISL_1372262, EPI_ISL_1372263, EPI_ISL_1372264, EPI_ISL_1372265, EPI_ISL_1372266, EPI_ISL_1372267 | | | |
| see above | Mayo Clinic & Mayo Clinic Laboratories | Minnesota Department of Health, Public Health Laboratory | Alexandra Lorentz, Jacob Garfin, Matt Plumb, and Xiong Wang |
| EPI_ISL_1372280, EPI_ISL_1372282 | Florida Bureau of Public Health Laboratories | Florida Bureau of Public Health Laboratories | Sarah Schmedes, Jason Blanton |
| EPI_ISL_1372464, EPI_ISL_1372465, EPI_ISL_1372466 | Mayo Clinic & Mayo Clinic Laboratories | Minnesota Department of Health, Public Health Laboratory | Alexandra Lorentz, Jacob Garfin, Matt Plumb, and Xiong Wang |
| EPI_ISL_1372467 | Minnesota Department of Health, Public Health Laboratory | Minnesota Department of Health, Public Health Laboratory | Alexandra Lorentz, Jacob Garfin, Matt Plumb, and Xiong Wang |
| EPI_ISL_1372468, EPI_ISL_1372469, EPI_ISL_1372470, EPI_ISL_1372471, EPI_ISL_1372472 | Mayo Clinic & Mayo Clinic Laboratories | Minnesota Department of Health, Public Health Laboratory | Alexandra Lorentz, Jacob Garfin, Matt Plumb, and Xiong Wang |
| EPI_ISL_1372473 | Vault Health | Minnesota Department of Health, Public Health Laboratory | Alexandra Lorentz, Jacob Garfin, Matt Plumb, and Xiong Wang |
| EPI_ISL_1372474, EPI_ISL_1372475 | Mayo Clinic & Mayo Clinic Laboratories | Minnesota Department of Health, Public Health Laboratory | Alexandra Lorentz, Jacob Garfin, Matt Plumb, and Xiong Wang |
| EPI_ISL_1372476 | M Health Fairview | Minnesota Department of Health, Public Health Laboratory | Alexandra Lorentz, Jacob Garfin, Matt Plumb, and Xiong Wang |
| EPI_ISL_1372477, EPI_ISL_1372478, EPI_ISL_1372479, EPI_ISL_1372480 | Mayo Clinic & Mayo Clinic Laboratories | Minnesota Department of Health, Public Health Laboratory | Alexandra Lorentz, Jacob Garfin, Matt Plumb, and Xiong Wang |
| EPI_ISL_1372481, EPI_ISL_1372482 | Vault Health | Minnesota Department of Health, Public Health Laboratory | Alexandra Lorentz, Jacob Garfin, Matt Plumb, and Xiong Wang |
| EPI_ISL_1372483 | Mayo Clinic & Mayo Clinic Laboratories | Minnesota Department of Health, Public Health Laboratory | Alexandra Lorentz, Jacob Garfin, Matt Plumb, and Xiong Wang |

| | | | |
|---|---|---|---|
| EPI_ISL_1372484, EPI_ISL_1372485 | Vault Health | Minnesota Department of Health, Public Health Laboratory | Alexandra Lorentz, Jacob Garfin, Matt Plumb, and Xiong Wang |
| EPI_ISL_1372486 | LAKE REGION HEALTHCARE | Minnesota Department of Health, Public Health Laboratory | Alexandra Lorentz, Jacob Garfin, Matt Plumb, and Xiong Wang |
| EPI_ISL_1372487 | M Health Fairview | Minnesota Department of Health, Public Health Laboratory | Alexandra Lorentz, Jacob Garfin, Matt Plumb, and Xiong Wang |
| EPI_ISL_1372488, EPI_ISL_1372489, EPI_ISL_1372490, EPI_ISL_1372491, EPI_ISL_1372492, EPI_ISL_1372493, EPI_ISL_1372494, EPI_ISL_1372495 | Mayo Clinic & Mayo Clinic Laboratories | Minnesota Department of Health, Public Health Laboratory | Alexandra Lorentz, Jacob Garfin, Matt Plumb, and Xiong Wang |
| EPI_ISL_1372496, EPI_ISL_1372497 | Minnesota Department of Health, Public Health Laboratory | Minnesota Department of Health, Public Health Laboratory | Alexandra Lorentz, Jacob Garfin, Matt Plumb, and Xiong Wang |
| EPI_ISL_1372498, EPI_ISL_1372499 | M Health Fairview | Minnesota Department of Health, Public Health Laboratory | Alexandra Lorentz, Jacob Garfin, Matt Plumb, and Xiong Wang |
| EPI_ISL_1372500 | Vault Health | Minnesota Department of Health, Public Health Laboratory | Alexandra Lorentz, Jacob Garfin, Matt Plumb, and Xiong Wang |
| EPI_ISL_1372510 | M Health Fairview | Minnesota Department of Health, Public Health Laboratory | Alexandra Lorentz, Jacob Garfin, Matt Plumb, and Xiong Wang |
| EPI_ISL_1372511 | SOUTH LAKE PEDIATRICS | Minnesota Department of Health, Public Health Laboratory | Alexandra Lorentz, Jacob Garfin, Matt Plumb, and Xiong Wang |
| EPI_ISL_1372512 | Minnesota Department of Health, Public Health Laboratory | Minnesota Department of Health, Public Health Laboratory | Alexandra Lorentz, Jacob Garfin, Matt Plumb, and Xiong Wang |
| EPI_ISL_1372513, EPI_ISL_1372514, EPI_ISL_1372515, EPI_ISL_1372516, EPI_ISL_1372517, EPI_ISL_1372518, EPI_ISL_1372519, EPI_ISL_1372520, EPI_ISL_1372521 | M Health Fairview | Minnesota Department of Health, Public Health Laboratory | Alexandra Lorentz, Jacob Garfin, Matt Plumb, and Xiong Wang |
| EPI_ISL_1372522, EPI_ISL_1372523, EPI_ISL_1372524, EPI_ISL_1372525, EPI_ISL_1372526, EPI_ISL_1372527, EPI_ISL_1372528, EPI_ISL_1372529, EPI_ISL_1372530, EPI_ISL_1372531, EPI_ISL_1372532, EPI_ISL_1372533, EPI_ISL_1372534, EPI_ISL_1372535 | | | |
| see above | Mayo Clinic & Mayo Clinic Laboratories | Minnesota Department of Health, Public Health Laboratory | Alexandra Lorentz, Jacob Garfin, Matt Plumb, and Xiong Wang |
| EPI_ISL_1372536, EPI_ISL_1372537, EPI_ISL_1372538, EPI_ISL_1372539 | Vault Health | Minnesota Department of Health, Public Health Laboratory | Alexandra Lorentz, Jacob Garfin, Matt Plumb, and Xiong Wang |
| EPI_ISL_1372572, EPI_ISL_1372575, EPI_ISL_1372577, EPI_ISL_1372580, EPI_ISL_1372581 | Wyoming Public Health Laboratory | Wyoming Public Health Laboratory | Jim Mildenberger, Wanda Manley, Noah Hull, Taylor Fearing, Lynette Gumbleton, Channing Weber, Ashley Norberg, Chayse Rowley, Marley Goetz, Brian Dominguez, Elliot Thomasson, Cari Sloma, and Rob Christensen |
| EPI_ISL_1372584, EPI_ISL_1372585, EPI_ISL_1372586, EPI_ISL_1372587, EPI_ISL_1372588, EPI_ISL_1372589, EPI_ISL_1372590, EPI_ISL_1372591, EPI_ISL_1372592, EPI_ISL_1372593, EPI_ISL_1372594, EPI_ISL_1372595, EPI_ISL_1372596, EPI_ISL_1372597, EPI_ISL_1372598, EPI_ISL_1372599, EPI_ISL_1372600, EPI_ISL_1372601, EPI_ISL_1372602, EPI_ISL_1372603, EPI_ISL_1372604, EPI_ISL_1372605, EPI_ISL_1372606, EPI_ISL_1372607, EPI_ISL_1372608, EPI_ISL_1372609, EPI_ISL_1372610, EPI_ISL_1372611, EPI_ISL_1372612, EPI_ISL_1372613, EPI_ISL_1372614, EPI_ISL_1372615, EPI_ISL_1372616, EPI_ISL_1372617, EPI_ISL_1372618, EPI_ISL_1372619, EPI_ISL_1372620, EPI_ISL_1372621, EPI_ISL_1372622, EPI_ISL_1372623, EPI_ISL_1372624, EPI_ISL_1372625, EPI_ISL_1372626, EPI_ISL_1372627, EPI_ISL_1372628, EPI_ISL_1372629, EPI_ISL_1372630, EPI_ISL_1372646, EPI_ISL_1372647, EPI_ISL_1372648, EPI_ISL_1372649, EPI_ISL_1372650, EPI_ISL_1372651, EPI_ISL_1372652, EPI_ISL_1372653, EPI_ISL_1372654, EPI_ISL_1372655, EPI_ISL_1372656, EPI_ISL_1372657, EPI_ISL_1372658, EPI_ISL_1372659, EPI_ISL_1372660, EPI_ISL_1372661, EPI_ISL_1372662, EPI_ISL_1372663, EPI_ISL_1372664, EPI_ISL_1372665, EPI_ISL_1372666, EPI_ISL_1372667, EPI_ISL_1372668, EPI_ISL_1372669, EPI_ISL_1372670, EPI_ISL_1372671, EPI_ISL_1372672, EPI_ISL_1372673, EPI_ISL_1372674, EPI_ISL_1372675, EPI_ISL_1372676, EPI_ISL_1372677, EPI_ISL_1372678, EPI_ISL_1372679, EPI_ISL_1372680, EPI_ISL_1372681, EPI_ISL_1372682, EPI_ISL_1372683, EPI_ISL_1372684, EPI_ISL_1372685, EPI_ISL_1372686, EPI_ISL_1372687, EPI_ISL_1372688, EPI_ISL_1372689, EPI_ISL_1372690, EPI_ISL_1372691, EPI_ISL_1372692, EPI_ISL_1372693, EPI_ISL_1372694, EPI_ISL_1372695, EPI_ISL_1372696, EPI_ISL_1372697, EPI_ISL_1372698, EPI_ISL_1372699, EPI_ISL_1372700, EPI_ISL_1372701, EPI_ISL_1372702, EPI_ISL_1372703, EPI_ISL_1372704, EPI_ISL_1372705, EPI_ISL_1372706, EPI_ISL_1372707, EPI_ISL_1372708, EPI_ISL_1372709, EPI_ISL_1372710, EPI_ISL_1372711, EPI_ISL_1372712, EPI_ISL_1372713, EPI_ISL_1372714, EPI_ISL_1372715, EPI_ISL_1372716, EPI_ISL_1372717, EPI_ISL_1372718, EPI_ISL_1372719, EPI_ISL_1372720, EPI_ISL_1372721, EPI_ISL_1372722, EPI_ISL_1372723, EPI_ISL_1372724, EPI_ISL_1372725, EPI_ISL_1372726, EPI_ISL_1372727, EPI_ISL_1372728, EPI_ISL_1372729, EPI_ISL_1372730, EPI_ISL_1372731, EPI_ISL_1372732, EPI_ISL_1372733, EPI_ISL_1372734, EPI_ISL_1372735, EPI_ISL_1372736, EPI_ISL_1372737, EPI_ISL_1372738, EPI_ISL_1372739, EPI_ISL_1372740, EPI_ISL_1372741, EPI_ISL_1372742, EPI_ISL_1372743, EPI_ISL_1372744, EPI_ISL_1372745, EPI_ISL_1372746, EPI_ISL_1372747, EPI_ISL_1372748, EPI_ISL_1372749, EPI_ISL_1372750, EPI_ISL_1372751, EPI_ISL_1372752, EPI_ISL_1372753, EPI_ISL_1372754, EPI_ISL_1372755, EPI_ISL_1372756, EPI_ISL_1372757, EPI_ISL_1372758, EPI_ISL_1372759, EPI_ISL_1372760, EPI_ISL_1372761, EPI_ISL_1372762, EPI_ISL_1372763, EPI_ISL_1372764, EPI_ISL_1372765, EPI_ISL_1372766, EPI_ISL_1372767, EPI_ISL_1372768, EPI_ISL_1372769, EPI_ISL_1372770, EPI_ISL_1372771, EPI_ISL_1372772, EPI_ISL_1372773, EPI_ISL_1372774, EPI_ISL_1372775, EPI_ISL_1372776, EPI_ISL_1372777, EPI_ISL_1372778, EPI_ISL_1372779, EPI_ISL_1372780, EPI_ISL_1372781, EPI_ISL_1372782, EPI_ISL_1372783, EPI_ISL_1372784, EPI_ISL_1372785, EPI_ISL_1372786, EPI_ISL_1372787, EPI_ISL_1372788, EPI_ISL_1372789, EPI_ISL_1372790, EPI_ISL_1372791, EPI_ISL_1372792, EPI_ISL_1372793, EPI_ISL_1372794, EPI_ISL_1372795, EPI_ISL_1372796, EPI_ISL_1372797, EPI_ISL_1372798, EPI_ISL_1372799, EPI_ISL_1372800, EPI_ISL_1372801, EPI_ISL_1372802, EPI_ISL_1372803, EPI_ISL_1372804, EPI_ISL_1372805, EPI_ISL_1372806, EPI_ISL_1372807, EPI_ISL_1372808, EPI_ISL_1372809, EPI_ISL_1372810, EPI_ISL_1372811, EPI_ISL_1372812, EPI_ISL_1372813, EPI_ISL_1372814, EPI_ISL_1372815, EPI_ISL_1372816, EPI_ISL_1372817, EPI_ISL_1372818, EPI_ISL_1372819, EPI_ISL_1372820, EPI_ISL_1372821, EPI_ISL_1372822, EPI_ISL_1372823, EPI_ISL_1372824, EPI_ISL_1372825, EPI_ISL_1372826, EPI_ISL_1372827, EPI_ISL_1372828, EPI_ISL_1372829, EPI_ISL_1372830, EPI_ISL_1372831, EPI_ISL_1372832, EPI_ISL_1372833, EPI_ISL_1372834, EPI_ISL_1372835, EPI_ISL_1372836, EPI_ISL_1372837, EPI_ISL_1372838, EPI_ISL_1372839, EPI_ISL_1372840, EPI_ISL_1372841, EPI_ISL_1372842, EPI_ISL_1372843, EPI_ISL_1372844, EPI_ISL_1372845, EPI_ISL_1372846, EPI_ISL_1372847, EPI_ISL_1372848, EPI_ISL_1372849, EPI_ISL_1372850, EPI_ISL_1372851, EPI_ISL_1372852, EPI_ISL_1372853, EPI_ISL_1372854, EPI_ISL_1372855, EPI_ISL_1372856, EPI_ISL_1372857, EPI_ISL_1372858, EPI_ISL_1372859, EPI_ISL_1372860, EPI_ISL_1372861, EPI_ISL_1372862, EPI_ISL_1372863, EPI_ISL_1372864, EPI_ISL_1372865, EPI_ISL_1372866, EPI_ISL_1372867, EPI_ISL_1372868, EPI_ISL_1372869, EPI_ISL_1372870, EPI_ISL_1372871, EPI_ISL_1372872, EPI_ISL_1372873, EPI_ISL_1372874, EPI_ISL_1372875, EPI_ISL_1372876, EPI_ISL_1372877, EPI_ISL_1372878, EPI_ISL_1372879, EPI_ISL_1372880, EPI_ISL_1372881, EPI_ISL_1372882, EPI_ISL_1372883, EPI_ISL_1372884, EPI_ISL_1372885, EPI_ISL_1372886, EPI_ISL_1372887, EPI_ISL_1372888, EPI_ISL_1372889, EPI_ISL_1372890, EPI_ISL_1372891, EPI_ISL_1372892, EPI_ISL_1372893, EPI_ISL_1372894, EPI_ISL_1372895, EPI_ISL_1372896, EPI_ISL_1372897 | | | |
| see above | Maine Health and Environmental Testing Laboratory | Tewhey Lab, The Jackson Laboratory | Matluk,N., Dewey,H., Iosue,F., Barter,M., Lynch,R., Munger,H. and Tewhey,R. |
| EPI_ISL_1372965, EPI_ISL_1372966, EPI_ISL_1372967, EPI_ISL_1372968, EPI_ISL_1372969, EPI_ISL_1372971, EPI_ISL_1372972, EPI_ISL_1372973, EPI_ISL_1372980, EPI_ISL_1372981, EPI_ISL_1372982, EPI_ISL_1372983, EPI_ISL_1372984, EPI_ISL_1372985, EPI_ISL_1372986, EPI_ISL_1372987, EPI_ISL_1372988, EPI_ISL_1372989, EPI_ISL_1372990, EPI_ISL_1372991, EPI_ISL_1372992, EPI_ISL_1372993, EPI_ISL_1372994, EPI_ISL_1372995, EPI_ISL_1372996, EPI_ISL_1372997, EPI_ISL_1372998, EPI_ISL_1372999, EPI_ISL_1373000, EPI_ISL_1373001, EPI_ISL_1373002, EPI_ISL_1373003, EPI_ISL_1373004, EPI_ISL_1373005, EPI_ISL_1373006, EPI_ISL_1373007, EPI_ISL_1373008, EPI_ISL_1373009, EPI_ISL_1373010, EPI_ISL_1373011, EPI_ISL_1373012, EPI_ISL_1373013 | | | |
| see above | US Air Force School of Aerospace Medicine | US Air Force School of Aerospace Medicine | Anthony Fries, Jennifer Meyer, William Gruner, William Buggele, Amanda Javorina, Sarah Purves, Clarise Starr, Elizabeth Macias |
| EPI_ISL_1373014, EPI_ISL_1373015, EPI_ISL_1373016, EPI_ISL_1373017, EPI_ISL_1373018, EPI_ISL_1373019, EPI_ISL_1373020, EPI_ISL_1373021, EPI_ISL_1373022, EPI_ISL_1373025, EPI_ISL_1373026, EPI_ISL_1373027, EPI_ISL_1373028, EPI_ISL_1373030, EPI_ISL_1373031, EPI_ISL_1373032, EPI_ISL_1373033, EPI_ISL_1373034, EPI_ISL_1373035, EPI_ISL_1373036 | | | |
| see above | Wyoming Public Health Laboratory | Wyoming Public Health Laboratory | Jim Mildenberger, Wanda Manley, Noah Hull, Taylor Fearing, Lynette Gumbleton, Channing Weber, Ashley Norberg, Chayse Rowley, Marley Goetz, Brian Dominguez, Elliot Thomasson, Cari Sloma, and Rob Christensen |
| EPI_ISL_1373135, EPI_ISL_1373136, EPI_ISL_1373137, EPI_ISL_1373138, EPI_ISL_1373139, EPI_ISL_1373141, EPI_ISL_1373142, EPI_ISL_1373143, EPI_ISL_1373144, EPI_ISL_1373145, EPI_ISL_1373147, EPI_ISL_1373148, EPI_ISL_1373149, EPI_ISL_1373150, EPI_ISL_1373151, EPI_ISL_1373152, EPI_ISL_1373153, EPI_ISL_1373155, EPI_ISL_1373158, EPI_ISL_1373159, EPI_ISL_1373160 | | | |
| see above | Alameda County Public Health Department | Alameda County Public Health Department | Kristna Hsieh, Maria Elma, Kevin Libuit |
| EPI_ISL_1373315, EPI_ISL_1373316, EPI_ISL_1373317, EPI_ISL_1373318, EPI_ISL_1373319, EPI_ISL_1373320, EPI_ISL_1373321, EPI_ISL_1373322, EPI_ISL_1373323, EPI_ISL_1373324, EPI_ISL_1373325, EPI_ISL_1373326, EPI_ISL_1373327, EPI_ISL_1373328, EPI_ISL_1373329, EPI_ISL_1373330, EPI_ISL_1373331, EPI_ISL_1373332, EPI_ISL_1373333, EPI_ISL_1373334, EPI_ISL_1373335, EPI_ISL_1373336, EPI_ISL_1373337, EPI_ISL_1373338, EPI_ISL_1373339, EPI_ISL_1373340, EPI_ISL_1373341, EPI_ISL_1373342, EPI_ISL_1373343, EPI_ISL_1373344, EPI_ISL_1373345, EPI_ISL_1373346, EPI_ISL_1373347, EPI_ISL_1373348, EPI_ISL_1373349, EPI_ISL_1373350, EPI_ISL_1373351, EPI_ISL_1373352, EPI_ISL_1373354, EPI_ISL_1373355, EPI_ISL_1373356, EPI_ISL_1373357, EPI_ISL_1373358 | | | |
| see above | MD PHL | MD PHL | Maryland Department of Health Laboratories Administration |
| EPI_ISL_1373376, EPI_ISL_1373377, EPI_ISL_1373378, EPI_ISL_1373379, EPI_ISL_1373380, EPI_ISL_1373381, EPI_ISL_1373382, EPI_ISL_1373383, EPI_ISL_1373384, EPI_ISL_1373386, EPI_ISL_1373387, EPI_ISL_1373388, EPI_ISL_1373389, EPI_ISL_1373390, EPI_ISL_1373391, EPI_ISL_1373392, EPI_ISL_1373393, EPI_ISL_1373394, EPI_ISL_1373395, EPI_ISL_1373396, EPI_ISL_1373398, EPI_ISL_1373399, EPI_ISL_1373400, EPI_ISL_1373401, EPI_ISL_1373402, EPI_ISL_1373403, EPI_ISL_1373404, EPI_ISL_1373405, EPI_ISL_1373406, EPI_ISL_1373407, EPI_ISL_1373408, EPI_ISL_1373409, EPI_ISL_1373410, EPI_ISL_1373411, EPI_ISL_1373412, EPI_ISL_1373413, EPI_ISL_1373414, EPI_ISL_1373415, EPI_ISL_1373416, EPI_ISL_1373417, EPI_ISL_1373418, EPI_ISL_1373419, EPI_ISL_1373420, EPI_ISL_1373421, EPI_ISL_1373422, EPI_ISL_1373423, EPI_ISL_1373424, EPI_ISL_1373425, EPI_ISL_1373426, EPI_ISL_1373427, EPI_ISL_1373428, EPI_ISL_1373429, EPI_ISL_1373430, EPI_ISL_1373431, EPI_ISL_1373432, EPI_ISL_1373433, EPI_ISL_1373434, EPI_ISL_1373435, EPI_ISL_1373436, EPI_ISL_1373437, EPI_ISL_1373438, EPI_ISL_1373439, EPI_ISL_1373440 | | | |
| see above | Wyoming Public Health Laboratory | Wyoming Public Health Laboratory | Jim Mildenberger, Wanda Manley, Noah Hull, Taylor Fearing, Lynette Gumbleton, Channing Weber, Ashley Norberg, Chayse Rowley, Marley Goetz, Brian Dominguez, Elliot Thomasson, Cari Sloma, and Rob Christensen |
| EPI_ISL_1373441, EPI_ISL_1373442, EPI_ISL_1373443, EPI_ISL_1373444, EPI_ISL_1373445, EPI_ISL_1373446, EPI_ISL_1373447, EPI_ISL_1373453, EPI_ISL_1373454, EPI_ISL_1373455, EPI_ISL_1373456, EPI_ISL_1373457, EPI_ISL_1373458, EPI_ISL_1373459, EPI_ISL_1373460 | | | |
| see above | UNC-CH COVID Surveillance Lab | Jeremy Wang | Jeremy Wang, Alexander Rubinsteyn, Melissa Miller, Corbin Jones, Amy James Loftis, Amir Barzin, Susan Fiscus |
| EPI_ISL_1373462, EPI_ISL_1373463, EPI_ISL_1373464, EPI_ISL_1373466, EPI_ISL_1373467, EPI_ISL_1373468, EPI_ISL_1373470, EPI_ISL_1373472, EPI_ISL_1373473, EPI_ISL_1373474, EPI_ISL_1373475, EPI_ISL_1373476, EPI_ISL_1373477, EPI_ISL_1373478, EPI_ISL_1373479, EPI_ISL_1373480, EPI_ISL_1373481, EPI_ISL_1373483, EPI_ISL_1373484, EPI_ISL_1373486, EPI_ISL_1373488, EPI_ISL_1373490, EPI_ISL_1373491, EPI_ISL_1373492, EPI_ISL_1373493, EPI_ISL_1373494, EPI_ISL_1373495, EPI_ISL_1373496, EPI_ISL_1373497, EPI_ISL_1373498, EPI_ISL_1373501, EPI_ISL_1373503, EPI_ISL_1373504, EPI_ISL_1373505, EPI_ISL_1373506, EPI_ISL_1373507, EPI_ISL_1373510, EPI_ISL_1373511 | | | |
| see above | Clinical Molecular Microbiology Laboratory, UNC Hospitals | Jeremy Wang | Jeremy Wang, Alexander Rubinsteyn, Colleen Rice, Jason Smedberg, Shawn Hawken, Melissa Miller, Corbin Jones, Robert Hagan |
| EPI_ISL_1373512, EPI_ISL_1373513, EPI_ISL_1373514, EPI_ISL_1373515, EPI_ISL_1373516, EPI_ISL_1373517, EPI_ISL_1373518, EPI_ISL_1373519, EPI_ISL_1373520, EPI_ISL_1373521, EPI_ISL_1373522, EPI_ISL_1373524, EPI_ISL_1373525, EPI_ISL_1373526, EPI_ISL_1373527, EPI_ISL_1373528, EPI_ISL_1373529, EPI_ISL_1373531, EPI_ISL_1373532, EPI_ISL_1373533, EPI_ISL_1373534, EPI_ISL_1373535, EPI_ISL_1373536, EPI_ISL_1373537, EPI_ISL_1373538, EPI_ISL_1373539, EPI_ISL_1373540, EPI_ISL_1373541, EPI_ISL_1373542, EPI_ISL_1373543, EPI_ISL_1373544, EPI_ISL_1373545, EPI_ISL_1373546, EPI_ISL_1373547, EPI_ISL_1373548, EPI_ISL_1373549, EPI_ISL_1373550, EPI_ISL_1373551, EPI_ISL_1373552, EPI_ISL_1373553, EPI_ISL_1373554, EPI_ISL_1373555, EPI_ISL_1373556, EPI_ISL_1373557, EPI_ISL_1373558, EPI_ISL_1373559, EPI_ISL_1373560, EPI_ISL_1373561, EPI_ISL_1373563, EPI_ISL_1373564, EPI_ISL_1373565, EPI_ISL_1373566, EPI_ISL_1373567, EPI_ISL_1373568, EPI_ISL_1373569, EPI_ISL_1373570, EPI_ISL_1373571, EPI_ISL_1373572, EPI_ISL_1373573, EPI_ISL_1373574, EPI_ISL_1373575, EPI_ISL_1373576, EPI_ISL_1373577, EPI_ISL_1373578, EPI_ISL_1373579, EPI_ISL_1373580, EPI_ISL_1373581, EPI_ISL_1373582, EPI_ISL_1373583 | | | |
| see above | Vault Health | Minnesota Department of Health, Public Health Laboratory | Alexandra Lorentz, Jacob Garfin, Matt Plumb, and Xiong Wang |
| EPI_ISL_1373584, EPI_ISL_1373585, EPI_ISL_1373586, EPI_ISL_1373587, | Mayo Clinic & Mayo Clinic Laboratories | Minnesota Department of Health, Public Health Laboratory | Alexandra Lorentz, Jacob Garfin, Matt Plumb, and Xiong Wang |

| | | | |
|---|---|---|---|
| EPI_ISL_1373588 | | | |
| EPI_ISL_1373602, EPI_ISL_1373603, EPI_ISL_1373604, EPI_ISL_1373605, EPI_ISL_1373606, EPI_ISL_1373607, EPI_ISL_1373608, EPI_ISL_1373609, EPI_ISL_1373610, EPI_ISL_1373611, EPI_ISL_1373612, EPI_ISL_1373613, EPI_ISL_1373614, EPI_ISL_1373615, EPI_ISL_1373616, EPI_ISL_1373617, EPI_ISL_1373618, EPI_ISL_1373619, EPI_ISL_1373620, EPI_ISL_1373621, EPI_ISL_1373622, EPI_ISL_1373623, EPI_ISL_1373624, EPI_ISL_1373625, EPI_ISL_1373626, EPI_ISL_1373627, EPI_ISL_1373628, EPI_ISL_1373629, EPI_ISL_1373630, EPI_ISL_1373631, EPI_ISL_1373632, EPI_ISL_1373633, EPI_ISL_1373634, EPI_ISL_1373635, EPI_ISL_1373636, EPI_ISL_1373637, EPI_ISL_1373638, EPI_ISL_1373639, EPI_ISL_1373640, EPI_ISL_1373641, EPI_ISL_1373642, EPI_ISL_1373643, EPI_ISL_1373644, EPI_ISL_1373645, EPI_ISL_1373646, EPI_ISL_1373647, EPI_ISL_1373648, EPI_ISL_1373649, EPI_ISL_1373650, EPI_ISL_1373651, EPI_ISL_1373652, EPI_ISL_1373653, EPI_ISL_1373654, EPI_ISL_1373655, EPI_ISL_1373656, EPI_ISL_1373657, EPI_ISL_1373658, EPI_ISL_1373659, EPI_ISL_1373660, EPI_ISL_1373661, EPI_ISL_1373662, EPI_ISL_1373663, EPI_ISL_1373664, EPI_ISL_1373665, EPI_ISL_1373666, EPI_ISL_1373667, EPI_ISL_1373668, EPI_ISL_1373669, EPI_ISL_1373670, EPI_ISL_1373671, EPI_ISL_1373672, EPI_ISL_1373673, EPI_ISL_1373674, EPI_ISL_1373675, EPI_ISL_1373676, EPI_ISL_1373677, EPI_ISL_1373678, EPI_ISL_1373679, EPI_ISL_1373680, EPI_ISL_1373681, EPI_ISL_1373682, EPI_ISL_1373683, EPI_ISL_1373684, EPI_ISL_1373685, EPI_ISL_1373686, EPI_ISL_1373687, EPI_ISL_1373688, EPI_ISL_1373689, EPI_ISL_1373690, EPI_ISL_1373691, EPI_ISL_1373692, EPI_ISL_1373693, EPI_ISL_1373694, EPI_ISL_1373695, EPI_ISL_1373696, EPI_ISL_1373697, EPI_ISL_1373698, EPI_ISL_1373699, EPI_ISL_1373700, EPI_ISL_1373701, EPI_ISL_1373702, EPI_ISL_1373703, EPI_ISL_1373704, EPI_ISL_1373705, EPI_ISL_1373706, EPI_ISL_1373707, EPI_ISL_1373708, EPI_ISL_1373709, EPI_ISL_1373710, EPI_ISL_1373711, EPI_ISL_1373712, EPI_ISL_1373713, EPI_ISL_1373714, EPI_ISL_1373715, EPI_ISL_1373716, EPI_ISL_1373717, EPI_ISL_1373718, EPI_ISL_1373719, EPI_ISL_1373720, EPI_ISL_1373721, EPI_ISL_1373722, EPI_ISL_1373723, EPI_ISL_1373724 | | | |
| see above | UPMC Clinical Microbiology Laboratory | Microbial Genome Sequencing Center; Microbial Genomic Epidemiology Laboratory, University of Pittsburgh | Lee H. Harrison, Jane W. Marsh, Marissa P. Griffith, Stephanie L. Mitchell, Vatsala R. Srinivasa, Kady D. Waggle, Daniel J. Snyder, Vaughn S. Cooper |
| EPI_ISL_1373725 | UPMC Children's Hospital of Pittsburgh | Genomics and Discovery, Respiratory Viruses Branch, Division of Viral Diseases, Centers for Disease Control and Prevention | Ying Tao, Jing Zhang, Yan Li, Brian Lynch, Anna Kelleher, Krista Queen, Anna Uehara, Peter Cook, Han Jia Justin Ng, Clinton R. Paden, Haibin Wang, Suxiang Tong |
| EPI_ISL_1373726 | CDPH, Viral and Rickettsial Disease Laboratory | Genomics and Discovery, Respiratory Viruses Branch, Division of Viral Diseases, Centers for Disease Control and Prevention | Ying Tao, Jing Zhang, Yan Li, Brian Lynch, Anna Kelleher, Krista Queen, Anna Uehara, Peter Cook, Han Jia Justin Ng, Clinton R. Paden, Haibin Wang, Suxiang Tong |
| EPI_ISL_1373727, EPI_ISL_1373728 | UPMC Children's Hospital of Pittsburgh | Genomics and Discovery, Respiratory Viruses Branch, Division of Viral Diseases, Centers for Disease Control and Prevention | Ying Tao, Jing Zhang, Yan Li, Brian Lynch, Anna Kelleher, Krista Queen, Anna Uehara, Peter Cook, Han Jia Justin Ng, Clinton R. Paden, Haibin Wang, Suxiang Tong |
| EPI_ISL_1373729 | CDPH, Viral and Rickettsial Disease Laboratory | Genomics and Discovery, Respiratory Viruses Branch, Division of Viral Diseases, Centers for Disease Control and Prevention | Ying Tao, Jing Zhang, Yan Li, Brian Lynch, Anna Kelleher, Krista Queen, Anna Uehara, Peter Cook, Han Jia Justin Ng, Clinton R. Paden, Haibin Wang, Suxiang Tong |
| EPI_ISL_1373730 | Boston Childrens (Overcoming COVID) | Genomics and Discovery, Respiratory Viruses Branch, Division of Viral Diseases, Centers for Disease Control and Prevention | Ying Tao, Jing Zhang, Yan Li, Brian Lynch, Anna Kelleher, Krista Queen, Anna Uehara, Peter Cook, Han Jia Justin Ng, Clinton R. Paden, Haibin Wang, Suxiang Tong |
| EPI_ISL_1373731 | UPMC Children's Hospital of Pittsburgh | Genomics and Discovery, Respiratory Viruses Branch, Division of Viral Diseases, Centers for Disease Control and Prevention | Ying Tao, Jing Zhang, Yan Li, Brian Lynch, Anna Kelleher, Krista Queen, Anna Uehara, Peter Cook, Han Jia Justin Ng, Clinton R. Paden, Haibin Wang, Suxiang Tong |
| EPI_ISL_1373732, EPI_ISL_1373733, EPI_ISL_1373734, EPI_ISL_1373735, EPI_ISL_1373736 | Washington University School of Medicine | Genomics and Discovery, Respiratory Viruses Branch, Division of Viral Diseases, Centers for Disease Control and Prevention | Ying Tao, Jing Zhang, Yan Li, Brian Lynch, Anna Kelleher, Krista Queen, Anna Uehara, Peter Cook, Han Jia Justin Ng, Clinton R. Paden, Haibin Wang, Suxiang Tong |
| EPI_ISL_1373737 | CDPH, Viral and Rickettsial Disease Laboratory | Genomics and Discovery, Respiratory Viruses Branch, Division of Viral Diseases, Centers for Disease Control and Prevention | Ying Tao, Jing Zhang, Yan Li, Brian Lynch, Anna Kelleher, Krista Queen, Anna Uehara, Peter Cook, Han Jia Justin Ng, Clinton R. Paden, Haibin Wang, Suxiang Tong |
| EPI_ISL_1373738 | UPMC Children's Hospital of Pittsburgh | Genomics and Discovery, Respiratory Viruses Branch, Division of Viral Diseases, Centers for Disease Control and Prevention | Ying Tao, Jing Zhang, Yan Li, Brian Lynch, Anna Kelleher, Krista Queen, Anna Uehara, Peter Cook, Han Jia Justin Ng, Clinton R. Paden, Haibin Wang, Suxiang Tong |
| EPI_ISL_1373739 | CDPH, Viral and Rickettsial Disease Laboratory | Genomics and Discovery, Respiratory Viruses Branch, Division of Viral Diseases, Centers for Disease Control and Prevention | Ying Tao, Jing Zhang, Yan Li, Brian Lynch, Anna Kelleher, Krista Queen, Anna Uehara, Peter Cook, Han Jia Justin Ng, Clinton R. Paden, Haibin Wang, Suxiang Tong |
| EPI_ISL_1373740 | Washington University School of Medicine | Genomics and Discovery, Respiratory Viruses Branch, Division of Viral Diseases, Centers for Disease Control and Prevention | Ying Tao, Jing Zhang, Yan Li, Brian Lynch, Anna Kelleher, Krista Queen, Anna Uehara, Peter Cook, Han Jia Justin Ng, Clinton R. Paden, Haibin Wang, Suxiang Tong |
| EPI_ISL_1373741 | CDPH, Viral and Rickettsial Disease Laboratory | Genomics and Discovery, Respiratory Viruses Branch, Division of Viral Diseases, Centers for Disease Control and Prevention | Ying Tao, Jing Zhang, Yan Li, Brian Lynch, Anna Kelleher, Krista Queen, Anna Uehara, Peter Cook, Han Jia Justin Ng, Clinton R. Paden, Haibin Wang, Suxiang Tong |
| EPI_ISL_1373742 | Washington University School of Medicine | Genomics and Discovery, Respiratory Viruses Branch, Division of Viral Diseases, Centers for Disease Control and Prevention | Ying Tao, Jing Zhang, Yan Li, Brian Lynch, Anna Kelleher, Krista Queen, Anna Uehara, Peter Cook, Han Jia Justin Ng, Clinton R. Paden, Haibin Wang, Suxiang Tong |
| EPI_ISL_1373743 | UPMC Children's Hospital of Pittsburgh | Genomics and Discovery, Respiratory Viruses Branch, Division of Viral Diseases, Centers for Disease Control and Prevention | Ying Tao, Jing Zhang, Yan Li, Brian Lynch, Anna Kelleher, Krista Queen, Anna Uehara, Peter Cook, Han Jia Justin Ng, Clinton R. Paden, Haibin Wang, Suxiang Tong |
| EPI_ISL_1373744, EPI_ISL_1373745, EPI_ISL_1373746, EPI_ISL_1373747 | Washington University School of Medicine | Genomics and Discovery, Respiratory Viruses Branch, Division of Viral Diseases, Centers for Disease Control and Prevention | Ying Tao, Jing Zhang, Yan Li, Brian Lynch, Anna Kelleher, Krista Queen, Anna Uehara, Peter Cook, Han Jia Justin Ng, Clinton R. Paden, Haibin Wang, Suxiang Tong |
| EPI_ISL_1373748, EPI_ISL_1373749 | UPMC Children's Hospital of Pittsburgh | Genomics and Discovery, Respiratory Viruses Branch, Division of Viral Diseases, Centers for Disease Control and Prevention | Ying Tao, Jing Zhang, Yan Li, Brian Lynch, Anna Kelleher, Krista Queen, Anna Uehara, Peter Cook, Han Jia Justin Ng, Clinton R. Paden, Haibin Wang, Suxiang Tong |
| EPI_ISL_1373750 | Washington University School of Medicine | Genomics and Discovery, Respiratory Viruses Branch, Division of Viral Diseases, Centers for Disease Control and Prevention | Ying Tao, Jing Zhang, Yan Li, Brian Lynch, Anna Kelleher, Krista Queen, Anna Uehara, Peter Cook, Han Jia Justin Ng, Clinton R. Paden, Haibin Wang, Suxiang Tong |
| EPI_ISL_1373751 | CDPH, Viral and Rickettsial Disease Laboratory | Genomics and Discovery, Respiratory Viruses Branch, Division of Viral Diseases, Centers for Disease Control and Prevention | Ying Tao, Jing Zhang, Yan Li, Brian Lynch, Anna Kelleher, Krista Queen, Anna Uehara, Peter Cook, Han Jia Justin Ng, Clinton R. Paden, Haibin Wang, Suxiang Tong |
| EPI_ISL_1373752 | Boston Childrens (Overcoming COVID) | Genomics and Discovery, Respiratory Viruses Branch, Division of Viral Diseases, Centers for Disease Control and Prevention | Ying Tao, Jing Zhang, Yan Li, Brian Lynch, Anna Kelleher, Krista Queen, Anna Uehara, Peter Cook, Han Jia Justin Ng, Clinton R. Paden, Haibin Wang, Suxiang Tong |
| EPI_ISL_1373753 | UPMC Children's Hospital of Pittsburgh | Genomics and Discovery, Respiratory Viruses Branch, Division of Viral Diseases, Centers for Disease Control and Prevention | Ying Tao, Jing Zhang, Yan Li, Brian Lynch, Anna Kelleher, Krista Queen, Anna Uehara, Peter Cook, Han Jia Justin Ng, Clinton R. Paden, Haibin Wang, Suxiang Tong |
| EPI_ISL_1373754 | Boston Childrens (Overcoming COVID) | Genomics and Discovery, Respiratory Viruses Branch, Division of Viral Diseases, Centers for Disease Control and Prevention | Ying Tao, Jing Zhang, Yan Li, Brian Lynch, Anna Kelleher, Krista Queen, Anna Uehara, Peter Cook, Han Jia Justin Ng, Clinton R. Paden, Haibin Wang, Suxiang Tong |
| EPI_ISL_1373755 | UPMC Children's Hospital of Pittsburgh | Genomics and Discovery, Respiratory Viruses Branch, Division of Viral Diseases, Centers for Disease Control and Prevention | Ying Tao, Jing Zhang, Yan Li, Brian Lynch, Anna Kelleher, Krista Queen, Anna Uehara, Peter Cook, Han Jia Justin Ng, Clinton R. Paden, Haibin Wang, Suxiang Tong |
| EPI_ISL_1373756 | Springfield-Greene Co. Health Dept. | Genomics and Discovery, Respiratory Viruses Branch, Division of Viral Diseases, Centers for Disease Control and Prevention | Ying Tao, Jing Zhang, Yan Li, Brian Lynch, Anna Kelleher, Krista Queen, Anna Uehara, Peter Cook, Han Jia Justin Ng, Clinton R. Paden, Haibin Wang, Suxiang Tong |
| EPI_ISL_1373757 | UPMC Children's Hospital of Pittsburgh | Genomics and Discovery, Respiratory Viruses Branch, Division of Viral Diseases, Centers for Disease Control and Prevention | Ying Tao, Jing Zhang, Yan Li, Brian Lynch, Anna Kelleher, Krista Queen, Anna Uehara, Peter Cook, Han Jia Justin Ng, Clinton R. Paden, Haibin Wang, Suxiang Tong |
| EPI_ISL_1373758, EPI_ISL_1373759 | Boston Childrens (Overcoming COVID) | Genomics and Discovery, Respiratory Viruses Branch, Division of Viral Diseases, Centers for Disease Control and Prevention | Ying Tao, Jing Zhang, Yan Li, Brian Lynch, Anna Kelleher, Krista Queen, Anna Uehara, Peter Cook, Han Jia Justin Ng, Clinton R. Paden, Haibin Wang, Suxiang Tong |
| EPI_ISL_1373760, EPI_ISL_1373761, EPI_ISL_1373762, EPI_ISL_1373763 | Washington University School of Medicine | Genomics and Discovery, Respiratory Viruses Branch, Division of Viral Diseases, Centers for Disease Control and Prevention | Ying Tao, Jing Zhang, Yan Li, Brian Lynch, Anna Kelleher, Krista Queen, Anna Uehara, Peter Cook, Han Jia Justin Ng, Clinton R. Paden, Haibin Wang, Suxiang Tong |
| EPI_ISL_1373764 | UPMC Children's Hospital of Pittsburgh | Genomics and Discovery, Respiratory Viruses Branch, Division of Viral Diseases, Centers for Disease Control and Prevention | Ying Tao, Jing Zhang, Yan Li, Brian Lynch, Anna Kelleher, Krista Queen, Anna Uehara, Peter Cook, Han Jia Justin Ng, Clinton R. Paden, Haibin Wang, Suxiang Tong |
| EPI_ISL_1373765 | Washington University School of | Genomics and Discovery, Respiratory | Ying Tao, Jing Zhang, Yan Li, Brian Lynch, Anna Kelleher, Krista Queen, Anna Uehara, Peter Cook, Han Jia Justin Ng, Clinton R. Paden, Haibin Wang, Suxiang Tong |

| | | | |
|---|---|---|---|
| | Medicine | Viruses Branch, Division of Viral Diseases, Centers for Disease Control and Prevention | |
| EPI_ISL_1373766, EPI_ISL_1373767 | UPMC Children's Hospital of Pittsburgh | Genomics and Discovery, Respiratory Viruses Branch, Division of Viral Diseases, Centers for Disease Control and Prevention | Ying Tao, Jing Zhang, Yan Li, Brian Lynch, Anna Kelleher, Krista Queen, Anna Uehara, Peter Cook, Han Jia Justin Ng, Clinton R. Paden, Haibin Wang, Suxiang Tong |
| EPI_ISL_1373768 | Boston Childrens (Overcoming COVID) | Genomics and Discovery, Respiratory Viruses Branch, Division of Viral Diseases, Centers for Disease Control and Prevention | Ying Tao, Jing Zhang, Yan Li, Brian Lynch, Anna Kelleher, Krista Queen, Anna Uehara, Peter Cook, Han Jia Justin Ng, Clinton R. Paden, Haibin Wang, Suxiang Tong |
| EPI_ISL_1373769 | Washington University School of Medicine | Genomics and Discovery, Respiratory Viruses Branch, Division of Viral Diseases, Centers for Disease Control and Prevention | Ying Tao, Jing Zhang, Yan Li, Brian Lynch, Anna Kelleher, Krista Queen, Anna Uehara, Peter Cook, Han Jia Justin Ng, Clinton R. Paden, Haibin Wang, Suxiang Tong |
| EPI_ISL_1373770 | CDPH, Viral and Rickettsial Disease Laboratory | Genomics and Discovery, Respiratory Viruses Branch, Division of Viral Diseases, Centers for Disease Control and Prevention | Ying Tao, Jing Zhang, Yan Li, Brian Lynch, Anna Kelleher, Krista Queen, Anna Uehara, Peter Cook, Han Jia Justin Ng, Clinton R. Paden, Haibin Wang, Suxiang Tong |
| EPI_ISL_1373771 | Washington University School of Medicine | Genomics and Discovery, Respiratory Viruses Branch, Division of Viral Diseases, Centers for Disease Control and Prevention | Ying Tao, Jing Zhang, Yan Li, Brian Lynch, Anna Kelleher, Krista Queen, Anna Uehara, Peter Cook, Han Jia Justin Ng, Clinton R. Paden, Haibin Wang, Suxiang Tong |
| EPI_ISL_1373772 | Boston Childrens (Overcoming COVID) | Genomics and Discovery, Respiratory Viruses Branch, Division of Viral Diseases, Centers for Disease Control and Prevention | Ying Tao, Jing Zhang, Yan Li, Brian Lynch, Anna Kelleher, Krista Queen, Anna Uehara, Peter Cook, Han Jia Justin Ng, Clinton R. Paden, Haibin Wang, Suxiang Tong |
| EPI_ISL_1373773 | Washington University School of Medicine | Genomics and Discovery, Respiratory Viruses Branch, Division of Viral Diseases, Centers for Disease Control and Prevention | Ying Tao, Jing Zhang, Yan Li, Brian Lynch, Anna Kelleher, Krista Queen, Anna Uehara, Peter Cook, Han Jia Justin Ng, Clinton R. Paden, Haibin Wang, Suxiang Tong |
| EPI_ISL_1373774 | UPMC Children's Hospital of Pittsburgh | Genomics and Discovery, Respiratory Viruses Branch, Division of Viral Diseases, Centers for Disease Control and Prevention | Ying Tao, Jing Zhang, Yan Li, Brian Lynch, Anna Kelleher, Krista Queen, Anna Uehara, Peter Cook, Han Jia Justin Ng, Clinton R. Paden, Haibin Wang, Suxiang Tong |
| EPI_ISL_1373775 | Washington University School of Medicine | Genomics and Discovery, Respiratory Viruses Branch, Division of Viral Diseases, Centers for Disease Control and Prevention | Ying Tao, Jing Zhang, Yan Li, Brian Lynch, Anna Kelleher, Krista Queen, Anna Uehara, Peter Cook, Han Jia Justin Ng, Clinton R. Paden, Haibin Wang, Suxiang Tong |
| EPI_ISL_1373776 | CDPH, Viral and Rickettsial Disease Laboratory | Genomics and Discovery, Respiratory Viruses Branch, Division of Viral Diseases, Centers for Disease Control and Prevention | Ying Tao, Jing Zhang, Yan Li, Brian Lynch, Anna Kelleher, Krista Queen, Anna Uehara, Peter Cook, Han Jia Justin Ng, Clinton R. Paden, Haibin Wang, Suxiang Tong |
| EPI_ISL_1373777 | Boston Childrens (Overcoming COVID) | Genomics and Discovery, Respiratory Viruses Branch, Division of Viral Diseases, Centers for Disease Control and Prevention | Ying Tao, Jing Zhang, Yan Li, Brian Lynch, Anna Kelleher, Krista Queen, Anna Uehara, Peter Cook, Han Jia Justin Ng, Clinton R. Paden, Haibin Wang, Suxiang Tong |
| EPI_ISL_1373778 | CDPH, Viral and Rickettsial Disease Laboratory | Genomics and Discovery, Respiratory Viruses Branch, Division of Viral Diseases, Centers for Disease Control and Prevention | Ying Tao, Jing Zhang, Yan Li, Brian Lynch, Anna Kelleher, Krista Queen, Anna Uehara, Peter Cook, Han Jia Justin Ng, Clinton R. Paden, Haibin Wang, Suxiang Tong |
| EPI_ISL_1373779 | Washington University School of Medicine | Genomics and Discovery, Respiratory Viruses Branch, Division of Viral Diseases, Centers for Disease Control and Prevention | Ying Tao, Jing Zhang, Yan Li, Brian Lynch, Anna Kelleher, Krista Queen, Anna Uehara, Peter Cook, Han Jia Justin Ng, Clinton R. Paden, Haibin Wang, Suxiang Tong |
| EPI_ISL_1373780 | CDPH, Viral and Rickettsial Disease Laboratory | Genomics and Discovery, Respiratory Viruses Branch, Division of Viral Diseases, Centers for Disease Control and Prevention | Ying Tao, Jing Zhang, Yan Li, Brian Lynch, Anna Kelleher, Krista Queen, Anna Uehara, Peter Cook, Han Jia Justin Ng, Clinton R. Paden, Haibin Wang, Suxiang Tong |
| EPI_ISL_1373781, EPI_ISL_1373782, EPI_ISL_1373783, EPI_ISL_1373784 | Boston Childrens (Overcoming COVID) | Genomics and Discovery, Respiratory Viruses Branch, Division of Viral Diseases, Centers for Disease Control and Prevention | Ying Tao, Jing Zhang, Yan Li, Brian Lynch, Anna Kelleher, Krista Queen, Anna Uehara, Peter Cook, Han Jia Justin Ng, Clinton R. Paden, Haibin Wang, Suxiang Tong |
| EPI_ISL_1373785 | UPMC Children's Hospital of Pittsburgh | Genomics and Discovery, Respiratory Viruses Branch, Division of Viral Diseases, Centers for Disease Control and Prevention | Ying Tao, Jing Zhang, Yan Li, Brian Lynch, Anna Kelleher, Krista Queen, Anna Uehara, Peter Cook, Han Jia Justin Ng, Clinton R. Paden, Haibin Wang, Suxiang Tong |
| EPI_ISL_1373786, EPI_ISL_1373787 | CDPH, Viral and Rickettsial Disease Laboratory | Genomics and Discovery, Respiratory Viruses Branch, Division of Viral Diseases, Centers for Disease Control and Prevention | Ying Tao, Jing Zhang, Yan Li, Brian Lynch, Anna Kelleher, Krista Queen, Anna Uehara, Peter Cook, Han Jia Justin Ng, Clinton R. Paden, Haibin Wang, Suxiang Tong |
| EPI_ISL_1373788, EPI_ISL_1373789, EPI_ISL_1373790, EPI_ISL_1373791 | UPMC Children's Hospital of Pittsburgh | Genomics and Discovery, Respiratory Viruses Branch, Division of Viral Diseases, Centers for Disease Control and Prevention | Ying Tao, Jing Zhang, Yan Li, Brian Lynch, Anna Kelleher, Krista Queen, Anna Uehara, Peter Cook, Han Jia Justin Ng, Clinton R. Paden, Haibin Wang, Suxiang Tong |
| EPI_ISL_1373792 | Washington University School of Medicine | Genomics and Discovery, Respiratory Viruses Branch, Division of Viral Diseases, Centers for Disease Control and Prevention | Ying Tao, Jing Zhang, Yan Li, Brian Lynch, Anna Kelleher, Krista Queen, Anna Uehara, Peter Cook, Han Jia Justin Ng, Clinton R. Paden, Haibin Wang, Suxiang Tong |
| EPI_ISL_1373793 | CDPH, Viral and Rickettsial Disease Laboratory | Genomics and Discovery, Respiratory Viruses Branch, Division of Viral Diseases, Centers for Disease Control and Prevention | Ying Tao, Jing Zhang, Yan Li, Brian Lynch, Anna Kelleher, Krista Queen, Anna Uehara, Peter Cook, Han Jia Justin Ng, Clinton R. Paden, Haibin Wang, Suxiang Tong |
| EPI_ISL_1373794, EPI_ISL_1373795, EPI_ISL_1373796 | Washington University School of Medicine | Genomics and Discovery, Respiratory Viruses Branch, Division of Viral Diseases, Centers for Disease Control and Prevention | Ying Tao, Jing Zhang, Yan Li, Brian Lynch, Anna Kelleher, Krista Queen, Anna Uehara, Peter Cook, Han Jia Justin Ng, Clinton R. Paden, Haibin Wang, Suxiang Tong |
| EPI_ISL_1373797 | Boston Childrens (Overcoming COVID) | Genomics and Discovery, Respiratory Viruses Branch, Division of Viral Diseases, Centers for Disease Control and Prevention | Ying Tao, Jing Zhang, Yan Li, Brian Lynch, Anna Kelleher, Krista Queen, Anna Uehara, Peter Cook, Han Jia Justin Ng, Clinton R. Paden, Haibin Wang, Suxiang Tong |
| EPI_ISL_1373798 | Washington University School of Medicine | Genomics and Discovery, Respiratory Viruses Branch, Division of Viral Diseases, Centers for Disease Control and Prevention | Ying Tao, Jing Zhang, Yan Li, Brian Lynch, Anna Kelleher, Krista Queen, Anna Uehara, Peter Cook, Han Jia Justin Ng, Clinton R. Paden, Haibin Wang, Suxiang Tong |
| EPI_ISL_1373799 | Boston Childrens (Overcoming COVID) | Genomics and Discovery, Respiratory Viruses Branch, Division of Viral Diseases, Centers for Disease Control and Prevention | Ying Tao, Jing Zhang, Yan Li, Brian Lynch, Anna Kelleher, Krista Queen, Anna Uehara, Peter Cook, Han Jia Justin Ng, Clinton R. Paden, Haibin Wang, Suxiang Tong |
| EPI_ISL_1373800 | Washington University School of Medicine | Genomics and Discovery, Respiratory Viruses Branch, Division of Viral Diseases, Centers for Disease Control and Prevention | Ying Tao, Jing Zhang, Yan Li, Brian Lynch, Anna Kelleher, Krista Queen, Anna Uehara, Peter Cook, Han Jia Justin Ng, Clinton R. Paden, Haibin Wang, Suxiang Tong |
| EPI_ISL_1373801 | CDPH, Viral and Rickettsial Disease Laboratory | Genomics and Discovery, Respiratory Viruses Branch, Division of Viral Diseases, Centers for Disease Control and Prevention | Ying Tao, Jing Zhang, Yan Li, Brian Lynch, Anna Kelleher, Krista Queen, Anna Uehara, Peter Cook, Han Jia Justin Ng, Clinton R. Paden, Haibin Wang, Suxiang Tong |
| EPI_ISL_1373802 | UPMC Children's Hospital of Pittsburgh | Genomics and Discovery, Respiratory Viruses Branch, Division of Viral Diseases, Centers for Disease Control and Prevention | Ying Tao, Jing Zhang, Yan Li, Brian Lynch, Anna Kelleher, Krista Queen, Anna Uehara, Peter Cook, Han Jia Justin Ng, Clinton R. Paden, Haibin Wang, Suxiang Tong |
| EPI_ISL_1373803 | Washington University School of Medicine | Genomics and Discovery, Respiratory Viruses Branch, Division of Viral Diseases, Centers for Disease Control and Prevention | Ying Tao, Jing Zhang, Yan Li, Brian Lynch, Anna Kelleher, Krista Queen, Anna Uehara, Peter Cook, Han Jia Justin Ng, Clinton R. Paden, Haibin Wang, Suxiang Tong |
| EPI_ISL_1373804 | CDPH, Viral and Rickettsial Disease Laboratory | Genomics and Discovery, Respiratory Viruses Branch, Division of Viral Diseases, Centers for Disease Control and Prevention | Ying Tao, Jing Zhang, Yan Li, Brian Lynch, Anna Kelleher, Krista Queen, Anna Uehara, Peter Cook, Han Jia Justin Ng, Clinton R. Paden, Haibin Wang, Suxiang Tong |
| EPI_ISL_1373805, EPI_ISL_1373806, EPI_ISL_1373807, EPI_ISL_1373808 | UPMC Children's Hospital of Pittsburgh | Genomics and Discovery, Respiratory Viruses Branch, Division of Viral Diseases, Centers for Disease Control and Prevention | Ying Tao, Jing Zhang, Yan Li, Brian Lynch, Anna Kelleher, Krista Queen, Anna Uehara, Peter Cook, Han Jia Justin Ng, Clinton R. Paden, Haibin Wang, Suxiang Tong |
| EPI_ISL_1373809 | Washington University School of Medicine | Genomics and Discovery, Respiratory Viruses Branch, Division of Viral Diseases, | Ying Tao, Jing Zhang, Yan Li, Brian Lynch, Anna Kelleher, Krista Queen, Anna Uehara, Peter Cook, Han Jia Justin Ng, Clinton R. Paden, Haibin Wang, Suxiang Tong |

| | | | |
|---|---|---|---|
| | | Centers for Disease Control and Prevention | |
| EPI_ISL_1373810 | UPMC Children's Hospital of Pittsburgh | Genomics and Discovery, Respiratory Viruses Branch, Division of Viral Diseases, Centers for Disease Control and Prevention | Ying Tao, Jing Zhang, Yan Li, Brian Lynch, Anna Kelleher, Krista Queen, Anna Uehara, Peter Cook, Han Jia Justin Ng, Clinton R. Paden, Haibin Wang, Suxiang Tong |
| EPI_ISL_1373811 | GA Department of Public Health Laboratory | Genomics and Discovery, Respiratory Viruses Branch, Division of Viral Diseases, Centers for Disease Control and Prevention | Ying Tao, Jing Zhang, Yan Li, Brian Lynch, Anna Kelleher, Krista Queen, Anna Uehara, Peter Cook, Han Jia Justin Ng, Clinton R. Paden, Haibin Wang, Suxiang Tong |
| EPI_ISL_1373812 | Boston Childrens (Overcoming COVID) | Genomics and Discovery, Respiratory Viruses Branch, Division of Viral Diseases, Centers for Disease Control and Prevention | Ying Tao, Jing Zhang, Yan Li, Brian Lynch, Anna Kelleher, Krista Queen, Anna Uehara, Peter Cook, Han Jia Justin Ng, Clinton R. Paden, Haibin Wang, Suxiang Tong |
| EPI_ISL_1373813, EPI_ISL_1373814 | CDPH, Viral and Rickettsial Disease Laboratory | Genomics and Discovery, Respiratory Viruses Branch, Division of Viral Diseases, Centers for Disease Control and Prevention | Ying Tao, Jing Zhang, Yan Li, Brian Lynch, Anna Kelleher, Krista Queen, Anna Uehara, Peter Cook, Han Jia Justin Ng, Clinton R. Paden, Haibin Wang, Suxiang Tong |
| EPI_ISL_1373815 | Washington University School of Medicine | Genomics and Discovery, Respiratory Viruses Branch, Division of Viral Diseases, Centers for Disease Control and Prevention | Ying Tao, Jing Zhang, Yan Li, Brian Lynch, Anna Kelleher, Krista Queen, Anna Uehara, Peter Cook, Han Jia Justin Ng, Clinton R. Paden, Haibin Wang, Suxiang Tong |
| EPI_ISL_1373816 | CDPH, Viral and Rickettsial Disease Laboratory | Genomics and Discovery, Respiratory Viruses Branch, Division of Viral Diseases, Centers for Disease Control and Prevention | Ying Tao, Jing Zhang, Yan Li, Brian Lynch, Anna Kelleher, Krista Queen, Anna Uehara, Peter Cook, Han Jia Justin Ng, Clinton R. Paden, Haibin Wang, Suxiang Tong |
| EPI_ISL_1373817, EPI_ISL_1373818 | UPMC Children's Hospital of Pittsburgh | Genomics and Discovery, Respiratory Viruses Branch, Division of Viral Diseases, Centers for Disease Control and Prevention | Ying Tao, Jing Zhang, Yan Li, Brian Lynch, Anna Kelleher, Krista Queen, Anna Uehara, Peter Cook, Han Jia Justin Ng, Clinton R. Paden, Haibin Wang, Suxiang Tong |
| EPI_ISL_1373819, EPI_ISL_1373820 | Washington University School of Medicine | Genomics and Discovery, Respiratory Viruses Branch, Division of Viral Diseases, Centers for Disease Control and Prevention | Ying Tao, Jing Zhang, Yan Li, Brian Lynch, Anna Kelleher, Krista Queen, Anna Uehara, Peter Cook, Han Jia Justin Ng, Clinton R. Paden, Haibin Wang, Suxiang Tong |
| EPI_ISL_1373821 | Boston Childrens (Overcoming COVID) | Genomics and Discovery, Respiratory Viruses Branch, Division of Viral Diseases, Centers for Disease Control and Prevention | Ying Tao, Jing Zhang, Yan Li, Brian Lynch, Anna Kelleher, Krista Queen, Anna Uehara, Peter Cook, Han Jia Justin Ng, Clinton R. Paden, Haibin Wang, Suxiang Tong |
| EPI_ISL_1373822, EPI_ISL_1373823, EPI_ISL_1373824, EPI_ISL_1373825, EPI_ISL_1373826 | Washington University School of Medicine | Genomics and Discovery, Respiratory Viruses Branch, Division of Viral Diseases, Centers for Disease Control and Prevention | Ying Tao, Jing Zhang, Yan Li, Brian Lynch, Anna Kelleher, Krista Queen, Anna Uehara, Peter Cook, Han Jia Justin Ng, Clinton R. Paden, Haibin Wang, Suxiang Tong |
| EPI_ISL_1373827 | UPMC Children's Hospital of Pittsburgh | Genomics and Discovery, Respiratory Viruses Branch, Division of Viral Diseases, Centers for Disease Control and Prevention | Ying Tao, Jing Zhang, Yan Li, Brian Lynch, Anna Kelleher, Krista Queen, Anna Uehara, Peter Cook, Han Jia Justin Ng, Clinton R. Paden, Haibin Wang, Suxiang Tong |
| EPI_ISL_1373828 | CDPH, Viral and Rickettsial Disease Laboratory | Genomics and Discovery, Respiratory Viruses Branch, Division of Viral Diseases, Centers for Disease Control and Prevention | Ying Tao, Jing Zhang, Yan Li, Brian Lynch, Anna Kelleher, Krista Queen, Anna Uehara, Peter Cook, Han Jia Justin Ng, Clinton R. Paden, Haibin Wang, Suxiang Tong |
| EPI_ISL_1373829, EPI_ISL_1373830, EPI_ISL_1373831 | Washington University School of Medicine | Genomics and Discovery, Respiratory Viruses Branch, Division of Viral Diseases, Centers for Disease Control and Prevention | Ying Tao, Jing Zhang, Yan Li, Brian Lynch, Anna Kelleher, Krista Queen, Anna Uehara, Peter Cook, Han Jia Justin Ng, Clinton R. Paden, Haibin Wang, Suxiang Tong |
| EPI_ISL_1373832 | UPMC Children's Hospital of Pittsburgh | Genomics and Discovery, Respiratory Viruses Branch, Division of Viral Diseases, Centers for Disease Control and Prevention | Ying Tao, Jing Zhang, Yan Li, Brian Lynch, Anna Kelleher, Krista Queen, Anna Uehara, Peter Cook, Han Jia Justin Ng, Clinton R. Paden, Haibin Wang, Suxiang Tong |
| EPI_ISL_1373833, EPI_ISL_1373834, EPI_ISL_1373835 | Washington University School of Medicine | Genomics and Discovery, Respiratory Viruses Branch, Division of Viral Diseases, Centers for Disease Control and Prevention | Ying Tao, Jing Zhang, Yan Li, Brian Lynch, Anna Kelleher, Krista Queen, Anna Uehara, Peter Cook, Han Jia Justin Ng, Clinton R. Paden, Haibin Wang, Suxiang Tong |
| EPI_ISL_1373836 | UPMC Children's Hospital of Pittsburgh | Genomics and Discovery, Respiratory Viruses Branch, Division of Viral Diseases, Centers for Disease Control and Prevention | Ying Tao, Jing Zhang, Yan Li, Brian Lynch, Anna Kelleher, Krista Queen, Anna Uehara, Peter Cook, Han Jia Justin Ng, Clinton R. Paden, Haibin Wang, Suxiang Tong |
| EPI_ISL_1373837 | CDPH, Viral and Rickettsial Disease Laboratory | Genomics and Discovery, Respiratory Viruses Branch, Division of Viral Diseases, Centers for Disease Control and Prevention | Ying Tao, Jing Zhang, Yan Li, Brian Lynch, Anna Kelleher, Krista Queen, Anna Uehara, Peter Cook, Han Jia Justin Ng, Clinton R. Paden, Haibin Wang, Suxiang Tong |
| EPI_ISL_1373838 | UPMC Children's Hospital of Pittsburgh | Genomics and Discovery, Respiratory Viruses Branch, Division of Viral Diseases, Centers for Disease Control and Prevention | Ying Tao, Jing Zhang, Yan Li, Brian Lynch, Anna Kelleher, Krista Queen, Anna Uehara, Peter Cook, Han Jia Justin Ng, Clinton R. Paden, Haibin Wang, Suxiang Tong |
| EPI_ISL_1373839, EPI_ISL_1373840, EPI_ISL_1373841, EPI_ISL_1373842, EPI_ISL_1373843, EPI_ISL_1373844, EPI_ISL_1373845, EPI_ISL_1373846 | CDPH, Viral and Rickettsial Disease Laboratory | Genomics and Discovery, Respiratory Viruses Branch, Division of Viral Diseases, Centers for Disease Control and Prevention | Ying Tao, Jing Zhang, Yan Li, Brian Lynch, Anna Kelleher, Krista Queen, Anna Uehara, Peter Cook, Han Jia Justin Ng, Clinton R. Paden, Haibin Wang, Suxiang Tong |
| EPI_ISL_1373847 | Boston Childrens (Overcoming COVID) | Genomics and Discovery, Respiratory Viruses Branch, Division of Viral Diseases, Centers for Disease Control and Prevention | Ying Tao, Jing Zhang, Yan Li, Brian Lynch, Anna Kelleher, Krista Queen, Anna Uehara, Peter Cook, Han Jia Justin Ng, Clinton R. Paden, Haibin Wang, Suxiang Tong |
| EPI_ISL_1373848, EPI_ISL_1373849 | CDPH, Viral and Rickettsial Disease Laboratory | Genomics and Discovery, Respiratory Viruses Branch, Division of Viral Diseases, Centers for Disease Control and Prevention | Ying Tao, Jing Zhang, Yan Li, Brian Lynch, Anna Kelleher, Krista Queen, Anna Uehara, Peter Cook, Han Jia Justin Ng, Clinton R. Paden, Haibin Wang, Suxiang Tong |
| EPI_ISL_1373850 | Boston Childrens (Overcoming COVID) | Genomics and Discovery, Respiratory Viruses Branch, Division of Viral Diseases, Centers for Disease Control and Prevention | Ying Tao, Jing Zhang, Yan Li, Brian Lynch, Anna Kelleher, Krista Queen, Anna Uehara, Peter Cook, Han Jia Justin Ng, Clinton R. Paden, Haibin Wang, Suxiang Tong |
| EPI_ISL_1373851, EPI_ISL_1373852 | CDPH, Viral and Rickettsial Disease Laboratory | Genomics and Discovery, Respiratory Viruses Branch, Division of Viral Diseases, Centers for Disease Control and Prevention | Ying Tao, Jing Zhang, Yan Li, Brian Lynch, Anna Kelleher, Krista Queen, Anna Uehara, Peter Cook, Han Jia Justin Ng, Clinton R. Paden, Haibin Wang, Suxiang Tong |
| EPI_ISL_1373853, EPI_ISL_1373854 | Boston Childrens (Overcoming COVID) | Genomics and Discovery, Respiratory Viruses Branch, Division of Viral Diseases, Centers for Disease Control and Prevention | Ying Tao, Jing Zhang, Yan Li, Brian Lynch, Anna Kelleher, Krista Queen, Anna Uehara, Peter Cook, Han Jia Justin Ng, Clinton R. Paden, Haibin Wang, Suxiang Tong |
| EPI_ISL_1373855, EPI_ISL_1373856, EPI_ISL_1373857 | CDPH, Viral and Rickettsial Disease Laboratory | Genomics and Discovery, Respiratory Viruses Branch, Division of Viral Diseases, Centers for Disease Control and Prevention | Ying Tao, Jing Zhang, Yan Li, Brian Lynch, Anna Kelleher, Krista Queen, Anna Uehara, Peter Cook, Han Jia Justin Ng, Clinton R. Paden, Haibin Wang, Suxiang Tong |
| EPI_ISL_1373858 | Boston Childrens (Overcoming COVID) | Genomics and Discovery, Respiratory Viruses Branch, Division of Viral Diseases, Centers for Disease Control and Prevention | Ying Tao, Jing Zhang, Yan Li, Brian Lynch, Anna Kelleher, Krista Queen, Anna Uehara, Peter Cook, Han Jia Justin Ng, Clinton R. Paden, Haibin Wang, Suxiang Tong |
| EPI_ISL_1373859, EPI_ISL_1373860, EPI_ISL_1373861 | CDPH, Viral and Rickettsial Disease Laboratory | Genomics and Discovery, Respiratory Viruses Branch, Division of Viral Diseases, Centers for Disease Control and Prevention | Ying Tao, Jing Zhang, Yan Li, Brian Lynch, Anna Kelleher, Krista Queen, Anna Uehara, Peter Cook, Han Jia Justin Ng, Clinton R. Paden, Haibin Wang, Suxiang Tong |
| EPI_ISL_1373862, EPI_ISL_1373863 | Boston Childrens (Overcoming COVID) | Genomics and Discovery, Respiratory Viruses Branch, Division of Viral Diseases, Centers for Disease Control and Prevention | Ying Tao, Jing Zhang, Yan Li, Brian Lynch, Anna Kelleher, Krista Queen, Anna Uehara, Peter Cook, Han Jia Justin Ng, Clinton R. Paden, Haibin Wang, Suxiang Tong |
| EPI_ISL_1373864 | CDPH, Viral and Rickettsial Disease Laboratory | Genomics and Discovery, Respiratory Viruses Branch, Division of Viral Diseases, Centers for Disease Control and Prevention | Ying Tao, Jing Zhang, Yan Li, Brian Lynch, Anna Kelleher, Krista Queen, Anna Uehara, Peter Cook, Han Jia Justin Ng, Clinton R. Paden, Haibin Wang, Suxiang Tong |
| EPI_ISL_1373865 | Boston Childrens (Overcoming COVID) | Genomics and Discovery, Respiratory Viruses Branch, Division of Viral Diseases, | Ying Tao, Jing Zhang, Yan Li, Brian Lynch, Anna Kelleher, Krista Queen, Anna Uehara, Peter Cook, Han Jia Justin Ng, Clinton R. Paden, Haibin Wang, Suxiang Tong |

| | | | |
|---|---|---|---|
| | | Centers for Disease Control and Prevention | |
| EPI_ISL_1373866, EPI_ISL_1373867, EPI_ISL_1373868, EPI_ISL_1373869, EPI_ISL_1373870, EPI_ISL_1373871 | CDPH, Viral and Rickettsial Disease Laboratory | Genomics and Discovery, Respiratory Viruses Branch, Division of Viral Diseases, Centers for Disease Control and Prevention | Ying Tao, Jing Zhang, Yan Li, Brian Lynch, Anna Kelleher, Krista Queen, Anna Uehara, Peter Cook, Han Jia Justin Ng, Clinton R. Paden, Haibin Wang, Suxiang Tong |
| EPI_ISL_1373872 | Boston Childrens (Overcoming COVID) | Genomics and Discovery, Respiratory Viruses Branch, Division of Viral Diseases, Centers for Disease Control and Prevention | Ying Tao, Jing Zhang, Yan Li, Brian Lynch, Anna Kelleher, Krista Queen, Anna Uehara, Peter Cook, Han Jia Justin Ng, Clinton R. Paden, Haibin Wang, Suxiang Tong |
| EPI_ISL_1373873, EPI_ISL_1373874, EPI_ISL_1373875, EPI_ISL_1373876 | CDPH, Viral and Rickettsial Disease Laboratory | Genomics and Discovery, Respiratory Viruses Branch, Division of Viral Diseases, Centers for Disease Control and Prevention | Ying Tao, Jing Zhang, Yan Li, Brian Lynch, Anna Kelleher, Krista Queen, Anna Uehara, Peter Cook, Han Jia Justin Ng, Clinton R. Paden, Haibin Wang, Suxiang Tong |
| EPI_ISL_1373877 | Boston Childrens (Overcoming COVID) | Genomics and Discovery, Respiratory Viruses Branch, Division of Viral Diseases, Centers for Disease Control and Prevention | Ying Tao, Jing Zhang, Yan Li, Brian Lynch, Anna Kelleher, Krista Queen, Anna Uehara, Peter Cook, Han Jia Justin Ng, Clinton R. Paden, Haibin Wang, Suxiang Tong |
| EPI_ISL_1373878 | CDPH, Viral and Rickettsial Disease Laboratory | Genomics and Discovery, Respiratory Viruses Branch, Division of Viral Diseases, Centers for Disease Control and Prevention | Ying Tao, Jing Zhang, Yan Li, Brian Lynch, Anna Kelleher, Krista Queen, Anna Uehara, Peter Cook, Han Jia Justin Ng, Clinton R. Paden, Haibin Wang, Suxiang Tong |
| EPI_ISL_1373879 | PA Department of Health, Bureau of Laboratories | Pathogen Discovery, Respiratory Viruses Branch, Division of Viral Diseases, Centers for Disease Control and Prevention | Ying Tao, Jing Zhang, Yan Li, Brian Lynch, Anna Kelleher, Krista Queen, Anna Uehara, Peter Cook, Han Jia Justin Ng, Clinton R. Paden, Haibin Wang, Suxiang Tong |
| EPI_ISL_1373880, EPI_ISL_1373881 | PA Dept of Heath, Bureau of Laboratories | Pathogen Discovery, Respiratory Viruses Branch, Division of Viral Diseases, Centers for Disease Control and Prevention | Yan Li, Jing Zhang, Ying Tao, Brian Lynch, Anna Kelleher, Krista Queen, Anna Uehara, Peter Cook, Han Jia Justin Ng, Clinton R. Paden, Haibin Wang, Suxiang Tong |
| EPI_ISL_1373882, EPI_ISL_1373883, EPI_ISL_1373884, EPI_ISL_1373885, EPI_ISL_1373886, EPI_ISL_1373887, EPI_ISL_1373888, EPI_ISL_1373889, EPI_ISL_1373890, EPI_ISL_1373891, EPI_ISL_1373892, EPI_ISL_1373893, EPI_ISL_1373894, EPI_ISL_1373895, EPI_ISL_1373896, EPI_ISL_1373897, EPI_ISL_1373898, EPI_ISL_1373899, EPI_ISL_1373900, EPI_ISL_1373901, EPI_ISL_1373902, EPI_ISL_1373903, EPI_ISL_1373904, EPI_ISL_1373905, EPI_ISL_1373906, EPI_ISL_1373907, EPI_ISL_1373908, EPI_ISL_1373909, EPI_ISL_1373910, EPI_ISL_1373911, EPI_ISL_1373912, EPI_ISL_1373913, EPI_ISL_1373914, EPI_ISL_1373915, EPI_ISL_1373916, EPI_ISL_1373917, EPI_ISL_1373918, EPI_ISL_1373919, EPI_ISL_1373920, EPI_ISL_1373921, EPI_ISL_1373922, EPI_ISL_1373923, EPI_ISL_1373924, EPI_ISL_1373925, EPI_ISL_1373926, EPI_ISL_1373927, EPI_ISL_1373928, EPI_ISL_1373929, EPI_ISL_1373930, EPI_ISL_1373931, EPI_ISL_1373932, EPI_ISL_1373933, EPI_ISL_1373934, EPI_ISL_1373935, EPI_ISL_1373936, EPI_ISL_1373937, EPI_ISL_1373938, EPI_ISL_1373939, EPI_ISL_1373940, EPI_ISL_1373941, EPI_ISL_1373942, EPI_ISL_1373943, EPI_ISL_1373944, EPI_ISL_1373945, EPI_ISL_1373946, EPI_ISL_1373947, EPI_ISL_1373948, EPI_ISL_1373949, EPI_ISL_1373950, EPI_ISL_1373951, EPI_ISL_1373952, EPI_ISL_1373953, EPI_ISL_1373954, EPI_ISL_1373955, EPI_ISL_1373956, EPI_ISL_1373957, EPI_ISL_1373958, EPI_ISL_1373959, EPI_ISL_1373960, EPI_ISL_1373961, EPI_ISL_1373962, EPI_ISL_1373963, EPI_ISL_1373964, EPI_ISL_1373965, EPI_ISL_1373966, EPI_ISL_1373967, EPI_ISL_1373968, EPI_ISL_1373969, EPI_ISL_1373970, EPI_ISL_1373971, EPI_ISL_1373972, EPI_ISL_1373973, EPI_ISL_1373974, EPI_ISL_1373975, EPI_ISL_1373976, EPI_ISL_1373977, EPI_ISL_1373978, EPI_ISL_1373979, EPI_ISL_1373980, EPI_ISL_1373981, EPI_ISL_1373982, EPI_ISL_1373983, EPI_ISL_1373984, EPI_ISL_1373985, EPI_ISL_1373986, EPI_ISL_1373987, EPI_ISL_1373988, EPI_ISL_1373989, EPI_ISL_1373990, EPI_ISL_1373991, EPI_ISL_1373992, EPI_ISL_1373993, EPI_ISL_1373994 | | | |
| see above | Vanderbilt University Medical Center | Pathogen Discovery, Respiratory Viruses Branch, Division of Viral Diseases, Centers for Disease Control and Prevention | Jing Zhang, Yan Li, Ying Tao, Brian Lynch, Anna Kelleher, Krista Queen, Anna Uehara, Peter Cook, Han Jia Justin Ng, Clinton R. Paden, Haibin Wang, Suxiang Tong |
| EPI_ISL_1373995, EPI_ISL_1373996, EPI_ISL_1373997, EPI_ISL_1373998, EPI_ISL_1373999, EPI_ISL_1374000, EPI_ISL_1374001, EPI_ISL_1374002, EPI_ISL_1374003, EPI_ISL_1374004, EPI_ISL_1374005, EPI_ISL_1374006, EPI_ISL_1374007, EPI_ISL_1374008, EPI_ISL_1374009, EPI_ISL_1374010, EPI_ISL_1374011, EPI_ISL_1374012, EPI_ISL_1374014, EPI_ISL_1374015, EPI_ISL_1374016 | | | |
| see above | MO State Public Health Laboratory | Pathogen Discovery, Respiratory Viruses Branch, Division of Viral Diseases, Centers for Disease Control and Prevention | Yan Li, Jing Zhang, Ying Tao, Brian Lynch, Anna Kelleher, Krista Queen, Anna Uehara, Peter Cook, Han Jia Justin Ng, Clinton R. Paden, Haibin Wang, Suxiang Tong |
| EPI_ISL_1374017, EPI_ISL_1374018, EPI_ISL_1374019, EPI_ISL_1374020, EPI_ISL_1374021, EPI_ISL_1374022, EPI_ISL_1374023, EPI_ISL_1374024, EPI_ISL_1374025, EPI_ISL_1374026, EPI_ISL_1374027, EPI_ISL_1374028, EPI_ISL_1374029, EPI_ISL_1374030, EPI_ISL_1374031, EPI_ISL_1374032, EPI_ISL_1374033, EPI_ISL_1374034, EPI_ISL_1374035, EPI_ISL_1374036, EPI_ISL_1374037, EPI_ISL_1374038, EPI_ISL_1374039, EPI_ISL_1374040, EPI_ISL_1374041, EPI_ISL_1374042, EPI_ISL_1374043, EPI_ISL_1374044, EPI_ISL_1374045, EPI_ISL_1374046, EPI_ISL_1374047, EPI_ISL_1374048, EPI_ISL_1374049, EPI_ISL_1374050 | | | |
| see above | Springfield-Greene Co. Health Dept. | Pathogen Discovery, Respiratory Viruses Branch, Division of Viral Diseases, Centers for Disease Control and Prevention | Jing Zhang, Yan Li, Ying Tao, Brian Lynch, Anna Kelleher, Krista Queen, Anna Uehara, Peter Cook, Han Jia Justin Ng, Clinton R. Paden, Haibin Wang, Suxiang Tong |
| EPI_ISL_1378643, EPI_ISL_1378645, EPI_ISL_1378647, EPI_ISL_1378648, EPI_ISL_1378649, EPI_ISL_1378650, EPI_ISL_1378651, EPI_ISL_1378652, EPI_ISL_1378653, EPI_ISL_1378654, EPI_ISL_1378656, EPI_ISL_1378657, EPI_ISL_1378658, EPI_ISL_1378660, EPI_ISL_1378661, EPI_ISL_1378662, EPI_ISL_1378663, EPI_ISL_1378664, EPI_ISL_1378665, EPI_ISL_1378668, EPI_ISL_1378669, EPI_ISL_1378670, EPI_ISL_1378671 | | | |
| see above | Nevada State Public Health Laboratory | Nevada State Public Health Laboratory | Andrew Gorzalski, Mark Pandori |
| EPI_ISL_1378673, EPI_ISL_1378674, EPI_ISL_1378675, EPI_ISL_1378676, EPI_ISL_1378677, EPI_ISL_1378678, EPI_ISL_1378679, EPI_ISL_1378680, EPI_ISL_1378681, EPI_ISL_1378683, EPI_ISL_1378684, EPI_ISL_1378685, EPI_ISL_1378686, EPI_ISL_1378687, EPI_ISL_1378688, EPI_ISL_1378689, EPI_ISL_1378690, EPI_ISL_1378691, EPI_ISL_1378692, EPI_ISL_1378693 | | | |
| see above | Yale Clinical Virology Lab | Grubaugh Lab - Yale School of Public Health | Joseph Fauver, Mallery Breban, Isabell Ott, Tara Alpert, Mary Petrone, Anderson Brito, Chantal Vogels, Annie Watkins, Chaney Kalinich, Jessica Rothman, Marie L. Landry, Nathan Grubaugh |
| EPI_ISL_1378694, EPI_ISL_1378695, EPI_ISL_1378696 | Tempus | Grubaugh Lab - Yale School of Public Health | Joseph Fauver, Mallery Breban, Isabell Ott, Tara Alpert, Mary Petrone, Anderson Brito, Chantal Vogels, Annie Watkins, Chaney Kalinich, Jessica Rothman, Matthew J. MacKay, Gaurav Khullar, Jessica Metti, Joel T. Dudley, Megan Nash, Nike Beaubier, Christopher E. Mason, Nathan Grubaugh |
| EPI_ISL_1378697, EPI_ISL_1378698, EPI_ISL_1378699, EPI_ISL_1378700, EPI_ISL_1378701, EPI_ISL_1378702, EPI_ISL_1378703, EPI_ISL_1378704, EPI_ISL_1378705, EPI_ISL_1378706, EPI_ISL_1378708, EPI_ISL_1378709, EPI_ISL_1378710, EPI_ISL_1378711, EPI_ISL_1378712, EPI_ISL_1378714, EPI_ISL_1378715, EPI_ISL_1378716, EPI_ISL_1378717, EPI_ISL_1378718, EPI_ISL_1378719, EPI_ISL_1378720, EPI_ISL_1378721, EPI_ISL_1378722, EPI_ISL_1378723, EPI_ISL_1378724, EPI_ISL_1378725 | | | |
| see above | Yale Clinical Virology Lab | Grubaugh Lab - Yale School of Public Health | Joseph Fauver, Mallery Breban, Isabell Ott, Tara Alpert, Mary Petrone, Anderson Brito, Chantal Vogels, Annie Watkins, Chaney Kalinich, Jessica Rothman, Marie L. Landry, Nathan Grubaugh |
| EPI_ISL_1378726 | Grubaugh Lab - Yale School of Public Health | Grubaugh Lab - Yale School of Public Health | Joseph Fauver, Mallery Breban, Isabell Ott, Tara Alpert, Mary Petrone, Anderson Brito, Chantal Vogels, Annie Watkins, Chaney Kalinich, Jessica Rothman, Nathan Grubaugh |
| EPI_ISL_1378727, EPI_ISL_1378728, EPI_ISL_1378729, EPI_ISL_1378730, EPI_ISL_1378731, EPI_ISL_1378733, EPI_ISL_1378734, EPI_ISL_1378735, EPI_ISL_1378736, EPI_ISL_1378737, EPI_ISL_1378738, EPI_ISL_1378739, EPI_ISL_1378740, EPI_ISL_1378741, EPI_ISL_1378742, EPI_ISL_1378743, EPI_ISL_1378744, EPI_ISL_1378746, EPI_ISL_1378747, EPI_ISL_1378748, EPI_ISL_1378749, EPI_ISL_1378750, EPI_ISL_1378751, EPI_ISL_1378752 | | | |
| see above | Yale Clinical Virology Lab | Grubaugh Lab - Yale School of Public Health | Joseph Fauver, Mallery Breban, Isabell Ott, Tara Alpert, Mary Petrone, Anderson Brito, Chantal Vogels, Annie Watkins, Chaney Kalinich, Jessica Rothman, Marie L. Landry, Nathan Grubaugh |
| EPI_ISL_1378753 | Grubaugh Lab - Yale School of Public Health | Grubaugh Lab - Yale School of Public Health | Joseph Fauver, Mallery Breban, Isabell Ott, Tara Alpert, Mary Petrone, Anderson Brito, Chantal Vogels, Annie Watkins, Chaney Kalinich, Jessica Rothman, Nathan Grubaugh |
| EPI_ISL_1378759, EPI_ISL_1378761, EPI_ISL_1378762, EPI_ISL_1378763, EPI_ISL_1378764 | Yale Clinical Virology Lab | Grubaugh Lab - Yale School of Public Health | Joseph Fauver, Mallery Breban, Isabell Ott, Tara Alpert, Mary Petrone, Anderson Brito, Chantal Vogels, Annie Watkins, Chaney Kalinich, Jessica Rothman, Marie L. Landry, Nathan Grubaugh |
| EPI_ISL_1378765, EPI_ISL_1378766, EPI_ISL_1378767, EPI_ISL_1378768, EPI_ISL_1378769, EPI_ISL_1378770, EPI_ISL_1378771, EPI_ISL_1378772, EPI_ISL_1378773, EPI_ISL_1378774, EPI_ISL_1378775, EPI_ISL_1378776, EPI_ISL_1378777, EPI_ISL_1378778, EPI_ISL_1378779, EPI_ISL_1378780, EPI_ISL_1378781, EPI_ISL_1378782, EPI_ISL_1378783, EPI_ISL_1378784, EPI_ISL_1378785, EPI_ISL_1378786, EPI_ISL_1378787, EPI_ISL_1378788, EPI_ISL_1378789, EPI_ISL_1378790, EPI_ISL_1378791, EPI_ISL_1378792, EPI_ISL_1378793, EPI_ISL_1378794, EPI_ISL_1378795, EPI_ISL_1378796, EPI_ISL_1378797, EPI_ISL_1378798, EPI_ISL_1378799, EPI_ISL_1378800, EPI_ISL_1378801, EPI_ISL_1378802, EPI_ISL_1378803, EPI_ISL_1378804, EPI_ISL_1378805, EPI_ISL_1378806, EPI_ISL_1378807, EPI_ISL_1378808, EPI_ISL_1378809, EPI_ISL_1378810, EPI_ISL_1378811, EPI_ISL_1378812 | | | |
| see above | Tempus | Grubaugh Lab - Yale School of Public Health | Joseph Fauver, Mallery Breban, Isabell Ott, Tara Alpert, Mary Petrone, Anderson Brito, Chantal Vogels, Annie Watkins, Chaney Kalinich, Jessica Rothman, Matthew J. MacKay, Gaurav Khullar, Jessica Metti, Joel T. Dudley, Megan Nash, Nike Beaubier, Christopher E. Mason, Nathan Grubaugh |
| EPI_ISL_1378813 | Oak Ridge | Grubaugh Lab - Yale School of Public Health | Joseph Fauver, Mallery Breban, Isabell Ott, Tara Alpert, Mary Petrone, Anderson Brito, Chantal Vogels, Annie Watkins, Chaney Kalinich, Jessica Rothman, Daniel Jones, Nathan Grubaugh |
| EPI_ISL_1378814, EPI_ISL_1378815, EPI_ISL_1378816, EPI_ISL_1378818, EPI_ISL_1378819, EPI_ISL_1378820, EPI_ISL_1378821 | Yale Pathology Lab | Grubaugh Lab - Yale School of Public Health | Joseph Fauver, Mallery Breban, Isabell Ott, Tara Alpert, Mary Petrone, Anderson Brito, Chantal Vogels, Annie Watkins, Chaney Kalinich, Jessica Rothman, Jianhui Wang, Chen Liu, Pei Hui, Nathan Grubaugh |
| EPI_ISL_1378822, EPI_ISL_1378823, EPI_ISL_1378824, EPI_ISL_1378825, EPI_ISL_1378826, EPI_ISL_1378827, EPI_ISL_1378829 | Murphy Medical Association | Grubaugh Lab - Yale School of Public Health | Joseph Fauver, Mallery Breban, Isabell Ott, Tara Alpert, Mary Petrone, Anderson Brito, Chantal Vogels, Annie Watkins, Chaney Kalinich, Jessica Rothman, Caleb Neal, Eva Laszlo, Steven Murphy, Nathan Grubaugh |
| EPI_ISL_1378831, EPI_ISL_1378832 | Grubaugh Lab - Yale School of Public Health | Grubaugh Lab - Yale School of Public Health | Joseph Fauver, Mallery Breban, Isabell Ott, Tara Alpert, Mary Petrone, Anderson Brito, Chantal Vogels, Annie Watkins, Chaney Kalinich, Jessica Rothman, Nathan Grubaugh |
| EPI_ISL_1378917, EPI_ISL_1378936 | Wyoming Public Health Laboratory | Wyoming Public Health Laboratory | Jim Mildenberger, Wanda Manley, Noah Hull, Taylor Fearing, Lynette Gumbleton, Channing Weber, Ashley Norberg, Chayse Rowley, Marley Goetz, Brian Dominguez, Elliot Thomasson, Cari Sloma, and Rob Christensen |
| EPI_ISL_1379198, EPI_ISL_1379199, EPI_ISL_1379200, EPI_ISL_1379201, EPI_ISL_1379202, EPI_ISL_1379203, EPI_ISL_1379204, EPI_ISL_1379205, EPI_ISL_1379206, EPI_ISL_1379207, EPI_ISL_1379208, EPI_ISL_1379209, EPI_ISL_1379210, EPI_ISL_1379211, EPI_ISL_1379212, EPI_ISL_1379213, EPI_ISL_1379214, EPI_ISL_1379215, EPI_ISL_1379216, EPI_ISL_1379217, EPI_ISL_1379218, EPI_ISL_1379219, EPI_ISL_1379220, EPI_ISL_1379221, EPI_ISL_1379222, EPI_ISL_1379223, EPI_ISL_1379224, EPI_ISL_1379225, EPI_ISL_1379226, EPI_ISL_1379227, EPI_ISL_1379228, EPI_ISL_1379229, EPI_ISL_1379230, EPI_ISL_1379231, EPI_ISL_1379232, EPI_ISL_1379233, EPI_ISL_1379234, EPI_ISL_1379235, EPI_ISL_1379236, EPI_ISL_1379237, EPI_ISL_1379238, EPI_ISL_1379239, EPI_ISL_1379240, EPI_ISL_1379241, EPI_ISL_1379242, EPI_ISL_1379243, EPI_ISL_1379244, EPI_ISL_1379245, EPI_ISL_1379246, EPI_ISL_1379247, EPI_ISL_1379248, EPI_ISL_1379249, EPI_ISL_1379250, EPI_ISL_1379251, EPI_ISL_1379252, EPI_ISL_1379253, EPI_ISL_1379254, EPI_ISL_1379255, EPI_ISL_1379256, EPI_ISL_1379257, EPI_ISL_1379258, EPI_ISL_1379259, EPI_ISL_1379260, EPI_ISL_1379261, EPI_ISL_1379262, EPI_ISL_1379263, EPI_ISL_1379264, EPI_ISL_1379265, EPI_ISL_1379266 | | | |

| | | | |
|---|---|---|---|
| see above | NYP-WCM | New York Genome Center | Michael Zody, Andre Corvelo, Dayna M. Oschwald, Samantha Fennessey, Tom Maniatis, Melissa Cushing, Olivier Elemento, Margaret Elizabeth Ross, Chris Mason, Priya Velu, Hanna Rennert, Arryn Craney, Lars F Westblade |
| EPI_ISL_1379267, EPI_ISL_1379268, EPI_ISL_1379269, EPI_ISL_1379270, EPI_ISL_1379271, EPI_ISL_1379272, EPI_ISL_1379273, EPI_ISL_1379274, EPI_ISL_1379275, EPI_ISL_1379276, EPI_ISL_1379277, EPI_ISL_1379278, EPI_ISL_1379279, EPI_ISL_1379280, EPI_ISL_1379281, EPI_ISL_1379282, EPI_ISL_1379283, EPI_ISL_1379284, EPI_ISL_1379285, EPI_ISL_1379286, EPI_ISL_1379287, EPI_ISL_1379288, EPI_ISL_1379289, EPI_ISL_1379290, EPI_ISL_1379291, EPI_ISL_1379292, EPI_ISL_1379293, EPI_ISL_1379294, EPI_ISL_1379295, EPI_ISL_1379296, EPI_ISL_1379297, EPI_ISL_1379298, EPI_ISL_1379299, EPI_ISL_1379300, EPI_ISL_1379301, EPI_ISL_1379302, EPI_ISL_1379303, EPI_ISL_1379304, EPI_ISL_1379305, EPI_ISL_1379306, EPI_ISL_1379307, EPI_ISL_1379308, EPI_ISL_1379309, EPI_ISL_1379310, EPI_ISL_1379311, EPI_ISL_1379312, EPI_ISL_1379313, EPI_ISL_1379314, EPI_ISL_1379315, EPI_ISL_1379316, EPI_ISL_1379317, EPI_ISL_1379318, EPI_ISL_1379319, EPI_ISL_1379320, EPI_ISL_1379321, EPI_ISL_1379322, EPI_ISL_1379323, EPI_ISL_1379324, EPI_ISL_1379325, EPI_ISL_1379326, EPI_ISL_1379327, EPI_ISL_1379328, EPI_ISL_1379329, EPI_ISL_1379330, EPI_ISL_1379331, EPI_ISL_1379332, EPI_ISL_1379333, EPI_ISL_1379334, EPI_ISL_1379335, EPI_ISL_1379336, EPI_ISL_1379337, EPI_ISL_1379338 | | | |
| see above | Hackensack Meridian Health | New York Genome Center | Michael Zody, Andre Corvelo, Dayna M. Oschwald, Samantha Fennessey, Tom Maniatis, Liang Chen, Jose Mediavilla, Marcus Cunningham, Kaelea Composto, Kar Chow, David Perlin, Barry Kreiswirth |
| EPI_ISL_1379339, EPI_ISL_1379340, EPI_ISL_1379341, EPI_ISL_1379342, EPI_ISL_1379343, EPI_ISL_1379344, EPI_ISL_1379345, EPI_ISL_1379346, EPI_ISL_1379347, EPI_ISL_1379348, EPI_ISL_1379349, EPI_ISL_1379350, EPI_ISL_1379351, EPI_ISL_1379352, EPI_ISL_1379353, EPI_ISL_1379354, EPI_ISL_1379355, EPI_ISL_1379356, EPI_ISL_1379357, EPI_ISL_1379358, EPI_ISL_1379359, EPI_ISL_1379360, EPI_ISL_1379361, EPI_ISL_1379362, EPI_ISL_1379363, EPI_ISL_1379364, EPI_ISL_1379365, EPI_ISL_1379366, EPI_ISL_1379367, EPI_ISL_1379368, EPI_ISL_1379369, EPI_ISL_1379370, EPI_ISL_1379371, EPI_ISL_1379372, EPI_ISL_1379373, EPI_ISL_1379374, EPI_ISL_1379375, EPI_ISL_1379376, EPI_ISL_1379377, EPI_ISL_1379378, EPI_ISL_1379379, EPI_ISL_1379380, EPI_ISL_1379381, EPI_ISL_1379382, EPI_ISL_1379383, EPI_ISL_1379384, EPI_ISL_1379385, EPI_ISL_1379386, EPI_ISL_1379387, EPI_ISL_1379388, EPI_ISL_1379389, EPI_ISL_1379390 | | | |
| see above | NYP-WCM | New York Genome Center | Michael Zody, Andre Corvelo, Dayna M. Oschwald, Samantha Fennessey, Tom Maniatis, Melissa Cushing, Olivier Elemento, Margaret Elizabeth Ross, Chris Mason, Priya Velu, Hanna Rennert, Arryn Craney, Lars F Westblade |
| EPI_ISL_1379444, EPI_ISL_1379445, EPI_ISL_1379446, EPI_ISL_1379447, EPI_ISL_1379448, EPI_ISL_1379449, EPI_ISL_1379450, EPI_ISL_1379451, EPI_ISL_1379452, EPI_ISL_1379453, EPI_ISL_1379454, EPI_ISL_1379455, EPI_ISL_1379456, EPI_ISL_1379457, EPI_ISL_1379458, EPI_ISL_1379459, EPI_ISL_1379460, EPI_ISL_1379461, EPI_ISL_1379462, EPI_ISL_1379463, EPI_ISL_1379464, EPI_ISL_1379465, EPI_ISL_1379466, EPI_ISL_1379467, EPI_ISL_1379468, EPI_ISL_1379469, EPI_ISL_1379470, EPI_ISL_1379471, EPI_ISL_1379472, EPI_ISL_1379473, EPI_ISL_1379474, EPI_ISL_1379475, EPI_ISL_1379476, EPI_ISL_1379477, EPI_ISL_1379478, EPI_ISL_1379479, EPI_ISL_1379480, EPI_ISL_1379481, EPI_ISL_1379482, EPI_ISL_1379483, EPI_ISL_1379484, EPI_ISL_1379485, EPI_ISL_1379486, EPI_ISL_1379487, EPI_ISL_1379488, EPI_ISL_1379489, EPI_ISL_1379490, EPI_ISL_1379491, EPI_ISL_1379492, EPI_ISL_1379493, EPI_ISL_1379494, EPI_ISL_1379495, EPI_ISL_1379496, EPI_ISL_1379497, EPI_ISL_1379498, EPI_ISL_1379499, EPI_ISL_1379500, EPI_ISL_1379501, EPI_ISL_1379502, EPI_ISL_1379503, EPI_ISL_1379504, EPI_ISL_1379505, EPI_ISL_1379506, EPI_ISL_1379507, EPI_ISL_1379508, EPI_ISL_1379509, EPI_ISL_1379510, EPI_ISL_1379511, EPI_ISL_1379512, EPI_ISL_1379513, EPI_ISL_1379514, EPI_ISL_1379515, EPI_ISL_1379516, EPI_ISL_1379517, EPI_ISL_1379518, EPI_ISL_1379519, EPI_ISL_1379520, EPI_ISL_1379521, EPI_ISL_1379522, EPI_ISL_1379523, EPI_ISL_1379524, EPI_ISL_1379525, EPI_ISL_1379526, EPI_ISL_1379527, EPI_ISL_1379528, EPI_ISL_1379529, EPI_ISL_1379530, EPI_ISL_1379531, EPI_ISL_1379532, EPI_ISL_1379533, EPI_ISL_1379534, EPI_ISL_1379535, EPI_ISL_1379536, EPI_ISL_1379537, EPI_ISL_1379538, EPI_ISL_1379539, EPI_ISL_1379540, EPI_ISL_1379541, EPI_ISL_1379542, EPI_ISL_1379543, EPI_ISL_1379544, EPI_ISL_1379545, EPI_ISL_1379546, EPI_ISL_1379547, EPI_ISL_1379548, EPI_ISL_1379549, EPI_ISL_1379550, EPI_ISL_1379551, EPI_ISL_1379552, EPI_ISL_1379553, EPI_ISL_1379554, EPI_ISL_1379555, EPI_ISL_1379556, EPI_ISL_1379557, EPI_ISL_1379558, EPI_ISL_1379559, EPI_ISL_1379560, EPI_ISL_1379561, EPI_ISL_1379562, EPI_ISL_1379563, EPI_ISL_1379564, EPI_ISL_1379565, EPI_ISL_1379566, EPI_ISL_1379567, EPI_ISL_1379568, EPI_ISL_1379569, EPI_ISL_1379570, EPI_ISL_1379571, EPI_ISL_1379572, EPI_ISL_1379573, EPI_ISL_1379574, EPI_ISL_1379575, EPI_ISL_1379576, EPI_ISL_1379577, EPI_ISL_1379578, EPI_ISL_1379579, EPI_ISL_1379580, EPI_ISL_1379581, EPI_ISL_1379582, EPI_ISL_1379583, EPI_ISL_1379584, EPI_ISL_1379585, EPI_ISL_1379586, EPI_ISL_1379587, EPI_ISL_1379588, EPI_ISL_1379589, EPI_ISL_1379590 | | | |
| see above | Tampa General Hospital Esoteric Lab | Tampa General Hospital Esoteric Research & Development Lab | Grant Vestal, Deanna Becker, Dominic Uy, Vicki Healer, Amorce Lima, Suzane Silbert |
| EPI_ISL_1379593, EPI_ISL_1379594, EPI_ISL_1379595, EPI_ISL_1379596, EPI_ISL_1379597, EPI_ISL_1379598, EPI_ISL_1379599, EPI_ISL_1379600, EPI_ISL_1379601, EPI_ISL_1379602, EPI_ISL_1379603, EPI_ISL_1379604, EPI_ISL_1379605, EPI_ISL_1379606, EPI_ISL_1379607, EPI_ISL_1379608, EPI_ISL_1379609, EPI_ISL_1379610, EPI_ISL_1379611, EPI_ISL_1379612, EPI_ISL_1379613, EPI_ISL_1379615, EPI_ISL_1379616, EPI_ISL_1379617, EPI_ISL_1379618, EPI_ISL_1379619, EPI_ISL_1379620, EPI_ISL_1379621, EPI_ISL_1379622, EPI_ISL_1379623, EPI_ISL_1379624, EPI_ISL_1379625, EPI_ISL_1379626, EPI_ISL_1379628, EPI_ISL_1379629, EPI_ISL_1379631, EPI_ISL_1379632, EPI_ISL_1379633, EPI_ISL_1379634 | | | |
| see above | Wyoming Public Health Laboratory | Wyoming Public Health Laboratory | Jim Mildenberger, Wanda Manley, Noah Hull, Taylor Fearing, Lynette Gumbleton, Channing Weber, Ashley Norberg, Chayse Rowley, Marley Goetz, Brian Dominguez, Elliot Thomasson, Cari Sloma, and Rob Christensen |
| EPI_ISL_1379635, EPI_ISL_1379636, EPI_ISL_1379638, EPI_ISL_1379639, EPI_ISL_1379640, EPI_ISL_1379641, EPI_ISL_1379642, EPI_ISL_1379643, EPI_ISL_1379644, EPI_ISL_1379645, EPI_ISL_1379646, EPI_ISL_1379647, EPI_ISL_1379648, EPI_ISL_1379649, EPI_ISL_1379650, EPI_ISL_1379651, EPI_ISL_1379652, EPI_ISL_1379653, EPI_ISL_1379654, EPI_ISL_1379655, EPI_ISL_1379656, EPI_ISL_1379657, EPI_ISL_1379658, EPI_ISL_1379659, EPI_ISL_1379660, EPI_ISL_1379661 | | | |
| see above | Springfield-Greene Co. Health Dept. | Pathogen Discovery, Respiratory Viruses Branch, Division of Viral Diseases, Centers for Disease Control and Prevention | Yan Li, Jing Zhang, Ying Tao, Brian Lynch, Anna Kelleher, Krista Queen, Anna Uehara, Peter Cook, Han Jia Justin Ng, Clinton R. Paden, Haibin Wang, Suxiang Tong |
| EPI_ISL_1379662 | Pathogen Discovery, Respiratory Viruses Branch, Division of Viral Diseases, Centers for Disease Control and Prevention | Pathogen Discovery, Respiratory Viruses Branch, Division of Viral Diseases, Centers for Disease Control and Prevention | Yan Li, Jing Zhang, Ying Tao, Brian Lynch, Anna Kelleher, Krista Queen, Anna Uehara, Peter Cook, Han Jia Justin Ng, Clinton R. Paden, Haibin Wang, Suxiang Tong |
| EPI_ISL_1379663, EPI_ISL_1379664 | Vanderbilt University | Pathogen Discovery, Respiratory Viruses Branch, Division of Viral Diseases, Centers for Disease Control and Prevention | Yan Li, Jing Zhang, Ying Tao, Brian Lynch, Anna Kelleher, Krista Queen, Anna Uehara, Peter Cook, Han Jia Justin Ng, Clinton R. Paden, Haibin Wang, Suxiang Tong |
| EPI_ISL_1379665, EPI_ISL_1379666, EPI_ISL_1379667, EPI_ISL_1379668 | Pathogen Discovery, Respiratory Viruses Branch, Division of Viral Diseases, Centers for Disease Control and Prevention | Pathogen Discovery, Respiratory Viruses Branch, Division of Viral Diseases, Centers for Disease Control and Prevention | Yan Li, Jing Zhang, Ying Tao, Brian Lynch, Anna Kelleher, Krista Queen, Anna Uehara, Peter Cook, Han Jia Justin Ng, Clinton R. Paden, Haibin Wang, Suxiang Tong |
| EPI_ISL_1379669, EPI_ISL_1379670, EPI_ISL_1379671 | Vanderbilt University | Pathogen Discovery, Respiratory Viruses Branch, Division of Viral Diseases, Centers for Disease Control and Prevention | Yan Li, Jing Zhang, Ying Tao, Brian Lynch, Anna Kelleher, Krista Queen, Anna Uehara, Peter Cook, Han Jia Justin Ng, Clinton R. Paden, Haibin Wang, Suxiang Tong |
| EPI_ISL_1379672, EPI_ISL_1379673, EPI_ISL_1379674, EPI_ISL_1379675 | Pathogen Discovery, Respiratory Viruses Branch, Division of Viral Diseases, Centers for Disease Control and Prevention | Pathogen Discovery, Respiratory Viruses Branch, Division of Viral Diseases, Centers for Disease Control and Prevention | Yan Li, Jing Zhang, Ying Tao, Brian Lynch, Anna Kelleher, Krista Queen, Anna Uehara, Peter Cook, Han Jia Justin Ng, Clinton R. Paden, Haibin Wang, Suxiang Tong |
| EPI_ISL_1379676 | Vanderbilt University | Pathogen Discovery, Respiratory Viruses Branch, Division of Viral Diseases, Centers for Disease Control and Prevention | Yan Li, Jing Zhang, Ying Tao, Brian Lynch, Anna Kelleher, Krista Queen, Anna Uehara, Peter Cook, Han Jia Justin Ng, Clinton R. Paden, Haibin Wang, Suxiang Tong |
| EPI_ISL_1379678, EPI_ISL_1379679, EPI_ISL_1379680 | Pathogen Discovery, Respiratory Viruses Branch, Division of Viral Diseases, Centers for Disease Control and Prevention | Pathogen Discovery, Respiratory Viruses Branch, Division of Viral Diseases, Centers for Disease Control and Prevention | Yan Li, Jing Zhang, Ying Tao, Brian Lynch, Anna Kelleher, Krista Queen, Anna Uehara, Peter Cook, Han Jia Justin Ng, Clinton R. Paden, Haibin Wang, Suxiang Tong |
| EPI_ISL_1379681 | Vanderbilt University | Pathogen Discovery, Respiratory Viruses Branch, Division of Viral Diseases, Centers for Disease Control and Prevention | Yan Li, Jing Zhang, Ying Tao, Brian Lynch, Anna Kelleher, Krista Queen, Anna Uehara, Peter Cook, Han Jia Justin Ng, Clinton R. Paden, Haibin Wang, Suxiang Tong |
| EPI_ISL_1379682 | Pathogen Discovery, Respiratory Viruses Branch, Division of Viral Diseases, Centers for Disease Control and Prevention | Pathogen Discovery, Respiratory Viruses Branch, Division of Viral Diseases, Centers for Disease Control and Prevention | Yan Li, Jing Zhang, Ying Tao, Brian Lynch, Anna Kelleher, Krista Queen, Anna Uehara, Peter Cook, Han Jia Justin Ng, Clinton R. Paden, Haibin Wang, Suxiang Tong |
| EPI_ISL_1379683, EPI_ISL_1379684, EPI_ISL_1379685 | Vanderbilt University | Pathogen Discovery, Respiratory Viruses Branch, Division of Viral Diseases, Centers for Disease Control and Prevention | Yan Li, Jing Zhang, Ying Tao, Brian Lynch, Anna Kelleher, Krista Queen, Anna Uehara, Peter Cook, Han Jia Justin Ng, Clinton R. Paden, Haibin Wang, Suxiang Tong |
| EPI_ISL_1379686 | PA Dept of Heath, Bureau of Laboratories | Pathogen Discovery, Respiratory Viruses Branch, Division of Viral Diseases, Centers for Disease Control and Prevention | Yan Li, Jing Zhang, Ying Tao, Brian Lynch, Anna Kelleher, Krista Queen, Anna Uehara, Peter Cook, Han Jia Justin Ng, Clinton R. Paden, Haibin Wang, Suxiang Tong |
| EPI_ISL_1379687, EPI_ISL_1379688 | Pathogen Discovery, Respiratory Viruses Branch, Division of Viral Diseases, Centers for Disease Control and Prevention | Pathogen Discovery, Respiratory Viruses Branch, Division of Viral Diseases, Centers for Disease Control and Prevention | Yan Li, Jing Zhang, Ying Tao, Brian Lynch, Anna Kelleher, Krista Queen, Anna Uehara, Peter Cook, Han Jia Justin Ng, Clinton R. Paden, Haibin Wang, Suxiang Tong |
| EPI_ISL_1379689 | Springfield-Greene Co. Health Dept. | Pathogen Discovery, Respiratory Viruses Branch, Division of Viral Diseases, Centers for Disease Control and Prevention | Yan Li, Jing Zhang, Ying Tao, Brian Lynch, Anna Kelleher, Krista Queen, Anna Uehara, Peter Cook, Han Jia Justin Ng, Clinton R. Paden, Haibin Wang, Suxiang Tong |
| EPI_ISL_1379690, EPI_ISL_1379691, EPI_ISL_1379692 | Pathogen Discovery, Respiratory Viruses Branch, Division of Viral Diseases, Centers for Disease Control and Prevention | Pathogen Discovery, Respiratory Viruses Branch, Division of Viral Diseases, Centers for Disease Control and Prevention | Yan Li, Jing Zhang, Ying Tao, Brian Lynch, Anna Kelleher, Krista Queen, Anna Uehara, Peter Cook, Han Jia Justin Ng, Clinton R. Paden, Haibin Wang, Suxiang Tong |
| EPI_ISL_1379693 | PA Dept of Heath, Bureau of Laboratories | Pathogen Discovery, Respiratory Viruses Branch, Division of Viral Diseases, Centers for Disease Control and Prevention | Yan Li, Jing Zhang, Ying Tao, Brian Lynch, Anna Kelleher, Krista Queen, Anna Uehara, Peter Cook, Han Jia Justin Ng, Clinton R. Paden, Haibin Wang, Suxiang Tong |
| EPI_ISL_1379694 | Vanderbilt University | Pathogen Discovery, Respiratory Viruses Branch, Division of Viral Diseases, Centers for Disease Control and Prevention | Yan Li, Jing Zhang, Ying Tao, Brian Lynch, Anna Kelleher, Krista Queen, Anna Uehara, Peter Cook, Han Jia Justin Ng, Clinton R. Paden, Haibin Wang, Suxiang Tong |
| EPI_ISL_1379695 | Pathogen Discovery, Respiratory Viruses Branch, Division of Viral Diseases, Centers for Disease Control and Prevention | Pathogen Discovery, Respiratory Viruses Branch, Division of Viral Diseases, Centers for Disease Control and Prevention | Yan Li, Jing Zhang, Ying Tao, Brian Lynch, Anna Kelleher, Krista Queen, Anna Uehara, Peter Cook, Han Jia Justin Ng, Clinton R. Paden, Haibin Wang, Suxiang Tong |
| EPI_ISL_1379696 | Vanderbilt University | Pathogen Discovery, Respiratory Viruses Branch, Division of Viral Diseases, Centers for Disease Control and Prevention | Yan Li, Jing Zhang, Ying Tao, Brian Lynch, Anna Kelleher, Krista Queen, Anna Uehara, Peter Cook, Han Jia Justin Ng, Clinton R. Paden, Haibin Wang, Suxiang Tong |
| EPI_ISL_1379697, EPI_ISL_1379698 | Pathogen Discovery, Respiratory Viruses Branch, Division of Viral Diseases, Centers for Disease Control and Prevention | Pathogen Discovery, Respiratory Viruses Branch, Division of Viral Diseases, Centers for Disease Control and Prevention | Yan Li, Jing Zhang, Ying Tao, Brian Lynch, Anna Kelleher, Krista Queen, Anna Uehara, Peter Cook, Han Jia Justin Ng, Clinton R. Paden, Haibin Wang, Suxiang Tong |
| EPI_ISL_1379699, EPI_ISL_1379700 | Vanderbilt University | Pathogen Discovery, Respiratory Viruses Branch, Division of Viral Diseases, Centers for Disease Control and Prevention | Yan Li, Jing Zhang, Ying Tao, Brian Lynch, Anna Kelleher, Krista Queen, Anna Uehara, Peter Cook, Han Jia Justin Ng, Clinton R. Paden, Haibin Wang, Suxiang Tong |

| | | | |
|---|---|---|---|
| EPI_ISL_1379701 | Pathogen Discovery, Respiratory Viruses Branch, Division of Viral Diseases, Centers for Disease Control and Prevention | Pathogen Discovery, Respiratory Viruses Branch, Division of Viral Diseases, Centers for Disease Control and Prevention | Yan Li, Jing Zhang, Ying Tao, Brian Lynch, Anna Kelleher, Krista Queen, Anna Uehara, Peter Cook, Han Jia Justin Ng, Clinton R. Paden, Haibin Wang, Suxiang Tong |
| EPI_ISL_1379702 | Vanderbilt University | Pathogen Discovery, Respiratory Viruses Branch, Division of Viral Diseases, Centers for Disease Control and Prevention | Yan Li, Jing Zhang, Ying Tao, Brian Lynch, Anna Kelleher, Krista Queen, Anna Uehara, Peter Cook, Han Jia Justin Ng, Clinton R. Paden, Haibin Wang, Suxiang Tong |
| EPI_ISL_1379703, EPI_ISL_1379704 | Pathogen Discovery, Respiratory Viruses Branch, Division of Viral Diseases, Centers for Disease Control and Prevention | Pathogen Discovery, Respiratory Viruses Branch, Division of Viral Diseases, Centers for Disease Control and Prevention | Yan Li, Jing Zhang, Ying Tao, Brian Lynch, Anna Kelleher, Krista Queen, Anna Uehara, Peter Cook, Han Jia Justin Ng, Clinton R. Paden, Haibin Wang, Suxiang Tong |
| EPI_ISL_1379705, EPI_ISL_1379706, EPI_ISL_1379707, EPI_ISL_1379708 | Vanderbilt University | Pathogen Discovery, Respiratory Viruses Branch, Division of Viral Diseases, Centers for Disease Control and Prevention | Yan Li, Jing Zhang, Ying Tao, Brian Lynch, Anna Kelleher, Krista Queen, Anna Uehara, Peter Cook, Han Jia Justin Ng, Clinton R. Paden, Haibin Wang, Suxiang Tong |
| EPI_ISL_1379709, EPI_ISL_1379710 | Pathogen Discovery, Respiratory Viruses Branch, Division of Viral Diseases, Centers for Disease Control and Prevention | Pathogen Discovery, Respiratory Viruses Branch, Division of Viral Diseases, Centers for Disease Control and Prevention | Yan Li, Jing Zhang, Ying Tao, Brian Lynch, Anna Kelleher, Krista Queen, Anna Uehara, Peter Cook, Han Jia Justin Ng, Clinton R. Paden, Haibin Wang, Suxiang Tong |
| EPI_ISL_1379711 | Vanderbilt University | Pathogen Discovery, Respiratory Viruses Branch, Division of Viral Diseases, Centers for Disease Control and Prevention | Yan Li, Jing Zhang, Ying Tao, Brian Lynch, Anna Kelleher, Krista Queen, Anna Uehara, Peter Cook, Han Jia Justin Ng, Clinton R. Paden, Haibin Wang, Suxiang Tong |
| EPI_ISL_1379712 | Pathogen Discovery, Respiratory Viruses Branch, Division of Viral Diseases, Centers for Disease Control and Prevention | Pathogen Discovery, Respiratory Viruses Branch, Division of Viral Diseases, Centers for Disease Control and Prevention | Yan Li, Jing Zhang, Ying Tao, Brian Lynch, Anna Kelleher, Krista Queen, Anna Uehara, Peter Cook, Han Jia Justin Ng, Clinton R. Paden, Haibin Wang, Suxiang Tong |
| EPI_ISL_1379713 | Vanderbilt University | Pathogen Discovery, Respiratory Viruses Branch, Division of Viral Diseases, Centers for Disease Control and Prevention | Yan Li, Jing Zhang, Ying Tao, Brian Lynch, Anna Kelleher, Krista Queen, Anna Uehara, Peter Cook, Han Jia Justin Ng, Clinton R. Paden, Haibin Wang, Suxiang Tong |
| EPI_ISL_1379714 | Pathogen Discovery, Respiratory Viruses Branch, Division of Viral Diseases, Centers for Disease Control and Prevention | Pathogen Discovery, Respiratory Viruses Branch, Division of Viral Diseases, Centers for Disease Control and Prevention | Yan Li, Jing Zhang, Ying Tao, Brian Lynch, Anna Kelleher, Krista Queen, Anna Uehara, Peter Cook, Han Jia Justin Ng, Clinton R. Paden, Haibin Wang, Suxiang Tong |
| EPI_ISL_1379715 | Vanderbilt University | Pathogen Discovery, Respiratory Viruses Branch, Division of Viral Diseases, Centers for Disease Control and Prevention | Yan Li, Jing Zhang, Ying Tao, Brian Lynch, Anna Kelleher, Krista Queen, Anna Uehara, Peter Cook, Han Jia Justin Ng, Clinton R. Paden, Haibin Wang, Suxiang Tong |
| EPI_ISL_1379716 | Springfield-Greene Co. Health Dept. | Pathogen Discovery, Respiratory Viruses Branch, Division of Viral Diseases, Centers for Disease Control and Prevention | Yan Li, Jing Zhang, Ying Tao, Brian Lynch, Anna Kelleher, Krista Queen, Anna Uehara, Peter Cook, Han Jia Justin Ng, Clinton R. Paden, Haibin Wang, Suxiang Tong |
| EPI_ISL_1379717, EPI_ISL_1379718 | Pathogen Discovery, Respiratory Viruses Branch, Division of Viral Diseases, Centers for Disease Control and Prevention | Pathogen Discovery, Respiratory Viruses Branch, Division of Viral Diseases, Centers for Disease Control and Prevention | Yan Li, Jing Zhang, Ying Tao, Brian Lynch, Anna Kelleher, Krista Queen, Anna Uehara, Peter Cook, Han Jia Justin Ng, Clinton R. Paden, Haibin Wang, Suxiang Tong |
| EPI_ISL_1379719, EPI_ISL_1379720, EPI_ISL_1379721, EPI_ISL_1379722, EPI_ISL_1379723, EPI_ISL_1379724, EPI_ISL_1379725, EPI_ISL_1379726, EPI_ISL_1379727, EPI_ISL_1379728, EPI_ISL_1379729, EPI_ISL_1379730, EPI_ISL_1379731, EPI_ISL_1379732, EPI_ISL_1379733, EPI_ISL_1379734, EPI_ISL_1379735, EPI_ISL_1379736, EPI_ISL_1379737, EPI_ISL_1379738, EPI_ISL_1379739, EPI_ISL_1379740, EPI_ISL_1379741, EPI_ISL_1379742, EPI_ISL_1379743, EPI_ISL_1379744, EPI_ISL_1379745, EPI_ISL_1379746, EPI_ISL_1379747, EPI_ISL_1379748, EPI_ISL_1379749, EPI_ISL_1379750, EPI_ISL_1379751, EPI_ISL_1379752, EPI_ISL_1379753, EPI_ISL_1379754, EPI_ISL_1379755, EPI_ISL_1379756, EPI_ISL_1379757, EPI_ISL_1379758, EPI_ISL_1379759, EPI_ISL_1379760, EPI_ISL_1379761, EPI_ISL_1379762, EPI_ISL_1379763, EPI_ISL_1379764, EPI_ISL_1379765, EPI_ISL_1379766, EPI_ISL_1379767, EPI_ISL_1379768, EPI_ISL_1379769, EPI_ISL_1379770, EPI_ISL_1379771, EPI_ISL_1379772, EPI_ISL_1379773, EPI_ISL_1379774, EPI_ISL_1379775, EPI_ISL_1379776, EPI_ISL_1379777, EPI_ISL_1379778 | | | |
| see above | Stanford Health Care | Stanford University School of Medicine, Clinical Virology Laboratory | Fumiko Yamamoto, Malaya K. Sahoo, Mamdouh Sibai, Daniel Solis, Michelle Vergese, ChunHong Huang, James Zehnder, and Benjamin A. Pinsky |
| EPI_ISL_1379779, EPI_ISL_1379780, EPI_ISL_1379781, EPI_ISL_1379782, EPI_ISL_1379783, EPI_ISL_1379784, EPI_ISL_1379785, EPI_ISL_1379786, EPI_ISL_1379787, EPI_ISL_1379788, EPI_ISL_1379789, EPI_ISL_1379790, EPI_ISL_1379791, EPI_ISL_1379792, EPI_ISL_1379793, EPI_ISL_1379794, EPI_ISL_1379795, EPI_ISL_1379796, EPI_ISL_1379797, EPI_ISL_1379798, EPI_ISL_1379799, EPI_ISL_1379800, EPI_ISL_1379801, EPI_ISL_1379802, EPI_ISL_1379803, EPI_ISL_1379804, EPI_ISL_1379805, EPI_ISL_1379806, EPI_ISL_1379807, EPI_ISL_1379808, EPI_ISL_1379809, EPI_ISL_1379810, EPI_ISL_1379811, EPI_ISL_1379812, EPI_ISL_1379813, EPI_ISL_1379814, EPI_ISL_1379815, EPI_ISL_1379816 | | | |
| see above | Stanford Health Care | Stanford University School of Medicine, Clinical Virology Laboratory | Daniel Solis, Mamdouh Sibai, Fumiko Yamamoto, Malaya K. Sahoo, Michelle Verghese, ChunHong Huang, James Zehnder, and Benjamin A. Pinsky |
| EPI_ISL_1379817, EPI_ISL_1379818, EPI_ISL_1379819, EPI_ISL_1379820, EPI_ISL_1379821, EPI_ISL_1379822, EPI_ISL_1379823, EPI_ISL_1379824, EPI_ISL_1379825, EPI_ISL_1379826, EPI_ISL_1379827, EPI_ISL_1379828, EPI_ISL_1379829, EPI_ISL_1379830, EPI_ISL_1379831, EPI_ISL_1379832, EPI_ISL_1379833, EPI_ISL_1379834, EPI_ISL_1379835, EPI_ISL_1379836, EPI_ISL_1379837, EPI_ISL_1379838, EPI_ISL_1379839, EPI_ISL_1379840, EPI_ISL_1379841, EPI_ISL_1379842, EPI_ISL_1379843, EPI_ISL_1379844, EPI_ISL_1379845, EPI_ISL_1379846, EPI_ISL_1379847, EPI_ISL_1379848, EPI_ISL_1379849, EPI_ISL_1379850, EPI_ISL_1379851, EPI_ISL_1379852, EPI_ISL_1379853, EPI_ISL_1379854, EPI_ISL_1379855, EPI_ISL_1379856, EPI_ISL_1379857, EPI_ISL_1379858, EPI_ISL_1379859, EPI_ISL_1379860, EPI_ISL_1379861, EPI_ISL_1379862, EPI_ISL_1379863, EPI_ISL_1379864, EPI_ISL_1379865, EPI_ISL_1379866 | | | |
| see above | Stanford Health Care | Stanford University School of Medicine, Clinical Virology Laboratory | Mamdouh Sibai, Daniel Solis, Fumiko Yamamoto, Malaya K. Sahoo, Michelle Verghese, ChunHong Huang, James Zehnder, and Benjamin A. Pinsky |
| EPI_ISL_1379867, EPI_ISL_1379868, EPI_ISL_1379869, EPI_ISL_1379870, EPI_ISL_1379871, EPI_ISL_1379872, EPI_ISL_1379873, EPI_ISL_1379874, EPI_ISL_1379875, EPI_ISL_1379876, EPI_ISL_1379877, EPI_ISL_1379878, EPI_ISL_1379879, EPI_ISL_1379880, EPI_ISL_1379881, EPI_ISL_1379882, EPI_ISL_1379883, EPI_ISL_1379884, EPI_ISL_1379885, EPI_ISL_1379886 | | | |
| see above | Stanford Health Care | Stanford University School of Medicine, Clinical Virology Laboratory | Michelle Verghese, ChunHong Huang, Mamdouh Sibai, Daniel Solis, Fumiko Yamamoto, Malaya K. Sahoo, James Zehnder, and Benjamin A. Pinsky |
| EPI_ISL_1379887, EPI_ISL_1379888 | Stanford Health Care | Stanford University School of Medicine, Clinical Virology Laboratory | Daniel Solis, Mamdouh Sibai, Fumiko Yamamoto, Becky Jiang, Michelle Verghese, ChunHong Huang, James Zehnder, Malaya K. Sahoo, and Benjamin A. Pinsky |
| EPI_ISL_1379889 | Stanford Health Care | Stanford University School of Medicine, Clinical Virology Laboratory | Michelle Verghese, Becky Jiang, Naomi Iwai, Marilyn Mar, Malaya K. Sahoo, Fumiko Yamamoto, Kenji O. Mfuh, Jacob Miller, Hannah Wang, James Zehnder, and Benjamin A. Pinsky |
| EPI_ISL_1379956, EPI_ISL_1379957 | University of Vermont Medical Center | UW Virology Lab | Pavitra Roychoudhury, Hong Xie, Lasata Shrestha, Shah Mohamed Bakhash, Michelle Lin, Margaret Mills, Noah R. Baker, Sean Ellis, Saraswathi Sathees, Emily A. Bruce, Jessica W. Crothers, Beth D. Kirkpatrick, Allison L. Unger, Marya P. Carmolli, Meei-Li Huang, Keith R. Jerome, Alexander Greninger |
| EPI_ISL_1379958, EPI_ISL_1379959, EPI_ISL_1379960, EPI_ISL_1379961, EPI_ISL_1379962, EPI_ISL_1379963, EPI_ISL_1379964, EPI_ISL_1379965, EPI_ISL_1379966, EPI_ISL_1379967, EPI_ISL_1379968, EPI_ISL_1379969, EPI_ISL_1379970, EPI_ISL_1379971, EPI_ISL_1379972, EPI_ISL_1379973, EPI_ISL_1379974, EPI_ISL_1379975 | | | |
| see above | UCLA Clinical Micro Lab | Los Angeles County Public Health Laboratories | P. Hemarajata et al. |
| EPI_ISL_1379977 | USC Clinical Lab | Los Angeles County Public Health Laboratories | P. Hemarajata et al. |
| EPI_ISL_1379978 | Flow Diagnostics Lab | Los Angeles County Public Health Laboratories | P. Hemarajata et al. |
| EPI_ISL_1380052, EPI_ISL_1380053, EPI_ISL_1380054, EPI_ISL_1380056, EPI_ISL_1380057, EPI_ISL_1380058 | Clinical Molecular Microbiology Laboratory, UNC Hospitals | Jeremy Wang | Jeremy Wang, Alexander Rubinsteyn, Colleen Rice, Jason Smedberg, Shawn Hawken, Melissa Miller, Corbin Jones, Robert Hagan |
| EPI_ISL_1380499 | University of Vermont Medical Center | UW Virology Lab | Pavitra Roychoudhury, Hong Xie, Lasata Shrestha, Shah Mohamed Bakhash, Michelle Lin, Margaret Mills, Noah R. Baker, Sean Ellis, Saraswathi Sathees, Emily A. Bruce, Jessica W. Crothers, Beth D. Kirkpatrick, Allison L. Unger, Marya P. Carmolli, Meei-Li Huang, Keith R. Jerome, Alexander Greninger |
| EPI_ISL_1381234, EPI_ISL_1381235, EPI_ISL_1381236, EPI_ISL_1381237, EPI_ISL_1381238, EPI_ISL_1381239, EPI_ISL_1381240, EPI_ISL_1381241, EPI_ISL_1381242, EPI_ISL_1381243, EPI_ISL_1381244, EPI_ISL_1381245, EPI_ISL_1381246, EPI_ISL_1381247, EPI_ISL_1381248, EPI_ISL_1381249, EPI_ISL_1381250, EPI_ISL_1381251, EPI_ISL_1381252, EPI_ISL_1381253, EPI_ISL_1381254, EPI_ISL_1381255, EPI_ISL_1381256, EPI_ISL_1381257, EPI_ISL_1381258 | | | |
| see above | Centro de Investigacion Biomedica del Noreste (CIBIN) | Unidad de Genomica Avanzada | Consorcio Mexicano de Vigilancia Genomica (CoViGen-Mex). Authors (in alphabetical order): Julio Elias Alvarado-Yaah, Carlos F. Arias, Santiago Avila-Rios, Victor Hugo Borja-Aburto, Celia Boukadida, Juan Bautista Chale-Dzul , Jose Antonio Enciso-Moreno, Gloria Elena Espinoza-Ayala, Fernando Fontove-Herrera, Concepcion Grajales-Muniz, Ricardo Grande, Alfredo Herrera-Estrella, Carla Ivon Herrera-Najera, Pavel Isa, Brenda Irasema Maldonado-Meza, Bernardo Martinez-Miguel, Margarita Matias-Florentino, Maria Guadalupe de Jesus Mireles-Rivera, Gloria Maria Molina-Salinas, Hector Montoya-Fuentes, Jose Esteban Munoz-Medina, Jose de Jesus Nunez-Contreras, Alicia Ocana-Mondragon, Luis Alberto Ochoa-Carrera, Hector Esteban Paz-Juarez, Francisco Pulido, Helen Haydee Fernanda Ramirez-Plascencia, Angel Gustavo Salas-Lais, Jorge Ivan Salinal-Nevarez, Alejandro Sanchez-Flores, Clara Esperanza Santacruz-Tinoco, Maria Guadalupe Santiago-Mauricio, Nelly Selem-Mojica, Blanca Taboada, Gloria Vazquez |
| EPI_ISL_1381880, EPI_ISL_1381881, EPI_ISL_1381882, EPI_ISL_1381883, EPI_ISL_1381884, EPI_ISL_1381885, EPI_ISL_1381886, EPI_ISL_1381887, EPI_ISL_1381888, EPI_ISL_1381889, EPI_ISL_1381890, EPI_ISL_1381891, EPI_ISL_1381892, EPI_ISL_1381893, EPI_ISL_1381894, EPI_ISL_1381895, EPI_ISL_1381896, EPI_ISL_1381897, EPI_ISL_1381898, EPI_ISL_1381899, EPI_ISL_1381900, EPI_ISL_1381901, EPI_ISL_1381902, EPI_ISL_1381903, EPI_ISL_1381904, EPI_ISL_1381905, EPI_ISL_1381906, EPI_ISL_1381907, EPI_ISL_1381908, EPI_ISL_1381909, EPI_ISL_1381910, EPI_ISL_1381911, EPI_ISL_1381912, EPI_ISL_1381913, EPI_ISL_1381914, EPI_ISL_1381915, EPI_ISL_1381916, EPI_ISL_1381917, EPI_ISL_1381918, EPI_ISL_1381919, EPI_ISL_1381924, EPI_ISL_1381925, EPI_ISL_1381926, EPI_ISL_1381927, EPI_ISL_1381928, EPI_ISL_1381929, EPI_ISL_1381930, EPI_ISL_1381931, EPI_ISL_1381932, EPI_ISL_1381933, EPI_ISL_1381934, EPI_ISL_1381935, EPI_ISL_1381936, EPI_ISL_1381937, EPI_ISL_1381938, EPI_ISL_1381939, EPI_ISL_1381940, EPI_ISL_1381941, EPI_ISL_1381942, EPI_ISL_1381943, EPI_ISL_1381944, EPI_ISL_1381945, EPI_ISL_1381946, EPI_ISL_1382078, EPI_ISL_1382079, EPI_ISL_1382080, EPI_ISL_1382081, EPI_ISL_1382082, EPI_ISL_1382083, EPI_ISL_1382084, EPI_ISL_1382085, EPI_ISL_1382086, EPI_ISL_1382087, EPI_ISL_1382088, EPI_ISL_1382089, EPI_ISL_1382090, EPI_ISL_1382091, EPI_ISL_1382092, EPI_ISL_1382093, EPI_ISL_1382094, EPI_ISL_1382095, EPI_ISL_1382096, EPI_ISL_1382097, EPI_ISL_1382098, EPI_ISL_1382099, EPI_ISL_1382100, EPI_ISL_1382102, EPI_ISL_1382103, EPI_ISL_1382104, EPI_ISL_1382105, EPI_ISL_1382106, EPI_ISL_1382107, EPI_ISL_1382108, EPI_ISL_1382109, EPI_ISL_1382110, EPI_ISL_1382111 | | | |
| see above | Virginia Division of Consolidated Laboratory Services | Virginia Division of Consolidated Laboratory Services | Virginia DCLS |
| EPI_ISL_1382112, EPI_ISL_1382115 | UNC-CH COVID Surveillance Lab | Jeremy Wang | Jeremy Wang, Alexander Rubinsteyn, Melissa Miller, Corbin Jones, Amy James Loftis, Amir Barzin, Susan Fiscus |

| | | | |
|---|---|---|---|
| EPI_ISL_1382116 | Clinical Molecular Microbiology Laboratory, UNC Hospitals | Jeremy Wang | Jeremy Wang, Alexander Rubinsteyn, Colleen Rice, Jason Smedberg, Shawn Hawken, Melissa Miller, Corbin Jones, Robert Hagan |
| EPI_ISL_1382118, EPI_ISL_1382119, EPI_ISL_1382120, EPI_ISL_1382121, EPI_ISL_1382122, EPI_ISL_1382123, EPI_ISL_1382124, EPI_ISL_1382125, EPI_ISL_1382126, EPI_ISL_1382127, EPI_ISL_1382128, EPI_ISL_1382129, EPI_ISL_1382130, EPI_ISL_1382131, EPI_ISL_1382132, EPI_ISL_1382133, EPI_ISL_1382134, EPI_ISL_1382135, EPI_ISL_1382136, EPI_ISL_1382137, EPI_ISL_1382138, EPI_ISL_1382139, EPI_ISL_1382140, EPI_ISL_1382141, EPI_ISL_1382142, EPI_ISL_1382143, EPI_ISL_1382144 | | | |
| see above | Gundersen Clinical Microbiology Laboratory | Kabara Cancer Research Institute | Craig S. Richmond, Paraic A. Kenny |
| EPI_ISL_1383233 | Noblis | Noblis, Inc. | Lauren Brinkac, Shiela Diepold, Shane Mitchell, June Kang, Lee F. Kolakowski, William M. Nelson, Katharine Jennings |
| EPI_ISL_1383234 | Bioinformatics and Life Sciences, Noblis | Bioinformatics and Life Sciences, Noblis | Brinkac,L.M., Diepold,S., Mitchell,S., Kang,J., Kolakowski,L.F.,Nelson,W.M. and Jennings,K. |
| EPI_ISL_1384885, EPI_ISL_1384886, EPI_ISL_1384887, EPI_ISL_1384888, EPI_ISL_1384889, EPI_ISL_1384890, EPI_ISL_1384891, EPI_ISL_1384892, EPI_ISL_1384893, EPI_ISL_1384894, EPI_ISL_1384895, EPI_ISL_1384896, EPI_ISL_1384897, EPI_ISL_1384898, EPI_ISL_1384899, EPI_ISL_1384900, EPI_ISL_1384901, EPI_ISL_1384902, EPI_ISL_1384903, EPI_ISL_1384904, EPI_ISL_1384905, EPI_ISL_1384906, EPI_ISL_1384907, EPI_ISL_1384908, EPI_ISL_1384909, EPI_ISL_1384910, EPI_ISL_1384911, EPI_ISL_1384912, EPI_ISL_1384913, EPI_ISL_1384914, EPI_ISL_1384915, EPI_ISL_1384916, EPI_ISL_1384917, EPI_ISL_1384918, EPI_ISL_1384919, EPI_ISL_1384920, EPI_ISL_1384921, EPI_ISL_1384922, EPI_ISL_1384923, EPI_ISL_1384924, EPI_ISL_1384925, EPI_ISL_1384926, EPI_ISL_1384927, EPI_ISL_1384928, EPI_ISL_1384929, EPI_ISL_1384930, EPI_ISL_1384931, EPI_ISL_1384932, EPI_ISL_1384933, EPI_ISL_1384934, EPI_ISL_1384935, EPI_ISL_1384936, EPI_ISL_1384937, EPI_ISL_1384938, EPI_ISL_1384940, EPI_ISL_1384941, EPI_ISL_1384942, EPI_ISL_1384943, EPI_ISL_1384944, EPI_ISL_1384945, EPI_ISL_1384947, EPI_ISL_1384949, EPI_ISL_1384950, EPI_ISL_1384951, EPI_ISL_1384952, EPI_ISL_1384953, EPI_ISL_1384954, EPI_ISL_1384955, EPI_ISL_1384956, EPI_ISL_1384957, EPI_ISL_1384958, EPI_ISL_1384959, EPI_ISL_1384960, EPI_ISL_1384961, EPI_ISL_1384962, EPI_ISL_1384963, EPI_ISL_1384964, EPI_ISL_1384965, EPI_ISL_1384966, EPI_ISL_1384967, EPI_ISL_1384968, EPI_ISL_1384969, EPI_ISL_1384970, EPI_ISL_1384971, EPI_ISL_1384972, EPI_ISL_1384973, EPI_ISL_1384974, EPI_ISL_1384975, EPI_ISL_1384976, EPI_ISL_1384978, EPI_ISL_1384979, EPI_ISL_1384980, EPI_ISL_1384982, EPI_ISL_1384983, EPI_ISL_1384984, EPI_ISL_1384985, EPI_ISL_1384986, EPI_ISL_1384987, EPI_ISL_1384988, EPI_ISL_1384989, EPI_ISL_1384990, EPI_ISL_1384991, EPI_ISL_1384992, EPI_ISL_1384993, EPI_ISL_1384994, EPI_ISL_1384997, EPI_ISL_1384999, EPI_ISL_1385001, EPI_ISL_1385002, EPI_ISL_1385003, EPI_ISL_1385004, EPI_ISL_1385005, EPI_ISL_1385008, EPI_ISL_1385010, EPI_ISL_1385011, EPI_ISL_1385012, EPI_ISL_1385013, EPI_ISL_1385014, EPI_ISL_1385015, EPI_ISL_1385016, EPI_ISL_1385017, EPI_ISL_1385018, EPI_ISL_1385019, EPI_ISL_1385020, EPI_ISL_1385021, EPI_ISL_1385022, EPI_ISL_1385023, EPI_ISL_1385024, EPI_ISL_1385025, EPI_ISL_1385026, EPI_ISL_1385027, EPI_ISL_1385028, EPI_ISL_1385029, EPI_ISL_1385030, EPI_ISL_1385031, EPI_ISL_1385032, EPI_ISL_1385033, EPI_ISL_1385034, EPI_ISL_1385035, EPI_ISL_1385038, EPI_ISL_1385039, EPI_ISL_1385040, EPI_ISL_1385041, EPI_ISL_1385044, EPI_ISL_1385045, EPI_ISL_1385046, EPI_ISL_1385047, EPI_ISL_1385048, EPI_ISL_1385049, EPI_ISL_1385050, EPI_ISL_1385051, EPI_ISL_1385052, EPI_ISL_1385053, EPI_ISL_1385054, EPI_ISL_1385056, EPI_ISL_1385057, EPI_ISL_1385058, EPI_ISL_1385059, EPI_ISL_1385060, EPI_ISL_1385061, EPI_ISL_1385062, EPI_ISL_1385063, EPI_ISL_1385064, EPI_ISL_1385065, EPI_ISL_1385066, EPI_ISL_1385067, EPI_ISL_1385068, EPI_ISL_1385069, EPI_ISL_1385070, EPI_ISL_1385071, EPI_ISL_1385072, EPI_ISL_1385074, EPI_ISL_1385075, EPI_ISL_1385076, EPI_ISL_1385077, EPI_ISL_1385078, EPI_ISL_1385079, EPI_ISL_1385080, EPI_ISL_1385081, EPI_ISL_1385082, EPI_ISL_1385083, EPI_ISL_1385084, EPI_ISL_1385085, EPI_ISL_1385086, EPI_ISL_1385087, EPI_ISL_1385089, EPI_ISL_1385090, EPI_ISL_1385091, EPI_ISL_1385092, EPI_ISL_1385093, EPI_ISL_1385094, EPI_ISL_1385095, EPI_ISL_1385096, EPI_ISL_1385097, EPI_ISL_1385098, EPI_ISL_1385099, EPI_ISL_1385100, EPI_ISL_1385101, EPI_ISL_1385102, EPI_ISL_1385103, EPI_ISL_1385104, EPI_ISL_1385105, EPI_ISL_1385106, EPI_ISL_1385107, EPI_ISL_1385109, EPI_ISL_1385110, EPI_ISL_1385111, EPI_ISL_1385112, EPI_ISL_1385113, EPI_ISL_1385114, EPI_ISL_1385115, EPI_ISL_1385116, EPI_ISL_1385117, EPI_ISL_1385118, EPI_ISL_1385119, EPI_ISL_1385120, EPI_ISL_1385121, EPI_ISL_1385123, EPI_ISL_1385124, EPI_ISL_1385125, EPI_ISL_1385126, EPI_ISL_1385127, EPI_ISL_1385128, EPI_ISL_1385129, EPI_ISL_1385130, EPI_ISL_1385131, EPI_ISL_1385132, EPI_ISL_1385133, EPI_ISL_1385135, EPI_ISL_1385137, EPI_ISL_1385138, EPI_ISL_1385139, EPI_ISL_1385140, EPI_ISL_1385141, EPI_ISL_1385142, EPI_ISL_1385143, EPI_ISL_1385144, EPI_ISL_1385145, EPI_ISL_1385146, EPI_ISL_1385147, EPI_ISL_1385148, EPI_ISL_1385149, EPI_ISL_1385150, EPI_ISL_1385151, EPI_ISL_1385152, EPI_ISL_1385153, EPI_ISL_1385154, EPI_ISL_1385155, EPI_ISL_1385156, EPI_ISL_1385157, EPI_ISL_1385158, EPI_ISL_1385159, EPI_ISL_1385160, EPI_ISL_1385161, EPI_ISL_1385162, EPI_ISL_1385163, EPI_ISL_1385164, EPI_ISL_1385165, EPI_ISL_1385166, EPI_ISL_1385167, EPI_ISL_1385168, EPI_ISL_1385170, EPI_ISL_1385171, EPI_ISL_1385172, EPI_ISL_1385173, EPI_ISL_1385174, EPI_ISL_1385175, EPI_ISL_1385176, EPI_ISL_1385177, EPI_ISL_1385178, EPI_ISL_1385179, EPI_ISL_1385180, EPI_ISL_1385181, EPI_ISL_1385182, EPI_ISL_1385183, EPI_ISL_1385184, EPI_ISL_1385186, EPI_ISL_1385187, EPI_ISL_1385188, EPI_ISL_1385189, EPI_ISL_1385190, EPI_ISL_1385191, EPI_ISL_1385192, EPI_ISL_1385193, EPI_ISL_1385194, EPI_ISL_1385195, EPI_ISL_1385196, EPI_ISL_1385197, EPI_ISL_1385198, EPI_ISL_1385199, EPI_ISL_1385200, EPI_ISL_1385201, EPI_ISL_1385202, EPI_ISL_1385203, EPI_ISL_1385204, EPI_ISL_1385205, EPI_ISL_1385206, EPI_ISL_1385207, EPI_ISL_1385209, EPI_ISL_1385210, EPI_ISL_1385211, EPI_ISL_1385212, EPI_ISL_1385213, EPI_ISL_1385215, EPI_ISL_1385216, EPI_ISL_1385217, EPI_ISL_1385218, EPI_ISL_1385219, EPI_ISL_1385220, EPI_ISL_1385221, EPI_ISL_1385222, EPI_ISL_1385223, EPI_ISL_1385224, EPI_ISL_1385225, EPI_ISL_1385226, EPI_ISL_1385227, EPI_ISL_1385228, EPI_ISL_1385229, EPI_ISL_1385230, EPI_ISL_1385234, EPI_ISL_1385235, EPI_ISL_1385236, EPI_ISL_1385237, EPI_ISL_1385238, EPI_ISL_1385239, EPI_ISL_1385240, EPI_ISL_1385241, EPI_ISL_1385243, EPI_ISL_1385244, EPI_ISL_1385245, EPI_ISL_1385246, EPI_ISL_1385247, EPI_ISL_1385249, EPI_ISL_1385250, EPI_ISL_1385251, EPI_ISL_1385252, EPI_ISL_1385253, EPI_ISL_1385254, EPI_ISL_1385255, EPI_ISL_1385256, EPI_ISL_1385257, EPI_ISL_1385258, EPI_ISL_1385259, EPI_ISL_1385260, EPI_ISL_1385261, EPI_ISL_1385262, EPI_ISL_1385263, EPI_ISL_1385266, EPI_ISL_1385267, EPI_ISL_1385268, EPI_ISL_1385269, EPI_ISL_1385270, EPI_ISL_1385271, EPI_ISL_1385272, EPI_ISL_1385273, EPI_ISL_1385274, EPI_ISL_1385275, EPI_ISL_1385276, EPI_ISL_1385277, EPI_ISL_1385278, EPI_ISL_1385279, EPI_ISL_1385280, EPI_ISL_1385281, EPI_ISL_1385282, EPI_ISL_1385283, EPI_ISL_1385285, EPI_ISL_1385286, EPI_ISL_1385287, EPI_ISL_1385289, EPI_ISL_1385290, EPI_ISL_1385291, EPI_ISL_1385292, EPI_ISL_1385293, EPI_ISL_1385294, EPI_ISL_1385295, EPI_ISL_1385297, EPI_ISL_1385298, EPI_ISL_1385299, EPI_ISL_1385300, EPI_ISL_1385301, EPI_ISL_1385302, EPI_ISL_1385303, EPI_ISL_1385304, EPI_ISL_1385305, EPI_ISL_1385306, EPI_ISL_1385307, EPI_ISL_1385308, EPI_ISL_1385309, EPI_ISL_1385310, EPI_ISL_1385311, EPI_ISL_1385312, EPI_ISL_1385313, EPI_ISL_1385315, EPI_ISL_1385317, EPI_ISL_1385318, EPI_ISL_1385319, EPI_ISL_1385320, EPI_ISL_1385321, EPI_ISL_1385322, EPI_ISL_1385323, EPI_ISL_1385324, EPI_ISL_1385325, EPI_ISL_1385326, EPI_ISL_1385327, EPI_ISL_1385328, EPI_ISL_1385329, EPI_ISL_1385330, EPI_ISL_1385331, EPI_ISL_1385332, EPI_ISL_1385333, EPI_ISL_1385335, EPI_ISL_1385336, EPI_ISL_1385337, EPI_ISL_1385338, EPI_ISL_1385339, EPI_ISL_1385340, EPI_ISL_1385341, EPI_ISL_1385342, EPI_ISL_1385343, EPI_ISL_1385345, EPI_ISL_1385346, EPI_ISL_1385347, EPI_ISL_1385348, EPI_ISL_1385350, EPI_ISL_1385351, EPI_ISL_1385353, EPI_ISL_1385354, EPI_ISL_1385355, EPI_ISL_1385356, EPI_ISL_1385357, EPI_ISL_1385358, EPI_ISL_1385359, EPI_ISL_1385360, EPI_ISL_1385361, EPI_ISL_1385363, EPI_ISL_1385364, EPI_ISL_1385365, EPI_ISL_1385366, EPI_ISL_1385367, EPI_ISL_1385368, EPI_ISL_1385369, EPI_ISL_1385370, EPI_ISL_1385371, EPI_ISL_1385372, EPI_ISL_1385374, EPI_ISL_1385375, EPI_ISL_1385376, EPI_ISL_1385377, EPI_ISL_1385378, EPI_ISL_1385379, EPI_ISL_1385380, EPI_ISL_1385382, EPI_ISL_1385383, EPI_ISL_1385384, EPI_ISL_1385385, EPI_ISL_1385387, EPI_ISL_1385388, EPI_ISL_1385389, EPI_ISL_1385390, EPI_ISL_1385391, EPI_ISL_1385392, EPI_ISL_1385394, EPI_ISL_1385395, EPI_ISL_1385396, EPI_ISL_1385397, EPI_ISL_1385398, EPI_ISL_1385399, EPI_ISL_1385400, EPI_ISL_1385401, EPI_ISL_1385402, EPI_ISL_1385403, EPI_ISL_1385404, EPI_ISL_1385406, EPI_ISL_1385407, EPI_ISL_1385408, EPI_ISL_1385410, EPI_ISL_1385411, EPI_ISL_1385412, EPI_ISL_1385413, EPI_ISL_1385414, EPI_ISL_1385415, EPI_ISL_1385416, EPI_ISL_1385417, EPI_ISL_1385418, EPI_ISL_1385419, EPI_ISL_1385420, EPI_ISL_1385421, EPI_ISL_1385422, EPI_ISL_1385423, EPI_ISL_1385424, EPI_ISL_1385425, EPI_ISL_1385426, EPI_ISL_1385428, EPI_ISL_1385430, EPI_ISL_1385432, EPI_ISL_1385433, EPI_ISL_1385434, EPI_ISL_1385436, EPI_ISL_1385437, EPI_ISL_1385439, EPI_ISL_1385440, EPI_ISL_1385442, EPI_ISL_1385443, EPI_ISL_1385445, EPI_ISL_1385447, EPI_ISL_1385448, EPI_ISL_1385450, EPI_ISL_1385451, EPI_ISL_1385452, EPI_ISL_1385453, EPI_ISL_1385454, EPI_ISL_1385455, EPI_ISL_1385456, EPI_ISL_1385457, EPI_ISL_1385458, EPI_ISL_1385459, EPI_ISL_1385460, EPI_ISL_1385461, EPI_ISL_1385462, EPI_ISL_1385463, EPI_ISL_1385464, EPI_ISL_1385465, EPI_ISL_1385466, EPI_ISL_1385467, EPI_ISL_1385468, EPI_ISL_1385469, EPI_ISL_1385470, EPI_ISL_1385471, EPI_ISL_1385472, EPI_ISL_1385473, EPI_ISL_1385474, EPI_ISL_1385476, EPI_ISL_1385477, EPI_ISL_1385479, EPI_ISL_1385480, EPI_ISL_1385481, EPI_ISL_1385482, EPI_ISL_1385483, EPI_ISL_1385484, EPI_ISL_1385485, EPI_ISL_1385487, EPI_ISL_1385488, EPI_ISL_1385489, EPI_ISL_1385490, EPI_ISL_1385491, EPI_ISL_1385492, EPI_ISL_1385493, EPI_ISL_1385494, EPI_ISL_1385495, EPI_ISL_1385496, EPI_ISL_1385497, EPI_ISL_1385498, EPI_ISL_1385499, EPI_ISL_1385500, EPI_ISL_1385501, EPI_ISL_1385502, EPI_ISL_1385503, EPI_ISL_1385504, EPI_ISL_1385505, EPI_ISL_1385506, EPI_ISL_1385507, EPI_ISL_1385508, EPI_ISL_1385509, EPI_ISL_1385511, EPI_ISL_1385514, EPI_ISL_1385515, EPI_ISL_1385516, EPI_ISL_1385517, EPI_ISL_1385518, EPI_ISL_1385519, EPI_ISL_1385520, EPI_ISL_1385521, EPI_ISL_1385522, EPI_ISL_1385523, EPI_ISL_1385525, EPI_ISL_1385526, EPI_ISL_1385527, EPI_ISL_1385528, EPI_ISL_1385529, EPI_ISL_1385530, EPI_ISL_1385531, EPI_ISL_1385532, EPI_ISL_1385533, EPI_ISL_1385535, EPI_ISL_1385536, EPI_ISL_1385537, EPI_ISL_1385538, EPI_ISL_1385539, EPI_ISL_1385540, EPI_ISL_1385541, EPI_ISL_1385543, EPI_ISL_1385544, EPI_ISL_1385545, EPI_ISL_1385546, EPI_ISL_1385547, EPI_ISL_1385548, EPI_ISL_1385549, EPI_ISL_1385550, EPI_ISL_1385551, EPI_ISL_1385552, EPI_ISL_1385553, EPI_ISL_1385554, EPI_ISL_1385555, EPI_ISL_1385556, EPI_ISL_1385557, EPI_ISL_1385558, EPI_ISL_1385559, EPI_ISL_1385560, EPI_ISL_1385561, EPI_ISL_1385562, EPI_ISL_1385563, EPI_ISL_1385564, EPI_ISL_1385566, EPI_ISL_1385567, EPI_ISL_1385569, EPI_ISL_1385570, EPI_ISL_1385571, EPI_ISL_1385572, EPI_ISL_1385573, EPI_ISL_1385574, EPI_ISL_1385575, EPI_ISL_1385576, EPI_ISL_1385577, EPI_ISL_1385578, EPI_ISL_1385579, EPI_ISL_1385580, EPI_ISL_1385581, EPI_ISL_1385582, EPI_ISL_1385583, EPI_ISL_1385584, EPI_ISL_1385585, EPI_ISL_1385586, EPI_ISL_1385587, EPI_ISL_1385588, EPI_ISL_1385589, EPI_ISL_1385590, EPI_ISL_1385591, EPI_ISL_1385592, EPI_ISL_1385593, EPI_ISL_1385594, EPI_ISL_1385595, EPI_ISL_1385597, EPI_ISL_1385598, EPI_ISL_1385599, EPI_ISL_1385600, EPI_ISL_1385601, EPI_ISL_1385602, EPI_ISL_1385603, EPI_ISL_1385604, EPI_ISL_1385605, EPI_ISL_1385606, EPI_ISL_1385607, EPI_ISL_1385608, EPI_ISL_1385609, EPI_ISL_1385611, EPI_ISL_1385612, EPI_ISL_1385613, EPI_ISL_1385614, EPI_ISL_1385615, EPI_ISL_1385616, EPI_ISL_1385617, EPI_ISL_1385618, EPI_ISL_1385619, EPI_ISL_1385620, EPI_ISL_1385622, EPI_ISL_1385623, EPI_ISL_1385624, EPI_ISL_1385625, EPI_ISL_1385627, EPI_ISL_1385628, EPI_ISL_1385629, EPI_ISL_1385630, EPI_ISL_1385631, EPI_ISL_1385632, EPI_ISL_1385633, EPI_ISL_1385634, EPI_ISL_1385635, EPI_ISL_1385636, EPI_ISL_1385637, EPI_ISL_1385638, EPI_ISL_1385640, EPI_ISL_1385641, EPI_ISL_1385642, EPI_ISL_1385643, EPI_ISL_1385644, EPI_ISL_1385645, EPI_ISL_1385646, EPI_ISL_1385647, EPI_ISL_1385648, EPI_ISL_1385649, EPI_ISL_1385650, EPI_ISL_1385651, EPI_ISL_1385652, EPI_ISL_1385653, EPI_ISL_1385654, EPI_ISL_1385655, EPI_ISL_1385656, EPI_ISL_1385658, EPI_ISL_1385659, EPI_ISL_1385660, EPI_ISL_1385661, EPI_ISL_1385662, EPI_ISL_1385663, EPI_ISL_1385664, EPI_ISL_1385665, EPI_ISL_1385666, EPI_ISL_1385667, EPI_ISL_1385668, EPI_ISL_1385669, EPI_ISL_1385670, EPI_ISL_1385671, EPI_ISL_1385672, EPI_ISL_1385673, EPI_ISL_1385674, EPI_ISL_1385675, EPI_ISL_1385676, EPI_ISL_1385678, EPI_ISL_1385680, EPI_ISL_1385681, EPI_ISL_1385682, EPI_ISL_1385683, EPI_ISL_1385684, EPI_ISL_1385686, EPI_ISL_1385688, EPI_ISL_1385689, EPI_ISL_1385690, EPI_ISL_1385691, EPI_ISL_1385692, EPI_ISL_1385693, EPI_ISL_1385695, EPI_ISL_1385696, EPI_ISL_1385697, EPI_ISL_1385698, EPI_ISL_1385700, EPI_ISL_1385701, EPI_ISL_1385703, EPI_ISL_1385704, EPI_ISL_1385705, EPI_ISL_1385706, EPI_ISL_1385708, EPI_ISL_1385709, EPI_ISL_1385711, EPI_ISL_1385712, EPI_ISL_1385713, EPI_ISL_1385714, EPI_ISL_1385715, EPI_ISL_1385716, EPI_ISL_1385717, EPI_ISL_1385718, EPI_ISL_1385719, EPI_ISL_1385720, EPI_ISL_1385721, EPI_ISL_1385722, EPI_ISL_1385723, EPI_ISL_1385724, EPI_ISL_1385725, EPI_ISL_1385726, EPI_ISL_1385727, EPI_ISL_1385728, EPI_ISL_1385729, EPI_ISL_1385730, EPI_ISL_1385731, EPI_ISL_1385733, EPI_ISL_1385734, EPI_ISL_1385735, EPI_ISL_1385736, EPI_ISL_1385737, EPI_ISL_1385738, EPI_ISL_1385739, EPI_ISL_1385740, EPI_ISL_1385741, EPI_ISL_1385742, EPI_ISL_1385743, EPI_ISL_1385744, EPI_ISL_1385745, EPI_ISL_1385746, EPI_ISL_1385747, EPI_ISL_1385748, EPI_ISL_1385749, EPI_ISL_1385750, EPI_ISL_1385751, EPI_ISL_1385752, EPI_ISL_1385753, EPI_ISL_1385754, EPI_ISL_1385755, EPI_ISL_1385757, EPI_ISL_1385758, EPI_ISL_1385759, EPI_ISL_1385760, EPI_ISL_1385762, EPI_ISL_1385763, EPI_ISL_1385764, EPI_ISL_1385765, EPI_ISL_1385766, EPI_ISL_1385767, EPI_ISL_1385768, EPI_ISL_1385769, EPI_ISL_1385770, EPI_ISL_1385771, EPI_ISL_1385772, EPI_ISL_1385773, EPI_ISL_1385774, EPI_ISL_1385775, EPI_ISL_1385776, EPI_ISL_1385777, EPI_ISL_1385778, EPI_ISL_1385779, EPI_ISL_1385780, EPI_ISL_1385781, EPI_ISL_1385782, EPI_ISL_1385783 | | | |
| see above | Pandemic Response Lab - NYC | Pandemic Response Lab, R&D | Henry Lee, Michael Hammerling, Melissa Hopkins, Cybill del Castillo, Shinyoung Clair Kang, William Ward, Pradeep Bugga, Sol Rey, Dylan Law, Haiping Hao, Jon Laurent |
| EPI_ISL_1386661, EPI_ISL_1389160, EPI_ISL_1389161, EPI_ISL_1389162, EPI_ISL_1389275, EPI_ISL_1389397, EPI_ISL_1389399, EPI_ISL_1390768 | Ohio Department of Health Laboratory | Ohio Department of Health Laboratory | Holmes, Jennifer; Eric Brandt, Keoni Omura, Glen McGillivary, Caitlin McDonnell, Jade Mowery, Stephanie Mccracken, Tyler Payne, Kirtana Ramadugu, Erica Leasure, Brent Lee, Kelsey Florek, Heather Blankenship, Allison Black, Quanta Brown, and Tammy Bannerman |
| EPI_ISL_1391363 | Ohio State University Wexner Medical Center | Polaris Molecular Laboratory | Wang H, Jones D , Chappell D, Tu H, Snyder P, Leber A |
| EPI_ISL_1391372, EPI_ISL_1391373, EPI_ISL_1391374, EPI_ISL_1391375, EPI_ISL_1391376, EPI_ISL_1391377, EPI_ISL_1391378, EPI_ISL_1391379, EPI_ISL_1391380, EPI_ISL_1391381, EPI_ISL_1391382, EPI_ISL_1391383, EPI_ISL_1391384, EPI_ISL_1391385, EPI_ISL_1391386, EPI_ISL_1391387, EPI_ISL_1391388, EPI_ISL_1391389, EPI_ISL_1391390, EPI_ISL_1391391, EPI_ISL_1391392 | | | |
| see above | DC Public Health Lab/ Dept. of Forensic Sciences | DC Public Health Lab/ Dept. of Forensic Sciences | Janis Doss, Scott Nguyen, Elizabeth Zelaya, Sarah Scott, Connie Maza, Monica Mann, Brittany Hamilton, David Payne, Jocelyn Hauser |
| EPI_ISL_1391413 | Baylor Scott & White-Irving | Baylor Scott & White-Temple | Ari Rao, Linden Morales, Kimberly Walker, Marcus Volz, Shelby Hendrickson, Caitlin Maloney |
| EPI_ISL_1391415 | Baylor Scott & White-Waxahachie | Baylor Scott & White-Temple | Ari Rao, Linden Morales, Kimberly Walker, Marcus Volz, Shelby Hendrickson, Caitlin Maloney |
| EPI_ISL_1391416 | Baylor Scott & White-Temple | Baylor Scott & White-Temple | Ari Rao, Linden Morales, Kimberly Walker, Marcus Volz, Shelby Hendrickson, Caitlin Maloney |
| EPI_ISL_1391417, EPI_ISL_1391418, EPI_ISL_1391419, EPI_ISL_1391420 | Quest Diagnostics Incorporated | Centers for Disease Control and Prevention Division of Viral Diseases, Pathogen Discovery | Peter W. Cook, Dakota Howard, Dhwani Batra, Ben L. Rambo-Martin, S. H. Rosenthal, A. Gerasimova, R. M. Kagan, B. Anderson, M. Hua, Y. Liu, L.E. Bernstein, K.E. Livingston, A. Perez, I. A. Shlyakhter, R. V. Rolando, R. Owen, P. Tanpaiboon, F. Lacbawan, Clinton R. Paden, Suxiang Tong, Duncan MacCannell |
| EPI_ISL_1391421 | Baylor Scott & White-Temple | Baylor Scott & White-Temple | Ari Rao, Linden Morales, Kimberly Walker, Marcus Volz, Shelby Hendrickson, Caitlin Maloney |
| EPI_ISL_1391422, EPI_ISL_1391423, EPI_ISL_1391424, EPI_ISL_1391425, EPI_ISL_1391426, EPI_ISL_1391427, EPI_ISL_1391428, EPI_ISL_1391429, EPI_ISL_1391430, EPI_ISL_1391431, EPI_ISL_1391432, EPI_ISL_1391433, EPI_ISL_1391434, EPI_ISL_1391435, EPI_ISL_1391436, EPI_ISL_1391437, EPI_ISL_1391438, EPI_ISL_1391439, EPI_ISL_1391440, EPI_ISL_1391441, EPI_ISL_1391442, EPI_ISL_1391443, EPI_ISL_1391444, EPI_ISL_1391445, EPI_ISL_1391446, EPI_ISL_1391447, EPI_ISL_1391448, EPI_ISL_1391449, EPI_ISL_1391450, EPI_ISL_1391451, EPI_ISL_1391452, EPI_ISL_1391453, EPI_ISL_1391454, EPI_ISL_1391455, EPI_ISL_1391456, EPI_ISL_1391457, EPI_ISL_1391458, EPI_ISL_1391459, EPI_ISL_1391460, EPI_ISL_1391461, EPI_ISL_1391462, EPI_ISL_1391463, EPI_ISL_1391464, EPI_ISL_1391465, EPI_ISL_1391466, EPI_ISL_1391467, EPI_ISL_1391468, EPI_ISL_1391469, EPI_ISL_1391470, EPI_ISL_1391471, EPI_ISL_1391472, EPI_ISL_1391473, EPI_ISL_1391474, EPI_ISL_1391475, EPI_ISL_1391476, EPI_ISL_1391477, EPI_ISL_1391478, EPI_ISL_1391479, EPI_ISL_1391480, EPI_ISL_1391481, EPI_ISL_1391482, EPI_ISL_1391483, EPI_ISL_1391484, EPI_ISL_1391485, EPI_ISL_1391486, EPI_ISL_1391487, EPI_ISL_1391488, EPI_ISL_1391489, EPI_ISL_1391490, EPI_ISL_1391491, EPI_ISL_1391492, EPI_ISL_1391493, EPI_ISL_1391494, EPI_ISL_1391495, EPI_ISL_1391496, EPI_ISL_1391497, EPI_ISL_1391498, EPI_ISL_1391499, EPI_ISL_1391500, EPI_ISL_1391501, EPI_ISL_1391502, EPI_ISL_1391503, EPI_ISL_1391504, EPI_ISL_1391505, EPI_ISL_1391506, EPI_ISL_1391507, EPI_ISL_1391508, EPI_ISL_1391509, EPI_ISL_1391510, EPI_ISL_1391511, EPI_ISL_1391512, EPI_ISL_1391513, EPI_ISL_1391514, EPI_ISL_1391515, EPI_ISL_1391516, EPI_ISL_1391517, EPI_ISL_1391518, EPI_ISL_1391519, EPI_ISL_1391520, EPI_ISL_1391521, EPI_ISL_1391522, EPI_ISL_1391523, EPI_ISL_1391524, EPI_ISL_1391525, EPI_ISL_1391526, EPI_ISL_1391527, EPI_ISL_1391528, EPI_ISL_1391529, EPI_ISL_1391530, EPI_ISL_1391531, EPI_ISL_1391532, EPI_ISL_1391533, EPI_ISL_1391534, EPI_ISL_1391535, EPI_ISL_1391536, EPI_ISL_1391537, EPI_ISL_1391538, EPI_ISL_1391539, EPI_ISL_1391540, EPI_ISL_1391541, EPI_ISL_1391542, EPI_ISL_1391543, EPI_ISL_1391544, EPI_ISL_1391545, EPI_ISL_1391546, EPI_ISL_1391547, EPI_ISL_1391548, EPI_ISL_1391549, EPI_ISL_1391550, EPI_ISL_1391551, EPI_ISL_1391552, EPI_ISL_1391553, EPI_ISL_1391554, EPI_ISL_1391555, EPI_ISL_1391556, EPI_ISL_1391557, EPI_ISL_1391558, EPI_ISL_1391559, EPI_ISL_1391560, EPI_ISL_1391561, EPI_ISL_1391562, EPI_ISL_1391563, EPI_ISL_1391564, EPI_ISL_1391565, EPI_ISL_1391566, EPI_ISL_1391567, EPI_ISL_1391568, EPI_ISL_1391569, EPI_ISL_1391570, EPI_ISL_1391571, EPI_ISL_1391572, EPI_ISL_1391573, EPI_ISL_1391574, EPI_ISL_1391575, EPI_ISL_1391576, EPI_ISL_1391577, EPI_ISL_1391578, EPI_ISL_1391579, EPI_ISL_1391580, EPI_ISL_1391581, EPI_ISL_1391582, EPI_ISL_1391583, EPI_ISL_1391584, EPI_ISL_1391585, EPI_ISL_1391586, EPI_ISL_1391587, EPI_ISL_1391588, EPI_ISL_1391589, EPI_ISL_1391590, EPI_ISL_1391591, EPI_ISL_1391592, EPI_ISL_1391593, EPI_ISL_1391594, EPI_ISL_1391595, EPI_ISL_1391596, EPI_ISL_1391597, EPI_ISL_1391598, EPI_ISL_1391599, EPI_ISL_1391600, EPI_ISL_1391601, EPI_ISL_1391602, EPI_ISL_1391603, EPI_ISL_1391604, EPI_ISL_1391605, EPI_ISL_1391606, EPI_ISL_1391607, EPI_ISL_1391608, EPI_ISL_1391609, EPI_ISL_1391610, EPI_ISL_1391611, EPI_ISL_1391612, EPI_ISL_1391613, EPI_ISL_1391614, EPI_ISL_1391615, EPI_ISL_1391616, EPI_ISL_1391617, EPI_ISL_1391618, EPI_ISL_1391619, EPI_ISL_1391620 | | | |
| see above | Quest Diagnostics Incorporated | Centers for Disease Control and Prevention Division of Viral Diseases, Pathogen Discovery | Peter W. Cook, Dakota Howard, Dhwani Batra, Ben L. Rambo-Martin, S. H. Rosenthal, A. Gerasimova, R. M. Kagan, B. Anderson, M. Hua, Y. Liu, L.E. Bernstein, K.E. Livingston, A. Perez, I. A. Shlyakhter, R. V. Rolando, R. Owen, P. Tanpaiboon, F. Lacbawan, Clinton R. Paden, Suxiang Tong, Duncan MacCannell |
| EPI_ISL_1391621 | Baylor Scott & White-Temple | Baylor Scott & White-Temple | Ari Rao, Linden Morales, Kimberly Walker, Marcus Volz, Shelby Hendrickson, Caitlin Maloney |
| EPI_ISL_1391622, EPI_ISL_1391623, EPI_ISL_1391624, EPI_ISL_1391625, EPI_ISL_1391626, EPI_ISL_1391627, EPI_ISL_1391628, EPI_ISL_1391629, EPI_ISL_1391630, EPI_ISL_1391631, EPI_ISL_1391632, EPI_ISL_1391633, EPI_ISL_1391634, EPI_ISL_1391635, EPI_ISL_1391636, EPI_ISL_1391637, EPI_ISL_1391638, EPI_ISL_1391639, EPI_ISL_1391640, EPI_ISL_1391641, EPI_ISL_1391642, EPI_ISL_1391643, EPI_ISL_1391644, EPI_ISL_1391645, EPI_ISL_1391646, EPI_ISL_1391647, EPI_ISL_1391648, EPI_ISL_1391649, EPI_ISL_1391650, EPI_ISL_1391651, EPI_ISL_1391652, EPI_ISL_1391653, EPI_ISL_1391654, EPI_ISL_1391655, EPI_ISL_1391656, EPI_ISL_1391657, EPI_ISL_1391658, EPI_ISL_1391659, EPI_ISL_1391660, EPI_ISL_1391661, EPI_ISL_1391662, EPI_ISL_1391663, EPI_ISL_1391664, EPI_ISL_1391665, EPI_ISL_1391666, EPI_ISL_1391667, EPI_ISL_1391668, EPI_ISL_1391669, EPI_ISL_1391670, EPI_ISL_1391671, EPI_ISL_1391672, EPI_ISL_1391673, EPI_ISL_1391674, EPI_ISL_1391675, EPI_ISL_1391676, EPI_ISL_1391677, EPI_ISL_1391678, EPI_ISL_1391679, EPI_ISL_1391680, EPI_ISL_1391681, EPI_ISL_1391682, EPI_ISL_1391683, EPI_ISL_1391684, EPI_ISL_1391685, EPI_ISL_1391686, EPI_ISL_1391687, EPI_ISL_1391688, EPI_ISL_1391689, EPI_ISL_1391690, EPI_ISL_1391691, EPI_ISL_1391692, EPI_ISL_1391693, EPI_ISL_1391694, EPI_ISL_1391695, EPI_ISL_1391696, EPI_ISL_1391697, EPI_ISL_1391698 | | | |
| see above | Quest Diagnostics Incorporated | Centers for Disease Control and Prevention Division of Viral Diseases, Pathogen Discovery | Peter W. Cook, Dakota Howard, Dhwani Batra, Ben L. Rambo-Martin, S. H. Rosenthal, A. Gerasimova, R. M. Kagan, B. Anderson, M. Hua, Y. Liu, L.E. Bernstein, K.E. Livingston, A. Perez, I. A. Shlyakhter, R. V. Rolando, R. Owen, P. Tanpaiboon, F. Lacbawan, Clinton R. Paden, Suxiang Tong, Duncan MacCannell |
| EPI_ISL_1391699, EPI_ISL_1391700, EPI_ISL_1391701, EPI_ISL_1391702, EPI_ISL_1391703, EPI_ISL_1391704, EPI_ISL_1391705, EPI_ISL_1391706, EPI_ISL_1391707, EPI_ISL_1391708, EPI_ISL_1391709, EPI_ISL_1391710, EPI_ISL_1391711, EPI_ISL_1391712, EPI_ISL_1391713, EPI_ISL_1391714, EPI_ISL_1391715, EPI_ISL_1391716, EPI_ISL_1391717, EPI_ISL_1391718, EPI_ISL_1391719, EPI_ISL_1391720, EPI_ISL_1391721, EPI_ISL_1391722, EPI_ISL_1391723, EPI_ISL_1391724, EPI_ISL_1391725, EPI_ISL_1391726, EPI_ISL_1391727, EPI_ISL_1391728, EPI_ISL_1391729, EPI_ISL_1391730, EPI_ISL_1391731, EPI_ISL_1391732, EPI_ISL_1391733, EPI_ISL_1391734, EPI_ISL_1391735, EPI_ISL_1391736, EPI_ISL_1391737, EPI_ISL_1391738, EPI_ISL_1391739, EPI_ISL_1391740, EPI_ISL_1391741, EPI_ISL_1391742, EPI_ISL_1391743, EPI_ISL_1391744, EPI_ISL_1391745, EPI_ISL_1391746, EPI_ISL_1391747, EPI_ISL_1391748, EPI_ISL_1391749, EPI_ISL_1391750, EPI_ISL_1391751, EPI_ISL_1391752, EPI_ISL_1391753, EPI_ISL_1391754, EPI_ISL_1391755, EPI_ISL_1391756, EPI_ISL_1391757, EPI_ISL_1391758, EPI_ISL_1391759, EPI_ISL_1391760, EPI_ISL_1391761, EPI_ISL_1391762, EPI_ISL_1391763, EPI_ISL_1391764, EPI_ISL_1391765, EPI_ISL_1391766, EPI_ISL_1391767, EPI_ISL_1391768, EPI_ISL_1391769, EPI_ISL_1391770, EPI_ISL_1391771, EPI_ISL_1391772, EPI_ISL_1391773, EPI_ISL_1391774, EPI_ISL_1391775, EPI_ISL_1391776, EPI_ISL_1391777, EPI_ISL_1391778, EPI_ISL_1391779, EPI_ISL_1391780, EPI_ISL_1391782, EPI_ISL_1391783, EPI_ISL_1391784, EPI_ISL_1391785, EPI_ISL_1391786, EPI_ISL_1391787, EPI_ISL_1391788, EPI_ISL_1391789, EPI_ISL_1391790, EPI_ISL_1391791, EPI_ISL_1391792, EPI_ISL_1391793, EPI_ISL_1391794, EPI_ISL_1391795, EPI_ISL_1391796, EPI_ISL_1391797, EPI_ISL_1391798, EPI_ISL_1391799, EPI_ISL_1391800, EPI_ISL_1391801, EPI_ISL_1391802, EPI_ISL_1391803, EPI_ISL_1391804, EPI_ISL_1391805, EPI_ISL_1391806, EPI_ISL_1391807, EPI_ISL_1391808, EPI_ISL_1391809, EPI_ISL_1391810, EPI_ISL_1391811, EPI_ISL_1391812, EPI_ISL_1391813, EPI_ISL_1391814, EPI_ISL_1391815, EPI_ISL_1391816, EPI_ISL_1391817, EPI_ISL_1391818, EPI_ISL_1391819, EPI_ISL_1391820, EPI_ISL_1391821, EPI_ISL_1391822, EPI_ISL_1391823, EPI_ISL_1391824, EPI_ISL_1391825, EPI_ISL_1391826, EPI_ISL_1391827, EPI_ISL_1391828, EPI_ISL_1391829, EPI_ISL_1391830, EPI_ISL_1391831, EPI_ISL_1391832, EPI_ISL_1391833, EPI_ISL_1391834, EPI_ISL_1391835, EPI_ISL_1391836, EPI_ISL_1391837, EPI_ISL_1391838, EPI_ISL_1391839, EPI_ISL_1391841, EPI_ISL_1391842, EPI_ISL_1391843, EPI_ISL_1391844, EPI_ISL_1391845, EPI_ISL_1391846, EPI_ISL_1391847, EPI_ISL_1391848, EPI_ISL_1391849, EPI_ISL_1391850, EPI_ISL_1391851, EPI_ISL_1391852, EPI_ISL_1391854, EPI_ISL_1391855, EPI_ISL_1391856, EPI_ISL_1391857, EPI_ISL_1391858, EPI_ISL_1391859, EPI_ISL_1391860, EPI_ISL_1391861, EPI_ISL_1391864, EPI_ISL_1391865, EPI_ISL_1391866, EPI_ISL_1391867, EPI_ISL_1391868, EPI_ISL_1391869, EPI_ISL_1391870, EPI_ISL_1391871, EPI_ISL_1391872, EPI_ISL_1391873, EPI_ISL_1391874, EPI_ISL_1391875, EPI_ISL_1391876, EPI_ISL_1391877, EPI_ISL_1391878, EPI_ISL_1391879, EPI_ISL_1391880, EPI_ISL_1391881, EPI_ISL_1391882, EPI_ISL_1391883, EPI_ISL_1391884, EPI_ISL_1391885, EPI_ISL_1391886, EPI_ISL_1391887, EPI_ISL_1391888, EPI_ISL_1391889, EPI_ISL_1391890, EPI_ISL_1391891, EPI_ISL_1391892, EPI_ISL_1391893, EPI_ISL_1391894, EPI_ISL_1391895, EPI_ISL_1391896, EPI_ISL_1391897, EPI_ISL_1391898, EPI_ISL_1391899, EPI_ISL_1391900, EPI_ISL_1391901, EPI_ISL_1391902, EPI_ISL_1391903, EPI_ISL_1391904, EPI_ISL_1391905, EPI_ISL_1391906, EPI_ISL_1391907, EPI_ISL_1391908, EPI_ISL_1391909, EPI_ISL_1391910, EPI_ISL_1391911, EPI_ISL_1391912, EPI_ISL_1391913, EPI_ISL_1391914, EPI_ISL_1391915, EPI_ISL_1391916, EPI_ISL_1391918, EPI_ISL_1391919, EPI_ISL_1391920, EPI_ISL_1391921, EPI_ISL_1391922, EPI_ISL_1391923, EPI_ISL_1391924, EPI_ISL_1391925, EPI_ISL_1391927, EPI_ISL_1391928, EPI_ISL_1391929, EPI_ISL_1391930, EPI_ISL_1391931, EPI_ISL_1391932, EPI_ISL_1391933, EPI_ISL_1391934, EPI_ISL_1391935, EPI_ISL_1391936, EPI_ISL_1391937, EPI_ISL_1391938, EPI_ISL_1391939, EPI_ISL_1391941, EPI_ISL_1391942, EPI_ISL_1391943, EPI_ISL_1391944, EPI_ISL_1391945, EPI_ISL_1391946, EPI_ISL_1391947, EPI_ISL_1391948, EPI_ISL_1391949, EPI_ISL_1391950, EPI_ISL_1391951, EPI_ISL_1391952, EPI_ISL_1391953, EPI_ISL_1391955, EPI_ISL_1391957, EPI_ISL_1391959, EPI_ISL_1391960, EPI_ISL_1391961, EPI_ISL_1391962, EPI_ISL_1391963, EPI_ISL_1391965, EPI_ISL_1391966, EPI_ISL_1391967, EPI_ISL_1391968, EPI_ISL_1391969, EPI_ISL_1391970, EPI_ISL_1391971, EPI_ISL_1391972, EPI_ISL_1391973, EPI_ISL_1391974, | | | |

| | | | |
|---|---|---|---|
| EPI_ISL_1391975, EPI_ISL_1391976, EPI_ISL_1391977, EPI_ISL_1391979, EPI_ISL_1391980, EPI_ISL_1391981, EPI_ISL_1391982, EPI_ISL_1391983, EPI_ISL_1391984, EPI_ISL_1391985, EPI_ISL_1391986, EPI_ISL_1391987, EPI_ISL_1391988, EPI_ISL_1391989, EPI_ISL_1391990, EPI_ISL_1391991, EPI_ISL_1391992, EPI_ISL_1391993, EPI_ISL_1391994, EPI_ISL_1391995, EPI_ISL_1391996, EPI_ISL_1391997, EPI_ISL_1391998, EPI_ISL_1391999 | | | |
| see above | Helix/Illumina | Centers for Disease Control and Prevention Division of Viral Diseases, Pathogen Discovery | Peter W. Cook, Dakota Howard, Dhwani Batra, Ben L. Rambo-Martin, Eileen de Feo, Jan Antico, Christine Tran, Matthew Tolentino, Shannon Wickline, Kim Gietzen, Brad Sickler, Jingtao Liu, Eric Allen, Phil Febbo, Summer Galloway, Nicole L. Washington, Simon White, Geraint Levan, Kelly Schiabor Barrett, Elizabeth Cirulli, Alexandre Bolze, Ary Ascencio, Charlotte Rivera-Garcia, Ryan Cho, Jason Nguyen, Sherry Wang, Jimmy Ramirez, Tyler Cassens, Efren Sandoval, Magnus Isaksson, William Lee, David Becker, Marc Laurent, James Lu, Clinton R. Paden, Suxiang Tong, Duncan MacCannell |
| EPI_ISL_1392000 | TGen North | TGen North | "Jolene Bowers, Heather Centner, Chris French, Hayley Yaglom, Ashlyn Pfeiffer, Darrin Lemmer, Dave Engelthaler, The Arizona COVID Genomics Union (ACGU)" |
| EPI_ISL_1392001, EPI_ISL_1392002, EPI_ISL_1392003, EPI_ISL_1392005, EPI_ISL_1392006, EPI_ISL_1392007, EPI_ISL_1392008, EPI_ISL_1392009, EPI_ISL_1392010, EPI_ISL_1392011, EPI_ISL_1392016, EPI_ISL_1392017 | | | |
| see above | Sonora Quest Laboratories | TGen North | "Jolene Bowers, Heather Centner, Chris French, Hayley Yaglom, Ashlyn Pfeiffer, Darrin Lemmer, Dave Engelthaler, The Arizona COVID Genomics Union (ACGU)" |
| EPI_ISL_1392019 | TGen North | TGen North | "Jolene Bowers, Heather Centner, Chris French, Hayley Yaglom, Ashlyn Pfeiffer, Darrin Lemmer, Dave Engelthaler, The Arizona COVID Genomics Union (ACGU)" |
| EPI_ISL_1392020 | Sonora Quest Laboratories | TGen North | "Jolene Bowers, Heather Centner, Chris French, Hayley Yaglom, Ashlyn Pfeiffer, Darrin Lemmer, Dave Engelthaler, The Arizona COVID Genomics Union (ACGU)" |
| EPI_ISL_1392021 | TGen North | TGen North | "Jolene Bowers, Heather Centner, Chris French, Hayley Yaglom, Ashlyn Pfeiffer, Darrin Lemmer, Dave Engelthaler, The Arizona COVID Genomics Union (ACGU)" |
| EPI_ISL_1392022, EPI_ISL_1392023, EPI_ISL_1392024, EPI_ISL_1392025, EPI_ISL_1392033, EPI_ISL_1392034, EPI_ISL_1392035, EPI_ISL_1392037, EPI_ISL_1392038, EPI_ISL_1392039, EPI_ISL_1392040, EPI_ISL_1392041, EPI_ISL_1392042, EPI_ISL_1392043, EPI_ISL_1392044, EPI_ISL_1392046, EPI_ISL_1392047, EPI_ISL_1392048, EPI_ISL_1392049, EPI_ISL_1392052, EPI_ISL_1392053, EPI_ISL_1392054, EPI_ISL_1392057, EPI_ISL_1392058, EPI_ISL_1392059, EPI_ISL_1392062, EPI_ISL_1392063, EPI_ISL_1392064, EPI_ISL_1392065, EPI_ISL_1392066, EPI_ISL_1392067, EPI_ISL_1392069, EPI_ISL_1392072, EPI_ISL_1392073, EPI_ISL_1392074, EPI_ISL_1392075, EPI_ISL_1392076, EPI_ISL_1392077, EPI_ISL_1392078, EPI_ISL_1392079, EPI_ISL_1392083, EPI_ISL_1392086, EPI_ISL_1392088, EPI_ISL_1392089, EPI_ISL_1392090, EPI_ISL_1392091, EPI_ISL_1392092, EPI_ISL_1392093, EPI_ISL_1392094, EPI_ISL_1392095, EPI_ISL_1392096, EPI_ISL_1392097, EPI_ISL_1392098, EPI_ISL_1392099, EPI_ISL_1392100, EPI_ISL_1392101, EPI_ISL_1392102, EPI_ISL_1392103, EPI_ISL_1392105, EPI_ISL_1392107, EPI_ISL_1392108, EPI_ISL_1392109, EPI_ISL_1392111, EPI_ISL_1392112, EPI_ISL_1392114, EPI_ISL_1392115, EPI_ISL_1392116, EPI_ISL_1392117, EPI_ISL_1392118, EPI_ISL_1392119, EPI_ISL_1392120, EPI_ISL_1392121, EPI_ISL_1392122, EPI_ISL_1392123, EPI_ISL_1392126, EPI_ISL_1392127, EPI_ISL_1392128, EPI_ISL_1392130, EPI_ISL_1392131, EPI_ISL_1392134, EPI_ISL_1392135, EPI_ISL_1392137, EPI_ISL_1392138 | | | |
| see above | Sonora Quest Laboratories | TGen North | "Jolene Bowers, Heather Centner, Chris French, Hayley Yaglom, Ashlyn Pfeiffer, Darrin Lemmer, Dave Engelthaler, The Arizona COVID Genomics Union (ACGU)" |
| EPI_ISL_1392140 | TGen North | TGen North | "Jolene Bowers, Heather Centner, Chris French, Hayley Yaglom, Ashlyn Pfeiffer, Darrin Lemmer, Dave Engelthaler, The Arizona COVID Genomics Union (ACGU)" |
| EPI_ISL_1392142, EPI_ISL_1392144, EPI_ISL_1392145, EPI_ISL_1392146, EPI_ISL_1392147, EPI_ISL_1392148, EPI_ISL_1392150, EPI_ISL_1392151, EPI_ISL_1392152, EPI_ISL_1392154, EPI_ISL_1392155 | | | |
| see above | Sonora Quest Laboratories | TGen North | "Jolene Bowers, Heather Centner, Chris French, Hayley Yaglom, Ashlyn Pfeiffer, Darrin Lemmer, Dave Engelthaler, The Arizona COVID Genomics Union (ACGU)" |
| EPI_ISL_1392156 | TGen North | TGen North | "Jolene Bowers, Heather Centner, Chris French, Hayley Yaglom, Ashlyn Pfeiffer, Darrin Lemmer, Dave Engelthaler, The Arizona COVID Genomics Union (ACGU)" |
| EPI_ISL_1392157, EPI_ISL_1392159, EPI_ISL_1392161 | Sonora Quest Laboratories | TGen North | "Jolene Bowers, Heather Centner, Chris French, Hayley Yaglom, Ashlyn Pfeiffer, Darrin Lemmer, Dave Engelthaler, The Arizona COVID Genomics Union (ACGU)" |
| EPI_ISL_1392164 | TGen North | TGen North | "Jolene Bowers, Heather Centner, Chris French, Hayley Yaglom, Ashlyn Pfeiffer, Darrin Lemmer, Dave Engelthaler, The Arizona COVID Genomics Union (ACGU)" |
| EPI_ISL_1392165, EPI_ISL_1392166, EPI_ISL_1392169, EPI_ISL_1392170, EPI_ISL_1392172 | Sonora Quest Laboratories | TGen North | "Jolene Bowers, Heather Centner, Chris French, Hayley Yaglom, Ashlyn Pfeiffer, Darrin Lemmer, Dave Engelthaler, The Arizona COVID Genomics Union (ACGU)" |
| EPI_ISL_1392173 | TGen North | TGen North | "Jolene Bowers, Heather Centner, Chris French, Hayley Yaglom, Ashlyn Pfeiffer, Darrin Lemmer, Dave Engelthaler, The Arizona COVID Genomics Union (ACGU)" |
| EPI_ISL_1392174, EPI_ISL_1392175, EPI_ISL_1392176 | Sonora Quest Laboratories | TGen North | "Jolene Bowers, Heather Centner, Chris French, Hayley Yaglom, Ashlyn Pfeiffer, Darrin Lemmer, Dave Engelthaler, The Arizona COVID Genomics Union (ACGU)" |
| EPI_ISL_1392177 | TGen North | TGen North | "Jolene Bowers, Heather Centner, Chris French, Hayley Yaglom, Ashlyn Pfeiffer, Darrin Lemmer, Dave Engelthaler, The Arizona COVID Genomics Union (ACGU)" |
| EPI_ISL_1392178, EPI_ISL_1392179, EPI_ISL_1392181, EPI_ISL_1392182, EPI_ISL_1392184, EPI_ISL_1392185, EPI_ISL_1392186, EPI_ISL_1392187, EPI_ISL_1392188, EPI_ISL_1392189, EPI_ISL_1392190, EPI_ISL_1392191, EPI_ISL_1392192, EPI_ISL_1392194, EPI_ISL_1392195, EPI_ISL_1392196, EPI_ISL_1392197, EPI_ISL_1392198, EPI_ISL_1392200, EPI_ISL_1392201, EPI_ISL_1392203, EPI_ISL_1392204, EPI_ISL_1392205, EPI_ISL_1392206, EPI_ISL_1392207, EPI_ISL_1392208, EPI_ISL_1392209, EPI_ISL_1392210, EPI_ISL_1392211, EPI_ISL_1392212, EPI_ISL_1392213, EPI_ISL_1392214, EPI_ISL_1392215, EPI_ISL_1392217, EPI_ISL_1392218, EPI_ISL_1392219, EPI_ISL_1392220, EPI_ISL_1392221, EPI_ISL_1392222, EPI_ISL_1392225, EPI_ISL_1392226, EPI_ISL_1392228, EPI_ISL_1392229, EPI_ISL_1392230, EPI_ISL_1392231, EPI_ISL_1392232, EPI_ISL_1392233, EPI_ISL_1392234, EPI_ISL_1392235 | | | |
| see above | Sonora Quest Laboratories | TGen North | "Jolene Bowers, Heather Centner, Chris French, Hayley Yaglom, Ashlyn Pfeiffer, Darrin Lemmer, Dave Engelthaler, The Arizona COVID Genomics Union (ACGU)" |
| EPI_ISL_1392237, EPI_ISL_1392239, EPI_ISL_1392240 | Quest Diagnostics Incorporated | Centers for Disease Control and Prevention Division of Viral Diseases, Pathogen Discovery | Peter W. Cook, Dakota Howard, Dhwani Batra, Ben L. Rambo-Martin, S. H. Rosenthal, A. Gerasimova, R. M. Kagan, B. Anderson, M. Hua, Y. Liu, L.E. Bernstein, K.E. Livingston, A. Perez, I. A. Shlyakhter, R. V. Rolando, R. Owen, P. Tanpaiboon, F. Lacbawan, Clinton R. Paden, Suxiang Tong, Duncan MacCannell |
| EPI_ISL_1392241 | Sonora Quest Laboratories | TGen North | "Jolene Bowers, Heather Centner, Chris French, Hayley Yaglom, Ashlyn Pfeiffer, Darrin Lemmer, Dave Engelthaler, The Arizona COVID Genomics Union (ACGU)" |
| EPI_ISL_1392242, EPI_ISL_1392244 | Quest Diagnostics Incorporated | Centers for Disease Control and Prevention Division of Viral Diseases, Pathogen Discovery | Peter W. Cook, Dakota Howard, Dhwani Batra, Ben L. Rambo-Martin, S. H. Rosenthal, A. Gerasimova, R. M. Kagan, B. Anderson, M. Hua, Y. Liu, L.E. Bernstein, K.E. Livingston, A. Perez, I. A. Shlyakhter, R. V. Rolando, R. Owen, P. Tanpaiboon, F. Lacbawan, Clinton R. Paden, Suxiang Tong, Duncan MacCannell |
| EPI_ISL_1392245 | Sonora Quest Laboratories | TGen North | "Jolene Bowers, Heather Centner, Chris French, Hayley Yaglom, Ashlyn Pfeiffer, Darrin Lemmer, Dave Engelthaler, The Arizona COVID Genomics Union (ACGU)" |
| EPI_ISL_1392246 | Quest Diagnostics Incorporated | Centers for Disease Control and Prevention Division of Viral Diseases, Pathogen Discovery | Peter W. Cook, Dakota Howard, Dhwani Batra, Ben L. Rambo-Martin, S. H. Rosenthal, A. Gerasimova, R. M. Kagan, B. Anderson, M. Hua, Y. Liu, L.E. Bernstein, K.E. Livingston, A. Perez, I. A. Shlyakhter, R. V. Rolando, R. Owen, P. Tanpaiboon, F. Lacbawan, Clinton R. Paden, Suxiang Tong, Duncan MacCannell |
| EPI_ISL_1392247 | Sonora Quest Laboratories | TGen North | "Jolene Bowers, Heather Centner, Chris French, Hayley Yaglom, Ashlyn Pfeiffer, Darrin Lemmer, Dave Engelthaler, The Arizona COVID Genomics Union (ACGU)" |
| EPI_ISL_1392248 | Quest Diagnostics Incorporated | Centers for Disease Control and Prevention Division of Viral Diseases, Pathogen Discovery | Peter W. Cook, Dakota Howard, Dhwani Batra, Ben L. Rambo-Martin, S. H. Rosenthal, A. Gerasimova, R. M. Kagan, B. Anderson, M. Hua, Y. Liu, L.E. Bernstein, K.E. Livingston, A. Perez, I. A. Shlyakhter, R. V. Rolando, R. Owen, P. Tanpaiboon, F. Lacbawan, Clinton R. Paden, Suxiang Tong, Duncan MacCannell |
| EPI_ISL_1392249 | Sonora Quest Laboratories | TGen North | "Jolene Bowers, Heather Centner, Chris French, Hayley Yaglom, Ashlyn Pfeiffer, Darrin Lemmer, Dave Engelthaler, The Arizona COVID Genomics Union (ACGU)" |
| EPI_ISL_1392250 | Quest Diagnostics Incorporated | Centers for Disease Control and Prevention Division of Viral Diseases, Pathogen Discovery | Peter W. Cook, Dakota Howard, Dhwani Batra, Ben L. Rambo-Martin, S. H. Rosenthal, A. Gerasimova, R. M. Kagan, B. Anderson, M. Hua, Y. Liu, L.E. Bernstein, K.E. Livingston, A. Perez, I. A. Shlyakhter, R. V. Rolando, R. Owen, P. Tanpaiboon, F. Lacbawan, Clinton R. Paden, Suxiang Tong, Duncan MacCannell |
| EPI_ISL_1392251 | Sonora Quest Laboratories | TGen North | "Jolene Bowers, Heather Centner, Chris French, Hayley Yaglom, Ashlyn Pfeiffer, Darrin Lemmer, Dave Engelthaler, The Arizona COVID Genomics Union (ACGU)" |
| EPI_ISL_1392252, EPI_ISL_1392254, EPI_ISL_1392255 | Quest Diagnostics Incorporated | Centers for Disease Control and Prevention Division of Viral Diseases, Pathogen Discovery | Peter W. Cook, Dakota Howard, Dhwani Batra, Ben L. Rambo-Martin, S. H. Rosenthal, A. Gerasimova, R. M. Kagan, B. Anderson, M. Hua, Y. Liu, L.E. Bernstein, K.E. Livingston, A. Perez, I. A. Shlyakhter, R. V. Rolando, R. Owen, P. Tanpaiboon, F. Lacbawan, Clinton R. Paden, Suxiang Tong, Duncan MacCannell |
| EPI_ISL_1392257 | Sonora Quest Laboratories | TGen North | "Jolene Bowers, Heather Centner, Chris French, Hayley Yaglom, Ashlyn Pfeiffer, Darrin Lemmer, Dave Engelthaler, The Arizona COVID Genomics Union (ACGU)" |
| EPI_ISL_1392258, EPI_ISL_1392260 | Quest Diagnostics Incorporated | Centers for Disease Control and Prevention Division of Viral Diseases, Pathogen Discovery | Peter W. Cook, Dakota Howard, Dhwani Batra, Ben L. Rambo-Martin, S. H. Rosenthal, A. Gerasimova, R. M. Kagan, B. Anderson, M. Hua, Y. Liu, L.E. Bernstein, K.E. Livingston, A. Perez, I. A. Shlyakhter, R. V. Rolando, R. Owen, P. Tanpaiboon, F. Lacbawan, Clinton R. Paden, Suxiang Tong, Duncan MacCannell |
| EPI_ISL_1392261 | Sonora Quest Laboratories | TGen North | "Jolene Bowers, Heather Centner, Chris French, Hayley Yaglom, Ashlyn Pfeiffer, Darrin Lemmer, Dave Engelthaler, The Arizona COVID Genomics Union (ACGU)" |
| EPI_ISL_1392262 | Quest Diagnostics Incorporated | Centers for Disease Control and Prevention Division of Viral Diseases, Pathogen Discovery | Peter W. Cook, Dakota Howard, Dhwani Batra, Ben L. Rambo-Martin, S. H. Rosenthal, A. Gerasimova, R. M. Kagan, B. Anderson, M. Hua, Y. Liu, L.E. Bernstein, K.E. Livingston, A. Perez, I. A. Shlyakhter, R. V. Rolando, R. Owen, P. Tanpaiboon, F. Lacbawan, Clinton R. Paden, Suxiang Tong, Duncan MacCannell |
| EPI_ISL_1392263 | Sonora Quest Laboratories | TGen North | "Jolene Bowers, Heather Centner, Chris French, Hayley Yaglom, Ashlyn Pfeiffer, Darrin Lemmer, Dave Engelthaler, The Arizona COVID Genomics Union (ACGU)" |
| EPI_ISL_1392264 | Quest Diagnostics Incorporated | Centers for Disease Control and Prevention Division of Viral Diseases, Pathogen Discovery | Peter W. Cook, Dakota Howard, Dhwani Batra, Ben L. Rambo-Martin, S. H. Rosenthal, A. Gerasimova, R. M. Kagan, B. Anderson, M. Hua, Y. Liu, L.E. Bernstein, K.E. Livingston, A. Perez, I. A. Shlyakhter, R. V. Rolando, R. Owen, P. Tanpaiboon, F. Lacbawan, Clinton R. Paden, Suxiang Tong, Duncan MacCannell |
| EPI_ISL_1392265 | Sonora Quest Laboratories | TGen North | "Jolene Bowers, Heather Centner, Chris French, Hayley Yaglom, Ashlyn Pfeiffer, Darrin Lemmer, Dave Engelthaler, The Arizona COVID Genomics Union (ACGU)" |
| EPI_ISL_1392266, EPI_ISL_1392268 | Quest Diagnostics Incorporated | Centers for Disease Control and Prevention Division of Viral Diseases, Pathogen Discovery | Peter W. Cook, Dakota Howard, Dhwani Batra, Ben L. Rambo-Martin, S. H. Rosenthal, A. Gerasimova, R. M. Kagan, B. Anderson, M. Hua, Y. Liu, L.E. Bernstein, K.E. Livingston, A. Perez, I. A. Shlyakhter, R. V. Rolando, R. Owen, P. Tanpaiboon, F. Lacbawan, Clinton R. Paden, Suxiang Tong, Duncan MacCannell |
| EPI_ISL_1392269 | Sonora Quest Laboratories | TGen North | "Jolene Bowers, Heather Centner, Chris French, Hayley Yaglom, Ashlyn Pfeiffer, Darrin Lemmer, Dave Engelthaler, The Arizona COVID Genomics Union (ACGU)" |
| EPI_ISL_1392270 | Quest Diagnostics Incorporated | Centers for Disease Control and Prevention Division of Viral Diseases, Pathogen Discovery | Peter W. Cook, Dakota Howard, Dhwani Batra, Ben L. Rambo-Martin, S. H. Rosenthal, A. Gerasimova, R. M. Kagan, B. Anderson, M. Hua, Y. Liu, L.E. Bernstein, K.E. Livingston, A. Perez, I. A. Shlyakhter, R. V. Rolando, R. Owen, P. Tanpaiboon, F. Lacbawan, Clinton R. Paden, Suxiang Tong, Duncan MacCannell |
| EPI_ISL_1392271 | Sonora Quest Laboratories | TGen North | "Jolene Bowers, Heather Centner, Chris French, Hayley Yaglom, Ashlyn Pfeiffer, Darrin Lemmer, Dave Engelthaler, The Arizona COVID Genomics Union (ACGU)" |
| EPI_ISL_1392272, EPI_ISL_1392274 | Quest Diagnostics Incorporated | Centers for Disease Control and Prevention Division of Viral Diseases, Pathogen Discovery | Peter W. Cook, Dakota Howard, Dhwani Batra, Ben L. Rambo-Martin, S. H. Rosenthal, A. Gerasimova, R. M. Kagan, B. Anderson, M. Hua, Y. Liu, L.E. Bernstein, K.E. Livingston, A. Perez, I. A. Shlyakhter, R. V. Rolando, R. Owen, P. Tanpaiboon, F. Lacbawan, Clinton R. Paden, Suxiang Tong, Duncan MacCannell |
| EPI_ISL_1392275 | Sonora Quest Laboratories | TGen North | "Jolene Bowers, Heather Centner, Chris French, Hayley Yaglom, Ashlyn Pfeiffer, Darrin Lemmer, Dave Engelthaler, The Arizona COVID Genomics Union (ACGU)" |
| EPI_ISL_1392276 | Quest Diagnostics Incorporated | Centers for Disease Control and Prevention Division of Viral Diseases, Pathogen Discovery | Peter W. Cook, Dakota Howard, Dhwani Batra, Ben L. Rambo-Martin, S. H. Rosenthal, A. Gerasimova, R. M. Kagan, B. Anderson, M. Hua, Y. Liu, L.E. Bernstein, K.E. Livingston, A. Perez, I. A. Shlyakhter, R. V. Rolando, R. Owen, P. Tanpaiboon, F. Lacbawan, Clinton R. Paden, Suxiang Tong, Duncan MacCannell |

| | | | |
|---|---|---|---|
| EPI_ISL_1392277 | TGen North | TGen North | "Jolene Bowers, Heather Centner, Chris French, Hayley Yaglom, Ashlyn Pfeiffer, Darrin Lemmer, Dave Engelthaler, The Arizona COVID Genomics Union (ACGU)" |
| EPI_ISL_1392278 | Quest Diagnostics Incorporated | Centers for Disease Control and Prevention Division of Viral Diseases, Pathogen Discovery | Peter W. Cook, Dakota Howard, Dhwani Batra, Ben L. Rambo-Martin, S. H. Rosenthal, A. Gerasimova, R. M. Kagan, B. Anderson, M. Hua, Y. Liu, L.E. Bernstein, K.E. Livingston, A. Perez, I. A. Shlyakhter, R. V. Rolando, R. Owen, P. Tanpaiboon, F. Lacbawan, Clinton R. Paden, Suxiang Tong, Duncan MacCannell |
| EPI_ISL_1392279 | Sonora Quest Laboratories | TGen North | "Jolene Bowers, Heather Centner, Chris French, Hayley Yaglom, Ashlyn Pfeiffer, Darrin Lemmer, Dave Engelthaler, The Arizona COVID Genomics Union (ACGU)" |
| EPI_ISL_1392280 | Quest Diagnostics Incorporated | Centers for Disease Control and Prevention Division of Viral Diseases, Pathogen Discovery | Peter W. Cook, Dakota Howard, Dhwani Batra, Ben L. Rambo-Martin, S. H. Rosenthal, A. Gerasimova, R. M. Kagan, B. Anderson, M. Hua, Y. Liu, L.E. Bernstein, K.E. Livingston, A. Perez, I. A. Shlyakhter, R. V. Rolando, R. Owen, P. Tanpaiboon, F. Lacbawan, Clinton R. Paden, Suxiang Tong, Duncan MacCannell |
| EPI_ISL_1392281 | Sonora Quest Laboratories | TGen North | "Jolene Bowers, Heather Centner, Chris French, Hayley Yaglom, Ashlyn Pfeiffer, Darrin Lemmer, Dave Engelthaler, The Arizona COVID Genomics Union (ACGU)" |
| EPI_ISL_1392282 | Quest Diagnostics Incorporated | Centers for Disease Control and Prevention Division of Viral Diseases, Pathogen Discovery | Peter W. Cook, Dakota Howard, Dhwani Batra, Ben L. Rambo-Martin, S. H. Rosenthal, A. Gerasimova, R. M. Kagan, B. Anderson, M. Hua, Y. Liu, L.E. Bernstein, K.E. Livingston, A. Perez, I. A. Shlyakhter, R. V. Rolando, R. Owen, P. Tanpaiboon, F. Lacbawan, Clinton R. Paden, Suxiang Tong, Duncan MacCannell |
| EPI_ISL_1392283 | Sonora Quest Laboratories | TGen North | "Jolene Bowers, Heather Centner, Chris French, Hayley Yaglom, Ashlyn Pfeiffer, Darrin Lemmer, Dave Engelthaler, The Arizona COVID Genomics Union (ACGU)" |
| EPI_ISL_1392284 | Quest Diagnostics Incorporated | Centers for Disease Control and Prevention Division of Viral Diseases, Pathogen Discovery | Peter W. Cook, Dakota Howard, Dhwani Batra, Ben L. Rambo-Martin, S. H. Rosenthal, A. Gerasimova, R. M. Kagan, B. Anderson, M. Hua, Y. Liu, L.E. Bernstein, K.E. Livingston, A. Perez, I. A. Shlyakhter, R. V. Rolando, R. Owen, P. Tanpaiboon, F. Lacbawan, Clinton R. Paden, Suxiang Tong, Duncan MacCannell |
| EPI_ISL_1392285 | Sonora Quest Laboratories | TGen North | "Jolene Bowers, Heather Centner, Chris French, Hayley Yaglom, Ashlyn Pfeiffer, Darrin Lemmer, Dave Engelthaler, The Arizona COVID Genomics Union (ACGU)" |
| EPI_ISL_1392286, EPI_ISL_1392288 | Quest Diagnostics Incorporated | Centers for Disease Control and Prevention Division of Viral Diseases, Pathogen Discovery | Peter W. Cook, Dakota Howard, Dhwani Batra, Ben L. Rambo-Martin, S. H. Rosenthal, A. Gerasimova, R. M. Kagan, B. Anderson, M. Hua, Y. Liu, L.E. Bernstein, K.E. Livingston, A. Perez, I. A. Shlyakhter, R. V. Rolando, R. Owen, P. Tanpaiboon, F. Lacbawan, Clinton R. Paden, Suxiang Tong, Duncan MacCannell |
| EPI_ISL_1392289 | Sonora Quest Laboratories | TGen North | "Jolene Bowers, Heather Centner, Chris French, Hayley Yaglom, Ashlyn Pfeiffer, Darrin Lemmer, Dave Engelthaler, The Arizona COVID Genomics Union (ACGU)" |
| EPI_ISL_1392290 | Quest Diagnostics Incorporated | Centers for Disease Control and Prevention Division of Viral Diseases, Pathogen Discovery | Peter W. Cook, Dakota Howard, Dhwani Batra, Ben L. Rambo-Martin, S. H. Rosenthal, A. Gerasimova, R. M. Kagan, B. Anderson, M. Hua, Y. Liu, L.E. Bernstein, K.E. Livingston, A. Perez, I. A. Shlyakhter, R. V. Rolando, R. Owen, P. Tanpaiboon, F. Lacbawan, Clinton R. Paden, Suxiang Tong, Duncan MacCannell |
| EPI_ISL_1392291 | Sonora Quest Laboratories | TGen North | "Jolene Bowers, Heather Centner, Chris French, Hayley Yaglom, Ashlyn Pfeiffer, Darrin Lemmer, Dave Engelthaler, The Arizona COVID Genomics Union (ACGU)" |
| EPI_ISL_1392292, EPI_ISL_1392294 | Quest Diagnostics Incorporated | Centers for Disease Control and Prevention Division of Viral Diseases, Pathogen Discovery | Peter W. Cook, Dakota Howard, Dhwani Batra, Ben L. Rambo-Martin, S. H. Rosenthal, A. Gerasimova, R. M. Kagan, B. Anderson, M. Hua, Y. Liu, L.E. Bernstein, K.E. Livingston, A. Perez, I. A. Shlyakhter, R. V. Rolando, R. Owen, P. Tanpaiboon, F. Lacbawan, Clinton R. Paden, Suxiang Tong, Duncan MacCannell |
| EPI_ISL_1392295 | Sonora Quest Laboratories | TGen North | "Jolene Bowers, Heather Centner, Chris French, Hayley Yaglom, Ashlyn Pfeiffer, Darrin Lemmer, Dave Engelthaler, The Arizona COVID Genomics Union (ACGU)" |
| EPI_ISL_1392296 | Quest Diagnostics Incorporated | Centers for Disease Control and Prevention Division of Viral Diseases, Pathogen Discovery | Peter W. Cook, Dakota Howard, Dhwani Batra, Ben L. Rambo-Martin, S. H. Rosenthal, A. Gerasimova, R. M. Kagan, B. Anderson, M. Hua, Y. Liu, L.E. Bernstein, K.E. Livingston, A. Perez, I. A. Shlyakhter, R. V. Rolando, R. Owen, P. Tanpaiboon, F. Lacbawan, Clinton R. Paden, Suxiang Tong, Duncan MacCannell |
| EPI_ISL_1392297 | Sonora Quest Laboratories | TGen North | "Jolene Bowers, Heather Centner, Chris French, Hayley Yaglom, Ashlyn Pfeiffer, Darrin Lemmer, Dave Engelthaler, The Arizona COVID Genomics Union (ACGU)" |
| EPI_ISL_1392298 | Quest Diagnostics Incorporated | Centers for Disease Control and Prevention Division of Viral Diseases, Pathogen Discovery | Peter W. Cook, Dakota Howard, Dhwani Batra, Ben L. Rambo-Martin, S. H. Rosenthal, A. Gerasimova, R. M. Kagan, B. Anderson, M. Hua, Y. Liu, L.E. Bernstein, K.E. Livingston, A. Perez, I. A. Shlyakhter, R. V. Rolando, R. Owen, P. Tanpaiboon, F. Lacbawan, Clinton R. Paden, Suxiang Tong, Duncan MacCannell |
| EPI_ISL_1392299 | Sonora Quest Laboratories | TGen North | "Jolene Bowers, Heather Centner, Chris French, Hayley Yaglom, Ashlyn Pfeiffer, Darrin Lemmer, Dave Engelthaler, The Arizona COVID Genomics Union (ACGU)" |
| EPI_ISL_1392300, EPI_ISL_1392302 | Quest Diagnostics Incorporated | Centers for Disease Control and Prevention Division of Viral Diseases, Pathogen Discovery | Peter W. Cook, Dakota Howard, Dhwani Batra, Ben L. Rambo-Martin, S. H. Rosenthal, A. Gerasimova, R. M. Kagan, B. Anderson, M. Hua, Y. Liu, L.E. Bernstein, K.E. Livingston, A. Perez, I. A. Shlyakhter, R. V. Rolando, R. Owen, P. Tanpaiboon, F. Lacbawan, Clinton R. Paden, Suxiang Tong, Duncan MacCannell |
| EPI_ISL_1392303 | Sonora Quest Laboratories | TGen North | "Jolene Bowers, Heather Centner, Chris French, Hayley Yaglom, Ashlyn Pfeiffer, Darrin Lemmer, Dave Engelthaler, The Arizona COVID Genomics Union (ACGU)" |
| EPI_ISL_1392304, EPI_ISL_1392305, EPI_ISL_1392307 | Quest Diagnostics Incorporated | Centers for Disease Control and Prevention Division of Viral Diseases, Pathogen Discovery | Peter W. Cook, Dakota Howard, Dhwani Batra, Ben L. Rambo-Martin, S. H. Rosenthal, A. Gerasimova, R. M. Kagan, B. Anderson, M. Hua, Y. Liu, L.E. Bernstein, K.E. Livingston, A. Perez, I. A. Shlyakhter, R. V. Rolando, R. Owen, P. Tanpaiboon, F. Lacbawan, Clinton R. Paden, Suxiang Tong, Duncan MacCannell |
| EPI_ISL_1392308 | Sonora Quest Laboratories | TGen North | "Jolene Bowers, Heather Centner, Chris French, Hayley Yaglom, Ashlyn Pfeiffer, Darrin Lemmer, Dave Engelthaler, The Arizona COVID Genomics Union (ACGU)" |
| EPI_ISL_1392309, EPI_ISL_1392311, EPI_ISL_1392313 | Quest Diagnostics Incorporated | Centers for Disease Control and Prevention Division of Viral Diseases, Pathogen Discovery | Peter W. Cook, Dakota Howard, Dhwani Batra, Ben L. Rambo-Martin, S. H. Rosenthal, A. Gerasimova, R. M. Kagan, B. Anderson, M. Hua, Y. Liu, L.E. Bernstein, K.E. Livingston, A. Perez, I. A. Shlyakhter, R. V. Rolando, R. Owen, P. Tanpaiboon, F. Lacbawan, Clinton R. Paden, Suxiang Tong, Duncan MacCannell |
| EPI_ISL_1392314 | Sonora Quest Laboratories | TGen North | "Jolene Bowers, Heather Centner, Chris French, Hayley Yaglom, Ashlyn Pfeiffer, Darrin Lemmer, Dave Engelthaler, The Arizona COVID Genomics Union (ACGU)" |
| EPI_ISL_1392315, EPI_ISL_1392317, EPI_ISL_1392319 | Quest Diagnostics Incorporated | Centers for Disease Control and Prevention Division of Viral Diseases, Pathogen Discovery | Peter W. Cook, Dakota Howard, Dhwani Batra, Ben L. Rambo-Martin, S. H. Rosenthal, A. Gerasimova, R. M. Kagan, B. Anderson, M. Hua, Y. Liu, L.E. Bernstein, K.E. Livingston, A. Perez, I. A. Shlyakhter, R. V. Rolando, R. Owen, P. Tanpaiboon, F. Lacbawan, Clinton R. Paden, Suxiang Tong, Duncan MacCannell |
| EPI_ISL_1392320 | Sonora Quest Laboratories | TGen North | "Jolene Bowers, Heather Centner, Chris French, Hayley Yaglom, Ashlyn Pfeiffer, Darrin Lemmer, Dave Engelthaler, The Arizona COVID Genomics Union (ACGU)" |
| EPI_ISL_1392321 | Quest Diagnostics Incorporated | Centers for Disease Control and Prevention Division of Viral Diseases, Pathogen Discovery | Peter W. Cook, Dakota Howard, Dhwani Batra, Ben L. Rambo-Martin, S. H. Rosenthal, A. Gerasimova, R. M. Kagan, B. Anderson, M. Hua, Y. Liu, L.E. Bernstein, K.E. Livingston, A. Perez, I. A. Shlyakhter, R. V. Rolando, R. Owen, P. Tanpaiboon, F. Lacbawan, Clinton R. Paden, Suxiang Tong, Duncan MacCannell |
| EPI_ISL_1392322 | Sonora Quest Laboratories | TGen North | "Jolene Bowers, Heather Centner, Chris French, Hayley Yaglom, Ashlyn Pfeiffer, Darrin Lemmer, Dave Engelthaler, The Arizona COVID Genomics Union (ACGU)" |
| EPI_ISL_1392323 | Quest Diagnostics Incorporated | Centers for Disease Control and Prevention Division of Viral Diseases, Pathogen Discovery | Peter W. Cook, Dakota Howard, Dhwani Batra, Ben L. Rambo-Martin, S. H. Rosenthal, A. Gerasimova, R. M. Kagan, B. Anderson, M. Hua, Y. Liu, L.E. Bernstein, K.E. Livingston, A. Perez, I. A. Shlyakhter, R. V. Rolando, R. Owen, P. Tanpaiboon, F. Lacbawan, Clinton R. Paden, Suxiang Tong, Duncan MacCannell |
| EPI_ISL_1392324 | Sonora Quest Laboratories | TGen North | "Jolene Bowers, Heather Centner, Chris French, Hayley Yaglom, Ashlyn Pfeiffer, Darrin Lemmer, Dave Engelthaler, The Arizona COVID Genomics Union (ACGU)" |
| EPI_ISL_1392325, EPI_ISL_1392327 | Quest Diagnostics Incorporated | Centers for Disease Control and Prevention Division of Viral Diseases, Pathogen Discovery | Peter W. Cook, Dakota Howard, Dhwani Batra, Ben L. Rambo-Martin, S. H. Rosenthal, A. Gerasimova, R. M. Kagan, B. Anderson, M. Hua, Y. Liu, L.E. Bernstein, K.E. Livingston, A. Perez, I. A. Shlyakhter, R. V. Rolando, R. Owen, P. Tanpaiboon, F. Lacbawan, Clinton R. Paden, Suxiang Tong, Duncan MacCannell |
| EPI_ISL_1392328 | Sonora Quest Laboratories | TGen North | "Jolene Bowers, Heather Centner, Chris French, Hayley Yaglom, Ashlyn Pfeiffer, Darrin Lemmer, Dave Engelthaler, The Arizona COVID Genomics Union (ACGU)" |
| EPI_ISL_1392329 | Quest Diagnostics Incorporated | Centers for Disease Control and Prevention Division of Viral Diseases, Pathogen Discovery | Peter W. Cook, Dakota Howard, Dhwani Batra, Ben L. Rambo-Martin, S. H. Rosenthal, A. Gerasimova, R. M. Kagan, B. Anderson, M. Hua, Y. Liu, L.E. Bernstein, K.E. Livingston, A. Perez, I. A. Shlyakhter, R. V. Rolando, R. Owen, P. Tanpaiboon, F. Lacbawan, Clinton R. Paden, Suxiang Tong, Duncan MacCannell |
| EPI_ISL_1392330 | Sonora Quest Laboratories | TGen North | "Jolene Bowers, Heather Centner, Chris French, Hayley Yaglom, Ashlyn Pfeiffer, Darrin Lemmer, Dave Engelthaler, The Arizona COVID Genomics Union (ACGU)" |
| EPI_ISL_1392331, EPI_ISL_1392333, EPI_ISL_1392335, EPI_ISL_1392336 | Quest Diagnostics Incorporated | Centers for Disease Control and Prevention Division of Viral Diseases, Pathogen Discovery | Peter W. Cook, Dakota Howard, Dhwani Batra, Ben L. Rambo-Martin, S. H. Rosenthal, A. Gerasimova, R. M. Kagan, B. Anderson, M. Hua, Y. Liu, L.E. Bernstein, K.E. Livingston, A. Perez, I. A. Shlyakhter, R. V. Rolando, R. Owen, P. Tanpaiboon, F. Lacbawan, Clinton R. Paden, Suxiang Tong, Duncan MacCannell |
| EPI_ISL_1392337 | Sonora Quest Laboratories | TGen North | "Jolene Bowers, Heather Centner, Chris French, Hayley Yaglom, Ashlyn Pfeiffer, Darrin Lemmer, Dave Engelthaler, The Arizona COVID Genomics Union (ACGU)" |
| EPI_ISL_1392338 | Quest Diagnostics Incorporated | Centers for Disease Control and Prevention Division of Viral Diseases, Pathogen Discovery | Peter W. Cook, Dakota Howard, Dhwani Batra, Ben L. Rambo-Martin, S. H. Rosenthal, A. Gerasimova, R. M. Kagan, B. Anderson, M. Hua, Y. Liu, L.E. Bernstein, K.E. Livingston, A. Perez, I. A. Shlyakhter, R. V. Rolando, R. Owen, P. Tanpaiboon, F. Lacbawan, Clinton R. Paden, Suxiang Tong, Duncan MacCannell |
| EPI_ISL_1392339 | Sonora Quest Laboratories | TGen North | "Jolene Bowers, Heather Centner, Chris French, Hayley Yaglom, Ashlyn Pfeiffer, Darrin Lemmer, Dave Engelthaler, The Arizona COVID Genomics Union (ACGU)" |
| EPI_ISL_1392340 | Quest Diagnostics Incorporated | Centers for Disease Control and Prevention Division of Viral Diseases, Pathogen Discovery | Peter W. Cook, Dakota Howard, Dhwani Batra, Ben L. Rambo-Martin, S. H. Rosenthal, A. Gerasimova, R. M. Kagan, B. Anderson, M. Hua, Y. Liu, L.E. Bernstein, K.E. Livingston, A. Perez, I. A. Shlyakhter, R. V. Rolando, R. Owen, P. Tanpaiboon, F. Lacbawan, Clinton R. Paden, Suxiang Tong, Duncan MacCannell |
| EPI_ISL_1392342 | Sonora Quest Laboratories | TGen North | "Jolene Bowers, Heather Centner, Chris French, Hayley Yaglom, Ashlyn Pfeiffer, Darrin Lemmer, Dave Engelthaler, The Arizona COVID Genomics Union (ACGU)" |
| EPI_ISL_1392343 | Quest Diagnostics Incorporated | Centers for Disease Control and Prevention Division of Viral Diseases, Pathogen Discovery | Peter W. Cook, Dakota Howard, Dhwani Batra, Ben L. Rambo-Martin, S. H. Rosenthal, A. Gerasimova, R. M. Kagan, B. Anderson, M. Hua, Y. Liu, L.E. Bernstein, K.E. Livingston, A. Perez, I. A. Shlyakhter, R. V. Rolando, R. Owen, P. Tanpaiboon, F. Lacbawan, Clinton R. Paden, Suxiang Tong, Duncan MacCannell |
| EPI_ISL_1392344 | Sonora Quest Laboratories | TGen North | "Jolene Bowers, Heather Centner, Chris French, Hayley Yaglom, Ashlyn Pfeiffer, Darrin Lemmer, Dave Engelthaler, The Arizona COVID Genomics Union (ACGU)" |
| EPI_ISL_1392345 | Quest Diagnostics Incorporated | Centers for Disease Control and Prevention Division of Viral Diseases, Pathogen Discovery | Peter W. Cook, Dakota Howard, Dhwani Batra, Ben L. Rambo-Martin, S. H. Rosenthal, A. Gerasimova, R. M. Kagan, B. Anderson, M. Hua, Y. Liu, L.E. Bernstein, K.E. Livingston, A. Perez, I. A. Shlyakhter, R. V. Rolando, R. Owen, P. Tanpaiboon, F. Lacbawan, Clinton R. Paden, Suxiang Tong, Duncan MacCannell |
| EPI_ISL_1392346 | Sonora Quest Laboratories | TGen North | "Jolene Bowers, Heather Centner, Chris French, Hayley Yaglom, Ashlyn Pfeiffer, Darrin Lemmer, Dave Engelthaler, The Arizona COVID Genomics Union (ACGU)" |

| | | | |
|---|---|---|---|
| EPI_ISL_1392347 | Quest Diagnostics Incorporated | Centers for Disease Control and Prevention Division of Viral Diseases, Pathogen Discovery | Peter W. Cook, Dakota Howard, Dhwani Batra, Ben L. Rambo-Martin, S. H. Rosenthal, A. Gerasimova, R. M. Kagan, B. Anderson, M. Hua, Y. Liu, L.E. Bernstein, K.E. Livingston, A. Perez, I. A. Shlyakhter, R. V. Rolando, R. Owen, P. Tanpaiboon, F. Lacbawan, Clinton R. Paden, Suxiang Tong, Duncan MacCannell |
| EPI_ISL_1392348 | Sonora Quest Laboratories | TGen North | "Jolene Bowers, Heather Centner, Chris French, Hayley Yaglom, Ashlyn Pfeiffer, Darrin Lemmer, Dave Engelthaler, The Arizona COVID Genomics Union (ACGU)" |
| EPI_ISL_1392349 | Quest Diagnostics Incorporated | Centers for Disease Control and Prevention Division of Viral Diseases, Pathogen Discovery | Peter W. Cook, Dakota Howard, Dhwani Batra, Ben L. Rambo-Martin, S. H. Rosenthal, A. Gerasimova, R. M. Kagan, B. Anderson, M. Hua, Y. Liu, L.E. Bernstein, K.E. Livingston, A. Perez, I. A. Shlyakhter, R. V. Rolando, R. Owen, P. Tanpaiboon, F. Lacbawan, Clinton R. Paden, Suxiang Tong, Duncan MacCannell |
| EPI_ISL_1392350 | Sonora Quest Laboratories | TGen North | "Jolene Bowers, Heather Centner, Chris French, Hayley Yaglom, Ashlyn Pfeiffer, Darrin Lemmer, Dave Engelthaler, The Arizona COVID Genomics Union (ACGU)" |
| EPI_ISL_1392351 | Quest Diagnostics Incorporated | Centers for Disease Control and Prevention Division of Viral Diseases, Pathogen Discovery | Peter W. Cook, Dakota Howard, Dhwani Batra, Ben L. Rambo-Martin, S. H. Rosenthal, A. Gerasimova, R. M. Kagan, B. Anderson, M. Hua, Y. Liu, L.E. Bernstein, K.E. Livingston, A. Perez, I. A. Shlyakhter, R. V. Rolando, R. Owen, P. Tanpaiboon, F. Lacbawan, Clinton R. Paden, Suxiang Tong, Duncan MacCannell |
| EPI_ISL_1392352 | Sonora Quest Laboratories | TGen North | "Jolene Bowers, Heather Centner, Chris French, Hayley Yaglom, Ashlyn Pfeiffer, Darrin Lemmer, Dave Engelthaler, The Arizona COVID Genomics Union (ACGU)" |
| EPI_ISL_1392353 | Quest Diagnostics Incorporated | Centers for Disease Control and Prevention Division of Viral Diseases, Pathogen Discovery | Peter W. Cook, Dakota Howard, Dhwani Batra, Ben L. Rambo-Martin, S. H. Rosenthal, A. Gerasimova, R. M. Kagan, B. Anderson, M. Hua, Y. Liu, L.E. Bernstein, K.E. Livingston, A. Perez, I. A. Shlyakhter, R. V. Rolando, R. Owen, P. Tanpaiboon, F. Lacbawan, Clinton R. Paden, Suxiang Tong, Duncan MacCannell |
| EPI_ISL_1392354 | Sonora Quest Laboratories | TGen North | "Jolene Bowers, Heather Centner, Chris French, Hayley Yaglom, Ashlyn Pfeiffer, Darrin Lemmer, Dave Engelthaler, The Arizona COVID Genomics Union (ACGU)" |
| EPI_ISL_1392355 | Quest Diagnostics Incorporated | Centers for Disease Control and Prevention Division of Viral Diseases, Pathogen Discovery | Peter W. Cook, Dakota Howard, Dhwani Batra, Ben L. Rambo-Martin, S. H. Rosenthal, A. Gerasimova, R. M. Kagan, B. Anderson, M. Hua, Y. Liu, L.E. Bernstein, K.E. Livingston, A. Perez, I. A. Shlyakhter, R. V. Rolando, R. Owen, P. Tanpaiboon, F. Lacbawan, Clinton R. Paden, Suxiang Tong, Duncan MacCannell |
| EPI_ISL_1392356 | Sonora Quest Laboratories | TGen North | "Jolene Bowers, Heather Centner, Chris French, Hayley Yaglom, Ashlyn Pfeiffer, Darrin Lemmer, Dave Engelthaler, The Arizona COVID Genomics Union (ACGU)" |
| EPI_ISL_1392357 | Quest Diagnostics Incorporated | Centers for Disease Control and Prevention Division of Viral Diseases, Pathogen Discovery | Peter W. Cook, Dakota Howard, Dhwani Batra, Ben L. Rambo-Martin, S. H. Rosenthal, A. Gerasimova, R. M. Kagan, B. Anderson, M. Hua, Y. Liu, L.E. Bernstein, K.E. Livingston, A. Perez, I. A. Shlyakhter, R. V. Rolando, R. Owen, P. Tanpaiboon, F. Lacbawan, Clinton R. Paden, Suxiang Tong, Duncan MacCannell |
| EPI_ISL_1392358 | Sonora Quest Laboratories | TGen North | "Jolene Bowers, Heather Centner, Chris French, Hayley Yaglom, Ashlyn Pfeiffer, Darrin Lemmer, Dave Engelthaler, The Arizona COVID Genomics Union (ACGU)" |
| EPI_ISL_1392359 | Quest Diagnostics Incorporated | Centers for Disease Control and Prevention Division of Viral Diseases, Pathogen Discovery | Peter W. Cook, Dakota Howard, Dhwani Batra, Ben L. Rambo-Martin, S. H. Rosenthal, A. Gerasimova, R. M. Kagan, B. Anderson, M. Hua, Y. Liu, L.E. Bernstein, K.E. Livingston, A. Perez, I. A. Shlyakhter, R. V. Rolando, R. Owen, P. Tanpaiboon, F. Lacbawan, Clinton R. Paden, Suxiang Tong, Duncan MacCannell |
| EPI_ISL_1392360 | Sonora Quest Laboratories | TGen North | "Jolene Bowers, Heather Centner, Chris French, Hayley Yaglom, Ashlyn Pfeiffer, Darrin Lemmer, Dave Engelthaler, The Arizona COVID Genomics Union (ACGU)" |
| EPI_ISL_1392361 | Quest Diagnostics Incorporated | Centers for Disease Control and Prevention Division of Viral Diseases, Pathogen Discovery | Peter W. Cook, Dakota Howard, Dhwani Batra, Ben L. Rambo-Martin, S. H. Rosenthal, A. Gerasimova, R. M. Kagan, B. Anderson, M. Hua, Y. Liu, L.E. Bernstein, K.E. Livingston, A. Perez, I. A. Shlyakhter, R. V. Rolando, R. Owen, P. Tanpaiboon, F. Lacbawan, Clinton R. Paden, Suxiang Tong, Duncan MacCannell |
| EPI_ISL_1392362 | Sonora Quest Laboratories | TGen North | "Jolene Bowers, Heather Centner, Chris French, Hayley Yaglom, Ashlyn Pfeiffer, Darrin Lemmer, Dave Engelthaler, The Arizona COVID Genomics Union (ACGU)" |
| EPI_ISL_1392363 | Quest Diagnostics Incorporated | Centers for Disease Control and Prevention Division of Viral Diseases, Pathogen Discovery | Peter W. Cook, Dakota Howard, Dhwani Batra, Ben L. Rambo-Martin, S. H. Rosenthal, A. Gerasimova, R. M. Kagan, B. Anderson, M. Hua, Y. Liu, L.E. Bernstein, K.E. Livingston, A. Perez, I. A. Shlyakhter, R. V. Rolando, R. Owen, P. Tanpaiboon, F. Lacbawan, Clinton R. Paden, Suxiang Tong, Duncan MacCannell |
| EPI_ISL_1392364 | Sonora Quest Laboratories | TGen North | "Jolene Bowers, Heather Centner, Chris French, Hayley Yaglom, Ashlyn Pfeiffer, Darrin Lemmer, Dave Engelthaler, The Arizona COVID Genomics Union (ACGU)" |
| EPI_ISL_1392365 | Quest Diagnostics Incorporated | Centers for Disease Control and Prevention Division of Viral Diseases, Pathogen Discovery | Peter W. Cook, Dakota Howard, Dhwani Batra, Ben L. Rambo-Martin, S. H. Rosenthal, A. Gerasimova, R. M. Kagan, B. Anderson, M. Hua, Y. Liu, L.E. Bernstein, K.E. Livingston, A. Perez, I. A. Shlyakhter, R. V. Rolando, R. Owen, P. Tanpaiboon, F. Lacbawan, Clinton R. Paden, Suxiang Tong, Duncan MacCannell |
| EPI_ISL_1392366 | Sonora Quest Laboratories | TGen North | "Jolene Bowers, Heather Centner, Chris French, Hayley Yaglom, Ashlyn Pfeiffer, Darrin Lemmer, Dave Engelthaler, The Arizona COVID Genomics Union (ACGU)" |
| EPI_ISL_1392367, EPI_ISL_1392368 | Quest Diagnostics Incorporated | Centers for Disease Control and Prevention Division of Viral Diseases, Pathogen Discovery | Peter W. Cook, Dakota Howard, Dhwani Batra, Ben L. Rambo-Martin, S. H. Rosenthal, A. Gerasimova, R. M. Kagan, B. Anderson, M. Hua, Y. Liu, L.E. Bernstein, K.E. Livingston, A. Perez, I. A. Shlyakhter, R. V. Rolando, R. Owen, P. Tanpaiboon, F. Lacbawan, Clinton R. Paden, Suxiang Tong, Duncan MacCannell |
| EPI_ISL_1392369 | Sonora Quest Laboratories | TGen North | "Jolene Bowers, Heather Centner, Chris French, Hayley Yaglom, Ashlyn Pfeiffer, Darrin Lemmer, Dave Engelthaler, The Arizona COVID Genomics Union (ACGU)" |
| EPI_ISL_1392370 | Quest Diagnostics Incorporated | Centers for Disease Control and Prevention Division of Viral Diseases, Pathogen Discovery | Peter W. Cook, Dakota Howard, Dhwani Batra, Ben L. Rambo-Martin, S. H. Rosenthal, A. Gerasimova, R. M. Kagan, B. Anderson, M. Hua, Y. Liu, L.E. Bernstein, K.E. Livingston, A. Perez, I. A. Shlyakhter, R. V. Rolando, R. Owen, P. Tanpaiboon, F. Lacbawan, Clinton R. Paden, Suxiang Tong, Duncan MacCannell |
| EPI_ISL_1392371 | Sonora Quest Laboratories | TGen North | "Jolene Bowers, Heather Centner, Chris French, Hayley Yaglom, Ashlyn Pfeiffer, Darrin Lemmer, Dave Engelthaler, The Arizona COVID Genomics Union (ACGU)" |
| EPI_ISL_1392372 | Quest Diagnostics Incorporated | Centers for Disease Control and Prevention Division of Viral Diseases, Pathogen Discovery | Peter W. Cook, Dakota Howard, Dhwani Batra, Ben L. Rambo-Martin, S. H. Rosenthal, A. Gerasimova, R. M. Kagan, B. Anderson, M. Hua, Y. Liu, L.E. Bernstein, K.E. Livingston, A. Perez, I. A. Shlyakhter, R. V. Rolando, R. Owen, P. Tanpaiboon, F. Lacbawan, Clinton R. Paden, Suxiang Tong, Duncan MacCannell |
| EPI_ISL_1392373 | Sonora Quest Laboratories | TGen North | "Jolene Bowers, Heather Centner, Chris French, Hayley Yaglom, Ashlyn Pfeiffer, Darrin Lemmer, Dave Engelthaler, The Arizona COVID Genomics Union (ACGU)" |
| EPI_ISL_1392374, EPI_ISL_1392376 | Quest Diagnostics Incorporated | Centers for Disease Control and Prevention Division of Viral Diseases, Pathogen Discovery | Peter W. Cook, Dakota Howard, Dhwani Batra, Ben L. Rambo-Martin, S. H. Rosenthal, A. Gerasimova, R. M. Kagan, B. Anderson, M. Hua, Y. Liu, L.E. Bernstein, K.E. Livingston, A. Perez, I. A. Shlyakhter, R. V. Rolando, R. Owen, P. Tanpaiboon, F. Lacbawan, Clinton R. Paden, Suxiang Tong, Duncan MacCannell |
| EPI_ISL_1392377 | Sonora Quest Laboratories | TGen North | "Jolene Bowers, Heather Centner, Chris French, Hayley Yaglom, Ashlyn Pfeiffer, Darrin Lemmer, Dave Engelthaler, The Arizona COVID Genomics Union (ACGU)" |
| EPI_ISL_1392378 | Quest Diagnostics Incorporated | Centers for Disease Control and Prevention Division of Viral Diseases, Pathogen Discovery | Peter W. Cook, Dakota Howard, Dhwani Batra, Ben L. Rambo-Martin, S. H. Rosenthal, A. Gerasimova, R. M. Kagan, B. Anderson, M. Hua, Y. Liu, L.E. Bernstein, K.E. Livingston, A. Perez, I. A. Shlyakhter, R. V. Rolando, R. Owen, P. Tanpaiboon, F. Lacbawan, Clinton R. Paden, Suxiang Tong, Duncan MacCannell |
| EPI_ISL_1392379 | Sonora Quest Laboratories | TGen North | "Jolene Bowers, Heather Centner, Chris French, Hayley Yaglom, Ashlyn Pfeiffer, Darrin Lemmer, Dave Engelthaler, The Arizona COVID Genomics Union (ACGU)" |
| EPI_ISL_1392380 | Quest Diagnostics Incorporated | Centers for Disease Control and Prevention Division of Viral Diseases, Pathogen Discovery | Peter W. Cook, Dakota Howard, Dhwani Batra, Ben L. Rambo-Martin, S. H. Rosenthal, A. Gerasimova, R. M. Kagan, B. Anderson, M. Hua, Y. Liu, L.E. Bernstein, K.E. Livingston, A. Perez, I. A. Shlyakhter, R. V. Rolando, R. Owen, P. Tanpaiboon, F. Lacbawan, Clinton R. Paden, Suxiang Tong, Duncan MacCannell |
| EPI_ISL_1392381 | Sonora Quest Laboratories | TGen North | "Jolene Bowers, Heather Centner, Chris French, Hayley Yaglom, Ashlyn Pfeiffer, Darrin Lemmer, Dave Engelthaler, The Arizona COVID Genomics Union (ACGU)" |
| EPI_ISL_1392382 | Quest Diagnostics Incorporated | Centers for Disease Control and Prevention Division of Viral Diseases, Pathogen Discovery | Peter W. Cook, Dakota Howard, Dhwani Batra, Ben L. Rambo-Martin, S. H. Rosenthal, A. Gerasimova, R. M. Kagan, B. Anderson, M. Hua, Y. Liu, L.E. Bernstein, K.E. Livingston, A. Perez, I. A. Shlyakhter, R. V. Rolando, R. Owen, P. Tanpaiboon, F. Lacbawan, Clinton R. Paden, Suxiang Tong, Duncan MacCannell |
| EPI_ISL_1392383 | Sonora Quest Laboratories | TGen North | "Jolene Bowers, Heather Centner, Chris French, Hayley Yaglom, Ashlyn Pfeiffer, Darrin Lemmer, Dave Engelthaler, The Arizona COVID Genomics Union (ACGU)" |
| EPI_ISL_1392384, EPI_ISL_1392386 | Quest Diagnostics Incorporated | Centers for Disease Control and Prevention Division of Viral Diseases, Pathogen Discovery | Peter W. Cook, Dakota Howard, Dhwani Batra, Ben L. Rambo-Martin, S. H. Rosenthal, A. Gerasimova, R. M. Kagan, B. Anderson, M. Hua, Y. Liu, L.E. Bernstein, K.E. Livingston, A. Perez, I. A. Shlyakhter, R. V. Rolando, R. Owen, P. Tanpaiboon, F. Lacbawan, Clinton R. Paden, Suxiang Tong, Duncan MacCannell |
| EPI_ISL_1392387 | Sonora Quest Laboratories | TGen North | "Jolene Bowers, Heather Centner, Chris French, Hayley Yaglom, Ashlyn Pfeiffer, Darrin Lemmer, Dave Engelthaler, The Arizona COVID Genomics Union (ACGU)" |
| EPI_ISL_1392388 | Quest Diagnostics Incorporated | Centers for Disease Control and Prevention Division of Viral Diseases, Pathogen Discovery | Peter W. Cook, Dakota Howard, Dhwani Batra, Ben L. Rambo-Martin, S. H. Rosenthal, A. Gerasimova, R. M. Kagan, B. Anderson, M. Hua, Y. Liu, L.E. Bernstein, K.E. Livingston, A. Perez, I. A. Shlyakhter, R. V. Rolando, R. Owen, P. Tanpaiboon, F. Lacbawan, Clinton R. Paden, Suxiang Tong, Duncan MacCannell |
| EPI_ISL_1392389 | Sonora Quest Laboratories | TGen North | "Jolene Bowers, Heather Centner, Chris French, Hayley Yaglom, Ashlyn Pfeiffer, Darrin Lemmer, Dave Engelthaler, The Arizona COVID Genomics Union (ACGU)" |
| EPI_ISL_1392390 | Quest Diagnostics Incorporated | Centers for Disease Control and Prevention Division of Viral Diseases, Pathogen Discovery | Peter W. Cook, Dakota Howard, Dhwani Batra, Ben L. Rambo-Martin, S. H. Rosenthal, A. Gerasimova, R. M. Kagan, B. Anderson, M. Hua, Y. Liu, L.E. Bernstein, K.E. Livingston, A. Perez, I. A. Shlyakhter, R. V. Rolando, R. Owen, P. Tanpaiboon, F. Lacbawan, Clinton R. Paden, Suxiang Tong, Duncan MacCannell |
| EPI_ISL_1392391 | Sonora Quest Laboratories | TGen North | "Jolene Bowers, Heather Centner, Chris French, Hayley Yaglom, Ashlyn Pfeiffer, Darrin Lemmer, Dave Engelthaler, The Arizona COVID Genomics Union (ACGU)" |
| EPI_ISL_1392392 | Quest Diagnostics Incorporated | Centers for Disease Control and Prevention Division of Viral Diseases, Pathogen Discovery | Peter W. Cook, Dakota Howard, Dhwani Batra, Ben L. Rambo-Martin, S. H. Rosenthal, A. Gerasimova, R. M. Kagan, B. Anderson, M. Hua, Y. Liu, L.E. Bernstein, K.E. Livingston, A. Perez, I. A. Shlyakhter, R. V. Rolando, R. Owen, P. Tanpaiboon, F. Lacbawan, Clinton R. Paden, Suxiang Tong, Duncan MacCannell |
| EPI_ISL_1392393 | Sonora Quest Laboratories | TGen North | "Jolene Bowers, Heather Centner, Chris French, Hayley Yaglom, Ashlyn Pfeiffer, Darrin Lemmer, Dave Engelthaler, The Arizona COVID Genomics Union (ACGU)" |
| EPI_ISL_1392394 | Quest Diagnostics Incorporated | Centers for Disease Control and Prevention Division of Viral Diseases, Pathogen Discovery | Peter W. Cook, Dakota Howard, Dhwani Batra, Ben L. Rambo-Martin, S. H. Rosenthal, A. Gerasimova, R. M. Kagan, B. Anderson, M. Hua, Y. Liu, L.E. Bernstein, K.E. Livingston, A. Perez, I. A. Shlyakhter, R. V. Rolando, R. Owen, P. Tanpaiboon, F. Lacbawan, Clinton R. Paden, Suxiang Tong, Duncan MacCannell |
| EPI_ISL_1392395 | Sonora Quest Laboratories | TGen North | "Jolene Bowers, Heather Centner, Chris French, Hayley Yaglom, Ashlyn Pfeiffer, Darrin Lemmer, Dave Engelthaler, The Arizona COVID Genomics Union (ACGU)" |
| EPI_ISL_1392396 | Quest Diagnostics Incorporated | Centers for Disease Control and Prevention Division of Viral Diseases, Pathogen | Peter W. Cook, Dakota Howard, Dhwani Batra, Ben L. Rambo-Martin, S. H. Rosenthal, A. Gerasimova, R. M. Kagan, B. Anderson, M. Hua, Y. Liu, L.E. Bernstein, K.E. Livingston, A. Perez, I. A. Shlyakhter, R. V. Rolando, R. Owen, P. Tanpaiboon, F. Lacbawan, Clinton R. Paden, Suxiang Tong, Duncan MacCannell |

| | | | |
|---|---|---|---|
| | | Discovery | |
| EPI_ISL_1392397 | Sonora Quest Laboratories | TGen North | "Jolene Bowers, Heather Centner, Chris French, Hayley Yaglom, Ashlyn Pfeiffer, Darrin Lemmer, Dave Engelthaler, The Arizona COVID Genomics Union (ACGU)" |
| EPI_ISL_1392398, EPI_ISL_1392400, EPI_ISL_1392402 | Quest Diagnostics Incorporated | Centers for Disease Control and Prevention Division of Viral Diseases, Pathogen Discovery | Peter W. Cook, Dakota Howard, Dhwani Batra, Ben L. Rambo-Martin, S. H. Rosenthal, A. Gerasimova, R. M. Kagan, B. Anderson, M. Hua, Y. Liu, L.E. Bernstein, K.E. Livingston, A. Perez, I. A. Shlyakhter, R. V. Rolando, R. Owen, P. Tanpaiboon, F. Lacbawan, Clinton R. Paden, Suxiang Tong, Duncan MacCannell |
| EPI_ISL_1392403 | Sonora Quest Laboratories | TGen North | "Jolene Bowers, Heather Centner, Chris French, Hayley Yaglom, Ashlyn Pfeiffer, Darrin Lemmer, Dave Engelthaler, The Arizona COVID Genomics Union (ACGU)" |
| EPI_ISL_1392404 | Quest Diagnostics Incorporated | Centers for Disease Control and Prevention Division of Viral Diseases, Pathogen Discovery | Peter W. Cook, Dakota Howard, Dhwani Batra, Ben L. Rambo-Martin, S. H. Rosenthal, A. Gerasimova, R. M. Kagan, B. Anderson, M. Hua, Y. Liu, L.E. Bernstein, K.E. Livingston, A. Perez, I. A. Shlyakhter, R. V. Rolando, R. Owen, P. Tanpaiboon, F. Lacbawan, Clinton R. Paden, Suxiang Tong, Duncan MacCannell |
| EPI_ISL_1392405 | Sonora Quest Laboratories | TGen North | "Jolene Bowers, Heather Centner, Chris French, Hayley Yaglom, Ashlyn Pfeiffer, Darrin Lemmer, Dave Engelthaler, The Arizona COVID Genomics Union (ACGU)" |
| EPI_ISL_1392406 | Quest Diagnostics Incorporated | Centers for Disease Control and Prevention Division of Viral Diseases, Pathogen Discovery | Peter W. Cook, Dakota Howard, Dhwani Batra, Ben L. Rambo-Martin, S. H. Rosenthal, A. Gerasimova, R. M. Kagan, B. Anderson, M. Hua, Y. Liu, L.E. Bernstein, K.E. Livingston, A. Perez, I. A. Shlyakhter, R. V. Rolando, R. Owen, P. Tanpaiboon, F. Lacbawan, Clinton R. Paden, Suxiang Tong, Duncan MacCannell |
| EPI_ISL_1392407 | Sonora Quest Laboratories | TGen North | "Jolene Bowers, Heather Centner, Chris French, Hayley Yaglom, Ashlyn Pfeiffer, Darrin Lemmer, Dave Engelthaler, The Arizona COVID Genomics Union (ACGU)" |
| EPI_ISL_1392408, EPI_ISL_1392409 | Quest Diagnostics Incorporated | Centers for Disease Control and Prevention Division of Viral Diseases, Pathogen Discovery | Peter W. Cook, Dakota Howard, Dhwani Batra, Ben L. Rambo-Martin, S. H. Rosenthal, A. Gerasimova, R. M. Kagan, B. Anderson, M. Hua, Y. Liu, L.E. Bernstein, K.E. Livingston, A. Perez, I. A. Shlyakhter, R. V. Rolando, R. Owen, P. Tanpaiboon, F. Lacbawan, Clinton R. Paden, Suxiang Tong, Duncan MacCannell |
| EPI_ISL_1392410, EPI_ISL_1392411 | Sonora Quest Laboratories | TGen North | "Jolene Bowers, Heather Centner, Chris French, Hayley Yaglom, Ashlyn Pfeiffer, Darrin Lemmer, Dave Engelthaler, The Arizona COVID Genomics Union (ACGU)" |
| EPI_ISL_1392412, EPI_ISL_1392414, EPI_ISL_1392416, EPI_ISL_1392417 | Quest Diagnostics Incorporated | Centers for Disease Control and Prevention Division of Viral Diseases, Pathogen Discovery | Peter W. Cook, Dakota Howard, Dhwani Batra, Ben L. Rambo-Martin, S. H. Rosenthal, A. Gerasimova, R. M. Kagan, B. Anderson, M. Hua, Y. Liu, L.E. Bernstein, K.E. Livingston, A. Perez, I. A. Shlyakhter, R. V. Rolando, R. Owen, P. Tanpaiboon, F. Lacbawan, Clinton R. Paden, Suxiang Tong, Duncan MacCannell |
| EPI_ISL_1392418, EPI_ISL_1392419 | Sonora Quest Laboratories | TGen North | "Jolene Bowers, Heather Centner, Chris French, Hayley Yaglom, Ashlyn Pfeiffer, Darrin Lemmer, Dave Engelthaler, The Arizona COVID Genomics Union (ACGU)" |
| EPI_ISL_1392420, EPI_ISL_1392422 | Quest Diagnostics Incorporated | Centers for Disease Control and Prevention Division of Viral Diseases, Pathogen Discovery | Peter W. Cook, Dakota Howard, Dhwani Batra, Ben L. Rambo-Martin, S. H. Rosenthal, A. Gerasimova, R. M. Kagan, B. Anderson, M. Hua, Y. Liu, L.E. Bernstein, K.E. Livingston, A. Perez, I. A. Shlyakhter, R. V. Rolando, R. Owen, P. Tanpaiboon, F. Lacbawan, Clinton R. Paden, Suxiang Tong, Duncan MacCannell |
| EPI_ISL_1392423 | Sonora Quest Laboratories | TGen North | "Jolene Bowers, Heather Centner, Chris French, Hayley Yaglom, Ashlyn Pfeiffer, Darrin Lemmer, Dave Engelthaler, The Arizona COVID Genomics Union (ACGU)" |
| EPI_ISL_1392424 | Quest Diagnostics Incorporated | Centers for Disease Control and Prevention Division of Viral Diseases, Pathogen Discovery | Peter W. Cook, Dakota Howard, Dhwani Batra, Ben L. Rambo-Martin, S. H. Rosenthal, A. Gerasimova, R. M. Kagan, B. Anderson, M. Hua, Y. Liu, L.E. Bernstein, K.E. Livingston, A. Perez, I. A. Shlyakhter, R. V. Rolando, R. Owen, P. Tanpaiboon, F. Lacbawan, Clinton R. Paden, Suxiang Tong, Duncan MacCannell |
| EPI_ISL_1392425 | Sonora Quest Laboratories | TGen North | "Jolene Bowers, Heather Centner, Chris French, Hayley Yaglom, Ashlyn Pfeiffer, Darrin Lemmer, Dave Engelthaler, The Arizona COVID Genomics Union (ACGU)" |
| EPI_ISL_1392426 | Quest Diagnostics Incorporated | Centers for Disease Control and Prevention Division of Viral Diseases, Pathogen Discovery | Peter W. Cook, Dakota Howard, Dhwani Batra, Ben L. Rambo-Martin, S. H. Rosenthal, A. Gerasimova, R. M. Kagan, B. Anderson, M. Hua, Y. Liu, L.E. Bernstein, K.E. Livingston, A. Perez, I. A. Shlyakhter, R. V. Rolando, R. Owen, P. Tanpaiboon, F. Lacbawan, Clinton R. Paden, Suxiang Tong, Duncan MacCannell |
| EPI_ISL_1392427 | Sonora Quest Laboratories | TGen North | "Jolene Bowers, Heather Centner, Chris French, Hayley Yaglom, Ashlyn Pfeiffer, Darrin Lemmer, Dave Engelthaler, The Arizona COVID Genomics Union (ACGU)" |
| EPI_ISL_1392428 | Quest Diagnostics Incorporated | Centers for Disease Control and Prevention Division of Viral Diseases, Pathogen Discovery | Peter W. Cook, Dakota Howard, Dhwani Batra, Ben L. Rambo-Martin, S. H. Rosenthal, A. Gerasimova, R. M. Kagan, B. Anderson, M. Hua, Y. Liu, L.E. Bernstein, K.E. Livingston, A. Perez, I. A. Shlyakhter, R. V. Rolando, R. Owen, P. Tanpaiboon, F. Lacbawan, Clinton R. Paden, Suxiang Tong, Duncan MacCannell |
| EPI_ISL_1392429 | Sonora Quest Laboratories | TGen North | "Jolene Bowers, Heather Centner, Chris French, Hayley Yaglom, Ashlyn Pfeiffer, Darrin Lemmer, Dave Engelthaler, The Arizona COVID Genomics Union (ACGU)" |
| EPI_ISL_1392430, EPI_ISL_1392431 | Quest Diagnostics Incorporated | Centers for Disease Control and Prevention Division of Viral Diseases, Pathogen Discovery | Peter W. Cook, Dakota Howard, Dhwani Batra, Ben L. Rambo-Martin, S. H. Rosenthal, A. Gerasimova, R. M. Kagan, B. Anderson, M. Hua, Y. Liu, L.E. Bernstein, K.E. Livingston, A. Perez, I. A. Shlyakhter, R. V. Rolando, R. Owen, P. Tanpaiboon, F. Lacbawan, Clinton R. Paden, Suxiang Tong, Duncan MacCannell |
| EPI_ISL_1392432, EPI_ISL_1392433 | Sonora Quest Laboratories | TGen North | "Jolene Bowers, Heather Centner, Chris French, Hayley Yaglom, Ashlyn Pfeiffer, Darrin Lemmer, Dave Engelthaler, The Arizona COVID Genomics Union (ACGU)" |
| EPI_ISL_1392434 | Quest Diagnostics Incorporated | Centers for Disease Control and Prevention Division of Viral Diseases, Pathogen Discovery | Peter W. Cook, Dakota Howard, Dhwani Batra, Ben L. Rambo-Martin, S. H. Rosenthal, A. Gerasimova, R. M. Kagan, B. Anderson, M. Hua, Y. Liu, L.E. Bernstein, K.E. Livingston, A. Perez, I. A. Shlyakhter, R. V. Rolando, R. Owen, P. Tanpaiboon, F. Lacbawan, Clinton R. Paden, Suxiang Tong, Duncan MacCannell |
| EPI_ISL_1392435 | Sonora Quest Laboratories | TGen North | "Jolene Bowers, Heather Centner, Chris French, Hayley Yaglom, Ashlyn Pfeiffer, Darrin Lemmer, Dave Engelthaler, The Arizona COVID Genomics Union (ACGU)" |
| EPI_ISL_1392436 | Quest Diagnostics Incorporated | Centers for Disease Control and Prevention Division of Viral Diseases, Pathogen Discovery | Peter W. Cook, Dakota Howard, Dhwani Batra, Ben L. Rambo-Martin, S. H. Rosenthal, A. Gerasimova, R. M. Kagan, B. Anderson, M. Hua, Y. Liu, L.E. Bernstein, K.E. Livingston, A. Perez, I. A. Shlyakhter, R. V. Rolando, R. Owen, P. Tanpaiboon, F. Lacbawan, Clinton R. Paden, Suxiang Tong, Duncan MacCannell |
| EPI_ISL_1392439, EPI_ISL_1392440, EPI_ISL_1392441, EPI_ISL_1392442, EPI_ISL_1392443, EPI_ISL_1392444, EPI_ISL_1392445, EPI_ISL_1392446 | Sonora Quest Laboratories | TGen North | "Jolene Bowers, Heather Centner, Chris French, Hayley Yaglom, Ashlyn Pfeiffer, Darrin Lemmer, Dave Engelthaler, The Arizona COVID Genomics Union (ACGU)" |
| EPI_ISL_1392447 | TGen North | TGen North | "Jolene Bowers, Heather Centner, Chris French, Hayley Yaglom, Ashlyn Pfeiffer, Darrin Lemmer, Dave Engelthaler, The Arizona COVID Genomics Union (ACGU)" |
| EPI_ISL_1392448, EPI_ISL_1392449, EPI_ISL_1392450, EPI_ISL_1392451, EPI_ISL_1392452, EPI_ISL_1392453, EPI_ISL_1392454, EPI_ISL_1392455, EPI_ISL_1392456, EPI_ISL_1392457, EPI_ISL_1392458, EPI_ISL_1392459, EPI_ISL_1392460, EPI_ISL_1392462, EPI_ISL_1392463 | | | |
| see above | Sonora Quest Laboratories | TGen North | "Jolene Bowers, Heather Centner, Chris French, Hayley Yaglom, Ashlyn Pfeiffer, Darrin Lemmer, Dave Engelthaler, The Arizona COVID Genomics Union (ACGU)" |
| EPI_ISL_1392464 | TGen North | TGen North | "Jolene Bowers, Heather Centner, Chris French, Hayley Yaglom, Ashlyn Pfeiffer, Darrin Lemmer, Dave Engelthaler, The Arizona COVID Genomics Union (ACGU)" |
| EPI_ISL_1392466, EPI_ISL_1392467, EPI_ISL_1392469, EPI_ISL_1392470, EPI_ISL_1392471, EPI_ISL_1392472, EPI_ISL_1392473, EPI_ISL_1392474, EPI_ISL_1392475, EPI_ISL_1392478, EPI_ISL_1392479, EPI_ISL_1392480, EPI_ISL_1392481, EPI_ISL_1392482, EPI_ISL_1392483, EPI_ISL_1392484, EPI_ISL_1392485, EPI_ISL_1392486, EPI_ISL_1392487, EPI_ISL_1392489, EPI_ISL_1392490, EPI_ISL_1392491, EPI_ISL_1392492, EPI_ISL_1392494, EPI_ISL_1392495, EPI_ISL_1392496, EPI_ISL_1392498, EPI_ISL_1392500, EPI_ISL_1392501, EPI_ISL_1392502, EPI_ISL_1392503, EPI_ISL_1392504, EPI_ISL_1392505, EPI_ISL_1392507, EPI_ISL_1392508, EPI_ISL_1392510, EPI_ISL_1392512, EPI_ISL_1392513, EPI_ISL_1392514, EPI_ISL_1392515, EPI_ISL_1392516, EPI_ISL_1392519, EPI_ISL_1392520, EPI_ISL_1392521, EPI_ISL_1392522, EPI_ISL_1392523, EPI_ISL_1392524, EPI_ISL_1392525, EPI_ISL_1392526, EPI_ISL_1392527, EPI_ISL_1392528, EPI_ISL_1392529, EPI_ISL_1392530, EPI_ISL_1392531, EPI_ISL_1392532, EPI_ISL_1392533, EPI_ISL_1392535, EPI_ISL_1392536, EPI_ISL_1392537, EPI_ISL_1392538, EPI_ISL_1392539, EPI_ISL_1392540 | | | |
| see above | Sonora Quest Laboratories | TGen North | "Jolene Bowers, Heather Centner, Chris French, Hayley Yaglom, Ashlyn Pfeiffer, Darrin Lemmer, Dave Engelthaler, The Arizona COVID Genomics Union (ACGU)" |
| EPI_ISL_1392541 | TGen North | TGen North | "Jolene Bowers, Heather Centner, Chris French, Hayley Yaglom, Ashlyn Pfeiffer, Darrin Lemmer, Dave Engelthaler, The Arizona COVID Genomics Union (ACGU)" |
| EPI_ISL_1392542, EPI_ISL_1392544, EPI_ISL_1392545, EPI_ISL_1392546, EPI_ISL_1392548, EPI_ISL_1392550, EPI_ISL_1392551, EPI_ISL_1392552, EPI_ISL_1392553, EPI_ISL_1392554, EPI_ISL_1392555, EPI_ISL_1392556, EPI_ISL_1392557, EPI_ISL_1392558, EPI_ISL_1392560, EPI_ISL_1392562, EPI_ISL_1392563, EPI_ISL_1392564, EPI_ISL_1392565, EPI_ISL_1392566, EPI_ISL_1392568, EPI_ISL_1392569, EPI_ISL_1392571, EPI_ISL_1392572 | | | |
| see above | Sonora Quest Laboratories | TGen North | "Jolene Bowers, Heather Centner, Chris French, Hayley Yaglom, Ashlyn Pfeiffer, Darrin Lemmer, Dave Engelthaler, The Arizona COVID Genomics Union (ACGU)" |
| EPI_ISL_1392573 | TGen North | TGen North | "Jolene Bowers, Heather Centner, Chris French, Hayley Yaglom, Ashlyn Pfeiffer, Darrin Lemmer, Dave Engelthaler, The Arizona COVID Genomics Union (ACGU)" |
| EPI_ISL_1392574, EPI_ISL_1392576, EPI_ISL_1392577, EPI_ISL_1392578, EPI_ISL_1392581, EPI_ISL_1392582, EPI_ISL_1392583, EPI_ISL_1392585, EPI_ISL_1392586, EPI_ISL_1392587, EPI_ISL_1392589, EPI_ISL_1392591, EPI_ISL_1392592, EPI_ISL_1392593, EPI_ISL_1392594, EPI_ISL_1392600, EPI_ISL_1392601, EPI_ISL_1392602, EPI_ISL_1392603, EPI_ISL_1392604, EPI_ISL_1392605, EPI_ISL_1392606, EPI_ISL_1392607, EPI_ISL_1392608, EPI_ISL_1392609, EPI_ISL_1392610, EPI_ISL_1392611, EPI_ISL_1392612, EPI_ISL_1392615, EPI_ISL_1392617, EPI_ISL_1392619, EPI_ISL_1392620, EPI_ISL_1392621, EPI_ISL_1392622, EPI_ISL_1392623, EPI_ISL_1392624, EPI_ISL_1392625, EPI_ISL_1392626, EPI_ISL_1392627, EPI_ISL_1392628, EPI_ISL_1392629, EPI_ISL_1392630, EPI_ISL_1392632 | | | |
| see above | Sonora Quest Laboratories | TGen North | "Jolene Bowers, Heather Centner, Chris French, Hayley Yaglom, Ashlyn Pfeiffer, Darrin Lemmer, Dave Engelthaler, The Arizona COVID Genomics Union (ACGU)" |
| EPI_ISL_1392633 | TGen North | TGen North | "Jolene Bowers, Heather Centner, Chris French, Hayley Yaglom, Ashlyn Pfeiffer, Darrin Lemmer, Dave Engelthaler, The Arizona COVID Genomics Union (ACGU)" |
| EPI_ISL_1392634, EPI_ISL_1392635, EPI_ISL_1392636, EPI_ISL_1392637, EPI_ISL_1392638 | Sonora Quest Laboratories | TGen North | "Jolene Bowers, Heather Centner, Chris French, Hayley Yaglom, Ashlyn Pfeiffer, Darrin Lemmer, Dave Engelthaler, The Arizona COVID Genomics Union (ACGU)" |
| EPI_ISL_1392715, EPI_ISL_1392716, EPI_ISL_1392717, EPI_ISL_1392718, EPI_ISL_1392721, EPI_ISL_1392722, EPI_ISL_1392723, EPI_ISL_1392724, EPI_ISL_1392725, EPI_ISL_1392726, EPI_ISL_1392728, EPI_ISL_1392729, EPI_ISL_1392730, EPI_ISL_1392731, EPI_ISL_1392732, EPI_ISL_1392733, EPI_ISL_1392734, EPI_ISL_1392735, EPI_ISL_1392736, EPI_ISL_1392737, EPI_ISL_1392738, EPI_ISL_1392739, EPI_ISL_1392740, EPI_ISL_1392741, EPI_ISL_1392742, EPI_ISL_1392743, EPI_ISL_1392744, EPI_ISL_1392745, EPI_ISL_1392746, EPI_ISL_1392747, EPI_ISL_1392748, EPI_ISL_1392749, EPI_ISL_1392750, EPI_ISL_1392751, EPI_ISL_1392752, EPI_ISL_1392754, EPI_ISL_1392755, EPI_ISL_1392756, EPI_ISL_1392757, EPI_ISL_1392758, EPI_ISL_1392759, EPI_ISL_1392760, EPI_ISL_1392761, EPI_ISL_1392762, EPI_ISL_1392763, EPI_ISL_1392764, EPI_ISL_1392765, EPI_ISL_1392766, EPI_ISL_1392767, EPI_ISL_1392768, EPI_ISL_1392769, EPI_ISL_1392770, EPI_ISL_1392771, EPI_ISL_1392772, EPI_ISL_1392773, EPI_ISL_1392774, EPI_ISL_1392775, EPI_ISL_1392776, EPI_ISL_1392777, EPI_ISL_1392778, EPI_ISL_1392779, EPI_ISL_1392780, EPI_ISL_1392781, EPI_ISL_1392782, EPI_ISL_1392783, EPI_ISL_1392784, EPI_ISL_1392785, EPI_ISL_1392786, EPI_ISL_1392787, EPI_ISL_1392788, EPI_ISL_1392790, EPI_ISL_1392791, EPI_ISL_1392792, EPI_ISL_1392793, EPI_ISL_1392796, EPI_ISL_1392797, EPI_ISL_1392798, EPI_ISL_1392799 | | | |
| see above | TXDSHS | TXDSHS | Rashmi Tuladhar, Bonnie Oh, Jenny Zhang, Maliha Rahman, Mayela Pedrueza, Anita Pokharel, Lorraine Rodriguez, Myong Koag, Chun Wang, Rachel Lee, Grace Kubin |
| EPI_ISL_1392807, EPI_ISL_1392808, EPI_ISL_1392809, EPI_ISL_1392810, EPI_ISL_1392811, EPI_ISL_1392812, EPI_ISL_1392813, EPI_ISL_1392814, EPI_ISL_1392815, EPI_ISL_1392816, EPI_ISL_1392817, EPI_ISL_1392818, EPI_ISL_1392819, EPI_ISL_1392821, EPI_ISL_1392822, EPI_ISL_1392823, EPI_ISL_1392824, EPI_ISL_1392825, EPI_ISL_1392826, EPI_ISL_1392827, EPI_ISL_1392828, EPI_ISL_1392829, EPI_ISL_1392830, EPI_ISL_1392831, EPI_ISL_1392832, EPI_ISL_1392833, EPI_ISL_1392834, EPI_ISL_1392835, EPI_ISL_1392836, EPI_ISL_1392837, EPI_ISL_1392838, EPI_ISL_1392840, EPI_ISL_1392841, EPI_ISL_1392842, EPI_ISL_1392843, EPI_ISL_1392844, EPI_ISL_1392845, EPI_ISL_1392846, EPI_ISL_1392847, EPI_ISL_1392848, EPI_ISL_1392849, EPI_ISL_1392850, EPI_ISL_1392851, EPI_ISL_1392852, EPI_ISL_1392853, EPI_ISL_1392854, EPI_ISL_1392855, EPI_ISL_1392856, EPI_ISL_1392857, EPI_ISL_1392858, EPI_ISL_1392859, EPI_ISL_1392860, EPI_ISL_1392861, EPI_ISL_1392862, EPI_ISL_1392863, EPI_ISL_1392864, EPI_ISL_1392865, EPI_ISL_1392866, EPI_ISL_1392867, EPI_ISL_1392868, EPI_ISL_1392869, EPI_ISL_1392870, EPI_ISL_1392871, EPI_ISL_1392872, EPI_ISL_1392873, EPI_ISL_1392874, EPI_ISL_1392875, EPI_ISL_1392876, EPI_ISL_1392877, EPI_ISL_1392878, EPI_ISL_1392879, EPI_ISL_1392880, EPI_ISL_1392881, EPI_ISL_1392882, EPI_ISL_1392883, EPI_ISL_1392884, EPI_ISL_1392885, EPI_ISL_1392886, EPI_ISL_1392887, EPI_ISL_1392888 | | | |

| | | | |
|---|---|---|---|
| see above | Missouri State Public Health Laboratory | Missouri State Public Health Laboratory | Matthew Sinn, Joshua Barry, Ashley New |
| EPI_ISL_1392907, EPI_ISL_1392908, EPI_ISL_1392909, EPI_ISL_1392910, EPI_ISL_1392911, EPI_ISL_1392912, EPI_ISL_1392913, EPI_ISL_1392914, EPI_ISL_1392915 | Cytocheck Laboratory | Kansas Health and Environmental Lab | Mike Grose, Jonathan Barnell, Ben Olsen, and Phil Adam |
| EPI_ISL_1392916, EPI_ISL_1392917, EPI_ISL_1392918, EPI_ISL_1392919, EPI_ISL_1392920, EPI_ISL_1392921 | Kansas Health and Environmental Lab | Kansas Health and Environmental Lab | Mike Grose, Jonathan Barnell, Ben Olsen, and Phil Adam |
| EPI_ISL_1392922 | Instituto Nacional de Medicina Genomica | Instituto Nacional de Medicina Genomica | Hidalgo-Miranda A, Mendoza-Vargas A, Reyes-Grajeda JP, Cedro-Tanda A, Alcaraz N, Gonzalez-Barrera D, Rangel-DeLeon D, Ramirez-Vega O, Herrera-Montalvo LA |
| EPI_ISL_1393629 | Oregon State Public Health Laboratory | Oregon State Public Health Laboratory | John Fontana, Eugene Yeboah, Vanda Makris, Rafia Razzaque |
| EPI_ISL_1394338 | Oregon State Public Health Laboratory | Oregon State Public Health Laboratory | Rafia Razzaque, Eugene Yeboah, Vanda Makris, John Fontana, Shane Sevey |
| EPI_ISL_1394593, EPI_ISL_1394594, EPI_ISL_1394595, EPI_ISL_1394596, EPI_ISL_1394597, EPI_ISL_1394598, EPI_ISL_1394599, EPI_ISL_1394600, EPI_ISL_1394601, EPI_ISL_1394602, EPI_ISL_1394603, EPI_ISL_1394604, EPI_ISL_1394605, EPI_ISL_1394606, EPI_ISL_1394607, EPI_ISL_1394608, EPI_ISL_1394609, EPI_ISL_1394610, EPI_ISL_1394611, EPI_ISL_1394612, EPI_ISL_1394613, EPI_ISL_1394614, EPI_ISL_1394615, EPI_ISL_1394616, EPI_ISL_1394617, EPI_ISL_1394618, EPI_ISL_1394619, EPI_ISL_1394620, EPI_ISL_1394621, EPI_ISL_1394622, EPI_ISL_1394623, EPI_ISL_1394624, EPI_ISL_1394625, EPI_ISL_1394626, EPI_ISL_1394627, EPI_ISL_1394628 | | | |
| see above | WESTCHESTER MEDICAL CENTER | Wadsworth Center, New York State Department of Health | Kirsten St. George, Daryl M. Lamson, Alexis Russel, Matthew Shudt, Melissa A Leisner, Jonathan Plitnick, Navjot Singh, John Kelly, Erasmus Schneider, Erica Lasek-Nesselquist |
| EPI_ISL_1394629, EPI_ISL_1394630, EPI_ISL_1394631, EPI_ISL_1394632, EPI_ISL_1394633, EPI_ISL_1394634, EPI_ISL_1394635, EPI_ISL_1394636, EPI_ISL_1394637, EPI_ISL_1394638, EPI_ISL_1394639, EPI_ISL_1394640, EPI_ISL_1394641 | | | |
| see above | URMC LABS | Wadsworth Center, New York State Department of Health | Kirsten St. George, Daryl M. Lamson, Alexis Russel, Matthew Shudt, Melissa A Leisner, Jonathan Plitnick, Navjot Singh, John Kelly, Erasmus Schneider, Erica Lasek-Nesselquist |
| EPI_ISL_1394642 | New York Presbyterian Hospital | Wadsworth Center, New York State Department of Health | Kirsten St. George, Daryl M. Lamson, Alexis Russel, Matthew Shudt, Melissa A Leisner, Jonathan Plitnick, Navjot Singh, John Kelly, Erasmus Schneider, Erica Lasek-Nesselquist |
| EPI_ISL_1394643 | URMC LABS | Wadsworth Center, New York State Department of Health | Kirsten St. George, Daryl M. Lamson, Alexis Russel, Matthew Shudt, Melissa A Leisner, Jonathan Plitnick, Navjot Singh, John Kelly, Erasmus Schneider, Erica Lasek-Nesselquist |
| EPI_ISL_1394644, EPI_ISL_1394645, EPI_ISL_1394646, EPI_ISL_1394647, EPI_ISL_1394648, EPI_ISL_1394649, EPI_ISL_1394650, EPI_ISL_1394651, EPI_ISL_1394652, EPI_ISL_1394653, EPI_ISL_1394654, EPI_ISL_1394655, EPI_ISL_1394656, EPI_ISL_1394657, EPI_ISL_1394658, EPI_ISL_1394659, EPI_ISL_1394660, EPI_ISL_1394661, EPI_ISL_1394662, EPI_ISL_1394663, EPI_ISL_1394664 | | | |
| see above | New York Presbyterian Hospital | Wadsworth Center, New York State Department of Health | Kirsten St. George, Daryl M. Lamson, Alexis Russel, Matthew Shudt, Melissa A Leisner, Jonathan Plitnick, Navjot Singh, John Kelly, Erasmus Schneider, Erica Lasek-Nesselquist |
| EPI_ISL_1394840, EPI_ISL_1394841, EPI_ISL_1394842, EPI_ISL_1394843, EPI_ISL_1394844, EPI_ISL_1394845, EPI_ISL_1394846, EPI_ISL_1394847, EPI_ISL_1394848, EPI_ISL_1394849, EPI_ISL_1394850, EPI_ISL_1394851, EPI_ISL_1394852, EPI_ISL_1394853, EPI_ISL_1394854, EPI_ISL_1394855, EPI_ISL_1394856, EPI_ISL_1394857 | | | |
| see above | IN State Department of Health Laboratory Services | IN State Department of Health Laboratory Services | Cassandra Campion, Jamie Yeadon, Brian Pope, Lixia Liu, Kyle Brownlee, Melissa Hindenlang, Ankita Kashikar, Mark Glazier |
| EPI_ISL_1394858, EPI_ISL_1394859, EPI_ISL_1394860, EPI_ISL_1394861, EPI_ISL_1394864, EPI_ISL_1394866, EPI_ISL_1394868 | Oregon State University TRACE Project | OSU Center for Genome Research and Biocomputing | Oregon State University TRACE Project |
| EPI_ISL_1395706 | NYC Pandemic Response Lab | Wadsworth Center, New York State Department of Health | Kirsten St. George, Daryl M. Lamson, Alexis Russel, Matthew Shudt, Melissa A Leisner, Jonathan Plitnick, Navjot Singh, John Kelly, Erasmus Schneider, Erica Lasek-Nesselquist |
| EPI_ISL_1395707 | NORTHWELL HEALTH LABORATORIES | Wadsworth Center, New York State Department of Health | Kirsten St. George, Daryl M. Lamson, Alexis Russel, Matthew Shudt, Melissa A Leisner, Jonathan Plitnick, Navjot Singh, John Kelly, Erasmus Schneider, Erica Lasek-Nesselquist |
| EPI_ISL_1395708, EPI_ISL_1395709 | NYC Pandemic Response Lab | Wadsworth Center, New York State Department of Health | Kirsten St. George, Daryl M. Lamson, Alexis Russel, Matthew Shudt, Melissa A Leisner, Jonathan Plitnick, Navjot Singh, John Kelly, Erasmus Schneider, Erica Lasek-Nesselquist |
| EPI_ISL_1395710, EPI_ISL_1395711, EPI_ISL_1395712, EPI_ISL_1395713, EPI_ISL_1395714, EPI_ISL_1395715, EPI_ISL_1395716, EPI_ISL_1395717, EPI_ISL_1395718, EPI_ISL_1395719, EPI_ISL_1395720, EPI_ISL_1395721, EPI_ISL_1395722, EPI_ISL_1395723, EPI_ISL_1395724, EPI_ISL_1395725 | | | |
| see above | MEMORIAL SLOAN KETTERING CANCER CENTER | Wadsworth Center, New York State Department of Health | Kirsten St. George, Daryl M. Lamson, Alexis Russel, Matthew Shudt, Melissa A Leisner, Jonathan Plitnick, Navjot Singh, John Kelly, Erasmus Schneider, Erica Lasek-Nesselquist |
| EPI_ISL_1395726, EPI_ISL_1395727, EPI_ISL_1395728, EPI_ISL_1395729, EPI_ISL_1395730, EPI_ISL_1395731, EPI_ISL_1395732, EPI_ISL_1395733, EPI_ISL_1395734, EPI_ISL_1395735, EPI_ISL_1395736, EPI_ISL_1395737, EPI_ISL_1395738, EPI_ISL_1395739, EPI_ISL_1395740, EPI_ISL_1395741, EPI_ISL_1395742, EPI_ISL_1395743, EPI_ISL_1395744, EPI_ISL_1395745, EPI_ISL_1395746, EPI_ISL_1395747, EPI_ISL_1395748, EPI_ISL_1395749 | | | |
| see above | Columbia University Irving Medical Center | Wadsworth Center, New York State Department of Health | Kirsten St. George, Daryl M. Lamson, Alexis Russel, Matthew Shudt, Melissa A Leisner, Jonathan Plitnick, Navjot Singh, John Kelly, Erasmus Schneider, Erica Lasek-Nesselquist |
| EPI_ISL_1395750 | NORTH SHORE UNIVERSITY HOSPITAL | Wadsworth Center, New York State Department of Health | Kirsten St. George, Daryl M. Lamson, Alexis Russel, Matthew Shudt, Melissa A Leisner, Jonathan Plitnick, Navjot Singh, John Kelly, Erasmus Schneider, Erica Lasek-Nesselquist |
| EPI_ISL_1395751, EPI_ISL_1395752 | SUNY UPSTATE MEDICAL UNIVERSITY | Wadsworth Center, New York State Department of Health | Kirsten St. George, Daryl M. Lamson, Alexis Russel, Matthew Shudt, Melissa A Leisner, Jonathan Plitnick, Navjot Singh, John Kelly, Erasmus Schneider, Erica Lasek-Nesselquist |
| EPI_ISL_1395753, EPI_ISL_1395754 | NORTH SHORE UNIVERSITY HOSPITAL | Wadsworth Center, New York State Department of Health | Kirsten St. George, Daryl M. Lamson, Alexis Russel, Matthew Shudt, Melissa A Leisner, Jonathan Plitnick, Navjot Singh, John Kelly, Erasmus Schneider, Erica Lasek-Nesselquist |
| EPI_ISL_1395755 | SUNY UPSTATE MEDICAL UNIVERSITY | Wadsworth Center, New York State Department of Health | Kirsten St. George, Daryl M. Lamson, Alexis Russel, Matthew Shudt, Melissa A Leisner, Jonathan Plitnick, Navjot Singh, John Kelly, Erasmus Schneider, Erica Lasek-Nesselquist |
| EPI_ISL_1395756, EPI_ISL_1395757, EPI_ISL_1395758 | NORTH SHORE UNIVERSITY HOSPITAL | Wadsworth Center, New York State Department of Health | Kirsten St. George, Daryl M. Lamson, Alexis Russel, Matthew Shudt, Melissa A Leisner, Jonathan Plitnick, Navjot Singh, John Kelly, Erasmus Schneider, Erica Lasek-Nesselquist |
| EPI_ISL_1395759 | SUNY UPSTATE MEDICAL UNIVERSITY | Wadsworth Center, New York State Department of Health | Kirsten St. George, Daryl M. Lamson, Alexis Russel, Matthew Shudt, Melissa A Leisner, Jonathan Plitnick, Navjot Singh, John Kelly, Erasmus Schneider, Erica Lasek-Nesselquist |
| EPI_ISL_1395760 | NORTH SHORE UNIVERSITY HOSPITAL | Wadsworth Center, New York State Department of Health | Kirsten St. George, Daryl M. Lamson, Alexis Russel, Matthew Shudt, Melissa A Leisner, Jonathan Plitnick, Navjot Singh, John Kelly, Erasmus Schneider, Erica Lasek-Nesselquist |
| EPI_ISL_1395761, EPI_ISL_1395762 | SUNY UPSTATE MEDICAL UNIVERSITY | Wadsworth Center, New York State Department of Health | Kirsten St. George, Daryl M. Lamson, Alexis Russel, Matthew Shudt, Melissa A Leisner, Jonathan Plitnick, Navjot Singh, John Kelly, Erasmus Schneider, Erica Lasek-Nesselquist |
| EPI_ISL_1395763 | NORTH SHORE UNIVERSITY HOSPITAL | Wadsworth Center, New York State Department of Health | Kirsten St. George, Daryl M. Lamson, Alexis Russel, Matthew Shudt, Melissa A Leisner, Jonathan Plitnick, Navjot Singh, John Kelly, Erasmus Schneider, Erica Lasek-Nesselquist |
| EPI_ISL_1395764, EPI_ISL_1395765 | SUNY UPSTATE MEDICAL UNIVERSITY | Wadsworth Center, New York State Department of Health | Kirsten St. George, Daryl M. Lamson, Alexis Russel, Matthew Shudt, Melissa A Leisner, Jonathan Plitnick, Navjot Singh, John Kelly, Erasmus Schneider, Erica Lasek-Nesselquist |
| EPI_ISL_1395766 | THE MARY IMOGENE BASSETT HOSPITAL | Wadsworth Center, New York State Department of Health | Kirsten St. George, Daryl M. Lamson, Alexis Russel, Matthew Shudt, Melissa A Leisner, Jonathan Plitnick, Navjot Singh, John Kelly, Erasmus Schneider, Erica Lasek-Nesselquist |
| EPI_ISL_1395767 | SUNY UPSTATE MEDICAL UNIVERSITY | Wadsworth Center, New York State Department of Health | Kirsten St. George, Daryl M. Lamson, Alexis Russel, Matthew Shudt, Melissa A Leisner, Jonathan Plitnick, Navjot Singh, John Kelly, Erasmus Schneider, Erica Lasek-Nesselquist |
| EPI_ISL_1395768, EPI_ISL_1395769 | THE MARY IMOGENE BASSETT HOSPITAL | Wadsworth Center, New York State Department of Health | Kirsten St. George, Daryl M. Lamson, Alexis Russel, Matthew Shudt, Melissa A Leisner, Jonathan Plitnick, Navjot Singh, John Kelly, Erasmus Schneider, Erica Lasek-Nesselquist |
| EPI_ISL_1395770 | SUNY UPSTATE MEDICAL UNIVERSITY | Wadsworth Center, New York State Department of Health | Kirsten St. George, Daryl M. Lamson, Alexis Russel, Matthew Shudt, Melissa A Leisner, Jonathan Plitnick, Navjot Singh, John Kelly, Erasmus Schneider, Erica Lasek-Nesselquist |
| EPI_ISL_1395771 | THE MARY IMOGENE BASSETT HOSPITAL | Wadsworth Center, New York State Department of Health | Kirsten St. George, Daryl M. Lamson, Alexis Russel, Matthew Shudt, Melissa A Leisner, Jonathan Plitnick, Navjot Singh, John Kelly, Erasmus Schneider, Erica Lasek-Nesselquist |
| EPI_ISL_1395772 | SUNY UPSTATE MEDICAL UNIVERSITY | Wadsworth Center, New York State Department of Health | Kirsten St. George, Daryl M. Lamson, Alexis Russel, Matthew Shudt, Melissa A Leisner, Jonathan Plitnick, Navjot Singh, John Kelly, Erasmus Schneider, Erica Lasek-Nesselquist |
| EPI_ISL_1395773 | THE MARY IMOGENE BASSETT HOSPITAL | Wadsworth Center, New York State Department of Health | Kirsten St. George, Daryl M. Lamson, Alexis Russel, Matthew Shudt, Melissa A Leisner, Jonathan Plitnick, Navjot Singh, John Kelly, Erasmus Schneider, Erica Lasek-Nesselquist |

| | | | |
|---|---|---|---|
| EPI_ISL_1395774, EPI_ISL_1395775, EPI_ISL_1395776 | SUNY UPSTATE MEDICAL UNIVERSITY | Wadsworth Center, New York State Department of Health | Kirsten St. George, Daryl M. Lamson, Alexis Russel, Matthew Shudt, Melissa A Leisner, Jonathan Plitnick, Navjot Singh, John Kelly, Erasmus Schneider, Erica Lasek-Nesselquist |
| EPI_ISL_1396407 | DC Public Health Lab/ Dept. of Forensic Sciences | DC Public Health Lab/ Dept. of Forensic Sciences | Janis Doss, Scott Nguyen, Elizabeth Zelaya, Sarah Scott, Connie Maza, Monica Mann, Brittany Hamilton, David Payne, Jocelyn Hauser |
| EPI_ISL_1396409, EPI_ISL_1396410 | Mayo Clinic & Mayo Clinic Laboratories | Minnesota Department of Health, Public Health Laboratory | Alexandra Lorentz, Jacob Garfin, Matt Plumb, and Xiong Wang |
| EPI_ISL_1396411 | M Health Fairview | Minnesota Department of Health, Public Health Laboratory | Alexandra Lorentz, Jacob Garfin, Matt Plumb, and Xiong Wang |
| EPI_ISL_1396554, EPI_ISL_1396555 | Cytocheck Laboratory | Kansas Health and Environmental Lab | Mike Grose, Jonathan Barnell, Ben Olsen, and Phil Adam |
| EPI_ISL_1396634, EPI_ISL_1396635, EPI_ISL_1396636, EPI_ISL_1396637, EPI_ISL_1396638, EPI_ISL_1396639, EPI_ISL_1396640, EPI_ISL_1396641, EPI_ISL_1396642, EPI_ISL_1396643, EPI_ISL_1396644, EPI_ISL_1396645, EPI_ISL_1396646 | | | |
| see above | Clinical Reference Laboratory | Kansas Health and Environmental Lab | Mike Grose, Jonathan Barnell, Ben Olsen, and Phil Adam |
| EPI_ISL_1396647, EPI_ISL_1396648, EPI_ISL_1396649 | Kansas Health and Environmental Lab | Kansas Health and Environmental Lab | Mike Grose, Jonathan Barnell, Ben Olsen, and Phil Adam |
| EPI_ISL_1396656, EPI_ISL_1396657, EPI_ISL_1396658, EPI_ISL_1396659, EPI_ISL_1396660, EPI_ISL_1396661, EPI_ISL_1396662, EPI_ISL_1396663, EPI_ISL_1396664, EPI_ISL_1396665, EPI_ISL_1396666 | | | |
| see above | IN State Department of Health Laboratory Services | IN State Department of Health Laboratory Services | Cassandra Campion, Jamie Yeadon, Brian Pope, Lixia Liu, Kyle Brownlee, Melissa Hindenlang, Ankita Kashikar, Mark Glazier |
| EPI_ISL_1397383, EPI_ISL_1397384, EPI_ISL_1397385, EPI_ISL_1397386, EPI_ISL_1397387, EPI_ISL_1397388 | MEMORIAL SLOAN KETTERING CANCER CENTER | Wadsworth Center, New York State Department of Health | Kirsten St. George, Daryl M. Lamson, Alexis Russel, Matthew Shudt, Melissa A Leisner, Jonathan Plitnick, Navjot Singh, John Kelly, Erasmus Schneider, Erica Lasek-Nesselquist |
| EPI_ISL_1397389, EPI_ISL_1397390, EPI_ISL_1397391, EPI_ISL_1397392 | SUNY UPSTATE MEDICAL UNIVERSITY | Wadsworth Center, New York State Department of Health | Kirsten St. George, Daryl M. Lamson, Alexis Russel, Matthew Shudt, Melissa A Leisner, Jonathan Plitnick, Navjot Singh, John Kelly, Erasmus Schneider, Erica Lasek-Nesselquist |
| EPI_ISL_1397393 | NORTH SHORE UNIVERSITY HOSPITAL | Wadsworth Center, New York State Department of Health | Kirsten St. George, Daryl M. Lamson, Alexis Russel, Matthew Shudt, Melissa A Leisner, Jonathan Plitnick, Navjot Singh, John Kelly, Erasmus Schneider, Erica Lasek-Nesselquist |
| EPI_ISL_1397394 | SUNY UPSTATE MEDICAL UNIVERSITY | Wadsworth Center, New York State Department of Health | Kirsten St. George, Daryl M. Lamson, Alexis Russel, Matthew Shudt, Melissa A Leisner, Jonathan Plitnick, Navjot Singh, John Kelly, Erasmus Schneider, Erica Lasek-Nesselquist |
| EPI_ISL_1397395 | NORTH SHORE UNIVERSITY HOSPITAL | Wadsworth Center, New York State Department of Health | Kirsten St. George, Daryl M. Lamson, Alexis Russel, Matthew Shudt, Melissa A Leisner, Jonathan Plitnick, Navjot Singh, John Kelly, Erasmus Schneider, Erica Lasek-Nesselquist |
| EPI_ISL_1397396, EPI_ISL_1397397, EPI_ISL_1397398, EPI_ISL_1397399, EPI_ISL_1397400, EPI_ISL_1397401, EPI_ISL_1397402 | SUNY UPSTATE MEDICAL UNIVERSITY | Wadsworth Center, New York State Department of Health | Kirsten St. George, Daryl M. Lamson, Alexis Russel, Matthew Shudt, Melissa A Leisner, Jonathan Plitnick, Navjot Singh, John Kelly, Erasmus Schneider, Erica Lasek-Nesselquist |
| EPI_ISL_1397403 | NORTH SHORE UNIVERSITY HOSPITAL | Wadsworth Center, New York State Department of Health | Kirsten St. George, Daryl M. Lamson, Alexis Russel, Matthew Shudt, Melissa A Leisner, Jonathan Plitnick, Navjot Singh, John Kelly, Erasmus Schneider, Erica Lasek-Nesselquist |
| EPI_ISL_1397404 | SUNY UPSTATE MEDICAL UNIVERSITY | Wadsworth Center, New York State Department of Health | Kirsten St. George, Daryl M. Lamson, Alexis Russel, Matthew Shudt, Melissa A Leisner, Jonathan Plitnick, Navjot Singh, John Kelly, Erasmus Schneider, Erica Lasek-Nesselquist |
| EPI_ISL_1397405 | NORTH SHORE UNIVERSITY HOSPITAL | Wadsworth Center, New York State Department of Health | Kirsten St. George, Daryl M. Lamson, Alexis Russel, Matthew Shudt, Melissa A Leisner, Jonathan Plitnick, Navjot Singh, John Kelly, Erasmus Schneider, Erica Lasek-Nesselquist |
| EPI_ISL_1397406 | SUNY UPSTATE MEDICAL UNIVERSITY | Wadsworth Center, New York State Department of Health | Kirsten St. George, Daryl M. Lamson, Alexis Russel, Matthew Shudt, Melissa A Leisner, Jonathan Plitnick, Navjot Singh, John Kelly, Erasmus Schneider, Erica Lasek-Nesselquist |
| EPI_ISL_1397407 | NORTH SHORE UNIVERSITY HOSPITAL | Wadsworth Center, New York State Department of Health | Kirsten St. George, Daryl M. Lamson, Alexis Russel, Matthew Shudt, Melissa A Leisner, Jonathan Plitnick, Navjot Singh, John Kelly, Erasmus Schneider, Erica Lasek-Nesselquist |
| EPI_ISL_1397408 | SUNY UPSTATE MEDICAL UNIVERSITY | Wadsworth Center, New York State Department of Health | Kirsten St. George, Daryl M. Lamson, Alexis Russel, Matthew Shudt, Melissa A Leisner, Jonathan Plitnick, Navjot Singh, John Kelly, Erasmus Schneider, Erica Lasek-Nesselquist |
| EPI_ISL_1397409 | NORTH SHORE UNIVERSITY HOSPITAL | Wadsworth Center, New York State Department of Health | Kirsten St. George, Daryl M. Lamson, Alexis Russel, Matthew Shudt, Melissa A Leisner, Jonathan Plitnick, Navjot Singh, John Kelly, Erasmus Schneider, Erica Lasek-Nesselquist |
| EPI_ISL_1397410, EPI_ISL_1397411, EPI_ISL_1397412, EPI_ISL_1397413, EPI_ISL_1397414, EPI_ISL_1397415, EPI_ISL_1397416, EPI_ISL_1397417, EPI_ISL_1397418, EPI_ISL_1397419, EPI_ISL_1397420, EPI_ISL_1397421, EPI_ISL_1397422, EPI_ISL_1397423 | | | |
| see above | SUNY UPSTATE MEDICAL UNIVERSITY | Wadsworth Center, New York State Department of Health | Kirsten St. George, Daryl M. Lamson, Alexis Russel, Matthew Shudt, Melissa A Leisner, Jonathan Plitnick, Navjot Singh, John Kelly, Erasmus Schneider, Erica Lasek-Nesselquist |
| EPI_ISL_1397424, EPI_ISL_1397425, EPI_ISL_1397426, EPI_ISL_1397427, EPI_ISL_1397428, EPI_ISL_1397429, EPI_ISL_1397430, EPI_ISL_1397431, EPI_ISL_1397432, EPI_ISL_1397433, EPI_ISL_1397434, EPI_ISL_1397435, EPI_ISL_1397436, EPI_ISL_1397437, EPI_ISL_1397438, EPI_ISL_1397439, EPI_ISL_1397440, EPI_ISL_1397441, EPI_ISL_1397442, EPI_ISL_1397443 | | | |
| see above | ADIRONDACK MEDICAL CENTER | Wadsworth Center, New York State Department of Health | Kirsten St. George, Daryl M. Lamson, Alexis Russel, Matthew Shudt, Melissa A Leisner, Jonathan Plitnick, Navjot Singh, John Kelly, Erasmus Schneider, Erica Lasek-Nesselquist |
| EPI_ISL_1397444, EPI_ISL_1397445, EPI_ISL_1397446, EPI_ISL_1397447, EPI_ISL_1397448, EPI_ISL_1397449, EPI_ISL_1397450, EPI_ISL_1397451, EPI_ISL_1397452, EPI_ISL_1397453, EPI_ISL_1397454, EPI_ISL_1397455, EPI_ISL_1397456, EPI_ISL_1397457, EPI_ISL_1397458 | | | |
| see above | WESTCHESTER MEDICAL CENTER | Wadsworth Center, New York State Department of Health | Kirsten St. George, Daryl M. Lamson, Alexis Russel, Matthew Shudt, Melissa A Leisner, Jonathan Plitnick, Navjot Singh, John Kelly, Erasmus Schneider, Erica Lasek-Nesselquist |
| EPI_ISL_1397459 | SUNY UPSTATE MEDICAL UNIVERSITY | Wadsworth Center, New York State Department of Health | Kirsten St. George, Daryl M. Lamson, Alexis Russel, Matthew Shudt, Melissa A Leisner, Jonathan Plitnick, Navjot Singh, John Kelly, Erasmus Schneider, Erica Lasek-Nesselquist |
| EPI_ISL_1397460, EPI_ISL_1397461, EPI_ISL_1397462, EPI_ISL_1397463, EPI_ISL_1397464, EPI_ISL_1397465, EPI_ISL_1397466, EPI_ISL_1397467, EPI_ISL_1397468, EPI_ISL_1397469, EPI_ISL_1397470, EPI_ISL_1397471, EPI_ISL_1397472, EPI_ISL_1397473, EPI_ISL_1397474, EPI_ISL_1397475, EPI_ISL_1397476, EPI_ISL_1397477, EPI_ISL_1397478, EPI_ISL_1397479, EPI_ISL_1397480, EPI_ISL_1397481, EPI_ISL_1397482, EPI_ISL_1397483, EPI_ISL_1397484, EPI_ISL_1397485, EPI_ISL_1397486, EPI_ISL_1397487, EPI_ISL_1397488, EPI_ISL_1397489, EPI_ISL_1397490, EPI_ISL_1397491, EPI_ISL_1397492, EPI_ISL_1397493, EPI_ISL_1397494, EPI_ISL_1397495, EPI_ISL_1397496, EPI_ISL_1397497, EPI_ISL_1397498, EPI_ISL_1397499, EPI_ISL_1397500, EPI_ISL_1397501, EPI_ISL_1397502, EPI_ISL_1397503, EPI_ISL_1397504, EPI_ISL_1397505, EPI_ISL_1397506, EPI_ISL_1397507, EPI_ISL_1397508, EPI_ISL_1397509, EPI_ISL_1397510, EPI_ISL_1397511, EPI_ISL_1397512, EPI_ISL_1397513, EPI_ISL_1397514, EPI_ISL_1397515, EPI_ISL_1397516, EPI_ISL_1397517, EPI_ISL_1397518, EPI_ISL_1397519, EPI_ISL_1397520, EPI_ISL_1397521, EPI_ISL_1397522, EPI_ISL_1397523, EPI_ISL_1397524, EPI_ISL_1397525, EPI_ISL_1397526, EPI_ISL_1397527, EPI_ISL_1397528, EPI_ISL_1397529, EPI_ISL_1397530, EPI_ISL_1397531, EPI_ISL_1397532, EPI_ISL_1397533, EPI_ISL_1397534, EPI_ISL_1397535, EPI_ISL_1397536, EPI_ISL_1397537, EPI_ISL_1397538, EPI_ISL_1397539, EPI_ISL_1397540, EPI_ISL_1397541 | | | |
| see above | New York Presbyterian Hospital | Wadsworth Center, New York State Department of Health | Kirsten St. George, Daryl M. Lamson, Alexis Russel, Matthew Shudt, Melissa A Leisner, Jonathan Plitnick, Navjot Singh, John Kelly, Erasmus Schneider, Erica Lasek-Nesselquist |
| EPI_ISL_1397542, EPI_ISL_1397543, EPI_ISL_1397544, EPI_ISL_1397545 | ALBANY MEDICAL CENTER | Wadsworth Center, New York State Department of Health | Kirsten St. George, Daryl M. Lamson, Alexis Russel, Matthew Shudt, Melissa A Leisner, Jonathan Plitnick, Navjot Singh, John Kelly, Erasmus Schneider, Erica Lasek-Nesselquist |
| EPI_ISL_1397546, EPI_ISL_1397547, EPI_ISL_1397548, EPI_ISL_1397549 | GLENS FALLS HOSPITAL LABORATORY | Wadsworth Center, New York State Department of Health | Kirsten St. George, Daryl M. Lamson, Alexis Russel, Matthew Shudt, Melissa A Leisner, Jonathan Plitnick, Navjot Singh, John Kelly, Erasmus Schneider, Erica Lasek-Nesselquist |
| EPI_ISL_1397550 | WESTCHESTER MEDICAL CENTER | Wadsworth Center, New York State Department of Health | Kirsten St. George, Daryl M. Lamson, Alexis Russel, Matthew Shudt, Melissa A Leisner, Jonathan Plitnick, Navjot Singh, John Kelly, Erasmus Schneider, Erica Lasek-Nesselquist |
| EPI_ISL_1397551 | ALBANY MEDICAL CENTER | Wadsworth Center, New York State Department of Health | Kirsten St. George, Daryl M. Lamson, Alexis Russel, Matthew Shudt, Melissa A Leisner, Jonathan Plitnick, Navjot Singh, John Kelly, Erasmus Schneider, Erica Lasek-Nesselquist |
| EPI_ISL_1397552 | URMC LABS | Wadsworth Center, New York State Department of Health | Kirsten St. George, Daryl M. Lamson, Alexis Russel, Matthew Shudt, Melissa A Leisner, Jonathan Plitnick, Navjot Singh, John Kelly, Erasmus Schneider, Erica Lasek-Nesselquist |
| EPI_ISL_1397553, EPI_ISL_1397554 | TEMPUS LABS INC | Wadsworth Center, New York State Department of Health | Kirsten St. George, Daryl M. Lamson, Alexis Russel, Matthew Shudt, Melissa A Leisner, Jonathan Plitnick, Navjot Singh, John Kelly, Erasmus Schneider, Erica Lasek-Nesselquist |
| EPI_ISL_1397555, EPI_ISL_1397556, EPI_ISL_1397557, EPI_ISL_1397558, EPI_ISL_1397559, EPI_ISL_1397560, EPI_ISL_1397561, EPI_ISL_1397562, EPI_ISL_1397563, EPI_ISL_1397564, EPI_ISL_1397565, EPI_ISL_1397566, EPI_ISL_1397567, EPI_ISL_1397568, EPI_ISL_1397569, EPI_ISL_1397570, EPI_ISL_1397571, EPI_ISL_1397572, EPI_ISL_1397573, EPI_ISL_1397574, EPI_ISL_1397575, EPI_ISL_1397576, EPI_ISL_1397577, EPI_ISL_1397578, EPI_ISL_1397579, EPI_ISL_1397580 | | | |
| see above | URMC LABS | Wadsworth Center, New York State | Kirsten St. George, Daryl M. Lamson, Alexis Russel, Matthew Shudt, Melissa A Leisner, Jonathan Plitnick, Navjot Singh, John Kelly, Erasmus Schneider, Erica Lasek-Nesselquist |

| | | | |
|---|---|---|---|
| | | Department of Health | |
| EPI_ISL_1397581, EPI_ISL_1397582, EPI_ISL_1397583, EPI_ISL_1397584, EPI_ISL_1397585, EPI_ISL_1397586, EPI_ISL_1397587, EPI_ISL_1397588, EPI_ISL_1397589, EPI_ISL_1397590, EPI_ISL_1397591 | | | |
| see above | TEMPUS LABS INC | Wadsworth Center, New York State Department of Health | Kirsten St. George, Daryl M. Lamson, Alexis Russel, Matthew Shudt, Melissa A Leisner, Jonathan Plitnick, Navjot Singh, John Kelly, Erasmus Schneider, Erica Lasek-Nesselquist |
| EPI_ISL_1397592, EPI_ISL_1397593, EPI_ISL_1397594, EPI_ISL_1397595, EPI_ISL_1397596, EPI_ISL_1397597, EPI_ISL_1397598, EPI_ISL_1397599, EPI_ISL_1397600, EPI_ISL_1397601, EPI_ISL_1397602, EPI_ISL_1397603, EPI_ISL_1397604, EPI_ISL_1397605, EPI_ISL_1397606, EPI_ISL_1397607, EPI_ISL_1397608, EPI_ISL_1397609, EPI_ISL_1397610, EPI_ISL_1397611 | | | |
| see above | SUNY UPSTATE MEDICAL UNIVERSITY | Wadsworth Center, New York State Department of Health | Kirsten St. George, Daryl M. Lamson, Alexis Russel, Matthew Shudt, Melissa A Leisner, Jonathan Plitnick, Navjot Singh, John Kelly, Erasmus Schneider, Erica Lasek-Nesselquist |
| EPI_ISL_1397612, EPI_ISL_1397613, EPI_ISL_1397614, EPI_ISL_1397615, EPI_ISL_1397616, EPI_ISL_1397617 | THE MARY IMOGENE BASSETT HOSPITAL | Wadsworth Center, New York State Department of Health | Kirsten St. George, Daryl M. Lamson, Alexis Russel, Matthew Shudt, Melissa A Leisner, Jonathan Plitnick, Navjot Singh, John Kelly, Erasmus Schneider, Erica Lasek-Nesselquist |
| EPI_ISL_1397618, EPI_ISL_1397619, EPI_ISL_1397620, EPI_ISL_1397621, EPI_ISL_1397622 | MONTEFIORE MEDICAL CENTER LABORATORIES | Wadsworth Center, New York State Department of Health | Kirsten St. George, Daryl M. Lamson, Alexis Russel, Matthew Shudt, Melissa A Leisner, Jonathan Plitnick, Navjot Singh, John Kelly, Erasmus Schneider, Erica Lasek-Nesselquist |
| EPI_ISL_1397623, EPI_ISL_1397624, EPI_ISL_1397625, EPI_ISL_1397626, EPI_ISL_1397627, EPI_ISL_1397628 | NYC Pandemic Response Lab | Wadsworth Center, New York State Department of Health | Kirsten St. George, Daryl M. Lamson, Alexis Russel, Matthew Shudt, Melissa A Leisner, Jonathan Plitnick, Navjot Singh, John Kelly, Erasmus Schneider, Erica Lasek-Nesselquist |
| EPI_ISL_1397629 | SUNY UPSTATE MEDICAL UNIVERSITY | Wadsworth Center, New York State Department of Health | Kirsten St. George, Daryl M. Lamson, Alexis Russel, Matthew Shudt, Melissa A Leisner, Jonathan Plitnick, Navjot Singh, John Kelly, Erasmus Schneider, Erica Lasek-Nesselquist |
| EPI_ISL_1397630, EPI_ISL_1397631, EPI_ISL_1397632, EPI_ISL_1397633, EPI_ISL_1397634, EPI_ISL_1397635 | URMC LABS | Wadsworth Center, New York State Department of Health | Kirsten St. George, Daryl M. Lamson, Alexis Russel, Matthew Shudt, Melissa A Leisner, Jonathan Plitnick, Navjot Singh, John Kelly, Erasmus Schneider, Erica Lasek-Nesselquist |
| EPI_ISL_1397636, EPI_ISL_1397637, EPI_ISL_1397638, EPI_ISL_1397639, EPI_ISL_1397640, EPI_ISL_1397641, EPI_ISL_1397642, EPI_ISL_1397643, EPI_ISL_1397644, EPI_ISL_1397645, EPI_ISL_1397646, EPI_ISL_1397647, EPI_ISL_1397648, EPI_ISL_1397649 | | | |
| see above | Columbia University Irving Medical Center | Wadsworth Center, New York State Department of Health | Kirsten St. George, Daryl M. Lamson, Alexis Russel, Matthew Shudt, Melissa A Leisner, Jonathan Plitnick, Navjot Singh, John Kelly, Erasmus Schneider, Erica Lasek-Nesselquist |
| EPI_ISL_1397650, EPI_ISL_1397651, EPI_ISL_1397652, EPI_ISL_1397653, EPI_ISL_1397654, EPI_ISL_1397655, EPI_ISL_1397656, EPI_ISL_1397657, EPI_ISL_1397658, EPI_ISL_1397659, EPI_ISL_1397660, EPI_ISL_1397661, EPI_ISL_1397662, EPI_ISL_1397663, EPI_ISL_1397664, EPI_ISL_1397665, EPI_ISL_1397666, EPI_ISL_1397667, EPI_ISL_1397668, EPI_ISL_1397669, EPI_ISL_1397670, EPI_ISL_1397671, EPI_ISL_1397672, EPI_ISL_1397673, EPI_ISL_1397674, EPI_ISL_1397675, EPI_ISL_1397676, EPI_ISL_1397677, EPI_ISL_1397678, EPI_ISL_1397679, EPI_ISL_1397680, EPI_ISL_1397681, EPI_ISL_1397682, EPI_ISL_1397683, EPI_ISL_1397684, EPI_ISL_1397685, EPI_ISL_1397686, EPI_ISL_1397687, EPI_ISL_1397688, EPI_ISL_1397689, EPI_ISL_1397690, EPI_ISL_1397691, EPI_ISL_1397692, EPI_ISL_1397693 | | | |
| see above | NORTH SHORE UNIVERSITY HOSPITAL | Wadsworth Center, New York State Department of Health | Kirsten St. George, Daryl M. Lamson, Alexis Russel, Matthew Shudt, Melissa A Leisner, Jonathan Plitnick, Navjot Singh, John Kelly, Erasmus Schneider, Erica Lasek-Nesselquist |
| EPI_ISL_1397694, EPI_ISL_1397695, EPI_ISL_1397696, EPI_ISL_1397697, EPI_ISL_1397698, EPI_ISL_1397699 | BIO-REFERENCE LABORATORIES | Wadsworth Center, New York State Department of Health | Kirsten St. George, Daryl M. Lamson, Alexis Russel, Matthew Shudt, Melissa A Leisner, Jonathan Plitnick, Navjot Singh, John Kelly, Erasmus Schneider, Erica Lasek-Nesselquist |
| EPI_ISL_1397700, EPI_ISL_1397701, EPI_ISL_1397702, EPI_ISL_1397703, EPI_ISL_1397704, EPI_ISL_1397705 | TEMPUS LABS INC | Wadsworth Center, New York State Department of Health | Kirsten St. George, Daryl M. Lamson, Alexis Russel, Matthew Shudt, Melissa A Leisner, Jonathan Plitnick, Navjot Singh, John Kelly, Erasmus Schneider, Erica Lasek-Nesselquist |
| EPI_ISL_1397706 | SUNY UPSTATE MEDICAL UNIVERSITY | Wadsworth Center, New York State Department of Health | Kirsten St. George, Daryl M. Lamson, Alexis Russel, Matthew Shudt, Melissa A Leisner, Jonathan Plitnick, Navjot Singh, John Kelly, Erasmus Schneider, Erica Lasek-Nesselquist |
| EPI_ISL_1397707, EPI_ISL_1397708, EPI_ISL_1397709, EPI_ISL_1397710 | ALBANY MEDICAL CENTER | Wadsworth Center, New York State Department of Health | Kirsten St. George, Daryl M. Lamson, Alexis Russel, Matthew Shudt, Melissa A Leisner, Jonathan Plitnick, Navjot Singh, John Kelly, Erasmus Schneider, Erica Lasek-Nesselquist |
| EPI_ISL_1397711 | ADIRONDACK MEDICAL CENTER | Wadsworth Center, New York State Department of Health | Kirsten St. George, Daryl M. Lamson, Alexis Russel, Matthew Shudt, Melissa A Leisner, Jonathan Plitnick, Navjot Singh, John Kelly, Erasmus Schneider, Erica Lasek-Nesselquist |
| EPI_ISL_1397712, EPI_ISL_1397713, EPI_ISL_1397714, EPI_ISL_1397715, EPI_ISL_1397716, EPI_ISL_1397717, EPI_ISL_1397718, EPI_ISL_1397719, EPI_ISL_1397720 | ALBANY MEDICAL CENTER | Wadsworth Center, New York State Department of Health | Kirsten St. George, Daryl M. Lamson, Alexis Russel, Matthew Shudt, Melissa A Leisner, Jonathan Plitnick, Navjot Singh, John Kelly, Erasmus Schneider, Erica Lasek-Nesselquist |
| EPI_ISL_1397721 | ADIRONDACK MEDICAL CENTER | Wadsworth Center, New York State Department of Health | Kirsten St. George, Daryl M. Lamson, Alexis Russel, Matthew Shudt, Melissa A Leisner, Jonathan Plitnick, Navjot Singh, John Kelly, Erasmus Schneider, Erica Lasek-Nesselquist |
| EPI_ISL_1397722 | ALBANY MEDICAL CENTER | Wadsworth Center, New York State Department of Health | Kirsten St. George, Daryl M. Lamson, Alexis Russel, Matthew Shudt, Melissa A Leisner, Jonathan Plitnick, Navjot Singh, John Kelly, Erasmus Schneider, Erica Lasek-Nesselquist |
| EPI_ISL_1397723 | ADIRONDACK MEDICAL CENTER | Wadsworth Center, New York State Department of Health | Kirsten St. George, Daryl M. Lamson, Alexis Russel, Matthew Shudt, Melissa A Leisner, Jonathan Plitnick, Navjot Singh, John Kelly, Erasmus Schneider, Erica Lasek-Nesselquist |
| EPI_ISL_1397724, EPI_ISL_1397725, EPI_ISL_1397726, EPI_ISL_1397727, EPI_ISL_1397728, EPI_ISL_1397729, EPI_ISL_1397730 | ALBANY MEDICAL CENTER | Wadsworth Center, New York State Department of Health | Kirsten St. George, Daryl M. Lamson, Alexis Russel, Matthew Shudt, Melissa A Leisner, Jonathan Plitnick, Navjot Singh, John Kelly, Erasmus Schneider, Erica Lasek-Nesselquist |
| EPI_ISL_1397731 | ADIRONDACK MEDICAL CENTER | Wadsworth Center, New York State Department of Health | Kirsten St. George, Daryl M. Lamson, Alexis Russel, Matthew Shudt, Melissa A Leisner, Jonathan Plitnick, Navjot Singh, John Kelly, Erasmus Schneider, Erica Lasek-Nesselquist |
| EPI_ISL_1397732 | ALBANY MEDICAL CENTER | Wadsworth Center, New York State Department of Health | Kirsten St. George, Daryl M. Lamson, Alexis Russel, Matthew Shudt, Melissa A Leisner, Jonathan Plitnick, Navjot Singh, John Kelly, Erasmus Schneider, Erica Lasek-Nesselquist |
| EPI_ISL_1397733, EPI_ISL_1397734, EPI_ISL_1397735 | ADIRONDACK MEDICAL CENTER | Wadsworth Center, New York State Department of Health | Kirsten St. George, Daryl M. Lamson, Alexis Russel, Matthew Shudt, Melissa A Leisner, Jonathan Plitnick, Navjot Singh, John Kelly, Erasmus Schneider, Erica Lasek-Nesselquist |
| EPI_ISL_1397736, EPI_ISL_1397737 | ALBANY MEDICAL CENTER | Wadsworth Center, New York State Department of Health | Kirsten St. George, Daryl M. Lamson, Alexis Russel, Matthew Shudt, Melissa A Leisner, Jonathan Plitnick, Navjot Singh, John Kelly, Erasmus Schneider, Erica Lasek-Nesselquist |
| EPI_ISL_1397738, EPI_ISL_1397739 | ADIRONDACK MEDICAL CENTER | Wadsworth Center, New York State Department of Health | Kirsten St. George, Daryl M. Lamson, Alexis Russel, Matthew Shudt, Melissa A Leisner, Jonathan Plitnick, Navjot Singh, John Kelly, Erasmus Schneider, Erica Lasek-Nesselquist |
| EPI_ISL_1397740 | ALBANY MEDICAL CENTER | Wadsworth Center, New York State Department of Health | Kirsten St. George, Daryl M. Lamson, Alexis Russel, Matthew Shudt, Melissa A Leisner, Jonathan Plitnick, Navjot Singh, John Kelly, Erasmus Schneider, Erica Lasek-Nesselquist |
| EPI_ISL_1397741 | ADIRONDACK MEDICAL CENTER | Wadsworth Center, New York State Department of Health | Kirsten St. George, Daryl M. Lamson, Alexis Russel, Matthew Shudt, Melissa A Leisner, Jonathan Plitnick, Navjot Singh, John Kelly, Erasmus Schneider, Erica Lasek-Nesselquist |

| | | | |
|---|---|---|---|
| EPI_ISL_1397742 | ALBANY MEDICAL CENTER | Wadsworth Center, New York State Department of Health | Kirsten St. George, Daryl M. Lamson, Alexis Russel, Matthew Shudt, Melissa A Leisner, Jonathan Plitnick, Navjot Singh, John Kelly, Erasmus Schneider, Erica Lasek-Nesselquist |
| EPI_ISL_1397743 | ADIRONDACK MEDICAL CENTER | Wadsworth Center, New York State Department of Health | Kirsten St. George, Daryl M. Lamson, Alexis Russel, Matthew Shudt, Melissa A Leisner, Jonathan Plitnick, Navjot Singh, John Kelly, Erasmus Schneider, Erica Lasek-Nesselquist |
| EPI_ISL_1397744 | ALBANY MEDICAL CENTER | Wadsworth Center, New York State Department of Health | Kirsten St. George, Daryl M. Lamson, Alexis Russel, Matthew Shudt, Melissa A Leisner, Jonathan Plitnick, Navjot Singh, John Kelly, Erasmus Schneider, Erica Lasek-Nesselquist |
| EPI_ISL_1397745, EPI_ISL_1397746, EPI_ISL_1397747, EPI_ISL_1397748, EPI_ISL_1397749, EPI_ISL_1397750, EPI_ISL_1397751 | ADIRONDACK MEDICAL CENTER | Wadsworth Center, New York State Department of Health | Kirsten St. George, Daryl M. Lamson, Alexis Russel, Matthew Shudt, Melissa A Leisner, Jonathan Plitnick, Navjot Singh, John Kelly, Erasmus Schneider, Erica Lasek-Nesselquist |
| EPI_ISL_1397752, EPI_ISL_1397753, EPI_ISL_1397754, EPI_ISL_1397755, EPI_ISL_1397756, EPI_ISL_1397757, EPI_ISL_1397758, EPI_ISL_1397759, EPI_ISL_1397760, EPI_ISL_1397761, EPI_ISL_1397762, EPI_ISL_1397763, EPI_ISL_1397764, EPI_ISL_1397765 | | | |
| see above | ALBANY MEDICAL CENTER | Wadsworth Center, New York State Department of Health | Kirsten St. George, Daryl M. Lamson, Alexis Russel, Matthew Shudt, Melissa A Leisner, Jonathan Plitnick, Navjot Singh, John Kelly, Erasmus Schneider, Erica Lasek-Nesselquist |
| EPI_ISL_1397766 | ADIRONDACK MEDICAL CENTER | Wadsworth Center, New York State Department of Health | Kirsten St. George, Daryl M. Lamson, Alexis Russel, Matthew Shudt, Melissa A Leisner, Jonathan Plitnick, Navjot Singh, John Kelly, Erasmus Schneider, Erica Lasek-Nesselquist |
| EPI_ISL_1397767 | ALBANY MEDICAL CENTER | Wadsworth Center, New York State Department of Health | Kirsten St. George, Daryl M. Lamson, Alexis Russel, Matthew Shudt, Melissa A Leisner, Jonathan Plitnick, Navjot Singh, John Kelly, Erasmus Schneider, Erica Lasek-Nesselquist |
| EPI_ISL_1397768, EPI_ISL_1397769, EPI_ISL_1397770, EPI_ISL_1397771, EPI_ISL_1397772, EPI_ISL_1397773, EPI_ISL_1397774, EPI_ISL_1397775, EPI_ISL_1397776, EPI_ISL_1397777, EPI_ISL_1397778, EPI_ISL_1397779, EPI_ISL_1397780, EPI_ISL_1397781, EPI_ISL_1397782 | | | |
| see above | WESTCHESTER MEDICAL CENTER | Wadsworth Center, New York State Department of Health | Kirsten St. George, Daryl M. Lamson, Alexis Russel, Matthew Shudt, Melissa A Leisner, Jonathan Plitnick, Navjot Singh, John Kelly, Erasmus Schneider, Erica Lasek-Nesselquist |
| EPI_ISL_1397783 | THE MARY IMOGENE BASSETT HOSPITAL | Wadsworth Center, New York State Department of Health | Kirsten St. George, Daryl M. Lamson, Alexis Russel, Matthew Shudt, Melissa A Leisner, Jonathan Plitnick, Navjot Singh, John Kelly, Erasmus Schneider, Erica Lasek-Nesselquist |
| EPI_ISL_1397784, EPI_ISL_1397785, EPI_ISL_1397786, EPI_ISL_1397787 | SUNY UPSTATE MEDICAL UNIVERSITY | Wadsworth Center, New York State Department of Health | Kirsten St. George, Daryl M. Lamson, Alexis Russel, Matthew Shudt, Melissa A Leisner, Jonathan Plitnick, Navjot Singh, John Kelly, Erasmus Schneider, Erica Lasek-Nesselquist |
| EPI_ISL_1397788, EPI_ISL_1397789, EPI_ISL_1397790, EPI_ISL_1397791, EPI_ISL_1397792, EPI_ISL_1397793, EPI_ISL_1397794, EPI_ISL_1397795, EPI_ISL_1397796, EPI_ISL_1397797, EPI_ISL_1397798, EPI_ISL_1397799, EPI_ISL_1397800, EPI_ISL_1397801, EPI_ISL_1397802, EPI_ISL_1397803, EPI_ISL_1397804, EPI_ISL_1397805, EPI_ISL_1397806, EPI_ISL_1397807, EPI_ISL_1397808, EPI_ISL_1397809 | | | |
| see above | MONTEFIORE MEDICAL CENTER LABORATORIES | Wadsworth Center, New York State Department of Health | Kirsten St. George, Daryl M. Lamson, Alexis Russel, Matthew Shudt, Melissa A Leisner, Jonathan Plitnick, Navjot Singh, John Kelly, Erasmus Schneider, Erica Lasek-Nesselquist |
| EPI_ISL_1397810, EPI_ISL_1397811, EPI_ISL_1397812, EPI_ISL_1397813, EPI_ISL_1397814, EPI_ISL_1397815, EPI_ISL_1397816, EPI_ISL_1397817, EPI_ISL_1397818, EPI_ISL_1397819, EPI_ISL_1397820, EPI_ISL_1397821, EPI_ISL_1397822, EPI_ISL_1397823 | | | |
| see above | URMC LABS | Wadsworth Center, New York State Department of Health | Kirsten St. George, Daryl M. Lamson, Alexis Russel, Matthew Shudt, Melissa A Leisner, Jonathan Plitnick, Navjot Singh, John Kelly, Erasmus Schneider, Erica Lasek-Nesselquist |
| EPI_ISL_1397824, EPI_ISL_1397825, EPI_ISL_1397826, EPI_ISL_1397827, EPI_ISL_1397828 | Columbia University Irving Medical Center | Wadsworth Center, New York State Department of Health | Kirsten St. George, Daryl M. Lamson, Alexis Russel, Matthew Shudt, Melissa A Leisner, Jonathan Plitnick, Navjot Singh, John Kelly, Erasmus Schneider, Erica Lasek-Nesselquist |
| EPI_ISL_1397829, EPI_ISL_1397830 | NORTHWELL HEALTH LABORATORIES | Wadsworth Center, New York State Department of Health | Kirsten St. George, Daryl M. Lamson, Alexis Russel, Matthew Shudt, Melissa A Leisner, Jonathan Plitnick, Navjot Singh, John Kelly, Erasmus Schneider, Erica Lasek-Nesselquist |
| EPI_ISL_1397831, EPI_ISL_1397832 | STONY BROOK UNIVERSITY HOSPITAL | Wadsworth Center, New York State Department of Health | Kirsten St. George, Daryl M. Lamson, Alexis Russel, Matthew Shudt, Melissa A Leisner, Jonathan Plitnick, Navjot Singh, John Kelly, Erasmus Schneider, Erica Lasek-Nesselquist |
| EPI_ISL_1397833 | Wadsworth Center, New York State Department of Health | Wadsworth Center, New York State Department of Health | Kirsten St. George, Daryl M. Lamson, Alexis Russel, Matthew Shudt, Melissa A Leisner, Jonathan Plitnick, Navjot Singh, John Kelly, Erasmus Schneider, Erica Lasek-Nesselquist |
| EPI_ISL_1397834, EPI_ISL_1397835 | ALBANY MEDICAL CENTER | Wadsworth Center, New York State Department of Health | Kirsten St. George, Daryl M. Lamson, Alexis Russel, Matthew Shudt, Melissa A Leisner, Jonathan Plitnick, Navjot Singh, John Kelly, Erasmus Schneider, Erica Lasek-Nesselquist |
| EPI_ISL_1397836 | TEMPUS LABS INC | Wadsworth Center, New York State Department of Health | Kirsten St. George, Daryl M. Lamson, Alexis Russel, Matthew Shudt, Melissa A Leisner, Jonathan Plitnick, Navjot Singh, John Kelly, Erasmus Schneider, Erica Lasek-Nesselquist |
| EPI_ISL_1397837 | ALBANY MEDICAL CENTER | Wadsworth Center, New York State Department of Health | Kirsten St. George, Daryl M. Lamson, Alexis Russel, Matthew Shudt, Melissa A Leisner, Jonathan Plitnick, Navjot Singh, John Kelly, Erasmus Schneider, Erica Lasek-Nesselquist |
| EPI_ISL_1397838, EPI_ISL_1397839, EPI_ISL_1397840, EPI_ISL_1397841, EPI_ISL_1397842 | TEMPUS LABS INC | Wadsworth Center, New York State Department of Health | Kirsten St. George, Daryl M. Lamson, Alexis Russel, Matthew Shudt, Melissa A Leisner, Jonathan Plitnick, Navjot Singh, John Kelly, Erasmus Schneider, Erica Lasek-Nesselquist |
| EPI_ISL_1397843 | ALBANY MEDICAL CENTER | Wadsworth Center, New York State Department of Health | Kirsten St. George, Daryl M. Lamson, Alexis Russel, Matthew Shudt, Melissa A Leisner, Jonathan Plitnick, Navjot Singh, John Kelly, Erasmus Schneider, Erica Lasek-Nesselquist |
| EPI_ISL_1397844, EPI_ISL_1397845, EPI_ISL_1397846, EPI_ISL_1397847, EPI_ISL_1397848, EPI_ISL_1397849, EPI_ISL_1397850, EPI_ISL_1397851, EPI_ISL_1397852 | TEMPUS LABS INC | Wadsworth Center, New York State Department of Health | Kirsten St. George, Daryl M. Lamson, Alexis Russel, Matthew Shudt, Melissa A Leisner, Jonathan Plitnick, Navjot Singh, John Kelly, Erasmus Schneider, Erica Lasek-Nesselquist |
| EPI_ISL_1397853, EPI_ISL_1397854, EPI_ISL_1397855, EPI_ISL_1397856 | ALBANY MEDICAL CENTER | Wadsworth Center, New York State Department of Health | Kirsten St. George, Daryl M. Lamson, Alexis Russel, Matthew Shudt, Melissa A Leisner, Jonathan Plitnick, Navjot Singh, John Kelly, Erasmus Schneider, Erica Lasek-Nesselquist |
| EPI_ISL_1397857 | TEMPUS LABS INC | Wadsworth Center, New York State Department of Health | Kirsten St. George, Daryl M. Lamson, Alexis Russel, Matthew Shudt, Melissa A Leisner, Jonathan Plitnick, Navjot Singh, John Kelly, Erasmus Schneider, Erica Lasek-Nesselquist |
| EPI_ISL_1397858 | ALBANY MEDICAL CENTER | Wadsworth Center, New York State Department of Health | Kirsten St. George, Daryl M. Lamson, Alexis Russel, Matthew Shudt, Melissa A Leisner, Jonathan Plitnick, Navjot Singh, John Kelly, Erasmus Schneider, Erica Lasek-Nesselquist |
| EPI_ISL_1397859, EPI_ISL_1397860 | TEMPUS LABS INC | Wadsworth Center, New York State Department of Health | Kirsten St. George, Daryl M. Lamson, Alexis Russel, Matthew Shudt, Melissa A Leisner, Jonathan Plitnick, Navjot Singh, John Kelly, Erasmus Schneider, Erica Lasek-Nesselquist |
| EPI_ISL_1397861 | ALBANY MEDICAL CENTER | Wadsworth Center, New York State Department of Health | Kirsten St. George, Daryl M. Lamson, Alexis Russel, Matthew Shudt, Melissa A Leisner, Jonathan Plitnick, Navjot Singh, John Kelly, Erasmus Schneider, Erica Lasek-Nesselquist |
| EPI_ISL_1397862, EPI_ISL_1397863, EPI_ISL_1397864, EPI_ISL_1397865 | TEMPUS LABS INC | Wadsworth Center, New York State Department of Health | Kirsten St. George, Daryl M. Lamson, Alexis Russel, Matthew Shudt, Melissa A Leisner, Jonathan Plitnick, Navjot Singh, John Kelly, Erasmus Schneider, Erica Lasek-Nesselquist |
| EPI_ISL_1397866, EPI_ISL_1397867, EPI_ISL_1397868, EPI_ISL_1397869 | ALBANY MEDICAL CENTER | Wadsworth Center, New York State Department of Health | Kirsten St. George, Daryl M. Lamson, Alexis Russel, Matthew Shudt, Melissa A Leisner, Jonathan Plitnick, Navjot Singh, John Kelly, Erasmus Schneider, Erica Lasek-Nesselquist |
| EPI_ISL_1397870, EPI_ISL_1397871, EPI_ISL_1397872, EPI_ISL_1397873, EPI_ISL_1397874, EPI_ISL_1397875, EPI_ISL_1397876, EPI_ISL_1397877, EPI_ISL_1397878, EPI_ISL_1397879, EPI_ISL_1397880, EPI_ISL_1397881, EPI_ISL_1397882, EPI_ISL_1397883, EPI_ISL_1397884, EPI_ISL_1397885 | | | |
| see above | MONTEFIORE MEDICAL CENTER LABORATORIES | Wadsworth Center, New York State Department of Health | Kirsten St. George, Daryl M. Lamson, Alexis Russel, Matthew Shudt, Melissa A Leisner, Jonathan Plitnick, Navjot Singh, John Kelly, Erasmus Schneider, Erica Lasek-Nesselquist |
| EPI_ISL_1397886, EPI_ISL_1397887, EPI_ISL_1397888, EPI_ISL_1397889, EPI_ISL_1397890, EPI_ISL_1397891, EPI_ISL_1397892, EPI_ISL_1397893, EPI_ISL_1397894, EPI_ISL_1397895, EPI_ISL_1397896, EPI_ISL_1397897, EPI_ISL_1397898, EPI_ISL_1397899, EPI_ISL_1397900, EPI_ISL_1397901, EPI_ISL_1397902, EPI_ISL_1397903, EPI_ISL_1397904, EPI_ISL_1397905, EPI_ISL_1397906 | | | |
| see above | Columbia University Irving Medical Center | Wadsworth Center, New York State Department of Health | Kirsten St. George, Daryl M. Lamson, Alexis Russel, Matthew Shudt, Melissa A Leisner, Jonathan Plitnick, Navjot Singh, John Kelly, Erasmus Schneider, Erica Lasek-Nesselquist |

| | | | |
|---|---|---|---|
| EPI_ISL_1397907, EPI_ISL_1397908, EPI_ISL_1397909, EPI_ISL_1397910, EPI_ISL_1397911 | NORTHWELL HEALTH LABORATORIES | Wadsworth Center, New York State Department of Health | Kirsten St. George, Daryl M. Lamson, Alexis Russel, Matthew Shudt, Melissa A Leisner, Jonathan Plitnick, Navjot Singh, John Kelly, Erasmus Schneider, Erica Lasek-Nesselquist |
| EPI_ISL_1397912, EPI_ISL_1397913 | BIO-REFERENCE LABORATORIES | Wadsworth Center, New York State Department of Health | Kirsten St. George, Daryl M. Lamson, Alexis Russel, Matthew Shudt, Melissa A Leisner, Jonathan Plitnick, Navjot Singh, John Kelly, Erasmus Schneider, Erica Lasek-Nesselquist |
| EPI_ISL_1397914, EPI_ISL_1397915, EPI_ISL_1397916, EPI_ISL_1397917, EPI_ISL_1397918, EPI_ISL_1397919, EPI_ISL_1397920, EPI_ISL_1397921, EPI_ISL_1397922, EPI_ISL_1397923, EPI_ISL_1397924, EPI_ISL_1397925, EPI_ISL_1397926, EPI_ISL_1397927, EPI_ISL_1397928, EPI_ISL_1397929, EPI_ISL_1397930, EPI_ISL_1397931, EPI_ISL_1397932, EPI_ISL_1397933, EPI_ISL_1397934, EPI_ISL_1397935, EPI_ISL_1397936, EPI_ISL_1397937, EPI_ISL_1397938, EPI_ISL_1397939, EPI_ISL_1397940, EPI_ISL_1397941, EPI_ISL_1397942, EPI_ISL_1397943, EPI_ISL_1397944, EPI_ISL_1397945, EPI_ISL_1397946, EPI_ISL_1397947, EPI_ISL_1397948, EPI_ISL_1397949, EPI_ISL_1397950, EPI_ISL_1397951, EPI_ISL_1397952 | | | |
| see above | WESTCHESTER MEDICAL CENTER | Wadsworth Center, New York State Department of Health | Kirsten St. George, Daryl M. Lamson, Alexis Russel, Matthew Shudt, Melissa A Leisner, Jonathan Plitnick, Navjot Singh, John Kelly, Erasmus Schneider, Erica Lasek-Nesselquist |
| EPI_ISL_1397961 | ALBANY MEDICAL CENTER | Wadsworth Center, New York State Department of Health | Kirsten St. George, Daryl M. Lamson, Alexis Russel, Matthew Shudt, Melissa A Leisner, Jonathan Plitnick, Navjot Singh, John Kelly, Erasmus Schneider, Erica Lasek-Nesselquist |
| EPI_ISL_1397962, EPI_ISL_1397963, EPI_ISL_1397964, EPI_ISL_1397965 | GLENS FALLS HOSPITAL LABORATORY | Wadsworth Center, New York State Department of Health | Kirsten St. George, Daryl M. Lamson, Alexis Russel, Matthew Shudt, Melissa A Leisner, Jonathan Plitnick, Navjot Singh, John Kelly, Erasmus Schneider, Erica Lasek-Nesselquist |
| EPI_ISL_1397966, EPI_ISL_1397967 | THE MARY IMOGENE BASSETT HOSPITAL | Wadsworth Center, New York State Department of Health | Kirsten St. George, Daryl M. Lamson, Alexis Russel, Matthew Shudt, Melissa A Leisner, Jonathan Plitnick, Navjot Singh, John Kelly, Erasmus Schneider, Erica Lasek-Nesselquist |
| EPI_ISL_1397968, EPI_ISL_1397969, EPI_ISL_1397970 | SUNY UPSTATE MEDICAL UNIVERSITY | Wadsworth Center, New York State Department of Health | Kirsten St. George, Daryl M. Lamson, Alexis Russel, Matthew Shudt, Melissa A Leisner, Jonathan Plitnick, Navjot Singh, John Kelly, Erasmus Schneider, Erica Lasek-Nesselquist |
| EPI_ISL_1397971, EPI_ISL_1397972, EPI_ISL_1397973, EPI_ISL_1397974, EPI_ISL_1397975, EPI_ISL_1397976, EPI_ISL_1397977, EPI_ISL_1397978, EPI_ISL_1397979, EPI_ISL_1397980, EPI_ISL_1397981, EPI_ISL_1397982 | | | |
| see above | URMC LABS | Wadsworth Center, New York State Department of Health | Kirsten St. George, Daryl M. Lamson, Alexis Russel, Matthew Shudt, Melissa A Leisner, Jonathan Plitnick, Navjot Singh, John Kelly, Erasmus Schneider, Erica Lasek-Nesselquist |
| EPI_ISL_1397983 | SUNY UPSTATE MEDICAL UNIVERSITY | Wadsworth Center, New York State Department of Health | Kirsten St. George, Daryl M. Lamson, Alexis Russel, Matthew Shudt, Melissa A Leisner, Jonathan Plitnick, Navjot Singh, John Kelly, Erasmus Schneider, Erica Lasek-Nesselquist |
| EPI_ISL_1397984, EPI_ISL_1397985, EPI_ISL_1397986, EPI_ISL_1397987 | URMC LABS | Wadsworth Center, New York State Department of Health | Kirsten St. George, Daryl M. Lamson, Alexis Russel, Matthew Shudt, Melissa A Leisner, Jonathan Plitnick, Navjot Singh, John Kelly, Erasmus Schneider, Erica Lasek-Nesselquist |
| EPI_ISL_1397988 | THE MARY IMOGENE BASSETT HOSPITAL | Wadsworth Center, New York State Department of Health | Kirsten St. George, Daryl M. Lamson, Alexis Russel, Matthew Shudt, Melissa A Leisner, Jonathan Plitnick, Navjot Singh, John Kelly, Erasmus Schneider, Erica Lasek-Nesselquist |
| EPI_ISL_1397989, EPI_ISL_1397990, EPI_ISL_1397991, EPI_ISL_1397992, EPI_ISL_1397993, EPI_ISL_1397994, EPI_ISL_1397995, EPI_ISL_1397996, EPI_ISL_1397997, EPI_ISL_1397998, EPI_ISL_1397999, EPI_ISL_1398000, EPI_ISL_1398001 | | | |
| see above | URMC LABS | Wadsworth Center, New York State Department of Health | Kirsten St. George, Daryl M. Lamson, Alexis Russel, Matthew Shudt, Melissa A Leisner, Jonathan Plitnick, Navjot Singh, John Kelly, Erasmus Schneider, Erica Lasek-Nesselquist |
| EPI_ISL_1398002, EPI_ISL_1398003, EPI_ISL_1398004 | SUNY UPSTATE MEDICAL UNIVERSITY | Wadsworth Center, New York State Department of Health | Kirsten St. George, Daryl M. Lamson, Alexis Russel, Matthew Shudt, Melissa A Leisner, Jonathan Plitnick, Navjot Singh, John Kelly, Erasmus Schneider, Erica Lasek-Nesselquist |
| EPI_ISL_1398005, EPI_ISL_1398006, EPI_ISL_1398007, EPI_ISL_1398008, EPI_ISL_1398009, EPI_ISL_1398010, EPI_ISL_1398011, EPI_ISL_1398012, EPI_ISL_1398013, EPI_ISL_1398014, EPI_ISL_1398015, EPI_ISL_1398016, EPI_ISL_1398017, EPI_ISL_1398018, EPI_ISL_1398019, EPI_ISL_1398020 | | | |
| see above | URMC LABS | Wadsworth Center, New York State Department of Health | Kirsten St. George, Daryl M. Lamson, Alexis Russel, Matthew Shudt, Melissa A Leisner, Jonathan Plitnick, Navjot Singh, John Kelly, Erasmus Schneider, Erica Lasek-Nesselquist |
| EPI_ISL_1398021, EPI_ISL_1398022, EPI_ISL_1398023, EPI_ISL_1398024, EPI_ISL_1398025, EPI_ISL_1398026, EPI_ISL_1398027 | ALBANY MEDICAL CENTER | Wadsworth Center, New York State Department of Health | Kirsten St. George, Daryl M. Lamson, Alexis Russel, Matthew Shudt, Melissa A Leisner, Jonathan Plitnick, Navjot Singh, John Kelly, Erasmus Schneider, Erica Lasek-Nesselquist |
| EPI_ISL_1398028, EPI_ISL_1398029, EPI_ISL_1398030, EPI_ISL_1398031, EPI_ISL_1398032, EPI_ISL_1398033 | WESTCHESTER MEDICAL CENTER | Wadsworth Center, New York State Department of Health | Kirsten St. George, Daryl M. Lamson, Alexis Russel, Matthew Shudt, Melissa A Leisner, Jonathan Plitnick, Navjot Singh, John Kelly, Erasmus Schneider, Erica Lasek-Nesselquist |
| EPI_ISL_1398034, EPI_ISL_1398035, EPI_ISL_1398036, EPI_ISL_1398037, EPI_ISL_1398038 | MONTEFIORE MEDICAL CENTER LABORATORIES | Wadsworth Center, New York State Department of Health | Kirsten St. George, Daryl M. Lamson, Alexis Russel, Matthew Shudt, Melissa A Leisner, Jonathan Plitnick, Navjot Singh, John Kelly, Erasmus Schneider, Erica Lasek-Nesselquist |
| EPI_ISL_1398040, EPI_ISL_1398041 | MEMORIAL SLOAN KETTERING CANCER CENTER | Wadsworth Center, New York State Department of Health | Kirsten St. George, Daryl M. Lamson, Alexis Russel, Matthew Shudt, Melissa A Leisner, Jonathan Plitnick, Navjot Singh, John Kelly, Erasmus Schneider, Erica Lasek-Nesselquist |
| EPI_ISL_1398042 | NORTHWELL HEALTH LABORATORIES | Wadsworth Center, New York State Department of Health | Kirsten St. George, Daryl M. Lamson, Alexis Russel, Matthew Shudt, Melissa A Leisner, Jonathan Plitnick, Navjot Singh, John Kelly, Erasmus Schneider, Erica Lasek-Nesselquist |
| EPI_ISL_1398043 | MEMORIAL SLOAN KETTERING CANCER CENTER | Wadsworth Center, New York State Department of Health | Kirsten St. George, Daryl M. Lamson, Alexis Russel, Matthew Shudt, Melissa A Leisner, Jonathan Plitnick, Navjot Singh, John Kelly, Erasmus Schneider, Erica Lasek-Nesselquist |
| EPI_ISL_1398044, EPI_ISL_1398045, EPI_ISL_1398046, EPI_ISL_1398047 | Columbia University Irving Medical Center | Wadsworth Center, New York State Department of Health | Kirsten St. George, Daryl M. Lamson, Alexis Russel, Matthew Shudt, Melissa A Leisner, Jonathan Plitnick, Navjot Singh, John Kelly, Erasmus Schneider, Erica Lasek-Nesselquist |
| EPI_ISL_1398048, EPI_ISL_1398049, EPI_ISL_1398050, EPI_ISL_1398051, EPI_ISL_1398052, EPI_ISL_1398053, EPI_ISL_1398054, EPI_ISL_1398055, EPI_ISL_1398056, EPI_ISL_1398057, EPI_ISL_1398058, EPI_ISL_1398059, EPI_ISL_1398060, EPI_ISL_1398061, EPI_ISL_1398062, EPI_ISL_1398063, EPI_ISL_1398064, EPI_ISL_1398065, EPI_ISL_1398066, EPI_ISL_1398067 | | | |
| see above | MONTEFIORE MEDICAL CENTER LABORATORIES | Wadsworth Center, New York State Department of Health | Kirsten St. George, Daryl M. Lamson, Alexis Russel, Matthew Shudt, Melissa A Leisner, Jonathan Plitnick, Navjot Singh, John Kelly, Erasmus Schneider, Erica Lasek-Nesselquist |
| EPI_ISL_1398068, EPI_ISL_1398069, EPI_ISL_1398070, EPI_ISL_1398071, EPI_ISL_1398072, EPI_ISL_1398073, EPI_ISL_1398074, EPI_ISL_1398075, EPI_ISL_1398076, EPI_ISL_1398077, EPI_ISL_1398078, EPI_ISL_1398079, EPI_ISL_1398080, EPI_ISL_1398081, EPI_ISL_1398082, EPI_ISL_1398083, EPI_ISL_1398084, EPI_ISL_1398085, EPI_ISL_1398086, EPI_ISL_1398087, EPI_ISL_1398088, EPI_ISL_1398089, EPI_ISL_1398090, EPI_ISL_1398091, EPI_ISL_1398092, EPI_ISL_1398093, EPI_ISL_1398094, EPI_ISL_1398095, EPI_ISL_1398096, EPI_ISL_1398097, EPI_ISL_1398098, EPI_ISL_1398099, EPI_ISL_1398100, EPI_ISL_1398101, EPI_ISL_1398102, EPI_ISL_1398103, EPI_ISL_1398104, EPI_ISL_1398105, EPI_ISL_1398106, EPI_ISL_1398107, EPI_ISL_1398108, EPI_ISL_1398109, EPI_ISL_1398110, EPI_ISL_1398111, EPI_ISL_1398112, EPI_ISL_1398113, EPI_ISL_1398114, EPI_ISL_1398115, EPI_ISL_1398116, EPI_ISL_1398117, EPI_ISL_1398118, EPI_ISL_1398119 | | | |
| see above | NORTHWELL HEALTH LABORATORIES | Wadsworth Center, New York State Department of Health | Kirsten St. George, Daryl M. Lamson, Alexis Russel, Matthew Shudt, Melissa A Leisner, Jonathan Plitnick, Navjot Singh, John Kelly, Erasmus Schneider, Erica Lasek-Nesselquist |
| EPI_ISL_1398120, EPI_ISL_1398121, EPI_ISL_1398122 | MONTEFIORE MEDICAL CENTER LABORATORIES | Wadsworth Center, New York State Department of Health | Kirsten St. George, Daryl M. Lamson, Alexis Russel, Matthew Shudt, Melissa A Leisner, Jonathan Plitnick, Navjot Singh, John Kelly, Erasmus Schneider, Erica Lasek-Nesselquist |
| EPI_ISL_1398123, EPI_ISL_1398124, EPI_ISL_1398125, EPI_ISL_1398126, EPI_ISL_1398127, EPI_ISL_1398128, EPI_ISL_1398129, EPI_ISL_1398130, EPI_ISL_1398131 | URMC LABS | Wadsworth Center, New York State Department of Health | Kirsten St. George, Daryl M. Lamson, Alexis Russel, Matthew Shudt, Melissa A Leisner, Jonathan Plitnick, Navjot Singh, John Kelly, Erasmus Schneider, Erica Lasek-Nesselquist |
| EPI_ISL_1398132, EPI_ISL_1398133, EPI_ISL_1398134, EPI_ISL_1398135, EPI_ISL_1398136 | MONTEFIORE MEDICAL CENTER LABORATORIES | Wadsworth Center, New York State Department of Health | Kirsten St. George, Daryl M. Lamson, Alexis Russel, Matthew Shudt, Melissa A Leisner, Jonathan Plitnick, Navjot Singh, John Kelly, Erasmus Schneider, Erica Lasek-Nesselquist |

| | | | |
|---|---|---|---|
| EPI_ISL_1398137, EPI_ISL_1398138, EPI_ISL_1398139, EPI_ISL_1398140, EPI_ISL_1398141, EPI_ISL_1398142, EPI_ISL_1398143 | URMC LABS | Wadsworth Center, New York State Department of Health | Kirsten St. George, Daryl M. Lamson, Alexis Russel, Matthew Shudt, Melissa A Leisner, Jonathan Plitnick, Navjot Singh, John Kelly, Erasmus Schneider, Erica Lasek-Nesselquist |
| EPI_ISL_1398144, EPI_ISL_1398145, EPI_ISL_1398146 | Columbia University Irving Medical Center | Wadsworth Center, New York State Department of Health | Kirsten St. George, Daryl M. Lamson, Alexis Russel, Matthew Shudt, Melissa A Leisner, Jonathan Plitnick, Navjot Singh, John Kelly, Erasmus Schneider, Erica Lasek-Nesselquist |
| EPI_ISL_1398147, EPI_ISL_1398148, EPI_ISL_1398149, EPI_ISL_1398150, EPI_ISL_1398151, EPI_ISL_1398152, EPI_ISL_1398153 | ALBANY MEDICAL CENTER | Wadsworth Center, New York State Department of Health | Kirsten St. George, Daryl M. Lamson, Alexis Russel, Matthew Shudt, Melissa A Leisner, Jonathan Plitnick, Navjot Singh, John Kelly, Erasmus Schneider, Erica Lasek-Nesselquist |
| EPI_ISL_1398154, EPI_ISL_1398155 | GLENS FALLS HOSPITAL LABORATORY | Wadsworth Center, New York State Department of Health | Kirsten St. George, Daryl M. Lamson, Alexis Russel, Matthew Shudt, Melissa A Leisner, Jonathan Plitnick, Navjot Singh, John Kelly, Erasmus Schneider, Erica Lasek-Nesselquist |
| EPI_ISL_1398156, EPI_ISL_1398157, EPI_ISL_1398158, EPI_ISL_1398159, EPI_ISL_1398160, EPI_ISL_1398161, EPI_ISL_1398162 | SUNY UPSTATE MEDICAL UNIVERSITY | Wadsworth Center, New York State Department of Health | Kirsten St. George, Daryl M. Lamson, Alexis Russel, Matthew Shudt, Melissa A Leisner, Jonathan Plitnick, Navjot Singh, John Kelly, Erasmus Schneider, Erica Lasek-Nesselquist |
| EPI_ISL_1398163, EPI_ISL_1398164, EPI_ISL_1398165, EPI_ISL_1398166, EPI_ISL_1398167, EPI_ISL_1398168, EPI_ISL_1398169, EPI_ISL_1398170, EPI_ISL_1398171, EPI_ISL_1398172, EPI_ISL_1398173, EPI_ISL_1398174, EPI_ISL_1398175, EPI_ISL_1398176, EPI_ISL_1398177, EPI_ISL_1398178, EPI_ISL_1398179, EPI_ISL_1398180, EPI_ISL_1398181, EPI_ISL_1398182, EPI_ISL_1398183, EPI_ISL_1398184, EPI_ISL_1398185, EPI_ISL_1398186, EPI_ISL_1398187, EPI_ISL_1398188, EPI_ISL_1398189, EPI_ISL_1398190, EPI_ISL_1398191, EPI_ISL_1398192, EPI_ISL_1398193, EPI_ISL_1398194, EPI_ISL_1398195, EPI_ISL_1398196, EPI_ISL_1398197, EPI_ISL_1398198, EPI_ISL_1398199, EPI_ISL_1398200, EPI_ISL_1398201, EPI_ISL_1398202, EPI_ISL_1398203 | | | |
| see above | Columbia University Irving Medical Center | Wadsworth Center, New York State Department of Health | Kirsten St. George, Daryl M. Lamson, Alexis Russel, Matthew Shudt, Melissa A Leisner, Jonathan Plitnick, Navjot Singh, John Kelly, Erasmus Schneider, Erica Lasek-Nesselquist |
| EPI_ISL_1398204, EPI_ISL_1398205, EPI_ISL_1398206, EPI_ISL_1398207, EPI_ISL_1398208 | LENOX HILL HOSPITAL | Wadsworth Center, New York State Department of Health | Kirsten St. George, Daryl M. Lamson, Alexis Russel, Matthew Shudt, Melissa A Leisner, Jonathan Plitnick, Navjot Singh, John Kelly, Erasmus Schneider, Erica Lasek-Nesselquist |
| EPI_ISL_1398209, EPI_ISL_1398210, EPI_ISL_1398211, EPI_ISL_1398212, EPI_ISL_1398213, EPI_ISL_1398214, EPI_ISL_1398215, EPI_ISL_1398216, EPI_ISL_1398217, EPI_ISL_1398218, EPI_ISL_1398219, EPI_ISL_1398220, EPI_ISL_1398221, EPI_ISL_1398222, EPI_ISL_1398223, EPI_ISL_1398224, EPI_ISL_1398225, EPI_ISL_1398226, EPI_ISL_1398227, EPI_ISL_1398228, EPI_ISL_1398229, EPI_ISL_1398230, EPI_ISL_1398231, EPI_ISL_1398232, EPI_ISL_1398233, EPI_ISL_1398234, EPI_ISL_1398235, EPI_ISL_1398236, EPI_ISL_1398237, EPI_ISL_1398238, EPI_ISL_1398239, EPI_ISL_1398240, EPI_ISL_1398241, EPI_ISL_1398242, EPI_ISL_1398243, EPI_ISL_1398244, EPI_ISL_1398245, EPI_ISL_1398246, EPI_ISL_1398247, EPI_ISL_1398248, EPI_ISL_1398249, EPI_ISL_1398250, EPI_ISL_1398251, EPI_ISL_1398252, EPI_ISL_1398253, EPI_ISL_1398254, EPI_ISL_1398255, EPI_ISL_1398256, EPI_ISL_1398257, EPI_ISL_1398258 | | | |
| see above | KALEIDA CENTER FOR LABORATORY MEDICINE | Wadsworth Center, New York State Department of Health | Kirsten St. George, Daryl M. Lamson, Alexis Russel, Matthew Shudt, Melissa A Leisner, Jonathan Plitnick, Navjot Singh, John Kelly, Erasmus Schneider, Erica Lasek-Nesselquist |
| EPI_ISL_1398259, EPI_ISL_1398260, EPI_ISL_1398261, EPI_ISL_1398262, EPI_ISL_1398263 | THE MARY IMOGENE BASSETT HOSPITAL | Wadsworth Center, New York State Department of Health | Kirsten St. George, Daryl M. Lamson, Alexis Russel, Matthew Shudt, Melissa A Leisner, Jonathan Plitnick, Navjot Singh, John Kelly, Erasmus Schneider, Erica Lasek-Nesselquist |
| EPI_ISL_1398268 | WHITE PLAINS HOSPITAL CENTER LABORATORY | Wadsworth Center, New York State Department of Health | Kirsten St. George, Daryl M. Lamson, Alexis Russel, Matthew Shudt, Melissa A Leisner, Jonathan Plitnick, Navjot Singh, John Kelly, Erasmus Schneider, Erica Lasek-Nesselquist |
| EPI_ISL_1398269 | SARATOGA HOSPITAL LABORATORY | Wadsworth Center, New York State Department of Health | Kirsten St. George, Daryl M. Lamson, Alexis Russel, Matthew Shudt, Melissa A Leisner, Jonathan Plitnick, Navjot Singh, John Kelly, Erasmus Schneider, Erica Lasek-Nesselquist |
| EPI_ISL_1398270 | GLENS FALLS HOSPITAL LABORATORY | Wadsworth Center, New York State Department of Health | Kirsten St. George, Daryl M. Lamson, Alexis Russel, Matthew Shudt, Melissa A Leisner, Jonathan Plitnick, Navjot Singh, John Kelly, Erasmus Schneider, Erica Lasek-Nesselquist |
| EPI_ISL_1398271, EPI_ISL_1398272, EPI_ISL_1398273 | URMC LABS | Wadsworth Center, New York State Department of Health | Kirsten St. George, Daryl M. Lamson, Alexis Russel, Matthew Shudt, Melissa A Leisner, Jonathan Plitnick, Navjot Singh, John Kelly, Erasmus Schneider, Erica Lasek-Nesselquist |
| EPI_ISL_1398274, EPI_ISL_1398275, EPI_ISL_1398276, EPI_ISL_1398277, EPI_ISL_1398278, EPI_ISL_1398279, EPI_ISL_1398280, EPI_ISL_1398281, EPI_ISL_1398282, EPI_ISL_1398283 | ALBANY MEDICAL CENTER | Wadsworth Center, New York State Department of Health | Kirsten St. George, Daryl M. Lamson, Alexis Russel, Matthew Shudt, Melissa A Leisner, Jonathan Plitnick, Navjot Singh, John Kelly, Erasmus Schneider, Erica Lasek-Nesselquist |
| EPI_ISL_1398284, EPI_ISL_1398285, EPI_ISL_1398286, EPI_ISL_1398287, EPI_ISL_1398288, EPI_ISL_1398289, EPI_ISL_1398290, EPI_ISL_1398291, EPI_ISL_1398292, EPI_ISL_1398293, EPI_ISL_1398294, EPI_ISL_1398295, EPI_ISL_1398296, EPI_ISL_1398297, EPI_ISL_1398298, EPI_ISL_1398299, EPI_ISL_1398300, EPI_ISL_1398301, EPI_ISL_1398302, EPI_ISL_1398303, EPI_ISL_1398304, EPI_ISL_1398305, EPI_ISL_1398306, EPI_ISL_1398307, EPI_ISL_1398308, EPI_ISL_1398309, EPI_ISL_1398310, EPI_ISL_1398311 | | | |
| see above | NYC Pandemic Response Lab | Wadsworth Center, New York State Department of Health | Kirsten St. George, Daryl M. Lamson, Alexis Russel, Matthew Shudt, Melissa A Leisner, Jonathan Plitnick, Navjot Singh, John Kelly, Erasmus Schneider, Erica Lasek-Nesselquist |
| EPI_ISL_1398312, EPI_ISL_1398313, EPI_ISL_1398314, EPI_ISL_1398315, EPI_ISL_1398316, EPI_ISL_1398317 | URMC LABS | Wadsworth Center, New York State Department of Health | Kirsten St. George, Daryl M. Lamson, Alexis Russel, Matthew Shudt, Melissa A Leisner, Jonathan Plitnick, Navjot Singh, John Kelly, Erasmus Schneider, Erica Lasek-Nesselquist |
| EPI_ISL_1398318, EPI_ISL_1398319, EPI_ISL_1398320, EPI_ISL_1398321, EPI_ISL_1398322, EPI_ISL_1398323, EPI_ISL_1398324, EPI_ISL_1398325, EPI_ISL_1398326, EPI_ISL_1398327, EPI_ISL_1398328, EPI_ISL_1398329, EPI_ISL_1398330 | | | |
| see above | NYC Pandemic Response Lab | Wadsworth Center, New York State Department of Health | Kirsten St. George, Daryl M. Lamson, Alexis Russel, Matthew Shudt, Melissa A Leisner, Jonathan Plitnick, Navjot Singh, John Kelly, Erasmus Schneider, Erica Lasek-Nesselquist |
| EPI_ISL_1398331, EPI_ISL_1398333, EPI_ISL_1398334, EPI_ISL_1398335, EPI_ISL_1398336, EPI_ISL_1398337, EPI_ISL_1398338, EPI_ISL_1398339 | MONTEFIORE MEDICAL CENTER LABORATORIES | Wadsworth Center, New York State Department of Health | Kirsten St. George, Daryl M. Lamson, Alexis Russel, Matthew Shudt, Melissa A Leisner, Jonathan Plitnick, Navjot Singh, John Kelly, Erasmus Schneider, Erica Lasek-Nesselquist |
| EPI_ISL_1398340, EPI_ISL_1398341, EPI_ISL_1398342, EPI_ISL_1398343, EPI_ISL_1398344, EPI_ISL_1398345, EPI_ISL_1398346, EPI_ISL_1398347, EPI_ISL_1398348, EPI_ISL_1398349, EPI_ISL_1398350, EPI_ISL_1398351, EPI_ISL_1398352 | | | |
| see above | NYC Pandemic Response Lab | Wadsworth Center, New York State Department of Health | Kirsten St. George, Daryl M. Lamson, Alexis Russel, Matthew Shudt, Melissa A Leisner, Jonathan Plitnick, Navjot Singh, John Kelly, Erasmus Schneider, Erica Lasek-Nesselquist |
| EPI_ISL_1398956, EPI_ISL_1398957, EPI_ISL_1398958, EPI_ISL_1398959, EPI_ISL_1398960, EPI_ISL_1398961, EPI_ISL_1398963, EPI_ISL_1398964, EPI_ISL_1398965, EPI_ISL_1398966, EPI_ISL_1398967, EPI_ISL_1398969, EPI_ISL_1398970, EPI_ISL_1398971, EPI_ISL_1398972, EPI_ISL_1398973, EPI_ISL_1398974, EPI_ISL_1398975, EPI_ISL_1398976, EPI_ISL_1398977, EPI_ISL_1398978, EPI_ISL_1398979 | | | |
| see above | Wyoming Public Health Laboratory | Wyoming Public Health Laboratory | Jim Mildenberger, Wanda Manley, Noah Hull, Taylor Fearing, Lynette Gumbleton, Channing Weber, Ashley Norberg, Chayse Rowley, Marley Goetz, Brian Dominguez, Elliot Thomasson, Cari Sloma, and Rob Christensen |
| EPI_ISL_1399162, EPI_ISL_1399163, EPI_ISL_1399164, EPI_ISL_1399165, EPI_ISL_1399166, EPI_ISL_1399167, EPI_ISL_1399168, EPI_ISL_1399169, EPI_ISL_1399170, EPI_ISL_1399171, EPI_ISL_1399172, EPI_ISL_1399173, EPI_ISL_1399174, EPI_ISL_1399177, EPI_ISL_1399178, EPI_ISL_1399179 | | | |
| see above | Nevada State Public Health Laboratory | Nevada State Public Health Laboratory | Andrew Gorzalski, Mark Pandori |
| EPI_ISL_1399181, EPI_ISL_1399184, EPI_ISL_1399185, EPI_ISL_1399186, EPI_ISL_1399187, EPI_ISL_1399188, EPI_ISL_1399189, EPI_ISL_1399190, EPI_ISL_1399191, EPI_ISL_1399192, EPI_ISL_1399193, EPI_ISL_1399194, EPI_ISL_1399195, EPI_ISL_1399196, EPI_ISL_1399198, EPI_ISL_1399199, EPI_ISL_1399200, EPI_ISL_1399201, EPI_ISL_1399202, EPI_ISL_1399203, EPI_ISL_1399204, EPI_ISL_1399205, EPI_ISL_1399206, EPI_ISL_1399207, EPI_ISL_1399208, EPI_ISL_1399209, EPI_ISL_1399211, EPI_ISL_1399212, EPI_ISL_1399213, EPI_ISL_1399214, EPI_ISL_1399215, EPI_ISL_1399216, EPI_ISL_1399217, EPI_ISL_1399218, EPI_ISL_1399219, EPI_ISL_1399220, EPI_ISL_1399222, EPI_ISL_1399223, EPI_ISL_1399224, EPI_ISL_1399225, EPI_ISL_1399226, EPI_ISL_1399227 | | | |
| see above | Wyoming Public Health Laboratory | Wyoming Public Health Laboratory | Jim Mildenberger, Wanda Manley, Noah Hull, Taylor Fearing, Lynette Gumbleton, Channing Weber, Ashley Norberg, Chayse Rowley, Marley Goetz, Brian Dominguez, Elliot Thomasson, Cari Sloma, and Rob Christensen |
| EPI_ISL_1399249, | Washington State Department of Health | Washington State Department of Health | Drew MacKellar, Philip Dykema, Denny Russell, Joenice Gonzalez, Geoff Melly, Vanessa De Los Santos, Darren Lucas, JohnAric Peterson, Avi Singh, Rebecca Cao |

| | | | |
|---|---|---|---|
| EPI_ISL_1399250, EPI_ISL_1399251, EPI_ISL_1399252, EPI_ISL_1399253, EPI_ISL_1399254, EPI_ISL_1399255, EPI_ISL_1399256 | Public Health Laboratories | Public Health Laboratories | |
| EPI_ISL_1399259 | LESP Jalisco | Instituto de Diagnostico y Referencia Epidemiologicos (INDRE) | Claudia Wong-Arambula, Abril Rodriguez-Maldonado, Vanessa Rivero-Arredondo, Ariadna Medina-Benitez, Joaquin Quiroz-Mercado, Sergio Rangel-Guerrero, Natividad Cruz-Ortiz, Tatiana Nunez-Garcia, Gisela Barrera-Badillo, Lucia Hernandez-Rivas, Irma Lopez-Martinez, Ernesto Ramirez-Gonzalez. |
| EPI_ISL_1399260, EPI_ISL_1399261 | LESP Guanajuato | Instituto de Diagnostico y Referencia Epidemiologicos (INDRE) | Claudia Wong-Arambula, Abril Rodriguez-Maldonado, Vanessa Rivero-Arredondo, Ariadna Medina-Benitez, Joaquin Quiroz-Mercado, Sergio Rangel-Guerrero, Natividad Cruz-Ortiz, Tatiana Nunez-Garcia, Gisela Barrera-Badillo, Lucia Hernandez-Rivas, Irma Lopez-Martinez, Ernesto Ramirez-Gonzalez. |
| EPI_ISL_1399262, EPI_ISL_1399263, EPI_ISL_1399264, EPI_ISL_1399265, EPI_ISL_1399266, EPI_ISL_1399267, EPI_ISL_1399268, EPI_ISL_1399269 | LESP Nuevo Leon | Instituto de Diagnostico y Referencia Epidemiologicos (INDRE) | Claudia Wong-Arambula, Abril Rodriguez-Maldonado, Vanessa Rivero-Arredondo, Ariadna Medina-Benitez, Joaquin Quiroz-Mercado, Sergio Rangel-Guerrero, Natividad Cruz-Ortiz, Tatiana Nunez-Garcia, Gisela Barrera-Badillo, Lucia Hernandez-Rivas, Irma Lopez-Martinez, Ernesto Ramirez-Gonzalez. |
| EPI_ISL_1399270 | LESP Jalisco | Instituto de Diagnostico y Referencia Epidemiologicos (INDRE) | Claudia Wong-Arambula, Abril Rodriguez-Maldonado, Vanessa Rivero-Arredondo, Ariadna Medina-Benitez, Joaquin Quiroz-Mercado, Sergio Rangel-Guerrero, Natividad Cruz-Ortiz, Tatiana Nunez-Garcia, Gisela Barrera-Badillo, Lucia Hernandez-Rivas, Irma Lopez-Martinez, Ernesto Ramirez-Gonzalez. |
| EPI_ISL_1399271 | LESP Chihuahua | Instituto de Diagnostico y Referencia Epidemiologicos (INDRE) | Claudia Wong-Arambula, Abril Rodriguez-Maldonado, Vanessa Rivero-Arredondo, Ariadna Medina-Benitez, Joaquin Quiroz-Mercado, Sergio Rangel-Guerrero, Natividad Cruz-Ortiz, Tatiana Nunez-Garcia, Gisela Barrera-Badillo, Lucia Hernandez-Rivas, Irma Lopez-Martinez, Ernesto Ramirez-Gonzalez. |
| EPI_ISL_1399272 | LESP Queretaro | Instituto de Diagnostico y Referencia Epidemiologicos (INDRE) | Claudia Wong-Arambula, Abril Rodriguez-Maldonado, Vanessa Rivero-Arredondo, Ariadna Medina-Benitez, Joaquin Quiroz-Mercado, Sergio Rangel-Guerrero, Natividad Cruz-Ortiz, Tatiana Nunez-Garcia, Gisela Barrera-Badillo, Lucia Hernandez-Rivas, Irma Lopez-Martinez, Ernesto Ramirez-Gonzalez. |
| EPI_ISL_1399273, EPI_ISL_1399274 | LESP Nayarit | Instituto de Diagnostico y Referencia Epidemiologicos (INDRE) | Claudia Wong-Arambula, Abril Rodriguez-Maldonado, Vanessa Rivero-Arredondo, Ariadna Medina-Benitez, Joaquin Quiroz-Mercado, Sergio Rangel-Guerrero, Natividad Cruz-Ortiz, Tatiana Nunez-Garcia, Gisela Barrera-Badillo, Lucia Hernandez-Rivas, Irma Lopez-Martinez, Ernesto Ramirez-Gonzalez. |
| EPI_ISL_1399275, EPI_ISL_1399276, EPI_ISL_1399277 | LESP Tamaulipas | Instituto de Diagnostico y Referencia Epidemiologicos (INDRE) | Claudia Wong-Arambula, Abril Rodriguez-Maldonado, Vanessa Rivero-Arredondo, Ariadna Medina-Benitez, Joaquin Quiroz-Mercado, Sergio Rangel-Guerrero, Natividad Cruz-Ortiz, Tatiana Nunez-Garcia, Gisela Barrera-Badillo, Lucia Hernandez-Rivas, Irma Lopez-Martinez, Ernesto Ramirez-Gonzalez. |
| EPI_ISL_1399278 | LESP Coahuila | Instituto de Diagnostico y Referencia Epidemiologicos (INDRE) | Claudia Wong-Arambula, Abril Rodriguez-Maldonado, Vanessa Rivero-Arredondo, Ariadna Medina-Benitez, Joaquin Quiroz-Mercado, Sergio Rangel-Guerrero, Natividad Cruz-Ortiz, Tatiana Nunez-Garcia, Gisela Barrera-Badillo, Lucia Hernandez-Rivas, Irma Lopez-Martinez, Ernesto Ramirez-Gonzalez. |
| EPI_ISL_1399279 | LESP Veracruz | Instituto de Diagnostico y Referencia Epidemiologicos (INDRE) | Claudia Wong-Arambula, Abril Rodriguez-Maldonado, Vanessa Rivero-Arredondo, Ariadna Medina-Benitez, Joaquin Quiroz-Mercado, Sergio Rangel-Guerrero, Natividad Cruz-Ortiz, Tatiana Nunez-Garcia, Gisela Barrera-Badillo, Lucia Hernandez-Rivas, Irma Lopez-Martinez, Ernesto Ramirez-Gonzalez. |
| EPI_ISL_1399280 | LESP Baja California | Instituto de Diagnostico y Referencia Epidemiologicos (INDRE) | Claudia Wong-Arambula, Abril Rodriguez-Maldonado, Vanessa Rivero-Arredondo, Ariadna Medina-Benitez, Joaquin Quiroz-Mercado, Sergio Rangel-Guerrero, Natividad Cruz-Ortiz, Tatiana Nunez-Garcia, Gisela Barrera-Badillo, Lucia Hernandez-Rivas, Irma Lopez-Martinez, Ernesto Ramirez-Gonzalez. |
| EPI_ISL_1399281 | LESP Ciudad de Mexico | Instituto de Diagnostico y Referencia Epidemiologicos (INDRE) | Claudia Wong-Arambula, Abril Rodriguez-Maldonado, Vanessa Rivero-Arredondo, Ariadna Medina-Benitez, Joaquin Quiroz-Mercado, Sergio Rangel-Guerrero, Natividad Cruz-Ortiz, Tatiana Nunez-Garcia, Gisela Barrera-Badillo, Lucia Hernandez-Rivas, Irma Lopez-Martinez, Ernesto Ramirez-Gonzalez. |
| EPI_ISL_1400298 | LESP Jalisco | Instituto de Diagnostico y Referencia Epidemiologicos (INDRE) | Claudia Wong-Arambula, Abril Rodriguez-Maldonado, Vanessa Rivero-Arredondo, Ariadna Medina-Benitez, Joaquin Quiroz-Mercado, Sergio Rangel-Guerrero, Natividad Cruz-Ortiz, Tatiana Nunez-Garcia, Gisela Barrera-Badillo, Lucia Hernandez-Rivas, Irma Lopez-Martinez, Ernesto Ramirez-Gonzalez. |
| EPI_ISL_1400299 | LESP Campeche | Instituto de Diagnostico y Referencia Epidemiologicos (INDRE) | Claudia Wong-Arambula, Abril Rodriguez-Maldonado, Vanessa Rivero-Arredondo, Ariadna Medina-Benitez, Joaquin Quiroz-Mercado, Sergio Rangel-Guerrero, Natividad Cruz-Ortiz, Tatiana Nunez-Garcia, Gisela Barrera-Badillo, Lucia Hernandez-Rivas, Irma Lopez-Martinez, Ernesto Ramirez-Gonzalez. |
| EPI_ISL_1400300, EPI_ISL_1400301, EPI_ISL_1400302, EPI_ISL_1400303 | LESP Morelos | Instituto de Diagnostico y Referencia Epidemiologicos (INDRE) | Claudia Wong-Arambula, Abril Rodriguez-Maldonado, Vanessa Rivero-Arredondo, Ariadna Medina-Benitez, Joaquin Quiroz-Mercado, Sergio Rangel-Guerrero, Natividad Cruz-Ortiz, Tatiana Nunez-Garcia, Gisela Barrera-Badillo, Lucia Hernandez-Rivas, Irma Lopez-Martinez, Ernesto Ramirez-Gonzalez. |
| EPI_ISL_1400304 | LESP Guanajuato | Instituto de Diagnostico y Referencia Epidemiologicos (INDRE) | Claudia Wong-Arambula, Abril Rodriguez-Maldonado, Vanessa Rivero-Arredondo, Ariadna Medina-Benitez, Joaquin Quiroz-Mercado, Sergio Rangel-Guerrero, Natividad Cruz-Ortiz, Tatiana Nunez-Garcia, Gisela Barrera-Badillo, Lucia Hernandez-Rivas, Irma Lopez-Martinez, Ernesto Ramirez-Gonzalez. |
| EPI_ISL_1400305, EPI_ISL_1400306, EPI_ISL_1400307, EPI_ISL_1400308, EPI_ISL_1400309 | LESP Morelos | Instituto de Diagnostico y Referencia Epidemiologicos (INDRE) | Claudia Wong-Arambula, Abril Rodriguez-Maldonado, Vanessa Rivero-Arredondo, Ariadna Medina-Benitez, Joaquin Quiroz-Mercado, Sergio Rangel-Guerrero, Natividad Cruz-Ortiz, Tatiana Nunez-Garcia, Gisela Barrera-Badillo, Lucia Hernandez-Rivas, Irma Lopez-Martinez, Ernesto Ramirez-Gonzalez. |
| EPI_ISL_1400310, EPI_ISL_1400311, EPI_ISL_1400312, EPI_ISL_1400313 | LESP Puebla | Instituto de Diagnostico y Referencia Epidemiologicos (INDRE) | Claudia Wong-Arambula, Abril Rodriguez-Maldonado, Vanessa Rivero-Arredondo, Ariadna Medina-Benitez, Joaquin Quiroz-Mercado, Sergio Rangel-Guerrero, Natividad Cruz-Ortiz, Tatiana Nunez-Garcia, Gisela Barrera-Badillo, Lucia Hernandez-Rivas, Irma Lopez-Martinez, Ernesto Ramirez-Gonzalez. |
| EPI_ISL_1400314, EPI_ISL_1400315, EPI_ISL_1400316, EPI_ISL_1400317, EPI_ISL_1400318, EPI_ISL_1400319, EPI_ISL_1400320, EPI_ISL_1400321, EPI_ISL_1400322, EPI_ISL_1400323, EPI_ISL_1400324, EPI_ISL_1400325, EPI_ISL_1400326, EPI_ISL_1400327, EPI_ISL_1400328, EPI_ISL_1400329, EPI_ISL_1400330, EPI_ISL_1400331, EPI_ISL_1400332, EPI_ISL_1400333, EPI_ISL_1400334, EPI_ISL_1400335, EPI_ISL_1400336, EPI_ISL_1400337, EPI_ISL_1400338, EPI_ISL_1400339, EPI_ISL_1400340, EPI_ISL_1400341, EPI_ISL_1400342, EPI_ISL_1400343, EPI_ISL_1400344, EPI_ISL_1400345, EPI_ISL_1400346, EPI_ISL_1400347, EPI_ISL_1400348, EPI_ISL_1400349 | | | |
| see above | LESP Guanajuato | Instituto de Diagnostico y Referencia Epidemiologicos (INDRE) | Claudia Wong-Arambula, Abril Rodriguez-Maldonado, Vanessa Rivero-Arredondo, Ariadna Medina-Benitez, Joaquin Quiroz-Mercado, Sergio Rangel-Guerrero, Natividad Cruz-Ortiz, Tatiana Nunez-Garcia, Gisela Barrera-Badillo, Lucia Hernandez-Rivas, Irma Lopez-Martinez, Ernesto Ramirez-Gonzalez. |
| EPI_ISL_1400350, EPI_ISL_1400351, EPI_ISL_1400352, EPI_ISL_1400353, EPI_ISL_1400354, EPI_ISL_1400355, EPI_ISL_1400356, EPI_ISL_1400357, EPI_ISL_1400358, EPI_ISL_1400359, EPI_ISL_1400360, EPI_ISL_1400361, EPI_ISL_1400362, EPI_ISL_1400363, EPI_ISL_1400364, EPI_ISL_1400365, EPI_ISL_1400366, EPI_ISL_1400367, EPI_ISL_1400368, EPI_ISL_1400369, EPI_ISL_1400370, EPI_ISL_1400371, EPI_ISL_1400372, EPI_ISL_1400373, EPI_ISL_1400374, EPI_ISL_1400375, EPI_ISL_1400376, EPI_ISL_1400377, EPI_ISL_1400378, EPI_ISL_1400379, EPI_ISL_1400380, EPI_ISL_1400381, EPI_ISL_1400382, EPI_ISL_1400383, EPI_ISL_1400384, EPI_ISL_1400385, EPI_ISL_1400386, EPI_ISL_1400387, EPI_ISL_1400388, EPI_ISL_1400389, EPI_ISL_1400390, EPI_ISL_1400391, EPI_ISL_1400392, EPI_ISL_1400393, EPI_ISL_1400394, EPI_ISL_1400395, EPI_ISL_1400396, EPI_ISL_1400397, EPI_ISL_1400398, EPI_ISL_1400399, EPI_ISL_1400400, EPI_ISL_1400401, EPI_ISL_1400402, EPI_ISL_1400403, EPI_ISL_1400404, EPI_ISL_1400405, EPI_ISL_1400406, EPI_ISL_1400407, EPI_ISL_1400408, EPI_ISL_1400409, EPI_ISL_1400410, EPI_ISL_1400411, EPI_ISL_1400412, EPI_ISL_1400413, EPI_ISL_1400414, EPI_ISL_1400415, EPI_ISL_1400416, EPI_ISL_1400417, EPI_ISL_1400418, EPI_ISL_1400419, EPI_ISL_1400420, EPI_ISL_1400421, EPI_ISL_1400422, EPI_ISL_1400423, EPI_ISL_1400424, EPI_ISL_1400425, EPI_ISL_1400426, EPI_ISL_1400427, EPI_ISL_1400428, EPI_ISL_1400429, EPI_ISL_1400430, EPI_ISL_1400431, EPI_ISL_1400432, EPI_ISL_1400433, EPI_ISL_1400434, EPI_ISL_1400435, EPI_ISL_1400436, EPI_ISL_1400437, EPI_ISL_1400438, EPI_ISL_1400439, EPI_ISL_1400440, EPI_ISL_1400441, EPI_ISL_1400442, EPI_ISL_1400443 | | | |
| see above | UW Virology Lab | UW Virology Lab | Pavitra Roychoudhury, Hong Xie, Lasata Shrestha, Shah Mohamed Bakhash, Michelle Lin, Noah R. Baker, Sean Ellis, Saraswathi Sathees, Meei-Li Huang, Keith R Jerome, Alexander Greninger |
| EPI_ISL_1400444, EPI_ISL_1400445, EPI_ISL_1400446, EPI_ISL_1400447, EPI_ISL_1400448, EPI_ISL_1400449, EPI_ISL_1400450, EPI_ISL_1400451, EPI_ISL_1400452, EPI_ISL_1400453, EPI_ISL_1400454, EPI_ISL_1400455, EPI_ISL_1400456, EPI_ISL_1400460 | | | |
| see above | IN State Department of Health Laboratory Services | IN State Department of Health Laboratory Services | Cassandra Campion, Jamie Yeadon, Brian Pope, Lixia Liu, Kyle Brownlee, Melissa Hindenlang, Ankita Kashikar, Mark Glazier |
| EPI_ISL_1400461, EPI_ISL_1400462, EPI_ISL_1400463, EPI_ISL_1400464, EPI_ISL_1400465, EPI_ISL_1400466, EPI_ISL_1400467, EPI_ISL_1400468, EPI_ISL_1400469, EPI_ISL_1400470, EPI_ISL_1400471 | | | |
| see above | Nebraska Public Health Laboratory | NPHL COVID-19 Response Team | NPHL COVID-19 Response Team |
| EPI_ISL_1400472, EPI_ISL_1400473, EPI_ISL_1400474, EPI_ISL_1400475, EPI_ISL_1400476, EPI_ISL_1400477, EPI_ISL_1400478, EPI_ISL_1400479, EPI_ISL_1400480, EPI_ISL_1400481, EPI_ISL_1400482, EPI_ISL_1400483, EPI_ISL_1400484, EPI_ISL_1400485, EPI_ISL_1400486, EPI_ISL_1400487, EPI_ISL_1400488 | | | |
| see above | IN State Department of Health Laboratory Services | IN State Department of Health Laboratory Services | Cassandra Campion, Jamie Yeadon, Brian Pope, Lixia Liu, Kyle Brownlee, Melissa Hindenlang, Ankita Kashikar, Mark Glazier |
| EPI_ISL_1400795, EPI_ISL_1400796, EPI_ISL_1400797, EPI_ISL_1400798, EPI_ISL_1400799 | Instituto Nacional de Medicina Genomica | Instituto Nacional de Medicina Genomica | Hidalgo-Miranda A, Mendoza-Vargas A, Reyes-Grajeda JP, Cedro-Tanda A, Alcaraz N, Gonzalez-Barrera D, Rangel-DeLeon D, Miranda-Ortíz H, Rosas-Escobar P, Canseco Mendez JC, Ramirez-Vega O, Munguia-Garza, Garcia-Cárdenas FJ, González-Woge MA, Herrera-Montalvo LA |
| EPI_ISL_1400800 | Baylor Esoteric + Molecular Lab | Baylor Esoteric + Molecular Lab | Elizabeth Forrester; Dawn Richards |
| EPI_ISL_1400801 | ETSU Clinical Lab | Baylor Esoteric + Molecular Lab | Elizabeth Forrester; Dawn Richards |
| EPI_ISL_1400802, EPI_ISL_1400803, EPI_ISL_1400804, EPI_ISL_1400805, EPI_ISL_1400806, EPI_ISL_1400807, EPI_ISL_1400808, EPI_ISL_1400809, EPI_ISL_1400811, EPI_ISL_1400813, EPI_ISL_1400814, EPI_ISL_1400815, EPI_ISL_1400816, EPI_ISL_1400817, EPI_ISL_1400818, EPI_ISL_1400819, EPI_ISL_1400820, EPI_ISL_1400821, EPI_ISL_1400822, EPI_ISL_1400823, EPI_ISL_1400824, EPI_ISL_1400825, EPI_ISL_1400826, EPI_ISL_1400827, EPI_ISL_1400828, EPI_ISL_1400829, EPI_ISL_1400830, EPI_ISL_1400831, EPI_ISL_1400832, EPI_ISL_1400833, EPI_ISL_1400834, EPI_ISL_1400835, EPI_ISL_1400836, EPI_ISL_1400837, EPI_ISL_1400838, EPI_ISL_1400839, EPI_ISL_1400840, EPI_ISL_1400841, EPI_ISL_1400842, EPI_ISL_1400843, EPI_ISL_1400844, EPI_ISL_1400846, EPI_ISL_1400847, EPI_ISL_1400848, EPI_ISL_1400849, EPI_ISL_1400850, EPI_ISL_1400852, EPI_ISL_1400853, EPI_ISL_1400854, EPI_ISL_1400855, EPI_ISL_1400856, EPI_ISL_1400857, EPI_ISL_1400858, EPI_ISL_1400859, EPI_ISL_1400860, EPI_ISL_1400861, EPI_ISL_1400862, EPI_ISL_1400863, EPI_ISL_1400864, EPI_ISL_1400865, EPI_ISL_1400866, EPI_ISL_1400867, EPI_ISL_1400868, EPI_ISL_1400869, EPI_ISL_1400870, EPI_ISL_1400871, EPI_ISL_1400872, EPI_ISL_1400873, EPI_ISL_1400874, EPI_ISL_1400875, EPI_ISL_1400876, EPI_ISL_1400877, EPI_ISL_1400878, EPI_ISL_1400879, EPI_ISL_1400880, EPI_ISL_1400881, EPI_ISL_1400882, EPI_ISL_1400883, EPI_ISL_1400884, EPI_ISL_1400885, EPI_ISL_1400886, EPI_ISL_1400887, EPI_ISL_1400888, EPI_ISL_1400889, EPI_ISL_1400890, EPI_ISL_1400891, EPI_ISL_1400892, EPI_ISL_1400893, EPI_ISL_1400894, EPI_ISL_1400895, EPI_ISL_1400896, EPI_ISL_1400897, EPI_ISL_1400898, EPI_ISL_1400899, EPI_ISL_1400900, EPI_ISL_1400901, EPI_ISL_1400902, EPI_ISL_1400903, EPI_ISL_1400904, EPI_ISL_1400905, EPI_ISL_1400906, EPI_ISL_1400907, EPI_ISL_1400908, EPI_ISL_1400909, EPI_ISL_1400910, EPI_ISL_1400911, EPI_ISL_1400912, EPI_ISL_1400913, EPI_ISL_1400914, EPI_ISL_1400915, EPI_ISL_1400916, EPI_ISL_1400917, EPI_ISL_1400918, EPI_ISL_1400919, EPI_ISL_1400920, EPI_ISL_1400921, EPI_ISL_1400922, EPI_ISL_1400923, EPI_ISL_1400924, EPI_ISL_1400925, EPI_ISL_1400928, EPI_ISL_1400929, EPI_ISL_1400930, EPI_ISL_1400931, EPI_ISL_1400932, EPI_ISL_1400933, EPI_ISL_1400935, EPI_ISL_1400936, EPI_ISL_1400937, EPI_ISL_1400938, EPI_ISL_1400939, EPI_ISL_1400941 | | | |
| see above | Maryland Genomics, Institute for Genome Sciences, University of Maryland School of Medicine | Maryland Genomics, Institute for Genome Sciences, University of Maryland School of Medicine | Tallon, Luke J; Sadzewicz, Lisa D; Humphrys, Mike; Ott, Sandra; Roussey, Holly; Mehta, Aditya; Vavikolanu, Kranthi; Fraser, Claire M; Ravel, Jacques |
| EPI_ISL_1400942, EPI_ISL_1400943, EPI_ISL_1400944, EPI_ISL_1400945, EPI_ISL_1400946, EPI_ISL_1400947, EPI_ISL_1400948, EPI_ISL_1400949, EPI_ISL_1400951, EPI_ISL_1400952, EPI_ISL_1400953, EPI_ISL_1400954, EPI_ISL_1400955, EPI_ISL_1400956, EPI_ISL_1400957, EPI_ISL_1400958, EPI_ISL_1400959, EPI_ISL_1400960, EPI_ISL_1400961, EPI_ISL_1400962, EPI_ISL_1400963 | | | |
| see above | New Mexico Department of Health Scientific Laboratory | New Mexico Department of Health Scientific Laboratory | Ellie Johnson, Anastacia Griego-Fisher, D'eldra Malone, Jennifer Benoit |

| | | | |
|---|---|---|---|
| EPI_ISL_1400966 | TriCore Reference Laboratory Inc. | New Mexico Department of Health Scientific Laboratory | Ellie Johnson, Anastacia Griego-Fisher, D'eldra Malone, Jennifer Benoit |
| EPI_ISL_1400967, EPI_ISL_1400968, EPI_ISL_1400969, EPI_ISL_1400970, EPI_ISL_1400971, EPI_ISL_1400972, EPI_ISL_1400973, EPI_ISL_1400975, EPI_ISL_1400976, EPI_ISL_1400977, EPI_ISL_1400978, EPI_ISL_1400979, EPI_ISL_1400981 | | | |
| see above | New Mexico Department of Health Scientific Laboratory | New Mexico Department of Health Scientific Laboratory | Ellie Johnson, Anastacia Griego-Fisher, D'eldra Malone, Jennifer Benoit |
| EPI_ISL_1400982, EPI_ISL_1400983 | San Juan Regional Medical Center | New Mexico Department of Health Scientific Laboratory | Ellie Johnson, Anastacia Griego-Fisher, D'eldra Malone, Jennifer Benoit |
| EPI_ISL_1400985, EPI_ISL_1400986, EPI_ISL_1400987, EPI_ISL_1400988, EPI_ISL_1400989, EPI_ISL_1400990, EPI_ISL_1400991, EPI_ISL_1400992, EPI_ISL_1400993, EPI_ISL_1400994, EPI_ISL_1400995, EPI_ISL_1400996, EPI_ISL_1400997, EPI_ISL_1400998, EPI_ISL_1400999, EPI_ISL_1401000, EPI_ISL_1401001, EPI_ISL_1401002, EPI_ISL_1401003, EPI_ISL_1401004, EPI_ISL_1401005, EPI_ISL_1401006, EPI_ISL_1401007, EPI_ISL_1401008, EPI_ISL_1401009, EPI_ISL_1401010, EPI_ISL_1401011, EPI_ISL_1401012, EPI_ISL_1401013, EPI_ISL_1401014, EPI_ISL_1401015, EPI_ISL_1401016, EPI_ISL_1401017, EPI_ISL_1401018, EPI_ISL_1401019, EPI_ISL_1401020, EPI_ISL_1401021, EPI_ISL_1401022, EPI_ISL_1401023, EPI_ISL_1401024, EPI_ISL_1401025, EPI_ISL_1401026, EPI_ISL_1401027, EPI_ISL_1401028, EPI_ISL_1401029, EPI_ISL_1401030, EPI_ISL_1401031, EPI_ISL_1401032, EPI_ISL_1401033, EPI_ISL_1401034, EPI_ISL_1401035, EPI_ISL_1401036, EPI_ISL_1401037, EPI_ISL_1401038, EPI_ISL_1401039, EPI_ISL_1401040, EPI_ISL_1401041, EPI_ISL_1401042, EPI_ISL_1401043, EPI_ISL_1401044, EPI_ISL_1401045, EPI_ISL_1401046, EPI_ISL_1401047, EPI_ISL_1401048, EPI_ISL_1401049, EPI_ISL_1401050, EPI_ISL_1401051, EPI_ISL_1401052, EPI_ISL_1401053, EPI_ISL_1401054, EPI_ISL_1401055, EPI_ISL_1401056, EPI_ISL_1401057, EPI_ISL_1401058, EPI_ISL_1401059, EPI_ISL_1401060, EPI_ISL_1401061, EPI_ISL_1401062, EPI_ISL_1401063, EPI_ISL_1401064, EPI_ISL_1401065, EPI_ISL_1401066, EPI_ISL_1401067, EPI_ISL_1401068, EPI_ISL_1401069 | | | |
| see above | Erie County Public Health (ECPHL) | University at Buffalo Genomics and Bioinformatics Core | Jonathan Bard, Natalie Lamb, Alyssa Pohlman, Brandon Marzullo, Amanda Boccolucci, Norma Nowak, Donald Yergeau, Jennifer Surtees |
| EPI_ISL_1401070, EPI_ISL_1401071, EPI_ISL_1401072, EPI_ISL_1401073, EPI_ISL_1401074, EPI_ISL_1401075, EPI_ISL_1401076, EPI_ISL_1401077, EPI_ISL_1401078, EPI_ISL_1401079, EPI_ISL_1401080, EPI_ISL_1401081, EPI_ISL_1401082, EPI_ISL_1401083, EPI_ISL_1401084, EPI_ISL_1401085, EPI_ISL_1401086, EPI_ISL_1401087, EPI_ISL_1401088, EPI_ISL_1401089, EPI_ISL_1401091, EPI_ISL_1401092, EPI_ISL_1401093, EPI_ISL_1401094, EPI_ISL_1401095, EPI_ISL_1401096, EPI_ISL_1401097, EPI_ISL_1401098 | | | |
| see above | South Dakota Public Health Laboratory | South Dakota Public Health Laboratory | Jacob Garfin and Chris Carlson |
| EPI_ISL_1401473, EPI_ISL_1401474, EPI_ISL_1401475, EPI_ISL_1401476, EPI_ISL_1401477, EPI_ISL_1401478, EPI_ISL_1401479, EPI_ISL_1401480, EPI_ISL_1401481, EPI_ISL_1401482, EPI_ISL_1401483, EPI_ISL_1401484, EPI_ISL_1401485, EPI_ISL_1401486, EPI_ISL_1401487, EPI_ISL_1401488, EPI_ISL_1401489, EPI_ISL_1401490, EPI_ISL_1401491, EPI_ISL_1401492, EPI_ISL_1401493, EPI_ISL_1401494, EPI_ISL_1401495, EPI_ISL_1401496, EPI_ISL_1401497, EPI_ISL_1401498, EPI_ISL_1401499, EPI_ISL_1401500, EPI_ISL_1401501, EPI_ISL_1401503, EPI_ISL_1401504, EPI_ISL_1401505, EPI_ISL_1401506, EPI_ISL_1401507, EPI_ISL_1401508, EPI_ISL_1401509, EPI_ISL_1401510, EPI_ISL_1401511, EPI_ISL_1401512, EPI_ISL_1401513, EPI_ISL_1401514, EPI_ISL_1401515, EPI_ISL_1401516, EPI_ISL_1401517, EPI_ISL_1401518, EPI_ISL_1401519, EPI_ISL_1401520, EPI_ISL_1401521, EPI_ISL_1401522, EPI_ISL_1401523, EPI_ISL_1401524, EPI_ISL_1401525, EPI_ISL_1401526, EPI_ISL_1401527, EPI_ISL_1401528, EPI_ISL_1401529, EPI_ISL_1401530, EPI_ISL_1401531, EPI_ISL_1401532, EPI_ISL_1401533, EPI_ISL_1401534, EPI_ISL_1401535, EPI_ISL_1401536, EPI_ISL_1401537, EPI_ISL_1401538, EPI_ISL_1401539, EPI_ISL_1401540, EPI_ISL_1401541, EPI_ISL_1401542, EPI_ISL_1401543, EPI_ISL_1401544, EPI_ISL_1401545, EPI_ISL_1401546, EPI_ISL_1401547, EPI_ISL_1401548, EPI_ISL_1401549, EPI_ISL_1401550, EPI_ISL_1401551, EPI_ISL_1401552, EPI_ISL_1401553, EPI_ISL_1401554, EPI_ISL_1401555, EPI_ISL_1401556, EPI_ISL_1401557, EPI_ISL_1401558, EPI_ISL_1401560, EPI_ISL_1401561, EPI_ISL_1401562, EPI_ISL_1401563, EPI_ISL_1401564, EPI_ISL_1401565 | | | |
| see above | The Jackson Laboratory | The Jackson Laboratory | Bergeron D, Uvalic J, Renzette N, Adams M, Omerza G, Kelly K, Li L |
| EPI_ISL_1401566 | Indiana State Health Department | Quantigen Biosciences | Paul J Childress |
| EPI_ISL_1401966, EPI_ISL_1401967, EPI_ISL_1401968, EPI_ISL_1401969, EPI_ISL_1401970, EPI_ISL_1401971, EPI_ISL_1401972, EPI_ISL_1401973, EPI_ISL_1401974, EPI_ISL_1401975, EPI_ISL_1401976, EPI_ISL_1401977, EPI_ISL_1401978, EPI_ISL_1401979, EPI_ISL_1401980, EPI_ISL_1401981, EPI_ISL_1401982, EPI_ISL_1401983, EPI_ISL_1401984, EPI_ISL_1401985 | | | |
| see above | Kansas Health and Environmental Lab | Kansas Health and Environmental Lab | Mike Grose, Jonathan Barnell, Ben Olsen, and Phil Adam |
| EPI_ISL_1402541 | Oregon State Public Health Laboratory | Oregon State Public Health Laboratory | Rafia Razzaque, Eugene Yeboah, Vanda Makris, John Fontana and Shane Sevey |
| EPI_ISL_1402587, EPI_ISL_1402588, EPI_ISL_1402589, EPI_ISL_1402590, EPI_ISL_1402591, EPI_ISL_1402592, EPI_ISL_1402593, EPI_ISL_1402594, EPI_ISL_1402595, EPI_ISL_1402596, EPI_ISL_1402597, EPI_ISL_1402598, EPI_ISL_1402599, EPI_ISL_1402600, EPI_ISL_1402601, EPI_ISL_1402602, EPI_ISL_1402603, EPI_ISL_1402604, EPI_ISL_1402605, EPI_ISL_1402606, EPI_ISL_1402607, EPI_ISL_1402608, EPI_ISL_1402609, EPI_ISL_1402610, EPI_ISL_1402611, EPI_ISL_1402612, EPI_ISL_1402613, EPI_ISL_1402614, EPI_ISL_1402615, EPI_ISL_1402617, EPI_ISL_1402618, EPI_ISL_1402620, EPI_ISL_1402621, EPI_ISL_1402622, EPI_ISL_1402623, EPI_ISL_1402624, EPI_ISL_1402625, EPI_ISL_1402626, EPI_ISL_1402627, EPI_ISL_1402628, EPI_ISL_1402629, EPI_ISL_1402630, EPI_ISL_1402631, EPI_ISL_1402632, EPI_ISL_1402633, EPI_ISL_1402634, EPI_ISL_1402635, EPI_ISL_1402636, EPI_ISL_1402637, EPI_ISL_1402638, EPI_ISL_1402639, EPI_ISL_1402640, EPI_ISL_1402641, EPI_ISL_1402642, EPI_ISL_1402643, EPI_ISL_1402644, EPI_ISL_1402645, EPI_ISL_1402646, EPI_ISL_1402648, EPI_ISL_1402649, EPI_ISL_1402650, EPI_ISL_1402651, EPI_ISL_1402652, EPI_ISL_1402653, EPI_ISL_1402654 | | | |
| see above | UCSC Genomics Institute | UCSC Genomics Institute | Mark Akeson, Ikenna Anigbogu, Eric Beraut, Isabel Bjork, Molly Cassatt-Johnstone, Terren Chang, Russell Corbett-Detig, Namrita Dhillon, Maximilian Haeussler, David Haussler, Angie Hinrichs, Miten Jain, Joshua Kapp, A. Marm Kilpatrick, Jakob McBroome, Hugh Olsen, Jeremy Sanford, Beth Shapiro, Michael Stone, Bryan Thornlow, Yatish Turakhia |
| EPI_ISL_1402707 | Oregon State Public Health Laboratory | Oregon State Public Health Laboratory | Rafia Razzaque, Eugene Yeboah, Vanda Makris, John Fontana and Shane Sevey |
| EPI_ISL_1402708, EPI_ISL_1402709, EPI_ISL_1402710, EPI_ISL_1402711, EPI_ISL_1402713, EPI_ISL_1402714, EPI_ISL_1402715, EPI_ISL_1402716, EPI_ISL_1402717, EPI_ISL_1402718, EPI_ISL_1402719, EPI_ISL_1402720, EPI_ISL_1402721, EPI_ISL_1402722, EPI_ISL_1402723, EPI_ISL_1402724 | | | |
| see above | Minnesota Department of Health, Public Health Laboratory | Minnesota Department of Health, Public Health Laboratory | Alexandra Lorentz, Jacob Garfin, Matt Plumb, and Xiong Wang |
| EPI_ISL_1402725 | M Health Fairview | Minnesota Department of Health, Public Health Laboratory | Alexandra Lorentz, Jacob Garfin, Matt Plumb, and Xiong Wang |
| EPI_ISL_1402726, EPI_ISL_1402727, EPI_ISL_1402728, EPI_ISL_1402729, EPI_ISL_1402730, EPI_ISL_1402731, EPI_ISL_1402732, EPI_ISL_1402733, EPI_ISL_1402734, EPI_ISL_1402735, EPI_ISL_1402736, EPI_ISL_1402737, EPI_ISL_1402738, EPI_ISL_1402739, EPI_ISL_1402740 | | | |
| see above | Minnesota Department of Health, Public Health Laboratory | Minnesota Department of Health, Public Health Laboratory | Alexandra Lorentz, Jacob Garfin, Matt Plumb, and Xiong Wang |
| EPI_ISL_1402741, EPI_ISL_1402742 | M Health Fairview | Minnesota Department of Health, Public Health Laboratory | Alexandra Lorentz, Jacob Garfin, Matt Plumb, and Xiong Wang |
| EPI_ISL_1402743 | Vault Health | Minnesota Department of Health, Public Health Laboratory | Alexandra Lorentz, Jacob Garfin, Matt Plumb, and Xiong Wang |
| EPI_ISL_1402744, EPI_ISL_1402745, EPI_ISL_1402746, EPI_ISL_1402747, EPI_ISL_1402748, EPI_ISL_1402749, EPI_ISL_1402750, EPI_ISL_1402751, EPI_ISL_1402752, EPI_ISL_1402753, EPI_ISL_1402754, EPI_ISL_1402755, EPI_ISL_1402756, EPI_ISL_1402757, EPI_ISL_1402758, EPI_ISL_1402759, EPI_ISL_1402760, EPI_ISL_1402761, EPI_ISL_1402762, EPI_ISL_1402763, EPI_ISL_1402764, EPI_ISL_1402765, EPI_ISL_1402766, EPI_ISL_1402767, EPI_ISL_1402768, EPI_ISL_1402769, EPI_ISL_1402770, EPI_ISL_1402771, EPI_ISL_1402772, EPI_ISL_1402773, EPI_ISL_1402774, EPI_ISL_1402775, EPI_ISL_1402776, EPI_ISL_1402777, EPI_ISL_1402778, EPI_ISL_1402779, EPI_ISL_1402780, EPI_ISL_1402781 | | | |
| see above | M Health Fairview | Minnesota Department of Health, Public Health Laboratory | Alexandra Lorentz, Jacob Garfin, Matt Plumb, and Xiong Wang |
| EPI_ISL_1402786, EPI_ISL_1402787, EPI_ISL_1402788, EPI_ISL_1402789, EPI_ISL_1402790, EPI_ISL_1402791, EPI_ISL_1402792, EPI_ISL_1402793, EPI_ISL_1402794, EPI_ISL_1402795, EPI_ISL_1402796, EPI_ISL_1402797, EPI_ISL_1402798, EPI_ISL_1402799, EPI_ISL_1402800, EPI_ISL_1402801, EPI_ISL_1402802, EPI_ISL_1402803, EPI_ISL_1402804, EPI_ISL_1402805, EPI_ISL_1402806, EPI_ISL_1402807, EPI_ISL_1402808, EPI_ISL_1402809, EPI_ISL_1402810, EPI_ISL_1402811, EPI_ISL_1402812, EPI_ISL_1402813, EPI_ISL_1402814, EPI_ISL_1402815, EPI_ISL_1402816, EPI_ISL_1402817, EPI_ISL_1402818, EPI_ISL_1402819, EPI_ISL_1402820, EPI_ISL_1402821, EPI_ISL_1402822, EPI_ISL_1402823, EPI_ISL_1402824, EPI_ISL_1402825, EPI_ISL_1402826, EPI_ISL_1402828, EPI_ISL_1402829, EPI_ISL_1402830, EPI_ISL_1402832, EPI_ISL_1402833, EPI_ISL_1402834, EPI_ISL_1402835, EPI_ISL_1402836, EPI_ISL_1402837, EPI_ISL_1402838, EPI_ISL_1402839, EPI_ISL_1402840, EPI_ISL_1402841, EPI_ISL_1402842, EPI_ISL_1402843, EPI_ISL_1402844, EPI_ISL_1402845, EPI_ISL_1402846, EPI_ISL_1402847, EPI_ISL_1402848, EPI_ISL_1402849, EPI_ISL_1402850, EPI_ISL_1402851, EPI_ISL_1402852, EPI_ISL_1402853, EPI_ISL_1402854, EPI_ISL_1402855, EPI_ISL_1402856, EPI_ISL_1402857, EPI_ISL_1402858, EPI_ISL_1402859, EPI_ISL_1402860, EPI_ISL_1402861, EPI_ISL_1402862, EPI_ISL_1402863, EPI_ISL_1402865, EPI_ISL_1402866, EPI_ISL_1402867, EPI_ISL_1402868, EPI_ISL_1402869, EPI_ISL_1402870, EPI_ISL_1402871, EPI_ISL_1402872, EPI_ISL_1402873, EPI_ISL_1402874, EPI_ISL_1402875, EPI_ISL_1402877, EPI_ISL_1402878, EPI_ISL_1402879, EPI_ISL_1402880, EPI_ISL_1402881, EPI_ISL_1402882, EPI_ISL_1402883, EPI_ISL_1402884, EPI_ISL_1402885, EPI_ISL_1402886, EPI_ISL_1402888, EPI_ISL_1402889, EPI_ISL_1402890, EPI_ISL_1402892, EPI_ISL_1402893, EPI_ISL_1402895, EPI_ISL_1402896, EPI_ISL_1402897, EPI_ISL_1402898, EPI_ISL_1402899, EPI_ISL_1402900, EPI_ISL_1402901, EPI_ISL_1402902, EPI_ISL_1402903, EPI_ISL_1402904, EPI_ISL_1402905, EPI_ISL_1402906, EPI_ISL_1402907, EPI_ISL_1402908, EPI_ISL_1402909, EPI_ISL_1402910, EPI_ISL_1402911, EPI_ISL_1402912, EPI_ISL_1402913, EPI_ISL_1402914, EPI_ISL_1402915, EPI_ISL_1402916, EPI_ISL_1402917, EPI_ISL_1402918, EPI_ISL_1402919, EPI_ISL_1402920, EPI_ISL_1402921, EPI_ISL_1402922, EPI_ISL_1402923, EPI_ISL_1402924, EPI_ISL_1402925, EPI_ISL_1402926, EPI_ISL_1402927, EPI_ISL_1402929, EPI_ISL_1402930, EPI_ISL_1402931 | | | |
| see above | Johns Hopkins Hospital Department of Pathology | Johns Hopkins Hospital Department of Pathology | C. Paul Morris, Chun Huai Luo, Adannaya Amadi, Matthew Schwartz, Heba H. Mostafa |
| EPI_ISL_1402977, EPI_ISL_1402978, EPI_ISL_1402979, EPI_ISL_1402980, EPI_ISL_1402981, EPI_ISL_1402982, EPI_ISL_1402983, EPI_ISL_1402984, EPI_ISL_1402985, EPI_ISL_1402986, EPI_ISL_1402987, EPI_ISL_1402988, EPI_ISL_1402989, EPI_ISL_1402990, EPI_ISL_1402991, EPI_ISL_1402992, EPI_ISL_1402993, EPI_ISL_1402994, EPI_ISL_1402995, EPI_ISL_1402996, EPI_ISL_1402997, EPI_ISL_1402998, EPI_ISL_1402999, EPI_ISL_1403000, EPI_ISL_1403001, EPI_ISL_1403002, EPI_ISL_1403003, EPI_ISL_1403004, EPI_ISL_1403005, EPI_ISL_1403006, EPI_ISL_1403007, EPI_ISL_1403008, EPI_ISL_1403009, EPI_ISL_1403010, EPI_ISL_1403011, EPI_ISL_1403012, EPI_ISL_1403013, EPI_ISL_1403014, EPI_ISL_1403015, EPI_ISL_1403016, EPI_ISL_1403017, EPI_ISL_1403018, EPI_ISL_1403019, EPI_ISL_1403020, EPI_ISL_1403021, EPI_ISL_1403022, EPI_ISL_1403023, EPI_ISL_1403024, EPI_ISL_1403025, EPI_ISL_1403026, EPI_ISL_1403027, EPI_ISL_1403028, EPI_ISL_1403029, EPI_ISL_1403030, EPI_ISL_1403031, EPI_ISL_1403032, EPI_ISL_1403033, EPI_ISL_1403034, EPI_ISL_1403035, EPI_ISL_1403036, EPI_ISL_1403037, EPI_ISL_1403038, EPI_ISL_1403039, EPI_ISL_1403040, EPI_ISL_1403041, EPI_ISL_1403042, EPI_ISL_1403043, EPI_ISL_1403044, EPI_ISL_1403045, EPI_ISL_1403046, EPI_ISL_1403047, EPI_ISL_1403048 | | | |
| see above | Hospital of the University of Pennsylvania Molecular Pathology Lab | Bushman Lab - University of Pennsylvania | John Everett, Kyle Rodino, Shantan Reddy, Pascha Hokama, Aoife M. Roche, Young Hwang, Abigail Glascock, Scott Sherrill-Mix, Samantha A. Whiteside, Jevon Graham-Wooten, Layla A. Khatib, Ayannah S. Fitzgerald, Arupa Ganguly, Mike Feldman, Brendan Kelly, Ronald G. Collman and Frederic Bushman |
| EPI_ISL_1403049, EPI_ISL_1403050 | Oregon State Public Health Laboratory | Oregon State Public Health Laboratory | Rafia Razzaque, Eugene Yeboah, Vanda Makris, John Fontana and Shane Sevey |
| EPI_ISL_1403051, EPI_ISL_1403052, EPI_ISL_1403054, EPI_ISL_1403055, EPI_ISL_1403056, EPI_ISL_1403057, EPI_ISL_1403058, EPI_ISL_1403059, EPI_ISL_1403060, EPI_ISL_1403061, EPI_ISL_1403062, EPI_ISL_1403063, EPI_ISL_1403064, EPI_ISL_1403066, EPI_ISL_1403067, EPI_ISL_1403068, EPI_ISL_1403069, EPI_ISL_1403070, EPI_ISL_1403071, EPI_ISL_1403073, EPI_ISL_1403075, EPI_ISL_1403076, EPI_ISL_1403077, EPI_ISL_1403078, EPI_ISL_1403079, EPI_ISL_1403080 | | | |
| see above | IN State Department of Health Laboratory Services | IN State Department of Health Laboratory Services | Cassandra Campion, Jamie Yeadon, Brian Pope, Lixia Liu, Kyle Brownlee, Melissa Hindenlang, Ankita Kashikar, Mark Glazier |
| EPI_ISL_1403385 | Arizona State University | Arizona State University | Rabia Maqsood, LaRinda A. Holland, Peter T. Skidmore, Emily A. Kaelin, Nicholas J. Mellor, Kristina Buss, Joy M. Blain, Neal W Woodbury, Valerie Harris, Joshua LaBaer, Vel Murugan, Efrem S. Lim |
| EPI_ISL_1403386 | Oregon State Public Health Laboratory | Oregon State Public Health Laboratory | Rafia Razzaque, Eugene Yeboah, Vanda Makris, John Fontana and Shane Sevey |
| EPI_ISL_1403613, EPI_ISL_1403614 | DC Public Health Lab/ Dept. of Forensic Sciences | DC Public Health Lab/ Dept. of Forensic Sciences | David Payne, Scott Nguyen, Janis Doss, Elizabeth Zelaya, Sarah Scott, Connie Maza, Monica Mann, Brittany Hamilton, Jocelyn Hauser |
| EPI_ISL_1403624, EPI_ISL_1403625, EPI_ISL_1403626, EPI_ISL_1403627, EPI_ISL_1403628, EPI_ISL_1403629, EPI_ISL_1403630, EPI_ISL_1403631, EPI_ISL_1403632, EPI_ISL_1403633, EPI_ISL_1403634, EPI_ISL_1403635, EPI_ISL_1403636, EPI_ISL_1403637, EPI_ISL_1403646, EPI_ISL_1403647, EPI_ISL_1403648, EPI_ISL_1403649, EPI_ISL_1403650, EPI_ISL_1403651, EPI_ISL_1403652, EPI_ISL_1403653, EPI_ISL_1403654, EPI_ISL_1403655, EPI_ISL_1403656, EPI_ISL_1403657, EPI_ISL_1403658, EPI_ISL_1403659, EPI_ISL_1403660, EPI_ISL_1403661, EPI_ISL_1403662, EPI_ISL_1403663, EPI_ISL_1403664 | | | |
| see above | University of Wisconsin-Madison AIDS Vaccine Research Laboratories | University of Wisconsin-Madison AIDS Vaccine Research Laboratories | Gage Moreno, Katarina Braun, et al. AIDS Vaccine Research Laboratories |
| EPI_ISL_1403666 | Oregon State Public Health Laboratory | Oregon State Public Health Laboratory | Rafia Razzaque, Eugene Yeboah, Vanda Makris, John Fontana and Shane Sevey |
| EPI_ISL_1403667, EPI_ISL_1403668 | M Health Fairview | Minnesota Department of Health, Public Health Laboratory | Alexandra Lorentz, Jacob Garfin, Matt Plumb, and Xiong Wang |
| EPI_ISL_1403669 | Oregon State Public Health Laboratory | Oregon State Public Health Laboratory | Rafia Razzaque, Eugene Yeboah, Vanda Makris, John Fontana and Shane Sevey |
| EPI_ISL_1403677, EPI_ISL_1403678, EPI_ISL_1403679, EPI_ISL_1403680, EPI_ISL_1403681, EPI_ISL_1403682, EPI_ISL_1403683, EPI_ISL_1403684, EPI_ISL_1403685 | Wyoming Public Health Laboratory | Wyoming Public Health Laboratory | Jim Mildenberger, Wanda Manley, Noah Hull, Taylor Fearing, Lynette Gumbleton, Channing Weber, Ashley Norberg, Chayse Rowley, Marley Goetz, Brian Dominguez, Elliot Thomasson, Cari Sloma, and Rob Christensen |
| EPI_ISL_1403781, | Oregon State Public Health Laboratory | Oregon State Public Health Laboratory | Rafia Razzaque, Eugene Yeboah, Vanda Makris, John Fontana and Shane Sevey |

| | | | |
|---|---|---|---|
| EPI_ISL_1403782, EPI_ISL_1403783 | | | |
| EPI_ISL_1403784, EPI_ISL_1403785, EPI_ISL_1403786, EPI_ISL_1403787, EPI_ISL_1403788, EPI_ISL_1403789, EPI_ISL_1403790, EPI_ISL_1403791, EPI_ISL_1403792, EPI_ISL_1403793, EPI_ISL_1403794, EPI_ISL_1403796, EPI_ISL_1403797, EPI_ISL_1403799, EPI_ISL_1403800, EPI_ISL_1403801, EPI_ISL_1403802, EPI_ISL_1403803, EPI_ISL_1403804, EPI_ISL_1403805, EPI_ISL_1403806, EPI_ISL_1403807, EPI_ISL_1403808 | | | |
| see above | Wyoming Public Health Laboratory | Wyoming Public Health Laboratory | Jim Mildenberger, Wanda Manley, Noah Hull, Taylor Fearing, Lynette Gumbleton, Channing Weber, Ashley Norberg, Chayse Rowley, Marley Goetz, Brian Dominguez, Elliot Thomasson, Cari Sloma, and Rob Christensen |
| EPI_ISL_1403809 | Oregon State Public Health Laboratory | Oregon State Public Health Laboratory | Rafia Razzaque, Eugene Yeboah, Vanda Makris, John Fontana and Shane Sevey |
| EPI_ISL_1403810, EPI_ISL_1403811, EPI_ISL_1403812, EPI_ISL_1403813, EPI_ISL_1403814, EPI_ISL_1403815, EPI_ISL_1403816, EPI_ISL_1403817, EPI_ISL_1403818, EPI_ISL_1403819, EPI_ISL_1403821, EPI_ISL_1403823, EPI_ISL_1403824, EPI_ISL_1403825, EPI_ISL_1403826 | | | |
| see above | University of Wisconsin-Madison AIDS Vaccine Research Laboratories | University of Wisconsin-Madison AIDS Vaccine Research Laboratories | Gage Moreno, Katarina Braun, et al. AIDS Vaccine Research Laboratories |
| EPI_ISL_1403827 | Oregon State Public Health Laboratory | Oregon State Public Health Laboratory | Rafia Razzaque, Eugene Yeboah, Vanda Makris, John Fontana and Shane Sevey |
| EPI_ISL_1403828, EPI_ISL_1403829, EPI_ISL_1403830, EPI_ISL_1403831, EPI_ISL_1403832, EPI_ISL_1403833, EPI_ISL_1403834, EPI_ISL_1403835, EPI_ISL_1403836, EPI_ISL_1403837, EPI_ISL_1403838, EPI_ISL_1403839, EPI_ISL_1403840, EPI_ISL_1403841, EPI_ISL_1403842, EPI_ISL_1403843, EPI_ISL_1403844, EPI_ISL_1403845, EPI_ISL_1403846, EPI_ISL_1403847, EPI_ISL_1403848, EPI_ISL_1403849, EPI_ISL_1403850, EPI_ISL_1403851, EPI_ISL_1403852, EPI_ISL_1403853, EPI_ISL_1403854, EPI_ISL_1403855, EPI_ISL_1403856, EPI_ISL_1403857, EPI_ISL_1403858, EPI_ISL_1403859, EPI_ISL_1403860, EPI_ISL_1403861, EPI_ISL_1403862, EPI_ISL_1403863, EPI_ISL_1403864, EPI_ISL_1403865, EPI_ISL_1403866, EPI_ISL_1403867, EPI_ISL_1403868, EPI_ISL_1403869, EPI_ISL_1403870, EPI_ISL_1403871, EPI_ISL_1403872, EPI_ISL_1403873, EPI_ISL_1403874, EPI_ISL_1403875, EPI_ISL_1403876, EPI_ISL_1403877, EPI_ISL_1403878, EPI_ISL_1403879, EPI_ISL_1403880, EPI_ISL_1403881, EPI_ISL_1403882, EPI_ISL_1403883, EPI_ISL_1403884, EPI_ISL_1403885, EPI_ISL_1403886, EPI_ISL_1403887, EPI_ISL_1403888, EPI_ISL_1403889, EPI_ISL_1403890, EPI_ISL_1403891, EPI_ISL_1403892, EPI_ISL_1403893, EPI_ISL_1403894, EPI_ISL_1403895, EPI_ISL_1403896, EPI_ISL_1403897, EPI_ISL_1403898, EPI_ISL_1403899, EPI_ISL_1403900, EPI_ISL_1403901, EPI_ISL_1403902, EPI_ISL_1403903, EPI_ISL_1403904, EPI_ISL_1403905, EPI_ISL_1403906, EPI_ISL_1403907, EPI_ISL_1403908, EPI_ISL_1403909, EPI_ISL_1403910, EPI_ISL_1403911, EPI_ISL_1403912, EPI_ISL_1403913, EPI_ISL_1403914, EPI_ISL_1403915, EPI_ISL_1403916, EPI_ISL_1403917, EPI_ISL_1403918, EPI_ISL_1403919, EPI_ISL_1403920, EPI_ISL_1403921, EPI_ISL_1403922, EPI_ISL_1403923, EPI_ISL_1403924, EPI_ISL_1403925, EPI_ISL_1403926, EPI_ISL_1403927, EPI_ISL_1403928, EPI_ISL_1403929, EPI_ISL_1403930, EPI_ISL_1403931, EPI_ISL_1403932, EPI_ISL_1403933, EPI_ISL_1403934, EPI_ISL_1403935, EPI_ISL_1403936, EPI_ISL_1403937, EPI_ISL_1403938, EPI_ISL_1403939, EPI_ISL_1403940, EPI_ISL_1403941, EPI_ISL_1403942, EPI_ISL_1403943, EPI_ISL_1403944, EPI_ISL_1403945, EPI_ISL_1403946, EPI_ISL_1403947, EPI_ISL_1403948, EPI_ISL_1403949, EPI_ISL_1403950, EPI_ISL_1403951, EPI_ISL_1403952, EPI_ISL_1403953, EPI_ISL_1403954, EPI_ISL_1403955, EPI_ISL_1403956, EPI_ISL_1403957, EPI_ISL_1403958, EPI_ISL_1403959, EPI_ISL_1403960, EPI_ISL_1403961, EPI_ISL_1403962, EPI_ISL_1403963, EPI_ISL_1403964, EPI_ISL_1403965, EPI_ISL_1403966, EPI_ISL_1403967, EPI_ISL_1403968, EPI_ISL_1403969, EPI_ISL_1403970, EPI_ISL_1403971, EPI_ISL_1403972, EPI_ISL_1403973, EPI_ISL_1403974, EPI_ISL_1403975, EPI_ISL_1403976, EPI_ISL_1403977, EPI_ISL_1403978, EPI_ISL_1403979, EPI_ISL_1403980, EPI_ISL_1403981, EPI_ISL_1403982, EPI_ISL_1403983, EPI_ISL_1403984, EPI_ISL_1403985, EPI_ISL_1403986, EPI_ISL_1403987, EPI_ISL_1403988, EPI_ISL_1403989, EPI_ISL_1403990, EPI_ISL_1403991, EPI_ISL_1403992, EPI_ISL_1403993, EPI_ISL_1403994, EPI_ISL_1403995, EPI_ISL_1403996, EPI_ISL_1403997, EPI_ISL_1403998, EPI_ISL_1403999, EPI_ISL_1404000, EPI_ISL_1404001, EPI_ISL_1404002, EPI_ISL_1404003, EPI_ISL_1404004, EPI_ISL_1404005, EPI_ISL_1404006, EPI_ISL_1404007, EPI_ISL_1404008, EPI_ISL_1404009, EPI_ISL_1404010, EPI_ISL_1404011, EPI_ISL_1404012, EPI_ISL_1404013, EPI_ISL_1404014, EPI_ISL_1404015, EPI_ISL_1404016, EPI_ISL_1404017, EPI_ISL_1404018, EPI_ISL_1404019, EPI_ISL_1404020, EPI_ISL_1404021, EPI_ISL_1404022, EPI_ISL_1404023, EPI_ISL_1404024, EPI_ISL_1404025, EPI_ISL_1404026, EPI_ISL_1404027, EPI_ISL_1404028, EPI_ISL_1404029, EPI_ISL_1404030, EPI_ISL_1404031, EPI_ISL_1404032, EPI_ISL_1404033, EPI_ISL_1404034, EPI_ISL_1404035, EPI_ISL_1404036, EPI_ISL_1404037, EPI_ISL_1404038, EPI_ISL_1404039, EPI_ISL_1404040, EPI_ISL_1404041, EPI_ISL_1404042, EPI_ISL_1404043, EPI_ISL_1404044, EPI_ISL_1404045, EPI_ISL_1404046, EPI_ISL_1404047, EPI_ISL_1404048, EPI_ISL_1404049, EPI_ISL_1404050, EPI_ISL_1404051, EPI_ISL_1404052, EPI_ISL_1404053, EPI_ISL_1404054, EPI_ISL_1404055, EPI_ISL_1404056, EPI_ISL_1404057, EPI_ISL_1404058, EPI_ISL_1404059, EPI_ISL_1404060, EPI_ISL_1404061, EPI_ISL_1404062, EPI_ISL_1404063, EPI_ISL_1404064, EPI_ISL_1404065, EPI_ISL_1404066, EPI_ISL_1404067, EPI_ISL_1404068, EPI_ISL_1404069, EPI_ISL_1404070, EPI_ISL_1404071, EPI_ISL_1404072, EPI_ISL_1404073, EPI_ISL_1404074, EPI_ISL_1404075, EPI_ISL_1404076, EPI_ISL_1404077, EPI_ISL_1404078, EPI_ISL_1404079, EPI_ISL_1404080, EPI_ISL_1404081, EPI_ISL_1404082, EPI_ISL_1404083, EPI_ISL_1404084, EPI_ISL_1404085, EPI_ISL_1404086, EPI_ISL_1404087, EPI_ISL_1404088, EPI_ISL_1404089, EPI_ISL_1404090, EPI_ISL_1404091, EPI_ISL_1404092, EPI_ISL_1404093, EPI_ISL_1404094, EPI_ISL_1404095, EPI_ISL_1404096, EPI_ISL_1404097, EPI_ISL_1404098, EPI_ISL_1404099, EPI_ISL_1404100, EPI_ISL_1404101, EPI_ISL_1404102, EPI_ISL_1404103, EPI_ISL_1404104, EPI_ISL_1404105 | | | |
| see above | Utah Public Health Laboratory | Utah Public Health Laboratory | Erin L. Young, Kelly F. Oakeson, Tara Gallagher |
| EPI_ISL_1404106, EPI_ISL_1404107, EPI_ISL_1404108, EPI_ISL_1404110, EPI_ISL_1404111, EPI_ISL_1404112, EPI_ISL_1404113, EPI_ISL_1404114, EPI_ISL_1404115, EPI_ISL_1404116, EPI_ISL_1404117, EPI_ISL_1404118, EPI_ISL_1404119, EPI_ISL_1404120, EPI_ISL_1404121, EPI_ISL_1404122, EPI_ISL_1404123, EPI_ISL_1404124, EPI_ISL_1404126, EPI_ISL_1404127, EPI_ISL_1404129 | | | |
| see above | University of Wisconsin-Madison AIDS Vaccine Research Laboratories | University of Wisconsin-Madison AIDS Vaccine Research Laboratories | Gage Moreno, Katarina Braun, et al. AIDS Vaccine Research Laboratories |
| EPI_ISL_1404130, EPI_ISL_1404143 | Oregon State Public Health Laboratory | Oregon State Public Health Laboratory | Rafia Razzaque, Eugene Yeboah, Vanda Makris, John Fontana and Shane Sevey |
| EPI_ISL_1404145, EPI_ISL_1404146, EPI_ISL_1404147, EPI_ISL_1404149, EPI_ISL_1404150, EPI_ISL_1404151, EPI_ISL_1404152, EPI_ISL_1404153, EPI_ISL_1404154, EPI_ISL_1404155, EPI_ISL_1404156, EPI_ISL_1404157, EPI_ISL_1404158, EPI_ISL_1404159, EPI_ISL_1404160, EPI_ISL_1404161, EPI_ISL_1404162, EPI_ISL_1404163, EPI_ISL_1404164, EPI_ISL_1404165, EPI_ISL_1404166, EPI_ISL_1404167, EPI_ISL_1404168 | | | |
| see above | IN State Department of Health Laboratory Services | IN State Department of Health Laboratory Services | Cassandra Campion, Jamie Yeadon, Brian Pope, Lixia Liu, Kyle Brownlee, Melissa Hindenlang, Ankita Kashikar, Mark Glazier |
| EPI_ISL_1404249, EPI_ISL_1404250, EPI_ISL_1404251, EPI_ISL_1404252, EPI_ISL_1404253, EPI_ISL_1404254, EPI_ISL_1404255, EPI_ISL_1404256, EPI_ISL_1404257, EPI_ISL_1404258, EPI_ISL_1404259, EPI_ISL_1404260, EPI_ISL_1404261, EPI_ISL_1404262, EPI_ISL_1404263, EPI_ISL_1404264, EPI_ISL_1404265, EPI_ISL_1404266, EPI_ISL_1404267, EPI_ISL_1404268, EPI_ISL_1404269, EPI_ISL_1404270, EPI_ISL_1404271, EPI_ISL_1404272, EPI_ISL_1404273, EPI_ISL_1404274, EPI_ISL_1404275, EPI_ISL_1404276, EPI_ISL_1404277, EPI_ISL_1404278, EPI_ISL_1404279, EPI_ISL_1404280, EPI_ISL_1404281, EPI_ISL_1404282, EPI_ISL_1404283, EPI_ISL_1404284, EPI_ISL_1404285, EPI_ISL_1404286, EPI_ISL_1404287, EPI_ISL_1404288, EPI_ISL_1404289, EPI_ISL_1404290, EPI_ISL_1404291, EPI_ISL_1404292, EPI_ISL_1404293, EPI_ISL_1404294, EPI_ISL_1404295, EPI_ISL_1404296, EPI_ISL_1404297, EPI_ISL_1404298, EPI_ISL_1404299, EPI_ISL_1404300, EPI_ISL_1404301, EPI_ISL_1404302, EPI_ISL_1404303, EPI_ISL_1404304, EPI_ISL_1404305, EPI_ISL_1404306, EPI_ISL_1404307, EPI_ISL_1404308, EPI_ISL_1404309, EPI_ISL_1404310, EPI_ISL_1404311, EPI_ISL_1404312, EPI_ISL_1404313, EPI_ISL_1404314, EPI_ISL_1404315, EPI_ISL_1404316, EPI_ISL_1404317, EPI_ISL_1404318, EPI_ISL_1404319, EPI_ISL_1404320, EPI_ISL_1404321, EPI_ISL_1404322, EPI_ISL_1404323, EPI_ISL_1404324, EPI_ISL_1404325, EPI_ISL_1404326, EPI_ISL_1404327, EPI_ISL_1404328, EPI_ISL_1404329, EPI_ISL_1404330, EPI_ISL_1404331, EPI_ISL_1404332, EPI_ISL_1404333, EPI_ISL_1404334, EPI_ISL_1404335, EPI_ISL_1404336, EPI_ISL_1404337, EPI_ISL_1404338, EPI_ISL_1404339, EPI_ISL_1404340, EPI_ISL_1404341, EPI_ISL_1404342, EPI_ISL_1404343, EPI_ISL_1404344, EPI_ISL_1404345, EPI_ISL_1404346, EPI_ISL_1404347, EPI_ISL_1404348, EPI_ISL_1404349, EPI_ISL_1404350, EPI_ISL_1404351, EPI_ISL_1404352, EPI_ISL_1404353, EPI_ISL_1404354, EPI_ISL_1404355, EPI_ISL_1404356, EPI_ISL_1404357, EPI_ISL_1404358, EPI_ISL_1404359, EPI_ISL_1404360, EPI_ISL_1404361, EPI_ISL_1404362, EPI_ISL_1404363, EPI_ISL_1404364, EPI_ISL_1404365, EPI_ISL_1404366, EPI_ISL_1404367, EPI_ISL_1404368, EPI_ISL_1404369, EPI_ISL_1404370, EPI_ISL_1404371, EPI_ISL_1404372, EPI_ISL_1404373 | | | |
| see above | University of Michigan Clinical Microbiology Laboratory | Lauring Lab, University of Michigan, Department of Microbiology and Immunology | Valesano |
| EPI_ISL_1404436, EPI_ISL_1404437, EPI_ISL_1404438, EPI_ISL_1404439, EPI_ISL_1404440, EPI_ISL_1404441, EPI_ISL_1404442, EPI_ISL_1404443, EPI_ISL_1404444, EPI_ISL_1404445, EPI_ISL_1404446, EPI_ISL_1404447, EPI_ISL_1404448, EPI_ISL_1404449, EPI_ISL_1404450 | | | |
| see above | IN State Department of Health Laboratory Services | IN State Department of Health Laboratory Services | Cassandra Campion, Jamie Yeadon, Brian Pope, Lixia Liu, Kyle Brownlee, Melissa Hindenlang, Ankita Kashikar, Mark Glazier |
| EPI_ISL_1404576, EPI_ISL_1404577, EPI_ISL_1404578, EPI_ISL_1404579, EPI_ISL_1404580, EPI_ISL_1404581, EPI_ISL_1404582, EPI_ISL_1404583, EPI_ISL_1404584, EPI_ISL_1404585, EPI_ISL_1404586, EPI_ISL_1404587 | | | |
| see above | DC Public Health Lab/ Dept. of Forensic Sciences | DC Public Health Lab/ Dept. of Forensic Sciences | Janis Doss, Scott Nguyen, Elizabeth Zelaya, Sarah Scott, Connie Maza, Monica Mann, Brittany Hamilton, David Payne, Jocelyn Hauser |
| EPI_ISL_1404590, EPI_ISL_1404591, EPI_ISL_1404592, EPI_ISL_1404593, EPI_ISL_1404594, EPI_ISL_1404595, EPI_ISL_1404596, EPI_ISL_1404597, EPI_ISL_1404598, EPI_ISL_1404599, EPI_ISL_1404600, EPI_ISL_1404601, EPI_ISL_1404602, EPI_ISL_1404603, EPI_ISL_1404604, EPI_ISL_1404605, EPI_ISL_1404606, EPI_ISL_1404607, EPI_ISL_1404608, EPI_ISL_1404609, EPI_ISL_1404610, EPI_ISL_1404611, EPI_ISL_1404612, EPI_ISL_1404613 | | | |
| see above | Nevada State Public Health Laboratory | Nevada State Public Health Laboratory | Andrew Gorzalski, Mark Pandori |
| EPI_ISL_1404616 | siParadigm LLC | siParadigm LLC | Kamfai Chan |
| EPI_ISL_1404617, EPI_ISL_1404618, EPI_ISL_1404620, EPI_ISL_1404621, EPI_ISL_1404622, EPI_ISL_1404623 | Massachusetts State Public Health Laboratory | Massachusetts State Public Health Laboratory | Andrew Lang, Timelia Fink, Glen Gallagher, Sandra Smole |
| EPI_ISL_1404651, EPI_ISL_1404652, EPI_ISL_1404653, EPI_ISL_1404654, EPI_ISL_1404655, EPI_ISL_1404657, EPI_ISL_1404658, EPI_ISL_1404659, EPI_ISL_1404660, EPI_ISL_1404661, EPI_ISL_1404662, EPI_ISL_1404663, EPI_ISL_1404664, EPI_ISL_1404665, EPI_ISL_1404666, EPI_ISL_1404667, EPI_ISL_1404668, EPI_ISL_1404669, EPI_ISL_1404670, EPI_ISL_1404671, EPI_ISL_1404672, EPI_ISL_1404673, EPI_ISL_1404674, EPI_ISL_1404675, EPI_ISL_1404676, EPI_ISL_1404677, EPI_ISL_1404679 | | | |
| see above | Oregon State University TRACE Project | OSU Center for Genome Research and Biocomputing | Oregon State University TRACE Project |
| EPI_ISL_1404923, EPI_ISL_1404924, EPI_ISL_1404925, EPI_ISL_1404926, EPI_ISL_1404927, EPI_ISL_1404928, EPI_ISL_1404929, EPI_ISL_1404930, EPI_ISL_1404931, EPI_ISL_1404932, EPI_ISL_1404933, EPI_ISL_1404934, EPI_ISL_1404935, EPI_ISL_1404936, EPI_ISL_1404937, EPI_ISL_1404938, EPI_ISL_1404939, EPI_ISL_1404940, EPI_ISL_1404941, EPI_ISL_1404942, EPI_ISL_1404943, EPI_ISL_1404944, EPI_ISL_1404945, EPI_ISL_1404946, EPI_ISL_1404947, EPI_ISL_1404948, EPI_ISL_1404949, EPI_ISL_1404950, EPI_ISL_1404951, EPI_ISL_1404952, EPI_ISL_1404953, EPI_ISL_1404954, EPI_ISL_1404955, EPI_ISL_1404956, EPI_ISL_1404957, EPI_ISL_1404958, EPI_ISL_1404959, EPI_ISL_1404960, EPI_ISL_1404961, EPI_ISL_1404962, EPI_ISL_1404963, EPI_ISL_1404964, EPI_ISL_1404965, EPI_ISL_1404966, EPI_ISL_1404967, EPI_ISL_1404968, EPI_ISL_1404969, EPI_ISL_1404970, EPI_ISL_1404971, EPI_ISL_1404972, EPI_ISL_1404973, EPI_ISL_1404974, EPI_ISL_1404975, EPI_ISL_1404976, EPI_ISL_1404977, EPI_ISL_1404978, EPI_ISL_1404979, EPI_ISL_1404980, EPI_ISL_1404981, EPI_ISL_1404982, EPI_ISL_1404983, EPI_ISL_1404984, EPI_ISL_1404985, EPI_ISL_1404986, EPI_ISL_1404987, EPI_ISL_1404988, EPI_ISL_1404989, EPI_ISL_1404990, EPI_ISL_1404991, EPI_ISL_1404992, EPI_ISL_1404993, EPI_ISL_1404994, EPI_ISL_1404995, EPI_ISL_1404996 | | | |
| see above | Eurofins Diatherix | Hudsonalpha Genome Sequencing Center | Jane Grimwood, Melissa Williams, Lori H. Handley, Joshua Stough, Leslie Malone, Stefan Brzezinski, Ada Stewart, Teresa Jones, Jenell Webber, John Lovell, Jennifer Cart, and Jeremy Schmutz |
| EPI_ISL_1405000, EPI_ISL_1405001, EPI_ISL_1405003, EPI_ISL_1405004, EPI_ISL_1405005, EPI_ISL_1405007, EPI_ISL_1405009, EPI_ISL_1405010, EPI_ISL_1405011, EPI_ISL_1405012, EPI_ISL_1405013, EPI_ISL_1405016, EPI_ISL_1405017, EPI_ISL_1405018, EPI_ISL_1405019, EPI_ISL_1405020, EPI_ISL_1405021, EPI_ISL_1405023, EPI_ISL_1405024, EPI_ISL_1405025, EPI_ISL_1405026, EPI_ISL_1405027, EPI_ISL_1405028, EPI_ISL_1405029, EPI_ISL_1405030, EPI_ISL_1405031, EPI_ISL_1405032, EPI_ISL_1405033, EPI_ISL_1405034, EPI_ISL_1405035, EPI_ISL_1405037, EPI_ISL_1405038, EPI_ISL_1405039, EPI_ISL_1405040, EPI_ISL_1405044, EPI_ISL_1405045, EPI_ISL_1405046, EPI_ISL_1405047, EPI_ISL_1405048 | | | |
| see above | NJDOH, Public Health and Environmental Laboratories | New Jersey Public Health and Environmental Laboratories (NJ PHEL) | Lindsey Bodnar, Shiv K. Verma, Dana Woell, Byeong Jeong |
| EPI_ISL_1405050, EPI_ISL_1405051, EPI_ISL_1405052, EPI_ISL_1405053, EPI_ISL_1405054, EPI_ISL_1405057, EPI_ISL_1405058, EPI_ISL_1405059, EPI_ISL_1405060 | UCSC Genomics Institute | UCSC Genomics Institute | Mark Akeson, Ikenna Anigbogu, Eric Beraut, Isabel Bjork, Molly Cassatt-Johnstone, Terren Chang, Russell Corbett-Detig, Namrita Dhillon, Maximilian Haeussler, David Haussler, Angie Hinrichs, Miten Jain, Joshua Kapp, A. Marm Kilpatrick, Jakob McBroome, Hugh Olsen, Jeremy Sanford, Beth Shapiro, Michael Stone, Bryan Thornlow, Yatish Turakhia |
| EPI_ISL_1405061, EPI_ISL_1405062, EPI_ISL_1405063, EPI_ISL_1405064, EPI_ISL_1405065, EPI_ISL_1405066, EPI_ISL_1405067, EPI_ISL_1405068, EPI_ISL_1405069, EPI_ISL_1405070, EPI_ISL_1405071, EPI_ISL_1405072, EPI_ISL_1405073, EPI_ISL_1405074, EPI_ISL_1405075, EPI_ISL_1405076, EPI_ISL_1405077, EPI_ISL_1405078, EPI_ISL_1405079, EPI_ISL_1405080, EPI_ISL_1405081, EPI_ISL_1405082, EPI_ISL_1405083, EPI_ISL_1405084, EPI_ISL_1405085, EPI_ISL_1405086, EPI_ISL_1405087, EPI_ISL_1405088, EPI_ISL_1405089, EPI_ISL_1405090, EPI_ISL_1405091, EPI_ISL_1405092, EPI_ISL_1405093, EPI_ISL_1405094, EPI_ISL_1405095, EPI_ISL_1405096, EPI_ISL_1405097, EPI_ISL_1405098, EPI_ISL_1405099, EPI_ISL_1405100, EPI_ISL_1405101, EPI_ISL_1405102, EPI_ISL_1405103, EPI_ISL_1405104, EPI_ISL_1405105, EPI_ISL_1405106, EPI_ISL_1405107, EPI_ISL_1405108, EPI_ISL_1405109, EPI_ISL_1405110, EPI_ISL_1405111, EPI_ISL_1405112, EPI_ISL_1405113, EPI_ISL_1405114, EPI_ISL_1405115, EPI_ISL_1405116, EPI_ISL_1405117, EPI_ISL_1405118, EPI_ISL_1405119, EPI_ISL_1405120, EPI_ISL_1405121, EPI_ISL_1405122, EPI_ISL_1405123, EPI_ISL_1405124, EPI_ISL_1405125, EPI_ISL_1405126, EPI_ISL_1405128, EPI_ISL_1405129, EPI_ISL_1405130, EPI_ISL_1405131, EPI_ISL_1405132, EPI_ISL_1405133, EPI_ISL_1405134, EPI_ISL_1405135, EPI_ISL_1405136, EPI_ISL_1405138, EPI_ISL_1405139, EPI_ISL_1405140, EPI_ISL_1405141, EPI_ISL_1405142, EPI_ISL_1405143, EPI_ISL_1405144, EPI_ISL_1405145, EPI_ISL_1405146, EPI_ISL_1405147, EPI_ISL_1405148, EPI_ISL_1405149, EPI_ISL_1405150, EPI_ISL_1405151, EPI_ISL_1405152, EPI_ISL_1405153, EPI_ISL_1405154, EPI_ISL_1405155, EPI_ISL_1405156, EPI_ISL_1405157, EPI_ISL_1405158, EPI_ISL_1405159, EPI_ISL_1405160, EPI_ISL_1405161, EPI_ISL_1405162, EPI_ISL_1405163, EPI_ISL_1405164, EPI_ISL_1405165, EPI_ISL_1405166, EPI_ISL_1405167, EPI_ISL_1405168, EPI_ISL_1405169, EPI_ISL_1405170, EPI_ISL_1405171, EPI_ISL_1405172, EPI_ISL_1405173, EPI_ISL_1405174, EPI_ISL_1405175, EPI_ISL_1405176, EPI_ISL_1405177, EPI_ISL_1405178, EPI_ISL_1405179, EPI_ISL_1405180, EPI_ISL_1405181, EPI_ISL_1405182, EPI_ISL_1405183, EPI_ISL_1405184, EPI_ISL_1405185, EPI_ISL_1405186, EPI_ISL_1405187, EPI_ISL_1405188, EPI_ISL_1405189, EPI_ISL_1405190, EPI_ISL_1405191, EPI_ISL_1405192, EPI_ISL_1405193, EPI_ISL_1405194, EPI_ISL_1405195, EPI_ISL_1405196, EPI_ISL_1405197, EPI_ISL_1405198, EPI_ISL_1405199, EPI_ISL_1405200, EPI_ISL_1405201, EPI_ISL_1405202, EPI_ISL_1405203, EPI_ISL_1405204, EPI_ISL_1405206, EPI_ISL_1405207, EPI_ISL_1405208, EPI_ISL_1405209, EPI_ISL_1405210, EPI_ISL_1405211, EPI_ISL_1405212, EPI_ISL_1405213, EPI_ISL_1405214, EPI_ISL_1405215, EPI_ISL_1405216, EPI_ISL_1405217, EPI_ISL_1405218, EPI_ISL_1405219, EPI_ISL_1405220, EPI_ISL_1405221, EPI_ISL_1405222, EPI_ISL_1405223, EPI_ISL_1405224, EPI_ISL_1405225, EPI_ISL_1405227, EPI_ISL_1405228, EPI_ISL_1405229, EPI_ISL_1405230, EPI_ISL_1405231, EPI_ISL_1405232, EPI_ISL_1405234, EPI_ISL_1405235, EPI_ISL_1405236, EPI_ISL_1405237, EPI_ISL_1405238, EPI_ISL_1405239, EPI_ISL_1405241, EPI_ISL_1405242, EPI_ISL_1405243, EPI_ISL_1405244, EPI_ISL_1405245, EPI_ISL_1405246, EPI_ISL_1405247, EPI_ISL_1405248, EPI_ISL_1405249, EPI_ISL_1405250, EPI_ISL_1405251, EPI_ISL_1405252, EPI_ISL_1405253, EPI_ISL_1405254, EPI_ISL_1405255, EPI_ISL_1405256, EPI_ISL_1405257, EPI_ISL_1405258, EPI_ISL_1405259, EPI_ISL_1405260, EPI_ISL_1405261, EPI_ISL_1405262, EPI_ISL_1405263, EPI_ISL_1405264, EPI_ISL_1405265, EPI_ISL_1405266, EPI_ISL_1405267, EPI_ISL_1405269, EPI_ISL_1405270, EPI_ISL_1405271, EPI_ISL_1405272, EPI_ISL_1405273, EPI_ISL_1405274, EPI_ISL_1405275, EPI_ISL_1405276, EPI_ISL_1405277, EPI_ISL_1405278, EPI_ISL_1405279, EPI_ISL_1405280, EPI_ISL_1405281, EPI_ISL_1405282, EPI_ISL_1405283, EPI_ISL_1405285, EPI_ISL_1405286, EPI_ISL_1405289, EPI_ISL_1405290, EPI_ISL_1405291, EPI_ISL_1405292, EPI_ISL_1405293, EPI_ISL_1405294, EPI_ISL_1405295, EPI_ISL_1405296, EPI_ISL_1405297, EPI_ISL_1405298, EPI_ISL_1405299, EPI_ISL_1405300, EPI_ISL_1405301, EPI_ISL_1405303, EPI_ISL_1405304, EPI_ISL_1405305, EPI_ISL_1405306, EPI_ISL_1405307, EPI_ISL_1405308, EPI_ISL_1405309, EPI_ISL_1405310, EPI_ISL_1405311, EPI_ISL_1405312, EPI_ISL_1405313, EPI_ISL_1405314, EPI_ISL_1405315, EPI_ISL_1405316, EPI_ISL_1405318, EPI_ISL_1405319, EPI_ISL_1405320, EPI_ISL_1405321, EPI_ISL_1405322, EPI_ISL_1405323, EPI_ISL_1405325, EPI_ISL_1405327, EPI_ISL_1405328, EPI_ISL_1405329, EPI_ISL_1405330, EPI_ISL_1405331, EPI_ISL_1405332, EPI_ISL_1405333, EPI_ISL_1405334, EPI_ISL_1405335, EPI_ISL_1405336, EPI_ISL_1405337, EPI_ISL_1405338, EPI_ISL_1405339, EPI_ISL_1405340, EPI_ISL_1405341, EPI_ISL_1405342, EPI_ISL_1405343, EPI_ISL_1405344, EPI_ISL_1405345, EPI_ISL_1405346, EPI_ISL_1405348, EPI_ISL_1405349, EPI_ISL_1405350, EPI_ISL_1405351, EPI_ISL_1405352, EPI_ISL_1405353, EPI_ISL_1405355, EPI_ISL_1405356, EPI_ISL_1405357, EPI_ISL_1405358, EPI_ISL_1405359, EPI_ISL_1405360, EPI_ISL_1405362, EPI_ISL_1405363, EPI_ISL_1405364, EPI_ISL_1405365, EPI_ISL_1405366, EPI_ISL_1405367, EPI_ISL_1405368, EPI_ISL_1405369, EPI_ISL_1405370, EPI_ISL_1405371, EPI_ISL_1405372, EPI_ISL_1405373, EPI_ISL_1405374, EPI_ISL_1405375, EPI_ISL_1405376, EPI_ISL_1405377, EPI_ISL_1405378, EPI_ISL_1405379, EPI_ISL_1405380, EPI_ISL_1405381, EPI_ISL_1405382, EPI_ISL_1405383, EPI_ISL_1405384, EPI_ISL_1405385, EPI_ISL_1405386, EPI_ISL_1405387, EPI_ISL_1405388, EPI_ISL_1405389, EPI_ISL_1405390, EPI_ISL_1405391, EPI_ISL_1405392, EPI_ISL_1405393, EPI_ISL_1405394, EPI_ISL_1405395, EPI_ISL_1405396, EPI_ISL_1405397, EPI_ISL_1405398, EPI_ISL_1405399, EPI_ISL_1405400, EPI_ISL_1405401, EPI_ISL_1405402, EPI_ISL_1405403 | | | |
| see above | UW Virology Lab | UW Virology Lab | Pavitra Roychoudhury, Hong Xie, Lasata Shrestha, Shah Mohamed Bakhash, Michelle Lin, Noah R. Baker, Sean Ellis, Saraswathi Sathees, Meei-Li Huang, Keith R Jerome, Alexander Greninger |
| EPI_ISL_1405466, EPI_ISL_1405467, EPI_ISL_1405468, EPI_ISL_1405469, EPI_ISL_1405470, EPI_ISL_1405471, EPI_ISL_1405472, EPI_ISL_1405473, EPI_ISL_1405474, EPI_ISL_1405483, EPI_ISL_1405485, EPI_ISL_1405487, EPI_ISL_1405488, EPI_ISL_1405489, EPI_ISL_1405490, EPI_ISL_1405491, EPI_ISL_1405492, EPI_ISL_1405500, EPI_ISL_1405506, EPI_ISL_1405510, EPI_ISL_1405515, EPI_ISL_1405516, | | | |

| | | | |
|---|---|---|---|
| EPI_ISL_1405522, EPI_ISL_1405523, EPI_ISL_1405524, EPI_ISL_1405526, EPI_ISL_1405527, EPI_ISL_1405528, EPI_ISL_1405529, EPI_ISL_1405530, EPI_ISL_1405531, EPI_ISL_1405532, EPI_ISL_1405533, EPI_ISL_1405534, EPI_ISL_1405535, EPI_ISL_1405536, EPI_ISL_1405537, EPI_ISL_1405538, EPI_ISL_1405539, EPI_ISL_1405540, EPI_ISL_1405541, EPI_ISL_1405542, EPI_ISL_1405543, EPI_ISL_1405544, EPI_ISL_1405545, EPI_ISL_1405546, EPI_ISL_1405547, EPI_ISL_1405548, EPI_ISL_1405549, EPI_ISL_1405550, EPI_ISL_1405551, EPI_ISL_1405552, EPI_ISL_1405553, EPI_ISL_1405554, EPI_ISL_1405555, EPI_ISL_1405556, EPI_ISL_1405557, EPI_ISL_1405558, EPI_ISL_1405559, EPI_ISL_1405560, EPI_ISL_1405561, EPI_ISL_1405562, EPI_ISL_1405563, EPI_ISL_1405564, EPI_ISL_1405565, EPI_ISL_1405566, EPI_ISL_1405567, EPI_ISL_1405568, EPI_ISL_1405569, EPI_ISL_1405570, EPI_ISL_1405571, EPI_ISL_1405572, EPI_ISL_1405573, EPI_ISL_1405574, EPI_ISL_1405575, EPI_ISL_1405576, EPI_ISL_1405577, EPI_ISL_1405578, EPI_ISL_1405579, EPI_ISL_1405580, EPI_ISL_1405581, EPI_ISL_1405582, EPI_ISL_1405583, EPI_ISL_1405585, EPI_ISL_1405586, EPI_ISL_1405588, EPI_ISL_1405589, EPI_ISL_1405590, EPI_ISL_1405591, EPI_ISL_1405592, EPI_ISL_1405595, EPI_ISL_1405596, EPI_ISL_1405597, EPI_ISL_1405598 | | | |
| see above | Johns Hopkins Hospital Department of Pathology | Johns Hopkins Hospital Department of Pathology | C. Paul Morris, Chun Huai Luo, Adannaya Amadi, Matthew Schwartz, Heba H. Mostafa |
| EPI_ISL_1405599, EPI_ISL_1405600, EPI_ISL_1405601, EPI_ISL_1405602, EPI_ISL_1405603, EPI_ISL_1405604, EPI_ISL_1405605, EPI_ISL_1405606, EPI_ISL_1405607, EPI_ISL_1405608, EPI_ISL_1405609, EPI_ISL_1405610, EPI_ISL_1405611, EPI_ISL_1405612, EPI_ISL_1405613, EPI_ISL_1405614, EPI_ISL_1405615, EPI_ISL_1405616, EPI_ISL_1405617, EPI_ISL_1405618, EPI_ISL_1405619, EPI_ISL_1405620, EPI_ISL_1405621, EPI_ISL_1405622, EPI_ISL_1405623, EPI_ISL_1405624, EPI_ISL_1405625, EPI_ISL_1405626, EPI_ISL_1405627, EPI_ISL_1405628, EPI_ISL_1405629, EPI_ISL_1405630, EPI_ISL_1405631, EPI_ISL_1405632, EPI_ISL_1405633, EPI_ISL_1405634, EPI_ISL_1405635, EPI_ISL_1405636, EPI_ISL_1405637, EPI_ISL_1405638, EPI_ISL_1405639, EPI_ISL_1405640, EPI_ISL_1405641, EPI_ISL_1405642, EPI_ISL_1405643, EPI_ISL_1405644, EPI_ISL_1405645, EPI_ISL_1405646, EPI_ISL_1405647, EPI_ISL_1405648, EPI_ISL_1405649, EPI_ISL_1405650, EPI_ISL_1405651, EPI_ISL_1405652, EPI_ISL_1405653, EPI_ISL_1405654, EPI_ISL_1405655, EPI_ISL_1405657, EPI_ISL_1405658, EPI_ISL_1405659, EPI_ISL_1405660, EPI_ISL_1405661, EPI_ISL_1405662, EPI_ISL_1405663, EPI_ISL_1405664, EPI_ISL_1405666, EPI_ISL_1405667, EPI_ISL_1405668, EPI_ISL_1405669, EPI_ISL_1405670, EPI_ISL_1405671, EPI_ISL_1405672, EPI_ISL_1405673, EPI_ISL_1405674, EPI_ISL_1405675, EPI_ISL_1405676, EPI_ISL_1405678, EPI_ISL_1405679 | | | |
| see above | UCSC Genomics Institute | UCSC Genomics Institute | Mark Akeson, Ikenna Anigbogu, Eric Beraut, Isabel Bjork, Molly Cassatt-Johnstone, Terren Chang, Russell Corbett-Detig, Namrita Dhillon, Maximilian Haeussler, David Haussler, Angie Hinrichs, Miten Jain, Joshua Kapp, A. Marm Kilpatrick, Jakob McBroome, Hugh Olsen, Jeremy Sanford, Beth Shapiro, Michael Stone, Bryan Thornlow, Yatish Turakhia |
| EPI_ISL_1405712, EPI_ISL_1405714, EPI_ISL_1405715, EPI_ISL_1405716, EPI_ISL_1405717, EPI_ISL_1405718, EPI_ISL_1405719, EPI_ISL_1405720, EPI_ISL_1405721, EPI_ISL_1405722, EPI_ISL_1405723, EPI_ISL_1405724, EPI_ISL_1405727, EPI_ISL_1405728 | | | |
| see above | Idaho Bureau of Laboratories | Idaho Bureau of Laboratories | R. Beukelman, Matthew Charles Burns, Aimee Ceniseros, Robert L. Voermans, Christopher Ball |
| EPI_ISL_1405729, EPI_ISL_1405730, EPI_ISL_1405731, EPI_ISL_1405732, EPI_ISL_1405733, EPI_ISL_1405734, EPI_ISL_1405735, EPI_ISL_1405736, EPI_ISL_1405737, EPI_ISL_1405738, EPI_ISL_1405739, EPI_ISL_1405740, EPI_ISL_1405741, EPI_ISL_1405742, EPI_ISL_1405743, EPI_ISL_1405745, EPI_ISL_1405746, EPI_ISL_1405747, EPI_ISL_1405748, EPI_ISL_1405749, EPI_ISL_1405750, EPI_ISL_1405751, EPI_ISL_1405752, EPI_ISL_1405753, EPI_ISL_1405754, EPI_ISL_1405755, EPI_ISL_1405756, EPI_ISL_1405757, EPI_ISL_1405759, EPI_ISL_1405760, EPI_ISL_1405761, EPI_ISL_1405762, EPI_ISL_1405763, EPI_ISL_1405764, EPI_ISL_1405765, EPI_ISL_1405766, EPI_ISL_1405767, EPI_ISL_1405768, EPI_ISL_1405769, EPI_ISL_1405770, EPI_ISL_1405771, EPI_ISL_1405772, EPI_ISL_1405773, EPI_ISL_1405774, EPI_ISL_1405775, EPI_ISL_1405776, EPI_ISL_1405777, EPI_ISL_1405778, EPI_ISL_1405779, EPI_ISL_1405780, EPI_ISL_1405782, EPI_ISL_1405783, EPI_ISL_1405784, EPI_ISL_1405785, EPI_ISL_1405787, EPI_ISL_1405788, EPI_ISL_1405789, EPI_ISL_1405790, EPI_ISL_1405791, EPI_ISL_1405792, EPI_ISL_1405793, EPI_ISL_1405794, EPI_ISL_1405795, EPI_ISL_1405796, EPI_ISL_1405797, EPI_ISL_1405798, EPI_ISL_1405799, EPI_ISL_1405800, EPI_ISL_1405801, EPI_ISL_1405802, EPI_ISL_1405803, EPI_ISL_1405804, EPI_ISL_1405805, EPI_ISL_1405806, EPI_ISL_1405807, EPI_ISL_1405808, EPI_ISL_1405809, EPI_ISL_1405810, EPI_ISL_1405811, EPI_ISL_1405812, EPI_ISL_1405813 | | | |
| see above | Texas Department of State Health Services (TXDSHS) | Texas Department of State Health Services (TXDSHS) | Rashmi Tuladhar, Bonnie Oh, Jenny Zhang, Maliha Rahman, Mayela Pedrueza, Anita Pokharel, Lorraine Rodriguez, Myong Koag, Chun Wang, Rachel Lee, Grace Kubin |
| EPI_ISL_1405814, EPI_ISL_1405815, EPI_ISL_1405816, EPI_ISL_1405817, EPI_ISL_1405818, EPI_ISL_1405819, EPI_ISL_1405820, EPI_ISL_1405821, EPI_ISL_1405822, EPI_ISL_1405823, EPI_ISL_1405824, EPI_ISL_1405825, EPI_ISL_1405826, EPI_ISL_1405827, EPI_ISL_1405828, EPI_ISL_1405829, EPI_ISL_1405830, EPI_ISL_1405831, EPI_ISL_1405832, EPI_ISL_1405833, EPI_ISL_1405834, EPI_ISL_1405835, EPI_ISL_1405836, EPI_ISL_1405837, EPI_ISL_1405838, EPI_ISL_1405840, EPI_ISL_1405841, EPI_ISL_1405842, EPI_ISL_1405843, EPI_ISL_1405844, EPI_ISL_1405845, EPI_ISL_1405847, EPI_ISL_1405849, EPI_ISL_1405850, EPI_ISL_1405851, EPI_ISL_1405852, EPI_ISL_1405853, EPI_ISL_1405854, EPI_ISL_1405855, EPI_ISL_1405856, EPI_ISL_1405857, EPI_ISL_1405858, EPI_ISL_1405859, EPI_ISL_1405861, EPI_ISL_1405862, EPI_ISL_1405863, EPI_ISL_1405864, EPI_ISL_1405865, EPI_ISL_1405866, EPI_ISL_1405867, EPI_ISL_1405868, EPI_ISL_1405869, EPI_ISL_1405870, EPI_ISL_1405871, EPI_ISL_1405872, EPI_ISL_1405873, EPI_ISL_1405874, EPI_ISL_1405875, EPI_ISL_1405876, EPI_ISL_1405877, EPI_ISL_1405878, EPI_ISL_1405879, EPI_ISL_1405880, EPI_ISL_1405881, EPI_ISL_1405882, EPI_ISL_1405883, EPI_ISL_1405884, EPI_ISL_1405885, EPI_ISL_1405886, EPI_ISL_1405887, EPI_ISL_1405888, EPI_ISL_1405889, EPI_ISL_1405890, EPI_ISL_1405891, EPI_ISL_1405892, EPI_ISL_1405893, EPI_ISL_1405894, EPI_ISL_1405895, EPI_ISL_1405896, EPI_ISL_1405897 | | | |
| see above | UCSC Genomics Institute | UCSC Genomics Institute | Mark Akeson, Ikenna Anigbogu, Eric Beraut, Isabel Bjork, Molly Cassatt-Johnstone, Terren Chang, Russell Corbett-Detig, Namrita Dhillon, Maximilian Haeussler, David Haussler, Angie Hinrichs, Miten Jain, Joshua Kapp, A. Marm Kilpatrick, Jakob McBroome, Hugh Olsen, Jeremy Sanford, Beth Shapiro, Michael Stone, Bryan Thornlow, Yatish Turakhia |
| EPI_ISL_1405899 | LESP Morelos | Instituto de Diagnostico y Referencia Epidemiologicos (INDRE) | Claudia Wong-Arambula, Abril Rodriguez-Maldonado, Vanessa Rivero-Arredondo, Ariadna Medina-Benitez, Joaquin Quiroz-Mercado, Sergio Rangel-Guerrero, Natividad Cruz-Ortiz, Tatiana Nunez-Garcia, Gisela Barrera-Badillo, Lucia Hernandez-Rivas, Irma Lopez-Martinez, Ernesto Ramirez-Gonzalez. |
| EPI_ISL_1405900, EPI_ISL_1405901, EPI_ISL_1405902, EPI_ISL_1405903, EPI_ISL_1405904 | LESP Guanajuato | Instituto de Diagnostico y Referencia Epidemiologicos (INDRE) | Claudia Wong-Arambula, Abril Rodriguez-Maldonado, Vanessa Rivero-Arredondo, Ariadna Medina-Benitez, Joaquin Quiroz-Mercado, Sergio Rangel-Guerrero, Natividad Cruz-Ortiz, Tatiana Nunez-Garcia, Gisela Barrera-Badillo, Lucia Hernandez-Rivas, Irma Lopez-Martinez, Ernesto Ramirez-Gonzalez. |
| EPI_ISL_1405905 | LESP Chihuahua | Instituto de Diagnostico y Referencia Epidemiologicos (INDRE) | Claudia Wong-Arambula, Abril Rodriguez-Maldonado, Vanessa Rivero-Arredondo, Ariadna Medina-Benitez, Joaquin Quiroz-Mercado, Sergio Rangel-Guerrero, Natividad Cruz-Ortiz, Tatiana Nunez-Garcia, Gisela Barrera-Badillo, Lucia Hernandez-Rivas, Irma Lopez-Martinez, Ernesto Ramirez-Gonzalez. |
| EPI_ISL_1405906, EPI_ISL_1405907 | LESP Guanajuato | Instituto de Diagnostico y Referencia Epidemiologicos (INDRE) | Claudia Wong-Arambula, Abril Rodriguez-Maldonado, Vanessa Rivero-Arredondo, Ariadna Medina-Benitez, Joaquin Quiroz-Mercado, Sergio Rangel-Guerrero, Natividad Cruz-Ortiz, Tatiana Nunez-Garcia, Gisela Barrera-Badillo, Lucia Hernandez-Rivas, Irma Lopez-Martinez, Ernesto Ramirez-Gonzalez. |
| EPI_ISL_1405908 | LESP Jalisco | Instituto de Diagnostico y Referencia Epidemiologicos (INDRE) | Claudia Wong-Arambula, Abril Rodriguez-Maldonado, Vanessa Rivero-Arredondo, Ariadna Medina-Benitez, Joaquin Quiroz-Mercado, Sergio Rangel-Guerrero, Natividad Cruz-Ortiz, Tatiana Nunez-Garcia, Gisela Barrera-Badillo, Lucia Hernandez-Rivas, Irma Lopez-Martinez, Ernesto Ramirez-Gonzalez. |
| EPI_ISL_1405909, EPI_ISL_1405910 | LESP Guanajuato | Instituto de Diagnostico y Referencia Epidemiologicos (INDRE) | Claudia Wong-Arambula, Abril Rodriguez-Maldonado, Vanessa Rivero-Arredondo, Ariadna Medina-Benitez, Joaquin Quiroz-Mercado, Sergio Rangel-Guerrero, Natividad Cruz-Ortiz, Tatiana Nunez-Garcia, Gisela Barrera-Badillo, Lucia Hernandez-Rivas, Irma Lopez-Martinez, Ernesto Ramirez-Gonzalez. |
| EPI_ISL_1405911 | LESP Jalisco | Instituto de Diagnostico y Referencia Epidemiologicos (INDRE) | Claudia Wong-Arambula, Abril Rodriguez-Maldonado, Vanessa Rivero-Arredondo, Ariadna Medina-Benitez, Joaquin Quiroz-Mercado, Sergio Rangel-Guerrero, Natividad Cruz-Ortiz, Tatiana Nunez-Garcia, Gisela Barrera-Badillo, Lucia Hernandez-Rivas, Irma Lopez-Martinez, Ernesto Ramirez-Gonzalez. |
| EPI_ISL_1405912 | LESP Guanajuato | Instituto de Diagnostico y Referencia Epidemiologicos (INDRE) | Claudia Wong-Arambula, Abril Rodriguez-Maldonado, Vanessa Rivero-Arredondo, Ariadna Medina-Benitez, Joaquin Quiroz-Mercado, Sergio Rangel-Guerrero, Natividad Cruz-Ortiz, Tatiana Nunez-Garcia, Gisela Barrera-Badillo, Lucia Hernandez-Rivas, Irma Lopez-Martinez, Ernesto Ramirez-Gonzalez. |
| EPI_ISL_1405913 | LESP Jalisco | Instituto de Diagnostico y Referencia Epidemiologicos (INDRE) | Claudia Wong-Arambula, Abril Rodriguez-Maldonado, Vanessa Rivero-Arredondo, Ariadna Medina-Benitez, Joaquin Quiroz-Mercado, Sergio Rangel-Guerrero, Natividad Cruz-Ortiz, Tatiana Nunez-Garcia, Gisela Barrera-Badillo, Lucia Hernandez-Rivas, Irma Lopez-Martinez, Ernesto Ramirez-Gonzalez. |
| EPI_ISL_1405914, EPI_ISL_1405915 | LESP Guanajuato | Instituto de Diagnostico y Referencia Epidemiologicos (INDRE) | Claudia Wong-Arambula, Abril Rodriguez-Maldonado, Vanessa Rivero-Arredondo, Ariadna Medina-Benitez, Joaquin Quiroz-Mercado, Sergio Rangel-Guerrero, Natividad Cruz-Ortiz, Tatiana Nunez-Garcia, Gisela Barrera-Badillo, Lucia Hernandez-Rivas, Irma Lopez-Martinez, Ernesto Ramirez-Gonzalez. |
| EPI_ISL_1405918, EPI_ISL_1405920, EPI_ISL_1405922 | Arizona State University | Arizona State University | Peter T. Skidmore, LaRinda A. Holland, Rabia Maqsood, Emily A. Kaelin, Nicholas J. Mellor, Kristina Buss, Joy M. Blain, Neal W Woodbury, Valerie Harris, Joshua LaBaer, Vel Murugan, Efrem S. Lim |
| EPI_ISL_1405940, EPI_ISL_1405941, EPI_ISL_1405942, EPI_ISL_1405943, EPI_ISL_1405944, EPI_ISL_1405945 | Oregon State University TRACE Project | OSU Center for Genome Research and Biocomputing | Oregon State University TRACE Project |
| EPI_ISL_1406083, EPI_ISL_1406084, EPI_ISL_1406085, EPI_ISL_1406087, EPI_ISL_1406088, EPI_ISL_1406089, EPI_ISL_1406090, EPI_ISL_1406091, EPI_ISL_1406092, EPI_ISL_1406093, EPI_ISL_1406094, EPI_ISL_1406095, EPI_ISL_1406096, EPI_ISL_1406097, EPI_ISL_1406098, EPI_ISL_1406099, EPI_ISL_1406100, EPI_ISL_1406102, EPI_ISL_1406103, EPI_ISL_1406104, EPI_ISL_1406105, EPI_ISL_1406107, EPI_ISL_1406108, EPI_ISL_1406109, EPI_ISL_1406110, EPI_ISL_1406111, EPI_ISL_1406112, EPI_ISL_1406113, EPI_ISL_1406114, EPI_ISL_1406116, EPI_ISL_1406117, EPI_ISL_1406118, EPI_ISL_1406119, EPI_ISL_1406120, EPI_ISL_1406121, EPI_ISL_1406122, EPI_ISL_1406123, EPI_ISL_1406124, EPI_ISL_1406125, EPI_ISL_1406126, EPI_ISL_1406127, EPI_ISL_1406128, EPI_ISL_1406129, EPI_ISL_1406130, EPI_ISL_1406131, EPI_ISL_1406132, EPI_ISL_1406133, EPI_ISL_1406134, EPI_ISL_1406135, EPI_ISL_1406136, EPI_ISL_1406137, EPI_ISL_1406138, EPI_ISL_1406139, EPI_ISL_1406140, EPI_ISL_1406141, EPI_ISL_1406142 | | | |
| see above | Instituto Nacional de Medicina Genomica | Instituto Nacional de Medicina Genomica | Hidalgo-Miranda A, Mendoza-Vargas A, Reyes-Grajeda JP, Cedro-Tanda A, Alcaraz N, Gonzalez-Barrera D, Rangel-DeLeon D, Miranda-Ortiz H, Rosas-Escobar P, Canseco Mendez JC, Ramirez-Vega O, Munguia-Garza P, Garcia-Cardenas FJ, Gonzalez-Woge MA, Herrera-Montalvo LA |
| EPI_ISL_1406155, EPI_ISL_1406156, EPI_ISL_1406157, EPI_ISL_1406158, EPI_ISL_1406159, EPI_ISL_1406160, EPI_ISL_1406161, EPI_ISL_1406162, EPI_ISL_1406163, EPI_ISL_1406164, EPI_ISL_1406165, EPI_ISL_1406166, EPI_ISL_1406167, EPI_ISL_1406168, EPI_ISL_1406169, EPI_ISL_1406170, EPI_ISL_1406171, EPI_ISL_1406172, EPI_ISL_1406173, EPI_ISL_1406174, EPI_ISL_1406175 | | | |
| see above | Santa Clara County Public Health Laboratory | Santa Clara County Public Health Laboratory | Santa Clara County Public Health Department |
| EPI_ISL_1406205, EPI_ISL_1406206, EPI_ISL_1406207, EPI_ISL_1406208, EPI_ISL_1406210, EPI_ISL_1406211, EPI_ISL_1406212, EPI_ISL_1406213, EPI_ISL_1406214, EPI_ISL_1406216, EPI_ISL_1406217, EPI_ISL_1406218, EPI_ISL_1406219, EPI_ISL_1406222, EPI_ISL_1406223, EPI_ISL_1406224, EPI_ISL_1406225, EPI_ISL_1406227, EPI_ISL_1406228, EPI_ISL_1406229, EPI_ISL_1406230, EPI_ISL_1406231, EPI_ISL_1406233, EPI_ISL_1406234, EPI_ISL_1406235, EPI_ISL_1406237, EPI_ISL_1406238 | | | |
| see above | Wyoming Public Health Laboratory | Wyoming Public Health Laboratory | Jim Mildenberger, Wanda Manley, Noah Hull, Taylor Fearing, Lynette Gumbleton, Channing Weber, Ashley Norberg, Chayse Rowley, Marley Goetz, Brian Dominguez, Elliot Thomasson, Cari Sloma, and Rob Christensen |
| EPI_ISL_1406251, EPI_ISL_1406252, EPI_ISL_1406253, EPI_ISL_1406254, EPI_ISL_1406255, EPI_ISL_1406256, EPI_ISL_1406257, EPI_ISL_1406258, EPI_ISL_1406259, EPI_ISL_1406260, EPI_ISL_1406261, EPI_ISL_1406262, EPI_ISL_1406263, EPI_ISL_1406264, EPI_ISL_1406265, EPI_ISL_1406266, EPI_ISL_1406267, EPI_ISL_1406268, EPI_ISL_1406269, EPI_ISL_1406270, EPI_ISL_1406271 | | | |
| see above | University of Michigan Clinical Microbiology Laboratory | Lauring Lab, University of Michigan, Department of Microbiology and Immunology | Valesano |
| EPI_ISL_1406697, EPI_ISL_1406698, EPI_ISL_1406699, EPI_ISL_1406700, EPI_ISL_1406701, EPI_ISL_1406702, EPI_ISL_1406704 | Massachusetts State Public Health Laboratory | Massachusetts State Public Health Laboratory | Andrew Lang, Timelia Fink, Glen Gallagher, Sandra Smole |
| EPI_ISL_1406705, EPI_ISL_1406706, EPI_ISL_1406707, EPI_ISL_1406708, EPI_ISL_1406709, EPI_ISL_1406710, EPI_ISL_1406711, EPI_ISL_1406712, EPI_ISL_1406713, EPI_ISL_1406715, EPI_ISL_1406716, EPI_ISL_1406717, EPI_ISL_1406718, EPI_ISL_1406719, EPI_ISL_1406720, EPI_ISL_1406721 | | | |
| see above | Wyoming Public Health Laboratory | Wyoming Public Health Laboratory | Jim Mildenberger, Wanda Manley, Noah Hull, Taylor Fearing, Lynette Gumbleton, Channing Weber, Ashley Norberg, Chayse Rowley, Marley Goetz, Brian Dominguez, Elliot Thomasson, Cari Sloma, and Rob Christensen |
| EPI_ISL_1406722, EPI_ISL_1406723, EPI_ISL_1406724, EPI_ISL_1406725, EPI_ISL_1406726, EPI_ISL_1406727, EPI_ISL_1406728, EPI_ISL_1406729, EPI_ISL_1406730, EPI_ISL_1406731, EPI_ISL_1406732, EPI_ISL_1406733, EPI_ISL_1406734, EPI_ISL_1406735, EPI_ISL_1406736, EPI_ISL_1406737, EPI_ISL_1406738, EPI_ISL_1406739, EPI_ISL_1406740, EPI_ISL_1406741, EPI_ISL_1406742, EPI_ISL_1406743, EPI_ISL_1406744, EPI_ISL_1406745, EPI_ISL_1406746, EPI_ISL_1406747, EPI_ISL_1406748, EPI_ISL_1406749, EPI_ISL_1406750, EPI_ISL_1406751, EPI_ISL_1406752, EPI_ISL_1406753, EPI_ISL_1406754, EPI_ISL_1406755, EPI_ISL_1406756, EPI_ISL_1406757, EPI_ISL_1406758, EPI_ISL_1406759, EPI_ISL_1406760, EPI_ISL_1406761, EPI_ISL_1406762, EPI_ISL_1406763, EPI_ISL_1406764, EPI_ISL_1406765, EPI_ISL_1406766, EPI_ISL_1406767, EPI_ISL_1406768, EPI_ISL_1406769, EPI_ISL_1406770, EPI_ISL_1406771, EPI_ISL_1406772, EPI_ISL_1406773, EPI_ISL_1406774, EPI_ISL_1406775, EPI_ISL_1406776, EPI_ISL_1406777 | | | |
| see above | Instituto Nacional de Medicina Genomica | Instituto Nacional de Enfermedades Respiratorias (INER), Centro de Investigación en Enfermedades Infecciosas (CIENI) | Matias-Florentino M, Hidalgo-Miranda A, Mendoza-Vargas A, Reyes-Grajeda JP, Cedro-Tanda A, Arriaga-Canon C, Herrera-Montalvo LA, Reyes-Teran G, Avila-Rios S, Boukadida C |
| EPI_ISL_1407256, EPI_ISL_1407257, EPI_ISL_1407258, EPI_ISL_1407259, EPI_ISL_1407260, EPI_ISL_1407261, EPI_ISL_1407262, EPI_ISL_1407263, EPI_ISL_1407264, EPI_ISL_1407265, EPI_ISL_1407267, EPI_ISL_1407268, EPI_ISL_1407269, EPI_ISL_1407270, EPI_ISL_1407272, EPI_ISL_1407273, EPI_ISL_1407274, EPI_ISL_1407275, EPI_ISL_1407276, EPI_ISL_1407277, EPI_ISL_1407278, EPI_ISL_1407279, EPI_ISL_1407280, EPI_ISL_1407281, EPI_ISL_1407282, EPI_ISL_1407283, EPI_ISL_1407284, EPI_ISL_1407285, EPI_ISL_1407286, EPI_ISL_1407287, EPI_ISL_1407288, EPI_ISL_1407289, EPI_ISL_1407290, EPI_ISL_1407292, EPI_ISL_1407293, EPI_ISL_1407294, EPI_ISL_1407295, EPI_ISL_1407296, EPI_ISL_1407298, EPI_ISL_1407299, EPI_ISL_1407300, EPI_ISL_1407301, EPI_ISL_1407302, EPI_ISL_1407303, EPI_ISL_1407304, EPI_ISL_1407305, EPI_ISL_1407306, EPI_ISL_1407307, EPI_ISL_1407308, EPI_ISL_1407309, EPI_ISL_1407310, EPI_ISL_1407311, EPI_ISL_1407312, EPI_ISL_1407313, EPI_ISL_1407314, EPI_ISL_1407315, EPI_ISL_1407316, EPI_ISL_1407317, EPI_ISL_1407318, EPI_ISL_1407319, EPI_ISL_1407320, EPI_ISL_1407321, EPI_ISL_1407322, EPI_ISL_1407323, EPI_ISL_1407325, EPI_ISL_1407326, EPI_ISL_1407327, EPI_ISL_1407328, EPI_ISL_1407329, EPI_ISL_1407330, EPI_ISL_1407331, EPI_ISL_1407332, EPI_ISL_1407333 | | | |
| see above | Broad Institute Clinical Research Sequencing Platform | Infectious Disease Program, Broad Institute of Harvard and MIT | Siddle,K.J., Adams,G., Pearlman,L., Gladden-Young,A., Vicente,G., Blumenstiel,B., DeFelice,M., Lee,M., McGovern,S., Lagerborg,K., Rudy,M., DeRuff,K., Carter,A., Normandin,E., Bauer,M., Reilly,S., Tomkins-Tinch,C., Loreth,C., Chaluvadi,S., Meldrim,J., Granger,B., Lemieux,J.E., Birren,B.W., Sabeti,P.C., Larkin,K., Dodge,S., Lennon,N., Madoff,L., Brown,C., Gallagher,G., Smole,S., Park,D.J., Gabriel,S., and MacInnis,B.L. |
| EPI_ISL_1407334, EPI_ISL_1407335, EPI_ISL_1407336, EPI_ISL_1407337, EPI_ISL_1407338, EPI_ISL_1407339, EPI_ISL_1407340, EPI_ISL_1407341, EPI_ISL_1407342, EPI_ISL_1407343, EPI_ISL_1407344, EPI_ISL_1407345, EPI_ISL_1407346, EPI_ISL_1407347, EPI_ISL_1407348, EPI_ISL_1407349, EPI_ISL_1407350, EPI_ISL_1407351, EPI_ISL_1407352, EPI_ISL_1407353, EPI_ISL_1407354, EPI_ISL_1407355, | | | |

| | | | |
|---|---|---|---|
| EPI_ISL_1407356, EPI_ISL_1407357, EPI_ISL_1407358, EPI_ISL_1407359, EPI_ISL_1407360, EPI_ISL_1407361, EPI_ISL_1407362, EPI_ISL_1407363, EPI_ISL_1407364, EPI_ISL_1407365, EPI_ISL_1407366, EPI_ISL_1407367, EPI_ISL_1407368, EPI_ISL_1407370, EPI_ISL_1407372, EPI_ISL_1407373, EPI_ISL_1407374, EPI_ISL_1407375, EPI_ISL_1407376, EPI_ISL_1407377, EPI_ISL_1407378, EPI_ISL_1407379, EPI_ISL_1407380, EPI_ISL_1407381, EPI_ISL_1407382, EPI_ISL_1407383, EPI_ISL_1407384, EPI_ISL_1407385, EPI_ISL_1407386, EPI_ISL_1407387, EPI_ISL_1407388, EPI_ISL_1407389, EPI_ISL_1407390, EPI_ISL_1407391, EPI_ISL_1407392, EPI_ISL_1407393, EPI_ISL_1407395, EPI_ISL_1407396, EPI_ISL_1407397, EPI_ISL_1407398, EPI_ISL_1407399, EPI_ISL_1407400, EPI_ISL_1407401, EPI_ISL_1407402, EPI_ISL_1407403, EPI_ISL_1407404, EPI_ISL_1407405, EPI_ISL_1407406, EPI_ISL_1407407, EPI_ISL_1407408, EPI_ISL_1407409, EPI_ISL_1407410, EPI_ISL_1407411, EPI_ISL_1407413, EPI_ISL_1407414, EPI_ISL_1407415, EPI_ISL_1407416, EPI_ISL_1407417, EPI_ISL_1407418, EPI_ISL_1407419, EPI_ISL_1407420, EPI_ISL_1407421, EPI_ISL_1407422, EPI_ISL_1407423, EPI_ISL_1407424, EPI_ISL_1407425, EPI_ISL_1407426, EPI_ISL_1407427, EPI_ISL_1407428, EPI_ISL_1407429, EPI_ISL_1407431, EPI_ISL_1407432, EPI_ISL_1407433, EPI_ISL_1407434, EPI_ISL_1407435, EPI_ISL_1407436, EPI_ISL_1407437, EPI_ISL_1407438, EPI_ISL_1407439, EPI_ISL_1407440, EPI_ISL_1407441, EPI_ISL_1407442, EPI_ISL_1407443, EPI_ISL_1407444, EPI_ISL_1407445, EPI_ISL_1407446, EPI_ISL_1407447, EPI_ISL_1407448, EPI_ISL_1407449, EPI_ISL_1407450, EPI_ISL_1407452, EPI_ISL_1407453, EPI_ISL_1407454, EPI_ISL_1407455, EPI_ISL_1407456, EPI_ISL_1407457 | | | |
| see above | Rhode Island Department of Health | Infectious Disease Program, Broad Institute of Harvard and MIT | Siddle,K.J., Azevedo,K., Miller,A., Adams,G., Pearlman,L., Gladden-Young,A., Lagerborg,K., Rudy,M., DeRuff,K., Carter,A., Normandin,E., Bauer,M., Reilly,S., Tomkins-Tinch,C., Loreth,C., Chaluvadi,S., Lemieux,J.E., Birren,B.W., Sabeti,P.C., Huard,R., King,E., Park,D.J., and MacInnis,B.L. |
| EPI_ISL_1408887 | MT Public Health Laboratory | Genomics and Discovery, Respiratory Viruses Branch, Division of Viral Diseases, Centers for Disease Control and Prevention | Ying Tao, Jing Zhang, Yan Li, Brian Lynch, Anna Kelleher, Krista Queen, Anna Uehara, Peter Cook, Han Jia Justin Ng, Clinton R. Paden, Haibin Wang, Suxiang Tong |
| EPI_ISL_1408888, EPI_ISL_1408889 | TN Division of Laboratory Services | Genomics and Discovery, Respiratory Viruses Branch, Division of Viral Diseases, Centers for Disease Control and Prevention | Ying Tao, Jing Zhang, Yan Li, Brian Lynch, Anna Kelleher, Krista Queen, Anna Uehara, Peter Cook, Han Jia Justin Ng, Clinton R. Paden, Haibin Wang, Suxiang Tong |
| EPI_ISL_1408890 | "PA Department of Health, Bureau of Laboratories" | Genomics and Discovery, Respiratory Viruses Branch, Division of Viral Diseases, Centers for Disease Control and Prevention | Ying Tao, Jing Zhang, Yan Li, Brian Lynch, Anna Kelleher, Krista Queen, Anna Uehara, Peter Cook, Han Jia Justin Ng, Clinton R. Paden, Haibin Wang, Suxiang Tong |
| EPI_ISL_1408891 | GA Department of Public Health Laboratory | Genomics and Discovery, Respiratory Viruses Branch, Division of Viral Diseases, Centers for Disease Control and Prevention | Ying Tao, Jing Zhang, Yan Li, Brian Lynch, Anna Kelleher, Krista Queen, Anna Uehara, Peter Cook, Han Jia Justin Ng, Clinton R. Paden, Haibin Wang, Suxiang Tong |
| EPI_ISL_1408892 | NH Dept. of Health and Human Services Public Health Labs | Genomics and Discovery, Respiratory Viruses Branch, Division of Viral Diseases, Centers for Disease Control and Prevention | Ying Tao, Jing Zhang, Yan Li, Brian Lynch, Anna Kelleher, Krista Queen, Anna Uehara, Peter Cook, Han Jia Justin Ng, Clinton R. Paden, Haibin Wang, Suxiang Tong |
| EPI_ISL_1408893 | MT Public Health Laboratory | Genomics and Discovery, Respiratory Viruses Branch, Division of Viral Diseases, Centers for Disease Control and Prevention | Ying Tao, Jing Zhang, Yan Li, Brian Lynch, Anna Kelleher, Krista Queen, Anna Uehara, Peter Cook, Han Jia Justin Ng, Clinton R. Paden, Haibin Wang, Suxiang Tong |
| EPI_ISL_1408894 | ME Health and Environmental Testing Laboratory | Genomics and Discovery, Respiratory Viruses Branch, Division of Viral Diseases, Centers for Disease Control and Prevention | Ying Tao, Jing Zhang, Yan Li, Brian Lynch, Anna Kelleher, Krista Queen, Anna Uehara, Peter Cook, Han Jia Justin Ng, Clinton R. Paden, Haibin Wang, Suxiang Tong |
| EPI_ISL_1408895 | GA Department of Public Health Laboratory | Genomics and Discovery, Respiratory Viruses Branch, Division of Viral Diseases, Centers for Disease Control and Prevention | Ying Tao, Jing Zhang, Yan Li, Brian Lynch, Anna Kelleher, Krista Queen, Anna Uehara, Peter Cook, Han Jia Justin Ng, Clinton R. Paden, Haibin Wang, Suxiang Tong |
| EPI_ISL_1408896 | IL Dept. of Public Health Springfield Laboratory | Genomics and Discovery, Respiratory Viruses Branch, Division of Viral Diseases, Centers for Disease Control and Prevention | Ying Tao, Jing Zhang, Yan Li, Brian Lynch, Anna Kelleher, Krista Queen, Anna Uehara, Peter Cook, Han Jia Justin Ng, Clinton R. Paden, Haibin Wang, Suxiang Tong |
| EPI_ISL_1408897 | MT Public Health Laboratory | Genomics and Discovery, Respiratory Viruses Branch, Division of Viral Diseases, Centers for Disease Control and Prevention | Ying Tao, Jing Zhang, Yan Li, Brian Lynch, Anna Kelleher, Krista Queen, Anna Uehara, Peter Cook, Han Jia Justin Ng, Clinton R. Paden, Haibin Wang, Suxiang Tong |
| EPI_ISL_1408898 | TN Division of Laboratory Services | Genomics and Discovery, Respiratory Viruses Branch, Division of Viral Diseases, Centers for Disease Control and Prevention | Ying Tao, Jing Zhang, Yan Li, Brian Lynch, Anna Kelleher, Krista Queen, Anna Uehara, Peter Cook, Han Jia Justin Ng, Clinton R. Paden, Haibin Wang, Suxiang Tong |
| EPI_ISL_1411004, EPI_ISL_1411005, EPI_ISL_1411006, EPI_ISL_1411007 | DC Public Health Lab/ Dept. of Forensic Sciences | DC Public Health Lab/ Dept. of Forensic Sciences | Janis Doss, Scott Nguyen, Elizabeth Zelaya, Sarah Scott, Connie Maza, Monica Mann, Brittany Hamilton, David Payne, Jocelyn Hauser |
| EPI_ISL_1412475, EPI_ISL_1412478, EPI_ISL_1412481, EPI_ISL_1412484, EPI_ISL_1412487, EPI_ISL_1412490, EPI_ISL_1412493, EPI_ISL_1412496, EPI_ISL_1412499, EPI_ISL_1412502, EPI_ISL_1412505, EPI_ISL_1412508, EPI_ISL_1412510, EPI_ISL_1412513, EPI_ISL_1412516, EPI_ISL_1412519, EPI_ISL_1412522, EPI_ISL_1412525, EPI_ISL_1412528, EPI_ISL_1412530 | | | |
| see above | Kansas Health and Environmental Lab | Kansas Health and Environmental Lab | Mike Grose, Jonathan Barnell, Ben Olsen, and Phil Adam |
| EPI_ISL_1413133, EPI_ISL_1413136, EPI_ISL_1413139 | Massachusetts General Hospital | Infectious Disease Program, Broad Institute of Harvard and MIT | Lemieux,J.E., Siddle,K.J., Shaw,B., Adams,G., Pierce,V., Turbett,S., Anahtar,M., Branda,J., Slater,D., Harris,J., Lin,A.E., Gladden-Young,A., Lagerborg,K., Rudy,M., DeRuff,K., Carter,A., Normandin,E., Bauer,M., Reilly,S., Tomkins-Tinch,C., Loreth,C., Chaluvadi,S., Neumann,A., Cusick,C., Chapman,S.B., Gnirke,A., Flowers,K., Cerrato,F., Birren,B.W., Gallagher,G., Smole,S., Park,D.J., MacInnis,B.L., Ryan,E., LaRocque,R., Rosenberg,E. and Sabeti,P.C. |
| EPI_ISL_1413150, EPI_ISL_1413154, EPI_ISL_1413157, EPI_ISL_1413160, EPI_ISL_1413168, EPI_ISL_1413171, EPI_ISL_1413176, EPI_ISL_1413179, EPI_ISL_1413181, EPI_ISL_1413187, EPI_ISL_1413191, EPI_ISL_1413193, EPI_ISL_1413199, EPI_ISL_1413202, EPI_ISL_1413204, EPI_ISL_1413207, EPI_ISL_1413209, EPI_ISL_1413215, EPI_ISL_1413218, EPI_ISL_1413221, EPI_ISL_1413224, EPI_ISL_1413226, EPI_ISL_1413229, EPI_ISL_1413232, EPI_ISL_1413235, EPI_ISL_1413238, EPI_ISL_1413241, EPI_ISL_1413243, EPI_ISL_1413246, EPI_ISL_1413249, EPI_ISL_1413251, EPI_ISL_1413254, EPI_ISL_1413257, EPI_ISL_1413260, EPI_ISL_1413263, EPI_ISL_1413266, EPI_ISL_1413269, EPI_ISL_1413272, EPI_ISL_1413280, EPI_ISL_1413282, EPI_ISL_1413285, EPI_ISL_1413287, EPI_ISL_1413290, EPI_ISL_1413293, EPI_ISL_1413296, EPI_ISL_1413304, EPI_ISL_1413307, EPI_ISL_1413309, EPI_ISL_1413311, EPI_ISL_1413314, EPI_ISL_1413316, EPI_ISL_1413322, EPI_ISL_1413325, EPI_ISL_1413328, EPI_ISL_1413330, EPI_ISL_1413333, EPI_ISL_1413336, EPI_ISL_1413339, EPI_ISL_1413341, EPI_ISL_1413344, EPI_ISL_1413347, EPI_ISL_1413350, EPI_ISL_1413352, EPI_ISL_1413355, EPI_ISL_1413358, EPI_ISL_1413361, EPI_ISL_1413363, EPI_ISL_1413366, EPI_ISL_1413369, EPI_ISL_1413372, EPI_ISL_1413374, EPI_ISL_1413377, EPI_ISL_1413380, EPI_ISL_1413382, EPI_ISL_1413385, EPI_ISL_1413388, EPI_ISL_1413390, EPI_ISL_1413393, EPI_ISL_1413396, EPI_ISL_1413399, EPI_ISL_1413401, EPI_ISL_1413404, EPI_ISL_1413407, EPI_ISL_1413410, EPI_ISL_1413413, EPI_ISL_1413416, EPI_ISL_1413418, EPI_ISL_1413421, EPI_ISL_1413424, EPI_ISL_1413426, EPI_ISL_1413429, EPI_ISL_1413431, EPI_ISL_1413434, EPI_ISL_1413437, EPI_ISL_1413439, EPI_ISL_1413442, EPI_ISL_1413445, EPI_ISL_1413447, EPI_ISL_1413450, EPI_ISL_1413452, EPI_ISL_1413455, EPI_ISL_1413458, EPI_ISL_1413463, EPI_ISL_1413465, EPI_ISL_1413468, EPI_ISL_1413471, EPI_ISL_1413473, EPI_ISL_1413476, EPI_ISL_1413479, EPI_ISL_1413482, EPI_ISL_1413485, EPI_ISL_1413488, EPI_ISL_1413490, EPI_ISL_1413493, EPI_ISL_1413496, EPI_ISL_1413498, EPI_ISL_1413501, EPI_ISL_1413504, EPI_ISL_1413506, EPI_ISL_1413511 | | | |
| see above | Broad Institute Clinical Research Sequencing Platform | Infectious Disease Program, Broad Institute of Harvard and MIT | Siddle,K.J., Adams,G., Pearlman,L., Gladden-Young,A., Vicente,G., Blumenstiel,B., DeFelice,M., Lee,M., McGovern,S., Lagerborg,K., Rudy,M., DeRuff,K., Carter,A., Normandin,E., Bauer,M., Reilly,S., Tomkins-Tinch,C., Loreth,C., Chaluvadi,S., Meldrim,J., Granger,B., Lemieux,J.E., Birren,B.W., Sabeti,P.C., Larkin,K., Dodge,S., Lennon,N., Madoff,L., Brown,C., Gallagher,G., Smole,S., Park,D.J., Gabriel,S., and MacInnis,B.L. |
| EPI_ISL_1413514, EPI_ISL_1413516, EPI_ISL_1413519, EPI_ISL_1413522, EPI_ISL_1413525, EPI_ISL_1413528, EPI_ISL_1413530, EPI_ISL_1413533, EPI_ISL_1413536, EPI_ISL_1413538, EPI_ISL_1413542, EPI_ISL_1413544, EPI_ISL_1413547, EPI_ISL_1413549, EPI_ISL_1413551, EPI_ISL_1413554, EPI_ISL_1413556, EPI_ISL_1413559, EPI_ISL_1413561, EPI_ISL_1413563, EPI_ISL_1413566, EPI_ISL_1413568, EPI_ISL_1413571, EPI_ISL_1413574, EPI_ISL_1413576, EPI_ISL_1413579, EPI_ISL_1413582, EPI_ISL_1413585, EPI_ISL_1413587, EPI_ISL_1413589, EPI_ISL_1413592, EPI_ISL_1413594, EPI_ISL_1413597, EPI_ISL_1413600, EPI_ISL_1413602, EPI_ISL_1413605, EPI_ISL_1413607, EPI_ISL_1413610, EPI_ISL_1413613, EPI_ISL_1413615, EPI_ISL_1413618, EPI_ISL_1413621, EPI_ISL_1413623, EPI_ISL_1413626, EPI_ISL_1413628, EPI_ISL_1413631, EPI_ISL_1413634, EPI_ISL_1413637, EPI_ISL_1413639, EPI_ISL_1413642, EPI_ISL_1413645, EPI_ISL_1413647, EPI_ISL_1413651, EPI_ISL_1413653, EPI_ISL_1413656, EPI_ISL_1413658, EPI_ISL_1413664, EPI_ISL_1413667, EPI_ISL_1413672, EPI_ISL_1413678, EPI_ISL_1413680, EPI_ISL_1413685, EPI_ISL_1413687, EPI_ISL_1413696, EPI_ISL_1413702, EPI_ISL_1413704, EPI_ISL_1413707, EPI_ISL_1413710, EPI_ISL_1413712, EPI_ISL_1413715, EPI_ISL_1413726, EPI_ISL_1413733, EPI_ISL_1413736, EPI_ISL_1413742, EPI_ISL_1413748, EPI_ISL_1413750, EPI_ISL_1413755, EPI_ISL_1413761 | | | |
| see above | Rhode Island Department of Health | Infectious Disease Program, Broad Institute of Harvard and MIT | Siddle,K.J., Azevedo,K., Miller,A., Adams,G., Pearlman,L., Gladden-Young,A., Lagerborg,K., Rudy,M., DeRuff,K., Carter,A., Normandin,E., Bauer,M., Reilly,S., Tomkins-Tinch,C., Loreth,C., Chaluvadi,S., Lemieux,J.E., Birren,B.W., Sabeti,P.C., Huard,R., King,E., Park,D.J., and MacInnis,B.L. |
| EPI_ISL_1413769, EPI_ISL_1413771, EPI_ISL_1413781, EPI_ISL_1413784, EPI_ISL_1413787 | Warrior Diagnostics Laboratory | Infectious Disease Program, Broad Institute of Harvard and MIT | Lemieux,J.E., Siddle,K.J., Marshall,J., ONeill,M., Bronson,A., Adams,G., Gladden-Young,A., Lagerborg,K., Rudy,M., DeRuff,K., Carter,A., Normandin,E., Bauer,M., Reilly,S., Tomkins-Tinch,C., Loreth,C., Chaluvadi,S., Birren,B.W., Gallagher,G., Smole,S., Park,D.J., MacInnis,B.L., and Sabeti,P.C. |
| EPI_ISL_1413789, EPI_ISL_1413794, EPI_ISL_1413797, EPI_ISL_1413799, EPI_ISL_1413803, EPI_ISL_1413805, EPI_ISL_1413807, EPI_ISL_1413810, EPI_ISL_1413813, EPI_ISL_1413815, EPI_ISL_1413818, EPI_ISL_1413821, EPI_ISL_1413823, EPI_ISL_1413826, EPI_ISL_1413829, EPI_ISL_1413831, EPI_ISL_1413835, EPI_ISL_1413837, EPI_ISL_1413840, EPI_ISL_1413842, EPI_ISL_1413845, EPI_ISL_1413848, EPI_ISL_1413851, EPI_ISL_1413853, EPI_ISL_1413856, EPI_ISL_1413859, EPI_ISL_1413861, EPI_ISL_1413864 | | | |
| see above | Broad Institute Clinical Research Sequencing Platform | Infectious Disease Program, Broad Institute of Harvard and MIT | Siddle,K.J., Adams,G., Pearlman,L., Gladden-Young,A., Vicente,G., Blumenstiel,B., DeFelice,M., Lee,M., McGovern,S., Lagerborg,K., Rudy,M., DeRuff,K., Carter,A., Normandin,E., Bauer,M., Reilly,S., Tomkins-Tinch,C., Loreth,C., Chaluvadi,S., Meldrim,J., Granger,B., Lemieux,J.E., Birren,B.W., Sabeti,P.C., Larkin,K., Dodge,S., Lennon,N., Madoff,L., Brown,C., Gallagher,G., Smole,S., Park,D.J., Gabriel,S., and MacInnis,B.L. |
| EPI_ISL_1415061, EPI_ISL_1415062, EPI_ISL_1415063, EPI_ISL_1415064, EPI_ISL_1415065, EPI_ISL_1415066, EPI_ISL_1415067, EPI_ISL_1415068, EPI_ISL_1415070, EPI_ISL_1415071, EPI_ISL_1415072, EPI_ISL_1415073, EPI_ISL_1415074, EPI_ISL_1415075, EPI_ISL_1415077, EPI_ISL_1415078, EPI_ISL_1415079, EPI_ISL_1415080, EPI_ISL_1415082, EPI_ISL_1415083, EPI_ISL_1415084, EPI_ISL_1415085, EPI_ISL_1416193, EPI_ISL_1416194, EPI_ISL_1416195, EPI_ISL_1416196, EPI_ISL_1416197, EPI_ISL_1416198, EPI_ISL_1416199, EPI_ISL_1416200, EPI_ISL_1416201, EPI_ISL_1416202, EPI_ISL_1416203, EPI_ISL_1416204, EPI_ISL_1416205, EPI_ISL_1416206, EPI_ISL_1416207, EPI_ISL_1416208, EPI_ISL_1416209, EPI_ISL_1416210, EPI_ISL_1416211, EPI_ISL_1416212, EPI_ISL_1416213, EPI_ISL_1416214, EPI_ISL_1416215, EPI_ISL_1416216, EPI_ISL_1416217, EPI_ISL_1416218, EPI_ISL_1416219, EPI_ISL_1416221, EPI_ISL_1416223, EPI_ISL_1416224, EPI_ISL_1416225, EPI_ISL_1416226, EPI_ISL_1416227, EPI_ISL_1416228, EPI_ISL_1416229, EPI_ISL_1416230, EPI_ISL_1416231, EPI_ISL_1416232, EPI_ISL_1416233, EPI_ISL_1416234, EPI_ISL_1416235, EPI_ISL_1416236, EPI_ISL_1416237, EPI_ISL_1416238, EPI_ISL_1416239, EPI_ISL_1416240, EPI_ISL_1416241, EPI_ISL_1416242, EPI_ISL_1416243, EPI_ISL_1416244, EPI_ISL_1416245, EPI_ISL_1416246, EPI_ISL_1416247, EPI_ISL_1416248, EPI_ISL_1416249, EPI_ISL_1416250, EPI_ISL_1416251, EPI_ISL_1416252, EPI_ISL_1416253, EPI_ISL_1416254, EPI_ISL_1416255 | | | |
| see above | IN State Department of Health Laboratory Services | IN State Department of Health Laboratory Services | Cassandra Campion, Jamie Yeadon, Brian Pope, Lixia Liu, Kyle Brownlee, Melissa Hindenlang, Ankita Kashikar, Mark Glazier |
| EPI_ISL_1416256, EPI_ISL_1416257, EPI_ISL_1416258, EPI_ISL_1416259, EPI_ISL_1416260, EPI_ISL_1416261, EPI_ISL_1416262, EPI_ISL_1416263, EPI_ISL_1416264, EPI_ISL_1416265, EPI_ISL_1416266, EPI_ISL_1416267, EPI_ISL_1416268, EPI_ISL_1416269, EPI_ISL_1416270, EPI_ISL_1416271, EPI_ISL_1416272, EPI_ISL_1416273, EPI_ISL_1416274, EPI_ISL_1416275, EPI_ISL_1416276, EPI_ISL_1416277, EPI_ISL_1416278, EPI_ISL_1416279, EPI_ISL_1416280, EPI_ISL_1416281, EPI_ISL_1416282, EPI_ISL_1416283, EPI_ISL_1416284, EPI_ISL_1416285, EPI_ISL_1416286, EPI_ISL_1416287, EPI_ISL_1416288, EPI_ISL_1416289, EPI_ISL_1416290, EPI_ISL_1416291, EPI_ISL_1416292, EPI_ISL_1416293, EPI_ISL_1416294, EPI_ISL_1416295, EPI_ISL_1416296, EPI_ISL_1416297, EPI_ISL_1416298, EPI_ISL_1416299, EPI_ISL_1416300, EPI_ISL_1416301, EPI_ISL_1416302, EPI_ISL_1416303, EPI_ISL_1416304, EPI_ISL_1416305, EPI_ISL_1416306, EPI_ISL_1416307, EPI_ISL_1416308, EPI_ISL_1416309, EPI_ISL_1416310, EPI_ISL_1416311 | | | |
| see above | Oregon State University TRACE Project | OSU Center for Genome Research and Biocomputing | Oregon State University TRACE Project |
| EPI_ISL_1416327, EPI_ISL_1416328 | Montana Public Health Laboratory | Montana Public Health Laboratory | Joy Ritter, Michelle Mozer, Carrie Biskupiak, Deborah Gibson |
| EPI_ISL_1416332, EPI_ISL_1416333, EPI_ISL_1416334, EPI_ISL_1416335, EPI_ISL_1416336, EPI_ISL_1416337, EPI_ISL_1416338, EPI_ISL_1416339, EPI_ISL_1416340, EPI_ISL_1416341, EPI_ISL_1416342, EPI_ISL_1416343, EPI_ISL_1416344, EPI_ISL_1416345, EPI_ISL_1416346, EPI_ISL_1416347, EPI_ISL_1416348, EPI_ISL_1416349, EPI_ISL_1416350, EPI_ISL_1416351, EPI_ISL_1416352, EPI_ISL_1416353, EPI_ISL_1416354, EPI_ISL_1416355, EPI_ISL_1416356, EPI_ISL_1416357, EPI_ISL_1416358, EPI_ISL_1416359, EPI_ISL_1416360, EPI_ISL_1416361, EPI_ISL_1416362, EPI_ISL_1416363, EPI_ISL_1416364, EPI_ISL_1416365, EPI_ISL_1416366, EPI_ISL_1416367, EPI_ISL_1416368, EPI_ISL_1416369, EPI_ISL_1416370, EPI_ISL_1416371, EPI_ISL_1416372, EPI_ISL_1416373, EPI_ISL_1416374, EPI_ISL_1416375, EPI_ISL_1416376, EPI_ISL_1416377, EPI_ISL_1416378, EPI_ISL_1416379, EPI_ISL_1416380, EPI_ISL_1416381, EPI_ISL_1416382, EPI_ISL_1416383, EPI_ISL_1416384, EPI_ISL_1416385, EPI_ISL_1416386, EPI_ISL_1416387, EPI_ISL_1416388, EPI_ISL_1416389, EPI_ISL_1416390, EPI_ISL_1416391, EPI_ISL_1416392, EPI_ISL_1416393, EPI_ISL_1416394, EPI_ISL_1416395, EPI_ISL_1416396, EPI_ISL_1416397, EPI_ISL_1416398, EPI_ISL_1416399, EPI_ISL_1416400, EPI_ISL_1416401, EPI_ISL_1416402, EPI_ISL_1416403, EPI_ISL_1416404, EPI_ISL_1416405, EPI_ISL_1416406, EPI_ISL_1416407, EPI_ISL_1416408, EPI_ISL_1416409, EPI_ISL_1416410, EPI_ISL_1416411, EPI_ISL_1416412, EPI_ISL_1416413, EPI_ISL_1416414, EPI_ISL_1416415, EPI_ISL_1416416, EPI_ISL_1416417, EPI_ISL_1416418, EPI_ISL_1416419, EPI_ISL_1416420, EPI_ISL_1416421, EPI_ISL_1416422, EPI_ISL_1416423, EPI_ISL_1416424, EPI_ISL_1416425, EPI_ISL_1416426, EPI_ISL_1416427, EPI_ISL_1416428, EPI_ISL_1416429, EPI_ISL_1416430, EPI_ISL_1416431, EPI_ISL_1416432, EPI_ISL_1416433, EPI_ISL_1416434, EPI_ISL_1416435, EPI_ISL_1416436, EPI_ISL_1416437, EPI_ISL_1416438, EPI_ISL_1416439, EPI_ISL_1416440, EPI_ISL_1416441, EPI_ISL_1416442, EPI_ISL_1416443, EPI_ISL_1416444, EPI_ISL_1416445, EPI_ISL_1416446, EPI_ISL_1416447, EPI_ISL_1416448, EPI_ISL_1416449, EPI_ISL_1416450, EPI_ISL_1416451, EPI_ISL_1416452, EPI_ISL_1416453, EPI_ISL_1416454, EPI_ISL_1416455, EPI_ISL_1416456, EPI_ISL_1416457, EPI_ISL_1416458, EPI_ISL_1416459, EPI_ISL_1416461, EPI_ISL_1416462, EPI_ISL_1416463, EPI_ISL_1416464, EPI_ISL_1416465, EPI_ISL_1416466, EPI_ISL_1416467, EPI_ISL_1416468, EPI_ISL_1416469, EPI_ISL_1416470, EPI_ISL_1416471, EPI_ISL_1416472, EPI_ISL_1416473, EPI_ISL_1416474, EPI_ISL_1416475, EPI_ISL_1416476, EPI_ISL_1416477, EPI_ISL_1416478, EPI_ISL_1416479, EPI_ISL_1416480, EPI_ISL_1416481, EPI_ISL_1416482, EPI_ISL_1416483, EPI_ISL_1416484, EPI_ISL_1416485, EPI_ISL_1416486, EPI_ISL_1416487, EPI_ISL_1416488, EPI_ISL_1416489, EPI_ISL_1416490, EPI_ISL_1416491, EPI_ISL_1416492, EPI_ISL_1416493, EPI_ISL_1416494, EPI_ISL_1416495, EPI_ISL_1416496, EPI_ISL_1416497, EPI_ISL_1416498, EPI_ISL_1416499, EPI_ISL_1416500, EPI_ISL_1416501, EPI_ISL_1416502, EPI_ISL_1416503, EPI_ISL_1416504, EPI_ISL_1416505, EPI_ISL_1416506, EPI_ISL_1416507, EPI_ISL_1416508, EPI_ISL_1416509, EPI_ISL_1416510, EPI_ISL_1416511, EPI_ISL_1416512, EPI_ISL_1416513, EPI_ISL_1416514, EPI_ISL_1416515, EPI_ISL_1416516, EPI_ISL_1416517, EPI_ISL_1416518, EPI_ISL_1416519, EPI_ISL_1416520, EPI_ISL_1416521, EPI_ISL_1416522, EPI_ISL_1416523, EPI_ISL_1416524, EPI_ISL_1416525, EPI_ISL_1416526, EPI_ISL_1416527, EPI_ISL_1416528, EPI_ISL_1416529, EPI_ISL_1416530, EPI_ISL_1416531, EPI_ISL_1416532, EPI_ISL_1416533, EPI_ISL_1416534, EPI_ISL_1416535, EPI_ISL_1416536, EPI_ISL_1416537, EPI_ISL_1416538, EPI_ISL_1416539, EPI_ISL_1416540, EPI_ISL_1416541, EPI_ISL_1416542, EPI_ISL_1416543, EPI_ISL_1416544, EPI_ISL_1416545, EPI_ISL_1416546, EPI_ISL_1416547, EPI_ISL_1416548, EPI_ISL_1416549, EPI_ISL_1416550, EPI_ISL_1416551, EPI_ISL_1416552, EPI_ISL_1416553, EPI_ISL_1416554, EPI_ISL_1416555, EPI_ISL_1416556, EPI_ISL_1416557, EPI_ISL_1416558, EPI_ISL_1416559, EPI_ISL_1416560, EPI_ISL_1416561, EPI_ISL_1416562, EPI_ISL_1416563, EPI_ISL_1416564, EPI_ISL_1416565, EPI_ISL_1416566, EPI_ISL_1416567, EPI_ISL_1416568, EPI_ISL_1416569, EPI_ISL_1416570, EPI_ISL_1416571, EPI_ISL_1416572, EPI_ISL_1416573, EPI_ISL_1416574, EPI_ISL_1416575, EPI_ISL_1416576, EPI_ISL_1416577, EPI_ISL_1416578, EPI_ISL_1416579, EPI_ISL_1416580, EPI_ISL_1416581, EPI_ISL_1416582, EPI_ISL_1416583, EPI_ISL_1416584, EPI_ISL_1416585, EPI_ISL_1416586, EPI_ISL_1416587, EPI_ISL_1416588, EPI_ISL_1416589, EPI_ISL_1416590, EPI_ISL_1416591, EPI_ISL_1416592, EPI_ISL_1416593, EPI_ISL_1416594, EPI_ISL_1416595, EPI_ISL_1416596, | | | |

| | | | |
|---|---|---|---|
| EPI_ISL_1416597, EPI_ISL_1416598, EPI_ISL_1416599, EPI_ISL_1416600, EPI_ISL_1416601, EPI_ISL_1416602, EPI_ISL_1416603, EPI_ISL_1416605, EPI_ISL_1416606, EPI_ISL_1416607, EPI_ISL_1416608, EPI_ISL_1416609, EPI_ISL_1416610, EPI_ISL_1416611, EPI_ISL_1416612, EPI_ISL_1416613, EPI_ISL_1416614, EPI_ISL_1416615, EPI_ISL_1416616, EPI_ISL_1416617, EPI_ISL_1416618, EPI_ISL_1416619, EPI_ISL_1416620, EPI_ISL_1416621, EPI_ISL_1416622, EPI_ISL_1416623, EPI_ISL_1416624, EPI_ISL_1416625, EPI_ISL_1416626, EPI_ISL_1416627, EPI_ISL_1416628, EPI_ISL_1416629, EPI_ISL_1416630, EPI_ISL_1416631, EPI_ISL_1416632, EPI_ISL_1416633, EPI_ISL_1416634, EPI_ISL_1416635, EPI_ISL_1416636, EPI_ISL_1416637, EPI_ISL_1416638, EPI_ISL_1416639, EPI_ISL_1416640, EPI_ISL_1416641, EPI_ISL_1416642, EPI_ISL_1416643, EPI_ISL_1416644, EPI_ISL_1416645, EPI_ISL_1416646, EPI_ISL_1416647, EPI_ISL_1416648, EPI_ISL_1416649, EPI_ISL_1416650, EPI_ISL_1416651, EPI_ISL_1416652, EPI_ISL_1416653, EPI_ISL_1416654, EPI_ISL_1416655, EPI_ISL_1416656, EPI_ISL_1416657, EPI_ISL_1416658, EPI_ISL_1416659, EPI_ISL_1416660, EPI_ISL_1416661, EPI_ISL_1416662, EPI_ISL_1416663, EPI_ISL_1416664, EPI_ISL_1416665, EPI_ISL_1416666, EPI_ISL_1416667, EPI_ISL_1416668, EPI_ISL_1416669, EPI_ISL_1416670, EPI_ISL_1416671, EPI_ISL_1416672, EPI_ISL_1416673, EPI_ISL_1416674, EPI_ISL_1416675, EPI_ISL_1416676, EPI_ISL_1416677, EPI_ISL_1416678, EPI_ISL_1416679, EPI_ISL_1416680, EPI_ISL_1416681, EPI_ISL_1416682 | | | |
| see above | Laboratorio Central de Epidemiología (LCE) | Instituto de Biotecnología de la UNAM | Consorcio Mexicano de Vigilancia Genómica (CoViGen-Mex). Authors (in alphabetical order): Julio Elias Alvarado-Yaah, Carlos F. Arias, Santiago Ávila-Ríos, Víctor Hugo Borja-Aburto, Celia Boukadida, Juan Bautista Chale-Dzul , José Antonio Enciso-Moreno, Gloria Elena Espinoza-Ayala, Fernando Fontove-Herrera, Concepción Grajales-Muñiz, Ricardo Grande, Alfredo Herrera-Estrella, Carla Ivón Herrera-Najera, Pavel Isa, Brenda Irasema Maldonado-Meza, Bernardo Martínez-Miguel, Margarita Matías-Florentino, María Guadalupe de Jesús Mireles-Rivera, Gloria María Molina-Salinas, Hector Montoya-Fuentes, José Esteban Muñoz-Medina, José de Jesús Nuñez-Contreras, Alicia Ocaña-Mondragón, Luis Alberto Ochoa-Carrera, Hector Esteban Paz-Juárez, Francisco Pulido, Helen Haydee Fernanda Ramírez-Plascencia, Angel Gustavo Salas-Lais, Jorge Ivan Salinal-Nevarez, Alejandro Sanchez-Flores, Clara Esperanza Santacruz-Tinoco, María Guadalupe Santiago-Mauricio , Nelly Sélem-Mojica, Blanca Taboada , Gloria Vazquez |
| EPI_ISL_1416873 | Instituto Nacional de Medicina Genomica | Instituto Nacional de Medicina Genomica | Hidalgo-Miranda A, Mendoza-Vargas A, Reyes-Grajeda JP, Cedro-Tanda A, Alcaraz N, Gonzalez-Barrera D, Rangel-DeLeon D, Miranda-Ortiz H, Rosas-Escobar P, Canseco Mendez JC, Ramirez-Vega O, Munguia-Garza P, Garcia-Cardenas FJ, Gonzalez-Woge MA, Herrera-Montalvo LA. |
| EPI_ISL_1417442 | Instituto Nacional de Medicina Genomica | Instituto Nacional de Medicina Genomica | Hidalgo-Miranda A, Mendoza-Vargas A, Reyes-Grajeda JP, Cedro-Tanda A, Alcaraz N, Gonzalez-Barrera D, Rangel-DeLeon D, Miranda-Ortiz H, Rosas-Escobar P, Canseco Mendez JC, Ramirez-Vega O, Munguia-Garza P, Garcia-Cardenas FJ, Gonzalez-Woge MA, Herrera-Montalvo LA |
| EPI_ISL_1420789, EPI_ISL_1420790, EPI_ISL_1420791, EPI_ISL_1420792, EPI_ISL_1420794, EPI_ISL_1420795, EPI_ISL_1420796, EPI_ISL_1420797, EPI_ISL_1420798, EPI_ISL_1420800, EPI_ISL_1420802, EPI_ISL_1420804, EPI_ISL_1420805, EPI_ISL_1420806, EPI_ISL_1420807, EPI_ISL_1420808, EPI_ISL_1420809, EPI_ISL_1420810, EPI_ISL_1420812, EPI_ISL_1420813, EPI_ISL_1420814 | | | |
| see above | Wyoming Public Health Laboratory | Wyoming Public Health Laboratory | Jim Mildenberger, Wanda Manley, Noah Hull, Taylor Fearing, Lynette Gumbleton, Channing Weber, Ashley Norberg, Chayse Rowley, Marley Goetz, Brian Dominguez, Elliot Thomasson, Cari Sloma, and Rob Christensen |
| EPI_ISL_1420823, EPI_ISL_1420824, EPI_ISL_1420825, EPI_ISL_1420828 | Montana Public Health Laboratory | Montana Public Health Laboratory | Joy Ritter, Michelle Mozer, Carrie Biskupiak, Deborah Gibson |
| EPI_ISL_1420833, EPI_ISL_1420834, EPI_ISL_1420835, EPI_ISL_1420836, EPI_ISL_1420837, EPI_ISL_1420838, EPI_ISL_1420839, EPI_ISL_1420840, EPI_ISL_1420841, EPI_ISL_1420842, EPI_ISL_1420843, EPI_ISL_1420844, EPI_ISL_1420845, EPI_ISL_1420846, EPI_ISL_1420847, EPI_ISL_1420849, EPI_ISL_1420850, EPI_ISL_1420851, EPI_ISL_1420852, EPI_ISL_1420853, EPI_ISL_1420854, EPI_ISL_1420855, EPI_ISL_1420856, EPI_ISL_1420857, EPI_ISL_1420858, EPI_ISL_1420859, EPI_ISL_1420860, EPI_ISL_1420861, EPI_ISL_1420862, EPI_ISL_1420863, EPI_ISL_1420864, EPI_ISL_1420865, EPI_ISL_1420866, EPI_ISL_1420867, EPI_ISL_1420868, EPI_ISL_1420869, EPI_ISL_1420870, EPI_ISL_1420871, EPI_ISL_1420872, EPI_ISL_1420873, EPI_ISL_1420874, EPI_ISL_1420875, EPI_ISL_1420876, EPI_ISL_1420877, EPI_ISL_1420878, EPI_ISL_1420879, EPI_ISL_1420880, EPI_ISL_1420881, EPI_ISL_1420882, EPI_ISL_1420883, EPI_ISL_1420884, EPI_ISL_1420886, EPI_ISL_1420887, EPI_ISL_1420888, EPI_ISL_1420889, EPI_ISL_1420890, EPI_ISL_1420891, EPI_ISL_1420892, EPI_ISL_1420893, EPI_ISL_1420894, EPI_ISL_1420895, EPI_ISL_1420896, EPI_ISL_1420897, EPI_ISL_1420898, EPI_ISL_1420899, EPI_ISL_1420900, EPI_ISL_1420901, EPI_ISL_1420902, EPI_ISL_1420903, EPI_ISL_1420904, EPI_ISL_1420905, EPI_ISL_1420906, EPI_ISL_1420907, EPI_ISL_1420908, EPI_ISL_1420909, EPI_ISL_1420910, EPI_ISL_1420911, EPI_ISL_1420912, EPI_ISL_1420913, EPI_ISL_1420914, EPI_ISL_1420915, EPI_ISL_1420916, EPI_ISL_1420917, EPI_ISL_1420918, EPI_ISL_1420919, EPI_ISL_1420920, EPI_ISL_1420921, EPI_ISL_1420922, EPI_ISL_1420923, EPI_ISL_1420924, EPI_ISL_1420925, EPI_ISL_1420926, EPI_ISL_1420927, EPI_ISL_1420928, EPI_ISL_1420929, EPI_ISL_1420930, EPI_ISL_1420931, EPI_ISL_1420932, EPI_ISL_1420933, EPI_ISL_1420934, EPI_ISL_1420935, EPI_ISL_1420936, EPI_ISL_1420937, EPI_ISL_1420938, EPI_ISL_1420939, EPI_ISL_1420940, EPI_ISL_1420941, EPI_ISL_1420942, EPI_ISL_1420943, EPI_ISL_1420944, EPI_ISL_1420945, EPI_ISL_1420946, EPI_ISL_1420947, EPI_ISL_1420948, EPI_ISL_1420949, EPI_ISL_1420950, EPI_ISL_1420951, EPI_ISL_1420952, EPI_ISL_1420953, EPI_ISL_1420954, EPI_ISL_1420955, EPI_ISL_1420956, EPI_ISL_1420957, EPI_ISL_1420958, EPI_ISL_1420959, EPI_ISL_1420960, EPI_ISL_1420961, EPI_ISL_1420962, EPI_ISL_1420963, EPI_ISL_1420964, EPI_ISL_1420965, EPI_ISL_1420966, EPI_ISL_1420967, EPI_ISL_1420968, EPI_ISL_1420969, EPI_ISL_1420970, EPI_ISL_1420971, EPI_ISL_1420972 | | | |
| see above | Helix/Illumina | Centers for Disease Control and Prevention Division of Viral Diseases, Pathogen Discovery | Peter W. Cook, Dakota Howard, Dhwani Batra, Ben L. Rambo-Martin, Eileen de Feo, Jan Antico, Christine Tran, Matthew Tolentino, Shannon Wickline, Kim Gietzen, Brad Sickler, Jingtao Liu, Eric Allen, Phil Febbo, Summer Galloway, Nicole L. Washington, Simon White, Geraint Levan, Kelly Schiabor Barrett, Elizabeth Cirulli, Alexandre Bolze, Ary Ascencio, Charlotte Rivera-Garcia, Ryan Cho, Jason Nguyen, Sherry Wang, Jimmy Ramirez, Tyler Cassens, Efren Sandoval, Magnus Isaksson, William Lee, David Becker, Marc Laurent, James Lu, Clinton R. Paden, Suxiang Tong, Duncan MacCannell |
| EPI_ISL_1420973 | Wisconsin State Laboratory of Hygiene Communicable Disease Division | Wisconsin State Laboratory of Hygiene Communicable Disease Division | Kelsey R. Florek, Abigail C. Shockey |
| EPI_ISL_1420974 | Helix/Illumina | Centers for Disease Control and Prevention Division of Viral Diseases, Pathogen Discovery | Peter W. Cook, Dakota Howard, Dhwani Batra, Ben L. Rambo-Martin, Eileen de Feo, Jan Antico, Christine Tran, Matthew Tolentino, Shannon Wickline, Kim Gietzen, Brad Sickler, Jingtao Liu, Eric Allen, Phil Febbo, Summer Galloway, Nicole L. Washington, Simon White, Geraint Levan, Kelly Schiabor Barrett, Elizabeth Cirulli, Alexandre Bolze, Ary Ascencio, Charlotte Rivera-Garcia, Ryan Cho, Jason Nguyen, Sherry Wang, Jimmy Ramirez, Tyler Cassens, Efren Sandoval, Magnus Isaksson, William Lee, David Becker, Marc Laurent, James Lu, Clinton R. Paden, Suxiang Tong, Duncan MacCannell |
| EPI_ISL_1420975, EPI_ISL_1420976 | Wisconsin State Laboratory of Hygiene Communicable Disease Division | Wisconsin State Laboratory of Hygiene Communicable Disease Division | Kelsey R. Florek, Abigail C. Shockey |
| EPI_ISL_1420977 | Helix/Illumina | Centers for Disease Control and Prevention Division of Viral Diseases, Pathogen Discovery | Peter W. Cook, Dakota Howard, Dhwani Batra, Ben L. Rambo-Martin, Eileen de Feo, Jan Antico, Christine Tran, Matthew Tolentino, Shannon Wickline, Kim Gietzen, Brad Sickler, Jingtao Liu, Eric Allen, Phil Febbo, Summer Galloway, Nicole L. Washington, Simon White, Geraint Levan, Kelly Schiabor Barrett, Elizabeth Cirulli, Alexandre Bolze, Ary Ascencio, Charlotte Rivera-Garcia, Ryan Cho, Jason Nguyen, Sherry Wang, Jimmy Ramirez, Tyler Cassens, Efren Sandoval, Magnus Isaksson, William Lee, David Becker, Marc Laurent, James Lu, Clinton R. Paden, Suxiang Tong, Duncan MacCannell |
| EPI_ISL_1420978, EPI_ISL_1420979 | Wisconsin State Laboratory of Hygiene Communicable Disease Division | Wisconsin State Laboratory of Hygiene Communicable Disease Division | Kelsey R. Florek, Abigail C. Shockey |
| EPI_ISL_1420980 | Helix/Illumina | Centers for Disease Control and Prevention Division of Viral Diseases, Pathogen Discovery | Peter W. Cook, Dakota Howard, Dhwani Batra, Ben L. Rambo-Martin, Eileen de Feo, Jan Antico, Christine Tran, Matthew Tolentino, Shannon Wickline, Kim Gietzen, Brad Sickler, Jingtao Liu, Eric Allen, Phil Febbo, Summer Galloway, Nicole L. Washington, Simon White, Geraint Levan, Kelly Schiabor Barrett, Elizabeth Cirulli, Alexandre Bolze, Ary Ascencio, Charlotte Rivera-Garcia, Ryan Cho, Jason Nguyen, Sherry Wang, Jimmy Ramirez, Tyler Cassens, Efren Sandoval, Magnus Isaksson, William Lee, David Becker, Marc Laurent, James Lu, Clinton R. Paden, Suxiang Tong, Duncan MacCannell |
| EPI_ISL_1420981 | Wisconsin State Laboratory of Hygiene Communicable Disease Division | Wisconsin State Laboratory of Hygiene Communicable Disease Division | Kelsey R. Florek, Abigail C. Shockey |
| EPI_ISL_1420982 | Helix/Illumina | Centers for Disease Control and Prevention Division of Viral Diseases, Pathogen Discovery | Peter W. Cook, Dakota Howard, Dhwani Batra, Ben L. Rambo-Martin, Eileen de Feo, Jan Antico, Christine Tran, Matthew Tolentino, Shannon Wickline, Kim Gietzen, Brad Sickler, Jingtao Liu, Eric Allen, Phil Febbo, Summer Galloway, Nicole L. Washington, Simon White, Geraint Levan, Kelly Schiabor Barrett, Elizabeth Cirulli, Alexandre Bolze, Ary Ascencio, Charlotte Rivera-Garcia, Ryan Cho, Jason Nguyen, Sherry Wang, Jimmy Ramirez, Tyler Cassens, Efren Sandoval, Magnus Isaksson, William Lee, David Becker, Marc Laurent, James Lu, Clinton R. Paden, Suxiang Tong, Duncan MacCannell |
| EPI_ISL_1420983, EPI_ISL_1420984 | Wisconsin State Laboratory of Hygiene Communicable Disease Division | Wisconsin State Laboratory of Hygiene Communicable Disease Division | Kelsey R. Florek, Abigail C. Shockey |
| EPI_ISL_1420985 | Helix/Illumina | Centers for Disease Control and Prevention Division of Viral Diseases, Pathogen Discovery | Peter W. Cook, Dakota Howard, Dhwani Batra, Ben L. Rambo-Martin, Eileen de Feo, Jan Antico, Christine Tran, Matthew Tolentino, Shannon Wickline, Kim Gietzen, Brad Sickler, Jingtao Liu, Eric Allen, Phil Febbo, Summer Galloway, Nicole L. Washington, Simon White, Geraint Levan, Kelly Schiabor Barrett, Elizabeth Cirulli, Alexandre Bolze, Ary Ascencio, Charlotte Rivera-Garcia, Ryan Cho, Jason Nguyen, Sherry Wang, Jimmy Ramirez, Tyler Cassens, Efren Sandoval, Magnus Isaksson, William Lee, David Becker, Marc Laurent, James Lu, Clinton R. Paden, Suxiang Tong, Duncan MacCannell |
| EPI_ISL_1420986, EPI_ISL_1420987, EPI_ISL_1420988 | Wisconsin State Laboratory of Hygiene Communicable Disease Division | Wisconsin State Laboratory of Hygiene Communicable Disease Division | Kelsey R. Florek, Abigail C. Shockey |
| EPI_ISL_1420989 | Helix/Illumina | Centers for Disease Control and Prevention Division of Viral Diseases, Pathogen Discovery | Peter W. Cook, Dakota Howard, Dhwani Batra, Ben L. Rambo-Martin, Eileen de Feo, Jan Antico, Christine Tran, Matthew Tolentino, Shannon Wickline, Kim Gietzen, Brad Sickler, Jingtao Liu, Eric Allen, Phil Febbo, Summer Galloway, Nicole L. Washington, Simon White, Geraint Levan, Kelly Schiabor Barrett, Elizabeth Cirulli, Alexandre Bolze, Ary Ascencio, Charlotte Rivera-Garcia, Ryan Cho, Jason Nguyen, Sherry Wang, Jimmy Ramirez, Tyler Cassens, Efren Sandoval, Magnus Isaksson, William Lee, David Becker, Marc Laurent, James Lu, Clinton R. Paden, Suxiang Tong, Duncan MacCannell |
| EPI_ISL_1420990, EPI_ISL_1420991 | Wisconsin State Laboratory of Hygiene Communicable Disease Division | Wisconsin State Laboratory of Hygiene Communicable Disease Division | Kelsey R. Florek, Abigail C. Shockey |
| EPI_ISL_1420992 | Helix/Illumina | Centers for Disease Control and Prevention Division of Viral Diseases, Pathogen Discovery | Peter W. Cook, Dakota Howard, Dhwani Batra, Ben L. Rambo-Martin, Eileen de Feo, Jan Antico, Christine Tran, Matthew Tolentino, Shannon Wickline, Kim Gietzen, Brad Sickler, Jingtao Liu, Eric Allen, Phil Febbo, Summer Galloway, Nicole L. Washington, Simon White, Geraint Levan, Kelly Schiabor Barrett, Elizabeth Cirulli, Alexandre Bolze, Ary Ascencio, Charlotte Rivera-Garcia, Ryan Cho, Jason Nguyen, Sherry Wang, Jimmy Ramirez, Tyler Cassens, Efren Sandoval, Magnus Isaksson, William Lee, David Becker, Marc Laurent, James Lu, Clinton R. Paden, Suxiang Tong, Duncan MacCannell |
| EPI_ISL_1420993, EPI_ISL_1420994 | Wisconsin State Laboratory of Hygiene Communicable Disease Division | Wisconsin State Laboratory of Hygiene Communicable Disease Division | Kelsey R. Florek, Abigail C. Shockey |
| EPI_ISL_1420995 | Helix/Illumina | Centers for Disease Control and Prevention Division of Viral Diseases, Pathogen Discovery | Peter W. Cook, Dakota Howard, Dhwani Batra, Ben L. Rambo-Martin, Eileen de Feo, Jan Antico, Christine Tran, Matthew Tolentino, Shannon Wickline, Kim Gietzen, Brad Sickler, Jingtao Liu, Eric Allen, Phil Febbo, Summer Galloway, Nicole L. Washington, Simon White, Geraint Levan, Kelly Schiabor Barrett, Elizabeth Cirulli, Alexandre Bolze, Ary Ascencio, Charlotte Rivera-Garcia, Ryan Cho, Jason Nguyen, Sherry Wang, Jimmy Ramirez, Tyler Cassens, Efren Sandoval, Magnus Isaksson, William Lee, David Becker, Marc Laurent, James Lu, Clinton R. Paden, Suxiang Tong, Duncan MacCannell |
| EPI_ISL_1420996, EPI_ISL_1420997 | Wisconsin State Laboratory of Hygiene Communicable Disease Division | Wisconsin State Laboratory of Hygiene Communicable Disease Division | Kelsey R. Florek, Abigail C. Shockey |
| EPI_ISL_1420998 | Helix/Illumina | Centers for Disease Control and Prevention Division of Viral Diseases, Pathogen Discovery | Peter W. Cook, Dakota Howard, Dhwani Batra, Ben L. Rambo-Martin, Eileen de Feo, Jan Antico, Christine Tran, Matthew Tolentino, Shannon Wickline, Kim Gietzen, Brad Sickler, Jingtao Liu, Eric Allen, Phil Febbo, Summer Galloway, Nicole L. Washington, Simon White, Geraint Levan, Kelly Schiabor Barrett, Elizabeth Cirulli, Alexandre Bolze, Ary Ascencio, Charlotte Rivera-Garcia, Ryan Cho, Jason Nguyen, Sherry Wang, Jimmy Ramirez, Tyler Cassens, Efren Sandoval, Magnus Isaksson, William Lee, David Becker, Marc Laurent, James Lu, Clinton R. Paden, Suxiang Tong, Duncan MacCannell |
| EPI_ISL_1420999, EPI_ISL_1421000, EPI_ISL_1421001 | Wisconsin State Laboratory of Hygiene Communicable Disease Division | Wisconsin State Laboratory of Hygiene Communicable Disease Division | Kelsey R. Florek, Abigail C. Shockey |
| EPI_ISL_1421002 | Helix/Illumina | Centers for Disease Control and Prevention Division of Viral Diseases, Pathogen Discovery | Peter W. Cook, Dakota Howard, Dhwani Batra, Ben L. Rambo-Martin, Eileen de Feo, Jan Antico, Christine Tran, Matthew Tolentino, Shannon Wickline, Kim Gietzen, Brad Sickler, Jingtao Liu, Eric Allen, Phil Febbo, Summer Galloway, Nicole L. Washington, Simon White, Geraint Levan, Kelly Schiabor Barrett, Elizabeth Cirulli, Alexandre Bolze, Ary Ascencio, Charlotte Rivera-Garcia, Ryan Cho, Jason Nguyen, Sherry Wang, Jimmy Ramirez, Tyler Cassens, Efren Sandoval, Magnus Isaksson, William Lee, David Becker, Marc Laurent, James Lu, Clinton R. Paden, Suxiang Tong, Duncan MacCannell |
| EPI_ISL_1421003, EPI_ISL_1421004 | Wisconsin State Laboratory of Hygiene Communicable Disease Division | Wisconsin State Laboratory of Hygiene Communicable Disease Division | Kelsey R. Florek, Abigail C. Shockey |
| EPI_ISL_1421005 | Helix/Illumina | Centers for Disease Control and Prevention Division of Viral Diseases, Pathogen Discovery | Peter W. Cook, Dakota Howard, Dhwani Batra, Ben L. Rambo-Martin, Eileen de Feo, Jan Antico, Christine Tran, Matthew Tolentino, Shannon Wickline, Kim Gietzen, Brad Sickler, Jingtao Liu, Eric Allen, Phil Febbo, Summer Galloway, Nicole L. Washington, Simon White, Geraint Levan, Kelly Schiabor Barrett, Elizabeth Cirulli, Alexandre Bolze, Ary Ascencio, Charlotte Rivera-Garcia, Ryan Cho, Jason Nguyen, Sherry Wang, Jimmy Ramirez, Tyler Cassens, Efren Sandoval, Magnus Isaksson, William Lee, David Becker, Marc Laurent, James Lu, Clinton R. Paden, Suxiang Tong, Duncan MacCannell |
| EPI_ISL_1421006, EPI_ISL_1421007 | Wisconsin State Laboratory of Hygiene Communicable Disease Division | Wisconsin State Laboratory of Hygiene Communicable Disease Division | Kelsey R. Florek, Abigail C. Shockey |
| EPI_ISL_1421008 | Helix/Illumina | Centers for Disease Control and Prevention Division of Viral Diseases, Pathogen Discovery | Peter W. Cook, Dakota Howard, Dhwani Batra, Ben L. Rambo-Martin, Eileen de Feo, Jan Antico, Christine Tran, Matthew Tolentino, Shannon Wickline, Kim Gietzen, Brad Sickler, Jingtao Liu, Eric Allen, Phil Febbo, Summer Galloway, Nicole L. Washington, Simon White, Geraint Levan, Kelly Schiabor Barrett, Elizabeth Cirulli, Alexandre Bolze, Ary Ascencio, Charlotte Rivera-Garcia, Ryan Cho, Jason Nguyen, Sherry Wang, Jimmy Ramirez, Tyler Cassens, Efren Sandoval, Magnus Isaksson, William Lee, David Becker, Marc Laurent, James Lu, Clinton R. Paden, Suxiang Tong, Duncan MacCannell |
| EPI_ISL_1421009, EPI_ISL_1421010 | Wisconsin State Laboratory of Hygiene Communicable Disease Division | Wisconsin State Laboratory of Hygiene Communicable Disease Division | Kelsey R. Florek, Abigail C. Shockey |
| EPI_ISL_1421011 | Helix/Illumina | Centers for Disease Control and Prevention Division of Viral Diseases, Pathogen Discovery | Peter W. Cook, Dakota Howard, Dhwani Batra, Ben L. Rambo-Martin, Eileen de Feo, Jan Antico, Christine Tran, Matthew Tolentino, Shannon Wickline, Kim Gietzen, Brad Sickler, Jingtao Liu, Eric Allen, Phil Febbo, Summer Galloway, Nicole L. Washington, Simon White, Geraint Levan, Kelly Schiabor Barrett, Elizabeth Cirulli, Alexandre Bolze, Ary Ascencio, Charlotte Rivera-Garcia, Ryan Cho, Jason Nguyen, Sherry Wang, Jimmy Ramirez, Tyler Cassens, Efren Sandoval, Magnus Isaksson, William Lee, David Becker, Marc Laurent, James Lu, Clinton R. Paden, Suxiang Tong, Duncan MacCannell |

| | | | |
|---|---|---|---|
| EPI_ISL_1421012, EPI_ISL_1421013 | Wisconsin State Laboratory of Hygiene Communicable Disease Division | Wisconsin State Laboratory of Hygiene Communicable Disease Division | Kelsey R. Florek, Abigail C. Shockey |
| EPI_ISL_1421014 | Helix/Illumina | Centers for Disease Control and Prevention Division of Viral Diseases, Pathogen Discovery | Peter W. Cook, Dakota Howard, Dhwani Batra, Ben L. Rambo-Martin, Eileen de Feo, Jan Antico, Christine Tran, Matthew Tolentino, Shannon Wickline, Kim Gietzen, Brad Sickler, Jingtao Liu, Eric Allen, Phil Febbo, Summer Galloway, Nicole L. Washington, Simon White, Geraint Levan, Kelly Schiabor Barrett, Elizabeth Cirulli, Alexandre Bolze, Ary Ascencio, Charlotte Rivera-Garcia, Ryan Cho, Jason Nguyen, Sherry Wang, Jimmy Ramirez, Tyler Cassens, Efren Sandoval, Magnus Isaksson, William Lee, David Becker, Marc Laurent, James Lu, Clinton R. Paden, Suxiang Tong, Duncan MacCannell |
| EPI_ISL_1421015 | Wisconsin State Laboratory of Hygiene Communicable Disease Division | Wisconsin State Laboratory of Hygiene Communicable Disease Division | Kelsey R. Florek, Abigail C. Shockey |
| EPI_ISL_1421017 | Helix/Illumina | Centers for Disease Control and Prevention Division of Viral Diseases, Pathogen Discovery | Peter W. Cook, Dakota Howard, Dhwani Batra, Ben L. Rambo-Martin, Eileen de Feo, Jan Antico, Christine Tran, Matthew Tolentino, Shannon Wickline, Kim Gietzen, Brad Sickler, Jingtao Liu, Eric Allen, Phil Febbo, Summer Galloway, Nicole L. Washington, Simon White, Geraint Levan, Kelly Schiabor Barrett, Elizabeth Cirulli, Alexandre Bolze, Ary Ascencio, Charlotte Rivera-Garcia, Ryan Cho, Jason Nguyen, Sherry Wang, Jimmy Ramirez, Tyler Cassens, Efren Sandoval, Magnus Isaksson, William Lee, David Becker, Marc Laurent, James Lu, Clinton R. Paden, Suxiang Tong, Duncan MacCannell |
| EPI_ISL_1421018, EPI_ISL_1421019 | Wisconsin State Laboratory of Hygiene Communicable Disease Division | Wisconsin State Laboratory of Hygiene Communicable Disease Division | Kelsey R. Florek, Abigail C. Shockey |
| EPI_ISL_1421020 | Helix/Illumina | Centers for Disease Control and Prevention Division of Viral Diseases, Pathogen Discovery | Peter W. Cook, Dakota Howard, Dhwani Batra, Ben L. Rambo-Martin, Eileen de Feo, Jan Antico, Christine Tran, Matthew Tolentino, Shannon Wickline, Kim Gietzen, Brad Sickler, Jingtao Liu, Eric Allen, Phil Febbo, Summer Galloway, Nicole L. Washington, Simon White, Geraint Levan, Kelly Schiabor Barrett, Elizabeth Cirulli, Alexandre Bolze, Ary Ascencio, Charlotte Rivera-Garcia, Ryan Cho, Jason Nguyen, Sherry Wang, Jimmy Ramirez, Tyler Cassens, Efren Sandoval, Magnus Isaksson, William Lee, David Becker, Marc Laurent, James Lu, Clinton R. Paden, Suxiang Tong, Duncan MacCannell |
| EPI_ISL_1421021, EPI_ISL_1421022, EPI_ISL_1421023 | Wisconsin State Laboratory of Hygiene Communicable Disease Division | Wisconsin State Laboratory of Hygiene Communicable Disease Division | Kelsey R. Florek, Abigail C. Shockey |
| EPI_ISL_1421024 | Helix/Illumina | Centers for Disease Control and Prevention Division of Viral Diseases, Pathogen Discovery | Peter W. Cook, Dakota Howard, Dhwani Batra, Ben L. Rambo-Martin, Eileen de Feo, Jan Antico, Christine Tran, Matthew Tolentino, Shannon Wickline, Kim Gietzen, Brad Sickler, Jingtao Liu, Eric Allen, Phil Febbo, Summer Galloway, Nicole L. Washington, Simon White, Geraint Levan, Kelly Schiabor Barrett, Elizabeth Cirulli, Alexandre Bolze, Ary Ascencio, Charlotte Rivera-Garcia, Ryan Cho, Jason Nguyen, Sherry Wang, Jimmy Ramirez, Tyler Cassens, Efren Sandoval, Magnus Isaksson, William Lee, David Becker, Marc Laurent, James Lu, Clinton R. Paden, Suxiang Tong, Duncan MacCannell |
| EPI_ISL_1421025, EPI_ISL_1421026 | Wisconsin State Laboratory of Hygiene Communicable Disease Division | Wisconsin State Laboratory of Hygiene Communicable Disease Division | Kelsey R. Florek, Abigail C. Shockey |
| EPI_ISL_1421027 | Helix/Illumina | Centers for Disease Control and Prevention Division of Viral Diseases, Pathogen Discovery | Peter W. Cook, Dakota Howard, Dhwani Batra, Ben L. Rambo-Martin, Eileen de Feo, Jan Antico, Christine Tran, Matthew Tolentino, Shannon Wickline, Kim Gietzen, Brad Sickler, Jingtao Liu, Eric Allen, Phil Febbo, Summer Galloway, Nicole L. Washington, Simon White, Geraint Levan, Kelly Schiabor Barrett, Elizabeth Cirulli, Alexandre Bolze, Ary Ascencio, Charlotte Rivera-Garcia, Ryan Cho, Jason Nguyen, Sherry Wang, Jimmy Ramirez, Tyler Cassens, Efren Sandoval, Magnus Isaksson, William Lee, David Becker, Marc Laurent, James Lu, Clinton R. Paden, Suxiang Tong, Duncan MacCannell |
| EPI_ISL_1421028, EPI_ISL_1421029 | Wisconsin State Laboratory of Hygiene Communicable Disease Division | Wisconsin State Laboratory of Hygiene Communicable Disease Division | Kelsey R. Florek, Abigail C. Shockey |
| EPI_ISL_1421030 | Helix/Illumina | Centers for Disease Control and Prevention Division of Viral Diseases, Pathogen Discovery | Peter W. Cook, Dakota Howard, Dhwani Batra, Ben L. Rambo-Martin, Eileen de Feo, Jan Antico, Christine Tran, Matthew Tolentino, Shannon Wickline, Kim Gietzen, Brad Sickler, Jingtao Liu, Eric Allen, Phil Febbo, Summer Galloway, Nicole L. Washington, Simon White, Geraint Levan, Kelly Schiabor Barrett, Elizabeth Cirulli, Alexandre Bolze, Ary Ascencio, Charlotte Rivera-Garcia, Ryan Cho, Jason Nguyen, Sherry Wang, Jimmy Ramirez, Tyler Cassens, Efren Sandoval, Magnus Isaksson, William Lee, David Becker, Marc Laurent, James Lu, Clinton R. Paden, Suxiang Tong, Duncan MacCannell |
| EPI_ISL_1421031, EPI_ISL_1421032 | Wisconsin State Laboratory of Hygiene Communicable Disease Division | Wisconsin State Laboratory of Hygiene Communicable Disease Division | Kelsey R. Florek, Abigail C. Shockey |
| EPI_ISL_1421033 | Helix/Illumina | Centers for Disease Control and Prevention Division of Viral Diseases, Pathogen Discovery | Peter W. Cook, Dakota Howard, Dhwani Batra, Ben L. Rambo-Martin, Eileen de Feo, Jan Antico, Christine Tran, Matthew Tolentino, Shannon Wickline, Kim Gietzen, Brad Sickler, Jingtao Liu, Eric Allen, Phil Febbo, Summer Galloway, Nicole L. Washington, Simon White, Geraint Levan, Kelly Schiabor Barrett, Elizabeth Cirulli, Alexandre Bolze, Ary Ascencio, Charlotte Rivera-Garcia, Ryan Cho, Jason Nguyen, Sherry Wang, Jimmy Ramirez, Tyler Cassens, Efren Sandoval, Magnus Isaksson, William Lee, David Becker, Marc Laurent, James Lu, Clinton R. Paden, Suxiang Tong, Duncan MacCannell |
| EPI_ISL_1421034, EPI_ISL_1421035 | Wisconsin State Laboratory of Hygiene Communicable Disease Division | Wisconsin State Laboratory of Hygiene Communicable Disease Division | Kelsey R. Florek, Abigail C. Shockey |
| EPI_ISL_1421036 | Helix/Illumina | Centers for Disease Control and Prevention Division of Viral Diseases, Pathogen Discovery | Peter W. Cook, Dakota Howard, Dhwani Batra, Ben L. Rambo-Martin, Eileen de Feo, Jan Antico, Christine Tran, Matthew Tolentino, Shannon Wickline, Kim Gietzen, Brad Sickler, Jingtao Liu, Eric Allen, Phil Febbo, Summer Galloway, Nicole L. Washington, Simon White, Geraint Levan, Kelly Schiabor Barrett, Elizabeth Cirulli, Alexandre Bolze, Ary Ascencio, Charlotte Rivera-Garcia, Ryan Cho, Jason Nguyen, Sherry Wang, Jimmy Ramirez, Tyler Cassens, Efren Sandoval, Magnus Isaksson, William Lee, David Becker, Marc Laurent, James Lu, Clinton R. Paden, Suxiang Tong, Duncan MacCannell |
| EPI_ISL_1421037, EPI_ISL_1421038 | Wisconsin State Laboratory of Hygiene Communicable Disease Division | Wisconsin State Laboratory of Hygiene Communicable Disease Division | Kelsey R. Florek, Abigail C. Shockey |
| EPI_ISL_1421039 | Helix/Illumina | Centers for Disease Control and Prevention Division of Viral Diseases, Pathogen Discovery | Peter W. Cook, Dakota Howard, Dhwani Batra, Ben L. Rambo-Martin, Eileen de Feo, Jan Antico, Christine Tran, Matthew Tolentino, Shannon Wickline, Kim Gietzen, Brad Sickler, Jingtao Liu, Eric Allen, Phil Febbo, Summer Galloway, Nicole L. Washington, Simon White, Geraint Levan, Kelly Schiabor Barrett, Elizabeth Cirulli, Alexandre Bolze, Ary Ascencio, Charlotte Rivera-Garcia, Ryan Cho, Jason Nguyen, Sherry Wang, Jimmy Ramirez, Tyler Cassens, Efren Sandoval, Magnus Isaksson, William Lee, David Becker, Marc Laurent, James Lu, Clinton R. Paden, Suxiang Tong, Duncan MacCannell |
| EPI_ISL_1421040, EPI_ISL_1421041 | Wisconsin State Laboratory of Hygiene Communicable Disease Division | Wisconsin State Laboratory of Hygiene Communicable Disease Division | Kelsey R. Florek, Abigail C. Shockey |
| EPI_ISL_1421042 | Helix/Illumina | Centers for Disease Control and Prevention Division of Viral Diseases, Pathogen Discovery | Peter W. Cook, Dakota Howard, Dhwani Batra, Ben L. Rambo-Martin, Eileen de Feo, Jan Antico, Christine Tran, Matthew Tolentino, Shannon Wickline, Kim Gietzen, Brad Sickler, Jingtao Liu, Eric Allen, Phil Febbo, Summer Galloway, Nicole L. Washington, Simon White, Geraint Levan, Kelly Schiabor Barrett, Elizabeth Cirulli, Alexandre Bolze, Ary Ascencio, Charlotte Rivera-Garcia, Ryan Cho, Jason Nguyen, Sherry Wang, Jimmy Ramirez, Tyler Cassens, Efren Sandoval, Magnus Isaksson, William Lee, David Becker, Marc Laurent, James Lu, Clinton R. Paden, Suxiang Tong, Duncan MacCannell |
| EPI_ISL_1421043, EPI_ISL_1421044 | Wisconsin State Laboratory of Hygiene Communicable Disease Division | Wisconsin State Laboratory of Hygiene Communicable Disease Division | Kelsey R. Florek, Abigail C. Shockey |
| EPI_ISL_1421045 | Helix/Illumina | Centers for Disease Control and Prevention Division of Viral Diseases, Pathogen Discovery | Peter W. Cook, Dakota Howard, Dhwani Batra, Ben L. Rambo-Martin, Eileen de Feo, Jan Antico, Christine Tran, Matthew Tolentino, Shannon Wickline, Kim Gietzen, Brad Sickler, Jingtao Liu, Eric Allen, Phil Febbo, Summer Galloway, Nicole L. Washington, Simon White, Geraint Levan, Kelly Schiabor Barrett, Elizabeth Cirulli, Alexandre Bolze, Ary Ascencio, Charlotte Rivera-Garcia, Ryan Cho, Jason Nguyen, Sherry Wang, Jimmy Ramirez, Tyler Cassens, Efren Sandoval, Magnus Isaksson, William Lee, David Becker, Marc Laurent, James Lu, Clinton R. Paden, Suxiang Tong, Duncan MacCannell |
| EPI_ISL_1421046 | Wisconsin State Laboratory of Hygiene Communicable Disease Division | Wisconsin State Laboratory of Hygiene Communicable Disease Division | Kelsey R. Florek, Abigail C. Shockey |
| EPI_ISL_1421047 | Helix/Illumina | Centers for Disease Control and Prevention Division of Viral Diseases, Pathogen Discovery | Peter W. Cook, Dakota Howard, Dhwani Batra, Ben L. Rambo-Martin, Eileen de Feo, Jan Antico, Christine Tran, Matthew Tolentino, Shannon Wickline, Kim Gietzen, Brad Sickler, Jingtao Liu, Eric Allen, Phil Febbo, Summer Galloway, Nicole L. Washington, Simon White, Geraint Levan, Kelly Schiabor Barrett, Elizabeth Cirulli, Alexandre Bolze, Ary Ascencio, Charlotte Rivera-Garcia, Ryan Cho, Jason Nguyen, Sherry Wang, Jimmy Ramirez, Tyler Cassens, Efren Sandoval, Magnus Isaksson, William Lee, David Becker, Marc Laurent, James Lu, Clinton R. Paden, Suxiang Tong, Duncan MacCannell |
| EPI_ISL_1421048, EPI_ISL_1421049 | Wisconsin State Laboratory of Hygiene Communicable Disease Division | Wisconsin State Laboratory of Hygiene Communicable Disease Division | Kelsey R. Florek, Abigail C. Shockey |
| EPI_ISL_1421050 | Helix/Illumina | Centers for Disease Control and Prevention Division of Viral Diseases, Pathogen Discovery | Peter W. Cook, Dakota Howard, Dhwani Batra, Ben L. Rambo-Martin, Eileen de Feo, Jan Antico, Christine Tran, Matthew Tolentino, Shannon Wickline, Kim Gietzen, Brad Sickler, Jingtao Liu, Eric Allen, Phil Febbo, Summer Galloway, Nicole L. Washington, Simon White, Geraint Levan, Kelly Schiabor Barrett, Elizabeth Cirulli, Alexandre Bolze, Ary Ascencio, Charlotte Rivera-Garcia, Ryan Cho, Jason Nguyen, Sherry Wang, Jimmy Ramirez, Tyler Cassens, Efren Sandoval, Magnus Isaksson, William Lee, David Becker, Marc Laurent, James Lu, Clinton R. Paden, Suxiang Tong, Duncan MacCannell |
| EPI_ISL_1421051, EPI_ISL_1421052, EPI_ISL_1421053 | Wisconsin State Laboratory of Hygiene Communicable Disease Division | Wisconsin State Laboratory of Hygiene Communicable Disease Division | Kelsey R. Florek, Abigail C. Shockey |
| EPI_ISL_1421054 | Helix/Illumina | Centers for Disease Control and Prevention Division of Viral Diseases, Pathogen Discovery | Peter W. Cook, Dakota Howard, Dhwani Batra, Ben L. Rambo-Martin, Eileen de Feo, Jan Antico, Christine Tran, Matthew Tolentino, Shannon Wickline, Kim Gietzen, Brad Sickler, Jingtao Liu, Eric Allen, Phil Febbo, Summer Galloway, Nicole L. Washington, Simon White, Geraint Levan, Kelly Schiabor Barrett, Elizabeth Cirulli, Alexandre Bolze, Ary Ascencio, Charlotte Rivera-Garcia, Ryan Cho, Jason Nguyen, Sherry Wang, Jimmy Ramirez, Tyler Cassens, Efren Sandoval, Magnus Isaksson, William Lee, David Becker, Marc Laurent, James Lu, Clinton R. Paden, Suxiang Tong, Duncan MacCannell |
| EPI_ISL_1421055, EPI_ISL_1421056 | Wisconsin State Laboratory of Hygiene Communicable Disease Division | Wisconsin State Laboratory of Hygiene Communicable Disease Division | Kelsey R. Florek, Abigail C. Shockey |
| EPI_ISL_1421057 | Helix/Illumina | Centers for Disease Control and Prevention Division of Viral Diseases, Pathogen Discovery | Peter W. Cook, Dakota Howard, Dhwani Batra, Ben L. Rambo-Martin, Eileen de Feo, Jan Antico, Christine Tran, Matthew Tolentino, Shannon Wickline, Kim Gietzen, Brad Sickler, Jingtao Liu, Eric Allen, Phil Febbo, Summer Galloway, Nicole L. Washington, Simon White, Geraint Levan, Kelly Schiabor Barrett, Elizabeth Cirulli, Alexandre Bolze, Ary Ascencio, Charlotte Rivera-Garcia, Ryan Cho, Jason Nguyen, Sherry Wang, Jimmy Ramirez, Tyler Cassens, Efren Sandoval, Magnus Isaksson, William Lee, David Becker, Marc Laurent, James Lu, Clinton R. Paden, Suxiang Tong, Duncan MacCannell |
| EPI_ISL_1421058, EPI_ISL_1421059 | Wisconsin State Laboratory of Hygiene Communicable Disease Division | Wisconsin State Laboratory of Hygiene Communicable Disease Division | Kelsey R. Florek, Abigail C. Shockey |
| EPI_ISL_1421060 | Helix/Illumina | Centers for Disease Control and Prevention Division of Viral Diseases, Pathogen Discovery | Peter W. Cook, Dakota Howard, Dhwani Batra, Ben L. Rambo-Martin, Eileen de Feo, Jan Antico, Christine Tran, Matthew Tolentino, Shannon Wickline, Kim Gietzen, Brad Sickler, Jingtao Liu, Eric Allen, Phil Febbo, Summer Galloway, Nicole L. Washington, Simon White, Geraint Levan, Kelly Schiabor Barrett, Elizabeth Cirulli, Alexandre Bolze, Ary Ascencio, Charlotte Rivera-Garcia, Ryan Cho, Jason Nguyen, Sherry Wang, Jimmy Ramirez, Tyler Cassens, Efren Sandoval, Magnus Isaksson, William Lee, David Becker, Marc Laurent, James Lu, Clinton R. Paden, Suxiang Tong, Duncan MacCannell |
| EPI_ISL_1421061, EPI_ISL_1421062 | Wisconsin State Laboratory of Hygiene Communicable Disease Division | Wisconsin State Laboratory of Hygiene Communicable Disease Division | Kelsey R. Florek, Abigail C. Shockey |
| EPI_ISL_1421063 | Helix/Illumina | Centers for Disease Control and Prevention Division of Viral Diseases, Pathogen Discovery | Peter W. Cook, Dakota Howard, Dhwani Batra, Ben L. Rambo-Martin, Eileen de Feo, Jan Antico, Christine Tran, Matthew Tolentino, Shannon Wickline, Kim Gietzen, Brad Sickler, Jingtao Liu, Eric Allen, Phil Febbo, Summer Galloway, Nicole L. Washington, Simon White, Geraint Levan, Kelly Schiabor Barrett, Elizabeth Cirulli, Alexandre Bolze, Ary Ascencio, Charlotte Rivera-Garcia, Ryan Cho, Jason Nguyen, Sherry Wang, Jimmy Ramirez, Tyler Cassens, Efren Sandoval, Magnus Isaksson, William Lee, David Becker, Marc Laurent, James Lu, Clinton R. Paden, Suxiang Tong, Duncan MacCannell |
| EPI_ISL_1421064, EPI_ISL_1421065 | Wisconsin State Laboratory of Hygiene Communicable Disease Division | Wisconsin State Laboratory of Hygiene Communicable Disease Division | Kelsey R. Florek, Abigail C. Shockey |
| EPI_ISL_1421066 | Helix/Illumina | Centers for Disease Control and Prevention Division of Viral Diseases, Pathogen Discovery | Peter W. Cook, Dakota Howard, Dhwani Batra, Ben L. Rambo-Martin, Eileen de Feo, Jan Antico, Christine Tran, Matthew Tolentino, Shannon Wickline, Kim Gietzen, Brad Sickler, Jingtao Liu, Eric Allen, Phil Febbo, Summer Galloway, Nicole L. Washington, Simon White, Geraint Levan, Kelly Schiabor Barrett, Elizabeth Cirulli, Alexandre Bolze, Ary Ascencio, Charlotte Rivera-Garcia, Ryan Cho, Jason Nguyen, Sherry Wang, Jimmy Ramirez, Tyler Cassens, Efren Sandoval, Magnus Isaksson, William Lee, David Becker, Marc Laurent, James Lu, Clinton R. Paden, Suxiang Tong, Duncan MacCannell |
| EPI_ISL_1421067, EPI_ISL_1421068 | Wisconsin State Laboratory of Hygiene Communicable Disease Division | Wisconsin State Laboratory of Hygiene Communicable Disease Division | Kelsey R. Florek, Abigail C. Shockey |

| | | | |
|---|---|---|---|
| EPI_ISL_1421069 | Helix/Illumina | Centers for Disease Control and Prevention Division of Viral Diseases, Pathogen Discovery | Peter W. Cook, Dakota Howard, Dhwani Batra, Ben L. Rambo-Martin, Eileen de Feo, Jan Antico, Christine Tran, Matthew Tolentino, Shannon Wickline, Kim Gietzen, Brad Sickler, Jingtao Liu, Eric Allen, Phil Febbo, Summer Galloway, Nicole L. Washington, Simon White, Geraint Levan, Kelly Schiabor Barrett, Elizabeth Cirulli, Alexandre Bolze, Ary Ascencio, Charlotte Rivera-Garcia, Ryan Cho, Jason Nguyen, Sherry Wang, Jimmy Ramirez, Tyler Cassens, Efren Sandoval, Magnus Isaksson, William Lee, David Becker, Marc Laurent, James Lu, Clinton R. Paden, Suxiang Tong, |
| EPI_ISL_1421070 | Wisconsin State Laboratory of Hygiene Communicable Disease Division | Wisconsin State Laboratory of Hygiene Communicable Disease Division | Kelsey R. Florek, Abigail C. Shockey |
| EPI_ISL_1421071 | Helix/Illumina | Centers for Disease Control and Prevention Division of Viral Diseases, Pathogen Discovery | Peter W. Cook, Dakota Howard, Dhwani Batra, Ben L. Rambo-Martin, Eileen de Feo, Jan Antico, Christine Tran, Matthew Tolentino, Shannon Wickline, Kim Gietzen, Brad Sickler, Jingtao Liu, Eric Allen, Phil Febbo, Summer Galloway, Nicole L. Washington, Simon White, Geraint Levan, Kelly Schiabor Barrett, Elizabeth Cirulli, Alexandre Bolze, Ary Ascencio, Charlotte Rivera-Garcia, Ryan Cho, Jason Nguyen, Sherry Wang, Jimmy Ramirez, Tyler Cassens, Efren Sandoval, Magnus Isaksson, William Lee, David Becker, Marc Laurent, James Lu, Clinton R. Paden, Suxiang Tong, |
| EPI_ISL_1421072, EPI_ISL_1421073, EPI_ISL_1421074 | Wisconsin State Laboratory of Hygiene Communicable Disease Division | Wisconsin State Laboratory of Hygiene Communicable Disease Division | Kelsey R. Florek, Abigail C. Shockey |
| EPI_ISL_1421075 | Helix/Illumina | Centers for Disease Control and Prevention Division of Viral Diseases, Pathogen Discovery | Peter W. Cook, Dakota Howard, Dhwani Batra, Ben L. Rambo-Martin, Eileen de Feo, Jan Antico, Christine Tran, Matthew Tolentino, Shannon Wickline, Kim Gietzen, Brad Sickler, Jingtao Liu, Eric Allen, Phil Febbo, Summer Galloway, Nicole L. Washington, Simon White, Geraint Levan, Kelly Schiabor Barrett, Elizabeth Cirulli, Alexandre Bolze, Ary Ascencio, Charlotte Rivera-Garcia, Ryan Cho, Jason Nguyen, Sherry Wang, Jimmy Ramirez, Tyler Cassens, Efren Sandoval, Magnus Isaksson, William Lee, David Becker, Marc Laurent, James Lu, Clinton R. Paden, Suxiang Tong, Duncan MacCannell |
| EPI_ISL_1421076, EPI_ISL_1421077 | Wisconsin State Laboratory of Hygiene Communicable Disease Division | Wisconsin State Laboratory of Hygiene Communicable Disease Division | Kelsey R. Florek, Abigail C. Shockey |
| EPI_ISL_1421078 | Helix/Illumina | Centers for Disease Control and Prevention Division of Viral Diseases, Pathogen Discovery | Peter W. Cook, Dakota Howard, Dhwani Batra, Ben L. Rambo-Martin, Eileen de Feo, Jan Antico, Christine Tran, Matthew Tolentino, Shannon Wickline, Kim Gietzen, Brad Sickler, Jingtao Liu, Eric Allen, Phil Febbo, Summer Galloway, Nicole L. Washington, Simon White, Geraint Levan, Kelly Schiabor Barrett, Elizabeth Cirulli, Alexandre Bolze, Ary Ascencio, Charlotte Rivera-Garcia, Ryan Cho, Jason Nguyen, Sherry Wang, Jimmy Ramirez, Tyler Cassens, Efren Sandoval, Magnus Isaksson, William Lee, David Becker, Marc Laurent, James Lu, Clinton R. Paden, Suxiang Tong, Duncan MacCannell |
| EPI_ISL_1421079, EPI_ISL_1421080 | Wisconsin State Laboratory of Hygiene Communicable Disease Division | Wisconsin State Laboratory of Hygiene Communicable Disease Division | Kelsey R. Florek, Abigail C. Shockey |
| EPI_ISL_1421081 | Helix/Illumina | Centers for Disease Control and Prevention Division of Viral Diseases, Pathogen Discovery | Peter W. Cook, Dakota Howard, Dhwani Batra, Ben L. Rambo-Martin, Eileen de Feo, Jan Antico, Christine Tran, Matthew Tolentino, Shannon Wickline, Kim Gietzen, Brad Sickler, Jingtao Liu, Eric Allen, Phil Febbo, Summer Galloway, Nicole L. Washington, Simon White, Geraint Levan, Kelly Schiabor Barrett, Elizabeth Cirulli, Alexandre Bolze, Ary Ascencio, Charlotte Rivera-Garcia, Ryan Cho, Jason Nguyen, Sherry Wang, Jimmy Ramirez, Tyler Cassens, Efren Sandoval, Magnus Isaksson, William Lee, David Becker, Marc Laurent, James Lu, Clinton R. Paden, Suxiang Tong, Duncan MacCannell |
| EPI_ISL_1421083 | Wisconsin State Laboratory of Hygiene Communicable Disease Division | Wisconsin State Laboratory of Hygiene Communicable Disease Division | Kelsey R. Florek, Abigail C. Shockey |
| EPI_ISL_1421084 | Helix/Illumina | Centers for Disease Control and Prevention Division of Viral Diseases, Pathogen Discovery | Peter W. Cook, Dakota Howard, Dhwani Batra, Ben L. Rambo-Martin, Eileen de Feo, Jan Antico, Christine Tran, Matthew Tolentino, Shannon Wickline, Kim Gietzen, Brad Sickler, Jingtao Liu, Eric Allen, Phil Febbo, Summer Galloway, Nicole L. Washington, Simon White, Geraint Levan, Kelly Schiabor Barrett, Elizabeth Cirulli, Alexandre Bolze, Ary Ascencio, Charlotte Rivera-Garcia, Ryan Cho, Jason Nguyen, Sherry Wang, Jimmy Ramirez, Tyler Cassens, Efren Sandoval, Magnus Isaksson, William Lee, David Becker, Marc Laurent, James Lu, Clinton R. Paden, Suxiang Tong, Duncan MacCannell |
| EPI_ISL_1421085, EPI_ISL_1421086 | Wisconsin State Laboratory of Hygiene Communicable Disease Division | Wisconsin State Laboratory of Hygiene Communicable Disease Division | Kelsey R. Florek, Abigail C. Shockey |
| EPI_ISL_1421087 | Helix/Illumina | Centers for Disease Control and Prevention Division of Viral Diseases, Pathogen Discovery | Peter W. Cook, Dakota Howard, Dhwani Batra, Ben L. Rambo-Martin, Eileen de Feo, Jan Antico, Christine Tran, Matthew Tolentino, Shannon Wickline, Kim Gietzen, Brad Sickler, Jingtao Liu, Eric Allen, Phil Febbo, Summer Galloway, Nicole L. Washington, Simon White, Geraint Levan, Kelly Schiabor Barrett, Elizabeth Cirulli, Alexandre Bolze, Ary Ascencio, Charlotte Rivera-Garcia, Ryan Cho, Jason Nguyen, Sherry Wang, Jimmy Ramirez, Tyler Cassens, Efren Sandoval, Magnus Isaksson, William Lee, David Becker, Marc Laurent, James Lu, Clinton R. Paden, Suxiang Tong, Duncan MacCannell |
| EPI_ISL_1421088, EPI_ISL_1421089 | Wisconsin State Laboratory of Hygiene Communicable Disease Division | Wisconsin State Laboratory of Hygiene Communicable Disease Division | Kelsey R. Florek, Abigail C. Shockey |
| EPI_ISL_1421090 | Helix/Illumina | Centers for Disease Control and Prevention Division of Viral Diseases, Pathogen Discovery | Peter W. Cook, Dakota Howard, Dhwani Batra, Ben L. Rambo-Martin, Eileen de Feo, Jan Antico, Christine Tran, Matthew Tolentino, Shannon Wickline, Kim Gietzen, Brad Sickler, Jingtao Liu, Eric Allen, Phil Febbo, Summer Galloway, Nicole L. Washington, Simon White, Geraint Levan, Kelly Schiabor Barrett, Elizabeth Cirulli, Alexandre Bolze, Ary Ascencio, Charlotte Rivera-Garcia, Ryan Cho, Jason Nguyen, Sherry Wang, Jimmy Ramirez, Tyler Cassens, Efren Sandoval, Magnus Isaksson, William Lee, David Becker, Marc Laurent, James Lu, Clinton R. Paden, Suxiang Tong, Duncan MacCannell |
| EPI_ISL_1421091, EPI_ISL_1421092 | Wisconsin State Laboratory of Hygiene Communicable Disease Division | Wisconsin State Laboratory of Hygiene Communicable Disease Division | Kelsey R. Florek, Abigail C. Shockey |
| EPI_ISL_1421093 | Helix/Illumina | Centers for Disease Control and Prevention Division of Viral Diseases, Pathogen Discovery | Peter W. Cook, Dakota Howard, Dhwani Batra, Ben L. Rambo-Martin, Eileen de Feo, Jan Antico, Christine Tran, Matthew Tolentino, Shannon Wickline, Kim Gietzen, Brad Sickler, Jingtao Liu, Eric Allen, Phil Febbo, Summer Galloway, Nicole L. Washington, Simon White, Geraint Levan, Kelly Schiabor Barrett, Elizabeth Cirulli, Alexandre Bolze, Ary Ascencio, Charlotte Rivera-Garcia, Ryan Cho, Jason Nguyen, Sherry Wang, Jimmy Ramirez, Tyler Cassens, Efren Sandoval, Magnus Isaksson, William Lee, David Becker, Marc Laurent, James Lu, Clinton R. Paden, Suxiang Tong, Duncan MacCannell |
| EPI_ISL_1421094, EPI_ISL_1421095 | Wisconsin State Laboratory of Hygiene Communicable Disease Division | Wisconsin State Laboratory of Hygiene Communicable Disease Division | Kelsey R. Florek, Abigail C. Shockey |
| EPI_ISL_1421096 | Helix/Illumina | Centers for Disease Control and Prevention Division of Viral Diseases, Pathogen Discovery | Peter W. Cook, Dakota Howard, Dhwani Batra, Ben L. Rambo-Martin, Eileen de Feo, Jan Antico, Christine Tran, Matthew Tolentino, Shannon Wickline, Kim Gietzen, Brad Sickler, Jingtao Liu, Eric Allen, Phil Febbo, Summer Galloway, Nicole L. Washington, Simon White, Geraint Levan, Kelly Schiabor Barrett, Elizabeth Cirulli, Alexandre Bolze, Ary Ascencio, Charlotte Rivera-Garcia, Ryan Cho, Jason Nguyen, Sherry Wang, Jimmy Ramirez, Tyler Cassens, Efren Sandoval, Magnus Isaksson, William Lee, David Becker, Marc Laurent, James Lu, Clinton R. Paden, Suxiang Tong, Duncan MacCannell |
| EPI_ISL_1421097, EPI_ISL_1421098 | Wisconsin State Laboratory of Hygiene Communicable Disease Division | Wisconsin State Laboratory of Hygiene Communicable Disease Division | Kelsey R. Florek, Abigail C. Shockey |
| EPI_ISL_1421099 | Helix/Illumina | Centers for Disease Control and Prevention Division of Viral Diseases, Pathogen Discovery | Peter W. Cook, Dakota Howard, Dhwani Batra, Ben L. Rambo-Martin, Eileen de Feo, Jan Antico, Christine Tran, Matthew Tolentino, Shannon Wickline, Kim Gietzen, Brad Sickler, Jingtao Liu, Eric Allen, Phil Febbo, Summer Galloway, Nicole L. Washington, Simon White, Geraint Levan, Kelly Schiabor Barrett, Elizabeth Cirulli, Alexandre Bolze, Ary Ascencio, Charlotte Rivera-Garcia, Ryan Cho, Jason Nguyen, Sherry Wang, Jimmy Ramirez, Tyler Cassens, Efren Sandoval, Magnus Isaksson, William Lee, David Becker, Marc Laurent, James Lu, Clinton R. Paden, Suxiang Tong, Duncan MacCannell |
| EPI_ISL_1421100, EPI_ISL_1421101 | Wisconsin State Laboratory of Hygiene Communicable Disease Division | Wisconsin State Laboratory of Hygiene Communicable Disease Division | Kelsey R. Florek, Abigail C. Shockey |
| EPI_ISL_1421102 | Helix/Illumina | Centers for Disease Control and Prevention Division of Viral Diseases, Pathogen Discovery | Peter W. Cook, Dakota Howard, Dhwani Batra, Ben L. Rambo-Martin, Eileen de Feo, Jan Antico, Christine Tran, Matthew Tolentino, Shannon Wickline, Kim Gietzen, Brad Sickler, Jingtao Liu, Eric Allen, Phil Febbo, Summer Galloway, Nicole L. Washington, Simon White, Geraint Levan, Kelly Schiabor Barrett, Elizabeth Cirulli, Alexandre Bolze, Ary Ascencio, Charlotte Rivera-Garcia, Ryan Cho, Jason Nguyen, Sherry Wang, Jimmy Ramirez, Tyler Cassens, Efren Sandoval, Magnus Isaksson, William Lee, David Becker, Marc Laurent, James Lu, Clinton R. Paden, Suxiang Tong, Duncan MacCannell |
| EPI_ISL_1421103, EPI_ISL_1421104 | Wisconsin State Laboratory of Hygiene Communicable Disease Division | Wisconsin State Laboratory of Hygiene Communicable Disease Division | Kelsey R. Florek, Abigail C. Shockey |
| EPI_ISL_1421105 | Helix/Illumina | Centers for Disease Control and Prevention Division of Viral Diseases, Pathogen Discovery | Peter W. Cook, Dakota Howard, Dhwani Batra, Ben L. Rambo-Martin, Eileen de Feo, Jan Antico, Christine Tran, Matthew Tolentino, Shannon Wickline, Kim Gietzen, Brad Sickler, Jingtao Liu, Eric Allen, Phil Febbo, Summer Galloway, Nicole L. Washington, Simon White, Geraint Levan, Kelly Schiabor Barrett, Elizabeth Cirulli, Alexandre Bolze, Ary Ascencio, Charlotte Rivera-Garcia, Ryan Cho, Jason Nguyen, Sherry Wang, Jimmy Ramirez, Tyler Cassens, Efren Sandoval, Magnus Isaksson, William Lee, David Becker, Marc Laurent, James Lu, Clinton R. Paden, Suxiang Tong, Duncan MacCannell |
| EPI_ISL_1421106, EPI_ISL_1421107 | Wisconsin State Laboratory of Hygiene Communicable Disease Division | Wisconsin State Laboratory of Hygiene Communicable Disease Division | Kelsey R. Florek, Abigail C. Shockey |
| EPI_ISL_1421108 | Helix/Illumina | Centers for Disease Control and Prevention Division of Viral Diseases, Pathogen Discovery | Peter W. Cook, Dakota Howard, Dhwani Batra, Ben L. Rambo-Martin, Eileen de Feo, Jan Antico, Christine Tran, Matthew Tolentino, Shannon Wickline, Kim Gietzen, Brad Sickler, Jingtao Liu, Eric Allen, Phil Febbo, Summer Galloway, Nicole L. Washington, Simon White, Geraint Levan, Kelly Schiabor Barrett, Elizabeth Cirulli, Alexandre Bolze, Ary Ascencio, Charlotte Rivera-Garcia, Ryan Cho, Jason Nguyen, Sherry Wang, Jimmy Ramirez, Tyler Cassens, Efren Sandoval, Magnus Isaksson, William Lee, David Becker, Marc Laurent, James Lu, Clinton R. Paden, Suxiang Tong, Duncan MacCannell |
| EPI_ISL_1421109, EPI_ISL_1421110 | Wisconsin State Laboratory of Hygiene Communicable Disease Division | Wisconsin State Laboratory of Hygiene Communicable Disease Division | Kelsey R. Florek, Abigail C. Shockey |
| EPI_ISL_1421111 | Helix/Illumina | Centers for Disease Control and Prevention Division of Viral Diseases, Pathogen Discovery | Peter W. Cook, Dakota Howard, Dhwani Batra, Ben L. Rambo-Martin, Eileen de Feo, Jan Antico, Christine Tran, Matthew Tolentino, Shannon Wickline, Kim Gietzen, Brad Sickler, Jingtao Liu, Eric Allen, Phil Febbo, Summer Galloway, Nicole L. Washington, Simon White, Geraint Levan, Kelly Schiabor Barrett, Elizabeth Cirulli, Alexandre Bolze, Ary Ascencio, Charlotte Rivera-Garcia, Ryan Cho, Jason Nguyen, Sherry Wang, Jimmy Ramirez, Tyler Cassens, Efren Sandoval, Magnus Isaksson, William Lee, David Becker, Marc Laurent, James Lu, Clinton R. Paden, Suxiang Tong, Duncan MacCannell |
| EPI_ISL_1421112, EPI_ISL_1421113 | Wisconsin State Laboratory of Hygiene Communicable Disease Division | Wisconsin State Laboratory of Hygiene Communicable Disease Division | Kelsey R. Florek, Abigail C. Shockey |
| EPI_ISL_1421114 | Helix/Illumina | Centers for Disease Control and Prevention Division of Viral Diseases, Pathogen Discovery | Peter W. Cook, Dakota Howard, Dhwani Batra, Ben L. Rambo-Martin, Eileen de Feo, Jan Antico, Christine Tran, Matthew Tolentino, Shannon Wickline, Kim Gietzen, Brad Sickler, Jingtao Liu, Eric Allen, Phil Febbo, Summer Galloway, Nicole L. Washington, Simon White, Geraint Levan, Kelly Schiabor Barrett, Elizabeth Cirulli, Alexandre Bolze, Ary Ascencio, Charlotte Rivera-Garcia, Ryan Cho, Jason Nguyen, Sherry Wang, Jimmy Ramirez, Tyler Cassens, Efren Sandoval, Magnus Isaksson, William Lee, David Becker, Marc Laurent, James Lu, Clinton R. Paden, Suxiang Tong, Duncan MacCannell |
| EPI_ISL_1421115, EPI_ISL_1421116 | Wisconsin State Laboratory of Hygiene Communicable Disease Division | Wisconsin State Laboratory of Hygiene Communicable Disease Division | Kelsey R. Florek, Abigail C. Shockey |
| EPI_ISL_1421117 | Helix/Illumina | Centers for Disease Control and Prevention Division of Viral Diseases, Pathogen Discovery | Peter W. Cook, Dakota Howard, Dhwani Batra, Ben L. Rambo-Martin, Eileen de Feo, Jan Antico, Christine Tran, Matthew Tolentino, Shannon Wickline, Kim Gietzen, Brad Sickler, Jingtao Liu, Eric Allen, Phil Febbo, Summer Galloway, Nicole L. Washington, Simon White, Geraint Levan, Kelly Schiabor Barrett, Elizabeth Cirulli, Alexandre Bolze, Ary Ascencio, Charlotte Rivera-Garcia, Ryan Cho, Jason Nguyen, Sherry Wang, Jimmy Ramirez, Tyler Cassens, Efren Sandoval, Magnus Isaksson, William Lee, David Becker, Marc Laurent, James Lu, Clinton R. Paden, Suxiang Tong, Duncan MacCannell |
| EPI_ISL_1421118, EPI_ISL_1421119 | Wisconsin State Laboratory of Hygiene Communicable Disease Division | Wisconsin State Laboratory of Hygiene Communicable Disease Division | Kelsey R. Florek, Abigail C. Shockey |
| EPI_ISL_1421120 | Helix/Illumina | Centers for Disease Control and Prevention Division of Viral Diseases, Pathogen Discovery | Peter W. Cook, Dakota Howard, Dhwani Batra, Ben L. Rambo-Martin, Eileen de Feo, Jan Antico, Christine Tran, Matthew Tolentino, Shannon Wickline, Kim Gietzen, Brad Sickler, Jingtao Liu, Eric Allen, Phil Febbo, Summer Galloway, Nicole L. Washington, Simon White, Geraint Levan, Kelly Schiabor Barrett, Elizabeth Cirulli, Alexandre Bolze, Ary Ascencio, Charlotte Rivera-Garcia, Ryan Cho, Jason Nguyen, Sherry Wang, Jimmy Ramirez, Tyler Cassens, Efren Sandoval, Magnus Isaksson, William Lee, David Becker, Marc Laurent, James Lu, Clinton R. Paden, Suxiang Tong, Duncan MacCannell |
| EPI_ISL_1421121, EPI_ISL_1421122 | Wisconsin State Laboratory of Hygiene Communicable Disease Division | Wisconsin State Laboratory of Hygiene Communicable Disease Division | Kelsey R. Florek, Abigail C. Shockey |
| EPI_ISL_1421123 | Helix/Illumina | Centers for Disease Control and Prevention Division of Viral Diseases, Pathogen Discovery | Peter W. Cook, Dakota Howard, Dhwani Batra, Ben L. Rambo-Martin, Eileen de Feo, Jan Antico, Christine Tran, Matthew Tolentino, Shannon Wickline, Kim Gietzen, Brad Sickler, Jingtao Liu, Eric Allen, Phil Febbo, Summer Galloway, Nicole L. Washington, Simon White, Geraint Levan, Kelly Schiabor Barrett, Elizabeth Cirulli, Alexandre Bolze, Ary Ascencio, Charlotte Rivera-Garcia, Ryan Cho, Jason Nguyen, Sherry Wang, Jimmy Ramirez, Tyler Cassens, Efren Sandoval, Magnus Isaksson, William Lee, David Becker, Marc Laurent, James Lu, Clinton R. Paden, Suxiang Tong, Duncan MacCannell |

| | | | |
|---|---|---|---|
| EPI_ISL_1421124, EPI_ISL_1421125 | Wisconsin State Laboratory of Hygiene Communicable Disease Division | Wisconsin State Laboratory of Hygiene Communicable Disease Division | Kelsey R. Florek, Abigail C. Shockey |
| EPI_ISL_1421126 | Helix/Illumina | Centers for Disease Control and Prevention Division of Viral Diseases, Pathogen Discovery | Peter W. Cook, Dakota Howard, Dhwani Batra, Ben L. Rambo-Martin, Eileen de Feo, Jan Antico, Christine Tran, Matthew Tolentino, Shannon Wickline, Kim Gietzen, Brad Sickler, Jingtao Liu, Eric Allen, Phil Febbo, Summer Galloway, Nicole L. Washington, Simon White, Geraint Levan, Kelly Schiabor Barrett, Elizabeth Cirulli, Alexandre Bolze, Ary Ascencio, Charlotte Rivera-Garcia, Ryan Cho, Jason Nguyen, Sherry Wang, Jimmy Ramirez, Tyler Cassens, Efren Sandoval, Magnus Isaksson, William Lee, David Becker, Marc Laurent, James Lu, Clinton R. Paden, Suxiang Tong, Duncan MacCannell |
| EPI_ISL_1421127 | Wisconsin State Laboratory of Hygiene Communicable Disease Division | Wisconsin State Laboratory of Hygiene Communicable Disease Division | Kelsey R. Florek, Abigail C. Shockey |
| EPI_ISL_1421128 | Helix/Illumina | Centers for Disease Control and Prevention Division of Viral Diseases, Pathogen Discovery | Peter W. Cook, Dakota Howard, Dhwani Batra, Ben L. Rambo-Martin, Eileen de Feo, Jan Antico, Christine Tran, Matthew Tolentino, Shannon Wickline, Kim Gietzen, Brad Sickler, Jingtao Liu, Eric Allen, Phil Febbo, Summer Galloway, Nicole L. Washington, Simon White, Geraint Levan, Kelly Schiabor Barrett, Elizabeth Cirulli, Alexandre Bolze, Ary Ascencio, Charlotte Rivera-Garcia, Ryan Cho, Jason Nguyen, Sherry Wang, Jimmy Ramirez, Tyler Cassens, Efren Sandoval, Magnus Isaksson, William Lee, David Becker, Marc Laurent, James Lu, Clinton R. Paden, Suxiang Tong, Duncan MacCannell |
| EPI_ISL_1421129, EPI_ISL_1421130 | Wisconsin State Laboratory of Hygiene Communicable Disease Division | Wisconsin State Laboratory of Hygiene Communicable Disease Division | Kelsey R. Florek, Abigail C. Shockey |
| EPI_ISL_1421131 | Helix/Illumina | Centers for Disease Control and Prevention Division of Viral Diseases, Pathogen Discovery | Peter W. Cook, Dakota Howard, Dhwani Batra, Ben L. Rambo-Martin, Eileen de Feo, Jan Antico, Christine Tran, Matthew Tolentino, Shannon Wickline, Kim Gietzen, Brad Sickler, Jingtao Liu, Eric Allen, Phil Febbo, Summer Galloway, Nicole L. Washington, Simon White, Geraint Levan, Kelly Schiabor Barrett, Elizabeth Cirulli, Alexandre Bolze, Ary Ascencio, Charlotte Rivera-Garcia, Ryan Cho, Jason Nguyen, Sherry Wang, Jimmy Ramirez, Tyler Cassens, Efren Sandoval, Magnus Isaksson, William Lee, David Becker, Marc Laurent, James Lu, Clinton R. Paden, Suxiang Tong, Duncan MacCannell |
| EPI_ISL_1421132, EPI_ISL_1421133 | Wisconsin State Laboratory of Hygiene Communicable Disease Division | Wisconsin State Laboratory of Hygiene Communicable Disease Division | Kelsey R. Florek, Abigail C. Shockey |
| EPI_ISL_1421134 | Helix/Illumina | Centers for Disease Control and Prevention Division of Viral Diseases, Pathogen Discovery | Peter W. Cook, Dakota Howard, Dhwani Batra, Ben L. Rambo-Martin, Eileen de Feo, Jan Antico, Christine Tran, Matthew Tolentino, Shannon Wickline, Kim Gietzen, Brad Sickler, Jingtao Liu, Eric Allen, Phil Febbo, Summer Galloway, Nicole L. Washington, Simon White, Geraint Levan, Kelly Schiabor Barrett, Elizabeth Cirulli, Alexandre Bolze, Ary Ascencio, Charlotte Rivera-Garcia, Ryan Cho, Jason Nguyen, Sherry Wang, Jimmy Ramirez, Tyler Cassens, Efren Sandoval, Magnus Isaksson, William Lee, David Becker, Marc Laurent, James Lu, Clinton R. Paden, Suxiang Tong, Duncan MacCannell |
| EPI_ISL_1421135, EPI_ISL_1421136, EPI_ISL_1421137 | Wisconsin State Laboratory of Hygiene Communicable Disease Division | Wisconsin State Laboratory of Hygiene Communicable Disease Division | Kelsey R. Florek, Abigail C. Shockey |
| EPI_ISL_1421138 | Helix/Illumina | Centers for Disease Control and Prevention Division of Viral Diseases, Pathogen Discovery | Peter W. Cook, Dakota Howard, Dhwani Batra, Ben L. Rambo-Martin, Eileen de Feo, Jan Antico, Christine Tran, Matthew Tolentino, Shannon Wickline, Kim Gietzen, Brad Sickler, Jingtao Liu, Eric Allen, Phil Febbo, Summer Galloway, Nicole L. Washington, Simon White, Geraint Levan, Kelly Schiabor Barrett, Elizabeth Cirulli, Alexandre Bolze, Ary Ascencio, Charlotte Rivera-Garcia, Ryan Cho, Jason Nguyen, Sherry Wang, Jimmy Ramirez, Tyler Cassens, Efren Sandoval, Magnus Isaksson, William Lee, David Becker, Marc Laurent, James Lu, Clinton R. Paden, Suxiang Tong, Duncan MacCannell |
| EPI_ISL_1421139 | Wisconsin State Laboratory of Hygiene Communicable Disease Division | Wisconsin State Laboratory of Hygiene Communicable Disease Division | Kelsey R. Florek, Abigail C. Shockey |
| EPI_ISL_1421140 | Helix/Illumina | Centers for Disease Control and Prevention Division of Viral Diseases, Pathogen Discovery | Peter W. Cook, Dakota Howard, Dhwani Batra, Ben L. Rambo-Martin, Eileen de Feo, Jan Antico, Christine Tran, Matthew Tolentino, Shannon Wickline, Kim Gietzen, Brad Sickler, Jingtao Liu, Eric Allen, Phil Febbo, Summer Galloway, Nicole L. Washington, Simon White, Geraint Levan, Kelly Schiabor Barrett, Elizabeth Cirulli, Alexandre Bolze, Ary Ascencio, Charlotte Rivera-Garcia, Ryan Cho, Jason Nguyen, Sherry Wang, Jimmy Ramirez, Tyler Cassens, Efren Sandoval, Magnus Isaksson, William Lee, David Becker, Marc Laurent, James Lu, Clinton R. Paden, Suxiang Tong, Duncan MacCannell |
| EPI_ISL_1421141, EPI_ISL_1421142 | Wisconsin State Laboratory of Hygiene Communicable Disease Division | Wisconsin State Laboratory of Hygiene Communicable Disease Division | Kelsey R. Florek, Abigail C. Shockey |
| EPI_ISL_1421143 | Helix/Illumina | Centers for Disease Control and Prevention Division of Viral Diseases, Pathogen Discovery | Peter W. Cook, Dakota Howard, Dhwani Batra, Ben L. Rambo-Martin, Eileen de Feo, Jan Antico, Christine Tran, Matthew Tolentino, Shannon Wickline, Kim Gietzen, Brad Sickler, Jingtao Liu, Eric Allen, Phil Febbo, Summer Galloway, Nicole L. Washington, Simon White, Geraint Levan, Kelly Schiabor Barrett, Elizabeth Cirulli, Alexandre Bolze, Ary Ascencio, Charlotte Rivera-Garcia, Ryan Cho, Jason Nguyen, Sherry Wang, Jimmy Ramirez, Tyler Cassens, Efren Sandoval, Magnus Isaksson, William Lee, David Becker, Marc Laurent, James Lu, Clinton R. Paden, Suxiang Tong, Duncan MacCannell |
| EPI_ISL_1421144, EPI_ISL_1421145 | Wisconsin State Laboratory of Hygiene Communicable Disease Division | Wisconsin State Laboratory of Hygiene Communicable Disease Division | Kelsey R. Florek, Abigail C. Shockey |
| EPI_ISL_1421146 | Helix/Illumina | Centers for Disease Control and Prevention Division of Viral Diseases, Pathogen Discovery | Peter W. Cook, Dakota Howard, Dhwani Batra, Ben L. Rambo-Martin, Eileen de Feo, Jan Antico, Christine Tran, Matthew Tolentino, Shannon Wickline, Kim Gietzen, Brad Sickler, Jingtao Liu, Eric Allen, Phil Febbo, Summer Galloway, Nicole L. Washington, Simon White, Geraint Levan, Kelly Schiabor Barrett, Elizabeth Cirulli, Alexandre Bolze, Ary Ascencio, Charlotte Rivera-Garcia, Ryan Cho, Jason Nguyen, Sherry Wang, Jimmy Ramirez, Tyler Cassens, Efren Sandoval, Magnus Isaksson, William Lee, David Becker, Marc Laurent, James Lu, Clinton R. Paden, Suxiang Tong, Duncan MacCannell |
| EPI_ISL_1421147 | Wisconsin State Laboratory of Hygiene Communicable Disease Division | Wisconsin State Laboratory of Hygiene Communicable Disease Division | Kelsey R. Florek, Abigail C. Shockey |
| EPI_ISL_1421148 | Helix/Illumina | Centers for Disease Control and Prevention Division of Viral Diseases, Pathogen Discovery | Peter W. Cook, Dakota Howard, Dhwani Batra, Ben L. Rambo-Martin, Eileen de Feo, Jan Antico, Christine Tran, Matthew Tolentino, Shannon Wickline, Kim Gietzen, Brad Sickler, Jingtao Liu, Eric Allen, Phil Febbo, Summer Galloway, Nicole L. Washington, Simon White, Geraint Levan, Kelly Schiabor Barrett, Elizabeth Cirulli, Alexandre Bolze, Ary Ascencio, Charlotte Rivera-Garcia, Ryan Cho, Jason Nguyen, Sherry Wang, Jimmy Ramirez, Tyler Cassens, Efren Sandoval, Magnus Isaksson, William Lee, David Becker, Marc Laurent, James Lu, Clinton R. Paden, Suxiang Tong, Duncan MacCannell |
| EPI_ISL_1421149, EPI_ISL_1421150 | Wisconsin State Laboratory of Hygiene Communicable Disease Division | Wisconsin State Laboratory of Hygiene Communicable Disease Division | Kelsey R. Florek, Abigail C. Shockey |
| EPI_ISL_1421151 | Helix/Illumina | Centers for Disease Control and Prevention Division of Viral Diseases, Pathogen Discovery | Peter W. Cook, Dakota Howard, Dhwani Batra, Ben L. Rambo-Martin, Eileen de Feo, Jan Antico, Christine Tran, Matthew Tolentino, Shannon Wickline, Kim Gietzen, Brad Sickler, Jingtao Liu, Eric Allen, Phil Febbo, Summer Galloway, Nicole L. Washington, Simon White, Geraint Levan, Kelly Schiabor Barrett, Elizabeth Cirulli, Alexandre Bolze, Ary Ascencio, Charlotte Rivera-Garcia, Ryan Cho, Jason Nguyen, Sherry Wang, Jimmy Ramirez, Tyler Cassens, Efren Sandoval, Magnus Isaksson, William Lee, David Becker, Marc Laurent, James Lu, Clinton R. Paden, Suxiang Tong, Duncan MacCannell |
| EPI_ISL_1421152, EPI_ISL_1421153 | Wisconsin State Laboratory of Hygiene Communicable Disease Division | Wisconsin State Laboratory of Hygiene Communicable Disease Division | Kelsey R. Florek, Abigail C. Shockey |
| EPI_ISL_1421154 | Helix/Illumina | Centers for Disease Control and Prevention Division of Viral Diseases, Pathogen Discovery | Peter W. Cook, Dakota Howard, Dhwani Batra, Ben L. Rambo-Martin, Eileen de Feo, Jan Antico, Christine Tran, Matthew Tolentino, Shannon Wickline, Kim Gietzen, Brad Sickler, Jingtao Liu, Eric Allen, Phil Febbo, Summer Galloway, Nicole L. Washington, Simon White, Geraint Levan, Kelly Schiabor Barrett, Elizabeth Cirulli, Alexandre Bolze, Ary Ascencio, Charlotte Rivera-Garcia, Ryan Cho, Jason Nguyen, Sherry Wang, Jimmy Ramirez, Tyler Cassens, Efren Sandoval, Magnus Isaksson, William Lee, David Becker, Marc Laurent, James Lu, Clinton R. Paden, Suxiang Tong, Duncan MacCannell |
| EPI_ISL_1421155, EPI_ISL_1421156, EPI_ISL_1421157 | Wisconsin State Laboratory of Hygiene Communicable Disease Division | Wisconsin State Laboratory of Hygiene Communicable Disease Division | Kelsey R. Florek, Abigail C. Shockey |
| EPI_ISL_1421158 | Helix/Illumina | Centers for Disease Control and Prevention Division of Viral Diseases, Pathogen Discovery | Peter W. Cook, Dakota Howard, Dhwani Batra, Ben L. Rambo-Martin, Eileen de Feo, Jan Antico, Christine Tran, Matthew Tolentino, Shannon Wickline, Kim Gietzen, Brad Sickler, Jingtao Liu, Eric Allen, Phil Febbo, Summer Galloway, Nicole L. Washington, Simon White, Geraint Levan, Kelly Schiabor Barrett, Elizabeth Cirulli, Alexandre Bolze, Ary Ascencio, Charlotte Rivera-Garcia, Ryan Cho, Jason Nguyen, Sherry Wang, Jimmy Ramirez, Tyler Cassens, Efren Sandoval, Magnus Isaksson, William Lee, David Becker, Marc Laurent, James Lu, Clinton R. Paden, Suxiang Tong, Duncan MacCannell |
| EPI_ISL_1421160 | Wisconsin State Laboratory of Hygiene Communicable Disease Division | Wisconsin State Laboratory of Hygiene Communicable Disease Division | Kelsey R. Florek, Abigail C. Shockey |
| EPI_ISL_1421161 | Helix/Illumina | Centers for Disease Control and Prevention Division of Viral Diseases, Pathogen Discovery | Peter W. Cook, Dakota Howard, Dhwani Batra, Ben L. Rambo-Martin, Eileen de Feo, Jan Antico, Christine Tran, Matthew Tolentino, Shannon Wickline, Kim Gietzen, Brad Sickler, Jingtao Liu, Eric Allen, Phil Febbo, Summer Galloway, Nicole L. Washington, Simon White, Geraint Levan, Kelly Schiabor Barrett, Elizabeth Cirulli, Alexandre Bolze, Ary Ascencio, Charlotte Rivera-Garcia, Ryan Cho, Jason Nguyen, Sherry Wang, Jimmy Ramirez, Tyler Cassens, Efren Sandoval, Magnus Isaksson, William Lee, David Becker, Marc Laurent, James Lu, Clinton R. Paden, Suxiang Tong, Duncan MacCannell |
| EPI_ISL_1421162 | Wisconsin State Laboratory of Hygiene Communicable Disease Division | Wisconsin State Laboratory of Hygiene Communicable Disease Division | Kelsey R. Florek, Abigail C. Shockey |
| EPI_ISL_1421163 | Helix/Illumina | Centers for Disease Control and Prevention Division of Viral Diseases, Pathogen Discovery | Peter W. Cook, Dakota Howard, Dhwani Batra, Ben L. Rambo-Martin, Eileen de Feo, Jan Antico, Christine Tran, Matthew Tolentino, Shannon Wickline, Kim Gietzen, Brad Sickler, Jingtao Liu, Eric Allen, Phil Febbo, Summer Galloway, Nicole L. Washington, Simon White, Geraint Levan, Kelly Schiabor Barrett, Elizabeth Cirulli, Alexandre Bolze, Ary Ascencio, Charlotte Rivera-Garcia, Ryan Cho, Jason Nguyen, Sherry Wang, Jimmy Ramirez, Tyler Cassens, Efren Sandoval, Magnus Isaksson, William Lee, David Becker, Marc Laurent, James Lu, Clinton R. Paden, Suxiang Tong, Duncan MacCannell |
| EPI_ISL_1421164, EPI_ISL_1421165 | Wisconsin State Laboratory of Hygiene Communicable Disease Division | Wisconsin State Laboratory of Hygiene Communicable Disease Division | Kelsey R. Florek, Abigail C. Shockey |
| EPI_ISL_1421166 | Helix/Illumina | Centers for Disease Control and Prevention Division of Viral Diseases, Pathogen Discovery | Peter W. Cook, Dakota Howard, Dhwani Batra, Ben L. Rambo-Martin, Eileen de Feo, Jan Antico, Christine Tran, Matthew Tolentino, Shannon Wickline, Kim Gietzen, Brad Sickler, Jingtao Liu, Eric Allen, Phil Febbo, Summer Galloway, Nicole L. Washington, Simon White, Geraint Levan, Kelly Schiabor Barrett, Elizabeth Cirulli, Alexandre Bolze, Ary Ascencio, Charlotte Rivera-Garcia, Ryan Cho, Jason Nguyen, Sherry Wang, Jimmy Ramirez, Tyler Cassens, Efren Sandoval, Magnus Isaksson, William Lee, David Becker, Marc Laurent, James Lu, Clinton R. Paden, Suxiang Tong, Duncan MacCannell |
| EPI_ISL_1421167, EPI_ISL_1421168 | Wisconsin State Laboratory of Hygiene Communicable Disease Division | Wisconsin State Laboratory of Hygiene Communicable Disease Division | Kelsey R. Florek, Abigail C. Shockey |
| EPI_ISL_1421169 | Helix/Illumina | Centers for Disease Control and Prevention Division of Viral Diseases, Pathogen Discovery | Peter W. Cook, Dakota Howard, Dhwani Batra, Ben L. Rambo-Martin, Eileen de Feo, Jan Antico, Christine Tran, Matthew Tolentino, Shannon Wickline, Kim Gietzen, Brad Sickler, Jingtao Liu, Eric Allen, Phil Febbo, Summer Galloway, Nicole L. Washington, Simon White, Geraint Levan, Kelly Schiabor Barrett, Elizabeth Cirulli, Alexandre Bolze, Ary Ascencio, Charlotte Rivera-Garcia, Ryan Cho, Jason Nguyen, Sherry Wang, Jimmy Ramirez, Tyler Cassens, Efren Sandoval, Magnus Isaksson, William Lee, David Becker, Marc Laurent, James Lu, Clinton R. Paden, Suxiang Tong, Duncan MacCannell |
| EPI_ISL_1421170, EPI_ISL_1421171 | Wisconsin State Laboratory of Hygiene Communicable Disease Division | Wisconsin State Laboratory of Hygiene Communicable Disease Division | Kelsey R. Florek, Abigail C. Shockey |
| EPI_ISL_1421172 | Helix/Illumina | Centers for Disease Control and Prevention Division of Viral Diseases, Pathogen Discovery | Peter W. Cook, Dakota Howard, Dhwani Batra, Ben L. Rambo-Martin, Eileen de Feo, Jan Antico, Christine Tran, Matthew Tolentino, Shannon Wickline, Kim Gietzen, Brad Sickler, Jingtao Liu, Eric Allen, Phil Febbo, Summer Galloway, Nicole L. Washington, Simon White, Geraint Levan, Kelly Schiabor Barrett, Elizabeth Cirulli, Alexandre Bolze, Ary Ascencio, Charlotte Rivera-Garcia, Ryan Cho, Jason Nguyen, Sherry Wang, Jimmy Ramirez, Tyler Cassens, Efren Sandoval, Magnus Isaksson, William Lee, David Becker, Marc Laurent, James Lu, Clinton R. Paden, Suxiang Tong, Duncan MacCannell |
| EPI_ISL_1421174 | Wisconsin State Laboratory of Hygiene Communicable Disease Division | Wisconsin State Laboratory of Hygiene Communicable Disease Division | Kelsey R. Florek, Abigail C. Shockey |
| EPI_ISL_1421175 | Helix/Illumina | Centers for Disease Control and Prevention Division of Viral Diseases, Pathogen Discovery | Peter W. Cook, Dakota Howard, Dhwani Batra, Ben L. Rambo-Martin, Eileen de Feo, Jan Antico, Christine Tran, Matthew Tolentino, Shannon Wickline, Kim Gietzen, Brad Sickler, Jingtao Liu, Eric Allen, Phil Febbo, Summer Galloway, Nicole L. Washington, Simon White, Geraint Levan, Kelly Schiabor Barrett, Elizabeth Cirulli, Alexandre Bolze, Ary Ascencio, Charlotte Rivera-Garcia, Ryan Cho, Jason Nguyen, Sherry Wang, Jimmy Ramirez, Tyler Cassens, Efren Sandoval, Magnus Isaksson, William Lee, David Becker, Marc Laurent, James Lu, Clinton R. Paden, Suxiang Tong, Duncan MacCannell |
| EPI_ISL_1421176, EPI_ISL_1421177, | Wisconsin State Laboratory of Hygiene Communicable Disease Division | Wisconsin State Laboratory of Hygiene Communicable Disease Division | Kelsey R. Florek, Abigail C. Shockey |

| | | | |
|---|---|---|---|
| EPI_ISL_1421178 | | | |
| EPI_ISL_1421179 | Helix/Illumina | Centers for Disease Control and Prevention Division of Viral Diseases, Pathogen Discovery | Peter W. Cook, Dakota Howard, Dhwani Batra, Ben L. Rambo-Martin, Eileen de Feo, Jan Antico, Christine Tran, Matthew Tolentino, Shannon Wickline, Kim Gietzen, Brad Sickler, Jingtao Liu, Eric Allen, Phil Febbo, Summer Galloway, Nicole L. Washington, Simon White, Geraint Levan, Kelly Schiabor Barrett, Elizabeth Cirulli, Alexandre Bolze, Ary Ascencio, Charlotte Rivera-Garcia, Ryan Cho, Jason Nguyen, Sherry Wang, Jimmy Ramirez, Tyler Cassens, Efren Sandoval, Magnus Isaksson, William Lee, David Becker, Marc Laurent, James Lu, Clinton R. Paden, Suxiang Tong, Duncan MacCannell |
| EPI_ISL_1421180, EPI_ISL_1421181 | Wisconsin State Laboratory of Hygiene Communicable Disease Division | Wisconsin State Laboratory of Hygiene Communicable Disease Division | Kelsey R. Florek, Abigail C. Shockey |
| EPI_ISL_1421182 | Helix/Illumina | Centers for Disease Control and Prevention Division of Viral Diseases, Pathogen Discovery | Peter W. Cook, Dakota Howard, Dhwani Batra, Ben L. Rambo-Martin, Eileen de Feo, Jan Antico, Christine Tran, Matthew Tolentino, Shannon Wickline, Kim Gietzen, Brad Sickler, Jingtao Liu, Eric Allen, Phil Febbo, Summer Galloway, Nicole L. Washington, Simon White, Geraint Levan, Kelly Schiabor Barrett, Elizabeth Cirulli, Alexandre Bolze, Ary Ascencio, Charlotte Rivera-Garcia, Ryan Cho, Jason Nguyen, Sherry Wang, Jimmy Ramirez, Tyler Cassens, Efren Sandoval, Magnus Isaksson, William Lee, David Becker, Marc Laurent, James Lu, Clinton R. Paden, Suxiang Tong, Duncan MacCannell |
| EPI_ISL_1421183, EPI_ISL_1421184 | Wisconsin State Laboratory of Hygiene Communicable Disease Division | Wisconsin State Laboratory of Hygiene Communicable Disease Division | Kelsey R. Florek, Abigail C. Shockey |
| EPI_ISL_1421185 | Helix/Illumina | Centers for Disease Control and Prevention Division of Viral Diseases, Pathogen Discovery | Peter W. Cook, Dakota Howard, Dhwani Batra, Ben L. Rambo-Martin, Eileen de Feo, Jan Antico, Christine Tran, Matthew Tolentino, Shannon Wickline, Kim Gietzen, Brad Sickler, Jingtao Liu, Eric Allen, Phil Febbo, Summer Galloway, Nicole L. Washington, Simon White, Geraint Levan, Kelly Schiabor Barrett, Elizabeth Cirulli, Alexandre Bolze, Ary Ascencio, Charlotte Rivera-Garcia, Ryan Cho, Jason Nguyen, Sherry Wang, Jimmy Ramirez, Tyler Cassens, Efren Sandoval, Magnus Isaksson, William Lee, David Becker, Marc Laurent, James Lu, Clinton R. Paden, Suxiang Tong, Duncan MacCannell |
| EPI_ISL_1421186 | Wisconsin State Laboratory of Hygiene Communicable Disease Division | Wisconsin State Laboratory of Hygiene Communicable Disease Division | Kelsey R. Florek, Abigail C. Shockey |
| EPI_ISL_1421187 | Helix/Illumina | Centers for Disease Control and Prevention Division of Viral Diseases, Pathogen Discovery | Peter W. Cook, Dakota Howard, Dhwani Batra, Ben L. Rambo-Martin, Eileen de Feo, Jan Antico, Christine Tran, Matthew Tolentino, Shannon Wickline, Kim Gietzen, Brad Sickler, Jingtao Liu, Eric Allen, Phil Febbo, Summer Galloway, Nicole L. Washington, Simon White, Geraint Levan, Kelly Schiabor Barrett, Elizabeth Cirulli, Alexandre Bolze, Ary Ascencio, Charlotte Rivera-Garcia, Ryan Cho, Jason Nguyen, Sherry Wang, Jimmy Ramirez, Tyler Cassens, Efren Sandoval, Magnus Isaksson, William Lee, David Becker, Marc Laurent, James Lu, Clinton R. Paden, Suxiang Tong, Duncan MacCannell |
| EPI_ISL_1421188, EPI_ISL_1421189 | Wisconsin State Laboratory of Hygiene Communicable Disease Division | Wisconsin State Laboratory of Hygiene Communicable Disease Division | Kelsey R. Florek, Abigail C. Shockey |
| EPI_ISL_1421190 | Helix/Illumina | Centers for Disease Control and Prevention Division of Viral Diseases, Pathogen Discovery | Peter W. Cook, Dakota Howard, Dhwani Batra, Ben L. Rambo-Martin, Eileen de Feo, Jan Antico, Christine Tran, Matthew Tolentino, Shannon Wickline, Kim Gietzen, Brad Sickler, Jingtao Liu, Eric Allen, Phil Febbo, Summer Galloway, Nicole L. Washington, Simon White, Geraint Levan, Kelly Schiabor Barrett, Elizabeth Cirulli, Alexandre Bolze, Ary Ascencio, Charlotte Rivera-Garcia, Ryan Cho, Jason Nguyen, Sherry Wang, Jimmy Ramirez, Tyler Cassens, Efren Sandoval, Magnus Isaksson, William Lee, David Becker, Marc Laurent, James Lu, Clinton R. Paden, Suxiang Tong, Duncan MacCannell |
| EPI_ISL_1421191, EPI_ISL_1421192 | Wisconsin State Laboratory of Hygiene Communicable Disease Division | Wisconsin State Laboratory of Hygiene Communicable Disease Division | Kelsey R. Florek, Abigail C. Shockey |
| EPI_ISL_1421193 | Helix/Illumina | Centers for Disease Control and Prevention Division of Viral Diseases, Pathogen Discovery | Peter W. Cook, Dakota Howard, Dhwani Batra, Ben L. Rambo-Martin, Eileen de Feo, Jan Antico, Christine Tran, Matthew Tolentino, Shannon Wickline, Kim Gietzen, Brad Sickler, Jingtao Liu, Eric Allen, Phil Febbo, Summer Galloway, Nicole L. Washington, Simon White, Geraint Levan, Kelly Schiabor Barrett, Elizabeth Cirulli, Alexandre Bolze, Ary Ascencio, Charlotte Rivera-Garcia, Ryan Cho, Jason Nguyen, Sherry Wang, Jimmy Ramirez, Tyler Cassens, Efren Sandoval, Magnus Isaksson, William Lee, David Becker, Marc Laurent, James Lu, Clinton R. Paden, Suxiang Tong, Duncan MacCannell |
| EPI_ISL_1421194, EPI_ISL_1421195 | Wisconsin State Laboratory of Hygiene Communicable Disease Division | Wisconsin State Laboratory of Hygiene Communicable Disease Division | Kelsey R. Florek, Abigail C. Shockey |
| EPI_ISL_1421196 | Helix/Illumina | Centers for Disease Control and Prevention Division of Viral Diseases, Pathogen Discovery | Peter W. Cook, Dakota Howard, Dhwani Batra, Ben L. Rambo-Martin, Eileen de Feo, Jan Antico, Christine Tran, Matthew Tolentino, Shannon Wickline, Kim Gietzen, Brad Sickler, Jingtao Liu, Eric Allen, Phil Febbo, Summer Galloway, Nicole L. Washington, Simon White, Geraint Levan, Kelly Schiabor Barrett, Elizabeth Cirulli, Alexandre Bolze, Ary Ascencio, Charlotte Rivera-Garcia, Ryan Cho, Jason Nguyen, Sherry Wang, Jimmy Ramirez, Tyler Cassens, Efren Sandoval, Magnus Isaksson, William Lee, David Becker, Marc Laurent, James Lu, Clinton R. Paden, Suxiang Tong, Duncan MacCannell |
| EPI_ISL_1421198 | Wisconsin State Laboratory of Hygiene Communicable Disease Division | Wisconsin State Laboratory of Hygiene Communicable Disease Division | Kelsey R. Florek, Abigail C. Shockey |
| EPI_ISL_1421199 | Helix/Illumina | Centers for Disease Control and Prevention Division of Viral Diseases, Pathogen Discovery | Peter W. Cook, Dakota Howard, Dhwani Batra, Ben L. Rambo-Martin, Eileen de Feo, Jan Antico, Christine Tran, Matthew Tolentino, Shannon Wickline, Kim Gietzen, Brad Sickler, Jingtao Liu, Eric Allen, Phil Febbo, Summer Galloway, Nicole L. Washington, Simon White, Geraint Levan, Kelly Schiabor Barrett, Elizabeth Cirulli, Alexandre Bolze, Ary Ascencio, Charlotte Rivera-Garcia, Ryan Cho, Jason Nguyen, Sherry Wang, Jimmy Ramirez, Tyler Cassens, Efren Sandoval, Magnus Isaksson, William Lee, David Becker, Marc Laurent, James Lu, Clinton R. Paden, Suxiang Tong, Duncan MacCannell |
| EPI_ISL_1421200, EPI_ISL_1421201 | Wisconsin State Laboratory of Hygiene Communicable Disease Division | Wisconsin State Laboratory of Hygiene Communicable Disease Division | Kelsey R. Florek, Abigail C. Shockey |
| EPI_ISL_1421202 | Helix/Illumina | Centers for Disease Control and Prevention Division of Viral Diseases, Pathogen Discovery | Peter W. Cook, Dakota Howard, Dhwani Batra, Ben L. Rambo-Martin, Eileen de Feo, Jan Antico, Christine Tran, Matthew Tolentino, Shannon Wickline, Kim Gietzen, Brad Sickler, Jingtao Liu, Eric Allen, Phil Febbo, Summer Galloway, Nicole L. Washington, Simon White, Geraint Levan, Kelly Schiabor Barrett, Elizabeth Cirulli, Alexandre Bolze, Ary Ascencio, Charlotte Rivera-Garcia, Ryan Cho, Jason Nguyen, Sherry Wang, Jimmy Ramirez, Tyler Cassens, Efren Sandoval, Magnus Isaksson, William Lee, David Becker, Marc Laurent, James Lu, Clinton R. Paden, Suxiang Tong, Duncan MacCannell |
| EPI_ISL_1421203, EPI_ISL_1421204 | Wisconsin State Laboratory of Hygiene Communicable Disease Division | Wisconsin State Laboratory of Hygiene Communicable Disease Division | Kelsey R. Florek, Abigail C. Shockey |
| EPI_ISL_1421205 | Helix/Illumina | Centers for Disease Control and Prevention Division of Viral Diseases, Pathogen Discovery | Peter W. Cook, Dakota Howard, Dhwani Batra, Ben L. Rambo-Martin, Eileen de Feo, Jan Antico, Christine Tran, Matthew Tolentino, Shannon Wickline, Kim Gietzen, Brad Sickler, Jingtao Liu, Eric Allen, Phil Febbo, Summer Galloway, Nicole L. Washington, Simon White, Geraint Levan, Kelly Schiabor Barrett, Elizabeth Cirulli, Alexandre Bolze, Ary Ascencio, Charlotte Rivera-Garcia, Ryan Cho, Jason Nguyen, Sherry Wang, Jimmy Ramirez, Tyler Cassens, Efren Sandoval, Magnus Isaksson, William Lee, David Becker, Marc Laurent, James Lu, Clinton R. Paden, Suxiang Tong, Duncan MacCannell |
| EPI_ISL_1421206, EPI_ISL_1421207 | Wisconsin State Laboratory of Hygiene Communicable Disease Division | Wisconsin State Laboratory of Hygiene Communicable Disease Division | Kelsey R. Florek, Abigail C. Shockey |
| EPI_ISL_1421208 | Helix/Illumina | Centers for Disease Control and Prevention Division of Viral Diseases, Pathogen Discovery | Peter W. Cook, Dakota Howard, Dhwani Batra, Ben L. Rambo-Martin, Eileen de Feo, Jan Antico, Christine Tran, Matthew Tolentino, Shannon Wickline, Kim Gietzen, Brad Sickler, Jingtao Liu, Eric Allen, Phil Febbo, Summer Galloway, Nicole L. Washington, Simon White, Geraint Levan, Kelly Schiabor Barrett, Elizabeth Cirulli, Alexandre Bolze, Ary Ascencio, Charlotte Rivera-Garcia, Ryan Cho, Jason Nguyen, Sherry Wang, Jimmy Ramirez, Tyler Cassens, Efren Sandoval, Magnus Isaksson, William Lee, David Becker, Marc Laurent, James Lu, Clinton R. Paden, Suxiang Tong, Duncan MacCannell |
| EPI_ISL_1421209, EPI_ISL_1421210 | Wisconsin State Laboratory of Hygiene Communicable Disease Division | Wisconsin State Laboratory of Hygiene Communicable Disease Division | Kelsey R. Florek, Abigail C. Shockey |
| EPI_ISL_1421211 | Helix/Illumina | Centers for Disease Control and Prevention Division of Viral Diseases, Pathogen Discovery | Peter W. Cook, Dakota Howard, Dhwani Batra, Ben L. Rambo-Martin, Eileen de Feo, Jan Antico, Christine Tran, Matthew Tolentino, Shannon Wickline, Kim Gietzen, Brad Sickler, Jingtao Liu, Eric Allen, Phil Febbo, Summer Galloway, Nicole L. Washington, Simon White, Geraint Levan, Kelly Schiabor Barrett, Elizabeth Cirulli, Alexandre Bolze, Ary Ascencio, Charlotte Rivera-Garcia, Ryan Cho, Jason Nguyen, Sherry Wang, Jimmy Ramirez, Tyler Cassens, Efren Sandoval, Magnus Isaksson, William Lee, David Becker, Marc Laurent, James Lu, Clinton R. Paden, Suxiang Tong, Duncan MacCannell |
| EPI_ISL_1421212, EPI_ISL_1421213 | Wisconsin State Laboratory of Hygiene Communicable Disease Division | Wisconsin State Laboratory of Hygiene Communicable Disease Division | Kelsey R. Florek, Abigail C. Shockey |
| EPI_ISL_1421214 | Helix/Illumina | Centers for Disease Control and Prevention Division of Viral Diseases, Pathogen Discovery | Peter W. Cook, Dakota Howard, Dhwani Batra, Ben L. Rambo-Martin, Eileen de Feo, Jan Antico, Christine Tran, Matthew Tolentino, Shannon Wickline, Kim Gietzen, Brad Sickler, Jingtao Liu, Eric Allen, Phil Febbo, Summer Galloway, Nicole L. Washington, Simon White, Geraint Levan, Kelly Schiabor Barrett, Elizabeth Cirulli, Alexandre Bolze, Ary Ascencio, Charlotte Rivera-Garcia, Ryan Cho, Jason Nguyen, Sherry Wang, Jimmy Ramirez, Tyler Cassens, Efren Sandoval, Magnus Isaksson, William Lee, David Becker, Marc Laurent, James Lu, Clinton R. Paden, Suxiang Tong, Duncan MacCannell |
| EPI_ISL_1421215, EPI_ISL_1421216 | Wisconsin State Laboratory of Hygiene Communicable Disease Division | Wisconsin State Laboratory of Hygiene Communicable Disease Division | Kelsey R. Florek, Abigail C. Shockey |
| EPI_ISL_1421217 | Helix/Illumina | Centers for Disease Control and Prevention Division of Viral Diseases, Pathogen Discovery | Peter W. Cook, Dakota Howard, Dhwani Batra, Ben L. Rambo-Martin, Eileen de Feo, Jan Antico, Christine Tran, Matthew Tolentino, Shannon Wickline, Kim Gietzen, Brad Sickler, Jingtao Liu, Eric Allen, Phil Febbo, Summer Galloway, Nicole L. Washington, Simon White, Geraint Levan, Kelly Schiabor Barrett, Elizabeth Cirulli, Alexandre Bolze, Ary Ascencio, Charlotte Rivera-Garcia, Ryan Cho, Jason Nguyen, Sherry Wang, Jimmy Ramirez, Tyler Cassens, Efren Sandoval, Magnus Isaksson, William Lee, David Becker, Marc Laurent, James Lu, Clinton R. Paden, Suxiang Tong, Duncan MacCannell |
| EPI_ISL_1421218, EPI_ISL_1421219 | Wisconsin State Laboratory of Hygiene Communicable Disease Division | Wisconsin State Laboratory of Hygiene Communicable Disease Division | Kelsey R. Florek, Abigail C. Shockey |
| EPI_ISL_1421220 | Helix/Illumina | Centers for Disease Control and Prevention Division of Viral Diseases, Pathogen Discovery | Peter W. Cook, Dakota Howard, Dhwani Batra, Ben L. Rambo-Martin, Eileen de Feo, Jan Antico, Christine Tran, Matthew Tolentino, Shannon Wickline, Kim Gietzen, Brad Sickler, Jingtao Liu, Eric Allen, Phil Febbo, Summer Galloway, Nicole L. Washington, Simon White, Geraint Levan, Kelly Schiabor Barrett, Elizabeth Cirulli, Alexandre Bolze, Ary Ascencio, Charlotte Rivera-Garcia, Ryan Cho, Jason Nguyen, Sherry Wang, Jimmy Ramirez, Tyler Cassens, Efren Sandoval, Magnus Isaksson, William Lee, David Becker, Marc Laurent, James Lu, Clinton R. Paden, Suxiang Tong, Duncan MacCannell |
| EPI_ISL_1421221, EPI_ISL_1421222 | Wisconsin State Laboratory of Hygiene Communicable Disease Division | Wisconsin State Laboratory of Hygiene Communicable Disease Division | Kelsey R. Florek, Abigail C. Shockey |
| EPI_ISL_1421223 | Helix/Illumina | Centers for Disease Control and Prevention Division of Viral Diseases, Pathogen Discovery | Peter W. Cook, Dakota Howard, Dhwani Batra, Ben L. Rambo-Martin, Eileen de Feo, Jan Antico, Christine Tran, Matthew Tolentino, Shannon Wickline, Kim Gietzen, Brad Sickler, Jingtao Liu, Eric Allen, Phil Febbo, Summer Galloway, Nicole L. Washington, Simon White, Geraint Levan, Kelly Schiabor Barrett, Elizabeth Cirulli, Alexandre Bolze, Ary Ascencio, Charlotte Rivera-Garcia, Ryan Cho, Jason Nguyen, Sherry Wang, Jimmy Ramirez, Tyler Cassens, Efren Sandoval, Magnus Isaksson, William Lee, David Becker, Marc Laurent, James Lu, Clinton R. Paden, Suxiang Tong, Duncan MacCannell |
| EPI_ISL_1421224, EPI_ISL_1421225 | Wisconsin State Laboratory of Hygiene Communicable Disease Division | Wisconsin State Laboratory of Hygiene Communicable Disease Division | Kelsey R. Florek, Abigail C. Shockey |
| EPI_ISL_1421226 | Helix/Illumina | Centers for Disease Control and Prevention Division of Viral Diseases, Pathogen Discovery | Peter W. Cook, Dakota Howard, Dhwani Batra, Ben L. Rambo-Martin, Eileen de Feo, Jan Antico, Christine Tran, Matthew Tolentino, Shannon Wickline, Kim Gietzen, Brad Sickler, Jingtao Liu, Eric Allen, Phil Febbo, Summer Galloway, Nicole L. Washington, Simon White, Geraint Levan, Kelly Schiabor Barrett, Elizabeth Cirulli, Alexandre Bolze, Ary Ascencio, Charlotte Rivera-Garcia, Ryan Cho, Jason Nguyen, Sherry Wang, Jimmy Ramirez, Tyler Cassens, Efren Sandoval, Magnus Isaksson, William Lee, David Becker, Marc Laurent, James Lu, Clinton R. Paden, Suxiang Tong, Duncan MacCannell |
| EPI_ISL_1421227 | Wisconsin State Laboratory of Hygiene Communicable Disease Division | Wisconsin State Laboratory of Hygiene Communicable Disease Division | Kelsey R. Florek, Abigail C. Shockey |
| EPI_ISL_1421228, EPI_ISL_1421229 | Helix/Illumina | Centers for Disease Control and Prevention Division of Viral Diseases, Pathogen Discovery | Peter W. Cook, Dakota Howard, Dhwani Batra, Ben L. Rambo-Martin, Eileen de Feo, Jan Antico, Christine Tran, Matthew Tolentino, Shannon Wickline, Kim Gietzen, Brad Sickler, Jingtao Liu, Eric Allen, Phil Febbo, Summer Galloway, Nicole L. Washington, Simon White, Geraint Levan, Kelly Schiabor Barrett, Elizabeth Cirulli, Alexandre Bolze, Ary Ascencio, Charlotte Rivera-Garcia, Ryan Cho, Jason Nguyen, Sherry Wang, Jimmy Ramirez, Tyler Cassens, Efren Sandoval, Magnus Isaksson, William Lee, David Becker, Marc Laurent, James Lu, Clinton R. Paden, Suxiang Tong, Duncan MacCannell |
| EPI_ISL_1421230, EPI_ISL_1421232 | Wisconsin State Laboratory of Hygiene Communicable Disease Division | Wisconsin State Laboratory of Hygiene Communicable Disease Division | Kelsey R. Florek, Abigail C. Shockey |
| EPI_ISL_1421233 | Helix/Illumina | Centers for Disease Control and Prevention Division of Viral Diseases, Pathogen Discovery | Peter W. Cook, Dakota Howard, Dhwani Batra, Ben L. Rambo-Martin, Eileen de Feo, Jan Antico, Christine Tran, Matthew Tolentino, Shannon Wickline, Kim Gietzen, Brad Sickler, Jingtao Liu, Eric Allen, Phil Febbo, Summer Galloway, Nicole L. Washington, Simon White, Geraint Levan, Kelly Schiabor Barrett, Elizabeth Cirulli, Alexandre Bolze, Ary Ascencio, Charlotte Rivera-Garcia, Ryan Cho, Jason Nguyen, Sherry Wang, Jimmy Ramirez, Tyler Cassens, Efren Sandoval, Magnus Isaksson, William Lee, David Becker, Marc Laurent, James Lu, Clinton R. Paden, Suxiang Tong, Duncan MacCannell |

| | | | |
|---|---|---|---|
| EPI_ISL_1421234, EPI_ISL_1421235 | Wisconsin State Laboratory of Hygiene Communicable Disease Division | Wisconsin State Laboratory of Hygiene Communicable Disease Division | Kelsey R. Florek, Abigail C. Shockey |
| EPI_ISL_1421236 | Helix/Illumina | Centers for Disease Control and Prevention Division of Viral Diseases, Pathogen Discovery | Peter W. Cook, Dakota Howard, Dhwani Batra, Ben L. Rambo-Martin, Eileen de Feo, Jan Antico, Christine Tran, Matthew Tolentino, Shannon Wickline, Kim Gietzen, Brad Sickler, Jingtao Liu, Eric Allen, Phil Febbo, Summer Galloway, Nicole L. Washington, Simon White, Geraint Levan, Kelly Schiabor Barrett, Elizabeth Cirulli, Alexandre Bolze, Ary Ascencio, Charlotte Rivera-Garcia, Ryan Cho, Jason Nguyen, Sherry Wang, Jimmy Ramirez, Tyler Cassens, Efren Sandoval, Magnus Isaksson, William Lee, David Becker, Marc Laurent, James Lu, Clinton R. Paden, Suxiang Tong, Duncan MacCannell |
| EPI_ISL_1421237, EPI_ISL_1421238 | Wisconsin State Laboratory of Hygiene Communicable Disease Division | Wisconsin State Laboratory of Hygiene Communicable Disease Division | Kelsey R. Florek, Abigail C. Shockey |
| EPI_ISL_1421239 | Helix/Illumina | Centers for Disease Control and Prevention Division of Viral Diseases, Pathogen Discovery | Peter W. Cook, Dakota Howard, Dhwani Batra, Ben L. Rambo-Martin, Eileen de Feo, Jan Antico, Christine Tran, Matthew Tolentino, Shannon Wickline, Kim Gietzen, Brad Sickler, Jingtao Liu, Eric Allen, Phil Febbo, Summer Galloway, Nicole L. Washington, Simon White, Geraint Levan, Kelly Schiabor Barrett, Elizabeth Cirulli, Alexandre Bolze, Ary Ascencio, Charlotte Rivera-Garcia, Ryan Cho, Jason Nguyen, Sherry Wang, Jimmy Ramirez, Tyler Cassens, Efren Sandoval, Magnus Isaksson, William Lee, David Becker, Marc Laurent, James Lu, Clinton R. Paden, Suxiang Tong, Duncan MacCannell |
| EPI_ISL_1421240, EPI_ISL_1421241 | Wisconsin State Laboratory of Hygiene Communicable Disease Division | Wisconsin State Laboratory of Hygiene Communicable Disease Division | Kelsey R. Florek, Abigail C. Shockey |
| EPI_ISL_1421242 | Helix/Illumina | Centers for Disease Control and Prevention Division of Viral Diseases, Pathogen Discovery | Peter W. Cook, Dakota Howard, Dhwani Batra, Ben L. Rambo-Martin, Eileen de Feo, Jan Antico, Christine Tran, Matthew Tolentino, Shannon Wickline, Kim Gietzen, Brad Sickler, Jingtao Liu, Eric Allen, Phil Febbo, Summer Galloway, Nicole L. Washington, Simon White, Geraint Levan, Kelly Schiabor Barrett, Elizabeth Cirulli, Alexandre Bolze, Ary Ascencio, Charlotte Rivera-Garcia, Ryan Cho, Jason Nguyen, Sherry Wang, Jimmy Ramirez, Tyler Cassens, Efren Sandoval, Magnus Isaksson, William Lee, David Becker, Marc Laurent, James Lu, Clinton R. Paden, Suxiang Tong, Duncan MacCannell |
| EPI_ISL_1421243, EPI_ISL_1421244 | Wisconsin State Laboratory of Hygiene Communicable Disease Division | Wisconsin State Laboratory of Hygiene Communicable Disease Division | Kelsey R. Florek, Abigail C. Shockey |
| EPI_ISL_1421245 | Helix/Illumina | Centers for Disease Control and Prevention Division of Viral Diseases, Pathogen Discovery | Peter W. Cook, Dakota Howard, Dhwani Batra, Ben L. Rambo-Martin, Eileen de Feo, Jan Antico, Christine Tran, Matthew Tolentino, Shannon Wickline, Kim Gietzen, Brad Sickler, Jingtao Liu, Eric Allen, Phil Febbo, Summer Galloway, Nicole L. Washington, Simon White, Geraint Levan, Kelly Schiabor Barrett, Elizabeth Cirulli, Alexandre Bolze, Ary Ascencio, Charlotte Rivera-Garcia, Ryan Cho, Jason Nguyen, Sherry Wang, Jimmy Ramirez, Tyler Cassens, Efren Sandoval, Magnus Isaksson, William Lee, David Becker, Marc Laurent, James Lu, Clinton R. Paden, Suxiang Tong, Duncan MacCannell |
| EPI_ISL_1421246, EPI_ISL_1421247 | Wisconsin State Laboratory of Hygiene Communicable Disease Division | Wisconsin State Laboratory of Hygiene Communicable Disease Division | Kelsey R. Florek, Abigail C. Shockey |
| EPI_ISL_1421248 | Helix/Illumina | Centers for Disease Control and Prevention Division of Viral Diseases, Pathogen Discovery | Peter W. Cook, Dakota Howard, Dhwani Batra, Ben L. Rambo-Martin, Eileen de Feo, Jan Antico, Christine Tran, Matthew Tolentino, Shannon Wickline, Kim Gietzen, Brad Sickler, Jingtao Liu, Eric Allen, Phil Febbo, Summer Galloway, Nicole L. Washington, Simon White, Geraint Levan, Kelly Schiabor Barrett, Elizabeth Cirulli, Alexandre Bolze, Ary Ascencio, Charlotte Rivera-Garcia, Ryan Cho, Jason Nguyen, Sherry Wang, Jimmy Ramirez, Tyler Cassens, Efren Sandoval, Magnus Isaksson, William Lee, David Becker, Marc Laurent, James Lu, Clinton R. Paden, Suxiang Tong, Duncan MacCannell |
| EPI_ISL_1421249, EPI_ISL_1421250 | Wisconsin State Laboratory of Hygiene Communicable Disease Division | Wisconsin State Laboratory of Hygiene Communicable Disease Division | Kelsey R. Florek, Abigail C. Shockey |
| EPI_ISL_1421251 | Helix/Illumina | Centers for Disease Control and Prevention Division of Viral Diseases, Pathogen Discovery | Peter W. Cook, Dakota Howard, Dhwani Batra, Ben L. Rambo-Martin, Eileen de Feo, Jan Antico, Christine Tran, Matthew Tolentino, Shannon Wickline, Kim Gietzen, Brad Sickler, Jingtao Liu, Eric Allen, Phil Febbo, Summer Galloway, Nicole L. Washington, Simon White, Geraint Levan, Kelly Schiabor Barrett, Elizabeth Cirulli, Alexandre Bolze, Ary Ascencio, Charlotte Rivera-Garcia, Ryan Cho, Jason Nguyen, Sherry Wang, Jimmy Ramirez, Tyler Cassens, Efren Sandoval, Magnus Isaksson, William Lee, David Becker, Marc Laurent, James Lu, Clinton R. Paden, Suxiang Tong, Duncan MacCannell |
| EPI_ISL_1421252, EPI_ISL_1421253 | Wisconsin State Laboratory of Hygiene Communicable Disease Division | Wisconsin State Laboratory of Hygiene Communicable Disease Division | Kelsey R. Florek, Abigail C. Shockey |
| EPI_ISL_1421254 | Helix/Illumina | Centers for Disease Control and Prevention Division of Viral Diseases, Pathogen Discovery | Peter W. Cook, Dakota Howard, Dhwani Batra, Ben L. Rambo-Martin, Eileen de Feo, Jan Antico, Christine Tran, Matthew Tolentino, Shannon Wickline, Kim Gietzen, Brad Sickler, Jingtao Liu, Eric Allen, Phil Febbo, Summer Galloway, Nicole L. Washington, Simon White, Geraint Levan, Kelly Schiabor Barrett, Elizabeth Cirulli, Alexandre Bolze, Ary Ascencio, Charlotte Rivera-Garcia, Ryan Cho, Jason Nguyen, Sherry Wang, Jimmy Ramirez, Tyler Cassens, Efren Sandoval, Magnus Isaksson, William Lee, David Becker, Marc Laurent, James Lu, Clinton R. Paden, Suxiang Tong, Duncan MacCannell |
| EPI_ISL_1421255, EPI_ISL_1421256, EPI_ISL_1421258 | Wisconsin State Laboratory of Hygiene Communicable Disease Division | Wisconsin State Laboratory of Hygiene Communicable Disease Division | Kelsey R. Florek, Abigail C. Shockey |
| EPI_ISL_1421259 | Helix/Illumina | Centers for Disease Control and Prevention Division of Viral Diseases, Pathogen Discovery | Peter W. Cook, Dakota Howard, Dhwani Batra, Ben L. Rambo-Martin, Eileen de Feo, Jan Antico, Christine Tran, Matthew Tolentino, Shannon Wickline, Kim Gietzen, Brad Sickler, Jingtao Liu, Eric Allen, Phil Febbo, Summer Galloway, Nicole L. Washington, Simon White, Geraint Levan, Kelly Schiabor Barrett, Elizabeth Cirulli, Alexandre Bolze, Ary Ascencio, Charlotte Rivera-Garcia, Ryan Cho, Jason Nguyen, Sherry Wang, Jimmy Ramirez, Tyler Cassens, Efren Sandoval, Magnus Isaksson, William Lee, David Becker, Marc Laurent, James Lu, Clinton R. Paden, Suxiang Tong, Duncan MacCannell |
| EPI_ISL_1421260 | Wisconsin State Laboratory of Hygiene Communicable Disease Division | Wisconsin State Laboratory of Hygiene Communicable Disease Division | Kelsey R. Florek, Abigail C. Shockey |
| EPI_ISL_1421261, EPI_ISL_1421262 | Helix/Illumina | Centers for Disease Control and Prevention Division of Viral Diseases, Pathogen Discovery | Peter W. Cook, Dakota Howard, Dhwani Batra, Ben L. Rambo-Martin, Eileen de Feo, Jan Antico, Christine Tran, Matthew Tolentino, Shannon Wickline, Kim Gietzen, Brad Sickler, Jingtao Liu, Eric Allen, Phil Febbo, Summer Galloway, Nicole L. Washington, Simon White, Geraint Levan, Kelly Schiabor Barrett, Elizabeth Cirulli, Alexandre Bolze, Ary Ascencio, Charlotte Rivera-Garcia, Ryan Cho, Jason Nguyen, Sherry Wang, Jimmy Ramirez, Tyler Cassens, Efren Sandoval, Magnus Isaksson, William Lee, David Becker, Marc Laurent, James Lu, Clinton R. Paden, Suxiang Tong, Duncan MacCannell |
| EPI_ISL_1421263 | Wisconsin State Laboratory of Hygiene Communicable Disease Division | Wisconsin State Laboratory of Hygiene Communicable Disease Division | Kelsey R. Florek, Abigail C. Shockey |
| EPI_ISL_1421264, EPI_ISL_1421266 | Helix/Illumina | Centers for Disease Control and Prevention Division of Viral Diseases, Pathogen Discovery | Peter W. Cook, Dakota Howard, Dhwani Batra, Ben L. Rambo-Martin, Eileen de Feo, Jan Antico, Christine Tran, Matthew Tolentino, Shannon Wickline, Kim Gietzen, Brad Sickler, Jingtao Liu, Eric Allen, Phil Febbo, Summer Galloway, Nicole L. Washington, Simon White, Geraint Levan, Kelly Schiabor Barrett, Elizabeth Cirulli, Alexandre Bolze, Ary Ascencio, Charlotte Rivera-Garcia, Ryan Cho, Jason Nguyen, Sherry Wang, Jimmy Ramirez, Tyler Cassens, Efren Sandoval, Magnus Isaksson, William Lee, David Becker, Marc Laurent, James Lu, Clinton R. Paden, Suxiang Tong, Duncan MacCannell |
| EPI_ISL_1421267, EPI_ISL_1421268 | Wisconsin State Laboratory of Hygiene Communicable Disease Division | Wisconsin State Laboratory of Hygiene Communicable Disease Division | Kelsey R. Florek, Abigail C. Shockey |
| EPI_ISL_1421269 | Helix/Illumina | Centers for Disease Control and Prevention Division of Viral Diseases, Pathogen Discovery | Peter W. Cook, Dakota Howard, Dhwani Batra, Ben L. Rambo-Martin, Eileen de Feo, Jan Antico, Christine Tran, Matthew Tolentino, Shannon Wickline, Kim Gietzen, Brad Sickler, Jingtao Liu, Eric Allen, Phil Febbo, Summer Galloway, Nicole L. Washington, Simon White, Geraint Levan, Kelly Schiabor Barrett, Elizabeth Cirulli, Alexandre Bolze, Ary Ascencio, Charlotte Rivera-Garcia, Ryan Cho, Jason Nguyen, Sherry Wang, Jimmy Ramirez, Tyler Cassens, Efren Sandoval, Magnus Isaksson, William Lee, David Becker, Marc Laurent, James Lu, Clinton R. Paden, Suxiang Tong, Duncan MacCannell |
| EPI_ISL_1421270, EPI_ISL_1421271 | Wisconsin State Laboratory of Hygiene Communicable Disease Division | Wisconsin State Laboratory of Hygiene Communicable Disease Division | Kelsey R. Florek, Abigail C. Shockey |
| EPI_ISL_1421272 | Helix/Illumina | Centers for Disease Control and Prevention Division of Viral Diseases, Pathogen Discovery | Peter W. Cook, Dakota Howard, Dhwani Batra, Ben L. Rambo-Martin, Eileen de Feo, Jan Antico, Christine Tran, Matthew Tolentino, Shannon Wickline, Kim Gietzen, Brad Sickler, Jingtao Liu, Eric Allen, Phil Febbo, Summer Galloway, Nicole L. Washington, Simon White, Geraint Levan, Kelly Schiabor Barrett, Elizabeth Cirulli, Alexandre Bolze, Ary Ascencio, Charlotte Rivera-Garcia, Ryan Cho, Jason Nguyen, Sherry Wang, Jimmy Ramirez, Tyler Cassens, Efren Sandoval, Magnus Isaksson, William Lee, David Becker, Marc Laurent, James Lu, Clinton R. Paden, Suxiang Tong, Duncan MacCannell |
| EPI_ISL_1421273, EPI_ISL_1421274 | Wisconsin State Laboratory of Hygiene Communicable Disease Division | Wisconsin State Laboratory of Hygiene Communicable Disease Division | Kelsey R. Florek, Abigail C. Shockey |
| EPI_ISL_1421275 | Helix/Illumina | Centers for Disease Control and Prevention Division of Viral Diseases, Pathogen Discovery | Peter W. Cook, Dakota Howard, Dhwani Batra, Ben L. Rambo-Martin, Eileen de Feo, Jan Antico, Christine Tran, Matthew Tolentino, Shannon Wickline, Kim Gietzen, Brad Sickler, Jingtao Liu, Eric Allen, Phil Febbo, Summer Galloway, Nicole L. Washington, Simon White, Geraint Levan, Kelly Schiabor Barrett, Elizabeth Cirulli, Alexandre Bolze, Ary Ascencio, Charlotte Rivera-Garcia, Ryan Cho, Jason Nguyen, Sherry Wang, Jimmy Ramirez, Tyler Cassens, Efren Sandoval, Magnus Isaksson, William Lee, David Becker, Marc Laurent, James Lu, Clinton R. Paden, Suxiang Tong, Duncan MacCannell |
| EPI_ISL_1421276, EPI_ISL_1421277 | Wisconsin State Laboratory of Hygiene Communicable Disease Division | Wisconsin State Laboratory of Hygiene Communicable Disease Division | Kelsey R. Florek, Abigail C. Shockey |
| EPI_ISL_1421278 | Helix/Illumina | Centers for Disease Control and Prevention Division of Viral Diseases, Pathogen Discovery | Peter W. Cook, Dakota Howard, Dhwani Batra, Ben L. Rambo-Martin, Eileen de Feo, Jan Antico, Christine Tran, Matthew Tolentino, Shannon Wickline, Kim Gietzen, Brad Sickler, Jingtao Liu, Eric Allen, Phil Febbo, Summer Galloway, Nicole L. Washington, Simon White, Geraint Levan, Kelly Schiabor Barrett, Elizabeth Cirulli, Alexandre Bolze, Ary Ascencio, Charlotte Rivera-Garcia, Ryan Cho, Jason Nguyen, Sherry Wang, Jimmy Ramirez, Tyler Cassens, Efren Sandoval, Magnus Isaksson, William Lee, David Becker, Marc Laurent, James Lu, Clinton R. Paden, Suxiang Tong, Duncan MacCannell |
| EPI_ISL_1421279, EPI_ISL_1421280 | Wisconsin State Laboratory of Hygiene Communicable Disease Division | Wisconsin State Laboratory of Hygiene Communicable Disease Division | Kelsey R. Florek, Abigail C. Shockey |
| EPI_ISL_1421281 | Helix/Illumina | Centers for Disease Control and Prevention Division of Viral Diseases, Pathogen Discovery | Peter W. Cook, Dakota Howard, Dhwani Batra, Ben L. Rambo-Martin, Eileen de Feo, Jan Antico, Christine Tran, Matthew Tolentino, Shannon Wickline, Kim Gietzen, Brad Sickler, Jingtao Liu, Eric Allen, Phil Febbo, Summer Galloway, Nicole L. Washington, Simon White, Geraint Levan, Kelly Schiabor Barrett, Elizabeth Cirulli, Alexandre Bolze, Ary Ascencio, Charlotte Rivera-Garcia, Ryan Cho, Jason Nguyen, Sherry Wang, Jimmy Ramirez, Tyler Cassens, Efren Sandoval, Magnus Isaksson, William Lee, David Becker, Marc Laurent, James Lu, Clinton R. Paden, Suxiang Tong, Duncan MacCannell |
| EPI_ISL_1421282, EPI_ISL_1421283 | Wisconsin State Laboratory of Hygiene Communicable Disease Division | Wisconsin State Laboratory of Hygiene Communicable Disease Division | Kelsey R. Florek, Abigail C. Shockey |
| EPI_ISL_1421284 | Helix/Illumina | Centers for Disease Control and Prevention Division of Viral Diseases, Pathogen Discovery | Peter W. Cook, Dakota Howard, Dhwani Batra, Ben L. Rambo-Martin, Eileen de Feo, Jan Antico, Christine Tran, Matthew Tolentino, Shannon Wickline, Kim Gietzen, Brad Sickler, Jingtao Liu, Eric Allen, Phil Febbo, Summer Galloway, Nicole L. Washington, Simon White, Geraint Levan, Kelly Schiabor Barrett, Elizabeth Cirulli, Alexandre Bolze, Ary Ascencio, Charlotte Rivera-Garcia, Ryan Cho, Jason Nguyen, Sherry Wang, Jimmy Ramirez, Tyler Cassens, Efren Sandoval, Magnus Isaksson, William Lee, David Becker, Marc Laurent, James Lu, Clinton R. Paden, Suxiang Tong, Duncan MacCannell |
| EPI_ISL_1421285, EPI_ISL_1421286 | Wisconsin State Laboratory of Hygiene Communicable Disease Division | Wisconsin State Laboratory of Hygiene Communicable Disease Division | Kelsey R. Florek, Abigail C. Shockey |
| EPI_ISL_1421287 | Helix/Illumina | Centers for Disease Control and Prevention Division of Viral Diseases, Pathogen Discovery | Peter W. Cook, Dakota Howard, Dhwani Batra, Ben L. Rambo-Martin, Eileen de Feo, Jan Antico, Christine Tran, Matthew Tolentino, Shannon Wickline, Kim Gietzen, Brad Sickler, Jingtao Liu, Eric Allen, Phil Febbo, Summer Galloway, Nicole L. Washington, Simon White, Geraint Levan, Kelly Schiabor Barrett, Elizabeth Cirulli, Alexandre Bolze, Ary Ascencio, Charlotte Rivera-Garcia, Ryan Cho, Jason Nguyen, Sherry Wang, Jimmy Ramirez, Tyler Cassens, Efren Sandoval, Magnus Isaksson, William Lee, David Becker, Marc Laurent, James Lu, Clinton R. Paden, Suxiang Tong, Duncan MacCannell |
| EPI_ISL_1421288, EPI_ISL_1421289 | Wisconsin State Laboratory of Hygiene Communicable Disease Division | Wisconsin State Laboratory of Hygiene Communicable Disease Division | Kelsey R. Florek, Abigail C. Shockey |
| EPI_ISL_1421290 | Helix/Illumina | Centers for Disease Control and Prevention Division of Viral Diseases, Pathogen Discovery | Peter W. Cook, Dakota Howard, Dhwani Batra, Ben L. Rambo-Martin, Eileen de Feo, Jan Antico, Christine Tran, Matthew Tolentino, Shannon Wickline, Kim Gietzen, Brad Sickler, Jingtao Liu, Eric Allen, Phil Febbo, Summer Galloway, Nicole L. Washington, Simon White, Geraint Levan, Kelly Schiabor Barrett, Elizabeth Cirulli, Alexandre Bolze, Ary Ascencio, Charlotte Rivera-Garcia, Ryan Cho, Jason Nguyen, Sherry Wang, Jimmy Ramirez, Tyler Cassens, Efren Sandoval, Magnus Isaksson, William Lee, David Becker, Marc Laurent, James Lu, Clinton R. Paden, Suxiang Tong, Duncan MacCannell |
| EPI_ISL_1421292 | Wisconsin State Laboratory of Hygiene Communicable Disease Division | Wisconsin State Laboratory of Hygiene Communicable Disease Division | Kelsey R. Florek, Abigail C. Shockey |
| EPI_ISL_1421293 | Helix/Illumina | Centers for Disease Control and Prevention | Peter W. Cook, Dakota Howard, Dhwani Batra, Ben L. Rambo-Martin, Eileen de Feo, Jan Antico, Christine Tran, Matthew Tolentino, Shannon Wickline, Kim Gietzen, Brad Sickler, Jingtao Liu, Eric Allen, Phil Febbo, Summer Galloway, Nicole L. Washington, Simon White, Geraint Levan, Kelly |

| | | | |
|---|---|---|---|
| | | Division of Viral Diseases, Pathogen Discovery | Schiabor Barrett, Elizabeth Cirulli, Alexandre Bolze, Ary Ascencio, Charlotte Rivera-Garcia, Ryan Cho, Jason Nguyen, Sherry Wang, Jimmy Ramirez, Tyler Cassens, Efren Sandoval, Magnus Isaksson, William Lee, David Becker, Marc Laurent, James Lu, Clinton R. Paden, Suxiang Tong, Duncan MacCannell |
| EPI_ISL_1421294, EPI_ISL_1421295 | Wisconsin State Laboratory of Hygiene Communicable Disease Division | Wisconsin State Laboratory of Hygiene Communicable Disease Division | Kelsey R. Florek, Abigail C. Shockey |
| EPI_ISL_1421296 | Helix/Illumina | Centers for Disease Control and Prevention Division of Viral Diseases, Pathogen Discovery | Peter W. Cook, Dakota Howard, Dhwani Batra, Ben L. Rambo-Martin, Eileen de Feo, Jan Antico, Christine Tran, Matthew Tolentino, Shannon Wickline, Kim Gietzen, Brad Sickler, Jingtao Liu, Eric Allen, Phil Febbo, Summer Galloway, Nicole L. Washington, Simon White, Geraint Levan, Kelly Schiabor Barrett, Elizabeth Cirulli, Alexandre Bolze, Ary Ascencio, Charlotte Rivera-Garcia, Ryan Cho, Jason Nguyen, Sherry Wang, Jimmy Ramirez, Tyler Cassens, Efren Sandoval, Magnus Isaksson, William Lee, David Becker, Marc Laurent, James Lu, Clinton R. Paden, Suxiang Tong, Duncan MacCannell |
| EPI_ISL_1421297, EPI_ISL_1421298 | Wisconsin State Laboratory of Hygiene Communicable Disease Division | Wisconsin State Laboratory of Hygiene Communicable Disease Division | Kelsey R. Florek, Abigail C. Shockey |
| EPI_ISL_1421299, EPI_ISL_1421300 | Helix/Illumina | Centers for Disease Control and Prevention Division of Viral Diseases, Pathogen Discovery | Peter W. Cook, Dakota Howard, Dhwani Batra, Ben L. Rambo-Martin, Eileen de Feo, Jan Antico, Christine Tran, Matthew Tolentino, Shannon Wickline, Kim Gietzen, Brad Sickler, Jingtao Liu, Eric Allen, Phil Febbo, Summer Galloway, Nicole L. Washington, Simon White, Geraint Levan, Kelly Schiabor Barrett, Elizabeth Cirulli, Alexandre Bolze, Ary Ascencio, Charlotte Rivera-Garcia, Ryan Cho, Jason Nguyen, Sherry Wang, Jimmy Ramirez, Tyler Cassens, Efren Sandoval, Magnus Isaksson, William Lee, David Becker, Marc Laurent, James Lu, Clinton R. Paden, Suxiang Tong, Duncan MacCannell |
| EPI_ISL_1421301, EPI_ISL_1421302 | Wisconsin State Laboratory of Hygiene Communicable Disease Division | Wisconsin State Laboratory of Hygiene Communicable Disease Division | Kelsey R. Florek, Abigail C. Shockey |
| EPI_ISL_1421303 | Helix/Illumina | Centers for Disease Control and Prevention Division of Viral Diseases, Pathogen Discovery | Peter W. Cook, Dakota Howard, Dhwani Batra, Ben L. Rambo-Martin, Eileen de Feo, Jan Antico, Christine Tran, Matthew Tolentino, Shannon Wickline, Kim Gietzen, Brad Sickler, Jingtao Liu, Eric Allen, Phil Febbo, Summer Galloway, Nicole L. Washington, Simon White, Geraint Levan, Kelly Schiabor Barrett, Elizabeth Cirulli, Alexandre Bolze, Ary Ascencio, Charlotte Rivera-Garcia, Ryan Cho, Jason Nguyen, Sherry Wang, Jimmy Ramirez, Tyler Cassens, Efren Sandoval, Magnus Isaksson, William Lee, David Becker, Marc Laurent, James Lu, Clinton R. Paden, Suxiang Tong, Duncan MacCannell |
| EPI_ISL_1421304, EPI_ISL_1421305 | Wisconsin State Laboratory of Hygiene Communicable Disease Division | Wisconsin State Laboratory of Hygiene Communicable Disease Division | Kelsey R. Florek, Abigail C. Shockey |
| EPI_ISL_1421306 | Helix/Illumina | Centers for Disease Control and Prevention Division of Viral Diseases, Pathogen Discovery | Peter W. Cook, Dakota Howard, Dhwani Batra, Ben L. Rambo-Martin, Eileen de Feo, Jan Antico, Christine Tran, Matthew Tolentino, Shannon Wickline, Kim Gietzen, Brad Sickler, Jingtao Liu, Eric Allen, Phil Febbo, Summer Galloway, Nicole L. Washington, Simon White, Geraint Levan, Kelly Schiabor Barrett, Elizabeth Cirulli, Alexandre Bolze, Ary Ascencio, Charlotte Rivera-Garcia, Ryan Cho, Jason Nguyen, Sherry Wang, Jimmy Ramirez, Tyler Cassens, Efren Sandoval, Magnus Isaksson, William Lee, David Becker, Marc Laurent, James Lu, Clinton R. Paden, Suxiang Tong, Duncan MacCannell |
| EPI_ISL_1421307, EPI_ISL_1421308 | Wisconsin State Laboratory of Hygiene Communicable Disease Division | Wisconsin State Laboratory of Hygiene Communicable Disease Division | Kelsey R. Florek, Abigail C. Shockey |
| EPI_ISL_1421309 | Helix/Illumina | Centers for Disease Control and Prevention Division of Viral Diseases, Pathogen Discovery | Peter W. Cook, Dakota Howard, Dhwani Batra, Ben L. Rambo-Martin, Eileen de Feo, Jan Antico, Christine Tran, Matthew Tolentino, Shannon Wickline, Kim Gietzen, Brad Sickler, Jingtao Liu, Eric Allen, Phil Febbo, Summer Galloway, Nicole L. Washington, Simon White, Geraint Levan, Kelly Schiabor Barrett, Elizabeth Cirulli, Alexandre Bolze, Ary Ascencio, Charlotte Rivera-Garcia, Ryan Cho, Jason Nguyen, Sherry Wang, Jimmy Ramirez, Tyler Cassens, Efren Sandoval, Magnus Isaksson, William Lee, David Becker, Marc Laurent, James Lu, Clinton R. Paden, Suxiang Tong, Duncan MacCannell |
| EPI_ISL_1421310, EPI_ISL_1421311 | Wisconsin State Laboratory of Hygiene Communicable Disease Division | Wisconsin State Laboratory of Hygiene Communicable Disease Division | Kelsey R. Florek, Abigail C. Shockey |
| EPI_ISL_1421312 | Helix/Illumina | Centers for Disease Control and Prevention Division of Viral Diseases, Pathogen Discovery | Peter W. Cook, Dakota Howard, Dhwani Batra, Ben L. Rambo-Martin, Eileen de Feo, Jan Antico, Christine Tran, Matthew Tolentino, Shannon Wickline, Kim Gietzen, Brad Sickler, Jingtao Liu, Eric Allen, Phil Febbo, Summer Galloway, Nicole L. Washington, Simon White, Geraint Levan, Kelly Schiabor Barrett, Elizabeth Cirulli, Alexandre Bolze, Ary Ascencio, Charlotte Rivera-Garcia, Ryan Cho, Jason Nguyen, Sherry Wang, Jimmy Ramirez, Tyler Cassens, Efren Sandoval, Magnus Isaksson, William Lee, David Becker, Marc Laurent, James Lu, Clinton R. Paden, Suxiang Tong, Duncan MacCannell |
| EPI_ISL_1421313, EPI_ISL_1421314 | Wisconsin State Laboratory of Hygiene Communicable Disease Division | Wisconsin State Laboratory of Hygiene Communicable Disease Division | Kelsey R. Florek, Abigail C. Shockey |
| EPI_ISL_1421315 | Helix/Illumina | Centers for Disease Control and Prevention Division of Viral Diseases, Pathogen Discovery | Peter W. Cook, Dakota Howard, Dhwani Batra, Ben L. Rambo-Martin, Eileen de Feo, Jan Antico, Christine Tran, Matthew Tolentino, Shannon Wickline, Kim Gietzen, Brad Sickler, Jingtao Liu, Eric Allen, Phil Febbo, Summer Galloway, Nicole L. Washington, Simon White, Geraint Levan, Kelly Schiabor Barrett, Elizabeth Cirulli, Alexandre Bolze, Ary Ascencio, Charlotte Rivera-Garcia, Ryan Cho, Jason Nguyen, Sherry Wang, Jimmy Ramirez, Tyler Cassens, Efren Sandoval, Magnus Isaksson, William Lee, David Becker, Marc Laurent, James Lu, Clinton R. Paden, Suxiang Tong, Duncan MacCannell |
| EPI_ISL_1421316, EPI_ISL_1421317, EPI_ISL_1421319, EPI_ISL_1421320, EPI_ISL_1421321, EPI_ISL_1421322, EPI_ISL_1421323, EPI_ISL_1421324, EPI_ISL_1421325, EPI_ISL_1421326 | Wisconsin State Laboratory of Hygiene Communicable Disease Division | Wisconsin State Laboratory of Hygiene Communicable Disease Division | Kelsey R. Florek, Abigail C. Shockey |
| EPI_ISL_1421327, EPI_ISL_1421328 | Helix/Illumina | Centers for Disease Control and Prevention Division of Viral Diseases, Pathogen Discovery | Peter W. Cook, Dakota Howard, Dhwani Batra, Ben L. Rambo-Martin, Eileen de Feo, Jan Antico, Christine Tran, Matthew Tolentino, Shannon Wickline, Kim Gietzen, Brad Sickler, Jingtao Liu, Eric Allen, Phil Febbo, Summer Galloway, Nicole L. Washington, Simon White, Geraint Levan, Kelly Schiabor Barrett, Elizabeth Cirulli, Alexandre Bolze, Ary Ascencio, Charlotte Rivera-Garcia, Ryan Cho, Jason Nguyen, Sherry Wang, Jimmy Ramirez, Tyler Cassens, Efren Sandoval, Magnus Isaksson, William Lee, David Becker, Marc Laurent, James Lu, Clinton R. Paden, Suxiang Tong, Duncan MacCannell |
| EPI_ISL_1421329 | Wisconsin State Laboratory of Hygiene Communicable Disease Division | Wisconsin State Laboratory of Hygiene Communicable Disease Division | Kelsey R. Florek, Abigail C. Shockey |
| EPI_ISL_1421330 | Helix/Illumina | Centers for Disease Control and Prevention Division of Viral Diseases, Pathogen Discovery | Peter W. Cook, Dakota Howard, Dhwani Batra, Ben L. Rambo-Martin, Eileen de Feo, Jan Antico, Christine Tran, Matthew Tolentino, Shannon Wickline, Kim Gietzen, Brad Sickler, Jingtao Liu, Eric Allen, Phil Febbo, Summer Galloway, Nicole L. Washington, Simon White, Geraint Levan, Kelly Schiabor Barrett, Elizabeth Cirulli, Alexandre Bolze, Ary Ascencio, Charlotte Rivera-Garcia, Ryan Cho, Jason Nguyen, Sherry Wang, Jimmy Ramirez, Tyler Cassens, Efren Sandoval, Magnus Isaksson, William Lee, David Becker, Marc Laurent, James Lu, Clinton R. Paden, Suxiang Tong, Duncan MacCannell |
| EPI_ISL_1421331 | Wisconsin State Laboratory of Hygiene Communicable Disease Division | Wisconsin State Laboratory of Hygiene Communicable Disease Division | Kelsey R. Florek, Abigail C. Shockey |
| EPI_ISL_1421332 | Helix/Illumina | Centers for Disease Control and Prevention Division of Viral Diseases, Pathogen Discovery | Peter W. Cook, Dakota Howard, Dhwani Batra, Ben L. Rambo-Martin, Eileen de Feo, Jan Antico, Christine Tran, Matthew Tolentino, Shannon Wickline, Kim Gietzen, Brad Sickler, Jingtao Liu, Eric Allen, Phil Febbo, Summer Galloway, Nicole L. Washington, Simon White, Geraint Levan, Kelly Schiabor Barrett, Elizabeth Cirulli, Alexandre Bolze, Ary Ascencio, Charlotte Rivera-Garcia, Ryan Cho, Jason Nguyen, Sherry Wang, Jimmy Ramirez, Tyler Cassens, Efren Sandoval, Magnus Isaksson, William Lee, David Becker, Marc Laurent, James Lu, Clinton R. Paden, Suxiang Tong, Duncan MacCannell |
| EPI_ISL_1421333 | Wisconsin State Laboratory of Hygiene Communicable Disease Division | Wisconsin State Laboratory of Hygiene Communicable Disease Division | Kelsey R. Florek, Abigail C. Shockey |
| EPI_ISL_1421334, EPI_ISL_1421335 | Helix/Illumina | Centers for Disease Control and Prevention Division of Viral Diseases, Pathogen Discovery | Peter W. Cook, Dakota Howard, Dhwani Batra, Ben L. Rambo-Martin, Eileen de Feo, Jan Antico, Christine Tran, Matthew Tolentino, Shannon Wickline, Kim Gietzen, Brad Sickler, Jingtao Liu, Eric Allen, Phil Febbo, Summer Galloway, Nicole L. Washington, Simon White, Geraint Levan, Kelly Schiabor Barrett, Elizabeth Cirulli, Alexandre Bolze, Ary Ascencio, Charlotte Rivera-Garcia, Ryan Cho, Jason Nguyen, Sherry Wang, Jimmy Ramirez, Tyler Cassens, Efren Sandoval, Magnus Isaksson, William Lee, David Becker, Marc Laurent, James Lu, Clinton R. Paden, Suxiang Tong, Duncan MacCannell |
| EPI_ISL_1421336 | Wisconsin State Laboratory of Hygiene Communicable Disease Division | Wisconsin State Laboratory of Hygiene Communicable Disease Division | Kelsey R. Florek, Abigail C. Shockey |
| EPI_ISL_1421337 | Helix/Illumina | Centers for Disease Control and Prevention Division of Viral Diseases, Pathogen Discovery | Peter W. Cook, Dakota Howard, Dhwani Batra, Ben L. Rambo-Martin, Eileen de Feo, Jan Antico, Christine Tran, Matthew Tolentino, Shannon Wickline, Kim Gietzen, Brad Sickler, Jingtao Liu, Eric Allen, Phil Febbo, Summer Galloway, Nicole L. Washington, Simon White, Geraint Levan, Kelly Schiabor Barrett, Elizabeth Cirulli, Alexandre Bolze, Ary Ascencio, Charlotte Rivera-Garcia, Ryan Cho, Jason Nguyen, Sherry Wang, Jimmy Ramirez, Tyler Cassens, Efren Sandoval, Magnus Isaksson, William Lee, David Becker, Marc Laurent, James Lu, Clinton R. Paden, Suxiang Tong, Duncan MacCannell |
| EPI_ISL_1421338 | Wisconsin State Laboratory of Hygiene Communicable Disease Division | Wisconsin State Laboratory of Hygiene Communicable Disease Division | Kelsey R. Florek, Abigail C. Shockey |
| EPI_ISL_1421339 | Helix/Illumina | Centers for Disease Control and Prevention Division of Viral Diseases, Pathogen Discovery | Peter W. Cook, Dakota Howard, Dhwani Batra, Ben L. Rambo-Martin, Eileen de Feo, Jan Antico, Christine Tran, Matthew Tolentino, Shannon Wickline, Kim Gietzen, Brad Sickler, Jingtao Liu, Eric Allen, Phil Febbo, Summer Galloway, Nicole L. Washington, Simon White, Geraint Levan, Kelly Schiabor Barrett, Elizabeth Cirulli, Alexandre Bolze, Ary Ascencio, Charlotte Rivera-Garcia, Ryan Cho, Jason Nguyen, Sherry Wang, Jimmy Ramirez, Tyler Cassens, Efren Sandoval, Magnus Isaksson, William Lee, David Becker, Marc Laurent, James Lu, Clinton R. Paden, Suxiang Tong, Duncan MacCannell |
| EPI_ISL_1421340 | Wisconsin State Laboratory of Hygiene Communicable Disease Division | Wisconsin State Laboratory of Hygiene Communicable Disease Division | Kelsey R. Florek, Abigail C. Shockey |
| EPI_ISL_1421341, EPI_ISL_1421342 | Helix/Illumina | Centers for Disease Control and Prevention Division of Viral Diseases, Pathogen Discovery | Peter W. Cook, Dakota Howard, Dhwani Batra, Ben L. Rambo-Martin, Eileen de Feo, Jan Antico, Christine Tran, Matthew Tolentino, Shannon Wickline, Kim Gietzen, Brad Sickler, Jingtao Liu, Eric Allen, Phil Febbo, Summer Galloway, Nicole L. Washington, Simon White, Geraint Levan, Kelly Schiabor Barrett, Elizabeth Cirulli, Alexandre Bolze, Ary Ascencio, Charlotte Rivera-Garcia, Ryan Cho, Jason Nguyen, Sherry Wang, Jimmy Ramirez, Tyler Cassens, Efren Sandoval, Magnus Isaksson, William Lee, David Becker, Marc Laurent, James Lu, Clinton R. Paden, Suxiang Tong, Duncan MacCannell |
| EPI_ISL_1421343, EPI_ISL_1421344 | Wisconsin State Laboratory of Hygiene Communicable Disease Division | Wisconsin State Laboratory of Hygiene Communicable Disease Division | Kelsey R. Florek, Abigail C. Shockey |
| EPI_ISL_1421345 | Helix/Illumina | Centers for Disease Control and Prevention Division of Viral Diseases, Pathogen Discovery | Peter W. Cook, Dakota Howard, Dhwani Batra, Ben L. Rambo-Martin, Eileen de Feo, Jan Antico, Christine Tran, Matthew Tolentino, Shannon Wickline, Kim Gietzen, Brad Sickler, Jingtao Liu, Eric Allen, Phil Febbo, Summer Galloway, Nicole L. Washington, Simon White, Geraint Levan, Kelly Schiabor Barrett, Elizabeth Cirulli, Alexandre Bolze, Ary Ascencio, Charlotte Rivera-Garcia, Ryan Cho, Jason Nguyen, Sherry Wang, Jimmy Ramirez, Tyler Cassens, Efren Sandoval, Magnus Isaksson, William Lee, David Becker, Marc Laurent, James Lu, Clinton R. Paden, Suxiang Tong, Duncan MacCannell |
| EPI_ISL_1421346 | Wisconsin State Laboratory of Hygiene Communicable Disease Division | Wisconsin State Laboratory of Hygiene Communicable Disease Division | Kelsey R. Florek, Abigail C. Shockey |
| EPI_ISL_1421347 | Helix/Illumina | Centers for Disease Control and Prevention Division of Viral Diseases, Pathogen Discovery | Peter W. Cook, Dakota Howard, Dhwani Batra, Ben L. Rambo-Martin, Eileen de Feo, Jan Antico, Christine Tran, Matthew Tolentino, Shannon Wickline, Kim Gietzen, Brad Sickler, Jingtao Liu, Eric Allen, Phil Febbo, Summer Galloway, Nicole L. Washington, Simon White, Geraint Levan, Kelly Schiabor Barrett, Elizabeth Cirulli, Alexandre Bolze, Ary Ascencio, Charlotte Rivera-Garcia, Ryan Cho, Jason Nguyen, Sherry Wang, Jimmy Ramirez, Tyler Cassens, Efren Sandoval, Magnus Isaksson, William Lee, David Becker, Marc Laurent, James Lu, Clinton R. Paden, Suxiang Tong, Duncan MacCannell |
| EPI_ISL_1421348 | Wisconsin State Laboratory of Hygiene Communicable Disease Division | Wisconsin State Laboratory of Hygiene Communicable Disease Division | Kelsey R. Florek, Abigail C. Shockey |
| EPI_ISL_1421349, EPI_ISL_1421350 | Helix/Illumina | Centers for Disease Control and Prevention Division of Viral Diseases, Pathogen | Peter W. Cook, Dakota Howard, Dhwani Batra, Ben L. Rambo-Martin, Eileen de Feo, Jan Antico, Christine Tran, Matthew Tolentino, Shannon Wickline, Kim Gietzen, Brad Sickler, Jingtao Liu, Eric Allen, Phil Febbo, Summer Galloway, Nicole L. Washington, Simon White, Geraint Levan, Kelly Schiabor Barrett, Elizabeth Cirulli, Alexandre Bolze, Ary Ascencio, Charlotte Rivera-Garcia, Ryan Cho, Jason Nguyen, Sherry Wang, Jimmy Ramirez, Tyler Cassens, Efren Sandoval, Magnus Isaksson, William Lee, David Becker, Marc Laurent, James Lu, Clinton R. Paden, Suxiang Tong, |

| | | | |
|---|---|---|---|
| | | Discovery | Duncan MacCannell |
| EPI_ISL_1421351, EPI_ISL_1421352 | Wisconsin State Laboratory of Hygiene Communicable Disease Division | Wisconsin State Laboratory of Hygiene Communicable Disease Division | Kelsey R. Florek, Abigail C. Shockey |
| EPI_ISL_1421353 | Helix/Illumina | Centers for Disease Control and Prevention Division of Viral Diseases, Pathogen Discovery | Peter W. Cook, Dakota Howard, Dhwani Batra, Ben L. Rambo-Martin, Eileen de Feo, Jan Antico, Christine Tran, Matthew Tolentino, Shannon Wickline, Kim Gietzen, Brad Sickler, Jingtao Liu, Eric Allen, Phil Febbo, Summer Galloway, Nicole L. Washington, Simon White, Geraint Levan, Kelly Schiabor Barrett, Elizabeth Cirulli, Alexandre Bolze, Ary Ascencio, Charlotte Rivera-Garcia, Ryan Cho, Jason Nguyen, Sherry Wang, Jimmy Ramirez, Tyler Cassens, Efren Sandoval, Magnus Isaksson, William Lee, David Becker, Marc Laurent, James Lu, Clinton R. Paden, Suxiang Tong, Duncan MacCannell |
| EPI_ISL_1421354, EPI_ISL_1421355 | Wisconsin State Laboratory of Hygiene Communicable Disease Division | Wisconsin State Laboratory of Hygiene Communicable Disease Division | Kelsey R. Florek, Abigail C. Shockey |
| EPI_ISL_1421356 | Helix/Illumina | Centers for Disease Control and Prevention Division of Viral Diseases, Pathogen Discovery | Peter W. Cook, Dakota Howard, Dhwani Batra, Ben L. Rambo-Martin, Eileen de Feo, Jan Antico, Christine Tran, Matthew Tolentino, Shannon Wickline, Kim Gietzen, Brad Sickler, Jingtao Liu, Eric Allen, Phil Febbo, Summer Galloway, Nicole L. Washington, Simon White, Geraint Levan, Kelly Schiabor Barrett, Elizabeth Cirulli, Alexandre Bolze, Ary Ascencio, Charlotte Rivera-Garcia, Ryan Cho, Jason Nguyen, Sherry Wang, Jimmy Ramirez, Tyler Cassens, Efren Sandoval, Magnus Isaksson, William Lee, David Becker, Marc Laurent, James Lu, Clinton R. Paden, Suxiang Tong, Duncan MacCannell |
| EPI_ISL_1421357 | Wisconsin State Laboratory of Hygiene Communicable Disease Division | Wisconsin State Laboratory of Hygiene Communicable Disease Division | Kelsey R. Florek, Abigail C. Shockey |
| EPI_ISL_1421358 | Helix/Illumina | Centers for Disease Control and Prevention Division of Viral Diseases, Pathogen Discovery | Peter W. Cook, Dakota Howard, Dhwani Batra, Ben L. Rambo-Martin, Eileen de Feo, Jan Antico, Christine Tran, Matthew Tolentino, Shannon Wickline, Kim Gietzen, Brad Sickler, Jingtao Liu, Eric Allen, Phil Febbo, Summer Galloway, Nicole L. Washington, Simon White, Geraint Levan, Kelly Schiabor Barrett, Elizabeth Cirulli, Alexandre Bolze, Ary Ascencio, Charlotte Rivera-Garcia, Ryan Cho, Jason Nguyen, Sherry Wang, Jimmy Ramirez, Tyler Cassens, Efren Sandoval, Magnus Isaksson, William Lee, David Becker, Marc Laurent, James Lu, Clinton R. Paden, Suxiang Tong, Duncan MacCannell |
| EPI_ISL_1421359, EPI_ISL_1421360 | Wisconsin State Laboratory of Hygiene Communicable Disease Division | Wisconsin State Laboratory of Hygiene Communicable Disease Division | Kelsey R. Florek, Abigail C. Shockey |
| EPI_ISL_1421361 | Helix/Illumina | Centers for Disease Control and Prevention Division of Viral Diseases, Pathogen Discovery | Peter W. Cook, Dakota Howard, Dhwani Batra, Ben L. Rambo-Martin, Eileen de Feo, Jan Antico, Christine Tran, Matthew Tolentino, Shannon Wickline, Kim Gietzen, Brad Sickler, Jingtao Liu, Eric Allen, Phil Febbo, Summer Galloway, Nicole L. Washington, Simon White, Geraint Levan, Kelly Schiabor Barrett, Elizabeth Cirulli, Alexandre Bolze, Ary Ascencio, Charlotte Rivera-Garcia, Ryan Cho, Jason Nguyen, Sherry Wang, Jimmy Ramirez, Tyler Cassens, Efren Sandoval, Magnus Isaksson, William Lee, David Becker, Marc Laurent, James Lu, Clinton R. Paden, Suxiang Tong, Duncan MacCannell |
| EPI_ISL_1421362, EPI_ISL_1421363 | Wisconsin State Laboratory of Hygiene Communicable Disease Division | Wisconsin State Laboratory of Hygiene Communicable Disease Division | Kelsey R. Florek, Abigail C. Shockey |
| EPI_ISL_1421364 | Helix/Illumina | Centers for Disease Control and Prevention Division of Viral Diseases, Pathogen Discovery | Peter W. Cook, Dakota Howard, Dhwani Batra, Ben L. Rambo-Martin, Eileen de Feo, Jan Antico, Christine Tran, Matthew Tolentino, Shannon Wickline, Kim Gietzen, Brad Sickler, Jingtao Liu, Eric Allen, Phil Febbo, Summer Galloway, Nicole L. Washington, Simon White, Geraint Levan, Kelly Schiabor Barrett, Elizabeth Cirulli, Alexandre Bolze, Ary Ascencio, Charlotte Rivera-Garcia, Ryan Cho, Jason Nguyen, Sherry Wang, Jimmy Ramirez, Tyler Cassens, Efren Sandoval, Magnus Isaksson, William Lee, David Becker, Marc Laurent, James Lu, Clinton R. Paden, Suxiang Tong, Duncan MacCannell |
| EPI_ISL_1421365, EPI_ISL_1421366 | Wisconsin State Laboratory of Hygiene Communicable Disease Division | Wisconsin State Laboratory of Hygiene Communicable Disease Division | Kelsey R. Florek, Abigail C. Shockey |
| EPI_ISL_1421367 | Helix/Illumina | Centers for Disease Control and Prevention Division of Viral Diseases, Pathogen Discovery | Peter W. Cook, Dakota Howard, Dhwani Batra, Ben L. Rambo-Martin, Eileen de Feo, Jan Antico, Christine Tran, Matthew Tolentino, Shannon Wickline, Kim Gietzen, Brad Sickler, Jingtao Liu, Eric Allen, Phil Febbo, Summer Galloway, Nicole L. Washington, Simon White, Geraint Levan, Kelly Schiabor Barrett, Elizabeth Cirulli, Alexandre Bolze, Ary Ascencio, Charlotte Rivera-Garcia, Ryan Cho, Jason Nguyen, Sherry Wang, Jimmy Ramirez, Tyler Cassens, Efren Sandoval, Magnus Isaksson, William Lee, David Becker, Marc Laurent, James Lu, Clinton R. Paden, Suxiang Tong, Duncan MacCannell |
| EPI_ISL_1421368, EPI_ISL_1421369, EPI_ISL_1421370 | Wisconsin State Laboratory of Hygiene Communicable Disease Division | Wisconsin State Laboratory of Hygiene Communicable Disease Division | Kelsey R. Florek, Abigail C. Shockey |
| EPI_ISL_1421371 | Helix/Illumina | Centers for Disease Control and Prevention Division of Viral Diseases, Pathogen Discovery | Peter W. Cook, Dakota Howard, Dhwani Batra, Ben L. Rambo-Martin, Eileen de Feo, Jan Antico, Christine Tran, Matthew Tolentino, Shannon Wickline, Kim Gietzen, Brad Sickler, Jingtao Liu, Eric Allen, Phil Febbo, Summer Galloway, Nicole L. Washington, Simon White, Geraint Levan, Kelly Schiabor Barrett, Elizabeth Cirulli, Alexandre Bolze, Ary Ascencio, Charlotte Rivera-Garcia, Ryan Cho, Jason Nguyen, Sherry Wang, Jimmy Ramirez, Tyler Cassens, Efren Sandoval, Magnus Isaksson, William Lee, David Becker, Marc Laurent, James Lu, Clinton R. Paden, Suxiang Tong, Duncan MacCannell |
| EPI_ISL_1421372, EPI_ISL_1421373 | Wisconsin State Laboratory of Hygiene Communicable Disease Division | Wisconsin State Laboratory of Hygiene Communicable Disease Division | Kelsey R. Florek, Abigail C. Shockey |
| EPI_ISL_1421374 | Helix/Illumina | Centers for Disease Control and Prevention Division of Viral Diseases, Pathogen Discovery | Peter W. Cook, Dakota Howard, Dhwani Batra, Ben L. Rambo-Martin, Eileen de Feo, Jan Antico, Christine Tran, Matthew Tolentino, Shannon Wickline, Kim Gietzen, Brad Sickler, Jingtao Liu, Eric Allen, Phil Febbo, Summer Galloway, Nicole L. Washington, Simon White, Geraint Levan, Kelly Schiabor Barrett, Elizabeth Cirulli, Alexandre Bolze, Ary Ascencio, Charlotte Rivera-Garcia, Ryan Cho, Jason Nguyen, Sherry Wang, Jimmy Ramirez, Tyler Cassens, Efren Sandoval, Magnus Isaksson, William Lee, David Becker, Marc Laurent, James Lu, Clinton R. Paden, Suxiang Tong, Duncan MacCannell |
| EPI_ISL_1421375, EPI_ISL_1421376 | Wisconsin State Laboratory of Hygiene Communicable Disease Division | Wisconsin State Laboratory of Hygiene Communicable Disease Division | Kelsey R. Florek, Abigail C. Shockey |
| EPI_ISL_1421377, EPI_ISL_1421378 | Helix/Illumina | Centers for Disease Control and Prevention Division of Viral Diseases, Pathogen Discovery | Peter W. Cook, Dakota Howard, Dhwani Batra, Ben L. Rambo-Martin, Eileen de Feo, Jan Antico, Christine Tran, Matthew Tolentino, Shannon Wickline, Kim Gietzen, Brad Sickler, Jingtao Liu, Eric Allen, Phil Febbo, Summer Galloway, Nicole L. Washington, Simon White, Geraint Levan, Kelly Schiabor Barrett, Elizabeth Cirulli, Alexandre Bolze, Ary Ascencio, Charlotte Rivera-Garcia, Ryan Cho, Jason Nguyen, Sherry Wang, Jimmy Ramirez, Tyler Cassens, Efren Sandoval, Magnus Isaksson, William Lee, David Becker, Marc Laurent, James Lu, Clinton R. Paden, Suxiang Tong, Duncan MacCannell |
| EPI_ISL_1421379 | Wisconsin State Laboratory of Hygiene Communicable Disease Division | Wisconsin State Laboratory of Hygiene Communicable Disease Division | Kelsey R. Florek, Abigail C. Shockey |
| EPI_ISL_1421380, EPI_ISL_1421381 | Helix/Illumina | Centers for Disease Control and Prevention Division of Viral Diseases, Pathogen Discovery | Peter W. Cook, Dakota Howard, Dhwani Batra, Ben L. Rambo-Martin, Eileen de Feo, Jan Antico, Christine Tran, Matthew Tolentino, Shannon Wickline, Kim Gietzen, Brad Sickler, Jingtao Liu, Eric Allen, Phil Febbo, Summer Galloway, Nicole L. Washington, Simon White, Geraint Levan, Kelly Schiabor Barrett, Elizabeth Cirulli, Alexandre Bolze, Ary Ascencio, Charlotte Rivera-Garcia, Ryan Cho, Jason Nguyen, Sherry Wang, Jimmy Ramirez, Tyler Cassens, Efren Sandoval, Magnus Isaksson, William Lee, David Becker, Marc Laurent, James Lu, Clinton R. Paden, Suxiang Tong, Duncan MacCannell |
| EPI_ISL_1421382 | Wisconsin State Laboratory of Hygiene Communicable Disease Division | Wisconsin State Laboratory of Hygiene Communicable Disease Division | Kelsey R. Florek, Abigail C. Shockey |
| EPI_ISL_1421383 | Helix/Illumina | Centers for Disease Control and Prevention Division of Viral Diseases, Pathogen Discovery | Peter W. Cook, Dakota Howard, Dhwani Batra, Ben L. Rambo-Martin, Eileen de Feo, Jan Antico, Christine Tran, Matthew Tolentino, Shannon Wickline, Kim Gietzen, Brad Sickler, Jingtao Liu, Eric Allen, Phil Febbo, Summer Galloway, Nicole L. Washington, Simon White, Geraint Levan, Kelly Schiabor Barrett, Elizabeth Cirulli, Alexandre Bolze, Ary Ascencio, Charlotte Rivera-Garcia, Ryan Cho, Jason Nguyen, Sherry Wang, Jimmy Ramirez, Tyler Cassens, Efren Sandoval, Magnus Isaksson, William Lee, David Becker, Marc Laurent, James Lu, Clinton R. Paden, Suxiang Tong, Duncan MacCannell |
| EPI_ISL_1421384 | Wisconsin State Laboratory of Hygiene Communicable Disease Division | Wisconsin State Laboratory of Hygiene Communicable Disease Division | Kelsey R. Florek, Abigail C. Shockey |
| EPI_ISL_1421385 | Helix/Illumina | Centers for Disease Control and Prevention Division of Viral Diseases, Pathogen Discovery | Peter W. Cook, Dakota Howard, Dhwani Batra, Ben L. Rambo-Martin, Eileen de Feo, Jan Antico, Christine Tran, Matthew Tolentino, Shannon Wickline, Kim Gietzen, Brad Sickler, Jingtao Liu, Eric Allen, Phil Febbo, Summer Galloway, Nicole L. Washington, Simon White, Geraint Levan, Kelly Schiabor Barrett, Elizabeth Cirulli, Alexandre Bolze, Ary Ascencio, Charlotte Rivera-Garcia, Ryan Cho, Jason Nguyen, Sherry Wang, Jimmy Ramirez, Tyler Cassens, Efren Sandoval, Magnus Isaksson, William Lee, David Becker, Marc Laurent, James Lu, Clinton R. Paden, Suxiang Tong, Duncan MacCannell |
| EPI_ISL_1421386 | Wisconsin State Laboratory of Hygiene Communicable Disease Division | Wisconsin State Laboratory of Hygiene Communicable Disease Division | Kelsey R. Florek, Abigail C. Shockey |
| EPI_ISL_1421387, EPI_ISL_1421388 | Helix/Illumina | Centers for Disease Control and Prevention Division of Viral Diseases, Pathogen Discovery | Peter W. Cook, Dakota Howard, Dhwani Batra, Ben L. Rambo-Martin, Eileen de Feo, Jan Antico, Christine Tran, Matthew Tolentino, Shannon Wickline, Kim Gietzen, Brad Sickler, Jingtao Liu, Eric Allen, Phil Febbo, Summer Galloway, Nicole L. Washington, Simon White, Geraint Levan, Kelly Schiabor Barrett, Elizabeth Cirulli, Alexandre Bolze, Ary Ascencio, Charlotte Rivera-Garcia, Ryan Cho, Jason Nguyen, Sherry Wang, Jimmy Ramirez, Tyler Cassens, Efren Sandoval, Magnus Isaksson, William Lee, David Becker, Marc Laurent, James Lu, Clinton R. Paden, Suxiang Tong, Duncan MacCannell |
| EPI_ISL_1421389 | Wisconsin State Laboratory of Hygiene Communicable Disease Division | Wisconsin State Laboratory of Hygiene Communicable Disease Division | Kelsey R. Florek, Abigail C. Shockey |
| EPI_ISL_1421390 | Helix/Illumina | Centers for Disease Control and Prevention Division of Viral Diseases, Pathogen Discovery | Peter W. Cook, Dakota Howard, Dhwani Batra, Ben L. Rambo-Martin, Eileen de Feo, Jan Antico, Christine Tran, Matthew Tolentino, Shannon Wickline, Kim Gietzen, Brad Sickler, Jingtao Liu, Eric Allen, Phil Febbo, Summer Galloway, Nicole L. Washington, Simon White, Geraint Levan, Kelly Schiabor Barrett, Elizabeth Cirulli, Alexandre Bolze, Ary Ascencio, Charlotte Rivera-Garcia, Ryan Cho, Jason Nguyen, Sherry Wang, Jimmy Ramirez, Tyler Cassens, Efren Sandoval, Magnus Isaksson, William Lee, David Becker, Marc Laurent, James Lu, Clinton R. Paden, Suxiang Tong, Duncan MacCannell |
| EPI_ISL_1421391 | Wisconsin State Laboratory of Hygiene Communicable Disease Division | Wisconsin State Laboratory of Hygiene Communicable Disease Division | Kelsey R. Florek, Abigail C. Shockey |
| EPI_ISL_1421392 | Helix/Illumina | Centers for Disease Control and Prevention Division of Viral Diseases, Pathogen Discovery | Peter W. Cook, Dakota Howard, Dhwani Batra, Ben L. Rambo-Martin, Eileen de Feo, Jan Antico, Christine Tran, Matthew Tolentino, Shannon Wickline, Kim Gietzen, Brad Sickler, Jingtao Liu, Eric Allen, Phil Febbo, Summer Galloway, Nicole L. Washington, Simon White, Geraint Levan, Kelly Schiabor Barrett, Elizabeth Cirulli, Alexandre Bolze, Ary Ascencio, Charlotte Rivera-Garcia, Ryan Cho, Jason Nguyen, Sherry Wang, Jimmy Ramirez, Tyler Cassens, Efren Sandoval, Magnus Isaksson, William Lee, David Becker, Marc Laurent, James Lu, Clinton R. Paden, Suxiang Tong, Duncan MacCannell |
| EPI_ISL_1421393 | Wisconsin State Laboratory of Hygiene Communicable Disease Division | Wisconsin State Laboratory of Hygiene Communicable Disease Division | Kelsey R. Florek, Abigail C. Shockey |
| EPI_ISL_1421394 | Helix/Illumina | Centers for Disease Control and Prevention Division of Viral Diseases, Pathogen Discovery | Peter W. Cook, Dakota Howard, Dhwani Batra, Ben L. Rambo-Martin, Eileen de Feo, Jan Antico, Christine Tran, Matthew Tolentino, Shannon Wickline, Kim Gietzen, Brad Sickler, Jingtao Liu, Eric Allen, Phil Febbo, Summer Galloway, Nicole L. Washington, Simon White, Geraint Levan, Kelly Schiabor Barrett, Elizabeth Cirulli, Alexandre Bolze, Ary Ascencio, Charlotte Rivera-Garcia, Ryan Cho, Jason Nguyen, Sherry Wang, Jimmy Ramirez, Tyler Cassens, Efren Sandoval, Magnus Isaksson, William Lee, David Becker, Marc Laurent, James Lu, Clinton R. Paden, Suxiang Tong, Duncan MacCannell |
| EPI_ISL_1421395 | Wisconsin State Laboratory of Hygiene Communicable Disease Division | Wisconsin State Laboratory of Hygiene Communicable Disease Division | Kelsey R. Florek, Abigail C. Shockey |
| EPI_ISL_1421396 | Helix/Illumina | Centers for Disease Control and Prevention Division of Viral Diseases, Pathogen Discovery | Peter W. Cook, Dakota Howard, Dhwani Batra, Ben L. Rambo-Martin, Eileen de Feo, Jan Antico, Christine Tran, Matthew Tolentino, Shannon Wickline, Kim Gietzen, Brad Sickler, Jingtao Liu, Eric Allen, Phil Febbo, Summer Galloway, Nicole L. Washington, Simon White, Geraint Levan, Kelly Schiabor Barrett, Elizabeth Cirulli, Alexandre Bolze, Ary Ascencio, Charlotte Rivera-Garcia, Ryan Cho, Jason Nguyen, Sherry Wang, Jimmy Ramirez, Tyler Cassens, Efren Sandoval, Magnus Isaksson, William Lee, David Becker, Marc Laurent, James Lu, Clinton R. Paden, Suxiang Tong, Duncan MacCannell |
| EPI_ISL_1421397, EPI_ISL_1421398, EPI_ISL_1421399 | Wisconsin State Laboratory of Hygiene Communicable Disease Division | Wisconsin State Laboratory of Hygiene Communicable Disease Division | Kelsey R. Florek, Abigail C. Shockey |
| EPI_ISL_1421400 | Helix/Illumina | Centers for Disease Control and Prevention Division of Viral Diseases, Pathogen Discovery | Peter W. Cook, Dakota Howard, Dhwani Batra, Ben L. Rambo-Martin, Eileen de Feo, Jan Antico, Christine Tran, Matthew Tolentino, Shannon Wickline, Kim Gietzen, Brad Sickler, Jingtao Liu, Eric Allen, Phil Febbo, Summer Galloway, Nicole L. Washington, Simon White, Geraint Levan, Kelly Schiabor Barrett, Elizabeth Cirulli, Alexandre Bolze, Ary Ascencio, Charlotte Rivera-Garcia, Ryan Cho, Jason Nguyen, Sherry Wang, Jimmy Ramirez, Tyler Cassens, Efren Sandoval, Magnus Isaksson, William Lee, David Becker, Marc Laurent, James Lu, Clinton R. Paden, Suxiang Tong, Duncan MacCannell |
| EPI_ISL_1421401, | Wisconsin State Laboratory of Hygiene | Wisconsin State Laboratory of Hygiene | Kelsey R. Florek, Abigail C. Shockey |

| | | | |
|---|---|---|---|
| EPI_ISL_1421402 | Communicable Disease Division | Communicable Disease Division | |
| EPI_ISL_1421403 | Helix/Illumina | Centers for Disease Control and Prevention Division of Viral Diseases, Pathogen Discovery | Peter W. Cook, Dakota Howard, Dhwani Batra, Ben L. Rambo-Martin, Eileen de Feo, Jan Antico, Christine Tran, Matthew Tolentino, Shannon Wickline, Kim Gietzen, Brad Sickler, Jingtao Liu, Eric Allen, Phil Febbo, Summer Galloway, Nicole L. Washington, Simon White, Geraint Levan, Kelly Schiabor Barrett, Elizabeth Cirulli, Alexandre Bolze, Ary Ascencio, Charlotte Rivera-Garcia, Ryan Cho, Jason Nguyen, Sherry Wang, Jimmy Ramirez, Tyler Cassens, Efren Sandoval, Magnus Isaksson, William Lee, David Becker, Marc Laurent, James Lu, Clinton R. Paden, Suxiang Tong, Duncan MacCannell |
| EPI_ISL_1421404, EPI_ISL_1421405 | Wisconsin State Laboratory of Hygiene Communicable Disease Division | Wisconsin State Laboratory of Hygiene Communicable Disease Division | Kelsey R. Florek, Abigail C. Shockey |
| EPI_ISL_1421406 | Helix/Illumina | Centers for Disease Control and Prevention Division of Viral Diseases, Pathogen Discovery | Peter W. Cook, Dakota Howard, Dhwani Batra, Ben L. Rambo-Martin, Eileen de Feo, Jan Antico, Christine Tran, Matthew Tolentino, Shannon Wickline, Kim Gietzen, Brad Sickler, Jingtao Liu, Eric Allen, Phil Febbo, Summer Galloway, Nicole L. Washington, Simon White, Geraint Levan, Kelly Schiabor Barrett, Elizabeth Cirulli, Alexandre Bolze, Ary Ascencio, Charlotte Rivera-Garcia, Ryan Cho, Jason Nguyen, Sherry Wang, Jimmy Ramirez, Tyler Cassens, Efren Sandoval, Magnus Isaksson, William Lee, David Becker, Marc Laurent, James Lu, Clinton R. Paden, Suxiang Tong, Duncan MacCannell |
| EPI_ISL_1421407, EPI_ISL_1421408 | Wisconsin State Laboratory of Hygiene Communicable Disease Division | Wisconsin State Laboratory of Hygiene Communicable Disease Division | Kelsey R. Florek, Abigail C. Shockey |
| EPI_ISL_1421409 | Helix/Illumina | Centers for Disease Control and Prevention Division of Viral Diseases, Pathogen Discovery | Peter W. Cook, Dakota Howard, Dhwani Batra, Ben L. Rambo-Martin, Eileen de Feo, Jan Antico, Christine Tran, Matthew Tolentino, Shannon Wickline, Kim Gietzen, Brad Sickler, Jingtao Liu, Eric Allen, Phil Febbo, Summer Galloway, Nicole L. Washington, Simon White, Geraint Levan, Kelly Schiabor Barrett, Elizabeth Cirulli, Alexandre Bolze, Ary Ascencio, Charlotte Rivera-Garcia, Ryan Cho, Jason Nguyen, Sherry Wang, Jimmy Ramirez, Tyler Cassens, Efren Sandoval, Magnus Isaksson, William Lee, David Becker, Marc Laurent, James Lu, Clinton R. Paden, Suxiang Tong, Duncan MacCannell |
| EPI_ISL_1421410, EPI_ISL_1421411 | Wisconsin State Laboratory of Hygiene Communicable Disease Division | Wisconsin State Laboratory of Hygiene Communicable Disease Division | Kelsey R. Florek, Abigail C. Shockey |
| EPI_ISL_1421412 | Helix/Illumina | Centers for Disease Control and Prevention Division of Viral Diseases, Pathogen Discovery | Peter W. Cook, Dakota Howard, Dhwani Batra, Ben L. Rambo-Martin, Eileen de Feo, Jan Antico, Christine Tran, Matthew Tolentino, Shannon Wickline, Kim Gietzen, Brad Sickler, Jingtao Liu, Eric Allen, Phil Febbo, Summer Galloway, Nicole L. Washington, Simon White, Geraint Levan, Kelly Schiabor Barrett, Elizabeth Cirulli, Alexandre Bolze, Ary Ascencio, Charlotte Rivera-Garcia, Ryan Cho, Jason Nguyen, Sherry Wang, Jimmy Ramirez, Tyler Cassens, Efren Sandoval, Magnus Isaksson, William Lee, David Becker, Marc Laurent, James Lu, Clinton R. Paden, Suxiang Tong, Duncan MacCannell |
| EPI_ISL_1421413, EPI_ISL_1421414 | Wisconsin State Laboratory of Hygiene Communicable Disease Division | Wisconsin State Laboratory of Hygiene Communicable Disease Division | Kelsey R. Florek, Abigail C. Shockey |
| EPI_ISL_1421415 | Helix/Illumina | Centers for Disease Control and Prevention Division of Viral Diseases, Pathogen Discovery | Peter W. Cook, Dakota Howard, Dhwani Batra, Ben L. Rambo-Martin, Eileen de Feo, Jan Antico, Christine Tran, Matthew Tolentino, Shannon Wickline, Kim Gietzen, Brad Sickler, Jingtao Liu, Eric Allen, Phil Febbo, Summer Galloway, Nicole L. Washington, Simon White, Geraint Levan, Kelly Schiabor Barrett, Elizabeth Cirulli, Alexandre Bolze, Ary Ascencio, Charlotte Rivera-Garcia, Ryan Cho, Jason Nguyen, Sherry Wang, Jimmy Ramirez, Tyler Cassens, Efren Sandoval, Magnus Isaksson, William Lee, David Becker, Marc Laurent, James Lu, Clinton R. Paden, Suxiang Tong, Duncan MacCannell |
| EPI_ISL_1421416 | Wisconsin State Laboratory of Hygiene Communicable Disease Division | Wisconsin State Laboratory of Hygiene Communicable Disease Division | Kelsey R. Florek, Abigail C. Shockey |
| EPI_ISL_1421418 | Helix/Illumina | Centers for Disease Control and Prevention Division of Viral Diseases, Pathogen Discovery | Peter W. Cook, Dakota Howard, Dhwani Batra, Ben L. Rambo-Martin, Eileen de Feo, Jan Antico, Christine Tran, Matthew Tolentino, Shannon Wickline, Kim Gietzen, Brad Sickler, Jingtao Liu, Eric Allen, Phil Febbo, Summer Galloway, Nicole L. Washington, Simon White, Geraint Levan, Kelly Schiabor Barrett, Elizabeth Cirulli, Alexandre Bolze, Ary Ascencio, Charlotte Rivera-Garcia, Ryan Cho, Jason Nguyen, Sherry Wang, Jimmy Ramirez, Tyler Cassens, Efren Sandoval, Magnus Isaksson, William Lee, David Becker, Marc Laurent, James Lu, Clinton R. Paden, Suxiang Tong, Duncan MacCannell |
| EPI_ISL_1421419, EPI_ISL_1421420 | Wisconsin State Laboratory of Hygiene Communicable Disease Division | Wisconsin State Laboratory of Hygiene Communicable Disease Division | Kelsey R. Florek, Abigail C. Shockey |
| EPI_ISL_1421421 | Helix/Illumina | Centers for Disease Control and Prevention Division of Viral Diseases, Pathogen Discovery | Peter W. Cook, Dakota Howard, Dhwani Batra, Ben L. Rambo-Martin, Eileen de Feo, Jan Antico, Christine Tran, Matthew Tolentino, Shannon Wickline, Kim Gietzen, Brad Sickler, Jingtao Liu, Eric Allen, Phil Febbo, Summer Galloway, Nicole L. Washington, Simon White, Geraint Levan, Kelly Schiabor Barrett, Elizabeth Cirulli, Alexandre Bolze, Ary Ascencio, Charlotte Rivera-Garcia, Ryan Cho, Jason Nguyen, Sherry Wang, Jimmy Ramirez, Tyler Cassens, Efren Sandoval, Magnus Isaksson, William Lee, David Becker, Marc Laurent, James Lu, Clinton R. Paden, Suxiang Tong, Duncan MacCannell |
| EPI_ISL_1421422, EPI_ISL_1421423 | Wisconsin State Laboratory of Hygiene Communicable Disease Division | Wisconsin State Laboratory of Hygiene Communicable Disease Division | Kelsey R. Florek, Abigail C. Shockey |
| EPI_ISL_1421424 | Helix/Illumina | Centers for Disease Control and Prevention Division of Viral Diseases, Pathogen Discovery | Peter W. Cook, Dakota Howard, Dhwani Batra, Ben L. Rambo-Martin, Eileen de Feo, Jan Antico, Christine Tran, Matthew Tolentino, Shannon Wickline, Kim Gietzen, Brad Sickler, Jingtao Liu, Eric Allen, Phil Febbo, Summer Galloway, Nicole L. Washington, Simon White, Geraint Levan, Kelly Schiabor Barrett, Elizabeth Cirulli, Alexandre Bolze, Ary Ascencio, Charlotte Rivera-Garcia, Ryan Cho, Jason Nguyen, Sherry Wang, Jimmy Ramirez, Tyler Cassens, Efren Sandoval, Magnus Isaksson, William Lee, David Becker, Marc Laurent, James Lu, Clinton R. Paden, Suxiang Tong, Duncan MacCannell |
| EPI_ISL_1421425, EPI_ISL_1421426 | Wisconsin State Laboratory of Hygiene Communicable Disease Division | Wisconsin State Laboratory of Hygiene Communicable Disease Division | Kelsey R. Florek, Abigail C. Shockey |
| EPI_ISL_1421427 | Helix/Illumina | Centers for Disease Control and Prevention Division of Viral Diseases, Pathogen Discovery | Peter W. Cook, Dakota Howard, Dhwani Batra, Ben L. Rambo-Martin, Eileen de Feo, Jan Antico, Christine Tran, Matthew Tolentino, Shannon Wickline, Kim Gietzen, Brad Sickler, Jingtao Liu, Eric Allen, Phil Febbo, Summer Galloway, Nicole L. Washington, Simon White, Geraint Levan, Kelly Schiabor Barrett, Elizabeth Cirulli, Alexandre Bolze, Ary Ascencio, Charlotte Rivera-Garcia, Ryan Cho, Jason Nguyen, Sherry Wang, Jimmy Ramirez, Tyler Cassens, Efren Sandoval, Magnus Isaksson, William Lee, David Becker, Marc Laurent, James Lu, Clinton R. Paden, Suxiang Tong, Duncan MacCannell |
| EPI_ISL_1421428, EPI_ISL_1421429 | Wisconsin State Laboratory of Hygiene Communicable Disease Division | Wisconsin State Laboratory of Hygiene Communicable Disease Division | Kelsey R. Florek, Abigail C. Shockey |
| EPI_ISL_1421430 | Helix/Illumina | Centers for Disease Control and Prevention Division of Viral Diseases, Pathogen Discovery | Peter W. Cook, Dakota Howard, Dhwani Batra, Ben L. Rambo-Martin, Eileen de Feo, Jan Antico, Christine Tran, Matthew Tolentino, Shannon Wickline, Kim Gietzen, Brad Sickler, Jingtao Liu, Eric Allen, Phil Febbo, Summer Galloway, Nicole L. Washington, Simon White, Geraint Levan, Kelly Schiabor Barrett, Elizabeth Cirulli, Alexandre Bolze, Ary Ascencio, Charlotte Rivera-Garcia, Ryan Cho, Jason Nguyen, Sherry Wang, Jimmy Ramirez, Tyler Cassens, Efren Sandoval, Magnus Isaksson, William Lee, David Becker, Marc Laurent, James Lu, Clinton R. Paden, Suxiang Tong, Duncan MacCannell |
| EPI_ISL_1421431, EPI_ISL_1421432 | Wisconsin State Laboratory of Hygiene Communicable Disease Division | Wisconsin State Laboratory of Hygiene Communicable Disease Division | Kelsey R. Florek, Abigail C. Shockey |
| EPI_ISL_1421434 | Helix/Illumina | Centers for Disease Control and Prevention Division of Viral Diseases, Pathogen Discovery | Peter W. Cook, Dakota Howard, Dhwani Batra, Ben L. Rambo-Martin, Eileen de Feo, Jan Antico, Christine Tran, Matthew Tolentino, Shannon Wickline, Kim Gietzen, Brad Sickler, Jingtao Liu, Eric Allen, Phil Febbo, Summer Galloway, Nicole L. Washington, Simon White, Geraint Levan, Kelly Schiabor Barrett, Elizabeth Cirulli, Alexandre Bolze, Ary Ascencio, Charlotte Rivera-Garcia, Ryan Cho, Jason Nguyen, Sherry Wang, Jimmy Ramirez, Tyler Cassens, Efren Sandoval, Magnus Isaksson, William Lee, David Becker, Marc Laurent, James Lu, Clinton R. Paden, Suxiang Tong, Duncan MacCannell |
| EPI_ISL_1421435, EPI_ISL_1421436 | Wisconsin State Laboratory of Hygiene Communicable Disease Division | Wisconsin State Laboratory of Hygiene Communicable Disease Division | Kelsey R. Florek, Abigail C. Shockey |
| EPI_ISL_1421437 | Helix/Illumina | Centers for Disease Control and Prevention Division of Viral Diseases, Pathogen Discovery | Peter W. Cook, Dakota Howard, Dhwani Batra, Ben L. Rambo-Martin, Eileen de Feo, Jan Antico, Christine Tran, Matthew Tolentino, Shannon Wickline, Kim Gietzen, Brad Sickler, Jingtao Liu, Eric Allen, Phil Febbo, Summer Galloway, Nicole L. Washington, Simon White, Geraint Levan, Kelly Schiabor Barrett, Elizabeth Cirulli, Alexandre Bolze, Ary Ascencio, Charlotte Rivera-Garcia, Ryan Cho, Jason Nguyen, Sherry Wang, Jimmy Ramirez, Tyler Cassens, Efren Sandoval, Magnus Isaksson, William Lee, David Becker, Marc Laurent, James Lu, Clinton R. Paden, Suxiang Tong, Duncan MacCannell |
| EPI_ISL_1421438, EPI_ISL_1421439 | Wisconsin State Laboratory of Hygiene Communicable Disease Division | Wisconsin State Laboratory of Hygiene Communicable Disease Division | Kelsey R. Florek, Abigail C. Shockey |
| EPI_ISL_1421440 | Helix/Illumina | Centers for Disease Control and Prevention Division of Viral Diseases, Pathogen Discovery | Peter W. Cook, Dakota Howard, Dhwani Batra, Ben L. Rambo-Martin, Eileen de Feo, Jan Antico, Christine Tran, Matthew Tolentino, Shannon Wickline, Kim Gietzen, Brad Sickler, Jingtao Liu, Eric Allen, Phil Febbo, Summer Galloway, Nicole L. Washington, Simon White, Geraint Levan, Kelly Schiabor Barrett, Elizabeth Cirulli, Alexandre Bolze, Ary Ascencio, Charlotte Rivera-Garcia, Ryan Cho, Jason Nguyen, Sherry Wang, Jimmy Ramirez, Tyler Cassens, Efren Sandoval, Magnus Isaksson, William Lee, David Becker, Marc Laurent, James Lu, Clinton R. Paden, Suxiang Tong, Duncan MacCannell |
| EPI_ISL_1421441, EPI_ISL_1421442 | Wisconsin State Laboratory of Hygiene Communicable Disease Division | Wisconsin State Laboratory of Hygiene Communicable Disease Division | Kelsey R. Florek, Abigail C. Shockey |
| EPI_ISL_1421443 | Helix/Illumina | Centers for Disease Control and Prevention Division of Viral Diseases, Pathogen Discovery | Peter W. Cook, Dakota Howard, Dhwani Batra, Ben L. Rambo-Martin, Eileen de Feo, Jan Antico, Christine Tran, Matthew Tolentino, Shannon Wickline, Kim Gietzen, Brad Sickler, Jingtao Liu, Eric Allen, Phil Febbo, Summer Galloway, Nicole L. Washington, Simon White, Geraint Levan, Kelly Schiabor Barrett, Elizabeth Cirulli, Alexandre Bolze, Ary Ascencio, Charlotte Rivera-Garcia, Ryan Cho, Jason Nguyen, Sherry Wang, Jimmy Ramirez, Tyler Cassens, Efren Sandoval, Magnus Isaksson, William Lee, David Becker, Marc Laurent, James Lu, Clinton R. Paden, Suxiang Tong, Duncan MacCannell |
| EPI_ISL_1421444, EPI_ISL_1421445 | Wisconsin State Laboratory of Hygiene Communicable Disease Division | Wisconsin State Laboratory of Hygiene Communicable Disease Division | Kelsey R. Florek, Abigail C. Shockey |
| EPI_ISL_1421446 | Helix/Illumina | Centers for Disease Control and Prevention Division of Viral Diseases, Pathogen Discovery | Peter W. Cook, Dakota Howard, Dhwani Batra, Ben L. Rambo-Martin, Eileen de Feo, Jan Antico, Christine Tran, Matthew Tolentino, Shannon Wickline, Kim Gietzen, Brad Sickler, Jingtao Liu, Eric Allen, Phil Febbo, Summer Galloway, Nicole L. Washington, Simon White, Geraint Levan, Kelly Schiabor Barrett, Elizabeth Cirulli, Alexandre Bolze, Ary Ascencio, Charlotte Rivera-Garcia, Ryan Cho, Jason Nguyen, Sherry Wang, Jimmy Ramirez, Tyler Cassens, Efren Sandoval, Magnus Isaksson, William Lee, David Becker, Marc Laurent, James Lu, Clinton R. Paden, Suxiang Tong, Duncan MacCannell |
| EPI_ISL_1421447, EPI_ISL_1421448 | Wisconsin State Laboratory of Hygiene Communicable Disease Division | Wisconsin State Laboratory of Hygiene Communicable Disease Division | Kelsey R. Florek, Abigail C. Shockey |
| EPI_ISL_1421449 | Helix/Illumina | Centers for Disease Control and Prevention Division of Viral Diseases, Pathogen Discovery | Peter W. Cook, Dakota Howard, Dhwani Batra, Ben L. Rambo-Martin, Eileen de Feo, Jan Antico, Christine Tran, Matthew Tolentino, Shannon Wickline, Kim Gietzen, Brad Sickler, Jingtao Liu, Eric Allen, Phil Febbo, Summer Galloway, Nicole L. Washington, Simon White, Geraint Levan, Kelly Schiabor Barrett, Elizabeth Cirulli, Alexandre Bolze, Ary Ascencio, Charlotte Rivera-Garcia, Ryan Cho, Jason Nguyen, Sherry Wang, Jimmy Ramirez, Tyler Cassens, Efren Sandoval, Magnus Isaksson, William Lee, David Becker, Marc Laurent, James Lu, Clinton R. Paden, Suxiang Tong, Duncan MacCannell |
| EPI_ISL_1421450 | Wisconsin State Laboratory of Hygiene Communicable Disease Division | Wisconsin State Laboratory of Hygiene Communicable Disease Division | Kelsey R. Florek, Abigail C. Shockey |
| EPI_ISL_1421452 | Helix/Illumina | Centers for Disease Control and Prevention Division of Viral Diseases, Pathogen Discovery | Peter W. Cook, Dakota Howard, Dhwani Batra, Ben L. Rambo-Martin, Eileen de Feo, Jan Antico, Christine Tran, Matthew Tolentino, Shannon Wickline, Kim Gietzen, Brad Sickler, Jingtao Liu, Eric Allen, Phil Febbo, Summer Galloway, Nicole L. Washington, Simon White, Geraint Levan, Kelly Schiabor Barrett, Elizabeth Cirulli, Alexandre Bolze, Ary Ascencio, Charlotte Rivera-Garcia, Ryan Cho, Jason Nguyen, Sherry Wang, Jimmy Ramirez, Tyler Cassens, Efren Sandoval, Magnus Isaksson, William Lee, David Becker, Marc Laurent, James Lu, Clinton R. Paden, Suxiang Tong, Duncan MacCannell |
| EPI_ISL_1421453, EPI_ISL_1421454 | Wisconsin State Laboratory of Hygiene Communicable Disease Division | Wisconsin State Laboratory of Hygiene Communicable Disease Division | Kelsey R. Florek, Abigail C. Shockey |
| EPI_ISL_1421455 | Helix/Illumina | Centers for Disease Control and Prevention Division of Viral Diseases, Pathogen Discovery | Peter W. Cook, Dakota Howard, Dhwani Batra, Ben L. Rambo-Martin, Eileen de Feo, Jan Antico, Christine Tran, Matthew Tolentino, Shannon Wickline, Kim Gietzen, Brad Sickler, Jingtao Liu, Eric Allen, Phil Febbo, Summer Galloway, Nicole L. Washington, Simon White, Geraint Levan, Kelly Schiabor Barrett, Elizabeth Cirulli, Alexandre Bolze, Ary Ascencio, Charlotte Rivera-Garcia, Ryan Cho, Jason Nguyen, Sherry Wang, Jimmy Ramirez, Tyler Cassens, Efren Sandoval, Magnus Isaksson, William Lee, David Becker, Marc Laurent, James Lu, Clinton R. Paden, Suxiang Tong, Duncan MacCannell |
| EPI_ISL_1421456, EPI_ISL_1421457 | Wisconsin State Laboratory of Hygiene Communicable Disease Division | Wisconsin State Laboratory of Hygiene Communicable Disease Division | Kelsey R. Florek, Abigail C. Shockey |
| EPI_ISL_1421458 | Helix/Illumina | Centers for Disease Control and Prevention Division of Viral Diseases, Pathogen Discovery | Peter W. Cook, Dakota Howard, Dhwani Batra, Ben L. Rambo-Martin, Eileen de Feo, Jan Antico, Christine Tran, Matthew Tolentino, Shannon Wickline, Kim Gietzen, Brad Sickler, Jingtao Liu, Eric Allen, Phil Febbo, Summer Galloway, Nicole L. Washington, Simon White, Geraint Levan, Kelly Schiabor Barrett, Elizabeth Cirulli, Alexandre Bolze, Ary Ascencio, Charlotte Rivera-Garcia, Ryan Cho, Jason Nguyen, Sherry Wang, Jimmy Ramirez, Tyler Cassens, Efren Sandoval, Magnus Isaksson, William Lee, David Becker, Marc Laurent, James Lu, Clinton R. Paden, Suxiang Tong, Duncan MacCannell |

| | | | |
|---|---|---|---|
| EPI_ISL_1421459, EPI_ISL_1421460 | Wisconsin State Laboratory of Hygiene Communicable Disease Division | Wisconsin State Laboratory of Hygiene Communicable Disease Division | Kelsey R. Florek, Abigail C. Shockey |
| EPI_ISL_1421461 | Helix/Illumina | Centers for Disease Control and Prevention Division of Viral Diseases, Pathogen Discovery | Peter W. Cook, Dakota Howard, Dhwani Batra, Ben L. Rambo-Martin, Eileen de Feo, Jan Antico, Christine Tran, Matthew Tolentino, Shannon Wickline, Kim Gietzen, Brad Sickler, Jingtao Liu, Eric Allen, Phil Febbo, Summer Galloway, Nicole L. Washington, Simon White, Geraint Levan, Kelly Schiabor Barrett, Elizabeth Cirulli, Alexandre Bolze, Ary Ascencio, Charlotte Rivera-Garcia, Ryan Cho, Jason Nguyen, Sherry Wang, Jimmy Ramirez, Tyler Cassens, Efren Sandoval, Magnus Isaksson, William Lee, David Becker, Marc Laurent, James Lu, Clinton R. Paden, Suxiang Tong, Duncan MacCannell |
| EPI_ISL_1421462, EPI_ISL_1421463 | Wisconsin State Laboratory of Hygiene Communicable Disease Division | Wisconsin State Laboratory of Hygiene Communicable Disease Division | Kelsey R. Florek, Abigail C. Shockey |
| EPI_ISL_1421464 | Helix/Illumina | Centers for Disease Control and Prevention Division of Viral Diseases, Pathogen Discovery | Peter W. Cook, Dakota Howard, Dhwani Batra, Ben L. Rambo-Martin, Eileen de Feo, Jan Antico, Christine Tran, Matthew Tolentino, Shannon Wickline, Kim Gietzen, Brad Sickler, Jingtao Liu, Eric Allen, Phil Febbo, Summer Galloway, Nicole L. Washington, Simon White, Geraint Levan, Kelly Schiabor Barrett, Elizabeth Cirulli, Alexandre Bolze, Ary Ascencio, Charlotte Rivera-Garcia, Ryan Cho, Jason Nguyen, Sherry Wang, Jimmy Ramirez, Tyler Cassens, Efren Sandoval, Magnus Isaksson, William Lee, David Becker, Marc Laurent, James Lu, Clinton R. Paden, Suxiang Tong, Duncan MacCannell |
| EPI_ISL_1421465, EPI_ISL_1421466 | Wisconsin State Laboratory of Hygiene Communicable Disease Division | Wisconsin State Laboratory of Hygiene Communicable Disease Division | Kelsey R. Florek, Abigail C. Shockey |
| EPI_ISL_1421467 | Helix/Illumina | Centers for Disease Control and Prevention Division of Viral Diseases, Pathogen Discovery | Peter W. Cook, Dakota Howard, Dhwani Batra, Ben L. Rambo-Martin, Eileen de Feo, Jan Antico, Christine Tran, Matthew Tolentino, Shannon Wickline, Kim Gietzen, Brad Sickler, Jingtao Liu, Eric Allen, Phil Febbo, Summer Galloway, Nicole L. Washington, Simon White, Geraint Levan, Kelly Schiabor Barrett, Elizabeth Cirulli, Alexandre Bolze, Ary Ascencio, Charlotte Rivera-Garcia, Ryan Cho, Jason Nguyen, Sherry Wang, Jimmy Ramirez, Tyler Cassens, Efren Sandoval, Magnus Isaksson, William Lee, David Becker, Marc Laurent, James Lu, Clinton R. Paden, Suxiang Tong, Duncan MacCannell |
| EPI_ISL_1421468, EPI_ISL_1421469 | Wisconsin State Laboratory of Hygiene Communicable Disease Division | Wisconsin State Laboratory of Hygiene Communicable Disease Division | Kelsey R. Florek, Abigail C. Shockey |
| EPI_ISL_1421470 | Helix/Illumina | Centers for Disease Control and Prevention Division of Viral Diseases, Pathogen Discovery | Peter W. Cook, Dakota Howard, Dhwani Batra, Ben L. Rambo-Martin, Eileen de Feo, Jan Antico, Christine Tran, Matthew Tolentino, Shannon Wickline, Kim Gietzen, Brad Sickler, Jingtao Liu, Eric Allen, Phil Febbo, Summer Galloway, Nicole L. Washington, Simon White, Geraint Levan, Kelly Schiabor Barrett, Elizabeth Cirulli, Alexandre Bolze, Ary Ascencio, Charlotte Rivera-Garcia, Ryan Cho, Jason Nguyen, Sherry Wang, Jimmy Ramirez, Tyler Cassens, Efren Sandoval, Magnus Isaksson, William Lee, David Becker, Marc Laurent, James Lu, Clinton R. Paden, Suxiang Tong, Duncan MacCannell |
| EPI_ISL_1421471, EPI_ISL_1421472 | Wisconsin State Laboratory of Hygiene Communicable Disease Division | Wisconsin State Laboratory of Hygiene Communicable Disease Division | Kelsey R. Florek, Abigail C. Shockey |
| EPI_ISL_1421473, EPI_ISL_1421474 | Helix/Illumina | Centers for Disease Control and Prevention Division of Viral Diseases, Pathogen Discovery | Peter W. Cook, Dakota Howard, Dhwani Batra, Ben L. Rambo-Martin, Eileen de Feo, Jan Antico, Christine Tran, Matthew Tolentino, Shannon Wickline, Kim Gietzen, Brad Sickler, Jingtao Liu, Eric Allen, Phil Febbo, Summer Galloway, Nicole L. Washington, Simon White, Geraint Levan, Kelly Schiabor Barrett, Elizabeth Cirulli, Alexandre Bolze, Ary Ascencio, Charlotte Rivera-Garcia, Ryan Cho, Jason Nguyen, Sherry Wang, Jimmy Ramirez, Tyler Cassens, Efren Sandoval, Magnus Isaksson, William Lee, David Becker, Marc Laurent, James Lu, Clinton R. Paden, Suxiang Tong, Duncan MacCannell |
| EPI_ISL_1421475 | Wisconsin State Laboratory of Hygiene Communicable Disease Division | Wisconsin State Laboratory of Hygiene Communicable Disease Division | Kelsey R. Florek, Abigail C. Shockey |
| EPI_ISL_1421476 | Helix/Illumina | Centers for Disease Control and Prevention Division of Viral Diseases, Pathogen Discovery | Peter W. Cook, Dakota Howard, Dhwani Batra, Ben L. Rambo-Martin, Eileen de Feo, Jan Antico, Christine Tran, Matthew Tolentino, Shannon Wickline, Kim Gietzen, Brad Sickler, Jingtao Liu, Eric Allen, Phil Febbo, Summer Galloway, Nicole L. Washington, Simon White, Geraint Levan, Kelly Schiabor Barrett, Elizabeth Cirulli, Alexandre Bolze, Ary Ascencio, Charlotte Rivera-Garcia, Ryan Cho, Jason Nguyen, Sherry Wang, Jimmy Ramirez, Tyler Cassens, Efren Sandoval, Magnus Isaksson, William Lee, David Becker, Marc Laurent, James Lu, Clinton R. Paden, Suxiang Tong, Duncan MacCannell |
| EPI_ISL_1421477 | Wisconsin State Laboratory of Hygiene Communicable Disease Division | Wisconsin State Laboratory of Hygiene Communicable Disease Division | Kelsey R. Florek, Abigail C. Shockey |
| EPI_ISL_1421479 | Helix/Illumina | Centers for Disease Control and Prevention Division of Viral Diseases, Pathogen Discovery | Peter W. Cook, Dakota Howard, Dhwani Batra, Ben L. Rambo-Martin, Eileen de Feo, Jan Antico, Christine Tran, Matthew Tolentino, Shannon Wickline, Kim Gietzen, Brad Sickler, Jingtao Liu, Eric Allen, Phil Febbo, Summer Galloway, Nicole L. Washington, Simon White, Geraint Levan, Kelly Schiabor Barrett, Elizabeth Cirulli, Alexandre Bolze, Ary Ascencio, Charlotte Rivera-Garcia, Ryan Cho, Jason Nguyen, Sherry Wang, Jimmy Ramirez, Tyler Cassens, Efren Sandoval, Magnus Isaksson, William Lee, David Becker, Marc Laurent, James Lu, Clinton R. Paden, Suxiang Tong, Duncan MacCannell |
| EPI_ISL_1421480 | Wisconsin State Laboratory of Hygiene Communicable Disease Division | Wisconsin State Laboratory of Hygiene Communicable Disease Division | Kelsey R. Florek, Abigail C. Shockey |
| EPI_ISL_1421481 | Helix/Illumina | Centers for Disease Control and Prevention Division of Viral Diseases, Pathogen Discovery | Peter W. Cook, Dakota Howard, Dhwani Batra, Ben L. Rambo-Martin, Eileen de Feo, Jan Antico, Christine Tran, Matthew Tolentino, Shannon Wickline, Kim Gietzen, Brad Sickler, Jingtao Liu, Eric Allen, Phil Febbo, Summer Galloway, Nicole L. Washington, Simon White, Geraint Levan, Kelly Schiabor Barrett, Elizabeth Cirulli, Alexandre Bolze, Ary Ascencio, Charlotte Rivera-Garcia, Ryan Cho, Jason Nguyen, Sherry Wang, Jimmy Ramirez, Tyler Cassens, Efren Sandoval, Magnus Isaksson, William Lee, David Becker, Marc Laurent, James Lu, Clinton R. Paden, Suxiang Tong, Duncan MacCannell |
| EPI_ISL_1421482 | Wisconsin State Laboratory of Hygiene Communicable Disease Division | Wisconsin State Laboratory of Hygiene Communicable Disease Division | Kelsey R. Florek, Abigail C. Shockey |
| EPI_ISL_1421483 | Helix/Illumina | Centers for Disease Control and Prevention Division of Viral Diseases, Pathogen Discovery | Peter W. Cook, Dakota Howard, Dhwani Batra, Ben L. Rambo-Martin, Eileen de Feo, Jan Antico, Christine Tran, Matthew Tolentino, Shannon Wickline, Kim Gietzen, Brad Sickler, Jingtao Liu, Eric Allen, Phil Febbo, Summer Galloway, Nicole L. Washington, Simon White, Geraint Levan, Kelly Schiabor Barrett, Elizabeth Cirulli, Alexandre Bolze, Ary Ascencio, Charlotte Rivera-Garcia, Ryan Cho, Jason Nguyen, Sherry Wang, Jimmy Ramirez, Tyler Cassens, Efren Sandoval, Magnus Isaksson, William Lee, David Becker, Marc Laurent, James Lu, Clinton R. Paden, Suxiang Tong, Duncan MacCannell |
| EPI_ISL_1421484 | Wisconsin State Laboratory of Hygiene Communicable Disease Division | Wisconsin State Laboratory of Hygiene Communicable Disease Division | Kelsey R. Florek, Abigail C. Shockey |
| EPI_ISL_1421485 | Helix/Illumina | Centers for Disease Control and Prevention Division of Viral Diseases, Pathogen Discovery | Peter W. Cook, Dakota Howard, Dhwani Batra, Ben L. Rambo-Martin, Eileen de Feo, Jan Antico, Christine Tran, Matthew Tolentino, Shannon Wickline, Kim Gietzen, Brad Sickler, Jingtao Liu, Eric Allen, Phil Febbo, Summer Galloway, Nicole L. Washington, Simon White, Geraint Levan, Kelly Schiabor Barrett, Elizabeth Cirulli, Alexandre Bolze, Ary Ascencio, Charlotte Rivera-Garcia, Ryan Cho, Jason Nguyen, Sherry Wang, Jimmy Ramirez, Tyler Cassens, Efren Sandoval, Magnus Isaksson, William Lee, David Becker, Marc Laurent, James Lu, Clinton R. Paden, Suxiang Tong, Duncan MacCannell |
| EPI_ISL_1421486, EPI_ISL_1421487 | Wisconsin State Laboratory of Hygiene Communicable Disease Division | Wisconsin State Laboratory of Hygiene Communicable Disease Division | Kelsey R. Florek, Abigail C. Shockey |
| EPI_ISL_1421488 | Helix/Illumina | Centers for Disease Control and Prevention Division of Viral Diseases, Pathogen Discovery | Peter W. Cook, Dakota Howard, Dhwani Batra, Ben L. Rambo-Martin, Eileen de Feo, Jan Antico, Christine Tran, Matthew Tolentino, Shannon Wickline, Kim Gietzen, Brad Sickler, Jingtao Liu, Eric Allen, Phil Febbo, Summer Galloway, Nicole L. Washington, Simon White, Geraint Levan, Kelly Schiabor Barrett, Elizabeth Cirulli, Alexandre Bolze, Ary Ascencio, Charlotte Rivera-Garcia, Ryan Cho, Jason Nguyen, Sherry Wang, Jimmy Ramirez, Tyler Cassens, Efren Sandoval, Magnus Isaksson, William Lee, David Becker, Marc Laurent, James Lu, Clinton R. Paden, Suxiang Tong, Duncan MacCannell |
| EPI_ISL_1421489, EPI_ISL_1421490 | Wisconsin State Laboratory of Hygiene Communicable Disease Division | Wisconsin State Laboratory of Hygiene Communicable Disease Division | Kelsey R. Florek, Abigail C. Shockey |
| EPI_ISL_1421491 | Helix/Illumina | Centers for Disease Control and Prevention Division of Viral Diseases, Pathogen Discovery | Peter W. Cook, Dakota Howard, Dhwani Batra, Ben L. Rambo-Martin, Eileen de Feo, Jan Antico, Christine Tran, Matthew Tolentino, Shannon Wickline, Kim Gietzen, Brad Sickler, Jingtao Liu, Eric Allen, Phil Febbo, Summer Galloway, Nicole L. Washington, Simon White, Geraint Levan, Kelly Schiabor Barrett, Elizabeth Cirulli, Alexandre Bolze, Ary Ascencio, Charlotte Rivera-Garcia, Ryan Cho, Jason Nguyen, Sherry Wang, Jimmy Ramirez, Tyler Cassens, Efren Sandoval, Magnus Isaksson, William Lee, David Becker, Marc Laurent, James Lu, Clinton R. Paden, Suxiang Tong, Duncan MacCannell |
| EPI_ISL_1421492, EPI_ISL_1421493 | Wisconsin State Laboratory of Hygiene Communicable Disease Division | Wisconsin State Laboratory of Hygiene Communicable Disease Division | Kelsey R. Florek, Abigail C. Shockey |
| EPI_ISL_1421494 | Helix/Illumina | Centers for Disease Control and Prevention Division of Viral Diseases, Pathogen Discovery | Peter W. Cook, Dakota Howard, Dhwani Batra, Ben L. Rambo-Martin, Eileen de Feo, Jan Antico, Christine Tran, Matthew Tolentino, Shannon Wickline, Kim Gietzen, Brad Sickler, Jingtao Liu, Eric Allen, Phil Febbo, Summer Galloway, Nicole L. Washington, Simon White, Geraint Levan, Kelly Schiabor Barrett, Elizabeth Cirulli, Alexandre Bolze, Ary Ascencio, Charlotte Rivera-Garcia, Ryan Cho, Jason Nguyen, Sherry Wang, Jimmy Ramirez, Tyler Cassens, Efren Sandoval, Magnus Isaksson, William Lee, David Becker, Marc Laurent, James Lu, Clinton R. Paden, Suxiang Tong, Duncan MacCannell |
| EPI_ISL_1421495, EPI_ISL_1421496 | Wisconsin State Laboratory of Hygiene Communicable Disease Division | Wisconsin State Laboratory of Hygiene Communicable Disease Division | Kelsey R. Florek, Abigail C. Shockey |
| EPI_ISL_1421497 | Helix/Illumina | Centers for Disease Control and Prevention Division of Viral Diseases, Pathogen Discovery | Peter W. Cook, Dakota Howard, Dhwani Batra, Ben L. Rambo-Martin, Eileen de Feo, Jan Antico, Christine Tran, Matthew Tolentino, Shannon Wickline, Kim Gietzen, Brad Sickler, Jingtao Liu, Eric Allen, Phil Febbo, Summer Galloway, Nicole L. Washington, Simon White, Geraint Levan, Kelly Schiabor Barrett, Elizabeth Cirulli, Alexandre Bolze, Ary Ascencio, Charlotte Rivera-Garcia, Ryan Cho, Jason Nguyen, Sherry Wang, Jimmy Ramirez, Tyler Cassens, Efren Sandoval, Magnus Isaksson, William Lee, David Becker, Marc Laurent, James Lu, Clinton R. Paden, Suxiang Tong, Duncan MacCannell |
| EPI_ISL_1421498, EPI_ISL_1421499 | Wisconsin State Laboratory of Hygiene Communicable Disease Division | Wisconsin State Laboratory of Hygiene Communicable Disease Division | Kelsey R. Florek, Abigail C. Shockey |
| EPI_ISL_1421500 | Helix/Illumina | Centers for Disease Control and Prevention Division of Viral Diseases, Pathogen Discovery | Peter W. Cook, Dakota Howard, Dhwani Batra, Ben L. Rambo-Martin, Eileen de Feo, Jan Antico, Christine Tran, Matthew Tolentino, Shannon Wickline, Kim Gietzen, Brad Sickler, Jingtao Liu, Eric Allen, Phil Febbo, Summer Galloway, Nicole L. Washington, Simon White, Geraint Levan, Kelly Schiabor Barrett, Elizabeth Cirulli, Alexandre Bolze, Ary Ascencio, Charlotte Rivera-Garcia, Ryan Cho, Jason Nguyen, Sherry Wang, Jimmy Ramirez, Tyler Cassens, Efren Sandoval, Magnus Isaksson, William Lee, David Becker, Marc Laurent, James Lu, Clinton R. Paden, Suxiang Tong, Duncan MacCannell |
| EPI_ISL_1421501, EPI_ISL_1421502 | Wisconsin State Laboratory of Hygiene Communicable Disease Division | Wisconsin State Laboratory of Hygiene Communicable Disease Division | Kelsey R. Florek, Abigail C. Shockey |
| EPI_ISL_1421503 | Helix/Illumina | Centers for Disease Control and Prevention Division of Viral Diseases, Pathogen Discovery | Peter W. Cook, Dakota Howard, Dhwani Batra, Ben L. Rambo-Martin, Eileen de Feo, Jan Antico, Christine Tran, Matthew Tolentino, Shannon Wickline, Kim Gietzen, Brad Sickler, Jingtao Liu, Eric Allen, Phil Febbo, Summer Galloway, Nicole L. Washington, Simon White, Geraint Levan, Kelly Schiabor Barrett, Elizabeth Cirulli, Alexandre Bolze, Ary Ascencio, Charlotte Rivera-Garcia, Ryan Cho, Jason Nguyen, Sherry Wang, Jimmy Ramirez, Tyler Cassens, Efren Sandoval, Magnus Isaksson, William Lee, David Becker, Marc Laurent, James Lu, Clinton R. Paden, Suxiang Tong, Duncan MacCannell |
| EPI_ISL_1421504, EPI_ISL_1421505 | Wisconsin State Laboratory of Hygiene Communicable Disease Division | Wisconsin State Laboratory of Hygiene Communicable Disease Division | Kelsey R. Florek, Abigail C. Shockey |
| EPI_ISL_1421506 | Helix/Illumina | Centers for Disease Control and Prevention Division of Viral Diseases, Pathogen Discovery | Peter W. Cook, Dakota Howard, Dhwani Batra, Ben L. Rambo-Martin, Eileen de Feo, Jan Antico, Christine Tran, Matthew Tolentino, Shannon Wickline, Kim Gietzen, Brad Sickler, Jingtao Liu, Eric Allen, Phil Febbo, Summer Galloway, Nicole L. Washington, Simon White, Geraint Levan, Kelly Schiabor Barrett, Elizabeth Cirulli, Alexandre Bolze, Ary Ascencio, Charlotte Rivera-Garcia, Ryan Cho, Jason Nguyen, Sherry Wang, Jimmy Ramirez, Tyler Cassens, Efren Sandoval, Magnus Isaksson, William Lee, David Becker, Marc Laurent, James Lu, Clinton R. Paden, Suxiang Tong, Duncan MacCannell |
| EPI_ISL_1421507, EPI_ISL_1421508 | Wisconsin State Laboratory of Hygiene Communicable Disease Division | Wisconsin State Laboratory of Hygiene Communicable Disease Division | Kelsey R. Florek, Abigail C. Shockey |
| EPI_ISL_1421509 | Helix/Illumina | Centers for Disease Control and Prevention Division of Viral Diseases, Pathogen Discovery | Peter W. Cook, Dakota Howard, Dhwani Batra, Ben L. Rambo-Martin, Eileen de Feo, Jan Antico, Christine Tran, Matthew Tolentino, Shannon Wickline, Kim Gietzen, Brad Sickler, Jingtao Liu, Eric Allen, Phil Febbo, Summer Galloway, Nicole L. Washington, Simon White, Geraint Levan, Kelly Schiabor Barrett, Elizabeth Cirulli, Alexandre Bolze, Ary Ascencio, Charlotte Rivera-Garcia, Ryan Cho, Jason Nguyen, Sherry Wang, Jimmy Ramirez, Tyler Cassens, Efren Sandoval, Magnus Isaksson, William Lee, David Becker, Marc Laurent, James Lu, Clinton R. Paden, Suxiang Tong, Duncan MacCannell |
| EPI_ISL_1421510 | Wisconsin State Laboratory of Hygiene Communicable Disease Division | Wisconsin State Laboratory of Hygiene Communicable Disease Division | Kelsey R. Florek, Abigail C. Shockey |

| | | | |
|---|---|---|---|
| EPI_ISL_1421511 | Helix/Illumina | Centers for Disease Control and Prevention Division of Viral Diseases, Pathogen Discovery | Peter W. Cook, Dakota Howard, Dhwani Batra, Ben L. Rambo-Martin, Eileen de Feo, Jan Antico, Christine Tran, Matthew Tolentino, Shannon Wickline, Kim Gietzen, Brad Sickler, Jingtao Liu, Eric Allen, Phil Febbo, Summer Galloway, Nicole L. Washington, Simon White, Geraint Levan, Kelly Schiabor Barrett, Elizabeth Cirulli, Alexandre Bolze, Ary Ascencio, Charlotte Rivera-Garcia, Ryan Cho, Jason Nguyen, Sherry Wang, Jimmy Ramirez, Tyler Cassens, Efren Sandoval, Magnus Isaksson, William Lee, David Becker, Marc Laurent, James Lu, Clinton R. Paden, Suxiang Tong, Duncan MacCannell |
| EPI_ISL_1421512, EPI_ISL_1421513 | Wisconsin State Laboratory of Hygiene Communicable Disease Division | Wisconsin State Laboratory of Hygiene Communicable Disease Division | Kelsey R. Florek, Abigail C. Shockey |
| EPI_ISL_1421514 | Helix/Illumina | Centers for Disease Control and Prevention Division of Viral Diseases, Pathogen Discovery | Peter W. Cook, Dakota Howard, Dhwani Batra, Ben L. Rambo-Martin, Eileen de Feo, Jan Antico, Christine Tran, Matthew Tolentino, Shannon Wickline, Kim Gietzen, Brad Sickler, Jingtao Liu, Eric Allen, Phil Febbo, Summer Galloway, Nicole L. Washington, Simon White, Geraint Levan, Kelly Schiabor Barrett, Elizabeth Cirulli, Alexandre Bolze, Ary Ascencio, Charlotte Rivera-Garcia, Ryan Cho, Jason Nguyen, Sherry Wang, Jimmy Ramirez, Tyler Cassens, Efren Sandoval, Magnus Isaksson, William Lee, David Becker, Marc Laurent, James Lu, Clinton R. Paden, Suxiang Tong, Duncan MacCannell |
| EPI_ISL_1421515, EPI_ISL_1421516 | Wisconsin State Laboratory of Hygiene Communicable Disease Division | Wisconsin State Laboratory of Hygiene Communicable Disease Division | Kelsey R. Florek, Abigail C. Shockey |
| EPI_ISL_1421517 | Helix/Illumina | Centers for Disease Control and Prevention Division of Viral Diseases, Pathogen Discovery | Peter W. Cook, Dakota Howard, Dhwani Batra, Ben L. Rambo-Martin, Eileen de Feo, Jan Antico, Christine Tran, Matthew Tolentino, Shannon Wickline, Kim Gietzen, Brad Sickler, Jingtao Liu, Eric Allen, Phil Febbo, Summer Galloway, Nicole L. Washington, Simon White, Geraint Levan, Kelly Schiabor Barrett, Elizabeth Cirulli, Alexandre Bolze, Ary Ascencio, Charlotte Rivera-Garcia, Ryan Cho, Jason Nguyen, Sherry Wang, Jimmy Ramirez, Tyler Cassens, Efren Sandoval, Magnus Isaksson, William Lee, David Becker, Marc Laurent, James Lu, Clinton R. Paden, Suxiang Tong, Duncan MacCannell |
| EPI_ISL_1421518, EPI_ISL_1421519, EPI_ISL_1421520 | Wisconsin State Laboratory of Hygiene Communicable Disease Division | Wisconsin State Laboratory of Hygiene Communicable Disease Division | Kelsey R. Florek, Abigail C. Shockey |
| EPI_ISL_1421521 | Helix/Illumina | Centers for Disease Control and Prevention Division of Viral Diseases, Pathogen Discovery | Peter W. Cook, Dakota Howard, Dhwani Batra, Ben L. Rambo-Martin, Eileen de Feo, Jan Antico, Christine Tran, Matthew Tolentino, Shannon Wickline, Kim Gietzen, Brad Sickler, Jingtao Liu, Eric Allen, Phil Febbo, Summer Galloway, Nicole L. Washington, Simon White, Geraint Levan, Kelly Schiabor Barrett, Elizabeth Cirulli, Alexandre Bolze, Ary Ascencio, Charlotte Rivera-Garcia, Ryan Cho, Jason Nguyen, Sherry Wang, Jimmy Ramirez, Tyler Cassens, Efren Sandoval, Magnus Isaksson, William Lee, David Becker, Marc Laurent, James Lu, Clinton R. Paden, Suxiang Tong, Duncan MacCannell |
| EPI_ISL_1421522 | Wisconsin State Laboratory of Hygiene Communicable Disease Division | Wisconsin State Laboratory of Hygiene Communicable Disease Division | Kelsey R. Florek, Abigail C. Shockey |
| EPI_ISL_1421523 | Helix/Illumina | Centers for Disease Control and Prevention Division of Viral Diseases, Pathogen Discovery | Peter W. Cook, Dakota Howard, Dhwani Batra, Ben L. Rambo-Martin, Eileen de Feo, Jan Antico, Christine Tran, Matthew Tolentino, Shannon Wickline, Kim Gietzen, Brad Sickler, Jingtao Liu, Eric Allen, Phil Febbo, Summer Galloway, Nicole L. Washington, Simon White, Geraint Levan, Kelly Schiabor Barrett, Elizabeth Cirulli, Alexandre Bolze, Ary Ascencio, Charlotte Rivera-Garcia, Ryan Cho, Jason Nguyen, Sherry Wang, Jimmy Ramirez, Tyler Cassens, Efren Sandoval, Magnus Isaksson, William Lee, David Becker, Marc Laurent, James Lu, Clinton R. Paden, Suxiang Tong, Duncan MacCannell |
| EPI_ISL_1421524, EPI_ISL_1421525 | Wisconsin State Laboratory of Hygiene Communicable Disease Division | Wisconsin State Laboratory of Hygiene Communicable Disease Division | Kelsey R. Florek, Abigail C. Shockey |
| EPI_ISL_1421526 | Helix/Illumina | Centers for Disease Control and Prevention Division of Viral Diseases, Pathogen Discovery | Peter W. Cook, Dakota Howard, Dhwani Batra, Ben L. Rambo-Martin, Eileen de Feo, Jan Antico, Christine Tran, Matthew Tolentino, Shannon Wickline, Kim Gietzen, Brad Sickler, Jingtao Liu, Eric Allen, Phil Febbo, Summer Galloway, Nicole L. Washington, Simon White, Geraint Levan, Kelly Schiabor Barrett, Elizabeth Cirulli, Alexandre Bolze, Ary Ascencio, Charlotte Rivera-Garcia, Ryan Cho, Jason Nguyen, Sherry Wang, Jimmy Ramirez, Tyler Cassens, Efren Sandoval, Magnus Isaksson, William Lee, David Becker, Marc Laurent, James Lu, Clinton R. Paden, Suxiang Tong, Duncan MacCannell |
| EPI_ISL_1421527 | Wisconsin State Laboratory of Hygiene Communicable Disease Division | Wisconsin State Laboratory of Hygiene Communicable Disease Division | Kelsey R. Florek, Abigail C. Shockey |
| EPI_ISL_1421529 | Helix/Illumina | Centers for Disease Control and Prevention Division of Viral Diseases, Pathogen Discovery | Peter W. Cook, Dakota Howard, Dhwani Batra, Ben L. Rambo-Martin, Eileen de Feo, Jan Antico, Christine Tran, Matthew Tolentino, Shannon Wickline, Kim Gietzen, Brad Sickler, Jingtao Liu, Eric Allen, Phil Febbo, Summer Galloway, Nicole L. Washington, Simon White, Geraint Levan, Kelly Schiabor Barrett, Elizabeth Cirulli, Alexandre Bolze, Ary Ascencio, Charlotte Rivera-Garcia, Ryan Cho, Jason Nguyen, Sherry Wang, Jimmy Ramirez, Tyler Cassens, Efren Sandoval, Magnus Isaksson, William Lee, David Becker, Marc Laurent, James Lu, Clinton R. Paden, Suxiang Tong, Duncan MacCannell |
| EPI_ISL_1421530, EPI_ISL_1421531 | Wisconsin State Laboratory of Hygiene Communicable Disease Division | Wisconsin State Laboratory of Hygiene Communicable Disease Division | Kelsey R. Florek, Abigail C. Shockey |
| EPI_ISL_1421532 | Helix/Illumina | Centers for Disease Control and Prevention Division of Viral Diseases, Pathogen Discovery | Peter W. Cook, Dakota Howard, Dhwani Batra, Ben L. Rambo-Martin, Eileen de Feo, Jan Antico, Christine Tran, Matthew Tolentino, Shannon Wickline, Kim Gietzen, Brad Sickler, Jingtao Liu, Eric Allen, Phil Febbo, Summer Galloway, Nicole L. Washington, Simon White, Geraint Levan, Kelly Schiabor Barrett, Elizabeth Cirulli, Alexandre Bolze, Ary Ascencio, Charlotte Rivera-Garcia, Ryan Cho, Jason Nguyen, Sherry Wang, Jimmy Ramirez, Tyler Cassens, Efren Sandoval, Magnus Isaksson, William Lee, David Becker, Marc Laurent, James Lu, Clinton R. Paden, Suxiang Tong, Duncan MacCannell |
| EPI_ISL_1421533, EPI_ISL_1421534 | Wisconsin State Laboratory of Hygiene Communicable Disease Division | Wisconsin State Laboratory of Hygiene Communicable Disease Division | Kelsey R. Florek, Abigail C. Shockey |
| EPI_ISL_1421535 | Helix/Illumina | Centers for Disease Control and Prevention Division of Viral Diseases, Pathogen Discovery | Peter W. Cook, Dakota Howard, Dhwani Batra, Ben L. Rambo-Martin, Eileen de Feo, Jan Antico, Christine Tran, Matthew Tolentino, Shannon Wickline, Kim Gietzen, Brad Sickler, Jingtao Liu, Eric Allen, Phil Febbo, Summer Galloway, Nicole L. Washington, Simon White, Geraint Levan, Kelly Schiabor Barrett, Elizabeth Cirulli, Alexandre Bolze, Ary Ascencio, Charlotte Rivera-Garcia, Ryan Cho, Jason Nguyen, Sherry Wang, Jimmy Ramirez, Tyler Cassens, Efren Sandoval, Magnus Isaksson, William Lee, David Becker, Marc Laurent, James Lu, Clinton R. Paden, Suxiang Tong, Duncan MacCannell |
| EPI_ISL_1421536, EPI_ISL_1421537, EPI_ISL_1421538 | Wisconsin State Laboratory of Hygiene Communicable Disease Division | Wisconsin State Laboratory of Hygiene Communicable Disease Division | Kelsey R. Florek, Abigail C. Shockey |
| EPI_ISL_1421539 | Helix/Illumina | Centers for Disease Control and Prevention Division of Viral Diseases, Pathogen Discovery | Peter W. Cook, Dakota Howard, Dhwani Batra, Ben L. Rambo-Martin, Eileen de Feo, Jan Antico, Christine Tran, Matthew Tolentino, Shannon Wickline, Kim Gietzen, Brad Sickler, Jingtao Liu, Eric Allen, Phil Febbo, Summer Galloway, Nicole L. Washington, Simon White, Geraint Levan, Kelly Schiabor Barrett, Elizabeth Cirulli, Alexandre Bolze, Ary Ascencio, Charlotte Rivera-Garcia, Ryan Cho, Jason Nguyen, Sherry Wang, Jimmy Ramirez, Tyler Cassens, Efren Sandoval, Magnus Isaksson, William Lee, David Becker, Marc Laurent, James Lu, Clinton R. Paden, Suxiang Tong, Duncan MacCannell |
| EPI_ISL_1421540, EPI_ISL_1421541 | Wisconsin State Laboratory of Hygiene Communicable Disease Division | Wisconsin State Laboratory of Hygiene Communicable Disease Division | Kelsey R. Florek, Abigail C. Shockey |
| EPI_ISL_1421542 | Helix/Illumina | Centers for Disease Control and Prevention Division of Viral Diseases, Pathogen Discovery | Peter W. Cook, Dakota Howard, Dhwani Batra, Ben L. Rambo-Martin, Eileen de Feo, Jan Antico, Christine Tran, Matthew Tolentino, Shannon Wickline, Kim Gietzen, Brad Sickler, Jingtao Liu, Eric Allen, Phil Febbo, Summer Galloway, Nicole L. Washington, Simon White, Geraint Levan, Kelly Schiabor Barrett, Elizabeth Cirulli, Alexandre Bolze, Ary Ascencio, Charlotte Rivera-Garcia, Ryan Cho, Jason Nguyen, Sherry Wang, Jimmy Ramirez, Tyler Cassens, Efren Sandoval, Magnus Isaksson, William Lee, David Becker, Marc Laurent, James Lu, Clinton R. Paden, Suxiang Tong, Duncan MacCannell |
| EPI_ISL_1421543, EPI_ISL_1421544 | Wisconsin State Laboratory of Hygiene Communicable Disease Division | Wisconsin State Laboratory of Hygiene Communicable Disease Division | Kelsey R. Florek, Abigail C. Shockey |
| EPI_ISL_1421545 | Helix/Illumina | Centers for Disease Control and Prevention Division of Viral Diseases, Pathogen Discovery | Peter W. Cook, Dakota Howard, Dhwani Batra, Ben L. Rambo-Martin, Eileen de Feo, Jan Antico, Christine Tran, Matthew Tolentino, Shannon Wickline, Kim Gietzen, Brad Sickler, Jingtao Liu, Eric Allen, Phil Febbo, Summer Galloway, Nicole L. Washington, Simon White, Geraint Levan, Kelly Schiabor Barrett, Elizabeth Cirulli, Alexandre Bolze, Ary Ascencio, Charlotte Rivera-Garcia, Ryan Cho, Jason Nguyen, Sherry Wang, Jimmy Ramirez, Tyler Cassens, Efren Sandoval, Magnus Isaksson, William Lee, David Becker, Marc Laurent, James Lu, Clinton R. Paden, Suxiang Tong, Duncan MacCannell |
| EPI_ISL_1421546, EPI_ISL_1421547 | Wisconsin State Laboratory of Hygiene Communicable Disease Division | Wisconsin State Laboratory of Hygiene Communicable Disease Division | Kelsey R. Florek, Abigail C. Shockey |
| EPI_ISL_1421548 | Helix/Illumina | Centers for Disease Control and Prevention Division of Viral Diseases, Pathogen Discovery | Peter W. Cook, Dakota Howard, Dhwani Batra, Ben L. Rambo-Martin, Eileen de Feo, Jan Antico, Christine Tran, Matthew Tolentino, Shannon Wickline, Kim Gietzen, Brad Sickler, Jingtao Liu, Eric Allen, Phil Febbo, Summer Galloway, Nicole L. Washington, Simon White, Geraint Levan, Kelly Schiabor Barrett, Elizabeth Cirulli, Alexandre Bolze, Ary Ascencio, Charlotte Rivera-Garcia, Ryan Cho, Jason Nguyen, Sherry Wang, Jimmy Ramirez, Tyler Cassens, Efren Sandoval, Magnus Isaksson, William Lee, David Becker, Marc Laurent, James Lu, Clinton R. Paden, Suxiang Tong, Duncan MacCannell |
| EPI_ISL_1421549, EPI_ISL_1421550 | Wisconsin State Laboratory of Hygiene Communicable Disease Division | Wisconsin State Laboratory of Hygiene Communicable Disease Division | Kelsey R. Florek, Abigail C. Shockey |
| EPI_ISL_1421551 | Helix/Illumina | Centers for Disease Control and Prevention Division of Viral Diseases, Pathogen Discovery | Peter W. Cook, Dakota Howard, Dhwani Batra, Ben L. Rambo-Martin, Eileen de Feo, Jan Antico, Christine Tran, Matthew Tolentino, Shannon Wickline, Kim Gietzen, Brad Sickler, Jingtao Liu, Eric Allen, Phil Febbo, Summer Galloway, Nicole L. Washington, Simon White, Geraint Levan, Kelly Schiabor Barrett, Elizabeth Cirulli, Alexandre Bolze, Ary Ascencio, Charlotte Rivera-Garcia, Ryan Cho, Jason Nguyen, Sherry Wang, Jimmy Ramirez, Tyler Cassens, Efren Sandoval, Magnus Isaksson, William Lee, David Becker, Marc Laurent, James Lu, Clinton R. Paden, Suxiang Tong, Duncan MacCannell |
| EPI_ISL_1421552, EPI_ISL_1421553 | Wisconsin State Laboratory of Hygiene Communicable Disease Division | Wisconsin State Laboratory of Hygiene Communicable Disease Division | Kelsey R. Florek, Abigail C. Shockey |
| EPI_ISL_1421554, EPI_ISL_1421555 | Helix/Illumina | Centers for Disease Control and Prevention Division of Viral Diseases, Pathogen Discovery | Peter W. Cook, Dakota Howard, Dhwani Batra, Ben L. Rambo-Martin, Eileen de Feo, Jan Antico, Christine Tran, Matthew Tolentino, Shannon Wickline, Kim Gietzen, Brad Sickler, Jingtao Liu, Eric Allen, Phil Febbo, Summer Galloway, Nicole L. Washington, Simon White, Geraint Levan, Kelly Schiabor Barrett, Elizabeth Cirulli, Alexandre Bolze, Ary Ascencio, Charlotte Rivera-Garcia, Ryan Cho, Jason Nguyen, Sherry Wang, Jimmy Ramirez, Tyler Cassens, Efren Sandoval, Magnus Isaksson, William Lee, David Becker, Marc Laurent, James Lu, Clinton R. Paden, Suxiang Tong, Duncan MacCannell |
| EPI_ISL_1421556 | Wisconsin State Laboratory of Hygiene Communicable Disease Division | Wisconsin State Laboratory of Hygiene Communicable Disease Division | Kelsey R. Florek, Abigail C. Shockey |
| EPI_ISL_1421558 | Helix/Illumina | Centers for Disease Control and Prevention Division of Viral Diseases, Pathogen Discovery | Peter W. Cook, Dakota Howard, Dhwani Batra, Ben L. Rambo-Martin, Eileen de Feo, Jan Antico, Christine Tran, Matthew Tolentino, Shannon Wickline, Kim Gietzen, Brad Sickler, Jingtao Liu, Eric Allen, Phil Febbo, Summer Galloway, Nicole L. Washington, Simon White, Geraint Levan, Kelly Schiabor Barrett, Elizabeth Cirulli, Alexandre Bolze, Ary Ascencio, Charlotte Rivera-Garcia, Ryan Cho, Jason Nguyen, Sherry Wang, Jimmy Ramirez, Tyler Cassens, Efren Sandoval, Magnus Isaksson, William Lee, David Becker, Marc Laurent, James Lu, Clinton R. Paden, Suxiang Tong, Duncan MacCannell |
| EPI_ISL_1421559, EPI_ISL_1421560 | Wisconsin State Laboratory of Hygiene Communicable Disease Division | Wisconsin State Laboratory of Hygiene Communicable Disease Division | Kelsey R. Florek, Abigail C. Shockey |
| EPI_ISL_1421561 | Helix/Illumina | Centers for Disease Control and Prevention Division of Viral Diseases, Pathogen Discovery | Peter W. Cook, Dakota Howard, Dhwani Batra, Ben L. Rambo-Martin, Eileen de Feo, Jan Antico, Christine Tran, Matthew Tolentino, Shannon Wickline, Kim Gietzen, Brad Sickler, Jingtao Liu, Eric Allen, Phil Febbo, Summer Galloway, Nicole L. Washington, Simon White, Geraint Levan, Kelly Schiabor Barrett, Elizabeth Cirulli, Alexandre Bolze, Ary Ascencio, Charlotte Rivera-Garcia, Ryan Cho, Jason Nguyen, Sherry Wang, Jimmy Ramirez, Tyler Cassens, Efren Sandoval, Magnus Isaksson, William Lee, David Becker, Marc Laurent, James Lu, Clinton R. Paden, Suxiang Tong, Duncan MacCannell |
| EPI_ISL_1421562, EPI_ISL_1421563 | Wisconsin State Laboratory of Hygiene Communicable Disease Division | Wisconsin State Laboratory of Hygiene Communicable Disease Division | Kelsey R. Florek, Abigail C. Shockey |
| EPI_ISL_1421564 | Helix/Illumina | Centers for Disease Control and Prevention Division of Viral Diseases, Pathogen Discovery | Peter W. Cook, Dakota Howard, Dhwani Batra, Ben L. Rambo-Martin, Eileen de Feo, Jan Antico, Christine Tran, Matthew Tolentino, Shannon Wickline, Kim Gietzen, Brad Sickler, Jingtao Liu, Eric Allen, Phil Febbo, Summer Galloway, Nicole L. Washington, Simon White, Geraint Levan, Kelly Schiabor Barrett, Elizabeth Cirulli, Alexandre Bolze, Ary Ascencio, Charlotte Rivera-Garcia, Ryan Cho, Jason Nguyen, Sherry Wang, Jimmy Ramirez, Tyler Cassens, Efren Sandoval, Magnus Isaksson, William Lee, David Becker, Marc Laurent, James Lu, Clinton R. Paden, Suxiang Tong, Duncan MacCannell |
| EPI_ISL_1421565, EPI_ISL_1421566 | Wisconsin State Laboratory of Hygiene Communicable Disease Division | Wisconsin State Laboratory of Hygiene Communicable Disease Division | Kelsey R. Florek, Abigail C. Shockey |
| EPI_ISL_1421567 | Helix/Illumina | Centers for Disease Control and Prevention Division of Viral Diseases, Pathogen | Peter W. Cook, Dakota Howard, Dhwani Batra, Ben L. Rambo-Martin, Eileen de Feo, Jan Antico, Christine Tran, Matthew Tolentino, Shannon Wickline, Kim Gietzen, Brad Sickler, Jingtao Liu, Eric Allen, Phil Febbo, Summer Galloway, Nicole L. Washington, Simon White, Geraint Levan, Kelly Schiabor Barrett, Elizabeth Cirulli, Alexandre Bolze, Ary Ascencio, Charlotte Rivera-Garcia, Ryan Cho, Jason Nguyen, Sherry Wang, Jimmy Ramirez, Tyler Cassens, Efren Sandoval, Magnus Isaksson, William Lee, David Becker, Marc Laurent, James Lu, Clinton R. Paden, Suxiang Tong, |

| | | | |
|---|---|---|---|
| | | Discovery | Duncan MacCannell |
| EPI_ISL_1421568 | Wisconsin State Laboratory of Hygiene Communicable Disease Division | Wisconsin State Laboratory of Hygiene Communicable Disease Division | Kelsey R. Florek, Abigail C. Shockey |
| EPI_ISL_1421569 | Helix/Illumina | Centers for Disease Control and Prevention Division of Viral Diseases, Pathogen Discovery | Peter W. Cook, Dakota Howard, Dhwani Batra, Ben L. Rambo-Martin, Eileen de Feo, Jan Antico, Christine Tran, Matthew Tolentino, Shannon Wickline, Kim Gietzen, Brad Sickler, Jingtao Liu, Eric Allen, Phil Febbo, Summer Galloway, Nicole L. Washington, Simon White, Geraint Levan, Kelly Schiabor Barrett, Elizabeth Cirulli, Alexandre Bolze, Ary Ascencio, Charlotte Rivera-Garcia, Ryan Cho, Jason Nguyen, Sherry Wang, Jimmy Ramirez, Tyler Cassens, Efren Sandoval, Magnus Isaksson, William Lee, David Becker, Marc Laurent, James Lu, Clinton R. Paden, Suxiang Tong, Duncan MacCannell |
| EPI_ISL_1421570, EPI_ISL_1421571 | Wisconsin State Laboratory of Hygiene Communicable Disease Division | Wisconsin State Laboratory of Hygiene Communicable Disease Division | Kelsey R. Florek, Abigail C. Shockey |
| EPI_ISL_1421572 | Helix/Illumina | Centers for Disease Control and Prevention Division of Viral Diseases, Pathogen Discovery | Peter W. Cook, Dakota Howard, Dhwani Batra, Ben L. Rambo-Martin, Eileen de Feo, Jan Antico, Christine Tran, Matthew Tolentino, Shannon Wickline, Kim Gietzen, Brad Sickler, Jingtao Liu, Eric Allen, Phil Febbo, Summer Galloway, Nicole L. Washington, Simon White, Geraint Levan, Kelly Schiabor Barrett, Elizabeth Cirulli, Alexandre Bolze, Ary Ascencio, Charlotte Rivera-Garcia, Ryan Cho, Jason Nguyen, Sherry Wang, Jimmy Ramirez, Tyler Cassens, Efren Sandoval, Magnus Isaksson, William Lee, David Becker, Marc Laurent, James Lu, Clinton R. Paden, Suxiang Tong, Duncan MacCannell |
| EPI_ISL_1421573, EPI_ISL_1421574 | Wisconsin State Laboratory of Hygiene Communicable Disease Division | Wisconsin State Laboratory of Hygiene Communicable Disease Division | Kelsey R. Florek, Abigail C. Shockey |
| EPI_ISL_1421575 | Helix/Illumina | Centers for Disease Control and Prevention Division of Viral Diseases, Pathogen Discovery | Peter W. Cook, Dakota Howard, Dhwani Batra, Ben L. Rambo-Martin, Eileen de Feo, Jan Antico, Christine Tran, Matthew Tolentino, Shannon Wickline, Kim Gietzen, Brad Sickler, Jingtao Liu, Eric Allen, Phil Febbo, Summer Galloway, Nicole L. Washington, Simon White, Geraint Levan, Kelly Schiabor Barrett, Elizabeth Cirulli, Alexandre Bolze, Ary Ascencio, Charlotte Rivera-Garcia, Ryan Cho, Jason Nguyen, Sherry Wang, Jimmy Ramirez, Tyler Cassens, Efren Sandoval, Magnus Isaksson, William Lee, David Becker, Marc Laurent, James Lu, Clinton R. Paden, Suxiang Tong, Duncan MacCannell |
| EPI_ISL_1421576, EPI_ISL_1421577 | Wisconsin State Laboratory of Hygiene Communicable Disease Division | Wisconsin State Laboratory of Hygiene Communicable Disease Division | Kelsey R. Florek, Abigail C. Shockey |
| EPI_ISL_1421578 | Helix/Illumina | Centers for Disease Control and Prevention Division of Viral Diseases, Pathogen Discovery | Peter W. Cook, Dakota Howard, Dhwani Batra, Ben L. Rambo-Martin, Eileen de Feo, Jan Antico, Christine Tran, Matthew Tolentino, Shannon Wickline, Kim Gietzen, Brad Sickler, Jingtao Liu, Eric Allen, Phil Febbo, Summer Galloway, Nicole L. Washington, Simon White, Geraint Levan, Kelly Schiabor Barrett, Elizabeth Cirulli, Alexandre Bolze, Ary Ascencio, Charlotte Rivera-Garcia, Ryan Cho, Jason Nguyen, Sherry Wang, Jimmy Ramirez, Tyler Cassens, Efren Sandoval, Magnus Isaksson, William Lee, David Becker, Marc Laurent, James Lu, Clinton R. Paden, Suxiang Tong, Duncan MacCannell |
| EPI_ISL_1421579, EPI_ISL_1421580 | Wisconsin State Laboratory of Hygiene Communicable Disease Division | Wisconsin State Laboratory of Hygiene Communicable Disease Division | Kelsey R. Florek, Abigail C. Shockey |
| EPI_ISL_1421581 | Helix/Illumina | Centers for Disease Control and Prevention Division of Viral Diseases, Pathogen Discovery | Peter W. Cook, Dakota Howard, Dhwani Batra, Ben L. Rambo-Martin, Eileen de Feo, Jan Antico, Christine Tran, Matthew Tolentino, Shannon Wickline, Kim Gietzen, Brad Sickler, Jingtao Liu, Eric Allen, Phil Febbo, Summer Galloway, Nicole L. Washington, Simon White, Geraint Levan, Kelly Schiabor Barrett, Elizabeth Cirulli, Alexandre Bolze, Ary Ascencio, Charlotte Rivera-Garcia, Ryan Cho, Jason Nguyen, Sherry Wang, Jimmy Ramirez, Tyler Cassens, Efren Sandoval, Magnus Isaksson, William Lee, David Becker, Marc Laurent, James Lu, Clinton R. Paden, Suxiang Tong, Duncan MacCannell |
| EPI_ISL_1421582, EPI_ISL_1421583 | Wisconsin State Laboratory of Hygiene Communicable Disease Division | Wisconsin State Laboratory of Hygiene Communicable Disease Division | Kelsey R. Florek, Abigail C. Shockey |
| EPI_ISL_1421584 | Helix/Illumina | Centers for Disease Control and Prevention Division of Viral Diseases, Pathogen Discovery | Peter W. Cook, Dakota Howard, Dhwani Batra, Ben L. Rambo-Martin, Eileen de Feo, Jan Antico, Christine Tran, Matthew Tolentino, Shannon Wickline, Kim Gietzen, Brad Sickler, Jingtao Liu, Eric Allen, Phil Febbo, Summer Galloway, Nicole L. Washington, Simon White, Geraint Levan, Kelly Schiabor Barrett, Elizabeth Cirulli, Alexandre Bolze, Ary Ascencio, Charlotte Rivera-Garcia, Ryan Cho, Jason Nguyen, Sherry Wang, Jimmy Ramirez, Tyler Cassens, Efren Sandoval, Magnus Isaksson, William Lee, David Becker, Marc Laurent, James Lu, Clinton R. Paden, Suxiang Tong, Duncan MacCannell |
| EPI_ISL_1421585 | Wisconsin State Laboratory of Hygiene Communicable Disease Division | Wisconsin State Laboratory of Hygiene Communicable Disease Division | Kelsey R. Florek, Abigail C. Shockey |
| EPI_ISL_1421587 | Helix/Illumina | Centers for Disease Control and Prevention Division of Viral Diseases, Pathogen Discovery | Peter W. Cook, Dakota Howard, Dhwani Batra, Ben L. Rambo-Martin, Eileen de Feo, Jan Antico, Christine Tran, Matthew Tolentino, Shannon Wickline, Kim Gietzen, Brad Sickler, Jingtao Liu, Eric Allen, Phil Febbo, Summer Galloway, Nicole L. Washington, Simon White, Geraint Levan, Kelly Schiabor Barrett, Elizabeth Cirulli, Alexandre Bolze, Ary Ascencio, Charlotte Rivera-Garcia, Ryan Cho, Jason Nguyen, Sherry Wang, Jimmy Ramirez, Tyler Cassens, Efren Sandoval, Magnus Isaksson, William Lee, David Becker, Marc Laurent, James Lu, Clinton R. Paden, Suxiang Tong, Duncan MacCannell |
| EPI_ISL_1421588, EPI_ISL_1421589 | Wisconsin State Laboratory of Hygiene Communicable Disease Division | Wisconsin State Laboratory of Hygiene Communicable Disease Division | Kelsey R. Florek, Abigail C. Shockey |
| EPI_ISL_1421590 | Helix/Illumina | Centers for Disease Control and Prevention Division of Viral Diseases, Pathogen Discovery | Peter W. Cook, Dakota Howard, Dhwani Batra, Ben L. Rambo-Martin, Eileen de Feo, Jan Antico, Christine Tran, Matthew Tolentino, Shannon Wickline, Kim Gietzen, Brad Sickler, Jingtao Liu, Eric Allen, Phil Febbo, Summer Galloway, Nicole L. Washington, Simon White, Geraint Levan, Kelly Schiabor Barrett, Elizabeth Cirulli, Alexandre Bolze, Ary Ascencio, Charlotte Rivera-Garcia, Ryan Cho, Jason Nguyen, Sherry Wang, Jimmy Ramirez, Tyler Cassens, Efren Sandoval, Magnus Isaksson, William Lee, David Becker, Marc Laurent, James Lu, Clinton R. Paden, Suxiang Tong, Duncan MacCannell |
| EPI_ISL_1421591, EPI_ISL_1421592 | Wisconsin State Laboratory of Hygiene Communicable Disease Division | Wisconsin State Laboratory of Hygiene Communicable Disease Division | Kelsey R. Florek, Abigail C. Shockey |
| EPI_ISL_1421593 | Helix/Illumina | Centers for Disease Control and Prevention Division of Viral Diseases, Pathogen Discovery | Peter W. Cook, Dakota Howard, Dhwani Batra, Ben L. Rambo-Martin, Eileen de Feo, Jan Antico, Christine Tran, Matthew Tolentino, Shannon Wickline, Kim Gietzen, Brad Sickler, Jingtao Liu, Eric Allen, Phil Febbo, Summer Galloway, Nicole L. Washington, Simon White, Geraint Levan, Kelly Schiabor Barrett, Elizabeth Cirulli, Alexandre Bolze, Ary Ascencio, Charlotte Rivera-Garcia, Ryan Cho, Jason Nguyen, Sherry Wang, Jimmy Ramirez, Tyler Cassens, Efren Sandoval, Magnus Isaksson, William Lee, David Becker, Marc Laurent, James Lu, Clinton R. Paden, Suxiang Tong, Duncan MacCannell |
| EPI_ISL_1421594, EPI_ISL_1421595 | Wisconsin State Laboratory of Hygiene Communicable Disease Division | Wisconsin State Laboratory of Hygiene Communicable Disease Division | Kelsey R. Florek, Abigail C. Shockey |
| EPI_ISL_1421596, EPI_ISL_1421597 | Helix/Illumina | Centers for Disease Control and Prevention Division of Viral Diseases, Pathogen Discovery | Peter W. Cook, Dakota Howard, Dhwani Batra, Ben L. Rambo-Martin, Eileen de Feo, Jan Antico, Christine Tran, Matthew Tolentino, Shannon Wickline, Kim Gietzen, Brad Sickler, Jingtao Liu, Eric Allen, Phil Febbo, Summer Galloway, Nicole L. Washington, Simon White, Geraint Levan, Kelly Schiabor Barrett, Elizabeth Cirulli, Alexandre Bolze, Ary Ascencio, Charlotte Rivera-Garcia, Ryan Cho, Jason Nguyen, Sherry Wang, Jimmy Ramirez, Tyler Cassens, Efren Sandoval, Magnus Isaksson, William Lee, David Becker, Marc Laurent, James Lu, Clinton R. Paden, Suxiang Tong, Duncan MacCannell |
| EPI_ISL_1421598, EPI_ISL_1421599 | Wisconsin State Laboratory of Hygiene Communicable Disease Division | Wisconsin State Laboratory of Hygiene Communicable Disease Division | Kelsey R. Florek, Abigail C. Shockey |
| EPI_ISL_1421600 | Helix/Illumina | Centers for Disease Control and Prevention Division of Viral Diseases, Pathogen Discovery | Peter W. Cook, Dakota Howard, Dhwani Batra, Ben L. Rambo-Martin, Eileen de Feo, Jan Antico, Christine Tran, Matthew Tolentino, Shannon Wickline, Kim Gietzen, Brad Sickler, Jingtao Liu, Eric Allen, Phil Febbo, Summer Galloway, Nicole L. Washington, Simon White, Geraint Levan, Kelly Schiabor Barrett, Elizabeth Cirulli, Alexandre Bolze, Ary Ascencio, Charlotte Rivera-Garcia, Ryan Cho, Jason Nguyen, Sherry Wang, Jimmy Ramirez, Tyler Cassens, Efren Sandoval, Magnus Isaksson, William Lee, David Becker, Marc Laurent, James Lu, Clinton R. Paden, Suxiang Tong, Duncan MacCannell |
| EPI_ISL_1421601, EPI_ISL_1421602 | Wisconsin State Laboratory of Hygiene Communicable Disease Division | Wisconsin State Laboratory of Hygiene Communicable Disease Division | Kelsey R. Florek, Abigail C. Shockey |
| EPI_ISL_1421603 | Helix/Illumina | Centers for Disease Control and Prevention Division of Viral Diseases, Pathogen Discovery | Peter W. Cook, Dakota Howard, Dhwani Batra, Ben L. Rambo-Martin, Eileen de Feo, Jan Antico, Christine Tran, Matthew Tolentino, Shannon Wickline, Kim Gietzen, Brad Sickler, Jingtao Liu, Eric Allen, Phil Febbo, Summer Galloway, Nicole L. Washington, Simon White, Geraint Levan, Kelly Schiabor Barrett, Elizabeth Cirulli, Alexandre Bolze, Ary Ascencio, Charlotte Rivera-Garcia, Ryan Cho, Jason Nguyen, Sherry Wang, Jimmy Ramirez, Tyler Cassens, Efren Sandoval, Magnus Isaksson, William Lee, David Becker, Marc Laurent, James Lu, Clinton R. Paden, Suxiang Tong, Duncan MacCannell |
| EPI_ISL_1421604 | Wisconsin State Laboratory of Hygiene Communicable Disease Division | Wisconsin State Laboratory of Hygiene Communicable Disease Division | Kelsey R. Florek, Abigail C. Shockey |
| EPI_ISL_1421605 | Helix/Illumina | Centers for Disease Control and Prevention Division of Viral Diseases, Pathogen Discovery | Peter W. Cook, Dakota Howard, Dhwani Batra, Ben L. Rambo-Martin, Eileen de Feo, Jan Antico, Christine Tran, Matthew Tolentino, Shannon Wickline, Kim Gietzen, Brad Sickler, Jingtao Liu, Eric Allen, Phil Febbo, Summer Galloway, Nicole L. Washington, Simon White, Geraint Levan, Kelly Schiabor Barrett, Elizabeth Cirulli, Alexandre Bolze, Ary Ascencio, Charlotte Rivera-Garcia, Ryan Cho, Jason Nguyen, Sherry Wang, Jimmy Ramirez, Tyler Cassens, Efren Sandoval, Magnus Isaksson, William Lee, David Becker, Marc Laurent, James Lu, Clinton R. Paden, Suxiang Tong, Duncan MacCannell |
| EPI_ISL_1421606, EPI_ISL_1421607, EPI_ISL_1421608 | Wisconsin State Laboratory of Hygiene Communicable Disease Division | Wisconsin State Laboratory of Hygiene Communicable Disease Division | Kelsey R. Florek, Abigail C. Shockey |
| EPI_ISL_1421609 | Helix/Illumina | Centers for Disease Control and Prevention Division of Viral Diseases, Pathogen Discovery | Peter W. Cook, Dakota Howard, Dhwani Batra, Ben L. Rambo-Martin, Eileen de Feo, Jan Antico, Christine Tran, Matthew Tolentino, Shannon Wickline, Kim Gietzen, Brad Sickler, Jingtao Liu, Eric Allen, Phil Febbo, Summer Galloway, Nicole L. Washington, Simon White, Geraint Levan, Kelly Schiabor Barrett, Elizabeth Cirulli, Alexandre Bolze, Ary Ascencio, Charlotte Rivera-Garcia, Ryan Cho, Jason Nguyen, Sherry Wang, Jimmy Ramirez, Tyler Cassens, Efren Sandoval, Magnus Isaksson, William Lee, David Becker, Marc Laurent, James Lu, Clinton R. Paden, Suxiang Tong, Duncan MacCannell |
| EPI_ISL_1421610 | Wisconsin State Laboratory of Hygiene Communicable Disease Division | Wisconsin State Laboratory of Hygiene Communicable Disease Division | Kelsey R. Florek, Abigail C. Shockey |
| EPI_ISL_1421611, EPI_ISL_1421612 | Helix/Illumina | Centers for Disease Control and Prevention Division of Viral Diseases, Pathogen Discovery | Peter W. Cook, Dakota Howard, Dhwani Batra, Ben L. Rambo-Martin, Eileen de Feo, Jan Antico, Christine Tran, Matthew Tolentino, Shannon Wickline, Kim Gietzen, Brad Sickler, Jingtao Liu, Eric Allen, Phil Febbo, Summer Galloway, Nicole L. Washington, Simon White, Geraint Levan, Kelly Schiabor Barrett, Elizabeth Cirulli, Alexandre Bolze, Ary Ascencio, Charlotte Rivera-Garcia, Ryan Cho, Jason Nguyen, Sherry Wang, Jimmy Ramirez, Tyler Cassens, Efren Sandoval, Magnus Isaksson, William Lee, David Becker, Marc Laurent, James Lu, Clinton R. Paden, Suxiang Tong, Duncan MacCannell |
| EPI_ISL_1421613 | Wisconsin State Laboratory of Hygiene Communicable Disease Division | Wisconsin State Laboratory of Hygiene Communicable Disease Division | Kelsey R. Florek, Abigail C. Shockey |
| EPI_ISL_1421614 | Helix/Illumina | Centers for Disease Control and Prevention Division of Viral Diseases, Pathogen Discovery | Peter W. Cook, Dakota Howard, Dhwani Batra, Ben L. Rambo-Martin, Eileen de Feo, Jan Antico, Christine Tran, Matthew Tolentino, Shannon Wickline, Kim Gietzen, Brad Sickler, Jingtao Liu, Eric Allen, Phil Febbo, Summer Galloway, Nicole L. Washington, Simon White, Geraint Levan, Kelly Schiabor Barrett, Elizabeth Cirulli, Alexandre Bolze, Ary Ascencio, Charlotte Rivera-Garcia, Ryan Cho, Jason Nguyen, Sherry Wang, Jimmy Ramirez, Tyler Cassens, Efren Sandoval, Magnus Isaksson, William Lee, David Becker, Marc Laurent, James Lu, Clinton R. Paden, Suxiang Tong, Duncan MacCannell |
| EPI_ISL_1421615 | Wisconsin State Laboratory of Hygiene Communicable Disease Division | Wisconsin State Laboratory of Hygiene Communicable Disease Division | Kelsey R. Florek, Abigail C. Shockey |
| EPI_ISL_1421616 | Helix/Illumina | Centers for Disease Control and Prevention Division of Viral Diseases, Pathogen Discovery | Peter W. Cook, Dakota Howard, Dhwani Batra, Ben L. Rambo-Martin, Eileen de Feo, Jan Antico, Christine Tran, Matthew Tolentino, Shannon Wickline, Kim Gietzen, Brad Sickler, Jingtao Liu, Eric Allen, Phil Febbo, Summer Galloway, Nicole L. Washington, Simon White, Geraint Levan, Kelly Schiabor Barrett, Elizabeth Cirulli, Alexandre Bolze, Ary Ascencio, Charlotte Rivera-Garcia, Ryan Cho, Jason Nguyen, Sherry Wang, Jimmy Ramirez, Tyler Cassens, Efren Sandoval, Magnus Isaksson, William Lee, David Becker, Marc Laurent, James Lu, Clinton R. Paden, Suxiang Tong, Duncan MacCannell |
| EPI_ISL_1421617, EPI_ISL_1421618 | Wisconsin State Laboratory of Hygiene Communicable Disease Division | Wisconsin State Laboratory of Hygiene Communicable Disease Division | Kelsey R. Florek, Abigail C. Shockey |
| EPI_ISL_1421619 | Helix/Illumina | Centers for Disease Control and Prevention Division of Viral Diseases, Pathogen Discovery | Peter W. Cook, Dakota Howard, Dhwani Batra, Ben L. Rambo-Martin, Eileen de Feo, Jan Antico, Christine Tran, Matthew Tolentino, Shannon Wickline, Kim Gietzen, Brad Sickler, Jingtao Liu, Eric Allen, Phil Febbo, Summer Galloway, Nicole L. Washington, Simon White, Geraint Levan, Kelly Schiabor Barrett, Elizabeth Cirulli, Alexandre Bolze, Ary Ascencio, Charlotte Rivera-Garcia, Ryan Cho, Jason Nguyen, Sherry Wang, Jimmy Ramirez, Tyler Cassens, Efren Sandoval, Magnus Isaksson, William Lee, David Becker, Marc Laurent, James Lu, Clinton R. Paden, Suxiang Tong, Duncan MacCannell |
| EPI_ISL_1421620, | Wisconsin State Laboratory of Hygiene | Wisconsin State Laboratory of Hygiene | Kelsey R. Florek, Abigail C. Shockey |

| | | | |
|---|---|---|---|
| EPI_ISL_1421621, EPI_ISL_1421622 | Communicable Disease Division | Communicable Disease Division | |
| EPI_ISL_1421623 | Helix/Illumina | Centers for Disease Control and Prevention Division of Viral Diseases, Pathogen Discovery | Peter W. Cook, Dakota Howard, Dhwani Batra, Ben L. Rambo-Martin, Eileen de Feo, Jan Antico, Christine Tran, Matthew Tolentino, Shannon Wickline, Kim Gietzen, Brad Sickler, Jingtao Liu, Eric Allen, Phil Febbo, Summer Galloway, Nicole L. Washington, Simon White, Geraint Levan, Kelly Schiabor Barrett, Elizabeth Cirulli, Alexandre Bolze, Ary Ascencio, Charlotte Rivera-Garcia, Ryan Cho, Jason Nguyen, Sherry Wang, Jimmy Ramirez, Tyler Cassens, Efren Sandoval, Magnus Isaksson, William Lee, David Becker, Marc Laurent, James Lu, Clinton R. Paden, Suxiang Tong, Duncan MacCannell |
| EPI_ISL_1421624, EPI_ISL_1421625 | Wisconsin State Laboratory of Hygiene Communicable Disease Division | Wisconsin State Laboratory of Hygiene Communicable Disease Division | Kelsey R. Florek, Abigail C. Shockey |
| EPI_ISL_1421626 | Helix/Illumina | Centers for Disease Control and Prevention Division of Viral Diseases, Pathogen Discovery | Peter W. Cook, Dakota Howard, Dhwani Batra, Ben L. Rambo-Martin, Eileen de Feo, Jan Antico, Christine Tran, Matthew Tolentino, Shannon Wickline, Kim Gietzen, Brad Sickler, Jingtao Liu, Eric Allen, Phil Febbo, Summer Galloway, Nicole L. Washington, Simon White, Geraint Levan, Kelly Schiabor Barrett, Elizabeth Cirulli, Alexandre Bolze, Ary Ascencio, Charlotte Rivera-Garcia, Ryan Cho, Jason Nguyen, Sherry Wang, Jimmy Ramirez, Tyler Cassens, Efren Sandoval, Magnus Isaksson, William Lee, David Becker, Marc Laurent, James Lu, Clinton R. Paden, Suxiang Tong, Duncan MacCannell |
| EPI_ISL_1421627, EPI_ISL_1421628 | Wisconsin State Laboratory of Hygiene Communicable Disease Division | Wisconsin State Laboratory of Hygiene Communicable Disease Division | Kelsey R. Florek, Abigail C. Shockey |
| EPI_ISL_1421629 | Helix/Illumina | Centers for Disease Control and Prevention Division of Viral Diseases, Pathogen Discovery | Peter W. Cook, Dakota Howard, Dhwani Batra, Ben L. Rambo-Martin, Eileen de Feo, Jan Antico, Christine Tran, Matthew Tolentino, Shannon Wickline, Kim Gietzen, Brad Sickler, Jingtao Liu, Eric Allen, Phil Febbo, Summer Galloway, Nicole L. Washington, Simon White, Geraint Levan, Kelly Schiabor Barrett, Elizabeth Cirulli, Alexandre Bolze, Ary Ascencio, Charlotte Rivera-Garcia, Ryan Cho, Jason Nguyen, Sherry Wang, Jimmy Ramirez, Tyler Cassens, Efren Sandoval, Magnus Isaksson, William Lee, David Becker, Marc Laurent, James Lu, Clinton R. Paden, Suxiang Tong, Duncan MacCannell |
| EPI_ISL_1421630, EPI_ISL_1421631 | Wisconsin State Laboratory of Hygiene Communicable Disease Division | Wisconsin State Laboratory of Hygiene Communicable Disease Division | Kelsey R. Florek, Abigail C. Shockey |
| EPI_ISL_1421632 | Helix/Illumina | Centers for Disease Control and Prevention Division of Viral Diseases, Pathogen Discovery | Peter W. Cook, Dakota Howard, Dhwani Batra, Ben L. Rambo-Martin, Eileen de Feo, Jan Antico, Christine Tran, Matthew Tolentino, Shannon Wickline, Kim Gietzen, Brad Sickler, Jingtao Liu, Eric Allen, Phil Febbo, Summer Galloway, Nicole L. Washington, Simon White, Geraint Levan, Kelly Schiabor Barrett, Elizabeth Cirulli, Alexandre Bolze, Ary Ascencio, Charlotte Rivera-Garcia, Ryan Cho, Jason Nguyen, Sherry Wang, Jimmy Ramirez, Tyler Cassens, Efren Sandoval, Magnus Isaksson, William Lee, David Becker, Marc Laurent, James Lu, Clinton R. Paden, Suxiang Tong, Duncan MacCannell |
| EPI_ISL_1421633 | Wisconsin State Laboratory of Hygiene Communicable Disease Division | Wisconsin State Laboratory of Hygiene Communicable Disease Division | Kelsey R. Florek, Abigail C. Shockey |
| EPI_ISL_1421634 | Helix/Illumina | Centers for Disease Control and Prevention Division of Viral Diseases, Pathogen Discovery | Peter W. Cook, Dakota Howard, Dhwani Batra, Ben L. Rambo-Martin, Eileen de Feo, Jan Antico, Christine Tran, Matthew Tolentino, Shannon Wickline, Kim Gietzen, Brad Sickler, Jingtao Liu, Eric Allen, Phil Febbo, Summer Galloway, Nicole L. Washington, Simon White, Geraint Levan, Kelly Schiabor Barrett, Elizabeth Cirulli, Alexandre Bolze, Ary Ascencio, Charlotte Rivera-Garcia, Ryan Cho, Jason Nguyen, Sherry Wang, Jimmy Ramirez, Tyler Cassens, Efren Sandoval, Magnus Isaksson, William Lee, David Becker, Marc Laurent, James Lu, Clinton R. Paden, Suxiang Tong, Duncan MacCannell |
| EPI_ISL_1421635, EPI_ISL_1421636, EPI_ISL_1421637 | Wisconsin State Laboratory of Hygiene Communicable Disease Division | Wisconsin State Laboratory of Hygiene Communicable Disease Division | Kelsey R. Florek, Abigail C. Shockey |
| EPI_ISL_1421638 | Helix/Illumina | Centers for Disease Control and Prevention Division of Viral Diseases, Pathogen Discovery | Peter W. Cook, Dakota Howard, Dhwani Batra, Ben L. Rambo-Martin, Eileen de Feo, Jan Antico, Christine Tran, Matthew Tolentino, Shannon Wickline, Kim Gietzen, Brad Sickler, Jingtao Liu, Eric Allen, Phil Febbo, Summer Galloway, Nicole L. Washington, Simon White, Geraint Levan, Kelly Schiabor Barrett, Elizabeth Cirulli, Alexandre Bolze, Ary Ascencio, Charlotte Rivera-Garcia, Ryan Cho, Jason Nguyen, Sherry Wang, Jimmy Ramirez, Tyler Cassens, Efren Sandoval, Magnus Isaksson, William Lee, David Becker, Marc Laurent, James Lu, Clinton R. Paden, Suxiang Tong, Duncan MacCannell |
| EPI_ISL_1421639, EPI_ISL_1421640 | Wisconsin State Laboratory of Hygiene Communicable Disease Division | Wisconsin State Laboratory of Hygiene Communicable Disease Division | Kelsey R. Florek, Abigail C. Shockey |
| EPI_ISL_1421641 | Helix/Illumina | Centers for Disease Control and Prevention Division of Viral Diseases, Pathogen Discovery | Peter W. Cook, Dakota Howard, Dhwani Batra, Ben L. Rambo-Martin, Eileen de Feo, Jan Antico, Christine Tran, Matthew Tolentino, Shannon Wickline, Kim Gietzen, Brad Sickler, Jingtao Liu, Eric Allen, Phil Febbo, Summer Galloway, Nicole L. Washington, Simon White, Geraint Levan, Kelly Schiabor Barrett, Elizabeth Cirulli, Alexandre Bolze, Ary Ascencio, Charlotte Rivera-Garcia, Ryan Cho, Jason Nguyen, Sherry Wang, Jimmy Ramirez, Tyler Cassens, Efren Sandoval, Magnus Isaksson, William Lee, David Becker, Marc Laurent, James Lu, Clinton R. Paden, Suxiang Tong, Duncan MacCannell |
| EPI_ISL_1421642, EPI_ISL_1421643 | Wisconsin State Laboratory of Hygiene Communicable Disease Division | Wisconsin State Laboratory of Hygiene Communicable Disease Division | Kelsey R. Florek, Abigail C. Shockey |
| EPI_ISL_1421644 | Helix/Illumina | Centers for Disease Control and Prevention Division of Viral Diseases, Pathogen Discovery | Peter W. Cook, Dakota Howard, Dhwani Batra, Ben L. Rambo-Martin, Eileen de Feo, Jan Antico, Christine Tran, Matthew Tolentino, Shannon Wickline, Kim Gietzen, Brad Sickler, Jingtao Liu, Eric Allen, Phil Febbo, Summer Galloway, Nicole L. Washington, Simon White, Geraint Levan, Kelly Schiabor Barrett, Elizabeth Cirulli, Alexandre Bolze, Ary Ascencio, Charlotte Rivera-Garcia, Ryan Cho, Jason Nguyen, Sherry Wang, Jimmy Ramirez, Tyler Cassens, Efren Sandoval, Magnus Isaksson, William Lee, David Becker, Marc Laurent, James Lu, Clinton R. Paden, Suxiang Tong, Duncan MacCannell |
| EPI_ISL_1421645 | Wisconsin State Laboratory of Hygiene Communicable Disease Division | Wisconsin State Laboratory of Hygiene Communicable Disease Division | Kelsey R. Florek, Abigail C. Shockey |
| EPI_ISL_1421646 | Helix/Illumina | Centers for Disease Control and Prevention Division of Viral Diseases, Pathogen Discovery | Peter W. Cook, Dakota Howard, Dhwani Batra, Ben L. Rambo-Martin, Eileen de Feo, Jan Antico, Christine Tran, Matthew Tolentino, Shannon Wickline, Kim Gietzen, Brad Sickler, Jingtao Liu, Eric Allen, Phil Febbo, Summer Galloway, Nicole L. Washington, Simon White, Geraint Levan, Kelly Schiabor Barrett, Elizabeth Cirulli, Alexandre Bolze, Ary Ascencio, Charlotte Rivera-Garcia, Ryan Cho, Jason Nguyen, Sherry Wang, Jimmy Ramirez, Tyler Cassens, Efren Sandoval, Magnus Isaksson, William Lee, David Becker, Marc Laurent, James Lu, Clinton R. Paden, Suxiang Tong, Duncan MacCannell |
| EPI_ISL_1421647, EPI_ISL_1421648 | Wisconsin State Laboratory of Hygiene Communicable Disease Division | Wisconsin State Laboratory of Hygiene Communicable Disease Division | Kelsey R. Florek, Abigail C. Shockey |
| EPI_ISL_1421649 | Helix/Illumina | Centers for Disease Control and Prevention Division of Viral Diseases, Pathogen Discovery | Peter W. Cook, Dakota Howard, Dhwani Batra, Ben L. Rambo-Martin, Eileen de Feo, Jan Antico, Christine Tran, Matthew Tolentino, Shannon Wickline, Kim Gietzen, Brad Sickler, Jingtao Liu, Eric Allen, Phil Febbo, Summer Galloway, Nicole L. Washington, Simon White, Geraint Levan, Kelly Schiabor Barrett, Elizabeth Cirulli, Alexandre Bolze, Ary Ascencio, Charlotte Rivera-Garcia, Ryan Cho, Jason Nguyen, Sherry Wang, Jimmy Ramirez, Tyler Cassens, Efren Sandoval, Magnus Isaksson, William Lee, David Becker, Marc Laurent, James Lu, Clinton R. Paden, Suxiang Tong, Duncan MacCannell |
| EPI_ISL_1421650, EPI_ISL_1421651 | Wisconsin State Laboratory of Hygiene Communicable Disease Division | Wisconsin State Laboratory of Hygiene Communicable Disease Division | Kelsey R. Florek, Abigail C. Shockey |
| EPI_ISL_1421652 | Helix/Illumina | Centers for Disease Control and Prevention Division of Viral Diseases, Pathogen Discovery | Peter W. Cook, Dakota Howard, Dhwani Batra, Ben L. Rambo-Martin, Eileen de Feo, Jan Antico, Christine Tran, Matthew Tolentino, Shannon Wickline, Kim Gietzen, Brad Sickler, Jingtao Liu, Eric Allen, Phil Febbo, Summer Galloway, Nicole L. Washington, Simon White, Geraint Levan, Kelly Schiabor Barrett, Elizabeth Cirulli, Alexandre Bolze, Ary Ascencio, Charlotte Rivera-Garcia, Ryan Cho, Jason Nguyen, Sherry Wang, Jimmy Ramirez, Tyler Cassens, Efren Sandoval, Magnus Isaksson, William Lee, David Becker, Marc Laurent, James Lu, Clinton R. Paden, Suxiang Tong, Duncan MacCannell |
| EPI_ISL_1421653, EPI_ISL_1421654 | Wisconsin State Laboratory of Hygiene Communicable Disease Division | Wisconsin State Laboratory of Hygiene Communicable Disease Division | Kelsey R. Florek, Abigail C. Shockey |
| EPI_ISL_1421655 | Helix/Illumina | Centers for Disease Control and Prevention Division of Viral Diseases, Pathogen Discovery | Peter W. Cook, Dakota Howard, Dhwani Batra, Ben L. Rambo-Martin, Eileen de Feo, Jan Antico, Christine Tran, Matthew Tolentino, Shannon Wickline, Kim Gietzen, Brad Sickler, Jingtao Liu, Eric Allen, Phil Febbo, Summer Galloway, Nicole L. Washington, Simon White, Geraint Levan, Kelly Schiabor Barrett, Elizabeth Cirulli, Alexandre Bolze, Ary Ascencio, Charlotte Rivera-Garcia, Ryan Cho, Jason Nguyen, Sherry Wang, Jimmy Ramirez, Tyler Cassens, Efren Sandoval, Magnus Isaksson, William Lee, David Becker, Marc Laurent, James Lu, Clinton R. Paden, Suxiang Tong, Duncan MacCannell |
| EPI_ISL_1421656, EPI_ISL_1421657 | Wisconsin State Laboratory of Hygiene Communicable Disease Division | Wisconsin State Laboratory of Hygiene Communicable Disease Division | Kelsey R. Florek, Abigail C. Shockey |
| EPI_ISL_1421658 | Helix/Illumina | Centers for Disease Control and Prevention Division of Viral Diseases, Pathogen Discovery | Peter W. Cook, Dakota Howard, Dhwani Batra, Ben L. Rambo-Martin, Eileen de Feo, Jan Antico, Christine Tran, Matthew Tolentino, Shannon Wickline, Kim Gietzen, Brad Sickler, Jingtao Liu, Eric Allen, Phil Febbo, Summer Galloway, Nicole L. Washington, Simon White, Geraint Levan, Kelly Schiabor Barrett, Elizabeth Cirulli, Alexandre Bolze, Ary Ascencio, Charlotte Rivera-Garcia, Ryan Cho, Jason Nguyen, Sherry Wang, Jimmy Ramirez, Tyler Cassens, Efren Sandoval, Magnus Isaksson, William Lee, David Becker, Marc Laurent, James Lu, Clinton R. Paden, Suxiang Tong, Duncan MacCannell |
| EPI_ISL_1421659, EPI_ISL_1421660 | Wisconsin State Laboratory of Hygiene Communicable Disease Division | Wisconsin State Laboratory of Hygiene Communicable Disease Division | Kelsey R. Florek, Abigail C. Shockey |
| EPI_ISL_1421661 | Helix/Illumina | Centers for Disease Control and Prevention Division of Viral Diseases, Pathogen Discovery | Peter W. Cook, Dakota Howard, Dhwani Batra, Ben L. Rambo-Martin, Eileen de Feo, Jan Antico, Christine Tran, Matthew Tolentino, Shannon Wickline, Kim Gietzen, Brad Sickler, Jingtao Liu, Eric Allen, Phil Febbo, Summer Galloway, Nicole L. Washington, Simon White, Geraint Levan, Kelly Schiabor Barrett, Elizabeth Cirulli, Alexandre Bolze, Ary Ascencio, Charlotte Rivera-Garcia, Ryan Cho, Jason Nguyen, Sherry Wang, Jimmy Ramirez, Tyler Cassens, Efren Sandoval, Magnus Isaksson, William Lee, David Becker, Marc Laurent, James Lu, Clinton R. Paden, Suxiang Tong, Duncan MacCannell |
| EPI_ISL_1421662, EPI_ISL_1421663 | Wisconsin State Laboratory of Hygiene Communicable Disease Division | Wisconsin State Laboratory of Hygiene Communicable Disease Division | Kelsey R. Florek, Abigail C. Shockey |
| EPI_ISL_1421664 | Helix/Illumina | Centers for Disease Control and Prevention Division of Viral Diseases, Pathogen Discovery | Peter W. Cook, Dakota Howard, Dhwani Batra, Ben L. Rambo-Martin, Eileen de Feo, Jan Antico, Christine Tran, Matthew Tolentino, Shannon Wickline, Kim Gietzen, Brad Sickler, Jingtao Liu, Eric Allen, Phil Febbo, Summer Galloway, Nicole L. Washington, Simon White, Geraint Levan, Kelly Schiabor Barrett, Elizabeth Cirulli, Alexandre Bolze, Ary Ascencio, Charlotte Rivera-Garcia, Ryan Cho, Jason Nguyen, Sherry Wang, Jimmy Ramirez, Tyler Cassens, Efren Sandoval, Magnus Isaksson, William Lee, David Becker, Marc Laurent, James Lu, Clinton R. Paden, Suxiang Tong, Duncan MacCannell |
| EPI_ISL_1421665, EPI_ISL_1421666 | Wisconsin State Laboratory of Hygiene Communicable Disease Division | Wisconsin State Laboratory of Hygiene Communicable Disease Division | Kelsey R. Florek, Abigail C. Shockey |
| EPI_ISL_1421667 | Helix/Illumina | Centers for Disease Control and Prevention Division of Viral Diseases, Pathogen Discovery | Peter W. Cook, Dakota Howard, Dhwani Batra, Ben L. Rambo-Martin, Eileen de Feo, Jan Antico, Christine Tran, Matthew Tolentino, Shannon Wickline, Kim Gietzen, Brad Sickler, Jingtao Liu, Eric Allen, Phil Febbo, Summer Galloway, Nicole L. Washington, Simon White, Geraint Levan, Kelly Schiabor Barrett, Elizabeth Cirulli, Alexandre Bolze, Ary Ascencio, Charlotte Rivera-Garcia, Ryan Cho, Jason Nguyen, Sherry Wang, Jimmy Ramirez, Tyler Cassens, Efren Sandoval, Magnus Isaksson, William Lee, David Becker, Marc Laurent, James Lu, Clinton R. Paden, Suxiang Tong, Duncan MacCannell |
| EPI_ISL_1421668, EPI_ISL_1421669 | Wisconsin State Laboratory of Hygiene Communicable Disease Division | Wisconsin State Laboratory of Hygiene Communicable Disease Division | Kelsey R. Florek, Abigail C. Shockey |
| EPI_ISL_1421670 | Helix/Illumina | Centers for Disease Control and Prevention Division of Viral Diseases, Pathogen Discovery | Peter W. Cook, Dakota Howard, Dhwani Batra, Ben L. Rambo-Martin, Eileen de Feo, Jan Antico, Christine Tran, Matthew Tolentino, Shannon Wickline, Kim Gietzen, Brad Sickler, Jingtao Liu, Eric Allen, Phil Febbo, Summer Galloway, Nicole L. Washington, Simon White, Geraint Levan, Kelly Schiabor Barrett, Elizabeth Cirulli, Alexandre Bolze, Ary Ascencio, Charlotte Rivera-Garcia, Ryan Cho, Jason Nguyen, Sherry Wang, Jimmy Ramirez, Tyler Cassens, Efren Sandoval, Magnus Isaksson, William Lee, David Becker, Marc Laurent, James Lu, Clinton R. Paden, Suxiang Tong, Duncan MacCannell |
| EPI_ISL_1421671, EPI_ISL_1421672 | Wisconsin State Laboratory of Hygiene Communicable Disease Division | Wisconsin State Laboratory of Hygiene Communicable Disease Division | Kelsey R. Florek, Abigail C. Shockey |
| EPI_ISL_1421673 | Helix/Illumina | Centers for Disease Control and Prevention Division of Viral Diseases, Pathogen Discovery | Peter W. Cook, Dakota Howard, Dhwani Batra, Ben L. Rambo-Martin, Eileen de Feo, Jan Antico, Christine Tran, Matthew Tolentino, Shannon Wickline, Kim Gietzen, Brad Sickler, Jingtao Liu, Eric Allen, Phil Febbo, Summer Galloway, Nicole L. Washington, Simon White, Geraint Levan, Kelly Schiabor Barrett, Elizabeth Cirulli, Alexandre Bolze, Ary Ascencio, Charlotte Rivera-Garcia, Ryan Cho, Jason Nguyen, Sherry Wang, Jimmy Ramirez, Tyler Cassens, Efren Sandoval, Magnus Isaksson, William Lee, David Becker, Marc Laurent, James Lu, Clinton R. Paden, Suxiang Tong, Duncan MacCannell |
| EPI_ISL_1421674, EPI_ISL_1421675 | Wisconsin State Laboratory of Hygiene Communicable Disease Division | Wisconsin State Laboratory of Hygiene Communicable Disease Division | Kelsey R. Florek, Abigail C. Shockey |
| EPI_ISL_1421676 | Helix/Illumina | Centers for Disease Control and Prevention | Peter W. Cook, Dakota Howard, Dhwani Batra, Ben L. Rambo-Martin, Eileen de Feo, Jan Antico, Christine Tran, Matthew Tolentino, Shannon Wickline, Kim Gietzen, Brad Sickler, Jingtao Liu, Eric Allen, Phil Febbo, Summer Galloway, Nicole L. Washington, Simon White, Geraint Levan, Kelly |

| | | | |
|---|---|---|---|
| | | Division of Viral Diseases, Pathogen Discovery | Schiabor Barrett, Elizabeth Cirulli, Alexandre Bolze, Ary Ascencio, Charlotte Rivera-Garcia, Ryan Cho, Jason Nguyen, Sherry Wang, Jimmy Ramirez, Tyler Cassens, Efren Sandoval, Magnus Isaksson, William Lee, David Becker, Marc Laurent, James Lu, Clinton R. Paden, Suxiang Tong, Duncan MacCannell |
| EPI_ISL_1421677, EPI_ISL_1421678 | Wisconsin State Laboratory of Hygiene Communicable Disease Division | Wisconsin State Laboratory of Hygiene Communicable Disease Division | Kelsey R. Florek, Abigail C. Shockey |
| EPI_ISL_1421679 | Helix/Illumina | Centers for Disease Control and Prevention Division of Viral Diseases, Pathogen Discovery | Peter W. Cook, Dakota Howard, Dhwani Batra, Ben L. Rambo-Martin, Eileen de Feo, Jan Antico, Christine Tran, Matthew Tolentino, Shannon Wickline, Kim Gietzen, Brad Sickler, Jingtao Liu, Eric Allen, Phil Febbo, Summer Galloway, Nicole L. Washington, Simon White, Geraint Levan, Kelly Schiabor Barrett, Elizabeth Cirulli, Alexandre Bolze, Ary Ascencio, Charlotte Rivera-Garcia, Ryan Cho, Jason Nguyen, Sherry Wang, Jimmy Ramirez, Tyler Cassens, Efren Sandoval, Magnus Isaksson, William Lee, David Becker, Marc Laurent, James Lu, Clinton R. Paden, Suxiang Tong, Duncan MacCannell |
| EPI_ISL_1421680, EPI_ISL_1421681 | Wisconsin State Laboratory of Hygiene Communicable Disease Division | Wisconsin State Laboratory of Hygiene Communicable Disease Division | Kelsey R. Florek, Abigail C. Shockey |
| EPI_ISL_1421682 | Helix/Illumina | Centers for Disease Control and Prevention Division of Viral Diseases, Pathogen Discovery | Peter W. Cook, Dakota Howard, Dhwani Batra, Ben L. Rambo-Martin, Eileen de Feo, Jan Antico, Christine Tran, Matthew Tolentino, Shannon Wickline, Kim Gietzen, Brad Sickler, Jingtao Liu, Eric Allen, Phil Febbo, Summer Galloway, Nicole L. Washington, Simon White, Geraint Levan, Kelly Schiabor Barrett, Elizabeth Cirulli, Alexandre Bolze, Ary Ascencio, Charlotte Rivera-Garcia, Ryan Cho, Jason Nguyen, Sherry Wang, Jimmy Ramirez, Tyler Cassens, Efren Sandoval, Magnus Isaksson, William Lee, David Becker, Marc Laurent, James Lu, Clinton R. Paden, Suxiang Tong, Duncan MacCannell |
| EPI_ISL_1421683, EPI_ISL_1421684 | Wisconsin State Laboratory of Hygiene Communicable Disease Division | Wisconsin State Laboratory of Hygiene Communicable Disease Division | Kelsey R. Florek, Abigail C. Shockey |
| EPI_ISL_1421685 | Helix/Illumina | Centers for Disease Control and Prevention Division of Viral Diseases, Pathogen Discovery | Peter W. Cook, Dakota Howard, Dhwani Batra, Ben L. Rambo-Martin, Eileen de Feo, Jan Antico, Christine Tran, Matthew Tolentino, Shannon Wickline, Kim Gietzen, Brad Sickler, Jingtao Liu, Eric Allen, Phil Febbo, Summer Galloway, Nicole L. Washington, Simon White, Geraint Levan, Kelly Schiabor Barrett, Elizabeth Cirulli, Alexandre Bolze, Ary Ascencio, Charlotte Rivera-Garcia, Ryan Cho, Jason Nguyen, Sherry Wang, Jimmy Ramirez, Tyler Cassens, Efren Sandoval, Magnus Isaksson, William Lee, David Becker, Marc Laurent, James Lu, Clinton R. Paden, Suxiang Tong, Duncan MacCannell |
| EPI_ISL_1421686, EPI_ISL_1421687 | Wisconsin State Laboratory of Hygiene Communicable Disease Division | Wisconsin State Laboratory of Hygiene Communicable Disease Division | Kelsey R. Florek, Abigail C. Shockey |
| EPI_ISL_1421688 | Helix/Illumina | Centers for Disease Control and Prevention Division of Viral Diseases, Pathogen Discovery | Peter W. Cook, Dakota Howard, Dhwani Batra, Ben L. Rambo-Martin, Eileen de Feo, Jan Antico, Christine Tran, Matthew Tolentino, Shannon Wickline, Kim Gietzen, Brad Sickler, Jingtao Liu, Eric Allen, Phil Febbo, Summer Galloway, Nicole L. Washington, Simon White, Geraint Levan, Kelly Schiabor Barrett, Elizabeth Cirulli, Alexandre Bolze, Ary Ascencio, Charlotte Rivera-Garcia, Ryan Cho, Jason Nguyen, Sherry Wang, Jimmy Ramirez, Tyler Cassens, Efren Sandoval, Magnus Isaksson, William Lee, David Becker, Marc Laurent, James Lu, Clinton R. Paden, Suxiang Tong, Duncan MacCannell |
| EPI_ISL_1421689, EPI_ISL_1421690 | Wisconsin State Laboratory of Hygiene Communicable Disease Division | Wisconsin State Laboratory of Hygiene Communicable Disease Division | Kelsey R. Florek, Abigail C. Shockey |
| EPI_ISL_1421691 | Helix/Illumina | Centers for Disease Control and Prevention Division of Viral Diseases, Pathogen Discovery | Peter W. Cook, Dakota Howard, Dhwani Batra, Ben L. Rambo-Martin, Eileen de Feo, Jan Antico, Christine Tran, Matthew Tolentino, Shannon Wickline, Kim Gietzen, Brad Sickler, Jingtao Liu, Eric Allen, Phil Febbo, Summer Galloway, Nicole L. Washington, Simon White, Geraint Levan, Kelly Schiabor Barrett, Elizabeth Cirulli, Alexandre Bolze, Ary Ascencio, Charlotte Rivera-Garcia, Ryan Cho, Jason Nguyen, Sherry Wang, Jimmy Ramirez, Tyler Cassens, Efren Sandoval, Magnus Isaksson, William Lee, David Becker, Marc Laurent, James Lu, Clinton R. Paden, Suxiang Tong, Duncan MacCannell |
| EPI_ISL_1421692, EPI_ISL_1421693 | Wisconsin State Laboratory of Hygiene Communicable Disease Division | Wisconsin State Laboratory of Hygiene Communicable Disease Division | Kelsey R. Florek, Abigail C. Shockey |
| EPI_ISL_1421694 | Helix/Illumina | Centers for Disease Control and Prevention Division of Viral Diseases, Pathogen Discovery | Peter W. Cook, Dakota Howard, Dhwani Batra, Ben L. Rambo-Martin, Eileen de Feo, Jan Antico, Christine Tran, Matthew Tolentino, Shannon Wickline, Kim Gietzen, Brad Sickler, Jingtao Liu, Eric Allen, Phil Febbo, Summer Galloway, Nicole L. Washington, Simon White, Geraint Levan, Kelly Schiabor Barrett, Elizabeth Cirulli, Alexandre Bolze, Ary Ascencio, Charlotte Rivera-Garcia, Ryan Cho, Jason Nguyen, Sherry Wang, Jimmy Ramirez, Tyler Cassens, Efren Sandoval, Magnus Isaksson, William Lee, David Becker, Marc Laurent, James Lu, Clinton R. Paden, Suxiang Tong, Duncan MacCannell |
| EPI_ISL_1421695, EPI_ISL_1421696 | Wisconsin State Laboratory of Hygiene Communicable Disease Division | Wisconsin State Laboratory of Hygiene Communicable Disease Division | Kelsey R. Florek, Abigail C. Shockey |
| EPI_ISL_1421697 | Helix/Illumina | Centers for Disease Control and Prevention Division of Viral Diseases, Pathogen Discovery | Peter W. Cook, Dakota Howard, Dhwani Batra, Ben L. Rambo-Martin, Eileen de Feo, Jan Antico, Christine Tran, Matthew Tolentino, Shannon Wickline, Kim Gietzen, Brad Sickler, Jingtao Liu, Eric Allen, Phil Febbo, Summer Galloway, Nicole L. Washington, Simon White, Geraint Levan, Kelly Schiabor Barrett, Elizabeth Cirulli, Alexandre Bolze, Ary Ascencio, Charlotte Rivera-Garcia, Ryan Cho, Jason Nguyen, Sherry Wang, Jimmy Ramirez, Tyler Cassens, Efren Sandoval, Magnus Isaksson, William Lee, David Becker, Marc Laurent, James Lu, Clinton R. Paden, Suxiang Tong, Duncan MacCannell |
| EPI_ISL_1421698, EPI_ISL_1421699 | Wisconsin State Laboratory of Hygiene Communicable Disease Division | Wisconsin State Laboratory of Hygiene Communicable Disease Division | Kelsey R. Florek, Abigail C. Shockey |
| EPI_ISL_1421700 | Helix/Illumina | Centers for Disease Control and Prevention Division of Viral Diseases, Pathogen Discovery | Peter W. Cook, Dakota Howard, Dhwani Batra, Ben L. Rambo-Martin, Eileen de Feo, Jan Antico, Christine Tran, Matthew Tolentino, Shannon Wickline, Kim Gietzen, Brad Sickler, Jingtao Liu, Eric Allen, Phil Febbo, Summer Galloway, Nicole L. Washington, Simon White, Geraint Levan, Kelly Schiabor Barrett, Elizabeth Cirulli, Alexandre Bolze, Ary Ascencio, Charlotte Rivera-Garcia, Ryan Cho, Jason Nguyen, Sherry Wang, Jimmy Ramirez, Tyler Cassens, Efren Sandoval, Magnus Isaksson, William Lee, David Becker, Marc Laurent, James Lu, Clinton R. Paden, Suxiang Tong, Duncan MacCannell |
| EPI_ISL_1421701, EPI_ISL_1421702, EPI_ISL_1421703 | Wisconsin State Laboratory of Hygiene Communicable Disease Division | Wisconsin State Laboratory of Hygiene Communicable Disease Division | Kelsey R. Florek, Abigail C. Shockey |
| EPI_ISL_1421704 | Helix/Illumina | Centers for Disease Control and Prevention Division of Viral Diseases, Pathogen Discovery | Peter W. Cook, Dakota Howard, Dhwani Batra, Ben L. Rambo-Martin, Eileen de Feo, Jan Antico, Christine Tran, Matthew Tolentino, Shannon Wickline, Kim Gietzen, Brad Sickler, Jingtao Liu, Eric Allen, Phil Febbo, Summer Galloway, Nicole L. Washington, Simon White, Geraint Levan, Kelly Schiabor Barrett, Elizabeth Cirulli, Alexandre Bolze, Ary Ascencio, Charlotte Rivera-Garcia, Ryan Cho, Jason Nguyen, Sherry Wang, Jimmy Ramirez, Tyler Cassens, Efren Sandoval, Magnus Isaksson, William Lee, David Becker, Marc Laurent, James Lu, Clinton R. Paden, Suxiang Tong, Duncan MacCannell |
| EPI_ISL_1421706 | Wisconsin State Laboratory of Hygiene Communicable Disease Division | Wisconsin State Laboratory of Hygiene Communicable Disease Division | Kelsey R. Florek, Abigail C. Shockey |
| EPI_ISL_1421707 | Helix/Illumina | Centers for Disease Control and Prevention Division of Viral Diseases, Pathogen Discovery | Peter W. Cook, Dakota Howard, Dhwani Batra, Ben L. Rambo-Martin, Eileen de Feo, Jan Antico, Christine Tran, Matthew Tolentino, Shannon Wickline, Kim Gietzen, Brad Sickler, Jingtao Liu, Eric Allen, Phil Febbo, Summer Galloway, Nicole L. Washington, Simon White, Geraint Levan, Kelly Schiabor Barrett, Elizabeth Cirulli, Alexandre Bolze, Ary Ascencio, Charlotte Rivera-Garcia, Ryan Cho, Jason Nguyen, Sherry Wang, Jimmy Ramirez, Tyler Cassens, Efren Sandoval, Magnus Isaksson, William Lee, David Becker, Marc Laurent, James Lu, Clinton R. Paden, Suxiang Tong, Duncan MacCannell |
| EPI_ISL_1421708, EPI_ISL_1421709 | Wisconsin State Laboratory of Hygiene Communicable Disease Division | Wisconsin State Laboratory of Hygiene Communicable Disease Division | Kelsey R. Florek, Abigail C. Shockey |
| EPI_ISL_1421710 | Helix/Illumina | Centers for Disease Control and Prevention Division of Viral Diseases, Pathogen Discovery | Peter W. Cook, Dakota Howard, Dhwani Batra, Ben L. Rambo-Martin, Eileen de Feo, Jan Antico, Christine Tran, Matthew Tolentino, Shannon Wickline, Kim Gietzen, Brad Sickler, Jingtao Liu, Eric Allen, Phil Febbo, Summer Galloway, Nicole L. Washington, Simon White, Geraint Levan, Kelly Schiabor Barrett, Elizabeth Cirulli, Alexandre Bolze, Ary Ascencio, Charlotte Rivera-Garcia, Ryan Cho, Jason Nguyen, Sherry Wang, Jimmy Ramirez, Tyler Cassens, Efren Sandoval, Magnus Isaksson, William Lee, David Becker, Marc Laurent, James Lu, Clinton R. Paden, Suxiang Tong, Duncan MacCannell |
| EPI_ISL_1421711, EPI_ISL_1421712 | Wisconsin State Laboratory of Hygiene Communicable Disease Division | Wisconsin State Laboratory of Hygiene Communicable Disease Division | Kelsey R. Florek, Abigail C. Shockey |
| EPI_ISL_1421713 | Helix/Illumina | Centers for Disease Control and Prevention Division of Viral Diseases, Pathogen Discovery | Peter W. Cook, Dakota Howard, Dhwani Batra, Ben L. Rambo-Martin, Eileen de Feo, Jan Antico, Christine Tran, Matthew Tolentino, Shannon Wickline, Kim Gietzen, Brad Sickler, Jingtao Liu, Eric Allen, Phil Febbo, Summer Galloway, Nicole L. Washington, Simon White, Geraint Levan, Kelly Schiabor Barrett, Elizabeth Cirulli, Alexandre Bolze, Ary Ascencio, Charlotte Rivera-Garcia, Ryan Cho, Jason Nguyen, Sherry Wang, Jimmy Ramirez, Tyler Cassens, Efren Sandoval, Magnus Isaksson, William Lee, David Becker, Marc Laurent, James Lu, Clinton R. Paden, Suxiang Tong, Duncan MacCannell |
| EPI_ISL_1421714, EPI_ISL_1421715 | Wisconsin State Laboratory of Hygiene Communicable Disease Division | Wisconsin State Laboratory of Hygiene Communicable Disease Division | Kelsey R. Florek, Abigail C. Shockey |
| EPI_ISL_1421716 | Helix/Illumina | Centers for Disease Control and Prevention Division of Viral Diseases, Pathogen Discovery | Peter W. Cook, Dakota Howard, Dhwani Batra, Ben L. Rambo-Martin, Eileen de Feo, Jan Antico, Christine Tran, Matthew Tolentino, Shannon Wickline, Kim Gietzen, Brad Sickler, Jingtao Liu, Eric Allen, Phil Febbo, Summer Galloway, Nicole L. Washington, Simon White, Geraint Levan, Kelly Schiabor Barrett, Elizabeth Cirulli, Alexandre Bolze, Ary Ascencio, Charlotte Rivera-Garcia, Ryan Cho, Jason Nguyen, Sherry Wang, Jimmy Ramirez, Tyler Cassens, Efren Sandoval, Magnus Isaksson, William Lee, David Becker, Marc Laurent, James Lu, Clinton R. Paden, Suxiang Tong, Duncan MacCannell |
| EPI_ISL_1421717 | Wisconsin State Laboratory of Hygiene Communicable Disease Division | Wisconsin State Laboratory of Hygiene Communicable Disease Division | Kelsey R. Florek, Abigail C. Shockey |
| EPI_ISL_1421718, EPI_ISL_1421720, EPI_ISL_1421721, EPI_ISL_1421722, EPI_ISL_1421723, EPI_ISL_1421724, EPI_ISL_1421725, EPI_ISL_1421726, EPI_ISL_1421727, EPI_ISL_1421728, EPI_ISL_1421729, EPI_ISL_1421730, EPI_ISL_1421731, EPI_ISL_1421732, EPI_ISL_1421733, EPI_ISL_1421734, EPI_ISL_1421735, EPI_ISL_1421736, EPI_ISL_1421737, EPI_ISL_1421738, EPI_ISL_1421739, EPI_ISL_1421740, EPI_ISL_1421741, EPI_ISL_1421742, EPI_ISL_1421744, EPI_ISL_1421745, EPI_ISL_1421746, EPI_ISL_1421747, EPI_ISL_1421748, EPI_ISL_1421749, EPI_ISL_1421750, EPI_ISL_1421751, EPI_ISL_1421752, EPI_ISL_1421753, EPI_ISL_1421754 | | | |
| see above | Helix/Illumina | Centers for Disease Control and Prevention Division of Viral Diseases, Pathogen Discovery | Peter W. Cook, Dakota Howard, Dhwani Batra, Ben L. Rambo-Martin, Eileen de Feo, Jan Antico, Christine Tran, Matthew Tolentino, Shannon Wickline, Kim Gietzen, Brad Sickler, Jingtao Liu, Eric Allen, Phil Febbo, Summer Galloway, Nicole L. Washington, Simon White, Geraint Levan, Kelly Schiabor Barrett, Elizabeth Cirulli, Alexandre Bolze, Ary Ascencio, Charlotte Rivera-Garcia, Ryan Cho, Jason Nguyen, Sherry Wang, Jimmy Ramirez, Tyler Cassens, Efren Sandoval, Magnus Isaksson, William Lee, David Becker, Marc Laurent, James Lu, Clinton R. Paden, Suxiang Tong, Duncan MacCannell |
| EPI_ISL_1421755, EPI_ISL_1421756, EPI_ISL_1421757, EPI_ISL_1421759, EPI_ISL_1421764, EPI_ISL_1421765, EPI_ISL_1421766, EPI_ISL_1421769, EPI_ISL_1421770, EPI_ISL_1421771, EPI_ISL_1421774, EPI_ISL_1421776, EPI_ISL_1421777, EPI_ISL_1421778, EPI_ISL_1421781, EPI_ISL_1421782, EPI_ISL_1421785, EPI_ISL_1421789, EPI_ISL_1421790, EPI_ISL_1421791, EPI_ISL_1421793, EPI_ISL_1421794, EPI_ISL_1421795, EPI_ISL_1421797, EPI_ISL_1421799, EPI_ISL_1421800, EPI_ISL_1421801, EPI_ISL_1421804, EPI_ISL_1421805, EPI_ISL_1421806, EPI_ISL_1421807, EPI_ISL_1421808, EPI_ISL_1421810, EPI_ISL_1421811, EPI_ISL_1421813, EPI_ISL_1421816, EPI_ISL_1421817, EPI_ISL_1421818, EPI_ISL_1421820, EPI_ISL_1421821, EPI_ISL_1421822, EPI_ISL_1421823, EPI_ISL_1421827, EPI_ISL_1421830, EPI_ISL_1421832, EPI_ISL_1421834, EPI_ISL_1421835, EPI_ISL_1421836, EPI_ISL_1421838, EPI_ISL_1421839, EPI_ISL_1421840, EPI_ISL_1421841, EPI_ISL_1421842, EPI_ISL_1421843, EPI_ISL_1421846, EPI_ISL_1421849, EPI_ISL_1421851, EPI_ISL_1421853, EPI_ISL_1421854, EPI_ISL_1421855, EPI_ISL_1421857, EPI_ISL_1421859, EPI_ISL_1421864, EPI_ISL_1421866, EPI_ISL_1421867, EPI_ISL_1421868, EPI_ISL_1421870, EPI_ISL_1421873, EPI_ISL_1421874, EPI_ISL_1421875, EPI_ISL_1421876, EPI_ISL_1421880, EPI_ISL_1421884, EPI_ISL_1421885, EPI_ISL_1421889, EPI_ISL_1421890, EPI_ISL_1421894, EPI_ISL_1421896, EPI_ISL_1421897, EPI_ISL_1421901, EPI_ISL_1421905, EPI_ISL_1421906, EPI_ISL_1421907, EPI_ISL_1421908, EPI_ISL_1421909, EPI_ISL_1421910, EPI_ISL_1421912, EPI_ISL_1421914, EPI_ISL_1421918, EPI_ISL_1421922, EPI_ISL_1421924, EPI_ISL_1421925, EPI_ISL_1421928, EPI_ISL_1421930, EPI_ISL_1421933, EPI_ISL_1421934, EPI_ISL_1421935, EPI_ISL_1421939, EPI_ISL_1421944, EPI_ISL_1421945, EPI_ISL_1421949, EPI_ISL_1421952, EPI_ISL_1421957, EPI_ISL_1421958, EPI_ISL_1421960, EPI_ISL_1421962, EPI_ISL_1421963, EPI_ISL_1421964, EPI_ISL_1421971, EPI_ISL_1421972, EPI_ISL_1421973, EPI_ISL_1421974, EPI_ISL_1421975, EPI_ISL_1421978, EPI_ISL_1421981, EPI_ISL_1421982, EPI_ISL_1421983, EPI_ISL_1421984, EPI_ISL_1421989, EPI_ISL_1421990, EPI_ISL_1421993, EPI_ISL_1421996, EPI_ISL_1421997, EPI_ISL_1421998, EPI_ISL_1422001, EPI_ISL_1422005, EPI_ISL_1422008, EPI_ISL_1422010, EPI_ISL_1422012, EPI_ISL_1422014, EPI_ISL_1422017, EPI_ISL_1422018, EPI_ISL_1422019, EPI_ISL_1422021, EPI_ISL_1422022, EPI_ISL_1422025, EPI_ISL_1422026, EPI_ISL_1422028, EPI_ISL_1422029, EPI_ISL_1422031, EPI_ISL_1422032, EPI_ISL_1422034, EPI_ISL_1422035, EPI_ISL_1422036, EPI_ISL_1422037, EPI_ISL_1422038, EPI_ISL_1422039, EPI_ISL_1422040, EPI_ISL_1422041, EPI_ISL_1422043, EPI_ISL_1422044, EPI_ISL_1422046, EPI_ISL_1422047, EPI_ISL_1422048, EPI_ISL_1422050, EPI_ISL_1422051, EPI_ISL_1422052, EPI_ISL_1422054, EPI_ISL_1422055, EPI_ISL_1422057, EPI_ISL_1422058, EPI_ISL_1422060, EPI_ISL_1422061, EPI_ISL_1422062, EPI_ISL_1422063, EPI_ISL_1422064, EPI_ISL_1422065, EPI_ISL_1422066, EPI_ISL_1422067, EPI_ISL_1422068, EPI_ISL_1422069, EPI_ISL_1422071, EPI_ISL_1422072, EPI_ISL_1422073, EPI_ISL_1422075, EPI_ISL_1422076, EPI_ISL_1422078, EPI_ISL_1422079, EPI_ISL_1422080, EPI_ISL_1422082, EPI_ISL_1422086, EPI_ISL_1422088, EPI_ISL_1422089, EPI_ISL_1422090, EPI_ISL_1422091, EPI_ISL_1422092, EPI_ISL_1422094, EPI_ISL_1422095, EPI_ISL_1422096, EPI_ISL_1422099, EPI_ISL_1422100, EPI_ISL_1422102, EPI_ISL_1422103, EPI_ISL_1422104, EPI_ISL_1422105, EPI_ISL_1422106, EPI_ISL_1422107, EPI_ISL_1422108, EPI_ISL_1422109, EPI_ISL_1422110, EPI_ISL_1422112, EPI_ISL_1422113, EPI_ISL_1422114, EPI_ISL_1422115, EPI_ISL_1422117, EPI_ISL_1422118, EPI_ISL_1422120, EPI_ISL_1422121, EPI_ISL_1422126, EPI_ISL_1422129, EPI_ISL_1422131, EPI_ISL_1422132, EPI_ISL_1422133, EPI_ISL_1422135, EPI_ISL_1422137, EPI_ISL_1422138, EPI_ISL_1422139, EPI_ISL_1422140, EPI_ISL_1422143, EPI_ISL_1422144, EPI_ISL_1422147, EPI_ISL_1422148, EPI_ISL_1422149, EPI_ISL_1422155, EPI_ISL_1422156, EPI_ISL_1422160, EPI_ISL_1422164, EPI_ISL_1422165, EPI_ISL_1422167, EPI_ISL_1422168, EPI_ISL_1422170, EPI_ISL_1422173, EPI_ISL_1422174, EPI_ISL_1422176, EPI_ISL_1422177, EPI_ISL_1422179, EPI_ISL_1422180, EPI_ISL_1422181, EPI_ISL_1422182, EPI_ISL_1422183, EPI_ISL_1422184, EPI_ISL_1422186, EPI_ISL_1422190, EPI_ISL_1422193, EPI_ISL_1422194, EPI_ISL_1422195, EPI_ISL_1422197, EPI_ISL_1422200, EPI_ISL_1422204, EPI_ISL_1422209, EPI_ISL_1422211, EPI_ISL_1422213, EPI_ISL_1422214 | | | |
| see above | Laboratory Corporation of America | Centers for Disease Control and Prevention Division of Viral Diseases, Pathogen Discovery | Peter W. Cook, Dakota Howard, Dhwani Batra, Ben L. Rambo-Martin, Minoo Agarwal, Eyad Almasri, Debbie Boles, Ayla Burns, Nuthawin Charoensri, Oren Cohen, Susan Countryman, Mary Ann Cristobal, Bobbi Croy, Suzanne Dale, Hrushikesh Deshmukh, Amanda Douglas, Vincent Drouillon, Marcia Eisenberg, Howard Engler, Rama Ghatti, Prashant Gupta, Susan Hicks, Jake Humphrey, Lax Iyer, Manoj Jain, Mohan Kolli, Brian Krueger, Tim Kuphal, Stanley Letovsky, Michael Levandoski, Craig Lukasik, Jonathan Meltzer, Brian Norvell, Mindy Nye, Scott Parker, Christos Petropoulos, John Pruitt, Steven Ragan, Scott Ryan, Mike Sapeta, Jana Schroth, Suresh Babu Selvaraju, Goran Stevovic, Amanda Suchanek, Andrea Throop, Lyndon Tilson, Thomas Urban, Joe Voshell, Kimberly Wagner, Jonathan Williams, Mary Williamson, Qian Zeng, Tricia Zwiefelhofer, Clinton R. Paden, Suxiang Tong, Duncan MacCannell |
| EPI_ISL_1422216, EPI_ISL_1422217, EPI_ISL_1422218, EPI_ISL_1422219, EPI_ISL_1422220, EPI_ISL_1422221, EPI_ISL_1422222, EPI_ISL_1422223, EPI_ISL_1422224, EPI_ISL_1422225, EPI_ISL_1422226, EPI_ISL_1422227, EPI_ISL_1422228, EPI_ISL_1422229, EPI_ISL_1422230, EPI_ISL_1422231, EPI_ISL_1422232, EPI_ISL_1422233, EPI_ISL_1422235, EPI_ISL_1422236 | | | |
| see above | University of Wisconsin-Madison AIDS Vaccine Research Laboratories | University of Wisconsin-Madison AIDS Vaccine Research Laboratories | Gage Moreno, Katarina Braun, et al. AIDS Vaccine Research Laboratories |

| | | | |
|---|---|---|---|
| EPI_ISL_1422238, EPI_ISL_1422239, EPI_ISL_1422240, EPI_ISL_1422241, EPI_ISL_1422242, EPI_ISL_1422243, EPI_ISL_1422244, EPI_ISL_1422245, EPI_ISL_1422246, EPI_ISL_1422248, EPI_ISL_1422249, EPI_ISL_1422250, EPI_ISL_1422251, EPI_ISL_1422252, EPI_ISL_1422253, EPI_ISL_1422254, EPI_ISL_1422255, EPI_ISL_1422256, EPI_ISL_1422257, EPI_ISL_1422258, EPI_ISL_1422259, EPI_ISL_1422260, EPI_ISL_1422261, EPI_ISL_1422262, EPI_ISL_1422263, EPI_ISL_1422264, EPI_ISL_1422265, EPI_ISL_1422266, EPI_ISL_1422267, EPI_ISL_1422268, EPI_ISL_1422269, EPI_ISL_1422270, EPI_ISL_1422272, EPI_ISL_1422273, EPI_ISL_1422274, EPI_ISL_1422275, EPI_ISL_1422276, EPI_ISL_1422277, EPI_ISL_1422278, EPI_ISL_1422279, EPI_ISL_1422280, EPI_ISL_1422281, EPI_ISL_1422282, EPI_ISL_1422283, EPI_ISL_1422284, EPI_ISL_1422285, EPI_ISL_1422286, EPI_ISL_1422287, EPI_ISL_1422288, EPI_ISL_1422289, EPI_ISL_1422290, EPI_ISL_1422291, EPI_ISL_1422292, EPI_ISL_1422293, EPI_ISL_1422294, EPI_ISL_1422295, EPI_ISL_1422296, EPI_ISL_1422297, EPI_ISL_1422298, EPI_ISL_1422299, EPI_ISL_1422300, EPI_ISL_1422301, EPI_ISL_1422302, EPI_ISL_1422303, EPI_ISL_1422304, EPI_ISL_1422305, EPI_ISL_1422306, EPI_ISL_1422307, EPI_ISL_1422308, EPI_ISL_1422309, EPI_ISL_1422310, EPI_ISL_1422311, EPI_ISL_1422312, EPI_ISL_1422313, EPI_ISL_1422314, EPI_ISL_1422315, EPI_ISL_1422316, EPI_ISL_1422317, EPI_ISL_1422318, EPI_ISL_1422319, EPI_ISL_1422320, EPI_ISL_1422321, EPI_ISL_1422322, EPI_ISL_1422323, EPI_ISL_1422324, EPI_ISL_1422325, EPI_ISL_1422326 | | | |
| see above | TXDSHS | TXDSHS | Rashmi Tuladhar, Bonnie Oh, Jenny Zhang, Maliha Rahman, Mayela Pedrueza, Anita Pokharel, Lorraine Rodriguez, Myong Koag, Chun Wang, Rachel Lee, Grace Kubin |
| EPI_ISL_1422327, EPI_ISL_1422328, EPI_ISL_1422329, EPI_ISL_1422330, EPI_ISL_1422331, EPI_ISL_1422332, EPI_ISL_1422333, EPI_ISL_1422334, EPI_ISL_1422335, EPI_ISL_1422336, EPI_ISL_1422337, EPI_ISL_1422338, EPI_ISL_1422339, EPI_ISL_1422340, EPI_ISL_1422341, EPI_ISL_1422342, EPI_ISL_1422343, EPI_ISL_1422344, EPI_ISL_1422345, EPI_ISL_1422346, EPI_ISL_1422347, EPI_ISL_1422348, EPI_ISL_1422349, EPI_ISL_1422350, EPI_ISL_1422351, EPI_ISL_1422352, EPI_ISL_1422353, EPI_ISL_1422354, EPI_ISL_1422355, EPI_ISL_1422356, EPI_ISL_1422357, EPI_ISL_1422358, EPI_ISL_1422359, EPI_ISL_1422360, EPI_ISL_1422361, EPI_ISL_1422362, EPI_ISL_1422363, EPI_ISL_1422364, EPI_ISL_1422365, EPI_ISL_1422366, EPI_ISL_1422367, EPI_ISL_1422368, EPI_ISL_1422369, EPI_ISL_1422370, EPI_ISL_1422371, EPI_ISL_1422372, EPI_ISL_1422373, EPI_ISL_1422374, EPI_ISL_1422375, EPI_ISL_1422376, EPI_ISL_1422377, EPI_ISL_1422378, EPI_ISL_1422379, EPI_ISL_1422380, EPI_ISL_1422381, EPI_ISL_1422382, EPI_ISL_1422383, EPI_ISL_1422384, EPI_ISL_1422385, EPI_ISL_1422386, EPI_ISL_1422387, EPI_ISL_1422388, EPI_ISL_1422389, EPI_ISL_1422390, EPI_ISL_1422391, EPI_ISL_1422392, EPI_ISL_1422393, EPI_ISL_1422394, EPI_ISL_1422395, EPI_ISL_1422396, EPI_ISL_1422397, EPI_ISL_1422398, EPI_ISL_1422399, EPI_ISL_1422400, EPI_ISL_1422401, EPI_ISL_1422402, EPI_ISL_1422403, EPI_ISL_1422404, EPI_ISL_1422405, EPI_ISL_1422406, EPI_ISL_1422407, EPI_ISL_1422408, EPI_ISL_1422409, EPI_ISL_1422410, EPI_ISL_1422411, EPI_ISL_1422412, EPI_ISL_1422413, EPI_ISL_1422414, EPI_ISL_1422415, EPI_ISL_1422416, EPI_ISL_1422417, EPI_ISL_1422418, EPI_ISL_1422419, EPI_ISL_1422420, EPI_ISL_1422421, EPI_ISL_1422422, EPI_ISL_1422423, EPI_ISL_1422424, EPI_ISL_1422425, EPI_ISL_1422426, EPI_ISL_1422427, EPI_ISL_1422428, EPI_ISL_1422429, EPI_ISL_1422430, EPI_ISL_1422431, EPI_ISL_1422432, EPI_ISL_1422433, EPI_ISL_1422434, EPI_ISL_1422435, EPI_ISL_1422436, EPI_ISL_1422437, EPI_ISL_1422438, EPI_ISL_1422439, EPI_ISL_1422440, EPI_ISL_1422441, EPI_ISL_1422442, EPI_ISL_1422443, EPI_ISL_1422444, EPI_ISL_1422445, EPI_ISL_1422446, EPI_ISL_1422447, EPI_ISL_1422448, EPI_ISL_1422449, EPI_ISL_1422450, EPI_ISL_1422451, EPI_ISL_1422452, EPI_ISL_1422453, EPI_ISL_1422454, EPI_ISL_1422455, EPI_ISL_1422456, EPI_ISL_1422457, EPI_ISL_1422458, EPI_ISL_1422459, EPI_ISL_1422460, EPI_ISL_1422461, EPI_ISL_1422462, EPI_ISL_1422463, EPI_ISL_1422464, EPI_ISL_1422465, EPI_ISL_1422466, EPI_ISL_1422467, EPI_ISL_1422468, EPI_ISL_1422469, EPI_ISL_1422470, EPI_ISL_1422471, EPI_ISL_1422472, EPI_ISL_1422473, EPI_ISL_1422474, EPI_ISL_1422475, EPI_ISL_1422476, EPI_ISL_1422477, EPI_ISL_1422478, EPI_ISL_1422479, EPI_ISL_1422480, EPI_ISL_1422481, EPI_ISL_1422482, EPI_ISL_1422483, EPI_ISL_1422484, EPI_ISL_1422485, EPI_ISL_1422486, EPI_ISL_1422487, EPI_ISL_1422488, EPI_ISL_1422489, EPI_ISL_1422490, EPI_ISL_1422491, EPI_ISL_1422492, EPI_ISL_1422493, EPI_ISL_1422494, EPI_ISL_1422495, EPI_ISL_1422496, EPI_ISL_1422497, EPI_ISL_1422498, EPI_ISL_1422499, EPI_ISL_1422500, EPI_ISL_1422501, EPI_ISL_1422502, EPI_ISL_1422503, EPI_ISL_1422504, EPI_ISL_1422505, EPI_ISL_1422506, EPI_ISL_1422507, EPI_ISL_1422508, EPI_ISL_1422509, EPI_ISL_1422510, EPI_ISL_1422511, EPI_ISL_1422512, EPI_ISL_1422513, EPI_ISL_1422514, EPI_ISL_1422515, EPI_ISL_1422516, EPI_ISL_1422517, EPI_ISL_1422518, EPI_ISL_1422519, EPI_ISL_1422520, EPI_ISL_1422521, EPI_ISL_1422522, EPI_ISL_1422523, EPI_ISL_1422524, EPI_ISL_1422525, EPI_ISL_1422526, EPI_ISL_1422527, EPI_ISL_1422528, EPI_ISL_1422529, EPI_ISL_1422530, EPI_ISL_1422531, EPI_ISL_1422532, EPI_ISL_1422533, EPI_ISL_1422534, EPI_ISL_1422535, EPI_ISL_1422536, EPI_ISL_1422537, EPI_ISL_1422538, EPI_ISL_1422539, EPI_ISL_1422540, EPI_ISL_1422541, EPI_ISL_1422542, EPI_ISL_1422543, EPI_ISL_1422544, EPI_ISL_1422545, EPI_ISL_1422546, EPI_ISL_1422547, EPI_ISL_1422548, EPI_ISL_1422549, EPI_ISL_1422550, EPI_ISL_1422551, EPI_ISL_1422552, EPI_ISL_1422553, EPI_ISL_1422554, EPI_ISL_1422555, EPI_ISL_1422556, EPI_ISL_1422557, EPI_ISL_1422558, EPI_ISL_1422559, EPI_ISL_1422560, EPI_ISL_1422561 | | | |
| see above | Quest Diagnostics Incorporated | Centers for Disease Control and Prevention Division of Viral Diseases, Pathogen Discovery | Peter W. Cook, Dakota Howard, Dhwani Batra, Ben L. Rambo-Martin, S. H. Rosenthal, A. Gerasimova, R. M. Kagan, B. Anderson, M. Hua, Y. Liu, L.E. Bernstein, K.E. Livingston, A. Perez, I. A. Shlyakhter, R. V. Rolando, R. Owen, P. Tanpaiboon, F. Lacbawan, Clinton R. Paden, Suxiang Tong, Duncan MacCannell |
| EPI_ISL_1422562, EPI_ISL_1422563 | GA Department of Public Health | GA Department of Public Health | Stacy Reeves, Jonathan Edwards, Cynthia Dixey, Tonia Parrott, Aliyah Fields, Taylor Smith |
| EPI_ISL_1422564 | Quest Diagnostics Incorporated | Centers for Disease Control and Prevention Division of Viral Diseases, Pathogen Discovery | Peter W. Cook, Dakota Howard, Dhwani Batra, Ben L. Rambo-Martin, S. H. Rosenthal, A. Gerasimova, R. M. Kagan, B. Anderson, M. Hua, Y. Liu, L.E. Bernstein, K.E. Livingston, A. Perez, I. A. Shlyakhter, R. V. Rolando, R. Owen, P. Tanpaiboon, F. Lacbawan, Clinton R. Paden, Suxiang Tong, Duncan MacCannell |
| EPI_ISL_1422565, EPI_ISL_1422566 | GA Department of Public Health | GA Department of Public Health | Stacy Reeves, Jonathan Edwards, Cynthia Dixey, Tonia Parrott, Aliyah Fields, Taylor Smith |
| EPI_ISL_1422567 | Quest Diagnostics Incorporated | Centers for Disease Control and Prevention Division of Viral Diseases, Pathogen Discovery | Peter W. Cook, Dakota Howard, Dhwani Batra, Ben L. Rambo-Martin, S. H. Rosenthal, A. Gerasimova, R. M. Kagan, B. Anderson, M. Hua, Y. Liu, L.E. Bernstein, K.E. Livingston, A. Perez, I. A. Shlyakhter, R. V. Rolando, R. Owen, P. Tanpaiboon, F. Lacbawan, Clinton R. Paden, Suxiang Tong, Duncan MacCannell |
| EPI_ISL_1422568, EPI_ISL_1422569 | GA Department of Public Health | GA Department of Public Health | Stacy Reeves, Jonathan Edwards, Cynthia Dixey, Tonia Parrott, Aliyah Fields, Taylor Smith |
| EPI_ISL_1422570 | Quest Diagnostics Incorporated | Centers for Disease Control and Prevention Division of Viral Diseases, Pathogen Discovery | Peter W. Cook, Dakota Howard, Dhwani Batra, Ben L. Rambo-Martin, S. H. Rosenthal, A. Gerasimova, R. M. Kagan, B. Anderson, M. Hua, Y. Liu, L.E. Bernstein, K.E. Livingston, A. Perez, I. A. Shlyakhter, R. V. Rolando, R. Owen, P. Tanpaiboon, F. Lacbawan, Clinton R. Paden, Suxiang Tong, Duncan MacCannell |
| EPI_ISL_1422571, EPI_ISL_1422572 | GA Department of Public Health | GA Department of Public Health | Stacy Reeves, Jonathan Edwards, Cynthia Dixey, Tonia Parrott, Aliyah Fields, Taylor Smith |
| EPI_ISL_1422573 | Quest Diagnostics Incorporated | Centers for Disease Control and Prevention Division of Viral Diseases, Pathogen Discovery | Peter W. Cook, Dakota Howard, Dhwani Batra, Ben L. Rambo-Martin, S. H. Rosenthal, A. Gerasimova, R. M. Kagan, B. Anderson, M. Hua, Y. Liu, L.E. Bernstein, K.E. Livingston, A. Perez, I. A. Shlyakhter, R. V. Rolando, R. Owen, P. Tanpaiboon, F. Lacbawan, Clinton R. Paden, Suxiang Tong, Duncan MacCannell |
| EPI_ISL_1422574, EPI_ISL_1422575 | GA Department of Public Health | GA Department of Public Health | Stacy Reeves, Jonathan Edwards, Cynthia Dixey, Tonia Parrott, Aliyah Fields, Taylor Smith |
| EPI_ISL_1422576 | Quest Diagnostics Incorporated | Centers for Disease Control and Prevention Division of Viral Diseases, Pathogen Discovery | Peter W. Cook, Dakota Howard, Dhwani Batra, Ben L. Rambo-Martin, S. H. Rosenthal, A. Gerasimova, R. M. Kagan, B. Anderson, M. Hua, Y. Liu, L.E. Bernstein, K.E. Livingston, A. Perez, I. A. Shlyakhter, R. V. Rolando, R. Owen, P. Tanpaiboon, F. Lacbawan, Clinton R. Paden, Suxiang Tong, Duncan MacCannell |
| EPI_ISL_1422577, EPI_ISL_1422578 | GA Department of Public Health | GA Department of Public Health | Stacy Reeves, Jonathan Edwards, Cynthia Dixey, Tonia Parrott, Aliyah Fields, Taylor Smith |
| EPI_ISL_1422579 | Quest Diagnostics Incorporated | Centers for Disease Control and Prevention Division of Viral Diseases, Pathogen Discovery | Peter W. Cook, Dakota Howard, Dhwani Batra, Ben L. Rambo-Martin, S. H. Rosenthal, A. Gerasimova, R. M. Kagan, B. Anderson, M. Hua, Y. Liu, L.E. Bernstein, K.E. Livingston, A. Perez, I. A. Shlyakhter, R. V. Rolando, R. Owen, P. Tanpaiboon, F. Lacbawan, Clinton R. Paden, Suxiang Tong, Duncan MacCannell |
| EPI_ISL_1422580, EPI_ISL_1422581 | GA Department of Public Health | GA Department of Public Health | Stacy Reeves, Jonathan Edwards, Cynthia Dixey, Tonia Parrott, Aliyah Fields, Taylor Smith |
| EPI_ISL_1422582 | Quest Diagnostics Incorporated | Centers for Disease Control and Prevention Division of Viral Diseases, Pathogen Discovery | Peter W. Cook, Dakota Howard, Dhwani Batra, Ben L. Rambo-Martin, S. H. Rosenthal, A. Gerasimova, R. M. Kagan, B. Anderson, M. Hua, Y. Liu, L.E. Bernstein, K.E. Livingston, A. Perez, I. A. Shlyakhter, R. V. Rolando, R. Owen, P. Tanpaiboon, F. Lacbawan, Clinton R. Paden, Suxiang Tong, Duncan MacCannell |
| EPI_ISL_1422583, EPI_ISL_1422584 | GA Department of Public Health | GA Department of Public Health | Stacy Reeves, Jonathan Edwards, Cynthia Dixey, Tonia Parrott, Aliyah Fields, Taylor Smith |
| EPI_ISL_1422585 | Quest Diagnostics Incorporated | Centers for Disease Control and Prevention Division of Viral Diseases, Pathogen Discovery | Peter W. Cook, Dakota Howard, Dhwani Batra, Ben L. Rambo-Martin, S. H. Rosenthal, A. Gerasimova, R. M. Kagan, B. Anderson, M. Hua, Y. Liu, L.E. Bernstein, K.E. Livingston, A. Perez, I. A. Shlyakhter, R. V. Rolando, R. Owen, P. Tanpaiboon, F. Lacbawan, Clinton R. Paden, Suxiang Tong, Duncan MacCannell |
| EPI_ISL_1422586, EPI_ISL_1422587 | GA Department of Public Health | GA Department of Public Health | Stacy Reeves, Jonathan Edwards, Cynthia Dixey, Tonia Parrott, Aliyah Fields, Taylor Smith |
| EPI_ISL_1422588 | Quest Diagnostics Incorporated | Centers for Disease Control and Prevention Division of Viral Diseases, Pathogen Discovery | Peter W. Cook, Dakota Howard, Dhwani Batra, Ben L. Rambo-Martin, S. H. Rosenthal, A. Gerasimova, R. M. Kagan, B. Anderson, M. Hua, Y. Liu, L.E. Bernstein, K.E. Livingston, A. Perez, I. A. Shlyakhter, R. V. Rolando, R. Owen, P. Tanpaiboon, F. Lacbawan, Clinton R. Paden, Suxiang Tong, Duncan MacCannell |
| EPI_ISL_1422589, EPI_ISL_1422590 | GA Department of Public Health | GA Department of Public Health | Stacy Reeves, Jonathan Edwards, Cynthia Dixey, Tonia Parrott, Aliyah Fields, Taylor Smith |
| EPI_ISL_1422591, EPI_ISL_1422592, EPI_ISL_1422593, EPI_ISL_1422594, EPI_ISL_1422595, EPI_ISL_1422596, EPI_ISL_1422597, EPI_ISL_1422598, EPI_ISL_1422599, EPI_ISL_1422600, EPI_ISL_1422601, EPI_ISL_1422602, EPI_ISL_1422603, EPI_ISL_1422604, EPI_ISL_1422605, EPI_ISL_1422606, EPI_ISL_1422607, EPI_ISL_1422608, EPI_ISL_1422609, EPI_ISL_1422610, EPI_ISL_1422611, EPI_ISL_1422612, EPI_ISL_1422613, EPI_ISL_1422614, EPI_ISL_1422615, EPI_ISL_1422616, EPI_ISL_1422617, EPI_ISL_1422618, EPI_ISL_1422619, EPI_ISL_1422620, EPI_ISL_1422621, EPI_ISL_1422622, EPI_ISL_1422623, EPI_ISL_1422624, EPI_ISL_1422625, EPI_ISL_1422626, EPI_ISL_1422627, EPI_ISL_1422628, EPI_ISL_1422629, EPI_ISL_1422630, EPI_ISL_1422631, EPI_ISL_1422632, EPI_ISL_1422633, EPI_ISL_1422634, EPI_ISL_1422635, EPI_ISL_1422636, EPI_ISL_1422637, EPI_ISL_1422638, EPI_ISL_1422639, EPI_ISL_1422640, EPI_ISL_1422641, EPI_ISL_1422642, EPI_ISL_1422643, EPI_ISL_1422644, EPI_ISL_1422645, EPI_ISL_1422646, EPI_ISL_1422647, EPI_ISL_1422648, EPI_ISL_1422649, EPI_ISL_1422650, EPI_ISL_1422651, EPI_ISL_1422652, EPI_ISL_1422653, EPI_ISL_1422654, EPI_ISL_1422655, EPI_ISL_1422656, EPI_ISL_1422657, EPI_ISL_1422658, EPI_ISL_1422659, EPI_ISL_1422660, EPI_ISL_1422661, EPI_ISL_1422662, EPI_ISL_1422663, EPI_ISL_1422664, EPI_ISL_1422665, EPI_ISL_1422666, EPI_ISL_1422667, EPI_ISL_1422668, EPI_ISL_1422669, EPI_ISL_1422670, EPI_ISL_1422671, EPI_ISL_1422672, EPI_ISL_1422673, EPI_ISL_1422674, EPI_ISL_1422675, EPI_ISL_1422676, EPI_ISL_1422677, EPI_ISL_1422678, EPI_ISL_1422679, EPI_ISL_1422680, EPI_ISL_1422681, EPI_ISL_1422682, EPI_ISL_1422683, EPI_ISL_1422684, EPI_ISL_1422685, EPI_ISL_1422686, EPI_ISL_1422687, EPI_ISL_1422688, EPI_ISL_1422689, EPI_ISL_1422690, EPI_ISL_1422691, EPI_ISL_1422692, EPI_ISL_1422693, EPI_ISL_1422694, EPI_ISL_1422695, EPI_ISL_1422696, EPI_ISL_1422697, EPI_ISL_1422698, EPI_ISL_1422699, EPI_ISL_1422700, EPI_ISL_1422701, EPI_ISL_1422702, EPI_ISL_1422703, EPI_ISL_1422704, EPI_ISL_1422705, EPI_ISL_1422706, EPI_ISL_1422707, EPI_ISL_1422708, EPI_ISL_1422709, EPI_ISL_1422710, EPI_ISL_1422711, EPI_ISL_1422712, EPI_ISL_1422713, EPI_ISL_1422714, EPI_ISL_1422715, EPI_ISL_1422716, EPI_ISL_1422717, EPI_ISL_1422718, EPI_ISL_1422719, EPI_ISL_1422720, EPI_ISL_1422721, EPI_ISL_1422722, EPI_ISL_1422723, EPI_ISL_1422724, EPI_ISL_1422725, EPI_ISL_1422726, EPI_ISL_1422727, EPI_ISL_1422728, EPI_ISL_1422729, EPI_ISL_1422730, EPI_ISL_1422731, EPI_ISL_1422732, EPI_ISL_1422733, EPI_ISL_1422734, EPI_ISL_1422735, EPI_ISL_1422736, EPI_ISL_1422737, EPI_ISL_1422738, EPI_ISL_1422739, EPI_ISL_1422740, EPI_ISL_1422741, EPI_ISL_1422742, EPI_ISL_1422743, EPI_ISL_1422744, EPI_ISL_1422745, EPI_ISL_1422746, EPI_ISL_1422747, EPI_ISL_1422748, EPI_ISL_1422749, EPI_ISL_1422750, EPI_ISL_1422751, EPI_ISL_1422752, EPI_ISL_1422753, EPI_ISL_1422754, EPI_ISL_1422755, EPI_ISL_1422756, EPI_ISL_1422757, EPI_ISL_1422758, EPI_ISL_1422759, EPI_ISL_1422760, EPI_ISL_1422761, EPI_ISL_1422762, EPI_ISL_1422763, EPI_ISL_1422764, EPI_ISL_1422765, EPI_ISL_1422766, EPI_ISL_1422767, EPI_ISL_1422768, EPI_ISL_1422769, EPI_ISL_1422770, EPI_ISL_1422771, EPI_ISL_1422772, EPI_ISL_1422773, EPI_ISL_1422774, EPI_ISL_1422775, EPI_ISL_1422776, EPI_ISL_1422777, EPI_ISL_1422778, EPI_ISL_1422779, EPI_ISL_1422780, EPI_ISL_1422781, EPI_ISL_1422782, EPI_ISL_1422783, EPI_ISL_1422784, EPI_ISL_1422785, EPI_ISL_1422786, EPI_ISL_1422787, EPI_ISL_1422788, EPI_ISL_1422789, EPI_ISL_1422790, EPI_ISL_1422791, EPI_ISL_1422792, EPI_ISL_1422793, EPI_ISL_1422794, EPI_ISL_1422795, EPI_ISL_1422796, EPI_ISL_1422797, EPI_ISL_1422798, EPI_ISL_1422799, EPI_ISL_1422800, EPI_ISL_1422801, EPI_ISL_1422802, EPI_ISL_1422803, EPI_ISL_1422804, EPI_ISL_1422805, EPI_ISL_1422806, EPI_ISL_1422807, EPI_ISL_1422808, EPI_ISL_1422809, EPI_ISL_1422810, EPI_ISL_1422811, EPI_ISL_1422812, EPI_ISL_1422813, EPI_ISL_1422814, EPI_ISL_1422815, EPI_ISL_1422816, EPI_ISL_1422817, EPI_ISL_1422818, EPI_ISL_1422819, EPI_ISL_1422820, EPI_ISL_1422821, EPI_ISL_1422822, EPI_ISL_1422823, EPI_ISL_1422824, EPI_ISL_1422825, EPI_ISL_1422826, EPI_ISL_1422827, EPI_ISL_1422828, EPI_ISL_1422829, EPI_ISL_1422830, EPI_ISL_1422831, EPI_ISL_1422832, EPI_ISL_1422833, EPI_ISL_1422834, EPI_ISL_1422835, EPI_ISL_1422836, EPI_ISL_1422837, EPI_ISL_1422838, EPI_ISL_1422839, EPI_ISL_1422840, EPI_ISL_1422841, EPI_ISL_1422842, EPI_ISL_1422843, EPI_ISL_1422844, EPI_ISL_1422845, EPI_ISL_1422846, EPI_ISL_1422847, EPI_ISL_1422848, EPI_ISL_1422849, EPI_ISL_1422850, EPI_ISL_1422851, EPI_ISL_1422852, EPI_ISL_1422853, EPI_ISL_1422854, EPI_ISL_1422855, EPI_ISL_1422856, EPI_ISL_1422857, EPI_ISL_1422858, EPI_ISL_1422859, EPI_ISL_1422860, EPI_ISL_1422861, EPI_ISL_1422862, EPI_ISL_1422863, EPI_ISL_1422864, EPI_ISL_1422865, EPI_ISL_1422866, EPI_ISL_1422867, EPI_ISL_1422868, EPI_ISL_1422869, EPI_ISL_1422870, EPI_ISL_1422871, EPI_ISL_1422872, EPI_ISL_1422873, EPI_ISL_1422874, EPI_ISL_1422875, EPI_ISL_1422876, EPI_ISL_1422877, EPI_ISL_1422878, EPI_ISL_1422879, EPI_ISL_1422880, EPI_ISL_1422881, EPI_ISL_1422882, EPI_ISL_1422883, EPI_ISL_1422884, EPI_ISL_1422885, EPI_ISL_1422886, EPI_ISL_1422887, EPI_ISL_1422888, EPI_ISL_1422889, EPI_ISL_1422890, EPI_ISL_1422891, EPI_ISL_1422892, EPI_ISL_1422893, EPI_ISL_1422894, EPI_ISL_1422895, EPI_ISL_1422896, EPI_ISL_1422897, EPI_ISL_1422898, EPI_ISL_1422899, EPI_ISL_1422900, EPI_ISL_1422901, EPI_ISL_1422902, EPI_ISL_1422903, EPI_ISL_1422904, EPI_ISL_1422905, EPI_ISL_1422906, EPI_ISL_1422907, EPI_ISL_1422908, EPI_ISL_1422909, EPI_ISL_1422910, EPI_ISL_1422911, EPI_ISL_1422912, EPI_ISL_1422913, EPI_ISL_1422914, EPI_ISL_1422915, EPI_ISL_1422916, EPI_ISL_1422917, EPI_ISL_1422918, EPI_ISL_1422919, EPI_ISL_1422920, EPI_ISL_1422921, EPI_ISL_1422922, EPI_ISL_1422923, EPI_ISL_1422924, EPI_ISL_1422925, EPI_ISL_1422926, EPI_ISL_1422927, EPI_ISL_1422928, EPI_ISL_1422929, EPI_ISL_1422930, EPI_ISL_1422931, EPI_ISL_1422932, EPI_ISL_1422933, EPI_ISL_1422934, EPI_ISL_1422935, EPI_ISL_1422936, EPI_ISL_1422937, EPI_ISL_1422938, EPI_ISL_1422939, EPI_ISL_1422940, EPI_ISL_1422941, EPI_ISL_1422942, EPI_ISL_1422943, EPI_ISL_1422944, EPI_ISL_1422945, EPI_ISL_1422946, EPI_ISL_1422947, EPI_ISL_1422948, EPI_ISL_1422949, EPI_ISL_1422950, EPI_ISL_1422951, EPI_ISL_1422952, EPI_ISL_1422953, EPI_ISL_1422954, EPI_ISL_1422955, EPI_ISL_1422956, EPI_ISL_1422957, EPI_ISL_1422958, EPI_ISL_1422959, EPI_ISL_1422960, EPI_ISL_1422961, EPI_ISL_1422962, EPI_ISL_1422963, EPI_ISL_1422964, EPI_ISL_1422965, EPI_ISL_1422966, EPI_ISL_1422967, EPI_ISL_1422968, EPI_ISL_1422969, EPI_ISL_1422970, EPI_ISL_1422971, EPI_ISL_1422972, EPI_ISL_1422973, EPI_ISL_1422974, EPI_ISL_1422975, EPI_ISL_1422976, EPI_ISL_1422977, EPI_ISL_1422978, EPI_ISL_1422979, EPI_ISL_1422980, EPI_ISL_1422981, EPI_ISL_1422982, EPI_ISL_1422983, EPI_ISL_1422984, EPI_ISL_1422985, EPI_ISL_1422986, EPI_ISL_1422987, EPI_ISL_1422988, EPI_ISL_1422989, EPI_ISL_1422990, EPI_ISL_1422991 | | | |
| see above | Quest Diagnostics Incorporated | Centers for Disease Control and Prevention Division of Viral Diseases, Pathogen Discovery | Peter W. Cook, Dakota Howard, Dhwani Batra, Ben L. Rambo-Martin, S. H. Rosenthal, A. Gerasimova, R. M. Kagan, B. Anderson, M. Hua, Y. Liu, L.E. Bernstein, K.E. Livingston, A. Perez, I. A. Shlyakhter, R. V. Rolando, R. Owen, P. Tanpaiboon, F. Lacbawan, Clinton R. Paden, Suxiang Tong, Duncan MacCannell |
| EPI_ISL_1422992, EPI_ISL_1422993, EPI_ISL_1422994, EPI_ISL_1422995, EPI_ISL_1422996, EPI_ISL_1422997, EPI_ISL_1422998, EPI_ISL_1422999, EPI_ISL_1423000, EPI_ISL_1423001, EPI_ISL_1423002, EPI_ISL_1423003, EPI_ISL_1423004, EPI_ISL_1423005, EPI_ISL_1423006, EPI_ISL_1423007 | | | |
| see above | Nevada State Public Health Laboratory | Nevada State Public Health Laboratory | Andrew Gorzalski, Mark Pandori |
| EPI_ISL_1423009 | Jefferson Medical Center | WVU and Marshall University Combined Genomics Core Facilities | "James Denvir, Peter Stoilov, Peter Perrotta, Wesley Kimble, Ryan Percifield" |
| EPI_ISL_1423010, EPI_ISL_1423012, EPI_ISL_1423013, EPI_ISL_1423014 | Berkeley Medical Center | WVU and Marshall University Combined Genomics Core Facilities | "James Denvir, Peter Stoilov, Peter Perrotta, Wesley Kimble, Ryan Percifield" |
| EPI_ISL_1423015, | QLabs | WVU and Marshall University Combined | "James Denvir, Peter Stoilov, Peter Perrotta, Wesley Kimble, Ryan Percifield" |

| | | | |
|---|---|---|---|
| EPI_ISL_1423016, EPI_ISL_1423017, EPI_ISL_1423018, EPI_ISL_1423019, EPI_ISL_1423020 | | Genomics Core Facilities | |
| EPI_ISL_1423021, EPI_ISL_1423022, EPI_ISL_1423023, EPI_ISL_1423024, EPI_ISL_1423025, EPI_ISL_1423026, EPI_ISL_1423027, EPI_ISL_1423028, EPI_ISL_1423029, EPI_ISL_1423030, EPI_ISL_1423031, EPI_ISL_1423032, EPI_ISL_1423033, EPI_ISL_1423034, EPI_ISL_1423035, EPI_ISL_1423036, EPI_ISL_1423037, EPI_ISL_1423038, EPI_ISL_1423039, EPI_ISL_1423040, EPI_ISL_1423041, EPI_ISL_1423042, EPI_ISL_1423043, EPI_ISL_1423044, EPI_ISL_1423045, EPI_ISL_1423046, EPI_ISL_1423047, EPI_ISL_1423048, EPI_ISL_1423049, EPI_ISL_1423050, EPI_ISL_1423051, EPI_ISL_1423052, EPI_ISL_1423053, EPI_ISL_1423054, EPI_ISL_1423055, EPI_ISL_1423056, EPI_ISL_1423057, EPI_ISL_1423058, EPI_ISL_1423059, EPI_ISL_1423060, EPI_ISL_1423061, EPI_ISL_1423062, EPI_ISL_1423063, EPI_ISL_1423064, EPI_ISL_1423065, EPI_ISL_1423066, EPI_ISL_1423067, EPI_ISL_1423068, EPI_ISL_1423069, EPI_ISL_1423070, EPI_ISL_1423071, EPI_ISL_1423072, EPI_ISL_1423073, EPI_ISL_1423074, EPI_ISL_1423075, EPI_ISL_1423076, EPI_ISL_1423077, EPI_ISL_1423078, EPI_ISL_1423079, EPI_ISL_1423080, EPI_ISL_1423081, EPI_ISL_1423082, EPI_ISL_1423083, EPI_ISL_1423084, EPI_ISL_1423085, EPI_ISL_1423086, EPI_ISL_1423087, EPI_ISL_1423088, EPI_ISL_1423089, EPI_ISL_1423090, EPI_ISL_1423091, EPI_ISL_1423092, EPI_ISL_1423093, EPI_ISL_1423094, EPI_ISL_1423095, EPI_ISL_1423096, EPI_ISL_1423097, EPI_ISL_1423098, EPI_ISL_1423099, EPI_ISL_1423100, EPI_ISL_1423101, EPI_ISL_1423102, EPI_ISL_1423103, EPI_ISL_1423104, EPI_ISL_1423105, EPI_ISL_1423106, EPI_ISL_1423107, EPI_ISL_1423108, EPI_ISL_1423109, EPI_ISL_1423110, EPI_ISL_1423111 | | | |
| see above | WVU Rapid Development Laboratory | WVU and Marshall University Combined Genomics Core Facilities | "James Denvir, Peter Stoilov, Peter Perrotta, Wesley Kimble, Ryan Percifield" |
| EPI_ISL_1423112, EPI_ISL_1423113, EPI_ISL_1423114, EPI_ISL_1423115, EPI_ISL_1423117, EPI_ISL_1423118, EPI_ISL_1423119, EPI_ISL_1423120, EPI_ISL_1423121, EPI_ISL_1423122, EPI_ISL_1423123, EPI_ISL_1423124 | | | |
| see above | Berkeley Medical Center | WVU and Marshall University Combined Genomics Core Facilities | "James Denvir, Peter Stoilov, Peter Perrotta, Wesley Kimble, Ryan Percifield" |
| EPI_ISL_1423126 | QLabs | WVU and Marshall University Combined Genomics Core Facilities | "James Denvir, Peter Stoilov, Peter Perrotta, Wesley Kimble, Ryan Percifield" |
| EPI_ISL_1423127 | Camden Clark Medical Center | WVU and Marshall University Combined Genomics Core Facilities | "James Denvir, Peter Stoilov, Peter Perrotta, Wesley Kimble, Ryan Percifield" |
| EPI_ISL_1423128, EPI_ISL_1423129, EPI_ISL_1423130 | QLabs | WVU and Marshall University Combined Genomics Core Facilities | "James Denvir, Peter Stoilov, Peter Perrotta, Wesley Kimble, Ryan Percifield" |
| EPI_ISL_1423131, EPI_ISL_1423132, EPI_ISL_1423134, EPI_ISL_1423135, EPI_ISL_1423136, EPI_ISL_1423137, EPI_ISL_1423139, EPI_ISL_1423140, EPI_ISL_1423143, EPI_ISL_1423144, EPI_ISL_1423145, EPI_ISL_1423147, EPI_ISL_1423150, EPI_ISL_1423151, EPI_ISL_1423152, EPI_ISL_1423154, EPI_ISL_1423155, EPI_ISL_1423156, EPI_ISL_1423157, EPI_ISL_1423158, EPI_ISL_1423160, EPI_ISL_1423161, EPI_ISL_1423162, EPI_ISL_1423163, EPI_ISL_1423164, EPI_ISL_1423165, EPI_ISL_1423166, EPI_ISL_1423167, EPI_ISL_1423168, EPI_ISL_1423169, EPI_ISL_1423170, EPI_ISL_1423171, EPI_ISL_1423172, EPI_ISL_1423173, EPI_ISL_1423174, EPI_ISL_1423175, EPI_ISL_1423176, EPI_ISL_1423177, EPI_ISL_1423178, EPI_ISL_1423179, EPI_ISL_1423180, EPI_ISL_1423181, EPI_ISL_1423182, EPI_ISL_1423183, EPI_ISL_1423184, EPI_ISL_1423185, EPI_ISL_1423186, EPI_ISL_1423187, EPI_ISL_1423188, EPI_ISL_1423189, EPI_ISL_1423190, EPI_ISL_1423191, EPI_ISL_1423192, EPI_ISL_1423193, EPI_ISL_1423194, EPI_ISL_1423195, EPI_ISL_1423196, EPI_ISL_1423198, EPI_ISL_1423199, EPI_ISL_1423200 | | | |
| see above | WVU Rapid Development Laboratory | WVU and Marshall University Combined Genomics Core Facilities | "James Denvir, Peter Stoilov, Peter Perrotta, Wesley Kimble, Ryan Percifield" |
| EPI_ISL_1423203, EPI_ISL_1423204, EPI_ISL_1423205, EPI_ISL_1423207, EPI_ISL_1423208, EPI_ISL_1423210 | Berkeley Medical Center | WVU and Marshall University Combined Genomics Core Facilities | "James Denvir, Peter Stoilov, Peter Perrotta, Wesley Kimble, Ryan Percifield" |
| EPI_ISL_1423211, EPI_ISL_1423212 | QLabs | WVU and Marshall University Combined Genomics Core Facilities | "James Denvir, Peter Stoilov, Peter Perrotta, Wesley Kimble, Ryan Percifield" |
| EPI_ISL_1423213, EPI_ISL_1423214, EPI_ISL_1423215, EPI_ISL_1423216, EPI_ISL_1423217, EPI_ISL_1423218, EPI_ISL_1423219, EPI_ISL_1423220, EPI_ISL_1423223, EPI_ISL_1423225, EPI_ISL_1423227 | | | |
| see above | Berkeley Medical Center | WVU and Marshall University Combined Genomics Core Facilities | "James Denvir, Peter Stoilov, Peter Perrotta, Wesley Kimble, Ryan Percifield" |
| EPI_ISL_1423228, EPI_ISL_1423229, EPI_ISL_1423230, EPI_ISL_1423231, EPI_ISL_1423232, EPI_ISL_1423233, EPI_ISL_1423234, EPI_ISL_1423235, EPI_ISL_1423236, EPI_ISL_1423237, EPI_ISL_1423238, EPI_ISL_1423239, EPI_ISL_1423241, EPI_ISL_1423242, EPI_ISL_1423243, EPI_ISL_1423244, EPI_ISL_1423245 | | | |
| see above | J.W. Ruby Memorial Hospital | WVU and Marshall University Combined Genomics Core Facilities | "James Denvir, Peter Stoilov, Peter Perrotta, Wesley Kimble, Ryan Percifield" |
| EPI_ISL_1423246, EPI_ISL_1423247, EPI_ISL_1423248, EPI_ISL_1423249, EPI_ISL_1423250, EPI_ISL_1423251, EPI_ISL_1423252, EPI_ISL_1423253 | Berkeley Medical Center | WVU and Marshall University Combined Genomics Core Facilities | "James Denvir, Peter Stoilov, Peter Perrotta, Wesley Kimble, Ryan Percifield" |
| EPI_ISL_1423258, EPI_ISL_1423259, EPI_ISL_1423260, EPI_ISL_1423262, EPI_ISL_1423263, EPI_ISL_1423264, EPI_ISL_1423267, EPI_ISL_1423268, EPI_ISL_1423269, EPI_ISL_1423270, EPI_ISL_1423271, EPI_ISL_1423272, EPI_ISL_1423273 | | | |
| see above | J.W. Ruby Memorial Hospital | WVU and Marshall University Combined Genomics Core Facilities | "James Denvir, Peter Stoilov, Peter Perrotta, Wesley Kimble, Ryan Percifield" |
| EPI_ISL_1423275, EPI_ISL_1423276 | LabCorp Dublin | WVU and Marshall University Combined Genomics Core Facilities | "James Denvir, Peter Stoilov, Peter Perrotta, Wesley Kimble, Ryan Percifield" |
| EPI_ISL_1423277, EPI_ISL_1423278, EPI_ISL_1423279, EPI_ISL_1423282, EPI_ISL_1423284, EPI_ISL_1423285, EPI_ISL_1423286, EPI_ISL_1423288, EPI_ISL_1423289, EPI_ISL_1423290, EPI_ISL_1423291, EPI_ISL_1423292, EPI_ISL_1423294, EPI_ISL_1423295, EPI_ISL_1423296, EPI_ISL_1423297, EPI_ISL_1423298, EPI_ISL_1423299, EPI_ISL_1423300, EPI_ISL_1423302, EPI_ISL_1423303, EPI_ISL_1423304, EPI_ISL_1423305, EPI_ISL_1423306, EPI_ISL_1423307, EPI_ISL_1423308, EPI_ISL_1423309, EPI_ISL_1423310, EPI_ISL_1423311, EPI_ISL_1423313, EPI_ISL_1423314, EPI_ISL_1423315, EPI_ISL_1423316, EPI_ISL_1423318, EPI_ISL_1423319, EPI_ISL_1423320, EPI_ISL_1423321, EPI_ISL_1423323, EPI_ISL_1423324, EPI_ISL_1423325 | | | |
| see above | J.W. Ruby Memorial Hospital | WVU and Marshall University Combined Genomics Core Facilities | "James Denvir, Peter Stoilov, Peter Perrotta, Wesley Kimble, Ryan Percifield" |
| EPI_ISL_1423326, EPI_ISL_1423327, EPI_ISL_1423328, EPI_ISL_1423329, EPI_ISL_1423330, EPI_ISL_1423331, EPI_ISL_1423332, EPI_ISL_1423333, EPI_ISL_1423334, EPI_ISL_1423335, EPI_ISL_1423336, EPI_ISL_1423337, EPI_ISL_1423338, EPI_ISL_1423339, EPI_ISL_1423340 | | | |
| see above | Berkeley Medical Center | WVU and Marshall University Combined Genomics Core Facilities | "James Denvir, Peter Stoilov, Peter Perrotta, Wesley Kimble, Ryan Percifield" |
| EPI_ISL_1423341 | Grafton City Hospital | WVU and Marshall University Combined Genomics Core Facilities | "James Denvir, Peter Stoilov, Peter Perrotta, Wesley Kimble, Ryan Percifield" |
| EPI_ISL_1423343, EPI_ISL_1423345, EPI_ISL_1423347, EPI_ISL_1423348, EPI_ISL_1423350, EPI_ISL_1423351, EPI_ISL_1423352, EPI_ISL_1423353, EPI_ISL_1423354, EPI_ISL_1423355, EPI_ISL_1423356, EPI_ISL_1423357, EPI_ISL_1423358, EPI_ISL_1423359, EPI_ISL_1423360, EPI_ISL_1423365, EPI_ISL_1423369, EPI_ISL_1423370 | | | |
| see above | Wisconsin State Laboratory of Hygiene Communicable Disease Division | Ginkgo Bioworks Clinical Laboratory | Malaika Mckenzie-Bennett, James McGann, Jim Griffin, Keith Robison, Alex Plocik, Becky Schilling, Martha Pierson, Rebecca Littlefield, Michelle Spencer, Amanda Riccardi, Birgitte Simen, Kelsey R. Florek, Abigail C. Shockey |
| EPI_ISL_1423378, EPI_ISL_1423381, EPI_ISL_1423384, EPI_ISL_1423385, EPI_ISL_1423386, EPI_ISL_1423387, EPI_ISL_1423388, EPI_ISL_1423389, EPI_ISL_1423390, EPI_ISL_1423391, EPI_ISL_1423392, EPI_ISL_1423393, EPI_ISL_1423395, EPI_ISL_1423396, EPI_ISL_1423398, EPI_ISL_1423399, EPI_ISL_1423402, EPI_ISL_1423403, EPI_ISL_1423404, EPI_ISL_1423405, EPI_ISL_1423407, EPI_ISL_1423409, EPI_ISL_1423410, EPI_ISL_1423411, EPI_ISL_1423414, EPI_ISL_1423415, EPI_ISL_1423416, EPI_ISL_1423417, EPI_ISL_1423418, EPI_ISL_1423419, EPI_ISL_1423420, EPI_ISL_1423421, EPI_ISL_1423422, EPI_ISL_1423423, EPI_ISL_1423424, EPI_ISL_1423425, EPI_ISL_1423426, EPI_ISL_1423427, EPI_ISL_1423428, EPI_ISL_1423429, EPI_ISL_1423430, EPI_ISL_1423431, EPI_ISL_1423432, EPI_ISL_1423433, EPI_ISL_1423434, EPI_ISL_1423435, EPI_ISL_1423436, EPI_ISL_1423437, EPI_ISL_1423438, EPI_ISL_1423439, EPI_ISL_1423440, EPI_ISL_1423441, EPI_ISL_1423442, EPI_ISL_1423443, EPI_ISL_1423444, EPI_ISL_1423445, EPI_ISL_1423446, EPI_ISL_1423447, EPI_ISL_1423448, EPI_ISL_1423450, EPI_ISL_1423451, EPI_ISL_1423452, EPI_ISL_1423453, EPI_ISL_1423454, EPI_ISL_1423455, EPI_ISL_1423456, EPI_ISL_1423457, EPI_ISL_1423458, EPI_ISL_1423459, EPI_ISL_1423460, EPI_ISL_1423461, EPI_ISL_1423462, EPI_ISL_1423463, EPI_ISL_1423464, EPI_ISL_1423465, EPI_ISL_1423466, EPI_ISL_1423467, EPI_ISL_1423468, EPI_ISL_1423469, EPI_ISL_1423470, EPI_ISL_1423471, EPI_ISL_1423472, EPI_ISL_1423473, EPI_ISL_1423474, EPI_ISL_1423475, EPI_ISL_1423476, EPI_ISL_1423477, EPI_ISL_1423479, EPI_ISL_1423480, EPI_ISL_1423481, EPI_ISL_1423482, EPI_ISL_1423483, EPI_ISL_1423484, EPI_ISL_1423485, EPI_ISL_1423486, EPI_ISL_1423487, EPI_ISL_1423488, EPI_ISL_1423489, EPI_ISL_1423490, EPI_ISL_1423491, EPI_ISL_1423492, EPI_ISL_1423493, EPI_ISL_1423494, EPI_ISL_1423495, EPI_ISL_1423496, EPI_ISL_1423497, EPI_ISL_1423498, EPI_ISL_1423499, EPI_ISL_1423500, EPI_ISL_1423501, EPI_ISL_1423502, EPI_ISL_1423503 | | | |
| see above | NYU Langone Health | Departments of Pathology and Medicine, New York University School of Medicine | Adriana Heguy, Dacia Dimartino, Emily Guzman, Christian Marier, Peter Meyn, Sitharam Ramaswami, Gael Westby, Paul Zappile, Yutong Zhang, Paolo Cotzia, Guiqing Wang |
| EPI_ISL_1423505 | Unidad de Patologia Clinica | Instituto de Diagnostico y Referencia Epidemiologicos (INDRE) | Claudia Wong-Arambula, Abril Rodriguez-Maldonado, Vanessa Rivero-Arredondo, Ariadna Medina-Benitez, Joaquin Quiroz-Mercado, Sergio Rangel-Guerrero, Natividad Cruz-Ortiz, Tatiana Nunez-Garcia, Gisela Barrera-Badillo, Lucia Hernandez-Rivas, Irma Lopez-Martinez, Ernesto Ramirez-Gonzalez. |
| EPI_ISL_1423506, EPI_ISL_1423507, EPI_ISL_1423508, EPI_ISL_1423509 | LESP Chihuahua | Instituto de Diagnostico y Referencia Epidemiologicos (INDRE) | Claudia Wong-Arambula, Abril Rodriguez-Maldonado, Vanessa Rivero-Arredondo, Ariadna Medina-Benitez, Joaquin Quiroz-Mercado, Sergio Rangel-Guerrero, Natividad Cruz-Ortiz, Tatiana Nunez-Garcia, Gisela Barrera-Badillo, Lucia Hernandez-Rivas, Irma Lopez-Martinez, Ernesto Ramirez-Gonzalez. |
| EPI_ISL_1423510 | LESP Ciudad de Mexico | Instituto de Diagnostico y Referencia Epidemiologicos (INDRE) | Claudia Wong-Arambula, Abril Rodriguez-Maldonado, Vanessa Rivero-Arredondo, Ariadna Medina-Benitez, Joaquin Quiroz-Mercado, Sergio Rangel-Guerrero, Natividad Cruz-Ortiz, Tatiana Nunez-Garcia, Gisela Barrera-Badillo, Lucia Hernandez-Rivas, Irma Lopez-Martinez, Ernesto Ramirez-Gonzalez. |
| EPI_ISL_1423511 | LESP Tamaulipas | Instituto de Diagnostico y Referencia Epidemiologicos (INDRE) | Claudia Wong-Arambula, Abril Rodriguez-Maldonado, Vanessa Rivero-Arredondo, Ariadna Medina-Benitez, Joaquin Quiroz-Mercado, Sergio Rangel-Guerrero, Natividad Cruz-Ortiz, Tatiana Nunez-Garcia, Gisela Barrera-Badillo, Lucia Hernandez-Rivas, Irma Lopez-Martinez, Ernesto Ramirez-Gonzalez. |
| EPI_ISL_1423512 | LESP Chihuahua | Instituto de Diagnostico y Referencia Epidemiologicos (INDRE) | Claudia Wong-Arambula, Abril Rodriguez-Maldonado, Vanessa Rivero-Arredondo, Ariadna Medina-Benitez, Joaquin Quiroz-Mercado, Sergio Rangel-Guerrero, Natividad Cruz-Ortiz, Tatiana Nunez-Garcia, Gisela Barrera-Badillo, Lucia Hernandez-Rivas, Irma Lopez-Martinez, Ernesto Ramirez-Gonzalez. |
| EPI_ISL_1423513 | LESP Queretaro | Instituto de Diagnostico y Referencia Epidemiologicos (INDRE) | Claudia Wong-Arambula, Abril Rodriguez-Maldonado, Vanessa Rivero-Arredondo, Ariadna Medina-Benitez, Joaquin Quiroz-Mercado, Sergio Rangel-Guerrero, Natividad Cruz-Ortiz, Tatiana Nunez-Garcia, Gisela Barrera-Badillo, Lucia Hernandez-Rivas, Irma Lopez-Martinez, Ernesto Ramirez-Gonzalez. |
| EPI_ISL_1423514 | LESP Yucatan | Instituto de Diagnostico y Referencia Epidemiologicos (INDRE) | Claudia Wong-Arambula, Abril Rodriguez-Maldonado, Vanessa Rivero-Arredondo, Ariadna Medina-Benitez, Joaquin Quiroz-Mercado, Sergio Rangel-Guerrero, Natividad Cruz-Ortiz, Tatiana Nunez-Garcia, Gisela Barrera-Badillo, Lucia Hernandez-Rivas, Irma Lopez-Martinez, Ernesto Ramirez-Gonzalez. |
| EPI_ISL_1423516, EPI_ISL_1423517, EPI_ISL_1423518, EPI_ISL_1423519, EPI_ISL_1423520, EPI_ISL_1423521, EPI_ISL_1423522, EPI_ISL_1423523, EPI_ISL_1423524, EPI_ISL_1423525, EPI_ISL_1423526, EPI_ISL_1423527, EPI_ISL_1423528, EPI_ISL_1423529, EPI_ISL_1423530, EPI_ISL_1423531, EPI_ISL_1423532, EPI_ISL_1423533 | | | |
| see above | Minnesota Department of Health, Public Health Laboratory | Minnesota Department of Health, Public Health Laboratory | Alexandra Lorentz, Jacob Garfin, Matt Plumb, and Xiong Wang |

| | | | |
|---|---|---|---|
| EPI_ISL_1423534, EPI_ISL_1423535, EPI_ISL_1423536, EPI_ISL_1423537, EPI_ISL_1423538, EPI_ISL_1423539, EPI_ISL_1423540, EPI_ISL_1423541, EPI_ISL_1423542, EPI_ISL_1423543, EPI_ISL_1423544, EPI_ISL_1423545, EPI_ISL_1423546, EPI_ISL_1423547, EPI_ISL_1423548, EPI_ISL_1423549, EPI_ISL_1423550, EPI_ISL_1423551, EPI_ISL_1423552, EPI_ISL_1423553, EPI_ISL_1423554, EPI_ISL_1423555, EPI_ISL_1423556, EPI_ISL_1423557, EPI_ISL_1423558, EPI_ISL_1423559, EPI_ISL_1423560, EPI_ISL_1423561, EPI_ISL_1423562, EPI_ISL_1423563, EPI_ISL_1423564, EPI_ISL_1423565, EPI_ISL_1423566, EPI_ISL_1423567, EPI_ISL_1423568, EPI_ISL_1423569, EPI_ISL_1423570, EPI_ISL_1423571, EPI_ISL_1423572, EPI_ISL_1423573, EPI_ISL_1423574, EPI_ISL_1423575, EPI_ISL_1423576, EPI_ISL_1423577, EPI_ISL_1423578, EPI_ISL_1423579, EPI_ISL_1423580, EPI_ISL_1423581, EPI_ISL_1423582, EPI_ISL_1423583, EPI_ISL_1423584, EPI_ISL_1423585, EPI_ISL_1423586, EPI_ISL_1423587, EPI_ISL_1423588, EPI_ISL_1423589, EPI_ISL_1423590, EPI_ISL_1423591, EPI_ISL_1423592 | | | |
| see above | M Health Fairview | Minnesota Department of Health, Public Health Laboratory | Alexandra Lorentz, Jacob Garfin, Matt Plumb, and Xiong Wang |
| EPI_ISL_1423934, EPI_ISL_1423935, EPI_ISL_1423936, EPI_ISL_1423937, EPI_ISL_1423938, EPI_ISL_1423939, EPI_ISL_1423940, EPI_ISL_1423941, EPI_ISL_1423944, EPI_ISL_1423945, EPI_ISL_1423947, EPI_ISL_1423948, EPI_ISL_1423949, EPI_ISL_1423950, EPI_ISL_1423951, EPI_ISL_1423952, EPI_ISL_1423956, EPI_ISL_1423957, EPI_ISL_1423958, EPI_ISL_1423959, EPI_ISL_1423960, EPI_ISL_1423961, EPI_ISL_1423962, EPI_ISL_1423964, EPI_ISL_1423965, EPI_ISL_1423966, EPI_ISL_1423967, EPI_ISL_1423968, EPI_ISL_1423969, EPI_ISL_1423971, EPI_ISL_1423972, EPI_ISL_1423973, EPI_ISL_1423974, EPI_ISL_1423975, EPI_ISL_1423976 | | | |
| see above | Arizona State University | Arizona State University | Peter T. Skidmore, LaRinda A. Holland, Rabia Maqsood, Emily A. Kaelin, Nicholas J. Mellor, Kristina Buss, Joy M. Blain, Neal W Woodbury, Valerie Harris, Joshua LaBaer, Vel Murugan, Efrem S. Lim |
| EPI_ISL_1423977, EPI_ISL_1423978, EPI_ISL_1423979, EPI_ISL_1423980, EPI_ISL_1423981, EPI_ISL_1423982, EPI_ISL_1423983 | Lafene Health Center | Kansas State Veterinary Diagnostic Laboratory | Tyler Doerksen, Andreea Lu, Lance Noll, Elizabeth Porter, Kasey Hogan, Emily Cox, Jianfa Bai, Jamie Henningson, Rachel Palinski |
| EPI_ISL_1423984, EPI_ISL_1423985, EPI_ISL_1423986, EPI_ISL_1423987, EPI_ISL_1423988, EPI_ISL_1423989, EPI_ISL_1423990, EPI_ISL_1423991, EPI_ISL_1423992, EPI_ISL_1423993 | Oregon State University TRACE Project | OSU Center for Genome Research and Biocomputing | Oregon State University TRACE Project |
| EPI_ISL_1423994, EPI_ISL_1423995, EPI_ISL_1423996, EPI_ISL_1423997, EPI_ISL_1423998, EPI_ISL_1423999, EPI_ISL_1424000, EPI_ISL_1424001 | Instituto Nacional de Medicina Genomica | Instituto Nacional de Enfermedades Respiratorias (INER), Centro de Investigación en Enfermedades Infecciosas (CIENI) | Matias-Florentino M, Hidalgo-Miranda A, Mendoza-Vargas A, Reyes-Grajeda JP, Cedro-Tanda A, Arriaga-Canon C, Herrera-Montalvo LA, Reyes-Teran G, Avila-Rios S, Boukadida C |
| EPI_ISL_1424007 | LESP Yucatan | Instituto de Diagnostico y Referencia Epidemiologicos (INDRE) | Claudia Wong-Arambula, Abril Rodriguez-Maldonado, Vanessa Rivero-Arredondo, Ariadna Medina-Benitez, Joaquin Quiroz-Mercado, Sergio Rangel-Guerrero, Natividad Cruz-Ortiz, Tatiana Nunez-Garcia, Gisela Barrera-Badillo, Lucia Hernandez-Rivas, Irma Lopez-Martinez, Ernesto Ramirez-Gonzalez. |
| EPI_ISL_1424008, EPI_ISL_1424009 | LESP Hidalgo | Instituto de Diagnostico y Referencia Epidemiologicos (INDRE) | Claudia Wong-Arambula, Abril Rodriguez-Maldonado, Vanessa Rivero-Arredondo, Ariadna Medina-Benitez, Joaquin Quiroz-Mercado, Sergio Rangel-Guerrero, Natividad Cruz-Ortiz, Tatiana Nunez-Garcia, Gisela Barrera-Badillo, Lucia Hernandez-Rivas, Irma Lopez-Martinez, Ernesto Ramirez-Gonzalez. |
| EPI_ISL_1424010, EPI_ISL_1424011, EPI_ISL_1424012 | LESP Guanajuato | Instituto de Diagnostico y Referencia Epidemiologicos (INDRE) | Claudia Wong-Arambula, Abril Rodriguez-Maldonado, Vanessa Rivero-Arredondo, Ariadna Medina-Benitez, Joaquin Quiroz-Mercado, Sergio Rangel-Guerrero, Natividad Cruz-Ortiz, Tatiana Nunez-Garcia, Gisela Barrera-Badillo, Lucia Hernandez-Rivas, Irma Lopez-Martinez, Ernesto Ramirez-Gonzalez. |
| EPI_ISL_1424013 | LESP Chiapas | Instituto de Diagnostico y Referencia Epidemiologicos (INDRE) | Claudia Wong-Arambula, Abril Rodriguez-Maldonado, Vanessa Rivero-Arredondo, Ariadna Medina-Benitez, Joaquin Quiroz-Mercado, Sergio Rangel-Guerrero, Natividad Cruz-Ortiz, Tatiana Nunez-Garcia, Gisela Barrera-Badillo, Lucia Hernandez-Rivas, Irma Lopez-Martinez, Ernesto Ramirez-Gonzalez. |
| EPI_ISL_1424014 | LESP Yucatan | Instituto de Diagnostico y Referencia Epidemiologicos (INDRE) | Claudia Wong-Arambula, Abril Rodriguez-Maldonado, Vanessa Rivero-Arredondo, Ariadna Medina-Benitez, Joaquin Quiroz-Mercado, Sergio Rangel-Guerrero, Natividad Cruz-Ortiz, Tatiana Nunez-Garcia, Gisela Barrera-Badillo, Lucia Hernandez-Rivas, Irma Lopez-Martinez, Ernesto Ramirez-Gonzalez. |
| EPI_ISL_1424015 | LESP Campeche | Instituto de Diagnostico y Referencia Epidemiologicos (INDRE) | Claudia Wong-Arambula, Abril Rodriguez-Maldonado, Vanessa Rivero-Arredondo, Ariadna Medina-Benitez, Joaquin Quiroz-Mercado, Sergio Rangel-Guerrero, Natividad Cruz-Ortiz, Tatiana Nunez-Garcia, Gisela Barrera-Badillo, Lucia Hernandez-Rivas, Irma Lopez-Martinez, Ernesto Ramirez-Gonzalez. |
| EPI_ISL_1424016 | LESP Queretaro | Instituto de Diagnostico y Referencia Epidemiologicos (INDRE) | Claudia Wong-Arambula, Abril Rodriguez-Maldonado, Vanessa Rivero-Arredondo, Ariadna Medina-Benitez, Joaquin Quiroz-Mercado, Sergio Rangel-Guerrero, Natividad Cruz-Ortiz, Tatiana Nunez-Garcia, Gisela Barrera-Badillo, Lucia Hernandez-Rivas, Irma Lopez-Martinez, Ernesto Ramirez-Gonzalez. |
| EPI_ISL_1424017 | LESP Morelos | Instituto de Diagnostico y Referencia Epidemiologicos (INDRE) | Claudia Wong-Arambula, Abril Rodriguez-Maldonado, Vanessa Rivero-Arredondo, Ariadna Medina-Benitez, Joaquin Quiroz-Mercado, Sergio Rangel-Guerrero, Natividad Cruz-Ortiz, Tatiana Nunez-Garcia, Gisela Barrera-Badillo, Lucia Hernandez-Rivas, Irma Lopez-Martinez, Ernesto Ramirez-Gonzalez. |
| EPI_ISL_1424018 | LESP Veracruz | Instituto de Diagnostico y Referencia Epidemiologicos (INDRE) | Claudia Wong-Arambula, Abril Rodriguez-Maldonado, Vanessa Rivero-Arredondo, Ariadna Medina-Benitez, Joaquin Quiroz-Mercado, Sergio Rangel-Guerrero, Natividad Cruz-Ortiz, Tatiana Nunez-Garcia, Gisela Barrera-Badillo, Lucia Hernandez-Rivas, Irma Lopez-Martinez, Ernesto Ramirez-Gonzalez. |
| EPI_ISL_1424019 | LESP Morelos | Instituto de Diagnostico y Referencia Epidemiologicos (INDRE) | Claudia Wong-Arambula, Abril Rodriguez-Maldonado, Vanessa Rivero-Arredondo, Ariadna Medina-Benitez, Joaquin Quiroz-Mercado, Sergio Rangel-Guerrero, Natividad Cruz-Ortiz, Tatiana Nunez-Garcia, Gisela Barrera-Badillo, Lucia Hernandez-Rivas, Irma Lopez-Martinez, Ernesto Ramirez-Gonzalez. |
| EPI_ISL_1424020 | LESP Chiapas | Instituto de Diagnostico y Referencia Epidemiologicos (INDRE) | Claudia Wong-Arambula, Abril Rodriguez-Maldonado, Vanessa Rivero-Arredondo, Ariadna Medina-Benitez, Joaquin Quiroz-Mercado, Sergio Rangel-Guerrero, Natividad Cruz-Ortiz, Tatiana Nunez-Garcia, Gisela Barrera-Badillo, Lucia Hernandez-Rivas, Irma Lopez-Martinez, Ernesto Ramirez-Gonzalez. |
| EPI_ISL_1424021, EPI_ISL_1424022, EPI_ISL_1424023, EPI_ISL_1424024, EPI_ISL_1424025, EPI_ISL_1424026, EPI_ISL_1424027, EPI_ISL_1424028, EPI_ISL_1424029, EPI_ISL_1424030, EPI_ISL_1424031, EPI_ISL_1424032, EPI_ISL_1424033 | | | |
| see above | LESP Queretaro | Instituto de Diagnostico y Referencia Epidemiologicos (INDRE) | Claudia Wong-Arambula, Abril Rodriguez-Maldonado, Vanessa Rivero-Arredondo, Ariadna Medina-Benitez, Joaquin Quiroz-Mercado, Sergio Rangel-Guerrero, Natividad Cruz-Ortiz, Tatiana Nunez-Garcia, Gisela Barrera-Badillo, Lucia Hernandez-Rivas, Irma Lopez-Martinez, Ernesto Ramirez-Gonzalez. |
| EPI_ISL_1424034 | LESP Guanajuato | Instituto de Diagnostico y Referencia Epidemiologicos (INDRE) | Claudia Wong-Arambula, Abril Rodriguez-Maldonado, Vanessa Rivero-Arredondo, Ariadna Medina-Benitez, Joaquin Quiroz-Mercado, Sergio Rangel-Guerrero, Natividad Cruz-Ortiz, Tatiana Nunez-Garcia, Gisela Barrera-Badillo, Lucia Hernandez-Rivas, Irma Lopez-Martinez, Ernesto Ramirez-Gonzalez. |
| EPI_ISL_1424035, EPI_ISL_1424036, EPI_ISL_1424037, EPI_ISL_1424038 | LESP Yucatan | Instituto de Diagnostico y Referencia Epidemiologicos (INDRE) | Claudia Wong-Arambula, Abril Rodriguez-Maldonado, Vanessa Rivero-Arredondo, Ariadna Medina-Benitez, Joaquin Quiroz-Mercado, Sergio Rangel-Guerrero, Natividad Cruz-Ortiz, Tatiana Nunez-Garcia, Gisela Barrera-Badillo, Lucia Hernandez-Rivas, Irma Lopez-Martinez, Ernesto Ramirez-Gonzalez. |
| EPI_ISL_1424039, EPI_ISL_1424040, EPI_ISL_1424041 | LESP Tamaulipas | Instituto de Diagnostico y Referencia Epidemiologicos (INDRE) | Claudia Wong-Arambula, Abril Rodriguez-Maldonado, Vanessa Rivero-Arredondo, Ariadna Medina-Benitez, Joaquin Quiroz-Mercado, Sergio Rangel-Guerrero, Natividad Cruz-Ortiz, Tatiana Nunez-Garcia, Gisela Barrera-Badillo, Lucia Hernandez-Rivas, Irma Lopez-Martinez, Ernesto Ramirez-Gonzalez. |
| EPI_ISL_1424042 | LESP Morelos | Instituto de Diagnostico y Referencia Epidemiologicos (INDRE) | Claudia Wong-Arambula, Abril Rodriguez-Maldonado, Vanessa Rivero-Arredondo, Ariadna Medina-Benitez, Joaquin Quiroz-Mercado, Sergio Rangel-Guerrero, Natividad Cruz-Ortiz, Tatiana Nunez-Garcia, Gisela Barrera-Badillo, Lucia Hernandez-Rivas, Irma Lopez-Martinez, Ernesto Ramirez-Gonzalez. |
| EPI_ISL_1424043 | LESP Guerrero | Instituto de Diagnostico y Referencia Epidemiologicos (INDRE) | Claudia Wong-Arambula, Abril Rodriguez-Maldonado, Vanessa Rivero-Arredondo, Ariadna Medina-Benitez, Joaquin Quiroz-Mercado, Sergio Rangel-Guerrero, Natividad Cruz-Ortiz, Tatiana Nunez-Garcia, Gisela Barrera-Badillo, Lucia Hernandez-Rivas, Irma Lopez-Martinez, Ernesto Ramirez-Gonzalez. |
| EPI_ISL_1424044, EPI_ISL_1424045 | LESP Coahuila | Instituto de Diagnostico y Referencia Epidemiologicos (INDRE) | Claudia Wong-Arambula, Abril Rodriguez-Maldonado, Vanessa Rivero-Arredondo, Ariadna Medina-Benitez, Joaquin Quiroz-Mercado, Sergio Rangel-Guerrero, Natividad Cruz-Ortiz, Tatiana Nunez-Garcia, Gisela Barrera-Badillo, Lucia Hernandez-Rivas, Irma Lopez-Martinez, Ernesto Ramirez-Gonzalez. |
| EPI_ISL_1424046, EPI_ISL_1424047, EPI_ISL_1424048, EPI_ISL_1424049 | LESP Veracruz | Instituto de Diagnostico y Referencia Epidemiologicos (INDRE) | Claudia Wong-Arambula, Abril Rodriguez-Maldonado, Vanessa Rivero-Arredondo, Ariadna Medina-Benitez, Joaquin Quiroz-Mercado, Sergio Rangel-Guerrero, Natividad Cruz-Ortiz, Tatiana Nunez-Garcia, Gisela Barrera-Badillo, Lucia Hernandez-Rivas, Irma Lopez-Martinez, Ernesto Ramirez-Gonzalez. |
| EPI_ISL_1424050 | LESP Baja California | Instituto de Diagnostico y Referencia Epidemiologicos (INDRE) | Claudia Wong-Arambula, Abril Rodriguez-Maldonado, Vanessa Rivero-Arredondo, Ariadna Medina-Benitez, Joaquin Quiroz-Mercado, Sergio Rangel-Guerrero, Natividad Cruz-Ortiz, Tatiana Nunez-Garcia, Gisela Barrera-Badillo, Lucia Hernandez-Rivas, Irma Lopez-Martinez, Ernesto Ramirez-Gonzalez. |
| EPI_ISL_1424051, EPI_ISL_1424052, EPI_ISL_1424053 | LESP Ciudad de Mexico | Instituto de Diagnostico y Referencia Epidemiologicos (INDRE) | Claudia Wong-Arambula, Abril Rodriguez-Maldonado, Vanessa Rivero-Arredondo, Ariadna Medina-Benitez, Joaquin Quiroz-Mercado, Sergio Rangel-Guerrero, Natividad Cruz-Ortiz, Tatiana Nunez-Garcia, Gisela Barrera-Badillo, Lucia Hernandez-Rivas, Irma Lopez-Martinez, Ernesto Ramirez-Gonzalez. |
| EPI_ISL_1424066, EPI_ISL_1424067, EPI_ISL_1424068, EPI_ISL_1424069, EPI_ISL_1424070, EPI_ISL_1424071, EPI_ISL_1424072, EPI_ISL_1424073, EPI_ISL_1424074, EPI_ISL_1424075, EPI_ISL_1424076, EPI_ISL_1424077, EPI_ISL_1424078, EPI_ISL_1424079, EPI_ISL_1424080 | | | |
| see above | Stanford Health Care | Stanford University School of Medicine, Clinical Virology Laboratory | Michelle Verghese, ChunHong Huang, Mamdouh Sibai, Daniel Solis, Fumiko Yamamoto, Malaya K. Sahoo, James Zehnder, and Benjamin A. Pinsky |
| EPI_ISL_1424081, EPI_ISL_1424082, EPI_ISL_1424083, EPI_ISL_1424084, EPI_ISL_1424085, EPI_ISL_1424086, EPI_ISL_1424087, EPI_ISL_1424088, EPI_ISL_1424089, EPI_ISL_1424090 | Stanford Health Care | Stanford University School of Medicine, Clinical Virology Laboratory | Malaya K. Sahoo, Fumiko Yamamoto, Mamdouh Sibai, Daniel Solis, Michelle Verghese, ChunHong Huang, James Zehnder, and Benjamin A. Pinsky |

| | | | |
|---|---|---|---|
| EPI_ISL_1424091, EPI_ISL_1424092, EPI_ISL_1424093, EPI_ISL_1424094, EPI_ISL_1424095, EPI_ISL_1424096, EPI_ISL_1424097, EPI_ISL_1424098, EPI_ISL_1424099, EPI_ISL_1424100, EPI_ISL_1424101 | | | |
| see above | Stanford Health Care | Stanford University School of Medicine, Clinical Virology Laboratory | Fumiko Yamamoto, Malaya K. Sahoo, Mamdouh Sibai, Daniel Solis, Michelle Verghese, ChunHong Huang, James Zehnder, and Benjamin A. Pinsky |
| EPI_ISL_1424102, EPI_ISL_1424103, EPI_ISL_1424104, EPI_ISL_1424105, EPI_ISL_1424106, EPI_ISL_1424107, EPI_ISL_1424108, EPI_ISL_1424109, EPI_ISL_1424110, EPI_ISL_1424111 | Stanford Health Care | Stanford University School of Medicine, Clinical Virology Laboratory | Daniel Solis, Mamdouh Sibai, Fumiko Yamamoto, Becky Jiang, Michelle Verghese, ChunHong Huang, James Zehnder, Malaya K. Sahoo, and Benjamin A. Pinsky |
| EPI_ISL_1424112, EPI_ISL_1424113, EPI_ISL_1424114, EPI_ISL_1424115, EPI_ISL_1424116, EPI_ISL_1424117, EPI_ISL_1424118, EPI_ISL_1424119, EPI_ISL_1424120, EPI_ISL_1424121 | Stanford Health Care | Stanford University School of Medicine, Clinical Virology Laboratory | Mamdouh Sibai, Daniel Solis, Becky Jiang, Fumiko Yamamoto, Michelle Verghese, ChunHong Huang, James Zehnder, Malaya K. Sahoo, and Benjamin A. Pinsky |
| EPI_ISL_1424122, EPI_ISL_1424123, EPI_ISL_1424124, EPI_ISL_1424125, EPI_ISL_1424126, EPI_ISL_1424127, EPI_ISL_1424128, EPI_ISL_1424129 | Stanford Health Care | Stanford University School of Medicine, Clinical Virology Laboratory | Becky Jiang, Mamdouh Sibai, Daniel Solis, Fumiko Yamamoto, Michelle Verghese, ChunHong Huang, James Zehnder, Malaya K. Sahoo, and Benjamin A. Pinsky |
| EPI_ISL_1424131, EPI_ISL_1424132, EPI_ISL_1424133, EPI_ISL_1424134, EPI_ISL_1424135, EPI_ISL_1424136, EPI_ISL_1424138, EPI_ISL_1424139, EPI_ISL_1424140, EPI_ISL_1424141, EPI_ISL_1424142 | | | |
| see above | Oregon State Public Health Laboratory | Oregon State Public Health Laboratory | LaDonna Grenz, Karim Morey, Joel R. Sevinksy, Kevin, G. Libuit, John Fontana, Shane Sevey |
| EPI_ISL_1424478, EPI_ISL_1424479, EPI_ISL_1424480, EPI_ISL_1424481, EPI_ISL_1424482, EPI_ISL_1424483, EPI_ISL_1424484, EPI_ISL_1424485, EPI_ISL_1424486, EPI_ISL_1424487, EPI_ISL_1424488, EPI_ISL_1424489, EPI_ISL_1424490, EPI_ISL_1424491, EPI_ISL_1424492, EPI_ISL_1424493, EPI_ISL_1424494, EPI_ISL_1424495, EPI_ISL_1424496, EPI_ISL_1424497, EPI_ISL_1424498, EPI_ISL_1424499, EPI_ISL_1424500, EPI_ISL_1424501, EPI_ISL_1424502 | | | |
| see above | COVID Outreach Wood River | UW Virology Lab | Pavitra Roychoudhury, Hong Xie, Lasata Shrestha, Shah Mohamed Bakhash, Michelle Lin, Tom Archie, Ryan Cole, Meei-Li Huang, Keith R. Jerome, Alexander Greninger |
| EPI_ISL_1443199, EPI_ISL_1443200, EPI_ISL_1443201, EPI_ISL_1443202, EPI_ISL_1443203, EPI_ISL_1443204, EPI_ISL_1443205, EPI_ISL_1443206, EPI_ISL_1443207, EPI_ISL_1443208, EPI_ISL_1443209, EPI_ISL_1443210, EPI_ISL_1443211, EPI_ISL_1443212, EPI_ISL_1443213, EPI_ISL_1443214, EPI_ISL_1443215, EPI_ISL_1443216, EPI_ISL_1443217, EPI_ISL_1443219, EPI_ISL_1443220, EPI_ISL_1443221, EPI_ISL_1443222, EPI_ISL_1443223, EPI_ISL_1443224, EPI_ISL_1443225, EPI_ISL_1443226, EPI_ISL_1443227, EPI_ISL_1443229, EPI_ISL_1443231, EPI_ISL_1443233, EPI_ISL_1443234, EPI_ISL_1443235, EPI_ISL_1443236, EPI_ISL_1443237, EPI_ISL_1443238, EPI_ISL_1443239, EPI_ISL_1443241, EPI_ISL_1443242, EPI_ISL_1443244, EPI_ISL_1443245, EPI_ISL_1443246, EPI_ISL_1443247, EPI_ISL_1443248, EPI_ISL_1443249, EPI_ISL_1443250, EPI_ISL_1443251, EPI_ISL_1443252, EPI_ISL_1443254, EPI_ISL_1443255, EPI_ISL_1443256, EPI_ISL_1443257, EPI_ISL_1443258, EPI_ISL_1443259, EPI_ISL_1443260, EPI_ISL_1443261, EPI_ISL_1443262, EPI_ISL_1443263, EPI_ISL_1443264, EPI_ISL_1443265, EPI_ISL_1443266, EPI_ISL_1443267, EPI_ISL_1443268, EPI_ISL_1443269, EPI_ISL_1443270, EPI_ISL_1443271, EPI_ISL_1443272, EPI_ISL_1443273, EPI_ISL_1443274, EPI_ISL_1443275, EPI_ISL_1443276, EPI_ISL_1443277, EPI_ISL_1443278, EPI_ISL_1443279, EPI_ISL_1443280, EPI_ISL_1443281, EPI_ISL_1443282, EPI_ISL_1443283, EPI_ISL_1443284, EPI_ISL_1443285, EPI_ISL_1443286, EPI_ISL_1443287, EPI_ISL_1443288, EPI_ISL_1443289, EPI_ISL_1443290, EPI_ISL_1443291, EPI_ISL_1443292 | | | |
| see above | Illinois Department of Public Health - Springfield Lab | Illinois Department of Public Health - Springfield Lab | Bryan Sim, Gordon McCall |
| EPI_ISL_1443293, EPI_ISL_1443294, EPI_ISL_1443295, EPI_ISL_1443296, EPI_ISL_1443297, EPI_ISL_1443298, EPI_ISL_1443299, EPI_ISL_1443300, EPI_ISL_1443301, EPI_ISL_1443302, EPI_ISL_1443303, EPI_ISL_1443304, EPI_ISL_1443305, EPI_ISL_1443306, EPI_ISL_1443307, EPI_ISL_1443308, EPI_ISL_1443309, EPI_ISL_1443310, EPI_ISL_1443311, EPI_ISL_1443312, EPI_ISL_1443313, EPI_ISL_1443314, EPI_ISL_1443315, EPI_ISL_1443316, EPI_ISL_1443317, EPI_ISL_1443318, EPI_ISL_1443319, EPI_ISL_1443320, EPI_ISL_1443321, EPI_ISL_1443322, EPI_ISL_1443323, EPI_ISL_1443324, EPI_ISL_1443325, EPI_ISL_1443326, EPI_ISL_1443327, EPI_ISL_1443328, EPI_ISL_1443329, EPI_ISL_1443330, EPI_ISL_1443331, EPI_ISL_1443332, EPI_ISL_1443333, EPI_ISL_1443334, EPI_ISL_1443335, EPI_ISL_1443336 | | | |
| see above | Clinical Molecular Microbiology Laboratory, UNC Hospital | Dirk Dittmer | Justin T. Landis , Razia Moorad , Brent A. Eason, Melissa B. Miller, Linda Pluta, Dirk Dittmer, Angelica Juarez, Cecilia Thompson, Shawn Hawken, Cameroon Grant, Evelyn Hoffman, Patricio Cano, Jason Wong, Carolina Caro-Vegas, Ryan McNamara, Blossom Damania. |
| EPI_ISL_1443344, EPI_ISL_1443512, EPI_ISL_1443601 | Baylor Esoteric and Molecular Lab | Baylor Esoteric and Molecular Lab | Elizabeth Forrester and Dawn Richards |
| EPI_ISL_1443602, EPI_ISL_1443603, EPI_ISL_1443605, EPI_ISL_1443606, EPI_ISL_1443607, EPI_ISL_1443608, EPI_ISL_1443609, EPI_ISL_1443610, EPI_ISL_1443611, EPI_ISL_1443612, EPI_ISL_1443613, EPI_ISL_1443615, EPI_ISL_1443616, EPI_ISL_1443617, EPI_ISL_1443618, EPI_ISL_1443619, EPI_ISL_1443620, EPI_ISL_1443621, EPI_ISL_1443622, EPI_ISL_1443623, EPI_ISL_1443624, EPI_ISL_1443625, EPI_ISL_1443626, EPI_ISL_1443627, EPI_ISL_1443628, EPI_ISL_1443629, EPI_ISL_1443630, EPI_ISL_1443631, EPI_ISL_1443632, EPI_ISL_1443634, EPI_ISL_1443635, EPI_ISL_1443636, EPI_ISL_1443637, EPI_ISL_1443638, EPI_ISL_1443639, EPI_ISL_1443640 | | | |
| see above | Wyoming Public Health Laboratory | Wyoming Public Health Laboratory | Jim Mildenberger, Wanda Manley, Noah Hull, Taylor Fearing, Lynette Gumbleton, Channing Weber, Ashley Norberg, Chayse Rowley, Marley Goetz, Brian Dominguez, Elliot Thomasson, Cari Sloma, and Rob Christensen |
| EPI_ISL_1443756, EPI_ISL_1443757, EPI_ISL_1443758, EPI_ISL_1443759, EPI_ISL_1443760, EPI_ISL_1443761, EPI_ISL_1443762, EPI_ISL_1443763, EPI_ISL_1443764, EPI_ISL_1443765, EPI_ISL_1443766, EPI_ISL_1443767, EPI_ISL_1443768, EPI_ISL_1443769, EPI_ISL_1443770, EPI_ISL_1443771, EPI_ISL_1443772, EPI_ISL_1443773, EPI_ISL_1443774, EPI_ISL_1443775, EPI_ISL_1443776, EPI_ISL_1443777, EPI_ISL_1443778 | | | |
| see above | Kansas Health and Environmental Lab | Kansas Health and Environmental Lab | Mike Grose, Jonathan Barnell, Ben Olsen, and Phil Adam |
| EPI_ISL_1443779, EPI_ISL_1443780, EPI_ISL_1443781, EPI_ISL_1443782, EPI_ISL_1443783, EPI_ISL_1443784, EPI_ISL_1443786, EPI_ISL_1443787, EPI_ISL_1443788, EPI_ISL_1443789, EPI_ISL_1443790, EPI_ISL_1443793, EPI_ISL_1443794, EPI_ISL_1443795, EPI_ISL_1443796, EPI_ISL_1443797, EPI_ISL_1443798, EPI_ISL_1443799, EPI_ISL_1443800, EPI_ISL_1443801 | | | |
| see above | Arizona State Public Health Laboratory | Arizona State Public Health Laboratory | Trung Huynh, Jessica Escobar, Katherine Fullerton, Nobuko Fukushima, Stacy White, Linda Getsinger, Victor Waddell |
| EPI_ISL_1443802, EPI_ISL_1443803, EPI_ISL_1443804, EPI_ISL_1443805, EPI_ISL_1443806, EPI_ISL_1443807, EPI_ISL_1443808, EPI_ISL_1443809, EPI_ISL_1443810, EPI_ISL_1443811, EPI_ISL_1443812, EPI_ISL_1443813, EPI_ISL_1443814, EPI_ISL_1443815, EPI_ISL_1443816, EPI_ISL_1443817, EPI_ISL_1443818 | | | |
| see above | GA Department of Public Health | GA Department of Public Health | Stacy Reeves, Jonathan Edwards, Cynthia Dixey, Tonia Parrott, Aliyah Fields, Taylor Smith |
| EPI_ISL_1443819, EPI_ISL_1443820, EPI_ISL_1443821, EPI_ISL_1443822, EPI_ISL_1443823, EPI_ISL_1443824, EPI_ISL_1443825, EPI_ISL_1443826, EPI_ISL_1443827, EPI_ISL_1443828, EPI_ISL_1443829, EPI_ISL_1443830, EPI_ISL_1443831, EPI_ISL_1443832, EPI_ISL_1443833, EPI_ISL_1443834, EPI_ISL_1443835, EPI_ISL_1443836, EPI_ISL_1443837, EPI_ISL_1443839, EPI_ISL_1443840, EPI_ISL_1443842, EPI_ISL_1443843, EPI_ISL_1443844, EPI_ISL_1443845, EPI_ISL_1443846, EPI_ISL_1443847, EPI_ISL_1443848, EPI_ISL_1443849, EPI_ISL_1443850, EPI_ISL_1443851, EPI_ISL_1443852, EPI_ISL_1443853, EPI_ISL_1443854, EPI_ISL_1443855, EPI_ISL_1443856, EPI_ISL_1443857, EPI_ISL_1443859, EPI_ISL_1443860, EPI_ISL_1443861, EPI_ISL_1443862, EPI_ISL_1443863, EPI_ISL_1443864, EPI_ISL_1443865, EPI_ISL_1443866, EPI_ISL_1443867, EPI_ISL_1443868, EPI_ISL_1443911, EPI_ISL_1443912, EPI_ISL_1443913, EPI_ISL_1443914, EPI_ISL_1443915, EPI_ISL_1443916, EPI_ISL_1443917, EPI_ISL_1443918, EPI_ISL_1443919, EPI_ISL_1443920, EPI_ISL_1443922, EPI_ISL_1443923, EPI_ISL_1443924, EPI_ISL_1443925, EPI_ISL_1443926, EPI_ISL_1443927, EPI_ISL_1443928, EPI_ISL_1443929, EPI_ISL_1443930, EPI_ISL_1443931, EPI_ISL_1443932, EPI_ISL_1443933, EPI_ISL_1443934, EPI_ISL_1443935, EPI_ISL_1443936, EPI_ISL_1443937, EPI_ISL_1443938, EPI_ISL_1443939, EPI_ISL_1443940, EPI_ISL_1443941, EPI_ISL_1443942, EPI_ISL_1443943, EPI_ISL_1443944, EPI_ISL_1443945, EPI_ISL_1443946, EPI_ISL_1443947, EPI_ISL_1443948, EPI_ISL_1443949, EPI_ISL_1443950, EPI_ISL_1443951, EPI_ISL_1443952, EPI_ISL_1443953, EPI_ISL_1443954, EPI_ISL_1443955, EPI_ISL_1443956, EPI_ISL_1443957, EPI_ISL_1443958, EPI_ISL_1443959, EPI_ISL_1443960, EPI_ISL_1443961, EPI_ISL_1443962, EPI_ISL_1443963, EPI_ISL_1443964, EPI_ISL_1443965, EPI_ISL_1443966, EPI_ISL_1443967, EPI_ISL_1443968, EPI_ISL_1443969, EPI_ISL_1443970, EPI_ISL_1443971, EPI_ISL_1443972, EPI_ISL_1443973, EPI_ISL_1443974, EPI_ISL_1443975, EPI_ISL_1443976, EPI_ISL_1443977, EPI_ISL_1443978, EPI_ISL_1443979, EPI_ISL_1443980, EPI_ISL_1443981, EPI_ISL_1443982, EPI_ISL_1443983, EPI_ISL_1443984, EPI_ISL_1443985, EPI_ISL_1443986, EPI_ISL_1443987, EPI_ISL_1443988, EPI_ISL_1443989, EPI_ISL_1443990, EPI_ISL_1443991, EPI_ISL_1443992, EPI_ISL_1443993, EPI_ISL_1443994, EPI_ISL_1443995, EPI_ISL_1443996, EPI_ISL_1443997, EPI_ISL_1443998, EPI_ISL_1443999, EPI_ISL_1444000, EPI_ISL_1444001, EPI_ISL_1444002, EPI_ISL_1444003, EPI_ISL_1444004, EPI_ISL_1444005, EPI_ISL_1444006, EPI_ISL_1444007, EPI_ISL_1444008, EPI_ISL_1444009, EPI_ISL_1444010, EPI_ISL_1444011, EPI_ISL_1444012, EPI_ISL_1444013, EPI_ISL_1444014, EPI_ISL_1444015, EPI_ISL_1444016, EPI_ISL_1444017, EPI_ISL_1444018, EPI_ISL_1444019, EPI_ISL_1444020, EPI_ISL_1444021, EPI_ISL_1444022, EPI_ISL_1444023, EPI_ISL_1444024, EPI_ISL_1444025, EPI_ISL_1444026, EPI_ISL_1444027, EPI_ISL_1444028, EPI_ISL_1444029, EPI_ISL_1444030, EPI_ISL_1444031, EPI_ISL_1444032, EPI_ISL_1444033, EPI_ISL_1444034, EPI_ISL_1444035, EPI_ISL_1444036, EPI_ISL_1444037, EPI_ISL_1444038, EPI_ISL_1444039, EPI_ISL_1444040, EPI_ISL_1444041, EPI_ISL_1444042, EPI_ISL_1444043, EPI_ISL_1444044, EPI_ISL_1444045, EPI_ISL_1444046, EPI_ISL_1444047, EPI_ISL_1444048, EPI_ISL_1444049, EPI_ISL_1444050, EPI_ISL_1444051, EPI_ISL_1444052, EPI_ISL_1444053, EPI_ISL_1444054, EPI_ISL_1444055, EPI_ISL_1444056, EPI_ISL_1444057, EPI_ISL_1444058, EPI_ISL_1444059, EPI_ISL_1444060, EPI_ISL_1444061, EPI_ISL_1444062, EPI_ISL_1444063, EPI_ISL_1444064, EPI_ISL_1444065, EPI_ISL_1444066, EPI_ISL_1444067, EPI_ISL_1444068, EPI_ISL_1444069, EPI_ISL_1444070, EPI_ISL_1444071, EPI_ISL_1444072, EPI_ISL_1444073, EPI_ISL_1444074, EPI_ISL_1444075, EPI_ISL_1444076, EPI_ISL_1444077, EPI_ISL_1444078, EPI_ISL_1444079, EPI_ISL_1444080, EPI_ISL_1444081, EPI_ISL_1444082, EPI_ISL_1444083, EPI_ISL_1444084, EPI_ISL_1444085, EPI_ISL_1444086, EPI_ISL_1444087, EPI_ISL_1444088, EPI_ISL_1444089, EPI_ISL_1444090, EPI_ISL_1444091, EPI_ISL_1444092, EPI_ISL_1444093, EPI_ISL_1444094, EPI_ISL_1444095, EPI_ISL_1444096, EPI_ISL_1444097, EPI_ISL_1444098, EPI_ISL_1444099, EPI_ISL_1444100, EPI_ISL_1444101, EPI_ISL_1444102, EPI_ISL_1444103, EPI_ISL_1444104, EPI_ISL_1444105, EPI_ISL_1444106, EPI_ISL_1444107, EPI_ISL_1444108, EPI_ISL_1444110, EPI_ISL_1444113, EPI_ISL_1444117, EPI_ISL_1444120, EPI_ISL_1444123, EPI_ISL_1444127, EPI_ISL_1444133, EPI_ISL_1444136, EPI_ISL_1444139, EPI_ISL_1444142, EPI_ISL_1444145, EPI_ISL_1444152, EPI_ISL_1444155, EPI_ISL_1444158, EPI_ISL_1444162, EPI_ISL_1444165, EPI_ISL_1444167, EPI_ISL_1444168, EPI_ISL_1444169, EPI_ISL_1444170, EPI_ISL_1444171, EPI_ISL_1444172, EPI_ISL_1444173, EPI_ISL_1444174, EPI_ISL_1444175, EPI_ISL_1444176, EPI_ISL_1444177, EPI_ISL_1444178, EPI_ISL_1444179, EPI_ISL_1444180, EPI_ISL_1444181, EPI_ISL_1444183, EPI_ISL_1444184, EPI_ISL_1444185, EPI_ISL_1444186, EPI_ISL_1444187, EPI_ISL_1444188, EPI_ISL_1444189, EPI_ISL_1444190, EPI_ISL_1444191, EPI_ISL_1444192, EPI_ISL_1444194, EPI_ISL_1444195, EPI_ISL_1444196, EPI_ISL_1444197, EPI_ISL_1444198, EPI_ISL_1444199, EPI_ISL_1444200, EPI_ISL_1444201, EPI_ISL_1444202, EPI_ISL_1444203, EPI_ISL_1444204, EPI_ISL_1444205, EPI_ISL_1444206, EPI_ISL_1444207, EPI_ISL_1444208, EPI_ISL_1444209, EPI_ISL_1444210, EPI_ISL_1444211, EPI_ISL_1444212, EPI_ISL_1444213, EPI_ISL_1444214, EPI_ISL_1444215, EPI_ISL_1444216, EPI_ISL_1444217, EPI_ISL_1444218, EPI_ISL_1444219, EPI_ISL_1444220, EPI_ISL_1444221, EPI_ISL_1444222, EPI_ISL_1444223, EPI_ISL_1444224, EPI_ISL_1444225, EPI_ISL_1444226, EPI_ISL_1444227, EPI_ISL_1444228, EPI_ISL_1444229, EPI_ISL_1444230, EPI_ISL_1444231, EPI_ISL_1444232, EPI_ISL_1444233, EPI_ISL_1444234, EPI_ISL_1444235, EPI_ISL_1444236, EPI_ISL_1444237, EPI_ISL_1444238, EPI_ISL_1444239, EPI_ISL_1444240, EPI_ISL_1444241, EPI_ISL_1444242, EPI_ISL_1444243, EPI_ISL_1444244, EPI_ISL_1444245, EPI_ISL_1444246, EPI_ISL_1444247, EPI_ISL_1444248, EPI_ISL_1444249, EPI_ISL_1444250, EPI_ISL_1444251, EPI_ISL_1444252, EPI_ISL_1444253, EPI_ISL_1444254, EPI_ISL_1444255, EPI_ISL_1444256, EPI_ISL_1444257, EPI_ISL_1444258, EPI_ISL_1444259, EPI_ISL_1444261 | | | |
| see above | Helix/Illumina | Centers for Disease Control and Prevention Division of Viral Diseases, Pathogen Discovery | Peter W. Cook, Dakota Howard, Dhwani Batra, Ben L. Rambo-Martin, Eileen de Feo, Jan Antico, Christine Tran, Matthew Tolentino, Shannon Wickline, Kim Gietzen, Brad Sickler, Jingtao Liu, Eric Allen, Phil Febbo, Summer Galloway, Nicole L. Washington, Simon White, Geraint Levan, Kelly Schiabor Barrett, Elizabeth Cirulli, Alexandre Bolze, Ary Ascencio, Charlotte Rivera-Garcia, Ryan Cho, Jason Nguyen, Sherry Wang, Jimmy Ramirez, Tyler Cassens, Efren Sandoval, Magnus Isaksson, William Lee, David Becker, Marc Laurent, James Lu, Clinton R. Paden, Suxiang Tong, Duncan MacCannell |
| EPI_ISL_1444262, EPI_ISL_1444263, EPI_ISL_1444264, EPI_ISL_1444265, EPI_ISL_1444266, EPI_ISL_1444267, EPI_ISL_1444268, EPI_ISL_1444269, EPI_ISL_1444270, EPI_ISL_1444271, EPI_ISL_1444272, EPI_ISL_1444273, EPI_ISL_1444274, EPI_ISL_1444275, EPI_ISL_1444276, EPI_ISL_1444277, EPI_ISL_1444278, EPI_ISL_1444279, EPI_ISL_1444280, EPI_ISL_1444282, EPI_ISL_1444283, EPI_ISL_1444284, EPI_ISL_1444285, EPI_ISL_1444286, EPI_ISL_1444287, EPI_ISL_1444288, EPI_ISL_1444289, EPI_ISL_1444290, EPI_ISL_1444291, EPI_ISL_1444292, EPI_ISL_1444293, EPI_ISL_1444294, EPI_ISL_1444295, EPI_ISL_1444296, EPI_ISL_1444297, EPI_ISL_1444298, EPI_ISL_1444299, EPI_ISL_1444300, EPI_ISL_1444301, EPI_ISL_1444302, EPI_ISL_1444303, EPI_ISL_1444304, EPI_ISL_1444306, EPI_ISL_1444307, EPI_ISL_1444308, EPI_ISL_1444309, EPI_ISL_1444310, EPI_ISL_1444311, EPI_ISL_1444312, EPI_ISL_1444313, EPI_ISL_1444314, EPI_ISL_1444315, EPI_ISL_1444316, EPI_ISL_1444317, EPI_ISL_1444318, EPI_ISL_1444319, EPI_ISL_1444320, EPI_ISL_1444321, EPI_ISL_1444322, EPI_ISL_1444323, EPI_ISL_1444324, EPI_ISL_1444325, EPI_ISL_1444326, EPI_ISL_1444327, EPI_ISL_1444328, EPI_ISL_1444329, EPI_ISL_1444330, EPI_ISL_1444331, EPI_ISL_1444333, EPI_ISL_1444334, EPI_ISL_1444335, EPI_ISL_1444336, EPI_ISL_1444337, EPI_ISL_1444340, EPI_ISL_1444342, EPI_ISL_1444343, EPI_ISL_1444344, EPI_ISL_1444345, EPI_ISL_1444346 | | | |
| see above | TXDSHS | TXDSHS | Rashmi Tuladhar, Bonnie Oh, Jenny Zhang, Maliha Rahman, Mayela Pedrueza, Anita Pokharel, Lorraine Rodriguez, Myong Koag, Chun Wang, Rachel Lee, Grace Kubin |
| EPI_ISL_1444347, EPI_ISL_1444348, EPI_ISL_1444349, EPI_ISL_1444350, EPI_ISL_1444351, EPI_ISL_1444352, EPI_ISL_1444353, EPI_ISL_1444354, EPI_ISL_1444355, EPI_ISL_1444356, EPI_ISL_1444357, EPI_ISL_1444358, EPI_ISL_1444359, EPI_ISL_1444360, EPI_ISL_1444361, EPI_ISL_1444362, EPI_ISL_1444363, EPI_ISL_1444364, EPI_ISL_1444365, EPI_ISL_1444366, EPI_ISL_1444367, EPI_ISL_1444368, EPI_ISL_1444369, EPI_ISL_1444370, EPI_ISL_1444371, EPI_ISL_1444372, EPI_ISL_1444373, EPI_ISL_1444374, EPI_ISL_1444375, EPI_ISL_1444376, EPI_ISL_1444377, EPI_ISL_1444378, EPI_ISL_1444379, EPI_ISL_1444380, EPI_ISL_1444381, EPI_ISL_1444382, EPI_ISL_1444383, EPI_ISL_1444384, EPI_ISL_1444385, EPI_ISL_1444386, EPI_ISL_1444387, EPI_ISL_1444388, EPI_ISL_1444389, EPI_ISL_1444390, EPI_ISL_1444391, EPI_ISL_1444392, EPI_ISL_1444393, EPI_ISL_1444394, EPI_ISL_1444395, EPI_ISL_1444396, EPI_ISL_1444397, EPI_ISL_1444398, EPI_ISL_1444399, EPI_ISL_1444400, EPI_ISL_1444401, EPI_ISL_1444402, EPI_ISL_1444403, EPI_ISL_1444404, EPI_ISL_1444405, EPI_ISL_1444406, EPI_ISL_1444407, EPI_ISL_1444408, EPI_ISL_1444409, EPI_ISL_1444410, EPI_ISL_1444411, EPI_ISL_1444412, EPI_ISL_1444413, EPI_ISL_1444414, EPI_ISL_1444415, EPI_ISL_1444416, EPI_ISL_1444417, EPI_ISL_1444418, EPI_ISL_1444419, EPI_ISL_1444420, EPI_ISL_1444421, EPI_ISL_1444422, EPI_ISL_1444423, EPI_ISL_1444424, EPI_ISL_1444425, EPI_ISL_1444426, EPI_ISL_1444427, EPI_ISL_1444428, EPI_ISL_1444429, EPI_ISL_1444430, EPI_ISL_1444431, EPI_ISL_1444432, EPI_ISL_1444433, EPI_ISL_1444434, EPI_ISL_1444435, EPI_ISL_1444436, EPI_ISL_1444437, EPI_ISL_1444438, EPI_ISL_1444439, EPI_ISL_1444440, EPI_ISL_1444441, EPI_ISL_1444442, EPI_ISL_1444443, EPI_ISL_1444444, EPI_ISL_1444445, EPI_ISL_1444446, EPI_ISL_1444447, EPI_ISL_1444448, EPI_ISL_1444449, EPI_ISL_1444450, EPI_ISL_1444451, EPI_ISL_1444452, EPI_ISL_1444453, EPI_ISL_1444454, EPI_ISL_1444455, EPI_ISL_1444456, EPI_ISL_1444457, EPI_ISL_1444458, EPI_ISL_1444459, EPI_ISL_1444460, EPI_ISL_1444461, EPI_ISL_1444462, EPI_ISL_1444463, EPI_ISL_1444464, EPI_ISL_1444465, EPI_ISL_1444466, EPI_ISL_1444467, EPI_ISL_1444468, EPI_ISL_1444469, EPI_ISL_1444470, EPI_ISL_1444471, EPI_ISL_1444472, EPI_ISL_1444473, EPI_ISL_1444474, EPI_ISL_1444475, EPI_ISL_1444476, EPI_ISL_1444477, EPI_ISL_1444478, EPI_ISL_1444479, EPI_ISL_1444480, EPI_ISL_1444481, EPI_ISL_1444482, EPI_ISL_1444483, EPI_ISL_1444484, EPI_ISL_1444485, EPI_ISL_1444486, EPI_ISL_1444487, EPI_ISL_1444488, EPI_ISL_1444489, EPI_ISL_1444490, EPI_ISL_1444491, EPI_ISL_1444492, EPI_ISL_1444493, EPI_ISL_1444494, EPI_ISL_1444495, EPI_ISL_1444496, EPI_ISL_1444497, EPI_ISL_1444498, EPI_ISL_1444499, EPI_ISL_1444500, EPI_ISL_1444501, EPI_ISL_1444502, EPI_ISL_1444503, EPI_ISL_1444504, EPI_ISL_1444505, EPI_ISL_1444506, EPI_ISL_1444507, EPI_ISL_1444508, EPI_ISL_1444509, EPI_ISL_1444510, EPI_ISL_1444511, EPI_ISL_1444512, EPI_ISL_1444513, EPI_ISL_1444514, EPI_ISL_1444515, EPI_ISL_1444516, EPI_ISL_1444517, EPI_ISL_1444518, EPI_ISL_1444519, EPI_ISL_1444520, EPI_ISL_1444521, EPI_ISL_1444522, EPI_ISL_1444523, EPI_ISL_1444524, EPI_ISL_1444525, EPI_ISL_1444526, EPI_ISL_1444527, EPI_ISL_1444528, EPI_ISL_1444529, EPI_ISL_1444530, EPI_ISL_1444531, EPI_ISL_1444532, EPI_ISL_1444533, EPI_ISL_1444534, EPI_ISL_1444535, EPI_ISL_1444536, EPI_ISL_1444537, EPI_ISL_1444538, EPI_ISL_1444539, EPI_ISL_1444540, EPI_ISL_1444541, EPI_ISL_1444542, EPI_ISL_1444543, EPI_ISL_1444544, EPI_ISL_1444545, EPI_ISL_1444546, EPI_ISL_1444547, EPI_ISL_1444548, EPI_ISL_1444549, EPI_ISL_1444550, EPI_ISL_1444551, EPI_ISL_1444552, EPI_ISL_1444553, EPI_ISL_1444554, EPI_ISL_1444555, EPI_ISL_1444556, EPI_ISL_1444557, EPI_ISL_1444558, EPI_ISL_1444559, EPI_ISL_1444560, EPI_ISL_1444561, EPI_ISL_1444562, EPI_ISL_1444563, EPI_ISL_1444564, EPI_ISL_1444565, EPI_ISL_1444566, EPI_ISL_1444567, EPI_ISL_1444568, EPI_ISL_1444569, EPI_ISL_1444570, EPI_ISL_1444571, EPI_ISL_1444572, EPI_ISL_1444573, EPI_ISL_1444574, EPI_ISL_1444575, EPI_ISL_1444576, EPI_ISL_1444577, EPI_ISL_1444578, EPI_ISL_1444579, EPI_ISL_1444580, EPI_ISL_1444581, EPI_ISL_1444582, EPI_ISL_1444583, EPI_ISL_1444584, EPI_ISL_1444585, EPI_ISL_1444586, EPI_ISL_1444587, EPI_ISL_1444588, EPI_ISL_1444589, EPI_ISL_1444590, EPI_ISL_1444591, EPI_ISL_1444592, EPI_ISL_1444593, EPI_ISL_1444594, EPI_ISL_1444595, EPI_ISL_1444596, EPI_ISL_1444597, EPI_ISL_1444598, EPI_ISL_1444599, EPI_ISL_1444600, EPI_ISL_1444601, EPI_ISL_1444602, EPI_ISL_1444603, EPI_ISL_1444604, EPI_ISL_1444605, EPI_ISL_1444606, EPI_ISL_1444607, EPI_ISL_1444608, EPI_ISL_1444609, EPI_ISL_1444610, EPI_ISL_1444611, EPI_ISL_1444612, EPI_ISL_1444613, EPI_ISL_1444614, EPI_ISL_1444615, EPI_ISL_1444616, EPI_ISL_1444617, EPI_ISL_1444618, EPI_ISL_1444619, EPI_ISL_1444620, EPI_ISL_1444621, EPI_ISL_1444622, EPI_ISL_1444623, EPI_ISL_1444624, EPI_ISL_1444625, EPI_ISL_1444626, EPI_ISL_1444627, EPI_ISL_1444628, EPI_ISL_1444629, EPI_ISL_1444630, EPI_ISL_1444631, EPI_ISL_1444632, EPI_ISL_1444633, EPI_ISL_1444634, EPI_ISL_1444635, EPI_ISL_1444636, EPI_ISL_1444637, EPI_ISL_1444638, EPI_ISL_1444639, EPI_ISL_1444640, EPI_ISL_1444641, EPI_ISL_1444642, EPI_ISL_1444643, EPI_ISL_1444644, EPI_ISL_1444645, EPI_ISL_1444646, EPI_ISL_1444647, EPI_ISL_1444648, EPI_ISL_1444649, EPI_ISL_1444650, EPI_ISL_1444651, EPI_ISL_1444652, EPI_ISL_1444653, EPI_ISL_1444654, EPI_ISL_1444655, EPI_ISL_1444656, EPI_ISL_1444657, EPI_ISL_1444658, EPI_ISL_1444659, EPI_ISL_1444660, EPI_ISL_1444661, EPI_ISL_1444662, EPI_ISL_1444663, EPI_ISL_1444664, EPI_ISL_1444665, EPI_ISL_1444666, EPI_ISL_1444667, EPI_ISL_1444668, EPI_ISL_1444669, EPI_ISL_1444670, EPI_ISL_1444671, EPI_ISL_1444672 | | | |

| | | | |
|---|---|---|---|
| see above | Quest Diagnostics Incorporated | Centers for Disease Control and Prevention Division of Viral Diseases, Pathogen Discovery | Dakota Howard, Dhwani Batra, Peter W. Cook, Kara Moser, Adrian Paskey, Jason Caravas, Benjamin Rambo-Martin, Shatavia Morrison, Christopher Gulvick, Scott Sammons, Yvette Unoarumhi, Darlene Wagner, Matthew Schmerer, S. H. Rosenthal, A. Gerasimova, R. M. Kagan, B. Anderson, M. Hua, Y. Liu, L.E. Bernstein, K.E. Livingston, A. Perez, I. A. Shlyakhter, R. V. Rolando, R. Owen, P. Tanpaiboon, F. Lacbawan, Clinton R. Paden, Duncan MacCannell |
| EPI_ISL_1444673 | Nebraska Public Health Laboratory | NPHL COVID-19 Response Team | NPHL COVID-19 Response Team |
| EPI_ISL_1444675 | Quest Diagnostics Incorporated | Centers for Disease Control and Prevention Division of Viral Diseases, Pathogen Discovery | Dakota Howard, Dhwani Batra, Peter W. Cook, Kara Moser, Adrian Paskey, Jason Caravas, Benjamin Rambo-Martin, Shatavia Morrison, Christopher Gulvick, Scott Sammons, Yvette Unoarumhi, Darlene Wagner, Matthew Schmerer, S. H. Rosenthal, A. Gerasimova, R. M. Kagan, B. Anderson, M. Hua, Y. Liu, L.E. Bernstein, K.E. Livingston, A. Perez, I. A. Shlyakhter, R. V. Rolando, R. Owen, P. Tanpaiboon, F. Lacbawan, Clinton R. Paden, Duncan MacCannell |
| EPI_ISL_1444676, EPI_ISL_1444677 | Nebraska Public Health Laboratory | NPHL COVID-19 Response Team | NPHL COVID-19 Response Team |
| EPI_ISL_1444678 | Quest Diagnostics Incorporated | Centers for Disease Control and Prevention Division of Viral Diseases, Pathogen Discovery | Dakota Howard, Dhwani Batra, Peter W. Cook, Kara Moser, Adrian Paskey, Jason Caravas, Benjamin Rambo-Martin, Shatavia Morrison, Christopher Gulvick, Scott Sammons, Yvette Unoarumhi, Darlene Wagner, Matthew Schmerer, S. H. Rosenthal, A. Gerasimova, R. M. Kagan, B. Anderson, M. Hua, Y. Liu, L.E. Bernstein, K.E. Livingston, A. Perez, I. A. Shlyakhter, R. V. Rolando, R. Owen, P. Tanpaiboon, F. Lacbawan, Clinton R. Paden, Duncan MacCannell |
| EPI_ISL_1444679, EPI_ISL_1444680 | Nebraska Public Health Laboratory | NPHL COVID-19 Response Team | NPHL COVID-19 Response Team |
| EPI_ISL_1444681 | Quest Diagnostics Incorporated | Centers for Disease Control and Prevention Division of Viral Diseases, Pathogen Discovery | Dakota Howard, Dhwani Batra, Peter W. Cook, Kara Moser, Adrian Paskey, Jason Caravas, Benjamin Rambo-Martin, Shatavia Morrison, Christopher Gulvick, Scott Sammons, Yvette Unoarumhi, Darlene Wagner, Matthew Schmerer, S. H. Rosenthal, A. Gerasimova, R. M. Kagan, B. Anderson, M. Hua, Y. Liu, L.E. Bernstein, K.E. Livingston, A. Perez, I. A. Shlyakhter, R. V. Rolando, R. Owen, P. Tanpaiboon, F. Lacbawan, Clinton R. Paden, Duncan MacCannell |
| EPI_ISL_1444682, EPI_ISL_1444683 | Nebraska Public Health Laboratory | NPHL COVID-19 Response Team | NPHL COVID-19 Response Team |
| EPI_ISL_1444684 | Quest Diagnostics Incorporated | Centers for Disease Control and Prevention Division of Viral Diseases, Pathogen Discovery | Dakota Howard, Dhwani Batra, Peter W. Cook, Kara Moser, Adrian Paskey, Jason Caravas, Benjamin Rambo-Martin, Shatavia Morrison, Christopher Gulvick, Scott Sammons, Yvette Unoarumhi, Darlene Wagner, Matthew Schmerer, S. H. Rosenthal, A. Gerasimova, R. M. Kagan, B. Anderson, M. Hua, Y. Liu, L.E. Bernstein, K.E. Livingston, A. Perez, I. A. Shlyakhter, R. V. Rolando, R. Owen, P. Tanpaiboon, F. Lacbawan, Clinton R. Paden, Duncan MacCannell |
| EPI_ISL_1444685, EPI_ISL_1444686 | Nebraska Public Health Laboratory | NPHL COVID-19 Response Team | NPHL COVID-19 Response Team |
| EPI_ISL_1444687 | Quest Diagnostics Incorporated | Centers for Disease Control and Prevention Division of Viral Diseases, Pathogen Discovery | Dakota Howard, Dhwani Batra, Peter W. Cook, Kara Moser, Adrian Paskey, Jason Caravas, Benjamin Rambo-Martin, Shatavia Morrison, Christopher Gulvick, Scott Sammons, Yvette Unoarumhi, Darlene Wagner, Matthew Schmerer, S. H. Rosenthal, A. Gerasimova, R. M. Kagan, B. Anderson, M. Hua, Y. Liu, L.E. Bernstein, K.E. Livingston, A. Perez, I. A. Shlyakhter, R. V. Rolando, R. Owen, P. Tanpaiboon, F. Lacbawan, Clinton R. Paden, Duncan MacCannell |
| EPI_ISL_1444688 | Nebraska Public Health Laboratory | NPHL COVID-19 Response Team | NPHL COVID-19 Response Team |
| EPI_ISL_1444689, EPI_ISL_1444690, EPI_ISL_1444691, EPI_ISL_1444692, EPI_ISL_1444693, EPI_ISL_1444694, EPI_ISL_1444695, EPI_ISL_1444696, EPI_ISL_1444697, EPI_ISL_1444698, EPI_ISL_1444699, EPI_ISL_1444700, EPI_ISL_1444701, EPI_ISL_1444702, EPI_ISL_1444703, EPI_ISL_1444704, EPI_ISL_1444705, EPI_ISL_1444706, EPI_ISL_1444707, EPI_ISL_1444708, EPI_ISL_1444709, EPI_ISL_1444710, EPI_ISL_1444711, EPI_ISL_1444712, EPI_ISL_1444713, EPI_ISL_1444714, EPI_ISL_1444715, EPI_ISL_1444716, EPI_ISL_1444717, EPI_ISL_1444718, EPI_ISL_1444719, EPI_ISL_1444720, EPI_ISL_1444721, EPI_ISL_1444722, EPI_ISL_1444723, EPI_ISL_1444724, EPI_ISL_1444725, EPI_ISL_1444726, EPI_ISL_1444727, EPI_ISL_1444728, EPI_ISL_1444729, EPI_ISL_1444730, EPI_ISL_1444731 | | | |
| see above | Quest Diagnostics Incorporated | Centers for Disease Control and Prevention Division of Viral Diseases, Pathogen Discovery | Dakota Howard, Dhwani Batra, Peter W. Cook, Kara Moser, Adrian Paskey, Jason Caravas, Benjamin Rambo-Martin, Shatavia Morrison, Christopher Gulvick, Scott Sammons, Yvette Unoarumhi, Darlene Wagner, Matthew Schmerer, S. H. Rosenthal, A. Gerasimova, R. M. Kagan, B. Anderson, M. Hua, Y. Liu, L.E. Bernstein, K.E. Livingston, A. Perez, I. A. Shlyakhter, R. V. Rolando, R. Owen, P. Tanpaiboon, F. Lacbawan, Clinton R. Paden, Duncan MacCannell |
| EPI_ISL_1444777, EPI_ISL_1444778, EPI_ISL_1444779, EPI_ISL_1444781, EPI_ISL_1444782, EPI_ISL_1444783, EPI_ISL_1444784, EPI_ISL_1444785, EPI_ISL_1444786, EPI_ISL_1444787, EPI_ISL_1444789, EPI_ISL_1444790, EPI_ISL_1444791, EPI_ISL_1444792, EPI_ISL_1444793, EPI_ISL_1444794, EPI_ISL_1444795, EPI_ISL_1444797, EPI_ISL_1444799, EPI_ISL_1444800, EPI_ISL_1444802, EPI_ISL_1444803, EPI_ISL_1444805, EPI_ISL_1444806, EPI_ISL_1444807, EPI_ISL_1444808, EPI_ISL_1444810, EPI_ISL_1444811, EPI_ISL_1444812, EPI_ISL_1444813, EPI_ISL_1444814, EPI_ISL_1444815, EPI_ISL_1444816, EPI_ISL_1444817, EPI_ISL_1444818, EPI_ISL_1444819, EPI_ISL_1444820, EPI_ISL_1444821, EPI_ISL_1444822, EPI_ISL_1444824, EPI_ISL_1444825, EPI_ISL_1444826, EPI_ISL_1444827, EPI_ISL_1444828, EPI_ISL_1444829, EPI_ISL_1444830, EPI_ISL_1444831, EPI_ISL_1444832, EPI_ISL_1444833, EPI_ISL_1444834, EPI_ISL_1444835, EPI_ISL_1444836, EPI_ISL_1444837, EPI_ISL_1444838, EPI_ISL_1444839, EPI_ISL_1444840, EPI_ISL_1444841, EPI_ISL_1444842, EPI_ISL_1444843, EPI_ISL_1444844, EPI_ISL_1444845, EPI_ISL_1444846, EPI_ISL_1444847, EPI_ISL_1444848, EPI_ISL_1444849, EPI_ISL_1444850, EPI_ISL_1444851, EPI_ISL_1444852, EPI_ISL_1444853, EPI_ISL_1444854, EPI_ISL_1444855, EPI_ISL_1444856, EPI_ISL_1444857, EPI_ISL_1444858, EPI_ISL_1444859, EPI_ISL_1444860, EPI_ISL_1444861, EPI_ISL_1444862, EPI_ISL_1444863, EPI_ISL_1444864, EPI_ISL_1444865, EPI_ISL_1444866, EPI_ISL_1444867, EPI_ISL_1444868, EPI_ISL_1444869, EPI_ISL_1444870, EPI_ISL_1444871, EPI_ISL_1444872, EPI_ISL_1444873, EPI_ISL_1444874, EPI_ISL_1444875, EPI_ISL_1444876, EPI_ISL_1444877, EPI_ISL_1444878, EPI_ISL_1444879, EPI_ISL_1444880, EPI_ISL_1444881, EPI_ISL_1444882, EPI_ISL_1444883, EPI_ISL_1444884, EPI_ISL_1444885, EPI_ISL_1444886, EPI_ISL_1444887, EPI_ISL_1444888, EPI_ISL_1444890, EPI_ISL_1444891, EPI_ISL_1444892, EPI_ISL_1444893, EPI_ISL_1444894, EPI_ISL_1444895, EPI_ISL_1444897, EPI_ISL_1444898, EPI_ISL_1444899, EPI_ISL_1444900, EPI_ISL_1444901, EPI_ISL_1444903, EPI_ISL_1444904, EPI_ISL_1444905, EPI_ISL_1444906, EPI_ISL_1444907, EPI_ISL_1444908 | | | |
| see above | Maryland Genomics, Institute for Genome Sciences, University of Maryland School of Medicine | Maryland Genomics, Institute for Genome Sciences, University of Maryland School of Medicine | Tallon, Luke J; Sadzewicz, Lisa D; Humphrys, Mike; Ott, Sandra; Roussey, Holly; Mehta, Aditya; Vavikolanu, Kranthi; Fraser, Claire M; Ravel, Jacques |
| EPI_ISL_1444914, EPI_ISL_1444915, EPI_ISL_1444916, EPI_ISL_1444917, EPI_ISL_1444918, EPI_ISL_1444919, EPI_ISL_1444920, EPI_ISL_1444921, EPI_ISL_1444922, EPI_ISL_1444923, EPI_ISL_1444924, EPI_ISL_1444925, EPI_ISL_1444926, EPI_ISL_1444927, EPI_ISL_1444928, EPI_ISL_1444929, EPI_ISL_1444930, EPI_ISL_1444931, EPI_ISL_1444932, EPI_ISL_1444933, EPI_ISL_1444934, EPI_ISL_1444935, EPI_ISL_1444936, EPI_ISL_1444937, EPI_ISL_1444938, EPI_ISL_1444939, EPI_ISL_1444940, EPI_ISL_1444941, EPI_ISL_1444942, EPI_ISL_1444943, EPI_ISL_1444944, EPI_ISL_1444945, EPI_ISL_1444946, EPI_ISL_1444947, EPI_ISL_1444948, EPI_ISL_1444949, EPI_ISL_1444950, EPI_ISL_1444951, EPI_ISL_1444952, EPI_ISL_1444953, EPI_ISL_1444954, EPI_ISL_1444955, EPI_ISL_1444956, EPI_ISL_1444957, EPI_ISL_1444958, EPI_ISL_1444959, EPI_ISL_1444960, EPI_ISL_1444961, EPI_ISL_1444962, EPI_ISL_1444963, EPI_ISL_1444964, EPI_ISL_1444965, EPI_ISL_1444966, EPI_ISL_1444967, EPI_ISL_1444968, EPI_ISL_1444969, EPI_ISL_1444970, EPI_ISL_1444971, EPI_ISL_1444972, EPI_ISL_1444973, EPI_ISL_1444974, EPI_ISL_1444975, EPI_ISL_1444976, EPI_ISL_1444977, EPI_ISL_1444978, EPI_ISL_1444979, EPI_ISL_1444980, EPI_ISL_1444981, EPI_ISL_1444982, EPI_ISL_1444983, EPI_ISL_1444984, EPI_ISL_1444985, EPI_ISL_1444986, EPI_ISL_1444987, EPI_ISL_1444988, EPI_ISL_1444989, EPI_ISL_1444990, EPI_ISL_1444991, EPI_ISL_1444992, EPI_ISL_1444993, EPI_ISL_1444994, EPI_ISL_1444995, EPI_ISL_1444996, EPI_ISL_1444997, EPI_ISL_1444998, EPI_ISL_1444999, EPI_ISL_1445000, EPI_ISL_1445001, EPI_ISL_1445002, EPI_ISL_1445003, EPI_ISL_1445004, EPI_ISL_1445005, EPI_ISL_1445006, EPI_ISL_1445007, EPI_ISL_1445008, EPI_ISL_1445009, EPI_ISL_1445010, EPI_ISL_1445011, EPI_ISL_1445012, EPI_ISL_1445013, EPI_ISL_1445014, EPI_ISL_1445015, EPI_ISL_1445016, EPI_ISL_1445017, EPI_ISL_1445018, EPI_ISL_1445019 | | | |
| see above | Erie County Public Health (ECPHL) | University at Buffalo Genomics and Bioinformatics Core | Jonathan Bard, Natalie Lamb, Alyssa Pohlman, Brandon Marzullo, Amanda Boccolucci, Norma Nowak, Donald Yergeau, Jennifer Surtees |
| EPI_ISL_1445020, EPI_ISL_1445021, EPI_ISL_1445022, EPI_ISL_1445023, EPI_ISL_1445024, EPI_ISL_1445025, EPI_ISL_1445026, EPI_ISL_1445027, EPI_ISL_1445028, EPI_ISL_1445030, EPI_ISL_1445031, EPI_ISL_1445032, EPI_ISL_1445033, EPI_ISL_1445034, EPI_ISL_1445035, EPI_ISL_1445036, EPI_ISL_1445039, EPI_ISL_1445040, EPI_ISL_1445041, EPI_ISL_1445043, EPI_ISL_1445045, EPI_ISL_1445046, EPI_ISL_1445049, EPI_ISL_1445054, EPI_ISL_1445055, EPI_ISL_1445056, EPI_ISL_1445057, EPI_ISL_1445058, EPI_ISL_1445059, EPI_ISL_1445060, EPI_ISL_1445061, EPI_ISL_1445062, EPI_ISL_1445063, EPI_ISL_1445064 | | | |
| see above | Virginia Division of Consolidated Laboratory Services | Virginia Division of Consolidated Laboratory Services | Virginia DCLS |
| EPI_ISL_1445276, EPI_ISL_1445277, EPI_ISL_1445278, EPI_ISL_1445279, EPI_ISL_1445280, EPI_ISL_1445281, EPI_ISL_1445282, EPI_ISL_1445283, EPI_ISL_1445284, EPI_ISL_1445285, EPI_ISL_1445286, EPI_ISL_1445287, EPI_ISL_1445288, EPI_ISL_1445289, EPI_ISL_1445290, EPI_ISL_1445291, EPI_ISL_1445292, EPI_ISL_1445293, EPI_ISL_1445294, EPI_ISL_1445295, EPI_ISL_1445296, EPI_ISL_1445297, EPI_ISL_1445298, EPI_ISL_1445299, EPI_ISL_1445300, EPI_ISL_1445301, EPI_ISL_1445302, EPI_ISL_1445303, EPI_ISL_1445304, EPI_ISL_1445305, EPI_ISL_1445306, EPI_ISL_1445307, EPI_ISL_1445308, EPI_ISL_1445309, EPI_ISL_1445310, EPI_ISL_1445311, EPI_ISL_1445312, EPI_ISL_1445313, EPI_ISL_1445314, EPI_ISL_1445315, EPI_ISL_1445316, EPI_ISL_1445317, EPI_ISL_1445318, EPI_ISL_1445319, EPI_ISL_1445320, EPI_ISL_1445321, EPI_ISL_1445322, EPI_ISL_1445323, EPI_ISL_1445324, EPI_ISL_1445325, EPI_ISL_1445326, EPI_ISL_1445327, EPI_ISL_1445328, EPI_ISL_1445329, EPI_ISL_1445330, EPI_ISL_1445331, EPI_ISL_1445332, EPI_ISL_1445333, EPI_ISL_1445334, EPI_ISL_1445335, EPI_ISL_1445336, EPI_ISL_1445337, EPI_ISL_1445338, EPI_ISL_1445339, EPI_ISL_1445340, EPI_ISL_1445341, EPI_ISL_1445342, EPI_ISL_1445343, EPI_ISL_1445344, EPI_ISL_1445345, EPI_ISL_1445346, EPI_ISL_1445347, EPI_ISL_1445348, EPI_ISL_1445349, EPI_ISL_1445350, EPI_ISL_1445351, EPI_ISL_1445352, EPI_ISL_1445353, EPI_ISL_1445354, EPI_ISL_1445355, EPI_ISL_1445356, EPI_ISL_1445357, EPI_ISL_1445358, EPI_ISL_1445359, EPI_ISL_1445360, EPI_ISL_1445361, EPI_ISL_1445362, EPI_ISL_1445363, EPI_ISL_1445364, EPI_ISL_1445365, EPI_ISL_1445366, EPI_ISL_1445367, EPI_ISL_1445368, EPI_ISL_1445369, EPI_ISL_1445370, EPI_ISL_1445371, EPI_ISL_1445372, EPI_ISL_1445373, EPI_ISL_1445374, EPI_ISL_1445375, EPI_ISL_1445376, EPI_ISL_1445377, EPI_ISL_1445378, EPI_ISL_1445379, EPI_ISL_1445380, EPI_ISL_1445381, EPI_ISL_1445382, EPI_ISL_1445383, EPI_ISL_1445384, EPI_ISL_1445385, EPI_ISL_1445386, EPI_ISL_1445387, EPI_ISL_1445388, EPI_ISL_1445389, EPI_ISL_1445390, EPI_ISL_1445391, EPI_ISL_1445392, EPI_ISL_1445393, EPI_ISL_1445394, EPI_ISL_1445395, EPI_ISL_1445396, EPI_ISL_1445397, EPI_ISL_1445398, EPI_ISL_1445399, EPI_ISL_1445400, EPI_ISL_1445401, EPI_ISL_1445402, EPI_ISL_1445403, EPI_ISL_1445404, EPI_ISL_1445405, EPI_ISL_1445406, EPI_ISL_1445407, EPI_ISL_1445408, EPI_ISL_1445409, EPI_ISL_1445410, EPI_ISL_1445411, EPI_ISL_1445412, EPI_ISL_1445413, EPI_ISL_1445414, EPI_ISL_1445415, EPI_ISL_1445416, EPI_ISL_1445417, EPI_ISL_1445418, EPI_ISL_1445419, EPI_ISL_1445420, EPI_ISL_1445421, EPI_ISL_1445422, EPI_ISL_1445423, EPI_ISL_1445424, EPI_ISL_1445425, EPI_ISL_1445426, EPI_ISL_1445427, EPI_ISL_1445428, EPI_ISL_1445429, EPI_ISL_1445430, EPI_ISL_1445431, EPI_ISL_1445432, EPI_ISL_1445433, EPI_ISL_1445434, EPI_ISL_1445435, EPI_ISL_1445436, EPI_ISL_1445437, EPI_ISL_1445438, EPI_ISL_1445439, EPI_ISL_1445440, EPI_ISL_1445441, EPI_ISL_1445442, EPI_ISL_1445443, EPI_ISL_1445444, EPI_ISL_1445445, EPI_ISL_1445446, EPI_ISL_1445447, EPI_ISL_1445448, EPI_ISL_1445449, EPI_ISL_1445450, EPI_ISL_1445451, EPI_ISL_1445452, EPI_ISL_1445453, EPI_ISL_1445454, EPI_ISL_1445455, EPI_ISL_1445456, EPI_ISL_1445457, EPI_ISL_1445458, EPI_ISL_1445459, EPI_ISL_1445460, EPI_ISL_1445461, EPI_ISL_1445462, EPI_ISL_1445463, EPI_ISL_1445464, EPI_ISL_1445465, EPI_ISL_1445466, EPI_ISL_1445467, EPI_ISL_1445468, EPI_ISL_1445469, EPI_ISL_1445470, EPI_ISL_1445471, EPI_ISL_1445472, EPI_ISL_1445473, EPI_ISL_1445474, EPI_ISL_1445475, EPI_ISL_1445476, EPI_ISL_1445477, EPI_ISL_1445478, EPI_ISL_1445479, EPI_ISL_1445480, EPI_ISL_1445481, EPI_ISL_1445482, EPI_ISL_1445483, EPI_ISL_1445484, EPI_ISL_1445485, EPI_ISL_1445486, EPI_ISL_1445487, EPI_ISL_1445488, EPI_ISL_1445489, EPI_ISL_1445490, EPI_ISL_1445491, EPI_ISL_1445492, EPI_ISL_1445493, EPI_ISL_1445494, EPI_ISL_1445495, EPI_ISL_1445496, EPI_ISL_1445497, EPI_ISL_1445498, EPI_ISL_1445499, EPI_ISL_1445500, EPI_ISL_1445501, EPI_ISL_1445502, EPI_ISL_1445503, EPI_ISL_1445504, EPI_ISL_1445505, EPI_ISL_1445506, EPI_ISL_1445507, EPI_ISL_1445508, EPI_ISL_1445509, EPI_ISL_1445510, EPI_ISL_1445511, EPI_ISL_1445512, EPI_ISL_1445513, EPI_ISL_1445514, EPI_ISL_1445515, EPI_ISL_1445516, EPI_ISL_1445517, EPI_ISL_1445518, EPI_ISL_1445519, EPI_ISL_1445520, EPI_ISL_1445521, EPI_ISL_1445522, EPI_ISL_1445523, EPI_ISL_1445524, EPI_ISL_1445525, EPI_ISL_1445526, EPI_ISL_1445527, EPI_ISL_1445528, EPI_ISL_1445529, EPI_ISL_1445530, EPI_ISL_1445531, EPI_ISL_1445532, EPI_ISL_1445533, EPI_ISL_1445534, EPI_ISL_1445535, EPI_ISL_1445536, EPI_ISL_1445537, EPI_ISL_1445538, EPI_ISL_1445539, EPI_ISL_1445540, EPI_ISL_1445541, EPI_ISL_1445542, EPI_ISL_1445543, EPI_ISL_1445544, EPI_ISL_1445545, EPI_ISL_1445546, EPI_ISL_1445547, EPI_ISL_1445548, EPI_ISL_1445549, EPI_ISL_1445550, EPI_ISL_1445551, EPI_ISL_1445552, EPI_ISL_1445553, EPI_ISL_1445554, EPI_ISL_1445555, EPI_ISL_1445556, EPI_ISL_1445557, EPI_ISL_1445558, EPI_ISL_1445559, EPI_ISL_1445560, EPI_ISL_1445561, EPI_ISL_1445562, EPI_ISL_1445563, EPI_ISL_1445564, EPI_ISL_1445565, EPI_ISL_1445566, EPI_ISL_1445567, EPI_ISL_1445568, EPI_ISL_1445569, EPI_ISL_1445570, EPI_ISL_1445571, EPI_ISL_1445572, EPI_ISL_1445573, EPI_ISL_1445574, EPI_ISL_1445575, EPI_ISL_1445577, EPI_ISL_1445578, EPI_ISL_1445579, EPI_ISL_1445581, EPI_ISL_1445582, EPI_ISL_1445583, EPI_ISL_1445584, EPI_ISL_1445585, EPI_ISL_1445586, EPI_ISL_1445587, EPI_ISL_1445588, EPI_ISL_1445591, EPI_ISL_1445592, EPI_ISL_1445593, EPI_ISL_1445594, EPI_ISL_1445595, EPI_ISL_1445597, EPI_ISL_1445598, EPI_ISL_1445599, EPI_ISL_1445600, EPI_ISL_1445601, EPI_ISL_1445602, EPI_ISL_1445603, EPI_ISL_1445604, EPI_ISL_1445605, EPI_ISL_1445606, EPI_ISL_1445607, EPI_ISL_1445608, EPI_ISL_1445609, EPI_ISL_1445610, EPI_ISL_1445611, EPI_ISL_1445612, EPI_ISL_1445613, EPI_ISL_1445615, EPI_ISL_1445616, EPI_ISL_1445617, EPI_ISL_1445618, EPI_ISL_1445620, EPI_ISL_1445621, EPI_ISL_1445622, EPI_ISL_1445623, EPI_ISL_1445624, EPI_ISL_1445625, EPI_ISL_1445626, EPI_ISL_1445628, EPI_ISL_1445629, EPI_ISL_1445630, EPI_ISL_1445631, EPI_ISL_1445633, EPI_ISL_1445634, EPI_ISL_1445635, EPI_ISL_1445636, EPI_ISL_1445637, EPI_ISL_1445638, EPI_ISL_1445640, EPI_ISL_1445641, EPI_ISL_1445642, EPI_ISL_1445644, EPI_ISL_1445645, EPI_ISL_1445648, EPI_ISL_1445649, EPI_ISL_1445650, EPI_ISL_1445651, EPI_ISL_1445652, EPI_ISL_1445653, EPI_ISL_1445654, EPI_ISL_1445655, EPI_ISL_1445656, EPI_ISL_1445658, EPI_ISL_1445659, EPI_ISL_1445662, EPI_ISL_1445663, EPI_ISL_1445664, EPI_ISL_1445665, EPI_ISL_1445666, EPI_ISL_1445667, EPI_ISL_1445668, EPI_ISL_1445669, EPI_ISL_1445670, EPI_ISL_1445671, EPI_ISL_1445672, EPI_ISL_1445673, EPI_ISL_1445674, EPI_ISL_1445675, EPI_ISL_1445676, EPI_ISL_1445677, EPI_ISL_1445678, EPI_ISL_1445679, EPI_ISL_1445680, EPI_ISL_1445681, EPI_ISL_1445682, EPI_ISL_1445684, EPI_ISL_1445685, EPI_ISL_1445687, EPI_ISL_1445688, EPI_ISL_1445689, EPI_ISL_1445690, EPI_ISL_1445691, EPI_ISL_1445692, EPI_ISL_1445693, EPI_ISL_1445695, EPI_ISL_1445696, EPI_ISL_1445697, EPI_ISL_1445699, EPI_ISL_1445700, EPI_ISL_1445701, EPI_ISL_1445702, EPI_ISL_1445703, EPI_ISL_1445704, EPI_ISL_1445705, EPI_ISL_1445706, EPI_ISL_1445707, EPI_ISL_1445708, EPI_ISL_1445709, EPI_ISL_1445710, EPI_ISL_1445711, EPI_ISL_1445712, EPI_ISL_1445714, EPI_ISL_1445715, EPI_ISL_1445716, EPI_ISL_1445717, EPI_ISL_1445718, EPI_ISL_1445719, EPI_ISL_1445720, EPI_ISL_1445721, EPI_ISL_1445722, EPI_ISL_1445723, EPI_ISL_1445724, EPI_ISL_1445725, EPI_ISL_1445726, EPI_ISL_1445727, EPI_ISL_1445728, EPI_ISL_1445729, EPI_ISL_1445730, EPI_ISL_1445731, EPI_ISL_1445732, EPI_ISL_1445733, EPI_ISL_1445734, EPI_ISL_1445735, EPI_ISL_1445736, EPI_ISL_1445737, EPI_ISL_1445738, EPI_ISL_1445739, EPI_ISL_1445740, EPI_ISL_1445741, EPI_ISL_1445742, EPI_ISL_1445744, EPI_ISL_1445745, EPI_ISL_1445746, EPI_ISL_1445747, EPI_ISL_1445748, EPI_ISL_1445749, EPI_ISL_1445750, EPI_ISL_1445751, EPI_ISL_1445752, EPI_ISL_1445753, EPI_ISL_1445755, EPI_ISL_1445756, EPI_ISL_1445757, EPI_ISL_1445758, EPI_ISL_1445760, EPI_ISL_1445761, EPI_ISL_1445762, EPI_ISL_1445763, EPI_ISL_1445765, EPI_ISL_1445766, EPI_ISL_1445768, EPI_ISL_1445769, EPI_ISL_1445770, EPI_ISL_1445771, EPI_ISL_1445772, EPI_ISL_1445773, EPI_ISL_1445774, EPI_ISL_1445775, EPI_ISL_1445776, EPI_ISL_1445777, EPI_ISL_1445778, EPI_ISL_1445779, EPI_ISL_1445780, EPI_ISL_1445781, EPI_ISL_1445782, EPI_ISL_1445783, EPI_ISL_1445785, EPI_ISL_1445786, EPI_ISL_1445787, EPI_ISL_1445788, EPI_ISL_1445789, EPI_ISL_1445790, EPI_ISL_1445791, EPI_ISL_1445792, EPI_ISL_1445793, EPI_ISL_1445794, EPI_ISL_1445795, EPI_ISL_1445796, EPI_ISL_1445799, EPI_ISL_1445800, EPI_ISL_1445801, EPI_ISL_1445802, EPI_ISL_1445803, EPI_ISL_1445804, EPI_ISL_1445805, EPI_ISL_1445806, EPI_ISL_1445807, EPI_ISL_1445808, EPI_ISL_1445809, EPI_ISL_1445812, EPI_ISL_1445813, EPI_ISL_1445815, EPI_ISL_1445817, EPI_ISL_1445818, EPI_ISL_1445819, EPI_ISL_1445820, EPI_ISL_1445821, EPI_ISL_1445822, EPI_ISL_1445823, EPI_ISL_1445824, EPI_ISL_1445825, EPI_ISL_1445826, EPI_ISL_1445827, EPI_ISL_1445828, EPI_ISL_1445829, EPI_ISL_1445830, EPI_ISL_1445831, EPI_ISL_1445832, EPI_ISL_1445833, EPI_ISL_1445834, EPI_ISL_1445835, EPI_ISL_1445836, EPI_ISL_1445837, EPI_ISL_1445838, EPI_ISL_1445839, EPI_ISL_1445840, EPI_ISL_1445841, EPI_ISL_1445842, EPI_ISL_1445843, EPI_ISL_1445844, EPI_ISL_1445845, EPI_ISL_1445846, EPI_ISL_1445847, EPI_ISL_1445848, EPI_ISL_1445849, EPI_ISL_1445850, EPI_ISL_1445851, EPI_ISL_1445852, EPI_ISL_1445854, EPI_ISL_1445856, EPI_ISL_1445857, EPI_ISL_1445859, EPI_ISL_1445860, EPI_ISL_1445861, EPI_ISL_1445862, EPI_ISL_1445863, EPI_ISL_1445864, EPI_ISL_1445865, EPI_ISL_1445866, EPI_ISL_1445867, EPI_ISL_1445868, EPI_ISL_1445869, EPI_ISL_1445870, EPI_ISL_1445872, EPI_ISL_1445874, EPI_ISL_1445875, EPI_ISL_1445876, EPI_ISL_1445877, EPI_ISL_1445878, EPI_ISL_1445879, EPI_ISL_1445880, EPI_ISL_1445881, EPI_ISL_1445882, EPI_ISL_1445883, EPI_ISL_1445884, EPI_ISL_1445886, EPI_ISL_1445887, EPI_ISL_1445888, EPI_ISL_1445889, EPI_ISL_1445890, EPI_ISL_1445891, EPI_ISL_1445892, EPI_ISL_1445893, EPI_ISL_1445894, EPI_ISL_1445896, EPI_ISL_1445897, EPI_ISL_1445898, EPI_ISL_1445899, EPI_ISL_1445900, EPI_ISL_1445901, EPI_ISL_1445902, EPI_ISL_1445903, EPI_ISL_1445904, EPI_ISL_1445906, EPI_ISL_1445907, EPI_ISL_1445908, EPI_ISL_1445909, EPI_ISL_1445910, EPI_ISL_1445911, EPI_ISL_1445912, EPI_ISL_1445913, EPI_ISL_1445914, EPI_ISL_1445915, EPI_ISL_1445916, EPI_ISL_1445917, EPI_ISL_1445919, EPI_ISL_1445920, EPI_ISL_1445921, EPI_ISL_1445922, EPI_ISL_1445923, EPI_ISL_1445924, EPI_ISL_1445925, EPI_ISL_1445926, EPI_ISL_1445927, EPI_ISL_1445928, EPI_ISL_1445929, EPI_ISL_1445931, EPI_ISL_1445932, EPI_ISL_1445933, EPI_ISL_1445934, EPI_ISL_1445935, EPI_ISL_1445936, EPI_ISL_1445937, EPI_ISL_1445938, EPI_ISL_1445939, EPI_ISL_1445940, EPI_ISL_1445941, EPI_ISL_1445942, EPI_ISL_1445944, EPI_ISL_1445945, EPI_ISL_1445946, EPI_ISL_1445947, EPI_ISL_1445948, EPI_ISL_1445950, EPI_ISL_1445951, EPI_ISL_1445953, EPI_ISL_1445954, EPI_ISL_1445955, EPI_ISL_1445956, EPI_ISL_1445957, EPI_ISL_1445958, EPI_ISL_1445959, EPI_ISL_1445960, EPI_ISL_1445961, EPI_ISL_1445962, EPI_ISL_1445963, EPI_ISL_1445964, EPI_ISL_1445965, EPI_ISL_1445966, EPI_ISL_1445967, EPI_ISL_1445968, EPI_ISL_1445969, EPI_ISL_1445970, EPI_ISL_1445971, EPI_ISL_1445972, EPI_ISL_1445973, EPI_ISL_1445974, EPI_ISL_1445975, EPI_ISL_1445976, EPI_ISL_1445977 | | | |
| see above | Aegis Sciences Corporation | Centers for Disease Control and Prevention Division of Viral Diseases, Pathogen Discovery | Dakota Howard, Dhwani Batra, Peter W. Cook, Kara Moser, Adrian Paskey, Jason Caravas, Benjamin Rambo-Martin, Shatavia Morrison, Christopher Gulvick, Scott Sammons, Yvette Unoarumhi, Darlene Wagner, Matthew Schmerer, Cyndi Clark, Patrick Campbell, Rob Case, Vikramsinha Ghorpade, Holly Houdeshell, Ola Kvalvaag, Dillon Nall, Ethan Sanders, Alec Vest, Shaun Westlund, Matthew Hardison, Clinton R. Paden, Duncan MacCannell |
| EPI_ISL_1445978 | Vault Health | Minnesota Department of Health, Public Health Laboratory | Alexandra Lorentz, Jacob Garfin, Matt Plumb, and Xiong Wang |
| EPI_ISL_1445979 | M Health Fairview | Minnesota Department of Health, Public Health Laboratory | Alexandra Lorentz, Jacob Garfin, Matt Plumb, and Xiong Wang |
| EPI_ISL_1445980 | Aegis Sciences Corporation | Centers for Disease Control and Prevention Division of Viral Diseases, Pathogen Discovery | Dakota Howard, Dhwani Batra, Peter W. Cook, Kara Moser, Adrian Paskey, Jason Caravas, Benjamin Rambo-Martin, Shatavia Morrison, Christopher Gulvick, Scott Sammons, Yvette Unoarumhi, Darlene Wagner, Matthew Schmerer, Cyndi Clark, Patrick Campbell, Rob Case, Vikramsinha Ghorpade, Holly Houdeshell, Ola Kvalvaag, Dillon Nall, Ethan Sanders, Alec Vest, Shaun Westlund, Matthew Hardison, Clinton R. Paden, Duncan MacCannell |
| EPI_ISL_1445981 | M Health Fairview | Minnesota Department of Health, Public Health Laboratory | Alexandra Lorentz, Jacob Garfin, Matt Plumb, and Xiong Wang |
| EPI_ISL_1445982 | Vault Health | Minnesota Department of Health, Public Health Laboratory | Alexandra Lorentz, Jacob Garfin, Matt Plumb, and Xiong Wang |

| | | | |
|---|---|---|---|
| EPI_ISL_1445983 | Aegis Sciences Corporation | Centers for Disease Control and Prevention Division of Viral Diseases, Pathogen Discovery | Dakota Howard, Dhwani Batra, Peter W. Cook, Kara Moser, Adrian Paskey, Jason Caravas, Benjamin Rambo-Martin, Shatavia Morrison, Christopher Gulvick, Scott Sammons, Yvette Unoarumhi, Darlene Wagner, Matthew Schmerer, Cyndi Clark, Patrick Campbell, Rob Case, Vikramsinha Ghorpade, Holly Houdeshell, Ola Kvalvaag, Dillon Nall, Ethan Sanders, Alec Vest, Shaun Westlund, Matthew Hardison, Clinton R. Paden, Duncan MacCannell |
| EPI_ISL_1445984, EPI_ISL_1445985, EPI_ISL_1445986 | M Health Fairview | Minnesota Department of Health, Public Health Laboratory | Alexandra Lorentz, Jacob Garfin, Matt Plumb, and Xiong Wang |
| EPI_ISL_1445987 | Aegis Sciences Corporation | Centers for Disease Control and Prevention Division of Viral Diseases, Pathogen Discovery | Dakota Howard, Dhwani Batra, Peter W. Cook, Kara Moser, Adrian Paskey, Jason Caravas, Benjamin Rambo-Martin, Shatavia Morrison, Christopher Gulvick, Scott Sammons, Yvette Unoarumhi, Darlene Wagner, Matthew Schmerer, Cyndi Clark, Patrick Campbell, Rob Case, Vikramsinha Ghorpade, Holly Houdeshell, Ola Kvalvaag, Dillon Nall, Ethan Sanders, Alec Vest, Shaun Westlund, Matthew Hardison, Clinton R. Paden, Duncan MacCannell |
| EPI_ISL_1445988 | Minnesota Department of Health, Public Health Laboratory | Minnesota Department of Health, Public Health Laboratory | Alexandra Lorentz, Jacob Garfin, Matt Plumb, and Xiong Wang |
| EPI_ISL_1445989 | Vault Health | Minnesota Department of Health, Public Health Laboratory | Alexandra Lorentz, Jacob Garfin, Matt Plumb, and Xiong Wang |
| EPI_ISL_1445990 | Aegis Sciences Corporation | Centers for Disease Control and Prevention Division of Viral Diseases, Pathogen Discovery | Dakota Howard, Dhwani Batra, Peter W. Cook, Kara Moser, Adrian Paskey, Jason Caravas, Benjamin Rambo-Martin, Shatavia Morrison, Christopher Gulvick, Scott Sammons, Yvette Unoarumhi, Darlene Wagner, Matthew Schmerer, Cyndi Clark, Patrick Campbell, Rob Case, Vikramsinha Ghorpade, Holly Houdeshell, Ola Kvalvaag, Dillon Nall, Ethan Sanders, Alec Vest, Shaun Westlund, Matthew Hardison, Clinton R. Paden, Duncan MacCannell |
| EPI_ISL_1445991 | M Health Fairview | Minnesota Department of Health, Public Health Laboratory | Alexandra Lorentz, Jacob Garfin, Matt Plumb, and Xiong Wang |
| EPI_ISL_1445992 | Vault Health | Minnesota Department of Health, Public Health Laboratory | Alexandra Lorentz, Jacob Garfin, Matt Plumb, and Xiong Wang |
| EPI_ISL_1445994 | Minnesota Department of Health, Public Health Laboratory | Minnesota Department of Health, Public Health Laboratory | Alexandra Lorentz, Jacob Garfin, Matt Plumb, and Xiong Wang |
| EPI_ISL_1445995 | M Health Fairview | Minnesota Department of Health, Public Health Laboratory | Alexandra Lorentz, Jacob Garfin, Matt Plumb, and Xiong Wang |
| EPI_ISL_1445996 | Aegis Sciences Corporation | Centers for Disease Control and Prevention Division of Viral Diseases, Pathogen Discovery | Dakota Howard, Dhwani Batra, Peter W. Cook, Kara Moser, Adrian Paskey, Jason Caravas, Benjamin Rambo-Martin, Shatavia Morrison, Christopher Gulvick, Scott Sammons, Yvette Unoarumhi, Darlene Wagner, Matthew Schmerer, Cyndi Clark, Patrick Campbell, Rob Case, Vikramsinha Ghorpade, Holly Houdeshell, Ola Kvalvaag, Dillon Nall, Ethan Sanders, Alec Vest, Shaun Westlund, Matthew Hardison, Clinton R. Paden, Duncan MacCannell |
| EPI_ISL_1445997 | Minnesota Department of Health, Public Health Laboratory | Minnesota Department of Health, Public Health Laboratory | Alexandra Lorentz, Jacob Garfin, Matt Plumb, and Xiong Wang |
| EPI_ISL_1445998 | M Health Fairview | Minnesota Department of Health, Public Health Laboratory | Alexandra Lorentz, Jacob Garfin, Matt Plumb, and Xiong Wang |
| EPI_ISL_1446000, EPI_ISL_1446001 | Vault Health | Minnesota Department of Health, Public Health Laboratory | Alexandra Lorentz, Jacob Garfin, Matt Plumb, and Xiong Wang |
| EPI_ISL_1446002 | Aegis Sciences Corporation | Centers for Disease Control and Prevention Division of Viral Diseases, Pathogen Discovery | Dakota Howard, Dhwani Batra, Peter W. Cook, Kara Moser, Adrian Paskey, Jason Caravas, Benjamin Rambo-Martin, Shatavia Morrison, Christopher Gulvick, Scott Sammons, Yvette Unoarumhi, Darlene Wagner, Matthew Schmerer, Cyndi Clark, Patrick Campbell, Rob Case, Vikramsinha Ghorpade, Holly Houdeshell, Ola Kvalvaag, Dillon Nall, Ethan Sanders, Alec Vest, Shaun Westlund, Matthew Hardison, Clinton R. Paden, Duncan MacCannell |
| EPI_ISL_1446003, EPI_ISL_1446004 | Minnesota Department of Health, Public Health Laboratory | Minnesota Department of Health, Public Health Laboratory | Alexandra Lorentz, Jacob Garfin, Matt Plumb, and Xiong Wang |
| EPI_ISL_1446005 | Aegis Sciences Corporation | Centers for Disease Control and Prevention Division of Viral Diseases, Pathogen Discovery | Dakota Howard, Dhwani Batra, Peter W. Cook, Kara Moser, Adrian Paskey, Jason Caravas, Benjamin Rambo-Martin, Shatavia Morrison, Christopher Gulvick, Scott Sammons, Yvette Unoarumhi, Darlene Wagner, Matthew Schmerer, Cyndi Clark, Patrick Campbell, Rob Case, Vikramsinha Ghorpade, Holly Houdeshell, Ola Kvalvaag, Dillon Nall, Ethan Sanders, Alec Vest, Shaun Westlund, Matthew Hardison, Clinton R. Paden, Duncan MacCannell |
| EPI_ISL_1446006 | Minnesota Department of Health, Public Health Laboratory | Minnesota Department of Health, Public Health Laboratory | Alexandra Lorentz, Jacob Garfin, Matt Plumb, and Xiong Wang |
| EPI_ISL_1446007 | Vault Health | Minnesota Department of Health, Public Health Laboratory | Alexandra Lorentz, Jacob Garfin, Matt Plumb, and Xiong Wang |
| EPI_ISL_1446008 | Aegis Sciences Corporation | Centers for Disease Control and Prevention Division of Viral Diseases, Pathogen Discovery | Dakota Howard, Dhwani Batra, Peter W. Cook, Kara Moser, Adrian Paskey, Jason Caravas, Benjamin Rambo-Martin, Shatavia Morrison, Christopher Gulvick, Scott Sammons, Yvette Unoarumhi, Darlene Wagner, Matthew Schmerer, Cyndi Clark, Patrick Campbell, Rob Case, Vikramsinha Ghorpade, Holly Houdeshell, Ola Kvalvaag, Dillon Nall, Ethan Sanders, Alec Vest, Shaun Westlund, Matthew Hardison, Clinton R. Paden, Duncan MacCannell |
| EPI_ISL_1446009, EPI_ISL_1446010 | M Health Fairview | Minnesota Department of Health, Public Health Laboratory | Alexandra Lorentz, Jacob Garfin, Matt Plumb, and Xiong Wang |
| EPI_ISL_1446011 | Aegis Sciences Corporation | Centers for Disease Control and Prevention Division of Viral Diseases, Pathogen Discovery | Dakota Howard, Dhwani Batra, Peter W. Cook, Kara Moser, Adrian Paskey, Jason Caravas, Benjamin Rambo-Martin, Shatavia Morrison, Christopher Gulvick, Scott Sammons, Yvette Unoarumhi, Darlene Wagner, Matthew Schmerer, Cyndi Clark, Patrick Campbell, Rob Case, Vikramsinha Ghorpade, Holly Houdeshell, Ola Kvalvaag, Dillon Nall, Ethan Sanders, Alec Vest, Shaun Westlund, Matthew Hardison, Clinton R. Paden, Duncan MacCannell |
| EPI_ISL_1446012 | M Health Fairview | Minnesota Department of Health, Public Health Laboratory | Alexandra Lorentz, Jacob Garfin, Matt Plumb, and Xiong Wang |
| EPI_ISL_1446013 | Aegis Sciences Corporation | Centers for Disease Control and Prevention Division of Viral Diseases, Pathogen Discovery | Dakota Howard, Dhwani Batra, Peter W. Cook, Kara Moser, Adrian Paskey, Jason Caravas, Benjamin Rambo-Martin, Shatavia Morrison, Christopher Gulvick, Scott Sammons, Yvette Unoarumhi, Darlene Wagner, Matthew Schmerer, Cyndi Clark, Patrick Campbell, Rob Case, Vikramsinha Ghorpade, Holly Houdeshell, Ola Kvalvaag, Dillon Nall, Ethan Sanders, Alec Vest, Shaun Westlund, Matthew Hardison, Clinton R. Paden, Duncan MacCannell |
| EPI_ISL_1446014 | Minnesota Department of Health, Public Health Laboratory | Minnesota Department of Health, Public Health Laboratory | Alexandra Lorentz, Jacob Garfin, Matt Plumb, and Xiong Wang |
| EPI_ISL_1446015 | Vault Health | Minnesota Department of Health, Public Health Laboratory | Alexandra Lorentz, Jacob Garfin, Matt Plumb, and Xiong Wang |
| EPI_ISL_1446016, EPI_ISL_1446019, EPI_ISL_1446022 | Aegis Sciences Corporation | Centers for Disease Control and Prevention Division of Viral Diseases, Pathogen Discovery | Dakota Howard, Dhwani Batra, Peter W. Cook, Kara Moser, Adrian Paskey, Jason Caravas, Benjamin Rambo-Martin, Shatavia Morrison, Christopher Gulvick, Scott Sammons, Yvette Unoarumhi, Darlene Wagner, Matthew Schmerer, Cyndi Clark, Patrick Campbell, Rob Case, Vikramsinha Ghorpade, Holly Houdeshell, Ola Kvalvaag, Dillon Nall, Ethan Sanders, Alec Vest, Shaun Westlund, Matthew Hardison, Clinton R. Paden, Duncan MacCannell |
| EPI_ISL_1446023, EPI_ISL_1446024 | M Health Fairview | Minnesota Department of Health, Public Health Laboratory | Alexandra Lorentz, Jacob Garfin, Matt Plumb, and Xiong Wang |
| EPI_ISL_1446025 | Aegis Sciences Corporation | Centers for Disease Control and Prevention Division of Viral Diseases, Pathogen Discovery | Dakota Howard, Dhwani Batra, Peter W. Cook, Kara Moser, Adrian Paskey, Jason Caravas, Benjamin Rambo-Martin, Shatavia Morrison, Christopher Gulvick, Scott Sammons, Yvette Unoarumhi, Darlene Wagner, Matthew Schmerer, Cyndi Clark, Patrick Campbell, Rob Case, Vikramsinha Ghorpade, Holly Houdeshell, Ola Kvalvaag, Dillon Nall, Ethan Sanders, Alec Vest, Shaun Westlund, Matthew Hardison, Clinton R. Paden, Duncan MacCannell |
| EPI_ISL_1446026 | Vault Health | Minnesota Department of Health, Public Health Laboratory | Alexandra Lorentz, Jacob Garfin, Matt Plumb, and Xiong Wang |
| EPI_ISL_1446027 | M Health Fairview | Minnesota Department of Health, Public Health Laboratory | Alexandra Lorentz, Jacob Garfin, Matt Plumb, and Xiong Wang |
| EPI_ISL_1446028 | Mayo Clinic & Mayo Clinic Laboratories | Minnesota Department of Health, Public Health Laboratory | Alexandra Lorentz, Jacob Garfin, Matt Plumb, and Xiong Wang |
| EPI_ISL_1446029 | Aegis Sciences Corporation | Centers for Disease Control and Prevention Division of Viral Diseases, Pathogen Discovery | Dakota Howard, Dhwani Batra, Peter W. Cook, Kara Moser, Adrian Paskey, Jason Caravas, Benjamin Rambo-Martin, Shatavia Morrison, Christopher Gulvick, Scott Sammons, Yvette Unoarumhi, Darlene Wagner, Matthew Schmerer, Cyndi Clark, Patrick Campbell, Rob Case, Vikramsinha Ghorpade, Holly Houdeshell, Ola Kvalvaag, Dillon Nall, Ethan Sanders, Alec Vest, Shaun Westlund, Matthew Hardison, Clinton R. Paden, Duncan MacCannell |
| EPI_ISL_1446030, EPI_ISL_1446031, EPI_ISL_1446033, EPI_ISL_1446034 | Minnesota Department of Health, Public Health Laboratory | Minnesota Department of Health, Public Health Laboratory | Alexandra Lorentz, Jacob Garfin, Matt Plumb, and Xiong Wang |
| EPI_ISL_1446035 | Aegis Sciences Corporation | Centers for Disease Control and Prevention Division of Viral Diseases, Pathogen Discovery | Dakota Howard, Dhwani Batra, Peter W. Cook, Kara Moser, Adrian Paskey, Jason Caravas, Benjamin Rambo-Martin, Shatavia Morrison, Christopher Gulvick, Scott Sammons, Yvette Unoarumhi, Darlene Wagner, Matthew Schmerer, Cyndi Clark, Patrick Campbell, Rob Case, Vikramsinha Ghorpade, Holly Houdeshell, Ola Kvalvaag, Dillon Nall, Ethan Sanders, Alec Vest, Shaun Westlund, Matthew Hardison, Clinton R. Paden, Duncan MacCannell |
| EPI_ISL_1446036, EPI_ISL_1446037 | Minnesota Department of Health, Public Health Laboratory | Minnesota Department of Health, Public Health Laboratory | Alexandra Lorentz, Jacob Garfin, Matt Plumb, and Xiong Wang |
| EPI_ISL_1446038 | Aegis Sciences Corporation | Centers for Disease Control and Prevention Division of Viral Diseases, Pathogen Discovery | Dakota Howard, Dhwani Batra, Peter W. Cook, Kara Moser, Adrian Paskey, Jason Caravas, Benjamin Rambo-Martin, Shatavia Morrison, Christopher Gulvick, Scott Sammons, Yvette Unoarumhi, Darlene Wagner, Matthew Schmerer, Cyndi Clark, Patrick Campbell, Rob Case, Vikramsinha Ghorpade, Holly Houdeshell, Ola Kvalvaag, Dillon Nall, Ethan Sanders, Alec Vest, Shaun Westlund, Matthew Hardison, Clinton R. Paden, Duncan MacCannell |
| EPI_ISL_1446039, EPI_ISL_1446040 | Minnesota Department of Health, Public Health Laboratory | Minnesota Department of Health, Public Health Laboratory | Alexandra Lorentz, Jacob Garfin, Matt Plumb, and Xiong Wang |

| | | | |
|---|---|---|---|
| EPI_ISL_1446041 | Aegis Sciences Corporation | Centers for Disease Control and Prevention Division of Viral Diseases, Pathogen Discovery | Dakota Howard, Dhwani Batra, Peter W. Cook, Kara Moser, Adrian Paskey, Jason Caravas, Benjamin Rambo-Martin, Shatavia Morrison, Christopher Gulvick, Scott Sammons, Yvette Unoarumhi, Darlene Wagner, Matthew Schmerer, Cyndi Clark, Patrick Campbell, Rob Case, Vikramsinha Ghorpade, Holly Houdeshell, Ola Kvalvaag, Dillon Nall, Ethan Sanders, Alec Vest, Shaun Westlund, Matthew Hardison, Clinton R. Paden, Duncan MacCannell |
| EPI_ISL_1446042, EPI_ISL_1446043 | Minnesota Department of Health, Public Health Laboratory | Minnesota Department of Health, Public Health Laboratory | Alexandra Lorentz, Jacob Garfin, Matt Plumb, and Xiong Wang |
| EPI_ISL_1446044 | Aegis Sciences Corporation | Centers for Disease Control and Prevention Division of Viral Diseases, Pathogen Discovery | Dakota Howard, Dhwani Batra, Peter W. Cook, Kara Moser, Adrian Paskey, Jason Caravas, Benjamin Rambo-Martin, Shatavia Morrison, Christopher Gulvick, Scott Sammons, Yvette Unoarumhi, Darlene Wagner, Matthew Schmerer, Cyndi Clark, Patrick Campbell, Rob Case, Vikramsinha Ghorpade, Holly Houdeshell, Ola Kvalvaag, Dillon Nall, Ethan Sanders, Alec Vest, Shaun Westlund, Matthew Hardison, Clinton R. Paden, Duncan MacCannell |
| EPI_ISL_1446045, EPI_ISL_1446046 | Minnesota Department of Health, Public Health Laboratory | Minnesota Department of Health, Public Health Laboratory | Alexandra Lorentz, Jacob Garfin, Matt Plumb, and Xiong Wang |
| EPI_ISL_1446047 | Aegis Sciences Corporation | Centers for Disease Control and Prevention Division of Viral Diseases, Pathogen Discovery | Dakota Howard, Dhwani Batra, Peter W. Cook, Kara Moser, Adrian Paskey, Jason Caravas, Benjamin Rambo-Martin, Shatavia Morrison, Christopher Gulvick, Scott Sammons, Yvette Unoarumhi, Darlene Wagner, Matthew Schmerer, Cyndi Clark, Patrick Campbell, Rob Case, Vikramsinha Ghorpade, Holly Houdeshell, Ola Kvalvaag, Dillon Nall, Ethan Sanders, Alec Vest, Shaun Westlund, Matthew Hardison, Clinton R. Paden, Duncan MacCannell |
| EPI_ISL_1446048, EPI_ISL_1446049 | Mayo Clinic & Mayo Clinic Laboratories | Minnesota Department of Health, Public Health Laboratory | Alexandra Lorentz, Jacob Garfin, Matt Plumb, and Xiong Wang |
| EPI_ISL_1446050 | Aegis Sciences Corporation | Centers for Disease Control and Prevention Division of Viral Diseases, Pathogen Discovery | Dakota Howard, Dhwani Batra, Peter W. Cook, Kara Moser, Adrian Paskey, Jason Caravas, Benjamin Rambo-Martin, Shatavia Morrison, Christopher Gulvick, Scott Sammons, Yvette Unoarumhi, Darlene Wagner, Matthew Schmerer, Cyndi Clark, Patrick Campbell, Rob Case, Vikramsinha Ghorpade, Holly Houdeshell, Ola Kvalvaag, Dillon Nall, Ethan Sanders, Alec Vest, Shaun Westlund, Matthew Hardison, Clinton R. Paden, Duncan MacCannell |
| EPI_ISL_1446051, EPI_ISL_1446052 | Mayo Clinic & Mayo Clinic Laboratories | Minnesota Department of Health, Public Health Laboratory | Alexandra Lorentz, Jacob Garfin, Matt Plumb, and Xiong Wang |
| EPI_ISL_1446053 | Aegis Sciences Corporation | Centers for Disease Control and Prevention Division of Viral Diseases, Pathogen Discovery | Dakota Howard, Dhwani Batra, Peter W. Cook, Kara Moser, Adrian Paskey, Jason Caravas, Benjamin Rambo-Martin, Shatavia Morrison, Christopher Gulvick, Scott Sammons, Yvette Unoarumhi, Darlene Wagner, Matthew Schmerer, Cyndi Clark, Patrick Campbell, Rob Case, Vikramsinha Ghorpade, Holly Houdeshell, Ola Kvalvaag, Dillon Nall, Ethan Sanders, Alec Vest, Shaun Westlund, Matthew Hardison, Clinton R. Paden, Duncan MacCannell |
| EPI_ISL_1446054, EPI_ISL_1446055 | Mayo Clinic & Mayo Clinic Laboratories | Minnesota Department of Health, Public Health Laboratory | Alexandra Lorentz, Jacob Garfin, Matt Plumb, and Xiong Wang |
| EPI_ISL_1446056 | Vault Health | Minnesota Department of Health, Public Health Laboratory | Alexandra Lorentz, Jacob Garfin, Matt Plumb, and Xiong Wang |
| EPI_ISL_1446057 | Aegis Sciences Corporation | Centers for Disease Control and Prevention Division of Viral Diseases, Pathogen Discovery | Dakota Howard, Dhwani Batra, Peter W. Cook, Kara Moser, Adrian Paskey, Jason Caravas, Benjamin Rambo-Martin, Shatavia Morrison, Christopher Gulvick, Scott Sammons, Yvette Unoarumhi, Darlene Wagner, Matthew Schmerer, Cyndi Clark, Patrick Campbell, Rob Case, Vikramsinha Ghorpade, Holly Houdeshell, Ola Kvalvaag, Dillon Nall, Ethan Sanders, Alec Vest, Shaun Westlund, Matthew Hardison, Clinton R. Paden, Duncan MacCannell |
| EPI_ISL_1446058, EPI_ISL_1446059 | Vault Health | Minnesota Department of Health, Public Health Laboratory | Alexandra Lorentz, Jacob Garfin, Matt Plumb, and Xiong Wang |
| EPI_ISL_1446061, EPI_ISL_1446062 | M Health Fairview | Minnesota Department of Health, Public Health Laboratory | Alexandra Lorentz, Jacob Garfin, Matt Plumb, and Xiong Wang |
| EPI_ISL_1446063 | Aegis Sciences Corporation | Centers for Disease Control and Prevention Division of Viral Diseases, Pathogen Discovery | Dakota Howard, Dhwani Batra, Peter W. Cook, Kara Moser, Adrian Paskey, Jason Caravas, Benjamin Rambo-Martin, Shatavia Morrison, Christopher Gulvick, Scott Sammons, Yvette Unoarumhi, Darlene Wagner, Matthew Schmerer, Cyndi Clark, Patrick Campbell, Rob Case, Vikramsinha Ghorpade, Holly Houdeshell, Ola Kvalvaag, Dillon Nall, Ethan Sanders, Alec Vest, Shaun Westlund, Matthew Hardison, Clinton R. Paden, Duncan MacCannell |
| EPI_ISL_1446064, EPI_ISL_1446065 | M Health Fairview | Minnesota Department of Health, Public Health Laboratory | Alexandra Lorentz, Jacob Garfin, Matt Plumb, and Xiong Wang |
| EPI_ISL_1446066 | Aegis Sciences Corporation | Centers for Disease Control and Prevention Division of Viral Diseases, Pathogen Discovery | Dakota Howard, Dhwani Batra, Peter W. Cook, Kara Moser, Adrian Paskey, Jason Caravas, Benjamin Rambo-Martin, Shatavia Morrison, Christopher Gulvick, Scott Sammons, Yvette Unoarumhi, Darlene Wagner, Matthew Schmerer, Cyndi Clark, Patrick Campbell, Rob Case, Vikramsinha Ghorpade, Holly Houdeshell, Ola Kvalvaag, Dillon Nall, Ethan Sanders, Alec Vest, Shaun Westlund, Matthew Hardison, Clinton R. Paden, Duncan MacCannell |
| EPI_ISL_1446067, EPI_ISL_1446068 | M Health Fairview | Minnesota Department of Health, Public Health Laboratory | Alexandra Lorentz, Jacob Garfin, Matt Plumb, and Xiong Wang |
| EPI_ISL_1446069 | Aegis Sciences Corporation | Centers for Disease Control and Prevention Division of Viral Diseases, Pathogen Discovery | Dakota Howard, Dhwani Batra, Peter W. Cook, Kara Moser, Adrian Paskey, Jason Caravas, Benjamin Rambo-Martin, Shatavia Morrison, Christopher Gulvick, Scott Sammons, Yvette Unoarumhi, Darlene Wagner, Matthew Schmerer, Cyndi Clark, Patrick Campbell, Rob Case, Vikramsinha Ghorpade, Holly Houdeshell, Ola Kvalvaag, Dillon Nall, Ethan Sanders, Alec Vest, Shaun Westlund, Matthew Hardison, Clinton R. Paden, Duncan MacCannell |
| EPI_ISL_1446070 | M Health Fairview | Minnesota Department of Health, Public Health Laboratory | Alexandra Lorentz, Jacob Garfin, Matt Plumb, and Xiong Wang |
| EPI_ISL_1446071 | Aegis Sciences Corporation | Centers for Disease Control and Prevention Division of Viral Diseases, Pathogen Discovery | Dakota Howard, Dhwani Batra, Peter W. Cook, Kara Moser, Adrian Paskey, Jason Caravas, Benjamin Rambo-Martin, Shatavia Morrison, Christopher Gulvick, Scott Sammons, Yvette Unoarumhi, Darlene Wagner, Matthew Schmerer, Cyndi Clark, Patrick Campbell, Rob Case, Vikramsinha Ghorpade, Holly Houdeshell, Ola Kvalvaag, Dillon Nall, Ethan Sanders, Alec Vest, Shaun Westlund, Matthew Hardison, Clinton R. Paden, Duncan MacCannell |
| EPI_ISL_1446072, EPI_ISL_1446073, EPI_ISL_1446074 | M Health Fairview | Minnesota Department of Health, Public Health Laboratory | Alexandra Lorentz, Jacob Garfin, Matt Plumb, and Xiong Wang |
| EPI_ISL_1446075 | Aegis Sciences Corporation | Centers for Disease Control and Prevention Division of Viral Diseases, Pathogen Discovery | Dakota Howard, Dhwani Batra, Peter W. Cook, Kara Moser, Adrian Paskey, Jason Caravas, Benjamin Rambo-Martin, Shatavia Morrison, Christopher Gulvick, Scott Sammons, Yvette Unoarumhi, Darlene Wagner, Matthew Schmerer, Cyndi Clark, Patrick Campbell, Rob Case, Vikramsinha Ghorpade, Holly Houdeshell, Ola Kvalvaag, Dillon Nall, Ethan Sanders, Alec Vest, Shaun Westlund, Matthew Hardison, Clinton R. Paden, Duncan MacCannell |
| EPI_ISL_1446076 | M Health Fairview | Minnesota Department of Health, Public Health Laboratory | Alexandra Lorentz, Jacob Garfin, Matt Plumb, and Xiong Wang |
| EPI_ISL_1446077 | Aegis Sciences Corporation | Centers for Disease Control and Prevention Division of Viral Diseases, Pathogen Discovery | Dakota Howard, Dhwani Batra, Peter W. Cook, Kara Moser, Adrian Paskey, Jason Caravas, Benjamin Rambo-Martin, Shatavia Morrison, Christopher Gulvick, Scott Sammons, Yvette Unoarumhi, Darlene Wagner, Matthew Schmerer, Cyndi Clark, Patrick Campbell, Rob Case, Vikramsinha Ghorpade, Holly Houdeshell, Ola Kvalvaag, Dillon Nall, Ethan Sanders, Alec Vest, Shaun Westlund, Matthew Hardison, Clinton R. Paden, Duncan MacCannell |
| EPI_ISL_1446078, EPI_ISL_1446079, EPI_ISL_1446080 | M Health Fairview | Minnesota Department of Health, Public Health Laboratory | Alexandra Lorentz, Jacob Garfin, Matt Plumb, and Xiong Wang |
| EPI_ISL_1446081, EPI_ISL_1446082, EPI_ISL_1446083, EPI_ISL_1446084, EPI_ISL_1446085, EPI_ISL_1446086, EPI_ISL_1446087, EPI_ISL_1446088, EPI_ISL_1446089, EPI_ISL_1446091, EPI_ISL_1446092, EPI_ISL_1446093, EPI_ISL_1446094, EPI_ISL_1446095, EPI_ISL_1446096, EPI_ISL_1446097, EPI_ISL_1446098, EPI_ISL_1446099, EPI_ISL_1446100, EPI_ISL_1446101, EPI_ISL_1446102, EPI_ISL_1446103, EPI_ISL_1446104, EPI_ISL_1446105, EPI_ISL_1446106, EPI_ISL_1446107, EPI_ISL_1446108, EPI_ISL_1446109, EPI_ISL_1446110, EPI_ISL_1446111, EPI_ISL_1446112, EPI_ISL_1446113, EPI_ISL_1446114, EPI_ISL_1446115, EPI_ISL_1446117, EPI_ISL_1446118, EPI_ISL_1446119, EPI_ISL_1446120, EPI_ISL_1446121, EPI_ISL_1446122, EPI_ISL_1446124, EPI_ISL_1446125, EPI_ISL_1446126, EPI_ISL_1446127, EPI_ISL_1446128, EPI_ISL_1446129, EPI_ISL_1446130, EPI_ISL_1446131, EPI_ISL_1446132, EPI_ISL_1446133, EPI_ISL_1446134, EPI_ISL_1446135, EPI_ISL_1446136, EPI_ISL_1446137, EPI_ISL_1446138, EPI_ISL_1446139, EPI_ISL_1446140, EPI_ISL_1446141, EPI_ISL_1446143, EPI_ISL_1446144, EPI_ISL_1446146, EPI_ISL_1446147, EPI_ISL_1446148, EPI_ISL_1446149, EPI_ISL_1446150, EPI_ISL_1446151, EPI_ISL_1446153, EPI_ISL_1446154, EPI_ISL_1446155, EPI_ISL_1446156, EPI_ISL_1446158, EPI_ISL_1446159, EPI_ISL_1446160, EPI_ISL_1446161, EPI_ISL_1446162, EPI_ISL_1446163, EPI_ISL_1446164, EPI_ISL_1446165, EPI_ISL_1446166, EPI_ISL_1446167, EPI_ISL_1446168, EPI_ISL_1446169, EPI_ISL_1446170, EPI_ISL_1446171, EPI_ISL_1446172, EPI_ISL_1446173, EPI_ISL_1446174, EPI_ISL_1446175, EPI_ISL_1446176, EPI_ISL_1446177, EPI_ISL_1446178, EPI_ISL_1446179, EPI_ISL_1446181, EPI_ISL_1446182, EPI_ISL_1446183, EPI_ISL_1446184, EPI_ISL_1446185, EPI_ISL_1446186, EPI_ISL_1446187, EPI_ISL_1446189, EPI_ISL_1446190, EPI_ISL_1446192, EPI_ISL_1446193 | | | |
| see above | Aegis Sciences Corporation | Centers for Disease Control and Prevention Division of Viral Diseases, Pathogen Discovery | Dakota Howard, Dhwani Batra, Peter W. Cook, Kara Moser, Adrian Paskey, Jason Caravas, Benjamin Rambo-Martin, Shatavia Morrison, Christopher Gulvick, Scott Sammons, Yvette Unoarumhi, Darlene Wagner, Matthew Schmerer, Cyndi Clark, Patrick Campbell, Rob Case, Vikramsinha Ghorpade, Holly Houdeshell, Ola Kvalvaag, Dillon Nall, Ethan Sanders, Alec Vest, Shaun Westlund, Matthew Hardison, Clinton R. Paden, Duncan MacCannell |
| EPI_ISL_1446195, EPI_ISL_1446196, EPI_ISL_1446197, EPI_ISL_1446198, EPI_ISL_1446199, EPI_ISL_1446200, EPI_ISL_1446201, EPI_ISL_1446202, EPI_ISL_1446204, EPI_ISL_1446205, EPI_ISL_1446206, EPI_ISL_1446208, EPI_ISL_1446209, EPI_ISL_1446210, EPI_ISL_1446211, EPI_ISL_1446212, EPI_ISL_1446213, EPI_ISL_1446214, EPI_ISL_1446215, EPI_ISL_1446216, EPI_ISL_1446217, EPI_ISL_1446218, EPI_ISL_1446219, EPI_ISL_1446220, EPI_ISL_1446221, EPI_ISL_1446226, EPI_ISL_1446227, EPI_ISL_1446228, EPI_ISL_1446229, EPI_ISL_1446230, EPI_ISL_1446236, EPI_ISL_1446237, EPI_ISL_1446238, EPI_ISL_1446239, EPI_ISL_1446240, EPI_ISL_1446241, EPI_ISL_1446242, EPI_ISL_1446244, EPI_ISL_1446245, EPI_ISL_1446246, EPI_ISL_1446247, EPI_ISL_1446249, EPI_ISL_1446251, EPI_ISL_1446252, EPI_ISL_1446253, EPI_ISL_1446254, EPI_ISL_1446255, EPI_ISL_1446256, EPI_ISL_1446257, EPI_ISL_1446260, EPI_ISL_1446261, EPI_ISL_1446262, EPI_ISL_1446263, EPI_ISL_1446264, EPI_ISL_1446265, EPI_ISL_1446266, EPI_ISL_1446267, EPI_ISL_1446268, EPI_ISL_1446269, EPI_ISL_1446272, EPI_ISL_1446273, EPI_ISL_1446274, EPI_ISL_1446275, EPI_ISL_1446276, EPI_ISL_1446277, EPI_ISL_1446278, EPI_ISL_1446279, EPI_ISL_1446281, EPI_ISL_1446282, EPI_ISL_1446283 | | | |
| see above | Texas Children's Hospital | Texas Children's Microbiome Center | Ruth Ann Luna, Jennifer K. Spinler, James Dunn, James Versalovic, Ila Singh |
| EPI_ISL_1446284 | TN Division of Laboratory Services | Centers for Disease Control and Prevention Division of Viral Diseases, Pathogen Discovery | Mili Sheth, Sarah Nobles, Jasmine Padilla, Mark Burroughs, Shoshona Le, Katie Dillon, Peter Cook, Clinton R. Paden, Dhwani Batra, Krista Queen, Kristen Knipe, Dakota Howard, Yvette Unoarumhi, Darlene Wagner, Matthew Schmerer, Ben L. Rambo-Martin, Kristine Lacek, Sam Shepard, Alison Laufer Halpin, Dave Wentworth, Vivien Dugan, Suxiang Tong, Justin Lee |
| EPI_ISL_1446285 | Texas Children's Hospital | Texas Children's Microbiome Center | Ruth Ann Luna, Jennifer K. Spinler, James Dunn, James Versalovic, Ila Singh |
| EPI_ISL_1446287 | TN Division of Laboratory Services | Centers for Disease Control and Prevention Division of Viral Diseases, Pathogen Discovery | Mili Sheth, Sarah Nobles, Jasmine Padilla, Mark Burroughs, Shoshona Le, Katie Dillon, Peter Cook, Clinton R. Paden, Dhwani Batra, Krista Queen, Kristen Knipe, Dakota Howard, Yvette Unoarumhi, Darlene Wagner, Matthew Schmerer, Ben L. Rambo-Martin, Kristine Lacek, Sam Shepard, Alison Laufer Halpin, Dave Wentworth, Vivien Dugan, Suxiang Tong, Justin Lee |
| EPI_ISL_1446290 | MS Public Health Laboratory | Centers for Disease Control and Prevention Division of Viral Diseases, Pathogen Discovery | Mili Sheth, Sarah Nobles, Jasmine Padilla, Mark Burroughs, Shoshona Le, Katie Dillon, Peter Cook, Clinton R. Paden, Dhwani Batra, Krista Queen, Kristen Knipe, Dakota Howard, Yvette Unoarumhi, Darlene Wagner, Matthew Schmerer, Ben L. Rambo-Martin, Kristine Lacek, Sam Shepard, Alison Laufer Halpin, Dave Wentworth, Vivien Dugan, Suxiang Tong, Justin Lee |
| EPI_ISL_1446291 | Texas Children's Hospital | Texas Children's Microbiome Center | Ruth Ann Luna, Jennifer K. Spinler, James Dunn, James Versalovic, Ila Singh |
| EPI_ISL_1446293, EPI_ISL_1446296 | MS Public Health Laboratory | Centers for Disease Control and Prevention Division of Viral Diseases, Pathogen Discovery | Mili Sheth, Sarah Nobles, Jasmine Padilla, Mark Burroughs, Shoshona Le, Katie Dillon, Peter Cook, Clinton R. Paden, Dhwani Batra, Krista Queen, Kristen Knipe, Dakota Howard, Yvette Unoarumhi, Darlene Wagner, Matthew Schmerer, Ben L. Rambo-Martin, Kristine Lacek, Sam Shepard, Alison Laufer Halpin, Dave Wentworth, Vivien Dugan, Suxiang Tong, Justin Lee |
| EPI_ISL_1446298 | Texas Children's Hospital | Texas Children's Microbiome Center | Ruth Ann Luna, Jennifer K. Spinler, James Dunn, James Versalovic, Ila Singh |

| | | | |
|---|---|---|---|
| EPI_ISL_1446299 | MO State Public Health Laboratory | Centers for Disease Control and Prevention Division of Viral Diseases, Pathogen Discovery | Mili Sheth, Sarah Nobles, Jasmine Padilla, Mark Burroughs, Shoshona Le, Katie Dillon, Peter Cook, Clinton R. Paden, Dhwani Batra, Krista Queen, Kristen Knipe, Dakota Howard, Yvette Unoarumhi, Darlene Wagner, Matthew Schmerer, Ben L. Rambo-Martin, Kristine Lacek, Sam Shepard, Alison Laufer Halpin, Dave Wentworth, Vivien Dugan, Suxiang Tong, Justin Lee |
| EPI_ISL_1446300, EPI_ISL_1446301 | Texas Children's Hospital | Texas Children's Microbiome Center | Ruth Ann Luna, Jennifer K. Spinler, James Dunn, James Versalovic, Ila Singh |
| EPI_ISL_1446302, EPI_ISL_1446304 | MS Public Health Laboratory | Centers for Disease Control and Prevention Division of Viral Diseases, Pathogen Discovery | Mili Sheth, Sarah Nobles, Jasmine Padilla, Mark Burroughs, Shoshona Le, Katie Dillon, Peter Cook, Clinton R. Paden, Dhwani Batra, Krista Queen, Kristen Knipe, Dakota Howard, Yvette Unoarumhi, Darlene Wagner, Matthew Schmerer, Ben L. Rambo-Martin, Kristine Lacek, Sam Shepard, Alison Laufer Halpin, Dave Wentworth, Vivien Dugan, Suxiang Tong, Justin Lee |
| EPI_ISL_1446305, EPI_ISL_1446306 | Texas Children's Hospital | Texas Children's Microbiome Center | Ruth Ann Luna, Jennifer K. Spinler, James Dunn, James Versalovic, Ila Singh |
| EPI_ISL_1446307 | MS Public Health Laboratory | Centers for Disease Control and Prevention Division of Viral Diseases, Pathogen Discovery | Mili Sheth, Sarah Nobles, Jasmine Padilla, Mark Burroughs, Shoshona Le, Katie Dillon, Peter Cook, Clinton R. Paden, Dhwani Batra, Krista Queen, Kristen Knipe, Dakota Howard, Yvette Unoarumhi, Darlene Wagner, Matthew Schmerer, Ben L. Rambo-Martin, Kristine Lacek, Sam Shepard, Alison Laufer Halpin, Dave Wentworth, Vivien Dugan, Suxiang Tong, Justin Lee |
| EPI_ISL_1446308, EPI_ISL_1446309 | Texas Children's Hospital | Texas Children's Microbiome Center | Ruth Ann Luna, Jennifer K. Spinler, James Dunn, James Versalovic, Ila Singh |
| EPI_ISL_1446310 | MS Public Health Laboratory | Centers for Disease Control and Prevention Division of Viral Diseases, Pathogen Discovery | Mili Sheth, Sarah Nobles, Jasmine Padilla, Mark Burroughs, Shoshona Le, Katie Dillon, Peter Cook, Clinton R. Paden, Dhwani Batra, Krista Queen, Kristen Knipe, Dakota Howard, Yvette Unoarumhi, Darlene Wagner, Matthew Schmerer, Ben L. Rambo-Martin, Kristine Lacek, Sam Shepard, Alison Laufer Halpin, Dave Wentworth, Vivien Dugan, Suxiang Tong, Justin Lee |
| EPI_ISL_1446311, EPI_ISL_1446312 | Texas Children's Hospital | Texas Children's Microbiome Center | Ruth Ann Luna, Jennifer K. Spinler, James Dunn, James Versalovic, Ila Singh |
| EPI_ISL_1446313 | MO State Public Health Laboratory | Centers for Disease Control and Prevention Division of Viral Diseases, Pathogen Discovery | Mili Sheth, Sarah Nobles, Jasmine Padilla, Mark Burroughs, Shoshona Le, Katie Dillon, Peter Cook, Clinton R. Paden, Dhwani Batra, Krista Queen, Kristen Knipe, Dakota Howard, Yvette Unoarumhi, Darlene Wagner, Matthew Schmerer, Ben L. Rambo-Martin, Kristine Lacek, Sam Shepard, Alison Laufer Halpin, Dave Wentworth, Vivien Dugan, Suxiang Tong, Justin Lee |
| EPI_ISL_1446314 | Texas Children's Hospital | Texas Children's Microbiome Center | Ruth Ann Luna, Jennifer K. Spinler, James Dunn, James Versalovic, Ila Singh |
| EPI_ISL_1446315 | MS Public Health Laboratory | Centers for Disease Control and Prevention Division of Viral Diseases, Pathogen Discovery | Mili Sheth, Sarah Nobles, Jasmine Padilla, Mark Burroughs, Shoshona Le, Katie Dillon, Peter Cook, Clinton R. Paden, Dhwani Batra, Krista Queen, Kristen Knipe, Dakota Howard, Yvette Unoarumhi, Darlene Wagner, Matthew Schmerer, Ben L. Rambo-Martin, Kristine Lacek, Sam Shepard, Alison Laufer Halpin, Dave Wentworth, Vivien Dugan, Suxiang Tong, Justin Lee |
| EPI_ISL_1446316, EPI_ISL_1446317 | Texas Children's Hospital | Texas Children's Microbiome Center | Ruth Ann Luna, Jennifer K. Spinler, James Dunn, James Versalovic, Ila Singh |
| EPI_ISL_1446318 | MS Public Health Laboratory | Centers for Disease Control and Prevention Division of Viral Diseases, Pathogen Discovery | Mili Sheth, Sarah Nobles, Jasmine Padilla, Mark Burroughs, Shoshona Le, Katie Dillon, Peter Cook, Clinton R. Paden, Dhwani Batra, Krista Queen, Kristen Knipe, Dakota Howard, Yvette Unoarumhi, Darlene Wagner, Matthew Schmerer, Ben L. Rambo-Martin, Kristine Lacek, Sam Shepard, Alison Laufer Halpin, Dave Wentworth, Vivien Dugan, Suxiang Tong, Justin Lee |
| EPI_ISL_1446319, EPI_ISL_1446320 | Texas Children's Hospital | Texas Children's Microbiome Center | Ruth Ann Luna, Jennifer K. Spinler, James Dunn, James Versalovic, Ila Singh |
| EPI_ISL_1446321 | MS Public Health Laboratory | Centers for Disease Control and Prevention Division of Viral Diseases, Pathogen Discovery | Mili Sheth, Sarah Nobles, Jasmine Padilla, Mark Burroughs, Shoshona Le, Katie Dillon, Peter Cook, Clinton R. Paden, Dhwani Batra, Krista Queen, Kristen Knipe, Dakota Howard, Yvette Unoarumhi, Darlene Wagner, Matthew Schmerer, Ben L. Rambo-Martin, Kristine Lacek, Sam Shepard, Alison Laufer Halpin, Dave Wentworth, Vivien Dugan, Suxiang Tong, Justin Lee |
| EPI_ISL_1446322 | Texas Children's Hospital | Texas Children's Microbiome Center | Ruth Ann Luna, Jennifer K. Spinler, James Dunn, James Versalovic, Ila Singh |
| EPI_ISL_1446324 | OR State PHL-Virology/Immunology Section | Centers for Disease Control and Prevention Division of Viral Diseases, Pathogen Discovery | Mili Sheth, Sarah Nobles, Jasmine Padilla, Mark Burroughs, Shoshona Le, Katie Dillon, Peter Cook, Clinton R. Paden, Dhwani Batra, Krista Queen, Kristen Knipe, Dakota Howard, Yvette Unoarumhi, Darlene Wagner, Matthew Schmerer, Ben L. Rambo-Martin, Kristine Lacek, Sam Shepard, Alison Laufer Halpin, Dave Wentworth, Vivien Dugan, Suxiang Tong, Justin Lee |
| EPI_ISL_1446326 | Texas Children's Hospital | Texas Children's Microbiome Center | Ruth Ann Luna, Jennifer K. Spinler, James Dunn, James Versalovic, Ila Singh |
| EPI_ISL_1446327 | OR State PHL-Virology/Immunology Section | Centers for Disease Control and Prevention Division of Viral Diseases, Pathogen Discovery | Mili Sheth, Sarah Nobles, Jasmine Padilla, Mark Burroughs, Shoshona Le, Katie Dillon, Peter Cook, Clinton R. Paden, Dhwani Batra, Krista Queen, Kristen Knipe, Dakota Howard, Yvette Unoarumhi, Darlene Wagner, Matthew Schmerer, Ben L. Rambo-Martin, Kristine Lacek, Sam Shepard, Alison Laufer Halpin, Dave Wentworth, Vivien Dugan, Suxiang Tong, Justin Lee |
| EPI_ISL_1446328, EPI_ISL_1446329 | Texas Children's Hospital | Texas Children's Microbiome Center | Ruth Ann Luna, Jennifer K. Spinler, James Dunn, James Versalovic, Ila Singh |
| EPI_ISL_1446330 | OR State PHL-Virology/Immunology Section | Centers for Disease Control and Prevention Division of Viral Diseases, Pathogen Discovery | Mili Sheth, Sarah Nobles, Jasmine Padilla, Mark Burroughs, Shoshona Le, Katie Dillon, Peter Cook, Clinton R. Paden, Dhwani Batra, Krista Queen, Kristen Knipe, Dakota Howard, Yvette Unoarumhi, Darlene Wagner, Matthew Schmerer, Ben L. Rambo-Martin, Kristine Lacek, Sam Shepard, Alison Laufer Halpin, Dave Wentworth, Vivien Dugan, Suxiang Tong, Justin Lee |
| EPI_ISL_1446331 | Texas Children's Hospital | Texas Children's Microbiome Center | Ruth Ann Luna, Jennifer K. Spinler, James Dunn, James Versalovic, Ila Singh |
| EPI_ISL_1446332 | OR State PHL-Virology/Immunology Section | Centers for Disease Control and Prevention Division of Viral Diseases, Pathogen Discovery | Mili Sheth, Sarah Nobles, Jasmine Padilla, Mark Burroughs, Shoshona Le, Katie Dillon, Peter Cook, Clinton R. Paden, Dhwani Batra, Krista Queen, Kristen Knipe, Dakota Howard, Yvette Unoarumhi, Darlene Wagner, Matthew Schmerer, Ben L. Rambo-Martin, Kristine Lacek, Sam Shepard, Alison Laufer Halpin, Dave Wentworth, Vivien Dugan, Suxiang Tong, Justin Lee |
| EPI_ISL_1446333, EPI_ISL_1446334 | Texas Children's Hospital | Texas Children's Microbiome Center | Ruth Ann Luna, Jennifer K. Spinler, James Dunn, James Versalovic, Ila Singh |
| EPI_ISL_1446335 | KY State Public Health Lab | Centers for Disease Control and Prevention Division of Viral Diseases, Pathogen Discovery | Mili Sheth, Sarah Nobles, Jasmine Padilla, Mark Burroughs, Shoshona Le, Katie Dillon, Peter Cook, Clinton R. Paden, Dhwani Batra, Krista Queen, Kristen Knipe, Dakota Howard, Yvette Unoarumhi, Darlene Wagner, Matthew Schmerer, Ben L. Rambo-Martin, Kristine Lacek, Sam Shepard, Alison Laufer Halpin, Dave Wentworth, Vivien Dugan, Suxiang Tong, Justin Lee |
| EPI_ISL_1446336 | Texas Children's Hospital | Texas Children's Microbiome Center | Ruth Ann Luna, Jennifer K. Spinler, James Dunn, James Versalovic, Ila Singh |
| EPI_ISL_1446338 | KY State Public Health Lab | Centers for Disease Control and Prevention Division of Viral Diseases, Pathogen Discovery | Mili Sheth, Sarah Nobles, Jasmine Padilla, Mark Burroughs, Shoshona Le, Katie Dillon, Peter Cook, Clinton R. Paden, Dhwani Batra, Krista Queen, Kristen Knipe, Dakota Howard, Yvette Unoarumhi, Darlene Wagner, Matthew Schmerer, Ben L. Rambo-Martin, Kristine Lacek, Sam Shepard, Alison Laufer Halpin, Dave Wentworth, Vivien Dugan, Suxiang Tong, Justin Lee |
| EPI_ISL_1446340 | Texas Children's Hospital | Texas Children's Microbiome Center | Ruth Ann Luna, Jennifer K. Spinler, James Dunn, James Versalovic, Ila Singh |
| EPI_ISL_1446341 | OR State PHL-Virology/Immunology Section | Centers for Disease Control and Prevention Division of Viral Diseases, Pathogen Discovery | Mili Sheth, Sarah Nobles, Jasmine Padilla, Mark Burroughs, Shoshona Le, Katie Dillon, Peter Cook, Clinton R. Paden, Dhwani Batra, Krista Queen, Kristen Knipe, Dakota Howard, Yvette Unoarumhi, Darlene Wagner, Matthew Schmerer, Ben L. Rambo-Martin, Kristine Lacek, Sam Shepard, Alison Laufer Halpin, Dave Wentworth, Vivien Dugan, Suxiang Tong, Justin Lee |
| EPI_ISL_1446342, EPI_ISL_1446343 | Texas Children's Hospital | Texas Children's Microbiome Center | Ruth Ann Luna, Jennifer K. Spinler, James Dunn, James Versalovic, Ila Singh |
| EPI_ISL_1446344 | KY State Public Health Lab | Centers for Disease Control and Prevention Division of Viral Diseases, Pathogen Discovery | Mili Sheth, Sarah Nobles, Jasmine Padilla, Mark Burroughs, Shoshona Le, Katie Dillon, Peter Cook, Clinton R. Paden, Dhwani Batra, Krista Queen, Kristen Knipe, Dakota Howard, Yvette Unoarumhi, Darlene Wagner, Matthew Schmerer, Ben L. Rambo-Martin, Kristine Lacek, Sam Shepard, Alison Laufer Halpin, Dave Wentworth, Vivien Dugan, Suxiang Tong, Justin Lee |
| EPI_ISL_1446345, EPI_ISL_1446346 | Texas Children's Hospital | Texas Children's Microbiome Center | Ruth Ann Luna, Jennifer K. Spinler, James Dunn, James Versalovic, Ila Singh |
| EPI_ISL_1446347 | KY State Public Health Lab | Centers for Disease Control and Prevention Division of Viral Diseases, Pathogen Discovery | Mili Sheth, Sarah Nobles, Jasmine Padilla, Mark Burroughs, Shoshona Le, Katie Dillon, Peter Cook, Clinton R. Paden, Dhwani Batra, Krista Queen, Kristen Knipe, Dakota Howard, Yvette Unoarumhi, Darlene Wagner, Matthew Schmerer, Ben L. Rambo-Martin, Kristine Lacek, Sam Shepard, Alison Laufer Halpin, Dave Wentworth, Vivien Dugan, Suxiang Tong, Justin Lee |
| EPI_ISL_1446348, EPI_ISL_1446349 | Texas Children's Hospital | Texas Children's Microbiome Center | Ruth Ann Luna, Jennifer K. Spinler, James Dunn, James Versalovic, Ila Singh |
| EPI_ISL_1446350, EPI_ISL_1446352, EPI_ISL_1446355 | KY State Public Health Lab | Centers for Disease Control and Prevention Division of Viral Diseases, Pathogen Discovery | Mili Sheth, Sarah Nobles, Jasmine Padilla, Mark Burroughs, Shoshona Le, Katie Dillon, Peter Cook, Clinton R. Paden, Dhwani Batra, Krista Queen, Kristen Knipe, Dakota Howard, Yvette Unoarumhi, Darlene Wagner, Matthew Schmerer, Ben L. Rambo-Martin, Kristine Lacek, Sam Shepard, Alison Laufer Halpin, Dave Wentworth, Vivien Dugan, Suxiang Tong, Justin Lee |
| EPI_ISL_1446357 | Texas Children's Hospital | Texas Children's Microbiome Center | Ruth Ann Luna, Jennifer K. Spinler, James Dunn, James Versalovic, Ila Singh |
| EPI_ISL_1446358 | KY State Public Health Lab | Centers for Disease Control and Prevention Division of Viral Diseases, Pathogen Discovery | Mili Sheth, Sarah Nobles, Jasmine Padilla, Mark Burroughs, Shoshona Le, Katie Dillon, Peter Cook, Clinton R. Paden, Dhwani Batra, Krista Queen, Kristen Knipe, Dakota Howard, Yvette Unoarumhi, Darlene Wagner, Matthew Schmerer, Ben L. Rambo-Martin, Kristine Lacek, Sam Shepard, Alison Laufer Halpin, Dave Wentworth, Vivien Dugan, Suxiang Tong, Justin Lee |
| EPI_ISL_1446359, EPI_ISL_1446360 | Texas Children's Hospital | Texas Children's Microbiome Center | Ruth Ann Luna, Jennifer K. Spinler, James Dunn, James Versalovic, Ila Singh |
| EPI_ISL_1446361 | KY State Public Health Lab | Centers for Disease Control and Prevention Division of Viral Diseases, Pathogen Discovery | Mili Sheth, Sarah Nobles, Jasmine Padilla, Mark Burroughs, Shoshona Le, Katie Dillon, Peter Cook, Clinton R. Paden, Dhwani Batra, Krista Queen, Kristen Knipe, Dakota Howard, Yvette Unoarumhi, Darlene Wagner, Matthew Schmerer, Ben L. Rambo-Martin, Kristine Lacek, Sam Shepard, Alison Laufer Halpin, Dave Wentworth, Vivien Dugan, Suxiang Tong, Justin Lee |
| EPI_ISL_1446362, EPI_ISL_1446363 | Texas Children's Hospital | Texas Children's Microbiome Center | Ruth Ann Luna, Jennifer K. Spinler, James Dunn, James Versalovic, Ila Singh |

| | | | |
|---|---|---|---|
| EPI_ISL_1446364 | KY State Public Health Lab | Centers for Disease Control and Prevention Division of Viral Diseases, Pathogen Discovery | Mili Sheth, Sarah Nobles, Jasmine Padilla, Mark Burroughs, Shoshona Le, Katie Dillon, Peter Cook, Clinton R. Paden, Dhwani Batra, Krista Queen, Kristen Knipe, Dakota Howard, Yvette Unoarumhi, Darlene Wagner, Matthew Schmerer, Ben L. Rambo-Martin, Kristine Lacek, Sam Shepard, Alison Laufer Halpin, Dave Wentworth, Vivien Dugan, Suxiang Tong, Justin Lee |
| EPI_ISL_1446365, EPI_ISL_1446366 | Texas Children's Hospital | Texas Children's Microbiome Center | Ruth Ann Luna, Jennifer K. Spinler, James Dunn, James Versalovic, Ila Singh |
| EPI_ISL_1446367 | WI State Laboratory of Hygiene | Centers for Disease Control and Prevention Division of Viral Diseases, Pathogen Discovery | Mili Sheth, Sarah Nobles, Jasmine Padilla, Mark Burroughs, Shoshona Le, Katie Dillon, Peter Cook, Clinton R. Paden, Dhwani Batra, Krista Queen, Kristen Knipe, Dakota Howard, Yvette Unoarumhi, Darlene Wagner, Matthew Schmerer, Ben L. Rambo-Martin, Kristine Lacek, Sam Shepard, Alison Laufer Halpin, Dave Wentworth, Vivien Dugan, Suxiang Tong, Justin Lee |
| EPI_ISL_1446370, EPI_ISL_1446371, EPI_ISL_1446372, EPI_ISL_1446375, EPI_ISL_1446376, EPI_ISL_1446377, EPI_ISL_1446378, EPI_ISL_1446379, EPI_ISL_1446381, EPI_ISL_1446382 | Texas Children's Hospital | Texas Children's Microbiome Center | Ruth Ann Luna, Jennifer K. Spinler, James Dunn, James Versalovic, Ila Singh |
| EPI_ISL_1446383 | WI State Laboratory of Hygiene | Centers for Disease Control and Prevention Division of Viral Diseases, Pathogen Discovery | Mili Sheth, Sarah Nobles, Jasmine Padilla, Mark Burroughs, Shoshona Le, Katie Dillon, Peter Cook, Clinton R. Paden, Dhwani Batra, Krista Queen, Kristen Knipe, Dakota Howard, Yvette Unoarumhi, Darlene Wagner, Matthew Schmerer, Ben L. Rambo-Martin, Kristine Lacek, Sam Shepard, Alison Laufer Halpin, Dave Wentworth, Vivien Dugan, Suxiang Tong, Justin Lee |
| EPI_ISL_1446384, EPI_ISL_1446385 | Texas Children's Hospital | Texas Children's Microbiome Center | Ruth Ann Luna, Jennifer K. Spinler, James Dunn, James Versalovic, Ila Singh |
| EPI_ISL_1446386 | "NM Dept. Health, Scientific Laboratory Division " | Centers for Disease Control and Prevention Division of Viral Diseases, Pathogen Discovery | Mili Sheth, Sarah Nobles, Jasmine Padilla, Mark Burroughs, Shoshona Le, Katie Dillon, Peter Cook, Clinton R. Paden, Dhwani Batra, Krista Queen, Kristen Knipe, Dakota Howard, Yvette Unoarumhi, Darlene Wagner, Matthew Schmerer, Ben L. Rambo-Martin, Kristine Lacek, Sam Shepard, Alison Laufer Halpin, Dave Wentworth, Vivien Dugan, Suxiang Tong, Justin Lee |
| EPI_ISL_1446387, EPI_ISL_1446388 | Texas Children's Hospital | Texas Children's Microbiome Center | Ruth Ann Luna, Jennifer K. Spinler, James Dunn, James Versalovic, Ila Singh |
| EPI_ISL_1446389 | "NM Dept. Health, Scientific Laboratory Division " | Centers for Disease Control and Prevention Division of Viral Diseases, Pathogen Discovery | Mili Sheth, Sarah Nobles, Jasmine Padilla, Mark Burroughs, Shoshona Le, Katie Dillon, Peter Cook, Clinton R. Paden, Dhwani Batra, Krista Queen, Kristen Knipe, Dakota Howard, Yvette Unoarumhi, Darlene Wagner, Matthew Schmerer, Ben L. Rambo-Martin, Kristine Lacek, Sam Shepard, Alison Laufer Halpin, Dave Wentworth, Vivien Dugan, Suxiang Tong, Justin Lee |
| EPI_ISL_1446390 | Texas Children's Hospital | Texas Children's Microbiome Center | Ruth Ann Luna, Jennifer K. Spinler, James Dunn, James Versalovic, Ila Singh |
| EPI_ISL_1446392, EPI_ISL_1446395, EPI_ISL_1446398 | "NM Dept. Health, Scientific Laboratory Division " | Centers for Disease Control and Prevention Division of Viral Diseases, Pathogen Discovery | Mili Sheth, Sarah Nobles, Jasmine Padilla, Mark Burroughs, Shoshona Le, Katie Dillon, Peter Cook, Clinton R. Paden, Dhwani Batra, Krista Queen, Kristen Knipe, Dakota Howard, Yvette Unoarumhi, Darlene Wagner, Matthew Schmerer, Ben L. Rambo-Martin, Kristine Lacek, Sam Shepard, Alison Laufer Halpin, Dave Wentworth, Vivien Dugan, Suxiang Tong, Justin Lee |
| EPI_ISL_1446399, EPI_ISL_1446400 | Texas Children's Hospital | Texas Children's Microbiome Center | Ruth Ann Luna, Jennifer K. Spinler, James Dunn, James Versalovic, Ila Singh |
| EPI_ISL_1446401 | "NM Dept. Health, Scientific Laboratory Division " | Centers for Disease Control and Prevention Division of Viral Diseases, Pathogen Discovery | Mili Sheth, Sarah Nobles, Jasmine Padilla, Mark Burroughs, Shoshona Le, Katie Dillon, Peter Cook, Clinton R. Paden, Dhwani Batra, Krista Queen, Kristen Knipe, Dakota Howard, Yvette Unoarumhi, Darlene Wagner, Matthew Schmerer, Ben L. Rambo-Martin, Kristine Lacek, Sam Shepard, Alison Laufer Halpin, Dave Wentworth, Vivien Dugan, Suxiang Tong, Justin Lee |
| EPI_ISL_1446402, EPI_ISL_1446403 | Texas Children's Hospital | Texas Children's Microbiome Center | Ruth Ann Luna, Jennifer K. Spinler, James Dunn, James Versalovic, Ila Singh |
| EPI_ISL_1446404 | "NM Dept. Health, Scientific Laboratory Division " | Centers for Disease Control and Prevention Division of Viral Diseases, Pathogen Discovery | Mili Sheth, Sarah Nobles, Jasmine Padilla, Mark Burroughs, Shoshona Le, Katie Dillon, Peter Cook, Clinton R. Paden, Dhwani Batra, Krista Queen, Kristen Knipe, Dakota Howard, Yvette Unoarumhi, Darlene Wagner, Matthew Schmerer, Ben L. Rambo-Martin, Kristine Lacek, Sam Shepard, Alison Laufer Halpin, Dave Wentworth, Vivien Dugan, Suxiang Tong, Justin Lee |
| EPI_ISL_1446405 | Texas Children's Hospital | Texas Children's Microbiome Center | Ruth Ann Luna, Jennifer K. Spinler, James Dunn, James Versalovic, Ila Singh |
| EPI_ISL_1446406 | "NM Dept. Health, Scientific Laboratory Division " | Centers for Disease Control and Prevention Division of Viral Diseases, Pathogen Discovery | Mili Sheth, Sarah Nobles, Jasmine Padilla, Mark Burroughs, Shoshona Le, Katie Dillon, Peter Cook, Clinton R. Paden, Dhwani Batra, Krista Queen, Kristen Knipe, Dakota Howard, Yvette Unoarumhi, Darlene Wagner, Matthew Schmerer, Ben L. Rambo-Martin, Kristine Lacek, Sam Shepard, Alison Laufer Halpin, Dave Wentworth, Vivien Dugan, Suxiang Tong, Justin Lee |
| EPI_ISL_1446407 | Texas Children's Hospital | Texas Children's Microbiome Center | Ruth Ann Luna, Jennifer K. Spinler, James Dunn, James Versalovic, Ila Singh |
| EPI_ISL_1446409 | "NM Dept. Health, Scientific Laboratory Division " | Centers for Disease Control and Prevention Division of Viral Diseases, Pathogen Discovery | Mili Sheth, Sarah Nobles, Jasmine Padilla, Mark Burroughs, Shoshona Le, Katie Dillon, Peter Cook, Clinton R. Paden, Dhwani Batra, Krista Queen, Kristen Knipe, Dakota Howard, Yvette Unoarumhi, Darlene Wagner, Matthew Schmerer, Ben L. Rambo-Martin, Kristine Lacek, Sam Shepard, Alison Laufer Halpin, Dave Wentworth, Vivien Dugan, Suxiang Tong, Justin Lee |
| EPI_ISL_1446412, EPI_ISL_1446415 | NJ Public Health and Environmental Laboratories | Centers for Disease Control and Prevention Division of Viral Diseases, Pathogen Discovery | Mili Sheth, Sarah Nobles, Jasmine Padilla, Mark Burroughs, Shoshona Le, Katie Dillon, Peter Cook, Clinton R. Paden, Dhwani Batra, Krista Queen, Kristen Knipe, Dakota Howard, Yvette Unoarumhi, Darlene Wagner, Matthew Schmerer, Ben L. Rambo-Martin, Kristine Lacek, Sam Shepard, Alison Laufer Halpin, Dave Wentworth, Vivien Dugan, Suxiang Tong, Justin Lee |
| EPI_ISL_1446417 | Texas Children's Hospital | Texas Children's Microbiome Center | Ruth Ann Luna, Jennifer K. Spinler, James Dunn, James Versalovic, Ila Singh |
| EPI_ISL_1446418 | WY Public Health Laboratory | Centers for Disease Control and Prevention Division of Viral Diseases, Pathogen Discovery | Mili Sheth, Sarah Nobles, Jasmine Padilla, Mark Burroughs, Shoshona Le, Katie Dillon, Peter Cook, Clinton R. Paden, Dhwani Batra, Krista Queen, Kristen Knipe, Dakota Howard, Yvette Unoarumhi, Darlene Wagner, Matthew Schmerer, Ben L. Rambo-Martin, Kristine Lacek, Sam Shepard, Alison Laufer Halpin, Dave Wentworth, Vivien Dugan, Suxiang Tong, Justin Lee |
| EPI_ISL_1446419 | Texas Children's Hospital | Texas Children's Microbiome Center | Ruth Ann Luna, Jennifer K. Spinler, James Dunn, James Versalovic, Ila Singh |
| EPI_ISL_1446420 | WY Public Health Laboratory | Centers for Disease Control and Prevention Division of Viral Diseases, Pathogen Discovery | Mili Sheth, Sarah Nobles, Jasmine Padilla, Mark Burroughs, Shoshona Le, Katie Dillon, Peter Cook, Clinton R. Paden, Dhwani Batra, Krista Queen, Kristen Knipe, Dakota Howard, Yvette Unoarumhi, Darlene Wagner, Matthew Schmerer, Ben L. Rambo-Martin, Kristine Lacek, Sam Shepard, Alison Laufer Halpin, Dave Wentworth, Vivien Dugan, Suxiang Tong, Justin Lee |
| EPI_ISL_1446422 | Texas Children's Hospital | Texas Children's Microbiome Center | Ruth Ann Luna, Jennifer K. Spinler, James Dunn, James Versalovic, Ila Singh |
| EPI_ISL_1446423 | WY Public Health Laboratory | Centers for Disease Control and Prevention Division of Viral Diseases, Pathogen Discovery | Mili Sheth, Sarah Nobles, Jasmine Padilla, Mark Burroughs, Shoshona Le, Katie Dillon, Peter Cook, Clinton R. Paden, Dhwani Batra, Krista Queen, Kristen Knipe, Dakota Howard, Yvette Unoarumhi, Darlene Wagner, Matthew Schmerer, Ben L. Rambo-Martin, Kristine Lacek, Sam Shepard, Alison Laufer Halpin, Dave Wentworth, Vivien Dugan, Suxiang Tong, Justin Lee |
| EPI_ISL_1446424 | Texas Children's Hospital | Texas Children's Microbiome Center | Ruth Ann Luna, Jennifer K. Spinler, James Dunn, James Versalovic, Ila Singh |
| EPI_ISL_1446425 | WY Public Health Laboratory | Centers for Disease Control and Prevention Division of Viral Diseases, Pathogen Discovery | Mili Sheth, Sarah Nobles, Jasmine Padilla, Mark Burroughs, Shoshona Le, Katie Dillon, Peter Cook, Clinton R. Paden, Dhwani Batra, Krista Queen, Kristen Knipe, Dakota Howard, Yvette Unoarumhi, Darlene Wagner, Matthew Schmerer, Ben L. Rambo-Martin, Kristine Lacek, Sam Shepard, Alison Laufer Halpin, Dave Wentworth, Vivien Dugan, Suxiang Tong, Justin Lee |
| EPI_ISL_1446426 | Texas Children's Hospital | Texas Children's Microbiome Center | Ruth Ann Luna, Jennifer K. Spinler, James Dunn, James Versalovic, Ila Singh |
| EPI_ISL_1446427, EPI_ISL_1446430 | WY Public Health Laboratory | Centers for Disease Control and Prevention Division of Viral Diseases, Pathogen Discovery | Mili Sheth, Sarah Nobles, Jasmine Padilla, Mark Burroughs, Shoshona Le, Katie Dillon, Peter Cook, Clinton R. Paden, Dhwani Batra, Krista Queen, Kristen Knipe, Dakota Howard, Yvette Unoarumhi, Darlene Wagner, Matthew Schmerer, Ben L. Rambo-Martin, Kristine Lacek, Sam Shepard, Alison Laufer Halpin, Dave Wentworth, Vivien Dugan, Suxiang Tong, Justin Lee |
| EPI_ISL_1446433 | UT-Unified State Labs: Public Health Utah DOH | Centers for Disease Control and Prevention Division of Viral Diseases, Pathogen Discovery | Mili Sheth, Sarah Nobles, Jasmine Padilla, Mark Burroughs, Shoshona Le, Katie Dillon, Peter Cook, Clinton R. Paden, Dhwani Batra, Krista Queen, Kristen Knipe, Dakota Howard, Yvette Unoarumhi, Darlene Wagner, Matthew Schmerer, Ben L. Rambo-Martin, Kristine Lacek, Sam Shepard, Alison Laufer Halpin, Dave Wentworth, Vivien Dugan, Suxiang Tong, Justin Lee |
| EPI_ISL_1446435 | Texas Children's Hospital | Texas Children's Microbiome Center | Ruth Ann Luna, Jennifer K. Spinler, James Dunn, James Versalovic, Ila Singh |
| EPI_ISL_1446436 | NJ Public Health and Environmental Laboratories | Centers for Disease Control and Prevention Division of Viral Diseases, Pathogen Discovery | Mili Sheth, Sarah Nobles, Jasmine Padilla, Mark Burroughs, Shoshona Le, Katie Dillon, Peter Cook, Clinton R. Paden, Dhwani Batra, Krista Queen, Kristen Knipe, Dakota Howard, Yvette Unoarumhi, Darlene Wagner, Matthew Schmerer, Ben L. Rambo-Martin, Kristine Lacek, Sam Shepard, Alison Laufer Halpin, Dave Wentworth, Vivien Dugan, Suxiang Tong, Justin Lee |
| EPI_ISL_1446437, EPI_ISL_1446438 | Texas Children's Hospital | Texas Children's Microbiome Center | Ruth Ann Luna, Jennifer K. Spinler, James Dunn, James Versalovic, Ila Singh |
| EPI_ISL_1446439 | UT-Unified State Labs: Public Health Utah DOH | Centers for Disease Control and Prevention Division of Viral Diseases, Pathogen Discovery | Mili Sheth, Sarah Nobles, Jasmine Padilla, Mark Burroughs, Shoshona Le, Katie Dillon, Peter Cook, Clinton R. Paden, Dhwani Batra, Krista Queen, Kristen Knipe, Dakota Howard, Yvette Unoarumhi, Darlene Wagner, Matthew Schmerer, Ben L. Rambo-Martin, Kristine Lacek, Sam Shepard, Alison Laufer Halpin, Dave Wentworth, Vivien Dugan, Suxiang Tong, Justin Lee |
| EPI_ISL_1446440 | Texas Children's Hospital | Texas Children's Microbiome Center | Ruth Ann Luna, Jennifer K. Spinler, James Dunn, James Versalovic, Ila Singh |
| EPI_ISL_1446441 | UT-Unified State Labs: Public Health Utah DOH | Centers for Disease Control and Prevention Division of Viral Diseases, Pathogen Discovery | Mili Sheth, Sarah Nobles, Jasmine Padilla, Mark Burroughs, Shoshona Le, Katie Dillon, Peter Cook, Clinton R. Paden, Dhwani Batra, Krista Queen, Kristen Knipe, Dakota Howard, Yvette Unoarumhi, Darlene Wagner, Matthew Schmerer, Ben L. Rambo-Martin, Kristine Lacek, Sam Shepard, Alison Laufer Halpin, Dave Wentworth, Vivien Dugan, Suxiang Tong, Justin Lee |
| EPI_ISL_1446442 | Texas Children's Hospital | Texas Children's Microbiome Center | Ruth Ann Luna, Jennifer K. Spinler, James Dunn, James Versalovic, Ila Singh |

| | | | |
|---|---|---|---|
| EPI_ISL_1446444 | NJ Public Health and Environmental Laboratories | Centers for Disease Control and Prevention Division of Viral Diseases, Pathogen Discovery | Mili Sheth, Sarah Nobles, Jasmine Padilla, Mark Burroughs, Shoshona Le, Katie Dillon, Peter Cook, Clinton R. Paden, Dhwani Batra, Krista Queen, Kristen Knipe, Dakota Howard, Yvette Unoarumhi, Darlene Wagner, Matthew Schmerer, Ben L. Rambo-Martin, Kristine Lacek, Sam Shepard, Alison Laufer Halpin, Dave Wentworth, Vivien Dugan, Suxiang Tong, Justin Lee |
| EPI_ISL_1446445, EPI_ISL_1446446 | Texas Children's Hospital | Texas Children's Microbiome Center | Ruth Ann Luna, Jennifer K. Spinler, James Dunn, James Versalovic, Ila Singh |
| EPI_ISL_1446447 | MT Public Health Laboratory | Centers for Disease Control and Prevention Division of Viral Diseases, Pathogen Discovery | Mili Sheth, Sarah Nobles, Jasmine Padilla, Mark Burroughs, Shoshona Le, Katie Dillon, Peter Cook, Clinton R. Paden, Dhwani Batra, Krista Queen, Kristen Knipe, Dakota Howard, Yvette Unoarumhi, Darlene Wagner, Matthew Schmerer, Ben L. Rambo-Martin, Kristine Lacek, Sam Shepard, Alison Laufer Halpin, Dave Wentworth, Vivien Dugan, Suxiang Tong, Justin Lee |
| EPI_ISL_1446448, EPI_ISL_1446449 | Texas Children's Hospital | Texas Children's Microbiome Center | Ruth Ann Luna, Jennifer K. Spinler, James Dunn, James Versalovic, Ila Singh |
| EPI_ISL_1446450 | MT Public Health Laboratory | Centers for Disease Control and Prevention Division of Viral Diseases, Pathogen Discovery | Mili Sheth, Sarah Nobles, Jasmine Padilla, Mark Burroughs, Shoshona Le, Katie Dillon, Peter Cook, Clinton R. Paden, Dhwani Batra, Krista Queen, Kristen Knipe, Dakota Howard, Yvette Unoarumhi, Darlene Wagner, Matthew Schmerer, Ben L. Rambo-Martin, Kristine Lacek, Sam Shepard, Alison Laufer Halpin, Dave Wentworth, Vivien Dugan, Suxiang Tong, Justin Lee |
| EPI_ISL_1446451 | Texas Children's Hospital | Texas Children's Microbiome Center | Ruth Ann Luna, Jennifer K. Spinler, James Dunn, James Versalovic, Ila Singh |
| EPI_ISL_1446453 | MT Public Health Laboratory | Centers for Disease Control and Prevention Division of Viral Diseases, Pathogen Discovery | Mili Sheth, Sarah Nobles, Jasmine Padilla, Mark Burroughs, Shoshona Le, Katie Dillon, Peter Cook, Clinton R. Paden, Dhwani Batra, Krista Queen, Kristen Knipe, Dakota Howard, Yvette Unoarumhi, Darlene Wagner, Matthew Schmerer, Ben L. Rambo-Martin, Kristine Lacek, Sam Shepard, Alison Laufer Halpin, Dave Wentworth, Vivien Dugan, Suxiang Tong, Justin Lee |
| EPI_ISL_1446456, EPI_ISL_1446459, EPI_ISL_1446462, EPI_ISL_1446465, EPI_ISL_1446468, EPI_ISL_1446471, EPI_ISL_1446473 | "AR Dept. of Health-PHL, Molecular Diagnostics" | Centers for Disease Control and Prevention Division of Viral Diseases, Pathogen Discovery | Mili Sheth, Sarah Nobles, Jasmine Padilla, Mark Burroughs, Shoshona Le, Katie Dillon, Peter Cook, Clinton R. Paden, Dhwani Batra, Krista Queen, Kristen Knipe, Dakota Howard, Yvette Unoarumhi, Darlene Wagner, Matthew Schmerer, Ben L. Rambo-Martin, Kristine Lacek, Sam Shepard, Alison Laufer Halpin, Dave Wentworth, Vivien Dugan, Suxiang Tong, Justin Lee |
| EPI_ISL_1446474, EPI_ISL_1446475 | Texas Children's Hospital | Texas Children's Microbiome Center | Ruth Ann Luna, Jennifer K. Spinler, James Dunn, James Versalovic, Ila Singh |
| EPI_ISL_1446476 | MT Public Health Laboratory | Centers for Disease Control and Prevention Division of Viral Diseases, Pathogen Discovery | Mili Sheth, Sarah Nobles, Jasmine Padilla, Mark Burroughs, Shoshona Le, Katie Dillon, Peter Cook, Clinton R. Paden, Dhwani Batra, Krista Queen, Kristen Knipe, Dakota Howard, Yvette Unoarumhi, Darlene Wagner, Matthew Schmerer, Ben L. Rambo-Martin, Kristine Lacek, Sam Shepard, Alison Laufer Halpin, Dave Wentworth, Vivien Dugan, Suxiang Tong, Justin Lee |
| EPI_ISL_1446477, EPI_ISL_1446478, EPI_ISL_1446479 | Texas Children's Hospital | Texas Children's Microbiome Center | Ruth Ann Luna, Jennifer K. Spinler, James Dunn, James Versalovic, Ila Singh |
| EPI_ISL_1446480 | MT Public Health Laboratory | Centers for Disease Control and Prevention Division of Viral Diseases, Pathogen Discovery | Mili Sheth, Sarah Nobles, Jasmine Padilla, Mark Burroughs, Shoshona Le, Katie Dillon, Peter Cook, Clinton R. Paden, Dhwani Batra, Krista Queen, Kristen Knipe, Dakota Howard, Yvette Unoarumhi, Darlene Wagner, Matthew Schmerer, Ben L. Rambo-Martin, Kristine Lacek, Sam Shepard, Alison Laufer Halpin, Dave Wentworth, Vivien Dugan, Suxiang Tong, Justin Lee |
| EPI_ISL_1446481 | Texas Children's Hospital | Texas Children's Microbiome Center | Ruth Ann Luna, Jennifer K. Spinler, James Dunn, James Versalovic, Ila Singh |
| EPI_ISL_1446482 | MT Public Health Laboratory | Centers for Disease Control and Prevention Division of Viral Diseases, Pathogen Discovery | Mili Sheth, Sarah Nobles, Jasmine Padilla, Mark Burroughs, Shoshona Le, Katie Dillon, Peter Cook, Clinton R. Paden, Dhwani Batra, Krista Queen, Kristen Knipe, Dakota Howard, Yvette Unoarumhi, Darlene Wagner, Matthew Schmerer, Ben L. Rambo-Martin, Kristine Lacek, Sam Shepard, Alison Laufer Halpin, Dave Wentworth, Vivien Dugan, Suxiang Tong, Justin Lee |
| EPI_ISL_1446483, EPI_ISL_1446484, EPI_ISL_1446485 | Texas Children's Hospital | Texas Children's Microbiome Center | Ruth Ann Luna, Jennifer K. Spinler, James Dunn, James Versalovic, Ila Singh |
| EPI_ISL_1446486 | MT Public Health Laboratory | Centers for Disease Control and Prevention Division of Viral Diseases, Pathogen Discovery | Mili Sheth, Sarah Nobles, Jasmine Padilla, Mark Burroughs, Shoshona Le, Katie Dillon, Peter Cook, Clinton R. Paden, Dhwani Batra, Krista Queen, Kristen Knipe, Dakota Howard, Yvette Unoarumhi, Darlene Wagner, Matthew Schmerer, Ben L. Rambo-Martin, Kristine Lacek, Sam Shepard, Alison Laufer Halpin, Dave Wentworth, Vivien Dugan, Suxiang Tong, Justin Lee |
| EPI_ISL_1446487, EPI_ISL_1446488 | Texas Children's Hospital | Texas Children's Microbiome Center | Ruth Ann Luna, Jennifer K. Spinler, James Dunn, James Versalovic, Ila Singh |
| EPI_ISL_1446489 | NC State Laboratory of Public Health | Centers for Disease Control and Prevention Division of Viral Diseases, Pathogen Discovery | Mili Sheth, Sarah Nobles, Jasmine Padilla, Mark Burroughs, Shoshona Le, Katie Dillon, Peter Cook, Clinton R. Paden, Dhwani Batra, Krista Queen, Kristen Knipe, Dakota Howard, Yvette Unoarumhi, Darlene Wagner, Matthew Schmerer, Ben L. Rambo-Martin, Kristine Lacek, Sam Shepard, Alison Laufer Halpin, Dave Wentworth, Vivien Dugan, Suxiang Tong, Justin Lee |
| EPI_ISL_1446490 | Texas Children's Hospital | Texas Children's Microbiome Center | Ruth Ann Luna, Jennifer K. Spinler, James Dunn, James Versalovic, Ila Singh |
| EPI_ISL_1446491, EPI_ISL_1446494 | NC State Laboratory of Public Health | Centers for Disease Control and Prevention Division of Viral Diseases, Pathogen Discovery | Mili Sheth, Sarah Nobles, Jasmine Padilla, Mark Burroughs, Shoshona Le, Katie Dillon, Peter Cook, Clinton R. Paden, Dhwani Batra, Krista Queen, Kristen Knipe, Dakota Howard, Yvette Unoarumhi, Darlene Wagner, Matthew Schmerer, Ben L. Rambo-Martin, Kristine Lacek, Sam Shepard, Alison Laufer Halpin, Dave Wentworth, Vivien Dugan, Suxiang Tong, Justin Lee |
| EPI_ISL_1446495, EPI_ISL_1446496 | Texas Children's Hospital | Texas Children's Microbiome Center | Ruth Ann Luna, Jennifer K. Spinler, James Dunn, James Versalovic, Ila Singh |
| EPI_ISL_1446497 | NC State Laboratory of Public Health | Centers for Disease Control and Prevention Division of Viral Diseases, Pathogen Discovery | Mili Sheth, Sarah Nobles, Jasmine Padilla, Mark Burroughs, Shoshona Le, Katie Dillon, Peter Cook, Clinton R. Paden, Dhwani Batra, Krista Queen, Kristen Knipe, Dakota Howard, Yvette Unoarumhi, Darlene Wagner, Matthew Schmerer, Ben L. Rambo-Martin, Kristine Lacek, Sam Shepard, Alison Laufer Halpin, Dave Wentworth, Vivien Dugan, Suxiang Tong, Justin Lee |
| EPI_ISL_1446498 | Texas Children's Hospital | Texas Children's Microbiome Center | Ruth Ann Luna, Jennifer K. Spinler, James Dunn, James Versalovic, Ila Singh |
| EPI_ISL_1446499 | NC State Laboratory of Public Health | Centers for Disease Control and Prevention Division of Viral Diseases, Pathogen Discovery | Mili Sheth, Sarah Nobles, Jasmine Padilla, Mark Burroughs, Shoshona Le, Katie Dillon, Peter Cook, Clinton R. Paden, Dhwani Batra, Krista Queen, Kristen Knipe, Dakota Howard, Yvette Unoarumhi, Darlene Wagner, Matthew Schmerer, Ben L. Rambo-Martin, Kristine Lacek, Sam Shepard, Alison Laufer Halpin, Dave Wentworth, Vivien Dugan, Suxiang Tong, Justin Lee |
| EPI_ISL_1446500 | Texas Children's Hospital | Texas Children's Microbiome Center | Ruth Ann Luna, Jennifer K. Spinler, James Dunn, James Versalovic, Ila Singh |
| EPI_ISL_1446502, EPI_ISL_1446505 | NC State Laboratory of Public Health | Centers for Disease Control and Prevention Division of Viral Diseases, Pathogen Discovery | Mili Sheth, Sarah Nobles, Jasmine Padilla, Mark Burroughs, Shoshona Le, Katie Dillon, Peter Cook, Clinton R. Paden, Dhwani Batra, Krista Queen, Kristen Knipe, Dakota Howard, Yvette Unoarumhi, Darlene Wagner, Matthew Schmerer, Ben L. Rambo-Martin, Kristine Lacek, Sam Shepard, Alison Laufer Halpin, Dave Wentworth, Vivien Dugan, Suxiang Tong, Justin Lee |
| EPI_ISL_1446506, EPI_ISL_1446507 | Texas Children's Hospital | Texas Children's Microbiome Center | Ruth Ann Luna, Jennifer K. Spinler, James Dunn, James Versalovic, Ila Singh |
| EPI_ISL_1446508 | NC State Laboratory of Public Health | Centers for Disease Control and Prevention Division of Viral Diseases, Pathogen Discovery | Mili Sheth, Sarah Nobles, Jasmine Padilla, Mark Burroughs, Shoshona Le, Katie Dillon, Peter Cook, Clinton R. Paden, Dhwani Batra, Krista Queen, Kristen Knipe, Dakota Howard, Yvette Unoarumhi, Darlene Wagner, Matthew Schmerer, Ben L. Rambo-Martin, Kristine Lacek, Sam Shepard, Alison Laufer Halpin, Dave Wentworth, Vivien Dugan, Suxiang Tong, Justin Lee |
| EPI_ISL_1446509, EPI_ISL_1446510 | Texas Children's Hospital | Texas Children's Microbiome Center | Ruth Ann Luna, Jennifer K. Spinler, James Dunn, James Versalovic, Ila Singh |
| EPI_ISL_1446511 | NC State Laboratory of Public Health | Centers for Disease Control and Prevention Division of Viral Diseases, Pathogen Discovery | Mili Sheth, Sarah Nobles, Jasmine Padilla, Mark Burroughs, Shoshona Le, Katie Dillon, Peter Cook, Clinton R. Paden, Dhwani Batra, Krista Queen, Kristen Knipe, Dakota Howard, Yvette Unoarumhi, Darlene Wagner, Matthew Schmerer, Ben L. Rambo-Martin, Kristine Lacek, Sam Shepard, Alison Laufer Halpin, Dave Wentworth, Vivien Dugan, Suxiang Tong, Justin Lee |
| EPI_ISL_1446512 | Texas Children's Hospital | Texas Children's Microbiome Center | Ruth Ann Luna, Jennifer K. Spinler, James Dunn, James Versalovic, Ila Singh |
| EPI_ISL_1446513 | NC State Laboratory of Public Health | Centers for Disease Control and Prevention Division of Viral Diseases, Pathogen Discovery | Mili Sheth, Sarah Nobles, Jasmine Padilla, Mark Burroughs, Shoshona Le, Katie Dillon, Peter Cook, Clinton R. Paden, Dhwani Batra, Krista Queen, Kristen Knipe, Dakota Howard, Yvette Unoarumhi, Darlene Wagner, Matthew Schmerer, Ben L. Rambo-Martin, Kristine Lacek, Sam Shepard, Alison Laufer Halpin, Dave Wentworth, Vivien Dugan, Suxiang Tong, Justin Lee |
| EPI_ISL_1446516 | IN State Department of Health Laboratory Services | Centers for Disease Control and Prevention Division of Viral Diseases, Pathogen Discovery | Mili Sheth, Sarah Nobles, Jasmine Padilla, Mark Burroughs, Shoshona Le, Katie Dillon, Peter Cook, Clinton R. Paden, Dhwani Batra, Krista Queen, Kristen Knipe, Dakota Howard, Yvette Unoarumhi, Darlene Wagner, Matthew Schmerer, Ben L. Rambo-Martin, Kristine Lacek, Sam Shepard, Alison Laufer Halpin, Dave Wentworth, Vivien Dugan, Suxiang Tong, Justin Lee |
| EPI_ISL_1446518 | Texas Children's Hospital | Texas Children's Microbiome Center | Ruth Ann Luna, Jennifer K. Spinler, James Dunn, James Versalovic, Ila Singh |
| EPI_ISL_1446519 | IN State Department of Health Laboratory Services | Centers for Disease Control and Prevention Division of Viral Diseases, Pathogen Discovery | Mili Sheth, Sarah Nobles, Jasmine Padilla, Mark Burroughs, Shoshona Le, Katie Dillon, Peter Cook, Clinton R. Paden, Dhwani Batra, Krista Queen, Kristen Knipe, Dakota Howard, Yvette Unoarumhi, Darlene Wagner, Matthew Schmerer, Ben L. Rambo-Martin, Kristine Lacek, Sam Shepard, Alison Laufer Halpin, Dave Wentworth, Vivien Dugan, Suxiang Tong, Justin Lee |
| EPI_ISL_1446520, EPI_ISL_1446521 | Texas Children's Hospital | Texas Children's Microbiome Center | Ruth Ann Luna, Jennifer K. Spinler, James Dunn, James Versalovic, Ila Singh |
| EPI_ISL_1446522, EPI_ISL_1446524 | IN State Department of Health Laboratory Services | Centers for Disease Control and Prevention Division of Viral Diseases, Pathogen Discovery | Mili Sheth, Sarah Nobles, Jasmine Padilla, Mark Burroughs, Shoshona Le, Katie Dillon, Peter Cook, Clinton R. Paden, Dhwani Batra, Krista Queen, Kristen Knipe, Dakota Howard, Yvette Unoarumhi, Darlene Wagner, Matthew Schmerer, Ben L. Rambo-Martin, Kristine Lacek, Sam Shepard, Alison Laufer Halpin, Dave Wentworth, Vivien Dugan, Suxiang Tong, Justin Lee |
| EPI_ISL_1446525, | Texas Children's Hospital | Texas Children's Microbiome Center | Ruth Ann Luna, Jennifer K. Spinler, James Dunn, James Versalovic, Ila Singh |

| | | | |
|---|---|---|---|
| EPI_ISL_1446526 | | | |
| EPI_ISL_1446527 | IN State Department of Health Laboratory Services | Centers for Disease Control and Prevention Division of Viral Diseases, Pathogen Discovery | Mili Sheth, Sarah Nobles, Jasmine Padilla, Mark Burroughs, Shoshona Le, Katie Dillon, Peter Cook, Clinton R. Paden, Dhwani Batra, Krista Queen, Kristen Knipe, Dakota Howard, Yvette Unoarumhi, Darlene Wagner, Matthew Schmerer, Ben L. Rambo-Martin, Kristine Lacek, Sam Shepard, Alison Laufer Halpin, Dave Wentworth, Vivien Dugan, Suxiang Tong, Justin Lee |
| EPI_ISL_1446528 | Texas Children's Hospital | Texas Children's Microbiome Center | Ruth Ann Luna, Jennifer K. Spinler, James Dunn, James Versalovic, Ila Singh |
| EPI_ISL_1446530 | IN State Department of Health Laboratory Services | Centers for Disease Control and Prevention Division of Viral Diseases, Pathogen Discovery | Mili Sheth, Sarah Nobles, Jasmine Padilla, Mark Burroughs, Shoshona Le, Katie Dillon, Peter Cook, Clinton R. Paden, Dhwani Batra, Krista Queen, Kristen Knipe, Dakota Howard, Yvette Unoarumhi, Darlene Wagner, Matthew Schmerer, Ben L. Rambo-Martin, Kristine Lacek, Sam Shepard, Alison Laufer Halpin, Dave Wentworth, Vivien Dugan, Suxiang Tong, Justin Lee |
| EPI_ISL_1446531, EPI_ISL_1446532 | Texas Children's Hospital | Texas Children's Microbiome Center | Ruth Ann Luna, Jennifer K. Spinler, James Dunn, James Versalovic, Ila Singh |
| EPI_ISL_1446533, EPI_ISL_1446535 | IN State Department of Health Laboratory Services | Centers for Disease Control and Prevention Division of Viral Diseases, Pathogen Discovery | Mili Sheth, Sarah Nobles, Jasmine Padilla, Mark Burroughs, Shoshona Le, Katie Dillon, Peter Cook, Clinton R. Paden, Dhwani Batra, Krista Queen, Kristen Knipe, Dakota Howard, Yvette Unoarumhi, Darlene Wagner, Matthew Schmerer, Ben L. Rambo-Martin, Kristine Lacek, Sam Shepard, Alison Laufer Halpin, Dave Wentworth, Vivien Dugan, Suxiang Tong, Justin Lee |
| EPI_ISL_1446536, EPI_ISL_1446537 | Texas Children's Hospital | Texas Children's Microbiome Center | Ruth Ann Luna, Jennifer K. Spinler, James Dunn, James Versalovic, Ila Singh |
| EPI_ISL_1446538 | IN State Department of Health Laboratory Services | Centers for Disease Control and Prevention Division of Viral Diseases, Pathogen Discovery | Mili Sheth, Sarah Nobles, Jasmine Padilla, Mark Burroughs, Shoshona Le, Katie Dillon, Peter Cook, Clinton R. Paden, Dhwani Batra, Krista Queen, Kristen Knipe, Dakota Howard, Yvette Unoarumhi, Darlene Wagner, Matthew Schmerer, Ben L. Rambo-Martin, Kristine Lacek, Sam Shepard, Alison Laufer Halpin, Dave Wentworth, Vivien Dugan, Suxiang Tong, Justin Lee |
| EPI_ISL_1446540 | Texas Children's Hospital | Texas Children's Microbiome Center | Ruth Ann Luna, Jennifer K. Spinler, James Dunn, James Versalovic, Ila Singh |
| EPI_ISL_1446541 | NC State Laboratory of Public Health | Centers for Disease Control and Prevention Division of Viral Diseases, Pathogen Discovery | Mili Sheth, Sarah Nobles, Jasmine Padilla, Mark Burroughs, Shoshona Le, Katie Dillon, Peter Cook, Clinton R. Paden, Dhwani Batra, Krista Queen, Kristen Knipe, Dakota Howard, Yvette Unoarumhi, Darlene Wagner, Matthew Schmerer, Ben L. Rambo-Martin, Kristine Lacek, Sam Shepard, Alison Laufer Halpin, Dave Wentworth, Vivien Dugan, Suxiang Tong, Justin Lee |
| EPI_ISL_1446543, EPI_ISL_1446544, EPI_ISL_1446545, EPI_ISL_1446546 | "OK Public Health Laboratory, Oklahoma State DOH" | Centers for Disease Control and Prevention Division of Viral Diseases, Pathogen Discovery | Mili Sheth, Sarah Nobles, Jasmine Padilla, Mark Burroughs, Shoshona Le, Katie Dillon, Peter Cook, Clinton R. Paden, Dhwani Batra, Krista Queen, Kristen Knipe, Dakota Howard, Yvette Unoarumhi, Darlene Wagner, Matthew Schmerer, Ben L. Rambo-Martin, Kristine Lacek, Sam Shepard, Alison Laufer Halpin, Dave Wentworth, Vivien Dugan, Suxiang Tong, Justin Lee |
| EPI_ISL_1446547, EPI_ISL_1446548, EPI_ISL_1446549, EPI_ISL_1446550, EPI_ISL_1446551, EPI_ISL_1446552, EPI_ISL_1446553, EPI_ISL_1446554, EPI_ISL_1446555, EPI_ISL_1446556, EPI_ISL_1446557, EPI_ISL_1446558, EPI_ISL_1446559, EPI_ISL_1446560, EPI_ISL_1446561, EPI_ISL_1446562, EPI_ISL_1446563, EPI_ISL_1446564, EPI_ISL_1446565, EPI_ISL_1446566, EPI_ISL_1446567, EPI_ISL_1446568, EPI_ISL_1446569, EPI_ISL_1446570, EPI_ISL_1446571 | | | |
| see above | NC State Laboratory of Public Health | Centers for Disease Control and Prevention Division of Viral Diseases, Pathogen Discovery | Mili Sheth, Sarah Nobles, Jasmine Padilla, Mark Burroughs, Shoshona Le, Katie Dillon, Peter Cook, Clinton R. Paden, Dhwani Batra, Krista Queen, Kristen Knipe, Dakota Howard, Yvette Unoarumhi, Darlene Wagner, Matthew Schmerer, Ben L. Rambo-Martin, Kristine Lacek, Sam Shepard, Alison Laufer Halpin, Dave Wentworth, Vivien Dugan, Suxiang Tong, Justin Lee |
| EPI_ISL_1446572, EPI_ISL_1446573, EPI_ISL_1446574, EPI_ISL_1446575 | RI State Health Laboratories | Centers for Disease Control and Prevention Division of Viral Diseases, Pathogen Discovery | Mili Sheth, Sarah Nobles, Jasmine Padilla, Mark Burroughs, Shoshona Le, Katie Dillon, Peter Cook, Clinton R. Paden, Dhwani Batra, Krista Queen, Kristen Knipe, Dakota Howard, Yvette Unoarumhi, Darlene Wagner, Matthew Schmerer, Ben L. Rambo-Martin, Kristine Lacek, Sam Shepard, Alison Laufer Halpin, Dave Wentworth, Vivien Dugan, Suxiang Tong, Justin Lee |
| EPI_ISL_1446576, EPI_ISL_1446577, EPI_ISL_1446578, EPI_ISL_1446579, EPI_ISL_1446580, EPI_ISL_1446581 | UT-Unified State Labs: Public Health Utah DOH | Centers for Disease Control and Prevention Division of Viral Diseases, Pathogen Discovery | Mili Sheth, Sarah Nobles, Jasmine Padilla, Mark Burroughs, Shoshona Le, Katie Dillon, Peter Cook, Clinton R. Paden, Dhwani Batra, Krista Queen, Kristen Knipe, Dakota Howard, Yvette Unoarumhi, Darlene Wagner, Matthew Schmerer, Ben L. Rambo-Martin, Kristine Lacek, Sam Shepard, Alison Laufer Halpin, Dave Wentworth, Vivien Dugan, Suxiang Tong, Justin Lee |
| EPI_ISL_1446582 | TN Division of Laboratory Services | Centers for Disease Control and Prevention Division of Viral Diseases, Pathogen Discovery | Mili Sheth, Sarah Nobles, Jasmine Padilla, Mark Burroughs, Shoshona Le, Katie Dillon, Peter Cook, Clinton R. Paden, Dhwani Batra, Krista Queen, Kristen Knipe, Dakota Howard, Yvette Unoarumhi, Darlene Wagner, Matthew Schmerer, Ben L. Rambo-Martin, Kristine Lacek, Sam Shepard, Alison Laufer Halpin, Dave Wentworth, Vivien Dugan, Suxiang Tong, Justin Lee |
| EPI_ISL_1446583, EPI_ISL_1446584, EPI_ISL_1446585, EPI_ISL_1446586, EPI_ISL_1446587, EPI_ISL_1446588, EPI_ISL_1446589, EPI_ISL_1446590 | IL Dept. of Public Health Springfield Laboratory | Centers for Disease Control and Prevention Division of Viral Diseases, Pathogen Discovery | Mili Sheth, Sarah Nobles, Jasmine Padilla, Mark Burroughs, Shoshona Le, Katie Dillon, Peter Cook, Clinton R. Paden, Dhwani Batra, Krista Queen, Kristen Knipe, Dakota Howard, Yvette Unoarumhi, Darlene Wagner, Matthew Schmerer, Ben L. Rambo-Martin, Kristine Lacek, Sam Shepard, Alison Laufer Halpin, Dave Wentworth, Vivien Dugan, Suxiang Tong, Justin Lee |
| EPI_ISL_1446591 | LA Office of Public Health Laboratories | Centers for Disease Control and Prevention Division of Viral Diseases, Pathogen Discovery | Mili Sheth, Sarah Nobles, Jasmine Padilla, Mark Burroughs, Shoshona Le, Katie Dillon, Peter Cook, Clinton R. Paden, Dhwani Batra, Krista Queen, Kristen Knipe, Dakota Howard, Yvette Unoarumhi, Darlene Wagner, Matthew Schmerer, Ben L. Rambo-Martin, Kristine Lacek, Sam Shepard, Alison Laufer Halpin, Dave Wentworth, Vivien Dugan, Suxiang Tong, Justin Lee |
| EPI_ISL_1446592 | TN Division of Laboratory Services | Centers for Disease Control and Prevention Division of Viral Diseases, Pathogen Discovery | Mili Sheth, Sarah Nobles, Jasmine Padilla, Mark Burroughs, Shoshona Le, Katie Dillon, Peter Cook, Clinton R. Paden, Dhwani Batra, Krista Queen, Kristen Knipe, Dakota Howard, Yvette Unoarumhi, Darlene Wagner, Matthew Schmerer, Ben L. Rambo-Martin, Kristine Lacek, Sam Shepard, Alison Laufer Halpin, Dave Wentworth, Vivien Dugan, Suxiang Tong, Justin Lee |
| EPI_ISL_1446593 | LA Office of Public Health Laboratories | Centers for Disease Control and Prevention Division of Viral Diseases, Pathogen Discovery | Mili Sheth, Sarah Nobles, Jasmine Padilla, Mark Burroughs, Shoshona Le, Katie Dillon, Peter Cook, Clinton R. Paden, Dhwani Batra, Krista Queen, Kristen Knipe, Dakota Howard, Yvette Unoarumhi, Darlene Wagner, Matthew Schmerer, Ben L. Rambo-Martin, Kristine Lacek, Sam Shepard, Alison Laufer Halpin, Dave Wentworth, Vivien Dugan, Suxiang Tong, Justin Lee |
| EPI_ISL_1446594 | ME Health and Environmental Testing Laboratory | Centers for Disease Control and Prevention Division of Viral Diseases, Pathogen Discovery | Mili Sheth, Sarah Nobles, Jasmine Padilla, Mark Burroughs, Shoshona Le, Katie Dillon, Peter Cook, Clinton R. Paden, Dhwani Batra, Krista Queen, Kristen Knipe, Dakota Howard, Yvette Unoarumhi, Darlene Wagner, Matthew Schmerer, Ben L. Rambo-Martin, Kristine Lacek, Sam Shepard, Alison Laufer Halpin, Dave Wentworth, Vivien Dugan, Suxiang Tong, Justin Lee |
| EPI_ISL_1446595, EPI_ISL_1446596, EPI_ISL_1446597 | AL Dept. of Public Health Bureau of Clinical Laboratories | Centers for Disease Control and Prevention Division of Viral Diseases, Pathogen Discovery | Mili Sheth, Sarah Nobles, Jasmine Padilla, Mark Burroughs, Shoshona Le, Katie Dillon, Peter Cook, Clinton R. Paden, Dhwani Batra, Krista Queen, Kristen Knipe, Dakota Howard, Yvette Unoarumhi, Darlene Wagner, Matthew Schmerer, Ben L. Rambo-Martin, Kristine Lacek, Sam Shepard, Alison Laufer Halpin, Dave Wentworth, Vivien Dugan, Suxiang Tong, Justin Lee |
| EPI_ISL_1446598, EPI_ISL_1446599, EPI_ISL_1446600 | ME Health and Environmental Testing Laboratory | Centers for Disease Control and Prevention Division of Viral Diseases, Pathogen Discovery | Mili Sheth, Sarah Nobles, Jasmine Padilla, Mark Burroughs, Shoshona Le, Katie Dillon, Peter Cook, Clinton R. Paden, Dhwani Batra, Krista Queen, Kristen Knipe, Dakota Howard, Yvette Unoarumhi, Darlene Wagner, Matthew Schmerer, Ben L. Rambo-Martin, Kristine Lacek, Sam Shepard, Alison Laufer Halpin, Dave Wentworth, Vivien Dugan, Suxiang Tong, Justin Lee |
| EPI_ISL_1446601 | AL Dept. of Public Health Bureau of Clinical Laboratories | Centers for Disease Control and Prevention Division of Viral Diseases, Pathogen Discovery | Mili Sheth, Sarah Nobles, Jasmine Padilla, Mark Burroughs, Shoshona Le, Katie Dillon, Peter Cook, Clinton R. Paden, Dhwani Batra, Krista Queen, Kristen Knipe, Dakota Howard, Yvette Unoarumhi, Darlene Wagner, Matthew Schmerer, Ben L. Rambo-Martin, Kristine Lacek, Sam Shepard, Alison Laufer Halpin, Dave Wentworth, Vivien Dugan, Suxiang Tong, Justin Lee |
| EPI_ISL_1446602, EPI_ISL_1446603, EPI_ISL_1446604, EPI_ISL_1446605, EPI_ISL_1446606 | IA State Hygienic Laboratory | Centers for Disease Control and Prevention Division of Viral Diseases, Pathogen Discovery | Mili Sheth, Sarah Nobles, Jasmine Padilla, Mark Burroughs, Shoshona Le, Katie Dillon, Peter Cook, Clinton R. Paden, Dhwani Batra, Krista Queen, Kristen Knipe, Dakota Howard, Yvette Unoarumhi, Darlene Wagner, Matthew Schmerer, Ben L. Rambo-Martin, Kristine Lacek, Sam Shepard, Alison Laufer Halpin, Dave Wentworth, Vivien Dugan, Suxiang Tong, Justin Lee |
| EPI_ISL_1446607, EPI_ISL_1446608, EPI_ISL_1446609, EPI_ISL_1446610, EPI_ISL_1446611, EPI_ISL_1446612, EPI_ISL_1446613, EPI_ISL_1446614, EPI_ISL_1446615, EPI_ISL_1446616, EPI_ISL_1446617, EPI_ISL_1446618, EPI_ISL_1446619 | | | |
| see above | DE Public Health Laboratory | Centers for Disease Control and Prevention Division of Viral Diseases, Pathogen Discovery | Mili Sheth, Sarah Nobles, Jasmine Padilla, Mark Burroughs, Shoshona Le, Katie Dillon, Peter Cook, Clinton R. Paden, Dhwani Batra, Krista Queen, Kristen Knipe, Dakota Howard, Yvette Unoarumhi, Darlene Wagner, Matthew Schmerer, Ben L. Rambo-Martin, Kristine Lacek, Sam Shepard, Alison Laufer Halpin, Dave Wentworth, Vivien Dugan, Suxiang Tong, Justin Lee |
| EPI_ISL_1446620 | CT-Dr. Katherine A. Kelley State Public Health Lab | Centers for Disease Control and Prevention Division of Viral Diseases, Pathogen Discovery | Mili Sheth, Sarah Nobles, Jasmine Padilla, Mark Burroughs, Shoshona Le, Katie Dillon, Peter Cook, Clinton R. Paden, Dhwani Batra, Krista Queen, Kristen Knipe, Dakota Howard, Yvette Unoarumhi, Darlene Wagner, Matthew Schmerer, Ben L. Rambo-Martin, Kristine Lacek, Sam Shepard, Alison Laufer Halpin, Dave Wentworth, Vivien Dugan, Suxiang Tong, Justin Lee |
| EPI_ISL_1446621, EPI_ISL_1446622, EPI_ISL_1446623, EPI_ISL_1446624, EPI_ISL_1446625, EPI_ISL_1446626, EPI_ISL_1446627, EPI_ISL_1446628, EPI_ISL_1446629 | OH Department of Health Laboratory | Centers for Disease Control and Prevention Division of Viral Diseases, Pathogen Discovery | Mili Sheth, Sarah Nobles, Jasmine Padilla, Mark Burroughs, Shoshona Le, Katie Dillon, Peter Cook, Clinton R. Paden, Dhwani Batra, Krista Queen, Kristen Knipe, Dakota Howard, Yvette Unoarumhi, Darlene Wagner, Matthew Schmerer, Ben L. Rambo-Martin, Kristine Lacek, Sam Shepard, Alison Laufer Halpin, Dave Wentworth, Vivien Dugan, Suxiang Tong, Justin Lee |
| EPI_ISL_1446630, EPI_ISL_1446631 | MD DOH Laboratories Administration | Centers for Disease Control and Prevention Division of Viral Diseases, Pathogen Discovery | Mili Sheth, Sarah Nobles, Jasmine Padilla, Mark Burroughs, Shoshona Le, Katie Dillon, Peter Cook, Clinton R. Paden, Dhwani Batra, Krista Queen, Kristen Knipe, Dakota Howard, Yvette Unoarumhi, Darlene Wagner, Matthew Schmerer, Ben L. Rambo-Martin, Kristine Lacek, Sam Shepard, Alison Laufer Halpin, Dave Wentworth, Vivien Dugan, Suxiang Tong, Justin Lee |

| | | | |
|---|---|---|---|
| EPI_ISL_1446632, EPI_ISL_1446633 | "NM Dept. Health, Scientific Laboratory Division " | Centers for Disease Control and Prevention Division of Viral Diseases, Pathogen Discovery | Mili Sheth, Sarah Nobles, Jasmine Padilla, Mark Burroughs, Shoshona Le, Katie Dillon, Peter Cook, Clinton R. Paden, Dhwani Batra, Krista Queen, Kristen Knipe, Dakota Howard, Yvette Unoarumhi, Darlene Wagner, Matthew Schmerer, Ben L. Rambo-Martin, Kristine Lacek, Sam Shepard, Alison Laufer Halpin, Dave Wentworth, Vivien Dugan, Suxiang Tong, Justin Lee |
| EPI_ISL_1446634 | UT-Unified State Labs: Public Health Utah DOH | Centers for Disease Control and Prevention Division of Viral Diseases, Pathogen Discovery | Mili Sheth, Sarah Nobles, Jasmine Padilla, Mark Burroughs, Shoshona Le, Katie Dillon, Peter Cook, Clinton R. Paden, Dhwani Batra, Krista Queen, Kristen Knipe, Dakota Howard, Yvette Unoarumhi, Darlene Wagner, Matthew Schmerer, Ben L. Rambo-Martin, Kristine Lacek, Sam Shepard, Alison Laufer Halpin, Dave Wentworth, Vivien Dugan, Suxiang Tong, Justin Lee |
| EPI_ISL_1446635, EPI_ISL_1446636, EPI_ISL_1446637, EPI_ISL_1446638, EPI_ISL_1446639 | "NM Dept. Health, Scientific Laboratory Division " | Centers for Disease Control and Prevention Division of Viral Diseases, Pathogen Discovery | Mili Sheth, Sarah Nobles, Jasmine Padilla, Mark Burroughs, Shoshona Le, Katie Dillon, Peter Cook, Clinton R. Paden, Dhwani Batra, Krista Queen, Kristen Knipe, Dakota Howard, Yvette Unoarumhi, Darlene Wagner, Matthew Schmerer, Ben L. Rambo-Martin, Kristine Lacek, Sam Shepard, Alison Laufer Halpin, Dave Wentworth, Vivien Dugan, Suxiang Tong, Justin Lee |
| EPI_ISL_1446640 | "MN PHL Division, Minnesota Department of Health" | Centers for Disease Control and Prevention Division of Viral Diseases, Pathogen Discovery | Mili Sheth, Sarah Nobles, Jasmine Padilla, Mark Burroughs, Shoshona Le, Katie Dillon, Peter Cook, Clinton R. Paden, Dhwani Batra, Krista Queen, Kristen Knipe, Dakota Howard, Yvette Unoarumhi, Darlene Wagner, Matthew Schmerer, Ben L. Rambo-Martin, Kristine Lacek, Sam Shepard, Alison Laufer Halpin, Dave Wentworth, Vivien Dugan, Suxiang Tong, Justin Lee |
| EPI_ISL_1446641, EPI_ISL_1446642, EPI_ISL_1446643, EPI_ISL_1446644, EPI_ISL_1446645, EPI_ISL_1446646, EPI_ISL_1446647, EPI_ISL_1446648, EPI_ISL_1446649, EPI_ISL_1446650 | "TX DSHS, Lab Services Section MC 1947" | Centers for Disease Control and Prevention Division of Viral Diseases, Pathogen Discovery | Mili Sheth, Sarah Nobles, Jasmine Padilla, Mark Burroughs, Shoshona Le, Katie Dillon, Peter Cook, Clinton R. Paden, Dhwani Batra, Krista Queen, Kristen Knipe, Dakota Howard, Yvette Unoarumhi, Darlene Wagner, Matthew Schmerer, Ben L. Rambo-Martin, Kristine Lacek, Sam Shepard, Alison Laufer Halpin, Dave Wentworth, Vivien Dugan, Suxiang Tong, Justin Lee |
| EPI_ISL_1446651, EPI_ISL_1446652, EPI_ISL_1446653, EPI_ISL_1446654, EPI_ISL_1446655, EPI_ISL_1446656, EPI_ISL_1446657, EPI_ISL_1446658, EPI_ISL_1446659, EPI_ISL_1446660, EPI_ISL_1446661, EPI_ISL_1446662, EPI_ISL_1446663, EPI_ISL_1446664, EPI_ISL_1446665, EPI_ISL_1446666 | | | |
| see above | "AK State Public Health Lab, State Health Department" | Centers for Disease Control and Prevention Division of Viral Diseases, Pathogen Discovery | Mili Sheth, Sarah Nobles, Jasmine Padilla, Mark Burroughs, Shoshona Le, Katie Dillon, Peter Cook, Clinton R. Paden, Dhwani Batra, Krista Queen, Kristen Knipe, Dakota Howard, Yvette Unoarumhi, Darlene Wagner, Matthew Schmerer, Ben L. Rambo-Martin, Kristine Lacek, Sam Shepard, Alison Laufer Halpin, Dave Wentworth, Vivien Dugan, Suxiang Tong, Justin Lee |
| EPI_ISL_1446667, EPI_ISL_1446668, EPI_ISL_1446669, EPI_ISL_1446670 | VT Dept. of Health Laboratory | Centers for Disease Control and Prevention Division of Viral Diseases, Pathogen Discovery | Mili Sheth, Sarah Nobles, Jasmine Padilla, Mark Burroughs, Shoshona Le, Katie Dillon, Peter Cook, Clinton R. Paden, Dhwani Batra, Krista Queen, Kristen Knipe, Dakota Howard, Yvette Unoarumhi, Darlene Wagner, Matthew Schmerer, Ben L. Rambo-Martin, Kristine Lacek, Sam Shepard, Alison Laufer Halpin, Dave Wentworth, Vivien Dugan, Suxiang Tong, Justin Lee |
| EPI_ISL_1446671 | "AK State Public Health Lab, State Health Department" | Centers for Disease Control and Prevention Division of Viral Diseases, Pathogen Discovery | Mili Sheth, Sarah Nobles, Jasmine Padilla, Mark Burroughs, Shoshona Le, Katie Dillon, Peter Cook, Clinton R. Paden, Dhwani Batra, Krista Queen, Kristen Knipe, Dakota Howard, Yvette Unoarumhi, Darlene Wagner, Matthew Schmerer, Ben L. Rambo-Martin, Kristine Lacek, Sam Shepard, Alison Laufer Halpin, Dave Wentworth, Vivien Dugan, Suxiang Tong, Justin Lee |
| EPI_ISL_1446672, EPI_ISL_1446673, EPI_ISL_1446674, EPI_ISL_1446675 | VT Dept. of Health Laboratory | Centers for Disease Control and Prevention Division of Viral Diseases, Pathogen Discovery | Mili Sheth, Sarah Nobles, Jasmine Padilla, Mark Burroughs, Shoshona Le, Katie Dillon, Peter Cook, Clinton R. Paden, Dhwani Batra, Krista Queen, Kristen Knipe, Dakota Howard, Yvette Unoarumhi, Darlene Wagner, Matthew Schmerer, Ben L. Rambo-Martin, Kristine Lacek, Sam Shepard, Alison Laufer Halpin, Dave Wentworth, Vivien Dugan, Suxiang Tong, Justin Lee |
| EPI_ISL_1446676, EPI_ISL_1446677 | "NYSDOH Wadsworth Center, Virology Lab" | Centers for Disease Control and Prevention Division of Viral Diseases, Pathogen Discovery | Mili Sheth, Sarah Nobles, Jasmine Padilla, Mark Burroughs, Shoshona Le, Katie Dillon, Peter Cook, Clinton R. Paden, Dhwani Batra, Krista Queen, Kristen Knipe, Dakota Howard, Yvette Unoarumhi, Darlene Wagner, Matthew Schmerer, Ben L. Rambo-Martin, Kristine Lacek, Sam Shepard, Alison Laufer Halpin, Dave Wentworth, Vivien Dugan, Suxiang Tong, Justin Lee |
| EPI_ISL_1446678, EPI_ISL_1446679, EPI_ISL_1446680 | MI - Michigan Department of Health and Human Services - Bureau of Laboratories | Centers for Disease Control and Prevention Division of Viral Diseases, Pathogen Discovery | Mili Sheth, Sarah Nobles, Jasmine Padilla, Mark Burroughs, Shoshona Le, Katie Dillon, Peter Cook, Clinton R. Paden, Dhwani Batra, Krista Queen, Kristen Knipe, Dakota Howard, Yvette Unoarumhi, Darlene Wagner, Matthew Schmerer, Ben L. Rambo-Martin, Kristine Lacek, Sam Shepard, Alison Laufer Halpin, Dave Wentworth, Vivien Dugan, Suxiang Tong, Justin Lee |
| EPI_ISL_1446681, EPI_ISL_1446682, EPI_ISL_1446683 | WA State Department of Health | Centers for Disease Control and Prevention Division of Viral Diseases, Pathogen Discovery | Mili Sheth, Sarah Nobles, Jasmine Padilla, Mark Burroughs, Shoshona Le, Katie Dillon, Peter Cook, Clinton R. Paden, Dhwani Batra, Krista Queen, Kristen Knipe, Dakota Howard, Yvette Unoarumhi, Darlene Wagner, Matthew Schmerer, Ben L. Rambo-Martin, Kristine Lacek, Sam Shepard, Alison Laufer Halpin, Dave Wentworth, Vivien Dugan, Suxiang Tong, Justin Lee |
| EPI_ISL_1446684, EPI_ISL_1446685, EPI_ISL_1446686, EPI_ISL_1446687, EPI_ISL_1446688, EPI_ISL_1446689, EPI_ISL_1446690, EPI_ISL_1446691 | MT Public Health Laboratory | Centers for Disease Control and Prevention Division of Viral Diseases, Pathogen Discovery | Mili Sheth, Sarah Nobles, Jasmine Padilla, Mark Burroughs, Shoshona Le, Katie Dillon, Peter Cook, Clinton R. Paden, Dhwani Batra, Krista Queen, Kristen Knipe, Dakota Howard, Yvette Unoarumhi, Darlene Wagner, Matthew Schmerer, Ben L. Rambo-Martin, Kristine Lacek, Sam Shepard, Alison Laufer Halpin, Dave Wentworth, Vivien Dugan, Suxiang Tong, Justin Lee |
| EPI_ISL_1446692, EPI_ISL_1446693, EPI_ISL_1446694, EPI_ISL_1446695, EPI_ISL_1446696, EPI_ISL_1446697, EPI_ISL_1446698, EPI_ISL_1446699, EPI_ISL_1446700 | WA State Department of Health | Centers for Disease Control and Prevention Division of Viral Diseases, Pathogen Discovery | Mili Sheth, Sarah Nobles, Jasmine Padilla, Mark Burroughs, Shoshona Le, Katie Dillon, Peter Cook, Clinton R. Paden, Dhwani Batra, Krista Queen, Kristen Knipe, Dakota Howard, Yvette Unoarumhi, Darlene Wagner, Matthew Schmerer, Ben L. Rambo-Martin, Kristine Lacek, Sam Shepard, Alison Laufer Halpin, Dave Wentworth, Vivien Dugan, Suxiang Tong, Justin Lee |
| EPI_ISL_1446701 | "PA Department of Health, Bureau of Laboratories" | Centers for Disease Control and Prevention Division of Viral Diseases, Pathogen Discovery | Mili Sheth, Sarah Nobles, Jasmine Padilla, Mark Burroughs, Shoshona Le, Katie Dillon, Peter Cook, Clinton R. Paden, Dhwani Batra, Krista Queen, Kristen Knipe, Dakota Howard, Yvette Unoarumhi, Darlene Wagner, Matthew Schmerer, Ben L. Rambo-Martin, Kristine Lacek, Sam Shepard, Alison Laufer Halpin, Dave Wentworth, Vivien Dugan, Suxiang Tong, Justin Lee |
| EPI_ISL_1446702, EPI_ISL_1446703, EPI_ISL_1446704, EPI_ISL_1446705 | SD Public Health Laboratory | Centers for Disease Control and Prevention Division of Viral Diseases, Pathogen Discovery | Mili Sheth, Sarah Nobles, Jasmine Padilla, Mark Burroughs, Shoshona Le, Katie Dillon, Peter Cook, Clinton R. Paden, Dhwani Batra, Krista Queen, Kristen Knipe, Dakota Howard, Yvette Unoarumhi, Darlene Wagner, Matthew Schmerer, Ben L. Rambo-Martin, Kristine Lacek, Sam Shepard, Alison Laufer Halpin, Dave Wentworth, Vivien Dugan, Suxiang Tong, Justin Lee |
| EPI_ISL_1446706, EPI_ISL_1446707, EPI_ISL_1446708, EPI_ISL_1446709, EPI_ISL_1446710, EPI_ISL_1446711 | SC Dept of Health and Env. Control-Bureau of Laboratories | Centers for Disease Control and Prevention Division of Viral Diseases, Pathogen Discovery | Mili Sheth, Sarah Nobles, Jasmine Padilla, Mark Burroughs, Shoshona Le, Katie Dillon, Peter Cook, Clinton R. Paden, Dhwani Batra, Krista Queen, Kristen Knipe, Dakota Howard, Yvette Unoarumhi, Darlene Wagner, Matthew Schmerer, Ben L. Rambo-Martin, Kristine Lacek, Sam Shepard, Alison Laufer Halpin, Dave Wentworth, Vivien Dugan, Suxiang Tong, Justin Lee |
| EPI_ISL_1446712, EPI_ISL_1446713 | NJ Public Health and Environmental Laboratories | Centers for Disease Control and Prevention Division of Viral Diseases, Pathogen Discovery | Mili Sheth, Sarah Nobles, Jasmine Padilla, Mark Burroughs, Shoshona Le, Katie Dillon, Peter Cook, Clinton R. Paden, Dhwani Batra, Krista Queen, Kristen Knipe, Dakota Howard, Yvette Unoarumhi, Darlene Wagner, Matthew Schmerer, Ben L. Rambo-Martin, Kristine Lacek, Sam Shepard, Alison Laufer Halpin, Dave Wentworth, Vivien Dugan, Suxiang Tong, Justin Lee |
| EPI_ISL_1446714, EPI_ISL_1446715 | CA-Los Angeles County Public Health Laboratory | Centers for Disease Control and Prevention Division of Viral Diseases, Pathogen Discovery | Mili Sheth, Sarah Nobles, Jasmine Padilla, Mark Burroughs, Shoshona Le, Katie Dillon, Peter Cook, Clinton R. Paden, Dhwani Batra, Krista Queen, Kristen Knipe, Dakota Howard, Yvette Unoarumhi, Darlene Wagner, Matthew Schmerer, Ben L. Rambo-Martin, Kristine Lacek, Sam Shepard, Alison Laufer Halpin, Dave Wentworth, Vivien Dugan, Suxiang Tong, Justin Lee |
| EPI_ISL_1446716, EPI_ISL_1446717 | VT Dept. of Health Laboratory | Centers for Disease Control and Prevention Division of Viral Diseases, Pathogen Discovery | Mili Sheth, Sarah Nobles, Jasmine Padilla, Mark Burroughs, Shoshona Le, Katie Dillon, Peter Cook, Clinton R. Paden, Dhwani Batra, Krista Queen, Kristen Knipe, Dakota Howard, Yvette Unoarumhi, Darlene Wagner, Matthew Schmerer, Ben L. Rambo-Martin, Kristine Lacek, Sam Shepard, Alison Laufer Halpin, Dave Wentworth, Vivien Dugan, Suxiang Tong, Justin Lee |
| EPI_ISL_1446718, EPI_ISL_1446719 | WVDHHR - Office of Laboratory Services | Centers for Disease Control and Prevention Division of Viral Diseases, Pathogen Discovery | Mili Sheth, Sarah Nobles, Jasmine Padilla, Mark Burroughs, Shoshona Le, Katie Dillon, Peter Cook, Clinton R. Paden, Dhwani Batra, Krista Queen, Kristen Knipe, Dakota Howard, Yvette Unoarumhi, Darlene Wagner, Matthew Schmerer, Ben L. Rambo-Martin, Kristine Lacek, Sam Shepard, Alison Laufer Halpin, Dave Wentworth, Vivien Dugan, Suxiang Tong, Justin Lee |
| EPI_ISL_1446720 | WA State Department of Health | Centers for Disease Control and Prevention Division of Viral Diseases, Pathogen Discovery | Mili Sheth, Sarah Nobles, Jasmine Padilla, Mark Burroughs, Shoshona Le, Katie Dillon, Peter Cook, Clinton R. Paden, Dhwani Batra, Krista Queen, Kristen Knipe, Dakota Howard, Yvette Unoarumhi, Darlene Wagner, Matthew Schmerer, Ben L. Rambo-Martin, Kristine Lacek, Sam Shepard, Alison Laufer Halpin, Dave Wentworth, Vivien Dugan, Suxiang Tong, Justin Lee |
| EPI_ISL_1446721, EPI_ISL_1446722 | "MN PHL Division, Minnesota Department of Health" | Centers for Disease Control and Prevention Division of Viral Diseases, Pathogen Discovery | Mili Sheth, Sarah Nobles, Jasmine Padilla, Mark Burroughs, Shoshona Le, Katie Dillon, Peter Cook, Clinton R. Paden, Dhwani Batra, Krista Queen, Kristen Knipe, Dakota Howard, Yvette Unoarumhi, Darlene Wagner, Matthew Schmerer, Ben L. Rambo-Martin, Kristine Lacek, Sam Shepard, Alison Laufer Halpin, Dave Wentworth, Vivien Dugan, Suxiang Tong, Justin Lee |
| EPI_ISL_1446723, EPI_ISL_1446724 | ND Dept. of Health Laboratory Services-Microbiology | Centers for Disease Control and Prevention Division of Viral Diseases, Pathogen Discovery | Mili Sheth, Sarah Nobles, Jasmine Padilla, Mark Burroughs, Shoshona Le, Katie Dillon, Peter Cook, Clinton R. Paden, Dhwani Batra, Krista Queen, Kristen Knipe, Dakota Howard, Yvette Unoarumhi, Darlene Wagner, Matthew Schmerer, Ben L. Rambo-Martin, Kristine Lacek, Sam Shepard, Alison Laufer Halpin, Dave Wentworth, Vivien Dugan, Suxiang Tong, Justin Lee |

| | | | |
|---|---|---|---|
| EPI_ISL_1446725, EPI_ISL_1446726, EPI_ISL_1446727 | UT-Unified State Labs: Public Health Utah DOH | Centers for Disease Control and Prevention Division of Viral Diseases, Pathogen Discovery | Mili Sheth, Sarah Nobles, Jasmine Padilla, Mark Burroughs, Shoshona Le, Katie Dillon, Peter Cook, Clinton R. Paden, Dhwani Batra, Krista Queen, Kristen Knipe, Dakota Howard, Yvette Unoarumhi, Darlene Wagner, Matthew Schmerer, Ben L. Rambo-Martin, Kristine Lacek, Sam Shepard, Alison Laufer Halpin, Dave Wentworth, Vivien Dugan, Suxiang Tong, Justin Lee |
| EPI_ISL_1446728 | IA State Hygienic Laboratory | Centers for Disease Control and Prevention Division of Viral Diseases, Pathogen Discovery | Mili Sheth, Sarah Nobles, Jasmine Padilla, Mark Burroughs, Shoshona Le, Katie Dillon, Peter Cook, Clinton R. Paden, Dhwani Batra, Krista Queen, Kristen Knipe, Dakota Howard, Yvette Unoarumhi, Darlene Wagner, Matthew Schmerer, Ben L. Rambo-Martin, Kristine Lacek, Sam Shepard, Alison Laufer Halpin, Dave Wentworth, Vivien Dugan, Suxiang Tong, Justin Lee |
| EPI_ISL_1446729, EPI_ISL_1446730 | VA-Division of Consolidated Laboratory Services | Centers for Disease Control and Prevention Division of Viral Diseases, Pathogen Discovery | Mili Sheth, Sarah Nobles, Jasmine Padilla, Mark Burroughs, Shoshona Le, Katie Dillon, Peter Cook, Clinton R. Paden, Dhwani Batra, Krista Queen, Kristen Knipe, Dakota Howard, Yvette Unoarumhi, Darlene Wagner, Matthew Schmerer, Ben L. Rambo-Martin, Kristine Lacek, Sam Shepard, Alison Laufer Halpin, Dave Wentworth, Vivien Dugan, Suxiang Tong, Justin Lee |
| EPI_ISL_1446731 | WVDHHR - Office of Laboratory Services | Centers for Disease Control and Prevention Division of Viral Diseases, Pathogen Discovery | Mili Sheth, Sarah Nobles, Jasmine Padilla, Mark Burroughs, Shoshona Le, Katie Dillon, Peter Cook, Clinton R. Paden, Dhwani Batra, Krista Queen, Kristen Knipe, Dakota Howard, Yvette Unoarumhi, Darlene Wagner, Matthew Schmerer, Ben L. Rambo-Martin, Kristine Lacek, Sam Shepard, Alison Laufer Halpin, Dave Wentworth, Vivien Dugan, Suxiang Tong, Justin Lee |
| EPI_ISL_1446732 | VA-Division of Consolidated Laboratory Services | Centers for Disease Control and Prevention Division of Viral Diseases, Pathogen Discovery | Mili Sheth, Sarah Nobles, Jasmine Padilla, Mark Burroughs, Shoshona Le, Katie Dillon, Peter Cook, Clinton R. Paden, Dhwani Batra, Krista Queen, Kristen Knipe, Dakota Howard, Yvette Unoarumhi, Darlene Wagner, Matthew Schmerer, Ben L. Rambo-Martin, Kristine Lacek, Sam Shepard, Alison Laufer Halpin, Dave Wentworth, Vivien Dugan, Suxiang Tong, Justin Lee |
| EPI_ISL_1446733 | VT Dept. of Health Laboratory | Centers for Disease Control and Prevention Division of Viral Diseases, Pathogen Discovery | Mili Sheth, Sarah Nobles, Jasmine Padilla, Mark Burroughs, Shoshona Le, Katie Dillon, Peter Cook, Clinton R. Paden, Dhwani Batra, Krista Queen, Kristen Knipe, Dakota Howard, Yvette Unoarumhi, Darlene Wagner, Matthew Schmerer, Ben L. Rambo-Martin, Kristine Lacek, Sam Shepard, Alison Laufer Halpin, Dave Wentworth, Vivien Dugan, Suxiang Tong, Justin Lee |
| EPI_ISL_1446734 | WVDHHR - Office of Laboratory Services | Centers for Disease Control and Prevention Division of Viral Diseases, Pathogen Discovery | Mili Sheth, Sarah Nobles, Jasmine Padilla, Mark Burroughs, Shoshona Le, Katie Dillon, Peter Cook, Clinton R. Paden, Dhwani Batra, Krista Queen, Kristen Knipe, Dakota Howard, Yvette Unoarumhi, Darlene Wagner, Matthew Schmerer, Ben L. Rambo-Martin, Kristine Lacek, Sam Shepard, Alison Laufer Halpin, Dave Wentworth, Vivien Dugan, Suxiang Tong, Justin Lee |
| EPI_ISL_1446735, EPI_ISL_1446736, EPI_ISL_1446737 | SC Dept of Health and Env. Control-Bureau of Laboratories | Centers for Disease Control and Prevention Division of Viral Diseases, Pathogen Discovery | Mili Sheth, Sarah Nobles, Jasmine Padilla, Mark Burroughs, Shoshona Le, Katie Dillon, Peter Cook, Clinton R. Paden, Dhwani Batra, Krista Queen, Kristen Knipe, Dakota Howard, Yvette Unoarumhi, Darlene Wagner, Matthew Schmerer, Ben L. Rambo-Martin, Kristine Lacek, Sam Shepard, Alison Laufer Halpin, Dave Wentworth, Vivien Dugan, Suxiang Tong, Justin Lee |
| EPI_ISL_1446738 | "TX DSHS, Lab Services Section MC 1947" | Centers for Disease Control and Prevention Division of Viral Diseases, Pathogen Discovery | Mili Sheth, Sarah Nobles, Jasmine Padilla, Mark Burroughs, Shoshona Le, Katie Dillon, Peter Cook, Clinton R. Paden, Dhwani Batra, Krista Queen, Kristen Knipe, Dakota Howard, Yvette Unoarumhi, Darlene Wagner, Matthew Schmerer, Ben L. Rambo-Martin, Kristine Lacek, Sam Shepard, Alison Laufer Halpin, Dave Wentworth, Vivien Dugan, Suxiang Tong, Justin Lee |
| EPI_ISL_1446739 | WA State Department of Health | Centers for Disease Control and Prevention Division of Viral Diseases, Pathogen Discovery | Mili Sheth, Sarah Nobles, Jasmine Padilla, Mark Burroughs, Shoshona Le, Katie Dillon, Peter Cook, Clinton R. Paden, Dhwani Batra, Krista Queen, Kristen Knipe, Dakota Howard, Yvette Unoarumhi, Darlene Wagner, Matthew Schmerer, Ben L. Rambo-Martin, Kristine Lacek, Sam Shepard, Alison Laufer Halpin, Dave Wentworth, Vivien Dugan, Suxiang Tong, Justin Lee |
| EPI_ISL_1446740 | TN Division of Laboratory Services | Centers for Disease Control and Prevention Division of Viral Diseases, Pathogen Discovery | Mili Sheth, Sarah Nobles, Jasmine Padilla, Mark Burroughs, Shoshona Le, Katie Dillon, Peter Cook, Clinton R. Paden, Dhwani Batra, Krista Queen, Kristen Knipe, Dakota Howard, Yvette Unoarumhi, Darlene Wagner, Matthew Schmerer, Ben L. Rambo-Martin, Kristine Lacek, Sam Shepard, Alison Laufer Halpin, Dave Wentworth, Vivien Dugan, Suxiang Tong, Justin Lee |
| EPI_ISL_1446741, EPI_ISL_1446742 | NC State Laboratory of Public Health | Centers for Disease Control and Prevention Division of Viral Diseases, Pathogen Discovery | Mili Sheth, Sarah Nobles, Jasmine Padilla, Mark Burroughs, Shoshona Le, Katie Dillon, Peter Cook, Clinton R. Paden, Dhwani Batra, Krista Queen, Kristen Knipe, Dakota Howard, Yvette Unoarumhi, Darlene Wagner, Matthew Schmerer, Ben L. Rambo-Martin, Kristine Lacek, Sam Shepard, Alison Laufer Halpin, Dave Wentworth, Vivien Dugan, Suxiang Tong, Justin Lee |
| EPI_ISL_1446743 | NV State Public Health Laboratory | Centers for Disease Control and Prevention Division of Viral Diseases, Pathogen Discovery | Mili Sheth, Sarah Nobles, Jasmine Padilla, Mark Burroughs, Shoshona Le, Katie Dillon, Peter Cook, Clinton R. Paden, Dhwani Batra, Krista Queen, Kristen Knipe, Dakota Howard, Yvette Unoarumhi, Darlene Wagner, Matthew Schmerer, Ben L. Rambo-Martin, Kristine Lacek, Sam Shepard, Alison Laufer Halpin, Dave Wentworth, Vivien Dugan, Suxiang Tong, Justin Lee |
| EPI_ISL_1446744 | IL Dept. of Public Health Springfield Laboratory | Centers for Disease Control and Prevention Division of Viral Diseases, Pathogen Discovery | Mili Sheth, Sarah Nobles, Jasmine Padilla, Mark Burroughs, Shoshona Le, Katie Dillon, Peter Cook, Clinton R. Paden, Dhwani Batra, Krista Queen, Kristen Knipe, Dakota Howard, Yvette Unoarumhi, Darlene Wagner, Matthew Schmerer, Ben L. Rambo-Martin, Kristine Lacek, Sam Shepard, Alison Laufer Halpin, Dave Wentworth, Vivien Dugan, Suxiang Tong, Justin Lee |
| EPI_ISL_1446745 | CT-Dr. Katherine A. Kelley State Public Health Lab | Centers for Disease Control and Prevention Division of Viral Diseases, Pathogen Discovery | Mili Sheth, Sarah Nobles, Jasmine Padilla, Mark Burroughs, Shoshona Le, Katie Dillon, Peter Cook, Clinton R. Paden, Dhwani Batra, Krista Queen, Kristen Knipe, Dakota Howard, Yvette Unoarumhi, Darlene Wagner, Matthew Schmerer, Ben L. Rambo-Martin, Kristine Lacek, Sam Shepard, Alison Laufer Halpin, Dave Wentworth, Vivien Dugan, Suxiang Tong, Justin Lee |
| EPI_ISL_1446746, EPI_ISL_1446747, EPI_ISL_1446748, EPI_ISL_1446749, EPI_ISL_1446750, EPI_ISL_1446751, EPI_ISL_1446752, EPI_ISL_1446753, EPI_ISL_1446754, EPI_ISL_1446755, EPI_ISL_1446756, EPI_ISL_1446757, EPI_ISL_1446758 | | | |
| see above | ND Dept. of Health Laboratory Services-Microbiology | Centers for Disease Control and Prevention Division of Viral Diseases, Pathogen Discovery | Mili Sheth, Sarah Nobles, Jasmine Padilla, Mark Burroughs, Shoshona Le, Katie Dillon, Peter Cook, Clinton R. Paden, Dhwani Batra, Krista Queen, Kristen Knipe, Dakota Howard, Yvette Unoarumhi, Darlene Wagner, Matthew Schmerer, Ben L. Rambo-Martin, Kristine Lacek, Sam Shepard, Alison Laufer Halpin, Dave Wentworth, Vivien Dugan, Suxiang Tong, Justin Lee |
| EPI_ISL_1446759, EPI_ISL_1446760, EPI_ISL_1446762, EPI_ISL_1446763, EPI_ISL_1446764, EPI_ISL_1446765 | OR State PHL-Virology/Immunology Section | Centers for Disease Control and Prevention Division of Viral Diseases, Pathogen Discovery | Mili Sheth, Sarah Nobles, Jasmine Padilla, Mark Burroughs, Shoshona Le, Katie Dillon, Peter Cook, Clinton R. Paden, Dhwani Batra, Krista Queen, Kristen Knipe, Dakota Howard, Yvette Unoarumhi, Darlene Wagner, Matthew Schmerer, Ben L. Rambo-Martin, Kristine Lacek, Sam Shepard, Alison Laufer Halpin, Dave Wentworth, Vivien Dugan, Suxiang Tong, Justin Lee |
| EPI_ISL_1446766, EPI_ISL_1446767, EPI_ISL_1446768, EPI_ISL_1446769, EPI_ISL_1446770, EPI_ISL_1446771, EPI_ISL_1446772 | MS Public Health Laboratory | Centers for Disease Control and Prevention Division of Viral Diseases, Pathogen Discovery | Mili Sheth, Sarah Nobles, Jasmine Padilla, Mark Burroughs, Shoshona Le, Katie Dillon, Peter Cook, Clinton R. Paden, Dhwani Batra, Krista Queen, Kristen Knipe, Dakota Howard, Yvette Unoarumhi, Darlene Wagner, Matthew Schmerer, Ben L. Rambo-Martin, Kristine Lacek, Sam Shepard, Alison Laufer Halpin, Dave Wentworth, Vivien Dugan, Suxiang Tong, Justin Lee |
| EPI_ISL_1446773, EPI_ISL_1446774, EPI_ISL_1446775, EPI_ISL_1446776, EPI_ISL_1446777, EPI_ISL_1446778, EPI_ISL_1446779, EPI_ISL_1446780, EPI_ISL_1446781, EPI_ISL_1446782, EPI_ISL_1446783, EPI_ISL_1446784, EPI_ISL_1446785, EPI_ISL_1446786 | | | |
| see above | "CDPH, Viral and Rickettsial Disease Laboratory" | Centers for Disease Control and Prevention Division of Viral Diseases, Pathogen Discovery | Mili Sheth, Sarah Nobles, Jasmine Padilla, Mark Burroughs, Shoshona Le, Katie Dillon, Peter Cook, Clinton R. Paden, Dhwani Batra, Krista Queen, Kristen Knipe, Dakota Howard, Yvette Unoarumhi, Darlene Wagner, Matthew Schmerer, Ben L. Rambo-Martin, Kristine Lacek, Sam Shepard, Alison Laufer Halpin, Dave Wentworth, Vivien Dugan, Suxiang Tong, Justin Lee |
| EPI_ISL_1446787 | MO State Public Health Laboratory | Centers for Disease Control and Prevention Division of Viral Diseases, Pathogen Discovery | Mili Sheth, Sarah Nobles, Jasmine Padilla, Mark Burroughs, Shoshona Le, Katie Dillon, Peter Cook, Clinton R. Paden, Dhwani Batra, Krista Queen, Kristen Knipe, Dakota Howard, Yvette Unoarumhi, Darlene Wagner, Matthew Schmerer, Ben L. Rambo-Martin, Kristine Lacek, Sam Shepard, Alison Laufer Halpin, Dave Wentworth, Vivien Dugan, Suxiang Tong, Justin Lee |
| EPI_ISL_1446788, EPI_ISL_1446789 | "CDPH, Viral and Rickettsial Disease Laboratory" | Centers for Disease Control and Prevention Division of Viral Diseases, Pathogen Discovery | Mili Sheth, Sarah Nobles, Jasmine Padilla, Mark Burroughs, Shoshona Le, Katie Dillon, Peter Cook, Clinton R. Paden, Dhwani Batra, Krista Queen, Kristen Knipe, Dakota Howard, Yvette Unoarumhi, Darlene Wagner, Matthew Schmerer, Ben L. Rambo-Martin, Kristine Lacek, Sam Shepard, Alison Laufer Halpin, Dave Wentworth, Vivien Dugan, Suxiang Tong, Justin Lee |
| EPI_ISL_1446790 | MO State Public Health Laboratory | Centers for Disease Control and Prevention Division of Viral Diseases, Pathogen Discovery | Mili Sheth, Sarah Nobles, Jasmine Padilla, Mark Burroughs, Shoshona Le, Katie Dillon, Peter Cook, Clinton R. Paden, Dhwani Batra, Krista Queen, Kristen Knipe, Dakota Howard, Yvette Unoarumhi, Darlene Wagner, Matthew Schmerer, Ben L. Rambo-Martin, Kristine Lacek, Sam Shepard, Alison Laufer Halpin, Dave Wentworth, Vivien Dugan, Suxiang Tong, Justin Lee |
| EPI_ISL_1446791, EPI_ISL_1446792, EPI_ISL_1446793, EPI_ISL_1446794, EPI_ISL_1446795, EPI_ISL_1446796, EPI_ISL_1446797, EPI_ISL_1446798, EPI_ISL_1446799, EPI_ISL_1446800, EPI_ISL_1446801, EPI_ISL_1446802 | | | |
| see above | "CDPH, Viral and Rickettsial Disease Laboratory" | Centers for Disease Control and Prevention Division of Viral Diseases, Pathogen Discovery | Mili Sheth, Sarah Nobles, Jasmine Padilla, Mark Burroughs, Shoshona Le, Katie Dillon, Peter Cook, Clinton R. Paden, Dhwani Batra, Krista Queen, Kristen Knipe, Dakota Howard, Yvette Unoarumhi, Darlene Wagner, Matthew Schmerer, Ben L. Rambo-Martin, Kristine Lacek, Sam Shepard, Alison Laufer Halpin, Dave Wentworth, Vivien Dugan, Suxiang Tong, Justin Lee |
| EPI_ISL_1446803, EPI_ISL_1446804, EPI_ISL_1446805, EPI_ISL_1446806 | ME Health and Environmental Testing Laboratory | Centers for Disease Control and Prevention Division of Viral Diseases, Pathogen Discovery | Mili Sheth, Sarah Nobles, Jasmine Padilla, Mark Burroughs, Shoshona Le, Katie Dillon, Peter Cook, Clinton R. Paden, Dhwani Batra, Krista Queen, Kristen Knipe, Dakota Howard, Yvette Unoarumhi, Darlene Wagner, Matthew Schmerer, Ben L. Rambo-Martin, Kristine Lacek, Sam Shepard, Alison Laufer Halpin, Dave Wentworth, Vivien Dugan, Suxiang Tong, Justin Lee |
| EPI_ISL_1446807, EPI_ISL_1446808 | "PA Department of Health, Bureau of Laboratories" | Centers for Disease Control and Prevention Division of Viral Diseases, Pathogen Discovery | Mili Sheth, Sarah Nobles, Jasmine Padilla, Mark Burroughs, Shoshona Le, Katie Dillon, Peter Cook, Clinton R. Paden, Dhwani Batra, Krista Queen, Kristen Knipe, Dakota Howard, Yvette Unoarumhi, Darlene Wagner, Matthew Schmerer, Ben L. Rambo-Martin, Kristine Lacek, Sam Shepard, Alison Laufer Halpin, Dave Wentworth, Vivien Dugan, Suxiang Tong, Justin Lee |
| EPI_ISL_1446809, EPI_ISL_1446810, EPI_ISL_1446811, EPI_ISL_1446812, EPI_ISL_1446813, EPI_ISL_1446814, EPI_ISL_1446815, EPI_ISL_1446816, EPI_ISL_1446817, EPI_ISL_1446818, EPI_ISL_1446819, EPI_ISL_1446820, EPI_ISL_1446821, EPI_ISL_1446822, EPI_ISL_1446823, EPI_ISL_1446824 | | | |
| see above | GA Department of Public Health Laboratory | Centers for Disease Control and Prevention Division of Viral Diseases, Pathogen Discovery | Mili Sheth, Sarah Nobles, Jasmine Padilla, Mark Burroughs, Shoshona Le, Katie Dillon, Peter Cook, Clinton R. Paden, Dhwani Batra, Krista Queen, Kristen Knipe, Dakota Howard, Yvette Unoarumhi, Darlene Wagner, Matthew Schmerer, Ben L. Rambo-Martin, Kristine Lacek, Sam Shepard, Alison Laufer Halpin, Dave Wentworth, Vivien Dugan, Suxiang Tong, Justin Lee |
| EPI_ISL_1446825, EPI_ISL_1446826, EPI_ISL_1446827, EPI_ISL_1446828, EPI_ISL_1446829, EPI_ISL_1446830, EPI_ISL_1446831, EPI_ISL_1446832, EPI_ISL_1446833, EPI_ISL_1446834, EPI_ISL_1446835, EPI_ISL_1446836, EPI_ISL_1446837, EPI_ISL_1446838, EPI_ISL_1446839 | | | |
| see above | NE Public Health Laboratory | Centers for Disease Control and Prevention Division of Viral Diseases, Pathogen Discovery | Mili Sheth, Sarah Nobles, Jasmine Padilla, Mark Burroughs, Shoshona Le, Katie Dillon, Peter Cook, Clinton R. Paden, Dhwani Batra, Krista Queen, Kristen Knipe, Dakota Howard, Yvette Unoarumhi, Darlene Wagner, Matthew Schmerer, Ben L. Rambo-Martin, Kristine Lacek, Sam Shepard, Alison Laufer Halpin, Dave Wentworth, Vivien Dugan, Suxiang Tong, Justin Lee |
| EPI_ISL_1446840, | "CDPH, Viral and Rickettsial Disease | Centers for Disease Control and Prevention | Mili Sheth, Sarah Nobles, Jasmine Padilla, Mark Burroughs, Shoshona Le, Katie Dillon, Peter Cook, Clinton R. Paden, Dhwani Batra, Krista Queen, Kristen Knipe, Dakota Howard, Yvette Unoarumhi, Darlene Wagner, Matthew Schmerer, Ben L. Rambo-Martin, Kristine Lacek, Sam Shepard, |

| | | | |
|---|---|---|---|
| EPI_ISL_1446841, EPI_ISL_1446842, EPI_ISL_1446843, EPI_ISL_1446844, EPI_ISL_1446845 | Laboratory" | Division of Viral Diseases, Pathogen Discovery | Alison Laufer Halpin, Dave Wentworth, Vivien Dugan, Suxiang Tong, Justin Lee |
| EPI_ISL_1446846, EPI_ISL_1446847, EPI_ISL_1446848, EPI_ISL_1446849, EPI_ISL_1446850, EPI_ISL_1446851, EPI_ISL_1446852, EPI_ISL_1446853, EPI_ISL_1446854 | IN State Department of Health Laboratory Services | Centers for Disease Control and Prevention Division of Viral Diseases, Pathogen Discovery | Mili Sheth, Sarah Nobles, Jasmine Padilla, Mark Burroughs, Shoshona Le, Katie Dillon, Peter Cook, Clinton R. Paden, Dhwani Batra, Krista Queen, Kristen Knipe, Dakota Howard, Yvette Unoarumhi, Darlene Wagner, Matthew Schmerer, Ben L. Rambo-Martin, Kristine Lacek, Sam Shepard, Alison Laufer Halpin, Dave Wentworth, Vivien Dugan, Suxiang Tong, Justin Lee |
| EPI_ISL_1446855, EPI_ISL_1446856 | AL Dept. of Public Health Bureau of Clinical Laboratories | Centers for Disease Control and Prevention Division of Viral Diseases, Pathogen Discovery | Mili Sheth, Sarah Nobles, Jasmine Padilla, Mark Burroughs, Shoshona Le, Katie Dillon, Peter Cook, Clinton R. Paden, Dhwani Batra, Krista Queen, Kristen Knipe, Dakota Howard, Yvette Unoarumhi, Darlene Wagner, Matthew Schmerer, Ben L. Rambo-Martin, Kristine Lacek, Sam Shepard, Alison Laufer Halpin, Dave Wentworth, Vivien Dugan, Suxiang Tong, Justin Lee |
| EPI_ISL_1446857 | MT Public Health Laboratory | Centers for Disease Control and Prevention Division of Viral Diseases, Pathogen Discovery | Mili Sheth, Sarah Nobles, Jasmine Padilla, Mark Burroughs, Shoshona Le, Katie Dillon, Peter Cook, Clinton R. Paden, Dhwani Batra, Krista Queen, Kristen Knipe, Dakota Howard, Yvette Unoarumhi, Darlene Wagner, Matthew Schmerer, Ben L. Rambo-Martin, Kristine Lacek, Sam Shepard, Alison Laufer Halpin, Dave Wentworth, Vivien Dugan, Suxiang Tong, Justin Lee |
| EPI_ISL_1446858 | OH Department of Health Laboratory | Centers for Disease Control and Prevention Division of Viral Diseases, Pathogen Discovery | Mili Sheth, Sarah Nobles, Jasmine Padilla, Mark Burroughs, Shoshona Le, Katie Dillon, Peter Cook, Clinton R. Paden, Dhwani Batra, Krista Queen, Kristen Knipe, Dakota Howard, Yvette Unoarumhi, Darlene Wagner, Matthew Schmerer, Ben L. Rambo-Martin, Kristine Lacek, Sam Shepard, Alison Laufer Halpin, Dave Wentworth, Vivien Dugan, Suxiang Tong, Justin Lee |
| EPI_ISL_1446859, EPI_ISL_1446860 | MD DOH Laboratories Administration | Centers for Disease Control and Prevention Division of Viral Diseases, Pathogen Discovery | Mili Sheth, Sarah Nobles, Jasmine Padilla, Mark Burroughs, Shoshona Le, Katie Dillon, Peter Cook, Clinton R. Paden, Dhwani Batra, Krista Queen, Kristen Knipe, Dakota Howard, Yvette Unoarumhi, Darlene Wagner, Matthew Schmerer, Ben L. Rambo-Martin, Kristine Lacek, Sam Shepard, Alison Laufer Halpin, Dave Wentworth, Vivien Dugan, Suxiang Tong, Justin Lee |
| EPI_ISL_1446861 | WI State Laboratory of Hygiene | Centers for Disease Control and Prevention Division of Viral Diseases, Pathogen Discovery | Mili Sheth, Sarah Nobles, Jasmine Padilla, Mark Burroughs, Shoshona Le, Katie Dillon, Peter Cook, Clinton R. Paden, Dhwani Batra, Krista Queen, Kristen Knipe, Dakota Howard, Yvette Unoarumhi, Darlene Wagner, Matthew Schmerer, Ben L. Rambo-Martin, Kristine Lacek, Sam Shepard, Alison Laufer Halpin, Dave Wentworth, Vivien Dugan, Suxiang Tong, Justin Lee |
| EPI_ISL_1446862 | MD DOH Laboratories Administration | Centers for Disease Control and Prevention Division of Viral Diseases, Pathogen Discovery | Mili Sheth, Sarah Nobles, Jasmine Padilla, Mark Burroughs, Shoshona Le, Katie Dillon, Peter Cook, Clinton R. Paden, Dhwani Batra, Krista Queen, Kristen Knipe, Dakota Howard, Yvette Unoarumhi, Darlene Wagner, Matthew Schmerer, Ben L. Rambo-Martin, Kristine Lacek, Sam Shepard, Alison Laufer Halpin, Dave Wentworth, Vivien Dugan, Suxiang Tong, Justin Lee |
| EPI_ISL_1446863 | WI State Laboratory of Hygiene | Centers for Disease Control and Prevention Division of Viral Diseases, Pathogen Discovery | Mili Sheth, Sarah Nobles, Jasmine Padilla, Mark Burroughs, Shoshona Le, Katie Dillon, Peter Cook, Clinton R. Paden, Dhwani Batra, Krista Queen, Kristen Knipe, Dakota Howard, Yvette Unoarumhi, Darlene Wagner, Matthew Schmerer, Ben L. Rambo-Martin, Kristine Lacek, Sam Shepard, Alison Laufer Halpin, Dave Wentworth, Vivien Dugan, Suxiang Tong, Justin Lee |
| EPI_ISL_1446864, EPI_ISL_1446865, EPI_ISL_1446866, EPI_ISL_1446867 | GA Department of Public Health Laboratory | Centers for Disease Control and Prevention Division of Viral Diseases, Pathogen Discovery | Mili Sheth, Sarah Nobles, Jasmine Padilla, Mark Burroughs, Shoshona Le, Katie Dillon, Peter Cook, Clinton R. Paden, Dhwani Batra, Krista Queen, Kristen Knipe, Dakota Howard, Yvette Unoarumhi, Darlene Wagner, Matthew Schmerer, Ben L. Rambo-Martin, Kristine Lacek, Sam Shepard, Alison Laufer Halpin, Dave Wentworth, Vivien Dugan, Suxiang Tong, Justin Lee |
| EPI_ISL_1446868 | "NM Dept. Health, Scientific Laboratory Division " | Centers for Disease Control and Prevention Division of Viral Diseases, Pathogen Discovery | Mili Sheth, Sarah Nobles, Jasmine Padilla, Mark Burroughs, Shoshona Le, Katie Dillon, Peter Cook, Clinton R. Paden, Dhwani Batra, Krista Queen, Kristen Knipe, Dakota Howard, Yvette Unoarumhi, Darlene Wagner, Matthew Schmerer, Ben L. Rambo-Martin, Kristine Lacek, Sam Shepard, Alison Laufer Halpin, Dave Wentworth, Vivien Dugan, Suxiang Tong, Justin Lee |
| EPI_ISL_1446869, EPI_ISL_1446870, EPI_ISL_1446871 | "MN PHL Division, Minnesota Department of Health" | Centers for Disease Control and Prevention Division of Viral Diseases, Pathogen Discovery | Mili Sheth, Sarah Nobles, Jasmine Padilla, Mark Burroughs, Shoshona Le, Katie Dillon, Peter Cook, Clinton R. Paden, Dhwani Batra, Krista Queen, Kristen Knipe, Dakota Howard, Yvette Unoarumhi, Darlene Wagner, Matthew Schmerer, Ben L. Rambo-Martin, Kristine Lacek, Sam Shepard, Alison Laufer Halpin, Dave Wentworth, Vivien Dugan, Suxiang Tong, Justin Lee |
| EPI_ISL_1446872 | GA Department of Public Health Laboratory | Centers for Disease Control and Prevention Division of Viral Diseases, Pathogen Discovery | Mili Sheth, Sarah Nobles, Jasmine Padilla, Mark Burroughs, Shoshona Le, Katie Dillon, Peter Cook, Clinton R. Paden, Dhwani Batra, Krista Queen, Kristen Knipe, Dakota Howard, Yvette Unoarumhi, Darlene Wagner, Matthew Schmerer, Ben L. Rambo-Martin, Kristine Lacek, Sam Shepard, Alison Laufer Halpin, Dave Wentworth, Vivien Dugan, Suxiang Tong, Justin Lee |
| EPI_ISL_1446873 | "MN PHL Division, Minnesota Department of Health" | Centers for Disease Control and Prevention Division of Viral Diseases, Pathogen Discovery | Mili Sheth, Sarah Nobles, Jasmine Padilla, Mark Burroughs, Shoshona Le, Katie Dillon, Peter Cook, Clinton R. Paden, Dhwani Batra, Krista Queen, Kristen Knipe, Dakota Howard, Yvette Unoarumhi, Darlene Wagner, Matthew Schmerer, Ben L. Rambo-Martin, Kristine Lacek, Sam Shepard, Alison Laufer Halpin, Dave Wentworth, Vivien Dugan, Suxiang Tong, Justin Lee |
| EPI_ISL_1446874 | "NM Dept. Health, Scientific Laboratory Division " | Centers for Disease Control and Prevention Division of Viral Diseases, Pathogen Discovery | Mili Sheth, Sarah Nobles, Jasmine Padilla, Mark Burroughs, Shoshona Le, Katie Dillon, Peter Cook, Clinton R. Paden, Dhwani Batra, Krista Queen, Kristen Knipe, Dakota Howard, Yvette Unoarumhi, Darlene Wagner, Matthew Schmerer, Ben L. Rambo-Martin, Kristine Lacek, Sam Shepard, Alison Laufer Halpin, Dave Wentworth, Vivien Dugan, Suxiang Tong, Justin Lee |
| EPI_ISL_1446875 | "MN PHL Division, Minnesota Department of Health" | Centers for Disease Control and Prevention Division of Viral Diseases, Pathogen Discovery | Mili Sheth, Sarah Nobles, Jasmine Padilla, Mark Burroughs, Shoshona Le, Katie Dillon, Peter Cook, Clinton R. Paden, Dhwani Batra, Krista Queen, Kristen Knipe, Dakota Howard, Yvette Unoarumhi, Darlene Wagner, Matthew Schmerer, Ben L. Rambo-Martin, Kristine Lacek, Sam Shepard, Alison Laufer Halpin, Dave Wentworth, Vivien Dugan, Suxiang Tong, Justin Lee |
| EPI_ISL_1446876 | "NM Dept. Health, Scientific Laboratory Division " | Centers for Disease Control and Prevention Division of Viral Diseases, Pathogen Discovery | Mili Sheth, Sarah Nobles, Jasmine Padilla, Mark Burroughs, Shoshona Le, Katie Dillon, Peter Cook, Clinton R. Paden, Dhwani Batra, Krista Queen, Kristen Knipe, Dakota Howard, Yvette Unoarumhi, Darlene Wagner, Matthew Schmerer, Ben L. Rambo-Martin, Kristine Lacek, Sam Shepard, Alison Laufer Halpin, Dave Wentworth, Vivien Dugan, Suxiang Tong, Justin Lee |
| EPI_ISL_1446877 | GA Department of Public Health Laboratory | Centers for Disease Control and Prevention Division of Viral Diseases, Pathogen Discovery | Mili Sheth, Sarah Nobles, Jasmine Padilla, Mark Burroughs, Shoshona Le, Katie Dillon, Peter Cook, Clinton R. Paden, Dhwani Batra, Krista Queen, Kristen Knipe, Dakota Howard, Yvette Unoarumhi, Darlene Wagner, Matthew Schmerer, Ben L. Rambo-Martin, Kristine Lacek, Sam Shepard, Alison Laufer Halpin, Dave Wentworth, Vivien Dugan, Suxiang Tong, Justin Lee |
| EPI_ISL_1446878 | "NM Dept. Health, Scientific Laboratory Division " | Centers for Disease Control and Prevention Division of Viral Diseases, Pathogen Discovery | Mili Sheth, Sarah Nobles, Jasmine Padilla, Mark Burroughs, Shoshona Le, Katie Dillon, Peter Cook, Clinton R. Paden, Dhwani Batra, Krista Queen, Kristen Knipe, Dakota Howard, Yvette Unoarumhi, Darlene Wagner, Matthew Schmerer, Ben L. Rambo-Martin, Kristine Lacek, Sam Shepard, Alison Laufer Halpin, Dave Wentworth, Vivien Dugan, Suxiang Tong, Justin Lee |
| EPI_ISL_1446879 | "MN PHL Division, Minnesota Department of Health" | Centers for Disease Control and Prevention Division of Viral Diseases, Pathogen Discovery | Mili Sheth, Sarah Nobles, Jasmine Padilla, Mark Burroughs, Shoshona Le, Katie Dillon, Peter Cook, Clinton R. Paden, Dhwani Batra, Krista Queen, Kristen Knipe, Dakota Howard, Yvette Unoarumhi, Darlene Wagner, Matthew Schmerer, Ben L. Rambo-Martin, Kristine Lacek, Sam Shepard, Alison Laufer Halpin, Dave Wentworth, Vivien Dugan, Suxiang Tong, Justin Lee |
| EPI_ISL_1446880 | GA Department of Public Health Laboratory | Centers for Disease Control and Prevention Division of Viral Diseases, Pathogen Discovery | Mili Sheth, Sarah Nobles, Jasmine Padilla, Mark Burroughs, Shoshona Le, Katie Dillon, Peter Cook, Clinton R. Paden, Dhwani Batra, Krista Queen, Kristen Knipe, Dakota Howard, Yvette Unoarumhi, Darlene Wagner, Matthew Schmerer, Ben L. Rambo-Martin, Kristine Lacek, Sam Shepard, Alison Laufer Halpin, Dave Wentworth, Vivien Dugan, Suxiang Tong, Justin Lee |
| EPI_ISL_1446881 | "MN PHL Division, Minnesota Department of Health" | Centers for Disease Control and Prevention Division of Viral Diseases, Pathogen Discovery | Mili Sheth, Sarah Nobles, Jasmine Padilla, Mark Burroughs, Shoshona Le, Katie Dillon, Peter Cook, Clinton R. Paden, Dhwani Batra, Krista Queen, Kristen Knipe, Dakota Howard, Yvette Unoarumhi, Darlene Wagner, Matthew Schmerer, Ben L. Rambo-Martin, Kristine Lacek, Sam Shepard, Alison Laufer Halpin, Dave Wentworth, Vivien Dugan, Suxiang Tong, Justin Lee |
| EPI_ISL_1446882 | GA Department of Public Health Laboratory | Centers for Disease Control and Prevention Division of Viral Diseases, Pathogen Discovery | Mili Sheth, Sarah Nobles, Jasmine Padilla, Mark Burroughs, Shoshona Le, Katie Dillon, Peter Cook, Clinton R. Paden, Dhwani Batra, Krista Queen, Kristen Knipe, Dakota Howard, Yvette Unoarumhi, Darlene Wagner, Matthew Schmerer, Ben L. Rambo-Martin, Kristine Lacek, Sam Shepard, Alison Laufer Halpin, Dave Wentworth, Vivien Dugan, Suxiang Tong, Justin Lee |
| EPI_ISL_1446883 | "MN PHL Division, Minnesota Department of Health" | Centers for Disease Control and Prevention Division of Viral Diseases, Pathogen Discovery | Mili Sheth, Sarah Nobles, Jasmine Padilla, Mark Burroughs, Shoshona Le, Katie Dillon, Peter Cook, Clinton R. Paden, Dhwani Batra, Krista Queen, Kristen Knipe, Dakota Howard, Yvette Unoarumhi, Darlene Wagner, Matthew Schmerer, Ben L. Rambo-Martin, Kristine Lacek, Sam Shepard, Alison Laufer Halpin, Dave Wentworth, Vivien Dugan, Suxiang Tong, Justin Lee |
| EPI_ISL_1446884, EPI_ISL_1446885, EPI_ISL_1446886 | "NM Dept. Health, Scientific Laboratory Division " | Centers for Disease Control and Prevention Division of Viral Diseases, Pathogen Discovery | Mili Sheth, Sarah Nobles, Jasmine Padilla, Mark Burroughs, Shoshona Le, Katie Dillon, Peter Cook, Clinton R. Paden, Dhwani Batra, Krista Queen, Kristen Knipe, Dakota Howard, Yvette Unoarumhi, Darlene Wagner, Matthew Schmerer, Ben L. Rambo-Martin, Kristine Lacek, Sam Shepard, Alison Laufer Halpin, Dave Wentworth, Vivien Dugan, Suxiang Tong, Justin Lee |
| EPI_ISL_1446887 | GA Department of Public Health Laboratory | Centers for Disease Control and Prevention Division of Viral Diseases, Pathogen Discovery | Mili Sheth, Sarah Nobles, Jasmine Padilla, Mark Burroughs, Shoshona Le, Katie Dillon, Peter Cook, Clinton R. Paden, Dhwani Batra, Krista Queen, Kristen Knipe, Dakota Howard, Yvette Unoarumhi, Darlene Wagner, Matthew Schmerer, Ben L. Rambo-Martin, Kristine Lacek, Sam Shepard, Alison Laufer Halpin, Dave Wentworth, Vivien Dugan, Suxiang Tong, Justin Lee |
| EPI_ISL_1446888 | "NM Dept. Health, Scientific Laboratory Division " | Centers for Disease Control and Prevention Division of Viral Diseases, Pathogen Discovery | Mili Sheth, Sarah Nobles, Jasmine Padilla, Mark Burroughs, Shoshona Le, Katie Dillon, Peter Cook, Clinton R. Paden, Dhwani Batra, Krista Queen, Kristen Knipe, Dakota Howard, Yvette Unoarumhi, Darlene Wagner, Matthew Schmerer, Ben L. Rambo-Martin, Kristine Lacek, Sam Shepard, Alison Laufer Halpin, Dave Wentworth, Vivien Dugan, Suxiang Tong, Justin Lee |
| EPI_ISL_1446889, EPI_ISL_1446890, EPI_ISL_1446891, EPI_ISL_1446892, EPI_ISL_1446893, EPI_ISL_1446894, EPI_ISL_1446895, EPI_ISL_1446896, EPI_ISL_1446897, EPI_ISL_1446898, EPI_ISL_1446899 | | | |
| see above | "MN PHL Division, Minnesota Department of Health" | Centers for Disease Control and Prevention Division of Viral Diseases, Pathogen Discovery | Mili Sheth, Sarah Nobles, Jasmine Padilla, Mark Burroughs, Shoshona Le, Katie Dillon, Peter Cook, Clinton R. Paden, Dhwani Batra, Krista Queen, Kristen Knipe, Dakota Howard, Yvette Unoarumhi, Darlene Wagner, Matthew Schmerer, Ben L. Rambo-Martin, Kristine Lacek, Sam Shepard, Alison Laufer Halpin, Dave Wentworth, Vivien Dugan, Suxiang Tong, Justin Lee |
| EPI_ISL_1446900 | GA Department of Public Health Laboratory | Centers for Disease Control and Prevention Division of Viral Diseases, Pathogen | Mili Sheth, Sarah Nobles, Jasmine Padilla, Mark Burroughs, Shoshona Le, Katie Dillon, Peter Cook, Clinton R. Paden, Dhwani Batra, Krista Queen, Kristen Knipe, Dakota Howard, Yvette Unoarumhi, Darlene Wagner, Matthew Schmerer, Ben L. Rambo-Martin, Kristine Lacek, Sam Shepard, Alison Laufer Halpin, Dave Wentworth, Vivien Dugan, Suxiang Tong, Justin Lee |

| | | | |
|---|---|---|---|
| | | Discovery | |
| EPI_ISL_1446901 | "NM Dept. Health, Scientific Laboratory Division " | Centers for Disease Control and Prevention Division of Viral Diseases, Pathogen Discovery | Mili Sheth, Sarah Nobles, Jasmine Padilla, Mark Burroughs, Shoshona Le, Katie Dillon, Peter Cook, Clinton R. Paden, Dhwani Batra, Krista Queen, Kristen Knipe, Dakota Howard, Yvette Unoarumhi, Darlene Wagner, Matthew Schmerer, Ben L. Rambo-Martin, Kristine Lacek, Sam Shepard, Alison Laufer Halpin, Dave Wentworth, Vivien Dugan, Suxiang Tong, Justin Lee |
| EPI_ISL_1446902 | "MN PHL Division, Minnesota Department of Health" | Centers for Disease Control and Prevention Division of Viral Diseases, Pathogen Discovery | Mili Sheth, Sarah Nobles, Jasmine Padilla, Mark Burroughs, Shoshona Le, Katie Dillon, Peter Cook, Clinton R. Paden, Dhwani Batra, Krista Queen, Kristen Knipe, Dakota Howard, Yvette Unoarumhi, Darlene Wagner, Matthew Schmerer, Ben L. Rambo-Martin, Kristine Lacek, Sam Shepard, Alison Laufer Halpin, Dave Wentworth, Vivien Dugan, Suxiang Tong, Justin Lee |
| EPI_ISL_1446903 | "NM Dept. Health, Scientific Laboratory Division " | Centers for Disease Control and Prevention Division of Viral Diseases, Pathogen Discovery | Mili Sheth, Sarah Nobles, Jasmine Padilla, Mark Burroughs, Shoshona Le, Katie Dillon, Peter Cook, Clinton R. Paden, Dhwani Batra, Krista Queen, Kristen Knipe, Dakota Howard, Yvette Unoarumhi, Darlene Wagner, Matthew Schmerer, Ben L. Rambo-Martin, Kristine Lacek, Sam Shepard, Alison Laufer Halpin, Dave Wentworth, Vivien Dugan, Suxiang Tong, Justin Lee |
| EPI_ISL_1446904, EPI_ISL_1446905 | GA Department of Public Health Laboratory | Centers for Disease Control and Prevention Division of Viral Diseases, Pathogen Discovery | Mili Sheth, Sarah Nobles, Jasmine Padilla, Mark Burroughs, Shoshona Le, Katie Dillon, Peter Cook, Clinton R. Paden, Dhwani Batra, Krista Queen, Kristen Knipe, Dakota Howard, Yvette Unoarumhi, Darlene Wagner, Matthew Schmerer, Ben L. Rambo-Martin, Kristine Lacek, Sam Shepard, Alison Laufer Halpin, Dave Wentworth, Vivien Dugan, Suxiang Tong, Justin Lee |
| EPI_ISL_1446906 | "MN PHL Division, Minnesota Department of Health" | Centers for Disease Control and Prevention Division of Viral Diseases, Pathogen Discovery | Mili Sheth, Sarah Nobles, Jasmine Padilla, Mark Burroughs, Shoshona Le, Katie Dillon, Peter Cook, Clinton R. Paden, Dhwani Batra, Krista Queen, Kristen Knipe, Dakota Howard, Yvette Unoarumhi, Darlene Wagner, Matthew Schmerer, Ben L. Rambo-Martin, Kristine Lacek, Sam Shepard, Alison Laufer Halpin, Dave Wentworth, Vivien Dugan, Suxiang Tong, Justin Lee |
| EPI_ISL_1446907, EPI_ISL_1446908 | "NM Dept. Health, Scientific Laboratory Division " | Centers for Disease Control and Prevention Division of Viral Diseases, Pathogen Discovery | Mili Sheth, Sarah Nobles, Jasmine Padilla, Mark Burroughs, Shoshona Le, Katie Dillon, Peter Cook, Clinton R. Paden, Dhwani Batra, Krista Queen, Kristen Knipe, Dakota Howard, Yvette Unoarumhi, Darlene Wagner, Matthew Schmerer, Ben L. Rambo-Martin, Kristine Lacek, Sam Shepard, Alison Laufer Halpin, Dave Wentworth, Vivien Dugan, Suxiang Tong, Justin Lee |
| EPI_ISL_1446909 | "MN PHL Division, Minnesota Department of Health" | Centers for Disease Control and Prevention Division of Viral Diseases, Pathogen Discovery | Mili Sheth, Sarah Nobles, Jasmine Padilla, Mark Burroughs, Shoshona Le, Katie Dillon, Peter Cook, Clinton R. Paden, Dhwani Batra, Krista Queen, Kristen Knipe, Dakota Howard, Yvette Unoarumhi, Darlene Wagner, Matthew Schmerer, Ben L. Rambo-Martin, Kristine Lacek, Sam Shepard, Alison Laufer Halpin, Dave Wentworth, Vivien Dugan, Suxiang Tong, Justin Lee |
| EPI_ISL_1446910, EPI_ISL_1446911 | GA Department of Public Health Laboratory | Centers for Disease Control and Prevention Division of Viral Diseases, Pathogen Discovery | Mili Sheth, Sarah Nobles, Jasmine Padilla, Mark Burroughs, Shoshona Le, Katie Dillon, Peter Cook, Clinton R. Paden, Dhwani Batra, Krista Queen, Kristen Knipe, Dakota Howard, Yvette Unoarumhi, Darlene Wagner, Matthew Schmerer, Ben L. Rambo-Martin, Kristine Lacek, Sam Shepard, Alison Laufer Halpin, Dave Wentworth, Vivien Dugan, Suxiang Tong, Justin Lee |
| EPI_ISL_1446912 | "NM Dept. Health, Scientific Laboratory Division " | Centers for Disease Control and Prevention Division of Viral Diseases, Pathogen Discovery | Mili Sheth, Sarah Nobles, Jasmine Padilla, Mark Burroughs, Shoshona Le, Katie Dillon, Peter Cook, Clinton R. Paden, Dhwani Batra, Krista Queen, Kristen Knipe, Dakota Howard, Yvette Unoarumhi, Darlene Wagner, Matthew Schmerer, Ben L. Rambo-Martin, Kristine Lacek, Sam Shepard, Alison Laufer Halpin, Dave Wentworth, Vivien Dugan, Suxiang Tong, Justin Lee |
| EPI_ISL_1446913, EPI_ISL_1446914, EPI_ISL_1446915, EPI_ISL_1446916, EPI_ISL_1446917, EPI_ISL_1446918, EPI_ISL_1446919, EPI_ISL_1446920, EPI_ISL_1446921, EPI_ISL_1446922 | NC State Laboratory of Public Health | Centers for Disease Control and Prevention Division of Viral Diseases, Pathogen Discovery | Mili Sheth, Sarah Nobles, Jasmine Padilla, Mark Burroughs, Shoshona Le, Katie Dillon, Peter Cook, Clinton R. Paden, Dhwani Batra, Krista Queen, Kristen Knipe, Dakota Howard, Yvette Unoarumhi, Darlene Wagner, Matthew Schmerer, Ben L. Rambo-Martin, Kristine Lacek, Sam Shepard, Alison Laufer Halpin, Dave Wentworth, Vivien Dugan, Suxiang Tong, Justin Lee |
| EPI_ISL_1446923, EPI_ISL_1446924, EPI_ISL_1446925, EPI_ISL_1446926, EPI_ISL_1446927 | MO State Public Health Laboratory | Centers for Disease Control and Prevention Division of Viral Diseases, Pathogen Discovery | Mili Sheth, Sarah Nobles, Jasmine Padilla, Mark Burroughs, Shoshona Le, Katie Dillon, Peter Cook, Clinton R. Paden, Dhwani Batra, Krista Queen, Kristen Knipe, Dakota Howard, Yvette Unoarumhi, Darlene Wagner, Matthew Schmerer, Ben L. Rambo-Martin, Kristine Lacek, Sam Shepard, Alison Laufer Halpin, Dave Wentworth, Vivien Dugan, Suxiang Tong, Justin Lee |
| EPI_ISL_1446928, EPI_ISL_1446929, EPI_ISL_1446930, EPI_ISL_1446931 | IL Dept. of Public Health Springfield Laboratory | Centers for Disease Control and Prevention Division of Viral Diseases, Pathogen Discovery | Mili Sheth, Sarah Nobles, Jasmine Padilla, Mark Burroughs, Shoshona Le, Katie Dillon, Peter Cook, Clinton R. Paden, Dhwani Batra, Krista Queen, Kristen Knipe, Dakota Howard, Yvette Unoarumhi, Darlene Wagner, Matthew Schmerer, Ben L. Rambo-Martin, Kristine Lacek, Sam Shepard, Alison Laufer Halpin, Dave Wentworth, Vivien Dugan, Suxiang Tong, Justin Lee |
| EPI_ISL_1446932, EPI_ISL_1446933, EPI_ISL_1446934, EPI_ISL_1446935, EPI_ISL_1446936 | MS Public Health Laboratory | Centers for Disease Control and Prevention Division of Viral Diseases, Pathogen Discovery | Mili Sheth, Sarah Nobles, Jasmine Padilla, Mark Burroughs, Shoshona Le, Katie Dillon, Peter Cook, Clinton R. Paden, Dhwani Batra, Krista Queen, Kristen Knipe, Dakota Howard, Yvette Unoarumhi, Darlene Wagner, Matthew Schmerer, Ben L. Rambo-Martin, Kristine Lacek, Sam Shepard, Alison Laufer Halpin, Dave Wentworth, Vivien Dugan, Suxiang Tong, Justin Lee |
| EPI_ISL_1446937 | NE Public Health Laboratory | Centers for Disease Control and Prevention Division of Viral Diseases, Pathogen Discovery | Mili Sheth, Sarah Nobles, Jasmine Padilla, Mark Burroughs, Shoshona Le, Katie Dillon, Peter Cook, Clinton R. Paden, Dhwani Batra, Krista Queen, Kristen Knipe, Dakota Howard, Yvette Unoarumhi, Darlene Wagner, Matthew Schmerer, Ben L. Rambo-Martin, Kristine Lacek, Sam Shepard, Alison Laufer Halpin, Dave Wentworth, Vivien Dugan, Suxiang Tong, Justin Lee |
| EPI_ISL_1446938, EPI_ISL_1446939, EPI_ISL_1446940, EPI_ISL_1446942, EPI_ISL_1446943 | KS Health and Environmental Laboratories | Centers for Disease Control and Prevention Division of Viral Diseases, Pathogen Discovery | Mili Sheth, Sarah Nobles, Jasmine Padilla, Mark Burroughs, Shoshona Le, Katie Dillon, Peter Cook, Clinton R. Paden, Dhwani Batra, Krista Queen, Kristen Knipe, Dakota Howard, Yvette Unoarumhi, Darlene Wagner, Matthew Schmerer, Ben L. Rambo-Martin, Kristine Lacek, Sam Shepard, Alison Laufer Halpin, Dave Wentworth, Vivien Dugan, Suxiang Tong, Justin Lee |
| EPI_ISL_1446944 | AL Dept. of Public Health Bureau of Clinical Laboratories | Centers for Disease Control and Prevention Division of Viral Diseases, Pathogen Discovery | Mili Sheth, Sarah Nobles, Jasmine Padilla, Mark Burroughs, Shoshona Le, Katie Dillon, Peter Cook, Clinton R. Paden, Dhwani Batra, Krista Queen, Kristen Knipe, Dakota Howard, Yvette Unoarumhi, Darlene Wagner, Matthew Schmerer, Ben L. Rambo-Martin, Kristine Lacek, Sam Shepard, Alison Laufer Halpin, Dave Wentworth, Vivien Dugan, Suxiang Tong, Justin Lee |
| EPI_ISL_1446945 | TN Division of Laboratory Services | Centers for Disease Control and Prevention Division of Viral Diseases, Pathogen Discovery | Mili Sheth, Sarah Nobles, Jasmine Padilla, Mark Burroughs, Shoshona Le, Katie Dillon, Peter Cook, Clinton R. Paden, Dhwani Batra, Krista Queen, Kristen Knipe, Dakota Howard, Yvette Unoarumhi, Darlene Wagner, Matthew Schmerer, Ben L. Rambo-Martin, Kristine Lacek, Sam Shepard, Alison Laufer Halpin, Dave Wentworth, Vivien Dugan, Suxiang Tong, Justin Lee |
| EPI_ISL_1446946, EPI_ISL_1446947, EPI_ISL_1446948, EPI_ISL_1446949, EPI_ISL_1446950, EPI_ISL_1446951, EPI_ISL_1446952, EPI_ISL_1446953, EPI_ISL_1446954, EPI_ISL_1446955, EPI_ISL_1446956, EPI_ISL_1446957, EPI_ISL_1446958, EPI_ISL_1446959, EPI_ISL_1446960, EPI_ISL_1446961, EPI_ISL_1446962, EPI_ISL_1446963, EPI_ISL_1446964, EPI_ISL_1446965, EPI_ISL_1446966, EPI_ISL_1446967, EPI_ISL_1446968, EPI_ISL_1446969 | | | |
| see above | MI - Michigan Department of Health and Human Services - Bureau of Laboratories | Centers for Disease Control and Prevention Division of Viral Diseases, Pathogen Discovery | Mili Sheth, Sarah Nobles, Jasmine Padilla, Mark Burroughs, Shoshona Le, Katie Dillon, Peter Cook, Clinton R. Paden, Dhwani Batra, Krista Queen, Kristen Knipe, Dakota Howard, Yvette Unoarumhi, Darlene Wagner, Matthew Schmerer, Ben L. Rambo-Martin, Kristine Lacek, Sam Shepard, Alison Laufer Halpin, Dave Wentworth, Vivien Dugan, Suxiang Tong, Justin Lee |
| EPI_ISL_1446970, EPI_ISL_1446971, EPI_ISL_1446972, EPI_ISL_1446973 | DC Public Health Lab/ Dept. of Forensic Sciences | Centers for Disease Control and Prevention Division of Viral Diseases, Pathogen Discovery | Mili Sheth, Sarah Nobles, Jasmine Padilla, Mark Burroughs, Shoshona Le, Katie Dillon, Peter Cook, Clinton R. Paden, Dhwani Batra, Krista Queen, Kristen Knipe, Dakota Howard, Yvette Unoarumhi, Darlene Wagner, Matthew Schmerer, Ben L. Rambo-Martin, Kristine Lacek, Sam Shepard, Alison Laufer Halpin, Dave Wentworth, Vivien Dugan, Suxiang Tong, Justin Lee |
| EPI_ISL_1446974, EPI_ISL_1446975 | NYC Department of Health and Mental Hygiene | Centers for Disease Control and Prevention Division of Viral Diseases, Pathogen Discovery | Mili Sheth, Sarah Nobles, Jasmine Padilla, Mark Burroughs, Shoshona Le, Katie Dillon, Peter Cook, Clinton R. Paden, Dhwani Batra, Krista Queen, Kristen Knipe, Dakota Howard, Yvette Unoarumhi, Darlene Wagner, Matthew Schmerer, Ben L. Rambo-Martin, Kristine Lacek, Sam Shepard, Alison Laufer Halpin, Dave Wentworth, Vivien Dugan, Suxiang Tong, Justin Lee |
| EPI_ISL_1446976 | SC Dept of Health and Env. Control- Bureau of Laboratories | Centers for Disease Control and Prevention Division of Viral Diseases, Pathogen Discovery | Mili Sheth, Sarah Nobles, Jasmine Padilla, Mark Burroughs, Shoshona Le, Katie Dillon, Peter Cook, Clinton R. Paden, Dhwani Batra, Krista Queen, Kristen Knipe, Dakota Howard, Yvette Unoarumhi, Darlene Wagner, Matthew Schmerer, Ben L. Rambo-Martin, Kristine Lacek, Sam Shepard, Alison Laufer Halpin, Dave Wentworth, Vivien Dugan, Suxiang Tong, Justin Lee |
| EPI_ISL_1446977, EPI_ISL_1446978 | ID Bureau of Laboratories | Centers for Disease Control and Prevention Division of Viral Diseases, Pathogen Discovery | Mili Sheth, Sarah Nobles, Jasmine Padilla, Mark Burroughs, Shoshona Le, Katie Dillon, Peter Cook, Clinton R. Paden, Dhwani Batra, Krista Queen, Kristen Knipe, Dakota Howard, Yvette Unoarumhi, Darlene Wagner, Matthew Schmerer, Ben L. Rambo-Martin, Kristine Lacek, Sam Shepard, Alison Laufer Halpin, Dave Wentworth, Vivien Dugan, Suxiang Tong, Justin Lee |
| EPI_ISL_1446979 | IL Department of Public Health Chicago Laboratory | Centers for Disease Control and Prevention Division of Viral Diseases, Pathogen Discovery | Mili Sheth, Sarah Nobles, Jasmine Padilla, Mark Burroughs, Shoshona Le, Katie Dillon, Peter Cook, Clinton R. Paden, Dhwani Batra, Krista Queen, Kristen Knipe, Dakota Howard, Yvette Unoarumhi, Darlene Wagner, Matthew Schmerer, Ben L. Rambo-Martin, Kristine Lacek, Sam Shepard, Alison Laufer Halpin, Dave Wentworth, Vivien Dugan, Suxiang Tong, Justin Lee |
| EPI_ISL_1446980, EPI_ISL_1446981, EPI_ISL_1446982, EPI_ISL_1446983, EPI_ISL_1446984 | TX-Houston Health Department Laboratory | Centers for Disease Control and Prevention Division of Viral Diseases, Pathogen Discovery | Mili Sheth, Sarah Nobles, Jasmine Padilla, Mark Burroughs, Shoshona Le, Katie Dillon, Peter Cook, Clinton R. Paden, Dhwani Batra, Krista Queen, Kristen Knipe, Dakota Howard, Yvette Unoarumhi, Darlene Wagner, Matthew Schmerer, Ben L. Rambo-Martin, Kristine Lacek, Sam Shepard, Alison Laufer Halpin, Dave Wentworth, Vivien Dugan, Suxiang Tong, Justin Lee |
| EPI_ISL_1446985 | ND Dept. of Health Laboratory Services- Microbiology | Centers for Disease Control and Prevention Division of Viral Diseases, Pathogen Discovery | Mili Sheth, Sarah Nobles, Jasmine Padilla, Mark Burroughs, Shoshona Le, Katie Dillon, Peter Cook, Clinton R. Paden, Dhwani Batra, Krista Queen, Kristen Knipe, Dakota Howard, Yvette Unoarumhi, Darlene Wagner, Matthew Schmerer, Ben L. Rambo-Martin, Kristine Lacek, Sam Shepard, Alison Laufer Halpin, Dave Wentworth, Vivien Dugan, Suxiang Tong, Justin Lee |
| EPI_ISL_1446986 | "MN PHL Division, Minnesota Department of Health" | Centers for Disease Control and Prevention Division of Viral Diseases, Pathogen Discovery | Mili Sheth, Sarah Nobles, Jasmine Padilla, Mark Burroughs, Shoshona Le, Katie Dillon, Peter Cook, Clinton R. Paden, Dhwani Batra, Krista Queen, Kristen Knipe, Dakota Howard, Yvette Unoarumhi, Darlene Wagner, Matthew Schmerer, Ben L. Rambo-Martin, Kristine Lacek, Sam Shepard, Alison Laufer Halpin, Dave Wentworth, Vivien Dugan, Suxiang Tong, Justin Lee |

| | | | |
|---|---|---|---|
| EPI_ISL_1446987, EPI_ISL_1446988 | FL Bureau of Public Health Laboratories-Tampa | Centers for Disease Control and Prevention Division of Viral Diseases, Pathogen Discovery | Mili Sheth, Sarah Nobles, Jasmine Padilla, Mark Burroughs, Shoshona Le, Katie Dillon, Peter Cook, Clinton R. Paden, Dhwani Batra, Krista Queen, Kristen Knipe, Dakota Howard, Yvette Unoarumhi, Darlene Wagner, Matthew Schmerer, Ben L. Rambo-Martin, Kristine Lacek, Sam Shepard, Alison Laufer Halpin, Dave Wentworth, Vivien Dugan, Suxiang Tong, Justin Lee |
| EPI_ISL_1446989, EPI_ISL_1446990, EPI_ISL_1446991 | SD Public Health Laboratory | Centers for Disease Control and Prevention Division of Viral Diseases, Pathogen Discovery | Mili Sheth, Sarah Nobles, Jasmine Padilla, Mark Burroughs, Shoshona Le, Katie Dillon, Peter Cook, Clinton R. Paden, Dhwani Batra, Krista Queen, Kristen Knipe, Dakota Howard, Yvette Unoarumhi, Darlene Wagner, Matthew Schmerer, Ben L. Rambo-Martin, Kristine Lacek, Sam Shepard, Alison Laufer Halpin, Dave Wentworth, Vivien Dugan, Suxiang Tong, Justin Lee |
| EPI_ISL_1446992, EPI_ISL_1446993, EPI_ISL_1446994, EPI_ISL_1446995, EPI_ISL_1446996, EPI_ISL_1446997, EPI_ISL_1446998 | MA State Public Health Laboratory | Centers for Disease Control and Prevention Division of Viral Diseases, Pathogen Discovery | Mili Sheth, Sarah Nobles, Jasmine Padilla, Mark Burroughs, Shoshona Le, Katie Dillon, Peter Cook, Clinton R. Paden, Dhwani Batra, Krista Queen, Kristen Knipe, Dakota Howard, Yvette Unoarumhi, Darlene Wagner, Matthew Schmerer, Ben L. Rambo-Martin, Kristine Lacek, Sam Shepard, Alison Laufer Halpin, Dave Wentworth, Vivien Dugan, Suxiang Tong, Justin Lee |
| EPI_ISL_1446999, EPI_ISL_1447000, EPI_ISL_1447001 | FL Bur. of Public Health Laboratories-Jacksonville | Centers for Disease Control and Prevention Division of Viral Diseases, Pathogen Discovery | Mili Sheth, Sarah Nobles, Jasmine Padilla, Mark Burroughs, Shoshona Le, Katie Dillon, Peter Cook, Clinton R. Paden, Dhwani Batra, Krista Queen, Kristen Knipe, Dakota Howard, Yvette Unoarumhi, Darlene Wagner, Matthew Schmerer, Ben L. Rambo-Martin, Kristine Lacek, Sam Shepard, Alison Laufer Halpin, Dave Wentworth, Vivien Dugan, Suxiang Tong, Justin Lee |
| EPI_ISL_1447002 | AL Dept. of Public Health Bureau of Clinical Laboratories | Centers for Disease Control and Prevention Division of Viral Diseases, Pathogen Discovery | Mili Sheth, Sarah Nobles, Jasmine Padilla, Mark Burroughs, Shoshona Le, Katie Dillon, Peter Cook, Clinton R. Paden, Dhwani Batra, Krista Queen, Kristen Knipe, Dakota Howard, Yvette Unoarumhi, Darlene Wagner, Matthew Schmerer, Ben L. Rambo-Martin, Kristine Lacek, Sam Shepard, Alison Laufer Halpin, Dave Wentworth, Vivien Dugan, Suxiang Tong, Justin Lee |
| EPI_ISL_1447003, EPI_ISL_1447004, EPI_ISL_1447005, EPI_ISL_1447006, EPI_ISL_1447007, EPI_ISL_1447008, EPI_ISL_1447009, EPI_ISL_1447010, EPI_ISL_1447011 | GA Department of Public Health Laboratory | Centers for Disease Control and Prevention Division of Viral Diseases, Pathogen Discovery | Mili Sheth, Sarah Nobles, Jasmine Padilla, Mark Burroughs, Shoshona Le, Katie Dillon, Peter Cook, Clinton R. Paden, Dhwani Batra, Krista Queen, Kristen Knipe, Dakota Howard, Yvette Unoarumhi, Darlene Wagner, Matthew Schmerer, Ben L. Rambo-Martin, Kristine Lacek, Sam Shepard, Alison Laufer Halpin, Dave Wentworth, Vivien Dugan, Suxiang Tong, Justin Lee |
| EPI_ISL_1447012 | NC State Laboratory of Public Health | Centers for Disease Control and Prevention Division of Viral Diseases, Pathogen Discovery | Mili Sheth, Sarah Nobles, Jasmine Padilla, Mark Burroughs, Shoshona Le, Katie Dillon, Peter Cook, Clinton R. Paden, Dhwani Batra, Krista Queen, Kristen Knipe, Dakota Howard, Yvette Unoarumhi, Darlene Wagner, Matthew Schmerer, Ben L. Rambo-Martin, Kristine Lacek, Sam Shepard, Alison Laufer Halpin, Dave Wentworth, Vivien Dugan, Suxiang Tong, Justin Lee |
| EPI_ISL_1447013, EPI_ISL_1447014, EPI_ISL_1447015, EPI_ISL_1447016, EPI_ISL_1447017 | LA Office of Public Health Laboratories | Centers for Disease Control and Prevention Division of Viral Diseases, Pathogen Discovery | Mili Sheth, Sarah Nobles, Jasmine Padilla, Mark Burroughs, Shoshona Le, Katie Dillon, Peter Cook, Clinton R. Paden, Dhwani Batra, Krista Queen, Kristen Knipe, Dakota Howard, Yvette Unoarumhi, Darlene Wagner, Matthew Schmerer, Ben L. Rambo-Martin, Kristine Lacek, Sam Shepard, Alison Laufer Halpin, Dave Wentworth, Vivien Dugan, Suxiang Tong, Justin Lee |
| EPI_ISL_1447018 | "CDPH, Viral and Rickettsial Disease Laboratory" | Centers for Disease Control and Prevention Division of Viral Diseases, Pathogen Discovery | Mili Sheth, Sarah Nobles, Jasmine Padilla, Mark Burroughs, Shoshona Le, Katie Dillon, Peter Cook, Clinton R. Paden, Dhwani Batra, Krista Queen, Kristen Knipe, Dakota Howard, Yvette Unoarumhi, Darlene Wagner, Matthew Schmerer, Ben L. Rambo-Martin, Kristine Lacek, Sam Shepard, Alison Laufer Halpin, Dave Wentworth, Vivien Dugan, Suxiang Tong, Justin Lee |
| EPI_ISL_1447019, EPI_ISL_1447020, EPI_ISL_1447021, EPI_ISL_1447022, EPI_ISL_1447023, EPI_ISL_1447024, EPI_ISL_1447025, EPI_ISL_1447026 | "PA Department of Health, Bureau of Laboratories" | Centers for Disease Control and Prevention Division of Viral Diseases, Pathogen Discovery | Mili Sheth, Sarah Nobles, Jasmine Padilla, Mark Burroughs, Shoshona Le, Katie Dillon, Peter Cook, Clinton R. Paden, Dhwani Batra, Krista Queen, Kristen Knipe, Dakota Howard, Yvette Unoarumhi, Darlene Wagner, Matthew Schmerer, Ben L. Rambo-Martin, Kristine Lacek, Sam Shepard, Alison Laufer Halpin, Dave Wentworth, Vivien Dugan, Suxiang Tong, Justin Lee |
| EPI_ISL_1447027, EPI_ISL_1447028 | NJ Public Health and Environmental Laboratories | Centers for Disease Control and Prevention Division of Viral Diseases, Pathogen Discovery | Mili Sheth, Sarah Nobles, Jasmine Padilla, Mark Burroughs, Shoshona Le, Katie Dillon, Peter Cook, Clinton R. Paden, Dhwani Batra, Krista Queen, Kristen Knipe, Dakota Howard, Yvette Unoarumhi, Darlene Wagner, Matthew Schmerer, Ben L. Rambo-Martin, Kristine Lacek, Sam Shepard, Alison Laufer Halpin, Dave Wentworth, Vivien Dugan, Suxiang Tong, Justin Lee |
| EPI_ISL_1447029 | OR State PHL-Virology/Immunology Section | Centers for Disease Control and Prevention Division of Viral Diseases, Pathogen Discovery | Mili Sheth, Sarah Nobles, Jasmine Padilla, Mark Burroughs, Shoshona Le, Katie Dillon, Peter Cook, Clinton R. Paden, Dhwani Batra, Krista Queen, Kristen Knipe, Dakota Howard, Yvette Unoarumhi, Darlene Wagner, Matthew Schmerer, Ben L. Rambo-Martin, Kristine Lacek, Sam Shepard, Alison Laufer Halpin, Dave Wentworth, Vivien Dugan, Suxiang Tong, Justin Lee |
| EPI_ISL_1447030, EPI_ISL_1447031, EPI_ISL_1447032, EPI_ISL_1447033, EPI_ISL_1447034, EPI_ISL_1447035, EPI_ISL_1447036, EPI_ISL_1447037, EPI_ISL_1447038, EPI_ISL_1447039, EPI_ISL_1447040, EPI_ISL_1447041, EPI_ISL_1447042, EPI_ISL_1447043, EPI_ISL_1447044 | | | |
| see above | MI - Michigan Department of Health and Human Services - Bureau of Laboratories | Centers for Disease Control and Prevention Division of Viral Diseases, Pathogen Discovery | Mili Sheth, Sarah Nobles, Jasmine Padilla, Mark Burroughs, Shoshona Le, Katie Dillon, Peter Cook, Clinton R. Paden, Dhwani Batra, Krista Queen, Kristen Knipe, Dakota Howard, Yvette Unoarumhi, Darlene Wagner, Matthew Schmerer, Ben L. Rambo-Martin, Kristine Lacek, Sam Shepard, Alison Laufer Halpin, Dave Wentworth, Vivien Dugan, Suxiang Tong, Justin Lee |
| EPI_ISL_1447045 | VA-Division of Consolidated Laboratory Services | Centers for Disease Control and Prevention Division of Viral Diseases, Pathogen Discovery | Mili Sheth, Sarah Nobles, Jasmine Padilla, Mark Burroughs, Shoshona Le, Katie Dillon, Peter Cook, Clinton R. Paden, Dhwani Batra, Krista Queen, Kristen Knipe, Dakota Howard, Yvette Unoarumhi, Darlene Wagner, Matthew Schmerer, Ben L. Rambo-Martin, Kristine Lacek, Sam Shepard, Alison Laufer Halpin, Dave Wentworth, Vivien Dugan, Suxiang Tong, Justin Lee |
| EPI_ISL_1447046 | TX-Houston Health Department Laboratory | Centers for Disease Control and Prevention Division of Viral Diseases, Pathogen Discovery | Mili Sheth, Sarah Nobles, Jasmine Padilla, Mark Burroughs, Shoshona Le, Katie Dillon, Peter Cook, Clinton R. Paden, Dhwani Batra, Krista Queen, Kristen Knipe, Dakota Howard, Yvette Unoarumhi, Darlene Wagner, Matthew Schmerer, Ben L. Rambo-Martin, Kristine Lacek, Sam Shepard, Alison Laufer Halpin, Dave Wentworth, Vivien Dugan, Suxiang Tong, Justin Lee |
| EPI_ISL_1447047 | FL Bur. of Public Health Laboratories-Jacksonville | Centers for Disease Control and Prevention Division of Viral Diseases, Pathogen Discovery | Mili Sheth, Sarah Nobles, Jasmine Padilla, Mark Burroughs, Shoshona Le, Katie Dillon, Peter Cook, Clinton R. Paden, Dhwani Batra, Krista Queen, Kristen Knipe, Dakota Howard, Yvette Unoarumhi, Darlene Wagner, Matthew Schmerer, Ben L. Rambo-Martin, Kristine Lacek, Sam Shepard, Alison Laufer Halpin, Dave Wentworth, Vivien Dugan, Suxiang Tong, Justin Lee |
| EPI_ISL_1447048 | DC Public Health Lab/ Dept. of Forensic Sciences | Centers for Disease Control and Prevention Division of Viral Diseases, Pathogen Discovery | Mili Sheth, Sarah Nobles, Jasmine Padilla, Mark Burroughs, Shoshona Le, Katie Dillon, Peter Cook, Clinton R. Paden, Dhwani Batra, Krista Queen, Kristen Knipe, Dakota Howard, Yvette Unoarumhi, Darlene Wagner, Matthew Schmerer, Ben L. Rambo-Martin, Kristine Lacek, Sam Shepard, Alison Laufer Halpin, Dave Wentworth, Vivien Dugan, Suxiang Tong, Justin Lee |
| EPI_ISL_1447049, EPI_ISL_1447050, EPI_ISL_1447051 | CA-Los Angeles County Public Health Laboratory | Centers for Disease Control and Prevention Division of Viral Diseases, Pathogen Discovery | Mili Sheth, Sarah Nobles, Jasmine Padilla, Mark Burroughs, Shoshona Le, Katie Dillon, Peter Cook, Clinton R. Paden, Dhwani Batra, Krista Queen, Kristen Knipe, Dakota Howard, Yvette Unoarumhi, Darlene Wagner, Matthew Schmerer, Ben L. Rambo-Martin, Kristine Lacek, Sam Shepard, Alison Laufer Halpin, Dave Wentworth, Vivien Dugan, Suxiang Tong, Justin Lee |
| EPI_ISL_1447052 | MD DOH Laboratories Administration | Centers for Disease Control and Prevention Division of Viral Diseases, Pathogen Discovery | Mili Sheth, Sarah Nobles, Jasmine Padilla, Mark Burroughs, Shoshona Le, Katie Dillon, Peter Cook, Clinton R. Paden, Dhwani Batra, Krista Queen, Kristen Knipe, Dakota Howard, Yvette Unoarumhi, Darlene Wagner, Matthew Schmerer, Ben L. Rambo-Martin, Kristine Lacek, Sam Shepard, Alison Laufer Halpin, Dave Wentworth, Vivien Dugan, Suxiang Tong, Justin Lee |
| EPI_ISL_1447053 | "NYSDOH Wadsworth Center, Virology Lab" | Centers for Disease Control and Prevention Division of Viral Diseases, Pathogen Discovery | Mili Sheth, Sarah Nobles, Jasmine Padilla, Mark Burroughs, Shoshona Le, Katie Dillon, Peter Cook, Clinton R. Paden, Dhwani Batra, Krista Queen, Kristen Knipe, Dakota Howard, Yvette Unoarumhi, Darlene Wagner, Matthew Schmerer, Ben L. Rambo-Martin, Kristine Lacek, Sam Shepard, Alison Laufer Halpin, Dave Wentworth, Vivien Dugan, Suxiang Tong, Justin Lee |
| EPI_ISL_1447054 | FL Bureau of Public Health Laboratories-Miami | Centers for Disease Control and Prevention Division of Viral Diseases, Pathogen Discovery | Mili Sheth, Sarah Nobles, Jasmine Padilla, Mark Burroughs, Shoshona Le, Katie Dillon, Peter Cook, Clinton R. Paden, Dhwani Batra, Krista Queen, Kristen Knipe, Dakota Howard, Yvette Unoarumhi, Darlene Wagner, Matthew Schmerer, Ben L. Rambo-Martin, Kristine Lacek, Sam Shepard, Alison Laufer Halpin, Dave Wentworth, Vivien Dugan, Suxiang Tong, Justin Lee |
| EPI_ISL_1447055, EPI_ISL_1447056, EPI_ISL_1447057, EPI_ISL_1447058 | GA Department of Public Health Laboratory | Centers for Disease Control and Prevention Division of Viral Diseases, Pathogen Discovery | Mili Sheth, Sarah Nobles, Jasmine Padilla, Mark Burroughs, Shoshona Le, Katie Dillon, Peter Cook, Clinton R. Paden, Dhwani Batra, Krista Queen, Kristen Knipe, Dakota Howard, Yvette Unoarumhi, Darlene Wagner, Matthew Schmerer, Ben L. Rambo-Martin, Kristine Lacek, Sam Shepard, Alison Laufer Halpin, Dave Wentworth, Vivien Dugan, Suxiang Tong, Justin Lee |
| EPI_ISL_1447059 | NH Dept. of Health and Human Services Public Health Labs | Centers for Disease Control and Prevention Division of Viral Diseases, Pathogen Discovery | Mili Sheth, Sarah Nobles, Jasmine Padilla, Mark Burroughs, Shoshona Le, Katie Dillon, Peter Cook, Clinton R. Paden, Dhwani Batra, Krista Queen, Kristen Knipe, Dakota Howard, Yvette Unoarumhi, Darlene Wagner, Matthew Schmerer, Ben L. Rambo-Martin, Kristine Lacek, Sam Shepard, Alison Laufer Halpin, Dave Wentworth, Vivien Dugan, Suxiang Tong, Justin Lee |
| EPI_ISL_1447060, EPI_ISL_1447061, EPI_ISL_1447062 | NYC Department of Health and Mental Hygiene | Centers for Disease Control and Prevention Division of Viral Diseases, Pathogen Discovery | Mili Sheth, Sarah Nobles, Jasmine Padilla, Mark Burroughs, Shoshona Le, Katie Dillon, Peter Cook, Clinton R. Paden, Dhwani Batra, Krista Queen, Kristen Knipe, Dakota Howard, Yvette Unoarumhi, Darlene Wagner, Matthew Schmerer, Ben L. Rambo-Martin, Kristine Lacek, Sam Shepard, Alison Laufer Halpin, Dave Wentworth, Vivien Dugan, Suxiang Tong, Justin Lee |
| EPI_ISL_1447063, EPI_ISL_1447070 | Arizona State Public Health Laboratory | Arizona State Public Health Laboratory | Trung Huynh, Jessica Escobar, Katherine Fullerton, Nobuko Fukushima, Stacy White, Linda Getsinger, Victor Waddell |
| EPI_ISL_1447074, EPI_ISL_1447075, EPI_ISL_1447076, EPI_ISL_1447077, EPI_ISL_1447078, EPI_ISL_1447079, | Kansas Health and Environmental Lab | Kansas Health and Environmental Lab | Mike Grose, Jonathan Barnell, Ben Olsen, and Phil Adam |

| | | | |
|---|---|---|---|
| EPI_ISL_1447080, EPI_ISL_1447081, EPI_ISL_1447082, EPI_ISL_1447083 | | | |
| EPI_ISL_1447084 | Cytocheck Laboratory | Kansas Health and Environmental Lab | Mike Grose, Jonathan Barnell, Ben Olsen, and Phil Adam |
| EPI_ISL_1447085, EPI_ISL_1447086 | Kansas Health and Environmental Lab | Kansas Health and Environmental Lab | Mike Grose, Jonathan Barnell, Ben Olsen, and Phil Adam |
| EPI_ISL_1447087 | DOHMH Morrisania | New York City Public Health Laboratory | Jade Wang, et al. |
| EPI_ISL_1447088 | DOHMH Corona | New York City Public Health Laboratory | Jade Wang, et al. |
| EPI_ISL_1447089 | DOHMH Crown Heights | New York City Public Health Laboratory | Jade Wang, et al. |
| EPI_ISL_1447090 | DOHMH Morrisania | New York City Public Health Laboratory | Jade Wang, et al. |
| EPI_ISL_1447091 | OCME Office Of Chief Medical Examiner | New York City Public Health Laboratory | Jade Wang, et al. |
| EPI_ISL_1447092 | DOHMH Jamaica | New York City Public Health Laboratory | Jade Wang, et al. |
| EPI_ISL_1447093 | DOHMH Chelsea | New York City Public Health Laboratory | Jade Wang, et al. |
| EPI_ISL_1447094 | DOHMH Jamaica | New York City Public Health Laboratory | Jade Wang, et al. |
| EPI_ISL_1447095 | DOHMH Corona | New York City Public Health Laboratory | Jade Wang, et al. |
| EPI_ISL_1447096 | DOHMH Chelsea | New York City Public Health Laboratory | Jade Wang, et al. |
| EPI_ISL_1447097 | DOHMH Morrisania | New York City Public Health Laboratory | Jade Wang, et al. |
| EPI_ISL_1447098 | DOHMH Fort Greene | New York City Public Health Laboratory | Jade Wang, et al. |
| EPI_ISL_1447099 | DOHMH PHL | New York City Public Health Laboratory | Jade Wang, et al. |
| EPI_ISL_1447100 | OCME Office Of Chief Medical Examiner | New York City Public Health Laboratory | Jade Wang, et al. |
| EPI_ISL_1447101 | DOHMH Central Harlem | New York City Public Health Laboratory | Jade Wang, et al. |
| EPI_ISL_1447102 | Department of Homeless Services | New York City Public Health Laboratory | Jade Wang, et al. |
| EPI_ISL_1447103 | DOHMH Jamaica | New York City Public Health Laboratory | Jade Wang, et al. |
| EPI_ISL_1447104 | OCME Office Of Chief Medical Examiner | New York City Public Health Laboratory | Jade Wang, et al. |
| EPI_ISL_1447105 | DOHMH Riverside | New York City Public Health Laboratory | Jade Wang, et al. |
| EPI_ISL_1447106 | DOHMH Jamaica | New York City Public Health Laboratory | Jade Wang, et al. |
| EPI_ISL_1447107 | DOHMH PHL | New York City Public Health Laboratory | Jade Wang, et al. |
| EPI_ISL_1447108 | DOHMH Morrisania | New York City Public Health Laboratory | Jade Wang, et al. |
| EPI_ISL_1447109 | DOHMH Central Harlem | New York City Public Health Laboratory | Jade Wang, et al. |
| EPI_ISL_1447110 | DOHMH Morrisania | New York City Public Health Laboratory | Jade Wang, et al. |
| EPI_ISL_1447111 | OCME Office Of Chief Medical Examiner | New York City Public Health Laboratory | Jade Wang, et al. |
| EPI_ISL_1447112 | DOHMH Fort Greene | New York City Public Health Laboratory | Jade Wang, et al. |
| EPI_ISL_1447113 | DOHMH Crown Heights | New York City Public Health Laboratory | Jade Wang, et al. |
| EPI_ISL_1447114 | OCME Office Of Chief Medical Examiner | New York City Public Health Laboratory | Jade Wang, et al. |
| EPI_ISL_1447115 | DOHMH Fort Greene | New York City Public Health Laboratory | Jade Wang, et al. |
| EPI_ISL_1447116 | DOHMH Central Harlem | New York City Public Health Laboratory | Jade Wang, et al. |
| EPI_ISL_1447117 | DOHMH Jamaica | New York City Public Health Laboratory | Jade Wang, et al. |
| EPI_ISL_1447118 | DOHMH Chelsea | New York City Public Health Laboratory | Jade Wang, et al. |
| EPI_ISL_1447119 | OCME Office Of Chief Medical Examiner | New York City Public Health Laboratory | Jade Wang, et al. |
| EPI_ISL_1447120, EPI_ISL_1447121, EPI_ISL_1447122 | DOHMH Jamaica | New York City Public Health Laboratory | Jade Wang, et al. |
| EPI_ISL_1447123 | DOHMH Chelsea | New York City Public Health Laboratory | Jade Wang, et al. |
| EPI_ISL_1447124, EPI_ISL_1447125 | DOHMH Jamaica | New York City Public Health Laboratory | Jade Wang, et al. |
| EPI_ISL_1447126 | DOHMH Corona | New York City Public Health Laboratory | Jade Wang, et al. |
| EPI_ISL_1447127 | DOHMH Fort Greene | New York City Public Health Laboratory | Jade Wang, et al. |
| EPI_ISL_1447128 | DOHMH Crown Heights | New York City Public Health Laboratory | Jade Wang, et al. |
| EPI_ISL_1447129, EPI_ISL_1447130 | DOHMH Central Harlem | New York City Public Health Laboratory | Jade Wang, et al. |
| EPI_ISL_1447131, EPI_ISL_1447132 | DOHMH Jamaica | New York City Public Health Laboratory | Jade Wang, et al. |
| EPI_ISL_1447133 | DOHMH Chelsea | New York City Public Health Laboratory | Jade Wang, et al. |
| EPI_ISL_1447134 | DOHMH Central Harlem | New York City Public Health Laboratory | Jade Wang, et al. |
| EPI_ISL_1447135 | DOHMH Fort Greene | New York City Public Health Laboratory | Jade Wang, et al. |
| EPI_ISL_1447136, EPI_ISL_1447137 | DOHMH Corona | New York City Public Health Laboratory | Jade Wang, et al. |
| EPI_ISL_1447138 | OCME Office Of Chief Medical Examiner | New York City Public Health Laboratory | Jade Wang, et al. |
| EPI_ISL_1447139, EPI_ISL_1447140, EPI_ISL_1447141, EPI_ISL_1447142 | DOHMH Corona | New York City Public Health Laboratory | Jade Wang, et al. |
| EPI_ISL_1447143 | Rockefeller University Hospital | New York City Public Health Laboratory | Jade Wang, et al. |
| EPI_ISL_1447144 | DOHMH Corona | New York City Public Health Laboratory | Jade Wang, et al. |
| EPI_ISL_1447145 | DOHMH PHL | New York City Public Health Laboratory | Jade Wang, et al. |
| EPI_ISL_1447146 | DOHMH Jamaica | New York City Public Health Laboratory | Jade Wang, et al. |
| EPI_ISL_1447147 | DOHMH Corona | New York City Public Health Laboratory | Jade Wang, et al. |
| EPI_ISL_1447148 | DOHMH Crown Heights | New York City Public Health Laboratory | Jade Wang, et al. |
| EPI_ISL_1447149, EPI_ISL_1447150 | DOHMH PHL | New York City Public Health Laboratory | Jade Wang, et al. |
| EPI_ISL_1447151, EPI_ISL_1447152, EPI_ISL_1447153 | DOHMH Jamaica | New York City Public Health Laboratory | Jade Wang, et al. |
| EPI_ISL_1447154 | DOHMH Riverside | New York City Public Health Laboratory | Jade Wang, et al. |
| EPI_ISL_1447155 | DOHMH Corona | New York City Public Health Laboratory | Jade Wang, et al. |
| EPI_ISL_1447156, EPI_ISL_1447157 | DOHMH Chelsea | New York City Public Health Laboratory | Jade Wang, et al. |
| EPI_ISL_1447158, EPI_ISL_1447159 | DOHMH Jamaica | New York City Public Health Laboratory | Jade Wang, et al. |
| EPI_ISL_1447160 | DOHMH Corona | New York City Public Health Laboratory | Jade Wang, et al. |
| EPI_ISL_1447161, | DOHMH Crown Heights | New York City Public Health Laboratory | Jade Wang, et al. |

| | | | |
|---|---|---|---|
| EPI_ISL_1447162 | | | |
| EPI_ISL_1447163, EPI_ISL_1447164 | DOHMH Jamaica | New York City Public Health Laboratory | Jade Wang, et al. |
| EPI_ISL_1447165 | DOHMH Corona | New York City Public Health Laboratory | Jade Wang, et al. |
| EPI_ISL_1447166 | DOHMH Crown Heights | New York City Public Health Laboratory | Jade Wang, et al. |
| EPI_ISL_1447167, EPI_ISL_1447168 | DOHMH Central Harlem | New York City Public Health Laboratory | Jade Wang, et al. |
| EPI_ISL_1447169 | DOHMH Chelsea | New York City Public Health Laboratory | Jade Wang, et al. |
| EPI_ISL_1447170 | DOHMH Crown Heights | New York City Public Health Laboratory | Jade Wang, et al. |
| EPI_ISL_1447171, EPI_ISL_1447172 | DOHMH Corona | New York City Public Health Laboratory | Jade Wang, et al. |
| EPI_ISL_1447173 | DOHMH Riverside | New York City Public Health Laboratory | Jade Wang, et al. |
| EPI_ISL_1447174 | Department of Homeless Services | New York City Public Health Laboratory | Jade Wang, et al. |
| EPI_ISL_1447175 | DOHMH Jamaica | New York City Public Health Laboratory | Jade Wang, et al. |
| EPI_ISL_1447176 | DOHMH Fort Greene | New York City Public Health Laboratory | Jade Wang, et al. |
| EPI_ISL_1447177, EPI_ISL_1447178 | OCME Office Of Chief Medical Examiner | New York City Public Health Laboratory | Jade Wang, et al. |
| EPI_ISL_1447179, EPI_ISL_1447180 | DOHMH Jamaica | New York City Public Health Laboratory | Jade Wang, et al. |
| EPI_ISL_1447181 | DOHMH Morrisania | New York City Public Health Laboratory | Jade Wang, et al. |
| EPI_ISL_1447182 | Department of Homeless Services | New York City Public Health Laboratory | Jade Wang, et al. |
| EPI_ISL_1447183 | OCME Office Of Chief Medical Examiner | New York City Public Health Laboratory | Jade Wang, et al. |
| EPI_ISL_1447184 | DOHMH PHL | New York City Public Health Laboratory | Jade Wang, et al. |
| EPI_ISL_1447185 | DOHMH Corona | New York City Public Health Laboratory | Jade Wang, et al. |
| EPI_ISL_1447186 | DOHMH Central Harlem | New York City Public Health Laboratory | Jade Wang, et al. |
| EPI_ISL_1447187 | DOHMH Fort Greene | New York City Public Health Laboratory | Jade Wang, et al. |
| EPI_ISL_1447188, EPI_ISL_1447189, EPI_ISL_1447190 | DOHMH Jamaica | New York City Public Health Laboratory | Jade Wang, et al. |
| EPI_ISL_1447191 | DOHMH Crown Heights | New York City Public Health Laboratory | Jade Wang, et al. |
| EPI_ISL_1447192 | DOHMH Jamaica | New York City Public Health Laboratory | Jade Wang, et al. |
| EPI_ISL_1447193 | DOHMH Chelsea | New York City Public Health Laboratory | Jade Wang, et al. |
| EPI_ISL_1447488, EPI_ISL_1447489, EPI_ISL_1447490, EPI_ISL_1447491, EPI_ISL_1447492, EPI_ISL_1447493, EPI_ISL_1447494, EPI_ISL_1447495, EPI_ISL_1447496, EPI_ISL_1447497, EPI_ISL_1447498, EPI_ISL_1447499, EPI_ISL_1447500, EPI_ISL_1447501, EPI_ISL_1447502, EPI_ISL_1447503, EPI_ISL_1447504, EPI_ISL_1447505, EPI_ISL_1447506, EPI_ISL_1447507, EPI_ISL_1447508, EPI_ISL_1447509 | | | |
| see above | Washington State Department of Health Public Health Laboratories | Washington State Department of Health Public Health Laboratories | Drew MacKellar, Philip Dykema, Denny Russell, Joenice Gonzalez, Hannah Gray, Geoff Melly, Vanessa De Los Santos, Darren Lucas, JohnAric Peterson, Avi Singh, Rebecca Cao |
| EPI_ISL_1447510, EPI_ISL_1447511, EPI_ISL_1447512, EPI_ISL_1447513, EPI_ISL_1447514, EPI_ISL_1447516, EPI_ISL_1447517, EPI_ISL_1447518, EPI_ISL_1447519, EPI_ISL_1447520, EPI_ISL_1447521, EPI_ISL_1447522, EPI_ISL_1447523, EPI_ISL_1447524, EPI_ISL_1447525, EPI_ISL_1447526, EPI_ISL_1447527, EPI_ISL_1447528, EPI_ISL_1447529, EPI_ISL_1447530, EPI_ISL_1447531, EPI_ISL_1447532, EPI_ISL_1447533, EPI_ISL_1447534, EPI_ISL_1447535, EPI_ISL_1447536, EPI_ISL_1447537, EPI_ISL_1447538, EPI_ISL_1447540, EPI_ISL_1447541, EPI_ISL_1447542, EPI_ISL_1447543, EPI_ISL_1447546, EPI_ISL_1447547, EPI_ISL_1447549, EPI_ISL_1447551, EPI_ISL_1447552, EPI_ISL_1447553, EPI_ISL_1447554, EPI_ISL_1447556, EPI_ISL_1447557, EPI_ISL_1447558, EPI_ISL_1447559, EPI_ISL_1447560, EPI_ISL_1447561, EPI_ISL_1447562, EPI_ISL_1447563, EPI_ISL_1447564, EPI_ISL_1447565, EPI_ISL_1447566, EPI_ISL_1447569, EPI_ISL_1447570, EPI_ISL_1447571, EPI_ISL_1447572, EPI_ISL_1447573, EPI_ISL_1447574, EPI_ISL_1447575, EPI_ISL_1447578, EPI_ISL_1447579, EPI_ISL_1447580, EPI_ISL_1447581, EPI_ISL_1447582, EPI_ISL_1447584, EPI_ISL_1447585, EPI_ISL_1447586, EPI_ISL_1447587, EPI_ISL_1447588, EPI_ISL_1447589, EPI_ISL_1447590, EPI_ISL_1447591, EPI_ISL_1447592, EPI_ISL_1447593, EPI_ISL_1447594, EPI_ISL_1447596, EPI_ISL_1447597, EPI_ISL_1447598, EPI_ISL_1447599, EPI_ISL_1447601, EPI_ISL_1447602, EPI_ISL_1447603, EPI_ISL_1447604, EPI_ISL_1447605, EPI_ISL_1447606, EPI_ISL_1447607, EPI_ISL_1447608, EPI_ISL_1447609, EPI_ISL_1447610, EPI_ISL_1447611, EPI_ISL_1447612, EPI_ISL_1447613, EPI_ISL_1447614, EPI_ISL_1447615, EPI_ISL_1447616, EPI_ISL_1447617, EPI_ISL_1447618, EPI_ISL_1447619, EPI_ISL_1447620, EPI_ISL_1447621, EPI_ISL_1447622, EPI_ISL_1447623, EPI_ISL_1447625, EPI_ISL_1447626, EPI_ISL_1447628, EPI_ISL_1447629, EPI_ISL_1447630, EPI_ISL_1447632, EPI_ISL_1447633, EPI_ISL_1447634, EPI_ISL_1447635, EPI_ISL_1447636, EPI_ISL_1447637, EPI_ISL_1447640, EPI_ISL_1447641, EPI_ISL_1447642, EPI_ISL_1447643, EPI_ISL_1447644, EPI_ISL_1447645, EPI_ISL_1447646, EPI_ISL_1447647, EPI_ISL_1447648, EPI_ISL_1447649, EPI_ISL_1447650, EPI_ISL_1447651, EPI_ISL_1447652, EPI_ISL_1447653, EPI_ISL_1447654, EPI_ISL_1447655, EPI_ISL_1447656, EPI_ISL_1447657, EPI_ISL_1447658, EPI_ISL_1447659, EPI_ISL_1447660, EPI_ISL_1447661, EPI_ISL_1447662, EPI_ISL_1447664, EPI_ISL_1447665, EPI_ISL_1447666, EPI_ISL_1447667, EPI_ISL_1447669, EPI_ISL_1447670 | | | |
| see above | Yale Clinical Virology Lab | Grubaugh Lab - Yale School of Public Health | Joseph Fauver, Mallery Breban, Isabel Ott, Tara Alpert, Mary Petrone, Anderson Brito, Chantal Vogels, Annie Watkins, Chaney Kalinich, Jessica Rothman, Marie L. Landry, Nathan Grubaugh |
| EPI_ISL_1447671 | Yale Pathology Lab | Grubaugh Lab - Yale School of Public Health | Joseph Fauver, Mallery Breban, Isabel Ott, Tara Alpert, Mary Petrone, Anderson Brito, Chantal Vogels, Annie Watkins, Chaney Kalinich, Jessica Rothman, Jianhui Wang, Chen Liu, Pei Hui, Nathan Grubaugh |
| EPI_ISL_1447675 | Grubaugh Lab - Yale School of Public Health | Grubaugh Lab - Yale School of Public Health | Joseph Fauver, Mallery Breban, Isabell Ott, Tara Alpert, Mary Petrone, Anderson Brito, Chantal Vogels, Annie Watkins, Chaney Kalinich, Jessica Rothman, Nathan Grubaugh |
| EPI_ISL_1448029, EPI_ISL_1448030, EPI_ISL_1448031, EPI_ISL_1448035, EPI_ISL_1448036, EPI_ISL_1448037, EPI_ISL_1448038, EPI_ISL_1448039, EPI_ISL_1448040, EPI_ISL_1448041, EPI_ISL_1448042 | | | |
| see above | Maryland Genomics, Institute for Genome Sciences, University of Maryland School of Medicine | Maryland Genomics, Institute for Genome Sciences, University of Maryland School of Medicine | Tallon, Luke J; Sadzewicz, Lisa D; Humphrys, Mike; Ott, Sandra; Roussey, Holly; Mehta, Aditya; Vavikolanu, Kranthi; Fraser, Claire M; Ravel, Jacques |
| EPI_ISL_1448051, EPI_ISL_1448052, EPI_ISL_1448053, EPI_ISL_1448054, EPI_ISL_1448055, EPI_ISL_1448056, EPI_ISL_1448057, EPI_ISL_1448058, EPI_ISL_1448059, EPI_ISL_1448060, EPI_ISL_1448061, EPI_ISL_1448062, EPI_ISL_1448063, EPI_ISL_1448064, EPI_ISL_1448065, EPI_ISL_1448066, EPI_ISL_1448067, EPI_ISL_1448068, EPI_ISL_1448069, EPI_ISL_1448070, EPI_ISL_1448071, EPI_ISL_1448072, EPI_ISL_1448073, EPI_ISL_1448074, EPI_ISL_1448075, EPI_ISL_1448076, EPI_ISL_1448077, EPI_ISL_1448078, EPI_ISL_1448079, EPI_ISL_1448080, EPI_ISL_1448081, EPI_ISL_1448082, EPI_ISL_1448083, EPI_ISL_1448084, EPI_ISL_1448085, EPI_ISL_1448086, EPI_ISL_1448087, EPI_ISL_1448088, EPI_ISL_1448090, EPI_ISL_1448092, EPI_ISL_1448093, EPI_ISL_1448094, EPI_ISL_1448095, EPI_ISL_1448096, EPI_ISL_1448097, EPI_ISL_1448098, EPI_ISL_1448099, EPI_ISL_1448100, EPI_ISL_1448101, EPI_ISL_1448102, EPI_ISL_1448103, EPI_ISL_1448104, EPI_ISL_1448105, EPI_ISL_1448106, EPI_ISL_1448107, EPI_ISL_1448108, EPI_ISL_1448109, EPI_ISL_1448110, EPI_ISL_1448111, EPI_ISL_1448112, EPI_ISL_1448113, EPI_ISL_1448114, EPI_ISL_1448115, EPI_ISL_1448116, EPI_ISL_1448117, EPI_ISL_1448118, EPI_ISL_1448119, EPI_ISL_1448120, EPI_ISL_1448121, EPI_ISL_1448122, EPI_ISL_1448123, EPI_ISL_1448124, EPI_ISL_1448126, EPI_ISL_1448127, EPI_ISL_1448128, EPI_ISL_1448130, EPI_ISL_1448131, EPI_ISL_1448132, EPI_ISL_1448133, EPI_ISL_1448134, EPI_ISL_1448135, EPI_ISL_1448136, EPI_ISL_1448137, EPI_ISL_1448138, EPI_ISL_1448139, EPI_ISL_1448140, EPI_ISL_1448141, EPI_ISL_1448142, EPI_ISL_1448143, EPI_ISL_1448144, EPI_ISL_1448145, EPI_ISL_1448146, EPI_ISL_1448147, EPI_ISL_1448148, EPI_ISL_1448149, EPI_ISL_1448150, EPI_ISL_1448151, EPI_ISL_1448152, EPI_ISL_1448153, EPI_ISL_1448154, EPI_ISL_1448155, EPI_ISL_1448156, EPI_ISL_1448157, EPI_ISL_1448158, EPI_ISL_1448159, EPI_ISL_1448160, EPI_ISL_1448161, EPI_ISL_1448162, EPI_ISL_1448163, EPI_ISL_1448164, EPI_ISL_1448165, EPI_ISL_1448166, EPI_ISL_1448167, EPI_ISL_1448168, EPI_ISL_1448169, EPI_ISL_1448170, EPI_ISL_1448171, EPI_ISL_1448172, EPI_ISL_1448173, EPI_ISL_1448174, EPI_ISL_1448175, EPI_ISL_1448176, EPI_ISL_1448177, EPI_ISL_1448178, EPI_ISL_1448179, EPI_ISL_1448180, EPI_ISL_1448181, EPI_ISL_1448182, EPI_ISL_1448183, EPI_ISL_1448184, EPI_ISL_1448185, EPI_ISL_1448186, EPI_ISL_1448187, EPI_ISL_1448188, EPI_ISL_1448189, EPI_ISL_1448190, EPI_ISL_1448191, EPI_ISL_1448192, EPI_ISL_1448194, EPI_ISL_1448195, EPI_ISL_1448196, EPI_ISL_1448197, EPI_ISL_1448198, EPI_ISL_1448199, EPI_ISL_1448200, EPI_ISL_1448201, EPI_ISL_1448202, EPI_ISL_1448203, EPI_ISL_1448204, EPI_ISL_1448205, EPI_ISL_1448206, EPI_ISL_1448207, EPI_ISL_1448208, EPI_ISL_1448209, EPI_ISL_1448210, EPI_ISL_1448211, EPI_ISL_1448212, EPI_ISL_1448213, EPI_ISL_1448214, EPI_ISL_1448215, EPI_ISL_1448216, EPI_ISL_1448217, EPI_ISL_1448218, EPI_ISL_1448219, EPI_ISL_1448221, EPI_ISL_1448223, EPI_ISL_1448224, EPI_ISL_1448225, EPI_ISL_1448226, EPI_ISL_1448227, EPI_ISL_1448228, EPI_ISL_1448229, EPI_ISL_1448230, EPI_ISL_1448231, EPI_ISL_1448232, EPI_ISL_1448233, EPI_ISL_1448234, EPI_ISL_1448235, EPI_ISL_1448236, EPI_ISL_1448237, EPI_ISL_1448238, EPI_ISL_1448239, EPI_ISL_1448240, EPI_ISL_1448241, EPI_ISL_1448242, EPI_ISL_1448243, EPI_ISL_1448244, EPI_ISL_1448245, EPI_ISL_1448246, EPI_ISL_1448247, EPI_ISL_1448248, EPI_ISL_1448249, EPI_ISL_1448250, EPI_ISL_1448251, EPI_ISL_1448252, EPI_ISL_1448253, EPI_ISL_1448254, EPI_ISL_1448256, EPI_ISL_1448257, EPI_ISL_1448258, EPI_ISL_1448259, EPI_ISL_1448260, EPI_ISL_1448261, EPI_ISL_1448262, EPI_ISL_1448263, EPI_ISL_1448264, EPI_ISL_1448265, EPI_ISL_1448266, EPI_ISL_1448267, EPI_ISL_1448268, EPI_ISL_1448269, EPI_ISL_1448270, EPI_ISL_1448271, EPI_ISL_1448272, EPI_ISL_1448273, EPI_ISL_1448274, EPI_ISL_1448275, EPI_ISL_1448276, EPI_ISL_1448277, EPI_ISL_1448278, EPI_ISL_1448279, EPI_ISL_1448280, EPI_ISL_1448281, EPI_ISL_1448282, EPI_ISL_1448283, EPI_ISL_1448284, EPI_ISL_1448285, EPI_ISL_1448287, EPI_ISL_1448288, EPI_ISL_1448289, EPI_ISL_1448290, EPI_ISL_1448291, EPI_ISL_1448292, EPI_ISL_1448293, EPI_ISL_1448294, EPI_ISL_1448295, EPI_ISL_1448296, EPI_ISL_1448297, EPI_ISL_1448298, EPI_ISL_1448299, EPI_ISL_1448300, EPI_ISL_1448301, EPI_ISL_1448302, EPI_ISL_1448303, EPI_ISL_1448304, EPI_ISL_1448305, EPI_ISL_1448306, EPI_ISL_1448307, EPI_ISL_1448308, EPI_ISL_1448309, EPI_ISL_1448310, EPI_ISL_1448311, EPI_ISL_1448312, EPI_ISL_1448313, EPI_ISL_1448314, EPI_ISL_1448315, EPI_ISL_1448316, EPI_ISL_1448317, EPI_ISL_1448318, EPI_ISL_1448319, EPI_ISL_1448320, EPI_ISL_1448321, EPI_ISL_1448322, EPI_ISL_1448323, EPI_ISL_1448324, EPI_ISL_1448325, EPI_ISL_1448326, EPI_ISL_1448327, EPI_ISL_1448328, EPI_ISL_1448329, EPI_ISL_1448330, EPI_ISL_1448331, EPI_ISL_1448332, EPI_ISL_1448333, EPI_ISL_1448334, EPI_ISL_1448335, EPI_ISL_1448336, EPI_ISL_1448337, EPI_ISL_1448338, EPI_ISL_1448339, EPI_ISL_1448341, EPI_ISL_1448342, EPI_ISL_1448343, EPI_ISL_1448344, EPI_ISL_1448345, EPI_ISL_1448346, EPI_ISL_1448347, EPI_ISL_1448348, EPI_ISL_1448349, EPI_ISL_1448350, EPI_ISL_1448351, EPI_ISL_1448352, EPI_ISL_1448353, EPI_ISL_1448354, EPI_ISL_1448355, EPI_ISL_1448356, EPI_ISL_1448357, EPI_ISL_1448358, EPI_ISL_1448359, EPI_ISL_1448360, EPI_ISL_1448361, EPI_ISL_1448362, EPI_ISL_1448363, EPI_ISL_1448364, EPI_ISL_1448365, EPI_ISL_1448366, EPI_ISL_1448367, EPI_ISL_1448368, EPI_ISL_1448369, EPI_ISL_1448370, EPI_ISL_1448371, EPI_ISL_1448372, EPI_ISL_1448373, EPI_ISL_1448374, EPI_ISL_1448375, EPI_ISL_1448376, EPI_ISL_1448377, EPI_ISL_1448378, EPI_ISL_1448379, EPI_ISL_1448380, EPI_ISL_1448381, EPI_ISL_1448382, EPI_ISL_1448383, EPI_ISL_1448385, EPI_ISL_1448386, EPI_ISL_1448387, EPI_ISL_1448388, EPI_ISL_1448389, EPI_ISL_1448390, EPI_ISL_1448391, EPI_ISL_1448392, EPI_ISL_1448393, EPI_ISL_1448394, EPI_ISL_1448395, EPI_ISL_1448396, EPI_ISL_1448397 | | | |
| see above | UW Virology Lab | UW Virology Lab | Pavitra Roychoudhury, Hong Xie, Lasata Shrestha, Shah Mohamed Bakhash, Michelle Lin, Noah R. Baker, Sean Ellis, Saraswathi Sathees, Meei-Li Huang, Keith R Jerome, Alexander Greninger |
| EPI_ISL_1448511 | IL Dept. of Public Health Springfield Laboratory | Genomics and Discovery, Respiratory Viruses Branch, Division of Viral Diseases, Centers for Disease Control and Prevention | Yan Li, Anna Kelleher, Ying Tao, Jing Zhang, Brian Lynch, Krista Queen, Anna Uehara, Peter Cook, Han Jia Justin Ng, Clinton R. Paden, Haibin Wang, Suxiang Tong |
| EPI_ISL_1448512 | NE Public Health Laboratory | Genomics and Discovery, Respiratory Viruses Branch, Division of Viral Diseases, Centers for Disease Control and Prevention | Yan Li, Anna Kelleher, Ying Tao, Jing Zhang, Brian Lynch, Krista Queen, Anna Uehara, Peter Cook, Han Jia Justin Ng, Clinton R. Paden, Haibin Wang, Suxiang Tong |
| EPI_ISL_1448513, EPI_ISL_1448514, EPI_ISL_1448515 | IL Dept. of Public Health Springfield Laboratory | Genomics and Discovery, Respiratory Viruses Branch, Division of Viral Diseases, Centers for Disease Control and Prevention | Yan Li, Anna Kelleher, Ying Tao, Jing Zhang, Brian Lynch, Krista Queen, Anna Uehara, Peter Cook, Han Jia Justin Ng, Clinton R. Paden, Haibin Wang, Suxiang Tong |
| EPI_ISL_1448516 | IN State Department of Health Laboratory Services | Genomics and Discovery, Respiratory Viruses Branch, Division of Viral Diseases, Centers for Disease Control and Prevention | Yan Li, Anna Kelleher, Ying Tao, Jing Zhang, Brian Lynch, Krista Queen, Anna Uehara, Peter Cook, Han Jia Justin Ng, Clinton R. Paden, Haibin Wang, Suxiang Tong |
| EPI_ISL_1448517 | IL Dept. of Public Health Springfield Laboratory | Genomics and Discovery, Respiratory Viruses Branch, Division of Viral Diseases, Centers for Disease Control and Prevention | Yan Li, Anna Kelleher, Ying Tao, Jing Zhang, Brian Lynch, Krista Queen, Anna Uehara, Peter Cook, Han Jia Justin Ng, Clinton R. Paden, Haibin Wang, Suxiang Tong |
| EPI_ISL_1448518 | TN Division of Laboratory Services | Genomics and Discovery, Respiratory Viruses Branch, Division of Viral Diseases, Centers for Disease Control and Prevention | Yan Li, Anna Kelleher, Ying Tao, Jing Zhang, Brian Lynch, Krista Queen, Anna Uehara, Peter Cook, Han Jia Justin Ng, Clinton R. Paden, Haibin Wang, Suxiang Tong |
| EPI_ISL_1448519 | NE Public Health Laboratory | Genomics and Discovery, Respiratory Viruses Branch, Division of Viral Diseases, Centers for Disease Control and Prevention | Yan Li, Anna Kelleher, Ying Tao, Jing Zhang, Brian Lynch, Krista Queen, Anna Uehara, Peter Cook, Han Jia Justin Ng, Clinton R. Paden, Haibin Wang, Suxiang Tong |
| EPI_ISL_1448520, EPI_ISL_1448521 | IL Dept. of Public Health Springfield Laboratory | Genomics and Discovery, Respiratory Viruses Branch, Division of Viral Diseases, Centers for Disease Control and Prevention | Yan Li, Anna Kelleher, Ying Tao, Jing Zhang, Brian Lynch, Krista Queen, Anna Uehara, Peter Cook, Han Jia Justin Ng, Clinton R. Paden, Haibin Wang, Suxiang Tong |

| | | | |
|---|---|---|---|
| EPI_ISL_1448522 | LA Office of Public Health Laboratories | Genomics and Discovery, Respiratory Viruses Branch, Division of Viral Diseases, Centers for Disease Control and Prevention | Yan Li, Anna Kelleher, Ying Tao, Jing Zhang, Brian Lynch, Krista Queen, Anna Uehara, Peter Cook, Han Jia Justin Ng, Clinton R. Paden, Haibin Wang, Suxiang Tong |
| EPI_ISL_1448523 | TN Division of Laboratory Services | Genomics and Discovery, Respiratory Viruses Branch, Division of Viral Diseases, Centers for Disease Control and Prevention | Yan Li, Anna Kelleher, Ying Tao, Jing Zhang, Brian Lynch, Krista Queen, Anna Uehara, Peter Cook, Han Jia Justin Ng, Clinton R. Paden, Haibin Wang, Suxiang Tong |
| EPI_ISL_1448524 | IN State Department of Health Laboratory Services | Genomics and Discovery, Respiratory Viruses Branch, Division of Viral Diseases, Centers for Disease Control and Prevention | Yan Li, Anna Kelleher, Ying Tao, Jing Zhang, Brian Lynch, Krista Queen, Anna Uehara, Peter Cook, Han Jia Justin Ng, Clinton R. Paden, Haibin Wang, Suxiang Tong |
| EPI_ISL_1448525 | "PA Department of Health, Bureau of Laboratories" | Genomics and Discovery, Respiratory Viruses Branch, Division of Viral Diseases, Centers for Disease Control and Prevention | Yan Li, Anna Kelleher, Ying Tao, Jing Zhang, Brian Lynch, Krista Queen, Anna Uehara, Peter Cook, Han Jia Justin Ng, Clinton R. Paden, Haibin Wang, Suxiang Tong |
| EPI_ISL_1448526 | IL Dept. of Public Health Springfield Laboratory | Genomics and Discovery, Respiratory Viruses Branch, Division of Viral Diseases, Centers for Disease Control and Prevention | Yan Li, Anna Kelleher, Ying Tao, Jing Zhang, Brian Lynch, Krista Queen, Anna Uehara, Peter Cook, Han Jia Justin Ng, Clinton R. Paden, Haibin Wang, Suxiang Tong |

EPI_ISL_1457792, EPI_ISL_1457793, EPI_ISL_1457794, EPI_ISL_1457795, EPI_ISL_1457796, EPI_ISL_1457797, EPI_ISL_1457798, EPI_ISL_1457799, EPI_ISL_1457800, EPI_ISL_1457801, EPI_ISL_1457802, EPI_ISL_1457803, EPI_ISL_1457804, EPI_ISL_1457805, EPI_ISL_1457806, EPI_ISL_1457808, EPI_ISL_1457809, EPI_ISL_1457810, EPI_ISL_1457811, EPI_ISL_1457812, EPI_ISL_1457813, EPI_ISL_1457814,
EPI_ISL_1457815, EPI_ISL_1457816, EPI_ISL_1457817, EPI_ISL_1457818, EPI_ISL_1457819, EPI_ISL_1457820, EPI_ISL_1457821, EPI_ISL_1457822, EPI_ISL_1457823, EPI_ISL_1457824, EPI_ISL_1457825, EPI_ISL_1457826, EPI_ISL_1457827, EPI_ISL_1457828, EPI_ISL_1457829, EPI_ISL_1457830, EPI_ISL_1457831, EPI_ISL_1457832, EPI_ISL_1457833, EPI_ISL_1457834, EPI_ISL_1457835, EPI_ISL_1457836,
EPI_ISL_1457837, EPI_ISL_1457838, EPI_ISL_1457839, EPI_ISL_1457840, EPI_ISL_1457841, EPI_ISL_1457842, EPI_ISL_1457843, EPI_ISL_1457844, EPI_ISL_1457845, EPI_ISL_1457846, EPI_ISL_1457847, EPI_ISL_1457848, EPI_ISL_1457849, EPI_ISL_1457850, EPI_ISL_1457851, EPI_ISL_1457852, EPI_ISL_1457853, EPI_ISL_1457854, EPI_ISL_1457857, EPI_ISL_1457858, EPI_ISL_1457859, EPI_ISL_1457860,
EPI_ISL_1457861, EPI_ISL_1457862, EPI_ISL_1457863, EPI_ISL_1457864, EPI_ISL_1457865, EPI_ISL_1457866, EPI_ISL_1457867, EPI_ISL_1457868, EPI_ISL_1457869, EPI_ISL_1457870, EPI_ISL_1457871, EPI_ISL_1457872, EPI_ISL_1457873, EPI_ISL_1457874, EPI_ISL_1457875, EPI_ISL_1457876, EPI_ISL_1457877, EPI_ISL_1457878, EPI_ISL_1457879, EPI_ISL_1457880, EPI_ISL_1457881, EPI_ISL_1457882,
EPI_ISL_1457883, EPI_ISL_1457884, EPI_ISL_1457885, EPI_ISL_1457886, EPI_ISL_1457887, EPI_ISL_1457888, EPI_ISL_1457889, EPI_ISL_1457890, EPI_ISL_1457891, EPI_ISL_1457892, EPI_ISL_1457893, EPI_ISL_1457894, EPI_ISL_1457895, EPI_ISL_1457896, EPI_ISL_1457897, EPI_ISL_1457898, EPI_ISL_1457899, EPI_ISL_1457900, EPI_ISL_1457901, EPI_ISL_1457902, EPI_ISL_1457903, EPI_ISL_1457904,
EPI_ISL_1457905, EPI_ISL_1457906, EPI_ISL_1457907, EPI_ISL_1457908, EPI_ISL_1457909, EPI_ISL_1457910, EPI_ISL_1457911, EPI_ISL_1457912, EPI_ISL_1457913, EPI_ISL_1457914, EPI_ISL_1457915, EPI_ISL_1457916, EPI_ISL_1457918, EPI_ISL_1457919, EPI_ISL_1457921, EPI_ISL_1457922, EPI_ISL_1457923, EPI_ISL_1457924, EPI_ISL_1457926, EPI_ISL_1457927, EPI_ISL_1457928, EPI_ISL_1457929,
EPI_ISL_1457930, EPI_ISL_1457931, EPI_ISL_1457932, EPI_ISL_1457934, EPI_ISL_1457935, EPI_ISL_1457936, EPI_ISL_1457937, EPI_ISL_1457938, EPI_ISL_1457939, EPI_ISL_1457940, EPI_ISL_1457941, EPI_ISL_1457942, EPI_ISL_1457943, EPI_ISL_1457945, EPI_ISL_1457946, EPI_ISL_1457947, EPI_ISL_1457949, EPI_ISL_1457950, EPI_ISL_1457951, EPI_ISL_1457952, EPI_ISL_1457953, EPI_ISL_1457954,
EPI_ISL_1457955, EPI_ISL_1457956, EPI_ISL_1457957, EPI_ISL_1457958, EPI_ISL_1457959, EPI_ISL_1457960, EPI_ISL_1457961, EPI_ISL_1457962, EPI_ISL_1457963, EPI_ISL_1457964, EPI_ISL_1457965, EPI_ISL_1457966, EPI_ISL_1457967, EPI_ISL_1457968, EPI_ISL_1457969, EPI_ISL_1457970, EPI_ISL_1457972, EPI_ISL_1457973, EPI_ISL_1457975, EPI_ISL_1457976, EPI_ISL_1457977, EPI_ISL_1457978,
EPI_ISL_1457979, EPI_ISL_1457980, EPI_ISL_1457981, EPI_ISL_1457982, EPI_ISL_1457983, EPI_ISL_1457984, EPI_ISL_1457986, EPI_ISL_1457987, EPI_ISL_1457988, EPI_ISL_1457989, EPI_ISL_1457990, EPI_ISL_1457991, EPI_ISL_1457992, EPI_ISL_1457993, EPI_ISL_1457994, EPI_ISL_1457995, EPI_ISL_1457996, EPI_ISL_1457997, EPI_ISL_1457998, EPI_ISL_1457999, EPI_ISL_1458000, EPI_ISL_1458001,
EPI_ISL_1458002, EPI_ISL_1458003, EPI_ISL_1458004, EPI_ISL_1458005, EPI_ISL_1458006, EPI_ISL_1458007, EPI_ISL_1458009, EPI_ISL_1458010, EPI_ISL_1458011, EPI_ISL_1458012, EPI_ISL_1458013, EPI_ISL_1458014, EPI_ISL_1458015, EPI_ISL_1458016, EPI_ISL_1458017, EPI_ISL_1458018, EPI_ISL_1458019, EPI_ISL_1458020, EPI_ISL_1458022, EPI_ISL_1458023, EPI_ISL_1458024, EPI_ISL_1458025,
EPI_ISL_1458026, EPI_ISL_1458027, EPI_ISL_1458028, EPI_ISL_1458029, EPI_ISL_1458030, EPI_ISL_1458031, EPI_ISL_1458032, EPI_ISL_1458034, EPI_ISL_1458035, EPI_ISL_1458036, EPI_ISL_1458037, EPI_ISL_1458038, EPI_ISL_1458040, EPI_ISL_1458041, EPI_ISL_1458042, EPI_ISL_1458043, EPI_ISL_1458044, EPI_ISL_1458045, EPI_ISL_1458046, EPI_ISL_1458047, EPI_ISL_1458048, EPI_ISL_1458049,
EPI_ISL_1458050, EPI_ISL_1458051, EPI_ISL_1458052, EPI_ISL_1458053, EPI_ISL_1458054, EPI_ISL_1458055, EPI_ISL_1458056, EPI_ISL_1458057, EPI_ISL_1458058, EPI_ISL_1458059, EPI_ISL_1458060, EPI_ISL_1458061, EPI_ISL_1458062, EPI_ISL_1458063, EPI_ISL_1458064, EPI_ISL_1458065, EPI_ISL_1458066, EPI_ISL_1458067, EPI_ISL_1458068, EPI_ISL_1458069, EPI_ISL_1458070, EPI_ISL_1458071,
EPI_ISL_1458072, EPI_ISL_1458073, EPI_ISL_1458074, EPI_ISL_1458075, EPI_ISL_1458076, EPI_ISL_1458077, EPI_ISL_1458079, EPI_ISL_1458080, EPI_ISL_1458082, EPI_ISL_1458083, EPI_ISL_1458084, EPI_ISL_1458085, EPI_ISL_1458086, EPI_ISL_1458087, EPI_ISL_1458089, EPI_ISL_1458090, EPI_ISL_1458091, EPI_ISL_1458092, EPI_ISL_1458093, EPI_ISL_1458094, EPI_ISL_1458095, EPI_ISL_1458096,
EPI_ISL_1458097, EPI_ISL_1458098, EPI_ISL_1458099, EPI_ISL_1458100, EPI_ISL_1458101, EPI_ISL_1458102, EPI_ISL_1458103, EPI_ISL_1458104, EPI_ISL_1458105, EPI_ISL_1458107, EPI_ISL_1458108, EPI_ISL_1458109, EPI_ISL_1458110, EPI_ISL_1458111, EPI_ISL_1458112, EPI_ISL_1458113, EPI_ISL_1458114, EPI_ISL_1458115, EPI_ISL_1458116, EPI_ISL_1458117, EPI_ISL_1458118, EPI_ISL_1458119,
EPI_ISL_1458120, EPI_ISL_1458121, EPI_ISL_1458122, EPI_ISL_1458123, EPI_ISL_1458124, EPI_ISL_1458125, EPI_ISL_1458126, EPI_ISL_1458127, EPI_ISL_1458128, EPI_ISL_1458129, EPI_ISL_1458130, EPI_ISL_1458131, EPI_ISL_1458132, EPI_ISL_1458133, EPI_ISL_1458134, EPI_ISL_1458135, EPI_ISL_1458136, EPI_ISL_1458137, EPI_ISL_1458138, EPI_ISL_1458139, EPI_ISL_1458140, EPI_ISL_1458141,
EPI_ISL_1458142, EPI_ISL_1458143, EPI_ISL_1458144, EPI_ISL_1458145, EPI_ISL_1458146, EPI_ISL_1458147, EPI_ISL_1458148, EPI_ISL_1458149, EPI_ISL_1458150, EPI_ISL_1458151, EPI_ISL_1458152, EPI_ISL_1458153, EPI_ISL_1458154, EPI_ISL_1458155, EPI_ISL_1458156, EPI_ISL_1458157, EPI_ISL_1458158, EPI_ISL_1458159, EPI_ISL_1458160, EPI_ISL_1458161, EPI_ISL_1458162, EPI_ISL_1458163,
EPI_ISL_1458164, EPI_ISL_1458165, EPI_ISL_1458166, EPI_ISL_1458167, EPI_ISL_1458168, EPI_ISL_1458169, EPI_ISL_1458170, EPI_ISL_1458171, EPI_ISL_1458172, EPI_ISL_1458173, EPI_ISL_1458174, EPI_ISL_1458175, EPI_ISL_1458176, EPI_ISL_1458177, EPI_ISL_1458178, EPI_ISL_1458179, EPI_ISL_1458180, EPI_ISL_1458181, EPI_ISL_1458182, EPI_ISL_1458183, EPI_ISL_1458184, EPI_ISL_1458185,
EPI_ISL_1458186, EPI_ISL_1458187, EPI_ISL_1458188, EPI_ISL_1458189, EPI_ISL_1458190, EPI_ISL_1458191, EPI_ISL_1458192, EPI_ISL_1458193, EPI_ISL_1458194, EPI_ISL_1458195, EPI_ISL_1458196, EPI_ISL_1458197, EPI_ISL_1458198, EPI_ISL_1458199, EPI_ISL_1458200, EPI_ISL_1458201, EPI_ISL_1458202, EPI_ISL_1458204, EPI_ISL_1458205, EPI_ISL_1458206, EPI_ISL_1458207, EPI_ISL_1458208,
EPI_ISL_1458209, EPI_ISL_1458210, EPI_ISL_1458211, EPI_ISL_1458212, EPI_ISL_1458213, EPI_ISL_1458214, EPI_ISL_1458215, EPI_ISL_1458217, EPI_ISL_1458218, EPI_ISL_1458219, EPI_ISL_1458220, EPI_ISL_1458221, EPI_ISL_1458222, EPI_ISL_1458223, EPI_ISL_1458224, EPI_ISL_1458225, EPI_ISL_1458226, EPI_ISL_1458227, EPI_ISL_1458228, EPI_ISL_1458229, EPI_ISL_1458230, EPI_ISL_1458231,
EPI_ISL_1458232, EPI_ISL_1458233, EPI_ISL_1458234, EPI_ISL_1458235, EPI_ISL_1458236, EPI_ISL_1458238, EPI_ISL_1458239, EPI_ISL_1458240, EPI_ISL_1458241, EPI_ISL_1458242, EPI_ISL_1458243, EPI_ISL_1458244, EPI_ISL_1458245, EPI_ISL_1458246, EPI_ISL_1458247, EPI_ISL_1458248, EPI_ISL_1458249, EPI_ISL_1458250, EPI_ISL_1458251, EPI_ISL_1458252, EPI_ISL_1458253, EPI_ISL_1458254,
EPI_ISL_1458255, EPI_ISL_1458256, EPI_ISL_1458257, EPI_ISL_1458258, EPI_ISL_1458259, EPI_ISL_1458260, EPI_ISL_1458261, EPI_ISL_1458262, EPI_ISL_1458263, EPI_ISL_1458264, EPI_ISL_1458265, EPI_ISL_1458267, EPI_ISL_1458268, EPI_ISL_1458269, EPI_ISL_1458270, EPI_ISL_1458271, EPI_ISL_1458272, EPI_ISL_1458274, EPI_ISL_1458275, EPI_ISL_1458276, EPI_ISL_1458277, EPI_ISL_1458278,
EPI_ISL_1458279, EPI_ISL_1458280, EPI_ISL_1458281, EPI_ISL_1458282, EPI_ISL_1458284, EPI_ISL_1458285, EPI_ISL_1458286, EPI_ISL_1458287, EPI_ISL_1458288, EPI_ISL_1458289, EPI_ISL_1458290, EPI_ISL_1458291, EPI_ISL_1458292, EPI_ISL_1458293, EPI_ISL_1458294, EPI_ISL_1458295, EPI_ISL_1458296, EPI_ISL_1458297, EPI_ISL_1458298, EPI_ISL_1458299, EPI_ISL_1458300, EPI_ISL_1458301,
EPI_ISL_1458302, EPI_ISL_1458303, EPI_ISL_1458304, EPI_ISL_1458305, EPI_ISL_1458306, EPI_ISL_1458307, EPI_ISL_1458308, EPI_ISL_1458309, EPI_ISL_1458311, EPI_ISL_1458312, EPI_ISL_1458313, EPI_ISL_1458314, EPI_ISL_1458315, EPI_ISL_1458316, EPI_ISL_1458317, EPI_ISL_1458318, EPI_ISL_1458319, EPI_ISL_1458320, EPI_ISL_1458321, EPI_ISL_1458322, EPI_ISL_1458323, EPI_ISL_1458324,
EPI_ISL_1458325, EPI_ISL_1458326, EPI_ISL_1458327, EPI_ISL_1458328, EPI_ISL_1458329, EPI_ISL_1458330, EPI_ISL_1458331, EPI_ISL_1458332, EPI_ISL_1458333, EPI_ISL_1458334, EPI_ISL_1458335, EPI_ISL_1458336, EPI_ISL_1458337, EPI_ISL_1458338, EPI_ISL_1458339, EPI_ISL_1458340, EPI_ISL_1458341, EPI_ISL_1458342, EPI_ISL_1458343, EPI_ISL_1458344, EPI_ISL_1458345, EPI_ISL_1458346,
EPI_ISL_1458347, EPI_ISL_1458348, EPI_ISL_1458349, EPI_ISL_1458350, EPI_ISL_1458351, EPI_ISL_1458352, EPI_ISL_1458353, EPI_ISL_1458354, EPI_ISL_1458355, EPI_ISL_1458356, EPI_ISL_1458357, EPI_ISL_1458358, EPI_ISL_1458359, EPI_ISL_1458360, EPI_ISL_1458361, EPI_ISL_1458362, EPI_ISL_1458363, EPI_ISL_1458364, EPI_ISL_1458365, EPI_ISL_1458366, EPI_ISL_1458367, EPI_ISL_1458368,
EPI_ISL_1458369, EPI_ISL_1458370, EPI_ISL_1458371, EPI_ISL_1458372, EPI_ISL_1458373, EPI_ISL_1458374, EPI_ISL_1458375, EPI_ISL_1458376, EPI_ISL_1458377, EPI_ISL_1458378, EPI_ISL_1458379, EPI_ISL_1458380, EPI_ISL_1458381, EPI_ISL_1458382, EPI_ISL_1458383, EPI_ISL_1458384, EPI_ISL_1458385, EPI_ISL_1458386, EPI_ISL_1458387, EPI_ISL_1458388, EPI_ISL_1458389, EPI_ISL_1458391,
EPI_ISL_1458392, EPI_ISL_1458393, EPI_ISL_1458394, EPI_ISL_1458395, EPI_ISL_1458396, EPI_ISL_1458397, EPI_ISL_1458398, EPI_ISL_1458399, EPI_ISL_1458400, EPI_ISL_1458402, EPI_ISL_1458403, EPI_ISL_1458404, EPI_ISL_1458405, EPI_ISL_1458406, EPI_ISL_1458407, EPI_ISL_1458408, EPI_ISL_1458409, EPI_ISL_1458410, EPI_ISL_1458411, EPI_ISL_1458412, EPI_ISL_1458413, EPI_ISL_1458414,
EPI_ISL_1458415, EPI_ISL_1458416, EPI_ISL_1458417, EPI_ISL_1458418, EPI_ISL_1458419, EPI_ISL_1458420, EPI_ISL_1458421, EPI_ISL_1458422, EPI_ISL_1458423, EPI_ISL_1458424, EPI_ISL_1458425, EPI_ISL_1458426, EPI_ISL_1458427, EPI_ISL_1458428, EPI_ISL_1458429, EPI_ISL_1458430, EPI_ISL_1458431, EPI_ISL_1458432, EPI_ISL_1458433, EPI_ISL_1458434, EPI_ISL_1458435, EPI_ISL_1458436,
EPI_ISL_1458437, EPI_ISL_1458438, EPI_ISL_1458439, EPI_ISL_1458440, EPI_ISL_1458441, EPI_ISL_1458442, EPI_ISL_1458443, EPI_ISL_1458444, EPI_ISL_1458446, EPI_ISL_1458447, EPI_ISL_1458448, EPI_ISL_1458449, EPI_ISL_1458450, EPI_ISL_1458451, EPI_ISL_1458452, EPI_ISL_1458453, EPI_ISL_1458454, EPI_ISL_1458455, EPI_ISL_1458456, EPI_ISL_1458457, EPI_ISL_1458458, EPI_ISL_1458460,
EPI_ISL_1458461, EPI_ISL_1458462, EPI_ISL_1458463, EPI_ISL_1458464, EPI_ISL_1458465, EPI_ISL_1458466, EPI_ISL_1458467, EPI_ISL_1458468, EPI_ISL_1458469, EPI_ISL_1458470, EPI_ISL_1458471, EPI_ISL_1458472, EPI_ISL_1458473, EPI_ISL_1458474, EPI_ISL_1458475, EPI_ISL_1458476, EPI_ISL_1458477, EPI_ISL_1458478, EPI_ISL_1458479, EPI_ISL_1458480, EPI_ISL_1458481, EPI_ISL_1458482,
EPI_ISL_1458483, EPI_ISL_1458485, EPI_ISL_1458486, EPI_ISL_1458488, EPI_ISL_1458489, EPI_ISL_1458490, EPI_ISL_1458491, EPI_ISL_1458492, EPI_ISL_1458493, EPI_ISL_1458494, EPI_ISL_1458495, EPI_ISL_1458496, EPI_ISL_1458498, EPI_ISL_1458499, EPI_ISL_1458500, EPI_ISL_1458502, EPI_ISL_1458503, EPI_ISL_1458504, EPI_ISL_1458505, EPI_ISL_1458506, EPI_ISL_1458507, EPI_ISL_1458508,
EPI_ISL_1458509, EPI_ISL_1458510, EPI_ISL_1458511, EPI_ISL_1458512, EPI_ISL_1458513, EPI_ISL_1458514, EPI_ISL_1458515, EPI_ISL_1458516, EPI_ISL_1458517, EPI_ISL_1458518, EPI_ISL_1458519, EPI_ISL_1458520, EPI_ISL_1458522, EPI_ISL_1458523, EPI_ISL_1458524, EPI_ISL_1458526, EPI_ISL_1458527, EPI_ISL_1458528, EPI_ISL_1458529, EPI_ISL_1458530, EPI_ISL_1458531, EPI_ISL_1458533,
EPI_ISL_1458534, EPI_ISL_1458536, EPI_ISL_1458538, EPI_ISL_1458540, EPI_ISL_1458542, EPI_ISL_1458543, EPI_ISL_1458544, EPI_ISL_1458545, EPI_ISL_1458546, EPI_ISL_1458548, EPI_ISL_1458549, EPI_ISL_1458550, EPI_ISL_1458551, EPI_ISL_1458552, EPI_ISL_1458553, EPI_ISL_1458554, EPI_ISL_1458556, EPI_ISL_1458557, EPI_ISL_1458558, EPI_ISL_1458559, EPI_ISL_1458560, EPI_ISL_1458561,
EPI_ISL_1458562, EPI_ISL_1458563, EPI_ISL_1458564, EPI_ISL_1458565, EPI_ISL_1458566, EPI_ISL_1458567, EPI_ISL_1458568, EPI_ISL_1458569, EPI_ISL_1458570, EPI_ISL_1458571, EPI_ISL_1458572, EPI_ISL_1458573, EPI_ISL_1458574, EPI_ISL_1458575, EPI_ISL_1458576, EPI_ISL_1458577, EPI_ISL_1458579, EPI_ISL_1458580, EPI_ISL_1458581, EPI_ISL_1458583, EPI_ISL_1458584, EPI_ISL_1458585,
EPI_ISL_1458586, EPI_ISL_1458588, EPI_ISL_1458589, EPI_ISL_1458590, EPI_ISL_1458591, EPI_ISL_1458592, EPI_ISL_1458593, EPI_ISL_1458594, EPI_ISL_1458595, EPI_ISL_1458596, EPI_ISL_1458597, EPI_ISL_1458598, EPI_ISL_1458599, EPI_ISL_1458601, EPI_ISL_1458602, EPI_ISL_1458603, EPI_ISL_1458604, EPI_ISL_1458605, EPI_ISL_1458606, EPI_ISL_1458607, EPI_ISL_1458608, EPI_ISL_1458609,
EPI_ISL_1458610, EPI_ISL_1458611, EPI_ISL_1458612, EPI_ISL_1458613, EPI_ISL_1458614, EPI_ISL_1458615, EPI_ISL_1458616, EPI_ISL_1458617, EPI_ISL_1458618, EPI_ISL_1458619, EPI_ISL_1458620, EPI_ISL_1458622, EPI_ISL_1458623, EPI_ISL_1458624, EPI_ISL_1458625, EPI_ISL_1458626, EPI_ISL_1458627, EPI_ISL_1458629, EPI_ISL_1458630, EPI_ISL_1458631, EPI_ISL_1458632, EPI_ISL_1458633,
EPI_ISL_1458634, EPI_ISL_1458635, EPI_ISL_1458636, EPI_ISL_1458637, EPI_ISL_1458638, EPI_ISL_1458639, EPI_ISL_1458640, EPI_ISL_1458641, EPI_ISL_1458642, EPI_ISL_1458643, EPI_ISL_1458644, EPI_ISL_1458645, EPI_ISL_1458646, EPI_ISL_1458647, EPI_ISL_1458648, EPI_ISL_1458649, EPI_ISL_1458650, EPI_ISL_1458651, EPI_ISL_1458652, EPI_ISL_1458653, EPI_ISL_1458654, EPI_ISL_1458655,
EPI_ISL_1458656, EPI_ISL_1458657, EPI_ISL_1458658, EPI_ISL_1458659, EPI_ISL_1458660, EPI_ISL_1458661, EPI_ISL_1458662, EPI_ISL_1458663, EPI_ISL_1458664, EPI_ISL_1458665, EPI_ISL_1458666, EPI_ISL_1458667, EPI_ISL_1458668, EPI_ISL_1458669, EPI_ISL_1458670, EPI_ISL_1458671, EPI_ISL_1458672, EPI_ISL_1458673, EPI_ISL_1458674, EPI_ISL_1458675, EPI_ISL_1458676, EPI_ISL_1458677,
EPI_ISL_1458678, EPI_ISL_1458679, EPI_ISL_1458680, EPI_ISL_1458681, EPI_ISL_1458683, EPI_ISL_1458684, EPI_ISL_1458685, EPI_ISL_1458686, EPI_ISL_1458687, EPI_ISL_1458688, EPI_ISL_1458689, EPI_ISL_1458690, EPI_ISL_1458691, EPI_ISL_1458692, EPI_ISL_1458693, EPI_ISL_1458694, EPI_ISL_1458695, EPI_ISL_1458696, EPI_ISL_1458697, EPI_ISL_1458698, EPI_ISL_1458699, EPI_ISL_1458700,
EPI_ISL_1458701, EPI_ISL_1458702, EPI_ISL_1458703, EPI_ISL_1458704, EPI_ISL_1458705, EPI_ISL_1458707, EPI_ISL_1458708, EPI_ISL_1458709, EPI_ISL_1458711, EPI_ISL_1458712, EPI_ISL_1458713, EPI_ISL_1458714, EPI_ISL_1458715, EPI_ISL_1458716, EPI_ISL_1458717, EPI_ISL_1458718, EPI_ISL_1458719, EPI_ISL_1458720, EPI_ISL_1458721, EPI_ISL_1458722, EPI_ISL_1458723, EPI_ISL_1458724,
EPI_ISL_1458725, EPI_ISL_1458726, EPI_ISL_1458728, EPI_ISL_1458729, EPI_ISL_1458730, EPI_ISL_1458731, EPI_ISL_1458732, EPI_ISL_1458733, EPI_ISL_1458734, EPI_ISL_1458735, EPI_ISL_1458736, EPI_ISL_1458738, EPI_ISL_1458739, EPI_ISL_1458741, EPI_ISL_1458742, EPI_ISL_1458743, EPI_ISL_1458744, EPI_ISL_1458745, EPI_ISL_1458746, EPI_ISL_1458747, EPI_ISL_1458748, EPI_ISL_1458749,
EPI_ISL_1458750, EPI_ISL_1458751, EPI_ISL_1458752, EPI_ISL_1458753, EPI_ISL_1458754, EPI_ISL_1458755, EPI_ISL_1458757, EPI_ISL_1458758, EPI_ISL_1458759, EPI_ISL_1458760, EPI_ISL_1458762, EPI_ISL_1458763, EPI_ISL_1458764, EPI_ISL_1458765, EPI_ISL_1458766, EPI_ISL_1458767, EPI_ISL_1458769, EPI_ISL_1458771, EPI_ISL_1458772, EPI_ISL_1458773, EPI_ISL_1458775, EPI_ISL_1458776,
EPI_ISL_1458777, EPI_ISL_1458778, EPI_ISL_1458779, EPI_ISL_1458780, EPI_ISL_1458781, EPI_ISL_1458782, EPI_ISL_1458784, EPI_ISL_1458785, EPI_ISL_1458786, EPI_ISL_1458787, EPI_ISL_1458788, EPI_ISL_1458790, EPI_ISL_1458791, EPI_ISL_1458793, EPI_ISL_1458794, EPI_ISL_1458795, EPI_ISL_1458796, EPI_ISL_1458797, EPI_ISL_1458798, EPI_ISL_1458799, EPI_ISL_1458800, EPI_ISL_1458801,
EPI_ISL_1458802, EPI_ISL_1458803, EPI_ISL_1458804, EPI_ISL_1458805, EPI_ISL_1458806, EPI_ISL_1458807, EPI_ISL_1458808, EPI_ISL_1458809, EPI_ISL_1458810, EPI_ISL_1458811, EPI_ISL_1458812, EPI_ISL_1458813, EPI_ISL_1458814, EPI_ISL_1458815, EPI_ISL_1458816, EPI_ISL_1458817, EPI_ISL_1458818, EPI_ISL_1458819, EPI_ISL_1458844, EPI_ISL_1458845, EPI_ISL_1458847, EPI_ISL_1458850,
EPI_ISL_1458851, EPI_ISL_1458852, EPI_ISL_1458853, EPI_ISL_1458854, EPI_ISL_1458855, EPI_ISL_1458856, EPI_ISL_1458857, EPI_ISL_1458858, EPI_ISL_1458859, EPI_ISL_1458900, EPI_ISL_1458901, EPI_ISL_1458902, EPI_ISL_1458903, EPI_ISL_1458904, EPI_ISL_1458905, EPI_ISL_1458906, EPI_ISL_1458907, EPI_ISL_1458908, EPI_ISL_1458909, EPI_ISL_1458910, EPI_ISL_1458911, EPI_ISL_1458912,
EPI_ISL_1458913, EPI_ISL_1458914, EPI_ISL_1458915, EPI_ISL_1458916, EPI_ISL_1458917, EPI_ISL_1458918, EPI_ISL_1458919, EPI_ISL_1458920, EPI_ISL_1458921, EPI_ISL_1458922, EPI_ISL_1458923, EPI_ISL_1458924, EPI_ISL_1458925, EPI_ISL_1458926, EPI_ISL_1458927, EPI_ISL_1458928, EPI_ISL_1458929, EPI_ISL_1458930, EPI_ISL_1458931, EPI_ISL_1458932, EPI_ISL_1458933, EPI_ISL_1458934,
EPI_ISL_1458935, EPI_ISL_1458936, EPI_ISL_1458937, EPI_ISL_1458938, EPI_ISL_1458939, EPI_ISL_1458965, EPI_ISL_1458968, EPI_ISL_1458970, EPI_ISL_1458971, EPI_ISL_1458972, EPI_ISL_1458973, EPI_ISL_1458974, EPI_ISL_1458975, EPI_ISL_1458976, EPI_ISL_1458977, EPI_ISL_1458979, EPI_ISL_1459012, EPI_ISL_1459013, EPI_ISL_1459016, EPI_ISL_1459018, EPI_ISL_1459020, EPI_ISL_1459021,
EPI_ISL_1459022, EPI_ISL_1459023, EPI_ISL_1459024, EPI_ISL_1459025, EPI_ISL_1459026, EPI_ISL_1459027, EPI_ISL_1459028, EPI_ISL_1459029, EPI_ISL_1459070, EPI_ISL_1459071, EPI_ISL_1459072, EPI_ISL_1459073, EPI_ISL_1459074, EPI_ISL_1459075, EPI_ISL_1459076, EPI_ISL_1459077, EPI_ISL_1459078, EPI_ISL_1459079, EPI_ISL_1459080, EPI_ISL_1459081, EPI_ISL_1459083, EPI_ISL_1459084,
EPI_ISL_1459085, EPI_ISL_1459086, EPI_ISL_1459087, EPI_ISL_1459088, EPI_ISL_1459089, EPI_ISL_1459090, EPI_ISL_1459091, EPI_ISL_1459092, EPI_ISL_1459093, EPI_ISL_1459094, EPI_ISL_1459095, EPI_ISL_1459096, EPI_ISL_1459097, EPI_ISL_1459098, EPI_ISL_1459099, EPI_ISL_1459101, EPI_ISL_1459102, EPI_ISL_1459103, EPI_ISL_1459104, EPI_ISL_1459106, EPI_ISL_1459107, EPI_ISL_1459108,
EPI_ISL_1459109, EPI_ISL_1459133, EPI_ISL_1459134, EPI_ISL_1459137, EPI_ISL_1459138, EPI_ISL_1459139, EPI_ISL_1459140, EPI_ISL_1459141, EPI_ISL_1459142, EPI_ISL_1459143, EPI_ISL_1459144, EPI_ISL_1459145, EPI_ISL_1459146, EPI_ISL_1459147, EPI_ISL_1459148, EPI_ISL_1459149, EPI_ISL_1459190, EPI_ISL_1459191, EPI_ISL_1459192, EPI_ISL_1459193, EPI_ISL_1459194, EPI_ISL_1459195,
EPI_ISL_1459196, EPI_ISL_1459197, EPI_ISL_1459198, EPI_ISL_1459199, EPI_ISL_1459200, EPI_ISL_1459201, EPI_ISL_1459203, EPI_ISL_1459204, EPI_ISL_1459205, EPI_ISL_1459206, EPI_ISL_1459207, EPI_ISL_1459208, EPI_ISL_1459209, EPI_ISL_1459210, EPI_ISL_1459211, EPI_ISL_1459212, EPI_ISL_1459213, EPI_ISL_1459214, EPI_ISL_1459215, EPI_ISL_1459216, EPI_ISL_1459217, EPI_ISL_1459218,
EPI_ISL_1459220, EPI_ISL_1459221, EPI_ISL_1459222, EPI_ISL_1459223, EPI_ISL_1459224, EPI_ISL_1459225, EPI_ISL_1459227, EPI_ISL_1459229, EPI_ISL_1459256, EPI_ISL_1459257, EPI_ISL_1459258, EPI_ISL_1459259, EPI_ISL_1459260, EPI_ISL_1459261, EPI_ISL_1459262, EPI_ISL_1459263, EPI_ISL_1459264, EPI_ISL_1459265, EPI_ISL_1459266, EPI_ISL_1459267, EPI_ISL_1459268, EPI_ISL_1459269,
EPI_ISL_1459292, EPI_ISL_1459293, EPI_ISL_1459294, EPI_ISL_1459295, EPI_ISL_1459296, EPI_ISL_1459297, EPI_ISL_1459298, EPI_ISL_1459299, EPI_ISL_1459300, EPI_ISL_1459301, EPI_ISL_1459302, EPI_ISL_1459303, EPI_ISL_1459304, EPI_ISL_1459305, EPI_ISL_1459306, EPI_ISL_1459307, EPI_ISL_1459308, EPI_ISL_1459309, EPI_ISL_1459310, EPI_ISL_1459311, EPI_ISL_1459312, EPI_ISL_1459313,
EPI_ISL_1459315, EPI_ISL_1459316, EPI_ISL_1459317, EPI_ISL_1459318, EPI_ISL_1459319, EPI_ISL_1459320, EPI_ISL_1459321, EPI_ISL_1459322, EPI_ISL_1459323, EPI_ISL_1459324, EPI_ISL_1459325, EPI_ISL_1459326, EPI_ISL_1459327, EPI_ISL_1459328, EPI_ISL_1459329, EPI_ISL_1459330, EPI_ISL_1459331, EPI_ISL_1459332, EPI_ISL_1459333, EPI_ISL_1459334, EPI_ISL_1459335, EPI_ISL_1459336,
EPI_ISL_1459337, EPI_ISL_1459338, EPI_ISL_1459339, EPI_ISL_1459340, EPI_ISL_1459341, EPI_ISL_1459342, EPI_ISL_1459343, EPI_ISL_1459344, EPI_ISL_1459346, EPI_ISL_1459347, EPI_ISL_1459348, EPI_ISL_1459349, EPI_ISL_1459350, EPI_ISL_1459351, EPI_ISL_1459352, EPI_ISL_1459353, EPI_ISL_1459355, EPI_ISL_1459356, EPI_ISL_1459357, EPI_ISL_1459358, EPI_ISL_1459359, EPI_ISL_1459360,
EPI_ISL_1459361, EPI_ISL_1459362, EPI_ISL_1459363, EPI_ISL_1459365, EPI_ISL_1459366, EPI_ISL_1459367, EPI_ISL_1459368, EPI_ISL_1459369, EPI_ISL_1459370, EPI_ISL_1459372, EPI_ISL_1459373, EPI_ISL_1459374, EPI_ISL_1459375, EPI_ISL_1459376, EPI_ISL_1459377, EPI_ISL_1459378, EPI_ISL_1459379, EPI_ISL_1459380, EPI_ISL_1459381, EPI_ISL_1459382, EPI_ISL_1459383, EPI_ISL_1459384,
EPI_ISL_1459385, EPI_ISL_1459386, EPI_ISL_1459387, EPI_ISL_1459388, EPI_ISL_1459389, EPI_ISL_1459390, EPI_ISL_1459391, EPI_ISL_1459393, EPI_ISL_1459394, EPI_ISL_1459395, EPI_ISL_1459396, EPI_ISL_1459398, EPI_ISL_1459399, EPI_ISL_1459400, EPI_ISL_1459401, EPI_ISL_1459402, EPI_ISL_1459403, EPI_ISL_1459404, EPI_ISL_1459405, EPI_ISL_1459406, EPI_ISL_1459407, EPI_ISL_1459410,
EPI_ISL_1459411, EPI_ISL_1459412, EPI_ISL_1459414, EPI_ISL_1459415, EPI_ISL_1459416, EPI_ISL_1459417, EPI_ISL_1459418, EPI_ISL_1459419, EPI_ISL_1459420, EPI_ISL_1459421, EPI_ISL_1459422, EPI_ISL_1459424, EPI_ISL_1459425, EPI_ISL_1459426, EPI_ISL_1459428, EPI_ISL_1459430, EPI_ISL_1459431, EPI_ISL_1459432, EPI_ISL_1459433, EPI_ISL_1459434, EPI_ISL_1459435, EPI_ISL_1459437,
EPI_ISL_1459438, EPI_ISL_1459440, EPI_ISL_1459441, EPI_ISL_1459442, EPI_ISL_1459443, EPI_ISL_1459445, EPI_ISL_1459447, EPI_ISL_1459448, EPI_ISL_1459449, EPI_ISL_1459450, EPI_ISL_1459451, EPI_ISL_1459452, EPI_ISL_1459453, EPI_ISL_1459455, EPI_ISL_1459456, EPI_ISL_1459457, EPI_ISL_1459458, EPI_ISL_1459459, EPI_ISL_1459460, EPI_ISL_1459461, EPI_ISL_1459462, EPI_ISL_1459463,
EPI_ISL_1459464, EPI_ISL_1459465, EPI_ISL_1459468, EPI_ISL_1459469, EPI_ISL_1459470, EPI_ISL_1459471, EPI_ISL_1459472, EPI_ISL_1459473, EPI_ISL_1459474, EPI_ISL_1459475, EPI_ISL_1459478, EPI_ISL_1459480, EPI_ISL_1459483, EPI_ISL_1459484, EPI_ISL_1459486, EPI_ISL_1459487, EPI_ISL_1459488, EPI_ISL_1459489, EPI_ISL_1459490, EPI_ISL_1459491, EPI_ISL_1459493, EPI_ISL_1459497,
EPI_ISL_1459498, EPI_ISL_1459499, EPI_ISL_1459500, EPI_ISL_1459501, EPI_ISL_1459502, EPI_ISL_1459503, EPI_ISL_1459504, EPI_ISL_1459506, EPI_ISL_1459507, EPI_ISL_1459508, EPI_ISL_1459529, EPI_ISL_1459530, EPI_ISL_1459536, EPI_ISL_1459537, EPI_ISL_1459538, EPI_ISL_1459539, EPI_ISL_1459540, EPI_ISL_1459541, EPI_ISL_1459542, EPI_ISL_1459544, EPI_ISL_1459547, EPI_ISL_1459549,
EPI_ISL_1459550, EPI_ISL_1459552, EPI_ISL_1459554, EPI_ISL_1459555, EPI_ISL_1459556, EPI_ISL_1459557, EPI_ISL_1459558, EPI_ISL_1459560, EPI_ISL_1459561, EPI_ISL_1459563, EPI_ISL_1459564, EPI_ISL_1459565, EPI_ISL_1459566, EPI_ISL_1459567, EPI_ISL_1459568, EPI_ISL_1459570, EPI_ISL_1459571, EPI_ISL_1459572, EPI_ISL_1459574, EPI_ISL_1459575, EPI_ISL_1459576, EPI_ISL_1459577,
EPI_ISL_1459578, EPI_ISL_1459579, EPI_ISL_1459580, EPI_ISL_1459581, EPI_ISL_1459582, EPI_ISL_1459583, EPI_ISL_1459584, EPI_ISL_1459587, EPI_ISL_1459588, EPI_ISL_1459590, EPI_ISL_1459592, EPI_ISL_1459593, EPI_ISL_1459596, EPI_ISL_1459597, EPI_ISL_1459599, EPI_ISL_1459600, EPI_ISL_1459601, EPI_ISL_1459602, EPI_ISL_1459603, EPI_ISL_1459604, EPI_ISL_1459606, EPI_ISL_1459607,
EPI_ISL_1459608, EPI_ISL_1459609, EPI_ISL_1459610, EPI_ISL_1459611, EPI_ISL_1459612, EPI_ISL_1459613, EPI_ISL_1459614, EPI_ISL_1459617, EPI_ISL_1459618, EPI_ISL_1459619, EPI_ISL_1459620, EPI_ISL_1459621, EPI_ISL_1459622, EPI_ISL_1459623, EPI_ISL_1459624, EPI_ISL_1459626, EPI_ISL_1459627, EPI_ISL_1459628, EPI_ISL_1459629, EPI_ISL_1459630, EPI_ISL_1459631, EPI_ISL_1459632,
EPI_ISL_1459634, EPI_ISL_1459636, EPI_ISL_1459637, EPI_ISL_1459639, EPI_ISL_1459640, EPI_ISL_1459641, EPI_ISL_1459642, EPI_ISL_1459643, EPI_ISL_1459644, EPI_ISL_1459645, EPI_ISL_1459647, EPI_ISL_1459648, EPI_ISL_1459649, EPI_ISL_1459652, EPI_ISL_1459653, EPI_ISL_1459654, EPI_ISL_1459657, EPI_ISL_1459658, EPI_ISL_1459660, EPI_ISL_1459661, EPI_ISL_1459662, EPI_ISL_1459663,
EPI_ISL_1459664, EPI_ISL_1459665, EPI_ISL_1459666, EPI_ISL_1459668, EPI_ISL_1459670, EPI_ISL_1459672, EPI_ISL_1459673, EPI_ISL_1459674, EPI_ISL_1459675, EPI_ISL_1459676, EPI_ISL_1459677, EPI_ISL_1459678, EPI_ISL_1459679, EPI_ISL_1459680, EPI_ISL_1459681, EPI_ISL_1459682, EPI_ISL_1459683, EPI_ISL_1459686, EPI_ISL_1459687, EPI_ISL_1459688, EPI_ISL_1459689, EPI_ISL_1459690,
EPI_ISL_1459691, EPI_ISL_1459693, EPI_ISL_1459694, EPI_ISL_1459696, EPI_ISL_1459697, EPI_ISL_1459698, EPI_ISL_1459699, EPI_ISL_1459700, EPI_ISL_1459701, EPI_ISL_1459702, EPI_ISL_1459703, EPI_ISL_1459705, EPI_ISL_1459708, EPI_ISL_1459710, EPI_ISL_1459711, EPI_ISL_1459713, EPI_ISL_1459714, EPI_ISL_1459715, EPI_ISL_1459716, EPI_ISL_1459717, EPI_ISL_1459718, EPI_ISL_1459719,
EPI_ISL_1459721, EPI_ISL_1459722, EPI_ISL_1459723, EPI_ISL_1459724, EPI_ISL_1459725, EPI_ISL_1459726, EPI_ISL_1459727, EPI_ISL_1459728, EPI_ISL_1459730, EPI_ISL_1459731, EPI_ISL_1459732, EPI_ISL_1459734, EPI_ISL_1459735, EPI_ISL_1459736, EPI_ISL_1459737, EPI_ISL_1459738, EPI_ISL_1459739, EPI_ISL_1459740, EPI_ISL_1459741, EPI_ISL_1459742, EPI_ISL_1459743, EPI_ISL_1459744,
EPI_ISL_1459745, EPI_ISL_1459746, EPI_ISL_1459747, EPI_ISL_1459748, EPI_ISL_1459749, EPI_ISL_1459750, EPI_ISL_1459751, EPI_ISL_1459752, EPI_ISL_1459753, EPI_ISL_1459754, EPI_ISL_1459755, EPI_ISL_1459756, EPI_ISL_1459757, EPI_ISL_1459758, EPI_ISL_1459759, EPI_ISL_1459760, EPI_ISL_1459761, EPI_ISL_1459762, EPI_ISL_1459763, EPI_ISL_1459764, EPI_ISL_1459765, EPI_ISL_1459767,
EPI_ISL_1459768, EPI_ISL_1459769, EPI_ISL_1459771, EPI_ISL_1459772, EPI_ISL_1459773, EPI_ISL_1459775, EPI_ISL_1459776, EPI_ISL_1459777, EPI_ISL_1459778, EPI_ISL_1459779, EPI_ISL_1459780, EPI_ISL_1459782, EPI_ISL_1459783, EPI_ISL_1459784, EPI_ISL_1459785, EPI_ISL_1459786, EPI_ISL_1459787, EPI_ISL_1459788, EPI_ISL_1459789, EPI_ISL_1459790, EPI_ISL_1459791, EPI_ISL_1459792,
EPI_ISL_1459793, EPI_ISL_1459794, EPI_ISL_1459795, EPI_ISL_1459796, EPI_ISL_1459797, EPI_ISL_1459798, EPI_ISL_1459799, EPI_ISL_1459800, EPI_ISL_1459801, EPI_ISL_1459802, EPI_ISL_1459803, EPI_ISL_1459805, EPI_ISL_1459806, EPI_ISL_1459807, EPI_ISL_1459808, EPI_ISL_1459809, EPI_ISL_1459810, EPI_ISL_1459811, EPI_ISL_1459812, EPI_ISL_1459813, EPI_ISL_1459814, EPI_ISL_1459815,
EPI_ISL_1459816, EPI_ISL_1459819, EPI_ISL_1459820, EPI_ISL_1459823, EPI_ISL_1459824, EPI_ISL_1459825, EPI_ISL_1459827, EPI_ISL_1459830, EPI_ISL_1459831, EPI_ISL_1459832, EPI_ISL_1459833, EPI_ISL_1459834, EPI_ISL_1459835, EPI_ISL_1459836, EPI_ISL_1459837, EPI_ISL_1459838, EPI_ISL_1459839, EPI_ISL_1459840, EPI_ISL_1459841, EPI_ISL_1459842, EPI_ISL_1459843, EPI_ISL_1459844,
EPI_ISL_1459845, EPI_ISL_1459846, EPI_ISL_1459847, EPI_ISL_1459848, EPI_ISL_1459849, EPI_ISL_1459850, EPI_ISL_1459852, EPI_ISL_1459854, EPI_ISL_1459855, EPI_ISL_1459856, EPI_ISL_1459857, EPI_ISL_1459858, EPI_ISL_1459859, EPI_ISL_1459860, EPI_ISL_1459861, EPI_ISL_1459862, EPI_ISL_1459863, EPI_ISL_1459864, EPI_ISL_1459865, EPI_ISL_1459866, EPI_ISL_1459867, EPI_ISL_1459868,
EPI_ISL_1459869, EPI_ISL_1459870, EPI_ISL_1459871, EPI_ISL_1459872, EPI_ISL_1459876, EPI_ISL_1459877, EPI_ISL_1459878, EPI_ISL_1459879, EPI_ISL_1459880, EPI_ISL_1459881, EPI_ISL_1459882, EPI_ISL_1459883, EPI_ISL_1459884, EPI_ISL_1459885, EPI_ISL_1459887, EPI_ISL_1459889, EPI_ISL_1459890, EPI_ISL_1459891, EPI_ISL_1459892, EPI_ISL_1459893, EPI_ISL_1459894, EPI_ISL_1459895,
EPI_ISL_1459896, EPI_ISL_1459897, EPI_ISL_1459898, EPI_ISL_1459899, EPI_ISL_1459900, EPI_ISL_1459901, EPI_ISL_1459902, EPI_ISL_1459903, EPI_ISL_1459904, EPI_ISL_1459905, EPI_ISL_1459906, EPI_ISL_1459907, EPI_ISL_1459908, EPI_ISL_1459910, EPI_ISL_1459911, EPI_ISL_1459914, EPI_ISL_1459915, EPI_ISL_1459916, EPI_ISL_1459917, EPI_ISL_1459918, EPI_ISL_1459919, EPI_ISL_1459921,
EPI_ISL_1459922, EPI_ISL_1459923, EPI_ISL_1459924, EPI_ISL_1459925, EPI_ISL_1459926, EPI_ISL_1459928, EPI_ISL_1459930, EPI_ISL_1459931, EPI_ISL_1459933, EPI_ISL_1459934, EPI_ISL_1459935, EPI_ISL_1459936, EPI_ISL_1459938, EPI_ISL_1459939, EPI_ISL_1459941, EPI_ISL_1459942, EPI_ISL_1459943, EPI_ISL_1459944, EPI_ISL_1459945, EPI_ISL_1459946, EPI_ISL_1459947, EPI_ISL_1459948,
EPI_ISL_1459949, EPI_ISL_1459950, EPI_ISL_1459951, EPI_ISL_1459952, EPI_ISL_1459953, EPI_ISL_1459954, EPI_ISL_1459955, EPI_ISL_1459956, EPI_ISL_1459957, EPI_ISL_1459958, EPI_ISL_1459959, EPI_ISL_1459960, EPI_ISL_1459961, EPI_ISL_1459962, EPI_ISL_1459963, EPI_ISL_1459964, EPI_ISL_1459965, EPI_ISL_1459966, EPI_ISL_1459967, EPI_ISL_1459968, EPI_ISL_1459970, EPI_ISL_1459971,
EPI_ISL_1459972, EPI_ISL_1459973, EPI_ISL_1459974, EPI_ISL_1459975, EPI_ISL_1459976, EPI_ISL_1459977, EPI_ISL_1459978, EPI_ISL_1459979, EPI_ISL_1459980, EPI_ISL_1459982, EPI_ISL_1459983, EPI_ISL_1459984, EPI_ISL_1459985, EPI_ISL_1459986, EPI_ISL_1459987, EPI_ISL_1459988, EPI_ISL_1459989, EPI_ISL_1459990, EPI_ISL_1459991, EPI_ISL_1459992, EPI_ISL_1459993, EPI_ISL_1459994,
EPI_ISL_1459995, EPI_ISL_1459996, EPI_ISL_1459997, EPI_ISL_1459998, EPI_ISL_1460000, EPI_ISL_1460001, EPI_ISL_1460002, EPI_ISL_1460003, EPI_ISL_1460004, EPI_ISL_1460005, EPI_ISL_1460006, EPI_ISL_1460007, EPI_ISL_1460008, EPI_ISL_1460009, EPI_ISL_1460010, EPI_ISL_1460011, EPI_ISL_1460012, EPI_ISL_1460013, EPI_ISL_1460014, EPI_ISL_1460015, EPI_ISL_1460016, EPI_ISL_1460017,
EPI_ISL_1460018, EPI_ISL_1460019, EPI_ISL_1460020, EPI_ISL_1460021, EPI_ISL_1460022, EPI_ISL_1460023, EPI_ISL_1460024, EPI_ISL_1460025, EPI_ISL_1460026, EPI_ISL_1460027, EPI_ISL_1460028, EPI_ISL_1460029, EPI_ISL_1460030, EPI_ISL_1460031, EPI_ISL_1460032, EPI_ISL_1460034, EPI_ISL_1460035, EPI_ISL_1460036, EPI_ISL_1460037, EPI_ISL_1460038, EPI_ISL_1460040, EPI_ISL_1460042,
EPI_ISL_1460043, EPI_ISL_1460044, EPI_ISL_1460045, EPI_ISL_1460046, EPI_ISL_1460047, EPI_ISL_1460048, EPI_ISL_1460049, EPI_ISL_1460050, EPI_ISL_1460051, EPI_ISL_1460052, EPI_ISL_1460053, EPI_ISL_1460054, EPI_ISL_1460055, EPI_ISL_1460056, EPI_ISL_1460057, EPI_ISL_1460058, EPI_ISL_1460059, EPI_ISL_1460060, EPI_ISL_1460061, EPI_ISL_1460062, EPI_ISL_1460063, EPI_ISL_1460064,
EPI_ISL_1460065, EPI_ISL_1460066, EPI_ISL_1460067, EPI_ISL_1460068, EPI_ISL_1460069, EPI_ISL_1460070, EPI_ISL_1460071, EPI_ISL_1460072, EPI_ISL_1460073, EPI_ISL_1460074, EPI_ISL_1460075, EPI_ISL_1460076, EPI_ISL_1460077, EPI_ISL_1460078, EPI_ISL_1460079, EPI_ISL_1460080, EPI_ISL_1460081, EPI_ISL_1460082, EPI_ISL_1460083, EPI_ISL_1460084, EPI_ISL_1460085, EPI_ISL_1460086,
EPI_ISL_1460087, EPI_ISL_1460088, EPI_ISL_1460089, EPI_ISL_1460090, EPI_ISL_1460091, EPI_ISL_1460092, EPI_ISL_1460093, EPI_ISL_1460094, EPI_ISL_1460095, EPI_ISL_1460096, EPI_ISL_1460097, EPI_ISL_1460098, EPI_ISL_1460099, EPI_ISL_1460100, EPI_ISL_1460101, EPI_ISL_1460102, EPI_ISL_1460103, EPI_ISL_1460104, EPI_ISL_1460105, EPI_ISL_1460106, EPI_ISL_1460107, EPI_ISL_1460108,
EPI_ISL_1460109, EPI_ISL_1460110, EPI_ISL_1460111, EPI_ISL_1460112, EPI_ISL_1460113, EPI_ISL_1460114, EPI_ISL_1460115, EPI_ISL_1460116, EPI_ISL_1460117, EPI_ISL_1460118, EPI_ISL_1460119, EPI_ISL_1460120, EPI_ISL_1460122, EPI_ISL_1460123, EPI_ISL_1460124, EPI_ISL_1460125, EPI_ISL_1460126, EPI_ISL_1460127, EPI_ISL_1460128, EPI_ISL_1460129, EPI_ISL_1460130, EPI_ISL_1460131,
EPI_ISL_1460132, EPI_ISL_1460133, EPI_ISL_1460135, EPI_ISL_1460136, EPI_ISL_1460137, EPI_ISL_1460138, EPI_ISL_1460139, EPI_ISL_1460140, EPI_ISL_1460141, EPI_ISL_1460143, EPI_ISL_1460144, EPI_ISL_1460145, EPI_ISL_1460146, EPI_ISL_1460147, EPI_ISL_1460148, EPI_ISL_1460149, EPI_ISL_1460150, EPI_ISL_1460151, EPI_ISL_1460152, EPI_ISL_1460153, EPI_ISL_1460154, EPI_ISL_1460155,
EPI_ISL_1460156, EPI_ISL_1460157, EPI_ISL_1460158, EPI_ISL_1460159, EPI_ISL_1460160, EPI_ISL_1460161, EPI_ISL_1460162, EPI_ISL_1460163, EPI_ISL_1460164, EPI_ISL_1460165, EPI_ISL_1460166, EPI_ISL_1460167, EPI_ISL_1460168, EPI_ISL_1460169, EPI_ISL_1460170, EPI_ISL_1460171, EPI_ISL_1460172, EPI_ISL_1460173, EPI_ISL_1460174, EPI_ISL_1460175, EPI_ISL_1460176, EPI_ISL_1460177,
EPI_ISL_1460178, EPI_ISL_1460179, EPI_ISL_1460180, EPI_ISL_1460181, EPI_ISL_1460182, EPI_ISL_1460183, EPI_ISL_1460184, EPI_ISL_1460185, EPI_ISL_1460186, EPI_ISL_1460187, EPI_ISL_1460188, EPI_ISL_1460190, EPI_ISL_1460191, EPI_ISL_1460193, EPI_ISL_1460194, EPI_ISL_1460195, EPI_ISL_1460196, EPI_ISL_1460197, EPI_ISL_1460198, EPI_ISL_1460199, EPI_ISL_1460200, EPI_ISL_1460201,
EPI_ISL_1460202, EPI_ISL_1460203, EPI_ISL_1460204, EPI_ISL_1460205, EPI_ISL_1460206, EPI_ISL_1460207, EPI_ISL_1460208, EPI_ISL_1460209, EPI_ISL_1460210, EPI_ISL_1460211, EPI_ISL_1460212, EPI_ISL_1460213, EPI_ISL_1460214, EPI_ISL_1460215, EPI_ISL_1460216, EPI_ISL_1460217, EPI_ISL_1460218, EPI_ISL_1460220, EPI_ISL_1460221, EPI_ISL_1460222, EPI_ISL_1460223, EPI_ISL_1460224,
EPI_ISL_1460225, EPI_ISL_1460226, EPI_ISL_1460227, EPI_ISL_1460228, EPI_ISL_1460229, EPI_ISL_1460230, EPI_ISL_1460231, EPI_ISL_1460232, EPI_ISL_1460233, EPI_ISL_1460234, EPI_ISL_1460235, EPI_ISL_1460236, EPI_ISL_1460237, EPI_ISL_1460238, EPI_ISL_1460239, EPI_ISL_1460240, EPI_ISL_1460241, EPI_ISL_1460242, EPI_ISL_1460243, EPI_ISL_1460244, EPI_ISL_1460245, EPI_ISL_1460246,
EPI_ISL_1460247, EPI_ISL_1460248, EPI_ISL_1460249, EPI_ISL_1460251, EPI_ISL_1460252, EPI_ISL_1460253, EPI_ISL_1460254, EPI_ISL_1460255, EPI_ISL_1460256, EPI_ISL_1460258, EPI_ISL_1460259, EPI_ISL_1460260, EPI_ISL_1460261, EPI_ISL_1460262, EPI_ISL_1460263, EPI_ISL_1460264, EPI_ISL_1460266, EPI_ISL_1460267, EPI_ISL_1460268, EPI_ISL_1460269, EPI_ISL_1460270, EPI_ISL_1460271,
EPI_ISL_1460272, EPI_ISL_1460274, EPI_ISL_1460275, EPI_ISL_1460277, EPI_ISL_1460278, EPI_ISL_1460279, EPI_ISL_1460280, EPI_ISL_1460281, EPI_ISL_1460282, EPI_ISL_1460283, EPI_ISL_1460285, EPI_ISL_1460286, EPI_ISL_1460287, EPI_ISL_1460288, EPI_ISL_1460290, EPI_ISL_1460291, EPI_ISL_1460292, EPI_ISL_1460293, EPI_ISL_1460294, EPI_ISL_1460295, EPI_ISL_1460296, EPI_ISL_1460297,
EPI_ISL_1460298, EPI_ISL_1460299, EPI_ISL_1460301, EPI_ISL_1460302, EPI_ISL_1460304, EPI_ISL_1460305, EPI_ISL_1460307, EPI_ISL_1460308, EPI_ISL_1460309, EPI_ISL_1460310, EPI_ISL_1460311, EPI_ISL_1460312, EPI_ISL_1460313, EPI_ISL_1460314, EPI_ISL_1460316, EPI_ISL_1460317, EPI_ISL_1460318, EPI_ISL_1460319, EPI_ISL_1460320, EPI_ISL_1460321, EPI_ISL_1460323, EPI_ISL_1460324,
EPI_ISL_1460325, EPI_ISL_1460326, EPI_ISL_1460327, EPI_ISL_1460328, EPI_ISL_1460329, EPI_ISL_1460330, EPI_ISL_1460331, EPI_ISL_1460332, EPI_ISL_1460333, EPI_ISL_1460335, EPI_ISL_1460337, EPI_ISL_1460338, EPI_ISL_1460342, EPI_ISL_1460343, EPI_ISL_1460344, EPI_ISL_1460345, EPI_ISL_1460346, EPI_ISL_1460347, EPI_ISL_1460348, EPI_ISL_1460349, EPI_ISL_1460350, EPI_ISL_1460351,

| | | | |
|---|---|---|---|
| EPI_ISL_1460352, EPI_ISL_1460353, EPI_ISL_1460354, EPI_ISL_1460355, EPI_ISL_1460356, EPI_ISL_1460357, EPI_ISL_1460358, EPI_ISL_1460359, EPI_ISL_1460360, EPI_ISL_1460361, EPI_ISL_1460362, EPI_ISL_1460363, EPI_ISL_1460364, EPI_ISL_1460365, EPI_ISL_1460367, EPI_ISL_1460368, EPI_ISL_1460369, EPI_ISL_1460370, EPI_ISL_1460371, EPI_ISL_1460372, EPI_ISL_1460373, EPI_ISL_1460374, EPI_ISL_1460375, EPI_ISL_1460376, EPI_ISL_1460377, EPI_ISL_1460378, EPI_ISL_1460379, EPI_ISL_1460380, EPI_ISL_1460381, EPI_ISL_1460382, EPI_ISL_1460383, EPI_ISL_1460384, EPI_ISL_1460385, EPI_ISL_1460386, EPI_ISL_1460387, EPI_ISL_1460388, EPI_ISL_1460389, EPI_ISL_1460390, EPI_ISL_1460391, EPI_ISL_1460392, EPI_ISL_1460393, EPI_ISL_1460394, EPI_ISL_1460395, EPI_ISL_1460396, EPI_ISL_1460397, EPI_ISL_1460398, EPI_ISL_1460399, EPI_ISL_1460400, EPI_ISL_1460401, EPI_ISL_1460402, EPI_ISL_1460403, EPI_ISL_1460404, EPI_ISL_1460405, EPI_ISL_1460406, EPI_ISL_1460407, EPI_ISL_1460408, EPI_ISL_1460409, EPI_ISL_1460410, EPI_ISL_1460411, EPI_ISL_1460412, EPI_ISL_1460413, EPI_ISL_1460414, EPI_ISL_1460415, EPI_ISL_1460416, EPI_ISL_1460417, EPI_ISL_1460418, EPI_ISL_1460420, EPI_ISL_1460421, EPI_ISL_1460422, EPI_ISL_1460423, EPI_ISL_1460424, EPI_ISL_1460425, EPI_ISL_1460427, EPI_ISL_1460428, EPI_ISL_1460429, EPI_ISL_1460430, EPI_ISL_1460431, EPI_ISL_1460434, EPI_ISL_1460435, EPI_ISL_1460436, EPI_ISL_1460437, EPI_ISL_1460438, EPI_ISL_1460440, EPI_ISL_1460442, EPI_ISL_1460443, EPI_ISL_1460444, EPI_ISL_1460445, EPI_ISL_1460446, EPI_ISL_1460447, EPI_ISL_1460449, EPI_ISL_1460450, EPI_ISL_1460451, EPI_ISL_1460453, EPI_ISL_1460454, EPI_ISL_1460455, EPI_ISL_1460456, EPI_ISL_1460457, EPI_ISL_1460458, EPI_ISL_1460459, EPI_ISL_1460460, EPI_ISL_1460461, EPI_ISL_1460463, EPI_ISL_1460464, EPI_ISL_1460465, EPI_ISL_1460466, EPI_ISL_1460467, EPI_ISL_1460468, EPI_ISL_1460470, EPI_ISL_1460471, EPI_ISL_1460472, EPI_ISL_1460474, EPI_ISL_1460475, EPI_ISL_1460477, EPI_ISL_1460478, EPI_ISL_1460480, EPI_ISL_1460481, EPI_ISL_1460482, EPI_ISL_1460483, EPI_ISL_1460484, EPI_ISL_1460485, EPI_ISL_1460486, EPI_ISL_1460488, EPI_ISL_1460489, EPI_ISL_1460490, EPI_ISL_1460491, EPI_ISL_1460493, EPI_ISL_1460494, EPI_ISL_1460495, EPI_ISL_1460496, EPI_ISL_1460497, EPI_ISL_1460498, EPI_ISL_1460499, EPI_ISL_1460500, EPI_ISL_1460501, EPI_ISL_1460502, EPI_ISL_1460503, EPI_ISL_1460504, EPI_ISL_1460505, EPI_ISL_1460506, EPI_ISL_1460507, EPI_ISL_1460508, EPI_ISL_1460509, EPI_ISL_1460510, EPI_ISL_1460511, EPI_ISL_1460512, EPI_ISL_1460514, EPI_ISL_1460515, EPI_ISL_1460516, EPI_ISL_1460517, EPI_ISL_1460518, EPI_ISL_1460519, EPI_ISL_1460520, EPI_ISL_1460521, EPI_ISL_1460522, EPI_ISL_1460523, EPI_ISL_1460524, EPI_ISL_1460525, EPI_ISL_1460526, EPI_ISL_1460527, EPI_ISL_1460528, EPI_ISL_1460529, EPI_ISL_1460530, EPI_ISL_1460531, EPI_ISL_1460532, EPI_ISL_1460533, EPI_ISL_1460534, EPI_ISL_1460535, EPI_ISL_1460536, EPI_ISL_1460537, EPI_ISL_1460538, EPI_ISL_1460539, EPI_ISL_1460540, EPI_ISL_1460541, EPI_ISL_1460542, EPI_ISL_1460543, EPI_ISL_1460544, EPI_ISL_1460545, EPI_ISL_1460547, EPI_ISL_1460548, EPI_ISL_1460549, EPI_ISL_1460550, EPI_ISL_1460551, EPI_ISL_1460552, EPI_ISL_1460554, EPI_ISL_1460555, EPI_ISL_1460556, EPI_ISL_1460557, EPI_ISL_1460558, EPI_ISL_1460559, EPI_ISL_1460560, EPI_ISL_1460561, EPI_ISL_1460562, EPI_ISL_1460563, EPI_ISL_1460565, EPI_ISL_1460566, EPI_ISL_1460567, EPI_ISL_1460568, EPI_ISL_1460569, EPI_ISL_1460571, EPI_ISL_1460572, EPI_ISL_1460573, EPI_ISL_1460574, EPI_ISL_1460575, EPI_ISL_1460576, EPI_ISL_1460577, EPI_ISL_1460578, EPI_ISL_1460579, EPI_ISL_1460580, EPI_ISL_1460581, EPI_ISL_1460582, EPI_ISL_1460583, EPI_ISL_1460584, EPI_ISL_1460585, EPI_ISL_1460587, EPI_ISL_1460589, EPI_ISL_1460590, EPI_ISL_1460591, EPI_ISL_1460592, EPI_ISL_1460593, EPI_ISL_1460594, EPI_ISL_1460595, EPI_ISL_1460596, EPI_ISL_1460597, EPI_ISL_1460598, EPI_ISL_1460599, EPI_ISL_1460600, EPI_ISL_1460601, EPI_ISL_1460602, EPI_ISL_1460603, EPI_ISL_1460604, EPI_ISL_1460605, EPI_ISL_1460606, EPI_ISL_1460607, EPI_ISL_1460608, EPI_ISL_1460610, EPI_ISL_1460611, EPI_ISL_1460613, EPI_ISL_1460614, EPI_ISL_1460615, EPI_ISL_1460616, EPI_ISL_1460617, EPI_ISL_1460618, EPI_ISL_1460619, EPI_ISL_1460620, EPI_ISL_1460621, EPI_ISL_1460622, EPI_ISL_1460624, EPI_ISL_1460625, EPI_ISL_1460627, EPI_ISL_1460628, EPI_ISL_1460629, EPI_ISL_1460630, EPI_ISL_1460631, EPI_ISL_1460632, EPI_ISL_1460633, EPI_ISL_1460635, EPI_ISL_1460636, EPI_ISL_1460637, EPI_ISL_1460638, EPI_ISL_1460639, EPI_ISL_1460640, EPI_ISL_1460641, EPI_ISL_1460642, EPI_ISL_1460643, EPI_ISL_1460644, EPI_ISL_1460645, EPI_ISL_1460646, EPI_ISL_1460647, EPI_ISL_1460648, EPI_ISL_1460649, EPI_ISL_1460650, EPI_ISL_1460651, EPI_ISL_1460652, EPI_ISL_1460653, EPI_ISL_1460654, EPI_ISL_1460655, EPI_ISL_1460656, EPI_ISL_1460657, EPI_ISL_1460659, EPI_ISL_1460660, EPI_ISL_1460661, EPI_ISL_1460662, EPI_ISL_1460663, EPI_ISL_1460664, EPI_ISL_1460665, EPI_ISL_1460667, EPI_ISL_1460668, EPI_ISL_1460669, EPI_ISL_1460670, EPI_ISL_1460671, EPI_ISL_1460672, EPI_ISL_1460673, EPI_ISL_1460675, EPI_ISL_1460676, EPI_ISL_1460677, EPI_ISL_1460678, EPI_ISL_1460679, EPI_ISL_1460680, EPI_ISL_1460681, EPI_ISL_1460682, EPI_ISL_1460684, EPI_ISL_1460685, EPI_ISL_1460687, EPI_ISL_1460688, EPI_ISL_1460689, EPI_ISL_1460690, EPI_ISL_1460691, EPI_ISL_1460692, EPI_ISL_1460694, EPI_ISL_1460695, EPI_ISL_1460696, EPI_ISL_1460697, EPI_ISL_1460698, EPI_ISL_1460699, EPI_ISL_1460700, EPI_ISL_1460701, EPI_ISL_1460702, EPI_ISL_1460703, EPI_ISL_1460704, EPI_ISL_1460705, EPI_ISL_1460706, EPI_ISL_1460707, EPI_ISL_1460708, EPI_ISL_1460709, EPI_ISL_1460710, EPI_ISL_1460711, EPI_ISL_1460712, EPI_ISL_1460713, EPI_ISL_1460714, EPI_ISL_1460715, EPI_ISL_1460717, EPI_ISL_1460718, EPI_ISL_1460719, EPI_ISL_1460720, EPI_ISL_1460721, EPI_ISL_1460722, EPI_ISL_1460723, EPI_ISL_1460724, EPI_ISL_1460725, EPI_ISL_1460727, EPI_ISL_1460729 | | | |
| see above | Fulgent Genetics | Centers for Disease Control and Prevention Division of Viral Diseases, Pathogen Discovery | Dakota Howard, Dhwani Batra, Peter W. Cook, Kara Moser, Adrian Paskey, Jason Caravas, Benjamin Rambo-Martin, Shatavia Morrison, Christopher Gulvick, Scott Sammons, Yvette Unoarumhi, Darlene Wagner, Matthew Schmerer, Harry Gao, Mickey Li, John Gao, Joseph Fierro, Benafsh Sapra, Becky Tsai, Yan Meng, Doreen Ng, James Xie, Clinton R. Paden, Duncan MacCannell |
| EPI_ISL_1460730, EPI_ISL_1460731, EPI_ISL_1460732, EPI_ISL_1460733, EPI_ISL_1460734, EPI_ISL_1460735, EPI_ISL_1460736, EPI_ISL_1460737, EPI_ISL_1460738, EPI_ISL_1460739, EPI_ISL_1460740, EPI_ISL_1460741, EPI_ISL_1460742, EPI_ISL_1460743, EPI_ISL_1460744, EPI_ISL_1460745, EPI_ISL_1460746, EPI_ISL_1460747, EPI_ISL_1460748, EPI_ISL_1460749, EPI_ISL_1460750, EPI_ISL_1460751, EPI_ISL_1460752, EPI_ISL_1460753, EPI_ISL_1460754, EPI_ISL_1460755, EPI_ISL_1460756, EPI_ISL_1460757, EPI_ISL_1460758, EPI_ISL_1460759, EPI_ISL_1460760, EPI_ISL_1460761, EPI_ISL_1460762, EPI_ISL_1460763, EPI_ISL_1460764, EPI_ISL_1460765, EPI_ISL_1460766, EPI_ISL_1460767, EPI_ISL_1460768, EPI_ISL_1460769, EPI_ISL_1460770, EPI_ISL_1460771, EPI_ISL_1460772, EPI_ISL_1460773, EPI_ISL_1460774, EPI_ISL_1460775, EPI_ISL_1460776, EPI_ISL_1460777, EPI_ISL_1460778, EPI_ISL_1460779, EPI_ISL_1460780, EPI_ISL_1460781, EPI_ISL_1460782, EPI_ISL_1460783, EPI_ISL_1460784, EPI_ISL_1460785 | | | |
| see above | Quest Diagnostics Incorporated | Centers for Disease Control and Prevention Division of Viral Diseases, Pathogen Discovery | Dakota Howard, Dhwani Batra, Peter W. Cook, Kara Moser, Adrian Paskey, Jason Caravas, Benjamin Rambo-Martin, Shatavia Morrison, Christopher Gulvick, Scott Sammons, Yvette Unoarumhi, Darlene Wagner, Matthew Schmerer, S. H. Rosenthal, A. Gerasimova, R. M. Kagan, B. Anderson, M. Hua, Y. Liu, L.E. Bernstein, K.E. Livingston, A. Perez, I. A. Shlyakhter, R. V. Rolando, R. Owen, P. Tanpaiboon, F. Lacbawan, Clinton R. Paden, Duncan MacCannell |
| EPI_ISL_1460786, EPI_ISL_1460787, EPI_ISL_1460788, EPI_ISL_1460789, EPI_ISL_1460790, EPI_ISL_1460791, EPI_ISL_1460792, EPI_ISL_1460793, EPI_ISL_1460794, EPI_ISL_1460795, EPI_ISL_1460796, EPI_ISL_1460797, EPI_ISL_1460798, EPI_ISL_1460799, EPI_ISL_1460800, EPI_ISL_1460801, EPI_ISL_1460802, EPI_ISL_1460803, EPI_ISL_1460804, EPI_ISL_1460805, EPI_ISL_1460806, EPI_ISL_1460807, EPI_ISL_1460808, EPI_ISL_1460809, EPI_ISL_1460810, EPI_ISL_1460811, EPI_ISL_1460812, EPI_ISL_1460813, EPI_ISL_1460814, EPI_ISL_1460815, EPI_ISL_1460816, EPI_ISL_1460817, EPI_ISL_1460818, EPI_ISL_1460819, EPI_ISL_1460820, EPI_ISL_1460821, EPI_ISL_1460822, EPI_ISL_1460823, EPI_ISL_1460824, EPI_ISL_1460825, EPI_ISL_1460826, EPI_ISL_1460827, EPI_ISL_1460828, EPI_ISL_1460829, EPI_ISL_1460830, EPI_ISL_1460831, EPI_ISL_1460832, EPI_ISL_1460833, EPI_ISL_1460834, EPI_ISL_1460835, EPI_ISL_1460836, EPI_ISL_1460837, EPI_ISL_1460838, EPI_ISL_1460839, EPI_ISL_1460840, EPI_ISL_1460841, EPI_ISL_1460842, EPI_ISL_1460843, EPI_ISL_1460844, EPI_ISL_1460845, EPI_ISL_1460846, EPI_ISL_1460847, EPI_ISL_1460848, EPI_ISL_1460849, EPI_ISL_1460850, EPI_ISL_1460851, EPI_ISL_1460852, EPI_ISL_1460853, EPI_ISL_1460854, EPI_ISL_1460855, EPI_ISL_1460856, EPI_ISL_1460857, EPI_ISL_1460858, EPI_ISL_1460859, EPI_ISL_1460860, EPI_ISL_1460861, EPI_ISL_1460862, EPI_ISL_1460863, EPI_ISL_1460864, EPI_ISL_1460865, EPI_ISL_1460866, EPI_ISL_1460867, EPI_ISL_1460868, EPI_ISL_1460869, EPI_ISL_1460870, EPI_ISL_1460871, EPI_ISL_1460872, EPI_ISL_1460873, EPI_ISL_1460874, EPI_ISL_1460875, EPI_ISL_1460876, EPI_ISL_1460877, EPI_ISL_1460878, EPI_ISL_1460879, EPI_ISL_1460880, EPI_ISL_1460881, EPI_ISL_1460882, EPI_ISL_1460883, EPI_ISL_1460884, EPI_ISL_1460885, EPI_ISL_1460886, EPI_ISL_1460887, EPI_ISL_1460888, EPI_ISL_1460889, EPI_ISL_1460890, EPI_ISL_1460891, EPI_ISL_1460892, EPI_ISL_1460893, EPI_ISL_1460894, EPI_ISL_1460895, EPI_ISL_1460896, EPI_ISL_1460897, EPI_ISL_1460898, EPI_ISL_1460899, EPI_ISL_1460900, EPI_ISL_1460901, EPI_ISL_1460902, EPI_ISL_1460903, EPI_ISL_1460904, EPI_ISL_1460905, EPI_ISL_1460906, EPI_ISL_1460907, EPI_ISL_1460908, EPI_ISL_1460909, EPI_ISL_1460910, EPI_ISL_1460911, EPI_ISL_1460912, EPI_ISL_1460913, EPI_ISL_1460914, EPI_ISL_1460915, EPI_ISL_1460916, EPI_ISL_1460917, EPI_ISL_1460918, EPI_ISL_1460919, EPI_ISL_1460920, EPI_ISL_1460921, EPI_ISL_1460922, EPI_ISL_1460923, EPI_ISL_1460924, EPI_ISL_1460925, EPI_ISL_1460926, EPI_ISL_1460927, EPI_ISL_1460928, EPI_ISL_1460929, EPI_ISL_1460930, EPI_ISL_1460931, EPI_ISL_1460932, EPI_ISL_1460933, EPI_ISL_1460934, EPI_ISL_1460935, EPI_ISL_1460936, EPI_ISL_1460937, EPI_ISL_1460938, EPI_ISL_1460939, EPI_ISL_1460940, EPI_ISL_1460941, EPI_ISL_1460942, EPI_ISL_1460943, EPI_ISL_1460944, EPI_ISL_1460945, EPI_ISL_1460946, EPI_ISL_1460947, EPI_ISL_1460948, EPI_ISL_1460949, EPI_ISL_1460950, EPI_ISL_1460951, EPI_ISL_1460952, EPI_ISL_1460953, EPI_ISL_1460954, EPI_ISL_1460955, EPI_ISL_1460956, EPI_ISL_1460957, EPI_ISL_1460958, EPI_ISL_1460959, EPI_ISL_1460960, EPI_ISL_1460961, EPI_ISL_1460962, EPI_ISL_1460963, EPI_ISL_1460964, EPI_ISL_1460965, EPI_ISL_1460966, EPI_ISL_1460967, EPI_ISL_1460968, EPI_ISL_1460969, EPI_ISL_1460970, EPI_ISL_1460971, EPI_ISL_1460972, EPI_ISL_1460973, EPI_ISL_1460974, EPI_ISL_1460975, EPI_ISL_1460976, EPI_ISL_1460978, EPI_ISL_1460979, EPI_ISL_1460980, EPI_ISL_1460981, EPI_ISL_1460982, EPI_ISL_1460983, EPI_ISL_1460984, EPI_ISL_1460985, EPI_ISL_1460986, EPI_ISL_1460987, EPI_ISL_1460988, EPI_ISL_1460989, EPI_ISL_1460990, EPI_ISL_1460991, EPI_ISL_1460992, EPI_ISL_1460993, EPI_ISL_1460994, EPI_ISL_1460995, EPI_ISL_1460996, EPI_ISL_1460997, EPI_ISL_1460998, EPI_ISL_1460999, EPI_ISL_1461000, EPI_ISL_1461001, EPI_ISL_1461002 | | | |
| see above | Helix/Illumina | Centers for Disease Control and Prevention Division of Viral Diseases, Pathogen Discovery | Dakota Howard, Dhwani Batra, Peter W. Cook, Kara Moser, Adrian Paskey, Jason Caravas, Benjamin Rambo-Martin, Shatavia Morrison, Christopher Gulvick, Scott Sammons, Yvette Unoarumhi, Darlene Wagner, Matthew Schmerer, Eileen de Feo, Jan Antico, Christine Tran, Matthew Tolentino, Shannon Wickline, Kim Gietzen, Brad Sickler, Jingtao Liu, Eric Allen, Phil Febbo, Nicole L. Washington, Simon White, Geraint Levan, Kelly Schiabor Barrett, Elizabeth Cirulli, Alexandre Bolze, Ary Ascencio, Charlotte Rivera-Garcia, Ryan Cho, Jason Nguyen, Sherry Wang, Jimmy Ramirez, Tyler Cassens, Efren Sandoval, Magnus Isaksson, William Lee, David Becker, Marc Laurent, James Lu, Clinton R. Paden, Duncan MacCannell |
| EPI_ISL_1461278, EPI_ISL_1461279, EPI_ISL_1461280, EPI_ISL_1461281, EPI_ISL_1461282, EPI_ISL_1461283, EPI_ISL_1461284, EPI_ISL_1461287, EPI_ISL_1461288, EPI_ISL_1461289 | Michigan Department of Health and Human Services, Bureau of Laboratories | Michigan Department of Health and Human Services, Bureau of Laboratories | Blankenship HM, Riner D, Soehnlen MK |
| EPI_ISL_1461290, EPI_ISL_1461291, EPI_ISL_1461292 | URMC LABS | Wadsworth Center, New York State Department of Health | Kirsten St. George, Daryl M. Lamson, Alexis Russell, Matthew Shudt, Melissa A Leisner, Jonathan Plitnick, Catharine Prussing, Navjot Singh, John Kelly, Erasmus Schneider, Erica Lasek-Nesselquist |
| EPI_ISL_1461293 | GLENS FALLS HOSPITAL LABORATORY | Wadsworth Center, New York State Department of Health | Kirsten St. George, Daryl M. Lamson, Alexis Russell, Matthew Shudt, Melissa A Leisner, Jonathan Plitnick, Catharine Prussing, Navjot Singh, John Kelly, Erasmus Schneider, Erica Lasek-Nesselquist |
| EPI_ISL_1461294 | SUNY UPSTATE MEDICAL UNIVERSITY | Wadsworth Center, New York State Department of Health | Kirsten St. George, Daryl M. Lamson, Alexis Russell, Matthew Shudt, Melissa A Leisner, Jonathan Plitnick, Catharine Prussing, Navjot Singh, John Kelly, Erasmus Schneider, Erica Lasek-Nesselquist |
| EPI_ISL_1461295, EPI_ISL_1461296, EPI_ISL_1461297, EPI_ISL_1461298, EPI_ISL_1461299 | ALBANY MEDICAL CENTER | Wadsworth Center, New York State Department of Health | Kirsten St. George, Daryl M. Lamson, Alexis Russell, Matthew Shudt, Melissa A Leisner, Jonathan Plitnick, Catharine Prussing, Navjot Singh, John Kelly, Erasmus Schneider, Erica Lasek-Nesselquist |
| EPI_ISL_1461300, EPI_ISL_1461301, EPI_ISL_1461302, EPI_ISL_1461303, EPI_ISL_1461304, EPI_ISL_1461305, EPI_ISL_1461306, EPI_ISL_1461307, EPI_ISL_1461308, EPI_ISL_1461309 | URMC LABS | Wadsworth Center, New York State Department of Health | Kirsten St. George, Daryl M. Lamson, Alexis Russell, Matthew Shudt, Melissa A Leisner, Jonathan Plitnick, Catharine Prussing, Navjot Singh, John Kelly, Erasmus Schneider, Erica Lasek-Nesselquist |
| EPI_ISL_1461310, EPI_ISL_1461311, EPI_ISL_1461312, EPI_ISL_1461313, EPI_ISL_1461314, EPI_ISL_1461315, EPI_ISL_1461316, EPI_ISL_1461317 | SUNY UPSTATE MEDICAL UNIVERSITY | Wadsworth Center, New York State Department of Health | Kirsten St. George, Daryl M. Lamson, Alexis Russell, Matthew Shudt, Melissa A Leisner, Jonathan Plitnick, Catharine Prussing, Navjot Singh, John Kelly, Erasmus Schneider, Erica Lasek-Nesselquist |
| EPI_ISL_1461318 | URMC LABS | Wadsworth Center, New York State Department of Health | Kirsten St. George, Daryl M. Lamson, Alexis Russell, Matthew Shudt, Melissa A Leisner, Jonathan Plitnick, Catharine Prussing, Navjot Singh, John Kelly, Erasmus Schneider, Erica Lasek-Nesselquist |
| EPI_ISL_1461319, EPI_ISL_1461320, EPI_ISL_1461321 | NYC Pandemic Response Lab | Wadsworth Center, New York State Department of Health | Kirsten St. George, Daryl M. Lamson, Alexis Russell, Matthew Shudt, Melissa A Leisner, Jonathan Plitnick, Catharine Prussing, Navjot Singh, John Kelly, Erasmus Schneider, Erica Lasek-Nesselquist |
| EPI_ISL_1461322 | Michigan Department of Health and Human Services, Bureau of Laboratories | Michigan Department of Health and Human Services, Bureau of Laboratories | Blankenship HM, Riner D, Soehnlen MK |
| EPI_ISL_1461323, EPI_ISL_1461324, EPI_ISL_1461325, EPI_ISL_1461326, EPI_ISL_1461327, EPI_ISL_1461328, EPI_ISL_1461329, EPI_ISL_1461330, EPI_ISL_1461331, EPI_ISL_1461332, EPI_ISL_1461333, EPI_ISL_1461334, EPI_ISL_1461335, EPI_ISL_1461336, EPI_ISL_1461337, EPI_ISL_1461338, EPI_ISL_1461339, EPI_ISL_1461340, EPI_ISL_1461341, EPI_ISL_1461342, EPI_ISL_1461343, EPI_ISL_1461344, EPI_ISL_1461345, EPI_ISL_1461346, EPI_ISL_1461347, EPI_ISL_1461348, EPI_ISL_1461349, EPI_ISL_1461350, EPI_ISL_1461351, EPI_ISL_1461352, EPI_ISL_1461353, EPI_ISL_1461354, EPI_ISL_1461355 | | | |
| see above | NYC Pandemic Response Lab | Wadsworth Center, New York State Department of Health | Kirsten St. George, Daryl M. Lamson, Alexis Russell, Matthew Shudt, Melissa A Leisner, Jonathan Plitnick, Catharine Prussing, Navjot Singh, John Kelly, Erasmus Schneider, Erica Lasek-Nesselquist |
| EPI_ISL_1461356, EPI_ISL_1461357, EPI_ISL_1461358, EPI_ISL_1461359, EPI_ISL_1461360, EPI_ISL_1461361, EPI_ISL_1461362, EPI_ISL_1461363, EPI_ISL_1461364, EPI_ISL_1461365, EPI_ISL_1461366, EPI_ISL_1461367, EPI_ISL_1461368, EPI_ISL_1461369, EPI_ISL_1461370, EPI_ISL_1461371, EPI_ISL_1461372, EPI_ISL_1461373, EPI_ISL_1461374, EPI_ISL_1461375, EPI_ISL_1461376, EPI_ISL_1461377 | | | |
| see above | TEMPUS LABS INC | Wadsworth Center, New York State Department of Health | Kirsten St. George, Daryl M. Lamson, Alexis Russell, Matthew Shudt, Melissa A Leisner, Jonathan Plitnick, Catharine Prussing, Navjot Singh, John Kelly, Erasmus Schneider, Erica Lasek-Nesselquist |
| EPI_ISL_1461378, EPI_ISL_1461379, EPI_ISL_1461383, EPI_ISL_1461385, EPI_ISL_1461386, EPI_ISL_1461387, EPI_ISL_1461391, EPI_ISL_1461392, EPI_ISL_1461393, EPI_ISL_1461397, EPI_ISL_1461399, EPI_ISL_1461400, EPI_ISL_1461402, EPI_ISL_1461408, EPI_ISL_1461409, EPI_ISL_1461410, EPI_ISL_1461411, EPI_ISL_1461415, EPI_ISL_1461417, EPI_ISL_1461418, EPI_ISL_1461419, EPI_ISL_1461423, EPI_ISL_1461424, EPI_ISL_1461425, EPI_ISL_1461426, EPI_ISL_1461427, EPI_ISL_1461428, EPI_ISL_1461429, EPI_ISL_1461435, EPI_ISL_1461437, EPI_ISL_1461439, EPI_ISL_1461440, EPI_ISL_1461441, EPI_ISL_1461442, EPI_ISL_1461443, EPI_ISL_1461445, EPI_ISL_1461447, EPI_ISL_1461449, EPI_ISL_1461452, EPI_ISL_1461453, EPI_ISL_1461455, EPI_ISL_1461456, EPI_ISL_1461457, EPI_ISL_1461459, EPI_ISL_1461460, EPI_ISL_1461461, EPI_ISL_1461462, EPI_ISL_1461463, EPI_ISL_1461464, EPI_ISL_1461465, EPI_ISL_1461466, EPI_ISL_1461467, EPI_ISL_1461468, EPI_ISL_1461469, EPI_ISL_1461470, EPI_ISL_1461471, EPI_ISL_1461473, EPI_ISL_1461474, EPI_ISL_1461475, EPI_ISL_1461476, EPI_ISL_1461477, EPI_ISL_1461478, EPI_ISL_1461479, EPI_ISL_1461480, EPI_ISL_1461481, EPI_ISL_1461482, EPI_ISL_1461483, EPI_ISL_1461484, EPI_ISL_1461485, EPI_ISL_1461486, EPI_ISL_1461488, EPI_ISL_1461495, EPI_ISL_1461497, EPI_ISL_1461498, EPI_ISL_1461501, EPI_ISL_1461502, EPI_ISL_1461504, EPI_ISL_1461505, EPI_ISL_1461506, EPI_ISL_1461507, EPI_ISL_1461510, EPI_ISL_1461514, EPI_ISL_1461518, EPI_ISL_1461519, EPI_ISL_1461520, EPI_ISL_1461524, EPI_ISL_1461526, EPI_ISL_1461527, EPI_ISL_1461528, EPI_ISL_1461529, EPI_ISL_1461532, EPI_ISL_1461534, EPI_ISL_1461535, EPI_ISL_1461536, EPI_ISL_1461537, EPI_ISL_1461538, EPI_ISL_1461540, EPI_ISL_1461541, EPI_ISL_1461542, EPI_ISL_1461543, EPI_ISL_1461545, EPI_ISL_1461546, EPI_ISL_1461547, EPI_ISL_1461548, EPI_ISL_1461550, EPI_ISL_1461553, EPI_ISL_1461554, EPI_ISL_1461555, EPI_ISL_1461556, EPI_ISL_1461557, EPI_ISL_1461558, EPI_ISL_1461559, EPI_ISL_1461560, EPI_ISL_1461561, EPI_ISL_1461563, EPI_ISL_1461564, EPI_ISL_1461565, EPI_ISL_1461567, EPI_ISL_1461568, EPI_ISL_1461569, EPI_ISL_1461570, EPI_ISL_1461572, EPI_ISL_1461574, EPI_ISL_1461575, EPI_ISL_1461576, EPI_ISL_1461577, EPI_ISL_1461578, EPI_ISL_1461579, EPI_ISL_1461580, EPI_ISL_1461581, EPI_ISL_1461582, EPI_ISL_1461583, EPI_ISL_1461584, EPI_ISL_1461586, EPI_ISL_1461587, EPI_ISL_1461590, EPI_ISL_1461591, EPI_ISL_1461592, EPI_ISL_1461593, EPI_ISL_1461594, EPI_ISL_1461595, EPI_ISL_1461596, EPI_ISL_1461597, EPI_ISL_1461598, EPI_ISL_1461599, EPI_ISL_1461600, EPI_ISL_1461601, EPI_ISL_1461603, EPI_ISL_1461604, EPI_ISL_1461605, EPI_ISL_1461606, EPI_ISL_1461607, EPI_ISL_1461608, EPI_ISL_1461609, EPI_ISL_1461610, EPI_ISL_1461612, EPI_ISL_1461613, EPI_ISL_1461614, EPI_ISL_1461615, EPI_ISL_1461616, EPI_ISL_1461617, EPI_ISL_1461618, EPI_ISL_1461619, EPI_ISL_1461620, EPI_ISL_1461622, EPI_ISL_1461623, EPI_ISL_1461624, EPI_ISL_1461625 | | | |
| see above | Michigan Department of Health and | Michigan Department of Health and Human | Blankenship HM, Riner D, Soehnlen MK |

| | | | |
|---|---|---|---|
| | Human Services, Bureau of Laboratories | Services, Bureau of Laboratories | |
| EPI_ISL_1461626, EPI_ISL_1461627, EPI_ISL_1461628, EPI_ISL_1461629, EPI_ISL_1461630, EPI_ISL_1461631, EPI_ISL_1461632, EPI_ISL_1461633 | Kansas Health and Environmental Lab | Kansas Health and Environmental Lab | Mike Grose, Jonathan Barnell, Ben Olsen, and Phil Adam |
| EPI_ISL_1461634, EPI_ISL_1461635, EPI_ISL_1461636, EPI_ISL_1461637 | Cytocheck Laboratory | Kansas Health and Environmental Lab | Mike Grose, Jonathan Barnell, Ben Olsen, and Phil Adam |
| EPI_ISL_1461638 | Kansas Health and Environmental Lab | Kansas Health and Environmental Lab | Mike Grose, Jonathan Barnell, Ben Olsen, and Phil Adam |
| EPI_ISL_1461649, EPI_ISL_1461650, EPI_ISL_1461651, EPI_ISL_1461652, EPI_ISL_1461653, EPI_ISL_1461654, EPI_ISL_1461655, EPI_ISL_1461656, EPI_ISL_1461657, EPI_ISL_1461658, EPI_ISL_1461659, EPI_ISL_1461660, EPI_ISL_1461661, EPI_ISL_1461662, EPI_ISL_1461663, EPI_ISL_1461664, EPI_ISL_1461665, EPI_ISL_1461666, EPI_ISL_1461667, EPI_ISL_1461668, EPI_ISL_1461669, EPI_ISL_1461670, EPI_ISL_1461671, EPI_ISL_1461672, EPI_ISL_1461673, EPI_ISL_1461674, EPI_ISL_1461675, EPI_ISL_1461676, EPI_ISL_1461677, EPI_ISL_1461678, EPI_ISL_1461679, EPI_ISL_1461680, EPI_ISL_1461681, EPI_ISL_1461682, EPI_ISL_1461683, EPI_ISL_1461684, EPI_ISL_1461685, EPI_ISL_1461686, EPI_ISL_1461687, EPI_ISL_1461688, EPI_ISL_1461689, EPI_ISL_1461690, EPI_ISL_1461691, EPI_ISL_1461692, EPI_ISL_1461693, EPI_ISL_1461694, EPI_ISL_1461695, EPI_ISL_1461696, EPI_ISL_1461697, EPI_ISL_1461698, EPI_ISL_1461699, EPI_ISL_1461700, EPI_ISL_1461701, EPI_ISL_1461702, EPI_ISL_1461703, EPI_ISL_1461704, EPI_ISL_1461705, EPI_ISL_1461706, EPI_ISL_1461707, EPI_ISL_1461709 | | | |
| see above | Virginia Division of Consolidated Laboratory Services | Virginia Division of Consolidated Laboratory Services | Virginia DCLS |
| EPI_ISL_1461711, EPI_ISL_1461714, EPI_ISL_1461716, EPI_ISL_1461720, EPI_ISL_1461730, EPI_ISL_1461731, EPI_ISL_1461734, EPI_ISL_1461747, EPI_ISL_1461751, EPI_ISL_1461753, EPI_ISL_1461755, EPI_ISL_1461760, EPI_ISL_1461762, EPI_ISL_1461763, EPI_ISL_1461764, EPI_ISL_1461766, EPI_ISL_1461770, EPI_ISL_1461772, EPI_ISL_1461775, EPI_ISL_1461776, EPI_ISL_1461790, EPI_ISL_1461791, EPI_ISL_1461803, EPI_ISL_1461811, EPI_ISL_1461812, EPI_ISL_1461818, EPI_ISL_1461822, EPI_ISL_1461825, EPI_ISL_1461826, EPI_ISL_1461828, EPI_ISL_1461830, EPI_ISL_1461833, EPI_ISL_1461835, EPI_ISL_1461843, EPI_ISL_1461845, EPI_ISL_1461847, EPI_ISL_1461848, EPI_ISL_1461849, EPI_ISL_1461850, EPI_ISL_1461858, EPI_ISL_1461863, EPI_ISL_1461864, EPI_ISL_1461868, EPI_ISL_1461870, EPI_ISL_1461872, EPI_ISL_1461874, EPI_ISL_1461876, EPI_ISL_1461878, EPI_ISL_1461880, EPI_ISL_1461881, EPI_ISL_1461883, EPI_ISL_1461884, EPI_ISL_1461885, EPI_ISL_1461886, EPI_ISL_1461887, EPI_ISL_1461888, EPI_ISL_1461889, EPI_ISL_1461890, EPI_ISL_1461894, EPI_ISL_1461895, EPI_ISL_1461896, EPI_ISL_1461897, EPI_ISL_1461898, EPI_ISL_1461899, EPI_ISL_1461901, EPI_ISL_1461903, EPI_ISL_1461904, EPI_ISL_1461905, EPI_ISL_1461908, EPI_ISL_1461910, EPI_ISL_1461911, EPI_ISL_1461913, EPI_ISL_1461914, EPI_ISL_1461915, EPI_ISL_1461916, EPI_ISL_1461918, EPI_ISL_1461919, EPI_ISL_1461921, EPI_ISL_1461922, EPI_ISL_1461923, EPI_ISL_1461924, EPI_ISL_1461925, EPI_ISL_1461926, EPI_ISL_1461931, EPI_ISL_1461932, EPI_ISL_1461937, EPI_ISL_1461938, EPI_ISL_1461939, EPI_ISL_1461941, EPI_ISL_1461942, EPI_ISL_1461943, EPI_ISL_1461945, EPI_ISL_1461948, EPI_ISL_1461950, EPI_ISL_1461951, EPI_ISL_1461952, EPI_ISL_1461954, EPI_ISL_1461955, EPI_ISL_1461960, EPI_ISL_1461961, EPI_ISL_1461962, EPI_ISL_1461964, EPI_ISL_1461966, EPI_ISL_1461968, EPI_ISL_1461970, EPI_ISL_1461971, EPI_ISL_1461976, EPI_ISL_1461977, EPI_ISL_1461978, EPI_ISL_1461981, EPI_ISL_1461988, EPI_ISL_1461989, EPI_ISL_1461990, EPI_ISL_1461991, EPI_ISL_1461994, EPI_ISL_1461997, EPI_ISL_1461998, EPI_ISL_1461999, EPI_ISL_1462000, EPI_ISL_1462004, EPI_ISL_1462008, EPI_ISL_1462009, EPI_ISL_1462010, EPI_ISL_1462011, EPI_ISL_1462012, EPI_ISL_1462014, EPI_ISL_1462017, EPI_ISL_1462020, EPI_ISL_1462021, EPI_ISL_1462022, EPI_ISL_1462023, EPI_ISL_1462027, EPI_ISL_1462029, EPI_ISL_1462031, EPI_ISL_1462032, EPI_ISL_1462033, EPI_ISL_1462036, EPI_ISL_1462037, EPI_ISL_1462039, EPI_ISL_1462040, EPI_ISL_1462041, EPI_ISL_1462045, EPI_ISL_1462047, EPI_ISL_1462048, EPI_ISL_1462051, EPI_ISL_1462054, EPI_ISL_1462055, EPI_ISL_1462056, EPI_ISL_1462057, EPI_ISL_1462058, EPI_ISL_1462061, EPI_ISL_1462062, EPI_ISL_1462063, EPI_ISL_1462065, EPI_ISL_1462067, EPI_ISL_1462072, EPI_ISL_1462074, EPI_ISL_1462078, EPI_ISL_1462079, EPI_ISL_1462080, EPI_ISL_1462083, EPI_ISL_1462084, EPI_ISL_1462086, EPI_ISL_1462088, EPI_ISL_1462089, EPI_ISL_1462091, EPI_ISL_1462092, EPI_ISL_1462093, EPI_ISL_1462095, EPI_ISL_1462096, EPI_ISL_1462098, EPI_ISL_1462099, EPI_ISL_1462100, EPI_ISL_1462103, EPI_ISL_1462104, EPI_ISL_1462107, EPI_ISL_1462108, EPI_ISL_1462110, EPI_ISL_1462113, EPI_ISL_1462114, EPI_ISL_1462116, EPI_ISL_1462117, EPI_ISL_1462118, EPI_ISL_1462121, EPI_ISL_1462122, EPI_ISL_1462123, EPI_ISL_1462125, EPI_ISL_1462126, EPI_ISL_1462127, EPI_ISL_1462130, EPI_ISL_1462132, EPI_ISL_1462134, EPI_ISL_1462137, EPI_ISL_1462142, EPI_ISL_1462143, EPI_ISL_1462144, EPI_ISL_1462145, EPI_ISL_1462147, EPI_ISL_1462148, EPI_ISL_1462150, EPI_ISL_1462151, EPI_ISL_1462152, EPI_ISL_1462153, EPI_ISL_1462155, EPI_ISL_1462156, EPI_ISL_1462157, EPI_ISL_1462159, EPI_ISL_1462160, EPI_ISL_1462161, EPI_ISL_1462162, EPI_ISL_1462163, EPI_ISL_1462165, EPI_ISL_1462166, EPI_ISL_1462168, EPI_ISL_1462173, EPI_ISL_1462175, EPI_ISL_1462177, EPI_ISL_1462178, EPI_ISL_1462180, EPI_ISL_1462183, EPI_ISL_1462184, EPI_ISL_1462185, EPI_ISL_1462186, EPI_ISL_1462187, EPI_ISL_1462189, EPI_ISL_1462192, EPI_ISL_1462196, EPI_ISL_1462200, EPI_ISL_1462202, EPI_ISL_1462203, EPI_ISL_1462205, EPI_ISL_1462207, EPI_ISL_1462210, EPI_ISL_1462215, EPI_ISL_1462216, EPI_ISL_1462218, EPI_ISL_1462219, EPI_ISL_1462221, EPI_ISL_1462222, EPI_ISL_1462223, EPI_ISL_1462224, EPI_ISL_1462226, EPI_ISL_1462227, EPI_ISL_1462229, EPI_ISL_1462230, EPI_ISL_1462231, EPI_ISL_1462232, EPI_ISL_1462235, EPI_ISL_1462241, EPI_ISL_1462246, EPI_ISL_1462250, EPI_ISL_1462255, EPI_ISL_1462258, EPI_ISL_1462260, EPI_ISL_1462264, EPI_ISL_1462266, EPI_ISL_1462267, EPI_ISL_1462268, EPI_ISL_1462270, EPI_ISL_1462271, EPI_ISL_1462272, EPI_ISL_1462273, EPI_ISL_1462274, EPI_ISL_1462276, EPI_ISL_1462280, EPI_ISL_1462282, EPI_ISL_1462285, EPI_ISL_1462286, EPI_ISL_1462290, EPI_ISL_1462291, EPI_ISL_1462296, EPI_ISL_1462297, EPI_ISL_1462298, EPI_ISL_1462300, EPI_ISL_1462302, EPI_ISL_1462303, EPI_ISL_1462307, EPI_ISL_1462308, EPI_ISL_1462309, EPI_ISL_1462310, EPI_ISL_1462315, EPI_ISL_1462316, EPI_ISL_1462326, EPI_ISL_1462332, EPI_ISL_1462333, EPI_ISL_1462336, EPI_ISL_1462337, EPI_ISL_1462338, EPI_ISL_1462340, EPI_ISL_1462345, EPI_ISL_1462348, EPI_ISL_1462349, EPI_ISL_1462353, EPI_ISL_1462354, EPI_ISL_1462356, EPI_ISL_1462358, EPI_ISL_1462359, EPI_ISL_1462361, EPI_ISL_1462362, EPI_ISL_1462363, EPI_ISL_1462364, EPI_ISL_1462366, EPI_ISL_1462369, EPI_ISL_1462372, EPI_ISL_1462374, EPI_ISL_1462379, EPI_ISL_1462381, EPI_ISL_1462386, EPI_ISL_1462387, EPI_ISL_1462388, EPI_ISL_1462394, EPI_ISL_1462405, EPI_ISL_1462406, EPI_ISL_1462408, EPI_ISL_1462410, EPI_ISL_1462413, EPI_ISL_1462415, EPI_ISL_1462423, EPI_ISL_1462424, EPI_ISL_1462425, EPI_ISL_1462427, EPI_ISL_1462431, EPI_ISL_1462433, EPI_ISL_1462436, EPI_ISL_1462441, EPI_ISL_1462443, EPI_ISL_1462446, EPI_ISL_1462448, EPI_ISL_1462449, EPI_ISL_1462451, EPI_ISL_1462453, EPI_ISL_1462456, EPI_ISL_1462462, EPI_ISL_1462465, EPI_ISL_1462466, EPI_ISL_1462467, EPI_ISL_1462470, EPI_ISL_1462471, EPI_ISL_1462474, EPI_ISL_1462479, EPI_ISL_1462481, EPI_ISL_1462483, EPI_ISL_1462494, EPI_ISL_1462497, EPI_ISL_1462498 | | | |
| see above | Laboratory Corporation of America | Centers for Disease Control and Prevention Division of Viral Diseases, Pathogen Discovery | Dakota Howard, Dhwani Batra, Peter W. Cook, Kara Moser, Adrian Paskey, Jason Caravas, Benjamin Rambo-Martin, Shatavia Morrison, Christopher Gulvick, Scott Sammons, Yvette Unoarumhi, Darlene Wagner, Matthew Schmerer, Minoo Agarwal, Eyad Almasri, Debbie Boles, Ayla Burns, Nuthawin Charoensri, Oren Cohen, Susan Countryman, Mary Ann Cristobal, Bobbi Croy, Suzanne Dale, Hrushikesh Deshmukh, Amanda Douglas, Vincent Drouillon, Marcia Eisenberg, Howard Engler, Rama Ghatti, Prashant Gupta, Susan Hicks, Jake Humphrey, Lax Iyer, Manoj Jain, Mohan Kolli, Brian Krueger, Tim Kuphal, Stanley Letovsky, Michael Levandoski, Craig Lukasik, Jonathan Meltzer, Brian Norvell, Mindy Nye, Scott Parker, Christos Petropoulos, John Pruitt, Steven Ragan, Scott Ryan, Mike Sapeta, Jana Schroth, Suresh Babu Selvaraju, Goran Stevovic, Amanda Suchanek, Andrea Throop, Lyndon Tilson, Thomas Urban, Joe Voshell, Kimberly Wagner, Jonathan Williams, Mary Williamson, Qian Zeng, Tricia Zwiefelhofer, Clinton R. Paden, Duncan MacCannell |
| EPI_ISL_1462500 | SUNY UPSTATE MEDICAL UNIVERSITY | Wadsworth Center, New York State Department of Health | Kirsten St. George, Daryl M. Lamson, Alexis Russell, Matthew Shudt, Melissa A Leisner, Jonathan Plitnick, Catharine Prussing, Navjot Singh, John Kelly, Erasmus Schneider, Erica Lasek-Nesselquist |
| EPI_ISL_1462501 | Laboratory Corporation of America | Centers for Disease Control and Prevention Division of Viral Diseases, Pathogen Discovery | Dakota Howard, Dhwani Batra, Peter W. Cook, Kara Moser, Adrian Paskey, Jason Caravas, Benjamin Rambo-Martin, Shatavia Morrison, Christopher Gulvick, Scott Sammons, Yvette Unoarumhi, Darlene Wagner, Matthew Schmerer, Minoo Agarwal, Eyad Almasri, Debbie Boles, Ayla Burns, Nuthawin Charoensri, Oren Cohen, Susan Countryman, Mary Ann Cristobal, Bobbi Croy, Suzanne Dale, Hrushikesh Deshmukh, Amanda Douglas, Vincent Drouillon, Marcia Eisenberg, Howard Engler, Rama Ghatti, Prashant Gupta, Susan Hicks, Jake Humphrey, Lax Iyer, Manoj Jain, Mohan Kolli, Brian Krueger, Tim Kuphal, Stanley Letovsky, Michael Levandoski, Craig Lukasik, Jonathan Meltzer, Brian Norvell, Mindy Nye, Scott Parker, Christos Petropoulos, John Pruitt, Steven Ragan, Scott Ryan, Mike Sapeta, Jana Schroth, Suresh Babu Selvaraju, Goran Stevovic, Amanda Suchanek, Andrea Throop, Lyndon Tilson, Thomas Urban, Joe Voshell, Kimberly Wagner, Jonathan Williams, Mary Williamson, Qian Zeng, Tricia Zwiefelhofer, Clinton R. Paden, Duncan MacCannell |
| EPI_ISL_1462502, EPI_ISL_1462503, EPI_ISL_1462505 | SUNY UPSTATE MEDICAL UNIVERSITY | Wadsworth Center, New York State Department of Health | Kirsten St. George, Daryl M. Lamson, Alexis Russell, Matthew Shudt, Melissa A Leisner, Jonathan Plitnick, Catharine Prussing, Navjot Singh, John Kelly, Erasmus Schneider, Erica Lasek-Nesselquist |
| EPI_ISL_1462506, EPI_ISL_1462508 | ALBANY MEDICAL CENTER | Wadsworth Center, New York State Department of Health | Kirsten St. George, Daryl M. Lamson, Alexis Russell, Matthew Shudt, Melissa A Leisner, Jonathan Plitnick, Catharine Prussing, Navjot Singh, John Kelly, Erasmus Schneider, Erica Lasek-Nesselquist |
| EPI_ISL_1462509, EPI_ISL_1462510, EPI_ISL_1462512, EPI_ISL_1462513 | URMC LABS | Wadsworth Center, New York State Department of Health | Kirsten St. George, Daryl M. Lamson, Alexis Russell, Matthew Shudt, Melissa A Leisner, Jonathan Plitnick, Catharine Prussing, Navjot Singh, John Kelly, Erasmus Schneider, Erica Lasek-Nesselquist |
| EPI_ISL_1462514 | Laboratory Corporation of America | Centers for Disease Control and Prevention Division of Viral Diseases, Pathogen Discovery | Dakota Howard, Dhwani Batra, Peter W. Cook, Kara Moser, Adrian Paskey, Jason Caravas, Benjamin Rambo-Martin, Shatavia Morrison, Christopher Gulvick, Scott Sammons, Yvette Unoarumhi, Darlene Wagner, Matthew Schmerer, Minoo Agarwal, Eyad Almasri, Debbie Boles, Ayla Burns, Nuthawin Charoensri, Oren Cohen, Susan Countryman, Mary Ann Cristobal, Bobbi Croy, Suzanne Dale, Hrushikesh Deshmukh, Amanda Douglas, Vincent Drouillon, Marcia Eisenberg, Howard Engler, Rama Ghatti, Prashant Gupta, Susan Hicks, Jake Humphrey, Lax Iyer, Manoj Jain, Mohan Kolli, Brian Krueger, Tim Kuphal, Stanley Letovsky, Michael Levandoski, Craig Lukasik, Jonathan Meltzer, Brian Norvell, Mindy Nye, Scott Parker, Christos Petropoulos, John Pruitt, Steven Ragan, Scott Ryan, Mike Sapeta, Jana Schroth, Suresh Babu Selvaraju, Goran Stevovic, Amanda Suchanek, Andrea Throop, Lyndon Tilson, Thomas Urban, Joe Voshell, Kimberly Wagner, Jonathan Williams, Mary Williamson, Qian Zeng, Tricia Zwiefelhofer, Clinton R. Paden, Duncan MacCannell |
| EPI_ISL_1462515, EPI_ISL_1462516, EPI_ISL_1462518, EPI_ISL_1462519, EPI_ISL_1462521 | URMC LABS | Wadsworth Center, New York State Department of Health | Kirsten St. George, Daryl M. Lamson, Alexis Russell, Matthew Shudt, Melissa A Leisner, Jonathan Plitnick, Catharine Prussing, Navjot Singh, John Kelly, Erasmus Schneider, Erica Lasek-Nesselquist |
| EPI_ISL_1462522 | Laboratory Corporation of America | Centers for Disease Control and Prevention Division of Viral Diseases, Pathogen Discovery | Dakota Howard, Dhwani Batra, Peter W. Cook, Kara Moser, Adrian Paskey, Jason Caravas, Benjamin Rambo-Martin, Shatavia Morrison, Christopher Gulvick, Scott Sammons, Yvette Unoarumhi, Darlene Wagner, Matthew Schmerer, Minoo Agarwal, Eyad Almasri, Debbie Boles, Ayla Burns, Nuthawin Charoensri, Oren Cohen, Susan Countryman, Mary Ann Cristobal, Bobbi Croy, Suzanne Dale, Hrushikesh Deshmukh, Amanda Douglas, Vincent Drouillon, Marcia Eisenberg, Howard Engler, Rama Ghatti, Prashant Gupta, Susan Hicks, Jake Humphrey, Lax Iyer, Manoj Jain, Mohan Kolli, Brian Krueger, Tim Kuphal, Stanley Letovsky, Michael Levandoski, Craig Lukasik, Jonathan Meltzer, Brian Norvell, Mindy Nye, Scott Parker, Christos Petropoulos, John Pruitt, Steven Ragan, Scott Ryan, Mike Sapeta, Jana Schroth, Suresh Babu Selvaraju, Goran Stevovic, Amanda Suchanek, Andrea Throop, Lyndon Tilson, Thomas Urban, Joe Voshell, Kimberly Wagner, Jonathan Williams, Mary Williamson, Qian Zeng, Tricia Zwiefelhofer, Clinton R. Paden, Duncan MacCannell |
| EPI_ISL_1462523, EPI_ISL_1462524, EPI_ISL_1462525, EPI_ISL_1462527, EPI_ISL_1462528 | URMC LABS | Wadsworth Center, New York State Department of Health | Kirsten St. George, Daryl M. Lamson, Alexis Russell, Matthew Shudt, Melissa A Leisner, Jonathan Plitnick, Catharine Prussing, Navjot Singh, John Kelly, Erasmus Schneider, Erica Lasek-Nesselquist |
| EPI_ISL_1462529 | Laboratory Corporation of America | Centers for Disease Control and Prevention Division of Viral Diseases, Pathogen Discovery | Dakota Howard, Dhwani Batra, Peter W. Cook, Kara Moser, Adrian Paskey, Jason Caravas, Benjamin Rambo-Martin, Shatavia Morrison, Christopher Gulvick, Scott Sammons, Yvette Unoarumhi, Darlene Wagner, Matthew Schmerer, Minoo Agarwal, Eyad Almasri, Debbie Boles, Ayla Burns, Nuthawin Charoensri, Oren Cohen, Susan Countryman, Mary Ann Cristobal, Bobbi Croy, Suzanne Dale, Hrushikesh Deshmukh, Amanda Douglas, Vincent Drouillon, Marcia Eisenberg, Howard Engler, Rama Ghatti, Prashant Gupta, Susan Hicks, Jake Humphrey, Lax Iyer, Manoj Jain, Mohan Kolli, Brian Krueger, Tim Kuphal, Stanley Letovsky, Michael Levandoski, Craig Lukasik, Jonathan Meltzer, Brian Norvell, Mindy Nye, Scott Parker, Christos Petropoulos, John Pruitt, Steven Ragan, Scott Ryan, Mike Sapeta, Jana Schroth, Suresh Babu Selvaraju, Goran Stevovic, Amanda Suchanek, Andrea Throop, Lyndon Tilson, Thomas Urban, Joe Voshell, Kimberly Wagner, Jonathan Williams, Mary Williamson, Qian Zeng, Tricia Zwiefelhofer, Clinton R. Paden, Duncan MacCannell |
| EPI_ISL_1462530, EPI_ISL_1462531, EPI_ISL_1462533, EPI_ISL_1462534 | URMC LABS | Wadsworth Center, New York State Department of Health | Kirsten St. George, Daryl M. Lamson, Alexis Russell, Matthew Shudt, Melissa A Leisner, Jonathan Plitnick, Catharine Prussing, Navjot Singh, John Kelly, Erasmus Schneider, Erica Lasek-Nesselquist |
| EPI_ISL_1462535 | Laboratory Corporation of America | Centers for Disease Control and Prevention Division of Viral Diseases, Pathogen Discovery | Dakota Howard, Dhwani Batra, Peter W. Cook, Kara Moser, Adrian Paskey, Jason Caravas, Benjamin Rambo-Martin, Shatavia Morrison, Christopher Gulvick, Scott Sammons, Yvette Unoarumhi, Darlene Wagner, Matthew Schmerer, Minoo Agarwal, Eyad Almasri, Debbie Boles, Ayla Burns, Nuthawin Charoensri, Oren Cohen, Susan Countryman, Mary Ann Cristobal, Bobbi Croy, Suzanne Dale, Hrushikesh Deshmukh, Amanda Douglas, Vincent Drouillon, Marcia Eisenberg, Howard Engler, Rama Ghatti, Prashant Gupta, Susan Hicks, Jake Humphrey, Lax Iyer, Manoj Jain, Mohan Kolli, Brian Krueger, Tim Kuphal, Stanley Letovsky, Michael Levandoski, Craig Lukasik, Jonathan Meltzer, Brian Norvell, Mindy Nye, Scott Parker, Christos Petropoulos, John Pruitt, Steven Ragan, Scott Ryan, Mike Sapeta, Jana Schroth, Suresh Babu Selvaraju, Goran Stevovic, Amanda Suchanek, Andrea Throop, Lyndon Tilson, Thomas Urban, Joe Voshell, Kimberly Wagner, Jonathan Williams, Mary Williamson, Qian Zeng, Tricia Zwiefelhofer, Clinton R. Paden, Duncan MacCannell |
| EPI_ISL_1462536, EPI_ISL_1462537 | SUNY UPSTATE MEDICAL UNIVERSITY | Wadsworth Center, New York State Department of Health | Kirsten St. George, Daryl M. Lamson, Alexis Russell, Matthew Shudt, Melissa A Leisner, Jonathan Plitnick, Catharine Prussing, Navjot Singh, John Kelly, Erasmus Schneider, Erica Lasek-Nesselquist |
| EPI_ISL_1462538 | Laboratory Corporation of America | Centers for Disease Control and Prevention Division of Viral Diseases, Pathogen Discovery | Dakota Howard, Dhwani Batra, Peter W. Cook, Kara Moser, Adrian Paskey, Jason Caravas, Benjamin Rambo-Martin, Shatavia Morrison, Christopher Gulvick, Scott Sammons, Yvette Unoarumhi, Darlene Wagner, Matthew Schmerer, Minoo Agarwal, Eyad Almasri, Debbie Boles, Ayla Burns, Nuthawin Charoensri, Oren Cohen, Susan Countryman, Mary Ann Cristobal, Bobbi Croy, Suzanne Dale, Hrushikesh Deshmukh, Amanda Douglas, Vincent Drouillon, Marcia Eisenberg, Howard Engler, Rama Ghatti, Prashant Gupta, Susan Hicks, Jake Humphrey, Lax Iyer, Manoj Jain, Mohan Kolli, Brian Krueger, Tim Kuphal, Stanley Letovsky, Michael Levandoski, Craig Lukasik, Jonathan Meltzer, Brian Norvell, Mindy Nye, Scott Parker, Christos Petropoulos, John Pruitt, Steven Ragan, Scott Ryan, Mike Sapeta, Jana Schroth, Suresh Babu Selvaraju, Goran Stevovic, Amanda Suchanek, Andrea Throop, Lyndon Tilson, Thomas Urban, Joe Voshell, Kimberly Wagner, Jonathan Williams, Mary Williamson, Qian Zeng, Tricia Zwiefelhofer, Clinton R. Paden, Duncan MacCannell |
| EPI_ISL_1462539, EPI_ISL_1462540 | SUNY UPSTATE MEDICAL UNIVERSITY | Wadsworth Center, New York State Department of Health | Kirsten St. George, Daryl M. Lamson, Alexis Russell, Matthew Shudt, Melissa A Leisner, Jonathan Plitnick, Catharine Prussing, Navjot Singh, John Kelly, Erasmus Schneider, Erica Lasek-Nesselquist |
| EPI_ISL_1462541 | Laboratory Corporation of America | Centers for Disease Control and Prevention Division of Viral Diseases, Pathogen Discovery | Dakota Howard, Dhwani Batra, Peter W. Cook, Kara Moser, Adrian Paskey, Jason Caravas, Benjamin Rambo-Martin, Shatavia Morrison, Christopher Gulvick, Scott Sammons, Yvette Unoarumhi, Darlene Wagner, Matthew Schmerer, Minoo Agarwal, Eyad Almasri, Debbie Boles, Ayla Burns, Nuthawin Charoensri, Oren Cohen, Susan Countryman, Mary Ann Cristobal, Bobbi Croy, Suzanne Dale, Hrushikesh Deshmukh, Amanda Douglas, Vincent Drouillon, Marcia Eisenberg, Howard Engler, Rama Ghatti, Prashant Gupta, Susan Hicks, Jake Humphrey, Lax Iyer, Manoj Jain, Mohan Kolli, Brian Krueger, Tim Kuphal, Stanley Letovsky, Michael Levandoski, Craig Lukasik, Jonathan Meltzer, Brian Norvell, Mindy Nye, Scott Parker, Christos Petropoulos, John Pruitt, Steven Ragan, Scott Ryan, Mike Sapeta, Jana Schroth, Suresh Babu Selvaraju, Goran Stevovic, Amanda Suchanek, Andrea Throop, Lyndon Tilson, Thomas Urban, Joe Voshell, Kimberly Wagner, Jonathan Williams, Mary Williamson, Qian Zeng, Tricia Zwiefelhofer, Clinton R. Paden, Duncan MacCannell |
| EPI_ISL_1462542, EPI_ISL_1462543 | SUNY UPSTATE MEDICAL UNIVERSITY | Wadsworth Center, New York State Department of Health | Kirsten St. George, Daryl M. Lamson, Alexis Russell, Matthew Shudt, Melissa A Leisner, Jonathan Plitnick, Catharine Prussing, Navjot Singh, John Kelly, Erasmus Schneider, Erica Lasek-Nesselquist |
| EPI_ISL_1462544 | Laboratory Corporation of America | Centers for Disease Control and Prevention | Dakota Howard, Dhwani Batra, Peter W. Cook, Kara Moser, Adrian Paskey, Jason Caravas, Benjamin Rambo-Martin, Shatavia Morrison, Christopher Gulvick, Scott Sammons, Yvette Unoarumhi, Darlene Wagner, Matthew Schmerer, Minoo Agarwal, Eyad Almasri, Debbie Boles, Ayla Burns, |

| | | | |
|---|---|---|---|
| | | Division of Viral Diseases, Pathogen Discovery | Nuthawin Charoensri, Oren Cohen, Susan Countryman, Mary Ann Cristobal, Bobbi Croy, Suzanne Dale, Hrushikesh Deshmukh, Amanda Douglas, Vincent Drouillon, Marcia Eisenberg, Howard Engler, Rama Ghatti, Prashant Gupta, Susan Hicks, Jake Humphrey, Lax Iyer, Manoj Jain, Mohan Kolli, Brian Krueger, Tim Kuphal, Stanley Letovsky, Michael Levandoski, Craig Lukasik, Jonathan Meltzer, Brian Norvell, Mindy Nye, Scott Parker, Christos Petropoulos, John Pruitt, Steven Ragan, Scott Ryan, Mike Sapeta, Jana Schroth, Suresh Babu Selvaraju, Goran Stevovic, Amanda Suchanek, Andrea Throop, Lyndon Tilson, Thomas Urban, Joe Voshell, Kimberly Wagner, Jonathan Williams, Mary Williamson, Qian Zeng, Tricia Zwiefelhofer, Clinton R. Paden, Duncan MacCannell |
| EPI_ISL_1462545, EPI_ISL_1462546, EPI_ISL_1462548, EPI_ISL_1462549 | SUNY UPSTATE MEDICAL UNIVERSITY | Wadsworth Center, New York State Department of Health | Kirsten St. George, Daryl M. Lamson, Alexis Russell, Matthew Shudt, Melissa A Leisner, Jonathan Plitnick, Catharine Prussing, Navjot Singh, John Kelly, Erasmus Schneider, Erica Lasek-Nesselquist |
| EPI_ISL_1462551 | MONTEFIORE MEDICAL CENTER LABORATORIES | Wadsworth Center, New York State Department of Health | Kirsten St. George, Daryl M. Lamson, Alexis Russell, Matthew Shudt, Melissa A Leisner, Jonathan Plitnick, Catharine Prussing, Navjot Singh, John Kelly, Erasmus Schneider, Erica Lasek-Nesselquist |
| EPI_ISL_1462552 | TEMPUS LABS INC | Wadsworth Center, New York State Department of Health | Kirsten St. George, Daryl M. Lamson, Alexis Russell, Matthew Shudt, Melissa A Leisner, Jonathan Plitnick, Catharine Prussing, Navjot Singh, John Kelly, Erasmus Schneider, Erica Lasek-Nesselquist |
| EPI_ISL_1462553, EPI_ISL_1462555 | WESTCHESTER MEDICAL CENTER | Wadsworth Center, New York State Department of Health | Kirsten St. George, Daryl M. Lamson, Alexis Russell, Matthew Shudt, Melissa A Leisner, Jonathan Plitnick, Catharine Prussing, Navjot Singh, John Kelly, Erasmus Schneider, Erica Lasek-Nesselquist |
| EPI_ISL_1462556 | NORTHWELL HEALTH LABORATORIES | Wadsworth Center, New York State Department of Health | Kirsten St. George, Daryl M. Lamson, Alexis Russell, Matthew Shudt, Melissa A Leisner, Jonathan Plitnick, Catharine Prussing, Navjot Singh, John Kelly, Erasmus Schneider, Erica Lasek-Nesselquist |
| EPI_ISL_1462557 | Laboratory Corporation of America | Centers for Disease Control and Prevention Division of Viral Diseases, Pathogen Discovery | Dakota Howard, Dhwani Batra, Peter W. Cook, Kara Moser, Adrian Paskey, Jason Caravas, Benjamin Rambo-Martin, Shatavia Morrison, Christopher Gulvick, Scott Sammons, Yvette Unoarumhi, Darlene Wagner, Matthew Schmerer, Minoo Agarwal, Eyad Almasri, Debbie Boles, Ayla Burns, Nuthawin Charoensri, Oren Cohen, Susan Countryman, Mary Ann Cristobal, Bobbi Croy, Suzanne Dale, Hrushikesh Deshmukh, Amanda Douglas, Vincent Drouillon, Marcia Eisenberg, Howard Engler, Rama Ghatti, Prashant Gupta, Susan Hicks, Jake Humphrey, Lax Iyer, Manoj Jain, Mohan Kolli, Brian Krueger, Tim Kuphal, Stanley Letovsky, Michael Levandoski, Craig Lukasik, Jonathan Meltzer, Brian Norvell, Mindy Nye, Scott Parker, Christos Petropoulos, John Pruitt, Steven Ragan, Scott Ryan, Mike Sapeta, Jana Schroth, Suresh Babu Selvaraju, Goran Stevovic, Amanda Suchanek, Andrea Throop, Lyndon Tilson, Thomas Urban, Joe Voshell, Kimberly Wagner, Jonathan Williams, Mary Williamson, Qian Zeng, Tricia Zwiefelhofer, Clinton R. Paden, Duncan MacCannell |
| EPI_ISL_1462558, EPI_ISL_1462559, EPI_ISL_1462561, EPI_ISL_1462563, EPI_ISL_1462564 | NORTHWELL HEALTH LABORATORIES | Wadsworth Center, New York State Department of Health | Kirsten St. George, Daryl M. Lamson, Alexis Russell, Matthew Shudt, Melissa A Leisner, Jonathan Plitnick, Catharine Prussing, Navjot Singh, John Kelly, Erasmus Schneider, Erica Lasek-Nesselquist |
| EPI_ISL_1462565 | Laboratory Corporation of America | Centers for Disease Control and Prevention Division of Viral Diseases, Pathogen Discovery | Dakota Howard, Dhwani Batra, Peter W. Cook, Kara Moser, Adrian Paskey, Jason Caravas, Benjamin Rambo-Martin, Shatavia Morrison, Christopher Gulvick, Scott Sammons, Yvette Unoarumhi, Darlene Wagner, Matthew Schmerer, Minoo Agarwal, Eyad Almasri, Debbie Boles, Ayla Burns, Nuthawin Charoensri, Oren Cohen, Susan Countryman, Mary Ann Cristobal, Bobbi Croy, Suzanne Dale, Hrushikesh Deshmukh, Amanda Douglas, Vincent Drouillon, Marcia Eisenberg, Howard Engler, Rama Ghatti, Prashant Gupta, Susan Hicks, Jake Humphrey, Lax Iyer, Manoj Jain, Mohan Kolli, Brian Krueger, Tim Kuphal, Stanley Letovsky, Michael Levandoski, Craig Lukasik, Jonathan Meltzer, Brian Norvell, Mindy Nye, Scott Parker, Christos Petropoulos, John Pruitt, Steven Ragan, Scott Ryan, Mike Sapeta, Jana Schroth, Suresh Babu Selvaraju, Goran Stevovic, Amanda Suchanek, Andrea Throop, Lyndon Tilson, Thomas Urban, Joe Voshell, Kimberly Wagner, Jonathan Williams, Mary Williamson, Qian Zeng, Tricia Zwiefelhofer, Clinton R. Paden, Duncan MacCannell |
| EPI_ISL_1462566, EPI_ISL_1462567 | NORTHWELL HEALTH LABORATORIES | Wadsworth Center, New York State Department of Health | Kirsten St. George, Daryl M. Lamson, Alexis Russell, Matthew Shudt, Melissa A Leisner, Jonathan Plitnick, Catharine Prussing, Navjot Singh, John Kelly, Erasmus Schneider, Erica Lasek-Nesselquist |
| EPI_ISL_1462568 | Laboratory Corporation of America | Centers for Disease Control and Prevention Division of Viral Diseases, Pathogen Discovery | Dakota Howard, Dhwani Batra, Peter W. Cook, Kara Moser, Adrian Paskey, Jason Caravas, Benjamin Rambo-Martin, Shatavia Morrison, Christopher Gulvick, Scott Sammons, Yvette Unoarumhi, Darlene Wagner, Matthew Schmerer, Minoo Agarwal, Eyad Almasri, Debbie Boles, Ayla Burns, Nuthawin Charoensri, Oren Cohen, Susan Countryman, Mary Ann Cristobal, Bobbi Croy, Suzanne Dale, Hrushikesh Deshmukh, Amanda Douglas, Vincent Drouillon, Marcia Eisenberg, Howard Engler, Rama Ghatti, Prashant Gupta, Susan Hicks, Jake Humphrey, Lax Iyer, Manoj Jain, Mohan Kolli, Brian Krueger, Tim Kuphal, Stanley Letovsky, Michael Levandoski, Craig Lukasik, Jonathan Meltzer, Brian Norvell, Mindy Nye, Scott Parker, Christos Petropoulos, John Pruitt, Steven Ragan, Scott Ryan, Mike Sapeta, Jana Schroth, Suresh Babu Selvaraju, Goran Stevovic, Amanda Suchanek, Andrea Throop, Lyndon Tilson, Thomas Urban, Joe Voshell, Kimberly Wagner, Jonathan Williams, Mary Williamson, Qian Zeng, Tricia Zwiefelhofer, Clinton R. Paden, Duncan MacCannell |
| EPI_ISL_1462569, EPI_ISL_1462570 | NORTHWELL HEALTH LABORATORIES | Wadsworth Center, New York State Department of Health | Kirsten St. George, Daryl M. Lamson, Alexis Russell, Matthew Shudt, Melissa A Leisner, Jonathan Plitnick, Catharine Prussing, Navjot Singh, John Kelly, Erasmus Schneider, Erica Lasek-Nesselquist |
| EPI_ISL_1462572, EPI_ISL_1462573, EPI_ISL_1462575 | THE MARY IMOGENE BASSETT HOSPITAL | Wadsworth Center, New York State Department of Health | Kirsten St. George, Daryl M. Lamson, Alexis Russell, Matthew Shudt, Melissa A Leisner, Jonathan Plitnick, Catharine Prussing, Navjot Singh, John Kelly, Erasmus Schneider, Erica Lasek-Nesselquist |
| EPI_ISL_1462576, EPI_ISL_1462578, EPI_ISL_1462579 | NORTHWELL HEALTH LABORATORIES | Wadsworth Center, New York State Department of Health | Kirsten St. George, Daryl M. Lamson, Alexis Russell, Matthew Shudt, Melissa A Leisner, Jonathan Plitnick, Catharine Prussing, Navjot Singh, John Kelly, Erasmus Schneider, Erica Lasek-Nesselquist |
| EPI_ISL_1462580 | Laboratory Corporation of America | Centers for Disease Control and Prevention Division of Viral Diseases, Pathogen Discovery | Dakota Howard, Dhwani Batra, Peter W. Cook, Kara Moser, Adrian Paskey, Jason Caravas, Benjamin Rambo-Martin, Shatavia Morrison, Christopher Gulvick, Scott Sammons, Yvette Unoarumhi, Darlene Wagner, Matthew Schmerer, Minoo Agarwal, Eyad Almasri, Debbie Boles, Ayla Burns, Nuthawin Charoensri, Oren Cohen, Susan Countryman, Mary Ann Cristobal, Bobbi Croy, Suzanne Dale, Hrushikesh Deshmukh, Amanda Douglas, Vincent Drouillon, Marcia Eisenberg, Howard Engler, Rama Ghatti, Prashant Gupta, Susan Hicks, Jake Humphrey, Lax Iyer, Manoj Jain, Mohan Kolli, Brian Krueger, Tim Kuphal, Stanley Letovsky, Michael Levandoski, Craig Lukasik, Jonathan Meltzer, Brian Norvell, Mindy Nye, Scott Parker, Christos Petropoulos, John Pruitt, Steven Ragan, Scott Ryan, Mike Sapeta, Jana Schroth, Suresh Babu Selvaraju, Goran Stevovic, Amanda Suchanek, Andrea Throop, Lyndon Tilson, Thomas Urban, Joe Voshell, Kimberly Wagner, Jonathan Williams, Mary Williamson, Qian Zeng, Tricia Zwiefelhofer, Clinton R. Paden, Duncan MacCannell |
| EPI_ISL_1462581, EPI_ISL_1462582 | NORTHWELL HEALTH LABORATORIES | Wadsworth Center, New York State Department of Health | Kirsten St. George, Daryl M. Lamson, Alexis Russell, Matthew Shudt, Melissa A Leisner, Jonathan Plitnick, Catharine Prussing, Navjot Singh, John Kelly, Erasmus Schneider, Erica Lasek-Nesselquist |
| EPI_ISL_1462583 | Laboratory Corporation of America | Centers for Disease Control and Prevention Division of Viral Diseases, Pathogen Discovery | Dakota Howard, Dhwani Batra, Peter W. Cook, Kara Moser, Adrian Paskey, Jason Caravas, Benjamin Rambo-Martin, Shatavia Morrison, Christopher Gulvick, Scott Sammons, Yvette Unoarumhi, Darlene Wagner, Matthew Schmerer, Minoo Agarwal, Eyad Almasri, Debbie Boles, Ayla Burns, Nuthawin Charoensri, Oren Cohen, Susan Countryman, Mary Ann Cristobal, Bobbi Croy, Suzanne Dale, Hrushikesh Deshmukh, Amanda Douglas, Vincent Drouillon, Marcia Eisenberg, Howard Engler, Rama Ghatti, Prashant Gupta, Susan Hicks, Jake Humphrey, Lax Iyer, Manoj Jain, Mohan Kolli, Brian Krueger, Tim Kuphal, Stanley Letovsky, Michael Levandoski, Craig Lukasik, Jonathan Meltzer, Brian Norvell, Mindy Nye, Scott Parker, Christos Petropoulos, John Pruitt, Steven Ragan, Scott Ryan, Mike Sapeta, Jana Schroth, Suresh Babu Selvaraju, Goran Stevovic, Amanda Suchanek, Andrea Throop, Lyndon Tilson, Thomas Urban, Joe Voshell, Kimberly Wagner, Jonathan Williams, Mary Williamson, Qian Zeng, Tricia Zwiefelhofer, Clinton R. Paden, Duncan MacCannell |
| EPI_ISL_1462584, EPI_ISL_1462585, EPI_ISL_1462587, EPI_ISL_1462588, EPI_ISL_1462589, EPI_ISL_1462591, EPI_ISL_1462593, EPI_ISL_1462594 | NORTHWELL HEALTH LABORATORIES | Wadsworth Center, New York State Department of Health | Kirsten St. George, Daryl M. Lamson, Alexis Russell, Matthew Shudt, Melissa A Leisner, Jonathan Plitnick, Catharine Prussing, Navjot Singh, John Kelly, Erasmus Schneider, Erica Lasek-Nesselquist |
| EPI_ISL_1462595 | Laboratory Corporation of America | Centers for Disease Control and Prevention Division of Viral Diseases, Pathogen Discovery | Dakota Howard, Dhwani Batra, Peter W. Cook, Kara Moser, Adrian Paskey, Jason Caravas, Benjamin Rambo-Martin, Shatavia Morrison, Christopher Gulvick, Scott Sammons, Yvette Unoarumhi, Darlene Wagner, Matthew Schmerer, Minoo Agarwal, Eyad Almasri, Debbie Boles, Ayla Burns, Nuthawin Charoensri, Oren Cohen, Susan Countryman, Mary Ann Cristobal, Bobbi Croy, Suzanne Dale, Hrushikesh Deshmukh, Amanda Douglas, Vincent Drouillon, Marcia Eisenberg, Howard Engler, Rama Ghatti, Prashant Gupta, Susan Hicks, Jake Humphrey, Lax Iyer, Manoj Jain, Mohan Kolli, Brian Krueger, Tim Kuphal, Stanley Letovsky, Michael Levandoski, Craig Lukasik, Jonathan Meltzer, Brian Norvell, Mindy Nye, Scott Parker, Christos Petropoulos, John Pruitt, Steven Ragan, Scott Ryan, Mike Sapeta, Jana Schroth, Suresh Babu Selvaraju, Goran Stevovic, Amanda Suchanek, Andrea Throop, Lyndon Tilson, Thomas Urban, Joe Voshell, Kimberly Wagner, Jonathan Williams, Mary Williamson, Qian Zeng, Tricia Zwiefelhofer, Clinton R. Paden, Duncan MacCannell |
| EPI_ISL_1462596, EPI_ISL_1462597 | NORTHWELL HEALTH LABORATORIES | Wadsworth Center, New York State Department of Health | Kirsten St. George, Daryl M. Lamson, Alexis Russell, Matthew Shudt, Melissa A Leisner, Jonathan Plitnick, Catharine Prussing, Navjot Singh, John Kelly, Erasmus Schneider, Erica Lasek-Nesselquist |
| EPI_ISL_1462598 | Laboratory Corporation of America | Centers for Disease Control and Prevention Division of Viral Diseases, Pathogen Discovery | Dakota Howard, Dhwani Batra, Peter W. Cook, Kara Moser, Adrian Paskey, Jason Caravas, Benjamin Rambo-Martin, Shatavia Morrison, Christopher Gulvick, Scott Sammons, Yvette Unoarumhi, Darlene Wagner, Matthew Schmerer, Minoo Agarwal, Eyad Almasri, Debbie Boles, Ayla Burns, Nuthawin Charoensri, Oren Cohen, Susan Countryman, Mary Ann Cristobal, Bobbi Croy, Suzanne Dale, Hrushikesh Deshmukh, Amanda Douglas, Vincent Drouillon, Marcia Eisenberg, Howard Engler, Rama Ghatti, Prashant Gupta, Susan Hicks, Jake Humphrey, Lax Iyer, Manoj Jain, Mohan Kolli, Brian Krueger, Tim Kuphal, Stanley Letovsky, Michael Levandoski, Craig Lukasik, Jonathan Meltzer, Brian Norvell, Mindy Nye, Scott Parker, Christos Petropoulos, John Pruitt, Steven Ragan, Scott Ryan, Mike Sapeta, Jana Schroth, Suresh Babu Selvaraju, Goran Stevovic, Amanda Suchanek, Andrea Throop, Lyndon Tilson, Thomas Urban, Joe Voshell, Kimberly Wagner, Jonathan Williams, Mary Williamson, Qian Zeng, Tricia Zwiefelhofer, Clinton R. Paden, Duncan MacCannell |
| EPI_ISL_1462599, EPI_ISL_1462600, EPI_ISL_1462601, EPI_ISL_1462603, EPI_ISL_1462604, EPI_ISL_1462606, EPI_ISL_1462607 | NORTHWELL HEALTH LABORATORIES | Wadsworth Center, New York State Department of Health | Kirsten St. George, Daryl M. Lamson, Alexis Russell, Matthew Shudt, Melissa A Leisner, Jonathan Plitnick, Catharine Prussing, Navjot Singh, John Kelly, Erasmus Schneider, Erica Lasek-Nesselquist |
| EPI_ISL_1462608 | Laboratory Corporation of America | Centers for Disease Control and Prevention Division of Viral Diseases, Pathogen Discovery | Dakota Howard, Dhwani Batra, Peter W. Cook, Kara Moser, Adrian Paskey, Jason Caravas, Benjamin Rambo-Martin, Shatavia Morrison, Christopher Gulvick, Scott Sammons, Yvette Unoarumhi, Darlene Wagner, Matthew Schmerer, Minoo Agarwal, Eyad Almasri, Debbie Boles, Ayla Burns, Nuthawin Charoensri, Oren Cohen, Susan Countryman, Mary Ann Cristobal, Bobbi Croy, Suzanne Dale, Hrushikesh Deshmukh, Amanda Douglas, Vincent Drouillon, Marcia Eisenberg, Howard Engler, Rama Ghatti, Prashant Gupta, Susan Hicks, Jake Humphrey, Lax Iyer, Manoj Jain, Mohan Kolli, Brian Krueger, Tim Kuphal, Stanley Letovsky, Michael Levandoski, Craig Lukasik, Jonathan Meltzer, Brian Norvell, Mindy Nye, Scott Parker, Christos Petropoulos, John Pruitt, Steven Ragan, Scott Ryan, Mike Sapeta, Jana Schroth, Suresh Babu Selvaraju, Goran Stevovic, Amanda Suchanek, Andrea Throop, Lyndon Tilson, Thomas Urban, Joe Voshell, Kimberly Wagner, Jonathan Williams, Mary Williamson, Qian Zeng, Tricia Zwiefelhofer, Clinton R. Paden, Duncan MacCannell |
| EPI_ISL_1462609, EPI_ISL_1462610 | NORTHWELL HEALTH LABORATORIES | Wadsworth Center, New York State Department of Health | Kirsten St. George, Daryl M. Lamson, Alexis Russell, Matthew Shudt, Melissa A Leisner, Jonathan Plitnick, Catharine Prussing, Navjot Singh, John Kelly, Erasmus Schneider, Erica Lasek-Nesselquist |
| EPI_ISL_1462611 | Laboratory Corporation of America | Centers for Disease Control and Prevention Division of Viral Diseases, Pathogen Discovery | Dakota Howard, Dhwani Batra, Peter W. Cook, Kara Moser, Adrian Paskey, Jason Caravas, Benjamin Rambo-Martin, Shatavia Morrison, Christopher Gulvick, Scott Sammons, Yvette Unoarumhi, Darlene Wagner, Matthew Schmerer, Minoo Agarwal, Eyad Almasri, Debbie Boles, Ayla Burns, Nuthawin Charoensri, Oren Cohen, Susan Countryman, Mary Ann Cristobal, Bobbi Croy, Suzanne Dale, Hrushikesh Deshmukh, Amanda Douglas, Vincent Drouillon, Marcia Eisenberg, Howard Engler, Rama Ghatti, Prashant Gupta, Susan Hicks, Jake Humphrey, Lax Iyer, Manoj Jain, Mohan Kolli, Brian Krueger, Tim Kuphal, Stanley Letovsky, Michael Levandoski, Craig Lukasik, Jonathan Meltzer, Brian Norvell, Mindy Nye, Scott Parker, Christos Petropoulos, John Pruitt, Steven Ragan, Scott Ryan, Mike Sapeta, Jana Schroth, Suresh Babu Selvaraju, Goran Stevovic, Amanda Suchanek, Andrea Throop, Lyndon Tilson, Thomas Urban, Joe Voshell, Kimberly Wagner, Jonathan Williams, Mary Williamson, Qian Zeng, Tricia Zwiefelhofer, Clinton R. Paden, Duncan MacCannell |
| EPI_ISL_1462612, EPI_ISL_1462613, EPI_ISL_1462615, EPI_ISL_1462617, EPI_ISL_1462618, EPI_ISL_1462620, EPI_ISL_1462621, EPI_ISL_1462623, EPI_ISL_1462624 | NORTHWELL HEALTH LABORATORIES | Wadsworth Center, New York State Department of Health | Kirsten St. George, Daryl M. Lamson, Alexis Russell, Matthew Shudt, Melissa A Leisner, Jonathan Plitnick, Catharine Prussing, Navjot Singh, John Kelly, Erasmus Schneider, Erica Lasek-Nesselquist |

EPI_ISL_1462626, EPI_ISL_1462630, EPI_ISL_1462631, EPI_ISL_1462632, EPI_ISL_1462636, EPI_ISL_1462641, EPI_ISL_1462643, EPI_ISL_1462645, EPI_ISL_1462648, EPI_ISL_1462650, EPI_ISL_1462651, EPI_ISL_1462662, EPI_ISL_1462668, EPI_ISL_1462670, EPI_ISL_1462671, EPI_ISL_1462672, EPI_ISL_1462678, EPI_ISL_1462679, EPI_ISL_1462680, EPI_ISL_1462681, EPI_ISL_1462691, EPI_ISL_1462692, EPI_ISL_1462693, EPI_ISL_1462694, EPI_ISL_1462695, EPI_ISL_1462698, EPI_ISL_1462699, EPI_ISL_1462700, EPI_ISL_1462705, EPI_ISL_1462706, EPI_ISL_1462707, EPI_ISL_1462708, EPI_ISL_1462709, EPI_ISL_1462714, EPI_ISL_1462717, EPI_ISL_1462718, EPI_ISL_1462722, EPI_ISL_1462724, EPI_ISL_1462725, EPI_ISL_1462729, EPI_ISL_1462731, EPI_ISL_1462733, EPI_ISL_1462736, EPI_ISL_1462737,

| | | | |
|---|---|---|---|
| EPI_ISL_1462740, EPI_ISL_1462742, EPI_ISL_1462745, EPI_ISL_1462746, EPI_ISL_1462747, EPI_ISL_1462753, EPI_ISL_1462754, EPI_ISL_1462759, EPI_ISL_1462762, EPI_ISL_1462767, EPI_ISL_1462768, EPI_ISL_1462773, EPI_ISL_1462779, EPI_ISL_1462780, EPI_ISL_1462783, EPI_ISL_1462785, EPI_ISL_1462786, EPI_ISL_1462788, EPI_ISL_1462789, EPI_ISL_1462794, EPI_ISL_1462796, EPI_ISL_1462799, EPI_ISL_1462803, EPI_ISL_1462813, EPI_ISL_1462814, EPI_ISL_1462816, EPI_ISL_1462821, EPI_ISL_1462822, EPI_ISL_1462823, EPI_ISL_1462824, EPI_ISL_1462825, EPI_ISL_1462826, EPI_ISL_1462830, EPI_ISL_1462835, EPI_ISL_1462836, EPI_ISL_1462837, EPI_ISL_1462843, EPI_ISL_1462844, EPI_ISL_1462851, EPI_ISL_1462853, EPI_ISL_1462854, EPI_ISL_1462855, EPI_ISL_1462857, EPI_ISL_1462858, EPI_ISL_1462862, EPI_ISL_1462864, EPI_ISL_1462870, EPI_ISL_1462872, EPI_ISL_1462873, EPI_ISL_1462876, EPI_ISL_1462877, EPI_ISL_1462882, EPI_ISL_1462883, EPI_ISL_1462886, EPI_ISL_1462888, EPI_ISL_1462889, EPI_ISL_1462890, EPI_ISL_1462891, EPI_ISL_1462892, EPI_ISL_1462894, EPI_ISL_1462896, EPI_ISL_1462897, EPI_ISL_1462898, EPI_ISL_1462899, EPI_ISL_1462901, EPI_ISL_1462903, EPI_ISL_1462904, EPI_ISL_1462907, EPI_ISL_1462908, EPI_ISL_1462914, EPI_ISL_1462916, EPI_ISL_1462919, EPI_ISL_1462924, EPI_ISL_1462932, EPI_ISL_1462933, EPI_ISL_1462948, EPI_ISL_1462951, EPI_ISL_1462952, EPI_ISL_1462953, EPI_ISL_1462955, EPI_ISL_1462956, EPI_ISL_1462970, EPI_ISL_1462992, EPI_ISL_1462993, EPI_ISL_1463008, EPI_ISL_1463009, EPI_ISL_1463010, EPI_ISL_1463011, EPI_ISL_1463013, EPI_ISL_1463014, EPI_ISL_1463023, EPI_ISL_1463026, EPI_ISL_1463028, EPI_ISL_1463031, EPI_ISL_1463034, EPI_ISL_1463037, EPI_ISL_1463039, EPI_ISL_1463046, EPI_ISL_1463047, EPI_ISL_1463055, EPI_ISL_1463059, EPI_ISL_1463061, EPI_ISL_1463077, EPI_ISL_1463086, EPI_ISL_1463088, EPI_ISL_1463090, EPI_ISL_1463092, EPI_ISL_1463097, EPI_ISL_1463105, EPI_ISL_1463108, EPI_ISL_1463110, EPI_ISL_1463111, EPI_ISL_1463117, EPI_ISL_1463124, EPI_ISL_1463126, EPI_ISL_1463132, EPI_ISL_1463142, EPI_ISL_1463147, EPI_ISL_1463150 | | | |
| see above | Laboratory Corporation of America | Centers for Disease Control and Prevention Division of Viral Diseases, Pathogen Discovery | Dakota Howard, Dhwani Batra, Peter W. Cook, Kara Moser, Adrian Paskey, Jason Caravas, Benjamin Rambo-Martin, Shatavia Morrison, Christopher Gulvick, Scott Sammons, Yvette Unoarumhi, Darlene Wagner, Matthew Schmerer, Minoo Agarwal, Eyad Almasri, Debbie Boles, Ayla Burns, Nuthawin Charoensri, Oren Cohen, Susan Countryman, Mary Ann Cristobal, Bobbi Croy, Suzanne Dale, Hrushikesh Deshmukh, Amanda Douglas, Vincent Drouillon, Marcia Eisenberg, Howard Engler, Rama Ghatti, Prashant Gupta, Susan Hicks, Jake Humphrey, Lax Iyer, Manoj Jain, Mohan Kolli, Brian Krueger, Tim Kuphal, Stanley Letovsky, Michael Levandoski, Craig Lukasik, Jonathan Meltzer, Brian Norvell, Mindy Nye, Scott Parker, Christos Petropoulos, John Pruitt, Steven Ragan, Scott Ryan, Mike Sapeta, Jana Schroth, Suresh Babu Selvaraju, Goran Stevovic, Amanda Suchanek, Andrea Throop, Lyndon Tilson, Thomas Urban, Joe Voshell, Kimberly Wagner, Jonathan Williams, Mary Williamson, Qian Zeng, Tricia Zwiefelhofer, Clinton R. Paden, Duncan MacCannell |
| EPI_ISL_1463151 | Ohio Department of Health Laboratory | Ohio Department of Health Laboratory | Holmes, Jennifer; Eric Brandt, Keoni Omura, Glen McGillivary, Caitlin McDonnell, Jade Mowery, Stephanie Mccracken, Tyler Payne, Kirtana Ramadugu, Erica Leasure, Brent Lee, Kelsey Florek, Heather Blankenship, Allison Black, Quanta Brown, and Tammy Bannerman |
| EPI_ISL_1463152, EPI_ISL_1463157, EPI_ISL_1463166, EPI_ISL_1463168, EPI_ISL_1463171, EPI_ISL_1463173, EPI_ISL_1463175, EPI_ISL_1463181, EPI_ISL_1463186, EPI_ISL_1463188, EPI_ISL_1463190, EPI_ISL_1463234, EPI_ISL_1463336, EPI_ISL_1463362, EPI_ISL_1463388, EPI_ISL_1463402, EPI_ISL_1463410, EPI_ISL_1463411 | | | |
| see above | Laboratory Corporation of America | Centers for Disease Control and Prevention Division of Viral Diseases, Pathogen Discovery | Dakota Howard, Dhwani Batra, Peter W. Cook, Kara Moser, Adrian Paskey, Jason Caravas, Benjamin Rambo-Martin, Shatavia Morrison, Christopher Gulvick, Scott Sammons, Yvette Unoarumhi, Darlene Wagner, Matthew Schmerer, Minoo Agarwal, Eyad Almasri, Debbie Boles, Ayla Burns, Nuthawin Charoensri, Oren Cohen, Susan Countryman, Mary Ann Cristobal, Bobbi Croy, Suzanne Dale, Hrushikesh Deshmukh, Amanda Douglas, Vincent Drouillon, Marcia Eisenberg, Howard Engler, Rama Ghatti, Prashant Gupta, Susan Hicks, Jake Humphrey, Lax Iyer, Manoj Jain, Mohan Kolli, Brian Krueger, Tim Kuphal, Stanley Letovsky, Michael Levandoski, Craig Lukasik, Jonathan Meltzer, Brian Norvell, Mindy Nye, Scott Parker, Christos Petropoulos, John Pruitt, Steven Ragan, Scott Ryan, Mike Sapeta, Jana Schroth, Suresh Babu Selvaraju, Goran Stevovic, Amanda Suchanek, Andrea Throop, Lyndon Tilson, Thomas Urban, Joe Voshell, Kimberly Wagner, Jonathan Williams, Mary Williamson, Qian Zeng, Tricia Zwiefelhofer, Clinton R. Paden, Duncan MacCannell |
| EPI_ISL_1463445, EPI_ISL_1463446, EPI_ISL_1463447, EPI_ISL_1463541, EPI_ISL_1463542, EPI_ISL_1463543 | Ohio Department of Health Laboratory | Ohio Department of Health Laboratory | Holmes, Jennifer; Eric Brandt, Keoni Omura, Glen McGillivary, Caitlin McDonnell, Jade Mowery, Stephanie Mccracken, Tyler Payne, Kirtana Ramadugu, Erica Leasure, Brent Lee, Kelsey Florek, Heather Blankenship, Allison Black, Quanta Brown, and Tammy Bannerman |
| EPI_ISL_1464680, EPI_ISL_1464681, EPI_ISL_1464682, EPI_ISL_1464685, EPI_ISL_1464686, EPI_ISL_1464687, EPI_ISL_1464688, EPI_ISL_1464689, EPI_ISL_1464692 | Sonora Quest Laboratories | TGen North | "Jolene Bowers, Heather Centner, Chris French, Hayley Yaglom, Ashlyn Pfeiffer, Darrin Lemmer, Dave Engelthaler, The Arizona COVID Genomics Union (ACGU)" |
| EPI_ISL_1464693 | TGen North | TGen North | "Jolene Bowers, Heather Centner, Chris French, Hayley Yaglom, Ashlyn Pfeiffer, Darrin Lemmer, Dave Engelthaler, The Arizona COVID Genomics Union (ACGU)" |
| EPI_ISL_1464694, EPI_ISL_1464696, EPI_ISL_1464697, EPI_ISL_1464698, EPI_ISL_1464699, EPI_ISL_1464701, EPI_ISL_1464702, EPI_ISL_1464703, EPI_ISL_1464704, EPI_ISL_1464705, EPI_ISL_1464706, EPI_ISL_1464708, EPI_ISL_1464709, EPI_ISL_1464710, EPI_ISL_1464713, EPI_ISL_1464714, EPI_ISL_1464717 | | | |
| see above | Sonora Quest Laboratories | TGen North | "Jolene Bowers, Heather Centner, Chris French, Hayley Yaglom, Ashlyn Pfeiffer, Darrin Lemmer, Dave Engelthaler, The Arizona COVID Genomics Union (ACGU)" |
| EPI_ISL_1464719 | TGen North | TGen North | "Jolene Bowers, Heather Centner, Chris French, Hayley Yaglom, Ashlyn Pfeiffer, Darrin Lemmer, Dave Engelthaler, The Arizona COVID Genomics Union (ACGU)" |
| EPI_ISL_1464721, EPI_ISL_1464723, EPI_ISL_1464724, EPI_ISL_1464725, EPI_ISL_1464727, EPI_ISL_1464728, EPI_ISL_1464729, EPI_ISL_1464730, EPI_ISL_1464731, EPI_ISL_1464733, EPI_ISL_1464734, EPI_ISL_1464737, EPI_ISL_1464739, EPI_ISL_1464742, EPI_ISL_1464743, EPI_ISL_1464747, EPI_ISL_1464748, EPI_ISL_1464749, EPI_ISL_1464752, EPI_ISL_1464755, EPI_ISL_1464756, EPI_ISL_1464757, EPI_ISL_1464758, EPI_ISL_1464759, EPI_ISL_1464762, EPI_ISL_1464763, EPI_ISL_1464764, EPI_ISL_1464765, EPI_ISL_1464767, EPI_ISL_1464768, EPI_ISL_1464769, EPI_ISL_1464770, EPI_ISL_1464772, EPI_ISL_1464775, EPI_ISL_1464776, EPI_ISL_1464777, EPI_ISL_1464779, EPI_ISL_1464781, EPI_ISL_1464783, EPI_ISL_1464785, EPI_ISL_1464787, EPI_ISL_1464789, EPI_ISL_1464790, EPI_ISL_1464791, EPI_ISL_1464793, EPI_ISL_1464794, EPI_ISL_1464795, EPI_ISL_1464798, EPI_ISL_1464801, EPI_ISL_1464802, EPI_ISL_1464803, EPI_ISL_1464804, EPI_ISL_1464805, EPI_ISL_1464806, EPI_ISL_1464807, EPI_ISL_1464808, EPI_ISL_1464810, EPI_ISL_1464811, EPI_ISL_1464812, EPI_ISL_1464813 | | | |
| see above | Sonora Quest Laboratories | TGen North | "Jolene Bowers, Heather Centner, Chris French, Hayley Yaglom, Ashlyn Pfeiffer, Darrin Lemmer, Dave Engelthaler, The Arizona COVID Genomics Union (ACGU)" |
| EPI_ISL_1464814 | TGen North | TGen North | "Jolene Bowers, Heather Centner, Chris French, Hayley Yaglom, Ashlyn Pfeiffer, Darrin Lemmer, Dave Engelthaler, The Arizona COVID Genomics Union (ACGU)" |
| EPI_ISL_1464815, EPI_ISL_1464816, EPI_ISL_1464817, EPI_ISL_1464818, EPI_ISL_1464820, EPI_ISL_1464821, EPI_ISL_1464823, EPI_ISL_1464824, EPI_ISL_1464825, EPI_ISL_1464828, EPI_ISL_1464829, EPI_ISL_1464830, EPI_ISL_1464831, EPI_ISL_1464832, EPI_ISL_1464833, EPI_ISL_1464834, EPI_ISL_1464835, EPI_ISL_1464836, EPI_ISL_1464837, EPI_ISL_1464838, EPI_ISL_1464839, EPI_ISL_1464842, EPI_ISL_1464843, EPI_ISL_1464844, EPI_ISL_1464846, EPI_ISL_1464847, EPI_ISL_1464849, EPI_ISL_1464851, EPI_ISL_1464852, EPI_ISL_1464853, EPI_ISL_1464855, EPI_ISL_1464857, EPI_ISL_1464858, EPI_ISL_1464859, EPI_ISL_1464860, EPI_ISL_1464862, EPI_ISL_1464866, EPI_ISL_1464867, EPI_ISL_1464869, EPI_ISL_1464871, EPI_ISL_1464872, EPI_ISL_1464873, EPI_ISL_1464877, EPI_ISL_1464878, EPI_ISL_1464879, EPI_ISL_1464880, EPI_ISL_1464882, EPI_ISL_1464883, EPI_ISL_1464885, EPI_ISL_1464888, EPI_ISL_1464892, EPI_ISL_1464893, EPI_ISL_1464894, EPI_ISL_1464895, EPI_ISL_1464896, EPI_ISL_1464897, EPI_ISL_1464900, EPI_ISL_1464901 | | | |
| see above | Sonora Quest Laboratories | TGen North | "Jolene Bowers, Heather Centner, Chris French, Hayley Yaglom, Ashlyn Pfeiffer, Darrin Lemmer, Dave Engelthaler, The Arizona COVID Genomics Union (ACGU)" |
| EPI_ISL_1464904 | TGen North | TGen North | "Jolene Bowers, Heather Centner, Chris French, Hayley Yaglom, Ashlyn Pfeiffer, Darrin Lemmer, Dave Engelthaler, The Arizona COVID Genomics Union (ACGU)" |
| EPI_ISL_1464905, EPI_ISL_1464906, EPI_ISL_1464907, EPI_ISL_1464908, EPI_ISL_1464909, EPI_ISL_1464910, EPI_ISL_1464912, EPI_ISL_1464914, EPI_ISL_1464917, EPI_ISL_1464918, EPI_ISL_1464919, EPI_ISL_1464920, EPI_ISL_1464922, EPI_ISL_1464924, EPI_ISL_1464925, EPI_ISL_1464926, EPI_ISL_1464928, EPI_ISL_1464929, EPI_ISL_1464930, EPI_ISL_1464931, EPI_ISL_1464932, EPI_ISL_1464933, EPI_ISL_1464936, EPI_ISL_1464937, EPI_ISL_1464938, EPI_ISL_1464939, EPI_ISL_1464941, EPI_ISL_1464942, EPI_ISL_1464944, EPI_ISL_1464945, EPI_ISL_1464947 | | | |
| see above | Sonora Quest Laboratories | TGen North | "Jolene Bowers, Heather Centner, Chris French, Hayley Yaglom, Ashlyn Pfeiffer, Darrin Lemmer, Dave Engelthaler, The Arizona COVID Genomics Union (ACGU)" |
| EPI_ISL_1464950 | TGen North | TGen North | "Jolene Bowers, Heather Centner, Chris French, Hayley Yaglom, Ashlyn Pfeiffer, Darrin Lemmer, Dave Engelthaler, The Arizona COVID Genomics Union (ACGU)" |
| EPI_ISL_1464951, EPI_ISL_1464952, EPI_ISL_1464953, EPI_ISL_1464955, EPI_ISL_1464956, EPI_ISL_1464957, EPI_ISL_1464958, EPI_ISL_1464959, EPI_ISL_1464960, EPI_ISL_1464961, EPI_ISL_1464963, EPI_ISL_1464965, EPI_ISL_1464966, EPI_ISL_1464968 | | | |
| see above | Sonora Quest Laboratories | TGen North | "Jolene Bowers, Heather Centner, Chris French, Hayley Yaglom, Ashlyn Pfeiffer, Darrin Lemmer, Dave Engelthaler, The Arizona COVID Genomics Union (ACGU)" |
| EPI_ISL_1464969 | TGen North | TGen North | "Jolene Bowers, Heather Centner, Chris French, Hayley Yaglom, Ashlyn Pfeiffer, Darrin Lemmer, Dave Engelthaler, The Arizona COVID Genomics Union (ACGU)" |
| EPI_ISL_1464971, EPI_ISL_1464973, EPI_ISL_1464974, EPI_ISL_1464975, EPI_ISL_1464978, EPI_ISL_1464980, EPI_ISL_1464982, EPI_ISL_1464984, EPI_ISL_1464986, EPI_ISL_1464987, EPI_ISL_1464988, EPI_ISL_1464989, EPI_ISL_1464990, EPI_ISL_1464992, EPI_ISL_1464993, EPI_ISL_1464995, EPI_ISL_1464996, EPI_ISL_1464997, EPI_ISL_1464998, EPI_ISL_1465004, EPI_ISL_1465006, EPI_ISL_1465007, EPI_ISL_1465008, EPI_ISL_1465009, EPI_ISL_1465010, EPI_ISL_1465012, EPI_ISL_1465013, EPI_ISL_1465014, EPI_ISL_1465018, EPI_ISL_1465019, EPI_ISL_1465021, EPI_ISL_1465022, EPI_ISL_1465027, EPI_ISL_1465028, EPI_ISL_1465029, EPI_ISL_1465030, EPI_ISL_1465032, EPI_ISL_1465035, EPI_ISL_1465037, EPI_ISL_1465038, EPI_ISL_1465040, EPI_ISL_1465041, EPI_ISL_1465042, EPI_ISL_1465044, EPI_ISL_1465045, EPI_ISL_1465046, EPI_ISL_1465048, EPI_ISL_1465049, EPI_ISL_1465050, EPI_ISL_1465052, EPI_ISL_1465053, EPI_ISL_1465054, EPI_ISL_1465055, EPI_ISL_1465057, EPI_ISL_1465058, EPI_ISL_1465059, EPI_ISL_1465060, EPI_ISL_1465061, EPI_ISL_1465064, EPI_ISL_1465065, EPI_ISL_1465066, EPI_ISL_1465068, EPI_ISL_1465070, EPI_ISL_1465071, EPI_ISL_1465072, EPI_ISL_1465075, EPI_ISL_1465076, EPI_ISL_1465077, EPI_ISL_1465078, EPI_ISL_1465079, EPI_ISL_1465080, EPI_ISL_1465081, EPI_ISL_1465085, EPI_ISL_1465086, EPI_ISL_1465087, EPI_ISL_1465089, EPI_ISL_1465090, EPI_ISL_1465092, EPI_ISL_1465093, EPI_ISL_1465094, EPI_ISL_1465096, EPI_ISL_1465097, EPI_ISL_1465099, EPI_ISL_1465101, EPI_ISL_1465102, EPI_ISL_1465103, EPI_ISL_1465104, EPI_ISL_1465107, EPI_ISL_1465108, EPI_ISL_1465109, EPI_ISL_1465112, EPI_ISL_1465114, EPI_ISL_1465115, EPI_ISL_1465118, EPI_ISL_1465120, EPI_ISL_1465125, EPI_ISL_1465126, EPI_ISL_1465127, EPI_ISL_1465129, EPI_ISL_1465130, EPI_ISL_1465133, EPI_ISL_1465134, EPI_ISL_1465135, EPI_ISL_1465137, EPI_ISL_1465139, EPI_ISL_1465140, EPI_ISL_1465141, EPI_ISL_1465143, EPI_ISL_1465144, EPI_ISL_1465145, EPI_ISL_1465146, EPI_ISL_1465147, EPI_ISL_1465148, EPI_ISL_1465149, EPI_ISL_1465151, EPI_ISL_1465152, EPI_ISL_1465156 | | | |
| see above | Sonora Quest Laboratories | TGen North | "Jolene Bowers, Heather Centner, Chris French, Hayley Yaglom, Ashlyn Pfeiffer, Darrin Lemmer, Dave Engelthaler, The Arizona COVID Genomics Union (ACGU)" |
| EPI_ISL_1465157 | TGen North | TGen North | "Jolene Bowers, Heather Centner, Chris French, Hayley Yaglom, Ashlyn Pfeiffer, Darrin Lemmer, Dave Engelthaler, The Arizona COVID Genomics Union (ACGU)" |
| EPI_ISL_1465158, EPI_ISL_1465159, EPI_ISL_1465160, EPI_ISL_1465161, EPI_ISL_1465162, EPI_ISL_1465164, EPI_ISL_1465165, EPI_ISL_1465171, EPI_ISL_1465172, EPI_ISL_1465173, EPI_ISL_1465174, EPI_ISL_1465175, EPI_ISL_1465176, EPI_ISL_1465177, EPI_ISL_1465180, EPI_ISL_1465181, EPI_ISL_1465182, EPI_ISL_1465183, EPI_ISL_1465184, EPI_ISL_1465187, EPI_ISL_1465190, EPI_ISL_1465197, EPI_ISL_1465204, EPI_ISL_1465207, EPI_ISL_1465211, EPI_ISL_1465214, EPI_ISL_1465218, EPI_ISL_1465227, EPI_ISL_1465230, EPI_ISL_1465233, EPI_ISL_1465240, EPI_ISL_1465244, EPI_ISL_1465247, EPI_ISL_1465256, EPI_ISL_1465263, EPI_ISL_1465266, EPI_ISL_1465269, EPI_ISL_1465272, EPI_ISL_1465276, EPI_ISL_1465280, EPI_ISL_1465282, EPI_ISL_1465283, EPI_ISL_1465284, EPI_ISL_1465288, EPI_ISL_1465289, EPI_ISL_1465290, EPI_ISL_1465292, EPI_ISL_1465294, EPI_ISL_1465295, EPI_ISL_1465298, EPI_ISL_1465299, EPI_ISL_1465301, EPI_ISL_1465302, EPI_ISL_1465304, EPI_ISL_1465305, EPI_ISL_1465306, EPI_ISL_1465308, EPI_ISL_1465309, EPI_ISL_1465310, EPI_ISL_1465311, EPI_ISL_1465312, EPI_ISL_1465313, EPI_ISL_1465314, EPI_ISL_1465317, EPI_ISL_1465318, EPI_ISL_1465319, EPI_ISL_1465320, EPI_ISL_1465321, EPI_ISL_1465322, EPI_ISL_1465323, EPI_ISL_1465325, EPI_ISL_1465326, EPI_ISL_1465327, EPI_ISL_1465328, EPI_ISL_1465329, EPI_ISL_1465330, EPI_ISL_1465331, EPI_ISL_1465332, EPI_ISL_1465333, EPI_ISL_1465334, EPI_ISL_1465336, EPI_ISL_1465338, EPI_ISL_1465339, EPI_ISL_1465340, EPI_ISL_1465341, EPI_ISL_1465344, EPI_ISL_1465345, EPI_ISL_1465346, EPI_ISL_1465347, EPI_ISL_1465348, EPI_ISL_1465350, EPI_ISL_1465354, EPI_ISL_1465355, EPI_ISL_1465356, EPI_ISL_1465357, EPI_ISL_1465359 | | | |
| see above | Sonora Quest Laboratories | TGen North | "Jolene Bowers, Heather Centner, Chris French, Hayley Yaglom, Ashlyn Pfeiffer, Darrin Lemmer, Dave Engelthaler, The Arizona COVID Genomics Union (ACGU)" |
| EPI_ISL_1465360 | TGen North | TGen North | "Jolene Bowers, Heather Centner, Chris French, Hayley Yaglom, Ashlyn Pfeiffer, Darrin Lemmer, Dave Engelthaler, The Arizona COVID Genomics Union (ACGU)" |
| EPI_ISL_1465361 | Sonora Quest Laboratories | TGen North | "Jolene Bowers, Heather Centner, Chris French, Hayley Yaglom, Ashlyn Pfeiffer, Darrin Lemmer, Dave Engelthaler, The Arizona COVID Genomics Union (ACGU)" |
| EPI_ISL_1465362 | TGen North | TGen North | "Jolene Bowers, Heather Centner, Chris French, Hayley Yaglom, Ashlyn Pfeiffer, Darrin Lemmer, Dave Engelthaler, The Arizona COVID Genomics Union (ACGU)" |
| EPI_ISL_1465363, EPI_ISL_1465365, EPI_ISL_1465366, EPI_ISL_1465368, EPI_ISL_1465369, EPI_ISL_1465370, EPI_ISL_1465371, EPI_ISL_1465374, EPI_ISL_1465375, EPI_ISL_1465376, EPI_ISL_1465379, EPI_ISL_1465383, EPI_ISL_1465384, EPI_ISL_1465385, EPI_ISL_1465387, EPI_ISL_1465389, EPI_ISL_1465390, EPI_ISL_1465391, EPI_ISL_1465392, EPI_ISL_1465393, EPI_ISL_1465394, EPI_ISL_1465395, EPI_ISL_1465398, EPI_ISL_1465399, EPI_ISL_1465400, EPI_ISL_1465402, EPI_ISL_1465403, EPI_ISL_1465404, EPI_ISL_1465406, EPI_ISL_1465407, EPI_ISL_1465408, EPI_ISL_1465409, EPI_ISL_1465411, EPI_ISL_1465414, EPI_ISL_1465416 | | | |
| see above | Sonora Quest Laboratories | TGen North | "Jolene Bowers, Heather Centner, Chris French, Hayley Yaglom, Ashlyn Pfeiffer, Darrin Lemmer, Dave Engelthaler, The Arizona COVID Genomics Union (ACGU)" |
| EPI_ISL_1465417 | TGen North | TGen North | "Jolene Bowers, Heather Centner, Chris French, Hayley Yaglom, Ashlyn Pfeiffer, Darrin Lemmer, Dave Engelthaler, The Arizona COVID Genomics Union (ACGU)" |
| EPI_ISL_1465420, EPI_ISL_1465421 | Sonora Quest Laboratories | TGen North | "Jolene Bowers, Heather Centner, Chris French, Hayley Yaglom, Ashlyn Pfeiffer, Darrin Lemmer, Dave Engelthaler, The Arizona COVID Genomics Union (ACGU)" |
| EPI_ISL_1465423 | TGen North | TGen North | "Jolene Bowers, Heather Centner, Chris French, Hayley Yaglom, Ashlyn Pfeiffer, Darrin Lemmer, Dave Engelthaler, The Arizona COVID Genomics Union (ACGU)" |
| EPI_ISL_1465424, EPI_ISL_1465425, EPI_ISL_1465426, EPI_ISL_1465428, EPI_ISL_1465429, EPI_ISL_1465430, EPI_ISL_1465432, EPI_ISL_1465433, EPI_ISL_1465436, EPI_ISL_1465437 | Sonora Quest Laboratories | TGen North | "Jolene Bowers, Heather Centner, Chris French, Hayley Yaglom, Ashlyn Pfeiffer, Darrin Lemmer, Dave Engelthaler, The Arizona COVID Genomics Union (ACGU)" |
| EPI_ISL_1465615, EPI_ISL_1465616 | ALBANY MEDICAL CENTER | Wadsworth Center, New York State Department of Health | Kirsten St. George, Daryl M. Lamson, Alexis Russell, Matthew Shudt, Melissa A Leisner, Jonathan Plitnick, Catharine Prussing, Navjot Singh, John Kelly, Erasmus Schneider, Erica Lasek-Nesselquist |
| EPI_ISL_1465617 | GLENS FALLS HOSPITAL LABORATORY | Wadsworth Center, New York State Department of Health | Kirsten St. George, Daryl M. Lamson, Alexis Russell, Matthew Shudt, Melissa A Leisner, Jonathan Plitnick, Catharine Prussing, Navjot Singh, John Kelly, Erasmus Schneider, Erica Lasek-Nesselquist |
| EPI_ISL_1465618, EPI_ISL_1465619, EPI_ISL_1465620, | URMC LABS | Wadsworth Center, New York State Department of Health | Kirsten St. George, Daryl M. Lamson, Alexis Russell, Matthew Shudt, Melissa A Leisner, Jonathan Plitnick, Catharine Prussing, Navjot Singh, John Kelly, Erasmus Schneider, Erica Lasek-Nesselquist |

| | | | |
|---|---|---|---|
| EPI_ISL_1465621 | | | |
| EPI_ISL_1465622 | WESTCHESTER MEDICAL CENTER | Wadsworth Center, New York State Department of Health | Kirsten St. George, Daryl M. Lamson, Alexis Russell, Matthew Shudt, Melissa A Leisner, Jonathan Plitnick, Catharine Prussing, Navjot Singh, John Kelly, Erasmus Schneider, Erica Lasek-Nesselquist |
| EPI_ISL_1465623, EPI_ISL_1465624, EPI_ISL_1465625, EPI_ISL_1465626, EPI_ISL_1465627, EPI_ISL_1465628, EPI_ISL_1465629, EPI_ISL_1465630, EPI_ISL_1465631, EPI_ISL_1465632 | URMC LABS | Wadsworth Center, New York State Department of Health | Kirsten St. George, Daryl M. Lamson, Alexis Russell, Matthew Shudt, Melissa A Leisner, Jonathan Plitnick, Catharine Prussing, Navjot Singh, John Kelly, Erasmus Schneider, Erica Lasek-Nesselquist |
| EPI_ISL_1465633 | Columbia University Irving Medical Center | Wadsworth Center, New York State Department of Health | Kirsten St. George, Daryl M. Lamson, Alexis Russell, Matthew Shudt, Melissa A Leisner, Jonathan Plitnick, Catharine Prussing, Navjot Singh, John Kelly, Erasmus Schneider, Erica Lasek-Nesselquist |
| EPI_ISL_1465634, EPI_ISL_1465635, EPI_ISL_1465636 | ALBANY MEDICAL CENTER | Wadsworth Center, New York State Department of Health | Kirsten St. George, Daryl M. Lamson, Alexis Russell, Matthew Shudt, Melissa A Leisner, Jonathan Plitnick, Catharine Prussing, Navjot Singh, John Kelly, Erasmus Schneider, Erica Lasek-Nesselquist |
| EPI_ISL_1465637 | WESTCHESTER MEDICAL CENTER | Wadsworth Center, New York State Department of Health | Kirsten St. George, Daryl M. Lamson, Alexis Russell, Matthew Shudt, Melissa A Leisner, Jonathan Plitnick, Catharine Prussing, Navjot Singh, John Kelly, Erasmus Schneider, Erica Lasek-Nesselquist |
| EPI_ISL_1465638 | SUNY UPSTATE MEDICAL UNIVERSITY | Wadsworth Center, New York State Department of Health | Kirsten St. George, Daryl M. Lamson, Alexis Russell, Matthew Shudt, Melissa A Leisner, Jonathan Plitnick, Catharine Prussing, Navjot Singh, John Kelly, Erasmus Schneider, Erica Lasek-Nesselquist |
| EPI_ISL_1465639 | ALBANY MEDICAL CENTER | Wadsworth Center, New York State Department of Health | Kirsten St. George, Daryl M. Lamson, Alexis Russell, Matthew Shudt, Melissa A Leisner, Jonathan Plitnick, Catharine Prussing, Navjot Singh, John Kelly, Erasmus Schneider, Erica Lasek-Nesselquist |
| EPI_ISL_1465640, EPI_ISL_1465641, EPI_ISL_1465642, EPI_ISL_1465643, EPI_ISL_1465644, EPI_ISL_1465645, EPI_ISL_1465646, EPI_ISL_1465647, EPI_ISL_1465648, EPI_ISL_1465649, EPI_ISL_1465650, EPI_ISL_1465651, EPI_ISL_1465652, EPI_ISL_1465653, EPI_ISL_1465654, EPI_ISL_1465655, EPI_ISL_1465656, EPI_ISL_1465657, EPI_ISL_1465658, EPI_ISL_1465659, EPI_ISL_1465660, EPI_ISL_1465661, EPI_ISL_1465662, EPI_ISL_1465663, EPI_ISL_1465664 | | | |
| see above | URMC LABS | Wadsworth Center, New York State Department of Health | Kirsten St. George, Daryl M. Lamson, Alexis Russell, Matthew Shudt, Melissa A Leisner, Jonathan Plitnick, Catharine Prussing, Navjot Singh, John Kelly, Erasmus Schneider, Erica Lasek-Nesselquist |
| EPI_ISL_1465665, EPI_ISL_1465666 | SUNY UPSTATE MEDICAL UNIVERSITY | Wadsworth Center, New York State Department of Health | Kirsten St. George, Daryl M. Lamson, Alexis Russell, Matthew Shudt, Melissa A Leisner, Jonathan Plitnick, Catharine Prussing, Navjot Singh, John Kelly, Erasmus Schneider, Erica Lasek-Nesselquist |
| EPI_ISL_1465667 | URMC LABS | Wadsworth Center, New York State Department of Health | Kirsten St. George, Daryl M. Lamson, Alexis Russell, Matthew Shudt, Melissa A Leisner, Jonathan Plitnick, Catharine Prussing, Navjot Singh, John Kelly, Erasmus Schneider, Erica Lasek-Nesselquist |
| EPI_ISL_1465668, EPI_ISL_1465669, EPI_ISL_1465670, EPI_ISL_1465671, EPI_ISL_1465672 | MONTEFIORE MEDICAL CENTER LABORATORIES | Wadsworth Center, New York State Department of Health | Kirsten St. George, Daryl M. Lamson, Alexis Russell, Matthew Shudt, Melissa A Leisner, Jonathan Plitnick, Catharine Prussing, Navjot Singh, John Kelly, Erasmus Schneider, Erica Lasek-Nesselquist |
| EPI_ISL_1465673, EPI_ISL_1465674, EPI_ISL_1465675, EPI_ISL_1465676, EPI_ISL_1465677 | THE MARY IMOGENE BASSETT HOSPITAL | Wadsworth Center, New York State Department of Health | Kirsten St. George, Daryl M. Lamson, Alexis Russell, Matthew Shudt, Melissa A Leisner, Jonathan Plitnick, Catharine Prussing, Navjot Singh, John Kelly, Erasmus Schneider, Erica Lasek-Nesselquist |
| EPI_ISL_1465678, EPI_ISL_1465679, EPI_ISL_1465680, EPI_ISL_1465681, EPI_ISL_1465682 | NORTHWELL HEALTH LABORATORIES | Wadsworth Center, New York State Department of Health | Kirsten St. George, Daryl M. Lamson, Alexis Russell, Matthew Shudt, Melissa A Leisner, Jonathan Plitnick, Catharine Prussing, Navjot Singh, John Kelly, Erasmus Schneider, Erica Lasek-Nesselquist |
| EPI_ISL_1465683, EPI_ISL_1465684, EPI_ISL_1465685, EPI_ISL_1465686, EPI_ISL_1465687, EPI_ISL_1465688 | Kansas Health and Environmental Lab | Kansas Health and Environmental Lab | Mike Grose, Jonathan Barnell, Ben Olsen, and Phil Adam |
| EPI_ISL_1465689, EPI_ISL_1465690, EPI_ISL_1465691, EPI_ISL_1465692 | SUNY UPSTATE MEDICAL UNIVERSITY | Wadsworth Center, New York State Department of Health | Kirsten St. George, Daryl M. Lamson, Alexis Russell, Matthew Shudt, Melissa A Leisner, Jonathan Plitnick, Catharine Prussing, Navjot Singh, John Kelly, Erasmus Schneider, Erica Lasek-Nesselquist |
| EPI_ISL_1465693, EPI_ISL_1465694, EPI_ISL_1465695 | Columbia University Irving Medical Center | Wadsworth Center, New York State Department of Health | Kirsten St. George, Daryl M. Lamson, Alexis Russell, Matthew Shudt, Melissa A Leisner, Jonathan Plitnick, Catharine Prussing, Navjot Singh, John Kelly, Erasmus Schneider, Erica Lasek-Nesselquist |
| EPI_ISL_1465696 | URMC LABS | Wadsworth Center, New York State Department of Health | Kirsten St. George, Daryl M. Lamson, Alexis Russell, Matthew Shudt, Melissa A Leisner, Jonathan Plitnick, Catharine Prussing, Navjot Singh, John Kelly, Erasmus Schneider, Erica Lasek-Nesselquist |
| EPI_ISL_1465697, EPI_ISL_1465698, EPI_ISL_1465699, EPI_ISL_1465700 | SUNY UPSTATE MEDICAL UNIVERSITY | Wadsworth Center, New York State Department of Health | Kirsten St. George, Daryl M. Lamson, Alexis Russell, Matthew Shudt, Melissa A Leisner, Jonathan Plitnick, Catharine Prussing, Navjot Singh, John Kelly, Erasmus Schneider, Erica Lasek-Nesselquist |
| EPI_ISL_1465701, EPI_ISL_1465702, EPI_ISL_1465703, EPI_ISL_1465704, EPI_ISL_1465705, EPI_ISL_1465706, EPI_ISL_1465707, EPI_ISL_1465708, EPI_ISL_1465709, EPI_ISL_1465710, EPI_ISL_1465711, EPI_ISL_1465712, EPI_ISL_1465713 | | | |
| see above | MONTEFIORE MEDICAL CENTER LABORATORIES | Wadsworth Center, New York State Department of Health | Kirsten St. George, Daryl M. Lamson, Alexis Russell, Matthew Shudt, Melissa A Leisner, Jonathan Plitnick, Catharine Prussing, Navjot Singh, John Kelly, Erasmus Schneider, Erica Lasek-Nesselquist |
| EPI_ISL_1465714, EPI_ISL_1465715, EPI_ISL_1465716, EPI_ISL_1465717, EPI_ISL_1465718, EPI_ISL_1465719, EPI_ISL_1465720, EPI_ISL_1465721, EPI_ISL_1465722 | NYC Pandemic Response Lab | Wadsworth Center, New York State Department of Health | Kirsten St. George, Daryl M. Lamson, Alexis Russell, Matthew Shudt, Melissa A Leisner, Jonathan Plitnick, Catharine Prussing, Navjot Singh, John Kelly, Erasmus Schneider, Erica Lasek-Nesselquist |
| EPI_ISL_1465723, EPI_ISL_1465724, EPI_ISL_1465725, EPI_ISL_1465726, EPI_ISL_1465727, EPI_ISL_1465728, EPI_ISL_1465729, EPI_ISL_1465730, EPI_ISL_1465731, EPI_ISL_1465732, EPI_ISL_1465733, EPI_ISL_1465734, EPI_ISL_1465735, EPI_ISL_1465736, EPI_ISL_1465737, EPI_ISL_1465738, EPI_ISL_1465739, EPI_ISL_1465740, EPI_ISL_1465741, EPI_ISL_1465742, EPI_ISL_1465743, EPI_ISL_1465744, EPI_ISL_1465745, EPI_ISL_1465746, EPI_ISL_1465747, EPI_ISL_1465748, EPI_ISL_1465749, EPI_ISL_1465750, EPI_ISL_1465751, EPI_ISL_1465752, EPI_ISL_1465753, EPI_ISL_1465754 | | | |
| see above | URMC LABS | Wadsworth Center, New York State Department of Health | Kirsten St. George, Daryl M. Lamson, Alexis Russell, Matthew Shudt, Melissa A Leisner, Jonathan Plitnick, Catharine Prussing, Navjot Singh, John Kelly, Erasmus Schneider, Erica Lasek-Nesselquist |
| EPI_ISL_1465755, EPI_ISL_1465756, EPI_ISL_1465757, EPI_ISL_1465758, EPI_ISL_1465759 | NORTHWELL HEALTH LABORATORIES | Wadsworth Center, New York State Department of Health | Kirsten St. George, Daryl M. Lamson, Alexis Russell, Matthew Shudt, Melissa A Leisner, Jonathan Plitnick, Catharine Prussing, Navjot Singh, John Kelly, Erasmus Schneider, Erica Lasek-Nesselquist |
| EPI_ISL_1465760 | THE MARY IMOGENE BASSETT HOSPITAL | Wadsworth Center, New York State Department of Health | Kirsten St. George, Daryl M. Lamson, Alexis Russell, Matthew Shudt, Melissa A Leisner, Jonathan Plitnick, Catharine Prussing, Navjot Singh, John Kelly, Erasmus Schneider, Erica Lasek-Nesselquist |
| EPI_ISL_1465761, EPI_ISL_1465762, EPI_ISL_1465763, EPI_ISL_1465764, EPI_ISL_1465765, EPI_ISL_1465766 | NORTHWELL HEALTH LABORATORIES | Wadsworth Center, New York State Department of Health | Kirsten St. George, Daryl M. Lamson, Alexis Russell, Matthew Shudt, Melissa A Leisner, Jonathan Plitnick, Catharine Prussing, Navjot Singh, John Kelly, Erasmus Schneider, Erica Lasek-Nesselquist |
| EPI_ISL_1465767, EPI_ISL_1465768, EPI_ISL_1465769, EPI_ISL_1465770, EPI_ISL_1465771, EPI_ISL_1465772, EPI_ISL_1465773, EPI_ISL_1465774, EPI_ISL_1465775, EPI_ISL_1465776, EPI_ISL_1465777, EPI_ISL_1465778, EPI_ISL_1465779, EPI_ISL_1465780, EPI_ISL_1465781, EPI_ISL_1465782, EPI_ISL_1465783, EPI_ISL_1465784, EPI_ISL_1465785, EPI_ISL_1465786, EPI_ISL_1465787, EPI_ISL_1465788, EPI_ISL_1465789, EPI_ISL_1465790, EPI_ISL_1465791, EPI_ISL_1465792, EPI_ISL_1465793 | | | |

| | | | |
|---|---|---|---|
| see above | Gundersen Clinical Microbiology Laboratory | Kabara Cancer Research Institute | Craig S. Richmond, Paraic A. Kenny |
| EPI_ISL_1465794, EPI_ISL_1465795, EPI_ISL_1465796, EPI_ISL_1465797, EPI_ISL_1465798, EPI_ISL_1465799, EPI_ISL_1465800, EPI_ISL_1465801 | MONTEFIORE MEDICAL CENTER LABORATORIES | Wadsworth Center, New York State Department of Health | Kirsten St. George, Daryl M. Lamson, Alexis Russell, Matthew Shudt, Melissa A Leisner, Jonathan Plitnick, Catharine Prussing, Navjot Singh, John Kelly, Erasmus Schneider, Erica Lasek-Nesselquist |
| EPI_ISL_1465802 | MONTEFIORE NEW ROCHELLE HOSPITAL LABORATORY | Wadsworth Center, New York State Department of Health | Kirsten St. George, Daryl M. Lamson, Alexis Russell, Matthew Shudt, Melissa A Leisner, Jonathan Plitnick, Catharine Prussing, Navjot Singh, John Kelly, Erasmus Schneider, Erica Lasek-Nesselquist |
| EPI_ISL_1465803, EPI_ISL_1465804 | Columbia University Irving Medical Center | Wadsworth Center, New York State Department of Health | Kirsten St. George, Daryl M. Lamson, Alexis Russell, Matthew Shudt, Melissa A Leisner, Jonathan Plitnick, Catharine Prussing, Navjot Singh, John Kelly, Erasmus Schneider, Erica Lasek-Nesselquist |
| EPI_ISL_1465805, EPI_ISL_1465806, EPI_ISL_1465807, EPI_ISL_1465808, EPI_ISL_1465809, EPI_ISL_1465810, EPI_ISL_1465811, EPI_ISL_1465812, EPI_ISL_1465813, EPI_ISL_1465814, EPI_ISL_1465815, EPI_ISL_1465816, EPI_ISL_1465817, EPI_ISL_1465818, EPI_ISL_1465819, EPI_ISL_1465820, EPI_ISL_1465821, EPI_ISL_1465822, EPI_ISL_1465823, EPI_ISL_1465824, EPI_ISL_1465825, EPI_ISL_1465826, EPI_ISL_1465827, EPI_ISL_1465828, EPI_ISL_1465829, EPI_ISL_1465830, EPI_ISL_1465831, EPI_ISL_1465832, EPI_ISL_1465833, EPI_ISL_1465834 | | | |
| see above | NYC Pandemic Response Lab | Wadsworth Center, New York State Department of Health | Kirsten St. George, Daryl M. Lamson, Alexis Russell, Matthew Shudt, Melissa A Leisner, Jonathan Plitnick, Catharine Prussing, Navjot Singh, John Kelly, Erasmus Schneider, Erica Lasek-Nesselquist |
| EPI_ISL_1465835, EPI_ISL_1465836, EPI_ISL_1465837 | NORTHWELL HEALTH LABORATORIES | Wadsworth Center, New York State Department of Health | Kirsten St. George, Daryl M. Lamson, Alexis Russell, Matthew Shudt, Melissa A Leisner, Jonathan Plitnick, Catharine Prussing, Navjot Singh, John Kelly, Erasmus Schneider, Erica Lasek-Nesselquist |
| EPI_ISL_1465838, EPI_ISL_1465839, EPI_ISL_1465840, EPI_ISL_1465841, EPI_ISL_1465842, EPI_ISL_1465843, EPI_ISL_1465844, EPI_ISL_1465845, EPI_ISL_1465846, EPI_ISL_1465847 | STONY BROOK UNIVERSITY HOSPITAL | Wadsworth Center, New York State Department of Health | Kirsten St. George, Daryl M. Lamson, Alexis Russell, Matthew Shudt, Melissa A Leisner, Jonathan Plitnick, Catharine Prussing, Navjot Singh, John Kelly, Erasmus Schneider, Erica Lasek-Nesselquist |
| EPI_ISL_1465848 | GOOD SAMARITAN HOSPITAL LABORATORY | Wadsworth Center, New York State Department of Health | Kirsten St. George, Daryl M. Lamson, Alexis Russell, Matthew Shudt, Melissa A Leisner, Jonathan Plitnick, Catharine Prussing, Navjot Singh, John Kelly, Erasmus Schneider, Erica Lasek-Nesselquist |
| EPI_ISL_1465849, EPI_ISL_1465850, EPI_ISL_1465851, EPI_ISL_1465852 | MONTEFIORE NEW ROCHELLE HOSPITAL LABORATORY | Wadsworth Center, New York State Department of Health | Kirsten St. George, Daryl M. Lamson, Alexis Russell, Matthew Shudt, Melissa A Leisner, Jonathan Plitnick, Catharine Prussing, Navjot Singh, John Kelly, Erasmus Schneider, Erica Lasek-Nesselquist |
| EPI_ISL_1465853, EPI_ISL_1465854, EPI_ISL_1465855, EPI_ISL_1465856 | GLENS FALLS HOSPITAL LABORATORY | Wadsworth Center, New York State Department of Health | Kirsten St. George, Daryl M. Lamson, Alexis Russell, Matthew Shudt, Melissa A Leisner, Jonathan Plitnick, Catharine Prussing, Navjot Singh, John Kelly, Erasmus Schneider, Erica Lasek-Nesselquist |
| EPI_ISL_1465857 | ITHACA COLLEGE LABORATORY HAMMOND HEALTH CENTER | Wadsworth Center, New York State Department of Health | Kirsten St. George, Daryl M. Lamson, Alexis Russell, Matthew Shudt, Melissa A Leisner, Jonathan Plitnick, Catharine Prussing, Navjot Singh, John Kelly, Erasmus Schneider, Erica Lasek-Nesselquist |
| EPI_ISL_1465858 | GOOD SAMARITAN HOSPITAL LABORATORY | Wadsworth Center, New York State Department of Health | Kirsten St. George, Daryl M. Lamson, Alexis Russell, Matthew Shudt, Melissa A Leisner, Jonathan Plitnick, Catharine Prussing, Navjot Singh, John Kelly, Erasmus Schneider, Erica Lasek-Nesselquist |
| EPI_ISL_1465859, EPI_ISL_1465860, EPI_ISL_1465861, EPI_ISL_1465862, EPI_ISL_1465863, EPI_ISL_1465864, EPI_ISL_1465865, EPI_ISL_1465866, EPI_ISL_1465867, EPI_ISL_1465868, EPI_ISL_1465869, EPI_ISL_1465870, EPI_ISL_1465871, EPI_ISL_1465872, EPI_ISL_1465873, EPI_ISL_1465874, EPI_ISL_1465875, EPI_ISL_1465876, EPI_ISL_1465877, EPI_ISL_1465878 | | | |
| see above | TEMPUS LABS INC | Wadsworth Center, New York State Department of Health | Kirsten St. George, Daryl M. Lamson, Alexis Russell, Matthew Shudt, Melissa A Leisner, Jonathan Plitnick, Catharine Prussing, Navjot Singh, John Kelly, Erasmus Schneider, Erica Lasek-Nesselquist |
| EPI_ISL_1468405, EPI_ISL_1468406, EPI_ISL_1468407, EPI_ISL_1468408, EPI_ISL_1468409, EPI_ISL_1468410 | Ohio Department of Health Laboratory | Ohio Department of Health Laboratory | Holmes, Jennifer; Eric Brandt, Keoni Omura, Glen McGillivary, Caitlin McDonnell, Jade Mowery, Stephanie Mccracken, Tyler Payne, Kirtana Ramadugu, Erica Leasure, Brent Lee, Kelsey Florek, Heather Blankenship, Allison Black, Quanta Brown, and Tammy Bannerman |
| EPI_ISL_1468532, EPI_ISL_1468533, EPI_ISL_1468534, EPI_ISL_1468535, EPI_ISL_1468536, EPI_ISL_1468537, EPI_ISL_1468538, EPI_ISL_1468539, EPI_ISL_1468540, EPI_ISL_1468541, EPI_ISL_1468542, EPI_ISL_1468543, EPI_ISL_1468544, EPI_ISL_1468545, EPI_ISL_1468546, EPI_ISL_1468547, EPI_ISL_1468548, EPI_ISL_1468549, EPI_ISL_1468550, EPI_ISL_1468551, EPI_ISL_1468552, EPI_ISL_1468553, EPI_ISL_1468554, EPI_ISL_1468555, EPI_ISL_1468556, EPI_ISL_1468557, EPI_ISL_1468558, EPI_ISL_1468559, EPI_ISL_1468560, EPI_ISL_1468561, EPI_ISL_1468562, EPI_ISL_1468563, EPI_ISL_1468564, EPI_ISL_1468565, EPI_ISL_1468566, EPI_ISL_1468567, EPI_ISL_1468568, EPI_ISL_1468569, EPI_ISL_1468570, EPI_ISL_1468571, EPI_ISL_1468572, EPI_ISL_1468573, EPI_ISL_1468574, EPI_ISL_1468575, EPI_ISL_1468577, EPI_ISL_1468578, EPI_ISL_1468579, EPI_ISL_1468580, EPI_ISL_1468581, EPI_ISL_1468582, EPI_ISL_1468583, EPI_ISL_1468584, EPI_ISL_1468585, EPI_ISL_1468586, EPI_ISL_1468587, EPI_ISL_1468588, EPI_ISL_1468589, EPI_ISL_1468590, EPI_ISL_1468591, EPI_ISL_1468592, EPI_ISL_1468593, EPI_ISL_1468594, EPI_ISL_1468595, EPI_ISL_1468596, EPI_ISL_1468597, EPI_ISL_1468598, EPI_ISL_1468599, EPI_ISL_1468600, EPI_ISL_1468601, EPI_ISL_1468602, EPI_ISL_1468603, EPI_ISL_1468604, EPI_ISL_1468605, EPI_ISL_1468606, EPI_ISL_1468607, EPI_ISL_1468608, EPI_ISL_1468609, EPI_ISL_1468610, EPI_ISL_1468611, EPI_ISL_1468612, EPI_ISL_1468613, EPI_ISL_1468614, EPI_ISL_1468615, EPI_ISL_1468616, EPI_ISL_1468617, EPI_ISL_1468618, EPI_ISL_1468619, EPI_ISL_1468620, EPI_ISL_1468621, EPI_ISL_1468622, EPI_ISL_1468623, EPI_ISL_1468624, EPI_ISL_1468625, EPI_ISL_1468627, EPI_ISL_1468628, EPI_ISL_1468629, EPI_ISL_1468630, EPI_ISL_1468631, EPI_ISL_1468632, EPI_ISL_1468633, EPI_ISL_1468634, EPI_ISL_1468635, EPI_ISL_1468636, EPI_ISL_1468637, EPI_ISL_1468638, EPI_ISL_1468639, EPI_ISL_1468640, EPI_ISL_1468641, EPI_ISL_1468642, EPI_ISL_1468643, EPI_ISL_1468644, EPI_ISL_1468645, EPI_ISL_1468646, EPI_ISL_1468647, EPI_ISL_1468648, EPI_ISL_1468649, EPI_ISL_1468650, EPI_ISL_1468651, EPI_ISL_1468652, EPI_ISL_1468653, EPI_ISL_1468654, EPI_ISL_1468655 | | | |
| see above | Johns Hopkins Hospital Department of Pathology | Johns Hopkins Hospital Department of Pathology | C. Paul Morris, Chun Huai Luo, Adannaya Amadi, Matthew Schwartz, Heba H. Mostafa |
| EPI_ISL_1468656, EPI_ISL_1468658, EPI_ISL_1468659, EPI_ISL_1468661, EPI_ISL_1468662, EPI_ISL_1468664, EPI_ISL_1468665 | Wyoming Public Health Laboratory | Wyoming Public Health Laboratory | Jim Mildenberger, Wanda Manley, Noah Hull, Taylor Fearing, Lynette Gumbleton, Channing Weber, Ashley Norberg, Chayse Rowley, Marley Goetz, Brian Dominguez, Elliot Thomasson, Cari Sloma, and Rob Christensen |
| EPI_ISL_1468667, EPI_ISL_1468668, EPI_ISL_1468669, EPI_ISL_1468670 | Altius Institute | Seattle Flu Study | Deborah A. Nickerson, Chris D. Frazar, Jover Lee, Benjamin Pelle, Erica Ryke, Matthew Richardson, Amanda Adler, Elisabeth Brandstetter, Peter D. Han, Kairsten Fay, Misja Ilcisin, Kirsten Lacombe, Thomas R. Sibley, Melissa Truong, Caitlin R. Wolf, Ryan Alexander, Daniel Bates, Rebecca Bruders, Stephanie DeBaun, Clem Green, Muhammad Halimun, Jessica Halow, Kneshay Harper, Matt Hartman, Andrew Meuser, Alex Nguyen, Truong Nguyen, Sofia Olsson, Sadie Patraw, Hannah Petersen, Tobias Ragoczy, Joshua Richards, Jacob Rodriguez, John Stamatoyannopoulos, Julia Wald, Olivia Waltner, Michael Boeckh, Janet A. Englund, Michael Famulare, Barry R. Lutz, Mark J. Rieder, Lea M. Starita, Matthew Thompson, Helen Y. Chu, Jay Shendure, Trevor Bedford |
| EPI_ISL_1468671, EPI_ISL_1468672 | Seattle Flu Study | Seattle Flu Study | Deborah A. Nickerson, Chris D. Frazar, Jover Lee, Benjamin Pelle, Erica Ryke, Matthew Richardson, Amanda Adler, Elisabeth Brandstetter, Peter D. Han, Kairsten Fay, Misja Ilcisin, Kirsten Lacombe, Thomas R. Sibley, Melissa Truong, Caitlin R. Wolf, Karen Cowgill, Stephanie Schrag, Jeff Duchin, Michael Boeckh, Janet A. Englund, Michael Famulare, Barry R. Lutz, Mark J. Rieder, Lea M. Starita, Matthew Thompson, Helen Y. Chu, Trevor Bedford, Jay Shendure |
| EPI_ISL_1468673, EPI_ISL_1468674, EPI_ISL_1468675 | Altius Institute | Seattle Flu Study | Deborah A. Nickerson, Chris D. Frazar, Jover Lee, Benjamin Pelle, Erica Ryke, Matthew Richardson, Amanda Adler, Elisabeth Brandstetter, Peter D. Han, Kairsten Fay, Misja Ilcisin, Kirsten Lacombe, Thomas R. Sibley, Melissa Truong, Caitlin R. Wolf, Ryan Alexander, Daniel Bates, Rebecca Bruders, Stephanie DeBaun, Clem Green, Muhammad Halimun, Jessica Halow, Kneshay Harper, Matt Hartman, Andrew Meuser, Alex Nguyen, Truong Nguyen, Sofia Olsson, Sadie Patraw, Hannah Petersen, Tobias Ragoczy, Joshua Richards, Jacob Rodriguez, John Stamatoyannopoulos, Julia Wald, Olivia Waltner, Michael Boeckh, Janet A. Englund, Michael Famulare, Barry R. Lutz, Mark J. Rieder, Lea M. Starita, Matthew Thompson, Helen Y. Chu, Jay Shendure, Trevor Bedford |
| EPI_ISL_1468676, EPI_ISL_1468677 | Seattle Flu Study | Seattle Flu Study | Deborah A. Nickerson, Chris D. Frazar, Jover Lee, Benjamin Pelle, Erica Ryke, Matthew Richardson, Amanda Adler, Elisabeth Brandstetter, Peter D. Han, Kairsten Fay, Misja Ilcisin, Kirsten Lacombe, Thomas R. Sibley, Melissa Truong, Caitlin R. Wolf, Michael Boeckh, Janet A. Englund, Michael Famulare, Barry R. Lutz, Mark J. Rieder, Lea M. Starita, Matthew Thompson, Jay Shendure, Trevor Bedford, Helen Y. Chu |
| EPI_ISL_1468678, EPI_ISL_1468679 | Seattle Flu Study | Seattle Flu Study | Deborah A. Nickerson, Chris D. Frazar, Jover Lee, Benjamin Pelle, Erica Ryke, Matthew Richardson, Amanda Adler, Elisabeth Brandstetter, Peter D. Han, Kairsten Fay, Misja Ilcisin, Kirsten Lacombe, Thomas R. Sibley, Melissa Truong, Caitlin R. Wolf, Karen Cowgill, Stephanie Schrag, Jeff Duchin, Michael Boeckh, Janet A. Englund, Michael Famulare, Barry R. Lutz, Mark J. Rieder, Lea M. Starita, Matthew Thompson, Helen Y. Chu, Trevor Bedford, Jay Shendure |
| EPI_ISL_1468680, EPI_ISL_1468681, EPI_ISL_1468682 | Altius Institute | Seattle Flu Study | Deborah A. Nickerson, Chris D. Frazar, Jover Lee, Benjamin Pelle, Erica Ryke, Matthew Richardson, Amanda Adler, Elisabeth Brandstetter, Peter D. Han, Kairsten Fay, Misja Ilcisin, Kirsten Lacombe, Thomas R. Sibley, Melissa Truong, Caitlin R. Wolf, Ryan Alexander, Daniel Bates, Rebecca Bruders, Stephanie DeBaun, Clem Green, Muhammad Halimun, Jessica Halow, Kneshay Harper, Matt Hartman, Andrew Meuser, Alex Nguyen, Truong Nguyen, Sofia Olsson, Sadie Patraw, Hannah Petersen, Tobias Ragoczy, Joshua Richards, Jacob Rodriguez, John Stamatoyannopoulos, Julia Wald, Olivia Waltner, Michael Boeckh, Janet A. Englund, Michael Famulare, Barry R. Lutz, Mark J. Rieder, Lea M. Starita, Matthew Thompson, Helen Y. Chu, Jay Shendure, Trevor Bedford |
| EPI_ISL_1468683 | Seattle Flu Study | Seattle Flu Study | Deborah A. Nickerson, Chris D. Frazar, Jover Lee, Benjamin Pelle, Erica Ryke, Matthew Richardson, Amanda Adler, Elisabeth Brandstetter, Peter D. Han, Kairsten Fay, Misja Ilcisin, Kirsten Lacombe, Thomas R. Sibley, Melissa Truong, Caitlin R. Wolf, Karen Cowgill, Stephanie Schrag, Jeff Duchin, Michael Boeckh, Janet A. Englund, Michael Famulare, Barry R. Lutz, Mark J. Rieder, Lea M. Starita, Matthew Thompson, Helen Y. Chu, Trevor Bedford, Jay Shendure |
| EPI_ISL_1468684 | Altius Institute | Seattle Flu Study | Deborah A. Nickerson, Chris D. Frazar, Jover Lee, Benjamin Pelle, Erica Ryke, Matthew Richardson, Amanda Adler, Elisabeth Brandstetter, Peter D. Han, Kairsten Fay, Misja Ilcisin, Kirsten Lacombe, Thomas R. Sibley, Melissa Truong, Caitlin R. Wolf, Ryan Alexander, Daniel Bates, Rebecca Bruders, Stephanie DeBaun, Clem Green, Muhammad Halimun, Jessica Halow, Kneshay Harper, Matt Hartman, Andrew Meuser, Alex Nguyen, Truong Nguyen, Sofia Olsson, Sadie Patraw, Hannah Petersen, Tobias Ragoczy, Joshua Richards, Jacob Rodriguez, John Stamatoyannopoulos, Julia Wald, Olivia Waltner, Michael Boeckh, Janet A. Englund, Michael Famulare, Barry R. Lutz, Mark J. Rieder, Lea M. Starita, Matthew Thompson, Helen Y. Chu, Jay Shendure, Trevor Bedford |
| EPI_ISL_1468685, EPI_ISL_1468686, EPI_ISL_1468687 | Seattle Flu Study | Seattle Flu Study | Deborah A. Nickerson, Chris D. Frazar, Jover Lee, Benjamin Pelle, Erica Ryke, Matthew Richardson, Amanda Adler, Elisabeth Brandstetter, Peter D. Han, Kairsten Fay, Misja Ilcisin, Kirsten Lacombe, Thomas R. Sibley, Melissa Truong, Caitlin R. Wolf, Karen Cowgill, Stephanie Schrag, Jeff Duchin, Michael Boeckh, Janet A. Englund, Michael Famulare, Barry R. Lutz, Mark J. Rieder, Lea M. Starita, Matthew Thompson, Helen Y. Chu, Trevor Bedford, Jay Shendure |
| EPI_ISL_1468688, EPI_ISL_1468689 | Altius Institute | Seattle Flu Study | Deborah A. Nickerson, Chris D. Frazar, Jover Lee, Benjamin Pelle, Erica Ryke, Matthew Richardson, Amanda Adler, Elisabeth Brandstetter, Peter D. Han, Kairsten Fay, Misja Ilcisin, Kirsten Lacombe, Thomas R. Sibley, Melissa Truong, Caitlin R. Wolf, Ryan Alexander, Daniel Bates, Rebecca Bruders, Stephanie DeBaun, Clem Green, Muhammad Halimun, Jessica Halow, Kneshay Harper, Matt Hartman, Andrew Meuser, Alex Nguyen, Truong Nguyen, Sofia Olsson, Sadie Patraw, Hannah Petersen, Tobias Ragoczy, Joshua Richards, Jacob Rodriguez, John Stamatoyannopoulos, Julia Wald, Olivia Waltner, Michael Boeckh, Janet A. Englund, Michael Famulare, Barry R. Lutz, Mark J. Rieder, Lea M. Starita, Matthew Thompson, Helen Y. Chu, Jay Shendure, Trevor Bedford |
| EPI_ISL_1468691 | Seattle Flu Study | Seattle Flu Study | Deborah A. Nickerson, Chris D. Frazar, Jover Lee, Benjamin Pelle, Erica Ryke, Matthew Richardson, Amanda Adler, Elisabeth Brandstetter, Peter D. Han, Kairsten Fay, Misja Ilcisin, Kirsten Lacombe, Thomas R. Sibley, Melissa Truong, Caitlin R. Wolf, Karen Cowgill, Stephanie Schrag, Jeff |

| | | | |
|---|---|---|---|
| | | | Duchin, Michael Boeckh, Janet A. Englund, Michael Famulare, Barry R. Lutz, Mark J. Rieder, Lea M. Starita, Matthew Thompson, Helen Y. Chu, Trevor Bedford, Jay Shendure |
| EPI_ISL_1468692, EPI_ISL_1468693 | Altius Institute | Seattle Flu Study | Deborah A. Nickerson, Chris D. Frazar, Jover Lee, Benjamin Pelle, Erica Ryke, Matthew Richardson, Amanda Adler, Elisabeth Brandstetter, Peter D. Han, Kairsten Fay, Misja Ilcisin, Kirsten Lacombe, Thomas R. Sibley, Melissa Truong, Caitlin R. Wolf, Ryan Alexander, Daniel Bates, Rebecca Bruders, Stephanie DeBaun, Clem Green, Muhammad Halimun, Jessica Halow, Kneshay Harper, Matt Hartman, Andrew Meuser, Alex Nguyen, Truong Nguyen, Sofia Olsson, Sadie Patraw, Hannah Petersen, Tobias Ragoczy, Joshua Richards, Jacob Rodriguez, John Stamatoyannopoulos, Julia Wald, Olivia Waltner, Michael Boeckh, Janet A. Englund, Michael Famulare, Barry R. Lutz, Mark J. Rieder, Lea M. Starita, Matthew Thompson, Helen Y. Chu, Jay Shendure, Trevor Bedford |
| EPI_ISL_1468694 | Seattle Flu Study | Seattle Flu Study | Deborah A. Nickerson, Chris D. Frazar, Jover Lee, Benjamin Pelle, Erica Ryke, Matthew Richardson, Amanda Adler, Elisabeth Brandstetter, Peter D. Han, Kairsten Fay, Misja Ilcisin, Kirsten Lacombe, Thomas R. Sibley, Melissa Truong, Caitlin R. Wolf, Karen Cowgill, Stephanie Schrag, Jeff Duchin, Michael Boeckh, Janet A. Englund, Michael Famulare, Barry R. Lutz, Mark J. Rieder, Lea M. Starita, Matthew Thompson, Helen Y. Chu, Trevor Bedford, Jay Shendure |
| EPI_ISL_1468695 | Altius Institute | Seattle Flu Study | Deborah A. Nickerson, Chris D. Frazar, Jover Lee, Benjamin Pelle, Erica Ryke, Matthew Richardson, Amanda Adler, Elisabeth Brandstetter, Peter D. Han, Kairsten Fay, Misja Ilcisin, Kirsten Lacombe, Thomas R. Sibley, Melissa Truong, Caitlin R. Wolf, Ryan Alexander, Daniel Bates, Rebecca Bruders, Stephanie DeBaun, Clem Green, Muhammad Halimun, Jessica Halow, Kneshay Harper, Matt Hartman, Andrew Meuser, Alex Nguyen, Truong Nguyen, Sofia Olsson, Sadie Patraw, Hannah Petersen, Tobias Ragoczy, Joshua Richards, Jacob Rodriguez, John Stamatoyannopoulos, Julia Wald, Olivia Waltner, Michael Boeckh, Janet A. Englund, Michael Famulare, Barry R. Lutz, Mark J. Rieder, Lea M. Starita, Matthew Thompson, Helen Y. Chu, Jay Shendure, Trevor Bedford |
| EPI_ISL_1468696 | Seattle Flu Study | Seattle Flu Study | Deborah A. Nickerson, Chris D. Frazar, Jover Lee, Benjamin Pelle, Erica Ryke, Matthew Richardson, Amanda Adler, Elisabeth Brandstetter, Peter D. Han, Kairsten Fay, Misja Ilcisin, Kirsten Lacombe, Thomas R. Sibley, Melissa Truong, Caitlin R. Wolf, Karen Cowgill, Stephanie Schrag, Jeff Duchin, Michael Boeckh, Janet A. Englund, Michael Famulare, Barry R. Lutz, Mark J. Rieder, Lea M. Starita, Matthew Thompson, Helen Y. Chu, Trevor Bedford, Jay Shendure |
| EPI_ISL_1468697, EPI_ISL_1468698, EPI_ISL_1468699 | Altius Institute | Seattle Flu Study | Deborah A. Nickerson, Chris D. Frazar, Jover Lee, Benjamin Pelle, Erica Ryke, Matthew Richardson, Amanda Adler, Elisabeth Brandstetter, Peter D. Han, Kairsten Fay, Misja Ilcisin, Kirsten Lacombe, Thomas R. Sibley, Melissa Truong, Caitlin R. Wolf, Ryan Alexander, Daniel Bates, Rebecca Bruders, Stephanie DeBaun, Clem Green, Muhammad Halimun, Jessica Halow, Kneshay Harper, Matt Hartman, Andrew Meuser, Alex Nguyen, Truong Nguyen, Sofia Olsson, Sadie Patraw, Hannah Petersen, Tobias Ragoczy, Joshua Richards, Jacob Rodriguez, John Stamatoyannopoulos, Julia Wald, Olivia Waltner, Michael Boeckh, Janet A. Englund, Michael Famulare, Barry R. Lutz, Mark J. Rieder, Lea M. Starita, Matthew Thompson, Helen Y. Chu, Jay Shendure, Trevor Bedford |
| EPI_ISL_1468700 | Seattle Flu Study | Seattle Flu Study | Deborah A. Nickerson, Chris D. Frazar, Jover Lee, Benjamin Pelle, Erica Ryke, Matthew Richardson, Amanda Adler, Elisabeth Brandstetter, Peter D. Han, Kairsten Fay, Misja Ilcisin, Kirsten Lacombe, Thomas R. Sibley, Melissa Truong, Caitlin R. Wolf, Michael Boeckh, Janet A. Englund, Michael Famulare, Barry R. Lutz, Mark J. Rieder, Lea M. Starita, Matthew Thompson, Jay Shendure, Trevor Bedford, Helen Y. Chu |
| EPI_ISL_1468701, EPI_ISL_1468702 | Altius Institute | Seattle Flu Study | Deborah A. Nickerson, Chris D. Frazar, Jover Lee, Benjamin Pelle, Erica Ryke, Matthew Richardson, Amanda Adler, Elisabeth Brandstetter, Peter D. Han, Kairsten Fay, Misja Ilcisin, Kirsten Lacombe, Thomas R. Sibley, Melissa Truong, Caitlin R. Wolf, Ryan Alexander, Daniel Bates, Rebecca Bruders, Stephanie DeBaun, Clem Green, Muhammad Halimun, Jessica Halow, Kneshay Harper, Matt Hartman, Andrew Meuser, Alex Nguyen, Truong Nguyen, Sofia Olsson, Sadie Patraw, Hannah Petersen, Tobias Ragoczy, Joshua Richards, Jacob Rodriguez, John Stamatoyannopoulos, Julia Wald, Olivia Waltner, Michael Boeckh, Janet A. Englund, Michael Famulare, Barry R. Lutz, Mark J. Rieder, Lea M. Starita, Matthew Thompson, Helen Y. Chu, Jay Shendure, Trevor Bedford |
| EPI_ISL_1468703 | Seattle Flu Study | Seattle Flu Study | Deborah A. Nickerson, Chris D. Frazar, Jover Lee, Benjamin Pelle, Erica Ryke, Matthew Richardson, Amanda Adler, Elisabeth Brandstetter, Peter D. Han, Kairsten Fay, Misja Ilcisin, Kirsten Lacombe, Thomas R. Sibley, Melissa Truong, Caitlin R. Wolf, Karen Cowgill, Stephanie Schrag, Jeff Duchin, Michael Boeckh, Janet A. Englund, Michael Famulare, Barry R. Lutz, Mark J. Rieder, Lea M. Starita, Matthew Thompson, Helen Y. Chu, Trevor Bedford, Jay Shendure |
| EPI_ISL_1468704 | Altius Institute | Seattle Flu Study | Deborah A. Nickerson, Chris D. Frazar, Jover Lee, Benjamin Pelle, Erica Ryke, Matthew Richardson, Amanda Adler, Elisabeth Brandstetter, Peter D. Han, Kairsten Fay, Misja Ilcisin, Kirsten Lacombe, Thomas R. Sibley, Melissa Truong, Caitlin R. Wolf, Ryan Alexander, Daniel Bates, Rebecca Bruders, Stephanie DeBaun, Clem Green, Muhammad Halimun, Jessica Halow, Kneshay Harper, Matt Hartman, Andrew Meuser, Alex Nguyen, Truong Nguyen, Sofia Olsson, Sadie Patraw, Hannah Petersen, Tobias Ragoczy, Joshua Richards, Jacob Rodriguez, John Stamatoyannopoulos, Julia Wald, Olivia Waltner, Michael Boeckh, Janet A. Englund, Michael Famulare, Barry R. Lutz, Mark J. Rieder, Lea M. Starita, Matthew Thompson, Helen Y. Chu, Jay Shendure, Trevor Bedford |
| EPI_ISL_1468705 | Seattle Flu Study | Seattle Flu Study | Deborah A. Nickerson, Chris D. Frazar, Jover Lee, Benjamin Pelle, Erica Ryke, Matthew Richardson, Amanda Adler, Elisabeth Brandstetter, Peter D. Han, Kairsten Fay, Misja Ilcisin, Kirsten Lacombe, Thomas R. Sibley, Melissa Truong, Caitlin R. Wolf, Karen Cowgill, Stephanie Schrag, Jeff Duchin, Michael Boeckh, Janet A. Englund, Michael Famulare, Barry R. Lutz, Mark J. Rieder, Lea M. Starita, Matthew Thompson, Helen Y. Chu, Trevor Bedford, Jay Shendure |
| EPI_ISL_1468706, EPI_ISL_1468707, EPI_ISL_1468708 | Altius Institute | Seattle Flu Study | Deborah A. Nickerson, Chris D. Frazar, Jover Lee, Benjamin Pelle, Erica Ryke, Matthew Richardson, Amanda Adler, Elisabeth Brandstetter, Peter D. Han, Kairsten Fay, Misja Ilcisin, Kirsten Lacombe, Thomas R. Sibley, Melissa Truong, Caitlin R. Wolf, Ryan Alexander, Daniel Bates, Rebecca Bruders, Stephanie DeBaun, Clem Green, Muhammad Halimun, Jessica Halow, Kneshay Harper, Matt Hartman, Andrew Meuser, Alex Nguyen, Truong Nguyen, Sofia Olsson, Sadie Patraw, Hannah Petersen, Tobias Ragoczy, Joshua Richards, Jacob Rodriguez, John Stamatoyannopoulos, Julia Wald, Olivia Waltner, Michael Boeckh, Janet A. Englund, Michael Famulare, Barry R. Lutz, Mark J. Rieder, Lea M. Starita, Matthew Thompson, Helen Y. Chu, Jay Shendure, Trevor Bedford |
| EPI_ISL_1468709 | Seattle Flu Study | Seattle Flu Study | Deborah A. Nickerson, Chris D. Frazar, Jover Lee, Benjamin Pelle, Erica Ryke, Matthew Richardson, Amanda Adler, Elisabeth Brandstetter, Peter D. Han, Kairsten Fay, Misja Ilcisin, Kirsten Lacombe, Thomas R. Sibley, Melissa Truong, Caitlin R. Wolf, Karen Cowgill, Stephanie Schrag, Jeff Duchin, Michael Boeckh, Janet A. Englund, Michael Famulare, Barry R. Lutz, Mark J. Rieder, Lea M. Starita, Matthew Thompson, Helen Y. Chu, Trevor Bedford, Jay Shendure |
| EPI_ISL_1468710 | Altius Institute | Seattle Flu Study | Deborah A. Nickerson, Chris D. Frazar, Jover Lee, Benjamin Pelle, Erica Ryke, Matthew Richardson, Amanda Adler, Elisabeth Brandstetter, Peter D. Han, Kairsten Fay, Misja Ilcisin, Kirsten Lacombe, Thomas R. Sibley, Melissa Truong, Caitlin R. Wolf, Ryan Alexander, Daniel Bates, Rebecca Bruders, Stephanie DeBaun, Clem Green, Muhammad Halimun, Jessica Halow, Kneshay Harper, Matt Hartman, Andrew Meuser, Alex Nguyen, Truong Nguyen, Sofia Olsson, Sadie Patraw, Hannah Petersen, Tobias Ragoczy, Joshua Richards, Jacob Rodriguez, John Stamatoyannopoulos, Julia Wald, Olivia Waltner, Michael Boeckh, Janet A. Englund, Michael Famulare, Barry R. Lutz, Mark J. Rieder, Lea M. Starita, Matthew Thompson, Helen Y. Chu, Jay Shendure, Trevor Bedford |
| EPI_ISL_1468711 | Seattle Flu Study | Seattle Flu Study | Deborah A. Nickerson, Chris D. Frazar, Jover Lee, Benjamin Pelle, Erica Ryke, Matthew Richardson, Amanda Adler, Elisabeth Brandstetter, Peter D. Han, Kairsten Fay, Misja Ilcisin, Kirsten Lacombe, Thomas R. Sibley, Melissa Truong, Caitlin R. Wolf, Karen Cowgill, Stephanie Schrag, Jeff Duchin, Michael Boeckh, Janet A. Englund, Michael Famulare, Barry R. Lutz, Mark J. Rieder, Lea M. Starita, Matthew Thompson, Helen Y. Chu, Trevor Bedford, Jay Shendure |
| EPI_ISL_1468712, EPI_ISL_1468713, EPI_ISL_1468714, EPI_ISL_1468715, EPI_ISL_1468716 | Altius Institute | Seattle Flu Study | Deborah A. Nickerson, Chris D. Frazar, Jover Lee, Benjamin Pelle, Erica Ryke, Matthew Richardson, Amanda Adler, Elisabeth Brandstetter, Peter D. Han, Kairsten Fay, Misja Ilcisin, Kirsten Lacombe, Thomas R. Sibley, Melissa Truong, Caitlin R. Wolf, Ryan Alexander, Daniel Bates, Rebecca Bruders, Stephanie DeBaun, Clem Green, Muhammad Halimun, Jessica Halow, Kneshay Harper, Matt Hartman, Andrew Meuser, Alex Nguyen, Truong Nguyen, Sofia Olsson, Sadie Patraw, Hannah Petersen, Tobias Ragoczy, Joshua Richards, Jacob Rodriguez, John Stamatoyannopoulos, Julia Wald, Olivia Waltner, Michael Boeckh, Janet A. Englund, Michael Famulare, Barry R. Lutz, Mark J. Rieder, Lea M. Starita, Matthew Thompson, Helen Y. Chu, Jay Shendure, Trevor Bedford |
| EPI_ISL_1468717 | WA State PHL | Seattle Flu Study | Deborah A. Nickerson, Chris D. Frazar, Jover Lee, Benjamin Pelle, Erica Ryke, Matthew Richardson, Amanda Adler, Elisabeth Brandstetter, Peter D. Han, Kairsten Fay, Misja Ilcisin, Kirsten Lacombe, Thomas R. Sibley, Melissa Truong, Caitlin R. Wolf, Romesh Gautom, Geoff Melly, Brian Hiatt, Philip Dykema, Scott Lindquist, Michael Boeckh, Janet A. Englund, Michael Famulare, Barry R. Lutz, Mark J. Rieder, Lea M. Starita, Matthew Thompson, Helen Y. Chu, Jay Shendure, Trevor Bedford |
| EPI_ISL_1468718 | Altius Institute | Seattle Flu Study | Deborah A. Nickerson, Chris D. Frazar, Jover Lee, Benjamin Pelle, Erica Ryke, Matthew Richardson, Amanda Adler, Elisabeth Brandstetter, Peter D. Han, Kairsten Fay, Misja Ilcisin, Kirsten Lacombe, Thomas R. Sibley, Melissa Truong, Caitlin R. Wolf, Ryan Alexander, Daniel Bates, Rebecca Bruders, Stephanie DeBaun, Clem Green, Muhammad Halimun, Jessica Halow, Kneshay Harper, Matt Hartman, Andrew Meuser, Alex Nguyen, Truong Nguyen, Sofia Olsson, Sadie Patraw, Hannah Petersen, Tobias Ragoczy, Joshua Richards, Jacob Rodriguez, John Stamatoyannopoulos, Julia Wald, Olivia Waltner, Michael Boeckh, Janet A. Englund, Michael Famulare, Barry R. Lutz, Mark J. Rieder, Lea M. Starita, Matthew Thompson, Helen Y. Chu, Jay Shendure, Trevor Bedford |
| EPI_ISL_1468719 | Seattle Flu Study | Seattle Flu Study | Deborah A. Nickerson, Chris D. Frazar, Jover Lee, Benjamin Pelle, Erica Ryke, Matthew Richardson, Amanda Adler, Elisabeth Brandstetter, Peter D. Han, Kairsten Fay, Misja Ilcisin, Kirsten Lacombe, Thomas R. Sibley, Melissa Truong, Caitlin R. Wolf, Karen Cowgill, Stephanie Schrag, Jeff Duchin, Michael Boeckh, Janet A. Englund, Michael Famulare, Barry R. Lutz, Mark J. Rieder, Lea M. Starita, Matthew Thompson, Helen Y. Chu, Trevor Bedford, Jay Shendure |
| EPI_ISL_1468720, EPI_ISL_1468721, EPI_ISL_1468723 | Altius Institute | Seattle Flu Study | Deborah A. Nickerson, Chris D. Frazar, Jover Lee, Benjamin Pelle, Erica Ryke, Matthew Richardson, Amanda Adler, Elisabeth Brandstetter, Peter D. Han, Kairsten Fay, Misja Ilcisin, Kirsten Lacombe, Thomas R. Sibley, Melissa Truong, Caitlin R. Wolf, Ryan Alexander, Daniel Bates, Rebecca Bruders, Stephanie DeBaun, Clem Green, Muhammad Halimun, Jessica Halow, Kneshay Harper, Matt Hartman, Andrew Meuser, Alex Nguyen, Truong Nguyen, Sofia Olsson, Sadie Patraw, Hannah Petersen, Tobias Ragoczy, Joshua Richards, Jacob Rodriguez, John Stamatoyannopoulos, Julia Wald, Olivia Waltner, Michael Boeckh, Janet A. Englund, Michael Famulare, Barry R. Lutz, Mark J. Rieder, Lea M. Starita, Matthew Thompson, Helen Y. Chu, Jay Shendure, Trevor Bedford |
| EPI_ISL_1468724 | Seattle Flu Study | Seattle Flu Study | Deborah A. Nickerson, Chris D. Frazar, Jover Lee, Benjamin Pelle, Erica Ryke, Matthew Richardson, Amanda Adler, Elisabeth Brandstetter, Peter D. Han, Kairsten Fay, Misja Ilcisin, Kirsten Lacombe, Thomas R. Sibley, Melissa Truong, Caitlin R. Wolf, Karen Cowgill, Stephanie Schrag, Jeff Duchin, Michael Boeckh, Janet A. Englund, Michael Famulare, Barry R. Lutz, Mark J. Rieder, Lea M. Starita, Matthew Thompson, Helen Y. Chu, Trevor Bedford, Jay Shendure |
| EPI_ISL_1468725 | WA State PHL | Seattle Flu Study | Deborah A. Nickerson, Chris D. Frazar, Jover Lee, Benjamin Pelle, Erica Ryke, Matthew Richardson, Amanda Adler, Elisabeth Brandstetter, Peter D. Han, Kairsten Fay, Misja Ilcisin, Kirsten Lacombe, Thomas R. Sibley, Melissa Truong, Caitlin R. Wolf, Romesh Gautom, Geoff Melly, Brian Hiatt, Philip Dykema, Scott Lindquist, Michael Boeckh, Janet A. Englund, Michael Famulare, Barry R. Lutz, Mark J. Rieder, Lea M. Starita, Matthew Thompson, Helen Y. Chu, Jay Shendure, Trevor Bedford |
| EPI_ISL_1468726, EPI_ISL_1468727, EPI_ISL_1468728, EPI_ISL_1468729 | Altius Institute | Seattle Flu Study | Deborah A. Nickerson, Chris D. Frazar, Jover Lee, Benjamin Pelle, Erica Ryke, Matthew Richardson, Amanda Adler, Elisabeth Brandstetter, Peter D. Han, Kairsten Fay, Misja Ilcisin, Kirsten Lacombe, Thomas R. Sibley, Melissa Truong, Caitlin R. Wolf, Ryan Alexander, Daniel Bates, Rebecca Bruders, Stephanie DeBaun, Clem Green, Muhammad Halimun, Jessica Halow, Kneshay Harper, Matt Hartman, Andrew Meuser, Alex Nguyen, Truong Nguyen, Sofia Olsson, Sadie Patraw, Hannah Petersen, Tobias Ragoczy, Joshua Richards, Jacob Rodriguez, John Stamatoyannopoulos, Julia Wald, Olivia Waltner, Michael Boeckh, Janet A. Englund, Michael Famulare, Barry R. Lutz, Mark J. Rieder, Lea M. Starita, Matthew Thompson, Helen Y. Chu, Jay Shendure, Trevor Bedford |
| EPI_ISL_1468730, EPI_ISL_1468731 | Seattle Flu Study | Seattle Flu Study | Deborah A. Nickerson, Chris D. Frazar, Jover Lee, Benjamin Pelle, Erica Ryke, Matthew Richardson, Amanda Adler, Elisabeth Brandstetter, Peter D. Han, Kairsten Fay, Misja Ilcisin, Kirsten Lacombe, Thomas R. Sibley, Melissa Truong, Caitlin R. Wolf, Karen Cowgill, Stephanie Schrag, Jeff Duchin, Michael Boeckh, Janet A. Englund, Michael Famulare, Barry R. Lutz, Mark J. Rieder, Lea M. Starita, Matthew Thompson, Helen Y. Chu, Trevor Bedford, Jay Shendure |
| EPI_ISL_1468732, EPI_ISL_1468733, EPI_ISL_1468734 | Altius Institute | Seattle Flu Study | Deborah A. Nickerson, Chris D. Frazar, Jover Lee, Benjamin Pelle, Erica Ryke, Matthew Richardson, Amanda Adler, Elisabeth Brandstetter, Peter D. Han, Kairsten Fay, Misja Ilcisin, Kirsten Lacombe, Thomas R. Sibley, Melissa Truong, Caitlin R. Wolf, Ryan Alexander, Daniel Bates, Rebecca Bruders, Stephanie DeBaun, Clem Green, Muhammad Halimun, Jessica Halow, Kneshay Harper, Matt Hartman, Andrew Meuser, Alex Nguyen, Truong Nguyen, Sofia Olsson, Sadie Patraw, Hannah Petersen, Tobias Ragoczy, Joshua Richards, Jacob Rodriguez, John Stamatoyannopoulos, Julia Wald, Olivia Waltner, Michael Boeckh, Janet A. Englund, Michael Famulare, Barry R. Lutz, Mark J. Rieder, Lea M. Starita, Matthew Thompson, Helen Y. Chu, Jay Shendure, Trevor Bedford |
| EPI_ISL_1468735 | Seattle Flu Study | Seattle Flu Study | Deborah A. Nickerson, Chris D. Frazar, Jover Lee, Benjamin Pelle, Erica Ryke, Matthew Richardson, Amanda Adler, Elisabeth Brandstetter, Peter D. Han, Kairsten Fay, Misja Ilcisin, Kirsten Lacombe, Thomas R. Sibley, Melissa Truong, Caitlin R. Wolf, Karen Cowgill, Stephanie Schrag, Jeff Duchin, Michael Boeckh, Janet A. Englund, Michael Famulare, Barry R. Lutz, Mark J. Rieder, Lea M. Starita, Matthew Thompson, Helen Y. Chu, Trevor Bedford, Jay Shendure |
| EPI_ISL_1468736 | WA State PHL | Seattle Flu Study | Deborah A. Nickerson, Chris D. Frazar, Jover Lee, Benjamin Pelle, Erica Ryke, Matthew Richardson, Amanda Adler, Elisabeth Brandstetter, Peter D. Han, Kairsten Fay, Misja Ilcisin, Kirsten Lacombe, Thomas R. Sibley, Melissa Truong, Caitlin R. Wolf, Romesh Gautom, Geoff Melly, Brian Hiatt, Philip Dykema, Scott Lindquist, Michael Boeckh, Janet A. Englund, Michael Famulare, Barry R. Lutz, Mark J. Rieder, Lea M. Starita, Matthew Thompson, Helen Y. Chu, Jay Shendure, Trevor Bedford |
| EPI_ISL_1468737 | Altius Institute | Seattle Flu Study | Deborah A. Nickerson, Chris D. Frazar, Jover Lee, Benjamin Pelle, Erica Ryke, Matthew Richardson, Amanda Adler, Elisabeth Brandstetter, Peter D. Han, Kairsten Fay, Misja Ilcisin, Kirsten Lacombe, Thomas R. Sibley, Melissa Truong, Caitlin R. Wolf, Ryan Alexander, Daniel Bates, Rebecca Bruders, Stephanie DeBaun, Clem Green, Muhammad Halimun, Jessica Halow, Kneshay Harper, Matt Hartman, Andrew Meuser, Alex Nguyen, Truong Nguyen, Sofia Olsson, Sadie Patraw, Hannah Petersen, Tobias Ragoczy, Joshua Richards, Jacob Rodriguez, John Stamatoyannopoulos, Julia Wald, Olivia Waltner, Michael Boeckh, Janet A. Englund, Michael Famulare, Barry R. Lutz, Mark J. Rieder, Lea M. Starita, Matthew Thompson, Helen Y. Chu, Jay Shendure, Trevor Bedford |
| EPI_ISL_1468738, EPI_ISL_1468739 | Seattle Flu Study | Seattle Flu Study | Deborah A. Nickerson, Chris D. Frazar, Jover Lee, Benjamin Pelle, Erica Ryke, Matthew Richardson, Amanda Adler, Elisabeth Brandstetter, Peter D. Han, Kairsten Fay, Misja Ilcisin, Kirsten Lacombe, Thomas R. Sibley, Melissa Truong, Caitlin R. Wolf, Karen Cowgill, Stephanie Schrag, Jeff Duchin, Michael Boeckh, Janet A. Englund, Michael Famulare, Barry R. Lutz, Mark J. Rieder, Lea M. Starita, Matthew Thompson, Helen Y. Chu, Trevor Bedford, Jay Shendure |
| EPI_ISL_1468740 | Altius Institute | Seattle Flu Study | Deborah A. Nickerson, Chris D. Frazar, Jover Lee, Benjamin Pelle, Erica Ryke, Matthew Richardson, Amanda Adler, Elisabeth Brandstetter, Peter D. Han, Kairsten Fay, Misja Ilcisin, Kirsten Lacombe, Thomas R. Sibley, Melissa Truong, Caitlin R. Wolf, Ryan Alexander, Daniel Bates, Rebecca Bruders, Stephanie DeBaun, Clem Green, Muhammad Halimun, Jessica Halow, Kneshay Harper, Matt Hartman, Andrew Meuser, Alex Nguyen, Truong Nguyen, Sofia Olsson, Sadie Patraw, Hannah Petersen, Tobias Ragoczy, Joshua Richards, Jacob Rodriguez, John Stamatoyannopoulos, Julia Wald, Olivia Waltner, Michael Boeckh, Janet A. Englund, Michael Famulare, Barry R. Lutz, Mark J. Rieder, Lea M. Starita, Matthew Thompson, Helen Y. Chu, Jay Shendure, Trevor Bedford |
| EPI_ISL_1468741, EPI_ISL_1468742 | Seattle Flu Study | Seattle Flu Study | Deborah A. Nickerson, Chris D. Frazar, Jover Lee, Benjamin Pelle, Erica Ryke, Matthew Richardson, Amanda Adler, Elisabeth Brandstetter, Peter D. Han, Kairsten Fay, Misja Ilcisin, Kirsten Lacombe, Thomas R. Sibley, Melissa Truong, Caitlin R. Wolf, Michael Boeckh, Janet A. Englund, Michael Famulare, Barry R. Lutz, Mark J. Rieder, Lea M. Starita, Matthew Thompson, Jay Shendure, Trevor Bedford, Helen Y. Chu |
| EPI_ISL_1468743 | Seattle Flu Study | Seattle Flu Study | Deborah A. Nickerson, Chris D. Frazar, Jover Lee, Benjamin Pelle, Erica Ryke, Matthew Richardson, Amanda Adler, Elisabeth Brandstetter, Peter D. Han, Kairsten Fay, Misja Ilcisin, Kirsten Lacombe, Thomas R. Sibley, Melissa Truong, Caitlin R. Wolf, Karen Cowgill, Stephanie Schrag, Jeff Duchin, Michael Boeckh, Janet A. Englund, Michael Famulare, Barry R. Lutz, Mark J. Rieder, Lea M. Starita, Matthew Thompson, Helen Y. Chu, Trevor Bedford, Jay Shendure |
| EPI_ISL_1468744, EPI_ISL_1468745, EPI_ISL_1468746 | WA State PHL | Seattle Flu Study | Deborah A. Nickerson, Chris D. Frazar, Jover Lee, Benjamin Pelle, Erica Ryke, Matthew Richardson, Amanda Adler, Elisabeth Brandstetter, Peter D. Han, Kairsten Fay, Misja Ilcisin, Kirsten Lacombe, Thomas R. Sibley, Melissa Truong, Caitlin R. Wolf, Romesh Gautom, Geoff Melly, Brian Hiatt, Philip Dykema, Scott Lindquist, Michael Boeckh, Janet A. Englund, Michael Famulare, Barry R. Lutz, Mark J. Rieder, Lea M. Starita, Matthew Thompson, Helen Y. Chu, Jay Shendure, Trevor Bedford |
| EPI_ISL_1468747 | Seattle Flu Study | Seattle Flu Study | Deborah A. Nickerson, Chris D. Frazar, Jover Lee, Benjamin Pelle, Erica Ryke, Matthew Richardson, Amanda Adler, Elisabeth Brandstetter, Peter D. Han, Kairsten Fay, Misja Ilcisin, Kirsten Lacombe, Thomas R. Sibley, Melissa Truong, Caitlin R. Wolf, Karen Cowgill, Stephanie Schrag, Jeff Duchin, Michael Boeckh, Janet A. Englund, Michael Famulare, Barry R. Lutz, Mark J. Rieder, Lea M. Starita, Matthew Thompson, Helen Y. Chu, Trevor Bedford, Jay Shendure |
| EPI_ISL_1468748, EPI_ISL_1468749 | Altius Institute | Seattle Flu Study | Deborah A. Nickerson, Chris D. Frazar, Jover Lee, Benjamin Pelle, Erica Ryke, Matthew Richardson, Amanda Adler, Elisabeth Brandstetter, Peter D. Han, Kairsten Fay, Misja Ilcisin, Kirsten Lacombe, Thomas R. Sibley, Melissa Truong, Caitlin R. Wolf, Ryan Alexander, Daniel Bates, Rebecca Bruders, Stephanie DeBaun, Clem Green, Muhammad Halimun, Jessica Halow, Kneshay Harper, Matt Hartman, Andrew Meuser, Alex Nguyen, Truong Nguyen, Sofia Olsson, Sadie Patraw, Hannah Petersen, Tobias Ragoczy, Joshua Richards, Jacob Rodriguez, John Stamatoyannopoulos, Julia Wald, Olivia Waltner, Michael Boeckh, Janet A. Englund, Michael Famulare, Barry R. Lutz, Mark J. Rieder, Lea M. Starita, Matthew Thompson, Helen Y. Chu, Jay Shendure, Trevor Bedford |
| EPI_ISL_1468750, EPI_ISL_1468751 | Seattle Flu Study | Seattle Flu Study | Deborah A. Nickerson, Chris D. Frazar, Jover Lee, Benjamin Pelle, Erica Ryke, Matthew Richardson, Amanda Adler, Elisabeth Brandstetter, Peter D. Han, Kairsten Fay, Misja Ilcisin, Kirsten Lacombe, Thomas R. Sibley, Melissa Truong, Caitlin R. Wolf, Karen Cowgill, Stephanie Schrag, Jeff Duchin, Michael Boeckh, Janet A. Englund, Michael Famulare, Barry R. Lutz, Mark J. Rieder, Lea M. Starita, Matthew Thompson, Helen Y. Chu, Trevor Bedford, Jay Shendure |
| EPI_ISL_1468752, EPI_ISL_1468753 | Altius Institute | Seattle Flu Study | Deborah A. Nickerson, Chris D. Frazar, Jover Lee, Benjamin Pelle, Erica Ryke, Matthew Richardson, Amanda Adler, Elisabeth Brandstetter, Peter D. Han, Kairsten Fay, Misja Ilcisin, Kirsten Lacombe, Thomas R. Sibley, Melissa Truong, Caitlin R. Wolf, Ryan Alexander, Daniel Bates, Rebecca Bruders, Stephanie DeBaun, Clem Green, Muhammad Halimun, Jessica Halow, Kneshay Harper, Matt Hartman, Andrew Meuser, Alex Nguyen, Truong Nguyen, Sofia Olsson, Sadie Patraw, Hannah Petersen, Tobias Ragoczy, Joshua Richards, Jacob Rodriguez, John Stamatoyannopoulos, Julia Wald, Olivia Waltner, Michael Boeckh, Janet A. Englund, Michael Famulare, Barry R. Lutz, Mark J. Rieder, Lea M. Starita, Matthew Thompson, Helen Y. Chu, Jay Shendure, Trevor Bedford |
| EPI_ISL_1468754, | WA State PHL | Seattle Flu Study | Deborah A. Nickerson, Chris D. Frazar, Jover Lee, Benjamin Pelle, Erica Ryke, Matthew Richardson, Amanda Adler, Elisabeth Brandstetter, Peter D. Han, Kairsten Fay, Misja Ilcisin, Kirsten Lacombe, Thomas R. Sibley, Melissa Truong, Caitlin R. Wolf, Romesh Gautom, Geoff Melly, Brian |

| | | | |
|---|---|---|---|
| EPI_ISL_1468755, EPI_ISL_1468756, EPI_ISL_1468757 | | | Hiatt, Philip Dykema, Scott Lindquist, Michael Boeckh, Janet A. Englund, Michael Famulare, Barry R. Lutz, Mark J. Rieder, Lea M. Starita, Matthew Thompson, Helen Y. Chu, Jay Shendure, Trevor Bedford |
| EPI_ISL_1468815, EPI_ISL_1468816 | Seattle Flu Study | Seattle Flu Study | Deborah A. Nickerson, Chris D. Frazar, Jover Lee, Benjamin Pelle, Erica Ryke, Matthew Richardson, Amanda Adler, Elisabeth Brandstetter, Peter D. Han, Kairsten Fay, Misja Ilcisin, Kirsten Lacombe, Thomas R. Sibley, Melissa Truong, Caitlin R. Wolf, Karen Cowgill, Stephanie Schrag, Jeff Duchin, Michael Boeckh, Janet A. Englund, Michael Famulare, Barry R. Lutz, Mark J. Rieder, Lea M. Starita, Matthew Thompson, Helen Y. Chu, Trevor Bedford, Jay Shendure |
| EPI_ISL_1468817 | Seattle Flu Study | Seattle Flu Study | Deborah A. Nickerson, Chris D. Frazar, Jover Lee, Benjamin Pelle, Erica Ryke, Matthew Richardson, Amanda Adler, Elisabeth Brandstetter, Peter D. Han, Kairsten Fay, Misja Ilcisin, Kirsten Lacombe, Thomas R. Sibley, Melissa Truong, Caitlin R. Wolf, Michael Boeckh, Janet A. Englund, Michael Famulare, Barry R. Lutz, Mark J. Rieder, Lea M. Starita, Matthew Thompson, Jay Shendure, Trevor Bedford, Helen Y. Chu |
| EPI_ISL_1468818, EPI_ISL_1468819 | Seattle Flu Study | Seattle Flu Study | Deborah A. Nickerson, Chris D. Frazar, Jover Lee, Benjamin Pelle, Erica Ryke, Matthew Richardson, Amanda Adler, Elisabeth Brandstetter, Peter D. Han, Kairsten Fay, Misja Ilcisin, Kirsten Lacombe, Thomas R. Sibley, Melissa Truong, Caitlin R. Wolf, Karen Cowgill, Stephanie Schrag, Jeff Duchin, Michael Boeckh, Janet A. Englund, Michael Famulare, Barry R. Lutz, Mark J. Rieder, Lea M. Starita, Matthew Thompson, Helen Y. Chu, Trevor Bedford, Jay Shendure |
| EPI_ISL_1468820, EPI_ISL_1468821, EPI_ISL_1468822, EPI_ISL_1468823, EPI_ISL_1468824, EPI_ISL_1468825, EPI_ISL_1468826, EPI_ISL_1468827, EPI_ISL_1468828 | Altius Institute | Seattle Flu Study | Deborah A. Nickerson, Chris D. Frazar, Jover Lee, Benjamin Pelle, Erica Ryke, Matthew Richardson, Amanda Adler, Elisabeth Brandstetter, Peter D. Han, Kairsten Fay, Misja Ilcisin, Kirsten Lacombe, Thomas R. Sibley, Melissa Truong, Caitlin R. Wolf, Ryan Alexander, Daniel Bates, Rebecca Bruders, Stephanie DeBaun, Clem Green, Muhammad Halimun, Jessica Halow, Kneshay Harper, Matt Hartman, Andrew Meuser, Alex Nguyen, Truong Nguyen, Sofia Olsson, Sadie Patraw, Hannah Petersen, Tobias Ragoczy, Joshua Richards, Jacob Rodriguez, John Stamatoyannopoulos, Julia Wald, Olivia Waltner, Michael Boeckh, Janet A. Englund, Michael Famulare, Barry R. Lutz, Mark J. Rieder, Lea M. Starita, Matthew Thompson, Helen Y. Chu, Jay Shendure, Trevor Bedford |
| EPI_ISL_1468829, EPI_ISL_1468830, EPI_ISL_1468831 | Sharp HealthCare Laboratory | Andersen lab at Scripps Research | SEARCH Alliance San Diego with Aaron Harding, Jacquelyn Berumen, Cathy Woerle, Liam McGinnis, Art Mendoza, Omid Bakhtar |
| EPI_ISL_1468832 | San Diego County Public Health Laboratory | Andersen lab at Scripps Research | SEARCH Alliance San Diego with Tracy Basler, Jovan Shephard, Brett Austin |
| EPI_ISL_1468833, EPI_ISL_1468834, EPI_ISL_1468835, EPI_ISL_1468836 | Sharp HealthCare Laboratory | Andersen lab at Scripps Research | SEARCH Alliance San Diego with Aaron Harding, Jacquelyn Berumen, Cathy Woerle, Liam McGinnis, Art Mendoza, Omid Bakhtar |
| EPI_ISL_1468837 | San Diego County Public Health Laboratory | Andersen lab at Scripps Research | SEARCH Alliance San Diego with Tracy Basler, Jovan Shephard, Brett Austin |
| EPI_ISL_1468838 | Sharp HealthCare Laboratory | Andersen lab at Scripps Research | SEARCH Alliance San Diego with Aaron Harding, Jacquelyn Berumen, Cathy Woerle, Liam McGinnis, Art Mendoza, Omid Bakhtar |
| EPI_ISL_1468839 | San Diego County Public Health Laboratory | Andersen lab at Scripps Research | SEARCH Alliance San Diego with Tracy Basler, Jovan Shephard, Brett Austin |
| EPI_ISL_1468840 | Sharp HealthCare Laboratory | Andersen lab at Scripps Research | SEARCH Alliance San Diego with Aaron Harding, Jacquelyn Berumen, Cathy Woerle, Liam McGinnis, Art Mendoza, Omid Bakhtar |
| EPI_ISL_1468841 | San Diego County Public Health Laboratory | Andersen lab at Scripps Research | SEARCH Alliance San Diego with Tracy Basler, Jovan Shephard, Brett Austin |
| EPI_ISL_1468842 | Scripps Medical Laboratory | Andersen lab at Scripps Research | SEARCH Alliance San Diego with Michael Quigley, Ellen Stefanski, Ian Mchardy |
| EPI_ISL_1468843, EPI_ISL_1468844 | San Diego County Public Health Laboratory | Andersen lab at Scripps Research | SEARCH Alliance San Diego with Tracy Basler, Jovan Shephard, Brett Austin |
| EPI_ISL_1468845, EPI_ISL_1468846 | Sharp HealthCare Laboratory | Andersen lab at Scripps Research | SEARCH Alliance San Diego with Aaron Harding, Jacquelyn Berumen, Cathy Woerle, Liam McGinnis, Art Mendoza, Omid Bakhtar |
| EPI_ISL_1468847 | San Diego County Public Health Laboratory | Andersen lab at Scripps Research | SEARCH Alliance San Diego with Tracy Basler, Jovan Shephard, Brett Austin |
| EPI_ISL_1468848 | Sharp HealthCare Laboratory | Andersen lab at Scripps Research | SEARCH Alliance San Diego with Aaron Harding, Jacquelyn Berumen, Cathy Woerle, Liam McGinnis, Art Mendoza, Omid Bakhtar |
| EPI_ISL_1468849 | Scripps Medical Laboratory | Andersen lab at Scripps Research | SEARCH Alliance San Diego with Michael Quigley, Ellen Stefanski, Ian Mchardy |
| EPI_ISL_1468850, EPI_ISL_1468851, EPI_ISL_1468852, EPI_ISL_1468853 | Sharp HealthCare Laboratory | Andersen lab at Scripps Research | SEARCH Alliance San Diego with Aaron Harding, Jacquelyn Berumen, Cathy Woerle, Liam McGinnis, Art Mendoza, Omid Bakhtar |
| EPI_ISL_1468854 | San Diego County Public Health Laboratory | Andersen lab at Scripps Research | SEARCH Alliance San Diego with Tracy Basler, Jovan Shephard, Brett Austin |
| EPI_ISL_1468855, EPI_ISL_1468856, EPI_ISL_1468857, EPI_ISL_1468858, EPI_ISL_1468859, EPI_ISL_1468860, EPI_ISL_1468861, EPI_ISL_1468862, EPI_ISL_1468863, EPI_ISL_1468864, EPI_ISL_1468865, EPI_ISL_1468866, EPI_ISL_1468867, EPI_ISL_1468868, EPI_ISL_1468869, EPI_ISL_1468870, EPI_ISL_1468871, EPI_ISL_1468872, EPI_ISL_1468873 | | | |
| see above | Sharp HealthCare Laboratory | Andersen lab at Scripps Research | SEARCH Alliance San Diego with Aaron Harding, Jacquelyn Berumen, Cathy Woerle, Liam McGinnis, Art Mendoza, Omid Bakhtar |
| EPI_ISL_1468874, EPI_ISL_1468875, EPI_ISL_1468876, EPI_ISL_1468877, EPI_ISL_1468878, EPI_ISL_1468879, EPI_ISL_1468880, EPI_ISL_1468881 | San Diego County Public Health Laboratory | Andersen lab at Scripps Research | SEARCH Alliance San Diego with Tracy Basler, Jovan Shephard, Brett Austin |
| EPI_ISL_1468882, EPI_ISL_1468883, EPI_ISL_1468884, EPI_ISL_1468885, EPI_ISL_1468886, EPI_ISL_1468887, EPI_ISL_1468888 | Scripps Medical Laboratory | Andersen lab at Scripps Research | SEARCH Alliance San Diego with Michael Quigley, Ellen Stefanski, Ian Mchardy |
| EPI_ISL_1468889, EPI_ISL_1468890, EPI_ISL_1468891, EPI_ISL_1468892, EPI_ISL_1468893, EPI_ISL_1468894, EPI_ISL_1468895, EPI_ISL_1468896, EPI_ISL_1468897, EPI_ISL_1468898, EPI_ISL_1468899, EPI_ISL_1468900, EPI_ISL_1468901, EPI_ISL_1468902, EPI_ISL_1468903, EPI_ISL_1468904, EPI_ISL_1468905 | | | |
| see above | Sharp HealthCare Laboratory | Andersen lab at Scripps Research | SEARCH Alliance San Diego with Aaron Harding, Jacquelyn Berumen, Cathy Woerle, Liam McGinnis, Art Mendoza, Omid Bakhtar |
| EPI_ISL_1468906, EPI_ISL_1468907, EPI_ISL_1468908, EPI_ISL_1468909 | San Diego County Public Health Laboratory | Andersen lab at Scripps Research | SEARCH Alliance San Diego with Tracy Basler, Jovan Shephard, Brett Austin |
| EPI_ISL_1468910, EPI_ISL_1468911, EPI_ISL_1468912, EPI_ISL_1468913 | Sharp HealthCare Laboratory | Andersen lab at Scripps Research | SEARCH Alliance San Diego with Aaron Harding, Jacquelyn Berumen, Cathy Woerle, Liam McGinnis, Art Mendoza, Omid Bakhtar |
| EPI_ISL_1468914, EPI_ISL_1468915, EPI_ISL_1468916 | San Diego County Public Health Laboratory | Andersen lab at Scripps Research | SEARCH Alliance San Diego with Tracy Basler, Jovan Shephard, Brett Austin |
| EPI_ISL_1468917 | Sharp HealthCare Laboratory | Andersen lab at Scripps Research | SEARCH Alliance San Diego with Aaron Harding, Jacquelyn Berumen, Cathy Woerle, Liam McGinnis, Art Mendoza, Omid Bakhtar |
| EPI_ISL_1468922, EPI_ISL_1468923, EPI_ISL_1468924, EPI_ISL_1468925, EPI_ISL_1468926, EPI_ISL_1468927, EPI_ISL_1468928, EPI_ISL_1468929, EPI_ISL_1468930, EPI_ISL_1468931, EPI_ISL_1468932, EPI_ISL_1468933, EPI_ISL_1468934, EPI_ISL_1468935, EPI_ISL_1468936, EPI_ISL_1468937, EPI_ISL_1468938, EPI_ISL_1468939, EPI_ISL_1468940, EPI_ISL_1468941, EPI_ISL_1468942, EPI_ISL_1468943, EPI_ISL_1468944, EPI_ISL_1468945, EPI_ISL_1468946 | | | |
| see above | San Diego County Public Health Laboratory | Andersen lab at Scripps Research | SEARCH Alliance San Diego with Tracy Basler, Jovan Shephard, Brett Austin |
| EPI_ISL_1468947 | Scripps Medical Laboratory | Andersen lab at Scripps Research | SEARCH Alliance San Diego with Michael Quigley, Ellen Stefanski, Ian Mchardy |
| EPI_ISL_1468948, EPI_ISL_1468949, EPI_ISL_1468950, EPI_ISL_1468951 | San Diego County Public Health Laboratory | Andersen lab at Scripps Research | SEARCH Alliance San Diego with Tracy Basler, Jovan Shephard, Brett Austin |
| EPI_ISL_1468952 | Sharp HealthCare Laboratory | Andersen lab at Scripps Research | SEARCH Alliance San Diego with Aaron Harding, Jacquelyn Berumen, Cathy Woerle, Liam McGinnis, Art Mendoza, Omid Bakhtar |
| EPI_ISL_1468953 | San Diego County Public Health Laboratory | Andersen lab at Scripps Research | SEARCH Alliance San Diego with Tracy Basler, Jovan Shephard, Brett Austin |

| | | | |
|---|---|---|---|
| EPI_ISL_1468954 | Sharp HealthCare Laboratory | Andersen lab at Scripps Research | SEARCH Alliance San Diego with Aaron Harding, Jacquelyn Berumen, Cathy Woerle, Liam McGinnis, Art Mendoza, Omid Bakhtar |
| EPI_ISL_1468955, EPI_ISL_1468956 | Scripps Medical Laboratory | Andersen lab at Scripps Research | SEARCH Alliance San Diego with Michael Quigley, Ellen Stefanski, Ian Mchardy |
| EPI_ISL_1468957, EPI_ISL_1468958 | Sharp HealthCare Laboratory | Andersen lab at Scripps Research | SEARCH Alliance San Diego with Aaron Harding, Jacquelyn Berumen, Cathy Woerle, Liam McGinnis, Art Mendoza, Omid Bakhtar |
| EPI_ISL_1468959 | San Diego County Public Health Laboratory | Andersen lab at Scripps Research | SEARCH Alliance San Diego with Tracy Basler, Jovan Shephard, Brett Austin |
| EPI_ISL_1468960, EPI_ISL_1468961, EPI_ISL_1468962, EPI_ISL_1468963, EPI_ISL_1468964, EPI_ISL_1468965, EPI_ISL_1468966, EPI_ISL_1468967, EPI_ISL_1468968, EPI_ISL_1468969, EPI_ISL_1468970, EPI_ISL_1468971, EPI_ISL_1468972, EPI_ISL_1468973, EPI_ISL_1468974, EPI_ISL_1468975, EPI_ISL_1468976, EPI_ISL_1468977, EPI_ISL_1468978 | | | |
| see above | Sharp HealthCare Laboratory | Andersen lab at Scripps Research | SEARCH Alliance San Diego with Aaron Harding, Jacquelyn Berumen, Cathy Woerle, Liam McGinnis, Art Mendoza, Omid Bakhtar |
| EPI_ISL_1468979, EPI_ISL_1468980, EPI_ISL_1468981, EPI_ISL_1468982, EPI_ISL_1468983, EPI_ISL_1468984, EPI_ISL_1468985, EPI_ISL_1468986, EPI_ISL_1468987, EPI_ISL_1468988, EPI_ISL_1468989, EPI_ISL_1468990, EPI_ISL_1468991, EPI_ISL_1468992, EPI_ISL_1468993, EPI_ISL_1468994, EPI_ISL_1468995, EPI_ISL_1468996, EPI_ISL_1468997, EPI_ISL_1468998, EPI_ISL_1468999, EPI_ISL_1469000, EPI_ISL_1469001, EPI_ISL_1469002, EPI_ISL_1469003, EPI_ISL_1469004, EPI_ISL_1469005, EPI_ISL_1469006, EPI_ISL_1469007, EPI_ISL_1469008, EPI_ISL_1469009, EPI_ISL_1469010, EPI_ISL_1469011, EPI_ISL_1469012, EPI_ISL_1469013, EPI_ISL_1469014, EPI_ISL_1469015, EPI_ISL_1469016, EPI_ISL_1469017, EPI_ISL_1469018, EPI_ISL_1469019, EPI_ISL_1469020, EPI_ISL_1469021, EPI_ISL_1469022, EPI_ISL_1469023, EPI_ISL_1469024, EPI_ISL_1469025, EPI_ISL_1469026, EPI_ISL_1469027, EPI_ISL_1469028, EPI_ISL_1469029, EPI_ISL_1469030, EPI_ISL_1469031, EPI_ISL_1469032, EPI_ISL_1469033, EPI_ISL_1469034, EPI_ISL_1469035, EPI_ISL_1469036, EPI_ISL_1469037, EPI_ISL_1469038, EPI_ISL_1469039, EPI_ISL_1469040, EPI_ISL_1469041, EPI_ISL_1469042 | | | |
| see above | San Diego County Public Health Laboratory | Andersen lab at Scripps Research | SEARCH Alliance San Diego with Tracy Basler, Jovan Shephard, Brett Austin |
| EPI_ISL_1469043, EPI_ISL_1469044, EPI_ISL_1469045, EPI_ISL_1469046, EPI_ISL_1469047, EPI_ISL_1469048, EPI_ISL_1469049, EPI_ISL_1469050, EPI_ISL_1469051, EPI_ISL_1469052, EPI_ISL_1469053, EPI_ISL_1469054, EPI_ISL_1469055, EPI_ISL_1469056, EPI_ISL_1469057, EPI_ISL_1469058, EPI_ISL_1469059, EPI_ISL_1469060 | | | |
| see above | Scripps Medical Laboratory | Andersen lab at Scripps Research | SEARCH Alliance San Diego with Michael Quigley, Ellen Stefanski, Ian Mchardy |
| EPI_ISL_1469061, EPI_ISL_1469062, EPI_ISL_1469063, EPI_ISL_1469064, EPI_ISL_1469065, EPI_ISL_1469066, EPI_ISL_1469067, EPI_ISL_1469068, EPI_ISL_1469069, EPI_ISL_1469070, EPI_ISL_1469071, EPI_ISL_1469072, EPI_ISL_1469073, EPI_ISL_1469074, EPI_ISL_1469075, EPI_ISL_1469076, EPI_ISL_1469077, EPI_ISL_1469078, EPI_ISL_1469079, EPI_ISL_1469080, EPI_ISL_1469081, EPI_ISL_1469082, EPI_ISL_1469083, EPI_ISL_1469084, EPI_ISL_1469085, EPI_ISL_1469086, EPI_ISL_1469087 | | | |
| see above | Sharp HealthCare Laboratory | Andersen lab at Scripps Research | SEARCH Alliance San Diego with Aaron Harding, Jacquelyn Berumen, Cathy Woerle, Liam McGinnis, Art Mendoza, Omid Bakhtar |
| EPI_ISL_1469108, EPI_ISL_1469109, EPI_ISL_1469110, EPI_ISL_1469111, EPI_ISL_1469112, EPI_ISL_1469113, EPI_ISL_1469114, EPI_ISL_1469115, EPI_ISL_1469116, EPI_ISL_1469117, EPI_ISL_1469119 | | | |
| see above | Centro de Investigacion en Ciencias de la Salud y Biomedicina | CINVESTAV | Sofia Bernal, Andreu Comas |
| EPI_ISL_1469120, EPI_ISL_1469121 | UCSC Genomics Institute | UCSC Genomics Institute | Mark Akeson, Ikenna Anigbogu, Eric Beraut, Isabel Bjork, Molly Cassatt-Johnstone, Terren Chang, Russell Corbett-Detig, Namrita Dhillon, Maximilian Haeussler, David Haussler, Angie Hinrichs, Miten Jain, Joshua Kapp, A. Marm Kilpatrick, Jakob McBroome, Hugh Olsen, Jeremy Sanford, Beth Shapiro, Michael Stone, Bryan Thornlow, Yatish Turakhia |
| EPI_ISL_1469292, EPI_ISL_1469293, EPI_ISL_1469294, EPI_ISL_1469295, EPI_ISL_1469296, EPI_ISL_1469310, EPI_ISL_1469311, EPI_ISL_1469312 | Maryland Genomics, Institute for Genome Sciences, University of Maryland School of Medicine | Maryland Genomics, Institute for Genome Sciences, University of Maryland School of Medicine | Tallon, Luke J; Sadzewicz, Lisa D; Humphrys, Mike; Ott, Sandra; Roussey, Holly; Mehta, Aditya; Vavikolanu, Kranthi; Fraser, Claire M; Ravel, Jacques |
| EPI_ISL_1469493, EPI_ISL_1469494, EPI_ISL_1469495, EPI_ISL_1469496, EPI_ISL_1469498, EPI_ISL_1469499, EPI_ISL_1469500, EPI_ISL_1469501, EPI_ISL_1469502, EPI_ISL_1469503, EPI_ISL_1469504, EPI_ISL_1469505, EPI_ISL_1469506, EPI_ISL_1469507, EPI_ISL_1469508, EPI_ISL_1469509, EPI_ISL_1469511, EPI_ISL_1469512, EPI_ISL_1469513, EPI_ISL_1469514, EPI_ISL_1469515, EPI_ISL_1469516, EPI_ISL_1469517, EPI_ISL_1469518, EPI_ISL_1469519, EPI_ISL_1469520, EPI_ISL_1469521, EPI_ISL_1469523, EPI_ISL_1469524, EPI_ISL_1469526, EPI_ISL_1469528, EPI_ISL_1469529, EPI_ISL_1469531, EPI_ISL_1469532, EPI_ISL_1469533, EPI_ISL_1469534, EPI_ISL_1469535, EPI_ISL_1469536, EPI_ISL_1469537, EPI_ISL_1469538, EPI_ISL_1469539, EPI_ISL_1469540, EPI_ISL_1469542, EPI_ISL_1469543, EPI_ISL_1469544, EPI_ISL_1469545, EPI_ISL_1469546, EPI_ISL_1469547, EPI_ISL_1469548 | | | |
| see above | Nevada State Public Health Laboratory | Nevada State Public Health Laboratory | Andrew Gorzalski, Mark Pandori |
| EPI_ISL_1469852, EPI_ISL_1469853, EPI_ISL_1469854, EPI_ISL_1469855, EPI_ISL_1469856, EPI_ISL_1469857, EPI_ISL_1469858, EPI_ISL_1469859, EPI_ISL_1469860, EPI_ISL_1469861, EPI_ISL_1469862, EPI_ISL_1469863, EPI_ISL_1469864, EPI_ISL_1469865, EPI_ISL_1469866, EPI_ISL_1469867, EPI_ISL_1469868, EPI_ISL_1469869, EPI_ISL_1469870, EPI_ISL_1469871, EPI_ISL_1469872, EPI_ISL_1469873, EPI_ISL_1469874, EPI_ISL_1469875, EPI_ISL_1469876, EPI_ISL_1469877, EPI_ISL_1469878, EPI_ISL_1469879, EPI_ISL_1469880, EPI_ISL_1469881, EPI_ISL_1469882, EPI_ISL_1469883, EPI_ISL_1469884, EPI_ISL_1469886, EPI_ISL_1469887, EPI_ISL_1469888, EPI_ISL_1469889, EPI_ISL_1469890, EPI_ISL_1469891, EPI_ISL_1469892, EPI_ISL_1469893, EPI_ISL_1469894, EPI_ISL_1469895, EPI_ISL_1469896, EPI_ISL_1469897, EPI_ISL_1469898, EPI_ISL_1469899, EPI_ISL_1469900, EPI_ISL_1469901, EPI_ISL_1469902, EPI_ISL_1469903, EPI_ISL_1469905, EPI_ISL_1469906, EPI_ISL_1469907, EPI_ISL_1469908, EPI_ISL_1469909, EPI_ISL_1469910, EPI_ISL_1469911, EPI_ISL_1469912, EPI_ISL_1469914, EPI_ISL_1469915, EPI_ISL_1469916, EPI_ISL_1469917, EPI_ISL_1469918, EPI_ISL_1469919, EPI_ISL_1469921, EPI_ISL_1469922, EPI_ISL_1469923, EPI_ISL_1469924, EPI_ISL_1469925, EPI_ISL_1469926, EPI_ISL_1469927, EPI_ISL_1469929, EPI_ISL_1469930, EPI_ISL_1469931, EPI_ISL_1469932, EPI_ISL_1469933, EPI_ISL_1469934, EPI_ISL_1469935, EPI_ISL_1469936 | | | |
| see above | UCSC Genomics Institute | UCSC Genomics Institute | Mark Akeson, Ikenna Anigbogu, Eric Beraut, Isabel Bjork, Molly Cassatt-Johnstone, Terren Chang, Russell Corbett-Detig, Namrita Dhillon, Maximilian Haeussler, David Haussler, Angie Hinrichs, Miten Jain, Joshua Kapp, A. Marm Kilpatrick, Jakob McBroome, Hugh Olsen, Jeremy Sanford, Beth Shapiro, Michael Stone, Bryan Thornlow, Yatish Turakhia |
| EPI_ISL_1469937 | LESP Jalisco | Instituto de Diagnostico y Referencia Epidemiologicos (INDRE) | Claudia Wong-Arambula, Abril Rodriguez-Maldonado, Vanessa Rivero-Arredondo, Ariadna Medina-Benitez, Joaquin Quiroz-Mercado, Sergio Rangel-Guerrero, Natividad Cruz-Ortiz, Tatiana Nunez-Garcia, Gisela Barrera-Badillo, Lucia Hernandez-Rivas, Irma Lopez-Martinez, Ernesto Ramirez-Gonzalez. |
| EPI_ISL_1469938, EPI_ISL_1469939 | LESP Nuevo Leon | Instituto de Diagnostico y Referencia Epidemiologicos (INDRE) | Claudia Wong-Arambula, Abril Rodriguez-Maldonado, Vanessa Rivero-Arredondo, Ariadna Medina-Benitez, Joaquin Quiroz-Mercado, Sergio Rangel-Guerrero, Natividad Cruz-Ortiz, Tatiana Nunez-Garcia, Gisela Barrera-Badillo, Lucia Hernandez-Rivas, Irma Lopez-Martinez, Ernesto Ramirez-Gonzalez. |
| EPI_ISL_1469940 | LESP Sinaloa | Instituto de Diagnostico y Referencia Epidemiologicos (INDRE) | Claudia Wong-Arambula, Abril Rodriguez-Maldonado, Vanessa Rivero-Arredondo, Ariadna Medina-Benitez, Joaquin Quiroz-Mercado, Sergio Rangel-Guerrero, Natividad Cruz-Ortiz, Tatiana Nunez-Garcia, Gisela Barrera-Badillo, Lucia Hernandez-Rivas, Irma Lopez-Martinez, Ernesto Ramirez-Gonzalez. |
| EPI_ISL_1469941 | LESP Ciudad de Mexico | Instituto de Diagnostico y Referencia Epidemiologicos (INDRE) | Claudia Wong-Arambula, Abril Rodriguez-Maldonado, Vanessa Rivero-Arredondo, Ariadna Medina-Benitez, Joaquin Quiroz-Mercado, Sergio Rangel-Guerrero, Natividad Cruz-Ortiz, Tatiana Nunez-Garcia, Gisela Barrera-Badillo, Lucia Hernandez-Rivas, Irma Lopez-Martinez, Ernesto Ramirez-Gonzalez. |
| EPI_ISL_1469942, EPI_ISL_1469943, EPI_ISL_1469944 | LESP Guanajuato | Instituto de Diagnostico y Referencia Epidemiologicos (INDRE) | Claudia Wong-Arambula, Abril Rodriguez-Maldonado, Vanessa Rivero-Arredondo, Ariadna Medina-Benitez, Joaquin Quiroz-Mercado, Sergio Rangel-Guerrero, Natividad Cruz-Ortiz, Tatiana Nunez-Garcia, Gisela Barrera-Badillo, Lucia Hernandez-Rivas, Irma Lopez-Martinez, Ernesto Ramirez-Gonzalez. |
| EPI_ISL_1469988 | LSUHS Emerging Viral Threat Laboratory | LSUHS Emerging Viral Threat Laboratory | Alexander J. Mijalis, Gregory L. Ware, Rona S. Scott, Jennifer L. Carroll, Jeremy P. Kamil, Maarten Van Diest, Andrew D. Yurochko, John A. Vanchiere, Christopher G. Kevil |
| EPI_ISL_1470375 | Gravity Diagnostics | Kentucky State Public Health Lab | Stephanie Lunn, Karim George, Joshua Tobias, William Grooms, Vaneet Arora, Matthew Johnson, Rachel Zinner, Rhonda Lucas |
| EPI_ISL_1470376, EPI_ISL_1470377, EPI_ISL_1470378, EPI_ISL_1470379, EPI_ISL_1470380, EPI_ISL_1470381, EPI_ISL_1470382, EPI_ISL_1470383, EPI_ISL_1470384, EPI_ISL_1470385, EPI_ISL_1470386, EPI_ISL_1470387, EPI_ISL_1470388, EPI_ISL_1470389, EPI_ISL_1470390, EPI_ISL_1470391, EPI_ISL_1470392, EPI_ISL_1470393, EPI_ISL_1470394 | | | |
| see above | Kentucky State Public Health Lab | Kentucky State Public Health Lab | Stephanie Lunn, Karim George, Joshua Tobias, William Grooms, Vaneet Arora, Matthew Johnson, Rachel Zinner, Rhonda Lucas |
| EPI_ISL_1470395 | Gravity Diagnostics | Kentucky State Public Health Lab | Stephanie Lunn, Karim George, Joshua Tobias, William Grooms, Vaneet Arora, Matthew Johnson, Rachel Zinner, Rhonda Lucas |
| EPI_ISL_1470396, EPI_ISL_1470397, EPI_ISL_1470398, EPI_ISL_1470399, EPI_ISL_1470400, EPI_ISL_1470401, EPI_ISL_1470402, EPI_ISL_1470403, EPI_ISL_1470404, EPI_ISL_1470405, EPI_ISL_1470406, EPI_ISL_1470407, EPI_ISL_1470408, EPI_ISL_1470409, EPI_ISL_1470410, EPI_ISL_1470411, EPI_ISL_1470412 | | | |
| see above | Kentucky State Public Health Lab | Kentucky State Public Health Lab | Stephanie Lunn, Karim George, Joshua Tobias, William Grooms, Vaneet Arora, Matthew Johnson, Rachel Zinner, Rhonda Lucas |
| EPI_ISL_1470560, EPI_ISL_1470561, EPI_ISL_1470562, EPI_ISL_1470563, EPI_ISL_1470564, EPI_ISL_1470565, EPI_ISL_1470566, EPI_ISL_1470567, EPI_ISL_1470568, EPI_ISL_1470569, EPI_ISL_1470570, EPI_ISL_1470571, EPI_ISL_1470572, EPI_ISL_1470573, EPI_ISL_1470574, EPI_ISL_1470575, EPI_ISL_1470576, EPI_ISL_1470577, EPI_ISL_1470578, EPI_ISL_1470579, EPI_ISL_1470580, EPI_ISL_1470581, EPI_ISL_1470582, EPI_ISL_1470583, EPI_ISL_1470584, EPI_ISL_1470585, EPI_ISL_1470586, EPI_ISL_1470587, EPI_ISL_1470588, EPI_ISL_1470589, EPI_ISL_1470590, EPI_ISL_1470591, EPI_ISL_1470592, EPI_ISL_1470593, EPI_ISL_1470594, EPI_ISL_1470595, EPI_ISL_1470596, EPI_ISL_1470597, EPI_ISL_1470598, EPI_ISL_1470599, EPI_ISL_1470600, EPI_ISL_1470601, EPI_ISL_1470602, EPI_ISL_1470603, EPI_ISL_1470604, EPI_ISL_1470605, EPI_ISL_1470606, EPI_ISL_1470607, EPI_ISL_1470608, EPI_ISL_1470609, EPI_ISL_1470610, EPI_ISL_1470611, EPI_ISL_1470612, EPI_ISL_1470613, EPI_ISL_1470614, EPI_ISL_1470615, EPI_ISL_1470616, EPI_ISL_1470617, EPI_ISL_1470618, EPI_ISL_1470619, EPI_ISL_1470620, EPI_ISL_1470621, EPI_ISL_1470622, EPI_ISL_1470623, EPI_ISL_1470624, EPI_ISL_1470625, EPI_ISL_1470626, EPI_ISL_1470627, EPI_ISL_1470628, EPI_ISL_1470629, EPI_ISL_1470630, EPI_ISL_1470631, EPI_ISL_1470632, EPI_ISL_1470633, EPI_ISL_1470634, EPI_ISL_1470635, EPI_ISL_1470636, EPI_ISL_1470637, EPI_ISL_1470638, EPI_ISL_1470639, EPI_ISL_1470640, EPI_ISL_1470641, EPI_ISL_1470642, EPI_ISL_1470643, EPI_ISL_1470644, EPI_ISL_1470645, EPI_ISL_1470646, EPI_ISL_1470647, EPI_ISL_1470648, EPI_ISL_1470649, EPI_ISL_1470650, EPI_ISL_1470651, EPI_ISL_1470653, EPI_ISL_1470654, EPI_ISL_1470655, EPI_ISL_1470656, EPI_ISL_1470657, EPI_ISL_1470658, EPI_ISL_1470659, EPI_ISL_1470660, EPI_ISL_1470661, EPI_ISL_1470662, EPI_ISL_1470663, EPI_ISL_1470664, EPI_ISL_1470665, EPI_ISL_1470666, EPI_ISL_1470667, EPI_ISL_1470668, EPI_ISL_1470669, EPI_ISL_1470670, EPI_ISL_1470671, EPI_ISL_1470672, EPI_ISL_1470673, EPI_ISL_1470674, EPI_ISL_1470675, EPI_ISL_1470676, EPI_ISL_1470677, EPI_ISL_1470678, EPI_ISL_1470679, EPI_ISL_1470680, EPI_ISL_1470681, EPI_ISL_1470682, EPI_ISL_1470683, EPI_ISL_1470684, EPI_ISL_1470685, EPI_ISL_1470686, EPI_ISL_1470687, EPI_ISL_1470688, EPI_ISL_1470689, EPI_ISL_1470690, EPI_ISL_1470691, EPI_ISL_1470692, EPI_ISL_1470693, EPI_ISL_1470694, EPI_ISL_1470695, EPI_ISL_1470696, EPI_ISL_1470697, EPI_ISL_1470698, EPI_ISL_1470699, EPI_ISL_1470700, EPI_ISL_1470701, EPI_ISL_1470702, EPI_ISL_1470703, EPI_ISL_1470704, EPI_ISL_1470705, EPI_ISL_1470706, EPI_ISL_1470707, EPI_ISL_1470708, EPI_ISL_1470709, EPI_ISL_1470710, EPI_ISL_1470711, EPI_ISL_1470712, EPI_ISL_1470713, EPI_ISL_1470714, EPI_ISL_1470715, EPI_ISL_1470716, EPI_ISL_1470717, EPI_ISL_1470718, EPI_ISL_1470719, EPI_ISL_1470720, EPI_ISL_1470721, EPI_ISL_1470722, EPI_ISL_1470723, EPI_ISL_1470724, EPI_ISL_1470725, EPI_ISL_1470726, EPI_ISL_1470727, EPI_ISL_1470728, EPI_ISL_1470729, EPI_ISL_1470730, EPI_ISL_1470732, EPI_ISL_1470733, EPI_ISL_1470734, EPI_ISL_1470735, EPI_ISL_1470736, EPI_ISL_1470737, EPI_ISL_1470738, EPI_ISL_1470739, EPI_ISL_1470740 | | | |
| see above | The Jackson Laboratory | The Jackson Laboratory | Bergeron D, Renzette N, Adams M, Omerza G, Kelly K, Li L |
| EPI_ISL_1470741, EPI_ISL_1470742, EPI_ISL_1470743, EPI_ISL_1470744, EPI_ISL_1470745, EPI_ISL_1470746, EPI_ISL_1470747, EPI_ISL_1470748, EPI_ISL_1470749, EPI_ISL_1470750, EPI_ISL_1470751, EPI_ISL_1470752, EPI_ISL_1470753, EPI_ISL_1470755, EPI_ISL_1470756, EPI_ISL_1470757, EPI_ISL_1470758, EPI_ISL_1470759, EPI_ISL_1470760, EPI_ISL_1470761, EPI_ISL_1470763, EPI_ISL_1470764, EPI_ISL_1470765, EPI_ISL_1470766, EPI_ISL_1470767, EPI_ISL_1470768, EPI_ISL_1470769, EPI_ISL_1470770, EPI_ISL_1470771, EPI_ISL_1470772, EPI_ISL_1470773, EPI_ISL_1470774, EPI_ISL_1470775, EPI_ISL_1470776, EPI_ISL_1470778, EPI_ISL_1470779, EPI_ISL_1470781, EPI_ISL_1470782, EPI_ISL_1470784, EPI_ISL_1470785, EPI_ISL_1470786, EPI_ISL_1470787, EPI_ISL_1470788, EPI_ISL_1470789, EPI_ISL_1470790, EPI_ISL_1470791, EPI_ISL_1470792, EPI_ISL_1470793, EPI_ISL_1470795, EPI_ISL_1470796, EPI_ISL_1470797, EPI_ISL_1470798, EPI_ISL_1470799, EPI_ISL_1470800, EPI_ISL_1470801, EPI_ISL_1470802, EPI_ISL_1470804, EPI_ISL_1470805, EPI_ISL_1470806, EPI_ISL_1470807, EPI_ISL_1470808, EPI_ISL_1470809, EPI_ISL_1470810, EPI_ISL_1470811, EPI_ISL_1470812, EPI_ISL_1470813, EPI_ISL_1470814, EPI_ISL_1470815, EPI_ISL_1470817, EPI_ISL_1470818, EPI_ISL_1470819, EPI_ISL_1470820, EPI_ISL_1470821, EPI_ISL_1470822, EPI_ISL_1470823, EPI_ISL_1470824, EPI_ISL_1470825, EPI_ISL_1470826, EPI_ISL_1470828, EPI_ISL_1470829, EPI_ISL_1470830, EPI_ISL_1470831, EPI_ISL_1470832, EPI_ISL_1470833, EPI_ISL_1470834, EPI_ISL_1470835, EPI_ISL_1470836, EPI_ISL_1470837, EPI_ISL_1470838, EPI_ISL_1470839, EPI_ISL_1470840, EPI_ISL_1470841, EPI_ISL_1470842, EPI_ISL_1470843, EPI_ISL_1470844, EPI_ISL_1470845, EPI_ISL_1470846, EPI_ISL_1470847, EPI_ISL_1470848, EPI_ISL_1470849, EPI_ISL_1470850, EPI_ISL_1470851, EPI_ISL_1470853, EPI_ISL_1470854, EPI_ISL_1470855, EPI_ISL_1470856, EPI_ISL_1470857, EPI_ISL_1470858, EPI_ISL_1470859, EPI_ISL_1470861, EPI_ISL_1470863, EPI_ISL_1470864, EPI_ISL_1470865, EPI_ISL_1470866, EPI_ISL_1470867, EPI_ISL_1470868, EPI_ISL_1470869, EPI_ISL_1470870, EPI_ISL_1470871, EPI_ISL_1470872, EPI_ISL_1470874, EPI_ISL_1470876, EPI_ISL_1470877, EPI_ISL_1470878, EPI_ISL_1470879, EPI_ISL_1470880, EPI_ISL_1470881, EPI_ISL_1470882, EPI_ISL_1470883, EPI_ISL_1470884, EPI_ISL_1470885, EPI_ISL_1470886, EPI_ISL_1470887, EPI_ISL_1470888, EPI_ISL_1470889, EPI_ISL_1470890, EPI_ISL_1470892, EPI_ISL_1470893, EPI_ISL_1470894, EPI_ISL_1470895, EPI_ISL_1470896, EPI_ISL_1470897, EPI_ISL_1470898, EPI_ISL_1470899, EPI_ISL_1470902, EPI_ISL_1470903, EPI_ISL_1470904, EPI_ISL_1470906, EPI_ISL_1470907, EPI_ISL_1470908, EPI_ISL_1470909, EPI_ISL_1470910, EPI_ISL_1470911, EPI_ISL_1470913, EPI_ISL_1470914, EPI_ISL_1470915, EPI_ISL_1470916, EPI_ISL_1470917, EPI_ISL_1470919, EPI_ISL_1470920, EPI_ISL_1470921, EPI_ISL_1470922, EPI_ISL_1470923, EPI_ISL_1470924, EPI_ISL_1470925, EPI_ISL_1470926, EPI_ISL_1470927, EPI_ISL_1470928, EPI_ISL_1470929, EPI_ISL_1470930, EPI_ISL_1470931, EPI_ISL_1470932, EPI_ISL_1470933, EPI_ISL_1470934, EPI_ISL_1470935, EPI_ISL_1470936, EPI_ISL_1470937, EPI_ISL_1470938, EPI_ISL_1470939, EPI_ISL_1470940, EPI_ISL_1470941, EPI_ISL_1470942, EPI_ISL_1470943, EPI_ISL_1470944, EPI_ISL_1470945, EPI_ISL_1470946, EPI_ISL_1470947, EPI_ISL_1470948, EPI_ISL_1470949, EPI_ISL_1470950, EPI_ISL_1470951, EPI_ISL_1470952, EPI_ISL_1470953, EPI_ISL_1470954, EPI_ISL_1470955, EPI_ISL_1470956, EPI_ISL_1470957, EPI_ISL_1470958, EPI_ISL_1470959, EPI_ISL_1470960, EPI_ISL_1470961, EPI_ISL_1470962, EPI_ISL_1470964, EPI_ISL_1470966, EPI_ISL_1470967, EPI_ISL_1470968, EPI_ISL_1470969, EPI_ISL_1470970, EPI_ISL_1470971, EPI_ISL_1470972, EPI_ISL_1470973, EPI_ISL_1470974, EPI_ISL_1470975, EPI_ISL_1470976, EPI_ISL_1470978, EPI_ISL_1470979, EPI_ISL_1470980, EPI_ISL_1470981, EPI_ISL_1470982, EPI_ISL_1470983, EPI_ISL_1470985, EPI_ISL_1470986, EPI_ISL_1470987, EPI_ISL_1470988, EPI_ISL_1470989, EPI_ISL_1470990, EPI_ISL_1470991, EPI_ISL_1470993, EPI_ISL_1470996, EPI_ISL_1470998, EPI_ISL_1471000, EPI_ISL_1471001, EPI_ISL_1471002, EPI_ISL_1471003, EPI_ISL_1471004, EPI_ISL_1471006, EPI_ISL_1471007, EPI_ISL_1471008, EPI_ISL_1471009, EPI_ISL_1471010, EPI_ISL_1471011, EPI_ISL_1471012, EPI_ISL_1471013, EPI_ISL_1471014, EPI_ISL_1471015, EPI_ISL_1471016, EPI_ISL_1471017, EPI_ISL_1471018, EPI_ISL_1471019, EPI_ISL_1471021, EPI_ISL_1471022, EPI_ISL_1471023, EPI_ISL_1471024, EPI_ISL_1471025, EPI_ISL_1471026, EPI_ISL_1471027, EPI_ISL_1471028, EPI_ISL_1471029, EPI_ISL_1471030, EPI_ISL_1471031, EPI_ISL_1471032, EPI_ISL_1471033, EPI_ISL_1471034, EPI_ISL_1471035, EPI_ISL_1471036, EPI_ISL_1471037, EPI_ISL_1471038, EPI_ISL_1471039, EPI_ISL_1471040, EPI_ISL_1471041, EPI_ISL_1471042, EPI_ISL_1471043, EPI_ISL_1471044, EPI_ISL_1471045, EPI_ISL_1471047, EPI_ISL_1471048, EPI_ISL_1471049, EPI_ISL_1471050, EPI_ISL_1471051, EPI_ISL_1471052, EPI_ISL_1471053, EPI_ISL_1471054, EPI_ISL_1471055, EPI_ISL_1471056, EPI_ISL_1471057, EPI_ISL_1471058, EPI_ISL_1471059, EPI_ISL_1471060, EPI_ISL_1471061, EPI_ISL_1471062, EPI_ISL_1471063, EPI_ISL_1471064, EPI_ISL_1471066, EPI_ISL_1471067, EPI_ISL_1471068, EPI_ISL_1471069, EPI_ISL_1471071, EPI_ISL_1471072, EPI_ISL_1471073, EPI_ISL_1471074, EPI_ISL_1471076, EPI_ISL_1471077, EPI_ISL_1471078, EPI_ISL_1471079, EPI_ISL_1471080, EPI_ISL_1471081, EPI_ISL_1471082, EPI_ISL_1471083, EPI_ISL_1471084, EPI_ISL_1471085, EPI_ISL_1471086, EPI_ISL_1471087, EPI_ISL_1471088, EPI_ISL_1471089, EPI_ISL_1471090, EPI_ISL_1471091, EPI_ISL_1471092, EPI_ISL_1471093, EPI_ISL_1471094, EPI_ISL_1471095, EPI_ISL_1471096, EPI_ISL_1471097, EPI_ISL_1471098, EPI_ISL_1471099, EPI_ISL_1471100, EPI_ISL_1471101, EPI_ISL_1471102, EPI_ISL_1471103, EPI_ISL_1471104, EPI_ISL_1471105, EPI_ISL_1471106, EPI_ISL_1471107, EPI_ISL_1471108, EPI_ISL_1471109, EPI_ISL_1471110, EPI_ISL_1471111, EPI_ISL_1471113, EPI_ISL_1471114, EPI_ISL_1471115, EPI_ISL_1471116, EPI_ISL_1471117, EPI_ISL_1471118, EPI_ISL_1471119, EPI_ISL_1471120, EPI_ISL_1471121, EPI_ISL_1471122, EPI_ISL_1471123, EPI_ISL_1471124, EPI_ISL_1471125, EPI_ISL_1471126, EPI_ISL_1471128, EPI_ISL_1471130, EPI_ISL_1471131, EPI_ISL_1471132, EPI_ISL_1471133, EPI_ISL_1471136, EPI_ISL_1471137, EPI_ISL_1471138, EPI_ISL_1471139, EPI_ISL_1471140, EPI_ISL_1471141, EPI_ISL_1471142, EPI_ISL_1471143, EPI_ISL_1471144, EPI_ISL_1471145, EPI_ISL_1471146, EPI_ISL_1471147, EPI_ISL_1471148, EPI_ISL_1471149, EPI_ISL_1471150, EPI_ISL_1471151, EPI_ISL_1471153, EPI_ISL_1471154, EPI_ISL_1471155, EPI_ISL_1471156, EPI_ISL_1471157, EPI_ISL_1471158, EPI_ISL_1471159, EPI_ISL_1471160, EPI_ISL_1471161, EPI_ISL_1471162, EPI_ISL_1471163, EPI_ISL_1471164, EPI_ISL_1471166, EPI_ISL_1471167, EPI_ISL_1471168, EPI_ISL_1471169, EPI_ISL_1471170, EPI_ISL_1471171, EPI_ISL_1471172, EPI_ISL_1471173, EPI_ISL_1471174, EPI_ISL_1471176, EPI_ISL_1471177, EPI_ISL_1471178, EPI_ISL_1471179, EPI_ISL_1471180, EPI_ISL_1471181, EPI_ISL_1471183, EPI_ISL_1471184, EPI_ISL_1471185, EPI_ISL_1471186, EPI_ISL_1471187, EPI_ISL_1471188, EPI_ISL_1471189, EPI_ISL_1471190, EPI_ISL_1471191, EPI_ISL_1471192, EPI_ISL_1471193, EPI_ISL_1471195, EPI_ISL_1471196, EPI_ISL_1471197, EPI_ISL_1471198, EPI_ISL_1471199, EPI_ISL_1471200, EPI_ISL_1471201, EPI_ISL_1471202, EPI_ISL_1471203, EPI_ISL_1471204, EPI_ISL_1471205, EPI_ISL_1471207, EPI_ISL_1471208, EPI_ISL_1471209, EPI_ISL_1471210, EPI_ISL_1471211, EPI_ISL_1471213, EPI_ISL_1471214, EPI_ISL_1471215, EPI_ISL_1471216, EPI_ISL_1471217, EPI_ISL_1471218, EPI_ISL_1471219, EPI_ISL_1471220, EPI_ISL_1471223, EPI_ISL_1471224, EPI_ISL_1471225, EPI_ISL_1471226, EPI_ISL_1471227, EPI_ISL_1471228, EPI_ISL_1471229, EPI_ISL_1471230, EPI_ISL_1471231, EPI_ISL_1471233, EPI_ISL_1471234, EPI_ISL_1471235, EPI_ISL_1471236, EPI_ISL_1471237, EPI_ISL_1471238, EPI_ISL_1471239, EPI_ISL_1471240, EPI_ISL_1471241, EPI_ISL_1471242, EPI_ISL_1471243, EPI_ISL_1471244, EPI_ISL_1471245, EPI_ISL_1471246, EPI_ISL_1471247, EPI_ISL_1471248, EPI_ISL_1471249, EPI_ISL_1471250, EPI_ISL_1471251, EPI_ISL_1471252, EPI_ISL_1471254, EPI_ISL_1471256, EPI_ISL_1471257, EPI_ISL_1471258, EPI_ISL_1471261, EPI_ISL_1471262, EPI_ISL_1471263, EPI_ISL_1471264, EPI_ISL_1471265, EPI_ISL_1471266, EPI_ISL_1471267, EPI_ISL_1471268, EPI_ISL_1471269, EPI_ISL_1471270, EPI_ISL_1471271, EPI_ISL_1471272, EPI_ISL_1471273, EPI_ISL_1471274, EPI_ISL_1471276, EPI_ISL_1471277, EPI_ISL_1471278, EPI_ISL_1471279, EPI_ISL_1471280, EPI_ISL_1471281, EPI_ISL_1471282, EPI_ISL_1471283, EPI_ISL_1471284, EPI_ISL_1471285, EPI_ISL_1471286, EPI_ISL_1471287, EPI_ISL_1471288, EPI_ISL_1471290, EPI_ISL_1471291, EPI_ISL_1471293, EPI_ISL_1471294, EPI_ISL_1471295, EPI_ISL_1471297, EPI_ISL_1471298, EPI_ISL_1471299, EPI_ISL_1471300, EPI_ISL_1471301, EPI_ISL_1471302, EPI_ISL_1471303, EPI_ISL_1471304, EPI_ISL_1471305, EPI_ISL_1471307, EPI_ISL_1471308, EPI_ISL_1471309, EPI_ISL_1471310, EPI_ISL_1471311, EPI_ISL_1471312, EPI_ISL_1471313, EPI_ISL_1471314, EPI_ISL_1471315, EPI_ISL_1471316, EPI_ISL_1471317, EPI_ISL_1471318, EPI_ISL_1471319, EPI_ISL_1471320, EPI_ISL_1471321, EPI_ISL_1471322, EPI_ISL_1471323, EPI_ISL_1471324, EPI_ISL_1471325, EPI_ISL_1471326, EPI_ISL_1471327, | | | |

| | | | |
|---|---|---|---|
| EPI_ISL_1471328, EPI_ISL_1471329, EPI_ISL_1471330, EPI_ISL_1471332, EPI_ISL_1471333, EPI_ISL_1471334, EPI_ISL_1471335, EPI_ISL_1471336, EPI_ISL_1471337, EPI_ISL_1471338, EPI_ISL_1471339, EPI_ISL_1471340, EPI_ISL_1471341, EPI_ISL_1471342, EPI_ISL_1471343, EPI_ISL_1471344, EPI_ISL_1471345, EPI_ISL_1471346, EPI_ISL_1471347, EPI_ISL_1471348, EPI_ISL_1471349, EPI_ISL_1471350, EPI_ISL_1471351, EPI_ISL_1471352, EPI_ISL_1471353, EPI_ISL_1471354, EPI_ISL_1471355, EPI_ISL_1471356, EPI_ISL_1471360, EPI_ISL_1471361, EPI_ISL_1471362, EPI_ISL_1471363, EPI_ISL_1471364, EPI_ISL_1471365, EPI_ISL_1471367, EPI_ISL_1471368, EPI_ISL_1471369, EPI_ISL_1471370, EPI_ISL_1471371, EPI_ISL_1471372, EPI_ISL_1471373, EPI_ISL_1471374, EPI_ISL_1471375, EPI_ISL_1471376, EPI_ISL_1471377, EPI_ISL_1471378, EPI_ISL_1471379, EPI_ISL_1471380, EPI_ISL_1471381, EPI_ISL_1471382, EPI_ISL_1471383, EPI_ISL_1471384, EPI_ISL_1471385, EPI_ISL_1471386, EPI_ISL_1471387, EPI_ISL_1471388, EPI_ISL_1471389, EPI_ISL_1471390, EPI_ISL_1471391, EPI_ISL_1471392, EPI_ISL_1471393, EPI_ISL_1471394, EPI_ISL_1471395, EPI_ISL_1471396, EPI_ISL_1471397, EPI_ISL_1471398, EPI_ISL_1471399, EPI_ISL_1471400, EPI_ISL_1471402, EPI_ISL_1471403, EPI_ISL_1471404, EPI_ISL_1471405, EPI_ISL_1471406, EPI_ISL_1471407, EPI_ISL_1471408, EPI_ISL_1471409, EPI_ISL_1471411, EPI_ISL_1471412, EPI_ISL_1471413, EPI_ISL_1471415, EPI_ISL_1471417, EPI_ISL_1471419, EPI_ISL_1471420, EPI_ISL_1471421, EPI_ISL_1471422, EPI_ISL_1471423, EPI_ISL_1471424, EPI_ISL_1471425, EPI_ISL_1471426, EPI_ISL_1471427, EPI_ISL_1471428, EPI_ISL_1471429, EPI_ISL_1471430, EPI_ISL_1471431, EPI_ISL_1471432, EPI_ISL_1471433, EPI_ISL_1471434, EPI_ISL_1471435, EPI_ISL_1471436, EPI_ISL_1471437, EPI_ISL_1471438, EPI_ISL_1471439, EPI_ISL_1471440, EPI_ISL_1471441, EPI_ISL_1471442, EPI_ISL_1471443, EPI_ISL_1471444, EPI_ISL_1471445, EPI_ISL_1471446, EPI_ISL_1471448, EPI_ISL_1471449, EPI_ISL_1471450, EPI_ISL_1471451, EPI_ISL_1471452, EPI_ISL_1471453, EPI_ISL_1471454, EPI_ISL_1471455, EPI_ISL_1471456, EPI_ISL_1471457, EPI_ISL_1471458, EPI_ISL_1471459, EPI_ISL_1471460, EPI_ISL_1471461, EPI_ISL_1471463, EPI_ISL_1471464, EPI_ISL_1471465, EPI_ISL_1471466, EPI_ISL_1471467, EPI_ISL_1471468, EPI_ISL_1471469, EPI_ISL_1471470, EPI_ISL_1471471, EPI_ISL_1471472, EPI_ISL_1471473, EPI_ISL_1471474, EPI_ISL_1471475, EPI_ISL_1471476, EPI_ISL_1471477, EPI_ISL_1471478, EPI_ISL_1471479, EPI_ISL_1471480, EPI_ISL_1471481, EPI_ISL_1471482, EPI_ISL_1471483, EPI_ISL_1471484, EPI_ISL_1471485, EPI_ISL_1471486, EPI_ISL_1471487, EPI_ISL_1471488, EPI_ISL_1471489, EPI_ISL_1471490, EPI_ISL_1471491, EPI_ISL_1471493, EPI_ISL_1471494, EPI_ISL_1471495, EPI_ISL_1471497, EPI_ISL_1471498, EPI_ISL_1471499, EPI_ISL_1471500, EPI_ISL_1471501, EPI_ISL_1471502, EPI_ISL_1471504, EPI_ISL_1471505, EPI_ISL_1471506, EPI_ISL_1471507, EPI_ISL_1471509, EPI_ISL_1471510, EPI_ISL_1471511, EPI_ISL_1471513, EPI_ISL_1471514, EPI_ISL_1471515, EPI_ISL_1471516, EPI_ISL_1471517, EPI_ISL_1471518, EPI_ISL_1471519, EPI_ISL_1471520, EPI_ISL_1471521, EPI_ISL_1471522, EPI_ISL_1471523, EPI_ISL_1471524, EPI_ISL_1471525, EPI_ISL_1471526, EPI_ISL_1471527, EPI_ISL_1471528, EPI_ISL_1471529, EPI_ISL_1471530, EPI_ISL_1471533, EPI_ISL_1471534, EPI_ISL_1471535, EPI_ISL_1471536, EPI_ISL_1471537, EPI_ISL_1471538, EPI_ISL_1471539, EPI_ISL_1471540, EPI_ISL_1471541, EPI_ISL_1471542, EPI_ISL_1471543, EPI_ISL_1471544, EPI_ISL_1471545, EPI_ISL_1471546, EPI_ISL_1471547, EPI_ISL_1471548, EPI_ISL_1471549, EPI_ISL_1471550, EPI_ISL_1471551, EPI_ISL_1471552, EPI_ISL_1471553, EPI_ISL_1471554, EPI_ISL_1471555, EPI_ISL_1471556, EPI_ISL_1471557, EPI_ISL_1471559, EPI_ISL_1471560, EPI_ISL_1471561, EPI_ISL_1471562, EPI_ISL_1471563, EPI_ISL_1471564, EPI_ISL_1471565, EPI_ISL_1471566, EPI_ISL_1471567, EPI_ISL_1471568, EPI_ISL_1471569, EPI_ISL_1471570, EPI_ISL_1471571, EPI_ISL_1471572, EPI_ISL_1471574, EPI_ISL_1471575, EPI_ISL_1471576, EPI_ISL_1471577, EPI_ISL_1471578, EPI_ISL_1471579, EPI_ISL_1471580, EPI_ISL_1471581, EPI_ISL_1471582, EPI_ISL_1471583, EPI_ISL_1471584, EPI_ISL_1471585, EPI_ISL_1471586, EPI_ISL_1471587, EPI_ISL_1471588, EPI_ISL_1471589, EPI_ISL_1471590, EPI_ISL_1471591, EPI_ISL_1471592, EPI_ISL_1471593, EPI_ISL_1471594, EPI_ISL_1471595, EPI_ISL_1471596, EPI_ISL_1471597, EPI_ISL_1471598, EPI_ISL_1471599, EPI_ISL_1471600, EPI_ISL_1471601, EPI_ISL_1471602, EPI_ISL_1471603, EPI_ISL_1471604, EPI_ISL_1471605, EPI_ISL_1471606, EPI_ISL_1471607, EPI_ISL_1471608, EPI_ISL_1471609, EPI_ISL_1471610, EPI_ISL_1471611, EPI_ISL_1471612, EPI_ISL_1471613, EPI_ISL_1471614, EPI_ISL_1471615, EPI_ISL_1471617, EPI_ISL_1471618, EPI_ISL_1471619, EPI_ISL_1471620, EPI_ISL_1471621, EPI_ISL_1471622, EPI_ISL_1471623, EPI_ISL_1471624, EPI_ISL_1471625, EPI_ISL_1471626, EPI_ISL_1471627, EPI_ISL_1471628, EPI_ISL_1471629, EPI_ISL_1471630, EPI_ISL_1471631, EPI_ISL_1471632, EPI_ISL_1471633, EPI_ISL_1471634, EPI_ISL_1471635, EPI_ISL_1471636, EPI_ISL_1471637, EPI_ISL_1471638, EPI_ISL_1471639, EPI_ISL_1471640, EPI_ISL_1471641, EPI_ISL_1471642, EPI_ISL_1471643, EPI_ISL_1471644, EPI_ISL_1471645, EPI_ISL_1471646, EPI_ISL_1471647, EPI_ISL_1471648, EPI_ISL_1471649, EPI_ISL_1471650, EPI_ISL_1471651, EPI_ISL_1471652, EPI_ISL_1471653, EPI_ISL_1471654, EPI_ISL_1471655, EPI_ISL_1471656, EPI_ISL_1471657, EPI_ISL_1471658, EPI_ISL_1471659, EPI_ISL_1471660, EPI_ISL_1471661, EPI_ISL_1471662, EPI_ISL_1471663, EPI_ISL_1471664, EPI_ISL_1471665, EPI_ISL_1471666, EPI_ISL_1471667, EPI_ISL_1471668, EPI_ISL_1471669, EPI_ISL_1471670, EPI_ISL_1471671, EPI_ISL_1471672, EPI_ISL_1471673, EPI_ISL_1471675, EPI_ISL_1471676, EPI_ISL_1471677, EPI_ISL_1471678, EPI_ISL_1471679, EPI_ISL_1471680, EPI_ISL_1471681, EPI_ISL_1471682, EPI_ISL_1471683, EPI_ISL_1471684, EPI_ISL_1471685, EPI_ISL_1471686, EPI_ISL_1471687, EPI_ISL_1471688, EPI_ISL_1471689, EPI_ISL_1471690, EPI_ISL_1471691, EPI_ISL_1471693, EPI_ISL_1471694, EPI_ISL_1471695, EPI_ISL_1471696, EPI_ISL_1471697, EPI_ISL_1471698, EPI_ISL_1471699, EPI_ISL_1471700, EPI_ISL_1471701, EPI_ISL_1471702, EPI_ISL_1471703, EPI_ISL_1471704, EPI_ISL_1471705, EPI_ISL_1471706, EPI_ISL_1471707, EPI_ISL_1471708, EPI_ISL_1471709, EPI_ISL_1471710, EPI_ISL_1471711, EPI_ISL_1471712, EPI_ISL_1471713, EPI_ISL_1471714, EPI_ISL_1471715, EPI_ISL_1471716, EPI_ISL_1471717, EPI_ISL_1471718, EPI_ISL_1471719, EPI_ISL_1471720, EPI_ISL_1471721, EPI_ISL_1471722, EPI_ISL_1471723, EPI_ISL_1471724, EPI_ISL_1471726, EPI_ISL_1471727, EPI_ISL_1471728, EPI_ISL_1471729, EPI_ISL_1471730, EPI_ISL_1471731, EPI_ISL_1471732, EPI_ISL_1471733, EPI_ISL_1471734, EPI_ISL_1471735, EPI_ISL_1471736, EPI_ISL_1471737, EPI_ISL_1471738, EPI_ISL_1471739, EPI_ISL_1471740, EPI_ISL_1471741, EPI_ISL_1471742, EPI_ISL_1471743, EPI_ISL_1471744, EPI_ISL_1471745, EPI_ISL_1471746, EPI_ISL_1471747, EPI_ISL_1471748, EPI_ISL_1471749, EPI_ISL_1471750, EPI_ISL_1471751, EPI_ISL_1471752, EPI_ISL_1471753, EPI_ISL_1471754, EPI_ISL_1471755, EPI_ISL_1471757, EPI_ISL_1471758, EPI_ISL_1471759, EPI_ISL_1471760, EPI_ISL_1471761, EPI_ISL_1471762, EPI_ISL_1471763, EPI_ISL_1471764, EPI_ISL_1471765, EPI_ISL_1471766, EPI_ISL_1471767, EPI_ISL_1471768, EPI_ISL_1471770, EPI_ISL_1471771, EPI_ISL_1471772, EPI_ISL_1471773, EPI_ISL_1471774, EPI_ISL_1471775, EPI_ISL_1471776, EPI_ISL_1471777, EPI_ISL_1471778, EPI_ISL_1471779, EPI_ISL_1471780, EPI_ISL_1471782, EPI_ISL_1471783, EPI_ISL_1471784, EPI_ISL_1471785, EPI_ISL_1471786, EPI_ISL_1471788, EPI_ISL_1471789, EPI_ISL_1471790, EPI_ISL_1471791, EPI_ISL_1471792, EPI_ISL_1471793, EPI_ISL_1471794, EPI_ISL_1471795, EPI_ISL_1471796, EPI_ISL_1471797, EPI_ISL_1471798, EPI_ISL_1471800, EPI_ISL_1471801, EPI_ISL_1471802, EPI_ISL_1471803, EPI_ISL_1471804, EPI_ISL_1471805, EPI_ISL_1471806, EPI_ISL_1471807, EPI_ISL_1471808, EPI_ISL_1471809, EPI_ISL_1471810, EPI_ISL_1471811, EPI_ISL_1471812, EPI_ISL_1471813, EPI_ISL_1471814, EPI_ISL_1471815, EPI_ISL_1471816, EPI_ISL_1471817, EPI_ISL_1471818, EPI_ISL_1471819, EPI_ISL_1471820, EPI_ISL_1471821, EPI_ISL_1471823, EPI_ISL_1471824, EPI_ISL_1471826, EPI_ISL_1471827, EPI_ISL_1471828, EPI_ISL_1471829, EPI_ISL_1471830, EPI_ISL_1471831, EPI_ISL_1471832, EPI_ISL_1471833, EPI_ISL_1471834, EPI_ISL_1471835, EPI_ISL_1471836, EPI_ISL_1471837, EPI_ISL_1471838, EPI_ISL_1471839, EPI_ISL_1471840, EPI_ISL_1471844, EPI_ISL_1471845, EPI_ISL_1471846, EPI_ISL_1471847, EPI_ISL_1471849, EPI_ISL_1471850, EPI_ISL_1471851, EPI_ISL_1471852, EPI_ISL_1471853, EPI_ISL_1471854, EPI_ISL_1471855, EPI_ISL_1471856, EPI_ISL_1471857, EPI_ISL_1471858, EPI_ISL_1471859, EPI_ISL_1471860, EPI_ISL_1471863, EPI_ISL_1471864, EPI_ISL_1471865, EPI_ISL_1471866, EPI_ISL_1471867, EPI_ISL_1471868, EPI_ISL_1471869, EPI_ISL_1471870, EPI_ISL_1471871, EPI_ISL_1471872, EPI_ISL_1471874, EPI_ISL_1471875, EPI_ISL_1471876, EPI_ISL_1471877, EPI_ISL_1471878, EPI_ISL_1471879, EPI_ISL_1471880, EPI_ISL_1471882, EPI_ISL_1471883, EPI_ISL_1471884, EPI_ISL_1471885, EPI_ISL_1471886, EPI_ISL_1471887, EPI_ISL_1471888, EPI_ISL_1471889, EPI_ISL_1471890, EPI_ISL_1471891, EPI_ISL_1471892, EPI_ISL_1471893, EPI_ISL_1471894, EPI_ISL_1471895, EPI_ISL_1471896, EPI_ISL_1471897, EPI_ISL_1471898, EPI_ISL_1471899, EPI_ISL_1471900, EPI_ISL_1471903, EPI_ISL_1471904, EPI_ISL_1471905, EPI_ISL_1471906, EPI_ISL_1471907, EPI_ISL_1471908, EPI_ISL_1471909, EPI_ISL_1471910, EPI_ISL_1471911, EPI_ISL_1471912, EPI_ISL_1471913, EPI_ISL_1471914, EPI_ISL_1471915, EPI_ISL_1471916, EPI_ISL_1471917, EPI_ISL_1471918, EPI_ISL_1471919, EPI_ISL_1471920, EPI_ISL_1471921, EPI_ISL_1471922, EPI_ISL_1471923, EPI_ISL_1471924, EPI_ISL_1471925, EPI_ISL_1471926, EPI_ISL_1471927, EPI_ISL_1471928, EPI_ISL_1471929, EPI_ISL_1471930, EPI_ISL_1471933, EPI_ISL_1471934, EPI_ISL_1471935, EPI_ISL_1471936, EPI_ISL_1471937, EPI_ISL_1471938, EPI_ISL_1471939, EPI_ISL_1471940, EPI_ISL_1471941, EPI_ISL_1471942, EPI_ISL_1471943, EPI_ISL_1471944, EPI_ISL_1471945, EPI_ISL_1471947, EPI_ISL_1471948, EPI_ISL_1471949, EPI_ISL_1471950, EPI_ISL_1471951, EPI_ISL_1471952, EPI_ISL_1471953, EPI_ISL_1471954, EPI_ISL_1471955, EPI_ISL_1471956, EPI_ISL_1471957, EPI_ISL_1471958, EPI_ISL_1471959, EPI_ISL_1471960, EPI_ISL_1471961, EPI_ISL_1471962, EPI_ISL_1471963, EPI_ISL_1471964, EPI_ISL_1471965, EPI_ISL_1471966, EPI_ISL_1471968, EPI_ISL_1471969, EPI_ISL_1471970, EPI_ISL_1471971, EPI_ISL_1471973, EPI_ISL_1471974, EPI_ISL_1471975, EPI_ISL_1471976, EPI_ISL_1471977, EPI_ISL_1471978, EPI_ISL_1471979, EPI_ISL_1471980, EPI_ISL_1471981, EPI_ISL_1471982, EPI_ISL_1471983, EPI_ISL_1471984, EPI_ISL_1471985, EPI_ISL_1471986, EPI_ISL_1471987, EPI_ISL_1471988, EPI_ISL_1471989, EPI_ISL_1471990, EPI_ISL_1471991, EPI_ISL_1471992, EPI_ISL_1471993, EPI_ISL_1471994, EPI_ISL_1471995, EPI_ISL_1471996, EPI_ISL_1471997, EPI_ISL_1471998, EPI_ISL_1471999, EPI_ISL_1472000, EPI_ISL_1472001, EPI_ISL_1472002, EPI_ISL_1472003, EPI_ISL_1472004, EPI_ISL_1472005, EPI_ISL_1472006, EPI_ISL_1472007, EPI_ISL_1472008, EPI_ISL_1472009, EPI_ISL_1472011, EPI_ISL_1472012, EPI_ISL_1472013, EPI_ISL_1472014, EPI_ISL_1472015, EPI_ISL_1472016, EPI_ISL_1472017, EPI_ISL_1472018, EPI_ISL_1472019, EPI_ISL_1472020, EPI_ISL_1472021, EPI_ISL_1472022, EPI_ISL_1472024, EPI_ISL_1472025, EPI_ISL_1472026, EPI_ISL_1472027, EPI_ISL_1472028, EPI_ISL_1472029, EPI_ISL_1472030, EPI_ISL_1472031, EPI_ISL_1472032, EPI_ISL_1472033, EPI_ISL_1472034, EPI_ISL_1472035, EPI_ISL_1472036, EPI_ISL_1472037, EPI_ISL_1472038, EPI_ISL_1472039, EPI_ISL_1472040, EPI_ISL_1472041, EPI_ISL_1472042, EPI_ISL_1472043, EPI_ISL_1472044, EPI_ISL_1472045, EPI_ISL_1472047, EPI_ISL_1472048, EPI_ISL_1472049, EPI_ISL_1472050, EPI_ISL_1472051, EPI_ISL_1472052, EPI_ISL_1472053, EPI_ISL_1472054, EPI_ISL_1472055, EPI_ISL_1472056, EPI_ISL_1472057, EPI_ISL_1472060, EPI_ISL_1472061, EPI_ISL_1472062, EPI_ISL_1472063, EPI_ISL_1472066, EPI_ISL_1472067, EPI_ISL_1472069, EPI_ISL_1472070, EPI_ISL_1472071, EPI_ISL_1472072, EPI_ISL_1472073, EPI_ISL_1472075, EPI_ISL_1472076 | | | |
| see above | Pandemic Response Lab - NYC | Pandemic Response Lab, R&D | Henry Lee, Michael Hammerling, Melissa Hopkins, Cybill del Castillo, Shinyoung Clair Kang, William Ward, Pradeep Bugga, Sol Rey, Dylan Law, Haiping Hao, Jon Laurent |
| EPI_ISL_1472386, EPI_ISL_1472387, EPI_ISL_1472388, EPI_ISL_1472389, EPI_ISL_1472390, EPI_ISL_1472391, EPI_ISL_1472392, EPI_ISL_1472393, EPI_ISL_1472394, EPI_ISL_1472395, EPI_ISL_1472396, EPI_ISL_1472397, EPI_ISL_1472398, EPI_ISL_1472399, EPI_ISL_1472400, EPI_ISL_1472401, EPI_ISL_1472402, EPI_ISL_1472403, EPI_ISL_1472404, EPI_ISL_1472405, EPI_ISL_1472406, EPI_ISL_1472407 | | | |
| see above | University Hospitals Translational Laboratory (UHTL), University Hospitals | University Hospitals Translational Laboratory (UHTL), University Hospitals | Sadri,N., Alouani,D., Song,X. |
| EPI_ISL_1477167, EPI_ISL_1477168 | CA DPH Viral and Rickettsial Disease Laboratory | Chan-Zuckerberg Biohub | CZB Cliahub Consortium |
| EPI_ISL_1477169 | Humboldt County Public Health Laboratory | Chan-Zuckerberg Biohub | CZB Cliahub Consortium |
| EPI_ISL_1477170 | Alameda County Public Health Lab | Chan-Zuckerberg Biohub | CZB Cliahub Consortium |
| EPI_ISL_1477171 | CA DPH Viral and Rickettsial Disease | Chan-Zuckerberg Biohub | CZB Cliahub Consortium |
| EPI_ISL_1477172, EPI_ISL_1477173, EPI_ISL_1477174 | Humboldt County Public Health Laboratory | Chan-Zuckerberg Biohub | CZB Cliahub Consortium |
| EPI_ISL_1477175 | Alameda County Public Health Lab | Chan-Zuckerberg Biohub | CZB Cliahub Consortium |
| EPI_ISL_1477176 | Humboldt County Public Health Laboratory | Chan-Zuckerberg Biohub | CZB Cliahub Consortium |
| EPI_ISL_1477177 | CA DPH Viral and Rickettsial Disease Laboratory | Chan-Zuckerberg Biohub | CZB Cliahub Consortium |
| EPI_ISL_1477178 | Alameda County Public Health Lab | Chan-Zuckerberg Biohub | CZB Cliahub Consortium |
| EPI_ISL_1477179 | CA DPH Viral and Rickettsial Disease Laboratory | Chan-Zuckerberg Biohub | CZB Cliahub Consortium |
| EPI_ISL_1477180 | Humboldt County Public Health Laboratory | Chan-Zuckerberg Biohub | CZB Cliahub Consortium |
| EPI_ISL_1477181, EPI_ISL_1477182 | CA DPH Viral and Rickettsial Disease Laboratory | Chan-Zuckerberg Biohub | CZB Cliahub Consortium |
| EPI_ISL_1477183 | Humboldt County Public Health Laboratory | Chan-Zuckerberg Biohub | CZB Cliahub Consortium |
| EPI_ISL_1477184, EPI_ISL_1477185, EPI_ISL_1477186, EPI_ISL_1477188 | CA DPH Viral and Rickettsial Disease Laboratory | Chan-Zuckerberg Biohub | CZB Cliahub Consortium |
| EPI_ISL_1477189 | Humboldt County Public Health Laboratory | Chan-Zuckerberg Biohub | CZB Cliahub Consortium |
| EPI_ISL_1477190, EPI_ISL_1477191, EPI_ISL_1477192, EPI_ISL_1477193, EPI_ISL_1477194 | CA DPH Viral and Rickettsial Disease Laboratory | Chan-Zuckerberg Biohub | CZB Cliahub Consortium |
| EPI_ISL_1477195 | Humboldt County Public Health Laboratory | Chan-Zuckerberg Biohub | CZB Cliahub Consortium |
| EPI_ISL_1477196, EPI_ISL_1477197, EPI_ISL_1477198 | CA DPH Viral and Rickettsial Disease Laboratory | Chan-Zuckerberg Biohub | CZB Cliahub Consortium |
| EPI_ISL_1477199 | Alameda County Public Health Lab | Chan-Zuckerberg Biohub | CZB Cliahub Consortium |
| EPI_ISL_1477200, EPI_ISL_1477201, EPI_ISL_1477202, EPI_ISL_1477203, EPI_ISL_1477204, EPI_ISL_1477205 | CA DPH Viral and Rickettsial Disease Laboratory | Chan-Zuckerberg Biohub | CZB Cliahub Consortium |
| EPI_ISL_1477206, EPI_ISL_1477207, EPI_ISL_1477208 | Humboldt County Public Health Laboratory | Chan-Zuckerberg Biohub | CZB Cliahub Consortium |
| EPI_ISL_1477209, EPI_ISL_1477210, EPI_ISL_1477211, EPI_ISL_1477212, EPI_ISL_1477213, EPI_ISL_1477214, EPI_ISL_1477215 | CA DPH Viral and Rickettsial Disease Laboratory | Chan-Zuckerberg Biohub | CZB Cliahub Consortium |
| EPI_ISL_1477216 | Alameda County Public Health Lab | Chan-Zuckerberg Biohub | CZB Cliahub Consortium |
| EPI_ISL_1477217 | CA DPH Viral and Rickettsial Disease Laboratory | Chan-Zuckerberg Biohub | CZB Cliahub Consortium |
| EPI_ISL_1477218 | Humboldt County Public Health | Chan-Zuckerberg Biohub | CZB Cliahub Consortium |

| | | | |
|---|---|---|---|
| | Laboratory | | |
| EPI_ISL_1477219 | CA DPH Viral and Rickettsial Disease Laboratory | Chan-Zuckerberg Biohub | CZB Cliahub Consortium |
| EPI_ISL_1477220 | Humboldt County Public Health Laboratory | Chan-Zuckerberg Biohub | CZB Cliahub Consortium |
| EPI_ISL_1477221, EPI_ISL_1477222, EPI_ISL_1477223 | CA DPH Viral and Rickettsial Disease Laboratory | Chan-Zuckerberg Biohub | CZB Cliahub Consortium |
| EPI_ISL_1477224 | Humboldt County Public Health Laboratory | Chan-Zuckerberg Biohub | CZB Cliahub Consortium |
| EPI_ISL_1477225 | CA DPH Viral and Rickettsial Disease Laboratory | Chan-Zuckerberg Biohub | CZB Cliahub Consortium |
| EPI_ISL_1477226, EPI_ISL_1477227 | Humboldt County Public Health Laboratory | Chan-Zuckerberg Biohub | CZB Cliahub Consortium |
| EPI_ISL_1477229, EPI_ISL_1477230, EPI_ISL_1477231, EPI_ISL_1477232, EPI_ISL_1477233 | CA DPH Viral and Rickettsial Disease Laboratory | Chan-Zuckerberg Biohub | CZB Cliahub Consortium |
| EPI_ISL_1477234 | Humboldt County Public Health Laboratory | Chan-Zuckerberg Biohub | CZB Cliahub Consortium |
| EPI_ISL_1477235 | CA DPH Viral and Rickettsial Disease Laboratory | Chan-Zuckerberg Biohub | CZB Cliahub Consortium |
| EPI_ISL_1477236 | Humboldt County Public Health Laboratory | Chan-Zuckerberg Biohub | CZB Cliahub Consortium |
| EPI_ISL_1477237, EPI_ISL_1477238 | CA DPH Viral and Rickettsial Disease Laboratory | Chan-Zuckerberg Biohub | CZB Cliahub Consortium |
| EPI_ISL_1477239 | Humboldt County Public Health Laboratory | Chan-Zuckerberg Biohub | CZB Cliahub Consortium |
| EPI_ISL_1477240, EPI_ISL_1477241, EPI_ISL_1477242 | CA DPH Viral and Rickettsial Disease Laboratory | Chan-Zuckerberg Biohub | CZB Cliahub Consortium |
| EPI_ISL_1477243, EPI_ISL_1477244 | Alameda County Public Health Lab | Chan-Zuckerberg Biohub | CZB Cliahub Consortium |
| EPI_ISL_1477245, EPI_ISL_1477246 | CA DPH Viral and Rickettsial Disease Laboratory | Chan-Zuckerberg Biohub | CZB Cliahub Consortium |
| EPI_ISL_1477247 | Humboldt County Public Health Laboratory | Chan-Zuckerberg Biohub | CZB Cliahub Consortium |
| EPI_ISL_1477248 | CA DPH Viral and Rickettsial Disease Laboratory | Chan-Zuckerberg Biohub | CZB Cliahub Consortium |
| EPI_ISL_1477249 | Humboldt County Public Health Laboratory | Chan-Zuckerberg Biohub | CZB Cliahub Consortium |
| EPI_ISL_1477250 | CA DPH Viral and Rickettsial Disease Laboratory | Chan-Zuckerberg Biohub | CZB Cliahub Consortium |
| EPI_ISL_1477251, EPI_ISL_1477252 | Alameda County Public Health Lab | Chan-Zuckerberg Biohub | CZB Cliahub Consortium |
| EPI_ISL_1477253, EPI_ISL_1477254 | CA DPH Viral and Rickettsial Disease Laboratory | Chan-Zuckerberg Biohub | CZB Cliahub Consortium |
| EPI_ISL_1477255 | Humboldt County Public Health Laboratory | Chan-Zuckerberg Biohub | CZB Cliahub Consortium |
| EPI_ISL_1477256 | CA DPH Viral and Rickettsial Disease Laboratory | Chan-Zuckerberg Biohub | CZB Cliahub Consortium |
| EPI_ISL_1477257 | Humboldt County Public Health Laboratory | Chan-Zuckerberg Biohub | CZB Cliahub Consortium |
| EPI_ISL_1477258 | CA DPH Viral and Rickettsial Disease Laboratory | Chan-Zuckerberg Biohub | CZB Cliahub Consortium |
| EPI_ISL_1477259, EPI_ISL_1477260 | Alameda County Public Health Lab | Chan-Zuckerberg Biohub | CZB Cliahub Consortium |
| EPI_ISL_1477261 | CA DPH Viral and Rickettsial Disease Laboratory | Chan-Zuckerberg Biohub | CZB Cliahub Consortium |
| EPI_ISL_1477262 | Alameda County Public Health Lab | Chan-Zuckerberg Biohub | CZB Cliahub Consortium |
| EPI_ISL_1477263 | CA DPH Viral and Rickettsial Disease Laboratory | Chan-Zuckerberg Biohub | CZB Cliahub Consortium |
| EPI_ISL_1477264 | Humboldt County Public Health Laboratory | Chan-Zuckerberg Biohub | CZB Cliahub Consortium |
| EPI_ISL_1477265, EPI_ISL_1477266, EPI_ISL_1477267 | CA DPH Viral and Rickettsial Disease Laboratory | Chan-Zuckerberg Biohub | CZB Cliahub Consortium |
| EPI_ISL_1477268, EPI_ISL_1477269, EPI_ISL_1477270, EPI_ISL_1477271 | Alameda County Public Health Lab | Chan-Zuckerberg Biohub | CZB Cliahub Consortium |
| EPI_ISL_1477272 | Humboldt County Public Health Laboratory | Chan-Zuckerberg Biohub | CZB Cliahub Consortium |
| EPI_ISL_1477273, EPI_ISL_1477274, EPI_ISL_1477275 | CA DPH Viral and Rickettsial Disease Laboratory | Chan-Zuckerberg Biohub | CZB Cliahub Consortium |
| EPI_ISL_1477276, EPI_ISL_1477277 | Humboldt County Public Health Laboratory | Chan-Zuckerberg Biohub | CZB Cliahub Consortium |
| EPI_ISL_1477278 | CA DPH Viral and Rickettsial Disease Laboratory | Chan-Zuckerberg Biohub | CZB Cliahub Consortium |
| EPI_ISL_1477279 | Alameda County Public Health Lab | Chan-Zuckerberg Biohub | CZB Cliahub Consortium |
| EPI_ISL_1477280 | Humboldt County Public Health Laboratory | Chan-Zuckerberg Biohub | CZB Cliahub Consortium |
| EPI_ISL_1477281 | CA DPH Viral and Rickettsial Disease Laboratory | Chan-Zuckerberg Biohub | CZB Cliahub Consortium |
| EPI_ISL_1477282 | Humboldt County Public Health Laboratory | Chan-Zuckerberg Biohub | CZB Cliahub Consortium |
| EPI_ISL_1477283 | CA DPH Viral and Rickettsial Disease Laboratory | Chan-Zuckerberg Biohub | CZB Cliahub Consortium |
| EPI_ISL_1477284 | Humboldt County Public Health | Chan-Zuckerberg Biohub | CZB Cliahub Consortium |

| | | | |
|---|---|---|---|
| | Laboratory | | |
| EPI_ISL_1477285, EPI_ISL_1477286 | CA DPH Viral and Rickettsial Disease Laboratory | Chan-Zuckerberg Biohub | CZB Cliahub Consortium |
| EPI_ISL_1477287, EPI_ISL_1477288 | Alameda County Public Health Lab | Chan-Zuckerberg Biohub | CZB Cliahub Consortium |
| EPI_ISL_1477289 | CA DPH Viral and Rickettsial Disease Laboratory | Chan-Zuckerberg Biohub | CZB Cliahub Consortium |
| EPI_ISL_1477290 | Humboldt County Public Health Laboratory | Chan-Zuckerberg Biohub | CZB Cliahub Consortium |
| EPI_ISL_1477291 | CA DPH Viral and Rickettsial Disease Laboratory | Chan-Zuckerberg Biohub | CZB Cliahub Consortium |
| EPI_ISL_1477292, EPI_ISL_1477293 | Alameda County Public Health Lab | Chan-Zuckerberg Biohub | CZB Cliahub Consortium |
| EPI_ISL_1477294 | Humboldt County Public Health Laboratory | Chan-Zuckerberg Biohub | CZB Cliahub Consortium |
| EPI_ISL_1477295 | Alameda County Public Health Lab | Chan-Zuckerberg Biohub | CZB Cliahub Consortium |
| EPI_ISL_1477296 | Humboldt County Public Health Laboratory | Chan-Zuckerberg Biohub | CZB Cliahub Consortium |
| EPI_ISL_1477297 | CA DPH Viral and Rickettsial Disease Laboratory | Chan-Zuckerberg Biohub | CZB Cliahub Consortium |
| EPI_ISL_1477299 | Humboldt County Public Health Laboratory | Chan-Zuckerberg Biohub | CZB Cliahub Consortium |
| EPI_ISL_1477300 | CA DPH Viral and Rickettsial Disease Laboratory | Chan-Zuckerberg Biohub | CZB Cliahub Consortium |
| EPI_ISL_1477301 | Alameda County Public Health Lab | Chan-Zuckerberg Biohub | CZB Cliahub Consortium |
| EPI_ISL_1477302, EPI_ISL_1477303, EPI_ISL_1477304, EPI_ISL_1477305 | CA DPH Viral and Rickettsial Disease Laboratory | Chan-Zuckerberg Biohub | CZB Cliahub Consortium |
| EPI_ISL_1477306 | Humboldt County Public Health Laboratory | Chan-Zuckerberg Biohub | CZB Cliahub Consortium |
| EPI_ISL_1477307 | CA DPH Viral and Rickettsial Disease Laboratory | Chan-Zuckerberg Biohub | CZB Cliahub Consortium |
| EPI_ISL_1477308, EPI_ISL_1477309 | Alameda County Public Health Lab | Chan-Zuckerberg Biohub | CZB Cliahub Consortium |
| EPI_ISL_1477310, EPI_ISL_1477311 | CA DPH Viral and Rickettsial Disease Laboratory | Chan-Zuckerberg Biohub | CZB Cliahub Consortium |
| EPI_ISL_1477312 | Humboldt County Public Health Laboratory | Chan-Zuckerberg Biohub | CZB Cliahub Consortium |
| EPI_ISL_1477313 | Alameda County Public Health Lab | Chan-Zuckerberg Biohub | CZB Cliahub Consortium |
| EPI_ISL_1477314, EPI_ISL_1477315, EPI_ISL_1477316, EPI_ISL_1477317 | CA DPH Viral and Rickettsial Disease Laboratory | Chan-Zuckerberg Biohub | CZB Cliahub Consortium |
| EPI_ISL_1477318 | Alameda County Public Health Lab | Chan-Zuckerberg Biohub | CZB Cliahub Consortium |
| EPI_ISL_1477319, EPI_ISL_1477320, EPI_ISL_1477321 | CA DPH Viral and Rickettsial Disease Laboratory | Chan-Zuckerberg Biohub | CZB Cliahub Consortium |
| EPI_ISL_1477322 | Alameda County Public Health Lab | Chan-Zuckerberg Biohub | CZB Cliahub Consortium |
| EPI_ISL_1477323, EPI_ISL_1477324 | CA DPH Viral and Rickettsial Disease Laboratory | Chan-Zuckerberg Biohub | CZB Cliahub Consortium |
| EPI_ISL_1477325, EPI_ISL_1477326 | Humboldt County Public Health Laboratory | Chan-Zuckerberg Biohub | CZB Cliahub Consortium |
| EPI_ISL_1477327 | CA DPH Viral and Rickettsial Disease Laboratory | Chan-Zuckerberg Biohub | CZB Cliahub Consortium |
| EPI_ISL_1477328 | Alameda County Public Health Lab | Chan-Zuckerberg Biohub | CZB Cliahub Consortium |
| EPI_ISL_1477330, EPI_ISL_1477331 | CA DPH Viral and Rickettsial Disease Laboratory | Chan-Zuckerberg Biohub | CZB Cliahub Consortium |
| EPI_ISL_1477332 | Humboldt County Public Health Laboratory | Chan-Zuckerberg Biohub | CZB Cliahub Consortium |
| EPI_ISL_1477333 | CA DPH Viral and Rickettsial Disease Laboratory | Chan-Zuckerberg Biohub | CZB Cliahub Consortium |
| EPI_ISL_1477334 | Humboldt County Public Health Laboratory | Chan-Zuckerberg Biohub | CZB Cliahub Consortium |
| EPI_ISL_1477335 | Alameda County Public Health Lab | Chan-Zuckerberg Biohub | CZB Cliahub Consortium |
| EPI_ISL_1477336 | CA DPH Viral and Rickettsial Disease Laboratory | Chan-Zuckerberg Biohub | CZB Cliahub Consortium |
| EPI_ISL_1477337 | Alameda County Public Health Lab | Chan-Zuckerberg Biohub | CZB Cliahub Consortium |
| EPI_ISL_1477338 | CA DPH Viral and Rickettsial Disease Laboratory | Chan-Zuckerberg Biohub | CZB Cliahub Consortium |
| EPI_ISL_1477339, EPI_ISL_1477340, EPI_ISL_1477341 | Humboldt County Public Health Laboratory | Chan-Zuckerberg Biohub | CZB Cliahub Consortium |
| EPI_ISL_1477342 | CA DPH Viral and Rickettsial Disease Laboratory | Chan-Zuckerberg Biohub | CZB Cliahub Consortium |
| EPI_ISL_1477343 | Alameda County Public Health Lab | Chan-Zuckerberg Biohub | CZB Cliahub Consortium |
| EPI_ISL_1477344, EPI_ISL_1477345 | CA DPH Viral and Rickettsial Disease Laboratory | Chan-Zuckerberg Biohub | CZB Cliahub Consortium |
| EPI_ISL_1477346 | Humboldt County Public Health Laboratory | Chan-Zuckerberg Biohub | CZB Cliahub Consortium |
| EPI_ISL_1477347 | CA DPH Viral and Rickettsial Disease Laboratory | Chan-Zuckerberg Biohub | CZB Cliahub Consortium |
| EPI_ISL_1477348 | Humboldt County Public Health Laboratory | Chan-Zuckerberg Biohub | CZB Cliahub Consortium |
| EPI_ISL_1477349, EPI_ISL_1477350, EPI_ISL_1477351 | CA DPH Viral and Rickettsial Disease Laboratory | Chan-Zuckerberg Biohub | CZB Cliahub Consortium |
| EPI_ISL_1477352 | Alameda County Public Health Lab | Chan-Zuckerberg Biohub | CZB Cliahub Consortium |
| EPI_ISL_1477353 | CA DPH Viral and Rickettsial Disease | Chan-Zuckerberg Biohub | CZB Cliahub Consortium |

| | | | |
|---|---|---|---|
| | Laboratory | | |
| EPI_ISL_1477354, EPI_ISL_1477355 | Humboldt County Public Health Laboratory | Chan-Zuckerberg Biohub | CZB Cliahub Consortium |
| EPI_ISL_1477356, EPI_ISL_1477357, EPI_ISL_1477358 | CA DPH Viral and Rickettsial Disease Laboratory | Chan-Zuckerberg Biohub | CZB Cliahub Consortium |
| EPI_ISL_1477359 | Alameda County Public Health Lab | Chan-Zuckerberg Biohub | CZB Cliahub Consortium |
| EPI_ISL_1477360, EPI_ISL_1477361, EPI_ISL_1477362, EPI_ISL_1477363 | CA DPH Viral and Rickettsial Disease Laboratory | Chan-Zuckerberg Biohub | CZB Cliahub Consortium |
| EPI_ISL_1477364 | Alameda County Public Health Lab | Chan-Zuckerberg Biohub | CZB Cliahub Consortium |
| EPI_ISL_1477365, EPI_ISL_1477366 | CA DPH Viral and Rickettsial Disease Laboratory | Chan-Zuckerberg Biohub | CZB Cliahub Consortium |
| EPI_ISL_1477367 | Alameda County Public Health Lab | Chan-Zuckerberg Biohub | CZB Cliahub Consortium |
| EPI_ISL_1477368, EPI_ISL_1477369, EPI_ISL_1477370 | CA DPH Viral and Rickettsial Disease Laboratory | Chan-Zuckerberg Biohub | CZB Cliahub Consortium |
| EPI_ISL_1477371 | Alameda County Public Health Lab | Chan-Zuckerberg Biohub | CZB Cliahub Consortium |
| EPI_ISL_1477372 | CA DPH Viral and Rickettsial Disease Laboratory | Chan-Zuckerberg Biohub | CZB Cliahub Consortium |
| EPI_ISL_1477373 | Alameda County Public Health Lab | Chan-Zuckerberg Biohub | CZB Cliahub Consortium |
| EPI_ISL_1477374 | CA DPH Viral and Rickettsial Disease Laboratory | Chan-Zuckerberg Biohub | CZB Cliahub Consortium |
| EPI_ISL_1477375, EPI_ISL_1477376 | Humboldt County Public Health Laboratory | Chan-Zuckerberg Biohub | CZB Cliahub Consortium |
| EPI_ISL_1477377 | Alameda County Public Health Lab | Chan-Zuckerberg Biohub | CZB Cliahub Consortium |
| EPI_ISL_1477378 | CA DPH Viral and Rickettsial Disease Laboratory | Chan-Zuckerberg Biohub | CZB Cliahub Consortium |
| EPI_ISL_1477379 | Humboldt County Public Health Laboratory | Chan-Zuckerberg Biohub | CZB Cliahub Consortium |
| EPI_ISL_1477380 | Alameda County Public Health Lab | Chan-Zuckerberg Biohub | CZB Cliahub Consortium |
| EPI_ISL_1477381 | Humboldt County Public Health Laboratory | Chan-Zuckerberg Biohub | CZB Cliahub Consortium |
| EPI_ISL_1477382 | Alameda County Public Health Lab | Chan-Zuckerberg Biohub | CZB Cliahub Consortium |
| EPI_ISL_1477383, EPI_ISL_1477384 | CA DPH Viral and Rickettsial Disease Laboratory | Chan-Zuckerberg Biohub | CZB Cliahub Consortium |
| EPI_ISL_1477385 | Alameda County Public Health Lab | Chan-Zuckerberg Biohub | CZB Cliahub Consortium |
| EPI_ISL_1477386, EPI_ISL_1477387 | CA DPH Viral and Rickettsial Disease Laboratory | Chan-Zuckerberg Biohub | CZB Cliahub Consortium |
| EPI_ISL_1477388 | Alameda County Public Health Lab | Chan-Zuckerberg Biohub | CZB Cliahub Consortium |
| EPI_ISL_1477389 | Humboldt County Public Health Laboratory | Chan-Zuckerberg Biohub | CZB Cliahub Consortium |
| EPI_ISL_1477390, EPI_ISL_1477391, EPI_ISL_1477392, EPI_ISL_1477393 | CA DPH Viral and Rickettsial Disease Laboratory | Chan-Zuckerberg Biohub | CZB Cliahub Consortium |
| EPI_ISL_1477394 | Humboldt County Public Health Laboratory | Chan-Zuckerberg Biohub | CZB Cliahub Consortium |
| EPI_ISL_1477395 | CA DPH Viral and Rickettsial Disease Laboratory | Chan-Zuckerberg Biohub | CZB Cliahub Consortium |
| EPI_ISL_1477396, EPI_ISL_1477397 | Humboldt County Public Health Laboratory | Chan-Zuckerberg Biohub | CZB Cliahub Consortium |
| EPI_ISL_1477398 | CA DPH Viral and Rickettsial Disease Laboratory | Chan-Zuckerberg Biohub | CZB Cliahub Consortium |
| EPI_ISL_1477399 | Humboldt County Public Health Laboratory | Chan-Zuckerberg Biohub | CZB Cliahub Consortium |
| EPI_ISL_1477400 | CA DPH Viral and Rickettsial Disease Laboratory | Chan-Zuckerberg Biohub | CZB Cliahub Consortium |
| EPI_ISL_1477401 | Humboldt County Public Health Laboratory | Chan-Zuckerberg Biohub | CZB Cliahub Consortium |
| EPI_ISL_1477402, EPI_ISL_1477403, EPI_ISL_1477404 | CA DPH Viral and Rickettsial Disease Laboratory | Chan-Zuckerberg Biohub | CZB Cliahub Consortium |
| EPI_ISL_1477405, EPI_ISL_1477406 | Alameda County Public Health Lab | Chan-Zuckerberg Biohub | CZB Cliahub Consortium |
| EPI_ISL_1477407 | CA DPH Viral and Rickettsial Disease Laboratory | Chan-Zuckerberg Biohub | CZB Cliahub Consortium |
| EPI_ISL_1477408 | Humboldt County Public Health Laboratory | Chan-Zuckerberg Biohub | CZB Cliahub Consortium |
| EPI_ISL_1477409 | CA DPH Viral and Rickettsial Disease Laboratory | Chan-Zuckerberg Biohub | CZB Cliahub Consortium |
| EPI_ISL_1477410, EPI_ISL_1477411 | Alameda County Public Health Lab | Chan-Zuckerberg Biohub | CZB Cliahub Consortium |
| EPI_ISL_1477412 | Humboldt County Public Health Laboratory | Chan-Zuckerberg Biohub | CZB Cliahub Consortium |
| EPI_ISL_1477413, EPI_ISL_1477414, EPI_ISL_1477415 | Alameda County Public Health Lab | Chan-Zuckerberg Biohub | CZB Cliahub Consortium |
| EPI_ISL_1477416 | CA DPH Viral and Rickettsial Disease Laboratory | Chan-Zuckerberg Biohub | CZB Cliahub Consortium |
| EPI_ISL_1477417 | Humboldt County Public Health Laboratory | Chan-Zuckerberg Biohub | CZB Cliahub Consortium |

EPI_ISL_1477418, EPI_ISL_1477419, EPI_ISL_1477420, EPI_ISL_1477421, EPI_ISL_1477422, EPI_ISL_1477423, EPI_ISL_1477424, EPI_ISL_1477425, EPI_ISL_1477426, EPI_ISL_1477427, EPI_ISL_1477428, EPI_ISL_1477429

| | | | |
|---|---|---|---|
| see above | CA DPH Viral and Rickettsial Disease Laboratory | Chan-Zuckerberg Biohub | CZB Cliahub Consortium |
| EPI_ISL_1477430 | Humboldt County Public Health Laboratory | Chan-Zuckerberg Biohub | CZB Cliahub Consortium |
| EPI_ISL_1477431 | CA DPH Viral and Rickettsial Disease Laboratory | Chan-Zuckerberg Biohub | CZB Cliahub Consortium |

| | | | |
|---|---|---|---|
| EPI_ISL_1477432, EPI_ISL_1477433 | Alameda County Public Health Lab | Chan-Zuckerberg Biohub | CZB Cliahub Consortium |
| EPI_ISL_1477434 | CA DPH Viral and Rickettsial Disease Laboratory | Chan-Zuckerberg Biohub | CZB Cliahub Consortium |
| EPI_ISL_1477435 | Alameda County Public Health Lab | Chan-Zuckerberg Biohub | CZB Cliahub Consortium |
| EPI_ISL_1477436 | County of San Luis Obispo Public Health Laboratory | Chan-Zuckerberg Biohub | CZB Cliahub Consortium |
| EPI_ISL_1477437, EPI_ISL_1477438, EPI_ISL_1477439, EPI_ISL_1477440 | CA DPH Viral and Rickettsial Disease Laboratory | Chan-Zuckerberg Biohub | CZB Cliahub Consortium |
| EPI_ISL_1477441 | Santa Clara County Public Health Laboratory | Chan-Zuckerberg Biohub | CZB Cliahub Consortium |
| EPI_ISL_1477442 | CA DPH Viral and Rickettsial Disease Laboratory | Chan-Zuckerberg Biohub | CZB Cliahub Consortium |
| EPI_ISL_1477443 | Orange County Public Health Lab | Chan-Zuckerberg Biohub | CZB Cliahub Consortium |
| EPI_ISL_1477444 | CA DPH Viral and Rickettsial Disease Laboratory | Chan-Zuckerberg Biohub | CZB Cliahub Consortium |
| EPI_ISL_1477446, EPI_ISL_1477447 | Alameda County Public Health Lab | Chan-Zuckerberg Biohub | CZB Cliahub Consortium |
| EPI_ISL_1477448, EPI_ISL_1477449, EPI_ISL_1477450, EPI_ISL_1477451, EPI_ISL_1477452, EPI_ISL_1477453 | CA DPH Viral and Rickettsial Disease Laboratory | Chan-Zuckerberg Biohub | CZB Cliahub Consortium |
| EPI_ISL_1477454 | County of San Luis Obispo Public Health Laboratory | Chan-Zuckerberg Biohub | CZB Cliahub Consortium |
| EPI_ISL_1477455 | Santa Clara County Public Health Laboratory | Chan-Zuckerberg Biohub | CZB Cliahub Consortium |
| EPI_ISL_1477456, EPI_ISL_1477457, EPI_ISL_1477458, EPI_ISL_1477460 | CA DPH Viral and Rickettsial Disease Laboratory | Chan-Zuckerberg Biohub | CZB Cliahub Consortium |
| EPI_ISL_1477461 | Santa Clara County Public Health Laboratory | Chan-Zuckerberg Biohub | CZB Cliahub Consortium |
| EPI_ISL_1477462 | Alameda County Public Health Lab | Chan-Zuckerberg Biohub | CZB Cliahub Consortium |
| EPI_ISL_1477463, EPI_ISL_1477464, EPI_ISL_1477465 | CA DPH Viral and Rickettsial Disease Laboratory | Chan-Zuckerberg Biohub | CZB Cliahub Consortium |
| EPI_ISL_1477466 | Alameda County Public Health Lab | Chan-Zuckerberg Biohub | CZB Cliahub Consortium |
| EPI_ISL_1477467 | CA DPH Viral and Rickettsial Disease Laboratory | Chan-Zuckerberg Biohub | CZB Cliahub Consortium |
| EPI_ISL_1477468 | Alameda County Public Health Lab | Chan-Zuckerberg Biohub | CZB Cliahub Consortium |
| EPI_ISL_1477469, EPI_ISL_1477470, EPI_ISL_1477471, EPI_ISL_1477472, EPI_ISL_1477473, EPI_ISL_1477474, EPI_ISL_1477475, EPI_ISL_1477476, EPI_ISL_1477477, EPI_ISL_1477478, EPI_ISL_1477479, EPI_ISL_1477480, EPI_ISL_1477481, EPI_ISL_1477482 | | | |
| see above | CA DPH Viral and Rickettsial Disease Laboratory | Chan-Zuckerberg Biohub | CZB Cliahub Consortium |
| EPI_ISL_1477483 | Alameda County Public Health Lab | Chan-Zuckerberg Biohub | CZB Cliahub Consortium |
| EPI_ISL_1477484 | Santa Clara County Public Health Laboratory | Chan-Zuckerberg Biohub | CZB Cliahub Consortium |
| EPI_ISL_1477485 | CA DPH Viral and Rickettsial Disease Laboratory | Chan-Zuckerberg Biohub | CZB Cliahub Consortium |
| EPI_ISL_1477486 | Santa Clara County Public Health Laboratory | Chan-Zuckerberg Biohub | CZB Cliahub Consortium |
| EPI_ISL_1477487 | Alameda County Public Health Lab | Chan-Zuckerberg Biohub | CZB Cliahub Consortium |
| EPI_ISL_1477488 | CA DPH Viral and Rickettsial Disease Laboratory | Chan-Zuckerberg Biohub | CZB Cliahub Consortium |
| EPI_ISL_1477489 | Alameda County Public Health Lab | Chan-Zuckerberg Biohub | CZB Cliahub Consortium |
| EPI_ISL_1477490 | CA DPH Viral and Rickettsial Disease Laboratory | Chan-Zuckerberg Biohub | CZB Cliahub Consortium |
| EPI_ISL_1477491 | Alameda County Public Health Lab | Chan-Zuckerberg Biohub | CZB Cliahub Consortium |
| EPI_ISL_1477492, EPI_ISL_1477493 | CA DPH Viral and Rickettsial Disease Laboratory | Chan-Zuckerberg Biohub | CZB Cliahub Consortium |
| EPI_ISL_1477494 | Santa Clara County Public Health Laboratory | Chan-Zuckerberg Biohub | CZB Cliahub Consortium |
| EPI_ISL_1477495 | Orange County Public Health Lab | Chan-Zuckerberg Biohub | CZB Cliahub Consortium |
| EPI_ISL_1477496 | Santa Clara County Public Health Laboratory | Chan-Zuckerberg Biohub | CZB Cliahub Consortium |
| EPI_ISL_1477497 | Alameda County Public Health Lab | Chan-Zuckerberg Biohub | CZB Cliahub Consortium |
| EPI_ISL_1477498 | Orange County Public Health Lab | Chan-Zuckerberg Biohub | CZB Cliahub Consortium |
| EPI_ISL_1477499 | Alameda County Public Health Lab | Chan-Zuckerberg Biohub | CZB Cliahub Consortium |
| EPI_ISL_1477500 | CA DPH Viral and Rickettsial Disease Laboratory | Chan-Zuckerberg Biohub | CZB Cliahub Consortium |
| EPI_ISL_1477501 | Santa Clara County Public Health Laboratory | Chan-Zuckerberg Biohub | CZB Cliahub Consortium |
| EPI_ISL_1477502, EPI_ISL_1477503, EPI_ISL_1477504, EPI_ISL_1477505, EPI_ISL_1477506 | CA DPH Viral and Rickettsial Disease Laboratory | Chan-Zuckerberg Biohub | CZB Cliahub Consortium |
| EPI_ISL_1477507 | Santa Clara County Public Health Laboratory | Chan-Zuckerberg Biohub | CZB Cliahub Consortium |
| EPI_ISL_1477508, EPI_ISL_1477509 | CA DPH Viral and Rickettsial Disease Laboratory | Chan-Zuckerberg Biohub | CZB Cliahub Consortium |
| EPI_ISL_1477510 | Santa Clara County Public Health Laboratory | Chan-Zuckerberg Biohub | CZB Cliahub Consortium |
| EPI_ISL_1477511, EPI_ISL_1477512, EPI_ISL_1477513, EPI_ISL_1477514, | CA DPH Viral and Rickettsial Disease Laboratory | Chan-Zuckerberg Biohub | CZB Cliahub Consortium |

| | | | | |
|---|---|---|---|---|
| EPI_ISL_1477515, EPI_ISL_1477516, EPI_ISL_1477517 | | | | |
| EPI_ISL_1477518, EPI_ISL_1477519 | Santa Clara County Public Health Laboratory | Chan-Zuckerberg Biohub | | CZB Cliahub Consortium |
| EPI_ISL_1477520 | Alameda County Public Health Lab | Chan-Zuckerberg Biohub | | CZB Cliahub Consortium |
| EPI_ISL_1477521, EPI_ISL_1477522, EPI_ISL_1477523, EPI_ISL_1477524, EPI_ISL_1477525, EPI_ISL_1477526, EPI_ISL_1477527, EPI_ISL_1477528 | CA DPH Viral and Rickettsial Disease Laboratory | Chan-Zuckerberg Biohub | | CZB Cliahub Consortium |
| EPI_ISL_1477529 | Santa Clara County Public Health Laboratory | Chan-Zuckerberg Biohub | | CZB Cliahub Consortium |
| EPI_ISL_1477530 | County of San Luis Obispo Public Health Laboratory | Chan-Zuckerberg Biohub | | CZB Cliahub Consortium |
| EPI_ISL_1477531, EPI_ISL_1477532, EPI_ISL_1477533 | CA DPH Viral and Rickettsial Disease Laboratory | Chan-Zuckerberg Biohub | | CZB Cliahub Consortium |
| EPI_ISL_1477534, EPI_ISL_1477535 | Santa Clara County Public Health Laboratory | Chan-Zuckerberg Biohub | | CZB Cliahub Consortium |
| EPI_ISL_1477536, EPI_ISL_1477537, EPI_ISL_1477538, EPI_ISL_1477539 | CA DPH Viral and Rickettsial Disease Laboratory | Chan-Zuckerberg Biohub | | CZB Cliahub Consortium |
| EPI_ISL_1477540, EPI_ISL_1477541 | Santa Clara County Public Health Laboratory | Chan-Zuckerberg Biohub | | CZB Cliahub Consortium |
| EPI_ISL_1477542, EPI_ISL_1477544 | CA DPH Viral and Rickettsial Disease Laboratory | Chan-Zuckerberg Biohub | | CZB Cliahub Consortium |
| EPI_ISL_1477545, EPI_ISL_1477546, EPI_ISL_1477547 | Alameda County Public Health Lab | Chan-Zuckerberg Biohub | | CZB Cliahub Consortium |
| EPI_ISL_1477548, EPI_ISL_1477549 | CA DPH Viral and Rickettsial Disease Laboratory | Chan-Zuckerberg Biohub | | CZB Cliahub Consortium |
| EPI_ISL_1477550 | County of San Luis Obispo Public Health Laboratory | Chan-Zuckerberg Biohub | | CZB Cliahub Consortium |
| EPI_ISL_1477551 | CA DPH Viral and Rickettsial Disease Laboratory | Chan-Zuckerberg Biohub | | CZB Cliahub Consortium |
| EPI_ISL_1477552 | Santa Clara County Public Health Laboratory | Chan-Zuckerberg Biohub | | CZB Cliahub Consortium |
| EPI_ISL_1477553 | CA DPH Viral and Rickettsial Disease Laboratory | Chan-Zuckerberg Biohub | | CZB Cliahub Consortium |
| EPI_ISL_1477554 | Alameda County Public Health Lab | Chan-Zuckerberg Biohub | | CZB Cliahub Consortium |
| EPI_ISL_1477555, EPI_ISL_1477556 | CA DPH Viral and Rickettsial Disease Laboratory | Chan-Zuckerberg Biohub | | CZB Cliahub Consortium |
| EPI_ISL_1477557 | Alameda County Public Health Lab | Chan-Zuckerberg Biohub | | CZB Cliahub Consortium |
| EPI_ISL_1477558 | Santa Clara County Public Health Laboratory | Chan-Zuckerberg Biohub | | CZB Cliahub Consortium |
| EPI_ISL_1477559, EPI_ISL_1477560, EPI_ISL_1477561, EPI_ISL_1477562, EPI_ISL_1477563 | CA DPH Viral and Rickettsial Disease Laboratory | Chan-Zuckerberg Biohub | | CZB Cliahub Consortium |
| EPI_ISL_1477564 | County of San Luis Obispo Public Health Laboratory | Chan-Zuckerberg Biohub | | CZB Cliahub Consortium |
| EPI_ISL_1477565 | Alameda County Public Health Lab | Chan-Zuckerberg Biohub | | CZB Cliahub Consortium |
| EPI_ISL_1477566 | Santa Clara County Public Health Laboratory | Chan-Zuckerberg Biohub | | CZB Cliahub Consortium |
| EPI_ISL_1477567, EPI_ISL_1477568 | CA DPH Viral and Rickettsial Disease Laboratory | Chan-Zuckerberg Biohub | | CZB Cliahub Consortium |
| EPI_ISL_1477569 | Alameda County Public Health Lab | Chan-Zuckerberg Biohub | | CZB Cliahub Consortium |
| EPI_ISL_1477570, EPI_ISL_1477571 | CA DPH Viral and Rickettsial Disease Laboratory | Chan-Zuckerberg Biohub | | CZB Cliahub Consortium |
| EPI_ISL_1477572 | Orange County Public Health Lab | Chan-Zuckerberg Biohub | | CZB Cliahub Consortium |
| EPI_ISL_1477573, EPI_ISL_1477574 | CA DPH Viral and Rickettsial Disease Laboratory | Chan-Zuckerberg Biohub | | CZB Cliahub Consortium |
| EPI_ISL_1477575 | Santa Clara County Public Health Laboratory | Chan-Zuckerberg Biohub | | CZB Cliahub Consortium |
| EPI_ISL_1477576, EPI_ISL_1477577 | CA DPH Viral and Rickettsial Disease Laboratory | Chan-Zuckerberg Biohub | | CZB Cliahub Consortium |
| EPI_ISL_1477578 | Santa Clara County Public Health Laboratory | Chan-Zuckerberg Biohub | | CZB Cliahub Consortium |
| EPI_ISL_1477579, EPI_ISL_1477580 | CA DPH Viral and Rickettsial Disease Laboratory | Chan-Zuckerberg Biohub | | CZB Cliahub Consortium |
| EPI_ISL_1477581 | Santa Clara County Public Health Laboratory | Chan-Zuckerberg Biohub | | CZB Cliahub Consortium |
| EPI_ISL_1477582 | Orange County Public Health Lab | Chan-Zuckerberg Biohub | | CZB Cliahub Consortium |
| EPI_ISL_1477583 | CA DPH Viral and Rickettsial Disease Laboratory | Chan-Zuckerberg Biohub | | CZB Cliahub Consortium |
| EPI_ISL_1477584 | Alameda County Public Health Lab | Chan-Zuckerberg Biohub | | CZB Cliahub Consortium |
| EPI_ISL_1477585, EPI_ISL_1477586, EPI_ISL_1477587, EPI_ISL_1477588, EPI_ISL_1477589 | CA DPH Viral and Rickettsial Disease Laboratory | Chan-Zuckerberg Biohub | | CZB Cliahub Consortium |
| EPI_ISL_1477590 | Santa Clara County Public Health Laboratory | Chan-Zuckerberg Biohub | | CZB Cliahub Consortium |
| EPI_ISL_1477591, EPI_ISL_1477592, EPI_ISL_1477593 | CA DPH Viral and Rickettsial Disease Laboratory | Chan-Zuckerberg Biohub | | CZB Cliahub Consortium |

| | | | |
|---|---|---|---|
| EPI_ISL_1477594, EPI_ISL_1477595 | Alameda County Public Health Lab | Chan-Zuckerberg Biohub | CZB Cliahub Consortium |
| EPI_ISL_1477596, EPI_ISL_1477597 | CA DPH Viral and Rickettsial Disease Laboratory | Chan-Zuckerberg Biohub | CZB Cliahub Consortium |
| EPI_ISL_1477598 | Santa Clara County Public Health Laboratory | Chan-Zuckerberg Biohub | CZB Cliahub Consortium |
| EPI_ISL_1477599, EPI_ISL_1477600, EPI_ISL_1477601, EPI_ISL_1477602 | CA DPH Viral and Rickettsial Disease Laboratory | Chan-Zuckerberg Biohub | CZB Cliahub Consortium |
| EPI_ISL_1477603 | Santa Clara County Public Health Laboratory | Chan-Zuckerberg Biohub | CZB Cliahub Consortium |
| EPI_ISL_1477604 | CA DPH Viral and Rickettsial Disease Laboratory | Chan-Zuckerberg Biohub | CZB Cliahub Consortium |
| EPI_ISL_1477605 | Alameda County Public Health Lab | Chan-Zuckerberg Biohub | CZB Cliahub Consortium |
| EPI_ISL_1477606, EPI_ISL_1477607, EPI_ISL_1477608, EPI_ISL_1477609, EPI_ISL_1477610 | CA DPH Viral and Rickettsial Disease Laboratory | Chan-Zuckerberg Biohub | CZB Cliahub Consortium |
| EPI_ISL_1477611 | Orange County Public Health Lab | Chan-Zuckerberg Biohub | CZB Cliahub Consortium |
| EPI_ISL_1477612, EPI_ISL_1477613, EPI_ISL_1477614, EPI_ISL_1477615, EPI_ISL_1477616 | CA DPH Viral and Rickettsial Disease Laboratory | Chan-Zuckerberg Biohub | CZB Cliahub Consortium |
| EPI_ISL_1477617 | Alameda County Public Health Lab | Chan-Zuckerberg Biohub | CZB Cliahub Consortium |
| EPI_ISL_1477618, EPI_ISL_1477619, EPI_ISL_1477620, EPI_ISL_1477621 | CA DPH Viral and Rickettsial Disease Laboratory | Chan-Zuckerberg Biohub | CZB Cliahub Consortium |
| EPI_ISL_1477622 | Santa Clara County Public Health Laboratory | Chan-Zuckerberg Biohub | CZB Cliahub Consortium |
| EPI_ISL_1477623, EPI_ISL_1477624, EPI_ISL_1477625 | CA DPH Viral and Rickettsial Disease Laboratory | Chan-Zuckerberg Biohub | CZB Cliahub Consortium |
| EPI_ISL_1477626 | County of San Luis Obispo Public Health Laboratory | Chan-Zuckerberg Biohub | CZB Cliahub Consortium |
| EPI_ISL_1477627 | Alameda County Public Health Lab | Chan-Zuckerberg Biohub | CZB Cliahub Consortium |
| EPI_ISL_1477628, EPI_ISL_1477629 | CA DPH Viral and Rickettsial Disease Laboratory | Chan-Zuckerberg Biohub | CZB Cliahub Consortium |
| EPI_ISL_1477630 | Alameda County Public Health Lab | Chan-Zuckerberg Biohub | CZB Cliahub Consortium |
| EPI_ISL_1477631 | CA DPH Viral and Rickettsial Disease Laboratory | Chan-Zuckerberg Biohub | CZB Cliahub Consortium |
| EPI_ISL_1477632 | Alameda County Public Health Lab | Chan-Zuckerberg Biohub | CZB Cliahub Consortium |
| EPI_ISL_1477633 | CA DPH Viral and Rickettsial Disease Laboratory | Chan-Zuckerberg Biohub | CZB Cliahub Consortium |
| EPI_ISL_1477634 | County of San Luis Obispo Public Health Laboratory | Chan-Zuckerberg Biohub | CZB Cliahub Consortium |
| EPI_ISL_1477635, EPI_ISL_1477636 | CA DPH Viral and Rickettsial Disease Laboratory | Chan-Zuckerberg Biohub | CZB Cliahub Consortium |
| EPI_ISL_1477637 | Santa Clara County Public Health Laboratory | Chan-Zuckerberg Biohub | CZB Cliahub Consortium |
| EPI_ISL_1477638 | CA DPH Viral and Rickettsial Disease Laboratory | Chan-Zuckerberg Biohub | CZB Cliahub Consortium |
| EPI_ISL_1477639 | Alameda County Public Health Lab | Chan-Zuckerberg Biohub | CZB Cliahub Consortium |
| EPI_ISL_1477640, EPI_ISL_1477641, EPI_ISL_1477642, EPI_ISL_1477643 | CA DPH Viral and Rickettsial Disease Laboratory | Chan-Zuckerberg Biohub | CZB Cliahub Consortium |
| EPI_ISL_1477644 | Santa Clara County Public Health Laboratory | Chan-Zuckerberg Biohub | CZB Cliahub Consortium |
| EPI_ISL_1477645, EPI_ISL_1477646 | CA DPH Viral and Rickettsial Disease Laboratory | Chan-Zuckerberg Biohub | CZB Cliahub Consortium |
| EPI_ISL_1477647 | Alameda County Public Health Lab | Chan-Zuckerberg Biohub | CZB Cliahub Consortium |
| EPI_ISL_1477648 | CA DPH Viral and Rickettsial Disease Laboratory | Chan-Zuckerberg Biohub | CZB Cliahub Consortium |
| EPI_ISL_1477649 | Alameda County Public Health Lab | Chan-Zuckerberg Biohub | CZB Cliahub Consortium |
| EPI_ISL_1477650, EPI_ISL_1477651, EPI_ISL_1477652, EPI_ISL_1477653, EPI_ISL_1477654 | CA DPH Viral and Rickettsial Disease Laboratory | Chan-Zuckerberg Biohub | CZB Cliahub Consortium |
| EPI_ISL_1477655 | Santa Clara County Public Health Laboratory | Chan-Zuckerberg Biohub | CZB Cliahub Consortium |
| EPI_ISL_1477656 | CA DPH Viral and Rickettsial Disease Laboratory | Chan-Zuckerberg Biohub | CZB Cliahub Consortium |
| EPI_ISL_1477657 | Alameda County Public Health Lab | Chan-Zuckerberg Biohub | CZB Cliahub Consortium |
| EPI_ISL_1477658, EPI_ISL_1477659, EPI_ISL_1477660, EPI_ISL_1477661, EPI_ISL_1477662 | CA DPH Viral and Rickettsial Disease Laboratory | Chan-Zuckerberg Biohub | CZB Cliahub Consortium |
| EPI_ISL_1477663 | Orange County Public Health Lab | Chan-Zuckerberg Biohub | CZB Cliahub Consortium |
| EPI_ISL_1477664 | Santa Clara County Public Health Laboratory | Chan-Zuckerberg Biohub | CZB Cliahub Consortium |
| EPI_ISL_1477665, EPI_ISL_1477666, EPI_ISL_1477667 | CA DPH Viral and Rickettsial Disease Laboratory | Chan-Zuckerberg Biohub | CZB Cliahub Consortium |
| EPI_ISL_1477668, EPI_ISL_1477669 | Alameda County Public Health Lab | Chan-Zuckerberg Biohub | CZB Cliahub Consortium |
| EPI_ISL_1477670 | Santa Clara County Public Health | Chan-Zuckerberg Biohub | CZB Cliahub Consortium |

| | | | |
|---|---|---|---|
| | Laboratory | | |
| EPI_ISL_1477671, EPI_ISL_1477672 | Alameda County Public Health Lab | Chan-Zuckerberg Biohub | CZB Cliahub Consortium |
| EPI_ISL_1477673 | CA DPH Viral and Rickettsial Disease Laboratory | Chan-Zuckerberg Biohub | CZB Cliahub Consortium |
| EPI_ISL_1477674 | Alameda County Public Health Lab | Chan-Zuckerberg Biohub | CZB Cliahub Consortium |
| EPI_ISL_1477675, EPI_ISL_1477676 | Contra Costa County Public Health Lab | Chan-Zuckerberg Biohub | CZB Cliahub Consortium |
| EPI_ISL_1477677 | Santa Clara County Public Health Laboratory | Chan-Zuckerberg Biohub | CZB Cliahub Consortium |
| EPI_ISL_1477678 | CA DPH Viral and Rickettsial Disease Laboratory | Chan-Zuckerberg Biohub | CZB Cliahub Consortium |
| EPI_ISL_1477679 | Contra Costa County Public Health Lab | Chan-Zuckerberg Biohub | CZB Cliahub Consortium |
| EPI_ISL_1477680 | CA DPH Viral and Rickettsial Disease Laboratory | Chan-Zuckerberg Biohub | CZB Cliahub Consortium |
| EPI_ISL_1477681, EPI_ISL_1477682 | Santa Clara County Public Health Laboratory | Chan-Zuckerberg Biohub | CZB Cliahub Consortium |
| EPI_ISL_1477683, EPI_ISL_1477684 | Alameda County Public Health Lab | Chan-Zuckerberg Biohub | CZB Cliahub Consortium |
| EPI_ISL_1477685 | Contra Costa County Public Health Lab | Chan-Zuckerberg Biohub | CZB Cliahub Consortium |
| EPI_ISL_1477686 | Alameda County Public Health Lab | Chan-Zuckerberg Biohub | CZB Cliahub Consortium |
| EPI_ISL_1477687 | CA DPH Viral and Rickettsial Disease Laboratory | Chan-Zuckerberg Biohub | CZB Cliahub Consortium |
| EPI_ISL_1477688 | Alameda County Public Health Lab | Chan-Zuckerberg Biohub | CZB Cliahub Consortium |
| EPI_ISL_1477689 | Contra Costa County Public Health Lab | Chan-Zuckerberg Biohub | CZB Cliahub Consortium |
| EPI_ISL_1477690 | Santa Clara County Public Health Laboratory | Chan-Zuckerberg Biohub | CZB Cliahub Consortium |
| EPI_ISL_1477691 | Alameda County Public Health Lab | Chan-Zuckerberg Biohub | CZB Cliahub Consortium |
| EPI_ISL_1477692, EPI_ISL_1477693, EPI_ISL_1477694 | CA DPH Viral and Rickettsial Disease Laboratory | Chan-Zuckerberg Biohub | CZB Cliahub Consortium |
| EPI_ISL_1477695 | Santa Clara County Public Health Laboratory | Chan-Zuckerberg Biohub | CZB Cliahub Consortium |
| EPI_ISL_1477696 | Alameda County Public Health Lab | Chan-Zuckerberg Biohub | CZB Cliahub Consortium |
| EPI_ISL_1477697, EPI_ISL_1477698 | Contra Costa County Public Health Lab | Chan-Zuckerberg Biohub | CZB Cliahub Consortium |
| EPI_ISL_1477699 | Santa Clara County Public Health Laboratory | Chan-Zuckerberg Biohub | CZB Cliahub Consortium |
| EPI_ISL_1477700 | CA DPH Viral and Rickettsial Disease Laboratory | Chan-Zuckerberg Biohub | CZB Cliahub Consortium |
| EPI_ISL_1477701, EPI_ISL_1477702, EPI_ISL_1477703 | Alameda County Public Health Lab | Chan-Zuckerberg Biohub | CZB Cliahub Consortium |
| EPI_ISL_1477704 | Contra Costa County Public Health Lab | Chan-Zuckerberg Biohub | CZB Cliahub Consortium |
| EPI_ISL_1477705 | CA DPH Viral and Rickettsial Disease Laboratory | Chan-Zuckerberg Biohub | CZB Cliahub Consortium |
| EPI_ISL_1477706 | Santa Clara County Public Health Laboratory | Chan-Zuckerberg Biohub | CZB Cliahub Consortium |
| EPI_ISL_1477707 | CA DPH Viral and Rickettsial Disease Laboratory | Chan-Zuckerberg Biohub | CZB Cliahub Consortium |
| EPI_ISL_1477708 | Alameda County Public Health Lab | Chan-Zuckerberg Biohub | CZB Cliahub Consortium |
| EPI_ISL_1477709 | CA DPH Viral and Rickettsial Disease Laboratory | Chan-Zuckerberg Biohub | CZB Cliahub Consortium |
| EPI_ISL_1477710 | Alameda County Public Health Lab | Chan-Zuckerberg Biohub | CZB Cliahub Consortium |
| EPI_ISL_1477711 | CA DPH Viral and Rickettsial Disease Laboratory | Chan-Zuckerberg Biohub | CZB Cliahub Consortium |
| EPI_ISL_1477712 | Alameda County Public Health Lab | Chan-Zuckerberg Biohub | CZB Cliahub Consortium |
| EPI_ISL_1477713, EPI_ISL_1477714 | Contra Costa County Public Health Lab | Chan-Zuckerberg Biohub | CZB Cliahub Consortium |
| EPI_ISL_1477715 | CA DPH Viral and Rickettsial Disease Laboratory | Chan-Zuckerberg Biohub | CZB Cliahub Consortium |
| EPI_ISL_1477716 | Contra Costa County Public Health Lab | Chan-Zuckerberg Biohub | CZB Cliahub Consortium |
| EPI_ISL_1477717, EPI_ISL_1477718, EPI_ISL_1477719 | CA DPH Viral and Rickettsial Disease Laboratory | Chan-Zuckerberg Biohub | CZB Cliahub Consortium |
| EPI_ISL_1477720 | Alameda County Public Health Lab | Chan-Zuckerberg Biohub | CZB Cliahub Consortium |
| EPI_ISL_1477721 | CA DPH Viral and Rickettsial Disease Laboratory | Chan-Zuckerberg Biohub | CZB Cliahub Consortium |
| EPI_ISL_1477722 | Santa Clara County Public Health Laboratory | Chan-Zuckerberg Biohub | CZB Cliahub Consortium |
| EPI_ISL_1477723 | CA DPH Viral and Rickettsial Disease Laboratory | Chan-Zuckerberg Biohub | CZB Cliahub Consortium |
| EPI_ISL_1477724 | Contra Costa County Public Health Lab | Chan-Zuckerberg Biohub | CZB Cliahub Consortium |
| EPI_ISL_1477725 | CA DPH Viral and Rickettsial Disease Laboratory | Chan-Zuckerberg Biohub | CZB Cliahub Consortium |
| EPI_ISL_1477726, EPI_ISL_1477727 | Santa Clara County Public Health Laboratory | Chan-Zuckerberg Biohub | CZB Cliahub Consortium |
| EPI_ISL_1477728 | CA DPH Viral and Rickettsial Disease Laboratory | Chan-Zuckerberg Biohub | CZB Cliahub Consortium |
| EPI_ISL_1477729, EPI_ISL_1477730, EPI_ISL_1477731 | Santa Clara County Public Health Laboratory | Chan-Zuckerberg Biohub | CZB Cliahub Consortium |
| EPI_ISL_1477732 | Contra Costa County Public Health Lab | Chan-Zuckerberg Biohub | CZB Cliahub Consortium |
| EPI_ISL_1477733 | Alameda County Public Health Lab | Chan-Zuckerberg Biohub | CZB Cliahub Consortium |
| EPI_ISL_1477734 | Santa Clara County Public Health Laboratory | Chan-Zuckerberg Biohub | CZB Cliahub Consortium |
| EPI_ISL_1477735 | Alameda County Public Health Lab | Chan-Zuckerberg Biohub | CZB Cliahub Consortium |
| EPI_ISL_1477736 | CA DPH Viral and Rickettsial Disease | Chan-Zuckerberg Biohub | CZB Cliahub Consortium |

| | | | |
|---|---|---|---|
| | Laboratory | | |
| EPI_ISL_1477737, EPI_ISL_1477738, EPI_ISL_1477739 | Contra Costa County Public Health Lab | Chan-Zuckerberg Biohub | CZB Cliahub Consortium |
| EPI_ISL_1477740 | CA DPH Viral and Rickettsial Disease Laboratory | Chan-Zuckerberg Biohub | CZB Cliahub Consortium |
| EPI_ISL_1477741 | Alameda County Public Health Lab | Chan-Zuckerberg Biohub | CZB Cliahub Consortium |
| EPI_ISL_1477742 | CA DPH Viral and Rickettsial Disease Laboratory | Chan-Zuckerberg Biohub | CZB Cliahub Consortium |
| EPI_ISL_1477743, EPI_ISL_1477744 | Contra Costa County Public Health Lab | Chan-Zuckerberg Biohub | CZB Cliahub Consortium |
| EPI_ISL_1477745 | Santa Clara County Public Health Laboratory | Chan-Zuckerberg Biohub | CZB Cliahub Consortium |
| EPI_ISL_1477746 | Contra Costa County Public Health Lab | Chan-Zuckerberg Biohub | CZB Cliahub Consortium |
| EPI_ISL_1477747 | Alameda County Public Health Lab | Chan-Zuckerberg Biohub | CZB Cliahub Consortium |
| EPI_ISL_1477748 | Contra Costa County Public Health Lab | Chan-Zuckerberg Biohub | CZB Cliahub Consortium |
| EPI_ISL_1477749, EPI_ISL_1477750 | Santa Clara County Public Health Laboratory | Chan-Zuckerberg Biohub | CZB Cliahub Consortium |
| EPI_ISL_1477751 | Alameda County Public Health Lab | Chan-Zuckerberg Biohub | CZB Cliahub Consortium |
| EPI_ISL_1477752 | Santa Clara County Public Health Laboratory | Chan-Zuckerberg Biohub | CZB Cliahub Consortium |
| EPI_ISL_1477753, EPI_ISL_1477754 | Alameda County Public Health Lab | Chan-Zuckerberg Biohub | CZB Cliahub Consortium |
| EPI_ISL_1477755 | Santa Clara County Public Health Laboratory | Chan-Zuckerberg Biohub | CZB Cliahub Consortium |
| EPI_ISL_1477756 | CA DPH Viral and Rickettsial Disease Laboratory | Chan-Zuckerberg Biohub | CZB Cliahub Consortium |
| EPI_ISL_1477757, EPI_ISL_1477758 | Alameda County Public Health Lab | Chan-Zuckerberg Biohub | CZB Cliahub Consortium |
| EPI_ISL_1477759 | CA DPH Viral and Rickettsial Disease Laboratory | Chan-Zuckerberg Biohub | CZB Cliahub Consortium |
| EPI_ISL_1477760, EPI_ISL_1477761 | Contra Costa County Public Health Lab | Chan-Zuckerberg Biohub | CZB Cliahub Consortium |
| EPI_ISL_1477762 | Santa Clara County Public Health Laboratory | Chan-Zuckerberg Biohub | CZB Cliahub Consortium |
| EPI_ISL_1477763 | CA DPH Viral and Rickettsial Disease Laboratory | Chan-Zuckerberg Biohub | CZB Cliahub Consortium |
| EPI_ISL_1477764, EPI_ISL_1477765 | Contra Costa County Public Health Lab | Chan-Zuckerberg Biohub | CZB Cliahub Consortium |
| EPI_ISL_1477766 | Alameda County Public Health Lab | Chan-Zuckerberg Biohub | CZB Cliahub Consortium |
| EPI_ISL_1477767, EPI_ISL_1477768 | Santa Clara County Public Health Laboratory | Chan-Zuckerberg Biohub | CZB Cliahub Consortium |
| EPI_ISL_1477769 | CA DPH Viral and Rickettsial Disease Laboratory | Chan-Zuckerberg Biohub | CZB Cliahub Consortium |
| EPI_ISL_1477770 | Contra Costa County Public Health Lab | Chan-Zuckerberg Biohub | CZB Cliahub Consortium |
| EPI_ISL_1477771, EPI_ISL_1477772, EPI_ISL_1477773 | Alameda County Public Health Lab | Chan-Zuckerberg Biohub | CZB Cliahub Consortium |
| EPI_ISL_1477774 | Contra Costa County Public Health Lab | Chan-Zuckerberg Biohub | CZB Cliahub Consortium |
| EPI_ISL_1477775 | Alameda County Public Health Lab | Chan-Zuckerberg Biohub | CZB Cliahub Consortium |
| EPI_ISL_1477776 | Santa Clara County Public Health Laboratory | Chan-Zuckerberg Biohub | CZB Cliahub Consortium |
| EPI_ISL_1477777, EPI_ISL_1477778 | CA DPH Viral and Rickettsial Disease Laboratory | Chan-Zuckerberg Biohub | CZB Cliahub Consortium |
| EPI_ISL_1477779 | Alameda County Public Health Lab | Chan-Zuckerberg Biohub | CZB Cliahub Consortium |
| EPI_ISL_1477780 | Santa Clara County Public Health Laboratory | Chan-Zuckerberg Biohub | CZB Cliahub Consortium |
| EPI_ISL_1477781, EPI_ISL_1477782 | Contra Costa County Public Health Lab | Chan-Zuckerberg Biohub | CZB Cliahub Consortium |
| EPI_ISL_1477783, EPI_ISL_1477784 | Alameda County Public Health Lab | Chan-Zuckerberg Biohub | CZB Cliahub Consortium |
| EPI_ISL_1477785 | Contra Costa County Public Health Lab | Chan-Zuckerberg Biohub | CZB Cliahub Consortium |
| EPI_ISL_1477786, EPI_ISL_1477787 | CA DPH Viral and Rickettsial Disease Laboratory | Chan-Zuckerberg Biohub | CZB Cliahub Consortium |
| EPI_ISL_1477788, EPI_ISL_1477789 | Contra Costa County Public Health Lab | Chan-Zuckerberg Biohub | CZB Cliahub Consortium |
| EPI_ISL_1477790 | Santa Clara County Public Health Laboratory | Chan-Zuckerberg Biohub | CZB Cliahub Consortium |
| EPI_ISL_1477791 | Alameda County Public Health Lab | Chan-Zuckerberg Biohub | CZB Cliahub Consortium |
| EPI_ISL_1477792 | CA DPH Viral and Rickettsial Disease Laboratory | Chan-Zuckerberg Biohub | CZB Cliahub Consortium |
| EPI_ISL_1477793 | Santa Clara County Public Health Laboratory | Chan-Zuckerberg Biohub | CZB Cliahub Consortium |
| EPI_ISL_1477794, EPI_ISL_1477795 | Contra Costa County Public Health Lab | Chan-Zuckerberg Biohub | CZB Cliahub Consortium |
| EPI_ISL_1477796 | CA DPH Viral and Rickettsial Disease Laboratory | Chan-Zuckerberg Biohub | CZB Cliahub Consortium |
| EPI_ISL_1477797 | Santa Clara County Public Health Laboratory | Chan-Zuckerberg Biohub | CZB Cliahub Consortium |
| EPI_ISL_1477798 | CA DPH Viral and Rickettsial Disease Laboratory | Chan-Zuckerberg Biohub | CZB Cliahub Consortium |
| EPI_ISL_1477799, EPI_ISL_1477800 | Santa Clara County Public Health Laboratory | Chan-Zuckerberg Biohub | CZB Cliahub Consortium |
| EPI_ISL_1477801 | Alameda County Public Health Lab | Chan-Zuckerberg Biohub | CZB Cliahub Consortium |
| EPI_ISL_1477802 | Contra Costa County Public Health Lab | Chan-Zuckerberg Biohub | CZB Cliahub Consortium |
| EPI_ISL_1477803 | CA DPH Viral and Rickettsial Disease Laboratory | Chan-Zuckerberg Biohub | CZB Cliahub Consortium |
| EPI_ISL_1477804, | Alameda County Public Health Lab | Chan-Zuckerberg Biohub | CZB Cliahub Consortium |

| | | | |
|---|---|---|---|
| EPI_ISL_1477805 | | | |
| EPI_ISL_1477806 | Contra Costa County Public Health Lab | Chan-Zuckerberg Biohub | CZB Cliahub Consortium |
| EPI_ISL_1477807 | Alameda County Public Health Lab | Chan-Zuckerberg Biohub | CZB Cliahub Consortium |
| EPI_ISL_1477808 | CA DPH Viral and Rickettsial Disease Laboratory | Chan-Zuckerberg Biohub | CZB Cliahub Consortium |
| EPI_ISL_1477809 | Alameda County Public Health Lab | Chan-Zuckerberg Biohub | CZB Cliahub Consortium |
| EPI_ISL_1477810 | Contra Costa County Public Health Lab | Chan-Zuckerberg Biohub | CZB Cliahub Consortium |
| EPI_ISL_1477811, EPI_ISL_1477812 | CA DPH Viral and Rickettsial Disease Laboratory | Chan-Zuckerberg Biohub | CZB Cliahub Consortium |
| EPI_ISL_1477813 | Contra Costa County Public Health Lab | Chan-Zuckerberg Biohub | CZB Cliahub Consortium |
| EPI_ISL_1477814 | Santa Clara County Public Health Laboratory | Chan-Zuckerberg Biohub | CZB Cliahub Consortium |
| EPI_ISL_1477815, EPI_ISL_1477816 | Contra Costa County Public Health Lab | Chan-Zuckerberg Biohub | CZB Cliahub Consortium |
| EPI_ISL_1477817, EPI_ISL_1477818 | Santa Clara County Public Health Laboratory | Chan-Zuckerberg Biohub | CZB Cliahub Consortium |
| EPI_ISL_1477819 | CA DPH Viral and Rickettsial Disease Laboratory | Chan-Zuckerberg Biohub | CZB Cliahub Consortium |
| EPI_ISL_1477820 | Alameda County Public Health Lab | Chan-Zuckerberg Biohub | CZB Cliahub Consortium |
| EPI_ISL_1477821 | Contra Costa County Public Health Lab | Chan-Zuckerberg Biohub | CZB Cliahub Consortium |
| EPI_ISL_1477822 | Santa Clara County Public Health Laboratory | Chan-Zuckerberg Biohub | CZB Cliahub Consortium |
| EPI_ISL_1477823, EPI_ISL_1477824, EPI_ISL_1477825 | Alameda County Public Health Lab | Chan-Zuckerberg Biohub | CZB Cliahub Consortium |
| EPI_ISL_1477826, EPI_ISL_1477827 | Contra Costa County Public Health Lab | Chan-Zuckerberg Biohub | CZB Cliahub Consortium |
| EPI_ISL_1477828 | Alameda County Public Health Lab | Chan-Zuckerberg Biohub | CZB Cliahub Consortium |
| EPI_ISL_1477829 | Contra Costa County Public Health Lab | Chan-Zuckerberg Biohub | CZB Cliahub Consortium |
| EPI_ISL_1477830, EPI_ISL_1477831 | CA DPH Viral and Rickettsial Disease Laboratory | Chan-Zuckerberg Biohub | CZB Cliahub Consortium |
| EPI_ISL_1477832 | Alameda County Public Health Lab | Chan-Zuckerberg Biohub | CZB Cliahub Consortium |
| EPI_ISL_1477833, EPI_ISL_1477834, EPI_ISL_1477835 | Contra Costa County Public Health Lab | Chan-Zuckerberg Biohub | CZB Cliahub Consortium |
| EPI_ISL_1477836 | Santa Clara County Public Health Laboratory | Chan-Zuckerberg Biohub | CZB Cliahub Consortium |
| EPI_ISL_1477837, EPI_ISL_1477838 | CA DPH Viral and Rickettsial Disease Laboratory | Chan-Zuckerberg Biohub | CZB Cliahub Consortium |
| EPI_ISL_1477839 | Santa Clara County Public Health Laboratory | Chan-Zuckerberg Biohub | CZB Cliahub Consortium |
| EPI_ISL_1477840 | CA DPH Viral and Rickettsial Disease Laboratory | Chan-Zuckerberg Biohub | CZB Cliahub Consortium |
| EPI_ISL_1477841 | Alameda County Public Health Lab | Chan-Zuckerberg Biohub | CZB Cliahub Consortium |
| EPI_ISL_1477842 | Santa Clara County Public Health Laboratory | Chan-Zuckerberg Biohub | CZB Cliahub Consortium |
| EPI_ISL_1477843, EPI_ISL_1477844 | Contra Costa County Public Health Lab | Chan-Zuckerberg Biohub | CZB Cliahub Consortium |
| EPI_ISL_1477845, EPI_ISL_1477846 | Santa Clara County Public Health Laboratory | Chan-Zuckerberg Biohub | CZB Cliahub Consortium |
| EPI_ISL_1477847 | Alameda County Public Health Lab | Chan-Zuckerberg Biohub | CZB Cliahub Consortium |
| EPI_ISL_1477848 | CA DPH Viral and Rickettsial Disease Laboratory | Chan-Zuckerberg Biohub | CZB Cliahub Consortium |
| EPI_ISL_1477849 | Contra Costa County Public Health Lab | Chan-Zuckerberg Biohub | CZB Cliahub Consortium |
| EPI_ISL_1477850 | Santa Clara County Public Health Laboratory | Chan-Zuckerberg Biohub | CZB Cliahub Consortium |
| EPI_ISL_1477851 | Contra Costa County Public Health Lab | Chan-Zuckerberg Biohub | CZB Cliahub Consortium |
| EPI_ISL_1477852, EPI_ISL_1477853 | CA DPH Viral and Rickettsial Disease Laboratory | Chan-Zuckerberg Biohub | CZB Cliahub Consortium |
| EPI_ISL_1477854 | Santa Clara County Public Health Laboratory | Chan-Zuckerberg Biohub | CZB Cliahub Consortium |
| EPI_ISL_1477855 | CA DPH Viral and Rickettsial Disease Laboratory | Chan-Zuckerberg Biohub | CZB Cliahub Consortium |
| EPI_ISL_1477856 | Santa Clara County Public Health Laboratory | Chan-Zuckerberg Biohub | CZB Cliahub Consortium |
| EPI_ISL_1477857, EPI_ISL_1477858 | Alameda County Public Health Lab | Chan-Zuckerberg Biohub | CZB Cliahub Consortium |
| EPI_ISL_1477859, EPI_ISL_1477860 | Santa Clara County Public Health Laboratory | Chan-Zuckerberg Biohub | CZB Cliahub Consortium |
| EPI_ISL_1477861 | Contra Costa County Public Health Lab | Chan-Zuckerberg Biohub | CZB Cliahub Consortium |
| EPI_ISL_1477862 | Alameda County Public Health Lab | Chan-Zuckerberg Biohub | CZB Cliahub Consortium |
| EPI_ISL_1477863 | Santa Clara County Public Health Laboratory | Chan-Zuckerberg Biohub | CZB Cliahub Consortium |
| EPI_ISL_1477864 | Contra Costa County Public Health Lab | Chan-Zuckerberg Biohub | CZB Cliahub Consortium |
| EPI_ISL_1477865 | Alameda County Public Health Lab | Chan-Zuckerberg Biohub | CZB Cliahub Consortium |
| EPI_ISL_1477866, EPI_ISL_1477867 | Contra Costa County Public Health Lab | Chan-Zuckerberg Biohub | CZB Cliahub Consortium |
| EPI_ISL_1477868 | CA DPH Viral and Rickettsial Disease Laboratory | Chan-Zuckerberg Biohub | CZB Cliahub Consortium |
| EPI_ISL_1477869, EPI_ISL_1477870 | Contra Costa County Public Health Lab | Chan-Zuckerberg Biohub | CZB Cliahub Consortium |
| EPI_ISL_1477871 | Santa Clara County Public Health Laboratory | Chan-Zuckerberg Biohub | CZB Cliahub Consortium |
| EPI_ISL_1477872 | Contra Costa County Public Health Lab | Chan-Zuckerberg Biohub | CZB Cliahub Consortium |
| EPI_ISL_1477873 | CA DPH Viral and Rickettsial Disease Laboratory | Chan-Zuckerberg Biohub | CZB Cliahub Consortium |
| EPI_ISL_1477874 | Santa Clara County Public Health | Chan-Zuckerberg Biohub | CZB Cliahub Consortium |

| | | | |
|---|---|---|---|
| | Laboratory | | |
| EPI_ISL_1477875 | CA DPH Viral and Rickettsial Disease Laboratory | Chan-Zuckerberg Biohub | CZB Cliahub Consortium |
| EPI_ISL_1477876 | Contra Costa County Public Health Lab | Chan-Zuckerberg Biohub | CZB Cliahub Consortium |
| EPI_ISL_1477877 | CA DPH Viral and Rickettsial Disease Laboratory | Chan-Zuckerberg Biohub | CZB Cliahub Consortium |
| EPI_ISL_1477878, EPI_ISL_1477879 | Santa Clara County Public Health Laboratory | Chan-Zuckerberg Biohub | CZB Cliahub Consortium |
| EPI_ISL_1477880 | Alameda County Public Health Lab | Chan-Zuckerberg Biohub | CZB Cliahub Consortium |
| EPI_ISL_1477881, EPI_ISL_1477882 | Santa Clara County Public Health Laboratory | Chan-Zuckerberg Biohub | CZB Cliahub Consortium |
| EPI_ISL_1477883, EPI_ISL_1477884 | CA DPH Viral and Rickettsial Disease Laboratory | Chan-Zuckerberg Biohub | CZB Cliahub Consortium |
| EPI_ISL_1477885 | Santa Clara County Public Health Laboratory | Chan-Zuckerberg Biohub | CZB Cliahub Consortium |
| EPI_ISL_1477886, EPI_ISL_1477887 | Alameda County Public Health Lab | Chan-Zuckerberg Biohub | CZB Cliahub Consortium |
| EPI_ISL_1477888 | Santa Clara County Public Health Laboratory | Chan-Zuckerberg Biohub | CZB Cliahub Consortium |
| EPI_ISL_1477889 | Alameda County Public Health Lab | Chan-Zuckerberg Biohub | CZB Cliahub Consortium |
| EPI_ISL_1477890, EPI_ISL_1477891 | CA DPH Viral and Rickettsial Disease Laboratory | Chan-Zuckerberg Biohub | CZB Cliahub Consortium |
| EPI_ISL_1477892, EPI_ISL_1477893 | Santa Clara County Public Health Laboratory | Chan-Zuckerberg Biohub | CZB Cliahub Consortium |
| EPI_ISL_1477894 | Contra Costa County Public Health Lab | Chan-Zuckerberg Biohub | CZB Cliahub Consortium |
| EPI_ISL_1477895 | Santa Clara County Public Health Laboratory | Chan-Zuckerberg Biohub | CZB Cliahub Consortium |
| EPI_ISL_1477896 | Alameda County Public Health Lab | Chan-Zuckerberg Biohub | CZB Cliahub Consortium |
| EPI_ISL_1477897 | CA DPH Viral and Rickettsial Disease Laboratory | Chan-Zuckerberg Biohub | CZB Cliahub Consortium |
| EPI_ISL_1477898 | Contra Costa County Public Health Lab | Chan-Zuckerberg Biohub | CZB Cliahub Consortium |
| EPI_ISL_1477899, EPI_ISL_1477900 | Alameda County Public Health Lab | Chan-Zuckerberg Biohub | CZB Cliahub Consortium |
| EPI_ISL_1477901 | CA DPH Viral and Rickettsial Disease Laboratory | Chan-Zuckerberg Biohub | CZB Cliahub Consortium |
| EPI_ISL_1477902 | Santa Clara County Public Health Laboratory | Chan-Zuckerberg Biohub | CZB Cliahub Consortium |
| EPI_ISL_1477903 | CA DPH Viral and Rickettsial Disease Laboratory | Chan-Zuckerberg Biohub | CZB Cliahub Consortium |
| EPI_ISL_1477904, EPI_ISL_1477905, EPI_ISL_1477906 | Alameda County Public Health Lab | Chan-Zuckerberg Biohub | CZB Cliahub Consortium |
| EPI_ISL_1477907 | Santa Clara County Public Health Laboratory | Chan-Zuckerberg Biohub | CZB Cliahub Consortium |
| EPI_ISL_1477908 | CA DPH Viral and Rickettsial Disease Laboratory | Chan-Zuckerberg Biohub | CZB Cliahub Consortium |
| EPI_ISL_1477909 | Santa Clara County Public Health Laboratory | Chan-Zuckerberg Biohub | CZB Cliahub Consortium |
| EPI_ISL_1477910, EPI_ISL_1477911 | Contra Costa County Public Health Lab | Chan-Zuckerberg Biohub | CZB Cliahub Consortium |
| EPI_ISL_1477912 | Santa Clara County Public Health Laboratory | Chan-Zuckerberg Biohub | CZB Cliahub Consortium |
| EPI_ISL_1477913 | CA DPH Viral and Rickettsial Disease Laboratory | Chan-Zuckerberg Biohub | CZB Cliahub Consortium |
| EPI_ISL_1477914, EPI_ISL_1477915 | Alameda County Public Health Lab | Chan-Zuckerberg Biohub | CZB Cliahub Consortium |
| EPI_ISL_1477916 | Contra Costa County Public Health Lab | Chan-Zuckerberg Biohub | CZB Cliahub Consortium |
| EPI_ISL_1477917 | CA DPH Viral and Rickettsial Disease Laboratory | Chan-Zuckerberg Biohub | CZB Cliahub Consortium |
| EPI_ISL_1477918 | Contra Costa County Public Health Lab | Chan-Zuckerberg Biohub | CZB Cliahub Consortium |
| EPI_ISL_1477919 | Alameda County Public Health Lab | Chan-Zuckerberg Biohub | CZB Cliahub Consortium |
| EPI_ISL_1477920, EPI_ISL_1477921 | CA DPH Viral and Rickettsial Disease Laboratory | Chan-Zuckerberg Biohub | CZB Cliahub Consortium |
| EPI_ISL_1477922 | Alameda County Public Health Lab | Chan-Zuckerberg Biohub | CZB Cliahub Consortium |
| EPI_ISL_1477923 | Santa Clara County Public Health Laboratory | Chan-Zuckerberg Biohub | CZB Cliahub Consortium |
| EPI_ISL_1477924 | CA DPH Viral and Rickettsial Disease Laboratory | Chan-Zuckerberg Biohub | CZB Cliahub Consortium |
| EPI_ISL_1477925, EPI_ISL_1477926 | Santa Clara County Public Health Laboratory | Chan-Zuckerberg Biohub | CZB Cliahub Consortium |
| EPI_ISL_1477927 | Contra Costa County Public Health Lab | Chan-Zuckerberg Biohub | CZB Cliahub Consortium |
| EPI_ISL_1477928 | Alameda County Public Health Lab | Chan-Zuckerberg Biohub | CZB Cliahub Consortium |
| EPI_ISL_1477929 | CA DPH Viral and Rickettsial Disease Laboratory | Chan-Zuckerberg Biohub | CZB Cliahub Consortium |
| EPI_ISL_1477930, EPI_ISL_1477931 | Contra Costa County Public Health Lab | Chan-Zuckerberg Biohub | CZB Cliahub Consortium |
| EPI_ISL_1477932, EPI_ISL_1477933 | Alameda County Public Health Lab | Chan-Zuckerberg Biohub | CZB Cliahub Consortium |
| EPI_ISL_1477934 | CA DPH Viral and Rickettsial Disease Laboratory | Chan-Zuckerberg Biohub | CZB Cliahub Consortium |
| EPI_ISL_1477935 | Santa Clara County Public Health Laboratory | Chan-Zuckerberg Biohub | CZB Cliahub Consortium |
| EPI_ISL_1477936 | Alameda County Public Health Lab | Chan-Zuckerberg Biohub | CZB Cliahub Consortium |
| EPI_ISL_1477937 | Santa Clara County Public Health Laboratory | Chan-Zuckerberg Biohub | CZB Cliahub Consortium |
| EPI_ISL_1477938 | CA DPH Viral and Rickettsial Disease Laboratory | Chan-Zuckerberg Biohub | CZB Cliahub Consortium |
| EPI_ISL_1477939 | Contra Costa County Public Health Lab | Chan-Zuckerberg Biohub | CZB Cliahub Consortium |

| | | | |
|---|---|---|---|
| EPI_ISL_1477940 | CA DPH Viral and Rickettsial Disease Laboratory | Chan-Zuckerberg Biohub | CZB Cliahub Consortium |
| EPI_ISL_1477941, EPI_ISL_1477942 | Alameda County Public Health Lab | Chan-Zuckerberg Biohub | CZB Cliahub Consortium |
| EPI_ISL_1477943 | Santa Clara County Public Health Laboratory | Chan-Zuckerberg Biohub | CZB Cliahub Consortium |
| EPI_ISL_1477944 | Contra Costa County Public Health Lab | Chan-Zuckerberg Biohub | CZB Cliahub Consortium |
| EPI_ISL_1477945 | Alameda County Public Health Lab | Chan-Zuckerberg Biohub | CZB Cliahub Consortium |
| EPI_ISL_1477946, EPI_ISL_1477947 | Contra Costa County Public Health Lab | Chan-Zuckerberg Biohub | CZB Cliahub Consortium |
| EPI_ISL_1477948 | Alameda County Public Health Lab | Chan-Zuckerberg Biohub | CZB Cliahub Consortium |
| EPI_ISL_1477949 | CA DPH Viral and Rickettsial Disease Laboratory | Chan-Zuckerberg Biohub | CZB Cliahub Consortium |
| EPI_ISL_1477950 | Contra Costa County Public Health Lab | Chan-Zuckerberg Biohub | CZB Cliahub Consortium |
| EPI_ISL_1477951 | Humboldt County Public Health Laboratory | Chan-Zuckerberg Biohub | CZB Cliahub Consortium |
| EPI_ISL_1477952, EPI_ISL_1477953 | Contra Costa County Public Health Lab | Chan-Zuckerberg Biohub | CZB Cliahub Consortium |
| EPI_ISL_1477954, EPI_ISL_1477955 | Humboldt County Public Health Laboratory | Chan-Zuckerberg Biohub | CZB Cliahub Consortium |
| EPI_ISL_1477956, EPI_ISL_1477957 | Contra Costa County Public Health Lab | Chan-Zuckerberg Biohub | CZB Cliahub Consortium |
| EPI_ISL_1477958 | Santa Clara County Public Health Laboratory | Chan-Zuckerberg Biohub | CZB Cliahub Consortium |
| EPI_ISL_1477959 | CA DPH Viral and Rickettsial Disease Laboratory | Chan-Zuckerberg Biohub | CZB Cliahub Consortium |
| EPI_ISL_1477960 | Contra Costa County Public Health Lab | Chan-Zuckerberg Biohub | CZB Cliahub Consortium |
| EPI_ISL_1477961 | Santa Clara County Public Health Laboratory | Chan-Zuckerberg Biohub | CZB Cliahub Consortium |
| EPI_ISL_1477962 | Contra Costa County Public Health Lab | Chan-Zuckerberg Biohub | CZB Cliahub Consortium |
| EPI_ISL_1477963 | Santa Clara County Public Health Laboratory | Chan-Zuckerberg Biohub | CZB Cliahub Consortium |
| EPI_ISL_1477964 | CA DPH Viral and Rickettsial Disease Laboratory | Chan-Zuckerberg Biohub | CZB Cliahub Consortium |
| EPI_ISL_1477965 | County of San Luis Obispo Public Health Laboratory | Chan-Zuckerberg Biohub | CZB Cliahub Consortium |
| EPI_ISL_1477966 | Contra Costa County Public Health Lab | Chan-Zuckerberg Biohub | CZB Cliahub Consortium |
| EPI_ISL_1477967 | Santa Clara County Public Health Laboratory | Chan-Zuckerberg Biohub | CZB Cliahub Consortium |
| EPI_ISL_1477968, EPI_ISL_1477969 | CA DPH Viral and Rickettsial Disease Laboratory | Chan-Zuckerberg Biohub | CZB Cliahub Consortium |
| EPI_ISL_1477970 | Humboldt County Public Health Laboratory | Chan-Zuckerberg Biohub | CZB Cliahub Consortium |
| EPI_ISL_1477971 | CA DPH Viral and Rickettsial Disease Laboratory | Chan-Zuckerberg Biohub | CZB Cliahub Consortium |
| EPI_ISL_1477972 | Contra Costa County Public Health Lab | Chan-Zuckerberg Biohub | CZB Cliahub Consortium |
| EPI_ISL_1477973 | Humboldt County Public Health Laboratory | Chan-Zuckerberg Biohub | CZB Cliahub Consortium |
| EPI_ISL_1477974 | Santa Clara County Public Health Laboratory | Chan-Zuckerberg Biohub | CZB Cliahub Consortium |
| EPI_ISL_1477975, EPI_ISL_1477976 | Humboldt County Public Health Laboratory | Chan-Zuckerberg Biohub | CZB Cliahub Consortium |
| EPI_ISL_1477977 | Contra Costa County Public Health Lab | Chan-Zuckerberg Biohub | CZB Cliahub Consortium |
| EPI_ISL_1477978 | CA DPH Viral and Rickettsial Disease Laboratory | Chan-Zuckerberg Biohub | CZB Cliahub Consortium |
| EPI_ISL_1477980 | Contra Costa County Public Health Lab | Chan-Zuckerberg Biohub | CZB Cliahub Consortium |
| EPI_ISL_1477981 | Humboldt County Public Health Laboratory | Chan-Zuckerberg Biohub | CZB Cliahub Consortium |
| EPI_ISL_1477983 | Santa Clara County Public Health Laboratory | Chan-Zuckerberg Biohub | CZB Cliahub Consortium |
| EPI_ISL_1477984, EPI_ISL_1477985 | CA DPH Viral and Rickettsial Disease Laboratory | Chan-Zuckerberg Biohub | CZB Cliahub Consortium |
| EPI_ISL_1477986 | Orange County Public Health Lab | Chan-Zuckerberg Biohub | CZB Cliahub Consortium |
| EPI_ISL_1477987 | Humboldt County Public Health Laboratory | Chan-Zuckerberg Biohub | CZB Cliahub Consortium |
| EPI_ISL_1477988, EPI_ISL_1477989 | CA DPH Viral and Rickettsial Disease Laboratory | Chan-Zuckerberg Biohub | CZB Cliahub Consortium |
| EPI_ISL_1477990 | Contra Costa County Public Health Lab | Chan-Zuckerberg Biohub | CZB Cliahub Consortium |
| EPI_ISL_1477991 | Humboldt County Public Health Laboratory | Chan-Zuckerberg Biohub | CZB Cliahub Consortium |
| EPI_ISL_1477992 | County of San Luis Obispo Public Health Laboratory | Chan-Zuckerberg Biohub | CZB Cliahub Consortium |
| EPI_ISL_1477993 | Contra Costa County Public Health Lab | Chan-Zuckerberg Biohub | CZB Cliahub Consortium |
| EPI_ISL_1477994 | CA DPH Viral and Rickettsial Disease Laboratory | Chan-Zuckerberg Biohub | CZB Cliahub Consortium |
| EPI_ISL_1477995 | Contra Costa County Public Health Lab | Chan-Zuckerberg Biohub | CZB Cliahub Consortium |
| EPI_ISL_1477996 | CA DPH Viral and Rickettsial Disease Laboratory | Chan-Zuckerberg Biohub | CZB Cliahub Consortium |
| EPI_ISL_1477997 | Humboldt County Public Health Laboratory | Chan-Zuckerberg Biohub | CZB Cliahub Consortium |
| EPI_ISL_1477998, EPI_ISL_1477999, EPI_ISL_1478000 | Contra Costa County Public Health Lab | Chan-Zuckerberg Biohub | CZB Cliahub Consortium |
| EPI_ISL_1478001 | CA DPH Viral and Rickettsial Disease Laboratory | Chan-Zuckerberg Biohub | CZB Cliahub Consortium |
| EPI_ISL_1478002, EPI_ISL_1478003 | Contra Costa County Public Health Lab | Chan-Zuckerberg Biohub | CZB Cliahub Consortium |

| | | | |
|---|---|---|---|
| EPI_ISL_1478004 | CA DPH Viral and Rickettsial Disease Laboratory | Chan-Zuckerberg Biohub | CZB Cliahub Consortium |
| EPI_ISL_1478005 | Contra Costa County Public Health Lab | Chan-Zuckerberg Biohub | CZB Cliahub Consortium |
| EPI_ISL_1478006 | County of San Luis Obispo Public Health Laboratory | Chan-Zuckerberg Biohub | CZB Cliahub Consortium |
| EPI_ISL_1478007, EPI_ISL_1478008 | Contra Costa County Public Health Lab | Chan-Zuckerberg Biohub | CZB Cliahub Consortium |
| EPI_ISL_1478009, EPI_ISL_1478010 | Humboldt County Public Health Laboratory | Chan-Zuckerberg Biohub | CZB Cliahub Consortium |
| EPI_ISL_1478011 | Contra Costa County Public Health Lab | Chan-Zuckerberg Biohub | CZB Cliahub Consortium |
| EPI_ISL_1478012 | Santa Clara County Public Health Laboratory | Chan-Zuckerberg Biohub | CZB Cliahub Consortium |
| EPI_ISL_1478014 | CA DPH Viral and Rickettsial Disease Laboratory | Chan-Zuckerberg Biohub | CZB Cliahub Consortium |
| EPI_ISL_1478015 | Humboldt County Public Health Laboratory | Chan-Zuckerberg Biohub | CZB Cliahub Consortium |
| EPI_ISL_1478016 | CA DPH Viral and Rickettsial Disease Laboratory | Chan-Zuckerberg Biohub | CZB Cliahub Consortium |
| EPI_ISL_1478017 | Contra Costa County Public Health Lab | Chan-Zuckerberg Biohub | CZB Cliahub Consortium |
| EPI_ISL_1478018 | County of San Luis Obispo Public Health Laboratory | Chan-Zuckerberg Biohub | CZB Cliahub Consortium |
| EPI_ISL_1478019 | Santa Clara County Public Health Laboratory | Chan-Zuckerberg Biohub | CZB Cliahub Consortium |
| EPI_ISL_1478020 | Contra Costa County Public Health Lab | Chan-Zuckerberg Biohub | CZB Cliahub Consortium |
| EPI_ISL_1478021 | CA DPH Viral and Rickettsial Disease Laboratory | Chan-Zuckerberg Biohub | CZB Cliahub Consortium |
| EPI_ISL_1478022, EPI_ISL_1478023 | Contra Costa County Public Health Lab | Chan-Zuckerberg Biohub | CZB Cliahub Consortium |
| EPI_ISL_1478024 | Orange County Public Health Lab | Chan-Zuckerberg Biohub | CZB Cliahub Consortium |
| EPI_ISL_1478026 | Humboldt County Public Health Laboratory | Chan-Zuckerberg Biohub | CZB Cliahub Consortium |
| EPI_ISL_1478027 | CA DPH Viral and Rickettsial Disease Laboratory | Chan-Zuckerberg Biohub | CZB Cliahub Consortium |
| EPI_ISL_1478028 | Santa Clara County Public Health Laboratory | Chan-Zuckerberg Biohub | CZB Cliahub Consortium |
| EPI_ISL_1478029 | CA DPH Viral and Rickettsial Disease Laboratory | Chan-Zuckerberg Biohub | CZB Cliahub Consortium |
| EPI_ISL_1478030 | Orange County Public Health Lab | Chan-Zuckerberg Biohub | CZB Cliahub Consortium |
| EPI_ISL_1478031 | Santa Clara County Public Health Laboratory | Chan-Zuckerberg Biohub | CZB Cliahub Consortium |
| EPI_ISL_1478032 | Contra Costa County Public Health Lab | Chan-Zuckerberg Biohub | CZB Cliahub Consortium |
| EPI_ISL_1478033 | CA DPH Viral and Rickettsial Disease Laboratory | Chan-Zuckerberg Biohub | CZB Cliahub Consortium |
| EPI_ISL_1478034, EPI_ISL_1478035, EPI_ISL_1478036, EPI_ISL_1478037 | Contra Costa County Public Health Lab | Chan-Zuckerberg Biohub | CZB Cliahub Consortium |
| EPI_ISL_1478038 | Santa Clara County Public Health Laboratory | Chan-Zuckerberg Biohub | CZB Cliahub Consortium |
| EPI_ISL_1478039, EPI_ISL_1478040 | Contra Costa County Public Health Lab | Chan-Zuckerberg Biohub | CZB Cliahub Consortium |
| EPI_ISL_1478041 | Humboldt County Public Health Laboratory | Chan-Zuckerberg Biohub | CZB Cliahub Consortium |
| EPI_ISL_1478042 | CA DPH Viral and Rickettsial Disease Laboratory | Chan-Zuckerberg Biohub | CZB Cliahub Consortium |
| EPI_ISL_1478043 | Contra Costa County Public Health Lab | Chan-Zuckerberg Biohub | CZB Cliahub Consortium |
| EPI_ISL_1478044, EPI_ISL_1478045 | County of San Luis Obispo Public Health Laboratory | Chan-Zuckerberg Biohub | CZB Cliahub Consortium |
| EPI_ISL_1478046 | Santa Clara County Public Health Laboratory | Chan-Zuckerberg Biohub | CZB Cliahub Consortium |
| EPI_ISL_1478047 | Contra Costa County Public Health Lab | Chan-Zuckerberg Biohub | CZB Cliahub Consortium |
| EPI_ISL_1478048 | Santa Clara County Public Health Laboratory | Chan-Zuckerberg Biohub | CZB Cliahub Consortium |
| EPI_ISL_1478049 | Contra Costa County Public Health Lab | Chan-Zuckerberg Biohub | CZB Cliahub Consortium |
| EPI_ISL_1478050 | Santa Clara County Public Health Laboratory | Chan-Zuckerberg Biohub | CZB Cliahub Consortium |
| EPI_ISL_1478051 | CA DPH Viral and Rickettsial Disease Laboratory | Chan-Zuckerberg Biohub | CZB Cliahub Consortium |
| EPI_ISL_1478052, EPI_ISL_1478053 | Contra Costa County Public Health Lab | Chan-Zuckerberg Biohub | CZB Cliahub Consortium |
| EPI_ISL_1478054, EPI_ISL_1478055 | Humboldt County Public Health Laboratory | Chan-Zuckerberg Biohub | CZB Cliahub Consortium |
| EPI_ISL_1478056 | CA DPH Viral and Rickettsial Disease Laboratory | Chan-Zuckerberg Biohub | CZB Cliahub Consortium |
| EPI_ISL_1478057 | Contra Costa County Public Health Lab | Chan-Zuckerberg Biohub | CZB Cliahub Consortium |
| EPI_ISL_1478058 | Humboldt County Public Health Laboratory | Chan-Zuckerberg Biohub | CZB Cliahub Consortium |
| EPI_ISL_1478060 | CA DPH Viral and Rickettsial Disease Laboratory | Chan-Zuckerberg Biohub | CZB Cliahub Consortium |
| EPI_ISL_1478061 | County of San Luis Obispo Public Health Laboratory | Chan-Zuckerberg Biohub | CZB Cliahub Consortium |
| EPI_ISL_1478062, EPI_ISL_1478063 | Humboldt County Public Health Laboratory | Chan-Zuckerberg Biohub | CZB Cliahub Consortium |
| EPI_ISL_1478064 | CA DPH Viral and Rickettsial Disease Laboratory | Chan-Zuckerberg Biohub | CZB Cliahub Consortium |
| EPI_ISL_1478065, EPI_ISL_1478066, EPI_ISL_1478067 | Humboldt County Public Health Laboratory | Chan-Zuckerberg Biohub | CZB Cliahub Consortium |

| | | | |
|---|---|---|---|
| EPI_ISL_1478068 | CA DPH Viral and Rickettsial Disease Laboratory | Chan-Zuckerberg Biohub | CZB Cliahub Consortium |
| EPI_ISL_1478069 | Contra Costa County Public Health Lab | Chan-Zuckerberg Biohub | CZB Cliahub Consortium |
| EPI_ISL_1478070 | County of San Luis Obispo Public Health Laboratory | Chan-Zuckerberg Biohub | CZB Cliahub Consortium |
| EPI_ISL_1478071 | CA DPH Viral and Rickettsial Disease Laboratory | Chan-Zuckerberg Biohub | CZB Cliahub Consortium |
| EPI_ISL_1478072 | Contra Costa County Public Health Lab | Chan-Zuckerberg Biohub | CZB Cliahub Consortium |
| EPI_ISL_1478073 | CA DPH Viral and Rickettsial Disease Laboratory | Chan-Zuckerberg Biohub | CZB Cliahub Consortium |
| EPI_ISL_1478074 | County of San Luis Obispo Public Health Laboratory | Chan-Zuckerberg Biohub | CZB Cliahub Consortium |
| EPI_ISL_1478075 | Humboldt County Public Health Laboratory | Chan-Zuckerberg Biohub | CZB Cliahub Consortium |
| EPI_ISL_1478076 | CA DPH Viral and Rickettsial Disease Laboratory | Chan-Zuckerberg Biohub | CZB Cliahub Consortium |
| EPI_ISL_1478077 | Humboldt County Public Health Laboratory | Chan-Zuckerberg Biohub | CZB Cliahub Consortium |
| EPI_ISL_1478078 | Contra Costa County Public Health Lab | Chan-Zuckerberg Biohub | CZB Cliahub Consortium |
| EPI_ISL_1478079 | CA DPH Viral and Rickettsial Disease Laboratory | Chan-Zuckerberg Biohub | CZB Cliahub Consortium |
| EPI_ISL_1478080 | County of San Luis Obispo Public Health Laboratory | Chan-Zuckerberg Biohub | CZB Cliahub Consortium |
| EPI_ISL_1478081 | Santa Clara County Public Health Laboratory | Chan-Zuckerberg Biohub | CZB Cliahub Consortium |
| EPI_ISL_1478082, EPI_ISL_1478083 | Humboldt County Public Health Laboratory | Chan-Zuckerberg Biohub | CZB Cliahub Consortium |
| EPI_ISL_1478084 | Santa Clara County Public Health Laboratory | Chan-Zuckerberg Biohub | CZB Cliahub Consortium |
| EPI_ISL_1478085 | CA DPH Viral and Rickettsial Disease Laboratory | Chan-Zuckerberg Biohub | CZB Cliahub Consortium |
| EPI_ISL_1478086 | Contra Costa County Public Health Lab | Chan-Zuckerberg Biohub | CZB Cliahub Consortium |
| EPI_ISL_1478087 | Santa Clara County Public Health Laboratory | Chan-Zuckerberg Biohub | CZB Cliahub Consortium |
| EPI_ISL_1478088 | Contra Costa County Public Health Lab | Chan-Zuckerberg Biohub | CZB Cliahub Consortium |
| EPI_ISL_1478089 | Santa Clara County Public Health Laboratory | Chan-Zuckerberg Biohub | CZB Cliahub Consortium |
| EPI_ISL_1478090, EPI_ISL_1478091 | Contra Costa County Public Health Lab | Chan-Zuckerberg Biohub | CZB Cliahub Consortium |
| EPI_ISL_1478092 | Santa Clara County Public Health Laboratory | Chan-Zuckerberg Biohub | CZB Cliahub Consortium |
| EPI_ISL_1478093, EPI_ISL_1478094 | Humboldt County Public Health Laboratory | Chan-Zuckerberg Biohub | CZB Cliahub Consortium |
| EPI_ISL_1478095 | Santa Clara County Public Health Laboratory | Chan-Zuckerberg Biohub | CZB Cliahub Consortium |
| EPI_ISL_1478096 | Humboldt County Public Health Laboratory | Chan-Zuckerberg Biohub | CZB Cliahub Consortium |
| EPI_ISL_1478097 | Santa Clara County Public Health Laboratory | Chan-Zuckerberg Biohub | CZB Cliahub Consortium |
| EPI_ISL_1478098, EPI_ISL_1478099 | Contra Costa County Public Health Lab | Chan-Zuckerberg Biohub | CZB Cliahub Consortium |
| EPI_ISL_1478100 | CA DPH Viral and Rickettsial Disease Laboratory | Chan-Zuckerberg Biohub | CZB Cliahub Consortium |
| EPI_ISL_1478101 | Contra Costa County Public Health Lab | Chan-Zuckerberg Biohub | CZB Cliahub Consortium |
| EPI_ISL_1478102, EPI_ISL_1478103 | Santa Clara County Public Health Laboratory | Chan-Zuckerberg Biohub | CZB Cliahub Consortium |
| EPI_ISL_1478104 | CA DPH Viral and Rickettsial Disease Laboratory | Chan-Zuckerberg Biohub | CZB Cliahub Consortium |
| EPI_ISL_1478105 | Humboldt County Public Health Laboratory | Chan-Zuckerberg Biohub | CZB Cliahub Consortium |
| EPI_ISL_1478106 | Orange County Public Health Lab | Chan-Zuckerberg Biohub | CZB Cliahub Consortium |
| EPI_ISL_1478107 | Humboldt County Public Health Laboratory | Chan-Zuckerberg Biohub | CZB Cliahub Consortium |
| EPI_ISL_1478108 | Contra Costa County Public Health Lab | Chan-Zuckerberg Biohub | CZB Cliahub Consortium |
| EPI_ISL_1478109 | CA DPH Viral and Rickettsial Disease Laboratory | Chan-Zuckerberg Biohub | CZB Cliahub Consortium |
| EPI_ISL_1478110 | Orange County Public Health Lab | Chan-Zuckerberg Biohub | CZB Cliahub Consortium |
| EPI_ISL_1478111 | Contra Costa County Public Health Lab | Chan-Zuckerberg Biohub | CZB Cliahub Consortium |
| EPI_ISL_1478112, EPI_ISL_1478113, EPI_ISL_1478114, EPI_ISL_1478115, EPI_ISL_1478116 | CA DPH Viral and Rickettsial Disease Laboratory | Chan-Zuckerberg Biohub | CZB Cliahub Consortium |
| EPI_ISL_1478117 | Contra Costa County Public Health Lab | Chan-Zuckerberg Biohub | CZB Cliahub Consortium |
| EPI_ISL_1478118 | County of San Luis Obispo Public Health Laboratory | Chan-Zuckerberg Biohub | CZB Cliahub Consortium |
| EPI_ISL_1478119 | CA DPH Viral and Rickettsial Disease Laboratory | Chan-Zuckerberg Biohub | CZB Cliahub Consortium |
| EPI_ISL_1478120 | Contra Costa County Public Health Lab | Chan-Zuckerberg Biohub | CZB Cliahub Consortium |
| EPI_ISL_1478121 | Humboldt County Public Health Laboratory | Chan-Zuckerberg Biohub | CZB Cliahub Consortium |
| EPI_ISL_1478122 | Contra Costa County Public Health Lab | Chan-Zuckerberg Biohub | CZB Cliahub Consortium |
| EPI_ISL_1478123, EPI_ISL_1478124 | Santa Clara County Public Health Laboratory | Chan-Zuckerberg Biohub | CZB Cliahub Consortium |
| EPI_ISL_1478125, EPI_ISL_1478126 | Contra Costa County Public Health Lab | Chan-Zuckerberg Biohub | CZB Cliahub Consortium |
| EPI_ISL_1478127 | CA DPH Viral and Rickettsial Disease Laboratory | Chan-Zuckerberg Biohub | CZB Cliahub Consortium |
| EPI_ISL_1478128 | Contra Costa County Public Health Lab | Chan-Zuckerberg Biohub | CZB Cliahub Consortium |

| | | | |
|---|---|---|---|
| EPI_ISL_1478129 | Santa Clara County Public Health Laboratory | Chan-Zuckerberg Biohub | CZB Cliahub Consortium |
| EPI_ISL_1478130 | CA DPH Viral and Rickettsial Disease Laboratory | Chan-Zuckerberg Biohub | CZB Cliahub Consortium |
| EPI_ISL_1478131 | Contra Costa County Public Health Lab | Chan-Zuckerberg Biohub | CZB Cliahub Consortium |
| EPI_ISL_1478132 | CA DPH Viral and Rickettsial Disease Laboratory | Chan-Zuckerberg Biohub | CZB Cliahub Consortium |
| EPI_ISL_1478133 | Contra Costa County Public Health Lab | Chan-Zuckerberg Biohub | CZB Cliahub Consortium |
| EPI_ISL_1478134, EPI_ISL_1478135 | Humboldt County Public Health Laboratory | Chan-Zuckerberg Biohub | CZB Cliahub Consortium |
| EPI_ISL_1478136 | County of San Luis Obispo Public Health Laboratory | Chan-Zuckerberg Biohub | CZB Cliahub Consortium |
| EPI_ISL_1478137 | CA DPH Viral and Rickettsial Disease Laboratory | Chan-Zuckerberg Biohub | CZB Cliahub Consortium |
| EPI_ISL_1478138 | Humboldt County Public Health Laboratory | Chan-Zuckerberg Biohub | CZB Cliahub Consortium |
| EPI_ISL_1478139 | Contra Costa County Public Health Lab | Chan-Zuckerberg Biohub | CZB Cliahub Consortium |
| EPI_ISL_1478140 | Humboldt County Public Health Laboratory | Chan-Zuckerberg Biohub | CZB Cliahub Consortium |
| EPI_ISL_1478141 | Contra Costa County Public Health Lab | Chan-Zuckerberg Biohub | CZB Cliahub Consortium |
| EPI_ISL_1478142 | CA DPH Viral and Rickettsial Disease Laboratory | Chan-Zuckerberg Biohub | CZB Cliahub Consortium |
| EPI_ISL_1478143 | Humboldt County Public Health Laboratory | Chan-Zuckerberg Biohub | CZB Cliahub Consortium |
| EPI_ISL_1478144 | CA DPH Viral and Rickettsial Disease Laboratory | Chan-Zuckerberg Biohub | CZB Cliahub Consortium |
| EPI_ISL_1478145 | Contra Costa County Public Health Lab | Chan-Zuckerberg Biohub | CZB Cliahub Consortium |
| EPI_ISL_1478146 | CA DPH Viral and Rickettsial Disease Laboratory | Chan-Zuckerberg Biohub | CZB Cliahub Consortium |
| EPI_ISL_1478147 | Contra Costa County Public Health Lab | Chan-Zuckerberg Biohub | CZB Cliahub Consortium |
| EPI_ISL_1478148 | CA DPH Viral and Rickettsial Disease Laboratory | Chan-Zuckerberg Biohub | CZB Cliahub Consortium |
| EPI_ISL_1478149 | Santa Clara County Public Health Laboratory | Chan-Zuckerberg Biohub | CZB Cliahub Consortium |
| EPI_ISL_1478150 | Humboldt County Public Health Laboratory | Chan-Zuckerberg Biohub | CZB Cliahub Consortium |
| EPI_ISL_1478152, EPI_ISL_1478153 | Santa Clara County Public Health Laboratory | Chan-Zuckerberg Biohub | CZB Cliahub Consortium |
| EPI_ISL_1478154, EPI_ISL_1478155 | Contra Costa County Public Health Lab | Chan-Zuckerberg Biohub | CZB Cliahub Consortium |
| EPI_ISL_1478156 | Humboldt County Public Health Laboratory | Chan-Zuckerberg Biohub | CZB Cliahub Consortium |
| EPI_ISL_1478157 | Contra Costa County Public Health Lab | Chan-Zuckerberg Biohub | CZB Cliahub Consortium |
| EPI_ISL_1478158 | CA DPH Viral and Rickettsial Disease Laboratory | Chan-Zuckerberg Biohub | CZB Cliahub Consortium |
| EPI_ISL_1478159 | Humboldt County Public Health Laboratory | Chan-Zuckerberg Biohub | CZB Cliahub Consortium |
| EPI_ISL_1478160 | Contra Costa County Public Health Lab | Chan-Zuckerberg Biohub | CZB Cliahub Consortium |
| EPI_ISL_1478161 | Santa Clara County Public Health Laboratory | Chan-Zuckerberg Biohub | CZB Cliahub Consortium |
| EPI_ISL_1478162 | CA DPH Viral and Rickettsial Disease Laboratory | Chan-Zuckerberg Biohub | CZB Cliahub Consortium |
| EPI_ISL_1478163, EPI_ISL_1478164, EPI_ISL_1478165 | Humboldt County Public Health Laboratory | Chan-Zuckerberg Biohub | CZB Cliahub Consortium |
| EPI_ISL_1478166, EPI_ISL_1478167, EPI_ISL_1478168 | CA DPH Viral and Rickettsial Disease Laboratory | Chan-Zuckerberg Biohub | CZB Cliahub Consortium |
| EPI_ISL_1478169 | Humboldt County Public Health Laboratory | Chan-Zuckerberg Biohub | CZB Cliahub Consortium |
| EPI_ISL_1478171 | Santa Clara County Public Health Laboratory | Chan-Zuckerberg Biohub | CZB Cliahub Consortium |
| EPI_ISL_1478172 | Humboldt County Public Health Laboratory | Chan-Zuckerberg Biohub | CZB Cliahub Consortium |
| EPI_ISL_1478173 | CA DPH Viral and Rickettsial Disease Laboratory | Chan-Zuckerberg Biohub | CZB Cliahub Consortium |
| EPI_ISL_1478174 | County of San Luis Obispo Public Health Laboratory | Chan-Zuckerberg Biohub | CZB Cliahub Consortium |
| EPI_ISL_1478175 | Orange County Public Health Lab | Chan-Zuckerberg Biohub | CZB Cliahub Consortium |
| EPI_ISL_1478176, EPI_ISL_1478177 | Humboldt County Public Health Laboratory | Chan-Zuckerberg Biohub | CZB Cliahub Consortium |
| EPI_ISL_1478178 | CA DPH Viral and Rickettsial Disease Laboratory | Chan-Zuckerberg Biohub | CZB Cliahub Consortium |
| EPI_ISL_1478179 | County of San Luis Obispo Public Health Laboratory | Chan-Zuckerberg Biohub | CZB Cliahub Consortium |
| EPI_ISL_1478180, EPI_ISL_1478181, EPI_ISL_1478182 | Contra Costa County Public Health Lab | Chan-Zuckerberg Biohub | CZB Cliahub Consortium |
| EPI_ISL_1478183, EPI_ISL_1478184, EPI_ISL_1478185 | CA DPH Viral and Rickettsial Disease Laboratory | Chan-Zuckerberg Biohub | CZB Cliahub Consortium |
| EPI_ISL_1478186 | Humboldt County Public Health Laboratory | Chan-Zuckerberg Biohub | CZB Cliahub Consortium |
| EPI_ISL_1478187, EPI_ISL_1478188, EPI_ISL_1478189 | Contra Costa County Public Health Lab | Chan-Zuckerberg Biohub | CZB Cliahub Consortium |
| EPI_ISL_1478190 | Humboldt County Public Health Laboratory | Chan-Zuckerberg Biohub | CZB Cliahub Consortium |
| EPI_ISL_1478191 | CA DPH Viral and Rickettsial Disease | Chan-Zuckerberg Biohub | CZB Cliahub Consortium |

| | | | |
|---|---|---|---|
| | Laboratory | | |
| EPI_ISL_1478192 | Santa Clara County Public Health Laboratory | Chan-Zuckerberg Biohub | CZB Cliahub Consortium |
| EPI_ISL_1478193 | Contra Costa County Public Health Lab | Chan-Zuckerberg Biohub | CZB Cliahub Consortium |
| EPI_ISL_1478194 | CA DPH Viral and Rickettsial Disease Laboratory | Chan-Zuckerberg Biohub | CZB Cliahub Consortium |
| EPI_ISL_1478195 | County of San Luis Obispo Public Health Laboratory | Chan-Zuckerberg Biohub | CZB Cliahub Consortium |
| EPI_ISL_1478196, EPI_ISL_1478197 | Contra Costa County Public Health Lab | Chan-Zuckerberg Biohub | CZB Cliahub Consortium |
| EPI_ISL_1478198 | County of San Luis Obispo Public Health Laboratory | Chan-Zuckerberg Biohub | CZB Cliahub Consortium |
| EPI_ISL_1478199 | Contra Costa County Public Health Lab | Chan-Zuckerberg Biohub | CZB Cliahub Consortium |
| EPI_ISL_1478200 | Santa Clara County Public Health Laboratory | Chan-Zuckerberg Biohub | CZB Cliahub Consortium |
| EPI_ISL_1478201 | CA DPH Viral and Rickettsial Disease Laboratory | Chan-Zuckerberg Biohub | CZB Cliahub Consortium |
| EPI_ISL_1478202 | NE Public Health Laboratory | Genomics and Discovery, Respiratory Viruses Branch, Division of Viral Diseases, Centers for Disease Control and Prevention | Ying Tao, Jing Zhang, Yan Li, Brian Lynch, Anna Kelleher, Krista Queen, Anna Uehara, Peter Cook, Han Jia Justin Ng, Clinton R. Paden, Haibin Wang, Suxiang Tong |
| EPI_ISL_1478844, EPI_ISL_1478845, EPI_ISL_1478846, EPI_ISL_1478847, EPI_ISL_1478848, EPI_ISL_1478849, EPI_ISL_1478850, EPI_ISL_1478851, EPI_ISL_1478852, EPI_ISL_1478853, EPI_ISL_1478854, EPI_ISL_1478855, EPI_ISL_1478856, EPI_ISL_1478857, EPI_ISL_1478858, EPI_ISL_1478859, EPI_ISL_1478860, EPI_ISL_1478861, EPI_ISL_1478862, EPI_ISL_1478863, EPI_ISL_1478864, EPI_ISL_1478865, EPI_ISL_1478866, EPI_ISL_1478867, EPI_ISL_1478868, EPI_ISL_1478869, EPI_ISL_1478870, EPI_ISL_1478871, EPI_ISL_1478873, EPI_ISL_1478874, EPI_ISL_1478875, EPI_ISL_1478876, EPI_ISL_1478877, EPI_ISL_1478878, EPI_ISL_1478879, EPI_ISL_1478880, EPI_ISL_1478881, EPI_ISL_1478882, EPI_ISL_1478883, EPI_ISL_1478884, EPI_ISL_1478885, EPI_ISL_1478886, EPI_ISL_1478887, EPI_ISL_1478888, EPI_ISL_1478889, EPI_ISL_1478890, EPI_ISL_1478891, EPI_ISL_1478892, EPI_ISL_1478893, EPI_ISL_1478894, EPI_ISL_1478895, EPI_ISL_1478896, EPI_ISL_1478897, EPI_ISL_1478898, EPI_ISL_1478899, EPI_ISL_1478900, EPI_ISL_1478901, EPI_ISL_1478902, EPI_ISL_1478903, EPI_ISL_1478904, EPI_ISL_1478905, EPI_ISL_1478906, EPI_ISL_1478907, EPI_ISL_1478908, EPI_ISL_1478909, EPI_ISL_1478910, EPI_ISL_1478911, EPI_ISL_1478912, EPI_ISL_1478913, EPI_ISL_1478914, EPI_ISL_1478915, EPI_ISL_1478916, EPI_ISL_1478917, EPI_ISL_1478918, EPI_ISL_1478919, EPI_ISL_1478920, EPI_ISL_1478921, EPI_ISL_1478922, EPI_ISL_1478923, EPI_ISL_1478924, EPI_ISL_1478925, EPI_ISL_1478926, EPI_ISL_1478927, EPI_ISL_1478929, EPI_ISL_1478930, EPI_ISL_1478931, EPI_ISL_1478932 | | | |
| see above | Illinois Department of Public Health - Springfield Lab | Illinois Department of Public Health - Springfield Lab | Bryan Sim, Gordon McCall |
| EPI_ISL_1478933, EPI_ISL_1478934, EPI_ISL_1478935, EPI_ISL_1478936, EPI_ISL_1478937, EPI_ISL_1478938, EPI_ISL_1478939, EPI_ISL_1478940, EPI_ISL_1478941, EPI_ISL_1478942, EPI_ISL_1478943, EPI_ISL_1478944, EPI_ISL_1478945, EPI_ISL_1478946, EPI_ISL_1478947, EPI_ISL_1478948, EPI_ISL_1478949, EPI_ISL_1478950, EPI_ISL_1478951, EPI_ISL_1478952, EPI_ISL_1478953, EPI_ISL_1478954, EPI_ISL_1478955, EPI_ISL_1478956, EPI_ISL_1478957, EPI_ISL_1478958, EPI_ISL_1478959, EPI_ISL_1478960, EPI_ISL_1478961, EPI_ISL_1478962, EPI_ISL_1478963, EPI_ISL_1478964, EPI_ISL_1478965, EPI_ISL_1478966, EPI_ISL_1478967, EPI_ISL_1478968, EPI_ISL_1478969, EPI_ISL_1478970, EPI_ISL_1478971, EPI_ISL_1478972, EPI_ISL_1478973, EPI_ISL_1478974, EPI_ISL_1478975, EPI_ISL_1478976, EPI_ISL_1478977, EPI_ISL_1478978, EPI_ISL_1478979, EPI_ISL_1478981, EPI_ISL_1478982, EPI_ISL_1478983, EPI_ISL_1478984, EPI_ISL_1478985, EPI_ISL_1478986, EPI_ISL_1478987, EPI_ISL_1478988, EPI_ISL_1478989, EPI_ISL_1478990, EPI_ISL_1478991, EPI_ISL_1478992, EPI_ISL_1478993, EPI_ISL_1478994, EPI_ISL_1478996, EPI_ISL_1478997, EPI_ISL_1478998, EPI_ISL_1478999, EPI_ISL_1479000, EPI_ISL_1479001, EPI_ISL_1479002, EPI_ISL_1479003, EPI_ISL_1479004, EPI_ISL_1479005, EPI_ISL_1479006, EPI_ISL_1479007, EPI_ISL_1479008, EPI_ISL_1479009, EPI_ISL_1479010, EPI_ISL_1479011, EPI_ISL_1479012, EPI_ISL_1479013, EPI_ISL_1479014, EPI_ISL_1479015 | | | |
| see above | TXDSHS | TXDSHS | Rashmi Tuladhar, Bonnie Oh, Jenny Zhang, Maliha Rahman, Mayela Pedrueza, Anita Pokharel, Lorraine Rodriguez, Myong Koag, Chun Wang, Rachel Lee, Grace Kubin |
| EPI_ISL_1479016, EPI_ISL_1479017 | M Health Fairview | Minnesota Department of Health, Public Health Laboratory | Alexandra Lorentz, Jacob Garfin, Matt Plumb, and Xiong Wang |
| EPI_ISL_1479019, EPI_ISL_1479020, EPI_ISL_1479021, EPI_ISL_1479022, EPI_ISL_1479023, EPI_ISL_1479026, EPI_ISL_1479028, EPI_ISL_1479029, EPI_ISL_1479030, EPI_ISL_1479031, EPI_ISL_1479032, EPI_ISL_1479033, EPI_ISL_1479034, EPI_ISL_1479035, EPI_ISL_1479036, EPI_ISL_1479037, EPI_ISL_1479038, EPI_ISL_1479039, EPI_ISL_1479040, EPI_ISL_1479041 | | | |
| see above | Minnesota Department of Health, Public Health Laboratory | Minnesota Department of Health, Public Health Laboratory | Alexandra Lorentz, Jacob Garfin, Matt Plumb, and Xiong Wang |
| EPI_ISL_1479042, EPI_ISL_1479043, EPI_ISL_1479045, EPI_ISL_1479046, EPI_ISL_1479047, EPI_ISL_1479048, EPI_ISL_1479050, EPI_ISL_1479051, EPI_ISL_1479052, EPI_ISL_1479053, EPI_ISL_1479055, EPI_ISL_1479056, EPI_ISL_1479057, EPI_ISL_1479058, EPI_ISL_1479059, EPI_ISL_1479060, EPI_ISL_1479061, EPI_ISL_1479062, EPI_ISL_1479063, EPI_ISL_1479064, EPI_ISL_1479065, EPI_ISL_1479066, EPI_ISL_1479068, EPI_ISL_1479069, EPI_ISL_1479070, EPI_ISL_1479071, EPI_ISL_1479073, EPI_ISL_1479075, EPI_ISL_1479076, EPI_ISL_1479077, EPI_ISL_1479078, EPI_ISL_1479080, EPI_ISL_1479082, EPI_ISL_1479084, EPI_ISL_1479085, EPI_ISL_1479086, EPI_ISL_1479087, EPI_ISL_1479088, EPI_ISL_1479089, EPI_ISL_1479090, EPI_ISL_1479091, EPI_ISL_1479092, EPI_ISL_1479093, EPI_ISL_1479094, EPI_ISL_1479096 | | | |
| see above | Mayo Clinic & Mayo Clinic Laboratories | Minnesota Department of Health, Public Health Laboratory | Alexandra Lorentz, Jacob Garfin, Matt Plumb, and Xiong Wang |
| EPI_ISL_1479097, EPI_ISL_1479098, EPI_ISL_1479099, EPI_ISL_1479100, EPI_ISL_1479101, EPI_ISL_1479102, EPI_ISL_1479103, EPI_ISL_1479104, EPI_ISL_1479105, EPI_ISL_1479106, EPI_ISL_1479107, EPI_ISL_1479108, EPI_ISL_1479109, EPI_ISL_1479110, EPI_ISL_1479111, EPI_ISL_1479112, EPI_ISL_1479113, EPI_ISL_1479114, EPI_ISL_1479115, EPI_ISL_1479116, EPI_ISL_1479117, EPI_ISL_1479118 | | | |
| see above | Houston Health Dept. | Houston Health Dept. | Ryker Penn, Pamela Brown, Adolpho Lara, Yanlai Lai |
| EPI_ISL_1479134, EPI_ISL_1479135, EPI_ISL_1479136 | Minnesota Department of Health, Public Health Laboratory | Minnesota Department of Health, Public Health Laboratory | Alexandra Lorentz, Jacob Garfin, Matt Plumb, and Xiong Wang |
| EPI_ISL_1479137, EPI_ISL_1479138 | M Health Fairview | Minnesota Department of Health, Public Health Laboratory | Alexandra Lorentz, Jacob Garfin, Matt Plumb, and Xiong Wang |
| EPI_ISL_1479139 | Essesntia Health- Holy Trinity Hospital | Minnesota Department of Health, Public Health Laboratory | Alexandra Lorentz, Jacob Garfin, Matt Plumb, and Xiong Wang |
| EPI_ISL_1479140, EPI_ISL_1479141, EPI_ISL_1479142, EPI_ISL_1479143, EPI_ISL_1479144, EPI_ISL_1479145, EPI_ISL_1479146, EPI_ISL_1479147, EPI_ISL_1479148, EPI_ISL_1479149, EPI_ISL_1479150, EPI_ISL_1479151, EPI_ISL_1479152, EPI_ISL_1479153, EPI_ISL_1479154, EPI_ISL_1479155, EPI_ISL_1479156, EPI_ISL_1479157, EPI_ISL_1479158, EPI_ISL_1479159 | | | |
| see above | Minnesota Department of Health, Public Health Laboratory | Minnesota Department of Health, Public Health Laboratory | Alexandra Lorentz, Jacob Garfin, Matt Plumb, and Xiong Wang |
| EPI_ISL_1479160, EPI_ISL_1479161, EPI_ISL_1479162, EPI_ISL_1479163, EPI_ISL_1479164, EPI_ISL_1479165, EPI_ISL_1479168, EPI_ISL_1479169, EPI_ISL_1479172, EPI_ISL_1479173, EPI_ISL_1479174, EPI_ISL_1479175, EPI_ISL_1479176, EPI_ISL_1479177, EPI_ISL_1479178, EPI_ISL_1479179, EPI_ISL_1479180, EPI_ISL_1479181, EPI_ISL_1479182, EPI_ISL_1479183, EPI_ISL_1479184, EPI_ISL_1479185, EPI_ISL_1479186, EPI_ISL_1479187, EPI_ISL_1479188, EPI_ISL_1479189, EPI_ISL_1479191, EPI_ISL_1479192, EPI_ISL_1479193, EPI_ISL_1479195, EPI_ISL_1479196, EPI_ISL_1479197, EPI_ISL_1479198, EPI_ISL_1479199, EPI_ISL_1479200, EPI_ISL_1479201 | | | |
| see above | Centracare Laboratory Services | Minnesota Department of Health, Public Health Laboratory | Alexandra Lorentz, Jacob Garfin, Matt Plumb, and Xiong Wang |
| EPI_ISL_1479202, EPI_ISL_1479203, EPI_ISL_1479204, EPI_ISL_1479205, EPI_ISL_1479206, EPI_ISL_1479207, EPI_ISL_1479208, EPI_ISL_1479209, EPI_ISL_1479210, EPI_ISL_1479211, EPI_ISL_1479212, EPI_ISL_1479213, EPI_ISL_1479214, EPI_ISL_1479215, EPI_ISL_1479216, EPI_ISL_1479217, EPI_ISL_1479218, EPI_ISL_1479219, EPI_ISL_1479220, EPI_ISL_1479221, EPI_ISL_1479222, EPI_ISL_1479223, EPI_ISL_1479224, EPI_ISL_1479225, EPI_ISL_1479226, EPI_ISL_1479227, EPI_ISL_1479228, EPI_ISL_1479229, EPI_ISL_1479230, EPI_ISL_1479231, EPI_ISL_1479232, EPI_ISL_1479233, EPI_ISL_1479234, EPI_ISL_1479235, EPI_ISL_1479236, EPI_ISL_1479237, EPI_ISL_1479238, EPI_ISL_1479239, EPI_ISL_1479240, EPI_ISL_1479241, EPI_ISL_1479242, EPI_ISL_1479243, EPI_ISL_1479244, EPI_ISL_1479245, EPI_ISL_1479246, EPI_ISL_1479247, EPI_ISL_1479248, EPI_ISL_1479249, EPI_ISL_1479250, EPI_ISL_1479251, EPI_ISL_1479252, EPI_ISL_1479253, EPI_ISL_1479254, EPI_ISL_1479255, EPI_ISL_1479256, EPI_ISL_1479257, EPI_ISL_1479258, EPI_ISL_1479259, EPI_ISL_1479260, EPI_ISL_1479261, EPI_ISL_1479262, EPI_ISL_1479263, EPI_ISL_1479264, EPI_ISL_1479265, EPI_ISL_1479266, EPI_ISL_1479267, EPI_ISL_1479268, EPI_ISL_1479269, EPI_ISL_1479270, EPI_ISL_1479271, EPI_ISL_1479272, EPI_ISL_1479273, EPI_ISL_1479274, EPI_ISL_1479275, EPI_ISL_1479276, EPI_ISL_1479277, EPI_ISL_1479278, EPI_ISL_1479279, EPI_ISL_1479280, EPI_ISL_1479281, EPI_ISL_1479282, EPI_ISL_1479283, EPI_ISL_1479284, EPI_ISL_1479285, EPI_ISL_1479286, EPI_ISL_1479287, EPI_ISL_1479288, EPI_ISL_1479289, EPI_ISL_1479290, EPI_ISL_1479291, EPI_ISL_1479292, EPI_ISL_1479293, EPI_ISL_1479294, EPI_ISL_1479295, EPI_ISL_1479296, EPI_ISL_1479297, EPI_ISL_1479298, EPI_ISL_1479299, EPI_ISL_1479300, EPI_ISL_1479301, EPI_ISL_1479302, EPI_ISL_1479303, EPI_ISL_1479304, EPI_ISL_1479305, EPI_ISL_1479306, EPI_ISL_1479307, EPI_ISL_1479308, EPI_ISL_1479309, EPI_ISL_1479310, EPI_ISL_1479311, EPI_ISL_1479312, EPI_ISL_1479313, EPI_ISL_1479314, EPI_ISL_1479315, EPI_ISL_1479316, EPI_ISL_1479317, EPI_ISL_1479318, EPI_ISL_1479319, EPI_ISL_1479320, EPI_ISL_1479321, EPI_ISL_1479322, EPI_ISL_1479323, EPI_ISL_1479324, EPI_ISL_1479325, EPI_ISL_1479326, EPI_ISL_1479327, EPI_ISL_1479328, EPI_ISL_1479329, EPI_ISL_1479330, EPI_ISL_1479331, EPI_ISL_1479332, EPI_ISL_1479333, EPI_ISL_1479334, EPI_ISL_1479335, EPI_ISL_1479336, EPI_ISL_1479337, EPI_ISL_1479338, EPI_ISL_1479339, EPI_ISL_1479340, EPI_ISL_1479341, EPI_ISL_1479342, EPI_ISL_1479343, EPI_ISL_1479344, EPI_ISL_1479345, EPI_ISL_1479346, EPI_ISL_1479347, EPI_ISL_1479348, EPI_ISL_1479349, EPI_ISL_1479350, EPI_ISL_1479351, EPI_ISL_1479352, EPI_ISL_1479353, EPI_ISL_1479354, EPI_ISL_1479355, EPI_ISL_1479356, EPI_ISL_1479357, EPI_ISL_1479358, EPI_ISL_1479359, EPI_ISL_1479360, EPI_ISL_1479361, EPI_ISL_1479362, EPI_ISL_1479363, EPI_ISL_1479364, EPI_ISL_1479365, EPI_ISL_1479366, EPI_ISL_1479367, EPI_ISL_1479368, EPI_ISL_1479369, EPI_ISL_1479370, EPI_ISL_1479371, EPI_ISL_1479372, EPI_ISL_1479373, EPI_ISL_1479374, EPI_ISL_1479375, EPI_ISL_1479376, EPI_ISL_1479377, EPI_ISL_1479378, EPI_ISL_1479379, EPI_ISL_1479380, EPI_ISL_1479381, EPI_ISL_1479382, EPI_ISL_1479383, EPI_ISL_1479384, EPI_ISL_1479385, EPI_ISL_1479386, EPI_ISL_1479387, EPI_ISL_1479388, EPI_ISL_1479389, EPI_ISL_1479390, EPI_ISL_1479391, EPI_ISL_1479392, EPI_ISL_1479393, EPI_ISL_1479394, EPI_ISL_1479395, EPI_ISL_1479396, EPI_ISL_1479397, EPI_ISL_1479398, EPI_ISL_1479399, EPI_ISL_1479400, EPI_ISL_1479401, EPI_ISL_1479402, EPI_ISL_1479403, EPI_ISL_1479404, EPI_ISL_1479405, EPI_ISL_1479406, EPI_ISL_1479407, EPI_ISL_1479408, EPI_ISL_1479409, EPI_ISL_1479410, EPI_ISL_1479411, EPI_ISL_1479412, EPI_ISL_1479413, EPI_ISL_1479414, EPI_ISL_1479415, EPI_ISL_1479416, EPI_ISL_1479417, EPI_ISL_1479418, EPI_ISL_1479419, EPI_ISL_1479420, EPI_ISL_1479421, EPI_ISL_1479422, EPI_ISL_1479423, EPI_ISL_1479424, EPI_ISL_1479425, EPI_ISL_1479426, EPI_ISL_1479427, EPI_ISL_1479428, EPI_ISL_1479429, EPI_ISL_1479430, EPI_ISL_1479431, EPI_ISL_1479432, EPI_ISL_1479433, EPI_ISL_1479434, EPI_ISL_1479435, EPI_ISL_1479436, EPI_ISL_1479437, EPI_ISL_1479438, EPI_ISL_1479439, EPI_ISL_1479440 | | | |
| see above | Helix/Illumina | Centers for Disease Control and Prevention Division of Viral Diseases, Pathogen Discovery | Dakota Howard, Dhwani Batra, Peter W. Cook, Kara Moser, Adrian Paskey, Jason Caravas, Benjamin Rambo-Martin, Shatavia Morrison, Christopher Gulvick, Scott Sammons, Yvette Unoarumhi, Darlene Wagner, Matthew Schmerer, Eileen de Feo, Jan Antico, Christine Tran, Matthew Tolentino, Shannon Wickline, Kim Gietzen, Brad Sickler, Jingtao Liu, Eric Allen, Phil Febbo, Nicole L. Washington, Simon White, Geraint Levan, Kelly Schiabor Barrett, Elizabeth Cirulli, Alexandre Bolze, Ary Ascencio, Charlotte Rivera-Garcia, Ryan Cho, Jason Nguyen, Sherry Wang, Jimmy Ramirez, Tyler Cassens, Efren Sandoval, Magnus Isaksson, William Lee, David Becker, Marc Laurent, James Lu, Clinton R. Paden, Duncan MacCannell |
| EPI_ISL_1479441, EPI_ISL_1479442, EPI_ISL_1479443, EPI_ISL_1479444, EPI_ISL_1479445, EPI_ISL_1479446, EPI_ISL_1479447, EPI_ISL_1479448, EPI_ISL_1479449, EPI_ISL_1479451, EPI_ISL_1479452, EPI_ISL_1479453, EPI_ISL_1479454, EPI_ISL_1479456, EPI_ISL_1479457, EPI_ISL_1479459, EPI_ISL_1479460, EPI_ISL_1479461, EPI_ISL_1479462, EPI_ISL_1479463, EPI_ISL_1479465, EPI_ISL_1479466, EPI_ISL_1479467, EPI_ISL_1479468, EPI_ISL_1479469, EPI_ISL_1479470, EPI_ISL_1479471, EPI_ISL_1479472, EPI_ISL_1479473, EPI_ISL_1479474, EPI_ISL_1479476, EPI_ISL_1479477, EPI_ISL_1479479, EPI_ISL_1479482, EPI_ISL_1479484, EPI_ISL_1479485, EPI_ISL_1479487, EPI_ISL_1479488, EPI_ISL_1479489, EPI_ISL_1479490, EPI_ISL_1479491, EPI_ISL_1479492, EPI_ISL_1479493, EPI_ISL_1479494, EPI_ISL_1479496, EPI_ISL_1479497, EPI_ISL_1479499, EPI_ISL_1479500, EPI_ISL_1479501, EPI_ISL_1479502, EPI_ISL_1479503, EPI_ISL_1479504, EPI_ISL_1479505, EPI_ISL_1479506, EPI_ISL_1479507, EPI_ISL_1479508, EPI_ISL_1479509, EPI_ISL_1479510, EPI_ISL_1479511, EPI_ISL_1479512, EPI_ISL_1479513, EPI_ISL_1479514, EPI_ISL_1479515, EPI_ISL_1479517, EPI_ISL_1479518, EPI_ISL_1479519, EPI_ISL_1479520, EPI_ISL_1479521, EPI_ISL_1479522, EPI_ISL_1479523, EPI_ISL_1479524, EPI_ISL_1479525, EPI_ISL_1479526, EPI_ISL_1479527, EPI_ISL_1479528, EPI_ISL_1479529, EPI_ISL_1479530, EPI_ISL_1479531, EPI_ISL_1479532, EPI_ISL_1479533, EPI_ISL_1479534, EPI_ISL_1479535, EPI_ISL_1479536, EPI_ISL_1479537, EPI_ISL_1479538, EPI_ISL_1479539, EPI_ISL_1479540, EPI_ISL_1479541, EPI_ISL_1479542, EPI_ISL_1479543, EPI_ISL_1479545, EPI_ISL_1479546, EPI_ISL_1479548, EPI_ISL_1479549, EPI_ISL_1479550, EPI_ISL_1479551, EPI_ISL_1479552, EPI_ISL_1479553, EPI_ISL_1479554, EPI_ISL_1479555, EPI_ISL_1479556, EPI_ISL_1479557, EPI_ISL_1479558, EPI_ISL_1479560, EPI_ISL_1479561, EPI_ISL_1479562, EPI_ISL_1479563, EPI_ISL_1479564, EPI_ISL_1479566, EPI_ISL_1479567, EPI_ISL_1479568, EPI_ISL_1479569, EPI_ISL_1479570, EPI_ISL_1479571, EPI_ISL_1479572, EPI_ISL_1479573, EPI_ISL_1479574, EPI_ISL_1479576, EPI_ISL_1479577, EPI_ISL_1479578, EPI_ISL_1479579, EPI_ISL_1479580, EPI_ISL_1479581, EPI_ISL_1479582, EPI_ISL_1479583, EPI_ISL_1479584, EPI_ISL_1479585, EPI_ISL_1479586, EPI_ISL_1479587, EPI_ISL_1479588, EPI_ISL_1479591, EPI_ISL_1479592, EPI_ISL_1479593, EPI_ISL_1479594, EPI_ISL_1479595, EPI_ISL_1479596, EPI_ISL_1479597, EPI_ISL_1479598, EPI_ISL_1479599, EPI_ISL_1479600, EPI_ISL_1479602, EPI_ISL_1479603, EPI_ISL_1479604, EPI_ISL_1479605, EPI_ISL_1479606, EPI_ISL_1479607, EPI_ISL_1479608, EPI_ISL_1479609, EPI_ISL_1479610, EPI_ISL_1479611, EPI_ISL_1479612, EPI_ISL_1479613, EPI_ISL_1479614, EPI_ISL_1479616, EPI_ISL_1479617, EPI_ISL_1479618, EPI_ISL_1479619, EPI_ISL_1479620, EPI_ISL_1479622, EPI_ISL_1479624, EPI_ISL_1479626, EPI_ISL_1479627, EPI_ISL_1479628, EPI_ISL_1479629, EPI_ISL_1479630, EPI_ISL_1479631, EPI_ISL_1479632, EPI_ISL_1479634, EPI_ISL_1479635, EPI_ISL_1479636, EPI_ISL_1479637, EPI_ISL_1479638, EPI_ISL_1479639, EPI_ISL_1479641, EPI_ISL_1479642, EPI_ISL_1479643, EPI_ISL_1479644, EPI_ISL_1479645, EPI_ISL_1479647, EPI_ISL_1479648, EPI_ISL_1479649, EPI_ISL_1479650, EPI_ISL_1479653, EPI_ISL_1479655, EPI_ISL_1479656, EPI_ISL_1479657, EPI_ISL_1479658, EPI_ISL_1479659, EPI_ISL_1479660, EPI_ISL_1479661, EPI_ISL_1479662, EPI_ISL_1479663, EPI_ISL_1479665, EPI_ISL_1479666, EPI_ISL_1479667, EPI_ISL_1479668, EPI_ISL_1479669, EPI_ISL_1479670, EPI_ISL_1479671, EPI_ISL_1479672, EPI_ISL_1479673, EPI_ISL_1479674, EPI_ISL_1479675, EPI_ISL_1479676, EPI_ISL_1479677, EPI_ISL_1479678, EPI_ISL_1479679, EPI_ISL_1479680, EPI_ISL_1479681, EPI_ISL_1479682 | | | |
| see above | Aegis Sciences Corporation | Centers for Disease Control and Prevention Division of Viral Diseases, Pathogen Discovery | Dakota Howard, Dhwani Batra, Peter W. Cook, Kara Moser, Adrian Paskey, Jason Caravas, Benjamin Rambo-Martin, Shatavia Morrison, Christopher Gulvick, Scott Sammons, Yvette Unoarumhi, Darlene Wagner, Matthew Schmerer, Cyndi Clark, Patrick Campbell, Rob Case, Vikramsinha Ghorpade, Holly Houdeshell, Ola Kvalvaag, Dillon Nall, Ethan Sanders, Alec Vest, Shaun Westlund, Matthew Hardison, Clinton R. Paden, Duncan MacCannell |
| EPI_ISL_1479683, EPI_ISL_1479684, EPI_ISL_1479685, EPI_ISL_1479686, EPI_ISL_1479687, EPI_ISL_1479688, EPI_ISL_1479689, EPI_ISL_1479690, EPI_ISL_1479691, EPI_ISL_1479692, EPI_ISL_1479693, EPI_ISL_1479694, EPI_ISL_1479695, EPI_ISL_1479696, EPI_ISL_1479697, EPI_ISL_1479698, EPI_ISL_1479699, EPI_ISL_1479700, EPI_ISL_1479701, EPI_ISL_1479702, EPI_ISL_1479703, EPI_ISL_1479704, EPI_ISL_1479705, EPI_ISL_1479706, EPI_ISL_1479707, EPI_ISL_1479708, EPI_ISL_1479709, EPI_ISL_1479710, EPI_ISL_1479711, EPI_ISL_1479712, EPI_ISL_1479713, EPI_ISL_1479714, EPI_ISL_1479715, EPI_ISL_1479716, EPI_ISL_1479717, EPI_ISL_1479718, EPI_ISL_1479719, EPI_ISL_1479720, EPI_ISL_1479721, EPI_ISL_1479722, EPI_ISL_1479723, EPI_ISL_1479724, EPI_ISL_1479725, EPI_ISL_1479726, EPI_ISL_1479727, EPI_ISL_1479728, EPI_ISL_1479729, EPI_ISL_1479730, EPI_ISL_1479731, EPI_ISL_1479732, EPI_ISL_1479733, EPI_ISL_1479734, EPI_ISL_1479735, EPI_ISL_1479736, EPI_ISL_1479737, EPI_ISL_1479738, EPI_ISL_1479739, EPI_ISL_1479740, EPI_ISL_1479741, EPI_ISL_1479742, EPI_ISL_1479743, EPI_ISL_1479744, EPI_ISL_1479745, EPI_ISL_1479746, EPI_ISL_1479747, EPI_ISL_1479748, EPI_ISL_1479749, EPI_ISL_1479750, EPI_ISL_1479751, EPI_ISL_1479752, EPI_ISL_1479753, EPI_ISL_1479754, EPI_ISL_1479755, EPI_ISL_1479756, EPI_ISL_1479757, EPI_ISL_1479758, EPI_ISL_1479759, EPI_ISL_1479760, EPI_ISL_1479761, EPI_ISL_1479762, EPI_ISL_1479763, EPI_ISL_1479764, EPI_ISL_1479765, EPI_ISL_1479766, EPI_ISL_1479767, EPI_ISL_1479768, EPI_ISL_1479769, EPI_ISL_1479770, EPI_ISL_1479771, EPI_ISL_1479772, EPI_ISL_1479773, EPI_ISL_1479774, EPI_ISL_1479775, EPI_ISL_1479776, EPI_ISL_1479777, EPI_ISL_1479778, EPI_ISL_1479779, EPI_ISL_1479780, EPI_ISL_1479781, EPI_ISL_1479782, EPI_ISL_1479783, EPI_ISL_1479784, EPI_ISL_1479785, EPI_ISL_1479786, EPI_ISL_1479787, EPI_ISL_1479788, EPI_ISL_1479789, EPI_ISL_1479790, EPI_ISL_1479791, EPI_ISL_1479792, EPI_ISL_1479793, EPI_ISL_1479794, EPI_ISL_1479795, EPI_ISL_1479796, EPI_ISL_1479797, EPI_ISL_1479798, EPI_ISL_1479799, EPI_ISL_1479800, EPI_ISL_1479801 | | | |
| see above | Quest Diagnostics Incorporated | Centers for Disease Control and Prevention Division of Viral Diseases, Pathogen Discovery | Dakota Howard, Dhwani Batra, Peter W. Cook, Kara Moser, Adrian Paskey, Jason Caravas, Benjamin Rambo-Martin, Shatavia Morrison, Christopher Gulvick, Scott Sammons, Yvette Unoarumhi, Darlene Wagner, Matthew Schmerer, S. H. Rosenthal, A. Gerasimova, R. M. Kagan, B. Anderson, M. Hua, Y. Liu, L.E. Bernstein, K.E. Livingston, A. Perez, I. A. Shlyakhter, R. V. Rolando, R. Owen, P. Tanpaiboon, F. Lacbawan, Clinton R. Paden, Duncan MacCannell |
| EPI_ISL_1479802, EPI_ISL_1479803, EPI_ISL_1479804, EPI_ISL_1479805, EPI_ISL_1479806, EPI_ISL_1479807, EPI_ISL_1479808, EPI_ISL_1479809, EPI_ISL_1479810, EPI_ISL_1479812, EPI_ISL_1479813, EPI_ISL_1479814, EPI_ISL_1479815, EPI_ISL_1479816, EPI_ISL_1479817, EPI_ISL_1479819, EPI_ISL_1479820, EPI_ISL_1479821, EPI_ISL_1479822, EPI_ISL_1479823, EPI_ISL_1479824, EPI_ISL_1479825, EPI_ISL_1479826, EPI_ISL_1479827, EPI_ISL_1479828, EPI_ISL_1479829, EPI_ISL_1479831, EPI_ISL_1479832, EPI_ISL_1479833, EPI_ISL_1479834, EPI_ISL_1479836, EPI_ISL_1479837, EPI_ISL_1479838, EPI_ISL_1479839, EPI_ISL_1479840, EPI_ISL_1479842, EPI_ISL_1479843, EPI_ISL_1479844, EPI_ISL_1479845, EPI_ISL_1479846, EPI_ISL_1479847, EPI_ISL_1479848, EPI_ISL_1479849, EPI_ISL_1479850, | | | |

| | | | |
|---|---|---|---|
| EPI_ISL_1479851, EPI_ISL_1479852, EPI_ISL_1479853, EPI_ISL_1479854, EPI_ISL_1479855, EPI_ISL_1479856, EPI_ISL_1479857, EPI_ISL_1479858, EPI_ISL_1479859, EPI_ISL_1479860, EPI_ISL_1479861, EPI_ISL_1479862, EPI_ISL_1479863, EPI_ISL_1479865, EPI_ISL_1479866, EPI_ISL_1479867, EPI_ISL_1479868, EPI_ISL_1479869 | | | |
| see above | Aegis Sciences Corporation | Centers for Disease Control and Prevention Division of Viral Diseases, Pathogen Discovery | Dakota Howard, Dhwani Batra, Peter W. Cook, Kara Moser, Adrian Paskey, Jason Caravas, Benjamin Rambo-Martin, Shatavia Morrison, Christopher Gulvick, Scott Sammons, Yvette Unoarumhi, Darlene Wagner, Matthew Schmerer, Cyndi Clark, Patrick Campbell, Rob Case, Vikramsinha Ghorpade, Holly Houdeshell, Ola Kvalvaag, Dillon Nall, Ethan Sanders, Alec Vest, Shaun Westlund, Matthew Hardison, Clinton R. Paden, Duncan MacCannell |
| EPI_ISL_1479870, EPI_ISL_1479871, EPI_ISL_1479872, EPI_ISL_1479873, EPI_ISL_1479874, EPI_ISL_1479875, EPI_ISL_1479876, EPI_ISL_1479877, EPI_ISL_1479878, EPI_ISL_1479879, EPI_ISL_1479880, EPI_ISL_1479881, EPI_ISL_1479882, EPI_ISL_1479883, EPI_ISL_1479884, EPI_ISL_1479885, EPI_ISL_1479886, EPI_ISL_1479887, EPI_ISL_1479888, EPI_ISL_1479889, EPI_ISL_1479890, EPI_ISL_1479891, EPI_ISL_1479892, EPI_ISL_1479893, EPI_ISL_1479894, EPI_ISL_1479895, EPI_ISL_1479896, EPI_ISL_1479897, EPI_ISL_1479898, EPI_ISL_1479899, EPI_ISL_1479900, EPI_ISL_1479901, EPI_ISL_1479902, EPI_ISL_1479903, EPI_ISL_1479904, EPI_ISL_1479905, EPI_ISL_1479906, EPI_ISL_1479907, EPI_ISL_1479908, EPI_ISL_1479909, EPI_ISL_1479910, EPI_ISL_1479911, EPI_ISL_1479912, EPI_ISL_1479913, EPI_ISL_1479914, EPI_ISL_1479915, EPI_ISL_1479916, EPI_ISL_1479917, EPI_ISL_1479918, EPI_ISL_1479919, EPI_ISL_1479920, EPI_ISL_1479921, EPI_ISL_1479922, EPI_ISL_1479923, EPI_ISL_1479924, EPI_ISL_1479925, EPI_ISL_1479926, EPI_ISL_1479927, EPI_ISL_1479928, EPI_ISL_1479929, EPI_ISL_1479930, EPI_ISL_1479931, EPI_ISL_1479932, EPI_ISL_1479933, EPI_ISL_1479934, EPI_ISL_1479935, EPI_ISL_1479936, EPI_ISL_1479937, EPI_ISL_1479938, EPI_ISL_1479939, EPI_ISL_1479940, EPI_ISL_1479941, EPI_ISL_1479942, EPI_ISL_1479943, EPI_ISL_1479944, EPI_ISL_1479945, EPI_ISL_1479946, EPI_ISL_1479947, EPI_ISL_1479948, EPI_ISL_1479949, EPI_ISL_1479950, EPI_ISL_1479951, EPI_ISL_1479952, EPI_ISL_1479953, EPI_ISL_1479954, EPI_ISL_1479955, EPI_ISL_1479956, EPI_ISL_1479957, EPI_ISL_1479958, EPI_ISL_1479959, EPI_ISL_1479960, EPI_ISL_1479961, EPI_ISL_1479962, EPI_ISL_1479963, EPI_ISL_1479964, EPI_ISL_1479965, EPI_ISL_1479966, EPI_ISL_1479967, EPI_ISL_1479968, EPI_ISL_1479969, EPI_ISL_1479970, EPI_ISL_1479971, EPI_ISL_1479972, EPI_ISL_1479973, EPI_ISL_1479974, EPI_ISL_1479975, EPI_ISL_1479976, EPI_ISL_1479977, EPI_ISL_1479978, EPI_ISL_1479979, EPI_ISL_1479980, EPI_ISL_1479981, EPI_ISL_1479982, EPI_ISL_1479983, EPI_ISL_1479984, EPI_ISL_1479985, EPI_ISL_1479986, EPI_ISL_1479987, EPI_ISL_1479988, EPI_ISL_1479989, EPI_ISL_1479990, EPI_ISL_1479991, EPI_ISL_1479992, EPI_ISL_1479993, EPI_ISL_1479994, EPI_ISL_1479995, EPI_ISL_1479996, EPI_ISL_1479997, EPI_ISL_1479998, EPI_ISL_1479999, EPI_ISL_1480000, EPI_ISL_1480001, EPI_ISL_1480002, EPI_ISL_1480003, EPI_ISL_1480004, EPI_ISL_1480005, EPI_ISL_1480006 | | | |
| see above | Quest Diagnostics Incorporated | Centers for Disease Control and Prevention Division of Viral Diseases, Pathogen Discovery | Dakota Howard, Dhwani Batra, Peter W. Cook, Kara Moser, Adrian Paskey, Jason Caravas, Benjamin Rambo-Martin, Shatavia Morrison, Christopher Gulvick, Scott Sammons, Yvette Unoarumhi, Darlene Wagner, Matthew Schmerer, S. H. Rosenthal, A. Gerasimova, R. M. Kagan, B. Anderson, M. Hua, Y. Liu, L.E. Bernstein, K.E. Livingston, A. Perez, I. A. Shlyakhter, R. V. Rolando, R. Owen, P. Tanpaiboon, F. Lacbawan, Clinton R. Paden, Duncan MacCannell |
| EPI_ISL_1480007, EPI_ISL_1480008, EPI_ISL_1480009, EPI_ISL_1480010, EPI_ISL_1480011, EPI_ISL_1480012, EPI_ISL_1480013, EPI_ISL_1480014, EPI_ISL_1480015, EPI_ISL_1480016, EPI_ISL_1480017, EPI_ISL_1480018, EPI_ISL_1480019, EPI_ISL_1480020, EPI_ISL_1480021, EPI_ISL_1480022, EPI_ISL_1480023, EPI_ISL_1480024, EPI_ISL_1480025, EPI_ISL_1480026, EPI_ISL_1480027, EPI_ISL_1480028, EPI_ISL_1480029, EPI_ISL_1480030, EPI_ISL_1480031, EPI_ISL_1480032, EPI_ISL_1480033, EPI_ISL_1480034, EPI_ISL_1480035, EPI_ISL_1480036, EPI_ISL_1480037, EPI_ISL_1480038, EPI_ISL_1480039, EPI_ISL_1480040, EPI_ISL_1480041, EPI_ISL_1480042, EPI_ISL_1480043, EPI_ISL_1480044, EPI_ISL_1480045, EPI_ISL_1480046, EPI_ISL_1480047, EPI_ISL_1480048, EPI_ISL_1480049, EPI_ISL_1480050, EPI_ISL_1480051, EPI_ISL_1480052, EPI_ISL_1480053, EPI_ISL_1480054, EPI_ISL_1480055, EPI_ISL_1480056, EPI_ISL_1480057, EPI_ISL_1480058, EPI_ISL_1480059, EPI_ISL_1480060, EPI_ISL_1480061, EPI_ISL_1480062, EPI_ISL_1480063, EPI_ISL_1480064, EPI_ISL_1480065, EPI_ISL_1480066, EPI_ISL_1480067, EPI_ISL_1480068, EPI_ISL_1480069, EPI_ISL_1480070, EPI_ISL_1480071, EPI_ISL_1480072, EPI_ISL_1480073, EPI_ISL_1480074, EPI_ISL_1480075, EPI_ISL_1480076, EPI_ISL_1480077, EPI_ISL_1480078, EPI_ISL_1480079, EPI_ISL_1480080, EPI_ISL_1480081, EPI_ISL_1480082, EPI_ISL_1480083, EPI_ISL_1480084, EPI_ISL_1480085, EPI_ISL_1480086, EPI_ISL_1480087, EPI_ISL_1480088, EPI_ISL_1480089, EPI_ISL_1480090, EPI_ISL_1480091, EPI_ISL_1480092, EPI_ISL_1480093, EPI_ISL_1480094, EPI_ISL_1480095, EPI_ISL_1480096, EPI_ISL_1480097, EPI_ISL_1480098, EPI_ISL_1480099, EPI_ISL_1480100, EPI_ISL_1480101, EPI_ISL_1480102, EPI_ISL_1480103, EPI_ISL_1480104, EPI_ISL_1480105, EPI_ISL_1480106, EPI_ISL_1480107, EPI_ISL_1480108, EPI_ISL_1480109, EPI_ISL_1480110, EPI_ISL_1480111, EPI_ISL_1480112, EPI_ISL_1480113, EPI_ISL_1480114, EPI_ISL_1480115, EPI_ISL_1480116, EPI_ISL_1480117, EPI_ISL_1480118, EPI_ISL_1480119, EPI_ISL_1480120, EPI_ISL_1480121, EPI_ISL_1480122, EPI_ISL_1480123, EPI_ISL_1480124, EPI_ISL_1480125, EPI_ISL_1480126, EPI_ISL_1480127, EPI_ISL_1480128, EPI_ISL_1480129, EPI_ISL_1480130, EPI_ISL_1480131, EPI_ISL_1480132, EPI_ISL_1480133, EPI_ISL_1480134, EPI_ISL_1480135, EPI_ISL_1480136, EPI_ISL_1480137, EPI_ISL_1480138, EPI_ISL_1480139, EPI_ISL_1480140, EPI_ISL_1480141, EPI_ISL_1480142, EPI_ISL_1480143, EPI_ISL_1480144, EPI_ISL_1480145, EPI_ISL_1480146, EPI_ISL_1480147, EPI_ISL_1480148, EPI_ISL_1480149, EPI_ISL_1480150, EPI_ISL_1480151, EPI_ISL_1480152, EPI_ISL_1480153, EPI_ISL_1480154, EPI_ISL_1480155, EPI_ISL_1480156, EPI_ISL_1480157, EPI_ISL_1480158, EPI_ISL_1480159, EPI_ISL_1480160, EPI_ISL_1480161, EPI_ISL_1480162, EPI_ISL_1480163, EPI_ISL_1480164, EPI_ISL_1480165, EPI_ISL_1480166, EPI_ISL_1480167, EPI_ISL_1480168, EPI_ISL_1480169, EPI_ISL_1480170, EPI_ISL_1480171, EPI_ISL_1480172, EPI_ISL_1480173, EPI_ISL_1480174, EPI_ISL_1480175, EPI_ISL_1480176, EPI_ISL_1480177, EPI_ISL_1480178, EPI_ISL_1480179, EPI_ISL_1480180, EPI_ISL_1480181, EPI_ISL_1480182, EPI_ISL_1480183, EPI_ISL_1480184, EPI_ISL_1480185, EPI_ISL_1480186, EPI_ISL_1480187, EPI_ISL_1480188, EPI_ISL_1480189, EPI_ISL_1480190, EPI_ISL_1480191, EPI_ISL_1480192, EPI_ISL_1480193, EPI_ISL_1480194, EPI_ISL_1480195, EPI_ISL_1480196, EPI_ISL_1480197, EPI_ISL_1480198, EPI_ISL_1480199, EPI_ISL_1480200, EPI_ISL_1480201, EPI_ISL_1480202, EPI_ISL_1480203, EPI_ISL_1480204, EPI_ISL_1480205, EPI_ISL_1480206, EPI_ISL_1480207, EPI_ISL_1480208, EPI_ISL_1480209, EPI_ISL_1480210, EPI_ISL_1480211, EPI_ISL_1480212, EPI_ISL_1480213, EPI_ISL_1480214, EPI_ISL_1480215, EPI_ISL_1480216, EPI_ISL_1480217, EPI_ISL_1480218, EPI_ISL_1480219, EPI_ISL_1480220, EPI_ISL_1480221, EPI_ISL_1480222, EPI_ISL_1480223, EPI_ISL_1480224, EPI_ISL_1480225, EPI_ISL_1480226, EPI_ISL_1480228, EPI_ISL_1480229, EPI_ISL_1480230, EPI_ISL_1480231, EPI_ISL_1480232, EPI_ISL_1480233, EPI_ISL_1480234, EPI_ISL_1480235, EPI_ISL_1480236, EPI_ISL_1480237, EPI_ISL_1480238, EPI_ISL_1480239, EPI_ISL_1480240, EPI_ISL_1480241, EPI_ISL_1480242, EPI_ISL_1480243, EPI_ISL_1480244, EPI_ISL_1480245, EPI_ISL_1480246, EPI_ISL_1480247, EPI_ISL_1480248, EPI_ISL_1480249, EPI_ISL_1480250, EPI_ISL_1480251, EPI_ISL_1480252, EPI_ISL_1480253, EPI_ISL_1480254, EPI_ISL_1480255, EPI_ISL_1480256, EPI_ISL_1480257, EPI_ISL_1480258, EPI_ISL_1480259, EPI_ISL_1480260, EPI_ISL_1480261, EPI_ISL_1480262, EPI_ISL_1480263, EPI_ISL_1480264, EPI_ISL_1480265, EPI_ISL_1480266, EPI_ISL_1480267, EPI_ISL_1480268, EPI_ISL_1480269, EPI_ISL_1480270, EPI_ISL_1480271, EPI_ISL_1480272, EPI_ISL_1480273, EPI_ISL_1480274, EPI_ISL_1480275, EPI_ISL_1480276, EPI_ISL_1480277, EPI_ISL_1480278, EPI_ISL_1480279, EPI_ISL_1480280, EPI_ISL_1480281, EPI_ISL_1480282, EPI_ISL_1480283, EPI_ISL_1480284, EPI_ISL_1480285, EPI_ISL_1480286, EPI_ISL_1480287, EPI_ISL_1480288, EPI_ISL_1480289, EPI_ISL_1480290, EPI_ISL_1480291, EPI_ISL_1480292, EPI_ISL_1480293, EPI_ISL_1480294, EPI_ISL_1480295, EPI_ISL_1480296, EPI_ISL_1480297, EPI_ISL_1480298, EPI_ISL_1480299, EPI_ISL_1480300, EPI_ISL_1480301, EPI_ISL_1480302, EPI_ISL_1480303, EPI_ISL_1480304, EPI_ISL_1480305, EPI_ISL_1480306, EPI_ISL_1480307, EPI_ISL_1480308, EPI_ISL_1480309, EPI_ISL_1480310, EPI_ISL_1480311, EPI_ISL_1480312, EPI_ISL_1480313, EPI_ISL_1480314, EPI_ISL_1480315, EPI_ISL_1480316, EPI_ISL_1480317, EPI_ISL_1480318, EPI_ISL_1480319, EPI_ISL_1480320, EPI_ISL_1480321, EPI_ISL_1480322, EPI_ISL_1480323, EPI_ISL_1480324, EPI_ISL_1480327, EPI_ISL_1480328, EPI_ISL_1480329, EPI_ISL_1480330, EPI_ISL_1480331, EPI_ISL_1480332, EPI_ISL_1480333, EPI_ISL_1480334, EPI_ISL_1480335, EPI_ISL_1480336, EPI_ISL_1480337, EPI_ISL_1480338, EPI_ISL_1480339, EPI_ISL_1480340, EPI_ISL_1480341, EPI_ISL_1480342, EPI_ISL_1480343, EPI_ISL_1480344, EPI_ISL_1480345, EPI_ISL_1480346, EPI_ISL_1480347, EPI_ISL_1480348, EPI_ISL_1480349, EPI_ISL_1480350, EPI_ISL_1480351, EPI_ISL_1480352, EPI_ISL_1480353, EPI_ISL_1480354, EPI_ISL_1480355, EPI_ISL_1480356, EPI_ISL_1480357, EPI_ISL_1480358, EPI_ISL_1480359, EPI_ISL_1480360, EPI_ISL_1480361, EPI_ISL_1480362, EPI_ISL_1480363, EPI_ISL_1480364, EPI_ISL_1480365, EPI_ISL_1480366, EPI_ISL_1480367, EPI_ISL_1480368, EPI_ISL_1480369, EPI_ISL_1480370, EPI_ISL_1480371, EPI_ISL_1480372, EPI_ISL_1480373, EPI_ISL_1480374, EPI_ISL_1480375, EPI_ISL_1480376, EPI_ISL_1480377, EPI_ISL_1480378, EPI_ISL_1480379, EPI_ISL_1480380, EPI_ISL_1480381, EPI_ISL_1480382, EPI_ISL_1480383, EPI_ISL_1480384, EPI_ISL_1480385, EPI_ISL_1480386, EPI_ISL_1480387, EPI_ISL_1480388, EPI_ISL_1480389, EPI_ISL_1480390, EPI_ISL_1480391, EPI_ISL_1480392, EPI_ISL_1480393, EPI_ISL_1480394, EPI_ISL_1480395, EPI_ISL_1480396, EPI_ISL_1480397, EPI_ISL_1480398, EPI_ISL_1480399, EPI_ISL_1480400, EPI_ISL_1480401, EPI_ISL_1480402, EPI_ISL_1480403, EPI_ISL_1480404, EPI_ISL_1480405, EPI_ISL_1480406, EPI_ISL_1480407, EPI_ISL_1480408, EPI_ISL_1480409, EPI_ISL_1480410, EPI_ISL_1480411, EPI_ISL_1480412, EPI_ISL_1480413, EPI_ISL_1480414, EPI_ISL_1480415, EPI_ISL_1480416, EPI_ISL_1480417, EPI_ISL_1480418, EPI_ISL_1480419, EPI_ISL_1480420, EPI_ISL_1480421, EPI_ISL_1480422, EPI_ISL_1480423, EPI_ISL_1480424, EPI_ISL_1480425, EPI_ISL_1480426, EPI_ISL_1480428, EPI_ISL_1480429, EPI_ISL_1480430, EPI_ISL_1480431, EPI_ISL_1480432, EPI_ISL_1480433, EPI_ISL_1480434, EPI_ISL_1480435, EPI_ISL_1480436, EPI_ISL_1480437, EPI_ISL_1480438, EPI_ISL_1480439, EPI_ISL_1480440, EPI_ISL_1480441, EPI_ISL_1480442, EPI_ISL_1480443, EPI_ISL_1480444, EPI_ISL_1480445, EPI_ISL_1480446, EPI_ISL_1480447, EPI_ISL_1480448, EPI_ISL_1480449, EPI_ISL_1480450, EPI_ISL_1480451, EPI_ISL_1480452, EPI_ISL_1480453, EPI_ISL_1480454, EPI_ISL_1480455, EPI_ISL_1480456, EPI_ISL_1480457, EPI_ISL_1480458, EPI_ISL_1480459, EPI_ISL_1480460, EPI_ISL_1480461, EPI_ISL_1480462, EPI_ISL_1480463, EPI_ISL_1480464, EPI_ISL_1480465, EPI_ISL_1480466, EPI_ISL_1480467, EPI_ISL_1480468, EPI_ISL_1480469, EPI_ISL_1480470, EPI_ISL_1480471, EPI_ISL_1480472, EPI_ISL_1480473, EPI_ISL_1480474, EPI_ISL_1480475, EPI_ISL_1480476, EPI_ISL_1480477, EPI_ISL_1480478, EPI_ISL_1480479, EPI_ISL_1480480, EPI_ISL_1480481, EPI_ISL_1480482, EPI_ISL_1480483, EPI_ISL_1480484, EPI_ISL_1480485, EPI_ISL_1480486, EPI_ISL_1480487, EPI_ISL_1480488, EPI_ISL_1480489, EPI_ISL_1480490, EPI_ISL_1480491, EPI_ISL_1480492, EPI_ISL_1480493, EPI_ISL_1480494, EPI_ISL_1480495, EPI_ISL_1480496, EPI_ISL_1480497, EPI_ISL_1480498, EPI_ISL_1480499, EPI_ISL_1480500, EPI_ISL_1480501, EPI_ISL_1480502, EPI_ISL_1480503, EPI_ISL_1480504, EPI_ISL_1480505, EPI_ISL_1480506, EPI_ISL_1480507, EPI_ISL_1480508, EPI_ISL_1480509, EPI_ISL_1480510, EPI_ISL_1480511, EPI_ISL_1480512, EPI_ISL_1480513, EPI_ISL_1480515, EPI_ISL_1480516, EPI_ISL_1480517, EPI_ISL_1480518, EPI_ISL_1480519, EPI_ISL_1480520 | | | |
| see above | Helix/Illumina | Centers for Disease Control and Prevention Division of Viral Diseases, Pathogen Discovery | Dakota Howard, Dhwani Batra, Peter W. Cook, Kara Moser, Adrian Paskey, Jason Caravas, Benjamin Rambo-Martin, Shatavia Morrison, Christopher Gulvick, Scott Sammons, Yvette Unoarumhi, Darlene Wagner, Matthew Schmerer, Eileen de Feo, Jan Antico, Christine Tran, Matthew Tolentino, Shannon Wickline, Kim Gietzen, Brad Sickler, Jingtao Liu, Eric Allen, Phil Febbo, Nicole L. Washington, Simon White, Geraint Levan, Kelly Schiabor Barrett, Elizabeth Cirulli, Alexandre Bolze, Ary Ascencio, Charlotte Rivera-Garcia, Ryan Cho, Jason Nguyen, Sherry Wang, Jimmy Ramirez, Tyler Cassens, Efren Sandoval, Magnus Isaksson, William Lee, David Becker, Marc Laurent, James Lu, Clinton R. Paden, Duncan MacCannell |
| EPI_ISL_1480521, EPI_ISL_1480522, EPI_ISL_1480523, EPI_ISL_1480524, EPI_ISL_1480525, EPI_ISL_1480526, EPI_ISL_1480527, EPI_ISL_1480528, EPI_ISL_1480529, EPI_ISL_1480530, EPI_ISL_1480531, EPI_ISL_1480532, EPI_ISL_1480533, EPI_ISL_1480534, EPI_ISL_1480535, EPI_ISL_1480536, EPI_ISL_1480537, EPI_ISL_1480538, EPI_ISL_1480539, EPI_ISL_1480540, EPI_ISL_1480541, EPI_ISL_1480542, EPI_ISL_1480543, EPI_ISL_1480544, EPI_ISL_1480545, EPI_ISL_1480546, EPI_ISL_1480547, EPI_ISL_1480548, EPI_ISL_1480549, EPI_ISL_1480550, EPI_ISL_1480551, EPI_ISL_1480552, EPI_ISL_1480553, EPI_ISL_1480554, EPI_ISL_1480555, EPI_ISL_1480556, EPI_ISL_1480557, EPI_ISL_1480558, EPI_ISL_1480559, EPI_ISL_1480560, EPI_ISL_1480561, EPI_ISL_1480562, EPI_ISL_1480563, EPI_ISL_1480564, EPI_ISL_1480565, EPI_ISL_1480566, EPI_ISL_1480567, EPI_ISL_1480568, EPI_ISL_1480569, EPI_ISL_1480570, EPI_ISL_1480571, EPI_ISL_1480572, EPI_ISL_1480573, EPI_ISL_1480574, EPI_ISL_1480575, EPI_ISL_1480576, EPI_ISL_1480577, EPI_ISL_1480578, EPI_ISL_1480579, EPI_ISL_1480580, EPI_ISL_1480581, EPI_ISL_1480582, EPI_ISL_1480583, EPI_ISL_1480584, EPI_ISL_1480585, EPI_ISL_1480586, EPI_ISL_1480587, EPI_ISL_1480588, EPI_ISL_1480589, EPI_ISL_1480590, EPI_ISL_1480591, EPI_ISL_1480592, EPI_ISL_1480593, EPI_ISL_1480594, EPI_ISL_1480595, EPI_ISL_1480596, EPI_ISL_1480597, EPI_ISL_1480598, EPI_ISL_1480599, EPI_ISL_1480600, EPI_ISL_1480601, EPI_ISL_1480602, EPI_ISL_1480603, EPI_ISL_1480604, EPI_ISL_1480605, EPI_ISL_1480606, EPI_ISL_1480607, EPI_ISL_1480608, EPI_ISL_1480609, EPI_ISL_1480610, EPI_ISL_1480611, EPI_ISL_1480612, EPI_ISL_1480613, EPI_ISL_1480614, EPI_ISL_1480615, EPI_ISL_1480616, EPI_ISL_1480617, EPI_ISL_1480618, EPI_ISL_1480619, EPI_ISL_1480620, EPI_ISL_1480621, EPI_ISL_1480622, EPI_ISL_1480623, EPI_ISL_1480624, EPI_ISL_1480625, EPI_ISL_1480626, EPI_ISL_1480627, EPI_ISL_1480628, EPI_ISL_1480629, EPI_ISL_1480630, EPI_ISL_1480631, EPI_ISL_1480632, EPI_ISL_1480633, EPI_ISL_1480634, EPI_ISL_1480635, EPI_ISL_1480636, EPI_ISL_1480637, EPI_ISL_1480638, EPI_ISL_1480639, EPI_ISL_1480640, EPI_ISL_1480641, EPI_ISL_1480642, EPI_ISL_1480643, EPI_ISL_1480644, EPI_ISL_1480645, EPI_ISL_1480646, EPI_ISL_1480647, EPI_ISL_1480648, EPI_ISL_1480649, EPI_ISL_1480650, EPI_ISL_1480651, EPI_ISL_1480652, EPI_ISL_1480653, EPI_ISL_1480654, EPI_ISL_1480655, EPI_ISL_1480656, EPI_ISL_1480657, EPI_ISL_1480658, EPI_ISL_1480659, EPI_ISL_1480660, EPI_ISL_1480661, EPI_ISL_1480662, EPI_ISL_1480663, EPI_ISL_1480664, EPI_ISL_1480665, EPI_ISL_1480666, EPI_ISL_1480667, EPI_ISL_1480668, EPI_ISL_1480669, EPI_ISL_1480670, EPI_ISL_1480671, EPI_ISL_1480672, EPI_ISL_1480673, EPI_ISL_1480674, EPI_ISL_1480675, EPI_ISL_1480676, EPI_ISL_1480677, EPI_ISL_1480678, EPI_ISL_1480679, EPI_ISL_1480680, EPI_ISL_1480681, EPI_ISL_1480682, EPI_ISL_1480683, EPI_ISL_1480684, EPI_ISL_1480685, EPI_ISL_1480686, EPI_ISL_1480687, EPI_ISL_1480688, EPI_ISL_1480689, EPI_ISL_1480690, EPI_ISL_1480691, EPI_ISL_1480692, EPI_ISL_1480693, EPI_ISL_1480694, EPI_ISL_1480695, EPI_ISL_1480696, EPI_ISL_1480697, EPI_ISL_1480698, EPI_ISL_1480699, EPI_ISL_1480700, EPI_ISL_1480701, EPI_ISL_1480702, EPI_ISL_1480703, EPI_ISL_1480704, EPI_ISL_1480705, EPI_ISL_1480706, EPI_ISL_1480707, EPI_ISL_1480708, EPI_ISL_1480709, EPI_ISL_1480710, EPI_ISL_1480711, EPI_ISL_1480712, EPI_ISL_1480713, EPI_ISL_1480714, EPI_ISL_1480715, EPI_ISL_1480716, EPI_ISL_1480717, EPI_ISL_1480718, EPI_ISL_1480719, EPI_ISL_1480720, EPI_ISL_1480721, EPI_ISL_1480722, EPI_ISL_1480723, EPI_ISL_1480724, EPI_ISL_1480725, EPI_ISL_1480726, EPI_ISL_1480727, EPI_ISL_1480728, EPI_ISL_1480729, EPI_ISL_1480730, EPI_ISL_1480731, EPI_ISL_1480732, EPI_ISL_1480733, EPI_ISL_1480734, EPI_ISL_1480735, EPI_ISL_1480736, EPI_ISL_1480737, EPI_ISL_1480738, EPI_ISL_1480739, EPI_ISL_1480740, EPI_ISL_1480741, EPI_ISL_1480742, EPI_ISL_1480743, EPI_ISL_1480744, EPI_ISL_1480745, EPI_ISL_1480746, EPI_ISL_1480747, EPI_ISL_1480748, EPI_ISL_1480749, EPI_ISL_1480750, EPI_ISL_1480751, EPI_ISL_1480752, EPI_ISL_1480753, EPI_ISL_1480754, EPI_ISL_1480755, EPI_ISL_1480756, EPI_ISL_1480757, EPI_ISL_1480758, EPI_ISL_1480759, EPI_ISL_1480760, EPI_ISL_1480761, EPI_ISL_1480762, EPI_ISL_1480763, EPI_ISL_1480764, EPI_ISL_1480765, EPI_ISL_1480766, EPI_ISL_1480767, EPI_ISL_1480768, EPI_ISL_1480769, EPI_ISL_1480770, EPI_ISL_1480771, EPI_ISL_1480772, EPI_ISL_1480773, EPI_ISL_1480774, EPI_ISL_1480775, EPI_ISL_1480776, EPI_ISL_1480777, EPI_ISL_1480778, EPI_ISL_1480779, EPI_ISL_1480780, EPI_ISL_1480781, EPI_ISL_1480782, EPI_ISL_1480783, EPI_ISL_1480784, EPI_ISL_1480785, EPI_ISL_1480786, EPI_ISL_1480787, EPI_ISL_1480788, EPI_ISL_1480789, EPI_ISL_1480790, EPI_ISL_1480791, EPI_ISL_1480792, EPI_ISL_1480793, EPI_ISL_1480794, EPI_ISL_1480795, EPI_ISL_1480796, EPI_ISL_1480797, EPI_ISL_1480798, EPI_ISL_1480799, EPI_ISL_1480800, EPI_ISL_1480801, EPI_ISL_1480802, EPI_ISL_1480803, EPI_ISL_1480804, EPI_ISL_1480805, EPI_ISL_1480806, EPI_ISL_1480807, EPI_ISL_1480808, EPI_ISL_1480809, EPI_ISL_1480810, EPI_ISL_1480811, EPI_ISL_1480812, EPI_ISL_1480813, EPI_ISL_1480814, EPI_ISL_1480815, EPI_ISL_1480816, EPI_ISL_1480817, EPI_ISL_1480818, EPI_ISL_1480819, EPI_ISL_1480820, EPI_ISL_1480821, EPI_ISL_1480822, EPI_ISL_1480823, EPI_ISL_1480824, EPI_ISL_1480825, EPI_ISL_1480826, EPI_ISL_1480827, EPI_ISL_1480828, EPI_ISL_1480829, EPI_ISL_1480830, EPI_ISL_1480831, EPI_ISL_1480832, EPI_ISL_1480833, EPI_ISL_1480834, EPI_ISL_1480835, EPI_ISL_1480836, EPI_ISL_1480837, EPI_ISL_1480838, EPI_ISL_1480839, EPI_ISL_1480840, EPI_ISL_1480841, EPI_ISL_1480842, EPI_ISL_1480843, EPI_ISL_1480844, EPI_ISL_1480845, EPI_ISL_1480846, EPI_ISL_1480847, EPI_ISL_1480848, EPI_ISL_1480849, EPI_ISL_1480850, EPI_ISL_1480851, EPI_ISL_1480852, EPI_ISL_1480853, EPI_ISL_1480854, EPI_ISL_1480855, EPI_ISL_1480856, EPI_ISL_1480857, EPI_ISL_1480858, EPI_ISL_1480859, EPI_ISL_1480860, EPI_ISL_1480861, EPI_ISL_1480862, EPI_ISL_1480863, EPI_ISL_1480864, EPI_ISL_1480865, EPI_ISL_1480866, EPI_ISL_1480867, EPI_ISL_1480868, EPI_ISL_1480869, EPI_ISL_1480870, EPI_ISL_1480871, EPI_ISL_1480872, EPI_ISL_1480873, EPI_ISL_1480874, EPI_ISL_1480875, EPI_ISL_1480876, EPI_ISL_1480877, EPI_ISL_1480878, EPI_ISL_1480879, EPI_ISL_1480880, EPI_ISL_1480881, EPI_ISL_1480882, EPI_ISL_1480883, EPI_ISL_1480884, EPI_ISL_1480885, EPI_ISL_1480886, EPI_ISL_1480887, EPI_ISL_1480888, EPI_ISL_1480889, EPI_ISL_1480890, EPI_ISL_1480891, EPI_ISL_1480892, EPI_ISL_1480893, EPI_ISL_1480894, EPI_ISL_1480895, EPI_ISL_1480896, EPI_ISL_1480897, EPI_ISL_1480898, EPI_ISL_1480899, EPI_ISL_1480900, EPI_ISL_1480901, EPI_ISL_1480902, EPI_ISL_1480903, EPI_ISL_1480904, EPI_ISL_1480905, EPI_ISL_1480906, EPI_ISL_1480907, EPI_ISL_1480908, EPI_ISL_1480909, EPI_ISL_1480910, EPI_ISL_1480911, EPI_ISL_1480912, EPI_ISL_1480913, EPI_ISL_1480914, EPI_ISL_1480915, EPI_ISL_1480916, EPI_ISL_1480917, EPI_ISL_1480918, EPI_ISL_1480919, EPI_ISL_1480920, EPI_ISL_1480921, EPI_ISL_1480922, EPI_ISL_1480923, EPI_ISL_1480924, EPI_ISL_1480925, EPI_ISL_1480926, EPI_ISL_1480927, EPI_ISL_1480928, EPI_ISL_1480929, EPI_ISL_1480930, EPI_ISL_1480931, EPI_ISL_1480932, EPI_ISL_1480933, EPI_ISL_1480934, EPI_ISL_1480935, EPI_ISL_1480936, EPI_ISL_1480937, EPI_ISL_1480938, EPI_ISL_1480939, EPI_ISL_1480940, EPI_ISL_1480941, EPI_ISL_1480942, EPI_ISL_1480943, EPI_ISL_1480944, EPI_ISL_1480945, EPI_ISL_1480946, EPI_ISL_1480947, EPI_ISL_1480948, EPI_ISL_1480949, EPI_ISL_1480950, EPI_ISL_1480951, EPI_ISL_1480952, EPI_ISL_1480953, EPI_ISL_1480954, EPI_ISL_1480955, EPI_ISL_1480956, EPI_ISL_1480957, EPI_ISL_1480958, EPI_ISL_1480959, EPI_ISL_1480960, EPI_ISL_1480961, EPI_ISL_1480962, EPI_ISL_1480963, EPI_ISL_1480964, EPI_ISL_1480965, EPI_ISL_1480966, EPI_ISL_1480967, EPI_ISL_1480968, EPI_ISL_1480969, EPI_ISL_1480970, EPI_ISL_1480971, EPI_ISL_1480972, EPI_ISL_1480973, EPI_ISL_1480974, EPI_ISL_1480975, EPI_ISL_1480976 | | | |
| see above | Quest Diagnostics Incorporated | Centers for Disease Control and Prevention Division of Viral Diseases, Pathogen Discovery | Dakota Howard, Dhwani Batra, Peter W. Cook, Kara Moser, Adrian Paskey, Jason Caravas, Benjamin Rambo-Martin, Shatavia Morrison, Christopher Gulvick, Scott Sammons, Yvette Unoarumhi, Darlene Wagner, Matthew Schmerer, S. H. Rosenthal, A. Gerasimova, R. M. Kagan, B. Anderson, M. Hua, Y. Liu, L.E. Bernstein, K.E. Livingston, A. Perez, I. A. Shlyakhter, R. V. Rolando, R. Owen, P. Tanpaiboon, F. Lacbawan, Clinton R. Paden, Duncan MacCannell |
| EPI_ISL_1480978, EPI_ISL_1480979, EPI_ISL_1480980, EPI_ISL_1480981, EPI_ISL_1480984, EPI_ISL_1480986, EPI_ISL_1480987, EPI_ISL_1480988, EPI_ISL_1480989, EPI_ISL_1480991, EPI_ISL_1480992, EPI_ISL_1480997, EPI_ISL_1480999, EPI_ISL_1481000, EPI_ISL_1481001, EPI_ISL_1481002, EPI_ISL_1481005, EPI_ISL_1481006, EPI_ISL_1481009, EPI_ISL_1481011, EPI_ISL_1481013, EPI_ISL_1481014, EPI_ISL_1481016, EPI_ISL_1481017, EPI_ISL_1481018, EPI_ISL_1481019, EPI_ISL_1481020, EPI_ISL_1481022, EPI_ISL_1481023, EPI_ISL_1481024, EPI_ISL_1481026, EPI_ISL_1481027, EPI_ISL_1481030, EPI_ISL_1481031, EPI_ISL_1481032, EPI_ISL_1481033, EPI_ISL_1481034, EPI_ISL_1481035, EPI_ISL_1481037, EPI_ISL_1481039, EPI_ISL_1481040, EPI_ISL_1481041, EPI_ISL_1481044, EPI_ISL_1481045, EPI_ISL_1481046, EPI_ISL_1481047, EPI_ISL_1481049, EPI_ISL_1481052, EPI_ISL_1481053, EPI_ISL_1481054, EPI_ISL_1481055, EPI_ISL_1481056, EPI_ISL_1481059, EPI_ISL_1481061, EPI_ISL_1481062, EPI_ISL_1481065, EPI_ISL_1481066, EPI_ISL_1481067, EPI_ISL_1481068, EPI_ISL_1481069, EPI_ISL_1481070, EPI_ISL_1481072, EPI_ISL_1481073, EPI_ISL_1481074, EPI_ISL_1481075, EPI_ISL_1481076, EPI_ISL_1481077, EPI_ISL_1481079, EPI_ISL_1481080, EPI_ISL_1481084, EPI_ISL_1481085, EPI_ISL_1481087, EPI_ISL_1481088, EPI_ISL_1481090, EPI_ISL_1481094, EPI_ISL_1481095, EPI_ISL_1481098, EPI_ISL_1481101, EPI_ISL_1481102, EPI_ISL_1481103, EPI_ISL_1481106, EPI_ISL_1481107, EPI_ISL_1481109, EPI_ISL_1481112, EPI_ISL_1481114, EPI_ISL_1481116, EPI_ISL_1481117, EPI_ISL_1481119, EPI_ISL_1481120, EPI_ISL_1481121, EPI_ISL_1481126, EPI_ISL_1481127, EPI_ISL_1481128, EPI_ISL_1481129, EPI_ISL_1481130, EPI_ISL_1481131, EPI_ISL_1481132, EPI_ISL_1481133, EPI_ISL_1481135, EPI_ISL_1481136, EPI_ISL_1481137, EPI_ISL_1481140, EPI_ISL_1481141, EPI_ISL_1481142, EPI_ISL_1481145, EPI_ISL_1481146, EPI_ISL_1481148, EPI_ISL_1481152, EPI_ISL_1481153, EPI_ISL_1481156, EPI_ISL_1481157, EPI_ISL_1481159, EPI_ISL_1481161, EPI_ISL_1481163, EPI_ISL_1481165, EPI_ISL_1481166, EPI_ISL_1481167, EPI_ISL_1481168, EPI_ISL_1481169, EPI_ISL_1481170, EPI_ISL_1481171, EPI_ISL_1481172, EPI_ISL_1481173, EPI_ISL_1481174, EPI_ISL_1481175, EPI_ISL_1481178, EPI_ISL_1481181, EPI_ISL_1481182, EPI_ISL_1481183, EPI_ISL_1481185, EPI_ISL_1481186, EPI_ISL_1481188, EPI_ISL_1481189, EPI_ISL_1481190, EPI_ISL_1481193, EPI_ISL_1481194, EPI_ISL_1481195, EPI_ISL_1481196, EPI_ISL_1481197, EPI_ISL_1481199, EPI_ISL_1481201, EPI_ISL_1481202, EPI_ISL_1481206, EPI_ISL_1481208, EPI_ISL_1481210, EPI_ISL_1481211, EPI_ISL_1481213, EPI_ISL_1481214, EPI_ISL_1481215, EPI_ISL_1481218, EPI_ISL_1481219, EPI_ISL_1481220, EPI_ISL_1481224, EPI_ISL_1481228, EPI_ISL_1481229, EPI_ISL_1481230, EPI_ISL_1481231, EPI_ISL_1481232, EPI_ISL_1481235, EPI_ISL_1481237, EPI_ISL_1481243, EPI_ISL_1481245, EPI_ISL_1481246, EPI_ISL_1481247, EPI_ISL_1481251, EPI_ISL_1481252, EPI_ISL_1481253, EPI_ISL_1481254, EPI_ISL_1481256, EPI_ISL_1481257, EPI_ISL_1481258, EPI_ISL_1481260, EPI_ISL_1481261, EPI_ISL_1481262, EPI_ISL_1481263, EPI_ISL_1481264, EPI_ISL_1481265, EPI_ISL_1481267, EPI_ISL_1481269, EPI_ISL_1481271, EPI_ISL_1481272, EPI_ISL_1481274, EPI_ISL_1481275, EPI_ISL_1481276, EPI_ISL_1481278, EPI_ISL_1481279, EPI_ISL_1481283, EPI_ISL_1481284, EPI_ISL_1481285, EPI_ISL_1481287, EPI_ISL_1481290, EPI_ISL_1481292, EPI_ISL_1481293, EPI_ISL_1481294, EPI_ISL_1481295, EPI_ISL_1481296, EPI_ISL_1481299, EPI_ISL_1481301, EPI_ISL_1481302, EPI_ISL_1481305, EPI_ISL_1481308, EPI_ISL_1481309, EPI_ISL_1481310, EPI_ISL_1481311, EPI_ISL_1481313, EPI_ISL_1481318, EPI_ISL_1481319, EPI_ISL_1481320, EPI_ISL_1481321, EPI_ISL_1481323, EPI_ISL_1481324, EPI_ISL_1481326, EPI_ISL_1481327, EPI_ISL_1481329, EPI_ISL_1481330, EPI_ISL_1481331, EPI_ISL_1481332, EPI_ISL_1481333, EPI_ISL_1481334, EPI_ISL_1481335, EPI_ISL_1481336, EPI_ISL_1481337, EPI_ISL_1481338, EPI_ISL_1481339, EPI_ISL_1481340, EPI_ISL_1481341, EPI_ISL_1481343, EPI_ISL_1481345, EPI_ISL_1481347, EPI_ISL_1481348, EPI_ISL_1481350, EPI_ISL_1481351, EPI_ISL_1481352, EPI_ISL_1481354, EPI_ISL_1481356, EPI_ISL_1481358, EPI_ISL_1481361, EPI_ISL_1481362, EPI_ISL_1481364, EPI_ISL_1481366, EPI_ISL_1481367, EPI_ISL_1481369, EPI_ISL_1481371, EPI_ISL_1481373, EPI_ISL_1481374, EPI_ISL_1481375, EPI_ISL_1481376, EPI_ISL_1481377, EPI_ISL_1481382, EPI_ISL_1481384, EPI_ISL_1481386, EPI_ISL_1481387, EPI_ISL_1481389, EPI_ISL_1481390, EPI_ISL_1481391, EPI_ISL_1481392, EPI_ISL_1481395, EPI_ISL_1481397, EPI_ISL_1481398, EPI_ISL_1481400, EPI_ISL_1481401, EPI_ISL_1481402, EPI_ISL_1481404, EPI_ISL_1481405, EPI_ISL_1481407, EPI_ISL_1481412, EPI_ISL_1481413, EPI_ISL_1481414, EPI_ISL_1481415, EPI_ISL_1481417, EPI_ISL_1481418, EPI_ISL_1481420, EPI_ISL_1481421, EPI_ISL_1481422, EPI_ISL_1481423, EPI_ISL_1481424, EPI_ISL_1481426, EPI_ISL_1481427, EPI_ISL_1481428, EPI_ISL_1481429, EPI_ISL_1481430, EPI_ISL_1481432, EPI_ISL_1481434, EPI_ISL_1481442, EPI_ISL_1481443, EPI_ISL_1481445, EPI_ISL_1481450, EPI_ISL_1481451, EPI_ISL_1481452, EPI_ISL_1481455, EPI_ISL_1481456, EPI_ISL_1481457, EPI_ISL_1481459, EPI_ISL_1481460, EPI_ISL_1481461, EPI_ISL_1481463, EPI_ISL_1481464, EPI_ISL_1481467, EPI_ISL_1481468, EPI_ISL_1481473, EPI_ISL_1481476, EPI_ISL_1481478, EPI_ISL_1481479, EPI_ISL_1481480, EPI_ISL_1481483, EPI_ISL_1481488, EPI_ISL_1481491, EPI_ISL_1481493, EPI_ISL_1481496, EPI_ISL_1481502, EPI_ISL_1481507, EPI_ISL_1481508, EPI_ISL_1481510, EPI_ISL_1481511, EPI_ISL_1481514, EPI_ISL_1481516, EPI_ISL_1481518, EPI_ISL_1481519, EPI_ISL_1481521, EPI_ISL_1481524, EPI_ISL_1481529, EPI_ISL_1481533, EPI_ISL_1481534, EPI_ISL_1481536, EPI_ISL_1481538, EPI_ISL_1481539, EPI_ISL_1481542, EPI_ISL_1481543, EPI_ISL_1481544, EPI_ISL_1481546 | | | |
| see above | Laboratory Corporation of America | Centers for Disease Control and Prevention Division of Viral Diseases, Pathogen Discovery | Dakota Howard, Dhwani Batra, Peter W. Cook, Kara Moser, Adrian Paskey, Jason Caravas, Benjamin Rambo-Martin, Shatavia Morrison, Christopher Gulvick, Scott Sammons, Yvette Unoarumhi, Darlene Wagner, Matthew Schmerer, Minoo Agarwal, Eyad Almasri, Debbie Boles, Ayla Burns, Nuthawin Charoensri, Oren Cohen, Susan Countryman, Mary Ann Cristobal, Bobbi Croy, Suzanne Dale, Hrushikesh Deshmukh, Amanda Douglas, Vincent Drouillon, Marcia Eisenberg, Howard Engler, Rama Ghatti, Prashant Gupta, Susan Hicks, Jake Humphrey, Lax Iyer, Manoj Jain, Mohan Kolli, Brian Krueger, Tim Kuphal, Stanley Letovsky, Michael Levandoski, Craig Lukasik, Jonathan Meltzer, Brian Norvell, Mindy Nye, Scott Parker, Christos Petropoulos, John Pruitt, Steven Ragan, Scott Ryan, Mike Sapeta, Jana Schroth, Suresh Babu Selvaraju, Goran Stevovic, Amanda Suchanek, Andrea Throop, Lyndon Tilson, Thomas Urban, Joe Voshell, Kimberly Wagner, Jonathan Williams, Mary Williamson, Qian Zeng, Tricia Zwiefelhofer, Clinton R. Paden, Duncan MacCannell |
| EPI_ISL_1481549, EPI_ISL_1481550, EPI_ISL_1481551, EPI_ISL_1481552, EPI_ISL_1481553, EPI_ISL_1481554, EPI_ISL_1481555, EPI_ISL_1481556, EPI_ISL_1481557, EPI_ISL_1481558, EPI_ISL_1481559, EPI_ISL_1481560, EPI_ISL_1481561, EPI_ISL_1481562, EPI_ISL_1481563, EPI_ISL_1481564, EPI_ISL_1481565, EPI_ISL_1481566, EPI_ISL_1481567, EPI_ISL_1481568, EPI_ISL_1481569, EPI_ISL_1481570, EPI_ISL_1481571, EPI_ISL_1481572, EPI_ISL_1481573, EPI_ISL_1481574, EPI_ISL_1481575, EPI_ISL_1481576, EPI_ISL_1481577, EPI_ISL_1481578, EPI_ISL_1481579, EPI_ISL_1481580, EPI_ISL_1481581, EPI_ISL_1481582, EPI_ISL_1481583, EPI_ISL_1481584, EPI_ISL_1481585, EPI_ISL_1481586, EPI_ISL_1481587, EPI_ISL_1481588, EPI_ISL_1481589, EPI_ISL_1481590, EPI_ISL_1481591, EPI_ISL_1481592, EPI_ISL_1481593, EPI_ISL_1481594, EPI_ISL_1481595, EPI_ISL_1481596, EPI_ISL_1481597, EPI_ISL_1481598, EPI_ISL_1481599, EPI_ISL_1481600, EPI_ISL_1481601, EPI_ISL_1481602, EPI_ISL_1481603, EPI_ISL_1481604, EPI_ISL_1481605, EPI_ISL_1481607, EPI_ISL_1481608, EPI_ISL_1481609, EPI_ISL_1481611, EPI_ISL_1481612, EPI_ISL_1481613, EPI_ISL_1481614, EPI_ISL_1481615, EPI_ISL_1481616, EPI_ISL_1481617, EPI_ISL_1481618, EPI_ISL_1481619, EPI_ISL_1481620, EPI_ISL_1481621, EPI_ISL_1481622, EPI_ISL_1481624, EPI_ISL_1481626, EPI_ISL_1481627, EPI_ISL_1481628, EPI_ISL_1481630, EPI_ISL_1481631, EPI_ISL_1481632, EPI_ISL_1481633, EPI_ISL_1481634, EPI_ISL_1481635, EPI_ISL_1481636, EPI_ISL_1481637, EPI_ISL_1481638, EPI_ISL_1481639, EPI_ISL_1481640, EPI_ISL_1481642, EPI_ISL_1481643, EPI_ISL_1481644, EPI_ISL_1481645, EPI_ISL_1481646, EPI_ISL_1481647, EPI_ISL_1481649, EPI_ISL_1481650, EPI_ISL_1481651, EPI_ISL_1481652, EPI_ISL_1481653, EPI_ISL_1481654, EPI_ISL_1481655, EPI_ISL_1481658, EPI_ISL_1481659, EPI_ISL_1481660, EPI_ISL_1481661, EPI_ISL_1481662, EPI_ISL_1481663, EPI_ISL_1481664, EPI_ISL_1481665, EPI_ISL_1481667, EPI_ISL_1481668, EPI_ISL_1481669, EPI_ISL_1481670, EPI_ISL_1481671, EPI_ISL_1481672, EPI_ISL_1481673, EPI_ISL_1481674, EPI_ISL_1481678, EPI_ISL_1481680, EPI_ISL_1481681, EPI_ISL_1481683, EPI_ISL_1481686, EPI_ISL_1481687, EPI_ISL_1481688, EPI_ISL_1481689, EPI_ISL_1481692, EPI_ISL_1481693, EPI_ISL_1481695, EPI_ISL_1481696, EPI_ISL_1481697, EPI_ISL_1481700, EPI_ISL_1481701, EPI_ISL_1481702, EPI_ISL_1481703, EPI_ISL_1481704, EPI_ISL_1481705, EPI_ISL_1481707, EPI_ISL_1481709, EPI_ISL_1481710, EPI_ISL_1481711, EPI_ISL_1481712, EPI_ISL_1481714, EPI_ISL_1481715, EPI_ISL_1481716, EPI_ISL_1481717, EPI_ISL_1481718, EPI_ISL_1481719, EPI_ISL_1481720, EPI_ISL_1481721, EPI_ISL_1481722, EPI_ISL_1481723, EPI_ISL_1481724, EPI_ISL_1481725, EPI_ISL_1481726, EPI_ISL_1481727, EPI_ISL_1481728, EPI_ISL_1481730, EPI_ISL_1481731, EPI_ISL_1481732, EPI_ISL_1481734, EPI_ISL_1481735, EPI_ISL_1481736, EPI_ISL_1481737, EPI_ISL_1481738, EPI_ISL_1481739, EPI_ISL_1481740, EPI_ISL_1481741, EPI_ISL_1481742, EPI_ISL_1481743, EPI_ISL_1481744, EPI_ISL_1481745, EPI_ISL_1481746, EPI_ISL_1481747, EPI_ISL_1481748, EPI_ISL_1481749, EPI_ISL_1481750, EPI_ISL_1481751, EPI_ISL_1481752, EPI_ISL_1481753, EPI_ISL_1481754, EPI_ISL_1481755, EPI_ISL_1481756, EPI_ISL_1481757, EPI_ISL_1481758, EPI_ISL_1481759, EPI_ISL_1481760, EPI_ISL_1481761, EPI_ISL_1481762, EPI_ISL_1481763, EPI_ISL_1481764, EPI_ISL_1481765, EPI_ISL_1481766, EPI_ISL_1481767, EPI_ISL_1481768, EPI_ISL_1481769, EPI_ISL_1481770, EPI_ISL_1481771, EPI_ISL_1481772, EPI_ISL_1481774, EPI_ISL_1481775, EPI_ISL_1481776, EPI_ISL_1481777, EPI_ISL_1481779, EPI_ISL_1481780, EPI_ISL_1481782, EPI_ISL_1481783, EPI_ISL_1481784, EPI_ISL_1481785, EPI_ISL_1481786, EPI_ISL_1481787, EPI_ISL_1481789, EPI_ISL_1481790, EPI_ISL_1481791, EPI_ISL_1481792, EPI_ISL_1481793, EPI_ISL_1481794, EPI_ISL_1481795, EPI_ISL_1481796, EPI_ISL_1481797, EPI_ISL_1481798, EPI_ISL_1481799, EPI_ISL_1481800, EPI_ISL_1481801, EPI_ISL_1481802, EPI_ISL_1481804, EPI_ISL_1481805, EPI_ISL_1481806, EPI_ISL_1481808, EPI_ISL_1481809, EPI_ISL_1481810, EPI_ISL_1481811, EPI_ISL_1481812, EPI_ISL_1481813, EPI_ISL_1481814, EPI_ISL_1481815, EPI_ISL_1481816, EPI_ISL_1481817, EPI_ISL_1481818, EPI_ISL_1481819, EPI_ISL_1481820, EPI_ISL_1481821, EPI_ISL_1481823, EPI_ISL_1481824, EPI_ISL_1481826, EPI_ISL_1481829, EPI_ISL_1481830, EPI_ISL_1481831, EPI_ISL_1481832, EPI_ISL_1481834, EPI_ISL_1481835, EPI_ISL_1481836, EPI_ISL_1481837, EPI_ISL_1481838, EPI_ISL_1481839, EPI_ISL_1481840, EPI_ISL_1481843, EPI_ISL_1481844, EPI_ISL_1481845, EPI_ISL_1481846, EPI_ISL_1481849, EPI_ISL_1481850, EPI_ISL_1481851, EPI_ISL_1481852, EPI_ISL_1481853, EPI_ISL_1481854, EPI_ISL_1481855, EPI_ISL_1481857, EPI_ISL_1481858, EPI_ISL_1481860, EPI_ISL_1481861, EPI_ISL_1481862, EPI_ISL_1481863, EPI_ISL_1481864, EPI_ISL_1481865, EPI_ISL_1481866, EPI_ISL_1481867, EPI_ISL_1481869, EPI_ISL_1481870, EPI_ISL_1481871, EPI_ISL_1481872, EPI_ISL_1481873, EPI_ISL_1481874, EPI_ISL_1481875, EPI_ISL_1481877, EPI_ISL_1481878, EPI_ISL_1481879, EPI_ISL_1481880, EPI_ISL_1481881, EPI_ISL_1481882, EPI_ISL_1481883, EPI_ISL_1481884, EPI_ISL_1481886, EPI_ISL_1481887, EPI_ISL_1481888, EPI_ISL_1481889, EPI_ISL_1481890, EPI_ISL_1481892, EPI_ISL_1481893, EPI_ISL_1481894, EPI_ISL_1481895, EPI_ISL_1481896, EPI_ISL_1481897, EPI_ISL_1481899, EPI_ISL_1481900, EPI_ISL_1481901, EPI_ISL_1481902, EPI_ISL_1481903, EPI_ISL_1481904, EPI_ISL_1481905, EPI_ISL_1481906, EPI_ISL_1481907, EPI_ISL_1481908, EPI_ISL_1481909, EPI_ISL_1481910, EPI_ISL_1481911, EPI_ISL_1481912, EPI_ISL_1481913, EPI_ISL_1481916, EPI_ISL_1481917, EPI_ISL_1481918, EPI_ISL_1481919, EPI_ISL_1481920, EPI_ISL_1481922, EPI_ISL_1481923, EPI_ISL_1481924, EPI_ISL_1481926, EPI_ISL_1481927, EPI_ISL_1481928, EPI_ISL_1481929, EPI_ISL_1481930, EPI_ISL_1481931, EPI_ISL_1481932, EPI_ISL_1481933, EPI_ISL_1481934, EPI_ISL_1481935, EPI_ISL_1481936, EPI_ISL_1481937, EPI_ISL_1481938, EPI_ISL_1481939, EPI_ISL_1481940, EPI_ISL_1481941, EPI_ISL_1481942, EPI_ISL_1481943, EPI_ISL_1481944, EPI_ISL_1481945, EPI_ISL_1481946, EPI_ISL_1481947, EPI_ISL_1481948, EPI_ISL_1481949, EPI_ISL_1481950, EPI_ISL_1481952, EPI_ISL_1481953, EPI_ISL_1481954, EPI_ISL_1481955, EPI_ISL_1481956, EPI_ISL_1481957, EPI_ISL_1481958, EPI_ISL_1481959, EPI_ISL_1481960, EPI_ISL_1481961, EPI_ISL_1481962, EPI_ISL_1481963, EPI_ISL_1481964, EPI_ISL_1481965, EPI_ISL_1481966, EPI_ISL_1481967, EPI_ISL_1481968, EPI_ISL_1481969, EPI_ISL_1481970, EPI_ISL_1481971, EPI_ISL_1481972, EPI_ISL_1481973, EPI_ISL_1481974, EPI_ISL_1481975, EPI_ISL_1481976, EPI_ISL_1481977, EPI_ISL_1481978, EPI_ISL_1481979, EPI_ISL_1481980, EPI_ISL_1481981, EPI_ISL_1481984, EPI_ISL_1481985, EPI_ISL_1481986, EPI_ISL_1481987, EPI_ISL_1481988, EPI_ISL_1481989, EPI_ISL_1481990, EPI_ISL_1481991, EPI_ISL_1481992, EPI_ISL_1481993, EPI_ISL_1481994, EPI_ISL_1481995, EPI_ISL_1481996, EPI_ISL_1481998, EPI_ISL_1482000, EPI_ISL_1482001, EPI_ISL_1482002, EPI_ISL_1482003, EPI_ISL_1482004, EPI_ISL_1482005, EPI_ISL_1482006, EPI_ISL_1482008, EPI_ISL_1482009, EPI_ISL_1482010, EPI_ISL_1482012, EPI_ISL_1482013, EPI_ISL_1482014, EPI_ISL_1482015, EPI_ISL_1482016, EPI_ISL_1482017, EPI_ISL_1482018, EPI_ISL_1482019, EPI_ISL_1482020, EPI_ISL_1482021, EPI_ISL_1482022, EPI_ISL_1482024, EPI_ISL_1482025, EPI_ISL_1482026, EPI_ISL_1482027, EPI_ISL_1482028, EPI_ISL_1482029, EPI_ISL_1482030, EPI_ISL_1482031, EPI_ISL_1482032, EPI_ISL_1482033, EPI_ISL_1482035, EPI_ISL_1482036, EPI_ISL_1482038, EPI_ISL_1482039, EPI_ISL_1482041, EPI_ISL_1482042, EPI_ISL_1482043, EPI_ISL_1482044, EPI_ISL_1482045, EPI_ISL_1482046, EPI_ISL_1482047, EPI_ISL_1482048, EPI_ISL_1482049, EPI_ISL_1482050, EPI_ISL_1482051, EPI_ISL_1482052, EPI_ISL_1482053, EPI_ISL_1482054, EPI_ISL_1482055, EPI_ISL_1482056, EPI_ISL_1482057, EPI_ISL_1482058, EPI_ISL_1482059, EPI_ISL_1482060, EPI_ISL_1482061, EPI_ISL_1482062, EPI_ISL_1482064, EPI_ISL_1482065, EPI_ISL_1482066, EPI_ISL_1482067, EPI_ISL_1482069, EPI_ISL_1482073, EPI_ISL_1482074, EPI_ISL_1482075, EPI_ISL_1482077, EPI_ISL_1482079, EPI_ISL_1482080, EPI_ISL_1482081, EPI_ISL_1482082, EPI_ISL_1482083, EPI_ISL_1482084, EPI_ISL_1482085, EPI_ISL_1482086, EPI_ISL_1482087, EPI_ISL_1482088, EPI_ISL_1482089, EPI_ISL_1482090, EPI_ISL_1482091, EPI_ISL_1482092, EPI_ISL_1482093, EPI_ISL_1482095, EPI_ISL_1482098, EPI_ISL_1482099, EPI_ISL_1482100, EPI_ISL_1482101, EPI_ISL_1482102, EPI_ISL_1482104, EPI_ISL_1482105, EPI_ISL_1482106, EPI_ISL_1482107, | | | |

| | | | |
|---|---|---|---|
| EPI_ISL_1482108, EPI_ISL_1482109, EPI_ISL_1482110, EPI_ISL_1482112, EPI_ISL_1482114, EPI_ISL_1482115, EPI_ISL_1482116, EPI_ISL_1482117, EPI_ISL_1482118, EPI_ISL_1482119, EPI_ISL_1482120, EPI_ISL_1482122, EPI_ISL_1482123, EPI_ISL_1482124, EPI_ISL_1482125, EPI_ISL_1482126, EPI_ISL_1482127, EPI_ISL_1482128, EPI_ISL_1482129, EPI_ISL_1482130, EPI_ISL_1482131, EPI_ISL_1482132, EPI_ISL_1482133, EPI_ISL_1482134, EPI_ISL_1482135, EPI_ISL_1482136, EPI_ISL_1482137, EPI_ISL_1482138, EPI_ISL_1482139, EPI_ISL_1482140, EPI_ISL_1482141, EPI_ISL_1482142, EPI_ISL_1482143, EPI_ISL_1482144, EPI_ISL_1482145, EPI_ISL_1482146, EPI_ISL_1482147, EPI_ISL_1482148, EPI_ISL_1482149, EPI_ISL_1482150, EPI_ISL_1482151, EPI_ISL_1482152, EPI_ISL_1482153, EPI_ISL_1482154, EPI_ISL_1482155, EPI_ISL_1482156, EPI_ISL_1482157, EPI_ISL_1482158, EPI_ISL_1482159, EPI_ISL_1482160, EPI_ISL_1482161, EPI_ISL_1482162, EPI_ISL_1482163, EPI_ISL_1482164, EPI_ISL_1482165, EPI_ISL_1482166, EPI_ISL_1482167, EPI_ISL_1482168, EPI_ISL_1482169, EPI_ISL_1482170, EPI_ISL_1482172, EPI_ISL_1482173, EPI_ISL_1482174, EPI_ISL_1482175, EPI_ISL_1482176, EPI_ISL_1482177, EPI_ISL_1482178, EPI_ISL_1482179, EPI_ISL_1482180, EPI_ISL_1482181, EPI_ISL_1482182, EPI_ISL_1482183, EPI_ISL_1482184, EPI_ISL_1482185, EPI_ISL_1482186, EPI_ISL_1482187, EPI_ISL_1482188, EPI_ISL_1482193, EPI_ISL_1482194, EPI_ISL_1482195, EPI_ISL_1482196, EPI_ISL_1482197, EPI_ISL_1482198, EPI_ISL_1482199, EPI_ISL_1482200, EPI_ISL_1482201, EPI_ISL_1482203, EPI_ISL_1482204, EPI_ISL_1482205, EPI_ISL_1482206, EPI_ISL_1482207, EPI_ISL_1482208, EPI_ISL_1482209, EPI_ISL_1482210, EPI_ISL_1482211, EPI_ISL_1482214, EPI_ISL_1482215, EPI_ISL_1482216, EPI_ISL_1482217, EPI_ISL_1482218, EPI_ISL_1482220, EPI_ISL_1482221, EPI_ISL_1482222, EPI_ISL_1482223, EPI_ISL_1482224, EPI_ISL_1482225, EPI_ISL_1482226, EPI_ISL_1482227, EPI_ISL_1482228, EPI_ISL_1482229, EPI_ISL_1482230, EPI_ISL_1482231, EPI_ISL_1482232, EPI_ISL_1482233, EPI_ISL_1482234, EPI_ISL_1482235, EPI_ISL_1482236, EPI_ISL_1482237, EPI_ISL_1482238, EPI_ISL_1482239, EPI_ISL_1482241, EPI_ISL_1482242, EPI_ISL_1482243, EPI_ISL_1482244, EPI_ISL_1482245, EPI_ISL_1482246, EPI_ISL_1482247, EPI_ISL_1482249, EPI_ISL_1482250, EPI_ISL_1482251, EPI_ISL_1482252, EPI_ISL_1482253, EPI_ISL_1482254, EPI_ISL_1482255, EPI_ISL_1482256, EPI_ISL_1482258, EPI_ISL_1482259, EPI_ISL_1482260, EPI_ISL_1482261, EPI_ISL_1482263, EPI_ISL_1482264, EPI_ISL_1482265, EPI_ISL_1482267, EPI_ISL_1482268, EPI_ISL_1482269, EPI_ISL_1482270, EPI_ISL_1482271, EPI_ISL_1482272, EPI_ISL_1482273 | | | |
| see above | Fulgent Genetics | Centers for Disease Control and Prevention Division of Viral Diseases, Pathogen Discovery | Dakota Howard, Dhwani Batra, Peter W. Cook, Kara Moser, Adrian Paskey, Jason Caravas, Benjamin Rambo-Martin, Shatavia Morrison, Christopher Gulvick, Scott Sammons, Yvette Unoarumhi, Darlene Wagner, Matthew Schmerer, Harry Gao, Mickey Li, John Gao, Joseph Fierro, Benafsh Sapra, Becky Tsai, Yan Meng, Doreen Ng, James Xie, Clinton R. Paden, Duncan MacCannell |
| EPI_ISL_1482274, EPI_ISL_1482275, EPI_ISL_1482276 | SUNY UPSTATE MEDICAL UNIVERSITY | Wadsworth Center, New York State Department of Health | Kirsten St. George, Daryl M. Lamson, Alexis Russell, Matthew Shudt, Melissa A Leisner, Jonathan Plitnick, Catharine Prussing, Navjot Singh, John Kelly, Erasmus Schneider, Erica Lasek-Nesselquist |
| EPI_ISL_1482277 | Columbia University Irving Medical Center | Wadsworth Center, New York State Department of Health | Kirsten St. George, Daryl M. Lamson, Alexis Russell, Matthew Shudt, Melissa A Leisner, Jonathan Plitnick, Catharine Prussing, Navjot Singh, John Kelly, Erasmus Schneider, Erica Lasek-Nesselquist |
| EPI_ISL_1482278, EPI_ISL_1482279, EPI_ISL_1482280, EPI_ISL_1482281, EPI_ISL_1482282, EPI_ISL_1482283, EPI_ISL_1482284, EPI_ISL_1482285, EPI_ISL_1482286 | URMC LABS | Wadsworth Center, New York State Department of Health | Kirsten St. George, Daryl M. Lamson, Alexis Russell, Matthew Shudt, Melissa A Leisner, Jonathan Plitnick, Catharine Prussing, Navjot Singh, John Kelly, Erasmus Schneider, Erica Lasek-Nesselquist |
| EPI_ISL_1482287, EPI_ISL_1482288, EPI_ISL_1482289, EPI_ISL_1482290, EPI_ISL_1482291, EPI_ISL_1482292, EPI_ISL_1482293, EPI_ISL_1482294, EPI_ISL_1482295 | NYC Pandemic Response Lab | Wadsworth Center, New York State Department of Health | Kirsten St. George, Daryl M. Lamson, Alexis Russell, Matthew Shudt, Melissa A Leisner, Jonathan Plitnick, Catharine Prussing, Navjot Singh, John Kelly, Erasmus Schneider, Erica Lasek-Nesselquist |
| EPI_ISL_1482296, EPI_ISL_1482297, EPI_ISL_1482298, EPI_ISL_1482299, EPI_ISL_1482300, EPI_ISL_1482301, EPI_ISL_1482302, EPI_ISL_1482303, EPI_ISL_1482304, EPI_ISL_1482305, EPI_ISL_1482306, EPI_ISL_1482307, EPI_ISL_1482308, EPI_ISL_1482309, EPI_ISL_1482310, EPI_ISL_1482311, EPI_ISL_1482312, EPI_ISL_1482313, EPI_ISL_1482314, EPI_ISL_1482315, EPI_ISL_1482316, EPI_ISL_1482317, EPI_ISL_1482318, EPI_ISL_1482319, EPI_ISL_1482320, EPI_ISL_1482321, EPI_ISL_1482322, EPI_ISL_1482323, EPI_ISL_1482324, EPI_ISL_1482325, EPI_ISL_1482326, EPI_ISL_1482327, EPI_ISL_1482328, EPI_ISL_1482329, EPI_ISL_1482330, EPI_ISL_1482331, EPI_ISL_1482332 | | | |
| see above | URMC LABS | Wadsworth Center, New York State Department of Health | Kirsten St. George, Daryl M. Lamson, Alexis Russell, Matthew Shudt, Melissa A Leisner, Jonathan Plitnick, Catharine Prussing, Navjot Singh, John Kelly, Erasmus Schneider, Erica Lasek-Nesselquist |
| EPI_ISL_1482333, EPI_ISL_1482334, EPI_ISL_1482335 | GLENS FALLS HOSPITAL LABORATORY | Wadsworth Center, New York State Department of Health | Kirsten St. George, Daryl M. Lamson, Alexis Russell, Matthew Shudt, Melissa A Leisner, Jonathan Plitnick, Catharine Prussing, Navjot Singh, John Kelly, Erasmus Schneider, Erica Lasek-Nesselquist |
| EPI_ISL_1482336, EPI_ISL_1482337, EPI_ISL_1482338, EPI_ISL_1482339, EPI_ISL_1482340, EPI_ISL_1482341, EPI_ISL_1482342, EPI_ISL_1482343, EPI_ISL_1482344, EPI_ISL_1482345 | MONTEFIORE NEW ROCHELLE HOSPITAL LABORATORY | Wadsworth Center, New York State Department of Health | Kirsten St. George, Daryl M. Lamson, Alexis Russell, Matthew Shudt, Melissa A Leisner, Jonathan Plitnick, Catharine Prussing, Navjot Singh, John Kelly, Erasmus Schneider, Erica Lasek-Nesselquist |
| EPI_ISL_1482346, EPI_ISL_1482347, EPI_ISL_1482348, EPI_ISL_1482349, EPI_ISL_1482350, EPI_ISL_1482351, EPI_ISL_1482352, EPI_ISL_1482353 | NORTH SHORE UNIVERSITY HOSPITAL | Wadsworth Center, New York State Department of Health | Kirsten St. George, Daryl M. Lamson, Alexis Russell, Matthew Shudt, Melissa A Leisner, Jonathan Plitnick, Catharine Prussing, Navjot Singh, John Kelly, Erasmus Schneider, Erica Lasek-Nesselquist |
| EPI_ISL_1482394, EPI_ISL_1482395, EPI_ISL_1482396, EPI_ISL_1482397, EPI_ISL_1482398, EPI_ISL_1482399, EPI_ISL_1482400, EPI_ISL_1482401, EPI_ISL_1482402, EPI_ISL_1482403, EPI_ISL_1482404, EPI_ISL_1482405 | | | |
| see above | WESTCHESTER MEDICAL CENTER | Wadsworth Center, New York State Department of Health | Kirsten St. George, Daryl M. Lamson, Alexis Russell, Matthew Shudt, Melissa A Leisner, Jonathan Plitnick, Catharine Prussing, Navjot Singh, John Kelly, Erasmus Schneider, Erica Lasek-Nesselquist |
| EPI_ISL_1482406, EPI_ISL_1482407 | SUNY UPSTATE MEDICAL UNIVERSITY | Wadsworth Center, New York State Department of Health | Kirsten St. George, Daryl M. Lamson, Alexis Russell, Matthew Shudt, Melissa A Leisner, Jonathan Plitnick, Catharine Prussing, Navjot Singh, John Kelly, Erasmus Schneider, Erica Lasek-Nesselquist |
| EPI_ISL_1482408, EPI_ISL_1482409 | BOSTON HEART DIAGNOSTICS CORP | Wadsworth Center, New York State Department of Health | Kirsten St. George, Daryl M. Lamson, Alexis Russell, Matthew Shudt, Melissa A Leisner, Jonathan Plitnick, Catharine Prussing, Navjot Singh, John Kelly, Erasmus Schneider, Erica Lasek-Nesselquist |
| EPI_ISL_1482410 | Columbia University Irving Medical Center | Wadsworth Center, New York State Department of Health | Kirsten St. George, Daryl M. Lamson, Alexis Russell, Matthew Shudt, Melissa A Leisner, Jonathan Plitnick, Catharine Prussing, Navjot Singh, John Kelly, Erasmus Schneider, Erica Lasek-Nesselquist |
| EPI_ISL_1482411, EPI_ISL_1482412, EPI_ISL_1482413, EPI_ISL_1482414, EPI_ISL_1482415, EPI_ISL_1482416, EPI_ISL_1482417, EPI_ISL_1482418, EPI_ISL_1482419, EPI_ISL_1482420, EPI_ISL_1482421, EPI_ISL_1482422, EPI_ISL_1482423, EPI_ISL_1482424, EPI_ISL_1482425, EPI_ISL_1482426, EPI_ISL_1482427, EPI_ISL_1482428, EPI_ISL_1482429, EPI_ISL_1482430, EPI_ISL_1482431, EPI_ISL_1482432, EPI_ISL_1482433, EPI_ISL_1482434, EPI_ISL_1482435, EPI_ISL_1482436, EPI_ISL_1482437, EPI_ISL_1482438, EPI_ISL_1482439, EPI_ISL_1482440, EPI_ISL_1482441, EPI_ISL_1482442, EPI_ISL_1482443, EPI_ISL_1482444, EPI_ISL_1482445 | | | |
| see above | WESTCHESTER MEDICAL CENTER | Wadsworth Center, New York State Department of Health | Kirsten St. George, Daryl M. Lamson, Alexis Russell, Matthew Shudt, Melissa A Leisner, Jonathan Plitnick, Catharine Prussing, Navjot Singh, John Kelly, Erasmus Schneider, Erica Lasek-Nesselquist |
| EPI_ISL_1482446, EPI_ISL_1482447, EPI_ISL_1482448, EPI_ISL_1482449, EPI_ISL_1482450, EPI_ISL_1482451, EPI_ISL_1482452, EPI_ISL_1482453 | BOSTON HEART DIAGNOSTICS CORP | Wadsworth Center, New York State Department of Health | Kirsten St. George, Daryl M. Lamson, Alexis Russell, Matthew Shudt, Melissa A Leisner, Jonathan Plitnick, Catharine Prussing, Navjot Singh, John Kelly, Erasmus Schneider, Erica Lasek-Nesselquist |
| EPI_ISL_1482454 | ST. FRANCIS HOSPITAL LABORATORY | Wadsworth Center, New York State Department of Health | Kirsten St. George, Daryl M. Lamson, Alexis Russell, Matthew Shudt, Melissa A Leisner, Jonathan Plitnick, Catharine Prussing, Navjot Singh, John Kelly, Erasmus Schneider, Erica Lasek-Nesselquist |
| EPI_ISL_1482455, EPI_ISL_1482456, EPI_ISL_1482457, EPI_ISL_1482458, EPI_ISL_1482459, EPI_ISL_1482460, EPI_ISL_1482461, EPI_ISL_1482462, EPI_ISL_1482463, EPI_ISL_1482464, EPI_ISL_1482465, EPI_ISL_1482466, EPI_ISL_1482467, EPI_ISL_1482468, EPI_ISL_1482469 | | | |
| see above | ALBANY MEDICAL CENTER | Wadsworth Center, New York State Department of Health | Kirsten St. George, Daryl M. Lamson, Alexis Russell, Matthew Shudt, Melissa A Leisner, Jonathan Plitnick, Catharine Prussing, Navjot Singh, John Kelly, Erasmus Schneider, Erica Lasek-Nesselquist |
| EPI_ISL_1482470 | Good Samaritan Hospital Laboratory | Wadsworth Center, New York State Department of Health | Kirsten St. George, Daryl M. Lamson, Alexis Russell, Matthew Shudt, Melissa A Leisner, Jonathan Plitnick, Catharine Prussing, Navjot Singh, John Kelly, Erasmus Schneider, Erica Lasek-Nesselquist |
| EPI_ISL_1482471, EPI_ISL_1482472, EPI_ISL_1482473, EPI_ISL_1482474, EPI_ISL_1482475, EPI_ISL_1482476, EPI_ISL_1482477, EPI_ISL_1482478, EPI_ISL_1482479, EPI_ISL_1482480, EPI_ISL_1482481, EPI_ISL_1482482, EPI_ISL_1482483, EPI_ISL_1482484, EPI_ISL_1482485, EPI_ISL_1482486, EPI_ISL_1482487, EPI_ISL_1482488, EPI_ISL_1482489, EPI_ISL_1482490, EPI_ISL_1482491, EPI_ISL_1482492, EPI_ISL_1482493, EPI_ISL_1482494, EPI_ISL_1482495, EPI_ISL_1482496, EPI_ISL_1482498, EPI_ISL_1482499, EPI_ISL_1482500, EPI_ISL_1482501, EPI_ISL_1482502, EPI_ISL_1482503, EPI_ISL_1482504, EPI_ISL_1482505, EPI_ISL_1482506, EPI_ISL_1482507, EPI_ISL_1482508, EPI_ISL_1482509, EPI_ISL_1482510, EPI_ISL_1482511, EPI_ISL_1482512, EPI_ISL_1482513, EPI_ISL_1482514, EPI_ISL_1482515, EPI_ISL_1482516, EPI_ISL_1482517, EPI_ISL_1482518, EPI_ISL_1482519, EPI_ISL_1482520, EPI_ISL_1482521, EPI_ISL_1482522, EPI_ISL_1482524, EPI_ISL_1482525, EPI_ISL_1482526, EPI_ISL_1482527, EPI_ISL_1482528, EPI_ISL_1482529, EPI_ISL_1482530, EPI_ISL_1482531, EPI_ISL_1482532, EPI_ISL_1482534, EPI_ISL_1482536 | | | |
| see above | Wyoming Public Health Laboratory | Wyoming Public Health Laboratory | Jim Mildenberger, Wanda Manley, Noah Hull, Taylor Fearing, Lynette Gumbleton, Channing Weber, Ashley Norberg, Chayse Rowley, Marley Goetz, Brian Dominguez, Elliot Thomasson, Cari Sloma, and Rob Christensen |
| EPI_ISL_1482539, EPI_ISL_1482540, EPI_ISL_1482541, EPI_ISL_1482542, EPI_ISL_1482543, EPI_ISL_1482544, EPI_ISL_1482545, EPI_ISL_1482546, EPI_ISL_1482547, EPI_ISL_1482548, EPI_ISL_1482549, EPI_ISL_1482550, EPI_ISL_1482551, EPI_ISL_1482552, EPI_ISL_1482553, EPI_ISL_1482554, EPI_ISL_1482555 | | | |
| see above | WESTCHESTER MEDICAL CENTER | Wadsworth Center, New York State Department of Health | Kirsten St. George, Daryl M. Lamson, Alexis Russell, Matthew Shudt, Melissa A Leisner, Jonathan Plitnick, Catharine Prussing, Navjot Singh, John Kelly, Erasmus Schneider, Erica Lasek-Nesselquist |
| EPI_ISL_1482556, EPI_ISL_1482557, EPI_ISL_1482558, EPI_ISL_1482559, EPI_ISL_1482560, EPI_ISL_1482561, EPI_ISL_1482562, EPI_ISL_1482563, EPI_ISL_1482564, EPI_ISL_1482565, EPI_ISL_1482566, EPI_ISL_1482567, EPI_ISL_1482568, EPI_ISL_1482569, EPI_ISL_1482570, EPI_ISL_1482571, EPI_ISL_1482572 | | | |
| see above | NORTHWELL HEALTH LABORATORIES | Wadsworth Center, New York State Department of Health | Kirsten St. George, Daryl M. Lamson, Alexis Russell, Matthew Shudt, Melissa A Leisner, Jonathan Plitnick, Catharine Prussing, Navjot Singh, John Kelly, Erasmus Schneider, Erica Lasek-Nesselquist |

| | | | |
|---|---|---|---|
| EPI_ISL_1482573, EPI_ISL_1482574, EPI_ISL_1482575, EPI_ISL_1482576, EPI_ISL_1482577, EPI_ISL_1482578, EPI_ISL_1482579 | WHITE PLAINS HOSPITAL CENTER LABORATORY | Wadsworth Center, New York State Department of Health | Kirsten St. George, Daryl M. Lamson, Alexis Russell, Matthew Shudt, Melissa A Leisner, Jonathan Plitnick, Catharine Prussing, Navjot Singh, John Kelly, Erasmus Schneider, Erica Lasek-Nesselquist |
| EPI_ISL_1482580, EPI_ISL_1482581, EPI_ISL_1482582, EPI_ISL_1482583, EPI_ISL_1482584, EPI_ISL_1482585, EPI_ISL_1482586, EPI_ISL_1482587, EPI_ISL_1482588, EPI_ISL_1482589, EPI_ISL_1482590, EPI_ISL_1482591, EPI_ISL_1482592, EPI_ISL_1482593, EPI_ISL_1482594, EPI_ISL_1482595, EPI_ISL_1482596, EPI_ISL_1482597, EPI_ISL_1482598, EPI_ISL_1482599, EPI_ISL_1482600, EPI_ISL_1482601, EPI_ISL_1482602, EPI_ISL_1482603, EPI_ISL_1482604, EPI_ISL_1482605, EPI_ISL_1482606, EPI_ISL_1482607, EPI_ISL_1482608, EPI_ISL_1482609, EPI_ISL_1482610, EPI_ISL_1482611, EPI_ISL_1482612, EPI_ISL_1482613, EPI_ISL_1482614, EPI_ISL_1482615, EPI_ISL_1482616 | | | |
| see above | NORTHWELL HEALTH LABORATORIES | Wadsworth Center, New York State Department of Health | Kirsten St. George, Daryl M. Lamson, Alexis Russell, Matthew Shudt, Melissa A Leisner, Jonathan Plitnick, Catharine Prussing, Navjot Singh, John Kelly, Erasmus Schneider, Erica Lasek-Nesselquist |
| EPI_ISL_1482617, EPI_ISL_1482618, EPI_ISL_1482619, EPI_ISL_1482620, EPI_ISL_1482621, EPI_ISL_1482622, EPI_ISL_1482623, EPI_ISL_1482624 | WESTCHESTER MEDICAL CENTER | Wadsworth Center, New York State Department of Health | Kirsten St. George, Daryl M. Lamson, Alexis Russell, Matthew Shudt, Melissa A Leisner, Jonathan Plitnick, Catharine Prussing, Navjot Singh, John Kelly, Erasmus Schneider, Erica Lasek-Nesselquist |
| EPI_ISL_1482625, EPI_ISL_1482626 | URMC LABS | Wadsworth Center, New York State Department of Health | Kirsten St. George, Daryl M. Lamson, Alexis Russell, Matthew Shudt, Melissa A Leisner, Jonathan Plitnick, Catharine Prussing, Navjot Singh, John Kelly, Erasmus Schneider, Erica Lasek-Nesselquist |
| EPI_ISL_1482627, EPI_ISL_1482629, EPI_ISL_1482630, EPI_ISL_1482631 | Hospital Sharp | Microbial Genomics Laboratory | Bruno Gomez-Gil, Julissa Enciso-Ibarra, Alejandra Garcia-Gasca, Daniel Fregoso-Rueda |
| EPI_ISL_1482632, EPI_ISL_1482634, EPI_ISL_1482635, EPI_ISL_1482638, EPI_ISL_1482639, EPI_ISL_1482640, EPI_ISL_1482641, EPI_ISL_1482642, EPI_ISL_1482643, EPI_ISL_1482644, EPI_ISL_1482645, EPI_ISL_1482646 | | | |
| see above | DC Public Health Lab/ Dept. of Forensic Sciences | DC Public Health Lab/ Dept. of Forensic Sciences | Janis Doss, Scott Nguyen, Elizabeth Zelaya, Sarah Scott, Connie Maza, Monica Mann, Brittany Hamilton, David Payne, Jocelyn Hauser |
| EPI_ISL_1482647 | Oregon State Public Health Laboratory | Oregon State Public Health Laboratory | Rafia Razzaque, Eugene Yeboah, Vanda Makris, Laura Tsaknaridis, John Fontana and Shane Sevey |
| EPI_ISL_1482648, EPI_ISL_1482649, EPI_ISL_1482651, EPI_ISL_1482652, EPI_ISL_1482653, EPI_ISL_1482654, EPI_ISL_1482655, EPI_ISL_1482656, EPI_ISL_1482657, EPI_ISL_1482658, EPI_ISL_1482659, EPI_ISL_1482660, EPI_ISL_1482661, EPI_ISL_1482662, EPI_ISL_1482663, EPI_ISL_1482664, EPI_ISL_1482666, EPI_ISL_1482667, EPI_ISL_1482668, EPI_ISL_1482669, EPI_ISL_1482670, EPI_ISL_1482672, EPI_ISL_1482673, EPI_ISL_1482674, EPI_ISL_1482675, EPI_ISL_1482676, EPI_ISL_1482677, EPI_ISL_1482678, EPI_ISL_1482679, EPI_ISL_1482680, EPI_ISL_1482681, EPI_ISL_1482682, EPI_ISL_1482684, EPI_ISL_1482685, EPI_ISL_1482687, EPI_ISL_1482688, EPI_ISL_1482689, EPI_ISL_1482690, EPI_ISL_1482691, EPI_ISL_1482692, EPI_ISL_1482693, EPI_ISL_1482694, EPI_ISL_1482695, EPI_ISL_1482696, EPI_ISL_1482697, EPI_ISL_1482698, EPI_ISL_1482699, EPI_ISL_1482701, EPI_ISL_1482702, EPI_ISL_1482703, EPI_ISL_1482704, EPI_ISL_1482707, EPI_ISL_1482708, EPI_ISL_1482710, EPI_ISL_1482711, EPI_ISL_1482712, EPI_ISL_1482713, EPI_ISL_1482714, EPI_ISL_1482715, EPI_ISL_1482716, EPI_ISL_1482717, EPI_ISL_1482718, EPI_ISL_1482719, EPI_ISL_1482720, EPI_ISL_1482722, EPI_ISL_1482723, EPI_ISL_1482724, EPI_ISL_1482725, EPI_ISL_1482727, EPI_ISL_1482729, EPI_ISL_1482730, EPI_ISL_1482731, EPI_ISL_1482733, EPI_ISL_1482734, EPI_ISL_1482735, EPI_ISL_1482736, EPI_ISL_1482737, EPI_ISL_1482738, EPI_ISL_1482740, EPI_ISL_1482741, EPI_ISL_1482742, EPI_ISL_1482743, EPI_ISL_1482744, EPI_ISL_1482745, EPI_ISL_1482746, EPI_ISL_1482747, EPI_ISL_1482748, EPI_ISL_1482749, EPI_ISL_1482751, EPI_ISL_1482752, EPI_ISL_1482755, EPI_ISL_1482756, EPI_ISL_1482757, EPI_ISL_1482758, EPI_ISL_1482759, EPI_ISL_1482760, EPI_ISL_1482762, EPI_ISL_1482764, EPI_ISL_1482765, EPI_ISL_1482767, EPI_ISL_1482768, EPI_ISL_1482769, EPI_ISL_1482770, EPI_ISL_1482772, EPI_ISL_1482773, EPI_ISL_1482774, EPI_ISL_1482775, EPI_ISL_1482776, EPI_ISL_1482777, EPI_ISL_1482779, EPI_ISL_1482781, EPI_ISL_1482782, EPI_ISL_1482783, EPI_ISL_1482784, EPI_ISL_1482785, EPI_ISL_1482786, EPI_ISL_1482787, EPI_ISL_1482788, EPI_ISL_1482791, EPI_ISL_1482792, EPI_ISL_1482793, EPI_ISL_1482794, EPI_ISL_1482795, EPI_ISL_1482796, EPI_ISL_1482797, EPI_ISL_1482798, EPI_ISL_1482799, EPI_ISL_1482800, EPI_ISL_1482801, EPI_ISL_1482802, EPI_ISL_1482803, EPI_ISL_1482804, EPI_ISL_1482805, EPI_ISL_1482806, EPI_ISL_1482807, EPI_ISL_1482809, EPI_ISL_1482810, EPI_ISL_1482811, EPI_ISL_1482812, EPI_ISL_1482813, EPI_ISL_1482814, EPI_ISL_1482817, EPI_ISL_1482818, EPI_ISL_1482820, EPI_ISL_1482821, EPI_ISL_1482822, EPI_ISL_1482823, EPI_ISL_1482824, EPI_ISL_1482825, EPI_ISL_1482826, EPI_ISL_1482827, EPI_ISL_1482828, EPI_ISL_1482829, EPI_ISL_1482830, EPI_ISL_1482831, EPI_ISL_1482833, EPI_ISL_1482834, EPI_ISL_1482837, EPI_ISL_1482838, EPI_ISL_1482839, EPI_ISL_1482840, EPI_ISL_1482841, EPI_ISL_1482842, EPI_ISL_1482843, EPI_ISL_1482844, EPI_ISL_1482845, EPI_ISL_1482846, EPI_ISL_1482851, EPI_ISL_1482852, EPI_ISL_1482853, EPI_ISL_1482854, EPI_ISL_1482855, EPI_ISL_1482856, EPI_ISL_1482857, EPI_ISL_1482858, EPI_ISL_1482860, EPI_ISL_1482861, EPI_ISL_1482862, EPI_ISL_1482863, EPI_ISL_1482864, EPI_ISL_1482865, EPI_ISL_1482866, EPI_ISL_1482867, EPI_ISL_1482869, EPI_ISL_1482870, EPI_ISL_1482871, EPI_ISL_1482872, EPI_ISL_1482873, EPI_ISL_1482874, EPI_ISL_1482875, EPI_ISL_1482876, EPI_ISL_1482877, EPI_ISL_1482878, EPI_ISL_1482879, EPI_ISL_1482880, EPI_ISL_1482881, EPI_ISL_1482882, EPI_ISL_1482883, EPI_ISL_1482884, EPI_ISL_1482885, EPI_ISL_1482886, EPI_ISL_1482887, EPI_ISL_1482888, EPI_ISL_1482889, EPI_ISL_1482890, EPI_ISL_1482891, EPI_ISL_1482892, EPI_ISL_1482893, EPI_ISL_1482894, EPI_ISL_1482895, EPI_ISL_1482896, EPI_ISL_1482897, EPI_ISL_1482898, EPI_ISL_1482899, EPI_ISL_1482900, EPI_ISL_1482901, EPI_ISL_1482902, EPI_ISL_1482903, EPI_ISL_1482904, EPI_ISL_1482905, EPI_ISL_1482906, EPI_ISL_1482907, EPI_ISL_1482908, EPI_ISL_1482909, EPI_ISL_1482910, EPI_ISL_1482911, EPI_ISL_1482912, EPI_ISL_1482913, EPI_ISL_1482914, EPI_ISL_1482915, EPI_ISL_1482916, EPI_ISL_1482917, EPI_ISL_1482918, EPI_ISL_1482919, EPI_ISL_1482920, EPI_ISL_1482921, EPI_ISL_1482922, EPI_ISL_1482923, EPI_ISL_1482924, EPI_ISL_1482925, EPI_ISL_1482926, EPI_ISL_1482927, EPI_ISL_1482928, EPI_ISL_1482929, EPI_ISL_1482930, EPI_ISL_1482931, EPI_ISL_1482932, EPI_ISL_1482933, EPI_ISL_1482934, EPI_ISL_1482935, EPI_ISL_1482936, EPI_ISL_1482937, EPI_ISL_1482938, EPI_ISL_1482939, EPI_ISL_1482940, EPI_ISL_1482941, EPI_ISL_1482942, EPI_ISL_1482943, EPI_ISL_1482944, EPI_ISL_1482945, EPI_ISL_1482946, EPI_ISL_1482947, EPI_ISL_1482948, EPI_ISL_1482949, EPI_ISL_1482950, EPI_ISL_1482951, EPI_ISL_1482952, EPI_ISL_1482953, EPI_ISL_1482954, EPI_ISL_1482955, EPI_ISL_1482956, EPI_ISL_1482957, EPI_ISL_1482958, EPI_ISL_1482959, EPI_ISL_1482960, EPI_ISL_1482961, EPI_ISL_1482962, EPI_ISL_1482963, EPI_ISL_1482964, EPI_ISL_1482965, EPI_ISL_1482966, EPI_ISL_1482967, EPI_ISL_1482968, EPI_ISL_1482969, EPI_ISL_1482970, EPI_ISL_1482971, EPI_ISL_1482972, EPI_ISL_1482973 | | | |
| see above | MUSC Molecular Pathology Laboratory | MUSC Molecular Pathology Laboratory | Julie W. Hirschhorn, W. Bailey Glen Jr, Dariusz Pytel, Jaclyn Dunne, Kristen Maurer, Frederick S. Nolte |
| EPI_ISL_1482974, EPI_ISL_1482975, EPI_ISL_1482976, EPI_ISL_1482977, EPI_ISL_1482978 | Oregon State Public Health Laboratory | Oregon State Public Health Laboratory | Rafia Razzaque, Eugene Yeboah, Vanda Makris, Laura Tsaknaridis, John Fontana and Shane Sevey |
| EPI_ISL_1482979 | Department of General Services Division of Consolidated Laboratory Services, Virginia Division of Consolidated Laboratory Services Sequencing Submission Group | Department of General Services Division of Consolidated Laboratory Services, Virginia Division of Consolidated Laboratory Services Sequencing Submission Group | Fink,L. |
| EPI_ISL_1482980, EPI_ISL_1482981, EPI_ISL_1482982, EPI_ISL_1482983, EPI_ISL_1482984, EPI_ISL_1482985, EPI_ISL_1482986, EPI_ISL_1482987, EPI_ISL_1482988, EPI_ISL_1482989, EPI_ISL_1482990, EPI_ISL_1482991, EPI_ISL_1482992, EPI_ISL_1482993, EPI_ISL_1482994, EPI_ISL_1482995, EPI_ISL_1482996, EPI_ISL_1482997, EPI_ISL_1482998, EPI_ISL_1482999, EPI_ISL_1483000, EPI_ISL_1483001, EPI_ISL_1483002, EPI_ISL_1483003, EPI_ISL_1483004, EPI_ISL_1483005, EPI_ISL_1483006, EPI_ISL_1483007, EPI_ISL_1483008, EPI_ISL_1483009, EPI_ISL_1483010, EPI_ISL_1483011, EPI_ISL_1483012, EPI_ISL_1483013, EPI_ISL_1483014, EPI_ISL_1483015, EPI_ISL_1483016, EPI_ISL_1483017, EPI_ISL_1483018, EPI_ISL_1483019, EPI_ISL_1483020, EPI_ISL_1483021, EPI_ISL_1483022, EPI_ISL_1483023, EPI_ISL_1483024, EPI_ISL_1483025 | | | |
| see above | Maryland Public Health Laboratory | Maryland Public Health Laboratory | Maryland Department of Health Laboratories Administration |
| EPI_ISL_1483026 | Public Health Laboratory - Infectious Disease Lab, Minnesota Department of Health Infectious Disease Laboratory Submission Group | Public Health Laboratory - Infectious Disease Lab, Minnesota Department of Health Infectious Disease Laboratory Submission Group | Lorentz,A., Garfin,J., Plumb,M. and Wang,X. |
| EPI_ISL_1483032, EPI_ISL_1483033, EPI_ISL_1483034, EPI_ISL_1483035, EPI_ISL_1483036, EPI_ISL_1483037 | LESP Nuevo Leon | Instituto de Diagnostico y Referencia Epidemiologicos (INDRE) | Claudia Wong-Arambula, Abril Rodriguez-Maldonado, Vanessa Rivero-Arredondo, Ariadna Medina-Benitez, Joaquin Quiroz-Mercado, Sergio Rangel-Guerrero, Natividad Cruz-Ortiz, Tatiana Nunez-Garcia, Gisela Barrera-Badillo, Lucia Hernandez-Rivas, Irma Lopez-Martinez, Ernesto Ramirez-Gonzalez. |
| EPI_ISL_1483038, EPI_ISL_1483039 | LESP Yucatan | Instituto de Diagnostico y Referencia Epidemiologicos (INDRE) | Claudia Wong-Arambula, Abril Rodriguez-Maldonado, Vanessa Rivero-Arredondo, Ariadna Medina-Benitez, Joaquin Quiroz-Mercado, Sergio Rangel-Guerrero, Natividad Cruz-Ortiz, Tatiana Nunez-Garcia, Gisela Barrera-Badillo, Lucia Hernandez-Rivas, Irma Lopez-Martinez, Ernesto Ramirez-Gonzalez. |
| EPI_ISL_1483040, EPI_ISL_1483041 | LESP Nayarit | Instituto de Diagnostico y Referencia Epidemiologicos (INDRE) | Claudia Wong-Arambula, Abril Rodriguez-Maldonado, Vanessa Rivero-Arredondo, Ariadna Medina-Benitez, Joaquin Quiroz-Mercado, Sergio Rangel-Guerrero, Natividad Cruz-Ortiz, Tatiana Nunez-Garcia, Gisela Barrera-Badillo, Lucia Hernandez-Rivas, Irma Lopez-Martinez, Ernesto Ramirez-Gonzalez. |
| EPI_ISL_1483042, EPI_ISL_1483043 | LESP Sinaloa | Instituto de Diagnostico y Referencia Epidemiologicos (INDRE) | Claudia Wong-Arambula, Abril Rodriguez-Maldonado, Vanessa Rivero-Arredondo, Ariadna Medina-Benitez, Joaquin Quiroz-Mercado, Sergio Rangel-Guerrero, Natividad Cruz-Ortiz, Tatiana Nunez-Garcia, Gisela Barrera-Badillo, Lucia Hernandez-Rivas, Irma Lopez-Martinez, Ernesto Ramirez-Gonzalez. |
| EPI_ISL_1483044, EPI_ISL_1483045, EPI_ISL_1483046, EPI_ISL_1483047 | LESP Queretaro | Instituto de Diagnostico y Referencia Epidemiologicos (INDRE) | Claudia Wong-Arambula, Abril Rodriguez-Maldonado, Vanessa Rivero-Arredondo, Ariadna Medina-Benitez, Joaquin Quiroz-Mercado, Sergio Rangel-Guerrero, Natividad Cruz-Ortiz, Tatiana Nunez-Garcia, Gisela Barrera-Badillo, Lucia Hernandez-Rivas, Irma Lopez-Martinez, Ernesto Ramirez-Gonzalez. |
| EPI_ISL_1483048 | LESP Guanajuato | Instituto de Diagnostico y Referencia Epidemiologicos (INDRE) | Claudia Wong-Arambula, Abril Rodriguez-Maldonado, Vanessa Rivero-Arredondo, Ariadna Medina-Benitez, Joaquin Quiroz-Mercado, Sergio Rangel-Guerrero, Natividad Cruz-Ortiz, Tatiana Nunez-Garcia, Gisela Barrera-Badillo, Lucia Hernandez-Rivas, Irma Lopez-Martinez, Ernesto Ramirez-Gonzalez. |
| EPI_ISL_1483049, EPI_ISL_1483050 | LESP Queretaro | Instituto de Diagnostico y Referencia Epidemiologicos (INDRE) | Claudia Wong-Arambula, Abril Rodriguez-Maldonado, Vanessa Rivero-Arredondo, Ariadna Medina-Benitez, Joaquin Quiroz-Mercado, Sergio Rangel-Guerrero, Natividad Cruz-Ortiz, Tatiana Nunez-Garcia, Gisela Barrera-Badillo, Lucia Hernandez-Rivas, Irma Lopez-Martinez, Ernesto Ramirez-Gonzalez. |
| EPI_ISL_1483051, EPI_ISL_1483052 | Gundersen Clinical Microbiology Laboratory | Kabara Cancer Research Institute | Craig S. Richmond, Paraic A. Kenny |
| EPI_ISL_1483053, EPI_ISL_1483054 | Altius Institute | Seattle Flu Study | Deborah A. Nickerson, Chris D. Frazar, Jover Lee, Benjamin Pelle, Erica Ryke, Matthew Richardson, Amanda Adler, Elisabeth Brandstetter, Peter D. Han, Kairsten Fay, Misja Ilcisin, Kirsten Lacombe, Thomas R. Sibley, Melissa Truong, Caitlin R. Wolf, Ryan Alexander, Daniel Bates, Rebecca Bruders, Stephanie DeBaun, Clem Green, Muhammad Halimun, Jessica Halow, Kneshay Harper, Matt Hartman, Andrew Meuser, Alex Nguyen, Truong Nguyen, Sofia Olsson, Sadie Patraw, Hannah Petersen, Tobias Ragoczy, Joshua Richards, Jacob Rodriguez, John Stamatoyannopoulos, Julia Wald, Olivia Waltner, Michael Boeckh, Janet A. Englund, Michael Famulare, Barry R. Lutz, Mark J. Rieder, Lea M. Starita, Matthew Thompson, Helen Y. Chu, Jay Shendure, Trevor Bedford |
| EPI_ISL_1483055, EPI_ISL_1483056, EPI_ISL_1483057 | LESP Veracruz | Instituto de Diagnostico y Referencia Epidemiologicos (INDRE) | Claudia Wong-Arambula, Abril Rodriguez-Maldonado, Vanessa Rivero-Arredondo, Ariadna Medina-Benitez, Joaquin Quiroz-Mercado, Sergio Rangel-Guerrero, Natividad Cruz-Ortiz, Tatiana Nunez-Garcia, Gisela Barrera-Badillo, Lucia Hernandez-Rivas, Irma Lopez-Martinez, Ernesto Ramirez-Gonzalez. |
| EPI_ISL_1483058 | LESP Hidalgo | Instituto de Diagnostico y Referencia Epidemiologicos (INDRE) | Claudia Wong-Arambula, Abril Rodriguez-Maldonado, Vanessa Rivero-Arredondo, Ariadna Medina-Benitez, Joaquin Quiroz-Mercado, Sergio Rangel-Guerrero, Natividad Cruz-Ortiz, Tatiana Nunez-Garcia, Gisela Barrera-Badillo, Lucia Hernandez-Rivas, Irma Lopez-Martinez, Ernesto Ramirez-Gonzalez. |
| EPI_ISL_1483059, EPI_ISL_1483060, EPI_ISL_1483061, EPI_ISL_1483062, EPI_ISL_1483063, EPI_ISL_1483064, | LESP Puebla | Instituto de Diagnostico y Referencia Epidemiologicos (INDRE) | Claudia Wong-Arambula, Abril Rodriguez-Maldonado, Vanessa Rivero-Arredondo, Ariadna Medina-Benitez, Joaquin Quiroz-Mercado, Sergio Rangel-Guerrero, Natividad Cruz-Ortiz, Tatiana Nunez-Garcia, Gisela Barrera-Badillo, Lucia Hernandez-Rivas, Irma Lopez-Martinez, Ernesto Ramirez-Gonzalez. |

| | | | |
|---|---|---|---|
| EPI_ISL_1483065 | | | |
| EPI_ISL_1483066, EPI_ISL_1483067 | LESP Campeche | Instituto de Diagnostico y Referencia Epidemiologicos (INDRE) | Claudia Wong-Arambula, Abril Rodriguez-Maldonado, Vanessa Rivero-Arredondo, Ariadna Medina-Benitez, Joaquin Quiroz-Mercado, Sergio Rangel-Guerrero, Natividad Cruz-Ortiz, Tatiana Nunez-Garcia, Gisela Barrera-Badillo, Lucia Hernandez-Rivas, Irma Lopez-Martinez, Ernesto Ramirez-Gonzalez. |
| EPI_ISL_1483068, EPI_ISL_1483069, EPI_ISL_1483070, EPI_ISL_1483071, EPI_ISL_1483072 | LESP Jalisco | Instituto de Diagnostico y Referencia Epidemiologicos (INDRE) | Claudia Wong-Arambula, Abril Rodriguez-Maldonado, Vanessa Rivero-Arredondo, Ariadna Medina-Benitez, Joaquin Quiroz-Mercado, Sergio Rangel-Guerrero, Natividad Cruz-Ortiz, Tatiana Nunez-Garcia, Gisela Barrera-Badillo, Lucia Hernandez-Rivas, Irma Lopez-Martinez, Ernesto Ramirez-Gonzalez. |
| EPI_ISL_1483073 | LESP Guerrero | Instituto de Diagnostico y Referencia Epidemiologicos (INDRE) | Claudia Wong-Arambula, Abril Rodriguez-Maldonado, Vanessa Rivero-Arredondo, Ariadna Medina-Benitez, Joaquin Quiroz-Mercado, Sergio Rangel-Guerrero, Natividad Cruz-Ortiz, Tatiana Nunez-Garcia, Gisela Barrera-Badillo, Lucia Hernandez-Rivas, Irma Lopez-Martinez, Ernesto Ramirez-Gonzalez. |
| EPI_ISL_1483074 | LESP Chihuahua | Instituto de Diagnostico y Referencia Epidemiologicos (INDRE) | Claudia Wong-Arambula, Abril Rodriguez-Maldonado, Vanessa Rivero-Arredondo, Ariadna Medina-Benitez, Joaquin Quiroz-Mercado, Sergio Rangel-Guerrero, Natividad Cruz-Ortiz, Tatiana Nunez-Garcia, Gisela Barrera-Badillo, Lucia Hernandez-Rivas, Irma Lopez-Martinez, Ernesto Ramirez-Gonzalez. |
| EPI_ISL_1483075, EPI_ISL_1483076, EPI_ISL_1483077, EPI_ISL_1483078, EPI_ISL_1483079, EPI_ISL_1483080, EPI_ISL_1483081, EPI_ISL_1483082, EPI_ISL_1483083, EPI_ISL_1483084, EPI_ISL_1483085, EPI_ISL_1483086, EPI_ISL_1483087, EPI_ISL_1483088, EPI_ISL_1483089, EPI_ISL_1483090, EPI_ISL_1483091, EPI_ISL_1483092, EPI_ISL_1483093, EPI_ISL_1483094, EPI_ISL_1483095, EPI_ISL_1483096, EPI_ISL_1483097, EPI_ISL_1483098, EPI_ISL_1483099, EPI_ISL_1483100, EPI_ISL_1483101, EPI_ISL_1483102, EPI_ISL_1483103, EPI_ISL_1483104, EPI_ISL_1483105, EPI_ISL_1483106, EPI_ISL_1483107, EPI_ISL_1483108, EPI_ISL_1483109, EPI_ISL_1483110, EPI_ISL_1483111, EPI_ISL_1483112, EPI_ISL_1483113, EPI_ISL_1483114, EPI_ISL_1483115, EPI_ISL_1483116, EPI_ISL_1483117, EPI_ISL_1483118, EPI_ISL_1483119, EPI_ISL_1483120, EPI_ISL_1483121, EPI_ISL_1483122, EPI_ISL_1483123, EPI_ISL_1483124, EPI_ISL_1483125, EPI_ISL_1483126, EPI_ISL_1483127, EPI_ISL_1483128, EPI_ISL_1483129, EPI_ISL_1483130, EPI_ISL_1483131, EPI_ISL_1483132, EPI_ISL_1483133, EPI_ISL_1483134, EPI_ISL_1483135, EPI_ISL_1483136, EPI_ISL_1483137, EPI_ISL_1483138, EPI_ISL_1483139, EPI_ISL_1483140, EPI_ISL_1483141, EPI_ISL_1483142, EPI_ISL_1483143, EPI_ISL_1483144, EPI_ISL_1483145, EPI_ISL_1483146, EPI_ISL_1483147, EPI_ISL_1483148, EPI_ISL_1483149, EPI_ISL_1483150, EPI_ISL_1483151, EPI_ISL_1483152, EPI_ISL_1483153, EPI_ISL_1483154, EPI_ISL_1483155, EPI_ISL_1483156, EPI_ISL_1483157, EPI_ISL_1483158, EPI_ISL_1483159, EPI_ISL_1483160, EPI_ISL_1483161, EPI_ISL_1483162, EPI_ISL_1483163, EPI_ISL_1483164, EPI_ISL_1483165, EPI_ISL_1483166, EPI_ISL_1483167, EPI_ISL_1483168, EPI_ISL_1483169, EPI_ISL_1483170, EPI_ISL_1483171, EPI_ISL_1483172, EPI_ISL_1483173, EPI_ISL_1483174, EPI_ISL_1483175, EPI_ISL_1483176, EPI_ISL_1483177, EPI_ISL_1483178, EPI_ISL_1483179, EPI_ISL_1483180, EPI_ISL_1483181, EPI_ISL_1483182, EPI_ISL_1483183, EPI_ISL_1483184, EPI_ISL_1483185, EPI_ISL_1483186, EPI_ISL_1483187, EPI_ISL_1483188, EPI_ISL_1483189, EPI_ISL_1483190, EPI_ISL_1483191, EPI_ISL_1483192, EPI_ISL_1483193, EPI_ISL_1483194, EPI_ISL_1483195, EPI_ISL_1483196, EPI_ISL_1483197, EPI_ISL_1483198, EPI_ISL_1483199, EPI_ISL_1483200, EPI_ISL_1483201, EPI_ISL_1483202, EPI_ISL_1483203, EPI_ISL_1483204, EPI_ISL_1483205, EPI_ISL_1483206 | | | |
| see above | Utah Public Health Laboratory | Utah Public Health Laboratory | Erin L. Young, Kelly F. Oakeson, Tara Gallagher |
| EPI_ISL_1483207, EPI_ISL_1483208 | Springfield-Greene Co. Health Dept. | Pathogen Discovery, Respiratory Viruses Branch, Division of Viral Diseases, Centers for Disease Control and Prevention | Yan Li, Jing Zhang, Ying Tao, Brian Lynch, Anna Kelleher, Krista Queen, Anna Uehara, Peter Cook, Han Jia Justin Ng, Clinton R. Paden, Haibin Wang, Suxiang Tong |
| EPI_ISL_1483210 | Pathogen Discovery, Respiratory Viruses Branch, Division of Viral Diseases, Centers for Disease Control and Prevention | Pathogen Discovery, Respiratory Viruses Branch, Division of Viral Diseases, Centers for Disease Control and Prevention | Yan Li, Jing Zhang, Ying Tao, Brian Lynch, Anna Kelleher, Krista Queen, Anna Uehara, Peter Cook, Han Jia Justin Ng, Clinton R. Paden, Haibin Wang, Suxiang Tong |
| EPI_ISL_1483211, EPI_ISL_1483212 | Vanderbilt University | Pathogen Discovery, Respiratory Viruses Branch, Division of Viral Diseases, Centers for Disease Control and Prevention | Yan Li, Jing Zhang, Ying Tao, Brian Lynch, Anna Kelleher, Krista Queen, Anna Uehara, Peter Cook, Han Jia Justin Ng, Clinton R. Paden, Haibin Wang, Suxiang Tong |
| EPI_ISL_1483213 | Pathogen Discovery, Respiratory Viruses Branch, Division of Viral Diseases, Centers for Disease Control and Prevention | Pathogen Discovery, Respiratory Viruses Branch, Division of Viral Diseases, Centers for Disease Control and Prevention | Yan Li, Jing Zhang, Ying Tao, Brian Lynch, Anna Kelleher, Krista Queen, Anna Uehara, Peter Cook, Han Jia Justin Ng, Clinton R. Paden, Haibin Wang, Suxiang Tong |
| EPI_ISL_1483214, EPI_ISL_1483215, EPI_ISL_1483216, EPI_ISL_1483217, EPI_ISL_1483218, EPI_ISL_1483219, EPI_ISL_1483220 | Vanderbilt University | Pathogen Discovery, Respiratory Viruses Branch, Division of Viral Diseases, Centers for Disease Control and Prevention | Yan Li, Jing Zhang, Ying Tao, Brian Lynch, Anna Kelleher, Krista Queen, Anna Uehara, Peter Cook, Han Jia Justin Ng, Clinton R. Paden, Haibin Wang, Suxiang Tong |
| EPI_ISL_1483221 | Pathogen Discovery, Respiratory Viruses Branch, Division of Viral Diseases, Centers for Disease Control and Prevention | Pathogen Discovery, Respiratory Viruses Branch, Division of Viral Diseases, Centers for Disease Control and Prevention | Yan Li, Jing Zhang, Ying Tao, Brian Lynch, Anna Kelleher, Krista Queen, Anna Uehara, Peter Cook, Han Jia Justin Ng, Clinton R. Paden, Haibin Wang, Suxiang Tong |
| EPI_ISL_1483222, EPI_ISL_1483223, EPI_ISL_1483224, EPI_ISL_1483225, EPI_ISL_1483226, EPI_ISL_1483227, EPI_ISL_1483228, EPI_ISL_1483229 | Vanderbilt University | Pathogen Discovery, Respiratory Viruses Branch, Division of Viral Diseases, Centers for Disease Control and Prevention | Yan Li, Jing Zhang, Ying Tao, Brian Lynch, Anna Kelleher, Krista Queen, Anna Uehara, Peter Cook, Han Jia Justin Ng, Clinton R. Paden, Haibin Wang, Suxiang Tong |
| EPI_ISL_1483230 | Pathogen Discovery, Respiratory Viruses Branch, Division of Viral Diseases, Centers for Disease Control and Prevention | Pathogen Discovery, Respiratory Viruses Branch, Division of Viral Diseases, Centers for Disease Control and Prevention | Yan Li, Jing Zhang, Ying Tao, Brian Lynch, Anna Kelleher, Krista Queen, Anna Uehara, Peter Cook, Han Jia Justin Ng, Clinton R. Paden, Haibin Wang, Suxiang Tong |
| EPI_ISL_1483232, EPI_ISL_1483233, EPI_ISL_1483234 | Springfield-Greene Co. Health Dept. | Pathogen Discovery, Respiratory Viruses Branch, Division of Viral Diseases, Centers for Disease Control and Prevention | Yan Li, Jing Zhang, Ying Tao, Brian Lynch, Anna Kelleher, Krista Queen, Anna Uehara, Peter Cook, Han Jia Justin Ng, Clinton R. Paden, Haibin Wang, Suxiang Tong |
| EPI_ISL_1483235 | Vanderbilt University | Pathogen Discovery, Respiratory Viruses Branch, Division of Viral Diseases, Centers for Disease Control and Prevention | Yan Li, Jing Zhang, Ying Tao, Brian Lynch, Anna Kelleher, Krista Queen, Anna Uehara, Peter Cook, Han Jia Justin Ng, Clinton R. Paden, Haibin Wang, Suxiang Tong |
| EPI_ISL_1483236, EPI_ISL_1483237, EPI_ISL_1483238 | Vanderbilt University Medical Center | Pathogen Discovery, Respiratory Viruses Branch, Division of Viral Diseases, Centers for Disease Control and Prevention | Yan Li, Jing Zhang, Ying Tao, Brian Lynch, Anna Kelleher, Krista Queen, Anna Uehara, Peter Cook, Han Jia Justin Ng, Clinton R. Paden, Haibin Wang, Suxiang Tong |
| EPI_ISL_1483242, EPI_ISL_1483243, EPI_ISL_1483244, EPI_ISL_1483245, EPI_ISL_1483246, EPI_ISL_1483247, EPI_ISL_1483248, EPI_ISL_1483249, EPI_ISL_1483250, EPI_ISL_1483251, EPI_ISL_1483253, EPI_ISL_1483254, EPI_ISL_1483255, EPI_ISL_1483256, EPI_ISL_1483257, EPI_ISL_1483259, EPI_ISL_1483260, EPI_ISL_1483261, EPI_ISL_1483262, EPI_ISL_1483263, EPI_ISL_1483264, EPI_ISL_1483265, EPI_ISL_1483266, EPI_ISL_1483267, EPI_ISL_1483268, EPI_ISL_1483269, EPI_ISL_1483270, EPI_ISL_1483271, EPI_ISL_1483272, EPI_ISL_1483273, EPI_ISL_1483274, EPI_ISL_1483275, EPI_ISL_1483276, EPI_ISL_1483277 | | | |
| see above | Nevada State Public Health Laboratory | Nevada State Public Health Laboratory | Andrew Gorzalski, Mark Pandori |
| EPI_ISL_1483279, EPI_ISL_1483280, EPI_ISL_1483281, EPI_ISL_1483282, EPI_ISL_1483283, EPI_ISL_1483284, EPI_ISL_1483285, EPI_ISL_1483286, EPI_ISL_1483287, EPI_ISL_1483288, EPI_ISL_1483289, EPI_ISL_1483290, EPI_ISL_1483291, EPI_ISL_1483292, EPI_ISL_1483293, EPI_ISL_1483294, EPI_ISL_1483295, EPI_ISL_1483296, EPI_ISL_1483297, EPI_ISL_1483298, EPI_ISL_1483299, EPI_ISL_1483300, EPI_ISL_1483301, EPI_ISL_1483302, EPI_ISL_1483303, EPI_ISL_1483304, EPI_ISL_1483305, EPI_ISL_1483306, EPI_ISL_1483307, EPI_ISL_1483308, EPI_ISL_1483309, EPI_ISL_1483310, EPI_ISL_1483311, EPI_ISL_1483312, EPI_ISL_1483313, EPI_ISL_1483314, EPI_ISL_1483315, EPI_ISL_1483316, EPI_ISL_1483317, EPI_ISL_1483318 | | | |
| see above | Tampa General Hospital Esoteric Lab | Tampa General Hospital Esoteric Research & Development Lab | Grant Vestal, Deanna Becker, Dominic Uy, Vicki Healer, Amorce Lima, Suzane Silbert |
| EPI_ISL_1483321, EPI_ISL_1483322, EPI_ISL_1483323, EPI_ISL_1483324, EPI_ISL_1483325, EPI_ISL_1483326, EPI_ISL_1483327, EPI_ISL_1483328, EPI_ISL_1483329, EPI_ISL_1483330, EPI_ISL_1483331, EPI_ISL_1483332, EPI_ISL_1483333, EPI_ISL_1483334, EPI_ISL_1483335, EPI_ISL_1483336, EPI_ISL_1483337, EPI_ISL_1483338, EPI_ISL_1483339, EPI_ISL_1483340, EPI_ISL_1483341, EPI_ISL_1483342, EPI_ISL_1483343, EPI_ISL_1483344, EPI_ISL_1483345, EPI_ISL_1483346, EPI_ISL_1483347, EPI_ISL_1483348, EPI_ISL_1483349, EPI_ISL_1483350, EPI_ISL_1483351, EPI_ISL_1483352, EPI_ISL_1483353, EPI_ISL_1483354, EPI_ISL_1483356, EPI_ISL_1483357, EPI_ISL_1483358, EPI_ISL_1483359, EPI_ISL_1483360, EPI_ISL_1483361, EPI_ISL_1483362 | | | |
| see above | Wyoming Public Health Laboratory | Wyoming Public Health Laboratory | Jim Mildenberger, Wanda Manley, Noah Hull, Taylor Fearing, Lynette Gumbleton, Channing Weber, Ashley Norberg, Chayse Rowley, Marley Goetz, Brian Dominguez, Elliot Thomasson, Cari Sloma, and Rob Christensen |
| EPI_ISL_1483573, EPI_ISL_1483574, EPI_ISL_1483575, EPI_ISL_1483576, EPI_ISL_1483577, EPI_ISL_1483578, EPI_ISL_1483579, EPI_ISL_1483580, EPI_ISL_1483581, EPI_ISL_1483582, EPI_ISL_1483583, EPI_ISL_1483584, EPI_ISL_1483585 | | | |
| see above | Arizona State University | Arizona State University | Rabia Maqsood, LaRinda A. Holland, Peter T. Skidmore, Nicholas J. Mellor, Joy M. Blain, Valerie Harris, Joshua LaBaer, Vel Murugan, Efrem S. Lim |
| EPI_ISL_1483586, EPI_ISL_1483587, EPI_ISL_1483588, EPI_ISL_1483589, EPI_ISL_1483590, EPI_ISL_1483591, EPI_ISL_1483592, EPI_ISL_1483593, EPI_ISL_1483594, EPI_ISL_1483595, EPI_ISL_1483596, EPI_ISL_1483597, EPI_ISL_1483598, EPI_ISL_1483599, EPI_ISL_1483600, EPI_ISL_1483601, EPI_ISL_1483602, EPI_ISL_1483603, EPI_ISL_1483604, EPI_ISL_1483605, EPI_ISL_1483606, EPI_ISL_1483607, EPI_ISL_1483608, EPI_ISL_1483609, EPI_ISL_1483610, EPI_ISL_1483611, EPI_ISL_1483612, EPI_ISL_1483613, EPI_ISL_1483614, EPI_ISL_1483615, EPI_ISL_1483616, EPI_ISL_1483617, EPI_ISL_1483618, EPI_ISL_1483619, EPI_ISL_1483620, EPI_ISL_1483621, EPI_ISL_1483622, EPI_ISL_1483623, EPI_ISL_1483624, EPI_ISL_1483625, EPI_ISL_1483626, EPI_ISL_1483627, EPI_ISL_1483628, EPI_ISL_1483629, EPI_ISL_1483630, EPI_ISL_1483631, EPI_ISL_1483632, EPI_ISL_1483633, EPI_ISL_1483634, EPI_ISL_1483635, EPI_ISL_1483636, EPI_ISL_1483637, EPI_ISL_1483638, EPI_ISL_1483639, EPI_ISL_1483640, EPI_ISL_1483641, EPI_ISL_1483642, EPI_ISL_1483643, EPI_ISL_1483644, EPI_ISL_1483645, EPI_ISL_1483646, EPI_ISL_1483647, EPI_ISL_1483648, EPI_ISL_1483649, EPI_ISL_1483650, EPI_ISL_1483651, EPI_ISL_1483652, EPI_ISL_1483653, EPI_ISL_1483654, EPI_ISL_1483655, EPI_ISL_1483656, EPI_ISL_1483657, EPI_ISL_1483658, EPI_ISL_1483659, EPI_ISL_1483660, EPI_ISL_1483661, EPI_ISL_1483662, EPI_ISL_1483663, EPI_ISL_1483664, EPI_ISL_1483665, EPI_ISL_1483666, EPI_ISL_1483667, EPI_ISL_1483668, EPI_ISL_1483669, EPI_ISL_1483670, EPI_ISL_1483671, EPI_ISL_1483672, EPI_ISL_1483673, EPI_ISL_1483674, EPI_ISL_1483675, EPI_ISL_1483676, EPI_ISL_1483677, EPI_ISL_1483678, EPI_ISL_1483679, EPI_ISL_1483680, EPI_ISL_1483681, EPI_ISL_1483682, EPI_ISL_1483683, EPI_ISL_1483684, EPI_ISL_1483685, EPI_ISL_1483686, EPI_ISL_1483687, EPI_ISL_1483688, EPI_ISL_1483689, EPI_ISL_1483690, EPI_ISL_1483691, EPI_ISL_1483692, EPI_ISL_1483693, EPI_ISL_1483694, EPI_ISL_1483695, EPI_ISL_1483696, EPI_ISL_1483697, EPI_ISL_1483698, EPI_ISL_1483699 | | | |
| see above | Alaska State Virology Laboratory | Alaska State Virology Laboratory | Stephanie DeRonde, Elva House, Lisa Smith, Ph.D., Jack Chen, Ph.D. |
| EPI_ISL_1483789, EPI_ISL_1483790, EPI_ISL_1483791, EPI_ISL_1483792, EPI_ISL_1483793, EPI_ISL_1483794 | El Camino Hospital | Santa Clara County Public Health Laboratory | Santa Clara County Public Health Department |
| EPI_ISL_1483795, EPI_ISL_1483796, EPI_ISL_1483797, EPI_ISL_1483798 | Stanford Clinical Virology Laboratory | Santa Clara County Public Health Laboratory | Santa Clara County Public Health Department |
| EPI_ISL_1483799 | Santa Clara Valley Medical Center | Santa Clara County Public Health Laboratory | Santa Clara County Public Health Department |
| EPI_ISL_1483800, EPI_ISL_1483801 | El Camino Hospital | Santa Clara County Public Health Laboratory | Santa Clara County Public Health Department |

| | | | |
|---|---|---|---|
| EPI_ISL_1483802 | Santa Clara County Public Health Laboratory | Santa Clara County Public Health Laboratory | Santa Clara County Public Health Department |
| EPI_ISL_1484314, EPI_ISL_1484316, EPI_ISL_1484319, EPI_ISL_1484322, EPI_ISL_1484324, EPI_ISL_1484327, EPI_ISL_1484330, EPI_ISL_1484333, EPI_ISL_1484336, EPI_ISL_1484339, EPI_ISL_1484342, EPI_ISL_1484345, EPI_ISL_1484347, EPI_ISL_1484350, EPI_ISL_1484353, EPI_ISL_1484357, EPI_ISL_1484360, EPI_ISL_1484362, EPI_ISL_1484368, EPI_ISL_1484371, EPI_ISL_1484374, EPI_ISL_1484381, EPI_ISL_1484384, EPI_ISL_1484387, EPI_ISL_1484389, EPI_ISL_1484392, EPI_ISL_1484395, EPI_ISL_1484398, EPI_ISL_1484401, EPI_ISL_1484403, EPI_ISL_1484406, EPI_ISL_1484409, EPI_ISL_1484412, EPI_ISL_1484415, EPI_ISL_1484418, EPI_ISL_1484421, EPI_ISL_1484424, EPI_ISL_1484427, EPI_ISL_1484430, EPI_ISL_1484433, EPI_ISL_1484435, EPI_ISL_1484438, EPI_ISL_1484441, EPI_ISL_1484444, EPI_ISL_1484447, EPI_ISL_1484450 | | | |
| see above | Wyoming Public Health Laboratory | Wyoming Public Health Laboratory | Jim Mildenberger, Wanda Manley, Noah Hull, Taylor Fearing, Lynette Gumbleton, Channing Weber, Ashley Norberg, Chayse Rowley, Marley Goetz, Brian Dominguez, Elliot Thomasson, Cari Sloma, and Rob Christensen |
| EPI_ISL_1489657, EPI_ISL_1489659, EPI_ISL_1489661, EPI_ISL_1489662, EPI_ISL_1489664, EPI_ISL_1489665, EPI_ISL_1489666, EPI_ISL_1489669, EPI_ISL_1489670, EPI_ISL_1489671, EPI_ISL_1489672, EPI_ISL_1489673, EPI_ISL_1489674, EPI_ISL_1489675, EPI_ISL_1489677, EPI_ISL_1489678, EPI_ISL_1489679, EPI_ISL_1489680, EPI_ISL_1489682, EPI_ISL_1489683, EPI_ISL_1489684, EPI_ISL_1489686, EPI_ISL_1489687, EPI_ISL_1489689, EPI_ISL_1489691, EPI_ISL_1489692, EPI_ISL_1489693, EPI_ISL_1489695, EPI_ISL_1489697, EPI_ISL_1489698, EPI_ISL_1489699, EPI_ISL_1489700, EPI_ISL_1489701, EPI_ISL_1489702, EPI_ISL_1489703, EPI_ISL_1489705, EPI_ISL_1489707, EPI_ISL_1489708, EPI_ISL_1489709, EPI_ISL_1489710, EPI_ISL_1489712, EPI_ISL_1489714, EPI_ISL_1489716 | | | |
| see above | NJDOH, Public Health and Environmental Laboratories | New Jersey Public Health and Environmental Laboratories (NJ PHEL) | Lindsey Bodnar, Shiv K. Verma, Dana Woell, Byeong Jeong |
| EPI_ISL_1489843, EPI_ISL_1489844, EPI_ISL_1489845, EPI_ISL_1489846, EPI_ISL_1489847, EPI_ISL_1489848, EPI_ISL_1489849, EPI_ISL_1489850, EPI_ISL_1489851, EPI_ISL_1489852, EPI_ISL_1489853, EPI_ISL_1489854, EPI_ISL_1489855, EPI_ISL_1489856, EPI_ISL_1489857, EPI_ISL_1489858, EPI_ISL_1489859, EPI_ISL_1489860, EPI_ISL_1489861, EPI_ISL_1489862, EPI_ISL_1489863, EPI_ISL_1489864, EPI_ISL_1489865, EPI_ISL_1489866, EPI_ISL_1489867, EPI_ISL_1489868, EPI_ISL_1489869, EPI_ISL_1489870, EPI_ISL_1489871, EPI_ISL_1489872, EPI_ISL_1489873, EPI_ISL_1489874, EPI_ISL_1489875, EPI_ISL_1489876, EPI_ISL_1489877, EPI_ISL_1489878, EPI_ISL_1489879, EPI_ISL_1489880, EPI_ISL_1489881, EPI_ISL_1489882, EPI_ISL_1489883, EPI_ISL_1489884, EPI_ISL_1489885, EPI_ISL_1489886, EPI_ISL_1489887, EPI_ISL_1489888, EPI_ISL_1489889, EPI_ISL_1489890, EPI_ISL_1489891, EPI_ISL_1489892, EPI_ISL_1489893, EPI_ISL_1489894, EPI_ISL_1489895, EPI_ISL_1489896, EPI_ISL_1489897, EPI_ISL_1489898, EPI_ISL_1489899, EPI_ISL_1489900, EPI_ISL_1489901, EPI_ISL_1489902, EPI_ISL_1489903, EPI_ISL_1489904, EPI_ISL_1489905, EPI_ISL_1489906, EPI_ISL_1489908, EPI_ISL_1489909, EPI_ISL_1489910, EPI_ISL_1489911, EPI_ISL_1489912, EPI_ISL_1489913, EPI_ISL_1489914, EPI_ISL_1489915, EPI_ISL_1489918, EPI_ISL_1489919, EPI_ISL_1489920, EPI_ISL_1489921, EPI_ISL_1489922, EPI_ISL_1489923 | | | |
| see above | Missouri State Public Health Laboratory | Missouri State Public Health Laboratory | Matthew Sinn, Joshua Barry, Ashley New |
| EPI_ISL_1489935, EPI_ISL_1489939, EPI_ISL_1490190, EPI_ISL_1490222, EPI_ISL_1490227, EPI_ISL_1490251 | Baylor Esoteric and Molecular Lab | Baylor Esoteric and Molecular Lab | Elizabeth Forrester and Dawn Richards |
| EPI_ISL_1490470, EPI_ISL_1490650, EPI_ISL_1490651 | Montana Public Health Laboratory | Montana Public Health Laboratory | Joy Ritter, Michelle Mozer, Carrie Biskupiak, Deborah Gibson |
| EPI_ISL_1490652 | Baylor Esoteric and Molecular Lab | Baylor Esoteric and Molecular Lab | Elizabeth Forrester and Dawn Richards |
| EPI_ISL_1490654, EPI_ISL_1490659 | Montana Public Health Laboratory | Montana Public Health Laboratory | Joy Ritter, Michelle Mozer, Carrie Biskupiak, Deborah Gibson |
| EPI_ISL_1490660 | Baylor Esoteric and Molecular Lab | Baylor Esoteric and Molecular Lab | Elizabeth Forrester and Dawn Richards |
| EPI_ISL_1490662, EPI_ISL_1490663, EPI_ISL_1490664, EPI_ISL_1490665 | Montana Public Health Laboratory | Montana Public Health Laboratory | Joy Ritter, Michelle Mozer, Carrie Biskupiak, Deborah Gibson |
| EPI_ISL_1490667 | Baylor Esoteric and Molecular Lab | Baylor Esoteric and Molecular Lab | Elizabeth Forrester and Dawn Richards |
| EPI_ISL_1490668, EPI_ISL_1490670, EPI_ISL_1490710 | Montana Public Health Laboratory | Montana Public Health Laboratory | Joy Ritter, Michelle Mozer, Carrie Biskupiak, Deborah Gibson |
| EPI_ISL_1490711 | Baylor Esoteric and Molecular Lab | Baylor Esoteric and Molecular Lab | Elizabeth Forrester and Dawn Richards |
| EPI_ISL_1490712, EPI_ISL_1490713, EPI_ISL_1490715 | Montana Public Health Laboratory | Montana Public Health Laboratory | Joy Ritter, Michelle Mozer, Carrie Biskupiak, Deborah Gibson |
| EPI_ISL_1490716, EPI_ISL_1490749 | Baylor Esoteric and Molecular Lab | Baylor Esoteric and Molecular Lab | Elizabeth Forrester and Dawn Richards |
| EPI_ISL_1490750, EPI_ISL_1490751, EPI_ISL_1490752, EPI_ISL_1490753, EPI_ISL_1490754, EPI_ISL_1490755, EPI_ISL_1490756, EPI_ISL_1490757, EPI_ISL_1490758, EPI_ISL_1490759, EPI_ISL_1490760, EPI_ISL_1490761, EPI_ISL_1490762, EPI_ISL_1490763, EPI_ISL_1490764, EPI_ISL_1490765, EPI_ISL_1490766, EPI_ISL_1490767, EPI_ISL_1490768, EPI_ISL_1490769, EPI_ISL_1490770, EPI_ISL_1490771, EPI_ISL_1490772, EPI_ISL_1490773, EPI_ISL_1490774, EPI_ISL_1490775, EPI_ISL_1490776, EPI_ISL_1490777, EPI_ISL_1490778, EPI_ISL_1490779, EPI_ISL_1490780, EPI_ISL_1490781, EPI_ISL_1490782, EPI_ISL_1490783, EPI_ISL_1490784, EPI_ISL_1490785, EPI_ISL_1490786, EPI_ISL_1490787, EPI_ISL_1490788, EPI_ISL_1490789, EPI_ISL_1490790, EPI_ISL_1490791 | | | |
| see above | IN State Department of Health Laboratory Services | IN State Department of Health Laboratory Services | Cassandra Campion, Jamie Yeadon, Brian Pope, Lixia Liu, Kyle Brownlee, Melissa Hindenlang, Ankita Kashikar, Mark Glazier |
| EPI_ISL_1490817 | Baylor Esoteric and Molecular Lab | Baylor Esoteric and Molecular Lab | Elizabeth Forrester and Dawn Richards |
| EPI_ISL_1490818, EPI_ISL_1490819, EPI_ISL_1490820, EPI_ISL_1490821, EPI_ISL_1490822, EPI_ISL_1490823, EPI_ISL_1490824, EPI_ISL_1490825, EPI_ISL_1490826, EPI_ISL_1490827, EPI_ISL_1490828, EPI_ISL_1490829, EPI_ISL_1490830, EPI_ISL_1490831, EPI_ISL_1490832, EPI_ISL_1490833, EPI_ISL_1490834, EPI_ISL_1490835, EPI_ISL_1490836, EPI_ISL_1490837, EPI_ISL_1490838, EPI_ISL_1490839, EPI_ISL_1490840, EPI_ISL_1490841, EPI_ISL_1490842, EPI_ISL_1490843, EPI_ISL_1490844, EPI_ISL_1490845, EPI_ISL_1490846, EPI_ISL_1490847, EPI_ISL_1490848, EPI_ISL_1490849, EPI_ISL_1490850, EPI_ISL_1490851, EPI_ISL_1490852, EPI_ISL_1490853, EPI_ISL_1490854, EPI_ISL_1490855, EPI_ISL_1490856, EPI_ISL_1490857, EPI_ISL_1490858, EPI_ISL_1490859, EPI_ISL_1490860 | | | |
| see above | IN State Department of Health Laboratory Services | IN State Department of Health Laboratory Services | Cassandra Campion, Jamie Yeadon, Brian Pope, Lixia Liu, Kyle Brownlee, Melissa Hindenlang, Ankita Kashikar, Mark Glazier |
| EPI_ISL_1490861, EPI_ISL_1490862, EPI_ISL_1490863, EPI_ISL_1490864, EPI_ISL_1490865, EPI_ISL_1490866, EPI_ISL_1490867, EPI_ISL_1490868, EPI_ISL_1490869, EPI_ISL_1490870, EPI_ISL_1490871, EPI_ISL_1490872, EPI_ISL_1490873, EPI_ISL_1490874, EPI_ISL_1490875, EPI_ISL_1490876, EPI_ISL_1490877, EPI_ISL_1490878, EPI_ISL_1490879, EPI_ISL_1490880, EPI_ISL_1490881, EPI_ISL_1490882, EPI_ISL_1490883, EPI_ISL_1490884, EPI_ISL_1490885, EPI_ISL_1490886, EPI_ISL_1490887, EPI_ISL_1490888, EPI_ISL_1490889, EPI_ISL_1490890, EPI_ISL_1490891, EPI_ISL_1490892, EPI_ISL_1490893, EPI_ISL_1490894, EPI_ISL_1490895, EPI_ISL_1490896 | | | |
| see above | UW Virology Lab | UW Virology Lab | Pavitra Roychoudhury, Hong Xie, Lasata Shrestha, Shah Mohamed Bakhash, Michelle Lin, Noah R. Baker, Sean Ellis, Saraswathi Sathees, Meei-Li Huang, Keith R Jerome, Alexander Greninger |
| EPI_ISL_1490897 | Baylor Esoteric and Molecular Lab | Baylor Esoteric and Molecular Lab | Elizabeth Forrester and Dawn Richards |
| EPI_ISL_1490899, EPI_ISL_1490900, EPI_ISL_1490901, EPI_ISL_1490903, EPI_ISL_1490904, EPI_ISL_1490905, EPI_ISL_1490907, EPI_ISL_1490908, EPI_ISL_1490909, EPI_ISL_1490910, EPI_ISL_1490911, EPI_ISL_1490912, EPI_ISL_1490913, EPI_ISL_1490914, EPI_ISL_1490915, EPI_ISL_1490916, EPI_ISL_1490918 | | | |
| see above | Wyoming Public Health Laboratory | Wyoming Public Health Laboratory | Jim Mildenberger, Wanda Manley, Noah Hull, Taylor Fearing, Lynette Gumbleton, Channing Weber, Ashley Norberg, Chayse Rowley, Marley Goetz, Brian Dominguez, Elliot Thomasson, Cari Sloma, and Rob Christensen |
| EPI_ISL_1491054 | Baylor Esoteric and Molecular Lab | Baylor Esoteric and Molecular Lab | Elizabeth Forrester and Dawn Richards |
| EPI_ISL_1491342 | Gravity Diagnostics | Kentucky State Public Health Lab | Stephanie Lunn, Karim George, Joshua Tobias, William Grooms, Vaneet Arora, Matthew Johnson, Rachel Zinner, Rhonda Lucas |
| EPI_ISL_1491343, EPI_ISL_1491344, EPI_ISL_1491345, EPI_ISL_1491346, EPI_ISL_1491347, EPI_ISL_1491348, EPI_ISL_1491349, EPI_ISL_1491350, EPI_ISL_1491351, EPI_ISL_1491352, EPI_ISL_1491353, EPI_ISL_1491354, EPI_ISL_1491355, EPI_ISL_1491356, EPI_ISL_1491357, EPI_ISL_1491358, EPI_ISL_1491359, EPI_ISL_1491360 | | | |
| see above | Kentucky State Public Health Lab | Kentucky State Public Health Lab | Stephanie Lunn, Karim George, Joshua Tobias, William Grooms, Vaneet Arora, Matthew Johnson, Rachel Zinner, Rhonda Lucas |
| EPI_ISL_1491361 | Baylor Esoteric and Molecular Lab | Baylor Esoteric and Molecular Lab | Elizabeth Forrester and Dawn Richards |
| EPI_ISL_1491362 | Imágenes Diagnósticas de Mazatlán | Microbial Genomics Laboratory | Bruno Gomez-Gil, Julissa Enciso-Ibarra, Alejandra Garcia-Gasca, Daniel Fregoso-Rueda |
| EPI_ISL_1491364 | Hospital Sharp | Microbial Genomics Laboratory | Bruno Gomez-Gil, Julissa Enciso-Ibarra, Alejandra Garcia-Gasca, Daniel Fregoso-Rueda |
| EPI_ISL_1491366 | Baylor Esoteric and Molecular Lab | Baylor Esoteric and Molecular Lab | Elizabeth Forrester and Dawn Richards |
| EPI_ISL_1491367 | Gravity Diagnostics | Kentucky State Public Health Lab | Stephanie Lunn, Karim George, Joshua Tobias, William Grooms, Vaneet Arora, Matthew Johnson, Rachel Zinner, Rhonda Lucas |
| EPI_ISL_1491368, EPI_ISL_1491369, EPI_ISL_1491370, EPI_ISL_1491371, EPI_ISL_1491372, EPI_ISL_1491373, EPI_ISL_1491374, EPI_ISL_1491375, EPI_ISL_1491376, EPI_ISL_1491377, EPI_ISL_1491378, EPI_ISL_1491379, EPI_ISL_1491380, EPI_ISL_1491381, EPI_ISL_1491382, EPI_ISL_1491384, EPI_ISL_1491385, EPI_ISL_1491386 | | | |
| see above | Kentucky State Public Health Lab | Kentucky State Public Health Lab | Stephanie Lunn, Karim George, Joshua Tobias, William Grooms, Vaneet Arora, Matthew Johnson, Rachel Zinner, Rhonda Lucas |
| EPI_ISL_1491387, EPI_ISL_1491388 | Baylor Esoteric and Molecular Lab | Baylor Esoteric and Molecular Lab | Elizabeth Forrester and Dawn Richards |
| EPI_ISL_1491390, EPI_ISL_1491391, EPI_ISL_1491392, EPI_ISL_1491393, EPI_ISL_1491394, EPI_ISL_1491395, EPI_ISL_1491396, EPI_ISL_1491397, EPI_ISL_1491398, EPI_ISL_1491399, EPI_ISL_1491400, EPI_ISL_1491401, EPI_ISL_1491402, EPI_ISL_1491403, EPI_ISL_1491404, EPI_ISL_1491405, EPI_ISL_1491406, EPI_ISL_1491407, EPI_ISL_1491408, EPI_ISL_1491409, EPI_ISL_1491410, EPI_ISL_1491411, EPI_ISL_1491412, EPI_ISL_1491413, EPI_ISL_1491414, EPI_ISL_1491415, EPI_ISL_1491416, EPI_ISL_1491417, EPI_ISL_1491418, EPI_ISL_1491419, EPI_ISL_1491420, EPI_ISL_1491421, EPI_ISL_1491422, EPI_ISL_1491423, EPI_ISL_1491424, EPI_ISL_1491425, EPI_ISL_1491426, EPI_ISL_1491427, EPI_ISL_1491428, EPI_ISL_1491429, EPI_ISL_1491430, EPI_ISL_1491431, EPI_ISL_1491432, EPI_ISL_1491433, EPI_ISL_1491434, EPI_ISL_1491435, EPI_ISL_1491436, EPI_ISL_1491437, EPI_ISL_1491438, EPI_ISL_1491439, EPI_ISL_1491440, EPI_ISL_1491441, EPI_ISL_1491442, EPI_ISL_1491443, EPI_ISL_1491444, EPI_ISL_1491445, EPI_ISL_1491446, EPI_ISL_1491447, EPI_ISL_1491448, EPI_ISL_1491449, EPI_ISL_1491450, EPI_ISL_1491451, EPI_ISL_1491452, EPI_ISL_1491453, EPI_ISL_1491454, EPI_ISL_1491455, EPI_ISL_1491456, EPI_ISL_1491457, EPI_ISL_1491458, EPI_ISL_1491459, EPI_ISL_1491460, EPI_ISL_1491461, EPI_ISL_1491462, EPI_ISL_1491463, EPI_ISL_1491464, EPI_ISL_1491465, EPI_ISL_1491466, EPI_ISL_1491467, EPI_ISL_1491468, EPI_ISL_1491469, EPI_ISL_1491470, EPI_ISL_1491471, EPI_ISL_1491472, EPI_ISL_1491473, EPI_ISL_1491474, EPI_ISL_1491475, EPI_ISL_1491476, EPI_ISL_1491477, EPI_ISL_1491478, EPI_ISL_1491479, EPI_ISL_1491480, EPI_ISL_1491481, EPI_ISL_1491482, EPI_ISL_1491483, EPI_ISL_1491484, EPI_ISL_1491485, EPI_ISL_1491486, EPI_ISL_1491487, EPI_ISL_1491488, EPI_ISL_1491489, EPI_ISL_1491490, EPI_ISL_1491491, EPI_ISL_1491492, EPI_ISL_1491493, EPI_ISL_1491494, EPI_ISL_1491495, EPI_ISL_1491496, EPI_ISL_1491497, EPI_ISL_1491498, EPI_ISL_1491499, EPI_ISL_1491500, EPI_ISL_1491501, EPI_ISL_1491502, EPI_ISL_1491503, EPI_ISL_1491504, EPI_ISL_1491505, EPI_ISL_1491506, EPI_ISL_1491507, EPI_ISL_1491508, EPI_ISL_1491509, EPI_ISL_1491510, EPI_ISL_1491511, EPI_ISL_1491512, EPI_ISL_1491513, EPI_ISL_1491514 | | | |
| see above | University of Michigan Clinical Microbiology Laboratory | Lauring Lab, University of Michigan, Department of Microbiology and Immunology | Valesano |
| EPI_ISL_1491515 | UPMC Children's Hospital of Pittsburgh | Genomics and Discovery, Respiratory Viruses Branch, Division of Viral Diseases, Centers for Disease Control and Prevention | Ying Tao, Jing Zhang, Yan Li, Brian Lynch, Anna Kelleher, Krista Queen, Anna Uehara, Peter Cook, Han Jia Justin Ng, Clinton R. Paden, Haibin Wang, Suxiang Tong |
| EPI_ISL_1491516, EPI_ISL_1491517 | "CDPH, Viral and Rickettsial Disease Laboratory" | Genomics and Discovery, Respiratory Viruses Branch, Division of Viral Diseases, Centers for Disease Control and Prevention | Ying Tao, Jing Zhang, Yan Li, Brian Lynch, Anna Kelleher, Krista Queen, Anna Uehara, Peter Cook, Han Jia Justin Ng, Clinton R. Paden, Haibin Wang, Suxiang Tong |
| EPI_ISL_1491518, EPI_ISL_1491519 | UPMC Children's Hospital of Pittsburgh | Genomics and Discovery, Respiratory Viruses Branch, Division of Viral Diseases, Centers for Disease Control and Prevention | Ying Tao, Jing Zhang, Yan Li, Brian Lynch, Anna Kelleher, Krista Queen, Anna Uehara, Peter Cook, Han Jia Justin Ng, Clinton R. Paden, Haibin Wang, Suxiang Tong |
| EPI_ISL_1491520 | Washington University School of Medicine | Genomics and Discovery, Respiratory Viruses Branch, Division of Viral Diseases, Centers for Disease Control and Prevention | Ying Tao, Jing Zhang, Yan Li, Brian Lynch, Anna Kelleher, Krista Queen, Anna Uehara, Peter Cook, Han Jia Justin Ng, Clinton R. Paden, Haibin Wang, Suxiang Tong |

| | | | |
|---|---|---|---|
| EPI_ISL_1491521 | UPMC Children's Hospital of Pittsburgh | Genomics and Discovery, Respiratory Viruses Branch, Division of Viral Diseases, Centers for Disease Control and Prevention | Ying Tao, Jing Zhang, Yan Li, Brian Lynch, Anna Kelleher, Krista Queen, Anna Uehara, Peter Cook, Han Jia Justin Ng, Clinton R. Paden, Haibin Wang, Suxiang Tong |
| EPI_ISL_1491522 | "CDPH, Viral and Rickettsial Disease Laboratory" | Genomics and Discovery, Respiratory Viruses Branch, Division of Viral Diseases, Centers for Disease Control and Prevention | Ying Tao, Jing Zhang, Yan Li, Brian Lynch, Anna Kelleher, Krista Queen, Anna Uehara, Peter Cook, Han Jia Justin Ng, Clinton R. Paden, Haibin Wang, Suxiang Tong |
| EPI_ISL_1491523, EPI_ISL_1491524 | DC Public Health Lab/ Dept. of Forensic Sciences | DC Public Health Lab/ Dept. of Forensic Sciences | Janis Doss, Scott Nguyen, Elizabeth Zelaya, Sarah Scott, Connie Maza, Monica Mann, Brittany Hamilton, David Payne, Jocelyn Hauser |
| EPI_ISL_1491734, EPI_ISL_1491735, EPI_ISL_1491736, EPI_ISL_1491737, EPI_ISL_1491738, EPI_ISL_1491739, EPI_ISL_1491740, EPI_ISL_1491741, EPI_ISL_1491742, EPI_ISL_1491743, EPI_ISL_1491744, EPI_ISL_1491745, EPI_ISL_1491746, EPI_ISL_1491747, EPI_ISL_1491748, EPI_ISL_1491749, EPI_ISL_1491750, EPI_ISL_1491751, EPI_ISL_1491752, EPI_ISL_1491753, EPI_ISL_1491754, EPI_ISL_1491755, EPI_ISL_1491756, EPI_ISL_1491757, EPI_ISL_1491758, EPI_ISL_1491760, EPI_ISL_1491761, EPI_ISL_1491762, EPI_ISL_1491763, EPI_ISL_1491764, EPI_ISL_1491765, EPI_ISL_1491766, EPI_ISL_1491767, EPI_ISL_1491768, EPI_ISL_1491769, EPI_ISL_1491770, EPI_ISL_1491771, EPI_ISL_1491772, EPI_ISL_1491773, EPI_ISL_1491774, EPI_ISL_1491775, EPI_ISL_1491776, EPI_ISL_1491777, EPI_ISL_1491778, EPI_ISL_1491779, EPI_ISL_1491780, EPI_ISL_1491781, EPI_ISL_1491782, EPI_ISL_1491783, EPI_ISL_1491784, EPI_ISL_1491785, EPI_ISL_1491786, EPI_ISL_1491787, EPI_ISL_1491788, EPI_ISL_1491789, EPI_ISL_1491790, EPI_ISL_1491791, EPI_ISL_1491792, EPI_ISL_1491793, EPI_ISL_1491794, EPI_ISL_1491795, EPI_ISL_1491796, EPI_ISL_1491797, EPI_ISL_1491798, EPI_ISL_1491799, EPI_ISL_1491800, EPI_ISL_1491801, EPI_ISL_1491802, EPI_ISL_1491803, EPI_ISL_1491804, EPI_ISL_1491805, EPI_ISL_1491806, EPI_ISL_1491808, EPI_ISL_1491809, EPI_ISL_1491811, EPI_ISL_1491812, EPI_ISL_1491813, EPI_ISL_1491814, EPI_ISL_1491815, EPI_ISL_1491816, EPI_ISL_1491817, EPI_ISL_1491818, EPI_ISL_1491819, EPI_ISL_1491820, EPI_ISL_1491821, EPI_ISL_1491822, EPI_ISL_1491823, EPI_ISL_1491824, EPI_ISL_1491825, EPI_ISL_1491826, EPI_ISL_1491827, EPI_ISL_1491828, EPI_ISL_1491829, EPI_ISL_1491830, EPI_ISL_1491831, EPI_ISL_1491832, EPI_ISL_1491833, EPI_ISL_1491834, EPI_ISL_1491835, EPI_ISL_1491836, EPI_ISL_1491837, EPI_ISL_1491838, EPI_ISL_1491839, EPI_ISL_1491840, EPI_ISL_1491841, EPI_ISL_1491842, EPI_ISL_1491843, EPI_ISL_1491844, EPI_ISL_1491845, EPI_ISL_1491846, EPI_ISL_1491847, EPI_ISL_1491848, EPI_ISL_1491849, EPI_ISL_1491850, EPI_ISL_1491851, EPI_ISL_1491852, EPI_ISL_1491853, EPI_ISL_1491854, EPI_ISL_1491855, EPI_ISL_1491856, EPI_ISL_1491857, EPI_ISL_1491858, EPI_ISL_1491859, EPI_ISL_1491860, EPI_ISL_1491861, EPI_ISL_1491862, EPI_ISL_1491863, EPI_ISL_1491864, EPI_ISL_1491866, EPI_ISL_1491867, EPI_ISL_1491868, EPI_ISL_1491869, EPI_ISL_1491870, EPI_ISL_1491871, EPI_ISL_1491872, EPI_ISL_1491873, EPI_ISL_1491874, EPI_ISL_1491875, EPI_ISL_1491876, EPI_ISL_1491877, EPI_ISL_1491878, EPI_ISL_1491879, EPI_ISL_1491880, EPI_ISL_1491881, EPI_ISL_1491882, EPI_ISL_1491883, EPI_ISL_1491884, EPI_ISL_1491885, EPI_ISL_1491886, EPI_ISL_1491887, EPI_ISL_1491888, EPI_ISL_1491889, EPI_ISL_1491890, EPI_ISL_1491891, EPI_ISL_1491892, EPI_ISL_1491894, EPI_ISL_1491895, EPI_ISL_1491896, EPI_ISL_1491897, EPI_ISL_1491898, EPI_ISL_1491899, EPI_ISL_1491900, EPI_ISL_1491901, EPI_ISL_1491902, EPI_ISL_1491904, EPI_ISL_1491905, EPI_ISL_1491907, EPI_ISL_1491908, EPI_ISL_1491909, EPI_ISL_1491910, EPI_ISL_1491913, EPI_ISL_1491914, EPI_ISL_1491915, EPI_ISL_1491916, EPI_ISL_1491917, EPI_ISL_1491918, EPI_ISL_1491919, EPI_ISL_1491920, EPI_ISL_1491921, EPI_ISL_1491922, EPI_ISL_1491923, EPI_ISL_1491924, EPI_ISL_1491925, EPI_ISL_1491926, EPI_ISL_1491927, EPI_ISL_1491928, EPI_ISL_1491929, EPI_ISL_1491930, EPI_ISL_1491931, EPI_ISL_1491933, EPI_ISL_1491934, EPI_ISL_1491935, EPI_ISL_1491936, EPI_ISL_1491937, EPI_ISL_1491938, EPI_ISL_1491939, EPI_ISL_1491940, EPI_ISL_1491941, EPI_ISL_1491942, EPI_ISL_1491943, EPI_ISL_1491944, EPI_ISL_1491945, EPI_ISL_1491946, EPI_ISL_1491947, EPI_ISL_1491948, EPI_ISL_1491949 | | | |
| see above | Aegis Sciences Corporation | Centers for Disease Control and Prevention Division of Viral Diseases, Pathogen Discovery | Dakota Howard, Dhwani Batra, Peter W. Cook, Kara Moser, Adrian Paskey, Jason Caravas, Benjamin Rambo-Martin, Shatavia Morrison, Christopher Gulvick, Scott Sammons, Yvette Unoarumhi, Darlene Wagner, Matthew Schmerer, Cyndi Clark, Patrick Campbell, Rob Case, Vikramsinha Ghorpade, Holly Houdeshell, Ola Kvalvaag, Dillon Nall, Ethan Sanders, Alec Vest, Shaun Westlund, Matthew Hardison, Clinton R. Paden, Duncan MacCannell |
| EPI_ISL_1491950, EPI_ISL_1491951, EPI_ISL_1491952, EPI_ISL_1491953, EPI_ISL_1491954, EPI_ISL_1491955, EPI_ISL_1491956, EPI_ISL_1491957, EPI_ISL_1491958, EPI_ISL_1491959, EPI_ISL_1491960, EPI_ISL_1491961, EPI_ISL_1491962, EPI_ISL_1491963, EPI_ISL_1491964, EPI_ISL_1491965, EPI_ISL_1491966, EPI_ISL_1491967, EPI_ISL_1491968, EPI_ISL_1491969, EPI_ISL_1491970, EPI_ISL_1491971, EPI_ISL_1491972, EPI_ISL_1491973, EPI_ISL_1491974, EPI_ISL_1491975, EPI_ISL_1491976, EPI_ISL_1491977, EPI_ISL_1491978, EPI_ISL_1491979, EPI_ISL_1491980, EPI_ISL_1491981, EPI_ISL_1491982, EPI_ISL_1491983, EPI_ISL_1491984, EPI_ISL_1491985, EPI_ISL_1491986, EPI_ISL_1491987, EPI_ISL_1491988, EPI_ISL_1491989, EPI_ISL_1491990, EPI_ISL_1491991, EPI_ISL_1491992, EPI_ISL_1491993, EPI_ISL_1491994, EPI_ISL_1491995, EPI_ISL_1491996, EPI_ISL_1491997, EPI_ISL_1491998, EPI_ISL_1491999, EPI_ISL_1492000, EPI_ISL_1492001, EPI_ISL_1492002, EPI_ISL_1492003, EPI_ISL_1492004, EPI_ISL_1492005, EPI_ISL_1492006, EPI_ISL_1492007, EPI_ISL_1492008, EPI_ISL_1492009, EPI_ISL_1492010, EPI_ISL_1492011, EPI_ISL_1492012, EPI_ISL_1492013, EPI_ISL_1492014, EPI_ISL_1492015, EPI_ISL_1492016, EPI_ISL_1492017, EPI_ISL_1492018, EPI_ISL_1492019, EPI_ISL_1492020, EPI_ISL_1492021, EPI_ISL_1492022, EPI_ISL_1492023, EPI_ISL_1492024, EPI_ISL_1492025, EPI_ISL_1492026, EPI_ISL_1492027, EPI_ISL_1492028, EPI_ISL_1492029, EPI_ISL_1492030, EPI_ISL_1492031, EPI_ISL_1492032, EPI_ISL_1492033, EPI_ISL_1492034, EPI_ISL_1492035, EPI_ISL_1492036, EPI_ISL_1492037, EPI_ISL_1492038, EPI_ISL_1492039, EPI_ISL_1492040, EPI_ISL_1492041, EPI_ISL_1492042, EPI_ISL_1492043, EPI_ISL_1492044, EPI_ISL_1492045, EPI_ISL_1492046, EPI_ISL_1492047, EPI_ISL_1492048, EPI_ISL_1492049, EPI_ISL_1492050, EPI_ISL_1492051, EPI_ISL_1492052, EPI_ISL_1492053, EPI_ISL_1492054, EPI_ISL_1492055, EPI_ISL_1492056, EPI_ISL_1492057, EPI_ISL_1492058, EPI_ISL_1492059, EPI_ISL_1492060, EPI_ISL_1492061, EPI_ISL_1492062, EPI_ISL_1492063, EPI_ISL_1492064, EPI_ISL_1492065, EPI_ISL_1492066, EPI_ISL_1492067, EPI_ISL_1492068, EPI_ISL_1492069, EPI_ISL_1492072, EPI_ISL_1492075, EPI_ISL_1492078, EPI_ISL_1492080, EPI_ISL_1492083, EPI_ISL_1492086, EPI_ISL_1492088, EPI_ISL_1492091, EPI_ISL_1492094, EPI_ISL_1492096, EPI_ISL_1492098, EPI_ISL_1492101, EPI_ISL_1492104, EPI_ISL_1492106, EPI_ISL_1492109, EPI_ISL_1492112, EPI_ISL_1492115, EPI_ISL_1492118, EPI_ISL_1492120, EPI_ISL_1492122, EPI_ISL_1492125, EPI_ISL_1492128, EPI_ISL_1492131, EPI_ISL_1492134, EPI_ISL_1492136, EPI_ISL_1492139, EPI_ISL_1492142, EPI_ISL_1492145 | | | |
| see above | Quest Diagnostics Incorporated | Centers for Disease Control and Prevention Division of Viral Diseases, Pathogen Discovery | Dakota Howard, Dhwani Batra, Peter W. Cook, Kara Moser, Adrian Paskey, Jason Caravas, Benjamin Rambo-Martin, Shatavia Morrison, Christopher Gulvick, Scott Sammons, Yvette Unoarumhi, Darlene Wagner, Matthew Schmerer, S. H. Rosenthal, A. Gerasimova, R. M. Kagan, B. Anderson, M. Hua, Y. Liu, L.E. Bernstein, K.E. Livingston, A. Perez, I. A. Shlyakhter, R. V. Rolando, R. Owen, P. Tanpaiboon, F. Lacbawan, Clinton R. Paden, Duncan MacCannell |
| EPI_ISL_1492444, EPI_ISL_1492445, EPI_ISL_1492446, EPI_ISL_1492447, EPI_ISL_1492449, EPI_ISL_1492450, EPI_ISL_1492451, EPI_ISL_1492452, EPI_ISL_1492454, EPI_ISL_1492455 | Tampa General Hospital Esoteric Lab | Tampa General Hospital Esoteric Research & Development Lab | Grant Vestal, Deanna Becker, Dominic Uy, Vicki Healer, Amorce Lima, Suzane Silbert |
| EPI_ISL_1492456 | LESP Jalisco | Instituto de diagnóstico y Referencia Epidemiologicos (INDRE) | Claudia Wong-Arambula, Abril Rodriguez-Maldonado, Vanessa Rivero-Arredondo, Ariadna Medina-Benitez, Joaquin Quiroz-Mercado, Sergio Rangel-Guerrero, Natividad Cruz-Ortiz, Tatiana Nunez-Garcia, Gisela Barrera-Badillo, Lucia Hernandez-Rivas, Irma Lopez-Martinez, Ernesto Ramirez-Gonzalez. |
| EPI_ISL_1492620, EPI_ISL_1492622, EPI_ISL_1492623, EPI_ISL_1492624, EPI_ISL_1492625, EPI_ISL_1492626, EPI_ISL_1492627, EPI_ISL_1492628, EPI_ISL_1492629, EPI_ISL_1492630, EPI_ISL_1492631, EPI_ISL_1492633, EPI_ISL_1492634, EPI_ISL_1492635, EPI_ISL_1492636 | | | |
| see above | Idaho Bureau of Laboratories | Idaho Bureau of Laboratories | "R. Beukelman, Matthew Charles Burns, Aimee Ceniseros, Robert L. Voermans, Christopher Ball" |
| EPI_ISL_1493040, EPI_ISL_1493041, EPI_ISL_1493042, EPI_ISL_1493043, EPI_ISL_1493044, EPI_ISL_1493045, EPI_ISL_1493046, EPI_ISL_1493047, EPI_ISL_1493048, EPI_ISL_1493049, EPI_ISL_1493050, EPI_ISL_1493051, EPI_ISL_1493052, EPI_ISL_1493053, EPI_ISL_1493054, EPI_ISL_1493055, EPI_ISL_1493056, EPI_ISL_1493057, EPI_ISL_1493058 | | | |
| see above | University of Michigan Clinical Microbiology Laboratory | Lauring Lab, University of Michigan, Department of Microbiology and Immunology | Valesano |
| EPI_ISL_1493060, EPI_ISL_1493061 | Oregon State Public Health Laboratory | Oregon State Public Health Laboratory | Rafia Razzaque, Eugene Yeboah, Vanda Makris, John Fontana and Shane Sevey |
| EPI_ISL_1493062, EPI_ISL_1493063, EPI_ISL_1493064 | Michigan Department of Health and Human Services, Bureau of Laboratories | Michigan Department of Health and Human Services, Bureau of Laboratories | Blankenship HM, Riner D, Soehnlen MK |
| EPI_ISL_1493065 | University of Wisconsin-Madison AIDS Vaccine Research Laboratories | University of Wisconsin-Madison AIDS Vaccine Research Laboratories | Gage Moreno, Katarina Braun, et al. AIDS Vaccine Research Laboratories |
| EPI_ISL_1493067, EPI_ISL_1493068 | Oregon State Public Health Laboratory | Oregon State Public Health Laboratory | Rafia Razzaque, Eugene Yeboah, Vanda Makris, John Fontana and Shane Sevey |
| EPI_ISL_1493070, EPI_ISL_1493072, EPI_ISL_1493073, EPI_ISL_1493074, EPI_ISL_1493078, EPI_ISL_1493079, EPI_ISL_1493080, EPI_ISL_1493083, EPI_ISL_1493084, EPI_ISL_1493085, EPI_ISL_1493087, EPI_ISL_1493088, EPI_ISL_1493089 | | | |
| see above | Michigan Department of Health and Human Services, Bureau of Laboratories | Michigan Department of Health and Human Services, Bureau of Laboratories | Blankenship HM, Riner D, Soehnlen MK |
| EPI_ISL_1493105, EPI_ISL_1493109 | Baylor Esoteric and Molecular Lab | Baylor Esoteric and Molecular Lab | Elizabeth Forrester and Dawn Richards |
| EPI_ISL_1493110, EPI_ISL_1493111, EPI_ISL_1493114, EPI_ISL_1493115, EPI_ISL_1493117, EPI_ISL_1493118, EPI_ISL_1493119, EPI_ISL_1493120, EPI_ISL_1493121, EPI_ISL_1493122 | University of Wisconsin-Madison AIDS Vaccine Research Laboratories | University of Wisconsin-Madison AIDS Vaccine Research Laboratories | Gage Moreno, Katarina Braun, et al. AIDS Vaccine Research Laboratories |
| EPI_ISL_1493123 | Baylor Esoteric and Molecular Lab | Baylor Esoteric and Molecular Lab | Elizabeth Forrester and Dawn Richards |
| EPI_ISL_1493124 | Oregon State Public Health Laboratory | Oregon State Public Health Laboratory | Rafia Razzaque, Eugene Yeboah, Vanda Makris, Laura Tsaknaridis, John Fontana and Shane Sevey |
| EPI_ISL_1493125, EPI_ISL_1493126, EPI_ISL_1493127, EPI_ISL_1493128, EPI_ISL_1493129, EPI_ISL_1493130, EPI_ISL_1493131, EPI_ISL_1493132, EPI_ISL_1493133, EPI_ISL_1493134, EPI_ISL_1493135, EPI_ISL_1493136, EPI_ISL_1493137, EPI_ISL_1493138, EPI_ISL_1493139, EPI_ISL_1493140, EPI_ISL_1493141, EPI_ISL_1493142, EPI_ISL_1493143, EPI_ISL_1493144 | | | |
| see above | University of Wisconsin-Madison AIDS Vaccine Research Laboratories | University of Wisconsin-Madison AIDS Vaccine Research Laboratories | Gage Moreno, Katarina Braun, et al. AIDS Vaccine Research Laboratories |
| EPI_ISL_1493145 | Oregon State Public Health Laboratory | Oregon State Public Health Laboratory | Rafia Razzaque, Eugene Yeboah, Vanda Makris, Laura Tsaknaridis, John Fontana and Shane Sevey |
| EPI_ISL_1493146, EPI_ISL_1493147, EPI_ISL_1493149, EPI_ISL_1493150, EPI_ISL_1493151, EPI_ISL_1493152, EPI_ISL_1493153, EPI_ISL_1493154, EPI_ISL_1493155, EPI_ISL_1493156, EPI_ISL_1493157, EPI_ISL_1493158, EPI_ISL_1493160, EPI_ISL_1493161, EPI_ISL_1493162, EPI_ISL_1493163, EPI_ISL_1493164 | | | |
| see above | Nevada State Public Health Laboratory | Nevada State Public Health Laboratory | Andrew Gorzalski, Mark Pandori |
| EPI_ISL_1493165, EPI_ISL_1493166, EPI_ISL_1493167 | Oregon State Public Health Laboratory | Oregon State Public Health Laboratory | Rafia Razzaque, Eugene Yeboah, Vanda Makris, Laura Tsaknaridis, John Fontana and Shane Sevey |
| EPI_ISL_1493168, EPI_ISL_1493169, EPI_ISL_1493170, EPI_ISL_1493171, EPI_ISL_1493172, EPI_ISL_1493173, EPI_ISL_1493174, EPI_ISL_1493175, EPI_ISL_1493176, EPI_ISL_1493177, EPI_ISL_1493178, EPI_ISL_1493179, EPI_ISL_1493180, EPI_ISL_1493181, EPI_ISL_1493182, EPI_ISL_1493183, EPI_ISL_1493184, EPI_ISL_1493185, EPI_ISL_1493186, EPI_ISL_1493187, EPI_ISL_1493188, EPI_ISL_1493189, EPI_ISL_1493190, EPI_ISL_1493191, EPI_ISL_1493192, EPI_ISL_1493193, EPI_ISL_1493194, EPI_ISL_1493195, EPI_ISL_1493196, EPI_ISL_1493197, EPI_ISL_1493198, EPI_ISL_1493199, EPI_ISL_1493200, EPI_ISL_1493201, EPI_ISL_1493202, EPI_ISL_1493203, EPI_ISL_1493204, EPI_ISL_1493205, EPI_ISL_1493206, EPI_ISL_1493207, EPI_ISL_1493208, EPI_ISL_1493209, EPI_ISL_1493210, EPI_ISL_1493211, EPI_ISL_1493212, EPI_ISL_1493213, EPI_ISL_1493214, EPI_ISL_1493215, EPI_ISL_1493216, EPI_ISL_1493217, EPI_ISL_1493218, EPI_ISL_1493219, EPI_ISL_1493220, EPI_ISL_1493221, EPI_ISL_1493222, EPI_ISL_1493223, EPI_ISL_1493224, EPI_ISL_1493225, EPI_ISL_1493226, EPI_ISL_1493227, EPI_ISL_1493228, EPI_ISL_1493229, EPI_ISL_1493230, EPI_ISL_1493231, EPI_ISL_1493232, EPI_ISL_1493233, EPI_ISL_1493234, EPI_ISL_1493235, EPI_ISL_1493236, EPI_ISL_1493237, EPI_ISL_1493238, EPI_ISL_1493239, EPI_ISL_1493240, EPI_ISL_1493241, EPI_ISL_1493242, EPI_ISL_1493243, EPI_ISL_1493244, EPI_ISL_1493245, EPI_ISL_1493246, EPI_ISL_1493247, EPI_ISL_1493248, EPI_ISL_1493249, EPI_ISL_1493250, EPI_ISL_1493251, EPI_ISL_1493252, EPI_ISL_1493253, EPI_ISL_1493254, EPI_ISL_1493255, EPI_ISL_1493256, EPI_ISL_1493257 | | | |
| see above | MD PHL | MD PHL | Maryland Department of Health Laboratories Administration |
| EPI_ISL_1493258, EPI_ISL_1493259, EPI_ISL_1493260, EPI_ISL_1493261, EPI_ISL_1493262, EPI_ISL_1493263, EPI_ISL_1493264, EPI_ISL_1493265, EPI_ISL_1493266, EPI_ISL_1493267, EPI_ISL_1493268, EPI_ISL_1493363, EPI_ISL_1493364, EPI_ISL_1493365 | | | |
| see above | Oregon State Public Health Laboratory | Oregon State Public Health Laboratory | Rafia Razzaque, Eugene Yeboah, Vanda Makris, Laura Tsaknaridis, John Fontana and Shane Sevey |
| EPI_ISL_1493366 | Willamette Valley Toxicology | Oregon State Public Health Laboratory | Rafia Razzaque, Eugene Yeboah, Vanda Makris, Laura Tsaknaridis, John Fontana and Shane Sevey |
| EPI_ISL_1493367, EPI_ISL_1493368, EPI_ISL_1493369, EPI_ISL_1493370, EPI_ISL_1493371, EPI_ISL_1493372, EPI_ISL_1493373, EPI_ISL_1493374, EPI_ISL_1493375, EPI_ISL_1493376, EPI_ISL_1493377, EPI_ISL_1493378, EPI_ISL_1493379, EPI_ISL_1493380, EPI_ISL_1493381, EPI_ISL_1493382, EPI_ISL_1493383, EPI_ISL_1493384, EPI_ISL_1493385, EPI_ISL_1493386, EPI_ISL_1493387, EPI_ISL_1493388 | | | |
| see above | City of Milwaukee Health Department | City of Milwaukee Health Department | Sanjib Bhattacharyya |

| | | | |
|---|---|---|---|
| | Laboratory | Laboratory | |
| EPI_ISL_1493389, EPI_ISL_1493390, EPI_ISL_1493391, EPI_ISL_1493392, EPI_ISL_1493393, EPI_ISL_1493394, EPI_ISL_1493395, EPI_ISL_1493396, EPI_ISL_1493397, EPI_ISL_1493398, EPI_ISL_1493400, EPI_ISL_1493401, EPI_ISL_1493402, EPI_ISL_1493403, EPI_ISL_1493404, EPI_ISL_1493405, EPI_ISL_1493406, EPI_ISL_1493407, EPI_ISL_1493408, EPI_ISL_1493409, EPI_ISL_1493410, EPI_ISL_1493411, EPI_ISL_1493412, EPI_ISL_1493413, EPI_ISL_1493414, EPI_ISL_1493415, EPI_ISL_1493416, EPI_ISL_1493417, EPI_ISL_1493418, EPI_ISL_1493419, EPI_ISL_1493420, EPI_ISL_1493421, EPI_ISL_1493422, EPI_ISL_1493423, EPI_ISL_1493424, EPI_ISL_1493425, EPI_ISL_1493426, EPI_ISL_1493427, EPI_ISL_1493428, EPI_ISL_1493429, EPI_ISL_1493430, EPI_ISL_1493431, EPI_ISL_1493432, EPI_ISL_1493433, EPI_ISL_1493434, EPI_ISL_1493435 | | | |
| see above | New Mexico Department of Health Scientific Laboratory | New Mexico Department of Health Scientific Laboratory | Ellie Johnson, Anastacia Griego-Fisher, D'eldra Malone, Jennifer Benoit |
| EPI_ISL_1493436, EPI_ISL_1493437, EPI_ISL_1493438, EPI_ISL_1493439 | MD PHL | MD PHL | Maryland Department of Health Laboratories Administration |
| EPI_ISL_1493440, EPI_ISL_1493441, EPI_ISL_1493442, EPI_ISL_1493443, EPI_ISL_1493444, EPI_ISL_1493445, EPI_ISL_1493446, EPI_ISL_1493447, EPI_ISL_1493448, EPI_ISL_1493449, EPI_ISL_1493450, EPI_ISL_1493451, EPI_ISL_1493452, EPI_ISL_1493453, EPI_ISL_1493454, EPI_ISL_1493455, EPI_ISL_1493456, EPI_ISL_1493457, EPI_ISL_1493458, EPI_ISL_1493460, EPI_ISL_1493461, EPI_ISL_1493462, EPI_ISL_1493464, EPI_ISL_1493465, EPI_ISL_1493466, EPI_ISL_1493469, EPI_ISL_1493471, EPI_ISL_1493474, EPI_ISL_1493475, EPI_ISL_1493476, EPI_ISL_1493479, EPI_ISL_1493482, EPI_ISL_1493483, EPI_ISL_1493484, EPI_ISL_1493485, EPI_ISL_1493486, EPI_ISL_1493487 | | | |
| see above | NJDOH, Public Health and Environmental Laboratories | NJ_PHEL | Lindsey Bodnar, Shiv K. Verma, Dana Woell, Byeong Jeong |
| EPI_ISL_1493494, EPI_ISL_1493495, EPI_ISL_1493496, EPI_ISL_1493497, EPI_ISL_1493498, EPI_ISL_1493499, EPI_ISL_1493500, EPI_ISL_1493501, EPI_ISL_1493502, EPI_ISL_1493503, EPI_ISL_1493504, EPI_ISL_1493505, EPI_ISL_1493506, EPI_ISL_1493507, EPI_ISL_1493508, EPI_ISL_1493509, EPI_ISL_1493510, EPI_ISL_1493511, EPI_ISL_1493512, EPI_ISL_1493513, EPI_ISL_1493514, EPI_ISL_1493515, EPI_ISL_1493516, EPI_ISL_1493517, EPI_ISL_1493518, EPI_ISL_1493519, EPI_ISL_1493520, EPI_ISL_1493521, EPI_ISL_1493522, EPI_ISL_1493523, EPI_ISL_1493524, EPI_ISL_1493525, EPI_ISL_1493526, EPI_ISL_1493527, EPI_ISL_1493528, EPI_ISL_1493529, EPI_ISL_1493530, EPI_ISL_1493531, EPI_ISL_1493532, EPI_ISL_1493533, EPI_ISL_1493534, EPI_ISL_1493535, EPI_ISL_1493536, EPI_ISL_1493537, EPI_ISL_1493538, EPI_ISL_1493539, EPI_ISL_1493540, EPI_ISL_1493541, EPI_ISL_1493542, EPI_ISL_1493543, EPI_ISL_1493544, EPI_ISL_1493545, EPI_ISL_1493546, EPI_ISL_1493547, EPI_ISL_1493548, EPI_ISL_1493549, EPI_ISL_1493550, EPI_ISL_1493551, EPI_ISL_1493552, EPI_ISL_1493553, EPI_ISL_1493554, EPI_ISL_1493555, EPI_ISL_1493556, EPI_ISL_1493557, EPI_ISL_1493558, EPI_ISL_1493559, EPI_ISL_1493560, EPI_ISL_1493561, EPI_ISL_1493562, EPI_ISL_1493563, EPI_ISL_1493564, EPI_ISL_1493565, EPI_ISL_1493566, EPI_ISL_1493567, EPI_ISL_1493568, EPI_ISL_1493569, EPI_ISL_1493570, EPI_ISL_1493571 | | | |
| see above | Aegis Sciences Corporation | Centers for Disease Control and Prevention Division of Viral Diseases, Pathogen Discovery | Dakota Howard, Dhwani Batra, Peter W. Cook, Kara Moser, Adrian Paskey, Jason Caravas, Benjamin Rambo-Martin, Shatavia Morrison, Christopher Gulvick, Scott Sammons, Yvette Unoarumhi, Darlene Wagner, Matthew Schmerer, Cyndi Clark, Patrick Campbell, Rob Case, Vikramsinha Ghorpade, Holly Houdeshell, Ola Kvalvaag, Dillon Nall, Ethan Sanders, Alec Vest, Shaun Westlund, Matthew Hardison, Clinton R. Paden, Duncan MacCannell |
| EPI_ISL_1493601, EPI_ISL_1493602, EPI_ISL_1493603, EPI_ISL_1493604, EPI_ISL_1493605, EPI_ISL_1493606, EPI_ISL_1493607, EPI_ISL_1493608, EPI_ISL_1493609, EPI_ISL_1493610, EPI_ISL_1493611, EPI_ISL_1493612, EPI_ISL_1493613, EPI_ISL_1493614, EPI_ISL_1493615, EPI_ISL_1493616, EPI_ISL_1493617, EPI_ISL_1493618, EPI_ISL_1493619, EPI_ISL_1493620, EPI_ISL_1493621, EPI_ISL_1493622, EPI_ISL_1493623, EPI_ISL_1493624, EPI_ISL_1493625, EPI_ISL_1493626, EPI_ISL_1493627, EPI_ISL_1493628, EPI_ISL_1493629, EPI_ISL_1493630, EPI_ISL_1493631, EPI_ISL_1493632, EPI_ISL_1493633, EPI_ISL_1493634, EPI_ISL_1493635, EPI_ISL_1493636, EPI_ISL_1493637, EPI_ISL_1493638, EPI_ISL_1493639, EPI_ISL_1493640, EPI_ISL_1493641, EPI_ISL_1493642, EPI_ISL_1493643, EPI_ISL_1493644, EPI_ISL_1493645, EPI_ISL_1493646, EPI_ISL_1493647, EPI_ISL_1493648, EPI_ISL_1493649, EPI_ISL_1493650, EPI_ISL_1493651, EPI_ISL_1493652, EPI_ISL_1493653, EPI_ISL_1493654, EPI_ISL_1493655, EPI_ISL_1493656, EPI_ISL_1493657, EPI_ISL_1493658, EPI_ISL_1493659, EPI_ISL_1493660, EPI_ISL_1493661, EPI_ISL_1493662, EPI_ISL_1493663, EPI_ISL_1493664, EPI_ISL_1493665, EPI_ISL_1493666, EPI_ISL_1493667, EPI_ISL_1493668, EPI_ISL_1493669, EPI_ISL_1493670, EPI_ISL_1493671, EPI_ISL_1493672, EPI_ISL_1493673, EPI_ISL_1493674, EPI_ISL_1493675, EPI_ISL_1493676, EPI_ISL_1493677, EPI_ISL_1493678, EPI_ISL_1493679, EPI_ISL_1493680, EPI_ISL_1493681, EPI_ISL_1493682, EPI_ISL_1493683, EPI_ISL_1493684, EPI_ISL_1493685, EPI_ISL_1493686, EPI_ISL_1493687, EPI_ISL_1493688, EPI_ISL_1493689, EPI_ISL_1493690, EPI_ISL_1493691, EPI_ISL_1493692, EPI_ISL_1493693, EPI_ISL_1493694, EPI_ISL_1493695, EPI_ISL_1493696, EPI_ISL_1493697, EPI_ISL_1493698, EPI_ISL_1493699, EPI_ISL_1493700, EPI_ISL_1493701, EPI_ISL_1493702, EPI_ISL_1493703, EPI_ISL_1493704, EPI_ISL_1493705, EPI_ISL_1493706, EPI_ISL_1493707, EPI_ISL_1493708, EPI_ISL_1493709, EPI_ISL_1493710, EPI_ISL_1493711, EPI_ISL_1493712, EPI_ISL_1493713, EPI_ISL_1493714, EPI_ISL_1493715, EPI_ISL_1493716, EPI_ISL_1493717, EPI_ISL_1493718, EPI_ISL_1493719, EPI_ISL_1493720, EPI_ISL_1493721 | | | |
| see above | Quest Diagnostics Incorporated | Centers for Disease Control and Prevention Division of Viral Diseases, Pathogen Discovery | Dakota Howard, Dhwani Batra, Peter W. Cook, Kara Moser, Adrian Paskey, Jason Caravas, Benjamin Rambo-Martin, Shatavia Morrison, Christopher Gulvick, Scott Sammons, Yvette Unoarumhi, Darlene Wagner, Matthew Schmerer, S. H. Rosenthal, A. Gerasimova, R. M. Kagan, B. Anderson, M. Hua, Y. Liu, L.E. Bernstein, K.E. Livingston, A. Perez, I. A. Shlyakhter, R. V. Rolando, R. Owen, P. Tanpaiboon, F. Lacbawan, Clinton R. Paden, Duncan MacCannell |
| EPI_ISL_1493722, EPI_ISL_1493723, EPI_ISL_1493724, EPI_ISL_1493725, EPI_ISL_1493726, EPI_ISL_1493727, EPI_ISL_1493728, EPI_ISL_1493729, EPI_ISL_1493730, EPI_ISL_1493731, EPI_ISL_1493732, EPI_ISL_1493733, EPI_ISL_1493734, EPI_ISL_1493735, EPI_ISL_1493736, EPI_ISL_1493737, EPI_ISL_1493738, EPI_ISL_1493739, EPI_ISL_1493740, EPI_ISL_1493741, EPI_ISL_1493742, EPI_ISL_1493743, EPI_ISL_1493744, EPI_ISL_1493745, EPI_ISL_1493746, EPI_ISL_1493747, EPI_ISL_1493748, EPI_ISL_1493749, EPI_ISL_1493750, EPI_ISL_1493751, EPI_ISL_1493752, EPI_ISL_1493753, EPI_ISL_1493754, EPI_ISL_1493755, EPI_ISL_1493756, EPI_ISL_1493757, EPI_ISL_1493758, EPI_ISL_1493759, EPI_ISL_1493760, EPI_ISL_1493761, EPI_ISL_1493762, EPI_ISL_1493763, EPI_ISL_1493764, EPI_ISL_1493765, EPI_ISL_1493766, EPI_ISL_1493767, EPI_ISL_1493768, EPI_ISL_1493769, EPI_ISL_1493770, EPI_ISL_1493771, EPI_ISL_1493772, EPI_ISL_1493773, EPI_ISL_1493774, EPI_ISL_1493775, EPI_ISL_1493776, EPI_ISL_1493777, EPI_ISL_1493778, EPI_ISL_1493779, EPI_ISL_1493780, EPI_ISL_1493781, EPI_ISL_1493782, EPI_ISL_1493783, EPI_ISL_1493784, EPI_ISL_1493785, EPI_ISL_1493786, EPI_ISL_1493787, EPI_ISL_1493788, EPI_ISL_1493789, EPI_ISL_1493790, EPI_ISL_1493791, EPI_ISL_1493792, EPI_ISL_1493793, EPI_ISL_1493794, EPI_ISL_1493795, EPI_ISL_1493796, EPI_ISL_1493797, EPI_ISL_1493798, EPI_ISL_1493799, EPI_ISL_1493800, EPI_ISL_1493801, EPI_ISL_1493802, EPI_ISL_1493803, EPI_ISL_1493804, EPI_ISL_1493806, EPI_ISL_1493807, EPI_ISL_1493808, EPI_ISL_1493809, EPI_ISL_1493810, EPI_ISL_1493811, EPI_ISL_1493812, EPI_ISL_1493813, EPI_ISL_1493814, EPI_ISL_1493815, EPI_ISL_1493816, EPI_ISL_1493817, EPI_ISL_1493818, EPI_ISL_1493819, EPI_ISL_1493820, EPI_ISL_1493821, EPI_ISL_1493822, EPI_ISL_1493823, EPI_ISL_1493824, EPI_ISL_1493825, EPI_ISL_1493826, EPI_ISL_1493827, EPI_ISL_1493828, EPI_ISL_1493829, EPI_ISL_1493830, EPI_ISL_1493831, EPI_ISL_1493832, EPI_ISL_1493833, EPI_ISL_1493835, EPI_ISL_1493836, EPI_ISL_1493837, EPI_ISL_1493838, EPI_ISL_1493839, EPI_ISL_1493840, EPI_ISL_1493841, EPI_ISL_1493842, EPI_ISL_1493843, EPI_ISL_1493845, EPI_ISL_1493846, EPI_ISL_1493847, EPI_ISL_1493848, EPI_ISL_1493849, EPI_ISL_1493850, EPI_ISL_1493851, EPI_ISL_1493852, EPI_ISL_1493853, EPI_ISL_1493854, EPI_ISL_1493855, EPI_ISL_1493856, EPI_ISL_1493857, EPI_ISL_1493858, EPI_ISL_1493859, EPI_ISL_1493860, EPI_ISL_1493861, EPI_ISL_1493862, EPI_ISL_1493863, EPI_ISL_1493864, EPI_ISL_1493865, EPI_ISL_1493866, EPI_ISL_1493867, EPI_ISL_1493868, EPI_ISL_1493869, EPI_ISL_1493870, EPI_ISL_1493871, EPI_ISL_1493872, EPI_ISL_1493873, EPI_ISL_1493874, EPI_ISL_1493875, EPI_ISL_1493876, EPI_ISL_1493877, EPI_ISL_1493878, EPI_ISL_1493879, EPI_ISL_1493880, EPI_ISL_1493881, EPI_ISL_1493882, EPI_ISL_1493884, EPI_ISL_1493885, EPI_ISL_1493886, EPI_ISL_1493887, EPI_ISL_1493888, EPI_ISL_1493889, EPI_ISL_1493890, EPI_ISL_1493892, EPI_ISL_1493893, EPI_ISL_1493894, EPI_ISL_1493895, EPI_ISL_1493896, EPI_ISL_1493897, EPI_ISL_1493898, EPI_ISL_1493899, EPI_ISL_1493900, EPI_ISL_1493901, EPI_ISL_1493903, EPI_ISL_1493904, EPI_ISL_1493905, EPI_ISL_1493907, EPI_ISL_1493908, EPI_ISL_1493909, EPI_ISL_1493910, EPI_ISL_1493911, EPI_ISL_1493912, EPI_ISL_1493913, EPI_ISL_1493914, EPI_ISL_1493915, EPI_ISL_1493916, EPI_ISL_1493917, EPI_ISL_1493918, EPI_ISL_1493919, EPI_ISL_1493920, EPI_ISL_1493921, EPI_ISL_1493922, EPI_ISL_1493923, EPI_ISL_1493924, EPI_ISL_1493925, EPI_ISL_1493926, EPI_ISL_1493927, EPI_ISL_1493928, EPI_ISL_1493929, EPI_ISL_1493930, EPI_ISL_1493931, EPI_ISL_1493932, EPI_ISL_1493933, EPI_ISL_1493934, EPI_ISL_1493935, EPI_ISL_1493936, EPI_ISL_1493937, EPI_ISL_1493938 | | | |
| see above | Helix/Illumina | Centers for Disease Control and Prevention Division of Viral Diseases, Pathogen Discovery | Dakota Howard, Dhwani Batra, Peter W. Cook, Kara Moser, Adrian Paskey, Jason Caravas, Benjamin Rambo-Martin, Shatavia Morrison, Christopher Gulvick, Scott Sammons, Yvette Unoarumhi, Darlene Wagner, Matthew Schmerer, Eileen de Feo, Jan Antico, Christine Tran, Matthew Tolentino, Shannon Wickline, Kim Gietzen, Brad Sickler, Jingtao Liu, Eric Allen, Phil Febbo, Nicole L. Washington, Simon White, Geraint Levan, Kelly Schiabor Barrett, Elizabeth Cirulli, Alexandre Bolze, Ary Ascencio, Charlotte Rivera-Garcia, Ryan Cho, Jason Nguyen, Sherry Wang, Jimmy Ramirez, Tyler Cassens, Efren Sandoval, Magnus Isaksson, William Lee, David Becker, Marc Laurent, James Lu, Clinton R. Paden, Duncan MacCannell |
| EPI_ISL_1493939, EPI_ISL_1493940, EPI_ISL_1493941, EPI_ISL_1493942, EPI_ISL_1493943, EPI_ISL_1493944, EPI_ISL_1493945, EPI_ISL_1493946, EPI_ISL_1493947, EPI_ISL_1493948, EPI_ISL_1493949, EPI_ISL_1493950, EPI_ISL_1493951, EPI_ISL_1493952, EPI_ISL_1493953, EPI_ISL_1493954, EPI_ISL_1493955, EPI_ISL_1493956, EPI_ISL_1493957, EPI_ISL_1493958, EPI_ISL_1493959, EPI_ISL_1493960, EPI_ISL_1493961, EPI_ISL_1493962, EPI_ISL_1493963, EPI_ISL_1493964, EPI_ISL_1493965, EPI_ISL_1493966, EPI_ISL_1493967, EPI_ISL_1493968, EPI_ISL_1493969, EPI_ISL_1493970, EPI_ISL_1493971, EPI_ISL_1493972, EPI_ISL_1493973, EPI_ISL_1493974, EPI_ISL_1493975, EPI_ISL_1493976, EPI_ISL_1493977, EPI_ISL_1493978, EPI_ISL_1493979, EPI_ISL_1493980, EPI_ISL_1493981, EPI_ISL_1493982, EPI_ISL_1493983, EPI_ISL_1493984, EPI_ISL_1493985, EPI_ISL_1493986, EPI_ISL_1493987, EPI_ISL_1493988, EPI_ISL_1493989, EPI_ISL_1493990, EPI_ISL_1493991, EPI_ISL_1493992, EPI_ISL_1493993, EPI_ISL_1493994, EPI_ISL_1493995, EPI_ISL_1493996, EPI_ISL_1493997, EPI_ISL_1493998, EPI_ISL_1493999, EPI_ISL_1494000, EPI_ISL_1494001, EPI_ISL_1494002, EPI_ISL_1494003, EPI_ISL_1494004, EPI_ISL_1494005, EPI_ISL_1494006, EPI_ISL_1494007, EPI_ISL_1494008, EPI_ISL_1494009, EPI_ISL_1494010, EPI_ISL_1494011, EPI_ISL_1494012, EPI_ISL_1494013, EPI_ISL_1494014, EPI_ISL_1494015, EPI_ISL_1494016, EPI_ISL_1494017, EPI_ISL_1494018, EPI_ISL_1494019, EPI_ISL_1494020, EPI_ISL_1494021, EPI_ISL_1494022, EPI_ISL_1494023, EPI_ISL_1494024, EPI_ISL_1494025, EPI_ISL_1494026, EPI_ISL_1494027, EPI_ISL_1494028, EPI_ISL_1494029, EPI_ISL_1494030, EPI_ISL_1494031 | | | |
| see above | Quest Diagnostics Incorporated | Centers for Disease Control and Prevention Division of Viral Diseases, Pathogen Discovery | Peter W. Cook, Dakota Howard, Dhwani Batra, Ben L. Rambo-Martin, S. H. Rosenthal, A. Gerasimova, R. M. Kagan, B. Anderson, M. Hua, Y. Liu, L.E. Bernstein, K.E. Livingston, A. Perez, I. A. Shlyakhter, R. V. Rolando, R. Owen, P. Tanpaiboon, F. Lacbawan, Clinton R. Paden, Suxiang Tong, Duncan MacCannell |
| EPI_ISL_1494032, EPI_ISL_1494035, EPI_ISL_1494036, EPI_ISL_1494037, EPI_ISL_1494038, EPI_ISL_1494039, EPI_ISL_1494040, EPI_ISL_1494041, EPI_ISL_1494042, EPI_ISL_1494043, EPI_ISL_1494044, EPI_ISL_1494045, EPI_ISL_1494046, EPI_ISL_1494047, EPI_ISL_1494049, EPI_ISL_1494051, EPI_ISL_1494052, EPI_ISL_1494053, EPI_ISL_1494054, EPI_ISL_1494055, EPI_ISL_1494056, EPI_ISL_1494057, EPI_ISL_1494058, EPI_ISL_1494059, EPI_ISL_1494060, EPI_ISL_1494061, EPI_ISL_1494062, EPI_ISL_1494064, EPI_ISL_1494067, EPI_ISL_1494068, EPI_ISL_1494070, EPI_ISL_1494071, EPI_ISL_1494072, EPI_ISL_1494073, EPI_ISL_1494074, EPI_ISL_1494075, EPI_ISL_1494076, EPI_ISL_1494077, EPI_ISL_1494078, EPI_ISL_1494079, EPI_ISL_1494080, EPI_ISL_1494081, EPI_ISL_1494082, EPI_ISL_1494084, EPI_ISL_1494085, EPI_ISL_1494087, EPI_ISL_1494088, EPI_ISL_1494089, EPI_ISL_1494090, EPI_ISL_1494091, EPI_ISL_1494092, EPI_ISL_1494094, EPI_ISL_1494096, EPI_ISL_1494097, EPI_ISL_1494098, EPI_ISL_1494099, EPI_ISL_1494100, EPI_ISL_1494101, EPI_ISL_1494102, EPI_ISL_1494104, EPI_ISL_1494105, EPI_ISL_1494106, EPI_ISL_1494107, EPI_ISL_1494108, EPI_ISL_1494109, EPI_ISL_1494110, EPI_ISL_1494111, EPI_ISL_1494112, EPI_ISL_1494113, EPI_ISL_1494114, EPI_ISL_1494115, EPI_ISL_1494116, EPI_ISL_1494117, EPI_ISL_1494118, EPI_ISL_1494119, EPI_ISL_1494120, EPI_ISL_1494121, EPI_ISL_1494123, EPI_ISL_1494124, EPI_ISL_1494125, EPI_ISL_1494126, EPI_ISL_1494127, EPI_ISL_1494128, EPI_ISL_1494129, EPI_ISL_1494130, EPI_ISL_1494131, EPI_ISL_1494132, EPI_ISL_1494133, EPI_ISL_1494134, EPI_ISL_1494135, EPI_ISL_1494136, EPI_ISL_1494140, EPI_ISL_1494141, EPI_ISL_1494142, EPI_ISL_1494143, EPI_ISL_1494144, EPI_ISL_1494146, EPI_ISL_1494147, EPI_ISL_1494148, EPI_ISL_1494150, EPI_ISL_1494151, EPI_ISL_1494152, EPI_ISL_1494154, EPI_ISL_1494155, EPI_ISL_1494157, EPI_ISL_1494158, EPI_ISL_1494159, EPI_ISL_1494160, EPI_ISL_1494162, EPI_ISL_1494164, EPI_ISL_1494165, EPI_ISL_1494166, EPI_ISL_1494167, EPI_ISL_1494168, EPI_ISL_1494169, EPI_ISL_1494170, EPI_ISL_1494171, EPI_ISL_1494172, EPI_ISL_1494174, EPI_ISL_1494175, EPI_ISL_1494176, EPI_ISL_1494177, EPI_ISL_1494178, EPI_ISL_1494180, EPI_ISL_1494181, EPI_ISL_1494182, EPI_ISL_1494183, EPI_ISL_1494184, EPI_ISL_1494185, EPI_ISL_1494186, EPI_ISL_1494187, EPI_ISL_1494188, EPI_ISL_1494189, EPI_ISL_1494191, EPI_ISL_1494192, EPI_ISL_1494193, EPI_ISL_1494195, EPI_ISL_1494197, EPI_ISL_1494198, EPI_ISL_1494199, EPI_ISL_1494200, EPI_ISL_1494201, EPI_ISL_1494202, EPI_ISL_1494203, EPI_ISL_1494204, EPI_ISL_1494205, EPI_ISL_1494207, EPI_ISL_1494208, EPI_ISL_1494209, EPI_ISL_1494210, EPI_ISL_1494211, EPI_ISL_1494212, EPI_ISL_1494213, EPI_ISL_1494214, EPI_ISL_1494215, EPI_ISL_1494216, EPI_ISL_1494217, EPI_ISL_1494218, EPI_ISL_1494219, EPI_ISL_1494220, EPI_ISL_1494221, EPI_ISL_1494222, EPI_ISL_1494224, EPI_ISL_1494225, EPI_ISL_1494226, EPI_ISL_1494227, EPI_ISL_1494228, EPI_ISL_1494229, EPI_ISL_1494230, EPI_ISL_1494231, EPI_ISL_1494233, EPI_ISL_1494234, EPI_ISL_1494235, EPI_ISL_1494236, EPI_ISL_1494237, EPI_ISL_1494238, EPI_ISL_1494239, EPI_ISL_1494240, EPI_ISL_1494241, EPI_ISL_1494242, EPI_ISL_1494243, EPI_ISL_1494244, EPI_ISL_1494245, EPI_ISL_1494246, EPI_ISL_1494247, EPI_ISL_1494248, EPI_ISL_1494250, EPI_ISL_1494251, EPI_ISL_1494252, EPI_ISL_1494253, EPI_ISL_1494254, EPI_ISL_1494255, EPI_ISL_1494256, EPI_ISL_1494257, EPI_ISL_1494258, EPI_ISL_1494259, EPI_ISL_1494260, EPI_ISL_1494261, EPI_ISL_1494262, EPI_ISL_1494263, EPI_ISL_1494264, EPI_ISL_1494265, EPI_ISL_1494266, EPI_ISL_1494267, EPI_ISL_1494268, EPI_ISL_1494269, EPI_ISL_1494270, EPI_ISL_1494271, EPI_ISL_1494272, EPI_ISL_1494273, EPI_ISL_1494274, EPI_ISL_1494275, EPI_ISL_1494276, EPI_ISL_1494277, EPI_ISL_1494278, EPI_ISL_1494279, EPI_ISL_1494282, EPI_ISL_1494283, EPI_ISL_1494284, EPI_ISL_1494285, EPI_ISL_1494286, EPI_ISL_1494287, EPI_ISL_1494288, EPI_ISL_1494289, EPI_ISL_1494290, EPI_ISL_1494291, EPI_ISL_1494292, EPI_ISL_1494293, EPI_ISL_1494294, EPI_ISL_1494295, EPI_ISL_1494296, EPI_ISL_1494297, EPI_ISL_1494299, EPI_ISL_1494300, EPI_ISL_1494301, EPI_ISL_1494303, EPI_ISL_1494304, EPI_ISL_1494305, EPI_ISL_1494306, EPI_ISL_1494308, EPI_ISL_1494309, EPI_ISL_1494310, EPI_ISL_1494313, EPI_ISL_1494314, EPI_ISL_1494315, EPI_ISL_1494316, EPI_ISL_1494317, EPI_ISL_1494321, EPI_ISL_1494322, EPI_ISL_1494324, EPI_ISL_1494325, EPI_ISL_1494327, EPI_ISL_1494328, EPI_ISL_1494329, EPI_ISL_1494331, EPI_ISL_1494332, EPI_ISL_1494333, EPI_ISL_1494334, EPI_ISL_1494335, EPI_ISL_1494336, EPI_ISL_1494337, EPI_ISL_1494338, EPI_ISL_1494339, EPI_ISL_1494340, EPI_ISL_1494341, EPI_ISL_1494342, EPI_ISL_1494343, EPI_ISL_1494344, EPI_ISL_1494345, EPI_ISL_1494346, EPI_ISL_1494347, EPI_ISL_1494348, EPI_ISL_1494350, EPI_ISL_1494351, EPI_ISL_1494353, EPI_ISL_1494354, EPI_ISL_1494355, EPI_ISL_1494356, EPI_ISL_1494357, EPI_ISL_1494358, EPI_ISL_1494359, EPI_ISL_1494360, EPI_ISL_1494361, EPI_ISL_1494362, EPI_ISL_1494363, EPI_ISL_1494364, EPI_ISL_1494367, EPI_ISL_1494368, EPI_ISL_1494369, EPI_ISL_1494370, EPI_ISL_1494371, EPI_ISL_1494372, EPI_ISL_1494373, EPI_ISL_1494374, EPI_ISL_1494375, EPI_ISL_1494376, EPI_ISL_1494377, EPI_ISL_1494378, EPI_ISL_1494379, EPI_ISL_1494380, EPI_ISL_1494381, EPI_ISL_1494382, EPI_ISL_1494383, EPI_ISL_1494384, EPI_ISL_1494385, EPI_ISL_1494386, EPI_ISL_1494387, EPI_ISL_1494388, EPI_ISL_1494389, EPI_ISL_1494390, EPI_ISL_1494391, EPI_ISL_1494392, EPI_ISL_1494393, EPI_ISL_1494394, EPI_ISL_1494395, EPI_ISL_1494396, EPI_ISL_1494397, EPI_ISL_1494398, EPI_ISL_1494401, EPI_ISL_1494402, EPI_ISL_1494403, EPI_ISL_1494404, EPI_ISL_1494405, EPI_ISL_1494406, EPI_ISL_1494407, EPI_ISL_1494408, EPI_ISL_1494409, EPI_ISL_1494410, EPI_ISL_1494411, EPI_ISL_1494412, EPI_ISL_1494413, EPI_ISL_1494414, EPI_ISL_1494415, EPI_ISL_1494416, EPI_ISL_1494418, EPI_ISL_1494419, EPI_ISL_1494420, EPI_ISL_1494421, EPI_ISL_1494422, EPI_ISL_1494423, EPI_ISL_1494424, EPI_ISL_1494425, EPI_ISL_1494427, EPI_ISL_1494428, EPI_ISL_1494431, EPI_ISL_1494432, EPI_ISL_1494433, EPI_ISL_1494434, EPI_ISL_1494435, EPI_ISL_1494436, EPI_ISL_1494437, EPI_ISL_1494438, EPI_ISL_1494440, EPI_ISL_1494441, EPI_ISL_1494442, EPI_ISL_1494443, EPI_ISL_1494444, EPI_ISL_1494445, EPI_ISL_1494447, EPI_ISL_1494448, EPI_ISL_1494449, EPI_ISL_1494450, EPI_ISL_1494451, EPI_ISL_1494452, EPI_ISL_1494453, EPI_ISL_1494454, EPI_ISL_1494455, EPI_ISL_1494456, EPI_ISL_1494457, EPI_ISL_1494458, EPI_ISL_1494459, EPI_ISL_1494460, EPI_ISL_1494461, EPI_ISL_1494462, EPI_ISL_1494463, EPI_ISL_1494465, EPI_ISL_1494466, EPI_ISL_1494467, EPI_ISL_1494468, EPI_ISL_1494469, EPI_ISL_1494471, EPI_ISL_1494472, EPI_ISL_1494473, EPI_ISL_1494474, EPI_ISL_1494475, EPI_ISL_1494476, EPI_ISL_1494477, EPI_ISL_1494478, EPI_ISL_1494479, EPI_ISL_1494480, EPI_ISL_1494481, EPI_ISL_1494482, EPI_ISL_1494483, EPI_ISL_1494484, EPI_ISL_1494485, EPI_ISL_1494486, EPI_ISL_1494487, EPI_ISL_1494489, EPI_ISL_1494490, EPI_ISL_1494491, EPI_ISL_1494492, EPI_ISL_1494493, EPI_ISL_1494494, EPI_ISL_1494495, EPI_ISL_1494496, EPI_ISL_1494497, EPI_ISL_1494498, EPI_ISL_1494499, EPI_ISL_1494500, EPI_ISL_1494501, EPI_ISL_1494502, EPI_ISL_1494503, EPI_ISL_1494504, EPI_ISL_1494505, EPI_ISL_1494506, EPI_ISL_1494507, EPI_ISL_1494508, EPI_ISL_1494509, EPI_ISL_1494510, EPI_ISL_1494511, EPI_ISL_1494512, EPI_ISL_1494513, EPI_ISL_1494514, EPI_ISL_1494515, EPI_ISL_1494516, EPI_ISL_1494517, EPI_ISL_1494518, EPI_ISL_1494519, EPI_ISL_1494520, EPI_ISL_1494521, EPI_ISL_1494522, EPI_ISL_1494523, EPI_ISL_1494524, EPI_ISL_1494525, EPI_ISL_1494526, EPI_ISL_1494527, EPI_ISL_1494528, EPI_ISL_1494529, EPI_ISL_1494530, EPI_ISL_1494531, EPI_ISL_1494532, EPI_ISL_1494533, EPI_ISL_1494534, EPI_ISL_1494535, EPI_ISL_1494537, EPI_ISL_1494538, EPI_ISL_1494539, EPI_ISL_1494540, EPI_ISL_1494541, EPI_ISL_1494542, EPI_ISL_1494543, EPI_ISL_1494545, EPI_ISL_1494546, EPI_ISL_1494547, EPI_ISL_1494548, EPI_ISL_1494549 | | | |
| see above | Illinois Department of Public Health | Gagnon Lab, Southern Illinois University | Keith Gagnon |
| EPI_ISL_1494551, EPI_ISL_1494552, EPI_ISL_1494553 | Kansas Health and Environmental Lab | Kansas Health and Environmental Lab | Mike Grose, Jonathan Barnell, Ben Olsen, and Phil Adam |
| EPI_ISL_1494554, EPI_ISL_1494555, EPI_ISL_1494556 | Cytocheck Laboratory | Kansas Health and Environmental Lab | Mike Grose, Jonathan Barnell, Ben Olsen, and Phil Adam |
| EPI_ISL_1494557, EPI_ISL_1494558, EPI_ISL_1494559, EPI_ISL_1494560, EPI_ISL_1494561, EPI_ISL_1494562, EPI_ISL_1494563, EPI_ISL_1494564, EPI_ISL_1494565, EPI_ISL_1494567, EPI_ISL_1494568, EPI_ISL_1494569, EPI_ISL_1494570, EPI_ISL_1494571, EPI_ISL_1494572, EPI_ISL_1494573, EPI_ISL_1494574, EPI_ISL_1494575, EPI_ISL_1494576, EPI_ISL_1494577, EPI_ISL_1494580, EPI_ISL_1494581, EPI_ISL_1494582, EPI_ISL_1494583, EPI_ISL_1494584, EPI_ISL_1494586, EPI_ISL_1494587, EPI_ISL_1494588, EPI_ISL_1494589, EPI_ISL_1494591, EPI_ISL_1494592, EPI_ISL_1494593, EPI_ISL_1494594, EPI_ISL_1494595, EPI_ISL_1494596, EPI_ISL_1494597, EPI_ISL_1494598, EPI_ISL_1494599, EPI_ISL_1494600, EPI_ISL_1494601, EPI_ISL_1494602, EPI_ISL_1494603, EPI_ISL_1494604, EPI_ISL_1494605, EPI_ISL_1494606, EPI_ISL_1494607, EPI_ISL_1494608, EPI_ISL_1494609, EPI_ISL_1494610, EPI_ISL_1494611, EPI_ISL_1494612, EPI_ISL_1494613, EPI_ISL_1494614, EPI_ISL_1494615, EPI_ISL_1494616, EPI_ISL_1494617, EPI_ISL_1494618, EPI_ISL_1494619, EPI_ISL_1494620, EPI_ISL_1494621, EPI_ISL_1494622, EPI_ISL_1494624, EPI_ISL_1494625 | | | |
| see above | Virginia Division of Consolidated Laboratory Services | Virginia Division of Consolidated Laboratory Services | Virginia DCLS |
| EPI_ISL_1494626, EPI_ISL_1494627, EPI_ISL_1494628, EPI_ISL_1494629, EPI_ISL_1494630 | LESP Chihuahua | Instituto de Diagnostico y Referencia Epidemiologicos (INDRE) | Claudia Wong-Arambula, Abril Rodriguez-Maldonado, Vanessa Rivero-Arredondo, Ariadna Medina-Benitez, Joaquin Quiroz-Mercado, Sergio Rangel-Guerrero, Natividad Cruz-Ortiz, Tatiana Nunez-Garcia, Gisela Barrera-Badillo, Lucia Hernandez-Rivas, Irma Lopez-Martinez, Ernesto Ramirez-Gonzalez. |
| EPI_ISL_1494631, EPI_ISL_1494632, EPI_ISL_1494633, EPI_ISL_1494634, EPI_ISL_1494635 | LESP Nuevo Leon | Instituto de Diagnostico y Referencia Epidemiologicos (INDRE) | Claudia Wong-Arambula, Abril Rodriguez-Maldonado, Vanessa Rivero-Arredondo, Ariadna Medina-Benitez, Joaquin Quiroz-Mercado, Sergio Rangel-Guerrero, Natividad Cruz-Ortiz, Tatiana Nunez-Garcia, Gisela Barrera-Badillo, Lucia Hernandez-Rivas, Irma Lopez-Martinez, Ernesto Ramirez-Gonzalez. |
| EPI_ISL_1494636, EPI_ISL_1494637, EPI_ISL_1494638, EPI_ISL_1494639, EPI_ISL_1494640, EPI_ISL_1494641, EPI_ISL_1494642, EPI_ISL_1494643, EPI_ISL_1494644, EPI_ISL_1494645, EPI_ISL_1494646, EPI_ISL_1494647 | | | |
| see above | LESP Ciudad de Mexico | Instituto de Diagnostico y Referencia Epidemiologicos (INDRE) | Claudia Wong-Arambula, Abril Rodriguez-Maldonado, Vanessa Rivero-Arredondo, Ariadna Medina-Benitez, Joaquin Quiroz-Mercado, Sergio Rangel-Guerrero, Natividad Cruz-Ortiz, Tatiana Nunez-Garcia, Gisela Barrera-Badillo, Lucia Hernandez-Rivas, Irma Lopez-Martinez, Ernesto Ramirez-Gonzalez. |
| EPI_ISL_1494648, EPI_ISL_1494649 | LESP Nuevo Leon | Instituto de Diagnostico y Referencia Epidemiologicos (INDRE) | Claudia Wong-Arambula, Abril Rodriguez-Maldonado, Vanessa Rivero-Arredondo, Ariadna Medina-Benitez, Joaquin Quiroz-Mercado, Sergio Rangel-Guerrero, Natividad Cruz-Ortiz, Tatiana Nunez-Garcia, Gisela Barrera-Badillo, Lucia Hernandez-Rivas, Irma Lopez-Martinez, Ernesto Ramirez-Gonzalez. |

| | | | |
|---|---|---|---|
| EPI_ISL_1494650, EPI_ISL_1494651, EPI_ISL_1494652, EPI_ISL_1494653, EPI_ISL_1494654 | LESP Tamaulipas | Instituto de Diagnostico y Referencia Epidemiologicos (INDRE) | Claudia Wong-Arambula, Abril Rodriguez-Maldonado, Vanessa Rivero-Arredondo, Ariadna Medina-Benitez, Joaquin Quiroz-Mercado, Sergio Rangel-Guerrero, Natividad Cruz-Ortiz, Tatiana Nunez-Garcia, Gisela Barrera-Badillo, Lucia Hernandez-Rivas, Irma Lopez-Martinez, Ernesto Ramirez-Gonzalez. |
| EPI_ISL_1494655, EPI_ISL_1494656 | LESP Hidalgo | Instituto de Diagnostico y Referencia Epidemiologicos (INDRE) | Claudia Wong-Arambula, Abril Rodriguez-Maldonado, Vanessa Rivero-Arredondo, Ariadna Medina-Benitez, Joaquin Quiroz-Mercado, Sergio Rangel-Guerrero, Natividad Cruz-Ortiz, Tatiana Nunez-Garcia, Gisela Barrera-Badillo, Lucia Hernandez-Rivas, Irma Lopez-Martinez, Ernesto Ramirez-Gonzalez. |
| EPI_ISL_1494657, EPI_ISL_1494658, EPI_ISL_1494659, EPI_ISL_1494660, EPI_ISL_1494661, EPI_ISL_1494662, EPI_ISL_1494663, EPI_ISL_1494664, EPI_ISL_1494665, EPI_ISL_1494666, EPI_ISL_1494667, EPI_ISL_1494668, EPI_ISL_1494669 | | | |
| see above | LESP Guanajuato | Instituto de Diagnostico y Referencia Epidemiologicos (INDRE) | Claudia Wong-Arambula, Abril Rodriguez-Maldonado, Vanessa Rivero-Arredondo, Ariadna Medina-Benitez, Joaquin Quiroz-Mercado, Sergio Rangel-Guerrero, Natividad Cruz-Ortiz, Tatiana Nunez-Garcia, Gisela Barrera-Badillo, Lucia Hernandez-Rivas, Irma Lopez-Martinez, Ernesto Ramirez-Gonzalez. |
| EPI_ISL_1494670, EPI_ISL_1494671, EPI_ISL_1494672 | LESP Sinaloa | Instituto de Diagnostico y Referencia Epidemiologicos (INDRE) | Claudia Wong-Arambula, Abril Rodriguez-Maldonado, Vanessa Rivero-Arredondo, Ariadna Medina-Benitez, Joaquin Quiroz-Mercado, Sergio Rangel-Guerrero, Natividad Cruz-Ortiz, Tatiana Nunez-Garcia, Gisela Barrera-Badillo, Lucia Hernandez-Rivas, Irma Lopez-Martinez, Ernesto Ramirez-Gonzalez. |
| EPI_ISL_1494673 | LESP Nuevo Leon | Instituto de Diagnostico y Referencia Epidemiologicos (INDRE) | Claudia Wong-Arambula, Abril Rodriguez-Maldonado, Vanessa Rivero-Arredondo, Ariadna Medina-Benitez, Joaquin Quiroz-Mercado, Sergio Rangel-Guerrero, Natividad Cruz-Ortiz, Tatiana Nunez-Garcia, Gisela Barrera-Badillo, Lucia Hernandez-Rivas, Irma Lopez-Martinez, Ernesto Ramirez-Gonzalez. |
| EPI_ISL_1494674, EPI_ISL_1494717 | Centro de Investigación en Ciencias de la Salud y Biomedicina, U.A.S.L.P | Centro de Investigación en Ciencias de la Salud y Biomedicina, U.A.S.L.P | MD PhD Sofía Bernal Silva |
| EPI_ISL_1494724, EPI_ISL_1494725, EPI_ISL_1494726, EPI_ISL_1494727, EPI_ISL_1494728, EPI_ISL_1494729, EPI_ISL_1494730 | Centro de Investigación en Ciencias de la Salud y Biomedicina, U.A.S.LP. | Centro de Investigación en Ciencias de la Salud y Biomedicina, U.A.S.LP. | MD PhD Sofía Bernal Silva |
| EPI_ISL_1494731, EPI_ISL_1494732, EPI_ISL_1494733, EPI_ISL_1494734, EPI_ISL_1494735, EPI_ISL_1494736, EPI_ISL_1494737, EPI_ISL_1494738, EPI_ISL_1494739, EPI_ISL_1494740, EPI_ISL_1494741, EPI_ISL_1494742, EPI_ISL_1494743, EPI_ISL_1494744, EPI_ISL_1494745, EPI_ISL_1494746, EPI_ISL_1494747, EPI_ISL_1494748, EPI_ISL_1494749, EPI_ISL_1494750, EPI_ISL_1494751, EPI_ISL_1494752, EPI_ISL_1494753, EPI_ISL_1494755, EPI_ISL_1494756 | | | |
| see above | City of Milwaukee Health Department Laboratory | City of Milwaukee Health Department Laboratory | Sanjib Bhattacharyya |
| EPI_ISL_1494925, EPI_ISL_1494926, EPI_ISL_1494927, EPI_ISL_1494928, EPI_ISL_1494929, EPI_ISL_1494930, EPI_ISL_1494931, EPI_ISL_1494932, EPI_ISL_1494933, EPI_ISL_1494934, EPI_ISL_1494935, EPI_ISL_1494936, EPI_ISL_1494937, EPI_ISL_1494938, EPI_ISL_1494939 | | | |
| see above | Oregon State University TRACE Project | OSU Center for Genome Research and Biocomputing | Oregon State University TRACE Project |
| EPI_ISL_1495983 | Maine Health and Environmental Testing Laboratory | Maine Health and Environmental Testing Laboratory | Matluk,N., Dewey,H., Martha,J., Barter,M., Lynch,R., Munger,H. and Tewhey,R. |
| EPI_ISL_1497525 | Pomona Valley Hospital Medical Center | Los Angeles County Public Health Laboratories | P. Hemarajata et al. |
| EPI_ISL_1497526, EPI_ISL_1497527, EPI_ISL_1497529, EPI_ISL_1497530, EPI_ISL_1497531, EPI_ISL_1497532, EPI_ISL_1497534, EPI_ISL_1497537, EPI_ISL_1497538, EPI_ISL_1497539, EPI_ISL_1497540 | | | |
| see above | USC Clinical Lab | Los Angeles County Public Health Laboratories | P. Hemarajata et al. |
| EPI_ISL_1497541 | UCLA Clinical Micro Lab | Los Angeles County Public Health Laboratories | P. Hemarajata et al. |
| EPI_ISL_1497542 | Flow Diagnostics Lab | Los Angeles County Public Health Laboratories | P. Hemarajata et al. |
| EPI_ISL_1497543 | Los Angeles County Public Health Laboratories | Los Angeles County Public Health Laboratories | P. Hemarajata et al. |
| EPI_ISL_1497544, EPI_ISL_1497546, EPI_ISL_1497547 | Huntington Memorial Hospital | Los Angeles County Public Health Laboratories | P. Hemarajata et al. |
| EPI_ISL_1497766 | Baylor Esoteric and Molecular Lab | Baylor Esoteric and Molecular Lab | Elizabeth Forrester and Dawn Richards |
| EPI_ISL_1497767, EPI_ISL_1497768, EPI_ISL_1497769, EPI_ISL_1497770, EPI_ISL_1497771, EPI_ISL_1497772, EPI_ISL_1497773, EPI_ISL_1497774, EPI_ISL_1497775, EPI_ISL_1497776, EPI_ISL_1497777, EPI_ISL_1497778, EPI_ISL_1497779, EPI_ISL_1497780, EPI_ISL_1497781, EPI_ISL_1497783, EPI_ISL_1497784, EPI_ISL_1497785, EPI_ISL_1497786, EPI_ISL_1497787, EPI_ISL_1497788, EPI_ISL_1497789, EPI_ISL_1497790, EPI_ISL_1497791, EPI_ISL_1497792, EPI_ISL_1497793, EPI_ISL_1497794, EPI_ISL_1497795, EPI_ISL_1497796, EPI_ISL_1497797, EPI_ISL_1497798, EPI_ISL_1497799, EPI_ISL_1497800, EPI_ISL_1497801, EPI_ISL_1497802, EPI_ISL_1497803, EPI_ISL_1497804, EPI_ISL_1497805, EPI_ISL_1497806, EPI_ISL_1497807, EPI_ISL_1497808, EPI_ISL_1497809, EPI_ISL_1497810, EPI_ISL_1497811, EPI_ISL_1497812, EPI_ISL_1497813, EPI_ISL_1497814, EPI_ISL_1497815, EPI_ISL_1497816, EPI_ISL_1497817, EPI_ISL_1497819, EPI_ISL_1497820, EPI_ISL_1497822, EPI_ISL_1497823, EPI_ISL_1497824, EPI_ISL_1497825, EPI_ISL_1497827, EPI_ISL_1497828, EPI_ISL_1497829, EPI_ISL_1497830, EPI_ISL_1497831, EPI_ISL_1497832, EPI_ISL_1497833, EPI_ISL_1497834, EPI_ISL_1497835, EPI_ISL_1497836, EPI_ISL_1497837, EPI_ISL_1497838, EPI_ISL_1497839, EPI_ISL_1497840, EPI_ISL_1497841, EPI_ISL_1497842, EPI_ISL_1497843, EPI_ISL_1497844, EPI_ISL_1497845, EPI_ISL_1497846, EPI_ISL_1497847, EPI_ISL_1497848, EPI_ISL_1497849, EPI_ISL_1497850, EPI_ISL_1497851, EPI_ISL_1497852, EPI_ISL_1497853, EPI_ISL_1497854, EPI_ISL_1497855, EPI_ISL_1497856, EPI_ISL_1497857, EPI_ISL_1497858, EPI_ISL_1497859, EPI_ISL_1497860, EPI_ISL_1497861, EPI_ISL_1497862, EPI_ISL_1497863, EPI_ISL_1497864, EPI_ISL_1497865, EPI_ISL_1497866, EPI_ISL_1497867, EPI_ISL_1497868, EPI_ISL_1497869, EPI_ISL_1497870, EPI_ISL_1497871, EPI_ISL_1497872, EPI_ISL_1497873, EPI_ISL_1497874, EPI_ISL_1497875, EPI_ISL_1497876, EPI_ISL_1497877, EPI_ISL_1497878, EPI_ISL_1497879, EPI_ISL_1497880, EPI_ISL_1497881, EPI_ISL_1497883, EPI_ISL_1497884, EPI_ISL_1497885, EPI_ISL_1497886, EPI_ISL_1497887, EPI_ISL_1497888, EPI_ISL_1497889, EPI_ISL_1497890, EPI_ISL_1497891, EPI_ISL_1497892, EPI_ISL_1497893, EPI_ISL_1497894, EPI_ISL_1497895, EPI_ISL_1497897, EPI_ISL_1497898, EPI_ISL_1497899, EPI_ISL_1497900, EPI_ISL_1497901, EPI_ISL_1497902, EPI_ISL_1497903, EPI_ISL_1497904, EPI_ISL_1497905, EPI_ISL_1497906, EPI_ISL_1497907, EPI_ISL_1497908, EPI_ISL_1497909, EPI_ISL_1497910, EPI_ISL_1497911, EPI_ISL_1497912, EPI_ISL_1497913, EPI_ISL_1497914, EPI_ISL_1497915, EPI_ISL_1497916, EPI_ISL_1497917, EPI_ISL_1497918, EPI_ISL_1497919, EPI_ISL_1497920, EPI_ISL_1497921, EPI_ISL_1497922, EPI_ISL_1497923, EPI_ISL_1497924, EPI_ISL_1497925, EPI_ISL_1497926, EPI_ISL_1497927, EPI_ISL_1497928, EPI_ISL_1497929, EPI_ISL_1497930, EPI_ISL_1497931, EPI_ISL_1497932, EPI_ISL_1497933, EPI_ISL_1497934, EPI_ISL_1497935, EPI_ISL_1497936, EPI_ISL_1497937, EPI_ISL_1497938, EPI_ISL_1497939, EPI_ISL_1497940, EPI_ISL_1497941, EPI_ISL_1497942, EPI_ISL_1497943, EPI_ISL_1497944, EPI_ISL_1497945, EPI_ISL_1497946, EPI_ISL_1497947, EPI_ISL_1497948, EPI_ISL_1497949, EPI_ISL_1497950, EPI_ISL_1497951, EPI_ISL_1497952, EPI_ISL_1497953, EPI_ISL_1497954, EPI_ISL_1497955, EPI_ISL_1497956, EPI_ISL_1497957, EPI_ISL_1497958, EPI_ISL_1497959, EPI_ISL_1497960, EPI_ISL_1497962, EPI_ISL_1497964, EPI_ISL_1497965, EPI_ISL_1497966, EPI_ISL_1497967, EPI_ISL_1497969, EPI_ISL_1497970, EPI_ISL_1497971, EPI_ISL_1497972, EPI_ISL_1497973, EPI_ISL_1497974, EPI_ISL_1497975, EPI_ISL_1497976, EPI_ISL_1497977, EPI_ISL_1497978, EPI_ISL_1497979, EPI_ISL_1497980, EPI_ISL_1497981, EPI_ISL_1497982, EPI_ISL_1497983, EPI_ISL_1497984, EPI_ISL_1497985, EPI_ISL_1497986, EPI_ISL_1497987, EPI_ISL_1497988, EPI_ISL_1497989, EPI_ISL_1497990, EPI_ISL_1497991, EPI_ISL_1497992, EPI_ISL_1497993, EPI_ISL_1497994, EPI_ISL_1497995, EPI_ISL_1497996, EPI_ISL_1497997, EPI_ISL_1497998, EPI_ISL_1497999, EPI_ISL_1498000, EPI_ISL_1498001, EPI_ISL_1498002, EPI_ISL_1498003, EPI_ISL_1498004, EPI_ISL_1498005, EPI_ISL_1498006, EPI_ISL_1498007, EPI_ISL_1498008, EPI_ISL_1498009, EPI_ISL_1498010, EPI_ISL_1498011, EPI_ISL_1498012, EPI_ISL_1498013, EPI_ISL_1498014, EPI_ISL_1498015, EPI_ISL_1498016, EPI_ISL_1498017, EPI_ISL_1498018, EPI_ISL_1498019, EPI_ISL_1498020, EPI_ISL_1498021, EPI_ISL_1498022, EPI_ISL_1498023, EPI_ISL_1498024, EPI_ISL_1498025, EPI_ISL_1498026, EPI_ISL_1498027, EPI_ISL_1498028, EPI_ISL_1498029, EPI_ISL_1498030, EPI_ISL_1498031, EPI_ISL_1498032, EPI_ISL_1498033, EPI_ISL_1498034, EPI_ISL_1498035, EPI_ISL_1498036, EPI_ISL_1498037, EPI_ISL_1498038, EPI_ISL_1498040, EPI_ISL_1498041, EPI_ISL_1498042, EPI_ISL_1498043, EPI_ISL_1498044, EPI_ISL_1498045, EPI_ISL_1498046, EPI_ISL_1498047, EPI_ISL_1498048, EPI_ISL_1498049, EPI_ISL_1498050, EPI_ISL_1498051, EPI_ISL_1498052, EPI_ISL_1498053, EPI_ISL_1498054, EPI_ISL_1498055, EPI_ISL_1498056, EPI_ISL_1498057, EPI_ISL_1498058, EPI_ISL_1498059, EPI_ISL_1498060, EPI_ISL_1498061, EPI_ISL_1498062, EPI_ISL_1498063, EPI_ISL_1498064, EPI_ISL_1498065, EPI_ISL_1498066, EPI_ISL_1498067, EPI_ISL_1498068, EPI_ISL_1498069, EPI_ISL_1498070, EPI_ISL_1498071, EPI_ISL_1498072, EPI_ISL_1498073, EPI_ISL_1498074, EPI_ISL_1498075, EPI_ISL_1498076, EPI_ISL_1498077, EPI_ISL_1498078, EPI_ISL_1498079, EPI_ISL_1498080, EPI_ISL_1498081, EPI_ISL_1498082, EPI_ISL_1498083, EPI_ISL_1498084, EPI_ISL_1498085, EPI_ISL_1498086, EPI_ISL_1498087, EPI_ISL_1498088, EPI_ISL_1498089, EPI_ISL_1498090, EPI_ISL_1498091, EPI_ISL_1498093 | | | |
| see above | UW Virology Lab | UW Virology Lab | Pavitra Roychoudhury, Hong Xie, Lasata Shrestha, Shah Mohamed Bakhash, Michelle Lin, Noah R. Baker, Sean Ellis, Saraswathi Sathees, Meei-Li Huang, Keith R Jerome, Alexander Greninger |
| EPI_ISL_1498094, EPI_ISL_1498096 | Baylor Esoteric and Molecular Lab | Baylor Esoteric and Molecular Lab | Elizabeth Forrester and Dawn Richards |
| EPI_ISL_1498121, EPI_ISL_1498918 | NCSLPH | NCSLPH | Chase K, Miller MC, Greene S, Glover W |
| EPI_ISL_1499106, EPI_ISL_1499107, EPI_ISL_1499108, EPI_ISL_1499109, EPI_ISL_1499110, EPI_ISL_1499111 | Lafene Health Center | Kansas State Veterinary Diagnostic Laboratory | Tyler Doerksen, Andreea Lu, Lance Noll, Elizabeth Porter, Kasey Hogan, Emily Cox, Jianfa Bai, Jamie Henningson, Rachel Palinski |
| EPI_ISL_1499112 | NCSLPH | NCSLPH | Chase K, Miller MC, Greene S, Glover W |
| EPI_ISL_1499364, EPI_ISL_1499365, EPI_ISL_1499366, EPI_ISL_1499367, EPI_ISL_1499368, EPI_ISL_1499369, EPI_ISL_1499370, EPI_ISL_1499371, EPI_ISL_1499372, EPI_ISL_1499373, EPI_ISL_1499374, EPI_ISL_1499375, EPI_ISL_1499376, EPI_ISL_1499377, EPI_ISL_1499379, EPI_ISL_1499380, EPI_ISL_1499381, EPI_ISL_1499382, EPI_ISL_1499383, EPI_ISL_1499384, EPI_ISL_1499385, EPI_ISL_1499386, EPI_ISL_1499387, EPI_ISL_1499388, EPI_ISL_1499389, EPI_ISL_1499390, EPI_ISL_1499391, EPI_ISL_1499392, EPI_ISL_1499393, EPI_ISL_1499394 | | | |
| see above | Wyoming Public Health Laboratory | Wyoming Public Health Laboratory | Jim Mildenberger, Wanda Manley, Noah Hull, Taylor Fearing, Lynette Gumbleton, Channing Weber, Ashley Norberg, Chayse Rowley, Marley Goetz, Brian Dominguez, Elliot Thomasson, Cari Sloma, and Rob Christensen |
| EPI_ISL_1499395, EPI_ISL_1499396, EPI_ISL_1499397, EPI_ISL_1499398, EPI_ISL_1499399, EPI_ISL_1499400, EPI_ISL_1499401, EPI_ISL_1499402, EPI_ISL_1499403, EPI_ISL_1499404, EPI_ISL_1499405, EPI_ISL_1499406, EPI_ISL_1499407, EPI_ISL_1499408, EPI_ISL_1499409, EPI_ISL_1499410, EPI_ISL_1499411, EPI_ISL_1499412, EPI_ISL_1499413, EPI_ISL_1499414, EPI_ISL_1499415, EPI_ISL_1499416, EPI_ISL_1499417, EPI_ISL_1499418, EPI_ISL_1499419, EPI_ISL_1499420, EPI_ISL_1499421, EPI_ISL_1499422, EPI_ISL_1499423, EPI_ISL_1499424, EPI_ISL_1499425, EPI_ISL_1499426, EPI_ISL_1499427, EPI_ISL_1499428, EPI_ISL_1499429, EPI_ISL_1499430, EPI_ISL_1499431, EPI_ISL_1499432, EPI_ISL_1499433, EPI_ISL_1499434, EPI_ISL_1499435, EPI_ISL_1499436 | | | |
| see above | URMC LABS | Wadsworth Center, New York State Department of Health | Kirsten St. George, Daryl M. Lamson, Alexis Russel, Matthew Shudt, Melissa A Leisner, Jonathan Plitnick, Navjot Singh, John Kelly, Erasmus Schneider, Erica Lasek-Nesselquist |
| EPI_ISL_1499437, EPI_ISL_1499438, EPI_ISL_1499439, EPI_ISL_1499440, EPI_ISL_1499441, EPI_ISL_1499442, EPI_ISL_1499443, EPI_ISL_1499444, EPI_ISL_1499445, EPI_ISL_1499446, EPI_ISL_1499447, EPI_ISL_1499448, EPI_ISL_1499449, EPI_ISL_1499450, EPI_ISL_1499451, EPI_ISL_1499452, EPI_ISL_1499453, EPI_ISL_1499454, EPI_ISL_1499455, EPI_ISL_1499456 | | | |
| see above | SUNY UPSTATE MEDICAL UNIVERSITY | Wadsworth Center, New York State Department of Health | Kirsten St. George, Daryl M. Lamson, Alexis Russel, Matthew Shudt, Melissa A Leisner, Jonathan Plitnick, Navjot Singh, John Kelly, Erasmus Schneider, Erica Lasek-Nesselquist |
| EPI_ISL_1499457, EPI_ISL_1499458, EPI_ISL_1499459, EPI_ISL_1499460, EPI_ISL_1499461, EPI_ISL_1499462, EPI_ISL_1499463, EPI_ISL_1499464, EPI_ISL_1499465, EPI_ISL_1499466, EPI_ISL_1499467, EPI_ISL_1499468, EPI_ISL_1499469, EPI_ISL_1499470, EPI_ISL_1499471, EPI_ISL_1499472, EPI_ISL_1499473 | | | |
| see above | ALBANY MEDICAL CENTER HOSPITAL CLINICAL LABORATORIES | Wadsworth Center, New York State Department of Health | Kirsten St. George, Daryl M. Lamson, Alexis Russel, Matthew Shudt, Melissa A Leisner, Jonathan Plitnick, Navjot Singh, John Kelly, Erasmus Schneider, Erica Lasek-Nesselquist |
| EPI_ISL_1499595, EPI_ISL_1499596, EPI_ISL_1499597, EPI_ISL_1499598, EPI_ISL_1499599, EPI_ISL_1499600, EPI_ISL_1499601, EPI_ISL_1499602, EPI_ISL_1499603, EPI_ISL_1499604, EPI_ISL_1499605, EPI_ISL_1499606, EPI_ISL_1499607, EPI_ISL_1499608, EPI_ISL_1499609, EPI_ISL_1499610, EPI_ISL_1499611, EPI_ISL_1499612, EPI_ISL_1499613, EPI_ISL_1499614, EPI_ISL_1499615, EPI_ISL_1499616, EPI_ISL_1499617, EPI_ISL_1499618, EPI_ISL_1499619, EPI_ISL_1499620 | | | |
| see above | Lafene Health Center | Kansas State Veterinary Diagnostic Laboratory | Tyler Doerksen, Andreea Lu, Lance Noll, Elizabeth Porter, Kasey Hogan, Emily Cox, Jianfa Bai, Jamie Henningson, Rachel Palinski |
| EPI_ISL_1499652, EPI_ISL_1499653, EPI_ISL_1499654, EPI_ISL_1499655, EPI_ISL_1499656, EPI_ISL_1499657, EPI_ISL_1499658, EPI_ISL_1499659, EPI_ISL_1499660, EPI_ISL_1499661, EPI_ISL_1499662, EPI_ISL_1499663, EPI_ISL_1499664, EPI_ISL_1499665, EPI_ISL_1499666, EPI_ISL_1499668, EPI_ISL_1499669, EPI_ISL_1499670, EPI_ISL_1499671, EPI_ISL_1499672, EPI_ISL_1499673, EPI_ISL_1499674, | | | |

| | | | |
|---|---|---|---|
| EPI_ISL_1499675, EPI_ISL_1499676, EPI_ISL_1499677, EPI_ISL_1499678, EPI_ISL_1499679, EPI_ISL_1499680, EPI_ISL_1499682, EPI_ISL_1499683, EPI_ISL_1499684, EPI_ISL_1499685 | | | |
| see above | Minnesota Department of Health, Public Health Laboratory | Minnesota Department of Health, Public Health Laboratory | Alexandra Lorentz, Jacob Garfin, Matt Plumb, and Xiong Wang |
| EPI_ISL_1499686, EPI_ISL_1499687, EPI_ISL_1499688 | Vault Health | Minnesota Department of Health, Public Health Laboratory | Alexandra Lorentz, Jacob Garfin, Matt Plumb, and Xiong Wang |
| EPI_ISL_1499689, EPI_ISL_1499690, EPI_ISL_1499691, EPI_ISL_1499692, EPI_ISL_1499693, EPI_ISL_1499694, EPI_ISL_1499695, EPI_ISL_1499696, EPI_ISL_1499697, EPI_ISL_1499698, EPI_ISL_1499699, EPI_ISL_1499700, EPI_ISL_1499701, EPI_ISL_1499702, EPI_ISL_1499703, EPI_ISL_1499704, EPI_ISL_1499705, EPI_ISL_1499706, EPI_ISL_1499707, EPI_ISL_1499708, EPI_ISL_1499709, EPI_ISL_1499710, EPI_ISL_1499711, EPI_ISL_1499712, EPI_ISL_1499713, EPI_ISL_1499714, EPI_ISL_1499715, EPI_ISL_1499716, EPI_ISL_1499717, EPI_ISL_1499718, EPI_ISL_1499719, EPI_ISL_1499720, EPI_ISL_1499721, EPI_ISL_1499722 | | | |
| see above | M Health Fairview | Minnesota Department of Health, Public Health Laboratory | Alexandra Lorentz, Jacob Garfin, Matt Plumb, and Xiong Wang |
| EPI_ISL_1500152, EPI_ISL_1500153, EPI_ISL_1500154, EPI_ISL_1500155, EPI_ISL_1500156, EPI_ISL_1500157, EPI_ISL_1500158, EPI_ISL_1500159, EPI_ISL_1500160, EPI_ISL_1500161, EPI_ISL_1500163, EPI_ISL_1500164, EPI_ISL_1500165, EPI_ISL_1500166, EPI_ISL_1500167, EPI_ISL_1500168, EPI_ISL_1500169, EPI_ISL_1500170, EPI_ISL_1500171, EPI_ISL_1500172, EPI_ISL_1500173, EPI_ISL_1500174, EPI_ISL_1500175, EPI_ISL_1500176, EPI_ISL_1500177, EPI_ISL_1500178, EPI_ISL_1500179, EPI_ISL_1500180, EPI_ISL_1500181, EPI_ISL_1500182, EPI_ISL_1500183, EPI_ISL_1500184, EPI_ISL_1500185, EPI_ISL_1500186, EPI_ISL_1500187, EPI_ISL_1500188, EPI_ISL_1500189, EPI_ISL_1500190, EPI_ISL_1500191, EPI_ISL_1500193, EPI_ISL_1500194, EPI_ISL_1500195, EPI_ISL_1500196, EPI_ISL_1500197, EPI_ISL_1500198, EPI_ISL_1500199, EPI_ISL_1500200, EPI_ISL_1500201, EPI_ISL_1500202, EPI_ISL_1500203, EPI_ISL_1500204, EPI_ISL_1500205, EPI_ISL_1500206, EPI_ISL_1500207, EPI_ISL_1500208, EPI_ISL_1500209, EPI_ISL_1500210, EPI_ISL_1500211, EPI_ISL_1500212, EPI_ISL_1500213, EPI_ISL_1500214, EPI_ISL_1500215, EPI_ISL_1500216, EPI_ISL_1500217, EPI_ISL_1500218, EPI_ISL_1500219, EPI_ISL_1500220, EPI_ISL_1500221, EPI_ISL_1500222, EPI_ISL_1500223, EPI_ISL_1500224, EPI_ISL_1500225, EPI_ISL_1500226, EPI_ISL_1500227, EPI_ISL_1500228, EPI_ISL_1500229, EPI_ISL_1500231, EPI_ISL_1500232, EPI_ISL_1500233, EPI_ISL_1500234, EPI_ISL_1500235, EPI_ISL_1500236, EPI_ISL_1500237, EPI_ISL_1500238, EPI_ISL_1500239, EPI_ISL_1500240, EPI_ISL_1500241, EPI_ISL_1500242, EPI_ISL_1500243, EPI_ISL_1500244, EPI_ISL_1500245 | | | |
| see above | Illinois Department of Public Health - Springfield Lab | Illinois Department of Public Health - Springfield Lab | Bryan Sim, Gordon McCall |
| EPI_ISL_1500246, EPI_ISL_1500247, EPI_ISL_1500250, EPI_ISL_1500251, EPI_ISL_1500253, EPI_ISL_1500254, EPI_ISL_1500255, EPI_ISL_1500256, EPI_ISL_1500257, EPI_ISL_1500258, EPI_ISL_1500259, EPI_ISL_1500260, EPI_ISL_1500261, EPI_ISL_1500262, EPI_ISL_1500263, EPI_ISL_1500264, EPI_ISL_1500265, EPI_ISL_1500266, EPI_ISL_1500267, EPI_ISL_1500268, EPI_ISL_1500269, EPI_ISL_1500270, EPI_ISL_1500271, EPI_ISL_1500272, EPI_ISL_1500273, EPI_ISL_1500274, EPI_ISL_1500275, EPI_ISL_1500276, EPI_ISL_1500277, EPI_ISL_1500278, EPI_ISL_1500279, EPI_ISL_1500280, EPI_ISL_1500281, EPI_ISL_1500282, EPI_ISL_1500283, EPI_ISL_1500284, EPI_ISL_1500285, EPI_ISL_1500286, EPI_ISL_1500287, EPI_ISL_1500288, EPI_ISL_1500289, EPI_ISL_1500290, EPI_ISL_1500291, EPI_ISL_1500292, EPI_ISL_1500294, EPI_ISL_1500295, EPI_ISL_1500296, EPI_ISL_1500297, EPI_ISL_1500298, EPI_ISL_1500299, EPI_ISL_1500300, EPI_ISL_1500301, EPI_ISL_1500302, EPI_ISL_1500303, EPI_ISL_1500304, EPI_ISL_1500305, EPI_ISL_1500306, EPI_ISL_1500307, EPI_ISL_1500308, EPI_ISL_1500309, EPI_ISL_1500310, EPI_ISL_1500311, EPI_ISL_1500312, EPI_ISL_1500313, EPI_ISL_1500314, EPI_ISL_1500315, EPI_ISL_1500316, EPI_ISL_1500317, EPI_ISL_1500318, EPI_ISL_1500319, EPI_ISL_1500320, EPI_ISL_1500321, EPI_ISL_1500322, EPI_ISL_1500323, EPI_ISL_1500324, EPI_ISL_1500325, EPI_ISL_1500326, EPI_ISL_1500327, EPI_ISL_1500328, EPI_ISL_1500329, EPI_ISL_1500330, EPI_ISL_1500331, EPI_ISL_1500332, EPI_ISL_1500333, EPI_ISL_1500334, EPI_ISL_1500335, EPI_ISL_1500336, EPI_ISL_1500337, EPI_ISL_1500338, EPI_ISL_1500339, EPI_ISL_1500340, EPI_ISL_1500341, EPI_ISL_1500342, EPI_ISL_1500343, EPI_ISL_1500344, EPI_ISL_1500345, EPI_ISL_1500346, EPI_ISL_1500347, EPI_ISL_1500348, EPI_ISL_1500349, EPI_ISL_1500350, EPI_ISL_1500351, EPI_ISL_1500352, EPI_ISL_1500353, EPI_ISL_1500354, EPI_ISL_1500355, EPI_ISL_1500356, EPI_ISL_1500357, EPI_ISL_1500358, EPI_ISL_1500359, EPI_ISL_1500360, EPI_ISL_1500361, EPI_ISL_1500362, EPI_ISL_1500363, EPI_ISL_1500364, EPI_ISL_1500365, EPI_ISL_1500366, EPI_ISL_1500367, EPI_ISL_1500368, EPI_ISL_1500369, EPI_ISL_1500370, EPI_ISL_1500371, EPI_ISL_1500372, EPI_ISL_1500373, EPI_ISL_1500374, EPI_ISL_1500375, EPI_ISL_1500376, EPI_ISL_1500377, EPI_ISL_1500378, EPI_ISL_1500379, EPI_ISL_1500380, EPI_ISL_1500381, EPI_ISL_1500382, EPI_ISL_1500383, EPI_ISL_1500384, EPI_ISL_1500385, EPI_ISL_1500386, EPI_ISL_1500387, EPI_ISL_1500388, EPI_ISL_1500389, EPI_ISL_1500390, EPI_ISL_1500391, EPI_ISL_1500392, EPI_ISL_1500393, EPI_ISL_1500394, EPI_ISL_1500395, EPI_ISL_1500396, EPI_ISL_1500397, EPI_ISL_1500398, EPI_ISL_1500400, EPI_ISL_1500401, EPI_ISL_1500402, EPI_ISL_1500403, EPI_ISL_1500404, EPI_ISL_1500405, EPI_ISL_1500406, EPI_ISL_1500407, EPI_ISL_1500408, EPI_ISL_1500409, EPI_ISL_1500410, EPI_ISL_1500411, EPI_ISL_1500412, EPI_ISL_1500413, EPI_ISL_1500414, EPI_ISL_1500415, EPI_ISL_1500416, EPI_ISL_1500417, EPI_ISL_1500418, EPI_ISL_1500419, EPI_ISL_1500420, EPI_ISL_1500421, EPI_ISL_1500422, EPI_ISL_1500423, EPI_ISL_1500424, EPI_ISL_1500425 | | | |
| see above | Maryland Genomics, Institute for Genome Sciences, University of Maryland School of Medicine | Maryland Genomics, Institute for Genome Sciences, University of Maryland School of Medicine | Tallon, Luke J; Sadzewicz, Lisa D; Humphrys, Mike; Ott, Sandra; Roussey, Holly; Mehta, Aditya; Vavikolanu, Kranthi; Fraser, Claire M; Ravel, Jacques |
| EPI_ISL_1500448, EPI_ISL_1500449, EPI_ISL_1500450, EPI_ISL_1500451, EPI_ISL_1500452, EPI_ISL_1500453, EPI_ISL_1500454, EPI_ISL_1500455, EPI_ISL_1500456, EPI_ISL_1500457, EPI_ISL_1500458, EPI_ISL_1500459, EPI_ISL_1500460, EPI_ISL_1500461, EPI_ISL_1500462, EPI_ISL_1500463, EPI_ISL_1500464, EPI_ISL_1500465, EPI_ISL_1500466, EPI_ISL_1500467, EPI_ISL_1500468, EPI_ISL_1500469, EPI_ISL_1500470 | | | |
| see above | Kansas Health and Environmental Lab | Kansas Health and Environmental Lab | Mike Grose, Jonathan Barnell, Ben Olsen, and Phil Adam |
| EPI_ISL_1500472, EPI_ISL_1500473, EPI_ISL_1500474, EPI_ISL_1500477, EPI_ISL_1500478, EPI_ISL_1500479, EPI_ISL_1500480, EPI_ISL_1500481, EPI_ISL_1500482, EPI_ISL_1500483, EPI_ISL_1500484, EPI_ISL_1500485, EPI_ISL_1500486, EPI_ISL_1500488, EPI_ISL_1500489, EPI_ISL_1500491, EPI_ISL_1500492, EPI_ISL_1500493, EPI_ISL_1500494, EPI_ISL_1500495, EPI_ISL_1500496, EPI_ISL_1500497, EPI_ISL_1500498, EPI_ISL_1500499, EPI_ISL_1500500, EPI_ISL_1500501, EPI_ISL_1500502, EPI_ISL_1500503, EPI_ISL_1500505, EPI_ISL_1500506, EPI_ISL_1500508, EPI_ISL_1500510, EPI_ISL_1500511, EPI_ISL_1500512, EPI_ISL_1500513, EPI_ISL_1500514, EPI_ISL_1500516, EPI_ISL_1500517, EPI_ISL_1500518, EPI_ISL_1500521, EPI_ISL_1500522, EPI_ISL_1500523, EPI_ISL_1500524, EPI_ISL_1500525, EPI_ISL_1500526, EPI_ISL_1500527, EPI_ISL_1500528, EPI_ISL_1500529, EPI_ISL_1500530, EPI_ISL_1500531, EPI_ISL_1500532, EPI_ISL_1500534, EPI_ISL_1500535, EPI_ISL_1500536, EPI_ISL_1500537, EPI_ISL_1500538, EPI_ISL_1500539, EPI_ISL_1500540, EPI_ISL_1500541, EPI_ISL_1500542, EPI_ISL_1500543, EPI_ISL_1500544, EPI_ISL_1500545, EPI_ISL_1500546, EPI_ISL_1500549, EPI_ISL_1500550, EPI_ISL_1500551, EPI_ISL_1500552, EPI_ISL_1500553, EPI_ISL_1500554, EPI_ISL_1500555, EPI_ISL_1500556, EPI_ISL_1500557, EPI_ISL_1500558, EPI_ISL_1500559, EPI_ISL_1500560, EPI_ISL_1500561, EPI_ISL_1500562, EPI_ISL_1500563, EPI_ISL_1500564, EPI_ISL_1500565, EPI_ISL_1500566, EPI_ISL_1500567, EPI_ISL_1500568, EPI_ISL_1500569, EPI_ISL_1500570, EPI_ISL_1500571, EPI_ISL_1500572, EPI_ISL_1500573, EPI_ISL_1500574, EPI_ISL_1500575, EPI_ISL_1500576, EPI_ISL_1500577, EPI_ISL_1500578, EPI_ISL_1500579, EPI_ISL_1500580, EPI_ISL_1500581, EPI_ISL_1500582, EPI_ISL_1500583, EPI_ISL_1500584, EPI_ISL_1500585, EPI_ISL_1500586, EPI_ISL_1500587, EPI_ISL_1500588, EPI_ISL_1500589, EPI_ISL_1500590, EPI_ISL_1500591, EPI_ISL_1500592, EPI_ISL_1500593, EPI_ISL_1500594, EPI_ISL_1500595, EPI_ISL_1500596, EPI_ISL_1500597, EPI_ISL_1500598, EPI_ISL_1500599, EPI_ISL_1500600, EPI_ISL_1500601, EPI_ISL_1500602, EPI_ISL_1500603, EPI_ISL_1500604, EPI_ISL_1500605, EPI_ISL_1500606, EPI_ISL_1500607, EPI_ISL_1500608, EPI_ISL_1500609, EPI_ISL_1500610, EPI_ISL_1500611, EPI_ISL_1500612, EPI_ISL_1500613, EPI_ISL_1500614, EPI_ISL_1500615, EPI_ISL_1500616, EPI_ISL_1500617, EPI_ISL_1500618, EPI_ISL_1500619, EPI_ISL_1500620, EPI_ISL_1500621, EPI_ISL_1500622, EPI_ISL_1500623, EPI_ISL_1500624, EPI_ISL_1500625, EPI_ISL_1500626, EPI_ISL_1500627, EPI_ISL_1500628, EPI_ISL_1500629, EPI_ISL_1500630, EPI_ISL_1500631, EPI_ISL_1500632, EPI_ISL_1500633, EPI_ISL_1500634, EPI_ISL_1500635, EPI_ISL_1500636, EPI_ISL_1500637 | | | |
| see above | Clinical Microbiology Laboratory, NewYork Presbyterian Hospital/Columbia University Irving Medical Center | Uhlemann Laboratory, Columbia University Irving Medical Center | Medini K. Annavajhala, Anne-Catrin Uhlemann, Hiroshi Mohri, David Ho |
| EPI_ISL_1500640 | Sonora Quest Laboratories | TGen North | "Jolene Bowers, Heather Centner, Chris French, Hayley Yaglom, Ashlyn Pfeiffer, Darrin Lemmer, Dave Engelthaler, The Arizona COVID Genomics Union (ACGU)" |
| EPI_ISL_1500642 | TGen North | TGen North | "Jolene Bowers, Heather Centner, Chris French, Hayley Yaglom, Ashlyn Pfeiffer, Darrin Lemmer, Dave Engelthaler, The Arizona COVID Genomics Union (ACGU)" |
| EPI_ISL_1500643, EPI_ISL_1500646, EPI_ISL_1500649, EPI_ISL_1500650, EPI_ISL_1500651, EPI_ISL_1500652, EPI_ISL_1500654, EPI_ISL_1500656, EPI_ISL_1500657, EPI_ISL_1500659, EPI_ISL_1500661, EPI_ISL_1500662, EPI_ISL_1500663 | | | |
| see above | Sonora Quest Laboratories | TGen North | "Jolene Bowers, Heather Centner, Chris French, Hayley Yaglom, Ashlyn Pfeiffer, Darrin Lemmer, Dave Engelthaler, The Arizona COVID Genomics Union (ACGU)" |
| EPI_ISL_1500666, EPI_ISL_1500667 | Lafene Health Center | Kansas State Veterinary Diagnostic Laboratory | Tyler Doerksen, Andreea Lu, Lance Noll, Elizabeth Porter, Kasey Hogan, Emily Cox, Jianfa Bai, Jamie Henningson, Rachel Palinski |
| EPI_ISL_1500668, EPI_ISL_1500669, EPI_ISL_1500670 | Sonora Quest Laboratories | TGen North | "Jolene Bowers, Heather Centner, Chris French, Hayley Yaglom, Ashlyn Pfeiffer, Darrin Lemmer, Dave Engelthaler, The Arizona COVID Genomics Union (ACGU)" |
| EPI_ISL_1500671 | TGen North | TGen North | "Jolene Bowers, Heather Centner, Chris French, Hayley Yaglom, Ashlyn Pfeiffer, Darrin Lemmer, Dave Engelthaler, The Arizona COVID Genomics Union (ACGU)" |
| EPI_ISL_1500672, EPI_ISL_1500673, EPI_ISL_1500674, EPI_ISL_1500677 | Sonora Quest Laboratories | TGen North | "Jolene Bowers, Heather Centner, Chris French, Hayley Yaglom, Ashlyn Pfeiffer, Darrin Lemmer, Dave Engelthaler, The Arizona COVID Genomics Union (ACGU)" |
| EPI_ISL_1500681 | TGen North | TGen North | "Jolene Bowers, Heather Centner, Chris French, Hayley Yaglom, Ashlyn Pfeiffer, Darrin Lemmer, Dave Engelthaler, The Arizona COVID Genomics Union (ACGU)" |
| EPI_ISL_1500682, EPI_ISL_1500687, EPI_ISL_1500688, EPI_ISL_1500692, EPI_ISL_1500693, EPI_ISL_1500694 | Sonora Quest Laboratories | TGen North | "Jolene Bowers, Heather Centner, Chris French, Hayley Yaglom, Ashlyn Pfeiffer, Darrin Lemmer, Dave Engelthaler, The Arizona COVID Genomics Union (ACGU)" |
| EPI_ISL_1500695 | TGen North | TGen North | "Jolene Bowers, Heather Centner, Chris French, Hayley Yaglom, Ashlyn Pfeiffer, Darrin Lemmer, Dave Engelthaler, The Arizona COVID Genomics Union (ACGU)" |
| EPI_ISL_1500696 | Sonora Quest Laboratories | TGen North | "Jolene Bowers, Heather Centner, Chris French, Hayley Yaglom, Ashlyn Pfeiffer, Darrin Lemmer, Dave Engelthaler, The Arizona COVID Genomics Union (ACGU)" |
| EPI_ISL_1500698 | TGen North | TGen North | "Jolene Bowers, Heather Centner, Chris French, Hayley Yaglom, Ashlyn Pfeiffer, Darrin Lemmer, Dave Engelthaler, The Arizona COVID Genomics Union (ACGU)" |
| EPI_ISL_1500702 | Sonora Quest Laboratories | TGen North | "Jolene Bowers, Heather Centner, Chris French, Hayley Yaglom, Ashlyn Pfeiffer, Darrin Lemmer, Dave Engelthaler, The Arizona COVID Genomics Union (ACGU)" |
| EPI_ISL_1500703 | TGen North | TGen North | "Jolene Bowers, Heather Centner, Chris French, Hayley Yaglom, Ashlyn Pfeiffer, Darrin Lemmer, Dave Engelthaler, The Arizona COVID Genomics Union (ACGU)" |
| EPI_ISL_1500704, EPI_ISL_1500706, EPI_ISL_1500708, EPI_ISL_1500710, EPI_ISL_1500712 | Sonora Quest Laboratories | TGen North | "Jolene Bowers, Heather Centner, Chris French, Hayley Yaglom, Ashlyn Pfeiffer, Darrin Lemmer, Dave Engelthaler, The Arizona COVID Genomics Union (ACGU)" |
| EPI_ISL_1500714 | TGen North | TGen North | "Jolene Bowers, Heather Centner, Chris French, Hayley Yaglom, Ashlyn Pfeiffer, Darrin Lemmer, Dave Engelthaler, The Arizona COVID Genomics Union (ACGU)" |
| EPI_ISL_1500715, EPI_ISL_1500716, EPI_ISL_1500720, EPI_ISL_1500721 | Sonora Quest Laboratories | TGen North | "Jolene Bowers, Heather Centner, Chris French, Hayley Yaglom, Ashlyn Pfeiffer, Darrin Lemmer, Dave Engelthaler, The Arizona COVID Genomics Union (ACGU)" |
| EPI_ISL_1500722 | TGen North | TGen North | "Jolene Bowers, Heather Centner, Chris French, Hayley Yaglom, Ashlyn Pfeiffer, Darrin Lemmer, Dave Engelthaler, The Arizona COVID Genomics Union (ACGU)" |
| EPI_ISL_1500723, EPI_ISL_1500725, EPI_ISL_1500727, EPI_ISL_1500730 | Sonora Quest Laboratories | TGen North | "Jolene Bowers, Heather Centner, Chris French, Hayley Yaglom, Ashlyn Pfeiffer, Darrin Lemmer, Dave Engelthaler, The Arizona COVID Genomics Union (ACGU)" |
| EPI_ISL_1500732 | TGen North | TGen North | "Jolene Bowers, Heather Centner, Chris French, Hayley Yaglom, Ashlyn Pfeiffer, Darrin Lemmer, Dave Engelthaler, The Arizona COVID Genomics Union (ACGU)" |
| EPI_ISL_1500733 | Sonora Quest Laboratories | TGen North | "Jolene Bowers, Heather Centner, Chris French, Hayley Yaglom, Ashlyn Pfeiffer, Darrin Lemmer, Dave Engelthaler, The Arizona COVID Genomics Union (ACGU)" |
| EPI_ISL_1500734 | TGen North | TGen North | "Jolene Bowers, Heather Centner, Chris French, Hayley Yaglom, Ashlyn Pfeiffer, Darrin Lemmer, Dave Engelthaler, The Arizona COVID Genomics Union (ACGU)" |
| EPI_ISL_1500737, EPI_ISL_1500738, EPI_ISL_1500740, EPI_ISL_1500742, EPI_ISL_1500744, EPI_ISL_1500746, EPI_ISL_1500747, EPI_ISL_1500748 | Sonora Quest Laboratories | TGen North | "Jolene Bowers, Heather Centner, Chris French, Hayley Yaglom, Ashlyn Pfeiffer, Darrin Lemmer, Dave Engelthaler, The Arizona COVID Genomics Union (ACGU)" |

| | | | |
|---|---|---|---|
| EPI_ISL_1500752 | TGen North | TGen North | "Jolene Bowers, Heather Centner, Chris French, Hayley Yaglom, Ashlyn Pfeiffer, Darrin Lemmer, Dave Engelthaler, The Arizona COVID Genomics Union (ACGU)" |
| EPI_ISL_1500755, EPI_ISL_1500756, EPI_ISL_1500757, EPI_ISL_1500758, EPI_ISL_1500759, EPI_ISL_1500760, EPI_ISL_1500761, EPI_ISL_1500766 | Sonora Quest Laboratories | TGen North | "Jolene Bowers, Heather Centner, Chris French, Hayley Yaglom, Ashlyn Pfeiffer, Darrin Lemmer, Dave Engelthaler, The Arizona COVID Genomics Union (ACGU)" |
| EPI_ISL_1500767 | TGen North | TGen North | "Jolene Bowers, Heather Centner, Chris French, Hayley Yaglom, Ashlyn Pfeiffer, Darrin Lemmer, Dave Engelthaler, The Arizona COVID Genomics Union (ACGU)" |
| EPI_ISL_1500768, EPI_ISL_1500770, EPI_ISL_1500771 | Sonora Quest Laboratories | TGen North | "Jolene Bowers, Heather Centner, Chris French, Hayley Yaglom, Ashlyn Pfeiffer, Darrin Lemmer, Dave Engelthaler, The Arizona COVID Genomics Union (ACGU)" |
| EPI_ISL_1500772, EPI_ISL_1500773 | TGen North | TGen North | "Jolene Bowers, Heather Centner, Chris French, Hayley Yaglom, Ashlyn Pfeiffer, Darrin Lemmer, Dave Engelthaler, The Arizona COVID Genomics Union (ACGU)" |
| EPI_ISL_1500774, EPI_ISL_1500775, EPI_ISL_1500776 | Sonora Quest Laboratories | TGen North | "Jolene Bowers, Heather Centner, Chris French, Hayley Yaglom, Ashlyn Pfeiffer, Darrin Lemmer, Dave Engelthaler, The Arizona COVID Genomics Union (ACGU)" |
| EPI_ISL_1500777, EPI_ISL_1500778 | TGen North | TGen North | "Jolene Bowers, Heather Centner, Chris French, Hayley Yaglom, Ashlyn Pfeiffer, Darrin Lemmer, Dave Engelthaler, The Arizona COVID Genomics Union (ACGU)" |
| EPI_ISL_1500779, EPI_ISL_1500781, EPI_ISL_1500782, EPI_ISL_1500783, EPI_ISL_1500784 | Sonora Quest Laboratories | TGen North | "Jolene Bowers, Heather Centner, Chris French, Hayley Yaglom, Ashlyn Pfeiffer, Darrin Lemmer, Dave Engelthaler, The Arizona COVID Genomics Union (ACGU)" |
| EPI_ISL_1500787 | TGen North | TGen North | "Jolene Bowers, Heather Centner, Chris French, Hayley Yaglom, Ashlyn Pfeiffer, Darrin Lemmer, Dave Engelthaler, The Arizona COVID Genomics Union (ACGU)" |
| EPI_ISL_1500788 | Sonora Quest Laboratories | TGen North | "Jolene Bowers, Heather Centner, Chris French, Hayley Yaglom, Ashlyn Pfeiffer, Darrin Lemmer, Dave Engelthaler, The Arizona COVID Genomics Union (ACGU)" |
| EPI_ISL_1500789 | TGen North | TGen North | "Jolene Bowers, Heather Centner, Chris French, Hayley Yaglom, Ashlyn Pfeiffer, Darrin Lemmer, Dave Engelthaler, The Arizona COVID Genomics Union (ACGU)" |
| EPI_ISL_1500791, EPI_ISL_1500792, EPI_ISL_1500794, EPI_ISL_1500796 | Sonora Quest Laboratories | TGen North | "Jolene Bowers, Heather Centner, Chris French, Hayley Yaglom, Ashlyn Pfeiffer, Darrin Lemmer, Dave Engelthaler, The Arizona COVID Genomics Union (ACGU)" |
| EPI_ISL_1500797 | TGen North | TGen North | "Jolene Bowers, Heather Centner, Chris French, Hayley Yaglom, Ashlyn Pfeiffer, Darrin Lemmer, Dave Engelthaler, The Arizona COVID Genomics Union (ACGU)" |
| EPI_ISL_1500800, EPI_ISL_1500803 | Sonora Quest Laboratories | TGen North | "Jolene Bowers, Heather Centner, Chris French, Hayley Yaglom, Ashlyn Pfeiffer, Darrin Lemmer, Dave Engelthaler, The Arizona COVID Genomics Union (ACGU)" |
| EPI_ISL_1500805 | TGen North | TGen North | "Jolene Bowers, Heather Centner, Chris French, Hayley Yaglom, Ashlyn Pfeiffer, Darrin Lemmer, Dave Engelthaler, The Arizona COVID Genomics Union (ACGU)" |
| EPI_ISL_1500806, EPI_ISL_1500808, EPI_ISL_1500809, EPI_ISL_1500811 | Sonora Quest Laboratories | TGen North | "Jolene Bowers, Heather Centner, Chris French, Hayley Yaglom, Ashlyn Pfeiffer, Darrin Lemmer, Dave Engelthaler, The Arizona COVID Genomics Union (ACGU)" |
| EPI_ISL_1500812, EPI_ISL_1500813 | TGen North | TGen North | "Jolene Bowers, Heather Centner, Chris French, Hayley Yaglom, Ashlyn Pfeiffer, Darrin Lemmer, Dave Engelthaler, The Arizona COVID Genomics Union (ACGU)" |
| EPI_ISL_1500814, EPI_ISL_1500817, EPI_ISL_1500820, EPI_ISL_1500821, EPI_ISL_1500824, EPI_ISL_1500829, EPI_ISL_1500831, EPI_ISL_1500832, EPI_ISL_1500833, EPI_ISL_1500835, EPI_ISL_1500837, EPI_ISL_1500839, EPI_ISL_1500840, EPI_ISL_1500844, EPI_ISL_1500845, EPI_ISL_1500846, EPI_ISL_1500848, EPI_ISL_1500850 | | | |
| see above | Sonora Quest Laboratories | TGen North | "Jolene Bowers, Heather Centner, Chris French, Hayley Yaglom, Ashlyn Pfeiffer, Darrin Lemmer, Dave Engelthaler, The Arizona COVID Genomics Union (ACGU)" |
| EPI_ISL_1500852 | TGen North | TGen North | "Jolene Bowers, Heather Centner, Chris French, Hayley Yaglom, Ashlyn Pfeiffer, Darrin Lemmer, Dave Engelthaler, The Arizona COVID Genomics Union (ACGU)" |
| EPI_ISL_1500853, EPI_ISL_1500857, EPI_ISL_1500858, EPI_ISL_1500859, EPI_ISL_1500861, EPI_ISL_1500862, EPI_ISL_1500863, EPI_ISL_1500864, EPI_ISL_1500865 | Sonora Quest Laboratories | TGen North | "Jolene Bowers, Heather Centner, Chris French, Hayley Yaglom, Ashlyn Pfeiffer, Darrin Lemmer, Dave Engelthaler, The Arizona COVID Genomics Union (ACGU)" |
| EPI_ISL_1500866 | Centro de Investigación en Ciencias de la Salud y Biomedicina, U.A.S.L.P. | Centro de Investigación en Ciencias de la Salud y Biomedicina, U.A.S.L.P. | MD PhD Sofía Bernal Silva |
| EPI_ISL_1500868, EPI_ISL_1500869, EPI_ISL_1500871, EPI_ISL_1500872, EPI_ISL_1500873, EPI_ISL_1500874, EPI_ISL_1500875, EPI_ISL_1500876, EPI_ISL_1500877 | Sonora Quest Laboratories | TGen North | "Jolene Bowers, Heather Centner, Chris French, Hayley Yaglom, Ashlyn Pfeiffer, Darrin Lemmer, Dave Engelthaler, The Arizona COVID Genomics Union (ACGU)" |
| EPI_ISL_1500878 | TGen North | TGen North | "Jolene Bowers, Heather Centner, Chris French, Hayley Yaglom, Ashlyn Pfeiffer, Darrin Lemmer, Dave Engelthaler, The Arizona COVID Genomics Union (ACGU)" |
| EPI_ISL_1500880, EPI_ISL_1500881, EPI_ISL_1500883, EPI_ISL_1500885, EPI_ISL_1500887, EPI_ISL_1500893, EPI_ISL_1500895 | Sonora Quest Laboratories | TGen North | "Jolene Bowers, Heather Centner, Chris French, Hayley Yaglom, Ashlyn Pfeiffer, Darrin Lemmer, Dave Engelthaler, The Arizona COVID Genomics Union (ACGU)" |
| EPI_ISL_1500896 | TGen North | TGen North | "Jolene Bowers, Heather Centner, Chris French, Hayley Yaglom, Ashlyn Pfeiffer, Darrin Lemmer, Dave Engelthaler, The Arizona COVID Genomics Union (ACGU)" |
| EPI_ISL_1500897, EPI_ISL_1500898, EPI_ISL_1500900, EPI_ISL_1500901, EPI_ISL_1500902, EPI_ISL_1500904, EPI_ISL_1500907, EPI_ISL_1500908 | Sonora Quest Laboratories | TGen North | "Jolene Bowers, Heather Centner, Chris French, Hayley Yaglom, Ashlyn Pfeiffer, Darrin Lemmer, Dave Engelthaler, The Arizona COVID Genomics Union (ACGU)" |
| EPI_ISL_1500910 | TGen North | TGen North | "Jolene Bowers, Heather Centner, Chris French, Hayley Yaglom, Ashlyn Pfeiffer, Darrin Lemmer, Dave Engelthaler, The Arizona COVID Genomics Union (ACGU)" |
| EPI_ISL_1500913, EPI_ISL_1500914 | Sonora Quest Laboratories | TGen North | "Jolene Bowers, Heather Centner, Chris French, Hayley Yaglom, Ashlyn Pfeiffer, Darrin Lemmer, Dave Engelthaler, The Arizona COVID Genomics Union (ACGU)" |
| EPI_ISL_1500915 | TGen North | TGen North | "Jolene Bowers, Heather Centner, Chris French, Hayley Yaglom, Ashlyn Pfeiffer, Darrin Lemmer, Dave Engelthaler, The Arizona COVID Genomics Union (ACGU)" |
| EPI_ISL_1500916, EPI_ISL_1500917, EPI_ISL_1500920 | Sonora Quest Laboratories | TGen North | "Jolene Bowers, Heather Centner, Chris French, Hayley Yaglom, Ashlyn Pfeiffer, Darrin Lemmer, Dave Engelthaler, The Arizona COVID Genomics Union (ACGU)" |
| EPI_ISL_1500921 | TGen North | TGen North | "Jolene Bowers, Heather Centner, Chris French, Hayley Yaglom, Ashlyn Pfeiffer, Darrin Lemmer, Dave Engelthaler, The Arizona COVID Genomics Union (ACGU)" |
| EPI_ISL_1500922, EPI_ISL_1500923, EPI_ISL_1500924, EPI_ISL_1500926, EPI_ISL_1500927, EPI_ISL_1500929, EPI_ISL_1500930, EPI_ISL_1500931, EPI_ISL_1500934, EPI_ISL_1500936, EPI_ISL_1500937 | | | |
| see above | Sonora Quest Laboratories | TGen North | "Jolene Bowers, Heather Centner, Chris French, Hayley Yaglom, Ashlyn Pfeiffer, Darrin Lemmer, Dave Engelthaler, The Arizona COVID Genomics Union (ACGU)" |
| EPI_ISL_1500939 | TGen North | TGen North | "Jolene Bowers, Heather Centner, Chris French, Hayley Yaglom, Ashlyn Pfeiffer, Darrin Lemmer, Dave Engelthaler, The Arizona COVID Genomics Union (ACGU)" |
| EPI_ISL_1500941, | Sonora Quest Laboratories | TGen North | "Jolene Bowers, Heather Centner, Chris French, Hayley Yaglom, Ashlyn Pfeiffer, Darrin Lemmer, Dave Engelthaler, The Arizona COVID Genomics Union (ACGU)" |

| | | | |
|---|---|---|---|
| EPI_ISL_1500942, EPI_ISL_1500943, EPI_ISL_1500947 | | | |
| EPI_ISL_1501675, EPI_ISL_1501676, EPI_ISL_1501677, EPI_ISL_1501678, EPI_ISL_1501679, EPI_ISL_1501680, EPI_ISL_1501681, EPI_ISL_1501682 | Nevada State Public Health Laboratory | Nevada State Public Health Laboratory | Andrew Gorzalski, Mark Pandori |
| EPI_ISL_1501684, EPI_ISL_1501685, EPI_ISL_1501686, EPI_ISL_1501687, EPI_ISL_1501688, EPI_ISL_1501689, EPI_ISL_1501690, EPI_ISL_1501691, EPI_ISL_1501692, EPI_ISL_1501693 | Northwestern Memorial Hospital | Northwestern University - Ozer Lab | Ramon Lorenzo-Redondo, Lacy M. Simons, Taylor J. Dean, Chad J. Achenbach, Lawrence J. Jennings, Chao Qi, Michael G. Ison, Judd F. Hultquist, Egon A. Ozer |
| EPI_ISL_1501695, EPI_ISL_1501696, EPI_ISL_1501697, EPI_ISL_1501698, EPI_ISL_1501699, EPI_ISL_1501700, EPI_ISL_1501701, EPI_ISL_1501702, EPI_ISL_1501703, EPI_ISL_1501704, EPI_ISL_1501705, EPI_ISL_1501706, EPI_ISL_1501707, EPI_ISL_1501708, EPI_ISL_1501709, EPI_ISL_1501710, EPI_ISL_1501711, EPI_ISL_1501712, EPI_ISL_1501714, EPI_ISL_1501715, EPI_ISL_1501716, EPI_ISL_1501717 | | | |
| see above | Lurie Children's Hospital of Chicago | Northwestern University - Ozer Lab | Ramon Lorenzo-Redondo, Lacy M. Simons, Taylor J. Dean, Michael G. Ison, Xiaotian, Zheng, William J. Muller, Larry K. Kociolek, Judd F. Hultquist, Egon A. Ozer |
| EPI_ISL_1501718, EPI_ISL_1501719 | Oregon State University TRACE Project | OSU Center for Genome Research and Biocomputing | Oregon State University TRACE Project |
| EPI_ISL_1501720 | Lurie Children's Hospital of Chicago | Northwestern University - Ozer Lab | Ramon Lorenzo-Redondo, Lacy M. Simons, Taylor J. Dean, Michael G. Ison, Xiaotian, Zheng, William J. Muller, Larry K. Kociolek, Judd F. Hultquist, Egon A. Ozer |
| EPI_ISL_1501721, EPI_ISL_1501722 | Oregon State University TRACE Project | OSU Center for Genome Research and Biocomputing | Oregon State University TRACE Project |
| EPI_ISL_1501723 | Lurie Children's Hospital of Chicago | Northwestern University - Ozer Lab | Ramon Lorenzo-Redondo, Lacy M. Simons, Taylor J. Dean, Michael G. Ison, Xiaotian, Zheng, William J. Muller, Larry K. Kociolek, Judd F. Hultquist, Egon A. Ozer |
| EPI_ISL_1501724, EPI_ISL_1501725 | Oregon State University TRACE Project | OSU Center for Genome Research and Biocomputing | Oregon State University TRACE Project |
| EPI_ISL_1501726 | Lurie Children's Hospital of Chicago | Northwestern University - Ozer Lab | Ramon Lorenzo-Redondo, Lacy M. Simons, Taylor J. Dean, Michael G. Ison, Xiaotian, Zheng, William J. Muller, Larry K. Kociolek, Judd F. Hultquist, Egon A. Ozer |
| EPI_ISL_1501727, EPI_ISL_1501728 | Oregon State University TRACE Project | OSU Center for Genome Research and Biocomputing | Oregon State University TRACE Project |
| EPI_ISL_1501729 | Lurie Children's Hospital of Chicago | Northwestern University - Ozer Lab | Ramon Lorenzo-Redondo, Lacy M. Simons, Taylor J. Dean, Michael G. Ison, Xiaotian, Zheng, William J. Muller, Larry K. Kociolek, Judd F. Hultquist, Egon A. Ozer |
| EPI_ISL_1501730, EPI_ISL_1501731 | Oregon State University TRACE Project | OSU Center for Genome Research and Biocomputing | Oregon State University TRACE Project |
| EPI_ISL_1501732 | Lurie Children's Hospital of Chicago | Northwestern University - Ozer Lab | Ramon Lorenzo-Redondo, Lacy M. Simons, Taylor J. Dean, Michael G. Ison, Xiaotian, Zheng, William J. Muller, Larry K. Kociolek, Judd F. Hultquist, Egon A. Ozer |
| EPI_ISL_1501733, EPI_ISL_1501734 | Oregon State University TRACE Project | OSU Center for Genome Research and Biocomputing | Oregon State University TRACE Project |
| EPI_ISL_1501735 | Lurie Children's Hospital of Chicago | Northwestern University - Ozer Lab | Ramon Lorenzo-Redondo, Lacy M. Simons, Taylor J. Dean, Michael G. Ison, Xiaotian, Zheng, William J. Muller, Larry K. Kociolek, Judd F. Hultquist, Egon A. Ozer |
| EPI_ISL_1501736, EPI_ISL_1501737, EPI_ISL_1501739, EPI_ISL_1501740, EPI_ISL_1501742, EPI_ISL_1501743 | Oregon State University TRACE Project | OSU Center for Genome Research and Biocomputing | Oregon State University TRACE Project |
| EPI_ISL_1501744 | Lurie Children's Hospital of Chicago | Northwestern University - Ozer Lab | Ramon Lorenzo-Redondo, Lacy M. Simons, Taylor J. Dean, Michael G. Ison, Xiaotian, Zheng, William J. Muller, Larry K. Kociolek, Judd F. Hultquist, Egon A. Ozer |
| EPI_ISL_1501745, EPI_ISL_1501746 | Oregon State University TRACE Project | OSU Center for Genome Research and Biocomputing | Oregon State University TRACE Project |
| EPI_ISL_1501747 | Lurie Children's Hospital of Chicago | Northwestern University - Ozer Lab | Ramon Lorenzo-Redondo, Lacy M. Simons, Taylor J. Dean, Michael G. Ison, Xiaotian, Zheng, William J. Muller, Larry K. Kociolek, Judd F. Hultquist, Egon A. Ozer |
| EPI_ISL_1501748 | Oregon State University TRACE Project | OSU Center for Genome Research and Biocomputing | Oregon State University TRACE Project |
| EPI_ISL_1501749 | Lurie Children's Hospital of Chicago | Northwestern University - Ozer Lab | Ramon Lorenzo-Redondo, Lacy M. Simons, Taylor J. Dean, Michael G. Ison, Xiaotian, Zheng, William J. Muller, Larry K. Kociolek, Judd F. Hultquist, Egon A. Ozer |
| EPI_ISL_1501750, EPI_ISL_1501751, EPI_ISL_1501752 | Oregon State University TRACE Project | OSU Center for Genome Research and Biocomputing | Oregon State University TRACE Project |
| EPI_ISL_1501753 | Lurie Children's Hospital of Chicago | Northwestern University - Ozer Lab | Ramon Lorenzo-Redondo, Lacy M. Simons, Taylor J. Dean, Michael G. Ison, Xiaotian, Zheng, William J. Muller, Larry K. Kociolek, Judd F. Hultquist, Egon A. Ozer |
| EPI_ISL_1501754, EPI_ISL_1501755 | Oregon State University TRACE Project | OSU Center for Genome Research and Biocomputing | Oregon State University TRACE Project |
| EPI_ISL_1501756 | Lurie Children's Hospital of Chicago | Northwestern University - Ozer Lab | Ramon Lorenzo-Redondo, Lacy M. Simons, Taylor J. Dean, Michael G. Ison, Xiaotian, Zheng, William J. Muller, Larry K. Kociolek, Judd F. Hultquist, Egon A. Ozer |
| EPI_ISL_1501757 | Oregon State University TRACE Project | OSU Center for Genome Research and Biocomputing | Oregon State University TRACE Project |
| EPI_ISL_1501758 | Lurie Children's Hospital of Chicago | Northwestern University - Ozer Lab | Ramon Lorenzo-Redondo, Lacy M. Simons, Taylor J. Dean, Michael G. Ison, Xiaotian, Zheng, William J. Muller, Larry K. Kociolek, Judd F. Hultquist, Egon A. Ozer |
| EPI_ISL_1501759, EPI_ISL_1501760 | Oregon State University TRACE Project | OSU Center for Genome Research and Biocomputing | Oregon State University TRACE Project |
| EPI_ISL_1501761 | Lurie Children's Hospital of Chicago | Northwestern University - Ozer Lab | Ramon Lorenzo-Redondo, Lacy M. Simons, Taylor J. Dean, Michael G. Ison, Xiaotian, Zheng, William J. Muller, Larry K. Kociolek, Judd F. Hultquist, Egon A. Ozer |
| EPI_ISL_1501762, EPI_ISL_1501763 | Oregon State University TRACE Project | OSU Center for Genome Research and Biocomputing | Oregon State University TRACE Project |
| EPI_ISL_1501764 | Lurie Children's Hospital of Chicago | Northwestern University - Ozer Lab | Ramon Lorenzo-Redondo, Lacy M. Simons, Taylor J. Dean, Michael G. Ison, Xiaotian, Zheng, William J. Muller, Larry K. Kociolek, Judd F. Hultquist, Egon A. Ozer |
| EPI_ISL_1501765, EPI_ISL_1501766 | Oregon State University TRACE Project | OSU Center for Genome Research and Biocomputing | Oregon State University TRACE Project |
| EPI_ISL_1501767 | Lurie Children's Hospital of Chicago | Northwestern University - Ozer Lab | Ramon Lorenzo-Redondo, Lacy M. Simons, Taylor J. Dean, Michael G. Ison, Xiaotian, Zheng, William J. Muller, Larry K. Kociolek, Judd F. Hultquist, Egon A. Ozer |
| EPI_ISL_1501768, EPI_ISL_1501769 | Oregon State University TRACE Project | OSU Center for Genome Research and Biocomputing | Oregon State University TRACE Project |
| EPI_ISL_1501770 | Lurie Children's Hospital of Chicago | Northwestern University - Ozer Lab | Ramon Lorenzo-Redondo, Lacy M. Simons, Taylor J. Dean, Michael G. Ison, Xiaotian, Zheng, William J. Muller, Larry K. Kociolek, Judd F. Hultquist, Egon A. Ozer |
| EPI_ISL_1501771, EPI_ISL_1501772 | Oregon State University TRACE Project | OSU Center for Genome Research and Biocomputing | Oregon State University TRACE Project |
| EPI_ISL_1501773 | Lurie Children's Hospital of Chicago | Northwestern University - Ozer Lab | Ramon Lorenzo-Redondo, Lacy M. Simons, Taylor J. Dean, Michael G. Ison, Xiaotian, Zheng, William J. Muller, Larry K. Kociolek, Judd F. Hultquist, Egon A. Ozer |
| EPI_ISL_1501774, EPI_ISL_1501775 | Oregon State University TRACE Project | OSU Center for Genome Research and Biocomputing | Oregon State University TRACE Project |
| EPI_ISL_1501776 | Lurie Children's Hospital of Chicago | Northwestern University - Ozer Lab | Ramon Lorenzo-Redondo, Lacy M. Simons, Taylor J. Dean, Michael G. Ison, Xiaotian, Zheng, William J. Muller, Larry K. Kociolek, Judd F. Hultquist, Egon A. Ozer |
| EPI_ISL_1501777, EPI_ISL_1501778 | Oregon State University TRACE Project | OSU Center for Genome Research and Biocomputing | Oregon State University TRACE Project |
| EPI_ISL_1501779 | Lurie Children's Hospital of Chicago | Northwestern University - Ozer Lab | Ramon Lorenzo-Redondo, Lacy M. Simons, Taylor J. Dean, Michael G. Ison, Xiaotian, Zheng, William J. Muller, Larry K. Kociolek, Judd F. Hultquist, Egon A. Ozer |
| EPI_ISL_1501780, EPI_ISL_1501781 | Oregon State University TRACE Project | OSU Center for Genome Research and Biocomputing | Oregon State University TRACE Project |
| EPI_ISL_1501782 | Lurie Children's Hospital of Chicago | Northwestern University - Ozer Lab | Ramon Lorenzo-Redondo, Lacy M. Simons, Taylor J. Dean, Michael G. Ison, Xiaotian, Zheng, William J. Muller, Larry K. Kociolek, Judd F. Hultquist, Egon A. Ozer |
| EPI_ISL_1501783, EPI_ISL_1501784 | Oregon State University TRACE Project | OSU Center for Genome Research and Biocomputing | Oregon State University TRACE Project |

| | | | |
|---|---|---|---|
| EPI_ISL_1501785 | Lurie Children's Hospital of Chicago | Northwestern University - Ozer Lab | Ramon Lorenzo-Redondo, Lacy M. Simons, Taylor J. Dean, Michael G. Ison, Xiaotian, Zheng, William J. Muller, Larry K. Kociolek, Judd F. Hultquist, Egon A. Ozer |
| EPI_ISL_1501786, EPI_ISL_1501787, EPI_ISL_1501789, EPI_ISL_1501790 | Oregon State University TRACE Project | OSU Center for Genome Research and Biocomputing | Oregon State University TRACE Project |
| EPI_ISL_1501791, EPI_ISL_1501792, EPI_ISL_1501793, EPI_ISL_1501794, EPI_ISL_1501795, EPI_ISL_1501796, EPI_ISL_1501797, EPI_ISL_1501798, EPI_ISL_1501799, EPI_ISL_1501801, EPI_ISL_1501802, EPI_ISL_1501803, EPI_ISL_1501804, EPI_ISL_1501805, EPI_ISL_1501807, EPI_ISL_1501808, EPI_ISL_1501809, EPI_ISL_1501810, EPI_ISL_1501811, EPI_ISL_1501812, EPI_ISL_1501813, EPI_ISL_1501814, EPI_ISL_1501815, EPI_ISL_1501816, EPI_ISL_1501819, EPI_ISL_1501820, EPI_ISL_1501822, EPI_ISL_1501823, EPI_ISL_1501824, EPI_ISL_1501825, EPI_ISL_1501826, EPI_ISL_1501827, EPI_ISL_1501828, EPI_ISL_1501829, EPI_ISL_1501830, EPI_ISL_1501831, EPI_ISL_1501832, EPI_ISL_1501833, EPI_ISL_1501834, EPI_ISL_1501835, EPI_ISL_1501836, EPI_ISL_1501837, EPI_ISL_1501838, EPI_ISL_1501839, EPI_ISL_1501840, EPI_ISL_1501841, EPI_ISL_1501842, EPI_ISL_1501843, EPI_ISL_1501844, EPI_ISL_1501845, EPI_ISL_1501846, EPI_ISL_1501847, EPI_ISL_1501848, EPI_ISL_1501849, EPI_ISL_1501850, EPI_ISL_1501851, EPI_ISL_1501852, EPI_ISL_1501853, EPI_ISL_1501854, EPI_ISL_1501855, EPI_ISL_1501856, EPI_ISL_1501857, EPI_ISL_1501858, EPI_ISL_1501859, EPI_ISL_1501860, EPI_ISL_1501861, EPI_ISL_1501862, EPI_ISL_1501863, EPI_ISL_1501864, EPI_ISL_1501865, EPI_ISL_1501866, EPI_ISL_1501867, EPI_ISL_1501868, EPI_ISL_1501869, EPI_ISL_1501870, EPI_ISL_1501871, EPI_ISL_1501872, EPI_ISL_1501873, EPI_ISL_1501874, EPI_ISL_1501875, EPI_ISL_1501876, EPI_ISL_1501877, EPI_ISL_1501878, EPI_ISL_1501879, EPI_ISL_1501880, EPI_ISL_1501881, EPI_ISL_1501882, EPI_ISL_1501883, EPI_ISL_1501884, EPI_ISL_1501885, EPI_ISL_1501886, EPI_ISL_1501887, EPI_ISL_1501888, EPI_ISL_1501889, EPI_ISL_1501890, EPI_ISL_1501891, EPI_ISL_1501892, EPI_ISL_1501893, EPI_ISL_1501894, EPI_ISL_1501895, EPI_ISL_1501896, EPI_ISL_1501897, EPI_ISL_1501898, EPI_ISL_1501899, EPI_ISL_1501900, EPI_ISL_1501901, EPI_ISL_1501902, EPI_ISL_1501903, EPI_ISL_1501904, EPI_ISL_1501905, EPI_ISL_1501906, EPI_ISL_1501907, EPI_ISL_1501908, EPI_ISL_1501909, EPI_ISL_1501910, EPI_ISL_1501911, EPI_ISL_1501912, EPI_ISL_1501913, EPI_ISL_1501914, EPI_ISL_1501915, EPI_ISL_1501916, EPI_ISL_1501917, EPI_ISL_1501918, EPI_ISL_1501919, EPI_ISL_1501920, EPI_ISL_1501921, EPI_ISL_1501922, EPI_ISL_1501923, EPI_ISL_1501924, EPI_ISL_1501925, EPI_ISL_1501926, EPI_ISL_1501927, EPI_ISL_1501928, EPI_ISL_1501929, EPI_ISL_1501930, EPI_ISL_1501931, EPI_ISL_1501932, EPI_ISL_1501933, EPI_ISL_1501934, EPI_ISL_1501935, EPI_ISL_1501936, EPI_ISL_1501937, EPI_ISL_1501938, EPI_ISL_1501939, EPI_ISL_1501940, EPI_ISL_1501941, EPI_ISL_1501942, EPI_ISL_1501943, EPI_ISL_1501944, EPI_ISL_1501945, EPI_ISL_1501946, EPI_ISL_1501947, EPI_ISL_1501948, EPI_ISL_1501949, EPI_ISL_1501950, EPI_ISL_1501951, EPI_ISL_1501952, EPI_ISL_1501953, EPI_ISL_1501954, EPI_ISL_1501955, EPI_ISL_1501956, EPI_ISL_1501957, EPI_ISL_1501958, EPI_ISL_1501959, EPI_ISL_1501960, EPI_ISL_1501961, EPI_ISL_1501962, EPI_ISL_1501963, EPI_ISL_1501964, EPI_ISL_1501965, EPI_ISL_1501966, EPI_ISL_1501967, EPI_ISL_1501968, EPI_ISL_1501969, EPI_ISL_1501970, EPI_ISL_1501971, EPI_ISL_1501972, EPI_ISL_1501973, EPI_ISL_1501974, EPI_ISL_1501975, EPI_ISL_1501976, EPI_ISL_1501977, EPI_ISL_1501978, EPI_ISL_1501979, EPI_ISL_1501980, EPI_ISL_1501981, EPI_ISL_1501982, EPI_ISL_1501983, EPI_ISL_1501984, EPI_ISL_1501985, EPI_ISL_1501986, EPI_ISL_1501987, EPI_ISL_1501988, EPI_ISL_1501989, EPI_ISL_1501990, EPI_ISL_1501991, EPI_ISL_1501992, EPI_ISL_1501993, EPI_ISL_1501994, EPI_ISL_1501995, EPI_ISL_1501996, EPI_ISL_1501997, EPI_ISL_1501998, EPI_ISL_1501999, EPI_ISL_1502000, EPI_ISL_1502001, EPI_ISL_1502002, EPI_ISL_1502003, EPI_ISL_1502004, EPI_ISL_1502005, EPI_ISL_1502006, EPI_ISL_1502007, EPI_ISL_1502008, EPI_ISL_1502009, EPI_ISL_1502010, EPI_ISL_1502011, EPI_ISL_1502012, EPI_ISL_1502013, EPI_ISL_1502014, EPI_ISL_1502015, EPI_ISL_1502016, EPI_ISL_1502017, EPI_ISL_1502018, EPI_ISL_1502019, EPI_ISL_1502020, EPI_ISL_1502021, EPI_ISL_1502022, EPI_ISL_1502023, EPI_ISL_1502024, EPI_ISL_1502025, EPI_ISL_1502026, EPI_ISL_1502027, EPI_ISL_1502028, EPI_ISL_1502029, EPI_ISL_1502030, EPI_ISL_1502031, EPI_ISL_1502032, EPI_ISL_1502033, EPI_ISL_1502034, EPI_ISL_1502035, EPI_ISL_1502036, EPI_ISL_1502037, EPI_ISL_1502038, EPI_ISL_1502039, EPI_ISL_1502040, EPI_ISL_1502041, EPI_ISL_1502042, EPI_ISL_1502043, EPI_ISL_1502044, EPI_ISL_1502045, EPI_ISL_1502046, EPI_ISL_1502047, EPI_ISL_1502048, EPI_ISL_1502049, EPI_ISL_1502050, EPI_ISL_1502051 | | | |
| see above | Lurie Children's Hospital of Chicago | Northwestern University - Ozer Lab | Ramon Lorenzo-Redondo, Lacy M. Simons, Taylor J. Dean, Michael G. Ison, Xiaotian, Zheng, William J. Muller, Larry K. Kociolek, Judd F. Hultquist, Egon A. Ozer |
| EPI_ISL_1502052, EPI_ISL_1502053, EPI_ISL_1502054, EPI_ISL_1502055, EPI_ISL_1502056, EPI_ISL_1502057, EPI_ISL_1502058, EPI_ISL_1502059, EPI_ISL_1502060, EPI_ISL_1502061, EPI_ISL_1502062, EPI_ISL_1502064, EPI_ISL_1502065, EPI_ISL_1502066, EPI_ISL_1502067, EPI_ISL_1502068, EPI_ISL_1502069, EPI_ISL_1502070, EPI_ISL_1502071, EPI_ISL_1502072, EPI_ISL_1502073, EPI_ISL_1502074 | | | |
| see above | Northwestern Memorial Hospital | Northwestern University - Ozer Lab | Ramon Lorenzo-Redondo, Lacy M. Simons, Taylor J. Dean, Chad J. Achenbach, Lawrence J. Jennings, Chao Qi, Michael G. Ison, Judd F. Hultquist, Egon A. Ozer |
| EPI_ISL_1502075, EPI_ISL_1502076, EPI_ISL_1502077, EPI_ISL_1502078, EPI_ISL_1502079, EPI_ISL_1502080, EPI_ISL_1502081, EPI_ISL_1502082, EPI_ISL_1502083, EPI_ISL_1502084, EPI_ISL_1502085, EPI_ISL_1502086 | | | |
| see above | DC Public Health Lab/ Dept. of Forensic Sciences | DC Public Health Lab/ Dept. of Forensic Sciences | Janis Doss, Scott Nguyen, Elizabeth Zelaya, Sarah Scott, Connie Maza, Monica Mann, Brittany Hamilton, David Payne, Jocelyn Hauser |
| EPI_ISL_1502087, EPI_ISL_1502088, EPI_ISL_1502089, EPI_ISL_1502090, EPI_ISL_1502091, EPI_ISL_1502092, EPI_ISL_1502093, EPI_ISL_1502094, EPI_ISL_1502095, EPI_ISL_1502096, EPI_ISL_1502097, EPI_ISL_1502098, EPI_ISL_1502099, EPI_ISL_1502100, EPI_ISL_1502101, EPI_ISL_1502102, EPI_ISL_1502103, EPI_ISL_1502104, EPI_ISL_1502105, EPI_ISL_1502106, EPI_ISL_1502107, EPI_ISL_1502108, EPI_ISL_1502110, EPI_ISL_1502111 | | | |
| see above | Maine HETL | Health and Environmental Testing Laboratory | Matluk,N., Dewey,H., Martha,J., Barter,M., Lynch,R., Munger,H. and Tewhey,R. |
| EPI_ISL_1502796, EPI_ISL_1502797, EPI_ISL_1502798, EPI_ISL_1502799, EPI_ISL_1502800, EPI_ISL_1502801, EPI_ISL_1502802, EPI_ISL_1502803 | Kansas Health and Environmental Lab | Kansas Health and Environmental Lab | Mike Grose, Jonathan Barnell, Ben Olsen, and Phil Adam |
| EPI_ISL_1502805, EPI_ISL_1502806, EPI_ISL_1502807, EPI_ISL_1502809 | MD Laboratories | Los Angeles County Public Health Laboratories | P. Hemarajata et al. |
| EPI_ISL_1502810, EPI_ISL_1502811 | USC Clinical Lab | Los Angeles County Public Health Laboratories | P. Hemarajata et al. |
| EPI_ISL_1502814, EPI_ISL_1502815, EPI_ISL_1502816, EPI_ISL_1502817, EPI_ISL_1503143 | Infectious Diseases Department , Instituto Nacional de Ciencias Medicas y Nutrición | Instituto Nacional de Ciencias Medicas y Nutricion Infectious Diseases | Guillermo M. Ruiz-Palacios, Pilar Ramos Cervantes, Violeta Ibarra Gonzalez, Fernando Ledesma Barrientos, Miriam Arciniega Fuentes , Fernando Arteaga Cabello, Diana Paola Davalos Martinez, Luis Alberto Garcia Andrade, Luz Elena Cervantes Villar |
| EPI_ISL_1503231 | Clinical Reference Laboratory | Kansas Health and Environmental Lab | Mike Grose, Jonathan Barnell, Ben Olsen, and Phil Adam |
| EPI_ISL_1503232, EPI_ISL_1503233, EPI_ISL_1503234, EPI_ISL_1503236, EPI_ISL_1503237, EPI_ISL_1503238, EPI_ISL_1503239, EPI_ISL_1503240, EPI_ISL_1503241, EPI_ISL_1503242, EPI_ISL_1503243, EPI_ISL_1503244, EPI_ISL_1503246 | | | |
| see above | Wichita State University - Molecular Diagnostics Lab | Kansas Health and Environmental Lab | Mike Grose, Jonathan Barnell, Ben Olsen, and Phil Adam |
| EPI_ISL_1503248, EPI_ISL_1503249, EPI_ISL_1503250, EPI_ISL_1503251, EPI_ISL_1503252, EPI_ISL_1503253, EPI_ISL_1503254, EPI_ISL_1503255, EPI_ISL_1503256, EPI_ISL_1503257, EPI_ISL_1503258, EPI_ISL_1503259, EPI_ISL_1503260, EPI_ISL_1503262, EPI_ISL_1503263, EPI_ISL_1503264, EPI_ISL_1503265, EPI_ISL_1503266, EPI_ISL_1503267, EPI_ISL_1503268, EPI_ISL_1503269, EPI_ISL_1503271, EPI_ISL_1503272, EPI_ISL_1503273, EPI_ISL_1503274, EPI_ISL_1503275, EPI_ISL_1503276, EPI_ISL_1503277, EPI_ISL_1503279, EPI_ISL_1503280, EPI_ISL_1503281, EPI_ISL_1503282, EPI_ISL_1503283, EPI_ISL_1503284, EPI_ISL_1503285, EPI_ISL_1503286, EPI_ISL_1503287, EPI_ISL_1503288, EPI_ISL_1503289, EPI_ISL_1503290, EPI_ISL_1503291, EPI_ISL_1503292, EPI_ISL_1503293, EPI_ISL_1503294, EPI_ISL_1503295, EPI_ISL_1503296, EPI_ISL_1503297, EPI_ISL_1503298 | | | |
| see above | Wyoming Public Health Laboratory | Wyoming Public Health Laboratory | Jim Mildenberger, Wanda Manley, Noah Hull, Taylor Fearing, Lynette Gumbleton, Channing Weber, Ashley Norberg, Chayse Rowley, Marley Goetz, Brian Dominguez, Elliot Thomasson, Cari Sloma, and Rob Christensen |
| EPI_ISL_1503299, EPI_ISL_1503300, EPI_ISL_1503301, EPI_ISL_1503302, EPI_ISL_1503303, EPI_ISL_1503304, EPI_ISL_1503307, EPI_ISL_1503308 | Arizona State Public Health Laboratory | Arizona State Public Health Laboratory | Trung Huynh, Jessica Escobar, Katherine Fullerton, Nobuko Fukushima, Stacy White, Linda Getsinger, Victor Waddell |
| EPI_ISL_1503309, EPI_ISL_1503310, EPI_ISL_1503311, EPI_ISL_1503312, EPI_ISL_1503313, EPI_ISL_1503314, EPI_ISL_1503315, EPI_ISL_1503316, EPI_ISL_1503317, EPI_ISL_1503318, EPI_ISL_1503319, EPI_ISL_1503320, EPI_ISL_1503321, EPI_ISL_1503322, EPI_ISL_1503324, EPI_ISL_1503325, EPI_ISL_1503326, EPI_ISL_1503327, EPI_ISL_1503328, EPI_ISL_1503329, EPI_ISL_1503330, EPI_ISL_1503331, EPI_ISL_1503332, EPI_ISL_1503333, EPI_ISL_1503334, EPI_ISL_1503335, EPI_ISL_1503336, EPI_ISL_1503337, EPI_ISL_1503338, EPI_ISL_1503339, EPI_ISL_1503340, EPI_ISL_1503341, EPI_ISL_1503342, EPI_ISL_1503343, EPI_ISL_1503344, EPI_ISL_1503345, EPI_ISL_1503346, EPI_ISL_1503347, EPI_ISL_1503348, EPI_ISL_1503349, EPI_ISL_1503350, EPI_ISL_1503351, EPI_ISL_1503352, EPI_ISL_1503354, EPI_ISL_1503355, EPI_ISL_1503356, EPI_ISL_1503357, EPI_ISL_1503359, EPI_ISL_1503360, EPI_ISL_1503361, EPI_ISL_1503362, EPI_ISL_1503363, EPI_ISL_1503364, EPI_ISL_1503365, EPI_ISL_1503366, EPI_ISL_1503367, EPI_ISL_1503368, EPI_ISL_1503369, EPI_ISL_1503373, EPI_ISL_1503374, EPI_ISL_1503377, EPI_ISL_1503379, EPI_ISL_1503380, EPI_ISL_1503381, EPI_ISL_1503382, EPI_ISL_1503383, EPI_ISL_1503384, EPI_ISL_1503385, EPI_ISL_1503386, EPI_ISL_1503387, EPI_ISL_1503388, EPI_ISL_1503389, EPI_ISL_1503391, EPI_ISL_1503392, EPI_ISL_1503393, EPI_ISL_1503394 | | | |
| see above | Texas Department of State Health Services (TXDSHS) | Texas Department of State Health Services (TXDSHS) | Rashmi Tuladhar, Bonnie Oh, Jenny Zhang, Maliha Rahman, Mayela Pedrueza, Anita Pokharel, Lorraine Rodriguez, Myong Koag, Chun Wang, Rachel Lee, Grace Kubin |
| EPI_ISL_1503395, EPI_ISL_1503397, EPI_ISL_1503398, EPI_ISL_1503399, EPI_ISL_1503402, EPI_ISL_1503404, EPI_ISL_1503405, EPI_ISL_1503406, EPI_ISL_1503407, EPI_ISL_1503409, EPI_ISL_1503410, EPI_ISL_1503411, EPI_ISL_1503413, EPI_ISL_1503414, EPI_ISL_1503415, EPI_ISL_1503416, EPI_ISL_1503418, EPI_ISL_1503425, EPI_ISL_1503427, EPI_ISL_1503430, EPI_ISL_1503432, EPI_ISL_1503435, EPI_ISL_1503436, EPI_ISL_1503439, EPI_ISL_1503440, EPI_ISL_1503441, EPI_ISL_1503444, EPI_ISL_1503445, EPI_ISL_1503447, EPI_ISL_1503450, EPI_ISL_1503451, EPI_ISL_1503452, EPI_ISL_1503453 | | | |
| see above | Seattle Flu Study | Seattle Flu Study | Deborah A. Nickerson, Chris D. Frazar, Jover Lee, Benjamin Pelle, Erica Ryke, Matthew Richardson, Amanda Adler, Elisabeth Brandstetter, Peter D. Han, Kairsten Fay, Misja Ilcisin, Kirsten Lacombe, Thomas R. Sibley, Melissa Truong, Caitlin R. Wolf, Michael Boeckh, Janet A. Englund, Michael Famulare, Barry R. Lutz, Mark J. Rieder, Lea M. Starita, Matthew Thompson, Jay Shendure, Trevor Bedford, Helen Y. Chu |
| EPI_ISL_1503461, EPI_ISL_1503462, EPI_ISL_1503463, EPI_ISL_1503464, EPI_ISL_1503466, EPI_ISL_1503467, EPI_ISL_1503470, EPI_ISL_1503471, EPI_ISL_1503472, EPI_ISL_1503473, EPI_ISL_1503474, EPI_ISL_1503475, EPI_ISL_1503476, EPI_ISL_1503477, EPI_ISL_1503478, EPI_ISL_1503479, EPI_ISL_1503480, EPI_ISL_1503481, EPI_ISL_1503482, EPI_ISL_1503483, EPI_ISL_1503484, EPI_ISL_1503485, EPI_ISL_1503486, EPI_ISL_1503487, EPI_ISL_1503488, EPI_ISL_1503489, EPI_ISL_1503490, EPI_ISL_1503491, EPI_ISL_1503492, EPI_ISL_1503493, EPI_ISL_1503494, EPI_ISL_1503495, EPI_ISL_1503496, EPI_ISL_1503497, EPI_ISL_1503498, EPI_ISL_1503500, EPI_ISL_1503501, EPI_ISL_1503502, EPI_ISL_1503503, EPI_ISL_1503504, EPI_ISL_1503505, EPI_ISL_1503506, EPI_ISL_1503507, EPI_ISL_1503509, EPI_ISL_1503510, EPI_ISL_1503511, EPI_ISL_1503512, EPI_ISL_1503513, EPI_ISL_1503514, EPI_ISL_1503515, EPI_ISL_1503517, EPI_ISL_1503518, EPI_ISL_1503521, EPI_ISL_1503522, EPI_ISL_1503523, EPI_ISL_1503524, EPI_ISL_1503525, EPI_ISL_1503527, EPI_ISL_1503528, EPI_ISL_1503529, EPI_ISL_1503530, EPI_ISL_1503531, EPI_ISL_1503533, EPI_ISL_1503534, EPI_ISL_1503535, EPI_ISL_1503538, EPI_ISL_1503541, EPI_ISL_1503542, EPI_ISL_1503547, EPI_ISL_1503548, EPI_ISL_1503549, EPI_ISL_1503551, EPI_ISL_1503552, EPI_ISL_1503554, EPI_ISL_1503555, EPI_ISL_1503557, EPI_ISL_1503559, EPI_ISL_1503560, EPI_ISL_1503561, EPI_ISL_1503562, EPI_ISL_1503563, EPI_ISL_1503564, EPI_ISL_1503565, EPI_ISL_1503566, EPI_ISL_1503567, EPI_ISL_1503570, EPI_ISL_1503579, EPI_ISL_1503580, EPI_ISL_1503581, EPI_ISL_1503582, EPI_ISL_1503583, EPI_ISL_1503584, EPI_ISL_1503585, EPI_ISL_1503586, EPI_ISL_1503587, EPI_ISL_1503588, EPI_ISL_1503590, EPI_ISL_1503592, EPI_ISL_1503593, EPI_ISL_1503594, EPI_ISL_1503595, EPI_ISL_1503596, EPI_ISL_1503597, EPI_ISL_1503598, EPI_ISL_1503599, EPI_ISL_1503602, EPI_ISL_1503603, EPI_ISL_1503604, EPI_ISL_1503605, EPI_ISL_1503606, EPI_ISL_1503607, EPI_ISL_1503608, EPI_ISL_1503609, EPI_ISL_1503610, EPI_ISL_1503611, EPI_ISL_1503612, EPI_ISL_1503613, EPI_ISL_1503614, EPI_ISL_1503615, EPI_ISL_1503616, EPI_ISL_1503617, EPI_ISL_1503618, EPI_ISL_1503619, EPI_ISL_1503620, EPI_ISL_1503621, EPI_ISL_1503623, EPI_ISL_1503624, EPI_ISL_1503625, EPI_ISL_1503626, EPI_ISL_1503627, EPI_ISL_1503628, EPI_ISL_1503629, EPI_ISL_1503630, EPI_ISL_1503631, EPI_ISL_1503632, EPI_ISL_1503633, EPI_ISL_1503635, EPI_ISL_1503636, EPI_ISL_1503637, EPI_ISL_1503638, EPI_ISL_1503640, EPI_ISL_1503641, EPI_ISL_1503642, EPI_ISL_1503643, EPI_ISL_1503644, EPI_ISL_1503645, EPI_ISL_1503646, EPI_ISL_1503647, EPI_ISL_1503648, EPI_ISL_1503649, EPI_ISL_1503652, EPI_ISL_1503653, EPI_ISL_1503654, EPI_ISL_1503655, EPI_ISL_1503656, EPI_ISL_1503657, EPI_ISL_1503658, EPI_ISL_1503660, EPI_ISL_1503661, EPI_ISL_1503662, EPI_ISL_1503664, EPI_ISL_1503665, EPI_ISL_1503666, EPI_ISL_1503667, EPI_ISL_1503668, EPI_ISL_1503669, EPI_ISL_1503670, EPI_ISL_1503671, EPI_ISL_1503672, EPI_ISL_1503673, EPI_ISL_1503674, EPI_ISL_1503675, EPI_ISL_1503677, EPI_ISL_1503678, EPI_ISL_1503679, EPI_ISL_1503680, EPI_ISL_1503681, EPI_ISL_1503682, EPI_ISL_1503683, EPI_ISL_1503684, EPI_ISL_1503685, EPI_ISL_1503686, EPI_ISL_1503687, EPI_ISL_1503689, EPI_ISL_1503691, EPI_ISL_1503692, EPI_ISL_1503693, EPI_ISL_1503694, EPI_ISL_1503695, EPI_ISL_1503696, EPI_ISL_1503697, EPI_ISL_1503698, EPI_ISL_1503699, EPI_ISL_1503700, EPI_ISL_1503701, EPI_ISL_1503702, EPI_ISL_1503703, EPI_ISL_1503704, EPI_ISL_1503705, EPI_ISL_1503706, EPI_ISL_1503707, EPI_ISL_1503708, EPI_ISL_1503709, EPI_ISL_1503711, EPI_ISL_1503712, EPI_ISL_1503713, EPI_ISL_1503714, EPI_ISL_1503715, EPI_ISL_1503718, EPI_ISL_1503720, EPI_ISL_1503721, EPI_ISL_1503722, EPI_ISL_1503723, EPI_ISL_1503724, EPI_ISL_1503726, EPI_ISL_1503727, EPI_ISL_1503728, EPI_ISL_1503729, EPI_ISL_1503731, EPI_ISL_1503732, EPI_ISL_1503733, EPI_ISL_1503734, EPI_ISL_1503735, EPI_ISL_1503736, EPI_ISL_1503738, EPI_ISL_1503739, EPI_ISL_1503740, EPI_ISL_1503741, EPI_ISL_1503742, EPI_ISL_1503743, EPI_ISL_1503744, EPI_ISL_1503745, EPI_ISL_1503746, EPI_ISL_1503747, EPI_ISL_1503748, EPI_ISL_1503749, EPI_ISL_1503750, EPI_ISL_1503752, EPI_ISL_1503753, EPI_ISL_1503755, EPI_ISL_1503756, EPI_ISL_1503757, EPI_ISL_1503758, EPI_ISL_1503759, EPI_ISL_1503760, EPI_ISL_1503761, EPI_ISL_1503762, EPI_ISL_1503763, EPI_ISL_1503764, EPI_ISL_1503765, EPI_ISL_1503766, EPI_ISL_1503767, EPI_ISL_1503768, EPI_ISL_1503769, EPI_ISL_1503770, EPI_ISL_1503771, EPI_ISL_1503772, EPI_ISL_1503773, EPI_ISL_1503774, EPI_ISL_1503775, EPI_ISL_1503776, EPI_ISL_1503778, EPI_ISL_1503779, EPI_ISL_1503780, EPI_ISL_1503781, EPI_ISL_1503782, EPI_ISL_1503783, EPI_ISL_1503784, EPI_ISL_1503785, EPI_ISL_1503786, EPI_ISL_1503787, EPI_ISL_1503788, EPI_ISL_1503789, EPI_ISL_1503790, EPI_ISL_1503791 | | | |
| see above | Massachusetts State Public Health Laboratory | Massachusetts State Public Health Laboratory | Andrew Lang, Timelia Fink, Glen Gallagher, Sandra Smole |
| EPI_ISL_1503793, EPI_ISL_1503794, EPI_ISL_1503795, EPI_ISL_1503796, EPI_ISL_1503797, EPI_ISL_1503798, EPI_ISL_1503799, EPI_ISL_1503800, EPI_ISL_1503801 | Nebraska Public Health Laboratory | NPHL COVID-19 Response Team | NPHL COVID-19 Response Team |
| EPI_ISL_1503921, EPI_ISL_1503923, EPI_ISL_1503924, EPI_ISL_1503925, EPI_ISL_1503926, EPI_ISL_1503927, EPI_ISL_1503928, EPI_ISL_1503929, EPI_ISL_1503930, EPI_ISL_1503931, EPI_ISL_1503932, EPI_ISL_1503933, EPI_ISL_1503934, EPI_ISL_1503935, EPI_ISL_1503936, EPI_ISL_1503937, EPI_ISL_1503939, EPI_ISL_1503940, EPI_ISL_1503941, EPI_ISL_1503942, EPI_ISL_1503943, EPI_ISL_1503944, EPI_ISL_1503945, EPI_ISL_1503946, EPI_ISL_1503947, EPI_ISL_1503948, EPI_ISL_1503949, EPI_ISL_1503950, EPI_ISL_1503951, EPI_ISL_1503952, EPI_ISL_1503953, EPI_ISL_1503954 | | | |

| | | | |
|---|---|---|---|
| see above | Grady Memorial Hospital, Clinical Microbiology Laboratory | Piantadosi Lab, Emory Department of Pathology | Yun F (Wayne) Wang, Anne Piantadosi |
| EPI_ISL_1503955, EPI_ISL_1503956, EPI_ISL_1503957, EPI_ISL_1503958, EPI_ISL_1503959, EPI_ISL_1503960, EPI_ISL_1503961, EPI_ISL_1503962, EPI_ISL_1503963, EPI_ISL_1503964, EPI_ISL_1503965, EPI_ISL_1503966, EPI_ISL_1503967, EPI_ISL_1503968, EPI_ISL_1503969, EPI_ISL_1503970, EPI_ISL_1503972, EPI_ISL_1503973, EPI_ISL_1503974, EPI_ISL_1503975, EPI_ISL_1503976, EPI_ISL_1503977, EPI_ISL_1503978 | | | |
| see above | Emory Molecular Diagnostics Laboratory, Emory Healthcare | Piantadosi Lab, Emory Department of Pathology | Ahmed Babiker, Anne Piantadosi |
| EPI_ISL_1503989, EPI_ISL_1503990 | USC Clinical Lab | Los Angeles County Public Health Laboratories | P. Hemarajata et al. |
| EPI_ISL_1503991 | Curative Labs | Los Angeles County Public Health Laboratories | P. Hemarajata et al. |
| EPI_ISL_1503992 | Los Angeles County Public Health Laboratories | Los Angeles County Public Health Laboratories | P. Hemarajata et al. |
| EPI_ISL_1503994 | USC Clinical Lab | Los Angeles County Public Health Laboratories | P. Hemarajata et al. |
| EPI_ISL_1503995 | Los Angeles County Public Health Laboratories | Los Angeles County Public Health Laboratories | P. Hemarajata et al. |
| EPI_ISL_1503996, EPI_ISL_1503997 | USC Clinical Lab | Los Angeles County Public Health Laboratories | P. Hemarajata et al. |
| EPI_ISL_1503999, EPI_ISL_1504000, EPI_ISL_1504001, EPI_ISL_1504002, EPI_ISL_1504003, EPI_ISL_1504004, EPI_ISL_1504005, EPI_ISL_1504006, EPI_ISL_1504007, EPI_ISL_1504008, EPI_ISL_1504009, EPI_ISL_1504011, EPI_ISL_1504012, EPI_ISL_1504013, EPI_ISL_1504014, EPI_ISL_1504015, EPI_ISL_1504016, EPI_ISL_1504017, EPI_ISL_1504018, EPI_ISL_1504019, EPI_ISL_1504020, EPI_ISL_1504021 | | | |
| see above | Wyoming Public Health Laboratory | Wyoming Public Health Laboratory | Jim Mildenberger, Wanda Manley, Noah Hull, Taylor Fearing, Lynette Gumbleton, Channing Weber, Ashley Norberg, Chayse Rowley, Marley Goetz, Brian Dominguez, Elliot Thomasson, Cari Sloma, and Rob Christensen |
| EPI_ISL_1504025 | USC Clinical Lab | Los Angeles County Public Health Laboratories | P. Hemarajata et al. |
| EPI_ISL_1504026 | VA Medical Center | Los Angeles County Public Health Laboratories | P. Hemarajata et al. |
| EPI_ISL_1504027 | Los Angeles County Public Health Laboratories | Los Angeles County Public Health Laboratories | P. Hemarajata et al. |
| EPI_ISL_1504029, EPI_ISL_1504030, EPI_ISL_1504032, EPI_ISL_1504033, EPI_ISL_1504034, EPI_ISL_1504035, EPI_ISL_1504036, EPI_ISL_1504037, EPI_ISL_1504038, EPI_ISL_1504039, EPI_ISL_1504040, EPI_ISL_1504041 | | | |
| see above | USC Clinical Lab | Los Angeles County Public Health Laboratories | P. Hemarajata et al. |
| EPI_ISL_1504043, EPI_ISL_1504044, EPI_ISL_1504045, EPI_ISL_1504046, EPI_ISL_1504047, EPI_ISL_1504048, EPI_ISL_1504050, EPI_ISL_1504052, EPI_ISL_1504053, EPI_ISL_1504054, EPI_ISL_1504055, EPI_ISL_1504056 | | | |
| see above | Oregon State University TRACE Project | OSU Center for Genome Research and Biocomputing | Oregon State University TRACE Project |
| EPI_ISL_1504058, EPI_ISL_1504059, EPI_ISL_1504060, EPI_ISL_1504061, EPI_ISL_1504062, EPI_ISL_1504063, EPI_ISL_1504064, EPI_ISL_1504065, EPI_ISL_1504066, EPI_ISL_1504067 | LESP Nuevo Leon | Instituto de Diagnostico y Referencia Epidemiologicos (INDRE) | Claudia Wong-Arambula, Abril Rodriguez-Maldonado, Vanessa Rivero-Arredondo, Ariadna Medina-Benitez, Joaquin Quiroz-Mercado, Sergio Rangel-Guerrero, Natividad Cruz-Ortiz, Tatiana Nunez-Garcia, Gisela Barrera-Badillo, Lucia Hernandez-Rivas, Irma Lopez-Martinez, Ernesto Ramirez-Gonzalez. |
| EPI_ISL_1504068, EPI_ISL_1504069, EPI_ISL_1504070, EPI_ISL_1504071, EPI_ISL_1504072, EPI_ISL_1504073, EPI_ISL_1504074, EPI_ISL_1504075, EPI_ISL_1504076, EPI_ISL_1504077, EPI_ISL_1504078, EPI_ISL_1504079, EPI_ISL_1504080, EPI_ISL_1504081, EPI_ISL_1504082 | | | |
| see above | LESP Morelos | Instituto de Diagnostico y Referencia Epidemiologicos (INDRE) | Claudia Wong-Arambula, Abril Rodriguez-Maldonado, Vanessa Rivero-Arredondo, Ariadna Medina-Benitez, Joaquin Quiroz-Mercado, Sergio Rangel-Guerrero, Natividad Cruz-Ortiz, Tatiana Nunez-Garcia, Gisela Barrera-Badillo, Lucia Hernandez-Rivas, Irma Lopez-Martinez, Ernesto Ramirez-Gonzalez. |
| EPI_ISL_1504083 | LESP Ciudad de Mexico | Instituto de Diagnostico y Referencia Epidemiologicos (INDRE) | Claudia Wong-Arambula, Abril Rodriguez-Maldonado, Vanessa Rivero-Arredondo, Ariadna Medina-Benitez, Joaquin Quiroz-Mercado, Sergio Rangel-Guerrero, Natividad Cruz-Ortiz, Tatiana Nunez-Garcia, Gisela Barrera-Badillo, Lucia Hernandez-Rivas, Irma Lopez-Martinez, Ernesto Ramirez-Gonzalez. |
| EPI_ISL_1504084, EPI_ISL_1504085, EPI_ISL_1504086, EPI_ISL_1504087, EPI_ISL_1504088 | LESP Chihuahua | Instituto de Diagnostico y Referencia Epidemiologicos (INDRE) | Claudia Wong-Arambula, Abril Rodriguez-Maldonado, Vanessa Rivero-Arredondo, Ariadna Medina-Benitez, Joaquin Quiroz-Mercado, Sergio Rangel-Guerrero, Natividad Cruz-Ortiz, Tatiana Nunez-Garcia, Gisela Barrera-Badillo, Lucia Hernandez-Rivas, Irma Lopez-Martinez, Ernesto Ramirez-Gonzalez. |
| EPI_ISL_1504089, EPI_ISL_1504090, EPI_ISL_1504091, EPI_ISL_1504092, EPI_ISL_1504093, EPI_ISL_1504094, EPI_ISL_1504095, EPI_ISL_1504096, EPI_ISL_1504097, EPI_ISL_1504098, EPI_ISL_1504099, EPI_ISL_1504100, EPI_ISL_1504101, EPI_ISL_1504102, EPI_ISL_1504103 | | | |
| see above | LESP Queretaro | Instituto de Diagnostico y Referencia Epidemiologicos (INDRE) | Claudia Wong-Arambula, Abril Rodriguez-Maldonado, Vanessa Rivero-Arredondo, Ariadna Medina-Benitez, Joaquin Quiroz-Mercado, Sergio Rangel-Guerrero, Natividad Cruz-Ortiz, Tatiana Nunez-Garcia, Gisela Barrera-Badillo, Lucia Hernandez-Rivas, Irma Lopez-Martinez, Ernesto Ramirez-Gonzalez. |
| EPI_ISL_1504104 | LESP Guerrero | Instituto de Diagnostico y Referencia Epidemiologicos (INDRE) | Claudia Wong-Arambula, Abril Rodriguez-Maldonado, Vanessa Rivero-Arredondo, Ariadna Medina-Benitez, Joaquin Quiroz-Mercado, Sergio Rangel-Guerrero, Natividad Cruz-Ortiz, Tatiana Nunez-Garcia, Gisela Barrera-Badillo, Lucia Hernandez-Rivas, Irma Lopez-Martinez, Ernesto Ramirez-Gonzalez. |
| EPI_ISL_1504105 | LESP Baja California Sur | Instituto de Diagnostico y Referencia Epidemiologicos (INDRE) | Claudia Wong-Arambula, Abril Rodriguez-Maldonado, Vanessa Rivero-Arredondo, Ariadna Medina-Benitez, Joaquin Quiroz-Mercado, Sergio Rangel-Guerrero, Natividad Cruz-Ortiz, Tatiana Nunez-Garcia, Gisela Barrera-Badillo, Lucia Hernandez-Rivas, Irma Lopez-Martinez, Ernesto Ramirez-Gonzalez. |
| EPI_ISL_1504106, EPI_ISL_1504107 | LESPZacatecas | Instituto de Diagnostico y Referencia Epidemiologicos (INDRE) | Claudia Wong-Arambula, Abril Rodriguez-Maldonado, Vanessa Rivero-Arredondo, Ariadna Medina-Benitez, Joaquin Quiroz-Mercado, Sergio Rangel-Guerrero, Natividad Cruz-Ortiz, Tatiana Nunez-Garcia, Gisela Barrera-Badillo, Lucia Hernandez-Rivas, Irma Lopez-Martinez, Ernesto Ramirez-Gonzalez. |
| EPI_ISL_1504108 | LESP Baja California | Instituto de Diagnostico y Referencia Epidemiologicos (INDRE) | Claudia Wong-Arambula, Abril Rodriguez-Maldonado, Vanessa Rivero-Arredondo, Ariadna Medina-Benitez, Joaquin Quiroz-Mercado, Sergio Rangel-Guerrero, Natividad Cruz-Ortiz, Tatiana Nunez-Garcia, Gisela Barrera-Badillo, Lucia Hernandez-Rivas, Irma Lopez-Martinez, Ernesto Ramirez-Gonzalez. |
| EPI_ISL_1504109 | LESP Jalisco | Instituto de Diagnostico y Referencia Epidemiologicos (INDRE) | Claudia Wong-Arambula, Abril Rodriguez-Maldonado, Vanessa Rivero-Arredondo, Ariadna Medina-Benitez, Joaquin Quiroz-Mercado, Sergio Rangel-Guerrero, Natividad Cruz-Ortiz, Tatiana Nunez-Garcia, Gisela Barrera-Badillo, Lucia Hernandez-Rivas, Irma Lopez-Martinez, Ernesto Ramirez-Gonzalez. |
| EPI_ISL_1504110, EPI_ISL_1504111 | LESP Queretaro | Instituto de Diagnostico y Referencia Epidemiologicos (INDRE) | Claudia Wong-Arambula, Abril Rodriguez-Maldonado, Vanessa Rivero-Arredondo, Ariadna Medina-Benitez, Joaquin Quiroz-Mercado, Sergio Rangel-Guerrero, Natividad Cruz-Ortiz, Tatiana Nunez-Garcia, Gisela Barrera-Badillo, Lucia Hernandez-Rivas, Irma Lopez-Martinez, Ernesto Ramirez-Gonzalez. |
| EPI_ISL_1506114 | UCLA Clinical Micro Lab | Los Angeles County Public Health Laboratories | P. Hemarajata et al. |
| EPI_ISL_1506115 | Pomona Valley Hospital Medical Center | Los Angeles County Public Health Laboratories | P. Hemarajata et al. |
| EPI_ISL_1506116 | Huntington Memorial Hospital | Los Angeles County Public Health Laboratories | P. Hemarajata et al. |
| EPI_ISL_1506117, EPI_ISL_1506118, EPI_ISL_1506119 | MD Laboratories | Los Angeles County Public Health Laboratories | P. Hemarajata et al. |
| EPI_ISL_1506120, EPI_ISL_1506121, EPI_ISL_1506122, EPI_ISL_1506123, EPI_ISL_1506124, EPI_ISL_1506125, EPI_ISL_1506126 | USC Clinical Lab | Los Angeles County Public Health Laboratories | P. Hemarajata et al. |
| EPI_ISL_1506127 | VA Medical Center | Los Angeles County Public Health Laboratories | P. Hemarajata et al. |
| EPI_ISL_1508853, EPI_ISL_1508854, EPI_ISL_1508855, EPI_ISL_1508856, EPI_ISL_1508857, EPI_ISL_1508858, EPI_ISL_1508859, EPI_ISL_1508860, EPI_ISL_1508861, EPI_ISL_1508862, EPI_ISL_1508863, EPI_ISL_1508864, EPI_ISL_1508865, EPI_ISL_1508866, EPI_ISL_1508867, EPI_ISL_1508868, EPI_ISL_1508869, EPI_ISL_1508870, EPI_ISL_1508871, EPI_ISL_1508872, EPI_ISL_1508873, EPI_ISL_1508874, EPI_ISL_1508875, EPI_ISL_1508876, EPI_ISL_1508877, EPI_ISL_1508878, EPI_ISL_1508879, EPI_ISL_1508880, EPI_ISL_1508881, EPI_ISL_1508882, EPI_ISL_1508883, EPI_ISL_1508884, EPI_ISL_1508885, EPI_ISL_1508886, EPI_ISL_1508887, EPI_ISL_1508888, EPI_ISL_1508889 | | | |
| see above | University of Oregon COVID-19 MAP Laboratory | University of Oregon Genomics and Cell Characterization Core Facility (GC3F) | Douglas Turnbull, Ariana White, Jeff Bishop, Jason Carriere |
| EPI_ISL_1508898, EPI_ISL_1508899, EPI_ISL_1508900 | Pomona Valley Hospital Medical Center | Los Angeles County Public Health Laboratories | P. Hemarajata et al. |

| | | | |
|---|---|---|---|
| EPI_ISL_1508903, EPI_ISL_1508904, EPI_ISL_1508905, EPI_ISL_1508906, EPI_ISL_1508908, EPI_ISL_1508911 | USC Clinical Lab | Los Angeles County Public Health Laboratories | P. Hemarajata et al. |
| EPI_ISL_1508917 | Kaiser Permanente West Los Angeles Medical Center | Los Angeles County Public Health Laboratories | P. Hemarajata et al. |
| EPI_ISL_1508918 | Kaiser Permanente Downey Medical Center | Los Angeles County Public Health Laboratories | P. Hemarajata et al. |
| EPI_ISL_1508919, EPI_ISL_1508920, EPI_ISL_1508921, EPI_ISL_1508922, EPI_ISL_1508923, EPI_ISL_1508924, EPI_ISL_1508925, EPI_ISL_1508926, EPI_ISL_1508927, EPI_ISL_1508928, EPI_ISL_1508929, EPI_ISL_1508930, EPI_ISL_1508931, EPI_ISL_1508932, EPI_ISL_1508934, EPI_ISL_1508935, EPI_ISL_1508936, EPI_ISL_1508937, EPI_ISL_1508938, EPI_ISL_1508939 | | | |
| see above | USC Clinical Lab | Los Angeles County Public Health Laboratories | P. Hemarajata et al. |
| EPI_ISL_1508963 | Stanford Clinical Virology Laboratory | Santa Clara County Public Health Laboratory | Santa Clara County Public Health Department |
| EPI_ISL_1508964 | Santa Clara County Public Health Laboratory | Santa Clara County Public Health Laboratory | Santa Clara County Public Health Department |
| EPI_ISL_1508967, EPI_ISL_1508968, EPI_ISL_1508969, EPI_ISL_1508971, EPI_ISL_1508972, EPI_ISL_1508973, EPI_ISL_1508974, EPI_ISL_1508975, EPI_ISL_1508976, EPI_ISL_1508977 | Kaiser Santa Clara | Santa Clara County Public Health Laboratory | Santa Clara County Public Health Department |
| EPI_ISL_1508978, EPI_ISL_1508981, EPI_ISL_1508982, EPI_ISL_1508983 | Good Samaritan Hospital | Santa Clara County Public Health Laboratory | Santa Clara County Public Health Department |
| EPI_ISL_1508984, EPI_ISL_1508985, EPI_ISL_1508986 | Kaiser Regional | Santa Clara County Public Health Laboratory | Santa Clara County Public Health Department |
| EPI_ISL_1508987 | Good Samaritan Hospital | Santa Clara County Public Health Laboratory | Santa Clara County Public Health Department |
| EPI_ISL_1509003, EPI_ISL_1509004, EPI_ISL_1509005, EPI_ISL_1509006, EPI_ISL_1509007, EPI_ISL_1509008, EPI_ISL_1509009, EPI_ISL_1509010, EPI_ISL_1509011, EPI_ISL_1509012, EPI_ISL_1509013, EPI_ISL_1509014, EPI_ISL_1509015, EPI_ISL_1509016, EPI_ISL_1509017, EPI_ISL_1509018, EPI_ISL_1509019, EPI_ISL_1509020, EPI_ISL_1509021, EPI_ISL_1509022, EPI_ISL_1509023, EPI_ISL_1509024, EPI_ISL_1509025, EPI_ISL_1509026, EPI_ISL_1509027, EPI_ISL_1509028, EPI_ISL_1509029, EPI_ISL_1509030, EPI_ISL_1509031, EPI_ISL_1509032, EPI_ISL_1509033, EPI_ISL_1509034, EPI_ISL_1509035, EPI_ISL_1509036, EPI_ISL_1509037, EPI_ISL_1509038, EPI_ISL_1509039, EPI_ISL_1509040, EPI_ISL_1509041, EPI_ISL_1509042, EPI_ISL_1509043, EPI_ISL_1509044, EPI_ISL_1509045, EPI_ISL_1509046, EPI_ISL_1509047, EPI_ISL_1509048, EPI_ISL_1509049, EPI_ISL_1509050, EPI_ISL_1509051, EPI_ISL_1509052, EPI_ISL_1509053, EPI_ISL_1509054, EPI_ISL_1509055, EPI_ISL_1509056, EPI_ISL_1509057, EPI_ISL_1509058, EPI_ISL_1509059, EPI_ISL_1509060, EPI_ISL_1509061, EPI_ISL_1509062, EPI_ISL_1509063, EPI_ISL_1509064, EPI_ISL_1509065, EPI_ISL_1509066, EPI_ISL_1509067, EPI_ISL_1509068, EPI_ISL_1509069, EPI_ISL_1509070, EPI_ISL_1509071, EPI_ISL_1509072, EPI_ISL_1509073, EPI_ISL_1509074, EPI_ISL_1509075, EPI_ISL_1509076, EPI_ISL_1509077, EPI_ISL_1509078, EPI_ISL_1509079, EPI_ISL_1509080, EPI_ISL_1509081, EPI_ISL_1509082, EPI_ISL_1509083, EPI_ISL_1509084, EPI_ISL_1509085, EPI_ISL_1509086, EPI_ISL_1509087, EPI_ISL_1509088, EPI_ISL_1509089, EPI_ISL_1509090, EPI_ISL_1509091, EPI_ISL_1509092, EPI_ISL_1509093, EPI_ISL_1509094 | | | |
| see above | Department of Laboratory Medicine, Clinical Center, National Institutes of Health | Laboratory of Parasitic Diseases, Systems Genomics Section, National Institute of Allergy and Infectious Diseases, National Institutes of Health | Allison Roder, Stephanie Banakis, Matthew Chung, Jung-ho Youn, Rachel Mercado, Wei Wang, Tara Palmore, Michael Bell, Heike Bailin, Jessica McCormick-Ell, Sanchita Das, Jennifer Kwan, Elodie Ghedin |
| EPI_ISL_1509104, EPI_ISL_1509105, EPI_ISL_1509106, EPI_ISL_1509107, EPI_ISL_1509108, EPI_ISL_1509109, EPI_ISL_1509110, EPI_ISL_1509112, EPI_ISL_1509113, EPI_ISL_1509114, EPI_ISL_1509115, EPI_ISL_1509116, EPI_ISL_1509117, EPI_ISL_1509118, EPI_ISL_1509119, EPI_ISL_1509120, EPI_ISL_1509121, EPI_ISL_1509122, EPI_ISL_1509123, EPI_ISL_1509124, EPI_ISL_1509126, EPI_ISL_1509128, EPI_ISL_1509131, EPI_ISL_1509132, EPI_ISL_1509133, EPI_ISL_1509135, EPI_ISL_1509136, EPI_ISL_1509137, EPI_ISL_1509140, EPI_ISL_1509141, EPI_ISL_1509142, EPI_ISL_1509143, EPI_ISL_1509144, EPI_ISL_1509145, EPI_ISL_1509146, EPI_ISL_1509147, EPI_ISL_1509148, EPI_ISL_1509149, EPI_ISL_1509150, EPI_ISL_1509151, EPI_ISL_1509152, EPI_ISL_1509153, EPI_ISL_1509154, EPI_ISL_1509156, EPI_ISL_1509157, EPI_ISL_1509158, EPI_ISL_1509163, EPI_ISL_1509164, EPI_ISL_1509165, EPI_ISL_1509166, EPI_ISL_1509167, EPI_ISL_1509168, EPI_ISL_1509169, EPI_ISL_1509170, EPI_ISL_1509171, EPI_ISL_1509172, EPI_ISL_1509173, EPI_ISL_1509174, EPI_ISL_1509175, EPI_ISL_1509176, EPI_ISL_1509177, EPI_ISL_1509178, EPI_ISL_1509179, EPI_ISL_1509180, EPI_ISL_1509181, EPI_ISL_1509182, EPI_ISL_1509183, EPI_ISL_1509184, EPI_ISL_1509185, EPI_ISL_1509186, EPI_ISL_1509187, EPI_ISL_1509188, EPI_ISL_1509189, EPI_ISL_1509191, EPI_ISL_1509192, EPI_ISL_1509193, EPI_ISL_1509194, EPI_ISL_1509196, EPI_ISL_1509197, EPI_ISL_1509198, EPI_ISL_1509199, EPI_ISL_1509200, EPI_ISL_1509201, EPI_ISL_1509202, EPI_ISL_1509203, EPI_ISL_1509204, EPI_ISL_1509205, EPI_ISL_1509206, EPI_ISL_1509207, EPI_ISL_1509208, EPI_ISL_1509209, EPI_ISL_1509210, EPI_ISL_1509211, EPI_ISL_1509213, EPI_ISL_1509214, EPI_ISL_1509215, EPI_ISL_1509216, EPI_ISL_1509217, EPI_ISL_1509218, EPI_ISL_1509219, EPI_ISL_1509220, EPI_ISL_1509222, EPI_ISL_1509223, EPI_ISL_1509224, EPI_ISL_1509225, EPI_ISL_1509226, EPI_ISL_1509227, EPI_ISL_1509228, EPI_ISL_1509229, EPI_ISL_1509230, EPI_ISL_1509231, EPI_ISL_1509232, EPI_ISL_1509234, EPI_ISL_1509235, EPI_ISL_1509236, EPI_ISL_1509237, EPI_ISL_1509238, EPI_ISL_1509239, EPI_ISL_1509240, EPI_ISL_1509241, EPI_ISL_1509242, EPI_ISL_1509243, EPI_ISL_1509245, EPI_ISL_1509246, EPI_ISL_1509247, EPI_ISL_1509248, EPI_ISL_1509249, EPI_ISL_1509250, EPI_ISL_1509251, EPI_ISL_1509252, EPI_ISL_1509253, EPI_ISL_1509254, EPI_ISL_1509255, EPI_ISL_1509258, EPI_ISL_1509259, EPI_ISL_1509260 | | | |
| see above | Yale Clinical Virology Lab | Grubaugh Lab - Yale School of Public Health | Joseph Fauver, Mallery Breban, Isabell Ott, Tara Alpert, Mary Petrone, Anderson Brito, Chantal Vogels, Annie Watkins, Chaney Kalinich, Jessica Rothman, Marie L. Landry, Nathan Grubaugh |
| EPI_ISL_1509263, EPI_ISL_1509264, EPI_ISL_1509265, EPI_ISL_1509266, EPI_ISL_1509267, EPI_ISL_1509268, EPI_ISL_1509269 | Murphy Medical Association | Grubaugh Lab - Yale School of Public Health | Joseph Fauver, Mallery Breban, Isabel Ott, Tara Alpert, Mary Petrone, Anderson Brito, Chantal Vogels, Annie Watkins, Chaney Kalinich, Jessica Rothman, Caleb Neal, Eva Laszlo, Steven Murphy, Nathan Grubaugh |
| EPI_ISL_1509271 | Grubaugh Lab - Yale School of Public Health | Grubaugh Lab - Yale School of Public Health | Joseph Fauver, Mallery Breban, Isabell Ott, Tara Alpert, Mary Petrone, Anderson Brito, Chantal Vogels, Annie Watkins, Chaney Kalinich, Jessica Rothman, Nathan Grubaugh |
| EPI_ISL_1509274, EPI_ISL_1509275, EPI_ISL_1509276, EPI_ISL_1509277, EPI_ISL_1509278, EPI_ISL_1509279, EPI_ISL_1509280, EPI_ISL_1509281, EPI_ISL_1509282, EPI_ISL_1509283, EPI_ISL_1509284, EPI_ISL_1509285, EPI_ISL_1509286, EPI_ISL_1509287, EPI_ISL_1509288, EPI_ISL_1509289, EPI_ISL_1509290, EPI_ISL_1509291, EPI_ISL_1509292, EPI_ISL_1509293, EPI_ISL_1509294, EPI_ISL_1509295, EPI_ISL_1509297 | | | |
| see above | USC Clinical Lab | Los Angeles County Public Health Laboratories | P. Hemarajata et al. |
| EPI_ISL_1509925 | IL Department of Public Health Chicago Laboratory | Genomics and Discovery, Respiratory Viruses Branch, Division of Viral Diseases, Centers for Disease Control and Prevention | Yan Li, Ying Tao, Jing Zhang, Brian Lynch, Anna Kelleher, Anna Montmayeur, Krista Queen, Anna Uehara, Peter Cook, Rachel Marine, Mark Burroughs, Han Jia Justin Ng, Clinton R. Paden, Haibin Wang, Justin Lee, Adam Retchless, Suxiang Tong |
| EPI_ISL_1509926 | MT Public Health Laboratory | Genomics and Discovery, Respiratory Viruses Branch, Division of Viral Diseases, Centers for Disease Control and Prevention | Yan Li, Ying Tao, Jing Zhang, Brian Lynch, Anna Kelleher, Anna Montmayeur, Krista Queen, Anna Uehara, Peter Cook, Rachel Marine, Mark Burroughs, Han Jia Justin Ng, Clinton R. Paden, Haibin Wang, Justin Lee, Adam Retchless, Suxiang Tong |
| EPI_ISL_1509927 | WVDHHR - Office of Laboratory Services | Genomics and Discovery, Respiratory Viruses Branch, Division of Viral Diseases, Centers for Disease Control and Prevention | Yan Li, Ying Tao, Jing Zhang, Brian Lynch, Anna Kelleher, Anna Montmayeur, Krista Queen, Anna Uehara, Peter Cook, Rachel Marine, Mark Burroughs, Han Jia Justin Ng, Clinton R. Paden, Haibin Wang, Justin Lee, Adam Retchless, Suxiang Tong |
| EPI_ISL_1509928, EPI_ISL_1509929 | "AR Dept. of Health-PHL, Molecular Diagnostics" | Genomics and Discovery, Respiratory Viruses Branch, Division of Viral Diseases, Centers for Disease Control and Prevention | Yan Li, Ying Tao, Jing Zhang, Brian Lynch, Anna Kelleher, Anna Montmayeur, Krista Queen, Anna Uehara, Peter Cook, Rachel Marine, Mark Burroughs, Han Jia Justin Ng, Clinton R. Paden, Haibin Wang, Justin Lee, Adam Retchless, Suxiang Tong |
| EPI_ISL_1509930 | NH Dept. of Health and Human Services Public Health Labs | Genomics and Discovery, Respiratory Viruses Branch, Division of Viral Diseases, Centers for Disease Control and Prevention | Yan Li, Ying Tao, Jing Zhang, Brian Lynch, Anna Kelleher, Anna Montmayeur, Krista Queen, Anna Uehara, Peter Cook, Rachel Marine, Mark Burroughs, Han Jia Justin Ng, Clinton R. Paden, Haibin Wang, Justin Lee, Adam Retchless, Suxiang Tong |
| EPI_ISL_1509931 | IL Dept. of Public Health Springfield Laboratory | Genomics and Discovery, Respiratory Viruses Branch, Division of Viral Diseases, Centers for Disease Control and Prevention | Yan Li, Ying Tao, Jing Zhang, Brian Lynch, Anna Kelleher, Anna Montmayeur, Krista Queen, Anna Uehara, Peter Cook, Rachel Marine, Mark Burroughs, Han Jia Justin Ng, Clinton R. Paden, Haibin Wang, Justin Lee, Adam Retchless, Suxiang Tong |
| EPI_ISL_1509932 | LA Office of Public Health Laboratories | Genomics and Discovery, Respiratory Viruses Branch, Division of Viral Diseases, Centers for Disease Control and Prevention | Yan Li, Ying Tao, Jing Zhang, Brian Lynch, Anna Kelleher, Anna Montmayeur, Krista Queen, Anna Uehara, Peter Cook, Rachel Marine, Mark Burroughs, Han Jia Justin Ng, Clinton R. Paden, Haibin Wang, Justin Lee, Adam Retchless, Suxiang Tong |
| EPI_ISL_1509934 | IL Department of Public Health Chicago Laboratory | Genomics and Discovery, Respiratory Viruses Branch, Division of Viral Diseases, Centers for Disease Control and Prevention | Yan Li, Ying Tao, Jing Zhang, Brian Lynch, Anna Kelleher, Anna Montmayeur, Krista Queen, Anna Uehara, Peter Cook, Rachel Marine, Mark Burroughs, Han Jia Justin Ng, Clinton R. Paden, Haibin Wang, Justin Lee, Adam Retchless, Suxiang Tong |
| EPI_ISL_1509935 | IL Dept. of Public Health Springfield Laboratory | Genomics and Discovery, Respiratory Viruses Branch, Division of Viral Diseases, Centers for Disease Control and Prevention | Yan Li, Ying Tao, Jing Zhang, Brian Lynch, Anna Kelleher, Anna Montmayeur, Krista Queen, Anna Uehara, Peter Cook, Rachel Marine, Mark Burroughs, Han Jia Justin Ng, Clinton R. Paden, Haibin Wang, Justin Lee, Adam Retchless, Suxiang Tong |
| EPI_ISL_1509936 | WVDHHR - Office of Laboratory Services | Genomics and Discovery, Respiratory Viruses Branch, Division of Viral Diseases, Centers for Disease Control and Prevention | Yan Li, Ying Tao, Jing Zhang, Brian Lynch, Anna Kelleher, Anna Montmayeur, Krista Queen, Anna Uehara, Peter Cook, Rachel Marine, Mark Burroughs, Han Jia Justin Ng, Clinton R. Paden, Haibin Wang, Justin Lee, Adam Retchless, Suxiang Tong |
| EPI_ISL_1509937 | "AR Dept. of Health-PHL, Molecular Diagnostics" | Genomics and Discovery, Respiratory Viruses Branch, Division of Viral Diseases, Centers for Disease Control and Prevention | Yan Li, Ying Tao, Jing Zhang, Brian Lynch, Anna Kelleher, Anna Montmayeur, Krista Queen, Anna Uehara, Peter Cook, Rachel Marine, Mark Burroughs, Han Jia Justin Ng, Clinton R. Paden, Haibin Wang, Justin Lee, Adam Retchless, Suxiang Tong |

| | | | |
|---|---|---|---|
| EPI_ISL_1509938 | IL Department of Public Health Chicago Laboratory | Genomics and Discovery, Respiratory Viruses Branch, Division of Viral Diseases, Centers for Disease Control and Prevention | Yan Li, Ying Tao, Jing Zhang, Brian Lynch, Anna Kelleher, Anna Montmayeur, Krista Queen, Anna Uehara, Peter Cook, Rachel Marine, Mark Burroughs, Han Jia Justin Ng, Clinton R. Paden, Haibin Wang, Justin Lee, Adam Retchless, Suxiang Tong |
| EPI_ISL_1509939 | IL Dept. of Public Health Springfield Laboratory | Genomics and Discovery, Respiratory Viruses Branch, Division of Viral Diseases, Centers for Disease Control and Prevention | Yan Li, Ying Tao, Jing Zhang, Brian Lynch, Anna Kelleher, Anna Montmayeur, Krista Queen, Anna Uehara, Peter Cook, Rachel Marine, Mark Burroughs, Han Jia Justin Ng, Clinton R. Paden, Haibin Wang, Justin Lee, Adam Retchless, Suxiang Tong |
| EPI_ISL_1509940 | IL Department of Public Health Chicago Laboratory | Genomics and Discovery, Respiratory Viruses Branch, Division of Viral Diseases, Centers for Disease Control and Prevention | Yan Li, Ying Tao, Jing Zhang, Brian Lynch, Anna Kelleher, Anna Montmayeur, Krista Queen, Anna Uehara, Peter Cook, Rachel Marine, Mark Burroughs, Han Jia Justin Ng, Clinton R. Paden, Haibin Wang, Justin Lee, Adam Retchless, Suxiang Tong |
| EPI_ISL_1509941 | WVDHHR - Office of Laboratory Services | Genomics and Discovery, Respiratory Viruses Branch, Division of Viral Diseases, Centers for Disease Control and Prevention | Yan Li, Ying Tao, Jing Zhang, Brian Lynch, Anna Kelleher, Anna Montmayeur, Krista Queen, Anna Uehara, Peter Cook, Rachel Marine, Mark Burroughs, Han Jia Justin Ng, Clinton R. Paden, Haibin Wang, Justin Lee, Adam Retchless, Suxiang Tong |
| EPI_ISL_1509942 | ME Health and Environmental Testing Laboratory | Genomics and Discovery, Respiratory Viruses Branch, Division of Viral Diseases, Centers for Disease Control and Prevention | Yan Li, Ying Tao, Jing Zhang, Brian Lynch, Anna Kelleher, Anna Montmayeur, Krista Queen, Anna Uehara, Peter Cook, Rachel Marine, Mark Burroughs, Han Jia Justin Ng, Clinton R. Paden, Haibin Wang, Justin Lee, Adam Retchless, Suxiang Tong |
| EPI_ISL_1509943 | "OK Public Health Laboratory, Oklahoma State DOH" | Genomics and Discovery, Respiratory Viruses Branch, Division of Viral Diseases, Centers for Disease Control and Prevention | Yan Li, Ying Tao, Jing Zhang, Brian Lynch, Anna Kelleher, Anna Montmayeur, Krista Queen, Anna Uehara, Peter Cook, Rachel Marine, Mark Burroughs, Han Jia Justin Ng, Clinton R. Paden, Haibin Wang, Justin Lee, Adam Retchless, Suxiang Tong |
| EPI_ISL_1509944, EPI_ISL_1509945 | IL Dept. of Public Health Springfield Laboratory | Genomics and Discovery, Respiratory Viruses Branch, Division of Viral Diseases, Centers for Disease Control and Prevention | Yan Li, Ying Tao, Jing Zhang, Brian Lynch, Anna Kelleher, Anna Montmayeur, Krista Queen, Anna Uehara, Peter Cook, Rachel Marine, Mark Burroughs, Han Jia Justin Ng, Clinton R. Paden, Haibin Wang, Justin Lee, Adam Retchless, Suxiang Tong |
| EPI_ISL_1509946 | MT Public Health Laboratory | Genomics and Discovery, Respiratory Viruses Branch, Division of Viral Diseases, Centers for Disease Control and Prevention | Yan Li, Ying Tao, Jing Zhang, Brian Lynch, Anna Kelleher, Anna Montmayeur, Krista Queen, Anna Uehara, Peter Cook, Rachel Marine, Mark Burroughs, Han Jia Justin Ng, Clinton R. Paden, Haibin Wang, Justin Lee, Adam Retchless, Suxiang Tong |
| EPI_ISL_1509947, EPI_ISL_1509948, EPI_ISL_1509949, EPI_ISL_1509950 | IL Department of Public Health Chicago Laboratory | Genomics and Discovery, Respiratory Viruses Branch, Division of Viral Diseases, Centers for Disease Control and Prevention | Yan Li, Ying Tao, Jing Zhang, Brian Lynch, Anna Kelleher, Anna Montmayeur, Krista Queen, Anna Uehara, Peter Cook, Rachel Marine, Mark Burroughs, Han Jia Justin Ng, Clinton R. Paden, Haibin Wang, Justin Lee, Adam Retchless, Suxiang Tong |
| EPI_ISL_1509951 | TN Division of Laboratory Services | Genomics and Discovery, Respiratory Viruses Branch, Division of Viral Diseases, Centers for Disease Control and Prevention | Yan Li, Ying Tao, Jing Zhang, Brian Lynch, Anna Kelleher, Anna Montmayeur, Krista Queen, Anna Uehara, Peter Cook, Rachel Marine, Mark Burroughs, Han Jia Justin Ng, Clinton R. Paden, Haibin Wang, Justin Lee, Adam Retchless, Suxiang Tong |
| EPI_ISL_1510260, EPI_ISL_1510261, EPI_ISL_1510262, EPI_ISL_1510263, EPI_ISL_1510264, EPI_ISL_1510265, EPI_ISL_1510266, EPI_ISL_1510267, EPI_ISL_1510268, EPI_ISL_1510269, EPI_ISL_1510270, EPI_ISL_1510271, EPI_ISL_1510272, EPI_ISL_1510273, EPI_ISL_1510274, EPI_ISL_1510275, EPI_ISL_1510276, EPI_ISL_1510277 | | | |
| see above | Idaho Bureau of Laboratories | Idaho Bureau of Laboratories | "R. Beukelman, Matthew Charles Burns, Aimee Ceniseros, Robert L. Voermans, Christopher Ball" |
| EPI_ISL_1510636, EPI_ISL_1510637, EPI_ISL_1510638, EPI_ISL_1510639, EPI_ISL_1510640 | Cytocheck Laboratory | Kansas Health and Environmental Lab | Mike Grose, Jonathan Barnell, Ben Olsen, and Phil Adam |
| EPI_ISL_1510641, EPI_ISL_1510642, EPI_ISL_1510643, EPI_ISL_1510644, EPI_ISL_1510645, EPI_ISL_1510646 | Kansas Health and Environmental Lab | Kansas Health and Environmental Lab | Mike Grose, Jonathan Barnell, Ben Olsen, and Phil Adam |
| EPI_ISL_1510647 | Cytocheck Laboratory | Kansas Health and Environmental Lab | Mike Grose, Jonathan Barnell, Ben Olsen, and Phil Adam |
| EPI_ISL_1510648, EPI_ISL_1510649, EPI_ISL_1510650 | Clincal Reference Laboratory | Kansas Health and Environmental Lab | Mike Grose, Jonathan Barnell, Ben Olsen, and Phil Adam |
| EPI_ISL_1511139, EPI_ISL_1511140, EPI_ISL_1511141, EPI_ISL_1511142, EPI_ISL_1511143, EPI_ISL_1511144, EPI_ISL_1511145, EPI_ISL_1511146, EPI_ISL_1511147, EPI_ISL_1511148, EPI_ISL_1511149, EPI_ISL_1511150, EPI_ISL_1511151, EPI_ISL_1511152, EPI_ISL_1511153, EPI_ISL_1511154, EPI_ISL_1511155, EPI_ISL_1511156, EPI_ISL_1511157, EPI_ISL_1511158, EPI_ISL_1511159, EPI_ISL_1511160, EPI_ISL_1511161, EPI_ISL_1511162, EPI_ISL_1511163, EPI_ISL_1511164, EPI_ISL_1511165, EPI_ISL_1511166, EPI_ISL_1511167, EPI_ISL_1511168, EPI_ISL_1511169, EPI_ISL_1511170, EPI_ISL_1511171, EPI_ISL_1511172, EPI_ISL_1511173, EPI_ISL_1511174, EPI_ISL_1511175, EPI_ISL_1511176, EPI_ISL_1511177, EPI_ISL_1511178, EPI_ISL_1511179, EPI_ISL_1511181, EPI_ISL_1511182, EPI_ISL_1511183, EPI_ISL_1511184, EPI_ISL_1511185, EPI_ISL_1511187, EPI_ISL_1511188, EPI_ISL_1511189, EPI_ISL_1511190, EPI_ISL_1511191, EPI_ISL_1511192, EPI_ISL_1511193, EPI_ISL_1511194, EPI_ISL_1511195, EPI_ISL_1511196, EPI_ISL_1511197 | | | |
| see above | DPHL | Delaware Public Health Lab | Rebecca Savage |
| EPI_ISL_1511238, EPI_ISL_1511239, EPI_ISL_1511241, EPI_ISL_1511242, EPI_ISL_1511243, EPI_ISL_1511244, EPI_ISL_1511245, EPI_ISL_1511246, EPI_ISL_1511247, EPI_ISL_1511248, EPI_ISL_1511249, EPI_ISL_1511250, EPI_ISL_1511251, EPI_ISL_1511252, EPI_ISL_1511253, EPI_ISL_1511254, EPI_ISL_1511255, EPI_ISL_1511256, EPI_ISL_1511257, EPI_ISL_1511258, EPI_ISL_1511259, EPI_ISL_1511260, EPI_ISL_1511261, EPI_ISL_1511262, EPI_ISL_1511263, EPI_ISL_1511264, EPI_ISL_1511265 | | | |
| see above | Wyoming Public Health Laboratory | Wyoming Public Health Laboratory | Jim Mildenberger, Wanda Manley, Noah Hull, Taylor Fearing, Lynette Gumbleton, Channing Weber, Ashley Norberg, Chayse Rowley, Marley Goetz, Brian Dominguez, Elliot Thomasson, Cari Sloma, and Rob Christensen |
| EPI_ISL_1511403, EPI_ISL_1511404, EPI_ISL_1511405, EPI_ISL_1511406, EPI_ISL_1511407, EPI_ISL_1511408, EPI_ISL_1511409, EPI_ISL_1511410, EPI_ISL_1511411, EPI_ISL_1511412, EPI_ISL_1511413, EPI_ISL_1511414, EPI_ISL_1511415, EPI_ISL_1511416, EPI_ISL_1511417, EPI_ISL_1511418, EPI_ISL_1511419, EPI_ISL_1511420, EPI_ISL_1511421, EPI_ISL_1511422, EPI_ISL_1511423, EPI_ISL_1511424, EPI_ISL_1511425, EPI_ISL_1511426, EPI_ISL_1511427, EPI_ISL_1511428, EPI_ISL_1511429, EPI_ISL_1511430, EPI_ISL_1511431, EPI_ISL_1511432, EPI_ISL_1511433, EPI_ISL_1511434, EPI_ISL_1511435, EPI_ISL_1511436, EPI_ISL_1511437, EPI_ISL_1511438, EPI_ISL_1511439, EPI_ISL_1511440, EPI_ISL_1511441, EPI_ISL_1511442, EPI_ISL_1511443, EPI_ISL_1511444, EPI_ISL_1511445, EPI_ISL_1511446, EPI_ISL_1511447, EPI_ISL_1511448, EPI_ISL_1511449, EPI_ISL_1511450, EPI_ISL_1511451, EPI_ISL_1511452, EPI_ISL_1511453, EPI_ISL_1511454, EPI_ISL_1511455, EPI_ISL_1511456, EPI_ISL_1511457, EPI_ISL_1511458, EPI_ISL_1511459, EPI_ISL_1511460, EPI_ISL_1511461, EPI_ISL_1511462, EPI_ISL_1511463, EPI_ISL_1511464, EPI_ISL_1511465, EPI_ISL_1511466, EPI_ISL_1511467, EPI_ISL_1511468, EPI_ISL_1511469, EPI_ISL_1511470, EPI_ISL_1511471, EPI_ISL_1511472, EPI_ISL_1511473, EPI_ISL_1511474, EPI_ISL_1511475, EPI_ISL_1511476, EPI_ISL_1511477, EPI_ISL_1511478, EPI_ISL_1511479, EPI_ISL_1511480, EPI_ISL_1511481, EPI_ISL_1511482, EPI_ISL_1511483, EPI_ISL_1511484, EPI_ISL_1511485, EPI_ISL_1511486, EPI_ISL_1511487, EPI_ISL_1511488, EPI_ISL_1511489, EPI_ISL_1511490, EPI_ISL_1511491, EPI_ISL_1511492, EPI_ISL_1511493, EPI_ISL_1511494, EPI_ISL_1511495, EPI_ISL_1511496, EPI_ISL_1511497, EPI_ISL_1511498, EPI_ISL_1511499, EPI_ISL_1511500, EPI_ISL_1511501, EPI_ISL_1511502, EPI_ISL_1511503, EPI_ISL_1511504, EPI_ISL_1511505, EPI_ISL_1511506, EPI_ISL_1511507, EPI_ISL_1511508, EPI_ISL_1511509, EPI_ISL_1511510, EPI_ISL_1511511, EPI_ISL_1511512, EPI_ISL_1511513, EPI_ISL_1511514, EPI_ISL_1511515, EPI_ISL_1511516, EPI_ISL_1511517, EPI_ISL_1511518, EPI_ISL_1511519, EPI_ISL_1511520, EPI_ISL_1511521, EPI_ISL_1511522, EPI_ISL_1511523, EPI_ISL_1511524, EPI_ISL_1511525, EPI_ISL_1511526, EPI_ISL_1511527, EPI_ISL_1511528, EPI_ISL_1511529, EPI_ISL_1511530, EPI_ISL_1511531, EPI_ISL_1511532, EPI_ISL_1511533, EPI_ISL_1511534, EPI_ISL_1511535, EPI_ISL_1511536, EPI_ISL_1511537, EPI_ISL_1511538, EPI_ISL_1511539, EPI_ISL_1511540, EPI_ISL_1511541, EPI_ISL_1511542, EPI_ISL_1511543, EPI_ISL_1511544, EPI_ISL_1511545, EPI_ISL_1511546, EPI_ISL_1511547, EPI_ISL_1511548, EPI_ISL_1511549, EPI_ISL_1511550, EPI_ISL_1511551, EPI_ISL_1511552, EPI_ISL_1511553, EPI_ISL_1511554, EPI_ISL_1511555, EPI_ISL_1511556, EPI_ISL_1511557, EPI_ISL_1511558, EPI_ISL_1511559, EPI_ISL_1511560, EPI_ISL_1511561, EPI_ISL_1511562, EPI_ISL_1511563, EPI_ISL_1511564, EPI_ISL_1511565, EPI_ISL_1511566, EPI_ISL_1511567, EPI_ISL_1511568, EPI_ISL_1511569, EPI_ISL_1511570, EPI_ISL_1511571, EPI_ISL_1511572, EPI_ISL_1511573, EPI_ISL_1511574, EPI_ISL_1511575, EPI_ISL_1511576, EPI_ISL_1511577, EPI_ISL_1511578, EPI_ISL_1511583, EPI_ISL_1511584, EPI_ISL_1511585, EPI_ISL_1511586, EPI_ISL_1511587, EPI_ISL_1511588, EPI_ISL_1511589, EPI_ISL_1511590, EPI_ISL_1511591, EPI_ISL_1511592, EPI_ISL_1511593, EPI_ISL_1511594, EPI_ISL_1511595, EPI_ISL_1511596, EPI_ISL_1511597, EPI_ISL_1511598, EPI_ISL_1511599, EPI_ISL_1511600, EPI_ISL_1511601, EPI_ISL_1511602 | | | |
| see above | Hospital of the University of Pennsylvania Molecular Pathology Lab | Bushman Lab - University of Pennsylvania | John Everett, Kyle Rodino, Shantan Reddy, Pascha Hokama, Aoife M. Roche, Young Hwang, Abigail Glascock, Scott Sherrill-Mix, Samantha A. Whiteside, Jevon Graham-Wooten, Layla A. Khatib, Ayannah S. Fitzgerald, Arupa Ganguly, Mike Feldman, Brendan Kelly, Ronald G. Collman and Frederic Bushman |
| EPI_ISL_1511631, EPI_ISL_1511632, EPI_ISL_1511634, EPI_ISL_1511637, EPI_ISL_1511638, EPI_ISL_1511639 | Nevada State Public Health Laboratory | Nevada State Public Health Laboratory | Andrew Gorzalski, Mark Pandori |
| EPI_ISL_1511640, EPI_ISL_1511642, EPI_ISL_1511645 | Oregon State Public Health Laboratory | Oregon State Public Health Laboratory | Rafia Razzaque, Eugene Yeboah, Vanda Makris, Laura Tsaknaridis, John Fontana and Shane Sevey |
| EPI_ISL_1511646, EPI_ISL_1511647, EPI_ISL_1511648, EPI_ISL_1511649, EPI_ISL_1511650, EPI_ISL_1511651, EPI_ISL_1511652, EPI_ISL_1511653, EPI_ISL_1511654, EPI_ISL_1511655, EPI_ISL_1511656, EPI_ISL_1511657, EPI_ISL_1511658, EPI_ISL_1511659, EPI_ISL_1511660, EPI_ISL_1511661, EPI_ISL_1511662, EPI_ISL_1511663, EPI_ISL_1511664, EPI_ISL_1511665, EPI_ISL_1511666, EPI_ISL_1511667, EPI_ISL_1511668, EPI_ISL_1511669, EPI_ISL_1511670, EPI_ISL_1511671, EPI_ISL_1511672, EPI_ISL_1511673, EPI_ISL_1511674, EPI_ISL_1511675, EPI_ISL_1511676, EPI_ISL_1511677, EPI_ISL_1511678, EPI_ISL_1511679, EPI_ISL_1511680, EPI_ISL_1511681, EPI_ISL_1511682, EPI_ISL_1511683, EPI_ISL_1511684, EPI_ISL_1511685, EPI_ISL_1511686, EPI_ISL_1511687, EPI_ISL_1511688, EPI_ISL_1511689, EPI_ISL_1511690, EPI_ISL_1511691, EPI_ISL_1511692, EPI_ISL_1511693, EPI_ISL_1511694, EPI_ISL_1511695, EPI_ISL_1511696 | | | |
| see above | Quest Diagnostics Incorporated | Centers for Disease Control and Prevention Division of Viral Diseases, Pathogen Discovery | Dakota Howard, Dhwani Batra, Peter W. Cook, Kara Moser, Adrian Paskey, Jason Caravas, Benjamin Rambo-Martin, Shatavia Morrison, Christopher Gulvick, Scott Sammons, Yvette Unoarumhi, Darlene Wagner, Matthew Schmerer, S. H. Rosenthal, A. Gerasimova, R. M. Kagan, B. Anderson, M. Hua, Y. Liu, L.E. Bernstein, K.E. Livingston, A. Perez, I. A. Shlyakhter, R. V. Rolando, R. Owen, P. Tanpaiboon, F. Lacbawan, Clinton R. Paden, Duncan MacCannell |
| EPI_ISL_1511697 | Oregon State Public Health Laboratory | Oregon State Public Health Laboratory | Rafia Razzaque, Eugene Yeboah, Vanda Makris, Laura Tsaknaridis, John Fontana and Shane Sevey |
| EPI_ISL_1511698, EPI_ISL_1511699, EPI_ISL_1511700, EPI_ISL_1511701, EPI_ISL_1511702, EPI_ISL_1511703, EPI_ISL_1511704, EPI_ISL_1511705, EPI_ISL_1511706, EPI_ISL_1511707, EPI_ISL_1511708, EPI_ISL_1511709, EPI_ISL_1511710, EPI_ISL_1511711, EPI_ISL_1511712, EPI_ISL_1511713, EPI_ISL_1511714, EPI_ISL_1511715, EPI_ISL_1511716, EPI_ISL_1511717, EPI_ISL_1511718, EPI_ISL_1511719, EPI_ISL_1511720, EPI_ISL_1511721, EPI_ISL_1511722, EPI_ISL_1511723, EPI_ISL_1511724, EPI_ISL_1511725, EPI_ISL_1511726, EPI_ISL_1511727, EPI_ISL_1511728, EPI_ISL_1511729, EPI_ISL_1511730, EPI_ISL_1511731, EPI_ISL_1511732, EPI_ISL_1511733, EPI_ISL_1511734, EPI_ISL_1511735, EPI_ISL_1511736, EPI_ISL_1511737, EPI_ISL_1511738, EPI_ISL_1511739, EPI_ISL_1511740, EPI_ISL_1511741, EPI_ISL_1511742, EPI_ISL_1511743, EPI_ISL_1511744, EPI_ISL_1511745, EPI_ISL_1511746, EPI_ISL_1511747, EPI_ISL_1511748, EPI_ISL_1511749, EPI_ISL_1511750, EPI_ISL_1511751, EPI_ISL_1511752, EPI_ISL_1511753, EPI_ISL_1511754, EPI_ISL_1511755, EPI_ISL_1511756, EPI_ISL_1511757, EPI_ISL_1511758, EPI_ISL_1511759, EPI_ISL_1511760, EPI_ISL_1511761, EPI_ISL_1511762, EPI_ISL_1511763, EPI_ISL_1511764, EPI_ISL_1511765, EPI_ISL_1511766, EPI_ISL_1511767, EPI_ISL_1511768, EPI_ISL_1511769, EPI_ISL_1511770, EPI_ISL_1511771, EPI_ISL_1511772, EPI_ISL_1511773, EPI_ISL_1511774, EPI_ISL_1511775, EPI_ISL_1511776, EPI_ISL_1511777, EPI_ISL_1511778, EPI_ISL_1511779, EPI_ISL_1511780, EPI_ISL_1511781, EPI_ISL_1511782, EPI_ISL_1511783, EPI_ISL_1511784, EPI_ISL_1511785, EPI_ISL_1511786, EPI_ISL_1511787, EPI_ISL_1511788, EPI_ISL_1511789, EPI_ISL_1511790, EPI_ISL_1511791, EPI_ISL_1511792, EPI_ISL_1511793, EPI_ISL_1511794, EPI_ISL_1511795, EPI_ISL_1511796, EPI_ISL_1511797, EPI_ISL_1511798, EPI_ISL_1511799, EPI_ISL_1511800, EPI_ISL_1511801, EPI_ISL_1511802, EPI_ISL_1511803, EPI_ISL_1511804, EPI_ISL_1511805, EPI_ISL_1511806, EPI_ISL_1511807, EPI_ISL_1511808, EPI_ISL_1511809, EPI_ISL_1511810, EPI_ISL_1511811, EPI_ISL_1511812, EPI_ISL_1511813, EPI_ISL_1511814, EPI_ISL_1511815, EPI_ISL_1511816, EPI_ISL_1511817, EPI_ISL_1511818, EPI_ISL_1511819, EPI_ISL_1511820, EPI_ISL_1511821, EPI_ISL_1511822, EPI_ISL_1511823, EPI_ISL_1511824, EPI_ISL_1511825, EPI_ISL_1511826, EPI_ISL_1511827, EPI_ISL_1511828, EPI_ISL_1511829, EPI_ISL_1511830, EPI_ISL_1511831, EPI_ISL_1511832, EPI_ISL_1511833, EPI_ISL_1511834, EPI_ISL_1511835, EPI_ISL_1511836, EPI_ISL_1511837, EPI_ISL_1511838, EPI_ISL_1511839, EPI_ISL_1511840, EPI_ISL_1511841, EPI_ISL_1511842, EPI_ISL_1511843, EPI_ISL_1511844, EPI_ISL_1511845, EPI_ISL_1511846, EPI_ISL_1511847, EPI_ISL_1511848, EPI_ISL_1511849, EPI_ISL_1511850, EPI_ISL_1511851, EPI_ISL_1511852, EPI_ISL_1511853, EPI_ISL_1511854, EPI_ISL_1511855, EPI_ISL_1511856, EPI_ISL_1511857, EPI_ISL_1511858, EPI_ISL_1511859 | | | |
| see above | Quest Diagnostics Incorporated | Centers for Disease Control and Prevention Division of Viral Diseases, Pathogen Discovery | Dakota Howard, Dhwani Batra, Peter W. Cook, Kara Moser, Adrian Paskey, Jason Caravas, Benjamin Rambo-Martin, Shatavia Morrison, Christopher Gulvick, Scott Sammons, Yvette Unoarumhi, Darlene Wagner, Matthew Schmerer, S. H. Rosenthal, A. Gerasimova, R. M. Kagan, B. Anderson, M. Hua, Y. Liu, L.E. Bernstein, K.E. Livingston, A. Perez, I. A. Shlyakhter, R. V. Rolando, R. Owen, P. Tanpaiboon, F. Lacbawan, Clinton R. Paden, Duncan MacCannell |
| EPI_ISL_1511860, EPI_ISL_1511861, EPI_ISL_1511862, EPI_ISL_1511863, EPI_ISL_1511864, EPI_ISL_1511865, EPI_ISL_1511866, EPI_ISL_1511867, EPI_ISL_1511868, EPI_ISL_1511869, EPI_ISL_1511870, EPI_ISL_1511871, EPI_ISL_1511872, EPI_ISL_1511873, EPI_ISL_1511874, EPI_ISL_1511875, EPI_ISL_1511877, EPI_ISL_1511878, EPI_ISL_1511879, EPI_ISL_1511880, EPI_ISL_1511881, EPI_ISL_1511882, EPI_ISL_1511883, EPI_ISL_1511884, EPI_ISL_1511885, EPI_ISL_1511886, EPI_ISL_1511887, EPI_ISL_1511889, EPI_ISL_1511890, EPI_ISL_1511891, EPI_ISL_1511892, EPI_ISL_1511893, EPI_ISL_1511894, EPI_ISL_1511895, EPI_ISL_1511896, EPI_ISL_1511897, EPI_ISL_1511898, EPI_ISL_1511899, EPI_ISL_1511900, EPI_ISL_1511901, EPI_ISL_1511902, EPI_ISL_1511903, EPI_ISL_1511904, EPI_ISL_1511905, EPI_ISL_1511906, EPI_ISL_1511907, EPI_ISL_1511908, EPI_ISL_1511909, EPI_ISL_1511910, EPI_ISL_1511911, EPI_ISL_1511912, EPI_ISL_1511913, EPI_ISL_1511914, EPI_ISL_1511915, EPI_ISL_1511916, EPI_ISL_1511917, EPI_ISL_1511918, EPI_ISL_1511919 | | | |

| | | | |
|---|---|---|---|
| see above | Helix/Illumina | Centers for Disease Control and Prevention Division of Viral Diseases, Pathogen Discovery | Dakota Howard, Dhwani Batra, Peter W. Cook, Kara Moser, Adrian Paskey, Jason Caravas, Benjamin Rambo-Martin, Shatavia Morrison, Christopher Gulvick, Scott Sammons, Yvette Unoarumhi, Darlene Wagner, Matthew Schmerer, Eileen de Feo, Jan Antico, Christine Tran, Matthew Tolentino, Shannon Wickline, Kim Gietzen, Brad Sickler, Jingtao Liu, Eric Allen, Phil Febbo, Nicole L. Washington, Simon White, Geraint Levan, Kelly Schiabor Barrett, Elizabeth Cirulli, Alexandre Bolze, Ary Ascencio, Charlotte Rivera-Garcia, Ryan Cho, Jason Nguyen, Sherry Wang, Jimmy Ramirez, Tyler Cassens, Efren Sandoval, Magnus Isaksson, William Lee, David Becker, Marc Laurent, James Lu, Clinton R. Paden, Duncan MacCannell |
| EPI_ISL_1511920 | Oregon State Public Health Laboratory | Oregon State Public Health Laboratory | Rafia Razzaque, Eugene Yeboah, Vanda Makris, Laura Tsaknaridis, John Fontana and Shane Sevey |
| EPI_ISL_1511921, EPI_ISL_1511922, EPI_ISL_1511923, EPI_ISL_1511924, EPI_ISL_1511925, EPI_ISL_1511926, EPI_ISL_1511927, EPI_ISL_1511928, EPI_ISL_1511929, EPI_ISL_1511930, EPI_ISL_1511931, EPI_ISL_1511932, EPI_ISL_1511933, EPI_ISL_1511934, EPI_ISL_1511935, EPI_ISL_1511936, EPI_ISL_1511937, EPI_ISL_1511938, EPI_ISL_1511940, EPI_ISL_1511941, EPI_ISL_1511942, EPI_ISL_1511943, EPI_ISL_1511944, EPI_ISL_1511945, EPI_ISL_1511946, EPI_ISL_1511947, EPI_ISL_1511948, EPI_ISL_1511949, EPI_ISL_1511950, EPI_ISL_1511951, EPI_ISL_1511952, EPI_ISL_1511953, EPI_ISL_1511954, EPI_ISL_1511955, EPI_ISL_1511956, EPI_ISL_1511957, EPI_ISL_1511958, EPI_ISL_1511959, EPI_ISL_1511960, EPI_ISL_1511961, EPI_ISL_1511962, EPI_ISL_1511963, EPI_ISL_1511964, EPI_ISL_1511965, EPI_ISL_1511966, EPI_ISL_1511967, EPI_ISL_1511968, EPI_ISL_1511969, EPI_ISL_1511971, EPI_ISL_1511972, EPI_ISL_1511973, EPI_ISL_1511974, EPI_ISL_1511975, EPI_ISL_1511976, EPI_ISL_1511977, EPI_ISL_1511978, EPI_ISL_1511979, EPI_ISL_1511980, EPI_ISL_1511981, EPI_ISL_1511982, EPI_ISL_1511983, EPI_ISL_1511984, EPI_ISL_1511985, EPI_ISL_1511986, EPI_ISL_1511987, EPI_ISL_1511988, EPI_ISL_1511989, EPI_ISL_1511990, EPI_ISL_1511991, EPI_ISL_1511992, EPI_ISL_1511993, EPI_ISL_1511994, EPI_ISL_1511995, EPI_ISL_1511996, EPI_ISL_1511997, EPI_ISL_1511998, EPI_ISL_1511999, EPI_ISL_1512000, EPI_ISL_1512001, EPI_ISL_1512002, EPI_ISL_1512004, EPI_ISL_1512005, EPI_ISL_1512006, EPI_ISL_1512007, EPI_ISL_1512008, EPI_ISL_1512010, EPI_ISL_1512011, EPI_ISL_1512012, EPI_ISL_1512013, EPI_ISL_1512014, EPI_ISL_1512015, EPI_ISL_1512016, EPI_ISL_1512017, EPI_ISL_1512018, EPI_ISL_1512019, EPI_ISL_1512020, EPI_ISL_1512021, EPI_ISL_1512022, EPI_ISL_1512023, EPI_ISL_1512024, EPI_ISL_1512025, EPI_ISL_1512026, EPI_ISL_1512027, EPI_ISL_1512028, EPI_ISL_1512029, EPI_ISL_1512030, EPI_ISL_1512031, EPI_ISL_1512032, EPI_ISL_1512033, EPI_ISL_1512034, EPI_ISL_1512035, EPI_ISL_1512036, EPI_ISL_1512037, EPI_ISL_1512038, EPI_ISL_1512039, EPI_ISL_1512040, EPI_ISL_1512041, EPI_ISL_1512042, EPI_ISL_1512043, EPI_ISL_1512044, EPI_ISL_1512045, EPI_ISL_1512046, EPI_ISL_1512047, EPI_ISL_1512048, EPI_ISL_1512049, EPI_ISL_1512050, EPI_ISL_1512051, EPI_ISL_1512052, EPI_ISL_1512053, EPI_ISL_1512054, EPI_ISL_1512055, EPI_ISL_1512056, EPI_ISL_1512057, EPI_ISL_1512058, EPI_ISL_1512059, EPI_ISL_1512060, EPI_ISL_1512061, EPI_ISL_1512062, EPI_ISL_1512063, EPI_ISL_1512064, EPI_ISL_1512065, EPI_ISL_1512066, EPI_ISL_1512067, EPI_ISL_1512068, EPI_ISL_1512069, EPI_ISL_1512070, EPI_ISL_1512071, EPI_ISL_1512072, EPI_ISL_1512073, EPI_ISL_1512074, EPI_ISL_1512075, EPI_ISL_1512076, EPI_ISL_1512077, EPI_ISL_1512078, EPI_ISL_1512079, EPI_ISL_1512080, EPI_ISL_1512081, EPI_ISL_1512082, EPI_ISL_1512083, EPI_ISL_1512084, EPI_ISL_1512085, EPI_ISL_1512086, EPI_ISL_1512087, EPI_ISL_1512088, EPI_ISL_1512089, EPI_ISL_1512090, EPI_ISL_1512091, EPI_ISL_1512092, EPI_ISL_1512093, EPI_ISL_1512094, EPI_ISL_1512095, EPI_ISL_1512096, EPI_ISL_1512097, EPI_ISL_1512098, EPI_ISL_1512099, EPI_ISL_1512100, EPI_ISL_1512101, EPI_ISL_1512102, EPI_ISL_1512103, EPI_ISL_1512104, EPI_ISL_1512105, EPI_ISL_1512106, EPI_ISL_1512107, EPI_ISL_1512108, EPI_ISL_1512109, EPI_ISL_1512110, EPI_ISL_1512111, EPI_ISL_1512112, EPI_ISL_1512113, EPI_ISL_1512114, EPI_ISL_1512115, EPI_ISL_1512116, EPI_ISL_1512117, EPI_ISL_1512118, EPI_ISL_1512119, EPI_ISL_1512120, EPI_ISL_1512121, EPI_ISL_1512122, EPI_ISL_1512123, EPI_ISL_1512124, EPI_ISL_1512125, EPI_ISL_1512126, EPI_ISL_1512127, EPI_ISL_1512128, EPI_ISL_1512129, EPI_ISL_1512130, EPI_ISL_1512131, EPI_ISL_1512132, EPI_ISL_1512133, EPI_ISL_1512134, EPI_ISL_1512135, EPI_ISL_1512136, EPI_ISL_1512137, EPI_ISL_1512138, EPI_ISL_1512139, EPI_ISL_1512140, EPI_ISL_1512141, EPI_ISL_1512142, EPI_ISL_1512143, EPI_ISL_1512144, EPI_ISL_1512145, EPI_ISL_1512146, EPI_ISL_1512147, EPI_ISL_1512148, EPI_ISL_1512149, EPI_ISL_1512150, EPI_ISL_1512151, EPI_ISL_1512152, EPI_ISL_1512153, EPI_ISL_1512154, EPI_ISL_1512155, EPI_ISL_1512156, EPI_ISL_1512157, EPI_ISL_1512158, EPI_ISL_1512159, EPI_ISL_1512160, EPI_ISL_1512161, EPI_ISL_1512162, EPI_ISL_1512163, EPI_ISL_1512164, EPI_ISL_1512165, EPI_ISL_1512166, EPI_ISL_1512167, EPI_ISL_1512168, EPI_ISL_1512169, EPI_ISL_1512170, EPI_ISL_1512171, EPI_ISL_1512172, EPI_ISL_1512173, EPI_ISL_1512174, EPI_ISL_1512175, EPI_ISL_1512176, EPI_ISL_1512177, EPI_ISL_1512178, EPI_ISL_1512179, EPI_ISL_1512180, EPI_ISL_1512181, EPI_ISL_1512182 | | | |
| see above | Helix/Illumina | Centers for Disease Control and Prevention Division of Viral Diseases, Pathogen Discovery | Dakota Howard, Dhwani Batra, Peter W. Cook, Kara Moser, Adrian Paskey, Jason Caravas, Benjamin Rambo-Martin, Shatavia Morrison, Christopher Gulvick, Scott Sammons, Yvette Unoarumhi, Darlene Wagner, Matthew Schmerer, Eileen de Feo, Jan Antico, Christine Tran, Matthew Tolentino, Shannon Wickline, Kim Gietzen, Brad Sickler, Jingtao Liu, Eric Allen, Phil Febbo, Nicole L. Washington, Simon White, Geraint Levan, Kelly Schiabor Barrett, Elizabeth Cirulli, Alexandre Bolze, Ary Ascencio, Charlotte Rivera-Garcia, Ryan Cho, Jason Nguyen, Sherry Wang, Jimmy Ramirez, Tyler Cassens, Efren Sandoval, Magnus Isaksson, William Lee, David Becker, Marc Laurent, James Lu, Clinton R. Paden, Duncan MacCannell |
| EPI_ISL_1512183 | Oregon State Public Health Laboratory | Oregon State Public Health Laboratory | Rafia Razzaque, Eugene Yeboah, Vanda Makris, Laura Tsaknaridis, John Fontana and Shane Sevey |
| EPI_ISL_1512184, EPI_ISL_1512185, EPI_ISL_1512186, EPI_ISL_1512187, EPI_ISL_1512188, EPI_ISL_1512189, EPI_ISL_1512190, EPI_ISL_1512191, EPI_ISL_1512192, EPI_ISL_1512193, EPI_ISL_1512194, EPI_ISL_1512195, EPI_ISL_1512196, EPI_ISL_1512197, EPI_ISL_1512198, EPI_ISL_1512199, EPI_ISL_1512200, EPI_ISL_1512201, EPI_ISL_1512202, EPI_ISL_1512203, EPI_ISL_1512204, EPI_ISL_1512205, EPI_ISL_1512206, EPI_ISL_1512207, EPI_ISL_1512208, EPI_ISL_1512209, EPI_ISL_1512210, EPI_ISL_1512211, EPI_ISL_1512212, EPI_ISL_1512213, EPI_ISL_1512214, EPI_ISL_1512215, EPI_ISL_1512216, EPI_ISL_1512217, EPI_ISL_1512218, EPI_ISL_1512219, EPI_ISL_1512220, EPI_ISL_1512221, EPI_ISL_1512222, EPI_ISL_1512223, EPI_ISL_1512224, EPI_ISL_1512225, EPI_ISL_1512226, EPI_ISL_1512227, EPI_ISL_1512228, EPI_ISL_1512229, EPI_ISL_1512230, EPI_ISL_1512231, EPI_ISL_1512232, EPI_ISL_1512233, EPI_ISL_1512234, EPI_ISL_1512235, EPI_ISL_1512236, EPI_ISL_1512237, EPI_ISL_1512238, EPI_ISL_1512239, EPI_ISL_1512240, EPI_ISL_1512241, EPI_ISL_1512242, EPI_ISL_1512243, EPI_ISL_1512244, EPI_ISL_1512245, EPI_ISL_1512246, EPI_ISL_1512247, EPI_ISL_1512248, EPI_ISL_1512249, EPI_ISL_1512250, EPI_ISL_1512251, EPI_ISL_1512252, EPI_ISL_1512253, EPI_ISL_1512254, EPI_ISL_1512255, EPI_ISL_1512256, EPI_ISL_1512257, EPI_ISL_1512258, EPI_ISL_1512259, EPI_ISL_1512260, EPI_ISL_1512261, EPI_ISL_1512262, EPI_ISL_1512263, EPI_ISL_1512264, EPI_ISL_1512265, EPI_ISL_1512266, EPI_ISL_1512267, EPI_ISL_1512268, EPI_ISL_1512269, EPI_ISL_1512270, EPI_ISL_1512271, EPI_ISL_1512272, EPI_ISL_1512273, EPI_ISL_1512274, EPI_ISL_1512275, EPI_ISL_1512276, EPI_ISL_1512277, EPI_ISL_1512278, EPI_ISL_1512279, EPI_ISL_1512280, EPI_ISL_1512281, EPI_ISL_1512282, EPI_ISL_1512283, EPI_ISL_1512284, EPI_ISL_1512285, EPI_ISL_1512286, EPI_ISL_1512287, EPI_ISL_1512288, EPI_ISL_1512289, EPI_ISL_1512290, EPI_ISL_1512291, EPI_ISL_1512292, EPI_ISL_1512293, EPI_ISL_1512294, EPI_ISL_1512295, EPI_ISL_1512296, EPI_ISL_1512297, EPI_ISL_1512298, EPI_ISL_1512299, EPI_ISL_1512300, EPI_ISL_1512301, EPI_ISL_1512302, EPI_ISL_1512303, EPI_ISL_1512304, EPI_ISL_1512305, EPI_ISL_1512306, EPI_ISL_1512307, EPI_ISL_1512308, EPI_ISL_1512309, EPI_ISL_1512310, EPI_ISL_1512311, EPI_ISL_1512312, EPI_ISL_1512313, EPI_ISL_1512314, EPI_ISL_1512315, EPI_ISL_1512317, EPI_ISL_1512318, EPI_ISL_1512319, EPI_ISL_1512320, EPI_ISL_1512321, EPI_ISL_1512322, EPI_ISL_1512323, EPI_ISL_1512324, EPI_ISL_1512325, EPI_ISL_1512326, EPI_ISL_1512327, EPI_ISL_1512328, EPI_ISL_1512329, EPI_ISL_1512330, EPI_ISL_1512331, EPI_ISL_1512332, EPI_ISL_1512333, EPI_ISL_1512334, EPI_ISL_1512335, EPI_ISL_1512336, EPI_ISL_1512337, EPI_ISL_1512338, EPI_ISL_1512339, EPI_ISL_1512340, EPI_ISL_1512341, EPI_ISL_1512342, EPI_ISL_1512343, EPI_ISL_1512344, EPI_ISL_1512346, EPI_ISL_1512347, EPI_ISL_1512348, EPI_ISL_1512349, EPI_ISL_1512350, EPI_ISL_1512351, EPI_ISL_1512352, EPI_ISL_1512353, EPI_ISL_1512354, EPI_ISL_1512355, EPI_ISL_1512356, EPI_ISL_1512357, EPI_ISL_1512358, EPI_ISL_1512359, EPI_ISL_1512360, EPI_ISL_1512361, EPI_ISL_1512362, EPI_ISL_1512363, EPI_ISL_1512364, EPI_ISL_1512365, EPI_ISL_1512366, EPI_ISL_1512367, EPI_ISL_1512368, EPI_ISL_1512369, EPI_ISL_1512370, EPI_ISL_1512371, EPI_ISL_1512372, EPI_ISL_1512373, EPI_ISL_1512374, EPI_ISL_1512375, EPI_ISL_1512376, EPI_ISL_1512378, EPI_ISL_1512379, EPI_ISL_1512380, EPI_ISL_1512381, EPI_ISL_1512382, EPI_ISL_1512383, EPI_ISL_1512384, EPI_ISL_1512385, EPI_ISL_1512386, EPI_ISL_1512387, EPI_ISL_1512388, EPI_ISL_1512389, EPI_ISL_1512390, EPI_ISL_1512391, EPI_ISL_1512392, EPI_ISL_1512393, EPI_ISL_1512394, EPI_ISL_1512395, EPI_ISL_1512396, EPI_ISL_1512397, EPI_ISL_1512398, EPI_ISL_1512399, EPI_ISL_1512400, EPI_ISL_1512401, EPI_ISL_1512402, EPI_ISL_1512403, EPI_ISL_1512404, EPI_ISL_1512405, EPI_ISL_1512406, EPI_ISL_1512407, EPI_ISL_1512408, EPI_ISL_1512409, EPI_ISL_1512410, EPI_ISL_1512411, EPI_ISL_1512412, EPI_ISL_1512413, EPI_ISL_1512414, EPI_ISL_1512415, EPI_ISL_1512416, EPI_ISL_1512417, EPI_ISL_1512418, EPI_ISL_1512419, EPI_ISL_1512420, EPI_ISL_1512421, EPI_ISL_1512422, EPI_ISL_1512423, EPI_ISL_1512424, EPI_ISL_1512425, EPI_ISL_1512426, EPI_ISL_1512427, EPI_ISL_1512428, EPI_ISL_1512429, EPI_ISL_1512430, EPI_ISL_1512431, EPI_ISL_1512432, EPI_ISL_1512433, EPI_ISL_1512434, EPI_ISL_1512435, EPI_ISL_1512436, EPI_ISL_1512437, EPI_ISL_1512438, EPI_ISL_1512439, EPI_ISL_1512440, EPI_ISL_1512441, EPI_ISL_1512442, EPI_ISL_1512443, EPI_ISL_1512444, EPI_ISL_1512445, EPI_ISL_1512446, EPI_ISL_1512447, EPI_ISL_1512448, EPI_ISL_1512449, EPI_ISL_1512450, EPI_ISL_1512451, EPI_ISL_1512452, EPI_ISL_1512453, EPI_ISL_1512454, EPI_ISL_1512455, EPI_ISL_1512456, EPI_ISL_1512457, EPI_ISL_1512458, EPI_ISL_1512459, EPI_ISL_1512460, EPI_ISL_1512461, EPI_ISL_1512462, EPI_ISL_1512463, EPI_ISL_1512464, EPI_ISL_1512465, EPI_ISL_1512466, EPI_ISL_1512467, EPI_ISL_1512468, EPI_ISL_1512469, EPI_ISL_1512470, EPI_ISL_1512471, EPI_ISL_1512472, EPI_ISL_1512473, EPI_ISL_1512474, EPI_ISL_1512475, EPI_ISL_1512476, EPI_ISL_1512477, EPI_ISL_1512478, EPI_ISL_1512479, EPI_ISL_1512480, EPI_ISL_1512481, EPI_ISL_1512482, EPI_ISL_1512483, EPI_ISL_1512484, EPI_ISL_1512485, EPI_ISL_1512486, EPI_ISL_1512487, EPI_ISL_1512488, EPI_ISL_1512489, EPI_ISL_1512490, EPI_ISL_1512491, EPI_ISL_1512492, EPI_ISL_1512493, EPI_ISL_1512494, EPI_ISL_1512495, EPI_ISL_1512496 | | | |
| see above | Helix/Illumina | Centers for Disease Control and Prevention Division of Viral Diseases, Pathogen Discovery | Dakota Howard, Dhwani Batra, Peter W. Cook, Kara Moser, Adrian Paskey, Jason Caravas, Benjamin Rambo-Martin, Shatavia Morrison, Christopher Gulvick, Scott Sammons, Yvette Unoarumhi, Darlene Wagner, Matthew Schmerer, Eileen de Feo, Jan Antico, Christine Tran, Matthew Tolentino, Shannon Wickline, Kim Gietzen, Brad Sickler, Jingtao Liu, Eric Allen, Phil Febbo, Nicole L. Washington, Simon White, Geraint Levan, Kelly Schiabor Barrett, Elizabeth Cirulli, Alexandre Bolze, Ary Ascencio, Charlotte Rivera-Garcia, Ryan Cho, Jason Nguyen, Sherry Wang, Jimmy Ramirez, Tyler Cassens, Efren Sandoval, Magnus Isaksson, William Lee, David Becker, Marc Laurent, James Lu, Clinton R. Paden, Duncan MacCannell |
| EPI_ISL_1512497 | Oregon State Public Health Laboratory | Oregon State Public Health Laboratory | Rafia Razzaque, Eugene Yeboah, Vanda Makris, Laura Tsaknaridis, John Fontana and Shane Sevey |
| EPI_ISL_1512498, EPI_ISL_1512499, EPI_ISL_1512500, EPI_ISL_1512501, EPI_ISL_1512502, EPI_ISL_1512503, EPI_ISL_1512504, EPI_ISL_1512505, EPI_ISL_1512506, EPI_ISL_1512507, EPI_ISL_1512508, EPI_ISL_1512509, EPI_ISL_1512510, EPI_ISL_1512511, EPI_ISL_1512512, EPI_ISL_1512513, EPI_ISL_1512514, EPI_ISL_1512515, EPI_ISL_1512516, EPI_ISL_1512517, EPI_ISL_1512518, EPI_ISL_1512519, EPI_ISL_1512520, EPI_ISL_1512521, EPI_ISL_1512522, EPI_ISL_1512523, EPI_ISL_1512524, EPI_ISL_1512525, EPI_ISL_1512526, EPI_ISL_1512527, EPI_ISL_1512528, EPI_ISL_1512529, EPI_ISL_1512530, EPI_ISL_1512531, EPI_ISL_1512532, EPI_ISL_1512533, EPI_ISL_1512534, EPI_ISL_1512535, EPI_ISL_1512536, EPI_ISL_1512537, EPI_ISL_1512538, EPI_ISL_1512539, EPI_ISL_1512540, EPI_ISL_1512541, EPI_ISL_1512542, EPI_ISL_1512543, EPI_ISL_1512544, EPI_ISL_1512545, EPI_ISL_1512546, EPI_ISL_1512547, EPI_ISL_1512548, EPI_ISL_1512549, EPI_ISL_1512550, EPI_ISL_1512551, EPI_ISL_1512552, EPI_ISL_1512553, EPI_ISL_1512554, EPI_ISL_1512555, EPI_ISL_1512556, EPI_ISL_1512557, EPI_ISL_1512558, EPI_ISL_1512559, EPI_ISL_1512560, EPI_ISL_1512561, EPI_ISL_1512562, EPI_ISL_1512563, EPI_ISL_1512564, EPI_ISL_1512565, EPI_ISL_1512566, EPI_ISL_1512567, EPI_ISL_1512568, EPI_ISL_1512569, EPI_ISL_1512570, EPI_ISL_1512571, EPI_ISL_1512572, EPI_ISL_1512573, EPI_ISL_1512574, EPI_ISL_1512575, EPI_ISL_1512576, EPI_ISL_1512577, EPI_ISL_1512578, EPI_ISL_1512579, EPI_ISL_1512580, EPI_ISL_1512581, EPI_ISL_1512582, EPI_ISL_1512583, EPI_ISL_1512584, EPI_ISL_1512585, EPI_ISL_1512586, EPI_ISL_1512587, EPI_ISL_1512588, EPI_ISL_1512589, EPI_ISL_1512590, EPI_ISL_1512591, EPI_ISL_1512592, EPI_ISL_1512593, EPI_ISL_1512594, EPI_ISL_1512595, EPI_ISL_1512596, EPI_ISL_1512597, EPI_ISL_1512598, EPI_ISL_1512599, EPI_ISL_1512600, EPI_ISL_1512601, EPI_ISL_1512602, EPI_ISL_1512603, EPI_ISL_1512604, EPI_ISL_1512605, EPI_ISL_1512606, EPI_ISL_1512607, EPI_ISL_1512608, EPI_ISL_1512609, EPI_ISL_1512610, EPI_ISL_1512611, EPI_ISL_1512612, EPI_ISL_1512613, EPI_ISL_1512614, EPI_ISL_1512615, EPI_ISL_1512616, EPI_ISL_1512617, EPI_ISL_1512618, EPI_ISL_1512619, EPI_ISL_1512620, EPI_ISL_1512621, EPI_ISL_1512622, EPI_ISL_1512623, EPI_ISL_1512624, EPI_ISL_1512625, EPI_ISL_1512626, EPI_ISL_1512627, EPI_ISL_1512628, EPI_ISL_1512629, EPI_ISL_1512630, EPI_ISL_1512631, EPI_ISL_1512632, EPI_ISL_1512633, EPI_ISL_1512634, EPI_ISL_1512635, EPI_ISL_1512636, EPI_ISL_1512637, EPI_ISL_1512638, EPI_ISL_1512639, EPI_ISL_1512640, EPI_ISL_1512641, EPI_ISL_1512642, EPI_ISL_1512643, EPI_ISL_1512644, EPI_ISL_1512645, EPI_ISL_1512646, EPI_ISL_1512647, EPI_ISL_1512648, EPI_ISL_1512649, EPI_ISL_1512650, EPI_ISL_1512651, EPI_ISL_1512652, EPI_ISL_1512653, EPI_ISL_1512654, EPI_ISL_1512655, EPI_ISL_1512656, EPI_ISL_1512657, EPI_ISL_1512658, EPI_ISL_1512659, EPI_ISL_1512660, EPI_ISL_1512661, EPI_ISL_1512662, EPI_ISL_1512663, EPI_ISL_1512664, EPI_ISL_1512665, EPI_ISL_1512666, EPI_ISL_1512667, EPI_ISL_1512668, EPI_ISL_1512669, EPI_ISL_1512670, EPI_ISL_1512671, EPI_ISL_1512672, EPI_ISL_1512673, EPI_ISL_1512674, EPI_ISL_1512675, EPI_ISL_1512676, EPI_ISL_1512677, EPI_ISL_1512678, EPI_ISL_1512679, EPI_ISL_1512680, EPI_ISL_1512681, EPI_ISL_1512682, EPI_ISL_1512683, EPI_ISL_1512684, EPI_ISL_1512685, EPI_ISL_1512686, EPI_ISL_1512687, EPI_ISL_1512688, EPI_ISL_1512689, EPI_ISL_1512690, EPI_ISL_1512691, EPI_ISL_1512692, EPI_ISL_1512693, EPI_ISL_1512694, EPI_ISL_1512695, EPI_ISL_1512696, EPI_ISL_1512697, EPI_ISL_1512698, EPI_ISL_1512699, EPI_ISL_1512700, EPI_ISL_1512701, EPI_ISL_1512702, EPI_ISL_1512703, EPI_ISL_1512705, EPI_ISL_1512706, EPI_ISL_1512707, EPI_ISL_1512708, EPI_ISL_1512709, EPI_ISL_1512710, EPI_ISL_1512712, EPI_ISL_1512713, EPI_ISL_1512714, EPI_ISL_1512715, EPI_ISL_1512716, EPI_ISL_1512717, EPI_ISL_1512718, EPI_ISL_1512719, EPI_ISL_1512720, EPI_ISL_1512721, EPI_ISL_1512722, EPI_ISL_1512723, EPI_ISL_1512724, EPI_ISL_1512725, EPI_ISL_1512726, EPI_ISL_1512727, EPI_ISL_1512728, EPI_ISL_1512729, EPI_ISL_1512730, EPI_ISL_1512731, EPI_ISL_1512732, EPI_ISL_1512733, EPI_ISL_1512734, EPI_ISL_1512735, EPI_ISL_1512736, EPI_ISL_1512737, EPI_ISL_1512738, EPI_ISL_1512740, EPI_ISL_1512741, EPI_ISL_1512742, EPI_ISL_1512743, EPI_ISL_1512744 | | | |
| see above | Helix/Illumina | Centers for Disease Control and Prevention Division of Viral Diseases, Pathogen Discovery | Dakota Howard, Dhwani Batra, Peter W. Cook, Kara Moser, Adrian Paskey, Jason Caravas, Benjamin Rambo-Martin, Shatavia Morrison, Christopher Gulvick, Scott Sammons, Yvette Unoarumhi, Darlene Wagner, Matthew Schmerer, Eileen de Feo, Jan Antico, Christine Tran, Matthew Tolentino, Shannon Wickline, Kim Gietzen, Brad Sickler, Jingtao Liu, Eric Allen, Phil Febbo, Nicole L. Washington, Simon White, Geraint Levan, Kelly Schiabor Barrett, Elizabeth Cirulli, Alexandre Bolze, Ary Ascencio, Charlotte Rivera-Garcia, Ryan Cho, Jason Nguyen, Sherry Wang, Jimmy Ramirez, Tyler Cassens, Efren Sandoval, Magnus Isaksson, William Lee, David Becker, Marc Laurent, James Lu, Clinton R. Paden, Duncan MacCannell |
| EPI_ISL_1512745 | Oregon State Public Health Laboratory | Oregon State Public Health Laboratory | Rafia Razzaque, Eugene Yeboah, Vanda Makris, Laura Tsaknaridis, John Fontana and Shane Sevey |
| EPI_ISL_1512746, EPI_ISL_1512747, EPI_ISL_1512749, EPI_ISL_1512751, EPI_ISL_1512752, EPI_ISL_1512754, EPI_ISL_1512755, EPI_ISL_1512756, EPI_ISL_1512758, EPI_ISL_1512759, EPI_ISL_1512760, EPI_ISL_1512761, EPI_ISL_1512762, EPI_ISL_1512763, EPI_ISL_1512764, EPI_ISL_1512765, EPI_ISL_1512766, EPI_ISL_1512767, EPI_ISL_1512768, EPI_ISL_1512769, EPI_ISL_1512770, EPI_ISL_1512771, EPI_ISL_1512772, EPI_ISL_1512773, EPI_ISL_1512774, EPI_ISL_1512775, EPI_ISL_1512776, EPI_ISL_1512777, EPI_ISL_1512778, EPI_ISL_1512779, EPI_ISL_1512782, EPI_ISL_1512783, EPI_ISL_1512784, EPI_ISL_1512785, EPI_ISL_1512786, EPI_ISL_1512787, EPI_ISL_1512788, EPI_ISL_1512789, EPI_ISL_1512790, EPI_ISL_1512792, EPI_ISL_1512793, EPI_ISL_1512794, EPI_ISL_1512795, EPI_ISL_1512796, EPI_ISL_1512797, EPI_ISL_1512798, EPI_ISL_1512799, EPI_ISL_1512800, EPI_ISL_1512801, EPI_ISL_1512802, EPI_ISL_1512803, EPI_ISL_1512804, EPI_ISL_1512805, EPI_ISL_1512807, EPI_ISL_1512808, EPI_ISL_1512809, EPI_ISL_1512810, EPI_ISL_1512811, EPI_ISL_1512812, EPI_ISL_1512813, EPI_ISL_1512814, EPI_ISL_1512815, EPI_ISL_1512816, EPI_ISL_1512817, EPI_ISL_1512818, EPI_ISL_1512821, EPI_ISL_1512822, EPI_ISL_1512824, EPI_ISL_1512825, EPI_ISL_1512829, EPI_ISL_1512830, EPI_ISL_1512831, EPI_ISL_1512832, EPI_ISL_1512833, EPI_ISL_1512834, EPI_ISL_1512835, EPI_ISL_1512837, EPI_ISL_1512838, EPI_ISL_1512839, EPI_ISL_1512840, EPI_ISL_1512841, EPI_ISL_1512842, EPI_ISL_1512843, EPI_ISL_1512845, EPI_ISL_1512846, EPI_ISL_1512847, EPI_ISL_1512848, EPI_ISL_1512849, EPI_ISL_1512850, EPI_ISL_1512851, EPI_ISL_1512852, EPI_ISL_1512853, EPI_ISL_1512855, EPI_ISL_1512856, EPI_ISL_1512857, EPI_ISL_1512858, EPI_ISL_1512859, EPI_ISL_1512860, EPI_ISL_1512861, EPI_ISL_1512862, EPI_ISL_1512863, EPI_ISL_1512864, EPI_ISL_1512865, EPI_ISL_1512867, EPI_ISL_1512868, EPI_ISL_1512869, EPI_ISL_1512870, EPI_ISL_1512871, EPI_ISL_1512872, EPI_ISL_1512873, EPI_ISL_1512874, EPI_ISL_1512875, EPI_ISL_1512876, EPI_ISL_1512877, EPI_ISL_1512878, EPI_ISL_1512879, EPI_ISL_1512880, EPI_ISL_1512882, EPI_ISL_1512883, EPI_ISL_1512884, EPI_ISL_1512885, EPI_ISL_1512886, EPI_ISL_1512887, EPI_ISL_1512889, EPI_ISL_1512890, EPI_ISL_1512891, EPI_ISL_1512892, EPI_ISL_1512893, EPI_ISL_1512894, EPI_ISL_1512895, EPI_ISL_1512896, EPI_ISL_1512898, EPI_ISL_1512899, EPI_ISL_1512900, EPI_ISL_1512901, EPI_ISL_1512903, EPI_ISL_1512904, EPI_ISL_1512906, EPI_ISL_1512907, EPI_ISL_1512908, EPI_ISL_1512909, EPI_ISL_1512910, EPI_ISL_1512911, EPI_ISL_1512912, EPI_ISL_1512913, EPI_ISL_1512914, EPI_ISL_1512915, EPI_ISL_1512916, EPI_ISL_1512917, EPI_ISL_1512919, EPI_ISL_1512920, EPI_ISL_1512921, EPI_ISL_1512922, EPI_ISL_1512923, EPI_ISL_1512925, EPI_ISL_1512926, EPI_ISL_1512927, EPI_ISL_1512929, EPI_ISL_1512930, EPI_ISL_1512931, EPI_ISL_1512932, EPI_ISL_1512933, EPI_ISL_1512934, EPI_ISL_1512936, EPI_ISL_1512937, EPI_ISL_1512938, EPI_ISL_1512940, EPI_ISL_1512941, EPI_ISL_1512942, EPI_ISL_1512944, EPI_ISL_1512945, EPI_ISL_1512946, EPI_ISL_1512948, EPI_ISL_1512950, EPI_ISL_1512951, EPI_ISL_1512952, EPI_ISL_1512954, EPI_ISL_1512956, EPI_ISL_1512960, EPI_ISL_1512961, EPI_ISL_1512963, EPI_ISL_1512964, EPI_ISL_1512965, EPI_ISL_1512966, EPI_ISL_1512967, EPI_ISL_1512969, EPI_ISL_1512970, EPI_ISL_1512971, EPI_ISL_1512972, EPI_ISL_1512973, EPI_ISL_1512974, EPI_ISL_1512975, EPI_ISL_1512976, EPI_ISL_1512977, EPI_ISL_1512978, EPI_ISL_1512979, EPI_ISL_1512980, EPI_ISL_1512981, EPI_ISL_1512982, EPI_ISL_1512983, EPI_ISL_1512984, EPI_ISL_1512985, EPI_ISL_1512986, EPI_ISL_1512987, EPI_ISL_1512988, EPI_ISL_1512989, EPI_ISL_1512990, EPI_ISL_1512991, EPI_ISL_1512992, EPI_ISL_1512993, EPI_ISL_1512995, EPI_ISL_1512996, EPI_ISL_1512997, EPI_ISL_1512998, EPI_ISL_1512999, EPI_ISL_1513001, EPI_ISL_1513002, EPI_ISL_1513003, EPI_ISL_1513004, EPI_ISL_1513005, EPI_ISL_1513006, EPI_ISL_1513007, EPI_ISL_1513008, EPI_ISL_1513009, EPI_ISL_1513010, EPI_ISL_1513012, EPI_ISL_1513014, EPI_ISL_1513015, EPI_ISL_1513016, EPI_ISL_1513017, EPI_ISL_1513018, EPI_ISL_1513019, EPI_ISL_1513020, EPI_ISL_1513021, EPI_ISL_1513022, EPI_ISL_1513023, EPI_ISL_1513024, EPI_ISL_1513026, EPI_ISL_1513027, EPI_ISL_1513028, EPI_ISL_1513029, EPI_ISL_1513031, EPI_ISL_1513032, EPI_ISL_1513033, EPI_ISL_1513034, EPI_ISL_1513035, EPI_ISL_1513036, EPI_ISL_1513037, EPI_ISL_1513038, EPI_ISL_1513039, EPI_ISL_1513040, EPI_ISL_1513043, EPI_ISL_1513044, EPI_ISL_1513045, EPI_ISL_1513046, EPI_ISL_1513047, EPI_ISL_1513048, EPI_ISL_1513049, EPI_ISL_1513050, EPI_ISL_1513051, EPI_ISL_1513052, EPI_ISL_1513053, EPI_ISL_1513054, EPI_ISL_1513055, EPI_ISL_1513056, EPI_ISL_1513057, EPI_ISL_1513058, EPI_ISL_1513059, EPI_ISL_1513060, EPI_ISL_1513061, EPI_ISL_1513062, EPI_ISL_1513063, EPI_ISL_1513064, EPI_ISL_1513065, EPI_ISL_1513066, EPI_ISL_1513067, EPI_ISL_1513068, EPI_ISL_1513070, EPI_ISL_1513071, EPI_ISL_1513072, EPI_ISL_1513073, EPI_ISL_1513074, EPI_ISL_1513075, EPI_ISL_1513076, EPI_ISL_1513077, EPI_ISL_1513078, EPI_ISL_1513079, EPI_ISL_1513080, EPI_ISL_1513081, EPI_ISL_1513082, EPI_ISL_1513083, EPI_ISL_1513084, EPI_ISL_1513085, EPI_ISL_1513086, EPI_ISL_1513087, EPI_ISL_1513088, EPI_ISL_1513089, EPI_ISL_1513090, EPI_ISL_1513091, EPI_ISL_1513092, EPI_ISL_1513093, EPI_ISL_1513094, EPI_ISL_1513095, EPI_ISL_1513096, EPI_ISL_1513097, EPI_ISL_1513098, EPI_ISL_1513099, EPI_ISL_1513100, EPI_ISL_1513101, EPI_ISL_1513102, EPI_ISL_1513103, EPI_ISL_1513104, EPI_ISL_1513105, EPI_ISL_1513107, EPI_ISL_1513108, EPI_ISL_1513109, EPI_ISL_1513110, EPI_ISL_1513111, EPI_ISL_1513112, EPI_ISL_1513114, EPI_ISL_1513115, EPI_ISL_1513116, EPI_ISL_1513117, EPI_ISL_1513118, EPI_ISL_1513119, EPI_ISL_1513121, EPI_ISL_1513122, EPI_ISL_1513123, EPI_ISL_1513124, EPI_ISL_1513125, EPI_ISL_1513126, EPI_ISL_1513127, EPI_ISL_1513128, EPI_ISL_1513129, EPI_ISL_1513131, EPI_ISL_1513132, EPI_ISL_1513133, EPI_ISL_1513135, EPI_ISL_1513136, EPI_ISL_1513137, EPI_ISL_1513138 | | | |
| see above | Infinity Biologix | Centers for Disease Control and Prevention Division of Viral Diseases, Pathogen Discovery | Dakota Howard, Dhwani Batra, Peter W. Cook, Kara Moser, Adrian Paskey, Jason Caravas, Benjamin Rambo-Martin, Shatavia Morrison, Christopher Gulvick, Scott Sammons, Yvette Unoarumhi, Darlene Wagner, Matthew Schmerer, Christian Bixby, Yihe Wang, Jonathan Schultz, Chirayu Goswami, Russ Hager, Robin Grimwood, Clinton R. Paden, Duncan MacCannell |
| EPI_ISL_1513139, EPI_ISL_1513140, EPI_ISL_1513141, EPI_ISL_1513142, EPI_ISL_1513143, EPI_ISL_1513144, EPI_ISL_1513145, EPI_ISL_1513146, EPI_ISL_1513147, EPI_ISL_1513148, EPI_ISL_1513149, EPI_ISL_1513150, EPI_ISL_1513151, EPI_ISL_1513152, EPI_ISL_1513153, EPI_ISL_1513154, EPI_ISL_1513155, EPI_ISL_1513156, EPI_ISL_1513157, EPI_ISL_1513159, EPI_ISL_1513160, EPI_ISL_1513161, EPI_ISL_1513163, EPI_ISL_1513164, EPI_ISL_1513165, EPI_ISL_1513166, EPI_ISL_1513167, EPI_ISL_1513168, EPI_ISL_1513169, EPI_ISL_1513170, EPI_ISL_1513171, EPI_ISL_1513172, EPI_ISL_1513173, EPI_ISL_1513176, EPI_ISL_1513177, EPI_ISL_1513178, EPI_ISL_1513180, EPI_ISL_1513181, EPI_ISL_1513182, EPI_ISL_1513184, EPI_ISL_1513185, EPI_ISL_1513186, EPI_ISL_1513187, EPI_ISL_1513188, EPI_ISL_1513189, EPI_ISL_1513190, EPI_ISL_1513192, EPI_ISL_1513193, EPI_ISL_1513194, EPI_ISL_1513195, EPI_ISL_1513197, EPI_ISL_1513198, EPI_ISL_1513199, EPI_ISL_1513200, EPI_ISL_1513201, EPI_ISL_1513202, EPI_ISL_1513203, EPI_ISL_1513206, EPI_ISL_1513207, EPI_ISL_1513208, EPI_ISL_1513209, EPI_ISL_1513210, EPI_ISL_1513211, EPI_ISL_1513212, EPI_ISL_1513213, EPI_ISL_1513214, EPI_ISL_1513215, EPI_ISL_1513216, EPI_ISL_1513217, EPI_ISL_1513218, EPI_ISL_1513219, EPI_ISL_1513220, EPI_ISL_1513221, EPI_ISL_1513222, EPI_ISL_1513223, EPI_ISL_1513224, EPI_ISL_1513225, EPI_ISL_1513226, EPI_ISL_1513227, EPI_ISL_1513228, EPI_ISL_1513229, EPI_ISL_1513230, EPI_ISL_1513231, EPI_ISL_1513232, EPI_ISL_1513233, EPI_ISL_1513234, EPI_ISL_1513235, EPI_ISL_1513236, EPI_ISL_1513237, EPI_ISL_1513238, EPI_ISL_1513239, EPI_ISL_1513240, EPI_ISL_1513241, EPI_ISL_1513242, EPI_ISL_1513243, EPI_ISL_1513244, EPI_ISL_1513245, EPI_ISL_1513246, EPI_ISL_1513247, EPI_ISL_1513248, EPI_ISL_1513250, EPI_ISL_1513251, EPI_ISL_1513252, EPI_ISL_1513253, EPI_ISL_1513254, EPI_ISL_1513255, EPI_ISL_1513256, EPI_ISL_1513257, EPI_ISL_1513258, EPI_ISL_1513259, EPI_ISL_1513260, EPI_ISL_1513261, EPI_ISL_1513262, EPI_ISL_1513263, EPI_ISL_1513264, EPI_ISL_1513266, EPI_ISL_1513267, EPI_ISL_1513268, EPI_ISL_1513269, EPI_ISL_1513270, EPI_ISL_1513271, EPI_ISL_1513272, EPI_ISL_1513273, EPI_ISL_1513274, EPI_ISL_1513275, EPI_ISL_1513276, EPI_ISL_1513277, EPI_ISL_1513278, EPI_ISL_1513279, EPI_ISL_1513280, EPI_ISL_1513281, EPI_ISL_1513282, EPI_ISL_1513283, EPI_ISL_1513284, EPI_ISL_1513285, EPI_ISL_1513286, EPI_ISL_1513287, EPI_ISL_1513288, EPI_ISL_1513289, EPI_ISL_1513290, EPI_ISL_1513291, EPI_ISL_1513292, EPI_ISL_1513293, EPI_ISL_1513295, EPI_ISL_1513297, EPI_ISL_1513298, EPI_ISL_1513299, EPI_ISL_1513300, EPI_ISL_1513301, EPI_ISL_1513302, EPI_ISL_1513303, EPI_ISL_1513304, EPI_ISL_1513305, EPI_ISL_1513306, EPI_ISL_1513307, EPI_ISL_1513308, EPI_ISL_1513309, EPI_ISL_1513311, EPI_ISL_1513312, EPI_ISL_1513313, EPI_ISL_1513314, EPI_ISL_1513315, EPI_ISL_1513316, EPI_ISL_1513318, EPI_ISL_1513319, EPI_ISL_1513320, EPI_ISL_1513321, EPI_ISL_1513322, EPI_ISL_1513323, EPI_ISL_1513324, EPI_ISL_1513325, EPI_ISL_1513326, EPI_ISL_1513327, EPI_ISL_1513328, EPI_ISL_1513329, EPI_ISL_1513330, EPI_ISL_1513331, EPI_ISL_1513332, EPI_ISL_1513333, EPI_ISL_1513334, EPI_ISL_1513335, EPI_ISL_1513336, EPI_ISL_1513337, EPI_ISL_1513338, EPI_ISL_1513339, EPI_ISL_1513340, EPI_ISL_1513341, EPI_ISL_1513343, EPI_ISL_1513344, EPI_ISL_1513345, EPI_ISL_1513347, EPI_ISL_1513348, EPI_ISL_1513349, EPI_ISL_1513350, EPI_ISL_1513351, EPI_ISL_1513352, EPI_ISL_1513353, EPI_ISL_1513354, EPI_ISL_1513355, EPI_ISL_1513356, EPI_ISL_1513357, EPI_ISL_1513358, EPI_ISL_1513360, EPI_ISL_1513362, EPI_ISL_1513363, EPI_ISL_1513365, EPI_ISL_1513366, EPI_ISL_1513367, EPI_ISL_1513368, EPI_ISL_1513369, EPI_ISL_1513370, EPI_ISL_1513371, EPI_ISL_1513372, EPI_ISL_1513373, EPI_ISL_1513374, EPI_ISL_1513375, EPI_ISL_1513376, EPI_ISL_1513377, EPI_ISL_1513378, EPI_ISL_1513379, EPI_ISL_1513380, EPI_ISL_1513381, EPI_ISL_1513382, EPI_ISL_1513383, EPI_ISL_1513384, EPI_ISL_1513385, EPI_ISL_1513386, EPI_ISL_1513387, EPI_ISL_1513388, EPI_ISL_1513389, EPI_ISL_1513390, EPI_ISL_1513391, EPI_ISL_1513392, EPI_ISL_1513394, EPI_ISL_1513395, EPI_ISL_1513396, EPI_ISL_1513397, EPI_ISL_1513398, EPI_ISL_1513399, EPI_ISL_1513400, EPI_ISL_1513403, EPI_ISL_1513404, EPI_ISL_1513405, EPI_ISL_1513406, EPI_ISL_1513407, EPI_ISL_1513408, EPI_ISL_1513409, EPI_ISL_1513410, EPI_ISL_1513411, EPI_ISL_1513412, EPI_ISL_1513413, EPI_ISL_1513414, EPI_ISL_1513415, EPI_ISL_1513416, EPI_ISL_1513417, EPI_ISL_1513418, EPI_ISL_1513419, EPI_ISL_1513420, EPI_ISL_1513421, EPI_ISL_1513422, EPI_ISL_1513423, EPI_ISL_1513424, EPI_ISL_1513425, EPI_ISL_1513426, EPI_ISL_1513427, EPI_ISL_1513428, EPI_ISL_1513429, EPI_ISL_1513430, EPI_ISL_1513431, EPI_ISL_1513432, EPI_ISL_1513433, EPI_ISL_1513434, EPI_ISL_1513435, EPI_ISL_1513436, EPI_ISL_1513437, EPI_ISL_1513438, EPI_ISL_1513439, EPI_ISL_1513441, EPI_ISL_1513442, EPI_ISL_1513443, EPI_ISL_1513444, EPI_ISL_1513445, EPI_ISL_1513446, EPI_ISL_1513447, EPI_ISL_1513448, EPI_ISL_1513449, EPI_ISL_1513450, EPI_ISL_1513451, EPI_ISL_1513452, EPI_ISL_1513453, EPI_ISL_1513454, EPI_ISL_1513455, EPI_ISL_1513456, EPI_ISL_1513457, EPI_ISL_1513458, EPI_ISL_1513459, EPI_ISL_1513460, EPI_ISL_1513461, EPI_ISL_1513462, EPI_ISL_1513463, EPI_ISL_1513464, EPI_ISL_1513465, EPI_ISL_1513466, EPI_ISL_1513467, EPI_ISL_1513468, EPI_ISL_1513469, EPI_ISL_1513470, EPI_ISL_1513471, EPI_ISL_1513472, EPI_ISL_1513473, EPI_ISL_1513474, EPI_ISL_1513475, EPI_ISL_1513476, EPI_ISL_1513477, EPI_ISL_1513478, EPI_ISL_1513479, EPI_ISL_1513480, EPI_ISL_1513481, EPI_ISL_1513482, EPI_ISL_1513483, EPI_ISL_1513484, EPI_ISL_1513485, EPI_ISL_1513486, EPI_ISL_1513487, EPI_ISL_1513488, EPI_ISL_1513489, EPI_ISL_1513490, EPI_ISL_1513492, EPI_ISL_1513493, EPI_ISL_1513494, EPI_ISL_1513495, EPI_ISL_1513496, EPI_ISL_1513497, EPI_ISL_1513498, EPI_ISL_1513499, EPI_ISL_1513500, EPI_ISL_1513503, EPI_ISL_1513504, EPI_ISL_1513505, EPI_ISL_1513506, EPI_ISL_1513507, EPI_ISL_1513508, EPI_ISL_1513509, EPI_ISL_1513510, EPI_ISL_1513511, EPI_ISL_1513512, EPI_ISL_1513513, EPI_ISL_1513514, EPI_ISL_1513515, EPI_ISL_1513516, EPI_ISL_1513517, EPI_ISL_1513519, EPI_ISL_1513521, EPI_ISL_1513522, EPI_ISL_1513523, EPI_ISL_1513524, EPI_ISL_1513525, EPI_ISL_1513526, EPI_ISL_1513527, EPI_ISL_1513528, EPI_ISL_1513529, EPI_ISL_1513530, EPI_ISL_1513531, EPI_ISL_1513532, EPI_ISL_1513533, EPI_ISL_1513534, EPI_ISL_1513535, EPI_ISL_1513536, EPI_ISL_1513537, EPI_ISL_1513538, EPI_ISL_1513539, EPI_ISL_1513540, EPI_ISL_1513541, EPI_ISL_1513542, EPI_ISL_1513543, EPI_ISL_1513544, EPI_ISL_1513545, EPI_ISL_1513546, EPI_ISL_1513547, EPI_ISL_1513548, EPI_ISL_1513549, EPI_ISL_1513550, EPI_ISL_1513551, EPI_ISL_1513552, EPI_ISL_1513553, EPI_ISL_1513554, EPI_ISL_1513555, EPI_ISL_1513556, EPI_ISL_1513557, EPI_ISL_1513558, EPI_ISL_1513559, EPI_ISL_1513560, EPI_ISL_1513561, EPI_ISL_1513562, EPI_ISL_1513563, EPI_ISL_1513564, EPI_ISL_1513565, EPI_ISL_1513566, EPI_ISL_1513567, EPI_ISL_1513568, EPI_ISL_1513569, EPI_ISL_1513570, EPI_ISL_1513571, EPI_ISL_1513572, EPI_ISL_1513573, EPI_ISL_1513574, EPI_ISL_1513575, EPI_ISL_1513576, EPI_ISL_1513577, EPI_ISL_1513578, EPI_ISL_1513579, EPI_ISL_1513580, EPI_ISL_1513581, EPI_ISL_1513582, EPI_ISL_1513583, EPI_ISL_1513584, EPI_ISL_1513585, EPI_ISL_1513586, EPI_ISL_1513587, EPI_ISL_1513588, EPI_ISL_1513589, EPI_ISL_1513590, EPI_ISL_1513591, EPI_ISL_1513593, EPI_ISL_1513594, EPI_ISL_1513595, EPI_ISL_1513596, EPI_ISL_1513597, EPI_ISL_1513598, EPI_ISL_1513599, EPI_ISL_1513600, EPI_ISL_1513601, EPI_ISL_1513602, EPI_ISL_1513604, EPI_ISL_1513605, EPI_ISL_1513606, EPI_ISL_1513607, EPI_ISL_1513608, EPI_ISL_1513609, EPI_ISL_1513610, EPI_ISL_1513611, EPI_ISL_1513612, EPI_ISL_1513613, EPI_ISL_1513614, EPI_ISL_1513615, EPI_ISL_1513616, EPI_ISL_1513617, EPI_ISL_1513618, EPI_ISL_1513619, EPI_ISL_1513620, EPI_ISL_1513621, EPI_ISL_1513622, EPI_ISL_1513623, EPI_ISL_1513624, EPI_ISL_1513625, EPI_ISL_1513626, EPI_ISL_1513627, EPI_ISL_1513628, EPI_ISL_1513629, EPI_ISL_1513630 | | | |
| see above | Aegis Sciences Corporation | Centers for Disease Control and Prevention Division of Viral Diseases, Pathogen Discovery | Dakota Howard, Dhwani Batra, Peter W. Cook, Kara Moser, Adrian Paskey, Jason Caravas, Benjamin Rambo-Martin, Shatavia Morrison, Christopher Gulvick, Scott Sammons, Yvette Unoarumhi, Darlene Wagner, Matthew Schmerer, Cyndi Clark, Patrick Campbell, Rob Case, Vikramsinha Ghorpade, Holly Houdeshell, Ola Kvalvaag, Dillon Nall, Ethan Sanders, Alec Vest, Shaun Westlund, Matthew Hardison, Clinton R. Paden, Duncan MacCannell |
| EPI_ISL_1513631, EPI_ISL_1513632, EPI_ISL_1513633, EPI_ISL_1513634, EPI_ISL_1513639, EPI_ISL_1513641, EPI_ISL_1513645, EPI_ISL_1513646, EPI_ISL_1513647, EPI_ISL_1513648, EPI_ISL_1513650, EPI_ISL_1513652, EPI_ISL_1513655, EPI_ISL_1513656, EPI_ISL_1513657, EPI_ISL_1513658, EPI_ISL_1513660, EPI_ISL_1513663, EPI_ISL_1513664, EPI_ISL_1513665, EPI_ISL_1513667, EPI_ISL_1513668, EPI_ISL_1513669, EPI_ISL_1513670, EPI_ISL_1513671, EPI_ISL_1513672, EPI_ISL_1513673, EPI_ISL_1513675, EPI_ISL_1513676, EPI_ISL_1513678, EPI_ISL_1513679, EPI_ISL_1513681, EPI_ISL_1513683, EPI_ISL_1513684, EPI_ISL_1513685, EPI_ISL_1513686, EPI_ISL_1513687, EPI_ISL_1513688, EPI_ISL_1513689, EPI_ISL_1513690, EPI_ISL_1513691, EPI_ISL_1513692, EPI_ISL_1513693, EPI_ISL_1513694, EPI_ISL_1513695, EPI_ISL_1513696, EPI_ISL_1513697, EPI_ISL_1513698, EPI_ISL_1513699, EPI_ISL_1513700, EPI_ISL_1513703, EPI_ISL_1513704, EPI_ISL_1513705, EPI_ISL_1513706, EPI_ISL_1513707, EPI_ISL_1513708, EPI_ISL_1513711, EPI_ISL_1513712, EPI_ISL_1513716, EPI_ISL_1513718, EPI_ISL_1513720, EPI_ISL_1513721, EPI_ISL_1513723, EPI_ISL_1513724, EPI_ISL_1513725, EPI_ISL_1513726, EPI_ISL_1513727, EPI_ISL_1513728, EPI_ISL_1513729, EPI_ISL_1513730, EPI_ISL_1513731, EPI_ISL_1513732, EPI_ISL_1513733, EPI_ISL_1513735, EPI_ISL_1513736, EPI_ISL_1513739, EPI_ISL_1513741, EPI_ISL_1513742, EPI_ISL_1513743, EPI_ISL_1513744, EPI_ISL_1513745, EPI_ISL_1513746, EPI_ISL_1513747, EPI_ISL_1513748, EPI_ISL_1513753, EPI_ISL_1513754, EPI_ISL_1513755, EPI_ISL_1513758, EPI_ISL_1513763, EPI_ISL_1513764, EPI_ISL_1513765, EPI_ISL_1513766, EPI_ISL_1513767, EPI_ISL_1513768, EPI_ISL_1513769, EPI_ISL_1513770, EPI_ISL_1513771, EPI_ISL_1513772, EPI_ISL_1513773, EPI_ISL_1513774, EPI_ISL_1513775, EPI_ISL_1513776, EPI_ISL_1513777, EPI_ISL_1513778, EPI_ISL_1513779, EPI_ISL_1513780, EPI_ISL_1513782, EPI_ISL_1513783, EPI_ISL_1513784, EPI_ISL_1513786, | | | |

| | | | |
|---|---|---|---|
| EPI_ISL_1513788, EPI_ISL_1513789, EPI_ISL_1513790, EPI_ISL_1513793, EPI_ISL_1513794, EPI_ISL_1513796, EPI_ISL_1513798, EPI_ISL_1513799, EPI_ISL_1513800, EPI_ISL_1513801, EPI_ISL_1513802, EPI_ISL_1513803, EPI_ISL_1513804, EPI_ISL_1513805, EPI_ISL_1513806, EPI_ISL_1513808, EPI_ISL_1513809, EPI_ISL_1513811, EPI_ISL_1513812, EPI_ISL_1513813, EPI_ISL_1513814, EPI_ISL_1513815, EPI_ISL_1513817, EPI_ISL_1513818, EPI_ISL_1513820, EPI_ISL_1513821, EPI_ISL_1513823, EPI_ISL_1513824, EPI_ISL_1513825, EPI_ISL_1513826, EPI_ISL_1513827, EPI_ISL_1513828, EPI_ISL_1513829, EPI_ISL_1513830, EPI_ISL_1513831, EPI_ISL_1513832, EPI_ISL_1513833, EPI_ISL_1513834, EPI_ISL_1513835, EPI_ISL_1513836, EPI_ISL_1513837, EPI_ISL_1513838, EPI_ISL_1513840, EPI_ISL_1513841, EPI_ISL_1513842, EPI_ISL_1513843, EPI_ISL_1513844, EPI_ISL_1513845, EPI_ISL_1513847, EPI_ISL_1513848, EPI_ISL_1513849, EPI_ISL_1513850, EPI_ISL_1513851, EPI_ISL_1513853, EPI_ISL_1513854, EPI_ISL_1513856, EPI_ISL_1513858, EPI_ISL_1513859, EPI_ISL_1513860, EPI_ISL_1513863, EPI_ISL_1513864, EPI_ISL_1513865, EPI_ISL_1513866, EPI_ISL_1513867, EPI_ISL_1513868, EPI_ISL_1513869, EPI_ISL_1513870, EPI_ISL_1513872, EPI_ISL_1513874, EPI_ISL_1513876, EPI_ISL_1513877, EPI_ISL_1513879, EPI_ISL_1513880, EPI_ISL_1513881, EPI_ISL_1513882, EPI_ISL_1513883, EPI_ISL_1513885, EPI_ISL_1513886, EPI_ISL_1513887, EPI_ISL_1513888, EPI_ISL_1513890, EPI_ISL_1513891, EPI_ISL_1513892, EPI_ISL_1513893, EPI_ISL_1513894, EPI_ISL_1513895, EPI_ISL_1513896, EPI_ISL_1513897, EPI_ISL_1513898, EPI_ISL_1513899, EPI_ISL_1513900, EPI_ISL_1513901, EPI_ISL_1513902, EPI_ISL_1513904, EPI_ISL_1513907, EPI_ISL_1513908, EPI_ISL_1513909, EPI_ISL_1513930, EPI_ISL_1513946, EPI_ISL_1513952, EPI_ISL_1513954, EPI_ISL_1513955, EPI_ISL_1513958, EPI_ISL_1513959, EPI_ISL_1513961, EPI_ISL_1513963, EPI_ISL_1513964, EPI_ISL_1513965, EPI_ISL_1513969, EPI_ISL_1513972, EPI_ISL_1513973, EPI_ISL_1514013, EPI_ISL_1514015, EPI_ISL_1514016, EPI_ISL_1514017, EPI_ISL_1514018, EPI_ISL_1514019, EPI_ISL_1514020, EPI_ISL_1514022, EPI_ISL_1514023, EPI_ISL_1514024, EPI_ISL_1514025, EPI_ISL_1514026, EPI_ISL_1514027, EPI_ISL_1514028, EPI_ISL_1514029, EPI_ISL_1514030, EPI_ISL_1514031, EPI_ISL_1514032, EPI_ISL_1514033, EPI_ISL_1514035, EPI_ISL_1514036, EPI_ISL_1514037, EPI_ISL_1514039, EPI_ISL_1514040, EPI_ISL_1514041, EPI_ISL_1514042, EPI_ISL_1514043, EPI_ISL_1514044, EPI_ISL_1514045, EPI_ISL_1514046, EPI_ISL_1514047, EPI_ISL_1514048, EPI_ISL_1514049, EPI_ISL_1514050, EPI_ISL_1514052, EPI_ISL_1514054, EPI_ISL_1514055, EPI_ISL_1514056, EPI_ISL_1514057, EPI_ISL_1514058, EPI_ISL_1514059, EPI_ISL_1514061, EPI_ISL_1514062, EPI_ISL_1514064, EPI_ISL_1514066, EPI_ISL_1514067, EPI_ISL_1514069, EPI_ISL_1514070, EPI_ISL_1514071, EPI_ISL_1514072, EPI_ISL_1514074, EPI_ISL_1514076, EPI_ISL_1514077, EPI_ISL_1514078, EPI_ISL_1514079, EPI_ISL_1514081, EPI_ISL_1514082, EPI_ISL_1514083, EPI_ISL_1514085, EPI_ISL_1514086, EPI_ISL_1514087, EPI_ISL_1514088, EPI_ISL_1514090, EPI_ISL_1514091, EPI_ISL_1514092, EPI_ISL_1514093, EPI_ISL_1514094, EPI_ISL_1514095, EPI_ISL_1514097, EPI_ISL_1514099, EPI_ISL_1514100, EPI_ISL_1514104, EPI_ISL_1514105, EPI_ISL_1514106, EPI_ISL_1514107, EPI_ISL_1514110, EPI_ISL_1514111, EPI_ISL_1514112, EPI_ISL_1514114, EPI_ISL_1514118, EPI_ISL_1514119, EPI_ISL_1514120, EPI_ISL_1514121, EPI_ISL_1514122, EPI_ISL_1514123, EPI_ISL_1514124, EPI_ISL_1514125, EPI_ISL_1514126, EPI_ISL_1514128, EPI_ISL_1514129, EPI_ISL_1514130, EPI_ISL_1514131, EPI_ISL_1514133, EPI_ISL_1514134, EPI_ISL_1514137, EPI_ISL_1514138, EPI_ISL_1514139, EPI_ISL_1514142, EPI_ISL_1514146, EPI_ISL_1514147, EPI_ISL_1514149, EPI_ISL_1514150, EPI_ISL_1514151, EPI_ISL_1514152, EPI_ISL_1514154, EPI_ISL_1514155, EPI_ISL_1514156, EPI_ISL_1514157, EPI_ISL_1514158, EPI_ISL_1514160, EPI_ISL_1514161, EPI_ISL_1514162, EPI_ISL_1514163, EPI_ISL_1514165, EPI_ISL_1514166, EPI_ISL_1514168, EPI_ISL_1514170, EPI_ISL_1514172, EPI_ISL_1514173, EPI_ISL_1514174, EPI_ISL_1514177, EPI_ISL_1514178, EPI_ISL_1514179, EPI_ISL_1514181, EPI_ISL_1514182, EPI_ISL_1514186, EPI_ISL_1514187, EPI_ISL_1514188, EPI_ISL_1514190, EPI_ISL_1514191, EPI_ISL_1514192, EPI_ISL_1514193, EPI_ISL_1514194, EPI_ISL_1514195, EPI_ISL_1514197, EPI_ISL_1514198, EPI_ISL_1514199, EPI_ISL_1514200, EPI_ISL_1514201, EPI_ISL_1514203, EPI_ISL_1514204, EPI_ISL_1514205, EPI_ISL_1514208, EPI_ISL_1514209, EPI_ISL_1514211, EPI_ISL_1514213, EPI_ISL_1514215, EPI_ISL_1514216, EPI_ISL_1514220, EPI_ISL_1514221, EPI_ISL_1514222, EPI_ISL_1514223, EPI_ISL_1514224, EPI_ISL_1514225, EPI_ISL_1514226, EPI_ISL_1514228, EPI_ISL_1514229, EPI_ISL_1514230, EPI_ISL_1514231, EPI_ISL_1514232, EPI_ISL_1514233, EPI_ISL_1514234, EPI_ISL_1514235, EPI_ISL_1514236, EPI_ISL_1514237, EPI_ISL_1514238, EPI_ISL_1514239, EPI_ISL_1514240, EPI_ISL_1514241, EPI_ISL_1514242, EPI_ISL_1514244, EPI_ISL_1514245, EPI_ISL_1514246, EPI_ISL_1514247, EPI_ISL_1514248, EPI_ISL_1514249, EPI_ISL_1514250, EPI_ISL_1514251, EPI_ISL_1514252, EPI_ISL_1514253, EPI_ISL_1514256, EPI_ISL_1514257, EPI_ISL_1514258, EPI_ISL_1514259, EPI_ISL_1514261, EPI_ISL_1514263, EPI_ISL_1514264, EPI_ISL_1514265, EPI_ISL_1514266, EPI_ISL_1514268, EPI_ISL_1514270, EPI_ISL_1514271, EPI_ISL_1514272, EPI_ISL_1514274, EPI_ISL_1514275, EPI_ISL_1514277, EPI_ISL_1514284, EPI_ISL_1514285, EPI_ISL_1514286, EPI_ISL_1514289, EPI_ISL_1514290, EPI_ISL_1514291, EPI_ISL_1514293, EPI_ISL_1514294, EPI_ISL_1514297, EPI_ISL_1514298, EPI_ISL_1514299, EPI_ISL_1514300, EPI_ISL_1514301, EPI_ISL_1514302, EPI_ISL_1514303, EPI_ISL_1514304, EPI_ISL_1514306, EPI_ISL_1514308, EPI_ISL_1514309, EPI_ISL_1514311, EPI_ISL_1514314, EPI_ISL_1514315, EPI_ISL_1514316, EPI_ISL_1514317, EPI_ISL_1514318, EPI_ISL_1514319, EPI_ISL_1514320, EPI_ISL_1514321, EPI_ISL_1514322, EPI_ISL_1514323, EPI_ISL_1514325, EPI_ISL_1514326, EPI_ISL_1514328, EPI_ISL_1514329, EPI_ISL_1514331, EPI_ISL_1514332, EPI_ISL_1514334, EPI_ISL_1514335, EPI_ISL_1514338, EPI_ISL_1514339, EPI_ISL_1514340, EPI_ISL_1514342, EPI_ISL_1514343, EPI_ISL_1514344, EPI_ISL_1514345, EPI_ISL_1514346, EPI_ISL_1514349, EPI_ISL_1514350, EPI_ISL_1514351, EPI_ISL_1514352, EPI_ISL_1514353, EPI_ISL_1514354, EPI_ISL_1514356, EPI_ISL_1514357, EPI_ISL_1514358, EPI_ISL_1514359, EPI_ISL_1514360, EPI_ISL_1514362, EPI_ISL_1514363, EPI_ISL_1514364, EPI_ISL_1514365, EPI_ISL_1514366, EPI_ISL_1514367, EPI_ISL_1514368, EPI_ISL_1514369, EPI_ISL_1514371, EPI_ISL_1514372, EPI_ISL_1514373, EPI_ISL_1514376, EPI_ISL_1514377, EPI_ISL_1514379, EPI_ISL_1514383, EPI_ISL_1514384, EPI_ISL_1514385, EPI_ISL_1514386, EPI_ISL_1514387, EPI_ISL_1514388, EPI_ISL_1514390, EPI_ISL_1514391, EPI_ISL_1514392, EPI_ISL_1514394, EPI_ISL_1514395, EPI_ISL_1514396, EPI_ISL_1514398, EPI_ISL_1514402, EPI_ISL_1514403, EPI_ISL_1514404, EPI_ISL_1514405, EPI_ISL_1514406, EPI_ISL_1514407, EPI_ISL_1514409, EPI_ISL_1514410, EPI_ISL_1514411, EPI_ISL_1514412, EPI_ISL_1514413, EPI_ISL_1514415, EPI_ISL_1514417, EPI_ISL_1514418, EPI_ISL_1514419, EPI_ISL_1514420, EPI_ISL_1514422, EPI_ISL_1514423, EPI_ISL_1514424, EPI_ISL_1514425, EPI_ISL_1514427, EPI_ISL_1514429, EPI_ISL_1514430, EPI_ISL_1514431, EPI_ISL_1514432, EPI_ISL_1514433, EPI_ISL_1514434, EPI_ISL_1514436, EPI_ISL_1514437, EPI_ISL_1514438, EPI_ISL_1514439, EPI_ISL_1514440, EPI_ISL_1514441, EPI_ISL_1514442, EPI_ISL_1514443, EPI_ISL_1514447, EPI_ISL_1514448, EPI_ISL_1514450, EPI_ISL_1514451, EPI_ISL_1514452, EPI_ISL_1514454, EPI_ISL_1514455, EPI_ISL_1514456, EPI_ISL_1514458, EPI_ISL_1514464, EPI_ISL_1514465, EPI_ISL_1514466, EPI_ISL_1514468, EPI_ISL_1514469, EPI_ISL_1514470, EPI_ISL_1514471, EPI_ISL_1514474, EPI_ISL_1514475, EPI_ISL_1514476, EPI_ISL_1514477, EPI_ISL_1514478, EPI_ISL_1514479, EPI_ISL_1514481, EPI_ISL_1514482, EPI_ISL_1514483, EPI_ISL_1514486, EPI_ISL_1514490, EPI_ISL_1514491, EPI_ISL_1514492, EPI_ISL_1514493, EPI_ISL_1514495, EPI_ISL_1514496, EPI_ISL_1514497, EPI_ISL_1514498, EPI_ISL_1514499, EPI_ISL_1514500, EPI_ISL_1514501, EPI_ISL_1514502, EPI_ISL_1514505, EPI_ISL_1514507, EPI_ISL_1514511, EPI_ISL_1514512, EPI_ISL_1514513, EPI_ISL_1514514, EPI_ISL_1514515, EPI_ISL_1514516, EPI_ISL_1514517, EPI_ISL_1514518, EPI_ISL_1514519, EPI_ISL_1514520, EPI_ISL_1514521, EPI_ISL_1514523, EPI_ISL_1514524, EPI_ISL_1514526, EPI_ISL_1514529, EPI_ISL_1514530, EPI_ISL_1514531, EPI_ISL_1514533, EPI_ISL_1514534, EPI_ISL_1514535, EPI_ISL_1514536, EPI_ISL_1514538, EPI_ISL_1514540, EPI_ISL_1514541, EPI_ISL_1514542, EPI_ISL_1514543, EPI_ISL_1514544, EPI_ISL_1514545, EPI_ISL_1514546, EPI_ISL_1514547, EPI_ISL_1514548, EPI_ISL_1514549, EPI_ISL_1514550, EPI_ISL_1514553, EPI_ISL_1514558, EPI_ISL_1514559, EPI_ISL_1514560, EPI_ISL_1514561, EPI_ISL_1514562, EPI_ISL_1514565, EPI_ISL_1514566, EPI_ISL_1514568, EPI_ISL_1514569, EPI_ISL_1514570, EPI_ISL_1514571, EPI_ISL_1514572, EPI_ISL_1514573, EPI_ISL_1514574, EPI_ISL_1514575, EPI_ISL_1514576, EPI_ISL_1514577, EPI_ISL_1514578, EPI_ISL_1514580, EPI_ISL_1514582, EPI_ISL_1514583, EPI_ISL_1514585, EPI_ISL_1514586, EPI_ISL_1514589, EPI_ISL_1514590, EPI_ISL_1514592, EPI_ISL_1514594, EPI_ISL_1514595, EPI_ISL_1514596, EPI_ISL_1514597, EPI_ISL_1514598, EPI_ISL_1514601, EPI_ISL_1514604, EPI_ISL_1514605, EPI_ISL_1514607, EPI_ISL_1514608, EPI_ISL_1514611, EPI_ISL_1514612, EPI_ISL_1514613, EPI_ISL_1514614, EPI_ISL_1514615, EPI_ISL_1514617, EPI_ISL_1514619, EPI_ISL_1514620, EPI_ISL_1514621, EPI_ISL_1514622, EPI_ISL_1514627, EPI_ISL_1514629, EPI_ISL_1514630, EPI_ISL_1514631, EPI_ISL_1514632, EPI_ISL_1514635, EPI_ISL_1514637, EPI_ISL_1514638, EPI_ISL_1514640, EPI_ISL_1514642, EPI_ISL_1514643, EPI_ISL_1514644, EPI_ISL_1514645, EPI_ISL_1514647, EPI_ISL_1514648, EPI_ISL_1514649, EPI_ISL_1514651, EPI_ISL_1514654, EPI_ISL_1514655, EPI_ISL_1514656, EPI_ISL_1514657, EPI_ISL_1514658, EPI_ISL_1514659, EPI_ISL_1514660, EPI_ISL_1514662, EPI_ISL_1514663, EPI_ISL_1514664, EPI_ISL_1514665, EPI_ISL_1514666, EPI_ISL_1514675, EPI_ISL_1514676, EPI_ISL_1514677, EPI_ISL_1514679, EPI_ISL_1514681, EPI_ISL_1514682, EPI_ISL_1514684, EPI_ISL_1514685, EPI_ISL_1514686, EPI_ISL_1514687, EPI_ISL_1514688, EPI_ISL_1514695, EPI_ISL_1514697, EPI_ISL_1514699, EPI_ISL_1514700, EPI_ISL_1514701, EPI_ISL_1514703, EPI_ISL_1514704, EPI_ISL_1514707, EPI_ISL_1514710, EPI_ISL_1514716, EPI_ISL_1514717, EPI_ISL_1514718, EPI_ISL_1514722, EPI_ISL_1514723, EPI_ISL_1514725, EPI_ISL_1514726, EPI_ISL_1514727, EPI_ISL_1514728, EPI_ISL_1514729, EPI_ISL_1514730, EPI_ISL_1514731, EPI_ISL_1514732, EPI_ISL_1514733, EPI_ISL_1514734, EPI_ISL_1514735, EPI_ISL_1514736, EPI_ISL_1514737, EPI_ISL_1514738, EPI_ISL_1514740, EPI_ISL_1514741, EPI_ISL_1514742, EPI_ISL_1514745, EPI_ISL_1514746, EPI_ISL_1514749, EPI_ISL_1514752, EPI_ISL_1514753, EPI_ISL_1514754, EPI_ISL_1514756, EPI_ISL_1514757, EPI_ISL_1514758, EPI_ISL_1514760, EPI_ISL_1514761, EPI_ISL_1514762, EPI_ISL_1514763, EPI_ISL_1514764, EPI_ISL_1514765, EPI_ISL_1514767, EPI_ISL_1514769, EPI_ISL_1514770, EPI_ISL_1514771, EPI_ISL_1514772, EPI_ISL_1514773, EPI_ISL_1514774, EPI_ISL_1514775, EPI_ISL_1514777, EPI_ISL_1514778, EPI_ISL_1514779, EPI_ISL_1514780, EPI_ISL_1514781, EPI_ISL_1514782, EPI_ISL_1514783, EPI_ISL_1514784, EPI_ISL_1514785, EPI_ISL_1514786, EPI_ISL_1514787, EPI_ISL_1514788, EPI_ISL_1514789, EPI_ISL_1514790, EPI_ISL_1514791, EPI_ISL_1514792, EPI_ISL_1514793, EPI_ISL_1514794, EPI_ISL_1514796, EPI_ISL_1514798, EPI_ISL_1514799, EPI_ISL_1514800, EPI_ISL_1514802, EPI_ISL_1514803, EPI_ISL_1514804, EPI_ISL_1514805, EPI_ISL_1514806, EPI_ISL_1514807, EPI_ISL_1514808, EPI_ISL_1514809, EPI_ISL_1514810, EPI_ISL_1514811, EPI_ISL_1514812, EPI_ISL_1514813, EPI_ISL_1514814, EPI_ISL_1514815, EPI_ISL_1514816, EPI_ISL_1514817, EPI_ISL_1514818, EPI_ISL_1514819, EPI_ISL_1514820, EPI_ISL_1514821, EPI_ISL_1514824, EPI_ISL_1514825, EPI_ISL_1514826, EPI_ISL_1514827, EPI_ISL_1514828, EPI_ISL_1514829, EPI_ISL_1514830, EPI_ISL_1514831, EPI_ISL_1514832, EPI_ISL_1514833, EPI_ISL_1514834, EPI_ISL_1514835, EPI_ISL_1514836, EPI_ISL_1514837, EPI_ISL_1514838, EPI_ISL_1514839, EPI_ISL_1514842, EPI_ISL_1514843, EPI_ISL_1514844, EPI_ISL_1514845, EPI_ISL_1514846, EPI_ISL_1514847, EPI_ISL_1514848, EPI_ISL_1514849, EPI_ISL_1514850, EPI_ISL_1514851, EPI_ISL_1514852, EPI_ISL_1514853, EPI_ISL_1514854, EPI_ISL_1514855, EPI_ISL_1514856, EPI_ISL_1514857, EPI_ISL_1514858, EPI_ISL_1514859, EPI_ISL_1514860, EPI_ISL_1514861, EPI_ISL_1514862, EPI_ISL_1514863, EPI_ISL_1514865, EPI_ISL_1514866, EPI_ISL_1514867, EPI_ISL_1514868, EPI_ISL_1514869, EPI_ISL_1514870, EPI_ISL_1514871, EPI_ISL_1514872, EPI_ISL_1514873, EPI_ISL_1514874, EPI_ISL_1514875, EPI_ISL_1514876, EPI_ISL_1514877, EPI_ISL_1514878, EPI_ISL_1514879, EPI_ISL_1514880, EPI_ISL_1514881, EPI_ISL_1514882, EPI_ISL_1514883, EPI_ISL_1514884, EPI_ISL_1514886, EPI_ISL_1514887, EPI_ISL_1514888, EPI_ISL_1514889, EPI_ISL_1514890, EPI_ISL_1514893, EPI_ISL_1514894, EPI_ISL_1514895, EPI_ISL_1514897, EPI_ISL_1514898, EPI_ISL_1514900, EPI_ISL_1514901, EPI_ISL_1514902, EPI_ISL_1514904, EPI_ISL_1514909, EPI_ISL_1514910, EPI_ISL_1514911, EPI_ISL_1514912, EPI_ISL_1514913, EPI_ISL_1514914, EPI_ISL_1514915, EPI_ISL_1514916, EPI_ISL_1514917, EPI_ISL_1514919, EPI_ISL_1514920, EPI_ISL_1514921, EPI_ISL_1514923, EPI_ISL_1514924, EPI_ISL_1514925, EPI_ISL_1514926, EPI_ISL_1514927, EPI_ISL_1514928, EPI_ISL_1514929, EPI_ISL_1514930, EPI_ISL_1514931, EPI_ISL_1514932, EPI_ISL_1514933, EPI_ISL_1514935, EPI_ISL_1514936, EPI_ISL_1514937, EPI_ISL_1514942, EPI_ISL_1514943, EPI_ISL_1514945, EPI_ISL_1514946, EPI_ISL_1514947, EPI_ISL_1514949, EPI_ISL_1514950, EPI_ISL_1514951, EPI_ISL_1514952, EPI_ISL_1514953, EPI_ISL_1514955, EPI_ISL_1514957, EPI_ISL_1514958, EPI_ISL_1514959, EPI_ISL_1514961, EPI_ISL_1514962, EPI_ISL_1514963, EPI_ISL_1514964, EPI_ISL_1514966, EPI_ISL_1514967, EPI_ISL_1514968, EPI_ISL_1514969, EPI_ISL_1514970, EPI_ISL_1514973, EPI_ISL_1514974, EPI_ISL_1514976, EPI_ISL_1514977, EPI_ISL_1514979, EPI_ISL_1514980, EPI_ISL_1514981, EPI_ISL_1514982, EPI_ISL_1514983, EPI_ISL_1514984, EPI_ISL_1514985, EPI_ISL_1514986, EPI_ISL_1514987, EPI_ISL_1514988, EPI_ISL_1514989, EPI_ISL_1514992, EPI_ISL_1514993, EPI_ISL_1514994, EPI_ISL_1514995, EPI_ISL_1514996, EPI_ISL_1514998, EPI_ISL_1514999, EPI_ISL_1515000, EPI_ISL_1515001, EPI_ISL_1515008, EPI_ISL_1515009, EPI_ISL_1515010, EPI_ISL_1515011, EPI_ISL_1515012, EPI_ISL_1515013, EPI_ISL_1515014, EPI_ISL_1515015, EPI_ISL_1515016, EPI_ISL_1515017, EPI_ISL_1515018, EPI_ISL_1515021, EPI_ISL_1515025, EPI_ISL_1515026, EPI_ISL_1515027, EPI_ISL_1515028, EPI_ISL_1515029, EPI_ISL_1515030, EPI_ISL_1515031, EPI_ISL_1515033, EPI_ISL_1515034, EPI_ISL_1515035, EPI_ISL_1515040, EPI_ISL_1515041, EPI_ISL_1515042, EPI_ISL_1515043, EPI_ISL_1515044, EPI_ISL_1515045, EPI_ISL_1515046, EPI_ISL_1515049, EPI_ISL_1515050, EPI_ISL_1515051, EPI_ISL_1515052, EPI_ISL_1515053, EPI_ISL_1515054, EPI_ISL_1515055, EPI_ISL_1515057, EPI_ISL_1515059, EPI_ISL_1515060, EPI_ISL_1515061, EPI_ISL_1515062, EPI_ISL_1515063, EPI_ISL_1515064, EPI_ISL_1515065, EPI_ISL_1515066, EPI_ISL_1515067, EPI_ISL_1515068, EPI_ISL_1515070, EPI_ISL_1515071, EPI_ISL_1515072, EPI_ISL_1515073, EPI_ISL_1515076, EPI_ISL_1515077, EPI_ISL_1515078, EPI_ISL_1515079, EPI_ISL_1515080, EPI_ISL_1515081, EPI_ISL_1515082, EPI_ISL_1515083, EPI_ISL_1515084, EPI_ISL_1515085, EPI_ISL_1515087, EPI_ISL_1515088, EPI_ISL_1515089, EPI_ISL_1515091, EPI_ISL_1515092, EPI_ISL_1515093, EPI_ISL_1515094, EPI_ISL_1515095, EPI_ISL_1515097, EPI_ISL_1515098, EPI_ISL_1515099, EPI_ISL_1515100, EPI_ISL_1515103, EPI_ISL_1515104, EPI_ISL_1515107, EPI_ISL_1515108, EPI_ISL_1515109, EPI_ISL_1515110, EPI_ISL_1515111, EPI_ISL_1515112, EPI_ISL_1515113, EPI_ISL_1515115, EPI_ISL_1515116, EPI_ISL_1515117, EPI_ISL_1515119, EPI_ISL_1515120, EPI_ISL_1515121, EPI_ISL_1515122, EPI_ISL_1515123, EPI_ISL_1515125, EPI_ISL_1515126, EPI_ISL_1515127, EPI_ISL_1515128, EPI_ISL_1515130, EPI_ISL_1515131, EPI_ISL_1515132, EPI_ISL_1515133, EPI_ISL_1515134, EPI_ISL_1515135, EPI_ISL_1515136, EPI_ISL_1515137, EPI_ISL_1515138, EPI_ISL_1515139, EPI_ISL_1515141, EPI_ISL_1515142, EPI_ISL_1515143, EPI_ISL_1515144, EPI_ISL_1515145, EPI_ISL_1515146, EPI_ISL_1515147, EPI_ISL_1515149, EPI_ISL_1515150, EPI_ISL_1515152, EPI_ISL_1515154, EPI_ISL_1515155, EPI_ISL_1515157, EPI_ISL_1515158, EPI_ISL_1515159, EPI_ISL_1515160, EPI_ISL_1515163, EPI_ISL_1515164, EPI_ISL_1515166, EPI_ISL_1515167, EPI_ISL_1515168, EPI_ISL_1515169, EPI_ISL_1515170, EPI_ISL_1515172, EPI_ISL_1515173, EPI_ISL_1515174, EPI_ISL_1515175, EPI_ISL_1515176, EPI_ISL_1515179, EPI_ISL_1515180, EPI_ISL_1515181, EPI_ISL_1515182, EPI_ISL_1515184, EPI_ISL_1515185, EPI_ISL_1515186, EPI_ISL_1515187, EPI_ISL_1515188, EPI_ISL_1515189, EPI_ISL_1515190, EPI_ISL_1515192, EPI_ISL_1515193, EPI_ISL_1515195, EPI_ISL_1515197, EPI_ISL_1515198, EPI_ISL_1515199, EPI_ISL_1515200, EPI_ISL_1515201, EPI_ISL_1515202, EPI_ISL_1515203, EPI_ISL_1515204, EPI_ISL_1515207, EPI_ISL_1515208, EPI_ISL_1515209, EPI_ISL_1515211, EPI_ISL_1515212, EPI_ISL_1515214, EPI_ISL_1515217, EPI_ISL_1515218, EPI_ISL_1515220, EPI_ISL_1515221, EPI_ISL_1515222, EPI_ISL_1515223, EPI_ISL_1515225, EPI_ISL_1515226, EPI_ISL_1515227, EPI_ISL_1515228, EPI_ISL_1515229, EPI_ISL_1515230, EPI_ISL_1515231, EPI_ISL_1515232, EPI_ISL_1515233, EPI_ISL_1515235, EPI_ISL_1515236, EPI_ISL_1515237, EPI_ISL_1515238, EPI_ISL_1515239, EPI_ISL_1515240, EPI_ISL_1515241, EPI_ISL_1515245, EPI_ISL_1515246, EPI_ISL_1515248, EPI_ISL_1515250, EPI_ISL_1515251, EPI_ISL_1515252, EPI_ISL_1515253, EPI_ISL_1515254, EPI_ISL_1515255, EPI_ISL_1515256, EPI_ISL_1515259, EPI_ISL_1515260, EPI_ISL_1515261, EPI_ISL_1515262, EPI_ISL_1515263, EPI_ISL_1515264, EPI_ISL_1515265, EPI_ISL_1515266, EPI_ISL_1515268, EPI_ISL_1515269, EPI_ISL_1515270, EPI_ISL_1515271, EPI_ISL_1515273, EPI_ISL_1515274, EPI_ISL_1515276, EPI_ISL_1515277, EPI_ISL_1515278, EPI_ISL_1515280, EPI_ISL_1515281, EPI_ISL_1515282, EPI_ISL_1515284, EPI_ISL_1515285, EPI_ISL_1515286, EPI_ISL_1515287, EPI_ISL_1515288, EPI_ISL_1515289, EPI_ISL_1515290, EPI_ISL_1515291, EPI_ISL_1515292, EPI_ISL_1515294, EPI_ISL_1515295, EPI_ISL_1515296, EPI_ISL_1515297, EPI_ISL_1515298, EPI_ISL_1515300, EPI_ISL_1515301, EPI_ISL_1515303, EPI_ISL_1515304, EPI_ISL_1515306, EPI_ISL_1515308, EPI_ISL_1515309, EPI_ISL_1515310, EPI_ISL_1515311, EPI_ISL_1515312, EPI_ISL_1515313, EPI_ISL_1515314, EPI_ISL_1515315, EPI_ISL_1515316, EPI_ISL_1515317, EPI_ISL_1515320, EPI_ISL_1515321, EPI_ISL_1515322, EPI_ISL_1515324, EPI_ISL_1515325, EPI_ISL_1515326, EPI_ISL_1515327, EPI_ISL_1515330, EPI_ISL_1515331, EPI_ISL_1515332, EPI_ISL_1515333, EPI_ISL_1515335, EPI_ISL_1515337, EPI_ISL_1515338, EPI_ISL_1515339, EPI_ISL_1515340, EPI_ISL_1515342, EPI_ISL_1515343, EPI_ISL_1515344, EPI_ISL_1515345, EPI_ISL_1515346, EPI_ISL_1515347, EPI_ISL_1515348, EPI_ISL_1515350, EPI_ISL_1515351, EPI_ISL_1515352, EPI_ISL_1515353, EPI_ISL_1515354, EPI_ISL_1515356, EPI_ISL_1515357, EPI_ISL_1515358, EPI_ISL_1515359, EPI_ISL_1515360, EPI_ISL_1515362, EPI_ISL_1515363, EPI_ISL_1515364, EPI_ISL_1515365, EPI_ISL_1515367, EPI_ISL_1515368, EPI_ISL_1515372, EPI_ISL_1515374, EPI_ISL_1515375, EPI_ISL_1515376, EPI_ISL_1515377, EPI_ISL_1515378, EPI_ISL_1515379, EPI_ISL_1515380, EPI_ISL_1515381, EPI_ISL_1515385, EPI_ISL_1515387, EPI_ISL_1515388, EPI_ISL_1515389, EPI_ISL_1515390, EPI_ISL_1515391, EPI_ISL_1515392, EPI_ISL_1515393, EPI_ISL_1515394, EPI_ISL_1515395, EPI_ISL_1515396, EPI_ISL_1515397, EPI_ISL_1515398, EPI_ISL_1515399, EPI_ISL_1515400, EPI_ISL_1515401, EPI_ISL_1515402, EPI_ISL_1515403, EPI_ISL_1515405, EPI_ISL_1515406, EPI_ISL_1515408, EPI_ISL_1515409, EPI_ISL_1515410, EPI_ISL_1515411, EPI_ISL_1515412, EPI_ISL_1515413, EPI_ISL_1515414, EPI_ISL_1515415, EPI_ISL_1515417, EPI_ISL_1515418, EPI_ISL_1515419, EPI_ISL_1515420, EPI_ISL_1515421, EPI_ISL_1515422, EPI_ISL_1515425, EPI_ISL_1515427, EPI_ISL_1515428, EPI_ISL_1515430, EPI_ISL_1515431, EPI_ISL_1515432, EPI_ISL_1515433, EPI_ISL_1515435, EPI_ISL_1515437, EPI_ISL_1515438, EPI_ISL_1515439, EPI_ISL_1515440, EPI_ISL_1515441, EPI_ISL_1515442, EPI_ISL_1515443, EPI_ISL_1515444, EPI_ISL_1515445, EPI_ISL_1515446, EPI_ISL_1515448, EPI_ISL_1515449, EPI_ISL_1515450, EPI_ISL_1515451, EPI_ISL_1515453, EPI_ISL_1515454, EPI_ISL_1515455, EPI_ISL_1515456, EPI_ISL_1515457, EPI_ISL_1515458, EPI_ISL_1515459, EPI_ISL_1515460, EPI_ISL_1515461, EPI_ISL_1515462, EPI_ISL_1515463, EPI_ISL_1515464, EPI_ISL_1515465, EPI_ISL_1515466, EPI_ISL_1515467, EPI_ISL_1515468, EPI_ISL_1515469, EPI_ISL_1515470, EPI_ISL_1515472, EPI_ISL_1515473, EPI_ISL_1515474, EPI_ISL_1515475, EPI_ISL_1515476, EPI_ISL_1515478, EPI_ISL_1515479, EPI_ISL_1515480, EPI_ISL_1515481, EPI_ISL_1515482, EPI_ISL_1515483, EPI_ISL_1515484, EPI_ISL_1515485, EPI_ISL_1515486, EPI_ISL_1515487, EPI_ISL_1515488, EPI_ISL_1515489, EPI_ISL_1515492, EPI_ISL_1515493, EPI_ISL_1515495, EPI_ISL_1515496, EPI_ISL_1515497, EPI_ISL_1515498, EPI_ISL_1515499, EPI_ISL_1515500, EPI_ISL_1515502, EPI_ISL_1515503, EPI_ISL_1515504, EPI_ISL_1515505, EPI_ISL_1515506, EPI_ISL_1515507, EPI_ISL_1515508, EPI_ISL_1515509, EPI_ISL_1515512, EPI_ISL_1515513, EPI_ISL_1515514, EPI_ISL_1515515, EPI_ISL_1515516, EPI_ISL_1515517, EPI_ISL_1515518, EPI_ISL_1515519, EPI_ISL_1515520, EPI_ISL_1515521, EPI_ISL_1515522, EPI_ISL_1515523, EPI_ISL_1515524, EPI_ISL_1515525, EPI_ISL_1515526, EPI_ISL_1515527, EPI_ISL_1515528, EPI_ISL_1515529, EPI_ISL_1515530, EPI_ISL_1515531, EPI_ISL_1515532, EPI_ISL_1515533, EPI_ISL_1515534, EPI_ISL_1515535, EPI_ISL_1515537, EPI_ISL_1515538, EPI_ISL_1515539, EPI_ISL_1515540, EPI_ISL_1515542, EPI_ISL_1515543, EPI_ISL_1515544, EPI_ISL_1515545, EPI_ISL_1515546, EPI_ISL_1515551, EPI_ISL_1515555, EPI_ISL_1515556, EPI_ISL_1515557, EPI_ISL_1515558, EPI_ISL_1515560, EPI_ISL_1515562, EPI_ISL_1515563, EPI_ISL_1515564, EPI_ISL_1515565, EPI_ISL_1515567, EPI_ISL_1515568, EPI_ISL_1515569, EPI_ISL_1515570, EPI_ISL_1515571, EPI_ISL_1515572, EPI_ISL_1515573, EPI_ISL_1515574, EPI_ISL_1515575, EPI_ISL_1515576, EPI_ISL_1515579, EPI_ISL_1515580, EPI_ISL_1515581, EPI_ISL_1515583, EPI_ISL_1515585, EPI_ISL_1515586, EPI_ISL_1515587, EPI_ISL_1515589, EPI_ISL_1515590, EPI_ISL_1515591, EPI_ISL_1515592, EPI_ISL_1515594, EPI_ISL_1515595, EPI_ISL_1515596, EPI_ISL_1515597, EPI_ISL_1515598, EPI_ISL_1515599, EPI_ISL_1515600, EPI_ISL_1515601, EPI_ISL_1515602, EPI_ISL_1515604, EPI_ISL_1515605, EPI_ISL_1515606, EPI_ISL_1515607, EPI_ISL_1515609, EPI_ISL_1515611, EPI_ISL_1515612, EPI_ISL_1515613, EPI_ISL_1515614, EPI_ISL_1515615, EPI_ISL_1515617, EPI_ISL_1515618, EPI_ISL_1515619, EPI_ISL_1515620, EPI_ISL_1515621, EPI_ISL_1515622, EPI_ISL_1515623, EPI_ISL_1515624, EPI_ISL_1515627, EPI_ISL_1515628, EPI_ISL_1515630, EPI_ISL_1515632, EPI_ISL_1515633, EPI_ISL_1515634, EPI_ISL_1515635, EPI_ISL_1515636, EPI_ISL_1515638, EPI_ISL_1515639, EPI_ISL_1515640, EPI_ISL_1515641, EPI_ISL_1515642, EPI_ISL_1515645, EPI_ISL_1515646, EPI_ISL_1515647, EPI_ISL_1515648, EPI_ISL_1515649, EPI_ISL_1515650, EPI_ISL_1515651, EPI_ISL_1515653, EPI_ISL_1515654, EPI_ISL_1515657, EPI_ISL_1515658, EPI_ISL_1515659, EPI_ISL_1515660, EPI_ISL_1515662, EPI_ISL_1515663, EPI_ISL_1515664, EPI_ISL_1515665, EPI_ISL_1515666, EPI_ISL_1515667, EPI_ISL_1515669, EPI_ISL_1515670, EPI_ISL_1515671, EPI_ISL_1515672, EPI_ISL_1515673, EPI_ISL_1515674, EPI_ISL_1515675, EPI_ISL_1515676, EPI_ISL_1515677, EPI_ISL_1515678, EPI_ISL_1515679, EPI_ISL_1515680, EPI_ISL_1515681, EPI_ISL_1515683, EPI_ISL_1515684, EPI_ISL_1515685, EPI_ISL_1515686, EPI_ISL_1515688, EPI_ISL_1515689, EPI_ISL_1515690, EPI_ISL_1515691, EPI_ISL_1515692, EPI_ISL_1515693, EPI_ISL_1515694, EPI_ISL_1515697, EPI_ISL_1515699, EPI_ISL_1515700, EPI_ISL_1515702, EPI_ISL_1515703, EPI_ISL_1515705, EPI_ISL_1515706, EPI_ISL_1515707, EPI_ISL_1515708, EPI_ISL_1515709, EPI_ISL_1515710, EPI_ISL_1515711, EPI_ISL_1515712, EPI_ISL_1515713, EPI_ISL_1515714, EPI_ISL_1515715, EPI_ISL_1515717, EPI_ISL_1515718, EPI_ISL_1515719, EPI_ISL_1515720, EPI_ISL_1515721, EPI_ISL_1515723, EPI_ISL_1515725, EPI_ISL_1515726, EPI_ISL_1515727, EPI_ISL_1515728, EPI_ISL_1515729, EPI_ISL_1515730, EPI_ISL_1515731, EPI_ISL_1515732, EPI_ISL_1515733, EPI_ISL_1515734, EPI_ISL_1515736, EPI_ISL_1515737, EPI_ISL_1515738, EPI_ISL_1515739, EPI_ISL_1515740, EPI_ISL_1515741, EPI_ISL_1515742, EPI_ISL_1515743, EPI_ISL_1515744, EPI_ISL_1515745, EPI_ISL_1515747, EPI_ISL_1515748, EPI_ISL_1515749, EPI_ISL_1515750, EPI_ISL_1515752, EPI_ISL_1515753, EPI_ISL_1515754, EPI_ISL_1515755, EPI_ISL_1515756, EPI_ISL_1515757, EPI_ISL_1515758, EPI_ISL_1515759, EPI_ISL_1515760, EPI_ISL_1515761 | | | |
| see above | Laboratory Corporation of America | Centers for Disease Control and Prevention Division of Viral Diseases, Pathogen Discovery | Dakota Howard, Dhwani Batra, Peter W. Cook, Kara Moser, Adrian Paskey, Jason Caravas, Benjamin Rambo-Martin, Shatavia Morrison, Christopher Gulvick, Scott Sammons, Yvette Unoarumhi, Darlene Wagner, Matthew Schmerer, Minoo Agarwal, Eyad Almasri, Debbie Boles, Ayla Burns, Nuthawin Charoenshri, Oren Cohen, Susan Countryman, Mary Ann Cristobal, Bobbi Croy, Suzanne Dale, Hrushikesh Deshmukh, Amanda Douglas, Vincent Drouillon, Marcia Eisenberg, Howard Engler, Rama Ghatti, Prashant Gupta, Susan Hicks, Jake Humphrey, Lax Iyer, Manoj Jain, Mohan Kolli, Brian Krueger, Tim Kuphal, Susan Letovsky, Michael Levandoski, Craig Lukasik, Jonathan Meltzer, Brian Norvell, Mindy Nye, Scott Parker, Christos Petropoulos, John Pruitt, Steven Ragan, Scott Ryan, Mike Sapeta, Jana Schroth, Suresh Babu Selvaraju, Goran Stevovic, Amanda Suchanek, Andrea Throop, Lyndon Tilson, Thomas Urban, Joe Voshell, Kimberly Wagner, Jonathan Williams, Mary Williamson, Qian Zeng, Tricia Zwiefelhofer, Clinton R. Paden, Duncan MacCannell |
| EPI_ISL_1515764, EPI_ISL_1515765, EPI_ISL_1515766, EPI_ISL_1515767, EPI_ISL_1515769, EPI_ISL_1515771, EPI_ISL_1515772, EPI_ISL_1515773, EPI_ISL_1515775, EPI_ISL_1515776, EPI_ISL_1515777, EPI_ISL_1515779, EPI_ISL_1515780, EPI_ISL_1515781, EPI_ISL_1515782, EPI_ISL_1515783, EPI_ISL_1515785, EPI_ISL_1515786, EPI_ISL_1515787, EPI_ISL_1515788, EPI_ISL_1515789 | | | |
| see above | Wyoming Public Health Laboratory | Wyoming Public Health Laboratory | Jim Mildenberger, Wanda Manley, Noah Hull, Taylor Fearing, Lynette Gumbleton, Channing Weber, Ashley Norberg, Chayse Rowley, Marley Goetz, Brian Dominguez, Elliot Thomasson, Cari Sloma, and Rob Christensen |
| EPI_ISL_1515790, EPI_ISL_1515791, EPI_ISL_1515792, EPI_ISL_1515793, EPI_ISL_1515794, EPI_ISL_1515795, EPI_ISL_1515796, EPI_ISL_1515797 | Lafene Health Center | Kansas State Veterinary Diagnostic Laboratory | Tyler Doerksen, Andreea Lu, Lance Noll, Elizabeth Porter, Kasey Hogan, Emily Cox, Jianfa Bai, Jamie Henningson, Rachel Palinski |
| EPI_ISL_1515798, EPI_ISL_1515799 | DC Public Health Lab/ Dept. of Forensic Sciences | DC Public Health Lab/ Dept. of Forensic Sciences | Janis Doss, Scott Nguyen, Elizabeth Zelaya, Sarah Scott, Connie Maza, Monica Mann, Brittany Hamilton, David Payne, Jocelyn Hauser |
| EPI_ISL_1515800 | USC Clinical Lab | Los Angeles County Public Health Laboratories | P. Hemarajata et al. |
| EPI_ISL_1515801, EPI_ISL_1515802, EPI_ISL_1515803, EPI_ISL_1515804, EPI_ISL_1515805, EPI_ISL_1515806, EPI_ISL_1515807, EPI_ISL_1515808, EPI_ISL_1515809, EPI_ISL_1515810, EPI_ISL_1515811, EPI_ISL_1515812, EPI_ISL_1515813, EPI_ISL_1515814, EPI_ISL_1515815, EPI_ISL_1515816, EPI_ISL_1515817, EPI_ISL_1515818, EPI_ISL_1515819, EPI_ISL_1515820, EPI_ISL_1515821, EPI_ISL_1515823, EPI_ISL_1515824, EPI_ISL_1515825, EPI_ISL_1515826, EPI_ISL_1515827, EPI_ISL_1515828, EPI_ISL_1515829, EPI_ISL_1515830, EPI_ISL_1515831, EPI_ISL_1515832, EPI_ISL_1515833, EPI_ISL_1515834, EPI_ISL_1515835, EPI_ISL_1515837, EPI_ISL_1515838, EPI_ISL_1515839, EPI_ISL_1515842, EPI_ISL_1515843, EPI_ISL_1515844, EPI_ISL_1515845, EPI_ISL_1515846, EPI_ISL_1515847, EPI_ISL_1515848, EPI_ISL_1515849, EPI_ISL_1515850, EPI_ISL_1515851, EPI_ISL_1515852, EPI_ISL_1515853, EPI_ISL_1515854, EPI_ISL_1515855, EPI_ISL_1515856, EPI_ISL_1515857, EPI_ISL_1515858, EPI_ISL_1515859, EPI_ISL_1515860, EPI_ISL_1515861, EPI_ISL_1515862, EPI_ISL_1515863, EPI_ISL_1515864, EPI_ISL_1515865, EPI_ISL_1515866, EPI_ISL_1515867, EPI_ISL_1515868, EPI_ISL_1515869, EPI_ISL_1515870, EPI_ISL_1515871, EPI_ISL_1515872, EPI_ISL_1515873, EPI_ISL_1515874, EPI_ISL_1515875, EPI_ISL_1515876, EPI_ISL_1515877, EPI_ISL_1515878, EPI_ISL_1515879, EPI_ISL_1515880, EPI_ISL_1515881, EPI_ISL_1515882, EPI_ISL_1515883, EPI_ISL_1515884, EPI_ISL_1515885, EPI_ISL_1515886, EPI_ISL_1515887, EPI_ISL_1515888, EPI_ISL_1515889, EPI_ISL_1515890, EPI_ISL_1515891, EPI_ISL_1515892, EPI_ISL_1515893, EPI_ISL_1515894, EPI_ISL_1515895, EPI_ISL_1515896, EPI_ISL_1515897, EPI_ISL_1515898, EPI_ISL_1515899, EPI_ISL_1515900, EPI_ISL_1515901, EPI_ISL_1515902, EPI_ISL_1515903, EPI_ISL_1515904, EPI_ISL_1515905, EPI_ISL_1515906, EPI_ISL_1515907, EPI_ISL_1515908, EPI_ISL_1515909, EPI_ISL_1515910, EPI_ISL_1515911, EPI_ISL_1515912, EPI_ISL_1515913, EPI_ISL_1515914, EPI_ISL_1515915, EPI_ISL_1515916, EPI_ISL_1515917, EPI_ISL_1515918, EPI_ISL_1515919, EPI_ISL_1515920, EPI_ISL_1515921, EPI_ISL_1515922, EPI_ISL_1515923, EPI_ISL_1515924, EPI_ISL_1515925, EPI_ISL_1515926, EPI_ISL_1515927, EPI_ISL_1515928, EPI_ISL_1515929, EPI_ISL_1515930, EPI_ISL_1515931, EPI_ISL_1515932, EPI_ISL_1515933, EPI_ISL_1515934, EPI_ISL_1515935, EPI_ISL_1515936, EPI_ISL_1515937, EPI_ISL_1515938, EPI_ISL_1515939, EPI_ISL_1515940, EPI_ISL_1515941, EPI_ISL_1515942, EPI_ISL_1515943, EPI_ISL_1515944, EPI_ISL_1515945, EPI_ISL_1515946, EPI_ISL_1515947, EPI_ISL_1515948, EPI_ISL_1515949, EPI_ISL_1515950, EPI_ISL_1515951, EPI_ISL_1515952, EPI_ISL_1515953, EPI_ISL_1515954, EPI_ISL_1515955, EPI_ISL_1515956, EPI_ISL_1515957, EPI_ISL_1515958, EPI_ISL_1515959, EPI_ISL_1515960, EPI_ISL_1515961, EPI_ISL_1515962, EPI_ISL_1515963, EPI_ISL_1515964, EPI_ISL_1515965, EPI_ISL_1515966, EPI_ISL_1515967, EPI_ISL_1515968, EPI_ISL_1515969, EPI_ISL_1515970, EPI_ISL_1515971, EPI_ISL_1515972, EPI_ISL_1515973, EPI_ISL_1515974, EPI_ISL_1515975, EPI_ISL_1515976, EPI_ISL_1515977, EPI_ISL_1515978, EPI_ISL_1515979, EPI_ISL_1515980, EPI_ISL_1515981, EPI_ISL_1515982, EPI_ISL_1515983, EPI_ISL_1515984, EPI_ISL_1515985, EPI_ISL_1515986, EPI_ISL_1515987, EPI_ISL_1515988, EPI_ISL_1515989, EPI_ISL_1515990, EPI_ISL_1515991, EPI_ISL_1515992, EPI_ISL_1515993, EPI_ISL_1515994, EPI_ISL_1515996, EPI_ISL_1515997, EPI_ISL_1515999, EPI_ISL_1516000, EPI_ISL_1516001, EPI_ISL_1516002, EPI_ISL_1516003, EPI_ISL_1516004, EPI_ISL_1516005, EPI_ISL_1516006 | | | |
| see above | State Laboratories Division, Hawaii State Department of Health | State Laboratories Division, Hawaii State Department of Health | Pamela O'Brien, Drew Kuwazaki, Ayana Garnet, Razvan Sultana, Edward Desmond |
| EPI_ISL_1516169, EPI_ISL_1516170, EPI_ISL_1516171 | SD Public Health Laboratory | Centers for Disease Control and Prevention Division of Viral Diseases, Pathogen Discovery | Mili Sheth, Sarah Nobles, Jasmine Padilla, Mark Burroughs, Shoshona Le, Katie Dillon, Peter Cook, Clinton R. Paden, Dhwani Batra, Krista Queen, Kristen Knipe, Dakota Howard, Yvette Unoarumhi, Darlene Wagner, Matthew Schmerer, Ben L. Rambo-Martin, Kristine Lacek, Sam Shepard, Alison Laufer Halpin, Dave Wentworth, Vivien Dugan, Suxiang Tong, Justin Lee |
| EPI_ISL_1516172, EPI_ISL_1516173, EPI_ISL_1516174, EPI_ISL_1516175, EPI_ISL_1516176, EPI_ISL_1516177, EPI_ISL_1516178, EPI_ISL_1516179, EPI_ISL_1516180, EPI_ISL_1516181, EPI_ISL_1516182, EPI_ISL_1516183, EPI_ISL_1516184, EPI_ISL_1516185, EPI_ISL_1516186, EPI_ISL_1516187 | | | |
| see above | NJ Public Health and Environmental Laboratories | Centers for Disease Control and Prevention Division of Viral Diseases, Pathogen Discovery | Mili Sheth, Sarah Nobles, Jasmine Padilla, Mark Burroughs, Shoshona Le, Katie Dillon, Peter Cook, Clinton R. Paden, Dhwani Batra, Krista Queen, Kristen Knipe, Dakota Howard, Yvette Unoarumhi, Darlene Wagner, Matthew Schmerer, Ben L. Rambo-Martin, Kristine Lacek, Sam Shepard, Alison Laufer Halpin, Dave Wentworth, Vivien Dugan, Suxiang Tong, Justin Lee |
| EPI_ISL_1516197 | TN Division of Laboratory Services | Centers for Disease Control and Prevention | Mili Sheth, Sarah Nobles, Jasmine Padilla, Mark Burroughs, Shoshona Le, Katie Dillon, Peter Cook, Clinton R. Paden, Dhwani Batra, Krista Queen, Kristen Knipe, Dakota Howard, Yvette Unoarumhi, Darlene Wagner, Matthew Schmerer, Ben L. Rambo-Martin, Kristine Lacek, Sam Shepard, |

| | | | |
|---|---|---|---|
| | | Division of Viral Diseases, Pathogen Discovery | Alison Laufer Halpin, Dave Wentworth, Vivien Dugan, Suxiang Tong, Justin Lee |
| EPI_ISL_1516198, EPI_ISL_1516199, EPI_ISL_1516200, EPI_ISL_1516201, EPI_ISL_1516202, EPI_ISL_1516203, EPI_ISL_1516204 | OR State PHL-Virology/Immunology Section | Centers for Disease Control and Prevention Division of Viral Diseases, Pathogen Discovery | Mili Sheth, Sarah Nobles, Jasmine Padilla, Mark Burroughs, Shoshona Le, Katie Dillon, Peter Cook, Clinton R. Paden, Dhwani Batra, Krista Queen, Kristen Knipe, Dakota Howard, Yvette Unoarumhi, Darlene Wagner, Matthew Schmerer, Ben L. Rambo-Martin, Kristine Lacek, Sam Shepard, Alison Laufer Halpin, Dave Wentworth, Vivien Dugan, Suxiang Tong, Justin Lee |
| EPI_ISL_1516205, EPI_ISL_1516206, EPI_ISL_1516207, EPI_ISL_1516208, EPI_ISL_1516209, EPI_ISL_1516210, EPI_ISL_1516211, EPI_ISL_1516212 | NYC Department of Health and Mental Hygiene | Centers for Disease Control and Prevention Division of Viral Diseases, Pathogen Discovery | Mili Sheth, Sarah Nobles, Jasmine Padilla, Mark Burroughs, Shoshona Le, Katie Dillon, Peter Cook, Clinton R. Paden, Dhwani Batra, Krista Queen, Kristen Knipe, Dakota Howard, Yvette Unoarumhi, Darlene Wagner, Matthew Schmerer, Ben L. Rambo-Martin, Kristine Lacek, Sam Shepard, Alison Laufer Halpin, Dave Wentworth, Vivien Dugan, Suxiang Tong, Justin Lee |
| EPI_ISL_1516213, EPI_ISL_1516214 | WA State Department of Health | Centers for Disease Control and Prevention Division of Viral Diseases, Pathogen Discovery | Mili Sheth, Sarah Nobles, Jasmine Padilla, Mark Burroughs, Shoshona Le, Katie Dillon, Peter Cook, Clinton R. Paden, Dhwani Batra, Krista Queen, Kristen Knipe, Dakota Howard, Yvette Unoarumhi, Darlene Wagner, Matthew Schmerer, Ben L. Rambo-Martin, Kristine Lacek, Sam Shepard, Alison Laufer Halpin, Dave Wentworth, Vivien Dugan, Suxiang Tong, Justin Lee |
| EPI_ISL_1516215, EPI_ISL_1516216, EPI_ISL_1516217, EPI_ISL_1516218, EPI_ISL_1516219, EPI_ISL_1516220, EPI_ISL_1516221, EPI_ISL_1516222, EPI_ISL_1516223, EPI_ISL_1516224, EPI_ISL_1516225, EPI_ISL_1516226, EPI_ISL_1516227, EPI_ISL_1516228, EPI_ISL_1516229, EPI_ISL_1516230, EPI_ISL_1516231, EPI_ISL_1516232, EPI_ISL_1516233, EPI_ISL_1516234, EPI_ISL_1516235 | | | |
| see above | WY Public Health Laboratory | Centers for Disease Control and Prevention Division of Viral Diseases, Pathogen Discovery | Mili Sheth, Sarah Nobles, Jasmine Padilla, Mark Burroughs, Shoshona Le, Katie Dillon, Peter Cook, Clinton R. Paden, Dhwani Batra, Krista Queen, Kristen Knipe, Dakota Howard, Yvette Unoarumhi, Darlene Wagner, Matthew Schmerer, Ben L. Rambo-Martin, Kristine Lacek, Sam Shepard, Alison Laufer Halpin, Dave Wentworth, Vivien Dugan, Suxiang Tong, Justin Lee |
| EPI_ISL_1516236, EPI_ISL_1516237, EPI_ISL_1516238, EPI_ISL_1516239, EPI_ISL_1516240, EPI_ISL_1516241, EPI_ISL_1516242, EPI_ISL_1516243, EPI_ISL_1516244 | NM Dept. Health, Scientific Laboratory Division | Centers for Disease Control and Prevention Division of Viral Diseases, Pathogen Discovery | Mili Sheth, Sarah Nobles, Jasmine Padilla, Mark Burroughs, Shoshona Le, Katie Dillon, Peter Cook, Clinton R. Paden, Dhwani Batra, Krista Queen, Kristen Knipe, Dakota Howard, Yvette Unoarumhi, Darlene Wagner, Matthew Schmerer, Ben L. Rambo-Martin, Kristine Lacek, Sam Shepard, Alison Laufer Halpin, Dave Wentworth, Vivien Dugan, Suxiang Tong, Justin Lee |
| EPI_ISL_1516245, EPI_ISL_1516247, EPI_ISL_1516248, EPI_ISL_1516249, EPI_ISL_1516250, EPI_ISL_1516251, EPI_ISL_1516252, EPI_ISL_1516253, EPI_ISL_1516254, EPI_ISL_1516255 | WI State Laboratory of Hygiene | Centers for Disease Control and Prevention Division of Viral Diseases, Pathogen Discovery | Mili Sheth, Sarah Nobles, Jasmine Padilla, Mark Burroughs, Shoshona Le, Katie Dillon, Peter Cook, Clinton R. Paden, Dhwani Batra, Krista Queen, Kristen Knipe, Dakota Howard, Yvette Unoarumhi, Darlene Wagner, Matthew Schmerer, Ben L. Rambo-Martin, Kristine Lacek, Sam Shepard, Alison Laufer Halpin, Dave Wentworth, Vivien Dugan, Suxiang Tong, Justin Lee |
| EPI_ISL_1516256, EPI_ISL_1516257, EPI_ISL_1516258, EPI_ISL_1516259, EPI_ISL_1516260, EPI_ISL_1516261 | ME Health and Environmental Testing Laboratory | Centers for Disease Control and Prevention Division of Viral Diseases, Pathogen Discovery | Mili Sheth, Sarah Nobles, Jasmine Padilla, Mark Burroughs, Shoshona Le, Katie Dillon, Peter Cook, Clinton R. Paden, Dhwani Batra, Krista Queen, Kristen Knipe, Dakota Howard, Yvette Unoarumhi, Darlene Wagner, Matthew Schmerer, Ben L. Rambo-Martin, Kristine Lacek, Sam Shepard, Alison Laufer Halpin, Dave Wentworth, Vivien Dugan, Suxiang Tong, Justin Lee |
| EPI_ISL_1516262, EPI_ISL_1516263 | PR Public Health Lab | Centers for Disease Control and Prevention Division of Viral Diseases, Pathogen Discovery | Mili Sheth, Sarah Nobles, Jasmine Padilla, Mark Burroughs, Shoshona Le, Katie Dillon, Peter Cook, Clinton R. Paden, Dhwani Batra, Krista Queen, Kristen Knipe, Dakota Howard, Yvette Unoarumhi, Darlene Wagner, Matthew Schmerer, Ben L. Rambo-Martin, Kristine Lacek, Sam Shepard, Alison Laufer Halpin, Dave Wentworth, Vivien Dugan, Suxiang Tong, Justin Lee |
| EPI_ISL_1516264, EPI_ISL_1516265, EPI_ISL_1516266, EPI_ISL_1516267, EPI_ISL_1516268 | WVDHHR - Office of Laboratory Services | Centers for Disease Control and Prevention Division of Viral Diseases, Pathogen Discovery | Mili Sheth, Sarah Nobles, Jasmine Padilla, Mark Burroughs, Shoshona Le, Katie Dillon, Peter Cook, Clinton R. Paden, Dhwani Batra, Krista Queen, Kristen Knipe, Dakota Howard, Yvette Unoarumhi, Darlene Wagner, Matthew Schmerer, Ben L. Rambo-Martin, Kristine Lacek, Sam Shepard, Alison Laufer Halpin, Dave Wentworth, Vivien Dugan, Suxiang Tong, Justin Lee |
| EPI_ISL_1516269, EPI_ISL_1516270, EPI_ISL_1516271, EPI_ISL_1516272, EPI_ISL_1516273, EPI_ISL_1516274, EPI_ISL_1516275 | SC Dept of Health and Env. Control-Bureau of Laboratories | Centers for Disease Control and Prevention Division of Viral Diseases, Pathogen Discovery | Mili Sheth, Sarah Nobles, Jasmine Padilla, Mark Burroughs, Shoshona Le, Katie Dillon, Peter Cook, Clinton R. Paden, Dhwani Batra, Krista Queen, Kristen Knipe, Dakota Howard, Yvette Unoarumhi, Darlene Wagner, Matthew Schmerer, Ben L. Rambo-Martin, Kristine Lacek, Sam Shepard, Alison Laufer Halpin, Dave Wentworth, Vivien Dugan, Suxiang Tong, Justin Lee |
| EPI_ISL_1516276, EPI_ISL_1516277, EPI_ISL_1516278, EPI_ISL_1516279, EPI_ISL_1516280, EPI_ISL_1516281, EPI_ISL_1516282, EPI_ISL_1516283, EPI_ISL_1516284, EPI_ISL_1516285, EPI_ISL_1516286, EPI_ISL_1516287, EPI_ISL_1516288, EPI_ISL_1516289, EPI_ISL_1516290, EPI_ISL_1516291, EPI_ISL_1516292, EPI_ISL_1516293, EPI_ISL_1516294, EPI_ISL_1516295 | | | |
| see above | PA Department of Health, Bureau of Laboratories | Centers for Disease Control and Prevention Division of Viral Diseases, Pathogen Discovery | Mili Sheth, Sarah Nobles, Jasmine Padilla, Mark Burroughs, Shoshona Le, Katie Dillon, Peter Cook, Clinton R. Paden, Dhwani Batra, Krista Queen, Kristen Knipe, Dakota Howard, Yvette Unoarumhi, Darlene Wagner, Matthew Schmerer, Ben L. Rambo-Martin, Kristine Lacek, Sam Shepard, Alison Laufer Halpin, Dave Wentworth, Vivien Dugan, Suxiang Tong, Justin Lee |
| EPI_ISL_1516296, EPI_ISL_1516297, EPI_ISL_1516298, EPI_ISL_1516299, EPI_ISL_1516300, EPI_ISL_1516301 | OK Public Health Laboratory, Oklahoma State DOH | Centers for Disease Control and Prevention Division of Viral Diseases, Pathogen Discovery | Mili Sheth, Sarah Nobles, Jasmine Padilla, Mark Burroughs, Shoshona Le, Katie Dillon, Peter Cook, Clinton R. Paden, Dhwani Batra, Krista Queen, Kristen Knipe, Dakota Howard, Yvette Unoarumhi, Darlene Wagner, Matthew Schmerer, Ben L. Rambo-Martin, Kristine Lacek, Sam Shepard, Alison Laufer Halpin, Dave Wentworth, Vivien Dugan, Suxiang Tong, Justin Lee |
| EPI_ISL_1516302, EPI_ISL_1516303, EPI_ISL_1516304 | MN PHL Division, Minnesota Department of Health | Centers for Disease Control and Prevention Division of Viral Diseases, Pathogen Discovery | Mili Sheth, Sarah Nobles, Jasmine Padilla, Mark Burroughs, Shoshona Le, Katie Dillon, Peter Cook, Clinton R. Paden, Dhwani Batra, Krista Queen, Kristen Knipe, Dakota Howard, Yvette Unoarumhi, Darlene Wagner, Matthew Schmerer, Ben L. Rambo-Martin, Kristine Lacek, Sam Shepard, Alison Laufer Halpin, Dave Wentworth, Vivien Dugan, Suxiang Tong, Justin Lee |
| EPI_ISL_1516305, EPI_ISL_1516306, EPI_ISL_1516307, EPI_ISL_1516308, EPI_ISL_1516309, EPI_ISL_1516310, EPI_ISL_1516311, EPI_ISL_1516312 | MT Public Health Laboratory | Centers for Disease Control and Prevention Division of Viral Diseases, Pathogen Discovery | Mili Sheth, Sarah Nobles, Jasmine Padilla, Mark Burroughs, Shoshona Le, Katie Dillon, Peter Cook, Clinton R. Paden, Dhwani Batra, Krista Queen, Kristen Knipe, Dakota Howard, Yvette Unoarumhi, Darlene Wagner, Matthew Schmerer, Ben L. Rambo-Martin, Kristine Lacek, Sam Shepard, Alison Laufer Halpin, Dave Wentworth, Vivien Dugan, Suxiang Tong, Justin Lee |
| EPI_ISL_1516313, EPI_ISL_1516314, EPI_ISL_1516315, EPI_ISL_1516316 | KS Health and Environmental Laboratories | Centers for Disease Control and Prevention Division of Viral Diseases, Pathogen Discovery | Mili Sheth, Sarah Nobles, Jasmine Padilla, Mark Burroughs, Shoshona Le, Katie Dillon, Peter Cook, Clinton R. Paden, Dhwani Batra, Krista Queen, Kristen Knipe, Dakota Howard, Yvette Unoarumhi, Darlene Wagner, Matthew Schmerer, Ben L. Rambo-Martin, Kristine Lacek, Sam Shepard, Alison Laufer Halpin, Dave Wentworth, Vivien Dugan, Suxiang Tong, Justin Lee |
| EPI_ISL_1516317, EPI_ISL_1516318, EPI_ISL_1516319, EPI_ISL_1516320, EPI_ISL_1516321 | AR Dept. of Health-PHL, Molecular Diagnostics | Centers for Disease Control and Prevention Division of Viral Diseases, Pathogen Discovery | Mili Sheth, Sarah Nobles, Jasmine Padilla, Mark Burroughs, Shoshona Le, Katie Dillon, Peter Cook, Clinton R. Paden, Dhwani Batra, Krista Queen, Kristen Knipe, Dakota Howard, Yvette Unoarumhi, Darlene Wagner, Matthew Schmerer, Ben L. Rambo-Martin, Kristine Lacek, Sam Shepard, Alison Laufer Halpin, Dave Wentworth, Vivien Dugan, Suxiang Tong, Justin Lee |
| EPI_ISL_1516322, EPI_ISL_1516323, EPI_ISL_1516324, EPI_ISL_1516325, EPI_ISL_1516326, EPI_ISL_1516327, EPI_ISL_1516328, EPI_ISL_1516329, EPI_ISL_1516330, EPI_ISL_1516331, EPI_ISL_1516332, EPI_ISL_1516333, EPI_ISL_1516334, EPI_ISL_1516335, EPI_ISL_1516336, EPI_ISL_1516337, EPI_ISL_1516338, EPI_ISL_1516339, EPI_ISL_1516340, EPI_ISL_1516341, EPI_ISL_1516342 | | | |
| see above | MD DOH Laboratories Administration | Centers for Disease Control and Prevention Division of Viral Diseases, Pathogen Discovery | Mili Sheth, Sarah Nobles, Jasmine Padilla, Mark Burroughs, Shoshona Le, Katie Dillon, Peter Cook, Clinton R. Paden, Dhwani Batra, Krista Queen, Kristen Knipe, Dakota Howard, Yvette Unoarumhi, Darlene Wagner, Matthew Schmerer, Ben L. Rambo-Martin, Kristine Lacek, Sam Shepard, Alison Laufer Halpin, Dave Wentworth, Vivien Dugan, Suxiang Tong, Justin Lee |
| EPI_ISL_1516343, EPI_ISL_1516344, EPI_ISL_1516345, | UT-Unified State Labs: Public Health Utah DOH | Centers for Disease Control and Prevention Division of Viral Diseases, Pathogen Discovery | Mili Sheth, Sarah Nobles, Jasmine Padilla, Mark Burroughs, Shoshona Le, Katie Dillon, Peter Cook, Clinton R. Paden, Dhwani Batra, Krista Queen, Kristen Knipe, Dakota Howard, Yvette Unoarumhi, Darlene Wagner, Matthew Schmerer, Ben L. Rambo-Martin, Kristine Lacek, Sam Shepard, Alison Laufer Halpin, Dave Wentworth, Vivien Dugan, Suxiang Tong, Justin Lee |

| | | | |
|---|---|---|---|
| EPI_ISL_1516346, EPI_ISL_1516347, EPI_ISL_1516348, EPI_ISL_1516349, EPI_ISL_1516350, EPI_ISL_1516351, EPI_ISL_1516352 | | | |
| EPI_ISL_1516353, EPI_ISL_1516354, EPI_ISL_1516355, EPI_ISL_1516356, EPI_ISL_1516357, EPI_ISL_1516358, EPI_ISL_1516359, EPI_ISL_1516360 | KY State Public Health Lab | Centers for Disease Control and Prevention Division of Viral Diseases, Pathogen Discovery | Mili Sheth, Sarah Nobles, Jasmine Padilla, Mark Burroughs, Shoshona Le, Katie Dillon, Peter Cook, Clinton R. Paden, Dhwani Batra, Krista Queen, Kristen Knipe, Dakota Howard, Yvette Unoarumhi, Darlene Wagner, Matthew Schmerer, Ben L. Rambo-Martin, Kristine Lacek, Sam Shepard, Alison Laufer Halpin, Dave Wentworth, Vivien Dugan, Suxiang Tong, Justin Lee |
| EPI_ISL_1516361 | CA-Los Angeles County Public Health Laboratory | Centers for Disease Control and Prevention Division of Viral Diseases, Pathogen Discovery | Mili Sheth, Sarah Nobles, Jasmine Padilla, Mark Burroughs, Shoshona Le, Katie Dillon, Peter Cook, Clinton R. Paden, Dhwani Batra, Krista Queen, Kristen Knipe, Dakota Howard, Yvette Unoarumhi, Darlene Wagner, Matthew Schmerer, Ben L. Rambo-Martin, Kristine Lacek, Sam Shepard, Alison Laufer Halpin, Dave Wentworth, Vivien Dugan, Suxiang Tong, Justin Lee |
| EPI_ISL_1516362, EPI_ISL_1516363 | MI - Michigan Department of Health and Human Services - Bureau of Laboratories | Centers for Disease Control and Prevention Division of Viral Diseases, Pathogen Discovery | Mili Sheth, Sarah Nobles, Jasmine Padilla, Mark Burroughs, Shoshona Le, Katie Dillon, Peter Cook, Clinton R. Paden, Dhwani Batra, Krista Queen, Kristen Knipe, Dakota Howard, Yvette Unoarumhi, Darlene Wagner, Matthew Schmerer, Ben L. Rambo-Martin, Kristine Lacek, Sam Shepard, Alison Laufer Halpin, Dave Wentworth, Vivien Dugan, Suxiang Tong, Justin Lee |
| EPI_ISL_1516364, EPI_ISL_1516365, EPI_ISL_1516366, EPI_ISL_1516367 | CT-Dr. Katherine A. Kelley State Public Health Lab | Centers for Disease Control and Prevention Division of Viral Diseases, Pathogen Discovery | Mili Sheth, Sarah Nobles, Jasmine Padilla, Mark Burroughs, Shoshona Le, Katie Dillon, Peter Cook, Clinton R. Paden, Dhwani Batra, Krista Queen, Kristen Knipe, Dakota Howard, Yvette Unoarumhi, Darlene Wagner, Matthew Schmerer, Ben L. Rambo-Martin, Kristine Lacek, Sam Shepard, Alison Laufer Halpin, Dave Wentworth, Vivien Dugan, Suxiang Tong, Justin Lee |
| EPI_ISL_1516368, EPI_ISL_1516369, EPI_ISL_1516370, EPI_ISL_1516371, EPI_ISL_1516372 | MS Public Health Laboratory | Centers for Disease Control and Prevention Division of Viral Diseases, Pathogen Discovery | Mili Sheth, Sarah Nobles, Jasmine Padilla, Mark Burroughs, Shoshona Le, Katie Dillon, Peter Cook, Clinton R. Paden, Dhwani Batra, Krista Queen, Kristen Knipe, Dakota Howard, Yvette Unoarumhi, Darlene Wagner, Matthew Schmerer, Ben L. Rambo-Martin, Kristine Lacek, Sam Shepard, Alison Laufer Halpin, Dave Wentworth, Vivien Dugan, Suxiang Tong, Justin Lee |
| EPI_ISL_1516373, EPI_ISL_1516374, EPI_ISL_1516375, EPI_ISL_1516376 | OH Department of Health Laboratory | Centers for Disease Control and Prevention Division of Viral Diseases, Pathogen Discovery | Mili Sheth, Sarah Nobles, Jasmine Padilla, Mark Burroughs, Shoshona Le, Katie Dillon, Peter Cook, Clinton R. Paden, Dhwani Batra, Krista Queen, Kristen Knipe, Dakota Howard, Yvette Unoarumhi, Darlene Wagner, Matthew Schmerer, Ben L. Rambo-Martin, Kristine Lacek, Sam Shepard, Alison Laufer Halpin, Dave Wentworth, Vivien Dugan, Suxiang Tong, Justin Lee |
| EPI_ISL_1516377, EPI_ISL_1516378, EPI_ISL_1516379, EPI_ISL_1516380, EPI_ISL_1516381, EPI_ISL_1516382, EPI_ISL_1516383, EPI_ISL_1516384, EPI_ISL_1516385, EPI_ISL_1516386, EPI_ISL_1516387 | | | |
| see above | RI State Health Laboratories | Centers for Disease Control and Prevention Division of Viral Diseases, Pathogen Discovery | Mili Sheth, Sarah Nobles, Jasmine Padilla, Mark Burroughs, Shoshona Le, Katie Dillon, Peter Cook, Clinton R. Paden, Dhwani Batra, Krista Queen, Kristen Knipe, Dakota Howard, Yvette Unoarumhi, Darlene Wagner, Matthew Schmerer, Ben L. Rambo-Martin, Kristine Lacek, Sam Shepard, Alison Laufer Halpin, Dave Wentworth, Vivien Dugan, Suxiang Tong, Justin Lee |
| EPI_ISL_1516388, EPI_ISL_1516389, EPI_ISL_1516390, EPI_ISL_1516391, EPI_ISL_1516392, EPI_ISL_1516393, EPI_ISL_1516394, EPI_ISL_1516395, EPI_ISL_1516396, EPI_ISL_1516397, EPI_ISL_1516398, EPI_ISL_1516399 | | | |
| see above | NC State Laboratory of Public Health | Centers for Disease Control and Prevention Division of Viral Diseases, Pathogen Discovery | Mili Sheth, Sarah Nobles, Jasmine Padilla, Mark Burroughs, Shoshona Le, Katie Dillon, Peter Cook, Clinton R. Paden, Dhwani Batra, Krista Queen, Kristen Knipe, Dakota Howard, Yvette Unoarumhi, Darlene Wagner, Matthew Schmerer, Ben L. Rambo-Martin, Kristine Lacek, Sam Shepard, Alison Laufer Halpin, Dave Wentworth, Vivien Dugan, Suxiang Tong, Justin Lee |
| EPI_ISL_1516400 | MO State Public Health Laboratory | Centers for Disease Control and Prevention Division of Viral Diseases, Pathogen Discovery | Mili Sheth, Sarah Nobles, Jasmine Padilla, Mark Burroughs, Shoshona Le, Katie Dillon, Peter Cook, Clinton R. Paden, Dhwani Batra, Krista Queen, Kristen Knipe, Dakota Howard, Yvette Unoarumhi, Darlene Wagner, Matthew Schmerer, Ben L. Rambo-Martin, Kristine Lacek, Sam Shepard, Alison Laufer Halpin, Dave Wentworth, Vivien Dugan, Suxiang Tong, Justin Lee |
| EPI_ISL_1516401, EPI_ISL_1516402, EPI_ISL_1516403, EPI_ISL_1516404, EPI_ISL_1516405, EPI_ISL_1516406, EPI_ISL_1516407, EPI_ISL_1516408, EPI_ISL_1516409, EPI_ISL_1516410, EPI_ISL_1516411, EPI_ISL_1516412, EPI_ISL_1516413, EPI_ISL_1516414, EPI_ISL_1516415, EPI_ISL_1516416, EPI_ISL_1516417, EPI_ISL_1516418, EPI_ISL_1516419, EPI_ISL_1516420, EPI_ISL_1516421, EPI_ISL_1516422, EPI_ISL_1516423 | | | |
| see above | LA Office of Public Health Laboratories | Centers for Disease Control and Prevention Division of Viral Diseases, Pathogen Discovery | Mili Sheth, Sarah Nobles, Jasmine Padilla, Mark Burroughs, Shoshona Le, Katie Dillon, Peter Cook, Clinton R. Paden, Dhwani Batra, Krista Queen, Kristen Knipe, Dakota Howard, Yvette Unoarumhi, Darlene Wagner, Matthew Schmerer, Ben L. Rambo-Martin, Kristine Lacek, Sam Shepard, Alison Laufer Halpin, Dave Wentworth, Vivien Dugan, Suxiang Tong, Justin Lee |
| EPI_ISL_1516424, EPI_ISL_1516425, EPI_ISL_1516426, EPI_ISL_1516427, EPI_ISL_1516428, EPI_ISL_1516429, EPI_ISL_1516430, EPI_ISL_1516431 | IA State Hygienic Laboratory | Centers for Disease Control and Prevention Division of Viral Diseases, Pathogen Discovery | Mili Sheth, Sarah Nobles, Jasmine Padilla, Mark Burroughs, Shoshona Le, Katie Dillon, Peter Cook, Clinton R. Paden, Dhwani Batra, Krista Queen, Kristen Knipe, Dakota Howard, Yvette Unoarumhi, Darlene Wagner, Matthew Schmerer, Ben L. Rambo-Martin, Kristine Lacek, Sam Shepard, Alison Laufer Halpin, Dave Wentworth, Vivien Dugan, Suxiang Tong, Justin Lee |
| EPI_ISL_1516432, EPI_ISL_1516433, EPI_ISL_1516434, EPI_ISL_1516435, EPI_ISL_1516436, EPI_ISL_1516437, EPI_ISL_1516438, EPI_ISL_1516439, EPI_ISL_1516440, EPI_ISL_1516441, EPI_ISL_1516442, EPI_ISL_1516443, EPI_ISL_1516444 | | | |
| see above | IN State Department of Health Laboratory Services | Centers for Disease Control and Prevention Division of Viral Diseases, Pathogen Discovery | Mili Sheth, Sarah Nobles, Jasmine Padilla, Mark Burroughs, Shoshona Le, Katie Dillon, Peter Cook, Clinton R. Paden, Dhwani Batra, Krista Queen, Kristen Knipe, Dakota Howard, Yvette Unoarumhi, Darlene Wagner, Matthew Schmerer, Ben L. Rambo-Martin, Kristine Lacek, Sam Shepard, Alison Laufer Halpin, Dave Wentworth, Vivien Dugan, Suxiang Tong, Justin Lee |
| EPI_ISL_1516446, EPI_ISL_1516447, EPI_ISL_1516448, EPI_ISL_1516449, EPI_ISL_1516450, EPI_ISL_1516451, EPI_ISL_1516452, EPI_ISL_1516453, EPI_ISL_1516454, EPI_ISL_1516455, EPI_ISL_1516456, EPI_ISL_1516457, EPI_ISL_1516458, EPI_ISL_1516459, EPI_ISL_1516460, EPI_ISL_1516461, EPI_ISL_1516462, EPI_ISL_1516463, EPI_ISL_1516464, EPI_ISL_1516465, EPI_ISL_1516466, EPI_ISL_1516467, EPI_ISL_1516468, EPI_ISL_1516469, EPI_ISL_1516470, EPI_ISL_1516471, EPI_ISL_1516472, EPI_ISL_1516473, EPI_ISL_1516474, EPI_ISL_1516475, EPI_ISL_1516476, EPI_ISL_1516477, EPI_ISL_1516478, EPI_ISL_1516479, EPI_ISL_1516480, EPI_ISL_1516481, EPI_ISL_1516482, EPI_ISL_1516483, EPI_ISL_1516484, EPI_ISL_1516485, EPI_ISL_1516486, EPI_ISL_1516487, EPI_ISL_1516488, EPI_ISL_1516489, EPI_ISL_1516490, EPI_ISL_1516491, EPI_ISL_1516492, EPI_ISL_1516493, EPI_ISL_1516494, EPI_ISL_1516495, EPI_ISL_1516496, EPI_ISL_1516497, EPI_ISL_1516498, EPI_ISL_1516499, EPI_ISL_1516500, EPI_ISL_1516501, EPI_ISL_1516502, EPI_ISL_1516503, EPI_ISL_1516504, EPI_ISL_1516505, EPI_ISL_1516506, EPI_ISL_1516507, EPI_ISL_1516508, EPI_ISL_1516509, EPI_ISL_1516510, EPI_ISL_1516511, EPI_ISL_1516512, EPI_ISL_1516513, EPI_ISL_1516514, EPI_ISL_1516515, EPI_ISL_1516516, EPI_ISL_1516517, EPI_ISL_1516518, EPI_ISL_1516519, EPI_ISL_1516520, EPI_ISL_1516521, EPI_ISL_1516522, EPI_ISL_1516523, EPI_ISL_1516524, EPI_ISL_1516525, EPI_ISL_1516526, EPI_ISL_1516527, EPI_ISL_1516528, EPI_ISL_1516529, EPI_ISL_1516530, EPI_ISL_1516531, EPI_ISL_1516532, EPI_ISL_1516533, EPI_ISL_1516534, EPI_ISL_1516535, EPI_ISL_1516536, EPI_ISL_1516537, EPI_ISL_1516538, EPI_ISL_1516539, EPI_ISL_1516540, EPI_ISL_1516541, EPI_ISL_1516542, EPI_ISL_1516543, EPI_ISL_1516544, EPI_ISL_1516545, EPI_ISL_1516546, EPI_ISL_1516547, EPI_ISL_1516548, EPI_ISL_1516549, EPI_ISL_1516550, EPI_ISL_1516551, EPI_ISL_1516552, EPI_ISL_1516553, EPI_ISL_1516554, EPI_ISL_1516555, EPI_ISL_1516556, EPI_ISL_1516557, EPI_ISL_1516558, EPI_ISL_1516559, EPI_ISL_1516560, EPI_ISL_1516561, EPI_ISL_1516562, EPI_ISL_1516563, EPI_ISL_1516564, EPI_ISL_1516565, EPI_ISL_1516566, EPI_ISL_1516567, EPI_ISL_1516568, EPI_ISL_1516569, EPI_ISL_1516570, EPI_ISL_1516571, EPI_ISL_1516572, EPI_ISL_1516573, EPI_ISL_1516574, EPI_ISL_1516575, EPI_ISL_1516576, EPI_ISL_1516577, EPI_ISL_1516578, EPI_ISL_1516579, EPI_ISL_1516580, EPI_ISL_1516581, EPI_ISL_1516582, EPI_ISL_1516583, EPI_ISL_1516584, EPI_ISL_1516585, EPI_ISL_1516586, EPI_ISL_1516587, EPI_ISL_1516588, EPI_ISL_1516589, EPI_ISL_1516590, EPI_ISL_1516591, EPI_ISL_1516592, EPI_ISL_1516593, EPI_ISL_1516594, EPI_ISL_1516595, EPI_ISL_1516596, EPI_ISL_1516597 | | | |
| see above | Broad Institute Clinical Research Sequencing Platform | Infectious Disease Program, Broad Institute of Harvard and MIT | Siddle,K.J., Adams,G., Pearlman,L., Gladden-Young,A., Vicente,G., Blumenstiel,B., DeFelice,M., Lee,M., McGovern,S., Lagerborg,K., Rudy,M., DeRuff,K., Carter,A., Normandin,E., Bauer,M., Reilly,S., Tomkins-Tinch,C., Loreth,C., Chaluvadi,S., Meldrim,J., Granger,B., Lemieux,J.E., Birren,B.W., Sabeti,P.C., Larkin,K., Dodge,S., Lennon,N., Madoff,L., Brown,C., Gallagher,G., Smole,S., Park,D.J., Gabriel,S., and MacInnis,B.L. |
| EPI_ISL_1516598, EPI_ISL_1516600, EPI_ISL_1516601, EPI_ISL_1516602, EPI_ISL_1516603, EPI_ISL_1516604, EPI_ISL_1516605, EPI_ISL_1516606, EPI_ISL_1516607, EPI_ISL_1516608, EPI_ISL_1516609, EPI_ISL_1516610, EPI_ISL_1516611, EPI_ISL_1516612, EPI_ISL_1516613, EPI_ISL_1516614, EPI_ISL_1516615, EPI_ISL_1516616, EPI_ISL_1516617, EPI_ISL_1516618, EPI_ISL_1516619, EPI_ISL_1516620, EPI_ISL_1516622, EPI_ISL_1516623, EPI_ISL_1516624, EPI_ISL_1516625, EPI_ISL_1516626, EPI_ISL_1516627, EPI_ISL_1516628, EPI_ISL_1516629, EPI_ISL_1516630, EPI_ISL_1516631, EPI_ISL_1516632, EPI_ISL_1516633, EPI_ISL_1516634, EPI_ISL_1516635, EPI_ISL_1516636, EPI_ISL_1516637, EPI_ISL_1516638, EPI_ISL_1516639, EPI_ISL_1516640, EPI_ISL_1516641, EPI_ISL_1516642, EPI_ISL_1516643, EPI_ISL_1516644, EPI_ISL_1516645, EPI_ISL_1516646, EPI_ISL_1516647, EPI_ISL_1516648, EPI_ISL_1516649, EPI_ISL_1516650, EPI_ISL_1516651, EPI_ISL_1516652, EPI_ISL_1516653, EPI_ISL_1516654, EPI_ISL_1516655, EPI_ISL_1516656, EPI_ISL_1516657, EPI_ISL_1516658, EPI_ISL_1516659, EPI_ISL_1516660, EPI_ISL_1516661, EPI_ISL_1516662, EPI_ISL_1516663, EPI_ISL_1516664 | | | |
| see above | Rhode Island Department of Health | Infectious Disease Program, Broad Institute of Harvard and MIT | Siddle,K.J., Azevedo,K., Miller,A., Adams,G., Pearlman,L., Gladden-Young,A., Lagerborg,K., Rudy,M., DeRuff,K., Carter,A., Normandin,E., Bauer,M., Reilly,S., Tomkins-Tinch,C., Loreth,C., Chaluvadi,S., Lemieux,J.E., Birren,B.W., Sabeti,P.C., Huard,R., King,E., Park,D.J., and MacInnis,B.L. |
| EPI_ISL_1516665, EPI_ISL_1516666, EPI_ISL_1516667, EPI_ISL_1516668, EPI_ISL_1516669, EPI_ISL_1516670, EPI_ISL_1516671, EPI_ISL_1516672, EPI_ISL_1516673, EPI_ISL_1516674, EPI_ISL_1516675, EPI_ISL_1516676, EPI_ISL_1516677, EPI_ISL_1516678, EPI_ISL_1516679, EPI_ISL_1516680, EPI_ISL_1516681, EPI_ISL_1516682, EPI_ISL_1516683, EPI_ISL_1516685, EPI_ISL_1516686, EPI_ISL_1516687, EPI_ISL_1516688, EPI_ISL_1516689, EPI_ISL_1516690, EPI_ISL_1516691, EPI_ISL_1516692, EPI_ISL_1516693, EPI_ISL_1516694, EPI_ISL_1516696, EPI_ISL_1516697, EPI_ISL_1516698, EPI_ISL_1516700, EPI_ISL_1516701, EPI_ISL_1516702, EPI_ISL_1516703, EPI_ISL_1516704, EPI_ISL_1516705, EPI_ISL_1516706, EPI_ISL_1516707, EPI_ISL_1516708, EPI_ISL_1516709, EPI_ISL_1516710, EPI_ISL_1516712, EPI_ISL_1516713, EPI_ISL_1516714, EPI_ISL_1516715, EPI_ISL_1516716, EPI_ISL_1516717, EPI_ISL_1516718, EPI_ISL_1516719, EPI_ISL_1516720, EPI_ISL_1516721, EPI_ISL_1516722, EPI_ISL_1516723, EPI_ISL_1516724, EPI_ISL_1516726, EPI_ISL_1516727, EPI_ISL_1516728, EPI_ISL_1516729, EPI_ISL_1516730, EPI_ISL_1516731, EPI_ISL_1516732, EPI_ISL_1516733, EPI_ISL_1516735, EPI_ISL_1516736, EPI_ISL_1516737, EPI_ISL_1516738, EPI_ISL_1516739, EPI_ISL_1516740, EPI_ISL_1516741, EPI_ISL_1516742, EPI_ISL_1516743, EPI_ISL_1516744, EPI_ISL_1516745 | | | |
| see above | Broad Institute Clinical Research Sequencing Platform | Infectious Disease Program, Broad Institute of Harvard and MIT | Siddle,K.J., Adams,G., Pearlman,L., Gladden-Young,A., Vicente,G., Blumenstiel,B., DeFelice,M., Lee,M., McGovern,S., Lagerborg,K., Rudy,M., DeRuff,K., Carter,A., Normandin,E., Bauer,M., Reilly,S., Tomkins-Tinch,C., Loreth,C., Chaluvadi,S., Meldrim,J., Granger,B., Lemieux,J.E., Birren,B.W., Sabeti,P.C., Larkin,K., Dodge,S., Lennon,N., Madoff,L., Brown,C., Gallagher,G., Smole,S., Park,D.J., Gabriel,S., and MacInnis,B.L. |
| EPI_ISL_1516746 | Flow Health | Infectious Disease Program, Broad Institute of Harvard and MIT | Lemieux,J.E., Siddle,K.J., Adams,G., Gladden-Young,A., Lagerborg,K., Rudy,M., DeRuff,K., Carter,A., Normandin,E., Bauer,M., Reilly,S., Tomkins-Tinch,C., Loreth,C., Chaluvadi,S., Birren,B.W., Gallagher,G., Smole,S., Park,D.J., MacInnis,B.L., and Sabeti,P.C. |
| EPI_ISL_1516749, EPI_ISL_1516750, EPI_ISL_1516751, EPI_ISL_1516752 | DC Public Health Lab/ Dept. of Forensic Sciences | DC Public Health Lab/ Dept. of Forensic Sciences | Janis Doss, Scott Nguyen, Elizabeth Zelaya, Sarah Scott, Connie Maza, Monica Mann, Brittany Hamilton, David Payne, Jocelyn Hauser |
| EPI_ISL_1516753 | Seattle Flu Study | Seattle Flu Study | Deborah A. Nickerson, Chris D. Frazar, Jover Lee, Benjamin Pelle, Erica Ryke, Matthew Richardson, Amanda Adler, Elisabeth Brandstetter, Peter D. Han, Kairsten Fay, Misja Ilcisin, Kirsten Lacombe, Thomas R. Sibley, Melissa Truong, Caitlin R. Wolf, Michael Boeckh, Janet A. Englund, Michael Famulare, Barry R. Lutz, Mark J. Rieder, Lea M. Starita, Matthew Thompson, Jay Shendure, Trevor Bedford, Helen Y. Chu |
| EPI_ISL_1516754, EPI_ISL_1516755, EPI_ISL_1516756, EPI_ISL_1516757, EPI_ISL_1516758, EPI_ISL_1516759 | Maryland Genomics, Institute for Genome Sciences, University of Maryland School of Medicine | Maryland Genomics, Institute for Genome Sciences, University of Maryland School of Medicine | Tallon, Luke J; Sadzewicz, Lisa D; Humphrys, Mike; Ott, Sandra; Roussey, Holly; Mehta, Aditya; Vavikolanu, Kranthi; Fraser, Claire M; Ravel, Jacques |
| EPI_ISL_1516762 | LESP Jalisco/UdeG | Instituto de Diagnostico y Referencia | Claudia Wong-Arambula, Abril Rodriguez-Maldonado, Vanessa Rivero-Arredondo, Ariadna Medina-Benitez, Joaquin Quiroz-Mercado, Sergio Rangel-Guerrero, Natividad Cruz-Ortiz, Tatiana Nunez-Garcia, Gisela Barrera-Badillo, Lucia Hernandez-Rivas, Irma Lopez-Martinez, Ernesto |

| | | | |
|---|---|---|---|
| | | Epidemiologicos (INDRE) | Ramirez-Gonzalez. |
| EPI_ISL_1516763 | LESP Hidalgo | Instituto de Diagnostico y Referencia Epidemiologicos (INDRE) | Claudia Wong-Arambula, Abril Rodriguez-Maldonado, Vanessa Rivero-Arredondo, Ariadna Medina-Benitez, Joaquin Quiroz-Mercado, Sergio Rangel-Guerrero, Natividad Cruz-Ortiz, Tatiana Nunez-Garcia, Gisela Barrera-Badillo, Lucia Hernandez-Rivas, Irma Lopez-Martinez, Ernesto Ramirez-Gonzalez. |
| EPI_ISL_1516764 | LESP Puebla | Instituto de Diagnostico y Referencia Epidemiologicos (INDRE) | Claudia Wong-Arambula, Abril Rodriguez-Maldonado, Vanessa Rivero-Arredondo, Ariadna Medina-Benitez, Joaquin Quiroz-Mercado, Sergio Rangel-Guerrero, Natividad Cruz-Ortiz, Tatiana Nunez-Garcia, Gisela Barrera-Badillo, Lucia Hernandez-Rivas, Irma Lopez-Martinez, Ernesto Ramirez-Gonzalez. |
| EPI_ISL_1516765 | LESP Jalisco | Instituto de Diagnostico y Referencia Epidemiologicos (INDRE) | Claudia Wong-Arambula, Abril Rodriguez-Maldonado, Vanessa Rivero-Arredondo, Ariadna Medina-Benitez, Joaquin Quiroz-Mercado, Sergio Rangel-Guerrero, Natividad Cruz-Ortiz, Tatiana Nunez-Garcia, Gisela Barrera-Badillo, Lucia Hernandez-Rivas, Irma Lopez-Martinez, Ernesto Ramirez-Gonzalez. |
| EPI_ISL_1516766, EPI_ISL_1516767, EPI_ISL_1516768 | LESP Chihuahua | Instituto de Diagnostico y Referencia Epidemiologicos (INDRE) | Claudia Wong-Arambula, Abril Rodriguez-Maldonado, Vanessa Rivero-Arredondo, Ariadna Medina-Benitez, Joaquin Quiroz-Mercado, Sergio Rangel-Guerrero, Natividad Cruz-Ortiz, Tatiana Nunez-Garcia, Gisela Barrera-Badillo, Lucia Hernandez-Rivas, Irma Lopez-Martinez, Ernesto Ramirez-Gonzalez. |
| EPI_ISL_1516769, EPI_ISL_1516770, EPI_ISL_1516771, EPI_ISL_1516772, EPI_ISL_1516773 | LESP Nuevo Leon | Instituto de Diagnostico y Referencia Epidemiologicos (INDRE) | Claudia Wong-Arambula, Abril Rodriguez-Maldonado, Vanessa Rivero-Arredondo, Ariadna Medina-Benitez, Joaquin Quiroz-Mercado, Sergio Rangel-Guerrero, Natividad Cruz-Ortiz, Tatiana Nunez-Garcia, Gisela Barrera-Badillo, Lucia Hernandez-Rivas, Irma Lopez-Martinez, Ernesto Ramirez-Gonzalez. |
| EPI_ISL_1516774 | LESP Yucatan | Instituto de Diagnostico y Referencia Epidemiologicos (INDRE) | Claudia Wong-Arambula, Abril Rodriguez-Maldonado, Vanessa Rivero-Arredondo, Ariadna Medina-Benitez, Joaquin Quiroz-Mercado, Sergio Rangel-Guerrero, Natividad Cruz-Ortiz, Tatiana Nunez-Garcia, Gisela Barrera-Badillo, Lucia Hernandez-Rivas, Irma Lopez-Martinez, Ernesto Ramirez-Gonzalez. |
| EPI_ISL_1516775 | LESP Queretaro | Instituto de Diagnostico y Referencia Epidemiologicos (INDRE) | Claudia Wong-Arambula, Abril Rodriguez-Maldonado, Vanessa Rivero-Arredondo, Ariadna Medina-Benitez, Joaquin Quiroz-Mercado, Sergio Rangel-Guerrero, Natividad Cruz-Ortiz, Tatiana Nunez-Garcia, Gisela Barrera-Badillo, Lucia Hernandez-Rivas, Irma Lopez-Martinez, Ernesto Ramirez-Gonzalez. |
| EPI_ISL_1516776 | LESP Tamaulipas | Instituto de Diagnostico y Referencia Epidemiologicos (INDRE) | Claudia Wong-Arambula, Abril Rodriguez-Maldonado, Vanessa Rivero-Arredondo, Ariadna Medina-Benitez, Joaquin Quiroz-Mercado, Sergio Rangel-Guerrero, Natividad Cruz-Ortiz, Tatiana Nunez-Garcia, Gisela Barrera-Badillo, Lucia Hernandez-Rivas, Irma Lopez-Martinez, Ernesto Ramirez-Gonzalez. |
| EPI_ISL_1516777, EPI_ISL_1516778, EPI_ISL_1516779, EPI_ISL_1516780, EPI_ISL_1516781, EPI_ISL_1516782, EPI_ISL_1516783, EPI_ISL_1516784, EPI_ISL_1516785, EPI_ISL_1516786 | LESP Aguascalientes | Instituto de Diagnostico y Referencia Epidemiologicos (INDRE) | Claudia Wong-Arambula, Abril Rodriguez-Maldonado, Vanessa Rivero-Arredondo, Ariadna Medina-Benitez, Joaquin Quiroz-Mercado, Sergio Rangel-Guerrero, Natividad Cruz-Ortiz, Tatiana Nunez-Garcia, Gisela Barrera-Badillo, Lucia Hernandez-Rivas, Irma Lopez-Martinez, Ernesto Ramirez-Gonzalez. |
| EPI_ISL_1516787, EPI_ISL_1516788 | LESP Ciudad de Mexico | Instituto de Diagnostico y Referencia Epidemiologicos (INDRE) | Claudia Wong-Arambula, Abril Rodriguez-Maldonado, Vanessa Rivero-Arredondo, Ariadna Medina-Benitez, Joaquin Quiroz-Mercado, Sergio Rangel-Guerrero, Natividad Cruz-Ortiz, Tatiana Nunez-Garcia, Gisela Barrera-Badillo, Lucia Hernandez-Rivas, Irma Lopez-Martinez, Ernesto Ramirez-Gonzalez. |
| EPI_ISL_1516789, EPI_ISL_1516790 | LESP Tamaulipas | Instituto de Diagnostico y Referencia Epidemiologicos (INDRE) | Claudia Wong-Arambula, Abril Rodriguez-Maldonado, Vanessa Rivero-Arredondo, Ariadna Medina-Benitez, Joaquin Quiroz-Mercado, Sergio Rangel-Guerrero, Natividad Cruz-Ortiz, Tatiana Nunez-Garcia, Gisela Barrera-Badillo, Lucia Hernandez-Rivas, Irma Lopez-Martinez, Ernesto Ramirez-Gonzalez. |
| EPI_ISL_1516791, EPI_ISL_1516792, EPI_ISL_1516793 | LESP Coahuila | Instituto de Diagnostico y Referencia Epidemiologicos (INDRE) | Claudia Wong-Arambula, Abril Rodriguez-Maldonado, Vanessa Rivero-Arredondo, Ariadna Medina-Benitez, Joaquin Quiroz-Mercado, Sergio Rangel-Guerrero, Natividad Cruz-Ortiz, Tatiana Nunez-Garcia, Gisela Barrera-Badillo, Lucia Hernandez-Rivas, Irma Lopez-Martinez, Ernesto Ramirez-Gonzalez. |
| EPI_ISL_1516794 | LESP Guanajuato | Instituto de Diagnostico y Referencia Epidemiologicos (INDRE) | Claudia Wong-Arambula, Abril Rodriguez-Maldonado, Vanessa Rivero-Arredondo, Ariadna Medina-Benitez, Joaquin Quiroz-Mercado, Sergio Rangel-Guerrero, Natividad Cruz-Ortiz, Tatiana Nunez-Garcia, Gisela Barrera-Badillo, Lucia Hernandez-Rivas, Irma Lopez-Martinez, Ernesto Ramirez-Gonzalez. |
| EPI_ISL_1516795 | LESP Jalisco | Instituto de Diagnostico y Referencia Epidemiologicos (INDRE) | Claudia Wong-Arambula, Abril Rodriguez-Maldonado, Vanessa Rivero-Arredondo, Ariadna Medina-Benitez, Joaquin Quiroz-Mercado, Sergio Rangel-Guerrero, Natividad Cruz-Ortiz, Tatiana Nunez-Garcia, Gisela Barrera-Badillo, Lucia Hernandez-Rivas, Irma Lopez-Martinez, Ernesto Ramirez-Gonzalez. |
| EPI_ISL_1516796 | LESP Guanajuato | Instituto de Diagnostico y Referencia Epidemiologicos (INDRE) | Claudia Wong-Arambula, Abril Rodriguez-Maldonado, Vanessa Rivero-Arredondo, Ariadna Medina-Benitez, Joaquin Quiroz-Mercado, Sergio Rangel-Guerrero, Natividad Cruz-Ortiz, Tatiana Nunez-Garcia, Gisela Barrera-Badillo, Lucia Hernandez-Rivas, Irma Lopez-Martinez, Ernesto Ramirez-Gonzalez. |
| EPI_ISL_1516797, EPI_ISL_1516798, EPI_ISL_1516799, EPI_ISL_1516800, EPI_ISL_1516801, EPI_ISL_1516802, EPI_ISL_1516803, EPI_ISL_1516804, EPI_ISL_1516805, EPI_ISL_1516806, EPI_ISL_1516807 | | | |
| see above | LESP Nuevo Leon | Instituto de Diagnostico y Referencia Epidemiologicos (INDRE) | Claudia Wong-Arambula, Abril Rodriguez-Maldonado, Vanessa Rivero-Arredondo, Ariadna Medina-Benitez, Joaquin Quiroz-Mercado, Sergio Rangel-Guerrero, Natividad Cruz-Ortiz, Tatiana Nunez-Garcia, Gisela Barrera-Badillo, Lucia Hernandez-Rivas, Irma Lopez-Martinez, Ernesto Ramirez-Gonzalez. |
| EPI_ISL_1516808, EPI_ISL_1516809, EPI_ISL_1516810, EPI_ISL_1516811, EPI_ISL_1516812, EPI_ISL_1516813, EPI_ISL_1516814, EPI_ISL_1516815 | LESP Morelos | Instituto de Diagnostico y Referencia Epidemiologicos (INDRE) | Claudia Wong-Arambula, Abril Rodriguez-Maldonado, Vanessa Rivero-Arredondo, Ariadna Medina-Benitez, Joaquin Quiroz-Mercado, Sergio Rangel-Guerrero, Natividad Cruz-Ortiz, Tatiana Nunez-Garcia, Gisela Barrera-Badillo, Lucia Hernandez-Rivas, Irma Lopez-Martinez, Ernesto Ramirez-Gonzalez. |
| EPI_ISL_1516816 | LESP Chihuahua | Instituto de Diagnostico y Referencia Epidemiologicos (INDRE) | Claudia Wong-Arambula, Abril Rodriguez-Maldonado, Vanessa Rivero-Arredondo, Ariadna Medina-Benitez, Joaquin Quiroz-Mercado, Sergio Rangel-Guerrero, Natividad Cruz-Ortiz, Tatiana Nunez-Garcia, Gisela Barrera-Badillo, Lucia Hernandez-Rivas, Irma Lopez-Martinez, Ernesto Ramirez-Gonzalez. |
| EPI_ISL_1516817 | LESP Ciudad de Mexico | Instituto de Diagnostico y Referencia Epidemiologicos (INDRE) | Claudia Wong-Arambula, Abril Rodriguez-Maldonado, Vanessa Rivero-Arredondo, Ariadna Medina-Benitez, Joaquin Quiroz-Mercado, Sergio Rangel-Guerrero, Natividad Cruz-Ortiz, Tatiana Nunez-Garcia, Gisela Barrera-Badillo, Lucia Hernandez-Rivas, Irma Lopez-Martinez, Ernesto Ramirez-Gonzalez. |
| EPI_ISL_1516818, EPI_ISL_1516819, EPI_ISL_1516820, EPI_ISL_1516821, EPI_ISL_1516822, EPI_ISL_1516823 | LESP Chihuahua | Instituto de Diagnostico y Referencia Epidemiologicos (INDRE) | Claudia Wong-Arambula, Abril Rodriguez-Maldonado, Vanessa Rivero-Arredondo, Ariadna Medina-Benitez, Joaquin Quiroz-Mercado, Sergio Rangel-Guerrero, Natividad Cruz-Ortiz, Tatiana Nunez-Garcia, Gisela Barrera-Badillo, Lucia Hernandez-Rivas, Irma Lopez-Martinez, Ernesto Ramirez-Gonzalez. |
| EPI_ISL_1516824, EPI_ISL_1516825, EPI_ISL_1516826 | LESP Queretaro | Instituto de Diagnostico y Referencia Epidemiologicos (INDRE) | Claudia Wong-Arambula, Abril Rodriguez-Maldonado, Vanessa Rivero-Arredondo, Ariadna Medina-Benitez, Joaquin Quiroz-Mercado, Sergio Rangel-Guerrero, Natividad Cruz-Ortiz, Tatiana Nunez-Garcia, Gisela Barrera-Badillo, Lucia Hernandez-Rivas, Irma Lopez-Martinez, Ernesto Ramirez-Gonzalez. |
| EPI_ISL_1520143 | Northwestern Medicine | Illinois Department of Public Health - Chicago Lab | Vineet K. Dhiman, Ira Heimler |
| EPI_ISL_1520144, EPI_ISL_1520145 | Quest Diagnostics | Illinois Department of Public Health - Chicago Lab | Vineet K. Dhiman, Ira Heimler |
| EPI_ISL_1520146, EPI_ISL_1520147, EPI_ISL_1520148, EPI_ISL_1520149, EPI_ISL_1520150, EPI_ISL_1520151, EPI_ISL_1520152, EPI_ISL_1520153, EPI_ISL_1520154 | Riverside Medical Center | Illinois Department of Public Health - Chicago Lab | Vineet K. Dhiman, Ira Heimler |
| EPI_ISL_1520155 | Alverno Laboratories | Illinois Department of Public Health - Chicago Lab | Vineet K. Dhiman, Ira Heimler |
| EPI_ISL_1520156, EPI_ISL_1520157 | Central DuPage Hospital | Illinois Department of Public Health - Chicago Lab | Vineet K. Dhiman, Ira Heimler |
| EPI_ISL_1520159, EPI_ISL_1520160, EPI_ISL_1520161, EPI_ISL_1520162, EPI_ISL_1520163, EPI_ISL_1520164 | The University of Chicago Medical Center | Illinois Department of Public Health - Chicago Lab | Vineet K. Dhiman, Ira Heimler |
| EPI_ISL_1520165 | Northwestern Medicine | Illinois Department of Public Health - Chicago Lab | Vineet K. Dhiman, Ira Heimler |
| EPI_ISL_1520166 | CGH Medical Center | Illinois Department of Public Health - Chicago Lab | Vineet K. Dhiman, Ira Heimler |
| EPI_ISL_1520167 | Northwest Community Hospital | Illinois Department of Public Health - Chicago | Vineet K. Dhiman, Ira Heimler |

| | | | |
|---|---|---|---|
| | | Lab | |
| EPI_ISL_1520168 | Pathnostics Laboratory | Illinois Department of Public Health - Chicago Lab | Vineet K. Dhiman, Ira Heimler |
| EPI_ISL_1522293, EPI_ISL_1522295, EPI_ISL_1522297, EPI_ISL_1522299, EPI_ISL_1522302, EPI_ISL_1522304, EPI_ISL_1522306, EPI_ISL_1522309, EPI_ISL_1522311, EPI_ISL_1522313, EPI_ISL_1522318, EPI_ISL_1522321, EPI_ISL_1522324, EPI_ISL_1522326, EPI_ISL_1522328, EPI_ISL_1522331, EPI_ISL_1522333, EPI_ISL_1522336, EPI_ISL_1522339, EPI_ISL_1522341, EPI_ISL_1522346, EPI_ISL_1522349, EPI_ISL_1522351, EPI_ISL_1522354, EPI_ISL_1522357, EPI_ISL_1522359, EPI_ISL_1522361, EPI_ISL_1522363, EPI_ISL_1522365, EPI_ISL_1522367, EPI_ISL_1522368, EPI_ISL_1522369, EPI_ISL_1522371, EPI_ISL_1522372, EPI_ISL_1522373, EPI_ISL_1522374, EPI_ISL_1522375, EPI_ISL_1522376, EPI_ISL_1522377, EPI_ISL_1522379, EPI_ISL_1522380, EPI_ISL_1522381 | | | |
| see above | Curative | New Mexico Department of Health Scientific Laboratory | Ellie Johnson, Anastacia Griego-Fisher, D'eldra Malone, Jennifer Benoit, Linda Salazar, Ratheesh Rajan |
| EPI_ISL_1523525, EPI_ISL_1523526, EPI_ISL_1523527, EPI_ISL_1523528, EPI_ISL_1523529, EPI_ISL_1523530, EPI_ISL_1523531, EPI_ISL_1523532, EPI_ISL_1523533, EPI_ISL_1523534, EPI_ISL_1523535, EPI_ISL_1523536, EPI_ISL_1523537, EPI_ISL_1523538, EPI_ISL_1523539, EPI_ISL_1523540, EPI_ISL_1523541, EPI_ISL_1523542, EPI_ISL_1523543 | | | |
| see above | Oregon State Public Health Laboratory | Oregon State Public Health Laboratory | LaDonna Grenz, Karim Morey, Joel R. Sevinksy, Kevin, G. Libuit, John Fontana, Shane Sevey |
| EPI_ISL_1523643, EPI_ISL_1523644, EPI_ISL_1523645, EPI_ISL_1523646, EPI_ISL_1523647, EPI_ISL_1523648, EPI_ISL_1523649, EPI_ISL_1523650, EPI_ISL_1523651, EPI_ISL_1523652, EPI_ISL_1523653, EPI_ISL_1523654, EPI_ISL_1523655, EPI_ISL_1523656, EPI_ISL_1523657, EPI_ISL_1523658, EPI_ISL_1523659, EPI_ISL_1523660, EPI_ISL_1523661, EPI_ISL_1523662, EPI_ISL_1523663, EPI_ISL_1523664, EPI_ISL_1523665, EPI_ISL_1523667, EPI_ISL_1523668, EPI_ISL_1523669, EPI_ISL_1523670, EPI_ISL_1523671, EPI_ISL_1523672, EPI_ISL_1523673, EPI_ISL_1523674, EPI_ISL_1523675, EPI_ISL_1523676, EPI_ISL_1523677, EPI_ISL_1523678, EPI_ISL_1523679, EPI_ISL_1523680, EPI_ISL_1523681, EPI_ISL_1523682, EPI_ISL_1523683, EPI_ISL_1523684, EPI_ISL_1523685, EPI_ISL_1523686, EPI_ISL_1523687, EPI_ISL_1523688, EPI_ISL_1523689, EPI_ISL_1523690, EPI_ISL_1523691, EPI_ISL_1523692, EPI_ISL_1523693, EPI_ISL_1523694, EPI_ISL_1523695, EPI_ISL_1523696, EPI_ISL_1523697, EPI_ISL_1523698, EPI_ISL_1523699, EPI_ISL_1523700, EPI_ISL_1523701, EPI_ISL_1523702, EPI_ISL_1523703, EPI_ISL_1523704, EPI_ISL_1523705, EPI_ISL_1523706, EPI_ISL_1523707, EPI_ISL_1523708, EPI_ISL_1523709, EPI_ISL_1523710, EPI_ISL_1523711, EPI_ISL_1523712, EPI_ISL_1523713, EPI_ISL_1523714, EPI_ISL_1523715, EPI_ISL_1523716, EPI_ISL_1523717, EPI_ISL_1523718, EPI_ISL_1523719, EPI_ISL_1523720, EPI_ISL_1523721, EPI_ISL_1523722, EPI_ISL_1523723, EPI_ISL_1523724, EPI_ISL_1523725, EPI_ISL_1523726, EPI_ISL_1523727, EPI_ISL_1523728, EPI_ISL_1523729, EPI_ISL_1523730, EPI_ISL_1523731, EPI_ISL_1523732, EPI_ISL_1523733, EPI_ISL_1523734, EPI_ISL_1523735, EPI_ISL_1523736, EPI_ISL_1523737, EPI_ISL_1523738, EPI_ISL_1523739, EPI_ISL_1523740, EPI_ISL_1523741, EPI_ISL_1523742, EPI_ISL_1523743, EPI_ISL_1523744, EPI_ISL_1523745, EPI_ISL_1523746, EPI_ISL_1523747, EPI_ISL_1523748, EPI_ISL_1523749, EPI_ISL_1523750, EPI_ISL_1523751, EPI_ISL_1523752, EPI_ISL_1523753, EPI_ISL_1523754, EPI_ISL_1523755, EPI_ISL_1523756, EPI_ISL_1523757, EPI_ISL_1523758, EPI_ISL_1523759, EPI_ISL_1523760, EPI_ISL_1523761, EPI_ISL_1523762, EPI_ISL_1523763, EPI_ISL_1523764, EPI_ISL_1523765, EPI_ISL_1523766, EPI_ISL_1523767, EPI_ISL_1523768, EPI_ISL_1523769, EPI_ISL_1523770, EPI_ISL_1523771, EPI_ISL_1523772, EPI_ISL_1523773, EPI_ISL_1523774, EPI_ISL_1523775, EPI_ISL_1523777, EPI_ISL_1523778, EPI_ISL_1523779, EPI_ISL_1523780, EPI_ISL_1523781, EPI_ISL_1523782, EPI_ISL_1523783, EPI_ISL_1523784, EPI_ISL_1523785, EPI_ISL_1523786, EPI_ISL_1523787, EPI_ISL_1523788, EPI_ISL_1523789, EPI_ISL_1523790, EPI_ISL_1523791, EPI_ISL_1523792 | | | |
| see above | Maine Health and Environmental Testing Laboratory | Tewhey Lab, The Jackson Laboratory | Matluk,N., Dewey,H., Iosue,F., Barter,M., Lynch,R., Munger,H. and Tewhey,R. |
| EPI_ISL_1523794, EPI_ISL_1523796, EPI_ISL_1523797, EPI_ISL_1523799, EPI_ISL_1523800, EPI_ISL_1523801, EPI_ISL_1523802, EPI_ISL_1523803, EPI_ISL_1523804, EPI_ISL_1523805, EPI_ISL_1523806, EPI_ISL_1523807, EPI_ISL_1523808, EPI_ISL_1523809, EPI_ISL_1523810, EPI_ISL_1523811, EPI_ISL_1523812, EPI_ISL_1523813, EPI_ISL_1523814, EPI_ISL_1523815, EPI_ISL_1523816, EPI_ISL_1523817, EPI_ISL_1523818, EPI_ISL_1523819, EPI_ISL_1523820, EPI_ISL_1523821, EPI_ISL_1523822, EPI_ISL_1523823, EPI_ISL_1523824, EPI_ISL_1523825, EPI_ISL_1523826, EPI_ISL_1523827, EPI_ISL_1523828, EPI_ISL_1523829, EPI_ISL_1523830, EPI_ISL_1523831, EPI_ISL_1523832, EPI_ISL_1523833, EPI_ISL_1523834, EPI_ISL_1523835, EPI_ISL_1523836, EPI_ISL_1523837, EPI_ISL_1523838, EPI_ISL_1523839, EPI_ISL_1523840, EPI_ISL_1523841, EPI_ISL_1523842, EPI_ISL_1523843, EPI_ISL_1523844, EPI_ISL_1523845, EPI_ISL_1523846, EPI_ISL_1523847, EPI_ISL_1523848, EPI_ISL_1523849, EPI_ISL_1523850, EPI_ISL_1523851, EPI_ISL_1523852, EPI_ISL_1523853, EPI_ISL_1523854, EPI_ISL_1523855, EPI_ISL_1523856, EPI_ISL_1523857, EPI_ISL_1523858, EPI_ISL_1523859, EPI_ISL_1523860, EPI_ISL_1523861, EPI_ISL_1523862, EPI_ISL_1523863, EPI_ISL_1523864, EPI_ISL_1523865, EPI_ISL_1523866, EPI_ISL_1523867, EPI_ISL_1523868, EPI_ISL_1523869, EPI_ISL_1523870, EPI_ISL_1523871, EPI_ISL_1523872, EPI_ISL_1523873, EPI_ISL_1523874, EPI_ISL_1523875, EPI_ISL_1523876, EPI_ISL_1523877, EPI_ISL_1523878, EPI_ISL_1523879, EPI_ISL_1523880, EPI_ISL_1523881, EPI_ISL_1523882, EPI_ISL_1523883, EPI_ISL_1523884, EPI_ISL_1523885, EPI_ISL_1523886, EPI_ISL_1523887, EPI_ISL_1523888, EPI_ISL_1523889, EPI_ISL_1523890, EPI_ISL_1523891, EPI_ISL_1523892, EPI_ISL_1523893, EPI_ISL_1523894, EPI_ISL_1523895, EPI_ISL_1523896, EPI_ISL_1523897, EPI_ISL_1523898, EPI_ISL_1523899, EPI_ISL_1523900, EPI_ISL_1523901, EPI_ISL_1523902, EPI_ISL_1523903, EPI_ISL_1523904, EPI_ISL_1523905, EPI_ISL_1523906, EPI_ISL_1523907, EPI_ISL_1523908, EPI_ISL_1523909, EPI_ISL_1523910, EPI_ISL_1523911, EPI_ISL_1523912, EPI_ISL_1523913, EPI_ISL_1523914, EPI_ISL_1523915, EPI_ISL_1523916, EPI_ISL_1523917, EPI_ISL_1523918, EPI_ISL_1523919, EPI_ISL_1523920, EPI_ISL_1523921, EPI_ISL_1523922, EPI_ISL_1523923, EPI_ISL_1523924, EPI_ISL_1523925, EPI_ISL_1523926, EPI_ISL_1523927, EPI_ISL_1523928, EPI_ISL_1523929, EPI_ISL_1523930, EPI_ISL_1523931, EPI_ISL_1523932, EPI_ISL_1523933, EPI_ISL_1523934, EPI_ISL_1523935, EPI_ISL_1523937, EPI_ISL_1523939, EPI_ISL_1523940, EPI_ISL_1523941, EPI_ISL_1523942, EPI_ISL_1523943, EPI_ISL_1523944, EPI_ISL_1523945, EPI_ISL_1523946, EPI_ISL_1523947, EPI_ISL_1523948, EPI_ISL_1523949, EPI_ISL_1523950, EPI_ISL_1523951, EPI_ISL_1523952, EPI_ISL_1523953, EPI_ISL_1523954, EPI_ISL_1523955, EPI_ISL_1523956, EPI_ISL_1523957, EPI_ISL_1523958, EPI_ISL_1523959, EPI_ISL_1523960, EPI_ISL_1523961, EPI_ISL_1523962, EPI_ISL_1523963, EPI_ISL_1523964, EPI_ISL_1523965, EPI_ISL_1523966, EPI_ISL_1523967, EPI_ISL_1523969, EPI_ISL_1523970, EPI_ISL_1523971, EPI_ISL_1523972, EPI_ISL_1523973, EPI_ISL_1523974, EPI_ISL_1523975, EPI_ISL_1523976, EPI_ISL_1523977, EPI_ISL_1523978, EPI_ISL_1523979, EPI_ISL_1523980, EPI_ISL_1523981, EPI_ISL_1523982, EPI_ISL_1523983, EPI_ISL_1523984, EPI_ISL_1523985, EPI_ISL_1523986, EPI_ISL_1523987, EPI_ISL_1523988, EPI_ISL_1523989, EPI_ISL_1523990, EPI_ISL_1523991, EPI_ISL_1523992, EPI_ISL_1523993, EPI_ISL_1523994, EPI_ISL_1523995, EPI_ISL_1523996, EPI_ISL_1523997, EPI_ISL_1523998, EPI_ISL_1523999, EPI_ISL_1524000, EPI_ISL_1524001, EPI_ISL_1524002, EPI_ISL_1524003, EPI_ISL_1524004, EPI_ISL_1524005, EPI_ISL_1524007, EPI_ISL_1524008, EPI_ISL_1524009, EPI_ISL_1524010, EPI_ISL_1524011, EPI_ISL_1524012, EPI_ISL_1524013, EPI_ISL_1524014 | | | |
| see above | UW Virology Lab | UW Virology Lab | Pavitra Roychoudhury, Hong Xie, Lasata Shrestha, Shah Mohamed Bakhash, Michelle Lin, Noah R. Baker, Sean Ellis, Saraswathi Sathees, Meei-Li Huang, Keith R Jerome, Alexander Greninger |
| EPI_ISL_1525002, EPI_ISL_1525003, EPI_ISL_1525004, EPI_ISL_1525005, EPI_ISL_1525006, EPI_ISL_1525007, EPI_ISL_1525008, EPI_ISL_1525009, EPI_ISL_1525010, EPI_ISL_1525011, EPI_ISL_1525012, EPI_ISL_1525013, EPI_ISL_1525014, EPI_ISL_1525015, EPI_ISL_1525016, EPI_ISL_1525017, EPI_ISL_1525018 | | | |
| see above | Indiana State Department of Health | Quantigen Biosciences | Paul J Childress |
| EPI_ISL_1525019, EPI_ISL_1525020, EPI_ISL_1525021, EPI_ISL_1525022, EPI_ISL_1525023, EPI_ISL_1525024, EPI_ISL_1525025, EPI_ISL_1525026, EPI_ISL_1525028, EPI_ISL_1525029, EPI_ISL_1525030, EPI_ISL_1525031, EPI_ISL_1525033, EPI_ISL_1525034, EPI_ISL_1525035, EPI_ISL_1525036, EPI_ISL_1525037, EPI_ISL_1525038, EPI_ISL_1525039, EPI_ISL_1525040, EPI_ISL_1525041, EPI_ISL_1525042, EPI_ISL_1525043, EPI_ISL_1525044, EPI_ISL_1525046, EPI_ISL_1525047, EPI_ISL_1525048, EPI_ISL_1525049, EPI_ISL_1525050, EPI_ISL_1525051, EPI_ISL_1525052, EPI_ISL_1525053, EPI_ISL_1525054, EPI_ISL_1525055, EPI_ISL_1525056, EPI_ISL_1525058, EPI_ISL_1525059, EPI_ISL_1525060, EPI_ISL_1525061, EPI_ISL_1525062, EPI_ISL_1525063, EPI_ISL_1525064, EPI_ISL_1525065, EPI_ISL_1525066, EPI_ISL_1525067, EPI_ISL_1525068, EPI_ISL_1525069, EPI_ISL_1525070, EPI_ISL_1525071, EPI_ISL_1525072, EPI_ISL_1525073, EPI_ISL_1525074, EPI_ISL_1525075, EPI_ISL_1525076, EPI_ISL_1525077, EPI_ISL_1525078, EPI_ISL_1525079, EPI_ISL_1525080, EPI_ISL_1525081, EPI_ISL_1525082, EPI_ISL_1525083, EPI_ISL_1525084, EPI_ISL_1525085, EPI_ISL_1525086, EPI_ISL_1525087, EPI_ISL_1525088, EPI_ISL_1525090, EPI_ISL_1525091, EPI_ISL_1525092, EPI_ISL_1525093, EPI_ISL_1525094, EPI_ISL_1525095, EPI_ISL_1525096, EPI_ISL_1525097, EPI_ISL_1525098, EPI_ISL_1525099, EPI_ISL_1525100, EPI_ISL_1525101, EPI_ISL_1525102, EPI_ISL_1525104, EPI_ISL_1525105, EPI_ISL_1525107, EPI_ISL_1525108, EPI_ISL_1525109, EPI_ISL_1525110, EPI_ISL_1525111, EPI_ISL_1525112, EPI_ISL_1525113, EPI_ISL_1525114, EPI_ISL_1525115, EPI_ISL_1525116, EPI_ISL_1525117, EPI_ISL_1525118, EPI_ISL_1525119, EPI_ISL_1525120, EPI_ISL_1525121, EPI_ISL_1525122, EPI_ISL_1525123, EPI_ISL_1525124, EPI_ISL_1525125, EPI_ISL_1525126, EPI_ISL_1525127, EPI_ISL_1525128, EPI_ISL_1525129, EPI_ISL_1525130, EPI_ISL_1525131, EPI_ISL_1525132, EPI_ISL_1525133, EPI_ISL_1525135, EPI_ISL_1525136, EPI_ISL_1525137, EPI_ISL_1525138, EPI_ISL_1525139, EPI_ISL_1525140, EPI_ISL_1525141, EPI_ISL_1525142, EPI_ISL_1525143, EPI_ISL_1525144, EPI_ISL_1525145, EPI_ISL_1525146, EPI_ISL_1525147, EPI_ISL_1525148, EPI_ISL_1525149, EPI_ISL_1525150, EPI_ISL_1525151, EPI_ISL_1525152, EPI_ISL_1525153, EPI_ISL_1525154, EPI_ISL_1525155, EPI_ISL_1525156, EPI_ISL_1525157, EPI_ISL_1525158, EPI_ISL_1525159, EPI_ISL_1525160, EPI_ISL_1525161, EPI_ISL_1525162, EPI_ISL_1525163, EPI_ISL_1525164, EPI_ISL_1525165, EPI_ISL_1525166, EPI_ISL_1525167, EPI_ISL_1525168, EPI_ISL_1525169, EPI_ISL_1525170, EPI_ISL_1525171, EPI_ISL_1525172, EPI_ISL_1525174, EPI_ISL_1525175, EPI_ISL_1525176, EPI_ISL_1525177, EPI_ISL_1525178, EPI_ISL_1525179, EPI_ISL_1525180, EPI_ISL_1525181, EPI_ISL_1525182, EPI_ISL_1525183, EPI_ISL_1525184, EPI_ISL_1525185, EPI_ISL_1525186, EPI_ISL_1525187, EPI_ISL_1525188, EPI_ISL_1525189, EPI_ISL_1525190, EPI_ISL_1525191, EPI_ISL_1525192, EPI_ISL_1525193, EPI_ISL_1525194, EPI_ISL_1525195, EPI_ISL_1525196, EPI_ISL_1525197, EPI_ISL_1525198, EPI_ISL_1525199, EPI_ISL_1525201, EPI_ISL_1525202, EPI_ISL_1525203, EPI_ISL_1525204, EPI_ISL_1525205, EPI_ISL_1525206, EPI_ISL_1525207, EPI_ISL_1525208, EPI_ISL_1525209, EPI_ISL_1525210, EPI_ISL_1525211, EPI_ISL_1525212, EPI_ISL_1525213, EPI_ISL_1525214, EPI_ISL_1525215, EPI_ISL_1525216, EPI_ISL_1525217, EPI_ISL_1525218, EPI_ISL_1525219, EPI_ISL_1525220, EPI_ISL_1525221, EPI_ISL_1525222, EPI_ISL_1525223, EPI_ISL_1525224, EPI_ISL_1525225, EPI_ISL_1525226, EPI_ISL_1525227, EPI_ISL_1525228, EPI_ISL_1525229, EPI_ISL_1525230, EPI_ISL_1525231, EPI_ISL_1525232, EPI_ISL_1525233, EPI_ISL_1525234, EPI_ISL_1525235, EPI_ISL_1525236, EPI_ISL_1525237, EPI_ISL_1525238, EPI_ISL_1525239, EPI_ISL_1525240, EPI_ISL_1525241, EPI_ISL_1525242, EPI_ISL_1525243, EPI_ISL_1525244, EPI_ISL_1525245, EPI_ISL_1525246, EPI_ISL_1525247, EPI_ISL_1525248 | | | |
| see above | Aegis Sciences Corporation | Centers for Disease Control and Prevention Division of Viral Diseases, Pathogen Discovery | Dakota Howard, Dhwani Batra, Peter W. Cook, Kara Moser, Adrian Paskey, Jason Caravas, Benjamin Rambo-Martin, Shatavia Morrison, Christopher Gulvick, Scott Sammons, Yvette Unoarumhi, Darlene Wagner, Matthew Schmerer, Cyndi Clark, Patrick Campbell, Rob Case, Vikramsinha Ghorpade, Holly Houdeshell, Ola Kvalvaag, Dillon Nall, Ethan Sanders, Alec Vest, Shaun Westlund, Matthew Hardison, Clinton R. Paden, Duncan MacCannell |
| EPI_ISL_1525267, EPI_ISL_1525268, EPI_ISL_1525269, EPI_ISL_1525270, EPI_ISL_1525271, EPI_ISL_1525272, EPI_ISL_1525273, EPI_ISL_1525274, EPI_ISL_1525275, EPI_ISL_1525276, EPI_ISL_1525277, EPI_ISL_1525278, EPI_ISL_1525279, EPI_ISL_1525280, EPI_ISL_1525281, EPI_ISL_1525282, EPI_ISL_1525283, EPI_ISL_1525284, EPI_ISL_1525285, EPI_ISL_1525286, EPI_ISL_1525287, EPI_ISL_1525288, EPI_ISL_1525289, EPI_ISL_1525290, EPI_ISL_1525291, EPI_ISL_1525292, EPI_ISL_1525293, EPI_ISL_1525294, EPI_ISL_1525295, EPI_ISL_1525296, EPI_ISL_1525297, EPI_ISL_1525298, EPI_ISL_1525299, EPI_ISL_1525300, EPI_ISL_1525301, EPI_ISL_1525302, EPI_ISL_1525304, EPI_ISL_1525305, EPI_ISL_1525306, EPI_ISL_1525308, EPI_ISL_1525309, EPI_ISL_1525310, EPI_ISL_1525311, EPI_ISL_1525312, EPI_ISL_1525314, EPI_ISL_1525315, EPI_ISL_1525316, EPI_ISL_1525317, EPI_ISL_1525319, EPI_ISL_1525320, EPI_ISL_1525321, EPI_ISL_1525322, EPI_ISL_1525323, EPI_ISL_1525324, EPI_ISL_1525325, EPI_ISL_1525326, EPI_ISL_1525327, EPI_ISL_1525328, EPI_ISL_1525329, EPI_ISL_1525330, EPI_ISL_1525331, EPI_ISL_1525332, EPI_ISL_1525333, EPI_ISL_1525334, EPI_ISL_1525335, EPI_ISL_1525336, EPI_ISL_1525337, EPI_ISL_1525338, EPI_ISL_1525339, EPI_ISL_1525340, EPI_ISL_1525341, EPI_ISL_1525342, EPI_ISL_1525343, EPI_ISL_1525344, EPI_ISL_1525345, EPI_ISL_1525346, EPI_ISL_1525347, EPI_ISL_1525348, EPI_ISL_1525349, EPI_ISL_1525350, EPI_ISL_1525351, EPI_ISL_1525352, EPI_ISL_1525353, EPI_ISL_1525355, EPI_ISL_1525356, EPI_ISL_1525357, EPI_ISL_1525358, EPI_ISL_1525359, EPI_ISL_1525360, EPI_ISL_1525361, EPI_ISL_1525362, EPI_ISL_1525363, EPI_ISL_1525364, EPI_ISL_1525365, EPI_ISL_1525366, EPI_ISL_1525367, EPI_ISL_1525368, EPI_ISL_1525369, EPI_ISL_1525370, EPI_ISL_1525371, EPI_ISL_1525372, EPI_ISL_1525374, EPI_ISL_1525375, EPI_ISL_1525376, EPI_ISL_1525377, EPI_ISL_1525378, EPI_ISL_1525379, EPI_ISL_1525381, EPI_ISL_1525382, EPI_ISL_1525383, EPI_ISL_1525384, EPI_ISL_1525385, EPI_ISL_1525386, EPI_ISL_1525387, EPI_ISL_1525388, EPI_ISL_1525389, EPI_ISL_1525390, EPI_ISL_1525391, EPI_ISL_1525392, EPI_ISL_1525393, EPI_ISL_1525395, EPI_ISL_1525396, EPI_ISL_1525397, EPI_ISL_1525398, EPI_ISL_1525400, EPI_ISL_1525401, EPI_ISL_1525402, EPI_ISL_1525403, EPI_ISL_1525404, EPI_ISL_1525405, EPI_ISL_1525406, EPI_ISL_1525407, EPI_ISL_1525408, EPI_ISL_1525410, EPI_ISL_1525411, EPI_ISL_1525412, EPI_ISL_1525413, EPI_ISL_1525414, EPI_ISL_1525415, EPI_ISL_1525416, EPI_ISL_1525417, EPI_ISL_1525418, EPI_ISL_1525419, EPI_ISL_1525420, EPI_ISL_1525421, EPI_ISL_1525422, EPI_ISL_1525423, EPI_ISL_1525424, EPI_ISL_1525425, EPI_ISL_1525426, EPI_ISL_1525427, EPI_ISL_1525428, EPI_ISL_1525429, EPI_ISL_1525432, EPI_ISL_1525433, EPI_ISL_1525434, EPI_ISL_1525435, EPI_ISL_1525436, EPI_ISL_1525437, EPI_ISL_1525438, EPI_ISL_1525439, EPI_ISL_1525440, EPI_ISL_1525441, EPI_ISL_1525442, EPI_ISL_1525443, EPI_ISL_1525444, EPI_ISL_1525445, EPI_ISL_1525446, EPI_ISL_1525447, EPI_ISL_1525448, EPI_ISL_1525449, EPI_ISL_1525450, EPI_ISL_1525451, EPI_ISL_1525452, EPI_ISL_1525454, EPI_ISL_1525455, EPI_ISL_1525456, EPI_ISL_1525457, EPI_ISL_1525458, EPI_ISL_1525459, EPI_ISL_1525460, EPI_ISL_1525461, EPI_ISL_1525462, EPI_ISL_1525463, EPI_ISL_1525464, EPI_ISL_1525465, EPI_ISL_1525466, EPI_ISL_1525467, EPI_ISL_1525468, EPI_ISL_1525469, EPI_ISL_1525470, EPI_ISL_1525471, EPI_ISL_1525472, EPI_ISL_1525474, EPI_ISL_1525475, EPI_ISL_1525476, EPI_ISL_1525477, EPI_ISL_1525478, EPI_ISL_1525479, EPI_ISL_1525480, EPI_ISL_1525481, EPI_ISL_1525482, EPI_ISL_1525483, EPI_ISL_1525484, EPI_ISL_1525485, EPI_ISL_1525486, EPI_ISL_1525487, EPI_ISL_1525488, EPI_ISL_1525489, EPI_ISL_1525490, EPI_ISL_1525491, EPI_ISL_1525492, EPI_ISL_1525493, EPI_ISL_1525494, EPI_ISL_1525495, EPI_ISL_1525496, EPI_ISL_1525497, EPI_ISL_1525498, EPI_ISL_1525500, EPI_ISL_1525501, EPI_ISL_1525502, EPI_ISL_1525503, EPI_ISL_1525504, EPI_ISL_1525505, EPI_ISL_1525506, EPI_ISL_1525507, EPI_ISL_1525508, EPI_ISL_1525509, EPI_ISL_1525510, EPI_ISL_1525511, EPI_ISL_1525513, EPI_ISL_1525514, EPI_ISL_1525515, EPI_ISL_1525516, EPI_ISL_1525517, EPI_ISL_1525518, EPI_ISL_1525519, EPI_ISL_1525521, EPI_ISL_1525522, EPI_ISL_1525523, EPI_ISL_1525524, EPI_ISL_1525525, EPI_ISL_1525526, EPI_ISL_1525527, EPI_ISL_1525528, EPI_ISL_1525529, EPI_ISL_1525530, EPI_ISL_1525531, EPI_ISL_1525532, EPI_ISL_1525533, EPI_ISL_1525534, EPI_ISL_1525535, EPI_ISL_1525536, EPI_ISL_1525538, EPI_ISL_1525539, EPI_ISL_1525540, EPI_ISL_1525541, EPI_ISL_1525542, EPI_ISL_1525543, EPI_ISL_1525544, EPI_ISL_1525545, EPI_ISL_1525546, EPI_ISL_1525547, EPI_ISL_1525548, EPI_ISL_1525549, EPI_ISL_1525550, EPI_ISL_1525551, EPI_ISL_1525552, EPI_ISL_1525553, EPI_ISL_1525554, EPI_ISL_1525555, EPI_ISL_1525556, EPI_ISL_1525557, EPI_ISL_1525558, EPI_ISL_1525559, EPI_ISL_1525560, EPI_ISL_1525561, EPI_ISL_1525562, EPI_ISL_1525563, EPI_ISL_1525565, EPI_ISL_1525566, EPI_ISL_1525567, EPI_ISL_1525568, EPI_ISL_1525569, EPI_ISL_1525570, EPI_ISL_1525571, EPI_ISL_1525572, EPI_ISL_1525573, EPI_ISL_1525574, EPI_ISL_1525575, EPI_ISL_1525577, EPI_ISL_1525578, EPI_ISL_1525579, EPI_ISL_1525580, EPI_ISL_1525581, EPI_ISL_1525582, EPI_ISL_1525583, EPI_ISL_1525584, EPI_ISL_1525585, EPI_ISL_1525586, EPI_ISL_1525587, EPI_ISL_1525588, EPI_ISL_1525589, EPI_ISL_1525590, EPI_ISL_1525591, EPI_ISL_1525592, EPI_ISL_1525593, EPI_ISL_1525594, EPI_ISL_1525595, EPI_ISL_1525596, EPI_ISL_1525597, EPI_ISL_1525598, EPI_ISL_1525599, EPI_ISL_1525600, EPI_ISL_1525601, EPI_ISL_1525602, EPI_ISL_1525603, EPI_ISL_1525604, EPI_ISL_1525605, EPI_ISL_1525607, EPI_ISL_1525608, EPI_ISL_1525609, EPI_ISL_1525610, EPI_ISL_1525611, EPI_ISL_1525612, EPI_ISL_1525613, EPI_ISL_1525614, EPI_ISL_1525615, EPI_ISL_1525616, EPI_ISL_1525618, EPI_ISL_1525619, EPI_ISL_1525621, EPI_ISL_1525622, EPI_ISL_1525623, EPI_ISL_1525624, EPI_ISL_1525625, EPI_ISL_1525626, EPI_ISL_1525627, EPI_ISL_1525628, EPI_ISL_1525629, EPI_ISL_1525630, EPI_ISL_1525631, EPI_ISL_1525632, EPI_ISL_1525633, EPI_ISL_1525635, EPI_ISL_1525636, EPI_ISL_1525637, EPI_ISL_1525638, EPI_ISL_1525639, EPI_ISL_1525640, EPI_ISL_1525641, EPI_ISL_1525642, EPI_ISL_1525643, EPI_ISL_1525644, EPI_ISL_1525645, EPI_ISL_1525646, EPI_ISL_1525647, EPI_ISL_1525648, EPI_ISL_1525649, EPI_ISL_1525650, EPI_ISL_1525651, EPI_ISL_1525652, EPI_ISL_1525653, EPI_ISL_1525654, EPI_ISL_1525655, EPI_ISL_1525656, EPI_ISL_1525657, EPI_ISL_1525659, EPI_ISL_1525660, EPI_ISL_1525661, EPI_ISL_1525662, EPI_ISL_1525663, EPI_ISL_1525664, EPI_ISL_1525666, EPI_ISL_1525667, EPI_ISL_1525668, EPI_ISL_1525669, EPI_ISL_1525670, EPI_ISL_1525671, EPI_ISL_1525672, EPI_ISL_1525673, EPI_ISL_1525674, EPI_ISL_1525675, EPI_ISL_1525676, EPI_ISL_1525677, EPI_ISL_1525678, EPI_ISL_1525679, EPI_ISL_1525680, EPI_ISL_1525681, EPI_ISL_1525682, EPI_ISL_1525683, EPI_ISL_1525684, EPI_ISL_1525685, EPI_ISL_1525686, EPI_ISL_1525687, EPI_ISL_1525688, EPI_ISL_1525689, EPI_ISL_1525690, EPI_ISL_1525691, EPI_ISL_1525692, EPI_ISL_1525693, EPI_ISL_1525694, EPI_ISL_1525695, EPI_ISL_1525696, EPI_ISL_1525697, EPI_ISL_1525698, EPI_ISL_1525699, EPI_ISL_1525700, EPI_ISL_1525701, EPI_ISL_1525702, EPI_ISL_1525703, EPI_ISL_1525704, EPI_ISL_1525705, EPI_ISL_1525706, EPI_ISL_1525707, EPI_ISL_1525708, EPI_ISL_1525709, EPI_ISL_1525710, EPI_ISL_1525711, EPI_ISL_1525712, EPI_ISL_1525713, EPI_ISL_1525714, EPI_ISL_1525715, EPI_ISL_1525716, EPI_ISL_1525717, EPI_ISL_1525718, EPI_ISL_1525719, EPI_ISL_1525720, EPI_ISL_1525721, EPI_ISL_1525722, EPI_ISL_1525724, EPI_ISL_1525725, EPI_ISL_1525726, EPI_ISL_1525727, EPI_ISL_1525728, EPI_ISL_1525729, EPI_ISL_1525730, EPI_ISL_1525731, EPI_ISL_1525732, EPI_ISL_1525733, EPI_ISL_1525734, EPI_ISL_1525735, EPI_ISL_1525736, EPI_ISL_1525737, EPI_ISL_1525739, EPI_ISL_1525740, EPI_ISL_1525741, EPI_ISL_1525742, EPI_ISL_1525743, EPI_ISL_1525744, EPI_ISL_1525745, EPI_ISL_1525746, EPI_ISL_1525747, EPI_ISL_1525748, EPI_ISL_1525749, EPI_ISL_1525750, EPI_ISL_1525751, EPI_ISL_1525752, EPI_ISL_1525753, EPI_ISL_1525754, EPI_ISL_1525755, EPI_ISL_1525756, EPI_ISL_1525757, EPI_ISL_1525758, EPI_ISL_1525759, EPI_ISL_1525760, EPI_ISL_1525761, EPI_ISL_1525762, EPI_ISL_1525763, EPI_ISL_1525765, EPI_ISL_1525766, EPI_ISL_1525767, EPI_ISL_1525768, EPI_ISL_1525769, EPI_ISL_1525770, EPI_ISL_1525771, EPI_ISL_1525772, EPI_ISL_1525773, EPI_ISL_1525774, EPI_ISL_1525775, EPI_ISL_1525776, EPI_ISL_1525777, EPI_ISL_1525778, EPI_ISL_1525779, EPI_ISL_1525780, EPI_ISL_1525781, EPI_ISL_1525782, EPI_ISL_1525784, EPI_ISL_1525785, EPI_ISL_1525786, EPI_ISL_1525787, EPI_ISL_1525788, EPI_ISL_1525789, EPI_ISL_1525790, EPI_ISL_1525791, EPI_ISL_1525792, EPI_ISL_1525793, EPI_ISL_1525794, EPI_ISL_1525795, EPI_ISL_1525796, EPI_ISL_1525797, EPI_ISL_1525798, EPI_ISL_1525799, EPI_ISL_1525800, EPI_ISL_1525802, EPI_ISL_1525803, EPI_ISL_1525804, EPI_ISL_1525805, EPI_ISL_1525806, EPI_ISL_1525807, EPI_ISL_1525808, EPI_ISL_1525809, EPI_ISL_1525811, EPI_ISL_1525812, EPI_ISL_1525813, EPI_ISL_1525815, EPI_ISL_1525816, EPI_ISL_1525817, EPI_ISL_1525818, EPI_ISL_1525819, EPI_ISL_1525820, EPI_ISL_1525821, EPI_ISL_1525822, EPI_ISL_1525823, EPI_ISL_1525824, EPI_ISL_1525825, EPI_ISL_1525826, EPI_ISL_1525827, EPI_ISL_1525829, EPI_ISL_1525830, EPI_ISL_1525831, EPI_ISL_1525832, EPI_ISL_1525834, EPI_ISL_1525835, EPI_ISL_1525836, EPI_ISL_1525838, EPI_ISL_1525840, EPI_ISL_1525841, EPI_ISL_1525842, EPI_ISL_1525843, EPI_ISL_1525844, EPI_ISL_1525845, EPI_ISL_1525846, EPI_ISL_1525847, EPI_ISL_1525848, EPI_ISL_1525849, EPI_ISL_1525850, EPI_ISL_1525851, EPI_ISL_1525852, EPI_ISL_1525854, EPI_ISL_1525855, EPI_ISL_1525857, EPI_ISL_1525858, EPI_ISL_1525859, EPI_ISL_1525860, EPI_ISL_1525861, EPI_ISL_1525862, EPI_ISL_1525863, EPI_ISL_1525865, EPI_ISL_1525866, EPI_ISL_1525868, EPI_ISL_1525869, EPI_ISL_1525870, EPI_ISL_1525871, EPI_ISL_1525872, EPI_ISL_1525873, EPI_ISL_1525874, EPI_ISL_1525875, EPI_ISL_1525876, EPI_ISL_1525877, EPI_ISL_1525878, EPI_ISL_1525879, EPI_ISL_1525880, EPI_ISL_1525881, EPI_ISL_1525882, EPI_ISL_1525883, EPI_ISL_1525884, EPI_ISL_1525885, EPI_ISL_1525886, EPI_ISL_1525887, EPI_ISL_1525888, EPI_ISL_1525889, EPI_ISL_1525890, EPI_ISL_1525891, EPI_ISL_1525892, EPI_ISL_1525893, EPI_ISL_1525894, EPI_ISL_1525895, EPI_ISL_1525896, EPI_ISL_1525897, EPI_ISL_1525898, EPI_ISL_1525899, EPI_ISL_1525900, EPI_ISL_1525901, EPI_ISL_1525902, EPI_ISL_1525903 | | | |
| see above | Fulgent Genetics | Centers for Disease Control and Prevention Division of Viral Diseases, Pathogen Discovery | Dakota Howard, Dhwani Batra, Peter W. Cook, Kara Moser, Adrian Paskey, Jason Caravas, Benjamin Rambo-Martin, Shatavia Morrison, Christopher Gulvick, Scott Sammons, Yvette Unoarumhi, Darlene Wagner, Matthew Schmerer, Harry Gao, Mickey Li, John Gao, Joseph Fierro, Benafsh Sapra, Becky Tsai, Yan Meng, Doreen Ng, James Xie, Clinton R. Paden, Duncan MacCannell |
| EPI_ISL_1525904, EPI_ISL_1525905, EPI_ISL_1525906, EPI_ISL_1525907, EPI_ISL_1525908, EPI_ISL_1525909, EPI_ISL_1525910, EPI_ISL_1525911, EPI_ISL_1525912, EPI_ISL_1525913, EPI_ISL_1525914, EPI_ISL_1525915, EPI_ISL_1525916, EPI_ISL_1525917, EPI_ISL_1525918, EPI_ISL_1525919, EPI_ISL_1525920, EPI_ISL_1525921, EPI_ISL_1525922, EPI_ISL_1525923, EPI_ISL_1525924, EPI_ISL_1525925, EPI_ISL_1525926, EPI_ISL_1525927, EPI_ISL_1525928, EPI_ISL_1525929, EPI_ISL_1525930, EPI_ISL_1525931, EPI_ISL_1525932, EPI_ISL_1525933, EPI_ISL_1525934, EPI_ISL_1525935, EPI_ISL_1525936, EPI_ISL_1525937, EPI_ISL_1525938, EPI_ISL_1525939, EPI_ISL_1525940, EPI_ISL_1525941, EPI_ISL_1525942, EPI_ISL_1525943, EPI_ISL_1525944, EPI_ISL_1525945, EPI_ISL_1525946, EPI_ISL_1525947, EPI_ISL_1525948, EPI_ISL_1525949, EPI_ISL_1525950, EPI_ISL_1525951, EPI_ISL_1525952, EPI_ISL_1525953, EPI_ISL_1525954, EPI_ISL_1525955, EPI_ISL_1525956, EPI_ISL_1525957, EPI_ISL_1525958, EPI_ISL_1525959, EPI_ISL_1525960, EPI_ISL_1525961, EPI_ISL_1525962, EPI_ISL_1525963, EPI_ISL_1525964, EPI_ISL_1525965, EPI_ISL_1525966, EPI_ISL_1525967, EPI_ISL_1525968, EPI_ISL_1525969, EPI_ISL_1525970, EPI_ISL_1525971, EPI_ISL_1525972, EPI_ISL_1525973, EPI_ISL_1525974, EPI_ISL_1525975, EPI_ISL_1525976, EPI_ISL_1525977, EPI_ISL_1525978, EPI_ISL_1525979, EPI_ISL_1525980, EPI_ISL_1525981, EPI_ISL_1525982, EPI_ISL_1525983, EPI_ISL_1525984, EPI_ISL_1525985, EPI_ISL_1525986, EPI_ISL_1525987, EPI_ISL_1525988, EPI_ISL_1525989, EPI_ISL_1525990, EPI_ISL_1525991, EPI_ISL_1525992, EPI_ISL_1525993, EPI_ISL_1525994, EPI_ISL_1525995, EPI_ISL_1525996, EPI_ISL_1525997, EPI_ISL_1525998, EPI_ISL_1525999, EPI_ISL_1526000, EPI_ISL_1526001, EPI_ISL_1526002, EPI_ISL_1526003, EPI_ISL_1526004, EPI_ISL_1526005, EPI_ISL_1526006, EPI_ISL_1526007, EPI_ISL_1526008, EPI_ISL_1526009, EPI_ISL_1526010, EPI_ISL_1526011, EPI_ISL_1526012, EPI_ISL_1526013, EPI_ISL_1526014, EPI_ISL_1526015, EPI_ISL_1526016, EPI_ISL_1526017, EPI_ISL_1526018, EPI_ISL_1526019, EPI_ISL_1526020, EPI_ISL_1526021, EPI_ISL_1526022, EPI_ISL_1526023, EPI_ISL_1526024, EPI_ISL_1526025, EPI_ISL_1526026, EPI_ISL_1526027, EPI_ISL_1526028, EPI_ISL_1526029, EPI_ISL_1526030, EPI_ISL_1526031, EPI_ISL_1526032, EPI_ISL_1526033, EPI_ISL_1526034, EPI_ISL_1526035, EPI_ISL_1526036, EPI_ISL_1526037, EPI_ISL_1526038, EPI_ISL_1526039, EPI_ISL_1526040, EPI_ISL_1526041, EPI_ISL_1526042, EPI_ISL_1526043, EPI_ISL_1526044, EPI_ISL_1526045, EPI_ISL_1526046, EPI_ISL_1526047, EPI_ISL_1526048, EPI_ISL_1526049, EPI_ISL_1526050, EPI_ISL_1526051, EPI_ISL_1526052, EPI_ISL_1526053, EPI_ISL_1526054, EPI_ISL_1526055, EPI_ISL_1526056, EPI_ISL_1526057, EPI_ISL_1526058, EPI_ISL_1526059, EPI_ISL_1526060, EPI_ISL_1526061, EPI_ISL_1526062, EPI_ISL_1526063, EPI_ISL_1526064, EPI_ISL_1526065, EPI_ISL_1526066, EPI_ISL_1526067, EPI_ISL_1526068, EPI_ISL_1526069, EPI_ISL_1526070 | | | |
| see above | Quest Diagnostics Incorporated | Centers for Disease Control and Prevention Division of Viral Diseases, Pathogen Discovery | Dakota Howard, Dhwani Batra, Peter W. Cook, Kara Moser, Adrian Paskey, Jason Caravas, Benjamin Rambo-Martin, Shatavia Morrison, Christopher Gulvick, Scott Sammons, Yvette Unoarumhi, Darlene Wagner, Matthew Schmerer, S. H. Rosenthal, A. Gerasimova, R. M. Kagan, B. Anderson, M. Hua, Y. Liu, L.E. Bernstein, K.E. Livingston, A. Perez, I. A. Shlyakhter, R. V. Rolando, R. Owen, P. Tanpaiboon, F. Lacbawan, Clinton R. Paden, Duncan MacCannell |
| EPI_ISL_1526795, EPI_ISL_1526796, EPI_ISL_1526797, EPI_ISL_1526798, EPI_ISL_1526799, EPI_ISL_1526800, EPI_ISL_1526801, EPI_ISL_1526802, EPI_ISL_1526803, EPI_ISL_1526804, EPI_ISL_1526805, EPI_ISL_1526806, EPI_ISL_1526807, EPI_ISL_1526808, EPI_ISL_1526809, EPI_ISL_1526810, EPI_ISL_1526811, EPI_ISL_1526812, EPI_ISL_1526813, EPI_ISL_1526814, EPI_ISL_1526815, EPI_ISL_1526816, EPI_ISL_1526817, EPI_ISL_1526818, EPI_ISL_1526819, EPI_ISL_1526820, EPI_ISL_1526821, EPI_ISL_1526822, EPI_ISL_1526823, EPI_ISL_1526824, EPI_ISL_1526825, EPI_ISL_1526826, EPI_ISL_1526827, EPI_ISL_1526828, EPI_ISL_1526829, EPI_ISL_1526830, EPI_ISL_1526831, EPI_ISL_1526832, EPI_ISL_1526833, EPI_ISL_1526834, EPI_ISL_1526835 | | | |
| see above | UW Virology Lab | UW Virology Lab | Pavitra Roychoudhury, Hong Xie, Lasata Shrestha, Shah Mohamed Bakhash, Michelle Lin, Margaret Mills, Noah Baker, Sean Ellis, Saraswathi Sathees, Meei-Li Huang, Keith R Jerome, Alexander Greninger |
| EPI_ISL_1526836, EPI_ISL_1526837, EPI_ISL_1526838, EPI_ISL_1526839, EPI_ISL_1526840, EPI_ISL_1526841, EPI_ISL_1526843, EPI_ISL_1526844, EPI_ISL_1526846, EPI_ISL_1526847, EPI_ISL_1526848, EPI_ISL_1526849, EPI_ISL_1526850, EPI_ISL_1526852, EPI_ISL_1526853, EPI_ISL_1526854, EPI_ISL_1526855, EPI_ISL_1526856, EPI_ISL_1526857, EPI_ISL_1526859, EPI_ISL_1526860, EPI_ISL_1526861, EPI_ISL_1526862, EPI_ISL_1526863, EPI_ISL_1526864, EPI_ISL_1526865, EPI_ISL_1526866, EPI_ISL_1526867, EPI_ISL_1526868, EPI_ISL_1526869, EPI_ISL_1526870, EPI_ISL_1526871, EPI_ISL_1526872, EPI_ISL_1526873, EPI_ISL_1526874, EPI_ISL_1526875, EPI_ISL_1526877, EPI_ISL_1526878, EPI_ISL_1526880, EPI_ISL_1526881, EPI_ISL_1526882, EPI_ISL_1526883, EPI_ISL_1526886, EPI_ISL_1526887, EPI_ISL_1526888, EPI_ISL_1526889, EPI_ISL_1526890, EPI_ISL_1526891, EPI_ISL_1526892, EPI_ISL_1526893, EPI_ISL_1526894, EPI_ISL_1526895, EPI_ISL_1526896, EPI_ISL_1526897, EPI_ISL_1526898, EPI_ISL_1526899, EPI_ISL_1526900, EPI_ISL_1526901, EPI_ISL_1526902, EPI_ISL_1526903, EPI_ISL_1526904, EPI_ISL_1526905, EPI_ISL_1526906, EPI_ISL_1526907, EPI_ISL_1526908, EPI_ISL_1526909 | | | |
| see above | Virginia Division of Consolidated Laboratory Services | Virginia Division of Consolidated Laboratory Services | Virginia DCLS |
| EPI_ISL_1526910, EPI_ISL_1526911, EPI_ISL_1526912, EPI_ISL_1526913, EPI_ISL_1526914, EPI_ISL_1526915, EPI_ISL_1526916, EPI_ISL_1526917, EPI_ISL_1526918, EPI_ISL_1526919, EPI_ISL_1526920, EPI_ISL_1526921, EPI_ISL_1526922, EPI_ISL_1526923, EPI_ISL_1526924, EPI_ISL_1526925, EPI_ISL_1526926, EPI_ISL_1526927, EPI_ISL_1526928, EPI_ISL_1526929, EPI_ISL_1526930, EPI_ISL_1526931, EPI_ISL_1526932, EPI_ISL_1526933, EPI_ISL_1526934, EPI_ISL_1526935, EPI_ISL_1526936, EPI_ISL_1526937, EPI_ISL_1526938, EPI_ISL_1526939, EPI_ISL_1526940, EPI_ISL_1526941, EPI_ISL_1526942, EPI_ISL_1526943, EPI_ISL_1526944, EPI_ISL_1526945, EPI_ISL_1526946, EPI_ISL_1526947, EPI_ISL_1526948, EPI_ISL_1526949, EPI_ISL_1526950, EPI_ISL_1526951, EPI_ISL_1526952, EPI_ISL_1526953, EPI_ISL_1526954, EPI_ISL_1526955, EPI_ISL_1526956, EPI_ISL_1526957, EPI_ISL_1526958, EPI_ISL_1526959, EPI_ISL_1526960, EPI_ISL_1526961, EPI_ISL_1526962, EPI_ISL_1526963, EPI_ISL_1526964, EPI_ISL_1526965, EPI_ISL_1526966, EPI_ISL_1526967, EPI_ISL_1526968, EPI_ISL_1526969, EPI_ISL_1526970, EPI_ISL_1526971, EPI_ISL_1526972, EPI_ISL_1526973, EPI_ISL_1526974, EPI_ISL_1526975, EPI_ISL_1526976, EPI_ISL_1526977, EPI_ISL_1526978, EPI_ISL_1526979, EPI_ISL_1526980, EPI_ISL_1526981, EPI_ISL_1526982, EPI_ISL_1526983, EPI_ISL_1526984, EPI_ISL_1526985, EPI_ISL_1526986, EPI_ISL_1526987, EPI_ISL_1526988, EPI_ISL_1526989 | | | |
| see above | UW Virology Lab | UW Virology Lab | Pavitra Roychoudhury, Hong Xie, Lasata Shrestha, Shah Mohamed Bakhash, Michelle Lin, Noah R. Baker, Sean Ellis, Saraswathi Sathees, Meei-Li Huang, Keith R Jerome, Alexander Greninger |

| | | | |
|---|---|---|---|
| EPI_ISL_1526990, EPI_ISL_1526991, EPI_ISL_1526992, EPI_ISL_1526993, EPI_ISL_1526994, EPI_ISL_1526995, EPI_ISL_1526997, EPI_ISL_1526998, EPI_ISL_1526999, EPI_ISL_1527000, EPI_ISL_1527001, EPI_ISL_1527002, EPI_ISL_1527003, EPI_ISL_1527004, EPI_ISL_1527005, EPI_ISL_1527006, EPI_ISL_1527007, EPI_ISL_1527008, EPI_ISL_1527009, EPI_ISL_1527010 | | | |
| see above | Washington State Department of Health Public Health Laboratories | Washington State Department of Health Public Health Laboratories | Drew MacKellar, Philip Dykema, Denny Russell, Joenice Gonzalez, Hannah Gray, Geoff Melly, Vanessa De Los Santos, Darren Lucas, JohnAric Peterson, Avi Singh, Rebecca Cao |
| EPI_ISL_1527029, EPI_ISL_1527030, EPI_ISL_1527031, EPI_ISL_1527032, EPI_ISL_1527033, EPI_ISL_1527034, EPI_ISL_1527035, EPI_ISL_1527036, EPI_ISL_1527037, EPI_ISL_1527038, EPI_ISL_1527039, EPI_ISL_1527040, EPI_ISL_1527042, EPI_ISL_1527043, EPI_ISL_1527044, EPI_ISL_1527045, EPI_ISL_1527047, EPI_ISL_1527048, EPI_ISL_1527049, EPI_ISL_1527050, EPI_ISL_1527051, EPI_ISL_1527052, EPI_ISL_1527054, EPI_ISL_1527056, EPI_ISL_1527057, EPI_ISL_1527058, EPI_ISL_1527059, EPI_ISL_1527060, EPI_ISL_1527061, EPI_ISL_1527062, EPI_ISL_1527063, EPI_ISL_1527064, EPI_ISL_1527065, EPI_ISL_1527066, EPI_ISL_1527067, EPI_ISL_1527068, EPI_ISL_1527069, EPI_ISL_1527070, EPI_ISL_1527071, EPI_ISL_1527072, EPI_ISL_1527073, EPI_ISL_1527074, EPI_ISL_1527075, EPI_ISL_1527076, EPI_ISL_1527077, EPI_ISL_1527078, EPI_ISL_1527079, EPI_ISL_1527081, EPI_ISL_1527082, EPI_ISL_1527083, EPI_ISL_1527084, EPI_ISL_1527085, EPI_ISL_1527086, EPI_ISL_1527087, EPI_ISL_1527089, EPI_ISL_1527090 | | | |
| see above | University of Mississippi Medical Center, Department of Pathology | University of Mississippi Medical Center, Molecular and Genomics Core Facility | Ashley C. Johnson, Ithiel J. Frame, Krishna K. Ayyalasomayajula, Michael R. Garrett, D. Ashley Robinson |
| EPI_ISL_1527091, EPI_ISL_1527092, EPI_ISL_1527093, EPI_ISL_1527094, EPI_ISL_1527095, EPI_ISL_1527096, EPI_ISL_1527097, EPI_ISL_1527098, EPI_ISL_1527099, EPI_ISL_1527100, EPI_ISL_1527101, EPI_ISL_1527102, EPI_ISL_1527103, EPI_ISL_1527104, EPI_ISL_1527105, EPI_ISL_1527106, EPI_ISL_1527107, EPI_ISL_1527108, EPI_ISL_1527109, EPI_ISL_1527110, EPI_ISL_1527111, EPI_ISL_1527112, EPI_ISL_1527113, EPI_ISL_1527114, EPI_ISL_1527115, EPI_ISL_1527116, EPI_ISL_1527117, EPI_ISL_1527118, EPI_ISL_1527119, EPI_ISL_1527120, EPI_ISL_1527121, EPI_ISL_1527122, EPI_ISL_1527123, EPI_ISL_1527124, EPI_ISL_1527125, EPI_ISL_1527126, EPI_ISL_1527127, EPI_ISL_1527128, EPI_ISL_1527129, EPI_ISL_1527130, EPI_ISL_1527131, EPI_ISL_1527132, EPI_ISL_1527133, EPI_ISL_1527134, EPI_ISL_1527135, EPI_ISL_1527136, EPI_ISL_1527137, EPI_ISL_1527138, EPI_ISL_1527139, EPI_ISL_1527140, EPI_ISL_1527141, EPI_ISL_1527142, EPI_ISL_1527143, EPI_ISL_1527144, EPI_ISL_1527145, EPI_ISL_1527146, EPI_ISL_1527147, EPI_ISL_1527148, EPI_ISL_1527149, EPI_ISL_1527150, EPI_ISL_1527151, EPI_ISL_1527152, EPI_ISL_1527153, EPI_ISL_1527154, EPI_ISL_1527155, EPI_ISL_1527156, EPI_ISL_1527157, EPI_ISL_1527158, EPI_ISL_1527159, EPI_ISL_1527160, EPI_ISL_1527161, EPI_ISL_1527162, EPI_ISL_1527163, EPI_ISL_1527164, EPI_ISL_1527165, EPI_ISL_1527166, EPI_ISL_1527167, EPI_ISL_1527168, EPI_ISL_1527169, EPI_ISL_1527170, EPI_ISL_1527171, EPI_ISL_1527172, EPI_ISL_1527173, EPI_ISL_1527174, EPI_ISL_1527175, EPI_ISL_1527176, EPI_ISL_1527177, EPI_ISL_1527178, EPI_ISL_1527179, EPI_ISL_1527180, EPI_ISL_1527181, EPI_ISL_1527182, EPI_ISL_1527183, EPI_ISL_1527184, EPI_ISL_1527185, EPI_ISL_1527186, EPI_ISL_1527187, EPI_ISL_1527188, EPI_ISL_1527189, EPI_ISL_1527190, EPI_ISL_1527191, EPI_ISL_1527192, EPI_ISL_1527193, EPI_ISL_1527194, EPI_ISL_1527195, EPI_ISL_1527196, EPI_ISL_1527197, EPI_ISL_1527198, EPI_ISL_1527199, EPI_ISL_1527200, EPI_ISL_1527217, EPI_ISL_1527218, EPI_ISL_1527219, EPI_ISL_1527220, EPI_ISL_1527221, EPI_ISL_1527222, EPI_ISL_1527223, EPI_ISL_1527224, EPI_ISL_1527225, EPI_ISL_1527226, EPI_ISL_1527227, EPI_ISL_1527228, EPI_ISL_1527229, EPI_ISL_1527230, EPI_ISL_1527231, EPI_ISL_1527232, EPI_ISL_1527233, EPI_ISL_1527234, EPI_ISL_1527235, EPI_ISL_1527236 | | | |
| see above | Massachusetts State Public Health Laboratory | Massachusetts State Public Health Laboratory | Andrew Lang, Timelia Fink, Glen Gallagher, Sandra Smole |
| EPI_ISL_1527237, EPI_ISL_1527238, EPI_ISL_1527239, EPI_ISL_1527240, EPI_ISL_1527241, EPI_ISL_1527242, EPI_ISL_1527244, EPI_ISL_1527245, EPI_ISL_1527246, EPI_ISL_1527247, EPI_ISL_1527249, EPI_ISL_1527250, EPI_ISL_1527253, EPI_ISL_1527254, EPI_ISL_1527255, EPI_ISL_1527256, EPI_ISL_1527257, EPI_ISL_1527259, EPI_ISL_1527261, EPI_ISL_1527262, EPI_ISL_1527263, EPI_ISL_1527264 | | | |
| see above | University of Mississippi Medical Center, Department of Pathology | University of Mississippi Medical Center, Molecular and Genomics Core Facility | Ashley C. Johnson, Ithiel J. Frame, Krishna K. Ayyalasomayajula, Michael R. Garrett, D. Ashley Robinson |
| EPI_ISL_1527265, EPI_ISL_1527266, EPI_ISL_1527267, EPI_ISL_1527268, EPI_ISL_1527269, EPI_ISL_1527270, EPI_ISL_1527271, EPI_ISL_1527272, EPI_ISL_1527273, EPI_ISL_1527274, EPI_ISL_1527275, EPI_ISL_1527276, EPI_ISL_1527277, EPI_ISL_1527278, EPI_ISL_1527279, EPI_ISL_1527280, EPI_ISL_1527281, EPI_ISL_1527282, EPI_ISL_1527283, EPI_ISL_1527284, EPI_ISL_1527285, EPI_ISL_1527286, EPI_ISL_1527287, EPI_ISL_1527288, EPI_ISL_1527289, EPI_ISL_1527290, EPI_ISL_1527291, EPI_ISL_1527292, EPI_ISL_1527293, EPI_ISL_1527294, EPI_ISL_1527295, EPI_ISL_1527296, EPI_ISL_1527297, EPI_ISL_1527298, EPI_ISL_1527299, EPI_ISL_1527300, EPI_ISL_1527301, EPI_ISL_1527302, EPI_ISL_1527303, EPI_ISL_1527304, EPI_ISL_1527305, EPI_ISL_1527306, EPI_ISL_1527307, EPI_ISL_1527308, EPI_ISL_1527309, EPI_ISL_1527310, EPI_ISL_1527311, EPI_ISL_1527312, EPI_ISL_1527313, EPI_ISL_1527314, EPI_ISL_1527315, EPI_ISL_1527316, EPI_ISL_1527317, EPI_ISL_1527318, EPI_ISL_1527319, EPI_ISL_1527320, EPI_ISL_1527321, EPI_ISL_1527322, EPI_ISL_1527323, EPI_ISL_1527324, EPI_ISL_1527325, EPI_ISL_1527326, EPI_ISL_1527327, EPI_ISL_1527328, EPI_ISL_1527329, EPI_ISL_1527330, EPI_ISL_1527331, EPI_ISL_1527334, EPI_ISL_1527335, EPI_ISL_1527336, EPI_ISL_1527337, EPI_ISL_1527338, EPI_ISL_1527339, EPI_ISL_1527340, EPI_ISL_1527341, EPI_ISL_1527342, EPI_ISL_1527343, EPI_ISL_1527344, EPI_ISL_1527345, EPI_ISL_1527346, EPI_ISL_1527347, EPI_ISL_1527348, EPI_ISL_1527349, EPI_ISL_1527350, EPI_ISL_1527351, EPI_ISL_1527352, EPI_ISL_1527353, EPI_ISL_1527354, EPI_ISL_1527355, EPI_ISL_1527356, EPI_ISL_1527357, EPI_ISL_1527358, EPI_ISL_1527359 | | | |
| see above | Washington State Department of Health Public Health Laboratories | Washington State Department of Health Public Health Laboratories | Drew MacKellar, Philip Dykema, Denny Russell, Joenice Gonzalez, Hannah Gray, Geoff Melly, Vanessa De Los Santos, Darren Lucas, JohnAric Peterson, Avi Singh, Rebecca Cao |
| EPI_ISL_1527400, EPI_ISL_1527402, EPI_ISL_1527403, EPI_ISL_1527404, EPI_ISL_1527405, EPI_ISL_1527406, EPI_ISL_1527411, EPI_ISL_1527412, EPI_ISL_1527415, EPI_ISL_1527416, EPI_ISL_1527417, EPI_ISL_1527418, EPI_ISL_1527420, EPI_ISL_1527422, EPI_ISL_1527424, EPI_ISL_1527425, EPI_ISL_1527427, EPI_ISL_1527428, EPI_ISL_1527429, EPI_ISL_1527430, EPI_ISL_1527432, EPI_ISL_1527433, EPI_ISL_1527435, EPI_ISL_1527436, EPI_ISL_1527439, EPI_ISL_1527440 | | | |
| see above | University of Mississippi Medical Center, Department of Pathology | University of Mississippi Medical Center, Molecular and Genomics Core Facility | Ashley C. Johnson, Ithiel J. Frame, Krishna K. Ayyalasomayajula, Michael R. Garrett, D. Ashley Robinson |
| EPI_ISL_1527444, EPI_ISL_1527445, EPI_ISL_1527446, EPI_ISL_1527447, EPI_ISL_1527448, EPI_ISL_1527449, EPI_ISL_1527450, EPI_ISL_1527451, EPI_ISL_1527452, EPI_ISL_1527453, EPI_ISL_1527454, EPI_ISL_1527455, EPI_ISL_1527456, EPI_ISL_1527457, EPI_ISL_1527458, EPI_ISL_1527459, EPI_ISL_1527460, EPI_ISL_1527461, EPI_ISL_1527462, EPI_ISL_1527463 | | | |
| see above | GA Department of Public Health | GA Department of Public Health | Stacy Reeves, Jonathan Edwards, Cynthia Dixey, Tonia Parrott, Aliyah Fields, Taylor Smith |
| EPI_ISL_1527464 | M Health Fairview | Minnesota Department of Health, Public Health Laboratory | Alexandra Lorentz, Jacob Garfin, Matt Plumb, and Xiong Wang |
| EPI_ISL_1527465 | Mayo Clinic & Mayo Clinic Laboratories | Minnesota Department of Health, Public Health Laboratory | Alexandra Lorentz, Jacob Garfin, Matt Plumb, and Xiong Wang |
| EPI_ISL_1527466 | Vault Health | Minnesota Department of Health, Public Health Laboratory | Alexandra Lorentz, Jacob Garfin, Matt Plumb, and Xiong Wang |
| EPI_ISL_1527467 | M Health Fairview | Minnesota Department of Health, Public Health Laboratory | Alexandra Lorentz, Jacob Garfin, Matt Plumb, and Xiong Wang |
| EPI_ISL_1527468 | Mayo Clinic & Mayo Clinic Laboratories | Minnesota Department of Health, Public Health Laboratory | Alexandra Lorentz, Jacob Garfin, Matt Plumb, and Xiong Wang |
| EPI_ISL_1527469 | M Health Fairview | Minnesota Department of Health, Public Health Laboratory | Alexandra Lorentz, Jacob Garfin, Matt Plumb, and Xiong Wang |
| EPI_ISL_1527470 | Mayo Clinic & Mayo Clinic Laboratories | Minnesota Department of Health, Public Health Laboratory | Alexandra Lorentz, Jacob Garfin, Matt Plumb, and Xiong Wang |
| EPI_ISL_1527471 | M Health Fairview | Minnesota Department of Health, Public Health Laboratory | Alexandra Lorentz, Jacob Garfin, Matt Plumb, and Xiong Wang |
| EPI_ISL_1527472 | Mayo Clinic & Mayo Clinic Laboratories | Minnesota Department of Health, Public Health Laboratory | Alexandra Lorentz, Jacob Garfin, Matt Plumb, and Xiong Wang |
| EPI_ISL_1527473 | Vault Health | Minnesota Department of Health, Public Health Laboratory | Alexandra Lorentz, Jacob Garfin, Matt Plumb, and Xiong Wang |
| EPI_ISL_1527474 | Mayo Clinic & Mayo Clinic Laboratories | Minnesota Department of Health, Public Health Laboratory | Alexandra Lorentz, Jacob Garfin, Matt Plumb, and Xiong Wang |
| EPI_ISL_1527475 | Vault Health | Minnesota Department of Health, Public Health Laboratory | Alexandra Lorentz, Jacob Garfin, Matt Plumb, and Xiong Wang |
| EPI_ISL_1527476, EPI_ISL_1527477, EPI_ISL_1527478, EPI_ISL_1527479, EPI_ISL_1527480, EPI_ISL_1527481, EPI_ISL_1527482, EPI_ISL_1527483, EPI_ISL_1527484, EPI_ISL_1527485, EPI_ISL_1527486, EPI_ISL_1527487 | | | |
| see above | Mayo Clinic & Mayo Clinic Laboratories | Minnesota Department of Health, Public Health Laboratory | Alexandra Lorentz, Jacob Garfin, Matt Plumb, and Xiong Wang |
| EPI_ISL_1527488, EPI_ISL_1527489, EPI_ISL_1527490, EPI_ISL_1527491 | Vault Health | Minnesota Department of Health, Public Health Laboratory | Alexandra Lorentz, Jacob Garfin, Matt Plumb, and Xiong Wang |
| EPI_ISL_1527492, EPI_ISL_1527493, EPI_ISL_1527494, EPI_ISL_1527495, EPI_ISL_1527496 | M Health Fairview | Minnesota Department of Health, Public Health Laboratory | Alexandra Lorentz, Jacob Garfin, Matt Plumb, and Xiong Wang |
| EPI_ISL_1527497 | Vault Health | Minnesota Department of Health, Public Health Laboratory | Alexandra Lorentz, Jacob Garfin, Matt Plumb, and Xiong Wang |
| EPI_ISL_1527498, EPI_ISL_1527499, EPI_ISL_1527500, EPI_ISL_1527501, EPI_ISL_1527502 | Mayo Clinic & Mayo Clinic Laboratories | Minnesota Department of Health, Public Health Laboratory | Alexandra Lorentz, Jacob Garfin, Matt Plumb, and Xiong Wang |
| EPI_ISL_1527503, EPI_ISL_1527504 | Vault Health | Minnesota Department of Health, Public Health Laboratory | Alexandra Lorentz, Jacob Garfin, Matt Plumb, and Xiong Wang |
| EPI_ISL_1527505, EPI_ISL_1527509 | M Health Fairview | Minnesota Department of Health, Public Health Laboratory | Alexandra Lorentz, Jacob Garfin, Matt Plumb, and Xiong Wang |
| EPI_ISL_1527510, EPI_ISL_1527511, EPI_ISL_1527512, EPI_ISL_1527513, EPI_ISL_1527514, EPI_ISL_1527515, EPI_ISL_1527516, EPI_ISL_1527517, EPI_ISL_1527518, EPI_ISL_1527519, EPI_ISL_1527520, EPI_ISL_1527521 | | | |
| see above | Mayo Clinic & Mayo Clinic Laboratories | Minnesota Department of Health, Public Health Laboratory | Alexandra Lorentz, Jacob Garfin, Matt Plumb, and Xiong Wang |
| EPI_ISL_1527522 | Vault Health | Minnesota Department of Health, Public Health Laboratory | Alexandra Lorentz, Jacob Garfin, Matt Plumb, and Xiong Wang |
| EPI_ISL_1527523, EPI_ISL_1527524, EPI_ISL_1527525, EPI_ISL_1527526, EPI_ISL_1527527 | M Health Fairview | Minnesota Department of Health, Public Health Laboratory | Alexandra Lorentz, Jacob Garfin, Matt Plumb, and Xiong Wang |
| EPI_ISL_1527528, EPI_ISL_1527529 | Vault Health | Minnesota Department of Health, Public Health Laboratory | Alexandra Lorentz, Jacob Garfin, Matt Plumb, and Xiong Wang |
| EPI_ISL_1527530, EPI_ISL_1527531, | School of Pharmacy, Shenandoah University | School of Pharmacy, Shenandoah University | Adams,S.M., Harralson,A.F., Kidd,R.S., Sawyer,G.W. |

| | | | |
|---|---|---|---|
| EPI_ISL_1527533, EPI_ISL_1527534, EPI_ISL_1527535, EPI_ISL_1527537 | | | |
| EPI_ISL_1527538, EPI_ISL_1527539, EPI_ISL_1527540, EPI_ISL_1527541, EPI_ISL_1527542, EPI_ISL_1527543, EPI_ISL_1527544, EPI_ISL_1527545, EPI_ISL_1527546, EPI_ISL_1527547, EPI_ISL_1527548, EPI_ISL_1527549, EPI_ISL_1527550, EPI_ISL_1527551, EPI_ISL_1527552, EPI_ISL_1527553, EPI_ISL_1527554, EPI_ISL_1527555, EPI_ISL_1527556, EPI_ISL_1527557, EPI_ISL_1527558, EPI_ISL_1527559, EPI_ISL_1527560, EPI_ISL_1527561, EPI_ISL_1527562, EPI_ISL_1527563, EPI_ISL_1527564, EPI_ISL_1527565, EPI_ISL_1527566, EPI_ISL_1527567, EPI_ISL_1527568, EPI_ISL_1527569, EPI_ISL_1527571, EPI_ISL_1527572, EPI_ISL_1527573, EPI_ISL_1527574, EPI_ISL_1527575, EPI_ISL_1527576, EPI_ISL_1527577, EPI_ISL_1527578, EPI_ISL_1527579, EPI_ISL_1527580, EPI_ISL_1527581, EPI_ISL_1527582, EPI_ISL_1527583, EPI_ISL_1527584, EPI_ISL_1527585, EPI_ISL_1527586, EPI_ISL_1527587, EPI_ISL_1527588, EPI_ISL_1527589, EPI_ISL_1527590, EPI_ISL_1527591, EPI_ISL_1527592, EPI_ISL_1527593, EPI_ISL_1527594, EPI_ISL_1527595, EPI_ISL_1527596, EPI_ISL_1527597, EPI_ISL_1527598, EPI_ISL_1527599, EPI_ISL_1527600, EPI_ISL_1527601, EPI_ISL_1527602, EPI_ISL_1527603, EPI_ISL_1527604, EPI_ISL_1527605, EPI_ISL_1527606, EPI_ISL_1527607, EPI_ISL_1527608, EPI_ISL_1527609, EPI_ISL_1527610, EPI_ISL_1527611, EPI_ISL_1527612, EPI_ISL_1527613, EPI_ISL_1527614, EPI_ISL_1527615, EPI_ISL_1527616, EPI_ISL_1527617, EPI_ISL_1527618, EPI_ISL_1527619, EPI_ISL_1527620, EPI_ISL_1527621, EPI_ISL_1527622, EPI_ISL_1527623, EPI_ISL_1527624, EPI_ISL_1527625, EPI_ISL_1527626, EPI_ISL_1527627, EPI_ISL_1527628, EPI_ISL_1527629 | | | |
| see above | TXDSHS | TXDSHS | Rashmi Tuladhar, Bonnie Oh, Jenny Zhang, Maliha Rahman, Mayela Pedrueza, Anita Pokharel, Lorraine Rodriguez, Myong Koag, Chun Wang, Rachel Lee, Grace Kubin |
| EPI_ISL_1527661, EPI_ISL_1527663, EPI_ISL_1527666, EPI_ISL_1527667, EPI_ISL_1527668, EPI_ISL_1527669, EPI_ISL_1527670, EPI_ISL_1527671, EPI_ISL_1527672, EPI_ISL_1527673, EPI_ISL_1527676, EPI_ISL_1527677, EPI_ISL_1527678, EPI_ISL_1527679, EPI_ISL_1527680, EPI_ISL_1527681, EPI_ISL_1527682, EPI_ISL_1527683, EPI_ISL_1527684, EPI_ISL_1527685, EPI_ISL_1527686, EPI_ISL_1527687, EPI_ISL_1527689, EPI_ISL_1527690, EPI_ISL_1527691, EPI_ISL_1527692, EPI_ISL_1527693, EPI_ISL_1527694, EPI_ISL_1527695, EPI_ISL_1527697, EPI_ISL_1527699, EPI_ISL_1527700, EPI_ISL_1527701, EPI_ISL_1527702, EPI_ISL_1527703, EPI_ISL_1527704, EPI_ISL_1527705, EPI_ISL_1527706, EPI_ISL_1527707, EPI_ISL_1527708, EPI_ISL_1527709, EPI_ISL_1527710, EPI_ISL_1527711, EPI_ISL_1527713, EPI_ISL_1527714, EPI_ISL_1527716, EPI_ISL_1527717, EPI_ISL_1527719, EPI_ISL_1527720, EPI_ISL_1527724, EPI_ISL_1527725, EPI_ISL_1527726, EPI_ISL_1527736, EPI_ISL_1527737, EPI_ISL_1527741, EPI_ISL_1527746, EPI_ISL_1527749, EPI_ISL_1527751, EPI_ISL_1527763, EPI_ISL_1527766, EPI_ISL_1527767, EPI_ISL_1527768, EPI_ISL_1527769, EPI_ISL_1527770, EPI_ISL_1527771, EPI_ISL_1527772, EPI_ISL_1527773, EPI_ISL_1527775, EPI_ISL_1527776, EPI_ISL_1527778, EPI_ISL_1527782, EPI_ISL_1527785, EPI_ISL_1527786, EPI_ISL_1527787, EPI_ISL_1527789, EPI_ISL_1527790, EPI_ISL_1527791, EPI_ISL_1527792, EPI_ISL_1527794, EPI_ISL_1527795, EPI_ISL_1527797, EPI_ISL_1527798, EPI_ISL_1527799, EPI_ISL_1527801, EPI_ISL_1527803, EPI_ISL_1527827, EPI_ISL_1527829, EPI_ISL_1527831, EPI_ISL_1527832, EPI_ISL_1527833, EPI_ISL_1527834, EPI_ISL_1527835, EPI_ISL_1527836, EPI_ISL_1527837, EPI_ISL_1527838, EPI_ISL_1527839, EPI_ISL_1527841, EPI_ISL_1527842, EPI_ISL_1527845, EPI_ISL_1527846, EPI_ISL_1527848, EPI_ISL_1527849, EPI_ISL_1527850, EPI_ISL_1527852, EPI_ISL_1527853, EPI_ISL_1527854, EPI_ISL_1527856, EPI_ISL_1527857, EPI_ISL_1527860, EPI_ISL_1527865, EPI_ISL_1527866, EPI_ISL_1527867, EPI_ISL_1527868, EPI_ISL_1527869, EPI_ISL_1527870, EPI_ISL_1527871, EPI_ISL_1527872, EPI_ISL_1527874, EPI_ISL_1527875, EPI_ISL_1527876, EPI_ISL_1527877, EPI_ISL_1527878, EPI_ISL_1527879, EPI_ISL_1527880, EPI_ISL_1527881, EPI_ISL_1527882, EPI_ISL_1527883, EPI_ISL_1527884, EPI_ISL_1527885, EPI_ISL_1527886, EPI_ISL_1527887, EPI_ISL_1527888, EPI_ISL_1527889, EPI_ISL_1527891, EPI_ISL_1527892, EPI_ISL_1527893, EPI_ISL_1527894, EPI_ISL_1527895, EPI_ISL_1527896, EPI_ISL_1527897, EPI_ISL_1527898, EPI_ISL_1527899, EPI_ISL_1527900, EPI_ISL_1527901, EPI_ISL_1527902, EPI_ISL_1527903, EPI_ISL_1527904, EPI_ISL_1527905, EPI_ISL_1527906, EPI_ISL_1527907, EPI_ISL_1527908, EPI_ISL_1527911, EPI_ISL_1527912, EPI_ISL_1527913, EPI_ISL_1527915, EPI_ISL_1527916, EPI_ISL_1527917, EPI_ISL_1527918, EPI_ISL_1527919, EPI_ISL_1527920, EPI_ISL_1527921, EPI_ISL_1527922, EPI_ISL_1527923, EPI_ISL_1527925, EPI_ISL_1527926, EPI_ISL_1527927, EPI_ISL_1527928, EPI_ISL_1527929, EPI_ISL_1527930, EPI_ISL_1527931, EPI_ISL_1527932, EPI_ISL_1527933, EPI_ISL_1527934, EPI_ISL_1527935, EPI_ISL_1527936, EPI_ISL_1527937, EPI_ISL_1527938, EPI_ISL_1527939, EPI_ISL_1527940, EPI_ISL_1527941 | | | |
| see above | Michigan Department of Health and Human Services, Bureau of Laboratories | Michigan Department of Health and Human Services, Bureau of Laboratories | Blankenship HM, Riner D, Soehnlen MK |
| EPI_ISL_1527942, EPI_ISL_1527943, EPI_ISL_1527944, EPI_ISL_1527945, EPI_ISL_1527946, EPI_ISL_1527947, EPI_ISL_1527948, EPI_ISL_1527949, EPI_ISL_1527950, EPI_ISL_1527951, EPI_ISL_1527952, EPI_ISL_1527953 | | | |
| see above | IN State Department of Health Laboratory Services | IN State Department of Health Laboratory Services | Cassandra Campion, Jamie Yeadon, Brian Pope, Lixia Liu, Kyle Brownlee, Melissa Hindenlang, Ankita Kashikar, Mark Glazier |
| EPI_ISL_1527954, EPI_ISL_1527956, EPI_ISL_1527958, EPI_ISL_1527959, EPI_ISL_1527961, EPI_ISL_1527962, EPI_ISL_1527964, EPI_ISL_1527965, EPI_ISL_1527967, EPI_ISL_1527968, EPI_ISL_1527970, EPI_ISL_1527971, EPI_ISL_1527973, EPI_ISL_1527974, EPI_ISL_1527975, EPI_ISL_1527978, EPI_ISL_1527979, EPI_ISL_1527980, EPI_ISL_1527982, EPI_ISL_1527983, EPI_ISL_1527984, EPI_ISL_1527986, EPI_ISL_1527987, EPI_ISL_1527988, EPI_ISL_1527989, EPI_ISL_1527992, EPI_ISL_1527993, EPI_ISL_1527995, EPI_ISL_1527996, EPI_ISL_1527997, EPI_ISL_1527998, EPI_ISL_1528000, EPI_ISL_1528002, EPI_ISL_1528003, EPI_ISL_1528004, EPI_ISL_1528005, EPI_ISL_1528006, EPI_ISL_1528007, EPI_ISL_1528009, EPI_ISL_1528010, EPI_ISL_1528012, EPI_ISL_1528013, EPI_ISL_1528014, EPI_ISL_1528016, EPI_ISL_1528017, EPI_ISL_1528018, EPI_ISL_1528019, EPI_ISL_1528020, EPI_ISL_1528021, EPI_ISL_1528022, EPI_ISL_1528023, EPI_ISL_1528024, EPI_ISL_1528025, EPI_ISL_1528026, EPI_ISL_1528027, EPI_ISL_1528028, EPI_ISL_1528029, EPI_ISL_1528030, EPI_ISL_1528032, EPI_ISL_1528033, EPI_ISL_1528034, EPI_ISL_1528035, EPI_ISL_1528036, EPI_ISL_1528038, EPI_ISL_1528039, EPI_ISL_1528040, EPI_ISL_1528041, EPI_ISL_1528042, EPI_ISL_1528043, EPI_ISL_1528044, EPI_ISL_1528045, EPI_ISL_1528046, EPI_ISL_1528047, EPI_ISL_1528048, EPI_ISL_1528049, EPI_ISL_1528050, EPI_ISL_1528051, EPI_ISL_1528052, EPI_ISL_1528053, EPI_ISL_1528054, EPI_ISL_1528055, EPI_ISL_1528056, EPI_ISL_1528057, EPI_ISL_1528058, EPI_ISL_1528059, EPI_ISL_1528061, EPI_ISL_1528062, EPI_ISL_1528063, EPI_ISL_1528064, EPI_ISL_1528065, EPI_ISL_1528066, EPI_ISL_1528067, EPI_ISL_1528068, EPI_ISL_1528069, EPI_ISL_1528070, EPI_ISL_1528071, EPI_ISL_1528072, EPI_ISL_1528073, EPI_ISL_1528074, EPI_ISL_1528075, EPI_ISL_1528076, EPI_ISL_1528077, EPI_ISL_1528078, EPI_ISL_1528079, EPI_ISL_1528080, EPI_ISL_1528081, EPI_ISL_1528082, EPI_ISL_1528083, EPI_ISL_1528084, EPI_ISL_1528085, EPI_ISL_1528087, EPI_ISL_1528088, EPI_ISL_1528090, EPI_ISL_1528091, EPI_ISL_1528092 | | | |
| see above | Michigan Department of Health and Human Services, Bureau of Laboratories | Michigan Department of Health and Human Services, Bureau of Laboratories | Blankenship HM, Riner D, Soehnlen MK |
| EPI_ISL_1528093, EPI_ISL_1528094, EPI_ISL_1528095, EPI_ISL_1528096, EPI_ISL_1528097, EPI_ISL_1528098, EPI_ISL_1528099, EPI_ISL_1528100, EPI_ISL_1528101, EPI_ISL_1528102, EPI_ISL_1528103, EPI_ISL_1528104, EPI_ISL_1528105, EPI_ISL_1528106, EPI_ISL_1528107, EPI_ISL_1528108, EPI_ISL_1528109, EPI_ISL_1528110, EPI_ISL_1528111, EPI_ISL_1528112, EPI_ISL_1528113, EPI_ISL_1528114, EPI_ISL_1528115, EPI_ISL_1528116, EPI_ISL_1528117, EPI_ISL_1528118, EPI_ISL_1528119, EPI_ISL_1528120, EPI_ISL_1528121, EPI_ISL_1528122, EPI_ISL_1528123, EPI_ISL_1528124, EPI_ISL_1528125, EPI_ISL_1528126, EPI_ISL_1528127, EPI_ISL_1528128, EPI_ISL_1528129, EPI_ISL_1528130, EPI_ISL_1528131, EPI_ISL_1528132, EPI_ISL_1528133, EPI_ISL_1528134, EPI_ISL_1528135, EPI_ISL_1528136, EPI_ISL_1528137, EPI_ISL_1528138, EPI_ISL_1528139, EPI_ISL_1528140, EPI_ISL_1528141, EPI_ISL_1528142, EPI_ISL_1528149, EPI_ISL_1528150, EPI_ISL_1528151, EPI_ISL_1528152, EPI_ISL_1528153, EPI_ISL_1528154, EPI_ISL_1528155, EPI_ISL_1528156, EPI_ISL_1528157, EPI_ISL_1528158, EPI_ISL_1528159 | | | |
| see above | Massachusetts State Public Health Laboratory | Massachusetts State Public Health Laboratory | Andrew Lang, Timelia Fink, Glen Gallagher, Sandra Smole |
| EPI_ISL_1528242, EPI_ISL_1528243, EPI_ISL_1528244, EPI_ISL_1528245, EPI_ISL_1528246, EPI_ISL_1528247, EPI_ISL_1528249, EPI_ISL_1528250, EPI_ISL_1528251, EPI_ISL_1528253, EPI_ISL_1528254, EPI_ISL_1528255, EPI_ISL_1528256, EPI_ISL_1528257, EPI_ISL_1528258, EPI_ISL_1528259, EPI_ISL_1528260, EPI_ISL_1528261, EPI_ISL_1528262, EPI_ISL_1528263, EPI_ISL_1528264, EPI_ISL_1528265 | | | |
| see above | IN State Department of Health Laboratory Services | IN State Department of Health Laboratory Services | Cassandra Campion, Jamie Yeadon, Brian Pope, Lixia Liu, Kyle Brownlee, Melissa Hindenlang, Ankita Kashikar, Mark Glazier |
| EPI_ISL_1528266, EPI_ISL_1528267, EPI_ISL_1528268, EPI_ISL_1528269 | SD Public Health Laboratory | Centers for Disease Control and Prevention Division of Viral Diseases, Pathogen Discovery | Mili Sheth, Sarah Nobles, Jasmine Padilla, Mark Burroughs, Shoshona Le, Katie Dillon, Peter Cook, Clinton R. Paden, Dhwani Batra, Krista Queen, Kristen Knipe, Dakota Howard, Yvette Unoarumhi, Darlene Wagner, Matthew Schmerer, Ben L. Rambo-Martin, Kristine Lacek, Sam Shepard, Alison Laufer Halpin, Dave Wentworth, Vivien Dugan, Suxiang Tong, Justin Lee |
| EPI_ISL_1528270, EPI_ISL_1528271 | SC Dept of Health and Env. Control-Bureau of Laboratories | Centers for Disease Control and Prevention Division of Viral Diseases, Pathogen Discovery | Mili Sheth, Sarah Nobles, Jasmine Padilla, Mark Burroughs, Shoshona Le, Katie Dillon, Peter Cook, Clinton R. Paden, Dhwani Batra, Krista Queen, Kristen Knipe, Dakota Howard, Yvette Unoarumhi, Darlene Wagner, Matthew Schmerer, Ben L. Rambo-Martin, Kristine Lacek, Sam Shepard, Alison Laufer Halpin, Dave Wentworth, Vivien Dugan, Suxiang Tong, Justin Lee |
| EPI_ISL_1528272, EPI_ISL_1528273, EPI_ISL_1528274 | "MN PHL Division, Minnesota Department of Health" | Centers for Disease Control and Prevention Division of Viral Diseases, Pathogen Discovery | Mili Sheth, Sarah Nobles, Jasmine Padilla, Mark Burroughs, Shoshona Le, Katie Dillon, Peter Cook, Clinton R. Paden, Dhwani Batra, Krista Queen, Kristen Knipe, Dakota Howard, Yvette Unoarumhi, Darlene Wagner, Matthew Schmerer, Ben L. Rambo-Martin, Kristine Lacek, Sam Shepard, Alison Laufer Halpin, Dave Wentworth, Vivien Dugan, Suxiang Tong, Justin Lee |
| EPI_ISL_1528275, EPI_ISL_1528276, EPI_ISL_1528277, EPI_ISL_1528278, EPI_ISL_1528279, EPI_ISL_1528280, EPI_ISL_1528281, EPI_ISL_1528282, EPI_ISL_1528283 | VA-Division of Consolidated Laboratory Services | Centers for Disease Control and Prevention Division of Viral Diseases, Pathogen Discovery | Mili Sheth, Sarah Nobles, Jasmine Padilla, Mark Burroughs, Shoshona Le, Katie Dillon, Peter Cook, Clinton R. Paden, Dhwani Batra, Krista Queen, Kristen Knipe, Dakota Howard, Yvette Unoarumhi, Darlene Wagner, Matthew Schmerer, Ben L. Rambo-Martin, Kristine Lacek, Sam Shepard, Alison Laufer Halpin, Dave Wentworth, Vivien Dugan, Suxiang Tong, Justin Lee |
| EPI_ISL_1528284, EPI_ISL_1528285 | FL Bureau of Public Health Laboratories-Tampa | Centers for Disease Control and Prevention Division of Viral Diseases, Pathogen Discovery | Mili Sheth, Sarah Nobles, Jasmine Padilla, Mark Burroughs, Shoshona Le, Katie Dillon, Peter Cook, Clinton R. Paden, Dhwani Batra, Krista Queen, Kristen Knipe, Dakota Howard, Yvette Unoarumhi, Darlene Wagner, Matthew Schmerer, Ben L. Rambo-Martin, Kristine Lacek, Sam Shepard, Alison Laufer Halpin, Dave Wentworth, Vivien Dugan, Suxiang Tong, Justin Lee |
| EPI_ISL_1528286, EPI_ISL_1528287, EPI_ISL_1528288, EPI_ISL_1528289, EPI_ISL_1528290 | NE Public Health Laboratory | Centers for Disease Control and Prevention Division of Viral Diseases, Pathogen Discovery | Mili Sheth, Sarah Nobles, Jasmine Padilla, Mark Burroughs, Shoshona Le, Katie Dillon, Peter Cook, Clinton R. Paden, Dhwani Batra, Krista Queen, Kristen Knipe, Dakota Howard, Yvette Unoarumhi, Darlene Wagner, Matthew Schmerer, Ben L. Rambo-Martin, Kristine Lacek, Sam Shepard, Alison Laufer Halpin, Dave Wentworth, Vivien Dugan, Suxiang Tong, Justin Lee |
| EPI_ISL_1528291, EPI_ISL_1528292, EPI_ISL_1528293 | "AR Dept. of Health-PHL, Molecular Diagnostics" | Centers for Disease Control and Prevention Division of Viral Diseases, Pathogen Discovery | Mili Sheth, Sarah Nobles, Jasmine Padilla, Mark Burroughs, Shoshona Le, Katie Dillon, Peter Cook, Clinton R. Paden, Dhwani Batra, Krista Queen, Kristen Knipe, Dakota Howard, Yvette Unoarumhi, Darlene Wagner, Matthew Schmerer, Ben L. Rambo-Martin, Kristine Lacek, Sam Shepard, Alison Laufer Halpin, Dave Wentworth, Vivien Dugan, Suxiang Tong, Justin Lee |
| EPI_ISL_1528294, EPI_ISL_1528295, EPI_ISL_1528296, EPI_ISL_1528297, EPI_ISL_1528298, EPI_ISL_1528299, EPI_ISL_1528300, EPI_ISL_1528301, EPI_ISL_1528302 | MT Public Health Laboratory | Centers for Disease Control and Prevention Division of Viral Diseases, Pathogen Discovery | Mili Sheth, Sarah Nobles, Jasmine Padilla, Mark Burroughs, Shoshona Le, Katie Dillon, Peter Cook, Clinton R. Paden, Dhwani Batra, Krista Queen, Kristen Knipe, Dakota Howard, Yvette Unoarumhi, Darlene Wagner, Matthew Schmerer, Ben L. Rambo-Martin, Kristine Lacek, Sam Shepard, Alison Laufer Halpin, Dave Wentworth, Vivien Dugan, Suxiang Tong, Justin Lee |
| EPI_ISL_1528303, EPI_ISL_1528304, EPI_ISL_1528305, EPI_ISL_1528306, EPI_ISL_1528307 | DE Public Health Laboratory | Centers for Disease Control and Prevention Division of Viral Diseases, Pathogen Discovery | Mili Sheth, Sarah Nobles, Jasmine Padilla, Mark Burroughs, Shoshona Le, Katie Dillon, Peter Cook, Clinton R. Paden, Dhwani Batra, Krista Queen, Kristen Knipe, Dakota Howard, Yvette Unoarumhi, Darlene Wagner, Matthew Schmerer, Ben L. Rambo-Martin, Kristine Lacek, Sam Shepard, Alison Laufer Halpin, Dave Wentworth, Vivien Dugan, Suxiang Tong, Justin Lee |
| EPI_ISL_1528308, EPI_ISL_1528309, EPI_ISL_1528310, EPI_ISL_1528311, EPI_ISL_1528312, EPI_ISL_1528313 | MI - Michigan Department of Health and Human Services - Bureau of Laboratories | Centers for Disease Control and Prevention Division of Viral Diseases, Pathogen Discovery | Mili Sheth, Sarah Nobles, Jasmine Padilla, Mark Burroughs, Shoshona Le, Katie Dillon, Peter Cook, Clinton R. Paden, Dhwani Batra, Krista Queen, Kristen Knipe, Dakota Howard, Yvette Unoarumhi, Darlene Wagner, Matthew Schmerer, Ben L. Rambo-Martin, Kristine Lacek, Sam Shepard, Alison Laufer Halpin, Dave Wentworth, Vivien Dugan, Suxiang Tong, Justin Lee |
| EPI_ISL_1528314, EPI_ISL_1528315, EPI_ISL_1528316, EPI_ISL_1528317, EPI_ISL_1528318, EPI_ISL_1528319, EPI_ISL_1528320 | FL Bur. of Public Health Laboratories-Jacksonville | Centers for Disease Control and Prevention Division of Viral Diseases, Pathogen Discovery | Mili Sheth, Sarah Nobles, Jasmine Padilla, Mark Burroughs, Shoshona Le, Katie Dillon, Peter Cook, Clinton R. Paden, Dhwani Batra, Krista Queen, Kristen Knipe, Dakota Howard, Yvette Unoarumhi, Darlene Wagner, Matthew Schmerer, Ben L. Rambo-Martin, Kristine Lacek, Sam Shepard, Alison Laufer Halpin, Dave Wentworth, Vivien Dugan, Suxiang Tong, Justin Lee |
| EPI_ISL_1528321, EPI_ISL_1528322, EPI_ISL_1528323, | OR State PHL-Virology/Immunology Section | Centers for Disease Control and Prevention Division of Viral Diseases, Pathogen Discovery | Mili Sheth, Sarah Nobles, Jasmine Padilla, Mark Burroughs, Shoshona Le, Katie Dillon, Peter Cook, Clinton R. Paden, Dhwani Batra, Krista Queen, Kristen Knipe, Dakota Howard, Yvette Unoarumhi, Darlene Wagner, Matthew Schmerer, Ben L. Rambo-Martin, Kristine Lacek, Sam Shepard, Alison Laufer Halpin, Dave Wentworth, Vivien Dugan, Suxiang Tong, Justin Lee |

| | | | |
|---|---|---|---|
| EPI_ISL_1528324, EPI_ISL_1528325 | | | |
| EPI_ISL_1528326, EPI_ISL_1528327, EPI_ISL_1528328, EPI_ISL_1528329, EPI_ISL_1528330, EPI_ISL_1528331, EPI_ISL_1528332, EPI_ISL_1528333, EPI_ISL_1528334, EPI_ISL_1528335, EPI_ISL_1528336, EPI_ISL_1528337, EPI_ISL_1528338 | | | |
| see above | NJ Public Health and Environmental Laboratories | Centers for Disease Control and Prevention Division of Viral Diseases, Pathogen Discovery | Mili Sheth, Sarah Nobles, Jasmine Padilla, Mark Burroughs, Shoshona Le, Katie Dillon, Peter Cook, Clinton R. Paden, Dhwani Batra, Krista Queen, Kristen Knipe, Dakota Howard, Yvette Unoarumhi, Darlene Wagner, Matthew Schmerer, Ben L. Rambo-Martin, Kristine Lacek, Sam Shepard, Alison Laufer Halpin, Dave Wentworth, Vivien Dugan, Suxiang Tong, Justin Lee |
| EPI_ISL_1528339, EPI_ISL_1528340, EPI_ISL_1528341, EPI_ISL_1528342, EPI_ISL_1528343, EPI_ISL_1528344, EPI_ISL_1528345, EPI_ISL_1528346, EPI_ISL_1528347, EPI_ISL_1528348, EPI_ISL_1528349, EPI_ISL_1528350, EPI_ISL_1528351, EPI_ISL_1528352, EPI_ISL_1528353, EPI_ISL_1528354, EPI_ISL_1528355, EPI_ISL_1528356, EPI_ISL_1528357, EPI_ISL_1528358, EPI_ISL_1528359, EPI_ISL_1528360, EPI_ISL_1528361, EPI_ISL_1528362 | | | |
| see above | "CO Dept. of Public Health and Environment, Lab Services Division" | Centers for Disease Control and Prevention Division of Viral Diseases, Pathogen Discovery | Mili Sheth, Sarah Nobles, Jasmine Padilla, Mark Burroughs, Shoshona Le, Katie Dillon, Peter Cook, Clinton R. Paden, Dhwani Batra, Krista Queen, Kristen Knipe, Dakota Howard, Yvette Unoarumhi, Darlene Wagner, Matthew Schmerer, Ben L. Rambo-Martin, Kristine Lacek, Sam Shepard, Alison Laufer Halpin, Dave Wentworth, Vivien Dugan, Suxiang Tong, Justin Lee |
| EPI_ISL_1528363, EPI_ISL_1528364 | ME Health and Environmental Testing Laboratory | Centers for Disease Control and Prevention Division of Viral Diseases, Pathogen Discovery | Mili Sheth, Sarah Nobles, Jasmine Padilla, Mark Burroughs, Shoshona Le, Katie Dillon, Peter Cook, Clinton R. Paden, Dhwani Batra, Krista Queen, Kristen Knipe, Dakota Howard, Yvette Unoarumhi, Darlene Wagner, Matthew Schmerer, Ben L. Rambo-Martin, Kristine Lacek, Sam Shepard, Alison Laufer Halpin, Dave Wentworth, Vivien Dugan, Suxiang Tong, Justin Lee |
| EPI_ISL_1528365, EPI_ISL_1528366, EPI_ISL_1528367, EPI_ISL_1528368, EPI_ISL_1528369, EPI_ISL_1528370, EPI_ISL_1528371, EPI_ISL_1528372, EPI_ISL_1528373 | IL Dept. of Public Health Springfield Laboratory | Centers for Disease Control and Prevention Division of Viral Diseases, Pathogen Discovery | Mili Sheth, Sarah Nobles, Jasmine Padilla, Mark Burroughs, Shoshona Le, Katie Dillon, Peter Cook, Clinton R. Paden, Dhwani Batra, Krista Queen, Kristen Knipe, Dakota Howard, Yvette Unoarumhi, Darlene Wagner, Matthew Schmerer, Ben L. Rambo-Martin, Kristine Lacek, Sam Shepard, Alison Laufer Halpin, Dave Wentworth, Vivien Dugan, Suxiang Tong, Justin Lee |
| EPI_ISL_1528374, EPI_ISL_1528375, EPI_ISL_1528376, EPI_ISL_1528377 | WA State Department of Health | Centers for Disease Control and Prevention Division of Viral Diseases, Pathogen Discovery | Mili Sheth, Sarah Nobles, Jasmine Padilla, Mark Burroughs, Shoshona Le, Katie Dillon, Peter Cook, Clinton R. Paden, Dhwani Batra, Krista Queen, Kristen Knipe, Dakota Howard, Yvette Unoarumhi, Darlene Wagner, Matthew Schmerer, Ben L. Rambo-Martin, Kristine Lacek, Sam Shepard, Alison Laufer Halpin, Dave Wentworth, Vivien Dugan, Suxiang Tong, Justin Lee |
| EPI_ISL_1528378, EPI_ISL_1528379, EPI_ISL_1528380, EPI_ISL_1528381, EPI_ISL_1528382, EPI_ISL_1528383, EPI_ISL_1528384, EPI_ISL_1528385 | "NM Dept. Health, Scientific Laboratory Division " | Centers for Disease Control and Prevention Division of Viral Diseases, Pathogen Discovery | Mili Sheth, Sarah Nobles, Jasmine Padilla, Mark Burroughs, Shoshona Le, Katie Dillon, Peter Cook, Clinton R. Paden, Dhwani Batra, Krista Queen, Kristen Knipe, Dakota Howard, Yvette Unoarumhi, Darlene Wagner, Matthew Schmerer, Ben L. Rambo-Martin, Kristine Lacek, Sam Shepard, Alison Laufer Halpin, Dave Wentworth, Vivien Dugan, Suxiang Tong, Justin Lee |
| EPI_ISL_1528386 | VT Dept. of Health Laboratory | Centers for Disease Control and Prevention Division of Viral Diseases, Pathogen Discovery | Mili Sheth, Sarah Nobles, Jasmine Padilla, Mark Burroughs, Shoshona Le, Katie Dillon, Peter Cook, Clinton R. Paden, Dhwani Batra, Krista Queen, Kristen Knipe, Dakota Howard, Yvette Unoarumhi, Darlene Wagner, Matthew Schmerer, Ben L. Rambo-Martin, Kristine Lacek, Sam Shepard, Alison Laufer Halpin, Dave Wentworth, Vivien Dugan, Suxiang Tong, Justin Lee |
| EPI_ISL_1528387, EPI_ISL_1528388, EPI_ISL_1528390, EPI_ISL_1528391, EPI_ISL_1528392, EPI_ISL_1528393, EPI_ISL_1528394, EPI_ISL_1528395, EPI_ISL_1528396 | IA State Hygienic Laboratory | Centers for Disease Control and Prevention Division of Viral Diseases, Pathogen Discovery | Mili Sheth, Sarah Nobles, Jasmine Padilla, Mark Burroughs, Shoshona Le, Katie Dillon, Peter Cook, Clinton R. Paden, Dhwani Batra, Krista Queen, Kristen Knipe, Dakota Howard, Yvette Unoarumhi, Darlene Wagner, Matthew Schmerer, Ben L. Rambo-Martin, Kristine Lacek, Sam Shepard, Alison Laufer Halpin, Dave Wentworth, Vivien Dugan, Suxiang Tong, Justin Lee |
| EPI_ISL_1528397, EPI_ISL_1528398 | "TX DSHS, Lab Services Section MC 1947" | Centers for Disease Control and Prevention Division of Viral Diseases, Pathogen Discovery | Mili Sheth, Sarah Nobles, Jasmine Padilla, Mark Burroughs, Shoshona Le, Katie Dillon, Peter Cook, Clinton R. Paden, Dhwani Batra, Krista Queen, Kristen Knipe, Dakota Howard, Yvette Unoarumhi, Darlene Wagner, Matthew Schmerer, Ben L. Rambo-Martin, Kristine Lacek, Sam Shepard, Alison Laufer Halpin, Dave Wentworth, Vivien Dugan, Suxiang Tong, Justin Lee |
| EPI_ISL_1528399, EPI_ISL_1528400, EPI_ISL_1528401, EPI_ISL_1528402, EPI_ISL_1528403 | KS Health and Environmental Laboratories | Centers for Disease Control and Prevention Division of Viral Diseases, Pathogen Discovery | Mili Sheth, Sarah Nobles, Jasmine Padilla, Mark Burroughs, Shoshona Le, Katie Dillon, Peter Cook, Clinton R. Paden, Dhwani Batra, Krista Queen, Kristen Knipe, Dakota Howard, Yvette Unoarumhi, Darlene Wagner, Matthew Schmerer, Ben L. Rambo-Martin, Kristine Lacek, Sam Shepard, Alison Laufer Halpin, Dave Wentworth, Vivien Dugan, Suxiang Tong, Justin Lee |
| EPI_ISL_1528404, EPI_ISL_1528405 | TN Division of Laboratory Services | Centers for Disease Control and Prevention Division of Viral Diseases, Pathogen Discovery | Mili Sheth, Sarah Nobles, Jasmine Padilla, Mark Burroughs, Shoshona Le, Katie Dillon, Peter Cook, Clinton R. Paden, Dhwani Batra, Krista Queen, Kristen Knipe, Dakota Howard, Yvette Unoarumhi, Darlene Wagner, Matthew Schmerer, Ben L. Rambo-Martin, Kristine Lacek, Sam Shepard, Alison Laufer Halpin, Dave Wentworth, Vivien Dugan, Suxiang Tong, Justin Lee |
| EPI_ISL_1528406, EPI_ISL_1528407, EPI_ISL_1528408, EPI_ISL_1528409, EPI_ISL_1528410 | WI State Laboratory of Hygiene | Centers for Disease Control and Prevention Division of Viral Diseases, Pathogen Discovery | Mili Sheth, Sarah Nobles, Jasmine Padilla, Mark Burroughs, Shoshona Le, Katie Dillon, Peter Cook, Clinton R. Paden, Dhwani Batra, Krista Queen, Kristen Knipe, Dakota Howard, Yvette Unoarumhi, Darlene Wagner, Matthew Schmerer, Ben L. Rambo-Martin, Kristine Lacek, Sam Shepard, Alison Laufer Halpin, Dave Wentworth, Vivien Dugan, Suxiang Tong, Justin Lee |
| EPI_ISL_1528411, EPI_ISL_1528412, EPI_ISL_1528413 | MS Public Health Laboratory | Centers for Disease Control and Prevention Division of Viral Diseases, Pathogen Discovery | Mili Sheth, Sarah Nobles, Jasmine Padilla, Mark Burroughs, Shoshona Le, Katie Dillon, Peter Cook, Clinton R. Paden, Dhwani Batra, Krista Queen, Kristen Knipe, Dakota Howard, Yvette Unoarumhi, Darlene Wagner, Matthew Schmerer, Ben L. Rambo-Martin, Kristine Lacek, Sam Shepard, Alison Laufer Halpin, Dave Wentworth, Vivien Dugan, Suxiang Tong, Justin Lee |
| EPI_ISL_1528414, EPI_ISL_1528415, EPI_ISL_1528416, EPI_ISL_1528417, EPI_ISL_1528418 | WY Public Health Laboratory | Centers for Disease Control and Prevention Division of Viral Diseases, Pathogen Discovery | Mili Sheth, Sarah Nobles, Jasmine Padilla, Mark Burroughs, Shoshona Le, Katie Dillon, Peter Cook, Clinton R. Paden, Dhwani Batra, Krista Queen, Kristen Knipe, Dakota Howard, Yvette Unoarumhi, Darlene Wagner, Matthew Schmerer, Ben L. Rambo-Martin, Kristine Lacek, Sam Shepard, Alison Laufer Halpin, Dave Wentworth, Vivien Dugan, Suxiang Tong, Justin Lee |
| EPI_ISL_1528419 | "AZ SPHL, Arizona Department of Health Services" | Centers for Disease Control and Prevention Division of Viral Diseases, Pathogen Discovery | Mili Sheth, Sarah Nobles, Jasmine Padilla, Mark Burroughs, Shoshona Le, Katie Dillon, Peter Cook, Clinton R. Paden, Dhwani Batra, Krista Queen, Kristen Knipe, Dakota Howard, Yvette Unoarumhi, Darlene Wagner, Matthew Schmerer, Ben L. Rambo-Martin, Kristine Lacek, Sam Shepard, Alison Laufer Halpin, Dave Wentworth, Vivien Dugan, Suxiang Tong, Justin Lee |
| EPI_ISL_1528420, EPI_ISL_1528421, EPI_ISL_1528422, EPI_ISL_1528423 | NV State Public Health Laboratory | Centers for Disease Control and Prevention Division of Viral Diseases, Pathogen Discovery | Mili Sheth, Sarah Nobles, Jasmine Padilla, Mark Burroughs, Shoshona Le, Katie Dillon, Peter Cook, Clinton R. Paden, Dhwani Batra, Krista Queen, Kristen Knipe, Dakota Howard, Yvette Unoarumhi, Darlene Wagner, Matthew Schmerer, Ben L. Rambo-Martin, Kristine Lacek, Sam Shepard, Alison Laufer Halpin, Dave Wentworth, Vivien Dugan, Suxiang Tong, Justin Lee |
| EPI_ISL_1528424, EPI_ISL_1528425, EPI_ISL_1528426, EPI_ISL_1528427, EPI_ISL_1528428, EPI_ISL_1528429, EPI_ISL_1528430, EPI_ISL_1528431 | IN State Department of Health Laboratory Services | Centers for Disease Control and Prevention Division of Viral Diseases, Pathogen Discovery | Mili Sheth, Sarah Nobles, Jasmine Padilla, Mark Burroughs, Shoshona Le, Katie Dillon, Peter Cook, Clinton R. Paden, Dhwani Batra, Krista Queen, Kristen Knipe, Dakota Howard, Yvette Unoarumhi, Darlene Wagner, Matthew Schmerer, Ben L. Rambo-Martin, Kristine Lacek, Sam Shepard, Alison Laufer Halpin, Dave Wentworth, Vivien Dugan, Suxiang Tong, Justin Lee |
| EPI_ISL_1528432, EPI_ISL_1528433, EPI_ISL_1528434, EPI_ISL_1528435, EPI_ISL_1528436, EPI_ISL_1528437, EPI_ISL_1528438, EPI_ISL_1528439, EPI_ISL_1528440, EPI_ISL_1528441 | UT-Unified State Labs: Public Health Utah DOH | Centers for Disease Control and Prevention Division of Viral Diseases, Pathogen Discovery | Mili Sheth, Sarah Nobles, Jasmine Padilla, Mark Burroughs, Shoshona Le, Katie Dillon, Peter Cook, Clinton R. Paden, Dhwani Batra, Krista Queen, Kristen Knipe, Dakota Howard, Yvette Unoarumhi, Darlene Wagner, Matthew Schmerer, Ben L. Rambo-Martin, Kristine Lacek, Sam Shepard, Alison Laufer Halpin, Dave Wentworth, Vivien Dugan, Suxiang Tong, Justin Lee |
| EPI_ISL_1528442, EPI_ISL_1528443 | DC Public Health Lab/ Dept. of Forensic Sciences | Centers for Disease Control and Prevention Division of Viral Diseases, Pathogen Discovery | Mili Sheth, Sarah Nobles, Jasmine Padilla, Mark Burroughs, Shoshona Le, Katie Dillon, Peter Cook, Clinton R. Paden, Dhwani Batra, Krista Queen, Kristen Knipe, Dakota Howard, Yvette Unoarumhi, Darlene Wagner, Matthew Schmerer, Ben L. Rambo-Martin, Kristine Lacek, Sam Shepard, Alison Laufer Halpin, Dave Wentworth, Vivien Dugan, Suxiang Tong, Justin Lee |
| EPI_ISL_1528444, EPI_ISL_1528445, | WVDHHR - Office of Laboratory Services | Centers for Disease Control and Prevention Division of Viral Diseases, Pathogen | Mili Sheth, Sarah Nobles, Jasmine Padilla, Mark Burroughs, Shoshona Le, Katie Dillon, Peter Cook, Clinton R. Paden, Dhwani Batra, Krista Queen, Kristen Knipe, Dakota Howard, Yvette Unoarumhi, Darlene Wagner, Matthew Schmerer, Ben L. Rambo-Martin, Kristine Lacek, Sam Shepard, Alison Laufer Halpin, Dave Wentworth, Vivien Dugan, Suxiang Tong, Justin Lee |

| | | | |
|---|---|---|---|
| EPI_ISL_1528446, EPI_ISL_1528447, EPI_ISL_1528448, EPI_ISL_1528449, EPI_ISL_1528450 | | Discovery | |
| EPI_ISL_1528451, EPI_ISL_1528452, EPI_ISL_1528454, EPI_ISL_1528455, EPI_ISL_1528456, EPI_ISL_1528457, EPI_ISL_1528458, EPI_ISL_1528459 | MD DOH Laboratories Administration | Centers for Disease Control and Prevention Division of Viral Diseases, Pathogen Discovery | Mili Sheth, Sarah Nobles, Jasmine Padilla, Mark Burroughs, Shoshona Le, Katie Dillon, Peter Cook, Clinton R. Paden, Dhwani Batra, Krista Queen, Kristen Knipe, Dakota Howard, Yvette Unoarumhi, Darlene Wagner, Matthew Schmerer, Ben L. Rambo-Martin, Kristine Lacek, Sam Shepard, Alison Laufer Halpin, Dave Wentworth, Vivien Dugan, Suxiang Tong, Justin Lee |
| EPI_ISL_1528460, EPI_ISL_1528461 | PR Public Health Lab | Centers for Disease Control and Prevention Division of Viral Diseases, Pathogen Discovery | Mili Sheth, Sarah Nobles, Jasmine Padilla, Mark Burroughs, Shoshona Le, Katie Dillon, Peter Cook, Clinton R. Paden, Dhwani Batra, Krista Queen, Kristen Knipe, Dakota Howard, Yvette Unoarumhi, Darlene Wagner, Matthew Schmerer, Ben L. Rambo-Martin, Kristine Lacek, Sam Shepard, Alison Laufer Halpin, Dave Wentworth, Vivien Dugan, Suxiang Tong, Justin Lee |
| EPI_ISL_1528462, EPI_ISL_1528463, EPI_ISL_1528464, EPI_ISL_1528465, EPI_ISL_1528466, EPI_ISL_1528467, EPI_ISL_1528468, EPI_ISL_1528469, EPI_ISL_1528470, EPI_ISL_1528471, EPI_ISL_1528472, EPI_ISL_1528473, EPI_ISL_1528474 | | | |
| see above | RI State Health Laboratories | Centers for Disease Control and Prevention Division of Viral Diseases, Pathogen Discovery | Mili Sheth, Sarah Nobles, Jasmine Padilla, Mark Burroughs, Shoshona Le, Katie Dillon, Peter Cook, Clinton R. Paden, Dhwani Batra, Krista Queen, Kristen Knipe, Dakota Howard, Yvette Unoarumhi, Darlene Wagner, Matthew Schmerer, Ben L. Rambo-Martin, Kristine Lacek, Sam Shepard, Alison Laufer Halpin, Dave Wentworth, Vivien Dugan, Suxiang Tong, Justin Lee |
| EPI_ISL_1528475, EPI_ISL_1528476, EPI_ISL_1528477 | LA Office of Public Health Laboratories | Centers for Disease Control and Prevention Division of Viral Diseases, Pathogen Discovery | Mili Sheth, Sarah Nobles, Jasmine Padilla, Mark Burroughs, Shoshona Le, Katie Dillon, Peter Cook, Clinton R. Paden, Dhwani Batra, Krista Queen, Kristen Knipe, Dakota Howard, Yvette Unoarumhi, Darlene Wagner, Matthew Schmerer, Ben L. Rambo-Martin, Kristine Lacek, Sam Shepard, Alison Laufer Halpin, Dave Wentworth, Vivien Dugan, Suxiang Tong, Justin Lee |
| EPI_ISL_1528478 | AL Dept. of Public Health Bureau of Clinical Laboratories | Centers for Disease Control and Prevention Division of Viral Diseases, Pathogen Discovery | Mili Sheth, Sarah Nobles, Jasmine Padilla, Mark Burroughs, Shoshona Le, Katie Dillon, Peter Cook, Clinton R. Paden, Dhwani Batra, Krista Queen, Kristen Knipe, Dakota Howard, Yvette Unoarumhi, Darlene Wagner, Matthew Schmerer, Ben L. Rambo-Martin, Kristine Lacek, Sam Shepard, Alison Laufer Halpin, Dave Wentworth, Vivien Dugan, Suxiang Tong, Justin Lee |
| EPI_ISL_1528479, EPI_ISL_1528480, EPI_ISL_1528481, EPI_ISL_1528482, EPI_ISL_1528483, EPI_ISL_1528484, EPI_ISL_1528485, EPI_ISL_1528486, EPI_ISL_1528487, EPI_ISL_1528488, EPI_ISL_1528489, EPI_ISL_1528490, EPI_ISL_1528491, EPI_ISL_1528492 | | | |
| see above | "AK State Public Health Lab, State Health Department" | Centers for Disease Control and Prevention Division of Viral Diseases, Pathogen Discovery | Mili Sheth, Sarah Nobles, Jasmine Padilla, Mark Burroughs, Shoshona Le, Katie Dillon, Peter Cook, Clinton R. Paden, Dhwani Batra, Krista Queen, Kristen Knipe, Dakota Howard, Yvette Unoarumhi, Darlene Wagner, Matthew Schmerer, Ben L. Rambo-Martin, Kristine Lacek, Sam Shepard, Alison Laufer Halpin, Dave Wentworth, Vivien Dugan, Suxiang Tong, Justin Lee |
| EPI_ISL_1528493, EPI_ISL_1528494, EPI_ISL_1528495, EPI_ISL_1528496 | NH Dept. of Health and Human Services Public Health Labs | Centers for Disease Control and Prevention Division of Viral Diseases, Pathogen Discovery | Mili Sheth, Sarah Nobles, Jasmine Padilla, Mark Burroughs, Shoshona Le, Katie Dillon, Peter Cook, Clinton R. Paden, Dhwani Batra, Krista Queen, Kristen Knipe, Dakota Howard, Yvette Unoarumhi, Darlene Wagner, Matthew Schmerer, Ben L. Rambo-Martin, Kristine Lacek, Sam Shepard, Alison Laufer Halpin, Dave Wentworth, Vivien Dugan, Suxiang Tong, Justin Lee |
| EPI_ISL_1528497, EPI_ISL_1528498, EPI_ISL_1528499, EPI_ISL_1528500, EPI_ISL_1528501, EPI_ISL_1528502 | OH Department of Health Laboratory | Centers for Disease Control and Prevention Division of Viral Diseases, Pathogen Discovery | Mili Sheth, Sarah Nobles, Jasmine Padilla, Mark Burroughs, Shoshona Le, Katie Dillon, Peter Cook, Clinton R. Paden, Dhwani Batra, Krista Queen, Kristen Knipe, Dakota Howard, Yvette Unoarumhi, Darlene Wagner, Matthew Schmerer, Ben L. Rambo-Martin, Kristine Lacek, Sam Shepard, Alison Laufer Halpin, Dave Wentworth, Vivien Dugan, Suxiang Tong, Justin Lee |
| EPI_ISL_1528503, EPI_ISL_1528504, EPI_ISL_1528505 | MO State Public Health Laboratory | Centers for Disease Control and Prevention Division of Viral Diseases, Pathogen Discovery | Mili Sheth, Sarah Nobles, Jasmine Padilla, Mark Burroughs, Shoshona Le, Katie Dillon, Peter Cook, Clinton R. Paden, Dhwani Batra, Krista Queen, Kristen Knipe, Dakota Howard, Yvette Unoarumhi, Darlene Wagner, Matthew Schmerer, Ben L. Rambo-Martin, Kristine Lacek, Sam Shepard, Alison Laufer Halpin, Dave Wentworth, Vivien Dugan, Suxiang Tong, Justin Lee |
| EPI_ISL_1528506, EPI_ISL_1528507 | "PA Department of Health, Bureau of Laboratories" | Centers for Disease Control and Prevention Division of Viral Diseases, Pathogen Discovery | Mili Sheth, Sarah Nobles, Jasmine Padilla, Mark Burroughs, Shoshona Le, Katie Dillon, Peter Cook, Clinton R. Paden, Dhwani Batra, Krista Queen, Kristen Knipe, Dakota Howard, Yvette Unoarumhi, Darlene Wagner, Matthew Schmerer, Ben L. Rambo-Martin, Kristine Lacek, Sam Shepard, Alison Laufer Halpin, Dave Wentworth, Vivien Dugan, Suxiang Tong, Justin Lee |
| EPI_ISL_1528508, EPI_ISL_1528509, EPI_ISL_1528510, EPI_ISL_1528511, EPI_ISL_1528512, EPI_ISL_1528513, EPI_ISL_1528514, EPI_ISL_1528515, EPI_ISL_1528516 | NC State Laboratory of Public Health | Centers for Disease Control and Prevention Division of Viral Diseases, Pathogen Discovery | Mili Sheth, Sarah Nobles, Jasmine Padilla, Mark Burroughs, Shoshona Le, Katie Dillon, Peter Cook, Clinton R. Paden, Dhwani Batra, Krista Queen, Kristen Knipe, Dakota Howard, Yvette Unoarumhi, Darlene Wagner, Matthew Schmerer, Ben L. Rambo-Martin, Kristine Lacek, Sam Shepard, Alison Laufer Halpin, Dave Wentworth, Vivien Dugan, Suxiang Tong, Justin Lee |
| EPI_ISL_1528517, EPI_ISL_1528518, EPI_ISL_1528519, EPI_ISL_1528520 | VI-US Virgin Islands Department of Health | Centers for Disease Control and Prevention Division of Viral Diseases, Pathogen Discovery | Mili Sheth, Sarah Nobles, Jasmine Padilla, Mark Burroughs, Shoshona Le, Katie Dillon, Peter Cook, Clinton R. Paden, Dhwani Batra, Krista Queen, Kristen Knipe, Dakota Howard, Yvette Unoarumhi, Darlene Wagner, Matthew Schmerer, Ben L. Rambo-Martin, Kristine Lacek, Sam Shepard, Alison Laufer Halpin, Dave Wentworth, Vivien Dugan, Suxiang Tong, Justin Lee |
| EPI_ISL_1528521, EPI_ISL_1528522, EPI_ISL_1528523, EPI_ISL_1528525, EPI_ISL_1528526, EPI_ISL_1528527, EPI_ISL_1528528, EPI_ISL_1528529, EPI_ISL_1528530 | GA Department of Public Health Laboratory | Centers for Disease Control and Prevention Division of Viral Diseases, Pathogen Discovery | Mili Sheth, Sarah Nobles, Jasmine Padilla, Mark Burroughs, Shoshona Le, Katie Dillon, Peter Cook, Clinton R. Paden, Dhwani Batra, Krista Queen, Kristen Knipe, Dakota Howard, Yvette Unoarumhi, Darlene Wagner, Matthew Schmerer, Ben L. Rambo-Martin, Kristine Lacek, Sam Shepard, Alison Laufer Halpin, Dave Wentworth, Vivien Dugan, Suxiang Tong, Justin Lee |
| EPI_ISL_1528531, EPI_ISL_1528532, EPI_ISL_1528533, EPI_ISL_1528534, EPI_ISL_1528535, EPI_ISL_1528536, EPI_ISL_1528537, EPI_ISL_1528538, EPI_ISL_1528539, EPI_ISL_1528540, EPI_ISL_1528541, EPI_ISL_1528542, EPI_ISL_1528543, EPI_ISL_1528544, EPI_ISL_1528545, EPI_ISL_1528546, EPI_ISL_1528547, EPI_ISL_1528548, EPI_ISL_1528549, EPI_ISL_1528551, EPI_ISL_1528552, EPI_ISL_1528553, EPI_ISL_1528554, EPI_ISL_1528555, EPI_ISL_1528556, EPI_ISL_1528557, EPI_ISL_1528558, EPI_ISL_1528559, EPI_ISL_1528560, EPI_ISL_1528561, EPI_ISL_1528562, EPI_ISL_1528564, EPI_ISL_1528565, EPI_ISL_1528566, EPI_ISL_1528567, EPI_ISL_1528568, EPI_ISL_1528569, EPI_ISL_1528570, EPI_ISL_1528571, EPI_ISL_1528572, EPI_ISL_1528573, EPI_ISL_1528574, EPI_ISL_1528575, EPI_ISL_1528576, EPI_ISL_1528577, EPI_ISL_1528578 | | | |
| see above | WVU Rapid Development Lab | WVU and Marshall University Combined Genomics Core Facilities | "James Denvir, Peter Stoilov, Peter Perrotta, Wesley Kimble, Ryan Percifield" |
| EPI_ISL_1528579, EPI_ISL_1528580, EPI_ISL_1528581, EPI_ISL_1528582, EPI_ISL_1528583, EPI_ISL_1528584, EPI_ISL_1528585, EPI_ISL_1528586, EPI_ISL_1528587, EPI_ISL_1528588, EPI_ISL_1528589, EPI_ISL_1528590, EPI_ISL_1528591, EPI_ISL_1528592, EPI_ISL_1528593, EPI_ISL_1528594, EPI_ISL_1528595, EPI_ISL_1528596, EPI_ISL_1528597, EPI_ISL_1528598, EPI_ISL_1528599, EPI_ISL_1528600 | | | |
| see above | Berkeley Medical Center | WVU and Marshall University Combined Genomics Core Facilities | "James Denvir, Peter Stoilov, Peter Perrotta, Wesley Kimble, Ryan Percifield" |
| EPI_ISL_1528601, EPI_ISL_1528602, EPI_ISL_1528603, EPI_ISL_1528604, EPI_ISL_1528605, EPI_ISL_1528606, EPI_ISL_1528607, EPI_ISL_1528608, EPI_ISL_1528609, EPI_ISL_1528610, EPI_ISL_1528611, EPI_ISL_1528612, EPI_ISL_1528613, EPI_ISL_1528614, EPI_ISL_1528615, EPI_ISL_1528616 | | | |
| see above | Wheeling Hospital | WVU and Marshall University Combined Genomics Core Facilities | "James Denvir, Peter Stoilov, Peter Perrotta, Wesley Kimble, Ryan Percifield" |
| EPI_ISL_1528617, EPI_ISL_1528618, EPI_ISL_1528619, EPI_ISL_1528620, EPI_ISL_1528621, EPI_ISL_1528622, EPI_ISL_1528623, EPI_ISL_1528624, EPI_ISL_1528625, EPI_ISL_1528626, EPI_ISL_1528627, EPI_ISL_1528628, EPI_ISL_1528629, EPI_ISL_1528630, EPI_ISL_1528631, EPI_ISL_1528632, EPI_ISL_1528633, EPI_ISL_1528634, EPI_ISL_1528635, EPI_ISL_1528636, EPI_ISL_1528637, EPI_ISL_1528638, EPI_ISL_1528639, EPI_ISL_1528640, EPI_ISL_1528641, EPI_ISL_1528642, EPI_ISL_1528643, EPI_ISL_1528644, EPI_ISL_1528645, EPI_ISL_1528646, EPI_ISL_1528647, EPI_ISL_1528648, EPI_ISL_1528649, EPI_ISL_1528650, EPI_ISL_1528651, EPI_ISL_1528652, EPI_ISL_1528653, EPI_ISL_1528654, EPI_ISL_1528655, EPI_ISL_1528656, EPI_ISL_1528657, EPI_ISL_1528658, EPI_ISL_1528660, EPI_ISL_1528661, EPI_ISL_1528663, EPI_ISL_1528664, EPI_ISL_1528665, EPI_ISL_1528666, EPI_ISL_1528667, EPI_ISL_1528668, EPI_ISL_1528669, EPI_ISL_1528670, EPI_ISL_1528671, EPI_ISL_1528672, EPI_ISL_1528673, EPI_ISL_1528674, EPI_ISL_1528675, EPI_ISL_1528676, EPI_ISL_1528677, EPI_ISL_1528678, EPI_ISL_1528679, EPI_ISL_1528680, EPI_ISL_1528681, EPI_ISL_1528683, EPI_ISL_1528684, EPI_ISL_1528685, EPI_ISL_1528686, EPI_ISL_1528687, EPI_ISL_1528688, EPI_ISL_1528689, EPI_ISL_1528690, EPI_ISL_1528691, EPI_ISL_1528692, EPI_ISL_1528694, EPI_ISL_1528695, EPI_ISL_1528696, EPI_ISL_1528697, EPI_ISL_1528698, EPI_ISL_1528699, EPI_ISL_1528700, EPI_ISL_1528701, EPI_ISL_1528702, EPI_ISL_1528703, EPI_ISL_1528704, EPI_ISL_1528706, EPI_ISL_1528707, EPI_ISL_1528708, EPI_ISL_1528709, EPI_ISL_1528710, EPI_ISL_1528711, EPI_ISL_1528712, EPI_ISL_1528713, EPI_ISL_1528714, EPI_ISL_1528715, EPI_ISL_1528716, EPI_ISL_1528717, EPI_ISL_1528718 | | | |
| see above | J.W. Ruby Memorial Hospital | WVU and Marshall University Combined Genomics Core Facilities | "James Denvir, Peter Stoilov, Peter Perrotta, Wesley Kimble, Ryan Percifield" |
| EPI_ISL_1528720, EPI_ISL_1528721, EPI_ISL_1528722, EPI_ISL_1528723, EPI_ISL_1528725, EPI_ISL_1528726, EPI_ISL_1528727, EPI_ISL_1528728, EPI_ISL_1528729, EPI_ISL_1528730, EPI_ISL_1528731, EPI_ISL_1528732, EPI_ISL_1528733, EPI_ISL_1528734, EPI_ISL_1528735, EPI_ISL_1528736 | | | |
| see above | QLabs | WVU and Marshall University Combined Genomics Core Facilities | "James Denvir, Peter Stoilov, Peter Perrotta, Wesley Kimble, Ryan Percifield" |
| EPI_ISL_1528737 | J.W. Ruby Memorial Hospital | WVU and Marshall University Combined Genomics Core Facilities | "James Denvir, Peter Stoilov, Peter Perrotta, Wesley Kimble, Ryan Percifield" |
| EPI_ISL_1528738, EPI_ISL_1528739 | QLabs | WVU and Marshall University Combined Genomics Core Facilities | "James Denvir, Peter Stoilov, Peter Perrotta, Wesley Kimble, Ryan Percifield" |
| EPI_ISL_1528740, EPI_ISL_1528741, EPI_ISL_1528742 | J.W. Ruby Memorial Hospital | WVU and Marshall University Combined Genomics Core Facilities | "James Denvir, Peter Stoilov, Peter Perrotta, Wesley Kimble, Ryan Percifield" |

| | | | |
|---|---|---|---|
| EPI_ISL_1528743 | QLabs | WVU and Marshall University Combined Genomics Core Facilities | "James Denvir, Peter Stoilov, Peter Perrotta, Wesley Kimble, Ryan Percifield" |
| EPI_ISL_1528744 | J.W. Ruby Memorial Hospital | WVU and Marshall University Combined Genomics Core Facilities | "James Denvir, Peter Stoilov, Peter Perrotta, Wesley Kimble, Ryan Percifield" |
| EPI_ISL_1528745, EPI_ISL_1528746, EPI_ISL_1528747, EPI_ISL_1528748, EPI_ISL_1528749, EPI_ISL_1528750, EPI_ISL_1528751, EPI_ISL_1528752, EPI_ISL_1528753, EPI_ISL_1528754, EPI_ISL_1528755 | | | |
| see above | Berkeley Medical Center | WVU and Marshall University Combined Genomics Core Facilities | "James Denvir, Peter Stoilov, Peter Perrotta, Wesley Kimble, Ryan Percifield" |
| EPI_ISL_1528756, EPI_ISL_1528757, EPI_ISL_1528758, EPI_ISL_1528759 | QLabs | WVU and Marshall University Combined Genomics Core Facilities | "James Denvir, Peter Stoilov, Peter Perrotta, Wesley Kimble, Ryan Percifield" |
| EPI_ISL_1528760, EPI_ISL_1528761, EPI_ISL_1528763, EPI_ISL_1528764, EPI_ISL_1528765, EPI_ISL_1528766, EPI_ISL_1528767, EPI_ISL_1528768, EPI_ISL_1528769, EPI_ISL_1528770, EPI_ISL_1528771, EPI_ISL_1528772, EPI_ISL_1528773, EPI_ISL_1528774, EPI_ISL_1528775, EPI_ISL_1528776, EPI_ISL_1528777, EPI_ISL_1528778, EPI_ISL_1528779, EPI_ISL_1528781, EPI_ISL_1528782, EPI_ISL_1528784, EPI_ISL_1528785, EPI_ISL_1528786, EPI_ISL_1528787, EPI_ISL_1528788, EPI_ISL_1528789, EPI_ISL_1528790, EPI_ISL_1528791, EPI_ISL_1528792, EPI_ISL_1528793, EPI_ISL_1528794, EPI_ISL_1528795, EPI_ISL_1528797, EPI_ISL_1528798, EPI_ISL_1528799, EPI_ISL_1528800, EPI_ISL_1528801, EPI_ISL_1528802, EPI_ISL_1528803, EPI_ISL_1528804, EPI_ISL_1528805, EPI_ISL_1528806, EPI_ISL_1528807, EPI_ISL_1528808, EPI_ISL_1528809, EPI_ISL_1528810, EPI_ISL_1528811, EPI_ISL_1528812, EPI_ISL_1528813, EPI_ISL_1528814, EPI_ISL_1528815, EPI_ISL_1528816 | | | |
| see above | J.W. Ruby Memorial Hospital | WVU and Marshall University Combined Genomics Core Facilities | "James Denvir, Peter Stoilov, Peter Perrotta, Wesley Kimble, Ryan Percifield" |
| EPI_ISL_1528817, EPI_ISL_1528818, EPI_ISL_1528819, EPI_ISL_1528820, EPI_ISL_1528821, EPI_ISL_1528823, EPI_ISL_1528824 | Wheeling Hospital | WVU and Marshall University Combined Genomics Core Facilities | "James Denvir, Peter Stoilov, Peter Perrotta, Wesley Kimble, Ryan Percifield" |
| EPI_ISL_1528826 | Global Health Research Complex (GHRC) | TxGen | Benjamin W. Neuman, Melissa M. Kahl-Mcdonagh, Kurt A. Zuelke, Charlie D. Johnson, Allison Ficht, Sankar P. Chaki, Sierra Guidry, Joshua Hill, Richard Metz, Marcel Brun |
| EPI_ISL_1528828, EPI_ISL_1528829, EPI_ISL_1528830, EPI_ISL_1528832, EPI_ISL_1528833 | WVU Rapid Development Lab | WVU and Marshall University Combined Genomics Core Facilities | "James Denvir, Peter Stoilov, Peter Perrotta, Wesley Kimble, Ryan Percifield" |
| EPI_ISL_1528834, EPI_ISL_1528835, EPI_ISL_1528836, EPI_ISL_1528837, EPI_ISL_1528838, EPI_ISL_1528839, EPI_ISL_1528840, EPI_ISL_1528842, EPI_ISL_1528843, EPI_ISL_1528844, EPI_ISL_1528845, EPI_ISL_1528846, EPI_ISL_1528847, EPI_ISL_1528848, EPI_ISL_1528849, EPI_ISL_1528850, EPI_ISL_1528851, EPI_ISL_1528852, EPI_ISL_1528853, EPI_ISL_1528854, EPI_ISL_1528856, EPI_ISL_1528857, EPI_ISL_1528858, EPI_ISL_1528859, EPI_ISL_1528860, EPI_ISL_1528861, EPI_ISL_1528862, EPI_ISL_1528863, EPI_ISL_1528864, EPI_ISL_1528865, EPI_ISL_1528866, EPI_ISL_1528867, EPI_ISL_1528868, EPI_ISL_1528869 | | | |
| see above | J.W. Ruby Memorial Hospital | WVU and Marshall University Combined Genomics Core Facilities | "James Denvir, Peter Stoilov, Peter Perrotta, Wesley Kimble, Ryan Percifield" |
| EPI_ISL_1528871 | 4926 MacCorkle Ave SE, Charleston, WV 25304 | WVU and Marshall University Combined Genomics Core Facilities | "James Denvir, Peter Stoilov, Peter Perrotta, Wesley Kimble, Ryan Percifield" |
| EPI_ISL_1528872 | 4945 MacCorkle Ave SE, Charleston, WV 25304 | WVU and Marshall University Combined Genomics Core Facilities | "James Denvir, Peter Stoilov, Peter Perrotta, Wesley Kimble, Ryan Percifield" |
| EPI_ISL_1528873 | 4935 MacCorkle Ave SE, Charleston, WV 25304 | WVU and Marshall University Combined Genomics Core Facilities | "James Denvir, Peter Stoilov, Peter Perrotta, Wesley Kimble, Ryan Percifield" |
| EPI_ISL_1528874 | 4944 MacCorkle Ave SE, Charleston, WV 25304 | WVU and Marshall University Combined Genomics Core Facilities | "James Denvir, Peter Stoilov, Peter Perrotta, Wesley Kimble, Ryan Percifield" |
| EPI_ISL_1528875 | 4954 MacCorkle Ave SE, Charleston, WV 25304 | WVU and Marshall University Combined Genomics Core Facilities | "James Denvir, Peter Stoilov, Peter Perrotta, Wesley Kimble, Ryan Percifield" |
| EPI_ISL_1528877 | 4932 MacCorkle Ave SE, Charleston, WV 25304 | WVU and Marshall University Combined Genomics Core Facilities | "James Denvir, Peter Stoilov, Peter Perrotta, Wesley Kimble, Ryan Percifield" |
| EPI_ISL_1528878, EPI_ISL_1528879, EPI_ISL_1528880, EPI_ISL_1528881, EPI_ISL_1528882, EPI_ISL_1528883, EPI_ISL_1528884 | J.W. Ruby Memorial Hospital | WVU and Marshall University Combined Genomics Core Facilities | "James Denvir, Peter Stoilov, Peter Perrotta, Wesley Kimble, Ryan Percifield" |
| EPI_ISL_1528885 | UHC | WVU and Marshall University Combined Genomics Core Facilities | "James Denvir, Peter Stoilov, Peter Perrotta, Wesley Kimble, Ryan Percifield" |
| EPI_ISL_1528886 | 4933 MacCorkle Ave SE, Charleston, WV 25304 | WVU and Marshall University Combined Genomics Core Facilities | "James Denvir, Peter Stoilov, Peter Perrotta, Wesley Kimble, Ryan Percifield" |
| EPI_ISL_1528887 | J.W. Ruby Memorial Hospital | WVU and Marshall University Combined Genomics Core Facilities | "James Denvir, Peter Stoilov, Peter Perrotta, Wesley Kimble, Ryan Percifield" |
| EPI_ISL_1528889 | 4958 MacCorkle Ave SE, Charleston, WV 25304 | WVU and Marshall University Combined Genomics Core Facilities | "James Denvir, Peter Stoilov, Peter Perrotta, Wesley Kimble, Ryan Percifield" |
| EPI_ISL_1528890, EPI_ISL_1528891 | J.W. Ruby Memorial Hospital | WVU and Marshall University Combined Genomics Core Facilities | "James Denvir, Peter Stoilov, Peter Perrotta, Wesley Kimble, Ryan Percifield" |
| EPI_ISL_1528892 | 4936 MacCorkle Ave SE, Charleston, WV 25304 | WVU and Marshall University Combined Genomics Core Facilities | "James Denvir, Peter Stoilov, Peter Perrotta, Wesley Kimble, Ryan Percifield" |
| EPI_ISL_1528893 | 4948 MacCorkle Ave SE, Charleston, WV 25304 | WVU and Marshall University Combined Genomics Core Facilities | "James Denvir, Peter Stoilov, Peter Perrotta, Wesley Kimble, Ryan Percifield" |
| EPI_ISL_1528894 | 4929 MacCorkle Ave SE, Charleston, WV 25304 | WVU and Marshall University Combined Genomics Core Facilities | "James Denvir, Peter Stoilov, Peter Perrotta, Wesley Kimble, Ryan Percifield" |
| EPI_ISL_1528895 | 4937 MacCorkle Ave SE, Charleston, WV 25304 | WVU and Marshall University Combined Genomics Core Facilities | "James Denvir, Peter Stoilov, Peter Perrotta, Wesley Kimble, Ryan Percifield" |
| EPI_ISL_1528896 | 4931 MacCorkle Ave SE, Charleston, WV 25304 | WVU and Marshall University Combined Genomics Core Facilities | "James Denvir, Peter Stoilov, Peter Perrotta, Wesley Kimble, Ryan Percifield" |
| EPI_ISL_1528897 | 4934 MacCorkle Ave SE, Charleston, WV 25304 | WVU and Marshall University Combined Genomics Core Facilities | "James Denvir, Peter Stoilov, Peter Perrotta, Wesley Kimble, Ryan Percifield" |
| EPI_ISL_1528898 | 4940 MacCorkle Ave SE, Charleston, WV 25304 | WVU and Marshall University Combined Genomics Core Facilities | "James Denvir, Peter Stoilov, Peter Perrotta, Wesley Kimble, Ryan Percifield" |
| EPI_ISL_1528900 | 4924 MacCorkle Ave SE, Charleston, WV 25304 | WVU and Marshall University Combined Genomics Core Facilities | "James Denvir, Peter Stoilov, Peter Perrotta, Wesley Kimble, Ryan Percifield" |
| EPI_ISL_1528901 | 4950 MacCorkle Ave SE, Charleston, WV 25304 | WVU and Marshall University Combined Genomics Core Facilities | "James Denvir, Peter Stoilov, Peter Perrotta, Wesley Kimble, Ryan Percifield" |
| EPI_ISL_1528903 | 4960 MacCorkle Ave SE, Charleston, WV 25304 | WVU and Marshall University Combined Genomics Core Facilities | "James Denvir, Peter Stoilov, Peter Perrotta, Wesley Kimble, Ryan Percifield" |
| EPI_ISL_1528904 | J.W. Ruby Memorial Hospital | WVU and Marshall University Combined Genomics Core Facilities | "James Denvir, Peter Stoilov, Peter Perrotta, Wesley Kimble, Ryan Percifield" |
| EPI_ISL_1528905 | 4959 MacCorkle Ave SE, Charleston, WV 25304 | WVU and Marshall University Combined Genomics Core Facilities | "James Denvir, Peter Stoilov, Peter Perrotta, Wesley Kimble, Ryan Percifield" |
| EPI_ISL_1528906 | J.W. Ruby Memorial Hospital | WVU and Marshall University Combined Genomics Core Facilities | "James Denvir, Peter Stoilov, Peter Perrotta, Wesley Kimble, Ryan Percifield" |
| EPI_ISL_1528907 | 4956 MacCorkle Ave SE, Charleston, WV 25304 | WVU and Marshall University Combined Genomics Core Facilities | "James Denvir, Peter Stoilov, Peter Perrotta, Wesley Kimble, Ryan Percifield" |
| EPI_ISL_1528908 | 4938 MacCorkle Ave SE, Charleston, WV 25304 | WVU and Marshall University Combined Genomics Core Facilities | "James Denvir, Peter Stoilov, Peter Perrotta, Wesley Kimble, Ryan Percifield" |

| | | | |
|---|---|---|---|
| EPI_ISL_1528909 | 4951 MacCorkle Ave SE, Charleston, WV 25304 | WVU and Marshall University Combined Genomics Core Facilities | "James Denvir, Peter Stoilov, Peter Perrotta, Wesley Kimble, Ryan Percifield" |
| EPI_ISL_1528910, EPI_ISL_1528911, EPI_ISL_1528912, EPI_ISL_1528913, EPI_ISL_1528914, EPI_ISL_1528915, EPI_ISL_1528917, EPI_ISL_1528918 | J.W. Ruby Memorial Hospital | WVU and Marshall University Combined Genomics Core Facilities | "James Denvir, Peter Stoilov, Peter Perrotta, Wesley Kimble, Ryan Percifield" |
| EPI_ISL_1528919 | NA | WVU and Marshall University Combined Genomics Core Facilities | "James Denvir, Peter Stoilov, Peter Perrotta, Wesley Kimble, Ryan Percifield" |
| EPI_ISL_1528920 | 4949 MacCorkle Ave SE, Charleston, WV 25304 | WVU and Marshall University Combined Genomics Core Facilities | "James Denvir, Peter Stoilov, Peter Perrotta, Wesley Kimble, Ryan Percifield" |
| EPI_ISL_1528921 | 4957 MacCorkle Ave SE, Charleston, WV 25304 | WVU and Marshall University Combined Genomics Core Facilities | "James Denvir, Peter Stoilov, Peter Perrotta, Wesley Kimble, Ryan Percifield" |
| EPI_ISL_1528922, EPI_ISL_1528923 | J.W. Ruby Memorial Hospital | WVU and Marshall University Combined Genomics Core Facilities | "James Denvir, Peter Stoilov, Peter Perrotta, Wesley Kimble, Ryan Percifield" |
| EPI_ISL_1528924 | NA | WVU and Marshall University Combined Genomics Core Facilities | "James Denvir, Peter Stoilov, Peter Perrotta, Wesley Kimble, Ryan Percifield" |
| EPI_ISL_1528926, EPI_ISL_1528927, EPI_ISL_1528928, EPI_ISL_1528929, EPI_ISL_1528930, EPI_ISL_1528931, EPI_ISL_1528932, EPI_ISL_1528933, EPI_ISL_1528934, EPI_ISL_1528935, EPI_ISL_1528936, EPI_ISL_1528937, EPI_ISL_1528939, EPI_ISL_1528940, EPI_ISL_1528941, EPI_ISL_1528942, EPI_ISL_1528943, EPI_ISL_1528944, EPI_ISL_1528945, EPI_ISL_1528946, EPI_ISL_1528947, EPI_ISL_1528948, EPI_ISL_1528949, EPI_ISL_1528950, EPI_ISL_1528951, EPI_ISL_1528952, EPI_ISL_1528953, EPI_ISL_1528954, EPI_ISL_1528955, EPI_ISL_1528956, EPI_ISL_1528957, EPI_ISL_1528958, EPI_ISL_1528959, EPI_ISL_1528960, EPI_ISL_1528961, EPI_ISL_1528962, EPI_ISL_1528963, EPI_ISL_1528964, EPI_ISL_1528965, EPI_ISL_1528966, EPI_ISL_1528967, EPI_ISL_1528968, EPI_ISL_1528969, EPI_ISL_1528970, EPI_ISL_1528971, EPI_ISL_1528972, EPI_ISL_1528973, EPI_ISL_1528974, EPI_ISL_1528975, EPI_ISL_1528976, EPI_ISL_1528977, EPI_ISL_1528978, EPI_ISL_1528979, EPI_ISL_1528981, EPI_ISL_1528982, EPI_ISL_1528983, EPI_ISL_1528984, EPI_ISL_1528985, EPI_ISL_1528986, EPI_ISL_1528987, EPI_ISL_1528988, EPI_ISL_1528989, EPI_ISL_1528990, EPI_ISL_1528991, EPI_ISL_1528992, EPI_ISL_1528993, EPI_ISL_1528994, EPI_ISL_1528995, EPI_ISL_1528996, EPI_ISL_1528997, EPI_ISL_1528998 | | | |
| see above | J.W. Ruby Memorial Hospital | WVU and Marshall University Combined Genomics Core Facilities | "James Denvir, Peter Stoilov, Peter Perrotta, Wesley Kimble, Ryan Percifield" |
| EPI_ISL_1528999 | Preston Taylor Community Health Center | WVU and Marshall University Combined Genomics Core Facilities | "James Denvir, Peter Stoilov, Peter Perrotta, Wesley Kimble, Ryan Percifield" |
| EPI_ISL_1529000, EPI_ISL_1529001, EPI_ISL_1529002 | NA | WVU and Marshall University Combined Genomics Core Facilities | "James Denvir, Peter Stoilov, Peter Perrotta, Wesley Kimble, Ryan Percifield" |
| EPI_ISL_1529003 | 4920 MacCorkle Ave SE, Charleston, WV 25304 | WVU and Marshall University Combined Genomics Core Facilities | "James Denvir, Peter Stoilov, Peter Perrotta, Wesley Kimble, Ryan Percifield" |
| EPI_ISL_1529004 | Mon Health Medical Center | WVU and Marshall University Combined Genomics Core Facilities | "James Denvir, Peter Stoilov, Peter Perrotta, Wesley Kimble, Ryan Percifield" |
| EPI_ISL_1529005 | 4919 MacCorkle Ave SE, Charleston, WV 25304 | WVU and Marshall University Combined Genomics Core Facilities | "James Denvir, Peter Stoilov, Peter Perrotta, Wesley Kimble, Ryan Percifield" |
| EPI_ISL_1529006, EPI_ISL_1529007, EPI_ISL_1529008, EPI_ISL_1529009, EPI_ISL_1529010, EPI_ISL_1529011, EPI_ISL_1529012, EPI_ISL_1529013, EPI_ISL_1529014, EPI_ISL_1529015, EPI_ISL_1529017, EPI_ISL_1529018, EPI_ISL_1529019, EPI_ISL_1529020, EPI_ISL_1529021, EPI_ISL_1529022, EPI_ISL_1529023, EPI_ISL_1529024, EPI_ISL_1529025, EPI_ISL_1529026, EPI_ISL_1529027, EPI_ISL_1529028 | | | |
| see above | J.W. Ruby Memorial Hospital | WVU and Marshall University Combined Genomics Core Facilities | "James Denvir, Peter Stoilov, Peter Perrotta, Wesley Kimble, Ryan Percifield" |
| EPI_ISL_1529029, EPI_ISL_1529030, EPI_ISL_1529031, EPI_ISL_1529032, EPI_ISL_1529033, EPI_ISL_1529034, EPI_ISL_1529036, EPI_ISL_1529037, EPI_ISL_1529038, EPI_ISL_1529039, EPI_ISL_1529040, EPI_ISL_1529041, EPI_ISL_1529042, EPI_ISL_1529043, EPI_ISL_1529044, EPI_ISL_1529045, EPI_ISL_1529046, EPI_ISL_1529047, EPI_ISL_1529048, EPI_ISL_1529049, EPI_ISL_1529050, EPI_ISL_1529051, EPI_ISL_1529052, EPI_ISL_1529053, EPI_ISL_1529054, EPI_ISL_1529055, EPI_ISL_1529056, EPI_ISL_1529057, EPI_ISL_1529058, EPI_ISL_1529059, EPI_ISL_1529060, EPI_ISL_1529061, EPI_ISL_1529063, EPI_ISL_1529064, EPI_ISL_1529065, EPI_ISL_1529066, EPI_ISL_1529067, EPI_ISL_1529068, EPI_ISL_1529069, EPI_ISL_1529070, EPI_ISL_1529071, EPI_ISL_1529072, EPI_ISL_1529073, EPI_ISL_1529074 | | | |
| see above | Berkeley Medical Center | WVU and Marshall University Combined Genomics Core Facilities | "James Denvir, Peter Stoilov, Peter Perrotta, Wesley Kimble, Ryan Percifield" |
| EPI_ISL_1529075, EPI_ISL_1529076, EPI_ISL_1529077, EPI_ISL_1529078, EPI_ISL_1529079, EPI_ISL_1529080, EPI_ISL_1529081, EPI_ISL_1529082, EPI_ISL_1529083, EPI_ISL_1529084, EPI_ISL_1529085, EPI_ISL_1529086, EPI_ISL_1529087, EPI_ISL_1529088, EPI_ISL_1529089, EPI_ISL_1529090, EPI_ISL_1529091, EPI_ISL_1529092, EPI_ISL_1529093, EPI_ISL_1529094, EPI_ISL_1529095, EPI_ISL_1529096, EPI_ISL_1529097, EPI_ISL_1529098 | | | |
| see above | Alaska State Virology Laboratory | Alaska State Virology Laboratory | Stephanie DeRonde, Elva House, Lisa Smith, Ph.D., Jack Chen, Ph.D. |
| EPI_ISL_1529099 | Global Health Research Complex (GHRC) | Texas A&M Institute for Genomic Sciences and Society (TIGSS) | Benjamin W. Neuman, Melissa M. Kahl-Mcdonagh, Kurt A. Zuelke, David Threadgill, Allison Ficht, Sankar P. Chaki, Sierra Guidry, Drew Hillhouse, Wesley Brashear |
| EPI_ISL_1529100 | CA DPH Viral and Rickettsial Disease Laboratory | Chan-Zuckerberg Biohub | CZB Cliahub Consortium |
| EPI_ISL_1529101, EPI_ISL_1529102 | Santa Clara County Public Health Laboratory | Chan-Zuckerberg Biohub | CZB Cliahub Consortium |
| EPI_ISL_1529103 | Contra Costa County Public Health Lab | Chan-Zuckerberg Biohub | CZB Cliahub Consortium |
| EPI_ISL_1529104, EPI_ISL_1529105 | CA DPH Viral and Rickettsial Disease Laboratory | Chan-Zuckerberg Biohub | CZB Cliahub Consortium |
| EPI_ISL_1529106, EPI_ISL_1529107, EPI_ISL_1529108, EPI_ISL_1529109, EPI_ISL_1529110 | Santa Clara County Public Health Laboratory | Chan-Zuckerberg Biohub | CZB Cliahub Consortium |
| EPI_ISL_1529111, EPI_ISL_1529112 | CA DPH Viral and Rickettsial Disease Laboratory | Chan-Zuckerberg Biohub | CZB Cliahub Consortium |
| EPI_ISL_1529113 | Santa Clara County Public Health Laboratory | Chan-Zuckerberg Biohub | CZB Cliahub Consortium |
| EPI_ISL_1529114, EPI_ISL_1529115, EPI_ISL_1529116, EPI_ISL_1529117, EPI_ISL_1529118 | CA DPH Viral and Rickettsial Disease Laboratory | Chan-Zuckerberg Biohub | CZB Cliahub Consortium |
| EPI_ISL_1529119 | Santa Clara County Public Health Laboratory | Chan-Zuckerberg Biohub | CZB Cliahub Consortium |
| EPI_ISL_1529120 | CA DPH Viral and Rickettsial Disease Laboratory | Chan-Zuckerberg Biohub | CZB Cliahub Consortium |
| EPI_ISL_1529121 | Santa Clara County Public Health Laboratory | Chan-Zuckerberg Biohub | CZB Cliahub Consortium |
| EPI_ISL_1529122, EPI_ISL_1529123, EPI_ISL_1529124 | Contra Costa County Public Health Lab | Chan-Zuckerberg Biohub | CZB Cliahub Consortium |
| EPI_ISL_1529125, EPI_ISL_1529126 | CA DPH Viral and Rickettsial Disease Laboratory | Chan-Zuckerberg Biohub | CZB Cliahub Consortium |
| EPI_ISL_1529127, EPI_ISL_1529128, EPI_ISL_1529129 | County of San Luis Obispo Public Health Laboratory | Chan-Zuckerberg Biohub | CZB Cliahub Consortium |
| EPI_ISL_1529130 | CA DPH Viral and Rickettsial Disease Laboratory | Chan-Zuckerberg Biohub | CZB Cliahub Consortium |
| EPI_ISL_1529131 | Contra Costa County Public Health Lab | Chan-Zuckerberg Biohub | CZB Cliahub Consortium |
| EPI_ISL_1529132 | CA DPH Viral and Rickettsial Disease Laboratory | Chan-Zuckerberg Biohub | CZB Cliahub Consortium |
| EPI_ISL_1529133, EPI_ISL_1529134, EPI_ISL_1529135 | Contra Costa County Public Health Lab | Chan-Zuckerberg Biohub | CZB Cliahub Consortium |
| EPI_ISL_1529136, | CA DPH Viral and Rickettsial Disease | Chan-Zuckerberg Biohub | CZB Cliahub Consortium |

| | | | |
|---|---|---|---|
| EPI_ISL_1529141, EPI_ISL_1529142, EPI_ISL_1529143 | Laboratory | | |
| EPI_ISL_1529144 | Contra Costa County Public Health Lab | Chan-Zuckerberg Biohub | CZB Cliahub Consortium |
| EPI_ISL_1529145, EPI_ISL_1529146 | CA DPH Viral and Rickettsial Disease Laboratory | Chan-Zuckerberg Biohub | CZB Cliahub Consortium |
| EPI_ISL_1529147 | County of San Luis Obispo Public Health Laboratory | Chan-Zuckerberg Biohub | CZB Cliahub Consortium |
| EPI_ISL_1529148 | Santa Clara County Public Health Laboratory | Chan-Zuckerberg Biohub | CZB Cliahub Consortium |
| EPI_ISL_1529149 | Contra Costa County Public Health Lab | Chan-Zuckerberg Biohub | CZB Cliahub Consortium |
| EPI_ISL_1529150 | CA DPH Viral and Rickettsial Disease Laboratory | Chan-Zuckerberg Biohub | CZB Cliahub Consortium |
| EPI_ISL_1529151, EPI_ISL_1529152 | Santa Clara County Public Health Laboratory | Chan-Zuckerberg Biohub | CZB Cliahub Consortium |
| EPI_ISL_1529153 | CA DPH Viral and Rickettsial Disease Laboratory | Chan-Zuckerberg Biohub | CZB Cliahub Consortium |
| EPI_ISL_1529154 | Santa Clara County Public Health Laboratory | Chan-Zuckerberg Biohub | CZB Cliahub Consortium |
| EPI_ISL_1529155 | Contra Costa County Public Health Lab | Chan-Zuckerberg Biohub | CZB Cliahub Consortium |
| EPI_ISL_1529156, EPI_ISL_1529157 | CA DPH Viral and Rickettsial Disease Laboratory | Chan-Zuckerberg Biohub | CZB Cliahub Consortium |
| EPI_ISL_1529158 | Orange County Public Health Lab | Chan-Zuckerberg Biohub | CZB Cliahub Consortium |
| EPI_ISL_1529159, EPI_ISL_1529160 | CA DPH Viral and Rickettsial Disease Laboratory | Chan-Zuckerberg Biohub | CZB Cliahub Consortium |
| EPI_ISL_1529161 | County of San Luis Obispo Public Health Laboratory | Chan-Zuckerberg Biohub | CZB Cliahub Consortium |
| EPI_ISL_1529162 | Santa Clara County Public Health Laboratory | Chan-Zuckerberg Biohub | CZB Cliahub Consortium |
| EPI_ISL_1529163, EPI_ISL_1529164, EPI_ISL_1529165, EPI_ISL_1529166 | Contra Costa County Public Health Lab | Chan-Zuckerberg Biohub | CZB Cliahub Consortium |
| EPI_ISL_1529167 | CA DPH Viral and Rickettsial Disease Laboratory | Chan-Zuckerberg Biohub | CZB Cliahub Consortium |
| EPI_ISL_1529168 | County of San Luis Obispo Public Health Laboratory | Chan-Zuckerberg Biohub | CZB Cliahub Consortium |
| EPI_ISL_1529169, EPI_ISL_1529170 | CA DPH Viral and Rickettsial Disease Laboratory | Chan-Zuckerberg Biohub | CZB Cliahub Consortium |
| EPI_ISL_1529171 | Contra Costa County Public Health Lab | Chan-Zuckerberg Biohub | CZB Cliahub Consortium |
| EPI_ISL_1529172 | CA DPH Viral and Rickettsial Disease Laboratory | Chan-Zuckerberg Biohub | CZB Cliahub Consortium |
| EPI_ISL_1529173 | Santa Clara County Public Health Laboratory | Chan-Zuckerberg Biohub | CZB Cliahub Consortium |
| EPI_ISL_1529174, EPI_ISL_1529175 | CA DPH Viral and Rickettsial Disease Laboratory | Chan-Zuckerberg Biohub | CZB Cliahub Consortium |
| EPI_ISL_1529176, EPI_ISL_1529177 | Contra Costa County Public Health Lab | Chan-Zuckerberg Biohub | CZB Cliahub Consortium |
| EPI_ISL_1529178 | CA DPH Viral and Rickettsial Disease Laboratory | Chan-Zuckerberg Biohub | CZB Cliahub Consortium |
| EPI_ISL_1529179 | Contra Costa County Public Health Lab | Chan-Zuckerberg Biohub | CZB Cliahub Consortium |
| EPI_ISL_1529180 | CA DPH Viral and Rickettsial Disease Laboratory | Chan-Zuckerberg Biohub | CZB Cliahub Consortium |
| EPI_ISL_1529181 | Contra Costa County Public Health Lab | Chan-Zuckerberg Biohub | CZB Cliahub Consortium |
| EPI_ISL_1529182 | CA DPH Viral and Rickettsial Disease Laboratory | Chan-Zuckerberg Biohub | CZB Cliahub Consortium |
| EPI_ISL_1529183 | Santa Clara County Public Health Laboratory | Chan-Zuckerberg Biohub | CZB Cliahub Consortium |
| EPI_ISL_1529184 | CA DPH Viral and Rickettsial Disease Laboratory | Chan-Zuckerberg Biohub | CZB Cliahub Consortium |
| EPI_ISL_1529185, EPI_ISL_1529186 | Contra Costa County Public Health Lab | Chan-Zuckerberg Biohub | CZB Cliahub Consortium |
| EPI_ISL_1529187 | CA DPH Viral and Rickettsial Disease Laboratory | Chan-Zuckerberg Biohub | CZB Cliahub Consortium |
| EPI_ISL_1529188 | Contra Costa County Public Health Lab | Chan-Zuckerberg Biohub | CZB Cliahub Consortium |
| EPI_ISL_1529189 | CA DPH Viral and Rickettsial Disease Laboratory | Chan-Zuckerberg Biohub | CZB Cliahub Consortium |
| EPI_ISL_1529190 | Contra Costa County Public Health Lab | Chan-Zuckerberg Biohub | CZB Cliahub Consortium |
| EPI_ISL_1529191 | Santa Clara County Public Health Laboratory | Chan-Zuckerberg Biohub | CZB Cliahub Consortium |
| EPI_ISL_1529192 | CA DPH Viral and Rickettsial Disease Laboratory | Chan-Zuckerberg Biohub | CZB Cliahub Consortium |
| EPI_ISL_1529193 | Santa Clara County Public Health Laboratory | Chan-Zuckerberg Biohub | CZB Cliahub Consortium |
| EPI_ISL_1529194, EPI_ISL_1529195, EPI_ISL_1529196 | Contra Costa County Public Health Lab | Chan-Zuckerberg Biohub | CZB Cliahub Consortium |
| EPI_ISL_1529197, EPI_ISL_1529198, EPI_ISL_1529199, EPI_ISL_1529200, EPI_ISL_1529201 | CA DPH Viral and Rickettsial Disease Laboratory | Chan-Zuckerberg Biohub | CZB Cliahub Consortium |
| EPI_ISL_1529202 | Contra Costa County Public Health Lab | Chan-Zuckerberg Biohub | CZB Cliahub Consortium |
| EPI_ISL_1529203 | CA DPH Viral and Rickettsial Disease Laboratory | Chan-Zuckerberg Biohub | CZB Cliahub Consortium |
| EPI_ISL_1529204 | Contra Costa County Public Health Lab | Chan-Zuckerberg Biohub | CZB Cliahub Consortium |
| EPI_ISL_1529205, EPI_ISL_1529206, EPI_ISL_1529207 | Santa Clara County Public Health Laboratory | Chan-Zuckerberg Biohub | CZB Cliahub Consortium |
| EPI_ISL_1529208 | County of San Luis Obispo Public Health | Chan-Zuckerberg Biohub | CZB Cliahub Consortium |

| | | | |
|---|---|---|---|
| | Laboratory | | |
| EPI_ISL_1529209, EPI_ISL_1529210, EPI_ISL_1529211, EPI_ISL_1529212 | Contra Costa County Public Health Lab | Chan-Zuckerberg Biohub | CZB Cliahub Consortium |
| EPI_ISL_1529213 | County of San Luis Obispo Public Health Laboratory | Chan-Zuckerberg Biohub | CZB Cliahub Consortium |
| EPI_ISL_1529214, EPI_ISL_1529215 | CA DPH Viral and Rickettsial Disease Laboratory | Chan-Zuckerberg Biohub | CZB Cliahub Consortium |
| EPI_ISL_1529216, EPI_ISL_1529217 | Contra Costa County Public Health Lab | Chan-Zuckerberg Biohub | CZB Cliahub Consortium |
| EPI_ISL_1529218 | Santa Clara County Public Health Laboratory | Chan-Zuckerberg Biohub | CZB Cliahub Consortium |
| EPI_ISL_1529219, EPI_ISL_1529220 | CA DPH Viral and Rickettsial Disease Laboratory | Chan-Zuckerberg Biohub | CZB Cliahub Consortium |
| EPI_ISL_1529221 | Santa Clara County Public Health Laboratory | Chan-Zuckerberg Biohub | CZB Cliahub Consortium |
| EPI_ISL_1529222, EPI_ISL_1529223 | CA DPH Viral and Rickettsial Disease Laboratory | Chan-Zuckerberg Biohub | CZB Cliahub Consortium |
| EPI_ISL_1529224 | Santa Clara County Public Health Laboratory | Chan-Zuckerberg Biohub | CZB Cliahub Consortium |
| EPI_ISL_1529225 | County of San Luis Obispo Public Health Laboratory | Chan-Zuckerberg Biohub | CZB Cliahub Consortium |
| EPI_ISL_1529226 | Santa Clara County Public Health Laboratory | Chan-Zuckerberg Biohub | CZB Cliahub Consortium |
| EPI_ISL_1529227, EPI_ISL_1529228, EPI_ISL_1529229 | Contra Costa County Public Health Lab | Chan-Zuckerberg Biohub | CZB Cliahub Consortium |
| EPI_ISL_1529230, EPI_ISL_1529231 | CA DPH Viral and Rickettsial Disease Laboratory | Chan-Zuckerberg Biohub | CZB Cliahub Consortium |
| EPI_ISL_1529232 | County of San Luis Obispo Public Health Laboratory | Chan-Zuckerberg Biohub | CZB Cliahub Consortium |
| EPI_ISL_1529233 | CA DPH Viral and Rickettsial Disease Laboratory | Chan-Zuckerberg Biohub | CZB Cliahub Consortium |
| EPI_ISL_1529234 | Santa Clara County Public Health Laboratory | Chan-Zuckerberg Biohub | CZB Cliahub Consortium |
| EPI_ISL_1529235, EPI_ISL_1529236 | Contra Costa County Public Health Lab | Chan-Zuckerberg Biohub | CZB Cliahub Consortium |
| EPI_ISL_1529237 | County of San Luis Obispo Public Health Laboratory | Chan-Zuckerberg Biohub | CZB Cliahub Consortium |
| EPI_ISL_1529238 | Santa Clara County Public Health Laboratory | Chan-Zuckerberg Biohub | CZB Cliahub Consortium |
| EPI_ISL_1529239 | Contra Costa County Public Health Lab | Chan-Zuckerberg Biohub | CZB Cliahub Consortium |
| EPI_ISL_1529240, EPI_ISL_1529241 | Santa Clara County Public Health Laboratory | Chan-Zuckerberg Biohub | CZB Cliahub Consortium |
| EPI_ISL_1529242, EPI_ISL_1529243, EPI_ISL_1529244 | CA DPH Viral and Rickettsial Disease Laboratory | Chan-Zuckerberg Biohub | CZB Cliahub Consortium |
| EPI_ISL_1529245 | Santa Clara County Public Health Laboratory | Chan-Zuckerberg Biohub | CZB Cliahub Consortium |
| EPI_ISL_1529246 | County of San Luis Obispo Public Health Laboratory | Chan-Zuckerberg Biohub | CZB Cliahub Consortium |
| EPI_ISL_1529247 | Contra Costa County Public Health Lab | Chan-Zuckerberg Biohub | CZB Cliahub Consortium |
| EPI_ISL_1529248 | County of San Luis Obispo Public Health Laboratory | Chan-Zuckerberg Biohub | CZB Cliahub Consortium |
| EPI_ISL_1529249 | CA DPH Viral and Rickettsial Disease Laboratory | Chan-Zuckerberg Biohub | CZB Cliahub Consortium |
| EPI_ISL_1529250, EPI_ISL_1529251 | Contra Costa County Public Health Lab | Chan-Zuckerberg Biohub | CZB Cliahub Consortium |
| EPI_ISL_1529252 | Santa Clara County Public Health Laboratory | Chan-Zuckerberg Biohub | CZB Cliahub Consortium |
| EPI_ISL_1529253 | Contra Costa County Public Health Lab | Chan-Zuckerberg Biohub | CZB Cliahub Consortium |
| EPI_ISL_1529254 | CA DPH Viral and Rickettsial Disease Laboratory | Chan-Zuckerberg Biohub | CZB Cliahub Consortium |
| EPI_ISL_1529255, EPI_ISL_1529256 | County of San Luis Obispo Public Health Laboratory | Chan-Zuckerberg Biohub | CZB Cliahub Consortium |
| EPI_ISL_1529257 | CA DPH Viral and Rickettsial Disease Laboratory | Chan-Zuckerberg Biohub | CZB Cliahub Consortium |
| EPI_ISL_1529258, EPI_ISL_1529259 | Santa Clara County Public Health Laboratory | Chan-Zuckerberg Biohub | CZB Cliahub Consortium |
| EPI_ISL_1529260 | CA DPH Viral and Rickettsial Disease Laboratory | Chan-Zuckerberg Biohub | CZB Cliahub Consortium |
| EPI_ISL_1529261 | Contra Costa County Public Health Lab | Chan-Zuckerberg Biohub | CZB Cliahub Consortium |
| EPI_ISL_1529262, EPI_ISL_1529263 | Santa Clara County Public Health Laboratory | Chan-Zuckerberg Biohub | CZB Cliahub Consortium |
| EPI_ISL_1529264, EPI_ISL_1529265, EPI_ISL_1529266 | CA DPH Viral and Rickettsial Disease Laboratory | Chan-Zuckerberg Biohub | CZB Cliahub Consortium |
| EPI_ISL_1529267 | Santa Clara County Public Health Laboratory | Chan-Zuckerberg Biohub | CZB Cliahub Consortium |
| EPI_ISL_1529268 | County of San Luis Obispo Public Health Laboratory | Chan-Zuckerberg Biohub | CZB Cliahub Consortium |
| EPI_ISL_1529269 | Contra Costa County Public Health Lab | Chan-Zuckerberg Biohub | CZB Cliahub Consortium |
| EPI_ISL_1529270 | County of San Luis Obispo Public Health Laboratory | Chan-Zuckerberg Biohub | CZB Cliahub Consortium |
| EPI_ISL_1529271, EPI_ISL_1529272 | Contra Costa County Public Health Lab | Chan-Zuckerberg Biohub | CZB Cliahub Consortium |
| EPI_ISL_1529273 | Santa Clara County Public Health Laboratory | Chan-Zuckerberg Biohub | CZB Cliahub Consortium |
| EPI_ISL_1529274 | CA DPH Viral and Rickettsial Disease | Chan-Zuckerberg Biohub | CZB Cliahub Consortium |

| | | | |
|---|---|---|---|
| | Laboratory | | |
| EPI_ISL_1529275 | Contra Costa County Public Health Lab | Chan-Zuckerberg Biohub | CZB Cliahub Consortium |
| EPI_ISL_1529276, EPI_ISL_1529277, EPI_ISL_1529278 | CA DPH Viral and Rickettsial Disease Laboratory | Chan-Zuckerberg Biohub | CZB Cliahub Consortium |
| EPI_ISL_1529279 | County of San Luis Obispo Public Health Laboratory | Chan-Zuckerberg Biohub | CZB Cliahub Consortium |
| EPI_ISL_1529280, EPI_ISL_1529281 | CA DPH Viral and Rickettsial Disease Laboratory | Chan-Zuckerberg Biohub | CZB Cliahub Consortium |
| EPI_ISL_1529282 | Contra Costa County Public Health Lab | Chan-Zuckerberg Biohub | CZB Cliahub Consortium |
| EPI_ISL_1529283, EPI_ISL_1529284 | Santa Clara County Public Health Laboratory | Chan-Zuckerberg Biohub | CZB Cliahub Consortium |
| EPI_ISL_1529285 | County of San Luis Obispo Public Health Laboratory | Chan-Zuckerberg Biohub | CZB Cliahub Consortium |
| EPI_ISL_1529286 | CA DPH Viral and Rickettsial Disease Laboratory | Chan-Zuckerberg Biohub | CZB Cliahub Consortium |
| EPI_ISL_1529287 | Santa Clara County Public Health Laboratory | Chan-Zuckerberg Biohub | CZB Cliahub Consortium |
| EPI_ISL_1529288, EPI_ISL_1529289, EPI_ISL_1529290, EPI_ISL_1529291 | CA DPH Viral and Rickettsial Disease Laboratory | Chan-Zuckerberg Biohub | CZB Cliahub Consortium |
| EPI_ISL_1529292, EPI_ISL_1529293 | County of San Luis Obispo Public Health Laboratory | Chan-Zuckerberg Biohub | CZB Cliahub Consortium |
| EPI_ISL_1529294 | CA DPH Viral and Rickettsial Disease Laboratory | Chan-Zuckerberg Biohub | CZB Cliahub Consortium |
| EPI_ISL_1529295 | Santa Clara County Public Health Laboratory | Chan-Zuckerberg Biohub | CZB Cliahub Consortium |
| EPI_ISL_1529296, EPI_ISL_1529297, EPI_ISL_1529298 | Contra Costa County Public Health Lab | Chan-Zuckerberg Biohub | CZB Cliahub Consortium |
| EPI_ISL_1529299 | CA DPH Viral and Rickettsial Disease Laboratory | Chan-Zuckerberg Biohub | CZB Cliahub Consortium |
| EPI_ISL_1529300, EPI_ISL_1529301, EPI_ISL_1529302 | Contra Costa County Public Health Lab | Chan-Zuckerberg Biohub | CZB Cliahub Consortium |
| EPI_ISL_1529303 | CA DPH Viral and Rickettsial Disease Laboratory | Chan-Zuckerberg Biohub | CZB Cliahub Consortium |
| EPI_ISL_1529304, EPI_ISL_1529305 | Contra Costa County Public Health Lab | Chan-Zuckerberg Biohub | CZB Cliahub Consortium |
| EPI_ISL_1529306, EPI_ISL_1529307 | CA DPH Viral and Rickettsial Disease Laboratory | Chan-Zuckerberg Biohub | CZB Cliahub Consortium |
| EPI_ISL_1529308 | Contra Costa County Public Health Lab | Chan-Zuckerberg Biohub | CZB Cliahub Consortium |
| EPI_ISL_1529309 | Santa Clara County Public Health Laboratory | Chan-Zuckerberg Biohub | CZB Cliahub Consortium |
| EPI_ISL_1529310 | Contra Costa County Public Health Lab | Chan-Zuckerberg Biohub | CZB Cliahub Consortium |
| EPI_ISL_1529311 | County of San Luis Obispo Public Health Laboratory | Chan-Zuckerberg Biohub | CZB Cliahub Consortium |
| EPI_ISL_1529312, EPI_ISL_1529313, EPI_ISL_1529314, EPI_ISL_1529315 | CA DPH Viral and Rickettsial Disease Laboratory | Chan-Zuckerberg Biohub | CZB Cliahub Consortium |
| EPI_ISL_1529316, EPI_ISL_1529317 | Santa Clara County Public Health Laboratory | Chan-Zuckerberg Biohub | CZB Cliahub Consortium |
| EPI_ISL_1529318 | Contra Costa County Public Health Lab | Chan-Zuckerberg Biohub | CZB Cliahub Consortium |
| EPI_ISL_1529319 | CA DPH Viral and Rickettsial Disease Laboratory | Chan-Zuckerberg Biohub | CZB Cliahub Consortium |
| EPI_ISL_1529320 | Contra Costa County Public Health Lab | Chan-Zuckerberg Biohub | CZB Cliahub Consortium |
| EPI_ISL_1529321 | CA DPH Viral and Rickettsial Disease Laboratory | Chan-Zuckerberg Biohub | CZB Cliahub Consortium |
| EPI_ISL_1529322, EPI_ISL_1529323 | Contra Costa County Public Health Lab | Chan-Zuckerberg Biohub | CZB Cliahub Consortium |
| EPI_ISL_1529324, EPI_ISL_1529325, EPI_ISL_1529326 | CA DPH Viral and Rickettsial Disease Laboratory | Chan-Zuckerberg Biohub | CZB Cliahub Consortium |
| EPI_ISL_1529327, EPI_ISL_1529328 | Contra Costa County Public Health Lab | Chan-Zuckerberg Biohub | CZB Cliahub Consortium |
| EPI_ISL_1529329 | County of San Luis Obispo Public Health Laboratory | Chan-Zuckerberg Biohub | CZB Cliahub Consortium |
| EPI_ISL_1529330, EPI_ISL_1529331 | Contra Costa County Public Health Lab | Chan-Zuckerberg Biohub | CZB Cliahub Consortium |
| EPI_ISL_1529332, EPI_ISL_1529333 | CA DPH Viral and Rickettsial Disease Laboratory | Chan-Zuckerberg Biohub | CZB Cliahub Consortium |
| EPI_ISL_1529334 | Contra Costa County Public Health Lab | Chan-Zuckerberg Biohub | CZB Cliahub Consortium |
| EPI_ISL_1529335 | CA DPH Viral and Rickettsial Disease Laboratory | Chan-Zuckerberg Biohub | CZB Cliahub Consortium |
| EPI_ISL_1529336 | Orange County Public Health Lab | Chan-Zuckerberg Biohub | CZB Cliahub Consortium |
| EPI_ISL_1529337, EPI_ISL_1529338, EPI_ISL_1529339 | CA DPH Viral and Rickettsial Disease Laboratory | Chan-Zuckerberg Biohub | CZB Cliahub Consortium |
| EPI_ISL_1529340 | Contra Costa County Public Health Lab | Chan-Zuckerberg Biohub | CZB Cliahub Consortium |
| EPI_ISL_1529341, EPI_ISL_1529342, EPI_ISL_1529343 | County of San Luis Obispo Public Health Laboratory | Chan-Zuckerberg Biohub | CZB Cliahub Consortium |
| EPI_ISL_1529344 | Santa Clara County Public Health Laboratory | Chan-Zuckerberg Biohub | CZB Cliahub Consortium |
| EPI_ISL_1529345, EPI_ISL_1529346 | Contra Costa County Public Health Lab | Chan-Zuckerberg Biohub | CZB Cliahub Consortium |
| EPI_ISL_1529347 | Santa Clara County Public Health Laboratory | Chan-Zuckerberg Biohub | CZB Cliahub Consortium |

| | | | |
|---|---|---|---|
| EPI_ISL_1529348 | Contra Costa County Public Health Lab | Chan-Zuckerberg Biohub | CZB Cliahub Consortium |
| EPI_ISL_1529349 | Santa Clara County Public Health Laboratory | Chan-Zuckerberg Biohub | CZB Cliahub Consortium |
| EPI_ISL_1529350 | Contra Costa County Public Health Lab | Chan-Zuckerberg Biohub | CZB Cliahub Consortium |
| EPI_ISL_1529351 | Santa Clara County Public Health Laboratory | Chan-Zuckerberg Biohub | CZB Cliahub Consortium |
| EPI_ISL_1529352, EPI_ISL_1529353, EPI_ISL_1529354, EPI_ISL_1529355 | Contra Costa County Public Health Lab | Chan-Zuckerberg Biohub | CZB Cliahub Consortium |
| EPI_ISL_1529356, EPI_ISL_1529357 | CA DPH Viral and Rickettsial Disease Laboratory | Chan-Zuckerberg Biohub | CZB Cliahub Consortium |
| EPI_ISL_1529358 | Contra Costa County Public Health Lab | Chan-Zuckerberg Biohub | CZB Cliahub Consortium |
| EPI_ISL_1529359, EPI_ISL_1529360, EPI_ISL_1529361, EPI_ISL_1529362, EPI_ISL_1529363 | CA DPH Viral and Rickettsial Disease Laboratory | Chan-Zuckerberg Biohub | CZB Cliahub Consortium |
| EPI_ISL_1529364, EPI_ISL_1529365 | Contra Costa County Public Health Lab | Chan-Zuckerberg Biohub | CZB Cliahub Consortium |
| EPI_ISL_1529366 | CA DPH Viral and Rickettsial Disease Laboratory | Chan-Zuckerberg Biohub | CZB Cliahub Consortium |
| EPI_ISL_1529367, EPI_ISL_1529368 | Contra Costa County Public Health Lab | Chan-Zuckerberg Biohub | CZB Cliahub Consortium |
| EPI_ISL_1529369 | CA DPH Viral and Rickettsial Disease Laboratory | Chan-Zuckerberg Biohub | CZB Cliahub Consortium |
| EPI_ISL_1529370, EPI_ISL_1529535, EPI_ISL_1529952 | Global Health Research Complex (GHRC) | Texas A&M Institute for Genomic Sciences and Society (TIGSS) | Benjamin W. Neuman, Melissa M. Kahl-Mcdonagh, Kurt A. Zuelke, David Threadgill, Allison Ficht, Sankar P. Chaki, Sierra Guidry, Drew Hillhouse, Wesley Brashear |
| EPI_ISL_1530555, EPI_ISL_1530558, EPI_ISL_1530559, EPI_ISL_1530560, EPI_ISL_1530561, EPI_ISL_1530562 | Sonora Quest Laboratories | TGen North | Jolene Bowers, Heather Centner, Chris French, Hayley Yaglom, Ashlyn Pfeiffer, Darrin Lemmer, Dave Engelthaler, The Arizona COVID Genomics Union (ACGU) |
| EPI_ISL_1530563 | TGen North | TGen North | Jolene Bowers, Heather Centner, Chris French, Hayley Yaglom, Ashlyn Pfeiffer, Darrin Lemmer, Dave Engelthaler, The Arizona COVID Genomics Union (ACGU) |
| EPI_ISL_1530564 | Sonora Quest Laboratories | TGen North | Jolene Bowers, Heather Centner, Chris French, Hayley Yaglom, Ashlyn Pfeiffer, Darrin Lemmer, Dave Engelthaler, The Arizona COVID Genomics Union (ACGU) |
| EPI_ISL_1530567, EPI_ISL_1530570 | TGen North | TGen North | Jolene Bowers, Heather Centner, Chris French, Hayley Yaglom, Ashlyn Pfeiffer, Darrin Lemmer, Dave Engelthaler, The Arizona COVID Genomics Union (ACGU) |
| EPI_ISL_1530573 | Sonora Quest Laboratories | TGen North | Jolene Bowers, Heather Centner, Chris French, Hayley Yaglom, Ashlyn Pfeiffer, Darrin Lemmer, Dave Engelthaler, The Arizona COVID Genomics Union (ACGU) |
| EPI_ISL_1530574 | TGen North | TGen North | Jolene Bowers, Heather Centner, Chris French, Hayley Yaglom, Ashlyn Pfeiffer, Darrin Lemmer, Dave Engelthaler, The Arizona COVID Genomics Union (ACGU) |
| EPI_ISL_1530575, EPI_ISL_1530577, EPI_ISL_1530578 | Sonora Quest Laboratories | TGen North | Jolene Bowers, Heather Centner, Chris French, Hayley Yaglom, Ashlyn Pfeiffer, Darrin Lemmer, Dave Engelthaler, The Arizona COVID Genomics Union (ACGU) |
| EPI_ISL_1530579 | TGen North | TGen North | Jolene Bowers, Heather Centner, Chris French, Hayley Yaglom, Ashlyn Pfeiffer, Darrin Lemmer, Dave Engelthaler, The Arizona COVID Genomics Union (ACGU) |
| EPI_ISL_1530580, EPI_ISL_1530581, EPI_ISL_1530583, EPI_ISL_1530584, EPI_ISL_1530585, EPI_ISL_1530586, EPI_ISL_1530588, EPI_ISL_1530591, EPI_ISL_1530594 | Sonora Quest Laboratories | TGen North | Jolene Bowers, Heather Centner, Chris French, Hayley Yaglom, Ashlyn Pfeiffer, Darrin Lemmer, Dave Engelthaler, The Arizona COVID Genomics Union (ACGU) |
| EPI_ISL_1530595, EPI_ISL_1530597 | TGen North | TGen North | Jolene Bowers, Heather Centner, Chris French, Hayley Yaglom, Ashlyn Pfeiffer, Darrin Lemmer, Dave Engelthaler, The Arizona COVID Genomics Union (ACGU) |
| EPI_ISL_1530598 | Sonora Quest Laboratories | TGen North | Jolene Bowers, Heather Centner, Chris French, Hayley Yaglom, Ashlyn Pfeiffer, Darrin Lemmer, Dave Engelthaler, The Arizona COVID Genomics Union (ACGU) |
| EPI_ISL_1530599, EPI_ISL_1530601, EPI_ISL_1530602, EPI_ISL_1530604 | TGen North | TGen North | Jolene Bowers, Heather Centner, Chris French, Hayley Yaglom, Ashlyn Pfeiffer, Darrin Lemmer, Dave Engelthaler, The Arizona COVID Genomics Union (ACGU) |
| EPI_ISL_1530605, EPI_ISL_1530606 | Sonora Quest Laboratories | TGen North | Jolene Bowers, Heather Centner, Chris French, Hayley Yaglom, Ashlyn Pfeiffer, Darrin Lemmer, Dave Engelthaler, The Arizona COVID Genomics Union (ACGU) |
| EPI_ISL_1530607 | TGen North | TGen North | Jolene Bowers, Heather Centner, Chris French, Hayley Yaglom, Ashlyn Pfeiffer, Darrin Lemmer, Dave Engelthaler, The Arizona COVID Genomics Union (ACGU) |
| EPI_ISL_1530611, EPI_ISL_1530612 | Sonora Quest Laboratories | TGen North | Jolene Bowers, Heather Centner, Chris French, Hayley Yaglom, Ashlyn Pfeiffer, Darrin Lemmer, Dave Engelthaler, The Arizona COVID Genomics Union (ACGU) |
| EPI_ISL_1530613 | TGen North | TGen North | Jolene Bowers, Heather Centner, Chris French, Hayley Yaglom, Ashlyn Pfeiffer, Darrin Lemmer, Dave Engelthaler, The Arizona COVID Genomics Union (ACGU) |
| EPI_ISL_1530615, EPI_ISL_1530616, EPI_ISL_1530617 | Sonora Quest Laboratories | TGen North | Jolene Bowers, Heather Centner, Chris French, Hayley Yaglom, Ashlyn Pfeiffer, Darrin Lemmer, Dave Engelthaler, The Arizona COVID Genomics Union (ACGU) |
| EPI_ISL_1530619 | TGen North | TGen North | Jolene Bowers, Heather Centner, Chris French, Hayley Yaglom, Ashlyn Pfeiffer, Darrin Lemmer, Dave Engelthaler, The Arizona COVID Genomics Union (ACGU) |
| EPI_ISL_1530621, EPI_ISL_1530622 | Sonora Quest Laboratories | TGen North | Jolene Bowers, Heather Centner, Chris French, Hayley Yaglom, Ashlyn Pfeiffer, Darrin Lemmer, Dave Engelthaler, The Arizona COVID Genomics Union (ACGU) |
| EPI_ISL_1530624 | TGen North | TGen North | Jolene Bowers, Heather Centner, Chris French, Hayley Yaglom, Ashlyn Pfeiffer, Darrin Lemmer, Dave Engelthaler, The Arizona COVID Genomics Union (ACGU) |
| EPI_ISL_1530626 | Sonora Quest Laboratories | TGen North | Jolene Bowers, Heather Centner, Chris French, Hayley Yaglom, Ashlyn Pfeiffer, Darrin Lemmer, Dave Engelthaler, The Arizona COVID Genomics Union (ACGU) |
| EPI_ISL_1530628 | TGen North | TGen North | Jolene Bowers, Heather Centner, Chris French, Hayley Yaglom, Ashlyn Pfeiffer, Darrin Lemmer, Dave Engelthaler, The Arizona COVID Genomics Union (ACGU) |
| EPI_ISL_1530631 | Sonora Quest Laboratories | TGen North | Jolene Bowers, Heather Centner, Chris French, Hayley Yaglom, Ashlyn Pfeiffer, Darrin Lemmer, Dave Engelthaler, The Arizona COVID Genomics Union (ACGU) |
| EPI_ISL_1530634 | TGen North | TGen North | Jolene Bowers, Heather Centner, Chris French, Hayley Yaglom, Ashlyn Pfeiffer, Darrin Lemmer, Dave Engelthaler, The Arizona COVID Genomics Union (ACGU) |
| EPI_ISL_1530636, EPI_ISL_1530637, EPI_ISL_1530639, EPI_ISL_1530641 | Sonora Quest Laboratories | TGen North | Jolene Bowers, Heather Centner, Chris French, Hayley Yaglom, Ashlyn Pfeiffer, Darrin Lemmer, Dave Engelthaler, The Arizona COVID Genomics Union (ACGU) |
| EPI_ISL_1530642 | TGen North | TGen North | Jolene Bowers, Heather Centner, Chris French, Hayley Yaglom, Ashlyn Pfeiffer, Darrin Lemmer, Dave Engelthaler, The Arizona COVID Genomics Union (ACGU) |
| EPI_ISL_1530643, EPI_ISL_1530644, EPI_ISL_1530645, EPI_ISL_1530646, EPI_ISL_1530650, EPI_ISL_1530652, EPI_ISL_1530654, EPI_ISL_1530659, EPI_ISL_1530660 | Sonora Quest Laboratories | TGen North | Jolene Bowers, Heather Centner, Chris French, Hayley Yaglom, Ashlyn Pfeiffer, Darrin Lemmer, Dave Engelthaler, The Arizona COVID Genomics Union (ACGU) |

| | | | |
|---|---|---|---|
| EPI_ISL_1530663 | TGen North | TGen North | Jolene Bowers, Heather Centner, Chris French, Hayley Yaglom, Ashlyn Pfeiffer, Darrin Lemmer, Dave Engelthaler, The Arizona COVID Genomics Union (ACGU) |
| EPI_ISL_1530664, EPI_ISL_1530665, EPI_ISL_1530669, EPI_ISL_1530670, EPI_ISL_1530672, EPI_ISL_1530673, EPI_ISL_1530674, EPI_ISL_1530675 | Sonora Quest Laboratories | TGen North | Jolene Bowers, Heather Centner, Chris French, Hayley Yaglom, Ashlyn Pfeiffer, Darrin Lemmer, Dave Engelthaler, The Arizona COVID Genomics Union (ACGU) |
| EPI_ISL_1530676 | TGen North | TGen North | Jolene Bowers, Heather Centner, Chris French, Hayley Yaglom, Ashlyn Pfeiffer, Darrin Lemmer, Dave Engelthaler, The Arizona COVID Genomics Union (ACGU) |
| EPI_ISL_1530678, EPI_ISL_1530679, EPI_ISL_1530681, EPI_ISL_1530683, EPI_ISL_1530684, EPI_ISL_1530685 | Sonora Quest Laboratories | TGen North | Jolene Bowers, Heather Centner, Chris French, Hayley Yaglom, Ashlyn Pfeiffer, Darrin Lemmer, Dave Engelthaler, The Arizona COVID Genomics Union (ACGU) |
| EPI_ISL_1530686 | TGen North | TGen North | Jolene Bowers, Heather Centner, Chris French, Hayley Yaglom, Ashlyn Pfeiffer, Darrin Lemmer, Dave Engelthaler, The Arizona COVID Genomics Union (ACGU) |
| EPI_ISL_1530687 | Sonora Quest Laboratories | TGen North | Jolene Bowers, Heather Centner, Chris French, Hayley Yaglom, Ashlyn Pfeiffer, Darrin Lemmer, Dave Engelthaler, The Arizona COVID Genomics Union (ACGU) |
| EPI_ISL_1530688 | TGen North | TGen North | Jolene Bowers, Heather Centner, Chris French, Hayley Yaglom, Ashlyn Pfeiffer, Darrin Lemmer, Dave Engelthaler, The Arizona COVID Genomics Union (ACGU) |
| EPI_ISL_1530690 | Sonora Quest Laboratories | TGen North | Jolene Bowers, Heather Centner, Chris French, Hayley Yaglom, Ashlyn Pfeiffer, Darrin Lemmer, Dave Engelthaler, The Arizona COVID Genomics Union (ACGU) |
| EPI_ISL_1530691 | TGen North | TGen North | Jolene Bowers, Heather Centner, Chris French, Hayley Yaglom, Ashlyn Pfeiffer, Darrin Lemmer, Dave Engelthaler, The Arizona COVID Genomics Union (ACGU) |
| EPI_ISL_1530692, EPI_ISL_1530694, EPI_ISL_1530695 | Sonora Quest Laboratories | TGen North | Jolene Bowers, Heather Centner, Chris French, Hayley Yaglom, Ashlyn Pfeiffer, Darrin Lemmer, Dave Engelthaler, The Arizona COVID Genomics Union (ACGU) |
| EPI_ISL_1530697 | TGen North | TGen North | Jolene Bowers, Heather Centner, Chris French, Hayley Yaglom, Ashlyn Pfeiffer, Darrin Lemmer, Dave Engelthaler, The Arizona COVID Genomics Union (ACGU) |
| EPI_ISL_1530698 | Sonora Quest Laboratories | TGen North | Jolene Bowers, Heather Centner, Chris French, Hayley Yaglom, Ashlyn Pfeiffer, Darrin Lemmer, Dave Engelthaler, The Arizona COVID Genomics Union (ACGU) |
| EPI_ISL_1530699 | TGen North | TGen North | Jolene Bowers, Heather Centner, Chris French, Hayley Yaglom, Ashlyn Pfeiffer, Darrin Lemmer, Dave Engelthaler, The Arizona COVID Genomics Union (ACGU) |
| EPI_ISL_1530700, EPI_ISL_1530701, EPI_ISL_1530703 | Sonora Quest Laboratories | TGen North | Jolene Bowers, Heather Centner, Chris French, Hayley Yaglom, Ashlyn Pfeiffer, Darrin Lemmer, Dave Engelthaler, The Arizona COVID Genomics Union (ACGU) |
| EPI_ISL_1530704 | TGen North | TGen North | Jolene Bowers, Heather Centner, Chris French, Hayley Yaglom, Ashlyn Pfeiffer, Darrin Lemmer, Dave Engelthaler, The Arizona COVID Genomics Union (ACGU) |
| EPI_ISL_1530705, EPI_ISL_1530706 | Sonora Quest Laboratories | TGen North | Jolene Bowers, Heather Centner, Chris French, Hayley Yaglom, Ashlyn Pfeiffer, Darrin Lemmer, Dave Engelthaler, The Arizona COVID Genomics Union (ACGU) |
| EPI_ISL_1530708 | TGen North | TGen North | Jolene Bowers, Heather Centner, Chris French, Hayley Yaglom, Ashlyn Pfeiffer, Darrin Lemmer, Dave Engelthaler, The Arizona COVID Genomics Union (ACGU) |
| EPI_ISL_1530709, EPI_ISL_1530710, EPI_ISL_1530712, EPI_ISL_1530713, EPI_ISL_1530714, EPI_ISL_1530715 | Sonora Quest Laboratories | TGen North | Jolene Bowers, Heather Centner, Chris French, Hayley Yaglom, Ashlyn Pfeiffer, Darrin Lemmer, Dave Engelthaler, The Arizona COVID Genomics Union (ACGU) |
| EPI_ISL_1530716, EPI_ISL_1530721, EPI_ISL_1530723, EPI_ISL_1530724 | TGen North | TGen North | Jolene Bowers, Heather Centner, Chris French, Hayley Yaglom, Ashlyn Pfeiffer, Darrin Lemmer, Dave Engelthaler, The Arizona COVID Genomics Union (ACGU) |
| EPI_ISL_1530727 | Sonora Quest Laboratories | TGen North | Jolene Bowers, Heather Centner, Chris French, Hayley Yaglom, Ashlyn Pfeiffer, Darrin Lemmer, Dave Engelthaler, The Arizona COVID Genomics Union (ACGU) |
| EPI_ISL_1530729, EPI_ISL_1530730 | TGen North | TGen North | Jolene Bowers, Heather Centner, Chris French, Hayley Yaglom, Ashlyn Pfeiffer, Darrin Lemmer, Dave Engelthaler, The Arizona COVID Genomics Union (ACGU) |
| EPI_ISL_1530731 | Sonora Quest Laboratories | TGen North | Jolene Bowers, Heather Centner, Chris French, Hayley Yaglom, Ashlyn Pfeiffer, Darrin Lemmer, Dave Engelthaler, The Arizona COVID Genomics Union (ACGU) |
| EPI_ISL_1530732 | TGen North | TGen North | Jolene Bowers, Heather Centner, Chris French, Hayley Yaglom, Ashlyn Pfeiffer, Darrin Lemmer, Dave Engelthaler, The Arizona COVID Genomics Union (ACGU) |
| EPI_ISL_1530733 | Sonora Quest Laboratories | TGen North | Jolene Bowers, Heather Centner, Chris French, Hayley Yaglom, Ashlyn Pfeiffer, Darrin Lemmer, Dave Engelthaler, The Arizona COVID Genomics Union (ACGU) |
| EPI_ISL_1530734 | TGen North | TGen North | Jolene Bowers, Heather Centner, Chris French, Hayley Yaglom, Ashlyn Pfeiffer, Darrin Lemmer, Dave Engelthaler, The Arizona COVID Genomics Union (ACGU) |
| EPI_ISL_1530735 | Sonora Quest Laboratories | TGen North | Jolene Bowers, Heather Centner, Chris French, Hayley Yaglom, Ashlyn Pfeiffer, Darrin Lemmer, Dave Engelthaler, The Arizona COVID Genomics Union (ACGU) |
| EPI_ISL_1530736 | TGen North | TGen North | Jolene Bowers, Heather Centner, Chris French, Hayley Yaglom, Ashlyn Pfeiffer, Darrin Lemmer, Dave Engelthaler, The Arizona COVID Genomics Union (ACGU) |
| EPI_ISL_1530737, EPI_ISL_1530738, EPI_ISL_1530739, EPI_ISL_1530740 | Sonora Quest Laboratories | TGen North | Jolene Bowers, Heather Centner, Chris French, Hayley Yaglom, Ashlyn Pfeiffer, Darrin Lemmer, Dave Engelthaler, The Arizona COVID Genomics Union (ACGU) |
| EPI_ISL_1530741, EPI_ISL_1530743 | TGen North | TGen North | Jolene Bowers, Heather Centner, Chris French, Hayley Yaglom, Ashlyn Pfeiffer, Darrin Lemmer, Dave Engelthaler, The Arizona COVID Genomics Union (ACGU) |
| EPI_ISL_1530746, EPI_ISL_1530748, EPI_ISL_1530750, EPI_ISL_1530751, EPI_ISL_1530753 | Sonora Quest Laboratories | TGen North | Jolene Bowers, Heather Centner, Chris French, Hayley Yaglom, Ashlyn Pfeiffer, Darrin Lemmer, Dave Engelthaler, The Arizona COVID Genomics Union (ACGU) |
| EPI_ISL_1530755 | TGen North | TGen North | Jolene Bowers, Heather Centner, Chris French, Hayley Yaglom, Ashlyn Pfeiffer, Darrin Lemmer, Dave Engelthaler, The Arizona COVID Genomics Union (ACGU) |
| EPI_ISL_1530756, EPI_ISL_1530757, EPI_ISL_1530759 | Sonora Quest Laboratories | TGen North | Jolene Bowers, Heather Centner, Chris French, Hayley Yaglom, Ashlyn Pfeiffer, Darrin Lemmer, Dave Engelthaler, The Arizona COVID Genomics Union (ACGU) |
| EPI_ISL_1530760 | TGen North | TGen North | Jolene Bowers, Heather Centner, Chris French, Hayley Yaglom, Ashlyn Pfeiffer, Darrin Lemmer, Dave Engelthaler, The Arizona COVID Genomics Union (ACGU) |
| EPI_ISL_1530761, EPI_ISL_1530762, EPI_ISL_1530765, EPI_ISL_1530766 | Sonora Quest Laboratories | TGen North | Jolene Bowers, Heather Centner, Chris French, Hayley Yaglom, Ashlyn Pfeiffer, Darrin Lemmer, Dave Engelthaler, The Arizona COVID Genomics Union (ACGU) |
| EPI_ISL_1530768 | TGen North | TGen North | Jolene Bowers, Heather Centner, Chris French, Hayley Yaglom, Ashlyn Pfeiffer, Darrin Lemmer, Dave Engelthaler, The Arizona COVID Genomics Union (ACGU) |
| EPI_ISL_1530771, EPI_ISL_1530772, EPI_ISL_1530773, EPI_ISL_1530774, EPI_ISL_1530777, EPI_ISL_1530778, EPI_ISL_1530780, EPI_ISL_1530781, EPI_ISL_1530782, EPI_ISL_1530784 | Sonora Quest Laboratories | TGen North | Jolene Bowers, Heather Centner, Chris French, Hayley Yaglom, Ashlyn Pfeiffer, Darrin Lemmer, Dave Engelthaler, The Arizona COVID Genomics Union (ACGU) |
| EPI_ISL_1530785 | TGen North | TGen North | Jolene Bowers, Heather Centner, Chris French, Hayley Yaglom, Ashlyn Pfeiffer, Darrin Lemmer, Dave Engelthaler, The Arizona COVID Genomics Union (ACGU) |
| EPI_ISL_1530787 | Sonora Quest Laboratories | TGen North | Jolene Bowers, Heather Centner, Chris French, Hayley Yaglom, Ashlyn Pfeiffer, Darrin Lemmer, Dave Engelthaler, The Arizona COVID Genomics Union (ACGU) |
| EPI_ISL_1530788 | TGen North | TGen North | Jolene Bowers, Heather Centner, Chris French, Hayley Yaglom, Ashlyn Pfeiffer, Darrin Lemmer, Dave Engelthaler, The Arizona COVID Genomics Union (ACGU) |
| EPI_ISL_1530790 | Sonora Quest Laboratories | TGen North | Jolene Bowers, Heather Centner, Chris French, Hayley Yaglom, Ashlyn Pfeiffer, Darrin Lemmer, Dave Engelthaler, The Arizona COVID Genomics Union (ACGU) |
| EPI_ISL_1530791, EPI_ISL_1530792 | TGen North | TGen North | Jolene Bowers, Heather Centner, Chris French, Hayley Yaglom, Ashlyn Pfeiffer, Darrin Lemmer, Dave Engelthaler, The Arizona COVID Genomics Union (ACGU) |
| EPI_ISL_1530793, EPI_ISL_1530795, | Sonora Quest Laboratories | TGen North | Jolene Bowers, Heather Centner, Chris French, Hayley Yaglom, Ashlyn Pfeiffer, Darrin Lemmer, Dave Engelthaler, The Arizona COVID Genomics Union (ACGU) |

| | | | |
|---|---|---|---|
| EPI_ISL_1530796, EPI_ISL_1530797, EPI_ISL_1530799, EPI_ISL_1530800, EPI_ISL_1530801 | | | |
| EPI_ISL_1530802 | TGen North | TGen North | Jolene Bowers, Heather Centner, Chris French, Hayley Yaglom, Ashlyn Pfeiffer, Darrin Lemmer, Dave Engelthaler, The Arizona COVID Genomics Union (ACGU) |
| EPI_ISL_1530804 | Sonora Quest Laboratories | TGen North | Jolene Bowers, Heather Centner, Chris French, Hayley Yaglom, Ashlyn Pfeiffer, Darrin Lemmer, Dave Engelthaler, The Arizona COVID Genomics Union (ACGU) |
| EPI_ISL_1530806, EPI_ISL_1530807, EPI_ISL_1530809 | TGen North | TGen North | Jolene Bowers, Heather Centner, Chris French, Hayley Yaglom, Ashlyn Pfeiffer, Darrin Lemmer, Dave Engelthaler, The Arizona COVID Genomics Union (ACGU) |
| EPI_ISL_1530812, EPI_ISL_1530815, EPI_ISL_1530819, EPI_ISL_1530820 | Sonora Quest Laboratories | TGen North | Jolene Bowers, Heather Centner, Chris French, Hayley Yaglom, Ashlyn Pfeiffer, Darrin Lemmer, Dave Engelthaler, The Arizona COVID Genomics Union (ACGU) |
| EPI_ISL_1530821, EPI_ISL_1530823, EPI_ISL_1530824, EPI_ISL_1530825 | TGen North | TGen North | Jolene Bowers, Heather Centner, Chris French, Hayley Yaglom, Ashlyn Pfeiffer, Darrin Lemmer, Dave Engelthaler, The Arizona COVID Genomics Union (ACGU) |
| EPI_ISL_1530828, EPI_ISL_1530829, EPI_ISL_1530830 | Sonora Quest Laboratories | TGen North | Jolene Bowers, Heather Centner, Chris French, Hayley Yaglom, Ashlyn Pfeiffer, Darrin Lemmer, Dave Engelthaler, The Arizona COVID Genomics Union (ACGU) |
| EPI_ISL_1530831 | TGen North | TGen North | Jolene Bowers, Heather Centner, Chris French, Hayley Yaglom, Ashlyn Pfeiffer, Darrin Lemmer, Dave Engelthaler, The Arizona COVID Genomics Union (ACGU) |
| EPI_ISL_1530832 | Sonora Quest Laboratories | TGen North | Jolene Bowers, Heather Centner, Chris French, Hayley Yaglom, Ashlyn Pfeiffer, Darrin Lemmer, Dave Engelthaler, The Arizona COVID Genomics Union (ACGU) |
| EPI_ISL_1530833 | TGen North | TGen North | Jolene Bowers, Heather Centner, Chris French, Hayley Yaglom, Ashlyn Pfeiffer, Darrin Lemmer, Dave Engelthaler, The Arizona COVID Genomics Union (ACGU) |
| EPI_ISL_1530834, EPI_ISL_1530835, EPI_ISL_1530837, EPI_ISL_1530838, EPI_ISL_1530840, EPI_ISL_1530841, EPI_ISL_1530844, EPI_ISL_1530845, EPI_ISL_1530847, EPI_ISL_1530848, EPI_ISL_1530849, EPI_ISL_1530850 | | | |
| see above | Sonora Quest Laboratories | TGen North | Jolene Bowers, Heather Centner, Chris French, Hayley Yaglom, Ashlyn Pfeiffer, Darrin Lemmer, Dave Engelthaler, The Arizona COVID Genomics Union (ACGU) |
| EPI_ISL_1530853 | TGen North | TGen North | Jolene Bowers, Heather Centner, Chris French, Hayley Yaglom, Ashlyn Pfeiffer, Darrin Lemmer, Dave Engelthaler, The Arizona COVID Genomics Union (ACGU) |
| EPI_ISL_1530854, EPI_ISL_1530856 | Sonora Quest Laboratories | TGen North | Jolene Bowers, Heather Centner, Chris French, Hayley Yaglom, Ashlyn Pfeiffer, Darrin Lemmer, Dave Engelthaler, The Arizona COVID Genomics Union (ACGU) |
| EPI_ISL_1530858 | TGen North | TGen North | Jolene Bowers, Heather Centner, Chris French, Hayley Yaglom, Ashlyn Pfeiffer, Darrin Lemmer, Dave Engelthaler, The Arizona COVID Genomics Union (ACGU) |
| EPI_ISL_1530860, EPI_ISL_1530861, EPI_ISL_1530863, EPI_ISL_1530864 | Sonora Quest Laboratories | TGen North | Jolene Bowers, Heather Centner, Chris French, Hayley Yaglom, Ashlyn Pfeiffer, Darrin Lemmer, Dave Engelthaler, The Arizona COVID Genomics Union (ACGU) |
| EPI_ISL_1530866, EPI_ISL_1530870, EPI_ISL_1530872 | TGen North | TGen North | Jolene Bowers, Heather Centner, Chris French, Hayley Yaglom, Ashlyn Pfeiffer, Darrin Lemmer, Dave Engelthaler, The Arizona COVID Genomics Union (ACGU) |
| EPI_ISL_1530873 | Sonora Quest Laboratories | TGen North | Jolene Bowers, Heather Centner, Chris French, Hayley Yaglom, Ashlyn Pfeiffer, Darrin Lemmer, Dave Engelthaler, The Arizona COVID Genomics Union (ACGU) |
| EPI_ISL_1530878, EPI_ISL_1530880, EPI_ISL_1530884, EPI_ISL_1530885, EPI_ISL_1530886, EPI_ISL_1530887, EPI_ISL_1530888, EPI_ISL_1530889, EPI_ISL_1530890, EPI_ISL_1530891, EPI_ISL_1530892, EPI_ISL_1530893, EPI_ISL_1530894, EPI_ISL_1530895, EPI_ISL_1530896, EPI_ISL_1530898, EPI_ISL_1530899, EPI_ISL_1530900, EPI_ISL_1530901, EPI_ISL_1530902, EPI_ISL_1530903, EPI_ISL_1530904, EPI_ISL_1530908, EPI_ISL_1530910, EPI_ISL_1530913, EPI_ISL_1530914, EPI_ISL_1530918, EPI_ISL_1530923, EPI_ISL_1530924, EPI_ISL_1530926, EPI_ISL_1530927, EPI_ISL_1530928, EPI_ISL_1530929, EPI_ISL_1530930, EPI_ISL_1530931, EPI_ISL_1530933, EPI_ISL_1530934, EPI_ISL_1530937, EPI_ISL_1530938, EPI_ISL_1530939, EPI_ISL_1530940, EPI_ISL_1530941, EPI_ISL_1530942, EPI_ISL_1530945 | | | |
| see above | NJDOH, Public Health and Environmental Laboratories | New Jersey Public Health and Environmental Laboratories (NJPHEL) | Lindsey Bodnar, Shiv K. Verma, Jacquelyn Deverell, Matthew Scarnati, Mohammad M Ali, Dana Woell, Byeong Jeong |
| EPI_ISL_1530946, EPI_ISL_1530947 | TriCore Reference Laboratories | New Mexico Department of Health Scientific Laboratory | Ellie Johnson, Anastacia Griego-Fisher, D'eldra Malone, Jennifer Benoit |
| EPI_ISL_1530948, EPI_ISL_1530949, EPI_ISL_1530950, EPI_ISL_1530951, EPI_ISL_1530952, EPI_ISL_1530953, EPI_ISL_1530954, EPI_ISL_1530955, EPI_ISL_1530956, EPI_ISL_1530957, EPI_ISL_1530958, EPI_ISL_1530959, EPI_ISL_1530960, EPI_ISL_1530961, EPI_ISL_1530962, EPI_ISL_1530963, EPI_ISL_1530964, EPI_ISL_1530965, EPI_ISL_1530966, EPI_ISL_1530967, EPI_ISL_1530968, EPI_ISL_1530969, EPI_ISL_1530970, EPI_ISL_1530971, EPI_ISL_1530972, EPI_ISL_1530973, EPI_ISL_1530974, EPI_ISL_1530975, EPI_ISL_1530976, EPI_ISL_1530977, EPI_ISL_1530978, EPI_ISL_1530980, EPI_ISL_1530981, EPI_ISL_1530982, EPI_ISL_1530983, EPI_ISL_1530984, EPI_ISL_1530985, EPI_ISL_1530986, EPI_ISL_1530987, EPI_ISL_1530988, EPI_ISL_1530989 | | | |
| see above | New Mexico Department of Health Scientific Laboratory | New Mexico Department of Health Scientific Laboratory | Ellie Johnson, Anastacia Griego-Fisher, D'eldra Malone, Jennifer Benoit |
| EPI_ISL_1530990, EPI_ISL_1530991, EPI_ISL_1530992, EPI_ISL_1530993, EPI_ISL_1530994, EPI_ISL_1530996, EPI_ISL_1530997, EPI_ISL_1530999, EPI_ISL_1531000, EPI_ISL_1531001, EPI_ISL_1531002, EPI_ISL_1531003, EPI_ISL_1531004, EPI_ISL_1531005, EPI_ISL_1531007, EPI_ISL_1531008, EPI_ISL_1531009, EPI_ISL_1531010, EPI_ISL_1531011, EPI_ISL_1531012, EPI_ISL_1531013, EPI_ISL_1531014, EPI_ISL_1531016, EPI_ISL_1531017, EPI_ISL_1531018, EPI_ISL_1531019, EPI_ISL_1531020, EPI_ISL_1531021, EPI_ISL_1531022, EPI_ISL_1531024, EPI_ISL_1531025, EPI_ISL_1531027, EPI_ISL_1531029, EPI_ISL_1531031, EPI_ISL_1531033, EPI_ISL_1531037, EPI_ISL_1531038, EPI_ISL_1531040, EPI_ISL_1531041, EPI_ISL_1531042, EPI_ISL_1531043, EPI_ISL_1531045, EPI_ISL_1531046, EPI_ISL_1531048, EPI_ISL_1531049, EPI_ISL_1531051, EPI_ISL_1531052, EPI_ISL_1531053, EPI_ISL_1531054, EPI_ISL_1531055, EPI_ISL_1531057, EPI_ISL_1531059, EPI_ISL_1531060, EPI_ISL_1531061, EPI_ISL_1531062, EPI_ISL_1531063, EPI_ISL_1531064, EPI_ISL_1531065, EPI_ISL_1531066, EPI_ISL_1531067, EPI_ISL_1531068, EPI_ISL_1531069, EPI_ISL_1531070, EPI_ISL_1531071, EPI_ISL_1531072, EPI_ISL_1531073, EPI_ISL_1531074, EPI_ISL_1531075, EPI_ISL_1531076, EPI_ISL_1531077, EPI_ISL_1531078, EPI_ISL_1531079, EPI_ISL_1531080, EPI_ISL_1531081, EPI_ISL_1531082, EPI_ISL_1531083, EPI_ISL_1531084, EPI_ISL_1531085, EPI_ISL_1531086, EPI_ISL_1531087, EPI_ISL_1531088, EPI_ISL_1531089, EPI_ISL_1531090, EPI_ISL_1531091, EPI_ISL_1531092, EPI_ISL_1531093, EPI_ISL_1531094, EPI_ISL_1531095, EPI_ISL_1531096, EPI_ISL_1531097, EPI_ISL_1531098, EPI_ISL_1531099, EPI_ISL_1531100, EPI_ISL_1531101, EPI_ISL_1531102, EPI_ISL_1531103, EPI_ISL_1531104, EPI_ISL_1531105, EPI_ISL_1531106, EPI_ISL_1531107, EPI_ISL_1531108, EPI_ISL_1531109, EPI_ISL_1531110, EPI_ISL_1531112, EPI_ISL_1531113, EPI_ISL_1531114, EPI_ISL_1531115, EPI_ISL_1531118, EPI_ISL_1531119, EPI_ISL_1531121, EPI_ISL_1531122, EPI_ISL_1531123, EPI_ISL_1531124, EPI_ISL_1531126, EPI_ISL_1531127, EPI_ISL_1531128, EPI_ISL_1531129, EPI_ISL_1531130, EPI_ISL_1531131, EPI_ISL_1531132, EPI_ISL_1531133, EPI_ISL_1531134, EPI_ISL_1531135, EPI_ISL_1531136, EPI_ISL_1531137, EPI_ISL_1531138, EPI_ISL_1531139, EPI_ISL_1531140, EPI_ISL_1531141, EPI_ISL_1531142, EPI_ISL_1531143, EPI_ISL_1531144, EPI_ISL_1531145, EPI_ISL_1531146, EPI_ISL_1531147, EPI_ISL_1531148, EPI_ISL_1531151, EPI_ISL_1531152, EPI_ISL_1531154, EPI_ISL_1531155, EPI_ISL_1531156, EPI_ISL_1531157, EPI_ISL_1531158, EPI_ISL_1531159, EPI_ISL_1531160, EPI_ISL_1531161, EPI_ISL_1531162, EPI_ISL_1531163, EPI_ISL_1531164, EPI_ISL_1531165, EPI_ISL_1531166, EPI_ISL_1531167, EPI_ISL_1531168, EPI_ISL_1531169, EPI_ISL_1531171, EPI_ISL_1531176, EPI_ISL_1531179, EPI_ISL_1531180, EPI_ISL_1531181, EPI_ISL_1531182, EPI_ISL_1531183, EPI_ISL_1531184, EPI_ISL_1531185, EPI_ISL_1531186, EPI_ISL_1531187, EPI_ISL_1531188, EPI_ISL_1531189, EPI_ISL_1531190, EPI_ISL_1531191, EPI_ISL_1531192, EPI_ISL_1531193, EPI_ISL_1531194, EPI_ISL_1531195, EPI_ISL_1531196, EPI_ISL_1531197, EPI_ISL_1531198, EPI_ISL_1531199, EPI_ISL_1531200, EPI_ISL_1531201, EPI_ISL_1531202, EPI_ISL_1531203, EPI_ISL_1531204, EPI_ISL_1531205, EPI_ISL_1531206, EPI_ISL_1531208, EPI_ISL_1531209, EPI_ISL_1531211, EPI_ISL_1531212, EPI_ISL_1531215, EPI_ISL_1531216, EPI_ISL_1531217, EPI_ISL_1531218, EPI_ISL_1531219, EPI_ISL_1531221, EPI_ISL_1531222, EPI_ISL_1531223, EPI_ISL_1531224, EPI_ISL_1531225, EPI_ISL_1531226, EPI_ISL_1531227, EPI_ISL_1531230, EPI_ISL_1531231, EPI_ISL_1531232, EPI_ISL_1531233, EPI_ISL_1531234, EPI_ISL_1531237, EPI_ISL_1531238, EPI_ISL_1531239, EPI_ISL_1531240, EPI_ISL_1531241, EPI_ISL_1531242, EPI_ISL_1531243, EPI_ISL_1531244, EPI_ISL_1531245, EPI_ISL_1531246, EPI_ISL_1531248, EPI_ISL_1531249, EPI_ISL_1531251, EPI_ISL_1531252, EPI_ISL_1531253, EPI_ISL_1531254, EPI_ISL_1531255, EPI_ISL_1531256, EPI_ISL_1531257, EPI_ISL_1531258, EPI_ISL_1531259, EPI_ISL_1531260, EPI_ISL_1531261, EPI_ISL_1531262, EPI_ISL_1531263, EPI_ISL_1531264, EPI_ISL_1531265, EPI_ISL_1531266, EPI_ISL_1531268, EPI_ISL_1531269, EPI_ISL_1531271, EPI_ISL_1531272, EPI_ISL_1531273, EPI_ISL_1531274, EPI_ISL_1531275, EPI_ISL_1531276, EPI_ISL_1531277, EPI_ISL_1531278, EPI_ISL_1531279, EPI_ISL_1531280, EPI_ISL_1531281, EPI_ISL_1531282, EPI_ISL_1531283, EPI_ISL_1531284, EPI_ISL_1531285, EPI_ISL_1531286, EPI_ISL_1531287, EPI_ISL_1531288, EPI_ISL_1531289, EPI_ISL_1531290, EPI_ISL_1531291, EPI_ISL_1531294, EPI_ISL_1531295, EPI_ISL_1531296, EPI_ISL_1531298, EPI_ISL_1531299, EPI_ISL_1531300, EPI_ISL_1531301, EPI_ISL_1531302, EPI_ISL_1531303, EPI_ISL_1531304, EPI_ISL_1531305, EPI_ISL_1531306, EPI_ISL_1531307, EPI_ISL_1531308, EPI_ISL_1531309, EPI_ISL_1531310, EPI_ISL_1531312, EPI_ISL_1531314, EPI_ISL_1531315, EPI_ISL_1531317, EPI_ISL_1531318, EPI_ISL_1531319, EPI_ISL_1531320, EPI_ISL_1531321, EPI_ISL_1531322, EPI_ISL_1531323, EPI_ISL_1531324, EPI_ISL_1531325, EPI_ISL_1531326, EPI_ISL_1531327, EPI_ISL_1531328, EPI_ISL_1531329, EPI_ISL_1531330, EPI_ISL_1531331, EPI_ISL_1531333, EPI_ISL_1531334, EPI_ISL_1531335, EPI_ISL_1531338, EPI_ISL_1531339, EPI_ISL_1531340, EPI_ISL_1531341, EPI_ISL_1531342, EPI_ISL_1531343, EPI_ISL_1531344, EPI_ISL_1531345, EPI_ISL_1531346, EPI_ISL_1531347, EPI_ISL_1531348, EPI_ISL_1531349, EPI_ISL_1531350, EPI_ISL_1531351, EPI_ISL_1531352, EPI_ISL_1531353, EPI_ISL_1531354, EPI_ISL_1531355, EPI_ISL_1531356, EPI_ISL_1531357, EPI_ISL_1531358, EPI_ISL_1531359, EPI_ISL_1531360, EPI_ISL_1531361, EPI_ISL_1531362, EPI_ISL_1531363, EPI_ISL_1531364, EPI_ISL_1531365, EPI_ISL_1531366, EPI_ISL_1531367, EPI_ISL_1531368, EPI_ISL_1531369, EPI_ISL_1531370, EPI_ISL_1531371, EPI_ISL_1531372, EPI_ISL_1531373, EPI_ISL_1531374, EPI_ISL_1531375, EPI_ISL_1531376, EPI_ISL_1531377, EPI_ISL_1531378, EPI_ISL_1531379, EPI_ISL_1531380, EPI_ISL_1531381, EPI_ISL_1531382, EPI_ISL_1531383, EPI_ISL_1531384, EPI_ISL_1531385, EPI_ISL_1531386, EPI_ISL_1531388, EPI_ISL_1531389, EPI_ISL_1531392, EPI_ISL_1531395, EPI_ISL_1531396, EPI_ISL_1531397, EPI_ISL_1531400, EPI_ISL_1531401, EPI_ISL_1531403, EPI_ISL_1531404, EPI_ISL_1531405, EPI_ISL_1531406, EPI_ISL_1531408, EPI_ISL_1531413, EPI_ISL_1531414, EPI_ISL_1531416 | | | |
| see above | Florida Bureau of Public Health Laboratories | Florida Bureau of Public Health Laboratories | Sarah Schmedes, Jason Blanton |
| EPI_ISL_1531417, EPI_ISL_1531418, EPI_ISL_1531419, EPI_ISL_1531421, EPI_ISL_1531422, EPI_ISL_1531423 | Wyoming Public Health Laboratory | Wyoming Public Health Laboratory | Jim Mildenberger, Wanda Manley, Noah Hull, Taylor Fearing, Lynette Gumbleton, Channing Weber, Ashley Norberg, Chayse Rowley, Marley Goetz, Brian Dominguez, Elliot Thomasson, Cari Sloma, and Rob Christensen |
| EPI_ISL_1531432, EPI_ISL_1531433, EPI_ISL_1531434, EPI_ISL_1531435, EPI_ISL_1531436, EPI_ISL_1531437, EPI_ISL_1531438, EPI_ISL_1531439, EPI_ISL_1531440, EPI_ISL_1531441, EPI_ISL_1531442, EPI_ISL_1531443, EPI_ISL_1531445, EPI_ISL_1531446, EPI_ISL_1531447, EPI_ISL_1531449, EPI_ISL_1531450, EPI_ISL_1531451, EPI_ISL_1531452, EPI_ISL_1531453, EPI_ISL_1531454, EPI_ISL_1531455, EPI_ISL_1531456, EPI_ISL_1531457, EPI_ISL_1531458, EPI_ISL_1531460, EPI_ISL_1531461, EPI_ISL_1531462, EPI_ISL_1531463, EPI_ISL_1531464, EPI_ISL_1531465, EPI_ISL_1531466, EPI_ISL_1531467, EPI_ISL_1531468, EPI_ISL_1531469, EPI_ISL_1531470, EPI_ISL_1531471, EPI_ISL_1531472, EPI_ISL_1531473, EPI_ISL_1531474, EPI_ISL_1531475, EPI_ISL_1531476, EPI_ISL_1531477, EPI_ISL_1531479, EPI_ISL_1531480, EPI_ISL_1531481, EPI_ISL_1531482, EPI_ISL_1531483, EPI_ISL_1531484, EPI_ISL_1531485, EPI_ISL_1531486, EPI_ISL_1531488, EPI_ISL_1531489, EPI_ISL_1531490, EPI_ISL_1531492, EPI_ISL_1531494, EPI_ISL_1531495, EPI_ISL_1531496, EPI_ISL_1531497, EPI_ISL_1531498, EPI_ISL_1531499, EPI_ISL_1531501, EPI_ISL_1531502, EPI_ISL_1531503, EPI_ISL_1531505, EPI_ISL_1531506, EPI_ISL_1531507, EPI_ISL_1531508, EPI_ISL_1531509, EPI_ISL_1531510, EPI_ISL_1531511, EPI_ISL_1531512, EPI_ISL_1531513, EPI_ISL_1531515, EPI_ISL_1531517, EPI_ISL_1531518, EPI_ISL_1531519, EPI_ISL_1531520, EPI_ISL_1531521, EPI_ISL_1531522, EPI_ISL_1531524, EPI_ISL_1531525, EPI_ISL_1531527, EPI_ISL_1531528, EPI_ISL_1531529, EPI_ISL_1531530 | | | |
| see above | US Air Force School of Aerospace Medicine | US Air Force School of Aerospace Medicine | Anthony Fries, Jennifer Meyer, William Gruner, William Buggele, Amanda Javorina, Sarah Purves, Clarise Starr, Elizabeth Macias |
| EPI_ISL_1531535, EPI_ISL_1531536, EPI_ISL_1531537, EPI_ISL_1531538, EPI_ISL_1531539, EPI_ISL_1531540, EPI_ISL_1531541, EPI_ISL_1531542, EPI_ISL_1531543, EPI_ISL_1531544, EPI_ISL_1531545, EPI_ISL_1531546, EPI_ISL_1531547, EPI_ISL_1531548, EPI_ISL_1531549, EPI_ISL_1531550, EPI_ISL_1531551, EPI_ISL_1531552 | | | |
| see above | Oregon State Public Health Laboratory | Oregon State Public Health Laboratory | John Fontana, Eugene Yeboah, Rafia Razzaque, Laura Tsaknadidis, Vanda Makris, Shane Sevey |
| EPI_ISL_1531574, EPI_ISL_1531575, EPI_ISL_1531576, EPI_ISL_1531577, EPI_ISL_1531578, EPI_ISL_1531579, EPI_ISL_1531580, EPI_ISL_1531581, EPI_ISL_1531582, EPI_ISL_1531583, EPI_ISL_1531584, EPI_ISL_1531585, EPI_ISL_1531586, EPI_ISL_1531587, EPI_ISL_1531588, EPI_ISL_1531589, EPI_ISL_1531590, EPI_ISL_1531592, EPI_ISL_1531593, EPI_ISL_1531594, EPI_ISL_1531595, EPI_ISL_1531596, EPI_ISL_1531597, EPI_ISL_1531598, EPI_ISL_1531599, EPI_ISL_1531600, EPI_ISL_1531601, EPI_ISL_1531602, EPI_ISL_1531603, EPI_ISL_1531604, EPI_ISL_1531605, EPI_ISL_1531606, EPI_ISL_1531607, EPI_ISL_1531608, EPI_ISL_1531609, EPI_ISL_1531610, EPI_ISL_1531611, EPI_ISL_1531612, EPI_ISL_1531613, EPI_ISL_1531614, EPI_ISL_1531615, EPI_ISL_1531616, EPI_ISL_1531617, EPI_ISL_1531618, EPI_ISL_1531619, EPI_ISL_1531620, EPI_ISL_1531621, EPI_ISL_1531622, EPI_ISL_1531623, EPI_ISL_1531624, EPI_ISL_1531625, EPI_ISL_1531627, EPI_ISL_1531628, EPI_ISL_1531629, EPI_ISL_1531630, EPI_ISL_1531631, EPI_ISL_1531633 | | | |
| see above | Minnesota Department of Health, Public Health Laboratory | Minnesota Department of Health, Public Health Laboratory | Alexandra Lorentz, Jacob Garfin, Matt Plumb, and Xiong Wang |
| EPI_ISL_1531634, EPI_ISL_1531635, EPI_ISL_1531636, EPI_ISL_1531637, EPI_ISL_1531638, EPI_ISL_1531640, EPI_ISL_1531641, EPI_ISL_1531642, EPI_ISL_1531643 | Vault Health | Minnesota Department of Health, Public Health Laboratory | Alexandra Lorentz, Jacob Garfin, Matt Plumb, and Xiong Wang |

| | | | |
|---|---|---|---|
| EPI_ISL_1531644, EPI_ISL_1531645, EPI_ISL_1531646, EPI_ISL_1531647, EPI_ISL_1531648, EPI_ISL_1531649, EPI_ISL_1531650, EPI_ISL_1531651, EPI_ISL_1531653, EPI_ISL_1531654, EPI_ISL_1531655, EPI_ISL_1531657, EPI_ISL_1531658, EPI_ISL_1531659, EPI_ISL_1531660, EPI_ISL_1531661, EPI_ISL_1531662, EPI_ISL_1531663, EPI_ISL_1531664, EPI_ISL_1531665, EPI_ISL_1531666, EPI_ISL_1531667, EPI_ISL_1531668, EPI_ISL_1531669, EPI_ISL_1531670, EPI_ISL_1531671, EPI_ISL_1531672, EPI_ISL_1531673, EPI_ISL_1531674, EPI_ISL_1531675, EPI_ISL_1531676, EPI_ISL_1531677, EPI_ISL_1531678, EPI_ISL_1531679, EPI_ISL_1531680, EPI_ISL_1531681, EPI_ISL_1531682, EPI_ISL_1531683, EPI_ISL_1531684, EPI_ISL_1531685, EPI_ISL_1531686, EPI_ISL_1531687, EPI_ISL_1531688, EPI_ISL_1531689, EPI_ISL_1531690, EPI_ISL_1531691, EPI_ISL_1531692, EPI_ISL_1531693, EPI_ISL_1531694, EPI_ISL_1531695, EPI_ISL_1531696, EPI_ISL_1531697, EPI_ISL_1531698, EPI_ISL_1531699, EPI_ISL_1531700, EPI_ISL_1531701, EPI_ISL_1531702, EPI_ISL_1531703, EPI_ISL_1531704, EPI_ISL_1531705, EPI_ISL_1531706, EPI_ISL_1531707, EPI_ISL_1531708, EPI_ISL_1531709, EPI_ISL_1531710, EPI_ISL_1531711, EPI_ISL_1531712, EPI_ISL_1531713, EPI_ISL_1531714, EPI_ISL_1531715, EPI_ISL_1531716, EPI_ISL_1531717, EPI_ISL_1531718, EPI_ISL_1531719, EPI_ISL_1531720, EPI_ISL_1531721, EPI_ISL_1531722, EPI_ISL_1531723, EPI_ISL_1531724, EPI_ISL_1531725, EPI_ISL_1531726, EPI_ISL_1531727, EPI_ISL_1531728, EPI_ISL_1531729, EPI_ISL_1531730, EPI_ISL_1531731, EPI_ISL_1531732, EPI_ISL_1531733 | | | |
| see above | M Health Fairview | Minnesota Department of Health, Public Health Laboratory | Alexandra Lorentz, Jacob Garfin, Matt Plumb, and Xiong Wang |
| EPI_ISL_1531736, EPI_ISL_1531739, EPI_ISL_1531741 | School of Pharmacy, Shenandoah University | School of Pharmacy, Shenandoah University | Adams,S.M., Harralson,A.F., Kidd,R.S., Sawyer,G.W. |
| EPI_ISL_1531745, EPI_ISL_1531746, EPI_ISL_1531747, EPI_ISL_1531748, EPI_ISL_1531749, EPI_ISL_1531750, EPI_ISL_1531751, EPI_ISL_1531752, EPI_ISL_1531753, EPI_ISL_1531754, EPI_ISL_1531755, EPI_ISL_1531756, EPI_ISL_1531757, EPI_ISL_1531758, EPI_ISL_1531759, EPI_ISL_1531760, EPI_ISL_1531761, EPI_ISL_1531762, EPI_ISL_1531763, EPI_ISL_1531764, EPI_ISL_1531765, EPI_ISL_1531766, EPI_ISL_1531767, EPI_ISL_1531768, EPI_ISL_1531769, EPI_ISL_1531770, EPI_ISL_1531771, EPI_ISL_1531772, EPI_ISL_1531773, EPI_ISL_1531774, EPI_ISL_1531775 | | | |
| see above | University of Oregon COVID-19 MAP Laboratory | University of Oregon Genomics and Cell Characterization Core Facility (GC3F) | Douglas Turnbull, Ariana White, Jeff Bishop, Jason Carriere |
| EPI_ISL_1531777, EPI_ISL_1531778, EPI_ISL_1531779, EPI_ISL_1531780, EPI_ISL_1531781, EPI_ISL_1531782, EPI_ISL_1531783, EPI_ISL_1531784, EPI_ISL_1531785, EPI_ISL_1531786, EPI_ISL_1531787, EPI_ISL_1531788, EPI_ISL_1531789, EPI_ISL_1531790, EPI_ISL_1531791, EPI_ISL_1531792, EPI_ISL_1531793, EPI_ISL_1531794, EPI_ISL_1531795, EPI_ISL_1531796, EPI_ISL_1531797, EPI_ISL_1531798, EPI_ISL_1531799, EPI_ISL_1531800, EPI_ISL_1531801, EPI_ISL_1531802, EPI_ISL_1531803, EPI_ISL_1531804, EPI_ISL_1531805, EPI_ISL_1531806 | | | |
| see above | Sharp HealthCare Laboratory | Andersen lab at Scripps Research | SEARCH Alliance San Diego with Aaron Harding, Jacquelyn Berumen, Cathy Woerle, Liam McGinnis, Art Mendoza, Omid Bakhtar |
| EPI_ISL_1531807, EPI_ISL_1531808, EPI_ISL_1531809, EPI_ISL_1531810, EPI_ISL_1531811, EPI_ISL_1531812, EPI_ISL_1531813, EPI_ISL_1531814, EPI_ISL_1531815, EPI_ISL_1531816, EPI_ISL_1531817, EPI_ISL_1531818, EPI_ISL_1531819, EPI_ISL_1531820, EPI_ISL_1531821, EPI_ISL_1531822, EPI_ISL_1531823, EPI_ISL_1531824, EPI_ISL_1531825, EPI_ISL_1531826, EPI_ISL_1531827, EPI_ISL_1531828, EPI_ISL_1531829, EPI_ISL_1531830, EPI_ISL_1531831, EPI_ISL_1531832, EPI_ISL_1531833, EPI_ISL_1531834, EPI_ISL_1531835, EPI_ISL_1531836, EPI_ISL_1531837, EPI_ISL_1531838, EPI_ISL_1531839, EPI_ISL_1531840, EPI_ISL_1531841, EPI_ISL_1531842, EPI_ISL_1531843, EPI_ISL_1531844, EPI_ISL_1531845, EPI_ISL_1531846, EPI_ISL_1531847, EPI_ISL_1531848, EPI_ISL_1531849, EPI_ISL_1531850, EPI_ISL_1531851, EPI_ISL_1531852, EPI_ISL_1531853, EPI_ISL_1531854, EPI_ISL_1531855, EPI_ISL_1531856, EPI_ISL_1531857, EPI_ISL_1531858, EPI_ISL_1531859, EPI_ISL_1531860, EPI_ISL_1531861, EPI_ISL_1531862, EPI_ISL_1531863, EPI_ISL_1531864, EPI_ISL_1531865, EPI_ISL_1531866, EPI_ISL_1531867, EPI_ISL_1531868, EPI_ISL_1531869, EPI_ISL_1531870, EPI_ISL_1531871, EPI_ISL_1531872, EPI_ISL_1531873, EPI_ISL_1531874, EPI_ISL_1531875, EPI_ISL_1531876, EPI_ISL_1531877, EPI_ISL_1531878, EPI_ISL_1531879, EPI_ISL_1531880, EPI_ISL_1531881, EPI_ISL_1531882, EPI_ISL_1531883, EPI_ISL_1531884, EPI_ISL_1531885, EPI_ISL_1531886, EPI_ISL_1531887, EPI_ISL_1531888, EPI_ISL_1531889, EPI_ISL_1531890, EPI_ISL_1531891, EPI_ISL_1531892, EPI_ISL_1531893, EPI_ISL_1531894, EPI_ISL_1531895, EPI_ISL_1531896, EPI_ISL_1531897, EPI_ISL_1531898, EPI_ISL_1531899, EPI_ISL_1531900, EPI_ISL_1531901, EPI_ISL_1531902, EPI_ISL_1531903, EPI_ISL_1531904, EPI_ISL_1531905, EPI_ISL_1531906, EPI_ISL_1531907, EPI_ISL_1531908, EPI_ISL_1531909, EPI_ISL_1531910, EPI_ISL_1531911, EPI_ISL_1531912, EPI_ISL_1531913, EPI_ISL_1531914, EPI_ISL_1531915, EPI_ISL_1531916, EPI_ISL_1531917, EPI_ISL_1531918, EPI_ISL_1531919, EPI_ISL_1531920, EPI_ISL_1531921, EPI_ISL_1531922, EPI_ISL_1531923, EPI_ISL_1531924, EPI_ISL_1531925, EPI_ISL_1531926, EPI_ISL_1531927, EPI_ISL_1531928, EPI_ISL_1531929, EPI_ISL_1531930, EPI_ISL_1531931, EPI_ISL_1531932, EPI_ISL_1531933, EPI_ISL_1531934, EPI_ISL_1531935, EPI_ISL_1531936, EPI_ISL_1531937, EPI_ISL_1531938, EPI_ISL_1531939, EPI_ISL_1531940, EPI_ISL_1531941 | | | |
| see above | Centro de Diagnostico COVID-19 UABC Tijuana | Andersen lab at Scripps Research | SEARCH Alliance San Diego with Idanya Rubi Serafin Higuera, Manuel Sanchez Alavez, Jorge Luis Jimenez Niebla, German Ibarra, Jonathan Vincent Baena, Oscar Efren Zazueta Fierro |
| EPI_ISL_1531942, EPI_ISL_1531943, EPI_ISL_1531944, EPI_ISL_1531945, EPI_ISL_1531946, EPI_ISL_1531947, EPI_ISL_1531948, EPI_ISL_1531949, EPI_ISL_1531950, EPI_ISL_1531951, EPI_ISL_1531952, EPI_ISL_1531953, EPI_ISL_1531954, EPI_ISL_1531955, EPI_ISL_1531956, EPI_ISL_1531957, EPI_ISL_1531958, EPI_ISL_1531959, EPI_ISL_1531960, EPI_ISL_1531961, EPI_ISL_1531962, EPI_ISL_1531963, EPI_ISL_1531964, EPI_ISL_1531965, EPI_ISL_1531966, EPI_ISL_1531967, EPI_ISL_1531968, EPI_ISL_1531969, EPI_ISL_1531970, EPI_ISL_1531971, EPI_ISL_1531972, EPI_ISL_1531973, EPI_ISL_1531974, EPI_ISL_1531975, EPI_ISL_1531976, EPI_ISL_1531977, EPI_ISL_1531978, EPI_ISL_1531979, EPI_ISL_1531980, EPI_ISL_1531981, EPI_ISL_1531982, EPI_ISL_1531983, EPI_ISL_1531984, EPI_ISL_1531985, EPI_ISL_1531986, EPI_ISL_1531987, EPI_ISL_1531988, EPI_ISL_1531989, EPI_ISL_1531990, EPI_ISL_1531991, EPI_ISL_1531992, EPI_ISL_1531993, EPI_ISL_1531994, EPI_ISL_1531995, EPI_ISL_1531996, EPI_ISL_1531997, EPI_ISL_1531998, EPI_ISL_1531999, EPI_ISL_1532000, EPI_ISL_1532001, EPI_ISL_1532002, EPI_ISL_1532003, EPI_ISL_1532004, EPI_ISL_1532005, EPI_ISL_1532006, EPI_ISL_1532007, EPI_ISL_1532008, EPI_ISL_1532009, EPI_ISL_1532010, EPI_ISL_1532011, EPI_ISL_1532012, EPI_ISL_1532013, EPI_ISL_1532014, EPI_ISL_1532015, EPI_ISL_1532016, EPI_ISL_1532017, EPI_ISL_1532018, EPI_ISL_1532019, EPI_ISL_1532020, EPI_ISL_1532021, EPI_ISL_1532022, EPI_ISL_1532023, EPI_ISL_1532024, EPI_ISL_1532025, EPI_ISL_1532026, EPI_ISL_1532027, EPI_ISL_1532028, EPI_ISL_1532029, EPI_ISL_1532030, EPI_ISL_1532031, EPI_ISL_1532032, EPI_ISL_1532033, EPI_ISL_1532034, EPI_ISL_1532035, EPI_ISL_1532036, EPI_ISL_1532037, EPI_ISL_1532038, EPI_ISL_1532039, EPI_ISL_1532040, EPI_ISL_1532041, EPI_ISL_1532042, EPI_ISL_1532043, EPI_ISL_1532044, EPI_ISL_1532045, EPI_ISL_1532046, EPI_ISL_1532047, EPI_ISL_1532048, EPI_ISL_1532049, EPI_ISL_1532050, EPI_ISL_1532051, EPI_ISL_1532052, EPI_ISL_1532053, EPI_ISL_1532054, EPI_ISL_1532055, EPI_ISL_1532056, EPI_ISL_1532057, EPI_ISL_1532058, EPI_ISL_1532059, EPI_ISL_1532060, EPI_ISL_1532061, EPI_ISL_1532062, EPI_ISL_1532063, EPI_ISL_1532064 | | | |
| see above | San Diego County Public Health Laboratory | Andersen lab at Scripps Research | SEARCH Alliance San Diego with Tracy Basler, Jovan Shephard, Brett Austin |
| EPI_ISL_1532066, EPI_ISL_1532067, EPI_ISL_1532068, EPI_ISL_1532069, EPI_ISL_1532071, EPI_ISL_1532072, EPI_ISL_1532073, EPI_ISL_1532074, EPI_ISL_1532075, EPI_ISL_1532076, EPI_ISL_1532077, EPI_ISL_1532078, EPI_ISL_1532079, EPI_ISL_1532080, EPI_ISL_1532081, EPI_ISL_1532082, EPI_ISL_1532083, EPI_ISL_1532084, EPI_ISL_1532085, EPI_ISL_1532086, EPI_ISL_1532087, EPI_ISL_1532088 | | | |
| see above | Idaho Bureau of Laboratories | Idaho Bureau of Laboratories | R. Beukelman, Matthew Charles Burns, Aimee Ceniseros, Robert L. Voermans, Christopher Ball |
| EPI_ISL_1532089, EPI_ISL_1532090, EPI_ISL_1532091, EPI_ISL_1532092, EPI_ISL_1532093, EPI_ISL_1532094, EPI_ISL_1532095, EPI_ISL_1532096, EPI_ISL_1532098, EPI_ISL_1532099, EPI_ISL_1532100, EPI_ISL_1532101, EPI_ISL_1532102, EPI_ISL_1532103, EPI_ISL_1532104, EPI_ISL_1532105, EPI_ISL_1532106, EPI_ISL_1532108, EPI_ISL_1532109, EPI_ISL_1532110, EPI_ISL_1532111, EPI_ISL_1532112, EPI_ISL_1532113, EPI_ISL_1532114, EPI_ISL_1532115, EPI_ISL_1532116, EPI_ISL_1532117 | | | |
| see above | Wyoming Public Health Laboratory | Wyoming Public Health Laboratory | Jim Mildenberger, Wanda Manley, Noah Hull, Taylor Fearing, Lynette Gumbleton, Channing Weber, Ashley Norberg, Chayse Rowley, Marley Goetz, Brian Dominguez, Elliot Thomasson, Cari Sloma, and Rob Christensen |
| EPI_ISL_1532118, EPI_ISL_1532120, EPI_ISL_1532121, EPI_ISL_1532122, EPI_ISL_1532123 | Good Samaritan Hospital | Santa Clara County Public Health Laboratory | Santa Clara County Public Health Department |
| EPI_ISL_1532124, EPI_ISL_1532125, EPI_ISL_1532126 | El Camino Hospital | Santa Clara County Public Health Laboratory | Santa Clara County Public Health Department |
| EPI_ISL_1532127, EPI_ISL_1532128, EPI_ISL_1532129, EPI_ISL_1532130, EPI_ISL_1532133, EPI_ISL_1532134, EPI_ISL_1532135, EPI_ISL_1532136, EPI_ISL_1532137 | Santa Clara Valley Medical Center | Santa Clara County Public Health Laboratory | Santa Clara County Public Health Department |
| EPI_ISL_1532139, EPI_ISL_1532140 | Santa Clara County Public Health Laboratory | Santa Clara County Public Health Laboratory | Santa Clara County Public Health Department |
| EPI_ISL_1532141, EPI_ISL_1532142, EPI_ISL_1532143 | Kaiser San Jose | Santa Clara County Public Health Laboratory | Santa Clara County Public Health Department |
| EPI_ISL_1532144 | Santa Clara County Public Health Laboratory | Santa Clara County Public Health Laboratory | Santa Clara County Public Health Department |
| EPI_ISL_1532236, EPI_ISL_1532237 | LESP Queretaro | Instituto de Diagnostico y Referencia Epidemiologicos (INDRE) | Claudia Wong-Arambula, Abril Rodriguez-Maldonado, Vanessa Rivero-Arredondo, Ariadna Medina-Benitez, Joaquin Quiroz-Mercado, Sergio Rangel-Guerrero, Natividad Cruz-Ortiz, Tatiana Nunez-Garcia, Gisela Barrera-Badillo, Lucia Hernandez-Rivas, Irma Lopez-Martinez, Ernesto Ramirez-Gonzalez. |
| EPI_ISL_1532238 | LESP Guanajuato | Instituto de Diagnostico y Referencia Epidemiologicos (INDRE) | Claudia Wong-Arambula, Abril Rodriguez-Maldonado, Vanessa Rivero-Arredondo, Ariadna Medina-Benitez, Joaquin Quiroz-Mercado, Sergio Rangel-Guerrero, Natividad Cruz-Ortiz, Tatiana Nunez-Garcia, Gisela Barrera-Badillo, Lucia Hernandez-Rivas, Irma Lopez-Martinez, Ernesto Ramirez-Gonzalez. |
| EPI_ISL_1532239 | LESP Queretaro | Instituto de Diagnostico y Referencia Epidemiologicos (INDRE) | Claudia Wong-Arambula, Abril Rodriguez-Maldonado, Vanessa Rivero-Arredondo, Ariadna Medina-Benitez, Joaquin Quiroz-Mercado, Sergio Rangel-Guerrero, Natividad Cruz-Ortiz, Tatiana Nunez-Garcia, Gisela Barrera-Badillo, Lucia Hernandez-Rivas, Irma Lopez-Martinez, Ernesto Ramirez-Gonzalez. |
| EPI_ISL_1532240, EPI_ISL_1532241, EPI_ISL_1532242 | LESP Guerrero | Instituto de Diagnostico y Referencia Epidemiologicos (INDRE) | Claudia Wong-Arambula, Abril Rodriguez-Maldonado, Vanessa Rivero-Arredondo, Ariadna Medina-Benitez, Joaquin Quiroz-Mercado, Sergio Rangel-Guerrero, Natividad Cruz-Ortiz, Tatiana Nunez-Garcia, Gisela Barrera-Badillo, Lucia Hernandez-Rivas, Irma Lopez-Martinez, Ernesto Ramirez-Gonzalez. |
| EPI_ISL_1532243, EPI_ISL_1532244, EPI_ISL_1532245 | LESP Baja California Sur | Instituto de Diagnostico y Referencia Epidemiologicos (INDRE) | Claudia Wong-Arambula, Abril Rodriguez-Maldonado, Vanessa Rivero-Arredondo, Ariadna Medina-Benitez, Joaquin Quiroz-Mercado, Sergio Rangel-Guerrero, Natividad Cruz-Ortiz, Tatiana Nunez-Garcia, Gisela Barrera-Badillo, Lucia Hernandez-Rivas, Irma Lopez-Martinez, Ernesto Ramirez-Gonzalez. |
| EPI_ISL_1532246, EPI_ISL_1532247 | LESP Chiapas | Instituto de Diagnostico y Referencia Epidemiologicos (INDRE) | Claudia Wong-Arambula, Abril Rodriguez-Maldonado, Vanessa Rivero-Arredondo, Ariadna Medina-Benitez, Joaquin Quiroz-Mercado, Sergio Rangel-Guerrero, Natividad Cruz-Ortiz, Tatiana Nunez-Garcia, Gisela Barrera-Badillo, Lucia Hernandez-Rivas, Irma Lopez-Martinez, Ernesto Ramirez-Gonzalez. |
| EPI_ISL_1532248 | LESP Yucatan | Instituto de Diagnostico y Referencia Epidemiologicos (INDRE) | Claudia Wong-Arambula, Abril Rodriguez-Maldonado, Vanessa Rivero-Arredondo, Ariadna Medina-Benitez, Joaquin Quiroz-Mercado, Sergio Rangel-Guerrero, Natividad Cruz-Ortiz, Tatiana Nunez-Garcia, Gisela Barrera-Badillo, Lucia Hernandez-Rivas, Irma Lopez-Martinez, Ernesto Ramirez-Gonzalez. |
| EPI_ISL_1532249 | LESP Estado de Mexico | Instituto de Diagnostico y Referencia Epidemiologicos (INDRE) | Claudia Wong-Arambula, Abril Rodriguez-Maldonado, Vanessa Rivero-Arredondo, Ariadna Medina-Benitez, Joaquin Quiroz-Mercado, Sergio Rangel-Guerrero, Natividad Cruz-Ortiz, Tatiana Nunez-Garcia, Gisela Barrera-Badillo, Lucia Hernandez-Rivas, Irma Lopez-Martinez, Ernesto Ramirez-Gonzalez. |
| EPI_ISL_1532250, EPI_ISL_1532251, EPI_ISL_1532252, EPI_ISL_1532253, EPI_ISL_1532254, EPI_ISL_1532255, EPI_ISL_1532256, EPI_ISL_1532257, EPI_ISL_1532258, EPI_ISL_1532259, EPI_ISL_1532260, EPI_ISL_1532261, EPI_ISL_1532262, EPI_ISL_1532263 | | | |
| see above | LESPZacatecas | Instituto de Diagnostico y Referencia Epidemiologicos (INDRE) | Claudia Wong-Arambula, Abril Rodriguez-Maldonado, Vanessa Rivero-Arredondo, Ariadna Medina-Benitez, Joaquin Quiroz-Mercado, Sergio Rangel-Guerrero, Natividad Cruz-Ortiz, Tatiana Nunez-Garcia, Gisela Barrera-Badillo, Lucia Hernandez-Rivas, Irma Lopez-Martinez, Ernesto Ramirez-Gonzalez. |
| EPI_ISL_1532264 | LESP Baja California | Instituto de Diagnostico y Referencia Epidemiologicos (INDRE) | Claudia Wong-Arambula, Abril Rodriguez-Maldonado, Vanessa Rivero-Arredondo, Ariadna Medina-Benitez, Joaquin Quiroz-Mercado, Sergio Rangel-Guerrero, Natividad Cruz-Ortiz, Tatiana Nunez-Garcia, Gisela Barrera-Badillo, Lucia Hernandez-Rivas, Irma Lopez-Martinez, Ernesto Ramirez-Gonzalez. |
| EPI_ISL_1532265 | LESP Ciudad de Mexico | Instituto de Diagnostico y Referencia Epidemiologicos (INDRE) | Claudia Wong-Arambula, Abril Rodriguez-Maldonado, Vanessa Rivero-Arredondo, Ariadna Medina-Benitez, Joaquin Quiroz-Mercado, Sergio Rangel-Guerrero, Natividad Cruz-Ortiz, Tatiana Nunez-Garcia, Gisela Barrera-Badillo, Lucia Hernandez-Rivas, Irma Lopez-Martinez, Ernesto Ramirez-Gonzalez. |
| EPI_ISL_1532266, EPI_ISL_1532267, EPI_ISL_1532268, EPI_ISL_1532269, EPI_ISL_1532270 | LESP Jalisco | Instituto de Diagnostico y Referencia Epidemiologicos (INDRE) | Claudia Wong-Arambula, Abril Rodriguez-Maldonado, Vanessa Rivero-Arredondo, Ariadna Medina-Benitez, Joaquin Quiroz-Mercado, Sergio Rangel-Guerrero, Natividad Cruz-Ortiz, Tatiana Nunez-Garcia, Gisela Barrera-Badillo, Lucia Hernandez-Rivas, Irma Lopez-Martinez, Ernesto Ramirez-Gonzalez. |
| EPI_ISL_1532271 | LESP Nuevo Leon | Instituto de Diagnostico y Referencia Epidemiologicos (INDRE) | Claudia Wong-Arambula, Abril Rodriguez-Maldonado, Vanessa Rivero-Arredondo, Ariadna Medina-Benitez, Joaquin Quiroz-Mercado, Sergio Rangel-Guerrero, Natividad Cruz-Ortiz, Tatiana Nunez-Garcia, Gisela Barrera-Badillo, Lucia Hernandez-Rivas, Irma Lopez-Martinez, Ernesto Ramirez-Gonzalez. |
| EPI_ISL_1532272, | LESP Queretaro | Instituto de Diagnostico y Referencia | Claudia Wong-Arambula, Abril Rodriguez-Maldonado, Vanessa Rivero-Arredondo, Ariadna Medina-Benitez, Joaquin Quiroz-Mercado, Sergio Rangel-Guerrero, Natividad Cruz-Ortiz, Tatiana Nunez-Garcia, Gisela Barrera-Badillo, Lucia Hernandez-Rivas, Irma Lopez-Martinez, Ernesto |

| | | | |
|---|---|---|---|
| EPI_ISL_1532273, EPI_ISL_1532274, EPI_ISL_1532275, EPI_ISL_1532276, EPI_ISL_1532277, EPI_ISL_1532278 | | Epidemiologicos (INDRE) | Ramirez-Gonzalez. |
| EPI_ISL_1532279 | LESP Ciudad de Mexico | Instituto de Diagnostico y Referencia Epidemiologicos (INDRE) | Claudia Wong-Arambula, Abril Rodriguez-Maldonado, Vanessa Rivero-Arredondo, Ariadna Medina-Benitez, Joaquin Quiroz-Mercado, Sergio Rangel-Guerrero, Natividad Cruz-Ortiz, Tatiana Nunez-Garcia, Gisela Barrera-Badillo, Lucia Hernandez-Rivas, Irma Lopez-Martinez, Ernesto Ramirez-Gonzalez. |
| EPI_ISL_1532280 | LESP Coahuila | Instituto de Diagnostico y Referencia Epidemiologicos (INDRE) | Claudia Wong-Arambula, Abril Rodriguez-Maldonado, Vanessa Rivero-Arredondo, Ariadna Medina-Benitez, Joaquin Quiroz-Mercado, Sergio Rangel-Guerrero, Natividad Cruz-Ortiz, Tatiana Nunez-Garcia, Gisela Barrera-Badillo, Lucia Hernandez-Rivas, Irma Lopez-Martinez, Ernesto Ramirez-Gonzalez. |
| EPI_ISL_1532281 | LESP Quintana Roo | Instituto de Diagnostico y Referencia Epidemiologicos (INDRE) | Claudia Wong-Arambula, Abril Rodriguez-Maldonado, Vanessa Rivero-Arredondo, Ariadna Medina-Benitez, Joaquin Quiroz-Mercado, Sergio Rangel-Guerrero, Natividad Cruz-Ortiz, Tatiana Nunez-Garcia, Gisela Barrera-Badillo, Lucia Hernandez-Rivas, Irma Lopez-Martinez, Ernesto Ramirez-Gonzalez. |
| EPI_ISL_1532306, EPI_ISL_1532307, EPI_ISL_1532308, EPI_ISL_1532309 | Arizona State University | Arizona State University | Rabia Maqsood, LaRinda A. Holland, Peter T. Skidmore, Nicholas J. Mellor, Joy M. Blain, Valerie Harris, Joshua LaBaer, Vel Murugan, Efrem S. Lim |
| EPI_ISL_1532312, EPI_ISL_1532313, EPI_ISL_1532314, EPI_ISL_1532315, EPI_ISL_1532316, EPI_ISL_1532317, EPI_ISL_1532318, EPI_ISL_1532319, EPI_ISL_1532320, EPI_ISL_1532321, EPI_ISL_1532322, EPI_ISL_1532323, EPI_ISL_1532324, EPI_ISL_1532325, EPI_ISL_1532326, EPI_ISL_1532327, EPI_ISL_1532328, EPI_ISL_1532329, EPI_ISL_1532330, EPI_ISL_1532331, EPI_ISL_1532332, EPI_ISL_1532333, EPI_ISL_1532334, EPI_ISL_1532335, EPI_ISL_1532336, EPI_ISL_1532337, EPI_ISL_1532338, EPI_ISL_1532339, EPI_ISL_1532340, EPI_ISL_1532341, EPI_ISL_1532342, EPI_ISL_1532343, EPI_ISL_1532344, EPI_ISL_1532345, EPI_ISL_1532346, EPI_ISL_1532347, EPI_ISL_1532348, EPI_ISL_1532349, EPI_ISL_1532350, EPI_ISL_1532351, EPI_ISL_1532352, EPI_ISL_1532353, EPI_ISL_1532354, EPI_ISL_1532355, EPI_ISL_1532356, EPI_ISL_1532357, EPI_ISL_1532358, EPI_ISL_1532359, EPI_ISL_1532360, EPI_ISL_1532361, EPI_ISL_1532362, EPI_ISL_1532363, EPI_ISL_1532364, EPI_ISL_1532365, EPI_ISL_1532366, EPI_ISL_1532367, EPI_ISL_1532368, EPI_ISL_1532369, EPI_ISL_1532370, EPI_ISL_1532371, EPI_ISL_1532372, EPI_ISL_1532373, EPI_ISL_1532374, EPI_ISL_1532375, EPI_ISL_1532376, EPI_ISL_1532377, EPI_ISL_1532378, EPI_ISL_1532379, EPI_ISL_1532380, EPI_ISL_1532381, EPI_ISL_1532382, EPI_ISL_1532383, EPI_ISL_1532384, EPI_ISL_1532385, EPI_ISL_1532386, EPI_ISL_1532387, EPI_ISL_1532388, EPI_ISL_1532389, EPI_ISL_1532390, EPI_ISL_1532391, EPI_ISL_1532392, EPI_ISL_1532393, EPI_ISL_1532394, EPI_ISL_1532395, EPI_ISL_1532396, EPI_ISL_1532397, EPI_ISL_1532398, EPI_ISL_1532399, EPI_ISL_1532400, EPI_ISL_1532401, EPI_ISL_1532402, EPI_ISL_1532403, EPI_ISL_1532527, EPI_ISL_1532528, EPI_ISL_1532529, EPI_ISL_1532530, EPI_ISL_1532531, EPI_ISL_1532532, EPI_ISL_1532533, EPI_ISL_1532534, EPI_ISL_1532535, EPI_ISL_1532536, EPI_ISL_1532537, EPI_ISL_1532538, EPI_ISL_1532539, EPI_ISL_1532540, EPI_ISL_1532541, EPI_ISL_1532542, EPI_ISL_1532543, EPI_ISL_1532544, EPI_ISL_1532545, EPI_ISL_1532546, EPI_ISL_1532547, EPI_ISL_1532548, EPI_ISL_1532549, EPI_ISL_1532550, EPI_ISL_1532551, EPI_ISL_1532552, EPI_ISL_1532553, EPI_ISL_1532554, EPI_ISL_1532555, EPI_ISL_1532556, EPI_ISL_1532557, EPI_ISL_1532558, EPI_ISL_1532559, EPI_ISL_1532560, EPI_ISL_1532561, EPI_ISL_1532562, EPI_ISL_1532563, EPI_ISL_1532564, EPI_ISL_1532565, EPI_ISL_1532566, EPI_ISL_1532567, EPI_ISL_1532568, EPI_ISL_1532569, EPI_ISL_1532570, EPI_ISL_1532571, EPI_ISL_1532572, EPI_ISL_1532573, EPI_ISL_1532574, EPI_ISL_1532575, EPI_ISL_1532576, EPI_ISL_1532577, EPI_ISL_1532578, EPI_ISL_1532579, EPI_ISL_1532580, EPI_ISL_1532581, EPI_ISL_1532582, EPI_ISL_1532583, EPI_ISL_1532584 | | | |
| see above | Arizona State University | Arizona State University | Peter T. Skidmore, LaRinda A. Holland, Rabia Maqsood, Emily A. Kaelin, Nicholas J. Mellor, Kristina Buss, Joy M. Blain, Neal W Woodbury, Valerie Harris, Joshua LaBaer, Vel Murugan, Efrem S. Lim |
| EPI_ISL_1532585, EPI_ISL_1532586, EPI_ISL_1532587, EPI_ISL_1532593, EPI_ISL_1532594, EPI_ISL_1532595, EPI_ISL_1532596, EPI_ISL_1532597, EPI_ISL_1532598, EPI_ISL_1532599, EPI_ISL_1532600, EPI_ISL_1532601, EPI_ISL_1532602, EPI_ISL_1532603, EPI_ISL_1532604, EPI_ISL_1532605, EPI_ISL_1532606, EPI_ISL_1532607, EPI_ISL_1532608, EPI_ISL_1532609, EPI_ISL_1532610, EPI_ISL_1532611, EPI_ISL_1532612, EPI_ISL_1532613, EPI_ISL_1532614, EPI_ISL_1532615, EPI_ISL_1532616, EPI_ISL_1532617, EPI_ISL_1532618, EPI_ISL_1532619, EPI_ISL_1532620, EPI_ISL_1532621, EPI_ISL_1532622, EPI_ISL_1532623, EPI_ISL_1532624, EPI_ISL_1532625, EPI_ISL_1532626, EPI_ISL_1532627, EPI_ISL_1532628, EPI_ISL_1532629, EPI_ISL_1532630, EPI_ISL_1532631, EPI_ISL_1532632, EPI_ISL_1532633, EPI_ISL_1532634, EPI_ISL_1532635, EPI_ISL_1532636, EPI_ISL_1532637, EPI_ISL_1532638, EPI_ISL_1532639, EPI_ISL_1532640, EPI_ISL_1532641, EPI_ISL_1532642, EPI_ISL_1532643, EPI_ISL_1532644, EPI_ISL_1532645, EPI_ISL_1532646, EPI_ISL_1532647, EPI_ISL_1532648, EPI_ISL_1532649, EPI_ISL_1532650, EPI_ISL_1532651 | | | |
| see above | Eurofins Diatherix | Hudsonalpha Genome Sequencing Center | Jane Grimwood, Melissa Williams, Lori H. Handley, Joshua Stough, Leslie Malone, Stefan Brzezinski, Ada Stewart, Teresa Jones, Jenell Webber, John Lovell, Jennifer Cart, and Jeremy Schmutz |
| EPI_ISL_1532695 | Wyoming Public Health Laboratory | Wyoming Public Health Laboratory | Jim Mildenberger, Wanda Manley, Noah Hull, Taylor Fearing, Lynette Gumbleton, Channing Weber, Ashley Norberg, Chayse Rowley, Marley Goetz, Brian Dominguez, Elliot Thomasson, Cari Sloma, and Rob Christensen |
| EPI_ISL_1532731, EPI_ISL_1532732, EPI_ISL_1532733, EPI_ISL_1532734, EPI_ISL_1532735, EPI_ISL_1532736, EPI_ISL_1532737, EPI_ISL_1532738, EPI_ISL_1532739, EPI_ISL_1532740, EPI_ISL_1532741, EPI_ISL_1532742, EPI_ISL_1532743, EPI_ISL_1532744, EPI_ISL_1532745, EPI_ISL_1532746, EPI_ISL_1532747, EPI_ISL_1532774, EPI_ISL_1532775, EPI_ISL_1532776, EPI_ISL_1532777, EPI_ISL_1532778, EPI_ISL_1532779, EPI_ISL_1532780, EPI_ISL_1532781, EPI_ISL_1532782, EPI_ISL_1532783, EPI_ISL_1532784, EPI_ISL_1532785 | | | |
| see above | NYU Langone Health | Departments of Pathology and Medicine, New York University School of Medicine | Adriana Heguy, Dacia Dimartino, Emily Guzman, Christian Marier, Peter Meyn, Sitharam Ramaswami, Gael Westby, Paul Zappile, Yutong Zhang, Paolo Cotzia, Guiqing Wang |
| EPI_ISL_1533424, EPI_ISL_1533425, EPI_ISL_1533428 | Ohio Department of Health Laboratory | Ohio Department of Health Laboratory | Holmes, Jennifer; Eric Brandt, Keoni Omura, Glen McGillivary, Caitlin McDonnell, Jade Mowery, Stephanie Mccracken, Tyler Payne, Kirtana Ramadugu, Erica Leasure, Brent Lee, Kelsey Florek, Heather Blankenship, Allison Black, Quanta Brown, and Tammy Bannerman |
| EPI_ISL_1533431, EPI_ISL_1533432, EPI_ISL_1533433, EPI_ISL_1533434, EPI_ISL_1533435 | Kaiser Permanente Washington Health Research Institute | Genomics and Discovery, Respiratory Viruses Branch, Division of Viral Diseases, Centers for Disease Control and Prevention | Yan Li, Ying Tao, Anna Kelleher, Jing Zhang, Anna Montmayeur, Brian Lynch, Krista Queen, Anna Uehara, Peter Cook, Han Jia Justin Ng, Rachel Marine, Clinton R. Paden, Haibin Wang, Mark Burroughs, Justin Lee, Adam Retchless, Suxiang Tong |
| EPI_ISL_1533436 | Atlanta VA Medical Center | Genomics and Discovery, Respiratory Viruses Branch, Division of Viral Diseases, Centers for Disease Control and Prevention | Yan Li, Ying Tao, Anna Kelleher, Jing Zhang, Anna Montmayeur, Brian Lynch, Krista Queen, Anna Uehara, Peter Cook, Han Jia Justin Ng, Rachel Marine, Clinton R. Paden, Haibin Wang, Mark Burroughs, Justin Lee, Adam Retchless, Suxiang Tong |
| EPI_ISL_1533437 | Kaiser Permanente Washington Health Research Institute | Genomics and Discovery, Respiratory Viruses Branch, Division of Viral Diseases, Centers for Disease Control and Prevention | Yan Li, Ying Tao, Anna Kelleher, Jing Zhang, Anna Montmayeur, Brian Lynch, Krista Queen, Anna Uehara, Peter Cook, Han Jia Justin Ng, Rachel Marine, Clinton R. Paden, Haibin Wang, Mark Burroughs, Justin Lee, Adam Retchless, Suxiang Tong |
| EPI_ISL_1533438 | Vanderbilt University Medical Center | Genomics and Discovery, Respiratory Viruses Branch, Division of Viral Diseases, Centers for Disease Control and Prevention | Yan Li, Ying Tao, Anna Kelleher, Jing Zhang, Anna Montmayeur, Brian Lynch, Krista Queen, Anna Uehara, Peter Cook, Han Jia Justin Ng, Rachel Marine, Clinton R. Paden, Haibin Wang, Mark Burroughs, Justin Lee, Adam Retchless, Suxiang Tong |
| EPI_ISL_1533439, EPI_ISL_1533440 | Kaiser Permanente Washington Health Research Institute | Genomics and Discovery, Respiratory Viruses Branch, Division of Viral Diseases, Centers for Disease Control and Prevention | Yan Li, Ying Tao, Anna Kelleher, Jing Zhang, Anna Montmayeur, Brian Lynch, Krista Queen, Anna Uehara, Peter Cook, Han Jia Justin Ng, Rachel Marine, Clinton R. Paden, Haibin Wang, Mark Burroughs, Justin Lee, Adam Retchless, Suxiang Tong |
| EPI_ISL_1533441, EPI_ISL_1533442 | Marshfield Clinic Research Institute | Genomics and Discovery, Respiratory Viruses Branch, Division of Viral Diseases, Centers for Disease Control and Prevention | Yan Li, Ying Tao, Anna Kelleher, Jing Zhang, Anna Montmayeur, Brian Lynch, Krista Queen, Anna Uehara, Peter Cook, Han Jia Justin Ng, Rachel Marine, Clinton R. Paden, Haibin Wang, Mark Burroughs, Justin Lee, Adam Retchless, Suxiang Tong |
| EPI_ISL_1533443, EPI_ISL_1533444 | Atlanta VA Medical Center | Genomics and Discovery, Respiratory Viruses Branch, Division of Viral Diseases, Centers for Disease Control and Prevention | Yan Li, Ying Tao, Anna Kelleher, Jing Zhang, Anna Montmayeur, Brian Lynch, Krista Queen, Anna Uehara, Peter Cook, Han Jia Justin Ng, Rachel Marine, Clinton R. Paden, Haibin Wang, Mark Burroughs, Justin Lee, Adam Retchless, Suxiang Tong |
| EPI_ISL_1533445 | Kaiser Permanente Washington Health Research Institute | Genomics and Discovery, Respiratory Viruses Branch, Division of Viral Diseases, Centers for Disease Control and Prevention | Yan Li, Ying Tao, Anna Kelleher, Jing Zhang, Anna Montmayeur, Brian Lynch, Krista Queen, Anna Uehara, Peter Cook, Han Jia Justin Ng, Rachel Marine, Clinton R. Paden, Haibin Wang, Mark Burroughs, Justin Lee, Adam Retchless, Suxiang Tong |
| EPI_ISL_1533446, EPI_ISL_1533447 | Atlanta VA Medical Center | Genomics and Discovery, Respiratory Viruses Branch, Division of Viral Diseases, Centers for Disease Control and Prevention | Yan Li, Ying Tao, Anna Kelleher, Jing Zhang, Anna Montmayeur, Brian Lynch, Krista Queen, Anna Uehara, Peter Cook, Han Jia Justin Ng, Rachel Marine, Clinton R. Paden, Haibin Wang, Mark Burroughs, Justin Lee, Adam Retchless, Suxiang Tong |
| EPI_ISL_1533448 | Kaiser Permanente Washington Health Research Institute | Genomics and Discovery, Respiratory Viruses Branch, Division of Viral Diseases, Centers for Disease Control and Prevention | Yan Li, Ying Tao, Anna Kelleher, Jing Zhang, Anna Montmayeur, Brian Lynch, Krista Queen, Anna Uehara, Peter Cook, Han Jia Justin Ng, Rachel Marine, Clinton R. Paden, Haibin Wang, Mark Burroughs, Justin Lee, Adam Retchless, Suxiang Tong |
| EPI_ISL_1533449, EPI_ISL_1533450 | Atlanta VA Medical Center | Genomics and Discovery, Respiratory Viruses Branch, Division of Viral Diseases, Centers for Disease Control and Prevention | Yan Li, Ying Tao, Anna Kelleher, Jing Zhang, Anna Montmayeur, Brian Lynch, Krista Queen, Anna Uehara, Peter Cook, Han Jia Justin Ng, Rachel Marine, Clinton R. Paden, Haibin Wang, Mark Burroughs, Justin Lee, Adam Retchless, Suxiang Tong |
| EPI_ISL_1533451 | Kaiser Permanente Washington Health Research Institute | Genomics and Discovery, Respiratory Viruses Branch, Division of Viral Diseases, Centers for Disease Control and Prevention | Yan Li, Ying Tao, Anna Kelleher, Jing Zhang, Anna Montmayeur, Brian Lynch, Krista Queen, Anna Uehara, Peter Cook, Han Jia Justin Ng, Rachel Marine, Clinton R. Paden, Haibin Wang, Mark Burroughs, Justin Lee, Adam Retchless, Suxiang Tong |
| EPI_ISL_1533452, EPI_ISL_1533453, EPI_ISL_1533454, EPI_ISL_1533455, EPI_ISL_1533456, EPI_ISL_1533457, EPI_ISL_1533458, EPI_ISL_1533459, EPI_ISL_1533460, EPI_ISL_1533461, EPI_ISL_1533462, EPI_ISL_1533463, EPI_ISL_1533464, EPI_ISL_1533465, EPI_ISL_1533466, EPI_ISL_1533467, EPI_ISL_1533468, EPI_ISL_1533469, EPI_ISL_1533470, EPI_ISL_1533471, EPI_ISL_1533472, EPI_ISL_1533473, EPI_ISL_1533474, EPI_ISL_1533475, EPI_ISL_1533476, EPI_ISL_1533477, EPI_ISL_1533478, EPI_ISL_1533479, EPI_ISL_1533480, EPI_ISL_1533481, EPI_ISL_1533482, EPI_ISL_1533483, EPI_ISL_1533484, EPI_ISL_1533485, EPI_ISL_1533486, EPI_ISL_1533487, EPI_ISL_1533488, EPI_ISL_1533489, EPI_ISL_1533490, EPI_ISL_1533491, EPI_ISL_1533492, EPI_ISL_1533493, EPI_ISL_1533494, EPI_ISL_1533495, EPI_ISL_1533496, EPI_ISL_1533497, EPI_ISL_1533498, EPI_ISL_1533499, EPI_ISL_1533500, EPI_ISL_1533501, EPI_ISL_1533502, EPI_ISL_1533503, EPI_ISL_1533504, EPI_ISL_1533505, EPI_ISL_1533506, EPI_ISL_1533507, EPI_ISL_1533508, EPI_ISL_1533509, EPI_ISL_1533510, EPI_ISL_1533511, EPI_ISL_1533512, EPI_ISL_1533513, EPI_ISL_1533514, EPI_ISL_1533515, EPI_ISL_1533516, EPI_ISL_1533517, EPI_ISL_1533518, EPI_ISL_1533519, EPI_ISL_1533520, EPI_ISL_1533521, EPI_ISL_1533522, EPI_ISL_1533523, EPI_ISL_1533524, EPI_ISL_1533525, EPI_ISL_1533526, EPI_ISL_1533527, EPI_ISL_1533528, EPI_ISL_1533529, EPI_ISL_1533530 | | | |
| see above | Atlanta VA Medical Center | Genomics and Discovery, Respiratory Viruses Branch, Division of Viral Diseases, Centers for Disease Control and Prevention | Yan Li, Ying Tao, Anna Kelleher, Jing Zhang, Anna Montmayeur, Brian Lynch, Krista Queen, Anna Uehara, Peter Cook, Han Jia Justin Ng, Rachel Marine, Clinton R. Paden, Haibin Wang, Mark Burroughs, Justin Lee, Adam Retchless, Suxiang Tong |
| EPI_ISL_1533531 | Kaiser Permanente Washington Health Research Institute | Genomics and Discovery, Respiratory Viruses Branch, Division of Viral Diseases, Centers for Disease Control and Prevention | Yan Li, Ying Tao, Anna Kelleher, Jing Zhang, Anna Montmayeur, Brian Lynch, Krista Queen, Anna Uehara, Peter Cook, Han Jia Justin Ng, Rachel Marine, Clinton R. Paden, Haibin Wang, Mark Burroughs, Justin Lee, Adam Retchless, Suxiang Tong |
| EPI_ISL_1533532 | Marshfield Clinic Research Institute | Genomics and Discovery, Respiratory Viruses Branch, Division of Viral Diseases, Centers for Disease Control and Prevention | Yan Li, Ying Tao, Anna Kelleher, Jing Zhang, Anna Montmayeur, Brian Lynch, Krista Queen, Anna Uehara, Peter Cook, Han Jia Justin Ng, Rachel Marine, Clinton R. Paden, Haibin Wang, Mark Burroughs, Justin Lee, Adam Retchless, Suxiang Tong |
| EPI_ISL_1533533 | Kaiser Permanente Washington Health Research Institute | Genomics and Discovery, Respiratory Viruses Branch, Division of Viral Diseases, Centers for Disease Control and Prevention | Yan Li, Ying Tao, Anna Kelleher, Jing Zhang, Anna Montmayeur, Brian Lynch, Krista Queen, Anna Uehara, Peter Cook, Han Jia Justin Ng, Rachel Marine, Clinton R. Paden, Haibin Wang, Mark Burroughs, Justin Lee, Adam Retchless, Suxiang Tong |
| EPI_ISL_1533534 | Marshfield Clinic Research Institute | Genomics and Discovery, Respiratory Viruses Branch, Division of Viral Diseases, Centers for Disease Control and Prevention | Yan Li, Ying Tao, Anna Kelleher, Jing Zhang, Anna Montmayeur, Brian Lynch, Krista Queen, Anna Uehara, Peter Cook, Han Jia Justin Ng, Rachel Marine, Clinton R. Paden, Haibin Wang, Mark Burroughs, Justin Lee, Adam Retchless, Suxiang Tong |
| EPI_ISL_1533535 | Kaiser Permanente Washington Health Research Institute | Genomics and Discovery, Respiratory Viruses Branch, Division of Viral Diseases, Centers for Disease Control and Prevention | Yan Li, Ying Tao, Anna Kelleher, Jing Zhang, Anna Montmayeur, Brian Lynch, Krista Queen, Anna Uehara, Peter Cook, Han Jia Justin Ng, Rachel Marine, Clinton R. Paden, Haibin Wang, Mark Burroughs, Justin Lee, Adam Retchless, Suxiang Tong |

| | | | |
|---|---|---|---|
| EPI_ISL_1533536, EPI_ISL_1533537, EPI_ISL_1533538 | Vanderbilt University Medical Center | Genomics and Discovery, Respiratory Viruses Branch, Division of Viral Diseases, Centers for Disease Control and Prevention | Yan Li, Ying Tao, Anna Kelleher, Jing Zhang, Anna Montmayeur, Brian Lynch, Krista Queen, Anna Uehara, Peter Cook, Han Jia Justin Ng, Rachel Marine, Clinton R. Paden, Haibin Wang, Mark Burroughs, Justin Lee, Adam Retchless, Suxiang Tong |

EPI_ISL_1533539, EPI_ISL_1533540, EPI_ISL_1533541, EPI_ISL_1533542, EPI_ISL_1533543, EPI_ISL_1533544, EPI_ISL_1533545, EPI_ISL_1533546, EPI_ISL_1533547, EPI_ISL_1533548, EPI_ISL_1533549, EPI_ISL_1533550, EPI_ISL_1533551, EPI_ISL_1533552, EPI_ISL_1533553, EPI_ISL_1533554, EPI_ISL_1533555, EPI_ISL_1533556, EPI_ISL_1533557, EPI_ISL_1533558, EPI_ISL_1533559, EPI_ISL_1533560, EPI_ISL_1533561, EPI_ISL_1533562, EPI_ISL_1533563

| | | | |
|---|---|---|---|
| see above | Atlanta VA Medical Center | Genomics and Discovery, Respiratory Viruses Branch, Division of Viral Diseases, Centers for Disease Control and Prevention | Yan Li, Ying Tao, Anna Kelleher, Jing Zhang, Anna Montmayeur, Brian Lynch, Krista Queen, Anna Uehara, Peter Cook, Han Jia Justin Ng, Rachel Marine, Clinton R. Paden, Haibin Wang, Mark Burroughs, Justin Lee, Adam Retchless, Suxiang Tong |

EPI_ISL_1533564, EPI_ISL_1533565, EPI_ISL_1533566, EPI_ISL_1533567, EPI_ISL_1533568, EPI_ISL_1533569, EPI_ISL_1533570, EPI_ISL_1533571, EPI_ISL_1533572, EPI_ISL_1533573, EPI_ISL_1533574, EPI_ISL_1533575, EPI_ISL_1533576, EPI_ISL_1533577, EPI_ISL_1533578, EPI_ISL_1533579, EPI_ISL_1533580, EPI_ISL_1533581, EPI_ISL_1533582, EPI_ISL_1533583, EPI_ISL_1533584, EPI_ISL_1533585, EPI_ISL_1533586, EPI_ISL_1533587, EPI_ISL_1533588, EPI_ISL_1533589, EPI_ISL_1533590, EPI_ISL_1533591, EPI_ISL_1533592, EPI_ISL_1533593, EPI_ISL_1533594, EPI_ISL_1533595, EPI_ISL_1533596, EPI_ISL_1533597, EPI_ISL_1533598, EPI_ISL_1533599, EPI_ISL_1533600, EPI_ISL_1533601, EPI_ISL_1533602, EPI_ISL_1533603, EPI_ISL_1533604, EPI_ISL_1533605, EPI_ISL_1533606, EPI_ISL_1533607, EPI_ISL_1533608

| | | | |
|---|---|---|---|
| see above | Kaiser Permanente Washington Health Research Institute | Genomics and Discovery, Respiratory Viruses Branch, Division of Viral Diseases, Centers for Disease Control and Prevention | Yan Li, Ying Tao, Anna Kelleher, Jing Zhang, Anna Montmayeur, Brian Lynch, Krista Queen, Anna Uehara, Peter Cook, Han Jia Justin Ng, Rachel Marine, Clinton R. Paden, Haibin Wang, Mark Burroughs, Justin Lee, Adam Retchless, Suxiang Tong |

EPI_ISL_1539244, EPI_ISL_1539245, EPI_ISL_1539252, EPI_ISL_1539264, EPI_ISL_1539271, EPI_ISL_1539273, EPI_ISL_1539274, EPI_ISL_1539276, EPI_ISL_1539277, EPI_ISL_1539278, EPI_ISL_1539291, EPI_ISL_1539292, EPI_ISL_1539294, EPI_ISL_1539300, EPI_ISL_1539307, EPI_ISL_1539315, EPI_ISL_1539317, EPI_ISL_1539320, EPI_ISL_1539321, EPI_ISL_1539337, EPI_ISL_1539345, EPI_ISL_1539348, EPI_ISL_1539350, EPI_ISL_1539351, EPI_ISL_1539352, EPI_ISL_1539355, EPI_ISL_1539356, EPI_ISL_1539361, EPI_ISL_1539365, EPI_ISL_1539366, EPI_ISL_1539367, EPI_ISL_1539369, EPI_ISL_1539375, EPI_ISL_1539376, EPI_ISL_1539378, EPI_ISL_1539382, EPI_ISL_1539386, EPI_ISL_1539387, EPI_ISL_1539389, EPI_ISL_1539395, EPI_ISL_1539397, EPI_ISL_1539398, EPI_ISL_1539399, EPI_ISL_1539400, EPI_ISL_1539402, EPI_ISL_1539403, EPI_ISL_1539405, EPI_ISL_1539406, EPI_ISL_1539408, EPI_ISL_1539409, EPI_ISL_1539410, EPI_ISL_1539411, EPI_ISL_1539412, EPI_ISL_1539414, EPI_ISL_1539415, EPI_ISL_1539416, EPI_ISL_1539418, EPI_ISL_1539420, EPI_ISL_1539421, EPI_ISL_1539422, EPI_ISL_1539425, EPI_ISL_1539426, EPI_ISL_1539427, EPI_ISL_1539431, EPI_ISL_1539432, EPI_ISL_1539433, EPI_ISL_1539435, EPI_ISL_1539437, EPI_ISL_1539439, EPI_ISL_1539440, EPI_ISL_1539442, EPI_ISL_1539443, EPI_ISL_1539445, EPI_ISL_1539447, EPI_ISL_1539448, EPI_ISL_1539449, EPI_ISL_1539450, EPI_ISL_1539451, EPI_ISL_1539452, EPI_ISL_1539453, EPI_ISL_1539454, EPI_ISL_1539455, EPI_ISL_1539456, EPI_ISL_1539458, EPI_ISL_1539459, EPI_ISL_1539460, EPI_ISL_1539461, EPI_ISL_1539462, EPI_ISL_1539466, EPI_ISL_1539467, EPI_ISL_1539468, EPI_ISL_1539469, EPI_ISL_1539470, EPI_ISL_1539471, EPI_ISL_1539472, EPI_ISL_1539473, EPI_ISL_1539474, EPI_ISL_1539475, EPI_ISL_1539476, EPI_ISL_1539477, EPI_ISL_1539478, EPI_ISL_1539479, EPI_ISL_1539480, EPI_ISL_1539481, EPI_ISL_1539482, EPI_ISL_1539483, EPI_ISL_1539484, EPI_ISL_1539485, EPI_ISL_1539486, EPI_ISL_1539487, EPI_ISL_1539488, EPI_ISL_1539489, EPI_ISL_1539490, EPI_ISL_1539491, EPI_ISL_1539492, EPI_ISL_1539493, EPI_ISL_1539494, EPI_ISL_1539495, EPI_ISL_1539496, EPI_ISL_1539498, EPI_ISL_1539500, EPI_ISL_1539501, EPI_ISL_1539502, EPI_ISL_1539503, EPI_ISL_1539504, EPI_ISL_1539505, EPI_ISL_1539508, EPI_ISL_1539509, EPI_ISL_1539510, EPI_ISL_1539511, EPI_ISL_1539512, EPI_ISL_1539513, EPI_ISL_1539514, EPI_ISL_1539515, EPI_ISL_1539516, EPI_ISL_1539517, EPI_ISL_1539518, EPI_ISL_1539519, EPI_ISL_1539520, EPI_ISL_1539521, EPI_ISL_1539522, EPI_ISL_1539523, EPI_ISL_1539524, EPI_ISL_1539525, EPI_ISL_1539526, EPI_ISL_1539527, EPI_ISL_1539528, EPI_ISL_1539529, EPI_ISL_1539530, EPI_ISL_1539531, EPI_ISL_1539532, EPI_ISL_1539534, EPI_ISL_1539535, EPI_ISL_1539536, EPI_ISL_1539537, EPI_ISL_1539539, EPI_ISL_1539540, EPI_ISL_1539542, EPI_ISL_1539543, EPI_ISL_1539544, EPI_ISL_1539545, EPI_ISL_1539546, EPI_ISL_1539547, EPI_ISL_1539548, EPI_ISL_1539549, EPI_ISL_1539550, EPI_ISL_1539551, EPI_ISL_1539553, EPI_ISL_1539555, EPI_ISL_1539556, EPI_ISL_1539557, EPI_ISL_1539558, EPI_ISL_1539559, EPI_ISL_1539560, EPI_ISL_1539561, EPI_ISL_1539562, EPI_ISL_1539563, EPI_ISL_1539564, EPI_ISL_1539565, EPI_ISL_1539566, EPI_ISL_1539567, EPI_ISL_1539568, EPI_ISL_1539569, EPI_ISL_1539570, EPI_ISL_1539571, EPI_ISL_1539572, EPI_ISL_1539573, EPI_ISL_1539574, EPI_ISL_1539575, EPI_ISL_1539576, EPI_ISL_1539577, EPI_ISL_1539578, EPI_ISL_1539579, EPI_ISL_1539580, EPI_ISL_1539581, EPI_ISL_1539582, EPI_ISL_1539583, EPI_ISL_1539584, EPI_ISL_1539585, EPI_ISL_1539586, EPI_ISL_1539587, EPI_ISL_1539588, EPI_ISL_1539589, EPI_ISL_1539590, EPI_ISL_1539591, EPI_ISL_1539592, EPI_ISL_1539593, EPI_ISL_1539594, EPI_ISL_1539595, EPI_ISL_1539596, EPI_ISL_1539597, EPI_ISL_1539598, EPI_ISL_1539599, EPI_ISL_1539600, EPI_ISL_1539601, EPI_ISL_1539602, EPI_ISL_1539603, EPI_ISL_1539604, EPI_ISL_1539605, EPI_ISL_1539606, EPI_ISL_1539607, EPI_ISL_1539608, EPI_ISL_1539609, EPI_ISL_1539610, EPI_ISL_1539613, EPI_ISL_1539614, EPI_ISL_1539615, EPI_ISL_1539616, EPI_ISL_1539617, EPI_ISL_1539618, EPI_ISL_1539619, EPI_ISL_1539620, EPI_ISL_1539621, EPI_ISL_1539623, EPI_ISL_1539624, EPI_ISL_1539625, EPI_ISL_1539626, EPI_ISL_1539627, EPI_ISL_1539629, EPI_ISL_1539630, EPI_ISL_1539631, EPI_ISL_1539632, EPI_ISL_1539633, EPI_ISL_1539634, EPI_ISL_1539635, EPI_ISL_1539637, EPI_ISL_1539645, EPI_ISL_1539646, EPI_ISL_1539647, EPI_ISL_1539648, EPI_ISL_1539649, EPI_ISL_1539650, EPI_ISL_1539651, EPI_ISL_1539652, EPI_ISL_1539653, EPI_ISL_1539654, EPI_ISL_1539655, EPI_ISL_1539656, EPI_ISL_1539657, EPI_ISL_1539658, EPI_ISL_1539659, EPI_ISL_1539660, EPI_ISL_1539661, EPI_ISL_1539662, EPI_ISL_1539663, EPI_ISL_1539664, EPI_ISL_1539665, EPI_ISL_1539667, EPI_ISL_1539668, EPI_ISL_1539669, EPI_ISL_1539672, EPI_ISL_1539673, EPI_ISL_1539674, EPI_ISL_1539675, EPI_ISL_1539676, EPI_ISL_1539678, EPI_ISL_1539679, EPI_ISL_1539680, EPI_ISL_1539681, EPI_ISL_1539682, EPI_ISL_1539683, EPI_ISL_1539685, EPI_ISL_1539687, EPI_ISL_1539688, EPI_ISL_1539690, EPI_ISL_1539692, EPI_ISL_1539693, EPI_ISL_1539694, EPI_ISL_1539698, EPI_ISL_1539699, EPI_ISL_1539701, EPI_ISL_1539703, EPI_ISL_1539704, EPI_ISL_1539705, EPI_ISL_1539707, EPI_ISL_1539708, EPI_ISL_1539709, EPI_ISL_1539710, EPI_ISL_1539711, EPI_ISL_1539712, EPI_ISL_1539713, EPI_ISL_1539715, EPI_ISL_1539716, EPI_ISL_1539717, EPI_ISL_1539718, EPI_ISL_1539719, EPI_ISL_1539722, EPI_ISL_1539723, EPI_ISL_1539724, EPI_ISL_1539725, EPI_ISL_1539726, EPI_ISL_1539727, EPI_ISL_1539728, EPI_ISL_1539730, EPI_ISL_1539731, EPI_ISL_1539732, EPI_ISL_1539733, EPI_ISL_1539734, EPI_ISL_1539735, EPI_ISL_1539736, EPI_ISL_1539737, EPI_ISL_1539738, EPI_ISL_1539739, EPI_ISL_1539740, EPI_ISL_1539741, EPI_ISL_1539742, EPI_ISL_1539743, EPI_ISL_1539744, EPI_ISL_1539745, EPI_ISL_1539746, EPI_ISL_1539749, EPI_ISL_1539750, EPI_ISL_1539751, EPI_ISL_1539752, EPI_ISL_1539753, EPI_ISL_1539754, EPI_ISL_1539755, EPI_ISL_1539756, EPI_ISL_1539757, EPI_ISL_1539758, EPI_ISL_1539760, EPI_ISL_1539762, EPI_ISL_1539764, EPI_ISL_1539765, EPI_ISL_1539766, EPI_ISL_1539767, EPI_ISL_1539769, EPI_ISL_1539770, EPI_ISL_1539771, EPI_ISL_1539774, EPI_ISL_1539775, EPI_ISL_1539776, EPI_ISL_1539777, EPI_ISL_1539778, EPI_ISL_1539779, EPI_ISL_1539780, EPI_ISL_1539781, EPI_ISL_1539783, EPI_ISL_1539785, EPI_ISL_1539786, EPI_ISL_1539787, EPI_ISL_1539789, EPI_ISL_1539790, EPI_ISL_1539791, EPI_ISL_1539792, EPI_ISL_1539793, EPI_ISL_1539794, EPI_ISL_1539795, EPI_ISL_1539796, EPI_ISL_1539797, EPI_ISL_1539798, EPI_ISL_1539799, EPI_ISL_1539802, EPI_ISL_1539803, EPI_ISL_1539804, EPI_ISL_1539806, EPI_ISL_1539807, EPI_ISL_1539808, EPI_ISL_1539809, EPI_ISL_1539810, EPI_ISL_1539811, EPI_ISL_1539812, EPI_ISL_1539813, EPI_ISL_1539814, EPI_ISL_1539815, EPI_ISL_1539817, EPI_ISL_1539818, EPI_ISL_1539819, EPI_ISL_1539821, EPI_ISL_1539822, EPI_ISL_1539823, EPI_ISL_1539824, EPI_ISL_1539825, EPI_ISL_1539826, EPI_ISL_1539827, EPI_ISL_1539828, EPI_ISL_1539829, EPI_ISL_1539831, EPI_ISL_1539832, EPI_ISL_1539834, EPI_ISL_1539835, EPI_ISL_1539836, EPI_ISL_1539837, EPI_ISL_1539838, EPI_ISL_1539841, EPI_ISL_1539842, EPI_ISL_1539843, EPI_ISL_1539844, EPI_ISL_1539845, EPI_ISL_1539846, EPI_ISL_1539847, EPI_ISL_1539849, EPI_ISL_1539850, EPI_ISL_1539851, EPI_ISL_1539852, EPI_ISL_1539853, EPI_ISL_1539854, EPI_ISL_1539855, EPI_ISL_1539856, EPI_ISL_1539857, EPI_ISL_1539859, EPI_ISL_1539860, EPI_ISL_1539861, EPI_ISL_1539862, EPI_ISL_1539863, EPI_ISL_1539865, EPI_ISL_1539866, EPI_ISL_1539867, EPI_ISL_1539868, EPI_ISL_1539869, EPI_ISL_1539871, EPI_ISL_1539872, EPI_ISL_1539873, EPI_ISL_1539874, EPI_ISL_1539875, EPI_ISL_1539876, EPI_ISL_1539877, EPI_ISL_1539878, EPI_ISL_1539879, EPI_ISL_1539880, EPI_ISL_1539881, EPI_ISL_1539882, EPI_ISL_1539883, EPI_ISL_1539884, EPI_ISL_1539886, EPI_ISL_1539887, EPI_ISL_1539888, EPI_ISL_1539890, EPI_ISL_1539891, EPI_ISL_1539892, EPI_ISL_1539893, EPI_ISL_1539894, EPI_ISL_1539895, EPI_ISL_1539896, EPI_ISL_1539897, EPI_ISL_1539898, EPI_ISL_1539899, EPI_ISL_1539900, EPI_ISL_1539902, EPI_ISL_1539903, EPI_ISL_1539905, EPI_ISL_1539906, EPI_ISL_1539907, EPI_ISL_1539908, EPI_ISL_1539910, EPI_ISL_1539912, EPI_ISL_1539913, EPI_ISL_1539915, EPI_ISL_1539917, EPI_ISL_1539918, EPI_ISL_1539919, EPI_ISL_1539921, EPI_ISL_1539922, EPI_ISL_1539923, EPI_ISL_1539924, EPI_ISL_1539925, EPI_ISL_1539926, EPI_ISL_1539927, EPI_ISL_1539928, EPI_ISL_1539929, EPI_ISL_1539930, EPI_ISL_1539931, EPI_ISL_1539932, EPI_ISL_1539933, EPI_ISL_1539934, EPI_ISL_1539935, EPI_ISL_1539936, EPI_ISL_1539937, EPI_ISL_1539939, EPI_ISL_1539940, EPI_ISL_1539941, EPI_ISL_1539942, EPI_ISL_1539944, EPI_ISL_1539945, EPI_ISL_1539947, EPI_ISL_1539948, EPI_ISL_1539949, EPI_ISL_1539951, EPI_ISL_1539952, EPI_ISL_1539954, EPI_ISL_1539955, EPI_ISL_1539956, EPI_ISL_1539957, EPI_ISL_1539958, EPI_ISL_1539959, EPI_ISL_1539960, EPI_ISL_1539961, EPI_ISL_1539962, EPI_ISL_1539963, EPI_ISL_1539964, EPI_ISL_1539965, EPI_ISL_1539966, EPI_ISL_1539967, EPI_ISL_1539968, EPI_ISL_1539969, EPI_ISL_1539970, EPI_ISL_1539971, EPI_ISL_1539972, EPI_ISL_1539973, EPI_ISL_1539974, EPI_ISL_1539975, EPI_ISL_1539976, EPI_ISL_1539977, EPI_ISL_1539978, EPI_ISL_1539979, EPI_ISL_1539980, EPI_ISL_1539981, EPI_ISL_1539982, EPI_ISL_1539983, EPI_ISL_1539984, EPI_ISL_1539985, EPI_ISL_1539986, EPI_ISL_1539987, EPI_ISL_1539988, EPI_ISL_1539989, EPI_ISL_1539990, EPI_ISL_1539991, EPI_ISL_1539992, EPI_ISL_1539993, EPI_ISL_1539994, EPI_ISL_1539995, EPI_ISL_1539996, EPI_ISL_1539997, EPI_ISL_1539999, EPI_ISL_1540000, EPI_ISL_1540001, EPI_ISL_1540002, EPI_ISL_1540003, EPI_ISL_1540004, EPI_ISL_1540005, EPI_ISL_1540006, EPI_ISL_1540007, EPI_ISL_1540008, EPI_ISL_1540009, EPI_ISL_1540010, EPI_ISL_1540011, EPI_ISL_1540012, EPI_ISL_1540013, EPI_ISL_1540014, EPI_ISL_1540015, EPI_ISL_1540016, EPI_ISL_1540017, EPI_ISL_1540018, EPI_ISL_1540019, EPI_ISL_1540020, EPI_ISL_1540023, EPI_ISL_1540024, EPI_ISL_1540025, EPI_ISL_1540026, EPI_ISL_1540081, EPI_ISL_1540082, EPI_ISL_1540083, EPI_ISL_1540085, EPI_ISL_1540086, EPI_ISL_1540087, EPI_ISL_1540088, EPI_ISL_1540089, EPI_ISL_1540090, EPI_ISL_1540091, EPI_ISL_1540092, EPI_ISL_1540094, EPI_ISL_1540095, EPI_ISL_1540096, EPI_ISL_1540097, EPI_ISL_1540098, EPI_ISL_1540101, EPI_ISL_1540103, EPI_ISL_1540104, EPI_ISL_1540105, EPI_ISL_1540106, EPI_ISL_1540107, EPI_ISL_1540108, EPI_ISL_1540110, EPI_ISL_1540111, EPI_ISL_1540112, EPI_ISL_1540113, EPI_ISL_1540114, EPI_ISL_1540115, EPI_ISL_1540116, EPI_ISL_1540117, EPI_ISL_1540118, EPI_ISL_1540127, EPI_ISL_1540128, EPI_ISL_1540129, EPI_ISL_1540130, EPI_ISL_1540131, EPI_ISL_1540133, EPI_ISL_1540134, EPI_ISL_1540135, EPI_ISL_1540136, EPI_ISL_1540137, EPI_ISL_1540138, EPI_ISL_1540139, EPI_ISL_1540140, EPI_ISL_1540141, EPI_ISL_1540142, EPI_ISL_1540143, EPI_ISL_1540144, EPI_ISL_1540145, EPI_ISL_1540146, EPI_ISL_1540147, EPI_ISL_1540148, EPI_ISL_1540149, EPI_ISL_1540150, EPI_ISL_1540151, EPI_ISL_1540152, EPI_ISL_1540153, EPI_ISL_1540154, EPI_ISL_1540155, EPI_ISL_1540156, EPI_ISL_1540157, EPI_ISL_1540158, EPI_ISL_1540159, EPI_ISL_1540160, EPI_ISL_1540161, EPI_ISL_1540162, EPI_ISL_1540164, EPI_ISL_1540165, EPI_ISL_1540167, EPI_ISL_1540168, EPI_ISL_1540169, EPI_ISL_1540170, EPI_ISL_1540172, EPI_ISL_1540173, EPI_ISL_1540174, EPI_ISL_1540175, EPI_ISL_1540176, EPI_ISL_1540177, EPI_ISL_1540179, EPI_ISL_1540180, EPI_ISL_1540182, EPI_ISL_1540184, EPI_ISL_1540186, EPI_ISL_1540188, EPI_ISL_1540192, EPI_ISL_1540194, EPI_ISL_1540195, EPI_ISL_1540197, EPI_ISL_1540199, EPI_ISL_1540200, EPI_ISL_1540201, EPI_ISL_1540202, EPI_ISL_1540204, EPI_ISL_1540207, EPI_ISL_1540208, EPI_ISL_1540209, EPI_ISL_1540210, EPI_ISL_1540211, EPI_ISL_1540212, EPI_ISL_1540213, EPI_ISL_1540214, EPI_ISL_1540215, EPI_ISL_1540216, EPI_ISL_1540217, EPI_ISL_1540220, EPI_ISL_1540221, EPI_ISL_1540222, EPI_ISL_1540223, EPI_ISL_1540224, EPI_ISL_1540226, EPI_ISL_1540227, EPI_ISL_1540228, EPI_ISL_1540229, EPI_ISL_1540230, EPI_ISL_1540231, EPI_ISL_1540233, EPI_ISL_1540234, EPI_ISL_1540235, EPI_ISL_1540236, EPI_ISL_1540238, EPI_ISL_1540240, EPI_ISL_1540241, EPI_ISL_1540242, EPI_ISL_1540243, EPI_ISL_1540245, EPI_ISL_1540246, EPI_ISL_1540247, EPI_ISL_1540248, EPI_ISL_1540249, EPI_ISL_1540250, EPI_ISL_1540251, EPI_ISL_1540252, EPI_ISL_1540253, EPI_ISL_1540254, EPI_ISL_1540255, EPI_ISL_1540256, EPI_ISL_1540257, EPI_ISL_1540258, EPI_ISL_1540259, EPI_ISL_1540260, EPI_ISL_1540261, EPI_ISL_1540262, EPI_ISL_1540263, EPI_ISL_1540264, EPI_ISL_1540265, EPI_ISL_1540266, EPI_ISL_1540267, EPI_ISL_1540268, EPI_ISL_1540269, EPI_ISL_1540270, EPI_ISL_1540272, EPI_ISL_1540273, EPI_ISL_1540274, EPI_ISL_1540275, EPI_ISL_1540276, EPI_ISL_1540277, EPI_ISL_1540278, EPI_ISL_1540279, EPI_ISL_1540281, EPI_ISL_1540283, EPI_ISL_1540284, EPI_ISL_1540285, EPI_ISL_1540286, EPI_ISL_1540287, EPI_ISL_1540288, EPI_ISL_1540289, EPI_ISL_1540290, EPI_ISL_1540291, EPI_ISL_1540292, EPI_ISL_1540293, EPI_ISL_1540294, EPI_ISL_1540295, EPI_ISL_1540296, EPI_ISL_1540297, EPI_ISL_1540298, EPI_ISL_1540299, EPI_ISL_1540300, EPI_ISL_1540301, EPI_ISL_1540303, EPI_ISL_1540304, EPI_ISL_1540305, EPI_ISL_1540307, EPI_ISL_1540308, EPI_ISL_1540309, EPI_ISL_1540310, EPI_ISL_1540311, EPI_ISL_1540313, EPI_ISL_1540315, EPI_ISL_1540316, EPI_ISL_1540317, EPI_ISL_1540318, EPI_ISL_1540320, EPI_ISL_1540321, EPI_ISL_1540322, EPI_ISL_1540323, EPI_ISL_1540324, EPI_ISL_1540325, EPI_ISL_1540326, EPI_ISL_1540327, EPI_ISL_1540328, EPI_ISL_1540329, EPI_ISL_1540330, EPI_ISL_1540331, EPI_ISL_1540333, EPI_ISL_1540334, EPI_ISL_1540335, EPI_ISL_1540337, EPI_ISL_1540338, EPI_ISL_1540339, EPI_ISL_1540340, EPI_ISL_1540341, EPI_ISL_1540342, EPI_ISL_1540343, EPI_ISL_1540344, EPI_ISL_1540347, EPI_ISL_1540348, EPI_ISL_1540349, EPI_ISL_1540350, EPI_ISL_1540351, EPI_ISL_1540352, EPI_ISL_1540353, EPI_ISL_1540354, EPI_ISL_1540355, EPI_ISL_1540356, EPI_ISL_1540357, EPI_ISL_1540358, EPI_ISL_1540359, EPI_ISL_1540360, EPI_ISL_1540361, EPI_ISL_1540362, EPI_ISL_1540364, EPI_ISL_1540365, EPI_ISL_1540366, EPI_ISL_1540367, EPI_ISL_1540368, EPI_ISL_1540369, EPI_ISL_1540370, EPI_ISL_1540373, EPI_ISL_1540375, EPI_ISL_1540376, EPI_ISL_1540378, EPI_ISL_1540379, EPI_ISL_1540380, EPI_ISL_1540382, EPI_ISL_1540383, EPI_ISL_1540384, EPI_ISL_1540385, EPI_ISL_1540387, EPI_ISL_1540389, EPI_ISL_1540390, EPI_ISL_1540391, EPI_ISL_1540393, EPI_ISL_1540394, EPI_ISL_1540396, EPI_ISL_1540397, EPI_ISL_1540398, EPI_ISL_1540399, EPI_ISL_1540400, EPI_ISL_1540402, EPI_ISL_1540403, EPI_ISL_1540404, EPI_ISL_1540405, EPI_ISL_1540406, EPI_ISL_1540407, EPI_ISL_1540408, EPI_ISL_1540409, EPI_ISL_1540410, EPI_ISL_1540411, EPI_ISL_1540413, EPI_ISL_1540415, EPI_ISL_1540416, EPI_ISL_1540417

| | | | |
|---|---|---|---|
| see above | Colorado Department of Public Health and Environment | Colorado Department of Public Health and Environment | Laura Bankers, Molly C. Hetherington-Rauth, Diana Ir, Shannon Ely, Shannon R. Matzinger, Sarah Elizabeth Totten, Emily A. Travanty |

EPI_ISL_1540418, EPI_ISL_1540420, EPI_ISL_1540421, EPI_ISL_1540422, EPI_ISL_1540424, EPI_ISL_1540425, EPI_ISL_1540426, EPI_ISL_1540427, EPI_ISL_1540429, EPI_ISL_1540430, EPI_ISL_1540431, EPI_ISL_1540432, EPI_ISL_1540433, EPI_ISL_1540434, EPI_ISL_1540435, EPI_ISL_1540436, EPI_ISL_1540437, EPI_ISL_1540438, EPI_ISL_1540439, EPI_ISL_1540440, EPI_ISL_1540441, EPI_ISL_1540442, EPI_ISL_1540443, EPI_ISL_1540444

| | | | |
|---|---|---|---|
| see above | Wyoming Public Health Laboratory | Wyoming Public Health Laboratory | Jim Mildenberger, Wanda Manley, Noah Hull, Taylor Fearing, Lynette Gumbleton, Channing Weber, Ashley Norberg, Chayse Rowley, Marley Goetz, Brian Dominguez, Elliot Thomasson, Cari Sloma, and Rob Christensen |

EPI_ISL_1540458, EPI_ISL_1540459, EPI_ISL_1540460, EPI_ISL_1540461, EPI_ISL_1540462, EPI_ISL_1540463, EPI_ISL_1540464, EPI_ISL_1540465, EPI_ISL_1540466, EPI_ISL_1540467, EPI_ISL_1540468, EPI_ISL_1540469, EPI_ISL_1540470, EPI_ISL_1540471, EPI_ISL_1540472, EPI_ISL_1540473, EPI_ISL_1540474, EPI_ISL_1540475, EPI_ISL_1540476, EPI_ISL_1540477, EPI_ISL_1540478, EPI_ISL_1540479, EPI_ISL_1540480, EPI_ISL_1540481, EPI_ISL_1540482, EPI_ISL_1540483, EPI_ISL_1540484, EPI_ISL_1540485, EPI_ISL_1540486, EPI_ISL_1540489, EPI_ISL_1540490, EPI_ISL_1540492, EPI_ISL_1540493, EPI_ISL_1540494, EPI_ISL_1540495, EPI_ISL_1540496, EPI_ISL_1540497, EPI_ISL_1540498, EPI_ISL_1540499, EPI_ISL_1540500

| | | | |
|---|---|---|---|
| see above | Virginia Division of Consolidated Laboratory Services | Virginia Division of Consolidated Laboratory Services | Virginia DCLS |

EPI_ISL_1540714, EPI_ISL_1540715, EPI_ISL_1540716, EPI_ISL_1540717, EPI_ISL_1540718, EPI_ISL_1540719, EPI_ISL_1540720, EPI_ISL_1540721, EPI_ISL_1540722, EPI_ISL_1540723, EPI_ISL_1540724, EPI_ISL_1540725, EPI_ISL_1540726, EPI_ISL_1540727, EPI_ISL_1540728, EPI_ISL_1540729, EPI_ISL_1540730, EPI_ISL_1540731, EPI_ISL_1540733, EPI_ISL_1540734, EPI_ISL_1540735, EPI_ISL_1540736, EPI_ISL_1540737, EPI_ISL_1540738, EPI_ISL_1540739, EPI_ISL_1540740, EPI_ISL_1540741, EPI_ISL_1540742, EPI_ISL_1540743, EPI_ISL_1540744, EPI_ISL_1540745, EPI_ISL_1540746, EPI_ISL_1540747, EPI_ISL_1540748, EPI_ISL_1540749, EPI_ISL_1540750, EPI_ISL_1540751, EPI_ISL_1540752, EPI_ISL_1540753, EPI_ISL_1540754, EPI_ISL_1540755, EPI_ISL_1540756, EPI_ISL_1540757, EPI_ISL_1540758, EPI_ISL_1540759, EPI_ISL_1540760, EPI_ISL_1540761, EPI_ISL_1540762, EPI_ISL_1540763, EPI_ISL_1540764, EPI_ISL_1540765, EPI_ISL_1540766, EPI_ISL_1540767, EPI_ISL_1540768, EPI_ISL_1540769, EPI_ISL_1540770, EPI_ISL_1540771, EPI_ISL_1540772, EPI_ISL_1540773, EPI_ISL_1540774, EPI_ISL_1540775, EPI_ISL_1540776, EPI_ISL_1540777, EPI_ISL_1540778, EPI_ISL_1540779, EPI_ISL_1540780, EPI_ISL_1540781, EPI_ISL_1540782, EPI_ISL_1540783, EPI_ISL_1540784, EPI_ISL_1540785, EPI_ISL_1540786, EPI_ISL_1540787, EPI_ISL_1540788, EPI_ISL_1540789, EPI_ISL_1540790, EPI_ISL_1540791, EPI_ISL_1540792, EPI_ISL_1540793, EPI_ISL_1540794, EPI_ISL_1540795, EPI_ISL_1540796, EPI_ISL_1540797, EPI_ISL_1540798, EPI_ISL_1540799, EPI_ISL_1540800, EPI_ISL_1540801, EPI_ISL_1540802, EPI_ISL_1540803, EPI_ISL_1540804, EPI_ISL_1540805, EPI_ISL_1540806, EPI_ISL_1540807, EPI_ISL_1540808, EPI_ISL_1540809, EPI_ISL_1540810, EPI_ISL_1540811, EPI_ISL_1540812, EPI_ISL_1540813, EPI_ISL_1540814, EPI_ISL_1540815, EPI_ISL_1540816, EPI_ISL_1540817, EPI_ISL_1540818, EPI_ISL_1540819, EPI_ISL_1540820, EPI_ISL_1540821, EPI_ISL_1540822, EPI_ISL_1540823, EPI_ISL_1540824, EPI_ISL_1540825, EPI_ISL_1540826, EPI_ISL_1540827, EPI_ISL_1540828, EPI_ISL_1540829, EPI_ISL_1540830, EPI_ISL_1540831, EPI_ISL_1540832, EPI_ISL_1540833, EPI_ISL_1540834, EPI_ISL_1540835, EPI_ISL_1540836, EPI_ISL_1540837, EPI_ISL_1540838, EPI_ISL_1540839, EPI_ISL_1540840, EPI_ISL_1540841, EPI_ISL_1540842, EPI_ISL_1540843, EPI_ISL_1540844, EPI_ISL_1540845, EPI_ISL_1540846, EPI_ISL_1540847, EPI_ISL_1540848, EPI_ISL_1540849, EPI_ISL_1540850, EPI_ISL_1540851, EPI_ISL_1540852

| | | | |
|---|---|---|---|
| see above | The Jackson Laboratory | The Jackson Laboratory | Bergeron D, Renzette N, Adams M, Omerza G, Kelly K, Long J, Li L |

EPI_ISL_1540853, EPI_ISL_1540854, EPI_ISL_1540855, EPI_ISL_1540856, EPI_ISL_1540857, EPI_ISL_1540858, EPI_ISL_1540859, EPI_ISL_1540860, EPI_ISL_1540861, EPI_ISL_1540862, EPI_ISL_1540863, EPI_ISL_1540864, EPI_ISL_1540865, EPI_ISL_1540866, EPI_ISL_1540867, EPI_ISL_1540868, EPI_ISL_1540869, EPI_ISL_1540871, EPI_ISL_1540872, EPI_ISL_1540873, EPI_ISL_1540874, EPI_ISL_1540875, EPI_ISL_1540877, EPI_ISL_1540878, EPI_ISL_1540879, EPI_ISL_1540880, EPI_ISL_1540882, EPI_ISL_1540883, EPI_ISL_1540884, EPI_ISL_1540885, EPI_ISL_1540886, EPI_ISL_1540887, EPI_ISL_1540888, EPI_ISL_1540889, EPI_ISL_1540890, EPI_ISL_1540892, EPI_ISL_1540893, EPI_ISL_1540894, EPI_ISL_1540895, EPI_ISL_1540896, EPI_ISL_1540897, EPI_ISL_1540898, EPI_ISL_1540899, EPI_ISL_1540900, EPI_ISL_1540901, EPI_ISL_1540903, EPI_ISL_1540904, EPI_ISL_1540905, EPI_ISL_1540906, EPI_ISL_1540907, EPI_ISL_1540908, EPI_ISL_1540909, EPI_ISL_1540910, EPI_ISL_1540911, EPI_ISL_1540913, EPI_ISL_1540915, EPI_ISL_1540916, EPI_ISL_1540917, EPI_ISL_1540921, EPI_ISL_1540922, EPI_ISL_1540924, EPI_ISL_1540925, EPI_ISL_1540926, EPI_ISL_1540927, EPI_ISL_1540931, EPI_ISL_1540932, EPI_ISL_1540934, EPI_ISL_1540936, EPI_ISL_1540938, EPI_ISL_1540939, EPI_ISL_1540940, EPI_ISL_1540941, EPI_ISL_1540942, EPI_ISL_1540943, EPI_ISL_1540944, EPI_ISL_1540945, EPI_ISL_1540946, EPI_ISL_1540947, EPI_ISL_1540948, EPI_ISL_1540950, EPI_ISL_1540951, EPI_ISL_1540952, EPI_ISL_1540953, EPI_ISL_1540954, EPI_ISL_1540955, EPI_ISL_1540956, EPI_ISL_1540957, EPI_ISL_1540958, EPI_ISL_1540960, EPI_ISL_1540961, EPI_ISL_1540962, EPI_ISL_1540963, EPI_ISL_1540964, EPI_ISL_1540965, EPI_ISL_1540966, EPI_ISL_1540967, EPI_ISL_1540968, EPI_ISL_1540969, EPI_ISL_1540970, EPI_ISL_1540972, EPI_ISL_1540973, EPI_ISL_1540976, EPI_ISL_1540978, EPI_ISL_1540979, EPI_ISL_1540980, EPI_ISL_1540981, EPI_ISL_1540984, EPI_ISL_1540986, EPI_ISL_1540988, EPI_ISL_1540990, EPI_ISL_1540992, EPI_ISL_1540994, EPI_ISL_1541073, EPI_ISL_1541074, EPI_ISL_1541078, EPI_ISL_1541080, EPI_ISL_1541081, EPI_ISL_1541082, EPI_ISL_1541083, EPI_ISL_1541084, EPI_ISL_1541085, EPI_ISL_1541087, EPI_ISL_1541088, EPI_ISL_1541089, EPI_ISL_1541090, EPI_ISL_1541091, EPI_ISL_1541092, EPI_ISL_1541093, EPI_ISL_1541094, EPI_ISL_1541095, EPI_ISL_1541097, EPI_ISL_1541098, EPI_ISL_1541099, EPI_ISL_1541100, EPI_ISL_1541101, EPI_ISL_1541102, EPI_ISL_1541103, EPI_ISL_1541104, EPI_ISL_1541105, EPI_ISL_1541106, EPI_ISL_1541107, EPI_ISL_1541108, EPI_ISL_1541109, EPI_ISL_1541111, EPI_ISL_1541112, EPI_ISL_1541113, EPI_ISL_1541114, EPI_ISL_1541115, EPI_ISL_1541116, EPI_ISL_1541117, EPI_ISL_1541118, EPI_ISL_1541119, EPI_ISL_1541120, EPI_ISL_1541121, EPI_ISL_1541122, EPI_ISL_1541123, EPI_ISL_1541124, EPI_ISL_1541125, EPI_ISL_1541126, EPI_ISL_1541127, EPI_ISL_1541128, EPI_ISL_1541130, EPI_ISL_1541132, EPI_ISL_1541133, EPI_ISL_1541134, EPI_ISL_1541135, EPI_ISL_1541136, EPI_ISL_1541137, EPI_ISL_1541138, EPI_ISL_1541139, EPI_ISL_1541140, EPI_ISL_1541143, EPI_ISL_1541144, EPI_ISL_1541146, EPI_ISL_1541147, EPI_ISL_1541148, EPI_ISL_1541150, EPI_ISL_1541151, EPI_ISL_1541152, EPI_ISL_1541153, EPI_ISL_1541154, EPI_ISL_1541155, EPI_ISL_1541156, EPI_ISL_1541157, EPI_ISL_1541158, EPI_ISL_1541159, EPI_ISL_1541160, EPI_ISL_1541161, EPI_ISL_1541162, EPI_ISL_1541163, EPI_ISL_1541164, EPI_ISL_1541165, EPI_ISL_1541166, EPI_ISL_1541167, EPI_ISL_1541168, EPI_ISL_1541169, EPI_ISL_1541170, EPI_ISL_1541171, EPI_ISL_1541172, EPI_ISL_1541173, EPI_ISL_1541174, EPI_ISL_1541175, EPI_ISL_1541176, EPI_ISL_1541177, EPI_ISL_1541178, EPI_ISL_1541179, EPI_ISL_1541180, EPI_ISL_1541181, EPI_ISL_1541182, EPI_ISL_1541183, EPI_ISL_1541184, EPI_ISL_1541185, EPI_ISL_1541189, EPI_ISL_1541190, EPI_ISL_1541191, EPI_ISL_1541192, EPI_ISL_1541193, EPI_ISL_1541194, EPI_ISL_1541196, EPI_ISL_1541197, EPI_ISL_1541198, EPI_ISL_1541199, EPI_ISL_1541200, EPI_ISL_1541201, EPI_ISL_1541202, EPI_ISL_1541203, EPI_ISL_1541205, EPI_ISL_1541206, EPI_ISL_1541208, EPI_ISL_1541209

| | | | |
|---|---|---|---|
| see above | Pathology and Laboratory Medicine Institute, Cleveland Clinic, Ohio, USA | Pathology and Laboratory Medicine Institute, Cleveland Clinic, Ohio, USA | Joy Nakitandwe, Zheng Jin Tu, Jay Brock, Yu-Wei Cheng, Gary Procop, Daniel Rhoads, Daniel H. Farkas, David Bosler |

EPI_ISL_1541210, EPI_ISL_1541211, EPI_ISL_1541212, EPI_ISL_1541213, EPI_ISL_1541215, EPI_ISL_1541216, EPI_ISL_1541217, EPI_ISL_1541218, EPI_ISL_1541219, EPI_ISL_1541220, EPI_ISL_1541221, EPI_ISL_1541223, EPI_ISL_1541224, EPI_ISL_1541226, EPI_ISL_1541230, EPI_ISL_1541231, EPI_ISL_1541232, EPI_ISL_1541233, EPI_ISL_1541234, EPI_ISL_1541235, EPI_ISL_1541236, EPI_ISL_1541237, EPI_ISL_1541238, EPI_ISL_1541239, EPI_ISL_1541240, EPI_ISL_1541241, EPI_ISL_1541243, EPI_ISL_1541244, EPI_ISL_1541245, EPI_ISL_1541246, EPI_ISL_1541249, EPI_ISL_1541250, EPI_ISL_1541251, EPI_ISL_1541252, EPI_ISL_1541253, EPI_ISL_1541254, EPI_ISL_1541255, EPI_ISL_1541256, EPI_ISL_1541257, EPI_ISL_1541258, EPI_ISL_1541259, EPI_ISL_1541260, EPI_ISL_1541261, EPI_ISL_1541262, EPI_ISL_1541265, EPI_ISL_1541266, EPI_ISL_1541270, EPI_ISL_1541271, EPI_ISL_1541272, EPI_ISL_1541273, EPI_ISL_1541274, EPI_ISL_1541277, EPI_ISL_1541278, EPI_ISL_1541279, EPI_ISL_1541280, EPI_ISL_1541281, EPI_ISL_1541282, EPI_ISL_1541283, EPI_ISL_1541284, EPI_ISL_1541285, EPI_ISL_1541286, EPI_ISL_1541287, EPI_ISL_1541288, EPI_ISL_1541289, EPI_ISL_1541290, EPI_ISL_1541291, EPI_ISL_1541292, EPI_ISL_1541293, EPI_ISL_1541294, EPI_ISL_1541295, EPI_ISL_1541296, EPI_ISL_1541298, EPI_ISL_1541300, EPI_ISL_1541301, EPI_ISL_1541302, EPI_ISL_1541303, EPI_ISL_1541304, EPI_ISL_1541305, EPI_ISL_1541306, EPI_ISL_1541307, EPI_ISL_1541308, EPI_ISL_1541309, EPI_ISL_1541310, EPI_ISL_1541311, EPI_ISL_1541312, EPI_ISL_1541313, EPI_ISL_1541314, EPI_ISL_1541315, EPI_ISL_1541318, EPI_ISL_1541319, EPI_ISL_1541320, EPI_ISL_1541322, EPI_ISL_1541324, EPI_ISL_1541325, EPI_ISL_1541326, EPI_ISL_1541327, EPI_ISL_1541328, EPI_ISL_1541329, EPI_ISL_1541330, EPI_ISL_1541331, EPI_ISL_1541332, EPI_ISL_1541333, EPI_ISL_1541334, EPI_ISL_1541336, EPI_ISL_1541337, EPI_ISL_1541339, EPI_ISL_1541340, EPI_ISL_1541342, EPI_ISL_1541344, EPI_ISL_1541345, EPI_ISL_1541346, EPI_ISL_1541347, EPI_ISL_1541348, EPI_ISL_1541349, EPI_ISL_1541351, EPI_ISL_1541352, EPI_ISL_1541353, EPI_ISL_1541354, EPI_ISL_1541355, EPI_ISL_1541356, EPI_ISL_1541357, EPI_ISL_1541358, EPI_ISL_1541359, EPI_ISL_1541360, EPI_ISL_1541362, EPI_ISL_1541363, EPI_ISL_1541364, EPI_ISL_1541365, EPI_ISL_1541366, EPI_ISL_1541367, EPI_ISL_1541368, EPI_ISL_1541369, EPI_ISL_1541370, EPI_ISL_1541371, EPI_ISL_1541372, EPI_ISL_1541373, EPI_ISL_1541375, EPI_ISL_1541377, EPI_ISL_1541378, EPI_ISL_1541379, EPI_ISL_1541380, EPI_ISL_1541382, EPI_ISL_1541383, EPI_ISL_1541384, EPI_ISL_1541385, EPI_ISL_1541386, EPI_ISL_1541387, EPI_ISL_1541388, EPI_ISL_1541389, EPI_ISL_1541391, EPI_ISL_1541392, EPI_ISL_1541393, EPI_ISL_1541394, EPI_ISL_1541395, EPI_ISL_1541396, EPI_ISL_1541397, EPI_ISL_1541398, EPI_ISL_1541400, EPI_ISL_1541401, EPI_ISL_1541402, EPI_ISL_1541403, EPI_ISL_1541404, EPI_ISL_1541405, EPI_ISL_1541406, EPI_ISL_1541407, EPI_ISL_1541408, EPI_ISL_1541409, EPI_ISL_1541410, EPI_ISL_1541411, EPI_ISL_1541412, EPI_ISL_1541413, EPI_ISL_1541414, EPI_ISL_1541416, EPI_ISL_1541417, EPI_ISL_1541418, EPI_ISL_1541419, EPI_ISL_1541420, EPI_ISL_1541422, EPI_ISL_1541423, EPI_ISL_1541424, EPI_ISL_1541426, EPI_ISL_1541427, EPI_ISL_1541428, EPI_ISL_1541429, EPI_ISL_1541430, EPI_ISL_1541431, EPI_ISL_1541432, EPI_ISL_1541433, EPI_ISL_1541434, EPI_ISL_1541435, EPI_ISL_1541436, EPI_ISL_1541437, EPI_ISL_1541438, EPI_ISL_1541439, EPI_ISL_1541440, EPI_ISL_1541441, EPI_ISL_1541442, EPI_ISL_1541443, EPI_ISL_1541444, EPI_ISL_1541445, EPI_ISL_1541446, EPI_ISL_1541447, EPI_ISL_1541448, EPI_ISL_1541449, EPI_ISL_1541450, EPI_ISL_1541452, EPI_ISL_1541453, EPI_ISL_1541454, EPI_ISL_1541456, EPI_ISL_1541457, EPI_ISL_1541458, EPI_ISL_1541459, EPI_ISL_1541460, EPI_ISL_1541461, EPI_ISL_1541462, EPI_ISL_1541463, EPI_ISL_1541464, EPI_ISL_1541465, EPI_ISL_1541466, EPI_ISL_1541468, EPI_ISL_1541469, EPI_ISL_1541470, EPI_ISL_1541471, EPI_ISL_1541472, EPI_ISL_1541473, EPI_ISL_1541474, EPI_ISL_1541475, EPI_ISL_1541476, EPI_ISL_1541477, EPI_ISL_1541478, EPI_ISL_1541479, EPI_ISL_1541481, EPI_ISL_1541482, EPI_ISL_1541483, EPI_ISL_1541484, EPI_ISL_1541485, EPI_ISL_1541486, EPI_ISL_1541487, EPI_ISL_1541488, EPI_ISL_1541490, EPI_ISL_1541491, EPI_ISL_1541492, EPI_ISL_1541493, EPI_ISL_1541494, EPI_ISL_1541496, EPI_ISL_1541498, EPI_ISL_1541499, EPI_ISL_1541500, EPI_ISL_1541501, EPI_ISL_1541503, EPI_ISL_1541504, EPI_ISL_1541505, EPI_ISL_1541506, EPI_ISL_1541507, EPI_ISL_1541508, EPI_ISL_1541509, EPI_ISL_1541510, EPI_ISL_1541511, EPI_ISL_1541512, EPI_ISL_1541513, EPI_ISL_1541514, EPI_ISL_1541515, EPI_ISL_1541516, EPI_ISL_1541518, EPI_ISL_1541519, EPI_ISL_1541520, EPI_ISL_1541521, EPI_ISL_1541522, EPI_ISL_1541523, EPI_ISL_1541524, EPI_ISL_1541525, EPI_ISL_1541526, EPI_ISL_1541527, EPI_ISL_1541529, EPI_ISL_1541530, EPI_ISL_1541531

| | | | |
|---|---|---|---|
| see above | OHSU Lab Services Molecular Microbiology Lab | Oregon SARS-CoV-2 Genome Sequencing Center | Brendan L. O'Connell, Sally Grindstaff, Kayla Carter, Sonia Acharya, Ruth V. Nichols, Alec J. Hirsch, Donna Hansel, Guang Fan, Xuan Qin, Daniel N. Streblow, William B. Messer, Andrew C. Adey, Benjamin N. Bimber, Brian J. O’Roak |
| EPI_ISL_1541532, EPI_ISL_1541533 | OHSU Lab Services Molecular Microbiology Lab | Oregon SARS-CoV-2 Genome Sequencing Center | Brendan L. O'Connell, Sally Grindstaff, Sonia Acharya, Cierra LaBlanc, Kayla Carter, Ruth V. Nichols, Alec J. Hirsch, Donna Hansel, Guang Fan, Xuan Qin, Daniel N. Streblow, William B. Messer, Andrew C. Adey, Benjamin N. Bimber, Brian J. O’Roak |

EPI_ISL_1541534, EPI_ISL_1541535, EPI_ISL_1541536, EPI_ISL_1541537, EPI_ISL_1541538, EPI_ISL_1541539, EPI_ISL_1541540, EPI_ISL_1541541, EPI_ISL_1541542, EPI_ISL_1541543, EPI_ISL_1541544, EPI_ISL_1541545, EPI_ISL_1541546, EPI_ISL_1541547, EPI_ISL_1541549, EPI_ISL_1541550, EPI_ISL_1541552, EPI_ISL_1541554, EPI_ISL_1541555, EPI_ISL_1541557, EPI_ISL_1541558, EPI_ISL_1541559, EPI_ISL_1541560, EPI_ISL_1541561, EPI_ISL_1541562, EPI_ISL_1541563, EPI_ISL_1541565, EPI_ISL_1541566, EPI_ISL_1541567, EPI_ISL_1541568, EPI_ISL_1541569, EPI_ISL_1541570, EPI_ISL_1541571, EPI_ISL_1541572, EPI_ISL_1541574, EPI_ISL_1541575, EPI_ISL_1541576, EPI_ISL_1541577, EPI_ISL_1541578, EPI_ISL_1541579, EPI_ISL_1541580, EPI_ISL_1541581, EPI_ISL_1541582, EPI_ISL_1541583,

| | | | |
|---|---|---|---|
| EPI_ISL_1541584, EPI_ISL_1541585, EPI_ISL_1541586, EPI_ISL_1541588, EPI_ISL_1541589, EPI_ISL_1541590, EPI_ISL_1541591, EPI_ISL_1541592, EPI_ISL_1541593, EPI_ISL_1541594, EPI_ISL_1541595, EPI_ISL_1541596, EPI_ISL_1541597, EPI_ISL_1541598, EPI_ISL_1541599, EPI_ISL_1541600, EPI_ISL_1541601, EPI_ISL_1541602, EPI_ISL_1541603, EPI_ISL_1541604, EPI_ISL_1541606, EPI_ISL_1541608, EPI_ISL_1541610, EPI_ISL_1541612, EPI_ISL_1541613, EPI_ISL_1541614, EPI_ISL_1541615, EPI_ISL_1541616, EPI_ISL_1541617, EPI_ISL_1541618, EPI_ISL_1541619, EPI_ISL_1541620, EPI_ISL_1541621, EPI_ISL_1541623, EPI_ISL_1541624, EPI_ISL_1541625, EPI_ISL_1541626, EPI_ISL_1541627, EPI_ISL_1541628, EPI_ISL_1541629, EPI_ISL_1541630, EPI_ISL_1541631, EPI_ISL_1541632, EPI_ISL_1541633, EPI_ISL_1541634, EPI_ISL_1541635, EPI_ISL_1541636, EPI_ISL_1541638, EPI_ISL_1541639, EPI_ISL_1541640, EPI_ISL_1541641, EPI_ISL_1541642, EPI_ISL_1541643, EPI_ISL_1541644, EPI_ISL_1541645, EPI_ISL_1541646, EPI_ISL_1541647, EPI_ISL_1541648, EPI_ISL_1541649, EPI_ISL_1541651, EPI_ISL_1541652, EPI_ISL_1541656, EPI_ISL_1541657, EPI_ISL_1541658, EPI_ISL_1541659, EPI_ISL_1541660, EPI_ISL_1541661, EPI_ISL_1541663, EPI_ISL_1541666, EPI_ISL_1541667, EPI_ISL_1541670, EPI_ISL_1541671, EPI_ISL_1541673, EPI_ISL_1541674, EPI_ISL_1541675, EPI_ISL_1541676, EPI_ISL_1541677, EPI_ISL_1541678, EPI_ISL_1541679, EPI_ISL_1541680, EPI_ISL_1541681, EPI_ISL_1541682, EPI_ISL_1541683, EPI_ISL_1541684, EPI_ISL_1541685, EPI_ISL_1541686, EPI_ISL_1541687, EPI_ISL_1541688, EPI_ISL_1541689, EPI_ISL_1541690, EPI_ISL_1541691, EPI_ISL_1541692, EPI_ISL_1541693, EPI_ISL_1541694, EPI_ISL_1541696, EPI_ISL_1541697, EPI_ISL_1541699, EPI_ISL_1541700, EPI_ISL_1541701, EPI_ISL_1541702, EPI_ISL_1541703, EPI_ISL_1541705, EPI_ISL_1541706, EPI_ISL_1541708, EPI_ISL_1541709, EPI_ISL_1541711, EPI_ISL_1541712, EPI_ISL_1541713, EPI_ISL_1541714, EPI_ISL_1541715, EPI_ISL_1541716, EPI_ISL_1541718, EPI_ISL_1541719, EPI_ISL_1541720, EPI_ISL_1541722, EPI_ISL_1541724, EPI_ISL_1541726, EPI_ISL_1541727, EPI_ISL_1541729, EPI_ISL_1541730, EPI_ISL_1541731, EPI_ISL_1541732, EPI_ISL_1541733, EPI_ISL_1541734, EPI_ISL_1541736, EPI_ISL_1541737, EPI_ISL_1541739, EPI_ISL_1541740, EPI_ISL_1541741, EPI_ISL_1541742, EPI_ISL_1541743, EPI_ISL_1541744, EPI_ISL_1541745, EPI_ISL_1541746, EPI_ISL_1541747, EPI_ISL_1541748, EPI_ISL_1541750, EPI_ISL_1541752, EPI_ISL_1541753, EPI_ISL_1541754, EPI_ISL_1541755, EPI_ISL_1541756, EPI_ISL_1541757, EPI_ISL_1541758, EPI_ISL_1541759, EPI_ISL_1541761, EPI_ISL_1541762, EPI_ISL_1541763, EPI_ISL_1541764, EPI_ISL_1541765, EPI_ISL_1541766, EPI_ISL_1541768, EPI_ISL_1541769, EPI_ISL_1541771, EPI_ISL_1541772, EPI_ISL_1541773, EPI_ISL_1541774, EPI_ISL_1541775, EPI_ISL_1541776, EPI_ISL_1541777, EPI_ISL_1541778, EPI_ISL_1541779, EPI_ISL_1541781, EPI_ISL_1541782, EPI_ISL_1541783, EPI_ISL_1541784, EPI_ISL_1541785, EPI_ISL_1541786, EPI_ISL_1541787, EPI_ISL_1541788, EPI_ISL_1541789, EPI_ISL_1541791, EPI_ISL_1541792, EPI_ISL_1541794, EPI_ISL_1541795, EPI_ISL_1541796, EPI_ISL_1541797, EPI_ISL_1541798, EPI_ISL_1541799, EPI_ISL_1541800, EPI_ISL_1541801, EPI_ISL_1541802, EPI_ISL_1541803, EPI_ISL_1541804, EPI_ISL_1541805, EPI_ISL_1541806, EPI_ISL_1541807, EPI_ISL_1541808, EPI_ISL_1541809, EPI_ISL_1541810, EPI_ISL_1541811, EPI_ISL_1541812, EPI_ISL_1541813, EPI_ISL_1541814, EPI_ISL_1541815, EPI_ISL_1541817, EPI_ISL_1541818, EPI_ISL_1541819, EPI_ISL_1541821 | | | |
| see above | OHSU Lab Services Molecular Microbiology Lab | Oregon SARS-CoV-2 Genome Sequencing Center | Brendan L. O'Connell, Sally Grindstaff, Kayla Carter, Sonia Acharya, Ruth V. Nichols, Alec J. Hirsch, Donna Hansel, Guang Fan, Xuan Qin, Daniel N. Streblow, William B. Messer, Andrew C. Adey, Benjamin N. Bimber, Brian J. O'Roak |
| EPI_ISL_1541822 | OHSU Lab Services Molecular Microbiology Lab | Oregon SARS-CoV-2 Genome Sequencing Center | Brendan L. O'Connell, Sally Grindstaff, Sonia Acharya, Cierra LaBlanc, Kayla Carter, Ruth V. Nichols, Alec J. Hirsch, Donna Hansel, Guang Fan, Xuan Qin, Daniel N. Streblow, William B. Messer, Andrew C. Adey, Benjamin N. Bimber, Brian J. O'Roak |
| EPI_ISL_1541823, EPI_ISL_1541824, EPI_ISL_1541825, EPI_ISL_1541826, EPI_ISL_1541827, EPI_ISL_1541828, EPI_ISL_1541829, EPI_ISL_1541830, EPI_ISL_1541831, EPI_ISL_1541833, EPI_ISL_1541834, EPI_ISL_1541835, EPI_ISL_1541836, EPI_ISL_1541837, EPI_ISL_1541838, EPI_ISL_1541839, EPI_ISL_1541840, EPI_ISL_1541841, EPI_ISL_1541842, EPI_ISL_1541843, EPI_ISL_1541844, EPI_ISL_1541845, EPI_ISL_1541846, EPI_ISL_1541847, EPI_ISL_1541848, EPI_ISL_1541849, EPI_ISL_1541851, EPI_ISL_1541852, EPI_ISL_1541854, EPI_ISL_1541855, EPI_ISL_1541856, EPI_ISL_1541858, EPI_ISL_1541859, EPI_ISL_1541860, EPI_ISL_1541861, EPI_ISL_1541862, EPI_ISL_1541863, EPI_ISL_1541864, EPI_ISL_1541865, EPI_ISL_1541867, EPI_ISL_1541868, EPI_ISL_1541869, EPI_ISL_1541870, EPI_ISL_1541871, EPI_ISL_1541872, EPI_ISL_1541873, EPI_ISL_1541874, EPI_ISL_1541875, EPI_ISL_1541876, EPI_ISL_1541877, EPI_ISL_1541878, EPI_ISL_1541879, EPI_ISL_1541880, EPI_ISL_1541881, EPI_ISL_1541882, EPI_ISL_1541885, EPI_ISL_1541886, EPI_ISL_1541887, EPI_ISL_1541888, EPI_ISL_1541889, EPI_ISL_1541891, EPI_ISL_1541892, EPI_ISL_1541894, EPI_ISL_1541895, EPI_ISL_1541896, EPI_ISL_1541897, EPI_ISL_1541898, EPI_ISL_1541899, EPI_ISL_1541900, EPI_ISL_1541901, EPI_ISL_1541903, EPI_ISL_1541904, EPI_ISL_1541905, EPI_ISL_1541906, EPI_ISL_1541907, EPI_ISL_1541908, EPI_ISL_1541909, EPI_ISL_1541910, EPI_ISL_1541911, EPI_ISL_1541913, EPI_ISL_1541914, EPI_ISL_1541915, EPI_ISL_1541916, EPI_ISL_1541917, EPI_ISL_1541918, EPI_ISL_1541919, EPI_ISL_1541920, EPI_ISL_1541921, EPI_ISL_1541922, EPI_ISL_1541923, EPI_ISL_1541924, EPI_ISL_1541925, EPI_ISL_1541926, EPI_ISL_1541928, EPI_ISL_1541929, EPI_ISL_1541930, EPI_ISL_1541931, EPI_ISL_1541932, EPI_ISL_1541933, EPI_ISL_1541934, EPI_ISL_1541935, EPI_ISL_1541936, EPI_ISL_1541938, EPI_ISL_1541939, EPI_ISL_1541940, EPI_ISL_1541941, EPI_ISL_1541942, EPI_ISL_1541943, EPI_ISL_1541944, EPI_ISL_1541945, EPI_ISL_1541947, EPI_ISL_1541948, EPI_ISL_1541949, EPI_ISL_1541950, EPI_ISL_1541951, EPI_ISL_1541953, EPI_ISL_1541954, EPI_ISL_1541955, EPI_ISL_1541956, EPI_ISL_1541958, EPI_ISL_1541959, EPI_ISL_1541961, EPI_ISL_1541963, EPI_ISL_1541964, EPI_ISL_1541965, EPI_ISL_1541966, EPI_ISL_1541967, EPI_ISL_1541968, EPI_ISL_1541969, EPI_ISL_1541970, EPI_ISL_1541971, EPI_ISL_1541972, EPI_ISL_1541973, EPI_ISL_1541974, EPI_ISL_1541975, EPI_ISL_1541976, EPI_ISL_1541978, EPI_ISL_1541979, EPI_ISL_1541980, EPI_ISL_1541981, EPI_ISL_1541982, EPI_ISL_1541983 | | | |
| see above | OHSU Lab Services Molecular Microbiology Lab | Oregon SARS-CoV-2 Genome Sequencing Center | Brendan L. O'Connell, Sally Grindstaff, Kayla Carter, Sonia Acharya, Ruth V. Nichols, Alec J. Hirsch, Donna Hansel, Guang Fan, Xuan Qin, Daniel N. Streblow, William B. Messer, Andrew C. Adey, Benjamin N. Bimber, Brian J. O'Roak |
| EPI_ISL_1541985 | OHSU Lab Services Molecular Microbiology Lab | Oregon SARS-CoV-2 Genome Sequencing Center | Brendan L. O'Connell, Sally Grindstaff, Sonia Acharya, Cierra LaBlanc, Kayla Carter, Ruth V. Nichols, Alec J. Hirsch, Donna Hansel, Guang Fan, Xuan Qin, Daniel N. Streblow, William B. Messer, Andrew C. Adey, Benjamin N. Bimber, Brian J. O'Roak |
| EPI_ISL_1541989, EPI_ISL_1541991, EPI_ISL_1541996, EPI_ISL_1541999, EPI_ISL_1542001, EPI_ISL_1542003, EPI_ISL_1542004, EPI_ISL_1542005, EPI_ISL_1542006, EPI_ISL_1542007, EPI_ISL_1542008, EPI_ISL_1542009, EPI_ISL_1542010, EPI_ISL_1542015, EPI_ISL_1542016, EPI_ISL_1542017, EPI_ISL_1542018, EPI_ISL_1542019, EPI_ISL_1542020, EPI_ISL_1542021, EPI_ISL_1542022, EPI_ISL_1542023, EPI_ISL_1542024, EPI_ISL_1542025, EPI_ISL_1542026, EPI_ISL_1542027, EPI_ISL_1542029, EPI_ISL_1542030, EPI_ISL_1542031, EPI_ISL_1542032, EPI_ISL_1542033, EPI_ISL_1542034, EPI_ISL_1542035, EPI_ISL_1542036, EPI_ISL_1542037, EPI_ISL_1542038, EPI_ISL_1542039, EPI_ISL_1542041, EPI_ISL_1542042, EPI_ISL_1542043, EPI_ISL_1542045, EPI_ISL_1542046, EPI_ISL_1542047, EPI_ISL_1542050, EPI_ISL_1542051, EPI_ISL_1542052, EPI_ISL_1542053, EPI_ISL_1542054, EPI_ISL_1542055, EPI_ISL_1542057, EPI_ISL_1542058, EPI_ISL_1542059, EPI_ISL_1542060, EPI_ISL_1542061, EPI_ISL_1542062, EPI_ISL_1542063, EPI_ISL_1542067, EPI_ISL_1542068, EPI_ISL_1542069, EPI_ISL_1542070, EPI_ISL_1542071, EPI_ISL_1542072 | | | |
| see above | OHSU Lab Services Molecular Microbiology Lab | Oregon SARS-CoV-2 Genome Sequencing Center | Brendan L. O'Connell, Sally Grindstaff, Kayla Carter, Sonia Acharya, Ruth V. Nichols, Alec J. Hirsch, Donna Hansel, Guang Fan, Xuan Qin, Daniel N. Streblow, William B. Messer, Andrew C. Adey, Benjamin N. Bimber, Brian J. O'Roak |
| EPI_ISL_1542074, EPI_ISL_1542075, EPI_ISL_1542076, EPI_ISL_1542077, EPI_ISL_1542080, EPI_ISL_1542081, EPI_ISL_1542082, EPI_ISL_1542083, EPI_ISL_1542084, EPI_ISL_1542085, EPI_ISL_1542086, EPI_ISL_1542087, EPI_ISL_1542088, EPI_ISL_1542089, EPI_ISL_1542090, EPI_ISL_1542091, EPI_ISL_1542092, EPI_ISL_1542093, EPI_ISL_1542094, EPI_ISL_1542095, EPI_ISL_1542096, EPI_ISL_1542098, EPI_ISL_1542099, EPI_ISL_1542100, EPI_ISL_1542101, EPI_ISL_1542102, EPI_ISL_1542103, EPI_ISL_1542104, EPI_ISL_1542105, EPI_ISL_1542106, EPI_ISL_1542108, EPI_ISL_1542109, EPI_ISL_1542110, EPI_ISL_1542111, EPI_ISL_1542112, EPI_ISL_1542113, EPI_ISL_1542114, EPI_ISL_1542115, EPI_ISL_1542116, EPI_ISL_1542117, EPI_ISL_1542118, EPI_ISL_1542119, EPI_ISL_1542120, EPI_ISL_1542121, EPI_ISL_1542122, EPI_ISL_1542123, EPI_ISL_1542126, EPI_ISL_1542127, EPI_ISL_1542128, EPI_ISL_1542129, EPI_ISL_1542130, EPI_ISL_1542131, EPI_ISL_1542132, EPI_ISL_1542133, EPI_ISL_1542134, EPI_ISL_1542135, EPI_ISL_1542136, EPI_ISL_1542137, EPI_ISL_1542138, EPI_ISL_1542139, EPI_ISL_1542140, EPI_ISL_1542141, EPI_ISL_1542143, EPI_ISL_1542144, EPI_ISL_1542146, EPI_ISL_1542147, EPI_ISL_1542148, EPI_ISL_1542149, EPI_ISL_1542150, EPI_ISL_1542151, EPI_ISL_1542152, EPI_ISL_1542153, EPI_ISL_1542155, EPI_ISL_1542157, EPI_ISL_1542158, EPI_ISL_1542160, EPI_ISL_1542161, EPI_ISL_1542162, EPI_ISL_1542163, EPI_ISL_1542164, EPI_ISL_1542165, EPI_ISL_1542166, EPI_ISL_1542167, EPI_ISL_1542168, EPI_ISL_1542169, EPI_ISL_1542170, EPI_ISL_1542171, EPI_ISL_1542172, EPI_ISL_1542173, EPI_ISL_1542174, EPI_ISL_1542175, EPI_ISL_1542176, EPI_ISL_1542177, EPI_ISL_1542178, EPI_ISL_1542179, EPI_ISL_1542180, EPI_ISL_1542181, EPI_ISL_1542182, EPI_ISL_1542183, EPI_ISL_1542184, EPI_ISL_1542185, EPI_ISL_1542188, EPI_ISL_1542189, EPI_ISL_1542190, EPI_ISL_1542191, EPI_ISL_1542192, EPI_ISL_1542193, EPI_ISL_1542194, EPI_ISL_1542195, EPI_ISL_1542196, EPI_ISL_1542197, EPI_ISL_1542198, EPI_ISL_1542199, EPI_ISL_1542200, EPI_ISL_1542201, EPI_ISL_1542202, EPI_ISL_1542203, EPI_ISL_1542204, EPI_ISL_1542205, EPI_ISL_1542206, EPI_ISL_1542207, EPI_ISL_1542208, EPI_ISL_1542209, EPI_ISL_1542210, EPI_ISL_1542211, EPI_ISL_1542212, EPI_ISL_1542215, EPI_ISL_1542216, EPI_ISL_1542217, EPI_ISL_1542218, EPI_ISL_1542220, EPI_ISL_1542221, EPI_ISL_1542222, EPI_ISL_1542223, EPI_ISL_1542224, EPI_ISL_1542225, EPI_ISL_1542226, EPI_ISL_1542227, EPI_ISL_1542228, EPI_ISL_1542229, EPI_ISL_1542230, EPI_ISL_1542231, EPI_ISL_1542232, EPI_ISL_1542233, EPI_ISL_1542234, EPI_ISL_1542235, EPI_ISL_1542236, EPI_ISL_1542237, EPI_ISL_1542238, EPI_ISL_1542239, EPI_ISL_1542240, EPI_ISL_1542242, EPI_ISL_1542243, EPI_ISL_1542244, EPI_ISL_1542245, EPI_ISL_1542246, EPI_ISL_1542247, EPI_ISL_1542248, EPI_ISL_1542249, EPI_ISL_1542250, EPI_ISL_1542251, EPI_ISL_1542253, EPI_ISL_1542254, EPI_ISL_1542255, EPI_ISL_1542256, EPI_ISL_1542257, EPI_ISL_1542258, EPI_ISL_1542259, EPI_ISL_1542260, EPI_ISL_1542261, EPI_ISL_1542262, EPI_ISL_1542263, EPI_ISL_1542264, EPI_ISL_1542265, EPI_ISL_1542266, EPI_ISL_1542267, EPI_ISL_1542268, EPI_ISL_1542269, EPI_ISL_1542270, EPI_ISL_1542271, EPI_ISL_1542272, EPI_ISL_1542273, EPI_ISL_1542275, EPI_ISL_1542276, EPI_ISL_1542277, EPI_ISL_1542279, EPI_ISL_1542280, EPI_ISL_1542281, EPI_ISL_1542282, EPI_ISL_1542283, EPI_ISL_1542284, EPI_ISL_1542285, EPI_ISL_1542286, EPI_ISL_1542287, EPI_ISL_1542288, EPI_ISL_1542289, EPI_ISL_1542290, EPI_ISL_1542291, EPI_ISL_1542293, EPI_ISL_1542294, EPI_ISL_1542295, EPI_ISL_1542296, EPI_ISL_1542297, EPI_ISL_1542298, EPI_ISL_1542300, EPI_ISL_1542301, EPI_ISL_1542302, EPI_ISL_1542303, EPI_ISL_1542304, EPI_ISL_1542305, EPI_ISL_1542307, EPI_ISL_1542308, EPI_ISL_1542309, EPI_ISL_1542312, EPI_ISL_1542313, EPI_ISL_1542314, EPI_ISL_1542315, EPI_ISL_1542316, EPI_ISL_1542317, EPI_ISL_1542318, EPI_ISL_1542319, EPI_ISL_1542320, EPI_ISL_1542321, EPI_ISL_1542322, EPI_ISL_1542323, EPI_ISL_1542324, EPI_ISL_1542325, EPI_ISL_1542326, EPI_ISL_1542327, EPI_ISL_1542328, EPI_ISL_1542329, EPI_ISL_1542330, EPI_ISL_1542331, EPI_ISL_1542332, EPI_ISL_1542333, EPI_ISL_1542334, EPI_ISL_1542337, EPI_ISL_1542340, EPI_ISL_1542341, EPI_ISL_1542342, EPI_ISL_1542343, EPI_ISL_1542345, EPI_ISL_1542346, EPI_ISL_1542347, EPI_ISL_1542349, EPI_ISL_1542351, EPI_ISL_1542352, EPI_ISL_1542353, EPI_ISL_1542354, EPI_ISL_1542355, EPI_ISL_1542356, EPI_ISL_1542357, EPI_ISL_1542358, EPI_ISL_1542359, EPI_ISL_1542360, EPI_ISL_1542361, EPI_ISL_1542362, EPI_ISL_1542363, EPI_ISL_1542364, EPI_ISL_1542365, EPI_ISL_1542366, EPI_ISL_1542367, EPI_ISL_1542368, EPI_ISL_1542369, EPI_ISL_1542370, EPI_ISL_1542371, EPI_ISL_1542372, EPI_ISL_1542373, EPI_ISL_1542374, EPI_ISL_1542375, EPI_ISL_1542377, EPI_ISL_1542378, EPI_ISL_1542379, EPI_ISL_1542380, EPI_ISL_1542381, EPI_ISL_1542382, EPI_ISL_1542383, EPI_ISL_1542384, EPI_ISL_1542385, EPI_ISL_1542386, EPI_ISL_1542387, EPI_ISL_1542388, EPI_ISL_1542389, EPI_ISL_1542390, EPI_ISL_1542391, EPI_ISL_1542392, EPI_ISL_1542393, EPI_ISL_1542394, EPI_ISL_1542396, EPI_ISL_1542397, EPI_ISL_1542398, EPI_ISL_1542399, EPI_ISL_1542400, EPI_ISL_1542401, EPI_ISL_1542402, EPI_ISL_1542403, EPI_ISL_1542404, EPI_ISL_1542405, EPI_ISL_1542406, EPI_ISL_1542408, EPI_ISL_1542409, EPI_ISL_1542411, EPI_ISL_1542413, EPI_ISL_1542414, EPI_ISL_1542415, EPI_ISL_1542416, EPI_ISL_1542417, EPI_ISL_1542418, EPI_ISL_1542419, EPI_ISL_1542420, EPI_ISL_1542421, EPI_ISL_1542422, EPI_ISL_1542423, EPI_ISL_1542424, EPI_ISL_1542425, EPI_ISL_1542426, EPI_ISL_1542427, EPI_ISL_1542428, EPI_ISL_1542429, EPI_ISL_1542430, EPI_ISL_1542431, EPI_ISL_1542432, EPI_ISL_1542433, EPI_ISL_1542434, EPI_ISL_1542435, EPI_ISL_1542436, EPI_ISL_1542437, EPI_ISL_1542439, EPI_ISL_1542440, EPI_ISL_1542441, EPI_ISL_1542442, EPI_ISL_1542443, EPI_ISL_1542444, EPI_ISL_1542445, EPI_ISL_1542446, EPI_ISL_1542447, EPI_ISL_1542448, EPI_ISL_1542449, EPI_ISL_1542450, EPI_ISL_1542451, EPI_ISL_1542452, EPI_ISL_1542454, EPI_ISL_1542455, EPI_ISL_1542456, EPI_ISL_1542457, EPI_ISL_1542458, EPI_ISL_1542461, EPI_ISL_1542462, EPI_ISL_1542463, EPI_ISL_1542464, EPI_ISL_1542465, EPI_ISL_1542466, EPI_ISL_1542467, EPI_ISL_1542468, EPI_ISL_1542469, EPI_ISL_1542471, EPI_ISL_1542472, EPI_ISL_1542473, EPI_ISL_1542474, EPI_ISL_1542475, EPI_ISL_1542476, EPI_ISL_1542478, EPI_ISL_1542479, EPI_ISL_1542480, EPI_ISL_1542481, EPI_ISL_1542482, EPI_ISL_1542483, EPI_ISL_1542484, EPI_ISL_1542486, EPI_ISL_1542487, EPI_ISL_1542488, EPI_ISL_1542489, EPI_ISL_1542490, EPI_ISL_1542491, EPI_ISL_1542492, EPI_ISL_1542493, EPI_ISL_1542494, EPI_ISL_1542495, EPI_ISL_1542496, EPI_ISL_1542497, EPI_ISL_1542499, EPI_ISL_1542500, EPI_ISL_1542503, EPI_ISL_1542504, EPI_ISL_1542505, EPI_ISL_1542506, EPI_ISL_1542509, EPI_ISL_1542510, EPI_ISL_1542511, EPI_ISL_1542512, EPI_ISL_1542513, EPI_ISL_1542515, EPI_ISL_1542517, EPI_ISL_1542518, EPI_ISL_1542519, EPI_ISL_1542520, EPI_ISL_1542521, EPI_ISL_1542522, EPI_ISL_1542523, EPI_ISL_1542524, EPI_ISL_1542526, EPI_ISL_1542527, EPI_ISL_1542528, EPI_ISL_1542529, EPI_ISL_1542530, EPI_ISL_1542531, EPI_ISL_1542532, EPI_ISL_1542533, EPI_ISL_1542535, EPI_ISL_1542536, EPI_ISL_1542537, EPI_ISL_1542538, EPI_ISL_1542539, EPI_ISL_1542541, EPI_ISL_1542542, EPI_ISL_1542543, EPI_ISL_1542544, EPI_ISL_1542545, EPI_ISL_1542546, EPI_ISL_1542547, EPI_ISL_1542548, EPI_ISL_1542549, EPI_ISL_1542550, EPI_ISL_1542551, EPI_ISL_1542552, EPI_ISL_1542553, EPI_ISL_1542554, EPI_ISL_1542555, EPI_ISL_1542556, EPI_ISL_1542557, EPI_ISL_1542558, EPI_ISL_1542559, EPI_ISL_1542560, EPI_ISL_1542562, EPI_ISL_1542563, EPI_ISL_1542564, EPI_ISL_1542565, EPI_ISL_1542566, EPI_ISL_1542567, EPI_ISL_1542568, EPI_ISL_1542569, EPI_ISL_1542570, EPI_ISL_1542571, EPI_ISL_1542572, EPI_ISL_1542573, EPI_ISL_1542574, EPI_ISL_1542575, EPI_ISL_1542576, EPI_ISL_1542577, EPI_ISL_1542578, EPI_ISL_1542579, EPI_ISL_1542580, EPI_ISL_1542581, EPI_ISL_1542582, EPI_ISL_1542583, EPI_ISL_1542585, EPI_ISL_1542586, EPI_ISL_1542587, EPI_ISL_1542588, EPI_ISL_1542589, EPI_ISL_1542590, EPI_ISL_1542591, EPI_ISL_1542592, EPI_ISL_1542593, EPI_ISL_1542594, EPI_ISL_1542595, EPI_ISL_1542596, EPI_ISL_1542597, EPI_ISL_1542598, EPI_ISL_1542599, EPI_ISL_1542600, EPI_ISL_1542601, EPI_ISL_1542602, EPI_ISL_1542603, EPI_ISL_1542604, EPI_ISL_1542605, EPI_ISL_1542607, EPI_ISL_1542608, EPI_ISL_1542609, EPI_ISL_1542610, EPI_ISL_1542611, EPI_ISL_1542612, EPI_ISL_1542613, EPI_ISL_1542614, EPI_ISL_1542615, EPI_ISL_1542616, EPI_ISL_1542617, EPI_ISL_1542618, EPI_ISL_1542619, EPI_ISL_1542620, EPI_ISL_1542621, EPI_ISL_1542622, EPI_ISL_1542623, EPI_ISL_1542624, EPI_ISL_1542625, EPI_ISL_1542626, EPI_ISL_1542627, EPI_ISL_1542628, EPI_ISL_1542629, EPI_ISL_1542630, EPI_ISL_1542631, EPI_ISL_1542632, EPI_ISL_1542633, EPI_ISL_1542634, EPI_ISL_1542635, EPI_ISL_1542636, EPI_ISL_1542637, EPI_ISL_1542638, EPI_ISL_1542639, EPI_ISL_1542640, EPI_ISL_1542641, EPI_ISL_1542642, EPI_ISL_1542643, EPI_ISL_1542644, EPI_ISL_1542645, EPI_ISL_1542646, EPI_ISL_1542647, EPI_ISL_1542649, EPI_ISL_1542650, EPI_ISL_1542652, EPI_ISL_1542653, EPI_ISL_1542654, EPI_ISL_1542655, EPI_ISL_1542656, EPI_ISL_1542657, EPI_ISL_1542658, EPI_ISL_1542659, EPI_ISL_1542660, EPI_ISL_1542661, EPI_ISL_1542663, EPI_ISL_1542664, EPI_ISL_1542665, EPI_ISL_1542666, EPI_ISL_1542667, EPI_ISL_1542668, EPI_ISL_1542669, EPI_ISL_1542670, EPI_ISL_1542671, EPI_ISL_1542672, EPI_ISL_1542673, EPI_ISL_1542675, EPI_ISL_1542676, EPI_ISL_1542677, EPI_ISL_1542678, EPI_ISL_1542679, EPI_ISL_1542680, EPI_ISL_1542682, EPI_ISL_1542683, EPI_ISL_1542684, EPI_ISL_1542685, EPI_ISL_1542687, EPI_ISL_1542688, EPI_ISL_1542689, EPI_ISL_1542690, EPI_ISL_1542691, EPI_ISL_1542692, EPI_ISL_1542693, EPI_ISL_1542694, EPI_ISL_1542695, EPI_ISL_1542696, EPI_ISL_1542697, EPI_ISL_1542698, EPI_ISL_1542700, EPI_ISL_1542701, EPI_ISL_1542702, EPI_ISL_1542703, EPI_ISL_1542704, EPI_ISL_1542705, EPI_ISL_1542706, EPI_ISL_1542707, EPI_ISL_1542709, EPI_ISL_1542711, EPI_ISL_1542713, EPI_ISL_1542714, EPI_ISL_1542715, EPI_ISL_1542716, EPI_ISL_1542717, EPI_ISL_1542718, EPI_ISL_1542719, EPI_ISL_1542720, EPI_ISL_1542721, EPI_ISL_1542722, EPI_ISL_1542723, EPI_ISL_1542724, EPI_ISL_1542725, EPI_ISL_1542726, EPI_ISL_1542727, EPI_ISL_1542728, EPI_ISL_1542729, EPI_ISL_1542730, EPI_ISL_1542731, EPI_ISL_1542732, EPI_ISL_1542733, EPI_ISL_1542734, EPI_ISL_1542735, EPI_ISL_1542736, EPI_ISL_1542737, EPI_ISL_1542739, EPI_ISL_1542740, EPI_ISL_1542741, EPI_ISL_1542742, EPI_ISL_1542743, EPI_ISL_1542744, EPI_ISL_1542745, EPI_ISL_1542746, EPI_ISL_1542748, EPI_ISL_1542749, EPI_ISL_1542750, EPI_ISL_1542751, EPI_ISL_1542752, EPI_ISL_1542754, EPI_ISL_1542755, EPI_ISL_1542756, EPI_ISL_1542757, EPI_ISL_1542758, EPI_ISL_1542759, EPI_ISL_1542760, EPI_ISL_1542761, EPI_ISL_1542762, EPI_ISL_1542763, EPI_ISL_1542764, EPI_ISL_1542765, EPI_ISL_1542766, EPI_ISL_1542767, EPI_ISL_1542768, EPI_ISL_1542769, EPI_ISL_1542771, EPI_ISL_1542772, EPI_ISL_1542773, EPI_ISL_1542774, EPI_ISL_1542775, EPI_ISL_1542776, EPI_ISL_1542777, EPI_ISL_1542779, EPI_ISL_1542781, EPI_ISL_1542782, EPI_ISL_1542783, EPI_ISL_1542784, EPI_ISL_1542785, EPI_ISL_1542787, EPI_ISL_1542788, EPI_ISL_1542789, EPI_ISL_1542790, EPI_ISL_1542791, EPI_ISL_1542792, EPI_ISL_1542793, EPI_ISL_1542794, EPI_ISL_1542795, EPI_ISL_1542796, EPI_ISL_1542797, EPI_ISL_1542798, EPI_ISL_1542799, EPI_ISL_1542800, EPI_ISL_1542801, EPI_ISL_1542802, EPI_ISL_1542803, EPI_ISL_1542804, EPI_ISL_1542805, EPI_ISL_1542806, EPI_ISL_1542807, EPI_ISL_1542808, EPI_ISL_1542809, EPI_ISL_1542810, EPI_ISL_1542811, EPI_ISL_1542812, EPI_ISL_1542813, EPI_ISL_1542814, EPI_ISL_1542815, EPI_ISL_1542816, EPI_ISL_1542817, EPI_ISL_1542818, EPI_ISL_1542819, EPI_ISL_1542820, EPI_ISL_1542821, EPI_ISL_1542822, EPI_ISL_1542824, EPI_ISL_1542825, EPI_ISL_1542826, EPI_ISL_1542827, EPI_ISL_1542829, EPI_ISL_1542830, EPI_ISL_1542832, EPI_ISL_1542833, EPI_ISL_1542834, EPI_ISL_1542835, EPI_ISL_1542837, EPI_ISL_1542838, EPI_ISL_1542839, EPI_ISL_1542840, EPI_ISL_1542841, EPI_ISL_1542842, EPI_ISL_1542843, EPI_ISL_1542844, EPI_ISL_1542845, EPI_ISL_1542846, EPI_ISL_1542847, EPI_ISL_1542848, EPI_ISL_1542849, EPI_ISL_1542850, EPI_ISL_1542852, EPI_ISL_1542853, EPI_ISL_1542854, EPI_ISL_1542855, EPI_ISL_1542856, EPI_ISL_1542857, EPI_ISL_1542858, EPI_ISL_1542859, EPI_ISL_1542860, EPI_ISL_1542861, EPI_ISL_1542862, EPI_ISL_1542864, EPI_ISL_1542865, EPI_ISL_1542866, EPI_ISL_1542868, EPI_ISL_1542869, EPI_ISL_1542870, EPI_ISL_1542871, EPI_ISL_1542872, EPI_ISL_1542873, EPI_ISL_1542875, EPI_ISL_1542876, EPI_ISL_1542877, EPI_ISL_1542878, EPI_ISL_1542879, EPI_ISL_1542880, EPI_ISL_1542881, EPI_ISL_1542883, EPI_ISL_1542884, EPI_ISL_1542886, EPI_ISL_1542887, EPI_ISL_1542888, EPI_ISL_1542890, EPI_ISL_1542891, EPI_ISL_1542892, EPI_ISL_1542893, EPI_ISL_1542895, EPI_ISL_1542897, EPI_ISL_1542898, EPI_ISL_1542899, EPI_ISL_1542900, EPI_ISL_1542901, EPI_ISL_1542902, EPI_ISL_1542904, EPI_ISL_1542905, EPI_ISL_1542906, EPI_ISL_1542907, EPI_ISL_1542908, EPI_ISL_1542909, EPI_ISL_1542910, EPI_ISL_1542911, EPI_ISL_1542912, EPI_ISL_1542913, EPI_ISL_1542915, EPI_ISL_1542916, EPI_ISL_1542917, EPI_ISL_1542919, EPI_ISL_1542920, EPI_ISL_1542921, EPI_ISL_1542923, EPI_ISL_1542924, EPI_ISL_1542925, EPI_ISL_1542927, EPI_ISL_1542928, EPI_ISL_1542929, EPI_ISL_1542930, EPI_ISL_1542931, EPI_ISL_1542932, EPI_ISL_1542933, EPI_ISL_1542934, EPI_ISL_1542935, EPI_ISL_1542936, EPI_ISL_1542937, EPI_ISL_1542938, EPI_ISL_1542939, EPI_ISL_1542940, EPI_ISL_1542941, EPI_ISL_1542942, EPI_ISL_1542943, EPI_ISL_1542944, EPI_ISL_1542945, EPI_ISL_1542946, EPI_ISL_1542947, EPI_ISL_1542948, EPI_ISL_1542949, EPI_ISL_1542950, EPI_ISL_1542952, EPI_ISL_1542953, EPI_ISL_1542954, EPI_ISL_1542955, EPI_ISL_1542957, EPI_ISL_1542958, EPI_ISL_1542959, EPI_ISL_1542960, EPI_ISL_1542961, EPI_ISL_1542962, EPI_ISL_1542963, EPI_ISL_1542964, EPI_ISL_1542965, EPI_ISL_1542966, EPI_ISL_1542967, EPI_ISL_1542968, EPI_ISL_1542969, EPI_ISL_1542971, EPI_ISL_1542972, EPI_ISL_1542973, EPI_ISL_1542974, EPI_ISL_1542975, EPI_ISL_1542976, EPI_ISL_1542977, EPI_ISL_1542978, EPI_ISL_1542979, EPI_ISL_1542980, EPI_ISL_1542981, EPI_ISL_1542982, EPI_ISL_1542983, EPI_ISL_1542984, EPI_ISL_1542985, EPI_ISL_1542986, EPI_ISL_1542987, EPI_ISL_1542988, EPI_ISL_1542989, EPI_ISL_1542990, EPI_ISL_1542991, EPI_ISL_1542992, EPI_ISL_1542993, EPI_ISL_1542994, EPI_ISL_1542995, EPI_ISL_1542996, EPI_ISL_1542997, EPI_ISL_1542998, EPI_ISL_1542999, EPI_ISL_1543000, EPI_ISL_1543001, EPI_ISL_1543002, EPI_ISL_1543003, EPI_ISL_1543004, EPI_ISL_1543005, EPI_ISL_1543006, EPI_ISL_1543008, EPI_ISL_1543009, EPI_ISL_1543010, EPI_ISL_1543011, EPI_ISL_1543012, EPI_ISL_1543013, EPI_ISL_1543014, EPI_ISL_1543015, EPI_ISL_1543016, EPI_ISL_1543017, EPI_ISL_1543018, EPI_ISL_1543019, EPI_ISL_1543020, EPI_ISL_1543021, EPI_ISL_1543022, EPI_ISL_1543023, EPI_ISL_1543024, EPI_ISL_1543025, EPI_ISL_1543026, EPI_ISL_1543027, EPI_ISL_1543028, EPI_ISL_1543029, EPI_ISL_1543030, EPI_ISL_1543031, EPI_ISL_1543032, EPI_ISL_1543033, EPI_ISL_1543034, EPI_ISL_1543035, EPI_ISL_1543036, EPI_ISL_1543037, EPI_ISL_1543038, EPI_ISL_1543039, EPI_ISL_1543040, EPI_ISL_1543041, EPI_ISL_1543042, EPI_ISL_1543043, EPI_ISL_1543044, EPI_ISL_1543045, EPI_ISL_1543046, EPI_ISL_1543047, EPI_ISL_1543048, EPI_ISL_1543049, EPI_ISL_1543050, EPI_ISL_1543051, EPI_ISL_1543052, EPI_ISL_1543053, EPI_ISL_1543054, EPI_ISL_1543055, EPI_ISL_1543057, EPI_ISL_1543059, EPI_ISL_1543060, EPI_ISL_1543061, EPI_ISL_1543062, EPI_ISL_1543063, EPI_ISL_1543064, EPI_ISL_1543065, EPI_ISL_1543066, EPI_ISL_1543067, EPI_ISL_1543069, EPI_ISL_1543070, EPI_ISL_1543071, EPI_ISL_1543072, EPI_ISL_1543073, EPI_ISL_1543074, EPI_ISL_1543075, EPI_ISL_1543076, EPI_ISL_1543077, EPI_ISL_1543078, EPI_ISL_1543079, EPI_ISL_1543080, EPI_ISL_1543081, EPI_ISL_1543082, EPI_ISL_1543083, EPI_ISL_1543084, EPI_ISL_1543086, EPI_ISL_1543087, EPI_ISL_1543088, EPI_ISL_1543089, EPI_ISL_1543090, EPI_ISL_1543091, EPI_ISL_1543093, EPI_ISL_1543094, EPI_ISL_1543096, EPI_ISL_1543097, EPI_ISL_1543098, EPI_ISL_1543099, EPI_ISL_1543100, EPI_ISL_1543101, EPI_ISL_1543102, EPI_ISL_1543103, EPI_ISL_1543104, EPI_ISL_1543105, EPI_ISL_1543107, EPI_ISL_1543108, EPI_ISL_1543109, EPI_ISL_1543110, EPI_ISL_1543111, EPI_ISL_1543112, EPI_ISL_1543113, EPI_ISL_1543114, EPI_ISL_1543115, EPI_ISL_1543116, EPI_ISL_1543117, EPI_ISL_1543118, EPI_ISL_1543119, EPI_ISL_1543120, EPI_ISL_1543121, EPI_ISL_1543122, EPI_ISL_1543123, EPI_ISL_1543124, EPI_ISL_1543125, EPI_ISL_1543126, EPI_ISL_1543128, EPI_ISL_1543129, EPI_ISL_1543130, EPI_ISL_1543131, EPI_ISL_1543132, EPI_ISL_1543134, EPI_ISL_1543135, EPI_ISL_1543136, EPI_ISL_1543137, EPI_ISL_1543138, EPI_ISL_1543139, EPI_ISL_1543140, EPI_ISL_1543141, EPI_ISL_1543142, EPI_ISL_1543143, EPI_ISL_1543144, EPI_ISL_1543145, EPI_ISL_1543146, EPI_ISL_1543147, EPI_ISL_1543148, EPI_ISL_1543149, EPI_ISL_1543150, EPI_ISL_1543151, EPI_ISL_1543152, EPI_ISL_1543153, EPI_ISL_1543155, EPI_ISL_1543156, EPI_ISL_1543157, EPI_ISL_1543158, EPI_ISL_1543159, EPI_ISL_1543160, EPI_ISL_1543161, EPI_ISL_1543162, EPI_ISL_1543163, EPI_ISL_1543164, EPI_ISL_1543165, EPI_ISL_1543166, EPI_ISL_1543167, EPI_ISL_1543168, EPI_ISL_1543169, EPI_ISL_1543171, EPI_ISL_1543172, EPI_ISL_1543173, EPI_ISL_1543174, EPI_ISL_1543175, EPI_ISL_1543176, EPI_ISL_1543177, EPI_ISL_1543178, EPI_ISL_1543180, EPI_ISL_1543181, EPI_ISL_1543183, EPI_ISL_1543184, EPI_ISL_1543185, EPI_ISL_1543186, EPI_ISL_1543187, EPI_ISL_1543188, EPI_ISL_1543189, EPI_ISL_1543190, EPI_ISL_1543191, EPI_ISL_1543192, EPI_ISL_1543193, EPI_ISL_1543194, EPI_ISL_1543195, EPI_ISL_1543196, EPI_ISL_1543197, EPI_ISL_1543198, EPI_ISL_1543199, EPI_ISL_1543200, EPI_ISL_1543201, EPI_ISL_1543202, EPI_ISL_1543203, EPI_ISL_1543204, EPI_ISL_1543205, EPI_ISL_1543206, EPI_ISL_1543207, EPI_ISL_1543209, EPI_ISL_1543210, EPI_ISL_1543211, EPI_ISL_1543212, EPI_ISL_1543213, EPI_ISL_1543214, EPI_ISL_1543215, EPI_ISL_1543216, EPI_ISL_1543217, EPI_ISL_1543218, EPI_ISL_1543219, EPI_ISL_1543220, EPI_ISL_1543221, EPI_ISL_1543222, EPI_ISL_1543223, EPI_ISL_1543224, EPI_ISL_1543225, EPI_ISL_1543226, EPI_ISL_1543227, EPI_ISL_1543228, EPI_ISL_1543229, EPI_ISL_1543230, EPI_ISL_1543231, EPI_ISL_1543232, EPI_ISL_1543233, EPI_ISL_1543234, EPI_ISL_1543235, EPI_ISL_1543236, EPI_ISL_1543237, EPI_ISL_1543238, EPI_ISL_1543239, EPI_ISL_1543240, EPI_ISL_1543241, EPI_ISL_1543242, EPI_ISL_1543243, EPI_ISL_1543244, EPI_ISL_1543245, EPI_ISL_1543246, EPI_ISL_1543247, EPI_ISL_1543248, EPI_ISL_1543249, EPI_ISL_1543250, EPI_ISL_1543251, EPI_ISL_1543252, EPI_ISL_1543253, EPI_ISL_1543255, EPI_ISL_1543256, EPI_ISL_1543257, EPI_ISL_1543258, EPI_ISL_1543259, EPI_ISL_1543260, EPI_ISL_1543261, EPI_ISL_1543262, EPI_ISL_1543263, EPI_ISL_1543264, EPI_ISL_1543265, EPI_ISL_1543266, EPI_ISL_1543267, EPI_ISL_1543268, EPI_ISL_1543269, EPI_ISL_1543271, EPI_ISL_1543272, EPI_ISL_1543273, EPI_ISL_1543274, EPI_ISL_1543275, EPI_ISL_1543276, EPI_ISL_1543277, EPI_ISL_1543278, EPI_ISL_1543279, EPI_ISL_1543280, EPI_ISL_1543281, EPI_ISL_1543282, EPI_ISL_1543283, EPI_ISL_1543285, EPI_ISL_1543286, EPI_ISL_1543287, EPI_ISL_1543288, EPI_ISL_1543290, EPI_ISL_1543291, EPI_ISL_1543293, EPI_ISL_1543294, EPI_ISL_1543296, EPI_ISL_1543297, EPI_ISL_1543298, EPI_ISL_1543299, EPI_ISL_1543300, EPI_ISL_1543301, EPI_ISL_1543302, EPI_ISL_1543303, EPI_ISL_1543304, EPI_ISL_1543305, EPI_ISL_1543306, EPI_ISL_1543307, EPI_ISL_1543308, EPI_ISL_1543309, EPI_ISL_1543310, EPI_ISL_1543311, EPI_ISL_1543312, EPI_ISL_1543313, EPI_ISL_1543314, EPI_ISL_1543315, EPI_ISL_1543316, EPI_ISL_1543317, EPI_ISL_1543318, EPI_ISL_1543319, EPI_ISL_1543320, EPI_ISL_1543321, EPI_ISL_1543322, EPI_ISL_1543323, EPI_ISL_1543324, EPI_ISL_1543325, EPI_ISL_1543326, EPI_ISL_1543327, EPI_ISL_1543328, EPI_ISL_1543329, EPI_ISL_1543330, EPI_ISL_1543331, EPI_ISL_1543332, EPI_ISL_1543333, EPI_ISL_1543334, EPI_ISL_1543335, EPI_ISL_1543336, EPI_ISL_1543337, EPI_ISL_1543338, EPI_ISL_1543339, EPI_ISL_1543340, EPI_ISL_1543341, EPI_ISL_1543342, EPI_ISL_1543343, EPI_ISL_1543344, EPI_ISL_1543345, EPI_ISL_1543346, EPI_ISL_1543347, EPI_ISL_1543348, EPI_ISL_1543349, EPI_ISL_1543350, EPI_ISL_1543351, EPI_ISL_1543352, EPI_ISL_1543353, EPI_ISL_1543354, EPI_ISL_1543355, EPI_ISL_1543356, EPI_ISL_1543357, EPI_ISL_1543359, EPI_ISL_1543361, EPI_ISL_1543362, EPI_ISL_1543363, EPI_ISL_1543364, EPI_ISL_1543365, EPI_ISL_1543366, EPI_ISL_1543367, EPI_ISL_1543368, EPI_ISL_1543370, EPI_ISL_1543371, EPI_ISL_1543372, EPI_ISL_1543373, EPI_ISL_1543374, EPI_ISL_1543375, EPI_ISL_1543376, EPI_ISL_1543377, EPI_ISL_1543378, EPI_ISL_1543379, EPI_ISL_1543380, EPI_ISL_1543381, EPI_ISL_1543382, EPI_ISL_1543383, EPI_ISL_1543384, EPI_ISL_1543386, EPI_ISL_1543387, EPI_ISL_1543388, EPI_ISL_1543389, EPI_ISL_1543390, EPI_ISL_1543391, EPI_ISL_1543392, EPI_ISL_1543393, EPI_ISL_1543395, EPI_ISL_1543396, EPI_ISL_1543398, EPI_ISL_1543399, EPI_ISL_1543400, EPI_ISL_1543401, EPI_ISL_1543402, EPI_ISL_1543403, EPI_ISL_1543404, EPI_ISL_1543405, EPI_ISL_1543406, EPI_ISL_1543409, EPI_ISL_1543410, EPI_ISL_1543411, EPI_ISL_1543413, EPI_ISL_1543414, EPI_ISL_1543415, EPI_ISL_1543417, EPI_ISL_1543418, EPI_ISL_1543419, EPI_ISL_1543420, EPI_ISL_1543423, EPI_ISL_1543424, EPI_ISL_1543425, EPI_ISL_1543426, EPI_ISL_1543427, EPI_ISL_1543428, EPI_ISL_1543429, EPI_ISL_1543430, EPI_ISL_1543431, EPI_ISL_1543433, EPI_ISL_1543436, EPI_ISL_1543438, EPI_ISL_1543439, EPI_ISL_1543440, EPI_ISL_1543442, EPI_ISL_1543444, EPI_ISL_1543445, EPI_ISL_1543446, EPI_ISL_1543447, EPI_ISL_1543448, EPI_ISL_1543450, EPI_ISL_1543451, EPI_ISL_1543452, EPI_ISL_1543453, EPI_ISL_1543455, EPI_ISL_1543456, EPI_ISL_1543458, EPI_ISL_1543459, EPI_ISL_1543460, EPI_ISL_1543461, EPI_ISL_1543463, EPI_ISL_1543464, EPI_ISL_1543465, EPI_ISL_1543466, EPI_ISL_1543467, EPI_ISL_1543468, EPI_ISL_1543469, EPI_ISL_1543470, EPI_ISL_1543472, EPI_ISL_1543473, EPI_ISL_1543474, EPI_ISL_1543475, EPI_ISL_1543476, EPI_ISL_1543477, EPI_ISL_1543478, EPI_ISL_1543479, EPI_ISL_1543481, EPI_ISL_1543482, EPI_ISL_1543483, EPI_ISL_1543485, EPI_ISL_1543486, EPI_ISL_1543487, EPI_ISL_1543488, EPI_ISL_1543489, EPI_ISL_1543490, EPI_ISL_1543491, EPI_ISL_1543492, EPI_ISL_1543493, EPI_ISL_1543494, EPI_ISL_1543495, EPI_ISL_1543496, EPI_ISL_1543497, EPI_ISL_1543498, EPI_ISL_1543499, EPI_ISL_1543500, EPI_ISL_1543501, EPI_ISL_1543502, EPI_ISL_1543503, EPI_ISL_1543504, EPI_ISL_1543505, EPI_ISL_1543506, EPI_ISL_1543507, EPI_ISL_1543508, EPI_ISL_1543509, EPI_ISL_1543510, EPI_ISL_1543511, EPI_ISL_1543512, EPI_ISL_1543513, EPI_ISL_1543514, EPI_ISL_1543515, EPI_ISL_1543516, EPI_ISL_1543517, EPI_ISL_1543518, EPI_ISL_1543519, EPI_ISL_1543521, EPI_ISL_1543523, EPI_ISL_1543526, EPI_ISL_1543527, EPI_ISL_1543528, EPI_ISL_1543529, EPI_ISL_1543530, EPI_ISL_1543532, EPI_ISL_1543533, EPI_ISL_1543534, EPI_ISL_1543535, EPI_ISL_1543536, EPI_ISL_1543537, EPI_ISL_1543539, EPI_ISL_1543540, EPI_ISL_1543541, EPI_ISL_1543543, EPI_ISL_1543544, EPI_ISL_1543545, EPI_ISL_1543546, EPI_ISL_1543547, EPI_ISL_1543548, EPI_ISL_1543549, EPI_ISL_1543550, EPI_ISL_1543551, EPI_ISL_1543552, EPI_ISL_1543553, EPI_ISL_1543554, EPI_ISL_1543555, EPI_ISL_1543556, EPI_ISL_1543557, EPI_ISL_1543558, EPI_ISL_1543561, EPI_ISL_1543562, EPI_ISL_1543563, EPI_ISL_1543564, EPI_ISL_1543565, EPI_ISL_1543566, EPI_ISL_1543567, EPI_ISL_1543568, EPI_ISL_1543569, EPI_ISL_1543572, EPI_ISL_1543573, EPI_ISL_1543574, EPI_ISL_1543575, EPI_ISL_1543576, EPI_ISL_1543577, EPI_ISL_1543578, EPI_ISL_1543579, EPI_ISL_1543580, EPI_ISL_1543582, EPI_ISL_1543583, EPI_ISL_1543584, EPI_ISL_1543586, EPI_ISL_1543587, EPI_ISL_1543588, EPI_ISL_1543589, EPI_ISL_1543590, EPI_ISL_1543591, EPI_ISL_1543592, EPI_ISL_1543593, EPI_ISL_1543594, EPI_ISL_1543595, EPI_ISL_1543596, EPI_ISL_1543597, EPI_ISL_1543598, EPI_ISL_1543599, EPI_ISL_1543600, EPI_ISL_1543601, EPI_ISL_1543602, EPI_ISL_1543603, EPI_ISL_1543604, EPI_ISL_1543605, EPI_ISL_1543607, EPI_ISL_1543608, EPI_ISL_1543609, EPI_ISL_1543610, EPI_ISL_1543612, EPI_ISL_1543613, EPI_ISL_1543614, EPI_ISL_1543615, EPI_ISL_1543616, EPI_ISL_1543617, EPI_ISL_1543620, EPI_ISL_1543621, EPI_ISL_1543623, EPI_ISL_1543624, EPI_ISL_1543626, EPI_ISL_1543628, EPI_ISL_1543629, EPI_ISL_1543630, EPI_ISL_1543631, EPI_ISL_1543632, EPI_ISL_1543633, EPI_ISL_1543635, EPI_ISL_1543636, EPI_ISL_1543637, EPI_ISL_1543638, EPI_ISL_1543639, EPI_ISL_1543640, EPI_ISL_1543641, EPI_ISL_1543642, EPI_ISL_1543643, EPI_ISL_1543644, EPI_ISL_1543646, EPI_ISL_1543647, EPI_ISL_1543648, EPI_ISL_1543649, EPI_ISL_1543651, EPI_ISL_1543652, EPI_ISL_1543654, EPI_ISL_1543655, EPI_ISL_1543656, EPI_ISL_1543657, EPI_ISL_1543658, EPI_ISL_1543659, EPI_ISL_1543660, EPI_ISL_1543662, EPI_ISL_1543663, EPI_ISL_1543664, EPI_ISL_1543665, EPI_ISL_1543669, EPI_ISL_1543670, EPI_ISL_1543671, EPI_ISL_1543673, EPI_ISL_1543675, EPI_ISL_1543676, EPI_ISL_1543677, EPI_ISL_1543678, EPI_ISL_1543679, EPI_ISL_1543680, EPI_ISL_1543681, EPI_ISL_1543682, EPI_ISL_1543683, EPI_ISL_1543685, EPI_ISL_1543687, EPI_ISL_1543688, EPI_ISL_1543689, EPI_ISL_1543690, EPI_ISL_1543691, EPI_ISL_1543692, EPI_ISL_1543693, EPI_ISL_1543694, EPI_ISL_1543696, EPI_ISL_1543697, EPI_ISL_1543698, EPI_ISL_1543699, EPI_ISL_1543700, EPI_ISL_1543701, EPI_ISL_1543702, EPI_ISL_1543703, EPI_ISL_1543704, EPI_ISL_1543705, EPI_ISL_1543706, EPI_ISL_1543707, EPI_ISL_1543708, EPI_ISL_1543711, EPI_ISL_1543712, EPI_ISL_1543713, EPI_ISL_1543714, EPI_ISL_1543715, EPI_ISL_1543716, EPI_ISL_1543717, EPI_ISL_1543718, EPI_ISL_1543719, EPI_ISL_1543720, EPI_ISL_1543721, EPI_ISL_1543722, EPI_ISL_1543723, EPI_ISL_1543724, EPI_ISL_1543725, EPI_ISL_1543726, EPI_ISL_1543727, EPI_ISL_1543728, EPI_ISL_1543730, EPI_ISL_1543731, EPI_ISL_1543732, EPI_ISL_1543733, EPI_ISL_1543736, EPI_ISL_1543737, EPI_ISL_1543739, EPI_ISL_1543740, EPI_ISL_1543741, EPI_ISL_1543742, EPI_ISL_1543743, EPI_ISL_1543745, EPI_ISL_1543746, EPI_ISL_1543748, EPI_ISL_1543749, EPI_ISL_1543750, EPI_ISL_1543751, EPI_ISL_1543752, EPI_ISL_1543753, EPI_ISL_1543754, EPI_ISL_1543755, EPI_ISL_1543756, EPI_ISL_1543757, EPI_ISL_1543758, EPI_ISL_1543760, EPI_ISL_1543761, EPI_ISL_1543762, EPI_ISL_1543764, EPI_ISL_1543765, EPI_ISL_1543766, EPI_ISL_1543767, EPI_ISL_1543768, EPI_ISL_1543769, EPI_ISL_1543772, EPI_ISL_1543773, EPI_ISL_1543774, EPI_ISL_1543775, EPI_ISL_1543776, EPI_ISL_1543777, EPI_ISL_1543778, EPI_ISL_1543780, EPI_ISL_1543781, EPI_ISL_1543782, EPI_ISL_1543783, EPI_ISL_1543785, EPI_ISL_1543786, EPI_ISL_1543787, EPI_ISL_1543789, EPI_ISL_1543790, EPI_ISL_1543791, EPI_ISL_1543792, EPI_ISL_1543794, EPI_ISL_1543795, EPI_ISL_1543796, EPI_ISL_1543798, EPI_ISL_1543799, EPI_ISL_1543800, EPI_ISL_1543802, EPI_ISL_1543803, EPI_ISL_1543805, EPI_ISL_1543807, EPI_ISL_1543808, EPI_ISL_1543809, EPI_ISL_1543810, EPI_ISL_1543811, EPI_ISL_1543812, EPI_ISL_1543814, EPI_ISL_1543815, EPI_ISL_1543817, EPI_ISL_1543818, EPI_ISL_1543819, EPI_ISL_1543820, EPI_ISL_1543821, EPI_ISL_1543822, EPI_ISL_1543823, EPI_ISL_1543825, EPI_ISL_1543826, EPI_ISL_1543829, EPI_ISL_1543830, EPI_ISL_1543831, EPI_ISL_1543833, EPI_ISL_1543835, EPI_ISL_1543836, EPI_ISL_1543837, EPI_ISL_1543838, EPI_ISL_1543839, EPI_ISL_1543840, EPI_ISL_1543841, EPI_ISL_1543844, EPI_ISL_1543845, EPI_ISL_1543846, EPI_ISL_1543847, EPI_ISL_1543848, EPI_ISL_1543850, EPI_ISL_1543851, EPI_ISL_1543852, EPI_ISL_1543853, EPI_ISL_1543854, EPI_ISL_1543855, EPI_ISL_1543856, EPI_ISL_1543857, EPI_ISL_1543859, EPI_ISL_1543861, EPI_ISL_1543862, EPI_ISL_1543863, EPI_ISL_1543866, EPI_ISL_1543867, EPI_ISL_1543868, EPI_ISL_1543869, EPI_ISL_1543870, EPI_ISL_1543871, EPI_ISL_1543872, EPI_ISL_1543873, EPI_ISL_1543874, EPI_ISL_1543875, EPI_ISL_1543876, EPI_ISL_1543877, EPI_ISL_1543878, EPI_ISL_1543879, EPI_ISL_1543880, EPI_ISL_1543881, EPI_ISL_1543882, EPI_ISL_1543883, EPI_ISL_1543884, EPI_ISL_1543885, EPI_ISL_1543886, EPI_ISL_1543887, EPI_ISL_1543888, EPI_ISL_1543889, EPI_ISL_1543890, EPI_ISL_1543891, EPI_ISL_1543892, EPI_ISL_1543893, EPI_ISL_1543895, EPI_ISL_1543896, EPI_ISL_1543897, EPI_ISL_1543898, EPI_ISL_1543899, EPI_ISL_1543900, EPI_ISL_1543901, EPI_ISL_1543902, EPI_ISL_1543903, EPI_ISL_1543904, EPI_ISL_1543905, EPI_ISL_1543906, EPI_ISL_1543907, EPI_ISL_1543908, EPI_ISL_1543909, EPI_ISL_1543910, EPI_ISL_1543911, EPI_ISL_1543912, EPI_ISL_1543914, EPI_ISL_1543915, EPI_ISL_1543918, EPI_ISL_1543919, EPI_ISL_1543920 | | | |

| | | | |
|---|---|---|---|
| see above | Pandemic Response Lab - NYC | Pandemic Response Lab, R&D | Henry Lee, Michael Hammerling, Melissa Hopkins, Cybill del Castillo, Shinyoung Clair Kang, William Ward, Pradeep Bugga, Sol Rey, Dylan Law, Katharine Nelson, Haiping Hao, Jon Laurent |
| EPI_ISL_1545442, EPI_ISL_1545443, EPI_ISL_1545444 | Ohio Department of Health Laboratory | Ohio Department of Health Laboratory | Holmes, Jennifer; Eric Brandt, Keoni Omura, Glen McGillivary, Caitlin McDonnell, Jade Mowery, Stephanie Mccracken, Tyler Payne, Kirtana Ramadugu, Erica Leasure, Brent Lee, Kelsey Florek, Heather Blankenship, Allison Black, Quanta Brown, and Tammy Bannerman |
| EPI_ISL_1547908, EPI_ISL_1547909, EPI_ISL_1547910, EPI_ISL_1547911, EPI_ISL_1547913, EPI_ISL_1547914, EPI_ISL_1547915, EPI_ISL_1547917, EPI_ISL_1547918, EPI_ISL_1547919, EPI_ISL_1547921, EPI_ISL_1547922, EPI_ISL_1547923, EPI_ISL_1547924, EPI_ISL_1547925, EPI_ISL_1547926, EPI_ISL_1547927, EPI_ISL_1547928, EPI_ISL_1547929, EPI_ISL_1547931, EPI_ISL_1547932, EPI_ISL_1547933, EPI_ISL_1547934, EPI_ISL_1547935, EPI_ISL_1547936, EPI_ISL_1547937, EPI_ISL_1547938, EPI_ISL_1547939, EPI_ISL_1547940, EPI_ISL_1547941, EPI_ISL_1547942, EPI_ISL_1547944, EPI_ISL_1547945, EPI_ISL_1547946, EPI_ISL_1547947, EPI_ISL_1547948, EPI_ISL_1547950, EPI_ISL_1547951, EPI_ISL_1547952, EPI_ISL_1547953, EPI_ISL_1547954, EPI_ISL_1547955, EPI_ISL_1547957, EPI_ISL_1547958, EPI_ISL_1547959, EPI_ISL_1547960, EPI_ISL_1547961, EPI_ISL_1547962, EPI_ISL_1547963, EPI_ISL_1547964, EPI_ISL_1547965, EPI_ISL_1547966, EPI_ISL_1547967, EPI_ISL_1547968, EPI_ISL_1547969, EPI_ISL_1547970, EPI_ISL_1547971, EPI_ISL_1547972, EPI_ISL_1547973, EPI_ISL_1547974, EPI_ISL_1547975, EPI_ISL_1547976, EPI_ISL_1547977, EPI_ISL_1547978, EPI_ISL_1547979, EPI_ISL_1547980, EPI_ISL_1547981, EPI_ISL_1547982, EPI_ISL_1547983, EPI_ISL_1547984, EPI_ISL_1547985, EPI_ISL_1547986, EPI_ISL_1547987, EPI_ISL_1547988, EPI_ISL_1547989, EPI_ISL_1547990, EPI_ISL_1547991, EPI_ISL_1547992, EPI_ISL_1547993, EPI_ISL_1547994, EPI_ISL_1547995, EPI_ISL_1547996, EPI_ISL_1547997, EPI_ISL_1547999, EPI_ISL_1548000, EPI_ISL_1548002, EPI_ISL_1548003, EPI_ISL_1548004, EPI_ISL_1548005, EPI_ISL_1548006, EPI_ISL_1548007, EPI_ISL_1548008, EPI_ISL_1548009, EPI_ISL_1548010, EPI_ISL_1548011, EPI_ISL_1548012, EPI_ISL_1548013, EPI_ISL_1548014, EPI_ISL_1548015, EPI_ISL_1548016, EPI_ISL_1548017, EPI_ISL_1548018, EPI_ISL_1548019, EPI_ISL_1548020, EPI_ISL_1548021, EPI_ISL_1548022, EPI_ISL_1548023, EPI_ISL_1548024, EPI_ISL_1548025, EPI_ISL_1548026, EPI_ISL_1548027, EPI_ISL_1548028, EPI_ISL_1548029, EPI_ISL_1548030, EPI_ISL_1548031, EPI_ISL_1548032, EPI_ISL_1548033, EPI_ISL_1548034, EPI_ISL_1548035, EPI_ISL_1548036, EPI_ISL_1548037, EPI_ISL_1548038 | | | |
| see above | Johns Hopkins Hospital Department of Pathology | Johns Hopkins Hospital Department of Pathology | C. Paul Morris, Chun Huai Luo, Adannaya Amadi, Matthew Schwartz, Heba H. Mostafa |
| EPI_ISL_1548039, EPI_ISL_1548042, EPI_ISL_1548043, EPI_ISL_1548073, EPI_ISL_1548075, EPI_ISL_1548076, EPI_ISL_1548077, EPI_ISL_1548111, EPI_ISL_1548112, EPI_ISL_1548113, EPI_ISL_1548114, EPI_ISL_1548115, EPI_ISL_1548116 | | | |
| see above | Montana Public Health Laboratory | Montana Public Health Laboratory | Joy Ritter, Michelle Mozer, Carrie Biskupiak, Deborah Gibson |
| EPI_ISL_1548118, EPI_ISL_1548119, EPI_ISL_1548120, EPI_ISL_1548125, EPI_ISL_1548126, EPI_ISL_1548129, EPI_ISL_1548130, EPI_ISL_1548132, EPI_ISL_1548133, EPI_ISL_1548136, EPI_ISL_1548138, EPI_ISL_1548141, EPI_ISL_1548143, EPI_ISL_1548145, EPI_ISL_1548146, EPI_ISL_1548151, EPI_ISL_1548152, EPI_ISL_1548154, EPI_ISL_1548155, EPI_ISL_1548156, EPI_ISL_1548158, EPI_ISL_1548160, EPI_ISL_1548162, EPI_ISL_1548167, EPI_ISL_1548168, EPI_ISL_1548169, EPI_ISL_1548171, EPI_ISL_1548174, EPI_ISL_1548175, EPI_ISL_1548177, EPI_ISL_1548178, EPI_ISL_1548179, EPI_ISL_1548182, EPI_ISL_1548183, EPI_ISL_1548184, EPI_ISL_1548185, EPI_ISL_1548186, EPI_ISL_1548187, EPI_ISL_1548190, EPI_ISL_1548191, EPI_ISL_1548192, EPI_ISL_1548193, EPI_ISL_1548194, EPI_ISL_1548195, EPI_ISL_1548196, EPI_ISL_1548197, EPI_ISL_1548198, EPI_ISL_1548199, EPI_ISL_1548200, EPI_ISL_1548201, EPI_ISL_1548202, EPI_ISL_1548203, EPI_ISL_1548204, EPI_ISL_1548205, EPI_ISL_1548206, EPI_ISL_1548207, EPI_ISL_1548208, EPI_ISL_1548209, EPI_ISL_1548212, EPI_ISL_1548216, EPI_ISL_1548223, EPI_ISL_1548224, EPI_ISL_1548226, EPI_ISL_1548230, EPI_ISL_1548231, EPI_ISL_1548232, EPI_ISL_1548233, EPI_ISL_1548234, EPI_ISL_1548235, EPI_ISL_1548236, EPI_ISL_1548239, EPI_ISL_1548240, EPI_ISL_1548241, EPI_ISL_1548242, EPI_ISL_1548246, EPI_ISL_1548249, EPI_ISL_1548251, EPI_ISL_1548253, EPI_ISL_1548256, EPI_ISL_1548257, EPI_ISL_1548259, EPI_ISL_1548261, EPI_ISL_1548263, EPI_ISL_1548264, EPI_ISL_1548265, EPI_ISL_1548270, EPI_ISL_1548277, EPI_ISL_1548278, EPI_ISL_1548279, EPI_ISL_1548280, EPI_ISL_1548285, EPI_ISL_1548286, EPI_ISL_1548287, EPI_ISL_1548288, EPI_ISL_1548289, EPI_ISL_1548290, EPI_ISL_1548291, EPI_ISL_1548293, EPI_ISL_1548294, EPI_ISL_1548297, EPI_ISL_1548299, EPI_ISL_1548300, EPI_ISL_1548302, EPI_ISL_1548303, EPI_ISL_1548304, EPI_ISL_1548305, EPI_ISL_1548306, EPI_ISL_1548307, EPI_ISL_1548308, EPI_ISL_1548310, EPI_ISL_1548311, EPI_ISL_1548314, EPI_ISL_1548315, EPI_ISL_1548320, EPI_ISL_1548321, EPI_ISL_1548322, EPI_ISL_1548323, EPI_ISL_1548324, EPI_ISL_1548325, EPI_ISL_1548326, EPI_ISL_1548329, EPI_ISL_1548331, EPI_ISL_1548333, EPI_ISL_1548334, EPI_ISL_1548335, EPI_ISL_1548336, EPI_ISL_1548337, EPI_ISL_1548339, EPI_ISL_1548340, EPI_ISL_1548341, EPI_ISL_1548342, EPI_ISL_1548343, EPI_ISL_1548344, EPI_ISL_1548347, EPI_ISL_1548348, EPI_ISL_1548354, EPI_ISL_1548356, EPI_ISL_1548360, EPI_ISL_1548363, EPI_ISL_1548364, EPI_ISL_1548365, EPI_ISL_1548366, EPI_ISL_1548367, EPI_ISL_1548378, EPI_ISL_1548380, EPI_ISL_1548382, EPI_ISL_1548383, EPI_ISL_1548387, EPI_ISL_1548391, EPI_ISL_1548392, EPI_ISL_1548395, EPI_ISL_1548396, EPI_ISL_1548401, EPI_ISL_1548402, EPI_ISL_1548403, EPI_ISL_1548405, EPI_ISL_1548406, EPI_ISL_1548408, EPI_ISL_1548409, EPI_ISL_1548410, EPI_ISL_1548412, EPI_ISL_1548414, EPI_ISL_1548416, EPI_ISL_1548417, EPI_ISL_1548419, EPI_ISL_1548421, EPI_ISL_1548426, EPI_ISL_1548430, EPI_ISL_1548431, EPI_ISL_1548432, EPI_ISL_1548433, EPI_ISL_1548434, EPI_ISL_1548436, EPI_ISL_1548437, EPI_ISL_1548440, EPI_ISL_1548441, EPI_ISL_1548443, EPI_ISL_1548445, EPI_ISL_1548447, EPI_ISL_1548450, EPI_ISL_1548455, EPI_ISL_1548456, EPI_ISL_1548457, EPI_ISL_1548458, EPI_ISL_1548461, EPI_ISL_1548462, EPI_ISL_1548463, EPI_ISL_1548464, EPI_ISL_1548465, EPI_ISL_1548468, EPI_ISL_1548469, EPI_ISL_1548470, EPI_ISL_1548471, EPI_ISL_1548472, EPI_ISL_1548474, EPI_ISL_1548475, EPI_ISL_1548476, EPI_ISL_1548477, EPI_ISL_1548478, EPI_ISL_1548479, EPI_ISL_1548483, EPI_ISL_1548484, EPI_ISL_1548485, EPI_ISL_1548489, EPI_ISL_1548492, EPI_ISL_1548493, EPI_ISL_1548494, EPI_ISL_1548495, EPI_ISL_1548496, EPI_ISL_1548497, EPI_ISL_1548502, EPI_ISL_1548503, EPI_ISL_1548506, EPI_ISL_1548507, EPI_ISL_1548508, EPI_ISL_1548510, EPI_ISL_1548513, EPI_ISL_1548514, EPI_ISL_1548516, EPI_ISL_1548517, EPI_ISL_1548518, EPI_ISL_1548519, EPI_ISL_1548520, EPI_ISL_1548523, EPI_ISL_1548524, EPI_ISL_1548526, EPI_ISL_1548527, EPI_ISL_1548528, EPI_ISL_1548530, EPI_ISL_1548531, EPI_ISL_1548532, EPI_ISL_1548533, EPI_ISL_1548535, EPI_ISL_1548536, EPI_ISL_1548537, EPI_ISL_1548538, EPI_ISL_1548539, EPI_ISL_1548540, EPI_ISL_1548541, EPI_ISL_1548542, EPI_ISL_1548544, EPI_ISL_1548545, EPI_ISL_1548546, EPI_ISL_1548547, EPI_ISL_1548549, EPI_ISL_1548550, EPI_ISL_1548551, EPI_ISL_1548552, EPI_ISL_1548554, EPI_ISL_1548556, EPI_ISL_1548557, EPI_ISL_1548558, EPI_ISL_1548560, EPI_ISL_1548562, EPI_ISL_1548563, EPI_ISL_1548565, EPI_ISL_1548566, EPI_ISL_1548567, EPI_ISL_1548568, EPI_ISL_1548569, EPI_ISL_1548570, EPI_ISL_1548571, EPI_ISL_1548573, EPI_ISL_1548574, EPI_ISL_1548575, EPI_ISL_1548577, EPI_ISL_1548578, EPI_ISL_1548579, EPI_ISL_1548581, EPI_ISL_1548582, EPI_ISL_1548583, EPI_ISL_1548585, EPI_ISL_1548586, EPI_ISL_1548587, EPI_ISL_1548592, EPI_ISL_1548593, EPI_ISL_1548595, EPI_ISL_1548596, EPI_ISL_1548597, EPI_ISL_1548598, EPI_ISL_1548599, EPI_ISL_1548601, EPI_ISL_1548602, EPI_ISL_1548603, EPI_ISL_1548604, EPI_ISL_1548605, EPI_ISL_1548607, EPI_ISL_1548608, EPI_ISL_1548609, EPI_ISL_1548611, EPI_ISL_1548612, EPI_ISL_1548613, EPI_ISL_1548616, EPI_ISL_1548617, EPI_ISL_1548619, EPI_ISL_1548621, EPI_ISL_1548626, EPI_ISL_1548627, EPI_ISL_1548629, EPI_ISL_1548630, EPI_ISL_1548632, EPI_ISL_1548633, EPI_ISL_1548634, EPI_ISL_1548636, EPI_ISL_1548641, EPI_ISL_1548642, EPI_ISL_1548643, EPI_ISL_1548644, EPI_ISL_1548645, EPI_ISL_1548646, EPI_ISL_1548648, EPI_ISL_1548649, EPI_ISL_1548650, EPI_ISL_1548652, EPI_ISL_1548653, EPI_ISL_1548654, EPI_ISL_1548655, EPI_ISL_1548658, EPI_ISL_1548661, EPI_ISL_1548662, EPI_ISL_1548663, EPI_ISL_1548664, EPI_ISL_1548666, EPI_ISL_1548667, EPI_ISL_1548668, EPI_ISL_1548669, EPI_ISL_1548670, EPI_ISL_1548671, EPI_ISL_1548672, EPI_ISL_1548673, EPI_ISL_1548674, EPI_ISL_1548675, EPI_ISL_1548676, EPI_ISL_1548679, EPI_ISL_1548680, EPI_ISL_1548682, EPI_ISL_1548683, EPI_ISL_1548685, EPI_ISL_1548689, EPI_ISL_1548690, EPI_ISL_1548692, EPI_ISL_1548693, EPI_ISL_1548694, EPI_ISL_1548697, EPI_ISL_1548698, EPI_ISL_1548699, EPI_ISL_1548700, EPI_ISL_1548701, EPI_ISL_1548703, EPI_ISL_1548704, EPI_ISL_1548706, EPI_ISL_1548707, EPI_ISL_1548708, EPI_ISL_1548709, EPI_ISL_1548711, EPI_ISL_1548712, EPI_ISL_1548713, EPI_ISL_1548718, EPI_ISL_1548723, EPI_ISL_1548725, EPI_ISL_1548726, EPI_ISL_1548727, EPI_ISL_1548728, EPI_ISL_1548729, EPI_ISL_1548731, EPI_ISL_1548736, EPI_ISL_1548737, EPI_ISL_1548738, EPI_ISL_1548739, EPI_ISL_1548741, EPI_ISL_1548743, EPI_ISL_1548745, EPI_ISL_1548746, EPI_ISL_1548748, EPI_ISL_1548750, EPI_ISL_1548751, EPI_ISL_1548753, EPI_ISL_1548755, EPI_ISL_1548757, EPI_ISL_1548759, EPI_ISL_1548760, EPI_ISL_1548761, EPI_ISL_1548762, EPI_ISL_1548763, EPI_ISL_1548764, EPI_ISL_1548768, EPI_ISL_1548769, EPI_ISL_1548770, EPI_ISL_1548772, EPI_ISL_1548774, EPI_ISL_1548777, EPI_ISL_1548778, EPI_ISL_1548779, EPI_ISL_1548780, EPI_ISL_1548781, EPI_ISL_1548782, EPI_ISL_1548784, EPI_ISL_1548786, EPI_ISL_1548787, EPI_ISL_1548789, EPI_ISL_1548790, EPI_ISL_1548791, EPI_ISL_1548792, EPI_ISL_1548793, EPI_ISL_1548794, EPI_ISL_1548797, EPI_ISL_1548798, EPI_ISL_1548799, EPI_ISL_1548800, EPI_ISL_1548802, EPI_ISL_1548803, EPI_ISL_1548804, EPI_ISL_1548806, EPI_ISL_1548807, EPI_ISL_1548809, EPI_ISL_1548811, EPI_ISL_1548812, EPI_ISL_1548813, EPI_ISL_1548814, EPI_ISL_1548815, EPI_ISL_1548816, EPI_ISL_1548817, EPI_ISL_1548818, EPI_ISL_1548819, EPI_ISL_1548823, EPI_ISL_1548825, EPI_ISL_1548826, EPI_ISL_1548828, EPI_ISL_1548829, EPI_ISL_1548830, EPI_ISL_1548832, EPI_ISL_1548834, EPI_ISL_1548835, EPI_ISL_1548837, EPI_ISL_1548838, EPI_ISL_1548839, EPI_ISL_1548840, EPI_ISL_1548841, EPI_ISL_1548842, EPI_ISL_1548843, EPI_ISL_1548845, EPI_ISL_1548846, EPI_ISL_1548847, EPI_ISL_1548848, EPI_ISL_1548850, EPI_ISL_1548851, EPI_ISL_1548853, EPI_ISL_1548855, EPI_ISL_1548856, EPI_ISL_1548857, EPI_ISL_1548858, EPI_ISL_1548859, EPI_ISL_1548861, EPI_ISL_1548862, EPI_ISL_1548863, EPI_ISL_1548865, EPI_ISL_1548867, EPI_ISL_1548869, EPI_ISL_1548871, EPI_ISL_1548872, EPI_ISL_1548874, EPI_ISL_1548877, EPI_ISL_1548878, EPI_ISL_1548879, EPI_ISL_1548881, EPI_ISL_1548883, EPI_ISL_1548887, EPI_ISL_1548888, EPI_ISL_1548890, EPI_ISL_1548891, EPI_ISL_1548892, EPI_ISL_1548894, EPI_ISL_1548895, EPI_ISL_1548899, EPI_ISL_1548900, EPI_ISL_1548901, EPI_ISL_1548903, EPI_ISL_1548904, EPI_ISL_1548905, EPI_ISL_1548906, EPI_ISL_1548907, EPI_ISL_1548908, EPI_ISL_1548909, EPI_ISL_1548910, EPI_ISL_1548911, EPI_ISL_1548917, EPI_ISL_1548918, EPI_ISL_1548919, EPI_ISL_1548922, EPI_ISL_1548923, EPI_ISL_1548924, EPI_ISL_1548925, EPI_ISL_1548927, EPI_ISL_1548928, EPI_ISL_1548929, EPI_ISL_1548931, EPI_ISL_1548932, EPI_ISL_1548934, EPI_ISL_1548935, EPI_ISL_1548936, EPI_ISL_1548938, EPI_ISL_1548939, EPI_ISL_1548941, EPI_ISL_1548942, EPI_ISL_1548944, EPI_ISL_1548945, EPI_ISL_1548947, EPI_ISL_1548948, EPI_ISL_1548949, EPI_ISL_1548953, EPI_ISL_1548954, EPI_ISL_1548957, EPI_ISL_1548958, EPI_ISL_1548959, EPI_ISL_1548960, EPI_ISL_1548966, EPI_ISL_1548967, EPI_ISL_1548968, EPI_ISL_1548975, EPI_ISL_1548977, EPI_ISL_1548978, EPI_ISL_1548979, EPI_ISL_1548980, EPI_ISL_1548981, EPI_ISL_1548983, EPI_ISL_1548985, EPI_ISL_1548986, EPI_ISL_1548987, EPI_ISL_1548988, EPI_ISL_1548989, EPI_ISL_1548990, EPI_ISL_1548993, EPI_ISL_1548994, EPI_ISL_1548995, EPI_ISL_1548996, EPI_ISL_1548997, EPI_ISL_1549000, EPI_ISL_1549001, EPI_ISL_1549002, EPI_ISL_1549004, EPI_ISL_1549005, EPI_ISL_1549006, EPI_ISL_1549008, EPI_ISL_1549009, EPI_ISL_1549012, EPI_ISL_1549014, EPI_ISL_1549015, EPI_ISL_1549020, EPI_ISL_1549021, EPI_ISL_1549022, EPI_ISL_1549024, EPI_ISL_1549025, EPI_ISL_1549026, EPI_ISL_1549027, EPI_ISL_1549029, EPI_ISL_1549040, EPI_ISL_1549050, EPI_ISL_1549053 | | | |
| see above | Laboratory Corporation of America | Centers for Disease Control and Prevention Division of Viral Diseases, Pathogen Discovery | Dakota Howard, Dhwani Batra, Peter W. Cook, Kara Moser, Adrian Paskey, Jason Caravas, Benjamin Rambo-Martin, Shatavia Morrison, Christopher Gulvick, Scott Sammons, Yvette Unoarumhi, Darlene Wagner, Matthew Schmerer, Minoo Agarwal, Eyad Almasri, Debbie Boles, Ayla Burns, Nuthawin Charoensri, Oren Cohen, Susan Countryman, Mary Ann Cristobal, Bobbi Croy, Suzanne Dale, Hrushikesh Deshmukh, Amanda Douglas, Vincent Drouillon, Marcia Eisenberg, Howard Engler, Rama Ghatti, Prashant Gupta, Susan Hicks, Jake Humphrey, Lax Iyer, Manoj Jain, Mohan Kolli, Brian Krueger, Tim Kuphal, Stanley Letovsky, Michael Levandoski, Craig Lukasik, Jonathan Meltzer, Brian Norvell, Mindy Nye, Scott Parker, Christos Petropoulos, John Pruitt, Steven Ragan, Scott Ryan, Mike Sapeta, Jana Schroth, Suresh Babu Selvaraju, Goran Stevovic, Amanda Suchanek, Andrea Throop, Lyndon Tilson, Thomas Urban, Joe Voshell, Kimberly Wagner, Jonathan Williams, Mary Williamson, Qian Zeng, Tricia Zwiefelhofer, Clinton R. Paden, Duncan MacCannell |
| EPI_ISL_1549139 | Montana Public Health Laboratory | Montana Public Health Laboratory | Joy Ritter, Michelle Mozer, Carrie Biskupiak, Deborah Gibson |
| EPI_ISL_1549143, EPI_ISL_1549144, EPI_ISL_1549145, EPI_ISL_1549146, EPI_ISL_1549147, EPI_ISL_1549148, EPI_ISL_1549150, EPI_ISL_1549153, EPI_ISL_1549154, EPI_ISL_1549155, EPI_ISL_1549156, EPI_ISL_1549157, EPI_ISL_1549158, EPI_ISL_1549159, EPI_ISL_1549160, EPI_ISL_1549161, EPI_ISL_1549162, EPI_ISL_1549163, EPI_ISL_1549164, EPI_ISL_1549165, EPI_ISL_1549166, EPI_ISL_1549167 | | | |
| see above | Laboratory Corporation of America | Centers for Disease Control and Prevention Division of Viral Diseases, Pathogen Discovery | Dakota Howard, Dhwani Batra, Peter W. Cook, Kara Moser, Adrian Paskey, Jason Caravas, Benjamin Rambo-Martin, Shatavia Morrison, Christopher Gulvick, Scott Sammons, Yvette Unoarumhi, Darlene Wagner, Matthew Schmerer, Minoo Agarwal, Eyad Almasri, Debbie Boles, Ayla Burns, Nuthawin Charoensri, Oren Cohen, Susan Countryman, Mary Ann Cristobal, Bobbi Croy, Suzanne Dale, Hrushikesh Deshmukh, Amanda Douglas, Vincent Drouillon, Marcia Eisenberg, Howard Engler, Rama Ghatti, Prashant Gupta, Susan Hicks, Jake Humphrey, Lax Iyer, Manoj Jain, Mohan Kolli, Brian Krueger, Tim Kuphal, Stanley Letovsky, Michael Levandoski, Craig Lukasik, Jonathan Meltzer, Brian Norvell, Mindy Nye, Scott Parker, Christos Petropoulos, John Pruitt, Steven Ragan, Scott Ryan, Mike Sapeta, Jana Schroth, Suresh Babu Selvaraju, Goran Stevovic, Amanda Suchanek, Andrea Throop, Lyndon Tilson, Thomas Urban, Joe Voshell, Kimberly Wagner, Jonathan Williams, Mary Williamson, Qian Zeng, Tricia Zwiefelhofer, Clinton R. Paden, Duncan MacCannell |
| EPI_ISL_1549168 | Montana Public Health Laboratory | Montana Public Health Laboratory | Joy Ritter, Michelle Mozer, Carrie Biskupiak, Deborah Gibson |
| EPI_ISL_1549170, EPI_ISL_1549171, EPI_ISL_1549172, EPI_ISL_1549173, EPI_ISL_1549175, EPI_ISL_1549176, EPI_ISL_1549177, EPI_ISL_1549178, EPI_ISL_1549179, EPI_ISL_1549180, EPI_ISL_1549181, EPI_ISL_1549182, EPI_ISL_1549183, EPI_ISL_1549184, EPI_ISL_1549185, EPI_ISL_1549186, EPI_ISL_1549187, EPI_ISL_1549188, EPI_ISL_1549189, EPI_ISL_1549190, EPI_ISL_1549191, EPI_ISL_1549193, EPI_ISL_1549194, EPI_ISL_1549195, EPI_ISL_1549196, EPI_ISL_1549197, EPI_ISL_1549198, EPI_ISL_1549199, EPI_ISL_1549200, EPI_ISL_1549201, EPI_ISL_1549202, EPI_ISL_1549203, EPI_ISL_1549204, EPI_ISL_1549205, EPI_ISL_1549207, EPI_ISL_1549208, EPI_ISL_1549212, EPI_ISL_1549213 | | | |
| see above | Laboratory Corporation of America | Centers for Disease Control and Prevention Division of Viral Diseases, Pathogen Discovery | Dakota Howard, Dhwani Batra, Peter W. Cook, Kara Moser, Adrian Paskey, Jason Caravas, Benjamin Rambo-Martin, Shatavia Morrison, Christopher Gulvick, Scott Sammons, Yvette Unoarumhi, Darlene Wagner, Matthew Schmerer, Minoo Agarwal, Eyad Almasri, Debbie Boles, Ayla Burns, Nuthawin Charoensri, Oren Cohen, Susan Countryman, Mary Ann Cristobal, Bobbi Croy, Suzanne Dale, Hrushikesh Deshmukh, Amanda Douglas, Vincent Drouillon, Marcia Eisenberg, Howard Engler, Rama Ghatti, Prashant Gupta, Susan Hicks, Jake Humphrey, Lax Iyer, Manoj Jain, Mohan Kolli, Brian Krueger, Tim Kuphal, Stanley Letovsky, Michael Levandoski, Craig Lukasik, Jonathan Meltzer, Brian Norvell, Mindy Nye, Scott Parker, Christos Petropoulos, John Pruitt, Steven Ragan, Scott Ryan, Mike Sapeta, Jana Schroth, Suresh Babu Selvaraju, Goran Stevovic, Amanda Suchanek, Andrea Throop, Lyndon Tilson, Thomas Urban, Joe Voshell, Kimberly Wagner, Jonathan Williams, Mary Williamson, Qian Zeng, Tricia Zwiefelhofer, Clinton R. Paden, Duncan MacCannell |
| EPI_ISL_1549214 | USA Health Care Authority | Synergy Laboratories | Megan Cornwell |
| EPI_ISL_1549219, EPI_ISL_1549220, EPI_ISL_1549222, EPI_ISL_1549224, EPI_ISL_1549225, EPI_ISL_1549226, EPI_ISL_1549227, EPI_ISL_1549228, EPI_ISL_1549229, EPI_ISL_1549230, EPI_ISL_1549231, EPI_ISL_1549233, EPI_ISL_1549234, EPI_ISL_1549236, EPI_ISL_1549237, EPI_ISL_1549239, EPI_ISL_1549240, EPI_ISL_1549241, EPI_ISL_1549242, EPI_ISL_1549243, EPI_ISL_1549244, EPI_ISL_1549245, EPI_ISL_1549246, EPI_ISL_1549247, EPI_ISL_1549248, EPI_ISL_1549251, EPI_ISL_1549253, EPI_ISL_1549340, EPI_ISL_1549342, EPI_ISL_1549344, EPI_ISL_1549347, EPI_ISL_1549352, EPI_ISL_1549355, EPI_ISL_1549356, EPI_ISL_1549357, EPI_ISL_1549391, EPI_ISL_1549392, EPI_ISL_1549393, EPI_ISL_1549394, EPI_ISL_1549395, EPI_ISL_1549396, EPI_ISL_1549397, EPI_ISL_1549399, EPI_ISL_1549400, EPI_ISL_1549401, EPI_ISL_1549402, EPI_ISL_1549404, EPI_ISL_1549405, EPI_ISL_1549406, EPI_ISL_1549407, EPI_ISL_1549408, EPI_ISL_1549409, EPI_ISL_1549410, EPI_ISL_1549411, EPI_ISL_1549412, EPI_ISL_1549413, EPI_ISL_1549414, EPI_ISL_1549415, EPI_ISL_1549416, EPI_ISL_1549417, EPI_ISL_1549418, EPI_ISL_1549419, EPI_ISL_1549420, EPI_ISL_1549421, EPI_ISL_1549422, EPI_ISL_1549423, EPI_ISL_1549424, EPI_ISL_1549425, EPI_ISL_1549426, EPI_ISL_1549427, EPI_ISL_1549429, EPI_ISL_1549430, EPI_ISL_1549431, EPI_ISL_1549432, EPI_ISL_1549433, EPI_ISL_1549434, EPI_ISL_1549435, EPI_ISL_1549436, EPI_ISL_1549437, EPI_ISL_1549438, EPI_ISL_1549439, EPI_ISL_1549440, EPI_ISL_1549441, EPI_ISL_1549442, EPI_ISL_1549443, EPI_ISL_1549444, EPI_ISL_1549445, EPI_ISL_1549446, EPI_ISL_1549447, EPI_ISL_1549448, EPI_ISL_1549449, EPI_ISL_1549450, EPI_ISL_1549451, EPI_ISL_1549452, EPI_ISL_1549453, EPI_ISL_1549454, EPI_ISL_1549455, EPI_ISL_1549456, EPI_ISL_1549457, EPI_ISL_1549458, EPI_ISL_1549459, EPI_ISL_1549460, EPI_ISL_1549461, EPI_ISL_1549462, EPI_ISL_1549463, EPI_ISL_1549464, EPI_ISL_1549465, EPI_ISL_1549466, EPI_ISL_1549467, EPI_ISL_1549468, EPI_ISL_1549470, EPI_ISL_1549471, EPI_ISL_1549472, EPI_ISL_1549473, EPI_ISL_1549474, EPI_ISL_1549475, EPI_ISL_1549476, EPI_ISL_1549477 | | | |
| see above | Laboratory Corporation of America | Centers for Disease Control and Prevention Division of Viral Diseases, Pathogen Discovery | Dakota Howard, Dhwani Batra, Peter W. Cook, Kara Moser, Adrian Paskey, Jason Caravas, Benjamin Rambo-Martin, Shatavia Morrison, Christopher Gulvick, Scott Sammons, Yvette Unoarumhi, Darlene Wagner, Matthew Schmerer, Minoo Agarwal, Eyad Almasri, Debbie Boles, Ayla Burns, Nuthawin Charoensri, Oren Cohen, Susan Countryman, Mary Ann Cristobal, Bobbi Croy, Suzanne Dale, Hrushikesh Deshmukh, Amanda Douglas, Vincent Drouillon, Marcia Eisenberg, Howard Engler, Rama Ghatti, Prashant Gupta, Susan Hicks, Jake Humphrey, Lax Iyer, Manoj Jain, Mohan Kolli, Brian Krueger, Tim Kuphal, Stanley Letovsky, Michael Levandoski, Craig Lukasik, Jonathan Meltzer, Brian Norvell, Mindy Nye, Scott Parker, Christos Petropoulos, John Pruitt, Steven Ragan, Scott Ryan, Mike Sapeta, Jana Schroth, Suresh Babu Selvaraju, Goran Stevovic, Amanda Suchanek, Andrea Throop, Lyndon Tilson, Thomas Urban, Joe Voshell, Kimberly Wagner, Jonathan Williams, Mary Williamson, Qian Zeng, Tricia Zwiefelhofer, Clinton R. Paden, Duncan MacCannell |
| EPI_ISL_1549478 | Montana Public Health Laboratory | Montana Public Health Laboratory | Joy Ritter, Michelle Mozer, Carrie Biskupiak, Deborah Gibson |
| EPI_ISL_1549479, EPI_ISL_1549480, EPI_ISL_1549481, EPI_ISL_1549482, EPI_ISL_1549483, EPI_ISL_1549484, EPI_ISL_1549485, EPI_ISL_1549486, EPI_ISL_1549487, EPI_ISL_1549488, EPI_ISL_1549489, EPI_ISL_1549490, EPI_ISL_1549491, EPI_ISL_1549492, EPI_ISL_1549493, EPI_ISL_1549494, EPI_ISL_1549495, EPI_ISL_1549496, EPI_ISL_1549497, EPI_ISL_1549498, EPI_ISL_1549499, EPI_ISL_1549500, EPI_ISL_1549501, EPI_ISL_1549502, EPI_ISL_1549503, EPI_ISL_1549504, EPI_ISL_1549505, EPI_ISL_1549506, EPI_ISL_1549507, EPI_ISL_1549508, EPI_ISL_1549509, EPI_ISL_1549510, EPI_ISL_1549511, EPI_ISL_1549512, EPI_ISL_1549513, EPI_ISL_1549514, EPI_ISL_1549515, EPI_ISL_1549516, EPI_ISL_1549517, EPI_ISL_1549518, EPI_ISL_1549519, EPI_ISL_1549520, EPI_ISL_1549521, EPI_ISL_1549522, EPI_ISL_1549523, EPI_ISL_1549524, EPI_ISL_1549525, EPI_ISL_1549526, EPI_ISL_1549527, EPI_ISL_1549528, EPI_ISL_1549529, EPI_ISL_1549530, EPI_ISL_1549531, EPI_ISL_1549533, EPI_ISL_1549534, EPI_ISL_1549535, EPI_ISL_1549536, EPI_ISL_1549537, EPI_ISL_1549538, EPI_ISL_1549540, EPI_ISL_1549541, EPI_ISL_1549542, EPI_ISL_1549543, EPI_ISL_1549544, EPI_ISL_1549545, EPI_ISL_1549546, EPI_ISL_1549547, EPI_ISL_1549548, EPI_ISL_1549549, EPI_ISL_1549550, EPI_ISL_1549551, EPI_ISL_1549552, EPI_ISL_1549553, EPI_ISL_1549554, EPI_ISL_1549555, EPI_ISL_1549556, EPI_ISL_1549557, EPI_ISL_1549558, EPI_ISL_1549559, EPI_ISL_1549560, EPI_ISL_1549561, EPI_ISL_1549562, EPI_ISL_1549563, EPI_ISL_1549564, EPI_ISL_1549565, EPI_ISL_1549566, EPI_ISL_1549567, EPI_ISL_1549568, EPI_ISL_1549569, EPI_ISL_1549571, EPI_ISL_1549574, EPI_ISL_1549577, EPI_ISL_1549580, EPI_ISL_1549583, EPI_ISL_1549585, EPI_ISL_1549589, EPI_ISL_1549590, EPI_ISL_1549591, EPI_ISL_1549592, EPI_ISL_1549593, EPI_ISL_1549594, EPI_ISL_1549595, EPI_ISL_1549596, EPI_ISL_1549597, EPI_ISL_1549598, EPI_ISL_1549599, EPI_ISL_1549600, EPI_ISL_1549601, EPI_ISL_1549602, EPI_ISL_1549603, EPI_ISL_1549604, EPI_ISL_1549606, EPI_ISL_1549607, EPI_ISL_1549608, EPI_ISL_1549609, EPI_ISL_1549610, EPI_ISL_1549611, EPI_ISL_1549612, EPI_ISL_1549613, EPI_ISL_1549614, EPI_ISL_1549615, EPI_ISL_1549616, EPI_ISL_1549617, EPI_ISL_1549619, EPI_ISL_1549620, EPI_ISL_1549622, EPI_ISL_1549623, EPI_ISL_1549624, EPI_ISL_1549625, EPI_ISL_1549626, EPI_ISL_1549627, EPI_ISL_1549628, EPI_ISL_1549629, EPI_ISL_1549631, EPI_ISL_1549632, EPI_ISL_1549633, EPI_ISL_1549634, EPI_ISL_1549635, EPI_ISL_1549636, EPI_ISL_1549637, EPI_ISL_1549638, EPI_ISL_1549639, EPI_ISL_1549640, EPI_ISL_1549641, EPI_ISL_1549642, EPI_ISL_1549643, EPI_ISL_1549644, EPI_ISL_1549645, EPI_ISL_1549646, EPI_ISL_1549647, EPI_ISL_1549648, EPI_ISL_1549649, EPI_ISL_1549651, EPI_ISL_1549652, EPI_ISL_1549653, EPI_ISL_1549654, EPI_ISL_1549655, EPI_ISL_1549656, EPI_ISL_1549657, EPI_ISL_1549658, EPI_ISL_1549659, EPI_ISL_1549660, EPI_ISL_1549661, EPI_ISL_1549663, EPI_ISL_1549664, EPI_ISL_1549665, EPI_ISL_1549666, EPI_ISL_1549667, EPI_ISL_1549668, EPI_ISL_1549670, EPI_ISL_1549671, EPI_ISL_1549672, EPI_ISL_1549673, EPI_ISL_1549674, EPI_ISL_1549675, EPI_ISL_1549676, EPI_ISL_1549677, EPI_ISL_1549679, EPI_ISL_1549680, EPI_ISL_1549681, EPI_ISL_1549682, EPI_ISL_1549683, EPI_ISL_1549684, EPI_ISL_1549685, EPI_ISL_1549686, EPI_ISL_1549687, EPI_ISL_1549688, EPI_ISL_1549689, EPI_ISL_1549690, EPI_ISL_1549691, EPI_ISL_1549692, EPI_ISL_1549693, EPI_ISL_1549694, EPI_ISL_1549695, EPI_ISL_1549696, EPI_ISL_1549697, EPI_ISL_1549698, EPI_ISL_1549699, EPI_ISL_1549700, EPI_ISL_1549701, EPI_ISL_1549702, EPI_ISL_1549703, EPI_ISL_1549704, EPI_ISL_1549705, EPI_ISL_1549706, EPI_ISL_1549707, EPI_ISL_1549708, EPI_ISL_1549709, EPI_ISL_1549710, EPI_ISL_1549711, EPI_ISL_1549712, EPI_ISL_1549714, EPI_ISL_1549715, EPI_ISL_1549716, EPI_ISL_1549717, EPI_ISL_1549718, EPI_ISL_1549719, EPI_ISL_1549721, EPI_ISL_1549722, EPI_ISL_1549723, EPI_ISL_1549724, EPI_ISL_1549725, EPI_ISL_1549726 | | | |
| see above | Laboratory Corporation of America | Centers for Disease Control and Prevention Division of Viral Diseases, Pathogen Discovery | Dakota Howard, Dhwani Batra, Peter W. Cook, Kara Moser, Adrian Paskey, Jason Caravas, Benjamin Rambo-Martin, Shatavia Morrison, Christopher Gulvick, Scott Sammons, Yvette Unoarumhi, Darlene Wagner, Matthew Schmerer, Minoo Agarwal, Eyad Almasri, Debbie Boles, Ayla Burns, Nuthawin Charoensri, Oren Cohen, Susan Countryman, Mary Ann Cristobal, Bobbi Croy, Suzanne Dale, Hrushikesh Deshmukh, Amanda Douglas, Vincent Drouillon, Marcia Eisenberg, Howard Engler, Rama Ghatti, Prashant Gupta, Susan Hicks, Jake Humphrey, Lax Iyer, Manoj Jain, Mohan Kolli, Brian Krueger, Tim Kuphal, Stanley Letovsky, Michael Levandoski, Craig Lukasik, Jonathan Meltzer, Brian Norvell, Mindy Nye, Scott Parker, Christos Petropoulos, John Pruitt, Steven Ragan, Scott Ryan, Mike Sapeta, Jana Schroth, Suresh Babu Selvaraju, Goran Stevovic, Amanda Suchanek, Andrea Throop, Lyndon Tilson, Thomas Urban, Joe Voshell, Kimberly Wagner, Jonathan Williams, Mary Williamson, Qian Zeng, Tricia Zwiefelhofer, Clinton R. Paden, Duncan MacCannell |
| EPI_ISL_1549727 | Montana Public Health Laboratory | Montana Public Health Laboratory | Joy Ritter, Michelle Mozer, Carrie Biskupiak, Deborah Gibson |
| EPI_ISL_1549728, EPI_ISL_1549729, EPI_ISL_1549730, EPI_ISL_1549731, EPI_ISL_1549732, EPI_ISL_1549733, EPI_ISL_1549734, EPI_ISL_1549735, EPI_ISL_1549736, EPI_ISL_1549737, EPI_ISL_1549738, EPI_ISL_1549739, EPI_ISL_1549740, EPI_ISL_1549741, EPI_ISL_1549742, EPI_ISL_1549743, EPI_ISL_1549745, EPI_ISL_1549746, EPI_ISL_1549747, EPI_ISL_1549748, EPI_ISL_1549749, EPI_ISL_1549750, EPI_ISL_1549751, EPI_ISL_1549752, EPI_ISL_1549755, EPI_ISL_1549756, EPI_ISL_1549758, EPI_ISL_1549759, EPI_ISL_1549760, EPI_ISL_1549761, EPI_ISL_1549762, EPI_ISL_1549763, EPI_ISL_1549764, EPI_ISL_1549765, EPI_ISL_1549766, EPI_ISL_1549767, EPI_ISL_1549768, EPI_ISL_1549769, EPI_ISL_1549770, EPI_ISL_1549771, EPI_ISL_1549772, EPI_ISL_1549773, EPI_ISL_1549774, EPI_ISL_1549775, EPI_ISL_1549776, EPI_ISL_1549777, EPI_ISL_1549778, EPI_ISL_1549779, EPI_ISL_1549780, EPI_ISL_1549781, EPI_ISL_1549782, EPI_ISL_1549783, EPI_ISL_1549784, EPI_ISL_1549785, EPI_ISL_1549786, EPI_ISL_1549787, EPI_ISL_1549788, EPI_ISL_1549789, EPI_ISL_1549790, EPI_ISL_1549791, EPI_ISL_1549792, EPI_ISL_1549793, EPI_ISL_1549794, EPI_ISL_1549795, EPI_ISL_1549796, EPI_ISL_1549798, EPI_ISL_1549801, EPI_ISL_1549802, EPI_ISL_1549803, EPI_ISL_1549804, EPI_ISL_1549805, EPI_ISL_1549806, EPI_ISL_1549807, EPI_ISL_1549808, EPI_ISL_1549811, EPI_ISL_1549812, EPI_ISL_1549813, EPI_ISL_1549814, EPI_ISL_1549815, EPI_ISL_1549816, EPI_ISL_1549818, EPI_ISL_1549819, EPI_ISL_1549820, EPI_ISL_1549821, EPI_ISL_1549822, EPI_ISL_1549825, EPI_ISL_1549826, EPI_ISL_1549827, EPI_ISL_1549828, EPI_ISL_1549829, EPI_ISL_1549830, EPI_ISL_1549831, EPI_ISL_1549832, EPI_ISL_1549833, EPI_ISL_1549834, EPI_ISL_1549837, EPI_ISL_1549838, EPI_ISL_1549839, EPI_ISL_1549840, EPI_ISL_1549841, EPI_ISL_1549843, EPI_ISL_1549844, EPI_ISL_1549847, EPI_ISL_1549848, EPI_ISL_1549849, EPI_ISL_1549850, EPI_ISL_1549853, EPI_ISL_1549854, EPI_ISL_1549857, EPI_ISL_1549858, EPI_ISL_1549859, EPI_ISL_1549860, EPI_ISL_1549863, EPI_ISL_1549864, EPI_ISL_1549865, EPI_ISL_1549866, EPI_ISL_1549867, EPI_ISL_1549869, EPI_ISL_1549870, EPI_ISL_1549872, EPI_ISL_1549873, EPI_ISL_1549877, EPI_ISL_1549878, EPI_ISL_1549879, EPI_ISL_1549881, EPI_ISL_1549882, EPI_ISL_1549883, EPI_ISL_1549885, EPI_ISL_1549887, EPI_ISL_1549888, EPI_ISL_1549889, EPI_ISL_1549890, EPI_ISL_1549891, EPI_ISL_1549892, EPI_ISL_1549893, EPI_ISL_1549894, EPI_ISL_1549895, EPI_ISL_1549897, EPI_ISL_1549898, EPI_ISL_1549901, EPI_ISL_1549903, EPI_ISL_1549904, EPI_ISL_1549905, EPI_ISL_1549906, EPI_ISL_1549907, EPI_ISL_1549908, EPI_ISL_1549909, EPI_ISL_1549910, EPI_ISL_1549911, EPI_ISL_1549912, EPI_ISL_1549913, EPI_ISL_1549914, EPI_ISL_1549915, EPI_ISL_1549916, EPI_ISL_1549917, EPI_ISL_1549919, EPI_ISL_1549920, EPI_ISL_1549921, EPI_ISL_1549923, EPI_ISL_1549925, EPI_ISL_1549926, EPI_ISL_1549928, EPI_ISL_1549930, EPI_ISL_1549931, EPI_ISL_1549932, EPI_ISL_1549933, EPI_ISL_1549935, EPI_ISL_1549936, EPI_ISL_1549937, EPI_ISL_1549938, EPI_ISL_1549939, EPI_ISL_1549940, EPI_ISL_1549941, EPI_ISL_1549942, EPI_ISL_1549943, EPI_ISL_1549945, EPI_ISL_1549946, EPI_ISL_1549947, EPI_ISL_1549949, EPI_ISL_1549951, EPI_ISL_1549952, EPI_ISL_1549953, EPI_ISL_1549954, EPI_ISL_1549956, EPI_ISL_1549959, EPI_ISL_1549960, EPI_ISL_1549961, EPI_ISL_1549962, EPI_ISL_1549963, EPI_ISL_1549964, EPI_ISL_1549965, EPI_ISL_1549966, EPI_ISL_1549967, EPI_ISL_1549968, EPI_ISL_1549969, EPI_ISL_1549970, EPI_ISL_1549972, EPI_ISL_1549973, EPI_ISL_1549974, EPI_ISL_1549976, EPI_ISL_1549977, EPI_ISL_1549978, EPI_ISL_1549979, EPI_ISL_1549981, EPI_ISL_1549982, EPI_ISL_1549983, EPI_ISL_1549984, EPI_ISL_1549985, EPI_ISL_1549986 | | | |
| see above | Laboratory Corporation of America | Centers for Disease Control and Prevention Division of Viral Diseases, Pathogen Discovery | Dakota Howard, Dhwani Batra, Peter W. Cook, Kara Moser, Adrian Paskey, Jason Caravas, Benjamin Rambo-Martin, Shatavia Morrison, Christopher Gulvick, Scott Sammons, Yvette Unoarumhi, Darlene Wagner, Matthew Schmerer, Minoo Agarwal, Eyad Almasri, Debbie Boles, Ayla Burns, Nuthawin Charoensri, Oren Cohen, Susan Countryman, Mary Ann Cristobal, Bobbi Croy, Suzanne Dale, Hrushikesh Deshmukh, Amanda Douglas, Vincent Drouillon, Marcia Eisenberg, Howard Engler, Rama Ghatti, Prashant Gupta, Susan Hicks, Jake Humphrey, Lax Iyer, Manoj Jain, Mohan Kolli, Brian Krueger, Tim Kuphal, Stanley Letovsky, Michael Levandoski, Craig Lukasik, Jonathan Meltzer, Brian Norvell, Mindy Nye, Scott Parker, Christos Petropoulos, John Pruitt, Steven Ragan, Scott Ryan, Mike Sapeta, Jana Schroth, Suresh Babu Selvaraju, Goran Stevovic, Amanda Suchanek, Andrea Throop, Lyndon Tilson, Thomas Urban, Joe Voshell, Kimberly Wagner, Jonathan Williams, Mary Williamson, Qian Zeng, Tricia Zwiefelhofer, Clinton R. Paden, Duncan MacCannell |
| EPI_ISL_1549987 | Montana Public Health Laboratory | Montana Public Health Laboratory | Joy Ritter, Michelle Mozer, Carrie Biskupiak, Deborah Gibson |

| | | | |
|---|---|---|---|
| EPI_ISL_1549988, EPI_ISL_1549989, EPI_ISL_1549990, EPI_ISL_1549991, EPI_ISL_1549992, EPI_ISL_1549993, EPI_ISL_1549994, EPI_ISL_1549995, EPI_ISL_1549997, EPI_ISL_1549998, EPI_ISL_1549999, EPI_ISL_1550000, EPI_ISL_1550001, EPI_ISL_1550003, EPI_ISL_1550004, EPI_ISL_1550005, EPI_ISL_1550006, EPI_ISL_1550008, EPI_ISL_1550009, EPI_ISL_1550010, EPI_ISL_1550011, EPI_ISL_1550013, EPI_ISL_1550014, EPI_ISL_1550015, EPI_ISL_1550016, EPI_ISL_1550017, EPI_ISL_1550018, EPI_ISL_1550019, EPI_ISL_1550020, EPI_ISL_1550022, EPI_ISL_1550023, EPI_ISL_1550024, EPI_ISL_1550025, EPI_ISL_1550027, EPI_ISL_1550028, EPI_ISL_1550029, EPI_ISL_1550030, EPI_ISL_1550031, EPI_ISL_1550032, EPI_ISL_1550033, EPI_ISL_1550035, EPI_ISL_1550036, EPI_ISL_1550038, EPI_ISL_1550040, EPI_ISL_1550041, EPI_ISL_1550042, EPI_ISL_1550044, EPI_ISL_1550045, EPI_ISL_1550046, EPI_ISL_1550047, EPI_ISL_1550049, EPI_ISL_1550051, EPI_ISL_1550053, EPI_ISL_1550054, EPI_ISL_1550056, EPI_ISL_1550057, EPI_ISL_1550058, EPI_ISL_1550063, EPI_ISL_1550065, EPI_ISL_1550066, EPI_ISL_1550067, EPI_ISL_1550068, EPI_ISL_1550070, EPI_ISL_1550071, EPI_ISL_1550072, EPI_ISL_1550073, EPI_ISL_1550074, EPI_ISL_1550075, EPI_ISL_1550076, EPI_ISL_1550078, EPI_ISL_1550079, EPI_ISL_1550080, EPI_ISL_1550081, EPI_ISL_1550082, EPI_ISL_1550085, EPI_ISL_1550086, EPI_ISL_1550087, EPI_ISL_1550088, EPI_ISL_1550089, EPI_ISL_1550091, EPI_ISL_1550092, EPI_ISL_1550093, EPI_ISL_1550094, EPI_ISL_1550095, EPI_ISL_1550096, EPI_ISL_1550097, EPI_ISL_1550099, EPI_ISL_1550100, EPI_ISL_1550101, EPI_ISL_1550102, EPI_ISL_1550104, EPI_ISL_1550107, EPI_ISL_1550108, EPI_ISL_1550109, EPI_ISL_1550110, EPI_ISL_1550112, EPI_ISL_1550114, EPI_ISL_1550115, EPI_ISL_1550116, EPI_ISL_1550117, EPI_ISL_1550118, EPI_ISL_1550119, EPI_ISL_1550120, EPI_ISL_1550122, EPI_ISL_1550123, EPI_ISL_1550124, EPI_ISL_1550125, EPI_ISL_1550126, EPI_ISL_1550128, EPI_ISL_1550130, EPI_ISL_1550131, EPI_ISL_1550132, EPI_ISL_1550133, EPI_ISL_1550134, EPI_ISL_1550135, EPI_ISL_1550136, EPI_ISL_1550137, EPI_ISL_1550139, EPI_ISL_1550141, EPI_ISL_1550142, EPI_ISL_1550143, EPI_ISL_1550144, EPI_ISL_1550145, EPI_ISL_1550146, EPI_ISL_1550147, EPI_ISL_1550148, EPI_ISL_1550149, EPI_ISL_1550150, EPI_ISL_1550151, EPI_ISL_1550152, EPI_ISL_1550153, EPI_ISL_1550154, EPI_ISL_1550155, EPI_ISL_1550156, EPI_ISL_1550157, EPI_ISL_1550158 | | | |
| see above | Laboratory Corporation of America | Centers for Disease Control and Prevention Division of Viral Diseases, Pathogen Discovery | Dakota Howard, Dhwani Batra, Peter W. Cook, Kara Moser, Adrian Paskey, Jason Caravas, Benjamin Rambo-Martin, Shatavia Morrison, Christopher Gulvick, Scott Sammons, Yvette Unoarumhi, Darlene Wagner, Matthew Schmerer, Minoo Agarwal, Eyad Almasri, Debbie Boles, Ayla Burns, Nuthawin Charoensri, Oren Cohen, Susan Countryman, Mary Ann Cristobal, Bobbi Croy, Suzanne Dale, Hrushikesh Deshmukh, Amanda Douglas, Vincent Drouillon, Marcia Eisenberg, Howard Engler, Rama Ghatti, Prashant Gupta, Susan Hicks, Jake Humphrey, Lax Iyer, Manoj Jain, Mohan Kolli, Brian Krueger, Tim Kuphal, Stanley Letovsky, Michael Levandoski, Craig Lukasik, Jonathan Meltzer, Brian Norvell, Mindy Nye, Scott Parker, Christos Petropoulos, John Pruitt, Steven Ragan, Scott Ryan, Mike Sapeta, Jana Schroth, Suresh Babu Selvaraju, Goran Stevovic, Amanda Suchanek, Andrea Throop, Lyndon Tilson, Thomas Urban, Joe Voshell, Kimberly Wagner, Jonathan Williams, Mary Williamson, Qian Zeng, Tricia Zwiefelhofer, Clinton R. Paden, Duncan MacCannell |
| EPI_ISL_1550159 | Montana Public Health Laboratory | Montana Public Health Laboratory | Joy Ritter, Michelle Mozer, Carrie Biskupiak, Deborah Gibson |
| EPI_ISL_1550160, EPI_ISL_1550161, EPI_ISL_1550162, EPI_ISL_1550163, EPI_ISL_1550165, EPI_ISL_1550166, EPI_ISL_1550167, EPI_ISL_1550168, EPI_ISL_1550170, EPI_ISL_1550173, EPI_ISL_1550174, EPI_ISL_1550175, EPI_ISL_1550176, EPI_ISL_1550177, EPI_ISL_1550179, EPI_ISL_1550180, EPI_ISL_1550181, EPI_ISL_1550182, EPI_ISL_1550184, EPI_ISL_1550186, EPI_ISL_1550188, EPI_ISL_1550189, EPI_ISL_1550190, EPI_ISL_1550191, EPI_ISL_1550194, EPI_ISL_1550195, EPI_ISL_1550196, EPI_ISL_1550197, EPI_ISL_1550198, EPI_ISL_1550199, EPI_ISL_1550201, EPI_ISL_1550202, EPI_ISL_1550203, EPI_ISL_1550206, EPI_ISL_1550207, EPI_ISL_1550209, EPI_ISL_1550210, EPI_ISL_1550211, EPI_ISL_1550212, EPI_ISL_1550214, EPI_ISL_1550216, EPI_ISL_1550217, EPI_ISL_1550218, EPI_ISL_1550221, EPI_ISL_1550223, EPI_ISL_1550224, EPI_ISL_1550225, EPI_ISL_1550228, EPI_ISL_1550229, EPI_ISL_1550230, EPI_ISL_1550232, EPI_ISL_1550233, EPI_ISL_1550234, EPI_ISL_1550235, EPI_ISL_1550236, EPI_ISL_1550237, EPI_ISL_1550238, EPI_ISL_1550239, EPI_ISL_1550240, EPI_ISL_1550242, EPI_ISL_1550243, EPI_ISL_1550245, EPI_ISL_1550246, EPI_ISL_1550247, EPI_ISL_1550248, EPI_ISL_1550249, EPI_ISL_1550251, EPI_ISL_1550252, EPI_ISL_1550253, EPI_ISL_1550254, EPI_ISL_1550255, EPI_ISL_1550256, EPI_ISL_1550258, EPI_ISL_1550259, EPI_ISL_1550260, EPI_ISL_1550263, EPI_ISL_1550264, EPI_ISL_1550265, EPI_ISL_1550266, EPI_ISL_1550267, EPI_ISL_1550268, EPI_ISL_1550269, EPI_ISL_1550270, EPI_ISL_1550271, EPI_ISL_1550272, EPI_ISL_1550273, EPI_ISL_1550274, EPI_ISL_1550275, EPI_ISL_1550276, EPI_ISL_1550277, EPI_ISL_1550278, EPI_ISL_1550279, EPI_ISL_1550280, EPI_ISL_1550281, EPI_ISL_1550282, EPI_ISL_1550283, EPI_ISL_1550285, EPI_ISL_1550288, EPI_ISL_1550289, EPI_ISL_1550290, EPI_ISL_1550291, EPI_ISL_1550292, EPI_ISL_1550293, EPI_ISL_1550294, EPI_ISL_1550296, EPI_ISL_1550297, EPI_ISL_1550298, EPI_ISL_1550299, EPI_ISL_1550300, EPI_ISL_1550301, EPI_ISL_1550302, EPI_ISL_1550303, EPI_ISL_1550304, EPI_ISL_1550305, EPI_ISL_1550306, EPI_ISL_1550307, EPI_ISL_1550308, EPI_ISL_1550309, EPI_ISL_1550310, EPI_ISL_1550311, EPI_ISL_1550313, EPI_ISL_1550314, EPI_ISL_1550315, EPI_ISL_1550317, EPI_ISL_1550318, EPI_ISL_1550319, EPI_ISL_1550320, EPI_ISL_1550321, EPI_ISL_1550324, EPI_ISL_1550325, EPI_ISL_1550326, EPI_ISL_1550327, EPI_ISL_1550328, EPI_ISL_1550329, EPI_ISL_1550330, EPI_ISL_1550331, EPI_ISL_1550332, EPI_ISL_1550333, EPI_ISL_1550334, EPI_ISL_1550335, EPI_ISL_1550336, EPI_ISL_1550337, EPI_ISL_1550338, EPI_ISL_1550339, EPI_ISL_1550340, EPI_ISL_1550341, EPI_ISL_1550343, EPI_ISL_1550344, EPI_ISL_1550345, EPI_ISL_1550346, EPI_ISL_1550347, EPI_ISL_1550348, EPI_ISL_1550349, EPI_ISL_1550350, EPI_ISL_1550351, EPI_ISL_1550352, EPI_ISL_1550353, EPI_ISL_1550355, EPI_ISL_1550356, EPI_ISL_1550357, EPI_ISL_1550358, EPI_ISL_1550359, EPI_ISL_1550360, EPI_ISL_1550362, EPI_ISL_1550363, EPI_ISL_1550364, EPI_ISL_1550365, EPI_ISL_1550366, EPI_ISL_1550367, EPI_ISL_1550368, EPI_ISL_1550369, EPI_ISL_1550370, EPI_ISL_1550371, EPI_ISL_1550372, EPI_ISL_1550373, EPI_ISL_1550374, EPI_ISL_1550375, EPI_ISL_1550378, EPI_ISL_1550379, EPI_ISL_1550380, EPI_ISL_1550381, EPI_ISL_1550382, EPI_ISL_1550383, EPI_ISL_1550384, EPI_ISL_1550385, EPI_ISL_1550386, EPI_ISL_1550387 | | | |
| see above | Laboratory Corporation of America | Centers for Disease Control and Prevention Division of Viral Diseases, Pathogen Discovery | Dakota Howard, Dhwani Batra, Peter W. Cook, Kara Moser, Adrian Paskey, Jason Caravas, Benjamin Rambo-Martin, Shatavia Morrison, Christopher Gulvick, Scott Sammons, Yvette Unoarumhi, Darlene Wagner, Matthew Schmerer, Minoo Agarwal, Eyad Almasri, Debbie Boles, Ayla Burns, Nuthawin Charoensri, Oren Cohen, Susan Countryman, Mary Ann Cristobal, Bobbi Croy, Suzanne Dale, Hrushikesh Deshmukh, Amanda Douglas, Vincent Drouillon, Marcia Eisenberg, Howard Engler, Rama Ghatti, Prashant Gupta, Susan Hicks, Jake Humphrey, Lax Iyer, Manoj Jain, Mohan Kolli, Brian Krueger, Tim Kuphal, Stanley Letovsky, Michael Levandoski, Craig Lukasik, Jonathan Meltzer, Brian Norvell, Mindy Nye, Scott Parker, Christos Petropoulos, John Pruitt, Steven Ragan, Scott Ryan, Mike Sapeta, Jana Schroth, Suresh Babu Selvaraju, Goran Stevovic, Amanda Suchanek, Andrea Throop, Lyndon Tilson, Thomas Urban, Joe Voshell, Kimberly Wagner, Jonathan Williams, Mary Williamson, Qian Zeng, Tricia Zwiefelhofer, Clinton R. Paden, Duncan MacCannell |
| EPI_ISL_1550389, EPI_ISL_1550391, EPI_ISL_1550392, EPI_ISL_1550393, EPI_ISL_1550394, EPI_ISL_1550395, EPI_ISL_1550396, EPI_ISL_1550397, EPI_ISL_1550398, EPI_ISL_1550400, EPI_ISL_1550401, EPI_ISL_1550402, EPI_ISL_1550403, EPI_ISL_1550404, EPI_ISL_1550405, EPI_ISL_1550406, EPI_ISL_1550407, EPI_ISL_1550408, EPI_ISL_1550411, EPI_ISL_1550413, EPI_ISL_1550414, EPI_ISL_1550415, EPI_ISL_1550416, EPI_ISL_1550417, EPI_ISL_1550418, EPI_ISL_1550419, EPI_ISL_1550420, EPI_ISL_1550421, EPI_ISL_1550422, EPI_ISL_1550423, EPI_ISL_1550424, EPI_ISL_1550425, EPI_ISL_1550426, EPI_ISL_1550427, EPI_ISL_1550428, EPI_ISL_1550429, EPI_ISL_1550430, EPI_ISL_1550431, EPI_ISL_1550432, EPI_ISL_1550433, EPI_ISL_1550434, EPI_ISL_1550435, EPI_ISL_1550436, EPI_ISL_1550437, EPI_ISL_1550439, EPI_ISL_1550441, EPI_ISL_1550442, EPI_ISL_1550443, EPI_ISL_1550444, EPI_ISL_1550445, EPI_ISL_1550446, EPI_ISL_1550447, EPI_ISL_1550448, EPI_ISL_1550449, EPI_ISL_1550450, EPI_ISL_1550451, EPI_ISL_1550452, EPI_ISL_1550453, EPI_ISL_1550454, EPI_ISL_1550456, EPI_ISL_1550457, EPI_ISL_1550458, EPI_ISL_1550459, EPI_ISL_1550460, EPI_ISL_1550461, EPI_ISL_1550462, EPI_ISL_1550463, EPI_ISL_1550464, EPI_ISL_1550465, EPI_ISL_1550466, EPI_ISL_1550467, EPI_ISL_1550468, EPI_ISL_1550469, EPI_ISL_1550470, EPI_ISL_1550471, EPI_ISL_1550472, EPI_ISL_1550473, EPI_ISL_1550474, EPI_ISL_1550475, EPI_ISL_1550476, EPI_ISL_1550477, EPI_ISL_1550478, EPI_ISL_1550479, EPI_ISL_1550480, EPI_ISL_1550481, EPI_ISL_1550482, EPI_ISL_1550483, EPI_ISL_1550484, EPI_ISL_1550485, EPI_ISL_1550486, EPI_ISL_1550487, EPI_ISL_1550488, EPI_ISL_1550489, EPI_ISL_1550490, EPI_ISL_1550491, EPI_ISL_1550492, EPI_ISL_1550494, EPI_ISL_1550495, EPI_ISL_1550497, EPI_ISL_1550498, EPI_ISL_1550499, EPI_ISL_1550500, EPI_ISL_1550501, EPI_ISL_1550502, EPI_ISL_1550503, EPI_ISL_1550504, EPI_ISL_1550505, EPI_ISL_1550507, EPI_ISL_1550508, EPI_ISL_1550509, EPI_ISL_1550510, EPI_ISL_1550511, EPI_ISL_1550512, EPI_ISL_1550513, EPI_ISL_1550514, EPI_ISL_1550515, EPI_ISL_1550516, EPI_ISL_1550517, EPI_ISL_1550518, EPI_ISL_1550519, EPI_ISL_1550520, EPI_ISL_1550521, EPI_ISL_1550522, EPI_ISL_1550523, EPI_ISL_1550524, EPI_ISL_1550525, EPI_ISL_1550526, EPI_ISL_1550527, EPI_ISL_1550528, EPI_ISL_1550529, EPI_ISL_1550530, EPI_ISL_1550531, EPI_ISL_1550532, EPI_ISL_1550534, EPI_ISL_1550535, EPI_ISL_1550536, EPI_ISL_1550537, EPI_ISL_1550538, EPI_ISL_1550539, EPI_ISL_1550540, EPI_ISL_1550541, EPI_ISL_1550542, EPI_ISL_1550543, EPI_ISL_1550544, EPI_ISL_1550545, EPI_ISL_1550546, EPI_ISL_1550547, EPI_ISL_1550548, EPI_ISL_1550549, EPI_ISL_1550550, EPI_ISL_1550551, EPI_ISL_1550552, EPI_ISL_1550553, EPI_ISL_1550554, EPI_ISL_1550555, EPI_ISL_1550556, EPI_ISL_1550557, EPI_ISL_1550558, EPI_ISL_1550559, EPI_ISL_1550560, EPI_ISL_1550561, EPI_ISL_1550563, EPI_ISL_1550564, EPI_ISL_1550565, EPI_ISL_1550566, EPI_ISL_1550567, EPI_ISL_1550568, EPI_ISL_1550569, EPI_ISL_1550570, EPI_ISL_1550571, EPI_ISL_1550572, EPI_ISL_1550573, EPI_ISL_1550574, EPI_ISL_1550575, EPI_ISL_1550576, EPI_ISL_1550577, EPI_ISL_1550578, EPI_ISL_1550579, EPI_ISL_1550580, EPI_ISL_1550581, EPI_ISL_1550585, EPI_ISL_1550586, EPI_ISL_1550587, EPI_ISL_1550589, EPI_ISL_1550590, EPI_ISL_1550592, EPI_ISL_1550593, EPI_ISL_1550594, EPI_ISL_1550595, EPI_ISL_1550596, EPI_ISL_1550597, EPI_ISL_1550598, EPI_ISL_1550599, EPI_ISL_1550600, EPI_ISL_1550601, EPI_ISL_1550602, EPI_ISL_1550603, EPI_ISL_1550604, EPI_ISL_1550605, EPI_ISL_1550606, EPI_ISL_1550607, EPI_ISL_1550608, EPI_ISL_1550609, EPI_ISL_1550610, EPI_ISL_1550611, EPI_ISL_1550612, EPI_ISL_1550613, EPI_ISL_1550614, EPI_ISL_1550615, EPI_ISL_1550616, EPI_ISL_1550617, EPI_ISL_1550618, EPI_ISL_1550619, EPI_ISL_1550620, EPI_ISL_1550621, EPI_ISL_1550622, EPI_ISL_1550623, EPI_ISL_1550624, EPI_ISL_1550625, EPI_ISL_1550626, EPI_ISL_1550627, EPI_ISL_1550628, EPI_ISL_1550629, EPI_ISL_1550630, EPI_ISL_1550631, EPI_ISL_1550632, EPI_ISL_1550633, EPI_ISL_1550634, EPI_ISL_1550635, EPI_ISL_1550636, EPI_ISL_1550637, EPI_ISL_1550638, EPI_ISL_1550639, EPI_ISL_1550640, EPI_ISL_1550641, EPI_ISL_1550642, EPI_ISL_1550643, EPI_ISL_1550644, EPI_ISL_1550645, EPI_ISL_1550646, EPI_ISL_1550648, EPI_ISL_1550649, EPI_ISL_1550650, EPI_ISL_1550651, EPI_ISL_1550652, EPI_ISL_1550654, EPI_ISL_1550655, EPI_ISL_1550656, EPI_ISL_1550658, EPI_ISL_1550659, EPI_ISL_1550660, EPI_ISL_1550661, EPI_ISL_1550662, EPI_ISL_1550664, EPI_ISL_1550665, EPI_ISL_1550666, EPI_ISL_1550667, EPI_ISL_1550668, EPI_ISL_1550669, EPI_ISL_1550670, EPI_ISL_1550671, EPI_ISL_1550672, EPI_ISL_1550673, EPI_ISL_1550674, EPI_ISL_1550675, EPI_ISL_1550676, EPI_ISL_1550677, EPI_ISL_1550678, EPI_ISL_1550679, EPI_ISL_1550680, EPI_ISL_1550681, EPI_ISL_1550682, EPI_ISL_1550683, EPI_ISL_1550684, EPI_ISL_1550685, EPI_ISL_1550686, EPI_ISL_1550687, EPI_ISL_1550688, EPI_ISL_1550690, EPI_ISL_1550691, EPI_ISL_1550693, EPI_ISL_1550694, EPI_ISL_1550695, EPI_ISL_1550696, EPI_ISL_1550698, EPI_ISL_1550699, EPI_ISL_1550700, EPI_ISL_1550701, EPI_ISL_1550702, EPI_ISL_1550703, EPI_ISL_1550704, EPI_ISL_1550705, EPI_ISL_1550706, EPI_ISL_1550707, EPI_ISL_1550708, EPI_ISL_1550709, EPI_ISL_1550710, EPI_ISL_1550711, EPI_ISL_1550712, EPI_ISL_1550713, EPI_ISL_1550714, EPI_ISL_1550715, EPI_ISL_1550716, EPI_ISL_1550717, EPI_ISL_1550718, EPI_ISL_1550719, EPI_ISL_1550720, EPI_ISL_1550721, EPI_ISL_1550722, EPI_ISL_1550723, EPI_ISL_1550724, EPI_ISL_1550725, EPI_ISL_1550726, EPI_ISL_1550727, EPI_ISL_1550728, EPI_ISL_1550729, EPI_ISL_1550730, EPI_ISL_1550731, EPI_ISL_1550732, EPI_ISL_1550733, EPI_ISL_1550734, EPI_ISL_1550735, EPI_ISL_1550736, EPI_ISL_1550737, EPI_ISL_1550738, EPI_ISL_1550739, EPI_ISL_1550740, EPI_ISL_1550741, EPI_ISL_1550742, EPI_ISL_1550743, EPI_ISL_1550744, EPI_ISL_1550745, EPI_ISL_1550746, EPI_ISL_1550747, EPI_ISL_1550748, EPI_ISL_1550749, EPI_ISL_1550750, EPI_ISL_1550751, EPI_ISL_1550752, EPI_ISL_1550753, EPI_ISL_1550754, EPI_ISL_1550755, EPI_ISL_1550756, EPI_ISL_1550757, EPI_ISL_1550758, EPI_ISL_1550759, EPI_ISL_1550760, EPI_ISL_1550761, EPI_ISL_1550762, EPI_ISL_1550763, EPI_ISL_1550764, EPI_ISL_1550765, EPI_ISL_1550766, EPI_ISL_1550767, EPI_ISL_1550768, EPI_ISL_1550770, EPI_ISL_1550771, EPI_ISL_1550772, EPI_ISL_1550773, EPI_ISL_1550774, EPI_ISL_1550775, EPI_ISL_1550777, EPI_ISL_1550778, EPI_ISL_1550779, EPI_ISL_1550780, EPI_ISL_1550781, EPI_ISL_1550782, EPI_ISL_1550783, EPI_ISL_1550784, EPI_ISL_1550786, EPI_ISL_1550787, EPI_ISL_1550788, EPI_ISL_1550789, EPI_ISL_1550790, EPI_ISL_1550791, EPI_ISL_1550792, EPI_ISL_1550793, EPI_ISL_1550794, EPI_ISL_1550795, EPI_ISL_1550796, EPI_ISL_1550797, EPI_ISL_1550798, EPI_ISL_1550799, EPI_ISL_1550800, EPI_ISL_1550801, EPI_ISL_1550802, EPI_ISL_1550803, EPI_ISL_1550804, EPI_ISL_1550805, EPI_ISL_1550806, EPI_ISL_1550807, EPI_ISL_1550808, EPI_ISL_1550809, EPI_ISL_1550810, EPI_ISL_1550811, EPI_ISL_1550812, EPI_ISL_1550813, EPI_ISL_1550814, EPI_ISL_1550815, EPI_ISL_1550817, EPI_ISL_1550818, EPI_ISL_1550819, EPI_ISL_1550820, EPI_ISL_1550821, EPI_ISL_1550822, EPI_ISL_1550823, EPI_ISL_1550824, EPI_ISL_1550825, EPI_ISL_1550826, EPI_ISL_1550827, EPI_ISL_1550828, EPI_ISL_1550829, EPI_ISL_1550830, EPI_ISL_1550831, EPI_ISL_1550832, EPI_ISL_1550833, EPI_ISL_1550834, EPI_ISL_1550835, EPI_ISL_1550836, EPI_ISL_1550837, EPI_ISL_1550838, EPI_ISL_1550839, EPI_ISL_1550840, EPI_ISL_1550841, EPI_ISL_1550843, EPI_ISL_1550844, EPI_ISL_1550845, EPI_ISL_1550846, EPI_ISL_1550847, EPI_ISL_1550848, EPI_ISL_1550849, EPI_ISL_1550850, EPI_ISL_1550851, EPI_ISL_1550852, EPI_ISL_1550853, EPI_ISL_1550854, EPI_ISL_1550855, EPI_ISL_1550856, EPI_ISL_1550857, EPI_ISL_1550858, EPI_ISL_1550859, EPI_ISL_1550860, EPI_ISL_1550861, EPI_ISL_1550862, EPI_ISL_1550863, EPI_ISL_1550864, EPI_ISL_1550865, EPI_ISL_1550866, EPI_ISL_1550867, EPI_ISL_1550868, EPI_ISL_1550869, EPI_ISL_1550870, EPI_ISL_1550871, EPI_ISL_1550872, EPI_ISL_1550874, EPI_ISL_1550875, EPI_ISL_1550876, EPI_ISL_1550877 | | | |
| see above | Aegis Sciences Corporation | Centers for Disease Control and Prevention Division of Viral Diseases, Pathogen Discovery | Dakota Howard, Dhwani Batra, Peter W. Cook, Kara Moser, Adrian Paskey, Jason Caravas, Benjamin Rambo-Martin, Shatavia Morrison, Christopher Gulvick, Scott Sammons, Yvette Unoarumhi, Darlene Wagner, Matthew Schmerer, Cyndi Clark, Patrick Campbell, Rob Case, Vikramsinha Ghorpade, Holly Houdeshell, Ola Kvalvaag, Dillon Nall, Ethan Sanders, Alec Vest, Shaun Westlund, Matthew Hardison, Clinton R. Paden, Duncan MacCannell |
| EPI_ISL_1550879 | Sonora Quest Laboratories | TGen North | "Jolene Bowers, Heather Centner, Chris French, Hayley Yaglom, Ashlyn Pfeiffer, Darrin Lemmer, Dave Engelthaler, The Arizona COVID Genomics Union (ACGU)" |
| EPI_ISL_1550882 | TGen North | TGen North | "Jolene Bowers, Heather Centner, Chris French, Hayley Yaglom, Ashlyn Pfeiffer, Darrin Lemmer, Dave Engelthaler, The Arizona COVID Genomics Union (ACGU)" |
| EPI_ISL_1550886, EPI_ISL_1550887, EPI_ISL_1550889, EPI_ISL_1550896, EPI_ISL_1550898 | Sonora Quest Laboratories | TGen North | "Jolene Bowers, Heather Centner, Chris French, Hayley Yaglom, Ashlyn Pfeiffer, Darrin Lemmer, Dave Engelthaler, The Arizona COVID Genomics Union (ACGU)" |
| EPI_ISL_1550899 | TGen North | TGen North | "Jolene Bowers, Heather Centner, Chris French, Hayley Yaglom, Ashlyn Pfeiffer, Darrin Lemmer, Dave Engelthaler, The Arizona COVID Genomics Union (ACGU)" |
| EPI_ISL_1550900 | Sonora Quest Laboratories | TGen North | "Jolene Bowers, Heather Centner, Chris French, Hayley Yaglom, Ashlyn Pfeiffer, Darrin Lemmer, Dave Engelthaler, The Arizona COVID Genomics Union (ACGU)" |
| EPI_ISL_1550901 | TGen North | TGen North | "Jolene Bowers, Heather Centner, Chris French, Hayley Yaglom, Ashlyn Pfeiffer, Darrin Lemmer, Dave Engelthaler, The Arizona COVID Genomics Union (ACGU)" |
| EPI_ISL_1550902, EPI_ISL_1550905, EPI_ISL_1550906, EPI_ISL_1550907 | Sonora Quest Laboratories | TGen North | "Jolene Bowers, Heather Centner, Chris French, Hayley Yaglom, Ashlyn Pfeiffer, Darrin Lemmer, Dave Engelthaler, The Arizona COVID Genomics Union (ACGU)" |
| EPI_ISL_1550963, EPI_ISL_1550964, EPI_ISL_1550965, EPI_ISL_1550966, EPI_ISL_1550967, EPI_ISL_1550968, EPI_ISL_1550969, EPI_ISL_1550970, EPI_ISL_1550971, EPI_ISL_1550973, EPI_ISL_1550974, EPI_ISL_1550976, EPI_ISL_1550977, EPI_ISL_1550978, EPI_ISL_1550979, EPI_ISL_1550980, EPI_ISL_1550981, EPI_ISL_1550982, EPI_ISL_1550983, EPI_ISL_1550984, EPI_ISL_1550985, EPI_ISL_1550986, EPI_ISL_1550987, EPI_ISL_1550988, EPI_ISL_1550989, EPI_ISL_1550990, EPI_ISL_1550991, EPI_ISL_1550992, EPI_ISL_1550993, EPI_ISL_1550994, EPI_ISL_1550995, EPI_ISL_1550996, EPI_ISL_1550997, EPI_ISL_1550998, EPI_ISL_1550999, EPI_ISL_1551000, EPI_ISL_1551002, EPI_ISL_1551003, EPI_ISL_1551004, EPI_ISL_1551005, EPI_ISL_1551006, EPI_ISL_1551007, EPI_ISL_1551008, EPI_ISL_1551009, EPI_ISL_1551010, EPI_ISL_1551011, EPI_ISL_1551012, EPI_ISL_1551014, EPI_ISL_1551015, EPI_ISL_1551016, EPI_ISL_1551017, EPI_ISL_1551018, EPI_ISL_1551019, EPI_ISL_1551020, EPI_ISL_1551021, EPI_ISL_1551022, EPI_ISL_1551023, EPI_ISL_1551024, EPI_ISL_1551025, EPI_ISL_1551026, EPI_ISL_1551027, EPI_ISL_1551029, EPI_ISL_1551030, EPI_ISL_1551031, EPI_ISL_1551032, EPI_ISL_1551033, EPI_ISL_1551034, EPI_ISL_1551035, EPI_ISL_1551037, EPI_ISL_1551038, EPI_ISL_1551039, EPI_ISL_1551040, EPI_ISL_1551041, EPI_ISL_1551042, EPI_ISL_1551043, EPI_ISL_1551044, EPI_ISL_1551045, EPI_ISL_1551046, EPI_ISL_1551047, EPI_ISL_1551048, EPI_ISL_1551049, EPI_ISL_1551050, EPI_ISL_1551051, EPI_ISL_1551052, EPI_ISL_1551053, EPI_ISL_1551054, EPI_ISL_1551055, EPI_ISL_1551056, EPI_ISL_1551057, EPI_ISL_1551058, EPI_ISL_1551059, EPI_ISL_1551060, EPI_ISL_1551061, EPI_ISL_1551062, EPI_ISL_1551063, EPI_ISL_1551064, EPI_ISL_1551065, EPI_ISL_1551066, EPI_ISL_1551067, EPI_ISL_1551068, EPI_ISL_1551069, EPI_ISL_1551070, EPI_ISL_1551071, EPI_ISL_1551072, EPI_ISL_1551073, EPI_ISL_1551074, EPI_ISL_1551075, EPI_ISL_1551076, EPI_ISL_1551077, EPI_ISL_1551078, EPI_ISL_1551079, EPI_ISL_1551080, EPI_ISL_1551081, EPI_ISL_1551082, EPI_ISL_1551083, EPI_ISL_1551084, EPI_ISL_1551085, EPI_ISL_1551086, EPI_ISL_1551087, EPI_ISL_1551088, EPI_ISL_1551089, EPI_ISL_1551090, EPI_ISL_1551091, EPI_ISL_1551092, EPI_ISL_1551093, EPI_ISL_1551094, EPI_ISL_1551095, EPI_ISL_1551096, EPI_ISL_1551097, EPI_ISL_1551098, EPI_ISL_1551099, EPI_ISL_1551100, EPI_ISL_1551101, EPI_ISL_1551102, EPI_ISL_1551103, EPI_ISL_1551104, EPI_ISL_1551105, EPI_ISL_1551106, EPI_ISL_1551107, EPI_ISL_1551108, EPI_ISL_1551109, EPI_ISL_1551110, EPI_ISL_1551111, EPI_ISL_1551112, EPI_ISL_1551113, EPI_ISL_1551114, EPI_ISL_1551115, EPI_ISL_1551116, EPI_ISL_1551117, EPI_ISL_1551118, EPI_ISL_1551120, EPI_ISL_1551121, EPI_ISL_1551122, EPI_ISL_1551123, EPI_ISL_1551124, EPI_ISL_1551125, EPI_ISL_1551126, EPI_ISL_1551127, EPI_ISL_1551128, EPI_ISL_1551129, EPI_ISL_1551130, EPI_ISL_1551131, EPI_ISL_1551132, EPI_ISL_1551133, EPI_ISL_1551134, EPI_ISL_1551135, EPI_ISL_1551136, EPI_ISL_1551137, EPI_ISL_1551138, EPI_ISL_1551139, EPI_ISL_1551140, EPI_ISL_1551141, EPI_ISL_1551142, EPI_ISL_1551143, EPI_ISL_1551144, EPI_ISL_1551145, EPI_ISL_1551146, EPI_ISL_1551147, EPI_ISL_1551148, EPI_ISL_1551149, EPI_ISL_1551150, EPI_ISL_1551151, EPI_ISL_1551152, EPI_ISL_1551153, EPI_ISL_1551154, EPI_ISL_1551155, EPI_ISL_1551157, EPI_ISL_1551158, EPI_ISL_1551159, EPI_ISL_1551160, EPI_ISL_1551161, EPI_ISL_1551162, EPI_ISL_1551163, EPI_ISL_1551164, EPI_ISL_1551165, EPI_ISL_1551166, EPI_ISL_1551167, EPI_ISL_1551168, EPI_ISL_1551169, EPI_ISL_1551170, EPI_ISL_1551171, EPI_ISL_1551172, EPI_ISL_1551173, EPI_ISL_1551174, EPI_ISL_1551175, EPI_ISL_1551176, EPI_ISL_1551177, EPI_ISL_1551178, EPI_ISL_1551179, EPI_ISL_1551180, EPI_ISL_1551181, EPI_ISL_1551182, EPI_ISL_1551183, EPI_ISL_1551184, EPI_ISL_1551185, EPI_ISL_1551187, EPI_ISL_1551188, EPI_ISL_1551189, EPI_ISL_1551190, EPI_ISL_1551191, EPI_ISL_1551192, EPI_ISL_1551193, EPI_ISL_1551194, EPI_ISL_1551195, EPI_ISL_1551196, EPI_ISL_1551197, EPI_ISL_1551198, EPI_ISL_1551199, EPI_ISL_1551200, EPI_ISL_1551201, EPI_ISL_1551202, EPI_ISL_1551203, EPI_ISL_1551204, EPI_ISL_1551205, EPI_ISL_1551206, EPI_ISL_1551208, EPI_ISL_1551209, EPI_ISL_1551210, EPI_ISL_1551211, EPI_ISL_1551212, EPI_ISL_1551213, EPI_ISL_1551214, EPI_ISL_1551215, EPI_ISL_1551216, EPI_ISL_1551217, EPI_ISL_1551218, EPI_ISL_1551219, EPI_ISL_1551220, EPI_ISL_1551221, EPI_ISL_1551222, EPI_ISL_1551223, EPI_ISL_1551224, EPI_ISL_1551225, EPI_ISL_1551226, EPI_ISL_1551227, EPI_ISL_1551229, EPI_ISL_1551230, EPI_ISL_1551231, EPI_ISL_1551232, EPI_ISL_1551233, EPI_ISL_1551234, EPI_ISL_1551235, EPI_ISL_1551236, EPI_ISL_1551237, EPI_ISL_1551238, EPI_ISL_1551239, EPI_ISL_1551242, EPI_ISL_1551243, EPI_ISL_1551244, EPI_ISL_1551245, EPI_ISL_1551246, EPI_ISL_1551247, EPI_ISL_1551248, EPI_ISL_1551249, EPI_ISL_1551250, EPI_ISL_1551251, EPI_ISL_1551252, EPI_ISL_1551254, EPI_ISL_1551255, EPI_ISL_1551256, EPI_ISL_1551257, EPI_ISL_1551258, EPI_ISL_1551259, EPI_ISL_1551260, EPI_ISL_1551261, EPI_ISL_1551262, EPI_ISL_1551264, EPI_ISL_1551265, EPI_ISL_1551266, EPI_ISL_1551267, EPI_ISL_1551268, EPI_ISL_1551269, EPI_ISL_1551270, EPI_ISL_1551271, EPI_ISL_1551272, EPI_ISL_1551273, EPI_ISL_1551274, EPI_ISL_1551275, EPI_ISL_1551276, EPI_ISL_1551277, EPI_ISL_1551278, EPI_ISL_1551279, EPI_ISL_1551280, EPI_ISL_1551281, EPI_ISL_1551282, EPI_ISL_1551283, EPI_ISL_1551284, EPI_ISL_1551285, EPI_ISL_1551286, EPI_ISL_1551287, EPI_ISL_1551288, EPI_ISL_1551289, EPI_ISL_1551290, EPI_ISL_1551291, EPI_ISL_1551292, EPI_ISL_1551293, EPI_ISL_1551294, EPI_ISL_1551295, EPI_ISL_1551296, EPI_ISL_1551297, EPI_ISL_1551298, EPI_ISL_1551299, EPI_ISL_1551301, EPI_ISL_1551302, EPI_ISL_1551303, EPI_ISL_1551304, EPI_ISL_1551305, EPI_ISL_1551306, EPI_ISL_1551308, EPI_ISL_1551309, EPI_ISL_1551310, EPI_ISL_1551311, EPI_ISL_1551312, EPI_ISL_1551313, EPI_ISL_1551314, EPI_ISL_1551315, EPI_ISL_1551316, EPI_ISL_1551317, EPI_ISL_1551318, EPI_ISL_1551319, EPI_ISL_1551320, EPI_ISL_1551321, EPI_ISL_1551322, EPI_ISL_1551323, EPI_ISL_1551324, EPI_ISL_1551325, EPI_ISL_1551326, EPI_ISL_1551327, EPI_ISL_1551328, EPI_ISL_1551329, EPI_ISL_1551330, EPI_ISL_1551331, EPI_ISL_1551332, EPI_ISL_1551333, EPI_ISL_1551334, EPI_ISL_1551335, EPI_ISL_1551336, EPI_ISL_1551337, EPI_ISL_1551338, EPI_ISL_1551339, EPI_ISL_1551340, EPI_ISL_1551341, EPI_ISL_1551342, EPI_ISL_1551343, EPI_ISL_1551344, EPI_ISL_1551345, EPI_ISL_1551346, EPI_ISL_1551347, EPI_ISL_1551348, EPI_ISL_1551349, EPI_ISL_1551350, EPI_ISL_1551351, EPI_ISL_1551352, EPI_ISL_1551353, EPI_ISL_1551354, EPI_ISL_1551355, EPI_ISL_1551356, EPI_ISL_1551357, EPI_ISL_1551358, EPI_ISL_1551359, EPI_ISL_1551360, EPI_ISL_1551361 | | | |
| see above | Aegis Sciences Corporation | Centers for Disease Control and Prevention Division of Viral Diseases, Pathogen Discovery | Dakota Howard, Dhwani Batra, Peter W. Cook, Kara Moser, Adrian Paskey, Jason Caravas, Benjamin Rambo-Martin, Shatavia Morrison, Christopher Gulvick, Scott Sammons, Yvette Unoarumhi, Darlene Wagner, Matthew Schmerer, Cyndi Clark, Patrick Campbell, Rob Case, Vikramsinha Ghorpade, Holly Houdeshell, Ola Kvalvaag, Dillon Nall, Ethan Sanders, Alec Vest, Shaun Westlund, Matthew Hardison, Clinton R. Paden, Duncan MacCannell |
| EPI_ISL_1551362 | DC Public Health Lab/ Dept. of Forensic Sciences | DC Public Health Lab/ Dept. of Forensic Sciences | Janis Doss, Scott Nguyen, Elizabeth Zelaya, Sarah Scott, Connie Maza, Monica Mann, Brittany Hamilton, David Payne, Jocelyn Hauser |
| EPI_ISL_1551363 | Aegis Sciences Corporation | Centers for Disease Control and Prevention Division of Viral Diseases, Pathogen Discovery | Dakota Howard, Dhwani Batra, Peter W. Cook, Kara Moser, Adrian Paskey, Jason Caravas, Benjamin Rambo-Martin, Shatavia Morrison, Christopher Gulvick, Scott Sammons, Yvette Unoarumhi, Darlene Wagner, Matthew Schmerer, Cyndi Clark, Patrick Campbell, Rob Case, Vikramsinha Ghorpade, Holly Houdeshell, Ola Kvalvaag, Dillon Nall, Ethan Sanders, Alec Vest, Shaun Westlund, Matthew Hardison, Clinton R. Paden, Duncan MacCannell |
| EPI_ISL_1551364, EPI_ISL_1551365 | DC Public Health Lab/ Dept. of Forensic Sciences | DC Public Health Lab/ Dept. of Forensic Sciences | Janis Doss, Scott Nguyen, Elizabeth Zelaya, Sarah Scott, Connie Maza, Monica Mann, Brittany Hamilton, David Payne, Jocelyn Hauser |
| EPI_ISL_1551366 | Aegis Sciences Corporation | Centers for Disease Control and Prevention Division of Viral Diseases, Pathogen Discovery | Dakota Howard, Dhwani Batra, Peter W. Cook, Kara Moser, Adrian Paskey, Jason Caravas, Benjamin Rambo-Martin, Shatavia Morrison, Christopher Gulvick, Scott Sammons, Yvette Unoarumhi, Darlene Wagner, Matthew Schmerer, Cyndi Clark, Patrick Campbell, Rob Case, Vikramsinha Ghorpade, Holly Houdeshell, Ola Kvalvaag, Dillon Nall, Ethan Sanders, Alec Vest, Shaun Westlund, Matthew Hardison, Clinton R. Paden, Duncan MacCannell |
| EPI_ISL_1551367, EPI_ISL_1551368 | DC Public Health Lab/ Dept. of Forensic Sciences | DC Public Health Lab/ Dept. of Forensic Sciences | Janis Doss, Scott Nguyen, Elizabeth Zelaya, Sarah Scott, Connie Maza, Monica Mann, Brittany Hamilton, David Payne, Jocelyn Hauser |
| EPI_ISL_1551369 | Aegis Sciences Corporation | Centers for Disease Control and Prevention Division of Viral Diseases, Pathogen Discovery | Dakota Howard, Dhwani Batra, Peter W. Cook, Kara Moser, Adrian Paskey, Jason Caravas, Benjamin Rambo-Martin, Shatavia Morrison, Christopher Gulvick, Scott Sammons, Yvette Unoarumhi, Darlene Wagner, Matthew Schmerer, Cyndi Clark, Patrick Campbell, Rob Case, Vikramsinha Ghorpade, Holly Houdeshell, Ola Kvalvaag, Dillon Nall, Ethan Sanders, Alec Vest, Shaun Westlund, Matthew Hardison, Clinton R. Paden, Duncan MacCannell |
| EPI_ISL_1551370, EPI_ISL_1551372, | DC Public Health Lab/ Dept. of Forensic Sciences | DC Public Health Lab/ Dept. of Forensic Sciences | Janis Doss, Scott Nguyen, Elizabeth Zelaya, Sarah Scott, Connie Maza, Monica Mann, Brittany Hamilton, David Payne, Jocelyn Hauser |

| | | | |
|---|---|---|---|
| EPI_ISL_1551373 | | | |
| EPI_ISL_1551374, EPI_ISL_1551375, EPI_ISL_1551376, EPI_ISL_1551377, EPI_ISL_1551378, EPI_ISL_1551379, EPI_ISL_1551380, EPI_ISL_1551381, EPI_ISL_1551382, EPI_ISL_1551383, EPI_ISL_1551384, EPI_ISL_1551385, EPI_ISL_1551386, EPI_ISL_1551387, EPI_ISL_1551388, EPI_ISL_1551389, EPI_ISL_1551390, EPI_ISL_1551391, EPI_ISL_1551392, EPI_ISL_1551393, EPI_ISL_1551394, EPI_ISL_1551395, EPI_ISL_1551396, EPI_ISL_1551397, EPI_ISL_1551398, EPI_ISL_1551399, EPI_ISL_1551400, EPI_ISL_1551401, EPI_ISL_1551402, EPI_ISL_1551403, EPI_ISL_1551404, EPI_ISL_1551405, EPI_ISL_1551406, EPI_ISL_1551407, EPI_ISL_1551408, EPI_ISL_1551409, EPI_ISL_1551410, EPI_ISL_1551412, EPI_ISL_1551413, EPI_ISL_1551414, EPI_ISL_1551415, EPI_ISL_1551416, EPI_ISL_1551417, EPI_ISL_1551418, EPI_ISL_1551419, EPI_ISL_1551420, EPI_ISL_1551421, EPI_ISL_1551422, EPI_ISL_1551423, EPI_ISL_1551424, EPI_ISL_1551425, EPI_ISL_1551426, EPI_ISL_1551427, EPI_ISL_1551428, EPI_ISL_1551429, EPI_ISL_1551431, EPI_ISL_1551432, EPI_ISL_1551433, EPI_ISL_1551434, EPI_ISL_1551435, EPI_ISL_1551436, EPI_ISL_1551438, EPI_ISL_1551439, EPI_ISL_1551440, EPI_ISL_1551441, EPI_ISL_1551442, EPI_ISL_1551443, EPI_ISL_1551444, EPI_ISL_1551445, EPI_ISL_1551446, EPI_ISL_1551447, EPI_ISL_1551448, EPI_ISL_1551449, EPI_ISL_1551450, EPI_ISL_1551451, EPI_ISL_1551452, EPI_ISL_1551453, EPI_ISL_1551454, EPI_ISL_1551455, EPI_ISL_1551456, EPI_ISL_1551457, EPI_ISL_1551458, EPI_ISL_1551459, EPI_ISL_1551460, EPI_ISL_1551461, EPI_ISL_1551462, EPI_ISL_1551463, EPI_ISL_1551464, EPI_ISL_1551465, EPI_ISL_1551466, EPI_ISL_1551467, EPI_ISL_1551468, EPI_ISL_1551469, EPI_ISL_1551470, EPI_ISL_1551471, EPI_ISL_1551472, EPI_ISL_1551473, EPI_ISL_1551474, EPI_ISL_1551475, EPI_ISL_1551476, EPI_ISL_1551477, EPI_ISL_1551478, EPI_ISL_1551479, EPI_ISL_1551480, EPI_ISL_1551481, EPI_ISL_1551482, EPI_ISL_1551483, EPI_ISL_1551484, EPI_ISL_1551485, EPI_ISL_1551486, EPI_ISL_1551487, EPI_ISL_1551488, EPI_ISL_1551489, EPI_ISL_1551490, EPI_ISL_1551491, EPI_ISL_1551492, EPI_ISL_1551493, EPI_ISL_1551494, EPI_ISL_1551496, EPI_ISL_1551497, EPI_ISL_1551499, EPI_ISL_1551500, EPI_ISL_1551501, EPI_ISL_1551502, EPI_ISL_1551503, EPI_ISL_1551504, EPI_ISL_1551505, EPI_ISL_1551506, EPI_ISL_1551507, EPI_ISL_1551508, EPI_ISL_1551509, EPI_ISL_1551510, EPI_ISL_1551511, EPI_ISL_1551512, EPI_ISL_1551513, EPI_ISL_1551514, EPI_ISL_1551515, EPI_ISL_1551516, EPI_ISL_1551517, EPI_ISL_1551518, EPI_ISL_1551519, EPI_ISL_1551520, EPI_ISL_1551521, EPI_ISL_1551523, EPI_ISL_1551524, EPI_ISL_1551525, EPI_ISL_1551526, EPI_ISL_1551527, EPI_ISL_1551529, EPI_ISL_1551530, EPI_ISL_1551531, EPI_ISL_1551532, EPI_ISL_1551533, EPI_ISL_1551534, EPI_ISL_1551535, EPI_ISL_1551536, EPI_ISL_1551537, EPI_ISL_1551538, EPI_ISL_1551540, EPI_ISL_1551541, EPI_ISL_1551542, EPI_ISL_1551543, EPI_ISL_1551544, EPI_ISL_1551545, EPI_ISL_1551546, EPI_ISL_1551547, EPI_ISL_1551548, EPI_ISL_1551549, EPI_ISL_1551550, EPI_ISL_1551551, EPI_ISL_1551552, EPI_ISL_1551553, EPI_ISL_1551554, EPI_ISL_1551555, EPI_ISL_1551556, EPI_ISL_1551557, EPI_ISL_1551558, EPI_ISL_1551559, EPI_ISL_1551560, EPI_ISL_1551561, EPI_ISL_1551562, EPI_ISL_1551563, EPI_ISL_1551564, EPI_ISL_1551565, EPI_ISL_1551566, EPI_ISL_1551568, EPI_ISL_1551569, EPI_ISL_1551571, EPI_ISL_1551572, EPI_ISL_1551574, EPI_ISL_1551575, EPI_ISL_1551576, EPI_ISL_1551577, EPI_ISL_1551578, EPI_ISL_1551579, EPI_ISL_1551580, EPI_ISL_1551581, EPI_ISL_1551582, EPI_ISL_1551583, EPI_ISL_1551584, EPI_ISL_1551585, EPI_ISL_1551586, EPI_ISL_1551587, EPI_ISL_1551588, EPI_ISL_1551589, EPI_ISL_1551590, EPI_ISL_1551591, EPI_ISL_1551592, EPI_ISL_1551593, EPI_ISL_1551595, EPI_ISL_1551596, EPI_ISL_1551597, EPI_ISL_1551599, EPI_ISL_1551600, EPI_ISL_1551601, EPI_ISL_1551602, EPI_ISL_1551603, EPI_ISL_1551604, EPI_ISL_1551605, EPI_ISL_1551606, EPI_ISL_1551607, EPI_ISL_1551608, EPI_ISL_1551609, EPI_ISL_1551610, EPI_ISL_1551611, EPI_ISL_1551612, EPI_ISL_1551613, EPI_ISL_1551614, EPI_ISL_1551615, EPI_ISL_1551616, EPI_ISL_1551617, EPI_ISL_1551618, EPI_ISL_1551619, EPI_ISL_1551620, EPI_ISL_1551621, EPI_ISL_1551622, EPI_ISL_1551623, EPI_ISL_1551624, EPI_ISL_1551625, EPI_ISL_1551626, EPI_ISL_1551627, EPI_ISL_1551628, EPI_ISL_1551629, EPI_ISL_1551630, EPI_ISL_1551631, EPI_ISL_1551632, EPI_ISL_1551633, EPI_ISL_1551634, EPI_ISL_1551635, EPI_ISL_1551636, EPI_ISL_1551638, EPI_ISL_1551639, EPI_ISL_1551640, EPI_ISL_1551641, EPI_ISL_1551642 | | | |
| see above | Aegis Sciences Corporation | Centers for Disease Control and Prevention Division of Viral Diseases, Pathogen Discovery | Dakota Howard, Dhwani Batra, Peter W. Cook, Kara Moser, Adrian Paskey, Jason Caravas, Benjamin Rambo-Martin, Shatavia Morrison, Christopher Gulvick, Scott Sammons, Yvette Unoarumhi, Darlene Wagner, Matthew Schmerer, Cyndi Clark, Patrick Campbell, Rob Case, Vikramsinha Ghorpade, Holly Houdeshell, Ola Kvalvaag, Dillon Nall, Ethan Sanders, Alec Vest, Shaun Westlund, Matthew Hardison, Clinton R. Paden, Duncan MacCannell |
| EPI_ISL_1551643, EPI_ISL_1551644, EPI_ISL_1551645, EPI_ISL_1551646, EPI_ISL_1551647, EPI_ISL_1551648, EPI_ISL_1551649, EPI_ISL_1551650, EPI_ISL_1551651, EPI_ISL_1551652, EPI_ISL_1551653, EPI_ISL_1551654, EPI_ISL_1551655, EPI_ISL_1551656, EPI_ISL_1551657, EPI_ISL_1551658, EPI_ISL_1551659, EPI_ISL_1551660, EPI_ISL_1551661, EPI_ISL_1551662, EPI_ISL_1551663, EPI_ISL_1551664, EPI_ISL_1551665, EPI_ISL_1551666, EPI_ISL_1551667, EPI_ISL_1551668, EPI_ISL_1551669, EPI_ISL_1551670, EPI_ISL_1551671, EPI_ISL_1551672, EPI_ISL_1551673, EPI_ISL_1551674, EPI_ISL_1551675, EPI_ISL_1551676, EPI_ISL_1551677, EPI_ISL_1551678, EPI_ISL_1551679, EPI_ISL_1551680, EPI_ISL_1551681, EPI_ISL_1551682, EPI_ISL_1551683, EPI_ISL_1551684, EPI_ISL_1551685, EPI_ISL_1551686, EPI_ISL_1551687, EPI_ISL_1551688, EPI_ISL_1551689, EPI_ISL_1551690, EPI_ISL_1551691, EPI_ISL_1551692, EPI_ISL_1551693 | | | |
| see above | Quest Diagnostics Incorporated | Centers for Disease Control and Prevention Division of Viral Diseases, Pathogen Discovery | Peter W. Cook, Dakota Howard, Dhwani Batra, Ben L. Rambo-Martin, S. H. Rosenthal, A. Gerasimova, R. M. Kagan, B. Anderson, M. Hua, Y. Liu, L.E. Bernstein, K.E. Livingston, A. Perez, I. A. Shlyakhter, R. V. Rolando, R. Owen, P. Tanpaiboon, F. Lacbawan, Clinton R. Paden, Suxiang Tong, Duncan MacCannell |
| EPI_ISL_1551694 | Montana Public Health Laboratory | Montana Public Health Laboratory | Joy Ritter, Michelle Mozer, Carrie Biskupiak, Deborah Gibson |
| EPI_ISL_1551695, EPI_ISL_1551696, EPI_ISL_1551697, EPI_ISL_1551698, EPI_ISL_1551699, EPI_ISL_1551700, EPI_ISL_1551701, EPI_ISL_1551702, EPI_ISL_1551703, EPI_ISL_1551704, EPI_ISL_1551705, EPI_ISL_1551706, EPI_ISL_1551707, EPI_ISL_1551708, EPI_ISL_1551709, EPI_ISL_1551710, EPI_ISL_1551711, EPI_ISL_1551712, EPI_ISL_1551713, EPI_ISL_1551714, EPI_ISL_1551715, EPI_ISL_1551716, EPI_ISL_1551717, EPI_ISL_1551718, EPI_ISL_1551719, EPI_ISL_1551720, EPI_ISL_1551721, EPI_ISL_1551722, EPI_ISL_1551723, EPI_ISL_1551724, EPI_ISL_1551725, EPI_ISL_1551726, EPI_ISL_1551727, EPI_ISL_1551728, EPI_ISL_1551729, EPI_ISL_1551730, EPI_ISL_1551731, EPI_ISL_1551732, EPI_ISL_1551733, EPI_ISL_1551734, EPI_ISL_1551735, EPI_ISL_1551736, EPI_ISL_1551737, EPI_ISL_1551738, EPI_ISL_1551739, EPI_ISL_1551740, EPI_ISL_1551741, EPI_ISL_1551742, EPI_ISL_1551743, EPI_ISL_1551744, EPI_ISL_1551745, EPI_ISL_1551746, EPI_ISL_1551747, EPI_ISL_1551748, EPI_ISL_1551749, EPI_ISL_1551750, EPI_ISL_1551751, EPI_ISL_1551752, EPI_ISL_1551753, EPI_ISL_1551754, EPI_ISL_1551755, EPI_ISL_1551756, EPI_ISL_1551757, EPI_ISL_1551758, EPI_ISL_1551759, EPI_ISL_1551760, EPI_ISL_1551761, EPI_ISL_1551762, EPI_ISL_1551763, EPI_ISL_1551764, EPI_ISL_1551765, EPI_ISL_1551766, EPI_ISL_1551767, EPI_ISL_1551768, EPI_ISL_1551769, EPI_ISL_1551770, EPI_ISL_1551771, EPI_ISL_1551772, EPI_ISL_1551773, EPI_ISL_1551774 | | | |
| see above | Quest Diagnostics Incorporated | Centers for Disease Control and Prevention Division of Viral Diseases, Pathogen Discovery | Peter W. Cook, Dakota Howard, Dhwani Batra, Ben L. Rambo-Martin, S. H. Rosenthal, A. Gerasimova, R. M. Kagan, B. Anderson, M. Hua, Y. Liu, L.E. Bernstein, K.E. Livingston, A. Perez, I. A. Shlyakhter, R. V. Rolando, R. Owen, P. Tanpaiboon, F. Lacbawan, Clinton R. Paden, Suxiang Tong, Duncan MacCannell |
| EPI_ISL_1551775, EPI_ISL_1551776, EPI_ISL_1551777, EPI_ISL_1551778, EPI_ISL_1551779, EPI_ISL_1551780, EPI_ISL_1551781, EPI_ISL_1551782, EPI_ISL_1551783, EPI_ISL_1551784, EPI_ISL_1551785, EPI_ISL_1551786, EPI_ISL_1551787, EPI_ISL_1551788, EPI_ISL_1551789, EPI_ISL_1551790, EPI_ISL_1551791, EPI_ISL_1551792, EPI_ISL_1551793, EPI_ISL_1551794, EPI_ISL_1551795, EPI_ISL_1551796, EPI_ISL_1551797, EPI_ISL_1551798, EPI_ISL_1551799, EPI_ISL_1551800, EPI_ISL_1551801, EPI_ISL_1551802, EPI_ISL_1551803, EPI_ISL_1551804, EPI_ISL_1551805, EPI_ISL_1551806, EPI_ISL_1551807, EPI_ISL_1551808, EPI_ISL_1551809, EPI_ISL_1551810, EPI_ISL_1551811, EPI_ISL_1551812, EPI_ISL_1551813, EPI_ISL_1551814, EPI_ISL_1551815, EPI_ISL_1551816, EPI_ISL_1551817, EPI_ISL_1551818, EPI_ISL_1551819, EPI_ISL_1551820, EPI_ISL_1551821, EPI_ISL_1551822, EPI_ISL_1551823, EPI_ISL_1551824, EPI_ISL_1551825, EPI_ISL_1551826, EPI_ISL_1551827, EPI_ISL_1551828, EPI_ISL_1551829, EPI_ISL_1551830, EPI_ISL_1551831, EPI_ISL_1551832, EPI_ISL_1551833, EPI_ISL_1551834, EPI_ISL_1551835, EPI_ISL_1551836, EPI_ISL_1551837, EPI_ISL_1551838, EPI_ISL_1551839, EPI_ISL_1551840, EPI_ISL_1551841, EPI_ISL_1551842, EPI_ISL_1551843, EPI_ISL_1551844, EPI_ISL_1551845, EPI_ISL_1551846, EPI_ISL_1551847, EPI_ISL_1551848, EPI_ISL_1551849, EPI_ISL_1551850, EPI_ISL_1551851, EPI_ISL_1551852, EPI_ISL_1551853, EPI_ISL_1551854, EPI_ISL_1551855, EPI_ISL_1551856, EPI_ISL_1551857, EPI_ISL_1551858, EPI_ISL_1551859, EPI_ISL_1551860, EPI_ISL_1551861, EPI_ISL_1551862, EPI_ISL_1551863, EPI_ISL_1551864, EPI_ISL_1551865, EPI_ISL_1551866, EPI_ISL_1551867, EPI_ISL_1551868, EPI_ISL_1551869, EPI_ISL_1551870, EPI_ISL_1551871, EPI_ISL_1551872, EPI_ISL_1551873, EPI_ISL_1551874, EPI_ISL_1551875, EPI_ISL_1551876, EPI_ISL_1551877, EPI_ISL_1551878, EPI_ISL_1551879, EPI_ISL_1551880, EPI_ISL_1551881, EPI_ISL_1551882, EPI_ISL_1551883, EPI_ISL_1551884, EPI_ISL_1551885, EPI_ISL_1551886, EPI_ISL_1551887, EPI_ISL_1551888, EPI_ISL_1551889, EPI_ISL_1551890, EPI_ISL_1551891, EPI_ISL_1551892, EPI_ISL_1551893, EPI_ISL_1551894, EPI_ISL_1551895, EPI_ISL_1551896, EPI_ISL_1551897, EPI_ISL_1551898, EPI_ISL_1551899, EPI_ISL_1551900, EPI_ISL_1551901, EPI_ISL_1551902, EPI_ISL_1551903, EPI_ISL_1551904, EPI_ISL_1551905, EPI_ISL_1551906, EPI_ISL_1551907, EPI_ISL_1551908, EPI_ISL_1551909, EPI_ISL_1551910, EPI_ISL_1551911, EPI_ISL_1551912, EPI_ISL_1551913, EPI_ISL_1551914, EPI_ISL_1551915, EPI_ISL_1551916, EPI_ISL_1551917, EPI_ISL_1551918, EPI_ISL_1551919, EPI_ISL_1551920, EPI_ISL_1551921, EPI_ISL_1551922, EPI_ISL_1551923, EPI_ISL_1551924, EPI_ISL_1551925, EPI_ISL_1551926, EPI_ISL_1551927, EPI_ISL_1551928, EPI_ISL_1551929, EPI_ISL_1551930, EPI_ISL_1551931, EPI_ISL_1551932, EPI_ISL_1551933, EPI_ISL_1551934, EPI_ISL_1551935, EPI_ISL_1551936, EPI_ISL_1551937, EPI_ISL_1551938, EPI_ISL_1551939, EPI_ISL_1551940, EPI_ISL_1551941, EPI_ISL_1551942, EPI_ISL_1551943, EPI_ISL_1551944, EPI_ISL_1551945, EPI_ISL_1551946, EPI_ISL_1551947, EPI_ISL_1551948, EPI_ISL_1551949, EPI_ISL_1551950, EPI_ISL_1551951 | | | |
| see above | Quest Diagnostics Incorporated | Centers for Disease Control and Prevention Division of Viral Diseases, Pathogen Discovery | Dakota Howard, Dhwani Batra, Peter W. Cook, Kara Moser, Adrian Paskey, Jason Caravas, Benjamin Rambo-Martin, Shatavia Morrison, Christopher Gulvick, Scott Sammons, Yvette Unoarumhi, Darlene Wagner, Matthew Schmerer, S. H. Rosenthal, A. Gerasimova, R. M. Kagan, B. Anderson, M. Hua, Y. Liu, L.E. Bernstein, K.E. Livingston, A. Perez, I. A. Shlyakhter, R. V. Rolando, R. Owen, P. Tanpaiboon, F. Lacbawan, Clinton R. Paden, Duncan MacCannell |
| EPI_ISL_1551952 | Montana Public Health Laboratory | Montana Public Health Laboratory | Joy Ritter, Michelle Mozer, Carrie Biskupiak, Deborah Gibson |
| EPI_ISL_1551953, EPI_ISL_1551954, EPI_ISL_1551955, EPI_ISL_1551956, EPI_ISL_1551957, EPI_ISL_1551958, EPI_ISL_1551959, EPI_ISL_1551960, EPI_ISL_1551961, EPI_ISL_1551962, EPI_ISL_1551963, EPI_ISL_1551964, EPI_ISL_1551965, EPI_ISL_1551966, EPI_ISL_1551967, EPI_ISL_1551968, EPI_ISL_1551969, EPI_ISL_1551970, EPI_ISL_1551971, EPI_ISL_1551972, EPI_ISL_1551973, EPI_ISL_1551974, EPI_ISL_1551975, EPI_ISL_1551976, EPI_ISL_1551977, EPI_ISL_1551978, EPI_ISL_1551979, EPI_ISL_1551980, EPI_ISL_1551981, EPI_ISL_1551982, EPI_ISL_1551983, EPI_ISL_1551984, EPI_ISL_1551985, EPI_ISL_1551986, EPI_ISL_1551987, EPI_ISL_1551988, EPI_ISL_1551989, EPI_ISL_1551990, EPI_ISL_1551991, EPI_ISL_1551992, EPI_ISL_1551993, EPI_ISL_1551994, EPI_ISL_1551995, EPI_ISL_1551996, EPI_ISL_1551997, EPI_ISL_1551998, EPI_ISL_1551999, EPI_ISL_1552000, EPI_ISL_1552001, EPI_ISL_1552002, EPI_ISL_1552003, EPI_ISL_1552004, EPI_ISL_1552005, EPI_ISL_1552006, EPI_ISL_1552007, EPI_ISL_1552008, EPI_ISL_1552009, EPI_ISL_1552010, EPI_ISL_1552011, EPI_ISL_1552012, EPI_ISL_1552013, EPI_ISL_1552014, EPI_ISL_1552015, EPI_ISL_1552016, EPI_ISL_1552017, EPI_ISL_1552018, EPI_ISL_1552019, EPI_ISL_1552020, EPI_ISL_1552021, EPI_ISL_1552022, EPI_ISL_1552023, EPI_ISL_1552024, EPI_ISL_1552025, EPI_ISL_1552026, EPI_ISL_1552027, EPI_ISL_1552028, EPI_ISL_1552029, EPI_ISL_1552030, EPI_ISL_1552031, EPI_ISL_1552032, EPI_ISL_1552033, EPI_ISL_1552034, EPI_ISL_1552035, EPI_ISL_1552036, EPI_ISL_1552037, EPI_ISL_1552038, EPI_ISL_1552039, EPI_ISL_1552040, EPI_ISL_1552041, EPI_ISL_1552042, EPI_ISL_1552043, EPI_ISL_1552044, EPI_ISL_1552045, EPI_ISL_1552046, EPI_ISL_1552047, EPI_ISL_1552048, EPI_ISL_1552049, EPI_ISL_1552050, EPI_ISL_1552051, EPI_ISL_1552052, EPI_ISL_1552053, EPI_ISL_1552054, EPI_ISL_1552055, EPI_ISL_1552056, EPI_ISL_1552057, EPI_ISL_1552058, EPI_ISL_1552059, EPI_ISL_1552060, EPI_ISL_1552061, EPI_ISL_1552062, EPI_ISL_1552063, EPI_ISL_1552064, EPI_ISL_1552065, EPI_ISL_1552066, EPI_ISL_1552067, EPI_ISL_1552068, EPI_ISL_1552069, EPI_ISL_1552070, EPI_ISL_1552071, EPI_ISL_1552072, EPI_ISL_1552073, EPI_ISL_1552074, EPI_ISL_1552075, EPI_ISL_1552076, EPI_ISL_1552077, EPI_ISL_1552078, EPI_ISL_1552079, EPI_ISL_1552080, EPI_ISL_1552081, EPI_ISL_1552082, EPI_ISL_1552083 | | | |
| see above | Quest Diagnostics Incorporated | Centers for Disease Control and Prevention Division of Viral Diseases, Pathogen Discovery | Dakota Howard, Dhwani Batra, Peter W. Cook, Kara Moser, Adrian Paskey, Jason Caravas, Benjamin Rambo-Martin, Shatavia Morrison, Christopher Gulvick, Scott Sammons, Yvette Unoarumhi, Darlene Wagner, Matthew Schmerer, S. H. Rosenthal, A. Gerasimova, R. M. Kagan, B. Anderson, M. Hua, Y. Liu, L.E. Bernstein, K.E. Livingston, A. Perez, I. A. Shlyakhter, R. V. Rolando, R. Owen, P. Tanpaiboon, F. Lacbawan, Clinton R. Paden, Duncan MacCannell |
| EPI_ISL_1552084 | Montana Public Health Laboratory | Montana Public Health Laboratory | Joy Ritter, Michelle Mozer, Carrie Biskupiak, Deborah Gibson |
| EPI_ISL_1552085, EPI_ISL_1552086, EPI_ISL_1552087, EPI_ISL_1552088, EPI_ISL_1552089, EPI_ISL_1552090, EPI_ISL_1552091, EPI_ISL_1552092, EPI_ISL_1552093, EPI_ISL_1552094, EPI_ISL_1552095, EPI_ISL_1552096, EPI_ISL_1552097, EPI_ISL_1552098, EPI_ISL_1552099, EPI_ISL_1552100, EPI_ISL_1552101, EPI_ISL_1552102, EPI_ISL_1552103, EPI_ISL_1552104, EPI_ISL_1552105, EPI_ISL_1552106, EPI_ISL_1552107, EPI_ISL_1552108, EPI_ISL_1552109, EPI_ISL_1552110, EPI_ISL_1552111, EPI_ISL_1552112, EPI_ISL_1552113, EPI_ISL_1552114, EPI_ISL_1552115, EPI_ISL_1552116, EPI_ISL_1552117, EPI_ISL_1552118, EPI_ISL_1552119, EPI_ISL_1552120, EPI_ISL_1552121, EPI_ISL_1552122, EPI_ISL_1552123, EPI_ISL_1552124, EPI_ISL_1552125, EPI_ISL_1552126, EPI_ISL_1552127, EPI_ISL_1552128, EPI_ISL_1552129, EPI_ISL_1552130, EPI_ISL_1552131, EPI_ISL_1552132, EPI_ISL_1552133, EPI_ISL_1552134, EPI_ISL_1552135, EPI_ISL_1552136, EPI_ISL_1552137, EPI_ISL_1552138, EPI_ISL_1552139, EPI_ISL_1552140, EPI_ISL_1552141, EPI_ISL_1552142, EPI_ISL_1552143, EPI_ISL_1552144, EPI_ISL_1552145, EPI_ISL_1552146, EPI_ISL_1552147, EPI_ISL_1552148, EPI_ISL_1552149, EPI_ISL_1552150, EPI_ISL_1552151, EPI_ISL_1552152, EPI_ISL_1552153, EPI_ISL_1552154, EPI_ISL_1552155, EPI_ISL_1552156, EPI_ISL_1552157, EPI_ISL_1552158, EPI_ISL_1552159, EPI_ISL_1552160, EPI_ISL_1552161, EPI_ISL_1552162, EPI_ISL_1552163, EPI_ISL_1552164, EPI_ISL_1552165, EPI_ISL_1552166, EPI_ISL_1552167, EPI_ISL_1552168, EPI_ISL_1552169, EPI_ISL_1552170, EPI_ISL_1552171, EPI_ISL_1552172, EPI_ISL_1552173, EPI_ISL_1552174, EPI_ISL_1552175, EPI_ISL_1552176, EPI_ISL_1552177, EPI_ISL_1552178, EPI_ISL_1552179, EPI_ISL_1552180, EPI_ISL_1552181, EPI_ISL_1552182, EPI_ISL_1552183, EPI_ISL_1552184, EPI_ISL_1552185, EPI_ISL_1552186, EPI_ISL_1552187, EPI_ISL_1552188, EPI_ISL_1552189, EPI_ISL_1552190, EPI_ISL_1552191, EPI_ISL_1552192, EPI_ISL_1552193, EPI_ISL_1552194, EPI_ISL_1552195, EPI_ISL_1552196, EPI_ISL_1552197, EPI_ISL_1552198, EPI_ISL_1552199, EPI_ISL_1552200, EPI_ISL_1552201, EPI_ISL_1552202, EPI_ISL_1552203, EPI_ISL_1552204, EPI_ISL_1552205, EPI_ISL_1552206, EPI_ISL_1552207, EPI_ISL_1552208, EPI_ISL_1552209, EPI_ISL_1552210, EPI_ISL_1552211, EPI_ISL_1552212, EPI_ISL_1552213, EPI_ISL_1552214, EPI_ISL_1552215, EPI_ISL_1552216, EPI_ISL_1552217, EPI_ISL_1552218, EPI_ISL_1552219, EPI_ISL_1552220, EPI_ISL_1552221, EPI_ISL_1552222, EPI_ISL_1552223, EPI_ISL_1552224, EPI_ISL_1552225, EPI_ISL_1552226, EPI_ISL_1552227, EPI_ISL_1552228, EPI_ISL_1552229, EPI_ISL_1552230, EPI_ISL_1552231, EPI_ISL_1552232, EPI_ISL_1552233, EPI_ISL_1552234, EPI_ISL_1552235, EPI_ISL_1552236, EPI_ISL_1552237, EPI_ISL_1552238, EPI_ISL_1552239, EPI_ISL_1552240, EPI_ISL_1552241, EPI_ISL_1552242, EPI_ISL_1552245, EPI_ISL_1552249, EPI_ISL_1552252, EPI_ISL_1552254, EPI_ISL_1552255, EPI_ISL_1552257, EPI_ISL_1552259, EPI_ISL_1552260, EPI_ISL_1552262, EPI_ISL_1552264, EPI_ISL_1552265, EPI_ISL_1552267, EPI_ISL_1552269, EPI_ISL_1552270, EPI_ISL_1552272, EPI_ISL_1552273, EPI_ISL_1552275, EPI_ISL_1552277, EPI_ISL_1552278, EPI_ISL_1552280, EPI_ISL_1552282, EPI_ISL_1552283, EPI_ISL_1552285, EPI_ISL_1552286, EPI_ISL_1552288, EPI_ISL_1552290, EPI_ISL_1552291, EPI_ISL_1552293, EPI_ISL_1552295, EPI_ISL_1552296, EPI_ISL_1552298, EPI_ISL_1552299, EPI_ISL_1552301, EPI_ISL_1552304, EPI_ISL_1552305, EPI_ISL_1552307, EPI_ISL_1552308, EPI_ISL_1552310, EPI_ISL_1552311, EPI_ISL_1552313, EPI_ISL_1552314, EPI_ISL_1552321, EPI_ISL_1552324, EPI_ISL_1552327, EPI_ISL_1552330, EPI_ISL_1552333, EPI_ISL_1552334, EPI_ISL_1552336, EPI_ISL_1552337, EPI_ISL_1552339, EPI_ISL_1552341, EPI_ISL_1552342, EPI_ISL_1552344, EPI_ISL_1552346, EPI_ISL_1552347, EPI_ISL_1552349, EPI_ISL_1552350, EPI_ISL_1552351, EPI_ISL_1552353, EPI_ISL_1552355, EPI_ISL_1552356, EPI_ISL_1552358, EPI_ISL_1552359, EPI_ISL_1552361, EPI_ISL_1552362, EPI_ISL_1552364, EPI_ISL_1552365, EPI_ISL_1552367, EPI_ISL_1552368, EPI_ISL_1552370, EPI_ISL_1552371, EPI_ISL_1552373, EPI_ISL_1552375, EPI_ISL_1552376, EPI_ISL_1552378, EPI_ISL_1552379, EPI_ISL_1552382, EPI_ISL_1552385, EPI_ISL_1552388, EPI_ISL_1552390, EPI_ISL_1552392, EPI_ISL_1552394, EPI_ISL_1552395, EPI_ISL_1552397, EPI_ISL_1552398, EPI_ISL_1552400, EPI_ISL_1552402, EPI_ISL_1552403, EPI_ISL_1552405, EPI_ISL_1552406, EPI_ISL_1552408, EPI_ISL_1552409, EPI_ISL_1552411, EPI_ISL_1552412, EPI_ISL_1552414, EPI_ISL_1552415, EPI_ISL_1552417, EPI_ISL_1552418, EPI_ISL_1552420, EPI_ISL_1552421, EPI_ISL_1552423, EPI_ISL_1552424, EPI_ISL_1552426, EPI_ISL_1552428, EPI_ISL_1552429, EPI_ISL_1552431, EPI_ISL_1552432, EPI_ISL_1552434, EPI_ISL_1552436, EPI_ISL_1552437, EPI_ISL_1552439, EPI_ISL_1552440, EPI_ISL_1552442, EPI_ISL_1552443, EPI_ISL_1552445, EPI_ISL_1552446, EPI_ISL_1552449, EPI_ISL_1552452, EPI_ISL_1552455, EPI_ISL_1552458, EPI_ISL_1552460, EPI_ISL_1552463, EPI_ISL_1552466, EPI_ISL_1552469, EPI_ISL_1552473, EPI_ISL_1552476, EPI_ISL_1552479, EPI_ISL_1552482, EPI_ISL_1552484, EPI_ISL_1552487, EPI_ISL_1552490, EPI_ISL_1552493, EPI_ISL_1552496, EPI_ISL_1552499, EPI_ISL_1552502, EPI_ISL_1552505, EPI_ISL_1552508, EPI_ISL_1552511, EPI_ISL_1552514, EPI_ISL_1552517, EPI_ISL_1552520, EPI_ISL_1552523, EPI_ISL_1552526, EPI_ISL_1552529, EPI_ISL_1552532, EPI_ISL_1552535, EPI_ISL_1552539, EPI_ISL_1552542, EPI_ISL_1552544, EPI_ISL_1552547, EPI_ISL_1552550, EPI_ISL_1552553, EPI_ISL_1552556, EPI_ISL_1552559, EPI_ISL_1552562, EPI_ISL_1552565, EPI_ISL_1552568, EPI_ISL_1552571, EPI_ISL_1552574, EPI_ISL_1552577, EPI_ISL_1552580, EPI_ISL_1552583, EPI_ISL_1552586, EPI_ISL_1552589, EPI_ISL_1552592, EPI_ISL_1552594, EPI_ISL_1552597, EPI_ISL_1552600, EPI_ISL_1552602, EPI_ISL_1552605, EPI_ISL_1552608, EPI_ISL_1552611, EPI_ISL_1552614, EPI_ISL_1552617, EPI_ISL_1552620, EPI_ISL_1552623, EPI_ISL_1552626, EPI_ISL_1552629, EPI_ISL_1552632, EPI_ISL_1552635, EPI_ISL_1552639, EPI_ISL_1552642, EPI_ISL_1552645, EPI_ISL_1552648, EPI_ISL_1552651, EPI_ISL_1552654, EPI_ISL_1552657, EPI_ISL_1552660, EPI_ISL_1552663, EPI_ISL_1552665, EPI_ISL_1552668, EPI_ISL_1552671, EPI_ISL_1552674, EPI_ISL_1552677, EPI_ISL_1552680, EPI_ISL_1552683, EPI_ISL_1552685, EPI_ISL_1552686, EPI_ISL_1552687, EPI_ISL_1552688, EPI_ISL_1552689, EPI_ISL_1552690, EPI_ISL_1552691, EPI_ISL_1552692, EPI_ISL_1552693, EPI_ISL_1552694, EPI_ISL_1552695, EPI_ISL_1552696, EPI_ISL_1552697, EPI_ISL_1552698, EPI_ISL_1552699, EPI_ISL_1552700, EPI_ISL_1552701, EPI_ISL_1552702, EPI_ISL_1552703, EPI_ISL_1552704, EPI_ISL_1552705, EPI_ISL_1552706, EPI_ISL_1552707, EPI_ISL_1552708, EPI_ISL_1552709, EPI_ISL_1552710, EPI_ISL_1552711, EPI_ISL_1552712, EPI_ISL_1552713, EPI_ISL_1552714, EPI_ISL_1552715, EPI_ISL_1552716, EPI_ISL_1552717, EPI_ISL_1552718, EPI_ISL_1552719, EPI_ISL_1552720, EPI_ISL_1552721, EPI_ISL_1552722, EPI_ISL_1552723, EPI_ISL_1552724, EPI_ISL_1552725, EPI_ISL_1552726, EPI_ISL_1552727, EPI_ISL_1552728, EPI_ISL_1552729, EPI_ISL_1552730, EPI_ISL_1552731, EPI_ISL_1552732, EPI_ISL_1552733, EPI_ISL_1552734, EPI_ISL_1552735, EPI_ISL_1552736, EPI_ISL_1552737, EPI_ISL_1552738, EPI_ISL_1552739, EPI_ISL_1552740, EPI_ISL_1552741, EPI_ISL_1552742, EPI_ISL_1552743, EPI_ISL_1552744, EPI_ISL_1552745, EPI_ISL_1552746, EPI_ISL_1552747, EPI_ISL_1552748, EPI_ISL_1552749, EPI_ISL_1552750, EPI_ISL_1552751, EPI_ISL_1552752, EPI_ISL_1552753, EPI_ISL_1552754, EPI_ISL_1552755, EPI_ISL_1552756, EPI_ISL_1552757, EPI_ISL_1552758, EPI_ISL_1552759, EPI_ISL_1552760, EPI_ISL_1552761, EPI_ISL_1552762, EPI_ISL_1552763, EPI_ISL_1552764, EPI_ISL_1552765, EPI_ISL_1552766, EPI_ISL_1552767, EPI_ISL_1552768, EPI_ISL_1552769, EPI_ISL_1552770, EPI_ISL_1552771, EPI_ISL_1552772, EPI_ISL_1552773, EPI_ISL_1552774, EPI_ISL_1552775, EPI_ISL_1552776, EPI_ISL_1552777, EPI_ISL_1552778, EPI_ISL_1552779, EPI_ISL_1552780, EPI_ISL_1552781, EPI_ISL_1552782, EPI_ISL_1552783, EPI_ISL_1552784, EPI_ISL_1552785, EPI_ISL_1552786, EPI_ISL_1552787, EPI_ISL_1552788, EPI_ISL_1552789, EPI_ISL_1552790, EPI_ISL_1552791, EPI_ISL_1552792, EPI_ISL_1552793, EPI_ISL_1552794, EPI_ISL_1552795, EPI_ISL_1552796, EPI_ISL_1552797, EPI_ISL_1552798, EPI_ISL_1552799, EPI_ISL_1552800, EPI_ISL_1552801, EPI_ISL_1552802, EPI_ISL_1552803, EPI_ISL_1552804, EPI_ISL_1552805, EPI_ISL_1552806, EPI_ISL_1552807, EPI_ISL_1552808, EPI_ISL_1552809, EPI_ISL_1552810, EPI_ISL_1552811, EPI_ISL_1552812, EPI_ISL_1552813, EPI_ISL_1552814, EPI_ISL_1552815, EPI_ISL_1552816, EPI_ISL_1552817, EPI_ISL_1552818, EPI_ISL_1552819, EPI_ISL_1552820, EPI_ISL_1552821, EPI_ISL_1552822, EPI_ISL_1552823, EPI_ISL_1552824, EPI_ISL_1552825, EPI_ISL_1552826, EPI_ISL_1552827, EPI_ISL_1552828, EPI_ISL_1552829, EPI_ISL_1552830, EPI_ISL_1552831, EPI_ISL_1552832, EPI_ISL_1552833, EPI_ISL_1552834, EPI_ISL_1552835, EPI_ISL_1552836, EPI_ISL_1552837, EPI_ISL_1552838, EPI_ISL_1552839, EPI_ISL_1552840, EPI_ISL_1552841, EPI_ISL_1552842, EPI_ISL_1552843, EPI_ISL_1552844, EPI_ISL_1552845, EPI_ISL_1552846, EPI_ISL_1552847, EPI_ISL_1552848, EPI_ISL_1552849, EPI_ISL_1552850, EPI_ISL_1552851, EPI_ISL_1552852, EPI_ISL_1552853, EPI_ISL_1552854, EPI_ISL_1552855, EPI_ISL_1552856, EPI_ISL_1552857, EPI_ISL_1552858, EPI_ISL_1552859, EPI_ISL_1552860, EPI_ISL_1552861, EPI_ISL_1552862, EPI_ISL_1552863, EPI_ISL_1552864, EPI_ISL_1552865, EPI_ISL_1552866, EPI_ISL_1552867, EPI_ISL_1552868, EPI_ISL_1552869, EPI_ISL_1552870, EPI_ISL_1552871, EPI_ISL_1552872, EPI_ISL_1552873, EPI_ISL_1552874, EPI_ISL_1552875, EPI_ISL_1552876, EPI_ISL_1552877, EPI_ISL_1552878, EPI_ISL_1552879, EPI_ISL_1552880, EPI_ISL_1552881, EPI_ISL_1552882, EPI_ISL_1552883, EPI_ISL_1552884, EPI_ISL_1552885, EPI_ISL_1552886, EPI_ISL_1552887, EPI_ISL_1552888, EPI_ISL_1552889, EPI_ISL_1552890, EPI_ISL_1552891, EPI_ISL_1552892, EPI_ISL_1552893, EPI_ISL_1552894, EPI_ISL_1552895, EPI_ISL_1552896, EPI_ISL_1552897, EPI_ISL_1552898, EPI_ISL_1552899, EPI_ISL_1552900, EPI_ISL_1552901, EPI_ISL_1552902, EPI_ISL_1552903, EPI_ISL_1552904, EPI_ISL_1552905, EPI_ISL_1552906, EPI_ISL_1552907, EPI_ISL_1552908, EPI_ISL_1552909, EPI_ISL_1552910, EPI_ISL_1552911, EPI_ISL_1552912, EPI_ISL_1552913, EPI_ISL_1552914, EPI_ISL_1552915, EPI_ISL_1552916, EPI_ISL_1552917, EPI_ISL_1552918, EPI_ISL_1552919, EPI_ISL_1552920, EPI_ISL_1552921, EPI_ISL_1552922, EPI_ISL_1552923, EPI_ISL_1552924, EPI_ISL_1552925, EPI_ISL_1552926, EPI_ISL_1552927, EPI_ISL_1552928, EPI_ISL_1552929, EPI_ISL_1552930, EPI_ISL_1552931, EPI_ISL_1552932, EPI_ISL_1552933, EPI_ISL_1552934, EPI_ISL_1552935, EPI_ISL_1552936, EPI_ISL_1552937, EPI_ISL_1552938, EPI_ISL_1552939, EPI_ISL_1552940, EPI_ISL_1552941, EPI_ISL_1552942, EPI_ISL_1552943, EPI_ISL_1552944, EPI_ISL_1552945, EPI_ISL_1552946, EPI_ISL_1552947, EPI_ISL_1552948, EPI_ISL_1552949, EPI_ISL_1552950, EPI_ISL_1552951, EPI_ISL_1552952, EPI_ISL_1552953, EPI_ISL_1552954, EPI_ISL_1552955, EPI_ISL_1552956, EPI_ISL_1552957, EPI_ISL_1552958, EPI_ISL_1552959, EPI_ISL_1552960, EPI_ISL_1552961, EPI_ISL_1552962, EPI_ISL_1552963, EPI_ISL_1552964, EPI_ISL_1552965, EPI_ISL_1552966, EPI_ISL_1552967, EPI_ISL_1552968, EPI_ISL_1552969, EPI_ISL_1552970, EPI_ISL_1552971, EPI_ISL_1552972, EPI_ISL_1552973, EPI_ISL_1552974, EPI_ISL_1552975, EPI_ISL_1552976, EPI_ISL_1552977, EPI_ISL_1552978, EPI_ISL_1552979, EPI_ISL_1552980, EPI_ISL_1552981, EPI_ISL_1552982, EPI_ISL_1552983, EPI_ISL_1552984, EPI_ISL_1552985, EPI_ISL_1552986, EPI_ISL_1552987, EPI_ISL_1552988, EPI_ISL_1552989, EPI_ISL_1552990, EPI_ISL_1552991, EPI_ISL_1552992, EPI_ISL_1552993, EPI_ISL_1552994, EPI_ISL_1552995, EPI_ISL_1552996, EPI_ISL_1552997, EPI_ISL_1552998, EPI_ISL_1552999, EPI_ISL_1553000, EPI_ISL_1553001, EPI_ISL_1553002, EPI_ISL_1553003, EPI_ISL_1553004, EPI_ISL_1553005, EPI_ISL_1553006, EPI_ISL_1553007, EPI_ISL_1553008, EPI_ISL_1553009, EPI_ISL_1553010, EPI_ISL_1553011, EPI_ISL_1553012, EPI_ISL_1553013, EPI_ISL_1553014, EPI_ISL_1553015, EPI_ISL_1553016, EPI_ISL_1553017, EPI_ISL_1553018, EPI_ISL_1553019, EPI_ISL_1553020, EPI_ISL_1553021, EPI_ISL_1553022, EPI_ISL_1553023, EPI_ISL_1553024, EPI_ISL_1553025, EPI_ISL_1553026, EPI_ISL_1553027, EPI_ISL_1553028, EPI_ISL_1553029, EPI_ISL_1553030, EPI_ISL_1553031, EPI_ISL_1553032, EPI_ISL_1553033, EPI_ISL_1553034, EPI_ISL_1553035, EPI_ISL_1553036, EPI_ISL_1553037, EPI_ISL_1553038, EPI_ISL_1553039, EPI_ISL_1553040, EPI_ISL_1553041, EPI_ISL_1553042, EPI_ISL_1553043, EPI_ISL_1553044, EPI_ISL_1553045, EPI_ISL_1553046, EPI_ISL_1553047, EPI_ISL_1553048, EPI_ISL_1553049, EPI_ISL_1553050, EPI_ISL_1553051, EPI_ISL_1553052, EPI_ISL_1553053, EPI_ISL_1553054, EPI_ISL_1553055, EPI_ISL_1553056, EPI_ISL_1553057, EPI_ISL_1553058, EPI_ISL_1553059, EPI_ISL_1553060, EPI_ISL_1553061, EPI_ISL_1553062, EPI_ISL_1553063, EPI_ISL_1553064, EPI_ISL_1553065, EPI_ISL_1553066, EPI_ISL_1553067, EPI_ISL_1553068, EPI_ISL_1553069, EPI_ISL_1553070, EPI_ISL_1553071, EPI_ISL_1553072, EPI_ISL_1553073, EPI_ISL_1553074, EPI_ISL_1553075, EPI_ISL_1553076, EPI_ISL_1553077, EPI_ISL_1553078, EPI_ISL_1553079, EPI_ISL_1553080, EPI_ISL_1553081, EPI_ISL_1553082, EPI_ISL_1553083, EPI_ISL_1553084, EPI_ISL_1553085, EPI_ISL_1553086, EPI_ISL_1553087, EPI_ISL_1553088, EPI_ISL_1553089, EPI_ISL_1553090, EPI_ISL_1553091, EPI_ISL_1553092, EPI_ISL_1553093, EPI_ISL_1553094, EPI_ISL_1553095, EPI_ISL_1553096, EPI_ISL_1553097, EPI_ISL_1553098, EPI_ISL_1553099, EPI_ISL_1553100, EPI_ISL_1553101, EPI_ISL_1553102, EPI_ISL_1553103, EPI_ISL_1553104, EPI_ISL_1553105, EPI_ISL_1553106, EPI_ISL_1553107, EPI_ISL_1553108, EPI_ISL_1553109, EPI_ISL_1553110, EPI_ISL_1553111, EPI_ISL_1553112, EPI_ISL_1553113, EPI_ISL_1553114, EPI_ISL_1553115, EPI_ISL_1553116, EPI_ISL_1553117, EPI_ISL_1553118, EPI_ISL_1553119, EPI_ISL_1553120, EPI_ISL_1553121, EPI_ISL_1553122, EPI_ISL_1553123, EPI_ISL_1553124, EPI_ISL_1553125, EPI_ISL_1553126, EPI_ISL_1553127, EPI_ISL_1553128, EPI_ISL_1553129, EPI_ISL_1553130, EPI_ISL_1553131, EPI_ISL_1553132, EPI_ISL_1553133, EPI_ISL_1553134, EPI_ISL_1553135, EPI_ISL_1553136, EPI_ISL_1553137, EPI_ISL_1553138, EPI_ISL_1553139, EPI_ISL_1553140, EPI_ISL_1553141, EPI_ISL_1553142, EPI_ISL_1553143, EPI_ISL_1553144, EPI_ISL_1553145, EPI_ISL_1553146, EPI_ISL_1553147, EPI_ISL_1553148, EPI_ISL_1553149, EPI_ISL_1553150, EPI_ISL_1553151, EPI_ISL_1553152, EPI_ISL_1553153, EPI_ISL_1553154, EPI_ISL_1553155, EPI_ISL_1553156, EPI_ISL_1553157, EPI_ISL_1553158, EPI_ISL_1553159, EPI_ISL_1553160, EPI_ISL_1553161, EPI_ISL_1553162, EPI_ISL_1553163, EPI_ISL_1553164, EPI_ISL_1553165, EPI_ISL_1553166, EPI_ISL_1553167, EPI_ISL_1553168, EPI_ISL_1553169, EPI_ISL_1553170, EPI_ISL_1553171, EPI_ISL_1553172 | | | |
| see above | Quest Diagnostics Incorporated | Centers for Disease Control and Prevention Division of Viral Diseases, Pathogen Discovery | Dakota Howard, Dhwani Batra, Peter W. Cook, Kara Moser, Adrian Paskey, Jason Caravas, Benjamin Rambo-Martin, Shatavia Morrison, Christopher Gulvick, Scott Sammons, Yvette Unoarumhi, Darlene Wagner, Matthew Schmerer, S. H. Rosenthal, A. Gerasimova, R. M. Kagan, B. Anderson, M. Hua, Y. Liu, L.E. Bernstein, K.E. Livingston, A. Perez, I. A. Shlyakhter, R. V. Rolando, R. Owen, P. Tanpaiboon, F. Lacbawan, Clinton R. Paden, Duncan MacCannell |
| EPI_ISL_1553173, EPI_ISL_1553174, EPI_ISL_1553175, EPI_ISL_1553176, EPI_ISL_1553177, EPI_ISL_1553178, EPI_ISL_1553179, EPI_ISL_1553181, EPI_ISL_1553182, EPI_ISL_1553183, EPI_ISL_1553184, EPI_ISL_1553187, EPI_ISL_1553189, EPI_ISL_1553190, EPI_ISL_1553191, EPI_ISL_1553192, EPI_ISL_1553193, EPI_ISL_1553194, EPI_ISL_1553195, EPI_ISL_1553196, EPI_ISL_1553200, EPI_ISL_1553201, EPI_ISL_1553202, EPI_ISL_1553203, EPI_ISL_1553204, EPI_ISL_1553205, EPI_ISL_1553206, EPI_ISL_1553207, EPI_ISL_1553208, EPI_ISL_1553209, EPI_ISL_1553210, EPI_ISL_1553211, EPI_ISL_1553213, EPI_ISL_1553214, EPI_ISL_1553215, EPI_ISL_1553216, EPI_ISL_1553217, EPI_ISL_1553218, EPI_ISL_1553219, EPI_ISL_1553220, EPI_ISL_1553221, EPI_ISL_1553222, EPI_ISL_1553223, EPI_ISL_1553224, EPI_ISL_1553225, EPI_ISL_1553226, EPI_ISL_1553227, EPI_ISL_1553228, EPI_ISL_1553229, EPI_ISL_1553230, EPI_ISL_1553231, EPI_ISL_1553232, EPI_ISL_1553233, EPI_ISL_1553235, EPI_ISL_1553236, EPI_ISL_1553237, EPI_ISL_1553238, EPI_ISL_1553239, EPI_ISL_1553240, EPI_ISL_1553241, EPI_ISL_1553242, EPI_ISL_1553243, EPI_ISL_1553244, EPI_ISL_1553245, EPI_ISL_1553246, EPI_ISL_1553247, EPI_ISL_1553248, EPI_ISL_1553251, EPI_ISL_1553252, EPI_ISL_1553253, EPI_ISL_1553254, EPI_ISL_1553255, EPI_ISL_1553256, EPI_ISL_1553257, EPI_ISL_1553258, EPI_ISL_1553259, EPI_ISL_1553260, EPI_ISL_1553262 | | | |
| see above | Illinois Department of Public Health - Springfield Lab | Illinois Department of Public Health - Springfield Lab | Bryan Sim, Gordon McCall |
| EPI_ISL_1553264, EPI_ISL_1553265, EPI_ISL_1553266, EPI_ISL_1553267, EPI_ISL_1553268, EPI_ISL_1553269, EPI_ISL_1553270, EPI_ISL_1553271, EPI_ISL_1553272, EPI_ISL_1553273, EPI_ISL_1553274, EPI_ISL_1553275, EPI_ISL_1553276, EPI_ISL_1553277, EPI_ISL_1553278, EPI_ISL_1553279, EPI_ISL_1553280, EPI_ISL_1553281, EPI_ISL_1553282, EPI_ISL_1553283, EPI_ISL_1553284, EPI_ISL_1553285, | | | |

EPI_ISL_1553286, EPI_ISL_1553287, EPI_ISL_1553288, EPI_ISL_1553289, EPI_ISL_1553290, EPI_ISL_1553291, EPI_ISL_1553292, EPI_ISL_1553293, EPI_ISL_1553294, EPI_ISL_1553295, EPI_ISL_1553296, EPI_ISL_1553297, EPI_ISL_1553298, EPI_ISL_1553299, EPI_ISL_1553300, EPI_ISL_1553301, EPI_ISL_1553302, EPI_ISL_1553303, EPI_ISL_1553304, EPI_ISL_1553305, EPI_ISL_1553306, EPI_ISL_1553307,
EPI_ISL_1553308, EPI_ISL_1553309, EPI_ISL_1553310, EPI_ISL_1553311, EPI_ISL_1553312, EPI_ISL_1553313, EPI_ISL_1553314, EPI_ISL_1553315, EPI_ISL_1553316, EPI_ISL_1553317, EPI_ISL_1553318, EPI_ISL_1553319, EPI_ISL_1553320, EPI_ISL_1553321, EPI_ISL_1553322, EPI_ISL_1553323, EPI_ISL_1553324, EPI_ISL_1553325, EPI_ISL_1553326, EPI_ISL_1553327, EPI_ISL_1553328, EPI_ISL_1553329,
EPI_ISL_1553330, EPI_ISL_1553331, EPI_ISL_1553332, EPI_ISL_1553333, EPI_ISL_1553334, EPI_ISL_1553335, EPI_ISL_1553336, EPI_ISL_1553337, EPI_ISL_1553338, EPI_ISL_1553340, EPI_ISL_1553341, EPI_ISL_1553342, EPI_ISL_1553343, EPI_ISL_1553344, EPI_ISL_1553345, EPI_ISL_1553346, EPI_ISL_1553347, EPI_ISL_1553348, EPI_ISL_1553349, EPI_ISL_1553350, EPI_ISL_1553351, EPI_ISL_1553352,
EPI_ISL_1553353, EPI_ISL_1553354, EPI_ISL_1553355, EPI_ISL_1553356, EPI_ISL_1553357, EPI_ISL_1553358, EPI_ISL_1553359, EPI_ISL_1553360, EPI_ISL_1553362, EPI_ISL_1553363, EPI_ISL_1553364, EPI_ISL_1553365, EPI_ISL_1553366, EPI_ISL_1553367, EPI_ISL_1553368, EPI_ISL_1553369, EPI_ISL_1553370, EPI_ISL_1553371, EPI_ISL_1553372, EPI_ISL_1553373, EPI_ISL_1553374, EPI_ISL_1553375,
EPI_ISL_1553376, EPI_ISL_1553377, EPI_ISL_1553378, EPI_ISL_1553379, EPI_ISL_1553380, EPI_ISL_1553381, EPI_ISL_1553382, EPI_ISL_1553383, EPI_ISL_1553384, EPI_ISL_1553385, EPI_ISL_1553386, EPI_ISL_1553387, EPI_ISL_1553388, EPI_ISL_1553389, EPI_ISL_1553390, EPI_ISL_1553391, EPI_ISL_1553392, EPI_ISL_1553393, EPI_ISL_1553394, EPI_ISL_1553395, EPI_ISL_1553396, EPI_ISL_1553397,
EPI_ISL_1553398, EPI_ISL_1553399, EPI_ISL_1553400, EPI_ISL_1553401, EPI_ISL_1553402, EPI_ISL_1553403, EPI_ISL_1553404, EPI_ISL_1553405, EPI_ISL_1553406, EPI_ISL_1553407, EPI_ISL_1553408, EPI_ISL_1553409, EPI_ISL_1553410, EPI_ISL_1553411, EPI_ISL_1553412, EPI_ISL_1553413, EPI_ISL_1553414, EPI_ISL_1553415, EPI_ISL_1553416, EPI_ISL_1553417, EPI_ISL_1553418, EPI_ISL_1553419,
EPI_ISL_1553420, EPI_ISL_1553421, EPI_ISL_1553422, EPI_ISL_1553423, EPI_ISL_1553424, EPI_ISL_1553425, EPI_ISL_1553426, EPI_ISL_1553427, EPI_ISL_1553428, EPI_ISL_1553429, EPI_ISL_1553430, EPI_ISL_1553431, EPI_ISL_1553432, EPI_ISL_1553433, EPI_ISL_1553434, EPI_ISL_1553435, EPI_ISL_1553436, EPI_ISL_1553437, EPI_ISL_1553438, EPI_ISL_1553439, EPI_ISL_1553440, EPI_ISL_1553441,
EPI_ISL_1553442, EPI_ISL_1553443, EPI_ISL_1553444, EPI_ISL_1553445, EPI_ISL_1553446, EPI_ISL_1553447, EPI_ISL_1553448, EPI_ISL_1553449, EPI_ISL_1553450, EPI_ISL_1553451, EPI_ISL_1553452, EPI_ISL_1553453, EPI_ISL_1553454, EPI_ISL_1553455, EPI_ISL_1553456, EPI_ISL_1553457, EPI_ISL_1553458, EPI_ISL_1553459, EPI_ISL_1553460, EPI_ISL_1553461, EPI_ISL_1553462, EPI_ISL_1553463,
EPI_ISL_1553464, EPI_ISL_1553465, EPI_ISL_1553466, EPI_ISL_1553467, EPI_ISL_1553468, EPI_ISL_1553469, EPI_ISL_1553470, EPI_ISL_1553471, EPI_ISL_1553472, EPI_ISL_1553473, EPI_ISL_1553474, EPI_ISL_1553475, EPI_ISL_1553476, EPI_ISL_1553477, EPI_ISL_1553478, EPI_ISL_1553479, EPI_ISL_1553480, EPI_ISL_1553481, EPI_ISL_1553482, EPI_ISL_1553483, EPI_ISL_1553484, EPI_ISL_1553485,
EPI_ISL_1553486, EPI_ISL_1553487, EPI_ISL_1553488, EPI_ISL_1553489, EPI_ISL_1553490, EPI_ISL_1553491, EPI_ISL_1553492, EPI_ISL_1553493, EPI_ISL_1553494, EPI_ISL_1553495, EPI_ISL_1553496, EPI_ISL_1553497, EPI_ISL_1553498, EPI_ISL_1553499, EPI_ISL_1553500, EPI_ISL_1553501, EPI_ISL_1553502, EPI_ISL_1553503, EPI_ISL_1553504, EPI_ISL_1553505, EPI_ISL_1553506, EPI_ISL_1553507,
EPI_ISL_1553508, EPI_ISL_1553509, EPI_ISL_1553510, EPI_ISL_1553511, EPI_ISL_1553512, EPI_ISL_1553513, EPI_ISL_1553514, EPI_ISL_1553515, EPI_ISL_1553516, EPI_ISL_1553517, EPI_ISL_1553518, EPI_ISL_1553519, EPI_ISL_1553521, EPI_ISL_1553522, EPI_ISL_1553523, EPI_ISL_1553524, EPI_ISL_1553525, EPI_ISL_1553526, EPI_ISL_1553527, EPI_ISL_1553528, EPI_ISL_1553529, EPI_ISL_1553530,
EPI_ISL_1553531, EPI_ISL_1553532, EPI_ISL_1553533, EPI_ISL_1553534, EPI_ISL_1553535, EPI_ISL_1553536, EPI_ISL_1553537, EPI_ISL_1553538, EPI_ISL_1553539, EPI_ISL_1553540, EPI_ISL_1553541, EPI_ISL_1553542, EPI_ISL_1553543, EPI_ISL_1553544, EPI_ISL_1553545, EPI_ISL_1553546, EPI_ISL_1553547, EPI_ISL_1553548, EPI_ISL_1553549, EPI_ISL_1553550, EPI_ISL_1553551, EPI_ISL_1553552,
EPI_ISL_1553553, EPI_ISL_1553554, EPI_ISL_1553555, EPI_ISL_1553556, EPI_ISL_1553557, EPI_ISL_1553558, EPI_ISL_1553559, EPI_ISL_1553560, EPI_ISL_1553561, EPI_ISL_1553562, EPI_ISL_1553563, EPI_ISL_1553564, EPI_ISL_1553565, EPI_ISL_1553566, EPI_ISL_1553567, EPI_ISL_1553568, EPI_ISL_1553569, EPI_ISL_1553570, EPI_ISL_1553571, EPI_ISL_1553572, EPI_ISL_1553573, EPI_ISL_1553574,
EPI_ISL_1553575, EPI_ISL_1553576, EPI_ISL_1553577, EPI_ISL_1553578, EPI_ISL_1553579, EPI_ISL_1553580, EPI_ISL_1553581, EPI_ISL_1553582, EPI_ISL_1553583, EPI_ISL_1553584, EPI_ISL_1553585, EPI_ISL_1553586, EPI_ISL_1553587, EPI_ISL_1553588, EPI_ISL_1553589, EPI_ISL_1553590, EPI_ISL_1553591, EPI_ISL_1553592, EPI_ISL_1553593, EPI_ISL_1553594, EPI_ISL_1553595, EPI_ISL_1553596,
EPI_ISL_1553597, EPI_ISL_1553598, EPI_ISL_1553599, EPI_ISL_1553600, EPI_ISL_1553601, EPI_ISL_1553602, EPI_ISL_1553603, EPI_ISL_1553604, EPI_ISL_1553605, EPI_ISL_1553606, EPI_ISL_1553607, EPI_ISL_1553608, EPI_ISL_1553609, EPI_ISL_1553610, EPI_ISL_1553611, EPI_ISL_1553612, EPI_ISL_1553613, EPI_ISL_1553614, EPI_ISL_1553615, EPI_ISL_1553616, EPI_ISL_1553617, EPI_ISL_1553618,
EPI_ISL_1553619, EPI_ISL_1553620, EPI_ISL_1553621, EPI_ISL_1553622, EPI_ISL_1553623, EPI_ISL_1553624, EPI_ISL_1553625, EPI_ISL_1553626, EPI_ISL_1553627, EPI_ISL_1553628, EPI_ISL_1553629, EPI_ISL_1553630, EPI_ISL_1553631, EPI_ISL_1553632, EPI_ISL_1553633, EPI_ISL_1553634, EPI_ISL_1553635, EPI_ISL_1553636, EPI_ISL_1553637, EPI_ISL_1553638, EPI_ISL_1553639, EPI_ISL_1553640,
EPI_ISL_1553641, EPI_ISL_1553642, EPI_ISL_1553643, EPI_ISL_1553644, EPI_ISL_1553645, EPI_ISL_1553646, EPI_ISL_1553647, EPI_ISL_1553648, EPI_ISL_1553649, EPI_ISL_1553650, EPI_ISL_1553651, EPI_ISL_1553652, EPI_ISL_1553653, EPI_ISL_1553654, EPI_ISL_1553655, EPI_ISL_1553656, EPI_ISL_1553657, EPI_ISL_1553658, EPI_ISL_1553659, EPI_ISL_1553660, EPI_ISL_1553661, EPI_ISL_1553662,
EPI_ISL_1553663, EPI_ISL_1553664, EPI_ISL_1553665, EPI_ISL_1553666, EPI_ISL_1553667, EPI_ISL_1553668, EPI_ISL_1553669, EPI_ISL_1553670, EPI_ISL_1553671, EPI_ISL_1553672, EPI_ISL_1553673, EPI_ISL_1553674, EPI_ISL_1553675, EPI_ISL_1553676, EPI_ISL_1553677, EPI_ISL_1553678, EPI_ISL_1553679, EPI_ISL_1553680, EPI_ISL_1553681, EPI_ISL_1553682, EPI_ISL_1553683, EPI_ISL_1553684,
EPI_ISL_1553685, EPI_ISL_1553686, EPI_ISL_1553687, EPI_ISL_1553688, EPI_ISL_1553689, EPI_ISL_1553690, EPI_ISL_1553691, EPI_ISL_1553692, EPI_ISL_1553693, EPI_ISL_1553694, EPI_ISL_1553695, EPI_ISL_1553696, EPI_ISL_1553697, EPI_ISL_1553698, EPI_ISL_1553699, EPI_ISL_1553700, EPI_ISL_1553701, EPI_ISL_1553702, EPI_ISL_1553703, EPI_ISL_1553704, EPI_ISL_1553705, EPI_ISL_1553706,
EPI_ISL_1553707, EPI_ISL_1553708, EPI_ISL_1553709, EPI_ISL_1553710, EPI_ISL_1553711, EPI_ISL_1553712, EPI_ISL_1553713, EPI_ISL_1553714, EPI_ISL_1553715, EPI_ISL_1553716, EPI_ISL_1553717, EPI_ISL_1553718, EPI_ISL_1553719, EPI_ISL_1553720, EPI_ISL_1553721, EPI_ISL_1553722, EPI_ISL_1553723, EPI_ISL_1553724, EPI_ISL_1553725, EPI_ISL_1553726, EPI_ISL_1553727, EPI_ISL_1553728,
EPI_ISL_1553729, EPI_ISL_1553730, EPI_ISL_1553731, EPI_ISL_1553732, EPI_ISL_1553733, EPI_ISL_1553734, EPI_ISL_1553735, EPI_ISL_1553736, EPI_ISL_1553737, EPI_ISL_1553738, EPI_ISL_1553739, EPI_ISL_1553740, EPI_ISL_1553741, EPI_ISL_1553742, EPI_ISL_1553743, EPI_ISL_1553744, EPI_ISL_1553745, EPI_ISL_1553746, EPI_ISL_1553747, EPI_ISL_1553748, EPI_ISL_1553749, EPI_ISL_1553750,
EPI_ISL_1553751, EPI_ISL_1553752, EPI_ISL_1553753, EPI_ISL_1553754, EPI_ISL_1553755, EPI_ISL_1553756, EPI_ISL_1553757, EPI_ISL_1553758, EPI_ISL_1553759, EPI_ISL_1553760, EPI_ISL_1553761, EPI_ISL_1553762, EPI_ISL_1553763, EPI_ISL_1553764, EPI_ISL_1553765, EPI_ISL_1553766, EPI_ISL_1553767, EPI_ISL_1553768, EPI_ISL_1553769, EPI_ISL_1553770, EPI_ISL_1553771, EPI_ISL_1553772,
EPI_ISL_1553773, EPI_ISL_1553774, EPI_ISL_1553775, EPI_ISL_1553776, EPI_ISL_1553777, EPI_ISL_1553778, EPI_ISL_1553779, EPI_ISL_1553780, EPI_ISL_1553781, EPI_ISL_1553782, EPI_ISL_1553783, EPI_ISL_1553784, EPI_ISL_1553785, EPI_ISL_1553786, EPI_ISL_1553787, EPI_ISL_1553788, EPI_ISL_1553789, EPI_ISL_1553790, EPI_ISL_1553791, EPI_ISL_1553792, EPI_ISL_1553793, EPI_ISL_1553794,
EPI_ISL_1553795, EPI_ISL_1553796, EPI_ISL_1553797, EPI_ISL_1553798, EPI_ISL_1553799, EPI_ISL_1553848, EPI_ISL_1553849, EPI_ISL_1553850, EPI_ISL_1553851, EPI_ISL_1553852, EPI_ISL_1553853, EPI_ISL_1553854, EPI_ISL_1553855, EPI_ISL_1553856, EPI_ISL_1553857, EPI_ISL_1553858, EPI_ISL_1553859, EPI_ISL_1553860, EPI_ISL_1553861, EPI_ISL_1553862, EPI_ISL_1553863, EPI_ISL_1553864,
EPI_ISL_1553865, EPI_ISL_1553866, EPI_ISL_1553867, EPI_ISL_1553868, EPI_ISL_1553869, EPI_ISL_1553870, EPI_ISL_1553871, EPI_ISL_1553872, EPI_ISL_1553873, EPI_ISL_1553874, EPI_ISL_1553875, EPI_ISL_1553876, EPI_ISL_1553877, EPI_ISL_1553878, EPI_ISL_1553879, EPI_ISL_1553880, EPI_ISL_1553881, EPI_ISL_1553882, EPI_ISL_1553884, EPI_ISL_1553885, EPI_ISL_1553886, EPI_ISL_1553887,
EPI_ISL_1553888, EPI_ISL_1553889, EPI_ISL_1553890, EPI_ISL_1553891, EPI_ISL_1553892, EPI_ISL_1553893, EPI_ISL_1553894, EPI_ISL_1553895, EPI_ISL_1553896, EPI_ISL_1553898, EPI_ISL_1553899, EPI_ISL_1553900, EPI_ISL_1553901, EPI_ISL_1553902, EPI_ISL_1553903, EPI_ISL_1553904, EPI_ISL_1553905, EPI_ISL_1553906, EPI_ISL_1553907, EPI_ISL_1553908, EPI_ISL_1553909, EPI_ISL_1553910,
EPI_ISL_1553911, EPI_ISL_1553912, EPI_ISL_1553913, EPI_ISL_1553914, EPI_ISL_1553915, EPI_ISL_1553916, EPI_ISL_1553917, EPI_ISL_1553918, EPI_ISL_1553919, EPI_ISL_1553920, EPI_ISL_1553921, EPI_ISL_1553922, EPI_ISL_1553923, EPI_ISL_1553924, EPI_ISL_1553925, EPI_ISL_1553926, EPI_ISL_1553927, EPI_ISL_1553928, EPI_ISL_1553929, EPI_ISL_1553930, EPI_ISL_1553931, EPI_ISL_1553932,
EPI_ISL_1553933, EPI_ISL_1553934, EPI_ISL_1553935, EPI_ISL_1553936, EPI_ISL_1553937, EPI_ISL_1553938, EPI_ISL_1553939, EPI_ISL_1553940, EPI_ISL_1553941, EPI_ISL_1553942, EPI_ISL_1553943, EPI_ISL_1553944, EPI_ISL_1553945, EPI_ISL_1553946, EPI_ISL_1553947, EPI_ISL_1553948, EPI_ISL_1553949, EPI_ISL_1553950, EPI_ISL_1553951, EPI_ISL_1553952, EPI_ISL_1553953, EPI_ISL_1553954,
EPI_ISL_1553955, EPI_ISL_1553956, EPI_ISL_1553957, EPI_ISL_1553958, EPI_ISL_1553959, EPI_ISL_1553960, EPI_ISL_1553961, EPI_ISL_1553963, EPI_ISL_1553964, EPI_ISL_1553965, EPI_ISL_1553966, EPI_ISL_1553967, EPI_ISL_1553968, EPI_ISL_1553969, EPI_ISL_1553970, EPI_ISL_1553971, EPI_ISL_1553972, EPI_ISL_1553973, EPI_ISL_1553974, EPI_ISL_1553975, EPI_ISL_1553976, EPI_ISL_1553978,
EPI_ISL_1553979, EPI_ISL_1553980, EPI_ISL_1553981, EPI_ISL_1553982, EPI_ISL_1553983, EPI_ISL_1553984, EPI_ISL_1553985, EPI_ISL_1553986, EPI_ISL_1553987, EPI_ISL_1553988, EPI_ISL_1553989, EPI_ISL_1553990, EPI_ISL_1553991, EPI_ISL_1553992, EPI_ISL_1553993, EPI_ISL_1553994, EPI_ISL_1553995, EPI_ISL_1553996, EPI_ISL_1553997, EPI_ISL_1553998, EPI_ISL_1553999, EPI_ISL_1554000,
EPI_ISL_1554001, EPI_ISL_1554002, EPI_ISL_1554003, EPI_ISL_1554004, EPI_ISL_1554005, EPI_ISL_1554006, EPI_ISL_1554007, EPI_ISL_1554008, EPI_ISL_1554009, EPI_ISL_1554010, EPI_ISL_1554011, EPI_ISL_1554012, EPI_ISL_1554013, EPI_ISL_1554014, EPI_ISL_1554015, EPI_ISL_1554016, EPI_ISL_1554017, EPI_ISL_1554018, EPI_ISL_1554019, EPI_ISL_1554020, EPI_ISL_1554021, EPI_ISL_1554022,
EPI_ISL_1554023, EPI_ISL_1554024, EPI_ISL_1554025, EPI_ISL_1554026, EPI_ISL_1554027, EPI_ISL_1554028, EPI_ISL_1554029, EPI_ISL_1554030, EPI_ISL_1554032, EPI_ISL_1554033, EPI_ISL_1554034, EPI_ISL_1554035, EPI_ISL_1554036, EPI_ISL_1554037, EPI_ISL_1554038, EPI_ISL_1554039, EPI_ISL_1554040, EPI_ISL_1554041, EPI_ISL_1554042, EPI_ISL_1554043, EPI_ISL_1554044, EPI_ISL_1554045,
EPI_ISL_1554046, EPI_ISL_1554047, EPI_ISL_1554048, EPI_ISL_1554049, EPI_ISL_1554050, EPI_ISL_1554051, EPI_ISL_1554052, EPI_ISL_1554053, EPI_ISL_1554054, EPI_ISL_1554055, EPI_ISL_1554056, EPI_ISL_1554057, EPI_ISL_1554058, EPI_ISL_1554059, EPI_ISL_1554060, EPI_ISL_1554061, EPI_ISL_1554062, EPI_ISL_1554063, EPI_ISL_1554064, EPI_ISL_1554065, EPI_ISL_1554066, EPI_ISL_1554067,
EPI_ISL_1554068, EPI_ISL_1554069, EPI_ISL_1554070, EPI_ISL_1554071, EPI_ISL_1554072, EPI_ISL_1554073, EPI_ISL_1554074, EPI_ISL_1554075, EPI_ISL_1554076, EPI_ISL_1554077, EPI_ISL_1554078, EPI_ISL_1554079, EPI_ISL_1554080, EPI_ISL_1554081, EPI_ISL_1554082, EPI_ISL_1554083, EPI_ISL_1554084, EPI_ISL_1554085, EPI_ISL_1554086, EPI_ISL_1554087, EPI_ISL_1554088, EPI_ISL_1554089,
EPI_ISL_1554090, EPI_ISL_1554091, EPI_ISL_1554092, EPI_ISL_1554093, EPI_ISL_1554094, EPI_ISL_1554095, EPI_ISL_1554096, EPI_ISL_1554097, EPI_ISL_1554098, EPI_ISL_1554099, EPI_ISL_1554100, EPI_ISL_1554101, EPI_ISL_1554102, EPI_ISL_1554103, EPI_ISL_1554104, EPI_ISL_1554105, EPI_ISL_1554106, EPI_ISL_1554107, EPI_ISL_1554108, EPI_ISL_1554109, EPI_ISL_1554110, EPI_ISL_1554111,
EPI_ISL_1554112, EPI_ISL_1554113, EPI_ISL_1554114, EPI_ISL_1554115, EPI_ISL_1554116, EPI_ISL_1554117, EPI_ISL_1554118, EPI_ISL_1554119, EPI_ISL_1554120, EPI_ISL_1554121, EPI_ISL_1554122, EPI_ISL_1554123, EPI_ISL_1554124, EPI_ISL_1554125, EPI_ISL_1554127, EPI_ISL_1554128, EPI_ISL_1554129, EPI_ISL_1554130, EPI_ISL_1554131, EPI_ISL_1554132, EPI_ISL_1554133, EPI_ISL_1554134,
EPI_ISL_1554135, EPI_ISL_1554136, EPI_ISL_1554137, EPI_ISL_1554138, EPI_ISL_1554139, EPI_ISL_1554140, EPI_ISL_1554141, EPI_ISL_1554142, EPI_ISL_1554143, EPI_ISL_1554144, EPI_ISL_1554145, EPI_ISL_1554146, EPI_ISL_1554147, EPI_ISL_1554148, EPI_ISL_1554149, EPI_ISL_1554150, EPI_ISL_1554151, EPI_ISL_1554152, EPI_ISL_1554153, EPI_ISL_1554154, EPI_ISL_1554155, EPI_ISL_1554156,
EPI_ISL_1554157, EPI_ISL_1554158, EPI_ISL_1554159, EPI_ISL_1554160, EPI_ISL_1554161, EPI_ISL_1554162, EPI_ISL_1554163, EPI_ISL_1554164, EPI_ISL_1554165, EPI_ISL_1554166, EPI_ISL_1554167, EPI_ISL_1554168, EPI_ISL_1554169, EPI_ISL_1554170, EPI_ISL_1554171, EPI_ISL_1554172, EPI_ISL_1554173, EPI_ISL_1554174, EPI_ISL_1554175, EPI_ISL_1554176, EPI_ISL_1554177, EPI_ISL_1554178,
EPI_ISL_1554179, EPI_ISL_1554180, EPI_ISL_1554181, EPI_ISL_1554182, EPI_ISL_1554183, EPI_ISL_1554184, EPI_ISL_1554185, EPI_ISL_1554186, EPI_ISL_1554187, EPI_ISL_1554188, EPI_ISL_1554189, EPI_ISL_1554190, EPI_ISL_1554191, EPI_ISL_1554192, EPI_ISL_1554193, EPI_ISL_1554194, EPI_ISL_1554195, EPI_ISL_1554196, EPI_ISL_1554197, EPI_ISL_1554198, EPI_ISL_1554199, EPI_ISL_1554200,
EPI_ISL_1554201, EPI_ISL_1554202, EPI_ISL_1554203, EPI_ISL_1554204, EPI_ISL_1554205, EPI_ISL_1554207, EPI_ISL_1554208, EPI_ISL_1554209, EPI_ISL_1554210, EPI_ISL_1554211, EPI_ISL_1554212, EPI_ISL_1554213, EPI_ISL_1554214, EPI_ISL_1554215, EPI_ISL_1554216, EPI_ISL_1554217, EPI_ISL_1554218, EPI_ISL_1554220, EPI_ISL_1554221, EPI_ISL_1554222, EPI_ISL_1554223, EPI_ISL_1554224,
EPI_ISL_1554225, EPI_ISL_1554226, EPI_ISL_1554227, EPI_ISL_1554228, EPI_ISL_1554229, EPI_ISL_1554230, EPI_ISL_1554231, EPI_ISL_1554232, EPI_ISL_1554233, EPI_ISL_1554234, EPI_ISL_1554235, EPI_ISL_1554236, EPI_ISL_1554237, EPI_ISL_1554238, EPI_ISL_1554239, EPI_ISL_1554240, EPI_ISL_1554241, EPI_ISL_1554242, EPI_ISL_1554243, EPI_ISL_1554244, EPI_ISL_1554245, EPI_ISL_1554246,
EPI_ISL_1554247, EPI_ISL_1554248, EPI_ISL_1554249, EPI_ISL_1554250, EPI_ISL_1554251, EPI_ISL_1554252, EPI_ISL_1554253, EPI_ISL_1554254, EPI_ISL_1554255, EPI_ISL_1554256, EPI_ISL_1554257, EPI_ISL_1554258, EPI_ISL_1554259, EPI_ISL_1554260, EPI_ISL_1554261, EPI_ISL_1554262, EPI_ISL_1554263, EPI_ISL_1554264, EPI_ISL_1554265, EPI_ISL_1554266, EPI_ISL_1554267, EPI_ISL_1554268,
EPI_ISL_1554269, EPI_ISL_1554270, EPI_ISL_1554271, EPI_ISL_1554272, EPI_ISL_1554273, EPI_ISL_1554274, EPI_ISL_1554275, EPI_ISL_1554276, EPI_ISL_1554277, EPI_ISL_1554278, EPI_ISL_1554279, EPI_ISL_1554280, EPI_ISL_1554281, EPI_ISL_1554282, EPI_ISL_1554283, EPI_ISL_1554284, EPI_ISL_1554285, EPI_ISL_1554286, EPI_ISL_1554287, EPI_ISL_1554288, EPI_ISL_1554289, EPI_ISL_1554290,
EPI_ISL_1554291, EPI_ISL_1554292, EPI_ISL_1554293, EPI_ISL_1554294, EPI_ISL_1554295, EPI_ISL_1554296, EPI_ISL_1554297, EPI_ISL_1554298, EPI_ISL_1554299, EPI_ISL_1554300, EPI_ISL_1554301, EPI_ISL_1554302, EPI_ISL_1554303, EPI_ISL_1554304, EPI_ISL_1554305, EPI_ISL_1554306, EPI_ISL_1554307, EPI_ISL_1554308, EPI_ISL_1554309, EPI_ISL_1554310, EPI_ISL_1554311, EPI_ISL_1554312,
EPI_ISL_1554313, EPI_ISL_1554314, EPI_ISL_1554315, EPI_ISL_1554316, EPI_ISL_1554317, EPI_ISL_1554318, EPI_ISL_1554319, EPI_ISL_1554320, EPI_ISL_1554321, EPI_ISL_1554322, EPI_ISL_1554323, EPI_ISL_1554324, EPI_ISL_1554325, EPI_ISL_1554326, EPI_ISL_1554328, EPI_ISL_1554329, EPI_ISL_1554330, EPI_ISL_1554331, EPI_ISL_1554332, EPI_ISL_1554333, EPI_ISL_1554334, EPI_ISL_1554335,
EPI_ISL_1554336, EPI_ISL_1554337, EPI_ISL_1554338, EPI_ISL_1554339, EPI_ISL_1554340, EPI_ISL_1554341, EPI_ISL_1554342, EPI_ISL_1554343, EPI_ISL_1554344, EPI_ISL_1554345, EPI_ISL_1554347, EPI_ISL_1554348, EPI_ISL_1554349, EPI_ISL_1554350, EPI_ISL_1554351, EPI_ISL_1554352, EPI_ISL_1554353, EPI_ISL_1554354, EPI_ISL_1554355, EPI_ISL_1554356, EPI_ISL_1554357, EPI_ISL_1554358,
EPI_ISL_1554359, EPI_ISL_1554360, EPI_ISL_1554361, EPI_ISL_1554362, EPI_ISL_1554363, EPI_ISL_1554364, EPI_ISL_1554365, EPI_ISL_1554366, EPI_ISL_1554367, EPI_ISL_1554368, EPI_ISL_1554369, EPI_ISL_1554370, EPI_ISL_1554371, EPI_ISL_1554372, EPI_ISL_1554373, EPI_ISL_1554374, EPI_ISL_1554375, EPI_ISL_1554376, EPI_ISL_1554377, EPI_ISL_1554378, EPI_ISL_1554379, EPI_ISL_1554380,
EPI_ISL_1554381, EPI_ISL_1554382, EPI_ISL_1554383, EPI_ISL_1554384, EPI_ISL_1554385, EPI_ISL_1554386, EPI_ISL_1554387, EPI_ISL_1554388, EPI_ISL_1554389, EPI_ISL_1554390, EPI_ISL_1554391, EPI_ISL_1554392, EPI_ISL_1554393, EPI_ISL_1554394, EPI_ISL_1554395, EPI_ISL_1554396, EPI_ISL_1554397, EPI_ISL_1554398, EPI_ISL_1554399, EPI_ISL_1554400, EPI_ISL_1554401, EPI_ISL_1554402,
EPI_ISL_1554403, EPI_ISL_1554404, EPI_ISL_1554405, EPI_ISL_1554406, EPI_ISL_1554407, EPI_ISL_1554408, EPI_ISL_1554409, EPI_ISL_1554410, EPI_ISL_1554411, EPI_ISL_1554412, EPI_ISL_1554413, EPI_ISL_1554414, EPI_ISL_1554415, EPI_ISL_1554416, EPI_ISL_1554417, EPI_ISL_1554418, EPI_ISL_1554419, EPI_ISL_1554420, EPI_ISL_1554421, EPI_ISL_1554422, EPI_ISL_1554423, EPI_ISL_1554424,
EPI_ISL_1554425, EPI_ISL_1554426, EPI_ISL_1554427, EPI_ISL_1554428, EPI_ISL_1554429, EPI_ISL_1554430, EPI_ISL_1554431, EPI_ISL_1554432, EPI_ISL_1554433, EPI_ISL_1554434, EPI_ISL_1554435, EPI_ISL_1554436, EPI_ISL_1554437, EPI_ISL_1554438, EPI_ISL_1554439, EPI_ISL_1554440, EPI_ISL_1554441, EPI_ISL_1554442, EPI_ISL_1554443, EPI_ISL_1554444, EPI_ISL_1554445, EPI_ISL_1554446,
EPI_ISL_1554447, EPI_ISL_1554448, EPI_ISL_1554449, EPI_ISL_1554450, EPI_ISL_1554451, EPI_ISL_1554452, EPI_ISL_1554453, EPI_ISL_1554454, EPI_ISL_1554455, EPI_ISL_1554456, EPI_ISL_1554457, EPI_ISL_1554458, EPI_ISL_1554459, EPI_ISL_1554461, EPI_ISL_1554462, EPI_ISL_1554463, EPI_ISL_1554464, EPI_ISL_1554465, EPI_ISL_1554466, EPI_ISL_1554467, EPI_ISL_1554468, EPI_ISL_1554469,
EPI_ISL_1554470, EPI_ISL_1554471, EPI_ISL_1554472, EPI_ISL_1554473, EPI_ISL_1554474, EPI_ISL_1554475, EPI_ISL_1554476, EPI_ISL_1554477, EPI_ISL_1554478, EPI_ISL_1554479, EPI_ISL_1554480, EPI_ISL_1554481, EPI_ISL_1554482, EPI_ISL_1554483, EPI_ISL_1554484, EPI_ISL_1554485, EPI_ISL_1554486, EPI_ISL_1554487, EPI_ISL_1554488, EPI_ISL_1554490, EPI_ISL_1554491, EPI_ISL_1554492,
EPI_ISL_1554493, EPI_ISL_1554494, EPI_ISL_1554495, EPI_ISL_1554496, EPI_ISL_1554497, EPI_ISL_1554498, EPI_ISL_1554499, EPI_ISL_1554500, EPI_ISL_1554501, EPI_ISL_1554502, EPI_ISL_1554503, EPI_ISL_1554504, EPI_ISL_1554505, EPI_ISL_1554506, EPI_ISL_1554507, EPI_ISL_1554508, EPI_ISL_1554509, EPI_ISL_1554510, EPI_ISL_1554511, EPI_ISL_1554512, EPI_ISL_1554513, EPI_ISL_1554514,
EPI_ISL_1554515, EPI_ISL_1554516, EPI_ISL_1554517, EPI_ISL_1554518, EPI_ISL_1554519, EPI_ISL_1554520, EPI_ISL_1554521, EPI_ISL_1554522, EPI_ISL_1554523, EPI_ISL_1554524, EPI_ISL_1554525, EPI_ISL_1554526, EPI_ISL_1554527, EPI_ISL_1554528, EPI_ISL_1554529, EPI_ISL_1554530, EPI_ISL_1554531, EPI_ISL_1554532, EPI_ISL_1554533, EPI_ISL_1554534, EPI_ISL_1554535, EPI_ISL_1554536,
EPI_ISL_1554537, EPI_ISL_1554538, EPI_ISL_1554539, EPI_ISL_1554540, EPI_ISL_1554541, EPI_ISL_1554542, EPI_ISL_1554543, EPI_ISL_1554544, EPI_ISL_1554545, EPI_ISL_1554546, EPI_ISL_1554547, EPI_ISL_1554548, EPI_ISL_1554549, EPI_ISL_1554550, EPI_ISL_1554551, EPI_ISL_1554552, EPI_ISL_1554553, EPI_ISL_1554554, EPI_ISL_1554555, EPI_ISL_1554556, EPI_ISL_1554557, EPI_ISL_1554558,
EPI_ISL_1554559, EPI_ISL_1554560, EPI_ISL_1554561, EPI_ISL_1554562, EPI_ISL_1554563, EPI_ISL_1554564, EPI_ISL_1554565, EPI_ISL_1554567, EPI_ISL_1554568, EPI_ISL_1554569, EPI_ISL_1554570, EPI_ISL_1554571, EPI_ISL_1554572, EPI_ISL_1554573, EPI_ISL_1554575, EPI_ISL_1554576, EPI_ISL_1554577, EPI_ISL_1554578, EPI_ISL_1554579, EPI_ISL_1554580, EPI_ISL_1554581, EPI_ISL_1554582,
EPI_ISL_1554583, EPI_ISL_1554584, EPI_ISL_1554585, EPI_ISL_1554586, EPI_ISL_1554587, EPI_ISL_1554588, EPI_ISL_1554589, EPI_ISL_1554590, EPI_ISL_1554591, EPI_ISL_1554592, EPI_ISL_1554593, EPI_ISL_1554594, EPI_ISL_1554595, EPI_ISL_1554596, EPI_ISL_1554597, EPI_ISL_1554598, EPI_ISL_1554599, EPI_ISL_1554600, EPI_ISL_1554601, EPI_ISL_1554602, EPI_ISL_1554603, EPI_ISL_1554604,
EPI_ISL_1554605, EPI_ISL_1554606, EPI_ISL_1554607, EPI_ISL_1554608, EPI_ISL_1554609, EPI_ISL_1554610, EPI_ISL_1554611, EPI_ISL_1554612, EPI_ISL_1554613, EPI_ISL_1554614, EPI_ISL_1554615, EPI_ISL_1554616, EPI_ISL_1554617, EPI_ISL_1554618, EPI_ISL_1554619, EPI_ISL_1554620, EPI_ISL_1554621, EPI_ISL_1554622, EPI_ISL_1554623, EPI_ISL_1554624, EPI_ISL_1554625, EPI_ISL_1554626,
EPI_ISL_1554627, EPI_ISL_1554628, EPI_ISL_1554629, EPI_ISL_1554630, EPI_ISL_1554631, EPI_ISL_1554632, EPI_ISL_1554633, EPI_ISL_1554634, EPI_ISL_1554635, EPI_ISL_1554636, EPI_ISL_1554637, EPI_ISL_1554638, EPI_ISL_1554639, EPI_ISL_1554640, EPI_ISL_1554641, EPI_ISL_1554642, EPI_ISL_1554643, EPI_ISL_1554644, EPI_ISL_1554645, EPI_ISL_1554646, EPI_ISL_1554647, EPI_ISL_1554648,
EPI_ISL_1554649, EPI_ISL_1554650, EPI_ISL_1554651, EPI_ISL_1554652, EPI_ISL_1554653, EPI_ISL_1554654, EPI_ISL_1554655, EPI_ISL_1554656, EPI_ISL_1554657, EPI_ISL_1554658, EPI_ISL_1554659, EPI_ISL_1554660, EPI_ISL_1554661, EPI_ISL_1554662, EPI_ISL_1554663, EPI_ISL_1554664, EPI_ISL_1554665, EPI_ISL_1554666, EPI_ISL_1554667, EPI_ISL_1554668, EPI_ISL_1554669, EPI_ISL_1554670,
EPI_ISL_1554671, EPI_ISL_1554672, EPI_ISL_1554673, EPI_ISL_1554674, EPI_ISL_1554675, EPI_ISL_1554676, EPI_ISL_1554677, EPI_ISL_1554678, EPI_ISL_1554679, EPI_ISL_1554680, EPI_ISL_1554681, EPI_ISL_1554682, EPI_ISL_1554683, EPI_ISL_1554684, EPI_ISL_1554685, EPI_ISL_1554686, EPI_ISL_1554687, EPI_ISL_1554688, EPI_ISL_1554689, EPI_ISL_1554690, EPI_ISL_1554691, EPI_ISL_1554692,
EPI_ISL_1554693, EPI_ISL_1554694, EPI_ISL_1554695, EPI_ISL_1554696, EPI_ISL_1554697, EPI_ISL_1554698, EPI_ISL_1554699, EPI_ISL_1554700, EPI_ISL_1554701, EPI_ISL_1554702, EPI_ISL_1554703, EPI_ISL_1554704, EPI_ISL_1554705, EPI_ISL_1554706, EPI_ISL_1554707, EPI_ISL_1554708, EPI_ISL_1554709, EPI_ISL_1554710, EPI_ISL_1554711, EPI_ISL_1554712, EPI_ISL_1554713, EPI_ISL_1554714,
EPI_ISL_1554715, EPI_ISL_1554716, EPI_ISL_1554717, EPI_ISL_1554718, EPI_ISL_1554719, EPI_ISL_1554720, EPI_ISL_1554721, EPI_ISL_1554722, EPI_ISL_1554723, EPI_ISL_1554724, EPI_ISL_1554725, EPI_ISL_1554726, EPI_ISL_1554727, EPI_ISL_1554728, EPI_ISL_1554729, EPI_ISL_1554730, EPI_ISL_1554731, EPI_ISL_1554732, EPI_ISL_1554733, EPI_ISL_1554734, EPI_ISL_1554735, EPI_ISL_1554736,
EPI_ISL_1554737, EPI_ISL_1554738, EPI_ISL_1554739, EPI_ISL_1554740, EPI_ISL_1554741, EPI_ISL_1554742, EPI_ISL_1554743, EPI_ISL_1554744, EPI_ISL_1554745, EPI_ISL_1554746, EPI_ISL_1554747, EPI_ISL_1554748, EPI_ISL_1554749, EPI_ISL_1554750, EPI_ISL_1554751, EPI_ISL_1554752, EPI_ISL_1554753, EPI_ISL_1554754, EPI_ISL_1554755, EPI_ISL_1554756, EPI_ISL_1554757, EPI_ISL_1554758,
EPI_ISL_1554759, EPI_ISL_1554760, EPI_ISL_1554761, EPI_ISL_1554762, EPI_ISL_1554763, EPI_ISL_1554764, EPI_ISL_1554765, EPI_ISL_1554766, EPI_ISL_1554767, EPI_ISL_1554768, EPI_ISL_1554769, EPI_ISL_1554770, EPI_ISL_1554771, EPI_ISL_1554773, EPI_ISL_1554774, EPI_ISL_1554775, EPI_ISL_1554776, EPI_ISL_1554777, EPI_ISL_1554779, EPI_ISL_1554780, EPI_ISL_1554781, EPI_ISL_1554782,
EPI_ISL_1554783, EPI_ISL_1554784, EPI_ISL_1554785, EPI_ISL_1554787, EPI_ISL_1554788, EPI_ISL_1554789, EPI_ISL_1554790, EPI_ISL_1554791, EPI_ISL_1554792, EPI_ISL_1554793, EPI_ISL_1554794, EPI_ISL_1554795, EPI_ISL_1554796, EPI_ISL_1554797, EPI_ISL_1554798, EPI_ISL_1554799, EPI_ISL_1554800, EPI_ISL_1554801, EPI_ISL_1554802, EPI_ISL_1554803, EPI_ISL_1554804, EPI_ISL_1554805,
EPI_ISL_1554806, EPI_ISL_1554807, EPI_ISL_1554808, EPI_ISL_1554809, EPI_ISL_1554810, EPI_ISL_1554811, EPI_ISL_1554812, EPI_ISL_1554813, EPI_ISL_1554814, EPI_ISL_1554815, EPI_ISL_1554816, EPI_ISL_1554817, EPI_ISL_1554818, EPI_ISL_1554819, EPI_ISL_1554820, EPI_ISL_1554821, EPI_ISL_1554822, EPI_ISL_1554823, EPI_ISL_1554824, EPI_ISL_1554825, EPI_ISL_1554826, EPI_ISL_1554827,
EPI_ISL_1554828, EPI_ISL_1554829, EPI_ISL_1554830, EPI_ISL_1554831, EPI_ISL_1554832, EPI_ISL_1554833, EPI_ISL_1554834, EPI_ISL_1554835, EPI_ISL_1554836, EPI_ISL_1554837, EPI_ISL_1554838, EPI_ISL_1554840, EPI_ISL_1554841, EPI_ISL_1554842, EPI_ISL_1554843, EPI_ISL_1554844, EPI_ISL_1554845, EPI_ISL_1554846, EPI_ISL_1554847, EPI_ISL_1554848, EPI_ISL_1554850, EPI_ISL_1554851,
EPI_ISL_1554852, EPI_ISL_1554853, EPI_ISL_1554854, EPI_ISL_1554855, EPI_ISL_1554856, EPI_ISL_1554857, EPI_ISL_1554858, EPI_ISL_1554859, EPI_ISL_1554860, EPI_ISL_1554861, EPI_ISL_1554862, EPI_ISL_1554863, EPI_ISL_1554864, EPI_ISL_1554865, EPI_ISL_1554866, EPI_ISL_1554867, EPI_ISL_1554868, EPI_ISL_1554869, EPI_ISL_1554870, EPI_ISL_1554871, EPI_ISL_1554872, EPI_ISL_1554873,
EPI_ISL_1554874, EPI_ISL_1554875, EPI_ISL_1554876, EPI_ISL_1554877, EPI_ISL_1554878, EPI_ISL_1554879, EPI_ISL_1554880, EPI_ISL_1554881, EPI_ISL_1554882, EPI_ISL_1554883, EPI_ISL_1554884, EPI_ISL_1554885, EPI_ISL_1554886

| | | | |
|---|---|---|---|
| see above | Helix/Illumina | Centers for Disease Control and Prevention Division of Viral Diseases, Pathogen Discovery | Dakota Howard, Dhwani Batra, Peter W. Cook, Kara Moser, Adrian Paskey, Jason Caravas, Benjamin Rambo-Martin, Shatavia Morrison, Christopher Gulvick, Scott Sammons, Yvette Unoarumhi, Darlene Wagner, Matthew Schmerer, Eileen de Feo, Jan Antico, Christine Tran, Matthew Tolentino, Shannon Wickline, Kim Gietzen, Brad Sickler, Jingtao Liu, Eric Allen, Phil Febbo, Nicole L. Washington, Simon White, Geraint Levan, Kelly Schiabor Barrett, Elizabeth Cirulli, Alexandre Bolze, Ary Ascencio, Charlotte Rivera-Garcia, Ryan Cho, Jason Nguyen, Sherry Wang, Jimmy Ramirez, Tyler Cassens, Efren Sandoval, Magnus Isaksson, William Lee, David Becker, Marc Laurent, James Lu, Clinton R. Paden, Duncan MacCannell |

EPI_ISL_1554950, EPI_ISL_1554951, EPI_ISL_1554953, EPI_ISL_1554954, EPI_ISL_1554956, EPI_ISL_1554957, EPI_ISL_1554959, EPI_ISL_1554960, EPI_ISL_1554962, EPI_ISL_1554963, EPI_ISL_1554964, EPI_ISL_1554965, EPI_ISL_1554966, EPI_ISL_1554967, EPI_ISL_1554968, EPI_ISL_1554969, EPI_ISL_1554970, EPI_ISL_1554971, EPI_ISL_1554972, EPI_ISL_1554973, EPI_ISL_1554974, EPI_ISL_1554975,
EPI_ISL_1554976, EPI_ISL_1554977, EPI_ISL_1554978, EPI_ISL_1554979, EPI_ISL_1554980, EPI_ISL_1554981, EPI_ISL_1554982, EPI_ISL_1554983, EPI_ISL_1554984, EPI_ISL_1554985, EPI_ISL_1554986, EPI_ISL_1554987, EPI_ISL_1554988, EPI_ISL_1554989, EPI_ISL_1554990, EPI_ISL_1554991, EPI_ISL_1554992, EPI_ISL_1554993, EPI_ISL_1554994, EPI_ISL_1554995, EPI_ISL_1554996, EPI_ISL_1554997,
EPI_ISL_1554998, EPI_ISL_1554999, EPI_ISL_1555000, EPI_ISL_1555001, EPI_ISL_1555002, EPI_ISL_1555003, EPI_ISL_1555004, EPI_ISL_1555005, EPI_ISL_1555006, EPI_ISL_1555007, EPI_ISL_1555008, EPI_ISL_1555009, EPI_ISL_1555010, EPI_ISL_1555011, EPI_ISL_1555012, EPI_ISL_1555013, EPI_ISL_1555014, EPI_ISL_1555015, EPI_ISL_1555016, EPI_ISL_1555017, EPI_ISL_1555018, EPI_ISL_1555019,
EPI_ISL_1555020, EPI_ISL_1555021, EPI_ISL_1555022, EPI_ISL_1555023, EPI_ISL_1555024, EPI_ISL_1555025, EPI_ISL_1555026, EPI_ISL_1555027, EPI_ISL_1555028, EPI_ISL_1555029, EPI_ISL_1555030, EPI_ISL_1555031, EPI_ISL_1555032, EPI_ISL_1555033, EPI_ISL_1555034, EPI_ISL_1555035, EPI_ISL_1555036, EPI_ISL_1555037, EPI_ISL_1555039, EPI_ISL_1555040, EPI_ISL_1555041, EPI_ISL_1555042,
EPI_ISL_1555043, EPI_ISL_1555044, EPI_ISL_1555045, EPI_ISL_1555046, EPI_ISL_1555047, EPI_ISL_1555048, EPI_ISL_1555049, EPI_ISL_1555050, EPI_ISL_1555051, EPI_ISL_1555053, EPI_ISL_1555054, EPI_ISL_1555055, EPI_ISL_1555056, EPI_ISL_1555057, EPI_ISL_1555058, EPI_ISL_1555059, EPI_ISL_1555060, EPI_ISL_1555061, EPI_ISL_1555062, EPI_ISL_1555063, EPI_ISL_1555064, EPI_ISL_1555065,
EPI_ISL_1555066, EPI_ISL_1555067, EPI_ISL_1555068, EPI_ISL_1555069, EPI_ISL_1555070, EPI_ISL_1555071, EPI_ISL_1555072, EPI_ISL_1555073, EPI_ISL_1555074, EPI_ISL_1555075, EPI_ISL_1555076, EPI_ISL_1555077, EPI_ISL_1555078, EPI_ISL_1555079, EPI_ISL_1555080, EPI_ISL_1555081, EPI_ISL_1555082, EPI_ISL_1555083, EPI_ISL_1555084, EPI_ISL_1555085, EPI_ISL_1555086, EPI_ISL_1555087,
EPI_ISL_1555088, EPI_ISL_1555089, EPI_ISL_1555090, EPI_ISL_1555091, EPI_ISL_1555092, EPI_ISL_1555093, EPI_ISL_1555094, EPI_ISL_1555095, EPI_ISL_1555096, EPI_ISL_1555097, EPI_ISL_1555098, EPI_ISL_1555099, EPI_ISL_1555100, EPI_ISL_1555101, EPI_ISL_1555102, EPI_ISL_1555103, EPI_ISL_1555104, EPI_ISL_1555105, EPI_ISL_1555106, EPI_ISL_1555107, EPI_ISL_1555108, EPI_ISL_1555109,
EPI_ISL_1555110, EPI_ISL_1555111, EPI_ISL_1555112, EPI_ISL_1555113, EPI_ISL_1555115, EPI_ISL_1555116, EPI_ISL_1555117, EPI_ISL_1555118, EPI_ISL_1555119, EPI_ISL_1555120, EPI_ISL_1555121, EPI_ISL_1555122, EPI_ISL_1555123, EPI_ISL_1555124, EPI_ISL_1555125, EPI_ISL_1555126, EPI_ISL_1555127, EPI_ISL_1555128, EPI_ISL_1555129, EPI_ISL_1555130, EPI_ISL_1555131, EPI_ISL_1555132,
EPI_ISL_1555133, EPI_ISL_1555134, EPI_ISL_1555135, EPI_ISL_1555136, EPI_ISL_1555137, EPI_ISL_1555138, EPI_ISL_1555139, EPI_ISL_1555140, EPI_ISL_1555141, EPI_ISL_1555142, EPI_ISL_1555143, EPI_ISL_1555144, EPI_ISL_1555145, EPI_ISL_1555146, EPI_ISL_1555148, EPI_ISL_1555149, EPI_ISL_1555150, EPI_ISL_1555151, EPI_ISL_1555152, EPI_ISL_1555153, EPI_ISL_1555154, EPI_ISL_1555155,
EPI_ISL_1555156, EPI_ISL_1555157, EPI_ISL_1555158, EPI_ISL_1555159, EPI_ISL_1555160, EPI_ISL_1555161, EPI_ISL_1555162, EPI_ISL_1555163, EPI_ISL_1555164, EPI_ISL_1555165, EPI_ISL_1555166, EPI_ISL_1555167, EPI_ISL_1555168, EPI_ISL_1555169, EPI_ISL_1555170, EPI_ISL_1555171, EPI_ISL_1555172, EPI_ISL_1555173, EPI_ISL_1555174, EPI_ISL_1555175, EPI_ISL_1555176, EPI_ISL_1555177,
EPI_ISL_1555178, EPI_ISL_1555179, EPI_ISL_1555180, EPI_ISL_1555181, EPI_ISL_1555182, EPI_ISL_1555183, EPI_ISL_1555184, EPI_ISL_1555185, EPI_ISL_1555186, EPI_ISL_1555187, EPI_ISL_1555188, EPI_ISL_1555190, EPI_ISL_1555191, EPI_ISL_1555192, EPI_ISL_1555193, EPI_ISL_1555194, EPI_ISL_1555195, EPI_ISL_1555196, EPI_ISL_1555197, EPI_ISL_1555198, EPI_ISL_1555199, EPI_ISL_1555200,
EPI_ISL_1555201, EPI_ISL_1555203, EPI_ISL_1555204, EPI_ISL_1555205, EPI_ISL_1555206, EPI_ISL_1555207, EPI_ISL_1555208, EPI_ISL_1555209, EPI_ISL_1555210, EPI_ISL_1555211, EPI_ISL_1555212, EPI_ISL_1555213, EPI_ISL_1555214, EPI_ISL_1555215, EPI_ISL_1555216, EPI_ISL_1555217, EPI_ISL_1555218, EPI_ISL_1555219, EPI_ISL_1555220, EPI_ISL_1555222, EPI_ISL_1555223, EPI_ISL_1555224,
EPI_ISL_1555225, EPI_ISL_1555226, EPI_ISL_1555227, EPI_ISL_1555228, EPI_ISL_1555229, EPI_ISL_1555230, EPI_ISL_1555231, EPI_ISL_1555232, EPI_ISL_1555233, EPI_ISL_1555234, EPI_ISL_1555235, EPI_ISL_1555236, EPI_ISL_1555237, EPI_ISL_1555238, EPI_ISL_1555239, EPI_ISL_1555240, EPI_ISL_1555241, EPI_ISL_1555242, EPI_ISL_1555243, EPI_ISL_1555244, EPI_ISL_1555245, EPI_ISL_1555246,
EPI_ISL_1555247, EPI_ISL_1555248, EPI_ISL_1555249, EPI_ISL_1555250, EPI_ISL_1555251, EPI_ISL_1555252, EPI_ISL_1555253, EPI_ISL_1555254, EPI_ISL_1555255, EPI_ISL_1555256, EPI_ISL_1555257, EPI_ISL_1555258, EPI_ISL_1555259, EPI_ISL_1555260, EPI_ISL_1555261, EPI_ISL_1555262, EPI_ISL_1555263, EPI_ISL_1555264, EPI_ISL_1555265, EPI_ISL_1555266, EPI_ISL_1555267, EPI_ISL_1555268,
EPI_ISL_1555269, EPI_ISL_1555270, EPI_ISL_1555271, EPI_ISL_1555272, EPI_ISL_1555273, EPI_ISL_1555274, EPI_ISL_1555275, EPI_ISL_1555276, EPI_ISL_1555277, EPI_ISL_1555278, EPI_ISL_1555279, EPI_ISL_1555280, EPI_ISL_1555281, EPI_ISL_1555282, EPI_ISL_1555283, EPI_ISL_1555284, EPI_ISL_1555285, EPI_ISL_1555286, EPI_ISL_1555287, EPI_ISL_1555288, EPI_ISL_1555289, EPI_ISL_1555290,
EPI_ISL_1555291, EPI_ISL_1555292, EPI_ISL_1555293, EPI_ISL_1555294, EPI_ISL_1555295, EPI_ISL_1555296, EPI_ISL_1555297, EPI_ISL_1555298, EPI_ISL_1555299, EPI_ISL_1555300, EPI_ISL_1555301, EPI_ISL_1555303, EPI_ISL_1555304, EPI_ISL_1555305, EPI_ISL_1555306, EPI_ISL_1555307, EPI_ISL_1555308, EPI_ISL_1555309, EPI_ISL_1555310, EPI_ISL_1555311, EPI_ISL_1555312, EPI_ISL_1555314,
EPI_ISL_1555315, EPI_ISL_1555316, EPI_ISL_1555317, EPI_ISL_1555318, EPI_ISL_1555319, EPI_ISL_1555320, EPI_ISL_1555321, EPI_ISL_1555322, EPI_ISL_1555323, EPI_ISL_1555324, EPI_ISL_1555325, EPI_ISL_1555326, EPI_ISL_1555327, EPI_ISL_1555328, EPI_ISL_1555329, EPI_ISL_1555330, EPI_ISL_1555331, EPI_ISL_1555332, EPI_ISL_1555333, EPI_ISL_1555334, EPI_ISL_1555335, EPI_ISL_1555336,
EPI_ISL_1555337, EPI_ISL_1555338, EPI_ISL_1555339, EPI_ISL_1555340, EPI_ISL_1555341, EPI_ISL_1555342, EPI_ISL_1555344, EPI_ISL_1555345, EPI_ISL_1555346, EPI_ISL_1555347, EPI_ISL_1555348, EPI_ISL_1555349, EPI_ISL_1555350, EPI_ISL_1555351, EPI_ISL_1555353, EPI_ISL_1555354, EPI_ISL_1555355, EPI_ISL_1555356, EPI_ISL_1555357, EPI_ISL_1555358, EPI_ISL_1555359, EPI_ISL_1555360,
EPI_ISL_1555361, EPI_ISL_1555362, EPI_ISL_1555364, EPI_ISL_1555365, EPI_ISL_1555366, EPI_ISL_1555367, EPI_ISL_1555368, EPI_ISL_1555369, EPI_ISL_1555370, EPI_ISL_1555371, EPI_ISL_1555372, EPI_ISL_1555373, EPI_ISL_1555374, EPI_ISL_1555375, EPI_ISL_1555376, EPI_ISL_1555377, EPI_ISL_1555378, EPI_ISL_1555379, EPI_ISL_1555380, EPI_ISL_1555381, EPI_ISL_1555382, EPI_ISL_1555383,
EPI_ISL_1555384, EPI_ISL_1555386, EPI_ISL_1555387, EPI_ISL_1555388, EPI_ISL_1555389, EPI_ISL_1555391, EPI_ISL_1555392, EPI_ISL_1555393, EPI_ISL_1555394, EPI_ISL_1555395, EPI_ISL_1555396, EPI_ISL_1555397, EPI_ISL_1555398, EPI_ISL_1555400, EPI_ISL_1555401, EPI_ISL_1555402, EPI_ISL_1555403, EPI_ISL_1555404, EPI_ISL_1555405, EPI_ISL_1555406, EPI_ISL_1555407, EPI_ISL_1555408,
EPI_ISL_1555409, EPI_ISL_1555410, EPI_ISL_1555411, EPI_ISL_1555412, EPI_ISL_1555413, EPI_ISL_1555414, EPI_ISL_1555415, EPI_ISL_1555416, EPI_ISL_1555417, EPI_ISL_1555418, EPI_ISL_1555419, EPI_ISL_1555420, EPI_ISL_1555421, EPI_ISL_1555422, EPI_ISL_1555423, EPI_ISL_1555424, EPI_ISL_1555425, EPI_ISL_1555426, EPI_ISL_1555427, EPI_ISL_1555428, EPI_ISL_1555429, EPI_ISL_1555430,
EPI_ISL_1555431, EPI_ISL_1555433, EPI_ISL_1555434, EPI_ISL_1555435, EPI_ISL_1555436, EPI_ISL_1555437, EPI_ISL_1555438, EPI_ISL_1555439, EPI_ISL_1555440, EPI_ISL_1555441, EPI_ISL_1555442, EPI_ISL_1555443, EPI_ISL_1555444, EPI_ISL_1555445, EPI_ISL_1555446, EPI_ISL_1555447, EPI_ISL_1555448, EPI_ISL_1555449, EPI_ISL_1555450, EPI_ISL_1555451, EPI_ISL_1555452, EPI_ISL_1555453,
EPI_ISL_1555454, EPI_ISL_1555455, EPI_ISL_1555456, EPI_ISL_1555457, EPI_ISL_1555458, EPI_ISL_1555459, EPI_ISL_1555460, EPI_ISL_1555461, EPI_ISL_1555462, EPI_ISL_1555463, EPI_ISL_1555464, EPI_ISL_1555465, EPI_ISL_1555466, EPI_ISL_1555467, EPI_ISL_1555468, EPI_ISL_1555469, EPI_ISL_1555470, EPI_ISL_1555471, EPI_ISL_1555472, EPI_ISL_1555473, EPI_ISL_1555474, EPI_ISL_1555475,
EPI_ISL_1555476, EPI_ISL_1555477, EPI_ISL_1555478, EPI_ISL_1555479, EPI_ISL_1555480, EPI_ISL_1555481, EPI_ISL_1555482, EPI_ISL_1555483, EPI_ISL_1555484, EPI_ISL_1555485, EPI_ISL_1555486, EPI_ISL_1555488, EPI_ISL_1555489, EPI_ISL_1555490, EPI_ISL_1555491, EPI_ISL_1555492, EPI_ISL_1555493, EPI_ISL_1555494, EPI_ISL_1555495, EPI_ISL_1555496, EPI_ISL_1555497, EPI_ISL_1555498,
EPI_ISL_1555499, EPI_ISL_1555500, EPI_ISL_1555501, EPI_ISL_1555502, EPI_ISL_1555503, EPI_ISL_1555504, EPI_ISL_1555505, EPI_ISL_1555506, EPI_ISL_1555508, EPI_ISL_1555509, EPI_ISL_1555510, EPI_ISL_1555511, EPI_ISL_1555512, EPI_ISL_1555513, EPI_ISL_1555514, EPI_ISL_1555515, EPI_ISL_1555516, EPI_ISL_1555517, EPI_ISL_1555518, EPI_ISL_1555519, EPI_ISL_1555520, EPI_ISL_1555521,
EPI_ISL_1555522, EPI_ISL_1555523, EPI_ISL_1555524, EPI_ISL_1555525, EPI_ISL_1555526, EPI_ISL_1555527, EPI_ISL_1555528, EPI_ISL_1555529, EPI_ISL_1555530, EPI_ISL_1555531, EPI_ISL_1555532, EPI_ISL_1555533, EPI_ISL_1555534, EPI_ISL_1555535, EPI_ISL_1555536, EPI_ISL_1555537, EPI_ISL_1555538, EPI_ISL_1555539, EPI_ISL_1555540, EPI_ISL_1555541, EPI_ISL_1555542, EPI_ISL_1555543,
EPI_ISL_1555544, EPI_ISL_1555545, EPI_ISL_1555546, EPI_ISL_1555547, EPI_ISL_1555548, EPI_ISL_1555549, EPI_ISL_1555550, EPI_ISL_1555551, EPI_ISL_1555552, EPI_ISL_1555553, EPI_ISL_1555554, EPI_ISL_1555555, EPI_ISL_1555556, EPI_ISL_1555557, EPI_ISL_1555558, EPI_ISL_1555559, EPI_ISL_1555560, EPI_ISL_1555561, EPI_ISL_1555562, EPI_ISL_1555563, EPI_ISL_1555564, EPI_ISL_1555565,
EPI_ISL_1555568, EPI_ISL_1555569, EPI_ISL_1555570, EPI_ISL_1555571, EPI_ISL_1555572, EPI_ISL_1555573, EPI_ISL_1555574, EPI_ISL_1555575, EPI_ISL_1555576, EPI_ISL_1555577, EPI_ISL_1555578, EPI_ISL_1555579, EPI_ISL_1555580, EPI_ISL_1555582, EPI_ISL_1555583, EPI_ISL_1555584, EPI_ISL_1555585, EPI_ISL_1555586, EPI_ISL_1555587, EPI_ISL_1555588, EPI_ISL_1555589, EPI_ISL_1555590,
EPI_ISL_1555591, EPI_ISL_1555592, EPI_ISL_1555593, EPI_ISL_1555594, EPI_ISL_1555595, EPI_ISL_1555596, EPI_ISL_1555597, EPI_ISL_1555598, EPI_ISL_1555599, EPI_ISL_1555600, EPI_ISL_1555601, EPI_ISL_1555602, EPI_ISL_1555603, EPI_ISL_1555604, EPI_ISL_1555606, EPI_ISL_1555607, EPI_ISL_1555608, EPI_ISL_1555609, EPI_ISL_1555610, EPI_ISL_1555611, EPI_ISL_1555612, EPI_ISL_1555613,
EPI_ISL_1555614, EPI_ISL_1555615, EPI_ISL_1555616, EPI_ISL_1555617, EPI_ISL_1555618, EPI_ISL_1555619, EPI_ISL_1555620, EPI_ISL_1555622, EPI_ISL_1555624, EPI_ISL_1555625, EPI_ISL_1555626, EPI_ISL_1555628, EPI_ISL_1555630, EPI_ISL_1555631, EPI_ISL_1555632, EPI_ISL_1555633, EPI_ISL_1555634, EPI_ISL_1555635, EPI_ISL_1555636, EPI_ISL_1555639, EPI_ISL_1555640, EPI_ISL_1555641,
EPI_ISL_1555642, EPI_ISL_1555645, EPI_ISL_1555646, EPI_ISL_1555647, EPI_ISL_1555648, EPI_ISL_1555649, EPI_ISL_1555650, EPI_ISL_1555651, EPI_ISL_1555652, EPI_ISL_1555654, EPI_ISL_1555655, EPI_ISL_1555656, EPI_ISL_1555657, EPI_ISL_1555658, EPI_ISL_1555659, EPI_ISL_1555660, EPI_ISL_1555661, EPI_ISL_1555662, EPI_ISL_1555663, EPI_ISL_1555664, EPI_ISL_1555665, EPI_ISL_1555666,
EPI_ISL_1555667, EPI_ISL_1555668, EPI_ISL_1555669, EPI_ISL_1555670, EPI_ISL_1555671, EPI_ISL_1555672, EPI_ISL_1555673, EPI_ISL_1555674, EPI_ISL_1555675, EPI_ISL_1555676, EPI_ISL_1555677, EPI_ISL_1555678, EPI_ISL_1555679, EPI_ISL_1555680, EPI_ISL_1555681, EPI_ISL_1555683, EPI_ISL_1555684, EPI_ISL_1555685, EPI_ISL_1555686, EPI_ISL_1555687, EPI_ISL_1555688, EPI_ISL_1555689,
EPI_ISL_1555690, EPI_ISL_1555691, EPI_ISL_1555692, EPI_ISL_1555693, EPI_ISL_1555694, EPI_ISL_1555695, EPI_ISL_1555696, EPI_ISL_1555697, EPI_ISL_1555698, EPI_ISL_1555699, EPI_ISL_1555700, EPI_ISL_1555701, EPI_ISL_1555702, EPI_ISL_1555703, EPI_ISL_1555704, EPI_ISL_1555705, EPI_ISL_1555706, EPI_ISL_1555707, EPI_ISL_1555708, EPI_ISL_1555709, EPI_ISL_1555710, EPI_ISL_1555711,
EPI_ISL_1555712, EPI_ISL_1555713, EPI_ISL_1555714, EPI_ISL_1555715, EPI_ISL_1555716, EPI_ISL_1555717, EPI_ISL_1555718, EPI_ISL_1555719, EPI_ISL_1555720, EPI_ISL_1555721, EPI_ISL_1555722, EPI_ISL_1555723, EPI_ISL_1555724, EPI_ISL_1555725, EPI_ISL_1555726, EPI_ISL_1555727, EPI_ISL_1555728, EPI_ISL_1555729, EPI_ISL_1555730, EPI_ISL_1555731, EPI_ISL_1555732, EPI_ISL_1555733,
EPI_ISL_1555734, EPI_ISL_1555735, EPI_ISL_1555736, EPI_ISL_1555737, EPI_ISL_1555738, EPI_ISL_1555740, EPI_ISL_1555741, EPI_ISL_1555742, EPI_ISL_1555743, EPI_ISL_1555744, EPI_ISL_1555745, EPI_ISL_1555746, EPI_ISL_1555747, EPI_ISL_1555748, EPI_ISL_1555749, EPI_ISL_1555750, EPI_ISL_1555751, EPI_ISL_1555752, EPI_ISL_1555753, EPI_ISL_1555754, EPI_ISL_1555755, EPI_ISL_1555756,
EPI_ISL_1555758, EPI_ISL_1555759, EPI_ISL_1555760, EPI_ISL_1555761, EPI_ISL_1555762, EPI_ISL_1555763, EPI_ISL_1555764, EPI_ISL_1555765, EPI_ISL_1555766, EPI_ISL_1555767, EPI_ISL_1555768, EPI_ISL_1555770, EPI_ISL_1555771, EPI_ISL_1555772, EPI_ISL_1555774, EPI_ISL_1555775, EPI_ISL_1555776, EPI_ISL_1555777, EPI_ISL_1555778, EPI_ISL_1555779, EPI_ISL_1555780, EPI_ISL_1555781,

EPI_ISL_1555782, EPI_ISL_1555783, EPI_ISL_1555784, EPI_ISL_1555785, EPI_ISL_1555786, EPI_ISL_1555787, EPI_ISL_1555788, EPI_ISL_1555789, EPI_ISL_1555790, EPI_ISL_1555791, EPI_ISL_1555792, EPI_ISL_1555794, EPI_ISL_1555795, EPI_ISL_1555796, EPI_ISL_1555797, EPI_ISL_1555798, EPI_ISL_1555799, EPI_ISL_1555800, EPI_ISL_1555801, EPI_ISL_1555802, EPI_ISL_1555803, EPI_ISL_1555804, EPI_ISL_1555805, EPI_ISL_1555806, EPI_ISL_1555807, EPI_ISL_1555808, EPI_ISL_1555809, EPI_ISL_1555811, EPI_ISL_1555812, EPI_ISL_1555813, EPI_ISL_1555814, EPI_ISL_1555815, EPI_ISL_1555816, EPI_ISL_1555817, EPI_ISL_1555818, EPI_ISL_1555819, EPI_ISL_1555820, EPI_ISL_1555821, EPI_ISL_1555822, EPI_ISL_1555823, EPI_ISL_1555824, EPI_ISL_1555825, EPI_ISL_1555826, EPI_ISL_1555827, EPI_ISL_1555828, EPI_ISL_1555829, EPI_ISL_1555830, EPI_ISL_1555831, EPI_ISL_1555832, EPI_ISL_1555833, EPI_ISL_1555834, EPI_ISL_1555835, EPI_ISL_1555836, EPI_ISL_1555837, EPI_ISL_1555838, EPI_ISL_1555839, EPI_ISL_1555840, EPI_ISL_1555841, EPI_ISL_1555842, EPI_ISL_1555844, EPI_ISL_1555845, EPI_ISL_1555846, EPI_ISL_1555847, EPI_ISL_1555848, EPI_ISL_1555849, EPI_ISL_1555850, EPI_ISL_1555851, EPI_ISL_1555852, EPI_ISL_1555853, EPI_ISL_1555854, EPI_ISL_1555855, EPI_ISL_1555856, EPI_ISL_1555857, EPI_ISL_1555858, EPI_ISL_1555859, EPI_ISL_1555860, EPI_ISL_1555862, EPI_ISL_1555863, EPI_ISL_1555865, EPI_ISL_1555866, EPI_ISL_1555867, EPI_ISL_1555868, EPI_ISL_1555869, EPI_ISL_1555870, EPI_ISL_1555871, EPI_ISL_1555872, EPI_ISL_1555873, EPI_ISL_1555874, EPI_ISL_1555875, EPI_ISL_1555876, EPI_ISL_1555877, EPI_ISL_1555878, EPI_ISL_1555879, EPI_ISL_1555880, EPI_ISL_1555881, EPI_ISL_1555882, EPI_ISL_1555883, EPI_ISL_1555884, EPI_ISL_1555885, EPI_ISL_1555886, EPI_ISL_1555888, EPI_ISL_1555889, EPI_ISL_1555890, EPI_ISL_1555891, EPI_ISL_1555892, EPI_ISL_1555893, EPI_ISL_1555894, EPI_ISL_1555895, EPI_ISL_1555896, EPI_ISL_1555897, EPI_ISL_1555898, EPI_ISL_1555899, EPI_ISL_1555900, EPI_ISL_1555901, EPI_ISL_1555902, EPI_ISL_1555903, EPI_ISL_1555904, EPI_ISL_1555905, EPI_ISL_1555906, EPI_ISL_1555907, EPI_ISL_1555908, EPI_ISL_1555909, EPI_ISL_1555910, EPI_ISL_1555911, EPI_ISL_1555912, EPI_ISL_1555913, EPI_ISL_1555914, EPI_ISL_1555915, EPI_ISL_1555916, EPI_ISL_1555917, EPI_ISL_1555918, EPI_ISL_1555920, EPI_ISL_1555922, EPI_ISL_1555923, EPI_ISL_1555924, EPI_ISL_1555925, EPI_ISL_1555926, EPI_ISL_1555927, EPI_ISL_1555928, EPI_ISL_1555929, EPI_ISL_1555930, EPI_ISL_1555931, EPI_ISL_1555932, EPI_ISL_1555933, EPI_ISL_1555934, EPI_ISL_1555935, EPI_ISL_1555936, EPI_ISL_1555937, EPI_ISL_1555938, EPI_ISL_1555939, EPI_ISL_1555940, EPI_ISL_1555941, EPI_ISL_1555942, EPI_ISL_1555943, EPI_ISL_1555944, EPI_ISL_1555945, EPI_ISL_1555946, EPI_ISL_1555947, EPI_ISL_1555948, EPI_ISL_1555949, EPI_ISL_1555950, EPI_ISL_1555951, EPI_ISL_1555952, EPI_ISL_1555953, EPI_ISL_1555954, EPI_ISL_1555955, EPI_ISL_1555957, EPI_ISL_1555958, EPI_ISL_1555959, EPI_ISL_1555960, EPI_ISL_1555961, EPI_ISL_1555962, EPI_ISL_1555963, EPI_ISL_1555964, EPI_ISL_1555965, EPI_ISL_1555966, EPI_ISL_1555967, EPI_ISL_1555968, EPI_ISL_1555970, EPI_ISL_1555971, EPI_ISL_1555972, EPI_ISL_1555973, EPI_ISL_1555974, EPI_ISL_1555976, EPI_ISL_1555977, EPI_ISL_1555978, EPI_ISL_1555979, EPI_ISL_1555980, EPI_ISL_1555981, EPI_ISL_1555982, EPI_ISL_1555983, EPI_ISL_1555984, EPI_ISL_1555985, EPI_ISL_1555986, EPI_ISL_1555987, EPI_ISL_1555988, EPI_ISL_1555989, EPI_ISL_1555990, EPI_ISL_1555991, EPI_ISL_1555992, EPI_ISL_1555993, EPI_ISL_1555994, EPI_ISL_1555995, EPI_ISL_1555996, EPI_ISL_1555997, EPI_ISL_1555998, EPI_ISL_1555999, EPI_ISL_1556000, EPI_ISL_1556001, EPI_ISL_1556002, EPI_ISL_1556003, EPI_ISL_1556004, EPI_ISL_1556005, EPI_ISL_1556006, EPI_ISL_1556008, EPI_ISL_1556009, EPI_ISL_1556010, EPI_ISL_1556011, EPI_ISL_1556012, EPI_ISL_1556013, EPI_ISL_1556014, EPI_ISL_1556015, EPI_ISL_1556016, EPI_ISL_1556017, EPI_ISL_1556018, EPI_ISL_1556019, EPI_ISL_1556020, EPI_ISL_1556021, EPI_ISL_1556022, EPI_ISL_1556023, EPI_ISL_1556024, EPI_ISL_1556025, EPI_ISL_1556026, EPI_ISL_1556029, EPI_ISL_1556030, EPI_ISL_1556031, EPI_ISL_1556032, EPI_ISL_1556033, EPI_ISL_1556035, EPI_ISL_1556036, EPI_ISL_1556037, EPI_ISL_1556038, EPI_ISL_1556039, EPI_ISL_1556040, EPI_ISL_1556041, EPI_ISL_1556043, EPI_ISL_1556044, EPI_ISL_1556045, EPI_ISL_1556046, EPI_ISL_1556047, EPI_ISL_1556049, EPI_ISL_1556051, EPI_ISL_1556052, EPI_ISL_1556053, EPI_ISL_1556054, EPI_ISL_1556055, EPI_ISL_1556056, EPI_ISL_1556057, EPI_ISL_1556058, EPI_ISL_1556060, EPI_ISL_1556061, EPI_ISL_1556062, EPI_ISL_1556063, EPI_ISL_1556064, EPI_ISL_1556066, EPI_ISL_1556067, EPI_ISL_1556068, EPI_ISL_1556069, EPI_ISL_1556070, EPI_ISL_1556071, EPI_ISL_1556072, EPI_ISL_1556073, EPI_ISL_1556074, EPI_ISL_1556075, EPI_ISL_1556076, EPI_ISL_1556077, EPI_ISL_1556078, EPI_ISL_1556079, EPI_ISL_1556080, EPI_ISL_1556081, EPI_ISL_1556082, EPI_ISL_1556083, EPI_ISL_1556084, EPI_ISL_1556085, EPI_ISL_1556086, EPI_ISL_1556087, EPI_ISL_1556088, EPI_ISL_1556089, EPI_ISL_1556090, EPI_ISL_1556091, EPI_ISL_1556092, EPI_ISL_1556093, EPI_ISL_1556094, EPI_ISL_1556095, EPI_ISL_1556096, EPI_ISL_1556100, EPI_ISL_1556101, EPI_ISL_1556103, EPI_ISL_1556104, EPI_ISL_1556105, EPI_ISL_1556106, EPI_ISL_1556107, EPI_ISL_1556108, EPI_ISL_1556109, EPI_ISL_1556110, EPI_ISL_1556111, EPI_ISL_1556112, EPI_ISL_1556113, EPI_ISL_1556115, EPI_ISL_1556116, EPI_ISL_1556117, EPI_ISL_1556118, EPI_ISL_1556119, EPI_ISL_1556120, EPI_ISL_1556121, EPI_ISL_1556122, EPI_ISL_1556123, EPI_ISL_1556124, EPI_ISL_1556125, EPI_ISL_1556128, EPI_ISL_1556129, EPI_ISL_1556130, EPI_ISL_1556131, EPI_ISL_1556132, EPI_ISL_1556134, EPI_ISL_1556135, EPI_ISL_1556136, EPI_ISL_1556137, EPI_ISL_1556138, EPI_ISL_1556139, EPI_ISL_1556140, EPI_ISL_1556141, EPI_ISL_1556142, EPI_ISL_1556143, EPI_ISL_1556144, EPI_ISL_1556145, EPI_ISL_1556146, EPI_ISL_1556148, EPI_ISL_1556149, EPI_ISL_1556150, EPI_ISL_1556151, EPI_ISL_1556153, EPI_ISL_1556154, EPI_ISL_1556155, EPI_ISL_1556156, EPI_ISL_1556157, EPI_ISL_1556158, EPI_ISL_1556160, EPI_ISL_1556161, EPI_ISL_1556162, EPI_ISL_1556163, EPI_ISL_1556164, EPI_ISL_1556165, EPI_ISL_1556166, EPI_ISL_1556168, EPI_ISL_1556169, EPI_ISL_1556170, EPI_ISL_1556171, EPI_ISL_1556173, EPI_ISL_1556174, EPI_ISL_1556175, EPI_ISL_1556176, EPI_ISL_1556178, EPI_ISL_1556179, EPI_ISL_1556180, EPI_ISL_1556181, EPI_ISL_1556182, EPI_ISL_1556184, EPI_ISL_1556185, EPI_ISL_1556186, EPI_ISL_1556187, EPI_ISL_1556188, EPI_ISL_1556189, EPI_ISL_1556190, EPI_ISL_1556191, EPI_ISL_1556192, EPI_ISL_1556193, EPI_ISL_1556194, EPI_ISL_1556195, EPI_ISL_1556196, EPI_ISL_1556197, EPI_ISL_1556198, EPI_ISL_1556199, EPI_ISL_1556200, EPI_ISL_1556201, EPI_ISL_1556202, EPI_ISL_1556203, EPI_ISL_1556204, EPI_ISL_1556205, EPI_ISL_1556206, EPI_ISL_1556207, EPI_ISL_1556209, EPI_ISL_1556210, EPI_ISL_1556211, EPI_ISL_1556212, EPI_ISL_1556213, EPI_ISL_1556214, EPI_ISL_1556215, EPI_ISL_1556217, EPI_ISL_1556218, EPI_ISL_1556219, EPI_ISL_1556220, EPI_ISL_1556221, EPI_ISL_1556222, EPI_ISL_1556223, EPI_ISL_1556224, EPI_ISL_1556225, EPI_ISL_1556226, EPI_ISL_1556227, EPI_ISL_1556228, EPI_ISL_1556229, EPI_ISL_1556230, EPI_ISL_1556231, EPI_ISL_1556233, EPI_ISL_1556234, EPI_ISL_1556235, EPI_ISL_1556236, EPI_ISL_1556237, EPI_ISL_1556238, EPI_ISL_1556239, EPI_ISL_1556240, EPI_ISL_1556242, EPI_ISL_1556243, EPI_ISL_1556244, EPI_ISL_1556245, EPI_ISL_1556246, EPI_ISL_1556247, EPI_ISL_1556248, EPI_ISL_1556249, EPI_ISL_1556250, EPI_ISL_1556251, EPI_ISL_1556253, EPI_ISL_1556255, EPI_ISL_1556256, EPI_ISL_1556258, EPI_ISL_1556259, EPI_ISL_1556260, EPI_ISL_1556261, EPI_ISL_1556263, EPI_ISL_1556264, EPI_ISL_1556265, EPI_ISL_1556267, EPI_ISL_1556268, EPI_ISL_1556269, EPI_ISL_1556270, EPI_ISL_1556271, EPI_ISL_1556272, EPI_ISL_1556274, EPI_ISL_1556275, EPI_ISL_1556277, EPI_ISL_1556278, EPI_ISL_1556279, EPI_ISL_1556280, EPI_ISL_1556282, EPI_ISL_1556283, EPI_ISL_1556284, EPI_ISL_1556285, EPI_ISL_1556286, EPI_ISL_1556287, EPI_ISL_1556288, EPI_ISL_1556289, EPI_ISL_1556291, EPI_ISL_1556292, EPI_ISL_1556293, EPI_ISL_1556294, EPI_ISL_1556295, EPI_ISL_1556296, EPI_ISL_1556297, EPI_ISL_1556298, EPI_ISL_1556299, EPI_ISL_1556300, EPI_ISL_1556301, EPI_ISL_1556302, EPI_ISL_1556303, EPI_ISL_1556304, EPI_ISL_1556305, EPI_ISL_1556306, EPI_ISL_1556307, EPI_ISL_1556309, EPI_ISL_1556310, EPI_ISL_1556311, EPI_ISL_1556312, EPI_ISL_1556313, EPI_ISL_1556314, EPI_ISL_1556315, EPI_ISL_1556316, EPI_ISL_1556317, EPI_ISL_1556318, EPI_ISL_1556319, EPI_ISL_1556320, EPI_ISL_1556321, EPI_ISL_1556324, EPI_ISL_1556325, EPI_ISL_1556326, EPI_ISL_1556328, EPI_ISL_1556330, EPI_ISL_1556331, EPI_ISL_1556332, EPI_ISL_1556333, EPI_ISL_1556334, EPI_ISL_1556335, EPI_ISL_1556336, EPI_ISL_1556338, EPI_ISL_1556340, EPI_ISL_1556341, EPI_ISL_1556342, EPI_ISL_1556343, EPI_ISL_1556344, EPI_ISL_1556345, EPI_ISL_1556346, EPI_ISL_1556347, EPI_ISL_1556348, EPI_ISL_1556349, EPI_ISL_1556350, EPI_ISL_1556351, EPI_ISL_1556352, EPI_ISL_1556353, EPI_ISL_1556354, EPI_ISL_1556355, EPI_ISL_1556356, EPI_ISL_1556357, EPI_ISL_1556358, EPI_ISL_1556359, EPI_ISL_1556360, EPI_ISL_1556361, EPI_ISL_1556362, EPI_ISL_1556363, EPI_ISL_1556365, EPI_ISL_1556366, EPI_ISL_1556368, EPI_ISL_1556369, EPI_ISL_1556370, EPI_ISL_1556371, EPI_ISL_1556373, EPI_ISL_1556374, EPI_ISL_1556375, EPI_ISL_1556376, EPI_ISL_1556377, EPI_ISL_1556378, EPI_ISL_1556380, EPI_ISL_1556381, EPI_ISL_1556382, EPI_ISL_1556383, EPI_ISL_1556384, EPI_ISL_1556386, EPI_ISL_1556387, EPI_ISL_1556388, EPI_ISL_1556389, EPI_ISL_1556390, EPI_ISL_1556391, EPI_ISL_1556392, EPI_ISL_1556393, EPI_ISL_1556394, EPI_ISL_1556395, EPI_ISL_1556396, EPI_ISL_1556397, EPI_ISL_1556398, EPI_ISL_1556399, EPI_ISL_1556400, EPI_ISL_1556401, EPI_ISL_1556402, EPI_ISL_1556403, EPI_ISL_1556404, EPI_ISL_1556405, EPI_ISL_1556406, EPI_ISL_1556407, EPI_ISL_1556408, EPI_ISL_1556409, EPI_ISL_1556410, EPI_ISL_1556411, EPI_ISL_1556412, EPI_ISL_1556413, EPI_ISL_1556414, EPI_ISL_1556415, EPI_ISL_1556417, EPI_ISL_1556418, EPI_ISL_1556419, EPI_ISL_1556420, EPI_ISL_1556421, EPI_ISL_1556423, EPI_ISL_1556424, EPI_ISL_1556425, EPI_ISL_1556426, EPI_ISL_1556427, EPI_ISL_1556428, EPI_ISL_1556429, EPI_ISL_1556430, EPI_ISL_1556431, EPI_ISL_1556432, EPI_ISL_1556433, EPI_ISL_1556434, EPI_ISL_1556435, EPI_ISL_1556436, EPI_ISL_1556437, EPI_ISL_1556438, EPI_ISL_1556439, EPI_ISL_1556440, EPI_ISL_1556441, EPI_ISL_1556442, EPI_ISL_1556443, EPI_ISL_1556444, EPI_ISL_1556445, EPI_ISL_1556446, EPI_ISL_1556447, EPI_ISL_1556448, EPI_ISL_1556449, EPI_ISL_1556450, EPI_ISL_1556451, EPI_ISL_1556452, EPI_ISL_1556453, EPI_ISL_1556454, EPI_ISL_1556455, EPI_ISL_1556456, EPI_ISL_1556457, EPI_ISL_1556459, EPI_ISL_1556460, EPI_ISL_1556461, EPI_ISL_1556462, EPI_ISL_1556463, EPI_ISL_1556464, EPI_ISL_1556465, EPI_ISL_1556468, EPI_ISL_1556469, EPI_ISL_1556470, EPI_ISL_1556471, EPI_ISL_1556472, EPI_ISL_1556473, EPI_ISL_1556474, EPI_ISL_1556475, EPI_ISL_1556476, EPI_ISL_1556477, EPI_ISL_1556478, EPI_ISL_1556479, EPI_ISL_1556480, EPI_ISL_1556481, EPI_ISL_1556482, EPI_ISL_1556483, EPI_ISL_1556484, EPI_ISL_1556485, EPI_ISL_1556486, EPI_ISL_1556487, EPI_ISL_1556488, EPI_ISL_1556489, EPI_ISL_1556490, EPI_ISL_1556491, EPI_ISL_1556492, EPI_ISL_1556493, EPI_ISL_1556494, EPI_ISL_1556495, EPI_ISL_1556496, EPI_ISL_1556497, EPI_ISL_1556498, EPI_ISL_1556499, EPI_ISL_1556500, EPI_ISL_1556501, EPI_ISL_1556502, EPI_ISL_1556504, EPI_ISL_1556505, EPI_ISL_1556506, EPI_ISL_1556507, EPI_ISL_1556509, EPI_ISL_1556510, EPI_ISL_1556511, EPI_ISL_1556512, EPI_ISL_1556513, EPI_ISL_1556514, EPI_ISL_1556515, EPI_ISL_1556517, EPI_ISL_1556518, EPI_ISL_1556519, EPI_ISL_1556520, EPI_ISL_1556522, EPI_ISL_1556523, EPI_ISL_1556524, EPI_ISL_1556525, EPI_ISL_1556526, EPI_ISL_1556527, EPI_ISL_1556528, EPI_ISL_1556529, EPI_ISL_1556530, EPI_ISL_1556531, EPI_ISL_1556532, EPI_ISL_1556533, EPI_ISL_1556534, EPI_ISL_1556535, EPI_ISL_1556537, EPI_ISL_1556538, EPI_ISL_1556539, EPI_ISL_1556540, EPI_ISL_1556541, EPI_ISL_1556543, EPI_ISL_1556544, EPI_ISL_1556545, EPI_ISL_1556546, EPI_ISL_1556548, EPI_ISL_1556549, EPI_ISL_1556550, EPI_ISL_1556551, EPI_ISL_1556552, EPI_ISL_1556553, EPI_ISL_1556555, EPI_ISL_1556556, EPI_ISL_1556558, EPI_ISL_1556559, EPI_ISL_1556560, EPI_ISL_1556561, EPI_ISL_1556562, EPI_ISL_1556563, EPI_ISL_1556565, EPI_ISL_1556566, EPI_ISL_1556567, EPI_ISL_1556568, EPI_ISL_1556569, EPI_ISL_1556570, EPI_ISL_1556571, EPI_ISL_1556572, EPI_ISL_1556573, EPI_ISL_1556574, EPI_ISL_1556575, EPI_ISL_1556576, EPI_ISL_1556577, EPI_ISL_1556578, EPI_ISL_1556579, EPI_ISL_1556580, EPI_ISL_1556581, EPI_ISL_1556582, EPI_ISL_1556583, EPI_ISL_1556584, EPI_ISL_1556585, EPI_ISL_1556586, EPI_ISL_1556587, EPI_ISL_1556589, EPI_ISL_1556590, EPI_ISL_1556591, EPI_ISL_1556592, EPI_ISL_1556593, EPI_ISL_1556594, EPI_ISL_1556595, EPI_ISL_1556596, EPI_ISL_1556597, EPI_ISL_1556599, EPI_ISL_1556600, EPI_ISL_1556601, EPI_ISL_1556603, EPI_ISL_1556605, EPI_ISL_1556606, EPI_ISL_1556607, EPI_ISL_1556608, EPI_ISL_1556609, EPI_ISL_1556610, EPI_ISL_1556611, EPI_ISL_1556612, EPI_ISL_1556613, EPI_ISL_1556614, EPI_ISL_1556615, EPI_ISL_1556616, EPI_ISL_1556617, EPI_ISL_1556618, EPI_ISL_1556620, EPI_ISL_1556621, EPI_ISL_1556622, EPI_ISL_1556623, EPI_ISL_1556625, EPI_ISL_1556626, EPI_ISL_1556628, EPI_ISL_1556629, EPI_ISL_1556630, EPI_ISL_1556631, EPI_ISL_1556632, EPI_ISL_1556633, EPI_ISL_1556634, EPI_ISL_1556635, EPI_ISL_1556636, EPI_ISL_1556638, EPI_ISL_1556639, EPI_ISL_1556640, EPI_ISL_1556641, EPI_ISL_1556643, EPI_ISL_1556644, EPI_ISL_1556645, EPI_ISL_1556646, EPI_ISL_1556647, EPI_ISL_1556648, EPI_ISL_1556649, EPI_ISL_1556650, EPI_ISL_1556651, EPI_ISL_1556653, EPI_ISL_1556654, EPI_ISL_1556655, EPI_ISL_1556656, EPI_ISL_1556657, EPI_ISL_1556658, EPI_ISL_1556659, EPI_ISL_1556660, EPI_ISL_1556661, EPI_ISL_1556663, EPI_ISL_1556664, EPI_ISL_1556665, EPI_ISL_1556666, EPI_ISL_1556667, EPI_ISL_1556668, EPI_ISL_1556669, EPI_ISL_1556670, EPI_ISL_1556671, EPI_ISL_1556672, EPI_ISL_1556673, EPI_ISL_1556674, EPI_ISL_1556676, EPI_ISL_1556677, EPI_ISL_1556678, EPI_ISL_1556679, EPI_ISL_1556680, EPI_ISL_1556681, EPI_ISL_1556682, EPI_ISL_1556683, EPI_ISL_1556684, EPI_ISL_1556685, EPI_ISL_1556686, EPI_ISL_1556687, EPI_ISL_1556688, EPI_ISL_1556690, EPI_ISL_1556691, EPI_ISL_1556692, EPI_ISL_1556693, EPI_ISL_1556694, EPI_ISL_1556695, EPI_ISL_1556696, EPI_ISL_1556697, EPI_ISL_1556698, EPI_ISL_1556699, EPI_ISL_1556700, EPI_ISL_1556701, EPI_ISL_1556702, EPI_ISL_1556703, EPI_ISL_1556704, EPI_ISL_1556705, EPI_ISL_1556706, EPI_ISL_1556708, EPI_ISL_1556709, EPI_ISL_1556710, EPI_ISL_1556712, EPI_ISL_1556713, EPI_ISL_1556714, EPI_ISL_1556715, EPI_ISL_1556716, EPI_ISL_1556717, EPI_ISL_1556718, EPI_ISL_1556719, EPI_ISL_1556720, EPI_ISL_1556721, EPI_ISL_1556722, EPI_ISL_1556723, EPI_ISL_1556724, EPI_ISL_1556725, EPI_ISL_1556727, EPI_ISL_1556728, EPI_ISL_1556729, EPI_ISL_1556730, EPI_ISL_1556731, EPI_ISL_1556733, EPI_ISL_1556734, EPI_ISL_1556735, EPI_ISL_1556736, EPI_ISL_1556737, EPI_ISL_1556739, EPI_ISL_1556740, EPI_ISL_1556741, EPI_ISL_1556742, EPI_ISL_1556743, EPI_ISL_1556744, EPI_ISL_1556745, EPI_ISL_1556746, EPI_ISL_1556747, EPI_ISL_1556748, EPI_ISL_1556749, EPI_ISL_1556750, EPI_ISL_1556751, EPI_ISL_1556752, EPI_ISL_1556753, EPI_ISL_1556754, EPI_ISL_1556755, EPI_ISL_1556756, EPI_ISL_1556757, EPI_ISL_1556758, EPI_ISL_1556759, EPI_ISL_1556760, EPI_ISL_1556761, EPI_ISL_1556762, EPI_ISL_1556763, EPI_ISL_1556764, EPI_ISL_1556765, EPI_ISL_1556767, EPI_ISL_1556768, EPI_ISL_1556769, EPI_ISL_1556770, EPI_ISL_1556771, EPI_ISL_1556772, EPI_ISL_1556773, EPI_ISL_1556774, EPI_ISL_1556775, EPI_ISL_1556776, EPI_ISL_1556777, EPI_ISL_1556779, EPI_ISL_1556780, EPI_ISL_1556781, EPI_ISL_1556782, EPI_ISL_1556783, EPI_ISL_1556784, EPI_ISL_1556785, EPI_ISL_1556786, EPI_ISL_1556787, EPI_ISL_1556788, EPI_ISL_1556789, EPI_ISL_1556790, EPI_ISL_1556791, EPI_ISL_1556792, EPI_ISL_1556793, EPI_ISL_1556794, EPI_ISL_1556795, EPI_ISL_1556796, EPI_ISL_1556797, EPI_ISL_1556798, EPI_ISL_1556799, EPI_ISL_1556800, EPI_ISL_1556802, EPI_ISL_1556803, EPI_ISL_1556804, EPI_ISL_1556805, EPI_ISL_1556806, EPI_ISL_1556807, EPI_ISL_1556811, EPI_ISL_1556812, EPI_ISL_1556813, EPI_ISL_1556814, EPI_ISL_1556815, EPI_ISL_1556816, EPI_ISL_1556817, EPI_ISL_1556818, EPI_ISL_1556819, EPI_ISL_1556821, EPI_ISL_1556822, EPI_ISL_1556823, EPI_ISL_1556824, EPI_ISL_1556825, EPI_ISL_1556826, EPI_ISL_1556827, EPI_ISL_1556828, EPI_ISL_1556829, EPI_ISL_1556830, EPI_ISL_1556831, EPI_ISL_1556832, EPI_ISL_1556834, EPI_ISL_1556835, EPI_ISL_1556836, EPI_ISL_1556837, EPI_ISL_1556838, EPI_ISL_1556839, EPI_ISL_1556840, EPI_ISL_1556841, EPI_ISL_1556842, EPI_ISL_1556843, EPI_ISL_1556844, EPI_ISL_1556845, EPI_ISL_1556846, EPI_ISL_1556847, EPI_ISL_1556848, EPI_ISL_1556849, EPI_ISL_1556850, EPI_ISL_1556851, EPI_ISL_1556852, EPI_ISL_1556854, EPI_ISL_1556855, EPI_ISL_1556856, EPI_ISL_1556857, EPI_ISL_1556858, EPI_ISL_1556859, EPI_ISL_1556860, EPI_ISL_1556861, EPI_ISL_1556862, EPI_ISL_1556863, EPI_ISL_1556864, EPI_ISL_1556865, EPI_ISL_1556866, EPI_ISL_1556867, EPI_ISL_1556868, EPI_ISL_1556869, EPI_ISL_1556870, EPI_ISL_1556871, EPI_ISL_1556872, EPI_ISL_1556873, EPI_ISL_1556874, EPI_ISL_1556875, EPI_ISL_1556876, EPI_ISL_1556877, EPI_ISL_1556878, EPI_ISL_1556879, EPI_ISL_1556880, EPI_ISL_1556881, EPI_ISL_1556882, EPI_ISL_1556883, EPI_ISL_1556884, EPI_ISL_1556885, EPI_ISL_1556886, EPI_ISL_1556887, EPI_ISL_1556888, EPI_ISL_1556889, EPI_ISL_1556890, EPI_ISL_1556891, EPI_ISL_1556892, EPI_ISL_1556893, EPI_ISL_1556894, EPI_ISL_1556895, EPI_ISL_1556896, EPI_ISL_1556897, EPI_ISL_1556898, EPI_ISL_1556899, EPI_ISL_1556901, EPI_ISL_1556902, EPI_ISL_1556903, EPI_ISL_1556904, EPI_ISL_1556905, EPI_ISL_1556906, EPI_ISL_1556907, EPI_ISL_1556908, EPI_ISL_1556909, EPI_ISL_1556910, EPI_ISL_1556911, EPI_ISL_1556912, EPI_ISL_1556913, EPI_ISL_1556914, EPI_ISL_1556915, EPI_ISL_1556916, EPI_ISL_1556917, EPI_ISL_1556918, EPI_ISL_1556919, EPI_ISL_1556921, EPI_ISL_1556922, EPI_ISL_1556924, EPI_ISL_1556925, EPI_ISL_1556926, EPI_ISL_1556927, EPI_ISL_1556928, EPI_ISL_1556929, EPI_ISL_1556930, EPI_ISL_1556931, EPI_ISL_1556932, EPI_ISL_1556933, EPI_ISL_1556934, EPI_ISL_1556935, EPI_ISL_1556936, EPI_ISL_1556937, EPI_ISL_1556938, EPI_ISL_1556939, EPI_ISL_1556940, EPI_ISL_1556941, EPI_ISL_1556942, EPI_ISL_1556943, EPI_ISL_1556944, EPI_ISL_1556945, EPI_ISL_1556946, EPI_ISL_1556947, EPI_ISL_1556948, EPI_ISL_1556949, EPI_ISL_1556950, EPI_ISL_1556951, EPI_ISL_1556952, EPI_ISL_1556953, EPI_ISL_1556954, EPI_ISL_1556955, EPI_ISL_1556956, EPI_ISL_1556957, EPI_ISL_1556958, EPI_ISL_1556959, EPI_ISL_1556960, EPI_ISL_1556961, EPI_ISL_1556962, EPI_ISL_1556963, EPI_ISL_1556964, EPI_ISL_1556965, EPI_ISL_1556966, EPI_ISL_1556967, EPI_ISL_1556968, EPI_ISL_1556969, EPI_ISL_1556971, EPI_ISL_1556972, EPI_ISL_1556973, EPI_ISL_1556975, EPI_ISL_1556976, EPI_ISL_1556977, EPI_ISL_1556978, EPI_ISL_1556979, EPI_ISL_1556980, EPI_ISL_1556981, EPI_ISL_1556982, EPI_ISL_1556983, EPI_ISL_1556984, EPI_ISL_1556985, EPI_ISL_1556987, EPI_ISL_1556988, EPI_ISL_1556989, EPI_ISL_1556990, EPI_ISL_1556991, EPI_ISL_1556992, EPI_ISL_1556993, EPI_ISL_1556994, EPI_ISL_1556995, EPI_ISL_1556996, EPI_ISL_1556997, EPI_ISL_1556998, EPI_ISL_1556999, EPI_ISL_1557000, EPI_ISL_1557001, EPI_ISL_1557002, EPI_ISL_1557003, EPI_ISL_1557004, EPI_ISL_1557005, EPI_ISL_1557006, EPI_ISL_1557007, EPI_ISL_1557008, EPI_ISL_1557009, EPI_ISL_1557010, EPI_ISL_1557011, EPI_ISL_1557012, EPI_ISL_1557013, EPI_ISL_1557014, EPI_ISL_1557015, EPI_ISL_1557016, EPI_ISL_1557017, EPI_ISL_1557018, EPI_ISL_1557019, EPI_ISL_1557020, EPI_ISL_1557021, EPI_ISL_1557022, EPI_ISL_1557024, EPI_ISL_1557025, EPI_ISL_1557027, EPI_ISL_1557028, EPI_ISL_1557029, EPI_ISL_1557030, EPI_ISL_1557031, EPI_ISL_1557032, EPI_ISL_1557033, EPI_ISL_1557034, EPI_ISL_1557035, EPI_ISL_1557036, EPI_ISL_1557037, EPI_ISL_1557038, EPI_ISL_1557039, EPI_ISL_1557040, EPI_ISL_1557041, EPI_ISL_1557042, EPI_ISL_1557043, EPI_ISL_1557044, EPI_ISL_1557045, EPI_ISL_1557046, EPI_ISL_1557048, EPI_ISL_1557049, EPI_ISL_1557050, EPI_ISL_1557051, EPI_ISL_1557053, EPI_ISL_1557054, EPI_ISL_1557055, EPI_ISL_1557056, EPI_ISL_1557057, EPI_ISL_1557058, EPI_ISL_1557059, EPI_ISL_1557060, EPI_ISL_1557061, EPI_ISL_1557062, EPI_ISL_1557063, EPI_ISL_1557064, EPI_ISL_1557065, EPI_ISL_1557066, EPI_ISL_1557067, EPI_ISL_1557068, EPI_ISL_1557069, EPI_ISL_1557070, EPI_ISL_1557071, EPI_ISL_1557072, EPI_ISL_1557073, EPI_ISL_1557074, EPI_ISL_1557075, EPI_ISL_1557076, EPI_ISL_1557077, EPI_ISL_1557078, EPI_ISL_1557079, EPI_ISL_1557080, EPI_ISL_1557083, EPI_ISL_1557084, EPI_ISL_1557085, EPI_ISL_1557086, EPI_ISL_1557087, EPI_ISL_1557088, EPI_ISL_1557089, EPI_ISL_1557092, EPI_ISL_1557093, EPI_ISL_1557094, EPI_ISL_1557095, EPI_ISL_1557096, EPI_ISL_1557097, EPI_ISL_1557098, EPI_ISL_1557099, EPI_ISL_1557100, EPI_ISL_1557101, EPI_ISL_1557102, EPI_ISL_1557103, EPI_ISL_1557104, EPI_ISL_1557106, EPI_ISL_1557107, EPI_ISL_1557108, EPI_ISL_1557109, EPI_ISL_1557110, EPI_ISL_1557112, EPI_ISL_1557113, EPI_ISL_1557114, EPI_ISL_1557115, EPI_ISL_1557116, EPI_ISL_1557117, EPI_ISL_1557118, EPI_ISL_1557120, EPI_ISL_1557122, EPI_ISL_1557123, EPI_ISL_1557124, EPI_ISL_1557125, EPI_ISL_1557126, EPI_ISL_1557127, EPI_ISL_1557128, EPI_ISL_1557129, EPI_ISL_1557130, EPI_ISL_1557131, EPI_ISL_1557132, EPI_ISL_1557133, EPI_ISL_1557134, EPI_ISL_1557135, EPI_ISL_1557137, EPI_ISL_1557140, EPI_ISL_1557141, EPI_ISL_1557142, EPI_ISL_1557143, EPI_ISL_1557144, EPI_ISL_1557145, EPI_ISL_1557146, EPI_ISL_1557147, EPI_ISL_1557148, EPI_ISL_1557149, EPI_ISL_1557150, EPI_ISL_1557151, EPI_ISL_1557152, EPI_ISL_1557153, EPI_ISL_1557154, EPI_ISL_1557155, EPI_ISL_1557156, EPI_ISL_1557157, EPI_ISL_1557158, EPI_ISL_1557159, EPI_ISL_1557160, EPI_ISL_1557161, EPI_ISL_1557162, EPI_ISL_1557163, EPI_ISL_1557164, EPI_ISL_1557165, EPI_ISL_1557166, EPI_ISL_1557167, EPI_ISL_1557168, EPI_ISL_1557169, EPI_ISL_1557170, EPI_ISL_1557171, EPI_ISL_1557172, EPI_ISL_1557173, EPI_ISL_1557174, EPI_ISL_1557175, EPI_ISL_1557176, EPI_ISL_1557177, EPI_ISL_1557179, EPI_ISL_1557180, EPI_ISL_1557181, EPI_ISL_1557182, EPI_ISL_1557183, EPI_ISL_1557184, EPI_ISL_1557185, EPI_ISL_1557186, EPI_ISL_1557187, EPI_ISL_1557188, EPI_ISL_1557189, EPI_ISL_1557190, EPI_ISL_1557191, EPI_ISL_1557192, EPI_ISL_1557193, EPI_ISL_1557194, EPI_ISL_1557195, EPI_ISL_1557196, EPI_ISL_1557197, EPI_ISL_1557198, EPI_ISL_1557199, EPI_ISL_1557200, EPI_ISL_1557201, EPI_ISL_1557202, EPI_ISL_1557203, EPI_ISL_1557204, EPI_ISL_1557205, EPI_ISL_1557206, EPI_ISL_1557207, EPI_ISL_1557208, EPI_ISL_1557209, EPI_ISL_1557210, EPI_ISL_1557211, EPI_ISL_1557212, EPI_ISL_1557213, EPI_ISL_1557214, EPI_ISL_1557215, EPI_ISL_1557216, EPI_ISL_1557217, EPI_ISL_1557218

| | | | |
|---|---|---|---|
| see above | Fulgent Genetics | Centers for Disease Control and Prevention Division of Viral Diseases, Pathogen Discovery | Dakota Howard, Dhwani Batra, Peter W. Cook, Kara Moser, Adrian Paskey, Jason Caravas, Benjamin Rambo-Martin, Shatavia Morrison, Christopher Gulvick, Scott Sammons, Yvette Unoarumhi, Darlene Wagner, Matthew Schmerer, Harry Gao, Mickey Li, John Gao, Joseph Fierro, Benafsh Sapra, Becky Tsai, Yan Meng, Doreen Ng, James Xie, Clinton R. Paden, Duncan MacCannell |

EPI_ISL_1558716, EPI_ISL_1558717, EPI_ISL_1558718, EPI_ISL_1558719, EPI_ISL_1558720, EPI_ISL_1558721, EPI_ISL_1558722, EPI_ISL_1558723, EPI_ISL_1558724, EPI_ISL_1558725, EPI_ISL_1558726, EPI_ISL_1558727, EPI_ISL_1558728, EPI_ISL_1558729, EPI_ISL_1558730, EPI_ISL_1558731, EPI_ISL_1558732, EPI_ISL_1558733, EPI_ISL_1558734, EPI_ISL_1558735, EPI_ISL_1558736, EPI_ISL_1558737, EPI_ISL_1558738, EPI_ISL_1558739, EPI_ISL_1558740, EPI_ISL_1558741, EPI_ISL_1558742, EPI_ISL_1558743, EPI_ISL_1558744, EPI_ISL_1558745, EPI_ISL_1558746, EPI_ISL_1558748, EPI_ISL_1558749, EPI_ISL_1558750, EPI_ISL_1558751, EPI_ISL_1558752, EPI_ISL_1558753, EPI_ISL_1558754, EPI_ISL_1558755, EPI_ISL_1558756, EPI_ISL_1558757, EPI_ISL_1558758, EPI_ISL_1558759, EPI_ISL_1558760, EPI_ISL_1558761, EPI_ISL_1558762, EPI_ISL_1558763, EPI_ISL_1558764, EPI_ISL_1558765, EPI_ISL_1558766, EPI_ISL_1558767, EPI_ISL_1558768, EPI_ISL_1558769, EPI_ISL_1558770, EPI_ISL_1558771, EPI_ISL_1558772, EPI_ISL_1558773, EPI_ISL_1558774, EPI_ISL_1558775, EPI_ISL_1558776, EPI_ISL_1558777, EPI_ISL_1558778, EPI_ISL_1558779, EPI_ISL_1558780, EPI_ISL_1558781, EPI_ISL_1558782, EPI_ISL_1558783, EPI_ISL_1558784, EPI_ISL_1558785, EPI_ISL_1558786, EPI_ISL_1558787, EPI_ISL_1558788, EPI_ISL_1558789, EPI_ISL_1558790

| | | | |
|---|---|---|---|
| see above | Pathology and Laboratory Medicine Institute, Cleveland Clinic, Ohio, USA | Pathology and Laboratory Medicine Institute, Cleveland Clinic, Ohio, USA | Joy Nakitandwe, Zheng Jin Tu, Jay Brock, Yu-Wei Cheng, Gary Procop, Daniel Rhoads, Daniel H. Farkas, David Bosler |
| EPI_ISL_1558791, EPI_ISL_1558792, EPI_ISL_1558793 | LESP Chihuahua | Instituto de Diagnostico y Referencia Epidemiologicos (INDRE) | Claudia Wong-Arambula, Abril Rodriguez-Maldonado, Vanessa Rivero-Arredondo, Ariadna Medina-Benitez, Joaquin Quiroz-Mercado, Sergio Rangel-Guerrero, Natividad Cruz-Ortiz, Tatiana Nunez-Garcia, Gisela Barrera-Badillo, Lucia Hernandez-Rivas, Irma Lopez-Martinez, Ernesto Ramirez-Gonzalez. |
| EPI_ISL_1558794, EPI_ISL_1558795 | LESP Queretaro | Instituto de Diagnostico y Referencia Epidemiologicos (INDRE) | Claudia Wong-Arambula, Abril Rodriguez-Maldonado, Vanessa Rivero-Arredondo, Ariadna Medina-Benitez, Joaquin Quiroz-Mercado, Sergio Rangel-Guerrero, Natividad Cruz-Ortiz, Tatiana Nunez-Garcia, Gisela Barrera-Badillo, Lucia Hernandez-Rivas, Irma Lopez-Martinez, Ernesto Ramirez-Gonzalez. |
| EPI_ISL_1558796, EPI_ISL_1558797 | LESPZacatecas | Instituto de Diagnostico y Referencia Epidemiologicos (INDRE) | Claudia Wong-Arambula, Abril Rodriguez-Maldonado, Vanessa Rivero-Arredondo, Ariadna Medina-Benitez, Joaquin Quiroz-Mercado, Sergio Rangel-Guerrero, Natividad Cruz-Ortiz, Tatiana Nunez-Garcia, Gisela Barrera-Badillo, Lucia Hernandez-Rivas, Irma Lopez-Martinez, Ernesto Ramirez-Gonzalez. |
| EPI_ISL_1558798 | LESP Baja California | Instituto de Diagnostico y Referencia Epidemiologicos (INDRE) | Claudia Wong-Arambula, Abril Rodriguez-Maldonado, Vanessa Rivero-Arredondo, Ariadna Medina-Benitez, Joaquin Quiroz-Mercado, Sergio Rangel-Guerrero, Natividad Cruz-Ortiz, Tatiana Nunez-Garcia, Gisela Barrera-Badillo, Lucia Hernandez-Rivas, Irma Lopez-Martinez, Ernesto Ramirez-Gonzalez. |
| EPI_ISL_1558799, EPI_ISL_1558800 | LESP Jalisco | Instituto de Diagnostico y Referencia Epidemiologicos (INDRE) | Claudia Wong-Arambula, Abril Rodriguez-Maldonado, Vanessa Rivero-Arredondo, Ariadna Medina-Benitez, Joaquin Quiroz-Mercado, Sergio Rangel-Guerrero, Natividad Cruz-Ortiz, Tatiana Nunez-Garcia, Gisela Barrera-Badillo, Lucia Hernandez-Rivas, Irma Lopez-Martinez, Ernesto Ramirez-Gonzalez. |
| EPI_ISL_1558801 | LESP Baja California Sur | Instituto de Diagnostico y Referencia Epidemiologicos (INDRE) | Claudia Wong-Arambula, Abril Rodriguez-Maldonado, Vanessa Rivero-Arredondo, Ariadna Medina-Benitez, Joaquin Quiroz-Mercado, Sergio Rangel-Guerrero, Natividad Cruz-Ortiz, Tatiana Nunez-Garcia, Gisela Barrera-Badillo, Lucia Hernandez-Rivas, Irma Lopez-Martinez, Ernesto Ramirez-Gonzalez. |

EPI_ISL_1558802, EPI_ISL_1558803, EPI_ISL_1558804, EPI_ISL_1558805, EPI_ISL_1558806, EPI_ISL_1558807, EPI_ISL_1558808, EPI_ISL_1558809, EPI_ISL_1558810, EPI_ISL_1558811, EPI_ISL_1558812, EPI_ISL_1558813, EPI_ISL_1558814, EPI_ISL_1558815, EPI_ISL_1558817, EPI_ISL_1558818, EPI_ISL_1558819, EPI_ISL_1558820, EPI_ISL_1558821, EPI_ISL_1558822, EPI_ISL_1558823, EPI_ISL_1558824, EPI_ISL_1558825, EPI_ISL_1558826, EPI_ISL_1558827

| | | | |
|---|---|---|---|
| see above | Instituto Nacional de Medicina Genomica | Instituto Nacional de Medicina Genomica | Hidalgo-Miranda A, Mendoza-Vargas A, Reyes-Grajeda JP, Cedro-Tanda A, Alcaraz N, Gonzalez-Barrera D, Rangel-DeLeon D, Miranda-Ortiz H, Rosas-Escobar P, Canseco Mendez JC, Ramirez-Vega O, Munguia-Garza P, Garcia-Cardenas FJ, Gonzalez-Woge MA, Herrera-Montalvo LA |
| EPI_ISL_1558828, EPI_ISL_1558829, EPI_ISL_1558830, EPI_ISL_1558831, EPI_ISL_1558832, EPI_ISL_1558833 | LESP Jalisco/UdeG | Instituto de Diagnostico y Referencia Epidemiologicos (INDRE) | Claudia Wong-Arambula, Abril Rodriguez-Maldonado, Vanessa Rivero-Arredondo, Ariadna Medina-Benitez, Joaquin Quiroz-Mercado, Sergio Rangel-Guerrero, Natividad Cruz-Ortiz, Tatiana Nunez-Garcia, Gisela Barrera-Badillo, Lucia Hernandez-Rivas, Irma Lopez-Martinez, Ernesto Ramirez-Gonzalez. |

EPI_ISL_1559333, EPI_ISL_1559335, EPI_ISL_1559336, EPI_ISL_1559337, EPI_ISL_1559338, EPI_ISL_1559339, EPI_ISL_1559340, EPI_ISL_1559341, EPI_ISL_1559343, EPI_ISL_1559344, EPI_ISL_1559345, EPI_ISL_1559346, EPI_ISL_1559347, EPI_ISL_1559348, EPI_ISL_1559349, EPI_ISL_1559350, EPI_ISL_1559351, EPI_ISL_1559354, EPI_ISL_1559355, EPI_ISL_1559359, EPI_ISL_1559360, EPI_ISL_1559361, EPI_ISL_1559362, EPI_ISL_1559363, EPI_ISL_1559364, EPI_ISL_1559365, EPI_ISL_1559366, EPI_ISL_1559367, EPI_ISL_1559368

| | | | |
|---|---|---|---|
| see above | NJDOH, Public Health and Environmental Laboratories | NJ_PHEL | Lindsey Bodnar, Shiv K. Verma, Jacquelyn Deverell, Matthew Scarnati, Mohammad M Ali, Dana Woell, Byeong Jeong |
| EPI_ISL_1559385 | Atlanta VA Medical Center | Genomics and Discovery, Respiratory Viruses Branch, Division of Viral Diseases, Centers for Disease Control and Prevention | Yan Li, Ying Tao, Anna Kelleher, Jing Zhang, Anna Montmayeur, Brian Lynch, Krista Queen, Anna Uehara, Peter Cook, Han Jia Justin Ng, Rachel Marine, Clinton R. Paden, Haibin Wang, Mark Burroughs, Justin Lee, Adam Retchless, Suxiang Tong |
| EPI_ISL_1559386, EPI_ISL_1559387 | Kaiser Permanente Washington Health Research Institute | Genomics and Discovery, Respiratory Viruses Branch, Division of Viral Diseases, Centers for Disease Control and Prevention | Yan Li, Ying Tao, Anna Kelleher, Jing Zhang, Anna Montmayeur, Brian Lynch, Krista Queen, Anna Uehara, Peter Cook, Han Jia Justin Ng, Rachel Marine, Clinton R. Paden, Haibin Wang, Mark Burroughs, Justin Lee, Adam Retchless, Suxiang Tong |

EPI_ISL_1559388, EPI_ISL_1559389, EPI_ISL_1559390, EPI_ISL_1559391, EPI_ISL_1559392, EPI_ISL_1559393, EPI_ISL_1559394, EPI_ISL_1559395, EPI_ISL_1559396, EPI_ISL_1559397, EPI_ISL_1559398, EPI_ISL_1559399, EPI_ISL_1559400, EPI_ISL_1559401, EPI_ISL_1559402, EPI_ISL_1559403, EPI_ISL_1559404, EPI_ISL_1559405, EPI_ISL_1559406, EPI_ISL_1559407, EPI_ISL_1559408, EPI_ISL_1559409,

EPI_ISL_1559410

| | | | |
|---|---|---|---|
| see above | Washington State Department of Health Public Health Laboratories | Washington State Department of Health Public Health Laboratories | Drew MacKellar, Philip Dykema, Denny Russell, Joenice Gonzalez, Hannah Gray, Geoff Melly, Vanessa De Los Santos, Darren Lucas, JohnAric Peterson, Avi Singh, Rebecca Cao |

EPI_ISL_1559411, EPI_ISL_1559412, EPI_ISL_1559413, EPI_ISL_1559414, EPI_ISL_1559415, EPI_ISL_1559416, EPI_ISL_1559417, EPI_ISL_1559418, EPI_ISL_1559419, EPI_ISL_1559420, EPI_ISL_1559421, EPI_ISL_1559422, EPI_ISL_1559423, EPI_ISL_1559424, EPI_ISL_1559425, EPI_ISL_1559426, EPI_ISL_1559427, EPI_ISL_1559428, EPI_ISL_1559429, EPI_ISL_1559430, EPI_ISL_1559431, EPI_ISL_1559432,
EPI_ISL_1559433, EPI_ISL_1559434, EPI_ISL_1559435, EPI_ISL_1559436, EPI_ISL_1559437, EPI_ISL_1559438, EPI_ISL_1559439, EPI_ISL_1559440, EPI_ISL_1559441, EPI_ISL_1559442, EPI_ISL_1559443, EPI_ISL_1559444, EPI_ISL_1559445, EPI_ISL_1559446, EPI_ISL_1559447, EPI_ISL_1559448, EPI_ISL_1559449, EPI_ISL_1559450, EPI_ISL_1559451, EPI_ISL_1559452, EPI_ISL_1559453, EPI_ISL_1559454,
EPI_ISL_1559455, EPI_ISL_1559456, EPI_ISL_1559457, EPI_ISL_1559458, EPI_ISL_1559459, EPI_ISL_1559460, EPI_ISL_1559461, EPI_ISL_1559462, EPI_ISL_1559463, EPI_ISL_1559464, EPI_ISL_1559465, EPI_ISL_1559466, EPI_ISL_1559467, EPI_ISL_1559468, EPI_ISL_1559469, EPI_ISL_1559470, EPI_ISL_1559471, EPI_ISL_1559472, EPI_ISL_1559473, EPI_ISL_1559474, EPI_ISL_1559475, EPI_ISL_1559476,
EPI_ISL_1559477, EPI_ISL_1559478, EPI_ISL_1559479, EPI_ISL_1559480, EPI_ISL_1559481, EPI_ISL_1559482, EPI_ISL_1559483, EPI_ISL_1559484, EPI_ISL_1559485, EPI_ISL_1559486, EPI_ISL_1559487, EPI_ISL_1559488, EPI_ISL_1559489, EPI_ISL_1559490, EPI_ISL_1559491, EPI_ISL_1559492, EPI_ISL_1559493, EPI_ISL_1559494, EPI_ISL_1559495, EPI_ISL_1559496, EPI_ISL_1559497, EPI_ISL_1559498,
EPI_ISL_1559499, EPI_ISL_1559500, EPI_ISL_1559501, EPI_ISL_1559502, EPI_ISL_1559503, EPI_ISL_1559504, EPI_ISL_1559505, EPI_ISL_1559506, EPI_ISL_1559507, EPI_ISL_1559508, EPI_ISL_1559509, EPI_ISL_1559510, EPI_ISL_1559511, EPI_ISL_1559512, EPI_ISL_1559513, EPI_ISL_1559514, EPI_ISL_1559515, EPI_ISL_1559516, EPI_ISL_1559517, EPI_ISL_1559518, EPI_ISL_1559519, EPI_ISL_1559520,
EPI_ISL_1559521, EPI_ISL_1559522, EPI_ISL_1559523, EPI_ISL_1559524, EPI_ISL_1559525, EPI_ISL_1559526, EPI_ISL_1559527, EPI_ISL_1559528, EPI_ISL_1559529, EPI_ISL_1559530, EPI_ISL_1559531, EPI_ISL_1559532, EPI_ISL_1559533, EPI_ISL_1559534, EPI_ISL_1559535, EPI_ISL_1559536, EPI_ISL_1559537, EPI_ISL_1559538, EPI_ISL_1559539, EPI_ISL_1559540, EPI_ISL_1559541, EPI_ISL_1559542,
EPI_ISL_1559543, EPI_ISL_1559544, EPI_ISL_1559545, EPI_ISL_1559546, EPI_ISL_1559547, EPI_ISL_1559548, EPI_ISL_1559549, EPI_ISL_1559550, EPI_ISL_1559551, EPI_ISL_1559552, EPI_ISL_1559553, EPI_ISL_1559554, EPI_ISL_1559555, EPI_ISL_1559556, EPI_ISL_1559557, EPI_ISL_1559558, EPI_ISL_1559559, EPI_ISL_1559560, EPI_ISL_1559561, EPI_ISL_1559562, EPI_ISL_1559563, EPI_ISL_1559564,
EPI_ISL_1559565, EPI_ISL_1559566, EPI_ISL_1559567, EPI_ISL_1559568, EPI_ISL_1559569, EPI_ISL_1559570, EPI_ISL_1559571, EPI_ISL_1559572, EPI_ISL_1559573, EPI_ISL_1559574, EPI_ISL_1559575, EPI_ISL_1559576, EPI_ISL_1559577, EPI_ISL_1559578, EPI_ISL_1559579, EPI_ISL_1559580, EPI_ISL_1559581, EPI_ISL_1559582, EPI_ISL_1559583, EPI_ISL_1559584, EPI_ISL_1559585, EPI_ISL_1559586,
EPI_ISL_1559587

| | | | |
|---|---|---|---|
| see above | Quest Diagnostics Incorporated | Centers for Disease Control and Prevention Division of Viral Diseases, Pathogen Discovery | Dakota Howard, Dhwani Batra, Peter W. Cook, Kara Moser, Adrian Paskey, Jason Caravas, Benjamin Rambo-Martin, Shatavia Morrison, Christopher Gulvick, Scott Sammons, Yvette Unoarumhi, Darlene Wagner, Matthew Schmerer, S. H. Rosenthal, A. Gerasimova, R. M. Kagan, B. Anderson, M. Hua, Y. Liu, L.E. Bernstein, K.E. Livingston, A. Perez, I. A. Shlyakhter, R. V. Rolando, R. Owen, P. Tanpaiboon, F. Lacbawan, Clinton R. Paden, Duncan MacCannell |
| EPI_ISL_1559588, EPI_ISL_1559590, EPI_ISL_1559591 | Florida Bureau of Public Health Laboratories | Florida Bureau of Public Health Laboratories | Sarah Schmedes, Jason Blanton |
| EPI_ISL_1559626 | Diagnostic Genomics Laboratory | Diagnostic Genomics Laboratory | Carolyn Banister, Fadi Abboodi, Ji (Emily) Hao, Diego Altomoare, Michael Shtutman, Phillip Buckhaults, Helmut Albrecht |

EPI_ISL_1559627, EPI_ISL_1559628, EPI_ISL_1559629, EPI_ISL_1559630, EPI_ISL_1559631, EPI_ISL_1559632, EPI_ISL_1559633, EPI_ISL_1559634, EPI_ISL_1559635, EPI_ISL_1559636, EPI_ISL_1559637, EPI_ISL_1559638, EPI_ISL_1559639, EPI_ISL_1559640, EPI_ISL_1559641, EPI_ISL_1559642, EPI_ISL_1559643, EPI_ISL_1559644, EPI_ISL_1559645, EPI_ISL_1559646, EPI_ISL_1559647, EPI_ISL_1559648,
EPI_ISL_1559649, EPI_ISL_1559650, EPI_ISL_1559651, EPI_ISL_1559652, EPI_ISL_1559653, EPI_ISL_1559654, EPI_ISL_1559655, EPI_ISL_1559656, EPI_ISL_1559657, EPI_ISL_1559658, EPI_ISL_1559659, EPI_ISL_1559660, EPI_ISL_1559661, EPI_ISL_1559662, EPI_ISL_1559663, EPI_ISL_1559664, EPI_ISL_1559665, EPI_ISL_1559666, EPI_ISL_1559667, EPI_ISL_1559668, EPI_ISL_1559669, EPI_ISL_1559670,
EPI_ISL_1559671, EPI_ISL_1559672, EPI_ISL_1559673, EPI_ISL_1559674, EPI_ISL_1559675, EPI_ISL_1559676, EPI_ISL_1559677, EPI_ISL_1559678, EPI_ISL_1559679, EPI_ISL_1559680, EPI_ISL_1559681, EPI_ISL_1559682, EPI_ISL_1559683, EPI_ISL_1559684, EPI_ISL_1559685, EPI_ISL_1559686, EPI_ISL_1559687, EPI_ISL_1559688, EPI_ISL_1559689, EPI_ISL_1559690, EPI_ISL_1559691, EPI_ISL_1559692,
EPI_ISL_1559693, EPI_ISL_1559694, EPI_ISL_1559695, EPI_ISL_1559696, EPI_ISL_1559697, EPI_ISL_1559698, EPI_ISL_1559699, EPI_ISL_1559700, EPI_ISL_1559701, EPI_ISL_1559702, EPI_ISL_1559703, EPI_ISL_1559704, EPI_ISL_1559705, EPI_ISL_1559706, EPI_ISL_1559707, EPI_ISL_1559708, EPI_ISL_1559709, EPI_ISL_1559710, EPI_ISL_1559711, EPI_ISL_1559712, EPI_ISL_1559713, EPI_ISL_1559714,
EPI_ISL_1559715, EPI_ISL_1559716, EPI_ISL_1559717, EPI_ISL_1559718, EPI_ISL_1559719, EPI_ISL_1559720, EPI_ISL_1559721, EPI_ISL_1559722, EPI_ISL_1559723, EPI_ISL_1559724, EPI_ISL_1559725, EPI_ISL_1559726, EPI_ISL_1559727, EPI_ISL_1559728, EPI_ISL_1559729, EPI_ISL_1559730, EPI_ISL_1559731, EPI_ISL_1559732, EPI_ISL_1559733, EPI_ISL_1559734, EPI_ISL_1559735, EPI_ISL_1559736,
EPI_ISL_1559737, EPI_ISL_1559738, EPI_ISL_1559739, EPI_ISL_1559741, EPI_ISL_1559742, EPI_ISL_1559743, EPI_ISL_1559744, EPI_ISL_1559745, EPI_ISL_1559746, EPI_ISL_1559747, EPI_ISL_1559748, EPI_ISL_1559749, EPI_ISL_1559750, EPI_ISL_1559751, EPI_ISL_1559752, EPI_ISL_1559753, EPI_ISL_1559754, EPI_ISL_1559755, EPI_ISL_1559756, EPI_ISL_1559757, EPI_ISL_1559758, EPI_ISL_1559759,
EPI_ISL_1559760, EPI_ISL_1559761, EPI_ISL_1559762, EPI_ISL_1559763, EPI_ISL_1559764, EPI_ISL_1559765, EPI_ISL_1559766, EPI_ISL_1559767, EPI_ISL_1559768, EPI_ISL_1559769, EPI_ISL_1559770, EPI_ISL_1559771, EPI_ISL_1559772, EPI_ISL_1559773, EPI_ISL_1559774, EPI_ISL_1559775, EPI_ISL_1559776, EPI_ISL_1559777, EPI_ISL_1559778, EPI_ISL_1559779, EPI_ISL_1559780, EPI_ISL_1559781,
EPI_ISL_1559782, EPI_ISL_1559783, EPI_ISL_1559784, EPI_ISL_1559785, EPI_ISL_1559786, EPI_ISL_1559787, EPI_ISL_1559788, EPI_ISL_1559789, EPI_ISL_1559790, EPI_ISL_1559791, EPI_ISL_1559792, EPI_ISL_1559793, EPI_ISL_1559794, EPI_ISL_1559795, EPI_ISL_1559796, EPI_ISL_1559797, EPI_ISL_1559798, EPI_ISL_1559799, EPI_ISL_1559800, EPI_ISL_1559801, EPI_ISL_1559802, EPI_ISL_1559803,
EPI_ISL_1559804, EPI_ISL_1559805, EPI_ISL_1559806, EPI_ISL_1559807, EPI_ISL_1559808, EPI_ISL_1559809, EPI_ISL_1559810, EPI_ISL_1559811, EPI_ISL_1559812, EPI_ISL_1559813, EPI_ISL_1559814, EPI_ISL_1559815, EPI_ISL_1559816, EPI_ISL_1559817, EPI_ISL_1559818, EPI_ISL_1559819, EPI_ISL_1559820, EPI_ISL_1559821, EPI_ISL_1559822, EPI_ISL_1559823, EPI_ISL_1559824, EPI_ISL_1559825,
EPI_ISL_1559826, EPI_ISL_1559827, EPI_ISL_1559828, EPI_ISL_1559829, EPI_ISL_1559830, EPI_ISL_1559831, EPI_ISL_1559832, EPI_ISL_1559833, EPI_ISL_1559834, EPI_ISL_1559835, EPI_ISL_1559836, EPI_ISL_1559837, EPI_ISL_1559838, EPI_ISL_1559839, EPI_ISL_1559840, EPI_ISL_1559841, EPI_ISL_1559842, EPI_ISL_1559843, EPI_ISL_1559844, EPI_ISL_1559845, EPI_ISL_1559846, EPI_ISL_1559847,
EPI_ISL_1559848, EPI_ISL_1559849, EPI_ISL_1559850, EPI_ISL_1559851, EPI_ISL_1559852, EPI_ISL_1559853, EPI_ISL_1559854, EPI_ISL_1559855, EPI_ISL_1559856, EPI_ISL_1559857, EPI_ISL_1559858, EPI_ISL_1559859, EPI_ISL_1559860, EPI_ISL_1559861, EPI_ISL_1559862, EPI_ISL_1559863, EPI_ISL_1559864, EPI_ISL_1559865, EPI_ISL_1559866, EPI_ISL_1559867, EPI_ISL_1559868, EPI_ISL_1559869,
EPI_ISL_1559870, EPI_ISL_1559871, EPI_ISL_1559872, EPI_ISL_1559873, EPI_ISL_1559874, EPI_ISL_1559875, EPI_ISL_1559876, EPI_ISL_1559877, EPI_ISL_1559878, EPI_ISL_1559879, EPI_ISL_1559880, EPI_ISL_1559881, EPI_ISL_1559882, EPI_ISL_1559883, EPI_ISL_1559884, EPI_ISL_1559885, EPI_ISL_1559886, EPI_ISL_1559887, EPI_ISL_1559888, EPI_ISL_1559889, EPI_ISL_1559890, EPI_ISL_1559891,
EPI_ISL_1559892, EPI_ISL_1559893, EPI_ISL_1559894, EPI_ISL_1559895, EPI_ISL_1559896, EPI_ISL_1559897, EPI_ISL_1559898, EPI_ISL_1559899, EPI_ISL_1559900, EPI_ISL_1559901, EPI_ISL_1559902, EPI_ISL_1559903, EPI_ISL_1559904, EPI_ISL_1559905, EPI_ISL_1559906, EPI_ISL_1559907, EPI_ISL_1559908, EPI_ISL_1559909, EPI_ISL_1559910, EPI_ISL_1559911, EPI_ISL_1559912, EPI_ISL_1559913,
EPI_ISL_1559914, EPI_ISL_1559915, EPI_ISL_1559916, EPI_ISL_1559917, EPI_ISL_1559918, EPI_ISL_1559919, EPI_ISL_1559920, EPI_ISL_1559921, EPI_ISL_1559922, EPI_ISL_1559923, EPI_ISL_1559924, EPI_ISL_1559925, EPI_ISL_1559926, EPI_ISL_1559927, EPI_ISL_1559928, EPI_ISL_1559929, EPI_ISL_1559930, EPI_ISL_1559931, EPI_ISL_1559932, EPI_ISL_1559933, EPI_ISL_1559934, EPI_ISL_1559935,
EPI_ISL_1559936, EPI_ISL_1559937, EPI_ISL_1559938, EPI_ISL_1559939, EPI_ISL_1559940, EPI_ISL_1559941, EPI_ISL_1559942, EPI_ISL_1559943, EPI_ISL_1559944, EPI_ISL_1559945, EPI_ISL_1559946, EPI_ISL_1559947, EPI_ISL_1559948, EPI_ISL_1559949, EPI_ISL_1559950, EPI_ISL_1559951, EPI_ISL_1559952, EPI_ISL_1559953, EPI_ISL_1559954, EPI_ISL_1559955, EPI_ISL_1559956, EPI_ISL_1559958,
EPI_ISL_1559959, EPI_ISL_1559960, EPI_ISL_1559961, EPI_ISL_1559962, EPI_ISL_1559963, EPI_ISL_1559964, EPI_ISL_1559965, EPI_ISL_1559967, EPI_ISL_1559968, EPI_ISL_1559969, EPI_ISL_1559970, EPI_ISL_1559971, EPI_ISL_1559972, EPI_ISL_1559973, EPI_ISL_1559974, EPI_ISL_1559975, EPI_ISL_1559976, EPI_ISL_1559977, EPI_ISL_1559978, EPI_ISL_1559979, EPI_ISL_1559980, EPI_ISL_1559981,
EPI_ISL_1559982, EPI_ISL_1559983, EPI_ISL_1559984, EPI_ISL_1559985, EPI_ISL_1559986, EPI_ISL_1559987, EPI_ISL_1559988, EPI_ISL_1559989, EPI_ISL_1559990, EPI_ISL_1559991, EPI_ISL_1559992, EPI_ISL_1559993, EPI_ISL_1559994, EPI_ISL_1559995, EPI_ISL_1559996, EPI_ISL_1559997, EPI_ISL_1559998, EPI_ISL_1559999, EPI_ISL_1560000, EPI_ISL_1560001, EPI_ISL_1560002, EPI_ISL_1560003,
EPI_ISL_1560004, EPI_ISL_1560005, EPI_ISL_1560007, EPI_ISL_1560008, EPI_ISL_1560009, EPI_ISL_1560010, EPI_ISL_1560011, EPI_ISL_1560012, EPI_ISL_1560013, EPI_ISL_1560014, EPI_ISL_1560015, EPI_ISL_1560016, EPI_ISL_1560017, EPI_ISL_1560018, EPI_ISL_1560019, EPI_ISL_1560020, EPI_ISL_1560021, EPI_ISL_1560022, EPI_ISL_1560023, EPI_ISL_1560024, EPI_ISL_1560025, EPI_ISL_1560026,
EPI_ISL_1560027, EPI_ISL_1560028, EPI_ISL_1560029, EPI_ISL_1560030, EPI_ISL_1560031, EPI_ISL_1560032, EPI_ISL_1560033, EPI_ISL_1560034, EPI_ISL_1560035, EPI_ISL_1560036, EPI_ISL_1560037, EPI_ISL_1560038, EPI_ISL_1560039, EPI_ISL_1560040, EPI_ISL_1560041, EPI_ISL_1560042, EPI_ISL_1560043, EPI_ISL_1560044, EPI_ISL_1560045, EPI_ISL_1560046, EPI_ISL_1560047, EPI_ISL_1560048,
EPI_ISL_1560049, EPI_ISL_1560050, EPI_ISL_1560051, EPI_ISL_1560052, EPI_ISL_1560053, EPI_ISL_1560054, EPI_ISL_1560055, EPI_ISL_1560056, EPI_ISL_1560057, EPI_ISL_1560058, EPI_ISL_1560059, EPI_ISL_1560060, EPI_ISL_1560061, EPI_ISL_1560062, EPI_ISL_1560063, EPI_ISL_1560064, EPI_ISL_1560065, EPI_ISL_1560066, EPI_ISL_1560067, EPI_ISL_1560068, EPI_ISL_1560069, EPI_ISL_1560070,
EPI_ISL_1560071, EPI_ISL_1560072, EPI_ISL_1560073, EPI_ISL_1560074, EPI_ISL_1560075, EPI_ISL_1560076, EPI_ISL_1560078, EPI_ISL_1560079, EPI_ISL_1560080, EPI_ISL_1560081, EPI_ISL_1560082, EPI_ISL_1560083, EPI_ISL_1560084, EPI_ISL_1560085, EPI_ISL_1560086, EPI_ISL_1560087, EPI_ISL_1560088, EPI_ISL_1560089, EPI_ISL_1560090, EPI_ISL_1560091, EPI_ISL_1560092, EPI_ISL_1560093,
EPI_ISL_1560094, EPI_ISL_1560095, EPI_ISL_1560096, EPI_ISL_1560097, EPI_ISL_1560098, EPI_ISL_1560099, EPI_ISL_1560100, EPI_ISL_1560101, EPI_ISL_1560102, EPI_ISL_1560104, EPI_ISL_1560105, EPI_ISL_1560106, EPI_ISL_1560107, EPI_ISL_1560108, EPI_ISL_1560109, EPI_ISL_1560110, EPI_ISL_1560111, EPI_ISL_1560112, EPI_ISL_1560114, EPI_ISL_1560115, EPI_ISL_1560116, EPI_ISL_1560117,
EPI_ISL_1560118, EPI_ISL_1560119, EPI_ISL_1560120, EPI_ISL_1560121, EPI_ISL_1560122, EPI_ISL_1560123, EPI_ISL_1560124, EPI_ISL_1560125, EPI_ISL_1560126, EPI_ISL_1560127, EPI_ISL_1560128, EPI_ISL_1560129, EPI_ISL_1560130, EPI_ISL_1560131, EPI_ISL_1560132, EPI_ISL_1560133, EPI_ISL_1560134, EPI_ISL_1560136, EPI_ISL_1560137, EPI_ISL_1560138, EPI_ISL_1560139, EPI_ISL_1560140,
EPI_ISL_1560141, EPI_ISL_1560142, EPI_ISL_1560143, EPI_ISL_1560144, EPI_ISL_1560145, EPI_ISL_1560146, EPI_ISL_1560147, EPI_ISL_1560148, EPI_ISL_1560149, EPI_ISL_1560150, EPI_ISL_1560151, EPI_ISL_1560152, EPI_ISL_1560153, EPI_ISL_1560154, EPI_ISL_1560155, EPI_ISL_1560156, EPI_ISL_1560157, EPI_ISL_1560158, EPI_ISL_1560159, EPI_ISL_1560160, EPI_ISL_1560161, EPI_ISL_1560163,
EPI_ISL_1560164, EPI_ISL_1560165, EPI_ISL_1560166, EPI_ISL_1560167, EPI_ISL_1560168, EPI_ISL_1560169, EPI_ISL_1560170, EPI_ISL_1560171, EPI_ISL_1560172, EPI_ISL_1560173, EPI_ISL_1560174, EPI_ISL_1560176, EPI_ISL_1560177, EPI_ISL_1560178, EPI_ISL_1560179, EPI_ISL_1560180, EPI_ISL_1560181, EPI_ISL_1560182, EPI_ISL_1560183, EPI_ISL_1560184, EPI_ISL_1560185, EPI_ISL_1560186,
EPI_ISL_1560187, EPI_ISL_1560188, EPI_ISL_1560189, EPI_ISL_1560190, EPI_ISL_1560191, EPI_ISL_1560192, EPI_ISL_1560193, EPI_ISL_1560194, EPI_ISL_1560195, EPI_ISL_1560196, EPI_ISL_1560197, EPI_ISL_1560198, EPI_ISL_1560199, EPI_ISL_1560200, EPI_ISL_1560201, EPI_ISL_1560202, EPI_ISL_1560203, EPI_ISL_1560204, EPI_ISL_1560205, EPI_ISL_1560206, EPI_ISL_1560207, EPI_ISL_1560208,
EPI_ISL_1560209, EPI_ISL_1560210, EPI_ISL_1560211, EPI_ISL_1560212, EPI_ISL_1560213, EPI_ISL_1560214, EPI_ISL_1560215, EPI_ISL_1560216, EPI_ISL_1560217, EPI_ISL_1560218, EPI_ISL_1560219, EPI_ISL_1560220, EPI_ISL_1560221, EPI_ISL_1560222, EPI_ISL_1560223, EPI_ISL_1560224, EPI_ISL_1560225, EPI_ISL_1560226, EPI_ISL_1560227, EPI_ISL_1560228, EPI_ISL_1560229, EPI_ISL_1560230,
EPI_ISL_1560231, EPI_ISL_1560232, EPI_ISL_1560233, EPI_ISL_1560234, EPI_ISL_1560235, EPI_ISL_1560236, EPI_ISL_1560237, EPI_ISL_1560238, EPI_ISL_1560239, EPI_ISL_1560240, EPI_ISL_1560241, EPI_ISL_1560242, EPI_ISL_1560243, EPI_ISL_1560244, EPI_ISL_1560245, EPI_ISL_1560246, EPI_ISL_1560247, EPI_ISL_1560248, EPI_ISL_1560249, EPI_ISL_1560250, EPI_ISL_1560251, EPI_ISL_1560252,
EPI_ISL_1560253, EPI_ISL_1560254, EPI_ISL_1560255, EPI_ISL_1560256, EPI_ISL_1560257, EPI_ISL_1560258, EPI_ISL_1560259, EPI_ISL_1560260, EPI_ISL_1560261, EPI_ISL_1560262, EPI_ISL_1560263, EPI_ISL_1560264, EPI_ISL_1560265, EPI_ISL_1560266, EPI_ISL_1560267, EPI_ISL_1560268, EPI_ISL_1560269, EPI_ISL_1560270, EPI_ISL_1560272, EPI_ISL_1560273, EPI_ISL_1560274, EPI_ISL_1560275,
EPI_ISL_1560276, EPI_ISL_1560277, EPI_ISL_1560278, EPI_ISL_1560279, EPI_ISL_1560280, EPI_ISL_1560281, EPI_ISL_1560283, EPI_ISL_1560284, EPI_ISL_1560285, EPI_ISL_1560286, EPI_ISL_1560287, EPI_ISL_1560288, EPI_ISL_1560289, EPI_ISL_1560291, EPI_ISL_1560292, EPI_ISL_1560293, EPI_ISL_1560294, EPI_ISL_1560295, EPI_ISL_1560296, EPI_ISL_1560297, EPI_ISL_1560298, EPI_ISL_1560299,
EPI_ISL_1560300, EPI_ISL_1560302, EPI_ISL_1560303, EPI_ISL_1560304, EPI_ISL_1560305, EPI_ISL_1560306, EPI_ISL_1560307, EPI_ISL_1560308, EPI_ISL_1560309, EPI_ISL_1560310, EPI_ISL_1560311, EPI_ISL_1560312, EPI_ISL_1560313, EPI_ISL_1560314, EPI_ISL_1560315, EPI_ISL_1560316, EPI_ISL_1560317, EPI_ISL_1560318, EPI_ISL_1560319, EPI_ISL_1560320, EPI_ISL_1560321, EPI_ISL_1560322,
EPI_ISL_1560323, EPI_ISL_1560324, EPI_ISL_1560325, EPI_ISL_1560326, EPI_ISL_1560327, EPI_ISL_1560328, EPI_ISL_1560329, EPI_ISL_1560330, EPI_ISL_1560331, EPI_ISL_1560332, EPI_ISL_1560333, EPI_ISL_1560334, EPI_ISL_1560335, EPI_ISL_1560336, EPI_ISL_1560337, EPI_ISL_1560338, EPI_ISL_1560339, EPI_ISL_1560340, EPI_ISL_1560341, EPI_ISL_1560342, EPI_ISL_1560343, EPI_ISL_1560344,
EPI_ISL_1560345, EPI_ISL_1560346, EPI_ISL_1560347, EPI_ISL_1560348, EPI_ISL_1560349, EPI_ISL_1560350, EPI_ISL_1560351, EPI_ISL_1560352, EPI_ISL_1560353, EPI_ISL_1560354, EPI_ISL_1560355, EPI_ISL_1560356, EPI_ISL_1560357, EPI_ISL_1560358, EPI_ISL_1560359, EPI_ISL_1560360, EPI_ISL_1560361, EPI_ISL_1560362, EPI_ISL_1560363, EPI_ISL_1560364, EPI_ISL_1560365, EPI_ISL_1560366,
EPI_ISL_1560367, EPI_ISL_1560368, EPI_ISL_1560369, EPI_ISL_1560370, EPI_ISL_1560371, EPI_ISL_1560372, EPI_ISL_1560373, EPI_ISL_1560374, EPI_ISL_1560375, EPI_ISL_1560376, EPI_ISL_1560377, EPI_ISL_1560378, EPI_ISL_1560379, EPI_ISL_1560380, EPI_ISL_1560381, EPI_ISL_1560382, EPI_ISL_1560383, EPI_ISL_1560384, EPI_ISL_1560385, EPI_ISL_1560386, EPI_ISL_1560387, EPI_ISL_1560388,
EPI_ISL_1560389, EPI_ISL_1560390, EPI_ISL_1560392, EPI_ISL_1560393, EPI_ISL_1560394, EPI_ISL_1560395, EPI_ISL_1560396, EPI_ISL_1560397, EPI_ISL_1560398, EPI_ISL_1560399, EPI_ISL_1560400, EPI_ISL_1560401, EPI_ISL_1560402, EPI_ISL_1560403, EPI_ISL_1560404, EPI_ISL_1560405, EPI_ISL_1560406, EPI_ISL_1560407, EPI_ISL_1560408, EPI_ISL_1560409, EPI_ISL_1560410, EPI_ISL_1560411,
EPI_ISL_1560412, EPI_ISL_1560413, EPI_ISL_1560414, EPI_ISL_1560415, EPI_ISL_1560416, EPI_ISL_1560417, EPI_ISL_1560418, EPI_ISL_1560419, EPI_ISL_1560420, EPI_ISL_1560421, EPI_ISL_1560422, EPI_ISL_1560423, EPI_ISL_1560424, EPI_ISL_1560425, EPI_ISL_1560426, EPI_ISL_1560427, EPI_ISL_1560428, EPI_ISL_1560429, EPI_ISL_1560430, EPI_ISL_1560431, EPI_ISL_1560432, EPI_ISL_1560434,
EPI_ISL_1560435, EPI_ISL_1560436, EPI_ISL_1560437, EPI_ISL_1560438, EPI_ISL_1560439, EPI_ISL_1560440, EPI_ISL_1560441, EPI_ISL_1560442, EPI_ISL_1560444, EPI_ISL_1560445, EPI_ISL_1560446, EPI_ISL_1560447, EPI_ISL_1560448, EPI_ISL_1560449, EPI_ISL_1560450, EPI_ISL_1560451, EPI_ISL_1560452, EPI_ISL_1560453, EPI_ISL_1560454, EPI_ISL_1560455, EPI_ISL_1560456, EPI_ISL_1560457,
EPI_ISL_1560458, EPI_ISL_1560459, EPI_ISL_1560460, EPI_ISL_1560461, EPI_ISL_1560462, EPI_ISL_1560463, EPI_ISL_1560464, EPI_ISL_1560465, EPI_ISL_1560466, EPI_ISL_1560467, EPI_ISL_1560468, EPI_ISL_1560469, EPI_ISL_1560470, EPI_ISL_1560471, EPI_ISL_1560472, EPI_ISL_1560473, EPI_ISL_1560474, EPI_ISL_1560475, EPI_ISL_1560476, EPI_ISL_1560477, EPI_ISL_1560478, EPI_ISL_1560479,
EPI_ISL_1560480, EPI_ISL_1560481, EPI_ISL_1560482, EPI_ISL_1560483, EPI_ISL_1560484, EPI_ISL_1560485, EPI_ISL_1560486, EPI_ISL_1560487, EPI_ISL_1560488, EPI_ISL_1560489, EPI_ISL_1560490, EPI_ISL_1560491, EPI_ISL_1560492, EPI_ISL_1560493, EPI_ISL_1560494, EPI_ISL_1560495, EPI_ISL_1560496, EPI_ISL_1560497, EPI_ISL_1560498, EPI_ISL_1560499, EPI_ISL_1560500, EPI_ISL_1560501,
EPI_ISL_1560502, EPI_ISL_1560503, EPI_ISL_1560504, EPI_ISL_1560505, EPI_ISL_1560506, EPI_ISL_1560507, EPI_ISL_1560508, EPI_ISL_1560509, EPI_ISL_1560510, EPI_ISL_1560511, EPI_ISL_1560512, EPI_ISL_1560513, EPI_ISL_1560514, EPI_ISL_1560515, EPI_ISL_1560516, EPI_ISL_1560517, EPI_ISL_1560518, EPI_ISL_1560519, EPI_ISL_1560520, EPI_ISL_1560521, EPI_ISL_1560522, EPI_ISL_1560523,
EPI_ISL_1560524, EPI_ISL_1560525, EPI_ISL_1560526, EPI_ISL_1560527, EPI_ISL_1560528, EPI_ISL_1560529, EPI_ISL_1560530, EPI_ISL_1560531, EPI_ISL_1560532, EPI_ISL_1560533, EPI_ISL_1560534, EPI_ISL_1560535, EPI_ISL_1560536, EPI_ISL_1560537, EPI_ISL_1560538, EPI_ISL_1560539, EPI_ISL_1560540, EPI_ISL_1560541, EPI_ISL_1560542, EPI_ISL_1560543, EPI_ISL_1560544, EPI_ISL_1560545,
EPI_ISL_1560546, EPI_ISL_1560547, EPI_ISL_1560548, EPI_ISL_1560549, EPI_ISL_1560550, EPI_ISL_1560551, EPI_ISL_1560552, EPI_ISL_1560553, EPI_ISL_1560554, EPI_ISL_1560555, EPI_ISL_1560556, EPI_ISL_1560557, EPI_ISL_1560558, EPI_ISL_1560559, EPI_ISL_1560560, EPI_ISL_1560561, EPI_ISL_1560562, EPI_ISL_1560563, EPI_ISL_1560564, EPI_ISL_1560565, EPI_ISL_1560566, EPI_ISL_1560567,
EPI_ISL_1560568, EPI_ISL_1560569, EPI_ISL_1560570, EPI_ISL_1560571, EPI_ISL_1560572, EPI_ISL_1560573, EPI_ISL_1560574, EPI_ISL_1560575, EPI_ISL_1560576, EPI_ISL_1560577, EPI_ISL_1560578, EPI_ISL_1560579, EPI_ISL_1560580, EPI_ISL_1560581, EPI_ISL_1560582, EPI_ISL_1560583, EPI_ISL_1560584, EPI_ISL_1560585, EPI_ISL_1560586, EPI_ISL_1560587, EPI_ISL_1560588, EPI_ISL_1560589,
EPI_ISL_1560590, EPI_ISL_1560591, EPI_ISL_1560592, EPI_ISL_1560593, EPI_ISL_1560594, EPI_ISL_1560595, EPI_ISL_1560596, EPI_ISL_1560597, EPI_ISL_1560598, EPI_ISL_1560599, EPI_ISL_1560600, EPI_ISL_1560601, EPI_ISL_1560602, EPI_ISL_1560603, EPI_ISL_1560604, EPI_ISL_1560605, EPI_ISL_1560606, EPI_ISL_1560607, EPI_ISL_1560608, EPI_ISL_1560609, EPI_ISL_1560610, EPI_ISL_1560611,
EPI_ISL_1560612, EPI_ISL_1560613, EPI_ISL_1560614, EPI_ISL_1560615, EPI_ISL_1560616, EPI_ISL_1560617, EPI_ISL_1560618, EPI_ISL_1560619, EPI_ISL_1560620, EPI_ISL_1560621, EPI_ISL_1560622, EPI_ISL_1560623, EPI_ISL_1560624, EPI_ISL_1560625, EPI_ISL_1560627, EPI_ISL_1560628, EPI_ISL_1560629, EPI_ISL_1560630, EPI_ISL_1560631, EPI_ISL_1560632, EPI_ISL_1560633, EPI_ISL_1560634,
EPI_ISL_1560635, EPI_ISL_1560636, EPI_ISL_1560637, EPI_ISL_1560638, EPI_ISL_1560639, EPI_ISL_1560640, EPI_ISL_1560641, EPI_ISL_1560642, EPI_ISL_1560643, EPI_ISL_1560644, EPI_ISL_1560645, EPI_ISL_1560646, EPI_ISL_1560647, EPI_ISL_1560648, EPI_ISL_1560649, EPI_ISL_1560650, EPI_ISL_1560651, EPI_ISL_1560652, EPI_ISL_1560653, EPI_ISL_1560654, EPI_ISL_1560655, EPI_ISL_1560656,
EPI_ISL_1560657, EPI_ISL_1560658, EPI_ISL_1560659, EPI_ISL_1560660, EPI_ISL_1560661, EPI_ISL_1560662, EPI_ISL_1560663, EPI_ISL_1560664, EPI_ISL_1560665, EPI_ISL_1560666, EPI_ISL_1560668, EPI_ISL_1560669, EPI_ISL_1560670, EPI_ISL_1560671, EPI_ISL_1560672, EPI_ISL_1560673, EPI_ISL_1560674, EPI_ISL_1560675, EPI_ISL_1560676, EPI_ISL_1560677, EPI_ISL_1560679, EPI_ISL_1560680,
EPI_ISL_1560681, EPI_ISL_1560682, EPI_ISL_1560683, EPI_ISL_1560684, EPI_ISL_1560685, EPI_ISL_1560686, EPI_ISL_1560687, EPI_ISL_1560688, EPI_ISL_1560689, EPI_ISL_1560690, EPI_ISL_1560691, EPI_ISL_1560692, EPI_ISL_1560693, EPI_ISL_1560694, EPI_ISL_1560695, EPI_ISL_1560696, EPI_ISL_1560697, EPI_ISL_1560698, EPI_ISL_1560699, EPI_ISL_1560700, EPI_ISL_1560701, EPI_ISL_1560702,
EPI_ISL_1560703, EPI_ISL_1560704, EPI_ISL_1560705, EPI_ISL_1560706, EPI_ISL_1560707, EPI_ISL_1560708, EPI_ISL_1560709, EPI_ISL_1560710, EPI_ISL_1560711, EPI_ISL_1560712, EPI_ISL_1560713, EPI_ISL_1560714, EPI_ISL_1560715, EPI_ISL_1560716, EPI_ISL_1560717, EPI_ISL_1560718, EPI_ISL_1560719, EPI_ISL_1560720, EPI_ISL_1560721, EPI_ISL_1560722, EPI_ISL_1560723, EPI_ISL_1560724,
EPI_ISL_1560725, EPI_ISL_1560726, EPI_ISL_1560727, EPI_ISL_1560728, EPI_ISL_1560729, EPI_ISL_1560730, EPI_ISL_1560731, EPI_ISL_1560732, EPI_ISL_1560733, EPI_ISL_1560734, EPI_ISL_1560735, EPI_ISL_1560736, EPI_ISL_1560737, EPI_ISL_1560738, EPI_ISL_1560739, EPI_ISL_1560740, EPI_ISL_1560741, EPI_ISL_1560742, EPI_ISL_1560743, EPI_ISL_1560744, EPI_ISL_1560745, EPI_ISL_1560746,
EPI_ISL_1560747, EPI_ISL_1560748, EPI_ISL_1560749, EPI_ISL_1560750, EPI_ISL_1560751, EPI_ISL_1560752, EPI_ISL_1560753, EPI_ISL_1560754, EPI_ISL_1560755, EPI_ISL_1560756, EPI_ISL_1560757, EPI_ISL_1560758, EPI_ISL_1560759, EPI_ISL_1560760, EPI_ISL_1560761, EPI_ISL_1560762, EPI_ISL_1560763, EPI_ISL_1560764, EPI_ISL_1560765, EPI_ISL_1560766, EPI_ISL_1560767, EPI_ISL_1560768,
EPI_ISL_1560769, EPI_ISL_1560770, EPI_ISL_1560771, EPI_ISL_1560772, EPI_ISL_1560773, EPI_ISL_1560774, EPI_ISL_1560775, EPI_ISL_1560776, EPI_ISL_1560777, EPI_ISL_1560778, EPI_ISL_1560779, EPI_ISL_1560780, EPI_ISL_1560781, EPI_ISL_1560782, EPI_ISL_1560783, EPI_ISL_1560784, EPI_ISL_1560785, EPI_ISL_1560786, EPI_ISL_1560787, EPI_ISL_1560788, EPI_ISL_1560789, EPI_ISL_1560790,
EPI_ISL_1560791, EPI_ISL_1560792, EPI_ISL_1560793, EPI_ISL_1560794, EPI_ISL_1560795, EPI_ISL_1560796, EPI_ISL_1560797, EPI_ISL_1560798, EPI_ISL_1560800, EPI_ISL_1560801, EPI_ISL_1560802, EPI_ISL_1560803, EPI_ISL_1560804, EPI_ISL_1560805, EPI_ISL_1560806, EPI_ISL_1560807, EPI_ISL_1560808, EPI_ISL_1560809, EPI_ISL_1560810, EPI_ISL_1560811, EPI_ISL_1560812, EPI_ISL_1560813,
EPI_ISL_1560814, EPI_ISL_1560815, EPI_ISL_1560816, EPI_ISL_1560817, EPI_ISL_1560818, EPI_ISL_1560819, EPI_ISL_1560820, EPI_ISL_1560821, EPI_ISL_1560822, EPI_ISL_1560823, EPI_ISL_1560824, EPI_ISL_1560825, EPI_ISL_1560826, EPI_ISL_1560827, EPI_ISL_1560828, EPI_ISL_1560829, EPI_ISL_1560830, EPI_ISL_1560831, EPI_ISL_1560833, EPI_ISL_1560834, EPI_ISL_1560835, EPI_ISL_1560836,
EPI_ISL_1560837, EPI_ISL_1560838, EPI_ISL_1560839, EPI_ISL_1560840, EPI_ISL_1560841, EPI_ISL_1560842, EPI_ISL_1560843, EPI_ISL_1560844, EPI_ISL_1560845, EPI_ISL_1560846, EPI_ISL_1560847, EPI_ISL_1560848, EPI_ISL_1560849, EPI_ISL_1560850, EPI_ISL_1560851, EPI_ISL_1560852, EPI_ISL_1560853, EPI_ISL_1560854, EPI_ISL_1560855, EPI_ISL_1560856, EPI_ISL_1560857, EPI_ISL_1560858,
EPI_ISL_1560859, EPI_ISL_1560860, EPI_ISL_1560861, EPI_ISL_1560862, EPI_ISL_1560863, EPI_ISL_1560865, EPI_ISL_1560866, EPI_ISL_1560867, EPI_ISL_1560868, EPI_ISL_1560869, EPI_ISL_1560870, EPI_ISL_1560871, EPI_ISL_1560872, EPI_ISL_1560873, EPI_ISL_1560874, EPI_ISL_1560875, EPI_ISL_1560876, EPI_ISL_1560877, EPI_ISL_1560878, EPI_ISL_1560879, EPI_ISL_1560880, EPI_ISL_1560881,
EPI_ISL_1560882, EPI_ISL_1560883, EPI_ISL_1560884, EPI_ISL_1560885, EPI_ISL_1560886, EPI_ISL_1560888, EPI_ISL_1560889, EPI_ISL_1560890, EPI_ISL_1560891, EPI_ISL_1560892, EPI_ISL_1560893, EPI_ISL_1560894, EPI_ISL_1560895, EPI_ISL_1560896, EPI_ISL_1560897, EPI_ISL_1560898, EPI_ISL_1560899, EPI_ISL_1560900, EPI_ISL_1560901, EPI_ISL_1560902, EPI_ISL_1560903, EPI_ISL_1560904,
EPI_ISL_1560905, EPI_ISL_1560906, EPI_ISL_1560907, EPI_ISL_1560908, EPI_ISL_1560909, EPI_ISL_1560910, EPI_ISL_1560911, EPI_ISL_1560912, EPI_ISL_1560913, EPI_ISL_1560914, EPI_ISL_1560915, EPI_ISL_1560916, EPI_ISL_1560917, EPI_ISL_1560918, EPI_ISL_1560919, EPI_ISL_1560920, EPI_ISL_1560921, EPI_ISL_1560922, EPI_ISL_1560923, EPI_ISL_1560924, EPI_ISL_1560925, EPI_ISL_1560926,
EPI_ISL_1560927, EPI_ISL_1560928, EPI_ISL_1560929, EPI_ISL_1560930, EPI_ISL_1560931, EPI_ISL_1560932, EPI_ISL_1560933, EPI_ISL_1560934, EPI_ISL_1560935, EPI_ISL_1560936, EPI_ISL_1560937, EPI_ISL_1560938, EPI_ISL_1560939, EPI_ISL_1560940, EPI_ISL_1560941, EPI_ISL_1560942, EPI_ISL_1560943, EPI_ISL_1560944, EPI_ISL_1560945, EPI_ISL_1560946, EPI_ISL_1560947, EPI_ISL_1560948,
EPI_ISL_1560950, EPI_ISL_1560951, EPI_ISL_1560952, EPI_ISL_1560953, EPI_ISL_1560954, EPI_ISL_1560955, EPI_ISL_1560956, EPI_ISL_1560957, EPI_ISL_1560958, EPI_ISL_1560959, EPI_ISL_1560960, EPI_ISL_1560961, EPI_ISL_1560962, EPI_ISL_1560963, EPI_ISL_1560964, EPI_ISL_1560965, EPI_ISL_1560966, EPI_ISL_1560967, EPI_ISL_1560968, EPI_ISL_1560969, EPI_ISL_1560970, EPI_ISL_1560971,
EPI_ISL_1560972, EPI_ISL_1560973, EPI_ISL_1560974, EPI_ISL_1560975, EPI_ISL_1560976, EPI_ISL_1560977, EPI_ISL_1560978, EPI_ISL_1560979, EPI_ISL_1560980, EPI_ISL_1560981, EPI_ISL_1560982, EPI_ISL_1560983, EPI_ISL_1560984, EPI_ISL_1560985, EPI_ISL_1560986, EPI_ISL_1560987, EPI_ISL_1560988, EPI_ISL_1560989, EPI_ISL_1560990, EPI_ISL_1560991, EPI_ISL_1560992, EPI_ISL_1560993,
EPI_ISL_1560994, EPI_ISL_1560995, EPI_ISL_1560996, EPI_ISL_1560997, EPI_ISL_1560998, EPI_ISL_1560999, EPI_ISL_1561000, EPI_ISL_1561001, EPI_ISL_1561002, EPI_ISL_1561003, EPI_ISL_1561004, EPI_ISL_1561005, EPI_ISL_1561007, EPI_ISL_1561008, EPI_ISL_1561009, EPI_ISL_1561010, EPI_ISL_1561011, EPI_ISL_1561012, EPI_ISL_1561013, EPI_ISL_1561014, EPI_ISL_1561015, EPI_ISL_1561016,
EPI_ISL_1561017, EPI_ISL_1561018, EPI_ISL_1561019, EPI_ISL_1561020, EPI_ISL_1561021, EPI_ISL_1561022, EPI_ISL_1561023, EPI_ISL_1561024, EPI_ISL_1561025, EPI_ISL_1561026, EPI_ISL_1561027, EPI_ISL_1561028, EPI_ISL_1561029, EPI_ISL_1561030, EPI_ISL_1561031, EPI_ISL_1561032, EPI_ISL_1561033, EPI_ISL_1561034, EPI_ISL_1561035, EPI_ISL_1561036, EPI_ISL_1561037, EPI_ISL_1561038,
EPI_ISL_1561039, EPI_ISL_1561040, EPI_ISL_1561041, EPI_ISL_1561042, EPI_ISL_1561043, EPI_ISL_1561044, EPI_ISL_1561045, EPI_ISL_1561046, EPI_ISL_1561047, EPI_ISL_1561048, EPI_ISL_1561049, EPI_ISL_1561050, EPI_ISL_1561051, EPI_ISL_1561052, EPI_ISL_1561053, EPI_ISL_1561054, EPI_ISL_1561055, EPI_ISL_1561056, EPI_ISL_1561057, EPI_ISL_1561058, EPI_ISL_1561059, EPI_ISL_1561060,
EPI_ISL_1561061, EPI_ISL_1561062, EPI_ISL_1561063, EPI_ISL_1561064, EPI_ISL_1561065, EPI_ISL_1561066, EPI_ISL_1561067, EPI_ISL_1561068, EPI_ISL_1561069, EPI_ISL_1561071, EPI_ISL_1561072, EPI_ISL_1561073, EPI_ISL_1561074, EPI_ISL_1561075, EPI_ISL_1561076, EPI_ISL_1561077, EPI_ISL_1561078, EPI_ISL_1561079, EPI_ISL_1561080, EPI_ISL_1561081, EPI_ISL_1561082, EPI_ISL_1561083,
EPI_ISL_1561084, EPI_ISL_1561085, EPI_ISL_1561086, EPI_ISL_1561087, EPI_ISL_1561088, EPI_ISL_1561089, EPI_ISL_1561090, EPI_ISL_1561091, EPI_ISL_1561092, EPI_ISL_1561093, EPI_ISL_1561094, EPI_ISL_1561095, EPI_ISL_1561096, EPI_ISL_1561097, EPI_ISL_1561098, EPI_ISL_1561099, EPI_ISL_1561100, EPI_ISL_1561101, EPI_ISL_1561102, EPI_ISL_1561103, EPI_ISL_1561104, EPI_ISL_1561105,
EPI_ISL_1561106, EPI_ISL_1561107, EPI_ISL_1561108, EPI_ISL_1561109, EPI_ISL_1561111, EPI_ISL_1561112, EPI_ISL_1561113, EPI_ISL_1561114, EPI_ISL_1561115, EPI_ISL_1561116, EPI_ISL_1561117, EPI_ISL_1561118, EPI_ISL_1561119, EPI_ISL_1561120, EPI_ISL_1561121, EPI_ISL_1561122, EPI_ISL_1561123, EPI_ISL_1561124, EPI_ISL_1561125, EPI_ISL_1561126, EPI_ISL_1561127, EPI_ISL_1561128,
EPI_ISL_1561129, EPI_ISL_1561130, EPI_ISL_1561131, EPI_ISL_1561132, EPI_ISL_1561133, EPI_ISL_1561134, EPI_ISL_1561135, EPI_ISL_1561136, EPI_ISL_1561137, EPI_ISL_1561138, EPI_ISL_1561139, EPI_ISL_1561140, EPI_ISL_1561141, EPI_ISL_1561142, EPI_ISL_1561143, EPI_ISL_1561144, EPI_ISL_1561145, EPI_ISL_1561146, EPI_ISL_1561147, EPI_ISL_1561148, EPI_ISL_1561150, EPI_ISL_1561151,
EPI_ISL_1561152, EPI_ISL_1561153, EPI_ISL_1561154, EPI_ISL_1561156, EPI_ISL_1561157, EPI_ISL_1561158, EPI_ISL_1561159, EPI_ISL_1561160, EPI_ISL_1561161, EPI_ISL_1561162, EPI_ISL_1561163, EPI_ISL_1561164, EPI_ISL_1561165, EPI_ISL_1561166, EPI_ISL_1561167, EPI_ISL_1561168, EPI_ISL_1561169, EPI_ISL_1561170, EPI_ISL_1561171, EPI_ISL_1561172, EPI_ISL_1561173, EPI_ISL_1561174,
EPI_ISL_1561175, EPI_ISL_1561176, EPI_ISL_1561177, EPI_ISL_1561178, EPI_ISL_1561179, EPI_ISL_1561180, EPI_ISL_1561181, EPI_ISL_1561182, EPI_ISL_1561183, EPI_ISL_1561184, EPI_ISL_1561185, EPI_ISL_1561186, EPI_ISL_1561187, EPI_ISL_1561188, EPI_ISL_1561189, EPI_ISL_1561190, EPI_ISL_1561191, EPI_ISL_1561192, EPI_ISL_1561193, EPI_ISL_1561194, EPI_ISL_1561195, EPI_ISL_1561196,
EPI_ISL_1561197, EPI_ISL_1561198, EPI_ISL_1561199, EPI_ISL_1561200, EPI_ISL_1561201, EPI_ISL_1561202, EPI_ISL_1561203, EPI_ISL_1561204, EPI_ISL_1561205, EPI_ISL_1561206, EPI_ISL_1561207, EPI_ISL_1561208, EPI_ISL_1561209, EPI_ISL_1561210, EPI_ISL_1561211, EPI_ISL_1561212, EPI_ISL_1561213, EPI_ISL_1561214, EPI_ISL_1561215, EPI_ISL_1561216, EPI_ISL_1561217, EPI_ISL_1561218,
EPI_ISL_1561219, EPI_ISL_1561220, EPI_ISL_1561221, EPI_ISL_1561222, EPI_ISL_1561223, EPI_ISL_1561224, EPI_ISL_1561225, EPI_ISL_1561226, EPI_ISL_1561227, EPI_ISL_1561228, EPI_ISL_1561229, EPI_ISL_1561230, EPI_ISL_1561231, EPI_ISL_1561232, EPI_ISL_1561233, EPI_ISL_1561234, EPI_ISL_1561235, EPI_ISL_1561236, EPI_ISL_1561237, EPI_ISL_1561238, EPI_ISL_1561239, EPI_ISL_1561240,
EPI_ISL_1561241, EPI_ISL_1561242, EPI_ISL_1561243, EPI_ISL_1561244, EPI_ISL_1561245, EPI_ISL_1561246, EPI_ISL_1561247, EPI_ISL_1561248, EPI_ISL_1561249, EPI_ISL_1561250, EPI_ISL_1561251, EPI_ISL_1561252, EPI_ISL_1561253, EPI_ISL_1561254, EPI_ISL_1561255, EPI_ISL_1561256, EPI_ISL_1561257, EPI_ISL_1561258, EPI_ISL_1561259, EPI_ISL_1561260, EPI_ISL_1561261, EPI_ISL_1561262,
EPI_ISL_1561263, EPI_ISL_1561264, EPI_ISL_1561265, EPI_ISL_1561266, EPI_ISL_1561267, EPI_ISL_1561268, EPI_ISL_1561269, EPI_ISL_1561270, EPI_ISL_1561271, EPI_ISL_1561272, EPI_ISL_1561273, EPI_ISL_1561274, EPI_ISL_1561275, EPI_ISL_1561276, EPI_ISL_1561277, EPI_ISL_1561278, EPI_ISL_1561279, EPI_ISL_1561280, EPI_ISL_1561281, EPI_ISL_1561282, EPI_ISL_1561283, EPI_ISL_1561284,
EPI_ISL_1561285, EPI_ISL_1561286, EPI_ISL_1561287, EPI_ISL_1561288, EPI_ISL_1561289, EPI_ISL_1561290, EPI_ISL_1561291, EPI_ISL_1561292, EPI_ISL_1561293, EPI_ISL_1561294, EPI_ISL_1561295, EPI_ISL_1561297, EPI_ISL_1561298, EPI_ISL_1561299, EPI_ISL_1561300, EPI_ISL_1561301, EPI_ISL_1561302, EPI_ISL_1561303, EPI_ISL_1561304, EPI_ISL_1561305, EPI_ISL_1561306, EPI_ISL_1561307,
EPI_ISL_1561308, EPI_ISL_1561309, EPI_ISL_1561310, EPI_ISL_1561311, EPI_ISL_1561312, EPI_ISL_1561313, EPI_ISL_1561314, EPI_ISL_1561315, EPI_ISL_1561316, EPI_ISL_1561317, EPI_ISL_1561318, EPI_ISL_1561319, EPI_ISL_1561320, EPI_ISL_1561321, EPI_ISL_1561322, EPI_ISL_1561323, EPI_ISL_1561324, EPI_ISL_1561325, EPI_ISL_1561326, EPI_ISL_1561327, EPI_ISL_1561328, EPI_ISL_1561329,
EPI_ISL_1561330, EPI_ISL_1561331, EPI_ISL_1561332, EPI_ISL_1561333, EPI_ISL_1561334, EPI_ISL_1561335, EPI_ISL_1561336, EPI_ISL_1561338, EPI_ISL_1561339, EPI_ISL_1561340, EPI_ISL_1561341, EPI_ISL_1561342, EPI_ISL_1561343, EPI_ISL_1561344, EPI_ISL_1561345, EPI_ISL_1561346, EPI_ISL_1561347, EPI_ISL_1561348, EPI_ISL_1561349, EPI_ISL_1561350, EPI_ISL_1561351, EPI_ISL_1561352,
EPI_ISL_1561353, EPI_ISL_1561354, EPI_ISL_1561355, EPI_ISL_1561356, EPI_ISL_1561357, EPI_ISL_1561358, EPI_ISL_1561359, EPI_ISL_1561360, EPI_ISL_1561361, EPI_ISL_1561362, EPI_ISL_1561363, EPI_ISL_1561364, EPI_ISL_1561365, EPI_ISL_1561366, EPI_ISL_1561367, EPI_ISL_1561368, EPI_ISL_1561369, EPI_ISL_1561370, EPI_ISL_1561371, EPI_ISL_1561372, EPI_ISL_1561373, EPI_ISL_1561374,
EPI_ISL_1561375, EPI_ISL_1561376, EPI_ISL_1561377, EPI_ISL_1561378, EPI_ISL_1561379, EPI_ISL_1561380, EPI_ISL_1561381, EPI_ISL_1561382, EPI_ISL_1561383, EPI_ISL_1561384, EPI_ISL_1561385, EPI_ISL_1561386, EPI_ISL_1561387, EPI_ISL_1561388, EPI_ISL_1561389, EPI_ISL_1561390, EPI_ISL_1561391, EPI_ISL_1561392, EPI_ISL_1561393, EPI_ISL_1561394, EPI_ISL_1561395, EPI_ISL_1561396,
EPI_ISL_1561397, EPI_ISL_1561398, EPI_ISL_1561399, EPI_ISL_1561400, EPI_ISL_1561401, EPI_ISL_1561402, EPI_ISL_1561403, EPI_ISL_1561404, EPI_ISL_1561405, EPI_ISL_1561406, EPI_ISL_1561407, EPI_ISL_1561408, EPI_ISL_1561409, EPI_ISL_1561410, EPI_ISL_1561411, EPI_ISL_1561412, EPI_ISL_1561413, EPI_ISL_1561414, EPI_ISL_1561415, EPI_ISL_1561416, EPI_ISL_1561417, EPI_ISL_1561418,
EPI_ISL_1561419, EPI_ISL_1561420, EPI_ISL_1561421, EPI_ISL_1561422, EPI_ISL_1561423, EPI_ISL_1561424, EPI_ISL_1561426, EPI_ISL_1561427, EPI_ISL_1561429, EPI_ISL_1561430, EPI_ISL_1561431, EPI_ISL_1561432, EPI_ISL_1561433, EPI_ISL_1561434, EPI_ISL_1561435, EPI_ISL_1561436, EPI_ISL_1561437, EPI_ISL_1561438, EPI_ISL_1561439, EPI_ISL_1561440, EPI_ISL_1561441, EPI_ISL_1561442,
EPI_ISL_1561443, EPI_ISL_1561444, EPI_ISL_1561445, EPI_ISL_1561446, EPI_ISL_1561447, EPI_ISL_1561448, EPI_ISL_1561449, EPI_ISL_1561451, EPI_ISL_1561452, EPI_ISL_1561453, EPI_ISL_1561454, EPI_ISL_1561455, EPI_ISL_1561456, EPI_ISL_1561457, EPI_ISL_1561458, EPI_ISL_1561459, EPI_ISL_1561460, EPI_ISL_1561461, EPI_ISL_1561462, EPI_ISL_1561463, EPI_ISL_1561464, EPI_ISL_1561465,
EPI_ISL_1561466, EPI_ISL_1561467, EPI_ISL_1561468, EPI_ISL_1561469, EPI_ISL_1561470, EPI_ISL_1561471, EPI_ISL_1561472, EPI_ISL_1561473, EPI_ISL_1561474, EPI_ISL_1561475, EPI_ISL_1561476, EPI_ISL_1561477, EPI_ISL_1561478, EPI_ISL_1561479, EPI_ISL_1561480, EPI_ISL_1561481, EPI_ISL_1561482, EPI_ISL_1561483, EPI_ISL_1561484, EPI_ISL_1561485, EPI_ISL_1561486, EPI_ISL_1561487,
EPI_ISL_1561488, EPI_ISL_1561489, EPI_ISL_1561490, EPI_ISL_1561491, EPI_ISL_1561492, EPI_ISL_1561493, EPI_ISL_1561494, EPI_ISL_1561495, EPI_ISL_1561496, EPI_ISL_1561497, EPI_ISL_1561498, EPI_ISL_1561499, EPI_ISL_1561500, EPI_ISL_1561501, EPI_ISL_1561502, EPI_ISL_1561503, EPI_ISL_1561504, EPI_ISL_1561505, EPI_ISL_1561506, EPI_ISL_1561507, EPI_ISL_1561508, EPI_ISL_1561510,
EPI_ISL_1561511, EPI_ISL_1561512, EPI_ISL_1561514, EPI_ISL_1561515, EPI_ISL_1561516, EPI_ISL_1561518, EPI_ISL_1561519, EPI_ISL_1561520, EPI_ISL_1561521, EPI_ISL_1561522, EPI_ISL_1561523, EPI_ISL_1561524, EPI_ISL_1561525, EPI_ISL_1561526, EPI_ISL_1561527, EPI_ISL_1561528, EPI_ISL_1561529, EPI_ISL_1561530, EPI_ISL_1561531, EPI_ISL_1561532, EPI_ISL_1561533, EPI_ISL_1561534,
EPI_ISL_1561535, EPI_ISL_1561536, EPI_ISL_1561537, EPI_ISL_1561538, EPI_ISL_1561539, EPI_ISL_1561540, EPI_ISL_1561541, EPI_ISL_1561542, EPI_ISL_1561543, EPI_ISL_1561544, EPI_ISL_1561545, EPI_ISL_1561546, EPI_ISL_1561547, EPI_ISL_1561548, EPI_ISL_1561549, EPI_ISL_1561550, EPI_ISL_1561551, EPI_ISL_1561552, EPI_ISL_1561553, EPI_ISL_1561554, EPI_ISL_1561555, EPI_ISL_1561556,
EPI_ISL_1561557, EPI_ISL_1561558, EPI_ISL_1561559, EPI_ISL_1561560, EPI_ISL_1561561, EPI_ISL_1561562, EPI_ISL_1561563, EPI_ISL_1561564, EPI_ISL_1561566, EPI_ISL_1561567, EPI_ISL_1561568, EPI_ISL_1561569, EPI_ISL_1561570, EPI_ISL_1561571, EPI_ISL_1561572, EPI_ISL_1561573, EPI_ISL_1561574, EPI_ISL_1561575, EPI_ISL_1561576, EPI_ISL_1561577, EPI_ISL_1561578, EPI_ISL_1561579,
EPI_ISL_1561580, EPI_ISL_1561581, EPI_ISL_1561582, EPI_ISL_1561583, EPI_ISL_1561584, EPI_ISL_1561585, EPI_ISL_1561586, EPI_ISL_1561587, EPI_ISL_1561588, EPI_ISL_1561589, EPI_ISL_1561590, EPI_ISL_1561591, EPI_ISL_1561592, EPI_ISL_1561593, EPI_ISL_1561594, EPI_ISL_1561595, EPI_ISL_1561596, EPI_ISL_1561597, EPI_ISL_1561598, EPI_ISL_1561599, EPI_ISL_1561600, EPI_ISL_1561601,

| | | | |
|---|---|---|---|
| EPI_ISL_1561602, EPI_ISL_1561603, EPI_ISL_1561605, EPI_ISL_1561606, EPI_ISL_1561607, EPI_ISL_1561608, EPI_ISL_1561609, EPI_ISL_1561610, EPI_ISL_1561611, EPI_ISL_1561612, EPI_ISL_1561613, EPI_ISL_1561614, EPI_ISL_1561615, EPI_ISL_1561616, EPI_ISL_1561617, EPI_ISL_1561618, EPI_ISL_1561619, EPI_ISL_1561620, EPI_ISL_1561621, EPI_ISL_1561622, EPI_ISL_1561624, EPI_ISL_1561625, EPI_ISL_1561626, EPI_ISL_1561627, EPI_ISL_1561628, EPI_ISL_1561629, EPI_ISL_1561630, EPI_ISL_1561631, EPI_ISL_1561632, EPI_ISL_1561633, EPI_ISL_1561634, EPI_ISL_1561635, EPI_ISL_1561636, EPI_ISL_1561637, EPI_ISL_1561638, EPI_ISL_1561639, EPI_ISL_1561640, EPI_ISL_1561641, EPI_ISL_1561642, EPI_ISL_1561643, EPI_ISL_1561644, EPI_ISL_1561645, EPI_ISL_1561646, EPI_ISL_1561647, EPI_ISL_1561648, EPI_ISL_1561649, EPI_ISL_1561650, EPI_ISL_1561651, EPI_ISL_1561652, EPI_ISL_1561653, EPI_ISL_1561654, EPI_ISL_1561655, EPI_ISL_1561656, EPI_ISL_1561657, EPI_ISL_1561658, EPI_ISL_1561659, EPI_ISL_1561660, EPI_ISL_1561661, EPI_ISL_1561662, EPI_ISL_1561663, EPI_ISL_1561664, EPI_ISL_1561665, EPI_ISL_1561666, EPI_ISL_1561667, EPI_ISL_1561668, EPI_ISL_1561669, EPI_ISL_1561670, EPI_ISL_1561671, EPI_ISL_1561672, EPI_ISL_1561673, EPI_ISL_1561674, EPI_ISL_1561675, EPI_ISL_1561676, EPI_ISL_1561677, EPI_ISL_1561678, EPI_ISL_1561679, EPI_ISL_1561680, EPI_ISL_1561681, EPI_ISL_1561682, EPI_ISL_1561683, EPI_ISL_1561684, EPI_ISL_1561685, EPI_ISL_1561686, EPI_ISL_1561687, EPI_ISL_1561688, EPI_ISL_1561689, EPI_ISL_1561690, EPI_ISL_1561691, EPI_ISL_1561692, EPI_ISL_1561693, EPI_ISL_1561694, EPI_ISL_1561695, EPI_ISL_1561696, EPI_ISL_1561697, EPI_ISL_1561698, EPI_ISL_1561699, EPI_ISL_1561700, EPI_ISL_1561701, EPI_ISL_1561702, EPI_ISL_1561703, EPI_ISL_1561704, EPI_ISL_1561705, EPI_ISL_1561706, EPI_ISL_1561707, EPI_ISL_1561708, EPI_ISL_1561709, EPI_ISL_1561710, EPI_ISL_1561711, EPI_ISL_1561712, EPI_ISL_1561713, EPI_ISL_1561714, EPI_ISL_1561715, EPI_ISL_1561716, EPI_ISL_1561717, EPI_ISL_1561718, EPI_ISL_1561719, EPI_ISL_1561720, EPI_ISL_1561721, EPI_ISL_1561722, EPI_ISL_1561723, EPI_ISL_1561724, EPI_ISL_1561725, EPI_ISL_1561726, EPI_ISL_1561727, EPI_ISL_1561728, EPI_ISL_1561729, EPI_ISL_1561730, EPI_ISL_1561731, EPI_ISL_1561732, EPI_ISL_1561733, EPI_ISL_1561735, EPI_ISL_1561736, EPI_ISL_1561737, EPI_ISL_1561739, EPI_ISL_1561740, EPI_ISL_1561741, EPI_ISL_1561742, EPI_ISL_1561743, EPI_ISL_1561744, EPI_ISL_1561745, EPI_ISL_1561747, EPI_ISL_1561748, EPI_ISL_1561749, EPI_ISL_1561750, EPI_ISL_1561751, EPI_ISL_1561752, EPI_ISL_1561753, EPI_ISL_1561754, EPI_ISL_1561755, EPI_ISL_1561756, EPI_ISL_1561757, EPI_ISL_1561758, EPI_ISL_1561759, EPI_ISL_1561760, EPI_ISL_1561761, EPI_ISL_1561762, EPI_ISL_1561763, EPI_ISL_1561765, EPI_ISL_1561766, EPI_ISL_1561768, EPI_ISL_1561769, EPI_ISL_1561770, EPI_ISL_1561771, EPI_ISL_1561772, EPI_ISL_1561773, EPI_ISL_1561775, EPI_ISL_1561776, EPI_ISL_1561777, EPI_ISL_1561778, EPI_ISL_1561779, EPI_ISL_1561780, EPI_ISL_1561781, EPI_ISL_1561782, EPI_ISL_1561783, EPI_ISL_1561784, EPI_ISL_1561785, EPI_ISL_1561786, EPI_ISL_1561787, EPI_ISL_1561788, EPI_ISL_1561789, EPI_ISL_1561791, EPI_ISL_1561792, EPI_ISL_1561794, EPI_ISL_1561795, EPI_ISL_1561796, EPI_ISL_1561797, EPI_ISL_1561798, EPI_ISL_1561799, EPI_ISL_1561800, EPI_ISL_1561801, EPI_ISL_1561802, EPI_ISL_1561803, EPI_ISL_1561804, EPI_ISL_1561805, EPI_ISL_1561806, EPI_ISL_1561807, EPI_ISL_1561808, EPI_ISL_1561809, EPI_ISL_1561810, EPI_ISL_1561811, EPI_ISL_1561812, EPI_ISL_1561813, EPI_ISL_1561814, EPI_ISL_1561815, EPI_ISL_1561816, EPI_ISL_1561817, EPI_ISL_1561818, EPI_ISL_1561819, EPI_ISL_1561820, EPI_ISL_1561821, EPI_ISL_1561822, EPI_ISL_1561823, EPI_ISL_1561824, EPI_ISL_1561825, EPI_ISL_1561826, EPI_ISL_1561827, EPI_ISL_1561828, EPI_ISL_1561829, EPI_ISL_1561830, EPI_ISL_1561831, EPI_ISL_1561832, EPI_ISL_1561833, EPI_ISL_1561834, EPI_ISL_1561835, EPI_ISL_1561836, EPI_ISL_1561837, EPI_ISL_1561838, EPI_ISL_1561840, EPI_ISL_1561841, EPI_ISL_1561842, EPI_ISL_1561843, EPI_ISL_1561844, EPI_ISL_1561845, EPI_ISL_1561846, EPI_ISL_1561847, EPI_ISL_1561848, EPI_ISL_1561850, EPI_ISL_1561851, EPI_ISL_1561852, EPI_ISL_1561853, EPI_ISL_1561854, EPI_ISL_1561855, EPI_ISL_1561856, EPI_ISL_1561857, EPI_ISL_1561858, EPI_ISL_1561859, EPI_ISL_1561860, EPI_ISL_1561861, EPI_ISL_1561862, EPI_ISL_1561863, EPI_ISL_1561864, EPI_ISL_1561865, EPI_ISL_1561867, EPI_ISL_1561868, EPI_ISL_1561869, EPI_ISL_1561870, EPI_ISL_1561871, EPI_ISL_1561872, EPI_ISL_1561873, EPI_ISL_1561874, EPI_ISL_1561875, EPI_ISL_1561876, EPI_ISL_1561877, EPI_ISL_1561878, EPI_ISL_1561879, EPI_ISL_1561880, EPI_ISL_1561881, EPI_ISL_1561882, EPI_ISL_1561883, EPI_ISL_1561884, EPI_ISL_1561885, EPI_ISL_1561886, EPI_ISL_1561887, EPI_ISL_1561888, EPI_ISL_1561889, EPI_ISL_1561890, EPI_ISL_1561891, EPI_ISL_1561892, EPI_ISL_1561893, EPI_ISL_1561894, EPI_ISL_1561895, EPI_ISL_1561897, EPI_ISL_1561898, EPI_ISL_1561899, EPI_ISL_1561900, EPI_ISL_1561901, EPI_ISL_1561902, EPI_ISL_1561903, EPI_ISL_1561904, EPI_ISL_1561905, EPI_ISL_1561906, EPI_ISL_1561907, EPI_ISL_1561908, EPI_ISL_1561909, EPI_ISL_1561910, EPI_ISL_1561911, EPI_ISL_1561912, EPI_ISL_1561913, EPI_ISL_1561914, EPI_ISL_1561915, EPI_ISL_1561916, EPI_ISL_1561917, EPI_ISL_1561918, EPI_ISL_1561919, EPI_ISL_1561920, EPI_ISL_1561921, EPI_ISL_1561922, EPI_ISL_1561924, EPI_ISL_1561925, EPI_ISL_1561926, EPI_ISL_1561927, EPI_ISL_1561928, EPI_ISL_1561929, EPI_ISL_1561930, EPI_ISL_1561931, EPI_ISL_1561932, EPI_ISL_1561933, EPI_ISL_1561934, EPI_ISL_1561935, EPI_ISL_1561936, EPI_ISL_1561937, EPI_ISL_1561938, EPI_ISL_1561939, EPI_ISL_1561940, EPI_ISL_1561941, EPI_ISL_1561942, EPI_ISL_1561943, EPI_ISL_1561944, EPI_ISL_1561945, EPI_ISL_1561946, EPI_ISL_1561947, EPI_ISL_1561948, EPI_ISL_1561949, EPI_ISL_1561950, EPI_ISL_1561951, EPI_ISL_1561952, EPI_ISL_1561953, EPI_ISL_1561954, EPI_ISL_1561955, EPI_ISL_1561956, EPI_ISL_1561957, EPI_ISL_1561958, EPI_ISL_1561959, EPI_ISL_1561960, EPI_ISL_1561961, EPI_ISL_1561962, EPI_ISL_1561963, EPI_ISL_1561964, EPI_ISL_1561965, EPI_ISL_1561966, EPI_ISL_1561967, EPI_ISL_1561968, EPI_ISL_1561969, EPI_ISL_1561970, EPI_ISL_1561972, EPI_ISL_1561973, EPI_ISL_1561974, EPI_ISL_1561975, EPI_ISL_1561976, EPI_ISL_1561977, EPI_ISL_1561978, EPI_ISL_1561979, EPI_ISL_1561980, EPI_ISL_1561981, EPI_ISL_1561982, EPI_ISL_1561983, EPI_ISL_1561984, EPI_ISL_1561985, EPI_ISL_1561986, EPI_ISL_1561987, EPI_ISL_1561988, EPI_ISL_1561989, EPI_ISL_1561990, EPI_ISL_1561991, EPI_ISL_1561992, EPI_ISL_1561994, EPI_ISL_1561996, EPI_ISL_1561997, EPI_ISL_1561998, EPI_ISL_1561999, EPI_ISL_1562000, EPI_ISL_1562001, EPI_ISL_1562002, EPI_ISL_1562003, EPI_ISL_1562004, EPI_ISL_1562005, EPI_ISL_1562006, EPI_ISL_1562007, EPI_ISL_1562008, EPI_ISL_1562009, EPI_ISL_1562010, EPI_ISL_1562011, EPI_ISL_1562012, EPI_ISL_1562013, EPI_ISL_1562014, EPI_ISL_1562015, EPI_ISL_1562016, EPI_ISL_1562017, EPI_ISL_1562018, EPI_ISL_1562019, EPI_ISL_1562020, EPI_ISL_1562021, EPI_ISL_1562022, EPI_ISL_1562023, EPI_ISL_1562024, EPI_ISL_1562025, EPI_ISL_1562026, EPI_ISL_1562027, EPI_ISL_1562028, EPI_ISL_1562029, EPI_ISL_1562030, EPI_ISL_1562031, EPI_ISL_1562032, EPI_ISL_1562033, EPI_ISL_1562034, EPI_ISL_1562036, EPI_ISL_1562037, EPI_ISL_1562038, EPI_ISL_1562041, EPI_ISL_1562042, EPI_ISL_1562043, EPI_ISL_1562044, EPI_ISL_1562045, EPI_ISL_1562046, EPI_ISL_1562047, EPI_ISL_1562049, EPI_ISL_1562050, EPI_ISL_1562051, EPI_ISL_1562052, EPI_ISL_1562054, EPI_ISL_1562055, EPI_ISL_1562056, EPI_ISL_1562058, EPI_ISL_1562059, EPI_ISL_1562060, EPI_ISL_1562061, EPI_ISL_1562062, EPI_ISL_1562063, EPI_ISL_1562064, EPI_ISL_1562065, EPI_ISL_1562066, EPI_ISL_1562067, EPI_ISL_1562068, EPI_ISL_1562069, EPI_ISL_1562070, EPI_ISL_1562071, EPI_ISL_1562072, EPI_ISL_1562073, EPI_ISL_1562074, EPI_ISL_1562075, EPI_ISL_1562076, EPI_ISL_1562077, EPI_ISL_1562078, EPI_ISL_1562079, EPI_ISL_1562080, EPI_ISL_1562081, EPI_ISL_1562082, EPI_ISL_1562083, EPI_ISL_1562085, EPI_ISL_1562086, EPI_ISL_1562087, EPI_ISL_1562088, EPI_ISL_1562089, EPI_ISL_1562090, EPI_ISL_1562091, EPI_ISL_1562092, EPI_ISL_1562093, EPI_ISL_1562094, EPI_ISL_1562095, EPI_ISL_1562096, EPI_ISL_1562097, EPI_ISL_1562098, EPI_ISL_1562099, EPI_ISL_1562100, EPI_ISL_1562101, EPI_ISL_1562102, EPI_ISL_1562103, EPI_ISL_1562104, EPI_ISL_1562106, EPI_ISL_1562108, EPI_ISL_1562109, EPI_ISL_1562110, EPI_ISL_1562111, EPI_ISL_1562112, EPI_ISL_1562113, EPI_ISL_1562114, EPI_ISL_1562115, EPI_ISL_1562116, EPI_ISL_1562117, EPI_ISL_1562118, EPI_ISL_1562119, EPI_ISL_1562120, EPI_ISL_1562121, EPI_ISL_1562122, EPI_ISL_1562123, EPI_ISL_1562124, EPI_ISL_1562125, EPI_ISL_1562126, EPI_ISL_1562127, EPI_ISL_1562128, EPI_ISL_1562129, EPI_ISL_1562130, EPI_ISL_1562131, EPI_ISL_1562132, EPI_ISL_1562133, EPI_ISL_1562134, EPI_ISL_1562135, EPI_ISL_1562136, EPI_ISL_1562137, EPI_ISL_1562138, EPI_ISL_1562139, EPI_ISL_1562140, EPI_ISL_1562141, EPI_ISL_1562142, EPI_ISL_1562143, EPI_ISL_1562144, EPI_ISL_1562145, EPI_ISL_1562146, EPI_ISL_1562147, EPI_ISL_1562148, EPI_ISL_1562149, EPI_ISL_1562150, EPI_ISL_1562151, EPI_ISL_1562152, EPI_ISL_1562153, EPI_ISL_1562155, EPI_ISL_1562156, EPI_ISL_1562157, EPI_ISL_1562158, EPI_ISL_1562159, EPI_ISL_1562160, EPI_ISL_1562162, EPI_ISL_1562164, EPI_ISL_1562165, EPI_ISL_1562166, EPI_ISL_1562167, EPI_ISL_1562168, EPI_ISL_1562169, EPI_ISL_1562170, EPI_ISL_1562171, EPI_ISL_1562172, EPI_ISL_1562173, EPI_ISL_1562174, EPI_ISL_1562175, EPI_ISL_1562176, EPI_ISL_1562177, EPI_ISL_1562179, EPI_ISL_1562180, EPI_ISL_1562181, EPI_ISL_1562183, EPI_ISL_1562184, EPI_ISL_1562185, EPI_ISL_1562186, EPI_ISL_1562187, EPI_ISL_1562188, EPI_ISL_1562189, EPI_ISL_1562190, EPI_ISL_1562192, EPI_ISL_1562193, EPI_ISL_1562194, EPI_ISL_1562195, EPI_ISL_1562196, EPI_ISL_1562197, EPI_ISL_1562198, EPI_ISL_1562199, EPI_ISL_1562200, EPI_ISL_1562201, EPI_ISL_1562202, EPI_ISL_1562203, EPI_ISL_1562204, EPI_ISL_1562205, EPI_ISL_1562206, EPI_ISL_1562207, EPI_ISL_1562208, EPI_ISL_1562209, EPI_ISL_1562211, EPI_ISL_1562213, EPI_ISL_1562214, EPI_ISL_1562216, EPI_ISL_1562217, EPI_ISL_1562218, EPI_ISL_1562219, EPI_ISL_1562220, EPI_ISL_1562221, EPI_ISL_1562222, EPI_ISL_1562223, EPI_ISL_1562224, EPI_ISL_1562225, EPI_ISL_1562226, EPI_ISL_1562228, EPI_ISL_1562229, EPI_ISL_1562230, EPI_ISL_1562231, EPI_ISL_1562232, EPI_ISL_1562233, EPI_ISL_1562234, EPI_ISL_1562235, EPI_ISL_1562236, EPI_ISL_1562237, EPI_ISL_1562238, EPI_ISL_1562239, EPI_ISL_1562240, EPI_ISL_1562241, EPI_ISL_1562242, EPI_ISL_1562243, EPI_ISL_1562244, EPI_ISL_1562245, EPI_ISL_1562246, EPI_ISL_1562247, EPI_ISL_1562248, EPI_ISL_1562249, EPI_ISL_1562250, EPI_ISL_1562251, EPI_ISL_1562252, EPI_ISL_1562253, EPI_ISL_1562254, EPI_ISL_1562255, EPI_ISL_1562259, EPI_ISL_1562260, EPI_ISL_1562261, EPI_ISL_1562262, EPI_ISL_1562264, EPI_ISL_1562265, EPI_ISL_1562266, EPI_ISL_1562267, EPI_ISL_1562268, EPI_ISL_1562269, EPI_ISL_1562270, EPI_ISL_1562271, EPI_ISL_1562272, EPI_ISL_1562273, EPI_ISL_1562274, EPI_ISL_1562275, EPI_ISL_1562276, EPI_ISL_1562277, EPI_ISL_1562278, EPI_ISL_1562279, EPI_ISL_1562280, EPI_ISL_1562281, EPI_ISL_1562282, EPI_ISL_1562283, EPI_ISL_1562284, EPI_ISL_1562286, EPI_ISL_1562287, EPI_ISL_1562288, EPI_ISL_1562289, EPI_ISL_1562290, EPI_ISL_1562291, EPI_ISL_1562292, EPI_ISL_1562293, EPI_ISL_1562294, EPI_ISL_1562295, EPI_ISL_1562296, EPI_ISL_1562297, EPI_ISL_1562298, EPI_ISL_1562299, EPI_ISL_1562300, EPI_ISL_1562301, EPI_ISL_1562302, EPI_ISL_1562303, EPI_ISL_1562305, EPI_ISL_1562306, EPI_ISL_1562307, EPI_ISL_1562308, EPI_ISL_1562309, EPI_ISL_1562311, EPI_ISL_1562312, EPI_ISL_1562313, EPI_ISL_1562314, EPI_ISL_1562315, EPI_ISL_1562316, EPI_ISL_1562317, EPI_ISL_1562318, EPI_ISL_1562321, EPI_ISL_1562322, EPI_ISL_1562323, EPI_ISL_1562324, EPI_ISL_1562325, EPI_ISL_1562326, EPI_ISL_1562327, EPI_ISL_1562329, EPI_ISL_1562330, EPI_ISL_1562331, EPI_ISL_1562332, EPI_ISL_1562333, EPI_ISL_1562334, EPI_ISL_1562335, EPI_ISL_1562336, EPI_ISL_1562337, EPI_ISL_1562338, EPI_ISL_1562339, EPI_ISL_1562341, EPI_ISL_1562342, EPI_ISL_1562343, EPI_ISL_1562344, EPI_ISL_1562345, EPI_ISL_1562346, EPI_ISL_1562347, EPI_ISL_1562348, EPI_ISL_1562349, EPI_ISL_1562350, EPI_ISL_1562351, EPI_ISL_1562352, EPI_ISL_1562353, EPI_ISL_1562354, EPI_ISL_1562355, EPI_ISL_1562356, EPI_ISL_1562358, EPI_ISL_1562359, EPI_ISL_1562360, EPI_ISL_1562361, EPI_ISL_1562362, EPI_ISL_1562363, EPI_ISL_1562364, EPI_ISL_1562365, EPI_ISL_1562366, EPI_ISL_1562367, EPI_ISL_1562368, EPI_ISL_1562369, EPI_ISL_1562370, EPI_ISL_1562371, EPI_ISL_1562372, EPI_ISL_1562373, EPI_ISL_1562374, EPI_ISL_1562375, EPI_ISL_1562376, EPI_ISL_1562377, EPI_ISL_1562378, EPI_ISL_1562379, EPI_ISL_1562380, EPI_ISL_1562381, EPI_ISL_1562382, EPI_ISL_1562383, EPI_ISL_1562384, EPI_ISL_1562385, EPI_ISL_1562386, EPI_ISL_1562387, EPI_ISL_1562388, EPI_ISL_1562389, EPI_ISL_1562390, EPI_ISL_1562391, EPI_ISL_1562392, EPI_ISL_1562393, EPI_ISL_1562394, EPI_ISL_1562395, EPI_ISL_1562396, EPI_ISL_1562397, EPI_ISL_1562398, EPI_ISL_1562399, EPI_ISL_1562400, EPI_ISL_1562401, EPI_ISL_1562402, EPI_ISL_1562403, EPI_ISL_1562404, EPI_ISL_1562405, EPI_ISL_1562406, EPI_ISL_1562407, EPI_ISL_1562408, EPI_ISL_1562409, EPI_ISL_1562411, EPI_ISL_1562412, EPI_ISL_1562413, EPI_ISL_1562414, EPI_ISL_1562415, EPI_ISL_1562416, EPI_ISL_1562417, EPI_ISL_1562418, EPI_ISL_1562419, EPI_ISL_1562421, EPI_ISL_1562422, EPI_ISL_1562423, EPI_ISL_1562424, EPI_ISL_1562425, EPI_ISL_1562426, EPI_ISL_1562427, EPI_ISL_1562428, EPI_ISL_1562429, EPI_ISL_1562431, EPI_ISL_1562432, EPI_ISL_1562433, EPI_ISL_1562434, EPI_ISL_1562435, EPI_ISL_1562436, EPI_ISL_1562437, EPI_ISL_1562438, EPI_ISL_1562439, EPI_ISL_1562440, EPI_ISL_1562441, EPI_ISL_1562442, EPI_ISL_1562444, EPI_ISL_1562445, EPI_ISL_1562446, EPI_ISL_1562447, EPI_ISL_1562448, EPI_ISL_1562449, EPI_ISL_1562450, EPI_ISL_1562451, EPI_ISL_1562452, EPI_ISL_1562453, EPI_ISL_1562454, EPI_ISL_1562455, EPI_ISL_1562456, EPI_ISL_1562457, EPI_ISL_1562458, EPI_ISL_1562459, EPI_ISL_1562460, EPI_ISL_1562461, EPI_ISL_1562462, EPI_ISL_1562463, EPI_ISL_1562464, EPI_ISL_1562465, EPI_ISL_1562466, EPI_ISL_1562467, EPI_ISL_1562468, EPI_ISL_1562469, EPI_ISL_1562470, EPI_ISL_1562472, EPI_ISL_1562473, EPI_ISL_1562474, EPI_ISL_1562475, EPI_ISL_1562476, EPI_ISL_1562477, EPI_ISL_1562478, EPI_ISL_1562479, EPI_ISL_1562480, EPI_ISL_1562482, EPI_ISL_1562483, EPI_ISL_1562484, EPI_ISL_1562485, EPI_ISL_1562487, EPI_ISL_1562488, EPI_ISL_1562489, EPI_ISL_1562490, EPI_ISL_1562491, EPI_ISL_1562492, EPI_ISL_1562493, EPI_ISL_1562494, EPI_ISL_1562495, EPI_ISL_1562496, EPI_ISL_1562497, EPI_ISL_1562499, EPI_ISL_1562500, EPI_ISL_1562501, EPI_ISL_1562502, EPI_ISL_1562503, EPI_ISL_1562504, EPI_ISL_1562505, EPI_ISL_1562506, EPI_ISL_1562507, EPI_ISL_1562508, EPI_ISL_1562509, EPI_ISL_1562510, EPI_ISL_1562511, EPI_ISL_1562512, EPI_ISL_1562513, EPI_ISL_1562515, EPI_ISL_1562516, EPI_ISL_1562517, EPI_ISL_1562518, EPI_ISL_1562519, EPI_ISL_1562520, EPI_ISL_1562521, EPI_ISL_1562522, EPI_ISL_1562523, EPI_ISL_1562524, EPI_ISL_1562525, EPI_ISL_1562526, EPI_ISL_1562527, EPI_ISL_1562528, EPI_ISL_1562530, EPI_ISL_1562531, EPI_ISL_1562532, EPI_ISL_1562533, EPI_ISL_1562534, EPI_ISL_1562535, EPI_ISL_1562536, EPI_ISL_1562537, EPI_ISL_1562538, EPI_ISL_1562539, EPI_ISL_1562540, EPI_ISL_1562541, EPI_ISL_1562542, EPI_ISL_1562543, EPI_ISL_1562544, EPI_ISL_1562545, EPI_ISL_1562547, EPI_ISL_1562548, EPI_ISL_1562549, EPI_ISL_1562550, EPI_ISL_1562551, EPI_ISL_1562552, EPI_ISL_1562553, EPI_ISL_1562554, EPI_ISL_1562556, EPI_ISL_1562557, EPI_ISL_1562558, EPI_ISL_1562559, EPI_ISL_1562560, EPI_ISL_1562561, EPI_ISL_1562562, EPI_ISL_1562563, EPI_ISL_1562564, EPI_ISL_1562565, EPI_ISL_1562566, EPI_ISL_1562567, EPI_ISL_1562568, EPI_ISL_1562569, EPI_ISL_1562570, EPI_ISL_1562571, EPI_ISL_1562572, EPI_ISL_1562574, EPI_ISL_1562576, EPI_ISL_1562577, EPI_ISL_1562578, EPI_ISL_1562579, EPI_ISL_1562580, EPI_ISL_1562581, EPI_ISL_1562582, EPI_ISL_1562583, EPI_ISL_1562584, EPI_ISL_1562585, EPI_ISL_1562586, EPI_ISL_1562587, EPI_ISL_1562588, EPI_ISL_1562589, EPI_ISL_1562590, EPI_ISL_1562591, EPI_ISL_1562592, EPI_ISL_1562594, EPI_ISL_1562595, EPI_ISL_1562596, EPI_ISL_1562597, EPI_ISL_1562598, EPI_ISL_1562599, EPI_ISL_1562600, EPI_ISL_1562602, EPI_ISL_1562603, EPI_ISL_1562604, EPI_ISL_1562605, EPI_ISL_1562606, EPI_ISL_1562607, EPI_ISL_1562608, EPI_ISL_1562609, EPI_ISL_1562610, EPI_ISL_1562612, EPI_ISL_1562613, EPI_ISL_1562614, EPI_ISL_1562615, EPI_ISL_1562616, EPI_ISL_1562617, EPI_ISL_1562618, EPI_ISL_1562619, EPI_ISL_1562620, EPI_ISL_1562621, EPI_ISL_1562622, EPI_ISL_1562623, EPI_ISL_1562624, EPI_ISL_1562625, EPI_ISL_1562626, EPI_ISL_1562628, EPI_ISL_1562629, EPI_ISL_1562630, EPI_ISL_1562631, EPI_ISL_1562632, EPI_ISL_1562633, EPI_ISL_1562634, EPI_ISL_1562635, EPI_ISL_1562636, EPI_ISL_1562637, EPI_ISL_1562638, EPI_ISL_1562639, EPI_ISL_1562640, EPI_ISL_1562641, EPI_ISL_1562642, EPI_ISL_1562643, EPI_ISL_1562644, EPI_ISL_1562645, EPI_ISL_1562646, EPI_ISL_1562647, EPI_ISL_1562648, EPI_ISL_1562649, EPI_ISL_1562650, EPI_ISL_1562651, EPI_ISL_1562652, EPI_ISL_1562653, EPI_ISL_1562654, EPI_ISL_1562655, EPI_ISL_1562657, EPI_ISL_1562658, EPI_ISL_1562661, EPI_ISL_1562662, EPI_ISL_1562663, EPI_ISL_1562664, EPI_ISL_1562665, EPI_ISL_1562666, EPI_ISL_1562667, EPI_ISL_1562668, EPI_ISL_1562669, EPI_ISL_1562670, EPI_ISL_1562671, EPI_ISL_1562673, EPI_ISL_1562675, EPI_ISL_1562676, EPI_ISL_1562677, EPI_ISL_1562678, EPI_ISL_1562679, EPI_ISL_1562680, EPI_ISL_1562681, EPI_ISL_1562682, EPI_ISL_1562683, EPI_ISL_1562684, EPI_ISL_1562685, EPI_ISL_1562686, EPI_ISL_1562687, EPI_ISL_1562688, EPI_ISL_1562689, EPI_ISL_1562690, EPI_ISL_1562691, EPI_ISL_1562692, EPI_ISL_1562694, EPI_ISL_1562695, EPI_ISL_1562696, EPI_ISL_1562697, EPI_ISL_1562698, EPI_ISL_1562699, EPI_ISL_1562700, EPI_ISL_1562701, EPI_ISL_1562702, EPI_ISL_1562703, EPI_ISL_1562704, EPI_ISL_1562705, EPI_ISL_1562706, EPI_ISL_1562707, EPI_ISL_1562708, EPI_ISL_1562709, EPI_ISL_1562710, EPI_ISL_1562711, EPI_ISL_1562712, EPI_ISL_1562713, EPI_ISL_1562714, EPI_ISL_1562715, EPI_ISL_1562716, EPI_ISL_1562717, EPI_ISL_1562718, EPI_ISL_1562719, EPI_ISL_1562720, EPI_ISL_1562721, EPI_ISL_1562722, EPI_ISL_1562723, EPI_ISL_1562724, EPI_ISL_1562725, EPI_ISL_1562726, EPI_ISL_1562727, EPI_ISL_1562728, EPI_ISL_1562729, EPI_ISL_1562730, EPI_ISL_1562731, EPI_ISL_1562732, EPI_ISL_1562733, EPI_ISL_1562734, EPI_ISL_1562735, EPI_ISL_1562736, EPI_ISL_1562737, EPI_ISL_1562738, EPI_ISL_1562739, EPI_ISL_1562740, EPI_ISL_1562741, EPI_ISL_1562742, EPI_ISL_1562743, EPI_ISL_1562744, EPI_ISL_1562745, EPI_ISL_1562747, EPI_ISL_1562748, EPI_ISL_1562749, EPI_ISL_1562750, EPI_ISL_1562751, EPI_ISL_1562752, EPI_ISL_1562753, EPI_ISL_1562754, EPI_ISL_1562755, EPI_ISL_1562756, EPI_ISL_1562757, EPI_ISL_1562758, EPI_ISL_1562759, EPI_ISL_1562760, EPI_ISL_1562762, EPI_ISL_1562763, EPI_ISL_1562764, EPI_ISL_1562765, EPI_ISL_1562766, EPI_ISL_1562767, EPI_ISL_1562768, EPI_ISL_1562769, EPI_ISL_1562770, EPI_ISL_1562771, EPI_ISL_1562772, EPI_ISL_1562773, EPI_ISL_1562774, EPI_ISL_1562775, EPI_ISL_1562776, EPI_ISL_1562778, EPI_ISL_1562779, EPI_ISL_1562780, EPI_ISL_1562781, EPI_ISL_1562782, EPI_ISL_1562783, EPI_ISL_1562784, EPI_ISL_1562785, EPI_ISL_1562786, EPI_ISL_1562787, EPI_ISL_1562788, EPI_ISL_1562789, EPI_ISL_1562790, EPI_ISL_1562791, EPI_ISL_1562792, EPI_ISL_1562793, EPI_ISL_1562794, EPI_ISL_1562795, EPI_ISL_1562796, EPI_ISL_1562797, EPI_ISL_1562798, EPI_ISL_1562799, EPI_ISL_1562800, EPI_ISL_1562801, EPI_ISL_1562802, EPI_ISL_1562803, EPI_ISL_1562804, EPI_ISL_1562805, EPI_ISL_1562806, EPI_ISL_1562807, EPI_ISL_1562808, EPI_ISL_1562810, EPI_ISL_1562811, EPI_ISL_1562812, EPI_ISL_1562813, EPI_ISL_1562814, EPI_ISL_1562815, EPI_ISL_1562816, EPI_ISL_1562817, EPI_ISL_1562818, EPI_ISL_1562819, EPI_ISL_1562820, EPI_ISL_1562821, EPI_ISL_1562822, EPI_ISL_1562823, EPI_ISL_1562824, EPI_ISL_1562825, EPI_ISL_1562826, EPI_ISL_1562827, EPI_ISL_1562828, EPI_ISL_1562829, EPI_ISL_1562830, EPI_ISL_1562831, EPI_ISL_1562832, EPI_ISL_1562834, EPI_ISL_1562835, EPI_ISL_1562836, EPI_ISL_1562837, EPI_ISL_1562838, EPI_ISL_1562839, EPI_ISL_1562840, EPI_ISL_1562841, EPI_ISL_1562843, EPI_ISL_1562844, EPI_ISL_1562845, EPI_ISL_1562846, EPI_ISL_1562847, EPI_ISL_1562848, EPI_ISL_1562849, EPI_ISL_1562850, EPI_ISL_1562851, EPI_ISL_1562852, EPI_ISL_1562853, EPI_ISL_1562854, EPI_ISL_1562855, EPI_ISL_1562856, EPI_ISL_1562858, EPI_ISL_1562859, EPI_ISL_1562860, EPI_ISL_1562861, EPI_ISL_1562862, EPI_ISL_1562864, EPI_ISL_1562865, EPI_ISL_1562866, EPI_ISL_1562867, EPI_ISL_1562868, EPI_ISL_1562869, EPI_ISL_1562870, EPI_ISL_1562871, EPI_ISL_1562873, EPI_ISL_1562874, EPI_ISL_1562875, EPI_ISL_1562876, EPI_ISL_1562877, EPI_ISL_1562878, EPI_ISL_1562879, EPI_ISL_1562880, EPI_ISL_1562881, EPI_ISL_1562882, EPI_ISL_1562883, EPI_ISL_1562884, EPI_ISL_1562885, EPI_ISL_1562886, EPI_ISL_1562887, EPI_ISL_1562888, EPI_ISL_1562889, EPI_ISL_1562890, EPI_ISL_1562891, EPI_ISL_1562892, EPI_ISL_1562893, EPI_ISL_1562894, EPI_ISL_1562895, EPI_ISL_1562896, EPI_ISL_1562897, EPI_ISL_1562898, EPI_ISL_1562899, EPI_ISL_1562900, EPI_ISL_1562901, EPI_ISL_1562902, EPI_ISL_1562904, EPI_ISL_1562905, EPI_ISL_1562906, EPI_ISL_1562907, EPI_ISL_1562908, EPI_ISL_1562909, EPI_ISL_1562911, EPI_ISL_1562912, EPI_ISL_1562913, EPI_ISL_1562914, EPI_ISL_1562915, EPI_ISL_1562916, EPI_ISL_1562917, EPI_ISL_1562918, EPI_ISL_1562919, EPI_ISL_1562920, EPI_ISL_1562921, EPI_ISL_1562922, EPI_ISL_1562923, EPI_ISL_1562924, EPI_ISL_1562925, EPI_ISL_1562926, EPI_ISL_1562927, EPI_ISL_1562928, EPI_ISL_1562929, EPI_ISL_1562930, EPI_ISL_1562931, EPI_ISL_1562932, EPI_ISL_1562933, EPI_ISL_1562934, EPI_ISL_1562935, EPI_ISL_1562936, EPI_ISL_1562937, EPI_ISL_1562938, EPI_ISL_1562939, EPI_ISL_1562940, EPI_ISL_1562941, EPI_ISL_1562942, EPI_ISL_1562944, EPI_ISL_1562945, EPI_ISL_1562946, EPI_ISL_1562947, EPI_ISL_1562948, EPI_ISL_1562949, EPI_ISL_1562950, EPI_ISL_1562951, EPI_ISL_1562953, EPI_ISL_1562954, EPI_ISL_1562955, EPI_ISL_1562956, EPI_ISL_1562957, EPI_ISL_1562959, EPI_ISL_1562960, EPI_ISL_1562961, EPI_ISL_1562963, EPI_ISL_1562964, EPI_ISL_1562965, EPI_ISL_1562967, EPI_ISL_1562968, EPI_ISL_1562969, EPI_ISL_1562970, EPI_ISL_1562971, EPI_ISL_1562972, EPI_ISL_1562973, EPI_ISL_1562974, EPI_ISL_1562975, EPI_ISL_1562976, EPI_ISL_1562977, EPI_ISL_1562978, EPI_ISL_1562979, EPI_ISL_1562980, EPI_ISL_1562981, EPI_ISL_1562982, EPI_ISL_1562983, EPI_ISL_1562984, EPI_ISL_1562985, EPI_ISL_1562986, EPI_ISL_1562987, EPI_ISL_1562989, EPI_ISL_1562990, EPI_ISL_1562991, EPI_ISL_1562992, EPI_ISL_1562994, EPI_ISL_1562995, EPI_ISL_1562996, EPI_ISL_1562997, EPI_ISL_1562998, EPI_ISL_1562999, EPI_ISL_1563000, EPI_ISL_1563001, EPI_ISL_1563002, EPI_ISL_1563003, EPI_ISL_1563004, EPI_ISL_1563005, EPI_ISL_1563006, EPI_ISL_1563007, EPI_ISL_1563008, EPI_ISL_1563010, EPI_ISL_1563011, EPI_ISL_1563012, EPI_ISL_1563013, EPI_ISL_1563014, EPI_ISL_1563015, EPI_ISL_1563016, EPI_ISL_1563017, EPI_ISL_1563018, EPI_ISL_1563019, EPI_ISL_1563020, EPI_ISL_1563021, EPI_ISL_1563022, EPI_ISL_1563023, EPI_ISL_1563024, EPI_ISL_1563025, EPI_ISL_1563026, EPI_ISL_1563027, EPI_ISL_1563028, EPI_ISL_1563029, EPI_ISL_1563030, EPI_ISL_1563031, EPI_ISL_1563032, EPI_ISL_1563033, EPI_ISL_1563034, EPI_ISL_1563035, EPI_ISL_1563036, EPI_ISL_1563037, EPI_ISL_1563038, EPI_ISL_1563039, EPI_ISL_1563040, EPI_ISL_1563041, EPI_ISL_1563042, EPI_ISL_1563044, EPI_ISL_1563045, EPI_ISL_1563046, EPI_ISL_1563047, EPI_ISL_1563049, EPI_ISL_1563050, EPI_ISL_1563051, EPI_ISL_1563052, EPI_ISL_1563053, EPI_ISL_1563054, EPI_ISL_1563056, EPI_ISL_1563057, EPI_ISL_1563058, EPI_ISL_1563059, EPI_ISL_1563060, EPI_ISL_1563061, EPI_ISL_1563062, EPI_ISL_1563063, EPI_ISL_1563064, EPI_ISL_1563065, EPI_ISL_1563067, EPI_ISL_1563068, EPI_ISL_1563069, EPI_ISL_1563070, EPI_ISL_1563071, EPI_ISL_1563072, EPI_ISL_1563073, EPI_ISL_1563074, EPI_ISL_1563076, EPI_ISL_1563077, EPI_ISL_1563078, EPI_ISL_1563079, EPI_ISL_1563080, EPI_ISL_1563081, EPI_ISL_1563082, EPI_ISL_1563083, EPI_ISL_1563085, EPI_ISL_1563086, EPI_ISL_1563087, EPI_ISL_1563088, EPI_ISL_1563089, EPI_ISL_1563090, EPI_ISL_1563091, EPI_ISL_1563092, EPI_ISL_1563093, EPI_ISL_1563094, EPI_ISL_1563095, EPI_ISL_1563096, EPI_ISL_1563097, EPI_ISL_1563098, EPI_ISL_1563099, EPI_ISL_1563100, EPI_ISL_1563101, EPI_ISL_1563102, EPI_ISL_1563103, EPI_ISL_1563104, EPI_ISL_1563110, EPI_ISL_1563111, EPI_ISL_1563112, EPI_ISL_1563113, EPI_ISL_1563114, EPI_ISL_1563115, EPI_ISL_1563117, EPI_ISL_1563118, EPI_ISL_1563119, EPI_ISL_1563120, EPI_ISL_1563121, EPI_ISL_1563122, EPI_ISL_1563123, EPI_ISL_1563124, EPI_ISL_1563125, EPI_ISL_1563126, EPI_ISL_1563127, EPI_ISL_1563128, EPI_ISL_1563129, EPI_ISL_1563130, EPI_ISL_1563131, EPI_ISL_1563132, EPI_ISL_1563133, EPI_ISL_1563134, EPI_ISL_1563135, EPI_ISL_1563136, EPI_ISL_1563137, EPI_ISL_1563138, EPI_ISL_1563139, EPI_ISL_1563140, EPI_ISL_1563141, EPI_ISL_1563142, EPI_ISL_1563143, EPI_ISL_1563144, EPI_ISL_1563145, EPI_ISL_1563146, EPI_ISL_1563147, EPI_ISL_1563148, EPI_ISL_1563149, EPI_ISL_1563150, EPI_ISL_1563151, EPI_ISL_1563152, EPI_ISL_1563153, EPI_ISL_1563154, EPI_ISL_1563155, EPI_ISL_1563156, EPI_ISL_1563157, EPI_ISL_1563158, EPI_ISL_1563159, EPI_ISL_1563160, EPI_ISL_1563161, EPI_ISL_1563162, EPI_ISL_1563163, EPI_ISL_1563164, EPI_ISL_1563165, EPI_ISL_1563166, EPI_ISL_1563167, EPI_ISL_1563168, EPI_ISL_1563169, EPI_ISL_1563170, EPI_ISL_1563171, EPI_ISL_1563172, EPI_ISL_1563173, EPI_ISL_1563174, EPI_ISL_1563175, EPI_ISL_1563176, EPI_ISL_1563177, EPI_ISL_1563178, EPI_ISL_1563179, EPI_ISL_1563180, EPI_ISL_1563181, EPI_ISL_1563182, EPI_ISL_1563183, EPI_ISL_1563184, EPI_ISL_1563185, EPI_ISL_1563186, EPI_ISL_1563187, EPI_ISL_1563188, EPI_ISL_1563189, EPI_ISL_1563190, EPI_ISL_1563191, EPI_ISL_1563192, EPI_ISL_1563193, EPI_ISL_1563194, EPI_ISL_1563195, EPI_ISL_1563196, EPI_ISL_1563197, EPI_ISL_1563198, EPI_ISL_1563199, EPI_ISL_1563201, EPI_ISL_1563202, EPI_ISL_1563203, EPI_ISL_1563204, EPI_ISL_1563205, EPI_ISL_1563206, EPI_ISL_1563207, EPI_ISL_1563208, EPI_ISL_1563209, EPI_ISL_1563210, EPI_ISL_1563211, EPI_ISL_1563212, EPI_ISL_1563213, EPI_ISL_1563214, EPI_ISL_1563215, EPI_ISL_1563216, EPI_ISL_1563217, EPI_ISL_1563218, EPI_ISL_1563219, EPI_ISL_1563220, EPI_ISL_1563221, EPI_ISL_1563222, EPI_ISL_1563223, EPI_ISL_1563224, EPI_ISL_1563225, EPI_ISL_1563226, EPI_ISL_1563227, EPI_ISL_1563228, EPI_ISL_1563229, EPI_ISL_1563230, EPI_ISL_1563231, EPI_ISL_1563232, EPI_ISL_1563233, EPI_ISL_1563234, EPI_ISL_1563235, EPI_ISL_1563236, EPI_ISL_1563238, EPI_ISL_1563239, EPI_ISL_1563240, EPI_ISL_1563241, EPI_ISL_1563243, EPI_ISL_1563244, EPI_ISL_1563245, EPI_ISL_1563246, EPI_ISL_1563247, EPI_ISL_1563248, EPI_ISL_1563249, EPI_ISL_1563250, EPI_ISL_1563251, EPI_ISL_1563252, EPI_ISL_1563253, EPI_ISL_1563254, EPI_ISL_1563255, EPI_ISL_1563256, EPI_ISL_1563257, EPI_ISL_1563258, EPI_ISL_1563259, EPI_ISL_1563260, EPI_ISL_1563261, EPI_ISL_1563262, EPI_ISL_1563263, EPI_ISL_1563264, EPI_ISL_1563265, EPI_ISL_1563266, EPI_ISL_1563267, EPI_ISL_1563268, EPI_ISL_1563269, EPI_ISL_1563270, EPI_ISL_1563271, EPI_ISL_1563272, EPI_ISL_1563273, EPI_ISL_1563274, EPI_ISL_1563275, EPI_ISL_1563276, EPI_ISL_1563277, EPI_ISL_1563278, EPI_ISL_1563279, EPI_ISL_1563280, EPI_ISL_1563281, EPI_ISL_1563282, EPI_ISL_1563283, EPI_ISL_1563284, EPI_ISL_1563285, EPI_ISL_1563286, EPI_ISL_1563287, EPI_ISL_1563288, EPI_ISL_1563289, EPI_ISL_1563290, EPI_ISL_1563291, EPI_ISL_1563293, EPI_ISL_1563294, EPI_ISL_1563295, EPI_ISL_1563296, EPI_ISL_1563297, EPI_ISL_1563298, EPI_ISL_1563299, EPI_ISL_1563300, EPI_ISL_1563301, EPI_ISL_1563303, EPI_ISL_1563304, EPI_ISL_1563305, EPI_ISL_1563306, EPI_ISL_1563307, EPI_ISL_1563308, EPI_ISL_1563309, EPI_ISL_1563310, EPI_ISL_1563311, EPI_ISL_1563312, EPI_ISL_1563313, EPI_ISL_1563314, EPI_ISL_1563315, EPI_ISL_1563316, EPI_ISL_1563318, EPI_ISL_1563320, EPI_ISL_1563321, EPI_ISL_1563322, EPI_ISL_1563323, EPI_ISL_1563324, EPI_ISL_1563326, EPI_ISL_1563327, EPI_ISL_1563329, EPI_ISL_1563331, EPI_ISL_1563332, EPI_ISL_1563333, EPI_ISL_1563334, EPI_ISL_1563335, EPI_ISL_1563336, EPI_ISL_1563337, EPI_ISL_1563339, EPI_ISL_1563340, EPI_ISL_1563341, EPI_ISL_1563342, EPI_ISL_1563343, EPI_ISL_1563344, EPI_ISL_1563345, EPI_ISL_1563347, EPI_ISL_1563349, EPI_ISL_1563350, EPI_ISL_1563351, EPI_ISL_1563352, EPI_ISL_1563354, EPI_ISL_1563355, EPI_ISL_1563356, EPI_ISL_1563357, EPI_ISL_1563358, EPI_ISL_1563359, EPI_ISL_1563360, EPI_ISL_1563361, EPI_ISL_1563362, EPI_ISL_1563363, EPI_ISL_1563364, EPI_ISL_1563366, EPI_ISL_1563367, EPI_ISL_1563369, EPI_ISL_1563371, EPI_ISL_1563372, EPI_ISL_1563373, EPI_ISL_1563374, EPI_ISL_1563375, EPI_ISL_1563376, EPI_ISL_1563377, EPI_ISL_1563378, EPI_ISL_1563380, EPI_ISL_1563381, EPI_ISL_1563382, EPI_ISL_1563383, EPI_ISL_1563384, EPI_ISL_1563385, EPI_ISL_1563386, EPI_ISL_1563387, EPI_ISL_1563388, EPI_ISL_1563389, EPI_ISL_1563390, EPI_ISL_1563391, EPI_ISL_1563392, EPI_ISL_1563393, EPI_ISL_1563394, EPI_ISL_1563395, EPI_ISL_1563396, EPI_ISL_1563397, EPI_ISL_1563398, EPI_ISL_1563399, EPI_ISL_1563400, EPI_ISL_1563402, EPI_ISL_1563403, EPI_ISL_1563404, EPI_ISL_1563405, EPI_ISL_1563406, EPI_ISL_1563408, EPI_ISL_1563409, EPI_ISL_1563410, EPI_ISL_1563411, EPI_ISL_1563412, EPI_ISL_1563413, EPI_ISL_1563414, EPI_ISL_1563415, EPI_ISL_1563416, EPI_ISL_1563418, EPI_ISL_1563419, EPI_ISL_1563420, EPI_ISL_1563421, EPI_ISL_1563422, EPI_ISL_1563423, EPI_ISL_1563424, EPI_ISL_1563425, EPI_ISL_1563426, EPI_ISL_1563428, EPI_ISL_1563429, EPI_ISL_1563430, EPI_ISL_1563431, EPI_ISL_1563432, EPI_ISL_1563433, EPI_ISL_1563434, EPI_ISL_1563435, EPI_ISL_1563436, EPI_ISL_1563437, EPI_ISL_1563438, EPI_ISL_1563439, EPI_ISL_1563440, EPI_ISL_1563441, EPI_ISL_1563443, EPI_ISL_1563444, EPI_ISL_1563445, EPI_ISL_1563447, EPI_ISL_1563448, EPI_ISL_1563449, EPI_ISL_1563450, EPI_ISL_1563451, EPI_ISL_1563452, EPI_ISL_1563453, EPI_ISL_1563455, EPI_ISL_1563456, EPI_ISL_1563457, EPI_ISL_1563459, EPI_ISL_1563460, EPI_ISL_1563461, EPI_ISL_1563462, EPI_ISL_1563463, EPI_ISL_1563464, EPI_ISL_1563465, EPI_ISL_1563466, EPI_ISL_1563468, EPI_ISL_1563469, EPI_ISL_1563470, EPI_ISL_1563471, EPI_ISL_1563472, EPI_ISL_1563473, EPI_ISL_1563474, EPI_ISL_1563476, EPI_ISL_1563477, EPI_ISL_1563478, EPI_ISL_1563479, EPI_ISL_1563480, EPI_ISL_1563481, EPI_ISL_1563482, EPI_ISL_1563483, EPI_ISL_1563484, EPI_ISL_1563485, EPI_ISL_1563486, EPI_ISL_1563487, EPI_ISL_1563488, EPI_ISL_1563489, EPI_ISL_1563490, EPI_ISL_1563491, EPI_ISL_1563492, EPI_ISL_1563493, EPI_ISL_1563494, EPI_ISL_1563495, EPI_ISL_1563496, EPI_ISL_1563497, EPI_ISL_1563498, EPI_ISL_1563499, EPI_ISL_1563500, EPI_ISL_1563501, EPI_ISL_1563502, EPI_ISL_1563503, EPI_ISL_1563504, EPI_ISL_1563505, EPI_ISL_1563506, EPI_ISL_1563508, EPI_ISL_1563509, EPI_ISL_1563510, EPI_ISL_1563511, EPI_ISL_1563512, EPI_ISL_1563513, EPI_ISL_1563514, EPI_ISL_1563515, EPI_ISL_1563516, EPI_ISL_1563517, EPI_ISL_1563518, EPI_ISL_1563519, EPI_ISL_1563520, EPI_ISL_1563521, EPI_ISL_1563522, EPI_ISL_1563523, EPI_ISL_1563524, EPI_ISL_1563525, EPI_ISL_1563526, EPI_ISL_1563527, EPI_ISL_1563529, EPI_ISL_1563531, EPI_ISL_1563532, EPI_ISL_1563533, EPI_ISL_1563534, EPI_ISL_1563535, EPI_ISL_1563536, EPI_ISL_1563537, EPI_ISL_1563538, EPI_ISL_1563539, EPI_ISL_1563540, EPI_ISL_1563541, EPI_ISL_1563542, EPI_ISL_1563543, EPI_ISL_1563544, EPI_ISL_1563545, EPI_ISL_1563546, EPI_ISL_1563547, EPI_ISL_1563548, EPI_ISL_1563549, EPI_ISL_1563550, EPI_ISL_1563551, EPI_ISL_1563552, EPI_ISL_1563553, EPI_ISL_1563554, EPI_ISL_1563556, EPI_ISL_1563557, EPI_ISL_1563558, EPI_ISL_1563560, EPI_ISL_1563561, EPI_ISL_1563562, EPI_ISL_1563563, EPI_ISL_1563564, EPI_ISL_1563565, EPI_ISL_1563566, EPI_ISL_1563567, EPI_ISL_1563568, EPI_ISL_1563569, EPI_ISL_1563570, EPI_ISL_1563571, EPI_ISL_1563572, EPI_ISL_1563574, EPI_ISL_1563575, EPI_ISL_1563576, EPI_ISL_1563577, EPI_ISL_1563578, EPI_ISL_1563579, EPI_ISL_1563580, EPI_ISL_1563581, EPI_ISL_1563583, EPI_ISL_1563584, EPI_ISL_1563585, EPI_ISL_1563586, EPI_ISL_1563587, EPI_ISL_1563588, EPI_ISL_1563589, EPI_ISL_1563590, EPI_ISL_1563591, EPI_ISL_1563592, EPI_ISL_1563593, EPI_ISL_1563594, EPI_ISL_1563595, EPI_ISL_1563596, EPI_ISL_1563597, EPI_ISL_1563598, EPI_ISL_1563599, EPI_ISL_1563600, EPI_ISL_1563601, EPI_ISL_1563602, EPI_ISL_1563603, EPI_ISL_1563604, EPI_ISL_1563606, EPI_ISL_1563607, EPI_ISL_1563608, EPI_ISL_1563609, EPI_ISL_1563610, EPI_ISL_1563611, EPI_ISL_1563612, EPI_ISL_1563613, EPI_ISL_1563614, EPI_ISL_1563616, EPI_ISL_1563617, EPI_ISL_1563619, EPI_ISL_1563620, EPI_ISL_1563621, EPI_ISL_1563622, EPI_ISL_1563623, EPI_ISL_1563624, EPI_ISL_1563625 | | | |
| see above | Aegis Sciences Corporation | Centers for Disease Control and Prevention Division of Viral Diseases, Pathogen Discovery | Dakota Howard, Dhwani Batra, Peter W. Cook, Kara Moser, Adrian Paskey, Jason Caravas, Benjamin Rambo-Martin, Shatavia Morrison, Christopher Gulvick, Scott Sammons, Yvette Unoarumhi, Darlene Wagner, Matthew Schmerer, Cyndi Clark, Patrick Campbell, Rob Case, Vikramsinha Ghorpade, Holly Houdeshell, Ola Kvalvaag, Dillon Nall, Ethan Sanders, Alec Vest, Shaun Westlund, Matthew Hardison, Clinton R. Paden, Duncan MacCannell |
| EPI_ISL_1563635, EPI_ISL_1563636, EPI_ISL_1563637, EPI_ISL_1563638, EPI_ISL_1563639, EPI_ISL_1563640, EPI_ISL_1563641, EPI_ISL_1563642, EPI_ISL_1563643, EPI_ISL_1563644, EPI_ISL_1563645, EPI_ISL_1563646, EPI_ISL_1563647, EPI_ISL_1563648, EPI_ISL_1563649, EPI_ISL_1563650, EPI_ISL_1563651, EPI_ISL_1563652, EPI_ISL_1563653, EPI_ISL_1563654, EPI_ISL_1563655, EPI_ISL_1563656, EPI_ISL_1563657, EPI_ISL_1563658, EPI_ISL_1563659, EPI_ISL_1563661, EPI_ISL_1563662, EPI_ISL_1563663, EPI_ISL_1563664, EPI_ISL_1563666, EPI_ISL_1563667, EPI_ISL_1563668, EPI_ISL_1563669, EPI_ISL_1563671, EPI_ISL_1563672, EPI_ISL_1563673, EPI_ISL_1563674, EPI_ISL_1563676, EPI_ISL_1563677 | | | |
| see above | IN State Department of Health Laboratory Services | IN State Department of Health Laboratory Services | Cassandra Campion, Jamie Yeadon, Brian Pope, Lixia Liu, Kyle Brownlee, Melissa Hindenlang, Ankita Kashikar, Mark Glazier |
| EPI_ISL_1563683, EPI_ISL_1563684, EPI_ISL_1563685, EPI_ISL_1563687, EPI_ISL_1563688, EPI_ISL_1563689, EPI_ISL_1563691, EPI_ISL_1563692, EPI_ISL_1563693, EPI_ISL_1563696, EPI_ISL_1563697, EPI_ISL_1563698, EPI_ISL_1563700, EPI_ISL_1563701, EPI_ISL_1563702, EPI_ISL_1563703, EPI_ISL_1563704, EPI_ISL_1563705, EPI_ISL_1563706, EPI_ISL_1563707, EPI_ISL_1563708, EPI_ISL_1563710, EPI_ISL_1563712, EPI_ISL_1563713, EPI_ISL_1563714, EPI_ISL_1563716, EPI_ISL_1563717, EPI_ISL_1563718, EPI_ISL_1563719, EPI_ISL_1563720, EPI_ISL_1563721, EPI_ISL_1563722, EPI_ISL_1563723, EPI_ISL_1563724, EPI_ISL_1563725, EPI_ISL_1563726, EPI_ISL_1563727, EPI_ISL_1563728, EPI_ISL_1563729, EPI_ISL_1563730, EPI_ISL_1563731, EPI_ISL_1563732, EPI_ISL_1563734, EPI_ISL_1563735, EPI_ISL_1563736 | | | |
| see above | Pathology and Laboratory Medicine Institute, Cleveland Clinic, Ohio, USA | Pathology and Laboratory Medicine Institute, Cleveland Clinic, Ohio, USA | Joy Nakitandwe, Zheng Jin Tu, Jay Brock, Yu-Wei Cheng, Gary Procop, Daniel Rhoads, Daniel H. Farkas, David Bosler |
| EPI_ISL_1570465, EPI_ISL_1570466, EPI_ISL_1570468, EPI_ISL_1570471, EPI_ISL_1570472, EPI_ISL_1570474, EPI_ISL_1570478, EPI_ISL_1570479, EPI_ISL_1570481, EPI_ISL_1570482, EPI_ISL_1570483, EPI_ISL_1570485, EPI_ISL_1570486, EPI_ISL_1570488, EPI_ISL_1570490, EPI_ISL_1570493, EPI_ISL_1570494, EPI_ISL_1570897, EPI_ISL_1570898, EPI_ISL_1570899, EPI_ISL_1570900, EPI_ISL_1570902, EPI_ISL_1570903, EPI_ISL_1570904, EPI_ISL_1570906, EPI_ISL_1570907, EPI_ISL_1570909, EPI_ISL_1570910, EPI_ISL_1570911, EPI_ISL_1570912, EPI_ISL_1570913, EPI_ISL_1570915, EPI_ISL_1570916, EPI_ISL_1570917, EPI_ISL_1570919, EPI_ISL_1570920, EPI_ISL_1570921, EPI_ISL_1570923, EPI_ISL_1570924, EPI_ISL_1570926, EPI_ISL_1570927, EPI_ISL_1570929, EPI_ISL_1570930, EPI_ISL_1570933, EPI_ISL_1570935, EPI_ISL_1571243, EPI_ISL_1571247, EPI_ISL_1571251, EPI_ISL_1571768, EPI_ISL_1571769, EPI_ISL_1571771, EPI_ISL_1571773, EPI_ISL_1571775, EPI_ISL_1571776, EPI_ISL_1571781, EPI_ISL_1571783, EPI_ISL_1571784, EPI_ISL_1571785, EPI_ISL_1571787, EPI_ISL_1571788, EPI_ISL_1571790, EPI_ISL_1571794, EPI_ISL_1571795, EPI_ISL_1571799, EPI_ISL_1571805, EPI_ISL_1571807, EPI_ISL_1571808, EPI_ISL_1571810, EPI_ISL_1571812, EPI_ISL_1571813, EPI_ISL_1571815, EPI_ISL_1571817, EPI_ISL_1571819, EPI_ISL_1571820, EPI_ISL_1571822, EPI_ISL_1571823, EPI_ISL_1571825, EPI_ISL_1571827, EPI_ISL_1571829, EPI_ISL_1571830, EPI_ISL_1572362, EPI_ISL_1572366, EPI_ISL_1572368, EPI_ISL_1572371, EPI_ISL_1572374, EPI_ISL_1572378, EPI_ISL_1572380, EPI_ISL_1572381, EPI_ISL_1572387, EPI_ISL_1572388, EPI_ISL_1572390, EPI_ISL_1572392, EPI_ISL_1572393, EPI_ISL_1572394, EPI_ISL_1572396 | | | |
| see above | Wyoming Public Health Laboratory | Wyoming Public Health Laboratory | Jim Mildenberger, Wanda Manley, Noah Hull, Taylor Fearing, Lynette Gumbleton, Channing Weber, Ashley Norberg, Chayse Rowley, Marley Goetz, Brian Dominguez, Elliot Thomasson, Cari Sloma, and Rob Christensen |
| EPI_ISL_1574966 | American Indian Health Service of Chicago | Illinois Department of Public Health - Chicago Lab | Vineet K. Dhiman, Ira Heimler |

| | | | |
|---|---|---|---|
| EPI_ISL_1574967 | Childrens Habilitation Center | Illinois Department of Public Health - Chicago Lab | Vineet K. Dhiman, Ira Heimler |
| EPI_ISL_1574968 | Near North Health Service | Illinois Department of Public Health - Chicago Lab | Vineet K. Dhiman, Ira Heimler |
| EPI_ISL_1574969 | Childrens Habilitation Center | Illinois Department of Public Health - Chicago Lab | Vineet K. Dhiman, Ira Heimler |
| EPI_ISL_1574970 | Lawndale Christian Health Center | Illinois Department of Public Health - Chicago Lab | Vineet K. Dhiman, Ira Heimler |
| EPI_ISL_1574971 | Loretto | Illinois Department of Public Health - Chicago Lab | Vineet K. Dhiman, Ira Heimler |
| EPI_ISL_1574972, EPI_ISL_1574973 | Near North Health Service | Illinois Department of Public Health - Chicago Lab | Vineet K. Dhiman, Ira Heimler |
| EPI_ISL_1574974 | Loretto | Illinois Department of Public Health - Chicago Lab | Vineet K. Dhiman, Ira Heimler |
| EPI_ISL_1574975, EPI_ISL_1574976 | Near North Health Service | Illinois Department of Public Health - Chicago Lab | Vineet K. Dhiman, Ira Heimler |
| EPI_ISL_1574977, EPI_ISL_1574978 | Loretto | Illinois Department of Public Health - Chicago Lab | Vineet K. Dhiman, Ira Heimler |
| EPI_ISL_1574979, EPI_ISL_1574980, EPI_ISL_1574981, EPI_ISL_1574982 | Near North Health Service | Illinois Department of Public Health - Chicago Lab | Vineet K. Dhiman, Ira Heimler |
| EPI_ISL_1574983 | Loretto | Illinois Department of Public Health - Chicago Lab | Vineet K. Dhiman, Ira Heimler |
| EPI_ISL_1574984 | Near North Health Service | Illinois Department of Public Health - Chicago Lab | Vineet K. Dhiman, Ira Heimler |
| EPI_ISL_1574985 | Loretto | Illinois Department of Public Health - Chicago Lab | Vineet K. Dhiman, Ira Heimler |
| EPI_ISL_1574986 | Childrens Habilitation Center | Illinois Department of Public Health - Chicago Lab | Vineet K. Dhiman, Ira Heimler |
| EPI_ISL_1574987 | Edward Hines Jr VA Hospital | Illinois Department of Public Health - Chicago Lab | Vineet K. Dhiman, Ira Heimler |
| EPI_ISL_1574988 | ARUP Laboratories | Illinois Department of Public Health - Chicago Lab | Vineet K. Dhiman, Ira Heimler |
| EPI_ISL_1574989 | Edward Hines Jr VA Hospital | Illinois Department of Public Health - Chicago Lab | Vineet K. Dhiman, Ira Heimler |
| EPI_ISL_1574990 | CentralDupage Hospital Microbiology Laboratory | Illinois Department of Public Health - Chicago Lab | Vineet K. Dhiman, Ira Heimler |
| EPI_ISL_1574991, EPI_ISL_1574992, EPI_ISL_1574993, EPI_ISL_1574994, EPI_ISL_1574995, EPI_ISL_1574996, EPI_ISL_1574997, EPI_ISL_1574998, EPI_ISL_1574999, EPI_ISL_1575000, EPI_ISL_1575001, EPI_ISL_1575002, EPI_ISL_1575003, EPI_ISL_1575004 | | | |
| see above | FHN Memorial Hospital | Illinois Department of Public Health - Chicago Lab | Vineet K. Dhiman, Ira Heimler |
| EPI_ISL_1575005 | Advocate Good Smaritian Hospital Laboratory | Illinois Department of Public Health - Chicago Lab | Vineet K. Dhiman, Ira Heimler |
| EPI_ISL_1575006 | Northwestern-Central Dupage Hospital | Illinois Department of Public Health - Chicago Lab | Vineet K. Dhiman, Ira Heimler |
| EPI_ISL_1575007 | Childrens Habilitation Center | Illinois Department of Public Health - Chicago Lab | Vineet K. Dhiman, Ira Heimler |
| EPI_ISL_1575008 | Loretto | Illinois Department of Public Health - Chicago Lab | Vineet K. Dhiman, Ira Heimler |
| EPI_ISL_1575009, EPI_ISL_1575010, EPI_ISL_1575011 | Near North Health Service | Illinois Department of Public Health - Chicago Lab | Vineet K. Dhiman, Ira Heimler |
| EPI_ISL_1575012 | Uptown Care Center | Illinois Department of Public Health - Chicago Lab | Vineet K. Dhiman, Ira Heimler |
| EPI_ISL_1575013, EPI_ISL_1575014 | Loretto | Illinois Department of Public Health - Chicago Lab | Vineet K. Dhiman, Ira Heimler |
| EPI_ISL_1575015 | Lydia Home Association | Illinois Department of Public Health - Chicago Lab | Vineet K. Dhiman, Ira Heimler |
| EPI_ISL_1575016, EPI_ISL_1575017, EPI_ISL_1575018 | Childrens Habilitation Center | Illinois Department of Public Health - Chicago Lab | Vineet K. Dhiman, Ira Heimler |
| EPI_ISL_1575019 | Near North Health Service | Illinois Department of Public Health - Chicago Lab | Vineet K. Dhiman, Ira Heimler |
| EPI_ISL_1575020, EPI_ISL_1575021, EPI_ISL_1575022 | Childrens Habilitation Center | Illinois Department of Public Health - Chicago Lab | Vineet K. Dhiman, Ira Heimler |
| EPI_ISL_1575023 | Loretto | Illinois Department of Public Health - Chicago Lab | Vineet K. Dhiman, Ira Heimler |
| EPI_ISL_1575024, EPI_ISL_1575025 | Childrens Habilitation Center | Illinois Department of Public Health - Chicago Lab | Vineet K. Dhiman, Ira Heimler |
| EPI_ISL_1575026, EPI_ISL_1575028 | Loretto | Illinois Department of Public Health - Chicago Lab | Vineet K. Dhiman, Ira Heimler |
| EPI_ISL_1575029 | Childrens Habilitation Center | Illinois Department of Public Health - Chicago Lab | Vineet K. Dhiman, Ira Heimler |
| EPI_ISL_1575030 | Loretto | Illinois Department of Public Health - Chicago Lab | Vineet K. Dhiman, Ira Heimler |
| EPI_ISL_1575031 | Near North Health Service | Illinois Department of Public Health - Chicago Lab | Vineet K. Dhiman, Ira Heimler |
| EPI_ISL_1575032 | Loretto | Illinois Department of Public Health - Chicago Lab | Vineet K. Dhiman, Ira Heimler |
| EPI_ISL_1575033 | Near North Health Service | Illinois Department of Public Health - Chicago Lab | Vineet K. Dhiman, Ira Heimler |
| EPI_ISL_1575034 | Loretto | Illinois Department of Public Health - Chicago Lab | Vineet K. Dhiman, Ira Heimler |
| EPI_ISL_1575036, EPI_ISL_1575037, EPI_ISL_1575038, EPI_ISL_1575039, EPI_ISL_1575040, EPI_ISL_1575041, EPI_ISL_1575042, EPI_ISL_1575043, EPI_ISL_1575044, EPI_ISL_1575045, EPI_ISL_1575046, EPI_ISL_1575047, EPI_ISL_1575048, EPI_ISL_1575049, EPI_ISL_1575050, EPI_ISL_1575051, EPI_ISL_1575052, EPI_ISL_1575053, EPI_ISL_1575054, EPI_ISL_1575055, EPI_ISL_1575056, EPI_ISL_1575057, EPI_ISL_1575058, EPI_ISL_1575059, EPI_ISL_1575060, EPI_ISL_1575061, EPI_ISL_1575062, EPI_ISL_1575063, EPI_ISL_1575064, EPI_ISL_1575065, EPI_ISL_1575066, EPI_ISL_1575067, EPI_ISL_1575068, EPI_ISL_1575069, EPI_ISL_1575070, EPI_ISL_1575071, EPI_ISL_1575072, EPI_ISL_1575073, EPI_ISL_1575074, EPI_ISL_1575075, EPI_ISL_1575076, EPI_ISL_1575077, EPI_ISL_1575078, EPI_ISL_1575079, EPI_ISL_1575081, EPI_ISL_1575082, EPI_ISL_1575083, EPI_ISL_1575084, EPI_ISL_1575085, EPI_ISL_1575086, EPI_ISL_1575087, EPI_ISL_1575088, EPI_ISL_1575089, EPI_ISL_1575090, EPI_ISL_1575091, EPI_ISL_1575092, EPI_ISL_1575093, EPI_ISL_1575094, EPI_ISL_1575095, EPI_ISL_1575096, EPI_ISL_1575097, EPI_ISL_1575098, EPI_ISL_1575099, EPI_ISL_1575100, EPI_ISL_1575101, EPI_ISL_1575102, EPI_ISL_1575103, EPI_ISL_1575104, EPI_ISL_1575105, EPI_ISL_1575106, EPI_ISL_1575107, EPI_ISL_1575108, EPI_ISL_1575109, EPI_ISL_1575110, EPI_ISL_1575111 | | | |
| see above | Alaska State Virology Laboratory | Alaska State Virology Laboratory | Stephanie DeRonde, Elva House, Lisa Smith, Ph.D., Jack Chen, Ph.D. |
| EPI_ISL_1575114, EPI_ISL_1575116, EPI_ISL_1575118, EPI_ISL_1575119, EPI_ISL_1575120, EPI_ISL_1575121, EPI_ISL_1575122, EPI_ISL_1575123, EPI_ISL_1575124, EPI_ISL_1575125, EPI_ISL_1575126 | | | |
| see above | Alameda County Public Health | Alameda County Public Health Department | Kristna Hsieh, Maria Elma, Kevin Lubuit |

| | | | |
|---|---|---|---|
| | Department | | |
| EPI_ISL_1575128 | Mayo Clinic & Mayo Clinic Laboratories | Minnesota Department of Health, Public Health Laboratory | Alexandra Lorentz, Jacob Garfin, Matt Plumb, and Xiong Wang |
| EPI_ISL_1575137, EPI_ISL_1575138, EPI_ISL_1575139, EPI_ISL_1575140, EPI_ISL_1575141, EPI_ISL_1575142, EPI_ISL_1575143, EPI_ISL_1575144, EPI_ISL_1575145, EPI_ISL_1575146, EPI_ISL_1575147, EPI_ISL_1575148, EPI_ISL_1575149, EPI_ISL_1575150, EPI_ISL_1575151, EPI_ISL_1575152, EPI_ISL_1575153, EPI_ISL_1575154, EPI_ISL_1575155, EPI_ISL_1575156, EPI_ISL_1575157, EPI_ISL_1575158, EPI_ISL_1575159, EPI_ISL_1575160, EPI_ISL_1575161, EPI_ISL_1575162, EPI_ISL_1575163, EPI_ISL_1575164, EPI_ISL_1575165, EPI_ISL_1575166, EPI_ISL_1575167, EPI_ISL_1575168, EPI_ISL_1575169, EPI_ISL_1575170, EPI_ISL_1575171, EPI_ISL_1575172, EPI_ISL_1575173, EPI_ISL_1575174, EPI_ISL_1575175, EPI_ISL_1575176, EPI_ISL_1575177, EPI_ISL_1575178, EPI_ISL_1575179, EPI_ISL_1575180, EPI_ISL_1575181, EPI_ISL_1575182, EPI_ISL_1575183 | | | |
| see above | MD PHL | MD PHL | Maryland Department of Health Laboratories Administration |
| EPI_ISL_1575184, EPI_ISL_1575185, EPI_ISL_1575187, EPI_ISL_1575188, EPI_ISL_1575190, EPI_ISL_1575191, EPI_ISL_1575193, EPI_ISL_1575196, EPI_ISL_1575199, EPI_ISL_1575201, EPI_ISL_1575202, EPI_ISL_1575203, EPI_ISL_1575204, EPI_ISL_1575205, EPI_ISL_1575207, EPI_ISL_1575209, EPI_ISL_1575212, EPI_ISL_1575215, EPI_ISL_1575216, EPI_ISL_1575218, EPI_ISL_1575219, EPI_ISL_1575220, EPI_ISL_1575221, EPI_ISL_1575222, EPI_ISL_1575223, EPI_ISL_1575224, EPI_ISL_1575225, EPI_ISL_1575226, EPI_ISL_1575228, EPI_ISL_1575229, EPI_ISL_1575230, EPI_ISL_1575231, EPI_ISL_1575232, EPI_ISL_1575233, EPI_ISL_1575235, EPI_ISL_1575236, EPI_ISL_1575239, EPI_ISL_1575243, EPI_ISL_1575244, EPI_ISL_1575245, EPI_ISL_1575246, EPI_ISL_1575248, EPI_ISL_1575249, EPI_ISL_1575250 | | | |
| see above | University of Texas, Genomic Sequencing and Analysis Facility | University of Texas, Genomic Sequencing and Analysis Facility | Jessica Podnar, Sylvie Beaudenon, Audrey Kelly, Zachary Carver, Anna Battenhouse, Andreas Matouschek |
| EPI_ISL_1575251, EPI_ISL_1575252, EPI_ISL_1575253, EPI_ISL_1575255, EPI_ISL_1575256, EPI_ISL_1575257, EPI_ISL_1575258, EPI_ISL_1575259, EPI_ISL_1575260, EPI_ISL_1575261, EPI_ISL_1575262, EPI_ISL_1575263, EPI_ISL_1575264, EPI_ISL_1575265, EPI_ISL_1575266, EPI_ISL_1575267, EPI_ISL_1575268, EPI_ISL_1575269, EPI_ISL_1575270, EPI_ISL_1575271, EPI_ISL_1575272, EPI_ISL_1575273, EPI_ISL_1575274, EPI_ISL_1575276, EPI_ISL_1575277, EPI_ISL_1575278, EPI_ISL_1575279, EPI_ISL_1575280, EPI_ISL_1575281, EPI_ISL_1575282, EPI_ISL_1575283, EPI_ISL_1575284, EPI_ISL_1575285, EPI_ISL_1575286, EPI_ISL_1575288, EPI_ISL_1575289, EPI_ISL_1575290, EPI_ISL_1575291, EPI_ISL_1575292, EPI_ISL_1575293, EPI_ISL_1575294 | | | |
| see above | Nevada State Public Health Laboratory | Nevada State Public Health Laboratory | Andrew Gorzalski, Mark Pandori |
| EPI_ISL_1575308, EPI_ISL_1575309, EPI_ISL_1575311, EPI_ISL_1575312, EPI_ISL_1575313, EPI_ISL_1575314, EPI_ISL_1575315, EPI_ISL_1575316, EPI_ISL_1575317, EPI_ISL_1575318, EPI_ISL_1575319, EPI_ISL_1575320, EPI_ISL_1575321, EPI_ISL_1575322, EPI_ISL_1575323, EPI_ISL_1575324, EPI_ISL_1575350, EPI_ISL_1575351, EPI_ISL_1575352, EPI_ISL_1575353, EPI_ISL_1575354, EPI_ISL_1575355, EPI_ISL_1575356, EPI_ISL_1575357, EPI_ISL_1575358, EPI_ISL_1575359, EPI_ISL_1575360, EPI_ISL_1575361, EPI_ISL_1575362, EPI_ISL_1575363, EPI_ISL_1575364, EPI_ISL_1575365, EPI_ISL_1575366, EPI_ISL_1575367, EPI_ISL_1575368, EPI_ISL_1575369, EPI_ISL_1575370, EPI_ISL_1575371, EPI_ISL_1575372, EPI_ISL_1575373, EPI_ISL_1575374, EPI_ISL_1575375, EPI_ISL_1575376, EPI_ISL_1575377, EPI_ISL_1575378, EPI_ISL_1575379, EPI_ISL_1575380, EPI_ISL_1575381, EPI_ISL_1575382, EPI_ISL_1575383, EPI_ISL_1575384, EPI_ISL_1575385, EPI_ISL_1575386, EPI_ISL_1575387, EPI_ISL_1575388, EPI_ISL_1575389, EPI_ISL_1575390, EPI_ISL_1575391, EPI_ISL_1575392, EPI_ISL_1575393, EPI_ISL_1575394, EPI_ISL_1575395, EPI_ISL_1575396, EPI_ISL_1575397, EPI_ISL_1575398, EPI_ISL_1575399, EPI_ISL_1575400, EPI_ISL_1575401, EPI_ISL_1575402, EPI_ISL_1575403, EPI_ISL_1575404, EPI_ISL_1575405, EPI_ISL_1575406, EPI_ISL_1575407, EPI_ISL_1575408, EPI_ISL_1575409, EPI_ISL_1575410, EPI_ISL_1575411, EPI_ISL_1575412, EPI_ISL_1575413, EPI_ISL_1575414 | | | |
| see above | Pathology and Laboratory Medicine Institute, Cleveland Clinic, Ohio, USA | Pathology and Laboratory Medicine Institute, Cleveland Clinic, Ohio, USA | Joy Nakitandwe, Zheng Jin Tu, Jay Brock, Yu-Wei Cheng, Gary Procop, Daniel Rhoads, Daniel H. Farkas, David Bosler |
| EPI_ISL_1575415, EPI_ISL_1575417 | Kansas Health and Environmental Lab | Kansas Health and Environmental Lab | Mike Grose, Jonathan Barnell, Ben Olsen, and Phil Adam |
| EPI_ISL_1575419 | Wichita State University - Molecular Diagnostics Lab | Kansas Health and Environmental Lab | Mike Grose, Jonathan Barnell, Ben Olsen, and Phil Adam |
| EPI_ISL_1575420, EPI_ISL_1575421, EPI_ISL_1575422, EPI_ISL_1575423, EPI_ISL_1575424, EPI_ISL_1575425, EPI_ISL_1575426, EPI_ISL_1575427, EPI_ISL_1575428, EPI_ISL_1575429, EPI_ISL_1575430, EPI_ISL_1575431, EPI_ISL_1575432, EPI_ISL_1575433, EPI_ISL_1575434 | | | |
| see above | Kansas Health and Environmental Lab | Kansas Health and Environmental Lab | Mike Grose, Jonathan Barnell, Ben Olsen, and Phil Adam |
| EPI_ISL_1575435 | Wichita State University - Molecular Diagnostics Lab | Kansas Health and Environmental Lab | Mike Grose, Jonathan Barnell, Ben Olsen, and Phil Adam |
| EPI_ISL_1575445 | SUNY UPSTATE MEDICAL UNIVERSITY | Wadsworth Center, New York State Department of Health | Kirsten St. George, Daryl M. Lamson, Alexis Russell, Matthew Shudt, Melissa A Leisner, Jonathan Plitnick, Catharine Prussing, Navjot Singh, John Kelly, Erasmus Schneider, Erica Lasek-Nesselquist |
| EPI_ISL_1575446, EPI_ISL_1575447, EPI_ISL_1575448 | URMC LABS | Wadsworth Center, New York State Department of Health | Kirsten St. George, Daryl M. Lamson, Alexis Russell, Matthew Shudt, Melissa A Leisner, Jonathan Plitnick, Catharine Prussing, Navjot Singh, John Kelly, Erasmus Schneider, Erica Lasek-Nesselquist |
| EPI_ISL_1575449 | NORTHWELL HEALTH LABORATORIES | Wadsworth Center, New York State Department of Health | Kirsten St. George, Daryl M. Lamson, Alexis Russell, Matthew Shudt, Melissa A Leisner, Jonathan Plitnick, Catharine Prussing, Navjot Singh, John Kelly, Erasmus Schneider, Erica Lasek-Nesselquist |
| EPI_ISL_1575450, EPI_ISL_1575451, EPI_ISL_1575452 | URMC LABS | Wadsworth Center, New York State Department of Health | Kirsten St. George, Daryl M. Lamson, Alexis Russell, Matthew Shudt, Melissa A Leisner, Jonathan Plitnick, Catharine Prussing, Navjot Singh, John Kelly, Erasmus Schneider, Erica Lasek-Nesselquist |
| EPI_ISL_1575453, EPI_ISL_1575454 | WESTCHESTER MEDICAL CENTER | Wadsworth Center, New York State Department of Health | Kirsten St. George, Daryl M. Lamson, Alexis Russell, Matthew Shudt, Melissa A Leisner, Jonathan Plitnick, Catharine Prussing, Navjot Singh, John Kelly, Erasmus Schneider, Erica Lasek-Nesselquist |
| EPI_ISL_1575455, EPI_ISL_1575456, EPI_ISL_1575457, EPI_ISL_1575458 | URMC LABS | Wadsworth Center, New York State Department of Health | Kirsten St. George, Daryl M. Lamson, Alexis Russell, Matthew Shudt, Melissa A Leisner, Jonathan Plitnick, Catharine Prussing, Navjot Singh, John Kelly, Erasmus Schneider, Erica Lasek-Nesselquist |
| EPI_ISL_1575459 | NYC Pandemic Response Lab | Wadsworth Center, New York State Department of Health | Kirsten St. George, Daryl M. Lamson, Alexis Russell, Matthew Shudt, Melissa A Leisner, Jonathan Plitnick, Catharine Prussing, Navjot Singh, John Kelly, Erasmus Schneider, Erica Lasek-Nesselquist |
| EPI_ISL_1575460, EPI_ISL_1575461, EPI_ISL_1575462, EPI_ISL_1575463, EPI_ISL_1575464, EPI_ISL_1575465, EPI_ISL_1575466, EPI_ISL_1575467, EPI_ISL_1575468 | URMC LABS | Wadsworth Center, New York State Department of Health | Kirsten St. George, Daryl M. Lamson, Alexis Russell, Matthew Shudt, Melissa A Leisner, Jonathan Plitnick, Catharine Prussing, Navjot Singh, John Kelly, Erasmus Schneider, Erica Lasek-Nesselquist |
| EPI_ISL_1575469 | NORTHWELL HEALTH LABORATORIES | Wadsworth Center, New York State Department of Health | Kirsten St. George, Daryl M. Lamson, Alexis Russell, Matthew Shudt, Melissa A Leisner, Jonathan Plitnick, Catharine Prussing, Navjot Singh, John Kelly, Erasmus Schneider, Erica Lasek-Nesselquist |
| EPI_ISL_1575470 | MONTEFIORE MEDICAL CENTER LABORATORIES | Wadsworth Center, New York State Department of Health | Kirsten St. George, Daryl M. Lamson, Alexis Russell, Matthew Shudt, Melissa A Leisner, Jonathan Plitnick, Catharine Prussing, Navjot Singh, John Kelly, Erasmus Schneider, Erica Lasek-Nesselquist |
| EPI_ISL_1575471 | TEMPUS LABS INC | Wadsworth Center, New York State Department of Health | Kirsten St. George, Daryl M. Lamson, Alexis Russell, Matthew Shudt, Melissa A Leisner, Jonathan Plitnick, Catharine Prussing, Navjot Singh, John Kelly, Erasmus Schneider, Erica Lasek-Nesselquist |
| EPI_ISL_1575472 | Columbia University Irving Medical Center | Wadsworth Center, New York State Department of Health | Kirsten St. George, Daryl M. Lamson, Alexis Russell, Matthew Shudt, Melissa A Leisner, Jonathan Plitnick, Catharine Prussing, Navjot Singh, John Kelly, Erasmus Schneider, Erica Lasek-Nesselquist |
| EPI_ISL_1575473, EPI_ISL_1575474, EPI_ISL_1575475, EPI_ISL_1575476 | URMC LABS | Wadsworth Center, New York State Department of Health | Kirsten St. George, Daryl M. Lamson, Alexis Russell, Matthew Shudt, Melissa A Leisner, Jonathan Plitnick, Catharine Prussing, Navjot Singh, John Kelly, Erasmus Schneider, Erica Lasek-Nesselquist |
| EPI_ISL_1575477 | BOSTON HEART DIAGNOSTICS CORP | Wadsworth Center, New York State Department of Health | Kirsten St. George, Daryl M. Lamson, Alexis Russell, Matthew Shudt, Melissa A Leisner, Jonathan Plitnick, Catharine Prussing, Navjot Singh, John Kelly, Erasmus Schneider, Erica Lasek-Nesselquist |
| EPI_ISL_1575478 | NORTHWELL HEALTH LABORATORIES | Wadsworth Center, New York State Department of Health | Kirsten St. George, Daryl M. Lamson, Alexis Russell, Matthew Shudt, Melissa A Leisner, Jonathan Plitnick, Catharine Prussing, Navjot Singh, John Kelly, Erasmus Schneider, Erica Lasek-Nesselquist |
| EPI_ISL_1575480, EPI_ISL_1575481, EPI_ISL_1575482, EPI_ISL_1575483, EPI_ISL_1575484, EPI_ISL_1575485, EPI_ISL_1575486, EPI_ISL_1575487, EPI_ISL_1575488, EPI_ISL_1575489, EPI_ISL_1575490, EPI_ISL_1575491, EPI_ISL_1575492, EPI_ISL_1575493, EPI_ISL_1575494, EPI_ISL_1575495, EPI_ISL_1575496, EPI_ISL_1575497, EPI_ISL_1575498, EPI_ISL_1575499, EPI_ISL_1575500, EPI_ISL_1575501, EPI_ISL_1575502, EPI_ISL_1575503, EPI_ISL_1575504, EPI_ISL_1575505, EPI_ISL_1575506, EPI_ISL_1575507, EPI_ISL_1575508, EPI_ISL_1575509, EPI_ISL_1575510, EPI_ISL_1575511, EPI_ISL_1575512, EPI_ISL_1575513, EPI_ISL_1575514, EPI_ISL_1575515, EPI_ISL_1575516, EPI_ISL_1575517, EPI_ISL_1575518, EPI_ISL_1575519, EPI_ISL_1575520, EPI_ISL_1575521, EPI_ISL_1575522, EPI_ISL_1575523, EPI_ISL_1575524, EPI_ISL_1575525, EPI_ISL_1575526, EPI_ISL_1575527, EPI_ISL_1575528, EPI_ISL_1575529, EPI_ISL_1575530, EPI_ISL_1575531, EPI_ISL_1575532, EPI_ISL_1575533, EPI_ISL_1575534, EPI_ISL_1575535, EPI_ISL_1575536, EPI_ISL_1575537, EPI_ISL_1575538, EPI_ISL_1575539, EPI_ISL_1575540, EPI_ISL_1575541, EPI_ISL_1575542, EPI_ISL_1575543, EPI_ISL_1575544, EPI_ISL_1575545, EPI_ISL_1575547, EPI_ISL_1575548, EPI_ISL_1575549, EPI_ISL_1575550, EPI_ISL_1575551, EPI_ISL_1575552, EPI_ISL_1575553, EPI_ISL_1575554, EPI_ISL_1575555, EPI_ISL_1575556, EPI_ISL_1575557, EPI_ISL_1575558, EPI_ISL_1575559, EPI_ISL_1575560, EPI_ISL_1575561, EPI_ISL_1575562, EPI_ISL_1575563, EPI_ISL_1575564, EPI_ISL_1575565, EPI_ISL_1575566, EPI_ISL_1575567, EPI_ISL_1575568, EPI_ISL_1575569, EPI_ISL_1575570, EPI_ISL_1575571, EPI_ISL_1575572, EPI_ISL_1575573, EPI_ISL_1575574, EPI_ISL_1575575, EPI_ISL_1575576, EPI_ISL_1575577, EPI_ISL_1575578, EPI_ISL_1575579, EPI_ISL_1575580, EPI_ISL_1575581, EPI_ISL_1575582, EPI_ISL_1575583, EPI_ISL_1575584, EPI_ISL_1575585, EPI_ISL_1575586, EPI_ISL_1575587, EPI_ISL_1575588, EPI_ISL_1575589, EPI_ISL_1575590, EPI_ISL_1575591, EPI_ISL_1575592, EPI_ISL_1575593, EPI_ISL_1575594, EPI_ISL_1575595, EPI_ISL_1575596, EPI_ISL_1575597, EPI_ISL_1575598, EPI_ISL_1575599, EPI_ISL_1575600, EPI_ISL_1575601, EPI_ISL_1575602, EPI_ISL_1575603, EPI_ISL_1575604, EPI_ISL_1575605, EPI_ISL_1575606, EPI_ISL_1575607, EPI_ISL_1575608, EPI_ISL_1575609, EPI_ISL_1575610, EPI_ISL_1575611, EPI_ISL_1575612, EPI_ISL_1575613, EPI_ISL_1575614, EPI_ISL_1575615, EPI_ISL_1575616, EPI_ISL_1575617, EPI_ISL_1575618, EPI_ISL_1575619, EPI_ISL_1575620, EPI_ISL_1575621, EPI_ISL_1575622, EPI_ISL_1575623, EPI_ISL_1575624, EPI_ISL_1575625, EPI_ISL_1575626, EPI_ISL_1575627, EPI_ISL_1575628, EPI_ISL_1575629, EPI_ISL_1575630, EPI_ISL_1575631, EPI_ISL_1575632, EPI_ISL_1575633, EPI_ISL_1575634, EPI_ISL_1575636, EPI_ISL_1575637, EPI_ISL_1575638, EPI_ISL_1575639, EPI_ISL_1575640, EPI_ISL_1575641, EPI_ISL_1575642, EPI_ISL_1575643, EPI_ISL_1575644, EPI_ISL_1575645, EPI_ISL_1575646, EPI_ISL_1575647, EPI_ISL_1575648, EPI_ISL_1575649, EPI_ISL_1575650, EPI_ISL_1575651, EPI_ISL_1575652, EPI_ISL_1575653, EPI_ISL_1575654, EPI_ISL_1575655, EPI_ISL_1575656, EPI_ISL_1575657, EPI_ISL_1575658, EPI_ISL_1575659, EPI_ISL_1575660, EPI_ISL_1575661, EPI_ISL_1575662, EPI_ISL_1575663, EPI_ISL_1575664, EPI_ISL_1575666, EPI_ISL_1575667, EPI_ISL_1575668, EPI_ISL_1575669, EPI_ISL_1575670, EPI_ISL_1575671, EPI_ISL_1575673, EPI_ISL_1575675, EPI_ISL_1575676, EPI_ISL_1575680, EPI_ISL_1575682, EPI_ISL_1575683, EPI_ISL_1575685, EPI_ISL_1575687, EPI_ISL_1575688, EPI_ISL_1575690, EPI_ISL_1575691, EPI_ISL_1575693, EPI_ISL_1575696, EPI_ISL_1575699, EPI_ISL_1575701, EPI_ISL_1575703, EPI_ISL_1575706, EPI_ISL_1575707, EPI_ISL_1575709, EPI_ISL_1575710, EPI_ISL_1575712, EPI_ISL_1575713, EPI_ISL_1575715, EPI_ISL_1575717, EPI_ISL_1575718, EPI_ISL_1575721, EPI_ISL_1575723, EPI_ISL_1575724, EPI_ISL_1575726, EPI_ISL_1575727, EPI_ISL_1575729, EPI_ISL_1575730, EPI_ISL_1575732, EPI_ISL_1575733, EPI_ISL_1575735, EPI_ISL_1575737, EPI_ISL_1575738, EPI_ISL_1575741, EPI_ISL_1575742, EPI_ISL_1575744, EPI_ISL_1575745, EPI_ISL_1575747, EPI_ISL_1575748, EPI_ISL_1575750, EPI_ISL_1575751, EPI_ISL_1575753, EPI_ISL_1575754, EPI_ISL_1575756, EPI_ISL_1575757, EPI_ISL_1575759, EPI_ISL_1575762, EPI_ISL_1575764, EPI_ISL_1575765, EPI_ISL_1575767, EPI_ISL_1575768, EPI_ISL_1575770, EPI_ISL_1575771, EPI_ISL_1575773, EPI_ISL_1575775, EPI_ISL_1575777, EPI_ISL_1575778, EPI_ISL_1575781, EPI_ISL_1575782, EPI_ISL_1575784, EPI_ISL_1575785, EPI_ISL_1575788, EPI_ISL_1575789, EPI_ISL_1575790, EPI_ISL_1575791, EPI_ISL_1575792, EPI_ISL_1575793, EPI_ISL_1575794, EPI_ISL_1575795, EPI_ISL_1575796, EPI_ISL_1575797, EPI_ISL_1575798, EPI_ISL_1575799, EPI_ISL_1575800, EPI_ISL_1575801, EPI_ISL_1575802, EPI_ISL_1575803, EPI_ISL_1575804, EPI_ISL_1575805, EPI_ISL_1575806, EPI_ISL_1575807, EPI_ISL_1575808, EPI_ISL_1575809, EPI_ISL_1575810, EPI_ISL_1575811, EPI_ISL_1575812, EPI_ISL_1575813, EPI_ISL_1575814, EPI_ISL_1575815, EPI_ISL_1575817, EPI_ISL_1575818, EPI_ISL_1575819, EPI_ISL_1575820, EPI_ISL_1575821, EPI_ISL_1575822, EPI_ISL_1575823, EPI_ISL_1575824, EPI_ISL_1575825, EPI_ISL_1575826, EPI_ISL_1575827, EPI_ISL_1575828, EPI_ISL_1575829, EPI_ISL_1575830, EPI_ISL_1575831, EPI_ISL_1575832, EPI_ISL_1575833, EPI_ISL_1575834, EPI_ISL_1575835, EPI_ISL_1575836, EPI_ISL_1575837, EPI_ISL_1575838, EPI_ISL_1575840, EPI_ISL_1575841, EPI_ISL_1575842, EPI_ISL_1575843, EPI_ISL_1575844, EPI_ISL_1575845, EPI_ISL_1575846, EPI_ISL_1575847, EPI_ISL_1575848, EPI_ISL_1575849, EPI_ISL_1575850, EPI_ISL_1575851, EPI_ISL_1575852, EPI_ISL_1575853, EPI_ISL_1575854, EPI_ISL_1575855, EPI_ISL_1575856, EPI_ISL_1575857, EPI_ISL_1575858, EPI_ISL_1575859, EPI_ISL_1575860, EPI_ISL_1575861, EPI_ISL_1575862, EPI_ISL_1575863, EPI_ISL_1575864, EPI_ISL_1575865, EPI_ISL_1575866, EPI_ISL_1575867, EPI_ISL_1575868, EPI_ISL_1575869, EPI_ISL_1575870, EPI_ISL_1575871, EPI_ISL_1575872, EPI_ISL_1575873, EPI_ISL_1575874, EPI_ISL_1575875, EPI_ISL_1575877, EPI_ISL_1575878, EPI_ISL_1575880, EPI_ISL_1575881, EPI_ISL_1575882, EPI_ISL_1575883, EPI_ISL_1575884, EPI_ISL_1575885, EPI_ISL_1575886, EPI_ISL_1575887, EPI_ISL_1575888, EPI_ISL_1575889, EPI_ISL_1575890, EPI_ISL_1575891, EPI_ISL_1575892, EPI_ISL_1575893, EPI_ISL_1575894, EPI_ISL_1575895, EPI_ISL_1575896, EPI_ISL_1575897, EPI_ISL_1575898, EPI_ISL_1575899, EPI_ISL_1575900, EPI_ISL_1575901, EPI_ISL_1575902, EPI_ISL_1575903, EPI_ISL_1575904, EPI_ISL_1575905, EPI_ISL_1575907, EPI_ISL_1575908, EPI_ISL_1575909, EPI_ISL_1575910, EPI_ISL_1575911, EPI_ISL_1575912, EPI_ISL_1575913, EPI_ISL_1575914, EPI_ISL_1575915, EPI_ISL_1575916, EPI_ISL_1575917, EPI_ISL_1575918, EPI_ISL_1575919, EPI_ISL_1575920, EPI_ISL_1575921, EPI_ISL_1575922, EPI_ISL_1575923, EPI_ISL_1575925, EPI_ISL_1575926, EPI_ISL_1575928, EPI_ISL_1575929, EPI_ISL_1575931, EPI_ISL_1575933, EPI_ISL_1575934, EPI_ISL_1575936, EPI_ISL_1575937, EPI_ISL_1575939, EPI_ISL_1575940, EPI_ISL_1575942, EPI_ISL_1575944, EPI_ISL_1575945, EPI_ISL_1575946, EPI_ISL_1575949, EPI_ISL_1575951, EPI_ISL_1575953, EPI_ISL_1575954, EPI_ISL_1575956, EPI_ISL_1575958, EPI_ISL_1575960, EPI_ISL_1575962, EPI_ISL_1575965, EPI_ISL_1575967, EPI_ISL_1575969, EPI_ISL_1575970, EPI_ISL_1575972, EPI_ISL_1575973, EPI_ISL_1575975, EPI_ISL_1575977, EPI_ISL_1575978, EPI_ISL_1575980, EPI_ISL_1575981, EPI_ISL_1575983, EPI_ISL_1575985, EPI_ISL_1575986, EPI_ISL_1575988, EPI_ISL_1575989, EPI_ISL_1575991, EPI_ISL_1575994, EPI_ISL_1575995, EPI_ISL_1575997, EPI_ISL_1575998, EPI_ISL_1576000, EPI_ISL_1576002, EPI_ISL_1576005, EPI_ISL_1576006, EPI_ISL_1576008, EPI_ISL_1576009, EPI_ISL_1576011, EPI_ISL_1576013, EPI_ISL_1576014, EPI_ISL_1576016, EPI_ISL_1576017, EPI_ISL_1576019, EPI_ISL_1576020, EPI_ISL_1576022, EPI_ISL_1576024, EPI_ISL_1576026, EPI_ISL_1576028, EPI_ISL_1576030, EPI_ISL_1576032, EPI_ISL_1576034, EPI_ISL_1576035, EPI_ISL_1576037, EPI_ISL_1576038, EPI_ISL_1576040, EPI_ISL_1576041, EPI_ISL_1576043, EPI_ISL_1576044, EPI_ISL_1576045, EPI_ISL_1576046, EPI_ISL_1576047, EPI_ISL_1576048, EPI_ISL_1576049, EPI_ISL_1576050, EPI_ISL_1576051, EPI_ISL_1576052, EPI_ISL_1576053, EPI_ISL_1576054, EPI_ISL_1576055, EPI_ISL_1576056, EPI_ISL_1576058, EPI_ISL_1576059, EPI_ISL_1576060, EPI_ISL_1576061, EPI_ISL_1576062, EPI_ISL_1576063, EPI_ISL_1576064, EPI_ISL_1576065, EPI_ISL_1576066, EPI_ISL_1576067, EPI_ISL_1576068, EPI_ISL_1576070, EPI_ISL_1576071, EPI_ISL_1576072, EPI_ISL_1576073, EPI_ISL_1576074, EPI_ISL_1576075, EPI_ISL_1576076, EPI_ISL_1576077, EPI_ISL_1576078, EPI_ISL_1576079, EPI_ISL_1576080, EPI_ISL_1576081, EPI_ISL_1576082, EPI_ISL_1576083, EPI_ISL_1576084, EPI_ISL_1576085, EPI_ISL_1576086, EPI_ISL_1576087, EPI_ISL_1576088, EPI_ISL_1576089, EPI_ISL_1576090, EPI_ISL_1576091, EPI_ISL_1576092, EPI_ISL_1576093, EPI_ISL_1576094, EPI_ISL_1576095, EPI_ISL_1576096, EPI_ISL_1576097, EPI_ISL_1576098, EPI_ISL_1576099, EPI_ISL_1576100 | | | |
| see above | Helix/Illumina | Centers for Disease Control and Prevention | Dakota Howard, Dhwani Batra, Peter W. Cook, Kara Moser, Adrian Paskey, Jason Caravas, Benjamin Rambo-Martin, Shatavia Morrison, Christopher Gulvick, Scott Sammons, Yvette Unoarumhi, Darlene Wagner, Matthew Schmerer, Eileen de Feo, Jan Antico, Christine Tran, Matthew |

| | | | |
|---|---|---|---|
| | | Division of Viral Diseases, Pathogen Discovery | Tolentino, Shannon Wickline, Kim Gietzen, Brad Sickler, Jingtao Liu, Eric Allen, Phil Febbo, Nicole L. Washington, Simon White, Geraint Levan, Kelly Schiabor Barrett, Elizabeth Cirulli, Alexandre Bolze, Ary Ascencio, Charlotte Rivera-Garcia, Ryan Cho, Jason Nguyen, Sherry Wang, Jimmy Ramirez, Tyler Cassens, Efren Sandoval, Magnus Isaksson, William Lee, David Becker, Marc Laurent, James Lu, Clinton R. Paden, Duncan MacCannell |
| EPI_ISL_1576101 | Montana Public Health Laboratory | Montana Public Health Laboratory | Joy Ritter, Michelle Mozer, Carrie Biskupiak, Deborah Gibson |
| EPI_ISL_1576102, EPI_ISL_1576103, EPI_ISL_1576104, EPI_ISL_1576105, EPI_ISL_1576106, EPI_ISL_1576107, EPI_ISL_1576108, EPI_ISL_1576109, EPI_ISL_1576110, EPI_ISL_1576111, EPI_ISL_1576112, EPI_ISL_1576113, EPI_ISL_1576114, EPI_ISL_1576115, EPI_ISL_1576116, EPI_ISL_1576117, EPI_ISL_1576118, EPI_ISL_1576119, EPI_ISL_1576120, EPI_ISL_1576121, EPI_ISL_1576122, EPI_ISL_1576123, EPI_ISL_1576124, EPI_ISL_1576125, EPI_ISL_1576126, EPI_ISL_1576127, EPI_ISL_1576128, EPI_ISL_1576129, EPI_ISL_1576130, EPI_ISL_1576131, EPI_ISL_1576132, EPI_ISL_1576133, EPI_ISL_1576134, EPI_ISL_1576135, EPI_ISL_1576136, EPI_ISL_1576137, EPI_ISL_1576138, EPI_ISL_1576139, EPI_ISL_1576140, EPI_ISL_1576141, EPI_ISL_1576142, EPI_ISL_1576143, EPI_ISL_1576144, EPI_ISL_1576145, EPI_ISL_1576146, EPI_ISL_1576147, EPI_ISL_1576148, EPI_ISL_1576149, EPI_ISL_1576150, EPI_ISL_1576151, EPI_ISL_1576152, EPI_ISL_1576153, EPI_ISL_1576154, EPI_ISL_1576155, EPI_ISL_1576156, EPI_ISL_1576157, EPI_ISL_1576158, EPI_ISL_1576159, EPI_ISL_1576160, EPI_ISL_1576161, EPI_ISL_1576162, EPI_ISL_1576163, EPI_ISL_1576164, EPI_ISL_1576165, EPI_ISL_1576166, EPI_ISL_1576167, EPI_ISL_1576168, EPI_ISL_1576169, EPI_ISL_1576170, EPI_ISL_1576171, EPI_ISL_1576172, EPI_ISL_1576173, EPI_ISL_1576174, EPI_ISL_1576175, EPI_ISL_1576176, EPI_ISL_1576177, EPI_ISL_1576178, EPI_ISL_1576179, EPI_ISL_1576180, EPI_ISL_1576181, EPI_ISL_1576182, EPI_ISL_1576183, EPI_ISL_1576184, EPI_ISL_1576185, EPI_ISL_1576186, EPI_ISL_1576187, EPI_ISL_1576188, EPI_ISL_1576189, EPI_ISL_1576190, EPI_ISL_1576191, EPI_ISL_1576192, EPI_ISL_1576193, EPI_ISL_1576194, EPI_ISL_1576195, EPI_ISL_1576196, EPI_ISL_1576197, EPI_ISL_1576198, EPI_ISL_1576199, EPI_ISL_1576200, EPI_ISL_1576201, EPI_ISL_1576202, EPI_ISL_1576203, EPI_ISL_1576204, EPI_ISL_1576205, EPI_ISL_1576206, EPI_ISL_1576207, EPI_ISL_1576208, EPI_ISL_1576209, EPI_ISL_1576210, EPI_ISL_1576211, EPI_ISL_1576212, EPI_ISL_1576213, EPI_ISL_1576214, EPI_ISL_1576215, EPI_ISL_1576216, EPI_ISL_1576217, EPI_ISL_1576218, EPI_ISL_1576219, EPI_ISL_1576220, EPI_ISL_1576221, EPI_ISL_1576222, EPI_ISL_1576223, EPI_ISL_1576224, EPI_ISL_1576225, EPI_ISL_1576226, EPI_ISL_1576227, EPI_ISL_1576228, EPI_ISL_1576229, EPI_ISL_1576230, EPI_ISL_1576231, EPI_ISL_1576232, EPI_ISL_1576233, EPI_ISL_1576234, EPI_ISL_1576235, EPI_ISL_1576236, EPI_ISL_1576237, EPI_ISL_1576238, EPI_ISL_1576239, EPI_ISL_1576240, EPI_ISL_1576241, EPI_ISL_1576242, EPI_ISL_1576243, EPI_ISL_1576244, EPI_ISL_1576245, EPI_ISL_1576246, EPI_ISL_1576247, EPI_ISL_1576248, EPI_ISL_1576249, EPI_ISL_1576250, EPI_ISL_1576251, EPI_ISL_1576252, EPI_ISL_1576253, EPI_ISL_1576254, EPI_ISL_1576255, EPI_ISL_1576256, EPI_ISL_1576257, EPI_ISL_1576258, EPI_ISL_1576259, EPI_ISL_1576260, EPI_ISL_1576261, EPI_ISL_1576262, EPI_ISL_1576263, EPI_ISL_1576264, EPI_ISL_1576265, EPI_ISL_1576266, EPI_ISL_1576267, EPI_ISL_1576268, EPI_ISL_1576269, EPI_ISL_1576270, EPI_ISL_1576271, EPI_ISL_1576272, EPI_ISL_1576273, EPI_ISL_1576274, EPI_ISL_1576275, EPI_ISL_1576276, EPI_ISL_1576277, EPI_ISL_1576278, EPI_ISL_1576279, EPI_ISL_1576280, EPI_ISL_1576281, EPI_ISL_1576282, EPI_ISL_1576283, EPI_ISL_1576284, EPI_ISL_1576285, EPI_ISL_1576286, EPI_ISL_1576287, EPI_ISL_1576288, EPI_ISL_1576289, EPI_ISL_1576290, EPI_ISL_1576291, EPI_ISL_1576292, EPI_ISL_1576293, EPI_ISL_1576294, EPI_ISL_1576295, EPI_ISL_1576296, EPI_ISL_1576297, EPI_ISL_1576298, EPI_ISL_1576299, EPI_ISL_1576300 | | | |
| see above | Helix/Illumina | Centers for Disease Control and Prevention Division of Viral Diseases, Pathogen Discovery | Dakota Howard, Dhwani Batra, Peter W. Cook, Kara Moser, Adrian Paskey, Jason Caravas, Benjamin Rambo-Martin, Shatavia Morrison, Christopher Gulvick, Scott Sammons, Yvette Unoarumhi, Darlene Wagner, Matthew Schmerer, Eileen de Feo, Jan Antico, Christine Tran, Matthew Tolentino, Shannon Wickline, Kim Gietzen, Brad Sickler, Jingtao Liu, Eric Allen, Phil Febbo, Nicole L. Washington, Simon White, Geraint Levan, Kelly Schiabor Barrett, Elizabeth Cirulli, Alexandre Bolze, Ary Ascencio, Charlotte Rivera-Garcia, Ryan Cho, Jason Nguyen, Sherry Wang, Jimmy Ramirez, Tyler Cassens, Efren Sandoval, Magnus Isaksson, William Lee, David Becker, Marc Laurent, James Lu, Clinton R. Paden, Duncan MacCannell |
| EPI_ISL_1576301 | Montana Public Health Laboratory | Montana Public Health Laboratory | Joy Ritter, Michelle Mozer, Carrie Biskupiak, Deborah Gibson |
| EPI_ISL_1576302, EPI_ISL_1576303, EPI_ISL_1576304, EPI_ISL_1576305, EPI_ISL_1576306, EPI_ISL_1576307, EPI_ISL_1576308, EPI_ISL_1576309, EPI_ISL_1576310, EPI_ISL_1576311, EPI_ISL_1576312, EPI_ISL_1576313, EPI_ISL_1576314, EPI_ISL_1576315, EPI_ISL_1576316, EPI_ISL_1576317, EPI_ISL_1576318, EPI_ISL_1576319, EPI_ISL_1576320, EPI_ISL_1576321, EPI_ISL_1576322, EPI_ISL_1576323, EPI_ISL_1576324, EPI_ISL_1576325, EPI_ISL_1576326, EPI_ISL_1576327, EPI_ISL_1576328, EPI_ISL_1576329, EPI_ISL_1576330, EPI_ISL_1576331, EPI_ISL_1576332, EPI_ISL_1576333, EPI_ISL_1576334, EPI_ISL_1576335, EPI_ISL_1576336, EPI_ISL_1576337, EPI_ISL_1576338, EPI_ISL_1576339, EPI_ISL_1576340, EPI_ISL_1576341, EPI_ISL_1576342, EPI_ISL_1576343, EPI_ISL_1576344, EPI_ISL_1576345, EPI_ISL_1576346, EPI_ISL_1576347, EPI_ISL_1576348, EPI_ISL_1576349, EPI_ISL_1576350, EPI_ISL_1576351, EPI_ISL_1576352, EPI_ISL_1576353, EPI_ISL_1576354, EPI_ISL_1576355, EPI_ISL_1576356, EPI_ISL_1576357, EPI_ISL_1576358, EPI_ISL_1576359, EPI_ISL_1576360, EPI_ISL_1576361, EPI_ISL_1576362, EPI_ISL_1576363, EPI_ISL_1576364, EPI_ISL_1576365, EPI_ISL_1576366, EPI_ISL_1576367, EPI_ISL_1576368, EPI_ISL_1576369, EPI_ISL_1576370, EPI_ISL_1576371, EPI_ISL_1576372, EPI_ISL_1576373, EPI_ISL_1576374, EPI_ISL_1576375, EPI_ISL_1576376, EPI_ISL_1576377, EPI_ISL_1576378, EPI_ISL_1576379, EPI_ISL_1576380, EPI_ISL_1576381, EPI_ISL_1576382, EPI_ISL_1576383, EPI_ISL_1576384, EPI_ISL_1576385, EPI_ISL_1576386, EPI_ISL_1576387, EPI_ISL_1576388, EPI_ISL_1576389, EPI_ISL_1576390, EPI_ISL_1576391, EPI_ISL_1576392, EPI_ISL_1576393, EPI_ISL_1576394, EPI_ISL_1576395, EPI_ISL_1576396, EPI_ISL_1576397, EPI_ISL_1576398, EPI_ISL_1576399, EPI_ISL_1576400, EPI_ISL_1576401, EPI_ISL_1576402, EPI_ISL_1576403, EPI_ISL_1576404, EPI_ISL_1576405, EPI_ISL_1576406, EPI_ISL_1576407, EPI_ISL_1576408, EPI_ISL_1576409, EPI_ISL_1576410, EPI_ISL_1576411, EPI_ISL_1576412, EPI_ISL_1576413, EPI_ISL_1576414, EPI_ISL_1576415, EPI_ISL_1576416, EPI_ISL_1576417, EPI_ISL_1576418, EPI_ISL_1576419, EPI_ISL_1576420, EPI_ISL_1576421, EPI_ISL_1576422, EPI_ISL_1576423, EPI_ISL_1576424, EPI_ISL_1576425, EPI_ISL_1576426, EPI_ISL_1576427, EPI_ISL_1576428, EPI_ISL_1576429, EPI_ISL_1576430, EPI_ISL_1576431, EPI_ISL_1576432, EPI_ISL_1576433, EPI_ISL_1576434, EPI_ISL_1576435, EPI_ISL_1576436, EPI_ISL_1576437, EPI_ISL_1576438, EPI_ISL_1576439, EPI_ISL_1576440, EPI_ISL_1576441, EPI_ISL_1576442, EPI_ISL_1576443, EPI_ISL_1576444, EPI_ISL_1576445, EPI_ISL_1576446, EPI_ISL_1576447, EPI_ISL_1576448, EPI_ISL_1576449, EPI_ISL_1576450, EPI_ISL_1576451, EPI_ISL_1576452, EPI_ISL_1576453, EPI_ISL_1576454, EPI_ISL_1576455, EPI_ISL_1576456, EPI_ISL_1576457, EPI_ISL_1576458, EPI_ISL_1576459, EPI_ISL_1576460, EPI_ISL_1576461, EPI_ISL_1576462, EPI_ISL_1576463, EPI_ISL_1576464, EPI_ISL_1576465, EPI_ISL_1576466, EPI_ISL_1576467, EPI_ISL_1576468, EPI_ISL_1576469, EPI_ISL_1576470, EPI_ISL_1576471, EPI_ISL_1576472, EPI_ISL_1576473, EPI_ISL_1576474, EPI_ISL_1576475 | | | |
| see above | Helix/Illumina | Centers for Disease Control and Prevention Division of Viral Diseases, Pathogen Discovery | Dakota Howard, Dhwani Batra, Peter W. Cook, Kara Moser, Adrian Paskey, Jason Caravas, Benjamin Rambo-Martin, Shatavia Morrison, Christopher Gulvick, Scott Sammons, Yvette Unoarumhi, Darlene Wagner, Matthew Schmerer, Eileen de Feo, Jan Antico, Christine Tran, Matthew Tolentino, Shannon Wickline, Kim Gietzen, Brad Sickler, Jingtao Liu, Eric Allen, Phil Febbo, Nicole L. Washington, Simon White, Geraint Levan, Kelly Schiabor Barrett, Elizabeth Cirulli, Alexandre Bolze, Ary Ascencio, Charlotte Rivera-Garcia, Ryan Cho, Jason Nguyen, Sherry Wang, Jimmy Ramirez, Tyler Cassens, Efren Sandoval, Magnus Isaksson, William Lee, David Becker, Marc Laurent, James Lu, Clinton R. Paden, Duncan MacCannell |
| EPI_ISL_1576476 | Montana Public Health Laboratory | Montana Public Health Laboratory | Joy Ritter, Michelle Mozer, Carrie Biskupiak, Deborah Gibson |
| EPI_ISL_1576477, EPI_ISL_1576478, EPI_ISL_1576479, EPI_ISL_1576480, EPI_ISL_1576481, EPI_ISL_1576482, EPI_ISL_1576483, EPI_ISL_1576484, EPI_ISL_1576485, EPI_ISL_1576486, EPI_ISL_1576487, EPI_ISL_1576488, EPI_ISL_1576489, EPI_ISL_1576490, EPI_ISL_1576491, EPI_ISL_1576492, EPI_ISL_1576493, EPI_ISL_1576494, EPI_ISL_1576495, EPI_ISL_1576496, EPI_ISL_1576497, EPI_ISL_1576498, EPI_ISL_1576499, EPI_ISL_1576500, EPI_ISL_1576501, EPI_ISL_1576502, EPI_ISL_1576503, EPI_ISL_1576504, EPI_ISL_1576505, EPI_ISL_1576506, EPI_ISL_1576507, EPI_ISL_1576508, EPI_ISL_1576509, EPI_ISL_1576510, EPI_ISL_1576511, EPI_ISL_1576512, EPI_ISL_1576513, EPI_ISL_1576514, EPI_ISL_1576515, EPI_ISL_1576516, EPI_ISL_1576517, EPI_ISL_1576518, EPI_ISL_1576519, EPI_ISL_1576520, EPI_ISL_1576521, EPI_ISL_1576522, EPI_ISL_1576523, EPI_ISL_1576524, EPI_ISL_1576525, EPI_ISL_1576526, EPI_ISL_1576527, EPI_ISL_1576528, EPI_ISL_1576529, EPI_ISL_1576530, EPI_ISL_1576531, EPI_ISL_1576532, EPI_ISL_1576533, EPI_ISL_1576534, EPI_ISL_1576535, EPI_ISL_1576536, EPI_ISL_1576537, EPI_ISL_1576538, EPI_ISL_1576539, EPI_ISL_1576540, EPI_ISL_1576541, EPI_ISL_1576542, EPI_ISL_1576543, EPI_ISL_1576544, EPI_ISL_1576545, EPI_ISL_1576546, EPI_ISL_1576547, EPI_ISL_1576548, EPI_ISL_1576549, EPI_ISL_1576550, EPI_ISL_1576551, EPI_ISL_1576552, EPI_ISL_1576553, EPI_ISL_1576554, EPI_ISL_1576555, EPI_ISL_1576556, EPI_ISL_1576557, EPI_ISL_1576558, EPI_ISL_1576559, EPI_ISL_1576560, EPI_ISL_1576561, EPI_ISL_1576562, EPI_ISL_1576563, EPI_ISL_1576564, EPI_ISL_1576565, EPI_ISL_1576566, EPI_ISL_1576567, EPI_ISL_1576568, EPI_ISL_1576569, EPI_ISL_1576570, EPI_ISL_1576571, EPI_ISL_1576572, EPI_ISL_1576573, EPI_ISL_1576574, EPI_ISL_1576575, EPI_ISL_1576576, EPI_ISL_1576577, EPI_ISL_1576578, EPI_ISL_1576579, EPI_ISL_1576580, EPI_ISL_1576581, EPI_ISL_1576582, EPI_ISL_1576583, EPI_ISL_1576584, EPI_ISL_1576585, EPI_ISL_1576586, EPI_ISL_1576587, EPI_ISL_1576588, EPI_ISL_1576589, EPI_ISL_1576590, EPI_ISL_1576591, EPI_ISL_1576592, EPI_ISL_1576593, EPI_ISL_1576594, EPI_ISL_1576595, EPI_ISL_1576596, EPI_ISL_1576597, EPI_ISL_1576598, EPI_ISL_1576599, EPI_ISL_1576600, EPI_ISL_1576601, EPI_ISL_1576602, EPI_ISL_1576603, EPI_ISL_1576604, EPI_ISL_1576605, EPI_ISL_1576606, EPI_ISL_1576607, EPI_ISL_1576608, EPI_ISL_1576609, EPI_ISL_1576610, EPI_ISL_1576611, EPI_ISL_1576612, EPI_ISL_1576613, EPI_ISL_1576614, EPI_ISL_1576615, EPI_ISL_1576616, EPI_ISL_1576617, EPI_ISL_1576618, EPI_ISL_1576619, EPI_ISL_1576620, EPI_ISL_1576621, EPI_ISL_1576622, EPI_ISL_1576623, EPI_ISL_1576624, EPI_ISL_1576625, EPI_ISL_1576626, EPI_ISL_1576627, EPI_ISL_1576628, EPI_ISL_1576629, EPI_ISL_1576630, EPI_ISL_1576631, EPI_ISL_1576632, EPI_ISL_1576633, EPI_ISL_1576634, EPI_ISL_1576635, EPI_ISL_1576636, EPI_ISL_1576637, EPI_ISL_1576638, EPI_ISL_1576639, EPI_ISL_1576640, EPI_ISL_1576641, EPI_ISL_1576642, EPI_ISL_1576643, EPI_ISL_1576644, EPI_ISL_1576645, EPI_ISL_1576646, EPI_ISL_1576647, EPI_ISL_1576648, EPI_ISL_1576649, EPI_ISL_1576650, EPI_ISL_1576651, EPI_ISL_1576652, EPI_ISL_1576653, EPI_ISL_1576654, EPI_ISL_1576655, EPI_ISL_1576656, EPI_ISL_1576657, EPI_ISL_1576658, EPI_ISL_1576659, EPI_ISL_1576660, EPI_ISL_1576661, EPI_ISL_1576662, EPI_ISL_1576663, EPI_ISL_1576664, EPI_ISL_1576665, EPI_ISL_1576666, EPI_ISL_1576667, EPI_ISL_1576668, EPI_ISL_1576669, EPI_ISL_1576670, EPI_ISL_1576671, EPI_ISL_1576672, EPI_ISL_1576673, EPI_ISL_1576674, EPI_ISL_1576675, EPI_ISL_1576676, EPI_ISL_1576677, EPI_ISL_1576678, EPI_ISL_1576679, EPI_ISL_1576680, EPI_ISL_1576681, EPI_ISL_1576682, EPI_ISL_1576683, EPI_ISL_1576684, EPI_ISL_1576685, EPI_ISL_1576686, EPI_ISL_1576687, EPI_ISL_1576688, EPI_ISL_1576689, EPI_ISL_1576690, EPI_ISL_1576691, EPI_ISL_1576692, EPI_ISL_1576693, EPI_ISL_1576694, EPI_ISL_1576695, EPI_ISL_1576696, EPI_ISL_1576697, EPI_ISL_1576698, EPI_ISL_1576699, EPI_ISL_1576700, EPI_ISL_1576701, EPI_ISL_1576702, EPI_ISL_1576703, EPI_ISL_1576704, EPI_ISL_1576705, EPI_ISL_1576706, EPI_ISL_1576707, EPI_ISL_1576708, EPI_ISL_1576709, EPI_ISL_1576710, EPI_ISL_1576711, EPI_ISL_1576712, EPI_ISL_1576713, EPI_ISL_1576714, EPI_ISL_1576715, EPI_ISL_1576716, EPI_ISL_1576717, EPI_ISL_1576718, EPI_ISL_1576719, EPI_ISL_1576720, EPI_ISL_1576721, EPI_ISL_1576722, EPI_ISL_1576723, EPI_ISL_1576724, EPI_ISL_1576725, EPI_ISL_1576726, EPI_ISL_1576727, EPI_ISL_1576728, EPI_ISL_1576729, EPI_ISL_1576730, EPI_ISL_1576731, EPI_ISL_1576732, EPI_ISL_1576733, EPI_ISL_1576734, EPI_ISL_1576735, EPI_ISL_1576736, EPI_ISL_1576737, EPI_ISL_1576738, EPI_ISL_1576739, EPI_ISL_1576740, EPI_ISL_1576741, EPI_ISL_1576742, EPI_ISL_1576743, EPI_ISL_1576744, EPI_ISL_1576745, EPI_ISL_1576746, EPI_ISL_1576747, EPI_ISL_1576748, EPI_ISL_1576749, EPI_ISL_1576750, EPI_ISL_1576751, EPI_ISL_1576752, EPI_ISL_1576753, EPI_ISL_1576754, EPI_ISL_1576755, EPI_ISL_1576756, EPI_ISL_1576757, EPI_ISL_1576758 | | | |
| see above | Helix/Illumina | Centers for Disease Control and Prevention Division of Viral Diseases, Pathogen Discovery | Dakota Howard, Dhwani Batra, Peter W. Cook, Kara Moser, Adrian Paskey, Jason Caravas, Benjamin Rambo-Martin, Shatavia Morrison, Christopher Gulvick, Scott Sammons, Yvette Unoarumhi, Darlene Wagner, Matthew Schmerer, Eileen de Feo, Jan Antico, Christine Tran, Matthew Tolentino, Shannon Wickline, Kim Gietzen, Brad Sickler, Jingtao Liu, Eric Allen, Phil Febbo, Nicole L. Washington, Simon White, Geraint Levan, Kelly Schiabor Barrett, Elizabeth Cirulli, Alexandre Bolze, Ary Ascencio, Charlotte Rivera-Garcia, Ryan Cho, Jason Nguyen, Sherry Wang, Jimmy Ramirez, Tyler Cassens, Efren Sandoval, Magnus Isaksson, William Lee, David Becker, Marc Laurent, James Lu, Clinton R. Paden, Duncan MacCannell |
| EPI_ISL_1576759, EPI_ISL_1576760, EPI_ISL_1576761, EPI_ISL_1576762 | Montana Public Health Laboratory | Montana Public Health Laboratory | Joy Ritter, Michelle Mozer, Carrie Biskupiak, Deborah Gibson |
| EPI_ISL_1576763, EPI_ISL_1576764, EPI_ISL_1576765, EPI_ISL_1576766, EPI_ISL_1576767, EPI_ISL_1576768, EPI_ISL_1576769, EPI_ISL_1576770, EPI_ISL_1576771, EPI_ISL_1576772, EPI_ISL_1576773, EPI_ISL_1576774, EPI_ISL_1576775, EPI_ISL_1576776, EPI_ISL_1576777, EPI_ISL_1576778, EPI_ISL_1576779, EPI_ISL_1576780, EPI_ISL_1576781, EPI_ISL_1576782, EPI_ISL_1576783, EPI_ISL_1576784, EPI_ISL_1576785, EPI_ISL_1576786, EPI_ISL_1576787, EPI_ISL_1576788, EPI_ISL_1576789, EPI_ISL_1576790, EPI_ISL_1576791, EPI_ISL_1576792, EPI_ISL_1576793, EPI_ISL_1576794, EPI_ISL_1576795, EPI_ISL_1576796, EPI_ISL_1576797, EPI_ISL_1576798, EPI_ISL_1576799, EPI_ISL_1576800, EPI_ISL_1576801, EPI_ISL_1576802, EPI_ISL_1576803, EPI_ISL_1576804, EPI_ISL_1576805, EPI_ISL_1576806, EPI_ISL_1576807, EPI_ISL_1576808, EPI_ISL_1576809 | | | |
| see above | MD PHL | MD PHL | Maryland Department of Health Laboratories Administration |
| EPI_ISL_1576810, EPI_ISL_1576811 | Montana Public Health Laboratory | Montana Public Health Laboratory | Joy Ritter, Michelle Mozer, Carrie Biskupiak, Deborah Gibson |
| EPI_ISL_1576812, EPI_ISL_1576813 | SUNY UPSTATE MEDICAL UNIVERSITY | Wadsworth Center, New York State Department of Health | Kirsten St. George, Daryl M. Lamson, Alexis Russell, Matthew Shudt, Melissa A Leisner, Jonathan Plitnick, Catharine Prussing, Navjot Singh, John Kelly, Erasmus Schneider, Erica Lasek-Nesselquist |
| EPI_ISL_1576814 | TEMPUS LABS INC | Wadsworth Center, New York State Department of Health | Kirsten St. George, Daryl M. Lamson, Alexis Russell, Matthew Shudt, Melissa A Leisner, Jonathan Plitnick, Catharine Prussing, Navjot Singh, John Kelly, Erasmus Schneider, Erica Lasek-Nesselquist |
| EPI_ISL_1576815 | NORTHWELL HEALTH LABORATORIES | Wadsworth Center, New York State Department of Health | Kirsten St. George, Daryl M. Lamson, Alexis Russell, Matthew Shudt, Melissa A Leisner, Jonathan Plitnick, Catharine Prussing, Navjot Singh, John Kelly, Erasmus Schneider, Erica Lasek-Nesselquist |
| EPI_ISL_1576816 | MONTEFIORE MEDICAL CENTER LABORATORIES | Wadsworth Center, New York State Department of Health | Kirsten St. George, Daryl M. Lamson, Alexis Russell, Matthew Shudt, Melissa A Leisner, Jonathan Plitnick, Catharine Prussing, Navjot Singh, John Kelly, Erasmus Schneider, Erica Lasek-Nesselquist |
| EPI_ISL_1576817, EPI_ISL_1576818 | URMC LABS | Wadsworth Center, New York State Department of Health | Kirsten St. George, Daryl M. Lamson, Alexis Russell, Matthew Shudt, Melissa A Leisner, Jonathan Plitnick, Catharine Prussing, Navjot Singh, John Kelly, Erasmus Schneider, Erica Lasek-Nesselquist |
| EPI_ISL_1576819 | MONTEFIORE MEDICAL CENTER LABORATORIES | Wadsworth Center, New York State Department of Health | Kirsten St. George, Daryl M. Lamson, Alexis Russell, Matthew Shudt, Melissa A Leisner, Jonathan Plitnick, Catharine Prussing, Navjot Singh, John Kelly, Erasmus Schneider, Erica Lasek-Nesselquist |
| EPI_ISL_1576820 | Columbia University Irving Medical Center | Wadsworth Center, New York State Department of Health | Kirsten St. George, Daryl M. Lamson, Alexis Russell, Matthew Shudt, Melissa A Leisner, Jonathan Plitnick, Catharine Prussing, Navjot Singh, John Kelly, Erasmus Schneider, Erica Lasek-Nesselquist |
| EPI_ISL_1576821, EPI_ISL_1576822 | TEMPUS LABS INC | Wadsworth Center, New York State Department of Health | Kirsten St. George, Daryl M. Lamson, Alexis Russell, Matthew Shudt, Melissa A Leisner, Jonathan Plitnick, Catharine Prussing, Navjot Singh, John Kelly, Erasmus Schneider, Erica Lasek-Nesselquist |
| EPI_ISL_1576823 | URMC LABS | Wadsworth Center, New York State Department of Health | Kirsten St. George, Daryl M. Lamson, Alexis Russell, Matthew Shudt, Melissa A Leisner, Jonathan Plitnick, Catharine Prussing, Navjot Singh, John Kelly, Erasmus Schneider, Erica Lasek-Nesselquist |
| EPI_ISL_1576824 | ALBANY MEDICAL CENTER | Wadsworth Center, New York State Department of Health | Kirsten St. George, Daryl M. Lamson, Alexis Russell, Matthew Shudt, Melissa A Leisner, Jonathan Plitnick, Catharine Prussing, Navjot Singh, John Kelly, Erasmus Schneider, Erica Lasek-Nesselquist |
| EPI_ISL_1576825, EPI_ISL_1576826 | NORTHWELL HEALTH LABORATORIES | Wadsworth Center, New York State Department of Health | Kirsten St. George, Daryl M. Lamson, Alexis Russell, Matthew Shudt, Melissa A Leisner, Jonathan Plitnick, Catharine Prussing, Navjot Singh, John Kelly, Erasmus Schneider, Erica Lasek-Nesselquist |
| EPI_ISL_1576828, EPI_ISL_1576829, EPI_ISL_1576830, EPI_ISL_1576831 | Montana Public Health Laboratory | Montana Public Health Laboratory | Joy Ritter, Michelle Mozer, Carrie Biskupiak, Deborah Gibson |
| EPI_ISL_1576836, EPI_ISL_1576837, EPI_ISL_1576838, EPI_ISL_1576839, EPI_ISL_1576840, EPI_ISL_1576841, EPI_ISL_1576842, EPI_ISL_1576843, EPI_ISL_1576844, EPI_ISL_1576845, EPI_ISL_1576846, EPI_ISL_1576847, EPI_ISL_1576848, EPI_ISL_1576849, EPI_ISL_1576850, EPI_ISL_1576851, EPI_ISL_1576852, EPI_ISL_1576853, EPI_ISL_1576854, EPI_ISL_1576855, EPI_ISL_1576856, EPI_ISL_1576857, EPI_ISL_1576858, EPI_ISL_1576859, EPI_ISL_1576860 | | | |
| see above | Houston Health Dept. | Houston Health Dept. | Ryker Penn, Pamela Brown, Adolpho Lara, Yanlai Lai |
| EPI_ISL_1576861 | Montana Public Health Laboratory | Montana Public Health Laboratory | Joy Ritter, Michelle Mozer, Carrie Biskupiak, Deborah Gibson |
| EPI_ISL_1576862, EPI_ISL_1576863, EPI_ISL_1576864, EPI_ISL_1576865, EPI_ISL_1576866, EPI_ISL_1576867, EPI_ISL_1576868, EPI_ISL_1576869, EPI_ISL_1576870, EPI_ISL_1576871, EPI_ISL_1576872, EPI_ISL_1576873, EPI_ISL_1576874, EPI_ISL_1576875, EPI_ISL_1576876, EPI_ISL_1576877, EPI_ISL_1576878, EPI_ISL_1576879, EPI_ISL_1576880, EPI_ISL_1576881, EPI_ISL_1576882, EPI_ISL_1576883, EPI_ISL_1576884, EPI_ISL_1576885, EPI_ISL_1576886, EPI_ISL_1576887, EPI_ISL_1576888, EPI_ISL_1576889, EPI_ISL_1576890, EPI_ISL_1576891, EPI_ISL_1576892, EPI_ISL_1576893, EPI_ISL_1576894, EPI_ISL_1576895, EPI_ISL_1576896, EPI_ISL_1576897, EPI_ISL_1576898, EPI_ISL_1576899, EPI_ISL_1576900, EPI_ISL_1576901, EPI_ISL_1576902, EPI_ISL_1576903, EPI_ISL_1576904, EPI_ISL_1576905, EPI_ISL_1576906, EPI_ISL_1576907, EPI_ISL_1576908, EPI_ISL_1576909, EPI_ISL_1576910, EPI_ISL_1576911, EPI_ISL_1576912, EPI_ISL_1576913, EPI_ISL_1576914, EPI_ISL_1576915, EPI_ISL_1576916, EPI_ISL_1576917, EPI_ISL_1576918, EPI_ISL_1576919, EPI_ISL_1576920, EPI_ISL_1576921, EPI_ISL_1576922, EPI_ISL_1576923, EPI_ISL_1576924, EPI_ISL_1576925, EPI_ISL_1576926, EPI_ISL_1576927, EPI_ISL_1576928, EPI_ISL_1576929, EPI_ISL_1576930, EPI_ISL_1576931, EPI_ISL_1576933, EPI_ISL_1576934, EPI_ISL_1576935, EPI_ISL_1576936, EPI_ISL_1576937, EPI_ISL_1576938, EPI_ISL_1576939, EPI_ISL_1576941, EPI_ISL_1576942 | | | |
| see above | NYU Langone Health | Departments of Pathology and Medicine, New York University School of Medicine | Adriana Heguy, Dacia Dimartino, Emily Guzman, Christian Marier, Peter Meyn, Sitharam Ramaswami, Gael Westby, Paul Zappile, Yutong Zhang, Paolo Cotzia, Guiqing Wang |
| EPI_ISL_1576947 | Montana Public Health Laboratory | Montana Public Health Laboratory | Joy Ritter, Michelle Mozer, Carrie Biskupiak, Deborah Gibson |
| EPI_ISL_1576991, EPI_ISL_1576992, EPI_ISL_1576993, EPI_ISL_1576994, EPI_ISL_1576995, EPI_ISL_1576996, EPI_ISL_1576997, EPI_ISL_1576998, EPI_ISL_1576999, EPI_ISL_1577000, EPI_ISL_1577001, EPI_ISL_1577002 | | | |
| see above | Alameda County Public Health Department | Alameda County Public Health Department | Kristna Hsieh, Maria Elma, Kevin Libuit |

| | | | |
|---|---|---|---|
| EPI_ISL_1577003, EPI_ISL_1577005 | Montana Public Health Laboratory | Montana Public Health Laboratory | Joy Ritter, Michelle Mozer, Carrie Biskupiak, Deborah Gibson |
| EPI_ISL_1577007, EPI_ISL_1577011, EPI_ISL_1577013, EPI_ISL_1577014, EPI_ISL_1577016, EPI_ISL_1577017, EPI_ISL_1577018, EPI_ISL_1577019, EPI_ISL_1577021, EPI_ISL_1577022, EPI_ISL_1577023, EPI_ISL_1577024, EPI_ISL_1577025 | | | |
| see above | UTGSAF | UTGSAF | Jessica Podnar, Sylvie Beaudenon, Audrey Kelly, Zachary Carver, Anna Battenhouse, Andreas Matouschek |
| EPI_ISL_1577211, EPI_ISL_1577212, EPI_ISL_1577213, EPI_ISL_1577220, EPI_ISL_1577329, EPI_ISL_1577330 | Montana Public Health Laboratory | Montana Public Health Laboratory | Joy Ritter, Michelle Mozer, Carrie Biskupiak, Deborah Gibson |
| EPI_ISL_1577656, EPI_ISL_1577657, EPI_ISL_1577658, EPI_ISL_1577659, EPI_ISL_1577660, EPI_ISL_1577661 | Pathology and Laboratory Medicine Institute, Cleveland Clinic, Ohio, USA | Pathology and Laboratory Medicine Institute, Cleveland Clinic, Ohio, USA | Joy Nakitandwe, Zheng Jin Tu, Jay Brock, Yu-Wei Cheng, Gary Procop, Daniel Rhoads, Daniel H. Farkas, David Bosler |
| EPI_ISL_1578039, EPI_ISL_1578040, EPI_ISL_1578041, EPI_ISL_1578042, EPI_ISL_1578043, EPI_ISL_1578044, EPI_ISL_1578045, EPI_ISL_1578046, EPI_ISL_1578047, EPI_ISL_1578048, EPI_ISL_1578049, EPI_ISL_1578050, EPI_ISL_1578051, EPI_ISL_1578052, EPI_ISL_1578053, EPI_ISL_1578054, EPI_ISL_1578055, EPI_ISL_1578056, EPI_ISL_1578057, EPI_ISL_1578058, EPI_ISL_1578059, EPI_ISL_1578060, EPI_ISL_1578061, EPI_ISL_1578062, EPI_ISL_1578063, EPI_ISL_1578064, EPI_ISL_1578065, EPI_ISL_1578066, EPI_ISL_1578067, EPI_ISL_1578068, EPI_ISL_1578069, EPI_ISL_1578070, EPI_ISL_1578071, EPI_ISL_1578072, EPI_ISL_1578073, EPI_ISL_1578074, EPI_ISL_1578075, EPI_ISL_1578078, EPI_ISL_1578079, EPI_ISL_1578080, EPI_ISL_1578081, EPI_ISL_1578082, EPI_ISL_1578083, EPI_ISL_1578084, EPI_ISL_1578085, EPI_ISL_1578086, EPI_ISL_1578087, EPI_ISL_1578088, EPI_ISL_1578089, EPI_ISL_1578090, EPI_ISL_1578091, EPI_ISL_1578092, EPI_ISL_1578093, EPI_ISL_1578094, EPI_ISL_1578095, EPI_ISL_1578096, EPI_ISL_1578097, EPI_ISL_1578098, EPI_ISL_1578099, EPI_ISL_1578100, EPI_ISL_1578101, EPI_ISL_1578102, EPI_ISL_1578103, EPI_ISL_1578104, EPI_ISL_1578105, EPI_ISL_1578106, EPI_ISL_1578107, EPI_ISL_1578108, EPI_ISL_1578109, EPI_ISL_1578110 | | | |
| see above | South Dakota Public Health Laboratory | South Dakota Public Health Laboratory | Jacob Garfin and Chris Carlson |
| EPI_ISL_1578133 | Broad Institute Clinical Research Sequencing Platform | Infectious Disease Program, Broad Institute of Harvard and MIT | Siddle,K.J., Adams,G., Pearlman,L., Gladden-Young,A., Vicente,G., Blumenstiel,B., DeFelice,M., Lee,M., McGovern,S., Lagerborg,K., Rudy,M., DeRuff,K., Carter,A., Normandin,E., Bauer,M., Reilly,S., Tomkins-Tinch,C., Loreth,C., Chaluvadi,S., Meldrim,J., Granger,B., Lemieux,J.E., Birren,B.W., Sabeti,P.C., Larkin,K., Dodge,S., Lennon,N., Madoff,L., Brown,C., Gallagher,G., Smole,S., Park,D.J., Gabriel,S., and MacInnis,B.L. |
| EPI_ISL_1578134, EPI_ISL_1578135 | Rhode Island Department of Health | Infectious Disease Program, Broad Institute of Harvard and MIT | Siddle,K.J., Azevedo,K., Miller,A., Adams,G., Pearlman,L., Gladden-Young,A., Lagerborg,K., Rudy,M., DeRuff,K., Carter,A., Normandin,E., Bauer,M., Reilly,S., Tomkins-Tinch,C., Loreth,C., Chaluvadi,S., Lemieux,J.E., Birren,B.W., Sabeti,P.C., Huard,R., King,E., Park,D.J., and MacInnis,B.L. |
| EPI_ISL_1578136 | Flow Health | Infectious Disease Program, Broad Institute of Harvard and MIT | Lemieux,J.E., Siddle,K.J., Adams,G., Gladden-Young,A., Lagerborg,K., Rudy,M., DeRuff,K., Carter,A., Normandin,E., Bauer,M., Reilly,S., Tomkins-Tinch,C., Loreth,C., Chaluvadi,S., Birren,B.W., Gallagher,G., Smole,S., Park,D.J., MacInnis,B.L., and Sabeti,P.C. |
| EPI_ISL_1578137, EPI_ISL_1578138 | Rhode Island Department of Health | Infectious Disease Program, Broad Institute of Harvard and MIT | Siddle,K.J., Azevedo,K., Miller,A., Adams,G., Pearlman,L., Gladden-Young,A., Lagerborg,K., Rudy,M., DeRuff,K., Carter,A., Normandin,E., Bauer,M., Reilly,S., Tomkins-Tinch,C., Loreth,C., Chaluvadi,S., Lemieux,J.E., Birren,B.W., Sabeti,P.C., Huard,R., King,E., Park,D.J., and MacInnis,B.L. |
| EPI_ISL_1578139 | Flow Health | Infectious Disease Program, Broad Institute of Harvard and MIT | Lemieux,J.E., Siddle,K.J., Adams,G., Gladden-Young,A., Lagerborg,K., Rudy,M., DeRuff,K., Carter,A., Normandin,E., Bauer,M., Reilly,S., Tomkins-Tinch,C., Loreth,C., Chaluvadi,S., Birren,B.W., Gallagher,G., Smole,S., Park,D.J., MacInnis,B.L., and Sabeti,P.C. |
| EPI_ISL_1578140, EPI_ISL_1578141, EPI_ISL_1578142, EPI_ISL_1578143 | Broad Institute Clinical Research Sequencing Platform | Infectious Disease Program, Broad Institute of Harvard and MIT | Siddle,K.J., Adams,G., Pearlman,L., Gladden-Young,A., Vicente,G., Blumenstiel,B., DeFelice,M., Lee,M., McGovern,S., Lagerborg,K., Rudy,M., DeRuff,K., Carter,A., Normandin,E., Bauer,M., Reilly,S., Tomkins-Tinch,C., Loreth,C., Chaluvadi,S., Meldrim,J., Granger,B., Lemieux,J.E., Birren,B.W., Sabeti,P.C., Larkin,K., Dodge,S., Lennon,N., Madoff,L., Brown,C., Gallagher,G., Smole,S., Park,D.J., Gabriel,S., and MacInnis,B.L. |
| EPI_ISL_1578144 | Flow Health | Infectious Disease Program, Broad Institute of Harvard and MIT | Lemieux,J.E., Siddle,K.J., Adams,G., Gladden-Young,A., Lagerborg,K., Rudy,M., DeRuff,K., Carter,A., Normandin,E., Bauer,M., Reilly,S., Tomkins-Tinch,C., Loreth,C., Chaluvadi,S., Birren,B.W., Gallagher,G., Smole,S., Park,D.J., MacInnis,B.L., and Sabeti,P.C. |
| EPI_ISL_1578145, EPI_ISL_1578146, EPI_ISL_1578147, EPI_ISL_1578148 | Rhode Island Department of Health | Infectious Disease Program, Broad Institute of Harvard and MIT | Siddle,K.J., Azevedo,K., Miller,A., Adams,G., Pearlman,L., Gladden-Young,A., Lagerborg,K., Rudy,M., DeRuff,K., Carter,A., Normandin,E., Bauer,M., Reilly,S., Tomkins-Tinch,C., Loreth,C., Chaluvadi,S., Lemieux,J.E., Birren,B.W., Sabeti,P.C., Huard,R., King,E., Park,D.J., and MacInnis,B.L. |
| EPI_ISL_1578149 | Broad Institute Clinical Research Sequencing Platform | Infectious Disease Program, Broad Institute of Harvard and MIT | Siddle,K.J., Adams,G., Pearlman,L., Gladden-Young,A., Vicente,G., Blumenstiel,B., DeFelice,M., Lee,M., McGovern,S., Lagerborg,K., Rudy,M., DeRuff,K., Carter,A., Normandin,E., Bauer,M., Reilly,S., Tomkins-Tinch,C., Loreth,C., Chaluvadi,S., Meldrim,J., Granger,B., Lemieux,J.E., Birren,B.W., Sabeti,P.C., Larkin,K., Dodge,S., Lennon,N., Madoff,L., Brown,C., Gallagher,G., Smole,S., Park,D.J., Gabriel,S., and MacInnis,B.L. |
| EPI_ISL_1578150, EPI_ISL_1578151 | Rhode Island Department of Health | Infectious Disease Program, Broad Institute of Harvard and MIT | Siddle,K.J., Azevedo,K., Miller,A., Adams,G., Pearlman,L., Gladden-Young,A., Lagerborg,K., Rudy,M., DeRuff,K., Carter,A., Normandin,E., Bauer,M., Reilly,S., Tomkins-Tinch,C., Loreth,C., Chaluvadi,S., Lemieux,J.E., Birren,B.W., Sabeti,P.C., Huard,R., King,E., Park,D.J., and MacInnis,B.L. |
| EPI_ISL_1578152 | Broad Institute Clinical Research Sequencing Platform | Infectious Disease Program, Broad Institute of Harvard and MIT | Siddle,K.J., Adams,G., Pearlman,L., Gladden-Young,A., Vicente,G., Blumenstiel,B., DeFelice,M., Lee,M., McGovern,S., Lagerborg,K., Rudy,M., DeRuff,K., Carter,A., Normandin,E., Bauer,M., Reilly,S., Tomkins-Tinch,C., Loreth,C., Chaluvadi,S., Meldrim,J., Granger,B., Lemieux,J.E., Birren,B.W., Sabeti,P.C., Larkin,K., Dodge,S., Lennon,N., Madoff,L., Brown,C., Gallagher,G., Smole,S., Park,D.J., Gabriel,S., and MacInnis,B.L. |
| EPI_ISL_1578153 | Flow Health | Infectious Disease Program, Broad Institute of Harvard and MIT | Lemieux,J.E., Siddle,K.J., Adams,G., Gladden-Young,A., Lagerborg,K., Rudy,M., DeRuff,K., Carter,A., Normandin,E., Bauer,M., Reilly,S., Tomkins-Tinch,C., Loreth,C., Chaluvadi,S., Birren,B.W., Gallagher,G., Smole,S., Park,D.J., MacInnis,B.L., and Sabeti,P.C. |
| EPI_ISL_1578154, EPI_ISL_1578155 | Rhode Island Department of Health | Infectious Disease Program, Broad Institute of Harvard and MIT | Siddle,K.J., Azevedo,K., Miller,A., Adams,G., Pearlman,L., Gladden-Young,A., Lagerborg,K., Rudy,M., DeRuff,K., Carter,A., Normandin,E., Bauer,M., Reilly,S., Tomkins-Tinch,C., Loreth,C., Chaluvadi,S., Lemieux,J.E., Birren,B.W., Sabeti,P.C., Huard,R., King,E., Park,D.J., and MacInnis,B.L. |
| EPI_ISL_1578156 | Broad Institute Clinical Research Sequencing Platform | Infectious Disease Program, Broad Institute of Harvard and MIT | Siddle,K.J., Adams,G., Pearlman,L., Gladden-Young,A., Vicente,G., Blumenstiel,B., DeFelice,M., Lee,M., McGovern,S., Lagerborg,K., Rudy,M., DeRuff,K., Carter,A., Normandin,E., Bauer,M., Reilly,S., Tomkins-Tinch,C., Loreth,C., Chaluvadi,S., Meldrim,J., Granger,B., Lemieux,J.E., Birren,B.W., Sabeti,P.C., Larkin,K., Dodge,S., Lennon,N., Madoff,L., Brown,C., Gallagher,G., Smole,S., Park,D.J., Gabriel,S., and MacInnis,B.L. |
| EPI_ISL_1578157 | Flow Health | Infectious Disease Program, Broad Institute of Harvard and MIT | Lemieux,J.E., Siddle,K.J., Adams,G., Gladden-Young,A., Lagerborg,K., Rudy,M., DeRuff,K., Carter,A., Normandin,E., Bauer,M., Reilly,S., Tomkins-Tinch,C., Loreth,C., Chaluvadi,S., Birren,B.W., Gallagher,G., Smole,S., Park,D.J., MacInnis,B.L., and Sabeti,P.C. |
| EPI_ISL_1578158 | Rhode Island Department of Health | Infectious Disease Program, Broad Institute of Harvard and MIT | Siddle,K.J., Azevedo,K., Miller,A., Adams,G., Pearlman,L., Gladden-Young,A., Lagerborg,K., Rudy,M., DeRuff,K., Carter,A., Normandin,E., Bauer,M., Reilly,S., Tomkins-Tinch,C., Loreth,C., Chaluvadi,S., Lemieux,J.E., Birren,B.W., Sabeti,P.C., Huard,R., King,E., Park,D.J., and MacInnis,B.L. |
| EPI_ISL_1578159, EPI_ISL_1578160, EPI_ISL_1578161 | Broad Institute Clinical Research Sequencing Platform | Infectious Disease Program, Broad Institute of Harvard and MIT | Siddle,K.J., Adams,G., Pearlman,L., Gladden-Young,A., Vicente,G., Blumenstiel,B., DeFelice,M., Lee,M., McGovern,S., Lagerborg,K., Rudy,M., DeRuff,K., Carter,A., Normandin,E., Bauer,M., Reilly,S., Tomkins-Tinch,C., Loreth,C., Chaluvadi,S., Meldrim,J., Granger,B., Lemieux,J.E., Birren,B.W., Sabeti,P.C., Larkin,K., Dodge,S., Lennon,N., Madoff,L., Brown,C., Gallagher,G., Smole,S., Park,D.J., Gabriel,S., and MacInnis,B.L. |
| EPI_ISL_1578162 | Flow Health | Infectious Disease Program, Broad Institute of Harvard and MIT | Lemieux,J.E., Siddle,K.J., Adams,G., Gladden-Young,A., Lagerborg,K., Rudy,M., DeRuff,K., Carter,A., Normandin,E., Bauer,M., Reilly,S., Tomkins-Tinch,C., Loreth,C., Chaluvadi,S., Birren,B.W., Gallagher,G., Smole,S., Park,D.J., MacInnis,B.L., and Sabeti,P.C. |
| EPI_ISL_1578163 | Rhode Island Department of Health | Infectious Disease Program, Broad Institute of Harvard and MIT | Siddle,K.J., Azevedo,K., Miller,A., Adams,G., Pearlman,L., Gladden-Young,A., Lagerborg,K., Rudy,M., DeRuff,K., Carter,A., Normandin,E., Bauer,M., Reilly,S., Tomkins-Tinch,C., Loreth,C., Chaluvadi,S., Lemieux,J.E., Birren,B.W., Sabeti,P.C., Huard,R., King,E., Park,D.J., and MacInnis,B.L. |
| EPI_ISL_1578164 | Broad Institute Clinical Research Sequencing Platform | Infectious Disease Program, Broad Institute of Harvard and MIT | Siddle,K.J., Adams,G., Pearlman,L., Gladden-Young,A., Vicente,G., Blumenstiel,B., DeFelice,M., Lee,M., McGovern,S., Lagerborg,K., Rudy,M., DeRuff,K., Carter,A., Normandin,E., Bauer,M., Reilly,S., Tomkins-Tinch,C., Loreth,C., Chaluvadi,S., Meldrim,J., Granger,B., Lemieux,J.E., Birren,B.W., Sabeti,P.C., Larkin,K., Dodge,S., Lennon,N., Madoff,L., Brown,C., Gallagher,G., Smole,S., Park,D.J., Gabriel,S., and MacInnis,B.L. |
| EPI_ISL_1578165 | Rhode Island Department of Health | Infectious Disease Program, Broad Institute of Harvard and MIT | Siddle,K.J., Azevedo,K., Miller,A., Adams,G., Pearlman,L., Gladden-Young,A., Lagerborg,K., Rudy,M., DeRuff,K., Carter,A., Normandin,E., Bauer,M., Reilly,S., Tomkins-Tinch,C., Loreth,C., Chaluvadi,S., Lemieux,J.E., Birren,B.W., Sabeti,P.C., Huard,R., King,E., Park,D.J., and MacInnis,B.L. |
| EPI_ISL_1578166, EPI_ISL_1578167, EPI_ISL_1578168, EPI_ISL_1578169, EPI_ISL_1578170, EPI_ISL_1578171, EPI_ISL_1578172, EPI_ISL_1578173, EPI_ISL_1578174 | Broad Institute Clinical Research Sequencing Platform | Infectious Disease Program, Broad Institute of Harvard and MIT | Siddle,K.J., Adams,G., Pearlman,L., Gladden-Young,A., Vicente,G., Blumenstiel,B., DeFelice,M., Lee,M., McGovern,S., Lagerborg,K., Rudy,M., DeRuff,K., Carter,A., Normandin,E., Bauer,M., Reilly,S., Tomkins-Tinch,C., Loreth,C., Chaluvadi,S., Meldrim,J., Granger,B., Lemieux,J.E., Birren,B.W., Sabeti,P.C., Larkin,K., Dodge,S., Lennon,N., Madoff,L., Brown,C., Gallagher,G., Smole,S., Park,D.J., Gabriel,S., and MacInnis,B.L. |
| EPI_ISL_1578175 | Rhode Island Department of Health | Infectious Disease Program, Broad Institute of Harvard and MIT | Siddle,K.J., Azevedo,K., Miller,A., Adams,G., Pearlman,L., Gladden-Young,A., Lagerborg,K., Rudy,M., DeRuff,K., Carter,A., Normandin,E., Bauer,M., Reilly,S., Tomkins-Tinch,C., Loreth,C., Chaluvadi,S., Lemieux,J.E., Birren,B.W., Sabeti,P.C., Huard,R., King,E., Park,D.J., and MacInnis,B.L. |
| EPI_ISL_1578176 | Flow Health | Infectious Disease Program, Broad Institute of Harvard and MIT | Lemieux,J.E., Siddle,K.J., Adams,G., Gladden-Young,A., Lagerborg,K., Rudy,M., DeRuff,K., Carter,A., Normandin,E., Bauer,M., Reilly,S., Tomkins-Tinch,C., Loreth,C., Chaluvadi,S., Birren,B.W., Gallagher,G., Smole,S., Park,D.J., MacInnis,B.L., and Sabeti,P.C. |
| EPI_ISL_1578177 | Rhode Island Department of Health | Infectious Disease Program, Broad Institute of Harvard and MIT | Siddle,K.J., Azevedo,K., Miller,A., Adams,G., Pearlman,L., Gladden-Young,A., Lagerborg,K., Rudy,M., DeRuff,K., Carter,A., Normandin,E., Bauer,M., Reilly,S., Tomkins-Tinch,C., Loreth,C., Chaluvadi,S., Lemieux,J.E., Birren,B.W., Sabeti,P.C., Huard,R., King,E., Park,D.J., and MacInnis,B.L. |
| EPI_ISL_1578178, EPI_ISL_1578179 | Broad Institute Clinical Research Sequencing Platform | Infectious Disease Program, Broad Institute of Harvard and MIT | Siddle,K.J., Adams,G., Pearlman,L., Gladden-Young,A., Vicente,G., Blumenstiel,B., DeFelice,M., Lee,M., McGovern,S., Lagerborg,K., Rudy,M., DeRuff,K., Carter,A., Normandin,E., Bauer,M., Reilly,S., Tomkins-Tinch,C., Loreth,C., Chaluvadi,S., Meldrim,J., Granger,B., Lemieux,J.E., Birren,B.W., Sabeti,P.C., Larkin,K., Dodge,S., Lennon,N., Madoff,L., Brown,C., Gallagher,G., Smole,S., Park,D.J., Gabriel,S., and MacInnis,B.L. |
| EPI_ISL_1578180 | Flow Health | Infectious Disease Program, Broad Institute of Harvard and MIT | Lemieux,J.E., Siddle,K.J., Adams,G., Gladden-Young,A., Lagerborg,K., Rudy,M., DeRuff,K., Carter,A., Normandin,E., Bauer,M., Reilly,S., Tomkins-Tinch,C., Loreth,C., Chaluvadi,S., Birren,B.W., Gallagher,G., Smole,S., Park,D.J., MacInnis,B.L., and Sabeti,P.C. |
| EPI_ISL_1578181, EPI_ISL_1578182, EPI_ISL_1578183, EPI_ISL_1578184, EPI_ISL_1578185, EPI_ISL_1578186, EPI_ISL_1578187 | Broad Institute Clinical Research Sequencing Platform | Infectious Disease Program, Broad Institute of Harvard and MIT | Siddle,K.J., Adams,G., Pearlman,L., Gladden-Young,A., Vicente,G., Blumenstiel,B., DeFelice,M., Lee,M., McGovern,S., Lagerborg,K., Rudy,M., DeRuff,K., Carter,A., Normandin,E., Bauer,M., Reilly,S., Tomkins-Tinch,C., Loreth,C., Chaluvadi,S., Meldrim,J., Granger,B., Lemieux,J.E., Birren,B.W., Sabeti,P.C., Larkin,K., Dodge,S., Lennon,N., Madoff,L., Brown,C., Gallagher,G., Smole,S., Park,D.J., Gabriel,S., and MacInnis,B.L. |

| | | | |
|---|---|---|---|
| EPI_ISL_1578188 | Rhode Island Department of Health | Infectious Disease Program, Broad Institute of Harvard and MIT | Siddle,K.J., Azevedo,K., Miller,A., Adams,G., Pearlman,L., Gladden-Young,A., Lagerborg,K., Rudy,M., DeRuff,K., Carter,A., Normandin,E., Bauer,M., Reilly,S., Tomkins-Tinch,C., Loreth,C., Chaluvadi,S., Lemieux,J.E., Birren,B.W., Sabeti,P.C., Huard,R., King,E., Park,D.J., and MacInnis,B.L. |
| EPI_ISL_1578189, EPI_ISL_1578190, EPI_ISL_1578191 | Broad Institute Clinical Research Sequencing Platform | Infectious Disease Program, Broad Institute of Harvard and MIT | Siddle,K.J., Adams,G., Pearlman,L., Gladden-Young,A., Vicente,G., Blumenstiel,B., DeFelice,M., Lee,M., McGovern,S., Lagerborg,K., Rudy,M., DeRuff,K., Carter,A., Normandin,E., Bauer,M., Reilly,S., Tomkins-Tinch,C., Loreth,C., Chaluvadi,S., Meldrim,J., Granger,B., Lemieux,J.E., Birren,B.W., Sabeti,P.C., Larkin,K., Dodge,S., Lennon,N., Madoff,L., Brown,C., Gallagher,G., Smole,S., Park,D.J., Gabriel,S., and MacInnis,B.L. |
| EPI_ISL_1578192 | Rhode Island Department of Health | Infectious Disease Program, Broad Institute of Harvard and MIT | Siddle,K.J., Azevedo,K., Miller,A., Adams,G., Pearlman,L., Gladden-Young,A., Lagerborg,K., Rudy,M., DeRuff,K., Carter,A., Normandin,E., Bauer,M., Reilly,S., Tomkins-Tinch,C., Loreth,C., Chaluvadi,S., Lemieux,J.E., Birren,B.W., Sabeti,P.C., Huard,R., King,E., Park,D.J., and MacInnis,B.L. |
| EPI_ISL_1578193, EPI_ISL_1578194, EPI_ISL_1578195, EPI_ISL_1578196, EPI_ISL_1578197, EPI_ISL_1578198, EPI_ISL_1578199, EPI_ISL_1578200, EPI_ISL_1578201, EPI_ISL_1578202 | Broad Institute Clinical Research Sequencing Platform | Infectious Disease Program, Broad Institute of Harvard and MIT | Siddle,K.J., Adams,G., Pearlman,L., Gladden-Young,A., Vicente,G., Blumenstiel,B., DeFelice,M., Lee,M., McGovern,S., Lagerborg,K., Rudy,M., DeRuff,K., Carter,A., Normandin,E., Bauer,M., Reilly,S., Tomkins-Tinch,C., Loreth,C., Chaluvadi,S., Meldrim,J., Granger,B., Lemieux,J.E., Birren,B.W., Sabeti,P.C., Larkin,K., Dodge,S., Lennon,N., Madoff,L., Brown,C., Gallagher,G., Smole,S., Park,D.J., Gabriel,S., and MacInnis,B.L. |
| EPI_ISL_1578203 | Rhode Island Department of Health | Infectious Disease Program, Broad Institute of Harvard and MIT | Siddle,K.J., Azevedo,K., Miller,A., Adams,G., Pearlman,L., Gladden-Young,A., Lagerborg,K., Rudy,M., DeRuff,K., Carter,A., Normandin,E., Bauer,M., Reilly,S., Tomkins-Tinch,C., Loreth,C., Chaluvadi,S., Lemieux,J.E., Birren,B.W., Sabeti,P.C., Huard,R., King,E., Park,D.J., and MacInnis,B.L. |
| EPI_ISL_1578204, EPI_ISL_1578205, EPI_ISL_1578206, EPI_ISL_1578207 | Broad Institute Clinical Research Sequencing Platform | Infectious Disease Program, Broad Institute of Harvard and MIT | Siddle,K.J., Adams,G., Pearlman,L., Gladden-Young,A., Vicente,G., Blumenstiel,B., DeFelice,M., Lee,M., McGovern,S., Lagerborg,K., Rudy,M., DeRuff,K., Carter,A., Normandin,E., Bauer,M., Reilly,S., Tomkins-Tinch,C., Loreth,C., Chaluvadi,S., Meldrim,J., Granger,B., Lemieux,J.E., Birren,B.W., Sabeti,P.C., Larkin,K., Dodge,S., Lennon,N., Madoff,L., Brown,C., Gallagher,G., Smole,S., Park,D.J., Gabriel,S., and MacInnis,B.L. |
| EPI_ISL_1578208 | Rhode Island Department of Health | Infectious Disease Program, Broad Institute of Harvard and MIT | Siddle,K.J., Azevedo,K., Miller,A., Adams,G., Pearlman,L., Gladden-Young,A., Lagerborg,K., Rudy,M., DeRuff,K., Carter,A., Normandin,E., Bauer,M., Reilly,S., Tomkins-Tinch,C., Loreth,C., Chaluvadi,S., Lemieux,J.E., Birren,B.W., Sabeti,P.C., Huard,R., King,E., Park,D.J., and MacInnis,B.L. |
| EPI_ISL_1578209 | Broad Institute Clinical Research Sequencing Platform | Infectious Disease Program, Broad Institute of Harvard and MIT | Siddle,K.J., Adams,G., Pearlman,L., Gladden-Young,A., Vicente,G., Blumenstiel,B., DeFelice,M., Lee,M., McGovern,S., Lagerborg,K., Rudy,M., DeRuff,K., Carter,A., Normandin,E., Bauer,M., Reilly,S., Tomkins-Tinch,C., Loreth,C., Chaluvadi,S., Meldrim,J., Granger,B., Lemieux,J.E., Birren,B.W., Sabeti,P.C., Larkin,K., Dodge,S., Lennon,N., Madoff,L., Brown,C., Gallagher,G., Smole,S., Park,D.J., Gabriel,S., and MacInnis,B.L. |
| EPI_ISL_1578210 | Rhode Island Department of Health | Infectious Disease Program, Broad Institute of Harvard and MIT | Siddle,K.J., Azevedo,K., Miller,A., Adams,G., Pearlman,L., Gladden-Young,A., Lagerborg,K., Rudy,M., DeRuff,K., Carter,A., Normandin,E., Bauer,M., Reilly,S., Tomkins-Tinch,C., Loreth,C., Chaluvadi,S., Lemieux,J.E., Birren,B.W., Sabeti,P.C., Huard,R., King,E., Park,D.J., and MacInnis,B.L. |
| EPI_ISL_1578211, EPI_ISL_1578212, EPI_ISL_1578213 | Broad Institute Clinical Research Sequencing Platform | Infectious Disease Program, Broad Institute of Harvard and MIT | Siddle,K.J., Adams,G., Pearlman,L., Gladden-Young,A., Vicente,G., Blumenstiel,B., DeFelice,M., Lee,M., McGovern,S., Lagerborg,K., Rudy,M., DeRuff,K., Carter,A., Normandin,E., Bauer,M., Reilly,S., Tomkins-Tinch,C., Loreth,C., Chaluvadi,S., Meldrim,J., Granger,B., Lemieux,J.E., Birren,B.W., Sabeti,P.C., Larkin,K., Dodge,S., Lennon,N., Madoff,L., Brown,C., Gallagher,G., Smole,S., Park,D.J., Gabriel,S., and MacInnis,B.L. |
| EPI_ISL_1578214 | Rhode Island Department of Health | Infectious Disease Program, Broad Institute of Harvard and MIT | Siddle,K.J., Azevedo,K., Miller,A., Adams,G., Pearlman,L., Gladden-Young,A., Lagerborg,K., Rudy,M., DeRuff,K., Carter,A., Normandin,E., Bauer,M., Reilly,S., Tomkins-Tinch,C., Loreth,C., Chaluvadi,S., Lemieux,J.E., Birren,B.W., Sabeti,P.C., Huard,R., King,E., Park,D.J., and MacInnis,B.L. |
| EPI_ISL_1578215 | Flow Health | Infectious Disease Program, Broad Institute of Harvard and MIT | Lemieux,J.E., Siddle,K.J., Adams,G., Gladden-Young,A., Lagerborg,K., Rudy,M., DeRuff,K., Carter,A., Normandin,E., Bauer,M., Reilly,S., Tomkins-Tinch,C., Loreth,C., Chaluvadi,S., Birren,B.W., Gallagher,G., Smole,S., Park,D.J., MacInnis,B.L., and Sabeti,P.C. |
| EPI_ISL_1578216 | Broad Institute Clinical Research Sequencing Platform | Infectious Disease Program, Broad Institute of Harvard and MIT | Siddle,K.J., Adams,G., Pearlman,L., Gladden-Young,A., Vicente,G., Blumenstiel,B., DeFelice,M., Lee,M., McGovern,S., Lagerborg,K., Rudy,M., DeRuff,K., Carter,A., Normandin,E., Bauer,M., Reilly,S., Tomkins-Tinch,C., Loreth,C., Chaluvadi,S., Meldrim,J., Granger,B., Lemieux,J.E., Birren,B.W., Sabeti,P.C., Larkin,K., Dodge,S., Lennon,N., Madoff,L., Brown,C., Gallagher,G., Smole,S., Park,D.J., Gabriel,S., and MacInnis,B.L. |
| EPI_ISL_1578217 | Rhode Island Department of Health | Infectious Disease Program, Broad Institute of Harvard and MIT | Siddle,K.J., Azevedo,K., Miller,A., Adams,G., Pearlman,L., Gladden-Young,A., Lagerborg,K., Rudy,M., DeRuff,K., Carter,A., Normandin,E., Bauer,M., Reilly,S., Tomkins-Tinch,C., Loreth,C., Chaluvadi,S., Lemieux,J.E., Birren,B.W., Sabeti,P.C., Huard,R., King,E., Park,D.J., and MacInnis,B.L. |
| EPI_ISL_1578218 | Broad Institute Clinical Research Sequencing Platform | Infectious Disease Program, Broad Institute of Harvard and MIT | Siddle,K.J., Adams,G., Pearlman,L., Gladden-Young,A., Vicente,G., Blumenstiel,B., DeFelice,M., Lee,M., McGovern,S., Lagerborg,K., Rudy,M., DeRuff,K., Carter,A., Normandin,E., Bauer,M., Reilly,S., Tomkins-Tinch,C., Loreth,C., Chaluvadi,S., Meldrim,J., Granger,B., Lemieux,J.E., Birren,B.W., Sabeti,P.C., Larkin,K., Dodge,S., Lennon,N., Madoff,L., Brown,C., Gallagher,G., Smole,S., Park,D.J., Gabriel,S., and MacInnis,B.L. |
| EPI_ISL_1578219 | Rhode Island Department of Health | Infectious Disease Program, Broad Institute of Harvard and MIT | Siddle,K.J., Azevedo,K., Miller,A., Adams,G., Pearlman,L., Gladden-Young,A., Lagerborg,K., Rudy,M., DeRuff,K., Carter,A., Normandin,E., Bauer,M., Reilly,S., Tomkins-Tinch,C., Loreth,C., Chaluvadi,S., Lemieux,J.E., Birren,B.W., Sabeti,P.C., Huard,R., King,E., Park,D.J., and MacInnis,B.L. |
| EPI_ISL_1578220 | Broad Institute Clinical Research Sequencing Platform | Infectious Disease Program, Broad Institute of Harvard and MIT | Siddle,K.J., Adams,G., Pearlman,L., Gladden-Young,A., Vicente,G., Blumenstiel,B., DeFelice,M., Lee,M., McGovern,S., Lagerborg,K., Rudy,M., DeRuff,K., Carter,A., Normandin,E., Bauer,M., Reilly,S., Tomkins-Tinch,C., Loreth,C., Chaluvadi,S., Meldrim,J., Granger,B., Lemieux,J.E., Birren,B.W., Sabeti,P.C., Larkin,K., Dodge,S., Lennon,N., Madoff,L., Brown,C., Gallagher,G., Smole,S., Park,D.J., Gabriel,S., and MacInnis,B.L. |
| EPI_ISL_1578221 | Rhode Island Department of Health | Infectious Disease Program, Broad Institute of Harvard and MIT | Siddle,K.J., Azevedo,K., Miller,A., Adams,G., Pearlman,L., Gladden-Young,A., Lagerborg,K., Rudy,M., DeRuff,K., Carter,A., Normandin,E., Bauer,M., Reilly,S., Tomkins-Tinch,C., Loreth,C., Chaluvadi,S., Lemieux,J.E., Birren,B.W., Sabeti,P.C., Huard,R., King,E., Park,D.J., and MacInnis,B.L. |
| EPI_ISL_1578222, EPI_ISL_1578223, EPI_ISL_1578224 | Broad Institute Clinical Research Sequencing Platform | Infectious Disease Program, Broad Institute of Harvard and MIT | Siddle,K.J., Adams,G., Pearlman,L., Gladden-Young,A., Vicente,G., Blumenstiel,B., DeFelice,M., Lee,M., McGovern,S., Lagerborg,K., Rudy,M., DeRuff,K., Carter,A., Normandin,E., Bauer,M., Reilly,S., Tomkins-Tinch,C., Loreth,C., Chaluvadi,S., Meldrim,J., Granger,B., Lemieux,J.E., Birren,B.W., Sabeti,P.C., Larkin,K., Dodge,S., Lennon,N., Madoff,L., Brown,C., Gallagher,G., Smole,S., Park,D.J., Gabriel,S., and MacInnis,B.L. |
| EPI_ISL_1578225 | Rhode Island Department of Health | Infectious Disease Program, Broad Institute of Harvard and MIT | Siddle,K.J., Azevedo,K., Miller,A., Adams,G., Pearlman,L., Gladden-Young,A., Lagerborg,K., Rudy,M., DeRuff,K., Carter,A., Normandin,E., Bauer,M., Reilly,S., Tomkins-Tinch,C., Loreth,C., Chaluvadi,S., Lemieux,J.E., Birren,B.W., Sabeti,P.C., Huard,R., King,E., Park,D.J., and MacInnis,B.L. |
| EPI_ISL_1578226 | Broad Institute Clinical Research Sequencing Platform | Infectious Disease Program, Broad Institute of Harvard and MIT | Siddle,K.J., Adams,G., Pearlman,L., Gladden-Young,A., Vicente,G., Blumenstiel,B., DeFelice,M., Lee,M., McGovern,S., Lagerborg,K., Rudy,M., DeRuff,K., Carter,A., Normandin,E., Bauer,M., Reilly,S., Tomkins-Tinch,C., Loreth,C., Chaluvadi,S., Meldrim,J., Granger,B., Lemieux,J.E., Birren,B.W., Sabeti,P.C., Larkin,K., Dodge,S., Lennon,N., Madoff,L., Brown,C., Gallagher,G., Smole,S., Park,D.J., Gabriel,S., and MacInnis,B.L. |
| EPI_ISL_1578227, EPI_ISL_1578228 | Rhode Island Department of Health | Infectious Disease Program, Broad Institute of Harvard and MIT | Siddle,K.J., Azevedo,K., Miller,A., Adams,G., Pearlman,L., Gladden-Young,A., Lagerborg,K., Rudy,M., DeRuff,K., Carter,A., Normandin,E., Bauer,M., Reilly,S., Tomkins-Tinch,C., Loreth,C., Chaluvadi,S., Lemieux,J.E., Birren,B.W., Sabeti,P.C., Huard,R., King,E., Park,D.J., and MacInnis,B.L. |
| EPI_ISL_1578229, EPI_ISL_1578230 | Broad Institute Clinical Research Sequencing Platform | Infectious Disease Program, Broad Institute of Harvard and MIT | Siddle,K.J., Adams,G., Pearlman,L., Gladden-Young,A., Vicente,G., Blumenstiel,B., DeFelice,M., Lee,M., McGovern,S., Lagerborg,K., Rudy,M., DeRuff,K., Carter,A., Normandin,E., Bauer,M., Reilly,S., Tomkins-Tinch,C., Loreth,C., Chaluvadi,S., Meldrim,J., Granger,B., Lemieux,J.E., Birren,B.W., Sabeti,P.C., Larkin,K., Dodge,S., Lennon,N., Madoff,L., Brown,C., Gallagher,G., Smole,S., Park,D.J., Gabriel,S., and MacInnis,B.L. |
| EPI_ISL_1578231 | Rhode Island Department of Health | Infectious Disease Program, Broad Institute of Harvard and MIT | Siddle,K.J., Azevedo,K., Miller,A., Adams,G., Pearlman,L., Gladden-Young,A., Lagerborg,K., Rudy,M., DeRuff,K., Carter,A., Normandin,E., Bauer,M., Reilly,S., Tomkins-Tinch,C., Loreth,C., Chaluvadi,S., Lemieux,J.E., Birren,B.W., Sabeti,P.C., Huard,R., King,E., Park,D.J., and MacInnis,B.L. |
| EPI_ISL_1578232, EPI_ISL_1578233, EPI_ISL_1578234 | Broad Institute Clinical Research Sequencing Platform | Infectious Disease Program, Broad Institute of Harvard and MIT | Siddle,K.J., Adams,G., Pearlman,L., Gladden-Young,A., Vicente,G., Blumenstiel,B., DeFelice,M., Lee,M., McGovern,S., Lagerborg,K., Rudy,M., DeRuff,K., Carter,A., Normandin,E., Bauer,M., Reilly,S., Tomkins-Tinch,C., Loreth,C., Chaluvadi,S., Meldrim,J., Granger,B., Lemieux,J.E., Birren,B.W., Sabeti,P.C., Larkin,K., Dodge,S., Lennon,N., Madoff,L., Brown,C., Gallagher,G., Smole,S., Park,D.J., Gabriel,S., and MacInnis,B.L. |
| EPI_ISL_1578235 | Rhode Island Department of Health | Infectious Disease Program, Broad Institute of Harvard and MIT | Siddle,K.J., Azevedo,K., Miller,A., Adams,G., Pearlman,L., Gladden-Young,A., Lagerborg,K., Rudy,M., DeRuff,K., Carter,A., Normandin,E., Bauer,M., Reilly,S., Tomkins-Tinch,C., Loreth,C., Chaluvadi,S., Lemieux,J.E., Birren,B.W., Sabeti,P.C., Huard,R., King,E., Park,D.J., and MacInnis,B.L. |
| EPI_ISL_1578236, EPI_ISL_1578237, EPI_ISL_1578238, EPI_ISL_1578239, EPI_ISL_1578240, EPI_ISL_1578241, EPI_ISL_1578242, EPI_ISL_1578243, EPI_ISL_1578244, EPI_ISL_1578245, EPI_ISL_1578246, EPI_ISL_1578247, EPI_ISL_1578248, EPI_ISL_1578249, EPI_ISL_1578250, EPI_ISL_1578251, EPI_ISL_1578252, EPI_ISL_1578253, EPI_ISL_1578254, EPI_ISL_1578255, EPI_ISL_1578256, EPI_ISL_1578257, EPI_ISL_1578258, EPI_ISL_1578259, EPI_ISL_1578260, EPI_ISL_1578261, EPI_ISL_1578262, EPI_ISL_1578263, EPI_ISL_1578264, EPI_ISL_1578265, EPI_ISL_1578266, EPI_ISL_1578267, EPI_ISL_1578268, EPI_ISL_1578269, EPI_ISL_1578270, EPI_ISL_1578271, EPI_ISL_1578272, EPI_ISL_1578273, EPI_ISL_1578274, EPI_ISL_1578275, EPI_ISL_1578276, EPI_ISL_1578277, EPI_ISL_1578278, EPI_ISL_1578279, EPI_ISL_1578280, EPI_ISL_1578281, EPI_ISL_1578282, EPI_ISL_1578283, EPI_ISL_1578284, EPI_ISL_1578285 | | | |
| see above | Broad Institute Clinical Research Sequencing Platform | Infectious Disease Program, Broad Institute of Harvard and MIT | Siddle,K.J., Adams,G., Pearlman,L., Gladden-Young,A., Vicente,G., Blumenstiel,B., DeFelice,M., Lee,M., McGovern,S., Lagerborg,K., Rudy,M., DeRuff,K., Carter,A., Normandin,E., Bauer,M., Reilly,S., Tomkins-Tinch,C., Loreth,C., Chaluvadi,S., Meldrim,J., Granger,B., Lemieux,J.E., Birren,B.W., Sabeti,P.C., Larkin,K., Dodge,S., Lennon,N., Madoff,L., Brown,C., Gallagher,G., Smole,S., Park,D.J., Gabriel,S., and MacInnis,B.L. |
| EPI_ISL_1578286, EPI_ISL_1578287, EPI_ISL_1578288, EPI_ISL_1578289, EPI_ISL_1578290, EPI_ISL_1578291, EPI_ISL_1578292, EPI_ISL_1578293, EPI_ISL_1578294, EPI_ISL_1578295, EPI_ISL_1578296, EPI_ISL_1578297, EPI_ISL_1578298 | | | |
| see above | Rhode Island Department of Health | Infectious Disease Program, Broad Institute of Harvard and MIT | Siddle,K.J., Azevedo,K., Miller,A., Adams,G., Pearlman,L., Gladden-Young,A., Lagerborg,K., Rudy,M., DeRuff,K., Carter,A., Normandin,E., Bauer,M., Reilly,S., Tomkins-Tinch,C., Loreth,C., Chaluvadi,S., Lemieux,J.E., Birren,B.W., Sabeti,P.C., Huard,R., King,E., Park,D.J., and MacInnis,B.L. |
| EPI_ISL_1578299 | Flow Health | Infectious Disease Program, Broad Institute of Harvard and MIT | Lemieux,J.E., Siddle,K.J., Adams,G., Gladden-Young,A., Lagerborg,K., Rudy,M., DeRuff,K., Carter,A., Normandin,E., Bauer,M., Reilly,S., Tomkins-Tinch,C., Loreth,C., Chaluvadi,S., Birren,B.W., Gallagher,G., Smole,S., Park,D.J., MacInnis,B.L., and Sabeti,P.C. |
| EPI_ISL_1578300, EPI_ISL_1578301, EPI_ISL_1578302, EPI_ISL_1578303, EPI_ISL_1578304, EPI_ISL_1578305, EPI_ISL_1578306, EPI_ISL_1578307, EPI_ISL_1578308, EPI_ISL_1578309, EPI_ISL_1578310, EPI_ISL_1578311, EPI_ISL_1578312, EPI_ISL_1578313, EPI_ISL_1578314, EPI_ISL_1578315, EPI_ISL_1578316, EPI_ISL_1578317, EPI_ISL_1578318, EPI_ISL_1578319, EPI_ISL_1578320, EPI_ISL_1578321, EPI_ISL_1578322 | | | |
| see above | Broad Institute Clinical Research Sequencing Platform | Infectious Disease Program, Broad Institute of Harvard and MIT | Siddle,K.J., Adams,G., Pearlman,L., Gladden-Young,A., Vicente,G., Blumenstiel,B., DeFelice,M., Lee,M., McGovern,S., Lagerborg,K., Rudy,M., DeRuff,K., Carter,A., Normandin,E., Bauer,M., Reilly,S., Tomkins-Tinch,C., Loreth,C., Chaluvadi,S., Meldrim,J., Granger,B., Lemieux,J.E., Birren,B.W., Sabeti,P.C., Larkin,K., Dodge,S., Lennon,N., Madoff,L., Brown,C., Gallagher,G., Smole,S., Park,D.J., Gabriel,S., and MacInnis,B.L. |
| EPI_ISL_1578323, EPI_ISL_1578324 | Rhode Island Department of Health | Infectious Disease Program, Broad Institute of Harvard and MIT | Siddle,K.J., Azevedo,K., Miller,A., Adams,G., Pearlman,L., Gladden-Young,A., Lagerborg,K., Rudy,M., DeRuff,K., Carter,A., Normandin,E., Bauer,M., Reilly,S., Tomkins-Tinch,C., Loreth,C., Chaluvadi,S., Lemieux,J.E., Birren,B.W., Sabeti,P.C., Huard,R., King,E., Park,D.J., and MacInnis,B.L. |
| EPI_ISL_1578325 | Broad Institute Clinical Research Sequencing Platform | Infectious Disease Program, Broad Institute of Harvard and MIT | Siddle,K.J., Adams,G., Pearlman,L., Gladden-Young,A., Vicente,G., Blumenstiel,B., DeFelice,M., Lee,M., McGovern,S., Lagerborg,K., Rudy,M., DeRuff,K., Carter,A., Normandin,E., Bauer,M., Reilly,S., Tomkins-Tinch,C., Loreth,C., Chaluvadi,S., Meldrim,J., Granger,B., Lemieux,J.E., Birren,B.W., Sabeti,P.C., Larkin,K., Dodge,S., Lennon,N., Madoff,L., Brown,C., Gallagher,G., Smole,S., Park,D.J., Gabriel,S., and MacInnis,B.L. |
| EPI_ISL_1578341 | Rhode Island Department of Health | Infectious Disease Program, Broad Institute of Harvard and MIT | Siddle,K.J., Azevedo,K., Miller,A., Adams,G., Pearlman,L., Gladden-Young,A., Lagerborg,K., Rudy,M., DeRuff,K., Carter,A., Normandin,E., Bauer,M., Reilly,S., Tomkins-Tinch,C., Loreth,C., Chaluvadi,S., Lemieux,J.E., Birren,B.W., Sabeti,P.C., Huard,R., King,E., Park,D.J., and MacInnis,B.L. |
| EPI_ISL_1578342, EPI_ISL_1578343, EPI_ISL_1578344, EPI_ISL_1578345, EPI_ISL_1578346, EPI_ISL_1578347, EPI_ISL_1578348 | Broad Institute Clinical Research Sequencing Platform | Infectious Disease Program, Broad Institute of Harvard and MIT | Siddle,K.J., Adams,G., Pearlman,L., Gladden-Young,A., Vicente,G., Blumenstiel,B., DeFelice,M., Lee,M., McGovern,S., Lagerborg,K., Rudy,M., DeRuff,K., Carter,A., Normandin,E., Bauer,M., Reilly,S., Tomkins-Tinch,C., Loreth,C., Chaluvadi,S., Meldrim,J., Granger,B., Lemieux,J.E., Birren,B.W., Sabeti,P.C., Larkin,K., Dodge,S., Lennon,N., Madoff,L., Brown,C., Gallagher,G., Smole,S., Park,D.J., Gabriel,S., and MacInnis,B.L. |
| EPI_ISL_1578349 | Rhode Island Department of Health | Infectious Disease Program, Broad Institute | Siddle,K.J., Azevedo,K., Miller,A., Adams,G., Pearlman,L., Gladden-Young,A., Lagerborg,K., Rudy,M., DeRuff,K., Carter,A., Normandin,E., Bauer,M., Reilly,S., Tomkins-Tinch,C., Loreth,C., Chaluvadi,S., Lemieux,J.E., Birren,B.W., Sabeti,P.C., Huard,R., King,E., Park,D.J., and MacInnis,B.L. |

| | | | |
|---|---|---|---|
| | | of Harvard and MIT | |
| EPI_ISL_1578350, EPI_ISL_1578351, EPI_ISL_1578352, EPI_ISL_1578353, EPI_ISL_1578354, EPI_ISL_1578355, EPI_ISL_1578356, EPI_ISL_1578357, EPI_ISL_1578358, EPI_ISL_1578359, EPI_ISL_1578360, EPI_ISL_1578361, EPI_ISL_1578362, EPI_ISL_1578363, EPI_ISL_1578364, EPI_ISL_1578365, EPI_ISL_1578366, EPI_ISL_1578367, EPI_ISL_1578368, EPI_ISL_1578369, EPI_ISL_1578370, EPI_ISL_1578371, EPI_ISL_1578372, EPI_ISL_1578373, EPI_ISL_1578374, EPI_ISL_1578375, EPI_ISL_1578376 | | | |
| see above | Broad Institute Clinical Research Sequencing Platform | Infectious Disease Program, Broad Institute of Harvard and MIT | Siddle,K.J., Adams,G., Pearlman,L., Gladden-Young,A., Vicente,G., Blumenstiel,B., DeFelice,M., Lee,M., McGovern,S., Lagerborg,K., Rudy,M., DeRuff,K., Carter,A., Normandin,E., Bauer,M., Reilly,S., Tomkins-Tinch,C., Loreth,C., Chaluvadi,S., Meldrim,J., Granger,B., Lemieux,J.E., Birren,B.W., Sabeti,P.C., Larkin,K., Dodge,S., Lennon,N., Madoff,L., Brown,C., Gallagher,G., Smole,S., Park,D.J., Gabriel,S., and MacInnis,B.L. |
| EPI_ISL_1578377 | Rhode Island Department of Health | Infectious Disease Program, Broad Institute of Harvard and MIT | Siddle,K.J., Azevedo,K., Miller,A., Adams,G., Pearlman,L., Gladden-Young,A., Lagerborg,K., Rudy,M., DeRuff,K., Carter,A., Normandin,E., Bauer,M., Reilly,S., Tomkins-Tinch,C., Loreth,C., Chaluvadi,S., Lemieux,J.E., Birren,B.W., Sabeti,P.C., Huard,R., King,E., Park,D.J., and MacInnis,B.L. |
| EPI_ISL_1578378, EPI_ISL_1578379, EPI_ISL_1578380, EPI_ISL_1578381, EPI_ISL_1578382, EPI_ISL_1578383 | Broad Institute Clinical Research Sequencing Platform | Infectious Disease Program, Broad Institute of Harvard and MIT | Siddle,K.J., Adams,G., Pearlman,L., Gladden-Young,A., Vicente,G., Blumenstiel,B., DeFelice,M., Lee,M., McGovern,S., Lagerborg,K., Rudy,M., DeRuff,K., Carter,A., Normandin,E., Bauer,M., Reilly,S., Tomkins-Tinch,C., Loreth,C., Chaluvadi,S., Meldrim,J., Granger,B., Lemieux,J.E., Birren,B.W., Sabeti,P.C., Larkin,K., Dodge,S., Lennon,N., Madoff,L., Brown,C., Gallagher,G., Smole,S., Park,D.J., Gabriel,S., and MacInnis,B.L. |
| EPI_ISL_1578384 | Rhode Island Department of Health | Infectious Disease Program, Broad Institute of Harvard and MIT | Siddle,K.J., Azevedo,K., Miller,A., Adams,G., Pearlman,L., Gladden-Young,A., Lagerborg,K., Rudy,M., DeRuff,K., Carter,A., Normandin,E., Bauer,M., Reilly,S., Tomkins-Tinch,C., Loreth,C., Chaluvadi,S., Lemieux,J.E., Birren,B.W., Sabeti,P.C., Huard,R., King,E., Park,D.J., and MacInnis,B.L. |
| EPI_ISL_1578385, EPI_ISL_1578386, EPI_ISL_1578387, EPI_ISL_1578388 | Broad Institute Clinical Research Sequencing Platform | Infectious Disease Program, Broad Institute of Harvard and MIT | Siddle,K.J., Adams,G., Pearlman,L., Gladden-Young,A., Vicente,G., Blumenstiel,B., DeFelice,M., Lee,M., McGovern,S., Lagerborg,K., Rudy,M., DeRuff,K., Carter,A., Normandin,E., Bauer,M., Reilly,S., Tomkins-Tinch,C., Loreth,C., Chaluvadi,S., Meldrim,J., Granger,B., Lemieux,J.E., Birren,B.W., Sabeti,P.C., Larkin,K., Dodge,S., Lennon,N., Madoff,L., Brown,C., Gallagher,G., Smole,S., Park,D.J., Gabriel,S., and MacInnis,B.L. |
| EPI_ISL_1578389, EPI_ISL_1578390 | Rhode Island Department of Health | Infectious Disease Program, Broad Institute of Harvard and MIT | Siddle,K.J., Azevedo,K., Miller,A., Adams,G., Pearlman,L., Gladden-Young,A., Lagerborg,K., Rudy,M., DeRuff,K., Carter,A., Normandin,E., Bauer,M., Reilly,S., Tomkins-Tinch,C., Loreth,C., Chaluvadi,S., Lemieux,J.E., Birren,B.W., Sabeti,P.C., Huard,R., King,E., Park,D.J., and MacInnis,B.L. |
| EPI_ISL_1578391, EPI_ISL_1578392, EPI_ISL_1578393, EPI_ISL_1578394, EPI_ISL_1578395, EPI_ISL_1578396, EPI_ISL_1578397, EPI_ISL_1578398, EPI_ISL_1578399, EPI_ISL_1578400, EPI_ISL_1578401, EPI_ISL_1578402, EPI_ISL_1578403, EPI_ISL_1578404, EPI_ISL_1578405, EPI_ISL_1578406 | | | |
| see above | Broad Institute Clinical Research Sequencing Platform | Infectious Disease Program, Broad Institute of Harvard and MIT | Siddle,K.J., Adams,G., Pearlman,L., Gladden-Young,A., Vicente,G., Blumenstiel,B., DeFelice,M., Lee,M., McGovern,S., Lagerborg,K., Rudy,M., DeRuff,K., Carter,A., Normandin,E., Bauer,M., Reilly,S., Tomkins-Tinch,C., Loreth,C., Chaluvadi,S., Meldrim,J., Granger,B., Lemieux,J.E., Birren,B.W., Sabeti,P.C., Larkin,K., Dodge,S., Lennon,N., Madoff,L., Brown,C., Gallagher,G., Smole,S., Park,D.J., Gabriel,S., and MacInnis,B.L. |
| EPI_ISL_1578407, EPI_ISL_1578408, EPI_ISL_1578409, EPI_ISL_1578410 | Rhode Island Department of Health | Infectious Disease Program, Broad Institute of Harvard and MIT | Siddle,K.J., Azevedo,K., Miller,A., Adams,G., Pearlman,L., Gladden-Young,A., Lagerborg,K., Rudy,M., DeRuff,K., Carter,A., Normandin,E., Bauer,M., Reilly,S., Tomkins-Tinch,C., Loreth,C., Chaluvadi,S., Lemieux,J.E., Birren,B.W., Sabeti,P.C., Huard,R., King,E., Park,D.J., and MacInnis,B.L. |
| EPI_ISL_1578411, EPI_ISL_1578412, EPI_ISL_1578413 | Broad Institute Clinical Research Sequencing Platform | Infectious Disease Program, Broad Institute of Harvard and MIT | Siddle,K.J., Adams,G., Pearlman,L., Gladden-Young,A., Vicente,G., Blumenstiel,B., DeFelice,M., Lee,M., McGovern,S., Lagerborg,K., Rudy,M., DeRuff,K., Carter,A., Normandin,E., Bauer,M., Reilly,S., Tomkins-Tinch,C., Loreth,C., Chaluvadi,S., Meldrim,J., Granger,B., Lemieux,J.E., Birren,B.W., Sabeti,P.C., Larkin,K., Dodge,S., Lennon,N., Madoff,L., Brown,C., Gallagher,G., Smole,S., Park,D.J., Gabriel,S., and MacInnis,B.L. |
| EPI_ISL_1578414 | Rhode Island Department of Health | Infectious Disease Program, Broad Institute of Harvard and MIT | Siddle,K.J., Azevedo,K., Miller,A., Adams,G., Pearlman,L., Gladden-Young,A., Lagerborg,K., Rudy,M., DeRuff,K., Carter,A., Normandin,E., Bauer,M., Reilly,S., Tomkins-Tinch,C., Loreth,C., Chaluvadi,S., Lemieux,J.E., Birren,B.W., Sabeti,P.C., Huard,R., King,E., Park,D.J., and MacInnis,B.L. |
| EPI_ISL_1578415, EPI_ISL_1578416, EPI_ISL_1578417, EPI_ISL_1578418, EPI_ISL_1578419, EPI_ISL_1578420, EPI_ISL_1578421, EPI_ISL_1578422, EPI_ISL_1578423, EPI_ISL_1578424, EPI_ISL_1578425, EPI_ISL_1578426, EPI_ISL_1578427, EPI_ISL_1578428, EPI_ISL_1578429, EPI_ISL_1578430, EPI_ISL_1578431, EPI_ISL_1578432, EPI_ISL_1578433, EPI_ISL_1578434, EPI_ISL_1578435, EPI_ISL_1578436, EPI_ISL_1578437, EPI_ISL_1578438, EPI_ISL_1578439, EPI_ISL_1578440, EPI_ISL_1578441, EPI_ISL_1578442 | | | |
| see above | Broad Institute Clinical Research Sequencing Platform | Infectious Disease Program, Broad Institute of Harvard and MIT | Siddle,K.J., Adams,G., Pearlman,L., Gladden-Young,A., Vicente,G., Blumenstiel,B., DeFelice,M., Lee,M., McGovern,S., Lagerborg,K., Rudy,M., DeRuff,K., Carter,A., Normandin,E., Bauer,M., Reilly,S., Tomkins-Tinch,C., Loreth,C., Chaluvadi,S., Meldrim,J., Granger,B., Lemieux,J.E., Birren,B.W., Sabeti,P.C., Larkin,K., Dodge,S., Lennon,N., Madoff,L., Brown,C., Gallagher,G., Smole,S., Park,D.J., Gabriel,S., and MacInnis,B.L. |
| EPI_ISL_1578443, EPI_ISL_1578444, EPI_ISL_1578445, EPI_ISL_1578446, EPI_ISL_1578447, EPI_ISL_1578448, EPI_ISL_1578449, EPI_ISL_1578450 | Rhode Island Department of Health | Infectious Disease Program, Broad Institute of Harvard and MIT | Siddle,K.J., Azevedo,K., Miller,A., Adams,G., Pearlman,L., Gladden-Young,A., Lagerborg,K., Rudy,M., DeRuff,K., Carter,A., Normandin,E., Bauer,M., Reilly,S., Tomkins-Tinch,C., Loreth,C., Chaluvadi,S., Lemieux,J.E., Birren,B.W., Sabeti,P.C., Huard,R., King,E., Park,D.J., and MacInnis,B.L. |
| EPI_ISL_1578451, EPI_ISL_1578452 | Flow Health | Infectious Disease Program, Broad Institute of Harvard and MIT | Lemieux,J.E., Siddle,K.J., Adams,G., Gladden-Young,A., Lagerborg,K., Rudy,M., DeRuff,K., Carter,A., Normandin,E., Bauer,M., Reilly,S., Tomkins-Tinch,C., Loreth,C., Chaluvadi,S., Birren,B.W., Gallagher,G., Smole,S., Park,D.J., MacInnis,B.L., and Sabeti,P.C. |
| EPI_ISL_1578536, EPI_ISL_1578537, EPI_ISL_1578538, EPI_ISL_1578539, EPI_ISL_1578540, EPI_ISL_1578541, EPI_ISL_1578542, EPI_ISL_1578543, EPI_ISL_1578545 | Minnesota Department of Health, Public Health Laboratory | Minnesota Department of Health, Public Health Laboratory | Alexandra Lorentz, Jacob Garfin, Matt Plumb, and Xiong Wang |
| EPI_ISL_1578546 | M Health Fairview | Minnesota Department of Health, Public Health Laboratory | Alexandra Lorentz, Jacob Garfin, Matt Plumb, and Xiong Wang |
| EPI_ISL_1578547, EPI_ISL_1578548, EPI_ISL_1578549, EPI_ISL_1578550, EPI_ISL_1578551, EPI_ISL_1578552, EPI_ISL_1578553, EPI_ISL_1578554, EPI_ISL_1578555, EPI_ISL_1578556, EPI_ISL_1578557, EPI_ISL_1578558, EPI_ISL_1578559, EPI_ISL_1578560, EPI_ISL_1578561, EPI_ISL_1578562, EPI_ISL_1578563, EPI_ISL_1578564, EPI_ISL_1578565, EPI_ISL_1578566, EPI_ISL_1578567 | | | |
| see above | Minnesota Department of Health, Public Health Laboratory | Minnesota Department of Health, Public Health Laboratory | Alexandra Lorentz, Jacob Garfin, Matt Plumb, and Xiong Wang |
| EPI_ISL_1578568 | M Health Fairview | Minnesota Department of Health, Public Health Laboratory | Alexandra Lorentz, Jacob Garfin, Matt Plumb, and Xiong Wang |
| EPI_ISL_1578569, EPI_ISL_1578570, EPI_ISL_1578571, EPI_ISL_1578572, EPI_ISL_1578573, EPI_ISL_1578574, EPI_ISL_1578575, EPI_ISL_1578576, EPI_ISL_1578577, EPI_ISL_1578578 | Minnesota Department of Health, Public Health Laboratory | Minnesota Department of Health, Public Health Laboratory | Alexandra Lorentz, Jacob Garfin, Matt Plumb, and Xiong Wang |
| EPI_ISL_1578579 | M Health Fairview | Minnesota Department of Health, Public Health Laboratory | Alexandra Lorentz, Jacob Garfin, Matt Plumb, and Xiong Wang |
| EPI_ISL_1578580, EPI_ISL_1578582, EPI_ISL_1578583, EPI_ISL_1578584, EPI_ISL_1578585, EPI_ISL_1578586, EPI_ISL_1578587, EPI_ISL_1578588, EPI_ISL_1578589, EPI_ISL_1578590, EPI_ISL_1578591, EPI_ISL_1578592, EPI_ISL_1578593, EPI_ISL_1578594, EPI_ISL_1578595 | | | |
| see above | Minnesota Department of Health, Public Health Laboratory | Minnesota Department of Health, Public Health Laboratory | Alexandra Lorentz, Jacob Garfin, Matt Plumb, and Xiong Wang |
| EPI_ISL_1578596, EPI_ISL_1578597, EPI_ISL_1578598, EPI_ISL_1578599, EPI_ISL_1578600, EPI_ISL_1578601, EPI_ISL_1578602, EPI_ISL_1578603, EPI_ISL_1578604, EPI_ISL_1578605, EPI_ISL_1578606, EPI_ISL_1578607 | | | |
| see above | M Health Fairview | Minnesota Department of Health, Public Health Laboratory | Alexandra Lorentz, Jacob Garfin, Matt Plumb, and Xiong Wang |
| EPI_ISL_1579230, EPI_ISL_1579231, EPI_ISL_1579232, EPI_ISL_1579233, EPI_ISL_1579234, EPI_ISL_1579235, EPI_ISL_1579236, EPI_ISL_1579237, EPI_ISL_1579238, EPI_ISL_1579239, EPI_ISL_1579240, EPI_ISL_1579241, EPI_ISL_1579242, EPI_ISL_1579243, EPI_ISL_1579244, EPI_ISL_1579245, EPI_ISL_1579246, EPI_ISL_1579247, EPI_ISL_1579248 | | | |
| see above | Nebraska Public Health Laboratory | NPHL COVID-19 Response Team | NPHL COVID-19 Response Team |
| EPI_ISL_1580353, EPI_ISL_1580355, EPI_ISL_1580356, EPI_ISL_1580357, EPI_ISL_1580358, EPI_ISL_1580359, EPI_ISL_1580360, EPI_ISL_1580361, EPI_ISL_1580362, EPI_ISL_1580365, EPI_ISL_1580366, EPI_ISL_1580367, EPI_ISL_1580368, EPI_ISL_1580369, EPI_ISL_1580370, EPI_ISL_1580371, EPI_ISL_1580372, EPI_ISL_1580373, EPI_ISL_1580374, EPI_ISL_1580375, EPI_ISL_1580376, EPI_ISL_1580377, EPI_ISL_1580378, EPI_ISL_1580379, EPI_ISL_1580380, EPI_ISL_1580381, EPI_ISL_1580382, EPI_ISL_1580383, EPI_ISL_1580384, EPI_ISL_1580385, EPI_ISL_1580386, EPI_ISL_1580387, EPI_ISL_1580388, EPI_ISL_1580389, EPI_ISL_1580390, EPI_ISL_1580391, EPI_ISL_1580392, EPI_ISL_1580393, EPI_ISL_1580395, EPI_ISL_1580396, EPI_ISL_1580397, EPI_ISL_1580398, EPI_ISL_1580399, EPI_ISL_1580400, EPI_ISL_1580401, EPI_ISL_1580402, EPI_ISL_1580403, EPI_ISL_1580404, EPI_ISL_1580405, EPI_ISL_1580406, EPI_ISL_1580407, EPI_ISL_1580408, EPI_ISL_1580409, EPI_ISL_1580410, EPI_ISL_1580411, EPI_ISL_1580412, EPI_ISL_1580413, EPI_ISL_1580414, EPI_ISL_1580415, EPI_ISL_1580416, EPI_ISL_1580417, EPI_ISL_1580418, EPI_ISL_1580419, EPI_ISL_1580420, EPI_ISL_1580421, EPI_ISL_1580422, EPI_ISL_1580424, EPI_ISL_1580425, EPI_ISL_1580426, EPI_ISL_1580427, EPI_ISL_1580428, EPI_ISL_1580429, EPI_ISL_1580430, EPI_ISL_1580431, EPI_ISL_1580432, EPI_ISL_1580433, EPI_ISL_1580434, EPI_ISL_1580435, EPI_ISL_1580436, EPI_ISL_1580437, EPI_ISL_1580438, EPI_ISL_1580439, EPI_ISL_1580440, EPI_ISL_1580441, EPI_ISL_1580442, EPI_ISL_1580443, EPI_ISL_1580444, EPI_ISL_1580445, EPI_ISL_1580446, EPI_ISL_1580448, EPI_ISL_1580449, EPI_ISL_1580450, EPI_ISL_1580451, EPI_ISL_1580452, EPI_ISL_1580453, EPI_ISL_1580454, EPI_ISL_1580455, EPI_ISL_1580456, EPI_ISL_1580457, EPI_ISL_1580458, EPI_ISL_1580459, EPI_ISL_1580460, EPI_ISL_1580461, EPI_ISL_1580463, EPI_ISL_1580464, EPI_ISL_1580465, EPI_ISL_1580466, EPI_ISL_1580467, EPI_ISL_1580468, EPI_ISL_1580469, EPI_ISL_1580470, EPI_ISL_1580471, EPI_ISL_1580472, EPI_ISL_1580473, EPI_ISL_1580474, EPI_ISL_1580475, EPI_ISL_1580476, EPI_ISL_1580477, EPI_ISL_1580479, EPI_ISL_1580480, EPI_ISL_1580481, EPI_ISL_1580482, EPI_ISL_1580483, EPI_ISL_1580484, EPI_ISL_1580485, EPI_ISL_1580486, EPI_ISL_1580487, EPI_ISL_1580488, EPI_ISL_1580489, EPI_ISL_1580490, EPI_ISL_1580491, EPI_ISL_1580492, EPI_ISL_1580493, EPI_ISL_1580494, EPI_ISL_1580495, EPI_ISL_1580496, EPI_ISL_1580497, EPI_ISL_1580498, EPI_ISL_1580499, EPI_ISL_1580500, EPI_ISL_1580501 | | | |
| see above | University of Michigan Clinical Microbiology Laboratory | Lauring Lab, University of Michigan, Department of Microbiology and Immunology | Valesano |
| EPI_ISL_1580667, EPI_ISL_1580668, EPI_ISL_1580669, EPI_ISL_1580670 | BOSTON HEART DIAGNOSTICS CORP | Wadsworth Center, New York State Department of Health | Kirsten St. George, Daryl M. Lamson, Alexis Russell, Matthew Shudt, Melissa A Leisner, Jonathan Plitnick, Catharine Prussing, Navjot Singh, John Kelly, Erasmus Schneider, Erica Lasek-Nesselquist |

| | | | |
|---|---|---|---|
| EPI_ISL_1580671, EPI_ISL_1580672 | URMC LABS | Wadsworth Center, New York State Department of Health | Kirsten St. George, Daryl M. Lamson, Alexis Russell, Matthew Shudt, Melissa A Leisner, Jonathan Plitnick, Catharine Prussing, Navjot Singh, John Kelly, Erasmus Schneider, Erica Lasek-Nesselquist |
| EPI_ISL_1580673, EPI_ISL_1580674, EPI_ISL_1580675, EPI_ISL_1580676, EPI_ISL_1580677 | WESTCHESTER MEDICAL CENTER | Wadsworth Center, New York State Department of Health | Kirsten St. George, Daryl M. Lamson, Alexis Russell, Matthew Shudt, Melissa A Leisner, Jonathan Plitnick, Catharine Prussing, Navjot Singh, John Kelly, Erasmus Schneider, Erica Lasek-Nesselquist |
| EPI_ISL_1580678, EPI_ISL_1580679, EPI_ISL_1580680, EPI_ISL_1580681, EPI_ISL_1580682, EPI_ISL_1580683, EPI_ISL_1580684, EPI_ISL_1580685 | SUNY UPSTATE MEDICAL UNIVERSITY | Wadsworth Center, New York State Department of Health | Kirsten St. George, Daryl M. Lamson, Alexis Russell, Matthew Shudt, Melissa A Leisner, Jonathan Plitnick, Catharine Prussing, Navjot Singh, John Kelly, Erasmus Schneider, Erica Lasek-Nesselquist |
| EPI_ISL_1580686 | MONTEFIORE MEDICAL CENTER LABORATORIES | Wadsworth Center, New York State Department of Health | Kirsten St. George, Daryl M. Lamson, Alexis Russell, Matthew Shudt, Melissa A Leisner, Jonathan Plitnick, Catharine Prussing, Navjot Singh, John Kelly, Erasmus Schneider, Erica Lasek-Nesselquist |
| EPI_ISL_1580687, EPI_ISL_1580688, EPI_ISL_1580689, EPI_ISL_1580690, EPI_ISL_1580691, EPI_ISL_1580692, EPI_ISL_1580693, EPI_ISL_1580694, EPI_ISL_1580695, EPI_ISL_1580696, EPI_ISL_1580697, EPI_ISL_1580698 | | | |
| see above | Columbia University Irving Medical Center | Wadsworth Center, New York State Department of Health | Kirsten St. George, Daryl M. Lamson, Alexis Russell, Matthew Shudt, Melissa A Leisner, Jonathan Plitnick, Catharine Prussing, Navjot Singh, John Kelly, Erasmus Schneider, Erica Lasek-Nesselquist |
| EPI_ISL_1580699 | WESTCHESTER MEDICAL CENTER | Wadsworth Center, New York State Department of Health | Kirsten St. George, Daryl M. Lamson, Alexis Russell, Matthew Shudt, Melissa A Leisner, Jonathan Plitnick, Catharine Prussing, Navjot Singh, John Kelly, Erasmus Schneider, Erica Lasek-Nesselquist |
| EPI_ISL_1580700 | ALBANY MEDICAL CENTER | Wadsworth Center, New York State Department of Health | Kirsten St. George, Daryl M. Lamson, Alexis Russell, Matthew Shudt, Melissa A Leisner, Jonathan Plitnick, Catharine Prussing, Navjot Singh, John Kelly, Erasmus Schneider, Erica Lasek-Nesselquist |
| EPI_ISL_1580701, EPI_ISL_1580702, EPI_ISL_1580703, EPI_ISL_1580704, EPI_ISL_1580705, EPI_ISL_1580706, EPI_ISL_1580707, EPI_ISL_1580708, EPI_ISL_1580709, EPI_ISL_1580710, EPI_ISL_1580711, EPI_ISL_1580712, EPI_ISL_1580713, EPI_ISL_1580714, EPI_ISL_1580715, EPI_ISL_1580716, EPI_ISL_1580717, EPI_ISL_1580718, EPI_ISL_1580719, EPI_ISL_1580720, EPI_ISL_1580721, EPI_ISL_1580722 | | | |
| see above | URMC LABS | Wadsworth Center, New York State Department of Health | Kirsten St. George, Daryl M. Lamson, Alexis Russell, Matthew Shudt, Melissa A Leisner, Jonathan Plitnick, Catharine Prussing, Navjot Singh, John Kelly, Erasmus Schneider, Erica Lasek-Nesselquist |
| EPI_ISL_1580723, EPI_ISL_1580724, EPI_ISL_1580725, EPI_ISL_1580726, EPI_ISL_1580727, EPI_ISL_1580728, EPI_ISL_1580729, EPI_ISL_1580730, EPI_ISL_1580731, EPI_ISL_1580732, EPI_ISL_1580733, EPI_ISL_1580734, EPI_ISL_1580735, EPI_ISL_1580736, EPI_ISL_1580737, EPI_ISL_1580738 | | | |
| see above | NORTHWELL HEALTH LABORATORIES | Wadsworth Center, New York State Department of Health | Kirsten St. George, Daryl M. Lamson, Alexis Russell, Matthew Shudt, Melissa A Leisner, Jonathan Plitnick, Catharine Prussing, Navjot Singh, John Kelly, Erasmus Schneider, Erica Lasek-Nesselquist |
| EPI_ISL_1580739, EPI_ISL_1580740 | SARATOGA HOSPITAL LABORATORY | Wadsworth Center, New York State Department of Health | Kirsten St. George, Daryl M. Lamson, Alexis Russell, Matthew Shudt, Melissa A Leisner, Jonathan Plitnick, Catharine Prussing, Navjot Singh, John Kelly, Erasmus Schneider, Erica Lasek-Nesselquist |
| EPI_ISL_1580741 | ALBANY MEDICAL CENTER | Wadsworth Center, New York State Department of Health | Kirsten St. George, Daryl M. Lamson, Alexis Russell, Matthew Shudt, Melissa A Leisner, Jonathan Plitnick, Catharine Prussing, Navjot Singh, John Kelly, Erasmus Schneider, Erica Lasek-Nesselquist |
| EPI_ISL_1580742, EPI_ISL_1580743, EPI_ISL_1580744, EPI_ISL_1580745, EPI_ISL_1580746, EPI_ISL_1580747 | Helix/Illumina | Centers for Disease Control and Prevention Division of Viral Diseases, Pathogen Discovery | Dakota Howard, Dhwani Batra, Peter W. Cook, Kara Moser, Adrian Paskey, Jason Caravas, Benjamin Rambo-Martin, Shatavia Morrison, Christopher Gulvick, Scott Sammons, Yvette Unoarumhi, Darlene Wagner, Matthew Schmerer, Eileen de Feo, Jan Antico, Christine Tran, Matthew Tolentino, Shannon Wickline, Kim Gietzen, Brad Sickler, Jingtao Liu, Eric Allen, Phil Febbo, Nicole L. Washington, Simon White, Geraint Levan, Kelly Schiabor Barrett, Elizabeth Cirulli, Alexandre Bolze, Ary Ascencio, Charlotte Rivera-Garcia, Ryan Cho, Jason Nguyen, Sherry Wang, Jimmy Ramirez, Tyler Cassens, Efren Sandoval, Magnus Isaksson, William Lee, David Becker, Marc Laurent, James Lu, Clinton R. Paden, Duncan MacCannell |
| EPI_ISL_1580748 | Reditus Laboratories | Reditus Laboratories | Joshua J. Geltz, Ph.D., Robert M. Sgambelluri, Ph.D., Cassy Philips, M.S., Alexa Eichelberger, M.S. |
| EPI_ISL_1580749 | Helix/Illumina | Centers for Disease Control and Prevention Division of Viral Diseases, Pathogen Discovery | Dakota Howard, Dhwani Batra, Peter W. Cook, Kara Moser, Adrian Paskey, Jason Caravas, Benjamin Rambo-Martin, Shatavia Morrison, Christopher Gulvick, Scott Sammons, Yvette Unoarumhi, Darlene Wagner, Matthew Schmerer, Eileen de Feo, Jan Antico, Christine Tran, Matthew Tolentino, Shannon Wickline, Kim Gietzen, Brad Sickler, Jingtao Liu, Eric Allen, Phil Febbo, Nicole L. Washington, Simon White, Geraint Levan, Kelly Schiabor Barrett, Elizabeth Cirulli, Alexandre Bolze, Ary Ascencio, Charlotte Rivera-Garcia, Ryan Cho, Jason Nguyen, Sherry Wang, Jimmy Ramirez, Tyler Cassens, Efren Sandoval, Magnus Isaksson, William Lee, David Becker, Marc Laurent, James Lu, Clinton R. Paden, Duncan MacCannell |
| EPI_ISL_1580750 | Reditus Laboratories | Reditus Laboratories | Joshua J. Geltz, Ph.D., Robert M. Sgambelluri, Ph.D., Cassy Philips, M.S., Alexa Eichelberger, M.S. |
| EPI_ISL_1580751 | Helix/Illumina | Centers for Disease Control and Prevention Division of Viral Diseases, Pathogen Discovery | Dakota Howard, Dhwani Batra, Peter W. Cook, Kara Moser, Adrian Paskey, Jason Caravas, Benjamin Rambo-Martin, Shatavia Morrison, Christopher Gulvick, Scott Sammons, Yvette Unoarumhi, Darlene Wagner, Matthew Schmerer, Eileen de Feo, Jan Antico, Christine Tran, Matthew Tolentino, Shannon Wickline, Kim Gietzen, Brad Sickler, Jingtao Liu, Eric Allen, Phil Febbo, Nicole L. Washington, Simon White, Geraint Levan, Kelly Schiabor Barrett, Elizabeth Cirulli, Alexandre Bolze, Ary Ascencio, Charlotte Rivera-Garcia, Ryan Cho, Jason Nguyen, Sherry Wang, Jimmy Ramirez, Tyler Cassens, Efren Sandoval, Magnus Isaksson, William Lee, David Becker, Marc Laurent, James Lu, Clinton R. Paden, Duncan MacCannell |
| EPI_ISL_1580752, EPI_ISL_1580753 | Reditus Laboratories | Reditus Laboratories | Joshua J. Geltz, Ph.D., Robert M. Sgambelluri, Ph.D., Cassy Philips, M.S., Alexa Eichelberger, M.S. |
| EPI_ISL_1580754 | Helix/Illumina | Centers for Disease Control and Prevention Division of Viral Diseases, Pathogen Discovery | Dakota Howard, Dhwani Batra, Peter W. Cook, Kara Moser, Adrian Paskey, Jason Caravas, Benjamin Rambo-Martin, Shatavia Morrison, Christopher Gulvick, Scott Sammons, Yvette Unoarumhi, Darlene Wagner, Matthew Schmerer, Eileen de Feo, Jan Antico, Christine Tran, Matthew Tolentino, Shannon Wickline, Kim Gietzen, Brad Sickler, Jingtao Liu, Eric Allen, Phil Febbo, Nicole L. Washington, Simon White, Geraint Levan, Kelly Schiabor Barrett, Elizabeth Cirulli, Alexandre Bolze, Ary Ascencio, Charlotte Rivera-Garcia, Ryan Cho, Jason Nguyen, Sherry Wang, Jimmy Ramirez, Tyler Cassens, Efren Sandoval, Magnus Isaksson, William Lee, David Becker, Marc Laurent, James Lu, Clinton R. Paden, Duncan MacCannell |
| EPI_ISL_1580755, EPI_ISL_1580756 | Reditus Laboratories | Reditus Laboratories | Joshua J. Geltz, Ph.D., Robert M. Sgambelluri, Ph.D., Cassy Philips, M.S., Alexa Eichelberger, M.S. |
| EPI_ISL_1580757 | Helix/Illumina | Centers for Disease Control and Prevention Division of Viral Diseases, Pathogen Discovery | Dakota Howard, Dhwani Batra, Peter W. Cook, Kara Moser, Adrian Paskey, Jason Caravas, Benjamin Rambo-Martin, Shatavia Morrison, Christopher Gulvick, Scott Sammons, Yvette Unoarumhi, Darlene Wagner, Matthew Schmerer, Eileen de Feo, Jan Antico, Christine Tran, Matthew Tolentino, Shannon Wickline, Kim Gietzen, Brad Sickler, Jingtao Liu, Eric Allen, Phil Febbo, Nicole L. Washington, Simon White, Geraint Levan, Kelly Schiabor Barrett, Elizabeth Cirulli, Alexandre Bolze, Ary Ascencio, Charlotte Rivera-Garcia, Ryan Cho, Jason Nguyen, Sherry Wang, Jimmy Ramirez, Tyler Cassens, Efren Sandoval, Magnus Isaksson, William Lee, David Becker, Marc Laurent, James Lu, Clinton R. Paden, Duncan MacCannell |
| EPI_ISL_1580758 | Reditus Laboratories | Reditus Laboratories | Joshua J. Geltz, Ph.D., Robert M. Sgambelluri, Ph.D., Cassy Philips, M.S., Alexa Eichelberger, M.S. |
| EPI_ISL_1580760 | Helix/Illumina | Centers for Disease Control and Prevention Division of Viral Diseases, Pathogen Discovery | Dakota Howard, Dhwani Batra, Peter W. Cook, Kara Moser, Adrian Paskey, Jason Caravas, Benjamin Rambo-Martin, Shatavia Morrison, Christopher Gulvick, Scott Sammons, Yvette Unoarumhi, Darlene Wagner, Matthew Schmerer, Eileen de Feo, Jan Antico, Christine Tran, Matthew Tolentino, Shannon Wickline, Kim Gietzen, Brad Sickler, Jingtao Liu, Eric Allen, Phil Febbo, Nicole L. Washington, Simon White, Geraint Levan, Kelly Schiabor Barrett, Elizabeth Cirulli, Alexandre Bolze, Ary Ascencio, Charlotte Rivera-Garcia, Ryan Cho, Jason Nguyen, Sherry Wang, Jimmy Ramirez, Tyler Cassens, Efren Sandoval, Magnus Isaksson, William Lee, David Becker, Marc Laurent, James Lu, Clinton R. Paden, Duncan MacCannell |
| EPI_ISL_1580761, EPI_ISL_1580762 | Reditus Laboratories | Reditus Laboratories | Joshua J. Geltz, Ph.D., Robert M. Sgambelluri, Ph.D., Cassy Philips, M.S., Alexa Eichelberger, M.S. |
| EPI_ISL_1580763 | Helix/Illumina | Centers for Disease Control and Prevention Division of Viral Diseases, Pathogen Discovery | Dakota Howard, Dhwani Batra, Peter W. Cook, Kara Moser, Adrian Paskey, Jason Caravas, Benjamin Rambo-Martin, Shatavia Morrison, Christopher Gulvick, Scott Sammons, Yvette Unoarumhi, Darlene Wagner, Matthew Schmerer, Eileen de Feo, Jan Antico, Christine Tran, Matthew Tolentino, Shannon Wickline, Kim Gietzen, Brad Sickler, Jingtao Liu, Eric Allen, Phil Febbo, Nicole L. Washington, Simon White, Geraint Levan, Kelly Schiabor Barrett, Elizabeth Cirulli, Alexandre Bolze, Ary Ascencio, Charlotte Rivera-Garcia, Ryan Cho, Jason Nguyen, Sherry Wang, Jimmy Ramirez, Tyler Cassens, Efren Sandoval, Magnus Isaksson, William Lee, David Becker, Marc Laurent, James Lu, Clinton R. Paden, Duncan MacCannell |
| EPI_ISL_1580764, EPI_ISL_1580765 | Reditus Laboratories | Reditus Laboratories | Joshua J. Geltz, Ph.D., Robert M. Sgambelluri, Ph.D., Cassy Philips, M.S., Alexa Eichelberger, M.S. |
| EPI_ISL_1580766 | Helix/Illumina | Centers for Disease Control and Prevention Division of Viral Diseases, Pathogen Discovery | Dakota Howard, Dhwani Batra, Peter W. Cook, Kara Moser, Adrian Paskey, Jason Caravas, Benjamin Rambo-Martin, Shatavia Morrison, Christopher Gulvick, Scott Sammons, Yvette Unoarumhi, Darlene Wagner, Matthew Schmerer, Eileen de Feo, Jan Antico, Christine Tran, Matthew Tolentino, Shannon Wickline, Kim Gietzen, Brad Sickler, Jingtao Liu, Eric Allen, Phil Febbo, Nicole L. Washington, Simon White, Geraint Levan, Kelly Schiabor Barrett, Elizabeth Cirulli, Alexandre Bolze, Ary Ascencio, Charlotte Rivera-Garcia, Ryan Cho, Jason Nguyen, Sherry Wang, Jimmy Ramirez, Tyler Cassens, Efren Sandoval, Magnus Isaksson, William Lee, David Becker, Marc Laurent, James Lu, Clinton R. Paden, Duncan MacCannell |
| EPI_ISL_1580767, EPI_ISL_1580768 | Reditus Laboratories | Reditus Laboratories | Joshua J. Geltz, Ph.D., Robert M. Sgambelluri, Ph.D., Cassy Philips, M.S., Alexa Eichelberger, M.S. |
| EPI_ISL_1580769 | Helix/Illumina | Centers for Disease Control and Prevention Division of Viral Diseases, Pathogen Discovery | Dakota Howard, Dhwani Batra, Peter W. Cook, Kara Moser, Adrian Paskey, Jason Caravas, Benjamin Rambo-Martin, Shatavia Morrison, Christopher Gulvick, Scott Sammons, Yvette Unoarumhi, Darlene Wagner, Matthew Schmerer, Eileen de Feo, Jan Antico, Christine Tran, Matthew Tolentino, Shannon Wickline, Kim Gietzen, Brad Sickler, Jingtao Liu, Eric Allen, Phil Febbo, Nicole L. Washington, Simon White, Geraint Levan, Kelly Schiabor Barrett, Elizabeth Cirulli, Alexandre Bolze, Ary Ascencio, Charlotte Rivera-Garcia, Ryan Cho, Jason Nguyen, Sherry Wang, Jimmy Ramirez, Tyler Cassens, Efren Sandoval, Magnus Isaksson, William Lee, David Becker, Marc Laurent, James Lu, Clinton R. Paden, Duncan MacCannell |
| EPI_ISL_1580770 | Reditus Laboratories | Reditus Laboratories | Joshua J. Geltz, Ph.D., Robert M. Sgambelluri, Ph.D., Cassy Philips, M.S., Alexa Eichelberger, M.S. |
| EPI_ISL_1580771 | Helix/Illumina | Centers for Disease Control and Prevention Division of Viral Diseases, Pathogen Discovery | Dakota Howard, Dhwani Batra, Peter W. Cook, Kara Moser, Adrian Paskey, Jason Caravas, Benjamin Rambo-Martin, Shatavia Morrison, Christopher Gulvick, Scott Sammons, Yvette Unoarumhi, Darlene Wagner, Matthew Schmerer, Eileen de Feo, Jan Antico, Christine Tran, Matthew Tolentino, Shannon Wickline, Kim Gietzen, Brad Sickler, Jingtao Liu, Eric Allen, Phil Febbo, Nicole L. Washington, Simon White, Geraint Levan, Kelly Schiabor Barrett, Elizabeth Cirulli, Alexandre Bolze, Ary Ascencio, Charlotte Rivera-Garcia, Ryan Cho, Jason Nguyen, Sherry Wang, Jimmy Ramirez, Tyler Cassens, Efren Sandoval, Magnus Isaksson, William Lee, David Becker, Marc Laurent, James Lu, Clinton R. Paden, Duncan MacCannell |
| EPI_ISL_1580772, EPI_ISL_1580773 | Reditus Laboratories | Reditus Laboratories | Joshua J. Geltz, Ph.D., Robert M. Sgambelluri, Ph.D., Cassy Philips, M.S., Alexa Eichelberger, M.S. |
| EPI_ISL_1580774 | Helix/Illumina | Centers for Disease Control and Prevention Division of Viral Diseases, Pathogen Discovery | Dakota Howard, Dhwani Batra, Peter W. Cook, Kara Moser, Adrian Paskey, Jason Caravas, Benjamin Rambo-Martin, Shatavia Morrison, Christopher Gulvick, Scott Sammons, Yvette Unoarumhi, Darlene Wagner, Matthew Schmerer, Eileen de Feo, Jan Antico, Christine Tran, Matthew Tolentino, Shannon Wickline, Kim Gietzen, Brad Sickler, Jingtao Liu, Eric Allen, Phil Febbo, Nicole L. Washington, Simon White, Geraint Levan, Kelly Schiabor Barrett, Elizabeth Cirulli, Alexandre Bolze, Ary Ascencio, Charlotte Rivera-Garcia, Ryan Cho, Jason Nguyen, Sherry Wang, Jimmy Ramirez, Tyler Cassens, Efren Sandoval, Magnus Isaksson, William Lee, David Becker, Marc Laurent, James Lu, Clinton R. Paden, Duncan MacCannell |
| EPI_ISL_1580775, EPI_ISL_1580776 | Reditus Laboratories | Reditus Laboratories | Joshua J. Geltz, Ph.D., Robert M. Sgambelluri, Ph.D., Cassy Philips, M.S., Alexa Eichelberger, M.S. |
| EPI_ISL_1580777 | Helix/Illumina | Centers for Disease Control and Prevention Division of Viral Diseases, Pathogen Discovery | Dakota Howard, Dhwani Batra, Peter W. Cook, Kara Moser, Adrian Paskey, Jason Caravas, Benjamin Rambo-Martin, Shatavia Morrison, Christopher Gulvick, Scott Sammons, Yvette Unoarumhi, Darlene Wagner, Matthew Schmerer, Eileen de Feo, Jan Antico, Christine Tran, Matthew Tolentino, Shannon Wickline, Kim Gietzen, Brad Sickler, Jingtao Liu, Eric Allen, Phil Febbo, Nicole L. Washington, Simon White, Geraint Levan, Kelly Schiabor Barrett, Elizabeth Cirulli, Alexandre Bolze, Ary Ascencio, Charlotte Rivera-Garcia, Ryan Cho, Jason Nguyen, Sherry Wang, Jimmy Ramirez, Tyler Cassens, Efren Sandoval, Magnus Isaksson, William Lee, David Becker, Marc Laurent, James Lu, Clinton R. Paden, Duncan MacCannell |
| EPI_ISL_1580778, | Reditus Laboratories | Reditus Laboratories | Joshua J. Geltz, Ph.D., Robert M. Sgambelluri, Ph.D., Cassy Philips, M.S., Alexa Eichelberger, M.S. |

| | | | |
|---|---|---|---|
| EPI_ISL_1580779 | | | |
| EPI_ISL_1580780 | Helix/Illumina | Centers for Disease Control and Prevention Division of Viral Diseases, Pathogen Discovery | Dakota Howard, Dhwani Batra, Peter W. Cook, Kara Moser, Adrian Paskey, Jason Caravas, Benjamin Rambo-Martin, Shatavia Morrison, Christopher Gulvick, Scott Sammons, Yvette Unoarumhi, Darlene Wagner, Matthew Schmerer, Eileen de Feo, Jan Antico, Christine Tran, Matthew Tolentino, Shannon Wickline, Kim Gietzen, Brad Sickler, Jingtao Liu, Eric Allen, Phil Febbo, Nicole L. Washington, Simon White, Geraint Levan, Kelly Schiabor Barrett, Elizabeth Cirulli, Alexandre Bolze, Ary Ascencio, Charlotte Rivera-Garcia, Ryan Cho, Jason Nguyen, Sherry Wang, Jimmy Ramirez, Tyler Cassens, Efren Sandoval, Magnus Isaksson, William Lee, David Becker, Marc Laurent, James Lu, Clinton R. Paden, Duncan MacCannell |
| EPI_ISL_1580781, EPI_ISL_1580782 | Reditus Laboratories | Reditus Laboratories | Joshua J. Geltz, Ph.D., Robert M. Sgambelluri, Ph.D., Cassy Philips, M.S., Alexa Eichelberger, M.S. |
| EPI_ISL_1580783 | Helix/Illumina | Centers for Disease Control and Prevention Division of Viral Diseases, Pathogen Discovery | Dakota Howard, Dhwani Batra, Peter W. Cook, Kara Moser, Adrian Paskey, Jason Caravas, Benjamin Rambo-Martin, Shatavia Morrison, Christopher Gulvick, Scott Sammons, Yvette Unoarumhi, Darlene Wagner, Matthew Schmerer, Eileen de Feo, Jan Antico, Christine Tran, Matthew Tolentino, Shannon Wickline, Kim Gietzen, Brad Sickler, Jingtao Liu, Eric Allen, Phil Febbo, Nicole L. Washington, Simon White, Geraint Levan, Kelly Schiabor Barrett, Elizabeth Cirulli, Alexandre Bolze, Ary Ascencio, Charlotte Rivera-Garcia, Ryan Cho, Jason Nguyen, Sherry Wang, Jimmy Ramirez, Tyler Cassens, Efren Sandoval, Magnus Isaksson, William Lee, David Becker, Marc Laurent, James Lu, Clinton R. Paden, Duncan MacCannell |
| EPI_ISL_1580784 | Reditus Laboratories | Reditus Laboratories | Joshua J. Geltz, Ph.D., Robert M. Sgambelluri, Ph.D., Cassy Philips, M.S., Alexa Eichelberger, M.S. |
| EPI_ISL_1580785 | Helix/Illumina | Centers for Disease Control and Prevention Division of Viral Diseases, Pathogen Discovery | Dakota Howard, Dhwani Batra, Peter W. Cook, Kara Moser, Adrian Paskey, Jason Caravas, Benjamin Rambo-Martin, Shatavia Morrison, Christopher Gulvick, Scott Sammons, Yvette Unoarumhi, Darlene Wagner, Matthew Schmerer, Eileen de Feo, Jan Antico, Christine Tran, Matthew Tolentino, Shannon Wickline, Kim Gietzen, Brad Sickler, Jingtao Liu, Eric Allen, Phil Febbo, Nicole L. Washington, Simon White, Geraint Levan, Kelly Schiabor Barrett, Elizabeth Cirulli, Alexandre Bolze, Ary Ascencio, Charlotte Rivera-Garcia, Ryan Cho, Jason Nguyen, Sherry Wang, Jimmy Ramirez, Tyler Cassens, Efren Sandoval, Magnus Isaksson, William Lee, David Becker, Marc Laurent, James Lu, Clinton R. Paden, Duncan MacCannell |
| EPI_ISL_1580786, EPI_ISL_1580787 | Reditus Laboratories | Reditus Laboratories | Joshua J. Geltz, Ph.D., Robert M. Sgambelluri, Ph.D., Cassy Philips, M.S., Alexa Eichelberger, M.S. |
| EPI_ISL_1580788 | Helix/Illumina | Centers for Disease Control and Prevention Division of Viral Diseases, Pathogen Discovery | Dakota Howard, Dhwani Batra, Peter W. Cook, Kara Moser, Adrian Paskey, Jason Caravas, Benjamin Rambo-Martin, Shatavia Morrison, Christopher Gulvick, Scott Sammons, Yvette Unoarumhi, Darlene Wagner, Matthew Schmerer, Eileen de Feo, Jan Antico, Christine Tran, Matthew Tolentino, Shannon Wickline, Kim Gietzen, Brad Sickler, Jingtao Liu, Eric Allen, Phil Febbo, Nicole L. Washington, Simon White, Geraint Levan, Kelly Schiabor Barrett, Elizabeth Cirulli, Alexandre Bolze, Ary Ascencio, Charlotte Rivera-Garcia, Ryan Cho, Jason Nguyen, Sherry Wang, Jimmy Ramirez, Tyler Cassens, Efren Sandoval, Magnus Isaksson, William Lee, David Becker, Marc Laurent, James Lu, Clinton R. Paden, Duncan MacCannell |
| EPI_ISL_1580789, EPI_ISL_1580790 | Reditus Laboratories | Reditus Laboratories | Joshua J. Geltz, Ph.D., Robert M. Sgambelluri, Ph.D., Cassy Philips, M.S., Alexa Eichelberger, M.S. |
| EPI_ISL_1580791 | Helix/Illumina | Centers for Disease Control and Prevention Division of Viral Diseases, Pathogen Discovery | Dakota Howard, Dhwani Batra, Peter W. Cook, Kara Moser, Adrian Paskey, Jason Caravas, Benjamin Rambo-Martin, Shatavia Morrison, Christopher Gulvick, Scott Sammons, Yvette Unoarumhi, Darlene Wagner, Matthew Schmerer, Eileen de Feo, Jan Antico, Christine Tran, Matthew Tolentino, Shannon Wickline, Kim Gietzen, Brad Sickler, Jingtao Liu, Eric Allen, Phil Febbo, Nicole L. Washington, Simon White, Geraint Levan, Kelly Schiabor Barrett, Elizabeth Cirulli, Alexandre Bolze, Ary Ascencio, Charlotte Rivera-Garcia, Ryan Cho, Jason Nguyen, Sherry Wang, Jimmy Ramirez, Tyler Cassens, Efren Sandoval, Magnus Isaksson, William Lee, David Becker, Marc Laurent, James Lu, Clinton R. Paden, Duncan MacCannell |
| EPI_ISL_1580792, EPI_ISL_1580793 | Reditus Laboratories | Reditus Laboratories | Joshua J. Geltz, Ph.D., Robert M. Sgambelluri, Ph.D., Cassy Philips, M.S., Alexa Eichelberger, M.S. |
| EPI_ISL_1580794 | Helix/Illumina | Centers for Disease Control and Prevention Division of Viral Diseases, Pathogen Discovery | Dakota Howard, Dhwani Batra, Peter W. Cook, Kara Moser, Adrian Paskey, Jason Caravas, Benjamin Rambo-Martin, Shatavia Morrison, Christopher Gulvick, Scott Sammons, Yvette Unoarumhi, Darlene Wagner, Matthew Schmerer, Eileen de Feo, Jan Antico, Christine Tran, Matthew Tolentino, Shannon Wickline, Kim Gietzen, Brad Sickler, Jingtao Liu, Eric Allen, Phil Febbo, Nicole L. Washington, Simon White, Geraint Levan, Kelly Schiabor Barrett, Elizabeth Cirulli, Alexandre Bolze, Ary Ascencio, Charlotte Rivera-Garcia, Ryan Cho, Jason Nguyen, Sherry Wang, Jimmy Ramirez, Tyler Cassens, Efren Sandoval, Magnus Isaksson, William Lee, David Becker, Marc Laurent, James Lu, Clinton R. Paden, Duncan MacCannell |
| EPI_ISL_1580795, EPI_ISL_1580796 | Reditus Laboratories | Reditus Laboratories | Joshua J. Geltz, Ph.D., Robert M. Sgambelluri, Ph.D., Cassy Philips, M.S., Alexa Eichelberger, M.S. |
| EPI_ISL_1580797, EPI_ISL_1580798, EPI_ISL_1580799, EPI_ISL_1580800, EPI_ISL_1580801, EPI_ISL_1580802, EPI_ISL_1580803, EPI_ISL_1580804, EPI_ISL_1580805, EPI_ISL_1580806, EPI_ISL_1580807, EPI_ISL_1580808, EPI_ISL_1580809, EPI_ISL_1580810, EPI_ISL_1580811, EPI_ISL_1580812, EPI_ISL_1580813, EPI_ISL_1580814, EPI_ISL_1580815, EPI_ISL_1580816, EPI_ISL_1580817, EPI_ISL_1580818, EPI_ISL_1580819, EPI_ISL_1580820, EPI_ISL_1580821, EPI_ISL_1580822, EPI_ISL_1580823, EPI_ISL_1580824, EPI_ISL_1580825, EPI_ISL_1580826, EPI_ISL_1580827, EPI_ISL_1580828, EPI_ISL_1580829, EPI_ISL_1580830, EPI_ISL_1580831, EPI_ISL_1580832, EPI_ISL_1580833, EPI_ISL_1580834, EPI_ISL_1580835, EPI_ISL_1580836, EPI_ISL_1580837, EPI_ISL_1580838, EPI_ISL_1580839, EPI_ISL_1580840, EPI_ISL_1580841, EPI_ISL_1580842, EPI_ISL_1580843, EPI_ISL_1580844, EPI_ISL_1580845, EPI_ISL_1580846, EPI_ISL_1580847, EPI_ISL_1580848, EPI_ISL_1580849, EPI_ISL_1580850, EPI_ISL_1580851, EPI_ISL_1580852, EPI_ISL_1580853, EPI_ISL_1580854, EPI_ISL_1580855, EPI_ISL_1580856, EPI_ISL_1580857, EPI_ISL_1580858, EPI_ISL_1580859, EPI_ISL_1580860, EPI_ISL_1580861, EPI_ISL_1580862, EPI_ISL_1580863, EPI_ISL_1580864, EPI_ISL_1580865, EPI_ISL_1580866, EPI_ISL_1580867, EPI_ISL_1580868, EPI_ISL_1580869, EPI_ISL_1580870, EPI_ISL_1580871, EPI_ISL_1580872, EPI_ISL_1580873, EPI_ISL_1580874, EPI_ISL_1580875, EPI_ISL_1580876, EPI_ISL_1580877, EPI_ISL_1580878, EPI_ISL_1580879, EPI_ISL_1580880, EPI_ISL_1580881, EPI_ISL_1580882, EPI_ISL_1580883, EPI_ISL_1580884, EPI_ISL_1580885, EPI_ISL_1580886, EPI_ISL_1580887, EPI_ISL_1580888, EPI_ISL_1580889, EPI_ISL_1580890, EPI_ISL_1580891, EPI_ISL_1580892, EPI_ISL_1580893, EPI_ISL_1580894, EPI_ISL_1580895, EPI_ISL_1580896, EPI_ISL_1580897, EPI_ISL_1580898, EPI_ISL_1580899, EPI_ISL_1580900, EPI_ISL_1580901, EPI_ISL_1580902, EPI_ISL_1580903, EPI_ISL_1580904, EPI_ISL_1580905, EPI_ISL_1580906, EPI_ISL_1580907, EPI_ISL_1580908, EPI_ISL_1580909, EPI_ISL_1580910, EPI_ISL_1580911, EPI_ISL_1580912, EPI_ISL_1580913, EPI_ISL_1580914, EPI_ISL_1580915, EPI_ISL_1580916, EPI_ISL_1580917, EPI_ISL_1580918, EPI_ISL_1580919, EPI_ISL_1580920, EPI_ISL_1580921, EPI_ISL_1580922, EPI_ISL_1580923, EPI_ISL_1580924, EPI_ISL_1580925, EPI_ISL_1580926, EPI_ISL_1580927, EPI_ISL_1580928, EPI_ISL_1580929, EPI_ISL_1580930, EPI_ISL_1580931, EPI_ISL_1580932, EPI_ISL_1580933, EPI_ISL_1580934, EPI_ISL_1580935, EPI_ISL_1580936, EPI_ISL_1580937, EPI_ISL_1580938, EPI_ISL_1580939, EPI_ISL_1580940, EPI_ISL_1580941, EPI_ISL_1580942, EPI_ISL_1580943, EPI_ISL_1580944, EPI_ISL_1580945, EPI_ISL_1580946, EPI_ISL_1580948, EPI_ISL_1580949, EPI_ISL_1580950, EPI_ISL_1580951, EPI_ISL_1580952, EPI_ISL_1580953, EPI_ISL_1580954, EPI_ISL_1580955, EPI_ISL_1580956, EPI_ISL_1580957, EPI_ISL_1580958, EPI_ISL_1580959, EPI_ISL_1580960, EPI_ISL_1580961, EPI_ISL_1580962, EPI_ISL_1580963, EPI_ISL_1580964, EPI_ISL_1580965, EPI_ISL_1580966, EPI_ISL_1580968, EPI_ISL_1580969, EPI_ISL_1580970, EPI_ISL_1580971, EPI_ISL_1580972, EPI_ISL_1580973, EPI_ISL_1580974, EPI_ISL_1580975, EPI_ISL_1580976, EPI_ISL_1580977, EPI_ISL_1580978, EPI_ISL_1580979, EPI_ISL_1580980, EPI_ISL_1580981, EPI_ISL_1580982, EPI_ISL_1580983, EPI_ISL_1580984, EPI_ISL_1580985, EPI_ISL_1580986, EPI_ISL_1580988, EPI_ISL_1580989, EPI_ISL_1580990, EPI_ISL_1580991, EPI_ISL_1580992, EPI_ISL_1580993, EPI_ISL_1580994, EPI_ISL_1580995, EPI_ISL_1580996, EPI_ISL_1580997, EPI_ISL_1580998, EPI_ISL_1580999, EPI_ISL_1581001, EPI_ISL_1581002, EPI_ISL_1581003, EPI_ISL_1581004, EPI_ISL_1581005, EPI_ISL_1581006, EPI_ISL_1581007, EPI_ISL_1581008, EPI_ISL_1581009, EPI_ISL_1581010, EPI_ISL_1581011, EPI_ISL_1581012, EPI_ISL_1581013, EPI_ISL_1581014, EPI_ISL_1581015, EPI_ISL_1581016, EPI_ISL_1581017, EPI_ISL_1581018, EPI_ISL_1581019, EPI_ISL_1581020, EPI_ISL_1581021, EPI_ISL_1581022, EPI_ISL_1581023, EPI_ISL_1581024, EPI_ISL_1581025, EPI_ISL_1581026, EPI_ISL_1581027, EPI_ISL_1581028, EPI_ISL_1581029, EPI_ISL_1581030, EPI_ISL_1581031, EPI_ISL_1581032, EPI_ISL_1581033, EPI_ISL_1581035, EPI_ISL_1581036, EPI_ISL_1581037, EPI_ISL_1581039, EPI_ISL_1581040, EPI_ISL_1581041, EPI_ISL_1581042, EPI_ISL_1581043, EPI_ISL_1581044, EPI_ISL_1581045, EPI_ISL_1581046, EPI_ISL_1581047, EPI_ISL_1581048, EPI_ISL_1581049, EPI_ISL_1581050, EPI_ISL_1581051, EPI_ISL_1581052, EPI_ISL_1581053, EPI_ISL_1581054, EPI_ISL_1581055, EPI_ISL_1581056, EPI_ISL_1581057, EPI_ISL_1581058, EPI_ISL_1581059, EPI_ISL_1581060, EPI_ISL_1581061, EPI_ISL_1581062, EPI_ISL_1581063, EPI_ISL_1581064, EPI_ISL_1581065, EPI_ISL_1581066, EPI_ISL_1581067, EPI_ISL_1581068, EPI_ISL_1581069, EPI_ISL_1581070, EPI_ISL_1581071, EPI_ISL_1581072, EPI_ISL_1581073, EPI_ISL_1581074, EPI_ISL_1581075, EPI_ISL_1581076, EPI_ISL_1581077, EPI_ISL_1581078, EPI_ISL_1581079, EPI_ISL_1581080, EPI_ISL_1581081, EPI_ISL_1581082, EPI_ISL_1581083, EPI_ISL_1581084, EPI_ISL_1581085, EPI_ISL_1581086, EPI_ISL_1581087, EPI_ISL_1581088, EPI_ISL_1581089, EPI_ISL_1581090, EPI_ISL_1581091, EPI_ISL_1581092, EPI_ISL_1581093, EPI_ISL_1581094, EPI_ISL_1581095, EPI_ISL_1581096, EPI_ISL_1581097, EPI_ISL_1581098, EPI_ISL_1581099, EPI_ISL_1581100, EPI_ISL_1581101, EPI_ISL_1581102, EPI_ISL_1581103, EPI_ISL_1581104, EPI_ISL_1581105, EPI_ISL_1581106, EPI_ISL_1581107, EPI_ISL_1581108, EPI_ISL_1581109, EPI_ISL_1581110, EPI_ISL_1581111, EPI_ISL_1581112, EPI_ISL_1581113, EPI_ISL_1581114, EPI_ISL_1581115, EPI_ISL_1581116, EPI_ISL_1581117, EPI_ISL_1581118, EPI_ISL_1581119, EPI_ISL_1581120, EPI_ISL_1581121, EPI_ISL_1581122, EPI_ISL_1581123, EPI_ISL_1581124, EPI_ISL_1581125, EPI_ISL_1581126, EPI_ISL_1581127, EPI_ISL_1581128, EPI_ISL_1581129, EPI_ISL_1581130, EPI_ISL_1581131, EPI_ISL_1581132, EPI_ISL_1581133, EPI_ISL_1581134, EPI_ISL_1581135, EPI_ISL_1581136, EPI_ISL_1581137, EPI_ISL_1581138, EPI_ISL_1581139, EPI_ISL_1581140, EPI_ISL_1581141, EPI_ISL_1581142, EPI_ISL_1581143, EPI_ISL_1581144, EPI_ISL_1581145, EPI_ISL_1581146, EPI_ISL_1581147, EPI_ISL_1581148, EPI_ISL_1581149, EPI_ISL_1581150, EPI_ISL_1581151, EPI_ISL_1581152, EPI_ISL_1581153, EPI_ISL_1581154, EPI_ISL_1581155, EPI_ISL_1581156, EPI_ISL_1581157, EPI_ISL_1581158, EPI_ISL_1581159, EPI_ISL_1581160, EPI_ISL_1581161, EPI_ISL_1581162, EPI_ISL_1581163, EPI_ISL_1581164, EPI_ISL_1581165, EPI_ISL_1581167, EPI_ISL_1581168, EPI_ISL_1581169, EPI_ISL_1581170, EPI_ISL_1581171, EPI_ISL_1581172, EPI_ISL_1581173, EPI_ISL_1581174, EPI_ISL_1581175, EPI_ISL_1581176, EPI_ISL_1581177, EPI_ISL_1581178, EPI_ISL_1581179, EPI_ISL_1581180, EPI_ISL_1581181, EPI_ISL_1581182, EPI_ISL_1581183, EPI_ISL_1581184, EPI_ISL_1581185, EPI_ISL_1581186, EPI_ISL_1581187, EPI_ISL_1581188, EPI_ISL_1581189, EPI_ISL_1581190, EPI_ISL_1581191, EPI_ISL_1581192, EPI_ISL_1581193, EPI_ISL_1581194, EPI_ISL_1581195, EPI_ISL_1581196, EPI_ISL_1581197, EPI_ISL_1581198, EPI_ISL_1581199, EPI_ISL_1581200, EPI_ISL_1581201, EPI_ISL_1581202, EPI_ISL_1581203, EPI_ISL_1581204, EPI_ISL_1581205, EPI_ISL_1581206, EPI_ISL_1581207, EPI_ISL_1581208, EPI_ISL_1581209, EPI_ISL_1581210, EPI_ISL_1581211, EPI_ISL_1581212, EPI_ISL_1581213, EPI_ISL_1581214, EPI_ISL_1581215, EPI_ISL_1581216, EPI_ISL_1581217, EPI_ISL_1581218, EPI_ISL_1581219, EPI_ISL_1581220, EPI_ISL_1581221, EPI_ISL_1581222, EPI_ISL_1581223, EPI_ISL_1581224, EPI_ISL_1581225, EPI_ISL_1581226, EPI_ISL_1581227, EPI_ISL_1581228, EPI_ISL_1581229, EPI_ISL_1581230, EPI_ISL_1581231, EPI_ISL_1581232, EPI_ISL_1581233, EPI_ISL_1581234, EPI_ISL_1581235, EPI_ISL_1581236, EPI_ISL_1581237, EPI_ISL_1581238, EPI_ISL_1581239, EPI_ISL_1581240, EPI_ISL_1581241, EPI_ISL_1581242, EPI_ISL_1581243, EPI_ISL_1581244, EPI_ISL_1581245, EPI_ISL_1581246, EPI_ISL_1581247, EPI_ISL_1581248, EPI_ISL_1581249, EPI_ISL_1581250, EPI_ISL_1581251, EPI_ISL_1581252, EPI_ISL_1581253, EPI_ISL_1581254, EPI_ISL_1581255, EPI_ISL_1581256, EPI_ISL_1581257, EPI_ISL_1581258, EPI_ISL_1581259, EPI_ISL_1581260, EPI_ISL_1581261, EPI_ISL_1581262, EPI_ISL_1581263, EPI_ISL_1581264, EPI_ISL_1581265, EPI_ISL_1581266, EPI_ISL_1581267, EPI_ISL_1581268, EPI_ISL_1581269, EPI_ISL_1581270, EPI_ISL_1581271, EPI_ISL_1581272, EPI_ISL_1581273, EPI_ISL_1581274, EPI_ISL_1581275, EPI_ISL_1581276, EPI_ISL_1581277, EPI_ISL_1581278, EPI_ISL_1581279, EPI_ISL_1581280, EPI_ISL_1581281, EPI_ISL_1581282, EPI_ISL_1581284, EPI_ISL_1581285, EPI_ISL_1581286, EPI_ISL_1581287, EPI_ISL_1581288, EPI_ISL_1581289, EPI_ISL_1581290, EPI_ISL_1581291, EPI_ISL_1581292, EPI_ISL_1581293, EPI_ISL_1581294, EPI_ISL_1581295, EPI_ISL_1581296, EPI_ISL_1581297, EPI_ISL_1581298, EPI_ISL_1581299, EPI_ISL_1581300, EPI_ISL_1581301, EPI_ISL_1581302, EPI_ISL_1581303, EPI_ISL_1581304, EPI_ISL_1581305, EPI_ISL_1581306, EPI_ISL_1581307, EPI_ISL_1581308, EPI_ISL_1581309, EPI_ISL_1581310, EPI_ISL_1581311, EPI_ISL_1581312, EPI_ISL_1581313, EPI_ISL_1581314, EPI_ISL_1581315, EPI_ISL_1581316, EPI_ISL_1581317, EPI_ISL_1581318, EPI_ISL_1581319, EPI_ISL_1581320, EPI_ISL_1581321, EPI_ISL_1581322, EPI_ISL_1581323, EPI_ISL_1581324, EPI_ISL_1581325, EPI_ISL_1581326, EPI_ISL_1581327, EPI_ISL_1581328, EPI_ISL_1581329, EPI_ISL_1581330, EPI_ISL_1581331, EPI_ISL_1581332, EPI_ISL_1581333, EPI_ISL_1581334, EPI_ISL_1581335, EPI_ISL_1581336, EPI_ISL_1581337, EPI_ISL_1581338, EPI_ISL_1581339, EPI_ISL_1581340, EPI_ISL_1581341, EPI_ISL_1581342, EPI_ISL_1581343, EPI_ISL_1581344, EPI_ISL_1581345, EPI_ISL_1581346, EPI_ISL_1581347, EPI_ISL_1581348, EPI_ISL_1581349, EPI_ISL_1581350, EPI_ISL_1581351, EPI_ISL_1581352, EPI_ISL_1581353, EPI_ISL_1581354, EPI_ISL_1581355, EPI_ISL_1581356, EPI_ISL_1581357, EPI_ISL_1581358, EPI_ISL_1581359, EPI_ISL_1581360, EPI_ISL_1581361, EPI_ISL_1581362, EPI_ISL_1581363, EPI_ISL_1581364, EPI_ISL_1581365, EPI_ISL_1581366, EPI_ISL_1581367, EPI_ISL_1581368, EPI_ISL_1581369, EPI_ISL_1581370, EPI_ISL_1581371, EPI_ISL_1581372, EPI_ISL_1581373, EPI_ISL_1581374, EPI_ISL_1581375, EPI_ISL_1581376, EPI_ISL_1581377, EPI_ISL_1581378, EPI_ISL_1581379, EPI_ISL_1581380, EPI_ISL_1581381, EPI_ISL_1581382, EPI_ISL_1581383, EPI_ISL_1581384, EPI_ISL_1581385, EPI_ISL_1581386, EPI_ISL_1581387, EPI_ISL_1581388, EPI_ISL_1581389, EPI_ISL_1581390, EPI_ISL_1581391, EPI_ISL_1581392, EPI_ISL_1581393, EPI_ISL_1581394, EPI_ISL_1581395, EPI_ISL_1581396, EPI_ISL_1581397, EPI_ISL_1581398, EPI_ISL_1581399, EPI_ISL_1581400, EPI_ISL_1581401, EPI_ISL_1581402, EPI_ISL_1581403, EPI_ISL_1581404, EPI_ISL_1581405, EPI_ISL_1581406, EPI_ISL_1581408, EPI_ISL_1581411, EPI_ISL_1581414, EPI_ISL_1581417, EPI_ISL_1581420, EPI_ISL_1581424, EPI_ISL_1581427, EPI_ISL_1581430, EPI_ISL_1581432, EPI_ISL_1581438, EPI_ISL_1581441, EPI_ISL_1581444, EPI_ISL_1581448, EPI_ISL_1581451, EPI_ISL_1581454, EPI_ISL_1581457, EPI_ISL_1581460, EPI_ISL_1581463, EPI_ISL_1581467, EPI_ISL_1581475, EPI_ISL_1581478, EPI_ISL_1581481, EPI_ISL_1581484, EPI_ISL_1581487, EPI_ISL_1581490, EPI_ISL_1581493, EPI_ISL_1581496, EPI_ISL_1581499, EPI_ISL_1581502, EPI_ISL_1581505, EPI_ISL_1581507, EPI_ISL_1581510, EPI_ISL_1581513, EPI_ISL_1581516, EPI_ISL_1581519, EPI_ISL_1581522, EPI_ISL_1581525, EPI_ISL_1581531, EPI_ISL_1581534, EPI_ISL_1581537, EPI_ISL_1581540, EPI_ISL_1581543, EPI_ISL_1581546, EPI_ISL_1581549, EPI_ISL_1581552, EPI_ISL_1581555, EPI_ISL_1581558, EPI_ISL_1581561, EPI_ISL_1581564, EPI_ISL_1581567, EPI_ISL_1581570, EPI_ISL_1581572, EPI_ISL_1581575, EPI_ISL_1581578, EPI_ISL_1581581, EPI_ISL_1581584, EPI_ISL_1581586, EPI_ISL_1581589, EPI_ISL_1581593, EPI_ISL_1581596, EPI_ISL_1581599, EPI_ISL_1581602, EPI_ISL_1581605, EPI_ISL_1581607, EPI_ISL_1581610, EPI_ISL_1581613, EPI_ISL_1581616, EPI_ISL_1581620, EPI_ISL_1581623, EPI_ISL_1581626, EPI_ISL_1581629, EPI_ISL_1581632, EPI_ISL_1581635, EPI_ISL_1581638, EPI_ISL_1581641, EPI_ISL_1581644, EPI_ISL_1581646, EPI_ISL_1581649, EPI_ISL_1581652, EPI_ISL_1581655, EPI_ISL_1581658, EPI_ISL_1581659, EPI_ISL_1581660, EPI_ISL_1581661, EPI_ISL_1581662, EPI_ISL_1581663, EPI_ISL_1581664, EPI_ISL_1581665, EPI_ISL_1581666, EPI_ISL_1581667, EPI_ISL_1581668, EPI_ISL_1581669, EPI_ISL_1581670, EPI_ISL_1581672, EPI_ISL_1581673, EPI_ISL_1581674, EPI_ISL_1581675, EPI_ISL_1581676, EPI_ISL_1581677, EPI_ISL_1581678, EPI_ISL_1581679, EPI_ISL_1581680, EPI_ISL_1581681, EPI_ISL_1581682, EPI_ISL_1581683, EPI_ISL_1581684, EPI_ISL_1581685, EPI_ISL_1581686, EPI_ISL_1581687, EPI_ISL_1581688, EPI_ISL_1581689, EPI_ISL_1581690, EPI_ISL_1581691, EPI_ISL_1581692, EPI_ISL_1581693, EPI_ISL_1581694, EPI_ISL_1581695, EPI_ISL_1581696, EPI_ISL_1581697, EPI_ISL_1581698, EPI_ISL_1581699, EPI_ISL_1581700, EPI_ISL_1581701, EPI_ISL_1581702, EPI_ISL_1581703, EPI_ISL_1581704, EPI_ISL_1581705, EPI_ISL_1581706, EPI_ISL_1581707, EPI_ISL_1581708, EPI_ISL_1581709, EPI_ISL_1581710, EPI_ISL_1581711, EPI_ISL_1581713, EPI_ISL_1581714, EPI_ISL_1581715, EPI_ISL_1581716, EPI_ISL_1581717, EPI_ISL_1581718, EPI_ISL_1581719, EPI_ISL_1581720, EPI_ISL_1581721, EPI_ISL_1581722, EPI_ISL_1581723, EPI_ISL_1581724, EPI_ISL_1581725, EPI_ISL_1581726, EPI_ISL_1581727, EPI_ISL_1581728, EPI_ISL_1581729, EPI_ISL_1581730, EPI_ISL_1581731, EPI_ISL_1581732, EPI_ISL_1581733, EPI_ISL_1581734, EPI_ISL_1581735, EPI_ISL_1581736, EPI_ISL_1581737, EPI_ISL_1581739, EPI_ISL_1581740, EPI_ISL_1581741, EPI_ISL_1581742, EPI_ISL_1581743, EPI_ISL_1581744, EPI_ISL_1581745, EPI_ISL_1581746, EPI_ISL_1581747, EPI_ISL_1581748, EPI_ISL_1581749, EPI_ISL_1581750, EPI_ISL_1581751, EPI_ISL_1581753, EPI_ISL_1581754, EPI_ISL_1581755, EPI_ISL_1581756, EPI_ISL_1581757, EPI_ISL_1581758, EPI_ISL_1581759, EPI_ISL_1581760, EPI_ISL_1581761, EPI_ISL_1581762, EPI_ISL_1581763, EPI_ISL_1581764, EPI_ISL_1581765, EPI_ISL_1581766, EPI_ISL_1581767, EPI_ISL_1581768, EPI_ISL_1581769, EPI_ISL_1581770, EPI_ISL_1581771, EPI_ISL_1581772, EPI_ISL_1581773, EPI_ISL_1581774, EPI_ISL_1581775, EPI_ISL_1581776, EPI_ISL_1581777, EPI_ISL_1581778, EPI_ISL_1581779, EPI_ISL_1581780, EPI_ISL_1581781, EPI_ISL_1581782, EPI_ISL_1581783, EPI_ISL_1581784, EPI_ISL_1581785, EPI_ISL_1581786, EPI_ISL_1581787, EPI_ISL_1581788, EPI_ISL_1581789, EPI_ISL_1581790, EPI_ISL_1581791, EPI_ISL_1581792, EPI_ISL_1581793, EPI_ISL_1581794, EPI_ISL_1581795, EPI_ISL_1581796, EPI_ISL_1581797, EPI_ISL_1581798, EPI_ISL_1581799, EPI_ISL_1581800, EPI_ISL_1581801, EPI_ISL_1581802, EPI_ISL_1581803, EPI_ISL_1581804, EPI_ISL_1581805, EPI_ISL_1581806, EPI_ISL_1581807, EPI_ISL_1581808, EPI_ISL_1581809, EPI_ISL_1581810, EPI_ISL_1581811, EPI_ISL_1581812, EPI_ISL_1581813, EPI_ISL_1581814, EPI_ISL_1581815, EPI_ISL_1581816, EPI_ISL_1581817, EPI_ISL_1581818, EPI_ISL_1581819, EPI_ISL_1581820, EPI_ISL_1581821, EPI_ISL_1581822, EPI_ISL_1581823, EPI_ISL_1581824, EPI_ISL_1581825, EPI_ISL_1581826, EPI_ISL_1581827, EPI_ISL_1581828, EPI_ISL_1581829, EPI_ISL_1581830, EPI_ISL_1581831, EPI_ISL_1581832, EPI_ISL_1581833, EPI_ISL_1581834, EPI_ISL_1581835, EPI_ISL_1581836, EPI_ISL_1581837, EPI_ISL_1581838, EPI_ISL_1581839, EPI_ISL_1581840, EPI_ISL_1581841, EPI_ISL_1581842, EPI_ISL_1581843, EPI_ISL_1581844, EPI_ISL_1581845, EPI_ISL_1581846, EPI_ISL_1581847, EPI_ISL_1581848, EPI_ISL_1581849, EPI_ISL_1581850, EPI_ISL_1581851, EPI_ISL_1581852, EPI_ISL_1581853, EPI_ISL_1581854 | | | |
| see above | Helix/Illumina | Centers for Disease Control and Prevention Division of Viral Diseases, Pathogen Discovery | Dakota Howard, Dhwani Batra, Peter W. Cook, Kara Moser, Adrian Paskey, Jason Caravas, Benjamin Rambo-Martin, Shatavia Morrison, Christopher Gulvick, Scott Sammons, Yvette Unoarumhi, Darlene Wagner, Matthew Schmerer, Eileen de Feo, Jan Antico, Christine Tran, Matthew Tolentino, Shannon Wickline, Kim Gietzen, Brad Sickler, Jingtao Liu, Eric Allen, Phil Febbo, Nicole L. Washington, Simon White, Geraint Levan, Kelly Schiabor Barrett, Elizabeth Cirulli, Alexandre Bolze, Ary Ascencio, Charlotte Rivera-Garcia, Ryan Cho, Jason Nguyen, Sherry Wang, Jimmy Ramirez, Tyler Cassens, Efren Sandoval, Magnus Isaksson, William Lee, David Becker, Marc Laurent, James Lu, Clinton R. Paden, Duncan MacCannell |
| EPI_ISL_1581855, EPI_ISL_1581856, EPI_ISL_1581857, EPI_ISL_1581858, EPI_ISL_1581859, EPI_ISL_1581860, EPI_ISL_1581861, EPI_ISL_1581862, EPI_ISL_1581863, EPI_ISL_1581864, EPI_ISL_1581865, EPI_ISL_1581866, EPI_ISL_1581867, EPI_ISL_1581868, EPI_ISL_1581869, EPI_ISL_1581870, EPI_ISL_1581871, EPI_ISL_1581872, EPI_ISL_1581873, EPI_ISL_1581874, EPI_ISL_1581875, EPI_ISL_1581876, EPI_ISL_1581877, EPI_ISL_1581878, EPI_ISL_1581879, EPI_ISL_1581880, EPI_ISL_1581881, EPI_ISL_1581882, EPI_ISL_1581883, EPI_ISL_1581884, EPI_ISL_1581885, EPI_ISL_1581886, EPI_ISL_1581887, EPI_ISL_1581888, EPI_ISL_1581889, EPI_ISL_1581890, EPI_ISL_1581891, EPI_ISL_1581892, EPI_ISL_1581893, EPI_ISL_1581894, EPI_ISL_1581895, EPI_ISL_1581896, EPI_ISL_1581897, EPI_ISL_1581898, EPI_ISL_1581899, EPI_ISL_1581900, EPI_ISL_1581901, EPI_ISL_1581902, EPI_ISL_1581903, EPI_ISL_1581904, EPI_ISL_1581905, EPI_ISL_1581906, EPI_ISL_1581907, EPI_ISL_1581908, EPI_ISL_1581909, EPI_ISL_1581910, EPI_ISL_1581911, EPI_ISL_1581912, EPI_ISL_1581913, EPI_ISL_1581914, EPI_ISL_1581915, EPI_ISL_1581916, EPI_ISL_1581917, EPI_ISL_1581918, EPI_ISL_1581919, EPI_ISL_1581920, EPI_ISL_1581921, EPI_ISL_1581922, EPI_ISL_1581923, EPI_ISL_1581924, EPI_ISL_1581925, EPI_ISL_1581926, EPI_ISL_1581927, EPI_ISL_1581928, EPI_ISL_1581929, EPI_ISL_1581930, EPI_ISL_1581931, EPI_ISL_1581932, EPI_ISL_1581933, EPI_ISL_1581934, EPI_ISL_1581935, EPI_ISL_1581936, EPI_ISL_1581937, EPI_ISL_1581938, EPI_ISL_1581939, EPI_ISL_1581940, EPI_ISL_1581941, EPI_ISL_1581942, EPI_ISL_1581943, EPI_ISL_1581944, EPI_ISL_1581945, EPI_ISL_1581946, EPI_ISL_1581947, EPI_ISL_1581948, EPI_ISL_1581949, EPI_ISL_1581950, EPI_ISL_1581951, EPI_ISL_1581952, EPI_ISL_1581953, EPI_ISL_1581954, EPI_ISL_1581955, EPI_ISL_1581956, EPI_ISL_1581957, EPI_ISL_1581958, EPI_ISL_1581959, EPI_ISL_1581960, EPI_ISL_1581961, EPI_ISL_1581962, EPI_ISL_1581963, EPI_ISL_1581964, EPI_ISL_1581965, EPI_ISL_1581966, EPI_ISL_1581967, EPI_ISL_1581968, EPI_ISL_1581969, EPI_ISL_1581970, EPI_ISL_1581971, EPI_ISL_1581972, EPI_ISL_1581973, EPI_ISL_1581974, EPI_ISL_1581975, EPI_ISL_1581976, EPI_ISL_1581977, EPI_ISL_1581978, EPI_ISL_1581979, EPI_ISL_1581980, EPI_ISL_1581981, EPI_ISL_1581982, EPI_ISL_1581983, EPI_ISL_1581984, EPI_ISL_1581985, EPI_ISL_1581986, EPI_ISL_1581987, EPI_ISL_1581988, EPI_ISL_1581989, EPI_ISL_1581990, EPI_ISL_1581991, EPI_ISL_1581992, EPI_ISL_1581993, EPI_ISL_1581994, EPI_ISL_1581995, EPI_ISL_1581996, EPI_ISL_1581997, EPI_ISL_1581998, EPI_ISL_1581999, EPI_ISL_1582000, EPI_ISL_1582001, EPI_ISL_1582002, EPI_ISL_1582003, EPI_ISL_1582004, EPI_ISL_1582005, EPI_ISL_1582006, EPI_ISL_1582007, EPI_ISL_1582008, EPI_ISL_1582009, EPI_ISL_1582010, EPI_ISL_1582011, EPI_ISL_1582012, EPI_ISL_1582013, EPI_ISL_1582014, EPI_ISL_1582015, EPI_ISL_1582016, EPI_ISL_1582017, EPI_ISL_1582018, EPI_ISL_1582019, EPI_ISL_1582020, EPI_ISL_1582021, EPI_ISL_1582022, EPI_ISL_1582023, EPI_ISL_1582024, EPI_ISL_1582025, EPI_ISL_1582026, EPI_ISL_1582027, EPI_ISL_1582028, EPI_ISL_1582029, EPI_ISL_1582030, EPI_ISL_1582031, EPI_ISL_1582032, EPI_ISL_1582033, EPI_ISL_1582034, EPI_ISL_1582035, EPI_ISL_1582036, EPI_ISL_1582037, EPI_ISL_1582038, EPI_ISL_1582039, EPI_ISL_1582040, EPI_ISL_1582041, EPI_ISL_1582042, EPI_ISL_1582043, EPI_ISL_1582044, EPI_ISL_1582045, EPI_ISL_1582046, EPI_ISL_1582047, EPI_ISL_1582048, EPI_ISL_1582049, EPI_ISL_1582050, EPI_ISL_1582051, EPI_ISL_1582052, EPI_ISL_1582053, EPI_ISL_1582054, EPI_ISL_1582055, EPI_ISL_1582056, EPI_ISL_1582057, EPI_ISL_1582058, EPI_ISL_1582059, EPI_ISL_1582060, EPI_ISL_1582061, EPI_ISL_1582062, EPI_ISL_1582063, EPI_ISL_1582064, EPI_ISL_1582065, EPI_ISL_1582066, EPI_ISL_1582067, EPI_ISL_1582068, EPI_ISL_1582069, EPI_ISL_1582070, EPI_ISL_1582071, EPI_ISL_1582072, EPI_ISL_1582073, EPI_ISL_1582074, EPI_ISL_1582075, EPI_ISL_1582076, EPI_ISL_1582077, EPI_ISL_1582078, EPI_ISL_1582079, EPI_ISL_1582080, EPI_ISL_1582081, EPI_ISL_1582082, EPI_ISL_1582083, EPI_ISL_1582084, EPI_ISL_1582085, EPI_ISL_1582086, EPI_ISL_1582087, EPI_ISL_1582088, EPI_ISL_1582089, EPI_ISL_1582090, EPI_ISL_1582091, EPI_ISL_1582092, EPI_ISL_1582093, EPI_ISL_1582094, EPI_ISL_1582095, EPI_ISL_1582096, EPI_ISL_1582097, EPI_ISL_1582098, EPI_ISL_1582099, EPI_ISL_1582100, EPI_ISL_1582101, EPI_ISL_1582102, EPI_ISL_1582103, EPI_ISL_1582104, EPI_ISL_1582105, EPI_ISL_1582106, EPI_ISL_1582107, EPI_ISL_1582108, EPI_ISL_1582109, EPI_ISL_1582110, EPI_ISL_1582111, EPI_ISL_1582112, EPI_ISL_1582113, EPI_ISL_1582114, EPI_ISL_1582115, EPI_ISL_1582116, EPI_ISL_1582117, EPI_ISL_1582118, EPI_ISL_1582119, EPI_ISL_1582120, EPI_ISL_1582121, EPI_ISL_1582122, EPI_ISL_1582123, EPI_ISL_1582124, EPI_ISL_1582125, EPI_ISL_1582126, EPI_ISL_1582127, EPI_ISL_1582128, EPI_ISL_1582129, EPI_ISL_1582130, EPI_ISL_1582131, EPI_ISL_1582132, EPI_ISL_1582133, EPI_ISL_1582134, EPI_ISL_1582135, EPI_ISL_1582136, EPI_ISL_1582137, EPI_ISL_1582138, EPI_ISL_1582139, EPI_ISL_1582140, EPI_ISL_1582141, EPI_ISL_1582142, EPI_ISL_1582143, EPI_ISL_1582144, EPI_ISL_1582145, EPI_ISL_1582146, EPI_ISL_1582147, EPI_ISL_1582148 | | | |
| see above | Quest Diagnostics Incorporated | Centers for Disease Control and Prevention Division of Viral Diseases, Pathogen Discovery | Dakota Howard, Dhwani Batra, Peter W. Cook, Kara Moser, Adrian Paskey, Jason Caravas, Benjamin Rambo-Martin, Shatavia Morrison, Christopher Gulvick, Scott Sammons, Yvette Unoarumhi, Darlene Wagner, Matthew Schmerer, S. H. Rosenthal, A. Gerasimova, R. M. Kagan, B. Anderson, M. Hua, Y. Liu, L.E. Bernstein, K.E. Livingston, A. Perez, I. A. Shlyakhter, R. V. Rolando, R. Owen, P. Tanpaiboon, F. Lacbawan, Clinton R. Paden, Duncan MacCannell |
| EPI_ISL_1582149 | SUNY UPSTATE MEDICAL UNIVERSITY | Wadsworth Center, New York State Department of Health | Kirsten St. George, Daryl M. Lamson, Alexis Russell, Matthew Shudt, Melissa A Leisner, Jonathan Plitnick, Catharine Prussing, Navjot Singh, John Kelly, Erasmus Schneider, Erica Lasek-Nesselquist |
| EPI_ISL_1582150 | Quest Diagnostics Incorporated | Centers for Disease Control and Prevention Division of Viral Diseases, Pathogen Discovery | Dakota Howard, Dhwani Batra, Peter W. Cook, Kara Moser, Adrian Paskey, Jason Caravas, Benjamin Rambo-Martin, Shatavia Morrison, Christopher Gulvick, Scott Sammons, Yvette Unoarumhi, Darlene Wagner, Matthew Schmerer, S. H. Rosenthal, A. Gerasimova, R. M. Kagan, B. Anderson, M. Hua, Y. Liu, L.E. Bernstein, K.E. Livingston, A. Perez, I. A. Shlyakhter, R. V. Rolando, R. Owen, P. Tanpaiboon, F. Lacbawan, Clinton R. Paden, Duncan MacCannell |
| EPI_ISL_1582151 | URMC LABS | Wadsworth Center, New York State Department of Health | Kirsten St. George, Daryl M. Lamson, Alexis Russell, Matthew Shudt, Melissa A Leisner, Jonathan Plitnick, Catharine Prussing, Navjot Singh, John Kelly, Erasmus Schneider, Erica Lasek-Nesselquist |
| EPI_ISL_1582152, EPI_ISL_1582153 | Quest Diagnostics Incorporated | Centers for Disease Control and Prevention Division of Viral Diseases, Pathogen Discovery | Dakota Howard, Dhwani Batra, Peter W. Cook, Kara Moser, Adrian Paskey, Jason Caravas, Benjamin Rambo-Martin, Shatavia Morrison, Christopher Gulvick, Scott Sammons, Yvette Unoarumhi, Darlene Wagner, Matthew Schmerer, S. H. Rosenthal, A. Gerasimova, R. M. Kagan, B. Anderson, M. Hua, Y. Liu, L.E. Bernstein, K.E. Livingston, A. Perez, I. A. Shlyakhter, R. V. Rolando, R. Owen, P. Tanpaiboon, F. Lacbawan, Clinton R. Paden, Duncan MacCannell |

| | | | |
|---|---|---|---|
| EPI_ISL_1582154 | ALBANY MEDICAL CENTER | Wadsworth Center, New York State Department of Health | Kirsten St. George, Daryl M. Lamson, Alexis Russell, Matthew Shudt, Melissa A Leisner, Jonathan Plitnick, Catharine Prussing, Navjot Singh, John Kelly, Erasmus Schneider, Erica Lasek-Nesselquist |
| EPI_ISL_1582155 | Quest Diagnostics Incorporated | Centers for Disease Control and Prevention Division of Viral Diseases, Pathogen Discovery | Dakota Howard, Dhwani Batra, Peter W. Cook, Kara Moser, Adrian Paskey, Jason Caravas, Benjamin Rambo-Martin, Shatavia Morrison, Christopher Gulvick, Scott Sammons, Yvette Unoarumhi, Darlene Wagner, Matthew Schmerer, S. H. Rosenthal, A. Gerasimova, R. M. Kagan, B. Anderson, M. Hua, Y. Liu, L.E. Bernstein, K.E. Livingston, A. Perez, I. A. Shlyakhter, R. V. Rolando, R. Owen, P. Tanpaiboon, F. Lacbawan, Clinton R. Paden, Duncan MacCannell |
| EPI_ISL_1582156 | NORTHWELL HEALTH LABORATORIES | Wadsworth Center, New York State Department of Health | Kirsten St. George, Daryl M. Lamson, Alexis Russell, Matthew Shudt, Melissa A Leisner, Jonathan Plitnick, Catharine Prussing, Navjot Singh, John Kelly, Erasmus Schneider, Erica Lasek-Nesselquist |
| EPI_ISL_1582157, EPI_ISL_1582158 | Quest Diagnostics Incorporated | Centers for Disease Control and Prevention Division of Viral Diseases, Pathogen Discovery | Dakota Howard, Dhwani Batra, Peter W. Cook, Kara Moser, Adrian Paskey, Jason Caravas, Benjamin Rambo-Martin, Shatavia Morrison, Christopher Gulvick, Scott Sammons, Yvette Unoarumhi, Darlene Wagner, Matthew Schmerer, S. H. Rosenthal, A. Gerasimova, R. M. Kagan, B. Anderson, M. Hua, Y. Liu, L.E. Bernstein, K.E. Livingston, A. Perez, I. A. Shlyakhter, R. V. Rolando, R. Owen, P. Tanpaiboon, F. Lacbawan, Clinton R. Paden, Duncan MacCannell |
| EPI_ISL_1582159 | MONTEFIORE MEDICAL CENTER LABORATORIES | Wadsworth Center, New York State Department of Health | Kirsten St. George, Daryl M. Lamson, Alexis Russell, Matthew Shudt, Melissa A Leisner, Jonathan Plitnick, Catharine Prussing, Navjot Singh, John Kelly, Erasmus Schneider, Erica Lasek-Nesselquist |
| EPI_ISL_1582160, EPI_ISL_1582161 | Quest Diagnostics Incorporated | Centers for Disease Control and Prevention Division of Viral Diseases, Pathogen Discovery | Dakota Howard, Dhwani Batra, Peter W. Cook, Kara Moser, Adrian Paskey, Jason Caravas, Benjamin Rambo-Martin, Shatavia Morrison, Christopher Gulvick, Scott Sammons, Yvette Unoarumhi, Darlene Wagner, Matthew Schmerer, S. H. Rosenthal, A. Gerasimova, R. M. Kagan, B. Anderson, M. Hua, Y. Liu, L.E. Bernstein, K.E. Livingston, A. Perez, I. A. Shlyakhter, R. V. Rolando, R. Owen, P. Tanpaiboon, F. Lacbawan, Clinton R. Paden, Duncan MacCannell |
| EPI_ISL_1582162 | MONTEFIORE MEDICAL CENTER LABORATORIES | Wadsworth Center, New York State Department of Health | Kirsten St. George, Daryl M. Lamson, Alexis Russell, Matthew Shudt, Melissa A Leisner, Jonathan Plitnick, Catharine Prussing, Navjot Singh, John Kelly, Erasmus Schneider, Erica Lasek-Nesselquist |
| EPI_ISL_1582163, EPI_ISL_1582164 | Quest Diagnostics Incorporated | Centers for Disease Control and Prevention Division of Viral Diseases, Pathogen Discovery | Dakota Howard, Dhwani Batra, Peter W. Cook, Kara Moser, Adrian Paskey, Jason Caravas, Benjamin Rambo-Martin, Shatavia Morrison, Christopher Gulvick, Scott Sammons, Yvette Unoarumhi, Darlene Wagner, Matthew Schmerer, S. H. Rosenthal, A. Gerasimova, R. M. Kagan, B. Anderson, M. Hua, Y. Liu, L.E. Bernstein, K.E. Livingston, A. Perez, I. A. Shlyakhter, R. V. Rolando, R. Owen, P. Tanpaiboon, F. Lacbawan, Clinton R. Paden, Duncan MacCannell |
| EPI_ISL_1582165 | MONTEFIORE MEDICAL CENTER LABORATORIES | Wadsworth Center, New York State Department of Health | Kirsten St. George, Daryl M. Lamson, Alexis Russell, Matthew Shudt, Melissa A Leisner, Jonathan Plitnick, Catharine Prussing, Navjot Singh, John Kelly, Erasmus Schneider, Erica Lasek-Nesselquist |
| EPI_ISL_1582166, EPI_ISL_1582167 | Quest Diagnostics Incorporated | Centers for Disease Control and Prevention Division of Viral Diseases, Pathogen Discovery | Dakota Howard, Dhwani Batra, Peter W. Cook, Kara Moser, Adrian Paskey, Jason Caravas, Benjamin Rambo-Martin, Shatavia Morrison, Christopher Gulvick, Scott Sammons, Yvette Unoarumhi, Darlene Wagner, Matthew Schmerer, S. H. Rosenthal, A. Gerasimova, R. M. Kagan, B. Anderson, M. Hua, Y. Liu, L.E. Bernstein, K.E. Livingston, A. Perez, I. A. Shlyakhter, R. V. Rolando, R. Owen, P. Tanpaiboon, F. Lacbawan, Clinton R. Paden, Duncan MacCannell |
| EPI_ISL_1582168 | MONTEFIORE MEDICAL CENTER LABORATORIES | Wadsworth Center, New York State Department of Health | Kirsten St. George, Daryl M. Lamson, Alexis Russell, Matthew Shudt, Melissa A Leisner, Jonathan Plitnick, Catharine Prussing, Navjot Singh, John Kelly, Erasmus Schneider, Erica Lasek-Nesselquist |
| EPI_ISL_1582169 | Quest Diagnostics Incorporated | Centers for Disease Control and Prevention Division of Viral Diseases, Pathogen Discovery | Dakota Howard, Dhwani Batra, Peter W. Cook, Kara Moser, Adrian Paskey, Jason Caravas, Benjamin Rambo-Martin, Shatavia Morrison, Christopher Gulvick, Scott Sammons, Yvette Unoarumhi, Darlene Wagner, Matthew Schmerer, S. H. Rosenthal, A. Gerasimova, R. M. Kagan, B. Anderson, M. Hua, Y. Liu, L.E. Bernstein, K.E. Livingston, A. Perez, I. A. Shlyakhter, R. V. Rolando, R. Owen, P. Tanpaiboon, F. Lacbawan, Clinton R. Paden, Duncan MacCannell |
| EPI_ISL_1582170 | MONTEFIORE MEDICAL CENTER LABORATORIES | Wadsworth Center, New York State Department of Health | Kirsten St. George, Daryl M. Lamson, Alexis Russell, Matthew Shudt, Melissa A Leisner, Jonathan Plitnick, Catharine Prussing, Navjot Singh, John Kelly, Erasmus Schneider, Erica Lasek-Nesselquist |
| EPI_ISL_1582171, EPI_ISL_1582172 | Quest Diagnostics Incorporated | Centers for Disease Control and Prevention Division of Viral Diseases, Pathogen Discovery | Dakota Howard, Dhwani Batra, Peter W. Cook, Kara Moser, Adrian Paskey, Jason Caravas, Benjamin Rambo-Martin, Shatavia Morrison, Christopher Gulvick, Scott Sammons, Yvette Unoarumhi, Darlene Wagner, Matthew Schmerer, S. H. Rosenthal, A. Gerasimova, R. M. Kagan, B. Anderson, M. Hua, Y. Liu, L.E. Bernstein, K.E. Livingston, A. Perez, I. A. Shlyakhter, R. V. Rolando, R. Owen, P. Tanpaiboon, F. Lacbawan, Clinton R. Paden, Duncan MacCannell |
| EPI_ISL_1582173, EPI_ISL_1582174, EPI_ISL_1582175, EPI_ISL_1582176, EPI_ISL_1582177, EPI_ISL_1582178 | MONTEFIORE MEDICAL CENTER LABORATORIES | Wadsworth Center, New York State Department of Health | Kirsten St. George, Daryl M. Lamson, Alexis Russell, Matthew Shudt, Melissa A Leisner, Jonathan Plitnick, Catharine Prussing, Navjot Singh, John Kelly, Erasmus Schneider, Erica Lasek-Nesselquist |
| EPI_ISL_1582179, EPI_ISL_1582180, EPI_ISL_1582181, EPI_ISL_1582182, EPI_ISL_1582183, EPI_ISL_1582184, EPI_ISL_1582185, EPI_ISL_1582186, EPI_ISL_1582187, EPI_ISL_1582188, EPI_ISL_1582189, EPI_ISL_1582190, EPI_ISL_1582191, EPI_ISL_1582192, EPI_ISL_1582193, EPI_ISL_1582194, EPI_ISL_1582195, EPI_ISL_1582196, EPI_ISL_1582197, EPI_ISL_1582198, EPI_ISL_1582199, EPI_ISL_1582200, EPI_ISL_1582201, EPI_ISL_1582202, EPI_ISL_1582203, EPI_ISL_1582204, EPI_ISL_1582205, EPI_ISL_1582206, EPI_ISL_1582207, EPI_ISL_1582208, EPI_ISL_1582209, EPI_ISL_1582210, EPI_ISL_1582211, EPI_ISL_1582212, EPI_ISL_1582213 | | | |
| see above | NYC Pandemic Response Lab | Wadsworth Center, New York State Department of Health | Kirsten St. George, Daryl M. Lamson, Alexis Russell, Matthew Shudt, Melissa A Leisner, Jonathan Plitnick, Catharine Prussing, Navjot Singh, John Kelly, Erasmus Schneider, Erica Lasek-Nesselquist |
| EPI_ISL_1582214, EPI_ISL_1582215, EPI_ISL_1582216, EPI_ISL_1582217 | Columbia University Irving Medical Center | Wadsworth Center, New York State Department of Health | Kirsten St. George, Daryl M. Lamson, Alexis Russell, Matthew Shudt, Melissa A Leisner, Jonathan Plitnick, Catharine Prussing, Navjot Singh, John Kelly, Erasmus Schneider, Erica Lasek-Nesselquist |
| EPI_ISL_1582218 | ALBANY MEDICAL CENTER | Wadsworth Center, New York State Department of Health | Kirsten St. George, Daryl M. Lamson, Alexis Russell, Matthew Shudt, Melissa A Leisner, Jonathan Plitnick, Catharine Prussing, Navjot Singh, John Kelly, Erasmus Schneider, Erica Lasek-Nesselquist |
| EPI_ISL_1582219, EPI_ISL_1582220, EPI_ISL_1582221, EPI_ISL_1582222, EPI_ISL_1582223, EPI_ISL_1582224 | URMC LABS | Wadsworth Center, New York State Department of Health | Kirsten St. George, Daryl M. Lamson, Alexis Russell, Matthew Shudt, Melissa A Leisner, Jonathan Plitnick, Catharine Prussing, Navjot Singh, John Kelly, Erasmus Schneider, Erica Lasek-Nesselquist |
| EPI_ISL_1582225, EPI_ISL_1582226 | NORTHWELL HEALTH LABORATORIES | Wadsworth Center, New York State Department of Health | Kirsten St. George, Daryl M. Lamson, Alexis Russell, Matthew Shudt, Melissa A Leisner, Jonathan Plitnick, Catharine Prussing, Navjot Singh, John Kelly, Erasmus Schneider, Erica Lasek-Nesselquist |
| EPI_ISL_1582227, EPI_ISL_1582228, EPI_ISL_1582229 | SARATOGA HOSPITAL LABORATORY | Wadsworth Center, New York State Department of Health | Kirsten St. George, Daryl M. Lamson, Alexis Russell, Matthew Shudt, Melissa A Leisner, Jonathan Plitnick, Catharine Prussing, Navjot Singh, John Kelly, Erasmus Schneider, Erica Lasek-Nesselquist |
| EPI_ISL_1582230 | NYC Pandemic Response Lab | Wadsworth Center, New York State Department of Health | Kirsten St. George, Daryl M. Lamson, Alexis Russell, Matthew Shudt, Melissa A Leisner, Jonathan Plitnick, Catharine Prussing, Navjot Singh, John Kelly, Erasmus Schneider, Erica Lasek-Nesselquist |
| EPI_ISL_1582231 | WESTCHESTER MEDICAL CENTER | Wadsworth Center, New York State Department of Health | Kirsten St. George, Daryl M. Lamson, Alexis Russell, Matthew Shudt, Melissa A Leisner, Jonathan Plitnick, Catharine Prussing, Navjot Singh, John Kelly, Erasmus Schneider, Erica Lasek-Nesselquist |
| EPI_ISL_1582232, EPI_ISL_1582233, EPI_ISL_1582234, EPI_ISL_1582235, EPI_ISL_1582236, EPI_ISL_1582237, EPI_ISL_1582238, EPI_ISL_1582239, EPI_ISL_1582240, EPI_ISL_1582241, EPI_ISL_1582242, EPI_ISL_1582243, EPI_ISL_1582244 | | | |
| see above | TEMPUS LABS INC | Wadsworth Center, New York State Department of Health | Kirsten St. George, Daryl M. Lamson, Alexis Russell, Matthew Shudt, Melissa A Leisner, Jonathan Plitnick, Catharine Prussing, Navjot Singh, John Kelly, Erasmus Schneider, Erica Lasek-Nesselquist |
| EPI_ISL_1582245, EPI_ISL_1582246, EPI_ISL_1582247, EPI_ISL_1582248, EPI_ISL_1582249, EPI_ISL_1582250, EPI_ISL_1582251, EPI_ISL_1582252, EPI_ISL_1582253, EPI_ISL_1582254 | STONY BROOK UNIVERSITY HOSPITAL | Wadsworth Center, New York State Department of Health | Kirsten St. George, Daryl M. Lamson, Alexis Russell, Matthew Shudt, Melissa A Leisner, Jonathan Plitnick, Catharine Prussing, Navjot Singh, John Kelly, Erasmus Schneider, Erica Lasek-Nesselquist |
| EPI_ISL_1582255 | GOOD SAMARITAN HOSPITAL LABORATORY | Wadsworth Center, New York State Department of Health | Kirsten St. George, Daryl M. Lamson, Alexis Russell, Matthew Shudt, Melissa A Leisner, Jonathan Plitnick, Catharine Prussing, Navjot Singh, John Kelly, Erasmus Schneider, Erica Lasek-Nesselquist |
| EPI_ISL_1582256, EPI_ISL_1582257, EPI_ISL_1582258, EPI_ISL_1582259 | NORTH SHORE UNIVERSITY HOSPITAL | Wadsworth Center, New York State Department of Health | Kirsten St. George, Daryl M. Lamson, Alexis Russell, Matthew Shudt, Melissa A Leisner, Jonathan Plitnick, Catharine Prussing, Navjot Singh, John Kelly, Erasmus Schneider, Erica Lasek-Nesselquist |
| EPI_ISL_1582260 | ALBANY MEDICAL CENTER | Wadsworth Center, New York State Department of Health | Kirsten St. George, Daryl M. Lamson, Alexis Russell, Matthew Shudt, Melissa A Leisner, Jonathan Plitnick, Catharine Prussing, Navjot Singh, John Kelly, Erasmus Schneider, Erica Lasek-Nesselquist |
| EPI_ISL_1582261, EPI_ISL_1582262, EPI_ISL_1582263, EPI_ISL_1582264, EPI_ISL_1582265, EPI_ISL_1582266, EPI_ISL_1582267, EPI_ISL_1582268, EPI_ISL_1582269, EPI_ISL_1582270, EPI_ISL_1582271 | | | |
| see above | Columbia University Irving Medical Center | Wadsworth Center, New York State Department of Health | Kirsten St. George, Daryl M. Lamson, Alexis Russell, Matthew Shudt, Melissa A Leisner, Jonathan Plitnick, Catharine Prussing, Navjot Singh, John Kelly, Erasmus Schneider, Erica Lasek-Nesselquist |
| EPI_ISL_1582272 | ST. FRANCIS HOSPITAL LABORATORY | Wadsworth Center, New York State Department of Health | Kirsten St. George, Daryl M. Lamson, Alexis Russell, Matthew Shudt, Melissa A Leisner, Jonathan Plitnick, Catharine Prussing, Navjot Singh, John Kelly, Erasmus Schneider, Erica Lasek-Nesselquist |
| EPI_ISL_1582273, EPI_ISL_1582274, EPI_ISL_1582275, EPI_ISL_1582276, | WHITE PLAINS HOSPITAL CENTER LABORATORY | Wadsworth Center, New York State Department of Health | Kirsten St. George, Daryl M. Lamson, Alexis Russell, Matthew Shudt, Melissa A Leisner, Jonathan Plitnick, Catharine Prussing, Navjot Singh, John Kelly, Erasmus Schneider, Erica Lasek-Nesselquist |

| | | | |
|---|---|---|---|
| EPI_ISL_1582277, EPI_ISL_1582278, EPI_ISL_1582279, EPI_ISL_1582280, EPI_ISL_1582281, EPI_ISL_1582282 | | | |
| EPI_ISL_1582283, EPI_ISL_1582284, EPI_ISL_1582285, EPI_ISL_1582286, EPI_ISL_1582287, EPI_ISL_1582288, EPI_ISL_1582289, EPI_ISL_1582290, EPI_ISL_1582291, EPI_ISL_1582292, EPI_ISL_1582293, EPI_ISL_1582294, EPI_ISL_1582295, EPI_ISL_1582296, EPI_ISL_1582297, EPI_ISL_1582298, EPI_ISL_1582299, EPI_ISL_1582300, EPI_ISL_1582301, EPI_ISL_1582302, EPI_ISL_1582303, EPI_ISL_1582304, EPI_ISL_1582305, EPI_ISL_1582306, EPI_ISL_1582307 | | | |
| see above | NORTHWELL HEALTH LABORATORIES | Wadsworth Center, New York State Department of Health | Kirsten St. George, Daryl M. Lamson, Alexis Russell, Matthew Shudt, Melissa A Leisner, Jonathan Plitnick, Catharine Prussing, Navjot Singh, John Kelly, Erasmus Schneider, Erica Lasek-Nesselquist |
| EPI_ISL_1582308, EPI_ISL_1582309, EPI_ISL_1582310, EPI_ISL_1582311, EPI_ISL_1582312, EPI_ISL_1582313, EPI_ISL_1582314, EPI_ISL_1582315, EPI_ISL_1582316 | WESTCHESTER MEDICAL CENTER | Wadsworth Center, New York State Department of Health | Kirsten St. George, Daryl M. Lamson, Alexis Russell, Matthew Shudt, Melissa A Leisner, Jonathan Plitnick, Catharine Prussing, Navjot Singh, John Kelly, Erasmus Schneider, Erica Lasek-Nesselquist |
| EPI_ISL_1582317 | NORTHWELL HEALTH LABORATORIES | Wadsworth Center, New York State Department of Health | Kirsten St. George, Daryl M. Lamson, Alexis Russell, Matthew Shudt, Melissa A Leisner, Jonathan Plitnick, Catharine Prussing, Navjot Singh, John Kelly, Erasmus Schneider, Erica Lasek-Nesselquist |
| EPI_ISL_1582318 | SUNY UPSTATE MEDICAL UNIVERSITY | Wadsworth Center, New York State Department of Health | Kirsten St. George, Daryl M. Lamson, Alexis Russell, Matthew Shudt, Melissa A Leisner, Jonathan Plitnick, Catharine Prussing, Navjot Singh, John Kelly, Erasmus Schneider, Erica Lasek-Nesselquist |
| EPI_ISL_1582319, EPI_ISL_1582320, EPI_ISL_1582321 | MONTEFIORE MEDICAL CENTER LABORATORIES | Wadsworth Center, New York State Department of Health | Kirsten St. George, Daryl M. Lamson, Alexis Russell, Matthew Shudt, Melissa A Leisner, Jonathan Plitnick, Catharine Prussing, Navjot Singh, John Kelly, Erasmus Schneider, Erica Lasek-Nesselquist |
| EPI_ISL_1582322, EPI_ISL_1582323, EPI_ISL_1582324, EPI_ISL_1582325, EPI_ISL_1582326, EPI_ISL_1582327, EPI_ISL_1582328, EPI_ISL_1582329, EPI_ISL_1582330, EPI_ISL_1582331, EPI_ISL_1582332, EPI_ISL_1582333, EPI_ISL_1582334, EPI_ISL_1582335, EPI_ISL_1582336 | | | |
| see above | NYC Pandemic Response Lab | Wadsworth Center, New York State Department of Health | Kirsten St. George, Daryl M. Lamson, Alexis Russell, Matthew Shudt, Melissa A Leisner, Jonathan Plitnick, Catharine Prussing, Navjot Singh, John Kelly, Erasmus Schneider, Erica Lasek-Nesselquist |
| EPI_ISL_1582337 | Columbia University Irving Medical Center | Wadsworth Center, New York State Department of Health | Kirsten St. George, Daryl M. Lamson, Alexis Russell, Matthew Shudt, Melissa A Leisner, Jonathan Plitnick, Catharine Prussing, Navjot Singh, John Kelly, Erasmus Schneider, Erica Lasek-Nesselquist |
| EPI_ISL_1582338 | WESTCHESTER MEDICAL CENTER | Wadsworth Center, New York State Department of Health | Kirsten St. George, Daryl M. Lamson, Alexis Russell, Matthew Shudt, Melissa A Leisner, Jonathan Plitnick, Catharine Prussing, Navjot Singh, John Kelly, Erasmus Schneider, Erica Lasek-Nesselquist |
| EPI_ISL_1582339, EPI_ISL_1582340, EPI_ISL_1582341, EPI_ISL_1582342, EPI_ISL_1582343, EPI_ISL_1582344, EPI_ISL_1582345, EPI_ISL_1582346, EPI_ISL_1582347, EPI_ISL_1582348, EPI_ISL_1582349, EPI_ISL_1582350, EPI_ISL_1582351, EPI_ISL_1582352, EPI_ISL_1582353 | | | |
| see above | URMC LABS | Wadsworth Center, New York State Department of Health | Kirsten St. George, Daryl M. Lamson, Alexis Russell, Matthew Shudt, Melissa A Leisner, Jonathan Plitnick, Catharine Prussing, Navjot Singh, John Kelly, Erasmus Schneider, Erica Lasek-Nesselquist |
| EPI_ISL_1582354, EPI_ISL_1582355, EPI_ISL_1582356 | NORTHWELL HEALTH LABORATORIES | Wadsworth Center, New York State Department of Health | Kirsten St. George, Daryl M. Lamson, Alexis Russell, Matthew Shudt, Melissa A Leisner, Jonathan Plitnick, Catharine Prussing, Navjot Singh, John Kelly, Erasmus Schneider, Erica Lasek-Nesselquist |
| EPI_ISL_1582357, EPI_ISL_1582358, EPI_ISL_1582359 | TEMPUS LABS INC | Wadsworth Center, New York State Department of Health | Kirsten St. George, Daryl M. Lamson, Alexis Russell, Matthew Shudt, Melissa A Leisner, Jonathan Plitnick, Catharine Prussing, Navjot Singh, John Kelly, Erasmus Schneider, Erica Lasek-Nesselquist |
| EPI_ISL_1582360, EPI_ISL_1582361, EPI_ISL_1582362, EPI_ISL_1582363, EPI_ISL_1582364, EPI_ISL_1582365, EPI_ISL_1582366, EPI_ISL_1582367 | SUNY UPSTATE MEDICAL UNIVERSITY | Wadsworth Center, New York State Department of Health | Kirsten St. George, Daryl M. Lamson, Alexis Russell, Matthew Shudt, Melissa A Leisner, Jonathan Plitnick, Catharine Prussing, Navjot Singh, John Kelly, Erasmus Schneider, Erica Lasek-Nesselquist |
| EPI_ISL_1582368 | Columbia University Irving Medical Center | Wadsworth Center, New York State Department of Health | Kirsten St. George, Daryl M. Lamson, Alexis Russell, Matthew Shudt, Melissa A Leisner, Jonathan Plitnick, Catharine Prussing, Navjot Singh, John Kelly, Erasmus Schneider, Erica Lasek-Nesselquist |
| EPI_ISL_1582369, EPI_ISL_1582370, EPI_ISL_1582371, EPI_ISL_1582372, EPI_ISL_1582373, EPI_ISL_1582374, EPI_ISL_1582375, EPI_ISL_1582376, EPI_ISL_1582377, EPI_ISL_1582378, EPI_ISL_1582379, EPI_ISL_1582380, EPI_ISL_1582381, EPI_ISL_1582382, EPI_ISL_1582383, EPI_ISL_1582384, EPI_ISL_1582385, EPI_ISL_1582386, EPI_ISL_1582387, EPI_ISL_1582388, EPI_ISL_1582389 | | | |
| see above | NORTHWELL HEALTH LABORATORIES | Wadsworth Center, New York State Department of Health | Kirsten St. George, Daryl M. Lamson, Alexis Russell, Matthew Shudt, Melissa A Leisner, Jonathan Plitnick, Catharine Prussing, Navjot Singh, John Kelly, Erasmus Schneider, Erica Lasek-Nesselquist |
| EPI_ISL_1582398 | URMC LABS | Wadsworth Center, New York State Department of Health | Kirsten St. George, Daryl M. Lamson, Alexis Russell, Matthew Shudt, Melissa A Leisner, Jonathan Plitnick, Catharine Prussing, Navjot Singh, John Kelly, Erasmus Schneider, Erica Lasek-Nesselquist |
| EPI_ISL_1582399 | SUNY UPSTATE MEDICAL UNIVERSITY | Wadsworth Center, New York State Department of Health | Kirsten St. George, Daryl M. Lamson, Alexis Russell, Matthew Shudt, Melissa A Leisner, Jonathan Plitnick, Catharine Prussing, Navjot Singh, John Kelly, Erasmus Schneider, Erica Lasek-Nesselquist |
| EPI_ISL_1582400, EPI_ISL_1582401, EPI_ISL_1582402 | NYC Pandemic Response Lab | Wadsworth Center, New York State Department of Health | Kirsten St. George, Daryl M. Lamson, Alexis Russell, Matthew Shudt, Melissa A Leisner, Jonathan Plitnick, Catharine Prussing, Navjot Singh, John Kelly, Erasmus Schneider, Erica Lasek-Nesselquist |
| EPI_ISL_1582403 | TEMPUS LABS INC | Wadsworth Center, New York State Department of Health | Kirsten St. George, Daryl M. Lamson, Alexis Russell, Matthew Shudt, Melissa A Leisner, Jonathan Plitnick, Catharine Prussing, Navjot Singh, John Kelly, Erasmus Schneider, Erica Lasek-Nesselquist |
| EPI_ISL_1582404 | URMC LABS | Wadsworth Center, New York State Department of Health | Kirsten St. George, Daryl M. Lamson, Alexis Russell, Matthew Shudt, Melissa A Leisner, Jonathan Plitnick, Catharine Prussing, Navjot Singh, John Kelly, Erasmus Schneider, Erica Lasek-Nesselquist |
| EPI_ISL_1582405 | NORTHWELL HEALTH LABORATORIES | Wadsworth Center, New York State Department of Health | Kirsten St. George, Daryl M. Lamson, Alexis Russell, Matthew Shudt, Melissa A Leisner, Jonathan Plitnick, Catharine Prussing, Navjot Singh, John Kelly, Erasmus Schneider, Erica Lasek-Nesselquist |
| EPI_ISL_1582406 | GLENS FALLS HOSPITAL LABORATORY | Wadsworth Center, New York State Department of Health | Kirsten St. George, Daryl M. Lamson, Alexis Russell, Matthew Shudt, Melissa A Leisner, Jonathan Plitnick, Catharine Prussing, Navjot Singh, John Kelly, Erasmus Schneider, Erica Lasek-Nesselquist |
| EPI_ISL_1582407 | ALBANY MEDICAL CENTER | Wadsworth Center, New York State Department of Health | Kirsten St. George, Daryl M. Lamson, Alexis Russell, Matthew Shudt, Melissa A Leisner, Jonathan Plitnick, Catharine Prussing, Navjot Singh, John Kelly, Erasmus Schneider, Erica Lasek-Nesselquist |
| EPI_ISL_1582425 | New Mexico Department of Health Scientific Laboratory | New Mexico Department of Health Scientific Laboratory | Ellie Johnson, Anastacia Griego-Fisher, D'eldra Malone, Jennifer Benoit |
| EPI_ISL_1582428, EPI_ISL_1582429, EPI_ISL_1582430, EPI_ISL_1582431, EPI_ISL_1582432, EPI_ISL_1582436, EPI_ISL_1582438, EPI_ISL_1582440, EPI_ISL_1582443 | Curative | New Mexico Department of Health Scientific Laboratory | Ellie Johnson, Anastacia Griego-Fisher, D'eldra Malone, Jennifer Benoit |
| EPI_ISL_1582444, EPI_ISL_1582445, EPI_ISL_1582446, EPI_ISL_1582447, EPI_ISL_1582448, EPI_ISL_1582449, EPI_ISL_1582450, EPI_ISL_1582451, EPI_ISL_1582452, EPI_ISL_1582453, EPI_ISL_1582454, EPI_ISL_1582455, EPI_ISL_1582457, EPI_ISL_1582458, EPI_ISL_1582459, EPI_ISL_1582461, EPI_ISL_1582462, EPI_ISL_1582463 | | | |
| see above | New Mexico Department of Health Scientific Laboratory | New Mexico Department of Health Scientific Laboratory | Ellie Johnson, Anastacia Griego-Fisher, D'eldra Malone, Jennifer Benoit |
| EPI_ISL_1582464, EPI_ISL_1582465, EPI_ISL_1582466, EPI_ISL_1582467 | TriCore Reference Laboratories | New Mexico Department of Health Scientific Laboratory | Ellie Johnson, Anastacia Griego-Fisher, D'eldra Malone, Jennifer Benoit |
| EPI_ISL_1582468, EPI_ISL_1582469 | New Mexico Department of Health Scientific Laboratory | New Mexico Department of Health Scientific Laboratory | Ellie Johnson, Anastacia Griego-Fisher, D'eldra Malone, Jennifer Benoit |
| EPI_ISL_1582470, EPI_ISL_1582471, EPI_ISL_1582472, EPI_ISL_1582473, EPI_ISL_1582474, EPI_ISL_1582475, EPI_ISL_1582476, EPI_ISL_1582477, EPI_ISL_1582478, EPI_ISL_1582479, EPI_ISL_1582480, EPI_ISL_1582481, EPI_ISL_1582482, EPI_ISL_1582483, EPI_ISL_1582484, EPI_ISL_1582485, EPI_ISL_1582486, EPI_ISL_1582487, EPI_ISL_1582488 | | | |
| see above | University of Wisconsin-Madison AIDS Vaccine Research Laboratories | University of Wisconsin-Madison AIDS Vaccine Research Laboratories | Gage Moreno, Katarina Braun, et al. AIDS Vaccine Research Laboratories |

| | | | |
|---|---|---|---|
| EPI_ISL_1582515 | University of Michigan Clinical Microbiology Laboratory | Lauring Lab, University of Michigan, Department of Microbiology and Immunology | Valesano |
| EPI_ISL_1582593, EPI_ISL_1582594, EPI_ISL_1582595, EPI_ISL_1582596, EPI_ISL_1582597, EPI_ISL_1582598, EPI_ISL_1582599, EPI_ISL_1582600, EPI_ISL_1582602, EPI_ISL_1582603, EPI_ISL_1582604, EPI_ISL_1582605, EPI_ISL_1582606, EPI_ISL_1582607, EPI_ISL_1582608, EPI_ISL_1582609, EPI_ISL_1582610, EPI_ISL_1582611, EPI_ISL_1582612, EPI_ISL_1582613, EPI_ISL_1582614, EPI_ISL_1582615, EPI_ISL_1582616, EPI_ISL_1582617, EPI_ISL_1582618, EPI_ISL_1582619, EPI_ISL_1582620, EPI_ISL_1582621, EPI_ISL_1582622, EPI_ISL_1582623, EPI_ISL_1582624, EPI_ISL_1582625, EPI_ISL_1582626, EPI_ISL_1582627, EPI_ISL_1582628, EPI_ISL_1582629, EPI_ISL_1582630, EPI_ISL_1582631, EPI_ISL_1582632, EPI_ISL_1582633, EPI_ISL_1582634, EPI_ISL_1582635, EPI_ISL_1582637, EPI_ISL_1582638 | | | |
| see above | University of Wisconsin-Madison AIDS Vaccine Research Laboratories | University of Wisconsin-Madison AIDS Vaccine Research Laboratories | Gage Moreno, Katarina Braun, et al. AIDS Vaccine Research Laboratories |
| EPI_ISL_1582759, EPI_ISL_1582761, EPI_ISL_1582763, EPI_ISL_1582764, EPI_ISL_1582765, EPI_ISL_1582766, EPI_ISL_1582767, EPI_ISL_1582768, EPI_ISL_1582769 | Wyoming Public Health Laboratory | Wyoming Public Health Laboratory | Jim Mildenberger, Wanda Manley, Noah Hull, Taylor Fearing, Lynette Gumbleton, Channing Weber, Ashley Norberg, Chayse Rowley, Marley Goetz, Brian Dominguez, Elliot Thomasson, Cari Sloma, and Rob Christensen |
| EPI_ISL_1582960, EPI_ISL_1582961, EPI_ISL_1582962, EPI_ISL_1582963, EPI_ISL_1582964, EPI_ISL_1582965, EPI_ISL_1582966, EPI_ISL_1582968, EPI_ISL_1582969, EPI_ISL_1582970, EPI_ISL_1582971, EPI_ISL_1582972, EPI_ISL_1582973, EPI_ISL_1582974, EPI_ISL_1582975, EPI_ISL_1582976 | | | |
| see above | Kansas Health and Environmental Lab | Kansas Health and Environmental Lab | Mike Grose, Jonathan Barnell, Ben Olsen, and Phil Adam |
| EPI_ISL_1582998, EPI_ISL_1583000, EPI_ISL_1583003, EPI_ISL_1583004, EPI_ISL_1583005 | Arizona State University | Arizona State University | Rabia Maqsood, LaRinda A. Holland, Peter T. Skidmore, Nicholas J. Mellor, Joy M. Blain, Valerie Harris, Joshua LaBaer, Vel Murugan, Efrem S. Lim |
| EPI_ISL_1584466, EPI_ISL_1584469, EPI_ISL_1584472, EPI_ISL_1584475, EPI_ISL_1584478, EPI_ISL_1584481, EPI_ISL_1584484, EPI_ISL_1584487, EPI_ISL_1584490, EPI_ISL_1584493 | GA Department of Public Health | GA Department of Public Health | Stacy Reeves, Jonathan Edwards, Cynthia Dixey, Tonia Parrott, Aliyah Fields, Taylor Smith |
| EPI_ISL_1584999, EPI_ISL_1585003, EPI_ISL_1585005, EPI_ISL_1585007, EPI_ISL_1585008, EPI_ISL_1585009, EPI_ISL_1585010, EPI_ISL_1585011, EPI_ISL_1585014, EPI_ISL_1585015, EPI_ISL_1585016, EPI_ISL_1585018, EPI_ISL_1585019, EPI_ISL_1585020, EPI_ISL_1585021, EPI_ISL_1585022, EPI_ISL_1585025, EPI_ISL_1585026, EPI_ISL_1585027, EPI_ISL_1585028, EPI_ISL_1585029, EPI_ISL_1585030, EPI_ISL_1585031, EPI_ISL_1585033, EPI_ISL_1585034, EPI_ISL_1585035, EPI_ISL_1585036, EPI_ISL_1585037, EPI_ISL_1585038, EPI_ISL_1585040, EPI_ISL_1585041 | | | |
| see above | North Dakota Department of Health, Public Health Laboratory | North Dakota Department of Health, Public Health Laboratory | Lisa Wingerter |
| EPI_ISL_1585045, EPI_ISL_1585046, EPI_ISL_1585048, EPI_ISL_1585050, EPI_ISL_1585051 | Lafene Health Center | Kansas State Veterinary Diagnostic Laboratory | Tyler Doerksen, Andreea Lu, Lance Noll, Elizabeth Porter, Kasey Hogan, Emily Cox, Jianfa Bai, Jamie Henningson, Rachel Palinski |
| EPI_ISL_1585052, EPI_ISL_1585053, EPI_ISL_1585054, EPI_ISL_1585055, EPI_ISL_1585056, EPI_ISL_1585057, EPI_ISL_1585058, EPI_ISL_1585059, EPI_ISL_1585060, EPI_ISL_1585061, EPI_ISL_1585062, EPI_ISL_1585063, EPI_ISL_1585064, EPI_ISL_1585065, EPI_ISL_1585066 | | | |
| see above | Oregon State University TRACE Project | OSU Center for Genome Research and Biocomputing | Oregon State University TRACE Project |
| EPI_ISL_1585067, EPI_ISL_1585068, EPI_ISL_1585069, EPI_ISL_1585070, EPI_ISL_1585073, EPI_ISL_1585074, EPI_ISL_1585075, EPI_ISL_1585076, EPI_ISL_1585077, EPI_ISL_1585078, EPI_ISL_1585079, EPI_ISL_1585080, EPI_ISL_1585081, EPI_ISL_1585082, EPI_ISL_1585083, EPI_ISL_1585084, EPI_ISL_1585085, EPI_ISL_1585086, EPI_ISL_1585087, EPI_ISL_1585088, EPI_ISL_1585089, EPI_ISL_1585090, EPI_ISL_1585092, EPI_ISL_1585093, EPI_ISL_1585094, EPI_ISL_1585095, EPI_ISL_1585096, EPI_ISL_1585097, EPI_ISL_1585098, EPI_ISL_1585099, EPI_ISL_1585100, EPI_ISL_1585101, EPI_ISL_1585102, EPI_ISL_1585103, EPI_ISL_1585104, EPI_ISL_1585105, EPI_ISL_1585106, EPI_ISL_1585108, EPI_ISL_1585109, EPI_ISL_1585110, EPI_ISL_1585111, EPI_ISL_1585112, EPI_ISL_1585113, EPI_ISL_1585114, EPI_ISL_1585115, EPI_ISL_1585116, EPI_ISL_1585117, EPI_ISL_1585118, EPI_ISL_1585119, EPI_ISL_1585120, EPI_ISL_1585121, EPI_ISL_1585122, EPI_ISL_1585124, EPI_ISL_1585125, EPI_ISL_1585126, EPI_ISL_1585127, EPI_ISL_1585128, EPI_ISL_1585129, EPI_ISL_1585130, EPI_ISL_1585131, EPI_ISL_1585132, EPI_ISL_1585133, EPI_ISL_1585134, EPI_ISL_1585135, EPI_ISL_1585136, EPI_ISL_1585137, EPI_ISL_1585138, EPI_ISL_1585139, EPI_ISL_1585140, EPI_ISL_1585141, EPI_ISL_1585142, EPI_ISL_1585143, EPI_ISL_1585144, EPI_ISL_1585145, EPI_ISL_1585146, EPI_ISL_1585148, EPI_ISL_1585149, EPI_ISL_1585150, EPI_ISL_1585151 | | | |
| see above | Arizona State University | Arizona State University | Rabia Maqsood, LaRinda A. Holland, Peter T. Skidmore, Nicholas J. Mellor, Joy M. Blain, Valerie Harris, Joshua LaBaer, Vel Murugan, Efrem S. Lim |
| EPI_ISL_1585152, EPI_ISL_1585154, EPI_ISL_1585155, EPI_ISL_1585156, EPI_ISL_1585157, EPI_ISL_1585158, EPI_ISL_1585159, EPI_ISL_1585160, EPI_ISL_1585161, EPI_ISL_1585162, EPI_ISL_1585163, EPI_ISL_1585164, EPI_ISL_1585165, EPI_ISL_1585166, EPI_ISL_1585167, EPI_ISL_1585168, EPI_ISL_1585169, EPI_ISL_1585170, EPI_ISL_1585171, EPI_ISL_1585172, EPI_ISL_1585173, EPI_ISL_1585174, EPI_ISL_1585175, EPI_ISL_1585176, EPI_ISL_1585177, EPI_ISL_1585178, EPI_ISL_1585179, EPI_ISL_1585180, EPI_ISL_1585181, EPI_ISL_1585182, EPI_ISL_1585183, EPI_ISL_1585184, EPI_ISL_1585185, EPI_ISL_1585186, EPI_ISL_1585187, EPI_ISL_1585188, EPI_ISL_1585189, EPI_ISL_1585190, EPI_ISL_1585191, EPI_ISL_1585192, EPI_ISL_1585193, EPI_ISL_1585194, EPI_ISL_1585195, EPI_ISL_1585196, EPI_ISL_1585197, EPI_ISL_1585198, EPI_ISL_1585199, EPI_ISL_1585200, EPI_ISL_1585201, EPI_ISL_1585202, EPI_ISL_1585203, EPI_ISL_1585204, EPI_ISL_1585205, EPI_ISL_1585206, EPI_ISL_1585207, EPI_ISL_1585208, EPI_ISL_1585209, EPI_ISL_1585210, EPI_ISL_1585211, EPI_ISL_1585212, EPI_ISL_1585213, EPI_ISL_1585214, EPI_ISL_1585215, EPI_ISL_1585216, EPI_ISL_1585217, EPI_ISL_1585218, EPI_ISL_1585219, EPI_ISL_1585220, EPI_ISL_1585221, EPI_ISL_1585222, EPI_ISL_1585223, EPI_ISL_1585224, EPI_ISL_1585226, EPI_ISL_1585227, EPI_ISL_1585228, EPI_ISL_1585229, EPI_ISL_1585230, EPI_ISL_1585231, EPI_ISL_1585232, EPI_ISL_1585233, EPI_ISL_1585234, EPI_ISL_1585235 | | | |
| see above | Texas Department of State Health Services (TXDSHS) | Texas Department of State Health Services (TXDSHS) | Rashmi Tuladhar, Bonnie Oh, Jenny Zhang, Maliha Rahman, Mayela Pedrueza, Anita Pokharel, Lorraine Rodriguez, Myong Koag, Chun Wang, Rachel Lee, Grace Kubin |
| EPI_ISL_1585236, EPI_ISL_1585237, EPI_ISL_1585238, EPI_ISL_1585239, EPI_ISL_1585240, EPI_ISL_1585241 | Nebraska Public Health Laboratory | Nebraska Public Health Laboratory COVID-19 Response Team | NPHL COVID-19 Response Team |
| EPI_ISL_1585383 | Lafene Health Center | Kansas State Veterinary Diagnostic Laboratory | Tyler Doerksen, Andreea Lu, Lance Noll, Elizabeth Porter, Kasey Hogan, Emily Cox, Jianfa Bai, Jamie Henningson, Rachel Palinski |
| EPI_ISL_1585384, EPI_ISL_1585385, EPI_ISL_1585386, EPI_ISL_1585387, EPI_ISL_1585388, EPI_ISL_1585389, EPI_ISL_1585390, EPI_ISL_1585391, EPI_ISL_1585392, EPI_ISL_1585393, EPI_ISL_1585394, EPI_ISL_1585395, EPI_ISL_1585396, EPI_ISL_1585397, EPI_ISL_1585398, EPI_ISL_1585399, EPI_ISL_1585400, EPI_ISL_1585401, EPI_ISL_1585402, EPI_ISL_1585403, EPI_ISL_1585404, EPI_ISL_1585405, EPI_ISL_1585406, EPI_ISL_1585407, EPI_ISL_1585408, EPI_ISL_1585409, EPI_ISL_1585410, EPI_ISL_1585411, EPI_ISL_1585412, EPI_ISL_1585413, EPI_ISL_1585414, EPI_ISL_1585415, EPI_ISL_1585416, EPI_ISL_1585417, EPI_ISL_1585418, EPI_ISL_1585419, EPI_ISL_1585420, EPI_ISL_1585421, EPI_ISL_1585422, EPI_ISL_1585423, EPI_ISL_1585424, EPI_ISL_1585425, EPI_ISL_1585426, EPI_ISL_1585427 | | | |
| see above | Centro de Investigación Biomédica del Noreste (CIBIN) | Instituto Nacional de Enfermedades Respiratorias (INER): Centro de Investigación en Enfermedades Infecciosas (CIENI) | Consorcio Mexicano de Vigilancia Genómica (CoViGen-Mex). Authors (in alphabetical order): Julio Elias Alvarado-Yaah, Carlos F. Arias, Santiago Ávila-Ríos, Víctor Hugo Borja-Aburto, Celia Boukadida, Juan Bautista Chale-Dzul, Célida Duque Molina, José Antonio Enciso-Moreno, Gloria Elena Espinosa-Ayala, Fernando Fontove-Herrera, Victor Eduardo Garcia-Arias, Concepción Grajales-Muñiz, Ricardo Grande, Alfredo Herrera-Estrella, Carla Ivón Herrera-Najera, Pavel Isa, Brenda Irasema Maldonado-Meza, Bernardo Martínez-Miguel, Margarita Matías-Florentino, María Guadalupe de Jesús Mireles-Rivera, Gloria María Molina-Salinas, Hector Montoya-Fuentes, José Esteban Muñoz-Medina, José de Jesús Nuñez-Contreras, Alicia Ocaña-Mondragón, Luis Alberto Ochoa-Carrera, Hector Esteban Paz-Juárez, Francisco Pulido, Helen Haydee Fernanda Ramírez-Plascencia, Angel Gustavo Salas-Lais, Alejandro Sanchez-Flores, Clara Esperanza Santacruz-Tinoco, María Guadalupe Santiago-Mauricio, Nelly Sélem-Mojica, Blanca Taboada, Gloria Vazquez |
| EPI_ISL_1585428, EPI_ISL_1585429, EPI_ISL_1585430, EPI_ISL_1585431, EPI_ISL_1585432, EPI_ISL_1585433, EPI_ISL_1585434, EPI_ISL_1585435, EPI_ISL_1585436, EPI_ISL_1585437, EPI_ISL_1585438, EPI_ISL_1585439, EPI_ISL_1585440, EPI_ISL_1585441, EPI_ISL_1585442, EPI_ISL_1585443, EPI_ISL_1585444, EPI_ISL_1585445, EPI_ISL_1585446, EPI_ISL_1585447, EPI_ISL_1585448, EPI_ISL_1585449, EPI_ISL_1585450, EPI_ISL_1585451, EPI_ISL_1585452, EPI_ISL_1585453, EPI_ISL_1585454 | | | |
| see above | Unidad de Investigación Médica de Yucatán (UIMY) | Instituto Nacional de Enfermedades Respiratorias (INER): Centro de Investigación en Enfermedades Infecciosas (CIENI) | Consorcio Mexicano de Vigilancia Genómica (CoViGen-Mex). Authors (in alphabetical order): Julio Elias Alvarado-Yaah, Carlos F. Arias, Santiago Ávila-Ríos, Víctor Hugo Borja-Aburto, Celia Boukadida, Juan Bautista Chale-Dzul, Célida Duque Molina, José Antonio Enciso-Moreno, Gloria Elena Espinosa-Ayala, Fernando Fontove-Herrera, Victor Eduardo Garcia-Arias, Concepción Grajales-Muñiz, Ricardo Grande, Alfredo Herrera-Estrella, Carla Ivón Herrera-Najera, Pavel Isa, Brenda Irasema Maldonado-Meza, Bernardo Martínez-Miguel, Margarita Matías-Florentino, María Guadalupe de Jesús Mireles-Rivera, Gloria María Molina-Salinas, Hector Montoya-Fuentes, José Esteban Muñoz-Medina, José de Jesús Nuñez-Contreras, Alicia Ocaña-Mondragón, Luis Alberto Ochoa-Carrera, Hector Esteban Paz-Juárez, Francisco Pulido, Helen Haydee Fernanda Ramírez-Plascencia, Angel Gustavo Salas-Lais, Alejandro Sanchez-Flores, Clara Esperanza Santacruz-Tinoco, María Guadalupe Santiago-Mauricio, Nelly Sélem-Mojica, Blanca Taboada, Gloria Vazquez |
| EPI_ISL_1585455, EPI_ISL_1585456, EPI_ISL_1585457, EPI_ISL_1585458, EPI_ISL_1585459, EPI_ISL_1585460, EPI_ISL_1585461, EPI_ISL_1585462, EPI_ISL_1585463, EPI_ISL_1585464, EPI_ISL_1585465, EPI_ISL_1585466, EPI_ISL_1585467, EPI_ISL_1585468, EPI_ISL_1585469, EPI_ISL_1585470, EPI_ISL_1585471, EPI_ISL_1585472, EPI_ISL_1585473, EPI_ISL_1585474, EPI_ISL_1585475, EPI_ISL_1585476, EPI_ISL_1585477, EPI_ISL_1585478, EPI_ISL_1585479 | | | |
| see above | Unidad de Investigación Biomédica de Zacatecas (UIBZ) | Instituto Nacional de Enfermedades Respiratorias (INER): Centro de Investigación en Enfermedades Infecciosas (CIENI) | Consorcio Mexicano de Vigilancia Genómica (CoViGen-Mex). Authors (in alphabetical order): Julio Elias Alvarado-Yaah, Carlos F. Arias, Santiago Ávila-Ríos, Víctor Hugo Borja-Aburto, Celia Boukadida, Juan Bautista Chale-Dzul, Célida Duque Molina, José Antonio Enciso-Moreno, Gloria Elena Espinosa-Ayala, Fernando Fontove-Herrera, Victor Eduardo Garcia-Arias, Concepción Grajales-Muñiz, Ricardo Grande, Alfredo Herrera-Estrella, Carla Ivón Herrera-Najera, Pavel Isa, Brenda Irasema Maldonado-Meza, Bernardo Martínez-Miguel, Margarita Matías-Florentino, María Guadalupe de Jesús Mireles-Rivera, Gloria María Molina-Salinas, Hector Montoya-Fuentes, José Esteban Muñoz-Medina, José de Jesús Nuñez-Contreras, Alicia Ocaña-Mondragón, Luis Alberto Ochoa-Carrera, Hector Esteban Paz-Juárez, Francisco Pulido, Helen Haydee Fernanda Ramírez-Plascencia, Angel Gustavo Salas-Lais, Alejandro Sanchez-Flores, Clara Esperanza Santacruz-Tinoco, María Guadalupe Santiago-Mauricio, Nelly Sélem-Mojica, Blanca Taboada, Gloria Vazquez |
| EPI_ISL_1585480, EPI_ISL_1585481, EPI_ISL_1585482, EPI_ISL_1585483, EPI_ISL_1585484, EPI_ISL_1585485, EPI_ISL_1585486, EPI_ISL_1585487, EPI_ISL_1585488, EPI_ISL_1585489, EPI_ISL_1585490, EPI_ISL_1585491, EPI_ISL_1585492, EPI_ISL_1585493, EPI_ISL_1585494, EPI_ISL_1585495, EPI_ISL_1585496, EPI_ISL_1585497, EPI_ISL_1585498, EPI_ISL_1585499, EPI_ISL_1585500, EPI_ISL_1585501, EPI_ISL_1585502, EPI_ISL_1585503, EPI_ISL_1585504, EPI_ISL_1585505, EPI_ISL_1585506, EPI_ISL_1585507, EPI_ISL_1585508, EPI_ISL_1585509, EPI_ISL_1585510, EPI_ISL_1585511, EPI_ISL_1585512, EPI_ISL_1585513, EPI_ISL_1585514, EPI_ISL_1585515, EPI_ISL_1585516, EPI_ISL_1585517, EPI_ISL_1585518, EPI_ISL_1585519, EPI_ISL_1585520, EPI_ISL_1585521, EPI_ISL_1585522, EPI_ISL_1585523, EPI_ISL_1585524, EPI_ISL_1585525, EPI_ISL_1585526, EPI_ISL_1585527, EPI_ISL_1585528, EPI_ISL_1585529, EPI_ISL_1585530, EPI_ISL_1585531, EPI_ISL_1585532, EPI_ISL_1585533, EPI_ISL_1585534, EPI_ISL_1585535, EPI_ISL_1585536, EPI_ISL_1585537, EPI_ISL_1585538, EPI_ISL_1585539, EPI_ISL_1585540, EPI_ISL_1585541, EPI_ISL_1585542, EPI_ISL_1585543, EPI_ISL_1585544, EPI_ISL_1585545, EPI_ISL_1585546, EPI_ISL_1585547, EPI_ISL_1585548, EPI_ISL_1585549, EPI_ISL_1585550, EPI_ISL_1585551, EPI_ISL_1585552, EPI_ISL_1585553, EPI_ISL_1585554, EPI_ISL_1585555, EPI_ISL_1585556, EPI_ISL_1585557, EPI_ISL_1585558, EPI_ISL_1585559, EPI_ISL_1585560, EPI_ISL_1585561, EPI_ISL_1585562, EPI_ISL_1585563 | | | |
| see above | Centro de Investigación Biomédica de Occidente (CIBO) | Instituto Nacional de Enfermedades Respiratorias (INER): Centro de Investigación en Enfermedades Infecciosas (CIENI) | Consorcio Mexicano de Vigilancia Genómica (CoViGen-Mex). Authors (in alphabetical order): Julio Elias Alvarado-Yaah, Carlos F. Arias, Santiago Ávila-Ríos, Víctor Hugo Borja-Aburto, Celia Boukadida, Juan Bautista Chale-Dzul, Célida Duque Molina, José Antonio Enciso-Moreno, Gloria Elena Espinosa-Ayala, Fernando Fontove-Herrera, Victor Eduardo Garcia-Arias, Concepción Grajales-Muñiz, Ricardo Grande, Alfredo Herrera-Estrella, Carla Ivón Herrera-Najera, Pavel Isa, Brenda Irasema Maldonado-Meza, Bernardo Martínez-Miguel, Margarita Matías-Florentino, María Guadalupe de Jesús Mireles-Rivera, Gloria María Molina-Salinas, Hector Montoya-Fuentes, José Esteban Muñoz-Medina, José de Jesús Nuñez-Contreras, Alicia Ocaña-Mondragón, Luis Alberto Ochoa-Carrera, Hector Esteban Paz-Juárez, Francisco Pulido, Helen Haydee Fernanda Ramírez-Plascencia, Angel Gustavo Salas-Lais, Alejandro Sanchez-Flores, Clara Esperanza Santacruz-Tinoco, María Guadalupe Santiago-Mauricio, Nelly Sélem-Mojica, Blanca Taboada, Gloria Vazquez |
| EPI_ISL_1585564, EPI_ISL_1585565, EPI_ISL_1585566, EPI_ISL_1585567, EPI_ISL_1585568, EPI_ISL_1585569, EPI_ISL_1585570, EPI_ISL_1585571, EPI_ISL_1585572, EPI_ISL_1585573, EPI_ISL_1585574, EPI_ISL_1585575, EPI_ISL_1585576, EPI_ISL_1585577, EPI_ISL_1585578, EPI_ISL_1585579, EPI_ISL_1585580, EPI_ISL_1585581, EPI_ISL_1585582, EPI_ISL_1585583, EPI_ISL_1585584, EPI_ISL_1585585, EPI_ISL_1585586, EPI_ISL_1585587, EPI_ISL_1585588, EPI_ISL_1585589, EPI_ISL_1585590, EPI_ISL_1585591, EPI_ISL_1585592, EPI_ISL_1585593, EPI_ISL_1585594, EPI_ISL_1585595, EPI_ISL_1585596, EPI_ISL_1585597, EPI_ISL_1585598, EPI_ISL_1585599, EPI_ISL_1585600, EPI_ISL_1585601, EPI_ISL_1585602, EPI_ISL_1585603, EPI_ISL_1585604, EPI_ISL_1585605, EPI_ISL_1585606, EPI_ISL_1585607, EPI_ISL_1585608, EPI_ISL_1585609, EPI_ISL_1585610, EPI_ISL_1585611, EPI_ISL_1585612, EPI_ISL_1585613, EPI_ISL_1585614, EPI_ISL_1585615, EPI_ISL_1585616, EPI_ISL_1585617, EPI_ISL_1585618, EPI_ISL_1585619, EPI_ISL_1585620, EPI_ISL_1585621, EPI_ISL_1585622, EPI_ISL_1585623, EPI_ISL_1585624, EPI_ISL_1585625, EPI_ISL_1585626, EPI_ISL_1585627, EPI_ISL_1585628, EPI_ISL_1585629, EPI_ISL_1585630, EPI_ISL_1585631, EPI_ISL_1585632, EPI_ISL_1585633, EPI_ISL_1585634, EPI_ISL_1585635, EPI_ISL_1585636, EPI_ISL_1585637, EPI_ISL_1585638, EPI_ISL_1585639, EPI_ISL_1585640, EPI_ISL_1585641, EPI_ISL_1585642, EPI_ISL_1585643, EPI_ISL_1585644, EPI_ISL_1585645, EPI_ISL_1585646, EPI_ISL_1585647, EPI_ISL_1585648, EPI_ISL_1585649, EPI_ISL_1585650, EPI_ISL_1585651, EPI_ISL_1585652, EPI_ISL_1585653, EPI_ISL_1585654, EPI_ISL_1585655, EPI_ISL_1585656, EPI_ISL_1585657, EPI_ISL_1585658, EPI_ISL_1585659, EPI_ISL_1585660, EPI_ISL_1585661, EPI_ISL_1585662, EPI_ISL_1585663, EPI_ISL_1585664, EPI_ISL_1585665, EPI_ISL_1585666, EPI_ISL_1585667, EPI_ISL_1585668, EPI_ISL_1585669, EPI_ISL_1585670 | | | |
| see above | Laboratorio Central de Epidemiología (LCE) | Instituto Nacional de Enfermedades Respiratorias (INER): Centro de Investigación en Enfermedades Infecciosas (CIENI) | Consorcio Mexicano de Vigilancia Genómica (CoViGen-Mex). Authors (in alphabetical order): Julio Elias Alvarado-Yaah, Carlos F. Arias, Santiago Ávila-Ríos, Víctor Hugo Borja-Aburto, Celia Boukadida, Juan Bautista Chale-Dzul, Célida Duque Molina, José Antonio Enciso-Moreno, Gloria Elena Espinosa-Ayala, Fernando Fontove-Herrera, Victor Eduardo Garcia-Arias, Concepción Grajales-Muñiz, Ricardo Grande, Alfredo Herrera-Estrella, Carla Ivón Herrera-Najera, Pavel Isa, Brenda Irasema Maldonado-Meza, Bernardo Martínez-Miguel, Margarita Matías-Florentino, María Guadalupe de Jesús Mireles-Rivera, Gloria María Molina-Salinas, Hector Montoya-Fuentes, José Esteban Muñoz-Medina, José de Jesús Nuñez-Contreras, Alicia Ocaña-Mondragón, Luis Alberto Ochoa-Carrera, Hector Esteban Paz-Juárez, Francisco Pulido, Helen Haydee Fernanda Ramírez- |

| | | | |
|---|---|---|---|
| | | | Plascencia, Angel Gustavo Salas-Lais, Alejandro Sanchez-Flores, Clara Esperanza Santacruz-Tinoco, María Guadalupe Santiago-Mauricio, Nelly Sélem-Mojica, Blanca Taboada, Gloria Vazquez |
| EPI_ISL_1585671, EPI_ISL_1585672, EPI_ISL_1585673, EPI_ISL_1585674, EPI_ISL_1585675, EPI_ISL_1585676, EPI_ISL_1585677, EPI_ISL_1585678, EPI_ISL_1585679, EPI_ISL_1585680, EPI_ISL_1585681, EPI_ISL_1585682, EPI_ISL_1585683, EPI_ISL_1585684 | | | |
| see above | Oregon State Public Health Laboratory | Oregon State Public Health Laboratory | Rafia Razzaque, Eugene Yeboah, Vanda Makris, Laura Tsaknaridis, John Fontana and Shane Sevey |
| EPI_ISL_1585892, EPI_ISL_1585893, EPI_ISL_1585894, EPI_ISL_1585895, EPI_ISL_1585896, EPI_ISL_1585897, EPI_ISL_1585898, EPI_ISL_1585899, EPI_ISL_1585900, EPI_ISL_1585901, EPI_ISL_1585902, EPI_ISL_1585903, EPI_ISL_1585904, EPI_ISL_1585905, EPI_ISL_1585906, EPI_ISL_1585907, EPI_ISL_1585908, EPI_ISL_1585909, EPI_ISL_1585910, EPI_ISL_1585911, EPI_ISL_1585912, EPI_ISL_1585913, EPI_ISL_1585914 | | | |
| see above | University of Michigan Clinical Microbiology Laboratory | Lauring Lab, University of Michigan, Department of Microbiology and Immunology | Valesano |
| EPI_ISL_1585916, EPI_ISL_1585917, EPI_ISL_1585918 | Instituto Nacional de Medicina Genomica | Instituto Nacional de Medicina Genomica | Hidalgo-Miranda A, Mendoza-Vargas A, Reyes-Grajeda JP, Cedro-Tanda A, Alcaraz N, Gonzalez-Barrera D, Rangel-DeLeon D, Miranda-Ortiz H, Rosas-Escobar P, Canseco Mendez JC, Ramirez-Vega O, Munguia-Garza P, Garcia-Cardenas FJ, Gonzalez-Woge MA, Herrera-Montalvo LA |
| EPI_ISL_1585920, EPI_ISL_1585921, EPI_ISL_1585923, EPI_ISL_1585924, EPI_ISL_1585925, EPI_ISL_1585926, EPI_ISL_1585927, EPI_ISL_1585930, EPI_ISL_1585931, EPI_ISL_1585932, EPI_ISL_1585933, EPI_ISL_1585934, EPI_ISL_1585936, EPI_ISL_1585937, EPI_ISL_1585938 | | | |
| see above | Nevada State Public Health Laboratory | Nevada State Public Health Laboratory | Andrew Gorzalski, Mark Pandori |
| EPI_ISL_1586955 | Seattle Flu Study | Seattle Flu Study | Deborah A. Nickerson, Chris D. Frazar, Jover Lee, Benjamin Pelle, Erica Ryke, Matthew Richardson, Amanda Adler, Elisabeth Brandstetter, Peter D. Han, Kairsten Fay, Misja Ilcisin, Kirsten Lacombe, Thomas R. Sibley, Melissa Truong, Caitlin R. Wolf, Karen Cowgill, Stephanie Schrag, Jeff Duchin, Michael Boeckh, Janet A. Englund, Michael Famulare, Barry R. Lutz, Mark J. Rieder, Lea M. Starita, Matthew Thompson, Helen Y. Chu, Trevor Bedford, Jay Shendure |
| EPI_ISL_1586958, EPI_ISL_1586960, EPI_ISL_1586961, EPI_ISL_1586963, EPI_ISL_1586966, EPI_ISL_1586967, EPI_ISL_1586968, EPI_ISL_1586969, EPI_ISL_1586970, EPI_ISL_1586972, EPI_ISL_1586974, EPI_ISL_1586975, EPI_ISL_1586977, EPI_ISL_1586979, EPI_ISL_1586981, EPI_ISL_1586982, EPI_ISL_1586983, EPI_ISL_1586985, EPI_ISL_1586986, EPI_ISL_1586987, EPI_ISL_1586988 | | | |
| see above | WA State PHL | Seattle Flu Study | Deborah A. Nickerson, Chris D. Frazar, Jover Lee, Benjamin Pelle, Erica Ryke, Matthew Richardson, Amanda Adler, Elisabeth Brandstetter, Peter D. Han, Kairsten Fay, Misja Ilcisin, Kirsten Lacombe, Thomas R. Sibley, Melissa Truong, Caitlin R. Wolf, Romesh Gautom, Geoff Melly, Brian Hiatt, Philip Dykema, Scott Lindquist, Michael Boeckh, Janet A. Englund, Michael Famulare, Barry R. Lutz, Mark J. Rieder, Lea M. Starita, Matthew Thompson, Helen Y. Chu, Jay Shendure, Trevor Bedford |
| EPI_ISL_1586990, EPI_ISL_1586992, EPI_ISL_1586993, EPI_ISL_1586994, EPI_ISL_1586995, EPI_ISL_1586996, EPI_ISL_1586997 | Seattle Flu Study | Seattle Flu Study | Deborah A. Nickerson, Chris D. Frazar, Jover Lee, Benjamin Pelle, Erica Ryke, Matthew Richardson, Amanda Adler, Elisabeth Brandstetter, Peter D. Han, Kairsten Fay, Misja Ilcisin, Kirsten Lacombe, Thomas R. Sibley, Melissa Truong, Caitlin R. Wolf, Karen Cowgill, Stephanie Schrag, Jeff Duchin, Michael Boeckh, Janet A. Englund, Michael Famulare, Barry R. Lutz, Mark J. Rieder, Lea M. Starita, Matthew Thompson, Helen Y. Chu, Trevor Bedford, Jay Shendure |
| EPI_ISL_1586998, EPI_ISL_1586999 | Seattle Flu Study | Seattle Flu Study | Deborah A. Nickerson, Chris D. Frazar, Jover Lee, Benjamin Pelle, Erica Ryke, Matthew Richardson, Amanda Adler, Elisabeth Brandstetter, Peter D. Han, Kairsten Fay, Misja Ilcisin, Kirsten Lacombe, Thomas R. Sibley, Melissa Truong, Caitlin R. Wolf, Michael Boeckh, Janet A. Englund, Michael Famulare, Barry R. Lutz, Mark J. Rieder, Lea M. Starita, Matthew Thompson, Jay Shendure, Trevor Bedford, Helen Y. Chu |
| EPI_ISL_1587000, EPI_ISL_1587001, EPI_ISL_1587003, EPI_ISL_1587004, EPI_ISL_1587006, EPI_ISL_1587007, EPI_ISL_1587008, EPI_ISL_1587009, EPI_ISL_1587010, EPI_ISL_1587012, EPI_ISL_1587013, EPI_ISL_1587014, EPI_ISL_1587015, EPI_ISL_1587016, EPI_ISL_1587017, EPI_ISL_1587019, EPI_ISL_1587020, EPI_ISL_1587022, EPI_ISL_1587024, EPI_ISL_1587025, EPI_ISL_1587027, EPI_ISL_1587028, EPI_ISL_1587029, EPI_ISL_1587031, EPI_ISL_1587033, EPI_ISL_1587034, EPI_ISL_1587035, EPI_ISL_1587036, EPI_ISL_1587037, EPI_ISL_1587038, EPI_ISL_1587039, EPI_ISL_1587040, EPI_ISL_1587044, EPI_ISL_1587048, EPI_ISL_1587049, EPI_ISL_1587050, EPI_ISL_1587051, EPI_ISL_1587052, EPI_ISL_1587053, EPI_ISL_1587054, EPI_ISL_1587055, EPI_ISL_1587056, EPI_ISL_1587057, EPI_ISL_1587058, EPI_ISL_1587059, EPI_ISL_1587060, EPI_ISL_1587061, EPI_ISL_1587066, EPI_ISL_1587067, EPI_ISL_1587069, EPI_ISL_1587070, EPI_ISL_1587071, EPI_ISL_1587072, EPI_ISL_1587073, EPI_ISL_1587074, EPI_ISL_1587075, EPI_ISL_1587076, EPI_ISL_1587077, EPI_ISL_1587079 | | | |
| see above | Altius Institute | Seattle Flu Study | Deborah A. Nickerson, Chris D. Frazar, Jover Lee, Benjamin Pelle, Erica Ryke, Matthew Richardson, Amanda Adler, Elisabeth Brandstetter, Peter D. Han, Kairsten Fay, Misja Ilcisin, Kirsten Lacombe, Thomas R. Sibley, Melissa Truong, Caitlin R. Wolf, Ryan Alexander, Daniel Bates, Rebecca Bruders, Stephanie DeBaun, Clem Green, Muhammad Halimun, Jessica Halow, Kneshay Harper, Matt Hartman, Andrew Meuser, Alex Nguyen, Truong Nguyen, Sofia Olsson, Sadie Patraw, Hannah Petersen, Tobias Ragoczy, Joshua Richards, Jacob Rodriguez, John Stamatoyannopoulos, Julia Wald, Olivia Waltner, Michael Boeckh, Janet A. Englund, Michael Famulare, Barry R. Lutz, Mark J. Rieder, Lea M. Starita, Matthew Thompson, Helen Y. Chu, Jay Shendure, Trevor Bedford |
| EPI_ISL_1587082, EPI_ISL_1587085, EPI_ISL_1587087, EPI_ISL_1587088, EPI_ISL_1587089, EPI_ISL_1587090 | Seattle Flu Study | Seattle Flu Study | Deborah A. Nickerson, Chris D. Frazar, Jover Lee, Benjamin Pelle, Erica Ryke, Matthew Richardson, Amanda Adler, Elisabeth Brandstetter, Peter D. Han, Kairsten Fay, Misja Ilcisin, Kirsten Lacombe, Thomas R. Sibley, Melissa Truong, Caitlin R. Wolf, Karen Cowgill, Stephanie Schrag, Jeff Duchin, Michael Boeckh, Janet A. Englund, Michael Famulare, Barry R. Lutz, Mark J. Rieder, Lea M. Starita, Matthew Thompson, Helen Y. Chu, Trevor Bedford, Jay Shendure |
| EPI_ISL_1587091 | Seattle Flu Study | Seattle Flu Study | Deborah A. Nickerson, Chris D. Frazar, Jover Lee, Benjamin Pelle, Erica Ryke, Matthew Richardson, Amanda Adler, Elisabeth Brandstetter, Peter D. Han, Kairsten Fay, Misja Ilcisin, Kirsten Lacombe, Thomas R. Sibley, Melissa Truong, Caitlin R. Wolf, Michael Boeckh, Janet A. Englund, Michael Famulare, Barry R. Lutz, Mark J. Rieder, Lea M. Starita, Matthew Thompson, Jay Shendure, Trevor Bedford, Helen Y. Chu |
| EPI_ISL_1587096, EPI_ISL_1587097, EPI_ISL_1587098 | Seattle Flu Study | Seattle Flu Study | Deborah A. Nickerson, Chris D. Frazar, Jover Lee, Benjamin Pelle, Erica Ryke, Matthew Richardson, Amanda Adler, Elisabeth Brandstetter, Peter D. Han, Kairsten Fay, Misja Ilcisin, Kirsten Lacombe, Thomas R. Sibley, Melissa Truong, Caitlin R. Wolf, Karen Cowgill, Stephanie Schrag, Jeff Duchin, Michael Boeckh, Janet A. Englund, Michael Famulare, Barry R. Lutz, Mark J. Rieder, Lea M. Starita, Matthew Thompson, Helen Y. Chu, Trevor Bedford, Jay Shendure |
| EPI_ISL_1587099 | Seattle Flu Study | Seattle Flu Study | Deborah A. Nickerson, Chris D. Frazar, Jover Lee, Benjamin Pelle, Erica Ryke, Matthew Richardson, Amanda Adler, Elisabeth Brandstetter, Peter D. Han, Kairsten Fay, Misja Ilcisin, Kirsten Lacombe, Thomas R. Sibley, Melissa Truong, Caitlin R. Wolf, Michael Boeckh, Janet A. Englund, Michael Famulare, Barry R. Lutz, Mark J. Rieder, Lea M. Starita, Matthew Thompson, Jay Shendure, Trevor Bedford, Helen Y. Chu |
| EPI_ISL_1587100, EPI_ISL_1587102, EPI_ISL_1587104, EPI_ISL_1587107 | Seattle Flu Study | Seattle Flu Study | Deborah A. Nickerson, Chris D. Frazar, Jover Lee, Benjamin Pelle, Erica Ryke, Matthew Richardson, Amanda Adler, Elisabeth Brandstetter, Peter D. Han, Kairsten Fay, Misja Ilcisin, Kirsten Lacombe, Thomas R. Sibley, Melissa Truong, Caitlin R. Wolf, Karen Cowgill, Stephanie Schrag, Jeff Duchin, Michael Boeckh, Janet A. Englund, Michael Famulare, Barry R. Lutz, Mark J. Rieder, Lea M. Starita, Matthew Thompson, Helen Y. Chu, Trevor Bedford, Jay Shendure |
| EPI_ISL_1587109, EPI_ISL_1587110, EPI_ISL_1587112 | Seattle Flu Study | Seattle Flu Study | Deborah A. Nickerson, Chris D. Frazar, Jover Lee, Benjamin Pelle, Erica Ryke, Matthew Richardson, Amanda Adler, Elisabeth Brandstetter, Peter D. Han, Kairsten Fay, Misja Ilcisin, Kirsten Lacombe, Thomas R. Sibley, Melissa Truong, Caitlin R. Wolf, Michael Boeckh, Janet A. Englund, Michael Famulare, Barry R. Lutz, Mark J. Rieder, Lea M. Starita, Matthew Thompson, Jay Shendure, Trevor Bedford, Helen Y. Chu |
| EPI_ISL_1587113 | Seattle Flu Study | Seattle Flu Study | Deborah A. Nickerson, Chris D. Frazar, Jover Lee, Benjamin Pelle, Erica Ryke, Matthew Richardson, Amanda Adler, Elisabeth Brandstetter, Peter D. Han, Kairsten Fay, Misja Ilcisin, Kirsten Lacombe, Thomas R. Sibley, Melissa Truong, Caitlin R. Wolf, Karen Cowgill, Stephanie Schrag, Jeff Duchin, Michael Boeckh, Janet A. Englund, Michael Famulare, Barry R. Lutz, Mark J. Rieder, Lea M. Starita, Matthew Thompson, Helen Y. Chu, Trevor Bedford, Jay Shendure |
| EPI_ISL_1587118, EPI_ISL_1587123, EPI_ISL_1587125, EPI_ISL_1587127 | Seattle Flu Study | Seattle Flu Study | Deborah A. Nickerson, Chris D. Frazar, Jover Lee, Benjamin Pelle, Erica Ryke, Matthew Richardson, Amanda Adler, Elisabeth Brandstetter, Peter D. Han, Kairsten Fay, Misja Ilcisin, Kirsten Lacombe, Thomas R. Sibley, Melissa Truong, Caitlin R. Wolf, Michael Boeckh, Janet A. Englund, Michael Famulare, Barry R. Lutz, Mark J. Rieder, Lea M. Starita, Matthew Thompson, Jay Shendure, Trevor Bedford, Helen Y. Chu |
| EPI_ISL_1587128, EPI_ISL_1587129, EPI_ISL_1587130 | Seattle Flu Study | Seattle Flu Study | Deborah A. Nickerson, Chris D. Frazar, Jover Lee, Benjamin Pelle, Erica Ryke, Matthew Richardson, Amanda Adler, Elisabeth Brandstetter, Peter D. Han, Kairsten Fay, Misja Ilcisin, Kirsten Lacombe, Thomas R. Sibley, Melissa Truong, Caitlin R. Wolf, Karen Cowgill, Stephanie Schrag, Jeff Duchin, Michael Boeckh, Janet A. Englund, Michael Famulare, Barry R. Lutz, Mark J. Rieder, Lea M. Starita, Matthew Thompson, Helen Y. Chu, Trevor Bedford, Jay Shendure |
| EPI_ISL_1587131, EPI_ISL_1587132, EPI_ISL_1587133, EPI_ISL_1587134, EPI_ISL_1587137, EPI_ISL_1587138, EPI_ISL_1587139, EPI_ISL_1587140, EPI_ISL_1587141, EPI_ISL_1587142, EPI_ISL_1587144, EPI_ISL_1587145, EPI_ISL_1587146, EPI_ISL_1587147, EPI_ISL_1587148, EPI_ISL_1587149, EPI_ISL_1587150, EPI_ISL_1587151, EPI_ISL_1587153, EPI_ISL_1587154, EPI_ISL_1587156, EPI_ISL_1587157, EPI_ISL_1587161 | | | |
| see above | Seattle Flu Study | Seattle Flu Study | Deborah A. Nickerson, Chris D. Frazar, Jover Lee, Benjamin Pelle, Erica Ryke, Matthew Richardson, Amanda Adler, Elisabeth Brandstetter, Peter D. Han, Kairsten Fay, Misja Ilcisin, Kirsten Lacombe, Thomas R. Sibley, Melissa Truong, Caitlin R. Wolf, Michael Boeckh, Janet A. Englund, Michael Famulare, Barry R. Lutz, Mark J. Rieder, Lea M. Starita, Matthew Thompson, Jay Shendure, Trevor Bedford, Helen Y. Chu |
| EPI_ISL_1587162, EPI_ISL_1587163, EPI_ISL_1587165, EPI_ISL_1587168, EPI_ISL_1587169, EPI_ISL_1587172, EPI_ISL_1587173 | Seattle Flu Study | Seattle Flu Study | Deborah A. Nickerson, Chris D. Frazar, Jover Lee, Benjamin Pelle, Erica Ryke, Matthew Richardson, Amanda Adler, Elisabeth Brandstetter, Peter D. Han, Kairsten Fay, Misja Ilcisin, Kirsten Lacombe, Thomas R. Sibley, Melissa Truong, Caitlin R. Wolf, Karen Cowgill, Stephanie Schrag, Jeff Duchin, Michael Boeckh, Janet A. Englund, Michael Famulare, Barry R. Lutz, Mark J. Rieder, Lea M. Starita, Matthew Thompson, Helen Y. Chu, Trevor Bedford, Jay Shendure |
| EPI_ISL_1587174, EPI_ISL_1587175, EPI_ISL_1587177, EPI_ISL_1587178, EPI_ISL_1587179, EPI_ISL_1587180, EPI_ISL_1587181, EPI_ISL_1587182, EPI_ISL_1587183, EPI_ISL_1587185, EPI_ISL_1587186, EPI_ISL_1587189, EPI_ISL_1587191, EPI_ISL_1587200, EPI_ISL_1587206, EPI_ISL_1587208, EPI_ISL_1587209, EPI_ISL_1587210, EPI_ISL_1587212, EPI_ISL_1587213, EPI_ISL_1587214, EPI_ISL_1587217, EPI_ISL_1587221, EPI_ISL_1587222, EPI_ISL_1587223, EPI_ISL_1587224, EPI_ISL_1587225, EPI_ISL_1587226, EPI_ISL_1587227, EPI_ISL_1587228, EPI_ISL_1587229, EPI_ISL_1587231, EPI_ISL_1587232, EPI_ISL_1587233, EPI_ISL_1587234, EPI_ISL_1587235, EPI_ISL_1587236, EPI_ISL_1587237, EPI_ISL_1587238, EPI_ISL_1587240, EPI_ISL_1587241, EPI_ISL_1587242, EPI_ISL_1587243, EPI_ISL_1587244, EPI_ISL_1587245, EPI_ISL_1587247, EPI_ISL_1587248, EPI_ISL_1587249, EPI_ISL_1587251, EPI_ISL_1587255 | | | |
| see above | Altius Institute | Seattle Flu Study | Deborah A. Nickerson, Chris D. Frazar, Jover Lee, Benjamin Pelle, Erica Ryke, Matthew Richardson, Amanda Adler, Elisabeth Brandstetter, Peter D. Han, Kairsten Fay, Misja Ilcisin, Kirsten Lacombe, Thomas R. Sibley, Melissa Truong, Caitlin R. Wolf, Ryan Alexander, Daniel Bates, Rebecca Bruders, Stephanie DeBaun, Clem Green, Muhammad Halimun, Jessica Halow, Kneshay Harper, Matt Hartman, Andrew Meuser, Alex Nguyen, Truong Nguyen, Sofia Olsson, Sadie Patraw, Hannah Petersen, Tobias Ragoczy, Joshua Richards, Jacob Rodriguez, John Stamatoyannopoulos, Julia Wald, Olivia Waltner, Michael Boeckh, Janet A. Englund, Michael Famulare, Barry R. Lutz, Mark J. Rieder, Lea M. Starita, Matthew Thompson, Helen Y. Chu, Jay Shendure, Trevor Bedford |
| EPI_ISL_1587261, EPI_ISL_1587262, EPI_ISL_1587263, EPI_ISL_1587264, EPI_ISL_1587265, EPI_ISL_1587266, EPI_ISL_1587267, EPI_ISL_1587268, EPI_ISL_1587269, EPI_ISL_1587270, EPI_ISL_1587271, EPI_ISL_1587272, EPI_ISL_1587273, EPI_ISL_1587274, EPI_ISL_1587275, EPI_ISL_1587276, EPI_ISL_1587278, EPI_ISL_1587281, EPI_ISL_1587282, EPI_ISL_1587283, EPI_ISL_1587284, EPI_ISL_1587285, EPI_ISL_1587286, EPI_ISL_1587289, EPI_ISL_1587290, EPI_ISL_1587291, EPI_ISL_1587292, EPI_ISL_1587293, EPI_ISL_1587294, EPI_ISL_1587295, EPI_ISL_1587296, EPI_ISL_1587298, EPI_ISL_1587300, EPI_ISL_1587301, EPI_ISL_1587302, EPI_ISL_1587303, EPI_ISL_1587304, EPI_ISL_1587305, EPI_ISL_1587306, EPI_ISL_1587307, EPI_ISL_1587308, EPI_ISL_1587309, EPI_ISL_1587310, EPI_ISL_1587311, EPI_ISL_1587312, EPI_ISL_1587313, EPI_ISL_1587314, EPI_ISL_1587315, EPI_ISL_1587316, EPI_ISL_1587317, EPI_ISL_1587319, EPI_ISL_1587321, EPI_ISL_1587322, EPI_ISL_1587324, EPI_ISL_1587325, EPI_ISL_1587326, EPI_ISL_1587327, EPI_ISL_1587328, EPI_ISL_1587329, EPI_ISL_1587330, EPI_ISL_1587332, EPI_ISL_1587333, EPI_ISL_1587334, EPI_ISL_1587335, EPI_ISL_1587336, EPI_ISL_1587337, EPI_ISL_1587338, EPI_ISL_1587339, EPI_ISL_1587340, EPI_ISL_1587341, EPI_ISL_1587342, EPI_ISL_1587343, EPI_ISL_1587344, EPI_ISL_1587345, EPI_ISL_1587346, EPI_ISL_1587347, EPI_ISL_1587348, EPI_ISL_1587349, EPI_ISL_1587350, EPI_ISL_1587351, EPI_ISL_1587352, EPI_ISL_1587353, EPI_ISL_1587354, EPI_ISL_1587355, EPI_ISL_1587356, EPI_ISL_1587357, EPI_ISL_1587358, EPI_ISL_1587359, EPI_ISL_1587360, EPI_ISL_1587361, EPI_ISL_1587362, EPI_ISL_1587364, EPI_ISL_1587365, EPI_ISL_1587367, EPI_ISL_1587368, EPI_ISL_1587369, EPI_ISL_1587370, EPI_ISL_1587371, EPI_ISL_1587372, EPI_ISL_1587373, EPI_ISL_1587374, EPI_ISL_1587375, EPI_ISL_1587376, EPI_ISL_1587377, EPI_ISL_1587378, EPI_ISL_1587379, EPI_ISL_1587381, EPI_ISL_1587385, EPI_ISL_1587386, EPI_ISL_1587388, EPI_ISL_1587389, EPI_ISL_1587390, EPI_ISL_1587392, EPI_ISL_1587393, EPI_ISL_1587394, EPI_ISL_1587395, EPI_ISL_1587396, EPI_ISL_1587398, EPI_ISL_1587399, EPI_ISL_1587401, EPI_ISL_1587402, EPI_ISL_1587403, EPI_ISL_1587404, EPI_ISL_1587405, EPI_ISL_1587406, EPI_ISL_1587408, EPI_ISL_1587409, EPI_ISL_1587410, EPI_ISL_1587411, EPI_ISL_1587412, EPI_ISL_1587413, EPI_ISL_1587414, EPI_ISL_1587415, EPI_ISL_1587416, EPI_ISL_1587417, EPI_ISL_1587418, EPI_ISL_1587419, EPI_ISL_1587420, EPI_ISL_1587421, EPI_ISL_1587422, EPI_ISL_1587423, EPI_ISL_1587424, EPI_ISL_1587425, EPI_ISL_1587426, EPI_ISL_1587427 | | | |
| see above | Yale Clinical Virology Lab | Grubaugh Lab - Yale School of Public Health | Joseph Fauver, Mallery Breban, Isabel Ott, Tara Alpert, Mary Petrone, Anderson Brito, Chantal Vogels, Annie Watkins, Chaney Kalinich, Jessica Rothman, Marie L. Landry, Nathan Grubaugh |
| EPI_ISL_1587450, EPI_ISL_1587451 | Oregon State University TRACE Project | OSU Center for Genome Research and Biocomputing | Oregon State University TRACE Project |
| EPI_ISL_1587583, EPI_ISL_1587584, EPI_ISL_1587585, EPI_ISL_1587586, EPI_ISL_1587587, EPI_ISL_1587588, EPI_ISL_1587589, EPI_ISL_1587590, EPI_ISL_1587591, EPI_ISL_1587592, EPI_ISL_1587593, EPI_ISL_1587594, EPI_ISL_1587595, EPI_ISL_1587596, EPI_ISL_1587597, EPI_ISL_1587598, EPI_ISL_1587599, EPI_ISL_1587600, EPI_ISL_1587601, EPI_ISL_1587602, EPI_ISL_1587603, EPI_ISL_1587604, EPI_ISL_1587605, EPI_ISL_1587607, EPI_ISL_1587608, EPI_ISL_1587609, EPI_ISL_1587610, EPI_ISL_1587611, EPI_ISL_1587612, EPI_ISL_1587613, EPI_ISL_1587614, EPI_ISL_1587615, EPI_ISL_1587616, EPI_ISL_1587617, EPI_ISL_1587618, EPI_ISL_1587619, EPI_ISL_1587620, EPI_ISL_1587621, EPI_ISL_1587622, EPI_ISL_1587623, EPI_ISL_1587624, EPI_ISL_1587625, EPI_ISL_1587626, EPI_ISL_1587628, EPI_ISL_1587629, EPI_ISL_1587630, EPI_ISL_1587632, EPI_ISL_1587633, EPI_ISL_1587634, EPI_ISL_1587635, EPI_ISL_1587636, EPI_ISL_1587637, EPI_ISL_1587639, EPI_ISL_1587640, EPI_ISL_1587641, EPI_ISL_1587642, EPI_ISL_1587643, EPI_ISL_1587644, EPI_ISL_1587645, EPI_ISL_1587646, EPI_ISL_1587647, EPI_ISL_1587648, EPI_ISL_1587650, EPI_ISL_1587651, EPI_ISL_1587652, EPI_ISL_1587654, EPI_ISL_1587655, EPI_ISL_1587656, EPI_ISL_1587657, EPI_ISL_1587658, EPI_ISL_1587659, EPI_ISL_1587660, EPI_ISL_1587661, EPI_ISL_1587662, EPI_ISL_1587663, EPI_ISL_1587664, EPI_ISL_1587665, EPI_ISL_1587666, EPI_ISL_1587667, EPI_ISL_1587668, EPI_ISL_1587669, EPI_ISL_1587670, EPI_ISL_1587671, EPI_ISL_1587672, EPI_ISL_1587673, EPI_ISL_1587674, EPI_ISL_1587675, EPI_ISL_1587676, EPI_ISL_1587677, EPI_ISL_1587678, EPI_ISL_1587679, EPI_ISL_1587680, EPI_ISL_1587681, EPI_ISL_1587682, EPI_ISL_1587683, EPI_ISL_1587684, EPI_ISL_1587685, EPI_ISL_1587686, EPI_ISL_1587687, EPI_ISL_1587688, EPI_ISL_1587689, EPI_ISL_1587690, EPI_ISL_1587691, EPI_ISL_1587692, EPI_ISL_1587693, EPI_ISL_1587694, EPI_ISL_1587695, EPI_ISL_1587696, EPI_ISL_1587697, EPI_ISL_1587698, EPI_ISL_1587699, EPI_ISL_1587700, EPI_ISL_1587701, EPI_ISL_1587702, EPI_ISL_1587703, EPI_ISL_1587705, EPI_ISL_1587706, EPI_ISL_1587707, EPI_ISL_1587708, EPI_ISL_1587709, EPI_ISL_1587710, EPI_ISL_1587711, EPI_ISL_1587713, EPI_ISL_1587714, EPI_ISL_1587715, EPI_ISL_1587716, EPI_ISL_1587717, EPI_ISL_1587718, EPI_ISL_1587719, EPI_ISL_1587721, EPI_ISL_1587722, EPI_ISL_1587723, EPI_ISL_1587724, EPI_ISL_1587725, EPI_ISL_1587726, EPI_ISL_1587727, EPI_ISL_1587728, EPI_ISL_1587729, EPI_ISL_1587730, EPI_ISL_1587732, EPI_ISL_1587733, EPI_ISL_1587734, EPI_ISL_1587735, EPI_ISL_1587736, EPI_ISL_1587737, EPI_ISL_1587739, EPI_ISL_1587740, EPI_ISL_1587741, EPI_ISL_1587742, EPI_ISL_1587743, EPI_ISL_1587744, EPI_ISL_1587745, EPI_ISL_1587746, EPI_ISL_1587747, EPI_ISL_1587748, EPI_ISL_1587749, EPI_ISL_1587750, EPI_ISL_1587751, EPI_ISL_1587752, EPI_ISL_1587753, EPI_ISL_1587754, EPI_ISL_1587755, EPI_ISL_1587756, EPI_ISL_1587757, EPI_ISL_1587758, EPI_ISL_1587759, EPI_ISL_1587761, EPI_ISL_1587762, EPI_ISL_1587763, EPI_ISL_1587764, EPI_ISL_1587765, EPI_ISL_1587766, EPI_ISL_1587767, EPI_ISL_1587768, EPI_ISL_1587769, EPI_ISL_1587770, | | | |

| | | | |
|---|---|---|---|
| EPI_ISL_1587771, EPI_ISL_1587772, EPI_ISL_1587773, EPI_ISL_1587774, EPI_ISL_1587775, EPI_ISL_1587776, EPI_ISL_1587777, EPI_ISL_1587778, EPI_ISL_1587779, EPI_ISL_1587780, EPI_ISL_1587781, EPI_ISL_1587782, EPI_ISL_1587783, EPI_ISL_1587784, EPI_ISL_1587785, EPI_ISL_1587786, EPI_ISL_1587787, EPI_ISL_1587789, EPI_ISL_1587790, EPI_ISL_1587791, EPI_ISL_1587792, EPI_ISL_1587793, EPI_ISL_1587795, EPI_ISL_1587798, EPI_ISL_1587801, EPI_ISL_1587804, EPI_ISL_1587807, EPI_ISL_1587810, EPI_ISL_1587813, EPI_ISL_1587816, EPI_ISL_1587819, EPI_ISL_1587826, EPI_ISL_1587829, EPI_ISL_1587832, EPI_ISL_1587835, EPI_ISL_1587838, EPI_ISL_1587846, EPI_ISL_1587849, EPI_ISL_1587852, EPI_ISL_1587855, EPI_ISL_1587858, EPI_ISL_1587861, EPI_ISL_1587864, EPI_ISL_1587871, EPI_ISL_1587874, EPI_ISL_1587877, EPI_ISL_1587880, EPI_ISL_1587883, EPI_ISL_1587886, EPI_ISL_1587889, EPI_ISL_1587892, EPI_ISL_1587895, EPI_ISL_1587898, EPI_ISL_1587901, EPI_ISL_1587904, EPI_ISL_1587907, EPI_ISL_1587911, EPI_ISL_1587913, EPI_ISL_1587914, EPI_ISL_1587915, EPI_ISL_1587916, EPI_ISL_1587917, EPI_ISL_1587918, EPI_ISL_1587919, EPI_ISL_1587920, EPI_ISL_1587921, EPI_ISL_1587922, EPI_ISL_1587923, EPI_ISL_1587924, EPI_ISL_1587925, EPI_ISL_1587926, EPI_ISL_1587927, EPI_ISL_1587928, EPI_ISL_1587929, EPI_ISL_1587930, EPI_ISL_1587931, EPI_ISL_1587932, EPI_ISL_1587933, EPI_ISL_1587934, EPI_ISL_1587935, EPI_ISL_1587936, EPI_ISL_1587937, EPI_ISL_1587938, EPI_ISL_1587939, EPI_ISL_1587940, EPI_ISL_1587941, EPI_ISL_1587942, EPI_ISL_1587943, EPI_ISL_1587944, EPI_ISL_1587945, EPI_ISL_1587946, EPI_ISL_1587947, EPI_ISL_1587948, EPI_ISL_1587949, EPI_ISL_1587950, EPI_ISL_1587951, EPI_ISL_1587952, EPI_ISL_1587953, EPI_ISL_1587954, EPI_ISL_1587955, EPI_ISL_1587956, EPI_ISL_1587958, EPI_ISL_1587959, EPI_ISL_1587960, EPI_ISL_1587961, EPI_ISL_1587962, EPI_ISL_1587963, EPI_ISL_1587964, EPI_ISL_1587965, EPI_ISL_1587966, EPI_ISL_1587967, EPI_ISL_1587968, EPI_ISL_1587969, EPI_ISL_1587970, EPI_ISL_1587972, EPI_ISL_1587973, EPI_ISL_1587974, EPI_ISL_1587975, EPI_ISL_1587976, EPI_ISL_1587978, EPI_ISL_1587980, EPI_ISL_1587981, EPI_ISL_1587982, EPI_ISL_1587983, EPI_ISL_1587984, EPI_ISL_1587985, EPI_ISL_1587986, EPI_ISL_1587987, EPI_ISL_1587988, EPI_ISL_1587989, EPI_ISL_1587990, EPI_ISL_1587991, EPI_ISL_1587993, EPI_ISL_1587994, EPI_ISL_1587995, EPI_ISL_1587997, EPI_ISL_1587998, EPI_ISL_1587999, EPI_ISL_1588000, EPI_ISL_1588001, EPI_ISL_1588003, EPI_ISL_1588004, EPI_ISL_1588005, EPI_ISL_1588006, EPI_ISL_1588007, EPI_ISL_1588008, EPI_ISL_1588009, EPI_ISL_1588010, EPI_ISL_1588011, EPI_ISL_1588012, EPI_ISL_1588013, EPI_ISL_1588014, EPI_ISL_1588015, EPI_ISL_1588016, EPI_ISL_1588017, EPI_ISL_1588018, EPI_ISL_1588019, EPI_ISL_1588020, EPI_ISL_1588021, EPI_ISL_1588022, EPI_ISL_1588023, EPI_ISL_1588024, EPI_ISL_1588025, EPI_ISL_1588026, EPI_ISL_1588027, EPI_ISL_1588028, EPI_ISL_1588029, EPI_ISL_1588030, EPI_ISL_1588031, EPI_ISL_1588032, EPI_ISL_1588033, EPI_ISL_1588034, EPI_ISL_1588035, EPI_ISL_1588037, EPI_ISL_1588038, EPI_ISL_1588039, EPI_ISL_1588041, EPI_ISL_1588042, EPI_ISL_1588043, EPI_ISL_1588044, EPI_ISL_1588045, EPI_ISL_1588046, EPI_ISL_1588047, EPI_ISL_1588048, EPI_ISL_1588049, EPI_ISL_1588050, EPI_ISL_1588051, EPI_ISL_1588052, EPI_ISL_1588053, EPI_ISL_1588054, EPI_ISL_1588055, EPI_ISL_1588056, EPI_ISL_1588057, EPI_ISL_1588058, EPI_ISL_1588059, EPI_ISL_1588060, EPI_ISL_1588061, EPI_ISL_1588062, EPI_ISL_1588065, EPI_ISL_1588068, EPI_ISL_1588072, EPI_ISL_1588075, EPI_ISL_1588079, EPI_ISL_1588082, EPI_ISL_1588087, EPI_ISL_1588090, EPI_ISL_1588093, EPI_ISL_1588096, EPI_ISL_1588099, EPI_ISL_1588103, EPI_ISL_1588105, EPI_ISL_1588108, EPI_ISL_1588112, EPI_ISL_1588115, EPI_ISL_1588118, EPI_ISL_1588121, EPI_ISL_1588124, EPI_ISL_1588127, EPI_ISL_1588130, EPI_ISL_1588133, EPI_ISL_1588136, EPI_ISL_1588139, EPI_ISL_1588142, EPI_ISL_1588145, EPI_ISL_1588148, EPI_ISL_1588151, EPI_ISL_1588155, EPI_ISL_1588158, EPI_ISL_1588161, EPI_ISL_1588164, EPI_ISL_1588171, EPI_ISL_1588174, EPI_ISL_1588177, EPI_ISL_1588180, EPI_ISL_1588183, EPI_ISL_1588187, EPI_ISL_1588193, EPI_ISL_1588195, EPI_ISL_1588199, EPI_ISL_1588202, EPI_ISL_1588205, EPI_ISL_1588208, EPI_ISL_1588211, EPI_ISL_1588214, EPI_ISL_1588217, EPI_ISL_1588223, EPI_ISL_1588226, EPI_ISL_1588229, EPI_ISL_1588232, EPI_ISL_1588236, EPI_ISL_1588241, EPI_ISL_1588245, EPI_ISL_1588248, EPI_ISL_1588251, EPI_ISL_1588254, EPI_ISL_1588256, EPI_ISL_1588259, EPI_ISL_1588262, EPI_ISL_1588266, EPI_ISL_1588268, EPI_ISL_1588271, EPI_ISL_1588274, EPI_ISL_1588277, EPI_ISL_1588280, EPI_ISL_1588283, EPI_ISL_1588286, EPI_ISL_1588288, EPI_ISL_1588289, EPI_ISL_1588291, EPI_ISL_1588292, EPI_ISL_1588294, EPI_ISL_1588295, EPI_ISL_1588297, EPI_ISL_1588298, EPI_ISL_1588299, EPI_ISL_1588300, EPI_ISL_1588301, EPI_ISL_1588302, EPI_ISL_1588305, EPI_ISL_1588306, EPI_ISL_1588307, EPI_ISL_1588308, EPI_ISL_1588310, EPI_ISL_1588311, EPI_ISL_1588312, EPI_ISL_1588313, EPI_ISL_1588314, EPI_ISL_1588315, EPI_ISL_1588316, EPI_ISL_1588317, EPI_ISL_1588318, EPI_ISL_1588319, EPI_ISL_1588320, EPI_ISL_1588321, EPI_ISL_1588323, EPI_ISL_1588324, EPI_ISL_1588325, EPI_ISL_1588326, EPI_ISL_1588327, EPI_ISL_1588328, EPI_ISL_1588329, EPI_ISL_1588331, EPI_ISL_1588332, EPI_ISL_1588333, EPI_ISL_1588334, EPI_ISL_1588335, EPI_ISL_1588337, EPI_ISL_1588339, EPI_ISL_1588340, EPI_ISL_1588341, EPI_ISL_1588343, EPI_ISL_1588344, EPI_ISL_1588345, EPI_ISL_1588346, EPI_ISL_1588348, EPI_ISL_1588349, EPI_ISL_1588350, EPI_ISL_1588351, EPI_ISL_1588352, EPI_ISL_1588353, EPI_ISL_1588354, EPI_ISL_1588356, EPI_ISL_1588357, EPI_ISL_1588358, EPI_ISL_1588359, EPI_ISL_1588361, EPI_ISL_1588362, EPI_ISL_1588363, EPI_ISL_1588365, EPI_ISL_1588366, EPI_ISL_1588367, EPI_ISL_1588369, EPI_ISL_1588370, EPI_ISL_1588371, EPI_ISL_1588372, EPI_ISL_1588373, EPI_ISL_1588374, EPI_ISL_1588375, EPI_ISL_1588376, EPI_ISL_1588377, EPI_ISL_1588378, EPI_ISL_1588379, EPI_ISL_1588380, EPI_ISL_1588381, EPI_ISL_1588382, EPI_ISL_1588383, EPI_ISL_1588384, EPI_ISL_1588385, EPI_ISL_1588386, EPI_ISL_1588387, EPI_ISL_1588388, EPI_ISL_1588389, EPI_ISL_1588390, EPI_ISL_1588391, EPI_ISL_1588392, EPI_ISL_1588393, EPI_ISL_1588394, EPI_ISL_1588395, EPI_ISL_1588396, EPI_ISL_1588397, EPI_ISL_1588398, EPI_ISL_1588399, EPI_ISL_1588400, EPI_ISL_1588401, EPI_ISL_1588402, EPI_ISL_1588403, EPI_ISL_1588404, EPI_ISL_1588405, EPI_ISL_1588407, EPI_ISL_1588409, EPI_ISL_1588411, EPI_ISL_1588412, EPI_ISL_1588413, EPI_ISL_1588414, EPI_ISL_1588415, EPI_ISL_1588416, EPI_ISL_1588417, EPI_ISL_1588418, EPI_ISL_1588419, EPI_ISL_1588421, EPI_ISL_1588422, EPI_ISL_1588423, EPI_ISL_1588424, EPI_ISL_1588425, EPI_ISL_1588426, EPI_ISL_1588427, EPI_ISL_1588428, EPI_ISL_1588429, EPI_ISL_1588431, EPI_ISL_1588432, EPI_ISL_1588433, EPI_ISL_1588434, EPI_ISL_1588435, EPI_ISL_1588436 | | | |
| see above | Infinity Biologix | Centers for Disease Control and Prevention Division of Viral Diseases, Pathogen Discovery | Dakota Howard, Dhwani Batra, Peter W. Cook, Kara Moser, Adrian Paskey, Jason Caravas, Benjamin Rambo-Martin, Shatavia Morrison, Christopher Gulvick, Scott Sammons, Yvette Unoarumhi, Darlene Wagner, Matthew Schmerer, Christian Bixby, Yihe Wang, Jonathan Schultz, Chirayu Goswami, Russ Hager, Robin Grimwood, Clinton R. Paden, Duncan MacCannell |
| EPI_ISL_1591225 | NE Public Health Laboratory | Pathogen Discovery, Respiratory Viruses Branch, Division of Viral Diseases, Centers for Disease Control and Prevention | Yan Li, Ying Tao, Anna Kelleher, Jing Zhang, Anna Montmayeur, Brian Lynch, Krista Queen, Anna Uehara, Peter Cook, Han Jia Justin Ng, Rachel Marine, Clinton R. Paden, Haibin Wang, Mark Burroughs, Justin Lee, Adam Retchless, Suxiang Tong |
| EPI_ISL_1591226, EPI_ISL_1591227, EPI_ISL_1591228, EPI_ISL_1591229 | Washington University School of Medicine | Pathogen Discovery, Respiratory Viruses Branch, Division of Viral Diseases, Centers for Disease Control and Prevention | Yan Li, Ying Tao, Anna Kelleher, Jing Zhang, Anna Montmayeur, Brian Lynch, Krista Queen, Anna Uehara, Peter Cook, Han Jia Justin Ng, Rachel Marine, Clinton R. Paden, Haibin Wang, Mark Burroughs, Justin Lee, Adam Retchless, Suxiang Tong |
| EPI_ISL_1591230, EPI_ISL_1591231 | MO State Public Health Laboratory | Pathogen Discovery, Respiratory Viruses Branch, Division of Viral Diseases, Centers for Disease Control and Prevention | Yan Li, Ying Tao, Anna Kelleher, Jing Zhang, Anna Montmayeur, Brian Lynch, Krista Queen, Anna Uehara, Peter Cook, Han Jia Justin Ng, Rachel Marine, Clinton R. Paden, Haibin Wang, Mark Burroughs, Justin Lee, Adam Retchless, Suxiang Tong |
| EPI_ISL_1591236, EPI_ISL_1591237, EPI_ISL_1591238, EPI_ISL_1591239 | Vanderbilt University Medical Center | Pathogen Discovery, Respiratory Viruses Branch, Division of Viral Diseases, Centers for Disease Control and Prevention | Yan Li, Ying Tao, Anna Kelleher, Jing Zhang, Anna Montmayeur, Brian Lynch, Krista Queen, Anna Uehara, Peter Cook, Han Jia Justin Ng, Rachel Marine, Clinton R. Paden, Haibin Wang, Mark Burroughs, Justin Lee, Adam Retchless, Suxiang Tong |
| EPI_ISL_1591240, EPI_ISL_1591241 | CDPH, Viral and Rickettsial Disease Laboratory | Pathogen Discovery, Respiratory Viruses Branch, Division of Viral Diseases, Centers for Disease Control and Prevention | Yan Li, Ying Tao, Anna Kelleher, Jing Zhang, Anna Montmayeur, Brian Lynch, Krista Queen, Anna Uehara, Peter Cook, Han Jia Justin Ng, Rachel Marine, Clinton R. Paden, Haibin Wang, Mark Burroughs, Justin Lee, Adam Retchless, Suxiang Tong |
| EPI_ISL_1591244, EPI_ISL_1591245, EPI_ISL_1591247 | Kaiser Permanente Washington Health Research Institute | Pathogen Discovery, Respiratory Viruses Branch, Division of Viral Diseases, Centers for Disease Control and Prevention | Yan Li, Ying Tao, Anna Kelleher, Jing Zhang, Anna Montmayeur, Brian Lynch, Krista Queen, Anna Uehara, Peter Cook, Han Jia Justin Ng, Rachel Marine, Clinton R. Paden, Haibin Wang, Mark Burroughs, Justin Lee, Adam Retchless, Suxiang Tong |
| EPI_ISL_1591248 | Atlanta VA Medical Center | Pathogen Discovery, Respiratory Viruses Branch, Division of Viral Diseases, Centers for Disease Control and Prevention | Yan Li, Ying Tao, Anna Kelleher, Jing Zhang, Anna Montmayeur, Brian Lynch, Krista Queen, Anna Uehara, Peter Cook, Han Jia Justin Ng, Rachel Marine, Clinton R. Paden, Haibin Wang, Mark Burroughs, Justin Lee, Adam Retchless, Suxiang Tong |
| EPI_ISL_1591460, EPI_ISL_1591461, EPI_ISL_1591462, EPI_ISL_1591463, EPI_ISL_1591464, EPI_ISL_1591465, EPI_ISL_1591466, EPI_ISL_1591467, EPI_ISL_1591468, EPI_ISL_1591469, EPI_ISL_1591470, EPI_ISL_1591471, EPI_ISL_1591472, EPI_ISL_1591473, EPI_ISL_1591474, EPI_ISL_1591475, EPI_ISL_1591476, EPI_ISL_1591477, EPI_ISL_1591478, EPI_ISL_1591480, EPI_ISL_1591481, EPI_ISL_1591483, EPI_ISL_1591484, EPI_ISL_1591486, EPI_ISL_1591487, EPI_ISL_1591488, EPI_ISL_1591489, EPI_ISL_1591490, EPI_ISL_1591491, EPI_ISL_1591492, EPI_ISL_1591493, EPI_ISL_1591494, EPI_ISL_1591495, EPI_ISL_1591496, EPI_ISL_1591497, EPI_ISL_1591498, EPI_ISL_1591499, EPI_ISL_1591500, EPI_ISL_1591501, EPI_ISL_1591502, EPI_ISL_1591503, EPI_ISL_1591504, EPI_ISL_1591505, EPI_ISL_1591506, EPI_ISL_1591507, EPI_ISL_1591508, EPI_ISL_1591509, EPI_ISL_1591510, EPI_ISL_1591511, EPI_ISL_1591512, EPI_ISL_1591513, EPI_ISL_1591514, EPI_ISL_1591515, EPI_ISL_1591516, EPI_ISL_1591517, EPI_ISL_1591519, EPI_ISL_1591520, EPI_ISL_1591521, EPI_ISL_1591522, EPI_ISL_1591523, EPI_ISL_1591524, EPI_ISL_1591525, EPI_ISL_1591526, EPI_ISL_1591527, EPI_ISL_1591528, EPI_ISL_1591529, EPI_ISL_1591530, EPI_ISL_1591531, EPI_ISL_1591532, EPI_ISL_1591533, EPI_ISL_1591534, EPI_ISL_1591535, EPI_ISL_1591536, EPI_ISL_1591537, EPI_ISL_1591539, EPI_ISL_1591540, EPI_ISL_1591541, EPI_ISL_1591542, EPI_ISL_1591543, EPI_ISL_1591544, EPI_ISL_1591545, EPI_ISL_1591546, EPI_ISL_1591547, EPI_ISL_1591548, EPI_ISL_1591549, EPI_ISL_1591550, EPI_ISL_1591551, EPI_ISL_1591552, EPI_ISL_1591553, EPI_ISL_1591554, EPI_ISL_1591555, EPI_ISL_1591556, EPI_ISL_1591557, EPI_ISL_1591558, EPI_ISL_1591559, EPI_ISL_1591560, EPI_ISL_1591561, EPI_ISL_1591562, EPI_ISL_1591563, EPI_ISL_1591564, EPI_ISL_1591565, EPI_ISL_1591566, EPI_ISL_1591567, EPI_ISL_1591568, EPI_ISL_1591569, EPI_ISL_1591570, EPI_ISL_1591571, EPI_ISL_1591572, EPI_ISL_1591573, EPI_ISL_1591574, EPI_ISL_1591575, EPI_ISL_1591576, EPI_ISL_1591577, EPI_ISL_1591578, EPI_ISL_1591579, EPI_ISL_1591580, EPI_ISL_1591581, EPI_ISL_1591582, EPI_ISL_1591583, EPI_ISL_1591584, EPI_ISL_1591585, EPI_ISL_1591586, EPI_ISL_1591587, EPI_ISL_1591588, EPI_ISL_1591589, EPI_ISL_1591590, EPI_ISL_1591591, EPI_ISL_1591593, EPI_ISL_1591594, EPI_ISL_1591595, EPI_ISL_1591596, EPI_ISL_1591597, EPI_ISL_1591598, EPI_ISL_1591599, EPI_ISL_1591601, EPI_ISL_1591602, EPI_ISL_1591603, EPI_ISL_1591604, EPI_ISL_1591605, EPI_ISL_1591606, EPI_ISL_1591609, EPI_ISL_1591610, EPI_ISL_1591611, EPI_ISL_1591612, EPI_ISL_1591613, EPI_ISL_1591614, EPI_ISL_1591616, EPI_ISL_1591617, EPI_ISL_1591618, EPI_ISL_1591619, EPI_ISL_1591620, EPI_ISL_1591621, EPI_ISL_1591622, EPI_ISL_1591623, EPI_ISL_1591624, EPI_ISL_1591625, EPI_ISL_1591626, EPI_ISL_1591627, EPI_ISL_1591628, EPI_ISL_1591629, EPI_ISL_1591630, EPI_ISL_1591631, EPI_ISL_1591632, EPI_ISL_1591633, EPI_ISL_1591634, EPI_ISL_1591635, EPI_ISL_1591637, EPI_ISL_1591638, EPI_ISL_1591639, EPI_ISL_1591640, EPI_ISL_1591641, EPI_ISL_1591643, EPI_ISL_1591645, EPI_ISL_1591646, EPI_ISL_1591647, EPI_ISL_1591648, EPI_ISL_1591649, EPI_ISL_1591651, EPI_ISL_1591653, EPI_ISL_1591654, EPI_ISL_1591655, EPI_ISL_1591656, EPI_ISL_1591657, EPI_ISL_1591659, EPI_ISL_1591661, EPI_ISL_1591662, EPI_ISL_1591663, EPI_ISL_1591664, EPI_ISL_1591665, EPI_ISL_1591666, EPI_ISL_1591667, EPI_ISL_1591669, EPI_ISL_1591670, EPI_ISL_1591671, EPI_ISL_1591672, EPI_ISL_1591673, EPI_ISL_1591675, EPI_ISL_1591676, EPI_ISL_1591677, EPI_ISL_1591678, EPI_ISL_1591679, EPI_ISL_1591680, EPI_ISL_1591682, EPI_ISL_1591683, EPI_ISL_1591684, EPI_ISL_1591685, EPI_ISL_1591686, EPI_ISL_1591687, EPI_ISL_1591689, EPI_ISL_1591691, EPI_ISL_1591692, EPI_ISL_1591694, EPI_ISL_1591696, EPI_ISL_1591697, EPI_ISL_1591698, EPI_ISL_1591699, EPI_ISL_1591700, EPI_ISL_1591702, EPI_ISL_1591703, EPI_ISL_1591705, EPI_ISL_1591706, EPI_ISL_1591707, EPI_ISL_1591709, EPI_ISL_1591710, EPI_ISL_1591711, EPI_ISL_1591712, EPI_ISL_1591713 | | | |
| see above | Instituto Nacional de Medicina Genomica | Instituto Nacional de Medicina Genomica | Hidalgo-Miranda A, Mendoza-Vargas A, Reyes-Grajeda JP, Cedro-Tanda A, Alcaraz N, Gonzalez-Barrera D, Rangel-DeLeon D, Miranda-Ortiz H, Rosas-Escobar P, Canseco Mendez JC, Ramirez-Vega O, Munguia-Garza P, Garcia-Cardenas FJ, Gonzalez-Woge MA, Herrera-Montalvo LA |
| EPI_ISL_1591724, EPI_ISL_1591725, EPI_ISL_1591730, EPI_ISL_1591732 | TGen North | TGen North | Jolene Bowers, Heather Centner, Chris French, Hayley Yaglom, Ashlyn Pfeiffer, Darrin Lemmer, Dave Engelthaler, The Arizona COVID Genomics Union (ACGU) |
| EPI_ISL_1591737 | Sonora Quest Laboratories | TGen North | Jolene Bowers, Heather Centner, Chris French, Hayley Yaglom, Ashlyn Pfeiffer, Darrin Lemmer, Dave Engelthaler, The Arizona COVID Genomics Union (ACGU) |
| EPI_ISL_1591738, EPI_ISL_1591741, EPI_ISL_1591742, EPI_ISL_1591744, EPI_ISL_1591746, EPI_ISL_1591749, EPI_ISL_1591754, EPI_ISL_1591755, EPI_ISL_1591760, EPI_ISL_1591763, EPI_ISL_1591764, EPI_ISL_1591765, EPI_ISL_1591773, EPI_ISL_1591782, EPI_ISL_1591786 | | | |
| see above | TGen North | TGen North | Jolene Bowers, Heather Centner, Chris French, Hayley Yaglom, Ashlyn Pfeiffer, Darrin Lemmer, Dave Engelthaler, The Arizona COVID Genomics Union (ACGU) |
| EPI_ISL_1591789 | Sonora Quest Laboratories | TGen North | Jolene Bowers, Heather Centner, Chris French, Hayley Yaglom, Ashlyn Pfeiffer, Darrin Lemmer, Dave Engelthaler, The Arizona COVID Genomics Union (ACGU) |
| EPI_ISL_1591790, EPI_ISL_1591791, EPI_ISL_1591792, EPI_ISL_1591793, EPI_ISL_1591794, EPI_ISL_1591795, EPI_ISL_1591798, EPI_ISL_1591808, EPI_ISL_1591809, EPI_ISL_1591818, EPI_ISL_1591822, EPI_ISL_1591832, EPI_ISL_1591835, EPI_ISL_1591839, EPI_ISL_1591840, EPI_ISL_1591842, EPI_ISL_1591843, EPI_ISL_1591844, EPI_ISL_1591852 | | | |
| see above | TGen North | TGen North | Jolene Bowers, Heather Centner, Chris French, Hayley Yaglom, Ashlyn Pfeiffer, Darrin Lemmer, Dave Engelthaler, The Arizona COVID Genomics Union (ACGU) |
| EPI_ISL_1591853 | Sonora Quest Laboratories | TGen North | Jolene Bowers, Heather Centner, Chris French, Hayley Yaglom, Ashlyn Pfeiffer, Darrin Lemmer, Dave Engelthaler, The Arizona COVID Genomics Union (ACGU) |
| EPI_ISL_1591857, EPI_ISL_1591858, EPI_ISL_1591863, EPI_ISL_1591865, EPI_ISL_1591866, EPI_ISL_1591868, EPI_ISL_1591874, EPI_ISL_1591879, EPI_ISL_1591883, EPI_ISL_1591888, EPI_ISL_1591889, EPI_ISL_1591891, EPI_ISL_1591892, EPI_ISL_1591901, EPI_ISL_1591903, EPI_ISL_1591912, EPI_ISL_1591913, EPI_ISL_1591916, EPI_ISL_1591917, EPI_ISL_1591918, EPI_ISL_1591920, EPI_ISL_1591922, EPI_ISL_1591923, EPI_ISL_1591924, EPI_ISL_1591926, EPI_ISL_1591928, EPI_ISL_1591933, EPI_ISL_1591935 | | | |
| see above | TGen North | TGen North | Jolene Bowers, Heather Centner, Chris French, Hayley Yaglom, Ashlyn Pfeiffer, Darrin Lemmer, Dave Engelthaler, The Arizona COVID Genomics Union (ACGU) |
| EPI_ISL_1591939 | Sonora Quest Laboratories | TGen North | Jolene Bowers, Heather Centner, Chris French, Hayley Yaglom, Ashlyn Pfeiffer, Darrin Lemmer, Dave Engelthaler, The Arizona COVID Genomics Union (ACGU) |
| EPI_ISL_1591947 | TGen North | TGen North | Jolene Bowers, Heather Centner, Chris French, Hayley Yaglom, Ashlyn Pfeiffer, Darrin Lemmer, Dave Engelthaler, The Arizona COVID Genomics Union (ACGU) |
| EPI_ISL_1591948 | Sonora Quest Laboratories | TGen North | Jolene Bowers, Heather Centner, Chris French, Hayley Yaglom, Ashlyn Pfeiffer, Darrin Lemmer, Dave Engelthaler, The Arizona COVID Genomics Union (ACGU) |
| EPI_ISL_1591949, EPI_ISL_1591950, EPI_ISL_1591951, EPI_ISL_1591955, EPI_ISL_1591957, EPI_ISL_1591964, EPI_ISL_1591965, EPI_ISL_1591971, EPI_ISL_1591978, EPI_ISL_1591979, EPI_ISL_1591986, EPI_ISL_1591997, EPI_ISL_1591998, EPI_ISL_1591999, EPI_ISL_1592000, EPI_ISL_1592005, EPI_ISL_1592011, EPI_ISL_1592027, EPI_ISL_1592030, EPI_ISL_1592032, EPI_ISL_1592033, EPI_ISL_1592034, EPI_ISL_1592036, EPI_ISL_1592043, EPI_ISL_1592044, EPI_ISL_1592048, EPI_ISL_1592050, EPI_ISL_1592051, EPI_ISL_1592056, EPI_ISL_1592057, EPI_ISL_1592058, EPI_ISL_1592059, EPI_ISL_1592071, EPI_ISL_1592072, EPI_ISL_1592073 | | | |
| see above | TGen North | TGen North | Jolene Bowers, Heather Centner, Chris French, Hayley Yaglom, Ashlyn Pfeiffer, Darrin Lemmer, Dave Engelthaler, The Arizona COVID Genomics Union (ACGU) |
| EPI_ISL_1592074 | Sonora Quest Laboratories | TGen North | Jolene Bowers, Heather Centner, Chris French, Hayley Yaglom, Ashlyn Pfeiffer, Darrin Lemmer, Dave Engelthaler, The Arizona COVID Genomics Union (ACGU) |
| EPI_ISL_1592079, EPI_ISL_1592083, EPI_ISL_1592093, EPI_ISL_1592097, EPI_ISL_1592098, EPI_ISL_1592099, EPI_ISL_1592102, EPI_ISL_1592104, EPI_ISL_1592107, EPI_ISL_1592112, EPI_ISL_1592113, EPI_ISL_1592119, EPI_ISL_1592123, EPI_ISL_1592124, EPI_ISL_1592126, EPI_ISL_1592129, EPI_ISL_1592132, EPI_ISL_1592139, EPI_ISL_1592140, EPI_ISL_1592142, EPI_ISL_1592148, EPI_ISL_1592150, EPI_ISL_1592153, EPI_ISL_1592154, EPI_ISL_1592168, EPI_ISL_1592170, EPI_ISL_1592180, EPI_ISL_1592184, EPI_ISL_1592186, EPI_ISL_1592187, EPI_ISL_1592188, EPI_ISL_1592191, EPI_ISL_1592194, EPI_ISL_1592198 | | | |
| see above | TGen North | TGen North | Jolene Bowers, Heather Centner, Chris French, Hayley Yaglom, Ashlyn Pfeiffer, Darrin Lemmer, Dave Engelthaler, The Arizona COVID Genomics Union (ACGU) |
| EPI_ISL_1592200 | Arizona State University | Arizona State University | Peter T. Skidmore, LaRinda A. Holland, Rabia Maqsood, Nicholas J. Mellor, Joy M. Blain, Valerie Harris, Joshua LaBaer, Vel Murugan, Efrem S. Lim |
| EPI_ISL_1592219 | Seattle Flu Study | Seattle Flu Study | Deborah A. Nickerson, Chris D. Frazar, Jover Lee, Benjamin Pelle, Erica Ryke, Matthew Richardson, Amanda Adler, Elisabeth Brandstetter, Peter D. Han, Kairsten Fay, Misja Ilcisin, Kirsten Lacombe, Thomas R. Sibley, Melissa Truong, Caitlin R. Wolf, Karen Cowgill, Stephanie Schrag, Jeff Duchin, Michael Boeckh, Janet A. Englund, Michael Famulare, Barry R. Lutz, Mark J. Rieder, Lea M. Starita, Matthew Thompson, Helen Y. Chu, Trevor Bedford, Jay Shendure |
| EPI_ISL_1592221, EPI_ISL_1592222, EPI_ISL_1592223, EPI_ISL_1592225, EPI_ISL_1592226 | Altius Institute | Seattle Flu Study | Deborah A. Nickerson, Chris D. Frazar, Jover Lee, Benjamin Pelle, Erica Ryke, Matthew Richardson, Amanda Adler, Elisabeth Brandstetter, Peter D. Han, Kairsten Fay, Misja Ilcisin, Kirsten Lacombe, Thomas R. Sibley, Melissa Truong, Caitlin R. Wolf, Ryan Alexander, Daniel Bates, Rebecca Bruders, Stephanie DeBaun, Clem Green, Muhammad Halimun, Jessica Halow, Kneshay Harper, Matt Hartman, Andrew Meuser, Alex Nguyen, Truong Nguyen, Sofia Olsson, Sadie Patraw, Hannah Petersen, Tobias Ragoczy, Joshua Richards, Jacob Rodriguez, John Stamatoyannopoulos, Julia Wald, Olivia Waltner, Michael Boeckh, Janet A. Englund, Michael Famulare, Barry R. Lutz, Mark J. Rieder, Lea M. Starita, Matthew Thompson, Helen Y. Chu, Jay Shendure, Trevor Bedford |
| EPI_ISL_1592231, EPI_ISL_1592232, EPI_ISL_1592233, EPI_ISL_1592234, EPI_ISL_1592235, EPI_ISL_1592236, EPI_ISL_1592237, EPI_ISL_1592238, EPI_ISL_1592239, EPI_ISL_1592241, EPI_ISL_1592242, EPI_ISL_1592244, EPI_ISL_1592245, EPI_ISL_1592246, EPI_ISL_1592247, EPI_ISL_1592248, EPI_ISL_1592249, EPI_ISL_1592250, EPI_ISL_1592251, EPI_ISL_1592252 | | | |
| see above | City of Milwaukee Health Department Laboratory | City of Milwaukee Health Department Laboratory | Sanjib Bhattacharyya |
| EPI_ISL_1592253, EPI_ISL_1592255, EPI_ISL_1592256, EPI_ISL_1592257, EPI_ISL_1592258, EPI_ISL_1592259, EPI_ISL_1592262, EPI_ISL_1592264, EPI_ISL_1592265, EPI_ISL_1592266, EPI_ISL_1592268, EPI_ISL_1592269, EPI_ISL_1592270, EPI_ISL_1592271, EPI_ISL_1592272, EPI_ISL_1592274, EPI_ISL_1592275, EPI_ISL_1592279, EPI_ISL_1592280, EPI_ISL_1592283, EPI_ISL_1592286, EPI_ISL_1592288, EPI_ISL_1592290, EPI_ISL_1592291, EPI_ISL_1592292, EPI_ISL_1592294, EPI_ISL_1592297, EPI_ISL_1592298, EPI_ISL_1592299, EPI_ISL_1592301, EPI_ISL_1592302, EPI_ISL_1592303, EPI_ISL_1592305, EPI_ISL_1592306, EPI_ISL_1592307, EPI_ISL_1592308, EPI_ISL_1592309, EPI_ISL_1592311, EPI_ISL_1592314, EPI_ISL_1592315, EPI_ISL_1592316, EPI_ISL_1592318, EPI_ISL_1592319, EPI_ISL_1592320, | | | |

| | | | |
|---|---|---|---|
| EPI_ISL_1592321, EPI_ISL_1592322, EPI_ISL_1592323, EPI_ISL_1592324, EPI_ISL_1592325, EPI_ISL_1592326, EPI_ISL_1592329, EPI_ISL_1592330, EPI_ISL_1592331, EPI_ISL_1592333, EPI_ISL_1592335, EPI_ISL_1592336, EPI_ISL_1592337, EPI_ISL_1592338, EPI_ISL_1592339, EPI_ISL_1592340, EPI_ISL_1592341, EPI_ISL_1592342, EPI_ISL_1592343, EPI_ISL_1592345, EPI_ISL_1592347, EPI_ISL_1592348, EPI_ISL_1592351, EPI_ISL_1592352, EPI_ISL_1592354, EPI_ISL_1592357, EPI_ISL_1592359, EPI_ISL_1592360, EPI_ISL_1592363, EPI_ISL_1592365, EPI_ISL_1592367, EPI_ISL_1592368, EPI_ISL_1592370, EPI_ISL_1592371, EPI_ISL_1592373, EPI_ISL_1592374, EPI_ISL_1592375, EPI_ISL_1592376, EPI_ISL_1592377, EPI_ISL_1592378, EPI_ISL_1592384, EPI_ISL_1592386, EPI_ISL_1592387, EPI_ISL_1592389, EPI_ISL_1592390, EPI_ISL_1592392, EPI_ISL_1592393, EPI_ISL_1592394, EPI_ISL_1592395, EPI_ISL_1592397, EPI_ISL_1592398, EPI_ISL_1592399, EPI_ISL_1592401, EPI_ISL_1592402, EPI_ISL_1592404 | | | |
| see above | Arizona State University | Arizona State University | Peter T. Skidmore, LaRinda A. Holland, Rabia Maqsood, Nicholas J. Mellor, Joy M. Blain, Valerie Harris, Joshua LaBaer, Vel Murugan, Efrem S. Lim |
| EPI_ISL_1592405, EPI_ISL_1592406, EPI_ISL_1592407, EPI_ISL_1592408, EPI_ISL_1592409, EPI_ISL_1592410, EPI_ISL_1592411, EPI_ISL_1592412, EPI_ISL_1592413, EPI_ISL_1592414, EPI_ISL_1592415, EPI_ISL_1592416, EPI_ISL_1592417, EPI_ISL_1592418, EPI_ISL_1592419, EPI_ISL_1592420, EPI_ISL_1592421, EPI_ISL_1592422, EPI_ISL_1592423, EPI_ISL_1592424, EPI_ISL_1592425, EPI_ISL_1592426, EPI_ISL_1592427, EPI_ISL_1592428, EPI_ISL_1592429, EPI_ISL_1592430, EPI_ISL_1592431, EPI_ISL_1592432, EPI_ISL_1592433, EPI_ISL_1592434, EPI_ISL_1592435, EPI_ISL_1592436, EPI_ISL_1592437, EPI_ISL_1592438, EPI_ISL_1592439, EPI_ISL_1592440, EPI_ISL_1592441, EPI_ISL_1592442, EPI_ISL_1592443, EPI_ISL_1592444, EPI_ISL_1592445, EPI_ISL_1592446, EPI_ISL_1592447, EPI_ISL_1592448, EPI_ISL_1592449, EPI_ISL_1592451, EPI_ISL_1592452, EPI_ISL_1592453, EPI_ISL_1592454, EPI_ISL_1592455, EPI_ISL_1592456, EPI_ISL_1592457, EPI_ISL_1592458, EPI_ISL_1592459, EPI_ISL_1592460, EPI_ISL_1592461, EPI_ISL_1592462, EPI_ISL_1592463, EPI_ISL_1592464, EPI_ISL_1592465, EPI_ISL_1592466, EPI_ISL_1592467, EPI_ISL_1592468, EPI_ISL_1592469, EPI_ISL_1592470, EPI_ISL_1592471, EPI_ISL_1592472, EPI_ISL_1592473, EPI_ISL_1592474, EPI_ISL_1592475, EPI_ISL_1592476, EPI_ISL_1592477, EPI_ISL_1592478, EPI_ISL_1592479, EPI_ISL_1592480, EPI_ISL_1592481, EPI_ISL_1592482, EPI_ISL_1592483, EPI_ISL_1592484, EPI_ISL_1592486, EPI_ISL_1592487, EPI_ISL_1592488, EPI_ISL_1592489, EPI_ISL_1592490, EPI_ISL_1592491, EPI_ISL_1592492, EPI_ISL_1592493, EPI_ISL_1592494, EPI_ISL_1592495, EPI_ISL_1592496, EPI_ISL_1592497, EPI_ISL_1592498, EPI_ISL_1592499, EPI_ISL_1592500, EPI_ISL_1592501, EPI_ISL_1592502, EPI_ISL_1592503, EPI_ISL_1592504, EPI_ISL_1592505, EPI_ISL_1592506, EPI_ISL_1592507, EPI_ISL_1592508, EPI_ISL_1592509, EPI_ISL_1592510, EPI_ISL_1592511, EPI_ISL_1592512, EPI_ISL_1592513, EPI_ISL_1592514, EPI_ISL_1592515, EPI_ISL_1592516, EPI_ISL_1592517, EPI_ISL_1592518, EPI_ISL_1592519, EPI_ISL_1592520, EPI_ISL_1592521, EPI_ISL_1592522, EPI_ISL_1592523, EPI_ISL_1592524, EPI_ISL_1592525, EPI_ISL_1592526, EPI_ISL_1592527, EPI_ISL_1592528, EPI_ISL_1592529, EPI_ISL_1592530, EPI_ISL_1592531, EPI_ISL_1592532, EPI_ISL_1592533, EPI_ISL_1592534, EPI_ISL_1592535, EPI_ISL_1592536, EPI_ISL_1592537, EPI_ISL_1592538, EPI_ISL_1592539, EPI_ISL_1592540, EPI_ISL_1592541, EPI_ISL_1592542, EPI_ISL_1592543, EPI_ISL_1592544, EPI_ISL_1592545, EPI_ISL_1592546, EPI_ISL_1592547, EPI_ISL_1592548, EPI_ISL_1592549, EPI_ISL_1592550, EPI_ISL_1592551, EPI_ISL_1592552, EPI_ISL_1592553, EPI_ISL_1592554, EPI_ISL_1592555, EPI_ISL_1592556, EPI_ISL_1592557, EPI_ISL_1592558, EPI_ISL_1592559, EPI_ISL_1592560, EPI_ISL_1592562, EPI_ISL_1592563, EPI_ISL_1592564, EPI_ISL_1592565, EPI_ISL_1592566, EPI_ISL_1592567, EPI_ISL_1592568, EPI_ISL_1592569, EPI_ISL_1592570, EPI_ISL_1592571, EPI_ISL_1592572, EPI_ISL_1592573, EPI_ISL_1592574, EPI_ISL_1592575, EPI_ISL_1592577, EPI_ISL_1592578, EPI_ISL_1592579, EPI_ISL_1592580, EPI_ISL_1592581, EPI_ISL_1592582, EPI_ISL_1592583, EPI_ISL_1592584, EPI_ISL_1592585, EPI_ISL_1592586, EPI_ISL_1592587, EPI_ISL_1592588, EPI_ISL_1592589, EPI_ISL_1592590, EPI_ISL_1592591, EPI_ISL_1592592, EPI_ISL_1592593, EPI_ISL_1592594, EPI_ISL_1592595, EPI_ISL_1592596, EPI_ISL_1592598, EPI_ISL_1592599, EPI_ISL_1592600, EPI_ISL_1592601, EPI_ISL_1592602, EPI_ISL_1592603, EPI_ISL_1592604, EPI_ISL_1592605, EPI_ISL_1592606, EPI_ISL_1592607, EPI_ISL_1592608, EPI_ISL_1592609, EPI_ISL_1592610, EPI_ISL_1592611, EPI_ISL_1592612, EPI_ISL_1592613, EPI_ISL_1592614, EPI_ISL_1592615, EPI_ISL_1592616, EPI_ISL_1592617, EPI_ISL_1592618, EPI_ISL_1592619, EPI_ISL_1592620, EPI_ISL_1592621, EPI_ISL_1592622, EPI_ISL_1592623, EPI_ISL_1592624, EPI_ISL_1592626, EPI_ISL_1592627, EPI_ISL_1592628, EPI_ISL_1592629, EPI_ISL_1592630, EPI_ISL_1592631, EPI_ISL_1592632, EPI_ISL_1592633, EPI_ISL_1592634, EPI_ISL_1592635, EPI_ISL_1592636, EPI_ISL_1592637, EPI_ISL_1592638, EPI_ISL_1592639, EPI_ISL_1592640, EPI_ISL_1592641, EPI_ISL_1592642, EPI_ISL_1592643, EPI_ISL_1592644, EPI_ISL_1592645, EPI_ISL_1592646, EPI_ISL_1592647, EPI_ISL_1592648, EPI_ISL_1592649, EPI_ISL_1592650, EPI_ISL_1592651, EPI_ISL_1592652, EPI_ISL_1592653, EPI_ISL_1592654, EPI_ISL_1592655, EPI_ISL_1592656, EPI_ISL_1592657, EPI_ISL_1592658, EPI_ISL_1592659, EPI_ISL_1592660, EPI_ISL_1592661, EPI_ISL_1592662, EPI_ISL_1592663, EPI_ISL_1592664, EPI_ISL_1592665, EPI_ISL_1592666, EPI_ISL_1592667, EPI_ISL_1592668, EPI_ISL_1592669, EPI_ISL_1592670, EPI_ISL_1592671, EPI_ISL_1592672, EPI_ISL_1592673, EPI_ISL_1592674, EPI_ISL_1592675, EPI_ISL_1592676, EPI_ISL_1592677, EPI_ISL_1592678, EPI_ISL_1592679, EPI_ISL_1592680, EPI_ISL_1592681, EPI_ISL_1592682, EPI_ISL_1592683, EPI_ISL_1592684, EPI_ISL_1592685, EPI_ISL_1592686, EPI_ISL_1592687, EPI_ISL_1592688, EPI_ISL_1592689, EPI_ISL_1592690, EPI_ISL_1592692, EPI_ISL_1592693, EPI_ISL_1592694, EPI_ISL_1592695, EPI_ISL_1592696, EPI_ISL_1592697, EPI_ISL_1592698, EPI_ISL_1592699, EPI_ISL_1592700, EPI_ISL_1592701, EPI_ISL_1592702, EPI_ISL_1592703, EPI_ISL_1592704, EPI_ISL_1592705, EPI_ISL_1592706, EPI_ISL_1592707, EPI_ISL_1592708, EPI_ISL_1592709, EPI_ISL_1592710, EPI_ISL_1592711, EPI_ISL_1592712, EPI_ISL_1592713, EPI_ISL_1592714, EPI_ISL_1592715, EPI_ISL_1592716, EPI_ISL_1592717, EPI_ISL_1592718, EPI_ISL_1592719, EPI_ISL_1592720, EPI_ISL_1592721, EPI_ISL_1592722, EPI_ISL_1592723, EPI_ISL_1592724, EPI_ISL_1592725, EPI_ISL_1592726, EPI_ISL_1592727, EPI_ISL_1592728, EPI_ISL_1592729, EPI_ISL_1592730, EPI_ISL_1592731, EPI_ISL_1592732, EPI_ISL_1592733, EPI_ISL_1592734, EPI_ISL_1592735, EPI_ISL_1592736, EPI_ISL_1592737, EPI_ISL_1592738, EPI_ISL_1592739, EPI_ISL_1592740, EPI_ISL_1592741, EPI_ISL_1592743, EPI_ISL_1592744, EPI_ISL_1592745, EPI_ISL_1592746, EPI_ISL_1592747, EPI_ISL_1592748, EPI_ISL_1592749, EPI_ISL_1592750, EPI_ISL_1592751, EPI_ISL_1592752, EPI_ISL_1592753, EPI_ISL_1592754, EPI_ISL_1592755, EPI_ISL_1592756, EPI_ISL_1592757, EPI_ISL_1592758, EPI_ISL_1592759, EPI_ISL_1592760, EPI_ISL_1592762, EPI_ISL_1592763, EPI_ISL_1592764, EPI_ISL_1592765, EPI_ISL_1592766, EPI_ISL_1592767, EPI_ISL_1592768, EPI_ISL_1592769, EPI_ISL_1592770, EPI_ISL_1592771, EPI_ISL_1592772, EPI_ISL_1592773, EPI_ISL_1592774, EPI_ISL_1592775, EPI_ISL_1592776, EPI_ISL_1592777, EPI_ISL_1592778, EPI_ISL_1592779, EPI_ISL_1592780, EPI_ISL_1592781, EPI_ISL_1592782, EPI_ISL_1592783, EPI_ISL_1592784, EPI_ISL_1592785, EPI_ISL_1592786, EPI_ISL_1592787, EPI_ISL_1592788, EPI_ISL_1592789, EPI_ISL_1592790, EPI_ISL_1592791, EPI_ISL_1592792, EPI_ISL_1592793, EPI_ISL_1592794, EPI_ISL_1592795, EPI_ISL_1592796, EPI_ISL_1592797, EPI_ISL_1592798, EPI_ISL_1592799, EPI_ISL_1592800, EPI_ISL_1592801, EPI_ISL_1592802, EPI_ISL_1592803, EPI_ISL_1592804, EPI_ISL_1592805, EPI_ISL_1592806, EPI_ISL_1592807, EPI_ISL_1592808, EPI_ISL_1592809, EPI_ISL_1592810, EPI_ISL_1592811, EPI_ISL_1592812, EPI_ISL_1592813, EPI_ISL_1592814, EPI_ISL_1592815, EPI_ISL_1592816, EPI_ISL_1592817, EPI_ISL_1592818, EPI_ISL_1592819, EPI_ISL_1592820, EPI_ISL_1592821, EPI_ISL_1592822, EPI_ISL_1592823, EPI_ISL_1592824, EPI_ISL_1592825, EPI_ISL_1592826, EPI_ISL_1592827, EPI_ISL_1592828, EPI_ISL_1592829, EPI_ISL_1592830, EPI_ISL_1592831, EPI_ISL_1592832, EPI_ISL_1592833, EPI_ISL_1592834, EPI_ISL_1592835, EPI_ISL_1592836, EPI_ISL_1592837, EPI_ISL_1592838, EPI_ISL_1592839, EPI_ISL_1592840, EPI_ISL_1592841, EPI_ISL_1592842, EPI_ISL_1592843, EPI_ISL_1592844, EPI_ISL_1592845, EPI_ISL_1592846, EPI_ISL_1592847, EPI_ISL_1592848, EPI_ISL_1592849, EPI_ISL_1592850, EPI_ISL_1592851, EPI_ISL_1592852, EPI_ISL_1592853, EPI_ISL_1592854, EPI_ISL_1592855, EPI_ISL_1592856, EPI_ISL_1592857, EPI_ISL_1592858, EPI_ISL_1592859, EPI_ISL_1592860, EPI_ISL_1592861, EPI_ISL_1592862, EPI_ISL_1592863, EPI_ISL_1592864, EPI_ISL_1592865, EPI_ISL_1592866, EPI_ISL_1592867, EPI_ISL_1592868, EPI_ISL_1592869, EPI_ISL_1592870, EPI_ISL_1592871, EPI_ISL_1592872, EPI_ISL_1592873, EPI_ISL_1592874, EPI_ISL_1592875, EPI_ISL_1592876, EPI_ISL_1592877, EPI_ISL_1592878, EPI_ISL_1592879, EPI_ISL_1592880, EPI_ISL_1592881, EPI_ISL_1592882, EPI_ISL_1592883, EPI_ISL_1592884, EPI_ISL_1592885, EPI_ISL_1592886, EPI_ISL_1592887, EPI_ISL_1592888, EPI_ISL_1592889, EPI_ISL_1592890, EPI_ISL_1592891, EPI_ISL_1592892, EPI_ISL_1592893, EPI_ISL_1592894, EPI_ISL_1592895, EPI_ISL_1592896, EPI_ISL_1592897, EPI_ISL_1592898, EPI_ISL_1592899, EPI_ISL_1592900, EPI_ISL_1592901, EPI_ISL_1592902, EPI_ISL_1592903, EPI_ISL_1592904, EPI_ISL_1592905, EPI_ISL_1592906, EPI_ISL_1592907, EPI_ISL_1592908, EPI_ISL_1592909, EPI_ISL_1592910, EPI_ISL_1592911, EPI_ISL_1592912, EPI_ISL_1592913, EPI_ISL_1592914, EPI_ISL_1592915, EPI_ISL_1592916, EPI_ISL_1592917, EPI_ISL_1592918, EPI_ISL_1592919, EPI_ISL_1592920, EPI_ISL_1592921, EPI_ISL_1592922, EPI_ISL_1592923, EPI_ISL_1592924, EPI_ISL_1592925, EPI_ISL_1592927, EPI_ISL_1592928, EPI_ISL_1592929, EPI_ISL_1592930, EPI_ISL_1592931, EPI_ISL_1592932, EPI_ISL_1592933, EPI_ISL_1592934, EPI_ISL_1592935, EPI_ISL_1592936, EPI_ISL_1592938, EPI_ISL_1592939, EPI_ISL_1592940, EPI_ISL_1592941, EPI_ISL_1592942, EPI_ISL_1592943, EPI_ISL_1592944, EPI_ISL_1592945, EPI_ISL_1592946, EPI_ISL_1592947, EPI_ISL_1592948, EPI_ISL_1592949, EPI_ISL_1592950, EPI_ISL_1592951, EPI_ISL_1592952, EPI_ISL_1592953, EPI_ISL_1592954, EPI_ISL_1592955, EPI_ISL_1592956, EPI_ISL_1592957, EPI_ISL_1592958, EPI_ISL_1592959, EPI_ISL_1592960, EPI_ISL_1592961, EPI_ISL_1592962, EPI_ISL_1592963, EPI_ISL_1592964, EPI_ISL_1592965, EPI_ISL_1592966, EPI_ISL_1592967, EPI_ISL_1592968, EPI_ISL_1592969, EPI_ISL_1592970, EPI_ISL_1592971, EPI_ISL_1592972, EPI_ISL_1592973, EPI_ISL_1592974, EPI_ISL_1592975, EPI_ISL_1592976, EPI_ISL_1592977, EPI_ISL_1592978, EPI_ISL_1592979, EPI_ISL_1592980, EPI_ISL_1592981, EPI_ISL_1592982, EPI_ISL_1592983, EPI_ISL_1592984, EPI_ISL_1592985, EPI_ISL_1592986, EPI_ISL_1592987, EPI_ISL_1592988, EPI_ISL_1592989, EPI_ISL_1592990, EPI_ISL_1592991, EPI_ISL_1592992, EPI_ISL_1592993, EPI_ISL_1592994, EPI_ISL_1592995, EPI_ISL_1592996, EPI_ISL_1592997, EPI_ISL_1592998, EPI_ISL_1592999, EPI_ISL_1593000, EPI_ISL_1593001, EPI_ISL_1593002, EPI_ISL_1593003, EPI_ISL_1593004, EPI_ISL_1593005, EPI_ISL_1593006, EPI_ISL_1593007, EPI_ISL_1593008, EPI_ISL_1593009, EPI_ISL_1593010, EPI_ISL_1593011, EPI_ISL_1593012, EPI_ISL_1593013, EPI_ISL_1593014, EPI_ISL_1593015, EPI_ISL_1593016, EPI_ISL_1593017, EPI_ISL_1593018, EPI_ISL_1593019, EPI_ISL_1593020, EPI_ISL_1593021, EPI_ISL_1593022, EPI_ISL_1593023, EPI_ISL_1593024, EPI_ISL_1593025, EPI_ISL_1593026, EPI_ISL_1593027, EPI_ISL_1593028, EPI_ISL_1593029, EPI_ISL_1593030, EPI_ISL_1593031, EPI_ISL_1593032, EPI_ISL_1593033, EPI_ISL_1593034, EPI_ISL_1593035, EPI_ISL_1593036, EPI_ISL_1593037, EPI_ISL_1593038, EPI_ISL_1593039, EPI_ISL_1593040, EPI_ISL_1593041, EPI_ISL_1593042, EPI_ISL_1593043, EPI_ISL_1593044, EPI_ISL_1593045, EPI_ISL_1593046, EPI_ISL_1593047, EPI_ISL_1593048, EPI_ISL_1593049, EPI_ISL_1593050, EPI_ISL_1593051, EPI_ISL_1593052, EPI_ISL_1593053, EPI_ISL_1593054, EPI_ISL_1593055, EPI_ISL_1593056, EPI_ISL_1593057, EPI_ISL_1593058, EPI_ISL_1593059, EPI_ISL_1593060, EPI_ISL_1593061 | | | |
| see above | Helix/Illumina | Centers for Disease Control and Prevention Division of Viral Diseases, Pathogen Discovery | Dakota Howard, Dhwani Batra, Peter W. Cook, Kara Moser, Adrian Paskey, Jason Caravas, Benjamin Rambo-Martin, Shatavia Morrison, Christopher Gulvick, Scott Sammons, Yvette Unoarumhi, Darlene Wagner, Matthew Schmerer, Eileen de Feo, Jan Antico, Christine Tran, Matthew Tolentino, Shannon Wickline, Kim Gietzen, Brad Sickler, Jingtao Liu, Eric Allen, Phil Febbo, Nicole L. Washington, Simon White, Geraint Levan, Kelly Schiabor Barrett, Elizabeth Cirulli, Alexandre Bolze, Ary Ascencio, Charlotte Rivera-Garcia, Ryan Cho, Jason Nguyen, Sherry Wang, Jimmy Ramirez, Tyler Cassens, Efren Sandoval, Magnus Isaksson, William Lee, David Becker, Marc Laurent, James Lu, Clinton R. Paden, Duncan MacCannell |
| EPI_ISL_1593062 | University of Texas at Austin Genome Sequencing and Analysis Facility (UTGSAF) | University of Texas at Austin Genome Sequencing and Analysis Facility (UTGSAF) | Jessica Podnar, Sylvie Beaudenon, Audrey Kelly, Zachary Carver, Anna Battenhouse, Andreas Matouschek |
| EPI_ISL_1593063 | Helix/Illumina | Centers for Disease Control and Prevention Division of Viral Diseases, Pathogen Discovery | Dakota Howard, Dhwani Batra, Peter W. Cook, Kara Moser, Adrian Paskey, Jason Caravas, Benjamin Rambo-Martin, Shatavia Morrison, Christopher Gulvick, Scott Sammons, Yvette Unoarumhi, Darlene Wagner, Matthew Schmerer, Eileen de Feo, Jan Antico, Christine Tran, Matthew Tolentino, Shannon Wickline, Kim Gietzen, Brad Sickler, Jingtao Liu, Eric Allen, Phil Febbo, Nicole L. Washington, Simon White, Geraint Levan, Kelly Schiabor Barrett, Elizabeth Cirulli, Alexandre Bolze, Ary Ascencio, Charlotte Rivera-Garcia, Ryan Cho, Jason Nguyen, Sherry Wang, Jimmy Ramirez, Tyler Cassens, Efren Sandoval, Magnus Isaksson, William Lee, David Becker, Marc Laurent, James Lu, Clinton R. Paden, Duncan MacCannell |
| EPI_ISL_1593064 | University of Texas at Austin Genome Sequencing and Analysis Facility (UTGSAF) | University of Texas at Austin Genome Sequencing and Analysis Facility (UTGSAF) | Jessica Podnar, Sylvie Beaudenon, Audrey Kelly, Zachary Carver, Anna Battenhouse, Andreas Matouschek |
| EPI_ISL_1593065 | Helix/Illumina | Centers for Disease Control and Prevention Division of Viral Diseases, Pathogen Discovery | Dakota Howard, Dhwani Batra, Peter W. Cook, Kara Moser, Adrian Paskey, Jason Caravas, Benjamin Rambo-Martin, Shatavia Morrison, Christopher Gulvick, Scott Sammons, Yvette Unoarumhi, Darlene Wagner, Matthew Schmerer, Eileen de Feo, Jan Antico, Christine Tran, Matthew Tolentino, Shannon Wickline, Kim Gietzen, Brad Sickler, Jingtao Liu, Eric Allen, Phil Febbo, Nicole L. Washington, Simon White, Geraint Levan, Kelly Schiabor Barrett, Elizabeth Cirulli, Alexandre Bolze, Ary Ascencio, Charlotte Rivera-Garcia, Ryan Cho, Jason Nguyen, Sherry Wang, Jimmy Ramirez, Tyler Cassens, Efren Sandoval, Magnus Isaksson, William Lee, David Becker, Marc Laurent, James Lu, Clinton R. Paden, Duncan MacCannell |
| EPI_ISL_1593066, EPI_ISL_1593067 | University of Texas at Austin Genome Sequencing and Analysis Facility (UTGSAF) | University of Texas at Austin Genome Sequencing and Analysis Facility (UTGSAF) | Jessica Podnar, Sylvie Beaudenon, Audrey Kelly, Zachary Carver, Anna Battenhouse, Andreas Matouschek |
| EPI_ISL_1593068 | Helix/Illumina | Centers for Disease Control and Prevention Division of Viral Diseases, Pathogen Discovery | Dakota Howard, Dhwani Batra, Peter W. Cook, Kara Moser, Adrian Paskey, Jason Caravas, Benjamin Rambo-Martin, Shatavia Morrison, Christopher Gulvick, Scott Sammons, Yvette Unoarumhi, Darlene Wagner, Matthew Schmerer, Eileen de Feo, Jan Antico, Christine Tran, Matthew Tolentino, Shannon Wickline, Kim Gietzen, Brad Sickler, Jingtao Liu, Eric Allen, Phil Febbo, Nicole L. Washington, Simon White, Geraint Levan, Kelly Schiabor Barrett, Elizabeth Cirulli, Alexandre Bolze, Ary Ascencio, Charlotte Rivera-Garcia, Ryan Cho, Jason Nguyen, Sherry Wang, Jimmy Ramirez, Tyler Cassens, Efren Sandoval, Magnus Isaksson, William Lee, David Becker, Marc Laurent, James Lu, Clinton R. Paden, Duncan MacCannell |
| EPI_ISL_1593070 | University of Texas at Austin Genome Sequencing and Analysis Facility (UTGSAF) | University of Texas at Austin Genome Sequencing and Analysis Facility (UTGSAF) | Jessica Podnar, Sylvie Beaudenon, Audrey Kelly, Zachary Carver, Anna Battenhouse, Andreas Matouschek |
| EPI_ISL_1593071 | Helix/Illumina | Centers for Disease Control and Prevention Division of Viral Diseases, Pathogen Discovery | Dakota Howard, Dhwani Batra, Peter W. Cook, Kara Moser, Adrian Paskey, Jason Caravas, Benjamin Rambo-Martin, Shatavia Morrison, Christopher Gulvick, Scott Sammons, Yvette Unoarumhi, Darlene Wagner, Matthew Schmerer, Eileen de Feo, Jan Antico, Christine Tran, Matthew Tolentino, Shannon Wickline, Kim Gietzen, Brad Sickler, Jingtao Liu, Eric Allen, Phil Febbo, Nicole L. Washington, Simon White, Geraint Levan, Kelly Schiabor Barrett, Elizabeth Cirulli, Alexandre Bolze, Ary Ascencio, Charlotte Rivera-Garcia, Ryan Cho, Jason Nguyen, Sherry Wang, Jimmy Ramirez, Tyler Cassens, Efren Sandoval, Magnus Isaksson, William Lee, David Becker, Marc Laurent, James Lu, Clinton R. Paden, Duncan MacCannell |
| EPI_ISL_1593073 | University of Texas at Austin Genome Sequencing and Analysis Facility (UTGSAF) | University of Texas at Austin Genome Sequencing and Analysis Facility (UTGSAF) | Jessica Podnar, Sylvie Beaudenon, Audrey Kelly, Zachary Carver, Anna Battenhouse, Andreas Matouschek |
| EPI_ISL_1593074 | Helix/Illumina | Centers for Disease Control and Prevention Division of Viral Diseases, Pathogen Discovery | Dakota Howard, Dhwani Batra, Peter W. Cook, Kara Moser, Adrian Paskey, Jason Caravas, Benjamin Rambo-Martin, Shatavia Morrison, Christopher Gulvick, Scott Sammons, Yvette Unoarumhi, Darlene Wagner, Matthew Schmerer, Eileen de Feo, Jan Antico, Christine Tran, Matthew Tolentino, Shannon Wickline, Kim Gietzen, Brad Sickler, Jingtao Liu, Eric Allen, Phil Febbo, Nicole L. Washington, Simon White, Geraint Levan, Kelly Schiabor Barrett, Elizabeth Cirulli, Alexandre Bolze, Ary Ascencio, Charlotte Rivera-Garcia, Ryan Cho, Jason Nguyen, Sherry Wang, Jimmy Ramirez, Tyler Cassens, Efren Sandoval, Magnus Isaksson, William Lee, David Becker, Marc Laurent, James Lu, Clinton R. Paden, Duncan MacCannell |
| EPI_ISL_1593076 | University of Texas at Austin Genome Sequencing and Analysis Facility (UTGSAF) | University of Texas at Austin Genome Sequencing and Analysis Facility (UTGSAF) | Jessica Podnar, Sylvie Beaudenon, Audrey Kelly, Zachary Carver, Anna Battenhouse, Andreas Matouschek |
| EPI_ISL_1593077, EPI_ISL_1593079 | Helix/Illumina | Centers for Disease Control and Prevention Division of Viral Diseases, Pathogen Discovery | Dakota Howard, Dhwani Batra, Peter W. Cook, Kara Moser, Adrian Paskey, Jason Caravas, Benjamin Rambo-Martin, Shatavia Morrison, Christopher Gulvick, Scott Sammons, Yvette Unoarumhi, Darlene Wagner, Matthew Schmerer, Eileen de Feo, Jan Antico, Christine Tran, Matthew Tolentino, Shannon Wickline, Kim Gietzen, Brad Sickler, Jingtao Liu, Eric Allen, Phil Febbo, Nicole L. Washington, Simon White, Geraint Levan, Kelly Schiabor Barrett, Elizabeth Cirulli, Alexandre Bolze, Ary Ascencio, Charlotte Rivera-Garcia, Ryan Cho, Jason Nguyen, Sherry Wang, Jimmy Ramirez, Tyler Cassens, Efren Sandoval, Magnus Isaksson, William Lee, David Becker, Marc Laurent, James Lu, Clinton R. Paden, Duncan MacCannell |
| EPI_ISL_1593080, EPI_ISL_1593081 | University of Texas at Austin Genome Sequencing and Analysis Facility (UTGSAF) | University of Texas at Austin Genome Sequencing and Analysis Facility (UTGSAF) | Jessica Podnar, Sylvie Beaudenon, Audrey Kelly, Zachary Carver, Anna Battenhouse, Andreas Matouschek |
| EPI_ISL_1593082 | Helix/Illumina | Centers for Disease Control and Prevention Division of Viral Diseases, Pathogen Discovery | Dakota Howard, Dhwani Batra, Peter W. Cook, Kara Moser, Adrian Paskey, Jason Caravas, Benjamin Rambo-Martin, Shatavia Morrison, Christopher Gulvick, Scott Sammons, Yvette Unoarumhi, Darlene Wagner, Matthew Schmerer, Eileen de Feo, Jan Antico, Christine Tran, Matthew Tolentino, Shannon Wickline, Kim Gietzen, Brad Sickler, Jingtao Liu, Eric Allen, Phil Febbo, Nicole L. Washington, Simon White, Geraint Levan, Kelly Schiabor Barrett, Elizabeth Cirulli, Alexandre Bolze, Ary Ascencio, Charlotte Rivera-Garcia, Ryan Cho, Jason Nguyen, Sherry Wang, Jimmy Ramirez, Tyler Cassens, Efren Sandoval, Magnus Isaksson, William Lee, David Becker, Marc Laurent, James Lu, Clinton R. Paden, Duncan MacCannell |
| EPI_ISL_1593083, EPI_ISL_1593084 | University of Texas at Austin Genome Sequencing and Analysis Facility (UTGSAF) | University of Texas at Austin Genome Sequencing and Analysis Facility (UTGSAF) | Jessica Podnar, Sylvie Beaudenon, Audrey Kelly, Zachary Carver, Anna Battenhouse, Andreas Matouschek |
| EPI_ISL_1593085 | Helix/Illumina | Centers for Disease Control and Prevention Division of Viral Diseases, Pathogen Discovery | Dakota Howard, Dhwani Batra, Peter W. Cook, Kara Moser, Adrian Paskey, Jason Caravas, Benjamin Rambo-Martin, Shatavia Morrison, Christopher Gulvick, Scott Sammons, Yvette Unoarumhi, Darlene Wagner, Matthew Schmerer, Eileen de Feo, Jan Antico, Christine Tran, Matthew Tolentino, Shannon Wickline, Kim Gietzen, Brad Sickler, Jingtao Liu, Eric Allen, Phil Febbo, Nicole L. Washington, Simon White, Geraint Levan, Kelly Schiabor Barrett, Elizabeth Cirulli, Alexandre Bolze, Ary Ascencio, Charlotte Rivera-Garcia, Ryan Cho, Jason Nguyen, Sherry Wang, Jimmy Ramirez, Tyler Cassens, Efren Sandoval, Magnus Isaksson, William Lee, David Becker, Marc Laurent, James Lu, Clinton R. Paden, Duncan MacCannell |
| EPI_ISL_1593086 | University of Texas at Austin Genome Sequencing and Analysis Facility (UTGSAF) | University of Texas at Austin Genome Sequencing and Analysis Facility (UTGSAF) | Jessica Podnar, Sylvie Beaudenon, Audrey Kelly, Zachary Carver, Anna Battenhouse, Andreas Matouschek |
| EPI_ISL_1593087 | Helix/Illumina | Centers for Disease Control and Prevention Division of Viral Diseases, Pathogen Discovery | Dakota Howard, Dhwani Batra, Peter W. Cook, Kara Moser, Adrian Paskey, Jason Caravas, Benjamin Rambo-Martin, Shatavia Morrison, Christopher Gulvick, Scott Sammons, Yvette Unoarumhi, Darlene Wagner, Matthew Schmerer, Eileen de Feo, Jan Antico, Christine Tran, Matthew Tolentino, Shannon Wickline, Kim Gietzen, Brad Sickler, Jingtao Liu, Eric Allen, Phil Febbo, Nicole L. Washington, Simon White, Geraint Levan, Kelly Schiabor Barrett, Elizabeth Cirulli, Alexandre Bolze, Ary Ascencio, Charlotte Rivera-Garcia, Ryan Cho, Jason Nguyen, Sherry Wang, Jimmy Ramirez, Tyler Cassens, Efren Sandoval, Magnus Isaksson, William Lee, David Becker, Marc Laurent, James Lu, Clinton R. Paden, Duncan MacCannell |
| EPI_ISL_1593088, EPI_ISL_1593089 | University of Texas at Austin Genome Sequencing and Analysis Facility (UTGSAF) | University of Texas at Austin Genome Sequencing and Analysis Facility (UTGSAF) | Jessica Podnar, Sylvie Beaudenon, Audrey Kelly, Zachary Carver, Anna Battenhouse, Andreas Matouschek |
| EPI_ISL_1593090, EPI_ISL_1593093 | Helix/Illumina | Centers for Disease Control and Prevention Division of Viral Diseases, Pathogen Discovery | Dakota Howard, Dhwani Batra, Peter W. Cook, Kara Moser, Adrian Paskey, Jason Caravas, Benjamin Rambo-Martin, Shatavia Morrison, Christopher Gulvick, Scott Sammons, Yvette Unoarumhi, Darlene Wagner, Matthew Schmerer, Eileen de Feo, Jan Antico, Christine Tran, Matthew Tolentino, Shannon Wickline, Kim Gietzen, Brad Sickler, Jingtao Liu, Eric Allen, Phil Febbo, Nicole L. Washington, Simon White, Geraint Levan, Kelly Schiabor Barrett, Elizabeth Cirulli, Alexandre Bolze, Ary Ascencio, Charlotte Rivera-Garcia, Ryan Cho, Jason Nguyen, Sherry Wang, Jimmy Ramirez, Tyler Cassens, Efren Sandoval, Magnus Isaksson, William Lee, David Becker, Marc Laurent, James Lu, Clinton R. Paden, Duncan MacCannell |
| EPI_ISL_1593094, EPI_ISL_1593095 | University of Texas at Austin Genome Sequencing and Analysis Facility (UTGSAF) | University of Texas at Austin Genome Sequencing and Analysis Facility (UTGSAF) | Jessica Podnar, Sylvie Beaudenon, Audrey Kelly, Zachary Carver, Anna Battenhouse, Andreas Matouschek |

| | | | |
|---|---|---|---|
| EPI_ISL_1593096 | Helix/Illumina | Centers for Disease Control and Prevention Division of Viral Diseases, Pathogen Discovery | Dakota Howard, Dhwani Batra, Peter W. Cook, Kara Moser, Adrian Paskey, Jason Caravas, Benjamin Rambo-Martin, Shatavia Morrison, Christopher Gulvick, Scott Sammons, Yvette Unoarumhi, Darlene Wagner, Matthew Schmerer, Eileen de Feo, Jan Antico, Christine Tran, Matthew Tolentino, Shannon Wickline, Kim Gietzen, Brad Sickler, Jingtao Liu, Eric Allen, Phil Febbo, Nicole L. Washington, Simon White, Geraint Levan, Kelly Schiabor Barrett, Elizabeth Cirulli, Alexandre Bolze, Ary Ascencio, Charlotte Rivera-Garcia, Ryan Cho, Jason Nguyen, Sherry Wang, Jimmy Ramirez, Tyler Cassens, Efren Sandoval, Magnus Isaksson, William Lee, David Becker, Marc Laurent, James Lu, Clinton R. Paden, Duncan MacCannell |
| EPI_ISL_1593097 | University of Texas at Austin Genome Sequencing and Analysis Facility (UTGSAF) | University of Texas at Austin Genome Sequencing and Analysis Facility (UTGSAF) | Jessica Podnar, Sylvie Beaudenon, Audrey Kelly, Zachary Carver, Anna Battenhouse, Andreas Matouschek |
| EPI_ISL_1593098 | Helix/Illumina | Centers for Disease Control and Prevention Division of Viral Diseases, Pathogen Discovery | Dakota Howard, Dhwani Batra, Peter W. Cook, Kara Moser, Adrian Paskey, Jason Caravas, Benjamin Rambo-Martin, Shatavia Morrison, Christopher Gulvick, Scott Sammons, Yvette Unoarumhi, Darlene Wagner, Matthew Schmerer, Eileen de Feo, Jan Antico, Christine Tran, Matthew Tolentino, Shannon Wickline, Kim Gietzen, Brad Sickler, Jingtao Liu, Eric Allen, Phil Febbo, Nicole L. Washington, Simon White, Geraint Levan, Kelly Schiabor Barrett, Elizabeth Cirulli, Alexandre Bolze, Ary Ascencio, Charlotte Rivera-Garcia, Ryan Cho, Jason Nguyen, Sherry Wang, Jimmy Ramirez, Tyler Cassens, Efren Sandoval, Magnus Isaksson, William Lee, David Becker, Marc Laurent, James Lu, Clinton R. Paden, Duncan MacCannell |
| EPI_ISL_1593099, EPI_ISL_1593100 | University of Texas at Austin Genome Sequencing and Analysis Facility (UTGSAF) | University of Texas at Austin Genome Sequencing and Analysis Facility (UTGSAF) | Jessica Podnar, Sylvie Beaudenon, Audrey Kelly, Zachary Carver, Anna Battenhouse, Andreas Matouschek |
| EPI_ISL_1593101 | Helix/Illumina | Centers for Disease Control and Prevention Division of Viral Diseases, Pathogen Discovery | Dakota Howard, Dhwani Batra, Peter W. Cook, Kara Moser, Adrian Paskey, Jason Caravas, Benjamin Rambo-Martin, Shatavia Morrison, Christopher Gulvick, Scott Sammons, Yvette Unoarumhi, Darlene Wagner, Matthew Schmerer, Eileen de Feo, Jan Antico, Christine Tran, Matthew Tolentino, Shannon Wickline, Kim Gietzen, Brad Sickler, Jingtao Liu, Eric Allen, Phil Febbo, Nicole L. Washington, Simon White, Geraint Levan, Kelly Schiabor Barrett, Elizabeth Cirulli, Alexandre Bolze, Ary Ascencio, Charlotte Rivera-Garcia, Ryan Cho, Jason Nguyen, Sherry Wang, Jimmy Ramirez, Tyler Cassens, Efren Sandoval, Magnus Isaksson, William Lee, David Becker, Marc Laurent, James Lu, Clinton R. Paden, Duncan MacCannell |
| EPI_ISL_1593102 | University of Texas at Austin Genome Sequencing and Analysis Facility (UTGSAF) | University of Texas at Austin Genome Sequencing and Analysis Facility (UTGSAF) | Jessica Podnar, Sylvie Beaudenon, Audrey Kelly, Zachary Carver, Anna Battenhouse, Andreas Matouschek |
| EPI_ISL_1593104 | Helix/Illumina | Centers for Disease Control and Prevention Division of Viral Diseases, Pathogen Discovery | Dakota Howard, Dhwani Batra, Peter W. Cook, Kara Moser, Adrian Paskey, Jason Caravas, Benjamin Rambo-Martin, Shatavia Morrison, Christopher Gulvick, Scott Sammons, Yvette Unoarumhi, Darlene Wagner, Matthew Schmerer, Eileen de Feo, Jan Antico, Christine Tran, Matthew Tolentino, Shannon Wickline, Kim Gietzen, Brad Sickler, Jingtao Liu, Eric Allen, Phil Febbo, Nicole L. Washington, Simon White, Geraint Levan, Kelly Schiabor Barrett, Elizabeth Cirulli, Alexandre Bolze, Ary Ascencio, Charlotte Rivera-Garcia, Ryan Cho, Jason Nguyen, Sherry Wang, Jimmy Ramirez, Tyler Cassens, Efren Sandoval, Magnus Isaksson, William Lee, David Becker, Marc Laurent, James Lu, Clinton R. Paden, Duncan MacCannell |
| EPI_ISL_1593105 | University of Texas at Austin Genome Sequencing and Analysis Facility (UTGSAF) | University of Texas at Austin Genome Sequencing and Analysis Facility (UTGSAF) | Jessica Podnar, Sylvie Beaudenon, Audrey Kelly, Zachary Carver, Anna Battenhouse, Andreas Matouschek |
| EPI_ISL_1593106 | Helix/Illumina | Centers for Disease Control and Prevention Division of Viral Diseases, Pathogen Discovery | Dakota Howard, Dhwani Batra, Peter W. Cook, Kara Moser, Adrian Paskey, Jason Caravas, Benjamin Rambo-Martin, Shatavia Morrison, Christopher Gulvick, Scott Sammons, Yvette Unoarumhi, Darlene Wagner, Matthew Schmerer, Eileen de Feo, Jan Antico, Christine Tran, Matthew Tolentino, Shannon Wickline, Kim Gietzen, Brad Sickler, Jingtao Liu, Eric Allen, Phil Febbo, Nicole L. Washington, Simon White, Geraint Levan, Kelly Schiabor Barrett, Elizabeth Cirulli, Alexandre Bolze, Ary Ascencio, Charlotte Rivera-Garcia, Ryan Cho, Jason Nguyen, Sherry Wang, Jimmy Ramirez, Tyler Cassens, Efren Sandoval, Magnus Isaksson, William Lee, David Becker, Marc Laurent, James Lu, Clinton R. Paden, Duncan MacCannell |
| EPI_ISL_1593107, EPI_ISL_1593108 | University of Texas at Austin Genome Sequencing and Analysis Facility (UTGSAF) | University of Texas at Austin Genome Sequencing and Analysis Facility (UTGSAF) | Jessica Podnar, Sylvie Beaudenon, Audrey Kelly, Zachary Carver, Anna Battenhouse, Andreas Matouschek |
| EPI_ISL_1593109 | Helix/Illumina | Centers for Disease Control and Prevention Division of Viral Diseases, Pathogen Discovery | Dakota Howard, Dhwani Batra, Peter W. Cook, Kara Moser, Adrian Paskey, Jason Caravas, Benjamin Rambo-Martin, Shatavia Morrison, Christopher Gulvick, Scott Sammons, Yvette Unoarumhi, Darlene Wagner, Matthew Schmerer, Eileen de Feo, Jan Antico, Christine Tran, Matthew Tolentino, Shannon Wickline, Kim Gietzen, Brad Sickler, Jingtao Liu, Eric Allen, Phil Febbo, Nicole L. Washington, Simon White, Geraint Levan, Kelly Schiabor Barrett, Elizabeth Cirulli, Alexandre Bolze, Ary Ascencio, Charlotte Rivera-Garcia, Ryan Cho, Jason Nguyen, Sherry Wang, Jimmy Ramirez, Tyler Cassens, Efren Sandoval, Magnus Isaksson, William Lee, David Becker, Marc Laurent, James Lu, Clinton R. Paden, Duncan MacCannell |
| EPI_ISL_1593110 | University of Texas at Austin Genome Sequencing and Analysis Facility (UTGSAF) | University of Texas at Austin Genome Sequencing and Analysis Facility (UTGSAF) | Jessica Podnar, Sylvie Beaudenon, Audrey Kelly, Zachary Carver, Anna Battenhouse, Andreas Matouschek |
| EPI_ISL_1593111 | Helix/Illumina | Centers for Disease Control and Prevention Division of Viral Diseases, Pathogen Discovery | Dakota Howard, Dhwani Batra, Peter W. Cook, Kara Moser, Adrian Paskey, Jason Caravas, Benjamin Rambo-Martin, Shatavia Morrison, Christopher Gulvick, Scott Sammons, Yvette Unoarumhi, Darlene Wagner, Matthew Schmerer, Eileen de Feo, Jan Antico, Christine Tran, Matthew Tolentino, Shannon Wickline, Kim Gietzen, Brad Sickler, Jingtao Liu, Eric Allen, Phil Febbo, Nicole L. Washington, Simon White, Geraint Levan, Kelly Schiabor Barrett, Elizabeth Cirulli, Alexandre Bolze, Ary Ascencio, Charlotte Rivera-Garcia, Ryan Cho, Jason Nguyen, Sherry Wang, Jimmy Ramirez, Tyler Cassens, Efren Sandoval, Magnus Isaksson, William Lee, David Becker, Marc Laurent, James Lu, Clinton R. Paden, Duncan MacCannell |
| EPI_ISL_1593113 | University of Texas at Austin Genome Sequencing and Analysis Facility (UTGSAF) | University of Texas at Austin Genome Sequencing and Analysis Facility (UTGSAF) | Jessica Podnar, Sylvie Beaudenon, Audrey Kelly, Zachary Carver, Anna Battenhouse, Andreas Matouschek |
| EPI_ISL_1593114 | Helix/Illumina | Centers for Disease Control and Prevention Division of Viral Diseases, Pathogen Discovery | Dakota Howard, Dhwani Batra, Peter W. Cook, Kara Moser, Adrian Paskey, Jason Caravas, Benjamin Rambo-Martin, Shatavia Morrison, Christopher Gulvick, Scott Sammons, Yvette Unoarumhi, Darlene Wagner, Matthew Schmerer, Eileen de Feo, Jan Antico, Christine Tran, Matthew Tolentino, Shannon Wickline, Kim Gietzen, Brad Sickler, Jingtao Liu, Eric Allen, Phil Febbo, Nicole L. Washington, Simon White, Geraint Levan, Kelly Schiabor Barrett, Elizabeth Cirulli, Alexandre Bolze, Ary Ascencio, Charlotte Rivera-Garcia, Ryan Cho, Jason Nguyen, Sherry Wang, Jimmy Ramirez, Tyler Cassens, Efren Sandoval, Magnus Isaksson, William Lee, David Becker, Marc Laurent, James Lu, Clinton R. Paden, Duncan MacCannell |
| EPI_ISL_1593115, EPI_ISL_1593116 | University of Texas at Austin Genome Sequencing and Analysis Facility (UTGSAF) | University of Texas at Austin Genome Sequencing and Analysis Facility (UTGSAF) | Jessica Podnar, Sylvie Beaudenon, Audrey Kelly, Zachary Carver, Anna Battenhouse, Andreas Matouschek |
| EPI_ISL_1593117 | Helix/Illumina | Centers for Disease Control and Prevention Division of Viral Diseases, Pathogen Discovery | Dakota Howard, Dhwani Batra, Peter W. Cook, Kara Moser, Adrian Paskey, Jason Caravas, Benjamin Rambo-Martin, Shatavia Morrison, Christopher Gulvick, Scott Sammons, Yvette Unoarumhi, Darlene Wagner, Matthew Schmerer, Eileen de Feo, Jan Antico, Christine Tran, Matthew Tolentino, Shannon Wickline, Kim Gietzen, Brad Sickler, Jingtao Liu, Eric Allen, Phil Febbo, Nicole L. Washington, Simon White, Geraint Levan, Kelly Schiabor Barrett, Elizabeth Cirulli, Alexandre Bolze, Ary Ascencio, Charlotte Rivera-Garcia, Ryan Cho, Jason Nguyen, Sherry Wang, Jimmy Ramirez, Tyler Cassens, Efren Sandoval, Magnus Isaksson, William Lee, David Becker, Marc Laurent, James Lu, Clinton R. Paden, Duncan MacCannell |
| EPI_ISL_1593119 | University of Texas at Austin Genome Sequencing and Analysis Facility (UTGSAF) | University of Texas at Austin Genome Sequencing and Analysis Facility (UTGSAF) | Jessica Podnar, Sylvie Beaudenon, Audrey Kelly, Zachary Carver, Anna Battenhouse, Andreas Matouschek |
| EPI_ISL_1593120 | Helix/Illumina | Centers for Disease Control and Prevention Division of Viral Diseases, Pathogen Discovery | Dakota Howard, Dhwani Batra, Peter W. Cook, Kara Moser, Adrian Paskey, Jason Caravas, Benjamin Rambo-Martin, Shatavia Morrison, Christopher Gulvick, Scott Sammons, Yvette Unoarumhi, Darlene Wagner, Matthew Schmerer, Eileen de Feo, Jan Antico, Christine Tran, Matthew Tolentino, Shannon Wickline, Kim Gietzen, Brad Sickler, Jingtao Liu, Eric Allen, Phil Febbo, Nicole L. Washington, Simon White, Geraint Levan, Kelly Schiabor Barrett, Elizabeth Cirulli, Alexandre Bolze, Ary Ascencio, Charlotte Rivera-Garcia, Ryan Cho, Jason Nguyen, Sherry Wang, Jimmy Ramirez, Tyler Cassens, Efren Sandoval, Magnus Isaksson, William Lee, David Becker, Marc Laurent, James Lu, Clinton R. Paden, Duncan MacCannell |
| EPI_ISL_1593121, EPI_ISL_1593122 | University of Texas at Austin Genome Sequencing and Analysis Facility (UTGSAF) | University of Texas at Austin Genome Sequencing and Analysis Facility (UTGSAF) | Jessica Podnar, Sylvie Beaudenon, Audrey Kelly, Zachary Carver, Anna Battenhouse, Andreas Matouschek |
| EPI_ISL_1593123 | Helix/Illumina | Centers for Disease Control and Prevention Division of Viral Diseases, Pathogen Discovery | Dakota Howard, Dhwani Batra, Peter W. Cook, Kara Moser, Adrian Paskey, Jason Caravas, Benjamin Rambo-Martin, Shatavia Morrison, Christopher Gulvick, Scott Sammons, Yvette Unoarumhi, Darlene Wagner, Matthew Schmerer, Eileen de Feo, Jan Antico, Christine Tran, Matthew Tolentino, Shannon Wickline, Kim Gietzen, Brad Sickler, Jingtao Liu, Eric Allen, Phil Febbo, Nicole L. Washington, Simon White, Geraint Levan, Kelly Schiabor Barrett, Elizabeth Cirulli, Alexandre Bolze, Ary Ascencio, Charlotte Rivera-Garcia, Ryan Cho, Jason Nguyen, Sherry Wang, Jimmy Ramirez, Tyler Cassens, Efren Sandoval, Magnus Isaksson, William Lee, David Becker, Marc Laurent, James Lu, Clinton R. Paden, Duncan MacCannell |
| EPI_ISL_1593124 | University of Texas at Austin Genome Sequencing and Analysis Facility (UTGSAF) | University of Texas at Austin Genome Sequencing and Analysis Facility (UTGSAF) | Jessica Podnar, Sylvie Beaudenon, Audrey Kelly, Zachary Carver, Anna Battenhouse, Andreas Matouschek |
| EPI_ISL_1593125 | Helix/Illumina | Centers for Disease Control and Prevention Division of Viral Diseases, Pathogen Discovery | Dakota Howard, Dhwani Batra, Peter W. Cook, Kara Moser, Adrian Paskey, Jason Caravas, Benjamin Rambo-Martin, Shatavia Morrison, Christopher Gulvick, Scott Sammons, Yvette Unoarumhi, Darlene Wagner, Matthew Schmerer, Eileen de Feo, Jan Antico, Christine Tran, Matthew Tolentino, Shannon Wickline, Kim Gietzen, Brad Sickler, Jingtao Liu, Eric Allen, Phil Febbo, Nicole L. Washington, Simon White, Geraint Levan, Kelly Schiabor Barrett, Elizabeth Cirulli, Alexandre Bolze, Ary Ascencio, Charlotte Rivera-Garcia, Ryan Cho, Jason Nguyen, Sherry Wang, Jimmy Ramirez, Tyler Cassens, Efren Sandoval, Magnus Isaksson, William Lee, David Becker, Marc Laurent, James Lu, Clinton R. Paden, Duncan MacCannell |
| EPI_ISL_1593126, EPI_ISL_1593127 | University of Texas at Austin Genome Sequencing and Analysis Facility (UTGSAF) | University of Texas at Austin Genome Sequencing and Analysis Facility (UTGSAF) | Jessica Podnar, Sylvie Beaudenon, Audrey Kelly, Zachary Carver, Anna Battenhouse, Andreas Matouschek |
| EPI_ISL_1593128 | Helix/Illumina | Centers for Disease Control and Prevention Division of Viral Diseases, Pathogen Discovery | Dakota Howard, Dhwani Batra, Peter W. Cook, Kara Moser, Adrian Paskey, Jason Caravas, Benjamin Rambo-Martin, Shatavia Morrison, Christopher Gulvick, Scott Sammons, Yvette Unoarumhi, Darlene Wagner, Matthew Schmerer, Eileen de Feo, Jan Antico, Christine Tran, Matthew Tolentino, Shannon Wickline, Kim Gietzen, Brad Sickler, Jingtao Liu, Eric Allen, Phil Febbo, Nicole L. Washington, Simon White, Geraint Levan, Kelly Schiabor Barrett, Elizabeth Cirulli, Alexandre Bolze, Ary Ascencio, Charlotte Rivera-Garcia, Ryan Cho, Jason Nguyen, Sherry Wang, Jimmy Ramirez, Tyler Cassens, Efren Sandoval, Magnus Isaksson, William Lee, David Becker, Marc Laurent, James Lu, Clinton R. Paden, Duncan MacCannell |
| EPI_ISL_1593129, EPI_ISL_1593130 | University of Texas at Austin Genome Sequencing and Analysis Facility (UTGSAF) | University of Texas at Austin Genome Sequencing and Analysis Facility (UTGSAF) | Jessica Podnar, Sylvie Beaudenon, Audrey Kelly, Zachary Carver, Anna Battenhouse, Andreas Matouschek |
| EPI_ISL_1593131 | Helix/Illumina | Centers for Disease Control and Prevention Division of Viral Diseases, Pathogen Discovery | Dakota Howard, Dhwani Batra, Peter W. Cook, Kara Moser, Adrian Paskey, Jason Caravas, Benjamin Rambo-Martin, Shatavia Morrison, Christopher Gulvick, Scott Sammons, Yvette Unoarumhi, Darlene Wagner, Matthew Schmerer, Eileen de Feo, Jan Antico, Christine Tran, Matthew Tolentino, Shannon Wickline, Kim Gietzen, Brad Sickler, Jingtao Liu, Eric Allen, Phil Febbo, Nicole L. Washington, Simon White, Geraint Levan, Kelly Schiabor Barrett, Elizabeth Cirulli, Alexandre Bolze, Ary Ascencio, Charlotte Rivera-Garcia, Ryan Cho, Jason Nguyen, Sherry Wang, Jimmy Ramirez, Tyler Cassens, Efren Sandoval, Magnus Isaksson, William Lee, David Becker, Marc Laurent, James Lu, Clinton R. Paden, Duncan MacCannell |
| EPI_ISL_1593132 | University of Texas at Austin Genome Sequencing and Analysis Facility (UTGSAF) | University of Texas at Austin Genome Sequencing and Analysis Facility (UTGSAF) | Jessica Podnar, Sylvie Beaudenon, Audrey Kelly, Zachary Carver, Anna Battenhouse, Andreas Matouschek |
| EPI_ISL_1593133 | Helix/Illumina | Centers for Disease Control and Prevention Division of Viral Diseases, Pathogen Discovery | Dakota Howard, Dhwani Batra, Peter W. Cook, Kara Moser, Adrian Paskey, Jason Caravas, Benjamin Rambo-Martin, Shatavia Morrison, Christopher Gulvick, Scott Sammons, Yvette Unoarumhi, Darlene Wagner, Matthew Schmerer, Eileen de Feo, Jan Antico, Christine Tran, Matthew Tolentino, Shannon Wickline, Kim Gietzen, Brad Sickler, Jingtao Liu, Eric Allen, Phil Febbo, Nicole L. Washington, Simon White, Geraint Levan, Kelly Schiabor Barrett, Elizabeth Cirulli, Alexandre Bolze, Ary Ascencio, Charlotte Rivera-Garcia, Ryan Cho, Jason Nguyen, Sherry Wang, Jimmy Ramirez, Tyler Cassens, Efren Sandoval, Magnus Isaksson, William Lee, David Becker, Marc Laurent, James Lu, Clinton R. Paden, Duncan MacCannell |
| EPI_ISL_1593134, EPI_ISL_1593135 | University of Texas at Austin Genome Sequencing and Analysis Facility (UTGSAF) | University of Texas at Austin Genome Sequencing and Analysis Facility (UTGSAF) | Jessica Podnar, Sylvie Beaudenon, Audrey Kelly, Zachary Carver, Anna Battenhouse, Andreas Matouschek |
| EPI_ISL_1593136 | Helix/Illumina | Centers for Disease Control and Prevention Division of Viral Diseases, Pathogen Discovery | Dakota Howard, Dhwani Batra, Peter W. Cook, Kara Moser, Adrian Paskey, Jason Caravas, Benjamin Rambo-Martin, Shatavia Morrison, Christopher Gulvick, Scott Sammons, Yvette Unoarumhi, Darlene Wagner, Matthew Schmerer, Eileen de Feo, Jan Antico, Christine Tran, Matthew Tolentino, Shannon Wickline, Kim Gietzen, Brad Sickler, Jingtao Liu, Eric Allen, Phil Febbo, Nicole L. Washington, Simon White, Geraint Levan, Kelly Schiabor Barrett, Elizabeth Cirulli, Alexandre Bolze, Ary Ascencio, Charlotte Rivera-Garcia, Ryan Cho, Jason Nguyen, Sherry Wang, Jimmy Ramirez, Tyler Cassens, Efren Sandoval, Magnus Isaksson, William Lee, David Becker, Marc Laurent, James Lu, Clinton R. Paden, Duncan MacCannell |
| EPI_ISL_1593137, EPI_ISL_1593138 | University of Texas at Austin Genome Sequencing and Analysis Facility (UTGSAF) | University of Texas at Austin Genome Sequencing and Analysis Facility (UTGSAF) | Jessica Podnar, Sylvie Beaudenon, Audrey Kelly, Zachary Carver, Anna Battenhouse, Andreas Matouschek |

| | | | |
|---|---|---|---|
| EPI_ISL_1593139 | Helix/Illumina | Centers for Disease Control and Prevention Division of Viral Diseases, Pathogen Discovery | Dakota Howard, Dhwani Batra, Peter W. Cook, Kara Moser, Adrian Paskey, Jason Caravas, Benjamin Rambo-Martin, Shatavia Morrison, Christopher Gulvick, Scott Sammons, Yvette Unoarumhi, Darlene Wagner, Matthew Schmerer, Eileen de Feo, Jan Antico, Christine Tran, Matthew Tolentino, Shannon Wickline, Kim Gietzen, Brad Sickler, Jingtao Liu, Eric Allen, Phil Febbo, Nicole L. Washington, Simon White, Geraint Levan, Kelly Schiabor Barrett, Elizabeth Cirulli, Alexandre Bolze, Ary Ascencio, Charlotte Rivera-Garcia, Ryan Cho, Jason Nguyen, Sherry Wang, Jimmy Ramirez, Tyler Cassens, Efren Sandoval, Magnus Isaksson, William Lee, David Becker, Marc Laurent, James Lu, Clinton R. Paden, Duncan MacCannell |
| EPI_ISL_1593140 | University of Texas at Austin Genome Sequencing and Analysis Facility (UTGSAF) | University of Texas at Austin Genome Sequencing and Analysis Facility (UTGSAF) | Jessica Podnar, Sylvie Beaudenon, Audrey Kelly, Zachary Carver, Anna Battenhouse, Andreas Matouschek |
| EPI_ISL_1593141 | Helix/Illumina | Centers for Disease Control and Prevention Division of Viral Diseases, Pathogen Discovery | Dakota Howard, Dhwani Batra, Peter W. Cook, Kara Moser, Adrian Paskey, Jason Caravas, Benjamin Rambo-Martin, Shatavia Morrison, Christopher Gulvick, Scott Sammons, Yvette Unoarumhi, Darlene Wagner, Matthew Schmerer, Eileen de Feo, Jan Antico, Christine Tran, Matthew Tolentino, Shannon Wickline, Kim Gietzen, Brad Sickler, Jingtao Liu, Eric Allen, Phil Febbo, Nicole L. Washington, Simon White, Geraint Levan, Kelly Schiabor Barrett, Elizabeth Cirulli, Alexandre Bolze, Ary Ascencio, Charlotte Rivera-Garcia, Ryan Cho, Jason Nguyen, Sherry Wang, Jimmy Ramirez, Tyler Cassens, Efren Sandoval, Magnus Isaksson, William Lee, David Becker, Marc Laurent, James Lu, Clinton R. Paden, Duncan MacCannell |
| EPI_ISL_1593143 | University of Texas at Austin Genome Sequencing and Analysis Facility (UTGSAF) | University of Texas at Austin Genome Sequencing and Analysis Facility (UTGSAF) | Jessica Podnar, Sylvie Beaudenon, Audrey Kelly, Zachary Carver, Anna Battenhouse, Andreas Matouschek |
| EPI_ISL_1593144 | Helix/Illumina | Centers for Disease Control and Prevention Division of Viral Diseases, Pathogen Discovery | Dakota Howard, Dhwani Batra, Peter W. Cook, Kara Moser, Adrian Paskey, Jason Caravas, Benjamin Rambo-Martin, Shatavia Morrison, Christopher Gulvick, Scott Sammons, Yvette Unoarumhi, Darlene Wagner, Matthew Schmerer, Eileen de Feo, Jan Antico, Christine Tran, Matthew Tolentino, Shannon Wickline, Kim Gietzen, Brad Sickler, Jingtao Liu, Eric Allen, Phil Febbo, Nicole L. Washington, Simon White, Geraint Levan, Kelly Schiabor Barrett, Elizabeth Cirulli, Alexandre Bolze, Ary Ascencio, Charlotte Rivera-Garcia, Ryan Cho, Jason Nguyen, Sherry Wang, Jimmy Ramirez, Tyler Cassens, Efren Sandoval, Magnus Isaksson, William Lee, David Becker, Marc Laurent, James Lu, Clinton R. Paden, Duncan MacCannell |
| EPI_ISL_1593145, EPI_ISL_1593147, EPI_ISL_1593148 | University of Texas at Austin Genome Sequencing and Analysis Facility (UTGSAF) | University of Texas at Austin Genome Sequencing and Analysis Facility (UTGSAF) | Jessica Podnar, Sylvie Beaudenon, Audrey Kelly, Zachary Carver, Anna Battenhouse, Andreas Matouschek |
| EPI_ISL_1593149 | Helix/Illumina | Centers for Disease Control and Prevention Division of Viral Diseases, Pathogen Discovery | Dakota Howard, Dhwani Batra, Peter W. Cook, Kara Moser, Adrian Paskey, Jason Caravas, Benjamin Rambo-Martin, Shatavia Morrison, Christopher Gulvick, Scott Sammons, Yvette Unoarumhi, Darlene Wagner, Matthew Schmerer, Eileen de Feo, Jan Antico, Christine Tran, Matthew Tolentino, Shannon Wickline, Kim Gietzen, Brad Sickler, Jingtao Liu, Eric Allen, Phil Febbo, Nicole L. Washington, Simon White, Geraint Levan, Kelly Schiabor Barrett, Elizabeth Cirulli, Alexandre Bolze, Ary Ascencio, Charlotte Rivera-Garcia, Ryan Cho, Jason Nguyen, Sherry Wang, Jimmy Ramirez, Tyler Cassens, Efren Sandoval, Magnus Isaksson, William Lee, David Becker, Marc Laurent, James Lu, Clinton R. Paden, Duncan MacCannell |
| EPI_ISL_1593150 | University of Texas at Austin Genome Sequencing and Analysis Facility (UTGSAF) | University of Texas at Austin Genome Sequencing and Analysis Facility (UTGSAF) | Jessica Podnar, Sylvie Beaudenon, Audrey Kelly, Zachary Carver, Anna Battenhouse, Andreas Matouschek |
| EPI_ISL_1593152 | Helix/Illumina | Centers for Disease Control and Prevention Division of Viral Diseases, Pathogen Discovery | Dakota Howard, Dhwani Batra, Peter W. Cook, Kara Moser, Adrian Paskey, Jason Caravas, Benjamin Rambo-Martin, Shatavia Morrison, Christopher Gulvick, Scott Sammons, Yvette Unoarumhi, Darlene Wagner, Matthew Schmerer, Eileen de Feo, Jan Antico, Christine Tran, Matthew Tolentino, Shannon Wickline, Kim Gietzen, Brad Sickler, Jingtao Liu, Eric Allen, Phil Febbo, Nicole L. Washington, Simon White, Geraint Levan, Kelly Schiabor Barrett, Elizabeth Cirulli, Alexandre Bolze, Ary Ascencio, Charlotte Rivera-Garcia, Ryan Cho, Jason Nguyen, Sherry Wang, Jimmy Ramirez, Tyler Cassens, Efren Sandoval, Magnus Isaksson, William Lee, David Becker, Marc Laurent, James Lu, Clinton R. Paden, Duncan MacCannell |
| EPI_ISL_1593153 | University of Texas at Austin Genome Sequencing and Analysis Facility (UTGSAF) | University of Texas at Austin Genome Sequencing and Analysis Facility (UTGSAF) | Jessica Podnar, Sylvie Beaudenon, Audrey Kelly, Zachary Carver, Anna Battenhouse, Andreas Matouschek |
| EPI_ISL_1593154 | Helix/Illumina | Centers for Disease Control and Prevention Division of Viral Diseases, Pathogen Discovery | Dakota Howard, Dhwani Batra, Peter W. Cook, Kara Moser, Adrian Paskey, Jason Caravas, Benjamin Rambo-Martin, Shatavia Morrison, Christopher Gulvick, Scott Sammons, Yvette Unoarumhi, Darlene Wagner, Matthew Schmerer, Eileen de Feo, Jan Antico, Christine Tran, Matthew Tolentino, Shannon Wickline, Kim Gietzen, Brad Sickler, Jingtao Liu, Eric Allen, Phil Febbo, Nicole L. Washington, Simon White, Geraint Levan, Kelly Schiabor Barrett, Elizabeth Cirulli, Alexandre Bolze, Ary Ascencio, Charlotte Rivera-Garcia, Ryan Cho, Jason Nguyen, Sherry Wang, Jimmy Ramirez, Tyler Cassens, Efren Sandoval, Magnus Isaksson, William Lee, David Becker, Marc Laurent, James Lu, Clinton R. Paden, Duncan MacCannell |
| EPI_ISL_1593155, EPI_ISL_1593156 | University of Texas at Austin Genome Sequencing and Analysis Facility (UTGSAF) | University of Texas at Austin Genome Sequencing and Analysis Facility (UTGSAF) | Jessica Podnar, Sylvie Beaudenon, Audrey Kelly, Zachary Carver, Anna Battenhouse, Andreas Matouschek |
| EPI_ISL_1593157 | Helix/Illumina | Centers for Disease Control and Prevention Division of Viral Diseases, Pathogen Discovery | Dakota Howard, Dhwani Batra, Peter W. Cook, Kara Moser, Adrian Paskey, Jason Caravas, Benjamin Rambo-Martin, Shatavia Morrison, Christopher Gulvick, Scott Sammons, Yvette Unoarumhi, Darlene Wagner, Matthew Schmerer, Eileen de Feo, Jan Antico, Christine Tran, Matthew Tolentino, Shannon Wickline, Kim Gietzen, Brad Sickler, Jingtao Liu, Eric Allen, Phil Febbo, Nicole L. Washington, Simon White, Geraint Levan, Kelly Schiabor Barrett, Elizabeth Cirulli, Alexandre Bolze, Ary Ascencio, Charlotte Rivera-Garcia, Ryan Cho, Jason Nguyen, Sherry Wang, Jimmy Ramirez, Tyler Cassens, Efren Sandoval, Magnus Isaksson, William Lee, David Becker, Marc Laurent, James Lu, Clinton R. Paden, Duncan MacCannell |
| EPI_ISL_1593158 | University of Texas at Austin Genome Sequencing and Analysis Facility (UTGSAF) | University of Texas at Austin Genome Sequencing and Analysis Facility (UTGSAF) | Jessica Podnar, Sylvie Beaudenon, Audrey Kelly, Zachary Carver, Anna Battenhouse, Andreas Matouschek |
| EPI_ISL_1593159 | Helix/Illumina | Centers for Disease Control and Prevention Division of Viral Diseases, Pathogen Discovery | Dakota Howard, Dhwani Batra, Peter W. Cook, Kara Moser, Adrian Paskey, Jason Caravas, Benjamin Rambo-Martin, Shatavia Morrison, Christopher Gulvick, Scott Sammons, Yvette Unoarumhi, Darlene Wagner, Matthew Schmerer, Eileen de Feo, Jan Antico, Christine Tran, Matthew Tolentino, Shannon Wickline, Kim Gietzen, Brad Sickler, Jingtao Liu, Eric Allen, Phil Febbo, Nicole L. Washington, Simon White, Geraint Levan, Kelly Schiabor Barrett, Elizabeth Cirulli, Alexandre Bolze, Ary Ascencio, Charlotte Rivera-Garcia, Ryan Cho, Jason Nguyen, Sherry Wang, Jimmy Ramirez, Tyler Cassens, Efren Sandoval, Magnus Isaksson, William Lee, David Becker, Marc Laurent, James Lu, Clinton R. Paden, Duncan MacCannell |
| EPI_ISL_1593160, EPI_ISL_1593161 | University of Texas at Austin Genome Sequencing and Analysis Facility (UTGSAF) | University of Texas at Austin Genome Sequencing and Analysis Facility (UTGSAF) | Jessica Podnar, Sylvie Beaudenon, Audrey Kelly, Zachary Carver, Anna Battenhouse, Andreas Matouschek |
| EPI_ISL_1593162, EPI_ISL_1593163, EPI_ISL_1593164, EPI_ISL_1593165, EPI_ISL_1593166, EPI_ISL_1593167, EPI_ISL_1593168, EPI_ISL_1593169, EPI_ISL_1593170, EPI_ISL_1593171, EPI_ISL_1593172, EPI_ISL_1593173, EPI_ISL_1593174, EPI_ISL_1593175, EPI_ISL_1593176, EPI_ISL_1593177, EPI_ISL_1593178, EPI_ISL_1593179, EPI_ISL_1593180, EPI_ISL_1593181, EPI_ISL_1593182, EPI_ISL_1593183, EPI_ISL_1593185, EPI_ISL_1593186, EPI_ISL_1593187, EPI_ISL_1593188, EPI_ISL_1593189, EPI_ISL_1593190, EPI_ISL_1593191, EPI_ISL_1593192, EPI_ISL_1593193, EPI_ISL_1593194, EPI_ISL_1593195, EPI_ISL_1593196, EPI_ISL_1593197, EPI_ISL_1593198, EPI_ISL_1593199, EPI_ISL_1593200, EPI_ISL_1593201 | | | |
| see above | Helix/Illumina | Centers for Disease Control and Prevention Division of Viral Diseases, Pathogen Discovery | Dakota Howard, Dhwani Batra, Peter W. Cook, Kara Moser, Adrian Paskey, Jason Caravas, Benjamin Rambo-Martin, Shatavia Morrison, Christopher Gulvick, Scott Sammons, Yvette Unoarumhi, Darlene Wagner, Matthew Schmerer, Eileen de Feo, Jan Antico, Christine Tran, Matthew Tolentino, Shannon Wickline, Kim Gietzen, Brad Sickler, Jingtao Liu, Eric Allen, Phil Febbo, Nicole L. Washington, Simon White, Geraint Levan, Kelly Schiabor Barrett, Elizabeth Cirulli, Alexandre Bolze, Ary Ascencio, Charlotte Rivera-Garcia, Ryan Cho, Jason Nguyen, Sherry Wang, Jimmy Ramirez, Tyler Cassens, Efren Sandoval, Magnus Isaksson, William Lee, David Becker, Marc Laurent, James Lu, Clinton R. Paden, Duncan MacCannell |
| EPI_ISL_1593202, EPI_ISL_1593203, EPI_ISL_1593204, EPI_ISL_1593205, EPI_ISL_1593206, EPI_ISL_1593207, EPI_ISL_1593208, EPI_ISL_1593209, EPI_ISL_1593211, EPI_ISL_1593212, EPI_ISL_1593213, EPI_ISL_1593214, EPI_ISL_1593215, EPI_ISL_1593216, EPI_ISL_1593217, EPI_ISL_1593219, EPI_ISL_1593220, EPI_ISL_1593221, EPI_ISL_1593222, EPI_ISL_1593223, EPI_ISL_1593224, EPI_ISL_1593225, EPI_ISL_1593228, EPI_ISL_1593229, EPI_ISL_1593230, EPI_ISL_1593354, EPI_ISL_1593355, EPI_ISL_1593358, EPI_ISL_1593359, EPI_ISL_1593361, EPI_ISL_1593362, EPI_ISL_1593368, EPI_ISL_1593370, EPI_ISL_1593372, EPI_ISL_1593373, EPI_ISL_1593375, EPI_ISL_1593376, EPI_ISL_1593377, EPI_ISL_1593380, EPI_ISL_1593381, EPI_ISL_1593382, EPI_ISL_1593383, EPI_ISL_1593384, EPI_ISL_1593385, EPI_ISL_1593387, EPI_ISL_1593388, EPI_ISL_1593389, EPI_ISL_1593390, EPI_ISL_1593393, EPI_ISL_1593394, EPI_ISL_1593395, EPI_ISL_1593396, EPI_ISL_1593399, EPI_ISL_1593400, EPI_ISL_1593402, EPI_ISL_1593403, EPI_ISL_1593404, EPI_ISL_1593405, EPI_ISL_1593406, EPI_ISL_1593408, EPI_ISL_1593409, EPI_ISL_1593411, EPI_ISL_1593413, EPI_ISL_1593414, EPI_ISL_1593415, EPI_ISL_1593416, EPI_ISL_1593417, EPI_ISL_1593418, EPI_ISL_1593419, EPI_ISL_1593420, EPI_ISL_1593421, EPI_ISL_1593423, EPI_ISL_1593425, EPI_ISL_1593426, EPI_ISL_1593427, EPI_ISL_1593428, EPI_ISL_1593429, EPI_ISL_1593435, EPI_ISL_1593436, EPI_ISL_1593437, EPI_ISL_1593438, EPI_ISL_1593439, EPI_ISL_1593440, EPI_ISL_1593441, EPI_ISL_1593442, EPI_ISL_1593443, EPI_ISL_1593444, EPI_ISL_1593446, EPI_ISL_1593447, EPI_ISL_1593448, EPI_ISL_1593449, EPI_ISL_1593450, EPI_ISL_1593451, EPI_ISL_1593452, EPI_ISL_1593453, EPI_ISL_1593454, EPI_ISL_1593455, EPI_ISL_1593457, EPI_ISL_1593458, EPI_ISL_1593459, EPI_ISL_1593460, EPI_ISL_1593461 | | | |
| see above | North Dakota Department of Health, Public Health Laboratory | North Dakota Department of Health, Public Health Laboratory | Lisa Wingerter |
| EPI_ISL_1593476, EPI_ISL_1593477, EPI_ISL_1593478, EPI_ISL_1593479, EPI_ISL_1593480, EPI_ISL_1593481, EPI_ISL_1593482, EPI_ISL_1593483, EPI_ISL_1593484, EPI_ISL_1593485, EPI_ISL_1593486, EPI_ISL_1593487, EPI_ISL_1593488, EPI_ISL_1593489, EPI_ISL_1593490, EPI_ISL_1593491, EPI_ISL_1593492, EPI_ISL_1593493, EPI_ISL_1593494, EPI_ISL_1593495, EPI_ISL_1593496, EPI_ISL_1593497, EPI_ISL_1593498, EPI_ISL_1593499, EPI_ISL_1593500, EPI_ISL_1593501, EPI_ISL_1593502, EPI_ISL_1593503, EPI_ISL_1593504, EPI_ISL_1593505, EPI_ISL_1593506, EPI_ISL_1593507, EPI_ISL_1593508, EPI_ISL_1593509, EPI_ISL_1593510, EPI_ISL_1593511, EPI_ISL_1593512, EPI_ISL_1593513, EPI_ISL_1593514, EPI_ISL_1593515, EPI_ISL_1593516, EPI_ISL_1593517, EPI_ISL_1593518, EPI_ISL_1593519, EPI_ISL_1593520, EPI_ISL_1593521, EPI_ISL_1593522, EPI_ISL_1593523, EPI_ISL_1593524, EPI_ISL_1593525, EPI_ISL_1593526, EPI_ISL_1593527, EPI_ISL_1593528, EPI_ISL_1593529, EPI_ISL_1593530, EPI_ISL_1593531, EPI_ISL_1593532, EPI_ISL_1593533, EPI_ISL_1593534, EPI_ISL_1593535, EPI_ISL_1593536, EPI_ISL_1593537, EPI_ISL_1593538, EPI_ISL_1593539, EPI_ISL_1593540, EPI_ISL_1593542, EPI_ISL_1593543, EPI_ISL_1593544, EPI_ISL_1593545, EPI_ISL_1593546, EPI_ISL_1593547, EPI_ISL_1593548, EPI_ISL_1593549, EPI_ISL_1593550, EPI_ISL_1593551, EPI_ISL_1593552, EPI_ISL_1593553, EPI_ISL_1593554, EPI_ISL_1593555, EPI_ISL_1593556, EPI_ISL_1593557, EPI_ISL_1593558, EPI_ISL_1593559, EPI_ISL_1593560, EPI_ISL_1593561, EPI_ISL_1593562, EPI_ISL_1593563, EPI_ISL_1593564, EPI_ISL_1593565, EPI_ISL_1593566, EPI_ISL_1593567, EPI_ISL_1593568, EPI_ISL_1593569, EPI_ISL_1593570, EPI_ISL_1593571, EPI_ISL_1593572, EPI_ISL_1593574, EPI_ISL_1593575, EPI_ISL_1593576, EPI_ISL_1593577, EPI_ISL_1593579, EPI_ISL_1593580, EPI_ISL_1593581, EPI_ISL_1593582, EPI_ISL_1593583, EPI_ISL_1593584, EPI_ISL_1593585, EPI_ISL_1593586, EPI_ISL_1593587, EPI_ISL_1593588, EPI_ISL_1593589, EPI_ISL_1593590, EPI_ISL_1593591, EPI_ISL_1593592, EPI_ISL_1593593, EPI_ISL_1593594, EPI_ISL_1593595, EPI_ISL_1593596, EPI_ISL_1593597, EPI_ISL_1593598, EPI_ISL_1593599, EPI_ISL_1593600, EPI_ISL_1593601, EPI_ISL_1593602, EPI_ISL_1593603, EPI_ISL_1593604, EPI_ISL_1593605, EPI_ISL_1593606, EPI_ISL_1593607, EPI_ISL_1593608, EPI_ISL_1593609, EPI_ISL_1593610, EPI_ISL_1593611, EPI_ISL_1593612, EPI_ISL_1593613, EPI_ISL_1593614, EPI_ISL_1593615, EPI_ISL_1593616, EPI_ISL_1593617, EPI_ISL_1593618, EPI_ISL_1593619, EPI_ISL_1593620, EPI_ISL_1593621, EPI_ISL_1593622, EPI_ISL_1593623, EPI_ISL_1593624, EPI_ISL_1593625, EPI_ISL_1593626, EPI_ISL_1593627, EPI_ISL_1593628, EPI_ISL_1593629, EPI_ISL_1593630, EPI_ISL_1593631, EPI_ISL_1593632, EPI_ISL_1593633, EPI_ISL_1593634, EPI_ISL_1593635, EPI_ISL_1593636, EPI_ISL_1593637, EPI_ISL_1593638, EPI_ISL_1593639, EPI_ISL_1593640, EPI_ISL_1593641 | | | |
| see above | Wyoming Public Health Laboratory | Wyoming Public Health Laboratory | Jim Mildenberger, Taylor Fearing, Channing Weber, Ashley Norberg, Chayse Rowley, Marley Goetz, Brian Dominguez, Elliot Thomasson, Brittany Oher, Sam Britz, Cari Sloma, and Rob Christensen |
| EPI_ISL_1593642, EPI_ISL_1593643, EPI_ISL_1593644, EPI_ISL_1593645, EPI_ISL_1593646, EPI_ISL_1593647, EPI_ISL_1593648, EPI_ISL_1593649, EPI_ISL_1593650, EPI_ISL_1593651, EPI_ISL_1593652, EPI_ISL_1593653, EPI_ISL_1593654, EPI_ISL_1593655, EPI_ISL_1593656, EPI_ISL_1593657, EPI_ISL_1593658, EPI_ISL_1593659, EPI_ISL_1593660, EPI_ISL_1593661, EPI_ISL_1593662, EPI_ISL_1593663, EPI_ISL_1593664, EPI_ISL_1593665, EPI_ISL_1593666, EPI_ISL_1593667, EPI_ISL_1593668, EPI_ISL_1593669, EPI_ISL_1593670, EPI_ISL_1593671, EPI_ISL_1593672, EPI_ISL_1593673, EPI_ISL_1593674, EPI_ISL_1593675, EPI_ISL_1593676, EPI_ISL_1593677, EPI_ISL_1593678, EPI_ISL_1593679, EPI_ISL_1593680, EPI_ISL_1593681, EPI_ISL_1593682, EPI_ISL_1593683, EPI_ISL_1593684, EPI_ISL_1593685, EPI_ISL_1593686, EPI_ISL_1593687, EPI_ISL_1593688, EPI_ISL_1593689, EPI_ISL_1593690, EPI_ISL_1593692, EPI_ISL_1593693, EPI_ISL_1593694, EPI_ISL_1593695, EPI_ISL_1593696, EPI_ISL_1593697, EPI_ISL_1593698, EPI_ISL_1593699, EPI_ISL_1593700, EPI_ISL_1593701, EPI_ISL_1593702, EPI_ISL_1593703, EPI_ISL_1593704, EPI_ISL_1593705, EPI_ISL_1593706, EPI_ISL_1593707, EPI_ISL_1593708, EPI_ISL_1593709, EPI_ISL_1593710, EPI_ISL_1593711, EPI_ISL_1593712, EPI_ISL_1593713, EPI_ISL_1593714, EPI_ISL_1593715, EPI_ISL_1593716, EPI_ISL_1593717, EPI_ISL_1593718, EPI_ISL_1593719 | | | |
| see above | Minnesota Department of Health, Public Health Laboratory | Minnesota Department of Health, Public Health Laboratory | Alexandra Lorentz, Jacob Garfin, Matt Plumb, and Xiong Wang |
| EPI_ISL_1593720 | DOHMH Central Harlem | New York City Public Health Laboratory | Jade Wang, et al. |
| EPI_ISL_1593721 | DOHMH Corona | New York City Public Health Laboratory | Jade Wang, et al. |
| EPI_ISL_1593722 | DOHMH Central Harlem | New York City Public Health Laboratory | Jade Wang, et al. |
| EPI_ISL_1593723 | Bellevue Hospital Center | New York City Public Health Laboratory | Jade Wang, et al. |
| EPI_ISL_1593724 | Brookdale University Hospital Medical Center | New York City Public Health Laboratory | Jade Wang, et al. |
| EPI_ISL_1593729 | DOHMH Crown Heights | New York City Public Health Laboratory | Jade Wang, et al. |
| EPI_ISL_1593730 | DOHMH Jamaica | New York City Public Health Laboratory | Jade Wang, et al. |
| EPI_ISL_1593731 | DOHMH PHL | New York City Public Health Laboratory | Jade Wang, et al. |
| EPI_ISL_1593732 | DOHMH Jamaica | New York City Public Health Laboratory | Jade Wang, et al. |
| EPI_ISL_1593733 | DOHMH Chelsea | New York City Public Health Laboratory | Jade Wang, et al. |
| EPI_ISL_1593734 | DOHMH PHL | New York City Public Health Laboratory | Jade Wang, et al. |
| EPI_ISL_1593735 | DOHMH Jamaica | New York City Public Health Laboratory | Jade Wang, et al. |
| EPI_ISL_1593736 | DOHMH Riverside | New York City Public Health Laboratory | Jade Wang, et al. |
| EPI_ISL_1593737 | DOHMH Corona | New York City Public Health Laboratory | Jade Wang, et al. |
| EPI_ISL_1593738, EPI_ISL_1593739 | DOHMH Jamaica | New York City Public Health Laboratory | Jade Wang, et al. |
| EPI_ISL_1593740 | DOHMH Fort Greene | New York City Public Health Laboratory | Jade Wang, et al. |

| | | | |
|---|---|---|---|
| EPI_ISL_1593741 | DOHMH Crown Heights | New York City Public Health Laboratory | Jade Wang, et al. |
| EPI_ISL_1593742, EPI_ISL_1593743, EPI_ISL_1593744, EPI_ISL_1593745 | DOHMH Central Harlem | New York City Public Health Laboratory | Jade Wang, et al. |
| EPI_ISL_1593746 | DOHMH Corona | New York City Public Health Laboratory | Jade Wang, et al. |
| EPI_ISL_1593747, EPI_ISL_1593748 | DOHMH Jamaica | New York City Public Health Laboratory | Jade Wang, et al. |
| EPI_ISL_1593749 | DOHMH Crown Heights | New York City Public Health Laboratory | Jade Wang, et al. |
| EPI_ISL_1593750 | DOHMH Fort Greene | New York City Public Health Laboratory | Jade Wang, et al. |
| EPI_ISL_1593751, EPI_ISL_1593752 | DOHMH Central Harlem | New York City Public Health Laboratory | Jade Wang, et al. |
| EPI_ISL_1593753 | DOHMH Morrisania | New York City Public Health Laboratory | Jade Wang, et al. |
| EPI_ISL_1593754, EPI_ISL_1593755 | DOHMH Chelsea | New York City Public Health Laboratory | Jade Wang, et al. |
| EPI_ISL_1593756, EPI_ISL_1593757 | OCME Office Of Chief Medical Examiner | New York City Public Health Laboratory | Jade Wang, et al. |
| EPI_ISL_1593758 | DOHMH Fort Greene | New York City Public Health Laboratory | Jade Wang, et al. |
| EPI_ISL_1593759, EPI_ISL_1593760, EPI_ISL_1593761 | DOHMH Corona | New York City Public Health Laboratory | Jade Wang, et al. |
| EPI_ISL_1593762 | DOHMH Jamaica | New York City Public Health Laboratory | Jade Wang, et al. |
| EPI_ISL_1593763, EPI_ISL_1593764, EPI_ISL_1593765, EPI_ISL_1593766, EPI_ISL_1593767 | DOHMH Central Harlem | New York City Public Health Laboratory | Jade Wang, et al. |
| EPI_ISL_1593768 | DOHMH Corona | New York City Public Health Laboratory | Jade Wang, et al. |
| EPI_ISL_1593769 | DOHMH Crown Heights | New York City Public Health Laboratory | Jade Wang, et al. |
| EPI_ISL_1593770 | DOHMH Fort Greene | New York City Public Health Laboratory | Jade Wang, et al. |
| EPI_ISL_1593771 | Department of Homeless Services | New York City Public Health Laboratory | Jade Wang, et al. |
| EPI_ISL_1593772 | OCME Office Of Chief Medical Examiner | New York City Public Health Laboratory | Jade Wang, et al. |
| EPI_ISL_1593773, EPI_ISL_1593774, EPI_ISL_1593775, EPI_ISL_1593776, EPI_ISL_1593777, EPI_ISL_1593778, EPI_ISL_1593779, EPI_ISL_1593781, EPI_ISL_1593782, EPI_ISL_1593783, EPI_ISL_1593784, EPI_ISL_1593785, EPI_ISL_1593786, EPI_ISL_1593787, EPI_ISL_1593788, EPI_ISL_1593789, EPI_ISL_1593790, EPI_ISL_1593791, EPI_ISL_1593792, EPI_ISL_1593793, EPI_ISL_1593794, EPI_ISL_1593795, EPI_ISL_1593796, EPI_ISL_1593797, EPI_ISL_1593798, EPI_ISL_1593799, EPI_ISL_1593800, EPI_ISL_1593801, EPI_ISL_1593802, EPI_ISL_1593803, EPI_ISL_1593805, EPI_ISL_1593806, EPI_ISL_1593807, EPI_ISL_1593808, EPI_ISL_1593809, EPI_ISL_1593810, EPI_ISL_1593811, EPI_ISL_1593812, EPI_ISL_1593813, EPI_ISL_1593814, EPI_ISL_1593815, EPI_ISL_1593816, EPI_ISL_1593817, EPI_ISL_1593818, EPI_ISL_1593819, EPI_ISL_1593820, EPI_ISL_1593821, EPI_ISL_1593822, EPI_ISL_1593823, EPI_ISL_1593824, EPI_ISL_1593825 | | | |
| see above | Wyoming Public Health Laboratory | Wyoming Public Health Laboratory | Jim Mildenberger, Wanda Manley, Noah Hull, Taylor Fearing, Lynette Gumbleton, Channing Weber, Ashley Norberg, Chayse Rowley, Marley Goetz, Brian Dominguez, Elliot Thomasson, Sam Britz, Cari Sloma, and Rob Christensen |
| EPI_ISL_1593864, EPI_ISL_1593865 | WESTCHESTER MEDICAL CENTER | Wadsworth Center, New York State Department of Health | Kirsten St. George, Daryl M. Lamson, Alexis Russell, Matthew Shudt, Melissa A Leisner, Jonathan Plitnick, Catharine Prussing, Navjot Singh, John Kelly, Erasmus Schneider, Erica Lasek-Nesselquist |
| EPI_ISL_1593866, EPI_ISL_1593867 | URMC LABS | Wadsworth Center, New York State Department of Health | Kirsten St. George, Daryl M. Lamson, Alexis Russell, Matthew Shudt, Melissa A Leisner, Jonathan Plitnick, Catharine Prussing, Navjot Singh, John Kelly, Erasmus Schneider, Erica Lasek-Nesselquist |
| EPI_ISL_1593868, EPI_ISL_1593869 | MONTEFIORE MEDICAL CENTER LABORATORIES | Wadsworth Center, New York State Department of Health | Kirsten St. George, Daryl M. Lamson, Alexis Russell, Matthew Shudt, Melissa A Leisner, Jonathan Plitnick, Catharine Prussing, Navjot Singh, John Kelly, Erasmus Schneider, Erica Lasek-Nesselquist |
| EPI_ISL_1593870, EPI_ISL_1593871 | NYC Pandemic Response Lab | Wadsworth Center, New York State Department of Health | Kirsten St. George, Daryl M. Lamson, Alexis Russell, Matthew Shudt, Melissa A Leisner, Jonathan Plitnick, Catharine Prussing, Navjot Singh, John Kelly, Erasmus Schneider, Erica Lasek-Nesselquist |
| EPI_ISL_1593872 | WESTCHESTER MEDICAL CENTER | Wadsworth Center, New York State Department of Health | Kirsten St. George, Daryl M. Lamson, Alexis Russell, Matthew Shudt, Melissa A Leisner, Jonathan Plitnick, Catharine Prussing, Navjot Singh, John Kelly, Erasmus Schneider, Erica Lasek-Nesselquist |
| EPI_ISL_1593873, EPI_ISL_1593874, EPI_ISL_1593875 | TEMPUS LABS INC | Wadsworth Center, New York State Department of Health | Kirsten St. George, Daryl M. Lamson, Alexis Russell, Matthew Shudt, Melissa A Leisner, Jonathan Plitnick, Catharine Prussing, Navjot Singh, John Kelly, Erasmus Schneider, Erica Lasek-Nesselquist |
| EPI_ISL_1593876, EPI_ISL_1593877 | NORTHWELL HEALTH LABORATORIES | Wadsworth Center, New York State Department of Health | Kirsten St. George, Daryl M. Lamson, Alexis Russell, Matthew Shudt, Melissa A Leisner, Jonathan Plitnick, Catharine Prussing, Navjot Singh, John Kelly, Erasmus Schneider, Erica Lasek-Nesselquist |
| EPI_ISL_1593878 | NYC Pandemic Response Lab | Wadsworth Center, New York State Department of Health | Kirsten St. George, Daryl M. Lamson, Alexis Russell, Matthew Shudt, Melissa A Leisner, Jonathan Plitnick, Catharine Prussing, Navjot Singh, John Kelly, Erasmus Schneider, Erica Lasek-Nesselquist |
| EPI_ISL_1593881 | MONTEFIORE MEDICAL CENTER LABORATORIES | Wadsworth Center, New York State Department of Health | Kirsten St. George, Daryl M. Lamson, Alexis Russell, Matthew Shudt, Melissa A Leisner, Jonathan Plitnick, Catharine Prussing, Navjot Singh, John Kelly, Erasmus Schneider, Erica Lasek-Nesselquist |
| EPI_ISL_1593882 | URMC LABS | Wadsworth Center, New York State Department of Health | Kirsten St. George, Daryl M. Lamson, Alexis Russell, Matthew Shudt, Melissa A Leisner, Jonathan Plitnick, Catharine Prussing, Navjot Singh, John Kelly, Erasmus Schneider, Erica Lasek-Nesselquist |
| EPI_ISL_1593883 | NYC Pandemic Response Lab | Wadsworth Center, New York State Department of Health | Kirsten St. George, Daryl M. Lamson, Alexis Russell, Matthew Shudt, Melissa A Leisner, Jonathan Plitnick, Catharine Prussing, Navjot Singh, John Kelly, Erasmus Schneider, Erica Lasek-Nesselquist |
| EPI_ISL_1593884, EPI_ISL_1593885 | TEMPUS LABS INC | Wadsworth Center, New York State Department of Health | Kirsten St. George, Daryl M. Lamson, Alexis Russell, Matthew Shudt, Melissa A Leisner, Jonathan Plitnick, Catharine Prussing, Navjot Singh, John Kelly, Erasmus Schneider, Erica Lasek-Nesselquist |
| EPI_ISL_1593886 | WHITE PLAINS HOSPITAL CENTER LABORATORY | Wadsworth Center, New York State Department of Health | Kirsten St. George, Daryl M. Lamson, Alexis Russell, Matthew Shudt, Melissa A Leisner, Jonathan Plitnick, Catharine Prussing, Navjot Singh, John Kelly, Erasmus Schneider, Erica Lasek-Nesselquist |
| EPI_ISL_1593887 | NORTHWELL HEALTH LABORATORIES | Wadsworth Center, New York State Department of Health | Kirsten St. George, Daryl M. Lamson, Alexis Russell, Matthew Shudt, Melissa A Leisner, Jonathan Plitnick, Catharine Prussing, Navjot Singh, John Kelly, Erasmus Schneider, Erica Lasek-Nesselquist |
| EPI_ISL_1593889, EPI_ISL_1593890, EPI_ISL_1593891, EPI_ISL_1593893 | Arizona State Public Health Laboratory | Arizona State Public Health Laboratory | Trung Huynh, Jessica Escobar, Katherine Fullerton, Nobuko Fukushima, Stacy White, Linda Getsinger, Victor Waddell |
| EPI_ISL_1593895, EPI_ISL_1593896, EPI_ISL_1593897, EPI_ISL_1593898, EPI_ISL_1593899, EPI_ISL_1593900, EPI_ISL_1593901, EPI_ISL_1593902, EPI_ISL_1593903, EPI_ISL_1593904, EPI_ISL_1593905, EPI_ISL_1593906, EPI_ISL_1593907, EPI_ISL_1593908, EPI_ISL_1593909, EPI_ISL_1593910, EPI_ISL_1593911, EPI_ISL_1593912, EPI_ISL_1593913, EPI_ISL_1593914, EPI_ISL_1593915, EPI_ISL_1593916, EPI_ISL_1593917, EPI_ISL_1593918, EPI_ISL_1593919, EPI_ISL_1593920, EPI_ISL_1593921, EPI_ISL_1593922, EPI_ISL_1593923, EPI_ISL_1593924, EPI_ISL_1593925, EPI_ISL_1593926, EPI_ISL_1593927, EPI_ISL_1593928, EPI_ISL_1593929, EPI_ISL_1593930, EPI_ISL_1593931, EPI_ISL_1593932, EPI_ISL_1593934, EPI_ISL_1593935, EPI_ISL_1593936, EPI_ISL_1593938, EPI_ISL_1593939, EPI_ISL_1593940, EPI_ISL_1593941, EPI_ISL_1593942, EPI_ISL_1593943, EPI_ISL_1593944, EPI_ISL_1593945, EPI_ISL_1593946, EPI_ISL_1593947, EPI_ISL_1593948, EPI_ISL_1593949, EPI_ISL_1593950, EPI_ISL_1593951, EPI_ISL_1593952, EPI_ISL_1593953, EPI_ISL_1593954, EPI_ISL_1593955, EPI_ISL_1593956, EPI_ISL_1593957, EPI_ISL_1593958, EPI_ISL_1593959, EPI_ISL_1593960, EPI_ISL_1593961, EPI_ISL_1593962, EPI_ISL_1593963, EPI_ISL_1593964, EPI_ISL_1593965, EPI_ISL_1593967, EPI_ISL_1593968, EPI_ISL_1593970, EPI_ISL_1593972, EPI_ISL_1593973, EPI_ISL_1593974 | | | |
| see above | Missouri State Public Health Laboratory | Missouri State Public Health Laboratory | Matthew Sinn, Joshua Barry, Ashley New |
| EPI_ISL_1594000, EPI_ISL_1594001, EPI_ISL_1594002, EPI_ISL_1594003, EPI_ISL_1594004, EPI_ISL_1594005, EPI_ISL_1594006, EPI_ISL_1594007, EPI_ISL_1594008, EPI_ISL_1594009, EPI_ISL_1594010, EPI_ISL_1594011, EPI_ISL_1594012, EPI_ISL_1594013, EPI_ISL_1594014, EPI_ISL_1594015, EPI_ISL_1594016, EPI_ISL_1594018, EPI_ISL_1594019, EPI_ISL_1594020, EPI_ISL_1594021, EPI_ISL_1594022, EPI_ISL_1594023, EPI_ISL_1594024, EPI_ISL_1594025, EPI_ISL_1594026, EPI_ISL_1594027, EPI_ISL_1594028 | | | |
| see above | University of Wisconsin-Madison AIDS Vaccine Research Laboratories | University of Wisconsin-Madison AIDS Vaccine Research Laboratories | Gage Moreno, Katarina Braun, et al. AIDS Vaccine Research Laboratories |
| EPI_ISL_1594356, EPI_ISL_1594357, EPI_ISL_1594358, EPI_ISL_1594359, EPI_ISL_1594360, EPI_ISL_1594361, EPI_ISL_1594362, EPI_ISL_1594363, EPI_ISL_1594364, EPI_ISL_1594365, EPI_ISL_1594366, EPI_ISL_1594367, EPI_ISL_1594368, EPI_ISL_1594369, EPI_ISL_1594370, EPI_ISL_1594371, EPI_ISL_1594372, EPI_ISL_1594373, EPI_ISL_1594374, EPI_ISL_1594375, EPI_ISL_1594376, EPI_ISL_1594377, EPI_ISL_1594378 | | | |
| see above | Instituto Nacional de Medicina Genomica | Instituto Nacional de Medicina Genomica | Hidalgo-Miranda A, Mendoza-Vargas A, Reyes-Grajeda JP, Cedro-Tanda A, Alcaraz N, Gonzalez-Barrera D, Rangel-DeLeon D, Miranda-Ortiz H, Rosas-Escobar P, Canseco Mendez JC, Ramirez-Vega O, Munguia-Garza P, Garcia-Cardenas FJ, Gonzalez-Woge MA, Herrera-Montalvo LA |
| EPI_ISL_1595509, EPI_ISL_1595510, EPI_ISL_1595511, EPI_ISL_1595512, EPI_ISL_1595513, EPI_ISL_1595514, EPI_ISL_1595515, EPI_ISL_1595516, EPI_ISL_1595517, EPI_ISL_1595518, EPI_ISL_1595519, EPI_ISL_1595520, EPI_ISL_1595521, EPI_ISL_1595522, EPI_ISL_1595523, EPI_ISL_1595524, EPI_ISL_1595525, EPI_ISL_1595527, EPI_ISL_1595528, EPI_ISL_1595529, EPI_ISL_1595530, EPI_ISL_1595531, EPI_ISL_1595532, EPI_ISL_1595533, EPI_ISL_1595534, EPI_ISL_1595535, EPI_ISL_1595536, EPI_ISL_1595537, EPI_ISL_1595538, EPI_ISL_1595539, EPI_ISL_1595540, EPI_ISL_1595541, EPI_ISL_1595542, EPI_ISL_1595543, EPI_ISL_1595544, EPI_ISL_1595545, EPI_ISL_1595546, EPI_ISL_1595547, EPI_ISL_1595548, EPI_ISL_1595549, EPI_ISL_1595550, EPI_ISL_1595551, EPI_ISL_1595552, EPI_ISL_1595553, EPI_ISL_1595554, EPI_ISL_1595555, EPI_ISL_1595556, EPI_ISL_1595557, EPI_ISL_1595558, EPI_ISL_1595559, EPI_ISL_1595560, EPI_ISL_1595561, EPI_ISL_1595563, EPI_ISL_1595564, EPI_ISL_1595565, EPI_ISL_1595566, EPI_ISL_1595567, EPI_ISL_1595568, EPI_ISL_1595569, EPI_ISL_1595571, EPI_ISL_1595572, EPI_ISL_1595573, EPI_ISL_1595575, EPI_ISL_1595576, EPI_ISL_1595577, EPI_ISL_1595578, EPI_ISL_1595579, EPI_ISL_1595580, EPI_ISL_1595581, EPI_ISL_1595582, EPI_ISL_1595583, EPI_ISL_1595584, EPI_ISL_1595585, EPI_ISL_1595586, EPI_ISL_1595587, EPI_ISL_1595588, EPI_ISL_1595589, EPI_ISL_1595590, EPI_ISL_1595591, EPI_ISL_1595592 | | | |
| see above | Illinois Department of Public Health - Springfield Lab | Illinois Department of Public Health - Springfield Lab | Bryan Sim, Gordon McCall |
| EPI_ISL_1595595, EPI_ISL_1595596, EPI_ISL_1595597, | Centro de Investigación Biomédica del Noreste (CIBIN) | Instituto Nacional de Enfermedades Respiratorias (INER): Centro de Investigación en Enfermedades Infecciosas (CIENI) | Consorcio Mexicano de Vigilancia Genómica (CoViGen-Mex). Authors (in alphabetical order): Julio Elias Alvarado-Yaah, Carlos F. Arias, Santiago Ávila-Ríos, Víctor Hugo Borja-Aburto, Celia Boukadida, Juan Bautista Chale-Dzul, Célida Duque Molina, José Antonio Enciso-Moreno, Gloria Elena Espinosa-Ayala, Fernando Fontove-Herrera, Victor Eduardo Garcia-Arias, Concepción Grajales-Muñiz, Ricardo Grande, Alfredo Herrera-Estrella, Carla Ivón Herrera-Najera, Pavel Isa, Brenda Irasema Maldonado-Meza, Bernardo Martínez-Miguel, Margarita Matías-Florentino, María Guadalupe de Jesús Mireles-Rivera, Gloria María Molina-Salinas, Hector Montoya-Fuentes, José Esteban Muñoz-Medina, José de Jesús Nuñez-Contreras, Alicia Ocaña-Mondragón, Luis Alberto Ochoa-Carrera, Hector Esteban Paz-Juárez, Francisco Pulido, Helen Haydee Fernanda Ramírez- |

| | | | |
|---|---|---|---|
| EPI_ISL_1595598 | | | Plascencia, Angel Gustavo Salas-Lais, Alejandro Sanchez-Flores, Clara Esperanza Santacruz-Tinoco, María Guadalupe Santiago-Mauricio, Nelly Sélem-Mojica, Blanca Taboada, Gloria Vazquez |
| EPI_ISL_1595599 | Unidad de Investigación Médica de Yucatán (UIMY) | Instituto Nacional de Enfermedades Respiratorias (INER): Centro de Investigación en Enfermedades Infecciosas (CIENI) | Consorcio Mexicano de Vigilancia Genómica (CoViGen-Mex). Authors (in alphabetical order): Julio Elias Alvarado-Yaah, Carlos F. Arias, Santiago Ávila-Ríos, Víctor Hugo Borja-Aburto, Celia Boukadida, Juan Bautista Chale-Dzul, Célida Duque Molina, José Antonio Enciso-Moreno, Gloria Elena Espinosa-Ayala, Fernando Fontove-Herrera, Victor Eduardo Garcia-Arias, Concepción Grajales-Muñiz, Ricardo Grande, Alfredo Herrera-Estrella, Carla Ivón Herrera-Najera, Pavel Isa, Brenda Irasema Maldonado-Meza, Bernardo Martínez-Miguel, Margarita Matías-Florentino, María Guadalupe de Jesús Mireles-Rivera, Gloria María Molina-Salinas, Hector Montoya-Fuentes, José Esteban Muñoz-Medina, José de Jesús Nuñez-Contreras, Alicia Ocaña-Mondragón, Luis Alberto Ochoa-Carrera, Hector Esteban Paz-Juárez, Francisco Pulido, Helen Haydee Fernanda Ramírez-Plascencia, Angel Gustavo Salas-Lais, Alejandro Sanchez-Flores, Clara Esperanza Santacruz-Tinoco, María Guadalupe Santiago-Mauricio, Nelly Sélem-Mojica, Blanca Taboada, Gloria Vazquez |
| EPI_ISL_1595600 | Unidad de Investigación Biomédica de Zacatecas (UIBZ) | Instituto Nacional de Enfermedades Respiratorias (INER): Centro de Investigación en Enfermedades Infecciosas (CIENI) | Consorcio Mexicano de Vigilancia Genómica (CoViGen-Mex). Authors (in alphabetical order): Julio Elias Alvarado-Yaah, Carlos F. Arias, Santiago Ávila-Ríos, Víctor Hugo Borja-Aburto, Celia Boukadida, Juan Bautista Chale-Dzul, Célida Duque Molina, José Antonio Enciso-Moreno, Gloria Elena Espinosa-Ayala, Fernando Fontove-Herrera, Victor Eduardo Garcia-Arias, Concepción Grajales-Muñiz, Ricardo Grande, Alfredo Herrera-Estrella, Carla Ivón Herrera-Najera, Pavel Isa, Brenda Irasema Maldonado-Meza, Bernardo Martínez-Miguel, Margarita Matías-Florentino, María Guadalupe de Jesús Mireles-Rivera, Gloria María Molina-Salinas, Hector Montoya-Fuentes, José Esteban Muñoz-Medina, José de Jesús Nuñez-Contreras, Alicia Ocaña-Mondragón, Luis Alberto Ochoa-Carrera, Hector Esteban Paz-Juárez, Francisco Pulido, Helen Haydee Fernanda Ramírez-Plascencia, Angel Gustavo Salas-Lais, Alejandro Sanchez-Flores, Clara Esperanza Santacruz-Tinoco, María Guadalupe Santiago-Mauricio, Nelly Sélem-Mojica, Blanca Taboada, Gloria Vazquez |
| EPI_ISL_1595601, EPI_ISL_1595602, EPI_ISL_1595603, EPI_ISL_1595604, EPI_ISL_1595605, EPI_ISL_1595606, EPI_ISL_1595607 | Centro de Investigación Biomédica de Occidente (CIBO) | Instituto Nacional de Enfermedades Respiratorias (INER): Centro de Investigación en Enfermedades Infecciosas (CIENI) | Consorcio Mexicano de Vigilancia Genómica (CoViGen-Mex). Authors (in alphabetical order): Julio Elias Alvarado-Yaah, Carlos F. Arias, Santiago Ávila-Ríos, Víctor Hugo Borja-Aburto, Celia Boukadida, Juan Bautista Chale-Dzul, Célida Duque Molina, José Antonio Enciso-Moreno, Gloria Elena Espinosa-Ayala, Fernando Fontove-Herrera, Victor Eduardo Garcia-Arias, Concepción Grajales-Muñiz, Ricardo Grande, Alfredo Herrera-Estrella, Carla Ivón Herrera-Najera, Pavel Isa, Brenda Irasema Maldonado-Meza, Bernardo Martínez-Miguel, Margarita Matías-Florentino, María Guadalupe de Jesús Mireles-Rivera, Gloria María Molina-Salinas, Hector Montoya-Fuentes, José Esteban Muñoz-Medina, José de Jesús Nuñez-Contreras, Alicia Ocaña-Mondragón, Luis Alberto Ochoa-Carrera, Hector Esteban Paz-Juárez, Francisco Pulido, Helen Haydee Fernanda Ramírez-Plascencia, Angel Gustavo Salas-Lais, Alejandro Sanchez-Flores, Clara Esperanza Santacruz-Tinoco, María Guadalupe Santiago-Mauricio, Nelly Sélem-Mojica, Blanca Taboada, Gloria Vazquez |
| EPI_ISL_1595608, EPI_ISL_1595609, EPI_ISL_1595610, EPI_ISL_1595611 | Laboratorio Central de Epidemiología (LCE) | Instituto Nacional de Enfermedades Respiratorias (INER): Centro de Investigación en Enfermedades Infecciosas (CIENI) | Consorcio Mexicano de Vigilancia Genómica (CoViGen-Mex). Authors (in alphabetical order): Julio Elias Alvarado-Yaah, Carlos F. Arias, Santiago Ávila-Ríos, Víctor Hugo Borja-Aburto, Celia Boukadida, Juan Bautista Chale-Dzul, Célida Duque Molina, José Antonio Enciso-Moreno, Gloria Elena Espinosa-Ayala, Fernando Fontove-Herrera, Victor Eduardo Garcia-Arias, Concepción Grajales-Muñiz, Ricardo Grande, Alfredo Herrera-Estrella, Carla Ivón Herrera-Najera, Pavel Isa, Brenda Irasema Maldonado-Meza, Bernardo Martínez-Miguel, Margarita Matías-Florentino, María Guadalupe de Jesús Mireles-Rivera, Gloria María Molina-Salinas, Hector Montoya-Fuentes, José Esteban Muñoz-Medina, José de Jesús Nuñez-Contreras, Alicia Ocaña-Mondragón, Luis Alberto Ochoa-Carrera, Hector Esteban Paz-Juárez, Francisco Pulido, Helen Haydee Fernanda Ramírez-Plascencia, Angel Gustavo Salas-Lais, Alejandro Sanchez-Flores, Clara Esperanza Santacruz-Tinoco, María Guadalupe Santiago-Mauricio, Nelly Sélem-Mojica, Blanca Taboada, Gloria Vazquez |
| EPI_ISL_1595736, EPI_ISL_1595737, EPI_ISL_1595738, EPI_ISL_1595739, EPI_ISL_1595740, EPI_ISL_1595741, EPI_ISL_1595742, EPI_ISL_1595743, EPI_ISL_1595744, EPI_ISL_1595745, EPI_ISL_1595746, EPI_ISL_1595747, EPI_ISL_1595748, EPI_ISL_1595749, EPI_ISL_1595750, EPI_ISL_1595751, EPI_ISL_1595752 | | | |
| see above | DC Public Health Lab/ Dept. of Forensic Sciences | DC Public Health Lab/ Dept. of Forensic Sciences | Janis Doss, Scott Nguyen, Elizabeth Zelaya, Sarah Scott, Connie Maza, Monica Mann, Brittany Hamilton, David Payne, Jocelyn Hauser |
| EPI_ISL_1595844 | Compass Laboratory Services, LLC | Compass Laboratory Services | William Budd; PhD |
| EPI_ISL_1597646, EPI_ISL_1597647, EPI_ISL_1597648, EPI_ISL_1597649, EPI_ISL_1597650, EPI_ISL_1597651, EPI_ISL_1597652, EPI_ISL_1597653, EPI_ISL_1597654, EPI_ISL_1597655, EPI_ISL_1597656, EPI_ISL_1597657, EPI_ISL_1597658, EPI_ISL_1597659, EPI_ISL_1597660, EPI_ISL_1597661, EPI_ISL_1597662, EPI_ISL_1597663, EPI_ISL_1597664, EPI_ISL_1597665, EPI_ISL_1597666, EPI_ISL_1597667, EPI_ISL_1597668, EPI_ISL_1597669, EPI_ISL_1597670, EPI_ISL_1597671, EPI_ISL_1597672, EPI_ISL_1597673, EPI_ISL_1597674, EPI_ISL_1597675, EPI_ISL_1597676, EPI_ISL_1597677, EPI_ISL_1597678, EPI_ISL_1597679, EPI_ISL_1597680, EPI_ISL_1597681, EPI_ISL_1597682, EPI_ISL_1597683, EPI_ISL_1597684, EPI_ISL_1597685, EPI_ISL_1597686, EPI_ISL_1597687, EPI_ISL_1597688, EPI_ISL_1597689, EPI_ISL_1597690, EPI_ISL_1597691, EPI_ISL_1597692, EPI_ISL_1597693, EPI_ISL_1597694, EPI_ISL_1597695 | | | |
| see above | Rush University Medical Center | Abbott | Mary Hayden, Stefan Green, Alan Landay, Cheryl Jennings, Ana Olivo, Ana Vallari, Barbara Harris, Mary Rodgers, Todd Meyer, Gavin Cloherty |
| EPI_ISL_1597696, EPI_ISL_1597697, EPI_ISL_1597698, EPI_ISL_1597699, EPI_ISL_1597700, EPI_ISL_1597701, EPI_ISL_1597702, EPI_ISL_1597703, EPI_ISL_1597704, EPI_ISL_1597705, EPI_ISL_1597706, EPI_ISL_1597707, EPI_ISL_1597708, EPI_ISL_1597709, EPI_ISL_1597710, EPI_ISL_1597711, EPI_ISL_1597712, EPI_ISL_1597713, EPI_ISL_1597714, EPI_ISL_1597715, EPI_ISL_1597716, EPI_ISL_1597717 | | | |
| see above | Montefiore Medical Center | Abbott | Yitz Goldstein, Amy Fox, Ana Olivo, Ana Vallari, Barbara Harris, Mary Rodgers, Todd Meyer, Gavin Cloherty |
| EPI_ISL_1597718, EPI_ISL_1597719, EPI_ISL_1597720, EPI_ISL_1597721, EPI_ISL_1597722, EPI_ISL_1597723, EPI_ISL_1597724 | Rush University Medical Center | Abbott | Mary Hayden, Stefan Green, Alan Landay, Cheryl Jennings, Ana Olivo, Ana Vallari, Barbara Harris, Mary Rodgers, Todd Meyer, Gavin Cloherty |
| EPI_ISL_1597725, EPI_ISL_1597726, EPI_ISL_1597727, EPI_ISL_1597728, EPI_ISL_1597729, EPI_ISL_1597730, EPI_ISL_1597731, EPI_ISL_1597732, EPI_ISL_1597733 | Montefiore Medical Center | Abbott | Yitz Goldstein, Amy Fox, Ana Olivo, Ana Vallari, Barbara Harris, Mary Rodgers, Todd Meyer, Gavin Cloherty |
| EPI_ISL_1597734, EPI_ISL_1597735, EPI_ISL_1597736, EPI_ISL_1597737, EPI_ISL_1597738, EPI_ISL_1597739, EPI_ISL_1597740, EPI_ISL_1597741, EPI_ISL_1597742, EPI_ISL_1597743, EPI_ISL_1597744 | | | |
| see above | Rush University Medical Center | Abbott | Mary Hayden, Stefan Green, Alan Landay, Cheryl Jennings, Ana Olivo, Ana Vallari, Barbara Harris, Mary Rodgers, Todd Meyer, Gavin Cloherty |
| EPI_ISL_1597745, EPI_ISL_1597746, EPI_ISL_1597747, EPI_ISL_1597748, EPI_ISL_1597749, EPI_ISL_1597750, EPI_ISL_1597751, EPI_ISL_1597752, EPI_ISL_1597753, EPI_ISL_1597754, EPI_ISL_1597755, EPI_ISL_1597756, EPI_ISL_1597757, EPI_ISL_1597758, EPI_ISL_1597759, EPI_ISL_1597760, EPI_ISL_1597761, EPI_ISL_1597762, EPI_ISL_1597763, EPI_ISL_1597764, EPI_ISL_1597765, EPI_ISL_1597766, EPI_ISL_1597767, EPI_ISL_1597768, EPI_ISL_1597769, EPI_ISL_1597770, EPI_ISL_1597771, EPI_ISL_1597772, EPI_ISL_1597773, EPI_ISL_1597774, EPI_ISL_1597775, EPI_ISL_1597776, EPI_ISL_1597777, EPI_ISL_1597778, EPI_ISL_1597779, EPI_ISL_1597780, EPI_ISL_1597781 | | | |
| see above | Montefiore Medical Center | Abbott | Yitz Goldstein, Amy Fox, Ana Olivo, Ana Vallari, Barbara Harris, Mary Rodgers, Todd Meyer, Gavin Cloherty |
| EPI_ISL_1597782 | University of Wisconsin-Madison AIDS Vaccine Research Laboratories | University of Wisconsin-Madison AIDS Vaccine Research Laboratories | Gage Moreno, Katarina Braun, et al. AIDS Vaccine Research Laboratories |
| EPI_ISL_1599134 | Rush University Medical Center | RIPHL at Rush University Medical Center | Laura Furtado, Stefan Green, Kevin Kunstman, Max Kolton, Marieta Hyde |
| EPI_ISL_1599505, EPI_ISL_1599506, EPI_ISL_1599507, EPI_ISL_1599508, EPI_ISL_1599510, EPI_ISL_1599512, EPI_ISL_1599514, EPI_ISL_1599516, EPI_ISL_1599518, EPI_ISL_1599521, EPI_ISL_1599523 | | | |
| see above | NCSLPH | NCSLPH | Chase K, Miller MC, Greene S, Glover W |
| EPI_ISL_1601275, EPI_ISL_1601277, EPI_ISL_1601279, EPI_ISL_1601280, EPI_ISL_1601282, EPI_ISL_1601284, EPI_ISL_1601286, EPI_ISL_1601288, EPI_ISL_1601289, EPI_ISL_1601291, EPI_ISL_1601293, EPI_ISL_1601295, EPI_ISL_1601297, EPI_ISL_1601298, EPI_ISL_1601300, EPI_ISL_1601302, EPI_ISL_1601304, EPI_ISL_1601306, EPI_ISL_1601308, EPI_ISL_1601310, EPI_ISL_1601311, EPI_ISL_1601313, EPI_ISL_1601315, EPI_ISL_1601317, EPI_ISL_1601319, EPI_ISL_1601321, EPI_ISL_1601323, EPI_ISL_1601325, EPI_ISL_1601327, EPI_ISL_1601329, EPI_ISL_1601331, EPI_ISL_1601332, EPI_ISL_1601334, EPI_ISL_1601336, EPI_ISL_1601338, EPI_ISL_1601340, EPI_ISL_1601342, EPI_ISL_1601344, EPI_ISL_1601346, EPI_ISL_1601348, EPI_ISL_1601349, EPI_ISL_1601351, EPI_ISL_1601353, EPI_ISL_1601355, EPI_ISL_1601357, EPI_ISL_1601358, EPI_ISL_1601360, EPI_ISL_1601362, EPI_ISL_1601364, EPI_ISL_1601366, EPI_ISL_1601368, EPI_ISL_1601370, EPI_ISL_1601372, EPI_ISL_1601373, EPI_ISL_1601375, EPI_ISL_1601377, EPI_ISL_1601379, EPI_ISL_1601380, EPI_ISL_1601382, EPI_ISL_1601384, EPI_ISL_1601386, EPI_ISL_1601387, EPI_ISL_1601390, EPI_ISL_1601391, EPI_ISL_1601393, EPI_ISL_1601395, EPI_ISL_1601397, EPI_ISL_1601398, EPI_ISL_1601400, EPI_ISL_1601402, EPI_ISL_1601404, EPI_ISL_1601405, EPI_ISL_1601407, EPI_ISL_1601409, EPI_ISL_1601411, EPI_ISL_1601413, EPI_ISL_1601415, EPI_ISL_1601417, EPI_ISL_1601419, EPI_ISL_1601421, EPI_ISL_1601422, EPI_ISL_1601424, EPI_ISL_1601426, EPI_ISL_1601428, EPI_ISL_1601430, EPI_ISL_1601432, EPI_ISL_1601434, EPI_ISL_1601435, EPI_ISL_1601437, EPI_ISL_1601439, EPI_ISL_1601441, EPI_ISL_1601443, EPI_ISL_1601445, EPI_ISL_1601446, EPI_ISL_1601448, EPI_ISL_1601450, EPI_ISL_1601452, EPI_ISL_1601454, EPI_ISL_1601456, EPI_ISL_1601458, EPI_ISL_1601459, EPI_ISL_1601461, EPI_ISL_1601463, EPI_ISL_1601465, EPI_ISL_1601467, EPI_ISL_1601469, EPI_ISL_1601471, EPI_ISL_1601472, EPI_ISL_1601474, EPI_ISL_1601476, EPI_ISL_1601478, EPI_ISL_1601479, EPI_ISL_1601481, EPI_ISL_1601483, EPI_ISL_1601485, EPI_ISL_1601487, EPI_ISL_1601488, EPI_ISL_1601490, EPI_ISL_1601492, EPI_ISL_1601494, EPI_ISL_1601495, EPI_ISL_1601497, EPI_ISL_1601499, EPI_ISL_1601501, EPI_ISL_1601502, EPI_ISL_1601504, EPI_ISL_1601506, EPI_ISL_1601508, EPI_ISL_1601510, EPI_ISL_1601511, EPI_ISL_1601513, EPI_ISL_1601515, EPI_ISL_1601517, EPI_ISL_1601519, EPI_ISL_1601521, EPI_ISL_1601522, EPI_ISL_1601524, EPI_ISL_1601526, EPI_ISL_1601528, EPI_ISL_1601530, EPI_ISL_1601532, EPI_ISL_1601534, EPI_ISL_1601535, EPI_ISL_1601537, EPI_ISL_1601539, EPI_ISL_1601541, EPI_ISL_1604932, EPI_ISL_1604934, EPI_ISL_1604936, EPI_ISL_1604938, EPI_ISL_1604939, EPI_ISL_1604941, EPI_ISL_1604943, EPI_ISL_1604945, EPI_ISL_1604946, EPI_ISL_1604948, EPI_ISL_1604950, EPI_ISL_1604952, EPI_ISL_1604953, EPI_ISL_1604955, EPI_ISL_1604957, EPI_ISL_1604959, EPI_ISL_1604962, EPI_ISL_1604966, EPI_ISL_1604968, EPI_ISL_1604969, EPI_ISL_1604971, EPI_ISL_1604973, EPI_ISL_1604975, EPI_ISL_1604977, EPI_ISL_1604979, EPI_ISL_1604981, EPI_ISL_1604983, EPI_ISL_1604984, EPI_ISL_1604986, EPI_ISL_1604988, EPI_ISL_1604990, EPI_ISL_1604992, EPI_ISL_1604994, EPI_ISL_1604996, EPI_ISL_1604998, EPI_ISL_1605006, EPI_ISL_1605010, EPI_ISL_1605016, EPI_ISL_1605018, EPI_ISL_1605020, EPI_ISL_1605022, EPI_ISL_1605023 | | | |
| see above | UW Virology Lab | UW Virology Lab | Pavitra Roychoudhury, Hong Xie, Lasata Shrestha, Shah Mohamed Bakhash, Michelle Lin, Noah R. Baker, Sean Ellis, Saraswathi Sathees, Meei-Li Huang, Keith R Jerome, Alexander Greninger |
| EPI_ISL_1605699 | TEMPUS LABS INC | Wadsworth Center, New York State Department of Health | Kirsten St. George, Daryl M. Lamson, Alexis Russell, Matthew Shudt, Melissa A Leisner, Jonathan Plitnick, Catharine Prussing, Navjot Singh, John Kelly, Erasmus Schneider, Erica Lasek-Nesselquist |
| EPI_ISL_1605704 | NYC Pandemic Response Lab | Wadsworth Center, New York State Department of Health | Kirsten St. George, Daryl M. Lamson, Alexis Russell, Matthew Shudt, Melissa A Leisner, Jonathan Plitnick, Catharine Prussing, Navjot Singh, John Kelly, Erasmus Schneider, Erica Lasek-Nesselquist |
| EPI_ISL_1607244, EPI_ISL_1607248, EPI_ISL_1607258, EPI_ISL_1607273, EPI_ISL_1607287, EPI_ISL_1607296, EPI_ISL_1607304, EPI_ISL_1607308, EPI_ISL_1607310, EPI_ISL_1607333, EPI_ISL_1607340, EPI_ISL_1607360, EPI_ISL_1607368, EPI_ISL_1607381 | | | |
| see above | North Dakota Department of Health, Public Health Laboratory | North Dakota Department of Health, Public Health Laboratory | Lisa Wingerter |
| EPI_ISL_1607871 | Clinical Reference Laboratory | Kansas Health and Environmental Lab | Mike Grose, Jonathan Barnell, Ben Olsen, and Phil Adam |
| EPI_ISL_1607872, EPI_ISL_1607873, EPI_ISL_1607874, EPI_ISL_1607875, EPI_ISL_1607876, EPI_ISL_1607877 | Kansas Health and Environmental Lab | Kansas Health and Environmental Lab | Mike Grose, Jonathan Barnell, Ben Olsen, and Phil Adam |
| EPI_ISL_1607878 | Clinical Reference Laboratory | Kansas Health and Environmental Lab | Mike Grose, Jonathan Barnell, Ben Olsen, and Phil Adam |
| EPI_ISL_1607880, EPI_ISL_1607881, EPI_ISL_1607882, EPI_ISL_1607883, EPI_ISL_1607884, EPI_ISL_1607885, EPI_ISL_1607886, EPI_ISL_1607887, EPI_ISL_1607888, EPI_ISL_1607889, EPI_ISL_1607890, EPI_ISL_1607891, EPI_ISL_1607892, EPI_ISL_1607893, EPI_ISL_1607894, EPI_ISL_1607895, EPI_ISL_1607896, EPI_ISL_1607897, EPI_ISL_1607898, EPI_ISL_1607899, EPI_ISL_1607900, EPI_ISL_1607901, EPI_ISL_1607902, EPI_ISL_1607903, EPI_ISL_1607904, EPI_ISL_1607905, EPI_ISL_1607906, EPI_ISL_1607907, EPI_ISL_1607908, EPI_ISL_1607911, EPI_ISL_1607912, EPI_ISL_1607913, EPI_ISL_1607914, EPI_ISL_1607915, EPI_ISL_1607916, EPI_ISL_1607917, EPI_ISL_1607918, EPI_ISL_1607919, EPI_ISL_1607920, EPI_ISL_1607921, EPI_ISL_1607922, EPI_ISL_1607923, EPI_ISL_1607924, EPI_ISL_1607925, EPI_ISL_1607926, EPI_ISL_1607927, EPI_ISL_1607928, EPI_ISL_1607929, EPI_ISL_1607930, EPI_ISL_1607931, EPI_ISL_1607932, EPI_ISL_1607933, EPI_ISL_1607934, EPI_ISL_1607935, EPI_ISL_1607936, EPI_ISL_1607937, EPI_ISL_1607938, EPI_ISL_1607939, EPI_ISL_1607940, EPI_ISL_1607941, EPI_ISL_1607942, EPI_ISL_1607943, EPI_ISL_1607944, EPI_ISL_1607945, EPI_ISL_1607946, EPI_ISL_1607947, EPI_ISL_1607948, EPI_ISL_1607949, EPI_ISL_1607950, EPI_ISL_1607951, EPI_ISL_1607952, EPI_ISL_1607953, EPI_ISL_1607954, EPI_ISL_1607955, EPI_ISL_1607956, EPI_ISL_1607957, EPI_ISL_1607958, EPI_ISL_1607959, EPI_ISL_1607960, EPI_ISL_1607961, EPI_ISL_1607962, EPI_ISL_1607963, EPI_ISL_1607964, EPI_ISL_1607965, EPI_ISL_1607966, EPI_ISL_1607967 | | | |
| see above | TXDSHS | TXDSHS | Rashmi Tuladhar, Bonnie Oh, Jenny Zhang, Maliha Rahman, Mayela Pedrueza, Anita Pokharel, Lorraine Rodriguez, Myong Koag, Chun Wang, Rachel Lee, Grace Kubin |
| EPI_ISL_1608062, EPI_ISL_1608063, EPI_ISL_1608065, EPI_ISL_1608066, EPI_ISL_1608067 | Arizona State University | Arizona State University | Rabia Maqsood, LaRinda A. Holland, Peter T. Skidmore, Nicholas J. Mellor, Joy M. Blain, Valerie Harris, Joshua LaBaer, Vel Murugan, Efrem S. Lim |
| EPI_ISL_1608098 | DC Public Health Lab/ Dept. of Forensic Sciences | DC Public Health Lab/ Dept. of Forensic Sciences | Janis Doss, Scott Nguyen, Elizabeth Zelaya, Sarah Scott, Connie Maza, Monica Mann, Brittany Hamilton, David Payne, Jocelyn Hauser |

EPI_ISL_1608140, EPI_ISL_1608141, EPI_ISL_1608142, EPI_ISL_1608143, EPI_ISL_1608144, EPI_ISL_1608145, EPI_ISL_1608146, EPI_ISL_1608147, EPI_ISL_1608148, EPI_ISL_1608149, EPI_ISL_1608150, EPI_ISL_1608151, EPI_ISL_1608152, EPI_ISL_1608153, EPI_ISL_1608154, EPI_ISL_1608155, EPI_ISL_1608156, EPI_ISL_1608157, EPI_ISL_1608158, EPI_ISL_1608159, EPI_ISL_1608160

| | | | |
|---|---|---|---|
| see above | City of Milwaukee Health Department Laboratory | City of Milwaukee Health Department Laboratory | Sanjib Bhattacharyya |

EPI_ISL_1608173, EPI_ISL_1608180, EPI_ISL_1608182, EPI_ISL_1608183, EPI_ISL_1608185, EPI_ISL_1608186, EPI_ISL_1608187, EPI_ISL_1608189, EPI_ISL_1608190, EPI_ISL_1608194, EPI_ISL_1608198, EPI_ISL_1608200, EPI_ISL_1608201, EPI_ISL_1608202, EPI_ISL_1608203, EPI_ISL_1608206, EPI_ISL_1608207, EPI_ISL_1608208, EPI_ISL_1608210, EPI_ISL_1608216, EPI_ISL_1608218, EPI_ISL_1608221, EPI_ISL_1608222, EPI_ISL_1608223

| | | | |
|---|---|---|---|
| see above | North Dakota Department of Health, Public Health Laboratory | North Dakota Department of Health, Public Health Laboratory | Lisa Wingerter |

EPI_ISL_1608582, EPI_ISL_1608583, EPI_ISL_1608584, EPI_ISL_1608585, EPI_ISL_1608586, EPI_ISL_1608587, EPI_ISL_1608588, EPI_ISL_1608589, EPI_ISL_1608590, EPI_ISL_1608591, EPI_ISL_1608592, EPI_ISL_1608593, EPI_ISL_1608594, EPI_ISL_1608595, EPI_ISL_1608596, EPI_ISL_1608597, EPI_ISL_1608598, EPI_ISL_1608599, EPI_ISL_1608600, EPI_ISL_1608601, EPI_ISL_1608602, EPI_ISL_1608603, EPI_ISL_1608604, EPI_ISL_1608605, EPI_ISL_1608606, EPI_ISL_1608607, EPI_ISL_1608609, EPI_ISL_1608610, EPI_ISL_1608611, EPI_ISL_1608612, EPI_ISL_1608613, EPI_ISL_1608614, EPI_ISL_1608615, EPI_ISL_1608616, EPI_ISL_1608617, EPI_ISL_1608618, EPI_ISL_1608620, EPI_ISL_1608621, EPI_ISL_1608622, EPI_ISL_1608624, EPI_ISL_1608625, EPI_ISL_1608626, EPI_ISL_1608627, EPI_ISL_1608628, EPI_ISL_1608629, EPI_ISL_1608630, EPI_ISL_1608631, EPI_ISL_1608632, EPI_ISL_1608633, EPI_ISL_1608634, EPI_ISL_1608635, EPI_ISL_1608636, EPI_ISL_1608637, EPI_ISL_1608638, EPI_ISL_1608639, EPI_ISL_1608640, EPI_ISL_1608641, EPI_ISL_1608642, EPI_ISL_1608643, EPI_ISL_1608644, EPI_ISL_1608645, EPI_ISL_1608646, EPI_ISL_1608647, EPI_ISL_1608648, EPI_ISL_1608649, EPI_ISL_1608650, EPI_ISL_1608651, EPI_ISL_1608652, EPI_ISL_1608653, EPI_ISL_1608654, EPI_ISL_1608655, EPI_ISL_1608656, EPI_ISL_1608657, EPI_ISL_1608658, EPI_ISL_1608659, EPI_ISL_1608660, EPI_ISL_1608661, EPI_ISL_1608662, EPI_ISL_1608663, EPI_ISL_1608664, EPI_ISL_1608665, EPI_ISL_1608666, EPI_ISL_1608668, EPI_ISL_1608669, EPI_ISL_1608672, EPI_ISL_1608673, EPI_ISL_1608674, EPI_ISL_1608675, EPI_ISL_1608676, EPI_ISL_1608677, EPI_ISL_1608678, EPI_ISL_1608679, EPI_ISL_1608680, EPI_ISL_1608681, EPI_ISL_1608682, EPI_ISL_1608683, EPI_ISL_1608684, EPI_ISL_1608685, EPI_ISL_1608686, EPI_ISL_1608687, EPI_ISL_1608688, EPI_ISL_1608689, EPI_ISL_1608690, EPI_ISL_1608691, EPI_ISL_1608692, EPI_ISL_1608693, EPI_ISL_1608694, EPI_ISL_1608695, EPI_ISL_1608696, EPI_ISL_1608697, EPI_ISL_1608698, EPI_ISL_1608699, EPI_ISL_1608700, EPI_ISL_1608701, EPI_ISL_1608702, EPI_ISL_1608703, EPI_ISL_1608704, EPI_ISL_1608705, EPI_ISL_1608706, EPI_ISL_1608707, EPI_ISL_1608708, EPI_ISL_1608709, EPI_ISL_1608710, EPI_ISL_1608711, EPI_ISL_1608712, EPI_ISL_1608713, EPI_ISL_1608714, EPI_ISL_1608715, EPI_ISL_1608716, EPI_ISL_1608717, EPI_ISL_1608718, EPI_ISL_1608719, EPI_ISL_1608720, EPI_ISL_1608721, EPI_ISL_1608722, EPI_ISL_1608723, EPI_ISL_1608724, EPI_ISL_1608725, EPI_ISL_1608726, EPI_ISL_1608727, EPI_ISL_1608728, EPI_ISL_1608729, EPI_ISL_1608730, EPI_ISL_1608731, EPI_ISL_1608732, EPI_ISL_1608733, EPI_ISL_1608734, EPI_ISL_1608735, EPI_ISL_1608736, EPI_ISL_1608737, EPI_ISL_1608738, EPI_ISL_1608739, EPI_ISL_1608740, EPI_ISL_1608741, EPI_ISL_1608742, EPI_ISL_1608743, EPI_ISL_1608744, EPI_ISL_1608745, EPI_ISL_1608746, EPI_ISL_1608747, EPI_ISL_1608748, EPI_ISL_1608749, EPI_ISL_1608750, EPI_ISL_1608751, EPI_ISL_1608752, EPI_ISL_1608753, EPI_ISL_1608754, EPI_ISL_1608755, EPI_ISL_1608756, EPI_ISL_1608758, EPI_ISL_1608759, EPI_ISL_1608760, EPI_ISL_1608761

| | | | |
|---|---|---|---|
| see above | Maryland Genomics, Institute for Genome Sciences, University of Maryland School of Medicine | Maryland Genomics, Institute for Genome Sciences, University of Maryland School of Medicine | Tallon, Luke J; Sadzewicz, Lisa D; Humphrys, Mike; Ott, Sandra; Roussey, Holly; Mehta, Aditya; Vavikolanu, Kranthi; Fraser, Claire M; Ravel, Jacques |

EPI_ISL_1608762, EPI_ISL_1608763, EPI_ISL_1608764, EPI_ISL_1608765, EPI_ISL_1608766, EPI_ISL_1608767, EPI_ISL_1608768, EPI_ISL_1608769, EPI_ISL_1608770, EPI_ISL_1608771, EPI_ISL_1608772, EPI_ISL_1608775, EPI_ISL_1608778, EPI_ISL_1608779, EPI_ISL_1608781, EPI_ISL_1608782, EPI_ISL_1608783, EPI_ISL_1608784, EPI_ISL_1608785, EPI_ISL_1608786, EPI_ISL_1608787, EPI_ISL_1608789, EPI_ISL_1608790, EPI_ISL_1608792, EPI_ISL_1608793, EPI_ISL_1608794, EPI_ISL_1608795, EPI_ISL_1608796, EPI_ISL_1608797, EPI_ISL_1608798, EPI_ISL_1608799, EPI_ISL_1608800, EPI_ISL_1608803, EPI_ISL_1608804, EPI_ISL_1608805, EPI_ISL_1608807, EPI_ISL_1608808, EPI_ISL_1608809, EPI_ISL_1608810, EPI_ISL_1608813, EPI_ISL_1608814, EPI_ISL_1608815, EPI_ISL_1608816, EPI_ISL_1608817, EPI_ISL_1608819, EPI_ISL_1608820, EPI_ISL_1608821, EPI_ISL_1608822, EPI_ISL_1608823, EPI_ISL_1608825, EPI_ISL_1608826, EPI_ISL_1608827, EPI_ISL_1608828, EPI_ISL_1608829, EPI_ISL_1608830, EPI_ISL_1608831, EPI_ISL_1608833, EPI_ISL_1608834, EPI_ISL_1608835, EPI_ISL_1608837, EPI_ISL_1608838, EPI_ISL_1608839, EPI_ISL_1608840, EPI_ISL_1608841, EPI_ISL_1608842, EPI_ISL_1608843, EPI_ISL_1608844, EPI_ISL_1608845, EPI_ISL_1608847, EPI_ISL_1608848, EPI_ISL_1608849, EPI_ISL_1608850, EPI_ISL_1608851, EPI_ISL_1608852, EPI_ISL_1608853, EPI_ISL_1608855, EPI_ISL_1608856, EPI_ISL_1608857, EPI_ISL_1608858, EPI_ISL_1608859, EPI_ISL_1608860, EPI_ISL_1608861, EPI_ISL_1608862, EPI_ISL_1608864, EPI_ISL_1608865, EPI_ISL_1608866, EPI_ISL_1608868, EPI_ISL_1608869, EPI_ISL_1608870, EPI_ISL_1608871, EPI_ISL_1608873, EPI_ISL_1608874, EPI_ISL_1608875, EPI_ISL_1608876, EPI_ISL_1608877, EPI_ISL_1608878, EPI_ISL_1608879, EPI_ISL_1608880, EPI_ISL_1608881, EPI_ISL_1608882, EPI_ISL_1608884, EPI_ISL_1608885, EPI_ISL_1608886, EPI_ISL_1608888, EPI_ISL_1608889, EPI_ISL_1608892, EPI_ISL_1608893, EPI_ISL_1608896, EPI_ISL_1608897, EPI_ISL_1608898, EPI_ISL_1608899, EPI_ISL_1608901, EPI_ISL_1608902, EPI_ISL_1608903, EPI_ISL_1608904, EPI_ISL_1608905, EPI_ISL_1608906, EPI_ISL_1608907, EPI_ISL_1608908, EPI_ISL_1608910, EPI_ISL_1608911, EPI_ISL_1608912, EPI_ISL_1608913, EPI_ISL_1608914, EPI_ISL_1608915, EPI_ISL_1608918, EPI_ISL_1608919, EPI_ISL_1608920, EPI_ISL_1608921, EPI_ISL_1608922, EPI_ISL_1608923, EPI_ISL_1608924, EPI_ISL_1608925, EPI_ISL_1608926, EPI_ISL_1608928, EPI_ISL_1608929, EPI_ISL_1608930, EPI_ISL_1608932, EPI_ISL_1608934, EPI_ISL_1608935, EPI_ISL_1608936, EPI_ISL_1608937, EPI_ISL_1608939, EPI_ISL_1608940, EPI_ISL_1608941, EPI_ISL_1608942, EPI_ISL_1608943, EPI_ISL_1608944, EPI_ISL_1608945, EPI_ISL_1608947, EPI_ISL_1608948, EPI_ISL_1608949, EPI_ISL_1608950, EPI_ISL_1608951, EPI_ISL_1608952, EPI_ISL_1608954, EPI_ISL_1608956, EPI_ISL_1608957, EPI_ISL_1608959, EPI_ISL_1608960, EPI_ISL_1608961, EPI_ISL_1608962, EPI_ISL_1608963, EPI_ISL_1608966, EPI_ISL_1608967, EPI_ISL_1608968, EPI_ISL_1608969, EPI_ISL_1608971, EPI_ISL_1608972, EPI_ISL_1608973, EPI_ISL_1608976, EPI_ISL_1608978, EPI_ISL_1608980, EPI_ISL_1608981, EPI_ISL_1608982, EPI_ISL_1608983, EPI_ISL_1608984, EPI_ISL_1608985, EPI_ISL_1608986, EPI_ISL_1608987, EPI_ISL_1608988, EPI_ISL_1608989, EPI_ISL_1608990, EPI_ISL_1608991, EPI_ISL_1608992, EPI_ISL_1608994, EPI_ISL_1608995, EPI_ISL_1608996, EPI_ISL_1608997, EPI_ISL_1608998, EPI_ISL_1608999, EPI_ISL_1609000, EPI_ISL_1609001, EPI_ISL_1609002, EPI_ISL_1609003, EPI_ISL_1609005, EPI_ISL_1609006, EPI_ISL_1609008, EPI_ISL_1609009, EPI_ISL_1609010, EPI_ISL_1609011, EPI_ISL_1609012, EPI_ISL_1609013, EPI_ISL_1609014, EPI_ISL_1609015, EPI_ISL_1609016, EPI_ISL_1609017, EPI_ISL_1609018, EPI_ISL_1609020, EPI_ISL_1609021, EPI_ISL_1609022, EPI_ISL_1609025, EPI_ISL_1609026, EPI_ISL_1609027, EPI_ISL_1609030, EPI_ISL_1609031, EPI_ISL_1609032, EPI_ISL_1609033, EPI_ISL_1609034, EPI_ISL_1609035, EPI_ISL_1609036, EPI_ISL_1609037, EPI_ISL_1609038, EPI_ISL_1609039, EPI_ISL_1609040, EPI_ISL_1609041, EPI_ISL_1609043, EPI_ISL_1609044, EPI_ISL_1609045, EPI_ISL_1609047, EPI_ISL_1609048, EPI_ISL_1609049, EPI_ISL_1609050, EPI_ISL_1609051, EPI_ISL_1609052, EPI_ISL_1609054, EPI_ISL_1609055, EPI_ISL_1609056, EPI_ISL_1609058, EPI_ISL_1609061, EPI_ISL_1609062, EPI_ISL_1609064, EPI_ISL_1609065, EPI_ISL_1609066, EPI_ISL_1609067, EPI_ISL_1609068, EPI_ISL_1609069, EPI_ISL_1609070, EPI_ISL_1609071, EPI_ISL_1609073, EPI_ISL_1609074, EPI_ISL_1609075, EPI_ISL_1609076, EPI_ISL_1609077, EPI_ISL_1609079, EPI_ISL_1609080, EPI_ISL_1609081, EPI_ISL_1609082, EPI_ISL_1609084, EPI_ISL_1609085, EPI_ISL_1609086, EPI_ISL_1609090, EPI_ISL_1609091, EPI_ISL_1609092, EPI_ISL_1609093, EPI_ISL_1609094, EPI_ISL_1609095, EPI_ISL_1609097, EPI_ISL_1609100, EPI_ISL_1609101, EPI_ISL_1609103, EPI_ISL_1609104, EPI_ISL_1609105, EPI_ISL_1609106, EPI_ISL_1609108, EPI_ISL_1609109, EPI_ISL_1609110, EPI_ISL_1609112, EPI_ISL_1609115, EPI_ISL_1609116, EPI_ISL_1609117, EPI_ISL_1609119, EPI_ISL_1609121, EPI_ISL_1609122, EPI_ISL_1609123, EPI_ISL_1609124, EPI_ISL_1609125, EPI_ISL_1609126, EPI_ISL_1609127, EPI_ISL_1609128, EPI_ISL_1609129, EPI_ISL_1609130, EPI_ISL_1609131, EPI_ISL_1609133, EPI_ISL_1609134, EPI_ISL_1609135, EPI_ISL_1609137, EPI_ISL_1609138, EPI_ISL_1609139, EPI_ISL_1609140, EPI_ISL_1609144, EPI_ISL_1609150, EPI_ISL_1609151, EPI_ISL_1609152, EPI_ISL_1609153, EPI_ISL_1609154, EPI_ISL_1609155, EPI_ISL_1609156, EPI_ISL_1609157, EPI_ISL_1609158, EPI_ISL_1609159, EPI_ISL_1609160, EPI_ISL_1609162, EPI_ISL_1609164, EPI_ISL_1609165, EPI_ISL_1609166, EPI_ISL_1609168, EPI_ISL_1609169, EPI_ISL_1609171, EPI_ISL_1609174, EPI_ISL_1609175, EPI_ISL_1609176, EPI_ISL_1609177, EPI_ISL_1609181, EPI_ISL_1609184, EPI_ISL_1609185, EPI_ISL_1609187, EPI_ISL_1609188, EPI_ISL_1609189, EPI_ISL_1609190, EPI_ISL_1609191, EPI_ISL_1609195, EPI_ISL_1609196, EPI_ISL_1609197, EPI_ISL_1609198, EPI_ISL_1609202, EPI_ISL_1609203, EPI_ISL_1609204, EPI_ISL_1609206, EPI_ISL_1609208, EPI_ISL_1609209, EPI_ISL_1609211, EPI_ISL_1609212, EPI_ISL_1609213, EPI_ISL_1609214, EPI_ISL_1609216, EPI_ISL_1609218, EPI_ISL_1609219, EPI_ISL_1609220, EPI_ISL_1609221, EPI_ISL_1609222, EPI_ISL_1609223, EPI_ISL_1609224, EPI_ISL_1609227, EPI_ISL_1609228, EPI_ISL_1609230, EPI_ISL_1609231, EPI_ISL_1609233, EPI_ISL_1609235, EPI_ISL_1609236, EPI_ISL_1609237, EPI_ISL_1609238, EPI_ISL_1609239, EPI_ISL_1609240, EPI_ISL_1609241, EPI_ISL_1609242, EPI_ISL_1609244, EPI_ISL_1609245, EPI_ISL_1609246, EPI_ISL_1609247, EPI_ISL_1609248, EPI_ISL_1609249, EPI_ISL_1609251, EPI_ISL_1609253, EPI_ISL_1609255, EPI_ISL_1609256, EPI_ISL_1609257, EPI_ISL_1609258, EPI_ISL_1609260, EPI_ISL_1609261, EPI_ISL_1609262, EPI_ISL_1609263, EPI_ISL_1609264, EPI_ISL_1609265, EPI_ISL_1609266, EPI_ISL_1609267, EPI_ISL_1609268, EPI_ISL_1609269, EPI_ISL_1609272, EPI_ISL_1609273, EPI_ISL_1609274, EPI_ISL_1609276, EPI_ISL_1609278, EPI_ISL_1609279, EPI_ISL_1609280, EPI_ISL_1609282, EPI_ISL_1609283, EPI_ISL_1609284, EPI_ISL_1609285, EPI_ISL_1609286, EPI_ISL_1609287, EPI_ISL_1609289, EPI_ISL_1609290, EPI_ISL_1609291, EPI_ISL_1609292, EPI_ISL_1609293, EPI_ISL_1609294, EPI_ISL_1609295, EPI_ISL_1609297, EPI_ISL_1609300, EPI_ISL_1609304, EPI_ISL_1609306, EPI_ISL_1609307, EPI_ISL_1609308, EPI_ISL_1609309, EPI_ISL_1609311, EPI_ISL_1609312, EPI_ISL_1609313, EPI_ISL_1609314, EPI_ISL_1609315, EPI_ISL_1609316, EPI_ISL_1609318, EPI_ISL_1609320, EPI_ISL_1609322, EPI_ISL_1609323, EPI_ISL_1609324, EPI_ISL_1609325, EPI_ISL_1609326, EPI_ISL_1609327, EPI_ISL_1609329, EPI_ISL_1609330, EPI_ISL_1609331, EPI_ISL_1609332, EPI_ISL_1609333, EPI_ISL_1609336, EPI_ISL_1609337, EPI_ISL_1609338, EPI_ISL_1609341, EPI_ISL_1609344, EPI_ISL_1609345, EPI_ISL_1609346, EPI_ISL_1609347, EPI_ISL_1609349, EPI_ISL_1609350, EPI_ISL_1609351, EPI_ISL_1609352, EPI_ISL_1609353, EPI_ISL_1609354, EPI_ISL_1609355, EPI_ISL_1609356, EPI_ISL_1609357, EPI_ISL_1609358, EPI_ISL_1609359, EPI_ISL_1609360, EPI_ISL_1609361, EPI_ISL_1609362, EPI_ISL_1609363, EPI_ISL_1609364, EPI_ISL_1609365, EPI_ISL_1609366, EPI_ISL_1609369, EPI_ISL_1609372, EPI_ISL_1609374, EPI_ISL_1609375, EPI_ISL_1609376, EPI_ISL_1609377, EPI_ISL_1609378, EPI_ISL_1609379, EPI_ISL_1609381, EPI_ISL_1609383, EPI_ISL_1609385, EPI_ISL_1609387, EPI_ISL_1609388, EPI_ISL_1609390, EPI_ISL_1609391, EPI_ISL_1609392, EPI_ISL_1609393, EPI_ISL_1609394, EPI_ISL_1609395, EPI_ISL_1609397, EPI_ISL_1609398, EPI_ISL_1609399, EPI_ISL_1609400, EPI_ISL_1609402, EPI_ISL_1609403, EPI_ISL_1609404, EPI_ISL_1609405, EPI_ISL_1609409, EPI_ISL_1609410, EPI_ISL_1609411, EPI_ISL_1609412, EPI_ISL_1609413, EPI_ISL_1609414, EPI_ISL_1609415, EPI_ISL_1609416, EPI_ISL_1609417, EPI_ISL_1609418, EPI_ISL_1609419, EPI_ISL_1609420, EPI_ISL_1609422, EPI_ISL_1609424, EPI_ISL_1609426, EPI_ISL_1609427, EPI_ISL_1609428, EPI_ISL_1609429, EPI_ISL_1609430, EPI_ISL_1609431, EPI_ISL_1609432, EPI_ISL_1609434, EPI_ISL_1609435, EPI_ISL_1609436, EPI_ISL_1609437, EPI_ISL_1609438, EPI_ISL_1609439, EPI_ISL_1609440, EPI_ISL_1609441, EPI_ISL_1609442, EPI_ISL_1609444, EPI_ISL_1609446, EPI_ISL_1609447, EPI_ISL_1609449, EPI_ISL_1609450, EPI_ISL_1609451, EPI_ISL_1609452, EPI_ISL_1609453, EPI_ISL_1609454, EPI_ISL_1609455, EPI_ISL_1609456, EPI_ISL_1609457, EPI_ISL_1609461, EPI_ISL_1609462, EPI_ISL_1609463, EPI_ISL_1609464, EPI_ISL_1609466, EPI_ISL_1609467, EPI_ISL_1609468, EPI_ISL_1609469, EPI_ISL_1609470, EPI_ISL_1609471, EPI_ISL_1609472, EPI_ISL_1609473, EPI_ISL_1609474, EPI_ISL_1609475, EPI_ISL_1609478, EPI_ISL_1609479, EPI_ISL_1609480, EPI_ISL_1609482, EPI_ISL_1609485, EPI_ISL_1609486, EPI_ISL_1609487, EPI_ISL_1609489, EPI_ISL_1609490, EPI_ISL_1609492, EPI_ISL_1609493, EPI_ISL_1609494, EPI_ISL_1609495, EPI_ISL_1609496, EPI_ISL_1609497, EPI_ISL_1609498, EPI_ISL_1609499, EPI_ISL_1609501, EPI_ISL_1609502, EPI_ISL_1609503, EPI_ISL_1609504, EPI_ISL_1609505, EPI_ISL_1609507, EPI_ISL_1609508, EPI_ISL_1609509, EPI_ISL_1609510, EPI_ISL_1609511, EPI_ISL_1609512, EPI_ISL_1609513, EPI_ISL_1609517, EPI_ISL_1609520, EPI_ISL_1609521, EPI_ISL_1609522, EPI_ISL_1609523, EPI_ISL_1609525, EPI_ISL_1609527, EPI_ISL_1609528, EPI_ISL_1609529, EPI_ISL_1609530, EPI_ISL_1609531, EPI_ISL_1609533, EPI_ISL_1609534, EPI_ISL_1609535, EPI_ISL_1609536, EPI_ISL_1609537, EPI_ISL_1609538, EPI_ISL_1609539, EPI_ISL_1609541, EPI_ISL_1609542, EPI_ISL_1609543, EPI_ISL_1609545, EPI_ISL_1609547, EPI_ISL_1609548, EPI_ISL_1609549, EPI_ISL_1609550, EPI_ISL_1609551, EPI_ISL_1609554, EPI_ISL_1609556, EPI_ISL_1609557, EPI_ISL_1609558, EPI_ISL_1609559, EPI_ISL_1609560, EPI_ISL_1609561, EPI_ISL_1609563, EPI_ISL_1609564, EPI_ISL_1609565, EPI_ISL_1609566, EPI_ISL_1609567, EPI_ISL_1609568, EPI_ISL_1609569, EPI_ISL_1609570, EPI_ISL_1609571, EPI_ISL_1609572, EPI_ISL_1609573, EPI_ISL_1609574, EPI_ISL_1609575, EPI_ISL_1609576, EPI_ISL_1609577, EPI_ISL_1609579, EPI_ISL_1609581, EPI_ISL_1609582, EPI_ISL_1609583, EPI_ISL_1609584, EPI_ISL_1609586, EPI_ISL_1609587, EPI_ISL_1609588, EPI_ISL_1609589, EPI_ISL_1609590, EPI_ISL_1609591, EPI_ISL_1609592, EPI_ISL_1609594, EPI_ISL_1609595, EPI_ISL_1609596, EPI_ISL_1609597, EPI_ISL_1609598, EPI_ISL_1609599, EPI_ISL_1609600, EPI_ISL_1609601, EPI_ISL_1609603, EPI_ISL_1609604, EPI_ISL_1609606, EPI_ISL_1609607, EPI_ISL_1609608, EPI_ISL_1609609, EPI_ISL_1609610, EPI_ISL_1609611, EPI_ISL_1609612, EPI_ISL_1609613, EPI_ISL_1609614, EPI_ISL_1609615, EPI_ISL_1609616, EPI_ISL_1609617, EPI_ISL_1609619, EPI_ISL_1609621, EPI_ISL_1609622, EPI_ISL_1609623, EPI_ISL_1609624, EPI_ISL_1609625, EPI_ISL_1609626, EPI_ISL_1609627, EPI_ISL_1609629, EPI_ISL_1609630, EPI_ISL_1609631, EPI_ISL_1609632, EPI_ISL_1609633, EPI_ISL_1609635, EPI_ISL_1609636, EPI_ISL_1609638, EPI_ISL_1609639, EPI_ISL_1609640, EPI_ISL_1609641, EPI_ISL_1609642, EPI_ISL_1609643, EPI_ISL_1609645, EPI_ISL_1609646, EPI_ISL_1609649, EPI_ISL_1609650, EPI_ISL_1609651, EPI_ISL_1609652, EPI_ISL_1609653, EPI_ISL_1609654, EPI_ISL_1609655, EPI_ISL_1609656, EPI_ISL_1609657, EPI_ISL_1609658, EPI_ISL_1609659, EPI_ISL_1609660, EPI_ISL_1609661, EPI_ISL_1609662, EPI_ISL_1609663, EPI_ISL_1609664, EPI_ISL_1609665, EPI_ISL_1609666, EPI_ISL_1609668, EPI_ISL_1609669, EPI_ISL_1609670, EPI_ISL_1609671, EPI_ISL_1609672, EPI_ISL_1609673, EPI_ISL_1609674, EPI_ISL_1609675, EPI_ISL_1609676, EPI_ISL_1609679, EPI_ISL_1609680, EPI_ISL_1609681, EPI_ISL_1609682, EPI_ISL_1609684, EPI_ISL_1609685, EPI_ISL_1609686, EPI_ISL_1609687, EPI_ISL_1609688, EPI_ISL_1609689, EPI_ISL_1609690, EPI_ISL_1609691, EPI_ISL_1609692, EPI_ISL_1609693, EPI_ISL_1609694, EPI_ISL_1609695, EPI_ISL_1609696, EPI_ISL_1609697, EPI_ISL_1609699, EPI_ISL_1609700, EPI_ISL_1609701, EPI_ISL_1609705, EPI_ISL_1609706, EPI_ISL_1609707, EPI_ISL_1609708, EPI_ISL_1609715, EPI_ISL_1609724, EPI_ISL_1609726, EPI_ISL_1609727, EPI_ISL_1609729, EPI_ISL_1609730, EPI_ISL_1609731, EPI_ISL_1609733, EPI_ISL_1609736, EPI_ISL_1609741, EPI_ISL_1609744, EPI_ISL_1609746, EPI_ISL_1609747, EPI_ISL_1609751, EPI_ISL_1609753, EPI_ISL_1609755, EPI_ISL_1609757, EPI_ISL_1609759, EPI_ISL_1609762, EPI_ISL_1609763, EPI_ISL_1609764, EPI_ISL_1609766, EPI_ISL_1609771, EPI_ISL_1609772, EPI_ISL_1609776, EPI_ISL_1609777, EPI_ISL_1609779, EPI_ISL_1609783, EPI_ISL_1609785, EPI_ISL_1609786, EPI_ISL_1609788, EPI_ISL_1609790, EPI_ISL_1609793, EPI_ISL_1609794, EPI_ISL_1609796, EPI_ISL_1609797, EPI_ISL_1609798, EPI_ISL_1609799, EPI_ISL_1609801, EPI_ISL_1609802, EPI_ISL_1609805, EPI_ISL_1609806, EPI_ISL_1609807, EPI_ISL_1609808, EPI_ISL_1609809, EPI_ISL_1609810, EPI_ISL_1609811, EPI_ISL_1609812, EPI_ISL_1609814, EPI_ISL_1609817, EPI_ISL_1609818, EPI_ISL_1609821, EPI_ISL_1609822, EPI_ISL_1609823, EPI_ISL_1609824, EPI_ISL_1609825, EPI_ISL_1609826, EPI_ISL_1609828, EPI_ISL_1609831, EPI_ISL_1609832, EPI_ISL_1609833, EPI_ISL_1609835, EPI_ISL_1609840, EPI_ISL_1609841, EPI_ISL_1609842, EPI_ISL_1609845, EPI_ISL_1609846, EPI_ISL_1609847, EPI_ISL_1609848, EPI_ISL_1609853, EPI_ISL_1609854, EPI_ISL_1609856, EPI_ISL_1609857, EPI_ISL_1609858, EPI_ISL_1609859, EPI_ISL_1609860, EPI_ISL_1609861, EPI_ISL_1609862, EPI_ISL_1609863, EPI_ISL_1609864, EPI_ISL_1609865, EPI_ISL_1609866, EPI_ISL_1609868, EPI_ISL_1609869, EPI_ISL_1609871, EPI_ISL_1609872, EPI_ISL_1609873, EPI_ISL_1609874, EPI_ISL_1609877, EPI_ISL_1609878, EPI_ISL_1609880, EPI_ISL_1609881, EPI_ISL_1609882, EPI_ISL_1609884, EPI_ISL_1609885, EPI_ISL_1609886, EPI_ISL_1609887, EPI_ISL_1609888, EPI_ISL_1609889, EPI_ISL_1609890, EPI_ISL_1609891, EPI_ISL_1609893, EPI_ISL_1609894, EPI_ISL_1609895, EPI_ISL_1609896, EPI_ISL_1609897, EPI_ISL_1609898, EPI_ISL_1609899, EPI_ISL_1609900, EPI_ISL_1609901, EPI_ISL_1609902, EPI_ISL_1609903, EPI_ISL_1609904, EPI_ISL_1609906, EPI_ISL_1609910, EPI_ISL_1609912, EPI_ISL_1609913, EPI_ISL_1609914, EPI_ISL_1609917, EPI_ISL_1609918, EPI_ISL_1609920, EPI_ISL_1609921, EPI_ISL_1609924, EPI_ISL_1609925, EPI_ISL_1609926, EPI_ISL_1609927, EPI_ISL_1609928, EPI_ISL_1609929, EPI_ISL_1609930, EPI_ISL_1609932, EPI_ISL_1609933, EPI_ISL_1609934, EPI_ISL_1609935, EPI_ISL_1609936, EPI_ISL_1609937, EPI_ISL_1609940, EPI_ISL_1609941, EPI_ISL_1609942, EPI_ISL_1609943, EPI_ISL_1609944, EPI_ISL_1609945, EPI_ISL_1609946, EPI_ISL_1609948, EPI_ISL_1609952, EPI_ISL_1609953, EPI_ISL_1609954, EPI_ISL_1609955, EPI_ISL_1609956, EPI_ISL_1609957, EPI_ISL_1609959, EPI_ISL_1609960, EPI_ISL_1609961, EPI_ISL_1609962, EPI_ISL_1609963, EPI_ISL_1609964, EPI_ISL_1609966, EPI_ISL_1609967, EPI_ISL_1609969, EPI_ISL_1609970, EPI_ISL_1609971, EPI_ISL_1609972, EPI_ISL_1609973, EPI_ISL_1609975, EPI_ISL_1609976, EPI_ISL_1609977, EPI_ISL_1609980, EPI_ISL_1609981, EPI_ISL_1609982, EPI_ISL_1609983, EPI_ISL_1609984, EPI_ISL_1609985, EPI_ISL_1609986, EPI_ISL_1609988, EPI_ISL_1609989, EPI_ISL_1609991, EPI_ISL_1609992, EPI_ISL_1609993, EPI_ISL_1609994, EPI_ISL_1609996, EPI_ISL_1609997, EPI_ISL_1609998, EPI_ISL_1609999, EPI_ISL_1610000, EPI_ISL_1610001, EPI_ISL_1610002, EPI_ISL_1610005, EPI_ISL_1610006, EPI_ISL_1610007, EPI_ISL_1610008, EPI_ISL_1610010, EPI_ISL_1610011, EPI_ISL_1610012, EPI_ISL_1610013, EPI_ISL_1610014, EPI_ISL_1610015, EPI_ISL_1610016, EPI_ISL_1610017, EPI_ISL_1610018, EPI_ISL_1610019, EPI_ISL_1610020, EPI_ISL_1610021, EPI_ISL_1610022, EPI_ISL_1610023, EPI_ISL_1610024, EPI_ISL_1610025, EPI_ISL_1610026, EPI_ISL_1610027, EPI_ISL_1610030, EPI_ISL_1610031, EPI_ISL_1610032, EPI_ISL_1610033, EPI_ISL_1610034, EPI_ISL_1610036, EPI_ISL_1610038, EPI_ISL_1610039, EPI_ISL_1610040, EPI_ISL_1610041, EPI_ISL_1610042, EPI_ISL_1610043, EPI_ISL_1610044, EPI_ISL_1610046, EPI_ISL_1610048, EPI_ISL_1610049, EPI_ISL_1610051, EPI_ISL_1610052, EPI_ISL_1610053, EPI_ISL_1610054, EPI_ISL_1610055, EPI_ISL_1610057, EPI_ISL_1610058, EPI_ISL_1610059, EPI_ISL_1610061, EPI_ISL_1610062, EPI_ISL_1610066, EPI_ISL_1610067, EPI_ISL_1610068, EPI_ISL_1610071, EPI_ISL_1610072, EPI_ISL_1610073, EPI_ISL_1610074, EPI_ISL_1610075, EPI_ISL_1610076, EPI_ISL_1610078, EPI_ISL_1610080, EPI_ISL_1610081, EPI_ISL_1610082, EPI_ISL_1610083, EPI_ISL_1610084, EPI_ISL_1610085, EPI_ISL_1610086, EPI_ISL_1610087, EPI_ISL_1610088, EPI_ISL_1610089, EPI_ISL_1610090, EPI_ISL_1610092, EPI_ISL_1610096, EPI_ISL_1610097, EPI_ISL_1610098, EPI_ISL_1610099, EPI_ISL_1610100, EPI_ISL_1610102, EPI_ISL_1610103, EPI_ISL_1610105, EPI_ISL_1610108, EPI_ISL_1610109, EPI_ISL_1610110, EPI_ISL_1610111, EPI_ISL_1610112, EPI_ISL_1610113, EPI_ISL_1610114, EPI_ISL_1610115, EPI_ISL_1610116, EPI_ISL_1610117, EPI_ISL_1610118, EPI_ISL_1610119, EPI_ISL_1610121, EPI_ISL_1610122, EPI_ISL_1610123, EPI_ISL_1610124, EPI_ISL_1610125, EPI_ISL_1610126, EPI_ISL_1610127, EPI_ISL_1610129, EPI_ISL_1610130, EPI_ISL_1610131, EPI_ISL_1610132, EPI_ISL_1610133, EPI_ISL_1610134, EPI_ISL_1610137, EPI_ISL_1610138, EPI_ISL_1610141, EPI_ISL_1610144, EPI_ISL_1610145, EPI_ISL_1610146, EPI_ISL_1610147, EPI_ISL_1610148, EPI_ISL_1610149, EPI_ISL_1610150, EPI_ISL_1610152, EPI_ISL_1610154, EPI_ISL_1610155, EPI_ISL_1610156, EPI_ISL_1610157, EPI_ISL_1610159, EPI_ISL_1610161, EPI_ISL_1610162, EPI_ISL_1610165, EPI_ISL_1610166, EPI_ISL_1610167, EPI_ISL_1610168, EPI_ISL_1610170, EPI_ISL_1610171, EPI_ISL_1610172, EPI_ISL_1610174, EPI_ISL_1610176, EPI_ISL_1610178, EPI_ISL_1610179, EPI_ISL_1610180, EPI_ISL_1610181, EPI_ISL_1610183, EPI_ISL_1610184, EPI_ISL_1610185, EPI_ISL_1610188, EPI_ISL_1610190, EPI_ISL_1610191, EPI_ISL_1610192, EPI_ISL_1610194, EPI_ISL_1610195, EPI_ISL_1610198, EPI_ISL_1610200, EPI_ISL_1610201, EPI_ISL_1610202, EPI_ISL_1610203, EPI_ISL_1610204, EPI_ISL_1610205, EPI_ISL_1610206, EPI_ISL_1610207, EPI_ISL_1610209, EPI_ISL_1610210, EPI_ISL_1610211, EPI_ISL_1610212, EPI_ISL_1610213, EPI_ISL_1610214, EPI_ISL_1610215, EPI_ISL_1610216, EPI_ISL_1610217, EPI_ISL_1610219, EPI_ISL_1610220, EPI_ISL_1610222, EPI_ISL_1610223, EPI_ISL_1610224, EPI_ISL_1610225, EPI_ISL_1610227, EPI_ISL_1610230, EPI_ISL_1610231, EPI_ISL_1610232, EPI_ISL_1610233, EPI_ISL_1610234, EPI_ISL_1610235, EPI_ISL_1610237, EPI_ISL_1610238, EPI_ISL_1610239, EPI_ISL_1610240, EPI_ISL_1610241, EPI_ISL_1610242, EPI_ISL_1610243, EPI_ISL_1610244, EPI_ISL_1610245, EPI_ISL_1610246, EPI_ISL_1610248, EPI_ISL_1610249, EPI_ISL_1610250, EPI_ISL_1610251, EPI_ISL_1610252, EPI_ISL_1610253, EPI_ISL_1610254, EPI_ISL_1610255, EPI_ISL_1610256, EPI_ISL_1610257, EPI_ISL_1610258, EPI_ISL_1610259, EPI_ISL_1610260, EPI_ISL_1610261, EPI_ISL_1610264, EPI_ISL_1610265, EPI_ISL_1610266, EPI_ISL_1610268, EPI_ISL_1610269, EPI_ISL_1610270, EPI_ISL_1610271, EPI_ISL_1610272, EPI_ISL_1610273, EPI_ISL_1610274, EPI_ISL_1610275, EPI_ISL_1610276, EPI_ISL_1610278, EPI_ISL_1610279, EPI_ISL_1610281, EPI_ISL_1610282, EPI_ISL_1610283, EPI_ISL_1610284, EPI_ISL_1610285, EPI_ISL_1610286, EPI_ISL_1610288, EPI_ISL_1610289, EPI_ISL_1610290, EPI_ISL_1610291, EPI_ISL_1610292, EPI_ISL_1610294, EPI_ISL_1610295, EPI_ISL_1610297, EPI_ISL_1610298, EPI_ISL_1610299, EPI_ISL_1610300, EPI_ISL_1610301, EPI_ISL_1610302, EPI_ISL_1610303, EPI_ISL_1610305, EPI_ISL_1610306, EPI_ISL_1610307, EPI_ISL_1610308, EPI_ISL_1610309, EPI_ISL_1610310, EPI_ISL_1610311, EPI_ISL_1610313, EPI_ISL_1610314, EPI_ISL_1610316, EPI_ISL_1610317, EPI_ISL_1610318, EPI_ISL_1610319, EPI_ISL_1610320, EPI_ISL_1610321, EPI_ISL_1610322, EPI_ISL_1610323, EPI_ISL_1610324, EPI_ISL_1610326, EPI_ISL_1610327, EPI_ISL_1610328, EPI_ISL_1610331, EPI_ISL_1610332, EPI_ISL_1610333, EPI_ISL_1610334, EPI_ISL_1610335, EPI_ISL_1610337, EPI_ISL_1610339, EPI_ISL_1610340, EPI_ISL_1610341, EPI_ISL_1610342, EPI_ISL_1610343, EPI_ISL_1610344, EPI_ISL_1610346, EPI_ISL_1610347, EPI_ISL_1610348, EPI_ISL_1610349, EPI_ISL_1610350, EPI_ISL_1610351, EPI_ISL_1610352, EPI_ISL_1610353, EPI_ISL_1610354, EPI_ISL_1610355, EPI_ISL_1610356, EPI_ISL_1610357, EPI_ISL_1610358, EPI_ISL_1610359, EPI_ISL_1610360, EPI_ISL_1610361, EPI_ISL_1610362, EPI_ISL_1610363, EPI_ISL_1610364, EPI_ISL_1610365, EPI_ISL_1610366, EPI_ISL_1610367, EPI_ISL_1610369, EPI_ISL_1610370, EPI_ISL_1610371, EPI_ISL_1610373, EPI_ISL_1610374, EPI_ISL_1610375, EPI_ISL_1610376, EPI_ISL_1610379, EPI_ISL_1610380, EPI_ISL_1610384, EPI_ISL_1610385, EPI_ISL_1610386, EPI_ISL_1610388, EPI_ISL_1610389, EPI_ISL_1610390, EPI_ISL_1610391, EPI_ISL_1610392, EPI_ISL_1610393, EPI_ISL_1610395, EPI_ISL_1610396, EPI_ISL_1610397, EPI_ISL_1610399, EPI_ISL_1610401, EPI_ISL_1610402, EPI_ISL_1610404, EPI_ISL_1610405, EPI_ISL_1610407, EPI_ISL_1610408, EPI_ISL_1610410, EPI_ISL_1610411, EPI_ISL_1610412, EPI_ISL_1610414, EPI_ISL_1610416, EPI_ISL_1610419, EPI_ISL_1610421, EPI_ISL_1610422, EPI_ISL_1610424, EPI_ISL_1610426, EPI_ISL_1610429, EPI_ISL_1610433, EPI_ISL_1610435, EPI_ISL_1610436, EPI_ISL_1610438, EPI_ISL_1610440, EPI_ISL_1610441, EPI_ISL_1610443, EPI_ISL_1610445, EPI_ISL_1610446, EPI_ISL_1610448, EPI_ISL_1610449, EPI_ISL_1610451, EPI_ISL_1610453, EPI_ISL_1610454, EPI_ISL_1610457, EPI_ISL_1610459, EPI_ISL_1610460, EPI_ISL_1610462, EPI_ISL_1610463, EPI_ISL_1610467, EPI_ISL_1610468, EPI_ISL_1610470, EPI_ISL_1610471, EPI_ISL_1610473, EPI_ISL_1610474, EPI_ISL_1610476, EPI_ISL_1610478, EPI_ISL_1610479, EPI_ISL_1610481, EPI_ISL_1610482, EPI_ISL_1610486, EPI_ISL_1610487, EPI_ISL_1610489, EPI_ISL_1610492, EPI_ISL_1610495, EPI_ISL_1610498, EPI_ISL_1610500, EPI_ISL_1610501, EPI_ISL_1610503, EPI_ISL_1610504, EPI_ISL_1610506, EPI_ISL_1610507, EPI_ISL_1610509, EPI_ISL_1610511, EPI_ISL_1610512, EPI_ISL_1610513, EPI_ISL_1610515, EPI_ISL_1610517, EPI_ISL_1610518, EPI_ISL_1610520, EPI_ISL_1610521, EPI_ISL_1610523, EPI_ISL_1610525, EPI_ISL_1610527, EPI_ISL_1610529, EPI_ISL_1610530, EPI_ISL_1610533, EPI_ISL_1610537, EPI_ISL_1610538, EPI_ISL_1610540, EPI_ISL_1610541, EPI_ISL_1610542, EPI_ISL_1610544, EPI_ISL_1610545, EPI_ISL_1610547, EPI_ISL_1610549, EPI_ISL_1610550, EPI_ISL_1610552, EPI_ISL_1610553, EPI_ISL_1610555, EPI_ISL_1610558, EPI_ISL_1610560, EPI_ISL_1610561, EPI_ISL_1610563, EPI_ISL_1610565, EPI_ISL_1610568, EPI_ISL_1610569, EPI_ISL_1610571, EPI_ISL_1610574, EPI_ISL_1610576, EPI_ISL_1610578, EPI_ISL_1610579, EPI_ISL_1610581, EPI_ISL_1610582, EPI_ISL_1610584, EPI_ISL_1610586, EPI_ISL_1610591, EPI_ISL_1610592, EPI_ISL_1610594, EPI_ISL_1610595, EPI_ISL_1610601, EPI_ISL_1610606, EPI_ISL_1610607, EPI_ISL_1610610, EPI_ISL_1610612, EPI_ISL_1610614, EPI_ISL_1610617, EPI_ISL_1610620, EPI_ISL_1610622, EPI_ISL_1610625, EPI_ISL_1610627, EPI_ISL_1610628, EPI_ISL_1610630, EPI_ISL_1610633, EPI_ISL_1610636, EPI_ISL_1610638, EPI_ISL_1610639, EPI_ISL_1610641, EPI_ISL_1610648, EPI_ISL_1610649, EPI_ISL_1610651, EPI_ISL_1610652, EPI_ISL_1610653, EPI_ISL_1610654, EPI_ISL_1610655, EPI_ISL_1610656, EPI_ISL_1610657, EPI_ISL_1610658, EPI_ISL_1610659, EPI_ISL_1610660, EPI_ISL_1610661, EPI_ISL_1610662, EPI_ISL_1610664, EPI_ISL_1610665, EPI_ISL_1610666, EPI_ISL_1610667, EPI_ISL_1610668, EPI_ISL_1610669, EPI_ISL_1610670, EPI_ISL_1610671, EPI_ISL_1610672, EPI_ISL_1610673, EPI_ISL_1610674, EPI_ISL_1610675, EPI_ISL_1610676, EPI_ISL_1610677, EPI_ISL_1610678, EPI_ISL_1610679, EPI_ISL_1610680, EPI_ISL_1610681, EPI_ISL_1610683, EPI_ISL_1610684, EPI_ISL_1610685, EPI_ISL_1610686, EPI_ISL_1610687, EPI_ISL_1610689, EPI_ISL_1610690, EPI_ISL_1610692, EPI_ISL_1610693, EPI_ISL_1610695, EPI_ISL_1610696, EPI_ISL_1610697, EPI_ISL_1610699, EPI_ISL_1610700, EPI_ISL_1610701, EPI_ISL_1610702, EPI_ISL_1610703, EPI_ISL_1610704, EPI_ISL_1610705, EPI_ISL_1610706, EPI_ISL_1610707, EPI_ISL_1610708, EPI_ISL_1610709, EPI_ISL_1610710, EPI_ISL_1610711, EPI_ISL_1610713, EPI_ISL_1610714, EPI_ISL_1610715, EPI_ISL_1610716, EPI_ISL_1610717, EPI_ISL_1610720, EPI_ISL_1610721, EPI_ISL_1610722, EPI_ISL_1610723, EPI_ISL_1610724, EPI_ISL_1610725, EPI_ISL_1610726, EPI_ISL_1610727, EPI_ISL_1610728, EPI_ISL_1610729, EPI_ISL_1610730, EPI_ISL_1610732, EPI_ISL_1610733, EPI_ISL_1610734, EPI_ISL_1610735, EPI_ISL_1610736, EPI_ISL_1610737, EPI_ISL_1610738, EPI_ISL_1610740, EPI_ISL_1610741, EPI_ISL_1610742, EPI_ISL_1610743, EPI_ISL_1610744, EPI_ISL_1610747, EPI_ISL_1610748, EPI_ISL_1610749, EPI_ISL_1610750, EPI_ISL_1610751, EPI_ISL_1610752, EPI_ISL_1610753, EPI_ISL_1610754, EPI_ISL_1610755, EPI_ISL_1610756, EPI_ISL_1610757, EPI_ISL_1610758, EPI_ISL_1610759, EPI_ISL_1610760, EPI_ISL_1610761, EPI_ISL_1610762, EPI_ISL_1610763, EPI_ISL_1610764, EPI_ISL_1610765, EPI_ISL_1610766, EPI_ISL_1610767, EPI_ISL_1610768, EPI_ISL_1610769, EPI_ISL_1610770, EPI_ISL_1610771, EPI_ISL_1610772, EPI_ISL_1610773, EPI_ISL_1610774, EPI_ISL_1610775, EPI_ISL_1610776, EPI_ISL_1610777, EPI_ISL_1610778, EPI_ISL_1610779, EPI_ISL_1610780, EPI_ISL_1610781, EPI_ISL_1610782, EPI_ISL_1610783, EPI_ISL_1610784, EPI_ISL_1610785, EPI_ISL_1610786, EPI_ISL_1610787, EPI_ISL_1610788, EPI_ISL_1610790, EPI_ISL_1610791, EPI_ISL_1610792, EPI_ISL_1610793, EPI_ISL_1610794, EPI_ISL_1610795, EPI_ISL_1610796, EPI_ISL_1610797, EPI_ISL_1610798, EPI_ISL_1610799, EPI_ISL_1610800, EPI_ISL_1610801, EPI_ISL_1610802, EPI_ISL_1610804, EPI_ISL_1610805, EPI_ISL_1610806, EPI_ISL_1610808, EPI_ISL_1610809, EPI_ISL_1610810, EPI_ISL_1610812, EPI_ISL_1610813, EPI_ISL_1610814, EPI_ISL_1610815, EPI_ISL_1610816, EPI_ISL_1610817, EPI_ISL_1610818, EPI_ISL_1610820, EPI_ISL_1610821, EPI_ISL_1610823, EPI_ISL_1610824, EPI_ISL_1610825, EPI_ISL_1610826, EPI_ISL_1610827, EPI_ISL_1610828, EPI_ISL_1610829, EPI_ISL_1610830, EPI_ISL_1610832, EPI_ISL_1610833, EPI_ISL_1610835, EPI_ISL_1610836, EPI_ISL_1610837, EPI_ISL_1610838, EPI_ISL_1610839, EPI_ISL_1610840, EPI_ISL_1610841, EPI_ISL_1610842, EPI_ISL_1610843, EPI_ISL_1610844, EPI_ISL_1610846, EPI_ISL_1610847, EPI_ISL_1610848, EPI_ISL_1610849, EPI_ISL_1610850, EPI_ISL_1610852, EPI_ISL_1610853, EPI_ISL_1610855, EPI_ISL_1610856, EPI_ISL_1610859, EPI_ISL_1610860, EPI_ISL_1610861, EPI_ISL_1610862, EPI_ISL_1610863, EPI_ISL_1610864, EPI_ISL_1610865, EPI_ISL_1610866, EPI_ISL_1610867, EPI_ISL_1610868, EPI_ISL_1610869, EPI_ISL_1610870, EPI_ISL_1610871, EPI_ISL_1610872, EPI_ISL_1610873, EPI_ISL_1610874, EPI_ISL_1610875, EPI_ISL_1610876, EPI_ISL_1610877, EPI_ISL_1610878, EPI_ISL_1610881, EPI_ISL_1610882, EPI_ISL_1610883, EPI_ISL_1610885, EPI_ISL_1610886, EPI_ISL_1610889, EPI_ISL_1610890, EPI_ISL_1610891, EPI_ISL_1610892, EPI_ISL_1610893, EPI_ISL_1610894, EPI_ISL_1610895, EPI_ISL_1610896, EPI_ISL_1610897, EPI_ISL_1610898, EPI_ISL_1610899, EPI_ISL_1610900, EPI_ISL_1610903, EPI_ISL_1610905, EPI_ISL_1610906, EPI_ISL_1610907, EPI_ISL_1610908, EPI_ISL_1610909, EPI_ISL_1610910, EPI_ISL_1610911, EPI_ISL_1610912, EPI_ISL_1610913, EPI_ISL_1610915, EPI_ISL_1610916, EPI_ISL_1610920, EPI_ISL_1610922, EPI_ISL_1610923, EPI_ISL_1610924, EPI_ISL_1610925, EPI_ISL_1610926, EPI_ISL_1610927, EPI_ISL_1610928, EPI_ISL_1610930, EPI_ISL_1610931, EPI_ISL_1610933, EPI_ISL_1610934, EPI_ISL_1610936, EPI_ISL_1610937, EPI_ISL_1610938, EPI_ISL_1610939, EPI_ISL_1610940, EPI_ISL_1610941, EPI_ISL_1610943, EPI_ISL_1610944, EPI_ISL_1610945, EPI_ISL_1610947, EPI_ISL_1610948, EPI_ISL_1610950, EPI_ISL_1610951, EPI_ISL_1610955, EPI_ISL_1610956, EPI_ISL_1610957, EPI_ISL_1610958, EPI_ISL_1610959, EPI_ISL_1610960, EPI_ISL_1610961, EPI_ISL_1610962, EPI_ISL_1610963, EPI_ISL_1610964, EPI_ISL_1610965, EPI_ISL_1610966, EPI_ISL_1610968, EPI_ISL_1610970, EPI_ISL_1610971, EPI_ISL_1610972, EPI_ISL_1610974, EPI_ISL_1610975, EPI_ISL_1610976, EPI_ISL_1610977, EPI_ISL_1610978, EPI_ISL_1610979, EPI_ISL_1610980, EPI_ISL_1610982, EPI_ISL_1610984, EPI_ISL_1610986, EPI_ISL_1610987, EPI_ISL_1610988, EPI_ISL_1610989, EPI_ISL_1610990, EPI_ISL_1610991, EPI_ISL_1610994, EPI_ISL_1610995, EPI_ISL_1610996, EPI_ISL_1610997, EPI_ISL_1610998, EPI_ISL_1610999, EPI_ISL_1611001, EPI_ISL_1611003, EPI_ISL_1611004, EPI_ISL_1611005, EPI_ISL_1611007, EPI_ISL_1611008, EPI_ISL_1611009, EPI_ISL_1611011, EPI_ISL_1611012, EPI_ISL_1611014, EPI_ISL_1611015, EPI_ISL_1611016, EPI_ISL_1611017, EPI_ISL_1611018, EPI_ISL_1611019, EPI_ISL_1611020, EPI_ISL_1611021, EPI_ISL_1611022, EPI_ISL_1611023, EPI_ISL_1611024, EPI_ISL_1611025, EPI_ISL_1611026, EPI_ISL_1611027, EPI_ISL_1611029, EPI_ISL_1611030, EPI_ISL_1611031, EPI_ISL_1611033, EPI_ISL_1611035, EPI_ISL_1611036, EPI_ISL_1611037, EPI_ISL_1611038, EPI_ISL_1611039, EPI_ISL_1611040, EPI_ISL_1611041, EPI_ISL_1611042, EPI_ISL_1611043, EPI_ISL_1611044, EPI_ISL_1611046, EPI_ISL_1611047, EPI_ISL_1611048, EPI_ISL_1611049, EPI_ISL_1611050, EPI_ISL_1611052, EPI_ISL_1611053, EPI_ISL_1611054, EPI_ISL_1611055, EPI_ISL_1611056, EPI_ISL_1611057, EPI_ISL_1611058, EPI_ISL_1611059, EPI_ISL_1611060, EPI_ISL_1611061, EPI_ISL_1611063, EPI_ISL_1611065, EPI_ISL_1611067, EPI_ISL_1611068, EPI_ISL_1611069, EPI_ISL_1611070, EPI_ISL_1611071, EPI_ISL_1611073, EPI_ISL_1611074, EPI_ISL_1611075, EPI_ISL_1611076, EPI_ISL_1611077, EPI_ISL_1611079, EPI_ISL_1611080, EPI_ISL_1611081, EPI_ISL_1611082, EPI_ISL_1611084, EPI_ISL_1611085, EPI_ISL_1611086, EPI_ISL_1611087, EPI_ISL_1611088, EPI_ISL_1611089, EPI_ISL_1611090, EPI_ISL_1611091, EPI_ISL_1611092, EPI_ISL_1611093, EPI_ISL_1611094, EPI_ISL_1611095, EPI_ISL_1611096, EPI_ISL_1611097, EPI_ISL_1611098, EPI_ISL_1611099, EPI_ISL_1611100, EPI_ISL_1611101, EPI_ISL_1611102, EPI_ISL_1611103, EPI_ISL_1611104, EPI_ISL_1611107, EPI_ISL_1611108, EPI_ISL_1611109, EPI_ISL_1611110, EPI_ISL_1611111, EPI_ISL_1611112, EPI_ISL_1611113, EPI_ISL_1611115, EPI_ISL_1611116, EPI_ISL_1611117, EPI_ISL_1611118, EPI_ISL_1611119, EPI_ISL_1611120, EPI_ISL_1611121, EPI_ISL_1611122, EPI_ISL_1611123, EPI_ISL_1611124, EPI_ISL_1611125, EPI_ISL_1611126, EPI_ISL_1611127, EPI_ISL_1611128, EPI_ISL_1611129, EPI_ISL_1611130, EPI_ISL_1611131, EPI_ISL_1611132, EPI_ISL_1611133, EPI_ISL_1611134, EPI_ISL_1611137, EPI_ISL_1611138, EPI_ISL_1611139, EPI_ISL_1611140, EPI_ISL_1611141, EPI_ISL_1611142, EPI_ISL_1611143, EPI_ISL_1611144, EPI_ISL_1611145, EPI_ISL_1611146, EPI_ISL_1611147, EPI_ISL_1611148, EPI_ISL_1611149, EPI_ISL_1611150, EPI_ISL_1611151, EPI_ISL_1611152, EPI_ISL_1611154, EPI_ISL_1611155, EPI_ISL_1611156, EPI_ISL_1611157, EPI_ISL_1611159, EPI_ISL_1611160, EPI_ISL_1611162, EPI_ISL_1611163, EPI_ISL_1611164, EPI_ISL_1611165, EPI_ISL_1611166, EPI_ISL_1611167, EPI_ISL_1611168, EPI_ISL_1611169, EPI_ISL_1611170, EPI_ISL_1611171, EPI_ISL_1611172, EPI_ISL_1611173, EPI_ISL_1611174, EPI_ISL_1611175, EPI_ISL_1611176, EPI_ISL_1611177, EPI_ISL_1611178, EPI_ISL_1611179, EPI_ISL_1611180, EPI_ISL_1611181, EPI_ISL_1611182, EPI_ISL_1611183, EPI_ISL_1611184, EPI_ISL_1611185, EPI_ISL_1611186, EPI_ISL_1611187, EPI_ISL_1611188, EPI_ISL_1611189, EPI_ISL_1611190, EPI_ISL_1611192, EPI_ISL_1611193, EPI_ISL_1611194, EPI_ISL_1611195, EPI_ISL_1611196, EPI_ISL_1611197, EPI_ISL_1611198, EPI_ISL_1611200, EPI_ISL_1611201, EPI_ISL_1611202, EPI_ISL_1611203, EPI_ISL_1611204, EPI_ISL_1611205, EPI_ISL_1611206, EPI_ISL_1611207, EPI_ISL_1611208, EPI_ISL_1611209, EPI_ISL_1611210, EPI_ISL_1611211, EPI_ISL_1611213, EPI_ISL_1611214, EPI_ISL_1611215, EPI_ISL_1611216, EPI_ISL_1611217, EPI_ISL_1611218, EPI_ISL_1611219, EPI_ISL_1611220, EPI_ISL_1611221, EPI_ISL_1611222, EPI_ISL_1611223, EPI_ISL_1611224, EPI_ISL_1611225, EPI_ISL_1611226, EPI_ISL_1611227, EPI_ISL_1611228, EPI_ISL_1611229, EPI_ISL_1611230, EPI_ISL_1611231, EPI_ISL_1611232, EPI_ISL_1611233, EPI_ISL_1611234, EPI_ISL_1611236, EPI_ISL_1611237, EPI_ISL_1611238, EPI_ISL_1611239, EPI_ISL_1611240, EPI_ISL_1611241, EPI_ISL_1611242, EPI_ISL_1611245, EPI_ISL_1611247, EPI_ISL_1611248, EPI_ISL_1611249, EPI_ISL_1611251, EPI_ISL_1611252, EPI_ISL_1611253, EPI_ISL_1611255, EPI_ISL_1611257, EPI_ISL_1611258, EPI_ISL_1611259, EPI_ISL_1611260, EPI_ISL_1611261, EPI_ISL_1611263, EPI_ISL_1611264, EPI_ISL_1611265, EPI_ISL_1611267, EPI_ISL_1611268, EPI_ISL_1611269, EPI_ISL_1611270, EPI_ISL_1611271, EPI_ISL_1611272, EPI_ISL_1611273, EPI_ISL_1611274, EPI_ISL_1611275, EPI_ISL_1611276, EPI_ISL_1611277, EPI_ISL_1611278, EPI_ISL_1611279, EPI_ISL_1611280, EPI_ISL_1611281, EPI_ISL_1611282, EPI_ISL_1611283, EPI_ISL_1611284, EPI_ISL_1611285, EPI_ISL_1611286, EPI_ISL_1611287, EPI_ISL_1611288, EPI_ISL_1611289, EPI_ISL_1611290, EPI_ISL_1611291, EPI_ISL_1611293, EPI_ISL_1611294, EPI_ISL_1611295, EPI_ISL_1611296, EPI_ISL_1611297, EPI_ISL_1611298, EPI_ISL_1611299, EPI_ISL_1611300, EPI_ISL_1611302, EPI_ISL_1611303, EPI_ISL_1611304, EPI_ISL_1611305, EPI_ISL_1611307, EPI_ISL_1611308, EPI_ISL_1611309, EPI_ISL_1611310, EPI_ISL_1611311, EPI_ISL_1611312, EPI_ISL_1611314, EPI_ISL_1611315, EPI_ISL_1611316, EPI_ISL_1611317, EPI_ISL_1611318, EPI_ISL_1611320, EPI_ISL_1611321, EPI_ISL_1611322, EPI_ISL_1611323, EPI_ISL_1611324, EPI_ISL_1611325, EPI_ISL_1611327, EPI_ISL_1611328, EPI_ISL_1611329, EPI_ISL_1611330, EPI_ISL_1611331, EPI_ISL_1611332, EPI_ISL_1611333, EPI_ISL_1611334, EPI_ISL_1611335, EPI_ISL_1611337, EPI_ISL_1611338, EPI_ISL_1611340, EPI_ISL_1611341, EPI_ISL_1611342, EPI_ISL_1611343, EPI_ISL_1611344, EPI_ISL_1611346, EPI_ISL_1611347, EPI_ISL_1611348, EPI_ISL_1611349, EPI_ISL_1611351, EPI_ISL_1611352, EPI_ISL_1611353, EPI_ISL_1611354, EPI_ISL_1611355, EPI_ISL_1611356, EPI_ISL_1611357, EPI_ISL_1611358, EPI_ISL_1611359, EPI_ISL_1611360, EPI_ISL_1611362, EPI_ISL_1611366, EPI_ISL_1611367, EPI_ISL_1611369, EPI_ISL_1611370, EPI_ISL_1611371, EPI_ISL_1611373, EPI_ISL_1611374, EPI_ISL_1611375, EPI_ISL_1611377,

EPI_ISL_1611378, EPI_ISL_1611380, EPI_ISL_1611382, EPI_ISL_1611383, EPI_ISL_1611385, EPI_ISL_1611386, EPI_ISL_1611387, EPI_ISL_1611388, EPI_ISL_1611390, EPI_ISL_1611392, EPI_ISL_1611394, EPI_ISL_1611395, EPI_ISL_1611397, EPI_ISL_1611398, EPI_ISL_1611399, EPI_ISL_1611400, EPI_ISL_1611401, EPI_ISL_1611402, EPI_ISL_1611403, EPI_ISL_1611405, EPI_ISL_1611407, EPI_ISL_1611408, EPI_ISL_1611409, EPI_ISL_1611410, EPI_ISL_1611412, EPI_ISL_1611413, EPI_ISL_1611414, EPI_ISL_1611415, EPI_ISL_1611416, EPI_ISL_1611417, EPI_ISL_1611418, EPI_ISL_1611419, EPI_ISL_1611422, EPI_ISL_1611423, EPI_ISL_1611424, EPI_ISL_1611426, EPI_ISL_1611427, EPI_ISL_1611428, EPI_ISL_1611429, EPI_ISL_1611431, EPI_ISL_1611432, EPI_ISL_1611433, EPI_ISL_1611434, EPI_ISL_1611437, EPI_ISL_1611438, EPI_ISL_1611439, EPI_ISL_1611440, EPI_ISL_1611441, EPI_ISL_1611443, EPI_ISL_1611445, EPI_ISL_1611446, EPI_ISL_1611447, EPI_ISL_1611448, EPI_ISL_1611449, EPI_ISL_1611450, EPI_ISL_1611453, EPI_ISL_1611454, EPI_ISL_1611455, EPI_ISL_1611456, EPI_ISL_1611457, EPI_ISL_1611459, EPI_ISL_1611460, EPI_ISL_1611461, EPI_ISL_1611462, EPI_ISL_1611463, EPI_ISL_1611464, EPI_ISL_1611465, EPI_ISL_1611466, EPI_ISL_1611467, EPI_ISL_1611468, EPI_ISL_1611469, EPI_ISL_1611470, EPI_ISL_1611471, EPI_ISL_1611472, EPI_ISL_1611473, EPI_ISL_1611474, EPI_ISL_1611476, EPI_ISL_1611479, EPI_ISL_1611480, EPI_ISL_1611481, EPI_ISL_1611483, EPI_ISL_1611484, EPI_ISL_1611485, EPI_ISL_1611486, EPI_ISL_1611487, EPI_ISL_1611488, EPI_ISL_1611489, EPI_ISL_1611490, EPI_ISL_1611491, EPI_ISL_1611492, EPI_ISL_1611493, EPI_ISL_1611494, EPI_ISL_1611495, EPI_ISL_1611496, EPI_ISL_1611497, EPI_ISL_1611498, EPI_ISL_1611500, EPI_ISL_1611502, EPI_ISL_1611503, EPI_ISL_1611504, EPI_ISL_1611505, EPI_ISL_1611506, EPI_ISL_1611507, EPI_ISL_1611508, EPI_ISL_1611509, EPI_ISL_1611510, EPI_ISL_1611511, EPI_ISL_1611513, EPI_ISL_1611515, EPI_ISL_1611516, EPI_ISL_1611517, EPI_ISL_1611518, EPI_ISL_1611519, EPI_ISL_1611520, EPI_ISL_1611521, EPI_ISL_1611522, EPI_ISL_1611523, EPI_ISL_1611524, EPI_ISL_1611525, EPI_ISL_1611526, EPI_ISL_1611527, EPI_ISL_1611528, EPI_ISL_1611529, EPI_ISL_1611531, EPI_ISL_1611532, EPI_ISL_1611533, EPI_ISL_1611534, EPI_ISL_1611541, EPI_ISL_1611542, EPI_ISL_1611543, EPI_ISL_1611544, EPI_ISL_1611545, EPI_ISL_1611546, EPI_ISL_1611547, EPI_ISL_1611548, EPI_ISL_1611549, EPI_ISL_1611550, EPI_ISL_1611551, EPI_ISL_1611552, EPI_ISL_1611553, EPI_ISL_1611554, EPI_ISL_1611555, EPI_ISL_1611557, EPI_ISL_1611558, EPI_ISL_1611559, EPI_ISL_1611562, EPI_ISL_1611563, EPI_ISL_1611564, EPI_ISL_1611565, EPI_ISL_1611566, EPI_ISL_1611567, EPI_ISL_1611568, EPI_ISL_1611569, EPI_ISL_1611570, EPI_ISL_1611571, EPI_ISL_1611572, EPI_ISL_1611573, EPI_ISL_1611574, EPI_ISL_1611575, EPI_ISL_1611576, EPI_ISL_1611577, EPI_ISL_1611578, EPI_ISL_1611579, EPI_ISL_1611580, EPI_ISL_1611581, EPI_ISL_1611582, EPI_ISL_1611583, EPI_ISL_1611586, EPI_ISL_1611587, EPI_ISL_1611588, EPI_ISL_1611589, EPI_ISL_1611590, EPI_ISL_1611591, EPI_ISL_1611592, EPI_ISL_1611593, EPI_ISL_1611594, EPI_ISL_1611595, EPI_ISL_1611597, EPI_ISL_1611598, EPI_ISL_1611599, EPI_ISL_1611600, EPI_ISL_1611602, EPI_ISL_1611603, EPI_ISL_1611604, EPI_ISL_1611605, EPI_ISL_1611606, EPI_ISL_1611608, EPI_ISL_1611609, EPI_ISL_1611610, EPI_ISL_1611611, EPI_ISL_1611612, EPI_ISL_1611613, EPI_ISL_1611614, EPI_ISL_1611615, EPI_ISL_1611617, EPI_ISL_1611618, EPI_ISL_1611619, EPI_ISL_1611620, EPI_ISL_1611621, EPI_ISL_1611622, EPI_ISL_1611623, EPI_ISL_1611624, EPI_ISL_1611625, EPI_ISL_1611626, EPI_ISL_1611627, EPI_ISL_1611628, EPI_ISL_1611629, EPI_ISL_1611630, EPI_ISL_1611631, EPI_ISL_1611632, EPI_ISL_1611633, EPI_ISL_1611634, EPI_ISL_1611635, EPI_ISL_1611636, EPI_ISL_1611637, EPI_ISL_1611638, EPI_ISL_1611639, EPI_ISL_1611640, EPI_ISL_1611641, EPI_ISL_1611642, EPI_ISL_1611643, EPI_ISL_1611644, EPI_ISL_1611645, EPI_ISL_1611646, EPI_ISL_1611647, EPI_ISL_1611648, EPI_ISL_1611650, EPI_ISL_1611651, EPI_ISL_1611652, EPI_ISL_1611653, EPI_ISL_1611655, EPI_ISL_1611657, EPI_ISL_1611659, EPI_ISL_1611660, EPI_ISL_1611661, EPI_ISL_1611662, EPI_ISL_1611663, EPI_ISL_1611664, EPI_ISL_1611665, EPI_ISL_1611666, EPI_ISL_1611667, EPI_ISL_1611668, EPI_ISL_1611670, EPI_ISL_1611671, EPI_ISL_1611672, EPI_ISL_1611673, EPI_ISL_1611675, EPI_ISL_1611676, EPI_ISL_1611677, EPI_ISL_1611678, EPI_ISL_1611679, EPI_ISL_1611681, EPI_ISL_1611682, EPI_ISL_1611683, EPI_ISL_1611684, EPI_ISL_1611685, EPI_ISL_1611686, EPI_ISL_1611687, EPI_ISL_1611688, EPI_ISL_1611689, EPI_ISL_1611690, EPI_ISL_1611691, EPI_ISL_1611694, EPI_ISL_1611695, EPI_ISL_1611697, EPI_ISL_1611698, EPI_ISL_1611702, EPI_ISL_1611703, EPI_ISL_1611704, EPI_ISL_1611706, EPI_ISL_1611708, EPI_ISL_1611709, EPI_ISL_1611710, EPI_ISL_1611711, EPI_ISL_1611712, EPI_ISL_1611713, EPI_ISL_1611714, EPI_ISL_1611715, EPI_ISL_1611717, EPI_ISL_1611718, EPI_ISL_1611719, EPI_ISL_1611720, EPI_ISL_1611721, EPI_ISL_1611722, EPI_ISL_1611723, EPI_ISL_1611724, EPI_ISL_1611725, EPI_ISL_1611727, EPI_ISL_1611729, EPI_ISL_1611730, EPI_ISL_1611731, EPI_ISL_1611732, EPI_ISL_1611734, EPI_ISL_1611735, EPI_ISL_1611736, EPI_ISL_1611737, EPI_ISL_1611738, EPI_ISL_1611739, EPI_ISL_1611741, EPI_ISL_1611742, EPI_ISL_1611743, EPI_ISL_1611744, EPI_ISL_1611745, EPI_ISL_1611747, EPI_ISL_1611749, EPI_ISL_1611750, EPI_ISL_1611751, EPI_ISL_1611752, EPI_ISL_1611753, EPI_ISL_1611754, EPI_ISL_1611755, EPI_ISL_1611756, EPI_ISL_1611757, EPI_ISL_1611758, EPI_ISL_1611759, EPI_ISL_1611760, EPI_ISL_1611761, EPI_ISL_1611762, EPI_ISL_1611763, EPI_ISL_1611764, EPI_ISL_1611765, EPI_ISL_1611767, EPI_ISL_1611768, EPI_ISL_1611769, EPI_ISL_1611770, EPI_ISL_1611771, EPI_ISL_1611772, EPI_ISL_1611773, EPI_ISL_1611776, EPI_ISL_1611778, EPI_ISL_1611779, EPI_ISL_1611780, EPI_ISL_1611781, EPI_ISL_1611782, EPI_ISL_1611783, EPI_ISL_1611784, EPI_ISL_1611785, EPI_ISL_1611786, EPI_ISL_1611787, EPI_ISL_1611788, EPI_ISL_1611790, EPI_ISL_1611791, EPI_ISL_1611792, EPI_ISL_1611793, EPI_ISL_1611794, EPI_ISL_1611795, EPI_ISL_1611796, EPI_ISL_1611797, EPI_ISL_1611799, EPI_ISL_1611800, EPI_ISL_1611802, EPI_ISL_1611803, EPI_ISL_1611804, EPI_ISL_1611805, EPI_ISL_1611808, EPI_ISL_1611809, EPI_ISL_1611810, EPI_ISL_1611811, EPI_ISL_1611813, EPI_ISL_1611815, EPI_ISL_1611816, EPI_ISL_1611817, EPI_ISL_1611818, EPI_ISL_1611819, EPI_ISL_1611820, EPI_ISL_1611821, EPI_ISL_1611823, EPI_ISL_1611824, EPI_ISL_1611825, EPI_ISL_1611828, EPI_ISL_1611829, EPI_ISL_1611830, EPI_ISL_1611831, EPI_ISL_1611833, EPI_ISL_1611834, EPI_ISL_1611836, EPI_ISL_1611837, EPI_ISL_1611838, EPI_ISL_1611839, EPI_ISL_1611840, EPI_ISL_1611842, EPI_ISL_1611843, EPI_ISL_1611844, EPI_ISL_1611845, EPI_ISL_1611846, EPI_ISL_1611847, EPI_ISL_1611848, EPI_ISL_1611849, EPI_ISL_1611850, EPI_ISL_1611851, EPI_ISL_1611852, EPI_ISL_1611853, EPI_ISL_1611854, EPI_ISL_1611855, EPI_ISL_1611856, EPI_ISL_1611857, EPI_ISL_1611859, EPI_ISL_1611860, EPI_ISL_1611861, EPI_ISL_1611862, EPI_ISL_1611864, EPI_ISL_1611865, EPI_ISL_1611866, EPI_ISL_1611867, EPI_ISL_1611868, EPI_ISL_1611869, EPI_ISL_1611870, EPI_ISL_1611871, EPI_ISL_1611872, EPI_ISL_1611874, EPI_ISL_1611876, EPI_ISL_1611877, EPI_ISL_1611879, EPI_ISL_1611880, EPI_ISL_1611881, EPI_ISL_1611882, EPI_ISL_1611883, EPI_ISL_1611884, EPI_ISL_1611885, EPI_ISL_1611886, EPI_ISL_1611887, EPI_ISL_1611888, EPI_ISL_1611889, EPI_ISL_1611890, EPI_ISL_1611891, EPI_ISL_1611892, EPI_ISL_1611893, EPI_ISL_1611894, EPI_ISL_1611895, EPI_ISL_1611896, EPI_ISL_1611897, EPI_ISL_1611898, EPI_ISL_1611899, EPI_ISL_1611900, EPI_ISL_1611901, EPI_ISL_1611905, EPI_ISL_1611906, EPI_ISL_1611907, EPI_ISL_1611908, EPI_ISL_1611909, EPI_ISL_1611912, EPI_ISL_1611913, EPI_ISL_1611914, EPI_ISL_1611915, EPI_ISL_1611916, EPI_ISL_1611917, EPI_ISL_1611918, EPI_ISL_1611919, EPI_ISL_1611920, EPI_ISL_1611921, EPI_ISL_1611923, EPI_ISL_1611924, EPI_ISL_1611925, EPI_ISL_1611926, EPI_ISL_1611927, EPI_ISL_1611928, EPI_ISL_1611929, EPI_ISL_1611930, EPI_ISL_1611931, EPI_ISL_1611932, EPI_ISL_1611933, EPI_ISL_1611934, EPI_ISL_1611936, EPI_ISL_1611938, EPI_ISL_1611939, EPI_ISL_1611940, EPI_ISL_1611941, EPI_ISL_1611942, EPI_ISL_1611943, EPI_ISL_1611945, EPI_ISL_1611946, EPI_ISL_1611947, EPI_ISL_1611948, EPI_ISL_1611950, EPI_ISL_1611951, EPI_ISL_1611952, EPI_ISL_1611953, EPI_ISL_1611954, EPI_ISL_1611955, EPI_ISL_1611956, EPI_ISL_1611957, EPI_ISL_1611958, EPI_ISL_1611960, EPI_ISL_1611962, EPI_ISL_1611963, EPI_ISL_1611966, EPI_ISL_1611967, EPI_ISL_1611968, EPI_ISL_1611969, EPI_ISL_1611971, EPI_ISL_1611972, EPI_ISL_1611973, EPI_ISL_1611974, EPI_ISL_1611975, EPI_ISL_1611976, EPI_ISL_1611977, EPI_ISL_1611978, EPI_ISL_1611979, EPI_ISL_1611980, EPI_ISL_1611982, EPI_ISL_1611983, EPI_ISL_1611984, EPI_ISL_1611985, EPI_ISL_1611986, EPI_ISL_1611987, EPI_ISL_1611988, EPI_ISL_1611990, EPI_ISL_1611992, EPI_ISL_1611993, EPI_ISL_1611994, EPI_ISL_1611995, EPI_ISL_1611996, EPI_ISL_1611997, EPI_ISL_1611998, EPI_ISL_1611999, EPI_ISL_1612000, EPI_ISL_1612002, EPI_ISL_1612003, EPI_ISL_1612004, EPI_ISL_1612005, EPI_ISL_1612007, EPI_ISL_1612008, EPI_ISL_1612009, EPI_ISL_1612010, EPI_ISL_1612011, EPI_ISL_1612012, EPI_ISL_1612013, EPI_ISL_1612015, EPI_ISL_1612016, EPI_ISL_1612017, EPI_ISL_1612019, EPI_ISL_1612021, EPI_ISL_1612022, EPI_ISL_1612023, EPI_ISL_1612024, EPI_ISL_1612025, EPI_ISL_1612026, EPI_ISL_1612027, EPI_ISL_1612028, EPI_ISL_1612029, EPI_ISL_1612030, EPI_ISL_1612031, EPI_ISL_1612032, EPI_ISL_1612034, EPI_ISL_1612035, EPI_ISL_1612037, EPI_ISL_1612039, EPI_ISL_1612040, EPI_ISL_1612041, EPI_ISL_1612042, EPI_ISL_1612043, EPI_ISL_1612044, EPI_ISL_1612045, EPI_ISL_1612046, EPI_ISL_1612047, EPI_ISL_1612048, EPI_ISL_1612049, EPI_ISL_1612050, EPI_ISL_1612051, EPI_ISL_1612052, EPI_ISL_1612053, EPI_ISL_1612054, EPI_ISL_1612057, EPI_ISL_1612058, EPI_ISL_1612059, EPI_ISL_1612060, EPI_ISL_1612061, EPI_ISL_1612062, EPI_ISL_1612063, EPI_ISL_1612064, EPI_ISL_1612065, EPI_ISL_1612067, EPI_ISL_1612069, EPI_ISL_1612070, EPI_ISL_1612071, EPI_ISL_1612072, EPI_ISL_1612073, EPI_ISL_1612074, EPI_ISL_1612075, EPI_ISL_1612076, EPI_ISL_1612077, EPI_ISL_1612078, EPI_ISL_1612079, EPI_ISL_1612080, EPI_ISL_1612081, EPI_ISL_1612082, EPI_ISL_1612083, EPI_ISL_1612085, EPI_ISL_1612086, EPI_ISL_1612087, EPI_ISL_1612088, EPI_ISL_1612090, EPI_ISL_1612091, EPI_ISL_1612092, EPI_ISL_1612093, EPI_ISL_1612094, EPI_ISL_1612095, EPI_ISL_1612096, EPI_ISL_1612097, EPI_ISL_1612098, EPI_ISL_1612100, EPI_ISL_1612101, EPI_ISL_1612102, EPI_ISL_1612104, EPI_ISL_1612107, EPI_ISL_1612108, EPI_ISL_1612111, EPI_ISL_1612112, EPI_ISL_1612115, EPI_ISL_1612116, EPI_ISL_1612117, EPI_ISL_1612118, EPI_ISL_1612119, EPI_ISL_1612120, EPI_ISL_1612121, EPI_ISL_1612123, EPI_ISL_1612124, EPI_ISL_1612125, EPI_ISL_1612126, EPI_ISL_1612127, EPI_ISL_1612129, EPI_ISL_1612130, EPI_ISL_1612131, EPI_ISL_1612132, EPI_ISL_1612134, EPI_ISL_1612135, EPI_ISL_1612136, EPI_ISL_1612137, EPI_ISL_1612138, EPI_ISL_1612139, EPI_ISL_1612142, EPI_ISL_1612143, EPI_ISL_1612144, EPI_ISL_1612145, EPI_ISL_1612146, EPI_ISL_1612147, EPI_ISL_1612148, EPI_ISL_1612149, EPI_ISL_1612152, EPI_ISL_1612153, EPI_ISL_1612154, EPI_ISL_1612155, EPI_ISL_1612157, EPI_ISL_1612158, EPI_ISL_1612160, EPI_ISL_1612161, EPI_ISL_1612162, EPI_ISL_1612163, EPI_ISL_1612164, EPI_ISL_1612165, EPI_ISL_1612166, EPI_ISL_1612168, EPI_ISL_1612169, EPI_ISL_1612170, EPI_ISL_1612171, EPI_ISL_1612172, EPI_ISL_1612173, EPI_ISL_1612174, EPI_ISL_1612175, EPI_ISL_1612176, EPI_ISL_1612177, EPI_ISL_1612178, EPI_ISL_1612179, EPI_ISL_1612180, EPI_ISL_1612181, EPI_ISL_1612182, EPI_ISL_1612183, EPI_ISL_1612184, EPI_ISL_1612185, EPI_ISL_1612186, EPI_ISL_1612187, EPI_ISL_1612188, EPI_ISL_1612189, EPI_ISL_1612190, EPI_ISL_1612192, EPI_ISL_1612193, EPI_ISL_1612194, EPI_ISL_1612197, EPI_ISL_1612198, EPI_ISL_1612201, EPI_ISL_1612202, EPI_ISL_1612203, EPI_ISL_1612204, EPI_ISL_1612205, EPI_ISL_1612206, EPI_ISL_1612207, EPI_ISL_1612209, EPI_ISL_1612210, EPI_ISL_1612211, EPI_ISL_1612212, EPI_ISL_1612213, EPI_ISL_1612214, EPI_ISL_1612215, EPI_ISL_1612216, EPI_ISL_1612217, EPI_ISL_1612218, EPI_ISL_1612219, EPI_ISL_1612220, EPI_ISL_1612221, EPI_ISL_1612222, EPI_ISL_1612223, EPI_ISL_1612224, EPI_ISL_1612225, EPI_ISL_1612226, EPI_ISL_1612227, EPI_ISL_1612228, EPI_ISL_1612229, EPI_ISL_1612230, EPI_ISL_1612231, EPI_ISL_1612233, EPI_ISL_1612234, EPI_ISL_1612235, EPI_ISL_1612236, EPI_ISL_1612237, EPI_ISL_1612239, EPI_ISL_1612240, EPI_ISL_1612241, EPI_ISL_1612242, EPI_ISL_1612243, EPI_ISL_1612244, EPI_ISL_1612245, EPI_ISL_1612246, EPI_ISL_1612247, EPI_ISL_1612248, EPI_ISL_1612249, EPI_ISL_1612250, EPI_ISL_1612251, EPI_ISL_1612252, EPI_ISL_1612253, EPI_ISL_1612254, EPI_ISL_1612255, EPI_ISL_1612256, EPI_ISL_1612259, EPI_ISL_1612260, EPI_ISL_1612261, EPI_ISL_1612262, EPI_ISL_1612263, EPI_ISL_1612264, EPI_ISL_1612265, EPI_ISL_1612267, EPI_ISL_1612268, EPI_ISL_1612269, EPI_ISL_1612270, EPI_ISL_1612272, EPI_ISL_1612273, EPI_ISL_1612274, EPI_ISL_1612275, EPI_ISL_1612276, EPI_ISL_1612277, EPI_ISL_1612278, EPI_ISL_1612279, EPI_ISL_1612280, EPI_ISL_1612282, EPI_ISL_1612283, EPI_ISL_1612285, EPI_ISL_1612286, EPI_ISL_1612289, EPI_ISL_1612290, EPI_ISL_1612291, EPI_ISL_1612292, EPI_ISL_1612293, EPI_ISL_1612295, EPI_ISL_1612296, EPI_ISL_1612299, EPI_ISL_1612300, EPI_ISL_1612301, EPI_ISL_1612302, EPI_ISL_1612303, EPI_ISL_1612304, EPI_ISL_1612308, EPI_ISL_1612309, EPI_ISL_1612310, EPI_ISL_1612311, EPI_ISL_1612312, EPI_ISL_1612314, EPI_ISL_1612315, EPI_ISL_1612316, EPI_ISL_1612317, EPI_ISL_1612318, EPI_ISL_1612320, EPI_ISL_1612321, EPI_ISL_1612322, EPI_ISL_1612323, EPI_ISL_1612324, EPI_ISL_1612325, EPI_ISL_1612326, EPI_ISL_1612327, EPI_ISL_1612328, EPI_ISL_1612330, EPI_ISL_1612331, EPI_ISL_1612332, EPI_ISL_1612333, EPI_ISL_1612334, EPI_ISL_1612335, EPI_ISL_1612336, EPI_ISL_1612337, EPI_ISL_1612338, EPI_ISL_1612339, EPI_ISL_1612340, EPI_ISL_1612341, EPI_ISL_1612343, EPI_ISL_1612344, EPI_ISL_1612345, EPI_ISL_1612349

| | | | |
|---|---|---|---|
| see above | Laboratory Corporation of America | Centers for Disease Control and Prevention Division of Viral Diseases, Pathogen Discovery | Dakota Howard, Dhwani Batra, Peter W. Cook, Kara Moser, Adrian Paskey, Jason Caravas, Benjamin Rambo-Martin, Shatavia Morrison, Christopher Gulvick, Scott Sammons, Yvette Unoarumhi, Darlene Wagner, Matthew Schmerer, Minoo Agarwal, Eyad Almasri, Debbie Boles, Ayla Burns, Nuthawin Charoensri, Oren Cohen, Susan Countryman, Mary Ann Cristobal, Bobbi Croy, Suzanne Dale, Hrushikesh Deshmukh, Amanda Douglas, Vincent Drouillon, Marcia Eisenberg, Howard Engler, Rama Ghatti, Prashant Gupta, Susan Hicks, Jake Humphrey, Lax Iyer, Manoj Jain, Mohan Kolli, Brian Krueger, Tim Kuphal, Stanley Letovsky, Michael Levandoski, Craig Lukasik, Jonathan Meltzer, Brian Norvell, Mindy Nye, Scott Parker, Christos Petropoulos, John Pruitt, Steven Ragan, Scott Ryan, Mike Sapeta, Jana Schroth, Suresh Babu Selvaraju, Goran Stevovic, Amanda Suchanek, Andrea Throop, Lyndon Tilson, Thomas Urban, Joe Voshell, Kimberly Wagner, Jonathan Williams, Mary Williamson, Qian Zeng, Tricia Zwiefelhofer, Clinton R. Paden, Duncan MacCannell |
| EPI_ISL_1612350, EPI_ISL_1612351 | North Dakota Department of Health, Public Health Laboratory | North Dakota Department of Health, Public Health Laboratory | Lisa Wingerter |
| EPI_ISL_1612353 | Laboratory Corporation of America | Centers for Disease Control and Prevention Division of Viral Diseases, Pathogen Discovery | Dakota Howard, Dhwani Batra, Peter W. Cook, Kara Moser, Adrian Paskey, Jason Caravas, Benjamin Rambo-Martin, Shatavia Morrison, Christopher Gulvick, Scott Sammons, Yvette Unoarumhi, Darlene Wagner, Matthew Schmerer, Minoo Agarwal, Eyad Almasri, Debbie Boles, Ayla Burns, Nuthawin Charoensri, Oren Cohen, Susan Countryman, Mary Ann Cristobal, Bobbi Croy, Suzanne Dale, Hrushikesh Deshmukh, Amanda Douglas, Vincent Drouillon, Marcia Eisenberg, Howard Engler, Rama Ghatti, Prashant Gupta, Susan Hicks, Jake Humphrey, Lax Iyer, Manoj Jain, Mohan Kolli, Brian Krueger, Tim Kuphal, Stanley Letovsky, Michael Levandoski, Craig Lukasik, Jonathan Meltzer, Brian Norvell, Mindy Nye, Scott Parker, Christos Petropoulos, John Pruitt, Steven Ragan, Scott Ryan, Mike Sapeta, Jana Schroth, Suresh Babu Selvaraju, Goran Stevovic, Amanda Suchanek, Andrea Throop, Lyndon Tilson, Thomas Urban, Joe Voshell, Kimberly Wagner, Jonathan Williams, Mary Williamson, Qian Zeng, Tricia Zwiefelhofer, Clinton R. Paden, Duncan MacCannell |
| EPI_ISL_1612354 | North Dakota Department of Health, Public Health Laboratory | North Dakota Department of Health, Public Health Laboratory | Lisa Wingerter |
| EPI_ISL_1612358, EPI_ISL_1612361 | Laboratory Corporation of America | Centers for Disease Control and Prevention Division of Viral Diseases, Pathogen Discovery | Dakota Howard, Dhwani Batra, Peter W. Cook, Kara Moser, Adrian Paskey, Jason Caravas, Benjamin Rambo-Martin, Shatavia Morrison, Christopher Gulvick, Scott Sammons, Yvette Unoarumhi, Darlene Wagner, Matthew Schmerer, Minoo Agarwal, Eyad Almasri, Debbie Boles, Ayla Burns, Nuthawin Charoensri, Oren Cohen, Susan Countryman, Mary Ann Cristobal, Bobbi Croy, Suzanne Dale, Hrushikesh Deshmukh, Amanda Douglas, Vincent Drouillon, Marcia Eisenberg, Howard Engler, Rama Ghatti, Prashant Gupta, Susan Hicks, Jake Humphrey, Lax Iyer, Manoj Jain, Mohan Kolli, Brian Krueger, Tim Kuphal, Stanley Letovsky, Michael Levandoski, Craig Lukasik, Jonathan Meltzer, Brian Norvell, Mindy Nye, Scott Parker, Christos Petropoulos, John Pruitt, Steven Ragan, Scott Ryan, Mike Sapeta, Jana Schroth, Suresh Babu Selvaraju, Goran Stevovic, Amanda Suchanek, Andrea Throop, Lyndon Tilson, Thomas Urban, Joe Voshell, Kimberly Wagner, Jonathan Williams, Mary Williamson, Qian Zeng, Tricia Zwiefelhofer, Clinton R. Paden, Duncan MacCannell |
| EPI_ISL_1612363 | North Dakota Department of Health, Public Health Laboratory | North Dakota Department of Health, Public Health Laboratory | Lisa Wingerter |
| EPI_ISL_1612364 | Laboratory Corporation of America | Centers for Disease Control and Prevention Division of Viral Diseases, Pathogen Discovery | Dakota Howard, Dhwani Batra, Peter W. Cook, Kara Moser, Adrian Paskey, Jason Caravas, Benjamin Rambo-Martin, Shatavia Morrison, Christopher Gulvick, Scott Sammons, Yvette Unoarumhi, Darlene Wagner, Matthew Schmerer, Minoo Agarwal, Eyad Almasri, Debbie Boles, Ayla Burns, Nuthawin Charoensri, Oren Cohen, Susan Countryman, Mary Ann Cristobal, Bobbi Croy, Suzanne Dale, Hrushikesh Deshmukh, Amanda Douglas, Vincent Drouillon, Marcia Eisenberg, Howard Engler, Rama Ghatti, Prashant Gupta, Susan Hicks, Jake Humphrey, Lax Iyer, Manoj Jain, Mohan Kolli, Brian Krueger, Tim Kuphal, Stanley Letovsky, Michael Levandoski, Craig Lukasik, Jonathan Meltzer, Brian Norvell, Mindy Nye, Scott Parker, Christos Petropoulos, John Pruitt, Steven Ragan, Scott Ryan, Mike Sapeta, Jana Schroth, Suresh Babu Selvaraju, Goran Stevovic, Amanda Suchanek, Andrea Throop, Lyndon Tilson, Thomas Urban, Joe Voshell, Kimberly Wagner, Jonathan Williams, Mary Williamson, Qian Zeng, Tricia Zwiefelhofer, Clinton R. Paden, Duncan MacCannell |
| EPI_ISL_1612367, EPI_ISL_1612369, EPI_ISL_1612370, EPI_ISL_1612372 | North Dakota Department of Health, Public Health Laboratory | North Dakota Department of Health, Public Health Laboratory | Lisa Wingerter |
| EPI_ISL_1612374 | Laboratory Corporation of America | Centers for Disease Control and Prevention Division of Viral Diseases, Pathogen Discovery | Dakota Howard, Dhwani Batra, Peter W. Cook, Kara Moser, Adrian Paskey, Jason Caravas, Benjamin Rambo-Martin, Shatavia Morrison, Christopher Gulvick, Scott Sammons, Yvette Unoarumhi, Darlene Wagner, Matthew Schmerer, Minoo Agarwal, Eyad Almasri, Debbie Boles, Ayla Burns, Nuthawin Charoensri, Oren Cohen, Susan Countryman, Mary Ann Cristobal, Bobbi Croy, Suzanne Dale, Hrushikesh Deshmukh, Amanda Douglas, Vincent Drouillon, Marcia Eisenberg, Howard Engler, Rama Ghatti, Prashant Gupta, Susan Hicks, Jake Humphrey, Lax Iyer, Manoj Jain, Mohan Kolli, Brian Krueger, Tim Kuphal, Stanley Letovsky, Michael Levandoski, Craig Lukasik, Jonathan Meltzer, Brian Norvell, Mindy Nye, Scott Parker, Christos Petropoulos, John Pruitt, Steven Ragan, Scott Ryan, Mike Sapeta, Jana Schroth, Suresh Babu Selvaraju, Goran Stevovic, Amanda Suchanek, Andrea Throop, Lyndon Tilson, Thomas Urban, Joe Voshell, Kimberly Wagner, Jonathan Williams, Mary Williamson, Qian Zeng, Tricia Zwiefelhofer, Clinton R. Paden, Duncan MacCannell |
| EPI_ISL_1612378 | North Dakota Department of Health, Public Health Laboratory | North Dakota Department of Health, Public Health Laboratory | Lisa Wingerter |
| EPI_ISL_1612379 | Laboratory Corporation of America | Centers for Disease Control and Prevention Division of Viral Diseases, Pathogen Discovery | Dakota Howard, Dhwani Batra, Peter W. Cook, Kara Moser, Adrian Paskey, Jason Caravas, Benjamin Rambo-Martin, Shatavia Morrison, Christopher Gulvick, Scott Sammons, Yvette Unoarumhi, Darlene Wagner, Matthew Schmerer, Minoo Agarwal, Eyad Almasri, Debbie Boles, Ayla Burns, Nuthawin Charoensri, Oren Cohen, Susan Countryman, Mary Ann Cristobal, Bobbi Croy, Suzanne Dale, Hrushikesh Deshmukh, Amanda Douglas, Vincent Drouillon, Marcia Eisenberg, Howard Engler, Rama Ghatti, Prashant Gupta, Susan Hicks, Jake Humphrey, Lax Iyer, Manoj Jain, Mohan Kolli, Brian Krueger, Tim Kuphal, Stanley Letovsky, Michael Levandoski, Craig Lukasik, Jonathan Meltzer, Brian Norvell, Mindy Nye, Scott Parker, Christos Petropoulos, John Pruitt, Steven Ragan, Scott Ryan, Mike Sapeta, Jana Schroth, Suresh Babu Selvaraju, Goran Stevovic, Amanda Suchanek, Andrea Throop, Lyndon Tilson, Thomas Urban, Joe Voshell, Kimberly Wagner, Jonathan Williams, Mary Williamson, Qian Zeng, Tricia Zwiefelhofer, Clinton R. Paden, Duncan MacCannell |
| EPI_ISL_1612380 | North Dakota Department of Health, Public Health Laboratory | North Dakota Department of Health, Public Health Laboratory | Lisa Wingerter |
| EPI_ISL_1612383, EPI_ISL_1612385 | Laboratory Corporation of America | Centers for Disease Control and Prevention Division of Viral Diseases, Pathogen Discovery | Dakota Howard, Dhwani Batra, Peter W. Cook, Kara Moser, Adrian Paskey, Jason Caravas, Benjamin Rambo-Martin, Shatavia Morrison, Christopher Gulvick, Scott Sammons, Yvette Unoarumhi, Darlene Wagner, Matthew Schmerer, Minoo Agarwal, Eyad Almasri, Debbie Boles, Ayla Burns, Nuthawin Charoensri, Oren Cohen, Susan Countryman, Mary Ann Cristobal, Bobbi Croy, Suzanne Dale, Hrushikesh Deshmukh, Amanda Douglas, Vincent Drouillon, Marcia Eisenberg, Howard Engler, Rama Ghatti, Prashant Gupta, Susan Hicks, Jake Humphrey, Lax Iyer, Manoj Jain, Mohan Kolli, Brian Krueger, Tim Kuphal, Stanley Letovsky, Michael Levandoski, Craig Lukasik, Jonathan Meltzer, Brian Norvell, Mindy Nye, Scott Parker, Christos Petropoulos, John Pruitt, Steven Ragan, Scott Ryan, Mike Sapeta, Jana Schroth, Suresh Babu Selvaraju, Goran Stevovic, Amanda Suchanek, Andrea Throop, Lyndon Tilson, Thomas Urban, Joe Voshell, Kimberly Wagner, Jonathan Williams, Mary Williamson, Qian Zeng, Tricia Zwiefelhofer, Clinton R. Paden, Duncan MacCannell |
| EPI_ISL_1612386 | North Dakota Department of Health, Public Health Laboratory | North Dakota Department of Health, Public Health Laboratory | Lisa Wingerter |

EPI_ISL_1612387, EPI_ISL_1612388, EPI_ISL_1612389, EPI_ISL_1612390, EPI_ISL_1612391, EPI_ISL_1612393, EPI_ISL_1612394, EPI_ISL_1612396, EPI_ISL_1612398, EPI_ISL_1612399, EPI_ISL_1612401, EPI_ISL_1612403, EPI_ISL_1612404, EPI_ISL_1612405, EPI_ISL_1612406, EPI_ISL_1612407, EPI_ISL_1612408, EPI_ISL_1612409, EPI_ISL_1612410, EPI_ISL_1612411, EPI_ISL_1612412, EPI_ISL_1612414, EPI_ISL_1612415, EPI_ISL_1612416, EPI_ISL_1612417, EPI_ISL_1612418, EPI_ISL_1612419, EPI_ISL_1612420, EPI_ISL_1612421, EPI_ISL_1612422, EPI_ISL_1612423, EPI_ISL_1612426, EPI_ISL_1612427, EPI_ISL_1612429, EPI_ISL_1612430, EPI_ISL_1612432, EPI_ISL_1612434, EPI_ISL_1612435, EPI_ISL_1612436, EPI_ISL_1612437, EPI_ISL_1612438, EPI_ISL_1612442, EPI_ISL_1612444, EPI_ISL_1612445, EPI_ISL_1612446, EPI_ISL_1612447, EPI_ISL_1612448, EPI_ISL_1612449, EPI_ISL_1612450, EPI_ISL_1612452, EPI_ISL_1612453, EPI_ISL_1612454, EPI_ISL_1612455, EPI_ISL_1612457, EPI_ISL_1612458, EPI_ISL_1612459, EPI_ISL_1612460, EPI_ISL_1612461, EPI_ISL_1612463, EPI_ISL_1612465, EPI_ISL_1612466, EPI_ISL_1612467, EPI_ISL_1612468, EPI_ISL_1612469, EPI_ISL_1612470, EPI_ISL_1612471, EPI_ISL_1612472, EPI_ISL_1612473, EPI_ISL_1612474, EPI_ISL_1612475, EPI_ISL_1612476, EPI_ISL_1612478, EPI_ISL_1612479, EPI_ISL_1612480, EPI_ISL_1612481, EPI_ISL_1612483, EPI_ISL_1612484, EPI_ISL_1612488, EPI_ISL_1612490, EPI_ISL_1612492, EPI_ISL_1612493, EPI_ISL_1612494, EPI_ISL_1612495, EPI_ISL_1612496, EPI_ISL_1612497, EPI_ISL_1612498, EPI_ISL_1612499, EPI_ISL_1612500, EPI_ISL_1612501, EPI_ISL_1612502, EPI_ISL_1612503, EPI_ISL_1612504, EPI_ISL_1612505, EPI_ISL_1612506, EPI_ISL_1612507, EPI_ISL_1612508, EPI_ISL_1612509, EPI_ISL_1612510, EPI_ISL_1612511, EPI_ISL_1612512, EPI_ISL_1612513, EPI_ISL_1612515, EPI_ISL_1612516, EPI_ISL_1612517, EPI_ISL_1612518, EPI_ISL_1612519, EPI_ISL_1612520, EPI_ISL_1612521, EPI_ISL_1612522, EPI_ISL_1612524, EPI_ISL_1612525, EPI_ISL_1612526, EPI_ISL_1612527, EPI_ISL_1612529, EPI_ISL_1612530, EPI_ISL_1612531, EPI_ISL_1612532, EPI_ISL_1612535, EPI_ISL_1612538, EPI_ISL_1612539, EPI_ISL_1612540, EPI_ISL_1612541, EPI_ISL_1612542, EPI_ISL_1612543, EPI_ISL_1612545, EPI_ISL_1612546, EPI_ISL_1612548, EPI_ISL_1612549, EPI_ISL_1612550, EPI_ISL_1612551, EPI_ISL_1612552, EPI_ISL_1612553, EPI_ISL_1612554, EPI_ISL_1612555, EPI_ISL_1612556, EPI_ISL_1612557, EPI_ISL_1612558, EPI_ISL_1612559, EPI_ISL_1612560, EPI_ISL_1612561, EPI_ISL_1612562, EPI_ISL_1612563, EPI_ISL_1612564, EPI_ISL_1612565, EPI_ISL_1612566, EPI_ISL_1612568, EPI_ISL_1612569, EPI_ISL_1612570, EPI_ISL_1612572, EPI_ISL_1612573, EPI_ISL_1612574, EPI_ISL_1612575, EPI_ISL_1612576, EPI_ISL_1612577, EPI_ISL_1612578, EPI_ISL_1612579, EPI_ISL_1612580, EPI_ISL_1612581, EPI_ISL_1612582, EPI_ISL_1612583, EPI_ISL_1612585, EPI_ISL_1612586, EPI_ISL_1612587, EPI_ISL_1612588, EPI_ISL_1612589, EPI_ISL_1612591, EPI_ISL_1612592, EPI_ISL_1612595, EPI_ISL_1612596, EPI_ISL_1612597, EPI_ISL_1612598, EPI_ISL_1612599, EPI_ISL_1612600, EPI_ISL_1612601, EPI_ISL_1612602, EPI_ISL_1612604, EPI_ISL_1612605, EPI_ISL_1612606, EPI_ISL_1612607, EPI_ISL_1612609, EPI_ISL_1612611, EPI_ISL_1612612, EPI_ISL_1612613, EPI_ISL_1612614, EPI_ISL_1612617, EPI_ISL_1612618, EPI_ISL_1612619, EPI_ISL_1612620, EPI_ISL_1612621, EPI_ISL_1612622, EPI_ISL_1612623, EPI_ISL_1612624, EPI_ISL_1612625, EPI_ISL_1612626, EPI_ISL_1612627, EPI_ISL_1612628, EPI_ISL_1612629, EPI_ISL_1612631, EPI_ISL_1612632, EPI_ISL_1612633, EPI_ISL_1612634, EPI_ISL_1612635, EPI_ISL_1612637, EPI_ISL_1612638, EPI_ISL_1612639, EPI_ISL_1612640, EPI_ISL_1612641, EPI_ISL_1612642, EPI_ISL_1612643, EPI_ISL_1612644, EPI_ISL_1612645, EPI_ISL_1612647, EPI_ISL_1612648, EPI_ISL_1612649, EPI_ISL_1612651, EPI_ISL_1612654, EPI_ISL_1612655, EPI_ISL_1612656, EPI_ISL_1612657, EPI_ISL_1612658, EPI_ISL_1612659, EPI_ISL_1612660, EPI_ISL_1612661, EPI_ISL_1612662, EPI_ISL_1612663, EPI_ISL_1612664, EPI_ISL_1612666, EPI_ISL_1612667, EPI_ISL_1612669, EPI_ISL_1612672, EPI_ISL_1612673, EPI_ISL_1612674, EPI_ISL_1612675, EPI_ISL_1612677, EPI_ISL_1612678, EPI_ISL_1612679, EPI_ISL_1612681, EPI_ISL_1612682, EPI_ISL_1612683, EPI_ISL_1612684, EPI_ISL_1612685, EPI_ISL_1612686, EPI_ISL_1612687, EPI_ISL_1612688, EPI_ISL_1612689, EPI_ISL_1612690, EPI_ISL_1612691, EPI_ISL_1612693, EPI_ISL_1612694, EPI_ISL_1612695, EPI_ISL_1612696, EPI_ISL_1612697, EPI_ISL_1612698, EPI_ISL_1612699, EPI_ISL_1612700, EPI_ISL_1612701, EPI_ISL_1612702, EPI_ISL_1612703, EPI_ISL_1612704, EPI_ISL_1612705, EPI_ISL_1612706, EPI_ISL_1612707, EPI_ISL_1612708, EPI_ISL_1612709, EPI_ISL_1612710, EPI_ISL_1612711, EPI_ISL_1612712, EPI_ISL_1612713, EPI_ISL_1612714, EPI_ISL_1612715, EPI_ISL_1612716, EPI_ISL_1612717, EPI_ISL_1612718, EPI_ISL_1612719, EPI_ISL_1612720, EPI_ISL_1612723, EPI_ISL_1612724, EPI_ISL_1612726, EPI_ISL_1612728, EPI_ISL_1612729, EPI_ISL_1612730, EPI_ISL_1612731, EPI_ISL_1612732, EPI_ISL_1612733, EPI_ISL_1612734, EPI_ISL_1612735, EPI_ISL_1612736, EPI_ISL_1612738, EPI_ISL_1612739, EPI_ISL_1612741, EPI_ISL_1612742, EPI_ISL_1612743, EPI_ISL_1612744, EPI_ISL_1612745, EPI_ISL_1612746, EPI_ISL_1612747, EPI_ISL_1612748, EPI_ISL_1612749, EPI_ISL_1612750, EPI_ISL_1612752, EPI_ISL_1612754, EPI_ISL_1612755, EPI_ISL_1612756, EPI_ISL_1612757, EPI_ISL_1612758, EPI_ISL_1612760, EPI_ISL_1612762, EPI_ISL_1612763, EPI_ISL_1612764, EPI_ISL_1612765, EPI_ISL_1612766, EPI_ISL_1612767, EPI_ISL_1612768, EPI_ISL_1612769, EPI_ISL_1612770, EPI_ISL_1612771, EPI_ISL_1612772, EPI_ISL_1612774, EPI_ISL_1612775, EPI_ISL_1612776, EPI_ISL_1612778, EPI_ISL_1612779, EPI_ISL_1612780, EPI_ISL_1612781, EPI_ISL_1612782, EPI_ISL_1612784, EPI_ISL_1612785, EPI_ISL_1612787, EPI_ISL_1612789, EPI_ISL_1612790, EPI_ISL_1612791, EPI_ISL_1612792, EPI_ISL_1612793, EPI_ISL_1612794, EPI_ISL_1612796, EPI_ISL_1612797, EPI_ISL_1612798, EPI_ISL_1612799, EPI_ISL_1612800, EPI_ISL_1612801, EPI_ISL_1612805, EPI_ISL_1612806, EPI_ISL_1612807, EPI_ISL_1612808, EPI_ISL_1612809, EPI_ISL_1612810, EPI_ISL_1612811, EPI_ISL_1612812, EPI_ISL_1612816, EPI_ISL_1612817, EPI_ISL_1612819, EPI_ISL_1612820, EPI_ISL_1612821, EPI_ISL_1612824

| | | | |
|---|---|---|---|
| see above | Laboratory Corporation of America | Centers for Disease Control and Prevention Division of Viral Diseases, Pathogen Discovery | Dakota Howard, Dhwani Batra, Peter W. Cook, Kara Moser, Adrian Paskey, Jason Caravas, Benjamin Rambo-Martin, Shatavia Morrison, Christopher Gulvick, Scott Sammons, Yvette Unoarumhi, Darlene Wagner, Matthew Schmerer, Minoo Agarwal, Eyad Almasri, Debbie Boles, Ayla Burns, Nuthawin Charoensri, Oren Cohen, Susan Countryman, Mary Ann Cristobal, Bobbi Croy, Suzanne Dale, Hrushikesh Deshmukh, Amanda Douglas, Vincent Drouillon, Marcia Eisenberg, Howard Engler, Rama Ghatti, Prashant Gupta, Susan Hicks, Jake Humphrey, Lax Iyer, Manoj Jain, Mohan Kolli, Brian Krueger, Tim Kuphal, Stanley Letovsky, Michael Levandoski, Craig Lukasik, Jonathan Meltzer, Brian Norvell, Mindy Nye, Scott Parker, Christos Petropoulos, John Pruitt, Steven Ragan, Scott Ryan, Mike Sapeta, Jana Schroth, Suresh Babu Selvaraju, Goran Stevovic, Amanda Suchanek, Andrea Throop, Lyndon Tilson, Thomas Urban, Joe Voshell, Kimberly Wagner, Jonathan Williams, Mary Williamson, Qian Zeng, Tricia Zwiefelhofer, Clinton R. Paden, Duncan MacCannell |

EPI_ISL_1612825, EPI_ISL_1612826, EPI_ISL_1612827, EPI_ISL_1612829, EPI_ISL_1612830, EPI_ISL_1612831, EPI_ISL_1612832, EPI_ISL_1612833, EPI_ISL_1612834, EPI_ISL_1612835, EPI_ISL_1612837, EPI_ISL_1612838, EPI_ISL_1612839, EPI_ISL_1612840, EPI_ISL_1612841, EPI_ISL_1612843, EPI_ISL_1612844, EPI_ISL_1612845, EPI_ISL_1612846, EPI_ISL_1612847, EPI_ISL_1612848, EPI_ISL_1612849, EPI_ISL_1612850, EPI_ISL_1612851, EPI_ISL_1612852, EPI_ISL_1612853, EPI_ISL_1612854, EPI_ISL_1612855, EPI_ISL_1612856, EPI_ISL_1612857, EPI_ISL_1612858, EPI_ISL_1612859, EPI_ISL_1612860, EPI_ISL_1612861, EPI_ISL_1612862, EPI_ISL_1612863, EPI_ISL_1612864, EPI_ISL_1612866, EPI_ISL_1612867, EPI_ISL_1612868, EPI_ISL_1612870, EPI_ISL_1612872, EPI_ISL_1612873, EPI_ISL_1612874, EPI_ISL_1612875, EPI_ISL_1612876, EPI_ISL_1612877, EPI_ISL_1612878, EPI_ISL_1612879, EPI_ISL_1612880, EPI_ISL_1612881, EPI_ISL_1612883, EPI_ISL_1612884, EPI_ISL_1612886, EPI_ISL_1612887, EPI_ISL_1612888, EPI_ISL_1612889, EPI_ISL_1612891, EPI_ISL_1612892, EPI_ISL_1612893, EPI_ISL_1612894, EPI_ISL_1612896, EPI_ISL_1612898, EPI_ISL_1612899, EPI_ISL_1612900, EPI_ISL_1612901, EPI_ISL_1612902, EPI_ISL_1612903, EPI_ISL_1612904, EPI_ISL_1612905, EPI_ISL_1612906, EPI_ISL_1612907, EPI_ISL_1612909, EPI_ISL_1612910, EPI_ISL_1612911, EPI_ISL_1612912, EPI_ISL_1612913, EPI_ISL_1612914, EPI_ISL_1612915, EPI_ISL_1612916, EPI_ISL_1612917, EPI_ISL_1612918, EPI_ISL_1612919, EPI_ISL_1612920, EPI_ISL_1612921, EPI_ISL_1612922, EPI_ISL_1612923, EPI_ISL_1612924, EPI_ISL_1612925, EPI_ISL_1612926, EPI_ISL_1612928, EPI_ISL_1612929, EPI_ISL_1612930, EPI_ISL_1612931, EPI_ISL_1612932, EPI_ISL_1612933, EPI_ISL_1612934, EPI_ISL_1612935, EPI_ISL_1612936, EPI_ISL_1612937, EPI_ISL_1612939, EPI_ISL_1612940, EPI_ISL_1612941, EPI_ISL_1612942, EPI_ISL_1612943, EPI_ISL_1612944, EPI_ISL_1612945, EPI_ISL_1612946, EPI_ISL_1612947, EPI_ISL_1612948, EPI_ISL_1612949, EPI_ISL_1612950, EPI_ISL_1612951, EPI_ISL_1612952, EPI_ISL_1612953, EPI_ISL_1612954, EPI_ISL_1612955, EPI_ISL_1612956, EPI_ISL_1612957, EPI_ISL_1612958, EPI_ISL_1612959, EPI_ISL_1612960, EPI_ISL_1612961, EPI_ISL_1612962, EPI_ISL_1612963, EPI_ISL_1612964, EPI_ISL_1612965, EPI_ISL_1612966, EPI_ISL_1612967, EPI_ISL_1612968, EPI_ISL_1612969, EPI_ISL_1612970, EPI_ISL_1612971, EPI_ISL_1612972, EPI_ISL_1612973, EPI_ISL_1612974, EPI_ISL_1612975, EPI_ISL_1612976, EPI_ISL_1612977, EPI_ISL_1612978, EPI_ISL_1612980, EPI_ISL_1612981, EPI_ISL_1612983, EPI_ISL_1612984, EPI_ISL_1612985, EPI_ISL_1612986, EPI_ISL_1612987, EPI_ISL_1612988, EPI_ISL_1612989, EPI_ISL_1612990, EPI_ISL_1612991, EPI_ISL_1612992, EPI_ISL_1612993, EPI_ISL_1612994, EPI_ISL_1612996, EPI_ISL_1612998, EPI_ISL_1612999, EPI_ISL_1613000, EPI_ISL_1613001, EPI_ISL_1613002, EPI_ISL_1613003, EPI_ISL_1613004, EPI_ISL_1613005, EPI_ISL_1613006, EPI_ISL_1613007, EPI_ISL_1613008, EPI_ISL_1613009, EPI_ISL_1613010, EPI_ISL_1613011, EPI_ISL_1613012, EPI_ISL_1613013, EPI_ISL_1613014, EPI_ISL_1613015, EPI_ISL_1613016, EPI_ISL_1613017, EPI_ISL_1613018, EPI_ISL_1613019, EPI_ISL_1613020, EPI_ISL_1613021, EPI_ISL_1613022, EPI_ISL_1613023, EPI_ISL_1613024, EPI_ISL_1613025, EPI_ISL_1613026, EPI_ISL_1613027, EPI_ISL_1613029, EPI_ISL_1613030, EPI_ISL_1613031, EPI_ISL_1613032, EPI_ISL_1613033, EPI_ISL_1613034, EPI_ISL_1613035, EPI_ISL_1613036, EPI_ISL_1613037, EPI_ISL_1613038, EPI_ISL_1613040, EPI_ISL_1613041, EPI_ISL_1613042, EPI_ISL_1613043, EPI_ISL_1613044, EPI_ISL_1613045, EPI_ISL_1613046, EPI_ISL_1613047, EPI_ISL_1613048, EPI_ISL_1613049, EPI_ISL_1613050, EPI_ISL_1613051, EPI_ISL_1613052, EPI_ISL_1613053, EPI_ISL_1613054, EPI_ISL_1613055, EPI_ISL_1613056, EPI_ISL_1613058, EPI_ISL_1613059, EPI_ISL_1613060, EPI_ISL_1613061, EPI_ISL_1613062, EPI_ISL_1613063, EPI_ISL_1613064, EPI_ISL_1613065, EPI_ISL_1613066, EPI_ISL_1613067, EPI_ISL_1613068, EPI_ISL_1613069, EPI_ISL_1613071, EPI_ISL_1613072, EPI_ISL_1613073, EPI_ISL_1613074, EPI_ISL_1613075, EPI_ISL_1613076, EPI_ISL_1613077, EPI_ISL_1613078, EPI_ISL_1613079, EPI_ISL_1613080, EPI_ISL_1613081, EPI_ISL_1613082, EPI_ISL_1613083, EPI_ISL_1613084, EPI_ISL_1613085, EPI_ISL_1613086, EPI_ISL_1613087, EPI_ISL_1613089, EPI_ISL_1613090, EPI_ISL_1613091, EPI_ISL_1613092, EPI_ISL_1613095, EPI_ISL_1613096, EPI_ISL_1613097, EPI_ISL_1613099, EPI_ISL_1613100, EPI_ISL_1613102, EPI_ISL_1613103, EPI_ISL_1613105, EPI_ISL_1613106, EPI_ISL_1613107, EPI_ISL_1613108, EPI_ISL_1613109, EPI_ISL_1613110, EPI_ISL_1613111, EPI_ISL_1613112, EPI_ISL_1613113, EPI_ISL_1613114, EPI_ISL_1613117, EPI_ISL_1613118, EPI_ISL_1613119, EPI_ISL_1613120, EPI_ISL_1613121, EPI_ISL_1613122, EPI_ISL_1613123, EPI_ISL_1613126, EPI_ISL_1613129, EPI_ISL_1613132, EPI_ISL_1613135, EPI_ISL_1613138, EPI_ISL_1613144, EPI_ISL_1613146, EPI_ISL_1613149, EPI_ISL_1613152, EPI_ISL_1613155, EPI_ISL_1613158, EPI_ISL_1613161, EPI_ISL_1613164, EPI_ISL_1613167, EPI_ISL_1613170, EPI_ISL_1613173, EPI_ISL_1613176, EPI_ISL_1613185, EPI_ISL_1613188, EPI_ISL_1613191, EPI_ISL_1613194, EPI_ISL_1613197, EPI_ISL_1613200, EPI_ISL_1613203, EPI_ISL_1613206, EPI_ISL_1613209, EPI_ISL_1613212, EPI_ISL_1613215, EPI_ISL_1613218, EPI_ISL_1613221, EPI_ISL_1613224, EPI_ISL_1613226, EPI_ISL_1613229, EPI_ISL_1613232, EPI_ISL_1613235, EPI_ISL_1613238, EPI_ISL_1613241, EPI_ISL_1613245, EPI_ISL_1613248, EPI_ISL_1613251, EPI_ISL_1613254, EPI_ISL_1613257, EPI_ISL_1613260, EPI_ISL_1613263, EPI_ISL_1613266, EPI_ISL_1613269, EPI_ISL_1613275, EPI_ISL_1613277, EPI_ISL_1613280, EPI_ISL_1613287, EPI_ISL_1613290, EPI_ISL_1613292, EPI_ISL_1613295, EPI_ISL_1613298, EPI_ISL_1613304, EPI_ISL_1613307, EPI_ISL_1613310, EPI_ISL_1613313, EPI_ISL_1613316, EPI_ISL_1613319, EPI_ISL_1613321, EPI_ISL_1613324, EPI_ISL_1613327, EPI_ISL_1613331, EPI_ISL_1613334, EPI_ISL_1613336, EPI_ISL_1613339, EPI_ISL_1613342, EPI_ISL_1613345, EPI_ISL_1613348, EPI_ISL_1613351, EPI_ISL_1613354, EPI_ISL_1613357, EPI_ISL_1613360, EPI_ISL_1613363, EPI_ISL_1613366, EPI_ISL_1613369, EPI_ISL_1613372, EPI_ISL_1613375, EPI_ISL_1613378, EPI_ISL_1613384, EPI_ISL_1613386, EPI_ISL_1613389, EPI_ISL_1613392, EPI_ISL_1613395, EPI_ISL_1613398, EPI_ISL_1613401, EPI_ISL_1613404, EPI_ISL_1613407, EPI_ISL_1613410, EPI_ISL_1613413, EPI_ISL_1613416, EPI_ISL_1613419, EPI_ISL_1613422, EPI_ISL_1613425, EPI_ISL_1613428, EPI_ISL_1613431, EPI_ISL_1613433, EPI_ISL_1613436, EPI_ISL_1613439, EPI_ISL_1613442, EPI_ISL_1613445, EPI_ISL_1613448, EPI_ISL_1613451, EPI_ISL_1613454, EPI_ISL_1613458, EPI_ISL_1613461, EPI_ISL_1613464, EPI_ISL_1613467, EPI_ISL_1613469, EPI_ISL_1613472, EPI_ISL_1613475, EPI_ISL_1613478, EPI_ISL_1613481, EPI_ISL_1613485, EPI_ISL_1613487, EPI_ISL_1613490, EPI_ISL_1613493, EPI_ISL_1613496, EPI_ISL_1613499, EPI_ISL_1613502, EPI_ISL_1613505, EPI_ISL_1613508, EPI_ISL_1613511, EPI_ISL_1613514, EPI_ISL_1613517, EPI_ISL_1613520, EPI_ISL_1613522, EPI_ISL_1613525, EPI_ISL_1613529, EPI_ISL_1613532, EPI_ISL_1613534, EPI_ISL_1613538, EPI_ISL_1613540, EPI_ISL_1613543, EPI_ISL_1613546, EPI_ISL_1613549, EPI_ISL_1613552, EPI_ISL_1613555, EPI_ISL_1613558, EPI_ISL_1613562, EPI_ISL_1613565, EPI_ISL_1613567, EPI_ISL_1613570, EPI_ISL_1613573, EPI_ISL_1613576, EPI_ISL_1613579, EPI_ISL_1613582, EPI_ISL_1613585, EPI_ISL_1613591, EPI_ISL_1613593, EPI_ISL_1613596, EPI_ISL_1613599, EPI_ISL_1613602, EPI_ISL_1613605, EPI_ISL_1613608, EPI_ISL_1613611, EPI_ISL_1613614, EPI_ISL_1613620, EPI_ISL_1613622, EPI_ISL_1613626, EPI_ISL_1613629, EPI_ISL_1613632, EPI_ISL_1613635, EPI_ISL_1613637, EPI_ISL_1613640, EPI_ISL_1613643, EPI_ISL_1613646, EPI_ISL_1613649, EPI_ISL_1613652, EPI_ISL_1613655, EPI_ISL_1613658, EPI_ISL_1613664, EPI_ISL_1613667, EPI_ISL_1613670, EPI_ISL_1613673, EPI_ISL_1613676, EPI_ISL_1613679, EPI_ISL_1613681, EPI_ISL_1613690, EPI_ISL_1613693, EPI_ISL_1613699, EPI_ISL_1613702, EPI_ISL_1613705, EPI_ISL_1613711, EPI_ISL_1613714, EPI_ISL_1613717, EPI_ISL_1613725, EPI_ISL_1613728, EPI_ISL_1613731, EPI_ISL_1613734, EPI_ISL_1613737, EPI_ISL_1613740, EPI_ISL_1613743, EPI_ISL_1613746, EPI_ISL_1613749, EPI_ISL_1613751, EPI_ISL_1613755, EPI_ISL_1613757, EPI_ISL_1613760, EPI_ISL_1613764, EPI_ISL_1613766, EPI_ISL_1613770, EPI_ISL_1613775, EPI_ISL_1613778, EPI_ISL_1613781, EPI_ISL_1613787, EPI_ISL_1613790, EPI_ISL_1613793, EPI_ISL_1613796, EPI_ISL_1613799, EPI_ISL_1613802, EPI_ISL_1613805, EPI_ISL_1613808, EPI_ISL_1613810, EPI_ISL_1613813, EPI_ISL_1613816, EPI_ISL_1613819, EPI_ISL_1613822, EPI_ISL_1613825, EPI_ISL_1613828, EPI_ISL_1613831, EPI_ISL_1613834, EPI_ISL_1613836, EPI_ISL_1613839, EPI_ISL_1613843, EPI_ISL_1613849, EPI_ISL_1613852, EPI_ISL_1613854, EPI_ISL_1613857, EPI_ISL_1613859, EPI_ISL_1613862, EPI_ISL_1613865, EPI_ISL_1613871, EPI_ISL_1613874, EPI_ISL_1613877, EPI_ISL_1613880, EPI_ISL_1613883, EPI_ISL_1613886, EPI_ISL_1613889, EPI_ISL_1613891, EPI_ISL_1613894, EPI_ISL_1613897, EPI_ISL_1613900, EPI_ISL_1613902, EPI_ISL_1613905, EPI_ISL_1613908, EPI_ISL_1613914, EPI_ISL_1613916, EPI_ISL_1613919, EPI_ISL_1613922, EPI_ISL_1613925, EPI_ISL_1613928, EPI_ISL_1613931, EPI_ISL_1613934, EPI_ISL_1613937, EPI_ISL_1613938, EPI_ISL_1613939, EPI_ISL_1613940, EPI_ISL_1613941, EPI_ISL_1613942, EPI_ISL_1613943, EPI_ISL_1613944, EPI_ISL_1613945, EPI_ISL_1613946, EPI_ISL_1613947, EPI_ISL_1613948, EPI_ISL_1613949, EPI_ISL_1613952, EPI_ISL_1613953, EPI_ISL_1613954, EPI_ISL_1613955, EPI_ISL_1613956, EPI_ISL_1613957, EPI_ISL_1613958, EPI_ISL_1613959, EPI_ISL_1613960, EPI_ISL_1613961, EPI_ISL_1613962, EPI_ISL_1613963, EPI_ISL_1613965, EPI_ISL_1613966, EPI_ISL_1613967, EPI_ISL_1613968, EPI_ISL_1613969, EPI_ISL_1613970, EPI_ISL_1613972, EPI_ISL_1613973, EPI_ISL_1613974, EPI_ISL_1613975, EPI_ISL_1613976, EPI_ISL_1613977, EPI_ISL_1613978, EPI_ISL_1613979, EPI_ISL_1613980, EPI_ISL_1613981, EPI_ISL_1613982, EPI_ISL_1613983, EPI_ISL_1613984, EPI_ISL_1613985, EPI_ISL_1613986, EPI_ISL_1613987, EPI_ISL_1613988, EPI_ISL_1613989, EPI_ISL_1613990, EPI_ISL_1613992, EPI_ISL_1613993, EPI_ISL_1613994, EPI_ISL_1613995, EPI_ISL_1613996, EPI_ISL_1613997, EPI_ISL_1613998, EPI_ISL_1613999, EPI_ISL_1614000, EPI_ISL_1614001, EPI_ISL_1614002, EPI_ISL_1614003, EPI_ISL_1614004, EPI_ISL_1614005, EPI_ISL_1614006, EPI_ISL_1614007, EPI_ISL_1614008, EPI_ISL_1614009, EPI_ISL_1614010, EPI_ISL_1614012, EPI_ISL_1614013, EPI_ISL_1614014, EPI_ISL_1614015, EPI_ISL_1614017, EPI_ISL_1614019, EPI_ISL_1614020, EPI_ISL_1614021, EPI_ISL_1614022, EPI_ISL_1614023, EPI_ISL_1614024, EPI_ISL_1614025, EPI_ISL_1614026, EPI_ISL_1614027, EPI_ISL_1614028, EPI_ISL_1614029, EPI_ISL_1614030, EPI_ISL_1614031, EPI_ISL_1614032, EPI_ISL_1614033, EPI_ISL_1614034, EPI_ISL_1614037, EPI_ISL_1614038, EPI_ISL_1614039, EPI_ISL_1614040, EPI_ISL_1614041, EPI_ISL_1614042, EPI_ISL_1614043, EPI_ISL_1614044, EPI_ISL_1614045, EPI_ISL_1614046, EPI_ISL_1614047, EPI_ISL_1614048, EPI_ISL_1614049, EPI_ISL_1614050, EPI_ISL_1614051, EPI_ISL_1614052, EPI_ISL_1614053, EPI_ISL_1614054, EPI_ISL_1614055, EPI_ISL_1614056, EPI_ISL_1614057, EPI_ISL_1614058, EPI_ISL_1614059, EPI_ISL_1614060, EPI_ISL_1614061, EPI_ISL_1614062, EPI_ISL_1614064, EPI_ISL_1614065, EPI_ISL_1614066, EPI_ISL_1614067, EPI_ISL_1614069, EPI_ISL_1614070, EPI_ISL_1614071, EPI_ISL_1614072, EPI_ISL_1614073, EPI_ISL_1614074, EPI_ISL_1614075, EPI_ISL_1614076, EPI_ISL_1614077, EPI_ISL_1614078, EPI_ISL_1614079, EPI_ISL_1614080, EPI_ISL_1614081, EPI_ISL_1614082, EPI_ISL_1614083, EPI_ISL_1614084, EPI_ISL_1614085, EPI_ISL_1614086, EPI_ISL_1614087, EPI_ISL_1614088, EPI_ISL_1614090, EPI_ISL_1614091, EPI_ISL_1614092, EPI_ISL_1614093, EPI_ISL_1614094, EPI_ISL_1614095, EPI_ISL_1614096, EPI_ISL_1614097, EPI_ISL_1614098, EPI_ISL_1614099, EPI_ISL_1614100, EPI_ISL_1614101, EPI_ISL_1614102, EPI_ISL_1614103, EPI_ISL_1614104, EPI_ISL_1614105, EPI_ISL_1614106, EPI_ISL_1614108, EPI_ISL_1614109, EPI_ISL_1614110, EPI_ISL_1614111, EPI_ISL_1614112, EPI_ISL_1614113, EPI_ISL_1614114, EPI_ISL_1614115, EPI_ISL_1614116, EPI_ISL_1614117, EPI_ISL_1614118, EPI_ISL_1614119, EPI_ISL_1614120, EPI_ISL_1614121, EPI_ISL_1614122, EPI_ISL_1614123, EPI_ISL_1614124, EPI_ISL_1614125, EPI_ISL_1614126, EPI_ISL_1614127, EPI_ISL_1614128, EPI_ISL_1614129, EPI_ISL_1614130, EPI_ISL_1614131, EPI_ISL_1614133, EPI_ISL_1614134, EPI_ISL_1614135, EPI_ISL_1614136, EPI_ISL_1614137, EPI_ISL_1614138, EPI_ISL_1614140, EPI_ISL_1614141, EPI_ISL_1614142, EPI_ISL_1614143, EPI_ISL_1614144, EPI_ISL_1614145, EPI_ISL_1614146, EPI_ISL_1614147, EPI_ISL_1614148, EPI_ISL_1614149, EPI_ISL_1614150, EPI_ISL_1614151, EPI_ISL_1614152, EPI_ISL_1614154, EPI_ISL_1614155, EPI_ISL_1614156, EPI_ISL_1614157, EPI_ISL_1614159, EPI_ISL_1614161, EPI_ISL_1614162, EPI_ISL_1614163, EPI_ISL_1614164, EPI_ISL_1614165, EPI_ISL_1614167, EPI_ISL_1614168, EPI_ISL_1614169, EPI_ISL_1614170, EPI_ISL_1614171, EPI_ISL_1614173, EPI_ISL_1614174, EPI_ISL_1614175, EPI_ISL_1614176, EPI_ISL_1614177, EPI_ISL_1614178, EPI_ISL_1614179, EPI_ISL_1614180, EPI_ISL_1614181, EPI_ISL_1614182, EPI_ISL_1614184, EPI_ISL_1614185, EPI_ISL_1614186, EPI_ISL_1614187, EPI_ISL_1614189, EPI_ISL_1614190, EPI_ISL_1614191, EPI_ISL_1614192, EPI_ISL_1614193, EPI_ISL_1614194, EPI_ISL_1614195, EPI_ISL_1614196, EPI_ISL_1614197, EPI_ISL_1614198, EPI_ISL_1614199, EPI_ISL_1614200, EPI_ISL_1614201, EPI_ISL_1614202, EPI_ISL_1614203, EPI_ISL_1614204, EPI_ISL_1614205, EPI_ISL_1614206, EPI_ISL_1614207, EPI_ISL_1614208, EPI_ISL_1614209, EPI_ISL_1614211, EPI_ISL_1614212, EPI_ISL_1614213, EPI_ISL_1614214, EPI_ISL_1614215, EPI_ISL_1614216, EPI_ISL_1614217, EPI_ISL_1614218, EPI_ISL_1614219, EPI_ISL_1614221, EPI_ISL_1614222, EPI_ISL_1614223, EPI_ISL_1614224, EPI_ISL_1614225, EPI_ISL_1614226, EPI_ISL_1614227, EPI_ISL_1614229, EPI_ISL_1614230, EPI_ISL_1614231, EPI_ISL_1614232, EPI_ISL_1614233, EPI_ISL_1614234, EPI_ISL_1614235, EPI_ISL_1614236, EPI_ISL_1614237, EPI_ISL_1614238, EPI_ISL_1614240, EPI_ISL_1614241, EPI_ISL_1614242, EPI_ISL_1614243, EPI_ISL_1614244, EPI_ISL_1614245, EPI_ISL_1614246, EPI_ISL_1614247, EPI_ISL_1614248, EPI_ISL_1614249, EPI_ISL_1614250, EPI_ISL_1614251, EPI_ISL_1614252, EPI_ISL_1614253, EPI_ISL_1614254, EPI_ISL_1614255, EPI_ISL_1614256, EPI_ISL_1614258, EPI_ISL_1614260, EPI_ISL_1614262, EPI_ISL_1614263, EPI_ISL_1614264, EPI_ISL_1614265, EPI_ISL_1614266, EPI_ISL_1614267, EPI_ISL_1614268, EPI_ISL_1614271, EPI_ISL_1614272, EPI_ISL_1614273, EPI_ISL_1614274, EPI_ISL_1614275, EPI_ISL_1614279, EPI_ISL_1614280, EPI_ISL_1614281, EPI_ISL_1614283, EPI_ISL_1614284, EPI_ISL_1614285, EPI_ISL_1614286, EPI_ISL_1614287, EPI_ISL_1614288, EPI_ISL_1614289, EPI_ISL_1614290, EPI_ISL_1614291, EPI_ISL_1614292, EPI_ISL_1614293, EPI_ISL_1614294, EPI_ISL_1614295, EPI_ISL_1614296, EPI_ISL_1614297, EPI_ISL_1614298, EPI_ISL_1614299, EPI_ISL_1614300, EPI_ISL_1614301, EPI_ISL_1614302, EPI_ISL_1614303, EPI_ISL_1614304, EPI_ISL_1614305, EPI_ISL_1614306, EPI_ISL_1614307, EPI_ISL_1614308, EPI_ISL_1614309, EPI_ISL_1614311, EPI_ISL_1614312, EPI_ISL_1614313, EPI_ISL_1614314, EPI_ISL_1614315, EPI_ISL_1614316, EPI_ISL_1614317, EPI_ISL_1614319, EPI_ISL_1614320, EPI_ISL_1614321, EPI_ISL_1614323, EPI_ISL_1614324, EPI_ISL_1614325, EPI_ISL_1614326, EPI_ISL_1614327, EPI_ISL_1614328, EPI_ISL_1614329, EPI_ISL_1614330, EPI_ISL_1614331, EPI_ISL_1614332, EPI_ISL_1614333, EPI_ISL_1614335, EPI_ISL_1614338, EPI_ISL_1614340, EPI_ISL_1614341, EPI_ISL_1614342, EPI_ISL_1614343, EPI_ISL_1614344, EPI_ISL_1614346, EPI_ISL_1614347, EPI_ISL_1614348, EPI_ISL_1614349, EPI_ISL_1614350, EPI_ISL_1614352, EPI_ISL_1614353, EPI_ISL_1614355, EPI_ISL_1614356, EPI_ISL_1614357, EPI_ISL_1614359, EPI_ISL_1614360, EPI_ISL_1614361, EPI_ISL_1614362, EPI_ISL_1614363, EPI_ISL_1614365, EPI_ISL_1614366, EPI_ISL_1614367, EPI_ISL_1614368, EPI_ISL_1614369, EPI_ISL_1614370, EPI_ISL_1614371, EPI_ISL_1614372, EPI_ISL_1614373, EPI_ISL_1614374, EPI_ISL_1614375, EPI_ISL_1614376, EPI_ISL_1614377, EPI_ISL_1614378, EPI_ISL_1614379, EPI_ISL_1614381, EPI_ISL_1614382, EPI_ISL_1614383, EPI_ISL_1614385, EPI_ISL_1614386, EPI_ISL_1614387, EPI_ISL_1614388, EPI_ISL_1614389, EPI_ISL_1614390, EPI_ISL_1614391, EPI_ISL_1614392, EPI_ISL_1614394, EPI_ISL_1614395, EPI_ISL_1614396, EPI_ISL_1614397, EPI_ISL_1614399, EPI_ISL_1614400, EPI_ISL_1614401, EPI_ISL_1614402, EPI_ISL_1614403, EPI_ISL_1614404, EPI_ISL_1614405, EPI_ISL_1614406, EPI_ISL_1614407, EPI_ISL_1614409, EPI_ISL_1614410, EPI_ISL_1614411, EPI_ISL_1614412, EPI_ISL_1614413, EPI_ISL_1614414, EPI_ISL_1614415, EPI_ISL_1614416, EPI_ISL_1614417, EPI_ISL_1614418, EPI_ISL_1614419, EPI_ISL_1614420, EPI_ISL_1614424, EPI_ISL_1614425, EPI_ISL_1614426, EPI_ISL_1614427, EPI_ISL_1614428, EPI_ISL_1614429, EPI_ISL_1614430, EPI_ISL_1614432, EPI_ISL_1614433, EPI_ISL_1614434, EPI_ISL_1614435, EPI_ISL_1614436, EPI_ISL_1614437, EPI_ISL_1614438, EPI_ISL_1614439, EPI_ISL_1614440, EPI_ISL_1614441, EPI_ISL_1614442, EPI_ISL_1614444, EPI_ISL_1614445, EPI_ISL_1614446, EPI_ISL_1614447, EPI_ISL_1614448, EPI_ISL_1614449, EPI_ISL_1614450, EPI_ISL_1614451, EPI_ISL_1614452, EPI_ISL_1614453, EPI_ISL_1614455, EPI_ISL_1614456, EPI_ISL_1614457, EPI_ISL_1614458, EPI_ISL_1614460, EPI_ISL_1614461, EPI_ISL_1614463, EPI_ISL_1614464, EPI_ISL_1614465, EPI_ISL_1614466, EPI_ISL_1614467, EPI_ISL_1614468, EPI_ISL_1614469, EPI_ISL_1614470, EPI_ISL_1614471, EPI_ISL_1614472, EPI_ISL_1614474, EPI_ISL_1614475, EPI_ISL_1614476, EPI_ISL_1614477, EPI_ISL_1614478, EPI_ISL_1614479, EPI_ISL_1614480, EPI_ISL_1614481, EPI_ISL_1614482, EPI_ISL_1614484, EPI_ISL_1614485, EPI_ISL_1614486, EPI_ISL_1614487, EPI_ISL_1614488, EPI_ISL_1614489, EPI_ISL_1614490, EPI_ISL_1614491, EPI_ISL_1614492, EPI_ISL_1614493, EPI_ISL_1614494, EPI_ISL_1614495, EPI_ISL_1614496, EPI_ISL_1614497, EPI_ISL_1614498, EPI_ISL_1614499, EPI_ISL_1614500, EPI_ISL_1614501, EPI_ISL_1614502, EPI_ISL_1614503, EPI_ISL_1614504, EPI_ISL_1614506, EPI_ISL_1614507, EPI_ISL_1614508, EPI_ISL_1614509, EPI_ISL_1614510, EPI_ISL_1614511, EPI_ISL_1614512, EPI_ISL_1614513, EPI_ISL_1614514, EPI_ISL_1614515, EPI_ISL_1614516, EPI_ISL_1614517, EPI_ISL_1614518, EPI_ISL_1614519, EPI_ISL_1614520, EPI_ISL_1614521, EPI_ISL_1614522, EPI_ISL_1614523, EPI_ISL_1614524, EPI_ISL_1614525, EPI_ISL_1614526, EPI_ISL_1614527, EPI_ISL_1614528, EPI_ISL_1614529, EPI_ISL_1614530, EPI_ISL_1614531, EPI_ISL_1614532, EPI_ISL_1614533, EPI_ISL_1614534, EPI_ISL_1614535, EPI_ISL_1614536, EPI_ISL_1614537, EPI_ISL_1614538, EPI_ISL_1614539, EPI_ISL_1614540, EPI_ISL_1614541, EPI_ISL_1614542, EPI_ISL_1614543, EPI_ISL_1614544, EPI_ISL_1614545, EPI_ISL_1614546, EPI_ISL_1614547, EPI_ISL_1614548, EPI_ISL_1614549, EPI_ISL_1614550, EPI_ISL_1614551, EPI_ISL_1614553, EPI_ISL_1614554, EPI_ISL_1614555, EPI_ISL_1614556, EPI_ISL_1614557, EPI_ISL_1614558, EPI_ISL_1614559, EPI_ISL_1614560, EPI_ISL_1614561, EPI_ISL_1614562, EPI_ISL_1614563, EPI_ISL_1614564, EPI_ISL_1614565, EPI_ISL_1614566, EPI_ISL_1614567, EPI_ISL_1614568, EPI_ISL_1614569, EPI_ISL_1614570, EPI_ISL_1614571, EPI_ISL_1614572, EPI_ISL_1614574, EPI_ISL_1614575, EPI_ISL_1614576, EPI_ISL_1614577, EPI_ISL_1614578, EPI_ISL_1614579, EPI_ISL_1614581, EPI_ISL_1614582, EPI_ISL_1614583, EPI_ISL_1614584, EPI_ISL_1614585, EPI_ISL_1614586, EPI_ISL_1614587, EPI_ISL_1614588, EPI_ISL_1614589, EPI_ISL_1614590, EPI_ISL_1614591, EPI_ISL_1614592, EPI_ISL_1614593, EPI_ISL_1614594, EPI_ISL_1614595, EPI_ISL_1614596, EPI_ISL_1614597, EPI_ISL_1614598, EPI_ISL_1614599, EPI_ISL_1614600, EPI_ISL_1614601, EPI_ISL_1614602, EPI_ISL_1614603, EPI_ISL_1614604, EPI_ISL_1614605, EPI_ISL_1614606, EPI_ISL_1614607, EPI_ISL_1614608, EPI_ISL_1614609, EPI_ISL_1614610, EPI_ISL_1614611, EPI_ISL_1614612, EPI_ISL_1614613, EPI_ISL_1614614, EPI_ISL_1614615, EPI_ISL_1614616, EPI_ISL_1614617, EPI_ISL_1614618, EPI_ISL_1614619, EPI_ISL_1614620, EPI_ISL_1614621, EPI_ISL_1614622, EPI_ISL_1614623, EPI_ISL_1614624, EPI_ISL_1614625, EPI_ISL_1614626, EPI_ISL_1614628, EPI_ISL_1614630, EPI_ISL_1614631, EPI_ISL_1614632, EPI_ISL_1614633, EPI_ISL_1614634, EPI_ISL_1614635, EPI_ISL_1614636, EPI_ISL_1614637, EPI_ISL_1614639, EPI_ISL_1614640, EPI_ISL_1614641, EPI_ISL_1614642, EPI_ISL_1614643, EPI_ISL_1614644, EPI_ISL_1614645, EPI_ISL_1614646, EPI_ISL_1614647, EPI_ISL_1614649, EPI_ISL_1614650, EPI_ISL_1614651, EPI_ISL_1614652, EPI_ISL_1614653, EPI_ISL_1614654, EPI_ISL_1614655, EPI_ISL_1614656, EPI_ISL_1614657, EPI_ISL_1614658, EPI_ISL_1614659, EPI_ISL_1614660, EPI_ISL_1614661, EPI_ISL_1614662, EPI_ISL_1614663, EPI_ISL_1614664, EPI_ISL_1614665, EPI_ISL_1614666, EPI_ISL_1614667, EPI_ISL_1614668, EPI_ISL_1614669, EPI_ISL_1614671, EPI_ISL_1614672, EPI_ISL_1614673, EPI_ISL_1614674, EPI_ISL_1614675, EPI_ISL_1614676, EPI_ISL_1614677, EPI_ISL_1614678, EPI_ISL_1614679, EPI_ISL_1614680, EPI_ISL_1614681, EPI_ISL_1614682, EPI_ISL_1614683, EPI_ISL_1614684, EPI_ISL_1614685, EPI_ISL_1614686, EPI_ISL_1614687, EPI_ISL_1614688, EPI_ISL_1614689, EPI_ISL_1614690, EPI_ISL_1614691, EPI_ISL_1614692, EPI_ISL_1614693, EPI_ISL_1614694, EPI_ISL_1614695, EPI_ISL_1614696, EPI_ISL_1614697, EPI_ISL_1614698, EPI_ISL_1614699, EPI_ISL_1614700, EPI_ISL_1614701, EPI_ISL_1614702, EPI_ISL_1614703, EPI_ISL_1614704, EPI_ISL_1614705, EPI_ISL_1614706, EPI_ISL_1614707, EPI_ISL_1614708, EPI_ISL_1614709, EPI_ISL_1614710, EPI_ISL_1614712, EPI_ISL_1614713, EPI_ISL_1614714, EPI_ISL_1614716, EPI_ISL_1614717, EPI_ISL_1614718, EPI_ISL_1614719, EPI_ISL_1614720, EPI_ISL_1614721, EPI_ISL_1614722, EPI_ISL_1614723, EPI_ISL_1614724, EPI_ISL_1614725, EPI_ISL_1614726, EPI_ISL_1614727, EPI_ISL_1614728, EPI_ISL_1614729, EPI_ISL_1614730, EPI_ISL_1614731, EPI_ISL_1614732, EPI_ISL_1614733, EPI_ISL_1614734, EPI_ISL_1614735, EPI_ISL_1614737, EPI_ISL_1614739, EPI_ISL_1614740, EPI_ISL_1614741, EPI_ISL_1614742, EPI_ISL_1614743, EPI_ISL_1614745, EPI_ISL_1614747, EPI_ISL_1614748, EPI_ISL_1614749, EPI_ISL_1614750, EPI_ISL_1614751, EPI_ISL_1614752, EPI_ISL_1614753, EPI_ISL_1614754, EPI_ISL_1614755, EPI_ISL_1614756, EPI_ISL_1614757, EPI_ISL_1614758, EPI_ISL_1614759, EPI_ISL_1614760, EPI_ISL_1614761, EPI_ISL_1614762, EPI_ISL_1614763, EPI_ISL_1614764, EPI_ISL_1614765, EPI_ISL_1614767, EPI_ISL_1614768, EPI_ISL_1614769, EPI_ISL_1614770, EPI_ISL_1614771, EPI_ISL_1614772, EPI_ISL_1614775, EPI_ISL_1614776, EPI_ISL_1614777, EPI_ISL_1614778, EPI_ISL_1614779, EPI_ISL_1614780, EPI_ISL_1614781, EPI_ISL_1614782, EPI_ISL_1614783, EPI_ISL_1614784, EPI_ISL_1614785, EPI_ISL_1614786, EPI_ISL_1614787, EPI_ISL_1614788, EPI_ISL_1614789, EPI_ISL_1614790, EPI_ISL_1614791, EPI_ISL_1614792, EPI_ISL_1614793, EPI_ISL_1614794, EPI_ISL_1614795, EPI_ISL_1614796, EPI_ISL_1614797, EPI_ISL_1614798, EPI_ISL_1614799, EPI_ISL_1614800, EPI_ISL_1614801, EPI_ISL_1614802, EPI_ISL_1614803, EPI_ISL_1614804, EPI_ISL_1614805, EPI_ISL_1614806, EPI_ISL_1614807, EPI_ISL_1614808, EPI_ISL_1614809, EPI_ISL_1614810, EPI_ISL_1614811, EPI_ISL_1614812, EPI_ISL_1614813, EPI_ISL_1614814, EPI_ISL_1614815, EPI_ISL_1614816, EPI_ISL_1614817, EPI_ISL_1614819, EPI_ISL_1614820, EPI_ISL_1614821, EPI_ISL_1614822, EPI_ISL_1614823, EPI_ISL_1614824, EPI_ISL_1614825, EPI_ISL_1614826, EPI_ISL_1614827, EPI_ISL_1614828, EPI_ISL_1614829, EPI_ISL_1614830, EPI_ISL_1614831, EPI_ISL_1614832, EPI_ISL_1614833, EPI_ISL_1614834, EPI_ISL_1614835, EPI_ISL_1614836, EPI_ISL_1614837, EPI_ISL_1614838, EPI_ISL_1614840, EPI_ISL_1614841, EPI_ISL_1614842, EPI_ISL_1614843, EPI_ISL_1614844, EPI_ISL_1614845, EPI_ISL_1614846, EPI_ISL_1614847, EPI_ISL_1614849, EPI_ISL_1614850, EPI_ISL_1614851, EPI_ISL_1614852, EPI_ISL_1614853, EPI_ISL_1614854, EPI_ISL_1614855, EPI_ISL_1614856, EPI_ISL_1614857, EPI_ISL_1614858, EPI_ISL_1614859, EPI_ISL_1614860, EPI_ISL_1614861, EPI_ISL_1614862, EPI_ISL_1614864, EPI_ISL_1614865, EPI_ISL_1614866, EPI_ISL_1614867, EPI_ISL_1614868, EPI_ISL_1614869, EPI_ISL_1614871, EPI_ISL_1614872, EPI_ISL_1614873, EPI_ISL_1614874, EPI_ISL_1614875, EPI_ISL_1614876, EPI_ISL_1614877, EPI_ISL_1614878, EPI_ISL_1614879, EPI_ISL_1614880, EPI_ISL_1614881, EPI_ISL_1614882, EPI_ISL_1614883, EPI_ISL_1614884, EPI_ISL_1614885, EPI_ISL_1614886, EPI_ISL_1614887, EPI_ISL_1614889, EPI_ISL_1614890, EPI_ISL_1614891, EPI_ISL_1614893, EPI_ISL_1614895, EPI_ISL_1614898, EPI_ISL_1614900, EPI_ISL_1614901, EPI_ISL_1614902, EPI_ISL_1614903, EPI_ISL_1614904, EPI_ISL_1614905

| | | | |
|---|---|---|---|
| see above | Fulgent Genetics | Centers for Disease Control and Prevention Division of Viral Diseases, Pathogen Discovery | Dakota Howard, Dhwani Batra, Peter W. Cook, Kara Moser, Adrian Paskey, Jason Caravas, Benjamin Rambo-Martin, Shatavia Morrison, Christopher Gulvick, Scott Sammons, Yvette Unoarumhi, Darlene Wagner, Matthew Schmerer, Harry Gao, Mickey Li, John Gao, Joseph Fierro, Benafsh Sapra, Becky Tsai, Yan Meng, Doreen Ng, James Xie, Clinton R. Paden, Duncan MacCannell |

EPI_ISL_1614907, EPI_ISL_1614908, EPI_ISL_1614909, EPI_ISL_1614910, EPI_ISL_1614911, EPI_ISL_1614912, EPI_ISL_1614913, EPI_ISL_1614914, EPI_ISL_1614915, EPI_ISL_1614916, EPI_ISL_1614917, EPI_ISL_1614918, EPI_ISL_1614919, EPI_ISL_1614920, EPI_ISL_1614921, EPI_ISL_1614922, EPI_ISL_1614923, EPI_ISL_1614924, EPI_ISL_1614925, EPI_ISL_1614926, EPI_ISL_1614927, EPI_ISL_1614928, EPI_ISL_1614929, EPI_ISL_1614930, EPI_ISL_1614931, EPI_ISL_1614932, EPI_ISL_1614933, EPI_ISL_1614934, EPI_ISL_1614935, EPI_ISL_1614936, EPI_ISL_1614937, EPI_ISL_1614938, EPI_ISL_1614939, EPI_ISL_1614940, EPI_ISL_1614941, EPI_ISL_1614942, EPI_ISL_1614943, EPI_ISL_1614944, EPI_ISL_1614945, EPI_ISL_1614946, EPI_ISL_1614947, EPI_ISL_1614948, EPI_ISL_1614949, EPI_ISL_1614950, EPI_ISL_1614951, EPI_ISL_1614952, EPI_ISL_1614953, EPI_ISL_1614954, EPI_ISL_1614955, EPI_ISL_1614956, EPI_ISL_1614957, EPI_ISL_1614958, EPI_ISL_1614959, EPI_ISL_1614960, EPI_ISL_1614961, EPI_ISL_1614962, EPI_ISL_1614963, EPI_ISL_1614964, EPI_ISL_1614965, EPI_ISL_1614966, EPI_ISL_1614967, EPI_ISL_1614968, EPI_ISL_1614969, EPI_ISL_1614970, EPI_ISL_1614971, EPI_ISL_1614972, EPI_ISL_1614973, EPI_ISL_1614974, EPI_ISL_1614975, EPI_ISL_1614976, EPI_ISL_1614977, EPI_ISL_1614978, EPI_ISL_1614979, EPI_ISL_1614980, EPI_ISL_1614981, EPI_ISL_1614982, EPI_ISL_1614983, EPI_ISL_1614984, EPI_ISL_1614985, EPI_ISL_1614986, EPI_ISL_1614987, EPI_ISL_1614988, EPI_ISL_1614989, EPI_ISL_1614990, EPI_ISL_1614991, EPI_ISL_1614992, EPI_ISL_1614993, EPI_ISL_1614994, EPI_ISL_1614995, EPI_ISL_1614996, EPI_ISL_1614997, EPI_ISL_1614998, EPI_ISL_1614999, EPI_ISL_1615000, EPI_ISL_1615001, EPI_ISL_1615002, EPI_ISL_1615003, EPI_ISL_1615004, EPI_ISL_1615005, EPI_ISL_1615006, EPI_ISL_1615007, EPI_ISL_1615008, EPI_ISL_1615009, EPI_ISL_1615010, EPI_ISL_1615011, EPI_ISL_1615012, EPI_ISL_1615013, EPI_ISL_1615014, EPI_ISL_1615015, EPI_ISL_1615016, EPI_ISL_1615017, EPI_ISL_1615018, EPI_ISL_1615019, EPI_ISL_1615020, EPI_ISL_1615021, EPI_ISL_1615022, EPI_ISL_1615023, EPI_ISL_1615024, EPI_ISL_1615025, EPI_ISL_1615026, EPI_ISL_1615027, EPI_ISL_1615028, EPI_ISL_1615029, EPI_ISL_1615030, EPI_ISL_1615031, EPI_ISL_1615032, EPI_ISL_1615033, EPI_ISL_1615034, EPI_ISL_1615035, EPI_ISL_1615036, EPI_ISL_1615037, EPI_ISL_1615038, EPI_ISL_1615039, EPI_ISL_1615040, EPI_ISL_1615041, EPI_ISL_1615042, EPI_ISL_1615043, EPI_ISL_1615044, EPI_ISL_1615045, EPI_ISL_1615046, EPI_ISL_1615047, EPI_ISL_1615048, EPI_ISL_1615049, EPI_ISL_1615050, EPI_ISL_1615051, EPI_ISL_1615052, EPI_ISL_1615053, EPI_ISL_1615054, EPI_ISL_1615055, EPI_ISL_1615056, EPI_ISL_1615057, EPI_ISL_1615058, EPI_ISL_1615059, EPI_ISL_1615060, EPI_ISL_1615061, EPI_ISL_1615062, EPI_ISL_1615063, EPI_ISL_1615064, EPI_ISL_1615065, EPI_ISL_1615066, EPI_ISL_1615067, EPI_ISL_1615068

| | | | |
|---|---|---|---|
| see above | Quest Diagnostics Incorporated | Centers for Disease Control and Prevention Division of Viral Diseases, Pathogen Discovery | Dakota Howard, Dhwani Batra, Peter W. Cook, Kara Moser, Adrian Paskey, Jason Caravas, Benjamin Rambo-Martin, Shatavia Morrison, Christopher Gulvick, Scott Sammons, Yvette Unoarumhi, Darlene Wagner, Matthew Schmerer, S. H. Rosenthal, A. Gerasimova, R. M. Kagan, B. Anderson, M. Hua, Y. Liu, L.E. Bernstein, K.E. Livingston, A. Perez, I. A. Shlyakhter, R. V. Rolando, R. Owen, P. Tanpaiboon, F. Lacbawan, Clinton R. Paden, Duncan MacCannell |

EPI_ISL_1615070, EPI_ISL_1615071, EPI_ISL_1615072, EPI_ISL_1615073, EPI_ISL_1615074, EPI_ISL_1615075, EPI_ISL_1615076, EPI_ISL_1615077, EPI_ISL_1615078, EPI_ISL_1615079, EPI_ISL_1615080, EPI_ISL_1615081, EPI_ISL_1615082, EPI_ISL_1615083, EPI_ISL_1615084, EPI_ISL_1615085, EPI_ISL_1615086, EPI_ISL_1615087, EPI_ISL_1615088, EPI_ISL_1615089, EPI_ISL_1615090, EPI_ISL_1615091, EPI_ISL_1615092, EPI_ISL_1615093, EPI_ISL_1615094, EPI_ISL_1615095, EPI_ISL_1615096, EPI_ISL_1615097, EPI_ISL_1615098, EPI_ISL_1615099, EPI_ISL_1615100, EPI_ISL_1615101, EPI_ISL_1615102, EPI_ISL_1615103, EPI_ISL_1615104, EPI_ISL_1615105, EPI_ISL_1615106, EPI_ISL_1615107, EPI_ISL_1615108, EPI_ISL_1615109, EPI_ISL_1615110, EPI_ISL_1615111, EPI_ISL_1615112, EPI_ISL_1615113, EPI_ISL_1615114, EPI_ISL_1615115, EPI_ISL_1615116, EPI_ISL_1615117, EPI_ISL_1615118, EPI_ISL_1615119, EPI_ISL_1615120, EPI_ISL_1615121, EPI_ISL_1615122, EPI_ISL_1615123, EPI_ISL_1615124, EPI_ISL_1615125, EPI_ISL_1615126, EPI_ISL_1615127, EPI_ISL_1615128, EPI_ISL_1615129, EPI_ISL_1615130, EPI_ISL_1615131, EPI_ISL_1615132, EPI_ISL_1615133, EPI_ISL_1615134, EPI_ISL_1615135, EPI_ISL_1615136, EPI_ISL_1615137, EPI_ISL_1615138, EPI_ISL_1615139, EPI_ISL_1615140, EPI_ISL_1615141, EPI_ISL_1615142, EPI_ISL_1615143, EPI_ISL_1615144, EPI_ISL_1615145, EPI_ISL_1615146, EPI_ISL_1615147, EPI_ISL_1615148, EPI_ISL_1615149, EPI_ISL_1615150, EPI_ISL_1615151, EPI_ISL_1615152, EPI_ISL_1615153, EPI_ISL_1615154, EPI_ISL_1615155, EPI_ISL_1615156, EPI_ISL_1615157, EPI_ISL_1615158, EPI_ISL_1615159, EPI_ISL_1615160, EPI_ISL_1615161, EPI_ISL_1615162, EPI_ISL_1615163, EPI_ISL_1615164, EPI_ISL_1615165, EPI_ISL_1615166, EPI_ISL_1615167, EPI_ISL_1615168, EPI_ISL_1615169, EPI_ISL_1615170, EPI_ISL_1615171, EPI_ISL_1615172, EPI_ISL_1615173, EPI_ISL_1615174, EPI_ISL_1615175, EPI_ISL_1615176, EPI_ISL_1615177, EPI_ISL_1615178, EPI_ISL_1615179, EPI_ISL_1615180, EPI_ISL_1615181, EPI_ISL_1615182, EPI_ISL_1615183, EPI_ISL_1615185, EPI_ISL_1615186, EPI_ISL_1615187, EPI_ISL_1615188, EPI_ISL_1615189, EPI_ISL_1615190, EPI_ISL_1615191, EPI_ISL_1615192, EPI_ISL_1615193, EPI_ISL_1615194, EPI_ISL_1615195, EPI_ISL_1615196, EPI_ISL_1615197, EPI_ISL_1615198, EPI_ISL_1615199, EPI_ISL_1615200, EPI_ISL_1615201, EPI_ISL_1615202, EPI_ISL_1615203, EPI_ISL_1615204, EPI_ISL_1615205, EPI_ISL_1615206, EPI_ISL_1615207, EPI_ISL_1615208, EPI_ISL_1615209, EPI_ISL_1615210, EPI_ISL_1615211, EPI_ISL_1615212, EPI_ISL_1615213, EPI_ISL_1615214, EPI_ISL_1615215, EPI_ISL_1615216, EPI_ISL_1615217, EPI_ISL_1615218, EPI_ISL_1615219, EPI_ISL_1615220, EPI_ISL_1615222, EPI_ISL_1615224, EPI_ISL_1615225, EPI_ISL_1615226, EPI_ISL_1615227, EPI_ISL_1615228, EPI_ISL_1615229, EPI_ISL_1615230, EPI_ISL_1615231, EPI_ISL_1615232, EPI_ISL_1615233, EPI_ISL_1615234, EPI_ISL_1615235, EPI_ISL_1615236, EPI_ISL_1615237, EPI_ISL_1615238, EPI_ISL_1615239, EPI_ISL_1615240, EPI_ISL_1615241, EPI_ISL_1615242, EPI_ISL_1615243, EPI_ISL_1615244, EPI_ISL_1615245, EPI_ISL_1615246, EPI_ISL_1615247, EPI_ISL_1615248, EPI_ISL_1615249, EPI_ISL_1615250, EPI_ISL_1615251, EPI_ISL_1615252, EPI_ISL_1615253, EPI_ISL_1615254, EPI_ISL_1615255, EPI_ISL_1615256, EPI_ISL_1615257, EPI_ISL_1615258, EPI_ISL_1615259, EPI_ISL_1615260, EPI_ISL_1615261, EPI_ISL_1615262, EPI_ISL_1615263, EPI_ISL_1615264, EPI_ISL_1615265, EPI_ISL_1615266, EPI_ISL_1615267, EPI_ISL_1615268, EPI_ISL_1615269, EPI_ISL_1615270, EPI_ISL_1615271, EPI_ISL_1615272, EPI_ISL_1615273, EPI_ISL_1615274, EPI_ISL_1615275, EPI_ISL_1615277, EPI_ISL_1615278, EPI_ISL_1615279, EPI_ISL_1615280, EPI_ISL_1615281, EPI_ISL_1615282, EPI_ISL_1615283, EPI_ISL_1615284, EPI_ISL_1615285, EPI_ISL_1615286, EPI_ISL_1615287, EPI_ISL_1615288, EPI_ISL_1615289, EPI_ISL_1615290, EPI_ISL_1615291, EPI_ISL_1615292, EPI_ISL_1615293, EPI_ISL_1615294, EPI_ISL_1615295, EPI_ISL_1615296, EPI_ISL_1615297, EPI_ISL_1615298, EPI_ISL_1615299, EPI_ISL_1615300, EPI_ISL_1615301, EPI_ISL_1615302, EPI_ISL_1615303, EPI_ISL_1615304, EPI_ISL_1615305, EPI_ISL_1615306, EPI_ISL_1615307, EPI_ISL_1615308, EPI_ISL_1615309, EPI_ISL_1615310, EPI_ISL_1615311, EPI_ISL_1615312, EPI_ISL_1615313, EPI_ISL_1615314, EPI_ISL_1615315, EPI_ISL_1615316, EPI_ISL_1615317, EPI_ISL_1615318, EPI_ISL_1615319, EPI_ISL_1615320, EPI_ISL_1615321, EPI_ISL_1615322, EPI_ISL_1615323, EPI_ISL_1615324, EPI_ISL_1615325, EPI_ISL_1615326, EPI_ISL_1615327, EPI_ISL_1615329, EPI_ISL_1615330, EPI_ISL_1615331, EPI_ISL_1615332, EPI_ISL_1615333, EPI_ISL_1615334, EPI_ISL_1615335, EPI_ISL_1615336, EPI_ISL_1615337, EPI_ISL_1615338, EPI_ISL_1615339, EPI_ISL_1615340, EPI_ISL_1615341, EPI_ISL_1615342, EPI_ISL_1615343, EPI_ISL_1615344, EPI_ISL_1615345, EPI_ISL_1615346, EPI_ISL_1615347, EPI_ISL_1615348, EPI_ISL_1615349, EPI_ISL_1615350, EPI_ISL_1615351, EPI_ISL_1615352, EPI_ISL_1615353, EPI_ISL_1615354, EPI_ISL_1615355, EPI_ISL_1615356, EPI_ISL_1615357, EPI_ISL_1615358, EPI_ISL_1615359, EPI_ISL_1615360, EPI_ISL_1615361, EPI_ISL_1615362, EPI_ISL_1615363, EPI_ISL_1615364, EPI_ISL_1615365, EPI_ISL_1615366, EPI_ISL_1615367, EPI_ISL_1615368, EPI_ISL_1615369, EPI_ISL_1615370, EPI_ISL_1615371, EPI_ISL_1615372, EPI_ISL_1615373, EPI_ISL_1615374, EPI_ISL_1615375, EPI_ISL_1615376, EPI_ISL_1615377, EPI_ISL_1615378, EPI_ISL_1615379, EPI_ISL_1615380, EPI_ISL_1615381, EPI_ISL_1615382, EPI_ISL_1615383, EPI_ISL_1615384, EPI_ISL_1615385, EPI_ISL_1615386, EPI_ISL_1615387, EPI_ISL_1615388, EPI_ISL_1615389, EPI_ISL_1615390, EPI_ISL_1615391, EPI_ISL_1615392, EPI_ISL_1615393, EPI_ISL_1615394, EPI_ISL_1615395, EPI_ISL_1615396, EPI_ISL_1615397, EPI_ISL_1615398, EPI_ISL_1615399, EPI_ISL_1615400, EPI_ISL_1615401, EPI_ISL_1615402, EPI_ISL_1615403, EPI_ISL_1615404, EPI_ISL_1615405, EPI_ISL_1615406, EPI_ISL_1615407, EPI_ISL_1615408, EPI_ISL_1615409, EPI_ISL_1615410, EPI_ISL_1615411, EPI_ISL_1615412, EPI_ISL_1615413, EPI_ISL_1615414, EPI_ISL_1615415, EPI_ISL_1615416, EPI_ISL_1615417, EPI_ISL_1615418, EPI_ISL_1615419, EPI_ISL_1615420, EPI_ISL_1615421, EPI_ISL_1615422, EPI_ISL_1615423, EPI_ISL_1615424, EPI_ISL_1615425, EPI_ISL_1615426, EPI_ISL_1615427, EPI_ISL_1615428, EPI_ISL_1615429, EPI_ISL_1615430, EPI_ISL_1615431, EPI_ISL_1615432, EPI_ISL_1615433, EPI_ISL_1615434, EPI_ISL_1615435, EPI_ISL_1615436, EPI_ISL_1615437, EPI_ISL_1615438, EPI_ISL_1615439, EPI_ISL_1615440, EPI_ISL_1615441, EPI_ISL_1615442, EPI_ISL_1615443, EPI_ISL_1615444, EPI_ISL_1615445, EPI_ISL_1615446, EPI_ISL_1615447, EPI_ISL_1615448, EPI_ISL_1615449, EPI_ISL_1615450, EPI_ISL_1615451, EPI_ISL_1615452, EPI_ISL_1615453, EPI_ISL_1615454, EPI_ISL_1615455, EPI_ISL_1615456, EPI_ISL_1615457, EPI_ISL_1615458, EPI_ISL_1615459, EPI_ISL_1615460, EPI_ISL_1615461, EPI_ISL_1615462, EPI_ISL_1615463, EPI_ISL_1615464, EPI_ISL_1615465, EPI_ISL_1615466, EPI_ISL_1615467, EPI_ISL_1615468, EPI_ISL_1615469, EPI_ISL_1615470, EPI_ISL_1615471, EPI_ISL_1615472, EPI_ISL_1615473, EPI_ISL_1615474, EPI_ISL_1615475, EPI_ISL_1615476, EPI_ISL_1615477, EPI_ISL_1615478, EPI_ISL_1615479, EPI_ISL_1615480, EPI_ISL_1615481, EPI_ISL_1615482, EPI_ISL_1615483, EPI_ISL_1615484, EPI_ISL_1615485, EPI_ISL_1615486, EPI_ISL_1615487, EPI_ISL_1615488, EPI_ISL_1615489, EPI_ISL_1615490, EPI_ISL_1615491, EPI_ISL_1615492, EPI_ISL_1615493, EPI_ISL_1615494, EPI_ISL_1615495, EPI_ISL_1615496, EPI_ISL_1615497, EPI_ISL_1615498, EPI_ISL_1615499, EPI_ISL_1615500, EPI_ISL_1615501, EPI_ISL_1615502, EPI_ISL_1615503, EPI_ISL_1615504, EPI_ISL_1615505, EPI_ISL_1615506, EPI_ISL_1615507, EPI_ISL_1615508, EPI_ISL_1615509, EPI_ISL_1615510, EPI_ISL_1615511, EPI_ISL_1615512, EPI_ISL_1615513, EPI_ISL_1615514, EPI_ISL_1615515, EPI_ISL_1615516, EPI_ISL_1615517, EPI_ISL_1615518, EPI_ISL_1615519, EPI_ISL_1615520, EPI_ISL_1615521, EPI_ISL_1615522, EPI_ISL_1615523, EPI_ISL_1615524, EPI_ISL_1615525, EPI_ISL_1615526, EPI_ISL_1615527, EPI_ISL_1615528, EPI_ISL_1615529, EPI_ISL_1615530, EPI_ISL_1615531, EPI_ISL_1615532, EPI_ISL_1615533, EPI_ISL_1615534, EPI_ISL_1615535, EPI_ISL_1615536, EPI_ISL_1615537, EPI_ISL_1615538, EPI_ISL_1615539, EPI_ISL_1615540, EPI_ISL_1615541, EPI_ISL_1615542, EPI_ISL_1615543, EPI_ISL_1615544, EPI_ISL_1615545, EPI_ISL_1615546, EPI_ISL_1615547, EPI_ISL_1615548, EPI_ISL_1615549, EPI_ISL_1615550, EPI_ISL_1615551, EPI_ISL_1615552, EPI_ISL_1615553, EPI_ISL_1615554, EPI_ISL_1615555

| | | | |
|---|---|---|---|
| see above | Helix/Illumina | Centers for Disease Control and Prevention Division of Viral Diseases, Pathogen Discovery | Dakota Howard, Dhwani Batra, Peter W. Cook, Kara Moser, Adrian Paskey, Jason Caravas, Benjamin Rambo-Martin, Shatavia Morrison, Christopher Gulvick, Scott Sammons, Yvette Unoarumhi, Darlene Wagner, Matthew Schmerer, Eileen de Feo, Jan Antico, Christine Tran, Matthew Tolentino, Shannon Wickline, Kim Gietzen, Brad Sickler, Jingtao Liu, Eric Allen, Phil Febbo, Nicole L. Washington, Simon White, Geraint Levan, Kelly Schiabor Barrett, Elizabeth Cirulli, Alexandre Bolze, Ary Ascencio, Charlotte Rivera-Garcia, Ryan Cho, Jason Nguyen, Sherry Wang, Jimmy Ramirez, Tyler Cassens, Efren Sandoval, Magnus Isaksson, William Lee, David Becker, Marc Laurent, James Lu, Clinton R. Paden, Duncan MacCannell |

EPI_ISL_1615559, EPI_ISL_1615560, EPI_ISL_1615561, EPI_ISL_1615562, EPI_ISL_1615563, EPI_ISL_1615564, EPI_ISL_1615565, EPI_ISL_1615566, EPI_ISL_1615567, EPI_ISL_1615568, EPI_ISL_1615569, EPI_ISL_1615570, EPI_ISL_1615571, EPI_ISL_1615572, EPI_ISL_1615573, EPI_ISL_1615574, EPI_ISL_1615575, EPI_ISL_1615576, EPI_ISL_1615577, EPI_ISL_1615579, EPI_ISL_1615580, EPI_ISL_1615581, EPI_ISL_1615582, EPI_ISL_1615583, EPI_ISL_1615584, EPI_ISL_1615585

| | | | |
|---|---|---|---|
| see above | Wyoming Public Health Laboratory | Wyoming Public Health Laboratory | Jim Mildenberger, Wanda Manley, Noah Hull, Taylor Fearing, Lynette Gumbleton, Channing Weber, Ashley Norberg, Chayse Rowley, Marley Goetz, Brian Dominguez, Elliot Thomasson, Sam Britz, Cari Sloma, and Rob Christensen |

| | | | |
|---|---|---|---|
| EPI_ISL_1615601, EPI_ISL_1615603, EPI_ISL_1615606 | Wyoming Public Health Laboratory | Wyoming Public Health Laboratory | Jim Mildenberger, Taylor Fearing, Channing Weber, Ashley Norberg, Chayse Rowley, Marley Goetz, Brian Dominguez, Elliot Thomasson, Brittany Oher, Sam Britz, Cari Sloma, and Rob Christensen |
| EPI_ISL_1615622, EPI_ISL_1615623, EPI_ISL_1615624, EPI_ISL_1615625, EPI_ISL_1615626, EPI_ISL_1615627, EPI_ISL_1615628, EPI_ISL_1615629, EPI_ISL_1615630, EPI_ISL_1615631, EPI_ISL_1615632, EPI_ISL_1615633, EPI_ISL_1615634, EPI_ISL_1615635, EPI_ISL_1615638, EPI_ISL_1615639, EPI_ISL_1615640, EPI_ISL_1615641, EPI_ISL_1615642, EPI_ISL_1615643, EPI_ISL_1615644, EPI_ISL_1615645, EPI_ISL_1615646, EPI_ISL_1615647, EPI_ISL_1615648, EPI_ISL_1615649, EPI_ISL_1615650, EPI_ISL_1615666, EPI_ISL_1615667, EPI_ISL_1615670, EPI_ISL_1615672, EPI_ISL_1615673, EPI_ISL_1615676, EPI_ISL_1615681, EPI_ISL_1615682 | | | |
| see above | Wyoming Public Health Laboratory | Wyoming Public Health Laboratory | Jim Mildenberger, Wanda Manley, Noah Hull, Taylor Fearing, Lynette Gumbleton, Channing Weber, Ashley Norberg, Chayse Rowley, Marley Goetz, Brian Dominguez, Elliot Thomasson, Sam Britz, Cari Sloma, and Rob Christensen |
| EPI_ISL_1616617, EPI_ISL_1616618, EPI_ISL_1616619, EPI_ISL_1616620, EPI_ISL_1616621, EPI_ISL_1616622, EPI_ISL_1616623, EPI_ISL_1616624, EPI_ISL_1616625, EPI_ISL_1616626, EPI_ISL_1616627, EPI_ISL_1616628, EPI_ISL_1616629, EPI_ISL_1616630, EPI_ISL_1616631, EPI_ISL_1616632, EPI_ISL_1616633, EPI_ISL_1616634, EPI_ISL_1616635, EPI_ISL_1616636, EPI_ISL_1616637, EPI_ISL_1616638, EPI_ISL_1616639, EPI_ISL_1616640, EPI_ISL_1616641, EPI_ISL_1616642, EPI_ISL_1616643, EPI_ISL_1616644, EPI_ISL_1616645, EPI_ISL_1616646, EPI_ISL_1616647, EPI_ISL_1616648, EPI_ISL_1616649, EPI_ISL_1616650, EPI_ISL_1616651, EPI_ISL_1616652, EPI_ISL_1616653, EPI_ISL_1616654, EPI_ISL_1616655, EPI_ISL_1616656, EPI_ISL_1616657, EPI_ISL_1616658, EPI_ISL_1616659, EPI_ISL_1616660, EPI_ISL_1616661, EPI_ISL_1616662, EPI_ISL_1616663, EPI_ISL_1616664, EPI_ISL_1616665, EPI_ISL_1616666, EPI_ISL_1616667, EPI_ISL_1616668, EPI_ISL_1616669, EPI_ISL_1616670, EPI_ISL_1616671, EPI_ISL_1616672, EPI_ISL_1616673, EPI_ISL_1616674, EPI_ISL_1616675, EPI_ISL_1616676, EPI_ISL_1616677, EPI_ISL_1616678, EPI_ISL_1616679, EPI_ISL_1616680, EPI_ISL_1616681, EPI_ISL_1616682, EPI_ISL_1616683, EPI_ISL_1616684, EPI_ISL_1616685, EPI_ISL_1616686, EPI_ISL_1616687, EPI_ISL_1616688, EPI_ISL_1616689, EPI_ISL_1616690, EPI_ISL_1616691, EPI_ISL_1616692, EPI_ISL_1616693, EPI_ISL_1616694, EPI_ISL_1616695, EPI_ISL_1616696, EPI_ISL_1616697, EPI_ISL_1616698, EPI_ISL_1616699, EPI_ISL_1616700, EPI_ISL_1616701, EPI_ISL_1616702, EPI_ISL_1616703, EPI_ISL_1616704, EPI_ISL_1616705, EPI_ISL_1616706, EPI_ISL_1616707, EPI_ISL_1616708, EPI_ISL_1616709, EPI_ISL_1616710, EPI_ISL_1616711, EPI_ISL_1616712, EPI_ISL_1616713, EPI_ISL_1616714, EPI_ISL_1616715, EPI_ISL_1616716, EPI_ISL_1616717, EPI_ISL_1616718, EPI_ISL_1616719, EPI_ISL_1616720, EPI_ISL_1616721, EPI_ISL_1616722, EPI_ISL_1616723, EPI_ISL_1616724, EPI_ISL_1616725, EPI_ISL_1616726, EPI_ISL_1616727, EPI_ISL_1616728, EPI_ISL_1616729, EPI_ISL_1616730, EPI_ISL_1616731, EPI_ISL_1616732, EPI_ISL_1616733, EPI_ISL_1616734, EPI_ISL_1616735, EPI_ISL_1616736, EPI_ISL_1616737, EPI_ISL_1616738, EPI_ISL_1616739, EPI_ISL_1616740, EPI_ISL_1616741, EPI_ISL_1616742, EPI_ISL_1616743, EPI_ISL_1616744, EPI_ISL_1616745, EPI_ISL_1616746, EPI_ISL_1616747, EPI_ISL_1616748, EPI_ISL_1616749, EPI_ISL_1616750, EPI_ISL_1616751, EPI_ISL_1616752, EPI_ISL_1616753, EPI_ISL_1616754, EPI_ISL_1616755, EPI_ISL_1616756, EPI_ISL_1616757, EPI_ISL_1616758, EPI_ISL_1616759, EPI_ISL_1616760, EPI_ISL_1616761, EPI_ISL_1616762, EPI_ISL_1616763, EPI_ISL_1616764, EPI_ISL_1616765, EPI_ISL_1616766, EPI_ISL_1616767, EPI_ISL_1616768, EPI_ISL_1616769, EPI_ISL_1616770, EPI_ISL_1616771, EPI_ISL_1616772, EPI_ISL_1616773, EPI_ISL_1616774, EPI_ISL_1616775, EPI_ISL_1616776, EPI_ISL_1616777, EPI_ISL_1616778, EPI_ISL_1616779, EPI_ISL_1616780, EPI_ISL_1616781, EPI_ISL_1616782, EPI_ISL_1616783, EPI_ISL_1616784, EPI_ISL_1616785, EPI_ISL_1616786, EPI_ISL_1616787, EPI_ISL_1616788, EPI_ISL_1616789, EPI_ISL_1616790, EPI_ISL_1616791, EPI_ISL_1616792, EPI_ISL_1616793, EPI_ISL_1616794, EPI_ISL_1616795, EPI_ISL_1616796, EPI_ISL_1616797, EPI_ISL_1616798, EPI_ISL_1616799, EPI_ISL_1616800, EPI_ISL_1616801, EPI_ISL_1616802, EPI_ISL_1616803, EPI_ISL_1616804, EPI_ISL_1616805, EPI_ISL_1616807, EPI_ISL_1616808, EPI_ISL_1616809, EPI_ISL_1616810, EPI_ISL_1616811, EPI_ISL_1616812, EPI_ISL_1616813, EPI_ISL_1616814, EPI_ISL_1616815, EPI_ISL_1616816, EPI_ISL_1616817, EPI_ISL_1616818, EPI_ISL_1616819, EPI_ISL_1616820, EPI_ISL_1616821, EPI_ISL_1616822, EPI_ISL_1616823, EPI_ISL_1616824, EPI_ISL_1616825, EPI_ISL_1616826, EPI_ISL_1616827, EPI_ISL_1616828, EPI_ISL_1616829, EPI_ISL_1616830, EPI_ISL_1616831, EPI_ISL_1616832, EPI_ISL_1616833 | | | |
| see above | UW Virology Lab | UW Virology Lab | Pavitra Roychoudhury, Hong Xie, Lasata Shrestha, Shah Mohamed Bakhash, Michelle Lin, Noah R. Baker, Sean Ellis, Saraswathi Sathees, Meei-Li Huang, Keith R Jerome, Alexander Greninger |
| EPI_ISL_1616840, EPI_ISL_1616842, EPI_ISL_1616844, EPI_ISL_1616845, EPI_ISL_1616847, EPI_ISL_1616854, EPI_ISL_1616856, EPI_ISL_1616857, EPI_ISL_1616859, EPI_ISL_1616860, EPI_ISL_1616862, EPI_ISL_1616864, EPI_ISL_1616866, EPI_ISL_1616868, EPI_ISL_1616869, EPI_ISL_1616870, EPI_ISL_1616872, EPI_ISL_1616873, EPI_ISL_1616875, EPI_ISL_1616877, EPI_ISL_1616878, EPI_ISL_1616879, EPI_ISL_1616880, EPI_ISL_1616881, EPI_ISL_1616883, EPI_ISL_1616884, EPI_ISL_1616885, EPI_ISL_1616886, EPI_ISL_1616888, EPI_ISL_1616889, EPI_ISL_1616890, EPI_ISL_1616892, EPI_ISL_1616893, EPI_ISL_1616894, EPI_ISL_1616896, EPI_ISL_1616898, EPI_ISL_1616899, EPI_ISL_1616905, EPI_ISL_1616907, EPI_ISL_1616908, EPI_ISL_1616910, EPI_ISL_1616912, EPI_ISL_1616913, EPI_ISL_1616914, EPI_ISL_1616915, EPI_ISL_1616916, EPI_ISL_1616918, EPI_ISL_1616919, EPI_ISL_1616921, EPI_ISL_1616922, EPI_ISL_1616924, EPI_ISL_1616928, EPI_ISL_1616929, EPI_ISL_1616930, EPI_ISL_1616934, EPI_ISL_1616936, EPI_ISL_1616938, EPI_ISL_1616941, EPI_ISL_1616943, EPI_ISL_1616944, EPI_ISL_1616945, EPI_ISL_1616948, EPI_ISL_1616949, EPI_ISL_1616951, EPI_ISL_1616952, EPI_ISL_1616953, EPI_ISL_1616956, EPI_ISL_1616957, EPI_ISL_1616958, EPI_ISL_1616959, EPI_ISL_1616961, EPI_ISL_1616962, EPI_ISL_1616963, EPI_ISL_1616964, EPI_ISL_1616966, EPI_ISL_1616968, EPI_ISL_1616971, EPI_ISL_1616972, EPI_ISL_1616977, EPI_ISL_1616979, EPI_ISL_1616980, EPI_ISL_1616986, EPI_ISL_1616988, EPI_ISL_1616989, EPI_ISL_1616991, EPI_ISL_1616992, EPI_ISL_1616994, EPI_ISL_1616997, EPI_ISL_1616999, EPI_ISL_1617003, EPI_ISL_1617004, EPI_ISL_1617008, EPI_ISL_1617009, EPI_ISL_1617010, EPI_ISL_1617015, EPI_ISL_1617016, EPI_ISL_1617020, EPI_ISL_1617021, EPI_ISL_1617022, EPI_ISL_1617023, EPI_ISL_1617024, EPI_ISL_1617025, EPI_ISL_1617027, EPI_ISL_1617029, EPI_ISL_1617034, EPI_ISL_1617039, EPI_ISL_1617047, EPI_ISL_1617049, EPI_ISL_1617053, EPI_ISL_1617056, EPI_ISL_1617061, EPI_ISL_1617064, EPI_ISL_1617065, EPI_ISL_1617070, EPI_ISL_1617071, EPI_ISL_1617072, EPI_ISL_1617079, EPI_ISL_1617081, EPI_ISL_1617082, EPI_ISL_1617083, EPI_ISL_1617089, EPI_ISL_1617090, EPI_ISL_1617091, EPI_ISL_1617092, EPI_ISL_1617093, EPI_ISL_1617095, EPI_ISL_1617102, EPI_ISL_1617104, EPI_ISL_1617108, EPI_ISL_1617110, EPI_ISL_1617115, EPI_ISL_1617116, EPI_ISL_1617117, EPI_ISL_1617118, EPI_ISL_1617120, EPI_ISL_1617121, EPI_ISL_1617122, EPI_ISL_1617123, EPI_ISL_1617125, EPI_ISL_1617126, EPI_ISL_1617130, EPI_ISL_1617131, EPI_ISL_1617133, EPI_ISL_1617134, EPI_ISL_1617136, EPI_ISL_1617137, EPI_ISL_1617140, EPI_ISL_1617141, EPI_ISL_1617143, EPI_ISL_1617144, EPI_ISL_1617145, EPI_ISL_1617147, EPI_ISL_1617152, EPI_ISL_1617153, EPI_ISL_1617154, EPI_ISL_1617156, EPI_ISL_1617157, EPI_ISL_1617159, EPI_ISL_1617161, EPI_ISL_1617164, EPI_ISL_1617170, EPI_ISL_1617171, EPI_ISL_1617172, EPI_ISL_1617173, EPI_ISL_1617176, EPI_ISL_1617177, EPI_ISL_1617178, EPI_ISL_1617179, EPI_ISL_1617181, EPI_ISL_1617182, EPI_ISL_1617184, EPI_ISL_1617185, EPI_ISL_1617186, EPI_ISL_1617187, EPI_ISL_1617191, EPI_ISL_1617197, EPI_ISL_1617199, EPI_ISL_1617202, EPI_ISL_1617203, EPI_ISL_1617204, EPI_ISL_1617205, EPI_ISL_1617206, EPI_ISL_1617207, EPI_ISL_1617209, EPI_ISL_1617211, EPI_ISL_1617215, EPI_ISL_1617217, EPI_ISL_1617218, EPI_ISL_1617225, EPI_ISL_1617235, EPI_ISL_1617236, EPI_ISL_1617237, EPI_ISL_1617238, EPI_ISL_1617239, EPI_ISL_1617242, EPI_ISL_1617243, EPI_ISL_1617246, EPI_ISL_1617247, EPI_ISL_1617248, EPI_ISL_1617252, EPI_ISL_1617253, EPI_ISL_1617254, EPI_ISL_1617256, EPI_ISL_1617263, EPI_ISL_1617265, EPI_ISL_1617266, EPI_ISL_1617267, EPI_ISL_1617268, EPI_ISL_1617269, EPI_ISL_1617270, EPI_ISL_1617272, EPI_ISL_1617274, EPI_ISL_1617275, EPI_ISL_1617276, EPI_ISL_1617279, EPI_ISL_1617284, EPI_ISL_1617285, EPI_ISL_1617286, EPI_ISL_1617288, EPI_ISL_1617291, EPI_ISL_1617292, EPI_ISL_1617293, EPI_ISL_1617294, EPI_ISL_1617295, EPI_ISL_1617297, EPI_ISL_1617298, EPI_ISL_1617299, EPI_ISL_1617304, EPI_ISL_1617305, EPI_ISL_1617306, EPI_ISL_1617308, EPI_ISL_1617309, EPI_ISL_1617310, EPI_ISL_1617311, EPI_ISL_1617313, EPI_ISL_1617314, EPI_ISL_1617315, EPI_ISL_1617317, EPI_ISL_1617318, EPI_ISL_1617321, EPI_ISL_1617324, EPI_ISL_1617325, EPI_ISL_1617326, EPI_ISL_1617327, EPI_ISL_1617328, EPI_ISL_1617329, EPI_ISL_1617331, EPI_ISL_1617332, EPI_ISL_1617333, EPI_ISL_1617335, EPI_ISL_1617336 | | | |
| see above | Colorado Department of Public Health and Environment | Colorado Department of Public Health and Environment | Laura Bankers, Molly C. Hetherington-Rauth, Diana Ir, Shannon Ely, Shannon R. Matzinger, Sarah Elizabeth Totten, Emily A. Travanty |
| EPI_ISL_1620239, EPI_ISL_1620240, EPI_ISL_1620241, EPI_ISL_1620242, EPI_ISL_1620243, EPI_ISL_1620244, EPI_ISL_1620245, EPI_ISL_1620246, EPI_ISL_1620247, EPI_ISL_1620248, EPI_ISL_1620249, EPI_ISL_1620250, EPI_ISL_1620251, EPI_ISL_1620252, EPI_ISL_1620253, EPI_ISL_1620254, EPI_ISL_1620261, EPI_ISL_1620262, EPI_ISL_1620263, EPI_ISL_1620264, EPI_ISL_1620265, EPI_ISL_1620266, EPI_ISL_1620267, EPI_ISL_1620268, EPI_ISL_1620269, EPI_ISL_1620270, EPI_ISL_1620271, EPI_ISL_1620272, EPI_ISL_1620273, EPI_ISL_1620274, EPI_ISL_1620275, EPI_ISL_1620276, EPI_ISL_1620277, EPI_ISL_1620278, EPI_ISL_1620279, EPI_ISL_1620280, EPI_ISL_1620281, EPI_ISL_1620282, EPI_ISL_1620283, EPI_ISL_1620284, EPI_ISL_1620285, EPI_ISL_1620286, EPI_ISL_1620287, EPI_ISL_1620288, EPI_ISL_1620289, EPI_ISL_1620290, EPI_ISL_1620291, EPI_ISL_1620292, EPI_ISL_1620293, EPI_ISL_1620294, EPI_ISL_1620295, EPI_ISL_1620296, EPI_ISL_1620297, EPI_ISL_1620298, EPI_ISL_1620299, EPI_ISL_1620300, EPI_ISL_1620301, EPI_ISL_1620302, EPI_ISL_1620303, EPI_ISL_1620304, EPI_ISL_1620305, EPI_ISL_1620306, EPI_ISL_1620307, EPI_ISL_1620308, EPI_ISL_1620309, EPI_ISL_1620310, EPI_ISL_1620311, EPI_ISL_1620312, EPI_ISL_1620313, EPI_ISL_1620314, EPI_ISL_1620315, EPI_ISL_1620316, EPI_ISL_1620317, EPI_ISL_1620318, EPI_ISL_1620319, EPI_ISL_1620320, EPI_ISL_1620321, EPI_ISL_1620322, EPI_ISL_1620323, EPI_ISL_1620324, EPI_ISL_1620325, EPI_ISL_1620326, EPI_ISL_1620327, EPI_ISL_1620328, EPI_ISL_1620329, EPI_ISL_1620330, EPI_ISL_1620331, EPI_ISL_1620332 | | | |
| see above | TXDSHS | TXDSHS | Rashmi Tuladhar, Bonnie Oh, Jenny Zhang, Maliha Rahman, Mayela Pedrueza, Anita Pokharel, Lorraine Rodriguez, Myong Koag, Chun Wang, Rachel Lee, Grace Kubin |
| EPI_ISL_1620455 | Stanford Clinical Virology Laboratory | Santa Clara County Public Health Laboratory | Santa Clara County Public Health Department |
| EPI_ISL_1620456 | Santa Clara Valley Medical Center | Santa Clara County Public Health Laboratory | Santa Clara County Public Health Department |
| EPI_ISL_1620457, EPI_ISL_1620458, EPI_ISL_1620459, EPI_ISL_1620462, EPI_ISL_1620463, EPI_ISL_1620464, EPI_ISL_1620465 | Stanford Clinical Virology Laboratory | Santa Clara County Public Health Laboratory | Santa Clara County Public Health Department |
| EPI_ISL_1620533, EPI_ISL_1620534 | Inno Diagnostics/Center for Research Resources, Ponce Medical School Foundation, Inc. | Centers for Disease Control and Prevention, Dengue Branch | Gilberto A. Santiago, Glenda Gonzalez, Betzabel Flores, Keyla Charriez, Andrea Arias-Garcia, Raphael Sanchez-Torres, Vanessa Rivera-Amill, Gabriela Paz-Bailey, Jorge L. Munoz-Jordan |
| EPI_ISL_1620535, EPI_ISL_1620536, EPI_ISL_1620537 | Puerto Rico Department of Health | Centers for Disease Control and Prevention, Dengue Branch | Gilberto A. Santiago, Glenda Gonzalez, Betzabel Flores, Keyla Charriez, Gabriela Paz-Bailey, Jorge L. Munoz-Jordan |
| EPI_ISL_1620538, EPI_ISL_1620539, EPI_ISL_1620540, EPI_ISL_1620541, EPI_ISL_1620542, EPI_ISL_1620543, EPI_ISL_1620544, EPI_ISL_1620545 | Inno Diagnostics/Center for Research Resources, Ponce Medical School Foundation, Inc. | Centers for Disease Control and Prevention, Dengue Branch | Gilberto A. Santiago, Glenda Gonzalez, Betzabel Flores, Keyla Charriez, Andrea Arias-Garcia, Raphael Sanchez-Torres, Vanessa Rivera-Amill, Gabriela Paz-Bailey, Jorge L. Munoz-Jordan |
| EPI_ISL_1620546, EPI_ISL_1620547, EPI_ISL_1620548, EPI_ISL_1620549, EPI_ISL_1620550, EPI_ISL_1620551, EPI_ISL_1620552, EPI_ISL_1620553, EPI_ISL_1620554 | Puerto Rico Department of Health | Centers for Disease Control and Prevention, Dengue Branch | Gilberto A. Santiago, Glenda Gonzalez, Betzabel Flores, Keyla Charriez, Gabriela Paz-Bailey, Jorge L. Munoz-Jordan |
| EPI_ISL_1620555, EPI_ISL_1620556, EPI_ISL_1620557, EPI_ISL_1620558, EPI_ISL_1620559, EPI_ISL_1620560, EPI_ISL_1620561, EPI_ISL_1620562, EPI_ISL_1620563, EPI_ISL_1620564, EPI_ISL_1620565, EPI_ISL_1620566, EPI_ISL_1620567, EPI_ISL_1620568, EPI_ISL_1620569, EPI_ISL_1620570, EPI_ISL_1620571, EPI_ISL_1620572, EPI_ISL_1620573 | | | |
| see above | Inno Diagnostics/Center for Research Resources, Ponce Medical School Foundation, Inc. | Centers for Disease Control and Prevention, Dengue Branch | Gilberto A. Santiago, Glenda Gonzalez, Betzabel Flores, Keyla Charriez, Andrea Arias-Garcia, Raphael Sanchez-Torres, Vanessa Rivera-Amill, Gabriela Paz-Bailey, Jorge L. Munoz-Jordan |
| EPI_ISL_1620574, EPI_ISL_1620575, EPI_ISL_1620576, EPI_ISL_1620577, EPI_ISL_1620578, EPI_ISL_1620579, EPI_ISL_1620580 | Puerto Rico Department of Health | Centers for Disease Control and Prevention, Dengue Branch | Gilberto A. Santiago, Glenda Gonzalez, Betzabel Flores, Keyla Charriez, Gabriela Paz-Bailey, Jorge L. Munoz-Jordan |
| EPI_ISL_1620581, EPI_ISL_1620582, EPI_ISL_1620583, EPI_ISL_1620584, EPI_ISL_1620585, EPI_ISL_1620586, EPI_ISL_1620587, EPI_ISL_1620588, EPI_ISL_1620589, EPI_ISL_1620590, EPI_ISL_1620591, EPI_ISL_1620592 | | | |
| see above | Inno Diagnostics/Center for Research Resources, Ponce Medical School Foundation, Inc. | Centers for Disease Control and Prevention, Dengue Branch | Gilberto A. Santiago, Glenda Gonzalez, Betzabel Flores, Keyla Charriez, Andrea Arias-Garcia, Raphael Sanchez-Torres, Vanessa Rivera-Amill, Gabriela Paz-Bailey, Jorge L. Munoz-Jordan |
| EPI_ISL_1620593, EPI_ISL_1620594, EPI_ISL_1620595, EPI_ISL_1620596, EPI_ISL_1620597, EPI_ISL_1620598, EPI_ISL_1620599, EPI_ISL_1620600, EPI_ISL_1620601, EPI_ISL_1620602, EPI_ISL_1620603, EPI_ISL_1620604, EPI_ISL_1620605, EPI_ISL_1620606, EPI_ISL_1620607 | | | |
| see above | Puerto Rico Department of Health | Centers for Disease Control and Prevention, Dengue Branch | Gilberto A. Santiago, Glenda Gonzalez, Betzabel Flores, Keyla Charriez, Gabriela Paz-Bailey, Jorge L. Munoz-Jordan |
| EPI_ISL_1620608 | Inno Diagnostics/Center for Research Resources, Ponce Medical School Foundation, Inc. | Centers for Disease Control and Prevention, Dengue Branch | Gilberto A. Santiago, Glenda Gonzalez, Betzabel Flores, Keyla Charriez, Andrea Arias-Garcia, Raphael Sanchez-Torres, Vanessa Rivera-Amill, Gabriela Paz-Bailey, Jorge L. Munoz-Jordan |
| EPI_ISL_1620609, EPI_ISL_1620610, EPI_ISL_1620611, EPI_ISL_1620612, EPI_ISL_1620613, EPI_ISL_1620614, EPI_ISL_1620615, EPI_ISL_1620616, EPI_ISL_1620617, EPI_ISL_1620618, EPI_ISL_1620619, EPI_ISL_1620620, EPI_ISL_1620621, EPI_ISL_1620622, EPI_ISL_1620623, EPI_ISL_1620624, EPI_ISL_1620625, EPI_ISL_1620626, EPI_ISL_1620627 | | | |
| see above | Puerto Rico Department of Health | Centers for Disease Control and Prevention, Dengue Branch | Gilberto A. Santiago, Glenda Gonzalez, Betzabel Flores, Keyla Charriez, Gabriela Paz-Bailey, Jorge L. Munoz-Jordan |
| EPI_ISL_1620629, EPI_ISL_1620630, EPI_ISL_1620631, EPI_ISL_1620632, EPI_ISL_1620633, EPI_ISL_1620634, EPI_ISL_1620635, | Nevada State Public Health Laboratory | Nevada State Public Health Laboratory | Andrew Gorzalski, Mark Pandori |

| | | | |
|---|---|---|---|
| EPI_ISL_1620636, EPI_ISL_1620637 | | | |
| EPI_ISL_1620640, EPI_ISL_1620641, EPI_ISL_1620642, EPI_ISL_1620643, EPI_ISL_1620644, EPI_ISL_1620645, EPI_ISL_1620646, EPI_ISL_1620647, EPI_ISL_1620648, EPI_ISL_1620649, EPI_ISL_1620650, EPI_ISL_1620651, EPI_ISL_1620652, EPI_ISL_1620653, EPI_ISL_1620654, EPI_ISL_1620655, EPI_ISL_1620656, EPI_ISL_1620657, EPI_ISL_1620658, EPI_ISL_1620659, EPI_ISL_1620660, EPI_ISL_1620661 | | | |
| see above | GA Department of Public Health | GA Department of Public Health | Stacy Reeves, Jonathan Edwards, Cynthia Dixey, Tonia Parrott, Aliyah Fields, Taylor Smith |
| EPI_ISL_1620672, EPI_ISL_1620673, EPI_ISL_1620674, EPI_ISL_1620675, EPI_ISL_1620677, EPI_ISL_1620678, EPI_ISL_1620679, EPI_ISL_1620680, EPI_ISL_1620681, EPI_ISL_1620682, EPI_ISL_1620685, EPI_ISL_1620686, EPI_ISL_1620688, EPI_ISL_1620690, EPI_ISL_1620691, EPI_ISL_1620692, EPI_ISL_1620693, EPI_ISL_1620694, EPI_ISL_1620698, EPI_ISL_1620699, EPI_ISL_1620700 | | | |
| see above | North Dakota Department of Health, Public Health Laboratory | North Dakota Department of Health, Public Health Laboratory | Lisa Wingerter |
| EPI_ISL_1620808, EPI_ISL_1620809, EPI_ISL_1620810, EPI_ISL_1620811, EPI_ISL_1620812, EPI_ISL_1620813, EPI_ISL_1620814, EPI_ISL_1620815, EPI_ISL_1620816, EPI_ISL_1620817, EPI_ISL_1620818, EPI_ISL_1620819, EPI_ISL_1620820, EPI_ISL_1620821, EPI_ISL_1620822, EPI_ISL_1620823, EPI_ISL_1620824, EPI_ISL_1620825, EPI_ISL_1620826, EPI_ISL_1620827, EPI_ISL_1620828, EPI_ISL_1620829, EPI_ISL_1620830, EPI_ISL_1620831, EPI_ISL_1620832, EPI_ISL_1620833, EPI_ISL_1620834, EPI_ISL_1620835, EPI_ISL_1620836, EPI_ISL_1620837, EPI_ISL_1620838, EPI_ISL_1620839, EPI_ISL_1620840, EPI_ISL_1620841, EPI_ISL_1620842, EPI_ISL_1620843, EPI_ISL_1620844, EPI_ISL_1620845, EPI_ISL_1620846, EPI_ISL_1620847, EPI_ISL_1620848, EPI_ISL_1620849, EPI_ISL_1620850, EPI_ISL_1620851, EPI_ISL_1620852, EPI_ISL_1620853, EPI_ISL_1620854, EPI_ISL_1620855, EPI_ISL_1620856, EPI_ISL_1620857, EPI_ISL_1620858, EPI_ISL_1620859, EPI_ISL_1620860, EPI_ISL_1620861, EPI_ISL_1620862, EPI_ISL_1620863, EPI_ISL_1620864, EPI_ISL_1620865, EPI_ISL_1620866, EPI_ISL_1620867, EPI_ISL_1620868, EPI_ISL_1620869, EPI_ISL_1620870, EPI_ISL_1620871, EPI_ISL_1620872, EPI_ISL_1620873, EPI_ISL_1620874, EPI_ISL_1620875, EPI_ISL_1620876, EPI_ISL_1620877, EPI_ISL_1620878, EPI_ISL_1620879, EPI_ISL_1620880, EPI_ISL_1620881, EPI_ISL_1620882, EPI_ISL_1620883, EPI_ISL_1620884, EPI_ISL_1620885, EPI_ISL_1620886, EPI_ISL_1620887, EPI_ISL_1620888, EPI_ISL_1620889, EPI_ISL_1620890, EPI_ISL_1620891, EPI_ISL_1620892, EPI_ISL_1620893, EPI_ISL_1620894, EPI_ISL_1620895, EPI_ISL_1620896, EPI_ISL_1620897, EPI_ISL_1620898, EPI_ISL_1620899, EPI_ISL_1620900, EPI_ISL_1620901, EPI_ISL_1620902, EPI_ISL_1620903, EPI_ISL_1620904, EPI_ISL_1620905, EPI_ISL_1620906, EPI_ISL_1620907, EPI_ISL_1620908, EPI_ISL_1620909, EPI_ISL_1620910, EPI_ISL_1620911, EPI_ISL_1620912, EPI_ISL_1620913, EPI_ISL_1620914, EPI_ISL_1620915, EPI_ISL_1620916, EPI_ISL_1620917, EPI_ISL_1620918, EPI_ISL_1620919, EPI_ISL_1620920, EPI_ISL_1620921, EPI_ISL_1620922, EPI_ISL_1620923, EPI_ISL_1620924, EPI_ISL_1620925, EPI_ISL_1620926, EPI_ISL_1620927, EPI_ISL_1620928, EPI_ISL_1620929, EPI_ISL_1620930, EPI_ISL_1620931, EPI_ISL_1620932, EPI_ISL_1620933, EPI_ISL_1620934, EPI_ISL_1620935, EPI_ISL_1620936, EPI_ISL_1620937, EPI_ISL_1620938, EPI_ISL_1620939, EPI_ISL_1620940, EPI_ISL_1620941, EPI_ISL_1620942, EPI_ISL_1620943, EPI_ISL_1620954, EPI_ISL_1620955, EPI_ISL_1620956, EPI_ISL_1620957, EPI_ISL_1620958, EPI_ISL_1620959, EPI_ISL_1620960, EPI_ISL_1620961, EPI_ISL_1620962, EPI_ISL_1620963, EPI_ISL_1620964, EPI_ISL_1620965, EPI_ISL_1620966, EPI_ISL_1620967, EPI_ISL_1620968, EPI_ISL_1620969, EPI_ISL_1620970, EPI_ISL_1620971, EPI_ISL_1620972, EPI_ISL_1620973, EPI_ISL_1620974, EPI_ISL_1620975, EPI_ISL_1620976, EPI_ISL_1620977, EPI_ISL_1620978, EPI_ISL_1620979, EPI_ISL_1620980, EPI_ISL_1620981, EPI_ISL_1620982, EPI_ISL_1620983, EPI_ISL_1620984, EPI_ISL_1620985, EPI_ISL_1620986, EPI_ISL_1620987, EPI_ISL_1620988, EPI_ISL_1620989, EPI_ISL_1620990, EPI_ISL_1620991, EPI_ISL_1620992, EPI_ISL_1620993, EPI_ISL_1620994, EPI_ISL_1620995, EPI_ISL_1620996, EPI_ISL_1620997, EPI_ISL_1620998, EPI_ISL_1620999, EPI_ISL_1621000, EPI_ISL_1621001, EPI_ISL_1621002, EPI_ISL_1621003, EPI_ISL_1621004, EPI_ISL_1621005, EPI_ISL_1621006, EPI_ISL_1621007, EPI_ISL_1621008, EPI_ISL_1621009, EPI_ISL_1621010, EPI_ISL_1621011, EPI_ISL_1621012, EPI_ISL_1621013, EPI_ISL_1621014, EPI_ISL_1621015, EPI_ISL_1621016, EPI_ISL_1621017, EPI_ISL_1621018, EPI_ISL_1621019, EPI_ISL_1621020, EPI_ISL_1621021, EPI_ISL_1621022, EPI_ISL_1621023, EPI_ISL_1621024, EPI_ISL_1621025, EPI_ISL_1621026, EPI_ISL_1621027, EPI_ISL_1621028, EPI_ISL_1621029, EPI_ISL_1621030, EPI_ISL_1621031, EPI_ISL_1621032, EPI_ISL_1621033, EPI_ISL_1621034, EPI_ISL_1621035, EPI_ISL_1621036, EPI_ISL_1621037, EPI_ISL_1621038, EPI_ISL_1621039, EPI_ISL_1621040, EPI_ISL_1621041, EPI_ISL_1621042, EPI_ISL_1621043, EPI_ISL_1621044, EPI_ISL_1621045, EPI_ISL_1621046, EPI_ISL_1621047, EPI_ISL_1621048, EPI_ISL_1621049, EPI_ISL_1621050, EPI_ISL_1621051, EPI_ISL_1621052, EPI_ISL_1621053, EPI_ISL_1621054, EPI_ISL_1621055, EPI_ISL_1621056, EPI_ISL_1621057, EPI_ISL_1621058, EPI_ISL_1621059, EPI_ISL_1621060, EPI_ISL_1621061, EPI_ISL_1621062, EPI_ISL_1621063, EPI_ISL_1621064, EPI_ISL_1621065, EPI_ISL_1621066, EPI_ISL_1621067, EPI_ISL_1621068, EPI_ISL_1621069, EPI_ISL_1621070, EPI_ISL_1621071, EPI_ISL_1621072, EPI_ISL_1621073, EPI_ISL_1621074, EPI_ISL_1621075, EPI_ISL_1621076, EPI_ISL_1621077, EPI_ISL_1621078, EPI_ISL_1621079, EPI_ISL_1621080, EPI_ISL_1621081, EPI_ISL_1621082, EPI_ISL_1621083, EPI_ISL_1621084, EPI_ISL_1621085, EPI_ISL_1621086, EPI_ISL_1621087, EPI_ISL_1621088, EPI_ISL_1621089, EPI_ISL_1621090, EPI_ISL_1621091, EPI_ISL_1621092, EPI_ISL_1621093, EPI_ISL_1621094, EPI_ISL_1621095, EPI_ISL_1621096, EPI_ISL_1621097, EPI_ISL_1621098, EPI_ISL_1621099, EPI_ISL_1621100, EPI_ISL_1621101, EPI_ISL_1621102, EPI_ISL_1621103, EPI_ISL_1621104, EPI_ISL_1621105, EPI_ISL_1621106, EPI_ISL_1621107, EPI_ISL_1621108, EPI_ISL_1621109, EPI_ISL_1621110, EPI_ISL_1621111, EPI_ISL_1621112, EPI_ISL_1621113, EPI_ISL_1621114, EPI_ISL_1621115, EPI_ISL_1621116, EPI_ISL_1621117, EPI_ISL_1621118, EPI_ISL_1621119, EPI_ISL_1621120, EPI_ISL_1621121, EPI_ISL_1621122, EPI_ISL_1621123, EPI_ISL_1621124, EPI_ISL_1621125, EPI_ISL_1621126, EPI_ISL_1621127, EPI_ISL_1621128, EPI_ISL_1621129, EPI_ISL_1621130, EPI_ISL_1621131, EPI_ISL_1621132, EPI_ISL_1621133, EPI_ISL_1621134, EPI_ISL_1621135, EPI_ISL_1621136, EPI_ISL_1621137, EPI_ISL_1621138, EPI_ISL_1621139, EPI_ISL_1621140, EPI_ISL_1621141, EPI_ISL_1621142, EPI_ISL_1621143, EPI_ISL_1621144, EPI_ISL_1621145, EPI_ISL_1621146, EPI_ISL_1621147, EPI_ISL_1621148, EPI_ISL_1621149, EPI_ISL_1621150, EPI_ISL_1621151, EPI_ISL_1621152, EPI_ISL_1621153, EPI_ISL_1621154, EPI_ISL_1621155, EPI_ISL_1621156 | | | |
| see above | UW Virology Lab | UW Virology Lab | Pavitra Roychoudhury, Hong Xie, Lasata Shrestha, Shah Mohamed Bakhash, Michelle Lin, Noah R. Baker, Sean Ellis, Saraswathi Sathees, Meei-Li Huang, Keith R Jerome, Alexander Greninger |
| EPI_ISL_1621157, EPI_ISL_1621158 | Nebraska Public Health Laboratory | NPHL COVID-19 Response Team | NPHL COVID-19 Response Team |
| EPI_ISL_1621159, EPI_ISL_1621160, EPI_ISL_1621161, EPI_ISL_1621162, EPI_ISL_1621163, EPI_ISL_1621164, EPI_ISL_1621165, EPI_ISL_1621166, EPI_ISL_1621167, EPI_ISL_1621168, EPI_ISL_1621169, EPI_ISL_1621170, EPI_ISL_1621171, EPI_ISL_1621172, EPI_ISL_1621173, EPI_ISL_1621174, EPI_ISL_1621175, EPI_ISL_1621176, EPI_ISL_1621177, EPI_ISL_1621178, EPI_ISL_1621179, EPI_ISL_1621180, EPI_ISL_1621181, EPI_ISL_1621182, EPI_ISL_1621183, EPI_ISL_1621184, EPI_ISL_1621185, EPI_ISL_1621186, EPI_ISL_1621187, EPI_ISL_1621188, EPI_ISL_1621189, EPI_ISL_1621190, EPI_ISL_1621191, EPI_ISL_1621192, EPI_ISL_1621193, EPI_ISL_1621194, EPI_ISL_1621195, EPI_ISL_1621196 | | | |
| see above | Alaska State Virology Laboratory | Alaska State Virology Laboratory | Stephanie DeRonde, Elva House, Lisa Smith, Ph.D., Jack Chen, Ph.D. |
| EPI_ISL_1621225, EPI_ISL_1621226, EPI_ISL_1621227, EPI_ISL_1621228, EPI_ISL_1621229, EPI_ISL_1621231, EPI_ISL_1621232, EPI_ISL_1621233, EPI_ISL_1621234, EPI_ISL_1621236, EPI_ISL_1621237, EPI_ISL_1621238, EPI_ISL_1621239, EPI_ISL_1621240, EPI_ISL_1621243, EPI_ISL_1621244, EPI_ISL_1621245, EPI_ISL_1621247, EPI_ISL_1621248, EPI_ISL_1621249, EPI_ISL_1621250, EPI_ISL_1621251, EPI_ISL_1621252, EPI_ISL_1621254, EPI_ISL_1621255, EPI_ISL_1621256, EPI_ISL_1621257, EPI_ISL_1621263, EPI_ISL_1621264, EPI_ISL_1621265, EPI_ISL_1621266, EPI_ISL_1621267, EPI_ISL_1621268, EPI_ISL_1621269, EPI_ISL_1621270, EPI_ISL_1621271, EPI_ISL_1621272, EPI_ISL_1621273, EPI_ISL_1621275, EPI_ISL_1621276, EPI_ISL_1621277, EPI_ISL_1621279, EPI_ISL_1621280, EPI_ISL_1621281, EPI_ISL_1621282, EPI_ISL_1621283, EPI_ISL_1621284, EPI_ISL_1621285, EPI_ISL_1621286, EPI_ISL_1621287, EPI_ISL_1621288, EPI_ISL_1621289, EPI_ISL_1621290 | | | |
| see above | Texas Children's Hospital | Texas Children's Microbiome Center | Ruth Ann Luna, Jennifer K. Spinler, James Dunn, James Versalovic, Ila Singh |
| EPI_ISL_1621327, EPI_ISL_1621328, EPI_ISL_1621329, EPI_ISL_1621330, EPI_ISL_1621331, EPI_ISL_1621332, EPI_ISL_1621333, EPI_ISL_1621334, EPI_ISL_1621335, EPI_ISL_1621336, EPI_ISL_1621337, EPI_ISL_1621338, EPI_ISL_1621339, EPI_ISL_1621340, EPI_ISL_1621341, EPI_ISL_1621342, EPI_ISL_1621343, EPI_ISL_1621344, EPI_ISL_1621345, EPI_ISL_1621346, EPI_ISL_1621347, EPI_ISL_1621348, EPI_ISL_1621349, EPI_ISL_1621350, EPI_ISL_1621351, EPI_ISL_1621352, EPI_ISL_1621353, EPI_ISL_1621354, EPI_ISL_1621355, EPI_ISL_1621356, EPI_ISL_1621357, EPI_ISL_1621358, EPI_ISL_1621359, EPI_ISL_1621360, EPI_ISL_1621361, EPI_ISL_1621362, EPI_ISL_1621363, EPI_ISL_1621364, EPI_ISL_1621365, EPI_ISL_1621366, EPI_ISL_1621367, EPI_ISL_1621368, EPI_ISL_1621369, EPI_ISL_1621370, EPI_ISL_1621371, EPI_ISL_1621372, EPI_ISL_1621373, EPI_ISL_1621374, EPI_ISL_1621375, EPI_ISL_1621376, EPI_ISL_1621377, EPI_ISL_1621378, EPI_ISL_1621379, EPI_ISL_1621380, EPI_ISL_1621381, EPI_ISL_1621382, EPI_ISL_1621383, EPI_ISL_1621384, EPI_ISL_1621385, EPI_ISL_1621386, EPI_ISL_1621387, EPI_ISL_1621388, EPI_ISL_1621389, EPI_ISL_1621390, EPI_ISL_1621391, EPI_ISL_1621392, EPI_ISL_1621393, EPI_ISL_1621394, EPI_ISL_1621395, EPI_ISL_1621396, EPI_ISL_1621397, EPI_ISL_1621398, EPI_ISL_1621399, EPI_ISL_1621400, EPI_ISL_1621401, EPI_ISL_1621402, EPI_ISL_1621403, EPI_ISL_1621404, EPI_ISL_1621405, EPI_ISL_1621406, EPI_ISL_1621407, EPI_ISL_1621408, EPI_ISL_1621409, EPI_ISL_1621410, EPI_ISL_1621411, EPI_ISL_1621412, EPI_ISL_1621413, EPI_ISL_1621414, EPI_ISL_1621415, EPI_ISL_1621416, EPI_ISL_1621417, EPI_ISL_1621418, EPI_ISL_1621419, EPI_ISL_1621420, EPI_ISL_1621421, EPI_ISL_1621422, EPI_ISL_1621423, EPI_ISL_1621424, EPI_ISL_1621425, EPI_ISL_1621426, EPI_ISL_1621427, EPI_ISL_1621428, EPI_ISL_1621429, EPI_ISL_1621430, EPI_ISL_1621431, EPI_ISL_1621432, EPI_ISL_1621433, EPI_ISL_1621434, EPI_ISL_1621435, EPI_ISL_1621436, EPI_ISL_1621437, EPI_ISL_1621438, EPI_ISL_1621439, EPI_ISL_1621440, EPI_ISL_1621441, EPI_ISL_1621442, EPI_ISL_1621443, EPI_ISL_1621444, EPI_ISL_1621445, EPI_ISL_1621446, EPI_ISL_1621447, EPI_ISL_1621448, EPI_ISL_1621449, EPI_ISL_1621450, EPI_ISL_1621451, EPI_ISL_1621452, EPI_ISL_1621453, EPI_ISL_1621454, EPI_ISL_1621455, EPI_ISL_1621456, EPI_ISL_1621457, EPI_ISL_1621458, EPI_ISL_1621459, EPI_ISL_1621460, EPI_ISL_1621461, EPI_ISL_1621462, EPI_ISL_1621463, EPI_ISL_1621464, EPI_ISL_1621465, EPI_ISL_1621466, EPI_ISL_1621467, EPI_ISL_1621468, EPI_ISL_1621469, EPI_ISL_1621470, EPI_ISL_1621471, EPI_ISL_1621472, EPI_ISL_1621473, EPI_ISL_1621474, EPI_ISL_1621475, EPI_ISL_1621476, EPI_ISL_1621477, EPI_ISL_1621478, EPI_ISL_1621479, EPI_ISL_1621480, EPI_ISL_1621481, EPI_ISL_1621482, EPI_ISL_1621483, EPI_ISL_1621484 | | | |
| see above | Hospital of the University of Pennsylvania Molecular Pathology Lab | Bushman Lab - University of Pennsylvania | John Everett, Kyle Rodino, Shantan Reddy, Pascha Hokama, Aoife M. Roche, Young Hwang, Abigail Glascock, Scott Sherrill-Mix, Samantha A. Whiteside, Jevon Graham-Wooten, Layla A. Khatib, Ayannah S. Fitzgerald, Arupa Ganguly, Mike Feldman, Brendan Kelly, Ronald G. Collman and Frederic Bushman |
| EPI_ISL_1621591, EPI_ISL_1621592, EPI_ISL_1621593, EPI_ISL_1621594, EPI_ISL_1621595, EPI_ISL_1621618, EPI_ISL_1621619, EPI_ISL_1621620, EPI_ISL_1621621, EPI_ISL_1621622, EPI_ISL_1621623 | | | |
| see above | Arizona State University | Arizona State University | Peter T. Skidmore, LaRinda A. Holland, Rabia Maqsood, Emily A. Kaelin, Nicholas J. Mellor, Kristina Buss, Joy M. Blain, Neal W Woodbury, Valerie Harris, Joshua LaBaer, Vel Murugan, Efrem S. Lim |
| EPI_ISL_1621723, EPI_ISL_1621726, EPI_ISL_1621729, EPI_ISL_1621732, EPI_ISL_1621735, EPI_ISL_1621738, EPI_ISL_1621741, EPI_ISL_1621744, EPI_ISL_1621747, EPI_ISL_1621749, EPI_ISL_1621752, EPI_ISL_1621755, EPI_ISL_1621758, EPI_ISL_1621761, EPI_ISL_1621763, EPI_ISL_1621767, EPI_ISL_1621769, EPI_ISL_1621772, EPI_ISL_1621775, EPI_ISL_1621777, EPI_ISL_1621780, EPI_ISL_1621783, EPI_ISL_1621786, EPI_ISL_1621789, EPI_ISL_1621792, EPI_ISL_1621795, EPI_ISL_1621798, EPI_ISL_1621801, EPI_ISL_1621803, EPI_ISL_1621807, EPI_ISL_1621809, EPI_ISL_1621812, EPI_ISL_1621815, EPI_ISL_1621818, EPI_ISL_1621821, EPI_ISL_1621824, EPI_ISL_1621827, EPI_ISL_1621830, EPI_ISL_1621833, EPI_ISL_1621836, EPI_ISL_1621839, EPI_ISL_1621842, EPI_ISL_1621844, EPI_ISL_1621847, EPI_ISL_1621850, EPI_ISL_1621851, EPI_ISL_1621852, EPI_ISL_1621853, EPI_ISL_1621854, EPI_ISL_1621855, EPI_ISL_1621856, EPI_ISL_1621857, EPI_ISL_1621858, EPI_ISL_1621859, EPI_ISL_1621860, EPI_ISL_1621861, EPI_ISL_1621862, EPI_ISL_1621863, EPI_ISL_1621864, EPI_ISL_1621865, EPI_ISL_1621866, EPI_ISL_1621867, EPI_ISL_1621868, EPI_ISL_1621869, EPI_ISL_1621870, EPI_ISL_1621871, EPI_ISL_1621872, EPI_ISL_1621873, EPI_ISL_1621874, EPI_ISL_1621875, EPI_ISL_1621876, EPI_ISL_1621877, EPI_ISL_1621878, EPI_ISL_1621879, EPI_ISL_1621880, EPI_ISL_1621881, EPI_ISL_1621882, EPI_ISL_1621883, EPI_ISL_1621884, EPI_ISL_1621885, EPI_ISL_1621886, EPI_ISL_1621887, EPI_ISL_1621888, EPI_ISL_1621889, EPI_ISL_1621890, EPI_ISL_1621891, EPI_ISL_1621892, EPI_ISL_1621893, EPI_ISL_1621894, EPI_ISL_1621895, EPI_ISL_1621896, EPI_ISL_1621897, EPI_ISL_1621898, EPI_ISL_1621899, EPI_ISL_1621900, EPI_ISL_1621901, EPI_ISL_1621902, EPI_ISL_1621903, EPI_ISL_1621904, EPI_ISL_1621905, EPI_ISL_1621906, EPI_ISL_1621907, EPI_ISL_1621908, EPI_ISL_1621909, EPI_ISL_1621910, EPI_ISL_1621911, EPI_ISL_1621912, EPI_ISL_1621913, EPI_ISL_1621914, EPI_ISL_1621915, EPI_ISL_1621916, EPI_ISL_1621917, EPI_ISL_1621918, EPI_ISL_1621919, EPI_ISL_1621920, EPI_ISL_1621921, EPI_ISL_1621922, EPI_ISL_1621923, EPI_ISL_1621924, EPI_ISL_1621925, EPI_ISL_1621926, EPI_ISL_1621927, EPI_ISL_1621928, EPI_ISL_1621929, EPI_ISL_1621930, EPI_ISL_1621931, EPI_ISL_1621932, EPI_ISL_1621933, EPI_ISL_1621934, EPI_ISL_1621935, EPI_ISL_1621936, EPI_ISL_1621937, EPI_ISL_1621938, EPI_ISL_1621939, EPI_ISL_1621940, EPI_ISL_1621941, EPI_ISL_1621942, EPI_ISL_1621943, EPI_ISL_1621944, EPI_ISL_1621945, EPI_ISL_1621946, EPI_ISL_1621947, EPI_ISL_1621948, EPI_ISL_1621949, EPI_ISL_1621950, EPI_ISL_1621951, EPI_ISL_1621952, EPI_ISL_1621953, EPI_ISL_1621954, EPI_ISL_1621955, EPI_ISL_1621956, EPI_ISL_1621957, EPI_ISL_1621958, EPI_ISL_1621959, EPI_ISL_1621960, EPI_ISL_1621961, EPI_ISL_1621962, EPI_ISL_1621963, EPI_ISL_1621964, EPI_ISL_1621965, EPI_ISL_1621966, EPI_ISL_1621967, EPI_ISL_1621968, EPI_ISL_1621969, EPI_ISL_1621970, EPI_ISL_1621971, EPI_ISL_1621972, EPI_ISL_1621973, EPI_ISL_1621974, EPI_ISL_1621975, EPI_ISL_1621976, EPI_ISL_1621977, EPI_ISL_1621978, EPI_ISL_1621979, EPI_ISL_1621980, EPI_ISL_1621981, EPI_ISL_1621982, EPI_ISL_1621983, EPI_ISL_1621984, EPI_ISL_1621985, EPI_ISL_1621986, EPI_ISL_1621987, EPI_ISL_1621988, EPI_ISL_1621989, EPI_ISL_1621990, EPI_ISL_1621991, EPI_ISL_1621992, EPI_ISL_1621993, EPI_ISL_1621994, EPI_ISL_1621995, EPI_ISL_1621996, EPI_ISL_1621997, EPI_ISL_1621998, EPI_ISL_1621999, EPI_ISL_1622000, EPI_ISL_1622001, EPI_ISL_1622002, EPI_ISL_1622003, EPI_ISL_1622004, EPI_ISL_1622005, EPI_ISL_1622006, EPI_ISL_1622007, EPI_ISL_1622008, EPI_ISL_1622009, EPI_ISL_1622010, EPI_ISL_1622011, EPI_ISL_1622012, EPI_ISL_1622013, EPI_ISL_1622014, EPI_ISL_1622015, EPI_ISL_1622016, EPI_ISL_1622017, EPI_ISL_1622018, EPI_ISL_1622019, EPI_ISL_1622020, EPI_ISL_1622021, EPI_ISL_1622022, EPI_ISL_1622023, EPI_ISL_1622024, EPI_ISL_1622025, EPI_ISL_1622026, EPI_ISL_1622027, EPI_ISL_1622028, EPI_ISL_1622029, EPI_ISL_1622030, EPI_ISL_1622031, EPI_ISL_1622032, EPI_ISL_1622033, EPI_ISL_1622034, EPI_ISL_1622035, EPI_ISL_1622036, EPI_ISL_1622037, EPI_ISL_1622038, EPI_ISL_1622039, EPI_ISL_1622040, EPI_ISL_1622041, EPI_ISL_1622042, EPI_ISL_1622043, EPI_ISL_1622044, EPI_ISL_1622045, EPI_ISL_1622046, EPI_ISL_1622047, EPI_ISL_1622048, EPI_ISL_1622049, EPI_ISL_1622050, EPI_ISL_1622051, EPI_ISL_1622052, EPI_ISL_1622053, EPI_ISL_1622054, EPI_ISL_1622055, EPI_ISL_1622056, EPI_ISL_1622057, EPI_ISL_1622058, EPI_ISL_1622059, EPI_ISL_1622060, EPI_ISL_1622061, EPI_ISL_1622062, EPI_ISL_1622063, EPI_ISL_1622064, EPI_ISL_1622065, EPI_ISL_1622066, EPI_ISL_1622067, EPI_ISL_1622068, EPI_ISL_1622069, EPI_ISL_1622070, EPI_ISL_1622071, EPI_ISL_1622072, EPI_ISL_1622073, EPI_ISL_1622074, EPI_ISL_1622075, EPI_ISL_1622076, EPI_ISL_1622077, EPI_ISL_1622078, EPI_ISL_1622079, EPI_ISL_1622080, EPI_ISL_1622081, EPI_ISL_1622082, EPI_ISL_1622083, EPI_ISL_1622084, EPI_ISL_1622085, EPI_ISL_1622086, EPI_ISL_1622087, EPI_ISL_1622088, EPI_ISL_1622089, EPI_ISL_1622090, EPI_ISL_1622091, EPI_ISL_1622092, EPI_ISL_1622093, EPI_ISL_1622094, EPI_ISL_1622095, EPI_ISL_1622096, EPI_ISL_1622097, EPI_ISL_1622098, EPI_ISL_1622099, EPI_ISL_1622100, EPI_ISL_1622101, EPI_ISL_1622102, EPI_ISL_1622103, EPI_ISL_1622104, EPI_ISL_1622105, EPI_ISL_1622106, EPI_ISL_1622107, EPI_ISL_1622108, EPI_ISL_1622109, EPI_ISL_1622110 | | | |
| see above | Quest Diagnostics Incorporated | Centers for Disease Control and Prevention Division of Viral Diseases, Pathogen Discovery | Dakota Howard, Dhwani Batra, Peter W. Cook, Kara Moser, Adrian Paskey, Jason Caravas, Benjamin Rambo-Martin, Shatavia Morrison, Christopher Gulvick, Scott Sammons, Yvette Unoarumhi, Darlene Wagner, Matthew Schmerer, S. H. Rosenthal, A. Gerasimova, R. M. Kagan, B. Anderson, M. Hua, Y. Liu, L.E. Bernstein, K.E. Livingston, A. Perez, I. A. Shlyakhter, R. V. Rolando, R. Owen, P. Tanpaiboon, F. Lacbawan, Clinton R. Paden, Duncan MacCannell |
| EPI_ISL_1622878, EPI_ISL_1622879, EPI_ISL_1622880, EPI_ISL_1622881, EPI_ISL_1622882, EPI_ISL_1622904, EPI_ISL_1622905, EPI_ISL_1622906 | Arizona State University | Arizona State University | Peter T. Skidmore, LaRinda A. Holland, Rabia Maqsood, Nicholas J. Mellor, Joy M. Blain, Valerie Harris, Joshua LaBaer, Vel Murugan, Efrem S. Lim |
| EPI_ISL_1623694, EPI_ISL_1623695, EPI_ISL_1623696, EPI_ISL_1623697, EPI_ISL_1623698, EPI_ISL_1623699, EPI_ISL_1623700, EPI_ISL_1623701, EPI_ISL_1623702, EPI_ISL_1623703, EPI_ISL_1623704, EPI_ISL_1623705, EPI_ISL_1623706, EPI_ISL_1623707, EPI_ISL_1623708, EPI_ISL_1623709, EPI_ISL_1623710 | | | |
| see above | Oregon State Public Health Laboratory | Oregon State Public Health Laboratory | Rafia Razzaque, Eugene Yeboah, Vanda Makris, Laura Tsaknaridis, John Fontana and Shane Sevey |
| EPI_ISL_1623711, EPI_ISL_1623712, EPI_ISL_1623713, EPI_ISL_1623714, EPI_ISL_1623715, EPI_ISL_1623716, EPI_ISL_1623717, EPI_ISL_1623718, EPI_ISL_1623719, EPI_ISL_1623720, EPI_ISL_1623721, EPI_ISL_1623722, EPI_ISL_1623723, EPI_ISL_1623724, EPI_ISL_1623725, EPI_ISL_1623726, EPI_ISL_1623727, EPI_ISL_1623728, EPI_ISL_1623729, EPI_ISL_1623730, EPI_ISL_1623731, EPI_ISL_1623732, EPI_ISL_1623733, EPI_ISL_1623734, EPI_ISL_1623735, EPI_ISL_1623736, EPI_ISL_1623737, EPI_ISL_1623738, EPI_ISL_1623739, EPI_ISL_1623740, EPI_ISL_1623741, EPI_ISL_1623742, EPI_ISL_1623743, EPI_ISL_1623744, EPI_ISL_1623745, EPI_ISL_1623748, EPI_ISL_1623749, EPI_ISL_1623750, EPI_ISL_1623751, EPI_ISL_1623752, EPI_ISL_1623753, EPI_ISL_1623754, EPI_ISL_1623755 | | | |
| see above | MD PHL | MD PHL | Maryland Department of Health Laboratories Administration |
| EPI_ISL_1623763, EPI_ISL_1623764, EPI_ISL_1623765, EPI_ISL_1623767, EPI_ISL_1623768, EPI_ISL_1623769, EPI_ISL_1623770, EPI_ISL_1623771, EPI_ISL_1623772, EPI_ISL_1623773, EPI_ISL_1623774, EPI_ISL_1623775, EPI_ISL_1623776, EPI_ISL_1623777, EPI_ISL_1623778, EPI_ISL_1623779, EPI_ISL_1623780, EPI_ISL_1623781, EPI_ISL_1623782, EPI_ISL_1623783, EPI_ISL_1623784, EPI_ISL_1623785, EPI_ISL_1623786, EPI_ISL_1623787, EPI_ISL_1623788, EPI_ISL_1623789, EPI_ISL_1623790, EPI_ISL_1623791, EPI_ISL_1623792, EPI_ISL_1623793, EPI_ISL_1623794, EPI_ISL_1623795, EPI_ISL_1623796, EPI_ISL_1623797, EPI_ISL_1623798, EPI_ISL_1623799, EPI_ISL_1623800, EPI_ISL_1623801, EPI_ISL_1623802, EPI_ISL_1623803, EPI_ISL_1623804, EPI_ISL_1623805, EPI_ISL_1623806, EPI_ISL_1623807, EPI_ISL_1623808, EPI_ISL_1623809, EPI_ISL_1623810, EPI_ISL_1623811, EPI_ISL_1623812, EPI_ISL_1623813, EPI_ISL_1623814, EPI_ISL_1623815, EPI_ISL_1623816, EPI_ISL_1623817, EPI_ISL_1623818, EPI_ISL_1623819, EPI_ISL_1623820, EPI_ISL_1623821, EPI_ISL_1623822, EPI_ISL_1623823, EPI_ISL_1623824, EPI_ISL_1623825, EPI_ISL_1623826, EPI_ISL_1623827, EPI_ISL_1623828, EPI_ISL_1623829, EPI_ISL_1623830, EPI_ISL_1623831 | | | |
| see above | Baylor Scott & White-Temple | Baylor Scott & White-Temple | Ari Rao, Linden Morales, Kimberly Walker, Marcus Volz, Shelby Hendrickson, Caitlin Maloney |
| EPI_ISL_1623832 | Baylor Scott & White-Grapevine | Baylor Scott & White-Temple | Ari Rao, Linden Morales, Kimberly Walker, Marcus Volz, Shelby Hendrickson, Caitlin Maloney |
| EPI_ISL_1623833, EPI_ISL_1623834, EPI_ISL_1623835 | Vanderbilt University Medical Center | Pathogen Discovery, Respiratory Viruses Branch, Division of Viral Diseases, Centers for Disease Control and Prevention | Yan Li, Ying Tao, Anna Kelleher, Jing Zhang, Anna Montmayeur, Brian Lynch, Krista Queen, Anna Uehara, Peter Cook, Han Jia Justin Ng, Rachel Marine, Clinton R. Paden, Haibin Wang, Mark Burroughs, Justin Lee, Adam Retchless, Suxiang Tong |
| EPI_ISL_1623836 | CDPH, Viral and Rickettsial Disease Laboratory | Pathogen Discovery, Respiratory Viruses Branch, Division of Viral Diseases, Centers for Disease Control and Prevention | Yan Li, Ying Tao, Anna Kelleher, Jing Zhang, Anna Montmayeur, Brian Lynch, Krista Queen, Anna Uehara, Peter Cook, Han Jia Justin Ng, Rachel Marine, Clinton R. Paden, Haibin Wang, Mark Burroughs, Justin Lee, Adam Retchless, Suxiang Tong |
| EPI_ISL_1623837 | Kaiser Permanente Washington Health Research Institute | Pathogen Discovery, Respiratory Viruses Branch, Division of Viral Diseases, Centers for Disease Control and Prevention | Yan Li, Ying Tao, Anna Kelleher, Jing Zhang, Anna Montmayeur, Brian Lynch, Krista Queen, Anna Uehara, Peter Cook, Han Jia Justin Ng, Rachel Marine, Clinton R. Paden, Haibin Wang, Mark Burroughs, Justin Lee, Adam Retchless, Suxiang Tong |
| EPI_ISL_1623917 | Arizona State University | Arizona State University | Peter T. Skidmore, LaRinda A. Holland, Rabia Maqsood, Emily A. Kaelin, Nicholas J. Mellor, Kristina Buss, Joy M. Blain, Neal W Woodbury, Valerie Harris, Joshua LaBaer, Vel Murugan, Efrem S. Lim |
| EPI_ISL_1623918 | NORTHWELL HEALTH LABORATORIES | Wadsworth Center, New York State Department of Health | Kirsten St. George, Daryl M. Lamson, Alexis Russell, Matthew Shudt, Melissa A Leisner, Jonathan Plitnick, Catharine Prussing, Navjot Singh, John Kelly, Erasmus Schneider, Erica Lasek-Nesselquist |
| EPI_ISL_1623919, EPI_ISL_1623920, EPI_ISL_1623921, EPI_ISL_1623922, EPI_ISL_1623923, EPI_ISL_1623924, EPI_ISL_1623925, EPI_ISL_1623926, EPI_ISL_1623927, EPI_ISL_1623928, EPI_ISL_1623929, EPI_ISL_1623930, EPI_ISL_1623931, EPI_ISL_1623932, EPI_ISL_1623933, EPI_ISL_1623934, EPI_ISL_1623935, EPI_ISL_1623936, EPI_ISL_1623937, EPI_ISL_1623938, EPI_ISL_1623939, EPI_ISL_1623940, EPI_ISL_1623941, EPI_ISL_1623942, EPI_ISL_1623943, EPI_ISL_1623944, EPI_ISL_1623945, EPI_ISL_1623946, EPI_ISL_1623947, EPI_ISL_1623948, EPI_ISL_1623949, EPI_ISL_1623950, EPI_ISL_1623951 | | | |
| see above | MEMORIAL SLOAN KETTERING CANCER | Wadsworth Center, New York State | Kirsten St. George, Daryl M. Lamson, Alexis Russell, Matthew Shudt, Melissa A Leisner, Jonathan Plitnick, Catharine Prussing, Navjot Singh, John Kelly, Erasmus Schneider, Erica Lasek-Nesselquist |

| | | | |
|---|---|---|---|
| | CENTER | Department of Health | |
| EPI_ISL_1623952 | Columbia University Irving Medical Center | Wadsworth Center, New York State Department of Health | Kirsten St. George, Daryl M. Lamson, Alexis Russell, Matthew Shudt, Melissa A Leisner, Jonathan Plitnick, Catharine Prussing, Navjot Singh, John Kelly, Erasmus Schneider, Erica Lasek-Nesselquist |
| EPI_ISL_1623953, EPI_ISL_1623954, EPI_ISL_1623955, EPI_ISL_1623956, EPI_ISL_1623957, EPI_ISL_1623958, EPI_ISL_1623959, EPI_ISL_1623960, EPI_ISL_1623961, EPI_ISL_1623962, EPI_ISL_1623963, EPI_ISL_1623964, EPI_ISL_1623965, EPI_ISL_1623966, EPI_ISL_1623967, EPI_ISL_1623968, EPI_ISL_1623969, EPI_ISL_1623970, EPI_ISL_1623971, EPI_ISL_1623972 | | | |
| see above | MONTEFIORE MEDICAL CENTER LABORATORIES | Wadsworth Center, New York State Department of Health | Kirsten St. George, Daryl M. Lamson, Alexis Russell, Matthew Shudt, Melissa A Leisner, Jonathan Plitnick, Catharine Prussing, Navjot Singh, John Kelly, Erasmus Schneider, Erica Lasek-Nesselquist |
| EPI_ISL_1623973, EPI_ISL_1623974 | NYC Pandemic Response Lab | Wadsworth Center, New York State Department of Health | Kirsten St. George, Daryl M. Lamson, Alexis Russell, Matthew Shudt, Melissa A Leisner, Jonathan Plitnick, Catharine Prussing, Navjot Singh, John Kelly, Erasmus Schneider, Erica Lasek-Nesselquist |
| EPI_ISL_1623975, EPI_ISL_1623976, EPI_ISL_1623977, EPI_ISL_1623978, EPI_ISL_1623979, EPI_ISL_1623980, EPI_ISL_1623981, EPI_ISL_1623982, EPI_ISL_1623983, EPI_ISL_1623984, EPI_ISL_1623985, EPI_ISL_1623986, EPI_ISL_1623987, EPI_ISL_1623988, EPI_ISL_1623989 | | | |
| see above | MONTEFIORE MEDICAL CENTER LABORATORIES | Wadsworth Center, New York State Department of Health | Kirsten St. George, Daryl M. Lamson, Alexis Russell, Matthew Shudt, Melissa A Leisner, Jonathan Plitnick, Catharine Prussing, Navjot Singh, John Kelly, Erasmus Schneider, Erica Lasek-Nesselquist |
| EPI_ISL_1623990, EPI_ISL_1623991, EPI_ISL_1623992, EPI_ISL_1623993, EPI_ISL_1623994, EPI_ISL_1623995, EPI_ISL_1623996, EPI_ISL_1623997, EPI_ISL_1623998, EPI_ISL_1623999, EPI_ISL_1624000, EPI_ISL_1624001, EPI_ISL_1624002 | | | |
| see above | MEMORIAL SLOAN KETTERING CANCER CENTER | Wadsworth Center, New York State Department of Health | Kirsten St. George, Daryl M. Lamson, Alexis Russell, Matthew Shudt, Melissa A Leisner, Jonathan Plitnick, Catharine Prussing, Navjot Singh, John Kelly, Erasmus Schneider, Erica Lasek-Nesselquist |
| EPI_ISL_1624003 | New York Presbyterian Hospital | Wadsworth Center, New York State Department of Health | Kirsten St. George, Daryl M. Lamson, Alexis Russell, Matthew Shudt, Melissa A Leisner, Jonathan Plitnick, Catharine Prussing, Navjot Singh, John Kelly, Erasmus Schneider, Erica Lasek-Nesselquist |
| EPI_ISL_1624004 | NORTHWELL HEALTH LABORATORIES | Wadsworth Center, New York State Department of Health | Kirsten St. George, Daryl M. Lamson, Alexis Russell, Matthew Shudt, Melissa A Leisner, Jonathan Plitnick, Catharine Prussing, Navjot Singh, John Kelly, Erasmus Schneider, Erica Lasek-Nesselquist |
| EPI_ISL_1624005, EPI_ISL_1624006, EPI_ISL_1624007, EPI_ISL_1624008, EPI_ISL_1624009 | MEMORIAL SLOAN KETTERING CANCER CENTER | Wadsworth Center, New York State Department of Health | Kirsten St. George, Daryl M. Lamson, Alexis Russell, Matthew Shudt, Melissa A Leisner, Jonathan Plitnick, Catharine Prussing, Navjot Singh, John Kelly, Erasmus Schneider, Erica Lasek-Nesselquist |
| EPI_ISL_1624010, EPI_ISL_1624011 | Columbia University Irving Medical Center | Wadsworth Center, New York State Department of Health | Kirsten St. George, Daryl M. Lamson, Alexis Russell, Matthew Shudt, Melissa A Leisner, Jonathan Plitnick, Catharine Prussing, Navjot Singh, John Kelly, Erasmus Schneider, Erica Lasek-Nesselquist |
| EPI_ISL_1624012, EPI_ISL_1624013, EPI_ISL_1624014, EPI_ISL_1624015, EPI_ISL_1624016, EPI_ISL_1624017, EPI_ISL_1624018, EPI_ISL_1624019, EPI_ISL_1624020, EPI_ISL_1624021, EPI_ISL_1624022, EPI_ISL_1624023, EPI_ISL_1624024, EPI_ISL_1624025, EPI_ISL_1624026, EPI_ISL_1624027, EPI_ISL_1624028, EPI_ISL_1624029, EPI_ISL_1624030, EPI_ISL_1624031, EPI_ISL_1624032, EPI_ISL_1624033, EPI_ISL_1624034, EPI_ISL_1624035, EPI_ISL_1624036, EPI_ISL_1624037, EPI_ISL_1624038, EPI_ISL_1624039, EPI_ISL_1624040, EPI_ISL_1624041, EPI_ISL_1624042, EPI_ISL_1624043, EPI_ISL_1624044, EPI_ISL_1624045, EPI_ISL_1624046, EPI_ISL_1624047, EPI_ISL_1624048, EPI_ISL_1624049, EPI_ISL_1624050, EPI_ISL_1624051, EPI_ISL_1624052, EPI_ISL_1624053, EPI_ISL_1624054, EPI_ISL_1624055, EPI_ISL_1624056, EPI_ISL_1624057, EPI_ISL_1624058, EPI_ISL_1624059, EPI_ISL_1624060, EPI_ISL_1624061, EPI_ISL_1624062, EPI_ISL_1624063, EPI_ISL_1624064, EPI_ISL_1624065, EPI_ISL_1624066, EPI_ISL_1624067, EPI_ISL_1624068, EPI_ISL_1624069, EPI_ISL_1624070, EPI_ISL_1624071, EPI_ISL_1624072, EPI_ISL_1624073, EPI_ISL_1624074, EPI_ISL_1624075, EPI_ISL_1624076, EPI_ISL_1624077, EPI_ISL_1624078, EPI_ISL_1624079, EPI_ISL_1624080, EPI_ISL_1624081, EPI_ISL_1624082 | | | |
| see above | New York Presbyterian Hospital | Wadsworth Center, New York State Department of Health | Kirsten St. George, Daryl M. Lamson, Alexis Russell, Matthew Shudt, Melissa A Leisner, Jonathan Plitnick, Catharine Prussing, Navjot Singh, John Kelly, Erasmus Schneider, Erica Lasek-Nesselquist |
| EPI_ISL_1624083, EPI_ISL_1624084, EPI_ISL_1624085, EPI_ISL_1624086, EPI_ISL_1624087, EPI_ISL_1624088 | NYC Pandemic Response Lab | Wadsworth Center, New York State Department of Health | Kirsten St. George, Daryl M. Lamson, Alexis Russell, Matthew Shudt, Melissa A Leisner, Jonathan Plitnick, Catharine Prussing, Navjot Singh, John Kelly, Erasmus Schneider, Erica Lasek-Nesselquist |
| EPI_ISL_1624089, EPI_ISL_1624090 | GOOD SAMARITAN HOSPITAL LABORATORY | Wadsworth Center, New York State Department of Health | Kirsten St. George, Daryl M. Lamson, Alexis Russell, Matthew Shudt, Melissa A Leisner, Jonathan Plitnick, Catharine Prussing, Navjot Singh, John Kelly, Erasmus Schneider, Erica Lasek-Nesselquist |
| EPI_ISL_1624091, EPI_ISL_1624092, EPI_ISL_1624093, EPI_ISL_1624094, EPI_ISL_1624095, EPI_ISL_1624096, EPI_ISL_1624097, EPI_ISL_1624098, EPI_ISL_1624099, EPI_ISL_1624100, EPI_ISL_1624101, EPI_ISL_1624102, EPI_ISL_1624103, EPI_ISL_1624104, EPI_ISL_1624105, EPI_ISL_1624106, EPI_ISL_1624107, EPI_ISL_1624109, EPI_ISL_1624110, EPI_ISL_1624111, EPI_ISL_1624112, EPI_ISL_1624114, EPI_ISL_1624115, EPI_ISL_1624116, EPI_ISL_1624117, EPI_ISL_1624118, EPI_ISL_1624119, EPI_ISL_1624120, EPI_ISL_1624121, EPI_ISL_1624122, EPI_ISL_1624123, EPI_ISL_1624124, EPI_ISL_1624126, EPI_ISL_1624127, EPI_ISL_1624128, EPI_ISL_1624129, EPI_ISL_1624130, EPI_ISL_1624131, EPI_ISL_1624132, EPI_ISL_1624133, EPI_ISL_1624134, EPI_ISL_1624135, EPI_ISL_1624136, EPI_ISL_1624137, EPI_ISL_1624138, EPI_ISL_1624139, EPI_ISL_1624140, EPI_ISL_1624141, EPI_ISL_1624142, EPI_ISL_1624143, EPI_ISL_1624145, EPI_ISL_1624146, EPI_ISL_1624147, EPI_ISL_1624148, EPI_ISL_1624149, EPI_ISL_1624150, EPI_ISL_1624151, EPI_ISL_1624152, EPI_ISL_1624153, EPI_ISL_1624154, EPI_ISL_1624155, EPI_ISL_1624156, EPI_ISL_1624157, EPI_ISL_1624158, EPI_ISL_1624159, EPI_ISL_1624160, EPI_ISL_1624161, EPI_ISL_1624162, EPI_ISL_1624163, EPI_ISL_1624165, EPI_ISL_1624166, EPI_ISL_1624167, EPI_ISL_1624168, EPI_ISL_1624170, EPI_ISL_1624172, EPI_ISL_1624173, EPI_ISL_1624174 | | | |
| see above | Illinois Department of Public Health - Springfield Lab | Illinois Department of Public Health - Springfield Lab | Bryan Sim, Gordon McCall |
| EPI_ISL_1624175 | MT Public Health Laboratory | Genomics and Discovery, Respiratory Viruses Branch, Division of Viral Diseases, Centers for Disease Control and Prevention | Ying Tao, Jing Zhang, Yan Li, Brian Lynch, Anna Kelleher, Krista Queen, Anna Uehara, Peter Cook, Han Jia Justin Ng, Clinton R. Paden, Haibin Wang, Suxiang Tong |
| EPI_ISL_1624176, EPI_ISL_1624177, EPI_ISL_1624178, EPI_ISL_1624179 | WVDHHR - Office of Laboratory Services | Genomics and Discovery, Respiratory Viruses Branch, Division of Viral Diseases, Centers for Disease Control and Prevention | Ying Tao, Jing Zhang, Yan Li, Brian Lynch, Anna Kelleher, Krista Queen, Anna Uehara, Peter Cook, Han Jia Justin Ng, Clinton R. Paden, Haibin Wang, Suxiang Tong |
| EPI_ISL_1624180, EPI_ISL_1624181 | "AR Dept. of Health-PHL, Molecular Diagnostics" | Genomics and Discovery, Respiratory Viruses Branch, Division of Viral Diseases, Centers for Disease Control and Prevention | Ying Tao, Jing Zhang, Yan Li, Brian Lynch, Anna Kelleher, Krista Queen, Anna Uehara, Peter Cook, Han Jia Justin Ng, Clinton R. Paden, Haibin Wang, Suxiang Tong |
| EPI_ISL_1624182, EPI_ISL_1624183 | LA Office of Public Health Laboratories | Genomics and Discovery, Respiratory Viruses Branch, Division of Viral Diseases, Centers for Disease Control and Prevention | Ying Tao, Jing Zhang, Yan Li, Brian Lynch, Anna Kelleher, Krista Queen, Anna Uehara, Peter Cook, Han Jia Justin Ng, Clinton R. Paden, Haibin Wang, Suxiang Tong |
| EPI_ISL_1624184, EPI_ISL_1624185, EPI_ISL_1624186 | IL Dept. of Public Health Springfield Laboratory | Genomics and Discovery, Respiratory Viruses Branch, Division of Viral Diseases, Centers for Disease Control and Prevention | Ying Tao, Jing Zhang, Yan Li, Brian Lynch, Anna Kelleher, Krista Queen, Anna Uehara, Peter Cook, Han Jia Justin Ng, Clinton R. Paden, Haibin Wang, Suxiang Tong |
| EPI_ISL_1624187, EPI_ISL_1624188, EPI_ISL_1624189, EPI_ISL_1624190, EPI_ISL_1624191, EPI_ISL_1624192, EPI_ISL_1624193, EPI_ISL_1624194, EPI_ISL_1624195 | "PA Department of Health, Bureau of Laboratories" | Genomics and Discovery, Respiratory Viruses Branch, Division of Viral Diseases, Centers for Disease Control and Prevention | Ying Tao, Jing Zhang, Yan Li, Brian Lynch, Anna Kelleher, Krista Queen, Anna Uehara, Peter Cook, Han Jia Justin Ng, Clinton R. Paden, Haibin Wang, Suxiang Tong |
| EPI_ISL_1624196 | NH Dept. of Health and Human Services Public Health Labs | Genomics and Discovery, Respiratory Viruses Branch, Division of Viral Diseases, Centers for Disease Control and Prevention | Ying Tao, Jing Zhang, Yan Li, Brian Lynch, Anna Kelleher, Krista Queen, Anna Uehara, Peter Cook, Han Jia Justin Ng, Clinton R. Paden, Haibin Wang, Suxiang Tong |
| EPI_ISL_1624197, EPI_ISL_1624198, EPI_ISL_1624199, EPI_ISL_1624200, EPI_ISL_1624201 | GA Department of Public Health Laboratory | Genomics and Discovery, Respiratory Viruses Branch, Division of Viral Diseases, Centers for Disease Control and Prevention | Ying Tao, Jing Zhang, Yan Li, Brian Lynch, Anna Kelleher, Krista Queen, Anna Uehara, Peter Cook, Han Jia Justin Ng, Clinton R. Paden, Haibin Wang, Suxiang Tong |
| EPI_ISL_1624202, EPI_ISL_1624203, EPI_ISL_1624204, EPI_ISL_1624205, EPI_ISL_1624206 | TN Division of Laboratory Services | Genomics and Discovery, Respiratory Viruses Branch, Division of Viral Diseases, Centers for Disease Control and Prevention | Ying Tao, Jing Zhang, Yan Li, Brian Lynch, Anna Kelleher, Krista Queen, Anna Uehara, Peter Cook, Han Jia Justin Ng, Clinton R. Paden, Haibin Wang, Suxiang Tong |
| EPI_ISL_1624207, EPI_ISL_1624208, EPI_ISL_1624209 | ME Health and Environmental Testing Laboratory | Genomics and Discovery, Respiratory Viruses Branch, Division of Viral Diseases, Centers for Disease Control and Prevention | Ying Tao, Jing Zhang, Yan Li, Brian Lynch, Anna Kelleher, Krista Queen, Anna Uehara, Peter Cook, Han Jia Justin Ng, Clinton R. Paden, Haibin Wang, Suxiang Tong |
| EPI_ISL_1624210 | Naval Health Clinic Annapolis | Naval Medical Research Center Biological Defense Research Directorate | Logan Voegtly, Catherine Arnold, Bishwo Adhikari, Francisco Malagon Bautista, Andrea Luquette, Gregory Rice, Andrew Bennett, Kyle Long, Lindsay Glang, Michael Deschenes, Regina Cer, Kimberly Bishop-Lilly |
| EPI_ISL_1624211, EPI_ISL_1624212, EPI_ISL_1624213, EPI_ISL_1624214 | Naval Infectious Diseases Diagnostic Laboratory | Naval Medical Research Center Biological Defense Research Directorate | Logan Voegtly, Catherine Arnold, Bishwo Adhikari, Francisco Malagon Bautista, Andrea Luquette, Gregory Rice, Andrew Bennett, Kyle Long, Lindsay Glang, Michael Deschenes, Megan Schilling, Victor Sugiharto, Regina Cer, Kimberly Bishop-Lilly |
| EPI_ISL_1624215, EPI_ISL_1624216, EPI_ISL_1624217, EPI_ISL_1624218, EPI_ISL_1624219, | Naval Health Clinic Annapolis | Naval Medical Research Center Biological Defense Research Directorate | Logan Voegtly, Catherine Arnold, Bishwo Adhikari, Francisco Malagon Bautista, Andrea Luquette, Gregory Rice, Andrew Bennett, Kyle Long, Lindsay Glang, Michael Deschenes, Regina Cer, Kimberly Bishop-Lilly |

| | | | |
|---|---|---|---|
| EPI_ISL_1624220, EPI_ISL_1624221, EPI_ISL_1624222 | | | |
| EPI_ISL_1624223, EPI_ISL_1624224, EPI_ISL_1624225 | Naval Infectious Diseases Diagnostic Laboratory | Naval Medical Research Center Biological Defense Research Directorate | Logan Voegtly, Bishwo Adhikari, Catherine Arnold, Lindsay Glang, Gregory Rice, Kyle Long, Andrew Bennett, Francisco Malgon Bautista, Megan Schilling, Victor Sugiharto, Regina Cer, Kimberly Bishop-Lilly |
| EPI_ISL_1624226 | Naval Health Clinic Annapolis | Naval Medical Research Center Biological Defense Research Directorate | Logan Voegtly, Catherine Arnold, Bishwo Adhikari, Francisco Malagon Bautista, Andrea Luquette, Gregory Rice, Andrew Bennett, Kyle Long, Lindsay Glang, Michael Deschenes, Regina Cer, Kimberly Bishop-Lilly |
| EPI_ISL_1624227, EPI_ISL_1624228, EPI_ISL_1624229 | Naval Infectious Diseases Diagnostic Laboratory | Naval Medical Research Center Biological Defense Research Directorate | Logan Voegtly, Catherine Arnold, Bishwo Adhikari, Francisco Malagon Bautista, Andrea Luquette, Gregory Rice, Andrew Bennett, Kyle Long, Lindsay Glang, Michael Deschenes, Megan Schilling, Victor Sugiharto, Regina Cer, Kimberly Bishop-Lilly |
| EPI_ISL_1624230, EPI_ISL_1624231, EPI_ISL_1624232, EPI_ISL_1624233, EPI_ISL_1624234, EPI_ISL_1624235, EPI_ISL_1624236 | Naval Health Clinic Annapolis | Naval Medical Research Center Biological Defense Research Directorate | Logan Voegtly, Catherine Arnold, Bishwo Adhikari, Francisco Malagon Bautista, Andrea Luquette, Gregory Rice, Andrew Bennett, Kyle Long, Lindsay Glang, Michael Deschenes, Regina Cer, Kimberly Bishop-Lilly |
| EPI_ISL_1624237, EPI_ISL_1624238, EPI_ISL_1624239, EPI_ISL_1624240, EPI_ISL_1624241, EPI_ISL_1624242, EPI_ISL_1624243, EPI_ISL_1624244, EPI_ISL_1624245 | Naval Health Research Center | Naval Medical Research Center Biological Defense Research Directorate | Logan Voegtly, Lindsay Glang, Francisco Malagon Bautista, Kyle Long, Regina Cer, Kimberly Bishop-Lilly |
| EPI_ISL_1624246, EPI_ISL_1624247, EPI_ISL_1624248, EPI_ISL_1624249, EPI_ISL_1624250, EPI_ISL_1624251, EPI_ISL_1624252, EPI_ISL_1624253, EPI_ISL_1624254, EPI_ISL_1624255 | Naval Infectious Diseases Diagnostic Laboratory | Naval Medical Research Center Biological Defense Research Directorate | Logan Voegtly, Catherine Arnold, Bishwo Adhikari, Francisco Malagon Bautista, Andrea Luquette, Gregory Rice, Andrew Bennett, Kyle Long, Lindsay Glang, Michael Deschenes, Megan Schilling, Victor Sugiharto, Regina Cer, Kimberly Bishop-Lilly |
| EPI_ISL_1624256, EPI_ISL_1624257, EPI_ISL_1624258, EPI_ISL_1624259, EPI_ISL_1624260, EPI_ISL_1624261, EPI_ISL_1624262, EPI_ISL_1624263, EPI_ISL_1624264, EPI_ISL_1624265, EPI_ISL_1624266 | | | |
| see above | Naval Health Clinic Annapolis | Naval Medical Research Center Biological Defense Research Directorate | Logan Voegtly, Catherine Arnold, Bishwo Adhikari, Francisco Malagon Bautista, Andrea Luquette, Gregory Rice, Andrew Bennett, Kyle Long, Lindsay Glang, Michael Deschenes, Regina Cer, Kimberly Bishop-Lilly |
| EPI_ISL_1624267, EPI_ISL_1624268, EPI_ISL_1624269, EPI_ISL_1624270, EPI_ISL_1624271, EPI_ISL_1624272 | Naval Infectious Diseases Diagnostic Laboratory | Naval Medical Research Center Biological Defense Research Directorate | Logan Voegtly, Bishwo Adhikari, Catherine Arnold, Lindsay Glang, Gregory Rice, Kyle Long, Andrew Bennett, Francisco Malgon Bautista, Megan Schilling, Victor Sugiharto, Regina Cer, Kimberly Bishop-Lilly |
| EPI_ISL_1624273, EPI_ISL_1624274, EPI_ISL_1624275 | USNS Comfort T-AH 20 | Naval Medical Research Center Biological Defense Research Directorate | Logan Voegtly, Lindsay Glang, Francisco Malagon Bautista, Kyle Long, Regina Cer, Kimberly Bishop-Lilly |
| EPI_ISL_1624276 | Naval Infectious Diseases Diagnostic Laboratory | Naval Medical Research Center Biological Defense Research Directorate | Logan Voegtly, Catherine Arnold, Bishwo Adhikari, Francisco Malagon Bautista, Andrea Luquette, Gregory Rice, Andrew Bennett, Kyle Long, Lindsay Glang, Michael Deschenes, Megan Schilling, Victor Sugiharto, Regina Cer, Kimberly Bishop-Lilly |
| EPI_ISL_1624277, EPI_ISL_1624278, EPI_ISL_1624279, EPI_ISL_1624280, EPI_ISL_1624281, EPI_ISL_1624282, EPI_ISL_1624283, EPI_ISL_1624284, EPI_ISL_1624285, EPI_ISL_1624286, EPI_ISL_1624287, EPI_ISL_1624288, EPI_ISL_1624289, EPI_ISL_1624290, EPI_ISL_1624291, EPI_ISL_1624292, EPI_ISL_1624293, EPI_ISL_1624294, EPI_ISL_1624295, EPI_ISL_1624296 | | | |
| see above | Naval Infectious Diseases Diagnostic Laboratory | Naval Medical Research Center Biological Defense Research Directorate | Logan Voegtly, Bishwo Adhikari, Catherine Arnold, Lindsay Glang, Gregory Rice, Kyle Long, Andrew Bennett, Francisco Malgon Bautista, Megan Schilling, Victor Sugiharto, Regina Cer, Kimberly Bishop-Lilly |
| EPI_ISL_1624297, EPI_ISL_1624298, EPI_ISL_1624299, EPI_ISL_1624300, EPI_ISL_1624301, EPI_ISL_1624302, EPI_ISL_1624303, EPI_ISL_1624304, EPI_ISL_1624305, EPI_ISL_1624306 | Naval Health Research Center | Naval Medical Research Center Biological Defense Research Directorate | Logan Voegtly, Lindsay Glang, Francisco Malagon Bautista, Kyle Long, Regina Cer, Kimberly Bishop-Lilly |
| EPI_ISL_1624307, EPI_ISL_1624308, EPI_ISL_1624309, EPI_ISL_1624310, EPI_ISL_1624311, EPI_ISL_1624312, EPI_ISL_1624313, EPI_ISL_1624314, EPI_ISL_1624315, EPI_ISL_1624316, EPI_ISL_1624317, EPI_ISL_1624318, EPI_ISL_1624319, EPI_ISL_1624320, EPI_ISL_1624321, EPI_ISL_1624322, EPI_ISL_1624323, EPI_ISL_1624324, EPI_ISL_1624325, EPI_ISL_1624326, EPI_ISL_1624327, EPI_ISL_1624328, EPI_ISL_1624329, EPI_ISL_1624330, EPI_ISL_1624331, EPI_ISL_1624332, EPI_ISL_1624333, EPI_ISL_1624334, EPI_ISL_1624335, EPI_ISL_1624336, EPI_ISL_1624337, EPI_ISL_1624338, EPI_ISL_1624339, EPI_ISL_1624340, EPI_ISL_1624341, EPI_ISL_1624342 | | | |
| see above | Naval Infectious Diseases Diagnostic Laboratory | Naval Medical Research Center Biological Defense Research Directorate | Logan Voegtly, Catherine Arnold, Bishwo Adhikari, Francisco Malagon Bautista, Andrea Luquette, Gregory Rice, Andrew Bennett, Kyle Long, Lindsay Glang, Michael Deschenes, Megan Schilling, Victor Sugiharto, Regina Cer, Kimberly Bishop-Lilly |
| EPI_ISL_1624343, EPI_ISL_1624344, EPI_ISL_1624345, EPI_ISL_1624346, EPI_ISL_1624347, EPI_ISL_1624348, EPI_ISL_1624349, EPI_ISL_1624350, EPI_ISL_1624351, EPI_ISL_1624352, EPI_ISL_1624353, EPI_ISL_1624354, EPI_ISL_1624355, EPI_ISL_1624356, EPI_ISL_1624357, EPI_ISL_1624358, EPI_ISL_1624359, EPI_ISL_1624360, EPI_ISL_1624361, EPI_ISL_1624362, EPI_ISL_1624363, EPI_ISL_1624364, EPI_ISL_1624365, EPI_ISL_1624366, EPI_ISL_1624367, EPI_ISL_1624368, EPI_ISL_1624369, EPI_ISL_1624370, EPI_ISL_1624371, EPI_ISL_1624372, EPI_ISL_1624373, EPI_ISL_1624374, EPI_ISL_1624375, EPI_ISL_1624376, EPI_ISL_1624377, EPI_ISL_1624378, EPI_ISL_1624379, EPI_ISL_1624380, EPI_ISL_1624381 | | | |
| see above | Naval Infectious Diseases Diagnostic Laboratory | Naval Medical Research Center Biological Defense Research Directorate | Logan Voegtly, Bishwo Adhikari, Catherine Arnold, Lindsay Glang, Gregory Rice, Kyle Long, Andrew Bennett, Francisco Malgon Bautista, Megan Schilling, Victor Sugiharto, Regina Cer, Kimberly Bishop-Lilly |
| EPI_ISL_1624382, EPI_ISL_1624383, EPI_ISL_1624384, EPI_ISL_1624385, EPI_ISL_1624386, EPI_ISL_1624387, EPI_ISL_1624388, EPI_ISL_1624389, EPI_ISL_1624390, EPI_ISL_1624391, EPI_ISL_1624392, EPI_ISL_1624393, EPI_ISL_1624394, EPI_ISL_1624395, EPI_ISL_1624396, EPI_ISL_1624397, EPI_ISL_1624398, EPI_ISL_1624399, EPI_ISL_1624400, EPI_ISL_1624401, EPI_ISL_1624402, EPI_ISL_1624403 | | | |
| see above | Naval Infectious Diseases Diagnostic Laboratory | Naval Medical Research Center Biological Defense Research Directorate | Logan Voegtly, Lindsay Glang, Francisco Malagon Bautista, Kyle Long, Megan Schilling, Victor Sugiharto, Mark Simons, Regina Cer, Kimberly Bishop-Lilly |
| EPI_ISL_1624404, EPI_ISL_1624405 | Columbia University Irving Medical Center | Wadsworth Center, New York State Department of Health | Kirsten St. George, Daryl M. Lamson, Alexis Russell, Matthew Shudt, Melissa A Leisner, Jonathan Plitnick, Catharine Prussing, Navjot Singh, John Kelly, Erasmus Schneider, Erica Lasek-Nesselquist |
| EPI_ISL_1624406, EPI_ISL_1624407, EPI_ISL_1624408 | SUNY UPSTATE MEDICAL UNIVERSITY | Wadsworth Center, New York State Department of Health | Kirsten St. George, Daryl M. Lamson, Alexis Russell, Matthew Shudt, Melissa A Leisner, Jonathan Plitnick, Catharine Prussing, Navjot Singh, John Kelly, Erasmus Schneider, Erica Lasek-Nesselquist |
| EPI_ISL_1624409 | BOSTON HEART DIAGNOSTICS CORP | Wadsworth Center, New York State Department of Health | Kirsten St. George, Daryl M. Lamson, Alexis Russell, Matthew Shudt, Melissa A Leisner, Jonathan Plitnick, Catharine Prussing, Navjot Singh, John Kelly, Erasmus Schneider, Erica Lasek-Nesselquist |
| EPI_ISL_1624410, EPI_ISL_1624411, EPI_ISL_1624412, EPI_ISL_1624413, EPI_ISL_1624414, EPI_ISL_1624415, EPI_ISL_1624416, EPI_ISL_1624417, EPI_ISL_1624418, EPI_ISL_1624419 | URMC LABS | Wadsworth Center, New York State Department of Health | Kirsten St. George, Daryl M. Lamson, Alexis Russell, Matthew Shudt, Melissa A Leisner, Jonathan Plitnick, Catharine Prussing, Navjot Singh, John Kelly, Erasmus Schneider, Erica Lasek-Nesselquist |
| EPI_ISL_1624420 | SUNY UPSTATE MEDICAL UNIVERSITY | Wadsworth Center, New York State Department of Health | Kirsten St. George, Daryl M. Lamson, Alexis Russell, Matthew Shudt, Melissa A Leisner, Jonathan Plitnick, Catharine Prussing, Navjot Singh, John Kelly, Erasmus Schneider, Erica Lasek-Nesselquist |
| EPI_ISL_1624421, EPI_ISL_1624422, EPI_ISL_1624423, EPI_ISL_1624424, EPI_ISL_1624425, EPI_ISL_1624426, EPI_ISL_1624427, EPI_ISL_1624428, EPI_ISL_1624429, EPI_ISL_1624430, EPI_ISL_1624431, EPI_ISL_1624432, EPI_ISL_1624433, EPI_ISL_1624434, EPI_ISL_1624435, EPI_ISL_1624436, EPI_ISL_1624437, EPI_ISL_1624438, EPI_ISL_1624439, EPI_ISL_1624440, EPI_ISL_1624441, EPI_ISL_1624442, EPI_ISL_1624443, EPI_ISL_1624444, EPI_ISL_1624445, EPI_ISL_1624446, EPI_ISL_1624447, EPI_ISL_1624448, EPI_ISL_1624449, EPI_ISL_1624450, EPI_ISL_1624451, EPI_ISL_1624452, EPI_ISL_1624453, EPI_ISL_1624454, EPI_ISL_1624455, EPI_ISL_1624456, EPI_ISL_1624457, EPI_ISL_1624458, EPI_ISL_1624459, EPI_ISL_1624460, EPI_ISL_1624461, EPI_ISL_1624462, EPI_ISL_1624463, EPI_ISL_1624464, EPI_ISL_1624465, EPI_ISL_1624466, EPI_ISL_1624467, EPI_ISL_1624468, EPI_ISL_1624469, EPI_ISL_1624470, EPI_ISL_1624471, EPI_ISL_1624472, EPI_ISL_1624473, EPI_ISL_1624474, EPI_ISL_1624475, EPI_ISL_1624476, EPI_ISL_1624477, EPI_ISL_1624478, EPI_ISL_1624479, EPI_ISL_1624480, EPI_ISL_1624481, EPI_ISL_1624482 | | | |
| see above | URMC LABS | Wadsworth Center, New York State | Kirsten St. George, Daryl M. Lamson, Alexis Russell, Matthew Shudt, Melissa A Leisner, Jonathan Plitnick, Catharine Prussing, Navjot Singh, John Kelly, Erasmus Schneider, Erica Lasek-Nesselquist |

| | | | |
|---|---|---|---|
| | | Department of Health | |
| EPI_ISL_1624483, EPI_ISL_1624484, EPI_ISL_1624485, EPI_ISL_1624486 | SARATOGA HOSPITAL LABORATORY | Wadsworth Center, New York State Department of Health | Kirsten St. George, Daryl M. Lamson, Alexis Russell, Matthew Shudt, Melissa A Leisner, Jonathan Plitnick, Catharine Prussing, Navjot Singh, John Kelly, Erasmus Schneider, Erica Lasek-Nesselquist |
| EPI_ISL_1624487 | CHAMPLAIN VALLEY PHYSICIANS HOSPITAL MEDICAL CENTER | Wadsworth Center, New York State Department of Health | Kirsten St. George, Daryl M. Lamson, Alexis Russell, Matthew Shudt, Melissa A Leisner, Jonathan Plitnick, Catharine Prussing, Navjot Singh, John Kelly, Erasmus Schneider, Erica Lasek-Nesselquist |
| EPI_ISL_1624489, EPI_ISL_1624490, EPI_ISL_1624491, EPI_ISL_1624492, EPI_ISL_1624493, EPI_ISL_1624494, EPI_ISL_1624495, EPI_ISL_1624496, EPI_ISL_1624497, EPI_ISL_1624498, EPI_ISL_1624499, EPI_ISL_1624500, EPI_ISL_1624501, EPI_ISL_1624502, EPI_ISL_1624503, EPI_ISL_1624504, EPI_ISL_1624505, EPI_ISL_1624506, EPI_ISL_1624507, EPI_ISL_1624508, EPI_ISL_1624509, EPI_ISL_1624510, EPI_ISL_1624511, EPI_ISL_1624512, EPI_ISL_1624513, EPI_ISL_1624514, EPI_ISL_1624515, EPI_ISL_1624516, EPI_ISL_1624517, EPI_ISL_1624518, EPI_ISL_1624519, EPI_ISL_1624520, EPI_ISL_1624521, EPI_ISL_1624522, EPI_ISL_1624523, EPI_ISL_1624524, EPI_ISL_1624525, EPI_ISL_1624526, EPI_ISL_1624527, EPI_ISL_1624528, EPI_ISL_1624529, EPI_ISL_1624530, EPI_ISL_1624531, EPI_ISL_1624532, EPI_ISL_1624533, EPI_ISL_1624534, EPI_ISL_1624535, EPI_ISL_1624536, EPI_ISL_1624537, EPI_ISL_1624538, EPI_ISL_1624539, EPI_ISL_1624540, EPI_ISL_1624541, EPI_ISL_1624542, EPI_ISL_1624543, EPI_ISL_1624544, EPI_ISL_1624545, EPI_ISL_1624546, EPI_ISL_1624547, EPI_ISL_1624548, EPI_ISL_1624549, EPI_ISL_1624550, EPI_ISL_1624551, EPI_ISL_1624552, EPI_ISL_1624553, EPI_ISL_1624554, EPI_ISL_1624555, EPI_ISL_1624556, EPI_ISL_1624557, EPI_ISL_1624558, EPI_ISL_1624559, EPI_ISL_1624560, EPI_ISL_1624561, EPI_ISL_1624562, EPI_ISL_1624563, EPI_ISL_1624564, EPI_ISL_1624565, EPI_ISL_1624566, EPI_ISL_1624567, EPI_ISL_1624568, EPI_ISL_1624569, EPI_ISL_1624570, EPI_ISL_1624571, EPI_ISL_1624572, EPI_ISL_1624573, EPI_ISL_1624574, EPI_ISL_1624575, EPI_ISL_1624576, EPI_ISL_1624577, EPI_ISL_1624578, EPI_ISL_1624579, EPI_ISL_1624580, EPI_ISL_1624581, EPI_ISL_1624582, EPI_ISL_1624583, EPI_ISL_1624584, EPI_ISL_1624585, EPI_ISL_1624586, EPI_ISL_1624587, EPI_ISL_1624588, EPI_ISL_1624589, EPI_ISL_1624590, EPI_ISL_1624591, EPI_ISL_1624592, EPI_ISL_1624593, EPI_ISL_1624594, EPI_ISL_1624595, EPI_ISL_1624596, EPI_ISL_1624597, EPI_ISL_1624598, EPI_ISL_1624599, EPI_ISL_1624600, EPI_ISL_1624601, EPI_ISL_1624602, EPI_ISL_1624603, EPI_ISL_1624604, EPI_ISL_1624605, EPI_ISL_1624606, EPI_ISL_1624607, EPI_ISL_1624608, EPI_ISL_1624609, EPI_ISL_1624610, EPI_ISL_1624611, EPI_ISL_1624612, EPI_ISL_1624613, EPI_ISL_1624614, EPI_ISL_1624615, EPI_ISL_1624616, EPI_ISL_1624617, EPI_ISL_1624618, EPI_ISL_1624619, EPI_ISL_1624620, EPI_ISL_1624621, EPI_ISL_1624622, EPI_ISL_1624623, EPI_ISL_1624624, EPI_ISL_1624625, EPI_ISL_1624626, EPI_ISL_1624627, EPI_ISL_1624628, EPI_ISL_1624629, EPI_ISL_1624630, EPI_ISL_1624631, EPI_ISL_1624632, EPI_ISL_1624633, EPI_ISL_1624634, EPI_ISL_1624635, EPI_ISL_1624636, EPI_ISL_1624637, EPI_ISL_1624638, EPI_ISL_1624639, EPI_ISL_1624640, EPI_ISL_1624641, EPI_ISL_1624642, EPI_ISL_1624643, EPI_ISL_1624644, EPI_ISL_1624645, EPI_ISL_1624646, EPI_ISL_1624647, EPI_ISL_1624648, EPI_ISL_1624649, EPI_ISL_1624650, EPI_ISL_1624651, EPI_ISL_1624652, EPI_ISL_1624653, EPI_ISL_1624654, EPI_ISL_1624655, EPI_ISL_1624656, EPI_ISL_1624657, EPI_ISL_1624658, EPI_ISL_1624659, EPI_ISL_1624660, EPI_ISL_1624661, EPI_ISL_1624662, EPI_ISL_1624663, EPI_ISL_1624664, EPI_ISL_1624665, EPI_ISL_1624666, EPI_ISL_1624667, EPI_ISL_1624668, EPI_ISL_1624669, EPI_ISL_1624670, EPI_ISL_1624671, EPI_ISL_1624672, EPI_ISL_1624673, EPI_ISL_1624690, EPI_ISL_1624697, EPI_ISL_1624698, EPI_ISL_1624699, EPI_ISL_1624700, EPI_ISL_1624701, EPI_ISL_1624702, EPI_ISL_1624703, EPI_ISL_1624704, EPI_ISL_1624705, EPI_ISL_1624706, EPI_ISL_1624707, EPI_ISL_1624708, EPI_ISL_1624709, EPI_ISL_1624710, EPI_ISL_1624711, EPI_ISL_1624712, EPI_ISL_1624713, EPI_ISL_1624714, EPI_ISL_1624715, EPI_ISL_1624716 | | | |
| see above | Massachusetts State Public Health Laboratory | Massachusetts State Public Health Laboratory | Andrew Lang, Timelia Fink, Glen Gallagher, Sandra Smole |
| EPI_ISL_1624717, EPI_ISL_1624718, EPI_ISL_1624719, EPI_ISL_1624720, EPI_ISL_1624721, EPI_ISL_1624722, EPI_ISL_1624723, EPI_ISL_1624724, EPI_ISL_1624725, EPI_ISL_1624726, EPI_ISL_1624727, EPI_ISL_1624728, EPI_ISL_1624729, EPI_ISL_1624730, EPI_ISL_1624731, EPI_ISL_1624732, EPI_ISL_1624733, EPI_ISL_1624734, EPI_ISL_1624735, EPI_ISL_1624736, EPI_ISL_1624737, EPI_ISL_1624738, EPI_ISL_1624739, EPI_ISL_1624740, EPI_ISL_1624741, EPI_ISL_1624742, EPI_ISL_1624743, EPI_ISL_1624744, EPI_ISL_1624745, EPI_ISL_1624746, EPI_ISL_1624747, EPI_ISL_1624748, EPI_ISL_1624749, EPI_ISL_1624750, EPI_ISL_1624751, EPI_ISL_1624752, EPI_ISL_1624753, EPI_ISL_1624754, EPI_ISL_1624755, EPI_ISL_1624756, EPI_ISL_1624757 | | | |
| see above | WVU Rapid Development Lab | WVU and Marshall University Combined Genomics Core Facilities | James Denvir, Peter Stoilov, Peter Perrotta, Wesley Kimble, Ryan Percifield |
| EPI_ISL_1624758, EPI_ISL_1624759, EPI_ISL_1624760, EPI_ISL_1624761, EPI_ISL_1624762, EPI_ISL_1624763, EPI_ISL_1624764, EPI_ISL_1624765, EPI_ISL_1624766, EPI_ISL_1624767, EPI_ISL_1624768, EPI_ISL_1624769, EPI_ISL_1624770, EPI_ISL_1624771, EPI_ISL_1624773, EPI_ISL_1624774, EPI_ISL_1624775, EPI_ISL_1624776, EPI_ISL_1624777, EPI_ISL_1624778 | | | |
| see above | J.W. Ruby Memorial Hospital | WVU and Marshall University Combined Genomics Core Facilities | James Denvir, Peter Stoilov, Peter Perrotta, Wesley Kimble, Ryan Percifield |
| EPI_ISL_1624779 | Berkeley Medical Center | WVU and Marshall University Combined Genomics Core Facilities | James Denvir, Peter Stoilov, Peter Perrotta, Wesley Kimble, Ryan Percifield |
| EPI_ISL_1624780, EPI_ISL_1624781, EPI_ISL_1624782, EPI_ISL_1624783, EPI_ISL_1624784, EPI_ISL_1624785, EPI_ISL_1624786, EPI_ISL_1624787, EPI_ISL_1624788, EPI_ISL_1624789, EPI_ISL_1624791, EPI_ISL_1624792, EPI_ISL_1624793, EPI_ISL_1624794, EPI_ISL_1624795, EPI_ISL_1624796, EPI_ISL_1624797, EPI_ISL_1624799, EPI_ISL_1624801, EPI_ISL_1624802, EPI_ISL_1624803 | | | |
| see above | J.W. Ruby Memorial Hospital | WVU and Marshall University Combined Genomics Core Facilities | James Denvir, Peter Stoilov, Peter Perrotta, Wesley Kimble, Ryan Percifield |
| EPI_ISL_1624804 | Berkeley Medical Center | WVU and Marshall University Combined Genomics Core Facilities | James Denvir, Peter Stoilov, Peter Perrotta, Wesley Kimble, Ryan Percifield |
| EPI_ISL_1624805, EPI_ISL_1624806, EPI_ISL_1624807, EPI_ISL_1624808, EPI_ISL_1624809, EPI_ISL_1624810, EPI_ISL_1624811, EPI_ISL_1624812, EPI_ISL_1624813, EPI_ISL_1624814, EPI_ISL_1624816, EPI_ISL_1624817, EPI_ISL_1624819, EPI_ISL_1624820, EPI_ISL_1624821, EPI_ISL_1624822 | | | |
| see above | J.W. Ruby Memorial Hospital | WVU and Marshall University Combined Genomics Core Facilities | James Denvir, Peter Stoilov, Peter Perrotta, Wesley Kimble, Ryan Percifield |
| EPI_ISL_1624823 | Berkeley Medical Center | WVU and Marshall University Combined Genomics Core Facilities | James Denvir, Peter Stoilov, Peter Perrotta, Wesley Kimble, Ryan Percifield |
| EPI_ISL_1624824, EPI_ISL_1624826 | J.W. Ruby Memorial Hospital | WVU and Marshall University Combined Genomics Core Facilities | James Denvir, Peter Stoilov, Peter Perrotta, Wesley Kimble, Ryan Percifield |
| EPI_ISL_1624827, EPI_ISL_1624828, EPI_ISL_1624829, EPI_ISL_1624830, EPI_ISL_1624831, EPI_ISL_1624832, EPI_ISL_1624833, EPI_ISL_1624834, EPI_ISL_1624835, EPI_ISL_1624836, EPI_ISL_1624837, EPI_ISL_1624838, EPI_ISL_1624839, EPI_ISL_1624840, EPI_ISL_1624841, EPI_ISL_1624842, EPI_ISL_1624843, EPI_ISL_1624844, EPI_ISL_1624845, EPI_ISL_1624846, EPI_ISL_1624847, EPI_ISL_1624848, EPI_ISL_1624849, EPI_ISL_1624850, EPI_ISL_1624851, EPI_ISL_1624852 | | | |
| see above | Berkeley Medical Center | WVU and Marshall University Combined Genomics Core Facilities | James Denvir, Peter Stoilov, Peter Perrotta, Wesley Kimble, Ryan Percifield |
| EPI_ISL_1624853, EPI_ISL_1624854, EPI_ISL_1624855, EPI_ISL_1624856, EPI_ISL_1624857, EPI_ISL_1624858, EPI_ISL_1624859, EPI_ISL_1624860, EPI_ISL_1624861, EPI_ISL_1624862, EPI_ISL_1624863, EPI_ISL_1624864, EPI_ISL_1624865, EPI_ISL_1624866, EPI_ISL_1624867, EPI_ISL_1624868, EPI_ISL_1624869, EPI_ISL_1624870 | | | |
| see above | QLabs | WVU and Marshall University Combined Genomics Core Facilities | James Denvir, Peter Stoilov, Peter Perrotta, Wesley Kimble, Ryan Percifield |
| EPI_ISL_1624871, EPI_ISL_1624872, EPI_ISL_1624873, EPI_ISL_1624874, EPI_ISL_1624875, EPI_ISL_1624876, EPI_ISL_1624877, EPI_ISL_1624878, EPI_ISL_1624879, EPI_ISL_1624880, EPI_ISL_1624881, EPI_ISL_1624882, EPI_ISL_1624883, EPI_ISL_1624884, EPI_ISL_1624885, EPI_ISL_1624886, EPI_ISL_1624887, EPI_ISL_1624888, EPI_ISL_1624889, EPI_ISL_1624890, EPI_ISL_1624891, EPI_ISL_1624892, EPI_ISL_1624893, EPI_ISL_1624894, EPI_ISL_1624895 | | | |
| see above | Berkeley Medical Center | WVU and Marshall University Combined Genomics Core Facilities | James Denvir, Peter Stoilov, Peter Perrotta, Wesley Kimble, Ryan Percifield |
| EPI_ISL_1624896, EPI_ISL_1624897, EPI_ISL_1624898, EPI_ISL_1624899, EPI_ISL_1624900, EPI_ISL_1624901, EPI_ISL_1624902, EPI_ISL_1624903, EPI_ISL_1624904, EPI_ISL_1624905, EPI_ISL_1624906, EPI_ISL_1624907, EPI_ISL_1624908, EPI_ISL_1624909, EPI_ISL_1624910, EPI_ISL_1624911, EPI_ISL_1624912, EPI_ISL_1624913, EPI_ISL_1624914, EPI_ISL_1624915, EPI_ISL_1624916, EPI_ISL_1624917, EPI_ISL_1624918, EPI_ISL_1624919, EPI_ISL_1624920, EPI_ISL_1624921, EPI_ISL_1624922, EPI_ISL_1624924, EPI_ISL_1624925, EPI_ISL_1624926, EPI_ISL_1624927, EPI_ISL_1624929, EPI_ISL_1624930, EPI_ISL_1624931, EPI_ISL_1624932, EPI_ISL_1624933, EPI_ISL_1624934, EPI_ISL_1624935, EPI_ISL_1624936, EPI_ISL_1624937, EPI_ISL_1624939, EPI_ISL_1624940, EPI_ISL_1624941, EPI_ISL_1624942, EPI_ISL_1624944, EPI_ISL_1624945, EPI_ISL_1624946, EPI_ISL_1624947 | | | |
| see above | QLabs | WVU and Marshall University Combined Genomics Core Facilities | James Denvir, Peter Stoilov, Peter Perrotta, Wesley Kimble, Ryan Percifield |
| EPI_ISL_1624948, EPI_ISL_1624949, EPI_ISL_1624950, EPI_ISL_1624951, EPI_ISL_1624952, EPI_ISL_1624953 | J.W. Ruby Memorial Hospital | WVU and Marshall University Combined Genomics Core Facilities | James Denvir, Peter Stoilov, Peter Perrotta, Wesley Kimble, Ryan Percifield |
| EPI_ISL_1624954, EPI_ISL_1624955, EPI_ISL_1624956, EPI_ISL_1624957, EPI_ISL_1624958, EPI_ISL_1624960, EPI_ISL_1624961, EPI_ISL_1624962, EPI_ISL_1624963, EPI_ISL_1624964, EPI_ISL_1624965, EPI_ISL_1624966, EPI_ISL_1624967, EPI_ISL_1624968, EPI_ISL_1624969, EPI_ISL_1624970, EPI_ISL_1624971, EPI_ISL_1624972, EPI_ISL_1624974, EPI_ISL_1624975, EPI_ISL_1624976, EPI_ISL_1624977, EPI_ISL_1624978, EPI_ISL_1624979, EPI_ISL_1624980, EPI_ISL_1624981 | | | |
| see above | Berkeley Medical Center | WVU and Marshall University Combined Genomics Core Facilities | James Denvir, Peter Stoilov, Peter Perrotta, Wesley Kimble, Ryan Percifield |
| EPI_ISL_1624982, EPI_ISL_1624983, EPI_ISL_1624984, EPI_ISL_1624985, EPI_ISL_1624986, EPI_ISL_1624987, EPI_ISL_1624988, EPI_ISL_1624989, EPI_ISL_1624990, EPI_ISL_1624991, EPI_ISL_1624992, EPI_ISL_1624993, EPI_ISL_1624994, EPI_ISL_1624995, EPI_ISL_1624996, EPI_ISL_1624997, EPI_ISL_1624998, EPI_ISL_1624999, EPI_ISL_1625000, EPI_ISL_1625002 | | | |
| see above | J.W. Ruby Memorial Hospital | WVU and Marshall University Combined Genomics Core Facilities | James Denvir, Peter Stoilov, Peter Perrotta, Wesley Kimble, Ryan Percifield |
| EPI_ISL_1625540, EPI_ISL_1625541, EPI_ISL_1625542, EPI_ISL_1625543, EPI_ISL_1625544, EPI_ISL_1625545, EPI_ISL_1625546, EPI_ISL_1625547, EPI_ISL_1625548, EPI_ISL_1625549, EPI_ISL_1625550, EPI_ISL_1625551, EPI_ISL_1625552, EPI_ISL_1625553, EPI_ISL_1625554, EPI_ISL_1625555, EPI_ISL_1625556, EPI_ISL_1625557, EPI_ISL_1625558, EPI_ISL_1625559, EPI_ISL_1625560, EPI_ISL_1625561, EPI_ISL_1625562, EPI_ISL_1625563, EPI_ISL_1625564, EPI_ISL_1625565, EPI_ISL_1625566, EPI_ISL_1625567, EPI_ISL_1625568, EPI_ISL_1625569, EPI_ISL_1625570, EPI_ISL_1625571, EPI_ISL_1625572, EPI_ISL_1625573, EPI_ISL_1625574, EPI_ISL_1625575, EPI_ISL_1625576, EPI_ISL_1625577, EPI_ISL_1625578, EPI_ISL_1625579, EPI_ISL_1625580, EPI_ISL_1625581, EPI_ISL_1625582, EPI_ISL_1625583, EPI_ISL_1625584, EPI_ISL_1625585, EPI_ISL_1625586, EPI_ISL_1625587, EPI_ISL_1625588, EPI_ISL_1625589, EPI_ISL_1625590, EPI_ISL_1625591, EPI_ISL_1625592, EPI_ISL_1625593, EPI_ISL_1625594, EPI_ISL_1625595, EPI_ISL_1625596, EPI_ISL_1625597, EPI_ISL_1625598, EPI_ISL_1625599, EPI_ISL_1625600, EPI_ISL_1625601, EPI_ISL_1625602, EPI_ISL_1625603, EPI_ISL_1625604, EPI_ISL_1625605, EPI_ISL_1625606, EPI_ISL_1625607, EPI_ISL_1625608, EPI_ISL_1625609, EPI_ISL_1625610, EPI_ISL_1625611, EPI_ISL_1625612, EPI_ISL_1625613, EPI_ISL_1625614, EPI_ISL_1625615, EPI_ISL_1625617, EPI_ISL_1625618, EPI_ISL_1625619, EPI_ISL_1625620, EPI_ISL_1625621, EPI_ISL_1625622, EPI_ISL_1625623, EPI_ISL_1625624, EPI_ISL_1625625, EPI_ISL_1625626, EPI_ISL_1625627, EPI_ISL_1625628, EPI_ISL_1625629, EPI_ISL_1625630, EPI_ISL_1625631, EPI_ISL_1625632, EPI_ISL_1625633, EPI_ISL_1625634, EPI_ISL_1625635, EPI_ISL_1625636, EPI_ISL_1625637, EPI_ISL_1625638, EPI_ISL_1625639, EPI_ISL_1625640, EPI_ISL_1625641, EPI_ISL_1625642, EPI_ISL_1625643, EPI_ISL_1625644, EPI_ISL_1625645, EPI_ISL_1625646, EPI_ISL_1625647, EPI_ISL_1625648, EPI_ISL_1625649, EPI_ISL_1625650, EPI_ISL_1625651, EPI_ISL_1625652, EPI_ISL_1625653, EPI_ISL_1625654, EPI_ISL_1625655, EPI_ISL_1625656, EPI_ISL_1625657, EPI_ISL_1625658, EPI_ISL_1625659, EPI_ISL_1625660, EPI_ISL_1625661, EPI_ISL_1625662, EPI_ISL_1625663, EPI_ISL_1625664, EPI_ISL_1625665, EPI_ISL_1625666, EPI_ISL_1625667, EPI_ISL_1625668, EPI_ISL_1625669, EPI_ISL_1625671, EPI_ISL_1625672, EPI_ISL_1625674, EPI_ISL_1625675, EPI_ISL_1625676, EPI_ISL_1625677, EPI_ISL_1625678, EPI_ISL_1625679, EPI_ISL_1625680, EPI_ISL_1625681 | | | |
| see above | The Jackson Laboratory | The Jackson Laboratory | Bergeron D, Renzette N, Adams M, Omerza G, Kelly K, Long J, Li L |
| EPI_ISL_1625682, EPI_ISL_1625683, EPI_ISL_1625684, EPI_ISL_1625685, EPI_ISL_1625686, EPI_ISL_1625687, EPI_ISL_1625688, EPI_ISL_1625689, EPI_ISL_1625690, EPI_ISL_1625691, EPI_ISL_1625692, EPI_ISL_1625693, EPI_ISL_1625694, EPI_ISL_1625695, EPI_ISL_1625696, EPI_ISL_1625697, EPI_ISL_1625698, EPI_ISL_1625699, EPI_ISL_1625700, EPI_ISL_1625701, EPI_ISL_1625702, EPI_ISL_1625703, EPI_ISL_1625704, EPI_ISL_1625705, EPI_ISL_1625706, EPI_ISL_1625707, EPI_ISL_1625708, EPI_ISL_1625709, EPI_ISL_1625710, EPI_ISL_1625711, EPI_ISL_1625712, EPI_ISL_1625713, EPI_ISL_1625714, EPI_ISL_1625715, EPI_ISL_1625716, EPI_ISL_1625717, EPI_ISL_1625718, EPI_ISL_1625719, EPI_ISL_1625720, EPI_ISL_1625721, EPI_ISL_1625722, EPI_ISL_1625723, EPI_ISL_1625724, EPI_ISL_1625725, EPI_ISL_1625726, EPI_ISL_1625727, EPI_ISL_1625728, EPI_ISL_1625729, EPI_ISL_1625730, EPI_ISL_1625731, EPI_ISL_1625732, EPI_ISL_1625733, EPI_ISL_1625734, EPI_ISL_1625735, EPI_ISL_1625736, EPI_ISL_1625737, EPI_ISL_1625738, EPI_ISL_1625739, EPI_ISL_1625740, EPI_ISL_1625741, EPI_ISL_1625742, EPI_ISL_1625743, EPI_ISL_1625744, EPI_ISL_1625745, EPI_ISL_1625746, EPI_ISL_1625747, EPI_ISL_1625748, EPI_ISL_1625749, EPI_ISL_1625750, EPI_ISL_1625751, EPI_ISL_1625752, EPI_ISL_1625753, EPI_ISL_1625754, EPI_ISL_1625755, EPI_ISL_1625756, EPI_ISL_1625757, EPI_ISL_1625758, EPI_ISL_1625759, EPI_ISL_1625760, EPI_ISL_1625761, EPI_ISL_1625762, EPI_ISL_1625763, EPI_ISL_1625764, EPI_ISL_1625765, EPI_ISL_1625766, EPI_ISL_1625767, EPI_ISL_1625768, EPI_ISL_1625769, EPI_ISL_1625770, EPI_ISL_1625771, EPI_ISL_1625780, EPI_ISL_1625781, EPI_ISL_1625782, EPI_ISL_1625783, EPI_ISL_1625784, EPI_ISL_1625785, EPI_ISL_1625786, EPI_ISL_1625787, EPI_ISL_1625788, EPI_ISL_1625789, EPI_ISL_1625790, EPI_ISL_1625791, EPI_ISL_1625792, EPI_ISL_1625793, EPI_ISL_1625794, EPI_ISL_1625795, EPI_ISL_1625796, EPI_ISL_1625797, EPI_ISL_1625798, EPI_ISL_1625799, EPI_ISL_1625800, EPI_ISL_1625801, EPI_ISL_1625802, EPI_ISL_1625803, EPI_ISL_1625804, EPI_ISL_1625805, EPI_ISL_1625806, EPI_ISL_1625807, EPI_ISL_1625808, EPI_ISL_1625809, EPI_ISL_1625810, EPI_ISL_1625811, EPI_ISL_1625812, EPI_ISL_1625813, EPI_ISL_1625814, EPI_ISL_1625815, EPI_ISL_1625816, EPI_ISL_1625817, EPI_ISL_1625818, EPI_ISL_1625819, EPI_ISL_1625820, EPI_ISL_1625821, EPI_ISL_1625822, EPI_ISL_1625823, EPI_ISL_1625824, EPI_ISL_1625825, EPI_ISL_1625826, EPI_ISL_1625827, EPI_ISL_1625828, EPI_ISL_1625829, EPI_ISL_1625830, EPI_ISL_1625831, EPI_ISL_1625832, EPI_ISL_1625833, EPI_ISL_1625834 | | | |
| see above | Pathology and Laboratory Medicine Institute, Cleveland Clinic, Ohio, USA | Pathology and Laboratory Medicine Institute, Cleveland Clinic, Ohio, USA | Jennifer Starbuck, Joy Nakitandwe, Kristen McDonnell, David Plunkett, Zheng Jin Tu, Jay Brock, Yu-Wei Cheng, Gary Procop, Daniel Rhoads, Daniel H. Farkas, David Bosler |
| EPI_ISL_1625839, EPI_ISL_1625840, EPI_ISL_1625841, EPI_ISL_1625843, EPI_ISL_1625844, EPI_ISL_1625845, EPI_ISL_1625846, EPI_ISL_1625847, EPI_ISL_1625849, EPI_ISL_1625850, EPI_ISL_1625851, EPI_ISL_1625852, EPI_ISL_1625853, EPI_ISL_1625854, EPI_ISL_1625855, EPI_ISL_1625858, EPI_ISL_1625859, EPI_ISL_1625860, EPI_ISL_1625861, EPI_ISL_1625862, EPI_ISL_1625863, EPI_ISL_1625865, EPI_ISL_1625866, EPI_ISL_1625867, EPI_ISL_1625868, EPI_ISL_1625869, EPI_ISL_1625870, EPI_ISL_1625871, EPI_ISL_1625873, EPI_ISL_1625874, EPI_ISL_1625875, EPI_ISL_1625876, EPI_ISL_1625877, EPI_ISL_1625878, EPI_ISL_1625879, EPI_ISL_1625880, EPI_ISL_1625882, EPI_ISL_1625883, EPI_ISL_1625885, EPI_ISL_1625886, EPI_ISL_1625887, EPI_ISL_1625888, EPI_ISL_1625889, EPI_ISL_1625890, EPI_ISL_1625891, EPI_ISL_1625893, EPI_ISL_1625894, EPI_ISL_1625895, EPI_ISL_1625896, EPI_ISL_1625897, EPI_ISL_1625898, EPI_ISL_1625899, EPI_ISL_1625902, EPI_ISL_1625903, EPI_ISL_1625904, EPI_ISL_1625906, EPI_ISL_1625907, EPI_ISL_1625908, EPI_ISL_1625909, EPI_ISL_1625910, EPI_ISL_1625911 | | | |
| see above | NJDOH, Public Health and Environmental Laboratories | NJ_PHEL | Lindsey Bodnar, Shiv K. Verma, Jacquelyn Deverell, Matthew Scarnati, Mohammad M Ali, Dana Woell, Byeong Jeong |
| EPI_ISL_1625912, EPI_ISL_1625914, EPI_ISL_1625915, EPI_ISL_1625916, EPI_ISL_1625917, EPI_ISL_1625919, EPI_ISL_1625921, EPI_ISL_1625922, EPI_ISL_1625926, EPI_ISL_1625927, EPI_ISL_1625928, EPI_ISL_1625929, EPI_ISL_1625930, EPI_ISL_1625931, EPI_ISL_1625935, EPI_ISL_1625936, EPI_ISL_1625937, EPI_ISL_1625938, EPI_ISL_1625939, EPI_ISL_1625940, EPI_ISL_1625942, EPI_ISL_1625943, EPI_ISL_1625948, EPI_ISL_1625949, EPI_ISL_1625951, EPI_ISL_1625952, EPI_ISL_1625955, EPI_ISL_1625956, EPI_ISL_1625957, EPI_ISL_1625958, EPI_ISL_1625959, EPI_ISL_1625960, EPI_ISL_1625961, EPI_ISL_1625962, EPI_ISL_1625963, EPI_ISL_1625965, EPI_ISL_1625966, EPI_ISL_1625967, EPI_ISL_1625968, EPI_ISL_1625969, EPI_ISL_1625970, EPI_ISL_1625971 | | | |
| see above | UTGSAF | UTGSAF | Jessica Podnar, Sylvie Beaudenon, Audrey Kelly, Zachary Carver, Anna Battenhouse, Andreas Matouschek |

| | | | |
|---|---|---|---|
| EPI_ISL_1626010, EPI_ISL_1626011, EPI_ISL_1626012, EPI_ISL_1626013, EPI_ISL_1626014, EPI_ISL_1626015, EPI_ISL_1626017 | Atlanta VA Medical Center | Pathogen Discovery, Respiratory Viruses Branch, Division of Viral Diseases, Centers for Disease Control and Prevention | Yan Li, Ying Tao, Anna Kelleher, Jing Zhang, Anna Montmayeur, Brian Lynch, Krista Queen, Anna Uehara, Peter Cook, Han Jia Justin Ng, Rachel Marine, Clinton R. Paden, Haibin Wang, Mark Burroughs, Justin Lee, Adam Retchless, Suxiang Tong |
| EPI_ISL_1626018, EPI_ISL_1626019, EPI_ISL_1626020, EPI_ISL_1626021, EPI_ISL_1626022, EPI_ISL_1626023, EPI_ISL_1626025, EPI_ISL_1626026, EPI_ISL_1626027, EPI_ISL_1626028, EPI_ISL_1626029, EPI_ISL_1626030, EPI_ISL_1626031, EPI_ISL_1626032, EPI_ISL_1626033, EPI_ISL_1626034, EPI_ISL_1626035, EPI_ISL_1626036, EPI_ISL_1626037, EPI_ISL_1626038, EPI_ISL_1626040, EPI_ISL_1626041, EPI_ISL_1626042, EPI_ISL_1626043, EPI_ISL_1626044, EPI_ISL_1626045, EPI_ISL_1626046, EPI_ISL_1626047, EPI_ISL_1626048, EPI_ISL_1626049, EPI_ISL_1626050, EPI_ISL_1626051, EPI_ISL_1626052, EPI_ISL_1626053, EPI_ISL_1626054, EPI_ISL_1626055, EPI_ISL_1626056, EPI_ISL_1626057, EPI_ISL_1626058, EPI_ISL_1626059, EPI_ISL_1626060, EPI_ISL_1626061, EPI_ISL_1626062, EPI_ISL_1626064, EPI_ISL_1626065, EPI_ISL_1626066, EPI_ISL_1626067, EPI_ISL_1626068, EPI_ISL_1626069, EPI_ISL_1626071, EPI_ISL_1626072, EPI_ISL_1626073, EPI_ISL_1626074, EPI_ISL_1626075, EPI_ISL_1626077, EPI_ISL_1626078, EPI_ISL_1626079, EPI_ISL_1626080, EPI_ISL_1626081, EPI_ISL_1626082, EPI_ISL_1626083, EPI_ISL_1626084, EPI_ISL_1626085, EPI_ISL_1626086, EPI_ISL_1626087, EPI_ISL_1626088, EPI_ISL_1626090, EPI_ISL_1626091, EPI_ISL_1626092, EPI_ISL_1626093, EPI_ISL_1626094, EPI_ISL_1626095, EPI_ISL_1626096, EPI_ISL_1626097, EPI_ISL_1626098, EPI_ISL_1626099, EPI_ISL_1626100, EPI_ISL_1626101, EPI_ISL_1626102, EPI_ISL_1626103, EPI_ISL_1626104, EPI_ISL_1626105, EPI_ISL_1626106, EPI_ISL_1626107, EPI_ISL_1626108, EPI_ISL_1626109, EPI_ISL_1626110, EPI_ISL_1626111, EPI_ISL_1626112, EPI_ISL_1626113, EPI_ISL_1626114, EPI_ISL_1626115, EPI_ISL_1626116, EPI_ISL_1626117, EPI_ISL_1626118, EPI_ISL_1626119, EPI_ISL_1626120, EPI_ISL_1626121, EPI_ISL_1626122, EPI_ISL_1626123, EPI_ISL_1626124, EPI_ISL_1626125, EPI_ISL_1626126, EPI_ISL_1626127, EPI_ISL_1626128, EPI_ISL_1626129, EPI_ISL_1626130, EPI_ISL_1626131, EPI_ISL_1626132, EPI_ISL_1626133, EPI_ISL_1626134, EPI_ISL_1626135, EPI_ISL_1626136, EPI_ISL_1626137, EPI_ISL_1626138, EPI_ISL_1626139, EPI_ISL_1626141, EPI_ISL_1626142, EPI_ISL_1626143, EPI_ISL_1626144, EPI_ISL_1626145, EPI_ISL_1626146, EPI_ISL_1626147, EPI_ISL_1626148, EPI_ISL_1626149, EPI_ISL_1626150, EPI_ISL_1626151, EPI_ISL_1626152, EPI_ISL_1626153, EPI_ISL_1626154, EPI_ISL_1626155, EPI_ISL_1626156, EPI_ISL_1626157, EPI_ISL_1626158, EPI_ISL_1626159, EPI_ISL_1626160, EPI_ISL_1626161, EPI_ISL_1626162, EPI_ISL_1626163, EPI_ISL_1626164, EPI_ISL_1626165, EPI_ISL_1626166, EPI_ISL_1626167, EPI_ISL_1626168, EPI_ISL_1626169, EPI_ISL_1626170, EPI_ISL_1626171, EPI_ISL_1626172, EPI_ISL_1626173, EPI_ISL_1626174, EPI_ISL_1626175, EPI_ISL_1626176, EPI_ISL_1626178, EPI_ISL_1626179, EPI_ISL_1626180, EPI_ISL_1626182, EPI_ISL_1626183, EPI_ISL_1626184, EPI_ISL_1626185, EPI_ISL_1626186, EPI_ISL_1626187, EPI_ISL_1626188, EPI_ISL_1626189, EPI_ISL_1626190, EPI_ISL_1626191, EPI_ISL_1626193, EPI_ISL_1626194, EPI_ISL_1626195, EPI_ISL_1626196, EPI_ISL_1626197, EPI_ISL_1626198, EPI_ISL_1626199, EPI_ISL_1626200, EPI_ISL_1626201, EPI_ISL_1626202, EPI_ISL_1626203, EPI_ISL_1626204, EPI_ISL_1626205, EPI_ISL_1626206, EPI_ISL_1626207, EPI_ISL_1626208, EPI_ISL_1626209, EPI_ISL_1626210, EPI_ISL_1626211, EPI_ISL_1626212, EPI_ISL_1626213, EPI_ISL_1626214, EPI_ISL_1626215, EPI_ISL_1626216, EPI_ISL_1626217, EPI_ISL_1626218, EPI_ISL_1626219, EPI_ISL_1626220, EPI_ISL_1626221, EPI_ISL_1626222, EPI_ISL_1626223, EPI_ISL_1626224, EPI_ISL_1626225, EPI_ISL_1626226, EPI_ISL_1626227, EPI_ISL_1626228, EPI_ISL_1626229, EPI_ISL_1626230, EPI_ISL_1626231, EPI_ISL_1626232, EPI_ISL_1626233, EPI_ISL_1626234, EPI_ISL_1626235, EPI_ISL_1626236, EPI_ISL_1626237, EPI_ISL_1626238, EPI_ISL_1626239, EPI_ISL_1626240, EPI_ISL_1626241, EPI_ISL_1626242, EPI_ISL_1626243, EPI_ISL_1626244, EPI_ISL_1626245, EPI_ISL_1626246, EPI_ISL_1626247, EPI_ISL_1626248, EPI_ISL_1626249, EPI_ISL_1626250, EPI_ISL_1626251, EPI_ISL_1626253, EPI_ISL_1626254, EPI_ISL_1626255, EPI_ISL_1626256, EPI_ISL_1626258, EPI_ISL_1626259, EPI_ISL_1626260, EPI_ISL_1626261, EPI_ISL_1626262, EPI_ISL_1626263, EPI_ISL_1626264, EPI_ISL_1626265, EPI_ISL_1626266, EPI_ISL_1626270, EPI_ISL_1626271, EPI_ISL_1626272, EPI_ISL_1626273, EPI_ISL_1626274, EPI_ISL_1626275, EPI_ISL_1626276, EPI_ISL_1626277, EPI_ISL_1626278, EPI_ISL_1626279, EPI_ISL_1626280, EPI_ISL_1626281, EPI_ISL_1626282, EPI_ISL_1626283, EPI_ISL_1626284, EPI_ISL_1626285, EPI_ISL_1626286, EPI_ISL_1626287, EPI_ISL_1626288, EPI_ISL_1626289, EPI_ISL_1626290, EPI_ISL_1626292, EPI_ISL_1626293, EPI_ISL_1626294, EPI_ISL_1626295, EPI_ISL_1626296, EPI_ISL_1626298, EPI_ISL_1626300, EPI_ISL_1626301, EPI_ISL_1626303, EPI_ISL_1626304, EPI_ISL_1626305, EPI_ISL_1626306, EPI_ISL_1626307, EPI_ISL_1626308, EPI_ISL_1626309, EPI_ISL_1626310, EPI_ISL_1626311, EPI_ISL_1626313, EPI_ISL_1626315, EPI_ISL_1626316, EPI_ISL_1626317, EPI_ISL_1626318, EPI_ISL_1626320, EPI_ISL_1626321, EPI_ISL_1626322, EPI_ISL_1626323, EPI_ISL_1626324, EPI_ISL_1626325, EPI_ISL_1626326, EPI_ISL_1626327, EPI_ISL_1626328, EPI_ISL_1626329, EPI_ISL_1626330, EPI_ISL_1626331, EPI_ISL_1626332, EPI_ISL_1626333, EPI_ISL_1626334, EPI_ISL_1626335, EPI_ISL_1626336, EPI_ISL_1626337, EPI_ISL_1626338, EPI_ISL_1626339, EPI_ISL_1626340, EPI_ISL_1626341, EPI_ISL_1626342, EPI_ISL_1626343, EPI_ISL_1626344, EPI_ISL_1626345, EPI_ISL_1626346, EPI_ISL_1626347, EPI_ISL_1626348, EPI_ISL_1626349, EPI_ISL_1626350, EPI_ISL_1626351, EPI_ISL_1626352, EPI_ISL_1626353, EPI_ISL_1626354, EPI_ISL_1626355, EPI_ISL_1626356, EPI_ISL_1626357, EPI_ISL_1626359, EPI_ISL_1626360, EPI_ISL_1626361, EPI_ISL_1626362, EPI_ISL_1626363, EPI_ISL_1626364, EPI_ISL_1626365, EPI_ISL_1626366, EPI_ISL_1626367, EPI_ISL_1626368, EPI_ISL_1626369, EPI_ISL_1626370, EPI_ISL_1626371, EPI_ISL_1626372, EPI_ISL_1626373, EPI_ISL_1626374, EPI_ISL_1626375, EPI_ISL_1626376, EPI_ISL_1626377, EPI_ISL_1626378, EPI_ISL_1626379, EPI_ISL_1626380, EPI_ISL_1626381, EPI_ISL_1626382, EPI_ISL_1626383, EPI_ISL_1626384, EPI_ISL_1626385, EPI_ISL_1626386, EPI_ISL_1626387, EPI_ISL_1626388, EPI_ISL_1626389, EPI_ISL_1626390, EPI_ISL_1626392, EPI_ISL_1626393, EPI_ISL_1626394, EPI_ISL_1626395, EPI_ISL_1626396, EPI_ISL_1626397, EPI_ISL_1626398, EPI_ISL_1626399, EPI_ISL_1626400, EPI_ISL_1626401, EPI_ISL_1626402, EPI_ISL_1626403, EPI_ISL_1626404, EPI_ISL_1626405, EPI_ISL_1626406, EPI_ISL_1626407, EPI_ISL_1626408, EPI_ISL_1626409, EPI_ISL_1626410, EPI_ISL_1626411, EPI_ISL_1626412, EPI_ISL_1626413, EPI_ISL_1626414, EPI_ISL_1626415, EPI_ISL_1626416, EPI_ISL_1626417, EPI_ISL_1626418, EPI_ISL_1626419, EPI_ISL_1626420, EPI_ISL_1626421, EPI_ISL_1626422, EPI_ISL_1626423, EPI_ISL_1626424, EPI_ISL_1626425, EPI_ISL_1626426, EPI_ISL_1626427, EPI_ISL_1626428, EPI_ISL_1626429, EPI_ISL_1626430, EPI_ISL_1626431, EPI_ISL_1626432, EPI_ISL_1626433, EPI_ISL_1626434, EPI_ISL_1626435, EPI_ISL_1626436, EPI_ISL_1626437, EPI_ISL_1626438, EPI_ISL_1626439, EPI_ISL_1626440, EPI_ISL_1626441, EPI_ISL_1626442, EPI_ISL_1626443, EPI_ISL_1626444, EPI_ISL_1626445, EPI_ISL_1626446, EPI_ISL_1626447, EPI_ISL_1626448, EPI_ISL_1626449, EPI_ISL_1626450, EPI_ISL_1626451, EPI_ISL_1626452, EPI_ISL_1626453, EPI_ISL_1626454, EPI_ISL_1626455, EPI_ISL_1626456, EPI_ISL_1626457, EPI_ISL_1626458, EPI_ISL_1626459, EPI_ISL_1626460, EPI_ISL_1626461, EPI_ISL_1626462, EPI_ISL_1626463, EPI_ISL_1626464, EPI_ISL_1626465, EPI_ISL_1626466, EPI_ISL_1626467, EPI_ISL_1626468, EPI_ISL_1626469, EPI_ISL_1626470, EPI_ISL_1626471, EPI_ISL_1626472, EPI_ISL_1626473, EPI_ISL_1626475, EPI_ISL_1626476, EPI_ISL_1626477, EPI_ISL_1626478, EPI_ISL_1626479, EPI_ISL_1626480, EPI_ISL_1626481, EPI_ISL_1626482 | | | |
| see above | Wisconsin State Laboratory of Hygiene Communicable Disease Division | Wisconsin State Laboratory of Hygiene Communicable Disease Division | Kelsey R. Florek, Abigail C. Shockey |
| EPI_ISL_1626528, EPI_ISL_1626529, EPI_ISL_1626530, EPI_ISL_1626531, EPI_ISL_1626532, EPI_ISL_1626533, EPI_ISL_1626534, EPI_ISL_1626535 | Rush University Medical Center | RIPHL at Rush University Medical Center | Stefan Green, Kevin Kunstman, Max Kolton, Marieta Hyde, Laura Furtado, Hannah Barbian |
| EPI_ISL_1626536, EPI_ISL_1626537, EPI_ISL_1626538, EPI_ISL_1626539 | IN State Department of Health Laboratory Services | IN State Department of Health Laboratory Services | Cassandra Campion, Jamie Yeadon, Brian Pope, Lixia Liu, Kyle Brownlee, Melissa Hindenlang, Ankita Kashikar, Mark Glazier |
| EPI_ISL_1626540, EPI_ISL_1626541, EPI_ISL_1626542, EPI_ISL_1626543, EPI_ISL_1626544, EPI_ISL_1626545, EPI_ISL_1626546, EPI_ISL_1626547, EPI_ISL_1626548, EPI_ISL_1626549, EPI_ISL_1626550, EPI_ISL_1626551, EPI_ISL_1626552, EPI_ISL_1626553, EPI_ISL_1626554, EPI_ISL_1626555, EPI_ISL_1626556, EPI_ISL_1626557, EPI_ISL_1626558 | | | |
| see above | Nebraska Public Health Laboratory | NPHL COVID-19 Response Team | NPHL COVID-19 Response Team |
| EPI_ISL_1626559, EPI_ISL_1626560, EPI_ISL_1626561, EPI_ISL_1626562, EPI_ISL_1626563, EPI_ISL_1626564, EPI_ISL_1626565, EPI_ISL_1626566, EPI_ISL_1626573, EPI_ISL_1626574, EPI_ISL_1626575, EPI_ISL_1626576, EPI_ISL_1626577, EPI_ISL_1626578, EPI_ISL_1626579, EPI_ISL_1626580, EPI_ISL_1626581, EPI_ISL_1626582, EPI_ISL_1626583, EPI_ISL_1626584, EPI_ISL_1626585, EPI_ISL_1626586, EPI_ISL_1626587, EPI_ISL_1626588, EPI_ISL_1626589, EPI_ISL_1626590, EPI_ISL_1626591, EPI_ISL_1626592, EPI_ISL_1626593, EPI_ISL_1626594, EPI_ISL_1626595, EPI_ISL_1626596, EPI_ISL_1626597 | | | |
| see above | IN State Department of Health Laboratory Services | IN State Department of Health Laboratory Services | Cassandra Campion, Jamie Yeadon, Brian Pope, Lixia Liu, Kyle Brownlee, Melissa Hindenlang, Ankita Kashikar, Mark Glazier |
| EPI_ISL_1626598, EPI_ISL_1626599, EPI_ISL_1626600, EPI_ISL_1626601, EPI_ISL_1626602, EPI_ISL_1626603, EPI_ISL_1626604, EPI_ISL_1626605 | GA Department of Public Health | GA Department of Public Health | Stacy Reeves, Jonathan Edwards, Cynthia Dixey, Tonia Parrott, Aliyah Fields, Taylor Smith |
| EPI_ISL_1626614 | Instituto Nacional de Medicina Genomica | Instituto Nacional de Medicina Genomica | Hidalgo-Miranda A, Mendoza-Vargas A, Reyes-Grajeda JP, Cedro-Tanda A, Alcaraz N, Gonzalez-Barrera D, Rangel-DeLeon D, Miranda-Ortiz H, Rosas-Escobar P, Canseco Mendez JC, Munguia-Garza P, Ramirez-Vega O, Garcia-Cardenas FJ, Gonzalez-Woge MA, Herrera-Montalvo LA |
| EPI_ISL_1626622, EPI_ISL_1626623, EPI_ISL_1626624, EPI_ISL_1626625, EPI_ISL_1626626, EPI_ISL_1626627, EPI_ISL_1626628, EPI_ISL_1626629, EPI_ISL_1626630, EPI_ISL_1626631, EPI_ISL_1626632, EPI_ISL_1626633, EPI_ISL_1626634, EPI_ISL_1626635, EPI_ISL_1626637, EPI_ISL_1626638, EPI_ISL_1626639, EPI_ISL_1626640, EPI_ISL_1626641, EPI_ISL_1626642, EPI_ISL_1626643, EPI_ISL_1626644, EPI_ISL_1626645, EPI_ISL_1626646, EPI_ISL_1626647, EPI_ISL_1626648, EPI_ISL_1626649, EPI_ISL_1626650, EPI_ISL_1626651, EPI_ISL_1626652, EPI_ISL_1626653, EPI_ISL_1626654, EPI_ISL_1626655, EPI_ISL_1626656, EPI_ISL_1626657, EPI_ISL_1626658, EPI_ISL_1626659, EPI_ISL_1626660, EPI_ISL_1626661, EPI_ISL_1626662, EPI_ISL_1626663, EPI_ISL_1626664, EPI_ISL_1626665, EPI_ISL_1626666, EPI_ISL_1626667, EPI_ISL_1626668, EPI_ISL_1626669, EPI_ISL_1626670, EPI_ISL_1626671, EPI_ISL_1626672, EPI_ISL_1626673, EPI_ISL_1626674, EPI_ISL_1626675, EPI_ISL_1626676, EPI_ISL_1626677, EPI_ISL_1626678, EPI_ISL_1626679, EPI_ISL_1626680, EPI_ISL_1626681, EPI_ISL_1626682, EPI_ISL_1626683, EPI_ISL_1626684, EPI_ISL_1626685, EPI_ISL_1626686, EPI_ISL_1626687, EPI_ISL_1626688, EPI_ISL_1626689, EPI_ISL_1626691, EPI_ISL_1626692, EPI_ISL_1626693, EPI_ISL_1626694, EPI_ISL_1626695, EPI_ISL_1626696, EPI_ISL_1626697, EPI_ISL_1626698, EPI_ISL_1626699, EPI_ISL_1626700, EPI_ISL_1626701, EPI_ISL_1626702, EPI_ISL_1626703, EPI_ISL_1626704, EPI_ISL_1626705, EPI_ISL_1626706, EPI_ISL_1626707, EPI_ISL_1626708, EPI_ISL_1626709, EPI_ISL_1626710, EPI_ISL_1626711 | | | |
| see above | Texas Department of State Health Services (TXDSHS) | Texas Department of State Health Services (TXDSHS) | Rashmi Tuladhar, Bonnie Oh, Jenny Zhang, Maliha Rahman, Mayela Pedrueza, Anita Pokharel, Lorraine Rodriguez, Myong Koag, Chun Wang, Rachel Lee, Grace Kubin |
| EPI_ISL_1626786, EPI_ISL_1626787, EPI_ISL_1626788, EPI_ISL_1626789, EPI_ISL_1626790, EPI_ISL_1626791 | Kansas Health and Environmental Lab | Kansas Health and Environmental Lab | Mike Grose, Jonathan Barnell, Ben Olsen, and Phil Adam |
| EPI_ISL_1626792 | Clinical Refernce Laboratory | Kansas Health and Environmental Lab | Mike Grose, Jonathan Barnell, Ben Olsen, and Phil Adam |
| EPI_ISL_1626793 | Kansas Health and Environmental Lab | Kansas Health and Environmental Lab | Mike Grose, Jonathan Barnell, Ben Olsen, and Phil Adam |
| EPI_ISL_1626794, EPI_ISL_1626795, EPI_ISL_1626796, EPI_ISL_1626797, EPI_ISL_1626798, EPI_ISL_1626799, EPI_ISL_1626800, EPI_ISL_1626801 | LESP Chihuahua | Instituto de Diagnostico y Referencia Epidemiologicos (INDRE) | Claudia Wong-Arambula, Abril Rodriguez-Maldonado, Vanessa Rivero-Arredondo, Ariadna Medina-Benitez, Joaquin Quiroz-Mercado, Sergio Rangel-Guerrero, Natividad Cruz-Ortiz, Tatiana Nunez-Garcia, Gisela Barrera-Badillo, Lucia Hernandez-Rivas, Irma Lopez-Martinez, Ernesto Ramirez-Gonzalez. |
| EPI_ISL_1626802 | LESP Yucatan | Instituto de Diagnostico y Referencia Epidemiologicos (INDRE) | Claudia Wong-Arambula, Abril Rodriguez-Maldonado, Vanessa Rivero-Arredondo, Ariadna Medina-Benitez, Joaquin Quiroz-Mercado, Sergio Rangel-Guerrero, Natividad Cruz-Ortiz, Tatiana Nunez-Garcia, Gisela Barrera-Badillo, Lucia Hernandez-Rivas, Irma Lopez-Martinez, Ernesto Ramirez-Gonzalez. |
| EPI_ISL_1626803, EPI_ISL_1626804, EPI_ISL_1626805 | LESP Chihuahua | Instituto de Diagnostico y Referencia Epidemiologicos (INDRE) | Claudia Wong-Arambula, Abril Rodriguez-Maldonado, Vanessa Rivero-Arredondo, Ariadna Medina-Benitez, Joaquin Quiroz-Mercado, Sergio Rangel-Guerrero, Natividad Cruz-Ortiz, Tatiana Nunez-Garcia, Gisela Barrera-Badillo, Lucia Hernandez-Rivas, Irma Lopez-Martinez, Ernesto Ramirez-Gonzalez. |
| EPI_ISL_1626806 | LESP Nuevo Leon | Instituto de Diagnostico y Referencia Epidemiologicos (INDRE) | Claudia Wong-Arambula, Abril Rodriguez-Maldonado, Vanessa Rivero-Arredondo, Ariadna Medina-Benitez, Joaquin Quiroz-Mercado, Sergio Rangel-Guerrero, Natividad Cruz-Ortiz, Tatiana Nunez-Garcia, Gisela Barrera-Badillo, Lucia Hernandez-Rivas, Irma Lopez-Martinez, Ernesto Ramirez-Gonzalez. |
| EPI_ISL_1626807 | LESP Ciudad de Mexico | Instituto de Diagnostico y Referencia Epidemiologicos (INDRE) | Claudia Wong-Arambula, Abril Rodriguez-Maldonado, Vanessa Rivero-Arredondo, Ariadna Medina-Benitez, Joaquin Quiroz-Mercado, Sergio Rangel-Guerrero, Natividad Cruz-Ortiz, Tatiana Nunez-Garcia, Gisela Barrera-Badillo, Lucia Hernandez-Rivas, Irma Lopez-Martinez, Ernesto Ramirez-Gonzalez. |
| EPI_ISL_1626808 | LESP Hidalgo | Instituto de Diagnostico y Referencia Epidemiologicos (INDRE) | Claudia Wong-Arambula, Abril Rodriguez-Maldonado, Vanessa Rivero-Arredondo, Ariadna Medina-Benitez, Joaquin Quiroz-Mercado, Sergio Rangel-Guerrero, Natividad Cruz-Ortiz, Tatiana Nunez-Garcia, Gisela Barrera-Badillo, Lucia Hernandez-Rivas, Irma Lopez-Martinez, Ernesto Ramirez-Gonzalez. |
| EPI_ISL_1626809 | LESP Guanajuato | Instituto de Diagnostico y Referencia Epidemiologicos (INDRE) | Claudia Wong-Arambula, Abril Rodriguez-Maldonado, Vanessa Rivero-Arredondo, Ariadna Medina-Benitez, Joaquin Quiroz-Mercado, Sergio Rangel-Guerrero, Natividad Cruz-Ortiz, Tatiana Nunez-Garcia, Gisela Barrera-Badillo, Lucia Hernandez-Rivas, Irma Lopez-Martinez, Ernesto Ramirez-Gonzalez. |
| EPI_ISL_1626810 | LESP Chiapas | Instituto de Diagnostico y Referencia Epidemiologicos (INDRE) | Claudia Wong-Arambula, Abril Rodriguez-Maldonado, Vanessa Rivero-Arredondo, Ariadna Medina-Benitez, Joaquin Quiroz-Mercado, Sergio Rangel-Guerrero, Natividad Cruz-Ortiz, Tatiana Nunez-Garcia, Gisela Barrera-Badillo, Lucia Hernandez-Rivas, Irma Lopez-Martinez, Ernesto Ramirez-Gonzalez. |
| EPI_ISL_1626811, EPI_ISL_1626812, EPI_ISL_1626813, EPI_ISL_1626814, EPI_ISL_1626815 | LESP Baja California Sur | Instituto de Diagnostico y Referencia Epidemiologicos (INDRE) | Claudia Wong-Arambula, Abril Rodriguez-Maldonado, Vanessa Rivero-Arredondo, Ariadna Medina-Benitez, Joaquin Quiroz-Mercado, Sergio Rangel-Guerrero, Natividad Cruz-Ortiz, Tatiana Nunez-Garcia, Gisela Barrera-Badillo, Lucia Hernandez-Rivas, Irma Lopez-Martinez, Ernesto Ramirez-Gonzalez. |

| | | | |
|---|---|---|---|
| EPI_ISL_1626816 | LESP Guanajuato | Instituto de Diagnostico y Referencia Epidemiologicos (INDRE) | Claudia Wong-Arambula, Abril Rodriguez-Maldonado, Vanessa Rivero-Arredondo, Ariadna Medina-Benitez, Joaquin Quiroz-Mercado, Sergio Rangel-Guerrero, Natividad Cruz-Ortiz, Tatiana Nunez-Garcia, Gisela Barrera-Badillo, Lucia Hernandez-Rivas, Irma Lopez-Martinez, Ernesto Ramirez-Gonzalez. |
| EPI_ISL_1626817 | LESP Jalisco | Instituto de Diagnostico y Referencia Epidemiologicos (INDRE) | Claudia Wong-Arambula, Abril Rodriguez-Maldonado, Vanessa Rivero-Arredondo, Ariadna Medina-Benitez, Joaquin Quiroz-Mercado, Sergio Rangel-Guerrero, Natividad Cruz-Ortiz, Tatiana Nunez-Garcia, Gisela Barrera-Badillo, Lucia Hernandez-Rivas, Irma Lopez-Martinez, Ernesto Ramirez-Gonzalez. |
| EPI_ISL_1626818 | LESP Ciudad de Mexico | Instituto de Diagnostico y Referencia Epidemiologicos (INDRE) | Claudia Wong-Arambula, Abril Rodriguez-Maldonado, Vanessa Rivero-Arredondo, Ariadna Medina-Benitez, Joaquin Quiroz-Mercado, Sergio Rangel-Guerrero, Natividad Cruz-Ortiz, Tatiana Nunez-Garcia, Gisela Barrera-Badillo, Lucia Hernandez-Rivas, Irma Lopez-Martinez, Ernesto Ramirez-Gonzalez. |
| EPI_ISL_1626819, EPI_ISL_1626820 | LESP Guanajuato | Instituto de Diagnostico y Referencia Epidemiologicos (INDRE) | Claudia Wong-Arambula, Abril Rodriguez-Maldonado, Vanessa Rivero-Arredondo, Ariadna Medina-Benitez, Joaquin Quiroz-Mercado, Sergio Rangel-Guerrero, Natividad Cruz-Ortiz, Tatiana Nunez-Garcia, Gisela Barrera-Badillo, Lucia Hernandez-Rivas, Irma Lopez-Martinez, Ernesto Ramirez-Gonzalez. |
| EPI_ISL_1626821 | LESP Quintana Roo | Instituto de Diagnostico y Referencia Epidemiologicos (INDRE) | Claudia Wong-Arambula, Abril Rodriguez-Maldonado, Vanessa Rivero-Arredondo, Ariadna Medina-Benitez, Joaquin Quiroz-Mercado, Sergio Rangel-Guerrero, Natividad Cruz-Ortiz, Tatiana Nunez-Garcia, Gisela Barrera-Badillo, Lucia Hernandez-Rivas, Irma Lopez-Martinez, Ernesto Ramirez-Gonzalez. |
| EPI_ISL_1626822, EPI_ISL_1626823, EPI_ISL_1626824, EPI_ISL_1626825, EPI_ISL_1626826, EPI_ISL_1626827, EPI_ISL_1626828, EPI_ISL_1626829, EPI_ISL_1626830, EPI_ISL_1626831, EPI_ISL_1626832, EPI_ISL_1626833, EPI_ISL_1626834, EPI_ISL_1626835, EPI_ISL_1626836, EPI_ISL_1626837, EPI_ISL_1626838, EPI_ISL_1626839, EPI_ISL_1626840, EPI_ISL_1626841, EPI_ISL_1626842, EPI_ISL_1626843, EPI_ISL_1626844, EPI_ISL_1626845, EPI_ISL_1626846, EPI_ISL_1626847, EPI_ISL_1626848, EPI_ISL_1626849, EPI_ISL_1626850, EPI_ISL_1626851, EPI_ISL_1626852, EPI_ISL_1626853, EPI_ISL_1626854, EPI_ISL_1626855, EPI_ISL_1626856, EPI_ISL_1626857, EPI_ISL_1626858, EPI_ISL_1626859, EPI_ISL_1626860, EPI_ISL_1626861, EPI_ISL_1626862, EPI_ISL_1626863, EPI_ISL_1626864, EPI_ISL_1626865, EPI_ISL_1626866, EPI_ISL_1626867, EPI_ISL_1626868, EPI_ISL_1626869, EPI_ISL_1626870, EPI_ISL_1626871, EPI_ISL_1626872, EPI_ISL_1626873, EPI_ISL_1626874, EPI_ISL_1626875, EPI_ISL_1626876, EPI_ISL_1626877, EPI_ISL_1626878, EPI_ISL_1626879, EPI_ISL_1626880, EPI_ISL_1626881, EPI_ISL_1626882, EPI_ISL_1626883, EPI_ISL_1626884, EPI_ISL_1626885, EPI_ISL_1626886, EPI_ISL_1626887, EPI_ISL_1626888, EPI_ISL_1626889, EPI_ISL_1626890, EPI_ISL_1626891, EPI_ISL_1626892, EPI_ISL_1626893, EPI_ISL_1626894, EPI_ISL_1626895, EPI_ISL_1626896, EPI_ISL_1626897, EPI_ISL_1626898, EPI_ISL_1626899, EPI_ISL_1626900, EPI_ISL_1626901, EPI_ISL_1626902, EPI_ISL_1626903, EPI_ISL_1626904, EPI_ISL_1626905, EPI_ISL_1626906, EPI_ISL_1626907, EPI_ISL_1626908, EPI_ISL_1626909, EPI_ISL_1626910, EPI_ISL_1626911, EPI_ISL_1626912, EPI_ISL_1626913, EPI_ISL_1626914, EPI_ISL_1626915, EPI_ISL_1626916 | | | |
| see above | WI State Laboratory of Hygiene | Genomics and Discovery, Respiratory Viruses Branch, Division of Viral Diseases, Centers for Disease Control and Prevention | Jing Zhang, Yan Li, Brian Lynch, Anna Kelleher, Krista Queen, Anna Uehara, Peter Cook, Han Jia Justin Ng, Ying Tao, Clinton R. Paden, Haibin Wang, Suxiang Tong |
| EPI_ISL_1626917, EPI_ISL_1626918, EPI_ISL_1626919, EPI_ISL_1626920, EPI_ISL_1626921, EPI_ISL_1626922, EPI_ISL_1626923, EPI_ISL_1626924, EPI_ISL_1626925, EPI_ISL_1626926, EPI_ISL_1626927, EPI_ISL_1626928, EPI_ISL_1626929, EPI_ISL_1626930, EPI_ISL_1626931, EPI_ISL_1626932, EPI_ISL_1626933, EPI_ISL_1626934, EPI_ISL_1626935, EPI_ISL_1626936, EPI_ISL_1626937, EPI_ISL_1626938, EPI_ISL_1626939, EPI_ISL_1626940, EPI_ISL_1626941, EPI_ISL_1626942, EPI_ISL_1626943, EPI_ISL_1626944, EPI_ISL_1626945, EPI_ISL_1626946, EPI_ISL_1626947, EPI_ISL_1626948, EPI_ISL_1626949, EPI_ISL_1626950, EPI_ISL_1626951, EPI_ISL_1626952, EPI_ISL_1626953, EPI_ISL_1626954 | | | |
| see above | Global Health Research Complex | TIGSS | Benjamin W. Neuman, Melissa M. Kahl-Mcdonagh, Kurt A. Zuelke, Charlie D. Johnson, Allison Ficht, Sankar P. Chaki, Sierra Guidry, Joshua Hill, Richard Metz, Marcel Brun |
| EPI_ISL_1626955, EPI_ISL_1626956, EPI_ISL_1626957, EPI_ISL_1626958, EPI_ISL_1626959, EPI_ISL_1626960, EPI_ISL_1626961, EPI_ISL_1626962, EPI_ISL_1626963, EPI_ISL_1626964, EPI_ISL_1626965, EPI_ISL_1626966, EPI_ISL_1626967, EPI_ISL_1626968, EPI_ISL_1626969, EPI_ISL_1626970, EPI_ISL_1626971, EPI_ISL_1626972, EPI_ISL_1626973, EPI_ISL_1626974, EPI_ISL_1626975, EPI_ISL_1626976, EPI_ISL_1626977, EPI_ISL_1626978, EPI_ISL_1626979, EPI_ISL_1626980, EPI_ISL_1626981, EPI_ISL_1626982, EPI_ISL_1626983, EPI_ISL_1626984, EPI_ISL_1626985, EPI_ISL_1626986, EPI_ISL_1626987, EPI_ISL_1626988 | | | |
| see above | Global Health Research Complex | TxGen | Benjamin W. Neuman, Melissa M. Kahl-Mcdonagh, Kurt A. Zuelke, Charlie D. Johnson, Allison Ficht, Sankar P. Chaki, Sierra Guidry, Joshua Hill, Richard Metz, Marcel Brun |
| EPI_ISL_1626989, EPI_ISL_1626990, EPI_ISL_1626991, EPI_ISL_1626992, EPI_ISL_1626993, EPI_ISL_1626994, EPI_ISL_1626995, EPI_ISL_1626996, EPI_ISL_1626997, EPI_ISL_1626998, EPI_ISL_1626999, EPI_ISL_1627000, EPI_ISL_1627001, EPI_ISL_1627002, EPI_ISL_1627003, EPI_ISL_1627004, EPI_ISL_1627005, EPI_ISL_1627006, EPI_ISL_1627008, EPI_ISL_1627009, EPI_ISL_1627010, EPI_ISL_1627011, EPI_ISL_1627012, EPI_ISL_1627013, EPI_ISL_1627014, EPI_ISL_1627015, EPI_ISL_1627016, EPI_ISL_1627017, EPI_ISL_1627018, EPI_ISL_1627019, EPI_ISL_1627020, EPI_ISL_1627021, EPI_ISL_1627022, EPI_ISL_1627023, EPI_ISL_1627024, EPI_ISL_1627025, EPI_ISL_1627026, EPI_ISL_1627027, EPI_ISL_1627028, EPI_ISL_1627029, EPI_ISL_1627031, EPI_ISL_1627032, EPI_ISL_1627033, EPI_ISL_1627034, EPI_ISL_1627035, EPI_ISL_1627036, EPI_ISL_1627037, EPI_ISL_1627038, EPI_ISL_1627039, EPI_ISL_1627040, EPI_ISL_1627041, EPI_ISL_1627042, EPI_ISL_1627043, EPI_ISL_1627044, EPI_ISL_1627045, EPI_ISL_1627046, EPI_ISL_1627047, EPI_ISL_1627048, EPI_ISL_1627049, EPI_ISL_1627050, EPI_ISL_1627051, EPI_ISL_1627052, EPI_ISL_1627053, EPI_ISL_1627054, EPI_ISL_1627055, EPI_ISL_1627056, EPI_ISL_1627057, EPI_ISL_1627058, EPI_ISL_1627059, EPI_ISL_1627060, EPI_ISL_1627061, EPI_ISL_1627062, EPI_ISL_1627063, EPI_ISL_1627064, EPI_ISL_1627065, EPI_ISL_1627066, EPI_ISL_1627067, EPI_ISL_1627068, EPI_ISL_1627069, EPI_ISL_1627070, EPI_ISL_1627071, EPI_ISL_1627072, EPI_ISL_1627073, EPI_ISL_1627074, EPI_ISL_1627075, EPI_ISL_1627076, EPI_ISL_1627077, EPI_ISL_1627078 | | | |
| see above | Wyoming Public Health Laboratory | Center for Global Health, University of New Mexico Health Sciences Center | Daryl Domman, Kurt Schwalm, Rob Christensen, Wanda Manley, Cari Sloma, Noah Hull, Valerie Morley, Darrell Dinwiddie |
| EPI_ISL_1627079, EPI_ISL_1627080 | Hospital Sharp | Microbial Genomics Laboratory | Bruno Gomez-Gil, Julissa Enciso-Ibarra, Alejandra Garcia-Gasca, Daniel Fregoso-Rueda |
| EPI_ISL_1627081 | Imágenes Diagnósticas de Mazatlán | Microbial Genomics Laboratory | Bruno Gomez-Gil, Julissa Enciso-Ibarra, Alejandra Garcia-Gasca, Daniel Fregoso-Rueda |
| EPI_ISL_1627082 | Hospital Sharp | Microbial Genomics Laboratory | Bruno Gomez-Gil, Julissa Enciso-Ibarra, Alejandra Garcia-Gasca, Daniel Fregoso-Rueda |
| EPI_ISL_1627083, EPI_ISL_1627084 | Hospital Margarita Maza de Juárez | Microbial Genomics Laboratory | Bruno Gomez-Gil, Julissa Enciso-Ibarra, Alejandra Garcia-Gasca, Daniel Fregoso-Rueda |
| EPI_ISL_1627085 | Hospital Sharp | Microbial Genomics Laboratory | Bruno Gomez-Gil, Julissa Enciso-Ibarra, Alejandra Garcia-Gasca, Daniel Fregoso-Rueda |
| EPI_ISL_1627111, EPI_ISL_1627112, EPI_ISL_1627113, EPI_ISL_1627114, EPI_ISL_1627115, EPI_ISL_1627116, EPI_ISL_1627117, EPI_ISL_1627118, EPI_ISL_1627119, EPI_ISL_1627120, EPI_ISL_1627121, EPI_ISL_1627122, EPI_ISL_1627123, EPI_ISL_1627124, EPI_ISL_1627125, EPI_ISL_1627126, EPI_ISL_1627127, EPI_ISL_1627128, EPI_ISL_1627129, EPI_ISL_1627130, EPI_ISL_1627131, EPI_ISL_1627132, EPI_ISL_1627133, EPI_ISL_1627134, EPI_ISL_1627135, EPI_ISL_1627136, EPI_ISL_1627137, EPI_ISL_1627138, EPI_ISL_1627139, EPI_ISL_1627140, EPI_ISL_1627141, EPI_ISL_1627142, EPI_ISL_1627143, EPI_ISL_1627144, EPI_ISL_1627145, EPI_ISL_1627146, EPI_ISL_1627147, EPI_ISL_1627148, EPI_ISL_1627149, EPI_ISL_1627150, EPI_ISL_1627151, EPI_ISL_1627152, EPI_ISL_1627153, EPI_ISL_1627154, EPI_ISL_1627155, EPI_ISL_1627156 | | | |
| see above | Wyoming Public Health Laboratory | Center for Global Health, University of New Mexico Health Sciences Center | Daryl Domman, Kurt Schwalm, Rob Christensen, Wanda Manley, Cari Sloma, Noah Hull, Valerie Morley, Darrell Dinwiddie |
| EPI_ISL_1627251, EPI_ISL_1627252, EPI_ISL_1627253, EPI_ISL_1627254, EPI_ISL_1627255, EPI_ISL_1627256, EPI_ISL_1627257, EPI_ISL_1627258, EPI_ISL_1627259, EPI_ISL_1627260, EPI_ISL_1627261, EPI_ISL_1627262, EPI_ISL_1627263, EPI_ISL_1627264, EPI_ISL_1627265, EPI_ISL_1627266, EPI_ISL_1627267, EPI_ISL_1627283, EPI_ISL_1627284 | | | |
| see above | North Dakota Department of Health, Public Health Laboratory | North Dakota Department of Health, Public Health Laboratory | Lisa Wingerter |
| EPI_ISL_1627285, EPI_ISL_1627287, EPI_ISL_1627288, EPI_ISL_1627292, EPI_ISL_1627293, EPI_ISL_1627294, EPI_ISL_1627295, EPI_ISL_1627297, EPI_ISL_1627300, EPI_ISL_1627301, EPI_ISL_1627302, EPI_ISL_1627303, EPI_ISL_1627304, EPI_ISL_1627305, EPI_ISL_1627306, EPI_ISL_1627307, EPI_ISL_1627308, EPI_ISL_1627310, EPI_ISL_1627311, EPI_ISL_1627312, EPI_ISL_1627313, EPI_ISL_1627314, EPI_ISL_1627315, EPI_ISL_1627316, EPI_ISL_1627317, EPI_ISL_1627318, EPI_ISL_1627319 | | | |
| see above | Idaho Bureau of Laboratories | Idaho Bureau of Laboratories | R. Beukelman, Matthew Charles Burns, Aimee Ceniseros, Robert L. Voermans, Christopher Ball |
| EPI_ISL_1627364, EPI_ISL_1627365, EPI_ISL_1627366, EPI_ISL_1627367, EPI_ISL_1627368, EPI_ISL_1627369, EPI_ISL_1627370, EPI_ISL_1627371, EPI_ISL_1627372, EPI_ISL_1627373, EPI_ISL_1627374, EPI_ISL_1627375, EPI_ISL_1627376, EPI_ISL_1627377, EPI_ISL_1627378, EPI_ISL_1627379, EPI_ISL_1627380, EPI_ISL_1627381, EPI_ISL_1627382, EPI_ISL_1627383, EPI_ISL_1627384, EPI_ISL_1627385, EPI_ISL_1627386, EPI_ISL_1627387, EPI_ISL_1627388, EPI_ISL_1627389, EPI_ISL_1627390, EPI_ISL_1627392, EPI_ISL_1627393, EPI_ISL_1627394 | | | |
| see above | North Dakota Department of Health, Public Health Laboratory | North Dakota Department of Health, Public Health Laboratory | Lisa Wingerter |
| EPI_ISL_1627395, EPI_ISL_1627396, EPI_ISL_1627397, EPI_ISL_1627398, EPI_ISL_1627399, EPI_ISL_1627400, EPI_ISL_1627401, EPI_ISL_1627402, EPI_ISL_1627403, EPI_ISL_1627404, EPI_ISL_1627405, EPI_ISL_1627406, EPI_ISL_1627407, EPI_ISL_1627408, EPI_ISL_1627409, EPI_ISL_1627410, EPI_ISL_1627411, EPI_ISL_1627412, EPI_ISL_1627413, EPI_ISL_1627414, EPI_ISL_1627415, EPI_ISL_1627416, EPI_ISL_1627417, EPI_ISL_1627418, EPI_ISL_1627419, EPI_ISL_1627420, EPI_ISL_1627421, EPI_ISL_1627422, EPI_ISL_1627423, EPI_ISL_1627424, EPI_ISL_1627425, EPI_ISL_1627426, EPI_ISL_1627427, EPI_ISL_1627428, EPI_ISL_1627429, EPI_ISL_1627430, EPI_ISL_1627431, EPI_ISL_1627432, EPI_ISL_1627433, EPI_ISL_1627434, EPI_ISL_1627435, EPI_ISL_1627436, EPI_ISL_1627437, EPI_ISL_1627438, EPI_ISL_1627439, EPI_ISL_1627440, EPI_ISL_1627441, EPI_ISL_1627442, EPI_ISL_1627443, EPI_ISL_1627444, EPI_ISL_1627445, EPI_ISL_1627446, EPI_ISL_1627447, EPI_ISL_1627448, EPI_ISL_1627449, EPI_ISL_1627450, EPI_ISL_1627451, EPI_ISL_1627452, EPI_ISL_1627453, EPI_ISL_1627454, EPI_ISL_1627455, EPI_ISL_1627456, EPI_ISL_1627457, EPI_ISL_1627458, EPI_ISL_1627459, EPI_ISL_1627460, EPI_ISL_1627461, EPI_ISL_1627462, EPI_ISL_1627463, EPI_ISL_1627464, EPI_ISL_1627465, EPI_ISL_1627466, EPI_ISL_1627467, EPI_ISL_1627468, EPI_ISL_1627469, EPI_ISL_1627470 | | | |
| see above | Baylor Scott & White-Temple | Baylor Scott & White-Temple | Ari Rao, Linden Morales, Kimberly Walker, Marcus Volz, Shelby Hendrickson, Caitlin Maloney |
| EPI_ISL_1627471, EPI_ISL_1627472, EPI_ISL_1627474, EPI_ISL_1627475, EPI_ISL_1627476, EPI_ISL_1627477, EPI_ISL_1627478, EPI_ISL_1627479, EPI_ISL_1627480, EPI_ISL_1627481, EPI_ISL_1627482, EPI_ISL_1627483 | | | |
| see above | Nevada State Public Health Laboratory | Nevada State Public Health Laboratory | Andrew Gorzalski, Mark Pandori |
| EPI_ISL_1627484, EPI_ISL_1627485, EPI_ISL_1627486, EPI_ISL_1627487, EPI_ISL_1627488, EPI_ISL_1627489, EPI_ISL_1627490, EPI_ISL_1627491, EPI_ISL_1627492, EPI_ISL_1627493, EPI_ISL_1627494, EPI_ISL_1627495, EPI_ISL_1627496, EPI_ISL_1627497, EPI_ISL_1627498, EPI_ISL_1627499, EPI_ISL_1627500, EPI_ISL_1627501, EPI_ISL_1627502, EPI_ISL_1627503, EPI_ISL_1627504, EPI_ISL_1627505, EPI_ISL_1627506, EPI_ISL_1627507, EPI_ISL_1627508, EPI_ISL_1627509, EPI_ISL_1627510, EPI_ISL_1627511, EPI_ISL_1627512, EPI_ISL_1627513, EPI_ISL_1627514, EPI_ISL_1627515, EPI_ISL_1627516, EPI_ISL_1627517, EPI_ISL_1627518, EPI_ISL_1627519, EPI_ISL_1627520, EPI_ISL_1627521, EPI_ISL_1627522, EPI_ISL_1627523, EPI_ISL_1627524, EPI_ISL_1627525, EPI_ISL_1627545, EPI_ISL_1627546, EPI_ISL_1627547, EPI_ISL_1627548, EPI_ISL_1627549, EPI_ISL_1627550, EPI_ISL_1627551, EPI_ISL_1627552, EPI_ISL_1627553, EPI_ISL_1627554, EPI_ISL_1627555, EPI_ISL_1627556, EPI_ISL_1627557, EPI_ISL_1627558, EPI_ISL_1627559 | | | |
| see above | UTGSAF | UTGSAF | Jessica Podnar, Sylvie Beaudenon, Audrey Kelly, Zachary Carver, Anna Battenhouse, Andreas Matouschek |
| EPI_ISL_1628009, EPI_ISL_1628010, EPI_ISL_1628011, EPI_ISL_1628012, EPI_ISL_1628013, EPI_ISL_1628014, EPI_ISL_1628015, EPI_ISL_1628016, EPI_ISL_1628017, EPI_ISL_1628018, EPI_ISL_1628019, EPI_ISL_1628020, EPI_ISL_1628021, EPI_ISL_1628022, EPI_ISL_1628023, EPI_ISL_1628024, EPI_ISL_1628025, EPI_ISL_1628026, EPI_ISL_1628027, EPI_ISL_1628028, EPI_ISL_1628029, EPI_ISL_1628030, EPI_ISL_1628031, EPI_ISL_1628032, EPI_ISL_1628033, EPI_ISL_1628034, EPI_ISL_1628035, EPI_ISL_1628036, EPI_ISL_1628037, EPI_ISL_1628038, EPI_ISL_1628039, EPI_ISL_1628040, EPI_ISL_1628041, EPI_ISL_1628042, EPI_ISL_1628043, EPI_ISL_1628044, EPI_ISL_1628045, EPI_ISL_1628046, EPI_ISL_1628047, EPI_ISL_1628048, EPI_ISL_1628049, EPI_ISL_1628050, EPI_ISL_1628051, EPI_ISL_1628052, EPI_ISL_1628053, EPI_ISL_1628054, EPI_ISL_1628055, EPI_ISL_1628056, EPI_ISL_1628057, EPI_ISL_1628058, EPI_ISL_1628059, EPI_ISL_1628060, EPI_ISL_1628061, EPI_ISL_1628062, EPI_ISL_1628063, EPI_ISL_1628064, EPI_ISL_1628065, EPI_ISL_1628066, EPI_ISL_1628067, EPI_ISL_1628068, EPI_ISL_1628069, EPI_ISL_1628070, EPI_ISL_1628071, EPI_ISL_1628072, EPI_ISL_1628073, EPI_ISL_1628074, EPI_ISL_1628075, EPI_ISL_1628076, EPI_ISL_1628077, EPI_ISL_1628078, EPI_ISL_1628079, EPI_ISL_1628080, EPI_ISL_1628081, EPI_ISL_1628082, EPI_ISL_1628083, EPI_ISL_1628084, EPI_ISL_1628085, EPI_ISL_1628086, EPI_ISL_1628087, EPI_ISL_1628088, EPI_ISL_1628089, EPI_ISL_1628090, EPI_ISL_1628091, EPI_ISL_1628092, EPI_ISL_1628093, EPI_ISL_1628094, EPI_ISL_1628095, EPI_ISL_1628096, EPI_ISL_1628097, EPI_ISL_1628098, EPI_ISL_1628099, EPI_ISL_1628100, EPI_ISL_1628101, EPI_ISL_1628102, EPI_ISL_1628103, EPI_ISL_1628104, EPI_ISL_1628105, EPI_ISL_1628106, EPI_ISL_1628107, EPI_ISL_1628108, EPI_ISL_1628109, EPI_ISL_1628110, EPI_ISL_1628111, EPI_ISL_1628112, EPI_ISL_1628113, EPI_ISL_1628114, EPI_ISL_1628115, EPI_ISL_1628116, EPI_ISL_1628117, EPI_ISL_1628118, EPI_ISL_1628119, EPI_ISL_1628120, EPI_ISL_1628121, EPI_ISL_1628122, EPI_ISL_1628123, EPI_ISL_1628124, EPI_ISL_1628125, EPI_ISL_1628126, EPI_ISL_1628127, EPI_ISL_1628128, EPI_ISL_1628129, EPI_ISL_1628130, EPI_ISL_1628131, EPI_ISL_1628132, EPI_ISL_1628133, EPI_ISL_1628134, EPI_ISL_1628135, EPI_ISL_1628136, EPI_ISL_1628137, EPI_ISL_1628138, EPI_ISL_1628139, EPI_ISL_1628140, EPI_ISL_1628141, EPI_ISL_1628142, EPI_ISL_1628143, EPI_ISL_1628144, EPI_ISL_1628145, EPI_ISL_1628146, EPI_ISL_1628147, EPI_ISL_1628148, EPI_ISL_1628163, EPI_ISL_1628164, EPI_ISL_1628165, EPI_ISL_1628166, EPI_ISL_1628167, EPI_ISL_1628168, EPI_ISL_1628169, EPI_ISL_1628170, EPI_ISL_1628171, EPI_ISL_1628172, EPI_ISL_1628173, EPI_ISL_1628174, EPI_ISL_1628175, EPI_ISL_1628176, EPI_ISL_1628177, EPI_ISL_1628178, EPI_ISL_1628179, EPI_ISL_1628180, EPI_ISL_1628181, EPI_ISL_1628182, EPI_ISL_1628183, EPI_ISL_1628184, EPI_ISL_1628185, EPI_ISL_1628186, EPI_ISL_1628187, EPI_ISL_1628188, EPI_ISL_1628189, EPI_ISL_1628190, EPI_ISL_1628191, EPI_ISL_1628192, EPI_ISL_1628193, EPI_ISL_1628194, EPI_ISL_1628195, EPI_ISL_1628196, EPI_ISL_1628197, EPI_ISL_1628198, EPI_ISL_1628199, EPI_ISL_1628200, EPI_ISL_1628201, EPI_ISL_1628202, EPI_ISL_1628203, EPI_ISL_1628204, EPI_ISL_1628205, EPI_ISL_1628206, EPI_ISL_1628207, EPI_ISL_1628208, EPI_ISL_1628209, EPI_ISL_1628210, EPI_ISL_1628211, EPI_ISL_1628212, EPI_ISL_1628213, EPI_ISL_1628214, EPI_ISL_1628215, EPI_ISL_1628216, EPI_ISL_1628217, EPI_ISL_1628218, EPI_ISL_1628219, EPI_ISL_1628220, EPI_ISL_1628221, EPI_ISL_1628222, EPI_ISL_1628223, EPI_ISL_1628224, EPI_ISL_1628225, EPI_ISL_1628226, EPI_ISL_1628227, EPI_ISL_1628228, EPI_ISL_1628229, EPI_ISL_1628230, EPI_ISL_1628231, EPI_ISL_1628232, EPI_ISL_1628233, EPI_ISL_1628234, EPI_ISL_1628235, EPI_ISL_1628236, EPI_ISL_1628237, EPI_ISL_1628238, EPI_ISL_1628239, EPI_ISL_1628240, EPI_ISL_1628241, EPI_ISL_1628242, EPI_ISL_1628243, EPI_ISL_1628244, EPI_ISL_1628245, EPI_ISL_1628246, EPI_ISL_1628247, EPI_ISL_1628248, EPI_ISL_1628249, EPI_ISL_1628250, EPI_ISL_1628251, EPI_ISL_1628252, EPI_ISL_1628253, EPI_ISL_1628254, EPI_ISL_1628255, EPI_ISL_1628256, EPI_ISL_1628257, EPI_ISL_1628258, EPI_ISL_1628259, EPI_ISL_1628260, EPI_ISL_1628261, EPI_ISL_1628262, EPI_ISL_1628263, EPI_ISL_1628264, EPI_ISL_1628265, EPI_ISL_1628266, EPI_ISL_1628267, EPI_ISL_1628268, EPI_ISL_1628269, EPI_ISL_1628270, EPI_ISL_1628271, EPI_ISL_1628272, EPI_ISL_1628273, EPI_ISL_1628274, EPI_ISL_1628275, EPI_ISL_1628276, EPI_ISL_1628277, EPI_ISL_1628278, EPI_ISL_1628279, EPI_ISL_1628280, EPI_ISL_1628281, EPI_ISL_1628282, EPI_ISL_1628283, EPI_ISL_1628284, EPI_ISL_1628285, EPI_ISL_1628286, EPI_ISL_1628287, EPI_ISL_1628288, EPI_ISL_1628289, EPI_ISL_1628290, EPI_ISL_1628291, EPI_ISL_1628292, EPI_ISL_1628293, EPI_ISL_1628294, EPI_ISL_1628295, EPI_ISL_1628296, EPI_ISL_1628297, EPI_ISL_1628298, EPI_ISL_1628299, EPI_ISL_1628300, EPI_ISL_1628301, EPI_ISL_1628302, EPI_ISL_1628303, EPI_ISL_1628304, EPI_ISL_1628305, EPI_ISL_1628306, EPI_ISL_1628307, EPI_ISL_1628308, EPI_ISL_1628309, EPI_ISL_1628310, EPI_ISL_1628311, EPI_ISL_1628312, EPI_ISL_1628313, EPI_ISL_1628314, EPI_ISL_1628315, EPI_ISL_1628316, EPI_ISL_1628317, EPI_ISL_1628318, EPI_ISL_1628319, EPI_ISL_1628320, EPI_ISL_1628321, EPI_ISL_1628322, EPI_ISL_1628323, EPI_ISL_1628324 | | | |
| see above | UW Virology Lab | UW Virology Lab | Pavitra Roychoudhury, Hong Xie, Lasata Shrestha, Shah Mohamed Bakhash, Michelle Lin, Noah R. Baker, Sean Ellis, Meei-Li Huang, Keith R Jerome, Alexander Greninger |
| EPI_ISL_1628475, EPI_ISL_1628476, EPI_ISL_1628477, EPI_ISL_1628478, EPI_ISL_1628479, EPI_ISL_1628482, EPI_ISL_1628483, EPI_ISL_1628484, EPI_ISL_1628485, EPI_ISL_1628486, EPI_ISL_1628487, EPI_ISL_1628488, EPI_ISL_1628489, EPI_ISL_1628490, EPI_ISL_1628491, EPI_ISL_1628492, EPI_ISL_1628493, EPI_ISL_1628494, EPI_ISL_1628495, EPI_ISL_1628496, EPI_ISL_1628497, EPI_ISL_1628498 | | | |
| see above | University of Mississippi Medical Center, Department of Pathology | University of Mississippi Medical Center, Molecular and Genomics Core Facility | Ashley C. Johnson, Ithiel J. Frame, Krishna K. Ayyalasomayajula, Michael R. Garrett, D. Ashley Robinson |
| EPI_ISL_1628500 | Nationwide Children's Hospital | James Molecular Lab | H Wang, D Jones, A Leber |
| EPI_ISL_1628520, EPI_ISL_1628521, EPI_ISL_1628522, EPI_ISL_1628523, EPI_ISL_1628524, EPI_ISL_1628525, EPI_ISL_1628526, EPI_ISL_1628527, EPI_ISL_1628529, EPI_ISL_1628531, EPI_ISL_1628532, EPI_ISL_1628533, EPI_ISL_1628534, EPI_ISL_1628535, EPI_ISL_1628536, EPI_ISL_1628537, EPI_ISL_1628540, EPI_ISL_1628541, EPI_ISL_1628542, EPI_ISL_1628543, EPI_ISL_1628544, EPI_ISL_1628547, EPI_ISL_1628548, EPI_ISL_1628549, EPI_ISL_1628550, EPI_ISL_1628552, EPI_ISL_1628553, EPI_ISL_1628554, EPI_ISL_1628556, EPI_ISL_1628557, EPI_ISL_1628558, EPI_ISL_1628559, EPI_ISL_1628560, EPI_ISL_1628562, EPI_ISL_1628563, EPI_ISL_1628564, EPI_ISL_1628565, EPI_ISL_1628566, EPI_ISL_1628568, EPI_ISL_1628569, EPI_ISL_1628571, EPI_ISL_1628572, EPI_ISL_1628574, EPI_ISL_1628575, EPI_ISL_1628577, EPI_ISL_1628578, EPI_ISL_1628579, EPI_ISL_1628580, EPI_ISL_1628581, EPI_ISL_1628582, EPI_ISL_1628583, EPI_ISL_1628584, EPI_ISL_1628585, EPI_ISL_1628586, EPI_ISL_1628587, EPI_ISL_1628588, EPI_ISL_1628590, EPI_ISL_1628591, EPI_ISL_1628592, EPI_ISL_1628593, EPI_ISL_1628594, EPI_ISL_1628595 | | | |
| see above | Instituto Nacional de Medicina Genomica | Instituto Nacional de Medicina Genomica | Hidalgo-Miranda A, Mendoza-Vargas A, Reyes-Grajeda JP, Cedro-Tanda A, Alcaraz N, Gonzalez-Barrera D, Rangel-DeLeon D, Miranda-Ortiz H, Rosas-Escobar P, Canseco Mendez JC, Munguia-Garza P, Ramirez-Vega O, Garcia-Cardenas FJ, Gonzalez-Woge MA, Herrera-Montalvo LA |
| EPI_ISL_1628596, EPI_ISL_1628597, EPI_ISL_1628598, EPI_ISL_1628599, EPI_ISL_1628600, EPI_ISL_1628601, EPI_ISL_1628602, EPI_ISL_1628603, EPI_ISL_1628604, EPI_ISL_1628605, EPI_ISL_1628606, EPI_ISL_1628607, EPI_ISL_1628608, EPI_ISL_1628609, EPI_ISL_1628610, EPI_ISL_1628611, EPI_ISL_1628612, EPI_ISL_1628613, EPI_ISL_1628614, EPI_ISL_1628615, EPI_ISL_1628616, EPI_ISL_1628617, EPI_ISL_1628618, EPI_ISL_1628619, EPI_ISL_1628620, EPI_ISL_1628621, EPI_ISL_1628622, EPI_ISL_1628623, EPI_ISL_1628624, EPI_ISL_1628625, EPI_ISL_1628626, EPI_ISL_1628627, EPI_ISL_1628628, EPI_ISL_1628629, EPI_ISL_1628630, EPI_ISL_1628631, EPI_ISL_1628632, EPI_ISL_1628633, EPI_ISL_1628634, EPI_ISL_1628635, EPI_ISL_1628636, EPI_ISL_1628637, EPI_ISL_1628638, EPI_ISL_1628639, EPI_ISL_1628640, EPI_ISL_1628642, EPI_ISL_1628643, EPI_ISL_1628644, EPI_ISL_1628645, EPI_ISL_1628646, EPI_ISL_1628647, EPI_ISL_1628648, EPI_ISL_1628649, EPI_ISL_1628650, EPI_ISL_1628651, EPI_ISL_1628652, EPI_ISL_1628653, EPI_ISL_1628654, EPI_ISL_1628656, EPI_ISL_1628657, EPI_ISL_1628658, EPI_ISL_1628659, EPI_ISL_1628660, EPI_ISL_1628661, EPI_ISL_1628662, EPI_ISL_1628663, EPI_ISL_1628664, EPI_ISL_1628665, EPI_ISL_1628666, EPI_ISL_1628667, EPI_ISL_1628668, EPI_ISL_1628669, EPI_ISL_1628670, EPI_ISL_1628671, EPI_ISL_1628672, EPI_ISL_1628673, EPI_ISL_1628674, EPI_ISL_1628675, EPI_ISL_1628676, EPI_ISL_1628677, EPI_ISL_1628678, EPI_ISL_1628679, EPI_ISL_1628680, EPI_ISL_1628681, EPI_ISL_1628682, EPI_ISL_1628683, EPI_ISL_1628684 | | | |
| see above | InDRE | Instituto Nacional de Medicina Genomica | Hidalgo-Miranda A, Mendoza-Vargas A, Reyes-Grajeda JP, Cedro-Tanda A, Alcaraz N, Gisela Barrera-Badillo, Irma Lopez-Martinez, Jose Ernesto Ramirez González, Gonzalez-Barrera D, Rangel-DeLeon D, Miranda-Ortiz H, Rosas-Escobar P, Canseco Mendez JC, Munguia-Garza P, Ramirez- |

| | | | |
|---|---|---|---|
| | | | Vega O, Garcia-Cardenas FJ, Gonzalez-Woge MA, Herrera-Montalvo LA. |
| EPI_ISL_1629709 | Infectious Disease Diagnostics laboratory at the Children's Hospital of Philadelphia | Planet Lab, Children's Hospital of Philadelphia | Ahmed M. Moustafa, Colleen Bianco, Lidiya Denu, Azad Ahmed, Rebecca M. Harris, Josh Chang Mell, Paul J. Planet |
| EPI_ISL_1629715 | UW Virology Lab | UW Virology Lab | Pavitra Roychoudhury, Hong Xie, Lasata Shrestha, Shah Mohamed Bakhash, Michelle Lin, Noah R. Baker, Sean Ellis, Saraswathi Sathees, Meei-Li Huang, Keith R Jerome, Alexander Greninger |
| EPI_ISL_1630159, EPI_ISL_1630160, EPI_ISL_1630161, EPI_ISL_1630162, EPI_ISL_1630164 | UW Virology Lab | UW Virology Lab | Pavitra Roychoudhury, Hong Xie, Lasata Shrestha, Shah Mohamed Bakhash, Michelle Lin, Noah R. Baker, Sean Ellis, Meei-Li Huang, Keith R Jerome, Alexander Greninger |
| EPI_ISL_1632535, EPI_ISL_1632536, EPI_ISL_1632537, EPI_ISL_1632538, EPI_ISL_1632539, EPI_ISL_1632540, EPI_ISL_1632541, EPI_ISL_1632542, EPI_ISL_1632543, EPI_ISL_1632544, EPI_ISL_1632545, EPI_ISL_1632546, EPI_ISL_1632547, EPI_ISL_1632548, EPI_ISL_1632550, EPI_ISL_1632552, EPI_ISL_1632553, EPI_ISL_1632554, EPI_ISL_1632555, EPI_ISL_1632556, EPI_ISL_1632557, EPI_ISL_1632558, EPI_ISL_1632559, EPI_ISL_1632560, EPI_ISL_1632561, EPI_ISL_1632562, EPI_ISL_1632563, EPI_ISL_1632564, EPI_ISL_1632565, EPI_ISL_1632566, EPI_ISL_1632567, EPI_ISL_1632568, EPI_ISL_1632569, EPI_ISL_1632570, EPI_ISL_1632571, EPI_ISL_1632572, EPI_ISL_1632573, EPI_ISL_1632574, EPI_ISL_1632575, EPI_ISL_1632576, EPI_ISL_1632577, EPI_ISL_1632578, EPI_ISL_1632579, EPI_ISL_1632580, EPI_ISL_1632581, EPI_ISL_1632582, EPI_ISL_1632583, EPI_ISL_1632584, EPI_ISL_1632585, EPI_ISL_1632586, EPI_ISL_1632587, EPI_ISL_1632588, EPI_ISL_1632589, EPI_ISL_1632590, EPI_ISL_1632591, EPI_ISL_1632594, EPI_ISL_1632595, EPI_ISL_1632596, EPI_ISL_1632597, EPI_ISL_1632598, EPI_ISL_1632599, EPI_ISL_1632600, EPI_ISL_1632601, EPI_ISL_1632602, EPI_ISL_1632603, EPI_ISL_1632604, EPI_ISL_1632605, EPI_ISL_1632606, EPI_ISL_1632607, EPI_ISL_1632608, EPI_ISL_1632609, EPI_ISL_1632610, EPI_ISL_1632611, EPI_ISL_1632612, EPI_ISL_1632613, EPI_ISL_1632614, EPI_ISL_1632615, EPI_ISL_1632616, EPI_ISL_1632617, EPI_ISL_1632618, EPI_ISL_1632621, EPI_ISL_1632622, EPI_ISL_1632623, EPI_ISL_1632624, EPI_ISL_1632625, EPI_ISL_1632626, EPI_ISL_1632627, EPI_ISL_1632628, EPI_ISL_1632629, EPI_ISL_1632631, EPI_ISL_1632633, EPI_ISL_1632634, EPI_ISL_1632636, EPI_ISL_1632637, EPI_ISL_1632638, EPI_ISL_1632639, EPI_ISL_1632640, EPI_ISL_1632641, EPI_ISL_1632642, EPI_ISL_1632643, EPI_ISL_1632644, EPI_ISL_1632645, EPI_ISL_1632646, EPI_ISL_1632647, EPI_ISL_1632649, EPI_ISL_1632650, EPI_ISL_1632651, EPI_ISL_1632652, EPI_ISL_1632653, EPI_ISL_1632654, EPI_ISL_1632659, EPI_ISL_1632660, EPI_ISL_1632661, EPI_ISL_1632662, EPI_ISL_1632664, EPI_ISL_1632665, EPI_ISL_1632666, EPI_ISL_1632667, EPI_ISL_1632668, EPI_ISL_1632669, EPI_ISL_1632670, EPI_ISL_1632671, EPI_ISL_1632672, EPI_ISL_1632673, EPI_ISL_1632674, EPI_ISL_1632675, EPI_ISL_1632676, EPI_ISL_1632677, EPI_ISL_1632678, EPI_ISL_1632679, EPI_ISL_1632680, EPI_ISL_1632681, EPI_ISL_1632682, EPI_ISL_1632683, EPI_ISL_1632684, EPI_ISL_1632685, EPI_ISL_1632686, EPI_ISL_1632687, EPI_ISL_1632688, EPI_ISL_1632689, EPI_ISL_1632690, EPI_ISL_1632691, EPI_ISL_1632692, EPI_ISL_1632693, EPI_ISL_1632694, EPI_ISL_1632695, EPI_ISL_1632696, EPI_ISL_1632697, EPI_ISL_1632698, EPI_ISL_1632699, EPI_ISL_1632700, EPI_ISL_1632701, EPI_ISL_1632702 | | | |
| see above | Virginia Division of Consolidated Laboratory Services | Virginia Division of Consolidated Laboratory Services | Virginia DCLS |
| EPI_ISL_1632709, EPI_ISL_1632710 | Gundersen Clinical Microbiology Laboratory | Kabara Cancer Research Institute | Craig S. Richmond, Paraic A. Kenny |
| EPI_ISL_1633124, EPI_ISL_1633125, EPI_ISL_1633127, EPI_ISL_1633128, EPI_ISL_1633129, EPI_ISL_1633130, EPI_ISL_1633131, EPI_ISL_1633132, EPI_ISL_1633133, EPI_ISL_1633134, EPI_ISL_1633135, EPI_ISL_1633136, EPI_ISL_1633139, EPI_ISL_1633140, EPI_ISL_1633141, EPI_ISL_1633142, EPI_ISL_1633144, EPI_ISL_1633145, EPI_ISL_1633146, EPI_ISL_1633147, EPI_ISL_1633148, EPI_ISL_1633149, EPI_ISL_1633150, EPI_ISL_1633151, EPI_ISL_1633153, EPI_ISL_1633154, EPI_ISL_1633155, EPI_ISL_1633156, EPI_ISL_1633157, EPI_ISL_1633158, EPI_ISL_1633159, EPI_ISL_1633160, EPI_ISL_1633161, EPI_ISL_1633162, EPI_ISL_1633163, EPI_ISL_1633164, EPI_ISL_1633165, EPI_ISL_1633166, EPI_ISL_1633167, EPI_ISL_1633168, EPI_ISL_1633169, EPI_ISL_1633170, EPI_ISL_1633171, EPI_ISL_1633172, EPI_ISL_1633173, EPI_ISL_1633174, EPI_ISL_1633175, EPI_ISL_1633176, EPI_ISL_1633177, EPI_ISL_1633178, EPI_ISL_1633179, EPI_ISL_1633180, EPI_ISL_1633181, EPI_ISL_1633182, EPI_ISL_1633183, EPI_ISL_1633185, EPI_ISL_1633186, EPI_ISL_1633187, EPI_ISL_1633188, EPI_ISL_1633189, EPI_ISL_1633190, EPI_ISL_1633191, EPI_ISL_1633192, EPI_ISL_1633193, EPI_ISL_1633194, EPI_ISL_1633195, EPI_ISL_1633196, EPI_ISL_1633197, EPI_ISL_1633198, EPI_ISL_1633199, EPI_ISL_1633200, EPI_ISL_1633201, EPI_ISL_1633202, EPI_ISL_1633203, EPI_ISL_1633204, EPI_ISL_1633205, EPI_ISL_1633206, EPI_ISL_1633207, EPI_ISL_1633208, EPI_ISL_1633209, EPI_ISL_1633210, EPI_ISL_1633212, EPI_ISL_1633213, EPI_ISL_1633214, EPI_ISL_1633216, EPI_ISL_1633217, EPI_ISL_1633218, EPI_ISL_1633219, EPI_ISL_1633221, EPI_ISL_1633222, EPI_ISL_1633223, EPI_ISL_1633224, EPI_ISL_1633225, EPI_ISL_1633226, EPI_ISL_1633227, EPI_ISL_1633228, EPI_ISL_1633229, EPI_ISL_1633230, EPI_ISL_1633231, EPI_ISL_1633232, EPI_ISL_1633233, EPI_ISL_1633234, EPI_ISL_1633235, EPI_ISL_1633236, EPI_ISL_1633237, EPI_ISL_1633238, EPI_ISL_1633239, EPI_ISL_1633240, EPI_ISL_1633241, EPI_ISL_1633242, EPI_ISL_1633243, EPI_ISL_1633244, EPI_ISL_1633245, EPI_ISL_1633246, EPI_ISL_1633247, EPI_ISL_1633249, EPI_ISL_1633250, EPI_ISL_1633251, EPI_ISL_1633252, EPI_ISL_1633253, EPI_ISL_1633254, EPI_ISL_1633255, EPI_ISL_1633256, EPI_ISL_1633257, EPI_ISL_1633258, EPI_ISL_1633259, EPI_ISL_1633262, EPI_ISL_1633263, EPI_ISL_1633264, EPI_ISL_1633265, EPI_ISL_1633266, EPI_ISL_1633267, EPI_ISL_1633269, EPI_ISL_1633271, EPI_ISL_1633272, EPI_ISL_1633273, EPI_ISL_1633274 | | | |
| see above | Pathology and Laboratory Medicine Institute, Cleveland Clinic, Ohio, USA | Pathology and Laboratory Medicine Institute, Cleveland Clinic, Ohio, USA | Concetta Peck, Joy Nakitandwe, Kristen McDonnell, David Plunkett, Zheng Jin Tu, Jay Brock, Yu-Wei Cheng, Gary Procop, Daniel Rhoads, Daniel H. Farkas, David Bosler |
| EPI_ISL_1633342, EPI_ISL_1633343, EPI_ISL_1633344, EPI_ISL_1633345, EPI_ISL_1633346 | Arizona State Public Health Laboratory | Arizona State Public Health Laboratory | Trung Huynh, Jessica Escobar, Katherine Fullerton, Nobuko Fukushima, Stacy White, Linda Getsinger, Victor Waddell |
| EPI_ISL_1633574, EPI_ISL_1633575, EPI_ISL_1633576, EPI_ISL_1633577, EPI_ISL_1633578, EPI_ISL_1633579, EPI_ISL_1633580, EPI_ISL_1633581, EPI_ISL_1633582, EPI_ISL_1633583, EPI_ISL_1633584, EPI_ISL_1633585, EPI_ISL_1633586, EPI_ISL_1633587, EPI_ISL_1633588, EPI_ISL_1633589, EPI_ISL_1633590, EPI_ISL_1633591, EPI_ISL_1633592, EPI_ISL_1633593, EPI_ISL_1633594, EPI_ISL_1633595, EPI_ISL_1633596, EPI_ISL_1633597, EPI_ISL_1633598, EPI_ISL_1633599, EPI_ISL_1633600, EPI_ISL_1633601, EPI_ISL_1633602, EPI_ISL_1633603, EPI_ISL_1633604, EPI_ISL_1633605, EPI_ISL_1633606, EPI_ISL_1633607, EPI_ISL_1633608, EPI_ISL_1633609, EPI_ISL_1633610, EPI_ISL_1633611, EPI_ISL_1633612, EPI_ISL_1633613, EPI_ISL_1633614, EPI_ISL_1633615, EPI_ISL_1633616, EPI_ISL_1633618, EPI_ISL_1633619, EPI_ISL_1633620, EPI_ISL_1633621, EPI_ISL_1633622, EPI_ISL_1633623, EPI_ISL_1633624, EPI_ISL_1633625, EPI_ISL_1633626, EPI_ISL_1633627, EPI_ISL_1633628, EPI_ISL_1633629, EPI_ISL_1633630, EPI_ISL_1633631, EPI_ISL_1633632, EPI_ISL_1633633, EPI_ISL_1633634, EPI_ISL_1633635, EPI_ISL_1633636, EPI_ISL_1633637, EPI_ISL_1633638, EPI_ISL_1633639, EPI_ISL_1633640, EPI_ISL_1633641, EPI_ISL_1633642, EPI_ISL_1633643, EPI_ISL_1633644, EPI_ISL_1633645, EPI_ISL_1633646, EPI_ISL_1633647, EPI_ISL_1633648, EPI_ISL_1633649, EPI_ISL_1633650, EPI_ISL_1633651, EPI_ISL_1633652, EPI_ISL_1633653, EPI_ISL_1633654, EPI_ISL_1633655 | | | |
| see above | UW Virology Lab | UW Virology Lab | Pavitra Roychoudhury, Hong Xie, Lasata Shrestha, Shah Mohamed Bakhash, Michelle Lin, Noah R. Baker, Sean Ellis, Saraswathi Sathees, Meei-Li Huang, Keith R Jerome, Alexander Greninger |
| EPI_ISL_1633685, EPI_ISL_1633686, EPI_ISL_1633687, EPI_ISL_1633689, EPI_ISL_1633690, EPI_ISL_1633692, EPI_ISL_1633693, EPI_ISL_1633694, EPI_ISL_1633695, EPI_ISL_1633696, EPI_ISL_1633697, EPI_ISL_1633698, EPI_ISL_1633699, EPI_ISL_1633700, EPI_ISL_1633702, EPI_ISL_1633703, EPI_ISL_1633704, EPI_ISL_1633705, EPI_ISL_1633706, EPI_ISL_1633707, EPI_ISL_1633708, EPI_ISL_1633709, EPI_ISL_1633710, EPI_ISL_1633711, EPI_ISL_1633712, EPI_ISL_1633713, EPI_ISL_1633714, EPI_ISL_1633715, EPI_ISL_1633716, EPI_ISL_1633717, EPI_ISL_1633718, EPI_ISL_1633719, EPI_ISL_1633720, EPI_ISL_1633721, EPI_ISL_1633722, EPI_ISL_1633723, EPI_ISL_1633724, EPI_ISL_1633726, EPI_ISL_1633727, EPI_ISL_1633728, EPI_ISL_1633731, EPI_ISL_1633732, EPI_ISL_1633733, EPI_ISL_1633734, EPI_ISL_1633737, EPI_ISL_1633738, EPI_ISL_1633739, EPI_ISL_1633740, EPI_ISL_1633743, EPI_ISL_1633744, EPI_ISL_1633745, EPI_ISL_1633746, EPI_ISL_1633747, EPI_ISL_1633748, EPI_ISL_1633749, EPI_ISL_1633750, EPI_ISL_1633751, EPI_ISL_1633752, EPI_ISL_1633753, EPI_ISL_1633754, EPI_ISL_1633755, EPI_ISL_1633756, EPI_ISL_1633757, EPI_ISL_1633758, EPI_ISL_1633759, EPI_ISL_1633760, EPI_ISL_1633761, EPI_ISL_1633763, EPI_ISL_1633764, EPI_ISL_1633765, EPI_ISL_1633766, EPI_ISL_1633767, EPI_ISL_1633768, EPI_ISL_1633769, EPI_ISL_1633770, EPI_ISL_1633771, EPI_ISL_1633772, EPI_ISL_1633773, EPI_ISL_1633774, EPI_ISL_1633775, EPI_ISL_1633776, EPI_ISL_1633777, EPI_ISL_1633778, EPI_ISL_1633779, EPI_ISL_1633780, EPI_ISL_1633781, EPI_ISL_1633782, EPI_ISL_1633783, EPI_ISL_1633784, EPI_ISL_1633785, EPI_ISL_1633786, EPI_ISL_1633787, EPI_ISL_1633788, EPI_ISL_1633789, EPI_ISL_1633790, EPI_ISL_1633791, EPI_ISL_1633792, EPI_ISL_1633793, EPI_ISL_1633794, EPI_ISL_1633795, EPI_ISL_1633796, EPI_ISL_1633797, EPI_ISL_1633799, EPI_ISL_1633801, EPI_ISL_1633802, EPI_ISL_1633803, EPI_ISL_1633804, EPI_ISL_1633805, EPI_ISL_1633806, EPI_ISL_1633808, EPI_ISL_1633809, EPI_ISL_1633810, EPI_ISL_1633812, EPI_ISL_1633813, EPI_ISL_1633814, EPI_ISL_1633815, EPI_ISL_1633816, EPI_ISL_1633817, EPI_ISL_1633818, EPI_ISL_1633819, EPI_ISL_1633821, EPI_ISL_1633822, EPI_ISL_1633823, EPI_ISL_1633824, EPI_ISL_1633826, EPI_ISL_1633827, EPI_ISL_1633828, EPI_ISL_1633829, EPI_ISL_1633830, EPI_ISL_1633831, EPI_ISL_1633832, EPI_ISL_1633833, EPI_ISL_1633834, EPI_ISL_1633835, EPI_ISL_1633836, EPI_ISL_1633837, EPI_ISL_1633838, EPI_ISL_1633839, EPI_ISL_1633840, EPI_ISL_1633841, EPI_ISL_1633842, EPI_ISL_1633843, EPI_ISL_1633844, EPI_ISL_1633845, EPI_ISL_1633846, EPI_ISL_1633847, EPI_ISL_1633848, EPI_ISL_1633849, EPI_ISL_1633850, EPI_ISL_1633852, EPI_ISL_1633853, EPI_ISL_1633854, EPI_ISL_1633855, EPI_ISL_1633856, EPI_ISL_1633857, EPI_ISL_1633858, EPI_ISL_1633859, EPI_ISL_1633861, EPI_ISL_1633862, EPI_ISL_1633863, EPI_ISL_1633865, EPI_ISL_1633866, EPI_ISL_1633867, EPI_ISL_1633869, EPI_ISL_1633870, EPI_ISL_1633871, EPI_ISL_1633872, EPI_ISL_1633873, EPI_ISL_1633874, EPI_ISL_1633875, EPI_ISL_1633876, EPI_ISL_1633877, EPI_ISL_1633878, EPI_ISL_1633879, EPI_ISL_1633880, EPI_ISL_1633881, EPI_ISL_1633882, EPI_ISL_1633883, EPI_ISL_1633885, EPI_ISL_1633886, EPI_ISL_1633887, EPI_ISL_1633888, EPI_ISL_1633889, EPI_ISL_1633890, EPI_ISL_1633892, EPI_ISL_1633893, EPI_ISL_1633894, EPI_ISL_1633895, EPI_ISL_1633896, EPI_ISL_1633897, EPI_ISL_1633899, EPI_ISL_1633902, EPI_ISL_1633903, EPI_ISL_1633905, EPI_ISL_1633906, EPI_ISL_1633907, EPI_ISL_1633910, EPI_ISL_1633911, EPI_ISL_1633913, EPI_ISL_1633914, EPI_ISL_1633916, EPI_ISL_1633917, EPI_ISL_1633918, EPI_ISL_1633920, EPI_ISL_1633921, EPI_ISL_1633922, EPI_ISL_1633924, EPI_ISL_1633925, EPI_ISL_1633927, EPI_ISL_1633929, EPI_ISL_1633931, EPI_ISL_1633932, EPI_ISL_1633934, EPI_ISL_1633935, EPI_ISL_1633937, EPI_ISL_1633939, EPI_ISL_1633940, EPI_ISL_1633942, EPI_ISL_1633943, EPI_ISL_1633944, EPI_ISL_1633946, EPI_ISL_1633947, EPI_ISL_1633949, EPI_ISL_1633951, EPI_ISL_1633953, EPI_ISL_1633954, EPI_ISL_1633956, EPI_ISL_1633958, EPI_ISL_1633959, EPI_ISL_1633960, EPI_ISL_1633962, EPI_ISL_1633963, EPI_ISL_1633964, EPI_ISL_1633966, EPI_ISL_1633967, EPI_ISL_1633968, EPI_ISL_1633970, EPI_ISL_1633971, EPI_ISL_1633972, EPI_ISL_1633973, EPI_ISL_1633975, EPI_ISL_1633977, EPI_ISL_1633979, EPI_ISL_1633980, EPI_ISL_1633981, EPI_ISL_1633983, EPI_ISL_1633984, EPI_ISL_1633985, EPI_ISL_1633986, EPI_ISL_1633988, EPI_ISL_1633989, EPI_ISL_1633990, EPI_ISL_1633992, EPI_ISL_1633993, EPI_ISL_1633994, EPI_ISL_1633996, EPI_ISL_1633997, EPI_ISL_1633998, EPI_ISL_1634000, EPI_ISL_1634001, EPI_ISL_1634002, EPI_ISL_1634004, EPI_ISL_1634007, EPI_ISL_1634008, EPI_ISL_1634009, EPI_ISL_1634011, EPI_ISL_1634012, EPI_ISL_1634014, EPI_ISL_1634018, EPI_ISL_1634019, EPI_ISL_1634021, EPI_ISL_1634022, EPI_ISL_1634023, EPI_ISL_1634025, EPI_ISL_1634026, EPI_ISL_1634028, EPI_ISL_1634030, EPI_ISL_1634032, EPI_ISL_1634033, EPI_ISL_1634034, EPI_ISL_1634036, EPI_ISL_1634037, EPI_ISL_1634038, EPI_ISL_1634040, EPI_ISL_1634041, EPI_ISL_1634042, EPI_ISL_1634044, EPI_ISL_1634045, EPI_ISL_1634046, EPI_ISL_1634047, EPI_ISL_1634049, EPI_ISL_1634051, EPI_ISL_1634052, EPI_ISL_1634053, EPI_ISL_1634054, EPI_ISL_1634056, EPI_ISL_1634058, EPI_ISL_1634060, EPI_ISL_1634061, EPI_ISL_1634062, EPI_ISL_1634064, EPI_ISL_1634065, EPI_ISL_1634068, EPI_ISL_1634069, EPI_ISL_1634071, EPI_ISL_1634072, EPI_ISL_1634074, EPI_ISL_1634076, EPI_ISL_1634077, EPI_ISL_1634079, EPI_ISL_1634080, EPI_ISL_1634082, EPI_ISL_1634083, EPI_ISL_1634084, EPI_ISL_1634086, EPI_ISL_1634088, EPI_ISL_1634089, EPI_ISL_1634091, EPI_ISL_1634092, EPI_ISL_1634093, EPI_ISL_1634096, EPI_ISL_1634097, EPI_ISL_1634099, EPI_ISL_1634100, EPI_ISL_1634101, EPI_ISL_1634103, EPI_ISL_1634104, EPI_ISL_1634107, EPI_ISL_1634108, EPI_ISL_1634110, EPI_ISL_1634113, EPI_ISL_1634114, EPI_ISL_1634116, EPI_ISL_1634117, EPI_ISL_1634118, EPI_ISL_1634120, EPI_ISL_1634121, EPI_ISL_1634123, EPI_ISL_1634124, EPI_ISL_1634125, EPI_ISL_1634126, EPI_ISL_1634128, EPI_ISL_1634129, EPI_ISL_1634132, EPI_ISL_1634133, EPI_ISL_1634135, EPI_ISL_1634136, EPI_ISL_1634137, EPI_ISL_1634140, EPI_ISL_1634141, EPI_ISL_1634143, EPI_ISL_1634150, EPI_ISL_1634152, EPI_ISL_1634153, EPI_ISL_1634155, EPI_ISL_1634157, EPI_ISL_1634158, EPI_ISL_1634159, EPI_ISL_1634161, EPI_ISL_1634162, EPI_ISL_1634163, EPI_ISL_1634165, EPI_ISL_1634166, EPI_ISL_1634169, EPI_ISL_1634170, EPI_ISL_1634171, EPI_ISL_1634173, EPI_ISL_1634174, EPI_ISL_1634175, EPI_ISL_1634176, EPI_ISL_1634178, EPI_ISL_1634179, EPI_ISL_1634180, EPI_ISL_1634181, EPI_ISL_1634182, EPI_ISL_1634183, EPI_ISL_1634184, EPI_ISL_1634185, EPI_ISL_1634186, EPI_ISL_1634187, EPI_ISL_1634188, EPI_ISL_1634189, EPI_ISL_1634190, EPI_ISL_1634191, EPI_ISL_1634192, EPI_ISL_1634193, EPI_ISL_1634194, EPI_ISL_1634196, EPI_ISL_1634198, EPI_ISL_1634199, EPI_ISL_1634201, EPI_ISL_1634202, EPI_ISL_1634203, EPI_ISL_1634204, EPI_ISL_1634205, EPI_ISL_1634206, EPI_ISL_1634207, EPI_ISL_1634208, EPI_ISL_1634210, EPI_ISL_1634211, EPI_ISL_1634212, EPI_ISL_1634213, EPI_ISL_1634214, EPI_ISL_1634219, EPI_ISL_1634220, EPI_ISL_1634221, EPI_ISL_1634222, EPI_ISL_1634223, EPI_ISL_1634225, EPI_ISL_1634226, EPI_ISL_1634228, EPI_ISL_1634229, EPI_ISL_1634230, EPI_ISL_1634232, EPI_ISL_1634233, EPI_ISL_1634234, EPI_ISL_1634235, EPI_ISL_1634236, EPI_ISL_1634237, EPI_ISL_1634238, EPI_ISL_1634240, EPI_ISL_1634241, EPI_ISL_1634242, EPI_ISL_1634244, EPI_ISL_1634245, EPI_ISL_1634246, EPI_ISL_1634247, EPI_ISL_1634248, EPI_ISL_1634249, EPI_ISL_1634250, EPI_ISL_1634251, EPI_ISL_1634252, EPI_ISL_1634255, EPI_ISL_1634256, EPI_ISL_1634257, EPI_ISL_1634258, EPI_ISL_1634259, EPI_ISL_1634260, EPI_ISL_1634261, EPI_ISL_1634262, EPI_ISL_1634263, EPI_ISL_1634264, EPI_ISL_1634265, EPI_ISL_1634266, EPI_ISL_1634267, EPI_ISL_1634268, EPI_ISL_1634269, EPI_ISL_1634270, EPI_ISL_1634271, EPI_ISL_1634272, EPI_ISL_1634273, EPI_ISL_1634274, EPI_ISL_1634276, EPI_ISL_1634277, EPI_ISL_1634278, EPI_ISL_1634279, EPI_ISL_1634280, EPI_ISL_1634281, EPI_ISL_1634282, EPI_ISL_1634283, EPI_ISL_1634284, EPI_ISL_1634285, EPI_ISL_1634286, EPI_ISL_1634287, EPI_ISL_1634289, EPI_ISL_1634291, EPI_ISL_1634292, EPI_ISL_1634293, EPI_ISL_1634294, EPI_ISL_1634295, EPI_ISL_1634296, EPI_ISL_1634297, EPI_ISL_1634298, EPI_ISL_1634299, EPI_ISL_1634300, EPI_ISL_1634301, EPI_ISL_1634303, EPI_ISL_1634304, EPI_ISL_1634305, EPI_ISL_1634306, EPI_ISL_1634307, EPI_ISL_1634308, EPI_ISL_1634309, EPI_ISL_1634310, EPI_ISL_1634311, EPI_ISL_1634312, EPI_ISL_1634314, EPI_ISL_1634315, EPI_ISL_1634316, EPI_ISL_1634317, EPI_ISL_1634318, EPI_ISL_1634320, EPI_ISL_1634321, EPI_ISL_1634322, EPI_ISL_1634323, EPI_ISL_1634324, EPI_ISL_1634325, EPI_ISL_1634326, EPI_ISL_1634327, EPI_ISL_1634328, EPI_ISL_1634329, EPI_ISL_1634330, EPI_ISL_1634331, EPI_ISL_1634332, EPI_ISL_1634333, EPI_ISL_1634334, EPI_ISL_1634336, EPI_ISL_1634337, EPI_ISL_1634338, EPI_ISL_1634339, EPI_ISL_1635437, EPI_ISL_1635438, EPI_ISL_1635439, EPI_ISL_1635440, EPI_ISL_1635441, EPI_ISL_1635442, EPI_ISL_1635443, EPI_ISL_1635444, EPI_ISL_1635445, EPI_ISL_1635446, EPI_ISL_1635447, EPI_ISL_1635448, EPI_ISL_1635449, EPI_ISL_1635450, EPI_ISL_1635451, EPI_ISL_1635452, EPI_ISL_1635453, EPI_ISL_1635454, EPI_ISL_1635455, EPI_ISL_1635456, EPI_ISL_1635457, EPI_ISL_1635459, EPI_ISL_1635460, EPI_ISL_1635461, EPI_ISL_1635462, EPI_ISL_1635463, EPI_ISL_1635464, EPI_ISL_1635465, EPI_ISL_1635467, EPI_ISL_1635468, EPI_ISL_1635470, EPI_ISL_1635472, EPI_ISL_1635473, EPI_ISL_1635474, EPI_ISL_1635475, EPI_ISL_1635476, EPI_ISL_1635478, EPI_ISL_1635479, EPI_ISL_1635480, EPI_ISL_1635481, EPI_ISL_1635482, EPI_ISL_1635484, EPI_ISL_1635485, EPI_ISL_1635486, EPI_ISL_1635487, EPI_ISL_1635488, EPI_ISL_1635489, EPI_ISL_1635490, EPI_ISL_1635491, EPI_ISL_1635492, EPI_ISL_1635494, EPI_ISL_1635495, EPI_ISL_1635496, EPI_ISL_1635497, EPI_ISL_1635498, EPI_ISL_1635501, EPI_ISL_1635502, EPI_ISL_1635503, EPI_ISL_1635504, EPI_ISL_1635505, EPI_ISL_1635506, EPI_ISL_1635507, EPI_ISL_1635508, EPI_ISL_1635510, EPI_ISL_1635511, EPI_ISL_1635512, EPI_ISL_1635513, EPI_ISL_1635514, EPI_ISL_1635515, EPI_ISL_1635516, EPI_ISL_1635517, EPI_ISL_1635518, EPI_ISL_1635520, EPI_ISL_1635522, EPI_ISL_1635523, EPI_ISL_1635524, EPI_ISL_1635525, EPI_ISL_1635526, EPI_ISL_1635529, EPI_ISL_1635530, EPI_ISL_1635531, EPI_ISL_1635532, EPI_ISL_1635533, EPI_ISL_1635534, EPI_ISL_1635536, EPI_ISL_1635537, EPI_ISL_1635539, EPI_ISL_1635540, EPI_ISL_1635541, EPI_ISL_1635542, EPI_ISL_1635543, EPI_ISL_1635544, EPI_ISL_1635546, EPI_ISL_1635547, EPI_ISL_1635548, EPI_ISL_1635549, EPI_ISL_1635550, EPI_ISL_1635551, EPI_ISL_1635552, EPI_ISL_1635553, EPI_ISL_1635554, EPI_ISL_1635556, EPI_ISL_1635557, EPI_ISL_1635559, EPI_ISL_1635560, EPI_ISL_1635561, EPI_ISL_1635562, EPI_ISL_1635563, EPI_ISL_1635564, EPI_ISL_1635565, EPI_ISL_1635566, EPI_ISL_1635567, EPI_ISL_1635568, EPI_ISL_1635569, EPI_ISL_1635572, EPI_ISL_1635573, EPI_ISL_1635574, EPI_ISL_1635575, EPI_ISL_1635576, EPI_ISL_1635577, EPI_ISL_1635578, EPI_ISL_1635579, EPI_ISL_1635580, EPI_ISL_1635581, EPI_ISL_1635583, EPI_ISL_1635584, EPI_ISL_1635585, EPI_ISL_1635586, EPI_ISL_1635587, EPI_ISL_1635588, EPI_ISL_1635589, EPI_ISL_1635590, EPI_ISL_1635591, EPI_ISL_1635592, EPI_ISL_1635593, EPI_ISL_1635594, EPI_ISL_1635595, EPI_ISL_1635596, EPI_ISL_1635598, EPI_ISL_1635599, EPI_ISL_1635600, EPI_ISL_1635601, EPI_ISL_1635602, EPI_ISL_1635603, EPI_ISL_1635605, EPI_ISL_1635606, EPI_ISL_1635607, EPI_ISL_1635609, EPI_ISL_1635610, EPI_ISL_1635612, EPI_ISL_1635613, EPI_ISL_1635614, EPI_ISL_1635615, EPI_ISL_1635616, EPI_ISL_1635617, EPI_ISL_1635618, EPI_ISL_1635619, EPI_ISL_1635622, EPI_ISL_1635623, EPI_ISL_1635624, EPI_ISL_1635625, EPI_ISL_1635626, EPI_ISL_1635627, EPI_ISL_1635628, EPI_ISL_1635629, EPI_ISL_1635630, EPI_ISL_1635631, EPI_ISL_1635632, EPI_ISL_1635633, EPI_ISL_1635634, EPI_ISL_1635635, EPI_ISL_1635636, EPI_ISL_1635637, EPI_ISL_1635638, EPI_ISL_1635639, EPI_ISL_1635640, EPI_ISL_1635642, EPI_ISL_1635644, EPI_ISL_1635645, EPI_ISL_1635647, EPI_ISL_1635648, EPI_ISL_1635649, EPI_ISL_1635650, EPI_ISL_1635651, EPI_ISL_1635652, EPI_ISL_1635653, EPI_ISL_1635654, EPI_ISL_1635655, EPI_ISL_1635656, EPI_ISL_1635657, EPI_ISL_1635658, EPI_ISL_1635659, EPI_ISL_1635660, EPI_ISL_1635661, EPI_ISL_1635662, EPI_ISL_1635663, EPI_ISL_1635664, EPI_ISL_1635665, EPI_ISL_1635667, EPI_ISL_1635668, EPI_ISL_1635669, EPI_ISL_1635670, EPI_ISL_1635671, EPI_ISL_1635672, EPI_ISL_1635673, EPI_ISL_1635676, EPI_ISL_1635678, EPI_ISL_1635679, EPI_ISL_1635680, EPI_ISL_1635681, EPI_ISL_1635682, EPI_ISL_1635683, EPI_ISL_1635684, EPI_ISL_1635685, EPI_ISL_1635686, EPI_ISL_1635687, EPI_ISL_1635688, EPI_ISL_1635689, EPI_ISL_1635690, EPI_ISL_1635691, EPI_ISL_1635692, EPI_ISL_1635693, EPI_ISL_1635694, EPI_ISL_1635695, EPI_ISL_1635697, EPI_ISL_1635700, EPI_ISL_1635701, EPI_ISL_1635702, EPI_ISL_1635703, EPI_ISL_1635704, EPI_ISL_1635705, EPI_ISL_1635706, EPI_ISL_1635707, EPI_ISL_1635709, EPI_ISL_1635710, EPI_ISL_1635711, EPI_ISL_1635712, EPI_ISL_1635713, EPI_ISL_1635714, EPI_ISL_1635715, EPI_ISL_1635716, EPI_ISL_1635717, EPI_ISL_1635718, EPI_ISL_1635720, EPI_ISL_1635721, EPI_ISL_1635722, EPI_ISL_1635723, EPI_ISL_1635724, EPI_ISL_1635725, EPI_ISL_1635727, EPI_ISL_1635728, EPI_ISL_1635730, EPI_ISL_1635731, EPI_ISL_1635732, EPI_ISL_1635733, EPI_ISL_1635734, EPI_ISL_1635735, EPI_ISL_1635736, EPI_ISL_1635737, EPI_ISL_1635738, EPI_ISL_1635739, EPI_ISL_1635740, EPI_ISL_1635741, EPI_ISL_1635742, EPI_ISL_1635743, EPI_ISL_1635744, EPI_ISL_1635745, EPI_ISL_1635746, EPI_ISL_1635747, EPI_ISL_1635748, EPI_ISL_1635749, EPI_ISL_1635752, EPI_ISL_1635755, EPI_ISL_1635756, EPI_ISL_1635757, EPI_ISL_1635758, EPI_ISL_1635759, EPI_ISL_1635760, EPI_ISL_1635761, EPI_ISL_1635762, EPI_ISL_1635763, EPI_ISL_1635764, EPI_ISL_1635767, EPI_ISL_1635769, EPI_ISL_1635770, EPI_ISL_1635771, EPI_ISL_1635772, EPI_ISL_1635773, EPI_ISL_1635774, EPI_ISL_1635775, EPI_ISL_1635776, EPI_ISL_1635777, EPI_ISL_1635778, EPI_ISL_1635779, EPI_ISL_1635780, EPI_ISL_1635781, EPI_ISL_1635782, EPI_ISL_1635783, EPI_ISL_1635784, EPI_ISL_1635785, EPI_ISL_1635786, EPI_ISL_1635788, EPI_ISL_1635789, EPI_ISL_1635790, EPI_ISL_1635791, EPI_ISL_1635793, EPI_ISL_1635794, EPI_ISL_1635796, EPI_ISL_1635797, EPI_ISL_1635798, EPI_ISL_1635799, EPI_ISL_1635800, EPI_ISL_1635802, EPI_ISL_1635803, EPI_ISL_1635804, EPI_ISL_1635805, EPI_ISL_1635806, EPI_ISL_1635807, EPI_ISL_1635808, EPI_ISL_1635810, EPI_ISL_1635811, EPI_ISL_1635812, EPI_ISL_1635813, EPI_ISL_1635814, EPI_ISL_1635815, EPI_ISL_1635816, EPI_ISL_1635817, EPI_ISL_1635818, EPI_ISL_1635819, EPI_ISL_1635820, EPI_ISL_1635821, EPI_ISL_1635822, EPI_ISL_1635823, EPI_ISL_1635824, EPI_ISL_1635825, EPI_ISL_1635827, EPI_ISL_1635828, EPI_ISL_1635829, EPI_ISL_1635830, EPI_ISL_1635831, EPI_ISL_1635832, EPI_ISL_1635833, EPI_ISL_1635834, EPI_ISL_1635835, EPI_ISL_1635837, EPI_ISL_1635839, EPI_ISL_1635840, EPI_ISL_1635843, EPI_ISL_1635844, EPI_ISL_1635845, EPI_ISL_1635846, EPI_ISL_1635847, EPI_ISL_1635851, EPI_ISL_1635852, EPI_ISL_1635854, EPI_ISL_1635855, EPI_ISL_1635856, EPI_ISL_1635857, EPI_ISL_1635858, EPI_ISL_1635859, EPI_ISL_1635860, EPI_ISL_1635862, EPI_ISL_1635863, EPI_ISL_1635864, EPI_ISL_1635865, EPI_ISL_1635866, EPI_ISL_1635868, EPI_ISL_1635869, EPI_ISL_1635871, EPI_ISL_1635873, EPI_ISL_1635874, EPI_ISL_1635875, EPI_ISL_1635876, EPI_ISL_1635879, EPI_ISL_1635880, EPI_ISL_1635881, EPI_ISL_1635883, EPI_ISL_1635884, EPI_ISL_1635885, EPI_ISL_1635886, EPI_ISL_1635888, EPI_ISL_1635889, EPI_ISL_1635890, EPI_ISL_1635891, EPI_ISL_1635892, EPI_ISL_1635893, EPI_ISL_1635894, EPI_ISL_1635896, EPI_ISL_1635897, EPI_ISL_1635898, EPI_ISL_1635899, EPI_ISL_1635900, EPI_ISL_1635901, EPI_ISL_1635903, EPI_ISL_1635904, EPI_ISL_1635905, EPI_ISL_1635906, EPI_ISL_1635907, EPI_ISL_1635908, EPI_ISL_1635909, EPI_ISL_1635910, EPI_ISL_1635911, EPI_ISL_1635912, EPI_ISL_1635913, EPI_ISL_1635914, EPI_ISL_1635915, EPI_ISL_1635916, EPI_ISL_1635917, EPI_ISL_1635919, EPI_ISL_1635920, EPI_ISL_1635921, EPI_ISL_1635922, EPI_ISL_1635923, EPI_ISL_1635924, EPI_ISL_1635925, EPI_ISL_1635926, EPI_ISL_1635927, EPI_ISL_1635928, EPI_ISL_1635929, EPI_ISL_1635930, EPI_ISL_1635931, EPI_ISL_1635932, EPI_ISL_1635933, EPI_ISL_1635934, EPI_ISL_1635935, EPI_ISL_1635936, EPI_ISL_1635938, EPI_ISL_1635939, EPI_ISL_1635940, EPI_ISL_1635941, EPI_ISL_1635942, EPI_ISL_1635943, EPI_ISL_1635944, EPI_ISL_1635945, EPI_ISL_1635946, EPI_ISL_1635947, EPI_ISL_1635948, EPI_ISL_1635949, EPI_ISL_1635950, EPI_ISL_1635951, EPI_ISL_1635952, EPI_ISL_1635953, EPI_ISL_1635954, EPI_ISL_1635955, EPI_ISL_1635956, EPI_ISL_1635957, EPI_ISL_1635959, EPI_ISL_1635960, EPI_ISL_1635962, EPI_ISL_1635963, EPI_ISL_1635965, EPI_ISL_1635966, EPI_ISL_1635967, EPI_ISL_1635968, EPI_ISL_1635970, EPI_ISL_1635971, EPI_ISL_1635972, EPI_ISL_1635973, EPI_ISL_1635974, EPI_ISL_1635975, EPI_ISL_1635976, EPI_ISL_1635977, EPI_ISL_1635978, EPI_ISL_1635979, EPI_ISL_1635981, EPI_ISL_1635983, EPI_ISL_1635985, EPI_ISL_1635986, EPI_ISL_1635987, EPI_ISL_1635988, EPI_ISL_1635989, EPI_ISL_1635990, EPI_ISL_1635991, EPI_ISL_1635993, EPI_ISL_1635994, EPI_ISL_1635995, EPI_ISL_1635996, EPI_ISL_1635998, EPI_ISL_1635999, EPI_ISL_1636000, EPI_ISL_1636001, EPI_ISL_1636002, EPI_ISL_1636003, EPI_ISL_1636004, EPI_ISL_1636005, EPI_ISL_1636006, EPI_ISL_1636007, EPI_ISL_1636008, EPI_ISL_1636009, EPI_ISL_1636010, EPI_ISL_1636011, EPI_ISL_1636012, EPI_ISL_1636013, EPI_ISL_1636014, EPI_ISL_1636015, EPI_ISL_1636016, EPI_ISL_1636018, EPI_ISL_1636019, EPI_ISL_1636021, EPI_ISL_1636022, EPI_ISL_1636023, EPI_ISL_1636024, EPI_ISL_1636025, EPI_ISL_1636026, EPI_ISL_1636027, EPI_ISL_1636028, EPI_ISL_1636030, EPI_ISL_1636031, EPI_ISL_1636032, EPI_ISL_1636033, EPI_ISL_1636034, EPI_ISL_1636035, EPI_ISL_1636036, EPI_ISL_1636037, EPI_ISL_1636041, EPI_ISL_1636042, EPI_ISL_1636043, EPI_ISL_1636044, EPI_ISL_1636045, EPI_ISL_1636046, EPI_ISL_1636047, EPI_ISL_1636048, EPI_ISL_1636049, EPI_ISL_1636050, EPI_ISL_1636051, EPI_ISL_1636052, EPI_ISL_1636053, EPI_ISL_1636054, EPI_ISL_1636056, EPI_ISL_1636057, EPI_ISL_1636059, EPI_ISL_1636061, EPI_ISL_1636062, EPI_ISL_1636063, EPI_ISL_1636064, EPI_ISL_1636065, EPI_ISL_1636067, EPI_ISL_1636068, EPI_ISL_1636069, EPI_ISL_1636071, EPI_ISL_1636073, EPI_ISL_1636074, EPI_ISL_1636076, EPI_ISL_1636077, EPI_ISL_1636078, EPI_ISL_1636079, EPI_ISL_1636080, EPI_ISL_1636081, EPI_ISL_1636082, EPI_ISL_1636083, EPI_ISL_1636085, EPI_ISL_1636086, EPI_ISL_1636087, EPI_ISL_1636088, EPI_ISL_1636089, EPI_ISL_1636090, EPI_ISL_1636091, EPI_ISL_1636092, EPI_ISL_1636094, EPI_ISL_1636096, EPI_ISL_1636098, EPI_ISL_1636100, EPI_ISL_1636101, EPI_ISL_1636102, EPI_ISL_1636103, EPI_ISL_1636104, EPI_ISL_1636105, EPI_ISL_1636109, EPI_ISL_1636111, EPI_ISL_1636112, EPI_ISL_1636114, EPI_ISL_1636115, EPI_ISL_1636116, EPI_ISL_1636117, EPI_ISL_1636118, EPI_ISL_1636119, EPI_ISL_1636120, EPI_ISL_1636121, EPI_ISL_1636122, EPI_ISL_1636123, EPI_ISL_1636124, EPI_ISL_1636125, EPI_ISL_1636126, EPI_ISL_1636127, EPI_ISL_1636128, EPI_ISL_1636129, EPI_ISL_1636130, EPI_ISL_1636132, EPI_ISL_1636133, EPI_ISL_1636134, EPI_ISL_1636135, EPI_ISL_1636136, EPI_ISL_1636137, EPI_ISL_1636138, EPI_ISL_1636139, EPI_ISL_1636140, EPI_ISL_1636141, EPI_ISL_1636142, EPI_ISL_1636143, EPI_ISL_1636144, EPI_ISL_1636145, EPI_ISL_1636146, EPI_ISL_1636147, EPI_ISL_1636148, EPI_ISL_1636149, EPI_ISL_1636150, EPI_ISL_1636151, EPI_ISL_1636153, EPI_ISL_1636154, EPI_ISL_1636155, EPI_ISL_1636156, EPI_ISL_1636158, EPI_ISL_1636159, EPI_ISL_1636160, EPI_ISL_1636162, EPI_ISL_1636163, EPI_ISL_1636164, EPI_ISL_1636165, EPI_ISL_1636166, EPI_ISL_1636167, EPI_ISL_1636168, EPI_ISL_1636169, EPI_ISL_1636170, EPI_ISL_1636171, EPI_ISL_1636172, EPI_ISL_1636173, EPI_ISL_1636174, EPI_ISL_1636175, EPI_ISL_1636177, EPI_ISL_1636178, EPI_ISL_1636179, EPI_ISL_1636180, EPI_ISL_1636181, EPI_ISL_1636182, EPI_ISL_1636183, EPI_ISL_1636184, EPI_ISL_1636185, EPI_ISL_1636186, EPI_ISL_1636187, EPI_ISL_1636188, EPI_ISL_1636189, EPI_ISL_1636190, EPI_ISL_1636191, EPI_ISL_1636193, EPI_ISL_1636194, EPI_ISL_1636195, EPI_ISL_1636196, EPI_ISL_1636197, EPI_ISL_1636198, EPI_ISL_1636199, EPI_ISL_1636201, EPI_ISL_1636202, EPI_ISL_1636203, EPI_ISL_1636206, EPI_ISL_1636207, EPI_ISL_1636208, EPI_ISL_1636209, EPI_ISL_1636210, EPI_ISL_1636211, EPI_ISL_1636212, EPI_ISL_1636213, EPI_ISL_1636214, EPI_ISL_1636215, EPI_ISL_1636217, EPI_ISL_1636218, EPI_ISL_1636219, EPI_ISL_1636220, EPI_ISL_1636221, EPI_ISL_1636222, EPI_ISL_1636223, EPI_ISL_1636225, EPI_ISL_1636226, EPI_ISL_1636227, EPI_ISL_1636228, EPI_ISL_1636229, EPI_ISL_1636231, EPI_ISL_1636232, EPI_ISL_1636233, EPI_ISL_1636234, EPI_ISL_1636235, EPI_ISL_1636237, EPI_ISL_1636238, EPI_ISL_1636239, EPI_ISL_1636240, EPI_ISL_1636241, EPI_ISL_1636242, EPI_ISL_1636243, EPI_ISL_1636244, EPI_ISL_1636245, EPI_ISL_1636247, EPI_ISL_1636249, EPI_ISL_1636250, EPI_ISL_1636252, EPI_ISL_1636253, EPI_ISL_1636254, EPI_ISL_1636255, EPI_ISL_1636256, EPI_ISL_1636257, EPI_ISL_1636258, EPI_ISL_1636259, EPI_ISL_1636260, EPI_ISL_1636261, EPI_ISL_1636262, EPI_ISL_1636263, EPI_ISL_1636264, EPI_ISL_1636265, EPI_ISL_1636266, EPI_ISL_1636267, EPI_ISL_1636268, EPI_ISL_1636269, EPI_ISL_1636270, EPI_ISL_1636271, EPI_ISL_1636272, EPI_ISL_1636273, EPI_ISL_1636274, EPI_ISL_1636275, EPI_ISL_1636276, EPI_ISL_1636277, EPI_ISL_1636279, EPI_ISL_1636280, EPI_ISL_1636281, EPI_ISL_1636283, EPI_ISL_1636284, EPI_ISL_1636285, EPI_ISL_1636286, EPI_ISL_1636287, EPI_ISL_1636288, EPI_ISL_1636289, EPI_ISL_1636290, EPI_ISL_1636291, EPI_ISL_1636292, EPI_ISL_1636293, EPI_ISL_1636294, EPI_ISL_1636295, EPI_ISL_1636296, EPI_ISL_1636297, EPI_ISL_1636298, EPI_ISL_1636299, EPI_ISL_1636300, EPI_ISL_1636301, EPI_ISL_1636302, EPI_ISL_1636303, EPI_ISL_1636304, EPI_ISL_1636305, EPI_ISL_1636306, EPI_ISL_1636307, EPI_ISL_1636308, EPI_ISL_1636309, EPI_ISL_1636310, EPI_ISL_1636311, EPI_ISL_1636312, EPI_ISL_1636313, EPI_ISL_1636314, EPI_ISL_1636315, EPI_ISL_1636317, EPI_ISL_1636318, EPI_ISL_1636319, EPI_ISL_1636320, EPI_ISL_1636321, EPI_ISL_1636322, EPI_ISL_1636323, EPI_ISL_1636324, EPI_ISL_1636325, EPI_ISL_1636326, EPI_ISL_1636327, EPI_ISL_1636328, EPI_ISL_1636329, EPI_ISL_1636330, EPI_ISL_1636331, EPI_ISL_1636332, EPI_ISL_1636333, EPI_ISL_1636334, EPI_ISL_1636335, EPI_ISL_1636336, EPI_ISL_1636337, EPI_ISL_1636338, EPI_ISL_1636339, EPI_ISL_1636340, EPI_ISL_1636342, EPI_ISL_1636345, EPI_ISL_1636346, EPI_ISL_1636347, EPI_ISL_1636348, EPI_ISL_1636350, EPI_ISL_1636351, EPI_ISL_1636352, EPI_ISL_1636353, EPI_ISL_1636354, EPI_ISL_1636355, EPI_ISL_1636356, EPI_ISL_1636357, EPI_ISL_1636358, EPI_ISL_1636359, EPI_ISL_1636360, EPI_ISL_1636361, EPI_ISL_1636362, EPI_ISL_1636363, EPI_ISL_1636364, EPI_ISL_1636366, EPI_ISL_1636367, EPI_ISL_1636368, EPI_ISL_1636369, EPI_ISL_1636370, EPI_ISL_1636371, EPI_ISL_1636372, EPI_ISL_1636373, EPI_ISL_1636374, EPI_ISL_1636376, EPI_ISL_1636378, EPI_ISL_1636379, EPI_ISL_1636380, EPI_ISL_1636381, EPI_ISL_1636383, EPI_ISL_1636384, EPI_ISL_1636385, EPI_ISL_1636386, EPI_ISL_1636387, EPI_ISL_1636388, EPI_ISL_1636389, EPI_ISL_1636390, EPI_ISL_1636391, EPI_ISL_1636392, EPI_ISL_1636393, EPI_ISL_1636394, EPI_ISL_1636395, EPI_ISL_1636396, EPI_ISL_1636397 | | | |
| see above | Pandemic Response Lab - NYC | Pandemic Response Lab, R&D | Henry Lee, Michael Hammerling, Melissa Hopkins, Cybill del Castillo, Shinyoung Clair Kang, William Ward, Pradeep Bugga, Sol Rey, Dylan Law, Katharine Nelson, Haiping Hao, Jon Laurent |
| EPI_ISL_1637205, | NCSLPH | NCSLPH | Chase K, Miller MC, Greene S, Glover W |

| | | | |
|---|---|---|---|
| EPI_ISL_1641625, EPI_ISL_1644757, EPI_ISL_1646684, EPI_ISL_1647027, EPI_ISL_1647028, EPI_ISL_1647033, EPI_ISL_1647034, EPI_ISL_1647049 | | | |
| EPI_ISL_1647870, EPI_ISL_1647871, EPI_ISL_1647872, EPI_ISL_1647873, EPI_ISL_1647875, EPI_ISL_1647876, EPI_ISL_1647877, EPI_ISL_1647878, EPI_ISL_1647879, EPI_ISL_1647880, EPI_ISL_1647882, EPI_ISL_1647883, EPI_ISL_1647884, EPI_ISL_1647885, EPI_ISL_1647886, EPI_ISL_1647887, EPI_ISL_1647888, EPI_ISL_1647889, EPI_ISL_1647890, EPI_ISL_1647891, EPI_ISL_1647892, EPI_ISL_1647893, EPI_ISL_1647894, EPI_ISL_1647896, EPI_ISL_1647897, EPI_ISL_1647898, EPI_ISL_1647899, EPI_ISL_1647900, EPI_ISL_1647901, EPI_ISL_1647902, EPI_ISL_1647903, EPI_ISL_1647904, EPI_ISL_1647905, EPI_ISL_1647906, EPI_ISL_1647907, EPI_ISL_1647908, EPI_ISL_1647909, EPI_ISL_1647911, EPI_ISL_1647912, EPI_ISL_1647913, EPI_ISL_1647914, EPI_ISL_1647915, EPI_ISL_1647916, EPI_ISL_1647919, EPI_ISL_1647920, EPI_ISL_1647921, EPI_ISL_1647922, EPI_ISL_1647923, EPI_ISL_1647924, EPI_ISL_1647926, EPI_ISL_1647927, EPI_ISL_1647928, EPI_ISL_1647929, EPI_ISL_1647930, EPI_ISL_1647931, EPI_ISL_1647932, EPI_ISL_1647934, EPI_ISL_1647935, EPI_ISL_1647936, EPI_ISL_1647937, EPI_ISL_1647938, EPI_ISL_1647939, EPI_ISL_1647940 | | | |
| see above | Wyoming Public Health Laboratory | Wyoming Public Health Laboratory | Jim Mildenberger, Wanda Manley, Noah Hull, Taylor Fearing, Lynette Gumbleton, Channing Weber, Ashley Norberg, Chayse Rowley, Marley Goetz, Brian Dominguez, Elliot Thomasson, Cari Sloma, and Rob Christensen |
| EPI_ISL_1648045, EPI_ISL_1648046, EPI_ISL_1648047, EPI_ISL_1648048, EPI_ISL_1648049, EPI_ISL_1648050, EPI_ISL_1648051, EPI_ISL_1648052, EPI_ISL_1648053, EPI_ISL_1648054, EPI_ISL_1648055, EPI_ISL_1648056, EPI_ISL_1648057, EPI_ISL_1648058, EPI_ISL_1648059, EPI_ISL_1648060, EPI_ISL_1648061, EPI_ISL_1648062, EPI_ISL_1648063, EPI_ISL_1648064, EPI_ISL_1648065, EPI_ISL_1648066, EPI_ISL_1648067, EPI_ISL_1648068, EPI_ISL_1648069, EPI_ISL_1648070, EPI_ISL_1648071, EPI_ISL_1648073, EPI_ISL_1648074, EPI_ISL_1648075, EPI_ISL_1648076, EPI_ISL_1648077, EPI_ISL_1648078, EPI_ISL_1648079, EPI_ISL_1648080, EPI_ISL_1648081, EPI_ISL_1648082, EPI_ISL_1648083, EPI_ISL_1648084, EPI_ISL_1648085 | | | |
| see above | IN State Department of Health Laboratory Services | IN State Department of Health Laboratory Services | Cassandra Campion, Jamie Yeadon, Brian Pope, Lixia Liu, Kyle Brownlee, Melissa Hindenlang, Ankita Kashikar, Mark Glazier |
| EPI_ISL_1648086, EPI_ISL_1648088, EPI_ISL_1648089, EPI_ISL_1648090, EPI_ISL_1648091, EPI_ISL_1648092, EPI_ISL_1648093, EPI_ISL_1648094, EPI_ISL_1648096, EPI_ISL_1648098, EPI_ISL_1648099, EPI_ISL_1648100, EPI_ISL_1648101, EPI_ISL_1648102, EPI_ISL_1648105, EPI_ISL_1648106, EPI_ISL_1648109, EPI_ISL_1648110, EPI_ISL_1648111, EPI_ISL_1648113, EPI_ISL_1648114, EPI_ISL_1648116, EPI_ISL_1648120, EPI_ISL_1648121, EPI_ISL_1648122 | | | |
| see above | NJDOH, Public Health and Environmental Laboratories | NJ_PHEL | Lindsey Bodnar, Shiv K. Verma, Jacquelyn Deverell, Matthew Scarnati, Dana Woell, Byeong Jeong |
| EPI_ISL_1648123, EPI_ISL_1648124, EPI_ISL_1648125, EPI_ISL_1648126, EPI_ISL_1648127, EPI_ISL_1648128, EPI_ISL_1648129, EPI_ISL_1648130, EPI_ISL_1648131, EPI_ISL_1648132, EPI_ISL_1648133, EPI_ISL_1648134, EPI_ISL_1648135, EPI_ISL_1648136, EPI_ISL_1648137, EPI_ISL_1648138 | | | |
| see above | Nebraska Public Health Laboratory | NPHL COVID-19 Response Team | NPHL COVID-19 Response Team |
| EPI_ISL_1648139, EPI_ISL_1648140, EPI_ISL_1648141, EPI_ISL_1648142, EPI_ISL_1648143, EPI_ISL_1648144, EPI_ISL_1648145, EPI_ISL_1648146, EPI_ISL_1648147, EPI_ISL_1648148, EPI_ISL_1648149, EPI_ISL_1648150, EPI_ISL_1648151, EPI_ISL_1648152, EPI_ISL_1648153, EPI_ISL_1648154, EPI_ISL_1648155, EPI_ISL_1648156, EPI_ISL_1648157, EPI_ISL_1648158 | | | |
| see above | Quest Diagnostics Incorporated | Centers for Disease Control and Prevention Division of Viral Diseases, Pathogen Discovery | Dakota Howard, Dhwani Batra, Peter W. Cook, Kara Moser, Adrian Paskey, Jason Caravas, Benjamin Rambo-Martin, Shatavia Morrison, Christopher Gulvick, Scott Sammons, Yvette Unoarumhi, Darlene Wagner, Matthew Schmerer, S. H. Rosenthal, A. Gerasimova, R. M. Kagan, B. Anderson, M. Hua, Y. Liu, L.E. Bernstein, K.E. Livingston, A. Perez, I. A. Shlyakhter, R. V. Rolando, R. Owen, P. Tanpaiboon, F. Lacbawan, Clinton R. Paden, Duncan MacCannell |
| EPI_ISL_1648159 | NCSLPH | NCSLPH | Chase K, Miller MC, Greene S, Glover W |
| EPI_ISL_1648160 | Quest Diagnostics Incorporated | Centers for Disease Control and Prevention Division of Viral Diseases, Pathogen Discovery | Dakota Howard, Dhwani Batra, Peter W. Cook, Kara Moser, Adrian Paskey, Jason Caravas, Benjamin Rambo-Martin, Shatavia Morrison, Christopher Gulvick, Scott Sammons, Yvette Unoarumhi, Darlene Wagner, Matthew Schmerer, S. H. Rosenthal, A. Gerasimova, R. M. Kagan, B. Anderson, M. Hua, Y. Liu, L.E. Bernstein, K.E. Livingston, A. Perez, I. A. Shlyakhter, R. V. Rolando, R. Owen, P. Tanpaiboon, F. Lacbawan, Clinton R. Paden, Duncan MacCannell |
| EPI_ISL_1648161, EPI_ISL_1648162 | NCSLPH | NCSLPH | Chase K, Miller MC, Greene S, Glover W |
| EPI_ISL_1648163 | Quest Diagnostics Incorporated | Centers for Disease Control and Prevention Division of Viral Diseases, Pathogen Discovery | Dakota Howard, Dhwani Batra, Peter W. Cook, Kara Moser, Adrian Paskey, Jason Caravas, Benjamin Rambo-Martin, Shatavia Morrison, Christopher Gulvick, Scott Sammons, Yvette Unoarumhi, Darlene Wagner, Matthew Schmerer, S. H. Rosenthal, A. Gerasimova, R. M. Kagan, B. Anderson, M. Hua, Y. Liu, L.E. Bernstein, K.E. Livingston, A. Perez, I. A. Shlyakhter, R. V. Rolando, R. Owen, P. Tanpaiboon, F. Lacbawan, Clinton R. Paden, Duncan MacCannell |
| EPI_ISL_1648164, EPI_ISL_1648165 | NCSLPH | NCSLPH | Chase K, Miller MC, Greene S, Glover W |
| EPI_ISL_1648166 | Quest Diagnostics Incorporated | Centers for Disease Control and Prevention Division of Viral Diseases, Pathogen Discovery | Dakota Howard, Dhwani Batra, Peter W. Cook, Kara Moser, Adrian Paskey, Jason Caravas, Benjamin Rambo-Martin, Shatavia Morrison, Christopher Gulvick, Scott Sammons, Yvette Unoarumhi, Darlene Wagner, Matthew Schmerer, S. H. Rosenthal, A. Gerasimova, R. M. Kagan, B. Anderson, M. Hua, Y. Liu, L.E. Bernstein, K.E. Livingston, A. Perez, I. A. Shlyakhter, R. V. Rolando, R. Owen, P. Tanpaiboon, F. Lacbawan, Clinton R. Paden, Duncan MacCannell |
| EPI_ISL_1648167, EPI_ISL_1648168 | NCSLPH | NCSLPH | Chase K, Miller MC, Greene S, Glover W |
| EPI_ISL_1648169 | Quest Diagnostics Incorporated | Centers for Disease Control and Prevention Division of Viral Diseases, Pathogen Discovery | Dakota Howard, Dhwani Batra, Peter W. Cook, Kara Moser, Adrian Paskey, Jason Caravas, Benjamin Rambo-Martin, Shatavia Morrison, Christopher Gulvick, Scott Sammons, Yvette Unoarumhi, Darlene Wagner, Matthew Schmerer, S. H. Rosenthal, A. Gerasimova, R. M. Kagan, B. Anderson, M. Hua, Y. Liu, L.E. Bernstein, K.E. Livingston, A. Perez, I. A. Shlyakhter, R. V. Rolando, R. Owen, P. Tanpaiboon, F. Lacbawan, Clinton R. Paden, Duncan MacCannell |
| EPI_ISL_1648170, EPI_ISL_1648171 | NCSLPH | NCSLPH | Chase K, Miller MC, Greene S, Glover W |
| EPI_ISL_1648172 | Quest Diagnostics Incorporated | Centers for Disease Control and Prevention Division of Viral Diseases, Pathogen Discovery | Dakota Howard, Dhwani Batra, Peter W. Cook, Kara Moser, Adrian Paskey, Jason Caravas, Benjamin Rambo-Martin, Shatavia Morrison, Christopher Gulvick, Scott Sammons, Yvette Unoarumhi, Darlene Wagner, Matthew Schmerer, S. H. Rosenthal, A. Gerasimova, R. M. Kagan, B. Anderson, M. Hua, Y. Liu, L.E. Bernstein, K.E. Livingston, A. Perez, I. A. Shlyakhter, R. V. Rolando, R. Owen, P. Tanpaiboon, F. Lacbawan, Clinton R. Paden, Duncan MacCannell |
| EPI_ISL_1648173 | NCSLPH | NCSLPH | Chase K, Miller MC, Greene S, Glover W |
| EPI_ISL_1648174, EPI_ISL_1648175, EPI_ISL_1648176, EPI_ISL_1648177, EPI_ISL_1648178, EPI_ISL_1648179, EPI_ISL_1648180, EPI_ISL_1648181, EPI_ISL_1648182, EPI_ISL_1648183, EPI_ISL_1648184, EPI_ISL_1648185, EPI_ISL_1648186, EPI_ISL_1648187, EPI_ISL_1648188, EPI_ISL_1648189, EPI_ISL_1648190, EPI_ISL_1648191, EPI_ISL_1648192, EPI_ISL_1648193, EPI_ISL_1648194, EPI_ISL_1648195, EPI_ISL_1648196, EPI_ISL_1648197, EPI_ISL_1648198, EPI_ISL_1648199, EPI_ISL_1648200, EPI_ISL_1648201, EPI_ISL_1648202, EPI_ISL_1648203, EPI_ISL_1648204, EPI_ISL_1648205, EPI_ISL_1648206, EPI_ISL_1648207, EPI_ISL_1648208, EPI_ISL_1648209, EPI_ISL_1648210, EPI_ISL_1648211, EPI_ISL_1648212, EPI_ISL_1648213, EPI_ISL_1648214, EPI_ISL_1648215, EPI_ISL_1648216, EPI_ISL_1648217, EPI_ISL_1648218, EPI_ISL_1648219, EPI_ISL_1648220, EPI_ISL_1648221, EPI_ISL_1648222, EPI_ISL_1648223, EPI_ISL_1648224, EPI_ISL_1648225, EPI_ISL_1648226, EPI_ISL_1648227, EPI_ISL_1648228, EPI_ISL_1648229, EPI_ISL_1648230, EPI_ISL_1648231, EPI_ISL_1648232, EPI_ISL_1648233, EPI_ISL_1648234, EPI_ISL_1648235, EPI_ISL_1648236, EPI_ISL_1648237, EPI_ISL_1648238, EPI_ISL_1648239, EPI_ISL_1648240, EPI_ISL_1648241, EPI_ISL_1648242, EPI_ISL_1648243, EPI_ISL_1648244, EPI_ISL_1648245, EPI_ISL_1648246, EPI_ISL_1648247, EPI_ISL_1648248, EPI_ISL_1648249, EPI_ISL_1648250, EPI_ISL_1648251, EPI_ISL_1648252, EPI_ISL_1648253, EPI_ISL_1648254, EPI_ISL_1648255, EPI_ISL_1648256, EPI_ISL_1648257, EPI_ISL_1648258, EPI_ISL_1648259, EPI_ISL_1648260, EPI_ISL_1648261, EPI_ISL_1648262, EPI_ISL_1648263, EPI_ISL_1648264, EPI_ISL_1648265, EPI_ISL_1648266, EPI_ISL_1648267, EPI_ISL_1648268, EPI_ISL_1648269, EPI_ISL_1648270, EPI_ISL_1648271, EPI_ISL_1648272, EPI_ISL_1648273, EPI_ISL_1648274, EPI_ISL_1648275, EPI_ISL_1648276, EPI_ISL_1648277, EPI_ISL_1648278, EPI_ISL_1648279, EPI_ISL_1648280, EPI_ISL_1648281, EPI_ISL_1648282, EPI_ISL_1648283, EPI_ISL_1648284, EPI_ISL_1648285, EPI_ISL_1648286, EPI_ISL_1648287, EPI_ISL_1648288, EPI_ISL_1648289, EPI_ISL_1648290, EPI_ISL_1648291, EPI_ISL_1648292, EPI_ISL_1648293, EPI_ISL_1648294, EPI_ISL_1648295, EPI_ISL_1648296, EPI_ISL_1648297, EPI_ISL_1648298, EPI_ISL_1648299, EPI_ISL_1648300, EPI_ISL_1648301, EPI_ISL_1648302, EPI_ISL_1648303, EPI_ISL_1648304, EPI_ISL_1648305, EPI_ISL_1648306, EPI_ISL_1648307, EPI_ISL_1648308, EPI_ISL_1648309, EPI_ISL_1648310, EPI_ISL_1648311, EPI_ISL_1648312, EPI_ISL_1648313, EPI_ISL_1648314, EPI_ISL_1648315, EPI_ISL_1648316, EPI_ISL_1648317, EPI_ISL_1648318, EPI_ISL_1648319, EPI_ISL_1648320, EPI_ISL_1648321, EPI_ISL_1648322, EPI_ISL_1648323, EPI_ISL_1648324, EPI_ISL_1648325, EPI_ISL_1648326, EPI_ISL_1648327, EPI_ISL_1648328, EPI_ISL_1648329, EPI_ISL_1648330, EPI_ISL_1648331, EPI_ISL_1648332, EPI_ISL_1648333, EPI_ISL_1648334, EPI_ISL_1648335, EPI_ISL_1648336, EPI_ISL_1648337, EPI_ISL_1648338, EPI_ISL_1648339, EPI_ISL_1648340, EPI_ISL_1648341, EPI_ISL_1648342, EPI_ISL_1648343, EPI_ISL_1648344, EPI_ISL_1648345, EPI_ISL_1648346, EPI_ISL_1648347, EPI_ISL_1648348, EPI_ISL_1648349, EPI_ISL_1648350, EPI_ISL_1648351, EPI_ISL_1648352, EPI_ISL_1648353, EPI_ISL_1648354, EPI_ISL_1648355, EPI_ISL_1648356, EPI_ISL_1648357, EPI_ISL_1648358, EPI_ISL_1648359, EPI_ISL_1648360, EPI_ISL_1648361, EPI_ISL_1648362, EPI_ISL_1648363, EPI_ISL_1648364, EPI_ISL_1648365, EPI_ISL_1648366, EPI_ISL_1648367, EPI_ISL_1648368, EPI_ISL_1648369, EPI_ISL_1648370, EPI_ISL_1648371, EPI_ISL_1648372, EPI_ISL_1648373, EPI_ISL_1648374, EPI_ISL_1648375, EPI_ISL_1648376, EPI_ISL_1648377, EPI_ISL_1648378, EPI_ISL_1648379, EPI_ISL_1648380, EPI_ISL_1648381, EPI_ISL_1648382, EPI_ISL_1648383, EPI_ISL_1648384, EPI_ISL_1648385, EPI_ISL_1648386, EPI_ISL_1648387, EPI_ISL_1648388, EPI_ISL_1648389, EPI_ISL_1648390, EPI_ISL_1648391, EPI_ISL_1648392, EPI_ISL_1648393, EPI_ISL_1648394, EPI_ISL_1648395, EPI_ISL_1648396, EPI_ISL_1648397, EPI_ISL_1648398, EPI_ISL_1648399, EPI_ISL_1648400, EPI_ISL_1648401, EPI_ISL_1648402, EPI_ISL_1648403, EPI_ISL_1648404, EPI_ISL_1648405, EPI_ISL_1648406, EPI_ISL_1648407, EPI_ISL_1648408, EPI_ISL_1648409, EPI_ISL_1648410, EPI_ISL_1648411, EPI_ISL_1648412, EPI_ISL_1648413, EPI_ISL_1648414, EPI_ISL_1648415, EPI_ISL_1648416, EPI_ISL_1648417, EPI_ISL_1648418, EPI_ISL_1648419, EPI_ISL_1648420, EPI_ISL_1648421, EPI_ISL_1648422, EPI_ISL_1648423, EPI_ISL_1648424, EPI_ISL_1648425, EPI_ISL_1648426, EPI_ISL_1648427, EPI_ISL_1648428, EPI_ISL_1648429, EPI_ISL_1648430, EPI_ISL_1648431, EPI_ISL_1648432, EPI_ISL_1648433, EPI_ISL_1648434, EPI_ISL_1648435, EPI_ISL_1648436, EPI_ISL_1648437, EPI_ISL_1648438, EPI_ISL_1648439, EPI_ISL_1648440, EPI_ISL_1648441, EPI_ISL_1648442, EPI_ISL_1648443, EPI_ISL_1648444, EPI_ISL_1648445, EPI_ISL_1648446, EPI_ISL_1648447, EPI_ISL_1648448, EPI_ISL_1648449, EPI_ISL_1648450, EPI_ISL_1648451, EPI_ISL_1648452, EPI_ISL_1648453, EPI_ISL_1648454, EPI_ISL_1648455, EPI_ISL_1648456, EPI_ISL_1648457, EPI_ISL_1648458, EPI_ISL_1648459, EPI_ISL_1648460, EPI_ISL_1648461, EPI_ISL_1648462, EPI_ISL_1648463, EPI_ISL_1648464, EPI_ISL_1648465, EPI_ISL_1648466, EPI_ISL_1648467, EPI_ISL_1648468, EPI_ISL_1648469, EPI_ISL_1648470, EPI_ISL_1648471, EPI_ISL_1648472, EPI_ISL_1648473, EPI_ISL_1648474, EPI_ISL_1648475, EPI_ISL_1648476, EPI_ISL_1648477, EPI_ISL_1648478, EPI_ISL_1648479, EPI_ISL_1648480, EPI_ISL_1648481, EPI_ISL_1648482, EPI_ISL_1648483, EPI_ISL_1648484, EPI_ISL_1648485, EPI_ISL_1648486, EPI_ISL_1648487, EPI_ISL_1648488, EPI_ISL_1648489, EPI_ISL_1648490, EPI_ISL_1648491, EPI_ISL_1648492, EPI_ISL_1648493, EPI_ISL_1648494, EPI_ISL_1648495, EPI_ISL_1648496, EPI_ISL_1648497, EPI_ISL_1648498, EPI_ISL_1648499, EPI_ISL_1648500, EPI_ISL_1648501, EPI_ISL_1648502, EPI_ISL_1648503, EPI_ISL_1648504, EPI_ISL_1648505, EPI_ISL_1648506, EPI_ISL_1648507, EPI_ISL_1648508, EPI_ISL_1648509, EPI_ISL_1648510, EPI_ISL_1648511, EPI_ISL_1648512, EPI_ISL_1648513, EPI_ISL_1648514, EPI_ISL_1648515, EPI_ISL_1648516, EPI_ISL_1648517, EPI_ISL_1648518, EPI_ISL_1648519, EPI_ISL_1648520, EPI_ISL_1648521, EPI_ISL_1648522, EPI_ISL_1648523, EPI_ISL_1648524, EPI_ISL_1648525, EPI_ISL_1648526, EPI_ISL_1648527, EPI_ISL_1648528, EPI_ISL_1648529, EPI_ISL_1648530, EPI_ISL_1648531, EPI_ISL_1648532, EPI_ISL_1648533, EPI_ISL_1648534, EPI_ISL_1648535, EPI_ISL_1648536, EPI_ISL_1648537, EPI_ISL_1648538, EPI_ISL_1648539, EPI_ISL_1648540, EPI_ISL_1648541, EPI_ISL_1648542, EPI_ISL_1648543, EPI_ISL_1648544, EPI_ISL_1648545, EPI_ISL_1648546, EPI_ISL_1648547, EPI_ISL_1648548, EPI_ISL_1648549, EPI_ISL_1648550, EPI_ISL_1648551, EPI_ISL_1648552, EPI_ISL_1648553, EPI_ISL_1648554, EPI_ISL_1648555, EPI_ISL_1648556, EPI_ISL_1648557, EPI_ISL_1648558, EPI_ISL_1648559, EPI_ISL_1648560, EPI_ISL_1648561, EPI_ISL_1648562, EPI_ISL_1648563, EPI_ISL_1648564, EPI_ISL_1648565, EPI_ISL_1648566, EPI_ISL_1648567, EPI_ISL_1648568, EPI_ISL_1648569, EPI_ISL_1648570, EPI_ISL_1648571, EPI_ISL_1648572, EPI_ISL_1648573, EPI_ISL_1648574, EPI_ISL_1648575, EPI_ISL_1648576, EPI_ISL_1648577, EPI_ISL_1648578, EPI_ISL_1648579, EPI_ISL_1648580, EPI_ISL_1648581, EPI_ISL_1648582, EPI_ISL_1648583, EPI_ISL_1648584, EPI_ISL_1648585, EPI_ISL_1648586, EPI_ISL_1648587, EPI_ISL_1648588, EPI_ISL_1648589, EPI_ISL_1648590, EPI_ISL_1648591, EPI_ISL_1648592, EPI_ISL_1648593, EPI_ISL_1648594, EPI_ISL_1648595, EPI_ISL_1648596, EPI_ISL_1648597, EPI_ISL_1648598, EPI_ISL_1648599, EPI_ISL_1648600, EPI_ISL_1648601, EPI_ISL_1648602, EPI_ISL_1648603, EPI_ISL_1648604, EPI_ISL_1648605, EPI_ISL_1648606, EPI_ISL_1648607, EPI_ISL_1648608, EPI_ISL_1648609, EPI_ISL_1648610, EPI_ISL_1648611, EPI_ISL_1648612, EPI_ISL_1648613, EPI_ISL_1648614, EPI_ISL_1648615, EPI_ISL_1648616, EPI_ISL_1648617, EPI_ISL_1648618, EPI_ISL_1648619, EPI_ISL_1648620, EPI_ISL_1648621, EPI_ISL_1648622, EPI_ISL_1648623, EPI_ISL_1648624, EPI_ISL_1648625, EPI_ISL_1648626, EPI_ISL_1648627, EPI_ISL_1648628, EPI_ISL_1648629, EPI_ISL_1648630, EPI_ISL_1648631, EPI_ISL_1648632, EPI_ISL_1648633, EPI_ISL_1648634, EPI_ISL_1648635, EPI_ISL_1648636, EPI_ISL_1648637, EPI_ISL_1648638, EPI_ISL_1648639, EPI_ISL_1648640, EPI_ISL_1648641, EPI_ISL_1648642, EPI_ISL_1648643, EPI_ISL_1648644, EPI_ISL_1648645, EPI_ISL_1648646, EPI_ISL_1648647, EPI_ISL_1648648, EPI_ISL_1648649, EPI_ISL_1648650, EPI_ISL_1648651, EPI_ISL_1648652, EPI_ISL_1648653, EPI_ISL_1648654, EPI_ISL_1648655, EPI_ISL_1648656, EPI_ISL_1648657, EPI_ISL_1648658, EPI_ISL_1648659, EPI_ISL_1648660, EPI_ISL_1648661, EPI_ISL_1648662, EPI_ISL_1648663, EPI_ISL_1648664, EPI_ISL_1648665, EPI_ISL_1648666, EPI_ISL_1648667, EPI_ISL_1648668, EPI_ISL_1648669, EPI_ISL_1648670, EPI_ISL_1648671, EPI_ISL_1648672, EPI_ISL_1648673, EPI_ISL_1648674, EPI_ISL_1648675, EPI_ISL_1648676, EPI_ISL_1648677, EPI_ISL_1648678, EPI_ISL_1648679, EPI_ISL_1648680, EPI_ISL_1648681, EPI_ISL_1648682, EPI_ISL_1648683, EPI_ISL_1648684, EPI_ISL_1648685, EPI_ISL_1648686, EPI_ISL_1648687, EPI_ISL_1648688, EPI_ISL_1648689, EPI_ISL_1648690, EPI_ISL_1648691, EPI_ISL_1648692, EPI_ISL_1648693, EPI_ISL_1648694, EPI_ISL_1648695, EPI_ISL_1648696, EPI_ISL_1648697, EPI_ISL_1648698, EPI_ISL_1648699, EPI_ISL_1648700, EPI_ISL_1648701, EPI_ISL_1648702, EPI_ISL_1648703, EPI_ISL_1648704, EPI_ISL_1648705, EPI_ISL_1648706, EPI_ISL_1648707, EPI_ISL_1648708, EPI_ISL_1648709, EPI_ISL_1648710, EPI_ISL_1648711, EPI_ISL_1648712, EPI_ISL_1648713, EPI_ISL_1648714, EPI_ISL_1648715, EPI_ISL_1648716, EPI_ISL_1648717, EPI_ISL_1648718, EPI_ISL_1648719, EPI_ISL_1648720, EPI_ISL_1648721, EPI_ISL_1648722, EPI_ISL_1648723, EPI_ISL_1648724, EPI_ISL_1648725, EPI_ISL_1648726, EPI_ISL_1648727, EPI_ISL_1648728, EPI_ISL_1648729, EPI_ISL_1648730, EPI_ISL_1648731, EPI_ISL_1648732, EPI_ISL_1648733, EPI_ISL_1648734, EPI_ISL_1648735, EPI_ISL_1648736, EPI_ISL_1648737, EPI_ISL_1648738, EPI_ISL_1648739, EPI_ISL_1648740, EPI_ISL_1648741, EPI_ISL_1648742, EPI_ISL_1648743, EPI_ISL_1648744, EPI_ISL_1648745, EPI_ISL_1648746, EPI_ISL_1648747, EPI_ISL_1648748, EPI_ISL_1648749, EPI_ISL_1648750, EPI_ISL_1648751, EPI_ISL_1648752, EPI_ISL_1648753, EPI_ISL_1648754, EPI_ISL_1648755, EPI_ISL_1648756, EPI_ISL_1648757, EPI_ISL_1648758, EPI_ISL_1648759, EPI_ISL_1648760, EPI_ISL_1648761, EPI_ISL_1648762, EPI_ISL_1648763, EPI_ISL_1648764, EPI_ISL_1648765, EPI_ISL_1648766, EPI_ISL_1648767, EPI_ISL_1648768, EPI_ISL_1648769, EPI_ISL_1648770, EPI_ISL_1648771, EPI_ISL_1648772, EPI_ISL_1648773, EPI_ISL_1648774, EPI_ISL_1648775, EPI_ISL_1648776, EPI_ISL_1648777, EPI_ISL_1648778, EPI_ISL_1648779, EPI_ISL_1648780, EPI_ISL_1648781, EPI_ISL_1648782, EPI_ISL_1648783, EPI_ISL_1648784, EPI_ISL_1648785, EPI_ISL_1648786, EPI_ISL_1648787, EPI_ISL_1648788, EPI_ISL_1648789, EPI_ISL_1648790, EPI_ISL_1648791, EPI_ISL_1648792, EPI_ISL_1648793, EPI_ISL_1648794, EPI_ISL_1648795, EPI_ISL_1648796, EPI_ISL_1648797, EPI_ISL_1648798, EPI_ISL_1648799, EPI_ISL_1648800, EPI_ISL_1648801, EPI_ISL_1648802, EPI_ISL_1648803, EPI_ISL_1648804, EPI_ISL_1648805, EPI_ISL_1648806, EPI_ISL_1648807, EPI_ISL_1648808, EPI_ISL_1648809, EPI_ISL_1648810, EPI_ISL_1648811, EPI_ISL_1648812, EPI_ISL_1648813, EPI_ISL_1648814, EPI_ISL_1648815, EPI_ISL_1648816, EPI_ISL_1648817, EPI_ISL_1648818, EPI_ISL_1648819, EPI_ISL_1648820, EPI_ISL_1648821, EPI_ISL_1648822, EPI_ISL_1648823, EPI_ISL_1648824, EPI_ISL_1648825, EPI_ISL_1648826, EPI_ISL_1648827, EPI_ISL_1648828, EPI_ISL_1648829, EPI_ISL_1648830, EPI_ISL_1648831, EPI_ISL_1648832, EPI_ISL_1648833, EPI_ISL_1648834, EPI_ISL_1648835, EPI_ISL_1648836, EPI_ISL_1648837, EPI_ISL_1648838, EPI_ISL_1648839, EPI_ISL_1648840, EPI_ISL_1648841, EPI_ISL_1648842, EPI_ISL_1648843, EPI_ISL_1648844, EPI_ISL_1648845, EPI_ISL_1648846, EPI_ISL_1648847, EPI_ISL_1648848, EPI_ISL_1648849, EPI_ISL_1648850, EPI_ISL_1648851, EPI_ISL_1648852, EPI_ISL_1648853, EPI_ISL_1648854, EPI_ISL_1648855, EPI_ISL_1648856, EPI_ISL_1648857, EPI_ISL_1648858, EPI_ISL_1648859, EPI_ISL_1648860, EPI_ISL_1648861, EPI_ISL_1648862, EPI_ISL_1648863, EPI_ISL_1648864, EPI_ISL_1648865, EPI_ISL_1648866, EPI_ISL_1648867, EPI_ISL_1648868, EPI_ISL_1648869, EPI_ISL_1648870, EPI_ISL_1648871, EPI_ISL_1648872, EPI_ISL_1648873, EPI_ISL_1648874, EPI_ISL_1648875, EPI_ISL_1648876, EPI_ISL_1648877, EPI_ISL_1648878, EPI_ISL_1648879, EPI_ISL_1648880, EPI_ISL_1648881, EPI_ISL_1648882, EPI_ISL_1648883, EPI_ISL_1648884, EPI_ISL_1648885, EPI_ISL_1648886, EPI_ISL_1648887, EPI_ISL_1648888, EPI_ISL_1648889, EPI_ISL_1648890, EPI_ISL_1648891, EPI_ISL_1648892, EPI_ISL_1648893, EPI_ISL_1648894, EPI_ISL_1648895, EPI_ISL_1648896, EPI_ISL_1648897, EPI_ISL_1648898, EPI_ISL_1648899, EPI_ISL_1648900, EPI_ISL_1648901, EPI_ISL_1648902, EPI_ISL_1648903, EPI_ISL_1648904, EPI_ISL_1648905, EPI_ISL_1648906, EPI_ISL_1648907, EPI_ISL_1648908, EPI_ISL_1648909, EPI_ISL_1648910, EPI_ISL_1648911, EPI_ISL_1648912, EPI_ISL_1648913, EPI_ISL_1648914, EPI_ISL_1648915, EPI_ISL_1648916, EPI_ISL_1648917, EPI_ISL_1648918, EPI_ISL_1648919, EPI_ISL_1648920, EPI_ISL_1648921, EPI_ISL_1648922, EPI_ISL_1648923 | | | |
| see above | Quest Diagnostics Incorporated | Centers for Disease Control and Prevention Division of Viral Diseases, Pathogen Discovery | Dakota Howard, Dhwani Batra, Peter W. Cook, Kara Moser, Adrian Paskey, Jason Caravas, Benjamin Rambo-Martin, Shatavia Morrison, Christopher Gulvick, Scott Sammons, Yvette Unoarumhi, Darlene Wagner, Matthew Schmerer, S. H. Rosenthal, A. Gerasimova, R. M. Kagan, B. Anderson, M. Hua, Y. Liu, L.E. Bernstein, K.E. Livingston, A. Perez, I. A. Shlyakhter, R. V. Rolando, R. Owen, P. Tanpaiboon, F. Lacbawan, Clinton R. Paden, Duncan MacCannell |
| EPI_ISL_1648944, EPI_ISL_1648945, EPI_ISL_1648946, EPI_ISL_1648953, EPI_ISL_1648954, EPI_ISL_1648956, EPI_ISL_1648957, EPI_ISL_1648963 | University of Michigan Clinical Microbiology Laboratory | Lauring Lab, University of Michigan, Department of Microbiology and Immunology | Valesano |
| EPI_ISL_1648966, EPI_ISL_1648967, EPI_ISL_1648968, EPI_ISL_1648969, EPI_ISL_1648970, EPI_ISL_1648971, EPI_ISL_1648972, EPI_ISL_1648973, EPI_ISL_1648974, EPI_ISL_1648975, EPI_ISL_1648976, EPI_ISL_1648977, EPI_ISL_1648978, EPI_ISL_1648979, EPI_ISL_1648980, EPI_ISL_1648982, EPI_ISL_1648983, EPI_ISL_1648984, EPI_ISL_1648985, EPI_ISL_1648987, EPI_ISL_1648988, EPI_ISL_1648989, | | | |

EPI_ISL_1648990, EPI_ISL_1648991, EPI_ISL_1648992, EPI_ISL_1648993, EPI_ISL_1648994, EPI_ISL_1648995, EPI_ISL_1648996, EPI_ISL_1648997, EPI_ISL_1648998, EPI_ISL_1648999, EPI_ISL_1649000, EPI_ISL_1649001, EPI_ISL_1649002, EPI_ISL_1649003, EPI_ISL_1649004, EPI_ISL_1649005, EPI_ISL_1649006, EPI_ISL_1649007, EPI_ISL_1649008, EPI_ISL_1649009, EPI_ISL_1649010, EPI_ISL_1649011, EPI_ISL_1649012, EPI_ISL_1649013, EPI_ISL_1649014, EPI_ISL_1649015, EPI_ISL_1649016, EPI_ISL_1649017, EPI_ISL_1649018, EPI_ISL_1649019, EPI_ISL_1649020, EPI_ISL_1649021, EPI_ISL_1649022, EPI_ISL_1649023, EPI_ISL_1649024, EPI_ISL_1649025, EPI_ISL_1649026, EPI_ISL_1649027, EPI_ISL_1649028, EPI_ISL_1649029, EPI_ISL_1649030, EPI_ISL_1649031, EPI_ISL_1649032, EPI_ISL_1649033, EPI_ISL_1649034, EPI_ISL_1649035, EPI_ISL_1649036, EPI_ISL_1649037, EPI_ISL_1649038, EPI_ISL_1649039, EPI_ISL_1649040, EPI_ISL_1649041, EPI_ISL_1649042, EPI_ISL_1649043, EPI_ISL_1649045, EPI_ISL_1649046, EPI_ISL_1649047, EPI_ISL_1649048, EPI_ISL_1649049, EPI_ISL_1649050, EPI_ISL_1649051, EPI_ISL_1649052, EPI_ISL_1649053, EPI_ISL_1649054, EPI_ISL_1649055, EPI_ISL_1649056, EPI_ISL_1649057, EPI_ISL_1649058, EPI_ISL_1649059, EPI_ISL_1649060, EPI_ISL_1649061, EPI_ISL_1649064, EPI_ISL_1649066, EPI_ISL_1649067, EPI_ISL_1649068, EPI_ISL_1649069, EPI_ISL_1649070, EPI_ISL_1649071, EPI_ISL_1649072, EPI_ISL_1649073, EPI_ISL_1649074, EPI_ISL_1649075, EPI_ISL_1649076, EPI_ISL_1649077, EPI_ISL_1649078, EPI_ISL_1649079, EPI_ISL_1649080, EPI_ISL_1649081, EPI_ISL_1649082, EPI_ISL_1649083, EPI_ISL_1649084, EPI_ISL_1649085, EPI_ISL_1649086, EPI_ISL_1649087, EPI_ISL_1649088, EPI_ISL_1649089, EPI_ISL_1649090, EPI_ISL_1649091, EPI_ISL_1649092, EPI_ISL_1649093, EPI_ISL_1649094, EPI_ISL_1649095, EPI_ISL_1649096, EPI_ISL_1649097, EPI_ISL_1649098, EPI_ISL_1649099, EPI_ISL_1649101, EPI_ISL_1649102, EPI_ISL_1649103, EPI_ISL_1649104, EPI_ISL_1649105, EPI_ISL_1649106, EPI_ISL_1649107, EPI_ISL_1649108, EPI_ISL_1649109, EPI_ISL_1649111, EPI_ISL_1649112, EPI_ISL_1649113, EPI_ISL_1649114, EPI_ISL_1649116, EPI_ISL_1649117, EPI_ISL_1649118, EPI_ISL_1649119, EPI_ISL_1649120, EPI_ISL_1649121, EPI_ISL_1649122, EPI_ISL_1649123, EPI_ISL_1649124, EPI_ISL_1649125, EPI_ISL_1649126, EPI_ISL_1649127, EPI_ISL_1649128, EPI_ISL_1649129, EPI_ISL_1649130, EPI_ISL_1649131, EPI_ISL_1649132, EPI_ISL_1649133, EPI_ISL_1649135, EPI_ISL_1649136, EPI_ISL_1649137, EPI_ISL_1649138, EPI_ISL_1649139, EPI_ISL_1649140, EPI_ISL_1649141, EPI_ISL_1649142, EPI_ISL_1649143, EPI_ISL_1649144, EPI_ISL_1649145, EPI_ISL_1649146, EPI_ISL_1649147, EPI_ISL_1649148, EPI_ISL_1649149, EPI_ISL_1649150, EPI_ISL_1649152, EPI_ISL_1649154, EPI_ISL_1649155, EPI_ISL_1649156, EPI_ISL_1649157, EPI_ISL_1649158, EPI_ISL_1649159, EPI_ISL_1649160, EPI_ISL_1649161, EPI_ISL_1649162, EPI_ISL_1649164, EPI_ISL_1649165, EPI_ISL_1649166, EPI_ISL_1649167, EPI_ISL_1649168, EPI_ISL_1649169, EPI_ISL_1649170, EPI_ISL_1649171, EPI_ISL_1649172, EPI_ISL_1649173, EPI_ISL_1649174, EPI_ISL_1649175, EPI_ISL_1649176, EPI_ISL_1649177, EPI_ISL_1649178, EPI_ISL_1649179, EPI_ISL_1649180, EPI_ISL_1649181, EPI_ISL_1649182, EPI_ISL_1649183, EPI_ISL_1649184, EPI_ISL_1649185, EPI_ISL_1649186, EPI_ISL_1649187, EPI_ISL_1649188, EPI_ISL_1649189, EPI_ISL_1649190, EPI_ISL_1649191, EPI_ISL_1649192, EPI_ISL_1649194, EPI_ISL_1649195, EPI_ISL_1649196, EPI_ISL_1649197, EPI_ISL_1649198, EPI_ISL_1649199, EPI_ISL_1649200, EPI_ISL_1649201, EPI_ISL_1649202, EPI_ISL_1649203, EPI_ISL_1649205, EPI_ISL_1649206, EPI_ISL_1649208, EPI_ISL_1649209, EPI_ISL_1649210, EPI_ISL_1649211, EPI_ISL_1649212, EPI_ISL_1649213, EPI_ISL_1649214, EPI_ISL_1649215, EPI_ISL_1649217, EPI_ISL_1649218, EPI_ISL_1649219, EPI_ISL_1649220, EPI_ISL_1649221, EPI_ISL_1649222, EPI_ISL_1649223, EPI_ISL_1649224, EPI_ISL_1649225, EPI_ISL_1649226, EPI_ISL_1649227, EPI_ISL_1649228, EPI_ISL_1649229, EPI_ISL_1649230, EPI_ISL_1649231, EPI_ISL_1649232, EPI_ISL_1649233, EPI_ISL_1649234, EPI_ISL_1649235, EPI_ISL_1649236, EPI_ISL_1649237, EPI_ISL_1649238, EPI_ISL_1649239, EPI_ISL_1649240, EPI_ISL_1649241, EPI_ISL_1649243, EPI_ISL_1649244, EPI_ISL_1649245, EPI_ISL_1649246, EPI_ISL_1649247, EPI_ISL_1649248, EPI_ISL_1649249, EPI_ISL_1649250, EPI_ISL_1649251, EPI_ISL_1649252, EPI_ISL_1649253, EPI_ISL_1649255, EPI_ISL_1649256, EPI_ISL_1649257, EPI_ISL_1649258, EPI_ISL_1649259, EPI_ISL_1649260, EPI_ISL_1649261, EPI_ISL_1649262, EPI_ISL_1649263, EPI_ISL_1649264, EPI_ISL_1649265, EPI_ISL_1649266, EPI_ISL_1649267, EPI_ISL_1649268, EPI_ISL_1649269, EPI_ISL_1649270, EPI_ISL_1649271, EPI_ISL_1649272, EPI_ISL_1649273, EPI_ISL_1649274, EPI_ISL_1649275, EPI_ISL_1649276, EPI_ISL_1649277, EPI_ISL_1649278, EPI_ISL_1649279, EPI_ISL_1649280, EPI_ISL_1649281, EPI_ISL_1649282, EPI_ISL_1649283, EPI_ISL_1649284, EPI_ISL_1649285, EPI_ISL_1649286, EPI_ISL_1649287, EPI_ISL_1649288, EPI_ISL_1649289, EPI_ISL_1649290, EPI_ISL_1649291, EPI_ISL_1649292, EPI_ISL_1649293, EPI_ISL_1649294, EPI_ISL_1649295, EPI_ISL_1649297, EPI_ISL_1649298, EPI_ISL_1649299, EPI_ISL_1649300, EPI_ISL_1649301, EPI_ISL_1649302, EPI_ISL_1649303, EPI_ISL_1649304, EPI_ISL_1649305, EPI_ISL_1649306, EPI_ISL_1649307, EPI_ISL_1649308, EPI_ISL_1649309, EPI_ISL_1649311, EPI_ISL_1649312, EPI_ISL_1649313, EPI_ISL_1649314, EPI_ISL_1649315, EPI_ISL_1649316, EPI_ISL_1649317, EPI_ISL_1649318, EPI_ISL_1649319, EPI_ISL_1649320, EPI_ISL_1649321, EPI_ISL_1649322, EPI_ISL_1649323, EPI_ISL_1649324, EPI_ISL_1649325, EPI_ISL_1649326, EPI_ISL_1649328, EPI_ISL_1649329, EPI_ISL_1649330, EPI_ISL_1649331, EPI_ISL_1649332, EPI_ISL_1649334, EPI_ISL_1649335, EPI_ISL_1649336, EPI_ISL_1649337, EPI_ISL_1649338, EPI_ISL_1649339, EPI_ISL_1649340, EPI_ISL_1649341, EPI_ISL_1649342, EPI_ISL_1649343, EPI_ISL_1649344, EPI_ISL_1649345, EPI_ISL_1649346, EPI_ISL_1649347, EPI_ISL_1649348, EPI_ISL_1649349, EPI_ISL_1649350, EPI_ISL_1649351, EPI_ISL_1649352, EPI_ISL_1649353, EPI_ISL_1649354, EPI_ISL_1649355, EPI_ISL_1649356, EPI_ISL_1649357, EPI_ISL_1649358, EPI_ISL_1649359, EPI_ISL_1649360, EPI_ISL_1649361, EPI_ISL_1649362, EPI_ISL_1649363, EPI_ISL_1649364, EPI_ISL_1649366, EPI_ISL_1649367, EPI_ISL_1649368, EPI_ISL_1649369, EPI_ISL_1649370, EPI_ISL_1649371, EPI_ISL_1649372, EPI_ISL_1649373, EPI_ISL_1649374, EPI_ISL_1649375, EPI_ISL_1649376, EPI_ISL_1649377, EPI_ISL_1649378, EPI_ISL_1649379, EPI_ISL_1649380, EPI_ISL_1649381, EPI_ISL_1649382, EPI_ISL_1649383, EPI_ISL_1649384, EPI_ISL_1649385, EPI_ISL_1649386, EPI_ISL_1649387, EPI_ISL_1649388, EPI_ISL_1649389, EPI_ISL_1649390, EPI_ISL_1649393, EPI_ISL_1649394, EPI_ISL_1649395, EPI_ISL_1649396, EPI_ISL_1649397, EPI_ISL_1649398, EPI_ISL_1649399, EPI_ISL_1649400, EPI_ISL_1649401, EPI_ISL_1649402, EPI_ISL_1649403, EPI_ISL_1649404, EPI_ISL_1649405, EPI_ISL_1649406, EPI_ISL_1649407, EPI_ISL_1649408, EPI_ISL_1649409, EPI_ISL_1649410, EPI_ISL_1649411, EPI_ISL_1649412, EPI_ISL_1649413, EPI_ISL_1649414, EPI_ISL_1649415, EPI_ISL_1649416, EPI_ISL_1649417, EPI_ISL_1649418, EPI_ISL_1649419, EPI_ISL_1649420, EPI_ISL_1649421, EPI_ISL_1649422, EPI_ISL_1649423, EPI_ISL_1649424, EPI_ISL_1649425, EPI_ISL_1649426, EPI_ISL_1649427, EPI_ISL_1649428, EPI_ISL_1649429, EPI_ISL_1649430, EPI_ISL_1649431, EPI_ISL_1649432, EPI_ISL_1649433, EPI_ISL_1649434, EPI_ISL_1649435, EPI_ISL_1649436, EPI_ISL_1649437, EPI_ISL_1649438, EPI_ISL_1649439, EPI_ISL_1649440, EPI_ISL_1649441, EPI_ISL_1649442, EPI_ISL_1649443, EPI_ISL_1649444, EPI_ISL_1649445, EPI_ISL_1649446, EPI_ISL_1649447, EPI_ISL_1649448, EPI_ISL_1649449, EPI_ISL_1649450, EPI_ISL_1649451, EPI_ISL_1649452, EPI_ISL_1649453, EPI_ISL_1649454, EPI_ISL_1649455, EPI_ISL_1649456, EPI_ISL_1649457, EPI_ISL_1649459, EPI_ISL_1649460, EPI_ISL_1649461, EPI_ISL_1649463, EPI_ISL_1649464, EPI_ISL_1649465, EPI_ISL_1649466, EPI_ISL_1649467, EPI_ISL_1649468, EPI_ISL_1649469, EPI_ISL_1649470, EPI_ISL_1649471, EPI_ISL_1649472, EPI_ISL_1649473, EPI_ISL_1649474, EPI_ISL_1649475, EPI_ISL_1649476, EPI_ISL_1649477, EPI_ISL_1649478, EPI_ISL_1649479, EPI_ISL_1649480, EPI_ISL_1649481, EPI_ISL_1649482, EPI_ISL_1649483, EPI_ISL_1649484, EPI_ISL_1649485, EPI_ISL_1649486, EPI_ISL_1649487, EPI_ISL_1649488, EPI_ISL_1649489, EPI_ISL_1649490, EPI_ISL_1649492, EPI_ISL_1649493, EPI_ISL_1649494, EPI_ISL_1649495, EPI_ISL_1649496, EPI_ISL_1649497, EPI_ISL_1649498, EPI_ISL_1649499, EPI_ISL_1649500, EPI_ISL_1649501, EPI_ISL_1649502, EPI_ISL_1649503, EPI_ISL_1649504, EPI_ISL_1649505, EPI_ISL_1649506, EPI_ISL_1649507, EPI_ISL_1649508, EPI_ISL_1649509, EPI_ISL_1649510, EPI_ISL_1649511, EPI_ISL_1649512, EPI_ISL_1649513, EPI_ISL_1649514, EPI_ISL_1649515, EPI_ISL_1649516, EPI_ISL_1649517, EPI_ISL_1649518, EPI_ISL_1649519, EPI_ISL_1649520, EPI_ISL_1649521, EPI_ISL_1649522, EPI_ISL_1649523, EPI_ISL_1649524, EPI_ISL_1649525, EPI_ISL_1649526, EPI_ISL_1649527, EPI_ISL_1649528, EPI_ISL_1649529, EPI_ISL_1649530, EPI_ISL_1649531, EPI_ISL_1649532, EPI_ISL_1649533, EPI_ISL_1649534, EPI_ISL_1649536, EPI_ISL_1649537, EPI_ISL_1649538, EPI_ISL_1649539, EPI_ISL_1649540, EPI_ISL_1649541, EPI_ISL_1649542, EPI_ISL_1649543, EPI_ISL_1649544, EPI_ISL_1649545, EPI_ISL_1649546, EPI_ISL_1649547, EPI_ISL_1649549, EPI_ISL_1649550, EPI_ISL_1649551, EPI_ISL_1649552, EPI_ISL_1649553, EPI_ISL_1649554, EPI_ISL_1649555, EPI_ISL_1649556, EPI_ISL_1649557, EPI_ISL_1649558, EPI_ISL_1649559, EPI_ISL_1649560, EPI_ISL_1649561, EPI_ISL_1649562, EPI_ISL_1649563, EPI_ISL_1649564, EPI_ISL_1649565, EPI_ISL_1649566, EPI_ISL_1649567, EPI_ISL_1649568, EPI_ISL_1649569, EPI_ISL_1649570, EPI_ISL_1649571, EPI_ISL_1649572, EPI_ISL_1649573, EPI_ISL_1649574, EPI_ISL_1649575, EPI_ISL_1649576, EPI_ISL_1649577, EPI_ISL_1649578, EPI_ISL_1649579, EPI_ISL_1649580, EPI_ISL_1649581, EPI_ISL_1649582, EPI_ISL_1649583, EPI_ISL_1649584, EPI_ISL_1649585, EPI_ISL_1649586, EPI_ISL_1649587, EPI_ISL_1649588, EPI_ISL_1649589, EPI_ISL_1649590, EPI_ISL_1649591, EPI_ISL_1649592, EPI_ISL_1649593, EPI_ISL_1649594, EPI_ISL_1649595, EPI_ISL_1649596, EPI_ISL_1649597, EPI_ISL_1649599, EPI_ISL_1649600, EPI_ISL_1649603, EPI_ISL_1649604, EPI_ISL_1649605, EPI_ISL_1649606, EPI_ISL_1649607, EPI_ISL_1649608, EPI_ISL_1649609, EPI_ISL_1649610, EPI_ISL_1649611, EPI_ISL_1649612, EPI_ISL_1649613, EPI_ISL_1649614, EPI_ISL_1649615, EPI_ISL_1649616, EPI_ISL_1649617, EPI_ISL_1649618, EPI_ISL_1649619, EPI_ISL_1649620, EPI_ISL_1649621, EPI_ISL_1649622, EPI_ISL_1649623, EPI_ISL_1649625, EPI_ISL_1649626, EPI_ISL_1649627, EPI_ISL_1649628, EPI_ISL_1649629, EPI_ISL_1649630, EPI_ISL_1649631, EPI_ISL_1649632, EPI_ISL_1649633, EPI_ISL_1649634, EPI_ISL_1649635, EPI_ISL_1649636, EPI_ISL_1649638, EPI_ISL_1649639, EPI_ISL_1649640, EPI_ISL_1649641, EPI_ISL_1649642, EPI_ISL_1649643, EPI_ISL_1649644, EPI_ISL_1649645, EPI_ISL_1649646, EPI_ISL_1649647, EPI_ISL_1649648, EPI_ISL_1649649, EPI_ISL_1649650, EPI_ISL_1649651, EPI_ISL_1649652, EPI_ISL_1649653, EPI_ISL_1649654, EPI_ISL_1649655, EPI_ISL_1649656, EPI_ISL_1649657, EPI_ISL_1649658, EPI_ISL_1649659, EPI_ISL_1649660, EPI_ISL_1649661, EPI_ISL_1649663, EPI_ISL_1649664, EPI_ISL_1649665, EPI_ISL_1649666, EPI_ISL_1649667, EPI_ISL_1649668, EPI_ISL_1649669, EPI_ISL_1649670, EPI_ISL_1649671, EPI_ISL_1649672, EPI_ISL_1649673, EPI_ISL_1649674, EPI_ISL_1649675, EPI_ISL_1649676, EPI_ISL_1649677, EPI_ISL_1649678, EPI_ISL_1649679, EPI_ISL_1649680, EPI_ISL_1649681, EPI_ISL_1649682, EPI_ISL_1649683, EPI_ISL_1649684, EPI_ISL_1649685, EPI_ISL_1649686, EPI_ISL_1649687, EPI_ISL_1649688, EPI_ISL_1649689, EPI_ISL_1649690, EPI_ISL_1649691, EPI_ISL_1649692, EPI_ISL_1649693, EPI_ISL_1649694, EPI_ISL_1649695, EPI_ISL_1649696, EPI_ISL_1649697, EPI_ISL_1649698, EPI_ISL_1649699, EPI_ISL_1649700, EPI_ISL_1649701, EPI_ISL_1649702, EPI_ISL_1649703, EPI_ISL_1649704, EPI_ISL_1649705, EPI_ISL_1649706, EPI_ISL_1649707, EPI_ISL_1649708, EPI_ISL_1649709, EPI_ISL_1649710, EPI_ISL_1649711, EPI_ISL_1649712, EPI_ISL_1649713, EPI_ISL_1649714, EPI_ISL_1649715, EPI_ISL_1649716, EPI_ISL_1649717, EPI_ISL_1649718, EPI_ISL_1649719, EPI_ISL_1649720, EPI_ISL_1649721, EPI_ISL_1649722, EPI_ISL_1649724, EPI_ISL_1649725, EPI_ISL_1649726, EPI_ISL_1649727, EPI_ISL_1649728, EPI_ISL_1649729, EPI_ISL_1649730, EPI_ISL_1649731, EPI_ISL_1649732, EPI_ISL_1649735, EPI_ISL_1649736, EPI_ISL_1649737, EPI_ISL_1649738, EPI_ISL_1649739, EPI_ISL_1649740, EPI_ISL_1649741, EPI_ISL_1649742, EPI_ISL_1649744, EPI_ISL_1649745, EPI_ISL_1649746, EPI_ISL_1649747, EPI_ISL_1649748, EPI_ISL_1649749, EPI_ISL_1649750, EPI_ISL_1649752, EPI_ISL_1649753, EPI_ISL_1649754, EPI_ISL_1649755, EPI_ISL_1649756, EPI_ISL_1649757, EPI_ISL_1649758, EPI_ISL_1649759, EPI_ISL_1649760, EPI_ISL_1649761, EPI_ISL_1649762, EPI_ISL_1649763, EPI_ISL_1649764, EPI_ISL_1649765, EPI_ISL_1649766, EPI_ISL_1649767, EPI_ISL_1649768, EPI_ISL_1649769, EPI_ISL_1649770, EPI_ISL_1649771, EPI_ISL_1649772, EPI_ISL_1649773, EPI_ISL_1649774, EPI_ISL_1649775, EPI_ISL_1649776, EPI_ISL_1649777, EPI_ISL_1649778, EPI_ISL_1649781, EPI_ISL_1649784, EPI_ISL_1649787, EPI_ISL_1649790, EPI_ISL_1649792, EPI_ISL_1649799, EPI_ISL_1649801, EPI_ISL_1649804, EPI_ISL_1649807, EPI_ISL_1649810, EPI_ISL_1649813, EPI_ISL_1649815, EPI_ISL_1649816, EPI_ISL_1649818, EPI_ISL_1649819, EPI_ISL_1649821, EPI_ISL_1649822, EPI_ISL_1649823, EPI_ISL_1649824, EPI_ISL_1649825, EPI_ISL_1649826, EPI_ISL_1649829, EPI_ISL_1649831, EPI_ISL_1649834, EPI_ISL_1649835, EPI_ISL_1649836, EPI_ISL_1649837, EPI_ISL_1649839, EPI_ISL_1649840, EPI_ISL_1649845, EPI_ISL_1649846, EPI_ISL_1649847, EPI_ISL_1649848, EPI_ISL_1649849, EPI_ISL_1649853, EPI_ISL_1649854, EPI_ISL_1649855, EPI_ISL_1649856, EPI_ISL_1649858, EPI_ISL_1649860, EPI_ISL_1649861, EPI_ISL_1649862, EPI_ISL_1649863, EPI_ISL_1649864, EPI_ISL_1649865, EPI_ISL_1649867, EPI_ISL_1649868, EPI_ISL_1649869, EPI_ISL_1649872, EPI_ISL_1649874, EPI_ISL_1649875, EPI_ISL_1649879, EPI_ISL_1649880, EPI_ISL_1649881, EPI_ISL_1649882, EPI_ISL_1649883, EPI_ISL_1649884, EPI_ISL_1649885, EPI_ISL_1649886, EPI_ISL_1649887, EPI_ISL_1649888, EPI_ISL_1649889, EPI_ISL_1649890, EPI_ISL_1649891, EPI_ISL_1649892, EPI_ISL_1649893, EPI_ISL_1649894, EPI_ISL_1649895, EPI_ISL_1649896, EPI_ISL_1649897, EPI_ISL_1649898, EPI_ISL_1649899, EPI_ISL_1649900, EPI_ISL_1649901, EPI_ISL_1649902, EPI_ISL_1649903, EPI_ISL_1649904, EPI_ISL_1649905, EPI_ISL_1649906, EPI_ISL_1649907, EPI_ISL_1649908, EPI_ISL_1649909, EPI_ISL_1649910, EPI_ISL_1649911, EPI_ISL_1649912, EPI_ISL_1649913, EPI_ISL_1649914, EPI_ISL_1649915, EPI_ISL_1649916, EPI_ISL_1649917, EPI_ISL_1649918, EPI_ISL_1649919, EPI_ISL_1649920, EPI_ISL_1649921, EPI_ISL_1649922, EPI_ISL_1649923, EPI_ISL_1649924, EPI_ISL_1649925, EPI_ISL_1649926, EPI_ISL_1649927, EPI_ISL_1649928, EPI_ISL_1649929, EPI_ISL_1649930, EPI_ISL_1649931, EPI_ISL_1649932, EPI_ISL_1649933, EPI_ISL_1649934, EPI_ISL_1649935, EPI_ISL_1649936, EPI_ISL_1649937, EPI_ISL_1649938, EPI_ISL_1649939, EPI_ISL_1649940, EPI_ISL_1649942, EPI_ISL_1649943, EPI_ISL_1649944, EPI_ISL_1649945, EPI_ISL_1649946, EPI_ISL_1649947, EPI_ISL_1649948, EPI_ISL_1649949, EPI_ISL_1649951, EPI_ISL_1649952, EPI_ISL_1649953, EPI_ISL_1649954, EPI_ISL_1649955, EPI_ISL_1649956, EPI_ISL_1649957, EPI_ISL_1649958, EPI_ISL_1649959, EPI_ISL_1649960, EPI_ISL_1649961, EPI_ISL_1649962, EPI_ISL_1649963, EPI_ISL_1649964, EPI_ISL_1649965, EPI_ISL_1649966, EPI_ISL_1649967, EPI_ISL_1649968, EPI_ISL_1649969, EPI_ISL_1649970, EPI_ISL_1649971, EPI_ISL_1649972, EPI_ISL_1649973, EPI_ISL_1649974, EPI_ISL_1649975, EPI_ISL_1649976, EPI_ISL_1649977, EPI_ISL_1649978, EPI_ISL_1649979, EPI_ISL_1649980, EPI_ISL_1649981, EPI_ISL_1649982, EPI_ISL_1649983, EPI_ISL_1649984, EPI_ISL_1649985, EPI_ISL_1649986, EPI_ISL_1649987, EPI_ISL_1649988, EPI_ISL_1649989, EPI_ISL_1649990, EPI_ISL_1649991, EPI_ISL_1649992, EPI_ISL_1649993, EPI_ISL_1649994, EPI_ISL_1649995, EPI_ISL_1649996, EPI_ISL_1649997, EPI_ISL_1649998, EPI_ISL_1649999, EPI_ISL_1650000, EPI_ISL_1650001, EPI_ISL_1650002, EPI_ISL_1650003, EPI_ISL_1650004, EPI_ISL_1650005, EPI_ISL_1650006, EPI_ISL_1650007, EPI_ISL_1650008, EPI_ISL_1650009, EPI_ISL_1650010, EPI_ISL_1650011, EPI_ISL_1650012, EPI_ISL_1650013, EPI_ISL_1650014, EPI_ISL_1650015, EPI_ISL_1650016, EPI_ISL_1650017, EPI_ISL_1650018, EPI_ISL_1650019, EPI_ISL_1650020, EPI_ISL_1650021, EPI_ISL_1650022, EPI_ISL_1650023, EPI_ISL_1650024, EPI_ISL_1650025, EPI_ISL_1650026, EPI_ISL_1650027, EPI_ISL_1650028, EPI_ISL_1650029, EPI_ISL_1650030, EPI_ISL_1650031, EPI_ISL_1650032, EPI_ISL_1650033, EPI_ISL_1650034, EPI_ISL_1650035, EPI_ISL_1650036, EPI_ISL_1650037, EPI_ISL_1650038, EPI_ISL_1650039, EPI_ISL_1650040, EPI_ISL_1650041, EPI_ISL_1650042, EPI_ISL_1650043, EPI_ISL_1650044, EPI_ISL_1650045, EPI_ISL_1650046, EPI_ISL_1650047, EPI_ISL_1650048, EPI_ISL_1650049, EPI_ISL_1650050, EPI_ISL_1650051, EPI_ISL_1650052, EPI_ISL_1650053, EPI_ISL_1650054, EPI_ISL_1650055, EPI_ISL_1650056, EPI_ISL_1650057, EPI_ISL_1650058, EPI_ISL_1650059, EPI_ISL_1650060, EPI_ISL_1650061, EPI_ISL_1650062, EPI_ISL_1650063, EPI_ISL_1650064, EPI_ISL_1650065, EPI_ISL_1650066, EPI_ISL_1650067, EPI_ISL_1650068, EPI_ISL_1650069, EPI_ISL_1650070, EPI_ISL_1650071, EPI_ISL_1650072, EPI_ISL_1650073, EPI_ISL_1650074, EPI_ISL_1650075, EPI_ISL_1650076, EPI_ISL_1650077, EPI_ISL_1650078, EPI_ISL_1650079, EPI_ISL_1650080, EPI_ISL_1650081, EPI_ISL_1650082, EPI_ISL_1650083, EPI_ISL_1650084, EPI_ISL_1650085, EPI_ISL_1650086, EPI_ISL_1650087, EPI_ISL_1650088, EPI_ISL_1650089, EPI_ISL_1650090, EPI_ISL_1650091, EPI_ISL_1650092, EPI_ISL_1650093, EPI_ISL_1650094, EPI_ISL_1650095, EPI_ISL_1650096, EPI_ISL_1650097, EPI_ISL_1650098, EPI_ISL_1650099, EPI_ISL_1650100, EPI_ISL_1650102, EPI_ISL_1650103, EPI_ISL_1650104, EPI_ISL_1650105, EPI_ISL_1650106, EPI_ISL_1650107, EPI_ISL_1650108, EPI_ISL_1650109, EPI_ISL_1650110, EPI_ISL_1650111, EPI_ISL_1650112, EPI_ISL_1650113, EPI_ISL_1650114, EPI_ISL_1650115, EPI_ISL_1650116, EPI_ISL_1650117, EPI_ISL_1650118, EPI_ISL_1650119, EPI_ISL_1650120, EPI_ISL_1650121, EPI_ISL_1650122, EPI_ISL_1650123, EPI_ISL_1650124, EPI_ISL_1650125, EPI_ISL_1650126, EPI_ISL_1650127, EPI_ISL_1650128, EPI_ISL_1650130, EPI_ISL_1650131, EPI_ISL_1650132, EPI_ISL_1650133, EPI_ISL_1650134, EPI_ISL_1650135, EPI_ISL_1650136, EPI_ISL_1650137, EPI_ISL_1650138, EPI_ISL_1650139, EPI_ISL_1650140, EPI_ISL_1650141, EPI_ISL_1650142, EPI_ISL_1650143, EPI_ISL_1650144, EPI_ISL_1650145, EPI_ISL_1650146, EPI_ISL_1650147, EPI_ISL_1650148, EPI_ISL_1650149, EPI_ISL_1650150, EPI_ISL_1650151, EPI_ISL_1650152, EPI_ISL_1650153, EPI_ISL_1650154, EPI_ISL_1650155, EPI_ISL_1650156, EPI_ISL_1650157, EPI_ISL_1650158, EPI_ISL_1650159, EPI_ISL_1650160, EPI_ISL_1650161, EPI_ISL_1650162, EPI_ISL_1650163, EPI_ISL_1650164, EPI_ISL_1650165, EPI_ISL_1650166, EPI_ISL_1650167, EPI_ISL_1650168, EPI_ISL_1650169, EPI_ISL_1650170, EPI_ISL_1650171, EPI_ISL_1650172, EPI_ISL_1650173, EPI_ISL_1650174, EPI_ISL_1650175, EPI_ISL_1650176, EPI_ISL_1650177, EPI_ISL_1650178, EPI_ISL_1650179, EPI_ISL_1650180, EPI_ISL_1650181, EPI_ISL_1650182, EPI_ISL_1650183, EPI_ISL_1650184, EPI_ISL_1650185, EPI_ISL_1650186, EPI_ISL_1650187, EPI_ISL_1650188, EPI_ISL_1650189, EPI_ISL_1650190, EPI_ISL_1650191, EPI_ISL_1650192, EPI_ISL_1650193, EPI_ISL_1650194, EPI_ISL_1650195, EPI_ISL_1650196, EPI_ISL_1650197, EPI_ISL_1650198, EPI_ISL_1650199, EPI_ISL_1650200, EPI_ISL_1650201, EPI_ISL_1650202, EPI_ISL_1650203, EPI_ISL_1650204, EPI_ISL_1650205, EPI_ISL_1650206, EPI_ISL_1650207, EPI_ISL_1650208, EPI_ISL_1650209, EPI_ISL_1650210, EPI_ISL_1650211, EPI_ISL_1650212, EPI_ISL_1650213, EPI_ISL_1650214, EPI_ISL_1650215, EPI_ISL_1650216, EPI_ISL_1650217, EPI_ISL_1650218, EPI_ISL_1650219, EPI_ISL_1650220, EPI_ISL_1650221, EPI_ISL_1650222, EPI_ISL_1650223, EPI_ISL_1650224, EPI_ISL_1650225, EPI_ISL_1650226, EPI_ISL_1650227, EPI_ISL_1650228, EPI_ISL_1650229, EPI_ISL_1650230, EPI_ISL_1650231, EPI_ISL_1650232, EPI_ISL_1650233, EPI_ISL_1650234, EPI_ISL_1650235, EPI_ISL_1650236, EPI_ISL_1650237, EPI_ISL_1650238, EPI_ISL_1650239, EPI_ISL_1650240, EPI_ISL_1650241, EPI_ISL_1650242, EPI_ISL_1650243, EPI_ISL_1650244, EPI_ISL_1650245, EPI_ISL_1650246, EPI_ISL_1650247, EPI_ISL_1650248, EPI_ISL_1650249, EPI_ISL_1650250, EPI_ISL_1650251, EPI_ISL_1650252, EPI_ISL_1650253, EPI_ISL_1650254, EPI_ISL_1650255, EPI_ISL_1650256, EPI_ISL_1650257, EPI_ISL_1650258, EPI_ISL_1650259, EPI_ISL_1650260, EPI_ISL_1650261, EPI_ISL_1650262, EPI_ISL_1650263, EPI_ISL_1650264, EPI_ISL_1650265, EPI_ISL_1650266, EPI_ISL_1650267, EPI_ISL_1650268, EPI_ISL_1650269, EPI_ISL_1650270, EPI_ISL_1650271, EPI_ISL_1650272, EPI_ISL_1650273, EPI_ISL_1650274, EPI_ISL_1650275, EPI_ISL_1650276, EPI_ISL_1650277, EPI_ISL_1650278, EPI_ISL_1650279, EPI_ISL_1650280, EPI_ISL_1650281, EPI_ISL_1650282, EPI_ISL_1650283, EPI_ISL_1650284, EPI_ISL_1650285, EPI_ISL_1650286, EPI_ISL_1650287, EPI_ISL_1650288, EPI_ISL_1650289, EPI_ISL_1650290, EPI_ISL_1650291, EPI_ISL_1650292, EPI_ISL_1650293, EPI_ISL_1650294, EPI_ISL_1650295, EPI_ISL_1650296, EPI_ISL_1650297, EPI_ISL_1650298, EPI_ISL_1650299, EPI_ISL_1650300, EPI_ISL_1650301, EPI_ISL_1650302, EPI_ISL_1650303, EPI_ISL_1650304, EPI_ISL_1650305, EPI_ISL_1650306, EPI_ISL_1650307, EPI_ISL_1650308, EPI_ISL_1650309, EPI_ISL_1650310, EPI_ISL_1650311, EPI_ISL_1650312, EPI_ISL_1650313, EPI_ISL_1650314, EPI_ISL_1650315, EPI_ISL_1650316, EPI_ISL_1650317, EPI_ISL_1650318, EPI_ISL_1650319, EPI_ISL_1650320, EPI_ISL_1650321, EPI_ISL_1650322, EPI_ISL_1650323, EPI_ISL_1650324, EPI_ISL_1650325, EPI_ISL_1650326, EPI_ISL_1650327, EPI_ISL_1650328, EPI_ISL_1650329, EPI_ISL_1650330, EPI_ISL_1650331, EPI_ISL_1650332, EPI_ISL_1650333, EPI_ISL_1650334, EPI_ISL_1650335, EPI_ISL_1650336, EPI_ISL_1650338, EPI_ISL_1650339, EPI_ISL_1650340, EPI_ISL_1650341, EPI_ISL_1650342, EPI_ISL_1650343, EPI_ISL_1650344, EPI_ISL_1650345, EPI_ISL_1650346, EPI_ISL_1650347, EPI_ISL_1650348, EPI_ISL_1650349, EPI_ISL_1650350, EPI_ISL_1650351, EPI_ISL_1650352, EPI_ISL_1650353, EPI_ISL_1650354, EPI_ISL_1650355, EPI_ISL_1650356, EPI_ISL_1650357, EPI_ISL_1650358, EPI_ISL_1650359, EPI_ISL_1650360, EPI_ISL_1650361, EPI_ISL_1650362, EPI_ISL_1650363, EPI_ISL_1650364, EPI_ISL_1650365, EPI_ISL_1650366, EPI_ISL_1650367, EPI_ISL_1650368, EPI_ISL_1650369, EPI_ISL_1650370, EPI_ISL_1650371, EPI_ISL_1650372, EPI_ISL_1650373, EPI_ISL_1650374, EPI_ISL_1650375, EPI_ISL_1650376, EPI_ISL_1650377, EPI_ISL_1650378, EPI_ISL_1650380, EPI_ISL_1650381, EPI_ISL_1650382, EPI_ISL_1650383, EPI_ISL_1650384, EPI_ISL_1650385, EPI_ISL_1650386, EPI_ISL_1650387, EPI_ISL_1650388, EPI_ISL_1650389, EPI_ISL_1650390, EPI_ISL_1650391, EPI_ISL_1650392, EPI_ISL_1650393, EPI_ISL_1650394, EPI_ISL_1650395, EPI_ISL_1650396, EPI_ISL_1650397, EPI_ISL_1650398, EPI_ISL_1650399, EPI_ISL_1650400, EPI_ISL_1650401, EPI_ISL_1650402, EPI_ISL_1650403, EPI_ISL_1650404, EPI_ISL_1650405, EPI_ISL_1650406, EPI_ISL_1650407, EPI_ISL_1650408, EPI_ISL_1650409, EPI_ISL_1650410, EPI_ISL_1650411, EPI_ISL_1650412, EPI_ISL_1650413, EPI_ISL_1650414, EPI_ISL_1650415, EPI_ISL_1650416, EPI_ISL_1650417, EPI_ISL_1650418, EPI_ISL_1650419, EPI_ISL_1650420, EPI_ISL_1650421, EPI_ISL_1650422, EPI_ISL_1650423, EPI_ISL_1650424, EPI_ISL_1650425, EPI_ISL_1650428, EPI_ISL_1650429, EPI_ISL_1650430, EPI_ISL_1650431, EPI_ISL_1650432, EPI_ISL_1650433, EPI_ISL_1650434, EPI_ISL_1650435, EPI_ISL_1650436, EPI_ISL_1650437, EPI_ISL_1650438, EPI_ISL_1650439, EPI_ISL_1650440, EPI_ISL_1650441, EPI_ISL_1650443, EPI_ISL_1650444, EPI_ISL_1650445, EPI_ISL_1650446, EPI_ISL_1650447, EPI_ISL_1650448, EPI_ISL_1650449, EPI_ISL_1650450, EPI_ISL_1650451, EPI_ISL_1650452, EPI_ISL_1650453, EPI_ISL_1650454, EPI_ISL_1650455, EPI_ISL_1650456, EPI_ISL_1650457, EPI_ISL_1650458, EPI_ISL_1650459, EPI_ISL_1650460, EPI_ISL_1650461, EPI_ISL_1650462, EPI_ISL_1650463, EPI_ISL_1650464, EPI_ISL_1650465, EPI_ISL_1650466, EPI_ISL_1650467, EPI_ISL_1650468, EPI_ISL_1650469, EPI_ISL_1650470, EPI_ISL_1650471, EPI_ISL_1650472, EPI_ISL_1650473, EPI_ISL_1650474, EPI_ISL_1650475, EPI_ISL_1650476, EPI_ISL_1650477, EPI_ISL_1650478, EPI_ISL_1650479, EPI_ISL_1650480, EPI_ISL_1650481, EPI_ISL_1650482, EPI_ISL_1650483, EPI_ISL_1650484, EPI_ISL_1650485, EPI_ISL_1650486, EPI_ISL_1650487, EPI_ISL_1650488, EPI_ISL_1650489, EPI_ISL_1650490, EPI_ISL_1650491, EPI_ISL_1650492, EPI_ISL_1650494, EPI_ISL_1650495, EPI_ISL_1650496, EPI_ISL_1650497, EPI_ISL_1650498, EPI_ISL_1650499, EPI_ISL_1650500, EPI_ISL_1650501, EPI_ISL_1650502, EPI_ISL_1650503, EPI_ISL_1650504, EPI_ISL_1650505, EPI_ISL_1650506, EPI_ISL_1650507, EPI_ISL_1650508, EPI_ISL_1650509, EPI_ISL_1650510, EPI_ISL_1650511, EPI_ISL_1650512, EPI_ISL_1650513, EPI_ISL_1650514, EPI_ISL_1650515, EPI_ISL_1650516, EPI_ISL_1650517, EPI_ISL_1650518, EPI_ISL_1650519, EPI_ISL_1650520, EPI_ISL_1650521, EPI_ISL_1650522, EPI_ISL_1650523, EPI_ISL_1650524, EPI_ISL_1650525, EPI_ISL_1650526, EPI_ISL_1650527, EPI_ISL_1650528, EPI_ISL_1650529, EPI_ISL_1650530, EPI_ISL_1650531, EPI_ISL_1650532, EPI_ISL_1650533, EPI_ISL_1650534, EPI_ISL_1650535, EPI_ISL_1650536, EPI_ISL_1650538, EPI_ISL_1650539, EPI_ISL_1650541, EPI_ISL_1650542, EPI_ISL_1650543, EPI_ISL_1650544, EPI_ISL_1650545, EPI_ISL_1650546, EPI_ISL_1650547, EPI_ISL_1650548, EPI_ISL_1650549, EPI_ISL_1650550, EPI_ISL_1650551, EPI_ISL_1650552, EPI_ISL_1650553, EPI_ISL_1650554, EPI_ISL_1650555, EPI_ISL_1650556, EPI_ISL_1650557, EPI_ISL_1650558, EPI_ISL_1650559, EPI_ISL_1650560, EPI_ISL_1650561, EPI_ISL_1650562, EPI_ISL_1650563, EPI_ISL_1650564, EPI_ISL_1650565, EPI_ISL_1650566, EPI_ISL_1650567, EPI_ISL_1650568, EPI_ISL_1650569, EPI_ISL_1650570, EPI_ISL_1650571, EPI_ISL_1650572, EPI_ISL_1650573, EPI_ISL_1650574, EPI_ISL_1650575, EPI_ISL_1650576, EPI_ISL_1650577, EPI_ISL_1650578, EPI_ISL_1650579, EPI_ISL_1650580, EPI_ISL_1650581, EPI_ISL_1650582, EPI_ISL_1650583, EPI_ISL_1650584, EPI_ISL_1650585, EPI_ISL_1650586, EPI_ISL_1650587, EPI_ISL_1650588, EPI_ISL_1650589, EPI_ISL_1650590, EPI_ISL_1650591, EPI_ISL_1650592, EPI_ISL_1650593, EPI_ISL_1650594, EPI_ISL_1650595, EPI_ISL_1650596, EPI_ISL_1650597, EPI_ISL_1650598, EPI_ISL_1650599, EPI_ISL_1650600, EPI_ISL_1650601, EPI_ISL_1650602, EPI_ISL_1650603, EPI_ISL_1650604, EPI_ISL_1650605, EPI_ISL_1650606, EPI_ISL_1650607, EPI_ISL_1650608, EPI_ISL_1650609, EPI_ISL_1650610, EPI_ISL_1650611, EPI_ISL_1650612, EPI_ISL_1650613, EPI_ISL_1650614, EPI_ISL_1650615, EPI_ISL_1650616, EPI_ISL_1650617, EPI_ISL_1650618, EPI_ISL_1650619, EPI_ISL_1650620, EPI_ISL_1650621, EPI_ISL_1650622, EPI_ISL_1650623, EPI_ISL_1650624, EPI_ISL_1650625, EPI_ISL_1650626, EPI_ISL_1650627, EPI_ISL_1650628, EPI_ISL_1650629, EPI_ISL_1650630, EPI_ISL_1650631, EPI_ISL_1650632, EPI_ISL_1650633, EPI_ISL_1650634, EPI_ISL_1650635, EPI_ISL_1650636, EPI_ISL_1650637, EPI_ISL_1650638, EPI_ISL_1650639, EPI_ISL_1650640, EPI_ISL_1650641, EPI_ISL_1650642, EPI_ISL_1650643, EPI_ISL_1650644, EPI_ISL_1650645, EPI_ISL_1650646, EPI_ISL_1650647, EPI_ISL_1650648, EPI_ISL_1650649, EPI_ISL_1650650, EPI_ISL_1650651, EPI_ISL_1650652, EPI_ISL_1650653, EPI_ISL_1650654, EPI_ISL_1650655, EPI_ISL_1650656, EPI_ISL_1650657, EPI_ISL_1650658, EPI_ISL_1650659, EPI_ISL_1650660, EPI_ISL_1650661, EPI_ISL_1650662, EPI_ISL_1650663, EPI_ISL_1650664, EPI_ISL_1650665, EPI_ISL_1650666, EPI_ISL_1650667, EPI_ISL_1650668, EPI_ISL_1650669, EPI_ISL_1650670, EPI_ISL_1650671, EPI_ISL_1650672, EPI_ISL_1650673, EPI_ISL_1650674, EPI_ISL_1650675, EPI_ISL_1650676, EPI_ISL_1650677, EPI_ISL_1650678, EPI_ISL_1650679, EPI_ISL_1650680, EPI_ISL_1650681, EPI_ISL_1650682, EPI_ISL_1650683, EPI_ISL_1650684, EPI_ISL_1650685, EPI_ISL_1650686, EPI_ISL_1650687, EPI_ISL_1650688, EPI_ISL_1650689, EPI_ISL_1650690, EPI_ISL_1650691, EPI_ISL_1650692, EPI_ISL_1650693, EPI_ISL_1650694, EPI_ISL_1650695, EPI_ISL_1650696, EPI_ISL_1650697, EPI_ISL_1650698, EPI_ISL_1650699, EPI_ISL_1650700, EPI_ISL_1650701, EPI_ISL_1650702, EPI_ISL_1650703, EPI_ISL_1650704, EPI_ISL_1650705, EPI_ISL_1650706, EPI_ISL_1650707, EPI_ISL_1650708, EPI_ISL_1650709, EPI_ISL_1650710, EPI_ISL_1650711, EPI_ISL_1650712, EPI_ISL_1650713, EPI_ISL_1650714, EPI_ISL_1650715, EPI_ISL_1650716, EPI_ISL_1650717, EPI_ISL_1650718, EPI_ISL_1650719, EPI_ISL_1650720, EPI_ISL_1650721, EPI_ISL_1650722, EPI_ISL_1650723, EPI_ISL_1650724, EPI_ISL_1650725, EPI_ISL_1650726, EPI_ISL_1650727, EPI_ISL_1650728, EPI_ISL_1650729, EPI_ISL_1650730, EPI_ISL_1650731, EPI_ISL_1650732, EPI_ISL_1650733, EPI_ISL_1650734, EPI_ISL_1650735, EPI_ISL_1650736, EPI_ISL_1650737, EPI_ISL_1650738, EPI_ISL_1650739, EPI_ISL_1650740, EPI_ISL_1650741, EPI_ISL_1650742, EPI_ISL_1650743, EPI_ISL_1650744, EPI_ISL_1650745, EPI_ISL_1650746, EPI_ISL_1650747, EPI_ISL_1650748, EPI_ISL_1650749, EPI_ISL_1650750, EPI_ISL_1650751, EPI_ISL_1650752, EPI_ISL_1650753, EPI_ISL_1650754, EPI_ISL_1650755, EPI_ISL_1650756, EPI_ISL_1650757, EPI_ISL_1650759, EPI_ISL_1650760, EPI_ISL_1650761, EPI_ISL_1650762, EPI_ISL_1650763, EPI_ISL_1650764, EPI_ISL_1650765, EPI_ISL_1650766, EPI_ISL_1650767, EPI_ISL_1650768, EPI_ISL_1650769, EPI_ISL_1650770, EPI_ISL_1650771, EPI_ISL_1650772, EPI_ISL_1650773, EPI_ISL_1650774, EPI_ISL_1650775, EPI_ISL_1650776, EPI_ISL_1650777, EPI_ISL_1650778, EPI_ISL_1650779, EPI_ISL_1650780, EPI_ISL_1650781, EPI_ISL_1650782, EPI_ISL_1650783, EPI_ISL_1650784, EPI_ISL_1650785, EPI_ISL_1650786, EPI_ISL_1650787, EPI_ISL_1650788, EPI_ISL_1650789, EPI_ISL_1650790, EPI_ISL_1650791, EPI_ISL_1650792, EPI_ISL_1650793, EPI_ISL_1650794, EPI_ISL_1650795, EPI_ISL_1650796, EPI_ISL_1650797, EPI_ISL_1650798, EPI_ISL_1650799, EPI_ISL_1650800, EPI_ISL_1650801, EPI_ISL_1650802, EPI_ISL_1650803, EPI_ISL_1650804, EPI_ISL_1650805, EPI_ISL_1650806, EPI_ISL_1650807, EPI_ISL_1650808, EPI_ISL_1650809, EPI_ISL_1650810, EPI_ISL_1650811, EPI_ISL_1650812, EPI_ISL_1650813, EPI_ISL_1650814, EPI_ISL_1650815, EPI_ISL_1650816, EPI_ISL_1650817, EPI_ISL_1650818, EPI_ISL_1650819, EPI_ISL_1650820, EPI_ISL_1650821, EPI_ISL_1650822, EPI_ISL_1650823, EPI_ISL_1650824, EPI_ISL_1650825, EPI_ISL_1650826, EPI_ISL_1650827, EPI_ISL_1650828, EPI_ISL_1650829, EPI_ISL_1650830, EPI_ISL_1650831, EPI_ISL_1650832, EPI_ISL_1650833, EPI_ISL_1650834, EPI_ISL_1650835, EPI_ISL_1650836, EPI_ISL_1650837, EPI_ISL_1650838, EPI_ISL_1650839, EPI_ISL_1650840, EPI_ISL_1650841, EPI_ISL_1650842, EPI_ISL_1650843, EPI_ISL_1650844, EPI_ISL_1650845, EPI_ISL_1650846, EPI_ISL_1650847, EPI_ISL_1650848, EPI_ISL_1650849, EPI_ISL_1650850, EPI_ISL_1650851, EPI_ISL_1650852, EPI_ISL_1650853, EPI_ISL_1650854, EPI_ISL_1650855, EPI_ISL_1650856, EPI_ISL_1650857, EPI_ISL_1650858, EPI_ISL_1650859, EPI_ISL_1650860, EPI_ISL_1650861, EPI_ISL_1650862, EPI_ISL_1650863, EPI_ISL_1650864, EPI_ISL_1650865, EPI_ISL_1650866, EPI_ISL_1650867, EPI_ISL_1650868, EPI_ISL_1650869, EPI_ISL_1650870, EPI_ISL_1650871, EPI_ISL_1650872, EPI_ISL_1650873, EPI_ISL_1650874, EPI_ISL_1650875, EPI_ISL_1650876, EPI_ISL_1650877, EPI_ISL_1650878, EPI_ISL_1650879, EPI_ISL_1650880, EPI_ISL_1650881, EPI_ISL_1650882, EPI_ISL_1650883, EPI_ISL_1650884, EPI_ISL_1650885, EPI_ISL_1650886, EPI_ISL_1650887, EPI_ISL_1650888, EPI_ISL_1650889, EPI_ISL_1650890, EPI_ISL_1650891, EPI_ISL_1650892, EPI_ISL_1650893, EPI_ISL_1650894, EPI_ISL_1650895, EPI_ISL_1650896, EPI_ISL_1650897, EPI_ISL_1650898, EPI_ISL_1650899, EPI_ISL_1650900, EPI_ISL_1650901, EPI_ISL_1650902, EPI_ISL_1650903, EPI_ISL_1650904, EPI_ISL_1650905, EPI_ISL_1650906, EPI_ISL_1650907, EPI_ISL_1650908, EPI_ISL_1650909, EPI_ISL_1650910, EPI_ISL_1650911, EPI_ISL_1650912, EPI_ISL_1650913, EPI_ISL_1650914, EPI_ISL_1650915, EPI_ISL_1650917, EPI_ISL_1650918, EPI_ISL_1650919, EPI_ISL_1650920, EPI_ISL_1650921, EPI_ISL_1650922, EPI_ISL_1650923, EPI_ISL_1650924, EPI_ISL_1650925, EPI_ISL_1650926, EPI_ISL_1650927, EPI_ISL_1650928, EPI_ISL_1650929, EPI_ISL_1650930, EPI_ISL_1650932, EPI_ISL_1650933, EPI_ISL_1650934, EPI_ISL_1650935, EPI_ISL_1650936, EPI_ISL_1650937, EPI_ISL_1650938, EPI_ISL_1650939, EPI_ISL_1650940, EPI_ISL_1650941, EPI_ISL_1650942, EPI_ISL_1650943, EPI_ISL_1650944, EPI_ISL_1650945, EPI_ISL_1650946, EPI_ISL_1650947, EPI_ISL_1650948, EPI_ISL_1650949, EPI_ISL_1650950, EPI_ISL_1650951, EPI_ISL_1650952, EPI_ISL_1650953, EPI_ISL_1650954, EPI_ISL_1650955, EPI_ISL_1650956, EPI_ISL_1650957, EPI_ISL_1650958, EPI_ISL_1650959, EPI_ISL_1650960, EPI_ISL_1650961, EPI_ISL_1650962, EPI_ISL_1650963, EPI_ISL_1650964, EPI_ISL_1650965, EPI_ISL_1650966, EPI_ISL_1650967, EPI_ISL_1650968, EPI_ISL_1650969, EPI_ISL_1650970, EPI_ISL_1650971, EPI_ISL_1650972, EPI_ISL_1650973, EPI_ISL_1650974, EPI_ISL_1650975, EPI_ISL_1650976, EPI_ISL_1650977, EPI_ISL_1650978, EPI_ISL_1650979, EPI_ISL_1650980, EPI_ISL_1650981, EPI_ISL_1650983, EPI_ISL_1650984, EPI_ISL_1650985, EPI_ISL_1650986, EPI_ISL_1650987, EPI_ISL_1650988, EPI_ISL_1650989, EPI_ISL_1650990, EPI_ISL_1650991, EPI_ISL_1650992, EPI_ISL_1650993, EPI_ISL_1650994, EPI_ISL_1650995, EPI_ISL_1650996, EPI_ISL_1650997, EPI_ISL_1650998, EPI_ISL_1651000, EPI_ISL_1651001, EPI_ISL_1651002, EPI_ISL_1651003, EPI_ISL_1651004, EPI_ISL_1651005, EPI_ISL_1651006, EPI_ISL_1651007, EPI_ISL_1651008, EPI_ISL_1651009, EPI_ISL_1651010, EPI_ISL_1651011, EPI_ISL_1651012, EPI_ISL_1651013, EPI_ISL_1651014, EPI_ISL_1651015, EPI_ISL_1651016, EPI_ISL_1651017, EPI_ISL_1651018, EPI_ISL_1651019, EPI_ISL_1651020, EPI_ISL_1651021, EPI_ISL_1651022, EPI_ISL_1651023, EPI_ISL_1651024, EPI_ISL_1651025, EPI_ISL_1651026, EPI_ISL_1651027, EPI_ISL_1651028, EPI_ISL_1651029, EPI_ISL_1651030, EPI_ISL_1651031, EPI_ISL_1651032, EPI_ISL_1651033, EPI_ISL_1651034, EPI_ISL_1651035, EPI_ISL_1651036, EPI_ISL_1651037, EPI_ISL_1651038, EPI_ISL_1651039, EPI_ISL_1651040, EPI_ISL_1651041, EPI_ISL_1651042, EPI_ISL_1651043, EPI_ISL_1651044, EPI_ISL_1651045, EPI_ISL_1651046, EPI_ISL_1651047, EPI_ISL_1651048, EPI_ISL_1651049, EPI_ISL_1651050, EPI_ISL_1651051, EPI_ISL_1651052, EPI_ISL_1651053, EPI_ISL_1651054, EPI_ISL_1651055, EPI_ISL_1651056, EPI_ISL_1651057, EPI_ISL_1651058, EPI_ISL_1651059, EPI_ISL_1651060, EPI_ISL_1651061, EPI_ISL_1651062, EPI_ISL_1651063, EPI_ISL_1651064, EPI_ISL_1651065, EPI_ISL_1651066, EPI_ISL_1651067, EPI_ISL_1651068, EPI_ISL_1651069, EPI_ISL_1651070, EPI_ISL_1651071, EPI_ISL_1651072, EPI_ISL_1651073, EPI_ISL_1651074, EPI_ISL_1651075, EPI_ISL_1651076, EPI_ISL_1651077, EPI_ISL_1651078, EPI_ISL_1651079, EPI_ISL_1651080, EPI_ISL_1651081, EPI_ISL_1651082, EPI_ISL_1651083, EPI_ISL_1651084, EPI_ISL_1651085, EPI_ISL_1651086, EPI_ISL_1651087, EPI_ISL_1651088, EPI_ISL_1651089, EPI_ISL_1651090, EPI_ISL_1651091, EPI_ISL_1651092, EPI_ISL_1651093, EPI_ISL_1651094, EPI_ISL_1651095, EPI_ISL_1651096, EPI_ISL_1651097, EPI_ISL_1651098, EPI_ISL_1651099, EPI_ISL_1651100, EPI_ISL_1651101, EPI_ISL_1651102, EPI_ISL_1651103, EPI_ISL_1651104, EPI_ISL_1651105, EPI_ISL_1651106, EPI_ISL_1651107, EPI_ISL_1651108, EPI_ISL_1651109, EPI_ISL_1651110, EPI_ISL_1651111, EPI_ISL_1651112, EPI_ISL_1651113, EPI_ISL_1651114, EPI_ISL_1651115, EPI_ISL_1651116, EPI_ISL_1651117, EPI_ISL_1651118, EPI_ISL_1651119, EPI_ISL_1651120, EPI_ISL_1651121, EPI_ISL_1651122, EPI_ISL_1651123, EPI_ISL_1651124, EPI_ISL_1651125, EPI_ISL_1651126, EPI_ISL_1651127, EPI_ISL_1651128, EPI_ISL_1651129, EPI_ISL_1651130, EPI_ISL_1651131, EPI_ISL_1651132, EPI_ISL_1651133, EPI_ISL_1651134, EPI_ISL_1651135, EPI_ISL_1651136, EPI_ISL_1651137, EPI_ISL_1651138, EPI_ISL_1651139, EPI_ISL_1651140, EPI_ISL_1651141, EPI_ISL_1651142, EPI_ISL_1651143, EPI_ISL_1651144, EPI_ISL_1651145, EPI_ISL_1651146, EPI_ISL_1651147, EPI_ISL_1651148, EPI_ISL_1651149, EPI_ISL_1651150, EPI_ISL_1651151, EPI_ISL_1651152, EPI_ISL_1651153, EPI_ISL_1651154, EPI_ISL_1651155, EPI_ISL_1651156, EPI_ISL_1651157, EPI_ISL_1651158, EPI_ISL_1651159, EPI_ISL_1651160, EPI_ISL_1651161, EPI_ISL_1651162, EPI_ISL_1651163, EPI_ISL_1651164, EPI_ISL_1651165, EPI_ISL_1651166, EPI_ISL_1651167, EPI_ISL_1651168, EPI_ISL_1651169, EPI_ISL_1651170, EPI_ISL_1651171, EPI_ISL_1651173, EPI_ISL_1651174, EPI_ISL_1651175, EPI_ISL_1651176, EPI_ISL_1651177, EPI_ISL_1651178, EPI_ISL_1651179, EPI_ISL_1651180, EPI_ISL_1651181, EPI_ISL_1651182, EPI_ISL_1651183, EPI_ISL_1651184, EPI_ISL_1651185, EPI_ISL_1651186, EPI_ISL_1651187

| | | | |
|---|---|---|---|
| see above | Aegis Sciences Corporation | Centers for Disease Control and Prevention Division of Viral Diseases, Pathogen Discovery | Dakota Howard, Dhwani Batra, Peter W. Cook, Kara Moser, Adrian Paskey, Jason Caravas, Benjamin Rambo-Martin, Shatavia Morrison, Christopher Gulvick, Scott Sammons, Yvette Unoarumhi, Darlene Wagner, Matthew Schmerer, Cyndi Clark, Patrick Campbell, Rob Case, Vikramsinha Ghorpade, Holly Houdeshell, Ola Kvalvaag, Dillon Nall, Ethan Sanders, Alec Vest, Shaun Westlund, Matthew Hardison, Clinton R. Paden, Duncan MacCannell |

EPI_ISL_1651194, EPI_ISL_1651196, EPI_ISL_1651197, EPI_ISL_1651198, EPI_ISL_1651201, EPI_ISL_1651204, EPI_ISL_1651205, EPI_ISL_1651206, EPI_ISL_1651207, EPI_ISL_1651208, EPI_ISL_1651209, EPI_ISL_1651210

| | | | |
|---|---|---|---|
| see above | Wyoming Public Health Laboratory | Wyoming Public Health Laboratory | Jim Mildenberger, Wanda Manley, Noah Hull, Taylor Fearing, Lynette Gumbleton, Channing Weber, Ashley Norberg, Chayse Rowley, Marley Goetz, Brian Dominguez, Elliot Thomasson, Cari Sloma, and Rob Christensen |
| EPI_ISL_1651216, EPI_ISL_1651217 | MEMORIAL SLOAN KETTERING CANCER CENTER | Wadsworth Center, New York State Department of Health | Kirsten St. George, Daryl M. Lamson, Alexis Russell, Matthew Shudt, Melissa A Leisner, Jonathan Plitnick, Catharine Prussing, Navjot Singh, John Kelly, Erasmus Schneider, Erica Lasek-Nesselquist |
| EPI_ISL_1651218 | MONTEFIORE MEDICAL CENTER LABORATORIES | Wadsworth Center, New York State Department of Health | Kirsten St. George, Daryl M. Lamson, Alexis Russell, Matthew Shudt, Melissa A Leisner, Jonathan Plitnick, Catharine Prussing, Navjot Singh, John Kelly, Erasmus Schneider, Erica Lasek-Nesselquist |
| EPI_ISL_1651219, EPI_ISL_1651220, | New York Presbyterian Hospital | Wadsworth Center, New York State Department of Health | Kirsten St. George, Daryl M. Lamson, Alexis Russell, Matthew Shudt, Melissa A Leisner, Jonathan Plitnick, Catharine Prussing, Navjot Singh, John Kelly, Erasmus Schneider, Erica Lasek-Nesselquist |

| | | | |
|---|---|---|---|
| EPI_ISL_1651221 | | | |
| EPI_ISL_1651222, EPI_ISL_1651223 | URMC LABS | Wadsworth Center, New York State Department of Health | Kirsten St. George, Daryl M. Lamson, Alexis Russell, Matthew Shudt, Melissa A Leisner, Jonathan Plitnick, Catharine Prussing, Navjot Singh, John Kelly, Erasmus Schneider, Erica Lasek-Nesselquist |
| EPI_ISL_1651224 | SARATOGA HOSPITAL LABORATORY | Wadsworth Center, New York State Department of Health | Kirsten St. George, Daryl M. Lamson, Alexis Russell, Matthew Shudt, Melissa A Leisner, Jonathan Plitnick, Catharine Prussing, Navjot Singh, John Kelly, Erasmus Schneider, Erica Lasek-Nesselquist |
| EPI_ISL_1651225 | HOSPITAL FOR SPECIAL SURGERY | Wadsworth Center, New York State Department of Health | Kirsten St. George, Daryl M. Lamson, Alexis Russell, Matthew Shudt, Melissa A Leisner, Jonathan Plitnick, Catharine Prussing, Navjot Singh, John Kelly, Erasmus Schneider, Erica Lasek-Nesselquist |
| EPI_ISL_1651226 | MEMORIAL SLOAN KETTERING CANCER CENTER | Wadsworth Center, New York State Department of Health | Kirsten St. George, Daryl M. Lamson, Alexis Russell, Matthew Shudt, Melissa A Leisner, Jonathan Plitnick, Catharine Prussing, Navjot Singh, John Kelly, Erasmus Schneider, Erica Lasek-Nesselquist |
| EPI_ISL_1651227, EPI_ISL_1651228 | URMC LABS | Wadsworth Center, New York State Department of Health | Kirsten St. George, Daryl M. Lamson, Alexis Russell, Matthew Shudt, Melissa A Leisner, Jonathan Plitnick, Catharine Prussing, Navjot Singh, John Kelly, Erasmus Schneider, Erica Lasek-Nesselquist |
| EPI_ISL_1651229, EPI_ISL_1651230 | LESP Chiapas | Instituto de Diagnostico y Referencia Epidemiologicos (INDRE) | Claudia Wong-Arambula, Abril Rodriguez-Maldonado, Vanessa Rivero-Arredondo, Ariadna Medina-Benitez, Joaquin Quiroz-Mercado, Sergio Rangel-Guerrero, Natividad Cruz-Ortiz, Tatiana Nunez-Garcia, Gisela Barrera-Badillo, Lucia Hernandez-Rivas, Irma Lopez-Martinez, Ernesto Ramirez-Gonzalez. |
| EPI_ISL_1651231, EPI_ISL_1651232, EPI_ISL_1651233, EPI_ISL_1651234 | LESP Puebla | Instituto de Diagnostico y Referencia Epidemiologicos (INDRE) | Claudia Wong-Arambula, Abril Rodriguez-Maldonado, Vanessa Rivero-Arredondo, Ariadna Medina-Benitez, Joaquin Quiroz-Mercado, Sergio Rangel-Guerrero, Natividad Cruz-Ortiz, Tatiana Nunez-Garcia, Gisela Barrera-Badillo, Lucia Hernandez-Rivas, Irma Lopez-Martinez, Ernesto Ramirez-Gonzalez. |
| EPI_ISL_1651235, EPI_ISL_1651236, EPI_ISL_1651237, EPI_ISL_1651238, EPI_ISL_1651239, EPI_ISL_1651240, EPI_ISL_1651241, EPI_ISL_1651242, EPI_ISL_1651243, EPI_ISL_1651244, EPI_ISL_1651245, EPI_ISL_1651246, EPI_ISL_1651247, EPI_ISL_1651248, EPI_ISL_1651249 | | | |
| see above | LESP Nuevo Leon | Instituto de Diagnostico y Referencia Epidemiologicos (INDRE) | Claudia Wong-Arambula, Abril Rodriguez-Maldonado, Vanessa Rivero-Arredondo, Ariadna Medina-Benitez, Joaquin Quiroz-Mercado, Sergio Rangel-Guerrero, Natividad Cruz-Ortiz, Tatiana Nunez-Garcia, Gisela Barrera-Badillo, Lucia Hernandez-Rivas, Irma Lopez-Martinez, Ernesto Ramirez-Gonzalez. |
| EPI_ISL_1651250, EPI_ISL_1651251 | LESP Jalisco | Instituto de Diagnostico y Referencia Epidemiologicos (INDRE) | Claudia Wong-Arambula, Abril Rodriguez-Maldonado, Vanessa Rivero-Arredondo, Ariadna Medina-Benitez, Joaquin Quiroz-Mercado, Sergio Rangel-Guerrero, Natividad Cruz-Ortiz, Tatiana Nunez-Garcia, Gisela Barrera-Badillo, Lucia Hernandez-Rivas, Irma Lopez-Martinez, Ernesto Ramirez-Gonzalez. |
| EPI_ISL_1651252 | LESP Oaxaca | Instituto de Diagnostico y Referencia Epidemiologicos (INDRE) | Claudia Wong-Arambula, Abril Rodriguez-Maldonado, Vanessa Rivero-Arredondo, Ariadna Medina-Benitez, Joaquin Quiroz-Mercado, Sergio Rangel-Guerrero, Natividad Cruz-Ortiz, Tatiana Nunez-Garcia, Gisela Barrera-Badillo, Lucia Hernandez-Rivas, Irma Lopez-Martinez, Ernesto Ramirez-Gonzalez. |
| EPI_ISL_1651253 | LESP Sinaloa | Instituto de Diagnostico y Referencia Epidemiologicos (INDRE) | Claudia Wong-Arambula, Abril Rodriguez-Maldonado, Vanessa Rivero-Arredondo, Ariadna Medina-Benitez, Joaquin Quiroz-Mercado, Sergio Rangel-Guerrero, Natividad Cruz-Ortiz, Tatiana Nunez-Garcia, Gisela Barrera-Badillo, Lucia Hernandez-Rivas, Irma Lopez-Martinez, Ernesto Ramirez-Gonzalez. |
| EPI_ISL_1651254 | LESP Sonora | Instituto de Diagnostico y Referencia Epidemiologicos (INDRE) | Claudia Wong-Arambula, Abril Rodriguez-Maldonado, Vanessa Rivero-Arredondo, Ariadna Medina-Benitez, Joaquin Quiroz-Mercado, Sergio Rangel-Guerrero, Natividad Cruz-Ortiz, Tatiana Nunez-Garcia, Gisela Barrera-Badillo, Lucia Hernandez-Rivas, Irma Lopez-Martinez, Ernesto Ramirez-Gonzalez. |
| EPI_ISL_1651255, EPI_ISL_1651256, EPI_ISL_1651257, EPI_ISL_1651258, EPI_ISL_1651259, EPI_ISL_1651260, EPI_ISL_1651261, EPI_ISL_1651262, EPI_ISL_1651263, EPI_ISL_1651264, EPI_ISL_1651265, EPI_ISL_1651266, EPI_ISL_1651267, EPI_ISL_1651268, EPI_ISL_1651269, EPI_ISL_1651270, EPI_ISL_1651271, EPI_ISL_1651272, EPI_ISL_1651273, EPI_ISL_1651274, EPI_ISL_1651275, EPI_ISL_1651276 | | | |
| see above | LESP Morelos | Instituto de Diagnostico y Referencia Epidemiologicos (INDRE) | Claudia Wong-Arambula, Abril Rodriguez-Maldonado, Vanessa Rivero-Arredondo, Ariadna Medina-Benitez, Joaquin Quiroz-Mercado, Sergio Rangel-Guerrero, Natividad Cruz-Ortiz, Tatiana Nunez-Garcia, Gisela Barrera-Badillo, Lucia Hernandez-Rivas, Irma Lopez-Martinez, Ernesto Ramirez-Gonzalez. |
| EPI_ISL_1651277, EPI_ISL_1651278, EPI_ISL_1651279, EPI_ISL_1651280, EPI_ISL_1651281, EPI_ISL_1651282, EPI_ISL_1651283, EPI_ISL_1651284, EPI_ISL_1651285, EPI_ISL_1651286, EPI_ISL_1651287 | | | |
| see above | LESP Queretaro | Instituto de Diagnostico y Referencia Epidemiologicos (INDRE) | Claudia Wong-Arambula, Abril Rodriguez-Maldonado, Vanessa Rivero-Arredondo, Ariadna Medina-Benitez, Joaquin Quiroz-Mercado, Sergio Rangel-Guerrero, Natividad Cruz-Ortiz, Tatiana Nunez-Garcia, Gisela Barrera-Badillo, Lucia Hernandez-Rivas, Irma Lopez-Martinez, Ernesto Ramirez-Gonzalez. |
| EPI_ISL_1651288, EPI_ISL_1651289, EPI_ISL_1651290, EPI_ISL_1651291, EPI_ISL_1651292, EPI_ISL_1651293, EPI_ISL_1651294, EPI_ISL_1651295, EPI_ISL_1651296, EPI_ISL_1651297, EPI_ISL_1651298, EPI_ISL_1651299, EPI_ISL_1651300, EPI_ISL_1651301, EPI_ISL_1651302, EPI_ISL_1651303, EPI_ISL_1651304, EPI_ISL_1651305, EPI_ISL_1651306 | | | |
| see above | GA Department of Public Health | GA Department of Public Health | Stacy Reeves, Jonathan Edwards, Cynthia Dixey, Tonia Parrott, Aliyah Fields, Taylor Smith |
| EPI_ISL_1651313, EPI_ISL_1651314, EPI_ISL_1651315, EPI_ISL_1651316, EPI_ISL_1651317, EPI_ISL_1651318, EPI_ISL_1651319, EPI_ISL_1651320, EPI_ISL_1651321, EPI_ISL_1651322, EPI_ISL_1651323, EPI_ISL_1651324, EPI_ISL_1651325, EPI_ISL_1651326, EPI_ISL_1651327, EPI_ISL_1651328, EPI_ISL_1651329, EPI_ISL_1651330, EPI_ISL_1651331, EPI_ISL_1651332, EPI_ISL_1651333, EPI_ISL_1651334, EPI_ISL_1651335, EPI_ISL_1651337, EPI_ISL_1651338, EPI_ISL_1651339, EPI_ISL_1651340, EPI_ISL_1651341, EPI_ISL_1651342, EPI_ISL_1651344, EPI_ISL_1651345, EPI_ISL_1651346, EPI_ISL_1651347, EPI_ISL_1651348, EPI_ISL_1651349, EPI_ISL_1651350, EPI_ISL_1651352, EPI_ISL_1651355, EPI_ISL_1651356, EPI_ISL_1651357, EPI_ISL_1651358, EPI_ISL_1651359, EPI_ISL_1651360, EPI_ISL_1651361, EPI_ISL_1651362, EPI_ISL_1651363, EPI_ISL_1651364, EPI_ISL_1651365, EPI_ISL_1651367, EPI_ISL_1651369, EPI_ISL_1651370, EPI_ISL_1651371, EPI_ISL_1651372, EPI_ISL_1651374, EPI_ISL_1651375, EPI_ISL_1651376, EPI_ISL_1651377, EPI_ISL_1651378, EPI_ISL_1651379, EPI_ISL_1651380, EPI_ISL_1651381, EPI_ISL_1651383, EPI_ISL_1651384, EPI_ISL_1651387, EPI_ISL_1651389, EPI_ISL_1651390, EPI_ISL_1651391, EPI_ISL_1651392, EPI_ISL_1651393, EPI_ISL_1651395, EPI_ISL_1651396, EPI_ISL_1651398, EPI_ISL_1651399, EPI_ISL_1651400, EPI_ISL_1651401, EPI_ISL_1651402, EPI_ISL_1651403, EPI_ISL_1651404, EPI_ISL_1651405, EPI_ISL_1651406, EPI_ISL_1651407, EPI_ISL_1651408, EPI_ISL_1651409, EPI_ISL_1651411, EPI_ISL_1651412, EPI_ISL_1651413, EPI_ISL_1651414, EPI_ISL_1651416, EPI_ISL_1651417, EPI_ISL_1651418, EPI_ISL_1651419, EPI_ISL_1651420, EPI_ISL_1651421, EPI_ISL_1651422, EPI_ISL_1651423, EPI_ISL_1651424, EPI_ISL_1651426, EPI_ISL_1651427, EPI_ISL_1651428, EPI_ISL_1651429, EPI_ISL_1651430, EPI_ISL_1651432, EPI_ISL_1651433, EPI_ISL_1651434, EPI_ISL_1651436, EPI_ISL_1651437, EPI_ISL_1651438, EPI_ISL_1651440, EPI_ISL_1651441, EPI_ISL_1651442, EPI_ISL_1651443, EPI_ISL_1651445, EPI_ISL_1651446, EPI_ISL_1651447, EPI_ISL_1651448, EPI_ISL_1651449, EPI_ISL_1651450, EPI_ISL_1651451, EPI_ISL_1651453, EPI_ISL_1651454, EPI_ISL_1651455, EPI_ISL_1651456, EPI_ISL_1651457, EPI_ISL_1651458, EPI_ISL_1651459, EPI_ISL_1651460, EPI_ISL_1651461, EPI_ISL_1651462, EPI_ISL_1651463, EPI_ISL_1651464, EPI_ISL_1651465, EPI_ISL_1651466, EPI_ISL_1651467, EPI_ISL_1651470, EPI_ISL_1651472, EPI_ISL_1651473, EPI_ISL_1651474, EPI_ISL_1651475, EPI_ISL_1651476, EPI_ISL_1651477, EPI_ISL_1651478, EPI_ISL_1651479, EPI_ISL_1651480, EPI_ISL_1651481, EPI_ISL_1651482, EPI_ISL_1651483, EPI_ISL_1651484, EPI_ISL_1651485, EPI_ISL_1651486, EPI_ISL_1651487, EPI_ISL_1651488, EPI_ISL_1651489, EPI_ISL_1651490, EPI_ISL_1651491, EPI_ISL_1651492, EPI_ISL_1651493, EPI_ISL_1651494, EPI_ISL_1651495, EPI_ISL_1651496, EPI_ISL_1651497, EPI_ISL_1651498, EPI_ISL_1651499, EPI_ISL_1651500, EPI_ISL_1651501, EPI_ISL_1651502, EPI_ISL_1651503, EPI_ISL_1651504, EPI_ISL_1651505, EPI_ISL_1651506, EPI_ISL_1651507, EPI_ISL_1651508, EPI_ISL_1651509, EPI_ISL_1651510, EPI_ISL_1651511, EPI_ISL_1651512, EPI_ISL_1651514, EPI_ISL_1651516, EPI_ISL_1651517, EPI_ISL_1651518, EPI_ISL_1651519, EPI_ISL_1651520, EPI_ISL_1651521, EPI_ISL_1651522, EPI_ISL_1651523, EPI_ISL_1651524, EPI_ISL_1651525, EPI_ISL_1651526, EPI_ISL_1651529, EPI_ISL_1651530, EPI_ISL_1651531, EPI_ISL_1651532, EPI_ISL_1651533, EPI_ISL_1651534, EPI_ISL_1651535, EPI_ISL_1651536, EPI_ISL_1651537, EPI_ISL_1651539, EPI_ISL_1651540, EPI_ISL_1651541, EPI_ISL_1651542, EPI_ISL_1651543, EPI_ISL_1651544, EPI_ISL_1651545, EPI_ISL_1651546 | | | |
| see above | Florida Bureau of Public Health Laboratories | Florida Bureau of Public Health Laboratories | Sarah Schmedes, Jason Blanton |
| EPI_ISL_1651547, EPI_ISL_1651548, EPI_ISL_1651549, EPI_ISL_1651550, EPI_ISL_1651551, EPI_ISL_1651552, EPI_ISL_1651553, EPI_ISL_1651554, EPI_ISL_1651555, EPI_ISL_1651556, EPI_ISL_1651557, EPI_ISL_1651558, EPI_ISL_1651559, EPI_ISL_1651560, EPI_ISL_1651561, EPI_ISL_1651562, EPI_ISL_1651563, EPI_ISL_1651564, EPI_ISL_1651565, EPI_ISL_1651566, EPI_ISL_1651567, EPI_ISL_1651568, EPI_ISL_1651569, EPI_ISL_1651570, EPI_ISL_1651571, EPI_ISL_1651572, EPI_ISL_1651573, EPI_ISL_1651574, EPI_ISL_1651575, EPI_ISL_1651576, EPI_ISL_1651577, EPI_ISL_1651578, EPI_ISL_1651579, EPI_ISL_1651580, EPI_ISL_1651581, EPI_ISL_1651582, EPI_ISL_1651583, EPI_ISL_1651584, EPI_ISL_1651585, EPI_ISL_1651586, EPI_ISL_1651587, EPI_ISL_1651588, EPI_ISL_1651589, EPI_ISL_1651590, EPI_ISL_1651591, EPI_ISL_1651592, EPI_ISL_1651593, EPI_ISL_1651594, EPI_ISL_1651595, EPI_ISL_1651597, EPI_ISL_1651598, EPI_ISL_1651599, EPI_ISL_1651600, EPI_ISL_1651601, EPI_ISL_1651602, EPI_ISL_1651603, EPI_ISL_1651604, EPI_ISL_1651606, EPI_ISL_1651607, EPI_ISL_1651608, EPI_ISL_1651609, EPI_ISL_1651610, EPI_ISL_1651611, EPI_ISL_1651612, EPI_ISL_1651613, EPI_ISL_1651615, EPI_ISL_1651616, EPI_ISL_1651617, EPI_ISL_1651618, EPI_ISL_1651619, EPI_ISL_1651620, EPI_ISL_1651621, EPI_ISL_1651622, EPI_ISL_1651623, EPI_ISL_1651624, EPI_ISL_1651625, EPI_ISL_1651626, EPI_ISL_1651627, EPI_ISL_1651628, EPI_ISL_1651629, EPI_ISL_1651630, EPI_ISL_1651631, EPI_ISL_1651632, EPI_ISL_1651633, EPI_ISL_1651634, EPI_ISL_1651635, EPI_ISL_1651636, EPI_ISL_1651637, EPI_ISL_1651638, EPI_ISL_1651639, EPI_ISL_1651640, EPI_ISL_1651641, EPI_ISL_1651642, EPI_ISL_1651643, EPI_ISL_1651644, EPI_ISL_1651645, EPI_ISL_1651646, EPI_ISL_1651647, EPI_ISL_1651648, EPI_ISL_1651649, EPI_ISL_1651650, EPI_ISL_1651651, EPI_ISL_1651652, EPI_ISL_1651653, EPI_ISL_1651654, EPI_ISL_1651655, EPI_ISL_1651656, EPI_ISL_1651657, EPI_ISL_1651658, EPI_ISL_1651659, EPI_ISL_1651660, EPI_ISL_1651661, EPI_ISL_1651662, EPI_ISL_1651663, EPI_ISL_1651664, EPI_ISL_1651665, EPI_ISL_1651666, EPI_ISL_1651667, EPI_ISL_1651668, EPI_ISL_1651669, EPI_ISL_1651670, EPI_ISL_1651671, EPI_ISL_1651672, EPI_ISL_1651673, EPI_ISL_1651674, EPI_ISL_1651675, EPI_ISL_1651676, EPI_ISL_1651677, EPI_ISL_1651678, EPI_ISL_1651679, EPI_ISL_1651680, EPI_ISL_1651681, EPI_ISL_1651682, EPI_ISL_1651683, EPI_ISL_1651684, EPI_ISL_1651685, EPI_ISL_1651686, EPI_ISL_1651687, EPI_ISL_1651688, EPI_ISL_1651689, EPI_ISL_1651690, EPI_ISL_1651691, EPI_ISL_1651692, EPI_ISL_1651693, EPI_ISL_1651694, EPI_ISL_1651695, EPI_ISL_1651696, EPI_ISL_1651697, EPI_ISL_1651698, EPI_ISL_1651699, EPI_ISL_1651700, EPI_ISL_1651701, EPI_ISL_1651702, EPI_ISL_1651703, EPI_ISL_1651704, EPI_ISL_1651705, EPI_ISL_1651706, EPI_ISL_1651707, EPI_ISL_1651708, EPI_ISL_1651709, EPI_ISL_1651710, EPI_ISL_1651711, EPI_ISL_1651712, EPI_ISL_1651713, EPI_ISL_1651714, EPI_ISL_1651715 | | | |
| see above | University of Michigan Clinical Microbiology Laboratory | Lauring Lab, University of Michigan, Department of Microbiology and Immunology | Valesano |
| EPI_ISL_1651831, EPI_ISL_1651832, EPI_ISL_1651833, EPI_ISL_1651834, EPI_ISL_1651835, EPI_ISL_1651837, EPI_ISL_1651838, EPI_ISL_1651839, EPI_ISL_1651840, EPI_ISL_1651841, EPI_ISL_1651842, EPI_ISL_1651843, EPI_ISL_1651844, EPI_ISL_1651845, EPI_ISL_1651846, EPI_ISL_1651847, EPI_ISL_1651848, EPI_ISL_1651849, EPI_ISL_1651850, EPI_ISL_1651851, EPI_ISL_1651853, EPI_ISL_1651854, EPI_ISL_1651855, EPI_ISL_1651856, EPI_ISL_1651857, EPI_ISL_1651858, EPI_ISL_1651859, EPI_ISL_1651860, EPI_ISL_1651861, EPI_ISL_1651862, EPI_ISL_1651863, EPI_ISL_1651864, EPI_ISL_1651865, EPI_ISL_1651866, EPI_ISL_1651867, EPI_ISL_1651868, EPI_ISL_1651869, EPI_ISL_1651870, EPI_ISL_1651871, EPI_ISL_1651872, EPI_ISL_1651873, EPI_ISL_1651874, EPI_ISL_1651875 | | | |
| see above | Wyoming Public Health Laboratory | Wyoming Public Health Laboratory | Jim Mildenberger, Wanda Manley, Noah Hull, Taylor Fearing, Lynette Gumbleton, Channing Weber, Ashley Norberg, Chayse Rowley, Marley Goetz, Brian Dominguez, Elliot Thomasson, Sam Britz, Cari Sloma, and Rob Christensen |
| EPI_ISL_1651876 | LESP Queretaro | Instituto de Diagnostico y Referencia Epidemiologicos (INDRE) | Claudia Wong-Arambula, Abril Rodriguez-Maldonado, Vanessa Rivero-Arredondo, Ariadna Medina-Benitez, Joaquin Quiroz-Mercado, Sergio Rangel-Guerrero, Natividad Cruz-Ortiz, Tatiana Nunez-Garcia, Gisela Barrera-Badillo, Lucia Hernandez-Rivas, Irma Lopez-Martinez, Ernesto Ramirez-Gonzalez. |
| EPI_ISL_1651877, EPI_ISL_1651878, EPI_ISL_1651879, EPI_ISL_1651880 | LESP Chihuahua | Instituto de Diagnostico y Referencia Epidemiologicos (INDRE) | Claudia Wong-Arambula, Abril Rodriguez-Maldonado, Vanessa Rivero-Arredondo, Ariadna Medina-Benitez, Joaquin Quiroz-Mercado, Sergio Rangel-Guerrero, Natividad Cruz-Ortiz, Tatiana Nunez-Garcia, Gisela Barrera-Badillo, Lucia Hernandez-Rivas, Irma Lopez-Martinez, Ernesto Ramirez-Gonzalez. |
| EPI_ISL_1651881, EPI_ISL_1651882 | LESP Veracruz | Instituto de Diagnostico y Referencia Epidemiologicos (INDRE) | Claudia Wong-Arambula, Abril Rodriguez-Maldonado, Vanessa Rivero-Arredondo, Ariadna Medina-Benitez, Joaquin Quiroz-Mercado, Sergio Rangel-Guerrero, Natividad Cruz-Ortiz, Tatiana Nunez-Garcia, Gisela Barrera-Badillo, Lucia Hernandez-Rivas, Irma Lopez-Martinez, Ernesto Ramirez-Gonzalez. |
| EPI_ISL_1651883, EPI_ISL_1651884, EPI_ISL_1651885, EPI_ISL_1651886 | LESP Yucatan | Instituto de Diagnostico y Referencia Epidemiologicos (INDRE) | Claudia Wong-Arambula, Abril Rodriguez-Maldonado, Vanessa Rivero-Arredondo, Ariadna Medina-Benitez, Joaquin Quiroz-Mercado, Sergio Rangel-Guerrero, Natividad Cruz-Ortiz, Tatiana Nunez-Garcia, Gisela Barrera-Badillo, Lucia Hernandez-Rivas, Irma Lopez-Martinez, Ernesto Ramirez-Gonzalez. |
| EPI_ISL_1651887 | LESP Tamaulipas | Instituto de Diagnostico y Referencia Epidemiologicos (INDRE) | Claudia Wong-Arambula, Abril Rodriguez-Maldonado, Vanessa Rivero-Arredondo, Ariadna Medina-Benitez, Joaquin Quiroz-Mercado, Sergio Rangel-Guerrero, Natividad Cruz-Ortiz, Tatiana Nunez-Garcia, Gisela Barrera-Badillo, Lucia Hernandez-Rivas, Irma Lopez-Martinez, Ernesto Ramirez-Gonzalez. |
| EPI_ISL_1651888 | LESP Ciudad de Mexico | Instituto de Diagnostico y Referencia Epidemiologicos (INDRE) | Claudia Wong-Arambula, Abril Rodriguez-Maldonado, Vanessa Rivero-Arredondo, Ariadna Medina-Benitez, Joaquin Quiroz-Mercado, Sergio Rangel-Guerrero, Natividad Cruz-Ortiz, Tatiana Nunez-Garcia, Gisela Barrera-Badillo, Lucia Hernandez-Rivas, Irma Lopez-Martinez, Ernesto Ramirez-Gonzalez. |
| EPI_ISL_1651889 | LESP Coahuila | Instituto de Diagnostico y Referencia Epidemiologicos (INDRE) | Claudia Wong-Arambula, Abril Rodriguez-Maldonado, Vanessa Rivero-Arredondo, Ariadna Medina-Benitez, Joaquin Quiroz-Mercado, Sergio Rangel-Guerrero, Natividad Cruz-Ortiz, Tatiana Nunez-Garcia, Gisela Barrera-Badillo, Lucia Hernandez-Rivas, Irma Lopez-Martinez, Ernesto Ramirez-Gonzalez. |
| EPI_ISL_1651890, EPI_ISL_1651891, EPI_ISL_1651892, EPI_ISL_1651893, EPI_ISL_1651894, EPI_ISL_1651895 | LESP Baja California Sur | Instituto de Diagnostico y Referencia Epidemiologicos (INDRE) | Claudia Wong-Arambula, Abril Rodriguez-Maldonado, Vanessa Rivero-Arredondo, Ariadna Medina-Benitez, Joaquin Quiroz-Mercado, Sergio Rangel-Guerrero, Natividad Cruz-Ortiz, Tatiana Nunez-Garcia, Gisela Barrera-Badillo, Lucia Hernandez-Rivas, Irma Lopez-Martinez, Ernesto Ramirez-Gonzalez. |
| EPI_ISL_1651896, EPI_ISL_1651897, EPI_ISL_1651898 | LESP Quintana Roo | Instituto de Diagnostico y Referencia Epidemiologicos (INDRE) | Claudia Wong-Arambula, Abril Rodriguez-Maldonado, Vanessa Rivero-Arredondo, Ariadna Medina-Benitez, Joaquin Quiroz-Mercado, Sergio Rangel-Guerrero, Natividad Cruz-Ortiz, Tatiana Nunez-Garcia, Gisela Barrera-Badillo, Lucia Hernandez-Rivas, Irma Lopez-Martinez, Ernesto Ramirez-Gonzalez. |
| EPI_ISL_1651899 | LESP Ciudad de Mexico | Instituto de Diagnostico y Referencia Epidemiologicos (INDRE) | Claudia Wong-Arambula, Abril Rodriguez-Maldonado, Vanessa Rivero-Arredondo, Ariadna Medina-Benitez, Joaquin Quiroz-Mercado, Sergio Rangel-Guerrero, Natividad Cruz-Ortiz, Tatiana Nunez-Garcia, Gisela Barrera-Badillo, Lucia Hernandez-Rivas, Irma Lopez-Martinez, Ernesto Ramirez-Gonzalez. |
| EPI_ISL_1651900 | LESP Baja California Sur | Instituto de Diagnostico y Referencia Epidemiologicos (INDRE) | Claudia Wong-Arambula, Abril Rodriguez-Maldonado, Vanessa Rivero-Arredondo, Ariadna Medina-Benitez, Joaquin Quiroz-Mercado, Sergio Rangel-Guerrero, Natividad Cruz-Ortiz, Tatiana Nunez-Garcia, Gisela Barrera-Badillo, Lucia Hernandez-Rivas, Irma Lopez-Martinez, Ernesto Ramirez-Gonzalez. |
| EPI_ISL_1651901, EPI_ISL_1651902 | LESP Ciudad de Mexico | Instituto de Diagnostico y Referencia Epidemiologicos (INDRE) | Claudia Wong-Arambula, Abril Rodriguez-Maldonado, Vanessa Rivero-Arredondo, Ariadna Medina-Benitez, Joaquin Quiroz-Mercado, Sergio Rangel-Guerrero, Natividad Cruz-Ortiz, Tatiana Nunez-Garcia, Gisela Barrera-Badillo, Lucia Hernandez-Rivas, Irma Lopez-Martinez, Ernesto Ramirez-Gonzalez. |

| | | | |
|---|---|---|---|
| EPI_ISL_1651903 | LESP Estado de Mexico | Instituto de Diagnostico y Referencia Epidemiologicos (INDRE) | Claudia Wong-Arambula, Abril Rodriguez-Maldonado, Vanessa Rivero-Arredondo, Ariadna Medina-Benitez, Joaquin Quiroz-Mercado, Sergio Rangel-Guerrero, Natividad Cruz-Ortiz, Tatiana Nunez-Garcia, Gisela Barrera-Badillo, Lucia Hernandez-Rivas, Irma Lopez-Martinez, Ernesto Ramirez-Gonzalez. |
| EPI_ISL_1651904, EPI_ISL_1651906, EPI_ISL_1651907, EPI_ISL_1651908 | LESP Morelos | Instituto de Diagnostico y Referencia Epidemiologicos (INDRE) | Claudia Wong-Arambula, Abril Rodriguez-Maldonado, Vanessa Rivero-Arredondo, Ariadna Medina-Benitez, Joaquin Quiroz-Mercado, Sergio Rangel-Guerrero, Natividad Cruz-Ortiz, Tatiana Nunez-Garcia, Gisela Barrera-Badillo, Lucia Hernandez-Rivas, Irma Lopez-Martinez, Ernesto Ramirez-Gonzalez. |
| EPI_ISL_1652083, EPI_ISL_1652084 | Kansas Health and Environmental Lab | Kansas Health and Environmental Lab | Mike Grose, Jonathan Barnell, Ben Olsen, and Phil Adam |
| EPI_ISL_1652085, EPI_ISL_1652086, EPI_ISL_1652087, EPI_ISL_1652088, EPI_ISL_1652089 | Wichita State University - Molecular Diagnostics Lab | Kansas Health and Environmental Lab | Mike Grose, Jonathan Barnell, Ben Olsen, and Phil Adam |
| EPI_ISL_1652090 | Kansas Health and Environmental Lab | Kansas Health and Environmental Lab | Mike Grose, Jonathan Barnell, Ben Olsen, and Phil Adam |
| EPI_ISL_1652124, EPI_ISL_1652126, EPI_ISL_1652127, EPI_ISL_1652128, EPI_ISL_1652129, EPI_ISL_1652130, EPI_ISL_1652131, EPI_ISL_1652132, EPI_ISL_1652134, EPI_ISL_1652135, EPI_ISL_1652136, EPI_ISL_1652137, EPI_ISL_1652138, EPI_ISL_1652139, EPI_ISL_1652140, EPI_ISL_1652141, EPI_ISL_1652142, EPI_ISL_1652143, EPI_ISL_1652145, EPI_ISL_1652146, EPI_ISL_1652147, EPI_ISL_1652148, EPI_ISL_1652149, EPI_ISL_1652150, EPI_ISL_1652151 | | | |
| see above | Wyoming Public Health Laboratory | Wyoming Public Health Laboratory | Jim Mildenberger, Wanda Manley, Noah Hull, Taylor Fearing, Lynette Gumbleton, Channing Weber, Ashley Norberg, Chayse Rowley, Marley Goetz, Brian Dominguez, Elliot Thomasson, Sam Britz, Cari Sloma, and Rob Christensen |
| EPI_ISL_1652153, EPI_ISL_1652154, EPI_ISL_1652155, EPI_ISL_1652156, EPI_ISL_1652157, EPI_ISL_1652158, EPI_ISL_1652159, EPI_ISL_1652160, EPI_ISL_1652161, EPI_ISL_1652162, EPI_ISL_1652163, EPI_ISL_1652164, EPI_ISL_1652165, EPI_ISL_1652166, EPI_ISL_1652167, EPI_ISL_1652168, EPI_ISL_1652169, EPI_ISL_1652170, EPI_ISL_1652171, EPI_ISL_1652172, EPI_ISL_1652173, EPI_ISL_1652174, EPI_ISL_1652175, EPI_ISL_1652176, EPI_ISL_1652179, EPI_ISL_1652181, EPI_ISL_1652182, EPI_ISL_1652183, EPI_ISL_1652184, EPI_ISL_1652185, EPI_ISL_1652186, EPI_ISL_1652187, EPI_ISL_1652188, EPI_ISL_1652189, EPI_ISL_1652192, EPI_ISL_1652193, EPI_ISL_1652194, EPI_ISL_1652195 | | | |
| see above | City of Milwaukee Health Department Laboratory | City of Milwaukee Health Department Laboratory | Sanjib Bhattacharyya |
| EPI_ISL_1652197 | PA Department of Health, Bureau of Laboratories | Pathogen Discovery, Respiratory Viruses Branch, Division of Viral Diseases, Centers for Disease Control and Prevention | Ying Tao, Yan Li, Anna Kelleher, Jing Zhang, Anna Montmayeur, Brian Lynch, Krista Queen, Anna Uehara, Peter Cook, Han Jia Justin Ng, Rachel Marine, Clinton R. Paden, Haibin Wang, Mark Burroughs, Justin Lee, Adam Retchless, Suxiang Tong |
| EPI_ISL_1652201, EPI_ISL_1652202, EPI_ISL_1652203, EPI_ISL_1652204, EPI_ISL_1652205, EPI_ISL_1652206, EPI_ISL_1652207, EPI_ISL_1652208, EPI_ISL_1652209, EPI_ISL_1652210, EPI_ISL_1652211, EPI_ISL_1652212, EPI_ISL_1652213, EPI_ISL_1652214, EPI_ISL_1652215, EPI_ISL_1652216, EPI_ISL_1652217, EPI_ISL_1652218, EPI_ISL_1652219, EPI_ISL_1652220, EPI_ISL_1652221, EPI_ISL_1652222, EPI_ISL_1652223, EPI_ISL_1652224, EPI_ISL_1652225, EPI_ISL_1652226, EPI_ISL_1652227, EPI_ISL_1652228, EPI_ISL_1652229, EPI_ISL_1652230, EPI_ISL_1652231, EPI_ISL_1652232, EPI_ISL_1652233, EPI_ISL_1652234, EPI_ISL_1652235, EPI_ISL_1652236 | | | |
| see above | Alaska State Virology Laboratory | Alaska State Virology Laboratory | Stephanie DeRonde, Elva House, Lisa Smith, Ph.D., Jack Chen, Ph.D. |
| EPI_ISL_1652240, EPI_ISL_1652241, EPI_ISL_1652243, EPI_ISL_1652244, EPI_ISL_1652245, EPI_ISL_1652246, EPI_ISL_1652248, EPI_ISL_1652249, EPI_ISL_1652250, EPI_ISL_1652251, EPI_ISL_1652252, EPI_ISL_1652253, EPI_ISL_1652254, EPI_ISL_1652255, EPI_ISL_1652256, EPI_ISL_1652257, EPI_ISL_1652258, EPI_ISL_1652260, EPI_ISL_1652261, EPI_ISL_1652262, EPI_ISL_1652263, EPI_ISL_1652265, EPI_ISL_1652267, EPI_ISL_1652268, EPI_ISL_1652269, EPI_ISL_1652272, EPI_ISL_1652274, EPI_ISL_1652275, EPI_ISL_1652276, EPI_ISL_1652277, EPI_ISL_1652278, EPI_ISL_1652280, EPI_ISL_1652281, EPI_ISL_1652283, EPI_ISL_1652284, EPI_ISL_1652285, EPI_ISL_1652287, EPI_ISL_1652288, EPI_ISL_1652289, EPI_ISL_1652292, EPI_ISL_1652293, EPI_ISL_1652294, EPI_ISL_1652295, EPI_ISL_1652297, EPI_ISL_1652299, EPI_ISL_1652300, EPI_ISL_1652302, EPI_ISL_1652304, EPI_ISL_1652305, EPI_ISL_1652306, EPI_ISL_1652308, EPI_ISL_1652309, EPI_ISL_1652310, EPI_ISL_1652311, EPI_ISL_1652312, EPI_ISL_1652313, EPI_ISL_1652314, EPI_ISL_1652315, EPI_ISL_1652316, EPI_ISL_1652319, EPI_ISL_1652320, EPI_ISL_1652322, EPI_ISL_1652323, EPI_ISL_1652324, EPI_ISL_1652325, EPI_ISL_1652326, EPI_ISL_1652327, EPI_ISL_1652328, EPI_ISL_1652329, EPI_ISL_1652330, EPI_ISL_1652331, EPI_ISL_1652332, EPI_ISL_1652333, EPI_ISL_1652334, EPI_ISL_1652335, EPI_ISL_1652336, EPI_ISL_1652337, EPI_ISL_1652338, EPI_ISL_1652339, EPI_ISL_1652340, EPI_ISL_1652341, EPI_ISL_1652342, EPI_ISL_1652343, EPI_ISL_1652345, EPI_ISL_1652346, EPI_ISL_1652348, EPI_ISL_1652349, EPI_ISL_1652350, EPI_ISL_1652351, EPI_ISL_1652352, EPI_ISL_1652353, EPI_ISL_1652354, EPI_ISL_1652355, EPI_ISL_1652356, EPI_ISL_1652358, EPI_ISL_1652359, EPI_ISL_1652360, EPI_ISL_1652364, EPI_ISL_1652365, EPI_ISL_1652366, EPI_ISL_1652368, EPI_ISL_1652369, EPI_ISL_1652370, EPI_ISL_1652371, EPI_ISL_1652372, EPI_ISL_1652373, EPI_ISL_1652374, EPI_ISL_1652376, EPI_ISL_1652378, EPI_ISL_1652380, EPI_ISL_1652381, EPI_ISL_1652382, EPI_ISL_1652383, EPI_ISL_1652384, EPI_ISL_1652385, EPI_ISL_1652386, EPI_ISL_1652387, EPI_ISL_1652390, EPI_ISL_1652392, EPI_ISL_1652393, EPI_ISL_1652394, EPI_ISL_1652395, EPI_ISL_1652396, EPI_ISL_1652397, EPI_ISL_1652399, EPI_ISL_1652400, EPI_ISL_1652401, EPI_ISL_1652402, EPI_ISL_1652403, EPI_ISL_1652404, EPI_ISL_1652406, EPI_ISL_1652407, EPI_ISL_1652408, EPI_ISL_1652409, EPI_ISL_1652410, EPI_ISL_1652411, EPI_ISL_1652412, EPI_ISL_1652413, EPI_ISL_1652414, EPI_ISL_1652415, EPI_ISL_1652416, EPI_ISL_1652417, EPI_ISL_1652418, EPI_ISL_1652419, EPI_ISL_1652422, EPI_ISL_1652423, EPI_ISL_1652424, EPI_ISL_1652425, EPI_ISL_1652426, EPI_ISL_1652427, EPI_ISL_1652428, EPI_ISL_1652429, EPI_ISL_1652431, EPI_ISL_1652432, EPI_ISL_1652433, EPI_ISL_1652434, EPI_ISL_1652435, EPI_ISL_1652436, EPI_ISL_1652437, EPI_ISL_1652438, EPI_ISL_1652440, EPI_ISL_1652442, EPI_ISL_1652444, EPI_ISL_1652446, EPI_ISL_1652447, EPI_ISL_1652448, EPI_ISL_1652449, EPI_ISL_1652450, EPI_ISL_1652451, EPI_ISL_1652452, EPI_ISL_1652453, EPI_ISL_1652454, EPI_ISL_1652455, EPI_ISL_1652456, EPI_ISL_1652457, EPI_ISL_1652461, EPI_ISL_1652462, EPI_ISL_1652463, EPI_ISL_1652464, EPI_ISL_1652465, EPI_ISL_1652466, EPI_ISL_1652467, EPI_ISL_1652468, EPI_ISL_1652469, EPI_ISL_1652471, EPI_ISL_1652472, EPI_ISL_1652475, EPI_ISL_1652476, EPI_ISL_1652477, EPI_ISL_1652478, EPI_ISL_1652479, EPI_ISL_1652480, EPI_ISL_1652481, EPI_ISL_1652482, EPI_ISL_1652483, EPI_ISL_1652484, EPI_ISL_1652485, EPI_ISL_1652486, EPI_ISL_1652487, EPI_ISL_1652488, EPI_ISL_1652489, EPI_ISL_1652490, EPI_ISL_1652491 | | | |
| see above | Illinois Department of Public Health | Gagnon Lab, Southern Illinois University | Keith Gagnon |
| EPI_ISL_1652493, EPI_ISL_1652494, EPI_ISL_1652495, EPI_ISL_1652496, EPI_ISL_1652497, EPI_ISL_1652498, EPI_ISL_1652499, EPI_ISL_1652500, EPI_ISL_1652502, EPI_ISL_1652503, EPI_ISL_1652504, EPI_ISL_1652505, EPI_ISL_1652506, EPI_ISL_1652508, EPI_ISL_1652509, EPI_ISL_1652510, EPI_ISL_1652511, EPI_ISL_1652512, EPI_ISL_1652513, EPI_ISL_1652514, EPI_ISL_1652515, EPI_ISL_1652516, EPI_ISL_1652517, EPI_ISL_1652518, EPI_ISL_1652519, EPI_ISL_1652520, EPI_ISL_1652521, EPI_ISL_1652522, EPI_ISL_1652523, EPI_ISL_1652524, EPI_ISL_1652525, EPI_ISL_1652526, EPI_ISL_1652527, EPI_ISL_1652528, EPI_ISL_1652529, EPI_ISL_1652530 | | | |
| see above | Simple Laboratories | Gagnon Lab, Southern Illinois University | Keith Gagnon |
| EPI_ISL_1653771, EPI_ISL_1653773, EPI_ISL_1653774, EPI_ISL_1653775, EPI_ISL_1653776, EPI_ISL_1653777, EPI_ISL_1653778, EPI_ISL_1653779, EPI_ISL_1653780, EPI_ISL_1653781, EPI_ISL_1653782, EPI_ISL_1653783, EPI_ISL_1653784, EPI_ISL_1653785, EPI_ISL_1653786, EPI_ISL_1653787, EPI_ISL_1653788, EPI_ISL_1653789, EPI_ISL_1653790, EPI_ISL_1653791, EPI_ISL_1653792, EPI_ISL_1653793, EPI_ISL_1653794, EPI_ISL_1653795, EPI_ISL_1653796, EPI_ISL_1653797, EPI_ISL_1653798, EPI_ISL_1653799, EPI_ISL_1653800, EPI_ISL_1653801, EPI_ISL_1653802, EPI_ISL_1653803, EPI_ISL_1653804, EPI_ISL_1653805, EPI_ISL_1653806, EPI_ISL_1653807, EPI_ISL_1653809, EPI_ISL_1653811, EPI_ISL_1653812, EPI_ISL_1653813 | | | |
| see above | Southern Nevada Public Health Laboratory | Southern Nevada Public Health Laboratory | Michael Picker |
| EPI_ISL_1653816, EPI_ISL_1653817, EPI_ISL_1653818, EPI_ISL_1653819, EPI_ISL_1653820, EPI_ISL_1653821, EPI_ISL_1653822, EPI_ISL_1653823, EPI_ISL_1653824, EPI_ISL_1653825, EPI_ISL_1653826, EPI_ISL_1653827, EPI_ISL_1653828, EPI_ISL_1653829, EPI_ISL_1653830, EPI_ISL_1653831, EPI_ISL_1653832, EPI_ISL_1653833, EPI_ISL_1653834, EPI_ISL_1653835, EPI_ISL_1653836, EPI_ISL_1653837, EPI_ISL_1653838, EPI_ISL_1653839, EPI_ISL_1653840, EPI_ISL_1653841, EPI_ISL_1653842, EPI_ISL_1653843, EPI_ISL_1653844, EPI_ISL_1653845, EPI_ISL_1653846, EPI_ISL_1653847, EPI_ISL_1653848, EPI_ISL_1653849, EPI_ISL_1653850, EPI_ISL_1653851, EPI_ISL_1653852, EPI_ISL_1653853, EPI_ISL_1653854, EPI_ISL_1653855, EPI_ISL_1653856, EPI_ISL_1653857, EPI_ISL_1653858, EPI_ISL_1653859, EPI_ISL_1653860, EPI_ISL_1653861, EPI_ISL_1653862, EPI_ISL_1653863, EPI_ISL_1653864, EPI_ISL_1653865, EPI_ISL_1653866, EPI_ISL_1653867, EPI_ISL_1653868, EPI_ISL_1653869, EPI_ISL_1653870, EPI_ISL_1653871, EPI_ISL_1653872, EPI_ISL_1653873, EPI_ISL_1653874, EPI_ISL_1653875, EPI_ISL_1653876, EPI_ISL_1653877, EPI_ISL_1653878, EPI_ISL_1653879, EPI_ISL_1653880, EPI_ISL_1653881, EPI_ISL_1653882, EPI_ISL_1653883, EPI_ISL_1653884, EPI_ISL_1653885, EPI_ISL_1653886, EPI_ISL_1653887, EPI_ISL_1653888, EPI_ISL_1653889, EPI_ISL_1653890, EPI_ISL_1653891, EPI_ISL_1653892 | | | |
| see above | Washington State Department of Health Public Health Laboratories | Washington State Department of Health Public Health Laboratories | Drew MacKellar, Philip Dykema, Denny Russell, Joenice Gonzalez, Hannah Gray, Geoff Melly, Vanessa De Los Santos, Darren Lucas, JohnAric Peterson, Avi Singh, Rebecca Cao |
| EPI_ISL_1653895, EPI_ISL_1653896, EPI_ISL_1653897, EPI_ISL_1653898, EPI_ISL_1653899, EPI_ISL_1653900, EPI_ISL_1653901, EPI_ISL_1653902, EPI_ISL_1653903, EPI_ISL_1653904, EPI_ISL_1653905, EPI_ISL_1653906, EPI_ISL_1653907, EPI_ISL_1653908, EPI_ISL_1653909, EPI_ISL_1653910, EPI_ISL_1653911, EPI_ISL_1653912, EPI_ISL_1653913, EPI_ISL_1653914, EPI_ISL_1653915, EPI_ISL_1653916, EPI_ISL_1653917, EPI_ISL_1653918, EPI_ISL_1653919 | | | |
| see above | Alaska State Virology Laboratory | Alaska State Virology Laboratory | Stephanie DeRonde, Elva House, Lisa Smith, Ph.D., Jack Chen, Ph.D. |
| EPI_ISL_1654487, EPI_ISL_1654488, EPI_ISL_1654489, EPI_ISL_1654490, EPI_ISL_1654491, EPI_ISL_1654492, EPI_ISL_1654493, EPI_ISL_1654494, EPI_ISL_1654495, EPI_ISL_1654496 | Mayo Clinic & Mayo Clinic Laboratories | Minnesota Department of Health, Public Health Laboratory | Alexandra Lorentz, Jacob Garfin, Matt Plumb, and Xiong Wang |
| EPI_ISL_1654498, EPI_ISL_1654499, EPI_ISL_1654500, EPI_ISL_1654501, EPI_ISL_1654502, EPI_ISL_1654503, EPI_ISL_1654504, EPI_ISL_1654505, EPI_ISL_1654506, EPI_ISL_1654507, EPI_ISL_1654508, EPI_ISL_1654509, EPI_ISL_1654510, EPI_ISL_1654511, EPI_ISL_1654512, EPI_ISL_1654513, EPI_ISL_1654514, EPI_ISL_1654515, EPI_ISL_1654516, EPI_ISL_1654517, EPI_ISL_1654518, EPI_ISL_1654519, EPI_ISL_1654520, EPI_ISL_1654521, EPI_ISL_1654522, EPI_ISL_1654523, EPI_ISL_1654524, EPI_ISL_1654525, EPI_ISL_1654526, EPI_ISL_1654527, EPI_ISL_1654528, EPI_ISL_1654529, EPI_ISL_1654530, EPI_ISL_1654531, EPI_ISL_1654532, EPI_ISL_1654533, EPI_ISL_1654534, EPI_ISL_1654535, EPI_ISL_1654536, EPI_ISL_1654537, EPI_ISL_1654538, EPI_ISL_1654539, EPI_ISL_1654540, EPI_ISL_1654542, EPI_ISL_1654543, EPI_ISL_1654544, EPI_ISL_1654545, EPI_ISL_1654546, EPI_ISL_1654547, EPI_ISL_1654548, EPI_ISL_1654549, EPI_ISL_1654550, EPI_ISL_1654551, EPI_ISL_1654552, EPI_ISL_1654553, EPI_ISL_1654554, EPI_ISL_1654555, EPI_ISL_1654556, EPI_ISL_1654557, EPI_ISL_1654558, EPI_ISL_1654559, EPI_ISL_1654560, EPI_ISL_1654561, EPI_ISL_1654562, EPI_ISL_1654563, EPI_ISL_1654564, EPI_ISL_1654565, EPI_ISL_1654566, EPI_ISL_1654567, EPI_ISL_1654568, EPI_ISL_1654569, EPI_ISL_1654570, EPI_ISL_1654571, EPI_ISL_1654572, EPI_ISL_1654573 | | | |
| see above | M Health Fairview | Minnesota Department of Health, Public Health Laboratory | Alexandra Lorentz, Jacob Garfin, Matt Plumb, and Xiong Wang |
| EPI_ISL_1654574, EPI_ISL_1654575, EPI_ISL_1654576, EPI_ISL_1654577, EPI_ISL_1654578, EPI_ISL_1654579, EPI_ISL_1654580, EPI_ISL_1654581, EPI_ISL_1654583, EPI_ISL_1654584, EPI_ISL_1654585, EPI_ISL_1654587, EPI_ISL_1654588, EPI_ISL_1654589, EPI_ISL_1654590, EPI_ISL_1654591, EPI_ISL_1654592, EPI_ISL_1654593, EPI_ISL_1654594, EPI_ISL_1654595, EPI_ISL_1654596, EPI_ISL_1654597, EPI_ISL_1654598, EPI_ISL_1654599, EPI_ISL_1654603, EPI_ISL_1654604, EPI_ISL_1654612, EPI_ISL_1654613, EPI_ISL_1654614, EPI_ISL_1654615, EPI_ISL_1654616, EPI_ISL_1654617, EPI_ISL_1654618, EPI_ISL_1654619, EPI_ISL_1654620, EPI_ISL_1654621, EPI_ISL_1654622, EPI_ISL_1654623, EPI_ISL_1654624, EPI_ISL_1654625, EPI_ISL_1654626, EPI_ISL_1654627, EPI_ISL_1654628, EPI_ISL_1654629, EPI_ISL_1654630, EPI_ISL_1654631, EPI_ISL_1654632, EPI_ISL_1654633, EPI_ISL_1654634, EPI_ISL_1654635, EPI_ISL_1654636, EPI_ISL_1654637, EPI_ISL_1654638, EPI_ISL_1654639, EPI_ISL_1654640, EPI_ISL_1654641, EPI_ISL_1654642, EPI_ISL_1654643, EPI_ISL_1654644, EPI_ISL_1654645, EPI_ISL_1654646 | | | |
| see above | NYU Langone Health | Departments of Pathology and Medicine, New York University School of Medicine | Adriana Heguy, Dacia Dimartino, Emily Guzman, Christian Marier, Peter Meyn, Sitharam Ramaswami, Gael Westby, Paul Zappile, Yutong Zhang, Paolo Cotzia, Guiqing Wang |
| EPI_ISL_1656442, EPI_ISL_1656443, EPI_ISL_1656444, EPI_ISL_1656445, EPI_ISL_1656446, EPI_ISL_1656447, EPI_ISL_1656448, EPI_ISL_1656450, EPI_ISL_1656451, EPI_ISL_1656452, EPI_ISL_1656453, EPI_ISL_1656454, EPI_ISL_1656455, EPI_ISL_1656456, EPI_ISL_1656457, EPI_ISL_1656458 | | | |
| see above | University of Michigan Clinical Microbiology Laboratory | Lauring Lab, University of Michigan, Department of Microbiology and Immunology | Valesano |
| EPI_ISL_1660376, EPI_ISL_1660377, EPI_ISL_1660378, EPI_ISL_1660379, EPI_ISL_1660380, EPI_ISL_1660381, EPI_ISL_1660382, EPI_ISL_1660383, EPI_ISL_1660384, EPI_ISL_1660385, EPI_ISL_1660386, EPI_ISL_1660387, EPI_ISL_1660388, EPI_ISL_1660389, EPI_ISL_1660390, EPI_ISL_1660391, EPI_ISL_1660392, EPI_ISL_1660393, EPI_ISL_1660394 | | | |
| see above | Houston Health Dept. | Houston Health Dept. | Ryker Penn, Pamela Brown, Adolpho Lara, Yanlai Lai |
| EPI_ISL_1660527 | ME Health and Environmental Testing Laboratory | Pathogen Discovery, Respiratory Viruses Branch, Division of Viral Diseases, Centers for Disease Control and Prevention | Ying Tao, Yan Li, Anna Kelleher, Jing Zhang, Anna Montmayeur, Brian Lynch, Krista Queen, Anna Uehara, Peter Cook, Han Jia Justin Ng, Rachel Marine, Clinton R. Paden, Haibin Wang, Mark Burroughs, Justin Lee, Adam Retchless, Suxiang Tong |
| EPI_ISL_1660528, EPI_ISL_1660529, EPI_ISL_1660530, EPI_ISL_1660531, EPI_ISL_1660532, EPI_ISL_1660533, EPI_ISL_1660534, EPI_ISL_1660535, EPI_ISL_1660536, EPI_ISL_1660537, EPI_ISL_1660538, EPI_ISL_1660539, EPI_ISL_1660540, EPI_ISL_1660541, EPI_ISL_1660542, EPI_ISL_1660543, EPI_ISL_1660544, EPI_ISL_1660545, EPI_ISL_1660546, EPI_ISL_1660547, EPI_ISL_1660548, EPI_ISL_1660549, EPI_ISL_1660550, EPI_ISL_1660551, EPI_ISL_1660552, EPI_ISL_1660553, EPI_ISL_1660554, EPI_ISL_1660555, EPI_ISL_1660556, EPI_ISL_1660557, EPI_ISL_1660558, EPI_ISL_1660559, EPI_ISL_1660560, EPI_ISL_1660561, EPI_ISL_1660562, EPI_ISL_1660563, EPI_ISL_1660564, EPI_ISL_1660565, EPI_ISL_1660566, EPI_ISL_1660567, EPI_ISL_1660568, EPI_ISL_1660569, EPI_ISL_1660570, EPI_ISL_1660571, EPI_ISL_1660572, EPI_ISL_1660573, EPI_ISL_1660574, EPI_ISL_1660575, EPI_ISL_1660576, EPI_ISL_1660577, EPI_ISL_1660578, EPI_ISL_1660579, EPI_ISL_1660580, EPI_ISL_1660582, EPI_ISL_1660583, EPI_ISL_1660584, EPI_ISL_1660585, EPI_ISL_1660587, EPI_ISL_1660588, EPI_ISL_1660589, EPI_ISL_1660590, EPI_ISL_1660591, EPI_ISL_1660592, EPI_ISL_1660593, EPI_ISL_1660594, EPI_ISL_1660595, EPI_ISL_1660596, EPI_ISL_1660597, EPI_ISL_1660598, EPI_ISL_1660599, EPI_ISL_1660600, EPI_ISL_1660601, EPI_ISL_1660602, EPI_ISL_1660603, EPI_ISL_1660604, EPI_ISL_1660606, EPI_ISL_1660607, EPI_ISL_1660610, EPI_ISL_1660612, EPI_ISL_1660613, EPI_ISL_1660614, EPI_ISL_1660615 | | | |
| see above | Texas Department of State Health Services (TXDSHS) | Texas Department of State Health Services (TXDSHS) | Rashmi Tuladhar, Bonnie Oh, Jenny Zhang, Maliha Rahman, Mayela Pedrueza, Anita Pokharel, Lorraine Rodriguez, Myong Koag, Chun Wang, Rachel Lee, Grace Kubin |
| EPI_ISL_1660616 | Hospital Sharp | Microbial Genomics Laboratory | Bruno Gomez-Gil, Julissa Enciso-Ibarra, Alejandra Garcia-Gasca, Daniel Fregoso-Rueda |
| EPI_ISL_1660617 | Hospital Margarita Maza de Juárez | Microbial Genomics Laboratory | Bruno Gomez-Gil, Julissa Enciso-Ibarra, Alejandra Garcia-Gasca, Daniel Fregoso-Rueda |
| EPI_ISL_1660618 | Hospital Sharp | Microbial Genomics Laboratory | Bruno Gomez-Gil, Julissa Enciso-Ibarra, Alejandra Garcia-Gasca, Daniel Fregoso-Rueda |
| EPI_ISL_1660619 | Imágenes Diagnósticas de Mazatlán | Microbial Genomics Laboratory | Bruno Gomez-Gil, Julissa Enciso-Ibarra, Alejandra Garcia-Gasca, Daniel Fregoso-Rueda |

| | | | |
|---|---|---|---|
| EPI_ISL_1660620, EPI_ISL_1660621, EPI_ISL_1660622 | Hospital Margarita Maza de Juárez | Microbial Genomics Laboratory | Bruno Gomez-Gil, Julissa Enciso-Ibarra, Alejandra Garcia-Gasca, Daniel Fregoso-Rueda |
| EPI_ISL_1660623 | Hospital Sharp | Microbial Genomics Laboratory | Bruno Gomez-Gil, Julissa Enciso-Ibarra, Alejandra Garcia-Gasca, Daniel Fregoso-Rueda |
| EPI_ISL_1660624 | Hospital Margarita Maza de Juárez | Microbial Genomics Laboratory | Bruno Gomez-Gil, Julissa Enciso-Ibarra, Alejandra Garcia-Gasca, Daniel Fregoso-Rueda |
| EPI_ISL_1660625, EPI_ISL_1660626 | Hospital Sharp | Microbial Genomics Laboratory | Bruno Gomez-Gil, Julissa Enciso-Ibarra, Alejandra Garcia-Gasca, Daniel Fregoso-Rueda |
| EPI_ISL_1660627 | Imágenes Diagnósticas de Mazatlán | Microbial Genomics Laboratory | Bruno Gomez-Gil, Julissa Enciso-Ibarra, Alejandra Garcia-Gasca, Daniel Fregoso-Rueda |
| EPI_ISL_1660628 | Hospital Margarita Maza de Juárez | Microbial Genomics Laboratory | Bruno Gomez-Gil, Julissa Enciso-Ibarra, Alejandra Garcia-Gasca, Daniel Fregoso-Rueda |
| EPI_ISL_1661253, EPI_ISL_1661254 | Instituto Nacional de Medicina Genomica | Instituto Nacional de Medicina Genomica | Hidalgo-Miranda A, Mendoza-Vargas A, Reyes-Grajeda JP, Cedro-Tanda A, Alcaraz N, Gonzalez-Barrera D, Rangel-DeLeon D, Miranda-Ortiz H, Rosas-Escobar P, Canseco-Mendez JC, Cisneros-Villanueva M, Munguia-Garza P, Ramirez-Vega O, Garcia-Cardenas FJ, Gonzalez-Woge MA, Herrera-Montalvo LA |
| EPI_ISL_1661474, EPI_ISL_1661475, EPI_ISL_1661476, EPI_ISL_1661477 | LESP Morelos | Instituto de Diagnostico y Referencia Epidemiologicos (INDRE) | Claudia Wong-Arambula, Abril Rodriguez-Maldonado, Vanessa Rivero-Arredondo, Ariadna Medina-Benitez, Joaquin Quiroz-Mercado, Sergio Rangel-Guerrero, Natividad Cruz-Ortiz, Tatiana Nunez-Garcia, Gisela Barrera-Badillo, Lucia Hernandez-Rivas, Irma Lopez-Martinez, Ernesto Ramirez-Gonzalez. |
| EPI_ISL_1661478 | LESP Puebla | Instituto de Diagnostico y Referencia Epidemiologicos (INDRE) | Claudia Wong-Arambula, Abril Rodriguez-Maldonado, Vanessa Rivero-Arredondo, Ariadna Medina-Benitez, Joaquin Quiroz-Mercado, Sergio Rangel-Guerrero, Natividad Cruz-Ortiz, Tatiana Nunez-Garcia, Gisela Barrera-Badillo, Lucia Hernandez-Rivas, Irma Lopez-Martinez, Ernesto Ramirez-Gonzalez. |
| EPI_ISL_1661479, EPI_ISL_1661480 | LESP Morelos | Instituto de Diagnostico y Referencia Epidemiologicos (INDRE) | Claudia Wong-Arambula, Abril Rodriguez-Maldonado, Vanessa Rivero-Arredondo, Ariadna Medina-Benitez, Joaquin Quiroz-Mercado, Sergio Rangel-Guerrero, Natividad Cruz-Ortiz, Tatiana Nunez-Garcia, Gisela Barrera-Badillo, Lucia Hernandez-Rivas, Irma Lopez-Martinez, Ernesto Ramirez-Gonzalez. |
| EPI_ISL_1661481, EPI_ISL_1661482, EPI_ISL_1661483, EPI_ISL_1661484, EPI_ISL_1661485, EPI_ISL_1661486, EPI_ISL_1661487, EPI_ISL_1661488, EPI_ISL_1661489 | LESP Chihuahua | Instituto de Diagnostico y Referencia Epidemiologicos (INDRE) | Claudia Wong-Arambula, Abril Rodriguez-Maldonado, Vanessa Rivero-Arredondo, Ariadna Medina-Benitez, Joaquin Quiroz-Mercado, Sergio Rangel-Guerrero, Natividad Cruz-Ortiz, Tatiana Nunez-Garcia, Gisela Barrera-Badillo, Lucia Hernandez-Rivas, Irma Lopez-Martinez, Ernesto Ramirez-Gonzalez. |
| EPI_ISL_1661490 | LESP Ciudad de Mexico | Instituto de Diagnostico y Referencia Epidemiologicos (INDRE) | Claudia Wong-Arambula, Abril Rodriguez-Maldonado, Vanessa Rivero-Arredondo, Ariadna Medina-Benitez, Joaquin Quiroz-Mercado, Sergio Rangel-Guerrero, Natividad Cruz-Ortiz, Tatiana Nunez-Garcia, Gisela Barrera-Badillo, Lucia Hernandez-Rivas, Irma Lopez-Martinez, Ernesto Ramirez-Gonzalez. |
| EPI_ISL_1661491, EPI_ISL_1661492 | LESP Estado de Mexico | Instituto de Diagnostico y Referencia Epidemiologicos (INDRE) | Claudia Wong-Arambula, Abril Rodriguez-Maldonado, Vanessa Rivero-Arredondo, Ariadna Medina-Benitez, Joaquin Quiroz-Mercado, Sergio Rangel-Guerrero, Natividad Cruz-Ortiz, Tatiana Nunez-Garcia, Gisela Barrera-Badillo, Lucia Hernandez-Rivas, Irma Lopez-Martinez, Ernesto Ramirez-Gonzalez. |
| EPI_ISL_1661493, EPI_ISL_1661494 | LESP Ciudad de Mexico | Instituto de Diagnostico y Referencia Epidemiologicos (INDRE) | Claudia Wong-Arambula, Abril Rodriguez-Maldonado, Vanessa Rivero-Arredondo, Ariadna Medina-Benitez, Joaquin Quiroz-Mercado, Sergio Rangel-Guerrero, Natividad Cruz-Ortiz, Tatiana Nunez-Garcia, Gisela Barrera-Badillo, Lucia Hernandez-Rivas, Irma Lopez-Martinez, Ernesto Ramirez-Gonzalez. |
| EPI_ISL_1661495, EPI_ISL_1661496, EPI_ISL_1661497, EPI_ISL_1661498, EPI_ISL_1661499, EPI_ISL_1661500, EPI_ISL_1661501, EPI_ISL_1661502, EPI_ISL_1661503, EPI_ISL_1661504, EPI_ISL_1661505, EPI_ISL_1661506, EPI_ISL_1661507, EPI_ISL_1661508, EPI_ISL_1661509, EPI_ISL_1661510, EPI_ISL_1661511, EPI_ISL_1661512, EPI_ISL_1661513, EPI_ISL_1661514, EPI_ISL_1661515, EPI_ISL_1661516, EPI_ISL_1661517, EPI_ISL_1661518, EPI_ISL_1661519, EPI_ISL_1661520 | | | |
| see above | LESP Guanajuato | Instituto de Diagnostico y Referencia Epidemiologicos (INDRE) | Claudia Wong-Arambula, Abril Rodriguez-Maldonado, Vanessa Rivero-Arredondo, Ariadna Medina-Benitez, Joaquin Quiroz-Mercado, Sergio Rangel-Guerrero, Natividad Cruz-Ortiz, Tatiana Nunez-Garcia, Gisela Barrera-Badillo, Lucia Hernandez-Rivas, Irma Lopez-Martinez, Ernesto Ramirez-Gonzalez. |
| EPI_ISL_1661521 | LESP Estado de Mexico | Instituto de Diagnostico y Referencia Epidemiologicos (INDRE) | Claudia Wong-Arambula, Abril Rodriguez-Maldonado, Vanessa Rivero-Arredondo, Ariadna Medina-Benitez, Joaquin Quiroz-Mercado, Sergio Rangel-Guerrero, Natividad Cruz-Ortiz, Tatiana Nunez-Garcia, Gisela Barrera-Badillo, Lucia Hernandez-Rivas, Irma Lopez-Martinez, Ernesto Ramirez-Gonzalez. |
| EPI_ISL_1661522, EPI_ISL_1661523, EPI_ISL_1661524, EPI_ISL_1661525, EPI_ISL_1661526, EPI_ISL_1661527, EPI_ISL_1661528, EPI_ISL_1661529, EPI_ISL_1661530, EPI_ISL_1661531, EPI_ISL_1661532, EPI_ISL_1661533 | | | |
| see above | LESP Queretaro | Instituto de Diagnostico y Referencia Epidemiologicos (INDRE) | Claudia Wong-Arambula, Abril Rodriguez-Maldonado, Vanessa Rivero-Arredondo, Ariadna Medina-Benitez, Joaquin Quiroz-Mercado, Sergio Rangel-Guerrero, Natividad Cruz-Ortiz, Tatiana Nunez-Garcia, Gisela Barrera-Badillo, Lucia Hernandez-Rivas, Irma Lopez-Martinez, Ernesto Ramirez-Gonzalez. |
| EPI_ISL_1661534 | LESP Hidalgo | Instituto de Diagnostico y Referencia Epidemiologicos (INDRE) | Claudia Wong-Arambula, Abril Rodriguez-Maldonado, Vanessa Rivero-Arredondo, Ariadna Medina-Benitez, Joaquin Quiroz-Mercado, Sergio Rangel-Guerrero, Natividad Cruz-Ortiz, Tatiana Nunez-Garcia, Gisela Barrera-Badillo, Lucia Hernandez-Rivas, Irma Lopez-Martinez, Ernesto Ramirez-Gonzalez. |
| EPI_ISL_1661535 | LESP Puebla | Instituto de Diagnostico y Referencia Epidemiologicos (INDRE) | Claudia Wong-Arambula, Abril Rodriguez-Maldonado, Vanessa Rivero-Arredondo, Ariadna Medina-Benitez, Joaquin Quiroz-Mercado, Sergio Rangel-Guerrero, Natividad Cruz-Ortiz, Tatiana Nunez-Garcia, Gisela Barrera-Badillo, Lucia Hernandez-Rivas, Irma Lopez-Martinez, Ernesto Ramirez-Gonzalez. |
| EPI_ISL_1661536, EPI_ISL_1661537, EPI_ISL_1661538 | LESPZacatecas | Instituto de Diagnostico y Referencia Epidemiologicos (INDRE) | Claudia Wong-Arambula, Abril Rodriguez-Maldonado, Vanessa Rivero-Arredondo, Ariadna Medina-Benitez, Joaquin Quiroz-Mercado, Sergio Rangel-Guerrero, Natividad Cruz-Ortiz, Tatiana Nunez-Garcia, Gisela Barrera-Badillo, Lucia Hernandez-Rivas, Irma Lopez-Martinez, Ernesto Ramirez-Gonzalez. |
| EPI_ISL_1661539, EPI_ISL_1661540, EPI_ISL_1661541 | LESP Hidalgo | Instituto de Diagnostico y Referencia Epidemiologicos (INDRE) | Claudia Wong-Arambula, Abril Rodriguez-Maldonado, Vanessa Rivero-Arredondo, Ariadna Medina-Benitez, Joaquin Quiroz-Mercado, Sergio Rangel-Guerrero, Natividad Cruz-Ortiz, Tatiana Nunez-Garcia, Gisela Barrera-Badillo, Lucia Hernandez-Rivas, Irma Lopez-Martinez, Ernesto Ramirez-Gonzalez. |
| EPI_ISL_1661542, EPI_ISL_1661543 | LESP Morelos | Instituto de Diagnostico y Referencia Epidemiologicos (INDRE) | Claudia Wong-Arambula, Abril Rodriguez-Maldonado, Vanessa Rivero-Arredondo, Ariadna Medina-Benitez, Joaquin Quiroz-Mercado, Sergio Rangel-Guerrero, Natividad Cruz-Ortiz, Tatiana Nunez-Garcia, Gisela Barrera-Badillo, Lucia Hernandez-Rivas, Irma Lopez-Martinez, Ernesto Ramirez-Gonzalez. |
| EPI_ISL_1661544, EPI_ISL_1661545, EPI_ISL_1661546, EPI_ISL_1661547 | LESPZacatecas | Instituto de Diagnostico y Referencia Epidemiologicos (INDRE) | Claudia Wong-Arambula, Abril Rodriguez-Maldonado, Vanessa Rivero-Arredondo, Ariadna Medina-Benitez, Joaquin Quiroz-Mercado, Sergio Rangel-Guerrero, Natividad Cruz-Ortiz, Tatiana Nunez-Garcia, Gisela Barrera-Badillo, Lucia Hernandez-Rivas, Irma Lopez-Martinez, Ernesto Ramirez-Gonzalez. |
| EPI_ISL_1661548 | LESP Michoacan | Instituto de Diagnostico y Referencia Epidemiologicos (INDRE) | Claudia Wong-Arambula, Abril Rodriguez-Maldonado, Vanessa Rivero-Arredondo, Ariadna Medina-Benitez, Joaquin Quiroz-Mercado, Sergio Rangel-Guerrero, Natividad Cruz-Ortiz, Tatiana Nunez-Garcia, Gisela Barrera-Badillo, Lucia Hernandez-Rivas, Irma Lopez-Martinez, Ernesto Ramirez-Gonzalez. |
| EPI_ISL_1661549 | PEMEX | Instituto de Diagnostico y Referencia Epidemiologicos (INDRE) | Claudia Wong-Arambula, Abril Rodriguez-Maldonado, Vanessa Rivero-Arredondo, Ariadna Medina-Benitez, Joaquin Quiroz-Mercado, Sergio Rangel-Guerrero, Natividad Cruz-Ortiz, Tatiana Nunez-Garcia, Gisela Barrera-Badillo, Lucia Hernandez-Rivas, Irma Lopez-Martinez, Ernesto Ramirez-Gonzalez. |
| EPI_ISL_1661550 | LESP Baja California | Instituto de Diagnostico y Referencia Epidemiologicos (INDRE) | Claudia Wong-Arambula, Abril Rodriguez-Maldonado, Vanessa Rivero-Arredondo, Ariadna Medina-Benitez, Joaquin Quiroz-Mercado, Sergio Rangel-Guerrero, Natividad Cruz-Ortiz, Tatiana Nunez-Garcia, Gisela Barrera-Badillo, Lucia Hernandez-Rivas, Irma Lopez-Martinez, Ernesto Ramirez-Gonzalez. |
| EPI_ISL_1661551, EPI_ISL_1661552 | LESP Hidalgo | Instituto de Diagnostico y Referencia Epidemiologicos (INDRE) | Claudia Wong-Arambula, Abril Rodriguez-Maldonado, Vanessa Rivero-Arredondo, Ariadna Medina-Benitez, Joaquin Quiroz-Mercado, Sergio Rangel-Guerrero, Natividad Cruz-Ortiz, Tatiana Nunez-Garcia, Gisela Barrera-Badillo, Lucia Hernandez-Rivas, Irma Lopez-Martinez, Ernesto Ramirez-Gonzalez. |
| EPI_ISL_1661553, EPI_ISL_1661554, EPI_ISL_1661555, EPI_ISL_1661556 | LESP Nuevo Leon | Instituto de Diagnostico y Referencia Epidemiologicos (INDRE) | Claudia Wong-Arambula, Abril Rodriguez-Maldonado, Vanessa Rivero-Arredondo, Ariadna Medina-Benitez, Joaquin Quiroz-Mercado, Sergio Rangel-Guerrero, Natividad Cruz-Ortiz, Tatiana Nunez-Garcia, Gisela Barrera-Badillo, Lucia Hernandez-Rivas, Irma Lopez-Martinez, Ernesto Ramirez-Gonzalez. |
| EPI_ISL_1661557 | LESP San Luis Potosi | Instituto de Diagnostico y Referencia Epidemiologicos (INDRE) | Claudia Wong-Arambula, Abril Rodriguez-Maldonado, Vanessa Rivero-Arredondo, Ariadna Medina-Benitez, Joaquin Quiroz-Mercado, Sergio Rangel-Guerrero, Natividad Cruz-Ortiz, Tatiana Nunez-Garcia, Gisela Barrera-Badillo, Lucia Hernandez-Rivas, Irma Lopez-Martinez, Ernesto Ramirez-Gonzalez. |
| EPI_ISL_1661558, EPI_ISL_1661559, EPI_ISL_1661560, EPI_ISL_1661561, EPI_ISL_1661562, EPI_ISL_1661563, EPI_ISL_1661564 | LESP Nuevo Leon | Instituto de Diagnostico y Referencia Epidemiologicos (INDRE) | Claudia Wong-Arambula, Abril Rodriguez-Maldonado, Vanessa Rivero-Arredondo, Ariadna Medina-Benitez, Joaquin Quiroz-Mercado, Sergio Rangel-Guerrero, Natividad Cruz-Ortiz, Tatiana Nunez-Garcia, Gisela Barrera-Badillo, Lucia Hernandez-Rivas, Irma Lopez-Martinez, Ernesto Ramirez-Gonzalez. |
| EPI_ISL_1661565, EPI_ISL_1661566, EPI_ISL_1661567, EPI_ISL_1661568, EPI_ISL_1661569 | LESP Guanajuato | Instituto de Diagnostico y Referencia Epidemiologicos (INDRE) | Claudia Wong-Arambula, Abril Rodriguez-Maldonado, Vanessa Rivero-Arredondo, Ariadna Medina-Benitez, Joaquin Quiroz-Mercado, Sergio Rangel-Guerrero, Natividad Cruz-Ortiz, Tatiana Nunez-Garcia, Gisela Barrera-Badillo, Lucia Hernandez-Rivas, Irma Lopez-Martinez, Ernesto Ramirez-Gonzalez. |
| EPI_ISL_1661570 | LESP Guerrero | Instituto de Diagnostico y Referencia Epidemiologicos (INDRE) | Claudia Wong-Arambula, Abril Rodriguez-Maldonado, Vanessa Rivero-Arredondo, Ariadna Medina-Benitez, Joaquin Quiroz-Mercado, Sergio Rangel-Guerrero, Natividad Cruz-Ortiz, Tatiana Nunez-Garcia, Gisela Barrera-Badillo, Lucia Hernandez-Rivas, Irma Lopez-Martinez, Ernesto Ramirez-Gonzalez. |
| EPI_ISL_1661571, EPI_ISL_1661572, EPI_ISL_1661573, EPI_ISL_1661574 | Florida Bureau of Public Health Laboratories | Florida Bureau of Public Health Laboratories | Sarah Schmedes, Jason Blanton |
| EPI_ISL_1661842, EPI_ISL_1661843, EPI_ISL_1661844, EPI_ISL_1661845, EPI_ISL_1661846, EPI_ISL_1661847, EPI_ISL_1661848, EPI_ISL_1661849, EPI_ISL_1661850, EPI_ISL_1661852, EPI_ISL_1661853, EPI_ISL_1661854, EPI_ISL_1661855, EPI_ISL_1661856, EPI_ISL_1661857, EPI_ISL_1661858, EPI_ISL_1661859, EPI_ISL_1661860, EPI_ISL_1661861, EPI_ISL_1661862, EPI_ISL_1661863, EPI_ISL_1661864, EPI_ISL_1661865, EPI_ISL_1661866, EPI_ISL_1661867, EPI_ISL_1661868, EPI_ISL_1661869, EPI_ISL_1661870, EPI_ISL_1661871, EPI_ISL_1661872, EPI_ISL_1661873, EPI_ISL_1661874, EPI_ISL_1661875, EPI_ISL_1661876, EPI_ISL_1661877, EPI_ISL_1661878, EPI_ISL_1661879, EPI_ISL_1661880 | | | |
| see above | Texas Department of State Health | Texas Department of State Health Services | Rashmi Tuladhar, Bonnie Oh, Jenny Zhang, Maliha Rahman, Mayela Pedrueza, Anita Pokharel, Lorraine Rodriguez, Myong Koag, Chun Wang, Rachel Lee, Grace Kubin |

| | | | |
|---|---|---|---|
| | Services (TXDSHS) | (TXDSHS) | |
| EPI_ISL_1661977, EPI_ISL_1661978 | Laboratorio Central de Epidemiologia (LCE) | Unidad de Genomica Avanzada | Consorcio Mexicano de Vigilancia Genomica (CoViGen-Mex). Authors (in alphabetical order): Julio Elias Alvarado-Yaah, Carlos F. Arias, Santiago Ávila-Rios, Victor Hugo Borja-Aburto, Celia Boukadida, Juan Bautista Chale-Dzul, Celida Duque Molina, Jose Antonio Enciso-Moreno, Gloria Elena Espinosa-Ayala, Fernando Fontove-Herrera, Victor Eduardo Garcia-Arias,Concepcion Grajales-Muniz, Ricardo Grande, Alfredo Herrera-Estrella, Carla Ivon Herrera-Najera, Pavel Isa, Brenda Irasema Maldonado-Meza, Bernardo Martinez-Miguel, Margarita Matias-Florentino, Maria Guadalupe de Jesus Mireles-Rivera, Gloria Maria Molina-Salinas, Hector Montoya-Fuentes, Jose Esteban Munoz-Medina, Jose de Jesus Nunez-Contreras, Alicia Ocana-Mondragon, Luis Alberto Ochoa-Carrera, Hector Esteban Paz-Juarez, Francisco Pulido, Helen Haydee Fernanda Ramirez-Plascencia, Angel Gustavo Salas-Lais, Alejandro Sanchez-Flores, Clara Esperanza Santacruz-Tinoco, Maria Guadalupe Santiago-Mauricio, Nelly Selem-Mojica, Blanca Taboada, Gloria Vazquez |
| EPI_ISL_1661979 | Centro de Investigacion Biomedica de Occidente (CIBO) | Unidad de Genomica Avanzada | Consorcio Mexicano de Vigilancia Genomica (CoViGen-Mex). Authors (in alphabetical order): Julio Elias Alvarado-Yaah, Carlos F. Arias, Santiago Ávila-Rios, Victor Hugo Borja-Aburto, Celia Boukadida, Juan Bautista Chale-Dzul, Celida Duque Molina, Jose Antonio Enciso-Moreno, Gloria Elena Espinosa-Ayala, Fernando Fontove-Herrera, Victor Eduardo Garcia-Arias,Concepcion Grajales-Muniz, Ricardo Grande, Alfredo Herrera-Estrella, Carla Ivon Herrera-Najera, Pavel Isa, Brenda Irasema Maldonado-Meza, Bernardo Martinez-Miguel, Margarita Matias-Florentino, Maria Guadalupe de Jesus Mireles-Rivera, Gloria Maria Molina-Salinas, Hector Montoya-Fuentes, Jose Esteban Munoz-Medina, Jose de Jesus Nunez-Contreras, Alicia Ocana-Mondragon, Luis Alberto Ochoa-Carrera, Hector Esteban Paz-Juarez, Francisco Pulido, Helen Haydee Fernanda Ramirez-Plascencia, Angel Gustavo Salas-Lais, Alejandro Sanchez-Flores, Clara Esperanza Santacruz-Tinoco, Maria Guadalupe Santiago-Mauricio, Nelly Selem-Mojica, Blanca Taboada, Gloria Vazquez |
| EPI_ISL_1661980, EPI_ISL_1661981 | Laboratorio Central de Epidemiologia (LCE) | Unidad de Genomica Avanzada | Consorcio Mexicano de Vigilancia Genomica (CoViGen-Mex). Authors (in alphabetical order): Julio Elias Alvarado-Yaah, Carlos F. Arias, Santiago Ávila-Rios, Victor Hugo Borja-Aburto, Celia Boukadida, Juan Bautista Chale-Dzul, Celida Duque Molina, Jose Antonio Enciso-Moreno, Gloria Elena Espinosa-Ayala, Fernando Fontove-Herrera, Victor Eduardo Garcia-Arias,Concepcion Grajales-Muniz, Ricardo Grande, Alfredo Herrera-Estrella, Carla Ivon Herrera-Najera, Pavel Isa, Brenda Irasema Maldonado-Meza, Bernardo Martinez-Miguel, Margarita Matias-Florentino, Maria Guadalupe de Jesus Mireles-Rivera, Gloria Maria Molina-Salinas, Hector Montoya-Fuentes, Jose Esteban Munoz-Medina, Jose de Jesus Nunez-Contreras, Alicia Ocana-Mondragon, Luis Alberto Ochoa-Carrera, Hector Esteban Paz-Juarez, Francisco Pulido, Helen Haydee Fernanda Ramirez-Plascencia, Angel Gustavo Salas-Lais, Alejandro Sanchez-Flores, Clara Esperanza Santacruz-Tinoco, Maria Guadalupe Santiago-Mauricio, Nelly Selem-Mojica, Blanca Taboada, Gloria Vazquez |
| EPI_ISL_1661982, EPI_ISL_1661983, EPI_ISL_1661984, EPI_ISL_1661985, EPI_ISL_1661986, EPI_ISL_1661987, EPI_ISL_1661988, EPI_ISL_1661989, EPI_ISL_1661990 | Centro de Investigacion Biomedica del Noreste (CIBIN) | Unidad de Genomica Avanzada | Consorcio Mexicano de Vigilancia Genomica (CoViGen-Mex). Authors (in alphabetical order): Julio Elias Alvarado-Yaah, Carlos F. Arias, Santiago Ávila-Rios, Victor Hugo Borja-Aburto, Celia Boukadida, Juan Bautista Chale-Dzul, Celida Duque Molina, Jose Antonio Enciso-Moreno, Gloria Elena Espinosa-Ayala, Fernando Fontove-Herrera, Victor Eduardo Garcia-Arias,Concepcion Grajales-Muniz, Ricardo Grande, Alfredo Herrera-Estrella, Carla Ivon Herrera-Najera, Pavel Isa, Brenda Irasema Maldonado-Meza, Bernardo Martinez-Miguel, Margarita Matias-Florentino, Maria Guadalupe de Jesus Mireles-Rivera, Gloria Maria Molina-Salinas, Hector Montoya-Fuentes, Jose Esteban Munoz-Medina, Jose de Jesus Nunez-Contreras, Alicia Ocana-Mondragon, Luis Alberto Ochoa-Carrera, Hector Esteban Paz-Juarez, Francisco Pulido, Helen Haydee Fernanda Ramirez-Plascencia, Angel Gustavo Salas-Lais, Alejandro Sanchez-Flores, Clara Esperanza Santacruz-Tinoco, Maria Guadalupe Santiago-Mauricio, Nelly Selem-Mojica, Blanca Taboada, Gloria Vazquez |
| EPI_ISL_1661991, EPI_ISL_1661992, EPI_ISL_1661993, EPI_ISL_1661994, EPI_ISL_1661995, EPI_ISL_1661996, EPI_ISL_1661997, EPI_ISL_1661998, EPI_ISL_1661999, EPI_ISL_1662000, EPI_ISL_1662001, EPI_ISL_1662002, EPI_ISL_1662003, EPI_ISL_1662004, EPI_ISL_1662005, EPI_ISL_1662006, EPI_ISL_1662007, EPI_ISL_1662008, EPI_ISL_1662009, EPI_ISL_1662010, EPI_ISL_1662011, EPI_ISL_1662012, EPI_ISL_1662013, EPI_ISL_1662014, EPI_ISL_1662015, EPI_ISL_1662016, EPI_ISL_1662017, EPI_ISL_1662018, EPI_ISL_1662019, EPI_ISL_1662020, EPI_ISL_1662021, EPI_ISL_1662022, EPI_ISL_1662023, EPI_ISL_1662024, EPI_ISL_1662025, EPI_ISL_1662026, EPI_ISL_1662027, EPI_ISL_1662028, EPI_ISL_1662029, EPI_ISL_1662030, EPI_ISL_1662031, EPI_ISL_1662032, EPI_ISL_1662033, EPI_ISL_1662034 | | | |
| see above | Laboratorio Central de Epidemiologia (LCE) | Unidad de Genomica Avanzada | Consorcio Mexicano de Vigilancia Genomica (CoViGen-Mex). Authors (in alphabetical order): Julio Elias Alvarado-Yaah, Carlos F. Arias, Santiago Ávila-Rios, Victor Hugo Borja-Aburto, Celia Boukadida, Juan Bautista Chale-Dzul, Celida Duque Molina, Jose Antonio Enciso-Moreno, Gloria Elena Espinosa-Ayala, Fernando Fontove-Herrera, Victor Eduardo Garcia-Arias,Concepcion Grajales-Muniz, Ricardo Grande, Alfredo Herrera-Estrella, Carla Ivon Herrera-Najera, Pavel Isa, Brenda Irasema Maldonado-Meza, Bernardo Martinez-Miguel, Margarita Matias-Florentino, Maria Guadalupe de Jesus Mireles-Rivera, Gloria Maria Molina-Salinas, Hector Montoya-Fuentes, Jose Esteban Munoz-Medina, Jose de Jesus Nunez-Contreras, Alicia Ocana-Mondragon, Luis Alberto Ochoa-Carrera, Hector Esteban Paz-Juarez, Francisco Pulido, Helen Haydee Fernanda Ramirez-Plascencia, Angel Gustavo Salas-Lais, Alejandro Sanchez-Flores, Clara Esperanza Santacruz-Tinoco, Maria Guadalupe Santiago-Mauricio, Nelly Selem-Mojica, Blanca Taboada, Gloria Vazquez |
| EPI_ISL_1662035, EPI_ISL_1662036, EPI_ISL_1662037, EPI_ISL_1662038, EPI_ISL_1662039, EPI_ISL_1662040, EPI_ISL_1662041, EPI_ISL_1662042, EPI_ISL_1662043, EPI_ISL_1662044, EPI_ISL_1662045, EPI_ISL_1662046, EPI_ISL_1662047, EPI_ISL_1662048, EPI_ISL_1662049, EPI_ISL_1662050, EPI_ISL_1662051, EPI_ISL_1662052, EPI_ISL_1662053, EPI_ISL_1662054, EPI_ISL_1662055, EPI_ISL_1662056, EPI_ISL_1662057 | | | |
| see above | Centro de Investigacion Biomedica del Noreste (CIBIN) | Unidad de Genomica Avanzada | Consorcio Mexicano de Vigilancia Genomica (CoViGen-Mex). Authors (in alphabetical order): Julio Elias Alvarado-Yaah, Carlos F. Arias, Santiago Ávila-Rios, Victor Hugo Borja-Aburto, Celia Boukadida, Juan Bautista Chale-Dzul, Celida Duque Molina, Jose Antonio Enciso-Moreno, Gloria Elena Espinosa-Ayala, Fernando Fontove-Herrera, Victor Eduardo Garcia-Arias,Concepcion Grajales-Muniz, Ricardo Grande, Alfredo Herrera-Estrella, Carla Ivon Herrera-Najera, Pavel Isa, Brenda Irasema Maldonado-Meza, Bernardo Martinez-Miguel, Margarita Matias-Florentino, Maria Guadalupe de Jesus Mireles-Rivera, Gloria Maria Molina-Salinas, Hector Montoya-Fuentes, Jose Esteban Munoz-Medina, Jose de Jesus Nunez-Contreras, Alicia Ocana-Mondragon, Luis Alberto Ochoa-Carrera, Hector Esteban Paz-Juarez, Francisco Pulido, Helen Haydee Fernanda Ramirez-Plascencia, Angel Gustavo Salas-Lais, Alejandro Sanchez-Flores, Clara Esperanza Santacruz-Tinoco, Maria Guadalupe Santiago-Mauricio, Nelly Selem-Mojica, Blanca Taboada, Gloria Vazquez |
| EPI_ISL_1662058, EPI_ISL_1662059, EPI_ISL_1662060, EPI_ISL_1662061 | Laboratorio Central de Epidemiologia (LCE) | Unidad de Genomica Avanzada | Consorcio Mexicano de Vigilancia Genomica (CoViGen-Mex). Authors (in alphabetical order): Julio Elias Alvarado-Yaah, Carlos F. Arias, Santiago Ávila-Rios, Victor Hugo Borja-Aburto, Celia Boukadida, Juan Bautista Chale-Dzul, Celida Duque Molina, Jose Antonio Enciso-Moreno, Gloria Elena Espinosa-Ayala, Fernando Fontove-Herrera, Victor Eduardo Garcia-Arias,Concepcion Grajales-Muniz, Ricardo Grande, Alfredo Herrera-Estrella, Carla Ivon Herrera-Najera, Pavel Isa, Brenda Irasema Maldonado-Meza, Bernardo Martinez-Miguel, Margarita Matias-Florentino, Maria Guadalupe de Jesus Mireles-Rivera, Gloria Maria Molina-Salinas, Hector Montoya-Fuentes, Jose Esteban Munoz-Medina, Jose de Jesus Nunez-Contreras, Alicia Ocana-Mondragon, Luis Alberto Ochoa-Carrera, Hector Esteban Paz-Juarez, Francisco Pulido, Helen Haydee Fernanda Ramirez-Plascencia, Angel Gustavo Salas-Lais, Alejandro Sanchez-Flores, Clara Esperanza Santacruz-Tinoco, Maria Guadalupe Santiago-Mauricio, Nelly Selem-Mojica, Blanca Taboada, Gloria Vazquez |
| EPI_ISL_1662062, EPI_ISL_1662063, EPI_ISL_1662064, EPI_ISL_1662065, EPI_ISL_1662066, EPI_ISL_1662067, EPI_ISL_1662068, EPI_ISL_1662069 | Centro de Investigacion Biomedica de Occidente (CIBO) | Unidad de Genomica Avanzada | Consorcio Mexicano de Vigilancia Genomica (CoViGen-Mex). Authors (in alphabetical order): Julio Elias Alvarado-Yaah, Carlos F. Arias, Santiago Ávila-Rios, Victor Hugo Borja-Aburto, Celia Boukadida, Juan Bautista Chale-Dzul, Celida Duque Molina, Jose Antonio Enciso-Moreno, Gloria Elena Espinosa-Ayala, Fernando Fontove-Herrera, Victor Eduardo Garcia-Arias,Concepcion Grajales-Muniz, Ricardo Grande, Alfredo Herrera-Estrella, Carla Ivon Herrera-Najera, Pavel Isa, Brenda Irasema Maldonado-Meza, Bernardo Martinez-Miguel, Margarita Matias-Florentino, Maria Guadalupe de Jesus Mireles-Rivera, Gloria Maria Molina-Salinas, Hector Montoya-Fuentes, Jose Esteban Munoz-Medina, Jose de Jesus Nunez-Contreras, Alicia Ocana-Mondragon, Luis Alberto Ochoa-Carrera, Hector Esteban Paz-Juarez, Francisco Pulido, Helen Haydee Fernanda Ramirez-Plascencia, Angel Gustavo Salas-Lais, Alejandro Sanchez-Flores, Clara Esperanza Santacruz-Tinoco, Maria Guadalupe Santiago-Mauricio, Nelly Selem-Mojica, Blanca Taboada, Gloria Vazquez |
| EPI_ISL_1662070, EPI_ISL_1662071, EPI_ISL_1662072, EPI_ISL_1662073, EPI_ISL_1662074 | Laboratorio Central de Epidemiologia (LCE) | Unidad de Genomica Avanzada | Consorcio Mexicano de Vigilancia Genomica (CoViGen-Mex). Authors (in alphabetical order): Julio Elias Alvarado-Yaah, Carlos F. Arias, Santiago Ávila-Rios, Victor Hugo Borja-Aburto, Celia Boukadida, Juan Bautista Chale-Dzul, Celida Duque Molina, Jose Antonio Enciso-Moreno, Gloria Elena Espinosa-Ayala, Fernando Fontove-Herrera, Victor Eduardo Garcia-Arias,Concepcion Grajales-Muniz, Ricardo Grande, Alfredo Herrera-Estrella, Carla Ivon Herrera-Najera, Pavel Isa, Brenda Irasema Maldonado-Meza, Bernardo Martinez-Miguel, Margarita Matias-Florentino, Maria Guadalupe de Jesus Mireles-Rivera, Gloria Maria Molina-Salinas, Hector Montoya-Fuentes, Jose Esteban Munoz-Medina, Jose de Jesus Nunez-Contreras, Alicia Ocana-Mondragon, Luis Alberto Ochoa-Carrera, Hector Esteban Paz-Juarez, Francisco Pulido, Helen Haydee Fernanda Ramirez-Plascencia, Angel Gustavo Salas-Lais, Alejandro Sanchez-Flores, Clara Esperanza Santacruz-Tinoco, Maria Guadalupe Santiago-Mauricio, Nelly Selem-Mojica, Blanca Taboada, Gloria Vazquez |
| EPI_ISL_1662075, EPI_ISL_1662076, EPI_ISL_1662077, EPI_ISL_1662078, EPI_ISL_1662079, EPI_ISL_1662080, EPI_ISL_1662081, EPI_ISL_1662082, EPI_ISL_1662083, EPI_ISL_1662084, EPI_ISL_1662085, EPI_ISL_1662086, EPI_ISL_1662087, EPI_ISL_1662088, EPI_ISL_1662089, EPI_ISL_1662090, EPI_ISL_1662091, EPI_ISL_1662092, EPI_ISL_1662093, EPI_ISL_1662094, EPI_ISL_1662095, EPI_ISL_1662096, EPI_ISL_1662097, EPI_ISL_1662098, EPI_ISL_1662099, EPI_ISL_1662100, EPI_ISL_1662101, EPI_ISL_1662102, EPI_ISL_1662103 | | | |
| see above | Centro de Investigacion Biomedica de Occidente (CIBO) | Unidad de Genomica Avanzada | Consorcio Mexicano de Vigilancia Genomica (CoViGen-Mex). Authors (in alphabetical order): Julio Elias Alvarado-Yaah, Carlos F. Arias, Santiago Ávila-Rios, Victor Hugo Borja-Aburto, Celia Boukadida, Juan Bautista Chale-Dzul, Celida Duque Molina, Jose Antonio Enciso-Moreno, Gloria Elena Espinosa-Ayala, Fernando Fontove-Herrera, Victor Eduardo Garcia-Arias,Concepcion Grajales-Muniz, Ricardo Grande, Alfredo Herrera-Estrella, Carla Ivon Herrera-Najera, Pavel Isa, Brenda Irasema Maldonado-Meza, Bernardo Martinez-Miguel, Margarita Matias-Florentino, Maria Guadalupe de Jesus Mireles-Rivera, Gloria Maria Molina-Salinas, Hector Montoya-Fuentes, Jose Esteban Munoz-Medina, Jose de Jesus Nunez-Contreras, Alicia Ocana-Mondragon, Luis Alberto Ochoa-Carrera, Hector Esteban Paz-Juarez, Francisco Pulido, Helen Haydee Fernanda Ramirez-Plascencia, Angel Gustavo Salas-Lais, Alejandro Sanchez-Flores, Clara Esperanza Santacruz-Tinoco, Maria Guadalupe Santiago-Mauricio, Nelly Selem-Mojica, Blanca Taboada, Gloria Vazquez |
| EPI_ISL_1662104, EPI_ISL_1662105, EPI_ISL_1662106, EPI_ISL_1662107, EPI_ISL_1662108, EPI_ISL_1662109, EPI_ISL_1662110, EPI_ISL_1662111, EPI_ISL_1662112, EPI_ISL_1662113, EPI_ISL_1662114, EPI_ISL_1662115, EPI_ISL_1662116, EPI_ISL_1662117, EPI_ISL_1662118, EPI_ISL_1662119, EPI_ISL_1662120, EPI_ISL_1662121, EPI_ISL_1662122 | | | |
| see above | Laboratorio Central de Epidemiologia (LCE) | Unidad de Genomica Avanzada | Consorcio Mexicano de Vigilancia Genomica (CoViGen-Mex). Authors (in alphabetical order): Julio Elias Alvarado-Yaah, Carlos F. Arias, Santiago Ávila-Rios, Victor Hugo Borja-Aburto, Celia Boukadida, Juan Bautista Chale-Dzul, Celida Duque Molina, Jose Antonio Enciso-Moreno, Gloria Elena Espinosa-Ayala, Fernando Fontove-Herrera, Victor Eduardo Garcia-Arias,Concepcion Grajales-Muniz, Ricardo Grande, Alfredo Herrera-Estrella, Carla Ivon Herrera-Najera, Pavel Isa, Brenda Irasema Maldonado-Meza, Bernardo Martinez-Miguel, Margarita Matias-Florentino, Maria Guadalupe de Jesus Mireles-Rivera, Gloria Maria Molina-Salinas, Hector Montoya-Fuentes, Jose Esteban Munoz-Medina, Jose de Jesus Nunez-Contreras, Alicia Ocana-Mondragon, Luis Alberto Ochoa-Carrera, Hector Esteban Paz-Juarez, Francisco Pulido, Helen Haydee Fernanda Ramirez-Plascencia, Angel Gustavo Salas-Lais, Alejandro Sanchez-Flores, Clara Esperanza Santacruz-Tinoco, Maria Guadalupe Santiago-Mauricio, Nelly Selem-Mojica, Blanca Taboada, Gloria Vazquez |
| EPI_ISL_1662123, EPI_ISL_1662124, EPI_ISL_1662125, EPI_ISL_1662126, EPI_ISL_1662127 | Centro de Investigacion Biomedica de Occidente (CIBO) | Unidad de Genomica Avanzada | Consorcio Mexicano de Vigilancia Genomica (CoViGen-Mex). Authors (in alphabetical order): Julio Elias Alvarado-Yaah, Carlos F. Arias, Santiago Ávila-Rios, Victor Hugo Borja-Aburto, Celia Boukadida, Juan Bautista Chale-Dzul, Celida Duque Molina, Jose Antonio Enciso-Moreno, Gloria Elena Espinosa-Ayala, Fernando Fontove-Herrera, Victor Eduardo Garcia-Arias,Concepcion Grajales-Muniz, Ricardo Grande, Alfredo Herrera-Estrella, Carla Ivon Herrera-Najera, Pavel Isa, Brenda Irasema Maldonado-Meza, Bernardo Martinez-Miguel, Margarita Matias-Florentino, Maria Guadalupe de Jesus Mireles-Rivera, Gloria Maria Molina-Salinas, Hector Montoya-Fuentes, Jose Esteban Munoz-Medina, Jose de Jesus Nunez-Contreras, Alicia Ocana-Mondragon, Luis Alberto Ochoa-Carrera, Hector Esteban Paz-Juarez, Francisco Pulido, Helen Haydee Fernanda Ramirez-Plascencia, Angel Gustavo Salas-Lais, Alejandro Sanchez-Flores, Clara Esperanza Santacruz-Tinoco, Maria Guadalupe Santiago-Mauricio, Nelly Selem-Mojica, Blanca Taboada, Gloria Vazquez |
| EPI_ISL_1662128, EPI_ISL_1662129, EPI_ISL_1662130 | Laboratorio Central de Epidemiologia (LCE) | Unidad de Genomica Avanzada | Consorcio Mexicano de Vigilancia Genomica (CoViGen-Mex). Authors (in alphabetical order): Julio Elias Alvarado-Yaah, Carlos F. Arias, Santiago Ávila-Rios, Victor Hugo Borja-Aburto, Celia Boukadida, Juan Bautista Chale-Dzul, Celida Duque Molina, Jose Antonio Enciso-Moreno, Gloria Elena Espinosa-Ayala, Fernando Fontove-Herrera, Victor Eduardo Garcia-Arias,Concepcion Grajales-Muniz, Ricardo Grande, Alfredo Herrera-Estrella, Carla Ivon Herrera-Najera, Pavel Isa, Brenda Irasema Maldonado-Meza, Bernardo Martinez-Miguel, Margarita Matias-Florentino, Maria Guadalupe de Jesus Mireles-Rivera, Gloria Maria Molina-Salinas, Hector Montoya-Fuentes, Jose Esteban Munoz-Medina, Jose de Jesus Nunez-Contreras, Alicia Ocana-Mondragon, Luis Alberto Ochoa-Carrera, Hector Esteban Paz-Juarez, Francisco Pulido, Helen Haydee Fernanda Ramirez-Plascencia, Angel Gustavo Salas-Lais, Alejandro Sanchez-Flores, Clara Esperanza Santacruz-Tinoco, Maria Guadalupe Santiago-Mauricio, Nelly Selem-Mojica, Blanca Taboada, Gloria Vazquez |
| EPI_ISL_1662131, EPI_ISL_1662132 | Centro de Investigacion Biomedica de Occidente (CIBO) | Unidad de Genomica Avanzada | Consorcio Mexicano de Vigilancia Genomica (CoViGen-Mex). Authors (in alphabetical order): Julio Elias Alvarado-Yaah, Carlos F. Arias, Santiago Ávila-Rios, Victor Hugo Borja-Aburto, Celia Boukadida, Juan Bautista Chale-Dzul, Celida Duque Molina, Jose Antonio Enciso-Moreno, Gloria Elena Espinosa-Ayala, Fernando Fontove-Herrera, Victor Eduardo Garcia-Arias,Concepcion Grajales-Muniz, Ricardo Grande, Alfredo Herrera-Estrella, Carla Ivon Herrera-Najera, Pavel Isa, Brenda Irasema Maldonado-Meza, Bernardo Martinez-Miguel, Margarita Matias-Florentino, Maria Guadalupe de Jesus Mireles-Rivera, Gloria Maria Molina-Salinas, Hector Montoya-Fuentes, Jose Esteban Munoz-Medina, Jose de Jesus Nunez-Contreras, Alicia Ocana-Mondragon, Luis Alberto Ochoa-Carrera, Hector Esteban Paz-Juarez, Francisco Pulido, Helen Haydee Fernanda Ramirez-Plascencia, Angel Gustavo Salas-Lais, Alejandro Sanchez-Flores, Clara Esperanza Santacruz-Tinoco, Maria Guadalupe Santiago-Mauricio, Nelly Selem-Mojica, Blanca Taboada, Gloria Vazquez |
| EPI_ISL_1662133, EPI_ISL_1662134, EPI_ISL_1662135, EPI_ISL_1662136, EPI_ISL_1662137, EPI_ISL_1662138, EPI_ISL_1662139, EPI_ISL_1662140, EPI_ISL_1662141, EPI_ISL_1662142, EPI_ISL_1662143, EPI_ISL_1662144 | | | |
| see above | Centro de Investigacion Biomedica del Noreste (CIBIN) | Unidad de Genomica Avanzada | Consorcio Mexicano de Vigilancia Genomica (CoViGen-Mex). Authors (in alphabetical order): Julio Elias Alvarado-Yaah, Carlos F. Arias, Santiago Ávila-Rios, Victor Hugo Borja-Aburto, Celia Boukadida, Juan Bautista Chale-Dzul, Celida Duque Molina, Jose Antonio Enciso-Moreno, Gloria Elena Espinosa-Ayala, Fernando Fontove-Herrera, Victor Eduardo Garcia-Arias,Concepcion Grajales-Muniz, Ricardo Grande, Alfredo Herrera-Estrella, Carla Ivon Herrera-Najera, Pavel Isa, Brenda Irasema Maldonado-Meza, Bernardo Martinez-Miguel, Margarita Matias-Florentino, Maria Guadalupe de Jesus Mireles-Rivera, Gloria Maria Molina-Salinas, Hector Montoya-Fuentes, Jose Esteban Munoz-Medina, Jose de Jesus Nunez-Contreras, Alicia Ocana-Mondragon, Luis Alberto Ochoa-Carrera, Hector Esteban Paz-Juarez, Francisco Pulido, Helen Haydee Fernanda Ramirez-Plascencia, Angel Gustavo Salas-Lais, Alejandro Sanchez-Flores, Clara Esperanza Santacruz-Tinoco, Maria Guadalupe Santiago-Mauricio, Nelly Selem-Mojica, Blanca Taboada, Gloria Vazquez |
| EPI_ISL_1662145, EPI_ISL_1662146, EPI_ISL_1662147, EPI_ISL_1662148, EPI_ISL_1662149, EPI_ISL_1662150, EPI_ISL_1662151, EPI_ISL_1662152, EPI_ISL_1662153, EPI_ISL_1662154, EPI_ISL_1662155, EPI_ISL_1662156, EPI_ISL_1662157, EPI_ISL_1662158, EPI_ISL_1662159, EPI_ISL_1662160, EPI_ISL_1662161, EPI_ISL_1662162, EPI_ISL_1662163, EPI_ISL_1662164, EPI_ISL_1662165, EPI_ISL_1662166, EPI_ISL_1662167, EPI_ISL_1662168, EPI_ISL_1662169, EPI_ISL_1662170, EPI_ISL_1662171, EPI_ISL_1662172, EPI_ISL_1662173, EPI_ISL_1662174, EPI_ISL_1662175, EPI_ISL_1662176, EPI_ISL_1662177, EPI_ISL_1662178, EPI_ISL_1662179, EPI_ISL_1662180, EPI_ISL_1662181, EPI_ISL_1662182, EPI_ISL_1662183 | | | |
| see above | Laboratorio Central de Epidemiologia (LCE) | Unidad de Genomica Avanzada | Consorcio Mexicano de Vigilancia Genomica (CoViGen-Mex). Authors (in alphabetical order): Julio Elias Alvarado-Yaah, Carlos F. Arias, Santiago Ávila-Rios, Victor Hugo Borja-Aburto, Celia Boukadida, Juan Bautista Chale-Dzul, Celida Duque Molina, Jose Antonio Enciso-Moreno, Gloria Elena Espinosa-Ayala, Fernando Fontove-Herrera, Victor Eduardo Garcia-Arias,Concepcion Grajales-Muniz, Ricardo Grande, Alfredo Herrera-Estrella, Carla Ivon Herrera-Najera, Pavel Isa, Brenda Irasema Maldonado-Meza, Bernardo Martinez-Miguel, Margarita Matias-Florentino, Maria Guadalupe de Jesus Mireles-Rivera, Gloria Maria Molina-Salinas, Hector Montoya-Fuentes, Jose Esteban Munoz-Medina, Jose de Jesus Nunez-Contreras, Alicia Ocana-Mondragon, Luis Alberto Ochoa-Carrera, Hector Esteban Paz-Juarez, Francisco Pulido, Helen Haydee Fernanda Ramirez-Plascencia, Angel Gustavo Salas-Lais, Alejandro Sanchez-Flores, Clara Esperanza Santacruz-Tinoco, Maria Guadalupe Santiago-Mauricio, Nelly Selem-Mojica, Blanca Taboada, Gloria Vazquez |
| EPI_ISL_1662184, EPI_ISL_1662185 | Unidad de Investigacion Medica de Yucatan (UIMY) | Unidad de Genomica Avanzada | Consorcio Mexicano de Vigilancia Genomica (CoViGen-Mex). Authors (in alphabetical order): Julio Elias Alvarado-Yaah, Carlos F. Arias, Santiago Ávila-Rios, Victor Hugo Borja-Aburto, Celia Boukadida, Juan Bautista Chale-Dzul, Celida Duque Molina, Jose Antonio Enciso-Moreno, Gloria Elena Espinosa-Ayala, Fernando Fontove-Herrera, Victor Eduardo Garcia-Arias,Concepcion Grajales-Muniz, Ricardo Grande, Alfredo Herrera-Estrella, Carla Ivon Herrera-Najera, Pavel Isa, Brenda Irasema Maldonado-Meza, Bernardo Martinez-Miguel, Margarita Matias-Florentino, Maria Guadalupe de Jesus Mireles-Rivera, Gloria Maria Molina-Salinas, Hector Montoya-Fuentes, Jose Esteban Munoz-Medina, Jose de Jesus Nunez-Contreras, Alicia Ocana-Mondragon, Luis Alberto Ochoa-Carrera, Hector Esteban Paz-Juarez, Francisco Pulido, Helen Haydee Fernanda Ramirez-Plascencia, Angel Gustavo Salas-Lais, Alejandro Sanchez-Flores, Clara Esperanza Santacruz-Tinoco, Maria Guadalupe Santiago-Mauricio, Nelly Selem-Mojica, Blanca Taboada, Gloria Vazquez |
| EPI_ISL_1662186, EPI_ISL_1662187, EPI_ISL_1662188, EPI_ISL_1662189, EPI_ISL_1662190, EPI_ISL_1662191, EPI_ISL_1662192, EPI_ISL_1662193 | Centro de Investigacion Biomedica de Occidente (CIBO) | Unidad de Genomica Avanzada | Consorcio Mexicano de Vigilancia Genomica (CoViGen-Mex). Authors (in alphabetical order): Julio Elias Alvarado-Yaah, Carlos F. Arias, Santiago Ávila-Rios, Victor Hugo Borja-Aburto, Celia Boukadida, Juan Bautista Chale-Dzul, Celida Duque Molina, Jose Antonio Enciso-Moreno, Gloria Elena Espinosa-Ayala, Fernando Fontove-Herrera, Victor Eduardo Garcia-Arias,Concepcion Grajales-Muniz, Ricardo Grande, Alfredo Herrera-Estrella, Carla Ivon Herrera-Najera, Pavel Isa, Brenda Irasema Maldonado-Meza, Bernardo Martinez-Miguel, Margarita Matias-Florentino, Maria Guadalupe de Jesus Mireles-Rivera, Gloria Maria Molina-Salinas, Hector Montoya-Fuentes, Jose Esteban Munoz-Medina, Jose de Jesus Nunez-Contreras, Alicia Ocana-Mondragon, Luis Alberto Ochoa-Carrera, Hector Esteban Paz-Juarez, Francisco Pulido, Helen Haydee Fernanda Ramirez-Plascencia, Angel Gustavo Salas-Lais, Alejandro Sanchez-Flores, Clara Esperanza Santacruz-Tinoco, Maria Guadalupe Santiago-Mauricio, Nelly Selem-Mojica, Blanca Taboada, Gloria Vazquez |
| EPI_ISL_1662194, | Laboratorio Central de Epidemiologia | Unidad de Genomica Avanzada | Consorcio Mexicano de Vigilancia Genomica (CoViGen-Mex). Authors (in alphabetical order): Julio Elias Alvarado-Yaah, Carlos F. Arias, Santiago Ávila-Rios, Victor Hugo Borja-Aburto, Celia Boukadida, Juan Bautista Chale-Dzul, Celida Duque Molina, Jose Antonio Enciso-Moreno, Gloria |

| | | | |
|---|---|---|---|
| EPI_ISL_1662195, EPI_ISL_1662196 | (LCE) | | Elena Espinosa-Ayala, Fernando Fontove-Herrera, Victor Eduardo Garcia-Arias,Concepcion Grajales-Muniz, Ricardo Grande, Alfredo Herrera-Estrella, Carla Ivon Herrera-Najera, Pavel Isa, Brenda Irasema Maldonado-Meza, Bernardo Martinez-Miguel, Margarita Matias-Florentino, Maria Guadalupe de Jesus Mireles-Rivera, Gloria Maria Molina-Salinas, Hector Montoya-Fuentes, Jose Esteban Munoz-Medina, Jose de Jesus Nunez-Contreras, Alicia Ocana-Mondragon, Luis Alberto Ochoa-Carrera, Hector Esteban Paz-Juarez, Francisco Pulido, Helen Haydee Fernanda Ramirez-Plascencia, Angel Gustavo Salas-Lais, Alejandro Sanchez-Flores, Clara Esperanza Santacruz-Tinoco, Maria Guadalupe Santiago-Mauricio, Nelly Selem-Mojica, Blanca Taboada, Gloria Vazquez |
| EPI_ISL_1662197, EPI_ISL_1662198 | Centro de Investigacion Biomedica de Occidente (CIBO) | Unidad de Genomica Avanzada | Consorcio Mexicano de Vigilancia Genomica (CoViGen-Mex). Authors (in alphabetical order): Julio Elias Alvarado-Yaah, Carlos F. Arias, Santiago Ávila-Rios, Victor Hugo Borja-Aburto, Celia Boukadida, Juan Bautista Chale-Dzul, Celida Duque Molina, Jose Antonio Enciso-Moreno, Gloria Elena Espinosa-Ayala, Fernando Fontove-Herrera, Victor Eduardo Garcia-Arias,Concepcion Grajales-Muniz, Ricardo Grande, Alfredo Herrera-Estrella, Carla Ivon Herrera-Najera, Pavel Isa, Brenda Irasema Maldonado-Meza, Bernardo Martinez-Miguel, Margarita Matias-Florentino, Maria Guadalupe de Jesus Mireles-Rivera, Gloria Maria Molina-Salinas, Hector Montoya-Fuentes, Jose Esteban Munoz-Medina, Jose de Jesus Nunez-Contreras, Alicia Ocana-Mondragon, Luis Alberto Ochoa-Carrera, Hector Esteban Paz-Juarez, Francisco Pulido, Helen Haydee Fernanda Ramirez-Plascencia, Angel Gustavo Salas-Lais, Alejandro Sanchez-Flores, Clara Esperanza Santacruz-Tinoco, Maria Guadalupe Santiago-Mauricio, Nelly Selem-Mojica, Blanca Taboada, Gloria Vazquez |
| EPI_ISL_1662199, EPI_ISL_1662200 | Centro de Investigacion Biomedica del Noreste (CIBIN) | Unidad de Genomica Avanzada | Consorcio Mexicano de Vigilancia Genomica (CoViGen-Mex). Authors (in alphabetical order): Julio Elias Alvarado-Yaah, Carlos F. Arias, Santiago Ávila-Rios, Victor Hugo Borja-Aburto, Celia Boukadida, Juan Bautista Chale-Dzul, Celida Duque Molina, Jose Antonio Enciso-Moreno, Gloria Elena Espinosa-Ayala, Fernando Fontove-Herrera, Victor Eduardo Garcia-Arias,Concepcion Grajales-Muniz, Ricardo Grande, Alfredo Herrera-Estrella, Carla Ivon Herrera-Najera, Pavel Isa, Brenda Irasema Maldonado-Meza, Bernardo Martinez-Miguel, Margarita Matias-Florentino, Maria Guadalupe de Jesus Mireles-Rivera, Gloria Maria Molina-Salinas, Hector Montoya-Fuentes, Jose Esteban Munoz-Medina, Jose de Jesus Nunez-Contreras, Alicia Ocana-Mondragon, Luis Alberto Ochoa-Carrera, Hector Esteban Paz-Juarez, Francisco Pulido, Helen Haydee Fernanda Ramirez-Plascencia, Angel Gustavo Salas-Lais, Alejandro Sanchez-Flores, Clara Esperanza Santacruz-Tinoco, Maria Guadalupe Santiago-Mauricio, Nelly Selem-Mojica, Blanca Taboada, Gloria Vazquez |
| EPI_ISL_1662201, EPI_ISL_1662202, EPI_ISL_1662203, EPI_ISL_1662204, EPI_ISL_1662205, EPI_ISL_1662206, EPI_ISL_1662207, EPI_ISL_1662208, EPI_ISL_1662209, EPI_ISL_1662210, EPI_ISL_1662211, EPI_ISL_1662212 | | | |
| see above | Laboratorio Central de Epidemiologia (LCE) | Unidad de Genomica Avanzada | Consorcio Mexicano de Vigilancia Genomica (CoViGen-Mex). Authors (in alphabetical order): Julio Elias Alvarado-Yaah, Carlos F. Arias, Santiago Ávila-Rios, Victor Hugo Borja-Aburto, Celia Boukadida, Juan Bautista Chale-Dzul, Celida Duque Molina, Jose Antonio Enciso-Moreno, Gloria Elena Espinosa-Ayala, Fernando Fontove-Herrera, Victor Eduardo Garcia-Arias,Concepcion Grajales-Muniz, Ricardo Grande, Alfredo Herrera-Estrella, Carla Ivon Herrera-Najera, Pavel Isa, Brenda Irasema Maldonado-Meza, Bernardo Martinez-Miguel, Margarita Matias-Florentino, Maria Guadalupe de Jesus Mireles-Rivera, Gloria Maria Molina-Salinas, Hector Montoya-Fuentes, Jose Esteban Munoz-Medina, Jose de Jesus Nunez-Contreras, Alicia Ocana-Mondragon, Luis Alberto Ochoa-Carrera, Hector Esteban Paz-Juarez, Francisco Pulido, Helen Haydee Fernanda Ramirez-Plascencia, Angel Gustavo Salas-Lais, Alejandro Sanchez-Flores, Clara Esperanza Santacruz-Tinoco, Maria Guadalupe Santiago-Mauricio, Nelly Selem-Mojica, Blanca Taboada, Gloria Vazquez |
| EPI_ISL_1662213, EPI_ISL_1662214, EPI_ISL_1662215, EPI_ISL_1662216, EPI_ISL_1662217, EPI_ISL_1662218, EPI_ISL_1662219, EPI_ISL_1662220 | Unidad de Investigacion Medica de Yucatan (UIMY) | Unidad de Genomica Avanzada | Consorcio Mexicano de Vigilancia Genomica (CoViGen-Mex). Authors (in alphabetical order): Julio Elias Alvarado-Yaah, Carlos F. Arias, Santiago Ávila-Rios, Victor Hugo Borja-Aburto, Celia Boukadida, Juan Bautista Chale-Dzul, Celida Duque Molina, Jose Antonio Enciso-Moreno, Gloria Elena Espinosa-Ayala, Fernando Fontove-Herrera, Victor Eduardo Garcia-Arias,Concepcion Grajales-Muniz, Ricardo Grande, Alfredo Herrera-Estrella, Carla Ivon Herrera-Najera, Pavel Isa, Brenda Irasema Maldonado-Meza, Bernardo Martinez-Miguel, Margarita Matias-Florentino, Maria Guadalupe de Jesus Mireles-Rivera, Gloria Maria Molina-Salinas, Hector Montoya-Fuentes, Jose Esteban Munoz-Medina, Jose de Jesus Nunez-Contreras, Alicia Ocana-Mondragon, Luis Alberto Ochoa-Carrera, Hector Esteban Paz-Juarez, Francisco Pulido, Helen Haydee Fernanda Ramirez-Plascencia, Angel Gustavo Salas-Lais, Alejandro Sanchez-Flores, Clara Esperanza Santacruz-Tinoco, Maria Guadalupe Santiago-Mauricio, Nelly Selem-Mojica, Blanca Taboada, Gloria Vazquez |
| EPI_ISL_1662221, EPI_ISL_1662222, EPI_ISL_1662223, EPI_ISL_1662224, EPI_ISL_1662225, EPI_ISL_1662226, EPI_ISL_1662227, EPI_ISL_1662228, EPI_ISL_1662229, EPI_ISL_1662230, EPI_ISL_1662231, EPI_ISL_1662232 | | | |
| see above | Unidad de Investigacion Biomedica de Zacatecas (UIBZ) | Unidad de Genomica Avanzada | Consorcio Mexicano de Vigilancia Genomica (CoViGen-Mex). Authors (in alphabetical order): Julio Elias Alvarado-Yaah, Carlos F. Arias, Santiago Ávila-Rios, Victor Hugo Borja-Aburto, Celia Boukadida, Juan Bautista Chale-Dzul, Celida Duque Molina, Jose Antonio Enciso-Moreno, Gloria Elena Espinosa-Ayala, Fernando Fontove-Herrera, Victor Eduardo Garcia-Arias,Concepcion Grajales-Muniz, Ricardo Grande, Alfredo Herrera-Estrella, Carla Ivon Herrera-Najera, Pavel Isa, Brenda Irasema Maldonado-Meza, Bernardo Martinez-Miguel, Margarita Matias-Florentino, Maria Guadalupe de Jesus Mireles-Rivera, Gloria Maria Molina-Salinas, Hector Montoya-Fuentes, Jose Esteban Munoz-Medina, Jose de Jesus Nunez-Contreras, Alicia Ocana-Mondragon, Luis Alberto Ochoa-Carrera, Hector Esteban Paz-Juarez, Francisco Pulido, Helen Haydee Fernanda Ramirez-Plascencia, Angel Gustavo Salas-Lais, Alejandro Sanchez-Flores, Clara Esperanza Santacruz-Tinoco, Maria Guadalupe Santiago-Mauricio, Nelly Selem-Mojica, Blanca Taboada, Gloria Vazquez |
| EPI_ISL_1662233 | WI State Laboratory of Hygiene | Pathogen Discovery, Respiratory Viruses Branch, Division of Viral Diseases, Centers for Disease Control and Prevention | Ying Tao, Yan Li, Anna Kelleher, Jing Zhang, Anna Montmayeur, Brian Lynch, Krista Queen, Anna Uehara, Peter Cook, Han Jia Justin Ng, Rachel Marine, Clinton R. Paden, Haibin Wang, Mark Burroughs, Justin Lee, Adam Retchless, Suxiang Tong |
| EPI_ISL_1662235 | WVDHHR - Office of Laboratory Services | Pathogen Discovery, Respiratory Viruses Branch, Division of Viral Diseases, Centers for Disease Control and Prevention | Ying Tao, Yan Li, Anna Kelleher, Jing Zhang, Anna Montmayeur, Brian Lynch, Krista Queen, Anna Uehara, Peter Cook, Han Jia Justin Ng, Rachel Marine, Clinton R. Paden, Haibin Wang, Mark Burroughs, Justin Lee, Adam Retchless, Suxiang Tong |
| EPI_ISL_1662236 | AR Dept. of Health-PHL, Molecular Diagnostics | Pathogen Discovery, Respiratory Viruses Branch, Division of Viral Diseases, Centers for Disease Control and Prevention | Ying Tao, Yan Li, Anna Kelleher, Jing Zhang, Anna Montmayeur, Brian Lynch, Krista Queen, Anna Uehara, Peter Cook, Han Jia Justin Ng, Rachel Marine, Clinton R. Paden, Haibin Wang, Mark Burroughs, Justin Lee, Adam Retchless, Suxiang Tong |
| EPI_ISL_1662237 | NC State Laboratory of Public Health | Pathogen Discovery, Respiratory Viruses Branch, Division of Viral Diseases, Centers for Disease Control and Prevention | Ying Tao, Yan Li, Anna Kelleher, Jing Zhang, Anna Montmayeur, Brian Lynch, Krista Queen, Anna Uehara, Peter Cook, Han Jia Justin Ng, Rachel Marine, Clinton R. Paden, Haibin Wang, Mark Burroughs, Justin Lee, Adam Retchless, Suxiang Tong |
| EPI_ISL_1662238, EPI_ISL_1662239 | GA Department of Public Health Laboratory | Pathogen Discovery, Respiratory Viruses Branch, Division of Viral Diseases, Centers for Disease Control and Prevention | Ying Tao, Yan Li, Anna Kelleher, Jing Zhang, Anna Montmayeur, Brian Lynch, Krista Queen, Anna Uehara, Peter Cook, Han Jia Justin Ng, Rachel Marine, Clinton R. Paden, Haibin Wang, Mark Burroughs, Justin Lee, Adam Retchless, Suxiang Tong |
| EPI_ISL_1662520, EPI_ISL_1662521, EPI_ISL_1662522, EPI_ISL_1662523, EPI_ISL_1662524, EPI_ISL_1662525, EPI_ISL_1662526 | WI State Laboratory of Hygiene | Pathogen Discovery, Respiratory Viruses Branch, Division of Viral Diseases, Centers for Disease Control and Prevention | Ying Tao, Yan Li, Anna Kelleher, Jing Zhang, Anna Montmayeur, Brian Lynch, Krista Queen, Anna Uehara, Peter Cook, Han Jia Justin Ng, Rachel Marine, Clinton R. Paden, Haibin Wang, Mark Burroughs, Justin Lee, Adam Retchless, Suxiang Tong |
| EPI_ISL_1662596, EPI_ISL_1662598, EPI_ISL_1662599, EPI_ISL_1662600, EPI_ISL_1662601, EPI_ISL_1662602, EPI_ISL_1662603, EPI_ISL_1662604, EPI_ISL_1662605, EPI_ISL_1662606, EPI_ISL_1662607, EPI_ISL_1662608, EPI_ISL_1662609, EPI_ISL_1662610, EPI_ISL_1662611, EPI_ISL_1662612, EPI_ISL_1662613, EPI_ISL_1662614, EPI_ISL_1662615, EPI_ISL_1662616, EPI_ISL_1662617, EPI_ISL_1662618 | | | |
| see above | Wyoming Public Health Laboratory | Wyoming Public Health Laboratory | Jim Mildenberger, Wanda Manley, Noah Hull, Taylor Fearing, Lynette Gumbleton, Channing Weber, Ashley Norberg, Chayse Rowley, Marley Goetz, Brian Dominguez, Elliot Thomasson, Sam Britz, Cari Sloma, and Rob Christensen |
| EPI_ISL_1662621, EPI_ISL_1662622, EPI_ISL_1662623, EPI_ISL_1662624, EPI_ISL_1662625, EPI_ISL_1662626, EPI_ISL_1662628, EPI_ISL_1662629, EPI_ISL_1662630, EPI_ISL_1662631, EPI_ISL_1662632, EPI_ISL_1662633, EPI_ISL_1662634, EPI_ISL_1662635, EPI_ISL_1662636, EPI_ISL_1662637, EPI_ISL_1662638, EPI_ISL_1662639, EPI_ISL_1662640, EPI_ISL_1662641, EPI_ISL_1662642, EPI_ISL_1662643, EPI_ISL_1662644, EPI_ISL_1662645, EPI_ISL_1662646, EPI_ISL_1662647, EPI_ISL_1662648, EPI_ISL_1662649, EPI_ISL_1662650, EPI_ISL_1662651, EPI_ISL_1662652, EPI_ISL_1662653, EPI_ISL_1662655, EPI_ISL_1662656, EPI_ISL_1662657, EPI_ISL_1662658, EPI_ISL_1662659, EPI_ISL_1662660, EPI_ISL_1662661, EPI_ISL_1662662, EPI_ISL_1662663, EPI_ISL_1662664, EPI_ISL_1662665, EPI_ISL_1662666, EPI_ISL_1662667, EPI_ISL_1662669, EPI_ISL_1662670, EPI_ISL_1662672, EPI_ISL_1662674, EPI_ISL_1662675, EPI_ISL_1662676, EPI_ISL_1662678, EPI_ISL_1662679, EPI_ISL_1662680, EPI_ISL_1662682, EPI_ISL_1662683, EPI_ISL_1662684, EPI_ISL_1662685, EPI_ISL_1662686, EPI_ISL_1662687, EPI_ISL_1662688, EPI_ISL_1662689, EPI_ISL_1662690, EPI_ISL_1662691, EPI_ISL_1662692, EPI_ISL_1662693, EPI_ISL_1662695, EPI_ISL_1662696, EPI_ISL_1662697, EPI_ISL_1662698, EPI_ISL_1662699, EPI_ISL_1662700 | | | |
| see above | Delaware Public Health Lab | Delaware Public Health Lab | Rebecca Savage |
| EPI_ISL_1662701, EPI_ISL_1662702, EPI_ISL_1662703, EPI_ISL_1662704, EPI_ISL_1662705, EPI_ISL_1662706, EPI_ISL_1662707, EPI_ISL_1662708, EPI_ISL_1662709, EPI_ISL_1662710, EPI_ISL_1662711, EPI_ISL_1662712, EPI_ISL_1662713, EPI_ISL_1662714, EPI_ISL_1662715, EPI_ISL_1662716, EPI_ISL_1662717, EPI_ISL_1662718, EPI_ISL_1662719, EPI_ISL_1662720, EPI_ISL_1662721, EPI_ISL_1662722, EPI_ISL_1662723, EPI_ISL_1662724, EPI_ISL_1662725, EPI_ISL_1662726, EPI_ISL_1662727, EPI_ISL_1662728, EPI_ISL_1662729, EPI_ISL_1662730, EPI_ISL_1662731, EPI_ISL_1662732, EPI_ISL_1662733, EPI_ISL_1662734, EPI_ISL_1662735, EPI_ISL_1662736, EPI_ISL_1662737, EPI_ISL_1662738, EPI_ISL_1662739, EPI_ISL_1662740, EPI_ISL_1662741, EPI_ISL_1662742, EPI_ISL_1662743, EPI_ISL_1662744, EPI_ISL_1662745, EPI_ISL_1662746, EPI_ISL_1662747, EPI_ISL_1662748, EPI_ISL_1662749, EPI_ISL_1662750, EPI_ISL_1662751, EPI_ISL_1662752, EPI_ISL_1662753, EPI_ISL_1662754, EPI_ISL_1662755, EPI_ISL_1662756, EPI_ISL_1662757, EPI_ISL_1662758, EPI_ISL_1662759 | | | |
| see above | Marshfield Labs | Pathogen Discovery, Respiratory Viruses Branch, Division of Viral Diseases, Centers for Disease Control and Prevention | Ying Tao, Yan Li, Anna Kelleher, Jing Zhang, Anna Montmayeur, Brian Lynch, Krista Queen, Anna Uehara, Peter Cook, Han Jia Justin Ng, Rachel Marine, Clinton R. Paden, Haibin Wang, Mark Burroughs, Justin Lee, Adam Retchless, Suxiang Tong |
| EPI_ISL_1662762, EPI_ISL_1662763, EPI_ISL_1662764, EPI_ISL_1662765, EPI_ISL_1662766, EPI_ISL_1662767, EPI_ISL_1662768, EPI_ISL_1662769, EPI_ISL_1662770, EPI_ISL_1662771, EPI_ISL_1662772, EPI_ISL_1662773, EPI_ISL_1662774, EPI_ISL_1662775, EPI_ISL_1662776, EPI_ISL_1662777, EPI_ISL_1662778, EPI_ISL_1662779, EPI_ISL_1662780, EPI_ISL_1662781, EPI_ISL_1662782, EPI_ISL_1662783, EPI_ISL_1662784, EPI_ISL_1662785, EPI_ISL_1662786, EPI_ISL_1662787, EPI_ISL_1662788, EPI_ISL_1662789, EPI_ISL_1662790, EPI_ISL_1662791, EPI_ISL_1662792, EPI_ISL_1662793, EPI_ISL_1662794, EPI_ISL_1662795, EPI_ISL_1662796, EPI_ISL_1662797, EPI_ISL_1662798, EPI_ISL_1662799, EPI_ISL_1662800, EPI_ISL_1662801, EPI_ISL_1662802, EPI_ISL_1662803, EPI_ISL_1662804, EPI_ISL_1662805, EPI_ISL_1662806, EPI_ISL_1662807, EPI_ISL_1662808, EPI_ISL_1662809, EPI_ISL_1662810, EPI_ISL_1662811, EPI_ISL_1662812, EPI_ISL_1662813, EPI_ISL_1662814, EPI_ISL_1662815, EPI_ISL_1662817, EPI_ISL_1662818, EPI_ISL_1662819, EPI_ISL_1662821, EPI_ISL_1662822, EPI_ISL_1662823, EPI_ISL_1662824, EPI_ISL_1662825, EPI_ISL_1662826, EPI_ISL_1662827, EPI_ISL_1662828, EPI_ISL_1662829, EPI_ISL_1662830, EPI_ISL_1662831, EPI_ISL_1662832, EPI_ISL_1662833, EPI_ISL_1662834, EPI_ISL_1662835, EPI_ISL_1662836, EPI_ISL_1662837, EPI_ISL_1662838, EPI_ISL_1662839, EPI_ISL_1662840, EPI_ISL_1662841, EPI_ISL_1662842, EPI_ISL_1662843, EPI_ISL_1662844, EPI_ISL_1662845, EPI_ISL_1662846, EPI_ISL_1662847, EPI_ISL_1662848, EPI_ISL_1662849, EPI_ISL_1662850, EPI_ISL_1662851, EPI_ISL_1662852, EPI_ISL_1662853, EPI_ISL_1662854, EPI_ISL_1662855, EPI_ISL_1662856, EPI_ISL_1662857, EPI_ISL_1662858, EPI_ISL_1662859, EPI_ISL_1662860, EPI_ISL_1662861, EPI_ISL_1662862, EPI_ISL_1662863, EPI_ISL_1662864, EPI_ISL_1662865, EPI_ISL_1662866, EPI_ISL_1662867, EPI_ISL_1662868, EPI_ISL_1662869, EPI_ISL_1662870, EPI_ISL_1662871, EPI_ISL_1662872, EPI_ISL_1662873, EPI_ISL_1662874, EPI_ISL_1662875, EPI_ISL_1662876, EPI_ISL_1662877, EPI_ISL_1662879, EPI_ISL_1662880, EPI_ISL_1662881, EPI_ISL_1662882, EPI_ISL_1662883, EPI_ISL_1662884, EPI_ISL_1662885, EPI_ISL_1662886, EPI_ISL_1662887, EPI_ISL_1662888, EPI_ISL_1662889, EPI_ISL_1662890, EPI_ISL_1662891, EPI_ISL_1662892, EPI_ISL_1662893, EPI_ISL_1662894, EPI_ISL_1662895, EPI_ISL_1662896, EPI_ISL_1662897, EPI_ISL_1662898, EPI_ISL_1662899, EPI_ISL_1662900, EPI_ISL_1662901, EPI_ISL_1662902, EPI_ISL_1662903, EPI_ISL_1662904, EPI_ISL_1662905, EPI_ISL_1662906, EPI_ISL_1662907, EPI_ISL_1662908, EPI_ISL_1662909, EPI_ISL_1662910, EPI_ISL_1662911, EPI_ISL_1662912, EPI_ISL_1662913, EPI_ISL_1662914, EPI_ISL_1662915, EPI_ISL_1662916, EPI_ISL_1662917, EPI_ISL_1662918, EPI_ISL_1662919, EPI_ISL_1662921, EPI_ISL_1662922, EPI_ISL_1662923, EPI_ISL_1662924, EPI_ISL_1662925, EPI_ISL_1662926, EPI_ISL_1662927, EPI_ISL_1662928, EPI_ISL_1662929, EPI_ISL_1662931, EPI_ISL_1662932, EPI_ISL_1662934, EPI_ISL_1662935, EPI_ISL_1662936, EPI_ISL_1662937, EPI_ISL_1662938, EPI_ISL_1662939, EPI_ISL_1662940, EPI_ISL_1662941, EPI_ISL_1662942, EPI_ISL_1662943, EPI_ISL_1662944, EPI_ISL_1662945, EPI_ISL_1662946, EPI_ISL_1662947, EPI_ISL_1662948, EPI_ISL_1662949, EPI_ISL_1662950, EPI_ISL_1662951, EPI_ISL_1662952, EPI_ISL_1662953, EPI_ISL_1662954, EPI_ISL_1662955, EPI_ISL_1662957, EPI_ISL_1662958, EPI_ISL_1662959, EPI_ISL_1662960, EPI_ISL_1662962, EPI_ISL_1662963, EPI_ISL_1662964, EPI_ISL_1662965, EPI_ISL_1662966, EPI_ISL_1662967, EPI_ISL_1662969, EPI_ISL_1662970, EPI_ISL_1662971, EPI_ISL_1662972, EPI_ISL_1662973, EPI_ISL_1662974, EPI_ISL_1662975, EPI_ISL_1662976, EPI_ISL_1662977, EPI_ISL_1662978, EPI_ISL_1662979, EPI_ISL_1662980, EPI_ISL_1662982, EPI_ISL_1662983, EPI_ISL_1662984, EPI_ISL_1662985, EPI_ISL_1662986, EPI_ISL_1662987, EPI_ISL_1662988, EPI_ISL_1662990, EPI_ISL_1662991, EPI_ISL_1662992, EPI_ISL_1662993, EPI_ISL_1662994, EPI_ISL_1662995, EPI_ISL_1662996, EPI_ISL_1662997, EPI_ISL_1662998, EPI_ISL_1662999, EPI_ISL_1663000, EPI_ISL_1663001, EPI_ISL_1663002, EPI_ISL_1663003, EPI_ISL_1663004, EPI_ISL_1663005, EPI_ISL_1663006, EPI_ISL_1663007, EPI_ISL_1663008, EPI_ISL_1663009, EPI_ISL_1663010, EPI_ISL_1663011, EPI_ISL_1663012, EPI_ISL_1663013, EPI_ISL_1663014 | | | |
| see above | Johns Hopkins Hospital Department of Pathology | Johns Hopkins Hospital Department of Pathology | C. Paul Morris, Chun Huai Luo, Adannaya Amadi, Matthew Schwartz, Heba H. Mostafa |
| EPI_ISL_1663019, EPI_ISL_1663021, EPI_ISL_1663022, EPI_ISL_1663025, EPI_ISL_1663027, EPI_ISL_1663029, EPI_ISL_1663030, EPI_ISL_1663031, EPI_ISL_1663032, EPI_ISL_1663034, EPI_ISL_1663035, EPI_ISL_1663036, EPI_ISL_1663037, EPI_ISL_1663038, EPI_ISL_1663039, EPI_ISL_1663040, EPI_ISL_1663041, EPI_ISL_1663042, EPI_ISL_1663043, EPI_ISL_1663046, EPI_ISL_1663047, EPI_ISL_1663052, EPI_ISL_1663053, EPI_ISL_1663058 | | | |
| see above | Indiana State Department of Health | Quantigen Biosciences | Paul J. Childress |
| EPI_ISL_1663068, EPI_ISL_1663071, EPI_ISL_1663072, EPI_ISL_1663073, EPI_ISL_1663074, EPI_ISL_1663075, EPI_ISL_1663076, EPI_ISL_1663077, EPI_ISL_1663078, EPI_ISL_1663079, EPI_ISL_1663080, EPI_ISL_1663081, EPI_ISL_1663083, EPI_ISL_1663084, EPI_ISL_1663085, EPI_ISL_1663086, EPI_ISL_1663087, EPI_ISL_1663089, EPI_ISL_1663090, EPI_ISL_1663091, EPI_ISL_1663092, EPI_ISL_1663093, EPI_ISL_1663094, EPI_ISL_1663095, EPI_ISL_1663096, EPI_ISL_1663097, EPI_ISL_1663098, EPI_ISL_1663099, EPI_ISL_1663100, EPI_ISL_1663101, EPI_ISL_1663102, EPI_ISL_1663103, EPI_ISL_1663104, EPI_ISL_1663105, EPI_ISL_1663106, EPI_ISL_1663107, EPI_ISL_1663108, EPI_ISL_1663109, EPI_ISL_1663110, EPI_ISL_1663111, EPI_ISL_1663112, EPI_ISL_1663113, EPI_ISL_1663114, EPI_ISL_1663115, EPI_ISL_1663116, EPI_ISL_1663117, EPI_ISL_1663118, EPI_ISL_1663119, EPI_ISL_1663120, EPI_ISL_1663121, EPI_ISL_1663122, EPI_ISL_1663123, EPI_ISL_1663124, EPI_ISL_1663125, EPI_ISL_1663126, EPI_ISL_1663127, EPI_ISL_1663128, EPI_ISL_1663131, EPI_ISL_1663132, EPI_ISL_1663133, EPI_ISL_1663134, EPI_ISL_1663135, EPI_ISL_1663136, EPI_ISL_1663137, EPI_ISL_1663138, EPI_ISL_1663139 | | | |
| see above | Illinois Department of Public Health | Illinois Department of Public Health - Chicago Lab | Vineet K. Dhiman, Ira Heimler |
| EPI_ISL_1663142, EPI_ISL_1663143, EPI_ISL_1663144, EPI_ISL_1663145, EPI_ISL_1663147, EPI_ISL_1663148, EPI_ISL_1663149, EPI_ISL_1663150, EPI_ISL_1663151, EPI_ISL_1663152, EPI_ISL_1663153, EPI_ISL_1663154, EPI_ISL_1663155, EPI_ISL_1663156, EPI_ISL_1663157, EPI_ISL_1663158, EPI_ISL_1663159, EPI_ISL_1663160, EPI_ISL_1663161, EPI_ISL_1663162, EPI_ISL_1663163, EPI_ISL_1663164, EPI_ISL_1663165, EPI_ISL_1663166, EPI_ISL_1663167, EPI_ISL_1663168, EPI_ISL_1663169, EPI_ISL_1663170, EPI_ISL_1663171, EPI_ISL_1663172, EPI_ISL_1663173, EPI_ISL_1663174, EPI_ISL_1663175, EPI_ISL_1663176, EPI_ISL_1663177, EPI_ISL_1663178, EPI_ISL_1663179, EPI_ISL_1663180, EPI_ISL_1663181, EPI_ISL_1663182, EPI_ISL_1663183, EPI_ISL_1663184, EPI_ISL_1663185, EPI_ISL_1663186 | | | |
| see above | M Health Fairview | Minnesota Department of Health, Public Health Laboratory | Alexandra Lorentz, Jacob Garfin, Matt Plumb, and Xiong Wang |
| EPI_ISL_1663187, EPI_ISL_1663188, EPI_ISL_1663189, EPI_ISL_1663190, EPI_ISL_1663191, EPI_ISL_1663192, EPI_ISL_1663193, EPI_ISL_1663194, EPI_ISL_1663195, EPI_ISL_1663196, EPI_ISL_1663197, EPI_ISL_1663198, EPI_ISL_1663199, EPI_ISL_1663200, EPI_ISL_1663201, EPI_ISL_1663202, EPI_ISL_1663203, EPI_ISL_1663204, EPI_ISL_1663205, EPI_ISL_1663206, EPI_ISL_1663207, EPI_ISL_1663208, EPI_ISL_1663209, EPI_ISL_1663210, EPI_ISL_1663211, EPI_ISL_1663212, EPI_ISL_1663213, EPI_ISL_1663214, EPI_ISL_1663215, EPI_ISL_1663216, EPI_ISL_1663217, EPI_ISL_1663218, EPI_ISL_1663219, EPI_ISL_1663220, EPI_ISL_1663221, EPI_ISL_1663222, EPI_ISL_1663223, EPI_ISL_1663224, EPI_ISL_1663225, EPI_ISL_1663226, EPI_ISL_1663227 | | | |
| see above | Mayo Clinic & Mayo Clinic Laboratories | Minnesota Department of Health, Public Health Laboratory | Alexandra Lorentz, Jacob Garfin, Matt Plumb, and Xiong Wang |
| EPI_ISL_1663724, EPI_ISL_1663725, EPI_ISL_1663733, EPI_ISL_1663734, | Wyoming Public Health Laboratory | Wyoming Public Health Laboratory | Jim Mildenberger, Wanda Manley, Noah Hull, Taylor Fearing, Lynette Gumbleton, Channing Weber, Ashley Norberg, Chayse Rowley, Marley Goetz, Brian Dominguez, Elliot Thomasson, Sam Britz, Cari Sloma, and Rob Christensen |

| | | | |
|---|---|---|---|
| EPI_ISL_1663736, EPI_ISL_1663738 | | | |
| EPI_ISL_1663741, EPI_ISL_1663742, EPI_ISL_1663743, EPI_ISL_1663747, EPI_ISL_1663748, EPI_ISL_1663749, EPI_ISL_1663750, EPI_ISL_1663751, EPI_ISL_1663752, EPI_ISL_1663753, EPI_ISL_1663754, EPI_ISL_1663755, EPI_ISL_1663756, EPI_ISL_1663757, EPI_ISL_1663758, EPI_ISL_1663759, EPI_ISL_1663760, EPI_ISL_1663761, EPI_ISL_1663762, EPI_ISL_1663763, EPI_ISL_1663764, EPI_ISL_1663765, EPI_ISL_1663767, EPI_ISL_1663768, EPI_ISL_1663769, EPI_ISL_1663770, EPI_ISL_1663771, EPI_ISL_1663773, EPI_ISL_1663774, EPI_ISL_1663777, EPI_ISL_1663778, EPI_ISL_1663780, EPI_ISL_1663782, EPI_ISL_1663783, EPI_ISL_1663784, EPI_ISL_1663785, EPI_ISL_1663786, EPI_ISL_1663787, EPI_ISL_1663788, EPI_ISL_1663790, EPI_ISL_1663791, EPI_ISL_1663794, EPI_ISL_1663795, EPI_ISL_1663796, EPI_ISL_1663797, EPI_ISL_1663798, EPI_ISL_1663799, EPI_ISL_1663800, EPI_ISL_1663801, EPI_ISL_1663802, EPI_ISL_1663804, EPI_ISL_1663805, EPI_ISL_1663806, EPI_ISL_1663807, EPI_ISL_1663808, EPI_ISL_1663809, EPI_ISL_1663810, EPI_ISL_1663811 | | | |
| see above | UTGSAF | UTGSAF | Jessica Podnar, Sylvie Beaudenon, Audrey Kelly, Zachary Carver, Anna Battenhouse, Andreas Matouschek |
| EPI_ISL_1663812, EPI_ISL_1663813, EPI_ISL_1663814, EPI_ISL_1663815, EPI_ISL_1663816 | Washington University | Pathogen Discovery, Respiratory Viruses Branch, Division of Viral Diseases, Centers for Disease Control and Prevention | Ying Tao, Yan Li, Anna Kelleher, Jing Zhang, Anna Montmayeur, Brian Lynch, Krista Queen, Anna Uehara, Peter Cook, Han Jia Justin Ng, Rachel Marine, Clinton R. Paden, Haibin Wang, Mark Burroughs, Justin Lee, Adam Retchless, Suxiang Tong |
| EPI_ISL_1663817, EPI_ISL_1663819, EPI_ISL_1663820, EPI_ISL_1663821, EPI_ISL_1663822, EPI_ISL_1663824, EPI_ISL_1663825, EPI_ISL_1663826, EPI_ISL_1663827, EPI_ISL_1663828, EPI_ISL_1663829, EPI_ISL_1663830, EPI_ISL_1663831, EPI_ISL_1663832, EPI_ISL_1663833, EPI_ISL_1663834, EPI_ISL_1663835, EPI_ISL_1663836, EPI_ISL_1663837, EPI_ISL_1663838, EPI_ISL_1663839, EPI_ISL_1663840, EPI_ISL_1663843, EPI_ISL_1663844, EPI_ISL_1663845, EPI_ISL_1663846, EPI_ISL_1663847, EPI_ISL_1663848, EPI_ISL_1663850, EPI_ISL_1663851, EPI_ISL_1663852, EPI_ISL_1663853, EPI_ISL_1663856, EPI_ISL_1663858, EPI_ISL_1663859, EPI_ISL_1663862, EPI_ISL_1663863, EPI_ISL_1663864 | | | |
| see above | CURATIVE | New Mexico Department of Health Scientific Laboratory | Ellie Johnson, Anastacia Griego-Fisher, D'eldra Malone, Jennifer Benoit |
| EPI_ISL_1663868, EPI_ISL_1663869 | Oregon State University TRACE Project | OSU Center for Genome Research and Biocomputing | Oregon State University TRACE Project |
| EPI_ISL_1663873, EPI_ISL_1663874, EPI_ISL_1663875, EPI_ISL_1663876, EPI_ISL_1663877, EPI_ISL_1663878, EPI_ISL_1663879, EPI_ISL_1663881, EPI_ISL_1663883, EPI_ISL_1663884, EPI_ISL_1663885, EPI_ISL_1663887, EPI_ISL_1663888, EPI_ISL_1663889, EPI_ISL_1663890, EPI_ISL_1663891, EPI_ISL_1663892, EPI_ISL_1663894, EPI_ISL_1663895, EPI_ISL_1663896, EPI_ISL_1663898, EPI_ISL_1663899, EPI_ISL_1663900, EPI_ISL_1663901, EPI_ISL_1663903, EPI_ISL_1663904, EPI_ISL_1663905, EPI_ISL_1663906, EPI_ISL_1663907, EPI_ISL_1663908, EPI_ISL_1663909, EPI_ISL_1663910 | | | |
| see above | Nevada State Public Health Laboratory | Nevada State Public Health Laboratory | Andrew Gorzalski, Mark Pandori |
| EPI_ISL_1663913, EPI_ISL_1663915, EPI_ISL_1663916, EPI_ISL_1663918, EPI_ISL_1663919, EPI_ISL_1663920 | Notre Dame COVID Response and Testing Laboratory | Notre Dame Genomics & Bioinformatics Core Facility | Michael Pfrender, Joseph Sarro, Melissa Stephens |
| EPI_ISL_1663921, EPI_ISL_1663922 | CA DPH Viral and Rickettsial Disease Laboratory | Chan-Zuckerberg Biohub | CZB Cliahub Consortium |
| EPI_ISL_1663923 | Contra Costa County Public Health Lab | Chan-Zuckerberg Biohub | CZB Cliahub Consortium |
| EPI_ISL_1663924 | CA DPH Viral and Rickettsial Disease Laboratory | Chan-Zuckerberg Biohub | CZB Cliahub Consortium |
| EPI_ISL_1663926 | Contra Costa County Public Health Lab | Chan-Zuckerberg Biohub | CZB Cliahub Consortium |
| EPI_ISL_1663927, EPI_ISL_1663928 | Alameda County Public Health Lab | Chan-Zuckerberg Biohub | CZB Cliahub Consortium |
| EPI_ISL_1663929 | CA DPH Viral and Rickettsial Disease Laboratory | Chan-Zuckerberg Biohub | CZB Cliahub Consortium |
| EPI_ISL_1663930 | Contra Costa County Public Health Lab | Chan-Zuckerberg Biohub | CZB Cliahub Consortium |
| EPI_ISL_1663931 | Santa Clara County Public Health Laboratory | Chan-Zuckerberg Biohub | CZB Cliahub Consortium |
| EPI_ISL_1663932 | CA DPH Viral and Rickettsial Disease Laboratory | Chan-Zuckerberg Biohub | CZB Cliahub Consortium |
| EPI_ISL_1663933 | Santa Clara County Public Health Laboratory | Chan-Zuckerberg Biohub | CZB Cliahub Consortium |
| EPI_ISL_1663934 | Contra Costa County Public Health Lab | Chan-Zuckerberg Biohub | CZB Cliahub Consortium |
| EPI_ISL_1663935 | Alameda County Public Health Lab | Chan-Zuckerberg Biohub | CZB Cliahub Consortium |
| EPI_ISL_1663936, EPI_ISL_1663937 | Contra Costa County Public Health Lab | Chan-Zuckerberg Biohub | CZB Cliahub Consortium |
| EPI_ISL_1663938 | Alameda County Public Health Lab | Chan-Zuckerberg Biohub | CZB Cliahub Consortium |
| EPI_ISL_1663939 | Santa Clara County Public Health Laboratory | Chan-Zuckerberg Biohub | CZB Cliahub Consortium |
| EPI_ISL_1663940 | Contra Costa County Public Health Lab | Chan-Zuckerberg Biohub | CZB Cliahub Consortium |
| EPI_ISL_1663941, EPI_ISL_1663942 | CA DPH Viral and Rickettsial Disease Laboratory | Chan-Zuckerberg Biohub | CZB Cliahub Consortium |
| EPI_ISL_1663943 | Contra Costa County Public Health Lab | Chan-Zuckerberg Biohub | CZB Cliahub Consortium |
| EPI_ISL_1663944 | CA DPH Viral and Rickettsial Disease Laboratory | Chan-Zuckerberg Biohub | CZB Cliahub Consortium |
| EPI_ISL_1663946 | Contra Costa County Public Health Lab | Chan-Zuckerberg Biohub | CZB Cliahub Consortium |
| EPI_ISL_1663947 | Alameda County Public Health Lab | Chan-Zuckerberg Biohub | CZB Cliahub Consortium |
| EPI_ISL_1663948 | Contra Costa County Public Health Lab | Chan-Zuckerberg Biohub | CZB Cliahub Consortium |
| EPI_ISL_1663949 | CA DPH Viral and Rickettsial Disease Laboratory | Chan-Zuckerberg Biohub | CZB Cliahub Consortium |
| EPI_ISL_1663950, EPI_ISL_1663951, EPI_ISL_1663953, EPI_ISL_1663954, EPI_ISL_1663955 | Contra Costa County Public Health Lab | Chan-Zuckerberg Biohub | CZB Cliahub Consortium |
| EPI_ISL_1663956 | Alameda County Public Health Lab | Chan-Zuckerberg Biohub | CZB Cliahub Consortium |
| EPI_ISL_1663957 | Contra Costa County Public Health Lab | Chan-Zuckerberg Biohub | CZB Cliahub Consortium |
| EPI_ISL_1663958, EPI_ISL_1663959, EPI_ISL_1663960 | Alameda County Public Health Lab | Chan-Zuckerberg Biohub | CZB Cliahub Consortium |
| EPI_ISL_1663961 | CA DPH Viral and Rickettsial Disease Laboratory | Chan-Zuckerberg Biohub | CZB Cliahub Consortium |
| EPI_ISL_1663962 | Contra Costa County Public Health Lab | Chan-Zuckerberg Biohub | CZB Cliahub Consortium |
| EPI_ISL_1663963 | Santa Clara County Public Health Laboratory | Chan-Zuckerberg Biohub | CZB Cliahub Consortium |
| EPI_ISL_1663964 | Contra Costa County Public Health Lab | Chan-Zuckerberg Biohub | CZB Cliahub Consortium |
| EPI_ISL_1663965, EPI_ISL_1663966, EPI_ISL_1663967 | CA DPH Viral and Rickettsial Disease Laboratory | Chan-Zuckerberg Biohub | CZB Cliahub Consortium |
| EPI_ISL_1663968 | Contra Costa County Public Health Lab | Chan-Zuckerberg Biohub | CZB Cliahub Consortium |
| EPI_ISL_1663969 | Alameda County Public Health Lab | Chan-Zuckerberg Biohub | CZB Cliahub Consortium |
| EPI_ISL_1663970, EPI_ISL_1663971 | Contra Costa County Public Health Lab | Chan-Zuckerberg Biohub | CZB Cliahub Consortium |
| EPI_ISL_1663972 | Alameda County Public Health Lab | Chan-Zuckerberg Biohub | CZB Cliahub Consortium |
| EPI_ISL_1663973 | CA DPH Viral and Rickettsial Disease Laboratory | Chan-Zuckerberg Biohub | CZB Cliahub Consortium |
| EPI_ISL_1663974, EPI_ISL_1663975, | Contra Costa County Public Health Lab | Chan-Zuckerberg Biohub | CZB Cliahub Consortium |

| | | | |
|---|---|---|---|
| EPI_ISL_1663976, EPI_ISL_1663977 | | | |
| EPI_ISL_1663978 | CA DPH Viral and Rickettsial Disease Laboratory | Chan-Zuckerberg Biohub | CZB Cliahub Consortium |
| EPI_ISL_1663979, EPI_ISL_1663980 | Santa Clara County Public Health Laboratory | Chan-Zuckerberg Biohub | CZB Cliahub Consortium |
| EPI_ISL_1663981 | CA DPH Viral and Rickettsial Disease Laboratory | Chan-Zuckerberg Biohub | CZB Cliahub Consortium |
| EPI_ISL_1663982 | Santa Clara County Public Health Laboratory | Chan-Zuckerberg Biohub | CZB Cliahub Consortium |
| EPI_ISL_1663983, EPI_ISL_1663985 | Contra Costa County Public Health Lab | Chan-Zuckerberg Biohub | CZB Cliahub Consortium |
| EPI_ISL_1663986 | Alameda County Public Health Lab | Chan-Zuckerberg Biohub | CZB Cliahub Consortium |
| EPI_ISL_1663987 | Contra Costa County Public Health Lab | Chan-Zuckerberg Biohub | CZB Cliahub Consortium |
| EPI_ISL_1663988 | CA DPH Viral and Rickettsial Disease Laboratory | Chan-Zuckerberg Biohub | CZB Cliahub Consortium |
| EPI_ISL_1663989 | Contra Costa County Public Health Lab | Chan-Zuckerberg Biohub | CZB Cliahub Consortium |
| EPI_ISL_1663990 | Alameda County Public Health Lab | Chan-Zuckerberg Biohub | CZB Cliahub Consortium |
| EPI_ISL_1663991 | CA DPH Viral and Rickettsial Disease Laboratory | Chan-Zuckerberg Biohub | CZB Cliahub Consortium |
| EPI_ISL_1663992 | Contra Costa County Public Health Lab | Chan-Zuckerberg Biohub | CZB Cliahub Consortium |
| EPI_ISL_1663993 | Santa Clara County Public Health Laboratory | Chan-Zuckerberg Biohub | CZB Cliahub Consortium |
| EPI_ISL_1663994, EPI_ISL_1663995, EPI_ISL_1663996 | Contra Costa County Public Health Lab | Chan-Zuckerberg Biohub | CZB Cliahub Consortium |
| EPI_ISL_1663997, EPI_ISL_1663998 | CA DPH Viral and Rickettsial Disease Laboratory | Chan-Zuckerberg Biohub | CZB Cliahub Consortium |
| EPI_ISL_1663999 | Contra Costa County Public Health Lab | Chan-Zuckerberg Biohub | CZB Cliahub Consortium |
| EPI_ISL_1664000, EPI_ISL_1664001, EPI_ISL_1664002 | CA DPH Viral and Rickettsial Disease Laboratory | Chan-Zuckerberg Biohub | CZB Cliahub Consortium |
| EPI_ISL_1664003 | Alameda County Public Health Lab | Chan-Zuckerberg Biohub | CZB Cliahub Consortium |
| EPI_ISL_1664004 | Santa Clara County Public Health Laboratory | Chan-Zuckerberg Biohub | CZB Cliahub Consortium |
| EPI_ISL_1664005 | Contra Costa County Public Health Lab | Chan-Zuckerberg Biohub | CZB Cliahub Consortium |
| EPI_ISL_1664006 | Alameda County Public Health Lab | Chan-Zuckerberg Biohub | CZB Cliahub Consortium |
| EPI_ISL_1664007 | CA DPH Viral and Rickettsial Disease Laboratory | Chan-Zuckerberg Biohub | CZB Cliahub Consortium |
| EPI_ISL_1664008 | Contra Costa County Public Health Lab | Chan-Zuckerberg Biohub | CZB Cliahub Consortium |
| EPI_ISL_1664010 | Alameda County Public Health Lab | Chan-Zuckerberg Biohub | CZB Cliahub Consortium |
| EPI_ISL_1664011, EPI_ISL_1664012 | CA DPH Viral and Rickettsial Disease Laboratory | Chan-Zuckerberg Biohub | CZB Cliahub Consortium |
| EPI_ISL_1664014 | Contra Costa County Public Health Lab | Chan-Zuckerberg Biohub | CZB Cliahub Consortium |
| EPI_ISL_1664015 | Alameda County Public Health Lab | Chan-Zuckerberg Biohub | CZB Cliahub Consortium |
| EPI_ISL_1664016 | Santa Clara County Public Health Laboratory | Chan-Zuckerberg Biohub | CZB Cliahub Consortium |
| EPI_ISL_1664017, EPI_ISL_1664018, EPI_ISL_1664019 | CA DPH Viral and Rickettsial Disease Laboratory | Chan-Zuckerberg Biohub | CZB Cliahub Consortium |
| EPI_ISL_1664020 | Contra Costa County Public Health Lab | Chan-Zuckerberg Biohub | CZB Cliahub Consortium |
| EPI_ISL_1664021 | Alameda County Public Health Lab | Chan-Zuckerberg Biohub | CZB Cliahub Consortium |
| EPI_ISL_1664022, EPI_ISL_1664023 | CA DPH Viral and Rickettsial Disease Laboratory | Chan-Zuckerberg Biohub | CZB Cliahub Consortium |
| EPI_ISL_1664024 | Contra Costa County Public Health Lab | Chan-Zuckerberg Biohub | CZB Cliahub Consortium |
| EPI_ISL_1664025 | Santa Clara County Public Health Laboratory | Chan-Zuckerberg Biohub | CZB Cliahub Consortium |
| EPI_ISL_1664026 | Contra Costa County Public Health Lab | Chan-Zuckerberg Biohub | CZB Cliahub Consortium |
| EPI_ISL_1664027, EPI_ISL_1664029 | CA DPH Viral and Rickettsial Disease Laboratory | Chan-Zuckerberg Biohub | CZB Cliahub Consortium |
| EPI_ISL_1664030 | Contra Costa County Public Health Lab | Chan-Zuckerberg Biohub | CZB Cliahub Consortium |
| EPI_ISL_1664031 | CA DPH Viral and Rickettsial Disease Laboratory | Chan-Zuckerberg Biohub | CZB Cliahub Consortium |
| EPI_ISL_1664032 | Contra Costa County Public Health Lab | Chan-Zuckerberg Biohub | CZB Cliahub Consortium |
| EPI_ISL_1664033, EPI_ISL_1664034 | CA DPH Viral and Rickettsial Disease Laboratory | Chan-Zuckerberg Biohub | CZB Cliahub Consortium |
| EPI_ISL_1664035 | Contra Costa County Public Health Lab | Chan-Zuckerberg Biohub | CZB Cliahub Consortium |
| EPI_ISL_1664036 | Santa Clara County Public Health Laboratory | Chan-Zuckerberg Biohub | CZB Cliahub Consortium |
| EPI_ISL_1664037 | Alameda County Public Health Lab | Chan-Zuckerberg Biohub | CZB Cliahub Consortium |
| EPI_ISL_1664038 | Contra Costa County Public Health Lab | Chan-Zuckerberg Biohub | CZB Cliahub Consortium |
| EPI_ISL_1664039, EPI_ISL_1664040 | Alameda County Public Health Lab | Chan-Zuckerberg Biohub | CZB Cliahub Consortium |
| EPI_ISL_1664041, EPI_ISL_1664042 | CA DPH Viral and Rickettsial Disease Laboratory | Chan-Zuckerberg Biohub | CZB Cliahub Consortium |
| EPI_ISL_1664043 | Santa Clara County Public Health Laboratory | Chan-Zuckerberg Biohub | CZB Cliahub Consortium |
| EPI_ISL_1664044 | CA DPH Viral and Rickettsial Disease Laboratory | Chan-Zuckerberg Biohub | CZB Cliahub Consortium |
| EPI_ISL_1664045, EPI_ISL_1664046, EPI_ISL_1664047, EPI_ISL_1664049, EPI_ISL_1664050 | Contra Costa County Public Health Lab | Chan-Zuckerberg Biohub | CZB Cliahub Consortium |
| EPI_ISL_1664051, EPI_ISL_1664052 | Alameda County Public Health Lab | Chan-Zuckerberg Biohub | CZB Cliahub Consortium |
| EPI_ISL_1664053, | Contra Costa County Public Health Lab | Chan-Zuckerberg Biohub | CZB Cliahub Consortium |

| | | | |
|---|---|---|---|
| EPI_ISL_1664054 | | | |
| EPI_ISL_1664055 | CA DPH Viral and Rickettsial Disease Laboratory | Chan-Zuckerberg Biohub | CZB Cliahub Consortium |
| EPI_ISL_1664056 | Alameda County Public Health Lab | Chan-Zuckerberg Biohub | CZB Cliahub Consortium |
| EPI_ISL_1664057, EPI_ISL_1664058 | Contra Costa County Public Health Lab | Chan-Zuckerberg Biohub | CZB Cliahub Consortium |
| EPI_ISL_1664059 | CA DPH Viral and Rickettsial Disease Laboratory | Chan-Zuckerberg Biohub | CZB Cliahub Consortium |
| EPI_ISL_1664060 | Contra Costa County Public Health Lab | Chan-Zuckerberg Biohub | CZB Cliahub Consortium |
| EPI_ISL_1664062 | Santa Clara County Public Health Laboratory | Chan-Zuckerberg Biohub | CZB Cliahub Consortium |
| EPI_ISL_1664063 | Contra Costa County Public Health Lab | Chan-Zuckerberg Biohub | CZB Cliahub Consortium |
| EPI_ISL_1664065 | Alameda County Public Health Lab | Chan-Zuckerberg Biohub | CZB Cliahub Consortium |
| EPI_ISL_1664066, EPI_ISL_1664067 | CA DPH Viral and Rickettsial Disease Laboratory | Chan-Zuckerberg Biohub | CZB Cliahub Consortium |
| EPI_ISL_1664068, EPI_ISL_1664069, EPI_ISL_1664070, EPI_ISL_1664071, EPI_ISL_1664072 | Contra Costa County Public Health Lab | Chan-Zuckerberg Biohub | CZB Cliahub Consortium |
| EPI_ISL_1664073 | Santa Clara County Public Health Laboratory | Chan-Zuckerberg Biohub | CZB Cliahub Consortium |
| EPI_ISL_1664074, EPI_ISL_1664075, EPI_ISL_1664076, EPI_ISL_1664077, EPI_ISL_1664078 | CA DPH Viral and Rickettsial Disease Laboratory | Chan-Zuckerberg Biohub | CZB Cliahub Consortium |
| EPI_ISL_1664079, EPI_ISL_1664080 | Santa Clara County Public Health Laboratory | Chan-Zuckerberg Biohub | CZB Cliahub Consortium |
| EPI_ISL_1664081, EPI_ISL_1664082 | Contra Costa County Public Health Lab | Chan-Zuckerberg Biohub | CZB Cliahub Consortium |
| EPI_ISL_1664083 | Santa Clara County Public Health Laboratory | Chan-Zuckerberg Biohub | CZB Cliahub Consortium |
| EPI_ISL_1664084, EPI_ISL_1664085, EPI_ISL_1664086, EPI_ISL_1664087, EPI_ISL_1664088 | Contra Costa County Public Health Lab | Chan-Zuckerberg Biohub | CZB Cliahub Consortium |
| EPI_ISL_1664089 | Santa Clara County Public Health Laboratory | Chan-Zuckerberg Biohub | CZB Cliahub Consortium |
| EPI_ISL_1664090 | Alameda County Public Health Lab | Chan-Zuckerberg Biohub | CZB Cliahub Consortium |
| EPI_ISL_1664091 | Contra Costa County Public Health Lab | Chan-Zuckerberg Biohub | CZB Cliahub Consortium |
| EPI_ISL_1664092 | CA DPH Viral and Rickettsial Disease Laboratory | Chan-Zuckerberg Biohub | CZB Cliahub Consortium |
| EPI_ISL_1664093 | Contra Costa County Public Health Lab | Chan-Zuckerberg Biohub | CZB Cliahub Consortium |
| EPI_ISL_1664094, EPI_ISL_1664095 | Santa Clara County Public Health Laboratory | Chan-Zuckerberg Biohub | CZB Cliahub Consortium |
| EPI_ISL_1664096, EPI_ISL_1664097, EPI_ISL_1664098 | Contra Costa County Public Health Lab | Chan-Zuckerberg Biohub | CZB Cliahub Consortium |
| EPI_ISL_1664099 | CA DPH Viral and Rickettsial Disease Laboratory | Chan-Zuckerberg Biohub | CZB Cliahub Consortium |
| EPI_ISL_1664100 | Contra Costa County Public Health Lab | Chan-Zuckerberg Biohub | CZB Cliahub Consortium |
| EPI_ISL_1664101 | Alameda County Public Health Lab | Chan-Zuckerberg Biohub | CZB Cliahub Consortium |
| EPI_ISL_1664102, EPI_ISL_1664103, EPI_ISL_1664104, EPI_ISL_1664105 | Contra Costa County Public Health Lab | Chan-Zuckerberg Biohub | CZB Cliahub Consortium |
| EPI_ISL_1664106 | Alameda County Public Health Lab | Chan-Zuckerberg Biohub | CZB Cliahub Consortium |
| EPI_ISL_1664107 | Santa Clara County Public Health Laboratory | Chan-Zuckerberg Biohub | CZB Cliahub Consortium |
| EPI_ISL_1664108, EPI_ISL_1664109, EPI_ISL_1664110, EPI_ISL_1664111, EPI_ISL_1664112 | Contra Costa County Public Health Lab | Chan-Zuckerberg Biohub | CZB Cliahub Consortium |

EPI_ISL_1664203, EPI_ISL_1664204, EPI_ISL_1664206, EPI_ISL_1664208, EPI_ISL_1664209, EPI_ISL_1664211, EPI_ISL_1664215, EPI_ISL_1664216, EPI_ISL_1664217, EPI_ISL_1664221, EPI_ISL_1664223, EPI_ISL_1664224, EPI_ISL_1664227

| | | | |
|---|---|---|---|
| see above | Notre Dame COVID Response and Testing Laboratory | Notre Dame Genomics & Bioinformatics Core Facility | Michael Pfrender, Joseph Sarro, Melissa Stephens |
| EPI_ISL_1664231, EPI_ISL_1664232, EPI_ISL_1664233, EPI_ISL_1664235, EPI_ISL_1664239, EPI_ISL_1664244, EPI_ISL_1664245, EPI_ISL_1664250, EPI_ISL_1664251 | Kaiser San Jose | Santa Clara County public Health Laboratory | Santa Clara County Public Health Department |
| EPI_ISL_1664269 | Kaiser Santa Clara | Santa Clara County public Health Laboratory | Santa Clara County Public Health Department |
| EPI_ISL_1664272, EPI_ISL_1664273, EPI_ISL_1664274 | El Camino Hospital | Santa Clara County public Health Laboratory | Santa Clara County Public Health Department |

EPI_ISL_1664276, EPI_ISL_1664278, EPI_ISL_1664279, EPI_ISL_1664280, EPI_ISL_1664281, EPI_ISL_1664282, EPI_ISL_1664283, EPI_ISL_1664284, EPI_ISL_1664285, EPI_ISL_1664286, EPI_ISL_1664287, EPI_ISL_1664288, EPI_ISL_1664289, EPI_ISL_1664290, EPI_ISL_1664291, EPI_ISL_1664292, EPI_ISL_1664294, EPI_ISL_1664295, EPI_ISL_1664296, EPI_ISL_1664297, EPI_ISL_1664298, EPI_ISL_1664299

| | | | |
|---|---|---|---|
| see above | Santa Clara Valley Medical Center | Santa Clara County public Health Laboratory | Santa Clara County Public Health Department |
| EPI_ISL_1664310, EPI_ISL_1664311, EPI_ISL_1664312, EPI_ISL_1664313, EPI_ISL_1664315, EPI_ISL_1664316, EPI_ISL_1664318 | UNC-CH COVID Surveillance Lab | Jeremy Wang | Jeremy Wang, Alexander Rubinsteyn, Melissa Miller, Corbin Jones, Amy James Loftis, Amir Barzin, Susan Fiscus |

EPI_ISL_1664319, EPI_ISL_1664320, EPI_ISL_1664321, EPI_ISL_1664322, EPI_ISL_1664323, EPI_ISL_1664325, EPI_ISL_1664326, EPI_ISL_1664327, EPI_ISL_1664328, EPI_ISL_1664329, EPI_ISL_1664330, EPI_ISL_1664331, EPI_ISL_1664332, EPI_ISL_1664333, EPI_ISL_1664334, EPI_ISL_1664335, EPI_ISL_1664336, EPI_ISL_1664337, EPI_ISL_1664338, EPI_ISL_1664339, EPI_ISL_1664340, EPI_ISL_1664343,

EPI_ISL_1664344, EPI_ISL_1664345, EPI_ISL_1664346, EPI_ISL_1664347, EPI_ISL_1664348, EPI_ISL_1664349, EPI_ISL_1664350, EPI_ISL_1664351, EPI_ISL_1664352, EPI_ISL_1664354, EPI_ISL_1664355, EPI_ISL_1664356, EPI_ISL_1664357, EPI_ISL_1664359, EPI_ISL_1664360, EPI_ISL_1664361, EPI_ISL_1664363, EPI_ISL_1664364, EPI_ISL_1664365, EPI_ISL_1664366, EPI_ISL_1664367, EPI_ISL_1664368, EPI_ISL_1664369, EPI_ISL_1664370, EPI_ISL_1664371, EPI_ISL_1664372, EPI_ISL_1664373, EPI_ISL_1664374, EPI_ISL_1664375, EPI_ISL_1664376, EPI_ISL_1664377, EPI_ISL_1664378, EPI_ISL_1664379, EPI_ISL_1664380, EPI_ISL_1664381, EPI_ISL_1664382, EPI_ISL_1664383, EPI_ISL_1664384, EPI_ISL_1664385, EPI_ISL_1664386, EPI_ISL_1664387, EPI_ISL_1664388, EPI_ISL_1664389, EPI_ISL_1664390, EPI_ISL_1664391, EPI_ISL_1664392, EPI_ISL_1664393, EPI_ISL_1664394, EPI_ISL_1664395, EPI_ISL_1664396, EPI_ISL_1664397, EPI_ISL_1664398, EPI_ISL_1664399, EPI_ISL_1664400, EPI_ISL_1664401, EPI_ISL_1664402, EPI_ISL_1664403, EPI_ISL_1664404, EPI_ISL_1664405, EPI_ISL_1664406, EPI_ISL_1664407, EPI_ISL_1664408, EPI_ISL_1664409, EPI_ISL_1664410, EPI_ISL_1664411, EPI_ISL_1664412, EPI_ISL_1664413, EPI_ISL_1664414, EPI_ISL_1664415, EPI_ISL_1664416, EPI_ISL_1664417, EPI_ISL_1664418, EPI_ISL_1664419, EPI_ISL_1664420, EPI_ISL_1664421, EPI_ISL_1664422, EPI_ISL_1664423, EPI_ISL_1664424, EPI_ISL_1664425, EPI_ISL_1664426, EPI_ISL_1664427, EPI_ISL_1664428, EPI_ISL_1664429, EPI_ISL_1664430, EPI_ISL_1664431, EPI_ISL_1664432, EPI_ISL_1664433, EPI_ISL_1664434, EPI_ISL_1664435, EPI_ISL_1664436, EPI_ISL_1664437, EPI_ISL_1664438, EPI_ISL_1664441, EPI_ISL_1664442, EPI_ISL_1664443, EPI_ISL_1664444, EPI_ISL_1664445, EPI_ISL_1664446, EPI_ISL_1664447, EPI_ISL_1664448, EPI_ISL_1664449, EPI_ISL_1664450, EPI_ISL_1664453, EPI_ISL_1664454, EPI_ISL_1664455, EPI_ISL_1664456, EPI_ISL_1664457, EPI_ISL_1664458, EPI_ISL_1664459, EPI_ISL_1664460, EPI_ISL_1664461, EPI_ISL_1664462, EPI_ISL_1664463, EPI_ISL_1664464, EPI_ISL_1664465, EPI_ISL_1664466, EPI_ISL_1664469, EPI_ISL_1664470, EPI_ISL_1664471, EPI_ISL_1664472, EPI_ISL_1664473, EPI_ISL_1664474, EPI_ISL_1664475, EPI_ISL_1664476, EPI_ISL_1664477

| | | | |
|---|---|---|---|
| see above | Clinical Molecular Microbiology Laboratory, UNC Hospitals | Jeremy Wang | Jeremy Wang, Alexander Rubinsteyn, Colleen Rice, Jason Smedberg, Shawn Hawken, Melissa Miller, Corbin Jones, Robert Hagan |
| EPI_ISL_1664478, EPI_ISL_1664479, EPI_ISL_1664480 | New Mexico Department of Health Scientific Laboratory | Center for Global Health, University of New Mexico Health Sciences Center | Daryl Domman, Kurt Schwalm, Twila Kunde, Joseph Hicks, Anastacia Griego, Michael Edwards, Valerie Morley, Darrell Dinwiddie |
| EPI_ISL_1664481 | Clinical Molecular Microbiology Laboratory, UNC Hospitals | Jeremy Wang | Jeremy Wang, Alexander Rubinsteyn, Colleen Rice, Jason Smedberg, Shawn Hawken, Melissa Miller, Corbin Jones, Robert Hagan |
| EPI_ISL_1664482 | New Mexico Department of Health Scientific Laboratory | Center for Global Health, University of New Mexico Health Sciences Center | Daryl Domman, Kurt Schwalm, Twila Kunde, Joseph Hicks, Anastacia Griego, Michael Edwards, Valerie Morley, Darrell Dinwiddie |
| EPI_ISL_1664483 | Clinical Molecular Microbiology Laboratory, UNC Hospitals | Jeremy Wang | Jeremy Wang, Alexander Rubinsteyn, Colleen Rice, Jason Smedberg, Shawn Hawken, Melissa Miller, Corbin Jones, Robert Hagan |
| EPI_ISL_1664484, EPI_ISL_1664485 | New Mexico Department of Health Scientific Laboratory | Center for Global Health, University of New Mexico Health Sciences Center | Daryl Domman, Kurt Schwalm, Twila Kunde, Joseph Hicks, Anastacia Griego, Michael Edwards, Valerie Morley, Darrell Dinwiddie |
| EPI_ISL_1664486 | Clinical Molecular Microbiology Laboratory, UNC Hospitals | Jeremy Wang | Jeremy Wang, Alexander Rubinsteyn, Colleen Rice, Jason Smedberg, Shawn Hawken, Melissa Miller, Corbin Jones, Robert Hagan |
| EPI_ISL_1664488 | New Mexico Department of Health Scientific Laboratory | Center for Global Health, University of New Mexico Health Sciences Center | Daryl Domman, Kurt Schwalm, Twila Kunde, Joseph Hicks, Anastacia Griego, Michael Edwards, Valerie Morley, Darrell Dinwiddie |
| EPI_ISL_1664489 | Clinical Molecular Microbiology Laboratory, UNC Hospitals | Jeremy Wang | Jeremy Wang, Alexander Rubinsteyn, Colleen Rice, Jason Smedberg, Shawn Hawken, Melissa Miller, Corbin Jones, Robert Hagan |
| EPI_ISL_1664490, EPI_ISL_1664491 | New Mexico Department of Health Scientific Laboratory | Center for Global Health, University of New Mexico Health Sciences Center | Daryl Domman, Kurt Schwalm, Twila Kunde, Joseph Hicks, Anastacia Griego, Michael Edwards, Valerie Morley, Darrell Dinwiddie |
| EPI_ISL_1664492 | Clinical Molecular Microbiology Laboratory, UNC Hospitals | Jeremy Wang | Jeremy Wang, Alexander Rubinsteyn, Colleen Rice, Jason Smedberg, Shawn Hawken, Melissa Miller, Corbin Jones, Robert Hagan |
| EPI_ISL_1664493, EPI_ISL_1664494 | New Mexico Department of Health Scientific Laboratory | Center for Global Health, University of New Mexico Health Sciences Center | Daryl Domman, Kurt Schwalm, Twila Kunde, Joseph Hicks, Anastacia Griego, Michael Edwards, Valerie Morley, Darrell Dinwiddie |
| EPI_ISL_1664495 | Clinical Molecular Microbiology Laboratory, UNC Hospitals | Jeremy Wang | Jeremy Wang, Alexander Rubinsteyn, Colleen Rice, Jason Smedberg, Shawn Hawken, Melissa Miller, Corbin Jones, Robert Hagan |
| EPI_ISL_1664496, EPI_ISL_1664497 | New Mexico Department of Health Scientific Laboratory | Center for Global Health, University of New Mexico Health Sciences Center | Daryl Domman, Kurt Schwalm, Twila Kunde, Joseph Hicks, Anastacia Griego, Michael Edwards, Valerie Morley, Darrell Dinwiddie |
| EPI_ISL_1664498 | Clinical Molecular Microbiology Laboratory, UNC Hospitals | Jeremy Wang | Jeremy Wang, Alexander Rubinsteyn, Colleen Rice, Jason Smedberg, Shawn Hawken, Melissa Miller, Corbin Jones, Robert Hagan |
| EPI_ISL_1664499, EPI_ISL_1664500 | New Mexico Department of Health Scientific Laboratory | Center for Global Health, University of New Mexico Health Sciences Center | Daryl Domman, Kurt Schwalm, Twila Kunde, Joseph Hicks, Anastacia Griego, Michael Edwards, Valerie Morley, Darrell Dinwiddie |
| EPI_ISL_1664501 | Clinical Molecular Microbiology Laboratory, UNC Hospitals | Jeremy Wang | Jeremy Wang, Alexander Rubinsteyn, Colleen Rice, Jason Smedberg, Shawn Hawken, Melissa Miller, Corbin Jones, Robert Hagan |
| EPI_ISL_1664502, EPI_ISL_1664503 | New Mexico Department of Health Scientific Laboratory | Center for Global Health, University of New Mexico Health Sciences Center | Daryl Domman, Kurt Schwalm, Twila Kunde, Joseph Hicks, Anastacia Griego, Michael Edwards, Valerie Morley, Darrell Dinwiddie |
| EPI_ISL_1664504 | Clinical Molecular Microbiology Laboratory, UNC Hospitals | Jeremy Wang | Jeremy Wang, Alexander Rubinsteyn, Colleen Rice, Jason Smedberg, Shawn Hawken, Melissa Miller, Corbin Jones, Robert Hagan |
| EPI_ISL_1664505 | New Mexico Department of Health Scientific Laboratory | Center for Global Health, University of New Mexico Health Sciences Center | Daryl Domman, Kurt Schwalm, Twila Kunde, Joseph Hicks, Anastacia Griego, Michael Edwards, Valerie Morley, Darrell Dinwiddie |
| EPI_ISL_1664506 | Clinical Molecular Microbiology Laboratory, UNC Hospitals | Jeremy Wang | Jeremy Wang, Alexander Rubinsteyn, Colleen Rice, Jason Smedberg, Shawn Hawken, Melissa Miller, Corbin Jones, Robert Hagan |
| EPI_ISL_1664507, EPI_ISL_1664508 | New Mexico Department of Health Scientific Laboratory | Center for Global Health, University of New Mexico Health Sciences Center | Daryl Domman, Kurt Schwalm, Twila Kunde, Joseph Hicks, Anastacia Griego, Michael Edwards, Valerie Morley, Darrell Dinwiddie |
| EPI_ISL_1664509 | Clinical Molecular Microbiology Laboratory, UNC Hospitals | Jeremy Wang | Jeremy Wang, Alexander Rubinsteyn, Colleen Rice, Jason Smedberg, Shawn Hawken, Melissa Miller, Corbin Jones, Robert Hagan |
| EPI_ISL_1664510, EPI_ISL_1664511 | New Mexico Department of Health Scientific Laboratory | Center for Global Health, University of New Mexico Health Sciences Center | Daryl Domman, Kurt Schwalm, Twila Kunde, Joseph Hicks, Anastacia Griego, Michael Edwards, Valerie Morley, Darrell Dinwiddie |
| EPI_ISL_1664512 | Clinical Molecular Microbiology Laboratory, UNC Hospitals | Jeremy Wang | Jeremy Wang, Alexander Rubinsteyn, Colleen Rice, Jason Smedberg, Shawn Hawken, Melissa Miller, Corbin Jones, Robert Hagan |
| EPI_ISL_1664513, EPI_ISL_1664514 | New Mexico Department of Health Scientific Laboratory | Center for Global Health, University of New Mexico Health Sciences Center | Daryl Domman, Kurt Schwalm, Twila Kunde, Joseph Hicks, Anastacia Griego, Michael Edwards, Valerie Morley, Darrell Dinwiddie |
| EPI_ISL_1664515 | Clinical Molecular Microbiology Laboratory, UNC Hospitals | Jeremy Wang | Jeremy Wang, Alexander Rubinsteyn, Colleen Rice, Jason Smedberg, Shawn Hawken, Melissa Miller, Corbin Jones, Robert Hagan |
| EPI_ISL_1664516, EPI_ISL_1664517 | New Mexico Department of Health Scientific Laboratory | Center for Global Health, University of New Mexico Health Sciences Center | Daryl Domman, Kurt Schwalm, Twila Kunde, Joseph Hicks, Anastacia Griego, Michael Edwards, Valerie Morley, Darrell Dinwiddie |
| EPI_ISL_1664518, EPI_ISL_1664520 | Clinical Molecular Microbiology Laboratory, UNC Hospitals | Jeremy Wang | Jeremy Wang, Alexander Rubinsteyn, Colleen Rice, Jason Smedberg, Shawn Hawken, Melissa Miller, Corbin Jones, Robert Hagan |
| EPI_ISL_1664521, EPI_ISL_1664522 | New Mexico Department of Health Scientific Laboratory | Center for Global Health, University of New Mexico Health Sciences Center | Daryl Domman, Kurt Schwalm, Twila Kunde, Joseph Hicks, Anastacia Griego, Michael Edwards, Valerie Morley, Darrell Dinwiddie |
| EPI_ISL_1664523 | Clinical Molecular Microbiology Laboratory, UNC Hospitals | Jeremy Wang | Jeremy Wang, Alexander Rubinsteyn, Colleen Rice, Jason Smedberg, Shawn Hawken, Melissa Miller, Corbin Jones, Robert Hagan |
| EPI_ISL_1664524, EPI_ISL_1664525 | New Mexico Department of Health Scientific Laboratory | Center for Global Health, University of New Mexico Health Sciences Center | Daryl Domman, Kurt Schwalm, Twila Kunde, Joseph Hicks, Anastacia Griego, Michael Edwards, Valerie Morley, Darrell Dinwiddie |
| EPI_ISL_1664526 | Clinical Molecular Microbiology Laboratory, UNC Hospitals | Jeremy Wang | Jeremy Wang, Alexander Rubinsteyn, Colleen Rice, Jason Smedberg, Shawn Hawken, Melissa Miller, Corbin Jones, Robert Hagan |
| EPI_ISL_1664527, EPI_ISL_1664528 | New Mexico Department of Health Scientific Laboratory | Center for Global Health, University of New Mexico Health Sciences Center | Daryl Domman, Kurt Schwalm, Twila Kunde, Joseph Hicks, Anastacia Griego, Michael Edwards, Valerie Morley, Darrell Dinwiddie |
| EPI_ISL_1664529 | Clinical Molecular Microbiology Laboratory, UNC Hospitals | Jeremy Wang | Jeremy Wang, Alexander Rubinsteyn, Colleen Rice, Jason Smedberg, Shawn Hawken, Melissa Miller, Corbin Jones, Robert Hagan |
| EPI_ISL_1664530, EPI_ISL_1664531 | New Mexico Department of Health Scientific Laboratory | Center for Global Health, University of New Mexico Health Sciences Center | Daryl Domman, Kurt Schwalm, Twila Kunde, Joseph Hicks, Anastacia Griego, Michael Edwards, Valerie Morley, Darrell Dinwiddie |
| EPI_ISL_1664532 | Clinical Molecular Microbiology Laboratory, UNC Hospitals | Jeremy Wang | Jeremy Wang, Alexander Rubinsteyn, Colleen Rice, Jason Smedberg, Shawn Hawken, Melissa Miller, Corbin Jones, Robert Hagan |
| EPI_ISL_1664533, EPI_ISL_1664534 | New Mexico Department of Health Scientific Laboratory | Center for Global Health, University of New Mexico Health Sciences Center | Daryl Domman, Kurt Schwalm, Twila Kunde, Joseph Hicks, Anastacia Griego, Michael Edwards, Valerie Morley, Darrell Dinwiddie |
| EPI_ISL_1664535 | Clinical Molecular Microbiology Laboratory, UNC Hospitals | Jeremy Wang | Jeremy Wang, Alexander Rubinsteyn, Colleen Rice, Jason Smedberg, Shawn Hawken, Melissa Miller, Corbin Jones, Robert Hagan |
| EPI_ISL_1664536, EPI_ISL_1664537 | New Mexico Department of Health Scientific Laboratory | Center for Global Health, University of New Mexico Health Sciences Center | Daryl Domman, Kurt Schwalm, Twila Kunde, Joseph Hicks, Anastacia Griego, Michael Edwards, Valerie Morley, Darrell Dinwiddie |
| EPI_ISL_1664538 | Clinical Molecular Microbiology Laboratory, UNC Hospitals | Jeremy Wang | Jeremy Wang, Alexander Rubinsteyn, Colleen Rice, Jason Smedberg, Shawn Hawken, Melissa Miller, Corbin Jones, Robert Hagan |
| EPI_ISL_1664539, EPI_ISL_1664540 | New Mexico Department of Health Scientific Laboratory | Center for Global Health, University of New Mexico Health Sciences Center | Daryl Domman, Kurt Schwalm, Twila Kunde, Joseph Hicks, Anastacia Griego, Michael Edwards, Valerie Morley, Darrell Dinwiddie |

| | | | |
|---|---|---|---|
| EPI_ISL_1664541 | Clinical Molecular Microbiology Laboratory, UNC Hospitals | Jeremy Wang | Jeremy Wang, Alexander Rubinsteyn, Colleen Rice, Jason Smedberg, Shawn Hawken, Melissa Miller, Corbin Jones, Robert Hagan |
| EPI_ISL_1664542, EPI_ISL_1664543 | New Mexico Department of Health Scientific Laboratory | Center for Global Health, University of New Mexico Health Sciences Center | Daryl Domman, Kurt Schwalm, Twila Kunde, Joseph Hicks, Anastacia Griego, Michael Edwards, Valerie Morley, Darrell Dinwiddie |
| EPI_ISL_1664544 | Clinical Molecular Microbiology Laboratory, UNC Hospitals | Jeremy Wang | Jeremy Wang, Alexander Rubinsteyn, Colleen Rice, Jason Smedberg, Shawn Hawken, Melissa Miller, Corbin Jones, Robert Hagan |
| EPI_ISL_1664545, EPI_ISL_1664546, EPI_ISL_1664548, EPI_ISL_1664549 | New Mexico Department of Health Scientific Laboratory | Center for Global Health, University of New Mexico Health Sciences Center | Daryl Domman, Kurt Schwalm, Twila Kunde, Joseph Hicks, Anastacia Griego, Michael Edwards, Valerie Morley, Darrell Dinwiddie |
| EPI_ISL_1664550 | Clinical Molecular Microbiology Laboratory, UNC Hospitals | Jeremy Wang | Jeremy Wang, Alexander Rubinsteyn, Colleen Rice, Jason Smedberg, Shawn Hawken, Melissa Miller, Corbin Jones, Robert Hagan |
| EPI_ISL_1664551, EPI_ISL_1664552 | New Mexico Department of Health Scientific Laboratory | Center for Global Health, University of New Mexico Health Sciences Center | Daryl Domman, Kurt Schwalm, Twila Kunde, Joseph Hicks, Anastacia Griego, Michael Edwards, Valerie Morley, Darrell Dinwiddie |
| EPI_ISL_1664553 | Clinical Molecular Microbiology Laboratory, UNC Hospitals | Jeremy Wang | Jeremy Wang, Alexander Rubinsteyn, Colleen Rice, Jason Smedberg, Shawn Hawken, Melissa Miller, Corbin Jones, Robert Hagan |
| EPI_ISL_1664554, EPI_ISL_1664555 | New Mexico Department of Health Scientific Laboratory | Center for Global Health, University of New Mexico Health Sciences Center | Daryl Domman, Kurt Schwalm, Twila Kunde, Joseph Hicks, Anastacia Griego, Michael Edwards, Valerie Morley, Darrell Dinwiddie |
| EPI_ISL_1664556 | Clinical Molecular Microbiology Laboratory, UNC Hospitals | Jeremy Wang | Jeremy Wang, Alexander Rubinsteyn, Colleen Rice, Jason Smedberg, Shawn Hawken, Melissa Miller, Corbin Jones, Robert Hagan |
| EPI_ISL_1664557, EPI_ISL_1664558 | New Mexico Department of Health Scientific Laboratory | Center for Global Health, University of New Mexico Health Sciences Center | Daryl Domman, Kurt Schwalm, Twila Kunde, Joseph Hicks, Anastacia Griego, Michael Edwards, Valerie Morley, Darrell Dinwiddie |
| EPI_ISL_1664559 | Clinical Molecular Microbiology Laboratory, UNC Hospitals | Jeremy Wang | Jeremy Wang, Alexander Rubinsteyn, Colleen Rice, Jason Smedberg, Shawn Hawken, Melissa Miller, Corbin Jones, Robert Hagan |
| EPI_ISL_1664560, EPI_ISL_1664561 | New Mexico Department of Health Scientific Laboratory | Center for Global Health, University of New Mexico Health Sciences Center | Daryl Domman, Kurt Schwalm, Twila Kunde, Joseph Hicks, Anastacia Griego, Michael Edwards, Valerie Morley, Darrell Dinwiddie |
| EPI_ISL_1664562 | Clinical Molecular Microbiology Laboratory, UNC Hospitals | Jeremy Wang | Jeremy Wang, Alexander Rubinsteyn, Colleen Rice, Jason Smedberg, Shawn Hawken, Melissa Miller, Corbin Jones, Robert Hagan |
| EPI_ISL_1664563, EPI_ISL_1664564 | New Mexico Department of Health Scientific Laboratory | Center for Global Health, University of New Mexico Health Sciences Center | Daryl Domman, Kurt Schwalm, Twila Kunde, Joseph Hicks, Anastacia Griego, Michael Edwards, Valerie Morley, Darrell Dinwiddie |
| EPI_ISL_1664565 | Clinical Molecular Microbiology Laboratory, UNC Hospitals | Jeremy Wang | Jeremy Wang, Alexander Rubinsteyn, Colleen Rice, Jason Smedberg, Shawn Hawken, Melissa Miller, Corbin Jones, Robert Hagan |
| EPI_ISL_1664566, EPI_ISL_1664567 | New Mexico Department of Health Scientific Laboratory | Center for Global Health, University of New Mexico Health Sciences Center | Daryl Domman, Kurt Schwalm, Twila Kunde, Joseph Hicks, Anastacia Griego, Michael Edwards, Valerie Morley, Darrell Dinwiddie |
| EPI_ISL_1664568 | Clinical Molecular Microbiology Laboratory, UNC Hospitals | Jeremy Wang | Jeremy Wang, Alexander Rubinsteyn, Colleen Rice, Jason Smedberg, Shawn Hawken, Melissa Miller, Corbin Jones, Robert Hagan |
| EPI_ISL_1664569 | New Mexico Department of Health Scientific Laboratory | Center for Global Health, University of New Mexico Health Sciences Center | Daryl Domman, Kurt Schwalm, Twila Kunde, Joseph Hicks, Anastacia Griego, Michael Edwards, Valerie Morley, Darrell Dinwiddie |
| EPI_ISL_1664570 | Clinical Molecular Microbiology Laboratory, UNC Hospitals | Jeremy Wang | Jeremy Wang, Alexander Rubinsteyn, Colleen Rice, Jason Smedberg, Shawn Hawken, Melissa Miller, Corbin Jones, Robert Hagan |
| EPI_ISL_1664571, EPI_ISL_1664572 | New Mexico Department of Health Scientific Laboratory | Center for Global Health, University of New Mexico Health Sciences Center | Daryl Domman, Kurt Schwalm, Twila Kunde, Joseph Hicks, Anastacia Griego, Michael Edwards, Valerie Morley, Darrell Dinwiddie |
| EPI_ISL_1664573 | Clinical Molecular Microbiology Laboratory, UNC Hospitals | Jeremy Wang | Jeremy Wang, Alexander Rubinsteyn, Colleen Rice, Jason Smedberg, Shawn Hawken, Melissa Miller, Corbin Jones, Robert Hagan |
| EPI_ISL_1664574, EPI_ISL_1664576, EPI_ISL_1664578 | New Mexico Department of Health Scientific Laboratory | Center for Global Health, University of New Mexico Health Sciences Center | Daryl Domman, Kurt Schwalm, Twila Kunde, Joseph Hicks, Anastacia Griego, Michael Edwards, Valerie Morley, Darrell Dinwiddie |
| EPI_ISL_1664579 | Clinical Molecular Microbiology Laboratory, UNC Hospitals | Jeremy Wang | Jeremy Wang, Alexander Rubinsteyn, Colleen Rice, Jason Smedberg, Shawn Hawken, Melissa Miller, Corbin Jones, Robert Hagan |
| EPI_ISL_1664580 | New Mexico Department of Health Scientific Laboratory | Center for Global Health, University of New Mexico Health Sciences Center | Daryl Domman, Kurt Schwalm, Twila Kunde, Joseph Hicks, Anastacia Griego, Michael Edwards, Valerie Morley, Darrell Dinwiddie |
| EPI_ISL_1664581 | Clinical Molecular Microbiology Laboratory, UNC Hospitals | Jeremy Wang | Jeremy Wang, Alexander Rubinsteyn, Colleen Rice, Jason Smedberg, Shawn Hawken, Melissa Miller, Corbin Jones, Robert Hagan |
| EPI_ISL_1664582, EPI_ISL_1664583, EPI_ISL_1664586 | New Mexico Department of Health Scientific Laboratory | Center for Global Health, University of New Mexico Health Sciences Center | Daryl Domman, Kurt Schwalm, Twila Kunde, Joseph Hicks, Anastacia Griego, Michael Edwards, Valerie Morley, Darrell Dinwiddie |
| EPI_ISL_1664587 | Clinical Molecular Microbiology Laboratory, UNC Hospitals | Jeremy Wang | Jeremy Wang, Alexander Rubinsteyn, Colleen Rice, Jason Smedberg, Shawn Hawken, Melissa Miller, Corbin Jones, Robert Hagan |
| EPI_ISL_1664588 | New Mexico Department of Health Scientific Laboratory | Center for Global Health, University of New Mexico Health Sciences Center | Daryl Domman, Kurt Schwalm, Twila Kunde, Joseph Hicks, Anastacia Griego, Michael Edwards, Valerie Morley, Darrell Dinwiddie |
| EPI_ISL_1664589, EPI_ISL_1664590, EPI_ISL_1664591, EPI_ISL_1664592, EPI_ISL_1664593 | Clinical Molecular Microbiology Laboratory, UNC Hospitals | Jeremy Wang | Jeremy Wang, Alexander Rubinsteyn, Colleen Rice, Jason Smedberg, Shawn Hawken, Melissa Miller, Corbin Jones, Robert Hagan |
| EPI_ISL_1664594, EPI_ISL_1664595, EPI_ISL_1664596, EPI_ISL_1664597, EPI_ISL_1664598, EPI_ISL_1664599, EPI_ISL_1664600, EPI_ISL_1664601, EPI_ISL_1664602, EPI_ISL_1664603, EPI_ISL_1664604, EPI_ISL_1664605, EPI_ISL_1664606, EPI_ISL_1664607, EPI_ISL_1664608, EPI_ISL_1664610, EPI_ISL_1664611, EPI_ISL_1664612, EPI_ISL_1664613, EPI_ISL_1664614, EPI_ISL_1664615, EPI_ISL_1664616, EPI_ISL_1664617, EPI_ISL_1664618, EPI_ISL_1664619 | | | |
| see above | Monterey County Public Health Laboratory | Monterey County Public Health Laboratory | Monterey County Public Health Laboratory |
| EPI_ISL_1665283, EPI_ISL_1665284, EPI_ISL_1665285, EPI_ISL_1665287, EPI_ISL_1665291, EPI_ISL_1665295, EPI_ISL_1665297, EPI_ISL_1665299, EPI_ISL_1665300, EPI_ISL_1665301, EPI_ISL_1665302, EPI_ISL_1665303, EPI_ISL_1665304, EPI_ISL_1665305, EPI_ISL_1665307, EPI_ISL_1665310, EPI_ISL_1665314, EPI_ISL_1665318, EPI_ISL_1665319, EPI_ISL_1665320, EPI_ISL_1665321, EPI_ISL_1665323, EPI_ISL_1665325, EPI_ISL_1665326, EPI_ISL_1665330, EPI_ISL_1665333, EPI_ISL_1665334, EPI_ISL_1665341, EPI_ISL_1665344, EPI_ISL_1665351, EPI_ISL_1665356, EPI_ISL_1665361, EPI_ISL_1665365, EPI_ISL_1665367, EPI_ISL_1665370, EPI_ISL_1665371, EPI_ISL_1665375, EPI_ISL_1665376, EPI_ISL_1665378, EPI_ISL_1665380, EPI_ISL_1665387, EPI_ISL_1665392, EPI_ISL_1665398, EPI_ISL_1665401, EPI_ISL_1665404, EPI_ISL_1665405, EPI_ISL_1665411, EPI_ISL_1665415, EPI_ISL_1665419, EPI_ISL_1665424, EPI_ISL_1665425, EPI_ISL_1665435, EPI_ISL_1665436, EPI_ISL_1665441, EPI_ISL_1665453, EPI_ISL_1665454, EPI_ISL_1665456, EPI_ISL_1665458, EPI_ISL_1665459, EPI_ISL_1665462, EPI_ISL_1665466, EPI_ISL_1665467, EPI_ISL_1665473, EPI_ISL_1665478, EPI_ISL_1665482, EPI_ISL_1665487, EPI_ISL_1665488, EPI_ISL_1665494, EPI_ISL_1665513, EPI_ISL_1665519, EPI_ISL_1665520, EPI_ISL_1665523, EPI_ISL_1665524, EPI_ISL_1665525, EPI_ISL_1665526, EPI_ISL_1665529, EPI_ISL_1665530, EPI_ISL_1665531, EPI_ISL_1665532, EPI_ISL_1665533, EPI_ISL_1665534, EPI_ISL_1665538, EPI_ISL_1665540, EPI_ISL_1665541, EPI_ISL_1665542, EPI_ISL_1665543, EPI_ISL_1665545, EPI_ISL_1665546, EPI_ISL_1665547, EPI_ISL_1665548, EPI_ISL_1665550, EPI_ISL_1665552, EPI_ISL_1665553, EPI_ISL_1665554, EPI_ISL_1665556, EPI_ISL_1665561, EPI_ISL_1665563, EPI_ISL_1665566, EPI_ISL_1665570, EPI_ISL_1665573, EPI_ISL_1665575, EPI_ISL_1665583, EPI_ISL_1665584, EPI_ISL_1665585, EPI_ISL_1665588, EPI_ISL_1665591, EPI_ISL_1665594, EPI_ISL_1665595, EPI_ISL_1665597, EPI_ISL_1665598, EPI_ISL_1665599, EPI_ISL_1665600, EPI_ISL_1665603, EPI_ISL_1665604, EPI_ISL_1665606, EPI_ISL_1665611, EPI_ISL_1665612, EPI_ISL_1665614, EPI_ISL_1665615, EPI_ISL_1665617, EPI_ISL_1665618, EPI_ISL_1665619, EPI_ISL_1665620, EPI_ISL_1665621, EPI_ISL_1665623, EPI_ISL_1665624, EPI_ISL_1665626, EPI_ISL_1665627, EPI_ISL_1665629, EPI_ISL_1665631, EPI_ISL_1665632, EPI_ISL_1665635, EPI_ISL_1665637, EPI_ISL_1665639, EPI_ISL_1665640, EPI_ISL_1665641, EPI_ISL_1665642, EPI_ISL_1665644, EPI_ISL_1665647, EPI_ISL_1665648, EPI_ISL_1665650, EPI_ISL_1665653, EPI_ISL_1665660, EPI_ISL_1665661, EPI_ISL_1665662, EPI_ISL_1665663, EPI_ISL_1665665, EPI_ISL_1665667, EPI_ISL_1665668, EPI_ISL_1665669, EPI_ISL_1665670, EPI_ISL_1665673, EPI_ISL_1665674, EPI_ISL_1665676, EPI_ISL_1665677, EPI_ISL_1665679, EPI_ISL_1665688, EPI_ISL_1665690, EPI_ISL_1665691, EPI_ISL_1665693, EPI_ISL_1665694, EPI_ISL_1665695, EPI_ISL_1665697, EPI_ISL_1665699, EPI_ISL_1665700, EPI_ISL_1665703, EPI_ISL_1665708, EPI_ISL_1665710, EPI_ISL_1665711, EPI_ISL_1665713, EPI_ISL_1665715, EPI_ISL_1665716, EPI_ISL_1665717, EPI_ISL_1665718, EPI_ISL_1665719, EPI_ISL_1665721, EPI_ISL_1665723, EPI_ISL_1665724, EPI_ISL_1665725, EPI_ISL_1665728, EPI_ISL_1665732, EPI_ISL_1665733, EPI_ISL_1665734, EPI_ISL_1665735, EPI_ISL_1665736, EPI_ISL_1665745, EPI_ISL_1665747, EPI_ISL_1665748, EPI_ISL_1665749, EPI_ISL_1665750, EPI_ISL_1665753, EPI_ISL_1665757, EPI_ISL_1665758, EPI_ISL_1665759, EPI_ISL_1665760, EPI_ISL_1665761, EPI_ISL_1665767, EPI_ISL_1665768, EPI_ISL_1665770, EPI_ISL_1665772, EPI_ISL_1665776, EPI_ISL_1665779, EPI_ISL_1665780, EPI_ISL_1665782, EPI_ISL_1665785, EPI_ISL_1665786, EPI_ISL_1665787, EPI_ISL_1665788, EPI_ISL_1665791, EPI_ISL_1665792, EPI_ISL_1665793, EPI_ISL_1665794, EPI_ISL_1665797, EPI_ISL_1665799, EPI_ISL_1665800, EPI_ISL_1665802, EPI_ISL_1665803, EPI_ISL_1665804, EPI_ISL_1665805, EPI_ISL_1665809, EPI_ISL_1665811, EPI_ISL_1665813, EPI_ISL_1665817, EPI_ISL_1665818, EPI_ISL_1665819, EPI_ISL_1665820, EPI_ISL_1665823, EPI_ISL_1665826, EPI_ISL_1665829, EPI_ISL_1665831, EPI_ISL_1665832, EPI_ISL_1665834, EPI_ISL_1665835, EPI_ISL_1665837, EPI_ISL_1665838, EPI_ISL_1665839, EPI_ISL_1665843, EPI_ISL_1665846, EPI_ISL_1665848, EPI_ISL_1665850, EPI_ISL_1665851, EPI_ISL_1665854, EPI_ISL_1665858, EPI_ISL_1665860, EPI_ISL_1665863, EPI_ISL_1665864, EPI_ISL_1665867, EPI_ISL_1665868, EPI_ISL_1665871, EPI_ISL_1665872, EPI_ISL_1665873, EPI_ISL_1665875, EPI_ISL_1665876, EPI_ISL_1665880, EPI_ISL_1665881, EPI_ISL_1665887, EPI_ISL_1665890, EPI_ISL_1665891, EPI_ISL_1665896 | | | |
| see above | Colorado Department of Public Health and Environment | Colorado Department of Public Health and Environment | Laura Bankers, Molly C. Hetherington-Rauth, Diana Ir, Shannon Ely, Shannon R. Matzinger, Sarah Elizabeth Totten, Emily A. Travanty |
| EPI_ISL_1665898, EPI_ISL_1665899, EPI_ISL_1665902, EPI_ISL_1665903, EPI_ISL_1665904, EPI_ISL_1665905, EPI_ISL_1665906, EPI_ISL_1665907, EPI_ISL_1665908, EPI_ISL_1665909, EPI_ISL_1665911, EPI_ISL_1665913, EPI_ISL_1665915, EPI_ISL_1665916, EPI_ISL_1665918, EPI_ISL_1665919, EPI_ISL_1665920, EPI_ISL_1665921, EPI_ISL_1665922, EPI_ISL_1665923, EPI_ISL_1665924, EPI_ISL_1665925, EPI_ISL_1665926, EPI_ISL_1665927, EPI_ISL_1665928, EPI_ISL_1665930, EPI_ISL_1665931, EPI_ISL_1665932, EPI_ISL_1665933, EPI_ISL_1665935, EPI_ISL_1665936, EPI_ISL_1665937, EPI_ISL_1665938, EPI_ISL_1665939, EPI_ISL_1665940, EPI_ISL_1665941, EPI_ISL_1665942, EPI_ISL_1665943, EPI_ISL_1665945, EPI_ISL_1665946, EPI_ISL_1665947, EPI_ISL_1665948, EPI_ISL_1665949, EPI_ISL_1665950, EPI_ISL_1665951, EPI_ISL_1665952, EPI_ISL_1665953, EPI_ISL_1665954, EPI_ISL_1665955, EPI_ISL_1665956, EPI_ISL_1665957, EPI_ISL_1665958, EPI_ISL_1665959, EPI_ISL_1665960, EPI_ISL_1665961, EPI_ISL_1665962, EPI_ISL_1665963, EPI_ISL_1665964, EPI_ISL_1665965, EPI_ISL_1665966, EPI_ISL_1665967, EPI_ISL_1665968, EPI_ISL_1665969, EPI_ISL_1665970, EPI_ISL_1665971, EPI_ISL_1665972, EPI_ISL_1665974, EPI_ISL_1665975, EPI_ISL_1665976, EPI_ISL_1665977, EPI_ISL_1665978, EPI_ISL_1665979, EPI_ISL_1665980, EPI_ISL_1665981, EPI_ISL_1665982, EPI_ISL_1665983, EPI_ISL_1665984, EPI_ISL_1665985, EPI_ISL_1665986, EPI_ISL_1665987, EPI_ISL_1665988, EPI_ISL_1665989, EPI_ISL_1665991, EPI_ISL_1665992, EPI_ISL_1665993, EPI_ISL_1665994, EPI_ISL_1665995, EPI_ISL_1665996, EPI_ISL_1665997, EPI_ISL_1665998, EPI_ISL_1665999, EPI_ISL_1666000, EPI_ISL_1666001, EPI_ISL_1666002, EPI_ISL_1666003, EPI_ISL_1666004, EPI_ISL_1666005, EPI_ISL_1666006, EPI_ISL_1666007, EPI_ISL_1666008, EPI_ISL_1666009, EPI_ISL_1666010, EPI_ISL_1666011, EPI_ISL_1666012, EPI_ISL_1666013, EPI_ISL_1666014, EPI_ISL_1666015, EPI_ISL_1666016, EPI_ISL_1666017, EPI_ISL_1666018, EPI_ISL_1666019, EPI_ISL_1666020, EPI_ISL_1666021, EPI_ISL_1666022, EPI_ISL_1666023, EPI_ISL_1666024, EPI_ISL_1666025, EPI_ISL_1666026, EPI_ISL_1666028, EPI_ISL_1666029, EPI_ISL_1666030, EPI_ISL_1666031, EPI_ISL_1666032, EPI_ISL_1666033, EPI_ISL_1666034, EPI_ISL_1666035, EPI_ISL_1666036, EPI_ISL_1666037, EPI_ISL_1666039, EPI_ISL_1666040, EPI_ISL_1666041, EPI_ISL_1666042, EPI_ISL_1666043, EPI_ISL_1666044, EPI_ISL_1666045, EPI_ISL_1666046, EPI_ISL_1666047, EPI_ISL_1666048, EPI_ISL_1666049, EPI_ISL_1666051, EPI_ISL_1666052, EPI_ISL_1666053, EPI_ISL_1666054, EPI_ISL_1666055, EPI_ISL_1666056, EPI_ISL_1666057, EPI_ISL_1666058, EPI_ISL_1666059, EPI_ISL_1666060, EPI_ISL_1666061, EPI_ISL_1666062, EPI_ISL_1666063, EPI_ISL_1666064, EPI_ISL_1666065, EPI_ISL_1666066, EPI_ISL_1666067, EPI_ISL_1666069, EPI_ISL_1666070, EPI_ISL_1666071, EPI_ISL_1666072, EPI_ISL_1666073, EPI_ISL_1666074, EPI_ISL_1666075, EPI_ISL_1666076, EPI_ISL_1666077, EPI_ISL_1666078, EPI_ISL_1666079, EPI_ISL_1666080, EPI_ISL_1666081, EPI_ISL_1666083, EPI_ISL_1666084, EPI_ISL_1666085, EPI_ISL_1666086, EPI_ISL_1666087, EPI_ISL_1666088, EPI_ISL_1666089, EPI_ISL_1666090, EPI_ISL_1666091, EPI_ISL_1666092, EPI_ISL_1666093, EPI_ISL_1666094, EPI_ISL_1666095, EPI_ISL_1666096, EPI_ISL_1666097, EPI_ISL_1666098, EPI_ISL_1666099, EPI_ISL_1666100, EPI_ISL_1666101, EPI_ISL_1666102, EPI_ISL_1666103, EPI_ISL_1666104, EPI_ISL_1666105, EPI_ISL_1666106, EPI_ISL_1666107, EPI_ISL_1666108, EPI_ISL_1666109, EPI_ISL_1666111, EPI_ISL_1666112, EPI_ISL_1666113, EPI_ISL_1666114, EPI_ISL_1666115, EPI_ISL_1666116, EPI_ISL_1666117, EPI_ISL_1666118, EPI_ISL_1666119, EPI_ISL_1666120, EPI_ISL_1666122, EPI_ISL_1666123, EPI_ISL_1666124, EPI_ISL_1666125, EPI_ISL_1666127, EPI_ISL_1666128, EPI_ISL_1666129, EPI_ISL_1666130, EPI_ISL_1666131, EPI_ISL_1666132, EPI_ISL_1666133, EPI_ISL_1666134, EPI_ISL_1666135, EPI_ISL_1666136, EPI_ISL_1666137, EPI_ISL_1666138, EPI_ISL_1666139, EPI_ISL_1666140, EPI_ISL_1666141, EPI_ISL_1666142, EPI_ISL_1666143, EPI_ISL_1666144, EPI_ISL_1666145, EPI_ISL_1666146, EPI_ISL_1666147, EPI_ISL_1666148, EPI_ISL_1666149, EPI_ISL_1666150, EPI_ISL_1666151, EPI_ISL_1666152, EPI_ISL_1666153, EPI_ISL_1666154, EPI_ISL_1666155, EPI_ISL_1666157, EPI_ISL_1666158, EPI_ISL_1666159, EPI_ISL_1666160, EPI_ISL_1666161, EPI_ISL_1666162, EPI_ISL_1666163, EPI_ISL_1666164, EPI_ISL_1666165, EPI_ISL_1666166, EPI_ISL_1666167, EPI_ISL_1666169, EPI_ISL_1666171, EPI_ISL_1666172, EPI_ISL_1666173, EPI_ISL_1666175, EPI_ISL_1666176, EPI_ISL_1666177, EPI_ISL_1666178, EPI_ISL_1666179, EPI_ISL_1666180, EPI_ISL_1666181, EPI_ISL_1666182, EPI_ISL_1666183, EPI_ISL_1666184, | | | |

| | | | |
|---|---|---|---|
| EPI_ISL_1666185, EPI_ISL_1666187, EPI_ISL_1666188, EPI_ISL_1666189, EPI_ISL_1666190, EPI_ISL_1666191, EPI_ISL_1666192, EPI_ISL_1666193, EPI_ISL_1666194, EPI_ISL_1666195, EPI_ISL_1666196, EPI_ISL_1666197, EPI_ISL_1666198, EPI_ISL_1666199, EPI_ISL_1666200, EPI_ISL_1666201, EPI_ISL_1666202, EPI_ISL_1666203, EPI_ISL_1666204, EPI_ISL_1666205, EPI_ISL_1666206, EPI_ISL_1666207, EPI_ISL_1666208, EPI_ISL_1666209, EPI_ISL_1666210, EPI_ISL_1666211, EPI_ISL_1666212, EPI_ISL_1666213, EPI_ISL_1666214, EPI_ISL_1666215, EPI_ISL_1666216, EPI_ISL_1666217, EPI_ISL_1666218, EPI_ISL_1666219, EPI_ISL_1666220, EPI_ISL_1666221, EPI_ISL_1666223, EPI_ISL_1666224, EPI_ISL_1666225, EPI_ISL_1666226, EPI_ISL_1666227, EPI_ISL_1666228, EPI_ISL_1666229, EPI_ISL_1666230, EPI_ISL_1666231, EPI_ISL_1666232, EPI_ISL_1666233, EPI_ISL_1666234, EPI_ISL_1666235, EPI_ISL_1666236, EPI_ISL_1666238, EPI_ISL_1666239, EPI_ISL_1666240, EPI_ISL_1666242, EPI_ISL_1666243, EPI_ISL_1666244, EPI_ISL_1666245, EPI_ISL_1666246, EPI_ISL_1666247, EPI_ISL_1666248, EPI_ISL_1666249, EPI_ISL_1666250, EPI_ISL_1666251, EPI_ISL_1666252, EPI_ISL_1666253, EPI_ISL_1666254, EPI_ISL_1666255, EPI_ISL_1666257, EPI_ISL_1666258, EPI_ISL_1666259, EPI_ISL_1666260, EPI_ISL_1666261, EPI_ISL_1666262, EPI_ISL_1666263, EPI_ISL_1666264, EPI_ISL_1666265, EPI_ISL_1666266, EPI_ISL_1666267, EPI_ISL_1666268, EPI_ISL_1666269, EPI_ISL_1666270, EPI_ISL_1666271, EPI_ISL_1666272, EPI_ISL_1666273, EPI_ISL_1666274, EPI_ISL_1666275, EPI_ISL_1666276, EPI_ISL_1666277, EPI_ISL_1666278, EPI_ISL_1666280, EPI_ISL_1666282, EPI_ISL_1666283, EPI_ISL_1666284, EPI_ISL_1666285, EPI_ISL_1666287, EPI_ISL_1666288, EPI_ISL_1666289, EPI_ISL_1666290, EPI_ISL_1666291, EPI_ISL_1666292, EPI_ISL_1666293, EPI_ISL_1666294, EPI_ISL_1666295, EPI_ISL_1666296, EPI_ISL_1666297, EPI_ISL_1666298, EPI_ISL_1666299, EPI_ISL_1666300, EPI_ISL_1666301, EPI_ISL_1666302, EPI_ISL_1666303, EPI_ISL_1666304, EPI_ISL_1666306, EPI_ISL_1666307, EPI_ISL_1666308, EPI_ISL_1666309, EPI_ISL_1666310, EPI_ISL_1666311, EPI_ISL_1666312, EPI_ISL_1666313, EPI_ISL_1666314, EPI_ISL_1666315, EPI_ISL_1666316, EPI_ISL_1666317, EPI_ISL_1666318, EPI_ISL_1666319, EPI_ISL_1666320, EPI_ISL_1666321, EPI_ISL_1666322, EPI_ISL_1666323, EPI_ISL_1666324, EPI_ISL_1666325, EPI_ISL_1666326, EPI_ISL_1666327, EPI_ISL_1666329, EPI_ISL_1666330, EPI_ISL_1666331, EPI_ISL_1666332, EPI_ISL_1666333, EPI_ISL_1666334, EPI_ISL_1666335, EPI_ISL_1666336, EPI_ISL_1666338, EPI_ISL_1666340, EPI_ISL_1666342, EPI_ISL_1666343, EPI_ISL_1666344, EPI_ISL_1666345, EPI_ISL_1666346, EPI_ISL_1666347, EPI_ISL_1666349, EPI_ISL_1666350, EPI_ISL_1666351, EPI_ISL_1666352, EPI_ISL_1666353, EPI_ISL_1666354, EPI_ISL_1666355, EPI_ISL_1666356, EPI_ISL_1666357, EPI_ISL_1666358, EPI_ISL_1666359, EPI_ISL_1666360, EPI_ISL_1666361, EPI_ISL_1666362, EPI_ISL_1666363, EPI_ISL_1666364, EPI_ISL_1666365, EPI_ISL_1666366, EPI_ISL_1666369, EPI_ISL_1666370, EPI_ISL_1666371, EPI_ISL_1666372, EPI_ISL_1666373, EPI_ISL_1666375, EPI_ISL_1666376, EPI_ISL_1666377, EPI_ISL_1666378, EPI_ISL_1666379, EPI_ISL_1666380, EPI_ISL_1666381, EPI_ISL_1666382, EPI_ISL_1666383, EPI_ISL_1666384, EPI_ISL_1666385, EPI_ISL_1666386, EPI_ISL_1666387, EPI_ISL_1666388, EPI_ISL_1666389, EPI_ISL_1666390, EPI_ISL_1666391, EPI_ISL_1666392, EPI_ISL_1666393, EPI_ISL_1666394, EPI_ISL_1666395, EPI_ISL_1666396, EPI_ISL_1666397, EPI_ISL_1666398, EPI_ISL_1666399, EPI_ISL_1666400, EPI_ISL_1666401, EPI_ISL_1666402, EPI_ISL_1666404, EPI_ISL_1666405, EPI_ISL_1666407, EPI_ISL_1666408, EPI_ISL_1666409, EPI_ISL_1666410, EPI_ISL_1666411, EPI_ISL_1666412, EPI_ISL_1666413, EPI_ISL_1666414, EPI_ISL_1666415, EPI_ISL_1666416, EPI_ISL_1666417, EPI_ISL_1666418, EPI_ISL_1666419, EPI_ISL_1666420, EPI_ISL_1666421, EPI_ISL_1666422, EPI_ISL_1666423, EPI_ISL_1666424, EPI_ISL_1666425, EPI_ISL_1666426, EPI_ISL_1666427, EPI_ISL_1666428, EPI_ISL_1666429, EPI_ISL_1666430, EPI_ISL_1666431, EPI_ISL_1666432, EPI_ISL_1666433, EPI_ISL_1666434, EPI_ISL_1666435, EPI_ISL_1666436, EPI_ISL_1666437, EPI_ISL_1666438, EPI_ISL_1666439, EPI_ISL_1666440, EPI_ISL_1666441, EPI_ISL_1666442, EPI_ISL_1666443, EPI_ISL_1666444, EPI_ISL_1666445, EPI_ISL_1666446, EPI_ISL_1666447, EPI_ISL_1666448, EPI_ISL_1666449, EPI_ISL_1666451, EPI_ISL_1666452, EPI_ISL_1666453, EPI_ISL_1666454, EPI_ISL_1666455, EPI_ISL_1666456, EPI_ISL_1666458, EPI_ISL_1666459, EPI_ISL_1666463, EPI_ISL_1666464, EPI_ISL_1666465, EPI_ISL_1666466, EPI_ISL_1666467, EPI_ISL_1666468, EPI_ISL_1666469, EPI_ISL_1666470, EPI_ISL_1666471, EPI_ISL_1666472, EPI_ISL_1666473, EPI_ISL_1666474, EPI_ISL_1666475, EPI_ISL_1666476, EPI_ISL_1666477, EPI_ISL_1666478, EPI_ISL_1666479, EPI_ISL_1666480, EPI_ISL_1666481, EPI_ISL_1666483, EPI_ISL_1666484, EPI_ISL_1666485, EPI_ISL_1666486, EPI_ISL_1666487, EPI_ISL_1666488, EPI_ISL_1666489, EPI_ISL_1666491, EPI_ISL_1666492, EPI_ISL_1666493, EPI_ISL_1666494, EPI_ISL_1666495, EPI_ISL_1666496, EPI_ISL_1666497, EPI_ISL_1666499, EPI_ISL_1666500, EPI_ISL_1666501, EPI_ISL_1666502, EPI_ISL_1666503, EPI_ISL_1666504, EPI_ISL_1666505, EPI_ISL_1666506, EPI_ISL_1666507, EPI_ISL_1666508, EPI_ISL_1666509, EPI_ISL_1666510, EPI_ISL_1666511, EPI_ISL_1666512, EPI_ISL_1666513, EPI_ISL_1666514, EPI_ISL_1666515, EPI_ISL_1666516, EPI_ISL_1666517, EPI_ISL_1666518, EPI_ISL_1666519, EPI_ISL_1666520, EPI_ISL_1666521, EPI_ISL_1666522, EPI_ISL_1666523, EPI_ISL_1666524, EPI_ISL_1666525, EPI_ISL_1666526, EPI_ISL_1666527, EPI_ISL_1666528, EPI_ISL_1666529, EPI_ISL_1666530, EPI_ISL_1666531, EPI_ISL_1666532, EPI_ISL_1666533, EPI_ISL_1666534, EPI_ISL_1666535, EPI_ISL_1666536, EPI_ISL_1666538, EPI_ISL_1666539, EPI_ISL_1666540, EPI_ISL_1666541, EPI_ISL_1666542, EPI_ISL_1666543, EPI_ISL_1666545, EPI_ISL_1666546, EPI_ISL_1666547, EPI_ISL_1666548, EPI_ISL_1666549, EPI_ISL_1666550, EPI_ISL_1666551, EPI_ISL_1666553, EPI_ISL_1666554, EPI_ISL_1666555, EPI_ISL_1666557, EPI_ISL_1666558, EPI_ISL_1666559, EPI_ISL_1666560, EPI_ISL_1666561, EPI_ISL_1666562, EPI_ISL_1666563, EPI_ISL_1666564, EPI_ISL_1666567, EPI_ISL_1666568, EPI_ISL_1666569, EPI_ISL_1666570, EPI_ISL_1666571, EPI_ISL_1666572, EPI_ISL_1666573, EPI_ISL_1666574, EPI_ISL_1666576, EPI_ISL_1666577, EPI_ISL_1666581, EPI_ISL_1666582, EPI_ISL_1666583, EPI_ISL_1666584, EPI_ISL_1666588, EPI_ISL_1666589, EPI_ISL_1666591, EPI_ISL_1666594, EPI_ISL_1666595, EPI_ISL_1666596, EPI_ISL_1666597, EPI_ISL_1666598, EPI_ISL_1666599, EPI_ISL_1666600, EPI_ISL_1666601, EPI_ISL_1666603, EPI_ISL_1666605, EPI_ISL_1666606, EPI_ISL_1666607, EPI_ISL_1666608, EPI_ISL_1666609, EPI_ISL_1666611, EPI_ISL_1666613, EPI_ISL_1666615, EPI_ISL_1666616, EPI_ISL_1666617, EPI_ISL_1666618, EPI_ISL_1666619, EPI_ISL_1666620, EPI_ISL_1666621, EPI_ISL_1666622, EPI_ISL_1666623, EPI_ISL_1666624, EPI_ISL_1666625, EPI_ISL_1666626, EPI_ISL_1666628, EPI_ISL_1666629, EPI_ISL_1666630, EPI_ISL_1666632, EPI_ISL_1666633, EPI_ISL_1666634, EPI_ISL_1666635, EPI_ISL_1666636, EPI_ISL_1666637, EPI_ISL_1666638, EPI_ISL_1666639, EPI_ISL_1666640, EPI_ISL_1666641, EPI_ISL_1666643, EPI_ISL_1666644, EPI_ISL_1666645, EPI_ISL_1666646, EPI_ISL_1666647, EPI_ISL_1666648, EPI_ISL_1666650, EPI_ISL_1666651, EPI_ISL_1666652, EPI_ISL_1666653, EPI_ISL_1666654, EPI_ISL_1666655, EPI_ISL_1666657, EPI_ISL_1666658, EPI_ISL_1666659, EPI_ISL_1666662, EPI_ISL_1666666, EPI_ISL_1666669, EPI_ISL_1666672, EPI_ISL_1666675, EPI_ISL_1666681, EPI_ISL_1666684, EPI_ISL_1666687, EPI_ISL_1666690, EPI_ISL_1666693, EPI_ISL_1666696, EPI_ISL_1666699, EPI_ISL_1666705, EPI_ISL_1666708, EPI_ISL_1666711, EPI_ISL_1666715, EPI_ISL_1666721, EPI_ISL_1666724, EPI_ISL_1666728, EPI_ISL_1666731, EPI_ISL_1666734, EPI_ISL_1666737, EPI_ISL_1666740, EPI_ISL_1666743, EPI_ISL_1666746, EPI_ISL_1666749, EPI_ISL_1666756, EPI_ISL_1666762, EPI_ISL_1666765, EPI_ISL_1666768, EPI_ISL_1666771, EPI_ISL_1666774, EPI_ISL_1666777, EPI_ISL_1666780, EPI_ISL_1666786, EPI_ISL_1666789, EPI_ISL_1666793, EPI_ISL_1666796, EPI_ISL_1666799, EPI_ISL_1666802, EPI_ISL_1666808, EPI_ISL_1666811, EPI_ISL_1666814, EPI_ISL_1666817, EPI_ISL_1666820, EPI_ISL_1666824, EPI_ISL_1666827, EPI_ISL_1666833, EPI_ISL_1666836, EPI_ISL_1666839, EPI_ISL_1666842, EPI_ISL_1666845, EPI_ISL_1666848, EPI_ISL_1666857, EPI_ISL_1666861, EPI_ISL_1666864, EPI_ISL_1666870, EPI_ISL_1666879, EPI_ISL_1666885, EPI_ISL_1666888, EPI_ISL_1666892, EPI_ISL_1666895, EPI_ISL_1666898, EPI_ISL_1666901, EPI_ISL_1666904, EPI_ISL_1666906, EPI_ISL_1666910, EPI_ISL_1666913, EPI_ISL_1666916, EPI_ISL_1666919, EPI_ISL_1666925, EPI_ISL_1666931, EPI_ISL_1666934, EPI_ISL_1666937, EPI_ISL_1666940, EPI_ISL_1666943, EPI_ISL_1666949, EPI_ISL_1666955, EPI_ISL_1666958, EPI_ISL_1666961, EPI_ISL_1666964, EPI_ISL_1666968, EPI_ISL_1666971, EPI_ISL_1666974, EPI_ISL_1666977, EPI_ISL_1666983, EPI_ISL_1666984, EPI_ISL_1666985, EPI_ISL_1666986, EPI_ISL_1666987, EPI_ISL_1666988, EPI_ISL_1666989, EPI_ISL_1666990, EPI_ISL_1666991, EPI_ISL_1666993, EPI_ISL_1666994, EPI_ISL_1666995, EPI_ISL_1666996, EPI_ISL_1666997, EPI_ISL_1666998, EPI_ISL_1666999, EPI_ISL_1667000, EPI_ISL_1667001, EPI_ISL_1667002, EPI_ISL_1667003, EPI_ISL_1667005, EPI_ISL_1667006, EPI_ISL_1667007, EPI_ISL_1667008, EPI_ISL_1667009, EPI_ISL_1667010, EPI_ISL_1667012, EPI_ISL_1667013, EPI_ISL_1667015, EPI_ISL_1667016, EPI_ISL_1667019, EPI_ISL_1667021, EPI_ISL_1667022, EPI_ISL_1667023, EPI_ISL_1667024, EPI_ISL_1667025, EPI_ISL_1667026, EPI_ISL_1667027, EPI_ISL_1667028, EPI_ISL_1667029, EPI_ISL_1667031, EPI_ISL_1667032, EPI_ISL_1667033, EPI_ISL_1667034, EPI_ISL_1667035, EPI_ISL_1667037, EPI_ISL_1667038, EPI_ISL_1667039, EPI_ISL_1667041, EPI_ISL_1667042, EPI_ISL_1667043, EPI_ISL_1667044, EPI_ISL_1667045, EPI_ISL_1667048, EPI_ISL_1667049, EPI_ISL_1667050, EPI_ISL_1667051, EPI_ISL_1667052, EPI_ISL_1667053, EPI_ISL_1667054, EPI_ISL_1667055, EPI_ISL_1667056, EPI_ISL_1667057, EPI_ISL_1667058, EPI_ISL_1667059, EPI_ISL_1667060, EPI_ISL_1667061, EPI_ISL_1667063, EPI_ISL_1667065, EPI_ISL_1667066, EPI_ISL_1667067, EPI_ISL_1667068, EPI_ISL_1667069, EPI_ISL_1667070, EPI_ISL_1667071, EPI_ISL_1667072, EPI_ISL_1667074, EPI_ISL_1667075, EPI_ISL_1667076, EPI_ISL_1667077, EPI_ISL_1667078, EPI_ISL_1667079, EPI_ISL_1667080, EPI_ISL_1667081, EPI_ISL_1667082, EPI_ISL_1667083, EPI_ISL_1667084, EPI_ISL_1667085, EPI_ISL_1667086, EPI_ISL_1667087, EPI_ISL_1667088, EPI_ISL_1667089, EPI_ISL_1667090, EPI_ISL_1667091, EPI_ISL_1667092, EPI_ISL_1667093, EPI_ISL_1667094, EPI_ISL_1667095, EPI_ISL_1667096, EPI_ISL_1667097, EPI_ISL_1667098, EPI_ISL_1667099, EPI_ISL_1667100, EPI_ISL_1667101, EPI_ISL_1667102, EPI_ISL_1667103, EPI_ISL_1667104, EPI_ISL_1667105, EPI_ISL_1667106, EPI_ISL_1667107, EPI_ISL_1667108, EPI_ISL_1667109, EPI_ISL_1667110, EPI_ISL_1667111, EPI_ISL_1667112, EPI_ISL_1667113, EPI_ISL_1667114, EPI_ISL_1667115, EPI_ISL_1667116, EPI_ISL_1667117, EPI_ISL_1667118, EPI_ISL_1667119, EPI_ISL_1667120, EPI_ISL_1667121, EPI_ISL_1667122, EPI_ISL_1667123, EPI_ISL_1667124, EPI_ISL_1667125, EPI_ISL_1667126, EPI_ISL_1667127, EPI_ISL_1667128, EPI_ISL_1667129, EPI_ISL_1667130, EPI_ISL_1667131, EPI_ISL_1667132, EPI_ISL_1667133, EPI_ISL_1667134, EPI_ISL_1667135, EPI_ISL_1667136, EPI_ISL_1667137, EPI_ISL_1667138, EPI_ISL_1667139, EPI_ISL_1667140, EPI_ISL_1667141, EPI_ISL_1667142, EPI_ISL_1667144, EPI_ISL_1667145, EPI_ISL_1667146, EPI_ISL_1667147, EPI_ISL_1667148, EPI_ISL_1667149, EPI_ISL_1667150, EPI_ISL_1667151, EPI_ISL_1667152, EPI_ISL_1667153, EPI_ISL_1667154, EPI_ISL_1667155, EPI_ISL_1667156, EPI_ISL_1667157, EPI_ISL_1667158, EPI_ISL_1667159, EPI_ISL_1667160, EPI_ISL_1667161, EPI_ISL_1667162, EPI_ISL_1667163, EPI_ISL_1667164, EPI_ISL_1667165, EPI_ISL_1667166, EPI_ISL_1667167, EPI_ISL_1667168, EPI_ISL_1667169, EPI_ISL_1667170, EPI_ISL_1667171, EPI_ISL_1667173, EPI_ISL_1667174, EPI_ISL_1667175, EPI_ISL_1667176, EPI_ISL_1667177, EPI_ISL_1667178, EPI_ISL_1667179, EPI_ISL_1667181, EPI_ISL_1667182, EPI_ISL_1667183, EPI_ISL_1667184, EPI_ISL_1667185, EPI_ISL_1667186, EPI_ISL_1667187, EPI_ISL_1667188, EPI_ISL_1667189, EPI_ISL_1667190, EPI_ISL_1667191, EPI_ISL_1667192, EPI_ISL_1667193, EPI_ISL_1667194, EPI_ISL_1667195, EPI_ISL_1667196, EPI_ISL_1667197, EPI_ISL_1667198, EPI_ISL_1667199, EPI_ISL_1667200 | | | |
| see above | Fulgent Genetics | Centers for Disease Control and Prevention Division of Viral Diseases, Pathogen Discovery | Dakota Howard, Dhwani Batra, Peter W. Cook, Kara Moser, Adrian Paskey, Jason Caravas, Benjamin Rambo-Martin, Shatavia Morrison, Christopher Gulvick, Scott Sammons, Yvette Unoarumhi, Darlene Wagner, Matthew Schmerer, Harry Gao, Mickey Li, John Gao, Joseph Fierro, Benafsh Sapra, Becky Tsai, Yan Meng, Doreen Ng, James Xie, Clinton R. Paden, Duncan MacCannell |
| EPI_ISL_1667201 | Helix | Helix | Nicole L. Washington, Simon White, Geraint Levan, Kelly Schiabor Barrett, Elizabeth Cirulli, Alexandre Bolze, Ary Ascencio, Charlotte Rivera-Garcia, Ryan Cho, Jason Nguyen, Sherry Wang, Jimmy Ramirez, Tyler Cassens, Efren Sandoval, Magnus Isaksson, William Lee, David Becker, Marc Laurent, James Lu, Justin Eagles, Andrew dei Rossi, Dana Wyman, Shishi Luo, Henrique Machado |
| EPI_ISL_1667202, EPI_ISL_1667203, EPI_ISL_1667204, EPI_ISL_1667205, EPI_ISL_1667206, EPI_ISL_1667207, EPI_ISL_1667208, EPI_ISL_1667210, EPI_ISL_1667211, EPI_ISL_1667212, EPI_ISL_1667213, EPI_ISL_1667214, EPI_ISL_1667215, EPI_ISL_1667216, EPI_ISL_1667219, EPI_ISL_1667220, EPI_ISL_1667221, EPI_ISL_1667222, EPI_ISL_1667223, EPI_ISL_1667224, EPI_ISL_1667225, EPI_ISL_1667226, EPI_ISL_1667227, EPI_ISL_1667228, EPI_ISL_1667229, EPI_ISL_1667231, EPI_ISL_1667232, EPI_ISL_1667233, EPI_ISL_1667234, EPI_ISL_1667235, EPI_ISL_1667236, EPI_ISL_1667237, EPI_ISL_1667238, EPI_ISL_1667239, EPI_ISL_1667240, EPI_ISL_1667241, EPI_ISL_1667242, EPI_ISL_1667243, EPI_ISL_1667244, EPI_ISL_1667245, EPI_ISL_1667246, EPI_ISL_1667247, EPI_ISL_1667249, EPI_ISL_1667250, EPI_ISL_1667251, EPI_ISL_1667252, EPI_ISL_1667254, EPI_ISL_1667256, EPI_ISL_1667257, EPI_ISL_1667258, EPI_ISL_1667259, EPI_ISL_1667260, EPI_ISL_1667261, EPI_ISL_1667262, EPI_ISL_1667263, EPI_ISL_1667264, EPI_ISL_1667265, EPI_ISL_1667266, EPI_ISL_1667267, EPI_ISL_1667268, EPI_ISL_1667269, EPI_ISL_1667270, EPI_ISL_1667271, EPI_ISL_1667272, EPI_ISL_1667273, EPI_ISL_1667274, EPI_ISL_1667275, EPI_ISL_1667276, EPI_ISL_1667277, EPI_ISL_1667278, EPI_ISL_1667279, EPI_ISL_1667280, EPI_ISL_1667281, EPI_ISL_1667282, EPI_ISL_1667283, EPI_ISL_1667284, EPI_ISL_1667285, EPI_ISL_1667286, EPI_ISL_1667287, EPI_ISL_1667288, EPI_ISL_1667289, EPI_ISL_1667290, EPI_ISL_1667291, EPI_ISL_1667292, EPI_ISL_1667293, EPI_ISL_1667294, EPI_ISL_1667295, EPI_ISL_1667296, EPI_ISL_1667297, EPI_ISL_1667298, EPI_ISL_1667299, EPI_ISL_1667300, EPI_ISL_1667301, EPI_ISL_1667302, EPI_ISL_1667303, EPI_ISL_1667304, EPI_ISL_1667305, EPI_ISL_1667306, EPI_ISL_1667307, EPI_ISL_1667308, EPI_ISL_1667309, EPI_ISL_1667310, EPI_ISL_1667311, EPI_ISL_1667312, EPI_ISL_1667313, EPI_ISL_1667314, EPI_ISL_1667315, EPI_ISL_1667316, EPI_ISL_1667317, EPI_ISL_1667318, EPI_ISL_1667319, EPI_ISL_1667320, EPI_ISL_1667321, EPI_ISL_1667322, EPI_ISL_1667323, EPI_ISL_1667324, EPI_ISL_1667325, EPI_ISL_1667326, EPI_ISL_1667327, EPI_ISL_1667328, EPI_ISL_1667329, EPI_ISL_1667330, EPI_ISL_1667331, EPI_ISL_1667332, EPI_ISL_1667333, EPI_ISL_1667334, EPI_ISL_1667335, EPI_ISL_1667336, EPI_ISL_1667337, EPI_ISL_1667338, EPI_ISL_1667339, EPI_ISL_1667340, EPI_ISL_1667341, EPI_ISL_1667342, EPI_ISL_1667343, EPI_ISL_1667344, EPI_ISL_1667345, EPI_ISL_1667346, EPI_ISL_1667347, EPI_ISL_1667348, EPI_ISL_1667349, EPI_ISL_1667350, EPI_ISL_1667351, EPI_ISL_1667352, EPI_ISL_1667353, EPI_ISL_1667354, EPI_ISL_1667355, EPI_ISL_1667356, EPI_ISL_1667357, EPI_ISL_1667358, EPI_ISL_1667359, EPI_ISL_1667360, EPI_ISL_1667361, EPI_ISL_1667362, EPI_ISL_1667363, EPI_ISL_1667364, EPI_ISL_1667365, EPI_ISL_1667366, EPI_ISL_1667367, EPI_ISL_1667368, EPI_ISL_1667369, EPI_ISL_1667370, EPI_ISL_1667371, EPI_ISL_1667372, EPI_ISL_1667373, EPI_ISL_1667374, EPI_ISL_1667375, EPI_ISL_1667376, EPI_ISL_1667377, EPI_ISL_1667378, EPI_ISL_1667379, EPI_ISL_1667380, EPI_ISL_1667381, EPI_ISL_1667382, EPI_ISL_1667383, EPI_ISL_1667384, EPI_ISL_1667385, EPI_ISL_1667386, EPI_ISL_1667387, EPI_ISL_1667388, EPI_ISL_1667389, EPI_ISL_1667390, EPI_ISL_1667391, EPI_ISL_1667392, EPI_ISL_1667393, EPI_ISL_1667394, EPI_ISL_1667395, EPI_ISL_1667396, EPI_ISL_1667397, EPI_ISL_1667398, EPI_ISL_1667399, EPI_ISL_1667400, EPI_ISL_1667401, EPI_ISL_1667402, EPI_ISL_1667403, EPI_ISL_1667404, EPI_ISL_1667405, EPI_ISL_1667406, EPI_ISL_1667407, EPI_ISL_1667408, EPI_ISL_1667409, EPI_ISL_1667410, EPI_ISL_1667411, EPI_ISL_1667412, EPI_ISL_1667413, EPI_ISL_1667414, EPI_ISL_1667415, EPI_ISL_1667416, EPI_ISL_1667417, EPI_ISL_1667418, EPI_ISL_1667419, EPI_ISL_1667420, EPI_ISL_1667421, EPI_ISL_1667422, EPI_ISL_1667423, EPI_ISL_1667424, EPI_ISL_1667425, EPI_ISL_1667426, EPI_ISL_1667427, EPI_ISL_1667428, EPI_ISL_1667430 | | | |
| see above | Fulgent Genetics | Centers for Disease Control and Prevention Division of Viral Diseases, Pathogen Discovery | Dakota Howard, Dhwani Batra, Peter W. Cook, Kara Moser, Adrian Paskey, Jason Caravas, Benjamin Rambo-Martin, Shatavia Morrison, Christopher Gulvick, Scott Sammons, Yvette Unoarumhi, Darlene Wagner, Matthew Schmerer, Harry Gao, Mickey Li, John Gao, Joseph Fierro, Benafsh Sapra, Becky Tsai, Yan Meng, Doreen Ng, James Xie, Clinton R. Paden, Duncan MacCannell |
| EPI_ISL_1667431, EPI_ISL_1667432, EPI_ISL_1667436, EPI_ISL_1667439, EPI_ISL_1667440, EPI_ISL_1667441, EPI_ISL_1667442, EPI_ISL_1667443, EPI_ISL_1667444, EPI_ISL_1667445, EPI_ISL_1667446, EPI_ISL_1667447, EPI_ISL_1667448, EPI_ISL_1667449, EPI_ISL_1667450, EPI_ISL_1667451, EPI_ISL_1667452, EPI_ISL_1667453, EPI_ISL_1667454, EPI_ISL_1667455, EPI_ISL_1667456, EPI_ISL_1667457, EPI_ISL_1667458, EPI_ISL_1667459, EPI_ISL_1667460, EPI_ISL_1667461, EPI_ISL_1667462, EPI_ISL_1667463, EPI_ISL_1667464, EPI_ISL_1667465, EPI_ISL_1667466, EPI_ISL_1667467, EPI_ISL_1667468, EPI_ISL_1667470, EPI_ISL_1667473, EPI_ISL_1667476, EPI_ISL_1667477, EPI_ISL_1667478, EPI_ISL_1667479, EPI_ISL_1667480, EPI_ISL_1667482, EPI_ISL_1667483, EPI_ISL_1667484, EPI_ISL_1667485, EPI_ISL_1667486, EPI_ISL_1667487, EPI_ISL_1667488, EPI_ISL_1667489, EPI_ISL_1667490, EPI_ISL_1667491, EPI_ISL_1667492, EPI_ISL_1667493, EPI_ISL_1667494, EPI_ISL_1667495, EPI_ISL_1667496, EPI_ISL_1667497, EPI_ISL_1667498, EPI_ISL_1667499, EPI_ISL_1667500, EPI_ISL_1667501, EPI_ISL_1667502, EPI_ISL_1667503, EPI_ISL_1667504, EPI_ISL_1667505, EPI_ISL_1667506, EPI_ISL_1667507, EPI_ISL_1667508, EPI_ISL_1667509, EPI_ISL_1667510, EPI_ISL_1667511, EPI_ISL_1667512, EPI_ISL_1667513, EPI_ISL_1667514, EPI_ISL_1667515, EPI_ISL_1667516, EPI_ISL_1667517, EPI_ISL_1667518, EPI_ISL_1667519, EPI_ISL_1667520, EPI_ISL_1667521, EPI_ISL_1667522, EPI_ISL_1667523, EPI_ISL_1667524, EPI_ISL_1667525, EPI_ISL_1667526, EPI_ISL_1667527, EPI_ISL_1667528, EPI_ISL_1667529, EPI_ISL_1667530, EPI_ISL_1667531, EPI_ISL_1667532, EPI_ISL_1667533, EPI_ISL_1667534, EPI_ISL_1667535, EPI_ISL_1667536, EPI_ISL_1667537, EPI_ISL_1667538, EPI_ISL_1667540, EPI_ISL_1667541, EPI_ISL_1667542, EPI_ISL_1667543, EPI_ISL_1667544, EPI_ISL_1667545, EPI_ISL_1667546, EPI_ISL_1667547, EPI_ISL_1667548, EPI_ISL_1667549, EPI_ISL_1667550, EPI_ISL_1667551, EPI_ISL_1667552, EPI_ISL_1667553, EPI_ISL_1667554, EPI_ISL_1667555, EPI_ISL_1667556, EPI_ISL_1667557, EPI_ISL_1667558, EPI_ISL_1667559, EPI_ISL_1667560, EPI_ISL_1667561, EPI_ISL_1667562, EPI_ISL_1667563, EPI_ISL_1667564, EPI_ISL_1667565, EPI_ISL_1667566, EPI_ISL_1667567, EPI_ISL_1667568, EPI_ISL_1667569, EPI_ISL_1667570, EPI_ISL_1667571, EPI_ISL_1667572, EPI_ISL_1667573, EPI_ISL_1667574, EPI_ISL_1667575, EPI_ISL_1667576, EPI_ISL_1667577, EPI_ISL_1667578, EPI_ISL_1667579, EPI_ISL_1667580, EPI_ISL_1667581, EPI_ISL_1667582, EPI_ISL_1667583, EPI_ISL_1667584, EPI_ISL_1667585, EPI_ISL_1667586, EPI_ISL_1667587, EPI_ISL_1667588, EPI_ISL_1667589, EPI_ISL_1667590, EPI_ISL_1667591, EPI_ISL_1667592, EPI_ISL_1667593, EPI_ISL_1667595, EPI_ISL_1667596, EPI_ISL_1667597, EPI_ISL_1667598, EPI_ISL_1667599, EPI_ISL_1667600, EPI_ISL_1667601, EPI_ISL_1667602, EPI_ISL_1667603, EPI_ISL_1667604, EPI_ISL_1667605, EPI_ISL_1667606, EPI_ISL_1667607, EPI_ISL_1667608, EPI_ISL_1667609, EPI_ISL_1667610, EPI_ISL_1667611, EPI_ISL_1667612, EPI_ISL_1667613, EPI_ISL_1667614, EPI_ISL_1667615, EPI_ISL_1667616, EPI_ISL_1667617, EPI_ISL_1667618, EPI_ISL_1667619, EPI_ISL_1667620, EPI_ISL_1667621, EPI_ISL_1667622, EPI_ISL_1667623, EPI_ISL_1667624, EPI_ISL_1667625, EPI_ISL_1667626, EPI_ISL_1667627, EPI_ISL_1667629, EPI_ISL_1667630, EPI_ISL_1667631, EPI_ISL_1667632, EPI_ISL_1667633, EPI_ISL_1667634, EPI_ISL_1667635, EPI_ISL_1667636, EPI_ISL_1667637, EPI_ISL_1667638, EPI_ISL_1667639, EPI_ISL_1667640, EPI_ISL_1667641, EPI_ISL_1667642, EPI_ISL_1667643, EPI_ISL_1667644, EPI_ISL_1667645, EPI_ISL_1667646, EPI_ISL_1667647, EPI_ISL_1667648, EPI_ISL_1667649, EPI_ISL_1667650, EPI_ISL_1667651, EPI_ISL_1667652, EPI_ISL_1667653, EPI_ISL_1667654, EPI_ISL_1667655, EPI_ISL_1667656, EPI_ISL_1667657, EPI_ISL_1667658, EPI_ISL_1667659, EPI_ISL_1667660, EPI_ISL_1667661, EPI_ISL_1667662, EPI_ISL_1667663, EPI_ISL_1667664, EPI_ISL_1667665, EPI_ISL_1667666, EPI_ISL_1667667, EPI_ISL_1667668, EPI_ISL_1667669, EPI_ISL_1667670, EPI_ISL_1667671, EPI_ISL_1667672, EPI_ISL_1667673, EPI_ISL_1667674, EPI_ISL_1667675, EPI_ISL_1667676, EPI_ISL_1667677, EPI_ISL_1667678, EPI_ISL_1667679, EPI_ISL_1667680, EPI_ISL_1667681, EPI_ISL_1667682, EPI_ISL_1667683, EPI_ISL_1667684, EPI_ISL_1667685, EPI_ISL_1667686, EPI_ISL_1667687, EPI_ISL_1667688, EPI_ISL_1667689, EPI_ISL_1667690, EPI_ISL_1667691, EPI_ISL_1667692, EPI_ISL_1667693, EPI_ISL_1667694, EPI_ISL_1667695, EPI_ISL_1667696, EPI_ISL_1667697, EPI_ISL_1667698, EPI_ISL_1667699, EPI_ISL_1667700, EPI_ISL_1667701, EPI_ISL_1667702, EPI_ISL_1667703, EPI_ISL_1667704, EPI_ISL_1667705, EPI_ISL_1667706, EPI_ISL_1667707, EPI_ISL_1667708, EPI_ISL_1667709, EPI_ISL_1667710, EPI_ISL_1667711, EPI_ISL_1667712, EPI_ISL_1667713, EPI_ISL_1667714, EPI_ISL_1667715, EPI_ISL_1667716, EPI_ISL_1667717, EPI_ISL_1667718, EPI_ISL_1667719, EPI_ISL_1667720, EPI_ISL_1667721, EPI_ISL_1667722, EPI_ISL_1667723, EPI_ISL_1667724, EPI_ISL_1667725, EPI_ISL_1667726, EPI_ISL_1667727, EPI_ISL_1667728, EPI_ISL_1667729, EPI_ISL_1667730, EPI_ISL_1667731, EPI_ISL_1667732, EPI_ISL_1667733, EPI_ISL_1667734, EPI_ISL_1667735, EPI_ISL_1667736, EPI_ISL_1667737, EPI_ISL_1667738, EPI_ISL_1667739, EPI_ISL_1667740, EPI_ISL_1667741, EPI_ISL_1667742, EPI_ISL_1667743, EPI_ISL_1667744, EPI_ISL_1667746, EPI_ISL_1667747, EPI_ISL_1667748, EPI_ISL_1667749, EPI_ISL_1667750, EPI_ISL_1667751, EPI_ISL_1667752, EPI_ISL_1667753, EPI_ISL_1667754, EPI_ISL_1667755, EPI_ISL_1667756, EPI_ISL_1667757, EPI_ISL_1667758, EPI_ISL_1667759, EPI_ISL_1667760, EPI_ISL_1667761, EPI_ISL_1667762, EPI_ISL_1667763, EPI_ISL_1667764, EPI_ISL_1667765, EPI_ISL_1667766, EPI_ISL_1667767, EPI_ISL_1667768, EPI_ISL_1667769, EPI_ISL_1667770, EPI_ISL_1667771, EPI_ISL_1667772, EPI_ISL_1667773, EPI_ISL_1667774, EPI_ISL_1667775, EPI_ISL_1667776, EPI_ISL_1667777, EPI_ISL_1667778, EPI_ISL_1667779, EPI_ISL_1667780, EPI_ISL_1667781, EPI_ISL_1667782, EPI_ISL_1667783, EPI_ISL_1667784, EPI_ISL_1667785, EPI_ISL_1667786, EPI_ISL_1667787, EPI_ISL_1667788, EPI_ISL_1667789, EPI_ISL_1667790, EPI_ISL_1667791, EPI_ISL_1667792, EPI_ISL_1667793, EPI_ISL_1667794, EPI_ISL_1667795, EPI_ISL_1667796, EPI_ISL_1667797, EPI_ISL_1667798, EPI_ISL_1667799, EPI_ISL_1667800, EPI_ISL_1667801, EPI_ISL_1667802, EPI_ISL_1667803, EPI_ISL_1667804, EPI_ISL_1667805, EPI_ISL_1667806, EPI_ISL_1667807, EPI_ISL_1667808, EPI_ISL_1667809, EPI_ISL_1667810, EPI_ISL_1667811, EPI_ISL_1667812, EPI_ISL_1667813, EPI_ISL_1667814, EPI_ISL_1667815, EPI_ISL_1667816, EPI_ISL_1667817, EPI_ISL_1667818, EPI_ISL_1667819, EPI_ISL_1667820, EPI_ISL_1667821, EPI_ISL_1667822, EPI_ISL_1667823, EPI_ISL_1667824, EPI_ISL_1667825, EPI_ISL_1667826, EPI_ISL_1667827, EPI_ISL_1667828, EPI_ISL_1667829, EPI_ISL_1667830, EPI_ISL_1667831, EPI_ISL_1667832, EPI_ISL_1667833, EPI_ISL_1667835, EPI_ISL_1667837, EPI_ISL_1667838, EPI_ISL_1667839, EPI_ISL_1667840, EPI_ISL_1667841, EPI_ISL_1667843, EPI_ISL_1667845, EPI_ISL_1667847, EPI_ISL_1667850, EPI_ISL_1667852, EPI_ISL_1667854, EPI_ISL_1667856, EPI_ISL_1667857, EPI_ISL_1667858, EPI_ISL_1667859, EPI_ISL_1667860, EPI_ISL_1667861, EPI_ISL_1667862, EPI_ISL_1667863, EPI_ISL_1667864, EPI_ISL_1667865, EPI_ISL_1667866, EPI_ISL_1667867, EPI_ISL_1667868, EPI_ISL_1667869, EPI_ISL_1667870, EPI_ISL_1667871, EPI_ISL_1667872, EPI_ISL_1667873, EPI_ISL_1667874, EPI_ISL_1667875, EPI_ISL_1667876, EPI_ISL_1667877, EPI_ISL_1667878, EPI_ISL_1667879, EPI_ISL_1667880, EPI_ISL_1667881, EPI_ISL_1667882, EPI_ISL_1667884, EPI_ISL_1667886, EPI_ISL_1667888, EPI_ISL_1667890, EPI_ISL_1667892, EPI_ISL_1667896, EPI_ISL_1667898, EPI_ISL_1667900, EPI_ISL_1667902, EPI_ISL_1667905, EPI_ISL_1667907, EPI_ISL_1667909, EPI_ISL_1667911, EPI_ISL_1667913, EPI_ISL_1667915, EPI_ISL_1667917, EPI_ISL_1667919, EPI_ISL_1667921, EPI_ISL_1667923, EPI_ISL_1667925, EPI_ISL_1667927, EPI_ISL_1667929, EPI_ISL_1667931, EPI_ISL_1667933, EPI_ISL_1667935, EPI_ISL_1667937, EPI_ISL_1667939, EPI_ISL_1667941, EPI_ISL_1667943, EPI_ISL_1667945, EPI_ISL_1667947, EPI_ISL_1667949, EPI_ISL_1667951, EPI_ISL_1667953, EPI_ISL_1667955, EPI_ISL_1667957, EPI_ISL_1667959, EPI_ISL_1667961, EPI_ISL_1667963, EPI_ISL_1667965, EPI_ISL_1667967, EPI_ISL_1667969, EPI_ISL_1667970, EPI_ISL_1667971, EPI_ISL_1667972, EPI_ISL_1667973, EPI_ISL_1667974, EPI_ISL_1667975, EPI_ISL_1667976, EPI_ISL_1667977, EPI_ISL_1667978, EPI_ISL_1667979, EPI_ISL_1667980, EPI_ISL_1667981, EPI_ISL_1667982, EPI_ISL_1667983, EPI_ISL_1667984, EPI_ISL_1667985, EPI_ISL_1667986, EPI_ISL_1667987, EPI_ISL_1667988, EPI_ISL_1667989, EPI_ISL_1667990, EPI_ISL_1667991, EPI_ISL_1667992, EPI_ISL_1667993, EPI_ISL_1667994, EPI_ISL_1667995, EPI_ISL_1667996, EPI_ISL_1667997, EPI_ISL_1667998, EPI_ISL_1667999, EPI_ISL_1668000, EPI_ISL_1668001, EPI_ISL_1668002, EPI_ISL_1668003, EPI_ISL_1668004, EPI_ISL_1668005, EPI_ISL_1668006, EPI_ISL_1668007, EPI_ISL_1668008, EPI_ISL_1668009, EPI_ISL_1668010, EPI_ISL_1668011, EPI_ISL_1668012, EPI_ISL_1668013, EPI_ISL_1668014, EPI_ISL_1668015, EPI_ISL_1668016, EPI_ISL_1668017, EPI_ISL_1668018, EPI_ISL_1668019, EPI_ISL_1668020, EPI_ISL_1668021, EPI_ISL_1668022, EPI_ISL_1668023, EPI_ISL_1668024, EPI_ISL_1668025, EPI_ISL_1668026, EPI_ISL_1668027, EPI_ISL_1668028, EPI_ISL_1668029, EPI_ISL_1668030, EPI_ISL_1668031, EPI_ISL_1668032, EPI_ISL_1668033, EPI_ISL_1668034, EPI_ISL_1668035, EPI_ISL_1668036, EPI_ISL_1668037, EPI_ISL_1668038, EPI_ISL_1668039, EPI_ISL_1668040, EPI_ISL_1668041, EPI_ISL_1668042, EPI_ISL_1668043, EPI_ISL_1668044, EPI_ISL_1668045, EPI_ISL_1668046, EPI_ISL_1668047, EPI_ISL_1668049, EPI_ISL_1668050, EPI_ISL_1668051, EPI_ISL_1668052, EPI_ISL_1668053, EPI_ISL_1668054, EPI_ISL_1668055, EPI_ISL_1668056, EPI_ISL_1668057, EPI_ISL_1668058, EPI_ISL_1668059, EPI_ISL_1668060, EPI_ISL_1668061, EPI_ISL_1668062, EPI_ISL_1668063, EPI_ISL_1668065, EPI_ISL_1668066, EPI_ISL_1668067, EPI_ISL_1668068, EPI_ISL_1668069, EPI_ISL_1668070, EPI_ISL_1668071, EPI_ISL_1668072, EPI_ISL_1668073, EPI_ISL_1668074, EPI_ISL_1668075, EPI_ISL_1668076, EPI_ISL_1668077, EPI_ISL_1668078, EPI_ISL_1668079, EPI_ISL_1668080, EPI_ISL_1668081, EPI_ISL_1668082, EPI_ISL_1668083, EPI_ISL_1668084, EPI_ISL_1668085, EPI_ISL_1668086, EPI_ISL_1668087, EPI_ISL_1668088, EPI_ISL_1668089, EPI_ISL_1668090, EPI_ISL_1668091, EPI_ISL_1668092, EPI_ISL_1668093, EPI_ISL_1668094, EPI_ISL_1668095, EPI_ISL_1668096, EPI_ISL_1668097, EPI_ISL_1668098, EPI_ISL_1668099, EPI_ISL_1668100, EPI_ISL_1668102, EPI_ISL_1668103, EPI_ISL_1668104, EPI_ISL_1668105, EPI_ISL_1668106, EPI_ISL_1668107, EPI_ISL_1668108, EPI_ISL_1668109, EPI_ISL_1668110, EPI_ISL_1668111, EPI_ISL_1668112, EPI_ISL_1668113, EPI_ISL_1668114, EPI_ISL_1668115, EPI_ISL_1668116, EPI_ISL_1668117, EPI_ISL_1668118, EPI_ISL_1668119, EPI_ISL_1668120, EPI_ISL_1668121, EPI_ISL_1668122, EPI_ISL_1668123, EPI_ISL_1668124, EPI_ISL_1668125, EPI_ISL_1668126, EPI_ISL_1668127, EPI_ISL_1668128, EPI_ISL_1668129, EPI_ISL_1668130, EPI_ISL_1668131, EPI_ISL_1668132, EPI_ISL_1668133, EPI_ISL_1668134, EPI_ISL_1668135, EPI_ISL_1668136, EPI_ISL_1668137, EPI_ISL_1668138, EPI_ISL_1668139, EPI_ISL_1668140, EPI_ISL_1668141, EPI_ISL_1668142, EPI_ISL_1668143, EPI_ISL_1668144, EPI_ISL_1668145, EPI_ISL_1668146, EPI_ISL_1668147, EPI_ISL_1668148, EPI_ISL_1668149, EPI_ISL_1668150, EPI_ISL_1668151, EPI_ISL_1668152, EPI_ISL_1668153, EPI_ISL_1668154, EPI_ISL_1668155, EPI_ISL_1668156, EPI_ISL_1668157, EPI_ISL_1668158, EPI_ISL_1668159, EPI_ISL_1668160, EPI_ISL_1668161, EPI_ISL_1668162, EPI_ISL_1668163, EPI_ISL_1668164, EPI_ISL_1668165, EPI_ISL_1668166, EPI_ISL_1668167, EPI_ISL_1668168, EPI_ISL_1668169, EPI_ISL_1668170, EPI_ISL_1668171, EPI_ISL_1668172, EPI_ISL_1668173, EPI_ISL_1668174, EPI_ISL_1668175, EPI_ISL_1668176, EPI_ISL_1668177, EPI_ISL_1668178, EPI_ISL_1668179, EPI_ISL_1668180, EPI_ISL_1668181, EPI_ISL_1668182, EPI_ISL_1668183, EPI_ISL_1668184, EPI_ISL_1668185, EPI_ISL_1668186, EPI_ISL_1668187, EPI_ISL_1668188, EPI_ISL_1668189, EPI_ISL_1668190, EPI_ISL_1668191, EPI_ISL_1668192, EPI_ISL_1668193, EPI_ISL_1668194, EPI_ISL_1668195, EPI_ISL_1668196, EPI_ISL_1668197, EPI_ISL_1668198, EPI_ISL_1668199, EPI_ISL_1668200, EPI_ISL_1668201, EPI_ISL_1668202, EPI_ISL_1668203, EPI_ISL_1668204, EPI_ISL_1668205, EPI_ISL_1668206, EPI_ISL_1668207, EPI_ISL_1668208, EPI_ISL_1668209, EPI_ISL_1668210, EPI_ISL_1668211, EPI_ISL_1668212, EPI_ISL_1668213, EPI_ISL_1668214, EPI_ISL_1668215, EPI_ISL_1668216, EPI_ISL_1668217, EPI_ISL_1668218, EPI_ISL_1668219, EPI_ISL_1668220, EPI_ISL_1668221, EPI_ISL_1668222, EPI_ISL_1668223, EPI_ISL_1668224, EPI_ISL_1668225, EPI_ISL_1668226, EPI_ISL_1668227, EPI_ISL_1668228, EPI_ISL_1668229, EPI_ISL_1668230, EPI_ISL_1668231, EPI_ISL_1668232, EPI_ISL_1668233, EPI_ISL_1668234, EPI_ISL_1668235, EPI_ISL_1668236, EPI_ISL_1668237, EPI_ISL_1668238, EPI_ISL_1668239, EPI_ISL_1668240, EPI_ISL_1668241, EPI_ISL_1668242, EPI_ISL_1668243, EPI_ISL_1668245, EPI_ISL_1668246, EPI_ISL_1668247, EPI_ISL_1668248, EPI_ISL_1668249, EPI_ISL_1668250, EPI_ISL_1668251, EPI_ISL_1668252, EPI_ISL_1668253, EPI_ISL_1668254, EPI_ISL_1668255, EPI_ISL_1668256, EPI_ISL_1668257, EPI_ISL_1668258, EPI_ISL_1668259, EPI_ISL_1668260, EPI_ISL_1668261, EPI_ISL_1668262, EPI_ISL_1668263, EPI_ISL_1668264, EPI_ISL_1668265, EPI_ISL_1668266, EPI_ISL_1668267, EPI_ISL_1668268, EPI_ISL_1668269, EPI_ISL_1668270, EPI_ISL_1668271, EPI_ISL_1668272, EPI_ISL_1668273, EPI_ISL_1668274, EPI_ISL_1668275, EPI_ISL_1668276, EPI_ISL_1668277, EPI_ISL_1668278, EPI_ISL_1668279, EPI_ISL_1668280, EPI_ISL_1668281, EPI_ISL_1668282, EPI_ISL_1668283, EPI_ISL_1668284, EPI_ISL_1668285, EPI_ISL_1668286, EPI_ISL_1668287, EPI_ISL_1668288, EPI_ISL_1668289, EPI_ISL_1668290, EPI_ISL_1668291, EPI_ISL_1668292, EPI_ISL_1668293, EPI_ISL_1668294, EPI_ISL_1668295, EPI_ISL_1668296, EPI_ISL_1668297, EPI_ISL_1668298, EPI_ISL_1668299, EPI_ISL_1668300, EPI_ISL_1668301, EPI_ISL_1668302, EPI_ISL_1668303, EPI_ISL_1668304, EPI_ISL_1668305, EPI_ISL_1668306, EPI_ISL_1668307, EPI_ISL_1668308, EPI_ISL_1668309, EPI_ISL_1668310, EPI_ISL_1668311, EPI_ISL_1668312, EPI_ISL_1668313, EPI_ISL_1668314, EPI_ISL_1668315, EPI_ISL_1668316, EPI_ISL_1668317, EPI_ISL_1668318, EPI_ISL_1668319, EPI_ISL_1668320, EPI_ISL_1668321, EPI_ISL_1668322, EPI_ISL_1668323, EPI_ISL_1668324, EPI_ISL_1668325, EPI_ISL_1668326, EPI_ISL_1668327, EPI_ISL_1668328, EPI_ISL_1668329, EPI_ISL_1668330, EPI_ISL_1668331, EPI_ISL_1668332, EPI_ISL_1668333, EPI_ISL_1668334, EPI_ISL_1668335, EPI_ISL_1668336, EPI_ISL_1668337, EPI_ISL_1668338, EPI_ISL_1668339, EPI_ISL_1668340, EPI_ISL_1668341, EPI_ISL_1668342, EPI_ISL_1668343, EPI_ISL_1668344, EPI_ISL_1668345, EPI_ISL_1668346, EPI_ISL_1668347, EPI_ISL_1668348, EPI_ISL_1668349, EPI_ISL_1668350, EPI_ISL_1668351, EPI_ISL_1668352, EPI_ISL_1668353, EPI_ISL_1668354, EPI_ISL_1668355, EPI_ISL_1668356, EPI_ISL_1668357, EPI_ISL_1668358, EPI_ISL_1668359, EPI_ISL_1668360, EPI_ISL_1668361, EPI_ISL_1668362, EPI_ISL_1668363, EPI_ISL_1668364, EPI_ISL_1668365, EPI_ISL_1668366, EPI_ISL_1668367, EPI_ISL_1668368, EPI_ISL_1668369, EPI_ISL_1668370, EPI_ISL_1668371, EPI_ISL_1668372, EPI_ISL_1668373, EPI_ISL_1668374, EPI_ISL_1668375, EPI_ISL_1668376, EPI_ISL_1668377, EPI_ISL_1668378, EPI_ISL_1668379, EPI_ISL_1668380, EPI_ISL_1668381, EPI_ISL_1668382, EPI_ISL_1668383, EPI_ISL_1668384, EPI_ISL_1668385, EPI_ISL_1668386, EPI_ISL_1668387, EPI_ISL_1668388, EPI_ISL_1668389, EPI_ISL_1668390, EPI_ISL_1668391, EPI_ISL_1668392, EPI_ISL_1668393, EPI_ISL_1668394, EPI_ISL_1668395, EPI_ISL_1668396, EPI_ISL_1668397, EPI_ISL_1668398, EPI_ISL_1668399, EPI_ISL_1668400, EPI_ISL_1668401, EPI_ISL_1668402, EPI_ISL_1668403, EPI_ISL_1668404, EPI_ISL_1668405, EPI_ISL_1668406, EPI_ISL_1668408, EPI_ISL_1668409, EPI_ISL_1668410, EPI_ISL_1668411, EPI_ISL_1668412, EPI_ISL_1668413, EPI_ISL_1668414, EPI_ISL_1668415, EPI_ISL_1668416, EPI_ISL_1668417, EPI_ISL_1668418, EPI_ISL_1668419, EPI_ISL_1668420, EPI_ISL_1668421, EPI_ISL_1668422, EPI_ISL_1668423, EPI_ISL_1668424, EPI_ISL_1668425, EPI_ISL_1668426, EPI_ISL_1668427, EPI_ISL_1668428, EPI_ISL_1668429, EPI_ISL_1668430, EPI_ISL_1668431, EPI_ISL_1668432, EPI_ISL_1668433, EPI_ISL_1668434, EPI_ISL_1668435, EPI_ISL_1668436, EPI_ISL_1668437, EPI_ISL_1668438, EPI_ISL_1668439, EPI_ISL_1668440, EPI_ISL_1668441, EPI_ISL_1668442, EPI_ISL_1668443, EPI_ISL_1668444, EPI_ISL_1668445, EPI_ISL_1668446, EPI_ISL_1668447, EPI_ISL_1668448, EPI_ISL_1668449, EPI_ISL_1668450, EPI_ISL_1668451, EPI_ISL_1668452 | | | |
| see above | Aegis Sciences Corporation | Centers for Disease Control and Prevention Division of Viral Diseases, Pathogen Discovery | Dakota Howard, Dhwani Batra, Peter W. Cook, Kara Moser, Adrian Paskey, Jason Caravas, Benjamin Rambo-Martin, Shatavia Morrison, Christopher Gulvick, Scott Sammons, Yvette Unoarumhi, Darlene Wagner, Matthew Schmerer, Cyndi Clark, Patrick Campbell, Rob Case, Vikramsinha Ghorpade, Holly Houdeshell, Ola Kvalvaag, Dillon Nall, Ethan Sanders, Alec Vest, Shaun Westlund, Matthew Hardison, Clinton R. Paden, Duncan MacCannell |
| EPI_ISL_1668453, EPI_ISL_1668454, EPI_ISL_1668455, EPI_ISL_1668456, EPI_ISL_1668457, EPI_ISL_1668458, EPI_ISL_1668459, EPI_ISL_1668460, EPI_ISL_1668461, EPI_ISL_1668462, EPI_ISL_1668463, EPI_ISL_1668464, EPI_ISL_1668465, EPI_ISL_1668466, EPI_ISL_1668467, EPI_ISL_1668468, EPI_ISL_1668469, EPI_ISL_1668470, EPI_ISL_1668471, EPI_ISL_1668472, EPI_ISL_1668473, EPI_ISL_1668474, EPI_ISL_1668475, EPI_ISL_1668476, EPI_ISL_1668477, EPI_ISL_1668478, EPI_ISL_1668479, EPI_ISL_1668480, EPI_ISL_1668481, EPI_ISL_1668482, EPI_ISL_1668483, EPI_ISL_1668484, EPI_ISL_1668485, EPI_ISL_1668486, EPI_ISL_1668487, EPI_ISL_1668488, EPI_ISL_1668489, EPI_ISL_1668490, EPI_ISL_1668491, EPI_ISL_1668492, EPI_ISL_1668493, EPI_ISL_1668494, EPI_ISL_1668495, EPI_ISL_1668496, EPI_ISL_1668497, EPI_ISL_1668498, EPI_ISL_1668499, EPI_ISL_1668500, EPI_ISL_1668501, EPI_ISL_1668502, EPI_ISL_1668503, EPI_ISL_1668504, EPI_ISL_1668505, EPI_ISL_1668506, EPI_ISL_1668507, EPI_ISL_1668508, EPI_ISL_1668510, EPI_ISL_1668511, EPI_ISL_1668512, EPI_ISL_1668513, EPI_ISL_1668514, EPI_ISL_1668515, EPI_ISL_1668516, EPI_ISL_1668517, EPI_ISL_1668518, EPI_ISL_1668519, EPI_ISL_1668520, EPI_ISL_1668521, EPI_ISL_1668522, EPI_ISL_1668523, EPI_ISL_1668524, EPI_ISL_1668525, EPI_ISL_1668526, EPI_ISL_1668527, EPI_ISL_1668528, EPI_ISL_1668529, EPI_ISL_1668530, EPI_ISL_1668533, EPI_ISL_1668536, EPI_ISL_1668539, EPI_ISL_1668543, EPI_ISL_1668545, EPI_ISL_1668548, EPI_ISL_1668551, EPI_ISL_1668554, EPI_ISL_1668557, EPI_ISL_1668558, EPI_ISL_1668562, EPI_ISL_1668565, EPI_ISL_1668568, EPI_ISL_1668570, EPI_ISL_1668573, EPI_ISL_1668576, EPI_ISL_1668579, EPI_ISL_1668580, EPI_ISL_1668581, EPI_ISL_1668582, EPI_ISL_1668583, EPI_ISL_1668584, EPI_ISL_1668585, EPI_ISL_1668586, EPI_ISL_1668587, EPI_ISL_1668588, EPI_ISL_1668589, EPI_ISL_1668590, EPI_ISL_1668591, EPI_ISL_1668592, EPI_ISL_1668593, EPI_ISL_1668594, EPI_ISL_1668595, EPI_ISL_1668596, EPI_ISL_1668597, EPI_ISL_1668598, EPI_ISL_1668599, EPI_ISL_1668600, EPI_ISL_1668601, EPI_ISL_1668602, EPI_ISL_1668603, EPI_ISL_1668604, EPI_ISL_1668605, EPI_ISL_1668606, EPI_ISL_1668607, EPI_ISL_1668608, EPI_ISL_1668609, EPI_ISL_1668610, EPI_ISL_1668611, EPI_ISL_1668612, EPI_ISL_1668613, EPI_ISL_1668614, EPI_ISL_1668615, EPI_ISL_1668616, EPI_ISL_1668617, EPI_ISL_1668618, EPI_ISL_1668619, EPI_ISL_1668620, EPI_ISL_1668621, EPI_ISL_1668623, EPI_ISL_1668624, EPI_ISL_1668625, EPI_ISL_1668626, EPI_ISL_1668627, EPI_ISL_1668628, EPI_ISL_1668629, EPI_ISL_1668630, EPI_ISL_1668631, EPI_ISL_1668632, EPI_ISL_1668633, EPI_ISL_1668634, EPI_ISL_1668635, EPI_ISL_1668636, EPI_ISL_1668637, EPI_ISL_1668638, EPI_ISL_1668639, EPI_ISL_1668640, EPI_ISL_1668641, EPI_ISL_1668642, EPI_ISL_1668643, EPI_ISL_1668644, EPI_ISL_1668645, EPI_ISL_1668646, EPI_ISL_1668647, EPI_ISL_1668648, EPI_ISL_1668649, EPI_ISL_1668650, EPI_ISL_1668651, EPI_ISL_1668652, EPI_ISL_1668653, EPI_ISL_1668654, EPI_ISL_1668655, EPI_ISL_1668656, EPI_ISL_1668657, EPI_ISL_1668658, EPI_ISL_1668659, EPI_ISL_1668660, | | | |

| | | | |
|---|---|---|---|
| EPI_ISL_1668661, EPI_ISL_1668662, EPI_ISL_1668663, EPI_ISL_1668664, EPI_ISL_1668665, EPI_ISL_1668666, EPI_ISL_1668667, EPI_ISL_1668668, EPI_ISL_1668669, EPI_ISL_1668670, EPI_ISL_1668671, EPI_ISL_1668672, EPI_ISL_1668673, EPI_ISL_1668674, EPI_ISL_1668675, EPI_ISL_1668676, EPI_ISL_1668677, EPI_ISL_1668678, EPI_ISL_1668679, EPI_ISL_1668680, EPI_ISL_1668681, EPI_ISL_1668682, EPI_ISL_1668683, EPI_ISL_1668684, EPI_ISL_1668685, EPI_ISL_1668686, EPI_ISL_1668687, EPI_ISL_1668688, EPI_ISL_1668689, EPI_ISL_1668690, EPI_ISL_1668691, EPI_ISL_1668692, EPI_ISL_1668693, EPI_ISL_1668694, EPI_ISL_1668695, EPI_ISL_1668696, EPI_ISL_1668697, EPI_ISL_1668698, EPI_ISL_1668699, EPI_ISL_1668700, EPI_ISL_1668701, EPI_ISL_1668702, EPI_ISL_1668703, EPI_ISL_1668704, EPI_ISL_1668705, EPI_ISL_1668706, EPI_ISL_1668707, EPI_ISL_1668708, EPI_ISL_1668709, EPI_ISL_1668710, EPI_ISL_1668711, EPI_ISL_1668712, EPI_ISL_1668713, EPI_ISL_1668714, EPI_ISL_1668715, EPI_ISL_1668716, EPI_ISL_1668717, EPI_ISL_1668718, EPI_ISL_1668719, EPI_ISL_1668720, EPI_ISL_1668721, EPI_ISL_1668722, EPI_ISL_1668723, EPI_ISL_1668724, EPI_ISL_1668725, EPI_ISL_1668726, EPI_ISL_1668727, EPI_ISL_1668728, EPI_ISL_1668729, EPI_ISL_1668730, EPI_ISL_1668731, EPI_ISL_1668732, EPI_ISL_1668733, EPI_ISL_1668734, EPI_ISL_1668735, EPI_ISL_1668736, EPI_ISL_1668737, EPI_ISL_1668738, EPI_ISL_1668739, EPI_ISL_1668740, EPI_ISL_1668741, EPI_ISL_1668742, EPI_ISL_1668743, EPI_ISL_1668744, EPI_ISL_1668745, EPI_ISL_1668746, EPI_ISL_1668747, EPI_ISL_1668748, EPI_ISL_1668749, EPI_ISL_1668750, EPI_ISL_1668751, EPI_ISL_1668752, EPI_ISL_1668753, EPI_ISL_1668754, EPI_ISL_1668755, EPI_ISL_1668756, EPI_ISL_1668757, EPI_ISL_1668758, EPI_ISL_1668759, EPI_ISL_1668760, EPI_ISL_1668761, EPI_ISL_1668762, EPI_ISL_1668763, EPI_ISL_1668764, EPI_ISL_1668765, EPI_ISL_1668766, EPI_ISL_1668767, EPI_ISL_1668768, EPI_ISL_1668769, EPI_ISL_1668770, EPI_ISL_1668771, EPI_ISL_1668772, EPI_ISL_1668773, EPI_ISL_1668774, EPI_ISL_1668775, EPI_ISL_1668776, EPI_ISL_1668777, EPI_ISL_1668778, EPI_ISL_1668779, EPI_ISL_1668780, EPI_ISL_1668781, EPI_ISL_1668782, EPI_ISL_1668783, EPI_ISL_1668784, EPI_ISL_1668785, EPI_ISL_1668786, EPI_ISL_1668787, EPI_ISL_1668788, EPI_ISL_1668789, EPI_ISL_1668790, EPI_ISL_1668791, EPI_ISL_1668792, EPI_ISL_1668793, EPI_ISL_1668794, EPI_ISL_1668795, EPI_ISL_1668796, EPI_ISL_1668797, EPI_ISL_1668798, EPI_ISL_1668800, EPI_ISL_1668803, EPI_ISL_1668806, EPI_ISL_1668808, EPI_ISL_1668811, EPI_ISL_1668814, EPI_ISL_1668818, EPI_ISL_1668843, EPI_ISL_1668844, EPI_ISL_1668845, EPI_ISL_1668846, EPI_ISL_1668847, EPI_ISL_1668848, EPI_ISL_1668849, EPI_ISL_1668850, EPI_ISL_1668851, EPI_ISL_1668852, EPI_ISL_1668853, EPI_ISL_1668854, EPI_ISL_1668855, EPI_ISL_1668856, EPI_ISL_1668857, EPI_ISL_1668858, EPI_ISL_1668859, EPI_ISL_1668860, EPI_ISL_1668861, EPI_ISL_1668862, EPI_ISL_1668863, EPI_ISL_1668864, EPI_ISL_1668865, EPI_ISL_1668866, EPI_ISL_1668867, EPI_ISL_1668868, EPI_ISL_1668869, EPI_ISL_1668870, EPI_ISL_1668871, EPI_ISL_1668872, EPI_ISL_1668873, EPI_ISL_1668874, EPI_ISL_1668875, EPI_ISL_1668876, EPI_ISL_1668877, EPI_ISL_1668878, EPI_ISL_1668879, EPI_ISL_1668880, EPI_ISL_1668881, EPI_ISL_1668882, EPI_ISL_1668883, EPI_ISL_1668884, EPI_ISL_1668885, EPI_ISL_1668886, EPI_ISL_1668887, EPI_ISL_1668888, EPI_ISL_1668889, EPI_ISL_1668890, EPI_ISL_1668891, EPI_ISL_1668892, EPI_ISL_1668893, EPI_ISL_1668894, EPI_ISL_1668895, EPI_ISL_1668896, EPI_ISL_1668897, EPI_ISL_1668898, EPI_ISL_1668899, EPI_ISL_1668900, EPI_ISL_1668901, EPI_ISL_1668902, EPI_ISL_1668903, EPI_ISL_1668904, EPI_ISL_1668905, EPI_ISL_1668906, EPI_ISL_1668907, EPI_ISL_1668908, EPI_ISL_1668909, EPI_ISL_1668910, EPI_ISL_1668911, EPI_ISL_1668912, EPI_ISL_1668913, EPI_ISL_1668914, EPI_ISL_1668915, EPI_ISL_1668916, EPI_ISL_1668917, EPI_ISL_1668918, EPI_ISL_1668919, EPI_ISL_1668920, EPI_ISL_1668921, EPI_ISL_1668922, EPI_ISL_1668923, EPI_ISL_1668924, EPI_ISL_1668925, EPI_ISL_1668926, EPI_ISL_1668927, EPI_ISL_1668928, EPI_ISL_1668929, EPI_ISL_1668930, EPI_ISL_1668931, EPI_ISL_1668932, EPI_ISL_1668933, EPI_ISL_1668934, EPI_ISL_1668935, EPI_ISL_1668936, EPI_ISL_1668937, EPI_ISL_1668938, EPI_ISL_1668939, EPI_ISL_1668940, EPI_ISL_1668941, EPI_ISL_1668942, EPI_ISL_1668943, EPI_ISL_1668944, EPI_ISL_1668945, EPI_ISL_1668946, EPI_ISL_1668947, EPI_ISL_1668948, EPI_ISL_1668949, EPI_ISL_1668950, EPI_ISL_1668951, EPI_ISL_1668952, EPI_ISL_1668953, EPI_ISL_1668954, EPI_ISL_1668955, EPI_ISL_1668956, EPI_ISL_1668957, EPI_ISL_1668958, EPI_ISL_1668959, EPI_ISL_1668960, EPI_ISL_1668961, EPI_ISL_1668962, EPI_ISL_1668963, EPI_ISL_1668964, EPI_ISL_1668965, EPI_ISL_1668966, EPI_ISL_1668968, EPI_ISL_1668969, EPI_ISL_1668970, EPI_ISL_1668971, EPI_ISL_1668972, EPI_ISL_1668973, EPI_ISL_1668974, EPI_ISL_1668975, EPI_ISL_1668976, EPI_ISL_1668977, EPI_ISL_1668978, EPI_ISL_1668979, EPI_ISL_1668980, EPI_ISL_1668981, EPI_ISL_1668982, EPI_ISL_1668983, EPI_ISL_1668984, EPI_ISL_1668985, EPI_ISL_1668986, EPI_ISL_1668987, EPI_ISL_1668988, EPI_ISL_1668989, EPI_ISL_1668990, EPI_ISL_1668991, EPI_ISL_1668992, EPI_ISL_1668993, EPI_ISL_1668994, EPI_ISL_1668995, EPI_ISL_1668996, EPI_ISL_1668997, EPI_ISL_1668998, EPI_ISL_1668999, EPI_ISL_1669000, EPI_ISL_1669001, EPI_ISL_1669002, EPI_ISL_1669003, EPI_ISL_1669004, EPI_ISL_1669005, EPI_ISL_1669006, EPI_ISL_1669007, EPI_ISL_1669008, EPI_ISL_1669009, EPI_ISL_1669010, EPI_ISL_1669011, EPI_ISL_1669012, EPI_ISL_1669013, EPI_ISL_1669014, EPI_ISL_1669015, EPI_ISL_1669016, EPI_ISL_1669017, EPI_ISL_1669018, EPI_ISL_1669019, EPI_ISL_1669020, EPI_ISL_1669021, EPI_ISL_1669022, EPI_ISL_1669023, EPI_ISL_1669024, EPI_ISL_1669025, EPI_ISL_1669026, EPI_ISL_1669027, EPI_ISL_1669028, EPI_ISL_1669029, EPI_ISL_1669030, EPI_ISL_1669031, EPI_ISL_1669032, EPI_ISL_1669033, EPI_ISL_1669034, EPI_ISL_1669035, EPI_ISL_1669036, EPI_ISL_1669037, EPI_ISL_1669038, EPI_ISL_1669039, EPI_ISL_1669040, EPI_ISL_1669041, EPI_ISL_1669042, EPI_ISL_1669043, EPI_ISL_1669044, EPI_ISL_1669045, EPI_ISL_1669046, EPI_ISL_1669047, EPI_ISL_1669048, EPI_ISL_1669049, EPI_ISL_1669050, EPI_ISL_1669051, EPI_ISL_1669052, EPI_ISL_1669053, EPI_ISL_1669054, EPI_ISL_1669055, EPI_ISL_1669056, EPI_ISL_1669057, EPI_ISL_1669058, EPI_ISL_1669059, EPI_ISL_1669060, EPI_ISL_1669061, EPI_ISL_1669062, EPI_ISL_1669063, EPI_ISL_1669064, EPI_ISL_1669065, EPI_ISL_1669066, EPI_ISL_1669067, EPI_ISL_1669068, EPI_ISL_1669069, EPI_ISL_1669070, EPI_ISL_1669071, EPI_ISL_1669072, EPI_ISL_1669073, EPI_ISL_1669074, EPI_ISL_1669075, EPI_ISL_1669076, EPI_ISL_1669077, EPI_ISL_1669078, EPI_ISL_1669079, EPI_ISL_1669080, EPI_ISL_1669081, EPI_ISL_1669082, EPI_ISL_1669083, EPI_ISL_1669084, EPI_ISL_1669085, EPI_ISL_1669086, EPI_ISL_1669087, EPI_ISL_1669088, EPI_ISL_1669089, EPI_ISL_1669090, EPI_ISL_1669091, EPI_ISL_1669092, EPI_ISL_1669093, EPI_ISL_1669094, EPI_ISL_1669095, EPI_ISL_1669096, EPI_ISL_1669097, EPI_ISL_1669098, EPI_ISL_1669099, EPI_ISL_1669100, EPI_ISL_1669101, EPI_ISL_1669102, EPI_ISL_1669103, EPI_ISL_1669104, EPI_ISL_1669105, EPI_ISL_1669106, EPI_ISL_1669107, EPI_ISL_1669108, EPI_ISL_1669109, EPI_ISL_1669110, EPI_ISL_1669111, EPI_ISL_1669112, EPI_ISL_1669113, EPI_ISL_1669114, EPI_ISL_1669127, EPI_ISL_1669128, EPI_ISL_1669129, EPI_ISL_1669130, EPI_ISL_1669131, EPI_ISL_1669132, EPI_ISL_1669133, EPI_ISL_1669134, EPI_ISL_1669135, EPI_ISL_1669136, EPI_ISL_1669137, EPI_ISL_1669138, EPI_ISL_1669139, EPI_ISL_1669140, EPI_ISL_1669141, EPI_ISL_1669142, EPI_ISL_1669143, EPI_ISL_1669144, EPI_ISL_1669145, EPI_ISL_1669146, EPI_ISL_1669147, EPI_ISL_1669148, EPI_ISL_1669149, EPI_ISL_1669150, EPI_ISL_1669151, EPI_ISL_1669152, EPI_ISL_1669153, EPI_ISL_1669154, EPI_ISL_1669155, EPI_ISL_1669156, EPI_ISL_1669157, EPI_ISL_1669158, EPI_ISL_1669159, EPI_ISL_1669160, EPI_ISL_1669161, EPI_ISL_1669162, EPI_ISL_1669163, EPI_ISL_1669164, EPI_ISL_1669165, EPI_ISL_1669166, EPI_ISL_1669167, EPI_ISL_1669168, EPI_ISL_1669169, EPI_ISL_1669170, EPI_ISL_1669171, EPI_ISL_1669172, EPI_ISL_1669173, EPI_ISL_1669174, EPI_ISL_1669175, EPI_ISL_1669176, EPI_ISL_1669177, EPI_ISL_1669178, EPI_ISL_1669179, EPI_ISL_1669180, EPI_ISL_1669181, EPI_ISL_1669182, EPI_ISL_1669183, EPI_ISL_1669184, EPI_ISL_1669185, EPI_ISL_1669186, EPI_ISL_1669187, EPI_ISL_1669188, EPI_ISL_1669189, EPI_ISL_1669190, EPI_ISL_1669191, EPI_ISL_1669192, EPI_ISL_1669193, EPI_ISL_1669194, EPI_ISL_1669195, EPI_ISL_1669196, EPI_ISL_1669197, EPI_ISL_1669198, EPI_ISL_1669199, EPI_ISL_1669200, EPI_ISL_1669201, EPI_ISL_1669202, EPI_ISL_1669203, EPI_ISL_1669204, EPI_ISL_1669205, EPI_ISL_1669206, EPI_ISL_1669207, EPI_ISL_1669208, EPI_ISL_1669209, EPI_ISL_1669210, EPI_ISL_1669211, EPI_ISL_1669212, EPI_ISL_1669213, EPI_ISL_1669214, EPI_ISL_1669215, EPI_ISL_1669216, EPI_ISL_1669217, EPI_ISL_1669218, EPI_ISL_1669219, EPI_ISL_1669220, EPI_ISL_1669221, EPI_ISL_1669222, EPI_ISL_1669223, EPI_ISL_1669224, EPI_ISL_1669225, EPI_ISL_1669226, EPI_ISL_1669227, EPI_ISL_1669228, EPI_ISL_1669229, EPI_ISL_1669230, EPI_ISL_1669231, EPI_ISL_1669232, EPI_ISL_1669233, EPI_ISL_1669234, EPI_ISL_1669235, EPI_ISL_1669236, EPI_ISL_1669237, EPI_ISL_1669238, EPI_ISL_1669239, EPI_ISL_1669240, EPI_ISL_1669241, EPI_ISL_1669242, EPI_ISL_1669243, EPI_ISL_1669244, EPI_ISL_1669245, EPI_ISL_1669246, EPI_ISL_1669247, EPI_ISL_1669248, EPI_ISL_1669249, EPI_ISL_1669250, EPI_ISL_1669251, EPI_ISL_1669252, EPI_ISL_1669253, EPI_ISL_1669254, EPI_ISL_1669255, EPI_ISL_1669256, EPI_ISL_1669257, EPI_ISL_1669258, EPI_ISL_1669259, EPI_ISL_1669260, EPI_ISL_1669261, EPI_ISL_1669262, EPI_ISL_1669263, EPI_ISL_1669264, EPI_ISL_1669265, EPI_ISL_1669266, EPI_ISL_1669267, EPI_ISL_1669268, EPI_ISL_1669269, EPI_ISL_1669270, EPI_ISL_1669271, EPI_ISL_1669272, EPI_ISL_1669273, EPI_ISL_1669274, EPI_ISL_1669275, EPI_ISL_1669276, EPI_ISL_1669277, EPI_ISL_1669278, EPI_ISL_1669279, EPI_ISL_1669280, EPI_ISL_1669281, EPI_ISL_1669282, EPI_ISL_1669283, EPI_ISL_1669284, EPI_ISL_1669285, EPI_ISL_1669286, EPI_ISL_1669287, EPI_ISL_1669288, EPI_ISL_1669289, EPI_ISL_1669290, EPI_ISL_1669291, EPI_ISL_1669292, EPI_ISL_1669293, EPI_ISL_1669294, EPI_ISL_1669295, EPI_ISL_1669296, EPI_ISL_1669297, EPI_ISL_1669298, EPI_ISL_1669299, EPI_ISL_1669300, EPI_ISL_1669301, EPI_ISL_1669302, EPI_ISL_1669303, EPI_ISL_1669304, EPI_ISL_1669305, EPI_ISL_1669306, EPI_ISL_1669307, EPI_ISL_1669308, EPI_ISL_1669309, EPI_ISL_1669310, EPI_ISL_1669311, EPI_ISL_1669312, EPI_ISL_1669313, EPI_ISL_1669314, EPI_ISL_1669315, EPI_ISL_1669316, EPI_ISL_1669317, EPI_ISL_1669318, EPI_ISL_1669319 | | | |
| see above | Quest Diagnostics Incorporated | Centers for Disease Control and Prevention Division of Viral Diseases, Pathogen Discovery | Dakota Howard, Dhwani Batra, Peter W. Cook, Kara Moser, Adrian Paskey, Jason Caravas, Benjamin Rambo-Martin, Shatavia Morrison, Christopher Gulvick, Scott Sammons, Yvette Unoarumhi, Darlene Wagner, Matthew Schmerer, S. H. Rosenthal, A. Gerasimova, R. M. Kagan, B. Anderson, M. Hua, Y. Liu, L.E. Bernstein, K.E. Livingston, A. Perez, I. A. Shlyakhter, R. V. Rolando, R. Owen, P. Tanpaiboon, F. Lacbawan, Clinton R. Paden, Duncan MacCannell |
| EPI_ISL_1669340 | Providence Little Company of Mary Medical Center | Los Angeles County Public Health Laboratories | P. Hemarajata et al. |
| EPI_ISL_1669341, EPI_ISL_1669342, EPI_ISL_1669343, EPI_ISL_1669344, EPI_ISL_1669346, EPI_ISL_1669347 | Flow Diagnostics Lab | Los Angeles County Public Health Laboratories | P. Hemarajata et al. |
| EPI_ISL_1669348 | USC Clinical Lab | Los Angeles County Public Health Laboratories | P. Hemarajata et al. |
| EPI_ISL_1669801, EPI_ISL_1669802, EPI_ISL_1669803 | NYU Langone Health | Departments of Pathology and Medicine, New York University School of Medicine | Adriana Heguy, Dacia Dimartino, Emily Guzman, Christian Marier, Peter Meyn, Sitharam Ramaswami, Gael Westby, Paul Zappile, Yutong Zhang, Paolo Cotzia, Guiqing Wang |
| EPI_ISL_1669862 | Monterey County Public Health Laboratory | Monterey County Public Health Laboratory | Monterey County Public Health Laboratory |
| EPI_ISL_1670710, EPI_ISL_1670711, EPI_ISL_1670712, EPI_ISL_1670713, EPI_ISL_1670714, EPI_ISL_1670715, EPI_ISL_1670716, EPI_ISL_1670717, EPI_ISL_1670718, EPI_ISL_1670719, EPI_ISL_1670720, EPI_ISL_1670721, EPI_ISL_1670722, EPI_ISL_1670723, EPI_ISL_1670724, EPI_ISL_1670725, EPI_ISL_1670726, EPI_ISL_1670727, EPI_ISL_1670728, EPI_ISL_1670729, EPI_ISL_1670730, EPI_ISL_1670731, EPI_ISL_1670732, EPI_ISL_1670733, EPI_ISL_1670734, EPI_ISL_1670735, EPI_ISL_1670736, EPI_ISL_1670737, EPI_ISL_1670738, EPI_ISL_1670739, EPI_ISL_1670740, EPI_ISL_1670741, EPI_ISL_1670742, EPI_ISL_1670743, EPI_ISL_1670744, EPI_ISL_1670745, EPI_ISL_1670746, EPI_ISL_1670747, EPI_ISL_1670748, EPI_ISL_1670749, EPI_ISL_1670750, EPI_ISL_1670751, EPI_ISL_1670752, EPI_ISL_1670753, EPI_ISL_1670754, EPI_ISL_1670755, EPI_ISL_1670756, EPI_ISL_1670757, EPI_ISL_1670758, EPI_ISL_1670759, EPI_ISL_1670760, EPI_ISL_1670761, EPI_ISL_1670762, EPI_ISL_1670763, EPI_ISL_1670764, EPI_ISL_1670765, EPI_ISL_1670766, EPI_ISL_1670767, EPI_ISL_1670768, EPI_ISL_1670769, EPI_ISL_1670770, EPI_ISL_1670771, EPI_ISL_1670772, EPI_ISL_1670773, EPI_ISL_1670774, EPI_ISL_1670775, EPI_ISL_1670776, EPI_ISL_1670777, EPI_ISL_1670778, EPI_ISL_1670779, EPI_ISL_1670780, EPI_ISL_1670781, EPI_ISL_1670782, EPI_ISL_1670783, EPI_ISL_1670784, EPI_ISL_1670785, EPI_ISL_1670786, EPI_ISL_1670787, EPI_ISL_1670788, EPI_ISL_1670789, EPI_ISL_1670790, EPI_ISL_1670791, EPI_ISL_1670792, EPI_ISL_1670793, EPI_ISL_1670794, EPI_ISL_1670795, EPI_ISL_1670796, EPI_ISL_1670797, EPI_ISL_1670798, EPI_ISL_1670799, EPI_ISL_1670800, EPI_ISL_1670801, EPI_ISL_1670802, EPI_ISL_1670803, EPI_ISL_1670804, EPI_ISL_1670805, EPI_ISL_1670806, EPI_ISL_1670807, EPI_ISL_1670808, EPI_ISL_1670809, EPI_ISL_1670810, EPI_ISL_1670811, EPI_ISL_1670812, EPI_ISL_1670813, EPI_ISL_1670814, EPI_ISL_1670815, EPI_ISL_1670816, EPI_ISL_1670817, EPI_ISL_1670818, EPI_ISL_1670819, EPI_ISL_1670820, EPI_ISL_1670821, EPI_ISL_1670822, EPI_ISL_1670823, EPI_ISL_1670824, EPI_ISL_1670825, EPI_ISL_1670826, EPI_ISL_1670827, EPI_ISL_1670828, EPI_ISL_1670829, EPI_ISL_1670830, EPI_ISL_1670831, EPI_ISL_1670832, EPI_ISL_1670833, EPI_ISL_1670834, EPI_ISL_1670835, EPI_ISL_1670836, EPI_ISL_1670837, EPI_ISL_1670838, EPI_ISL_1670839, EPI_ISL_1670840, EPI_ISL_1670841, EPI_ISL_1670842, EPI_ISL_1670843, EPI_ISL_1670844, EPI_ISL_1670845, EPI_ISL_1670846, EPI_ISL_1670847, EPI_ISL_1670848, EPI_ISL_1670849, EPI_ISL_1670850, EPI_ISL_1670851, EPI_ISL_1670852, EPI_ISL_1670853, EPI_ISL_1670854, EPI_ISL_1670855, EPI_ISL_1670856, EPI_ISL_1670857, EPI_ISL_1670858, EPI_ISL_1670859, EPI_ISL_1670860, EPI_ISL_1670861, EPI_ISL_1670862, EPI_ISL_1670863, EPI_ISL_1670864, EPI_ISL_1670865, EPI_ISL_1670866, EPI_ISL_1670867, EPI_ISL_1670868, EPI_ISL_1670869, EPI_ISL_1670870, EPI_ISL_1670871, EPI_ISL_1670872, EPI_ISL_1670873, EPI_ISL_1670874, EPI_ISL_1670875, EPI_ISL_1670876, EPI_ISL_1670877, EPI_ISL_1670878, EPI_ISL_1670879, EPI_ISL_1670880, EPI_ISL_1670881, EPI_ISL_1670882 | | | |
| see above | UW Virology Lab | UW Virology Lab | Pavitra Roychoudhury, Hong Xie, Lasata Shrestha, Shah Mohamed Bakhash, Michelle Lin, Noah R. Baker, Sean Ellis, Meei-Li Huang, Keith R Jerome, Alexander Greninger |
| EPI_ISL_1671137 | MEMORIAL SLOAN KETTERING CANCER CENTER | Wadsworth Center, New York State Department of Health | Kirsten St. George, Daryl M. Lamson, Alexis Russell, Matthew Shudt, Melissa A Leisner, Jonathan Plitnick, Catharine Prussing, Navjot Singh, John Kelly, Erasmus Schneider, Erica Lasek-Nesselquist |
| EPI_ISL_1671138, EPI_ISL_1671139, EPI_ISL_1671140 | New York Presbyterian Hospital | Wadsworth Center, New York State Department of Health | Kirsten St. George, Daryl M. Lamson, Alexis Russell, Matthew Shudt, Melissa A Leisner, Jonathan Plitnick, Catharine Prussing, Navjot Singh, John Kelly, Erasmus Schneider, Erica Lasek-Nesselquist |
| EPI_ISL_1671141, EPI_ISL_1671142 | NYC Pandemic Response Lab | Wadsworth Center, New York State Department of Health | Kirsten St. George, Daryl M. Lamson, Alexis Russell, Matthew Shudt, Melissa A Leisner, Jonathan Plitnick, Catharine Prussing, Navjot Singh, John Kelly, Erasmus Schneider, Erica Lasek-Nesselquist |
| EPI_ISL_1671143, EPI_ISL_1671144 | WESTCHESTER MEDICAL CENTER | Wadsworth Center, New York State Department of Health | Kirsten St. George, Daryl M. Lamson, Alexis Russell, Matthew Shudt, Melissa A Leisner, Jonathan Plitnick, Catharine Prussing, Navjot Singh, John Kelly, Erasmus Schneider, Erica Lasek-Nesselquist |
| EPI_ISL_1671145 | MONTEFIORE MEDICAL CENTER LABORATORIES | Wadsworth Center, New York State Department of Health | Kirsten St. George, Daryl M. Lamson, Alexis Russell, Matthew Shudt, Melissa A Leisner, Jonathan Plitnick, Catharine Prussing, Navjot Singh, John Kelly, Erasmus Schneider, Erica Lasek-Nesselquist |
| EPI_ISL_1671146 | New York Presbyterian Hospital | Wadsworth Center, New York State Department of Health | Kirsten St. George, Daryl M. Lamson, Alexis Russell, Matthew Shudt, Melissa A Leisner, Jonathan Plitnick, Catharine Prussing, Navjot Singh, John Kelly, Erasmus Schneider, Erica Lasek-Nesselquist |
| EPI_ISL_1671147 | MEMORIAL SLOAN KETTERING CANCER CENTER | Wadsworth Center, New York State Department of Health | Kirsten St. George, Daryl M. Lamson, Alexis Russell, Matthew Shudt, Melissa A Leisner, Jonathan Plitnick, Catharine Prussing, Navjot Singh, John Kelly, Erasmus Schneider, Erica Lasek-Nesselquist |
| EPI_ISL_1671148, EPI_ISL_1671149, EPI_ISL_1671150, EPI_ISL_1671151, EPI_ISL_1671152, EPI_ISL_1671153, EPI_ISL_1671154, EPI_ISL_1671155 | NYC Pandemic Response Lab | Wadsworth Center, New York State Department of Health | Kirsten St. George, Daryl M. Lamson, Alexis Russell, Matthew Shudt, Melissa A Leisner, Jonathan Plitnick, Catharine Prussing, Navjot Singh, John Kelly, Erasmus Schneider, Erica Lasek-Nesselquist |
| EPI_ISL_1671156 | MONTEFIORE MEDICAL CENTER LABORATORIES | Wadsworth Center, New York State Department of Health | Kirsten St. George, Daryl M. Lamson, Alexis Russell, Matthew Shudt, Melissa A Leisner, Jonathan Plitnick, Catharine Prussing, Navjot Singh, John Kelly, Erasmus Schneider, Erica Lasek-Nesselquist |
| EPI_ISL_1671157 | NYC Pandemic Response Lab | Wadsworth Center, New York State Department of Health | Kirsten St. George, Daryl M. Lamson, Alexis Russell, Matthew Shudt, Melissa A Leisner, Jonathan Plitnick, Catharine Prussing, Navjot Singh, John Kelly, Erasmus Schneider, Erica Lasek-Nesselquist |
| EPI_ISL_1671158, EPI_ISL_1671159 | MONTEFIORE MEDICAL CENTER LABORATORIES | Wadsworth Center, New York State Department of Health | Kirsten St. George, Daryl M. Lamson, Alexis Russell, Matthew Shudt, Melissa A Leisner, Jonathan Plitnick, Catharine Prussing, Navjot Singh, John Kelly, Erasmus Schneider, Erica Lasek-Nesselquist |
| EPI_ISL_1671160 | Columbia University Irving Medical Center | Wadsworth Center, New York State Department of Health | Kirsten St. George, Daryl M. Lamson, Alexis Russell, Matthew Shudt, Melissa A Leisner, Jonathan Plitnick, Catharine Prussing, Navjot Singh, John Kelly, Erasmus Schneider, Erica Lasek-Nesselquist |
| EPI_ISL_1671161 | WESTCHESTER MEDICAL CENTER | Wadsworth Center, New York State Department of Health | Kirsten St. George, Daryl M. Lamson, Alexis Russell, Matthew Shudt, Melissa A Leisner, Jonathan Plitnick, Catharine Prussing, Navjot Singh, John Kelly, Erasmus Schneider, Erica Lasek-Nesselquist |
| EPI_ISL_1671162 | Columbia University Irving Medical Center | Wadsworth Center, New York State Department of Health | Kirsten St. George, Daryl M. Lamson, Alexis Russell, Matthew Shudt, Melissa A Leisner, Jonathan Plitnick, Catharine Prussing, Navjot Singh, John Kelly, Erasmus Schneider, Erica Lasek-Nesselquist |
| EPI_ISL_1671163, EPI_ISL_1671164 | MONTEFIORE MEDICAL CENTER LABORATORIES | Wadsworth Center, New York State Department of Health | Kirsten St. George, Daryl M. Lamson, Alexis Russell, Matthew Shudt, Melissa A Leisner, Jonathan Plitnick, Catharine Prussing, Navjot Singh, John Kelly, Erasmus Schneider, Erica Lasek-Nesselquist |
| EPI_ISL_1671165 | MEMORIAL SLOAN KETTERING CANCER CENTER | Wadsworth Center, New York State Department of Health | Kirsten St. George, Daryl M. Lamson, Alexis Russell, Matthew Shudt, Melissa A Leisner, Jonathan Plitnick, Catharine Prussing, Navjot Singh, John Kelly, Erasmus Schneider, Erica Lasek-Nesselquist |
| EPI_ISL_1671166 | New York Presbyterian Hospital | Wadsworth Center, New York State Department of Health | Kirsten St. George, Daryl M. Lamson, Alexis Russell, Matthew Shudt, Melissa A Leisner, Jonathan Plitnick, Catharine Prussing, Navjot Singh, John Kelly, Erasmus Schneider, Erica Lasek-Nesselquist |
| EPI_ISL_1671167, EPI_ISL_1671168 | NYC Pandemic Response Lab | Wadsworth Center, New York State Department of Health | Kirsten St. George, Daryl M. Lamson, Alexis Russell, Matthew Shudt, Melissa A Leisner, Jonathan Plitnick, Catharine Prussing, Navjot Singh, John Kelly, Erasmus Schneider, Erica Lasek-Nesselquist |

| | | | |
|---|---|---|---|
| EPI_ISL_1671169, EPI_ISL_1671170, EPI_ISL_1671171 | MONTEFIORE MEDICAL CENTER LABORATORIES | Wadsworth Center, New York State Department of Health | Kirsten St. George, Daryl M. Lamson, Alexis Russell, Matthew Shudt, Melissa A Leisner, Jonathan Plitnick, Catharine Prussing, Navjot Singh, John Kelly, Erasmus Schneider, Erica Lasek-Nesselquist |
| EPI_ISL_1671172, EPI_ISL_1671173 | NYC Pandemic Response Lab | Wadsworth Center, New York State Department of Health | Kirsten St. George, Daryl M. Lamson, Alexis Russell, Matthew Shudt, Melissa A Leisner, Jonathan Plitnick, Catharine Prussing, Navjot Singh, John Kelly, Erasmus Schneider, Erica Lasek-Nesselquist |
| EPI_ISL_1671174, EPI_ISL_1671175 | MONTEFIORE MEDICAL CENTER LABORATORIES | Wadsworth Center, New York State Department of Health | Kirsten St. George, Daryl M. Lamson, Alexis Russell, Matthew Shudt, Melissa A Leisner, Jonathan Plitnick, Catharine Prussing, Navjot Singh, John Kelly, Erasmus Schneider, Erica Lasek-Nesselquist |
| EPI_ISL_1671176 | Columbia University Irving Medical Center | Wadsworth Center, New York State Department of Health | Kirsten St. George, Daryl M. Lamson, Alexis Russell, Matthew Shudt, Melissa A Leisner, Jonathan Plitnick, Catharine Prussing, Navjot Singh, John Kelly, Erasmus Schneider, Erica Lasek-Nesselquist |
| EPI_ISL_1671177, EPI_ISL_1671178, EPI_ISL_1671179, EPI_ISL_1671180 | NYC Pandemic Response Lab | Wadsworth Center, New York State Department of Health | Kirsten St. George, Daryl M. Lamson, Alexis Russell, Matthew Shudt, Melissa A Leisner, Jonathan Plitnick, Catharine Prussing, Navjot Singh, John Kelly, Erasmus Schneider, Erica Lasek-Nesselquist |
| EPI_ISL_1671181, EPI_ISL_1671182 | MONTEFIORE MEDICAL CENTER LABORATORIES | Wadsworth Center, New York State Department of Health | Kirsten St. George, Daryl M. Lamson, Alexis Russell, Matthew Shudt, Melissa A Leisner, Jonathan Plitnick, Catharine Prussing, Navjot Singh, John Kelly, Erasmus Schneider, Erica Lasek-Nesselquist |
| EPI_ISL_1671183, EPI_ISL_1671184 | NYC Pandemic Response Lab | Wadsworth Center, New York State Department of Health | Kirsten St. George, Daryl M. Lamson, Alexis Russell, Matthew Shudt, Melissa A Leisner, Jonathan Plitnick, Catharine Prussing, Navjot Singh, John Kelly, Erasmus Schneider, Erica Lasek-Nesselquist |
| EPI_ISL_1671185, EPI_ISL_1671186, EPI_ISL_1671187, EPI_ISL_1671188, EPI_ISL_1671189, EPI_ISL_1671190, EPI_ISL_1671191, EPI_ISL_1671192 | MONTEFIORE MEDICAL CENTER LABORATORIES | Wadsworth Center, New York State Department of Health | Kirsten St. George, Daryl M. Lamson, Alexis Russell, Matthew Shudt, Melissa A Leisner, Jonathan Plitnick, Catharine Prussing, Navjot Singh, John Kelly, Erasmus Schneider, Erica Lasek-Nesselquist |
| EPI_ISL_1671193 | MEMORIAL SLOAN KETTERING CANCER CENTER | Wadsworth Center, New York State Department of Health | Kirsten St. George, Daryl M. Lamson, Alexis Russell, Matthew Shudt, Melissa A Leisner, Jonathan Plitnick, Catharine Prussing, Navjot Singh, John Kelly, Erasmus Schneider, Erica Lasek-Nesselquist |
| EPI_ISL_1671194 | New York Presbyterian Hospital | Wadsworth Center, New York State Department of Health | Kirsten St. George, Daryl M. Lamson, Alexis Russell, Matthew Shudt, Melissa A Leisner, Jonathan Plitnick, Catharine Prussing, Navjot Singh, John Kelly, Erasmus Schneider, Erica Lasek-Nesselquist |
| EPI_ISL_1671195, EPI_ISL_1671196, EPI_ISL_1671197, EPI_ISL_1671198, EPI_ISL_1671199, EPI_ISL_1671200 | NYC Pandemic Response Lab | Wadsworth Center, New York State Department of Health | Kirsten St. George, Daryl M. Lamson, Alexis Russell, Matthew Shudt, Melissa A Leisner, Jonathan Plitnick, Catharine Prussing, Navjot Singh, John Kelly, Erasmus Schneider, Erica Lasek-Nesselquist |
| EPI_ISL_1671201, EPI_ISL_1671202 | MEMORIAL SLOAN KETTERING CANCER CENTER | Wadsworth Center, New York State Department of Health | Kirsten St. George, Daryl M. Lamson, Alexis Russell, Matthew Shudt, Melissa A Leisner, Jonathan Plitnick, Catharine Prussing, Navjot Singh, John Kelly, Erasmus Schneider, Erica Lasek-Nesselquist |
| EPI_ISL_1671203, EPI_ISL_1671204 | MONTEFIORE MEDICAL CENTER LABORATORIES | Wadsworth Center, New York State Department of Health | Kirsten St. George, Daryl M. Lamson, Alexis Russell, Matthew Shudt, Melissa A Leisner, Jonathan Plitnick, Catharine Prussing, Navjot Singh, John Kelly, Erasmus Schneider, Erica Lasek-Nesselquist |
| EPI_ISL_1671205, EPI_ISL_1671206, EPI_ISL_1671207 | NYC Pandemic Response Lab | Wadsworth Center, New York State Department of Health | Kirsten St. George, Daryl M. Lamson, Alexis Russell, Matthew Shudt, Melissa A Leisner, Jonathan Plitnick, Catharine Prussing, Navjot Singh, John Kelly, Erasmus Schneider, Erica Lasek-Nesselquist |
| EPI_ISL_1671208 | MONTEFIORE MEDICAL CENTER LABORATORIES | Wadsworth Center, New York State Department of Health | Kirsten St. George, Daryl M. Lamson, Alexis Russell, Matthew Shudt, Melissa A Leisner, Jonathan Plitnick, Catharine Prussing, Navjot Singh, John Kelly, Erasmus Schneider, Erica Lasek-Nesselquist |
| EPI_ISL_1671209 | NORTHWELL HEALTH LABORATORIES | Wadsworth Center, New York State Department of Health | Kirsten St. George, Daryl M. Lamson, Alexis Russell, Matthew Shudt, Melissa A Leisner, Jonathan Plitnick, Catharine Prussing, Navjot Singh, John Kelly, Erasmus Schneider, Erica Lasek-Nesselquist |
| EPI_ISL_1671210, EPI_ISL_1671211, EPI_ISL_1671212 | Columbia University Irving Medical Center | Wadsworth Center, New York State Department of Health | Kirsten St. George, Daryl M. Lamson, Alexis Russell, Matthew Shudt, Melissa A Leisner, Jonathan Plitnick, Catharine Prussing, Navjot Singh, John Kelly, Erasmus Schneider, Erica Lasek-Nesselquist |
| EPI_ISL_1671213, EPI_ISL_1671214, EPI_ISL_1671215, EPI_ISL_1671216 | New York Presbyterian Hospital | Wadsworth Center, New York State Department of Health | Kirsten St. George, Daryl M. Lamson, Alexis Russell, Matthew Shudt, Melissa A Leisner, Jonathan Plitnick, Catharine Prussing, Navjot Singh, John Kelly, Erasmus Schneider, Erica Lasek-Nesselquist |
| EPI_ISL_1671217 | Columbia University Irving Medical Center | Wadsworth Center, New York State Department of Health | Kirsten St. George, Daryl M. Lamson, Alexis Russell, Matthew Shudt, Melissa A Leisner, Jonathan Plitnick, Catharine Prussing, Navjot Singh, John Kelly, Erasmus Schneider, Erica Lasek-Nesselquist |
| EPI_ISL_1671218, EPI_ISL_1671219, EPI_ISL_1671220 | WESTCHESTER MEDICAL CENTER | Wadsworth Center, New York State Department of Health | Kirsten St. George, Daryl M. Lamson, Alexis Russell, Matthew Shudt, Melissa A Leisner, Jonathan Plitnick, Catharine Prussing, Navjot Singh, John Kelly, Erasmus Schneider, Erica Lasek-Nesselquist |
| EPI_ISL_1671312 | TEMPUS LABS INC | Wadsworth Center, New York State Department of Health | Kirsten St. George, Daryl M. Lamson, Alexis Russell, Matthew Shudt, Melissa A Leisner, Jonathan Plitnick, Catharine Prussing, Navjot Singh, John Kelly, Erasmus Schneider, Erica Lasek-Nesselquist |
| EPI_ISL_1671313 | ALBANY MEDICAL CENTER | Wadsworth Center, New York State Department of Health | Kirsten St. George, Daryl M. Lamson, Alexis Russell, Matthew Shudt, Melissa A Leisner, Jonathan Plitnick, Catharine Prussing, Navjot Singh, John Kelly, Erasmus Schneider, Erica Lasek-Nesselquist |
| EPI_ISL_1671314 | TEMPUS LABS INC | Wadsworth Center, New York State Department of Health | Kirsten St. George, Daryl M. Lamson, Alexis Russell, Matthew Shudt, Melissa A Leisner, Jonathan Plitnick, Catharine Prussing, Navjot Singh, John Kelly, Erasmus Schneider, Erica Lasek-Nesselquist |
| EPI_ISL_1671315 | ALBANY MEDICAL CENTER | Wadsworth Center, New York State Department of Health | Kirsten St. George, Daryl M. Lamson, Alexis Russell, Matthew Shudt, Melissa A Leisner, Jonathan Plitnick, Catharine Prussing, Navjot Singh, John Kelly, Erasmus Schneider, Erica Lasek-Nesselquist |
| EPI_ISL_1671316 | STONY BROOK UNIVERSITY HOSPITAL | Wadsworth Center, New York State Department of Health | Kirsten St. George, Daryl M. Lamson, Alexis Russell, Matthew Shudt, Melissa A Leisner, Jonathan Plitnick, Catharine Prussing, Navjot Singh, John Kelly, Erasmus Schneider, Erica Lasek-Nesselquist |
| EPI_ISL_1671317, EPI_ISL_1671318, EPI_ISL_1671319, EPI_ISL_1671320, EPI_ISL_1671321 | ALBANY MEDICAL CENTER | Wadsworth Center, New York State Department of Health | Kirsten St. George, Daryl M. Lamson, Alexis Russell, Matthew Shudt, Melissa A Leisner, Jonathan Plitnick, Catharine Prussing, Navjot Singh, John Kelly, Erasmus Schneider, Erica Lasek-Nesselquist |
| EPI_ISL_1671322 | STONY BROOK UNIVERSITY HOSPITAL | Wadsworth Center, New York State Department of Health | Kirsten St. George, Daryl M. Lamson, Alexis Russell, Matthew Shudt, Melissa A Leisner, Jonathan Plitnick, Catharine Prussing, Navjot Singh, John Kelly, Erasmus Schneider, Erica Lasek-Nesselquist |
| EPI_ISL_1671323, EPI_ISL_1671324, EPI_ISL_1671325, EPI_ISL_1671326, EPI_ISL_1671327 | ALBANY MEDICAL CENTER | Wadsworth Center, New York State Department of Health | Kirsten St. George, Daryl M. Lamson, Alexis Russell, Matthew Shudt, Melissa A Leisner, Jonathan Plitnick, Catharine Prussing, Navjot Singh, John Kelly, Erasmus Schneider, Erica Lasek-Nesselquist |
| EPI_ISL_1671328, EPI_ISL_1671329, EPI_ISL_1671330 | STONY BROOK UNIVERSITY HOSPITAL | Wadsworth Center, New York State Department of Health | Kirsten St. George, Daryl M. Lamson, Alexis Russell, Matthew Shudt, Melissa A Leisner, Jonathan Plitnick, Catharine Prussing, Navjot Singh, John Kelly, Erasmus Schneider, Erica Lasek-Nesselquist |
| EPI_ISL_1671331, EPI_ISL_1671332, EPI_ISL_1671333, EPI_ISL_1671334, EPI_ISL_1671335 | TEMPUS LABS INC | Wadsworth Center, New York State Department of Health | Kirsten St. George, Daryl M. Lamson, Alexis Russell, Matthew Shudt, Melissa A Leisner, Jonathan Plitnick, Catharine Prussing, Navjot Singh, John Kelly, Erasmus Schneider, Erica Lasek-Nesselquist |
| EPI_ISL_1671336 | STONY BROOK UNIVERSITY HOSPITAL | Wadsworth Center, New York State Department of Health | Kirsten St. George, Daryl M. Lamson, Alexis Russell, Matthew Shudt, Melissa A Leisner, Jonathan Plitnick, Catharine Prussing, Navjot Singh, John Kelly, Erasmus Schneider, Erica Lasek-Nesselquist |
| EPI_ISL_1671337 | ALBANY MEDICAL CENTER | Wadsworth Center, New York State Department of Health | Kirsten St. George, Daryl M. Lamson, Alexis Russell, Matthew Shudt, Melissa A Leisner, Jonathan Plitnick, Catharine Prussing, Navjot Singh, John Kelly, Erasmus Schneider, Erica Lasek-Nesselquist |
| EPI_ISL_1671338, EPI_ISL_1671339, EPI_ISL_1671340 | TEMPUS LABS INC | Wadsworth Center, New York State Department of Health | Kirsten St. George, Daryl M. Lamson, Alexis Russell, Matthew Shudt, Melissa A Leisner, Jonathan Plitnick, Catharine Prussing, Navjot Singh, John Kelly, Erasmus Schneider, Erica Lasek-Nesselquist |
| EPI_ISL_1671341 | ALBANY MEDICAL CENTER | Wadsworth Center, New York State | Kirsten St. George, Daryl M. Lamson, Alexis Russell, Matthew Shudt, Melissa A Leisner, Jonathan Plitnick, Catharine Prussing, Navjot Singh, John Kelly, Erasmus Schneider, Erica Lasek-Nesselquist |

| | | | |
|---|---|---|---|
| | | Department of Health | |
| EPI_ISL_1671342, EPI_ISL_1671343, EPI_ISL_1671344, EPI_ISL_1671345, EPI_ISL_1671346, EPI_ISL_1671347, EPI_ISL_1671348 | TEMPUS LABS INC | Wadsworth Center, New York State Department of Health | Kirsten St. George, Daryl M. Lamson, Alexis Russell, Matthew Shudt, Melissa A Leisner, Jonathan Plitnick, Catharine Prussing, Navjot Singh, John Kelly, Erasmus Schneider, Erica Lasek-Nesselquist |
| EPI_ISL_1671349 | STONY BROOK UNIVERSITY HOSPITAL | Wadsworth Center, New York State Department of Health | Kirsten St. George, Daryl M. Lamson, Alexis Russell, Matthew Shudt, Melissa A Leisner, Jonathan Plitnick, Catharine Prussing, Navjot Singh, John Kelly, Erasmus Schneider, Erica Lasek-Nesselquist |
| EPI_ISL_1671350, EPI_ISL_1671351 | ALBANY MEDICAL CENTER | Wadsworth Center, New York State Department of Health | Kirsten St. George, Daryl M. Lamson, Alexis Russell, Matthew Shudt, Melissa A Leisner, Jonathan Plitnick, Catharine Prussing, Navjot Singh, John Kelly, Erasmus Schneider, Erica Lasek-Nesselquist |
| EPI_ISL_1671352, EPI_ISL_1671353 | TEMPUS LABS INC | Wadsworth Center, New York State Department of Health | Kirsten St. George, Daryl M. Lamson, Alexis Russell, Matthew Shudt, Melissa A Leisner, Jonathan Plitnick, Catharine Prussing, Navjot Singh, John Kelly, Erasmus Schneider, Erica Lasek-Nesselquist |
| EPI_ISL_1671354 | ALBANY MEDICAL CENTER | Wadsworth Center, New York State Department of Health | Kirsten St. George, Daryl M. Lamson, Alexis Russell, Matthew Shudt, Melissa A Leisner, Jonathan Plitnick, Catharine Prussing, Navjot Singh, John Kelly, Erasmus Schneider, Erica Lasek-Nesselquist |
| EPI_ISL_1671355 | TEMPUS LABS INC | Wadsworth Center, New York State Department of Health | Kirsten St. George, Daryl M. Lamson, Alexis Russell, Matthew Shudt, Melissa A Leisner, Jonathan Plitnick, Catharine Prussing, Navjot Singh, John Kelly, Erasmus Schneider, Erica Lasek-Nesselquist |
| EPI_ISL_1671356 | STONY BROOK UNIVERSITY HOSPITAL | Wadsworth Center, New York State Department of Health | Kirsten St. George, Daryl M. Lamson, Alexis Russell, Matthew Shudt, Melissa A Leisner, Jonathan Plitnick, Catharine Prussing, Navjot Singh, John Kelly, Erasmus Schneider, Erica Lasek-Nesselquist |
| EPI_ISL_1671357, EPI_ISL_1671358, EPI_ISL_1671359 | ALBANY MEDICAL CENTER | Wadsworth Center, New York State Department of Health | Kirsten St. George, Daryl M. Lamson, Alexis Russell, Matthew Shudt, Melissa A Leisner, Jonathan Plitnick, Catharine Prussing, Navjot Singh, John Kelly, Erasmus Schneider, Erica Lasek-Nesselquist |
| EPI_ISL_1671444, EPI_ISL_1671445, EPI_ISL_1671446, EPI_ISL_1671447 | BOSTON HEART DIAGNOSTICS CORP | Wadsworth Center, New York State Department of Health | Kirsten St. George, Daryl M. Lamson, Alexis Russell, Matthew Shudt, Melissa A Leisner, Jonathan Plitnick, Catharine Prussing, Navjot Singh, John Kelly, Erasmus Schneider, Erica Lasek-Nesselquist |
| EPI_ISL_1671448 | NORTH SHORE UNIVERSITY HOSPITAL | Wadsworth Center, New York State Department of Health | Kirsten St. George, Daryl M. Lamson, Alexis Russell, Matthew Shudt, Melissa A Leisner, Jonathan Plitnick, Catharine Prussing, Navjot Singh, John Kelly, Erasmus Schneider, Erica Lasek-Nesselquist |
| EPI_ISL_1671449 | WESTCHESTER MEDICAL CENTER | Wadsworth Center, New York State Department of Health | Kirsten St. George, Daryl M. Lamson, Alexis Russell, Matthew Shudt, Melissa A Leisner, Jonathan Plitnick, Catharine Prussing, Navjot Singh, John Kelly, Erasmus Schneider, Erica Lasek-Nesselquist |
| EPI_ISL_1671450, EPI_ISL_1671451 | ALBANY MEDICAL CENTER | Wadsworth Center, New York State Department of Health | Kirsten St. George, Daryl M. Lamson, Alexis Russell, Matthew Shudt, Melissa A Leisner, Jonathan Plitnick, Catharine Prussing, Navjot Singh, John Kelly, Erasmus Schneider, Erica Lasek-Nesselquist |
| EPI_ISL_1671452, EPI_ISL_1671453 | NORTH SHORE UNIVERSITY HOSPITAL | Wadsworth Center, New York State Department of Health | Kirsten St. George, Daryl M. Lamson, Alexis Russell, Matthew Shudt, Melissa A Leisner, Jonathan Plitnick, Catharine Prussing, Navjot Singh, John Kelly, Erasmus Schneider, Erica Lasek-Nesselquist |
| EPI_ISL_1671454, EPI_ISL_1671455, EPI_ISL_1671456, EPI_ISL_1671457 | BOSTON HEART DIAGNOSTICS CORP | Wadsworth Center, New York State Department of Health | Kirsten St. George, Daryl M. Lamson, Alexis Russell, Matthew Shudt, Melissa A Leisner, Jonathan Plitnick, Catharine Prussing, Navjot Singh, John Kelly, Erasmus Schneider, Erica Lasek-Nesselquist |
| EPI_ISL_1671458, EPI_ISL_1671459, EPI_ISL_1671460, EPI_ISL_1671461 | WESTCHESTER MEDICAL CENTER | Wadsworth Center, New York State Department of Health | Kirsten St. George, Daryl M. Lamson, Alexis Russell, Matthew Shudt, Melissa A Leisner, Jonathan Plitnick, Catharine Prussing, Navjot Singh, John Kelly, Erasmus Schneider, Erica Lasek-Nesselquist |
| EPI_ISL_1671462 | ALBANY MEDICAL CENTER | Wadsworth Center, New York State Department of Health | Kirsten St. George, Daryl M. Lamson, Alexis Russell, Matthew Shudt, Melissa A Leisner, Jonathan Plitnick, Catharine Prussing, Navjot Singh, John Kelly, Erasmus Schneider, Erica Lasek-Nesselquist |
| EPI_ISL_1671463, EPI_ISL_1671464, EPI_ISL_1671465 | SUNY UPSTATE MEDICAL UNIVERSITY | Wadsworth Center, New York State Department of Health | Kirsten St. George, Daryl M. Lamson, Alexis Russell, Matthew Shudt, Melissa A Leisner, Jonathan Plitnick, Catharine Prussing, Navjot Singh, John Kelly, Erasmus Schneider, Erica Lasek-Nesselquist |
| EPI_ISL_1671466 | MONTEFIORE MEDICAL CENTER LABORATORIES | Wadsworth Center, New York State Department of Health | Kirsten St. George, Daryl M. Lamson, Alexis Russell, Matthew Shudt, Melissa A Leisner, Jonathan Plitnick, Catharine Prussing, Navjot Singh, John Kelly, Erasmus Schneider, Erica Lasek-Nesselquist |
| EPI_ISL_1671467 | WESTCHESTER MEDICAL CENTER | Wadsworth Center, New York State Department of Health | Kirsten St. George, Daryl M. Lamson, Alexis Russell, Matthew Shudt, Melissa A Leisner, Jonathan Plitnick, Catharine Prussing, Navjot Singh, John Kelly, Erasmus Schneider, Erica Lasek-Nesselquist |
| EPI_ISL_1671468 | ST. FRANCIS HOSPITAL LABORATORY | Wadsworth Center, New York State Department of Health | Kirsten St. George, Daryl M. Lamson, Alexis Russell, Matthew Shudt, Melissa A Leisner, Jonathan Plitnick, Catharine Prussing, Navjot Singh, John Kelly, Erasmus Schneider, Erica Lasek-Nesselquist |
| EPI_ISL_1671469, EPI_ISL_1671470, EPI_ISL_1671471, EPI_ISL_1671472, EPI_ISL_1671473, EPI_ISL_1671474, EPI_ISL_1671475 | WESTCHESTER MEDICAL CENTER | Wadsworth Center, New York State Department of Health | Kirsten St. George, Daryl M. Lamson, Alexis Russell, Matthew Shudt, Melissa A Leisner, Jonathan Plitnick, Catharine Prussing, Navjot Singh, John Kelly, Erasmus Schneider, Erica Lasek-Nesselquist |
| EPI_ISL_1671476 | BOSTON HEART DIAGNOSTICS CORP | Wadsworth Center, New York State Department of Health | Kirsten St. George, Daryl M. Lamson, Alexis Russell, Matthew Shudt, Melissa A Leisner, Jonathan Plitnick, Catharine Prussing, Navjot Singh, John Kelly, Erasmus Schneider, Erica Lasek-Nesselquist |
| EPI_ISL_1671477, EPI_ISL_1671478, EPI_ISL_1671479 | MONTEFIORE MEDICAL CENTER LABORATORIES | Wadsworth Center, New York State Department of Health | Kirsten St. George, Daryl M. Lamson, Alexis Russell, Matthew Shudt, Melissa A Leisner, Jonathan Plitnick, Catharine Prussing, Navjot Singh, John Kelly, Erasmus Schneider, Erica Lasek-Nesselquist |
| EPI_ISL_1671480, EPI_ISL_1671481, EPI_ISL_1671482, EPI_ISL_1671483 | WESTCHESTER MEDICAL CENTER | Wadsworth Center, New York State Department of Health | Kirsten St. George, Daryl M. Lamson, Alexis Russell, Matthew Shudt, Melissa A Leisner, Jonathan Plitnick, Catharine Prussing, Navjot Singh, John Kelly, Erasmus Schneider, Erica Lasek-Nesselquist |
| EPI_ISL_1671484 | ST. FRANCIS HOSPITAL LABORATORY | Wadsworth Center, New York State Department of Health | Kirsten St. George, Daryl M. Lamson, Alexis Russell, Matthew Shudt, Melissa A Leisner, Jonathan Plitnick, Catharine Prussing, Navjot Singh, John Kelly, Erasmus Schneider, Erica Lasek-Nesselquist |
| EPI_ISL_1671485, EPI_ISL_1671486, EPI_ISL_1671487, EPI_ISL_1671488, EPI_ISL_1671489, EPI_ISL_1671490, EPI_ISL_1671491, EPI_ISL_1671492 | WESTCHESTER MEDICAL CENTER | Wadsworth Center, New York State Department of Health | Kirsten St. George, Daryl M. Lamson, Alexis Russell, Matthew Shudt, Melissa A Leisner, Jonathan Plitnick, Catharine Prussing, Navjot Singh, John Kelly, Erasmus Schneider, Erica Lasek-Nesselquist |
| EPI_ISL_1671493 | NORTH SHORE UNIVERSITY HOSPITAL | Wadsworth Center, New York State Department of Health | Kirsten St. George, Daryl M. Lamson, Alexis Russell, Matthew Shudt, Melissa A Leisner, Jonathan Plitnick, Catharine Prussing, Navjot Singh, John Kelly, Erasmus Schneider, Erica Lasek-Nesselquist |
| EPI_ISL_1671494 | SUNY UPSTATE MEDICAL UNIVERSITY | Wadsworth Center, New York State Department of Health | Kirsten St. George, Daryl M. Lamson, Alexis Russell, Matthew Shudt, Melissa A Leisner, Jonathan Plitnick, Catharine Prussing, Navjot Singh, John Kelly, Erasmus Schneider, Erica Lasek-Nesselquist |
| EPI_ISL_1671495, EPI_ISL_1671496 | BOSTON HEART DIAGNOSTICS CORP | Wadsworth Center, New York State Department of Health | Kirsten St. George, Daryl M. Lamson, Alexis Russell, Matthew Shudt, Melissa A Leisner, Jonathan Plitnick, Catharine Prussing, Navjot Singh, John Kelly, Erasmus Schneider, Erica Lasek-Nesselquist |
| EPI_ISL_1671497, EPI_ISL_1671498, EPI_ISL_1671499 | MONTEFIORE MEDICAL CENTER LABORATORIES | Wadsworth Center, New York State Department of Health | Kirsten St. George, Daryl M. Lamson, Alexis Russell, Matthew Shudt, Melissa A Leisner, Jonathan Plitnick, Catharine Prussing, Navjot Singh, John Kelly, Erasmus Schneider, Erica Lasek-Nesselquist |
| EPI_ISL_1671500, EPI_ISL_1671501, EPI_ISL_1671502, EPI_ISL_1671503, EPI_ISL_1671504, EPI_ISL_1671505, EPI_ISL_1671506, EPI_ISL_1671507, EPI_ISL_1671508, EPI_ISL_1671509, EPI_ISL_1671510, EPI_ISL_1671511 | | | |
| see above | WESTCHESTER MEDICAL CENTER | Wadsworth Center, New York State Department of Health | Kirsten St. George, Daryl M. Lamson, Alexis Russell, Matthew Shudt, Melissa A Leisner, Jonathan Plitnick, Catharine Prussing, Navjot Singh, John Kelly, Erasmus Schneider, Erica Lasek-Nesselquist |
| EPI_ISL_1671512, EPI_ISL_1671513, EPI_ISL_1671514 | SUNY UPSTATE MEDICAL UNIVERSITY | Wadsworth Center, New York State Department of Health | Kirsten St. George, Daryl M. Lamson, Alexis Russell, Matthew Shudt, Melissa A Leisner, Jonathan Plitnick, Catharine Prussing, Navjot Singh, John Kelly, Erasmus Schneider, Erica Lasek-Nesselquist |

| | | | |
|---|---|---|---|
| EPI_ISL_1671596 | BOSTON HEART DIAGNOSTICS CORP | Wadsworth Center, New York State Department of Health | Kirsten St. George, Daryl M. Lamson, Alexis Russell, Matthew Shudt, Melissa A Leisner, Jonathan Plitnick, Catharine Prussing, Navjot Singh, John Kelly, Erasmus Schneider, Erica Lasek-Nesselquist |
| EPI_ISL_1671597 | WESTCHESTER MEDICAL CENTER | Wadsworth Center, New York State Department of Health | Kirsten St. George, Daryl M. Lamson, Alexis Russell, Matthew Shudt, Melissa A Leisner, Jonathan Plitnick, Catharine Prussing, Navjot Singh, John Kelly, Erasmus Schneider, Erica Lasek-Nesselquist |
| EPI_ISL_1671598 | WHITE PLAINS HOSPITAL CENTER LABORATORY | Wadsworth Center, New York State Department of Health | Kirsten St. George, Daryl M. Lamson, Alexis Russell, Matthew Shudt, Melissa A Leisner, Jonathan Plitnick, Catharine Prussing, Navjot Singh, John Kelly, Erasmus Schneider, Erica Lasek-Nesselquist |
| EPI_ISL_1671599 | ALBANY MEDICAL CENTER | Wadsworth Center, New York State Department of Health | Kirsten St. George, Daryl M. Lamson, Alexis Russell, Matthew Shudt, Melissa A Leisner, Jonathan Plitnick, Catharine Prussing, Navjot Singh, John Kelly, Erasmus Schneider, Erica Lasek-Nesselquist |
| EPI_ISL_1671600 | WHITE PLAINS HOSPITAL CENTER LABORATORY | Wadsworth Center, New York State Department of Health | Kirsten St. George, Daryl M. Lamson, Alexis Russell, Matthew Shudt, Melissa A Leisner, Jonathan Plitnick, Catharine Prussing, Navjot Singh, John Kelly, Erasmus Schneider, Erica Lasek-Nesselquist |
| EPI_ISL_1671601, EPI_ISL_1671602, EPI_ISL_1671603, EPI_ISL_1671604, EPI_ISL_1671605, EPI_ISL_1671606 | NORTHWELL HEALTH LABORATORIES | Wadsworth Center, New York State Department of Health | Kirsten St. George, Daryl M. Lamson, Alexis Russell, Matthew Shudt, Melissa A Leisner, Jonathan Plitnick, Catharine Prussing, Navjot Singh, John Kelly, Erasmus Schneider, Erica Lasek-Nesselquist |
| EPI_ISL_1671607 | WHITE PLAINS HOSPITAL CENTER LABORATORY | Wadsworth Center, New York State Department of Health | Kirsten St. George, Daryl M. Lamson, Alexis Russell, Matthew Shudt, Melissa A Leisner, Jonathan Plitnick, Catharine Prussing, Navjot Singh, John Kelly, Erasmus Schneider, Erica Lasek-Nesselquist |
| EPI_ISL_1671608, EPI_ISL_1671609 | NORTHWELL HEALTH LABORATORIES | Wadsworth Center, New York State Department of Health | Kirsten St. George, Daryl M. Lamson, Alexis Russell, Matthew Shudt, Melissa A Leisner, Jonathan Plitnick, Catharine Prussing, Navjot Singh, John Kelly, Erasmus Schneider, Erica Lasek-Nesselquist |
| EPI_ISL_1671610, EPI_ISL_1671611 | WHITE PLAINS HOSPITAL CENTER LABORATORY | Wadsworth Center, New York State Department of Health | Kirsten St. George, Daryl M. Lamson, Alexis Russell, Matthew Shudt, Melissa A Leisner, Jonathan Plitnick, Catharine Prussing, Navjot Singh, John Kelly, Erasmus Schneider, Erica Lasek-Nesselquist |
| EPI_ISL_1671612 | SUNY UPSTATE MEDICAL UNIVERSITY | Wadsworth Center, New York State Department of Health | Kirsten St. George, Daryl M. Lamson, Alexis Russell, Matthew Shudt, Melissa A Leisner, Jonathan Plitnick, Catharine Prussing, Navjot Singh, John Kelly, Erasmus Schneider, Erica Lasek-Nesselquist |
| EPI_ISL_1671613 | URMC LABS | Wadsworth Center, New York State Department of Health | Kirsten St. George, Daryl M. Lamson, Alexis Russell, Matthew Shudt, Melissa A Leisner, Jonathan Plitnick, Catharine Prussing, Navjot Singh, John Kelly, Erasmus Schneider, Erica Lasek-Nesselquist |
| EPI_ISL_1671614, EPI_ISL_1671615, EPI_ISL_1671616, EPI_ISL_1671617 | NORTHWELL HEALTH LABORATORIES | Wadsworth Center, New York State Department of Health | Kirsten St. George, Daryl M. Lamson, Alexis Russell, Matthew Shudt, Melissa A Leisner, Jonathan Plitnick, Catharine Prussing, Navjot Singh, John Kelly, Erasmus Schneider, Erica Lasek-Nesselquist |
| EPI_ISL_1671618 | BOSTON HEART DIAGNOSTICS CORP | Wadsworth Center, New York State Department of Health | Kirsten St. George, Daryl M. Lamson, Alexis Russell, Matthew Shudt, Melissa A Leisner, Jonathan Plitnick, Catharine Prussing, Navjot Singh, John Kelly, Erasmus Schneider, Erica Lasek-Nesselquist |
| EPI_ISL_1671619 | WESTCHESTER MEDICAL CENTER | Wadsworth Center, New York State Department of Health | Kirsten St. George, Daryl M. Lamson, Alexis Russell, Matthew Shudt, Melissa A Leisner, Jonathan Plitnick, Catharine Prussing, Navjot Singh, John Kelly, Erasmus Schneider, Erica Lasek-Nesselquist |
| EPI_ISL_1671620, EPI_ISL_1671621 | NORTHWELL HEALTH LABORATORIES | Wadsworth Center, New York State Department of Health | Kirsten St. George, Daryl M. Lamson, Alexis Russell, Matthew Shudt, Melissa A Leisner, Jonathan Plitnick, Catharine Prussing, Navjot Singh, John Kelly, Erasmus Schneider, Erica Lasek-Nesselquist |
| EPI_ISL_1671622 | WHITE PLAINS HOSPITAL CENTER LABORATORY | Wadsworth Center, New York State Department of Health | Kirsten St. George, Daryl M. Lamson, Alexis Russell, Matthew Shudt, Melissa A Leisner, Jonathan Plitnick, Catharine Prussing, Navjot Singh, John Kelly, Erasmus Schneider, Erica Lasek-Nesselquist |
| EPI_ISL_1671623 | ALBANY MEDICAL CENTER | Wadsworth Center, New York State Department of Health | Kirsten St. George, Daryl M. Lamson, Alexis Russell, Matthew Shudt, Melissa A Leisner, Jonathan Plitnick, Catharine Prussing, Navjot Singh, John Kelly, Erasmus Schneider, Erica Lasek-Nesselquist |
| EPI_ISL_1671624, EPI_ISL_1671625 | WESTCHESTER MEDICAL CENTER | Wadsworth Center, New York State Department of Health | Kirsten St. George, Daryl M. Lamson, Alexis Russell, Matthew Shudt, Melissa A Leisner, Jonathan Plitnick, Catharine Prussing, Navjot Singh, John Kelly, Erasmus Schneider, Erica Lasek-Nesselquist |
| EPI_ISL_1671626, EPI_ISL_1671627 | URMC LABS | Wadsworth Center, New York State Department of Health | Kirsten St. George, Daryl M. Lamson, Alexis Russell, Matthew Shudt, Melissa A Leisner, Jonathan Plitnick, Catharine Prussing, Navjot Singh, John Kelly, Erasmus Schneider, Erica Lasek-Nesselquist |
| EPI_ISL_1671628, EPI_ISL_1671629, EPI_ISL_1671630, EPI_ISL_1671631, EPI_ISL_1671632, EPI_ISL_1671633 | NORTHWELL HEALTH LABORATORIES | Wadsworth Center, New York State Department of Health | Kirsten St. George, Daryl M. Lamson, Alexis Russell, Matthew Shudt, Melissa A Leisner, Jonathan Plitnick, Catharine Prussing, Navjot Singh, John Kelly, Erasmus Schneider, Erica Lasek-Nesselquist |
| EPI_ISL_1671634 | URMC LABS | Wadsworth Center, New York State Department of Health | Kirsten St. George, Daryl M. Lamson, Alexis Russell, Matthew Shudt, Melissa A Leisner, Jonathan Plitnick, Catharine Prussing, Navjot Singh, John Kelly, Erasmus Schneider, Erica Lasek-Nesselquist |
| EPI_ISL_1671635, EPI_ISL_1671636, EPI_ISL_1671637 | WESTCHESTER MEDICAL CENTER | Wadsworth Center, New York State Department of Health | Kirsten St. George, Daryl M. Lamson, Alexis Russell, Matthew Shudt, Melissa A Leisner, Jonathan Plitnick, Catharine Prussing, Navjot Singh, John Kelly, Erasmus Schneider, Erica Lasek-Nesselquist |
| EPI_ISL_1671638 | NORTHWELL HEALTH LABORATORIES | Wadsworth Center, New York State Department of Health | Kirsten St. George, Daryl M. Lamson, Alexis Russell, Matthew Shudt, Melissa A Leisner, Jonathan Plitnick, Catharine Prussing, Navjot Singh, John Kelly, Erasmus Schneider, Erica Lasek-Nesselquist |
| EPI_ISL_1671639 | WESTCHESTER MEDICAL CENTER | Wadsworth Center, New York State Department of Health | Kirsten St. George, Daryl M. Lamson, Alexis Russell, Matthew Shudt, Melissa A Leisner, Jonathan Plitnick, Catharine Prussing, Navjot Singh, John Kelly, Erasmus Schneider, Erica Lasek-Nesselquist |
| EPI_ISL_1671640, EPI_ISL_1671641 | NORTHWELL HEALTH LABORATORIES | Wadsworth Center, New York State Department of Health | Kirsten St. George, Daryl M. Lamson, Alexis Russell, Matthew Shudt, Melissa A Leisner, Jonathan Plitnick, Catharine Prussing, Navjot Singh, John Kelly, Erasmus Schneider, Erica Lasek-Nesselquist |
| EPI_ISL_1671642 | MONTEFIORE MEDICAL CENTER LABORATORIES | Wadsworth Center, New York State Department of Health | Kirsten St. George, Daryl M. Lamson, Alexis Russell, Matthew Shudt, Melissa A Leisner, Jonathan Plitnick, Catharine Prussing, Navjot Singh, John Kelly, Erasmus Schneider, Erica Lasek-Nesselquist |
| EPI_ISL_1671643 | TEMPUS LABS INC | Wadsworth Center, New York State Department of Health | Kirsten St. George, Daryl M. Lamson, Alexis Russell, Matthew Shudt, Melissa A Leisner, Jonathan Plitnick, Catharine Prussing, Navjot Singh, John Kelly, Erasmus Schneider, Erica Lasek-Nesselquist |
| EPI_ISL_1671644 | WHITE PLAINS HOSPITAL CENTER LABORATORY | Wadsworth Center, New York State Department of Health | Kirsten St. George, Daryl M. Lamson, Alexis Russell, Matthew Shudt, Melissa A Leisner, Jonathan Plitnick, Catharine Prussing, Navjot Singh, John Kelly, Erasmus Schneider, Erica Lasek-Nesselquist |
| EPI_ISL_1671645 | ALBANY MEDICAL CENTER | Wadsworth Center, New York State Department of Health | Kirsten St. George, Daryl M. Lamson, Alexis Russell, Matthew Shudt, Melissa A Leisner, Jonathan Plitnick, Catharine Prussing, Navjot Singh, John Kelly, Erasmus Schneider, Erica Lasek-Nesselquist |
| EPI_ISL_1671646, EPI_ISL_1671647 | WESTCHESTER MEDICAL CENTER | Wadsworth Center, New York State Department of Health | Kirsten St. George, Daryl M. Lamson, Alexis Russell, Matthew Shudt, Melissa A Leisner, Jonathan Plitnick, Catharine Prussing, Navjot Singh, John Kelly, Erasmus Schneider, Erica Lasek-Nesselquist |
| EPI_ISL_1671648, EPI_ISL_1671649, EPI_ISL_1671650, EPI_ISL_1671651, EPI_ISL_1671652, EPI_ISL_1671653 | NORTHWELL HEALTH LABORATORIES | Wadsworth Center, New York State Department of Health | Kirsten St. George, Daryl M. Lamson, Alexis Russell, Matthew Shudt, Melissa A Leisner, Jonathan Plitnick, Catharine Prussing, Navjot Singh, John Kelly, Erasmus Schneider, Erica Lasek-Nesselquist |
| EPI_ISL_1671654 | MONTEFIORE MEDICAL CENTER LABORATORIES | Wadsworth Center, New York State Department of Health | Kirsten St. George, Daryl M. Lamson, Alexis Russell, Matthew Shudt, Melissa A Leisner, Jonathan Plitnick, Catharine Prussing, Navjot Singh, John Kelly, Erasmus Schneider, Erica Lasek-Nesselquist |
| EPI_ISL_1671655, EPI_ISL_1671656 | WHITE PLAINS HOSPITAL CENTER LABORATORY | Wadsworth Center, New York State Department of Health | Kirsten St. George, Daryl M. Lamson, Alexis Russell, Matthew Shudt, Melissa A Leisner, Jonathan Plitnick, Catharine Prussing, Navjot Singh, John Kelly, Erasmus Schneider, Erica Lasek-Nesselquist |
| EPI_ISL_1671657, EPI_ISL_1671658, EPI_ISL_1671659, EPI_ISL_1671660 | WESTCHESTER MEDICAL CENTER | Wadsworth Center, New York State Department of Health | Kirsten St. George, Daryl M. Lamson, Alexis Russell, Matthew Shudt, Melissa A Leisner, Jonathan Plitnick, Catharine Prussing, Navjot Singh, John Kelly, Erasmus Schneider, Erica Lasek-Nesselquist |
| EPI_ISL_1671661, EPI_ISL_1671662, EPI_ISL_1671663 | NORTHWELL HEALTH LABORATORIES | Wadsworth Center, New York State Department of Health | Kirsten St. George, Daryl M. Lamson, Alexis Russell, Matthew Shudt, Melissa A Leisner, Jonathan Plitnick, Catharine Prussing, Navjot Singh, John Kelly, Erasmus Schneider, Erica Lasek-Nesselquist |
| EPI_ISL_1671664 | MONTEFIORE MEDICAL CENTER LABORATORIES | Wadsworth Center, New York State Department of Health | Kirsten St. George, Daryl M. Lamson, Alexis Russell, Matthew Shudt, Melissa A Leisner, Jonathan Plitnick, Catharine Prussing, Navjot Singh, John Kelly, Erasmus Schneider, Erica Lasek-Nesselquist |
| EPI_ISL_1671665 | WHITE PLAINS HOSPITAL CENTER LABORATORY | Wadsworth Center, New York State Department of Health | Kirsten St. George, Daryl M. Lamson, Alexis Russell, Matthew Shudt, Melissa A Leisner, Jonathan Plitnick, Catharine Prussing, Navjot Singh, John Kelly, Erasmus Schneider, Erica Lasek-Nesselquist |
| EPI_ISL_1671666 | WESTCHESTER MEDICAL CENTER | Wadsworth Center, New York State | Kirsten St. George, Daryl M. Lamson, Alexis Russell, Matthew Shudt, Melissa A Leisner, Jonathan Plitnick, Catharine Prussing, Navjot Singh, John Kelly, Erasmus Schneider, Erica Lasek-Nesselquist |

| | | | |
|---|---|---|---|
| | | Department of Health | |
| EPI_ISL_1671667 | URMC LABS | Wadsworth Center, New York State Department of Health | Kirsten St. George, Daryl M. Lamson, Alexis Russell, Matthew Shudt, Melissa A Leisner, Jonathan Plitnick, Catharine Prussing, Navjot Singh, John Kelly, Erasmus Schneider, Erica Lasek-Nesselquist |
| EPI_ISL_1671668, EPI_ISL_1671669, EPI_ISL_1671670, EPI_ISL_1671671, EPI_ISL_1671672, EPI_ISL_1671673, EPI_ISL_1671674, EPI_ISL_1671675, EPI_ISL_1671676 | NORTHWELL HEALTH LABORATORIES | Wadsworth Center, New York State Department of Health | Kirsten St. George, Daryl M. Lamson, Alexis Russell, Matthew Shudt, Melissa A Leisner, Jonathan Plitnick, Catharine Prussing, Navjot Singh, John Kelly, Erasmus Schneider, Erica Lasek-Nesselquist |
| EPI_ISL_1671677 | SARATOGA HOSPITAL LABORATORY | Wadsworth Center, New York State Department of Health | Kirsten St. George, Daryl M. Lamson, Alexis Russell, Matthew Shudt, Melissa A Leisner, Jonathan Plitnick, Catharine Prussing, Navjot Singh, John Kelly, Erasmus Schneider, Erica Lasek-Nesselquist |
| EPI_ISL_1671840, EPI_ISL_1671841, EPI_ISL_1671842, EPI_ISL_1671843, EPI_ISL_1671844, EPI_ISL_1671845, EPI_ISL_1671846, EPI_ISL_1671847, EPI_ISL_1671848, EPI_ISL_1671849, EPI_ISL_1671850, EPI_ISL_1671851, EPI_ISL_1671852, EPI_ISL_1671853, EPI_ISL_1671854, EPI_ISL_1671855, EPI_ISL_1671856, EPI_ISL_1671857, EPI_ISL_1671858, EPI_ISL_1671859, EPI_ISL_1671860, EPI_ISL_1671861, EPI_ISL_1671862, EPI_ISL_1671863, EPI_ISL_1671864, EPI_ISL_1671865, EPI_ISL_1671866, EPI_ISL_1671867, EPI_ISL_1671868, EPI_ISL_1671869, EPI_ISL_1671870, EPI_ISL_1671871, EPI_ISL_1671872, EPI_ISL_1671873, EPI_ISL_1671874, EPI_ISL_1671875, EPI_ISL_1671876, EPI_ISL_1671877, EPI_ISL_1671878, EPI_ISL_1671879, EPI_ISL_1671880, EPI_ISL_1671881, EPI_ISL_1671882, EPI_ISL_1671883, EPI_ISL_1671884, EPI_ISL_1671885, EPI_ISL_1671886, EPI_ISL_1671887, EPI_ISL_1671888, EPI_ISL_1671889, EPI_ISL_1671890, EPI_ISL_1671891, EPI_ISL_1671892, EPI_ISL_1671893, EPI_ISL_1671894, EPI_ISL_1671895, EPI_ISL_1671896, EPI_ISL_1671897, EPI_ISL_1671898, EPI_ISL_1671899, EPI_ISL_1671907, EPI_ISL_1671908, EPI_ISL_1671909, EPI_ISL_1671910, EPI_ISL_1671911, EPI_ISL_1671912, EPI_ISL_1671913, EPI_ISL_1671914, EPI_ISL_1671915, EPI_ISL_1671916, EPI_ISL_1671917, EPI_ISL_1671918, EPI_ISL_1671919, EPI_ISL_1671920, EPI_ISL_1671921 | | | |
| see above | Minnesota Department of Health, Public Health Laboratory | Minnesota Department of Health, Public Health Laboratory | Alexandra Lorentz, Jacob Garfin, Matt Plumb, and Xiong Wang |
| EPI_ISL_1672290, EPI_ISL_1672291 | M Health Fairview | Minnesota Department of Health, Public Health Laboratory | Alexandra Lorentz, Jacob Garfin, Matt Plumb, and Xiong Wang |
| EPI_ISL_1672292, EPI_ISL_1672293, EPI_ISL_1672294, EPI_ISL_1672295, EPI_ISL_1672296, EPI_ISL_1672297, EPI_ISL_1672298, EPI_ISL_1672299, EPI_ISL_1672300, EPI_ISL_1672301, EPI_ISL_1672302, EPI_ISL_1672303, EPI_ISL_1672304, EPI_ISL_1672305, EPI_ISL_1672306, EPI_ISL_1672307, EPI_ISL_1672308, EPI_ISL_1672309, EPI_ISL_1672310, EPI_ISL_1672311, EPI_ISL_1672312, EPI_ISL_1672313, EPI_ISL_1672314, EPI_ISL_1672315, EPI_ISL_1672316, EPI_ISL_1672317, EPI_ISL_1672318, EPI_ISL_1672319, EPI_ISL_1672320, EPI_ISL_1672321, EPI_ISL_1672322, EPI_ISL_1672323, EPI_ISL_1672324, EPI_ISL_1672325, EPI_ISL_1672326, EPI_ISL_1672327, EPI_ISL_1672328, EPI_ISL_1672329, EPI_ISL_1672330, EPI_ISL_1672331, EPI_ISL_1672332, EPI_ISL_1672333, EPI_ISL_1672334, EPI_ISL_1672335, EPI_ISL_1672336, EPI_ISL_1672337, EPI_ISL_1672338, EPI_ISL_1672339, EPI_ISL_1672340, EPI_ISL_1672341, EPI_ISL_1672342, EPI_ISL_1672343, EPI_ISL_1672344, EPI_ISL_1672345, EPI_ISL_1672346, EPI_ISL_1672347, EPI_ISL_1672348, EPI_ISL_1672349, EPI_ISL_1672350, EPI_ISL_1672351, EPI_ISL_1672352, EPI_ISL_1672353, EPI_ISL_1672354, EPI_ISL_1672355, EPI_ISL_1672356, EPI_ISL_1672357, EPI_ISL_1672358, EPI_ISL_1672359, EPI_ISL_1672360, EPI_ISL_1672361, EPI_ISL_1672362, EPI_ISL_1672363, EPI_ISL_1672364, EPI_ISL_1672365, EPI_ISL_1672366, EPI_ISL_1672367, EPI_ISL_1672368, EPI_ISL_1672369, EPI_ISL_1672370, EPI_ISL_1672371, EPI_ISL_1672372, EPI_ISL_1672373, EPI_ISL_1672374, EPI_ISL_1672375, EPI_ISL_1672376, EPI_ISL_1672377, EPI_ISL_1672378, EPI_ISL_1672379, EPI_ISL_1672380, EPI_ISL_1672381, EPI_ISL_1672382, EPI_ISL_1672383, EPI_ISL_1672384, EPI_ISL_1672385, EPI_ISL_1672386 | | | |
| see above | MD PHL | MD PHL | Maryland Department of Health Laboratories Administration |
| EPI_ISL_1672559 | Centracare Laboratory Services | Minnesota Department of Health, Public Health Laboratory | Alexandra Lorentz, Jacob Garfin, Matt Plumb, and Xiong Wang |
| EPI_ISL_1672560 | Minnesota Department of Health, Public Health Laboratory | Minnesota Department of Health, Public Health Laboratory | Alexandra Lorentz, Jacob Garfin, Matt Plumb, and Xiong Wang |
| EPI_ISL_1672561, EPI_ISL_1672562 | Centracare Laboratory Services | Minnesota Department of Health, Public Health Laboratory | Alexandra Lorentz, Jacob Garfin, Matt Plumb, and Xiong Wang |
| EPI_ISL_1672563, EPI_ISL_1672564, EPI_ISL_1672565, EPI_ISL_1672566, EPI_ISL_1672567, EPI_ISL_1672569, EPI_ISL_1672570, EPI_ISL_1672571, EPI_ISL_1672572, EPI_ISL_1672573, EPI_ISL_1672575, EPI_ISL_1672576, EPI_ISL_1672577, EPI_ISL_1672578, EPI_ISL_1672579, EPI_ISL_1672580, EPI_ISL_1672581, EPI_ISL_1672582, EPI_ISL_1672583, EPI_ISL_1672584, EPI_ISL_1672585, EPI_ISL_1672586, EPI_ISL_1672587, EPI_ISL_1672588, EPI_ISL_1672589, EPI_ISL_1672590, EPI_ISL_1672591, EPI_ISL_1672592, EPI_ISL_1672593, EPI_ISL_1672594, EPI_ISL_1672595, EPI_ISL_1672596, EPI_ISL_1672597, EPI_ISL_1672598, EPI_ISL_1672599, EPI_ISL_1672600, EPI_ISL_1672601, EPI_ISL_1672602, EPI_ISL_1672603, EPI_ISL_1672604, EPI_ISL_1672605, EPI_ISL_1672606, EPI_ISL_1672607, EPI_ISL_1672608, EPI_ISL_1672609, EPI_ISL_1672610, EPI_ISL_1672611, EPI_ISL_1672612, EPI_ISL_1672613, EPI_ISL_1672614, EPI_ISL_1672615, EPI_ISL_1672616, EPI_ISL_1672617, EPI_ISL_1672618, EPI_ISL_1672619, EPI_ISL_1672620, EPI_ISL_1672622, EPI_ISL_1672623, EPI_ISL_1672624, EPI_ISL_1672625, EPI_ISL_1672626, EPI_ISL_1672627, EPI_ISL_1672628, EPI_ISL_1672629, EPI_ISL_1672630, EPI_ISL_1672631, EPI_ISL_1672632 | | | |
| see above | Minnesota Department of Health, Public Health Laboratory | Minnesota Department of Health, Public Health Laboratory | Alexandra Lorentz, Jacob Garfin, Matt Plumb, and Xiong Wang |
| EPI_ISL_1672849 | Biologia molecular de enfermedades emergentes y EPOC, Instituto Nacional de Enfermedades Respiratorias | Biologia molecular de enfermedades emergentes y EPOC, Instituto Nacional de Enfermedades Respiratorias | Vazquez-Perez Joel Armando, Mejía-Nepomuceno Fidencio, Hernández-Teran Alejandra, Serna Ricardo, Salas-Hernández Jorge, Mario Mújica-Sánchez, Eduardo Becerril-Vargas, José Arturo Martínez-Orozco, Perez-Padilla Rogelio, |
| EPI_ISL_1672852, EPI_ISL_1672853, EPI_ISL_1672854, EPI_ISL_1672855, EPI_ISL_1672856, EPI_ISL_1672857, EPI_ISL_1672858, EPI_ISL_1672859, EPI_ISL_1672860, EPI_ISL_1672861, EPI_ISL_1672862, EPI_ISL_1672863 | | | |
| see above | Noblis | Noblis | Lauren Brinkac, Shiela Diepold, Shane Mitchell, June Kang, Lee F. Kolakowski, William M. Nelson, Katharine Jennings |
| EPI_ISL_1672864, EPI_ISL_1672865, EPI_ISL_1672866 | Biologia molecular de enfermedades emergentes y EPOC, Instituto Nacional de Enfermedades Respiratorias | Biologia molecular de enfermedades emergentes y EPOC, Instituto Nacional de Enfermedades Respiratorias | Vazquez-Perez Joel Armando, Mejía-Nepomuceno Fidencio, Hernández-Teran Alejandra, Serna Ricardo, Salas-Hernández Jorge, Mario Mújica-Sánchez, Eduardo Becerril-Vargas, José Arturo Martínez-Orozco, Perez-Padilla Rogelio |
| EPI_ISL_1672868, EPI_ISL_1672869, EPI_ISL_1672870, EPI_ISL_1672871, EPI_ISL_1672872, EPI_ISL_1672873, EPI_ISL_1672874, EPI_ISL_1672875, EPI_ISL_1672876, EPI_ISL_1672877, EPI_ISL_1672878, EPI_ISL_1672879, EPI_ISL_1672880, EPI_ISL_1672881, EPI_ISL_1672882, EPI_ISL_1672883, EPI_ISL_1672884, EPI_ISL_1672885 | | | |
| see above | Houston Health Dept. | Houston Health Dept. | Ryker Penn, Pamela Brown, Adolpho Lara, Yanlai Lai |
| EPI_ISL_1672888, EPI_ISL_1672889, EPI_ISL_1672890, EPI_ISL_1672892, EPI_ISL_1672894, EPI_ISL_1672895, EPI_ISL_1672896, EPI_ISL_1672897, EPI_ISL_1672898, EPI_ISL_1672899, EPI_ISL_1672900, EPI_ISL_1672901, EPI_ISL_1672903, EPI_ISL_1672904, EPI_ISL_1672905, EPI_ISL_1672906, EPI_ISL_1672907, EPI_ISL_1672908, EPI_ISL_1672909, EPI_ISL_1672910, EPI_ISL_1672911, EPI_ISL_1672912, EPI_ISL_1672913, EPI_ISL_1672914, EPI_ISL_1672915, EPI_ISL_1672916, EPI_ISL_1672917, EPI_ISL_1672918, EPI_ISL_1672919, EPI_ISL_1672920, EPI_ISL_1672921, EPI_ISL_1672926, EPI_ISL_1672927, EPI_ISL_1672929, EPI_ISL_1672930, EPI_ISL_1672932, EPI_ISL_1672934, EPI_ISL_1672935, EPI_ISL_1672937, EPI_ISL_1672938, EPI_ISL_1672939, EPI_ISL_1672940, EPI_ISL_1672941, EPI_ISL_1672942, EPI_ISL_1672943, EPI_ISL_1672944, EPI_ISL_1672945, EPI_ISL_1672947, EPI_ISL_1672949, EPI_ISL_1672951, EPI_ISL_1672952, EPI_ISL_1672954, EPI_ISL_1672955, EPI_ISL_1672956 | | | |
| see above | Wyoming Public Health Laboratory | Wyoming Public Health Laboratory | Jim Mildenberger, Wanda Manley, Noah Hull, Taylor Fearing, Lynette Gumbleton, Channing Weber, Ashley Norberg, Chayse Rowley, Marley Goetz, Brian Dominguez, Elliot Thomasson, Sam Britz, Cari Sloma, and Rob Christensen |
| EPI_ISL_1672958, EPI_ISL_1672959, EPI_ISL_1672960, EPI_ISL_1672961, EPI_ISL_1672962, EPI_ISL_1672963, EPI_ISL_1672964, EPI_ISL_1672965, EPI_ISL_1672966 | Nebraska Public Health Laboratory | NPHL COVID-19 Response Team | NPHL COVID-19 Response Team |
| EPI_ISL_1673263 | LESP Sinaloa | Instituto de Diagnostico y Referencia Epidemiologicos (INDRE) | Claudia Wong-Arambula, Abril Rodriguez-Maldonado, Vanessa Rivero-Arredondo, Ariadna Medina-Benitez, Joaquin Quiroz-Mercado, Sergio Rangel-Guerrero, Natividad Cruz-Ortiz, Tatiana Nunez-Garcia, Gisela Barrera-Badillo, Lucia Hernandez-Rivas, Irma Lopez-Martinez, Ernesto Ramirez-Gonzalez. |
| EPI_ISL_1673264 | LESP Queretaro | Instituto de Diagnostico y Referencia Epidemiologicos (INDRE) | Claudia Wong-Arambula, Abril Rodriguez-Maldonado, Vanessa Rivero-Arredondo, Ariadna Medina-Benitez, Joaquin Quiroz-Mercado, Sergio Rangel-Guerrero, Natividad Cruz-Ortiz, Tatiana Nunez-Garcia, Gisela Barrera-Badillo, Lucia Hernandez-Rivas, Irma Lopez-Martinez, Ernesto Ramirez-Gonzalez. |
| EPI_ISL_1673265 | LESP Guerrero | Instituto de Diagnostico y Referencia Epidemiologicos (INDRE) | Claudia Wong-Arambula, Abril Rodriguez-Maldonado, Vanessa Rivero-Arredondo, Ariadna Medina-Benitez, Joaquin Quiroz-Mercado, Sergio Rangel-Guerrero, Natividad Cruz-Ortiz, Tatiana Nunez-Garcia, Gisela Barrera-Badillo, Lucia Hernandez-Rivas, Irma Lopez-Martinez, Ernesto Ramirez-Gonzalez. |
| EPI_ISL_1673266 | LESP Morelos | Instituto de Diagnostico y Referencia Epidemiologicos (INDRE) | Claudia Wong-Arambula, Abril Rodriguez-Maldonado, Vanessa Rivero-Arredondo, Ariadna Medina-Benitez, Joaquin Quiroz-Mercado, Sergio Rangel-Guerrero, Natividad Cruz-Ortiz, Tatiana Nunez-Garcia, Gisela Barrera-Badillo, Lucia Hernandez-Rivas, Irma Lopez-Martinez, Ernesto Ramirez-Gonzalez. |
| EPI_ISL_1673267, EPI_ISL_1673268 | LESP Chihuahua | Instituto de Diagnostico y Referencia Epidemiologicos (INDRE) | Claudia Wong-Arambula, Abril Rodriguez-Maldonado, Vanessa Rivero-Arredondo, Ariadna Medina-Benitez, Joaquin Quiroz-Mercado, Sergio Rangel-Guerrero, Natividad Cruz-Ortiz, Tatiana Nunez-Garcia, Gisela Barrera-Badillo, Lucia Hernandez-Rivas, Irma Lopez-Martinez, Ernesto Ramirez-Gonzalez. |
| EPI_ISL_1673269, EPI_ISL_1673270, EPI_ISL_1673271, EPI_ISL_1673272, EPI_ISL_1673273 | LESP Guanajuato | Instituto de Diagnostico y Referencia Epidemiologicos (INDRE) | Claudia Wong-Arambula, Abril Rodriguez-Maldonado, Vanessa Rivero-Arredondo, Ariadna Medina-Benitez, Joaquin Quiroz-Mercado, Sergio Rangel-Guerrero, Natividad Cruz-Ortiz, Tatiana Nunez-Garcia, Gisela Barrera-Badillo, Lucia Hernandez-Rivas, Irma Lopez-Martinez, Ernesto Ramirez-Gonzalez. |
| EPI_ISL_1673274 | LESP Queretaro | Instituto de Diagnostico y Referencia Epidemiologicos (INDRE) | Claudia Wong-Arambula, Abril Rodriguez-Maldonado, Vanessa Rivero-Arredondo, Ariadna Medina-Benitez, Joaquin Quiroz-Mercado, Sergio Rangel-Guerrero, Natividad Cruz-Ortiz, Tatiana Nunez-Garcia, Gisela Barrera-Badillo, Lucia Hernandez-Rivas, Irma Lopez-Martinez, Ernesto Ramirez-Gonzalez. |
| EPI_ISL_1673275 | LESP Ciudad de Mexico/DIAGNOMOL | Instituto de Diagnostico y Referencia Epidemiologicos (INDRE) | Claudia Wong-Arambula, Abril Rodriguez-Maldonado, Vanessa Rivero-Arredondo, Ariadna Medina-Benitez, Joaquin Quiroz-Mercado, Sergio Rangel-Guerrero, Natividad Cruz-Ortiz, Tatiana Nunez-Garcia, Gisela Barrera-Badillo, Lucia Hernandez-Rivas, Irma Lopez-Martinez, Ernesto Ramirez-Gonzalez. |
| EPI_ISL_1673276 | LESP Jalisco | Instituto de Diagnostico y Referencia Epidemiologicos (INDRE) | Claudia Wong-Arambula, Abril Rodriguez-Maldonado, Vanessa Rivero-Arredondo, Ariadna Medina-Benitez, Joaquin Quiroz-Mercado, Sergio Rangel-Guerrero, Natividad Cruz-Ortiz, Tatiana Nunez-Garcia, Gisela Barrera-Badillo, Lucia Hernandez-Rivas, Irma Lopez-Martinez, Ernesto Ramirez-Gonzalez. |
| EPI_ISL_1673676, EPI_ISL_1673677, EPI_ISL_1673678, EPI_ISL_1673679 | Maryland Genomics, Institute for Genome Sciences, University of Maryland School of Medicine | Maryland Genomics, Institute for Genome Sciences, University of Maryland School of Medicine | Tallon, Luke J; Sadzewicz, Lisa D; Humphrys, Mike; Ott, Sandra; Roussey, Holly; Mehta, Aditya; Vavikolanu, Kranthi; Fraser, Claire M; Ravel, Jacques |
| EPI_ISL_1673686, EPI_ISL_1673687, EPI_ISL_1673689, EPI_ISL_1673690, EPI_ISL_1673691, EPI_ISL_1673692, EPI_ISL_1673693, EPI_ISL_1673694, EPI_ISL_1673695, EPI_ISL_1673696, EPI_ISL_1673697, EPI_ISL_1673698, EPI_ISL_1673699, EPI_ISL_1673700, EPI_ISL_1673701, EPI_ISL_1673702, EPI_ISL_1673703, EPI_ISL_1673704, EPI_ISL_1673705, EPI_ISL_1673706, EPI_ISL_1673707, EPI_ISL_1673708, EPI_ISL_1673709, EPI_ISL_1673710, EPI_ISL_1673711, EPI_ISL_1673712, EPI_ISL_1673713, EPI_ISL_1673714, EPI_ISL_1673715, EPI_ISL_1673716, EPI_ISL_1673717, EPI_ISL_1673718, EPI_ISL_1673719, EPI_ISL_1673720, EPI_ISL_1673721, EPI_ISL_1673722, EPI_ISL_1673723, EPI_ISL_1673724, EPI_ISL_1673725, EPI_ISL_1673726, EPI_ISL_1673727, EPI_ISL_1673728, EPI_ISL_1673729, EPI_ISL_1673730, EPI_ISL_1673732 | | | |
| see above | Wyoming Public Health Laboratory | Center for Global Health, University of New Mexico Health Sciences Center | Daryl Domman, Kurt Schwalm, Rob Christensen, Wanda Manley, Cari Sloma, Noah Hull, Valerie Morley, Darrell Dinwiddie |
| EPI_ISL_1674535, EPI_ISL_1674537, EPI_ISL_1674538, EPI_ISL_1674540, EPI_ISL_1674541, EPI_ISL_1674543, EPI_ISL_1674546, EPI_ISL_1674549, EPI_ISL_1674550, EPI_ISL_1674553, EPI_ISL_1674555, EPI_ISL_1674556, EPI_ISL_1674558, EPI_ISL_1674559, EPI_ISL_1674560, EPI_ISL_1674561, EPI_ISL_1674562, EPI_ISL_1674563, EPI_ISL_1674564, EPI_ISL_1674566, EPI_ISL_1674569, EPI_ISL_1674570, | | | |

| | | | |
|---|---|---|---|
| EPI_ISL_1674571, EPI_ISL_1674572, EPI_ISL_1674573, EPI_ISL_1674574, EPI_ISL_1674575, EPI_ISL_1674576, EPI_ISL_1674577, EPI_ISL_1674578, EPI_ISL_1674580, EPI_ISL_1674582, EPI_ISL_1674583, EPI_ISL_1674584, EPI_ISL_1674585, EPI_ISL_1674586, EPI_ISL_1674587, EPI_ISL_1674591, EPI_ISL_1674592, EPI_ISL_1674593, EPI_ISL_1674594, EPI_ISL_1674595, EPI_ISL_1674596, EPI_ISL_1674597, EPI_ISL_1674598, EPI_ISL_1674599, EPI_ISL_1674601, EPI_ISL_1674602, EPI_ISL_1674603, EPI_ISL_1674604, EPI_ISL_1674606, EPI_ISL_1674607, EPI_ISL_1674608, EPI_ISL_1674609, EPI_ISL_1674610, EPI_ISL_1674611, EPI_ISL_1674612, EPI_ISL_1674613, EPI_ISL_1674615, EPI_ISL_1674616, EPI_ISL_1674617, EPI_ISL_1674620, EPI_ISL_1674621, EPI_ISL_1674622, EPI_ISL_1674624, EPI_ISL_1674625, EPI_ISL_1674626, EPI_ISL_1674627, EPI_ISL_1674628, EPI_ISL_1674629, EPI_ISL_1674630, EPI_ISL_1674631, EPI_ISL_1674632, EPI_ISL_1674633, EPI_ISL_1674634, EPI_ISL_1674635, EPI_ISL_1674636, EPI_ISL_1674637, EPI_ISL_1674639, EPI_ISL_1674641, EPI_ISL_1674642, EPI_ISL_1674645, EPI_ISL_1674647, EPI_ISL_1674648, EPI_ISL_1674649, EPI_ISL_1674650, EPI_ISL_1674651, EPI_ISL_1674652, EPI_ISL_1674653, EPI_ISL_1674654, EPI_ISL_1674655, EPI_ISL_1674656, EPI_ISL_1674657, EPI_ISL_1674658, EPI_ISL_1674659, EPI_ISL_1674660, EPI_ISL_1674661, EPI_ISL_1674663, EPI_ISL_1674664, EPI_ISL_1674665, EPI_ISL_1674666, EPI_ISL_1674667, EPI_ISL_1674668, EPI_ISL_1674670, EPI_ISL_1674671, EPI_ISL_1674672, EPI_ISL_1674675, EPI_ISL_1674676, EPI_ISL_1674677, EPI_ISL_1674678, EPI_ISL_1674679, EPI_ISL_1674680, EPI_ISL_1674681, EPI_ISL_1674682, EPI_ISL_1674683, EPI_ISL_1674684, EPI_ISL_1674685, EPI_ISL_1674686, EPI_ISL_1674689, EPI_ISL_1674690, EPI_ISL_1674692, EPI_ISL_1674693, EPI_ISL_1674694, EPI_ISL_1674695, EPI_ISL_1674696 | | | |
| see above | Yale Clinical Virology Lab | Grubaugh Lab - Yale School of Public Health | Joseph Fauver, Mallery Breban, Isabel Ott, Tara Alpert, Mary Petrone, Anderson Brito, Chantal Vogels, Annie Watkins, Chaney Kalinich, Jessica Rothman, Marie L. Landry, Nathan Grubaugh |
| EPI_ISL_1674698, EPI_ISL_1674699, EPI_ISL_1674700, EPI_ISL_1674701, EPI_ISL_1674702, EPI_ISL_1674703, EPI_ISL_1674704, EPI_ISL_1674705 | Murphy Medical Association | Grubaugh Lab - Yale School of Public Health | Joseph Fauver, Mallery Breban, Isabel Ott, Tara Alpert, Mary Petrone, Anderson Brito, Chantal Vogels, Annie Watkins, Chaney Kalinich, Jessica Rothman, Caleb Neal, Eva Laszlo, Steven Murphy, Nathan Grubaugh |
| EPI_ISL_1674706, EPI_ISL_1674707, EPI_ISL_1674708 | Grubaugh Lab - Yale School of Public Health | Grubaugh Lab - Yale School of Public Health | Joseph Fauver, Mallery Breban, Isabel Ott, Tara Alpert, Mary Petrone, Anderson Brito, Chantal Vogels, Annie Watkins, Chaney Kalinich, Jessica Rothman, Nathan Grubaugh |
| EPI_ISL_1674711, EPI_ISL_1674712, EPI_ISL_1674713, EPI_ISL_1674714, EPI_ISL_1674715, EPI_ISL_1674716, EPI_ISL_1674717, EPI_ISL_1674718, EPI_ISL_1674719, EPI_ISL_1674720, EPI_ISL_1674721, EPI_ISL_1674722, EPI_ISL_1674723, EPI_ISL_1674724, EPI_ISL_1674725, EPI_ISL_1674726, EPI_ISL_1674727, EPI_ISL_1674729, EPI_ISL_1674730, EPI_ISL_1674731, EPI_ISL_1674732, EPI_ISL_1674733, EPI_ISL_1674734, EPI_ISL_1674735, EPI_ISL_1674736, EPI_ISL_1674737, EPI_ISL_1674738, EPI_ISL_1674739, EPI_ISL_1674740, EPI_ISL_1674741, EPI_ISL_1674742, EPI_ISL_1674743, EPI_ISL_1674744, EPI_ISL_1674745, EPI_ISL_1674746, EPI_ISL_1674747, EPI_ISL_1674748, EPI_ISL_1674749, EPI_ISL_1674750, EPI_ISL_1674751, EPI_ISL_1674752, EPI_ISL_1674753, EPI_ISL_1674754, EPI_ISL_1674755, EPI_ISL_1674756, EPI_ISL_1674757, EPI_ISL_1674758, EPI_ISL_1674759, EPI_ISL_1674760, EPI_ISL_1674761, EPI_ISL_1674762, EPI_ISL_1674763, EPI_ISL_1674764, EPI_ISL_1674765, EPI_ISL_1674766, EPI_ISL_1674767, EPI_ISL_1674768, EPI_ISL_1674770, EPI_ISL_1674771, EPI_ISL_1674772, EPI_ISL_1674773, EPI_ISL_1674774, EPI_ISL_1674775, EPI_ISL_1674776, EPI_ISL_1674777, EPI_ISL_1674778, EPI_ISL_1674779, EPI_ISL_1674780, EPI_ISL_1674782, EPI_ISL_1674783, EPI_ISL_1674784, EPI_ISL_1674785, EPI_ISL_1674787, EPI_ISL_1674788, EPI_ISL_1674789, EPI_ISL_1674790, EPI_ISL_1674791, EPI_ISL_1674792, EPI_ISL_1674793, EPI_ISL_1674794, EPI_ISL_1674795, EPI_ISL_1674798, EPI_ISL_1674799, EPI_ISL_1674800, EPI_ISL_1674801, EPI_ISL_1674802, EPI_ISL_1674803, EPI_ISL_1674804, EPI_ISL_1674805, EPI_ISL_1674806, EPI_ISL_1674807, EPI_ISL_1674808, EPI_ISL_1674809, EPI_ISL_1674810, EPI_ISL_1674811, EPI_ISL_1674812, EPI_ISL_1674813, EPI_ISL_1674814, EPI_ISL_1674815, EPI_ISL_1674816, EPI_ISL_1674817, EPI_ISL_1674818, EPI_ISL_1674819, EPI_ISL_1674820, EPI_ISL_1674821, EPI_ISL_1674822, EPI_ISL_1674823, EPI_ISL_1674824, EPI_ISL_1674825, EPI_ISL_1674826, EPI_ISL_1674827, EPI_ISL_1674828, EPI_ISL_1674829, EPI_ISL_1674831, EPI_ISL_1674832, EPI_ISL_1674833, EPI_ISL_1674834, EPI_ISL_1674835, EPI_ISL_1674836, EPI_ISL_1674837, EPI_ISL_1674838, EPI_ISL_1674839, EPI_ISL_1674840, EPI_ISL_1674841, EPI_ISL_1674842, EPI_ISL_1674843, EPI_ISL_1674844, EPI_ISL_1674845, EPI_ISL_1674846, EPI_ISL_1674847, EPI_ISL_1674848, EPI_ISL_1674849, EPI_ISL_1674850, EPI_ISL_1674851, EPI_ISL_1674853, EPI_ISL_1674855, EPI_ISL_1674856, EPI_ISL_1674857, EPI_ISL_1674858, EPI_ISL_1674859, EPI_ISL_1674860, EPI_ISL_1674861, EPI_ISL_1674862, EPI_ISL_1674863, EPI_ISL_1674864, EPI_ISL_1674865, EPI_ISL_1674866, EPI_ISL_1674868, EPI_ISL_1674869, EPI_ISL_1674870, EPI_ISL_1674871, EPI_ISL_1674872, EPI_ISL_1674873, EPI_ISL_1674874, EPI_ISL_1674875, EPI_ISL_1674876, EPI_ISL_1674878, EPI_ISL_1674879, EPI_ISL_1674880, EPI_ISL_1674881, EPI_ISL_1674882, EPI_ISL_1674883, EPI_ISL_1674884, EPI_ISL_1674885, EPI_ISL_1674886, EPI_ISL_1674887, EPI_ISL_1674888, EPI_ISL_1674890, EPI_ISL_1674891, EPI_ISL_1674892, EPI_ISL_1674893, EPI_ISL_1674894, EPI_ISL_1674895, EPI_ISL_1674896, EPI_ISL_1674897, EPI_ISL_1674898, EPI_ISL_1674899, EPI_ISL_1674900, EPI_ISL_1674901, EPI_ISL_1674902, EPI_ISL_1674903, EPI_ISL_1674904, EPI_ISL_1674905, EPI_ISL_1674906, EPI_ISL_1674907, EPI_ISL_1674908, EPI_ISL_1674909, EPI_ISL_1674910, EPI_ISL_1674911, EPI_ISL_1674912, EPI_ISL_1674913, EPI_ISL_1674914, EPI_ISL_1674915, EPI_ISL_1674916, EPI_ISL_1674917, EPI_ISL_1674918, EPI_ISL_1674919, EPI_ISL_1674920, EPI_ISL_1674921, EPI_ISL_1674922, EPI_ISL_1674923, EPI_ISL_1674924, EPI_ISL_1674925, EPI_ISL_1674926, EPI_ISL_1674927, EPI_ISL_1674928, EPI_ISL_1674929, EPI_ISL_1674930, EPI_ISL_1674931, EPI_ISL_1674933, EPI_ISL_1674934, EPI_ISL_1674935, EPI_ISL_1674936, EPI_ISL_1674938, EPI_ISL_1674939, EPI_ISL_1674940, EPI_ISL_1674941, EPI_ISL_1674942, EPI_ISL_1674943, EPI_ISL_1674944, EPI_ISL_1674945, EPI_ISL_1674946, EPI_ISL_1674947, EPI_ISL_1674948, EPI_ISL_1674949, EPI_ISL_1674950, EPI_ISL_1674951, EPI_ISL_1674952, EPI_ISL_1674953, EPI_ISL_1674954, EPI_ISL_1674955, EPI_ISL_1674956, EPI_ISL_1674957, EPI_ISL_1674958, EPI_ISL_1674959, EPI_ISL_1674960, EPI_ISL_1674961, EPI_ISL_1674962, EPI_ISL_1674963, EPI_ISL_1674964, EPI_ISL_1674965, EPI_ISL_1674966, EPI_ISL_1674967, EPI_ISL_1674968, EPI_ISL_1674969, EPI_ISL_1674970, EPI_ISL_1674972, EPI_ISL_1674973, EPI_ISL_1674974, EPI_ISL_1674975, EPI_ISL_1674976, EPI_ISL_1674977, EPI_ISL_1674978, EPI_ISL_1674979, EPI_ISL_1674980, EPI_ISL_1674981, EPI_ISL_1674982, EPI_ISL_1674983, EPI_ISL_1674984, EPI_ISL_1674985, EPI_ISL_1674986, EPI_ISL_1674987, EPI_ISL_1674988, EPI_ISL_1674990, EPI_ISL_1674991, EPI_ISL_1674992, EPI_ISL_1674993, EPI_ISL_1674994, EPI_ISL_1674995, EPI_ISL_1674997, EPI_ISL_1674998, EPI_ISL_1674999, EPI_ISL_1675000, EPI_ISL_1675002, EPI_ISL_1675003, EPI_ISL_1675004, EPI_ISL_1675005 | | | |
| see above | Florida Bureau of Public Health Laboratories | Florida Bureau of Public Health Laboratories | Sarah Schmedes, Jason Blanton |
| EPI_ISL_1675096, EPI_ISL_1675097, EPI_ISL_1675098, EPI_ISL_1675099, EPI_ISL_1675100, EPI_ISL_1675101, EPI_ISL_1675102, EPI_ISL_1675103, EPI_ISL_1675104, EPI_ISL_1675105, EPI_ISL_1675106, EPI_ISL_1675107, EPI_ISL_1675108, EPI_ISL_1675109, EPI_ISL_1675110, EPI_ISL_1675111, EPI_ISL_1675112, EPI_ISL_1675113, EPI_ISL_1675114, EPI_ISL_1675115, EPI_ISL_1675116, EPI_ISL_1675117, EPI_ISL_1675118, EPI_ISL_1675119, EPI_ISL_1675120, EPI_ISL_1675121, EPI_ISL_1675122, EPI_ISL_1675123, EPI_ISL_1675124, EPI_ISL_1675125, EPI_ISL_1675126, EPI_ISL_1675127, EPI_ISL_1675128, EPI_ISL_1675129, EPI_ISL_1675130, EPI_ISL_1675131 | | | |
| see above | Alaska State Virology Laboratory | Alaska State Virology Laboratory | Stephanie DeRonde, Elva House, Lisa Smith, Ph.D., Jack Chen, Ph.D. |
| EPI_ISL_1675145 | Oregon State University TRACE Project | OSU Center for Genome Research and Biocomputing | Oregon State University TRACE Project |
| EPI_ISL_1675148, EPI_ISL_1675149, EPI_ISL_1675150, EPI_ISL_1675151, EPI_ISL_1675152, EPI_ISL_1675153, EPI_ISL_1675154, EPI_ISL_1675155, EPI_ISL_1675156, EPI_ISL_1675157, EPI_ISL_1675159, EPI_ISL_1675160, EPI_ISL_1675161, EPI_ISL_1675162 | | | |
| see above | Nevada State Public Health Laboratory | Nevada State Public Health Laboratory | Andrew Gorzalski, Mark Pandori |
| EPI_ISL_1675164, EPI_ISL_1675165, EPI_ISL_1675166, EPI_ISL_1675167, EPI_ISL_1675168, EPI_ISL_1675169, EPI_ISL_1675170, EPI_ISL_1675171, EPI_ISL_1675172, EPI_ISL_1675173, EPI_ISL_1675174, EPI_ISL_1675175, EPI_ISL_1675176, EPI_ISL_1675177, EPI_ISL_1675178, EPI_ISL_1675179, EPI_ISL_1675180, EPI_ISL_1675181, EPI_ISL_1675182, EPI_ISL_1675183, EPI_ISL_1675184, EPI_ISL_1675185, EPI_ISL_1675186, EPI_ISL_1675187, EPI_ISL_1675188, EPI_ISL_1675189, EPI_ISL_1675190, EPI_ISL_1675191, EPI_ISL_1675192, EPI_ISL_1675193, EPI_ISL_1675194, EPI_ISL_1675195, EPI_ISL_1675196, EPI_ISL_1675197, EPI_ISL_1675198, EPI_ISL_1675199, EPI_ISL_1675200, EPI_ISL_1675202, EPI_ISL_1675204, EPI_ISL_1675205, EPI_ISL_1675207, EPI_ISL_1675208, EPI_ISL_1675209, EPI_ISL_1675210, EPI_ISL_1675211, EPI_ISL_1675212, EPI_ISL_1675213 | | | |
| see above | Department of Laboratory Medicine, Clinical Center, National Institutes of Health | Laboratory of Parasitic Diseases, Systems Genomics Section, National Institute of Allergy and Infectious Diseases, National Institutes of Health | Allison Roder, Stephanie Banakis, Matthew Chung, Jung-ho Youn, Rachel Mercado, Wei Wang, Tara Palmore, Michael Bell, Heike Bailin, Jessica McCormick-Ell, Sanchita Das, Jennifer Kwan, Elodie Ghedin |
| EPI_ISL_1675215, EPI_ISL_1675216, EPI_ISL_1675217, EPI_ISL_1675218 | Wyoming Public Health Laboratory | Wyoming Public Health Laboratory | Jim Mildenberger, Taylor Fearing, Channing Weber, Ashley Norberg, Chayse Rowley, Marley Goetz, Brian Dominguez, Elliot Thomasson, Brittany Oher, Sam Britz, Cari Sloma, and Rob Christensen |
| EPI_ISL_1675223, EPI_ISL_1675224, EPI_ISL_1675225 | Stanford Health Care | Stanford University School of Medicine, Clinical Virology Laboratory | Michelle Verghese, Becky Jiang, Naomi Iwai, Marilyn Mar, Malaya K. Sahoo, Daniel Solis, Mamdouh Sibai, Fumiko Yamamoto, Kenji O. Mfuh, Jacob Miller, Hannah Wang, James Zehnder, and Benjamin A. Pinsky |
| EPI_ISL_1675226, EPI_ISL_1675227 | Stanford Health Care | Stanford University School of Medicine, Clinical Virology Laboratory | Mamdouh Sibai, Daniel Solis, Michelle Verghese, Becky Jiang, Fumiko Yamamoto, Jacob Miller, Hannah Wang, Marilyn Mar, Naomi Iwai, Kenji O. Mfuh, James Zehnder, Malaya K. Sahoo, and Benjamin A. Pinsky |
| EPI_ISL_1675230, EPI_ISL_1675232, EPI_ISL_1675233, EPI_ISL_1675234, EPI_ISL_1675235, EPI_ISL_1675236, EPI_ISL_1675237, EPI_ISL_1675238, EPI_ISL_1675239, EPI_ISL_1675240, EPI_ISL_1675241 | | | |
| see above | Oregon State University TRACE Project | OSU Center for Genome Research and Biocomputing | Oregon State University TRACE Project |
| EPI_ISL_1677791 | Monterey County Public Health Laboratory | Monterey County Public Health Laboratory | Monterey County Public Health Laboratory |
| EPI_ISL_1678025, EPI_ISL_1678026, EPI_ISL_1678027, EPI_ISL_1678028, EPI_ISL_1678029, EPI_ISL_1678030 | "AR Dept. of Health-PHL, Molecular Diagnostics" | Centers for Disease Control and Prevention Division of Viral Diseases, Pathogen Discovery | Mili Sheth, Sarah Nobles, Jasmine Padilla, Mark Burroughs, Shoshona Le, Katie Dillon, Peter Cook, Clinton R. Paden, Dhwani Batra, Krista Queen, Kristen Knipe, Dakota Howard, Yvette Unoarumhi, Darlene Wagner, Matthew Schmerer, Ben L. Rambo-Martin, Kristine Lacek, Sam Shepard, Alison Laufer Halpin, Dave Wentworth, Vivien Dugan, Suxiang Tong, Justin Lee |
| EPI_ISL_1678031, EPI_ISL_1678032, EPI_ISL_1678033, EPI_ISL_1678034, EPI_ISL_1678035, EPI_ISL_1678036 | VA-Division of Consolidated Laboratory Services | Centers for Disease Control and Prevention Division of Viral Diseases, Pathogen Discovery | Mili Sheth, Sarah Nobles, Jasmine Padilla, Mark Burroughs, Shoshona Le, Katie Dillon, Peter Cook, Clinton R. Paden, Dhwani Batra, Krista Queen, Kristen Knipe, Dakota Howard, Yvette Unoarumhi, Darlene Wagner, Matthew Schmerer, Ben L. Rambo-Martin, Kristine Lacek, Sam Shepard, Alison Laufer Halpin, Dave Wentworth, Vivien Dugan, Suxiang Tong, Justin Lee |
| EPI_ISL_1678037, EPI_ISL_1678038 | "AZ SPHL, Arizona Department of Health Services" | Centers for Disease Control and Prevention Division of Viral Diseases, Pathogen Discovery | Mili Sheth, Sarah Nobles, Jasmine Padilla, Mark Burroughs, Shoshona Le, Katie Dillon, Peter Cook, Clinton R. Paden, Dhwani Batra, Krista Queen, Kristen Knipe, Dakota Howard, Yvette Unoarumhi, Darlene Wagner, Matthew Schmerer, Ben L. Rambo-Martin, Kristine Lacek, Sam Shepard, Alison Laufer Halpin, Dave Wentworth, Vivien Dugan, Suxiang Tong, Justin Lee |
| EPI_ISL_1678039, EPI_ISL_1678040, EPI_ISL_1678041, EPI_ISL_1678042, EPI_ISL_1678043, EPI_ISL_1678044, EPI_ISL_1678045 | MS Public Health Laboratory | Centers for Disease Control and Prevention Division of Viral Diseases, Pathogen Discovery | Mili Sheth, Sarah Nobles, Jasmine Padilla, Mark Burroughs, Shoshona Le, Katie Dillon, Peter Cook, Clinton R. Paden, Dhwani Batra, Krista Queen, Kristen Knipe, Dakota Howard, Yvette Unoarumhi, Darlene Wagner, Matthew Schmerer, Ben L. Rambo-Martin, Kristine Lacek, Sam Shepard, Alison Laufer Halpin, Dave Wentworth, Vivien Dugan, Suxiang Tong, Justin Lee |
| EPI_ISL_1678046, EPI_ISL_1678047, EPI_ISL_1678048 | WVDHHR - Office of Laboratory Services | Centers for Disease Control and Prevention Division of Viral Diseases, Pathogen Discovery | Mili Sheth, Sarah Nobles, Jasmine Padilla, Mark Burroughs, Shoshona Le, Katie Dillon, Peter Cook, Clinton R. Paden, Dhwani Batra, Krista Queen, Kristen Knipe, Dakota Howard, Yvette Unoarumhi, Darlene Wagner, Matthew Schmerer, Ben L. Rambo-Martin, Kristine Lacek, Sam Shepard, Alison Laufer Halpin, Dave Wentworth, Vivien Dugan, Suxiang Tong, Justin Lee |
| EPI_ISL_1678049 | IA State Hygienic Laboratory | Centers for Disease Control and Prevention Division of Viral Diseases, Pathogen Discovery | Mili Sheth, Sarah Nobles, Jasmine Padilla, Mark Burroughs, Shoshona Le, Katie Dillon, Peter Cook, Clinton R. Paden, Dhwani Batra, Krista Queen, Kristen Knipe, Dakota Howard, Yvette Unoarumhi, Darlene Wagner, Matthew Schmerer, Ben L. Rambo-Martin, Kristine Lacek, Sam Shepard, Alison Laufer Halpin, Dave Wentworth, Vivien Dugan, Suxiang Tong, Justin Lee |
| EPI_ISL_1678050, EPI_ISL_1678051, EPI_ISL_1678052, EPI_ISL_1678053, EPI_ISL_1678054, EPI_ISL_1678055 | KS Health and Environmental Laboratories | Centers for Disease Control and Prevention Division of Viral Diseases, Pathogen Discovery | Mili Sheth, Sarah Nobles, Jasmine Padilla, Mark Burroughs, Shoshona Le, Katie Dillon, Peter Cook, Clinton R. Paden, Dhwani Batra, Krista Queen, Kristen Knipe, Dakota Howard, Yvette Unoarumhi, Darlene Wagner, Matthew Schmerer, Ben L. Rambo-Martin, Kristine Lacek, Sam Shepard, Alison Laufer Halpin, Dave Wentworth, Vivien Dugan, Suxiang Tong, Justin Lee |
| EPI_ISL_1678056, EPI_ISL_1678057, EPI_ISL_1678058, EPI_ISL_1678059, EPI_ISL_1678060, EPI_ISL_1678061, EPI_ISL_1678062, EPI_ISL_1678063, EPI_ISL_1678064, EPI_ISL_1678065, EPI_ISL_1678066, EPI_ISL_1678067, EPI_ISL_1678068, EPI_ISL_1678069, EPI_ISL_1678070, EPI_ISL_1678071, EPI_ISL_1678072, EPI_ISL_1678073, EPI_ISL_1678074, EPI_ISL_1678075, EPI_ISL_1678076, EPI_ISL_1678077, EPI_ISL_1678078, EPI_ISL_1678079, EPI_ISL_1678080 | | | |
| see above | NYC Department of Health and Mental Hygiene | Centers for Disease Control and Prevention Division of Viral Diseases, Pathogen | Mili Sheth, Sarah Nobles, Jasmine Padilla, Mark Burroughs, Shoshona Le, Katie Dillon, Peter Cook, Clinton R. Paden, Dhwani Batra, Krista Queen, Kristen Knipe, Dakota Howard, Yvette Unoarumhi, Darlene Wagner, Matthew Schmerer, Ben L. Rambo-Martin, Kristine Lacek, Sam Shepard, Alison Laufer Halpin, Dave Wentworth, Vivien Dugan, Suxiang Tong, Justin Lee |

| | | | |
|---|---|---|---|
| | | Discovery | |
| EPI_ISL_1678081, EPI_ISL_1678082 | "NM Dept. Health, Scientific Laboratory Division " | Centers for Disease Control and Prevention Division of Viral Diseases, Pathogen Discovery | Mili Sheth, Sarah Nobles, Jasmine Padilla, Mark Burroughs, Shoshona Le, Katie Dillon, Peter Cook, Clinton R. Paden, Dhwani Batra, Krista Queen, Kristen Knipe, Dakota Howard, Yvette Unoarumhi, Darlene Wagner, Matthew Schmerer, Ben L. Rambo-Martin, Kristine Lacek, Sam Shepard, Alison Laufer Halpin, Dave Wentworth, Vivien Dugan, Suxiang Tong, Justin Lee |
| EPI_ISL_1678083 | "OK Public Health Laboratory, Oklahoma State DOH" | Centers for Disease Control and Prevention Division of Viral Diseases, Pathogen Discovery | Mili Sheth, Sarah Nobles, Jasmine Padilla, Mark Burroughs, Shoshona Le, Katie Dillon, Peter Cook, Clinton R. Paden, Dhwani Batra, Krista Queen, Kristen Knipe, Dakota Howard, Yvette Unoarumhi, Darlene Wagner, Matthew Schmerer, Ben L. Rambo-Martin, Kristine Lacek, Sam Shepard, Alison Laufer Halpin, Dave Wentworth, Vivien Dugan, Suxiang Tong, Justin Lee |
| EPI_ISL_1678084 | MI - Michigan Department of Health and Human Services - Bureau of Laboratories | Centers for Disease Control and Prevention Division of Viral Diseases, Pathogen Discovery | Mili Sheth, Sarah Nobles, Jasmine Padilla, Mark Burroughs, Shoshona Le, Katie Dillon, Peter Cook, Clinton R. Paden, Dhwani Batra, Krista Queen, Kristen Knipe, Dakota Howard, Yvette Unoarumhi, Darlene Wagner, Matthew Schmerer, Ben L. Rambo-Martin, Kristine Lacek, Sam Shepard, Alison Laufer Halpin, Dave Wentworth, Vivien Dugan, Suxiang Tong, Justin Lee |
| EPI_ISL_1678085, EPI_ISL_1678086, EPI_ISL_1678087, EPI_ISL_1678088, EPI_ISL_1678089, EPI_ISL_1678090, EPI_ISL_1678091, EPI_ISL_1678092, EPI_ISL_1678093, EPI_ISL_1678094, EPI_ISL_1678095 | | | |
| see above | CA-Los Angeles County Public Health Laboratory | Centers for Disease Control and Prevention Division of Viral Diseases, Pathogen Discovery | Mili Sheth, Sarah Nobles, Jasmine Padilla, Mark Burroughs, Shoshona Le, Katie Dillon, Peter Cook, Clinton R. Paden, Dhwani Batra, Krista Queen, Kristen Knipe, Dakota Howard, Yvette Unoarumhi, Darlene Wagner, Matthew Schmerer, Ben L. Rambo-Martin, Kristine Lacek, Sam Shepard, Alison Laufer Halpin, Dave Wentworth, Vivien Dugan, Suxiang Tong, Justin Lee |
| EPI_ISL_1678096, EPI_ISL_1678097, EPI_ISL_1678098, EPI_ISL_1678099 | UT-Unified State Labs: Public Health Utah DOH | Centers for Disease Control and Prevention Division of Viral Diseases, Pathogen Discovery | Mili Sheth, Sarah Nobles, Jasmine Padilla, Mark Burroughs, Shoshona Le, Katie Dillon, Peter Cook, Clinton R. Paden, Dhwani Batra, Krista Queen, Kristen Knipe, Dakota Howard, Yvette Unoarumhi, Darlene Wagner, Matthew Schmerer, Ben L. Rambo-Martin, Kristine Lacek, Sam Shepard, Alison Laufer Halpin, Dave Wentworth, Vivien Dugan, Suxiang Tong, Justin Lee |
| EPI_ISL_1678100, EPI_ISL_1678101, EPI_ISL_1678102, EPI_ISL_1678103, EPI_ISL_1678104 | DC Public Health Lab/ Dept. of Forensic Sciences | Centers for Disease Control and Prevention Division of Viral Diseases, Pathogen Discovery | Mili Sheth, Sarah Nobles, Jasmine Padilla, Mark Burroughs, Shoshona Le, Katie Dillon, Peter Cook, Clinton R. Paden, Dhwani Batra, Krista Queen, Kristen Knipe, Dakota Howard, Yvette Unoarumhi, Darlene Wagner, Matthew Schmerer, Ben L. Rambo-Martin, Kristine Lacek, Sam Shepard, Alison Laufer Halpin, Dave Wentworth, Vivien Dugan, Suxiang Tong, Justin Lee |
| EPI_ISL_1678105, EPI_ISL_1678106, EPI_ISL_1678107, EPI_ISL_1678108, EPI_ISL_1678109, EPI_ISL_1678110, EPI_ISL_1678111 | MT Public Health Laboratory | Centers for Disease Control and Prevention Division of Viral Diseases, Pathogen Discovery | Mili Sheth, Sarah Nobles, Jasmine Padilla, Mark Burroughs, Shoshona Le, Katie Dillon, Peter Cook, Clinton R. Paden, Dhwani Batra, Krista Queen, Kristen Knipe, Dakota Howard, Yvette Unoarumhi, Darlene Wagner, Matthew Schmerer, Ben L. Rambo-Martin, Kristine Lacek, Sam Shepard, Alison Laufer Halpin, Dave Wentworth, Vivien Dugan, Suxiang Tong, Justin Lee |
| EPI_ISL_1678112, EPI_ISL_1678113, EPI_ISL_1678114, EPI_ISL_1678115, EPI_ISL_1678116, EPI_ISL_1678117, EPI_ISL_1678118, EPI_ISL_1678119, EPI_ISL_1678120, EPI_ISL_1678121, EPI_ISL_1678122, EPI_ISL_1678123, EPI_ISL_1678124, EPI_ISL_1678125, EPI_ISL_1678126, EPI_ISL_1678127 | | | |
| see above | FL Bur. of Public Health Laboratories- Jacksonville | Centers for Disease Control and Prevention Division of Viral Diseases, Pathogen Discovery | Mili Sheth, Sarah Nobles, Jasmine Padilla, Mark Burroughs, Shoshona Le, Katie Dillon, Peter Cook, Clinton R. Paden, Dhwani Batra, Krista Queen, Kristen Knipe, Dakota Howard, Yvette Unoarumhi, Darlene Wagner, Matthew Schmerer, Ben L. Rambo-Martin, Kristine Lacek, Sam Shepard, Alison Laufer Halpin, Dave Wentworth, Vivien Dugan, Suxiang Tong, Justin Lee |
| EPI_ISL_1678128, EPI_ISL_1678129, EPI_ISL_1678130, EPI_ISL_1678131, EPI_ISL_1678132 | ME Health and Environmental Testing Laboratory | Centers for Disease Control and Prevention Division of Viral Diseases, Pathogen Discovery | Mili Sheth, Sarah Nobles, Jasmine Padilla, Mark Burroughs, Shoshona Le, Katie Dillon, Peter Cook, Clinton R. Paden, Dhwani Batra, Krista Queen, Kristen Knipe, Dakota Howard, Yvette Unoarumhi, Darlene Wagner, Matthew Schmerer, Ben L. Rambo-Martin, Kristine Lacek, Sam Shepard, Alison Laufer Halpin, Dave Wentworth, Vivien Dugan, Suxiang Tong, Justin Lee |
| EPI_ISL_1678133, EPI_ISL_1678134, EPI_ISL_1678135, EPI_ISL_1678136, EPI_ISL_1678137, EPI_ISL_1678138, EPI_ISL_1678139, EPI_ISL_1678140, EPI_ISL_1678141, EPI_ISL_1678142 | SC Dept of Health and Env. Control- Bureau of Laboratories | Centers for Disease Control and Prevention Division of Viral Diseases, Pathogen Discovery | Mili Sheth, Sarah Nobles, Jasmine Padilla, Mark Burroughs, Shoshona Le, Katie Dillon, Peter Cook, Clinton R. Paden, Dhwani Batra, Krista Queen, Kristen Knipe, Dakota Howard, Yvette Unoarumhi, Darlene Wagner, Matthew Schmerer, Ben L. Rambo-Martin, Kristine Lacek, Sam Shepard, Alison Laufer Halpin, Dave Wentworth, Vivien Dugan, Suxiang Tong, Justin Lee |
| EPI_ISL_1678143, EPI_ISL_1678144, EPI_ISL_1678145, EPI_ISL_1678146, EPI_ISL_1678147, EPI_ISL_1678148, EPI_ISL_1678149, EPI_ISL_1678150, EPI_ISL_1678151, EPI_ISL_1678152, EPI_ISL_1678153, EPI_ISL_1678154, EPI_ISL_1678155, EPI_ISL_1678156, EPI_ISL_1678157, EPI_ISL_1678158, EPI_ISL_1678159, EPI_ISL_1678160, EPI_ISL_1678161, EPI_ISL_1678162 | | | |
| see above | "CDPH, Viral and Rickettsial Disease Laboratory" | Centers for Disease Control and Prevention Division of Viral Diseases, Pathogen Discovery | Mili Sheth, Sarah Nobles, Jasmine Padilla, Mark Burroughs, Shoshona Le, Katie Dillon, Peter Cook, Clinton R. Paden, Dhwani Batra, Krista Queen, Kristen Knipe, Dakota Howard, Yvette Unoarumhi, Darlene Wagner, Matthew Schmerer, Ben L. Rambo-Martin, Kristine Lacek, Sam Shepard, Alison Laufer Halpin, Dave Wentworth, Vivien Dugan, Suxiang Tong, Justin Lee |
| EPI_ISL_1678163, EPI_ISL_1678164, EPI_ISL_1678165, EPI_ISL_1678166, EPI_ISL_1678167, EPI_ISL_1678168, EPI_ISL_1678169 | OR State PHL-Virology/Immunology Section | Centers for Disease Control and Prevention Division of Viral Diseases, Pathogen Discovery | Mili Sheth, Sarah Nobles, Jasmine Padilla, Mark Burroughs, Shoshona Le, Katie Dillon, Peter Cook, Clinton R. Paden, Dhwani Batra, Krista Queen, Kristen Knipe, Dakota Howard, Yvette Unoarumhi, Darlene Wagner, Matthew Schmerer, Ben L. Rambo-Martin, Kristine Lacek, Sam Shepard, Alison Laufer Halpin, Dave Wentworth, Vivien Dugan, Suxiang Tong, Justin Lee |
| EPI_ISL_1678170 | ID Bureau of Laboratories | Centers for Disease Control and Prevention Division of Viral Diseases, Pathogen Discovery | Mili Sheth, Sarah Nobles, Jasmine Padilla, Mark Burroughs, Shoshona Le, Katie Dillon, Peter Cook, Clinton R. Paden, Dhwani Batra, Krista Queen, Kristen Knipe, Dakota Howard, Yvette Unoarumhi, Darlene Wagner, Matthew Schmerer, Ben L. Rambo-Martin, Kristine Lacek, Sam Shepard, Alison Laufer Halpin, Dave Wentworth, Vivien Dugan, Suxiang Tong, Justin Lee |
| EPI_ISL_1678171, EPI_ISL_1678172, EPI_ISL_1678173, EPI_ISL_1678174, EPI_ISL_1678175, EPI_ISL_1678176, EPI_ISL_1678177, EPI_ISL_1678178, EPI_ISL_1678179 | ND Dept. of Health Laboratory Services- Microbiology | Centers for Disease Control and Prevention Division of Viral Diseases, Pathogen Discovery | Mili Sheth, Sarah Nobles, Jasmine Padilla, Mark Burroughs, Shoshona Le, Katie Dillon, Peter Cook, Clinton R. Paden, Dhwani Batra, Krista Queen, Kristen Knipe, Dakota Howard, Yvette Unoarumhi, Darlene Wagner, Matthew Schmerer, Ben L. Rambo-Martin, Kristine Lacek, Sam Shepard, Alison Laufer Halpin, Dave Wentworth, Vivien Dugan, Suxiang Tong, Justin Lee |
| EPI_ISL_1678180, EPI_ISL_1678181, EPI_ISL_1678182, EPI_ISL_1678183, EPI_ISL_1678184 | "PA Department of Health, Bureau of Laboratories" | Centers for Disease Control and Prevention Division of Viral Diseases, Pathogen Discovery | Mili Sheth, Sarah Nobles, Jasmine Padilla, Mark Burroughs, Shoshona Le, Katie Dillon, Peter Cook, Clinton R. Paden, Dhwani Batra, Krista Queen, Kristen Knipe, Dakota Howard, Yvette Unoarumhi, Darlene Wagner, Matthew Schmerer, Ben L. Rambo-Martin, Kristine Lacek, Sam Shepard, Alison Laufer Halpin, Dave Wentworth, Vivien Dugan, Suxiang Tong, Justin Lee |
| EPI_ISL_1678185, EPI_ISL_1678186, EPI_ISL_1678187, EPI_ISL_1678188, EPI_ISL_1678189, EPI_ISL_1678190, EPI_ISL_1678191, EPI_ISL_1678192, EPI_ISL_1678193, EPI_ISL_1678194, EPI_ISL_1678195, EPI_ISL_1678196, EPI_ISL_1678197, EPI_ISL_1678198, EPI_ISL_1678199, EPI_ISL_1678200, EPI_ISL_1678201, EPI_ISL_1678202, EPI_ISL_1678203, EPI_ISL_1678204, EPI_ISL_1678205, EPI_ISL_1678206, EPI_ISL_1678207, EPI_ISL_1678208, EPI_ISL_1678209, EPI_ISL_1678210, EPI_ISL_1678211, EPI_ISL_1678212 | | | |
| see above | "HI Dept. of Health, State Laboratories Division" | Centers for Disease Control and Prevention Division of Viral Diseases, Pathogen Discovery | Mili Sheth, Sarah Nobles, Jasmine Padilla, Mark Burroughs, Shoshona Le, Katie Dillon, Peter Cook, Clinton R. Paden, Dhwani Batra, Krista Queen, Kristen Knipe, Dakota Howard, Yvette Unoarumhi, Darlene Wagner, Matthew Schmerer, Ben L. Rambo-Martin, Kristine Lacek, Sam Shepard, Alison Laufer Halpin, Dave Wentworth, Vivien Dugan, Suxiang Tong, Justin Lee |
| EPI_ISL_1678213, EPI_ISL_1678214 | IN State Department of Health Laboratory Services | Centers for Disease Control and Prevention Division of Viral Diseases, Pathogen Discovery | Mili Sheth, Sarah Nobles, Jasmine Padilla, Mark Burroughs, Shoshona Le, Katie Dillon, Peter Cook, Clinton R. Paden, Dhwani Batra, Krista Queen, Kristen Knipe, Dakota Howard, Yvette Unoarumhi, Darlene Wagner, Matthew Schmerer, Ben L. Rambo-Martin, Kristine Lacek, Sam Shepard, Alison Laufer Halpin, Dave Wentworth, Vivien Dugan, Suxiang Tong, Justin Lee |
| EPI_ISL_1678215, EPI_ISL_1678216, EPI_ISL_1678217, EPI_ISL_1678218, EPI_ISL_1678219, EPI_ISL_1678220, EPI_ISL_1678221, EPI_ISL_1678222, EPI_ISL_1678223 | NC State Laboratory of Public Health | Centers for Disease Control and Prevention Division of Viral Diseases, Pathogen Discovery | Mili Sheth, Sarah Nobles, Jasmine Padilla, Mark Burroughs, Shoshona Le, Katie Dillon, Peter Cook, Clinton R. Paden, Dhwani Batra, Krista Queen, Kristen Knipe, Dakota Howard, Yvette Unoarumhi, Darlene Wagner, Matthew Schmerer, Ben L. Rambo-Martin, Kristine Lacek, Sam Shepard, Alison Laufer Halpin, Dave Wentworth, Vivien Dugan, Suxiang Tong, Justin Lee |
| EPI_ISL_1678224, EPI_ISL_1678225, EPI_ISL_1678226, EPI_ISL_1678227, EPI_ISL_1678228, EPI_ISL_1678229, | GA Department of Public Health Laboratory | Centers for Disease Control and Prevention Division of Viral Diseases, Pathogen Discovery | Mili Sheth, Sarah Nobles, Jasmine Padilla, Mark Burroughs, Shoshona Le, Katie Dillon, Peter Cook, Clinton R. Paden, Dhwani Batra, Krista Queen, Kristen Knipe, Dakota Howard, Yvette Unoarumhi, Darlene Wagner, Matthew Schmerer, Ben L. Rambo-Martin, Kristine Lacek, Sam Shepard, Alison Laufer Halpin, Dave Wentworth, Vivien Dugan, Suxiang Tong, Justin Lee |

| | | | |
|---|---|---|---|
| EPI_ISL_1678230, EPI_ISL_1678231, EPI_ISL_1678232 | | | |
| EPI_ISL_1678233, EPI_ISL_1678234, EPI_ISL_1678235, EPI_ISL_1678236, EPI_ISL_1678237, EPI_ISL_1678238 | NH Dept. of Health and Human Services Public Health Labs | Centers for Disease Control and Prevention Division of Viral Diseases, Pathogen Discovery | Mili Sheth, Sarah Nobles, Jasmine Padilla, Mark Burroughs, Shoshona Le, Katie Dillon, Peter Cook, Clinton R. Paden, Dhwani Batra, Krista Queen, Kristen Knipe, Dakota Howard, Yvette Unoarumhi, Darlene Wagner, Matthew Schmerer, Ben L. Rambo-Martin, Kristine Lacek, Sam Shepard, Alison Laufer Halpin, Dave Wentworth, Vivien Dugan, Suxiang Tong, Justin Lee |
| EPI_ISL_1678239, EPI_ISL_1678240, EPI_ISL_1678241, EPI_ISL_1678242, EPI_ISL_1678243, EPI_ISL_1678244 | "AR Dept. of Health-PHL, Molecular Diagnostics" | Centers for Disease Control and Prevention Division of Viral Diseases, Pathogen Discovery | Mili Sheth, Sarah Nobles, Jasmine Padilla, Mark Burroughs, Shoshona Le, Katie Dillon, Peter Cook, Clinton R. Paden, Dhwani Batra, Krista Queen, Kristen Knipe, Dakota Howard, Yvette Unoarumhi, Darlene Wagner, Matthew Schmerer, Ben L. Rambo-Martin, Kristine Lacek, Sam Shepard, Alison Laufer Halpin, Dave Wentworth, Vivien Dugan, Suxiang Tong, Justin Lee |
| EPI_ISL_1678245 | "MN PHL Division, Minnesota Department of Health" | Centers for Disease Control and Prevention Division of Viral Diseases, Pathogen Discovery | Mili Sheth, Sarah Nobles, Jasmine Padilla, Mark Burroughs, Shoshona Le, Katie Dillon, Peter Cook, Clinton R. Paden, Dhwani Batra, Krista Queen, Kristen Knipe, Dakota Howard, Yvette Unoarumhi, Darlene Wagner, Matthew Schmerer, Ben L. Rambo-Martin, Kristine Lacek, Sam Shepard, Alison Laufer Halpin, Dave Wentworth, Vivien Dugan, Suxiang Tong, Justin Lee |
| EPI_ISL_1678246, EPI_ISL_1678247, EPI_ISL_1678248, EPI_ISL_1678249 | MS Public Health Laboratory | Centers for Disease Control and Prevention Division of Viral Diseases, Pathogen Discovery | Mili Sheth, Sarah Nobles, Jasmine Padilla, Mark Burroughs, Shoshona Le, Katie Dillon, Peter Cook, Clinton R. Paden, Dhwani Batra, Krista Queen, Kristen Knipe, Dakota Howard, Yvette Unoarumhi, Darlene Wagner, Matthew Schmerer, Ben L. Rambo-Martin, Kristine Lacek, Sam Shepard, Alison Laufer Halpin, Dave Wentworth, Vivien Dugan, Suxiang Tong, Justin Lee |
| EPI_ISL_1678250 | WVDHHR - Office of Laboratory Services | Centers for Disease Control and Prevention Division of Viral Diseases, Pathogen Discovery | Mili Sheth, Sarah Nobles, Jasmine Padilla, Mark Burroughs, Shoshona Le, Katie Dillon, Peter Cook, Clinton R. Paden, Dhwani Batra, Krista Queen, Kristen Knipe, Dakota Howard, Yvette Unoarumhi, Darlene Wagner, Matthew Schmerer, Ben L. Rambo-Martin, Kristine Lacek, Sam Shepard, Alison Laufer Halpin, Dave Wentworth, Vivien Dugan, Suxiang Tong, Justin Lee |
| EPI_ISL_1678251, EPI_ISL_1678252, EPI_ISL_1678253 | IA State Hygienic Laboratory | Centers for Disease Control and Prevention Division of Viral Diseases, Pathogen Discovery | Mili Sheth, Sarah Nobles, Jasmine Padilla, Mark Burroughs, Shoshona Le, Katie Dillon, Peter Cook, Clinton R. Paden, Dhwani Batra, Krista Queen, Kristen Knipe, Dakota Howard, Yvette Unoarumhi, Darlene Wagner, Matthew Schmerer, Ben L. Rambo-Martin, Kristine Lacek, Sam Shepard, Alison Laufer Halpin, Dave Wentworth, Vivien Dugan, Suxiang Tong, Justin Lee |
| EPI_ISL_1678254, EPI_ISL_1678255, EPI_ISL_1678256 | KS Health and Environmental Laboratories | Centers for Disease Control and Prevention Division of Viral Diseases, Pathogen Discovery | Mili Sheth, Sarah Nobles, Jasmine Padilla, Mark Burroughs, Shoshona Le, Katie Dillon, Peter Cook, Clinton R. Paden, Dhwani Batra, Krista Queen, Kristen Knipe, Dakota Howard, Yvette Unoarumhi, Darlene Wagner, Matthew Schmerer, Ben L. Rambo-Martin, Kristine Lacek, Sam Shepard, Alison Laufer Halpin, Dave Wentworth, Vivien Dugan, Suxiang Tong, Justin Lee |
| EPI_ISL_1678257, EPI_ISL_1678258, EPI_ISL_1678259, EPI_ISL_1678260, EPI_ISL_1678261, EPI_ISL_1678262, EPI_ISL_1678263, EPI_ISL_1678264, EPI_ISL_1678265 | NYC Department of Health and Mental Hygiene | Centers for Disease Control and Prevention Division of Viral Diseases, Pathogen Discovery | Mili Sheth, Sarah Nobles, Jasmine Padilla, Mark Burroughs, Shoshona Le, Katie Dillon, Peter Cook, Clinton R. Paden, Dhwani Batra, Krista Queen, Kristen Knipe, Dakota Howard, Yvette Unoarumhi, Darlene Wagner, Matthew Schmerer, Ben L. Rambo-Martin, Kristine Lacek, Sam Shepard, Alison Laufer Halpin, Dave Wentworth, Vivien Dugan, Suxiang Tong, Justin Lee |
| EPI_ISL_1678266, EPI_ISL_1678267, EPI_ISL_1678268, EPI_ISL_1678269, EPI_ISL_1678270 | "NM Dept. Health, Scientific Laboratory Division " | Centers for Disease Control and Prevention Division of Viral Diseases, Pathogen Discovery | Mili Sheth, Sarah Nobles, Jasmine Padilla, Mark Burroughs, Shoshona Le, Katie Dillon, Peter Cook, Clinton R. Paden, Dhwani Batra, Krista Queen, Kristen Knipe, Dakota Howard, Yvette Unoarumhi, Darlene Wagner, Matthew Schmerer, Ben L. Rambo-Martin, Kristine Lacek, Sam Shepard, Alison Laufer Halpin, Dave Wentworth, Vivien Dugan, Suxiang Tong, Justin Lee |
| EPI_ISL_1678271, EPI_ISL_1678272, EPI_ISL_1678273 | "OK Public Health Laboratory, Oklahoma State DOH" | Centers for Disease Control and Prevention Division of Viral Diseases, Pathogen Discovery | Mili Sheth, Sarah Nobles, Jasmine Padilla, Mark Burroughs, Shoshona Le, Katie Dillon, Peter Cook, Clinton R. Paden, Dhwani Batra, Krista Queen, Kristen Knipe, Dakota Howard, Yvette Unoarumhi, Darlene Wagner, Matthew Schmerer, Ben L. Rambo-Martin, Kristine Lacek, Sam Shepard, Alison Laufer Halpin, Dave Wentworth, Vivien Dugan, Suxiang Tong, Justin Lee |
| EPI_ISL_1678274, EPI_ISL_1678275 | MI - Michigan Department of Health and Human Services - Bureau of Laboratories | Centers for Disease Control and Prevention Division of Viral Diseases, Pathogen Discovery | Mili Sheth, Sarah Nobles, Jasmine Padilla, Mark Burroughs, Shoshona Le, Katie Dillon, Peter Cook, Clinton R. Paden, Dhwani Batra, Krista Queen, Kristen Knipe, Dakota Howard, Yvette Unoarumhi, Darlene Wagner, Matthew Schmerer, Ben L. Rambo-Martin, Kristine Lacek, Sam Shepard, Alison Laufer Halpin, Dave Wentworth, Vivien Dugan, Suxiang Tong, Justin Lee |
| EPI_ISL_1678276, EPI_ISL_1678277, EPI_ISL_1678278, EPI_ISL_1678279, EPI_ISL_1678280, EPI_ISL_1678281, EPI_ISL_1678282, EPI_ISL_1678283, EPI_ISL_1678284, EPI_ISL_1678285, EPI_ISL_1678286, EPI_ISL_1678287 | | | |
| see above | CA-Los Angeles County Public Health Laboratory | Centers for Disease Control and Prevention Division of Viral Diseases, Pathogen Discovery | Mili Sheth, Sarah Nobles, Jasmine Padilla, Mark Burroughs, Shoshona Le, Katie Dillon, Peter Cook, Clinton R. Paden, Dhwani Batra, Krista Queen, Kristen Knipe, Dakota Howard, Yvette Unoarumhi, Darlene Wagner, Matthew Schmerer, Ben L. Rambo-Martin, Kristine Lacek, Sam Shepard, Alison Laufer Halpin, Dave Wentworth, Vivien Dugan, Suxiang Tong, Justin Lee |
| EPI_ISL_1678288 | UT-Unified State Labs: Public Health Utah DOH | Centers for Disease Control and Prevention Division of Viral Diseases, Pathogen Discovery | Mili Sheth, Sarah Nobles, Jasmine Padilla, Mark Burroughs, Shoshona Le, Katie Dillon, Peter Cook, Clinton R. Paden, Dhwani Batra, Krista Queen, Kristen Knipe, Dakota Howard, Yvette Unoarumhi, Darlene Wagner, Matthew Schmerer, Ben L. Rambo-Martin, Kristine Lacek, Sam Shepard, Alison Laufer Halpin, Dave Wentworth, Vivien Dugan, Suxiang Tong, Justin Lee |
| EPI_ISL_1678289, EPI_ISL_1678290, EPI_ISL_1678291, EPI_ISL_1678292 | DC Public Health Lab/ Dept. of Forensic Sciences | Centers for Disease Control and Prevention Division of Viral Diseases, Pathogen Discovery | Mili Sheth, Sarah Nobles, Jasmine Padilla, Mark Burroughs, Shoshona Le, Katie Dillon, Peter Cook, Clinton R. Paden, Dhwani Batra, Krista Queen, Kristen Knipe, Dakota Howard, Yvette Unoarumhi, Darlene Wagner, Matthew Schmerer, Ben L. Rambo-Martin, Kristine Lacek, Sam Shepard, Alison Laufer Halpin, Dave Wentworth, Vivien Dugan, Suxiang Tong, Justin Lee |
| EPI_ISL_1678293, EPI_ISL_1678294, EPI_ISL_1678295, EPI_ISL_1678296, EPI_ISL_1678297, EPI_ISL_1678298, EPI_ISL_1678299, EPI_ISL_1678300, EPI_ISL_1678301, EPI_ISL_1678302, EPI_ISL_1678303, EPI_ISL_1678304, EPI_ISL_1678305, EPI_ISL_1678306 | | | |
| see above | FL Bur. of Public Health Laboratories-Jacksonville | Centers for Disease Control and Prevention Division of Viral Diseases, Pathogen Discovery | Mili Sheth, Sarah Nobles, Jasmine Padilla, Mark Burroughs, Shoshona Le, Katie Dillon, Peter Cook, Clinton R. Paden, Dhwani Batra, Krista Queen, Kristen Knipe, Dakota Howard, Yvette Unoarumhi, Darlene Wagner, Matthew Schmerer, Ben L. Rambo-Martin, Kristine Lacek, Sam Shepard, Alison Laufer Halpin, Dave Wentworth, Vivien Dugan, Suxiang Tong, Justin Lee |
| EPI_ISL_1678307 | ME Health and Environmental Testing Laboratory | Centers for Disease Control and Prevention Division of Viral Diseases, Pathogen Discovery | Mili Sheth, Sarah Nobles, Jasmine Padilla, Mark Burroughs, Shoshona Le, Katie Dillon, Peter Cook, Clinton R. Paden, Dhwani Batra, Krista Queen, Kristen Knipe, Dakota Howard, Yvette Unoarumhi, Darlene Wagner, Matthew Schmerer, Ben L. Rambo-Martin, Kristine Lacek, Sam Shepard, Alison Laufer Halpin, Dave Wentworth, Vivien Dugan, Suxiang Tong, Justin Lee |
| EPI_ISL_1678308, EPI_ISL_1678309, EPI_ISL_1678310, EPI_ISL_1678311, EPI_ISL_1678312 | SC Dept of Health and Env. Control-Bureau of Laboratories | Centers for Disease Control and Prevention Division of Viral Diseases, Pathogen Discovery | Mili Sheth, Sarah Nobles, Jasmine Padilla, Mark Burroughs, Shoshona Le, Katie Dillon, Peter Cook, Clinton R. Paden, Dhwani Batra, Krista Queen, Kristen Knipe, Dakota Howard, Yvette Unoarumhi, Darlene Wagner, Matthew Schmerer, Ben L. Rambo-Martin, Kristine Lacek, Sam Shepard, Alison Laufer Halpin, Dave Wentworth, Vivien Dugan, Suxiang Tong, Justin Lee |
| EPI_ISL_1678313, EPI_ISL_1678314, EPI_ISL_1678315 | "CDPH, Viral and Rickettsial Disease Laboratory" | Centers for Disease Control and Prevention Division of Viral Diseases, Pathogen Discovery | Mili Sheth, Sarah Nobles, Jasmine Padilla, Mark Burroughs, Shoshona Le, Katie Dillon, Peter Cook, Clinton R. Paden, Dhwani Batra, Krista Queen, Kristen Knipe, Dakota Howard, Yvette Unoarumhi, Darlene Wagner, Matthew Schmerer, Ben L. Rambo-Martin, Kristine Lacek, Sam Shepard, Alison Laufer Halpin, Dave Wentworth, Vivien Dugan, Suxiang Tong, Justin Lee |
| EPI_ISL_1678316, EPI_ISL_1678317 | OR State PHL-Virology/Immunology Section | Centers for Disease Control and Prevention Division of Viral Diseases, Pathogen Discovery | Mili Sheth, Sarah Nobles, Jasmine Padilla, Mark Burroughs, Shoshona Le, Katie Dillon, Peter Cook, Clinton R. Paden, Dhwani Batra, Krista Queen, Kristen Knipe, Dakota Howard, Yvette Unoarumhi, Darlene Wagner, Matthew Schmerer, Ben L. Rambo-Martin, Kristine Lacek, Sam Shepard, Alison Laufer Halpin, Dave Wentworth, Vivien Dugan, Suxiang Tong, Justin Lee |
| EPI_ISL_1678318, EPI_ISL_1678319, EPI_ISL_1678320, EPI_ISL_1678321, EPI_ISL_1678322, EPI_ISL_1678323, EPI_ISL_1678324, EPI_ISL_1678325, EPI_ISL_1678326, EPI_ISL_1678327, EPI_ISL_1678328, EPI_ISL_1678329, EPI_ISL_1678330, EPI_ISL_1678331, EPI_ISL_1678332, EPI_ISL_1678333, EPI_ISL_1678334 | | | |
| see above | ID Bureau of Laboratories | Centers for Disease Control and Prevention Division of Viral Diseases, Pathogen Discovery | Mili Sheth, Sarah Nobles, Jasmine Padilla, Mark Burroughs, Shoshona Le, Katie Dillon, Peter Cook, Clinton R. Paden, Dhwani Batra, Krista Queen, Kristen Knipe, Dakota Howard, Yvette Unoarumhi, Darlene Wagner, Matthew Schmerer, Ben L. Rambo-Martin, Kristine Lacek, Sam Shepard, Alison Laufer Halpin, Dave Wentworth, Vivien Dugan, Suxiang Tong, Justin Lee |
| EPI_ISL_1678335, EPI_ISL_1678336, EPI_ISL_1678337, EPI_ISL_1678338, EPI_ISL_1678339, EPI_ISL_1678340, EPI_ISL_1678341, EPI_ISL_1678342, EPI_ISL_1678343, EPI_ISL_1678344, EPI_ISL_1678345, EPI_ISL_1678346 | | | |
| see above | "PA Department of Health, Bureau of Laboratories" | Centers for Disease Control and Prevention Division of Viral Diseases, Pathogen Discovery | Mili Sheth, Sarah Nobles, Jasmine Padilla, Mark Burroughs, Shoshona Le, Katie Dillon, Peter Cook, Clinton R. Paden, Dhwani Batra, Krista Queen, Kristen Knipe, Dakota Howard, Yvette Unoarumhi, Darlene Wagner, Matthew Schmerer, Ben L. Rambo-Martin, Kristine Lacek, Sam Shepard, Alison Laufer Halpin, Dave Wentworth, Vivien Dugan, Suxiang Tong, Justin Lee |
| EPI_ISL_1678347, EPI_ISL_1678348, EPI_ISL_1678349 | "HI Dept. of Health, State Laboratories Division" | Centers for Disease Control and Prevention Division of Viral Diseases, Pathogen Discovery | Mili Sheth, Sarah Nobles, Jasmine Padilla, Mark Burroughs, Shoshona Le, Katie Dillon, Peter Cook, Clinton R. Paden, Dhwani Batra, Krista Queen, Kristen Knipe, Dakota Howard, Yvette Unoarumhi, Darlene Wagner, Matthew Schmerer, Ben L. Rambo-Martin, Kristine Lacek, Sam Shepard, Alison Laufer Halpin, Dave Wentworth, Vivien Dugan, Suxiang Tong, Justin Lee |
| EPI_ISL_1678350, EPI_ISL_1678351, EPI_ISL_1678352, EPI_ISL_1678353, EPI_ISL_1678354, EPI_ISL_1678355, EPI_ISL_1678356 | IN State Department of Health Laboratory Services | Centers for Disease Control and Prevention Division of Viral Diseases, Pathogen Discovery | Mili Sheth, Sarah Nobles, Jasmine Padilla, Mark Burroughs, Shoshona Le, Katie Dillon, Peter Cook, Clinton R. Paden, Dhwani Batra, Krista Queen, Kristen Knipe, Dakota Howard, Yvette Unoarumhi, Darlene Wagner, Matthew Schmerer, Ben L. Rambo-Martin, Kristine Lacek, Sam Shepard, Alison Laufer Halpin, Dave Wentworth, Vivien Dugan, Suxiang Tong, Justin Lee |

| | | | |
|---|---|---|---|
| EPI_ISL_1678357 | LA Office of Public Health Laboratories | Centers for Disease Control and Prevention Division of Viral Diseases, Pathogen Discovery | Mili Sheth, Sarah Nobles, Jasmine Padilla, Mark Burroughs, Shoshona Le, Katie Dillon, Peter Cook, Clinton R. Paden, Dhwani Batra, Krista Queen, Kristen Knipe, Dakota Howard, Yvette Unoarumhi, Darlene Wagner, Matthew Schmerer, Ben L. Rambo-Martin, Kristine Lacek, Sam Shepard, Alison Laufer Halpin, Dave Wentworth, Vivien Dugan, Suxiang Tong, Justin Lee |
| EPI_ISL_1678358, EPI_ISL_1678359, EPI_ISL_1678360 | NC State Laboratory of Public Health | Centers for Disease Control and Prevention Division of Viral Diseases, Pathogen Discovery | Mili Sheth, Sarah Nobles, Jasmine Padilla, Mark Burroughs, Shoshona Le, Katie Dillon, Peter Cook, Clinton R. Paden, Dhwani Batra, Krista Queen, Kristen Knipe, Dakota Howard, Yvette Unoarumhi, Darlene Wagner, Matthew Schmerer, Ben L. Rambo-Martin, Kristine Lacek, Sam Shepard, Alison Laufer Halpin, Dave Wentworth, Vivien Dugan, Suxiang Tong, Justin Lee |
| EPI_ISL_1678361, EPI_ISL_1678362, EPI_ISL_1678363, EPI_ISL_1678364, EPI_ISL_1678365, EPI_ISL_1678366, EPI_ISL_1678367, EPI_ISL_1678368 | GA Department of Public Health Laboratory | Centers for Disease Control and Prevention Division of Viral Diseases, Pathogen Discovery | Mili Sheth, Sarah Nobles, Jasmine Padilla, Mark Burroughs, Shoshona Le, Katie Dillon, Peter Cook, Clinton R. Paden, Dhwani Batra, Krista Queen, Kristen Knipe, Dakota Howard, Yvette Unoarumhi, Darlene Wagner, Matthew Schmerer, Ben L. Rambo-Martin, Kristine Lacek, Sam Shepard, Alison Laufer Halpin, Dave Wentworth, Vivien Dugan, Suxiang Tong, Justin Lee |
| EPI_ISL_1678369, EPI_ISL_1678370, EPI_ISL_1678371, EPI_ISL_1678372, EPI_ISL_1678373, EPI_ISL_1678374, EPI_ISL_1678375 | NH Dept. of Health and Human Services Public Health Labs | Centers for Disease Control and Prevention Division of Viral Diseases, Pathogen Discovery | Mili Sheth, Sarah Nobles, Jasmine Padilla, Mark Burroughs, Shoshona Le, Katie Dillon, Peter Cook, Clinton R. Paden, Dhwani Batra, Krista Queen, Kristen Knipe, Dakota Howard, Yvette Unoarumhi, Darlene Wagner, Matthew Schmerer, Ben L. Rambo-Martin, Kristine Lacek, Sam Shepard, Alison Laufer Halpin, Dave Wentworth, Vivien Dugan, Suxiang Tong, Justin Lee |

EPI_ISL_1678875, EPI_ISL_1678876, EPI_ISL_1678877, EPI_ISL_1678878, EPI_ISL_1678879, EPI_ISL_1678880, EPI_ISL_1678881, EPI_ISL_1678882, EPI_ISL_1678883, EPI_ISL_1678884, EPI_ISL_1678885, EPI_ISL_1678886, EPI_ISL_1678887, EPI_ISL_1678888, EPI_ISL_1678889, EPI_ISL_1678890, EPI_ISL_1678891, EPI_ISL_1678892, EPI_ISL_1678893, EPI_ISL_1678894, EPI_ISL_1678895, EPI_ISL_1678896, EPI_ISL_1678897, EPI_ISL_1678899, EPI_ISL_1678900, EPI_ISL_1678901, EPI_ISL_1678902, EPI_ISL_1678903, EPI_ISL_1678904, EPI_ISL_1678905, EPI_ISL_1678906, EPI_ISL_1678907, EPI_ISL_1678908, EPI_ISL_1678909, EPI_ISL_1678910, EPI_ISL_1678911, EPI_ISL_1678912, EPI_ISL_1678913, EPI_ISL_1678914, EPI_ISL_1678915, EPI_ISL_1678916, EPI_ISL_1678917, EPI_ISL_1678918, EPI_ISL_1678919, EPI_ISL_1678920, EPI_ISL_1678921, EPI_ISL_1678922, EPI_ISL_1678923, EPI_ISL_1678924, EPI_ISL_1678925, EPI_ISL_1678926, EPI_ISL_1678927, EPI_ISL_1678928, EPI_ISL_1678929, EPI_ISL_1678930, EPI_ISL_1678931, EPI_ISL_1678933, EPI_ISL_1678935, EPI_ISL_1678936, EPI_ISL_1678937, EPI_ISL_1678938, EPI_ISL_1678939, EPI_ISL_1678940, EPI_ISL_1678941, EPI_ISL_1678942, EPI_ISL_1678943, EPI_ISL_1678944, EPI_ISL_1678945, EPI_ISL_1678946, EPI_ISL_1678947, EPI_ISL_1678948, EPI_ISL_1678949, EPI_ISL_1678950, EPI_ISL_1678951, EPI_ISL_1678952, EPI_ISL_1678953, EPI_ISL_1678954, EPI_ISL_1678955, EPI_ISL_1678956, EPI_ISL_1678957, EPI_ISL_1678958, EPI_ISL_1678960, EPI_ISL_1678961, EPI_ISL_1678962, EPI_ISL_1678963, EPI_ISL_1678964, EPI_ISL_1678965, EPI_ISL_1678966, EPI_ISL_1678967, EPI_ISL_1678968, EPI_ISL_1678969, EPI_ISL_1678970, EPI_ISL_1678971, EPI_ISL_1678972, EPI_ISL_1678973, EPI_ISL_1678974, EPI_ISL_1678976, EPI_ISL_1678977, EPI_ISL_1678978, EPI_ISL_1678979, EPI_ISL_1678980, EPI_ISL_1678981, EPI_ISL_1678982, EPI_ISL_1678983, EPI_ISL_1678984, EPI_ISL_1678985, EPI_ISL_1678986, EPI_ISL_1678987, EPI_ISL_1678988, EPI_ISL_1678989, EPI_ISL_1678990, EPI_ISL_1678991, EPI_ISL_1678992, EPI_ISL_1678993, EPI_ISL_1678994, EPI_ISL_1678995, EPI_ISL_1678996, EPI_ISL_1678997, EPI_ISL_1678999, EPI_ISL_1679000, EPI_ISL_1679001, EPI_ISL_1679002, EPI_ISL_1679003, EPI_ISL_1679004, EPI_ISL_1679005, EPI_ISL_1679006, EPI_ISL_1679007, EPI_ISL_1679008, EPI_ISL_1679009, EPI_ISL_1679010, EPI_ISL_1679011, EPI_ISL_1679012, EPI_ISL_1679013, EPI_ISL_1679014, EPI_ISL_1679015, EPI_ISL_1679016, EPI_ISL_1679017, EPI_ISL_1679018, EPI_ISL_1679019, EPI_ISL_1679020, EPI_ISL_1679021, EPI_ISL_1679022, EPI_ISL_1679023, EPI_ISL_1679024, EPI_ISL_1679025, EPI_ISL_1679026, EPI_ISL_1679027, EPI_ISL_1679029, EPI_ISL_1679030, EPI_ISL_1679031, EPI_ISL_1679032, EPI_ISL_1679033, EPI_ISL_1679034, EPI_ISL_1679035, EPI_ISL_1679036, EPI_ISL_1679037, EPI_ISL_1679038, EPI_ISL_1679039, EPI_ISL_1679040, EPI_ISL_1679041, EPI_ISL_1679042, EPI_ISL_1679044, EPI_ISL_1679045, EPI_ISL_1679046, EPI_ISL_1679047, EPI_ISL_1679048, EPI_ISL_1679049, EPI_ISL_1679050, EPI_ISL_1679051, EPI_ISL_1679052, EPI_ISL_1679053, EPI_ISL_1679054, EPI_ISL_1679055, EPI_ISL_1679056, EPI_ISL_1679057, EPI_ISL_1679058, EPI_ISL_1679059, EPI_ISL_1679061, EPI_ISL_1679062, EPI_ISL_1679063, EPI_ISL_1679064, EPI_ISL_1679065, EPI_ISL_1679066, EPI_ISL_1679067, EPI_ISL_1679068, EPI_ISL_1679069, EPI_ISL_1679070, EPI_ISL_1679071, EPI_ISL_1679072, EPI_ISL_1679073, EPI_ISL_1679074, EPI_ISL_1679075, EPI_ISL_1679076, EPI_ISL_1679077, EPI_ISL_1679078, EPI_ISL_1679079, EPI_ISL_1679080, EPI_ISL_1679081, EPI_ISL_1679082, EPI_ISL_1679083, EPI_ISL_1679084, EPI_ISL_1679085, EPI_ISL_1679086, EPI_ISL_1679087, EPI_ISL_1679088, EPI_ISL_1679089, EPI_ISL_1679090, EPI_ISL_1679091, EPI_ISL_1679092, EPI_ISL_1679093, EPI_ISL_1679094, EPI_ISL_1679095, EPI_ISL_1679096, EPI_ISL_1679097, EPI_ISL_1679098, EPI_ISL_1679099, EPI_ISL_1679100, EPI_ISL_1679101, EPI_ISL_1679102, EPI_ISL_1679103, EPI_ISL_1679104, EPI_ISL_1679105, EPI_ISL_1679106, EPI_ISL_1679107, EPI_ISL_1679108, EPI_ISL_1679109, EPI_ISL_1679110, EPI_ISL_1679111, EPI_ISL_1679112, EPI_ISL_1679113, EPI_ISL_1679114, EPI_ISL_1679115, EPI_ISL_1679116, EPI_ISL_1679117, EPI_ISL_1679118, EPI_ISL_1679119, EPI_ISL_1679120, EPI_ISL_1679121, EPI_ISL_1679122, EPI_ISL_1679123, EPI_ISL_1679124, EPI_ISL_1679125, EPI_ISL_1679126, EPI_ISL_1679127, EPI_ISL_1679128, EPI_ISL_1679129, EPI_ISL_1679130, EPI_ISL_1679131, EPI_ISL_1679132, EPI_ISL_1679133, EPI_ISL_1679134, EPI_ISL_1679135, EPI_ISL_1679136, EPI_ISL_1679137, EPI_ISL_1679138, EPI_ISL_1679139, EPI_ISL_1679140, EPI_ISL_1679141, EPI_ISL_1679142, EPI_ISL_1679143, EPI_ISL_1679144, EPI_ISL_1679145, EPI_ISL_1679146, EPI_ISL_1679147, EPI_ISL_1679148, EPI_ISL_1679149, EPI_ISL_1679150, EPI_ISL_1679151, EPI_ISL_1679152, EPI_ISL_1679153, EPI_ISL_1679154, EPI_ISL_1679155, EPI_ISL_1679156, EPI_ISL_1679157, EPI_ISL_1679158, EPI_ISL_1679159, EPI_ISL_1679160, EPI_ISL_1679161, EPI_ISL_1679162, EPI_ISL_1679163, EPI_ISL_1679164, EPI_ISL_1679165, EPI_ISL_1679166, EPI_ISL_1679167, EPI_ISL_1679168, EPI_ISL_1679169, EPI_ISL_1679170, EPI_ISL_1679171, EPI_ISL_1679172, EPI_ISL_1679173, EPI_ISL_1679174, EPI_ISL_1679175, EPI_ISL_1679176, EPI_ISL_1679177, EPI_ISL_1679178, EPI_ISL_1679179, EPI_ISL_1679180, EPI_ISL_1679181, EPI_ISL_1679182, EPI_ISL_1679183, EPI_ISL_1679184, EPI_ISL_1679185, EPI_ISL_1679186, EPI_ISL_1679187, EPI_ISL_1679188, EPI_ISL_1679189, EPI_ISL_1679190, EPI_ISL_1679191, EPI_ISL_1679192, EPI_ISL_1679193, EPI_ISL_1679194, EPI_ISL_1679195, EPI_ISL_1679196, EPI_ISL_1679197, EPI_ISL_1679198, EPI_ISL_1679199, EPI_ISL_1679200, EPI_ISL_1679201, EPI_ISL_1679202, EPI_ISL_1679203, EPI_ISL_1679204, EPI_ISL_1679205, EPI_ISL_1679206, EPI_ISL_1679207, EPI_ISL_1679208, EPI_ISL_1679209, EPI_ISL_1679210, EPI_ISL_1679211, EPI_ISL_1679212, EPI_ISL_1679213, EPI_ISL_1679214, EPI_ISL_1679215, EPI_ISL_1679216, EPI_ISL_1679217, EPI_ISL_1679218, EPI_ISL_1679219, EPI_ISL_1679220, EPI_ISL_1679221, EPI_ISL_1679222, EPI_ISL_1679223, EPI_ISL_1679224, EPI_ISL_1679225, EPI_ISL_1679226, EPI_ISL_1679227, EPI_ISL_1679228, EPI_ISL_1679229, EPI_ISL_1679230, EPI_ISL_1679231, EPI_ISL_1679232, EPI_ISL_1679233, EPI_ISL_1679234, EPI_ISL_1679235, EPI_ISL_1679236, EPI_ISL_1679237, EPI_ISL_1679238, EPI_ISL_1679239, EPI_ISL_1679240, EPI_ISL_1679241, EPI_ISL_1679242, EPI_ISL_1679243, EPI_ISL_1679244, EPI_ISL_1679245, EPI_ISL_1679246, EPI_ISL_1679247, EPI_ISL_1679248, EPI_ISL_1679249, EPI_ISL_1679250, EPI_ISL_1679251, EPI_ISL_1679252, EPI_ISL_1679253, EPI_ISL_1679254, EPI_ISL_1679255, EPI_ISL_1679256, EPI_ISL_1679257, EPI_ISL_1679258, EPI_ISL_1679259, EPI_ISL_1679260, EPI_ISL_1679261, EPI_ISL_1679262, EPI_ISL_1679263, EPI_ISL_1679264, EPI_ISL_1679265, EPI_ISL_1679266, EPI_ISL_1679267, EPI_ISL_1679268, EPI_ISL_1679269, EPI_ISL_1679270, EPI_ISL_1679271, EPI_ISL_1679272, EPI_ISL_1679273, EPI_ISL_1679274, EPI_ISL_1679275, EPI_ISL_1679276, EPI_ISL_1679277, EPI_ISL_1679278, EPI_ISL_1679279, EPI_ISL_1679280, EPI_ISL_1679281, EPI_ISL_1679282, EPI_ISL_1679283, EPI_ISL_1679284, EPI_ISL_1679285, EPI_ISL_1679286, EPI_ISL_1679287, EPI_ISL_1679288, EPI_ISL_1679289, EPI_ISL_1679290, EPI_ISL_1679291, EPI_ISL_1679292, EPI_ISL_1679293, EPI_ISL_1679294, EPI_ISL_1679295, EPI_ISL_1679296, EPI_ISL_1679297, EPI_ISL_1679298, EPI_ISL_1679299, EPI_ISL_1679300, EPI_ISL_1679301, EPI_ISL_1679302, EPI_ISL_1679303, EPI_ISL_1679304, EPI_ISL_1679305, EPI_ISL_1679306, EPI_ISL_1679307, EPI_ISL_1679308, EPI_ISL_1679309, EPI_ISL_1679310, EPI_ISL_1679311, EPI_ISL_1679312, EPI_ISL_1679313, EPI_ISL_1679314, EPI_ISL_1679315, EPI_ISL_1679316, EPI_ISL_1679317, EPI_ISL_1679318, EPI_ISL_1679319, EPI_ISL_1679320, EPI_ISL_1679321, EPI_ISL_1679322, EPI_ISL_1679323, EPI_ISL_1679324, EPI_ISL_1679325, EPI_ISL_1679326, EPI_ISL_1679327, EPI_ISL_1679328, EPI_ISL_1679329, EPI_ISL_1679330, EPI_ISL_1679331, EPI_ISL_1679332, EPI_ISL_1679333, EPI_ISL_1679334, EPI_ISL_1679335, EPI_ISL_1679336, EPI_ISL_1679337, EPI_ISL_1679338, EPI_ISL_1679339, EPI_ISL_1679340, EPI_ISL_1679341, EPI_ISL_1679342, EPI_ISL_1679343, EPI_ISL_1679344, EPI_ISL_1679345, EPI_ISL_1679346, EPI_ISL_1679347, EPI_ISL_1679348, EPI_ISL_1679349, EPI_ISL_1679350, EPI_ISL_1679351, EPI_ISL_1679352, EPI_ISL_1679353, EPI_ISL_1679354, EPI_ISL_1679355, EPI_ISL_1679356, EPI_ISL_1679357, EPI_ISL_1679358, EPI_ISL_1679359, EPI_ISL_1679360, EPI_ISL_1679361, EPI_ISL_1679362, EPI_ISL_1679363, EPI_ISL_1679364, EPI_ISL_1679365, EPI_ISL_1679366, EPI_ISL_1679367, EPI_ISL_1679368, EPI_ISL_1679369, EPI_ISL_1679370, EPI_ISL_1679371, EPI_ISL_1679372, EPI_ISL_1679373, EPI_ISL_1679374, EPI_ISL_1679375, EPI_ISL_1679376, EPI_ISL_1679377, EPI_ISL_1679378, EPI_ISL_1679379, EPI_ISL_1679380, EPI_ISL_1679381, EPI_ISL_1679382, EPI_ISL_1679383, EPI_ISL_1679384, EPI_ISL_1679385, EPI_ISL_1679386, EPI_ISL_1679387, EPI_ISL_1679388, EPI_ISL_1679389, EPI_ISL_1679390, EPI_ISL_1679391, EPI_ISL_1679392, EPI_ISL_1679393, EPI_ISL_1679394, EPI_ISL_1679395, EPI_ISL_1679396, EPI_ISL_1679397, EPI_ISL_1679398, EPI_ISL_1679399, EPI_ISL_1679400, EPI_ISL_1679401, EPI_ISL_1679402, EPI_ISL_1679403, EPI_ISL_1679404, EPI_ISL_1679405, EPI_ISL_1679406, EPI_ISL_1679407, EPI_ISL_1679408, EPI_ISL_1679409, EPI_ISL_1679410, EPI_ISL_1679411, EPI_ISL_1679412, EPI_ISL_1679413, EPI_ISL_1679414, EPI_ISL_1679415, EPI_ISL_1679416, EPI_ISL_1679417, EPI_ISL_1679418, EPI_ISL_1679419, EPI_ISL_1679420, EPI_ISL_1679421, EPI_ISL_1679422, EPI_ISL_1679423, EPI_ISL_1679424, EPI_ISL_1679425, EPI_ISL_1679426, EPI_ISL_1679427, EPI_ISL_1679428, EPI_ISL_1679429, EPI_ISL_1679430, EPI_ISL_1679431, EPI_ISL_1679432, EPI_ISL_1679433, EPI_ISL_1679434, EPI_ISL_1679435, EPI_ISL_1679436, EPI_ISL_1679437, EPI_ISL_1679438, EPI_ISL_1679439, EPI_ISL_1679440, EPI_ISL_1679441, EPI_ISL_1679442, EPI_ISL_1679443, EPI_ISL_1679444, EPI_ISL_1679445, EPI_ISL_1679446, EPI_ISL_1679447, EPI_ISL_1679448, EPI_ISL_1679449, EPI_ISL_1679450, EPI_ISL_1679451, EPI_ISL_1679452, EPI_ISL_1679453, EPI_ISL_1679454, EPI_ISL_1679455, EPI_ISL_1679456, EPI_ISL_1679457, EPI_ISL_1679458, EPI_ISL_1679460, EPI_ISL_1679461, EPI_ISL_1679462, EPI_ISL_1679463, EPI_ISL_1679464, EPI_ISL_1679465, EPI_ISL_1679466, EPI_ISL_1679467, EPI_ISL_1679468, EPI_ISL_1679469, EPI_ISL_1679470, EPI_ISL_1679471, EPI_ISL_1679472, EPI_ISL_1679473, EPI_ISL_1679474, EPI_ISL_1679475, EPI_ISL_1679476, EPI_ISL_1679477, EPI_ISL_1679478, EPI_ISL_1679479, EPI_ISL_1679480, EPI_ISL_1679481, EPI_ISL_1679482, EPI_ISL_1679483, EPI_ISL_1679484, EPI_ISL_1679485, EPI_ISL_1679486, EPI_ISL_1679487, EPI_ISL_1679488, EPI_ISL_1679489, EPI_ISL_1679490, EPI_ISL_1679491, EPI_ISL_1679492, EPI_ISL_1679493, EPI_ISL_1679494, EPI_ISL_1679495, EPI_ISL_1679496, EPI_ISL_1679497, EPI_ISL_1679498, EPI_ISL_1679499, EPI_ISL_1679500, EPI_ISL_1679501, EPI_ISL_1679502, EPI_ISL_1679503, EPI_ISL_1679504, EPI_ISL_1679505, EPI_ISL_1679506, EPI_ISL_1679507, EPI_ISL_1679508, EPI_ISL_1679509, EPI_ISL_1679510, EPI_ISL_1679511, EPI_ISL_1679512, EPI_ISL_1679513, EPI_ISL_1679514, EPI_ISL_1679515, EPI_ISL_1679516, EPI_ISL_1679517, EPI_ISL_1679518, EPI_ISL_1679519, EPI_ISL_1679520, EPI_ISL_1679521, EPI_ISL_1679522, EPI_ISL_1679523, EPI_ISL_1679524, EPI_ISL_1679525, EPI_ISL_1679526, EPI_ISL_1679527, EPI_ISL_1679528, EPI_ISL_1679529, EPI_ISL_1679530, EPI_ISL_1679531, EPI_ISL_1679532, EPI_ISL_1679533, EPI_ISL_1679534, EPI_ISL_1679535, EPI_ISL_1679536, EPI_ISL_1679537, EPI_ISL_1679538, EPI_ISL_1679539, EPI_ISL_1679540, EPI_ISL_1679541, EPI_ISL_1679542, EPI_ISL_1679543, EPI_ISL_1679544, EPI_ISL_1679545, EPI_ISL_1679546, EPI_ISL_1679547, EPI_ISL_1679548, EPI_ISL_1679549, EPI_ISL_1679550, EPI_ISL_1679551, EPI_ISL_1679552, EPI_ISL_1679553, EPI_ISL_1679554, EPI_ISL_1679555, EPI_ISL_1679556, EPI_ISL_1679557, EPI_ISL_1679558, EPI_ISL_1679559, EPI_ISL_1679560, EPI_ISL_1679561, EPI_ISL_1679562, EPI_ISL_1679563, EPI_ISL_1679564, EPI_ISL_1679565, EPI_ISL_1679566, EPI_ISL_1679567, EPI_ISL_1679568, EPI_ISL_1679569, EPI_ISL_1679570, EPI_ISL_1679571, EPI_ISL_1679572, EPI_ISL_1679573, EPI_ISL_1679574, EPI_ISL_1679575, EPI_ISL_1679576, EPI_ISL_1679577, EPI_ISL_1679578, EPI_ISL_1679579, EPI_ISL_1679580, EPI_ISL_1679581, EPI_ISL_1679582, EPI_ISL_1679583, EPI_ISL_1679584, EPI_ISL_1679585, EPI_ISL_1679586, EPI_ISL_1679587, EPI_ISL_1679588, EPI_ISL_1679589, EPI_ISL_1679590, EPI_ISL_1679591, EPI_ISL_1679592, EPI_ISL_1679593, EPI_ISL_1679594, EPI_ISL_1679595, EPI_ISL_1679596, EPI_ISL_1679597, EPI_ISL_1679598, EPI_ISL_1679599, EPI_ISL_1679600, EPI_ISL_1679601, EPI_ISL_1679602, EPI_ISL_1679603, EPI_ISL_1679604, EPI_ISL_1679605, EPI_ISL_1679606, EPI_ISL_1679607, EPI_ISL_1679608, EPI_ISL_1679609, EPI_ISL_1679610, EPI_ISL_1679611, EPI_ISL_1679612, EPI_ISL_1679613, EPI_ISL_1679614, EPI_ISL_1679615, EPI_ISL_1679616, EPI_ISL_1679617, EPI_ISL_1679618, EPI_ISL_1679619, EPI_ISL_1679620, EPI_ISL_1679621, EPI_ISL_1679622, EPI_ISL_1679623, EPI_ISL_1679624, EPI_ISL_1679625, EPI_ISL_1679626, EPI_ISL_1679627, EPI_ISL_1679628, EPI_ISL_1679629, EPI_ISL_1679630, EPI_ISL_1679631, EPI_ISL_1679632, EPI_ISL_1679633, EPI_ISL_1679634, EPI_ISL_1679635, EPI_ISL_1679636, EPI_ISL_1679637, EPI_ISL_1679638, EPI_ISL_1679639, EPI_ISL_1679640, EPI_ISL_1679641, EPI_ISL_1679642, EPI_ISL_1679643, EPI_ISL_1679644, EPI_ISL_1679645, EPI_ISL_1679646, EPI_ISL_1679647, EPI_ISL_1679648, EPI_ISL_1679649, EPI_ISL_1679650, EPI_ISL_1679651, EPI_ISL_1679652, EPI_ISL_1679653, EPI_ISL_1679654, EPI_ISL_1679655, EPI_ISL_1679656, EPI_ISL_1679657, EPI_ISL_1679658, EPI_ISL_1679659, EPI_ISL_1679660, EPI_ISL_1679661, EPI_ISL_1679662, EPI_ISL_1679663, EPI_ISL_1679664, EPI_ISL_1679665, EPI_ISL_1679666, EPI_ISL_1679667, EPI_ISL_1679668, EPI_ISL_1679669, EPI_ISL_1679670, EPI_ISL_1679671, EPI_ISL_1679672, EPI_ISL_1679673, EPI_ISL_1679674, EPI_ISL_1679675, EPI_ISL_1679676, EPI_ISL_1679677, EPI_ISL_1679678, EPI_ISL_1679679, EPI_ISL_1679680, EPI_ISL_1679681, EPI_ISL_1679682, EPI_ISL_1679683, EPI_ISL_1679684, EPI_ISL_1679685, EPI_ISL_1679686, EPI_ISL_1679687, EPI_ISL_1679688, EPI_ISL_1679689, EPI_ISL_1679690, EPI_ISL_1679691, EPI_ISL_1679692, EPI_ISL_1679693, EPI_ISL_1679694, EPI_ISL_1679695, EPI_ISL_1679696, EPI_ISL_1679697, EPI_ISL_1679698, EPI_ISL_1679699, EPI_ISL_1679700, EPI_ISL_1679701, EPI_ISL_1679702, EPI_ISL_1679703, EPI_ISL_1679704, EPI_ISL_1679705, EPI_ISL_1679706, EPI_ISL_1679707, EPI_ISL_1679708, EPI_ISL_1679709, EPI_ISL_1679710, EPI_ISL_1679711, EPI_ISL_1679712, EPI_ISL_1679713, EPI_ISL_1679714, EPI_ISL_1679715, EPI_ISL_1679716, EPI_ISL_1679717, EPI_ISL_1679718, EPI_ISL_1679719, EPI_ISL_1679720, EPI_ISL_1679721, EPI_ISL_1679722, EPI_ISL_1679723, EPI_ISL_1679724, EPI_ISL_1679725, EPI_ISL_1679726, EPI_ISL_1679727, EPI_ISL_1679728, EPI_ISL_1679729, EPI_ISL_1679730, EPI_ISL_1679731, EPI_ISL_1679732, EPI_ISL_1679733, EPI_ISL_1679734, EPI_ISL_1679735, EPI_ISL_1679736, EPI_ISL_1679737, EPI_ISL_1679738, EPI_ISL_1679739, EPI_ISL_1679740, EPI_ISL_1679741, EPI_ISL_1679742, EPI_ISL_1679743, EPI_ISL_1679744, EPI_ISL_1679745, EPI_ISL_1679746, EPI_ISL_1679747, EPI_ISL_1679748, EPI_ISL_1679749, EPI_ISL_1679750, EPI_ISL_1679751, EPI_ISL_1679752, EPI_ISL_1679753, EPI_ISL_1679754, EPI_ISL_1679755, EPI_ISL_1679756, EPI_ISL_1679757, EPI_ISL_1679758, EPI_ISL_1679759, EPI_ISL_1679760, EPI_ISL_1679761, EPI_ISL_1679762, EPI_ISL_1679763, EPI_ISL_1679764, EPI_ISL_1679765, EPI_ISL_1679766, EPI_ISL_1679767, EPI_ISL_1679768, EPI_ISL_1679769, EPI_ISL_1679770, EPI_ISL_1679771, EPI_ISL_1679772, EPI_ISL_1679773, EPI_ISL_1679774, EPI_ISL_1679775, EPI_ISL_1679776, EPI_ISL_1679777, EPI_ISL_1679778, EPI_ISL_1679779, EPI_ISL_1679780, EPI_ISL_1679781, EPI_ISL_1679782, EPI_ISL_1679783, EPI_ISL_1679784, EPI_ISL_1679785, EPI_ISL_1679786, EPI_ISL_1679787, EPI_ISL_1679788, EPI_ISL_1679789, EPI_ISL_1679790, EPI_ISL_1679791, EPI_ISL_1679792, EPI_ISL_1679793, EPI_ISL_1679794, EPI_ISL_1679795, EPI_ISL_1679796, EPI_ISL_1679797, EPI_ISL_1679798, EPI_ISL_1679799, EPI_ISL_1679800, EPI_ISL_1679801, EPI_ISL_1679802, EPI_ISL_1679803, EPI_ISL_1679804, EPI_ISL_1679805, EPI_ISL_1679806, EPI_ISL_1679807, EPI_ISL_1679808, EPI_ISL_1679809

| | | | |
|---|---|---|---|
| see above | Helix/Illumina | Centers for Disease Control and Prevention Division of Viral Diseases, Pathogen Discovery | Dakota Howard, Dhwani Batra, Peter W. Cook, Kara Moser, Adrian Paskey, Jason Caravas, Benjamin Rambo-Martin, Shatavia Morrison, Christopher Gulvick, Scott Sammons, Yvette Unoarumhi, Darlene Wagner, Matthew Schmerer, Eileen de Feo, Jan Antico, Christine Tran, Matthew Tolentino, Shannon Wickline, Kim Gietzen, Brad Sickler, Jingtao Liu, Eric Allen, Phil Febbo, Nicole L. Washington, Simon White, Geraint Levan, Kelly Schiabor Barrett, Elizabeth Cirulli, Alexandre Bolze, Ary Ascencio, Charlotte Rivera-Garcia, Ryan Cho, Jason Nguyen, Sherry Wang, Jimmy Ramirez, Tyler Cassens, Efren Sandoval, Magnus Isaksson, William Lee, David Becker, Marc Laurent, James Lu, Clinton R. Paden, Duncan MacCannell |
| EPI_ISL_1679810 | UW Virology Lab | UW Virology Lab | Pavitra Roychoudhury, Hong Xie, Lasata Shrestha, Shah Mohamed Bakhash, Michelle Lin, Noah R. Baker, Sean Ellis, Meei-Li Huang, Keith R Jerome, Alexander Greninger |
| EPI_ISL_1679811 | Helix/Illumina | Centers for Disease Control and Prevention Division of Viral Diseases, Pathogen Discovery | Dakota Howard, Dhwani Batra, Peter W. Cook, Kara Moser, Adrian Paskey, Jason Caravas, Benjamin Rambo-Martin, Shatavia Morrison, Christopher Gulvick, Scott Sammons, Yvette Unoarumhi, Darlene Wagner, Matthew Schmerer, Eileen de Feo, Jan Antico, Christine Tran, Matthew Tolentino, Shannon Wickline, Kim Gietzen, Brad Sickler, Jingtao Liu, Eric Allen, Phil Febbo, Nicole L. Washington, Simon White, Geraint Levan, Kelly Schiabor Barrett, Elizabeth Cirulli, Alexandre Bolze, Ary Ascencio, Charlotte Rivera-Garcia, Ryan Cho, Jason Nguyen, Sherry Wang, Jimmy Ramirez, Tyler Cassens, Efren Sandoval, Magnus Isaksson, William Lee, David Becker, Marc Laurent, James Lu, Clinton R. Paden, Duncan MacCannell |
| EPI_ISL_1679812, EPI_ISL_1679813 | UW Virology Lab | UW Virology Lab | Pavitra Roychoudhury, Hong Xie, Lasata Shrestha, Shah Mohamed Bakhash, Michelle Lin, Noah R. Baker, Sean Ellis, Meei-Li Huang, Keith R Jerome, Alexander Greninger |
| EPI_ISL_1679814 | Helix/Illumina | Centers for Disease Control and Prevention Division of Viral Diseases, Pathogen Discovery | Dakota Howard, Dhwani Batra, Peter W. Cook, Kara Moser, Adrian Paskey, Jason Caravas, Benjamin Rambo-Martin, Shatavia Morrison, Christopher Gulvick, Scott Sammons, Yvette Unoarumhi, Darlene Wagner, Matthew Schmerer, Eileen de Feo, Jan Antico, Christine Tran, Matthew Tolentino, Shannon Wickline, Kim Gietzen, Brad Sickler, Jingtao Liu, Eric Allen, Phil Febbo, Nicole L. Washington, Simon White, Geraint Levan, Kelly Schiabor Barrett, Elizabeth Cirulli, Alexandre Bolze, Ary Ascencio, Charlotte Rivera-Garcia, Ryan Cho, Jason Nguyen, Sherry Wang, Jimmy Ramirez, Tyler Cassens, Efren Sandoval, Magnus Isaksson, William Lee, David Becker, Marc Laurent, James Lu, Clinton R. Paden, Duncan MacCannell |
| EPI_ISL_1679815, EPI_ISL_1679816 | UW Virology Lab | UW Virology Lab | Pavitra Roychoudhury, Hong Xie, Lasata Shrestha, Shah Mohamed Bakhash, Michelle Lin, Noah R. Baker, Sean Ellis, Meei-Li Huang, Keith R Jerome, Alexander Greninger |
| EPI_ISL_1679817 | Helix/Illumina | Centers for Disease Control and Prevention Division of Viral Diseases, Pathogen Discovery | Dakota Howard, Dhwani Batra, Peter W. Cook, Kara Moser, Adrian Paskey, Jason Caravas, Benjamin Rambo-Martin, Shatavia Morrison, Christopher Gulvick, Scott Sammons, Yvette Unoarumhi, Darlene Wagner, Matthew Schmerer, Eileen de Feo, Jan Antico, Christine Tran, Matthew Tolentino, Shannon Wickline, Kim Gietzen, Brad Sickler, Jingtao Liu, Eric Allen, Phil Febbo, Nicole L. Washington, Simon White, Geraint Levan, Kelly Schiabor Barrett, Elizabeth Cirulli, Alexandre Bolze, Ary Ascencio, Charlotte Rivera-Garcia, Ryan Cho, Jason Nguyen, Sherry Wang, Jimmy Ramirez, Tyler Cassens, Efren Sandoval, Magnus Isaksson, William Lee, David Becker, Marc Laurent, James Lu, Clinton R. Paden, Duncan MacCannell |
| EPI_ISL_1679818, EPI_ISL_1679819 | UW Virology Lab | UW Virology Lab | Pavitra Roychoudhury, Hong Xie, Lasata Shrestha, Shah Mohamed Bakhash, Michelle Lin, Noah R. Baker, Sean Ellis, Meei-Li Huang, Keith R Jerome, Alexander Greninger |
| EPI_ISL_1679820 | Helix/Illumina | Centers for Disease Control and Prevention Division of Viral Diseases, Pathogen Discovery | Dakota Howard, Dhwani Batra, Peter W. Cook, Kara Moser, Adrian Paskey, Jason Caravas, Benjamin Rambo-Martin, Shatavia Morrison, Christopher Gulvick, Scott Sammons, Yvette Unoarumhi, Darlene Wagner, Matthew Schmerer, Eileen de Feo, Jan Antico, Christine Tran, Matthew Tolentino, Shannon Wickline, Kim Gietzen, Brad Sickler, Jingtao Liu, Eric Allen, Phil Febbo, Nicole L. Washington, Simon White, Geraint Levan, Kelly Schiabor Barrett, Elizabeth Cirulli, Alexandre Bolze, Ary Ascencio, Charlotte Rivera-Garcia, Ryan Cho, Jason Nguyen, Sherry Wang, Jimmy Ramirez, Tyler Cassens, Efren Sandoval, Magnus Isaksson, William Lee, David Becker, Marc Laurent, James Lu, Clinton R. Paden, Duncan MacCannell |
| EPI_ISL_1679821, EPI_ISL_1679822 | UW Virology Lab | UW Virology Lab | Pavitra Roychoudhury, Hong Xie, Lasata Shrestha, Shah Mohamed Bakhash, Michelle Lin, Noah R. Baker, Sean Ellis, Meei-Li Huang, Keith R Jerome, Alexander Greninger |
| EPI_ISL_1679823 | Helix/Illumina | Centers for Disease Control and Prevention Division of Viral Diseases, Pathogen Discovery | Dakota Howard, Dhwani Batra, Peter W. Cook, Kara Moser, Adrian Paskey, Jason Caravas, Benjamin Rambo-Martin, Shatavia Morrison, Christopher Gulvick, Scott Sammons, Yvette Unoarumhi, Darlene Wagner, Matthew Schmerer, Eileen de Feo, Jan Antico, Christine Tran, Matthew Tolentino, Shannon Wickline, Kim Gietzen, Brad Sickler, Jingtao Liu, Eric Allen, Phil Febbo, Nicole L. Washington, Simon White, Geraint Levan, Kelly Schiabor Barrett, Elizabeth Cirulli, Alexandre Bolze, Ary Ascencio, Charlotte Rivera-Garcia, Ryan Cho, Jason Nguyen, Sherry Wang, Jimmy Ramirez, Tyler Cassens, Efren Sandoval, Magnus Isaksson, William Lee, David Becker, Marc Laurent, James Lu, Clinton R. Paden, Duncan MacCannell |
| EPI_ISL_1679824, EPI_ISL_1679825 | UW Virology Lab | UW Virology Lab | Pavitra Roychoudhury, Hong Xie, Lasata Shrestha, Shah Mohamed Bakhash, Michelle Lin, Noah R. Baker, Sean Ellis, Meei-Li Huang, Keith R Jerome, Alexander Greninger |
| EPI_ISL_1679826 | Helix/Illumina | Centers for Disease Control and Prevention Division of Viral Diseases, Pathogen Discovery | Dakota Howard, Dhwani Batra, Peter W. Cook, Kara Moser, Adrian Paskey, Jason Caravas, Benjamin Rambo-Martin, Shatavia Morrison, Christopher Gulvick, Scott Sammons, Yvette Unoarumhi, Darlene Wagner, Matthew Schmerer, Eileen de Feo, Jan Antico, Christine Tran, Matthew Tolentino, Shannon Wickline, Kim Gietzen, Brad Sickler, Jingtao Liu, Eric Allen, Phil Febbo, Nicole L. Washington, Simon White, Geraint Levan, Kelly Schiabor Barrett, Elizabeth Cirulli, Alexandre Bolze, Ary Ascencio, Charlotte Rivera-Garcia, Ryan Cho, Jason Nguyen, Sherry Wang, Jimmy Ramirez, Tyler Cassens, Efren Sandoval, Magnus Isaksson, William Lee, David Becker, Marc Laurent, James Lu, Clinton R. Paden, Duncan MacCannell |
| EPI_ISL_1679827, EPI_ISL_1679828 | UW Virology Lab | UW Virology Lab | Pavitra Roychoudhury, Hong Xie, Lasata Shrestha, Shah Mohamed Bakhash, Michelle Lin, Noah R. Baker, Sean Ellis, Meei-Li Huang, Keith R Jerome, Alexander Greninger |
| EPI_ISL_1679829 | Helix/Illumina | Centers for Disease Control and Prevention Division of Viral Diseases, Pathogen Discovery | Dakota Howard, Dhwani Batra, Peter W. Cook, Kara Moser, Adrian Paskey, Jason Caravas, Benjamin Rambo-Martin, Shatavia Morrison, Christopher Gulvick, Scott Sammons, Yvette Unoarumhi, Darlene Wagner, Matthew Schmerer, Eileen de Feo, Jan Antico, Christine Tran, Matthew Tolentino, Shannon Wickline, Kim Gietzen, Brad Sickler, Jingtao Liu, Eric Allen, Phil Febbo, Nicole L. Washington, Simon White, Geraint Levan, Kelly Schiabor Barrett, Elizabeth Cirulli, Alexandre Bolze, Ary Ascencio, Charlotte Rivera-Garcia, Ryan Cho, Jason Nguyen, Sherry Wang, Jimmy Ramirez, Tyler Cassens, Efren Sandoval, Magnus Isaksson, William Lee, David Becker, Marc Laurent, James Lu, Clinton R. Paden, Duncan MacCannell |

| | | | |
|---|---|---|---|
| EPI_ISL_1679830 | UW Virology Lab | UW Virology Lab | Pavitra Roychoudhury, Hong Xie, Lasata Shrestha, Shah Mohamed Bakhash, Michelle Lin, Noah R. Baker, Sean Ellis, Meei-Li Huang, Keith R Jerome, Alexander Greninger |
| EPI_ISL_1679832 | Helix/Illumina | Centers for Disease Control and Prevention Division of Viral Diseases, Pathogen Discovery | Dakota Howard, Dhwani Batra, Peter W. Cook, Kara Moser, Adrian Paskey, Jason Caravas, Benjamin Rambo-Martin, Shatavia Morrison, Christopher Gulvick, Scott Sammons, Yvette Unoarumhi, Darlene Wagner, Matthew Schmerer, Eileen de Feo, Jan Antico, Christine Tran, Matthew Tolentino, Shannon Wickline, Kim Gietzen, Brad Sickler, Jingtao Liu, Eric Allen, Phil Febbo, Nicole L. Washington, Simon White, Geraint Levan, Kelly Schiabor Barrett, Elizabeth Cirulli, Alexandre Bolze, Ary Ascencio, Charlotte Rivera-Garcia, Ryan Cho, Jason Nguyen, Sherry Wang, Jimmy Ramirez, Tyler Cassens, Efren Sandoval, Magnus Isaksson, William Lee, David Becker, Marc Laurent, James Lu, Clinton R. Paden, Duncan MacCannell |
| EPI_ISL_1679833, EPI_ISL_1679834 | UW Virology Lab | UW Virology Lab | Pavitra Roychoudhury, Hong Xie, Lasata Shrestha, Shah Mohamed Bakhash, Michelle Lin, Noah R. Baker, Sean Ellis, Meei-Li Huang, Keith R Jerome, Alexander Greninger |
| EPI_ISL_1679835 | Helix/Illumina | Centers for Disease Control and Prevention Division of Viral Diseases, Pathogen Discovery | Dakota Howard, Dhwani Batra, Peter W. Cook, Kara Moser, Adrian Paskey, Jason Caravas, Benjamin Rambo-Martin, Shatavia Morrison, Christopher Gulvick, Scott Sammons, Yvette Unoarumhi, Darlene Wagner, Matthew Schmerer, Eileen de Feo, Jan Antico, Christine Tran, Matthew Tolentino, Shannon Wickline, Kim Gietzen, Brad Sickler, Jingtao Liu, Eric Allen, Phil Febbo, Nicole L. Washington, Simon White, Geraint Levan, Kelly Schiabor Barrett, Elizabeth Cirulli, Alexandre Bolze, Ary Ascencio, Charlotte Rivera-Garcia, Ryan Cho, Jason Nguyen, Sherry Wang, Jimmy Ramirez, Tyler Cassens, Efren Sandoval, Magnus Isaksson, William Lee, David Becker, Marc Laurent, James Lu, Clinton R. Paden, Duncan MacCannell |
| EPI_ISL_1679836, EPI_ISL_1679837 | UW Virology Lab | UW Virology Lab | Pavitra Roychoudhury, Hong Xie, Lasata Shrestha, Shah Mohamed Bakhash, Michelle Lin, Noah R. Baker, Sean Ellis, Meei-Li Huang, Keith R Jerome, Alexander Greninger |
| EPI_ISL_1679838 | Helix/Illumina | Centers for Disease Control and Prevention Division of Viral Diseases, Pathogen Discovery | Dakota Howard, Dhwani Batra, Peter W. Cook, Kara Moser, Adrian Paskey, Jason Caravas, Benjamin Rambo-Martin, Shatavia Morrison, Christopher Gulvick, Scott Sammons, Yvette Unoarumhi, Darlene Wagner, Matthew Schmerer, Eileen de Feo, Jan Antico, Christine Tran, Matthew Tolentino, Shannon Wickline, Kim Gietzen, Brad Sickler, Jingtao Liu, Eric Allen, Phil Febbo, Nicole L. Washington, Simon White, Geraint Levan, Kelly Schiabor Barrett, Elizabeth Cirulli, Alexandre Bolze, Ary Ascencio, Charlotte Rivera-Garcia, Ryan Cho, Jason Nguyen, Sherry Wang, Jimmy Ramirez, Tyler Cassens, Efren Sandoval, Magnus Isaksson, William Lee, David Becker, Marc Laurent, James Lu, Clinton R. Paden, Duncan MacCannell |
| EPI_ISL_1679839, EPI_ISL_1679840 | UW Virology Lab | UW Virology Lab | Pavitra Roychoudhury, Hong Xie, Lasata Shrestha, Shah Mohamed Bakhash, Michelle Lin, Noah R. Baker, Sean Ellis, Meei-Li Huang, Keith R Jerome, Alexander Greninger |
| EPI_ISL_1679841 | Helix/Illumina | Centers for Disease Control and Prevention Division of Viral Diseases, Pathogen Discovery | Dakota Howard, Dhwani Batra, Peter W. Cook, Kara Moser, Adrian Paskey, Jason Caravas, Benjamin Rambo-Martin, Shatavia Morrison, Christopher Gulvick, Scott Sammons, Yvette Unoarumhi, Darlene Wagner, Matthew Schmerer, Eileen de Feo, Jan Antico, Christine Tran, Matthew Tolentino, Shannon Wickline, Kim Gietzen, Brad Sickler, Jingtao Liu, Eric Allen, Phil Febbo, Nicole L. Washington, Simon White, Geraint Levan, Kelly Schiabor Barrett, Elizabeth Cirulli, Alexandre Bolze, Ary Ascencio, Charlotte Rivera-Garcia, Ryan Cho, Jason Nguyen, Sherry Wang, Jimmy Ramirez, Tyler Cassens, Efren Sandoval, Magnus Isaksson, William Lee, David Becker, Marc Laurent, James Lu, Clinton R. Paden, Duncan MacCannell |
| EPI_ISL_1679842, EPI_ISL_1679843 | UW Virology Lab | UW Virology Lab | Pavitra Roychoudhury, Hong Xie, Lasata Shrestha, Shah Mohamed Bakhash, Michelle Lin, Noah R. Baker, Sean Ellis, Meei-Li Huang, Keith R Jerome, Alexander Greninger |
| EPI_ISL_1679844 | Helix/Illumina | Centers for Disease Control and Prevention Division of Viral Diseases, Pathogen Discovery | Dakota Howard, Dhwani Batra, Peter W. Cook, Kara Moser, Adrian Paskey, Jason Caravas, Benjamin Rambo-Martin, Shatavia Morrison, Christopher Gulvick, Scott Sammons, Yvette Unoarumhi, Darlene Wagner, Matthew Schmerer, Eileen de Feo, Jan Antico, Christine Tran, Matthew Tolentino, Shannon Wickline, Kim Gietzen, Brad Sickler, Jingtao Liu, Eric Allen, Phil Febbo, Nicole L. Washington, Simon White, Geraint Levan, Kelly Schiabor Barrett, Elizabeth Cirulli, Alexandre Bolze, Ary Ascencio, Charlotte Rivera-Garcia, Ryan Cho, Jason Nguyen, Sherry Wang, Jimmy Ramirez, Tyler Cassens, Efren Sandoval, Magnus Isaksson, William Lee, David Becker, Marc Laurent, James Lu, Clinton R. Paden, Duncan MacCannell |
| EPI_ISL_1679845, EPI_ISL_1679846 | UW Virology Lab | UW Virology Lab | Pavitra Roychoudhury, Hong Xie, Lasata Shrestha, Shah Mohamed Bakhash, Michelle Lin, Noah R. Baker, Sean Ellis, Meei-Li Huang, Keith R Jerome, Alexander Greninger |
| EPI_ISL_1679847 | Helix/Illumina | Centers for Disease Control and Prevention Division of Viral Diseases, Pathogen Discovery | Dakota Howard, Dhwani Batra, Peter W. Cook, Kara Moser, Adrian Paskey, Jason Caravas, Benjamin Rambo-Martin, Shatavia Morrison, Christopher Gulvick, Scott Sammons, Yvette Unoarumhi, Darlene Wagner, Matthew Schmerer, Eileen de Feo, Jan Antico, Christine Tran, Matthew Tolentino, Shannon Wickline, Kim Gietzen, Brad Sickler, Jingtao Liu, Eric Allen, Phil Febbo, Nicole L. Washington, Simon White, Geraint Levan, Kelly Schiabor Barrett, Elizabeth Cirulli, Alexandre Bolze, Ary Ascencio, Charlotte Rivera-Garcia, Ryan Cho, Jason Nguyen, Sherry Wang, Jimmy Ramirez, Tyler Cassens, Efren Sandoval, Magnus Isaksson, William Lee, David Becker, Marc Laurent, James Lu, Clinton R. Paden, Duncan MacCannell |
| EPI_ISL_1679848, EPI_ISL_1679849 | UW Virology Lab | UW Virology Lab | Pavitra Roychoudhury, Hong Xie, Lasata Shrestha, Shah Mohamed Bakhash, Michelle Lin, Noah R. Baker, Sean Ellis, Meei-Li Huang, Keith R Jerome, Alexander Greninger |
| EPI_ISL_1679850, EPI_ISL_1679853 | Helix/Illumina | Centers for Disease Control and Prevention Division of Viral Diseases, Pathogen Discovery | Dakota Howard, Dhwani Batra, Peter W. Cook, Kara Moser, Adrian Paskey, Jason Caravas, Benjamin Rambo-Martin, Shatavia Morrison, Christopher Gulvick, Scott Sammons, Yvette Unoarumhi, Darlene Wagner, Matthew Schmerer, Eileen de Feo, Jan Antico, Christine Tran, Matthew Tolentino, Shannon Wickline, Kim Gietzen, Brad Sickler, Jingtao Liu, Eric Allen, Phil Febbo, Nicole L. Washington, Simon White, Geraint Levan, Kelly Schiabor Barrett, Elizabeth Cirulli, Alexandre Bolze, Ary Ascencio, Charlotte Rivera-Garcia, Ryan Cho, Jason Nguyen, Sherry Wang, Jimmy Ramirez, Tyler Cassens, Efren Sandoval, Magnus Isaksson, William Lee, David Becker, Marc Laurent, James Lu, Clinton R. Paden, Duncan MacCannell |
| EPI_ISL_1679854, EPI_ISL_1679855 | UW Virology Lab | UW Virology Lab | Pavitra Roychoudhury, Hong Xie, Lasata Shrestha, Shah Mohamed Bakhash, Michelle Lin, Noah R. Baker, Sean Ellis, Meei-Li Huang, Keith R Jerome, Alexander Greninger |
| EPI_ISL_1679856 | Helix/Illumina | Centers for Disease Control and Prevention Division of Viral Diseases, Pathogen Discovery | Dakota Howard, Dhwani Batra, Peter W. Cook, Kara Moser, Adrian Paskey, Jason Caravas, Benjamin Rambo-Martin, Shatavia Morrison, Christopher Gulvick, Scott Sammons, Yvette Unoarumhi, Darlene Wagner, Matthew Schmerer, Eileen de Feo, Jan Antico, Christine Tran, Matthew Tolentino, Shannon Wickline, Kim Gietzen, Brad Sickler, Jingtao Liu, Eric Allen, Phil Febbo, Nicole L. Washington, Simon White, Geraint Levan, Kelly Schiabor Barrett, Elizabeth Cirulli, Alexandre Bolze, Ary Ascencio, Charlotte Rivera-Garcia, Ryan Cho, Jason Nguyen, Sherry Wang, Jimmy Ramirez, Tyler Cassens, Efren Sandoval, Magnus Isaksson, William Lee, David Becker, Marc Laurent, James Lu, Clinton R. Paden, Duncan MacCannell |
| EPI_ISL_1679857, EPI_ISL_1679858 | UW Virology Lab | UW Virology Lab | Pavitra Roychoudhury, Hong Xie, Lasata Shrestha, Shah Mohamed Bakhash, Michelle Lin, Noah R. Baker, Sean Ellis, Meei-Li Huang, Keith R Jerome, Alexander Greninger |
| EPI_ISL_1679859 | Helix/Illumina | Centers for Disease Control and Prevention Division of Viral Diseases, Pathogen Discovery | Dakota Howard, Dhwani Batra, Peter W. Cook, Kara Moser, Adrian Paskey, Jason Caravas, Benjamin Rambo-Martin, Shatavia Morrison, Christopher Gulvick, Scott Sammons, Yvette Unoarumhi, Darlene Wagner, Matthew Schmerer, Eileen de Feo, Jan Antico, Christine Tran, Matthew Tolentino, Shannon Wickline, Kim Gietzen, Brad Sickler, Jingtao Liu, Eric Allen, Phil Febbo, Nicole L. Washington, Simon White, Geraint Levan, Kelly Schiabor Barrett, Elizabeth Cirulli, Alexandre Bolze, Ary Ascencio, Charlotte Rivera-Garcia, Ryan Cho, Jason Nguyen, Sherry Wang, Jimmy Ramirez, Tyler Cassens, Efren Sandoval, Magnus Isaksson, William Lee, David Becker, Marc Laurent, James Lu, Clinton R. Paden, Duncan MacCannell |
| EPI_ISL_1679860, EPI_ISL_1679861 | UW Virology Lab | UW Virology Lab | Pavitra Roychoudhury, Hong Xie, Lasata Shrestha, Shah Mohamed Bakhash, Michelle Lin, Noah R. Baker, Sean Ellis, Meei-Li Huang, Keith R Jerome, Alexander Greninger |
| EPI_ISL_1679862 | Helix/Illumina | Centers for Disease Control and Prevention Division of Viral Diseases, Pathogen Discovery | Dakota Howard, Dhwani Batra, Peter W. Cook, Kara Moser, Adrian Paskey, Jason Caravas, Benjamin Rambo-Martin, Shatavia Morrison, Christopher Gulvick, Scott Sammons, Yvette Unoarumhi, Darlene Wagner, Matthew Schmerer, Eileen de Feo, Jan Antico, Christine Tran, Matthew Tolentino, Shannon Wickline, Kim Gietzen, Brad Sickler, Jingtao Liu, Eric Allen, Phil Febbo, Nicole L. Washington, Simon White, Geraint Levan, Kelly Schiabor Barrett, Elizabeth Cirulli, Alexandre Bolze, Ary Ascencio, Charlotte Rivera-Garcia, Ryan Cho, Jason Nguyen, Sherry Wang, Jimmy Ramirez, Tyler Cassens, Efren Sandoval, Magnus Isaksson, William Lee, David Becker, Marc Laurent, James Lu, Clinton R. Paden, Duncan MacCannell |
| EPI_ISL_1679863, EPI_ISL_1679864 | UW Virology Lab | UW Virology Lab | Pavitra Roychoudhury, Hong Xie, Lasata Shrestha, Shah Mohamed Bakhash, Michelle Lin, Noah R. Baker, Sean Ellis, Meei-Li Huang, Keith R Jerome, Alexander Greninger |
| EPI_ISL_1679865 | Helix/Illumina | Centers for Disease Control and Prevention Division of Viral Diseases, Pathogen Discovery | Dakota Howard, Dhwani Batra, Peter W. Cook, Kara Moser, Adrian Paskey, Jason Caravas, Benjamin Rambo-Martin, Shatavia Morrison, Christopher Gulvick, Scott Sammons, Yvette Unoarumhi, Darlene Wagner, Matthew Schmerer, Eileen de Feo, Jan Antico, Christine Tran, Matthew Tolentino, Shannon Wickline, Kim Gietzen, Brad Sickler, Jingtao Liu, Eric Allen, Phil Febbo, Nicole L. Washington, Simon White, Geraint Levan, Kelly Schiabor Barrett, Elizabeth Cirulli, Alexandre Bolze, Ary Ascencio, Charlotte Rivera-Garcia, Ryan Cho, Jason Nguyen, Sherry Wang, Jimmy Ramirez, Tyler Cassens, Efren Sandoval, Magnus Isaksson, William Lee, David Becker, Marc Laurent, James Lu, Clinton R. Paden, Duncan MacCannell |
| EPI_ISL_1679867 | UW Virology Lab | UW Virology Lab | Pavitra Roychoudhury, Hong Xie, Lasata Shrestha, Shah Mohamed Bakhash, Michelle Lin, Noah R. Baker, Sean Ellis, Meei-Li Huang, Keith R Jerome, Alexander Greninger |
| EPI_ISL_1679868 | Helix/Illumina | Centers for Disease Control and Prevention Division of Viral Diseases, Pathogen Discovery | Dakota Howard, Dhwani Batra, Peter W. Cook, Kara Moser, Adrian Paskey, Jason Caravas, Benjamin Rambo-Martin, Shatavia Morrison, Christopher Gulvick, Scott Sammons, Yvette Unoarumhi, Darlene Wagner, Matthew Schmerer, Eileen de Feo, Jan Antico, Christine Tran, Matthew Tolentino, Shannon Wickline, Kim Gietzen, Brad Sickler, Jingtao Liu, Eric Allen, Phil Febbo, Nicole L. Washington, Simon White, Geraint Levan, Kelly Schiabor Barrett, Elizabeth Cirulli, Alexandre Bolze, Ary Ascencio, Charlotte Rivera-Garcia, Ryan Cho, Jason Nguyen, Sherry Wang, Jimmy Ramirez, Tyler Cassens, Efren Sandoval, Magnus Isaksson, William Lee, David Becker, Marc Laurent, James Lu, Clinton R. Paden, Duncan MacCannell |
| EPI_ISL_1679869, EPI_ISL_1679870 | UW Virology Lab | UW Virology Lab | Pavitra Roychoudhury, Hong Xie, Lasata Shrestha, Shah Mohamed Bakhash, Michelle Lin, Noah R. Baker, Sean Ellis, Meei-Li Huang, Keith R Jerome, Alexander Greninger |
| EPI_ISL_1679871 | Helix/Illumina | Centers for Disease Control and Prevention Division of Viral Diseases, Pathogen Discovery | Dakota Howard, Dhwani Batra, Peter W. Cook, Kara Moser, Adrian Paskey, Jason Caravas, Benjamin Rambo-Martin, Shatavia Morrison, Christopher Gulvick, Scott Sammons, Yvette Unoarumhi, Darlene Wagner, Matthew Schmerer, Eileen de Feo, Jan Antico, Christine Tran, Matthew Tolentino, Shannon Wickline, Kim Gietzen, Brad Sickler, Jingtao Liu, Eric Allen, Phil Febbo, Nicole L. Washington, Simon White, Geraint Levan, Kelly Schiabor Barrett, Elizabeth Cirulli, Alexandre Bolze, Ary Ascencio, Charlotte Rivera-Garcia, Ryan Cho, Jason Nguyen, Sherry Wang, Jimmy Ramirez, Tyler Cassens, Efren Sandoval, Magnus Isaksson, William Lee, David Becker, Marc Laurent, James Lu, Clinton R. Paden, Duncan MacCannell |
| EPI_ISL_1679872, EPI_ISL_1679873 | UW Virology Lab | UW Virology Lab | Pavitra Roychoudhury, Hong Xie, Lasata Shrestha, Shah Mohamed Bakhash, Michelle Lin, Noah R. Baker, Sean Ellis, Meei-Li Huang, Keith R Jerome, Alexander Greninger |
| EPI_ISL_1679874 | Helix/Illumina | Centers for Disease Control and Prevention Division of Viral Diseases, Pathogen Discovery | Dakota Howard, Dhwani Batra, Peter W. Cook, Kara Moser, Adrian Paskey, Jason Caravas, Benjamin Rambo-Martin, Shatavia Morrison, Christopher Gulvick, Scott Sammons, Yvette Unoarumhi, Darlene Wagner, Matthew Schmerer, Eileen de Feo, Jan Antico, Christine Tran, Matthew Tolentino, Shannon Wickline, Kim Gietzen, Brad Sickler, Jingtao Liu, Eric Allen, Phil Febbo, Nicole L. Washington, Simon White, Geraint Levan, Kelly Schiabor Barrett, Elizabeth Cirulli, Alexandre Bolze, Ary Ascencio, Charlotte Rivera-Garcia, Ryan Cho, Jason Nguyen, Sherry Wang, Jimmy Ramirez, Tyler Cassens, Efren Sandoval, Magnus Isaksson, William Lee, David Becker, Marc Laurent, James Lu, Clinton R. Paden, Duncan MacCannell |
| EPI_ISL_1679875, EPI_ISL_1679876, EPI_ISL_1679877 | UW Virology Lab | UW Virology Lab | Pavitra Roychoudhury, Hong Xie, Lasata Shrestha, Shah Mohamed Bakhash, Michelle Lin, Noah R. Baker, Sean Ellis, Meei-Li Huang, Keith R Jerome, Alexander Greninger |
| EPI_ISL_1679878 | Helix/Illumina | Centers for Disease Control and Prevention Division of Viral Diseases, Pathogen Discovery | Dakota Howard, Dhwani Batra, Peter W. Cook, Kara Moser, Adrian Paskey, Jason Caravas, Benjamin Rambo-Martin, Shatavia Morrison, Christopher Gulvick, Scott Sammons, Yvette Unoarumhi, Darlene Wagner, Matthew Schmerer, Eileen de Feo, Jan Antico, Christine Tran, Matthew Tolentino, Shannon Wickline, Kim Gietzen, Brad Sickler, Jingtao Liu, Eric Allen, Phil Febbo, Nicole L. Washington, Simon White, Geraint Levan, Kelly Schiabor Barrett, Elizabeth Cirulli, Alexandre Bolze, Ary Ascencio, Charlotte Rivera-Garcia, Ryan Cho, Jason Nguyen, Sherry Wang, Jimmy Ramirez, Tyler Cassens, Efren Sandoval, Magnus Isaksson, William Lee, David Becker, Marc Laurent, James Lu, Clinton R. Paden, Duncan MacCannell |
| EPI_ISL_1679879, EPI_ISL_1679880 | UW Virology Lab | UW Virology Lab | Pavitra Roychoudhury, Hong Xie, Lasata Shrestha, Shah Mohamed Bakhash, Michelle Lin, Noah R. Baker, Sean Ellis, Meei-Li Huang, Keith R Jerome, Alexander Greninger |
| EPI_ISL_1679881 | Helix/Illumina | Centers for Disease Control and Prevention Division of Viral Diseases, Pathogen Discovery | Dakota Howard, Dhwani Batra, Peter W. Cook, Kara Moser, Adrian Paskey, Jason Caravas, Benjamin Rambo-Martin, Shatavia Morrison, Christopher Gulvick, Scott Sammons, Yvette Unoarumhi, Darlene Wagner, Matthew Schmerer, Eileen de Feo, Jan Antico, Christine Tran, Matthew Tolentino, Shannon Wickline, Kim Gietzen, Brad Sickler, Jingtao Liu, Eric Allen, Phil Febbo, Nicole L. Washington, Simon White, Geraint Levan, Kelly Schiabor Barrett, Elizabeth Cirulli, Alexandre Bolze, Ary Ascencio, Charlotte Rivera-Garcia, Ryan Cho, Jason Nguyen, Sherry Wang, Jimmy Ramirez, Tyler Cassens, Efren Sandoval, Magnus Isaksson, William Lee, David Becker, Marc Laurent, James Lu, Clinton R. Paden, Duncan MacCannell |
| EPI_ISL_1679882, EPI_ISL_1679883 | UW Virology Lab | UW Virology Lab | Pavitra Roychoudhury, Hong Xie, Lasata Shrestha, Shah Mohamed Bakhash, Michelle Lin, Noah R. Baker, Sean Ellis, Meei-Li Huang, Keith R Jerome, Alexander Greninger |
| EPI_ISL_1679884 | Helix/Illumina | Centers for Disease Control and Prevention Division of Viral Diseases, Pathogen Discovery | Dakota Howard, Dhwani Batra, Peter W. Cook, Kara Moser, Adrian Paskey, Jason Caravas, Benjamin Rambo-Martin, Shatavia Morrison, Christopher Gulvick, Scott Sammons, Yvette Unoarumhi, Darlene Wagner, Matthew Schmerer, Eileen de Feo, Jan Antico, Christine Tran, Matthew Tolentino, Shannon Wickline, Kim Gietzen, Brad Sickler, Jingtao Liu, Eric Allen, Phil Febbo, Nicole L. Washington, Simon White, Geraint Levan, Kelly Schiabor Barrett, Elizabeth Cirulli, Alexandre Bolze, Ary Ascencio, Charlotte Rivera-Garcia, Ryan Cho, Jason Nguyen, Sherry Wang, Jimmy Ramirez, Tyler Cassens, Efren Sandoval, Magnus Isaksson, William Lee, David Becker, Marc Laurent, James Lu, Clinton R. Paden, Duncan MacCannell |
| EPI_ISL_1679885, EPI_ISL_1679886 | UW Virology Lab | UW Virology Lab | Pavitra Roychoudhury, Hong Xie, Lasata Shrestha, Shah Mohamed Bakhash, Michelle Lin, Noah R. Baker, Sean Ellis, Meei-Li Huang, Keith R Jerome, Alexander Greninger |
| EPI_ISL_1679887 | Helix/Illumina | Centers for Disease Control and Prevention Division of Viral Diseases, Pathogen Discovery | Dakota Howard, Dhwani Batra, Peter W. Cook, Kara Moser, Adrian Paskey, Jason Caravas, Benjamin Rambo-Martin, Shatavia Morrison, Christopher Gulvick, Scott Sammons, Yvette Unoarumhi, Darlene Wagner, Matthew Schmerer, Eileen de Feo, Jan Antico, Christine Tran, Matthew Tolentino, Shannon Wickline, Kim Gietzen, Brad Sickler, Jingtao Liu, Eric Allen, Phil Febbo, Nicole L. Washington, Simon White, Geraint Levan, Kelly Schiabor Barrett, Elizabeth Cirulli, Alexandre Bolze, Ary Ascencio, Charlotte Rivera-Garcia, Ryan Cho, Jason Nguyen, Sherry Wang, Jimmy Ramirez, Tyler Cassens, Efren Sandoval, Magnus Isaksson, William Lee, David Becker, Marc Laurent, James Lu, Clinton R. Paden, Duncan MacCannell |
| EPI_ISL_1679888, EPI_ISL_1679889 | UW Virology Lab | UW Virology Lab | Pavitra Roychoudhury, Hong Xie, Lasata Shrestha, Shah Mohamed Bakhash, Michelle Lin, Noah R. Baker, Sean Ellis, Meei-Li Huang, Keith R Jerome, Alexander Greninger |
| EPI_ISL_1679890, EPI_ISL_1679891 | Helix/Illumina | Centers for Disease Control and Prevention Division of Viral Diseases, Pathogen | Dakota Howard, Dhwani Batra, Peter W. Cook, Kara Moser, Adrian Paskey, Jason Caravas, Benjamin Rambo-Martin, Shatavia Morrison, Christopher Gulvick, Scott Sammons, Yvette Unoarumhi, Darlene Wagner, Matthew Schmerer, Eileen de Feo, Jan Antico, Christine Tran, Matthew Tolentino, Shannon Wickline, Kim Gietzen, Brad Sickler, Jingtao Liu, Eric Allen, Phil Febbo, Nicole L. Washington, Simon White, Geraint Levan, Kelly Schiabor Barrett, Elizabeth Cirulli, Alexandre Bolze, Ary Ascencio, Charlotte Rivera-Garcia, Ryan Cho, Jason Nguyen, Sherry Wang, Jimmy |

| | | | |
|---|---|---|---|
| | | Discovery | Ramirez, Tyler Cassens, Efren Sandoval, Magnus Isaksson, William Lee, David Becker, Marc Laurent, James Lu, Clinton R. Paden, Duncan MacCannell |
| EPI_ISL_1679892 | UW Virology Lab | UW Virology Lab | Pavitra Roychoudhury, Hong Xie, Lasata Shrestha, Shah Mohamed Bakhash, Michelle Lin, Noah R. Baker, Sean Ellis, Meei-Li Huang, Keith R Jerome, Alexander Greninger |
| EPI_ISL_1679893, EPI_ISL_1679894 | Helix/Illumina | Centers for Disease Control and Prevention Division of Viral Diseases, Pathogen Discovery | Dakota Howard, Dhwani Batra, Peter W. Cook, Kara Moser, Adrian Paskey, Jason Caravas, Benjamin Rambo-Martin, Shatavia Morrison, Christopher Gulvick, Scott Sammons, Yvette Unoarumhi, Darlene Wagner, Matthew Schmerer, Eileen de Feo, Jan Antico, Christine Tran, Matthew Tolentino, Shannon Wickline, Kim Gietzen, Brad Sickler, Jingtao Liu, Eric Allen, Phil Febbo, Nicole L. Washington, Simon White, Geraint Levan, Kelly Schiabor Barrett, Elizabeth Cirulli, Alexandre Bolze, Ary Ascencio, Charlotte Rivera-Garcia, Ryan Cho, Jason Nguyen, Sherry Wang, Jimmy Ramirez, Tyler Cassens, Efren Sandoval, Magnus Isaksson, William Lee, David Becker, Marc Laurent, James Lu, Clinton R. Paden, Duncan MacCannell |
| EPI_ISL_1679895 | UW Virology Lab | UW Virology Lab | Pavitra Roychoudhury, Hong Xie, Lasata Shrestha, Shah Mohamed Bakhash, Michelle Lin, Noah R. Baker, Sean Ellis, Meei-Li Huang, Keith R Jerome, Alexander Greninger |
| EPI_ISL_1679896, EPI_ISL_1679897 | Helix/Illumina | Centers for Disease Control and Prevention Division of Viral Diseases, Pathogen Discovery | Dakota Howard, Dhwani Batra, Peter W. Cook, Kara Moser, Adrian Paskey, Jason Caravas, Benjamin Rambo-Martin, Shatavia Morrison, Christopher Gulvick, Scott Sammons, Yvette Unoarumhi, Darlene Wagner, Matthew Schmerer, Eileen de Feo, Jan Antico, Christine Tran, Matthew Tolentino, Shannon Wickline, Kim Gietzen, Brad Sickler, Jingtao Liu, Eric Allen, Phil Febbo, Nicole L. Washington, Simon White, Geraint Levan, Kelly Schiabor Barrett, Elizabeth Cirulli, Alexandre Bolze, Ary Ascencio, Charlotte Rivera-Garcia, Ryan Cho, Jason Nguyen, Sherry Wang, Jimmy Ramirez, Tyler Cassens, Efren Sandoval, Magnus Isaksson, William Lee, David Becker, Marc Laurent, James Lu, Clinton R. Paden, Duncan MacCannell |
| EPI_ISL_1679898 | UW Virology Lab | UW Virology Lab | Pavitra Roychoudhury, Hong Xie, Lasata Shrestha, Shah Mohamed Bakhash, Michelle Lin, Noah R. Baker, Sean Ellis, Meei-Li Huang, Keith R Jerome, Alexander Greninger |
| EPI_ISL_1679899, EPI_ISL_1679900 | Helix/Illumina | Centers for Disease Control and Prevention Division of Viral Diseases, Pathogen Discovery | Dakota Howard, Dhwani Batra, Peter W. Cook, Kara Moser, Adrian Paskey, Jason Caravas, Benjamin Rambo-Martin, Shatavia Morrison, Christopher Gulvick, Scott Sammons, Yvette Unoarumhi, Darlene Wagner, Matthew Schmerer, Eileen de Feo, Jan Antico, Christine Tran, Matthew Tolentino, Shannon Wickline, Kim Gietzen, Brad Sickler, Jingtao Liu, Eric Allen, Phil Febbo, Nicole L. Washington, Simon White, Geraint Levan, Kelly Schiabor Barrett, Elizabeth Cirulli, Alexandre Bolze, Ary Ascencio, Charlotte Rivera-Garcia, Ryan Cho, Jason Nguyen, Sherry Wang, Jimmy Ramirez, Tyler Cassens, Efren Sandoval, Magnus Isaksson, William Lee, David Becker, Marc Laurent, James Lu, Clinton R. Paden, Duncan MacCannell |
| EPI_ISL_1679901 | UW Virology Lab | UW Virology Lab | Pavitra Roychoudhury, Hong Xie, Lasata Shrestha, Shah Mohamed Bakhash, Michelle Lin, Noah R. Baker, Sean Ellis, Meei-Li Huang, Keith R Jerome, Alexander Greninger |
| EPI_ISL_1679902, EPI_ISL_1679903 | Helix/Illumina | Centers for Disease Control and Prevention Division of Viral Diseases, Pathogen Discovery | Dakota Howard, Dhwani Batra, Peter W. Cook, Kara Moser, Adrian Paskey, Jason Caravas, Benjamin Rambo-Martin, Shatavia Morrison, Christopher Gulvick, Scott Sammons, Yvette Unoarumhi, Darlene Wagner, Matthew Schmerer, Eileen de Feo, Jan Antico, Christine Tran, Matthew Tolentino, Shannon Wickline, Kim Gietzen, Brad Sickler, Jingtao Liu, Eric Allen, Phil Febbo, Nicole L. Washington, Simon White, Geraint Levan, Kelly Schiabor Barrett, Elizabeth Cirulli, Alexandre Bolze, Ary Ascencio, Charlotte Rivera-Garcia, Ryan Cho, Jason Nguyen, Sherry Wang, Jimmy Ramirez, Tyler Cassens, Efren Sandoval, Magnus Isaksson, William Lee, David Becker, Marc Laurent, James Lu, Clinton R. Paden, Duncan MacCannell |
| EPI_ISL_1679904 | UW Virology Lab | UW Virology Lab | Pavitra Roychoudhury, Hong Xie, Lasata Shrestha, Shah Mohamed Bakhash, Michelle Lin, Noah R. Baker, Sean Ellis, Meei-Li Huang, Keith R Jerome, Alexander Greninger |
| EPI_ISL_1679905, EPI_ISL_1679906 | Helix/Illumina | Centers for Disease Control and Prevention Division of Viral Diseases, Pathogen Discovery | Dakota Howard, Dhwani Batra, Peter W. Cook, Kara Moser, Adrian Paskey, Jason Caravas, Benjamin Rambo-Martin, Shatavia Morrison, Christopher Gulvick, Scott Sammons, Yvette Unoarumhi, Darlene Wagner, Matthew Schmerer, Eileen de Feo, Jan Antico, Christine Tran, Matthew Tolentino, Shannon Wickline, Kim Gietzen, Brad Sickler, Jingtao Liu, Eric Allen, Phil Febbo, Nicole L. Washington, Simon White, Geraint Levan, Kelly Schiabor Barrett, Elizabeth Cirulli, Alexandre Bolze, Ary Ascencio, Charlotte Rivera-Garcia, Ryan Cho, Jason Nguyen, Sherry Wang, Jimmy Ramirez, Tyler Cassens, Efren Sandoval, Magnus Isaksson, William Lee, David Becker, Marc Laurent, James Lu, Clinton R. Paden, Duncan MacCannell |
| EPI_ISL_1679907 | UW Virology Lab | UW Virology Lab | Pavitra Roychoudhury, Hong Xie, Lasata Shrestha, Shah Mohamed Bakhash, Michelle Lin, Noah R. Baker, Sean Ellis, Meei-Li Huang, Keith R Jerome, Alexander Greninger |
| EPI_ISL_1679908, EPI_ISL_1679909 | Helix/Illumina | Centers for Disease Control and Prevention Division of Viral Diseases, Pathogen Discovery | Dakota Howard, Dhwani Batra, Peter W. Cook, Kara Moser, Adrian Paskey, Jason Caravas, Benjamin Rambo-Martin, Shatavia Morrison, Christopher Gulvick, Scott Sammons, Yvette Unoarumhi, Darlene Wagner, Matthew Schmerer, Eileen de Feo, Jan Antico, Christine Tran, Matthew Tolentino, Shannon Wickline, Kim Gietzen, Brad Sickler, Jingtao Liu, Eric Allen, Phil Febbo, Nicole L. Washington, Simon White, Geraint Levan, Kelly Schiabor Barrett, Elizabeth Cirulli, Alexandre Bolze, Ary Ascencio, Charlotte Rivera-Garcia, Ryan Cho, Jason Nguyen, Sherry Wang, Jimmy Ramirez, Tyler Cassens, Efren Sandoval, Magnus Isaksson, William Lee, David Becker, Marc Laurent, James Lu, Clinton R. Paden, Duncan MacCannell |
| EPI_ISL_1679910 | UW Virology Lab | UW Virology Lab | Pavitra Roychoudhury, Hong Xie, Lasata Shrestha, Shah Mohamed Bakhash, Michelle Lin, Noah R. Baker, Sean Ellis, Meei-Li Huang, Keith R Jerome, Alexander Greninger |
| EPI_ISL_1679911, EPI_ISL_1679912 | Helix/Illumina | Centers for Disease Control and Prevention Division of Viral Diseases, Pathogen Discovery | Dakota Howard, Dhwani Batra, Peter W. Cook, Kara Moser, Adrian Paskey, Jason Caravas, Benjamin Rambo-Martin, Shatavia Morrison, Christopher Gulvick, Scott Sammons, Yvette Unoarumhi, Darlene Wagner, Matthew Schmerer, Eileen de Feo, Jan Antico, Christine Tran, Matthew Tolentino, Shannon Wickline, Kim Gietzen, Brad Sickler, Jingtao Liu, Eric Allen, Phil Febbo, Nicole L. Washington, Simon White, Geraint Levan, Kelly Schiabor Barrett, Elizabeth Cirulli, Alexandre Bolze, Ary Ascencio, Charlotte Rivera-Garcia, Ryan Cho, Jason Nguyen, Sherry Wang, Jimmy Ramirez, Tyler Cassens, Efren Sandoval, Magnus Isaksson, William Lee, David Becker, Marc Laurent, James Lu, Clinton R. Paden, Duncan MacCannell |
| EPI_ISL_1679913 | UW Virology Lab | UW Virology Lab | Pavitra Roychoudhury, Hong Xie, Lasata Shrestha, Shah Mohamed Bakhash, Michelle Lin, Noah R. Baker, Sean Ellis, Meei-Li Huang, Keith R Jerome, Alexander Greninger |
| EPI_ISL_1679914, EPI_ISL_1679915 | Helix/Illumina | Centers for Disease Control and Prevention Division of Viral Diseases, Pathogen Discovery | Dakota Howard, Dhwani Batra, Peter W. Cook, Kara Moser, Adrian Paskey, Jason Caravas, Benjamin Rambo-Martin, Shatavia Morrison, Christopher Gulvick, Scott Sammons, Yvette Unoarumhi, Darlene Wagner, Matthew Schmerer, Eileen de Feo, Jan Antico, Christine Tran, Matthew Tolentino, Shannon Wickline, Kim Gietzen, Brad Sickler, Jingtao Liu, Eric Allen, Phil Febbo, Nicole L. Washington, Simon White, Geraint Levan, Kelly Schiabor Barrett, Elizabeth Cirulli, Alexandre Bolze, Ary Ascencio, Charlotte Rivera-Garcia, Ryan Cho, Jason Nguyen, Sherry Wang, Jimmy Ramirez, Tyler Cassens, Efren Sandoval, Magnus Isaksson, William Lee, David Becker, Marc Laurent, James Lu, Clinton R. Paden, Duncan MacCannell |
| EPI_ISL_1679916 | UW Virology Lab | UW Virology Lab | Pavitra Roychoudhury, Hong Xie, Lasata Shrestha, Shah Mohamed Bakhash, Michelle Lin, Noah R. Baker, Sean Ellis, Meei-Li Huang, Keith R Jerome, Alexander Greninger |
| EPI_ISL_1679917, EPI_ISL_1679918, EPI_ISL_1679919 | Helix/Illumina | Centers for Disease Control and Prevention Division of Viral Diseases, Pathogen Discovery | Dakota Howard, Dhwani Batra, Peter W. Cook, Kara Moser, Adrian Paskey, Jason Caravas, Benjamin Rambo-Martin, Shatavia Morrison, Christopher Gulvick, Scott Sammons, Yvette Unoarumhi, Darlene Wagner, Matthew Schmerer, Eileen de Feo, Jan Antico, Christine Tran, Matthew Tolentino, Shannon Wickline, Kim Gietzen, Brad Sickler, Jingtao Liu, Eric Allen, Phil Febbo, Nicole L. Washington, Simon White, Geraint Levan, Kelly Schiabor Barrett, Elizabeth Cirulli, Alexandre Bolze, Ary Ascencio, Charlotte Rivera-Garcia, Ryan Cho, Jason Nguyen, Sherry Wang, Jimmy Ramirez, Tyler Cassens, Efren Sandoval, Magnus Isaksson, William Lee, David Becker, Marc Laurent, James Lu, Clinton R. Paden, Duncan MacCannell |
| EPI_ISL_1679920 | UW Virology Lab | UW Virology Lab | Pavitra Roychoudhury, Hong Xie, Lasata Shrestha, Shah Mohamed Bakhash, Michelle Lin, Noah R. Baker, Sean Ellis, Meei-Li Huang, Keith R Jerome, Alexander Greninger |
| EPI_ISL_1679922 | Helix/Illumina | Centers for Disease Control and Prevention Division of Viral Diseases, Pathogen Discovery | Dakota Howard, Dhwani Batra, Peter W. Cook, Kara Moser, Adrian Paskey, Jason Caravas, Benjamin Rambo-Martin, Shatavia Morrison, Christopher Gulvick, Scott Sammons, Yvette Unoarumhi, Darlene Wagner, Matthew Schmerer, Eileen de Feo, Jan Antico, Christine Tran, Matthew Tolentino, Shannon Wickline, Kim Gietzen, Brad Sickler, Jingtao Liu, Eric Allen, Phil Febbo, Nicole L. Washington, Simon White, Geraint Levan, Kelly Schiabor Barrett, Elizabeth Cirulli, Alexandre Bolze, Ary Ascencio, Charlotte Rivera-Garcia, Ryan Cho, Jason Nguyen, Sherry Wang, Jimmy Ramirez, Tyler Cassens, Efren Sandoval, Magnus Isaksson, William Lee, David Becker, Marc Laurent, James Lu, Clinton R. Paden, Duncan MacCannell |
| EPI_ISL_1679923 | UW Virology Lab | UW Virology Lab | Pavitra Roychoudhury, Hong Xie, Lasata Shrestha, Shah Mohamed Bakhash, Michelle Lin, Noah R. Baker, Sean Ellis, Meei-Li Huang, Keith R Jerome, Alexander Greninger |
| EPI_ISL_1679924, EPI_ISL_1679925 | Helix/Illumina | Centers for Disease Control and Prevention Division of Viral Diseases, Pathogen Discovery | Dakota Howard, Dhwani Batra, Peter W. Cook, Kara Moser, Adrian Paskey, Jason Caravas, Benjamin Rambo-Martin, Shatavia Morrison, Christopher Gulvick, Scott Sammons, Yvette Unoarumhi, Darlene Wagner, Matthew Schmerer, Eileen de Feo, Jan Antico, Christine Tran, Matthew Tolentino, Shannon Wickline, Kim Gietzen, Brad Sickler, Jingtao Liu, Eric Allen, Phil Febbo, Nicole L. Washington, Simon White, Geraint Levan, Kelly Schiabor Barrett, Elizabeth Cirulli, Alexandre Bolze, Ary Ascencio, Charlotte Rivera-Garcia, Ryan Cho, Jason Nguyen, Sherry Wang, Jimmy Ramirez, Tyler Cassens, Efren Sandoval, Magnus Isaksson, William Lee, David Becker, Marc Laurent, James Lu, Clinton R. Paden, Duncan MacCannell |
| EPI_ISL_1679926 | UW Virology Lab | UW Virology Lab | Pavitra Roychoudhury, Hong Xie, Lasata Shrestha, Shah Mohamed Bakhash, Michelle Lin, Noah R. Baker, Sean Ellis, Meei-Li Huang, Keith R Jerome, Alexander Greninger |
| EPI_ISL_1679927, EPI_ISL_1679928 | Helix/Illumina | Centers for Disease Control and Prevention Division of Viral Diseases, Pathogen Discovery | Dakota Howard, Dhwani Batra, Peter W. Cook, Kara Moser, Adrian Paskey, Jason Caravas, Benjamin Rambo-Martin, Shatavia Morrison, Christopher Gulvick, Scott Sammons, Yvette Unoarumhi, Darlene Wagner, Matthew Schmerer, Eileen de Feo, Jan Antico, Christine Tran, Matthew Tolentino, Shannon Wickline, Kim Gietzen, Brad Sickler, Jingtao Liu, Eric Allen, Phil Febbo, Nicole L. Washington, Simon White, Geraint Levan, Kelly Schiabor Barrett, Elizabeth Cirulli, Alexandre Bolze, Ary Ascencio, Charlotte Rivera-Garcia, Ryan Cho, Jason Nguyen, Sherry Wang, Jimmy Ramirez, Tyler Cassens, Efren Sandoval, Magnus Isaksson, William Lee, David Becker, Marc Laurent, James Lu, Clinton R. Paden, Duncan MacCannell |
| EPI_ISL_1679929 | UW Virology Lab | UW Virology Lab | Pavitra Roychoudhury, Hong Xie, Lasata Shrestha, Shah Mohamed Bakhash, Michelle Lin, Noah R. Baker, Sean Ellis, Meei-Li Huang, Keith R Jerome, Alexander Greninger |
| EPI_ISL_1679930, EPI_ISL_1679931 | Helix/Illumina | Centers for Disease Control and Prevention Division of Viral Diseases, Pathogen Discovery | Dakota Howard, Dhwani Batra, Peter W. Cook, Kara Moser, Adrian Paskey, Jason Caravas, Benjamin Rambo-Martin, Shatavia Morrison, Christopher Gulvick, Scott Sammons, Yvette Unoarumhi, Darlene Wagner, Matthew Schmerer, Eileen de Feo, Jan Antico, Christine Tran, Matthew Tolentino, Shannon Wickline, Kim Gietzen, Brad Sickler, Jingtao Liu, Eric Allen, Phil Febbo, Nicole L. Washington, Simon White, Geraint Levan, Kelly Schiabor Barrett, Elizabeth Cirulli, Alexandre Bolze, Ary Ascencio, Charlotte Rivera-Garcia, Ryan Cho, Jason Nguyen, Sherry Wang, Jimmy Ramirez, Tyler Cassens, Efren Sandoval, Magnus Isaksson, William Lee, David Becker, Marc Laurent, James Lu, Clinton R. Paden, Duncan MacCannell |
| EPI_ISL_1679932 | UW Virology Lab | UW Virology Lab | Pavitra Roychoudhury, Hong Xie, Lasata Shrestha, Shah Mohamed Bakhash, Michelle Lin, Noah R. Baker, Sean Ellis, Meei-Li Huang, Keith R Jerome, Alexander Greninger |
| EPI_ISL_1679933, EPI_ISL_1679934 | Helix/Illumina | Centers for Disease Control and Prevention Division of Viral Diseases, Pathogen Discovery | Dakota Howard, Dhwani Batra, Peter W. Cook, Kara Moser, Adrian Paskey, Jason Caravas, Benjamin Rambo-Martin, Shatavia Morrison, Christopher Gulvick, Scott Sammons, Yvette Unoarumhi, Darlene Wagner, Matthew Schmerer, Eileen de Feo, Jan Antico, Christine Tran, Matthew Tolentino, Shannon Wickline, Kim Gietzen, Brad Sickler, Jingtao Liu, Eric Allen, Phil Febbo, Nicole L. Washington, Simon White, Geraint Levan, Kelly Schiabor Barrett, Elizabeth Cirulli, Alexandre Bolze, Ary Ascencio, Charlotte Rivera-Garcia, Ryan Cho, Jason Nguyen, Sherry Wang, Jimmy Ramirez, Tyler Cassens, Efren Sandoval, Magnus Isaksson, William Lee, David Becker, Marc Laurent, James Lu, Clinton R. Paden, Duncan MacCannell |
| EPI_ISL_1679935 | UW Virology Lab | UW Virology Lab | Pavitra Roychoudhury, Hong Xie, Lasata Shrestha, Shah Mohamed Bakhash, Michelle Lin, Noah R. Baker, Sean Ellis, Meei-Li Huang, Keith R Jerome, Alexander Greninger |
| EPI_ISL_1679936, EPI_ISL_1679937 | Helix/Illumina | Centers for Disease Control and Prevention Division of Viral Diseases, Pathogen Discovery | Dakota Howard, Dhwani Batra, Peter W. Cook, Kara Moser, Adrian Paskey, Jason Caravas, Benjamin Rambo-Martin, Shatavia Morrison, Christopher Gulvick, Scott Sammons, Yvette Unoarumhi, Darlene Wagner, Matthew Schmerer, Eileen de Feo, Jan Antico, Christine Tran, Matthew Tolentino, Shannon Wickline, Kim Gietzen, Brad Sickler, Jingtao Liu, Eric Allen, Phil Febbo, Nicole L. Washington, Simon White, Geraint Levan, Kelly Schiabor Barrett, Elizabeth Cirulli, Alexandre Bolze, Ary Ascencio, Charlotte Rivera-Garcia, Ryan Cho, Jason Nguyen, Sherry Wang, Jimmy Ramirez, Tyler Cassens, Efren Sandoval, Magnus Isaksson, William Lee, David Becker, Marc Laurent, James Lu, Clinton R. Paden, Duncan MacCannell |
| EPI_ISL_1679938 | UW Virology Lab | UW Virology Lab | Pavitra Roychoudhury, Hong Xie, Lasata Shrestha, Shah Mohamed Bakhash, Michelle Lin, Noah R. Baker, Sean Ellis, Meei-Li Huang, Keith R Jerome, Alexander Greninger |
| EPI_ISL_1679939, EPI_ISL_1679940 | Helix/Illumina | Centers for Disease Control and Prevention Division of Viral Diseases, Pathogen Discovery | Dakota Howard, Dhwani Batra, Peter W. Cook, Kara Moser, Adrian Paskey, Jason Caravas, Benjamin Rambo-Martin, Shatavia Morrison, Christopher Gulvick, Scott Sammons, Yvette Unoarumhi, Darlene Wagner, Matthew Schmerer, Eileen de Feo, Jan Antico, Christine Tran, Matthew Tolentino, Shannon Wickline, Kim Gietzen, Brad Sickler, Jingtao Liu, Eric Allen, Phil Febbo, Nicole L. Washington, Simon White, Geraint Levan, Kelly Schiabor Barrett, Elizabeth Cirulli, Alexandre Bolze, Ary Ascencio, Charlotte Rivera-Garcia, Ryan Cho, Jason Nguyen, Sherry Wang, Jimmy Ramirez, Tyler Cassens, Efren Sandoval, Magnus Isaksson, William Lee, David Becker, Marc Laurent, James Lu, Clinton R. Paden, Duncan MacCannell |
| EPI_ISL_1679941 | UW Virology Lab | UW Virology Lab | Pavitra Roychoudhury, Hong Xie, Lasata Shrestha, Shah Mohamed Bakhash, Michelle Lin, Noah R. Baker, Sean Ellis, Meei-Li Huang, Keith R Jerome, Alexander Greninger |
| EPI_ISL_1679942, EPI_ISL_1679943 | Helix/Illumina | Centers for Disease Control and Prevention Division of Viral Diseases, Pathogen Discovery | Dakota Howard, Dhwani Batra, Peter W. Cook, Kara Moser, Adrian Paskey, Jason Caravas, Benjamin Rambo-Martin, Shatavia Morrison, Christopher Gulvick, Scott Sammons, Yvette Unoarumhi, Darlene Wagner, Matthew Schmerer, Eileen de Feo, Jan Antico, Christine Tran, Matthew Tolentino, Shannon Wickline, Kim Gietzen, Brad Sickler, Jingtao Liu, Eric Allen, Phil Febbo, Nicole L. Washington, Simon White, Geraint Levan, Kelly Schiabor Barrett, Elizabeth Cirulli, Alexandre Bolze, Ary Ascencio, Charlotte Rivera-Garcia, Ryan Cho, Jason Nguyen, Sherry Wang, Jimmy Ramirez, Tyler Cassens, Efren Sandoval, Magnus Isaksson, William Lee, David Becker, Marc Laurent, James Lu, Clinton R. Paden, Duncan MacCannell |
| EPI_ISL_1679944 | UW Virology Lab | UW Virology Lab | Pavitra Roychoudhury, Hong Xie, Lasata Shrestha, Shah Mohamed Bakhash, Michelle Lin, Noah R. Baker, Sean Ellis, Meei-Li Huang, Keith R Jerome, Alexander Greninger |
| EPI_ISL_1679945, EPI_ISL_1679946 | Helix/Illumina | Centers for Disease Control and Prevention Division of Viral Diseases, Pathogen Discovery | Dakota Howard, Dhwani Batra, Peter W. Cook, Kara Moser, Adrian Paskey, Jason Caravas, Benjamin Rambo-Martin, Shatavia Morrison, Christopher Gulvick, Scott Sammons, Yvette Unoarumhi, Darlene Wagner, Matthew Schmerer, Eileen de Feo, Jan Antico, Christine Tran, Matthew Tolentino, Shannon Wickline, Kim Gietzen, Brad Sickler, Jingtao Liu, Eric Allen, Phil Febbo, Nicole L. Washington, Simon White, Geraint Levan, Kelly Schiabor Barrett, Elizabeth Cirulli, Alexandre Bolze, Ary Ascencio, Charlotte Rivera-Garcia, Ryan Cho, Jason Nguyen, Sherry Wang, Jimmy Ramirez, Tyler Cassens, Efren Sandoval, Magnus Isaksson, William Lee, David Becker, Marc Laurent, James Lu, Clinton R. Paden, Duncan MacCannell |
| EPI_ISL_1679947, EPI_ISL_1679948 | UW Virology Lab | UW Virology Lab | Pavitra Roychoudhury, Hong Xie, Lasata Shrestha, Shah Mohamed Bakhash, Michelle Lin, Noah R. Baker, Sean Ellis, Meei-Li Huang, Keith R Jerome, Alexander Greninger |
| EPI_ISL_1679949, EPI_ISL_1679950 | Helix/Illumina | Centers for Disease Control and Prevention Division of Viral Diseases, Pathogen Discovery | Dakota Howard, Dhwani Batra, Peter W. Cook, Kara Moser, Adrian Paskey, Jason Caravas, Benjamin Rambo-Martin, Shatavia Morrison, Christopher Gulvick, Scott Sammons, Yvette Unoarumhi, Darlene Wagner, Matthew Schmerer, Eileen de Feo, Jan Antico, Christine Tran, Matthew Tolentino, Shannon Wickline, Kim Gietzen, Brad Sickler, Jingtao Liu, Eric Allen, Phil Febbo, Nicole L. Washington, Simon White, Geraint Levan, Kelly Schiabor Barrett, Elizabeth Cirulli, Alexandre Bolze, Ary Ascencio, Charlotte Rivera-Garcia, Ryan Cho, Jason Nguyen, Sherry Wang, Jimmy Ramirez, Tyler Cassens, Efren Sandoval, Magnus Isaksson, William Lee, David Becker, Marc Laurent, James Lu, Clinton R. Paden, Duncan MacCannell |
| EPI_ISL_1679951 | UW Virology Lab | UW Virology Lab | Pavitra Roychoudhury, Hong Xie, Lasata Shrestha, Shah Mohamed Bakhash, Michelle Lin, Noah R. Baker, Sean Ellis, Meei-Li Huang, Keith R Jerome, Alexander Greninger |
| EPI_ISL_1679952, EPI_ISL_1679953 | Helix/Illumina | Centers for Disease Control and Prevention Division of Viral Diseases, Pathogen Discovery | Dakota Howard, Dhwani Batra, Peter W. Cook, Kara Moser, Adrian Paskey, Jason Caravas, Benjamin Rambo-Martin, Shatavia Morrison, Christopher Gulvick, Scott Sammons, Yvette Unoarumhi, Darlene Wagner, Matthew Schmerer, Eileen de Feo, Jan Antico, Christine Tran, Matthew Tolentino, Shannon Wickline, Kim Gietzen, Brad Sickler, Jingtao Liu, Eric Allen, Phil Febbo, Nicole L. Washington, Simon White, Geraint Levan, Kelly Schiabor Barrett, Elizabeth Cirulli, Alexandre Bolze, Ary Ascencio, Charlotte Rivera-Garcia, Ryan Cho, Jason Nguyen, Sherry Wang, Jimmy Ramirez, Tyler Cassens, Efren Sandoval, Magnus Isaksson, William Lee, David Becker, Marc Laurent, James Lu, Clinton R. Paden, Duncan MacCannell |
| EPI_ISL_1679954 | UW Virology Lab | UW Virology Lab | Pavitra Roychoudhury, Hong Xie, Lasata Shrestha, Shah Mohamed Bakhash, Michelle Lin, Noah R. Baker, Sean Ellis, Meei-Li Huang, Keith R Jerome, Alexander Greninger |
| EPI_ISL_1679955, EPI_ISL_1679956 | Helix/Illumina | Centers for Disease Control and Prevention Division of Viral Diseases, Pathogen Discovery | Dakota Howard, Dhwani Batra, Peter W. Cook, Kara Moser, Adrian Paskey, Jason Caravas, Benjamin Rambo-Martin, Shatavia Morrison, Christopher Gulvick, Scott Sammons, Yvette Unoarumhi, Darlene Wagner, Matthew Schmerer, Eileen de Feo, Jan Antico, Christine Tran, Matthew Tolentino, Shannon Wickline, Kim Gietzen, Brad Sickler, Jingtao Liu, Eric Allen, Phil Febbo, Nicole L. Washington, Simon White, Geraint Levan, Kelly Schiabor Barrett, Elizabeth Cirulli, Alexandre Bolze, Ary Ascencio, Charlotte Rivera-Garcia, Ryan Cho, Jason Nguyen, Sherry Wang, Jimmy Ramirez, Tyler Cassens, Efren Sandoval, Magnus Isaksson, William Lee, David Becker, Marc Laurent, James Lu, Clinton R. Paden, Duncan MacCannell |
| EPI_ISL_1679957 | UW Virology Lab | UW Virology Lab | Pavitra Roychoudhury, Hong Xie, Lasata Shrestha, Shah Mohamed Bakhash, Michelle Lin, Noah R. Baker, Sean Ellis, Meei-Li Huang, Keith R Jerome, Alexander Greninger |
| EPI_ISL_1679958, EPI_ISL_1679959 | Helix/Illumina | Centers for Disease Control and Prevention Division of Viral Diseases, Pathogen Discovery | Dakota Howard, Dhwani Batra, Peter W. Cook, Kara Moser, Adrian Paskey, Jason Caravas, Benjamin Rambo-Martin, Shatavia Morrison, Christopher Gulvick, Scott Sammons, Yvette Unoarumhi, Darlene Wagner, Matthew Schmerer, Eileen de Feo, Jan Antico, Christine Tran, Matthew Tolentino, Shannon Wickline, Kim Gietzen, Brad Sickler, Jingtao Liu, Eric Allen, Phil Febbo, Nicole L. Washington, Simon White, Geraint Levan, Kelly Schiabor Barrett, Elizabeth Cirulli, Alexandre Bolze, Ary Ascencio, Charlotte Rivera-Garcia, Ryan Cho, Jason Nguyen, Sherry Wang, Jimmy Ramirez, Tyler Cassens, Efren Sandoval, Magnus Isaksson, William Lee, David Becker, Marc Laurent, James Lu, Clinton R. Paden, Duncan MacCannell |

| | | | |
|---|---|---|---|
| EPI_ISL_1679960 | UW Virology Lab | UW Virology Lab | Pavitra Roychoudhury, Hong Xie, Lasata Shrestha, Shah Mohamed Bakhash, Michelle Lin, Noah R. Baker, Sean Ellis, Meei-Li Huang, Keith R Jerome, Alexander Greninger |
| EPI_ISL_1679961, EPI_ISL_1679962 | Helix/Illumina | Centers for Disease Control and Prevention Division of Viral Diseases, Pathogen Discovery | Dakota Howard, Dhwani Batra, Peter W. Cook, Kara Moser, Adrian Paskey, Jason Caravas, Benjamin Rambo-Martin, Shatavia Morrison, Christopher Gulvick, Scott Sammons, Yvette Unoarumhi, Darlene Wagner, Matthew Schmerer, Eileen de Feo, Jan Antico, Christine Tran, Matthew Tolentino, Shannon Wickline, Kim Gietzen, Brad Sickler, Jingtao Liu, Eric Allen, Phil Febbo, Nicole L. Washington, Simon White, Geraint Levan, Kelly Schiabor Barrett, Elizabeth Cirulli, Alexandre Bolze, Ary Ascencio, Charlotte Rivera-Garcia, Ryan Cho, Jason Nguyen, Sherry Wang, Jimmy Ramirez, Tyler Cassens, Efren Sandoval, Magnus Isaksson, William Lee, David Becker, Marc Laurent, James Lu, Clinton R. Paden, Duncan MacCannell |
| EPI_ISL_1679963 | UW Virology Lab | UW Virology Lab | Pavitra Roychoudhury, Hong Xie, Lasata Shrestha, Shah Mohamed Bakhash, Michelle Lin, Noah R. Baker, Sean Ellis, Meei-Li Huang, Keith R Jerome, Alexander Greninger |
| EPI_ISL_1679964, EPI_ISL_1679965 | Helix/Illumina | Centers for Disease Control and Prevention Division of Viral Diseases, Pathogen Discovery | Dakota Howard, Dhwani Batra, Peter W. Cook, Kara Moser, Adrian Paskey, Jason Caravas, Benjamin Rambo-Martin, Shatavia Morrison, Christopher Gulvick, Scott Sammons, Yvette Unoarumhi, Darlene Wagner, Matthew Schmerer, Eileen de Feo, Jan Antico, Christine Tran, Matthew Tolentino, Shannon Wickline, Kim Gietzen, Brad Sickler, Jingtao Liu, Eric Allen, Phil Febbo, Nicole L. Washington, Simon White, Geraint Levan, Kelly Schiabor Barrett, Elizabeth Cirulli, Alexandre Bolze, Ary Ascencio, Charlotte Rivera-Garcia, Ryan Cho, Jason Nguyen, Sherry Wang, Jimmy Ramirez, Tyler Cassens, Efren Sandoval, Magnus Isaksson, William Lee, David Becker, Marc Laurent, James Lu, Clinton R. Paden, Duncan MacCannell |
| EPI_ISL_1679966 | UW Virology Lab | UW Virology Lab | Pavitra Roychoudhury, Hong Xie, Lasata Shrestha, Shah Mohamed Bakhash, Michelle Lin, Noah R. Baker, Sean Ellis, Meei-Li Huang, Keith R Jerome, Alexander Greninger |
| EPI_ISL_1679967, EPI_ISL_1679968 | Helix/Illumina | Centers for Disease Control and Prevention Division of Viral Diseases, Pathogen Discovery | Dakota Howard, Dhwani Batra, Peter W. Cook, Kara Moser, Adrian Paskey, Jason Caravas, Benjamin Rambo-Martin, Shatavia Morrison, Christopher Gulvick, Scott Sammons, Yvette Unoarumhi, Darlene Wagner, Matthew Schmerer, Eileen de Feo, Jan Antico, Christine Tran, Matthew Tolentino, Shannon Wickline, Kim Gietzen, Brad Sickler, Jingtao Liu, Eric Allen, Phil Febbo, Nicole L. Washington, Simon White, Geraint Levan, Kelly Schiabor Barrett, Elizabeth Cirulli, Alexandre Bolze, Ary Ascencio, Charlotte Rivera-Garcia, Ryan Cho, Jason Nguyen, Sherry Wang, Jimmy Ramirez, Tyler Cassens, Efren Sandoval, Magnus Isaksson, William Lee, David Becker, Marc Laurent, James Lu, Clinton R. Paden, Duncan MacCannell |
| EPI_ISL_1679969 | UW Virology Lab | UW Virology Lab | Pavitra Roychoudhury, Hong Xie, Lasata Shrestha, Shah Mohamed Bakhash, Michelle Lin, Noah R. Baker, Sean Ellis, Meei-Li Huang, Keith R Jerome, Alexander Greninger |
| EPI_ISL_1679970, EPI_ISL_1679971 | Helix/Illumina | Centers for Disease Control and Prevention Division of Viral Diseases, Pathogen Discovery | Dakota Howard, Dhwani Batra, Peter W. Cook, Kara Moser, Adrian Paskey, Jason Caravas, Benjamin Rambo-Martin, Shatavia Morrison, Christopher Gulvick, Scott Sammons, Yvette Unoarumhi, Darlene Wagner, Matthew Schmerer, Eileen de Feo, Jan Antico, Christine Tran, Matthew Tolentino, Shannon Wickline, Kim Gietzen, Brad Sickler, Jingtao Liu, Eric Allen, Phil Febbo, Nicole L. Washington, Simon White, Geraint Levan, Kelly Schiabor Barrett, Elizabeth Cirulli, Alexandre Bolze, Ary Ascencio, Charlotte Rivera-Garcia, Ryan Cho, Jason Nguyen, Sherry Wang, Jimmy Ramirez, Tyler Cassens, Efren Sandoval, Magnus Isaksson, William Lee, David Becker, Marc Laurent, James Lu, Clinton R. Paden, Duncan MacCannell |
| EPI_ISL_1679972 | UW Virology Lab | UW Virology Lab | Pavitra Roychoudhury, Hong Xie, Lasata Shrestha, Shah Mohamed Bakhash, Michelle Lin, Noah R. Baker, Sean Ellis, Meei-Li Huang, Keith R Jerome, Alexander Greninger |
| EPI_ISL_1679973, EPI_ISL_1679974 | Helix/Illumina | Centers for Disease Control and Prevention Division of Viral Diseases, Pathogen Discovery | Dakota Howard, Dhwani Batra, Peter W. Cook, Kara Moser, Adrian Paskey, Jason Caravas, Benjamin Rambo-Martin, Shatavia Morrison, Christopher Gulvick, Scott Sammons, Yvette Unoarumhi, Darlene Wagner, Matthew Schmerer, Eileen de Feo, Jan Antico, Christine Tran, Matthew Tolentino, Shannon Wickline, Kim Gietzen, Brad Sickler, Jingtao Liu, Eric Allen, Phil Febbo, Nicole L. Washington, Simon White, Geraint Levan, Kelly Schiabor Barrett, Elizabeth Cirulli, Alexandre Bolze, Ary Ascencio, Charlotte Rivera-Garcia, Ryan Cho, Jason Nguyen, Sherry Wang, Jimmy Ramirez, Tyler Cassens, Efren Sandoval, Magnus Isaksson, William Lee, David Becker, Marc Laurent, James Lu, Clinton R. Paden, Duncan MacCannell |
| EPI_ISL_1679975 | UW Virology Lab | UW Virology Lab | Pavitra Roychoudhury, Hong Xie, Lasata Shrestha, Shah Mohamed Bakhash, Michelle Lin, Noah R. Baker, Sean Ellis, Meei-Li Huang, Keith R Jerome, Alexander Greninger |
| EPI_ISL_1679976, EPI_ISL_1679977 | Helix/Illumina | Centers for Disease Control and Prevention Division of Viral Diseases, Pathogen Discovery | Dakota Howard, Dhwani Batra, Peter W. Cook, Kara Moser, Adrian Paskey, Jason Caravas, Benjamin Rambo-Martin, Shatavia Morrison, Christopher Gulvick, Scott Sammons, Yvette Unoarumhi, Darlene Wagner, Matthew Schmerer, Eileen de Feo, Jan Antico, Christine Tran, Matthew Tolentino, Shannon Wickline, Kim Gietzen, Brad Sickler, Jingtao Liu, Eric Allen, Phil Febbo, Nicole L. Washington, Simon White, Geraint Levan, Kelly Schiabor Barrett, Elizabeth Cirulli, Alexandre Bolze, Ary Ascencio, Charlotte Rivera-Garcia, Ryan Cho, Jason Nguyen, Sherry Wang, Jimmy Ramirez, Tyler Cassens, Efren Sandoval, Magnus Isaksson, William Lee, David Becker, Marc Laurent, James Lu, Clinton R. Paden, Duncan MacCannell |
| EPI_ISL_1679978 | UW Virology Lab | UW Virology Lab | Pavitra Roychoudhury, Hong Xie, Lasata Shrestha, Shah Mohamed Bakhash, Michelle Lin, Noah R. Baker, Sean Ellis, Meei-Li Huang, Keith R Jerome, Alexander Greninger |
| EPI_ISL_1679979, EPI_ISL_1679980 | Helix/Illumina | Centers for Disease Control and Prevention Division of Viral Diseases, Pathogen Discovery | Dakota Howard, Dhwani Batra, Peter W. Cook, Kara Moser, Adrian Paskey, Jason Caravas, Benjamin Rambo-Martin, Shatavia Morrison, Christopher Gulvick, Scott Sammons, Yvette Unoarumhi, Darlene Wagner, Matthew Schmerer, Eileen de Feo, Jan Antico, Christine Tran, Matthew Tolentino, Shannon Wickline, Kim Gietzen, Brad Sickler, Jingtao Liu, Eric Allen, Phil Febbo, Nicole L. Washington, Simon White, Geraint Levan, Kelly Schiabor Barrett, Elizabeth Cirulli, Alexandre Bolze, Ary Ascencio, Charlotte Rivera-Garcia, Ryan Cho, Jason Nguyen, Sherry Wang, Jimmy Ramirez, Tyler Cassens, Efren Sandoval, Magnus Isaksson, William Lee, David Becker, Marc Laurent, James Lu, Clinton R. Paden, Duncan MacCannell |
| EPI_ISL_1679981 | UW Virology Lab | UW Virology Lab | Pavitra Roychoudhury, Hong Xie, Lasata Shrestha, Shah Mohamed Bakhash, Michelle Lin, Noah R. Baker, Sean Ellis, Meei-Li Huang, Keith R Jerome, Alexander Greninger |
| EPI_ISL_1679982, EPI_ISL_1679983 | Helix/Illumina | Centers for Disease Control and Prevention Division of Viral Diseases, Pathogen Discovery | Dakota Howard, Dhwani Batra, Peter W. Cook, Kara Moser, Adrian Paskey, Jason Caravas, Benjamin Rambo-Martin, Shatavia Morrison, Christopher Gulvick, Scott Sammons, Yvette Unoarumhi, Darlene Wagner, Matthew Schmerer, Eileen de Feo, Jan Antico, Christine Tran, Matthew Tolentino, Shannon Wickline, Kim Gietzen, Brad Sickler, Jingtao Liu, Eric Allen, Phil Febbo, Nicole L. Washington, Simon White, Geraint Levan, Kelly Schiabor Barrett, Elizabeth Cirulli, Alexandre Bolze, Ary Ascencio, Charlotte Rivera-Garcia, Ryan Cho, Jason Nguyen, Sherry Wang, Jimmy Ramirez, Tyler Cassens, Efren Sandoval, Magnus Isaksson, William Lee, David Becker, Marc Laurent, James Lu, Clinton R. Paden, Duncan MacCannell |
| EPI_ISL_1679984 | UW Virology Lab | UW Virology Lab | Pavitra Roychoudhury, Hong Xie, Lasata Shrestha, Shah Mohamed Bakhash, Michelle Lin, Noah R. Baker, Sean Ellis, Meei-Li Huang, Keith R Jerome, Alexander Greninger |
| EPI_ISL_1679985, EPI_ISL_1679986, EPI_ISL_1679987 | Helix/Illumina | Centers for Disease Control and Prevention Division of Viral Diseases, Pathogen Discovery | Dakota Howard, Dhwani Batra, Peter W. Cook, Kara Moser, Adrian Paskey, Jason Caravas, Benjamin Rambo-Martin, Shatavia Morrison, Christopher Gulvick, Scott Sammons, Yvette Unoarumhi, Darlene Wagner, Matthew Schmerer, Eileen de Feo, Jan Antico, Christine Tran, Matthew Tolentino, Shannon Wickline, Kim Gietzen, Brad Sickler, Jingtao Liu, Eric Allen, Phil Febbo, Nicole L. Washington, Simon White, Geraint Levan, Kelly Schiabor Barrett, Elizabeth Cirulli, Alexandre Bolze, Ary Ascencio, Charlotte Rivera-Garcia, Ryan Cho, Jason Nguyen, Sherry Wang, Jimmy Ramirez, Tyler Cassens, Efren Sandoval, Magnus Isaksson, William Lee, David Becker, Marc Laurent, James Lu, Clinton R. Paden, Duncan MacCannell |
| EPI_ISL_1679988 | UW Virology Lab | UW Virology Lab | Pavitra Roychoudhury, Hong Xie, Lasata Shrestha, Shah Mohamed Bakhash, Michelle Lin, Noah R. Baker, Sean Ellis, Meei-Li Huang, Keith R Jerome, Alexander Greninger |
| EPI_ISL_1679989, EPI_ISL_1679990 | Helix/Illumina | Centers for Disease Control and Prevention Division of Viral Diseases, Pathogen Discovery | Dakota Howard, Dhwani Batra, Peter W. Cook, Kara Moser, Adrian Paskey, Jason Caravas, Benjamin Rambo-Martin, Shatavia Morrison, Christopher Gulvick, Scott Sammons, Yvette Unoarumhi, Darlene Wagner, Matthew Schmerer, Eileen de Feo, Jan Antico, Christine Tran, Matthew Tolentino, Shannon Wickline, Kim Gietzen, Brad Sickler, Jingtao Liu, Eric Allen, Phil Febbo, Nicole L. Washington, Simon White, Geraint Levan, Kelly Schiabor Barrett, Elizabeth Cirulli, Alexandre Bolze, Ary Ascencio, Charlotte Rivera-Garcia, Ryan Cho, Jason Nguyen, Sherry Wang, Jimmy Ramirez, Tyler Cassens, Efren Sandoval, Magnus Isaksson, William Lee, David Becker, Marc Laurent, James Lu, Clinton R. Paden, Duncan MacCannell |
| EPI_ISL_1679991 | UW Virology Lab | UW Virology Lab | Pavitra Roychoudhury, Hong Xie, Lasata Shrestha, Shah Mohamed Bakhash, Michelle Lin, Noah R. Baker, Sean Ellis, Meei-Li Huang, Keith R Jerome, Alexander Greninger |
| EPI_ISL_1679992, EPI_ISL_1679993 | Helix/Illumina | Centers for Disease Control and Prevention Division of Viral Diseases, Pathogen Discovery | Dakota Howard, Dhwani Batra, Peter W. Cook, Kara Moser, Adrian Paskey, Jason Caravas, Benjamin Rambo-Martin, Shatavia Morrison, Christopher Gulvick, Scott Sammons, Yvette Unoarumhi, Darlene Wagner, Matthew Schmerer, Eileen de Feo, Jan Antico, Christine Tran, Matthew Tolentino, Shannon Wickline, Kim Gietzen, Brad Sickler, Jingtao Liu, Eric Allen, Phil Febbo, Nicole L. Washington, Simon White, Geraint Levan, Kelly Schiabor Barrett, Elizabeth Cirulli, Alexandre Bolze, Ary Ascencio, Charlotte Rivera-Garcia, Ryan Cho, Jason Nguyen, Sherry Wang, Jimmy Ramirez, Tyler Cassens, Efren Sandoval, Magnus Isaksson, William Lee, David Becker, Marc Laurent, James Lu, Clinton R. Paden, Duncan MacCannell |
| EPI_ISL_1679994 | UW Virology Lab | UW Virology Lab | Pavitra Roychoudhury, Hong Xie, Lasata Shrestha, Shah Mohamed Bakhash, Michelle Lin, Noah R. Baker, Sean Ellis, Meei-Li Huang, Keith R Jerome, Alexander Greninger |
| EPI_ISL_1679995, EPI_ISL_1679996 | Helix/Illumina | Centers for Disease Control and Prevention Division of Viral Diseases, Pathogen Discovery | Dakota Howard, Dhwani Batra, Peter W. Cook, Kara Moser, Adrian Paskey, Jason Caravas, Benjamin Rambo-Martin, Shatavia Morrison, Christopher Gulvick, Scott Sammons, Yvette Unoarumhi, Darlene Wagner, Matthew Schmerer, Eileen de Feo, Jan Antico, Christine Tran, Matthew Tolentino, Shannon Wickline, Kim Gietzen, Brad Sickler, Jingtao Liu, Eric Allen, Phil Febbo, Nicole L. Washington, Simon White, Geraint Levan, Kelly Schiabor Barrett, Elizabeth Cirulli, Alexandre Bolze, Ary Ascencio, Charlotte Rivera-Garcia, Ryan Cho, Jason Nguyen, Sherry Wang, Jimmy Ramirez, Tyler Cassens, Efren Sandoval, Magnus Isaksson, William Lee, David Becker, Marc Laurent, James Lu, Clinton R. Paden, Duncan MacCannell |
| EPI_ISL_1679997 | UW Virology Lab | UW Virology Lab | Pavitra Roychoudhury, Hong Xie, Lasata Shrestha, Shah Mohamed Bakhash, Michelle Lin, Noah R. Baker, Sean Ellis, Meei-Li Huang, Keith R Jerome, Alexander Greninger |
| EPI_ISL_1679998, EPI_ISL_1679999, EPI_ISL_1680000 | Helix/Illumina | Centers for Disease Control and Prevention Division of Viral Diseases, Pathogen Discovery | Dakota Howard, Dhwani Batra, Peter W. Cook, Kara Moser, Adrian Paskey, Jason Caravas, Benjamin Rambo-Martin, Shatavia Morrison, Christopher Gulvick, Scott Sammons, Yvette Unoarumhi, Darlene Wagner, Matthew Schmerer, Eileen de Feo, Jan Antico, Christine Tran, Matthew Tolentino, Shannon Wickline, Kim Gietzen, Brad Sickler, Jingtao Liu, Eric Allen, Phil Febbo, Nicole L. Washington, Simon White, Geraint Levan, Kelly Schiabor Barrett, Elizabeth Cirulli, Alexandre Bolze, Ary Ascencio, Charlotte Rivera-Garcia, Ryan Cho, Jason Nguyen, Sherry Wang, Jimmy Ramirez, Tyler Cassens, Efren Sandoval, Magnus Isaksson, William Lee, David Becker, Marc Laurent, James Lu, Clinton R. Paden, Duncan MacCannell |
| EPI_ISL_1680001 | UW Virology Lab | UW Virology Lab | Pavitra Roychoudhury, Hong Xie, Lasata Shrestha, Shah Mohamed Bakhash, Michelle Lin, Noah R. Baker, Sean Ellis, Meei-Li Huang, Keith R Jerome, Alexander Greninger |
| EPI_ISL_1680002, EPI_ISL_1680003 | Helix/Illumina | Centers for Disease Control and Prevention Division of Viral Diseases, Pathogen Discovery | Dakota Howard, Dhwani Batra, Peter W. Cook, Kara Moser, Adrian Paskey, Jason Caravas, Benjamin Rambo-Martin, Shatavia Morrison, Christopher Gulvick, Scott Sammons, Yvette Unoarumhi, Darlene Wagner, Matthew Schmerer, Eileen de Feo, Jan Antico, Christine Tran, Matthew Tolentino, Shannon Wickline, Kim Gietzen, Brad Sickler, Jingtao Liu, Eric Allen, Phil Febbo, Nicole L. Washington, Simon White, Geraint Levan, Kelly Schiabor Barrett, Elizabeth Cirulli, Alexandre Bolze, Ary Ascencio, Charlotte Rivera-Garcia, Ryan Cho, Jason Nguyen, Sherry Wang, Jimmy Ramirez, Tyler Cassens, Efren Sandoval, Magnus Isaksson, William Lee, David Becker, Marc Laurent, James Lu, Clinton R. Paden, Duncan MacCannell |
| EPI_ISL_1680004 | UW Virology Lab | UW Virology Lab | Pavitra Roychoudhury, Hong Xie, Lasata Shrestha, Shah Mohamed Bakhash, Michelle Lin, Noah R. Baker, Sean Ellis, Meei-Li Huang, Keith R Jerome, Alexander Greninger |
| EPI_ISL_1680005, EPI_ISL_1680006 | Helix/Illumina | Centers for Disease Control and Prevention Division of Viral Diseases, Pathogen Discovery | Dakota Howard, Dhwani Batra, Peter W. Cook, Kara Moser, Adrian Paskey, Jason Caravas, Benjamin Rambo-Martin, Shatavia Morrison, Christopher Gulvick, Scott Sammons, Yvette Unoarumhi, Darlene Wagner, Matthew Schmerer, Eileen de Feo, Jan Antico, Christine Tran, Matthew Tolentino, Shannon Wickline, Kim Gietzen, Brad Sickler, Jingtao Liu, Eric Allen, Phil Febbo, Nicole L. Washington, Simon White, Geraint Levan, Kelly Schiabor Barrett, Elizabeth Cirulli, Alexandre Bolze, Ary Ascencio, Charlotte Rivera-Garcia, Ryan Cho, Jason Nguyen, Sherry Wang, Jimmy Ramirez, Tyler Cassens, Efren Sandoval, Magnus Isaksson, William Lee, David Becker, Marc Laurent, James Lu, Clinton R. Paden, Duncan MacCannell |
| EPI_ISL_1680007 | UW Virology Lab | UW Virology Lab | Pavitra Roychoudhury, Hong Xie, Lasata Shrestha, Shah Mohamed Bakhash, Michelle Lin, Noah R. Baker, Sean Ellis, Meei-Li Huang, Keith R Jerome, Alexander Greninger |
| EPI_ISL_1680008, EPI_ISL_1680009 | Helix/Illumina | Centers for Disease Control and Prevention Division of Viral Diseases, Pathogen Discovery | Dakota Howard, Dhwani Batra, Peter W. Cook, Kara Moser, Adrian Paskey, Jason Caravas, Benjamin Rambo-Martin, Shatavia Morrison, Christopher Gulvick, Scott Sammons, Yvette Unoarumhi, Darlene Wagner, Matthew Schmerer, Eileen de Feo, Jan Antico, Christine Tran, Matthew Tolentino, Shannon Wickline, Kim Gietzen, Brad Sickler, Jingtao Liu, Eric Allen, Phil Febbo, Nicole L. Washington, Simon White, Geraint Levan, Kelly Schiabor Barrett, Elizabeth Cirulli, Alexandre Bolze, Ary Ascencio, Charlotte Rivera-Garcia, Ryan Cho, Jason Nguyen, Sherry Wang, Jimmy Ramirez, Tyler Cassens, Efren Sandoval, Magnus Isaksson, William Lee, David Becker, Marc Laurent, James Lu, Clinton R. Paden, Duncan MacCannell |
| EPI_ISL_1680010 | UW Virology Lab | UW Virology Lab | Pavitra Roychoudhury, Hong Xie, Lasata Shrestha, Shah Mohamed Bakhash, Michelle Lin, Noah R. Baker, Sean Ellis, Meei-Li Huang, Keith R Jerome, Alexander Greninger |
| EPI_ISL_1680011, EPI_ISL_1680012 | Helix/Illumina | Centers for Disease Control and Prevention Division of Viral Diseases, Pathogen Discovery | Dakota Howard, Dhwani Batra, Peter W. Cook, Kara Moser, Adrian Paskey, Jason Caravas, Benjamin Rambo-Martin, Shatavia Morrison, Christopher Gulvick, Scott Sammons, Yvette Unoarumhi, Darlene Wagner, Matthew Schmerer, Eileen de Feo, Jan Antico, Christine Tran, Matthew Tolentino, Shannon Wickline, Kim Gietzen, Brad Sickler, Jingtao Liu, Eric Allen, Phil Febbo, Nicole L. Washington, Simon White, Geraint Levan, Kelly Schiabor Barrett, Elizabeth Cirulli, Alexandre Bolze, Ary Ascencio, Charlotte Rivera-Garcia, Ryan Cho, Jason Nguyen, Sherry Wang, Jimmy Ramirez, Tyler Cassens, Efren Sandoval, Magnus Isaksson, William Lee, David Becker, Marc Laurent, James Lu, Clinton R. Paden, Duncan MacCannell |
| EPI_ISL_1680013 | UW Virology Lab | UW Virology Lab | Pavitra Roychoudhury, Hong Xie, Lasata Shrestha, Shah Mohamed Bakhash, Michelle Lin, Noah R. Baker, Sean Ellis, Meei-Li Huang, Keith R Jerome, Alexander Greninger |
| EPI_ISL_1680014, EPI_ISL_1680015 | Helix/Illumina | Centers for Disease Control and Prevention Division of Viral Diseases, Pathogen Discovery | Dakota Howard, Dhwani Batra, Peter W. Cook, Kara Moser, Adrian Paskey, Jason Caravas, Benjamin Rambo-Martin, Shatavia Morrison, Christopher Gulvick, Scott Sammons, Yvette Unoarumhi, Darlene Wagner, Matthew Schmerer, Eileen de Feo, Jan Antico, Christine Tran, Matthew Tolentino, Shannon Wickline, Kim Gietzen, Brad Sickler, Jingtao Liu, Eric Allen, Phil Febbo, Nicole L. Washington, Simon White, Geraint Levan, Kelly Schiabor Barrett, Elizabeth Cirulli, Alexandre Bolze, Ary Ascencio, Charlotte Rivera-Garcia, Ryan Cho, Jason Nguyen, Sherry Wang, Jimmy Ramirez, Tyler Cassens, Efren Sandoval, Magnus Isaksson, William Lee, David Becker, Marc Laurent, James Lu, Clinton R. Paden, Duncan MacCannell |
| EPI_ISL_1680016 | UW Virology Lab | UW Virology Lab | Pavitra Roychoudhury, Hong Xie, Lasata Shrestha, Shah Mohamed Bakhash, Michelle Lin, Noah R. Baker, Sean Ellis, Meei-Li Huang, Keith R Jerome, Alexander Greninger |
| EPI_ISL_1680017, EPI_ISL_1680018, EPI_ISL_1680019 | Helix/Illumina | Centers for Disease Control and Prevention Division of Viral Diseases, Pathogen Discovery | Dakota Howard, Dhwani Batra, Peter W. Cook, Kara Moser, Adrian Paskey, Jason Caravas, Benjamin Rambo-Martin, Shatavia Morrison, Christopher Gulvick, Scott Sammons, Yvette Unoarumhi, Darlene Wagner, Matthew Schmerer, Eileen de Feo, Jan Antico, Christine Tran, Matthew Tolentino, Shannon Wickline, Kim Gietzen, Brad Sickler, Jingtao Liu, Eric Allen, Phil Febbo, Nicole L. Washington, Simon White, Geraint Levan, Kelly Schiabor Barrett, Elizabeth Cirulli, Alexandre Bolze, Ary Ascencio, Charlotte Rivera-Garcia, Ryan Cho, Jason Nguyen, Sherry Wang, Jimmy Ramirez, Tyler Cassens, Efren Sandoval, Magnus Isaksson, William Lee, David Becker, Marc Laurent, James Lu, Clinton R. Paden, Duncan MacCannell |
| EPI_ISL_1680020 | UW Virology Lab | UW Virology Lab | Pavitra Roychoudhury, Hong Xie, Lasata Shrestha, Shah Mohamed Bakhash, Michelle Lin, Noah R. Baker, Sean Ellis, Meei-Li Huang, Keith R Jerome, Alexander Greninger |
| EPI_ISL_1680021, EPI_ISL_1680022 | Helix/Illumina | Centers for Disease Control and Prevention Division of Viral Diseases, Pathogen Discovery | Dakota Howard, Dhwani Batra, Peter W. Cook, Kara Moser, Adrian Paskey, Jason Caravas, Benjamin Rambo-Martin, Shatavia Morrison, Christopher Gulvick, Scott Sammons, Yvette Unoarumhi, Darlene Wagner, Matthew Schmerer, Eileen de Feo, Jan Antico, Christine Tran, Matthew Tolentino, Shannon Wickline, Kim Gietzen, Brad Sickler, Jingtao Liu, Eric Allen, Phil Febbo, Nicole L. Washington, Simon White, Geraint Levan, Kelly Schiabor Barrett, Elizabeth Cirulli, Alexandre Bolze, Ary Ascencio, Charlotte Rivera-Garcia, Ryan Cho, Jason Nguyen, Sherry Wang, Jimmy Ramirez, Tyler Cassens, Efren Sandoval, Magnus Isaksson, William Lee, David Becker, Marc Laurent, James Lu, Clinton R. Paden, Duncan MacCannell |
| EPI_ISL_1680023 | UW Virology Lab | UW Virology Lab | Pavitra Roychoudhury, Hong Xie, Lasata Shrestha, Shah Mohamed Bakhash, Michelle Lin, Noah R. Baker, Sean Ellis, Meei-Li Huang, Keith R Jerome, Alexander Greninger |
| EPI_ISL_1680024, EPI_ISL_1680025 | Helix/Illumina | Centers for Disease Control and Prevention Division of Viral Diseases, Pathogen Discovery | Dakota Howard, Dhwani Batra, Peter W. Cook, Kara Moser, Adrian Paskey, Jason Caravas, Benjamin Rambo-Martin, Shatavia Morrison, Christopher Gulvick, Scott Sammons, Yvette Unoarumhi, Darlene Wagner, Matthew Schmerer, Eileen de Feo, Jan Antico, Christine Tran, Matthew Tolentino, Shannon Wickline, Kim Gietzen, Brad Sickler, Jingtao Liu, Eric Allen, Phil Febbo, Nicole L. Washington, Simon White, Geraint Levan, Kelly Schiabor Barrett, Elizabeth Cirulli, Alexandre Bolze, Ary Ascencio, Charlotte Rivera-Garcia, Ryan Cho, Jason Nguyen, Sherry Wang, Jimmy Ramirez, Tyler Cassens, Efren Sandoval, Magnus Isaksson, William Lee, David Becker, Marc Laurent, James Lu, Clinton R. Paden, Duncan MacCannell |
| EPI_ISL_1680026 | UW Virology Lab | UW Virology Lab | Pavitra Roychoudhury, Hong Xie, Lasata Shrestha, Shah Mohamed Bakhash, Michelle Lin, Noah R. Baker, Sean Ellis, Meei-Li Huang, Keith R Jerome, Alexander Greninger |
| EPI_ISL_1680027, EPI_ISL_1680028 | Helix/Illumina | Centers for Disease Control and Prevention Division of Viral Diseases, Pathogen Discovery | Dakota Howard, Dhwani Batra, Peter W. Cook, Kara Moser, Adrian Paskey, Jason Caravas, Benjamin Rambo-Martin, Shatavia Morrison, Christopher Gulvick, Scott Sammons, Yvette Unoarumhi, Darlene Wagner, Matthew Schmerer, Eileen de Feo, Jan Antico, Christine Tran, Matthew Tolentino, Shannon Wickline, Kim Gietzen, Brad Sickler, Jingtao Liu, Eric Allen, Phil Febbo, Nicole L. Washington, Simon White, Geraint Levan, Kelly Schiabor Barrett, Elizabeth Cirulli, Alexandre Bolze, Ary Ascencio, Charlotte Rivera-Garcia, Ryan Cho, Jason Nguyen, Sherry Wang, Jimmy Ramirez, Tyler Cassens, Efren Sandoval, Magnus Isaksson, William Lee, David Becker, Marc Laurent, James Lu, Clinton R. Paden, Duncan MacCannell |
| EPI_ISL_1680029 | UW Virology Lab | UW Virology Lab | Pavitra Roychoudhury, Hong Xie, Lasata Shrestha, Shah Mohamed Bakhash, Michelle Lin, Noah R. Baker, Sean Ellis, Meei-Li Huang, Keith R Jerome, Alexander Greninger |

| | | | |
|---|---|---|---|
| EPI_ISL_1680030, EPI_ISL_1680031, EPI_ISL_1680032 | Helix/Illumina | Centers for Disease Control and Prevention Division of Viral Diseases, Pathogen Discovery | Dakota Howard, Dhwani Batra, Peter W. Cook, Kara Moser, Adrian Paskey, Jason Caravas, Benjamin Rambo-Martin, Shatavia Morrison, Christopher Gulvick, Scott Sammons, Yvette Unoarumhi, Darlene Wagner, Matthew Schmerer, Eileen de Feo, Jan Antico, Christine Tran, Matthew Tolentino, Shannon Wickline, Kim Gietzen, Brad Sickler, Jingtao Liu, Eric Allen, Phil Febbo, Nicole L. Washington, Simon White, Geraint Levan, Kelly Schiabor Barrett, Elizabeth Cirulli, Alexandre Bolze, Ary Ascencio, Charlotte Rivera-Garcia, Ryan Cho, Jason Nguyen, Sherry Wang, Jimmy Ramirez, Tyler Cassens, Efren Sandoval, Magnus Isaksson, William Lee, David Becker, Marc Laurent, James Lu, Clinton R. Paden, Duncan MacCannell |
| EPI_ISL_1680033 | UW Virology Lab | UW Virology Lab | Pavitra Roychoudhury, Hong Xie, Lasata Shrestha, Shah Mohamed Bakhash, Michelle Lin, Noah R. Baker, Sean Ellis, Meei-Li Huang, Keith R Jerome, Alexander Greninger |
| EPI_ISL_1680034, EPI_ISL_1680035 | Helix/Illumina | Centers for Disease Control and Prevention Division of Viral Diseases, Pathogen Discovery | Dakota Howard, Dhwani Batra, Peter W. Cook, Kara Moser, Adrian Paskey, Jason Caravas, Benjamin Rambo-Martin, Shatavia Morrison, Christopher Gulvick, Scott Sammons, Yvette Unoarumhi, Darlene Wagner, Matthew Schmerer, Eileen de Feo, Jan Antico, Christine Tran, Matthew Tolentino, Shannon Wickline, Kim Gietzen, Brad Sickler, Jingtao Liu, Eric Allen, Phil Febbo, Nicole L. Washington, Simon White, Geraint Levan, Kelly Schiabor Barrett, Elizabeth Cirulli, Alexandre Bolze, Ary Ascencio, Charlotte Rivera-Garcia, Ryan Cho, Jason Nguyen, Sherry Wang, Jimmy Ramirez, Tyler Cassens, Efren Sandoval, Magnus Isaksson, William Lee, David Becker, Marc Laurent, James Lu, Clinton R. Paden, Duncan MacCannell |
| EPI_ISL_1680036 | UW Virology Lab | UW Virology Lab | Pavitra Roychoudhury, Hong Xie, Lasata Shrestha, Shah Mohamed Bakhash, Michelle Lin, Noah R. Baker, Sean Ellis, Meei-Li Huang, Keith R Jerome, Alexander Greninger |
| EPI_ISL_1680037 | Helix/Illumina | Centers for Disease Control and Prevention Division of Viral Diseases, Pathogen Discovery | Dakota Howard, Dhwani Batra, Peter W. Cook, Kara Moser, Adrian Paskey, Jason Caravas, Benjamin Rambo-Martin, Shatavia Morrison, Christopher Gulvick, Scott Sammons, Yvette Unoarumhi, Darlene Wagner, Matthew Schmerer, Eileen de Feo, Jan Antico, Christine Tran, Matthew Tolentino, Shannon Wickline, Kim Gietzen, Brad Sickler, Jingtao Liu, Eric Allen, Phil Febbo, Nicole L. Washington, Simon White, Geraint Levan, Kelly Schiabor Barrett, Elizabeth Cirulli, Alexandre Bolze, Ary Ascencio, Charlotte Rivera-Garcia, Ryan Cho, Jason Nguyen, Sherry Wang, Jimmy Ramirez, Tyler Cassens, Efren Sandoval, Magnus Isaksson, William Lee, David Becker, Marc Laurent, James Lu, Clinton R. Paden, Duncan MacCannell |
| EPI_ISL_1680038 | Ohio Department of Health Laboratory | Ohio Department of Health Laboratory | Holmes, Jennifer; Eric Brandt, Keoni Omura, Glen McGillivary, Caitlin McDonnell, Jade Mowery, Stephanie Mccracken, Tyler Payne, Kirtana Ramadugu, Erica Leasure, Brent Lee, Kelsey Florek, Heather Blankenship, Allison Black, Quanta Brown, and Tammy Bannerman |
| EPI_ISL_1680039 | Helix/Illumina | Centers for Disease Control and Prevention Division of Viral Diseases, Pathogen Discovery | Dakota Howard, Dhwani Batra, Peter W. Cook, Kara Moser, Adrian Paskey, Jason Caravas, Benjamin Rambo-Martin, Shatavia Morrison, Christopher Gulvick, Scott Sammons, Yvette Unoarumhi, Darlene Wagner, Matthew Schmerer, Eileen de Feo, Jan Antico, Christine Tran, Matthew Tolentino, Shannon Wickline, Kim Gietzen, Brad Sickler, Jingtao Liu, Eric Allen, Phil Febbo, Nicole L. Washington, Simon White, Geraint Levan, Kelly Schiabor Barrett, Elizabeth Cirulli, Alexandre Bolze, Ary Ascencio, Charlotte Rivera-Garcia, Ryan Cho, Jason Nguyen, Sherry Wang, Jimmy Ramirez, Tyler Cassens, Efren Sandoval, Magnus Isaksson, William Lee, David Becker, Marc Laurent, James Lu, Clinton R. Paden, Duncan MacCannell |
| EPI_ISL_1680040 | UW Virology Lab | UW Virology Lab | Pavitra Roychoudhury, Hong Xie, Lasata Shrestha, Shah Mohamed Bakhash, Michelle Lin, Noah R. Baker, Sean Ellis, Meei-Li Huang, Keith R Jerome, Alexander Greninger |
| EPI_ISL_1680041, EPI_ISL_1680042 | Helix/Illumina | Centers for Disease Control and Prevention Division of Viral Diseases, Pathogen Discovery | Dakota Howard, Dhwani Batra, Peter W. Cook, Kara Moser, Adrian Paskey, Jason Caravas, Benjamin Rambo-Martin, Shatavia Morrison, Christopher Gulvick, Scott Sammons, Yvette Unoarumhi, Darlene Wagner, Matthew Schmerer, Eileen de Feo, Jan Antico, Christine Tran, Matthew Tolentino, Shannon Wickline, Kim Gietzen, Brad Sickler, Jingtao Liu, Eric Allen, Phil Febbo, Nicole L. Washington, Simon White, Geraint Levan, Kelly Schiabor Barrett, Elizabeth Cirulli, Alexandre Bolze, Ary Ascencio, Charlotte Rivera-Garcia, Ryan Cho, Jason Nguyen, Sherry Wang, Jimmy Ramirez, Tyler Cassens, Efren Sandoval, Magnus Isaksson, William Lee, David Becker, Marc Laurent, James Lu, Clinton R. Paden, Duncan MacCannell |
| EPI_ISL_1680043 | UW Virology Lab | UW Virology Lab | Pavitra Roychoudhury, Hong Xie, Lasata Shrestha, Shah Mohamed Bakhash, Michelle Lin, Noah R. Baker, Sean Ellis, Meei-Li Huang, Keith R Jerome, Alexander Greninger |
| EPI_ISL_1680044, EPI_ISL_1680045, EPI_ISL_1680046 | Helix/Illumina | Centers for Disease Control and Prevention Division of Viral Diseases, Pathogen Discovery | Dakota Howard, Dhwani Batra, Peter W. Cook, Kara Moser, Adrian Paskey, Jason Caravas, Benjamin Rambo-Martin, Shatavia Morrison, Christopher Gulvick, Scott Sammons, Yvette Unoarumhi, Darlene Wagner, Matthew Schmerer, Eileen de Feo, Jan Antico, Christine Tran, Matthew Tolentino, Shannon Wickline, Kim Gietzen, Brad Sickler, Jingtao Liu, Eric Allen, Phil Febbo, Nicole L. Washington, Simon White, Geraint Levan, Kelly Schiabor Barrett, Elizabeth Cirulli, Alexandre Bolze, Ary Ascencio, Charlotte Rivera-Garcia, Ryan Cho, Jason Nguyen, Sherry Wang, Jimmy Ramirez, Tyler Cassens, Efren Sandoval, Magnus Isaksson, William Lee, David Becker, Marc Laurent, James Lu, Clinton R. Paden, Duncan MacCannell |
| EPI_ISL_1680047 | UW Virology Lab | UW Virology Lab | Pavitra Roychoudhury, Hong Xie, Lasata Shrestha, Shah Mohamed Bakhash, Michelle Lin, Noah R. Baker, Sean Ellis, Meei-Li Huang, Keith R Jerome, Alexander Greninger |
| EPI_ISL_1680048, EPI_ISL_1680049 | Helix/Illumina | Centers for Disease Control and Prevention Division of Viral Diseases, Pathogen Discovery | Dakota Howard, Dhwani Batra, Peter W. Cook, Kara Moser, Adrian Paskey, Jason Caravas, Benjamin Rambo-Martin, Shatavia Morrison, Christopher Gulvick, Scott Sammons, Yvette Unoarumhi, Darlene Wagner, Matthew Schmerer, Eileen de Feo, Jan Antico, Christine Tran, Matthew Tolentino, Shannon Wickline, Kim Gietzen, Brad Sickler, Jingtao Liu, Eric Allen, Phil Febbo, Nicole L. Washington, Simon White, Geraint Levan, Kelly Schiabor Barrett, Elizabeth Cirulli, Alexandre Bolze, Ary Ascencio, Charlotte Rivera-Garcia, Ryan Cho, Jason Nguyen, Sherry Wang, Jimmy Ramirez, Tyler Cassens, Efren Sandoval, Magnus Isaksson, William Lee, David Becker, Marc Laurent, James Lu, Clinton R. Paden, Duncan MacCannell |
| EPI_ISL_1680050 | UW Virology Lab | UW Virology Lab | Pavitra Roychoudhury, Hong Xie, Lasata Shrestha, Shah Mohamed Bakhash, Michelle Lin, Noah R. Baker, Sean Ellis, Meei-Li Huang, Keith R Jerome, Alexander Greninger |
| EPI_ISL_1680051, EPI_ISL_1680052, EPI_ISL_1680054, EPI_ISL_1680055 | Helix/Illumina | Centers for Disease Control and Prevention Division of Viral Diseases, Pathogen Discovery | Dakota Howard, Dhwani Batra, Peter W. Cook, Kara Moser, Adrian Paskey, Jason Caravas, Benjamin Rambo-Martin, Shatavia Morrison, Christopher Gulvick, Scott Sammons, Yvette Unoarumhi, Darlene Wagner, Matthew Schmerer, Eileen de Feo, Jan Antico, Christine Tran, Matthew Tolentino, Shannon Wickline, Kim Gietzen, Brad Sickler, Jingtao Liu, Eric Allen, Phil Febbo, Nicole L. Washington, Simon White, Geraint Levan, Kelly Schiabor Barrett, Elizabeth Cirulli, Alexandre Bolze, Ary Ascencio, Charlotte Rivera-Garcia, Ryan Cho, Jason Nguyen, Sherry Wang, Jimmy Ramirez, Tyler Cassens, Efren Sandoval, Magnus Isaksson, William Lee, David Becker, Marc Laurent, James Lu, Clinton R. Paden, Duncan MacCannell |
| EPI_ISL_1680056 | UW Virology Lab | UW Virology Lab | Pavitra Roychoudhury, Hong Xie, Lasata Shrestha, Shah Mohamed Bakhash, Michelle Lin, Noah R. Baker, Sean Ellis, Meei-Li Huang, Keith R Jerome, Alexander Greninger |
| EPI_ISL_1680057, EPI_ISL_1680058 | Helix/Illumina | Centers for Disease Control and Prevention Division of Viral Diseases, Pathogen Discovery | Dakota Howard, Dhwani Batra, Peter W. Cook, Kara Moser, Adrian Paskey, Jason Caravas, Benjamin Rambo-Martin, Shatavia Morrison, Christopher Gulvick, Scott Sammons, Yvette Unoarumhi, Darlene Wagner, Matthew Schmerer, Eileen de Feo, Jan Antico, Christine Tran, Matthew Tolentino, Shannon Wickline, Kim Gietzen, Brad Sickler, Jingtao Liu, Eric Allen, Phil Febbo, Nicole L. Washington, Simon White, Geraint Levan, Kelly Schiabor Barrett, Elizabeth Cirulli, Alexandre Bolze, Ary Ascencio, Charlotte Rivera-Garcia, Ryan Cho, Jason Nguyen, Sherry Wang, Jimmy Ramirez, Tyler Cassens, Efren Sandoval, Magnus Isaksson, William Lee, David Becker, Marc Laurent, James Lu, Clinton R. Paden, Duncan MacCannell |
| EPI_ISL_1680059 | UW Virology Lab | UW Virology Lab | Pavitra Roychoudhury, Hong Xie, Lasata Shrestha, Shah Mohamed Bakhash, Michelle Lin, Noah R. Baker, Sean Ellis, Meei-Li Huang, Keith R Jerome, Alexander Greninger |
| EPI_ISL_1680060, EPI_ISL_1680061, EPI_ISL_1680062 | Helix/Illumina | Centers for Disease Control and Prevention Division of Viral Diseases, Pathogen Discovery | Dakota Howard, Dhwani Batra, Peter W. Cook, Kara Moser, Adrian Paskey, Jason Caravas, Benjamin Rambo-Martin, Shatavia Morrison, Christopher Gulvick, Scott Sammons, Yvette Unoarumhi, Darlene Wagner, Matthew Schmerer, Eileen de Feo, Jan Antico, Christine Tran, Matthew Tolentino, Shannon Wickline, Kim Gietzen, Brad Sickler, Jingtao Liu, Eric Allen, Phil Febbo, Nicole L. Washington, Simon White, Geraint Levan, Kelly Schiabor Barrett, Elizabeth Cirulli, Alexandre Bolze, Ary Ascencio, Charlotte Rivera-Garcia, Ryan Cho, Jason Nguyen, Sherry Wang, Jimmy Ramirez, Tyler Cassens, Efren Sandoval, Magnus Isaksson, William Lee, David Becker, Marc Laurent, James Lu, Clinton R. Paden, Duncan MacCannell |
| EPI_ISL_1680063 | UW Virology Lab | UW Virology Lab | Pavitra Roychoudhury, Hong Xie, Lasata Shrestha, Shah Mohamed Bakhash, Michelle Lin, Noah R. Baker, Sean Ellis, Meei-Li Huang, Keith R Jerome, Alexander Greninger |
| EPI_ISL_1680064 | Helix/Illumina | Centers for Disease Control and Prevention Division of Viral Diseases, Pathogen Discovery | Dakota Howard, Dhwani Batra, Peter W. Cook, Kara Moser, Adrian Paskey, Jason Caravas, Benjamin Rambo-Martin, Shatavia Morrison, Christopher Gulvick, Scott Sammons, Yvette Unoarumhi, Darlene Wagner, Matthew Schmerer, Eileen de Feo, Jan Antico, Christine Tran, Matthew Tolentino, Shannon Wickline, Kim Gietzen, Brad Sickler, Jingtao Liu, Eric Allen, Phil Febbo, Nicole L. Washington, Simon White, Geraint Levan, Kelly Schiabor Barrett, Elizabeth Cirulli, Alexandre Bolze, Ary Ascencio, Charlotte Rivera-Garcia, Ryan Cho, Jason Nguyen, Sherry Wang, Jimmy Ramirez, Tyler Cassens, Efren Sandoval, Magnus Isaksson, William Lee, David Becker, Marc Laurent, James Lu, Clinton R. Paden, Duncan MacCannell |
| EPI_ISL_1680065 | UW Virology Lab | UW Virology Lab | Pavitra Roychoudhury, Hong Xie, Lasata Shrestha, Shah Mohamed Bakhash, Michelle Lin, Noah R. Baker, Sean Ellis, Meei-Li Huang, Keith R Jerome, Alexander Greninger |
| EPI_ISL_1680066, EPI_ISL_1680067 | Helix/Illumina | Centers for Disease Control and Prevention Division of Viral Diseases, Pathogen Discovery | Dakota Howard, Dhwani Batra, Peter W. Cook, Kara Moser, Adrian Paskey, Jason Caravas, Benjamin Rambo-Martin, Shatavia Morrison, Christopher Gulvick, Scott Sammons, Yvette Unoarumhi, Darlene Wagner, Matthew Schmerer, Eileen de Feo, Jan Antico, Christine Tran, Matthew Tolentino, Shannon Wickline, Kim Gietzen, Brad Sickler, Jingtao Liu, Eric Allen, Phil Febbo, Nicole L. Washington, Simon White, Geraint Levan, Kelly Schiabor Barrett, Elizabeth Cirulli, Alexandre Bolze, Ary Ascencio, Charlotte Rivera-Garcia, Ryan Cho, Jason Nguyen, Sherry Wang, Jimmy Ramirez, Tyler Cassens, Efren Sandoval, Magnus Isaksson, William Lee, David Becker, Marc Laurent, James Lu, Clinton R. Paden, Duncan MacCannell |
| EPI_ISL_1680068 | UW Virology Lab | UW Virology Lab | Pavitra Roychoudhury, Hong Xie, Lasata Shrestha, Shah Mohamed Bakhash, Michelle Lin, Noah R. Baker, Sean Ellis, Meei-Li Huang, Keith R Jerome, Alexander Greninger |
| EPI_ISL_1680069, EPI_ISL_1680070 | Helix/Illumina | Centers for Disease Control and Prevention Division of Viral Diseases, Pathogen Discovery | Dakota Howard, Dhwani Batra, Peter W. Cook, Kara Moser, Adrian Paskey, Jason Caravas, Benjamin Rambo-Martin, Shatavia Morrison, Christopher Gulvick, Scott Sammons, Yvette Unoarumhi, Darlene Wagner, Matthew Schmerer, Eileen de Feo, Jan Antico, Christine Tran, Matthew Tolentino, Shannon Wickline, Kim Gietzen, Brad Sickler, Jingtao Liu, Eric Allen, Phil Febbo, Nicole L. Washington, Simon White, Geraint Levan, Kelly Schiabor Barrett, Elizabeth Cirulli, Alexandre Bolze, Ary Ascencio, Charlotte Rivera-Garcia, Ryan Cho, Jason Nguyen, Sherry Wang, Jimmy Ramirez, Tyler Cassens, Efren Sandoval, Magnus Isaksson, William Lee, David Becker, Marc Laurent, James Lu, Clinton R. Paden, Duncan MacCannell |
| EPI_ISL_1680071 | UW Virology Lab | UW Virology Lab | Pavitra Roychoudhury, Hong Xie, Lasata Shrestha, Shah Mohamed Bakhash, Michelle Lin, Noah R. Baker, Sean Ellis, Meei-Li Huang, Keith R Jerome, Alexander Greninger |
| EPI_ISL_1680072, EPI_ISL_1680073 | Helix/Illumina | Centers for Disease Control and Prevention Division of Viral Diseases, Pathogen Discovery | Dakota Howard, Dhwani Batra, Peter W. Cook, Kara Moser, Adrian Paskey, Jason Caravas, Benjamin Rambo-Martin, Shatavia Morrison, Christopher Gulvick, Scott Sammons, Yvette Unoarumhi, Darlene Wagner, Matthew Schmerer, Eileen de Feo, Jan Antico, Christine Tran, Matthew Tolentino, Shannon Wickline, Kim Gietzen, Brad Sickler, Jingtao Liu, Eric Allen, Phil Febbo, Nicole L. Washington, Simon White, Geraint Levan, Kelly Schiabor Barrett, Elizabeth Cirulli, Alexandre Bolze, Ary Ascencio, Charlotte Rivera-Garcia, Ryan Cho, Jason Nguyen, Sherry Wang, Jimmy Ramirez, Tyler Cassens, Efren Sandoval, Magnus Isaksson, William Lee, David Becker, Marc Laurent, James Lu, Clinton R. Paden, Duncan MacCannell |
| EPI_ISL_1680074 | UW Virology Lab | UW Virology Lab | Pavitra Roychoudhury, Hong Xie, Lasata Shrestha, Shah Mohamed Bakhash, Michelle Lin, Noah R. Baker, Sean Ellis, Meei-Li Huang, Keith R Jerome, Alexander Greninger |
| EPI_ISL_1680075, EPI_ISL_1680076 | Helix/Illumina | Centers for Disease Control and Prevention Division of Viral Diseases, Pathogen Discovery | Dakota Howard, Dhwani Batra, Peter W. Cook, Kara Moser, Adrian Paskey, Jason Caravas, Benjamin Rambo-Martin, Shatavia Morrison, Christopher Gulvick, Scott Sammons, Yvette Unoarumhi, Darlene Wagner, Matthew Schmerer, Eileen de Feo, Jan Antico, Christine Tran, Matthew Tolentino, Shannon Wickline, Kim Gietzen, Brad Sickler, Jingtao Liu, Eric Allen, Phil Febbo, Nicole L. Washington, Simon White, Geraint Levan, Kelly Schiabor Barrett, Elizabeth Cirulli, Alexandre Bolze, Ary Ascencio, Charlotte Rivera-Garcia, Ryan Cho, Jason Nguyen, Sherry Wang, Jimmy Ramirez, Tyler Cassens, Efren Sandoval, Magnus Isaksson, William Lee, David Becker, Marc Laurent, James Lu, Clinton R. Paden, Duncan MacCannell |
| EPI_ISL_1680077 | UW Virology Lab | UW Virology Lab | Pavitra Roychoudhury, Hong Xie, Lasata Shrestha, Shah Mohamed Bakhash, Michelle Lin, Noah R. Baker, Sean Ellis, Meei-Li Huang, Keith R Jerome, Alexander Greninger |
| EPI_ISL_1680078, EPI_ISL_1680079 | Helix/Illumina | Centers for Disease Control and Prevention Division of Viral Diseases, Pathogen Discovery | Dakota Howard, Dhwani Batra, Peter W. Cook, Kara Moser, Adrian Paskey, Jason Caravas, Benjamin Rambo-Martin, Shatavia Morrison, Christopher Gulvick, Scott Sammons, Yvette Unoarumhi, Darlene Wagner, Matthew Schmerer, Eileen de Feo, Jan Antico, Christine Tran, Matthew Tolentino, Shannon Wickline, Kim Gietzen, Brad Sickler, Jingtao Liu, Eric Allen, Phil Febbo, Nicole L. Washington, Simon White, Geraint Levan, Kelly Schiabor Barrett, Elizabeth Cirulli, Alexandre Bolze, Ary Ascencio, Charlotte Rivera-Garcia, Ryan Cho, Jason Nguyen, Sherry Wang, Jimmy Ramirez, Tyler Cassens, Efren Sandoval, Magnus Isaksson, William Lee, David Becker, Marc Laurent, James Lu, Clinton R. Paden, Duncan MacCannell |
| EPI_ISL_1680080 | UW Virology Lab | UW Virology Lab | Pavitra Roychoudhury, Hong Xie, Lasata Shrestha, Shah Mohamed Bakhash, Michelle Lin, Noah R. Baker, Sean Ellis, Meei-Li Huang, Keith R Jerome, Alexander Greninger |
| EPI_ISL_1680081, EPI_ISL_1680082 | Helix/Illumina | Centers for Disease Control and Prevention Division of Viral Diseases, Pathogen Discovery | Dakota Howard, Dhwani Batra, Peter W. Cook, Kara Moser, Adrian Paskey, Jason Caravas, Benjamin Rambo-Martin, Shatavia Morrison, Christopher Gulvick, Scott Sammons, Yvette Unoarumhi, Darlene Wagner, Matthew Schmerer, Eileen de Feo, Jan Antico, Christine Tran, Matthew Tolentino, Shannon Wickline, Kim Gietzen, Brad Sickler, Jingtao Liu, Eric Allen, Phil Febbo, Nicole L. Washington, Simon White, Geraint Levan, Kelly Schiabor Barrett, Elizabeth Cirulli, Alexandre Bolze, Ary Ascencio, Charlotte Rivera-Garcia, Ryan Cho, Jason Nguyen, Sherry Wang, Jimmy Ramirez, Tyler Cassens, Efren Sandoval, Magnus Isaksson, William Lee, David Becker, Marc Laurent, James Lu, Clinton R. Paden, Duncan MacCannell |
| EPI_ISL_1680083 | UW Virology Lab | UW Virology Lab | Pavitra Roychoudhury, Hong Xie, Lasata Shrestha, Shah Mohamed Bakhash, Michelle Lin, Noah R. Baker, Sean Ellis, Meei-Li Huang, Keith R Jerome, Alexander Greninger |
| EPI_ISL_1680084, EPI_ISL_1680085, EPI_ISL_1680087, EPI_ISL_1680088, EPI_ISL_1680089 | Helix/Illumina | Centers for Disease Control and Prevention Division of Viral Diseases, Pathogen Discovery | Dakota Howard, Dhwani Batra, Peter W. Cook, Kara Moser, Adrian Paskey, Jason Caravas, Benjamin Rambo-Martin, Shatavia Morrison, Christopher Gulvick, Scott Sammons, Yvette Unoarumhi, Darlene Wagner, Matthew Schmerer, Eileen de Feo, Jan Antico, Christine Tran, Matthew Tolentino, Shannon Wickline, Kim Gietzen, Brad Sickler, Jingtao Liu, Eric Allen, Phil Febbo, Nicole L. Washington, Simon White, Geraint Levan, Kelly Schiabor Barrett, Elizabeth Cirulli, Alexandre Bolze, Ary Ascencio, Charlotte Rivera-Garcia, Ryan Cho, Jason Nguyen, Sherry Wang, Jimmy Ramirez, Tyler Cassens, Efren Sandoval, Magnus Isaksson, William Lee, David Becker, Marc Laurent, James Lu, Clinton R. Paden, Duncan MacCannell |
| EPI_ISL_1680090 | UW Virology Lab | UW Virology Lab | Pavitra Roychoudhury, Hong Xie, Lasata Shrestha, Shah Mohamed Bakhash, Michelle Lin, Noah R. Baker, Sean Ellis, Meei-Li Huang, Keith R Jerome, Alexander Greninger |
| EPI_ISL_1680091, EPI_ISL_1680092 | Helix/Illumina | Centers for Disease Control and Prevention Division of Viral Diseases, Pathogen Discovery | Dakota Howard, Dhwani Batra, Peter W. Cook, Kara Moser, Adrian Paskey, Jason Caravas, Benjamin Rambo-Martin, Shatavia Morrison, Christopher Gulvick, Scott Sammons, Yvette Unoarumhi, Darlene Wagner, Matthew Schmerer, Eileen de Feo, Jan Antico, Christine Tran, Matthew Tolentino, Shannon Wickline, Kim Gietzen, Brad Sickler, Jingtao Liu, Eric Allen, Phil Febbo, Nicole L. Washington, Simon White, Geraint Levan, Kelly Schiabor Barrett, Elizabeth Cirulli, Alexandre Bolze, Ary Ascencio, Charlotte Rivera-Garcia, Ryan Cho, Jason Nguyen, Sherry Wang, Jimmy Ramirez, Tyler Cassens, Efren Sandoval, Magnus Isaksson, William Lee, David Becker, Marc Laurent, James Lu, Clinton R. Paden, Duncan MacCannell |
| EPI_ISL_1680093 | UW Virology Lab | UW Virology Lab | Pavitra Roychoudhury, Hong Xie, Lasata Shrestha, Shah Mohamed Bakhash, Michelle Lin, Noah R. Baker, Sean Ellis, Meei-Li Huang, Keith R Jerome, Alexander Greninger |
| EPI_ISL_1680094, EPI_ISL_1680095 | Helix/Illumina | Centers for Disease Control and Prevention Division of Viral Diseases, Pathogen Discovery | Dakota Howard, Dhwani Batra, Peter W. Cook, Kara Moser, Adrian Paskey, Jason Caravas, Benjamin Rambo-Martin, Shatavia Morrison, Christopher Gulvick, Scott Sammons, Yvette Unoarumhi, Darlene Wagner, Matthew Schmerer, Eileen de Feo, Jan Antico, Christine Tran, Matthew Tolentino, Shannon Wickline, Kim Gietzen, Brad Sickler, Jingtao Liu, Eric Allen, Phil Febbo, Nicole L. Washington, Simon White, Geraint Levan, Kelly Schiabor Barrett, Elizabeth Cirulli, Alexandre Bolze, Ary Ascencio, Charlotte Rivera-Garcia, Ryan Cho, Jason Nguyen, Sherry Wang, Jimmy Ramirez, Tyler Cassens, Efren Sandoval, Magnus Isaksson, William Lee, David Becker, Marc Laurent, James Lu, Clinton R. Paden, Duncan MacCannell |
| EPI_ISL_1680096 | UW Virology Lab | UW Virology Lab | Pavitra Roychoudhury, Hong Xie, Lasata Shrestha, Shah Mohamed Bakhash, Michelle Lin, Noah R. Baker, Sean Ellis, Meei-Li Huang, Keith R Jerome, Alexander Greninger |
| EPI_ISL_1680097, EPI_ISL_1680098, EPI_ISL_1680099 | Helix/Illumina | Centers for Disease Control and Prevention Division of Viral Diseases, Pathogen Discovery | Dakota Howard, Dhwani Batra, Peter W. Cook, Kara Moser, Adrian Paskey, Jason Caravas, Benjamin Rambo-Martin, Shatavia Morrison, Christopher Gulvick, Scott Sammons, Yvette Unoarumhi, Darlene Wagner, Matthew Schmerer, Eileen de Feo, Jan Antico, Christine Tran, Matthew Tolentino, Shannon Wickline, Kim Gietzen, Brad Sickler, Jingtao Liu, Eric Allen, Phil Febbo, Nicole L. Washington, Simon White, Geraint Levan, Kelly Schiabor Barrett, Elizabeth Cirulli, Alexandre Bolze, Ary Ascencio, Charlotte Rivera-Garcia, Ryan Cho, Jason Nguyen, Sherry Wang, Jimmy Ramirez, Tyler Cassens, Efren Sandoval, Magnus Isaksson, William Lee, David Becker, Marc Laurent, James Lu, Clinton R. Paden, Duncan MacCannell |
| EPI_ISL_1680100 | UW Virology Lab | UW Virology Lab | Pavitra Roychoudhury, Hong Xie, Lasata Shrestha, Shah Mohamed Bakhash, Michelle Lin, Noah R. Baker, Sean Ellis, Meei-Li Huang, Keith R Jerome, Alexander Greninger |
| EPI_ISL_1680101, EPI_ISL_1680102 | Helix/Illumina | Centers for Disease Control and Prevention Division of Viral Diseases, Pathogen Discovery | Dakota Howard, Dhwani Batra, Peter W. Cook, Kara Moser, Adrian Paskey, Jason Caravas, Benjamin Rambo-Martin, Shatavia Morrison, Christopher Gulvick, Scott Sammons, Yvette Unoarumhi, Darlene Wagner, Matthew Schmerer, Eileen de Feo, Jan Antico, Christine Tran, Matthew Tolentino, Shannon Wickline, Kim Gietzen, Brad Sickler, Jingtao Liu, Eric Allen, Phil Febbo, Nicole L. Washington, Simon White, Geraint Levan, Kelly Schiabor Barrett, Elizabeth Cirulli, Alexandre Bolze, Ary Ascencio, Charlotte Rivera-Garcia, Ryan Cho, Jason Nguyen, Sherry Wang, Jimmy Ramirez, Tyler Cassens, Efren Sandoval, Magnus Isaksson, William Lee, David Becker, Marc Laurent, James Lu, Clinton R. Paden, Duncan MacCannell |
| EPI_ISL_1680103 | UW Virology Lab | UW Virology Lab | Pavitra Roychoudhury, Hong Xie, Lasata Shrestha, Shah Mohamed Bakhash, Michelle Lin, Noah R. Baker, Sean Ellis, Meei-Li Huang, Keith R Jerome, Alexander Greninger |

| | | | |
|---|---|---|---|
| EPI_ISL_1680104, EPI_ISL_1680105, EPI_ISL_1680106 | Helix/Illumina | Centers for Disease Control and Prevention Division of Viral Diseases, Pathogen Discovery | Dakota Howard, Dhwani Batra, Peter W. Cook, Kara Moser, Adrian Paskey, Jason Caravas, Benjamin Rambo-Martin, Shatavia Morrison, Christopher Gulvick, Scott Sammons, Yvette Unoarumhi, Darlene Wagner, Matthew Schmerer, Eileen de Feo, Jan Antico, Christine Tran, Matthew Tolentino, Shannon Wickline, Kim Gietzen, Brad Sickler, Jingtao Liu, Eric Allen, Phil Febbo, Nicole L. Washington, Simon White, Geraint Levan, Kelly Schiabor Barrett, Elizabeth Cirulli, Alexandre Bolze, Ary Ascencio, Charlotte Rivera-Garcia, Ryan Cho, Jason Nguyen, Sherry Wang, Jimmy Ramirez, Tyler Cassens, Efren Sandoval, Magnus Isaksson, William Lee, David Becker, Marc Laurent, James Lu, Clinton R. Paden, Duncan MacCannell |
| EPI_ISL_1680107 | UW Virology Lab | UW Virology Lab | Pavitra Roychoudhury, Hong Xie, Lasata Shrestha, Shah Mohamed Bakhash, Michelle Lin, Noah R. Baker, Sean Ellis, Meei-Li Huang, Keith R Jerome, Alexander Greninger |
| EPI_ISL_1680108 | Helix/Illumina | Centers for Disease Control and Prevention Division of Viral Diseases, Pathogen Discovery | Dakota Howard, Dhwani Batra, Peter W. Cook, Kara Moser, Adrian Paskey, Jason Caravas, Benjamin Rambo-Martin, Shatavia Morrison, Christopher Gulvick, Scott Sammons, Yvette Unoarumhi, Darlene Wagner, Matthew Schmerer, Eileen de Feo, Jan Antico, Christine Tran, Matthew Tolentino, Shannon Wickline, Kim Gietzen, Brad Sickler, Jingtao Liu, Eric Allen, Phil Febbo, Nicole L. Washington, Simon White, Geraint Levan, Kelly Schiabor Barrett, Elizabeth Cirulli, Alexandre Bolze, Ary Ascencio, Charlotte Rivera-Garcia, Ryan Cho, Jason Nguyen, Sherry Wang, Jimmy Ramirez, Tyler Cassens, Efren Sandoval, Magnus Isaksson, William Lee, David Becker, Marc Laurent, James Lu, Clinton R. Paden, Duncan MacCannell |
| EPI_ISL_1680109 | UW Virology Lab | UW Virology Lab | Pavitra Roychoudhury, Hong Xie, Lasata Shrestha, Shah Mohamed Bakhash, Michelle Lin, Noah R. Baker, Sean Ellis, Meei-Li Huang, Keith R Jerome, Alexander Greninger |
| EPI_ISL_1680110, EPI_ISL_1680111 | Helix/Illumina | Centers for Disease Control and Prevention Division of Viral Diseases, Pathogen Discovery | Dakota Howard, Dhwani Batra, Peter W. Cook, Kara Moser, Adrian Paskey, Jason Caravas, Benjamin Rambo-Martin, Shatavia Morrison, Christopher Gulvick, Scott Sammons, Yvette Unoarumhi, Darlene Wagner, Matthew Schmerer, Eileen de Feo, Jan Antico, Christine Tran, Matthew Tolentino, Shannon Wickline, Kim Gietzen, Brad Sickler, Jingtao Liu, Eric Allen, Phil Febbo, Nicole L. Washington, Simon White, Geraint Levan, Kelly Schiabor Barrett, Elizabeth Cirulli, Alexandre Bolze, Ary Ascencio, Charlotte Rivera-Garcia, Ryan Cho, Jason Nguyen, Sherry Wang, Jimmy Ramirez, Tyler Cassens, Efren Sandoval, Magnus Isaksson, William Lee, David Becker, Marc Laurent, James Lu, Clinton R. Paden, Duncan MacCannell |
| EPI_ISL_1680112 | UW Virology Lab | UW Virology Lab | Pavitra Roychoudhury, Hong Xie, Lasata Shrestha, Shah Mohamed Bakhash, Michelle Lin, Noah R. Baker, Sean Ellis, Meei-Li Huang, Keith R Jerome, Alexander Greninger |
| EPI_ISL_1680113, EPI_ISL_1680114 | Helix/Illumina | Centers for Disease Control and Prevention Division of Viral Diseases, Pathogen Discovery | Dakota Howard, Dhwani Batra, Peter W. Cook, Kara Moser, Adrian Paskey, Jason Caravas, Benjamin Rambo-Martin, Shatavia Morrison, Christopher Gulvick, Scott Sammons, Yvette Unoarumhi, Darlene Wagner, Matthew Schmerer, Eileen de Feo, Jan Antico, Christine Tran, Matthew Tolentino, Shannon Wickline, Kim Gietzen, Brad Sickler, Jingtao Liu, Eric Allen, Phil Febbo, Nicole L. Washington, Simon White, Geraint Levan, Kelly Schiabor Barrett, Elizabeth Cirulli, Alexandre Bolze, Ary Ascencio, Charlotte Rivera-Garcia, Ryan Cho, Jason Nguyen, Sherry Wang, Jimmy Ramirez, Tyler Cassens, Efren Sandoval, Magnus Isaksson, William Lee, David Becker, Marc Laurent, James Lu, Clinton R. Paden, Duncan MacCannell |
| EPI_ISL_1680115 | UW Virology Lab | UW Virology Lab | Pavitra Roychoudhury, Hong Xie, Lasata Shrestha, Shah Mohamed Bakhash, Michelle Lin, Noah R. Baker, Sean Ellis, Meei-Li Huang, Keith R Jerome, Alexander Greninger |
| EPI_ISL_1680116, EPI_ISL_1680117 | Helix/Illumina | Centers for Disease Control and Prevention Division of Viral Diseases, Pathogen Discovery | Dakota Howard, Dhwani Batra, Peter W. Cook, Kara Moser, Adrian Paskey, Jason Caravas, Benjamin Rambo-Martin, Shatavia Morrison, Christopher Gulvick, Scott Sammons, Yvette Unoarumhi, Darlene Wagner, Matthew Schmerer, Eileen de Feo, Jan Antico, Christine Tran, Matthew Tolentino, Shannon Wickline, Kim Gietzen, Brad Sickler, Jingtao Liu, Eric Allen, Phil Febbo, Nicole L. Washington, Simon White, Geraint Levan, Kelly Schiabor Barrett, Elizabeth Cirulli, Alexandre Bolze, Ary Ascencio, Charlotte Rivera-Garcia, Ryan Cho, Jason Nguyen, Sherry Wang, Jimmy Ramirez, Tyler Cassens, Efren Sandoval, Magnus Isaksson, William Lee, David Becker, Marc Laurent, James Lu, Clinton R. Paden, Duncan MacCannell |
| EPI_ISL_1680118 | UW Virology Lab | UW Virology Lab | Pavitra Roychoudhury, Hong Xie, Lasata Shrestha, Shah Mohamed Bakhash, Michelle Lin, Noah R. Baker, Sean Ellis, Meei-Li Huang, Keith R Jerome, Alexander Greninger |
| EPI_ISL_1680119, EPI_ISL_1680120, EPI_ISL_1680121 | Helix/Illumina | Centers for Disease Control and Prevention Division of Viral Diseases, Pathogen Discovery | Dakota Howard, Dhwani Batra, Peter W. Cook, Kara Moser, Adrian Paskey, Jason Caravas, Benjamin Rambo-Martin, Shatavia Morrison, Christopher Gulvick, Scott Sammons, Yvette Unoarumhi, Darlene Wagner, Matthew Schmerer, Eileen de Feo, Jan Antico, Christine Tran, Matthew Tolentino, Shannon Wickline, Kim Gietzen, Brad Sickler, Jingtao Liu, Eric Allen, Phil Febbo, Nicole L. Washington, Simon White, Geraint Levan, Kelly Schiabor Barrett, Elizabeth Cirulli, Alexandre Bolze, Ary Ascencio, Charlotte Rivera-Garcia, Ryan Cho, Jason Nguyen, Sherry Wang, Jimmy Ramirez, Tyler Cassens, Efren Sandoval, Magnus Isaksson, William Lee, David Becker, Marc Laurent, James Lu, Clinton R. Paden, Duncan MacCannell |
| EPI_ISL_1680122 | UW Virology Lab | UW Virology Lab | Pavitra Roychoudhury, Hong Xie, Lasata Shrestha, Shah Mohamed Bakhash, Michelle Lin, Noah R. Baker, Sean Ellis, Meei-Li Huang, Keith R Jerome, Alexander Greninger |
| EPI_ISL_1680123, EPI_ISL_1680124 | Helix/Illumina | Centers for Disease Control and Prevention Division of Viral Diseases, Pathogen Discovery | Dakota Howard, Dhwani Batra, Peter W. Cook, Kara Moser, Adrian Paskey, Jason Caravas, Benjamin Rambo-Martin, Shatavia Morrison, Christopher Gulvick, Scott Sammons, Yvette Unoarumhi, Darlene Wagner, Matthew Schmerer, Eileen de Feo, Jan Antico, Christine Tran, Matthew Tolentino, Shannon Wickline, Kim Gietzen, Brad Sickler, Jingtao Liu, Eric Allen, Phil Febbo, Nicole L. Washington, Simon White, Geraint Levan, Kelly Schiabor Barrett, Elizabeth Cirulli, Alexandre Bolze, Ary Ascencio, Charlotte Rivera-Garcia, Ryan Cho, Jason Nguyen, Sherry Wang, Jimmy Ramirez, Tyler Cassens, Efren Sandoval, Magnus Isaksson, William Lee, David Becker, Marc Laurent, James Lu, Clinton R. Paden, Duncan MacCannell |
| EPI_ISL_1680125 | UW Virology Lab | UW Virology Lab | Pavitra Roychoudhury, Hong Xie, Lasata Shrestha, Shah Mohamed Bakhash, Michelle Lin, Noah R. Baker, Sean Ellis, Meei-Li Huang, Keith R Jerome, Alexander Greninger |
| EPI_ISL_1680126, EPI_ISL_1680127, EPI_ISL_1680128 | Helix/Illumina | Centers for Disease Control and Prevention Division of Viral Diseases, Pathogen Discovery | Dakota Howard, Dhwani Batra, Peter W. Cook, Kara Moser, Adrian Paskey, Jason Caravas, Benjamin Rambo-Martin, Shatavia Morrison, Christopher Gulvick, Scott Sammons, Yvette Unoarumhi, Darlene Wagner, Matthew Schmerer, Eileen de Feo, Jan Antico, Christine Tran, Matthew Tolentino, Shannon Wickline, Kim Gietzen, Brad Sickler, Jingtao Liu, Eric Allen, Phil Febbo, Nicole L. Washington, Simon White, Geraint Levan, Kelly Schiabor Barrett, Elizabeth Cirulli, Alexandre Bolze, Ary Ascencio, Charlotte Rivera-Garcia, Ryan Cho, Jason Nguyen, Sherry Wang, Jimmy Ramirez, Tyler Cassens, Efren Sandoval, Magnus Isaksson, William Lee, David Becker, Marc Laurent, James Lu, Clinton R. Paden, Duncan MacCannell |
| EPI_ISL_1680129 | UW Virology Lab | UW Virology Lab | Pavitra Roychoudhury, Hong Xie, Lasata Shrestha, Shah Mohamed Bakhash, Michelle Lin, Noah R. Baker, Sean Ellis, Meei-Li Huang, Keith R Jerome, Alexander Greninger |
| EPI_ISL_1680130, EPI_ISL_1680131 | Helix/Illumina | Centers for Disease Control and Prevention Division of Viral Diseases, Pathogen Discovery | Dakota Howard, Dhwani Batra, Peter W. Cook, Kara Moser, Adrian Paskey, Jason Caravas, Benjamin Rambo-Martin, Shatavia Morrison, Christopher Gulvick, Scott Sammons, Yvette Unoarumhi, Darlene Wagner, Matthew Schmerer, Eileen de Feo, Jan Antico, Christine Tran, Matthew Tolentino, Shannon Wickline, Kim Gietzen, Brad Sickler, Jingtao Liu, Eric Allen, Phil Febbo, Nicole L. Washington, Simon White, Geraint Levan, Kelly Schiabor Barrett, Elizabeth Cirulli, Alexandre Bolze, Ary Ascencio, Charlotte Rivera-Garcia, Ryan Cho, Jason Nguyen, Sherry Wang, Jimmy Ramirez, Tyler Cassens, Efren Sandoval, Magnus Isaksson, William Lee, David Becker, Marc Laurent, James Lu, Clinton R. Paden, Duncan MacCannell |
| EPI_ISL_1680132, EPI_ISL_1680133 | UW Virology Lab | UW Virology Lab | Pavitra Roychoudhury, Hong Xie, Lasata Shrestha, Shah Mohamed Bakhash, Michelle Lin, Noah R. Baker, Sean Ellis, Meei-Li Huang, Keith R Jerome, Alexander Greninger |
| EPI_ISL_1680134 | Helix/Illumina | Centers for Disease Control and Prevention Division of Viral Diseases, Pathogen Discovery | Dakota Howard, Dhwani Batra, Peter W. Cook, Kara Moser, Adrian Paskey, Jason Caravas, Benjamin Rambo-Martin, Shatavia Morrison, Christopher Gulvick, Scott Sammons, Yvette Unoarumhi, Darlene Wagner, Matthew Schmerer, Eileen de Feo, Jan Antico, Christine Tran, Matthew Tolentino, Shannon Wickline, Kim Gietzen, Brad Sickler, Jingtao Liu, Eric Allen, Phil Febbo, Nicole L. Washington, Simon White, Geraint Levan, Kelly Schiabor Barrett, Elizabeth Cirulli, Alexandre Bolze, Ary Ascencio, Charlotte Rivera-Garcia, Ryan Cho, Jason Nguyen, Sherry Wang, Jimmy Ramirez, Tyler Cassens, Efren Sandoval, Magnus Isaksson, William Lee, David Becker, Marc Laurent, James Lu, Clinton R. Paden, Duncan MacCannell |
| EPI_ISL_1680135, EPI_ISL_1680136 | UW Virology Lab | UW Virology Lab | Pavitra Roychoudhury, Hong Xie, Lasata Shrestha, Shah Mohamed Bakhash, Michelle Lin, Noah R. Baker, Sean Ellis, Meei-Li Huang, Keith R Jerome, Alexander Greninger |
| EPI_ISL_1680137 | Helix/Illumina | Centers for Disease Control and Prevention Division of Viral Diseases, Pathogen Discovery | Dakota Howard, Dhwani Batra, Peter W. Cook, Kara Moser, Adrian Paskey, Jason Caravas, Benjamin Rambo-Martin, Shatavia Morrison, Christopher Gulvick, Scott Sammons, Yvette Unoarumhi, Darlene Wagner, Matthew Schmerer, Eileen de Feo, Jan Antico, Christine Tran, Matthew Tolentino, Shannon Wickline, Kim Gietzen, Brad Sickler, Jingtao Liu, Eric Allen, Phil Febbo, Nicole L. Washington, Simon White, Geraint Levan, Kelly Schiabor Barrett, Elizabeth Cirulli, Alexandre Bolze, Ary Ascencio, Charlotte Rivera-Garcia, Ryan Cho, Jason Nguyen, Sherry Wang, Jimmy Ramirez, Tyler Cassens, Efren Sandoval, Magnus Isaksson, William Lee, David Becker, Marc Laurent, James Lu, Clinton R. Paden, Duncan MacCannell |
| EPI_ISL_1680138 | UW Virology Lab | UW Virology Lab | Pavitra Roychoudhury, Hong Xie, Lasata Shrestha, Shah Mohamed Bakhash, Michelle Lin, Noah R. Baker, Sean Ellis, Meei-Li Huang, Keith R Jerome, Alexander Greninger |
| EPI_ISL_1680139, EPI_ISL_1680140, EPI_ISL_1680141 | Helix/Illumina | Centers for Disease Control and Prevention Division of Viral Diseases, Pathogen Discovery | Dakota Howard, Dhwani Batra, Peter W. Cook, Kara Moser, Adrian Paskey, Jason Caravas, Benjamin Rambo-Martin, Shatavia Morrison, Christopher Gulvick, Scott Sammons, Yvette Unoarumhi, Darlene Wagner, Matthew Schmerer, Eileen de Feo, Jan Antico, Christine Tran, Matthew Tolentino, Shannon Wickline, Kim Gietzen, Brad Sickler, Jingtao Liu, Eric Allen, Phil Febbo, Nicole L. Washington, Simon White, Geraint Levan, Kelly Schiabor Barrett, Elizabeth Cirulli, Alexandre Bolze, Ary Ascencio, Charlotte Rivera-Garcia, Ryan Cho, Jason Nguyen, Sherry Wang, Jimmy Ramirez, Tyler Cassens, Efren Sandoval, Magnus Isaksson, William Lee, David Becker, Marc Laurent, James Lu, Clinton R. Paden, Duncan MacCannell |
| EPI_ISL_1680142 | UW Virology Lab | UW Virology Lab | Pavitra Roychoudhury, Hong Xie, Lasata Shrestha, Shah Mohamed Bakhash, Michelle Lin, Noah R. Baker, Sean Ellis, Meei-Li Huang, Keith R Jerome, Alexander Greninger |
| EPI_ISL_1680143, EPI_ISL_1680144 | Helix/Illumina | Centers for Disease Control and Prevention Division of Viral Diseases, Pathogen Discovery | Dakota Howard, Dhwani Batra, Peter W. Cook, Kara Moser, Adrian Paskey, Jason Caravas, Benjamin Rambo-Martin, Shatavia Morrison, Christopher Gulvick, Scott Sammons, Yvette Unoarumhi, Darlene Wagner, Matthew Schmerer, Eileen de Feo, Jan Antico, Christine Tran, Matthew Tolentino, Shannon Wickline, Kim Gietzen, Brad Sickler, Jingtao Liu, Eric Allen, Phil Febbo, Nicole L. Washington, Simon White, Geraint Levan, Kelly Schiabor Barrett, Elizabeth Cirulli, Alexandre Bolze, Ary Ascencio, Charlotte Rivera-Garcia, Ryan Cho, Jason Nguyen, Sherry Wang, Jimmy Ramirez, Tyler Cassens, Efren Sandoval, Magnus Isaksson, William Lee, David Becker, Marc Laurent, James Lu, Clinton R. Paden, Duncan MacCannell |
| EPI_ISL_1680145 | UW Virology Lab | UW Virology Lab | Pavitra Roychoudhury, Hong Xie, Lasata Shrestha, Shah Mohamed Bakhash, Michelle Lin, Noah R. Baker, Sean Ellis, Meei-Li Huang, Keith R Jerome, Alexander Greninger |
| EPI_ISL_1680146, EPI_ISL_1680147 | Helix/Illumina | Centers for Disease Control and Prevention Division of Viral Diseases, Pathogen Discovery | Dakota Howard, Dhwani Batra, Peter W. Cook, Kara Moser, Adrian Paskey, Jason Caravas, Benjamin Rambo-Martin, Shatavia Morrison, Christopher Gulvick, Scott Sammons, Yvette Unoarumhi, Darlene Wagner, Matthew Schmerer, Eileen de Feo, Jan Antico, Christine Tran, Matthew Tolentino, Shannon Wickline, Kim Gietzen, Brad Sickler, Jingtao Liu, Eric Allen, Phil Febbo, Nicole L. Washington, Simon White, Geraint Levan, Kelly Schiabor Barrett, Elizabeth Cirulli, Alexandre Bolze, Ary Ascencio, Charlotte Rivera-Garcia, Ryan Cho, Jason Nguyen, Sherry Wang, Jimmy Ramirez, Tyler Cassens, Efren Sandoval, Magnus Isaksson, William Lee, David Becker, Marc Laurent, James Lu, Clinton R. Paden, Duncan MacCannell |
| EPI_ISL_1680148 | UW Virology Lab | UW Virology Lab | Pavitra Roychoudhury, Hong Xie, Lasata Shrestha, Shah Mohamed Bakhash, Michelle Lin, Noah R. Baker, Sean Ellis, Meei-Li Huang, Keith R Jerome, Alexander Greninger |
| EPI_ISL_1680149, EPI_ISL_1680150 | Helix/Illumina | Centers for Disease Control and Prevention Division of Viral Diseases, Pathogen Discovery | Dakota Howard, Dhwani Batra, Peter W. Cook, Kara Moser, Adrian Paskey, Jason Caravas, Benjamin Rambo-Martin, Shatavia Morrison, Christopher Gulvick, Scott Sammons, Yvette Unoarumhi, Darlene Wagner, Matthew Schmerer, Eileen de Feo, Jan Antico, Christine Tran, Matthew Tolentino, Shannon Wickline, Kim Gietzen, Brad Sickler, Jingtao Liu, Eric Allen, Phil Febbo, Nicole L. Washington, Simon White, Geraint Levan, Kelly Schiabor Barrett, Elizabeth Cirulli, Alexandre Bolze, Ary Ascencio, Charlotte Rivera-Garcia, Ryan Cho, Jason Nguyen, Sherry Wang, Jimmy Ramirez, Tyler Cassens, Efren Sandoval, Magnus Isaksson, William Lee, David Becker, Marc Laurent, James Lu, Clinton R. Paden, Duncan MacCannell |
| EPI_ISL_1680151 | UW Virology Lab | UW Virology Lab | Pavitra Roychoudhury, Hong Xie, Lasata Shrestha, Shah Mohamed Bakhash, Michelle Lin, Noah R. Baker, Sean Ellis, Meei-Li Huang, Keith R Jerome, Alexander Greninger |
| EPI_ISL_1680152, EPI_ISL_1680153 | Helix/Illumina | Centers for Disease Control and Prevention Division of Viral Diseases, Pathogen Discovery | Dakota Howard, Dhwani Batra, Peter W. Cook, Kara Moser, Adrian Paskey, Jason Caravas, Benjamin Rambo-Martin, Shatavia Morrison, Christopher Gulvick, Scott Sammons, Yvette Unoarumhi, Darlene Wagner, Matthew Schmerer, Eileen de Feo, Jan Antico, Christine Tran, Matthew Tolentino, Shannon Wickline, Kim Gietzen, Brad Sickler, Jingtao Liu, Eric Allen, Phil Febbo, Nicole L. Washington, Simon White, Geraint Levan, Kelly Schiabor Barrett, Elizabeth Cirulli, Alexandre Bolze, Ary Ascencio, Charlotte Rivera-Garcia, Ryan Cho, Jason Nguyen, Sherry Wang, Jimmy Ramirez, Tyler Cassens, Efren Sandoval, Magnus Isaksson, William Lee, David Becker, Marc Laurent, James Lu, Clinton R. Paden, Duncan MacCannell |
| EPI_ISL_1680154 | UW Virology Lab | UW Virology Lab | Pavitra Roychoudhury, Hong Xie, Lasata Shrestha, Shah Mohamed Bakhash, Michelle Lin, Noah R. Baker, Sean Ellis, Meei-Li Huang, Keith R Jerome, Alexander Greninger |
| EPI_ISL_1680155, EPI_ISL_1680156 | Helix/Illumina | Centers for Disease Control and Prevention Division of Viral Diseases, Pathogen Discovery | Dakota Howard, Dhwani Batra, Peter W. Cook, Kara Moser, Adrian Paskey, Jason Caravas, Benjamin Rambo-Martin, Shatavia Morrison, Christopher Gulvick, Scott Sammons, Yvette Unoarumhi, Darlene Wagner, Matthew Schmerer, Eileen de Feo, Jan Antico, Christine Tran, Matthew Tolentino, Shannon Wickline, Kim Gietzen, Brad Sickler, Jingtao Liu, Eric Allen, Phil Febbo, Nicole L. Washington, Simon White, Geraint Levan, Kelly Schiabor Barrett, Elizabeth Cirulli, Alexandre Bolze, Ary Ascencio, Charlotte Rivera-Garcia, Ryan Cho, Jason Nguyen, Sherry Wang, Jimmy Ramirez, Tyler Cassens, Efren Sandoval, Magnus Isaksson, William Lee, David Becker, Marc Laurent, James Lu, Clinton R. Paden, Duncan MacCannell |
| EPI_ISL_1680157 | UW Virology Lab | UW Virology Lab | Pavitra Roychoudhury, Hong Xie, Lasata Shrestha, Shah Mohamed Bakhash, Michelle Lin, Noah R. Baker, Sean Ellis, Meei-Li Huang, Keith R Jerome, Alexander Greninger |
| EPI_ISL_1680158, EPI_ISL_1680159, EPI_ISL_1680160 | Helix/Illumina | Centers for Disease Control and Prevention Division of Viral Diseases, Pathogen Discovery | Dakota Howard, Dhwani Batra, Peter W. Cook, Kara Moser, Adrian Paskey, Jason Caravas, Benjamin Rambo-Martin, Shatavia Morrison, Christopher Gulvick, Scott Sammons, Yvette Unoarumhi, Darlene Wagner, Matthew Schmerer, Eileen de Feo, Jan Antico, Christine Tran, Matthew Tolentino, Shannon Wickline, Kim Gietzen, Brad Sickler, Jingtao Liu, Eric Allen, Phil Febbo, Nicole L. Washington, Simon White, Geraint Levan, Kelly Schiabor Barrett, Elizabeth Cirulli, Alexandre Bolze, Ary Ascencio, Charlotte Rivera-Garcia, Ryan Cho, Jason Nguyen, Sherry Wang, Jimmy Ramirez, Tyler Cassens, Efren Sandoval, Magnus Isaksson, William Lee, David Becker, Marc Laurent, James Lu, Clinton R. Paden, Duncan MacCannell |
| EPI_ISL_1680161 | UW Virology Lab | UW Virology Lab | Pavitra Roychoudhury, Hong Xie, Lasata Shrestha, Shah Mohamed Bakhash, Michelle Lin, Noah R. Baker, Sean Ellis, Meei-Li Huang, Keith R Jerome, Alexander Greninger |
| EPI_ISL_1680162, EPI_ISL_1680163 | Helix/Illumina | Centers for Disease Control and Prevention Division of Viral Diseases, Pathogen Discovery | Dakota Howard, Dhwani Batra, Peter W. Cook, Kara Moser, Adrian Paskey, Jason Caravas, Benjamin Rambo-Martin, Shatavia Morrison, Christopher Gulvick, Scott Sammons, Yvette Unoarumhi, Darlene Wagner, Matthew Schmerer, Eileen de Feo, Jan Antico, Christine Tran, Matthew Tolentino, Shannon Wickline, Kim Gietzen, Brad Sickler, Jingtao Liu, Eric Allen, Phil Febbo, Nicole L. Washington, Simon White, Geraint Levan, Kelly Schiabor Barrett, Elizabeth Cirulli, Alexandre Bolze, Ary Ascencio, Charlotte Rivera-Garcia, Ryan Cho, Jason Nguyen, Sherry Wang, Jimmy Ramirez, Tyler Cassens, Efren Sandoval, Magnus Isaksson, William Lee, David Becker, Marc Laurent, James Lu, Clinton R. Paden, Duncan MacCannell |
| EPI_ISL_1680164 | UW Virology Lab | UW Virology Lab | Pavitra Roychoudhury, Hong Xie, Lasata Shrestha, Shah Mohamed Bakhash, Michelle Lin, Noah R. Baker, Sean Ellis, Meei-Li Huang, Keith R Jerome, Alexander Greninger |
| EPI_ISL_1680165, EPI_ISL_1680166 | Helix/Illumina | Centers for Disease Control and Prevention Division of Viral Diseases, Pathogen Discovery | Dakota Howard, Dhwani Batra, Peter W. Cook, Kara Moser, Adrian Paskey, Jason Caravas, Benjamin Rambo-Martin, Shatavia Morrison, Christopher Gulvick, Scott Sammons, Yvette Unoarumhi, Darlene Wagner, Matthew Schmerer, Eileen de Feo, Jan Antico, Christine Tran, Matthew Tolentino, Shannon Wickline, Kim Gietzen, Brad Sickler, Jingtao Liu, Eric Allen, Phil Febbo, Nicole L. Washington, Simon White, Geraint Levan, Kelly Schiabor Barrett, Elizabeth Cirulli, Alexandre Bolze, Ary Ascencio, Charlotte Rivera-Garcia, Ryan Cho, Jason Nguyen, Sherry Wang, Jimmy Ramirez, Tyler Cassens, Efren Sandoval, Magnus Isaksson, William Lee, David Becker, Marc Laurent, James Lu, Clinton R. Paden, Duncan MacCannell |
| EPI_ISL_1680167 | UW Virology Lab | UW Virology Lab | Pavitra Roychoudhury, Hong Xie, Lasata Shrestha, Shah Mohamed Bakhash, Michelle Lin, Noah R. Baker, Sean Ellis, Meei-Li Huang, Keith R Jerome, Alexander Greninger |
| EPI_ISL_1680168, EPI_ISL_1680169 | Helix/Illumina | Centers for Disease Control and Prevention Division of Viral Diseases, Pathogen Discovery | Dakota Howard, Dhwani Batra, Peter W. Cook, Kara Moser, Adrian Paskey, Jason Caravas, Benjamin Rambo-Martin, Shatavia Morrison, Christopher Gulvick, Scott Sammons, Yvette Unoarumhi, Darlene Wagner, Matthew Schmerer, Eileen de Feo, Jan Antico, Christine Tran, Matthew Tolentino, Shannon Wickline, Kim Gietzen, Brad Sickler, Jingtao Liu, Eric Allen, Phil Febbo, Nicole L. Washington, Simon White, Geraint Levan, Kelly Schiabor Barrett, Elizabeth Cirulli, Alexandre Bolze, Ary Ascencio, Charlotte Rivera-Garcia, Ryan Cho, Jason Nguyen, Sherry Wang, Jimmy Ramirez, Tyler Cassens, Efren Sandoval, Magnus Isaksson, William Lee, David Becker, Marc Laurent, James Lu, Clinton R. Paden, Duncan MacCannell |
| EPI_ISL_1680170 | UW Virology Lab | UW Virology Lab | Pavitra Roychoudhury, Hong Xie, Lasata Shrestha, Shah Mohamed Bakhash, Michelle Lin, Noah R. Baker, Sean Ellis, Meei-Li Huang, Keith R Jerome, Alexander Greninger |
| EPI_ISL_1680171, EPI_ISL_1680172, EPI_ISL_1680173 | Helix/Illumina | Centers for Disease Control and Prevention Division of Viral Diseases, Pathogen Discovery | Dakota Howard, Dhwani Batra, Peter W. Cook, Kara Moser, Adrian Paskey, Jason Caravas, Benjamin Rambo-Martin, Shatavia Morrison, Christopher Gulvick, Scott Sammons, Yvette Unoarumhi, Darlene Wagner, Matthew Schmerer, Eileen de Feo, Jan Antico, Christine Tran, Matthew Tolentino, Shannon Wickline, Kim Gietzen, Brad Sickler, Jingtao Liu, Eric Allen, Phil Febbo, Nicole L. Washington, Simon White, Geraint Levan, Kelly Schiabor Barrett, Elizabeth Cirulli, Alexandre Bolze, Ary Ascencio, Charlotte Rivera-Garcia, Ryan Cho, Jason Nguyen, Sherry Wang, Jimmy Ramirez, Tyler Cassens, Efren Sandoval, Magnus Isaksson, William Lee, David Becker, Marc Laurent, James Lu, Clinton R. Paden, Duncan MacCannell |
| EPI_ISL_1680174 | UW Virology Lab | UW Virology Lab | Pavitra Roychoudhury, Hong Xie, Lasata Shrestha, Shah Mohamed Bakhash, Michelle Lin, Noah R. Baker, Sean Ellis, Meei-Li Huang, Keith R Jerome, Alexander Greninger |
| EPI_ISL_1680175, | Helix/Illumina | Centers for Disease Control and Prevention | Dakota Howard, Dhwani Batra, Peter W. Cook, Kara Moser, Adrian Paskey, Jason Caravas, Benjamin Rambo-Martin, Shatavia Morrison, Christopher Gulvick, Scott Sammons, Yvette Unoarumhi, Darlene Wagner, Matthew Schmerer, Eileen de Feo, Jan Antico, Christine Tran, Matthew |

| | | | |
|---|---|---|---|
| EPI_ISL_1680176 | | Division of Viral Diseases, Pathogen Discovery | Tolentino, Shannon Wickline, Kim Gietzen, Brad Sickler, Jingtao Liu, Eric Allen, Phil Febbo, Nicole L. Washington, Simon White, Geraint Levan, Kelly Schiabor Barrett, Elizabeth Cirulli, Alexandre Bolze, Ary Ascencio, Charlotte Rivera-Garcia, Ryan Cho, Jason Nguyen, Sherry Wang, Jimmy Ramirez, Tyler Cassens, Efren Sandoval, Magnus Isaksson, William Lee, David Becker, Marc Laurent, James Lu, Clinton R. Paden, Duncan MacCannell |
| EPI_ISL_1680177 | UW Virology Lab | UW Virology Lab | Pavitra Roychoudhury, Hong Xie, Lasata Shrestha, Shah Mohamed Bakhash, Michelle Lin, Noah R. Baker, Sean Ellis, Meei-Li Huang, Keith R Jerome, Alexander Greninger |
| EPI_ISL_1680178, EPI_ISL_1680179, EPI_ISL_1680180 | Helix/Illumina | Centers for Disease Control and Prevention Division of Viral Diseases, Pathogen Discovery | Dakota Howard, Dhwani Batra, Peter W. Cook, Kara Moser, Adrian Paskey, Jason Caravas, Benjamin Rambo-Martin, Shatavia Morrison, Christopher Gulvick, Scott Sammons, Yvette Unoarumhi, Darlene Wagner, Matthew Schmerer, Eileen de Feo, Jan Antico, Christine Tran, Matthew Tolentino, Shannon Wickline, Kim Gietzen, Brad Sickler, Jingtao Liu, Eric Allen, Phil Febbo, Nicole L. Washington, Simon White, Geraint Levan, Kelly Schiabor Barrett, Elizabeth Cirulli, Alexandre Bolze, Ary Ascencio, Charlotte Rivera-Garcia, Ryan Cho, Jason Nguyen, Sherry Wang, Jimmy Ramirez, Tyler Cassens, Efren Sandoval, Magnus Isaksson, William Lee, David Becker, Marc Laurent, James Lu, Clinton R. Paden, Duncan MacCannell |
| EPI_ISL_1680181 | UW Virology Lab | UW Virology Lab | Pavitra Roychoudhury, Hong Xie, Lasata Shrestha, Shah Mohamed Bakhash, Michelle Lin, Noah R. Baker, Sean Ellis, Meei-Li Huang, Keith R Jerome, Alexander Greninger |
| EPI_ISL_1680182, EPI_ISL_1680183 | Helix/Illumina | Centers for Disease Control and Prevention Division of Viral Diseases, Pathogen Discovery | Dakota Howard, Dhwani Batra, Peter W. Cook, Kara Moser, Adrian Paskey, Jason Caravas, Benjamin Rambo-Martin, Shatavia Morrison, Christopher Gulvick, Scott Sammons, Yvette Unoarumhi, Darlene Wagner, Matthew Schmerer, Eileen de Feo, Jan Antico, Christine Tran, Matthew Tolentino, Shannon Wickline, Kim Gietzen, Brad Sickler, Jingtao Liu, Eric Allen, Phil Febbo, Nicole L. Washington, Simon White, Geraint Levan, Kelly Schiabor Barrett, Elizabeth Cirulli, Alexandre Bolze, Ary Ascencio, Charlotte Rivera-Garcia, Ryan Cho, Jason Nguyen, Sherry Wang, Jimmy Ramirez, Tyler Cassens, Efren Sandoval, Magnus Isaksson, William Lee, David Becker, Marc Laurent, James Lu, Clinton R. Paden, Duncan MacCannell |
| EPI_ISL_1680184 | UW Virology Lab | UW Virology Lab | Pavitra Roychoudhury, Hong Xie, Lasata Shrestha, Shah Mohamed Bakhash, Michelle Lin, Noah R. Baker, Sean Ellis, Meei-Li Huang, Keith R Jerome, Alexander Greninger |
| EPI_ISL_1680185, EPI_ISL_1680186 | Helix/Illumina | Centers for Disease Control and Prevention Division of Viral Diseases, Pathogen Discovery | Dakota Howard, Dhwani Batra, Peter W. Cook, Kara Moser, Adrian Paskey, Jason Caravas, Benjamin Rambo-Martin, Shatavia Morrison, Christopher Gulvick, Scott Sammons, Yvette Unoarumhi, Darlene Wagner, Matthew Schmerer, Eileen de Feo, Jan Antico, Christine Tran, Matthew Tolentino, Shannon Wickline, Kim Gietzen, Brad Sickler, Jingtao Liu, Eric Allen, Phil Febbo, Nicole L. Washington, Simon White, Geraint Levan, Kelly Schiabor Barrett, Elizabeth Cirulli, Alexandre Bolze, Ary Ascencio, Charlotte Rivera-Garcia, Ryan Cho, Jason Nguyen, Sherry Wang, Jimmy Ramirez, Tyler Cassens, Efren Sandoval, Magnus Isaksson, William Lee, David Becker, Marc Laurent, James Lu, Clinton R. Paden, Duncan MacCannell |
| EPI_ISL_1680187 | UW Virology Lab | UW Virology Lab | Pavitra Roychoudhury, Hong Xie, Lasata Shrestha, Shah Mohamed Bakhash, Michelle Lin, Noah R. Baker, Sean Ellis, Meei-Li Huang, Keith R Jerome, Alexander Greninger |
| EPI_ISL_1680188 | Helix/Illumina | Centers for Disease Control and Prevention Division of Viral Diseases, Pathogen Discovery | Dakota Howard, Dhwani Batra, Peter W. Cook, Kara Moser, Adrian Paskey, Jason Caravas, Benjamin Rambo-Martin, Shatavia Morrison, Christopher Gulvick, Scott Sammons, Yvette Unoarumhi, Darlene Wagner, Matthew Schmerer, Eileen de Feo, Jan Antico, Christine Tran, Matthew Tolentino, Shannon Wickline, Kim Gietzen, Brad Sickler, Jingtao Liu, Eric Allen, Phil Febbo, Nicole L. Washington, Simon White, Geraint Levan, Kelly Schiabor Barrett, Elizabeth Cirulli, Alexandre Bolze, Ary Ascencio, Charlotte Rivera-Garcia, Ryan Cho, Jason Nguyen, Sherry Wang, Jimmy Ramirez, Tyler Cassens, Efren Sandoval, Magnus Isaksson, William Lee, David Becker, Marc Laurent, James Lu, Clinton R. Paden, Duncan MacCannell |
| EPI_ISL_1680189, EPI_ISL_1680190 | UW Virology Lab | UW Virology Lab | Pavitra Roychoudhury, Hong Xie, Lasata Shrestha, Shah Mohamed Bakhash, Michelle Lin, Noah R. Baker, Sean Ellis, Meei-Li Huang, Keith R Jerome, Alexander Greninger |
| EPI_ISL_1680191 | Helix/Illumina | Centers for Disease Control and Prevention Division of Viral Diseases, Pathogen Discovery | Dakota Howard, Dhwani Batra, Peter W. Cook, Kara Moser, Adrian Paskey, Jason Caravas, Benjamin Rambo-Martin, Shatavia Morrison, Christopher Gulvick, Scott Sammons, Yvette Unoarumhi, Darlene Wagner, Matthew Schmerer, Eileen de Feo, Jan Antico, Christine Tran, Matthew Tolentino, Shannon Wickline, Kim Gietzen, Brad Sickler, Jingtao Liu, Eric Allen, Phil Febbo, Nicole L. Washington, Simon White, Geraint Levan, Kelly Schiabor Barrett, Elizabeth Cirulli, Alexandre Bolze, Ary Ascencio, Charlotte Rivera-Garcia, Ryan Cho, Jason Nguyen, Sherry Wang, Jimmy Ramirez, Tyler Cassens, Efren Sandoval, Magnus Isaksson, William Lee, David Becker, Marc Laurent, James Lu, Clinton R. Paden, Duncan MacCannell |
| EPI_ISL_1680192, EPI_ISL_1680193 | UW Virology Lab | UW Virology Lab | Pavitra Roychoudhury, Hong Xie, Lasata Shrestha, Shah Mohamed Bakhash, Michelle Lin, Noah R. Baker, Sean Ellis, Meei-Li Huang, Keith R Jerome, Alexander Greninger |
| EPI_ISL_1680194 | Helix/Illumina | Centers for Disease Control and Prevention Division of Viral Diseases, Pathogen Discovery | Dakota Howard, Dhwani Batra, Peter W. Cook, Kara Moser, Adrian Paskey, Jason Caravas, Benjamin Rambo-Martin, Shatavia Morrison, Christopher Gulvick, Scott Sammons, Yvette Unoarumhi, Darlene Wagner, Matthew Schmerer, Eileen de Feo, Jan Antico, Christine Tran, Matthew Tolentino, Shannon Wickline, Kim Gietzen, Brad Sickler, Jingtao Liu, Eric Allen, Phil Febbo, Nicole L. Washington, Simon White, Geraint Levan, Kelly Schiabor Barrett, Elizabeth Cirulli, Alexandre Bolze, Ary Ascencio, Charlotte Rivera-Garcia, Ryan Cho, Jason Nguyen, Sherry Wang, Jimmy Ramirez, Tyler Cassens, Efren Sandoval, Magnus Isaksson, William Lee, David Becker, Marc Laurent, James Lu, Clinton R. Paden, Duncan MacCannell |
| EPI_ISL_1680196 | UW Virology Lab | UW Virology Lab | Pavitra Roychoudhury, Hong Xie, Lasata Shrestha, Shah Mohamed Bakhash, Michelle Lin, Noah R. Baker, Sean Ellis, Meei-Li Huang, Keith R Jerome, Alexander Greninger |
| EPI_ISL_1680197 | Helix/Illumina | Centers for Disease Control and Prevention Division of Viral Diseases, Pathogen Discovery | Dakota Howard, Dhwani Batra, Peter W. Cook, Kara Moser, Adrian Paskey, Jason Caravas, Benjamin Rambo-Martin, Shatavia Morrison, Christopher Gulvick, Scott Sammons, Yvette Unoarumhi, Darlene Wagner, Matthew Schmerer, Eileen de Feo, Jan Antico, Christine Tran, Matthew Tolentino, Shannon Wickline, Kim Gietzen, Brad Sickler, Jingtao Liu, Eric Allen, Phil Febbo, Nicole L. Washington, Simon White, Geraint Levan, Kelly Schiabor Barrett, Elizabeth Cirulli, Alexandre Bolze, Ary Ascencio, Charlotte Rivera-Garcia, Ryan Cho, Jason Nguyen, Sherry Wang, Jimmy Ramirez, Tyler Cassens, Efren Sandoval, Magnus Isaksson, William Lee, David Becker, Marc Laurent, James Lu, Clinton R. Paden, Duncan MacCannell |
| EPI_ISL_1680198, EPI_ISL_1680199 | UW Virology Lab | UW Virology Lab | Pavitra Roychoudhury, Hong Xie, Lasata Shrestha, Shah Mohamed Bakhash, Michelle Lin, Noah R. Baker, Sean Ellis, Meei-Li Huang, Keith R Jerome, Alexander Greninger |
| EPI_ISL_1680200 | Helix/Illumina | Centers for Disease Control and Prevention Division of Viral Diseases, Pathogen Discovery | Dakota Howard, Dhwani Batra, Peter W. Cook, Kara Moser, Adrian Paskey, Jason Caravas, Benjamin Rambo-Martin, Shatavia Morrison, Christopher Gulvick, Scott Sammons, Yvette Unoarumhi, Darlene Wagner, Matthew Schmerer, Eileen de Feo, Jan Antico, Christine Tran, Matthew Tolentino, Shannon Wickline, Kim Gietzen, Brad Sickler, Jingtao Liu, Eric Allen, Phil Febbo, Nicole L. Washington, Simon White, Geraint Levan, Kelly Schiabor Barrett, Elizabeth Cirulli, Alexandre Bolze, Ary Ascencio, Charlotte Rivera-Garcia, Ryan Cho, Jason Nguyen, Sherry Wang, Jimmy Ramirez, Tyler Cassens, Efren Sandoval, Magnus Isaksson, William Lee, David Becker, Marc Laurent, James Lu, Clinton R. Paden, Duncan MacCannell |
| EPI_ISL_1680202 | UW Virology Lab | UW Virology Lab | Pavitra Roychoudhury, Hong Xie, Lasata Shrestha, Shah Mohamed Bakhash, Michelle Lin, Noah R. Baker, Sean Ellis, Meei-Li Huang, Keith R Jerome, Alexander Greninger |
| EPI_ISL_1680203 | Helix/Illumina | Centers for Disease Control and Prevention Division of Viral Diseases, Pathogen Discovery | Dakota Howard, Dhwani Batra, Peter W. Cook, Kara Moser, Adrian Paskey, Jason Caravas, Benjamin Rambo-Martin, Shatavia Morrison, Christopher Gulvick, Scott Sammons, Yvette Unoarumhi, Darlene Wagner, Matthew Schmerer, Eileen de Feo, Jan Antico, Christine Tran, Matthew Tolentino, Shannon Wickline, Kim Gietzen, Brad Sickler, Jingtao Liu, Eric Allen, Phil Febbo, Nicole L. Washington, Simon White, Geraint Levan, Kelly Schiabor Barrett, Elizabeth Cirulli, Alexandre Bolze, Ary Ascencio, Charlotte Rivera-Garcia, Ryan Cho, Jason Nguyen, Sherry Wang, Jimmy Ramirez, Tyler Cassens, Efren Sandoval, Magnus Isaksson, William Lee, David Becker, Marc Laurent, James Lu, Clinton R. Paden, Duncan MacCannell |
| EPI_ISL_1680204, EPI_ISL_1680205 | UW Virology Lab | UW Virology Lab | Pavitra Roychoudhury, Hong Xie, Lasata Shrestha, Shah Mohamed Bakhash, Michelle Lin, Noah R. Baker, Sean Ellis, Meei-Li Huang, Keith R Jerome, Alexander Greninger |
| EPI_ISL_1680206 | Helix/Illumina | Centers for Disease Control and Prevention Division of Viral Diseases, Pathogen Discovery | Dakota Howard, Dhwani Batra, Peter W. Cook, Kara Moser, Adrian Paskey, Jason Caravas, Benjamin Rambo-Martin, Shatavia Morrison, Christopher Gulvick, Scott Sammons, Yvette Unoarumhi, Darlene Wagner, Matthew Schmerer, Eileen de Feo, Jan Antico, Christine Tran, Matthew Tolentino, Shannon Wickline, Kim Gietzen, Brad Sickler, Jingtao Liu, Eric Allen, Phil Febbo, Nicole L. Washington, Simon White, Geraint Levan, Kelly Schiabor Barrett, Elizabeth Cirulli, Alexandre Bolze, Ary Ascencio, Charlotte Rivera-Garcia, Ryan Cho, Jason Nguyen, Sherry Wang, Jimmy Ramirez, Tyler Cassens, Efren Sandoval, Magnus Isaksson, William Lee, David Becker, Marc Laurent, James Lu, Clinton R. Paden, Duncan MacCannell |
| EPI_ISL_1680207, EPI_ISL_1680208 | UW Virology Lab | UW Virology Lab | Pavitra Roychoudhury, Hong Xie, Lasata Shrestha, Shah Mohamed Bakhash, Michelle Lin, Noah R. Baker, Sean Ellis, Meei-Li Huang, Keith R Jerome, Alexander Greninger |
| EPI_ISL_1680209 | Helix/Illumina | Centers for Disease Control and Prevention Division of Viral Diseases, Pathogen Discovery | Dakota Howard, Dhwani Batra, Peter W. Cook, Kara Moser, Adrian Paskey, Jason Caravas, Benjamin Rambo-Martin, Shatavia Morrison, Christopher Gulvick, Scott Sammons, Yvette Unoarumhi, Darlene Wagner, Matthew Schmerer, Eileen de Feo, Jan Antico, Christine Tran, Matthew Tolentino, Shannon Wickline, Kim Gietzen, Brad Sickler, Jingtao Liu, Eric Allen, Phil Febbo, Nicole L. Washington, Simon White, Geraint Levan, Kelly Schiabor Barrett, Elizabeth Cirulli, Alexandre Bolze, Ary Ascencio, Charlotte Rivera-Garcia, Ryan Cho, Jason Nguyen, Sherry Wang, Jimmy Ramirez, Tyler Cassens, Efren Sandoval, Magnus Isaksson, William Lee, David Becker, Marc Laurent, James Lu, Clinton R. Paden, Duncan MacCannell |
| EPI_ISL_1680210 | UW Virology Lab | UW Virology Lab | Pavitra Roychoudhury, Hong Xie, Lasata Shrestha, Shah Mohamed Bakhash, Michelle Lin, Noah R. Baker, Sean Ellis, Meei-Li Huang, Keith R Jerome, Alexander Greninger |
| EPI_ISL_1680211 | Helix/Illumina | Centers for Disease Control and Prevention Division of Viral Diseases, Pathogen Discovery | Dakota Howard, Dhwani Batra, Peter W. Cook, Kara Moser, Adrian Paskey, Jason Caravas, Benjamin Rambo-Martin, Shatavia Morrison, Christopher Gulvick, Scott Sammons, Yvette Unoarumhi, Darlene Wagner, Matthew Schmerer, Eileen de Feo, Jan Antico, Christine Tran, Matthew Tolentino, Shannon Wickline, Kim Gietzen, Brad Sickler, Jingtao Liu, Eric Allen, Phil Febbo, Nicole L. Washington, Simon White, Geraint Levan, Kelly Schiabor Barrett, Elizabeth Cirulli, Alexandre Bolze, Ary Ascencio, Charlotte Rivera-Garcia, Ryan Cho, Jason Nguyen, Sherry Wang, Jimmy Ramirez, Tyler Cassens, Efren Sandoval, Magnus Isaksson, William Lee, David Becker, Marc Laurent, James Lu, Clinton R. Paden, Duncan MacCannell |
| EPI_ISL_1680212, EPI_ISL_1680213, EPI_ISL_1680214 | UW Virology Lab | UW Virology Lab | Pavitra Roychoudhury, Hong Xie, Lasata Shrestha, Shah Mohamed Bakhash, Michelle Lin, Noah R. Baker, Sean Ellis, Meei-Li Huang, Keith R Jerome, Alexander Greninger |
| EPI_ISL_1680215 | Helix/Illumina | Centers for Disease Control and Prevention Division of Viral Diseases, Pathogen Discovery | Dakota Howard, Dhwani Batra, Peter W. Cook, Kara Moser, Adrian Paskey, Jason Caravas, Benjamin Rambo-Martin, Shatavia Morrison, Christopher Gulvick, Scott Sammons, Yvette Unoarumhi, Darlene Wagner, Matthew Schmerer, Eileen de Feo, Jan Antico, Christine Tran, Matthew Tolentino, Shannon Wickline, Kim Gietzen, Brad Sickler, Jingtao Liu, Eric Allen, Phil Febbo, Nicole L. Washington, Simon White, Geraint Levan, Kelly Schiabor Barrett, Elizabeth Cirulli, Alexandre Bolze, Ary Ascencio, Charlotte Rivera-Garcia, Ryan Cho, Jason Nguyen, Sherry Wang, Jimmy Ramirez, Tyler Cassens, Efren Sandoval, Magnus Isaksson, William Lee, David Becker, Marc Laurent, James Lu, Clinton R. Paden, Duncan MacCannell |
| EPI_ISL_1680216 | UW Virology Lab | UW Virology Lab | Pavitra Roychoudhury, Hong Xie, Lasata Shrestha, Shah Mohamed Bakhash, Michelle Lin, Noah R. Baker, Sean Ellis, Meei-Li Huang, Keith R Jerome, Alexander Greninger |
| EPI_ISL_1680217 | Helix/Illumina | Centers for Disease Control and Prevention Division of Viral Diseases, Pathogen Discovery | Dakota Howard, Dhwani Batra, Peter W. Cook, Kara Moser, Adrian Paskey, Jason Caravas, Benjamin Rambo-Martin, Shatavia Morrison, Christopher Gulvick, Scott Sammons, Yvette Unoarumhi, Darlene Wagner, Matthew Schmerer, Eileen de Feo, Jan Antico, Christine Tran, Matthew Tolentino, Shannon Wickline, Kim Gietzen, Brad Sickler, Jingtao Liu, Eric Allen, Phil Febbo, Nicole L. Washington, Simon White, Geraint Levan, Kelly Schiabor Barrett, Elizabeth Cirulli, Alexandre Bolze, Ary Ascencio, Charlotte Rivera-Garcia, Ryan Cho, Jason Nguyen, Sherry Wang, Jimmy Ramirez, Tyler Cassens, Efren Sandoval, Magnus Isaksson, William Lee, David Becker, Marc Laurent, James Lu, Clinton R. Paden, Duncan MacCannell |
| EPI_ISL_1680218, EPI_ISL_1680219 | UW Virology Lab | UW Virology Lab | Pavitra Roychoudhury, Hong Xie, Lasata Shrestha, Shah Mohamed Bakhash, Michelle Lin, Noah R. Baker, Sean Ellis, Meei-Li Huang, Keith R Jerome, Alexander Greninger |
| EPI_ISL_1680220 | Helix/Illumina | Centers for Disease Control and Prevention Division of Viral Diseases, Pathogen Discovery | Dakota Howard, Dhwani Batra, Peter W. Cook, Kara Moser, Adrian Paskey, Jason Caravas, Benjamin Rambo-Martin, Shatavia Morrison, Christopher Gulvick, Scott Sammons, Yvette Unoarumhi, Darlene Wagner, Matthew Schmerer, Eileen de Feo, Jan Antico, Christine Tran, Matthew Tolentino, Shannon Wickline, Kim Gietzen, Brad Sickler, Jingtao Liu, Eric Allen, Phil Febbo, Nicole L. Washington, Simon White, Geraint Levan, Kelly Schiabor Barrett, Elizabeth Cirulli, Alexandre Bolze, Ary Ascencio, Charlotte Rivera-Garcia, Ryan Cho, Jason Nguyen, Sherry Wang, Jimmy Ramirez, Tyler Cassens, Efren Sandoval, Magnus Isaksson, William Lee, David Becker, Marc Laurent, James Lu, Clinton R. Paden, Duncan MacCannell |
| EPI_ISL_1680221, EPI_ISL_1680222, EPI_ISL_1680223 | UW Virology Lab | UW Virology Lab | Pavitra Roychoudhury, Hong Xie, Lasata Shrestha, Shah Mohamed Bakhash, Michelle Lin, Noah R. Baker, Sean Ellis, Meei-Li Huang, Keith R Jerome, Alexander Greninger |
| EPI_ISL_1680224 | Helix/Illumina | Centers for Disease Control and Prevention Division of Viral Diseases, Pathogen Discovery | Dakota Howard, Dhwani Batra, Peter W. Cook, Kara Moser, Adrian Paskey, Jason Caravas, Benjamin Rambo-Martin, Shatavia Morrison, Christopher Gulvick, Scott Sammons, Yvette Unoarumhi, Darlene Wagner, Matthew Schmerer, Eileen de Feo, Jan Antico, Christine Tran, Matthew Tolentino, Shannon Wickline, Kim Gietzen, Brad Sickler, Jingtao Liu, Eric Allen, Phil Febbo, Nicole L. Washington, Simon White, Geraint Levan, Kelly Schiabor Barrett, Elizabeth Cirulli, Alexandre Bolze, Ary Ascencio, Charlotte Rivera-Garcia, Ryan Cho, Jason Nguyen, Sherry Wang, Jimmy Ramirez, Tyler Cassens, Efren Sandoval, Magnus Isaksson, William Lee, David Becker, Marc Laurent, James Lu, Clinton R. Paden, Duncan MacCannell |
| EPI_ISL_1680225 | UW Virology Lab | UW Virology Lab | Pavitra Roychoudhury, Hong Xie, Lasata Shrestha, Shah Mohamed Bakhash, Michelle Lin, Noah R. Baker, Sean Ellis, Meei-Li Huang, Keith R Jerome, Alexander Greninger |
| EPI_ISL_1680226, EPI_ISL_1680227 | Helix/Illumina | Centers for Disease Control and Prevention Division of Viral Diseases, Pathogen Discovery | Dakota Howard, Dhwani Batra, Peter W. Cook, Kara Moser, Adrian Paskey, Jason Caravas, Benjamin Rambo-Martin, Shatavia Morrison, Christopher Gulvick, Scott Sammons, Yvette Unoarumhi, Darlene Wagner, Matthew Schmerer, Eileen de Feo, Jan Antico, Christine Tran, Matthew Tolentino, Shannon Wickline, Kim Gietzen, Brad Sickler, Jingtao Liu, Eric Allen, Phil Febbo, Nicole L. Washington, Simon White, Geraint Levan, Kelly Schiabor Barrett, Elizabeth Cirulli, Alexandre Bolze, Ary Ascencio, Charlotte Rivera-Garcia, Ryan Cho, Jason Nguyen, Sherry Wang, Jimmy Ramirez, Tyler Cassens, Efren Sandoval, Magnus Isaksson, William Lee, David Becker, Marc Laurent, James Lu, Clinton R. Paden, Duncan MacCannell |
| EPI_ISL_1680228 | UW Virology Lab | UW Virology Lab | Pavitra Roychoudhury, Hong Xie, Lasata Shrestha, Shah Mohamed Bakhash, Michelle Lin, Noah R. Baker, Sean Ellis, Meei-Li Huang, Keith R Jerome, Alexander Greninger |
| EPI_ISL_1680229, EPI_ISL_1680230 | Helix/Illumina | Centers for Disease Control and Prevention Division of Viral Diseases, Pathogen Discovery | Dakota Howard, Dhwani Batra, Peter W. Cook, Kara Moser, Adrian Paskey, Jason Caravas, Benjamin Rambo-Martin, Shatavia Morrison, Christopher Gulvick, Scott Sammons, Yvette Unoarumhi, Darlene Wagner, Matthew Schmerer, Eileen de Feo, Jan Antico, Christine Tran, Matthew Tolentino, Shannon Wickline, Kim Gietzen, Brad Sickler, Jingtao Liu, Eric Allen, Phil Febbo, Nicole L. Washington, Simon White, Geraint Levan, Kelly Schiabor Barrett, Elizabeth Cirulli, Alexandre Bolze, Ary Ascencio, Charlotte Rivera-Garcia, Ryan Cho, Jason Nguyen, Sherry Wang, Jimmy Ramirez, Tyler Cassens, Efren Sandoval, Magnus Isaksson, William Lee, David Becker, Marc Laurent, James Lu, Clinton R. Paden, Duncan MacCannell |
| EPI_ISL_1680231 | UW Virology Lab | UW Virology Lab | Pavitra Roychoudhury, Hong Xie, Lasata Shrestha, Shah Mohamed Bakhash, Michelle Lin, Noah R. Baker, Sean Ellis, Meei-Li Huang, Keith R Jerome, Alexander Greninger |
| EPI_ISL_1680232, EPI_ISL_1680233 | Helix/Illumina | Centers for Disease Control and Prevention Division of Viral Diseases, Pathogen Discovery | Dakota Howard, Dhwani Batra, Peter W. Cook, Kara Moser, Adrian Paskey, Jason Caravas, Benjamin Rambo-Martin, Shatavia Morrison, Christopher Gulvick, Scott Sammons, Yvette Unoarumhi, Darlene Wagner, Matthew Schmerer, Eileen de Feo, Jan Antico, Christine Tran, Matthew Tolentino, Shannon Wickline, Kim Gietzen, Brad Sickler, Jingtao Liu, Eric Allen, Phil Febbo, Nicole L. Washington, Simon White, Geraint Levan, Kelly Schiabor Barrett, Elizabeth Cirulli, Alexandre Bolze, Ary Ascencio, Charlotte Rivera-Garcia, Ryan Cho, Jason Nguyen, Sherry Wang, Jimmy Ramirez, Tyler Cassens, Efren Sandoval, Magnus Isaksson, William Lee, David Becker, Marc Laurent, James Lu, Clinton R. Paden, Duncan MacCannell |
| EPI_ISL_1680234 | UW Virology Lab | UW Virology Lab | Pavitra Roychoudhury, Hong Xie, Lasata Shrestha, Shah Mohamed Bakhash, Michelle Lin, Noah R. Baker, Sean Ellis, Meei-Li Huang, Keith R Jerome, Alexander Greninger |
| EPI_ISL_1680235, EPI_ISL_1680236 | Helix/Illumina | Centers for Disease Control and Prevention Division of Viral Diseases, Pathogen Discovery | Dakota Howard, Dhwani Batra, Peter W. Cook, Kara Moser, Adrian Paskey, Jason Caravas, Benjamin Rambo-Martin, Shatavia Morrison, Christopher Gulvick, Scott Sammons, Yvette Unoarumhi, Darlene Wagner, Matthew Schmerer, Eileen de Feo, Jan Antico, Christine Tran, Matthew Tolentino, Shannon Wickline, Kim Gietzen, Brad Sickler, Jingtao Liu, Eric Allen, Phil Febbo, Nicole L. Washington, Simon White, Geraint Levan, Kelly Schiabor Barrett, Elizabeth Cirulli, Alexandre Bolze, Ary Ascencio, Charlotte Rivera-Garcia, Ryan Cho, Jason Nguyen, Sherry Wang, Jimmy Ramirez, Tyler Cassens, Efren Sandoval, Magnus Isaksson, William Lee, David Becker, Marc Laurent, James Lu, Clinton R. Paden, Duncan MacCannell |

| | | | |
|---|---|---|---|
| EPI_ISL_1680237 | UW Virology Lab | UW Virology Lab | Pavitra Roychoudhury, Hong Xie, Lasata Shrestha, Shah Mohamed Bakhash, Michelle Lin, Noah R. Baker, Sean Ellis, Meei-Li Huang, Keith R Jerome, Alexander Greninger |
| EPI_ISL_1680238, EPI_ISL_1680239 | Helix/Illumina | Centers for Disease Control and Prevention Division of Viral Diseases, Pathogen Discovery | Dakota Howard, Dhwani Batra, Peter W. Cook, Kara Moser, Adrian Paskey, Jason Caravas, Benjamin Rambo-Martin, Shatavia Morrison, Christopher Gulvick, Scott Sammons, Yvette Unoarumhi, Darlene Wagner, Matthew Schmerer, Eileen de Feo, Jan Antico, Christine Tran, Matthew Tolentino, Shannon Wickline, Kim Gietzen, Brad Sickler, Jingtao Liu, Eric Allen, Phil Febbo, Nicole L. Washington, Simon White, Geraint Levan, Kelly Schiabor Barrett, Elizabeth Cirulli, Alexandre Bolze, Ary Ascencio, Charlotte Rivera-Garcia, Ryan Cho, Jason Nguyen, Sherry Wang, Jimmy Ramirez, Tyler Cassens, Efren Sandoval, Magnus Isaksson, William Lee, David Becker, Marc Laurent, James Lu, Clinton R. Paden, Duncan MacCannell |
| EPI_ISL_1680240 | UW Virology Lab | UW Virology Lab | Pavitra Roychoudhury, Hong Xie, Lasata Shrestha, Shah Mohamed Bakhash, Michelle Lin, Noah R. Baker, Sean Ellis, Meei-Li Huang, Keith R Jerome, Alexander Greninger |
| EPI_ISL_1680241, EPI_ISL_1680242 | Helix/Illumina | Centers for Disease Control and Prevention Division of Viral Diseases, Pathogen Discovery | Dakota Howard, Dhwani Batra, Peter W. Cook, Kara Moser, Adrian Paskey, Jason Caravas, Benjamin Rambo-Martin, Shatavia Morrison, Christopher Gulvick, Scott Sammons, Yvette Unoarumhi, Darlene Wagner, Matthew Schmerer, Eileen de Feo, Jan Antico, Christine Tran, Matthew Tolentino, Shannon Wickline, Kim Gietzen, Brad Sickler, Jingtao Liu, Eric Allen, Phil Febbo, Nicole L. Washington, Simon White, Geraint Levan, Kelly Schiabor Barrett, Elizabeth Cirulli, Alexandre Bolze, Ary Ascencio, Charlotte Rivera-Garcia, Ryan Cho, Jason Nguyen, Sherry Wang, Jimmy Ramirez, Tyler Cassens, Efren Sandoval, Magnus Isaksson, William Lee, David Becker, Marc Laurent, James Lu, Clinton R. Paden, Duncan MacCannell |
| EPI_ISL_1680243 | UW Virology Lab | UW Virology Lab | Pavitra Roychoudhury, Hong Xie, Lasata Shrestha, Shah Mohamed Bakhash, Michelle Lin, Noah R. Baker, Sean Ellis, Meei-Li Huang, Keith R Jerome, Alexander Greninger |
| EPI_ISL_1680244, EPI_ISL_1680245 | Helix/Illumina | Centers for Disease Control and Prevention Division of Viral Diseases, Pathogen Discovery | Dakota Howard, Dhwani Batra, Peter W. Cook, Kara Moser, Adrian Paskey, Jason Caravas, Benjamin Rambo-Martin, Shatavia Morrison, Christopher Gulvick, Scott Sammons, Yvette Unoarumhi, Darlene Wagner, Matthew Schmerer, Eileen de Feo, Jan Antico, Christine Tran, Matthew Tolentino, Shannon Wickline, Kim Gietzen, Brad Sickler, Jingtao Liu, Eric Allen, Phil Febbo, Nicole L. Washington, Simon White, Geraint Levan, Kelly Schiabor Barrett, Elizabeth Cirulli, Alexandre Bolze, Ary Ascencio, Charlotte Rivera-Garcia, Ryan Cho, Jason Nguyen, Sherry Wang, Jimmy Ramirez, Tyler Cassens, Efren Sandoval, Magnus Isaksson, William Lee, David Becker, Marc Laurent, James Lu, Clinton R. Paden, Duncan MacCannell |
| EPI_ISL_1680246 | UW Virology Lab | UW Virology Lab | Pavitra Roychoudhury, Hong Xie, Lasata Shrestha, Shah Mohamed Bakhash, Michelle Lin, Noah R. Baker, Sean Ellis, Meei-Li Huang, Keith R Jerome, Alexander Greninger |
| EPI_ISL_1680247, EPI_ISL_1680248, EPI_ISL_1680249 | Helix/Illumina | Centers for Disease Control and Prevention Division of Viral Diseases, Pathogen Discovery | Dakota Howard, Dhwani Batra, Peter W. Cook, Kara Moser, Adrian Paskey, Jason Caravas, Benjamin Rambo-Martin, Shatavia Morrison, Christopher Gulvick, Scott Sammons, Yvette Unoarumhi, Darlene Wagner, Matthew Schmerer, Eileen de Feo, Jan Antico, Christine Tran, Matthew Tolentino, Shannon Wickline, Kim Gietzen, Brad Sickler, Jingtao Liu, Eric Allen, Phil Febbo, Nicole L. Washington, Simon White, Geraint Levan, Kelly Schiabor Barrett, Elizabeth Cirulli, Alexandre Bolze, Ary Ascencio, Charlotte Rivera-Garcia, Ryan Cho, Jason Nguyen, Sherry Wang, Jimmy Ramirez, Tyler Cassens, Efren Sandoval, Magnus Isaksson, William Lee, David Becker, Marc Laurent, James Lu, Clinton R. Paden, Duncan MacCannell |
| EPI_ISL_1680250 | UW Virology Lab | UW Virology Lab | Pavitra Roychoudhury, Hong Xie, Lasata Shrestha, Shah Mohamed Bakhash, Michelle Lin, Noah R. Baker, Sean Ellis, Meei-Li Huang, Keith R Jerome, Alexander Greninger |
| EPI_ISL_1680251, EPI_ISL_1680252, EPI_ISL_1680254, EPI_ISL_1680255 | Helix/Illumina | Centers for Disease Control and Prevention Division of Viral Diseases, Pathogen Discovery | Dakota Howard, Dhwani Batra, Peter W. Cook, Kara Moser, Adrian Paskey, Jason Caravas, Benjamin Rambo-Martin, Shatavia Morrison, Christopher Gulvick, Scott Sammons, Yvette Unoarumhi, Darlene Wagner, Matthew Schmerer, Eileen de Feo, Jan Antico, Christine Tran, Matthew Tolentino, Shannon Wickline, Kim Gietzen, Brad Sickler, Jingtao Liu, Eric Allen, Phil Febbo, Nicole L. Washington, Simon White, Geraint Levan, Kelly Schiabor Barrett, Elizabeth Cirulli, Alexandre Bolze, Ary Ascencio, Charlotte Rivera-Garcia, Ryan Cho, Jason Nguyen, Sherry Wang, Jimmy Ramirez, Tyler Cassens, Efren Sandoval, Magnus Isaksson, William Lee, David Becker, Marc Laurent, James Lu, Clinton R. Paden, Duncan MacCannell |
| EPI_ISL_1680256 | UW Virology Lab | UW Virology Lab | Pavitra Roychoudhury, Hong Xie, Lasata Shrestha, Shah Mohamed Bakhash, Michelle Lin, Noah R. Baker, Sean Ellis, Meei-Li Huang, Keith R Jerome, Alexander Greninger |
| EPI_ISL_1680257, EPI_ISL_1680258 | Helix/Illumina | Centers for Disease Control and Prevention Division of Viral Diseases, Pathogen Discovery | Dakota Howard, Dhwani Batra, Peter W. Cook, Kara Moser, Adrian Paskey, Jason Caravas, Benjamin Rambo-Martin, Shatavia Morrison, Christopher Gulvick, Scott Sammons, Yvette Unoarumhi, Darlene Wagner, Matthew Schmerer, Eileen de Feo, Jan Antico, Christine Tran, Matthew Tolentino, Shannon Wickline, Kim Gietzen, Brad Sickler, Jingtao Liu, Eric Allen, Phil Febbo, Nicole L. Washington, Simon White, Geraint Levan, Kelly Schiabor Barrett, Elizabeth Cirulli, Alexandre Bolze, Ary Ascencio, Charlotte Rivera-Garcia, Ryan Cho, Jason Nguyen, Sherry Wang, Jimmy Ramirez, Tyler Cassens, Efren Sandoval, Magnus Isaksson, William Lee, David Becker, Marc Laurent, James Lu, Clinton R. Paden, Duncan MacCannell |
| EPI_ISL_1680259 | UW Virology Lab | UW Virology Lab | Pavitra Roychoudhury, Hong Xie, Lasata Shrestha, Shah Mohamed Bakhash, Michelle Lin, Noah R. Baker, Sean Ellis, Meei-Li Huang, Keith R Jerome, Alexander Greninger |
| EPI_ISL_1680260, EPI_ISL_1680261, EPI_ISL_1680263, EPI_ISL_1680264 | Helix/Illumina | Centers for Disease Control and Prevention Division of Viral Diseases, Pathogen Discovery | Dakota Howard, Dhwani Batra, Peter W. Cook, Kara Moser, Adrian Paskey, Jason Caravas, Benjamin Rambo-Martin, Shatavia Morrison, Christopher Gulvick, Scott Sammons, Yvette Unoarumhi, Darlene Wagner, Matthew Schmerer, Eileen de Feo, Jan Antico, Christine Tran, Matthew Tolentino, Shannon Wickline, Kim Gietzen, Brad Sickler, Jingtao Liu, Eric Allen, Phil Febbo, Nicole L. Washington, Simon White, Geraint Levan, Kelly Schiabor Barrett, Elizabeth Cirulli, Alexandre Bolze, Ary Ascencio, Charlotte Rivera-Garcia, Ryan Cho, Jason Nguyen, Sherry Wang, Jimmy Ramirez, Tyler Cassens, Efren Sandoval, Magnus Isaksson, William Lee, David Becker, Marc Laurent, James Lu, Clinton R. Paden, Duncan MacCannell |
| EPI_ISL_1680265 | UW Virology Lab | UW Virology Lab | Pavitra Roychoudhury, Hong Xie, Lasata Shrestha, Shah Mohamed Bakhash, Michelle Lin, Noah R. Baker, Sean Ellis, Meei-Li Huang, Keith R Jerome, Alexander Greninger |
| EPI_ISL_1680266, EPI_ISL_1680267, EPI_ISL_1680268 | Helix/Illumina | Centers for Disease Control and Prevention Division of Viral Diseases, Pathogen Discovery | Dakota Howard, Dhwani Batra, Peter W. Cook, Kara Moser, Adrian Paskey, Jason Caravas, Benjamin Rambo-Martin, Shatavia Morrison, Christopher Gulvick, Scott Sammons, Yvette Unoarumhi, Darlene Wagner, Matthew Schmerer, Eileen de Feo, Jan Antico, Christine Tran, Matthew Tolentino, Shannon Wickline, Kim Gietzen, Brad Sickler, Jingtao Liu, Eric Allen, Phil Febbo, Nicole L. Washington, Simon White, Geraint Levan, Kelly Schiabor Barrett, Elizabeth Cirulli, Alexandre Bolze, Ary Ascencio, Charlotte Rivera-Garcia, Ryan Cho, Jason Nguyen, Sherry Wang, Jimmy Ramirez, Tyler Cassens, Efren Sandoval, Magnus Isaksson, William Lee, David Becker, Marc Laurent, James Lu, Clinton R. Paden, Duncan MacCannell |
| EPI_ISL_1680269 | UW Virology Lab | UW Virology Lab | Pavitra Roychoudhury, Hong Xie, Lasata Shrestha, Shah Mohamed Bakhash, Michelle Lin, Noah R. Baker, Sean Ellis, Meei-Li Huang, Keith R Jerome, Alexander Greninger |
| EPI_ISL_1680270, EPI_ISL_1680271 | Helix/Illumina | Centers for Disease Control and Prevention Division of Viral Diseases, Pathogen Discovery | Dakota Howard, Dhwani Batra, Peter W. Cook, Kara Moser, Adrian Paskey, Jason Caravas, Benjamin Rambo-Martin, Shatavia Morrison, Christopher Gulvick, Scott Sammons, Yvette Unoarumhi, Darlene Wagner, Matthew Schmerer, Eileen de Feo, Jan Antico, Christine Tran, Matthew Tolentino, Shannon Wickline, Kim Gietzen, Brad Sickler, Jingtao Liu, Eric Allen, Phil Febbo, Nicole L. Washington, Simon White, Geraint Levan, Kelly Schiabor Barrett, Elizabeth Cirulli, Alexandre Bolze, Ary Ascencio, Charlotte Rivera-Garcia, Ryan Cho, Jason Nguyen, Sherry Wang, Jimmy Ramirez, Tyler Cassens, Efren Sandoval, Magnus Isaksson, William Lee, David Becker, Marc Laurent, James Lu, Clinton R. Paden, Duncan MacCannell |
| EPI_ISL_1680272 | UW Virology Lab | UW Virology Lab | Pavitra Roychoudhury, Hong Xie, Lasata Shrestha, Shah Mohamed Bakhash, Michelle Lin, Noah R. Baker, Sean Ellis, Meei-Li Huang, Keith R Jerome, Alexander Greninger |
| EPI_ISL_1680273, EPI_ISL_1680274 | Helix/Illumina | Centers for Disease Control and Prevention Division of Viral Diseases, Pathogen Discovery | Dakota Howard, Dhwani Batra, Peter W. Cook, Kara Moser, Adrian Paskey, Jason Caravas, Benjamin Rambo-Martin, Shatavia Morrison, Christopher Gulvick, Scott Sammons, Yvette Unoarumhi, Darlene Wagner, Matthew Schmerer, Eileen de Feo, Jan Antico, Christine Tran, Matthew Tolentino, Shannon Wickline, Kim Gietzen, Brad Sickler, Jingtao Liu, Eric Allen, Phil Febbo, Nicole L. Washington, Simon White, Geraint Levan, Kelly Schiabor Barrett, Elizabeth Cirulli, Alexandre Bolze, Ary Ascencio, Charlotte Rivera-Garcia, Ryan Cho, Jason Nguyen, Sherry Wang, Jimmy Ramirez, Tyler Cassens, Efren Sandoval, Magnus Isaksson, William Lee, David Becker, Marc Laurent, James Lu, Clinton R. Paden, Duncan MacCannell |
| EPI_ISL_1680275 | UW Virology Lab | UW Virology Lab | Pavitra Roychoudhury, Hong Xie, Lasata Shrestha, Shah Mohamed Bakhash, Michelle Lin, Noah R. Baker, Sean Ellis, Meei-Li Huang, Keith R Jerome, Alexander Greninger |
| EPI_ISL_1680276, EPI_ISL_1680277 | Helix/Illumina | Centers for Disease Control and Prevention Division of Viral Diseases, Pathogen Discovery | Dakota Howard, Dhwani Batra, Peter W. Cook, Kara Moser, Adrian Paskey, Jason Caravas, Benjamin Rambo-Martin, Shatavia Morrison, Christopher Gulvick, Scott Sammons, Yvette Unoarumhi, Darlene Wagner, Matthew Schmerer, Eileen de Feo, Jan Antico, Christine Tran, Matthew Tolentino, Shannon Wickline, Kim Gietzen, Brad Sickler, Jingtao Liu, Eric Allen, Phil Febbo, Nicole L. Washington, Simon White, Geraint Levan, Kelly Schiabor Barrett, Elizabeth Cirulli, Alexandre Bolze, Ary Ascencio, Charlotte Rivera-Garcia, Ryan Cho, Jason Nguyen, Sherry Wang, Jimmy Ramirez, Tyler Cassens, Efren Sandoval, Magnus Isaksson, William Lee, David Becker, Marc Laurent, James Lu, Clinton R. Paden, Duncan MacCannell |
| EPI_ISL_1680278 | UW Virology Lab | UW Virology Lab | Pavitra Roychoudhury, Hong Xie, Lasata Shrestha, Shah Mohamed Bakhash, Michelle Lin, Noah R. Baker, Sean Ellis, Meei-Li Huang, Keith R Jerome, Alexander Greninger |
| EPI_ISL_1680279, EPI_ISL_1680280 | Helix/Illumina | Centers for Disease Control and Prevention Division of Viral Diseases, Pathogen Discovery | Dakota Howard, Dhwani Batra, Peter W. Cook, Kara Moser, Adrian Paskey, Jason Caravas, Benjamin Rambo-Martin, Shatavia Morrison, Christopher Gulvick, Scott Sammons, Yvette Unoarumhi, Darlene Wagner, Matthew Schmerer, Eileen de Feo, Jan Antico, Christine Tran, Matthew Tolentino, Shannon Wickline, Kim Gietzen, Brad Sickler, Jingtao Liu, Eric Allen, Phil Febbo, Nicole L. Washington, Simon White, Geraint Levan, Kelly Schiabor Barrett, Elizabeth Cirulli, Alexandre Bolze, Ary Ascencio, Charlotte Rivera-Garcia, Ryan Cho, Jason Nguyen, Sherry Wang, Jimmy Ramirez, Tyler Cassens, Efren Sandoval, Magnus Isaksson, William Lee, David Becker, Marc Laurent, James Lu, Clinton R. Paden, Duncan MacCannell |
| EPI_ISL_1680281 | UW Virology Lab | UW Virology Lab | Pavitra Roychoudhury, Hong Xie, Lasata Shrestha, Shah Mohamed Bakhash, Michelle Lin, Noah R. Baker, Sean Ellis, Meei-Li Huang, Keith R Jerome, Alexander Greninger |
| EPI_ISL_1680282, EPI_ISL_1680283 | Helix/Illumina | Centers for Disease Control and Prevention Division of Viral Diseases, Pathogen Discovery | Dakota Howard, Dhwani Batra, Peter W. Cook, Kara Moser, Adrian Paskey, Jason Caravas, Benjamin Rambo-Martin, Shatavia Morrison, Christopher Gulvick, Scott Sammons, Yvette Unoarumhi, Darlene Wagner, Matthew Schmerer, Eileen de Feo, Jan Antico, Christine Tran, Matthew Tolentino, Shannon Wickline, Kim Gietzen, Brad Sickler, Jingtao Liu, Eric Allen, Phil Febbo, Nicole L. Washington, Simon White, Geraint Levan, Kelly Schiabor Barrett, Elizabeth Cirulli, Alexandre Bolze, Ary Ascencio, Charlotte Rivera-Garcia, Ryan Cho, Jason Nguyen, Sherry Wang, Jimmy Ramirez, Tyler Cassens, Efren Sandoval, Magnus Isaksson, William Lee, David Becker, Marc Laurent, James Lu, Clinton R. Paden, Duncan MacCannell |
| EPI_ISL_1680284 | UW Virology Lab | UW Virology Lab | Pavitra Roychoudhury, Hong Xie, Lasata Shrestha, Shah Mohamed Bakhash, Michelle Lin, Noah R. Baker, Sean Ellis, Meei-Li Huang, Keith R Jerome, Alexander Greninger |
| EPI_ISL_1680285, EPI_ISL_1680286 | Helix/Illumina | Centers for Disease Control and Prevention Division of Viral Diseases, Pathogen Discovery | Dakota Howard, Dhwani Batra, Peter W. Cook, Kara Moser, Adrian Paskey, Jason Caravas, Benjamin Rambo-Martin, Shatavia Morrison, Christopher Gulvick, Scott Sammons, Yvette Unoarumhi, Darlene Wagner, Matthew Schmerer, Eileen de Feo, Jan Antico, Christine Tran, Matthew Tolentino, Shannon Wickline, Kim Gietzen, Brad Sickler, Jingtao Liu, Eric Allen, Phil Febbo, Nicole L. Washington, Simon White, Geraint Levan, Kelly Schiabor Barrett, Elizabeth Cirulli, Alexandre Bolze, Ary Ascencio, Charlotte Rivera-Garcia, Ryan Cho, Jason Nguyen, Sherry Wang, Jimmy Ramirez, Tyler Cassens, Efren Sandoval, Magnus Isaksson, William Lee, David Becker, Marc Laurent, James Lu, Clinton R. Paden, Duncan MacCannell |
| EPI_ISL_1680287 | UW Virology Lab | UW Virology Lab | Pavitra Roychoudhury, Hong Xie, Lasata Shrestha, Shah Mohamed Bakhash, Michelle Lin, Noah R. Baker, Sean Ellis, Meei-Li Huang, Keith R Jerome, Alexander Greninger |
| EPI_ISL_1680288, EPI_ISL_1680289 | Helix/Illumina | Centers for Disease Control and Prevention Division of Viral Diseases, Pathogen Discovery | Dakota Howard, Dhwani Batra, Peter W. Cook, Kara Moser, Adrian Paskey, Jason Caravas, Benjamin Rambo-Martin, Shatavia Morrison, Christopher Gulvick, Scott Sammons, Yvette Unoarumhi, Darlene Wagner, Matthew Schmerer, Eileen de Feo, Jan Antico, Christine Tran, Matthew Tolentino, Shannon Wickline, Kim Gietzen, Brad Sickler, Jingtao Liu, Eric Allen, Phil Febbo, Nicole L. Washington, Simon White, Geraint Levan, Kelly Schiabor Barrett, Elizabeth Cirulli, Alexandre Bolze, Ary Ascencio, Charlotte Rivera-Garcia, Ryan Cho, Jason Nguyen, Sherry Wang, Jimmy Ramirez, Tyler Cassens, Efren Sandoval, Magnus Isaksson, William Lee, David Becker, Marc Laurent, James Lu, Clinton R. Paden, Duncan MacCannell |
| EPI_ISL_1680290 | UW Virology Lab | UW Virology Lab | Pavitra Roychoudhury, Hong Xie, Lasata Shrestha, Shah Mohamed Bakhash, Michelle Lin, Noah R. Baker, Sean Ellis, Meei-Li Huang, Keith R Jerome, Alexander Greninger |
| EPI_ISL_1680291, EPI_ISL_1680292 | Helix/Illumina | Centers for Disease Control and Prevention Division of Viral Diseases, Pathogen Discovery | Dakota Howard, Dhwani Batra, Peter W. Cook, Kara Moser, Adrian Paskey, Jason Caravas, Benjamin Rambo-Martin, Shatavia Morrison, Christopher Gulvick, Scott Sammons, Yvette Unoarumhi, Darlene Wagner, Matthew Schmerer, Eileen de Feo, Jan Antico, Christine Tran, Matthew Tolentino, Shannon Wickline, Kim Gietzen, Brad Sickler, Jingtao Liu, Eric Allen, Phil Febbo, Nicole L. Washington, Simon White, Geraint Levan, Kelly Schiabor Barrett, Elizabeth Cirulli, Alexandre Bolze, Ary Ascencio, Charlotte Rivera-Garcia, Ryan Cho, Jason Nguyen, Sherry Wang, Jimmy Ramirez, Tyler Cassens, Efren Sandoval, Magnus Isaksson, William Lee, David Becker, Marc Laurent, James Lu, Clinton R. Paden, Duncan MacCannell |
| EPI_ISL_1680293, EPI_ISL_1680294 | UW Virology Lab | UW Virology Lab | Pavitra Roychoudhury, Hong Xie, Lasata Shrestha, Shah Mohamed Bakhash, Michelle Lin, Noah R. Baker, Sean Ellis, Meei-Li Huang, Keith R Jerome, Alexander Greninger |
| EPI_ISL_1680295 | Helix/Illumina | Centers for Disease Control and Prevention Division of Viral Diseases, Pathogen Discovery | Dakota Howard, Dhwani Batra, Peter W. Cook, Kara Moser, Adrian Paskey, Jason Caravas, Benjamin Rambo-Martin, Shatavia Morrison, Christopher Gulvick, Scott Sammons, Yvette Unoarumhi, Darlene Wagner, Matthew Schmerer, Eileen de Feo, Jan Antico, Christine Tran, Matthew Tolentino, Shannon Wickline, Kim Gietzen, Brad Sickler, Jingtao Liu, Eric Allen, Phil Febbo, Nicole L. Washington, Simon White, Geraint Levan, Kelly Schiabor Barrett, Elizabeth Cirulli, Alexandre Bolze, Ary Ascencio, Charlotte Rivera-Garcia, Ryan Cho, Jason Nguyen, Sherry Wang, Jimmy Ramirez, Tyler Cassens, Efren Sandoval, Magnus Isaksson, William Lee, David Becker, Marc Laurent, James Lu, Clinton R. Paden, Duncan MacCannell |
| EPI_ISL_1680296 | UW Virology Lab | UW Virology Lab | Pavitra Roychoudhury, Hong Xie, Lasata Shrestha, Shah Mohamed Bakhash, Michelle Lin, Noah R. Baker, Sean Ellis, Meei-Li Huang, Keith R Jerome, Alexander Greninger |
| EPI_ISL_1680298 | Helix/Illumina | Centers for Disease Control and Prevention Division of Viral Diseases, Pathogen Discovery | Dakota Howard, Dhwani Batra, Peter W. Cook, Kara Moser, Adrian Paskey, Jason Caravas, Benjamin Rambo-Martin, Shatavia Morrison, Christopher Gulvick, Scott Sammons, Yvette Unoarumhi, Darlene Wagner, Matthew Schmerer, Eileen de Feo, Jan Antico, Christine Tran, Matthew Tolentino, Shannon Wickline, Kim Gietzen, Brad Sickler, Jingtao Liu, Eric Allen, Phil Febbo, Nicole L. Washington, Simon White, Geraint Levan, Kelly Schiabor Barrett, Elizabeth Cirulli, Alexandre Bolze, Ary Ascencio, Charlotte Rivera-Garcia, Ryan Cho, Jason Nguyen, Sherry Wang, Jimmy Ramirez, Tyler Cassens, Efren Sandoval, Magnus Isaksson, William Lee, David Becker, Marc Laurent, James Lu, Clinton R. Paden, Duncan MacCannell |
| EPI_ISL_1680299, EPI_ISL_1680300 | UW Virology Lab | UW Virology Lab | Pavitra Roychoudhury, Hong Xie, Lasata Shrestha, Shah Mohamed Bakhash, Michelle Lin, Noah R. Baker, Sean Ellis, Meei-Li Huang, Keith R Jerome, Alexander Greninger |
| EPI_ISL_1680301 | Helix/Illumina | Centers for Disease Control and Prevention Division of Viral Diseases, Pathogen Discovery | Dakota Howard, Dhwani Batra, Peter W. Cook, Kara Moser, Adrian Paskey, Jason Caravas, Benjamin Rambo-Martin, Shatavia Morrison, Christopher Gulvick, Scott Sammons, Yvette Unoarumhi, Darlene Wagner, Matthew Schmerer, Eileen de Feo, Jan Antico, Christine Tran, Matthew Tolentino, Shannon Wickline, Kim Gietzen, Brad Sickler, Jingtao Liu, Eric Allen, Phil Febbo, Nicole L. Washington, Simon White, Geraint Levan, Kelly Schiabor Barrett, Elizabeth Cirulli, Alexandre Bolze, Ary Ascencio, Charlotte Rivera-Garcia, Ryan Cho, Jason Nguyen, Sherry Wang, Jimmy Ramirez, Tyler Cassens, Efren Sandoval, Magnus Isaksson, William Lee, David Becker, Marc Laurent, James Lu, Clinton R. Paden, Duncan MacCannell |
| EPI_ISL_1680303 | UW Virology Lab | UW Virology Lab | Pavitra Roychoudhury, Hong Xie, Lasata Shrestha, Shah Mohamed Bakhash, Michelle Lin, Noah R. Baker, Sean Ellis, Meei-Li Huang, Keith R Jerome, Alexander Greninger |
| EPI_ISL_1680304 | Helix/Illumina | Centers for Disease Control and Prevention Division of Viral Diseases, Pathogen Discovery | Dakota Howard, Dhwani Batra, Peter W. Cook, Kara Moser, Adrian Paskey, Jason Caravas, Benjamin Rambo-Martin, Shatavia Morrison, Christopher Gulvick, Scott Sammons, Yvette Unoarumhi, Darlene Wagner, Matthew Schmerer, Eileen de Feo, Jan Antico, Christine Tran, Matthew Tolentino, Shannon Wickline, Kim Gietzen, Brad Sickler, Jingtao Liu, Eric Allen, Phil Febbo, Nicole L. Washington, Simon White, Geraint Levan, Kelly Schiabor Barrett, Elizabeth Cirulli, Alexandre Bolze, Ary Ascencio, Charlotte Rivera-Garcia, Ryan Cho, Jason Nguyen, Sherry Wang, Jimmy Ramirez, Tyler Cassens, Efren Sandoval, Magnus Isaksson, William Lee, David Becker, Marc Laurent, James Lu, Clinton R. Paden, Duncan MacCannell |
| EPI_ISL_1680305, EPI_ISL_1680306 | UW Virology Lab | UW Virology Lab | Pavitra Roychoudhury, Hong Xie, Lasata Shrestha, Shah Mohamed Bakhash, Michelle Lin, Noah R. Baker, Sean Ellis, Meei-Li Huang, Keith R Jerome, Alexander Greninger |
| EPI_ISL_1680307 | Helix/Illumina | Centers for Disease Control and Prevention Division of Viral Diseases, Pathogen Discovery | Dakota Howard, Dhwani Batra, Peter W. Cook, Kara Moser, Adrian Paskey, Jason Caravas, Benjamin Rambo-Martin, Shatavia Morrison, Christopher Gulvick, Scott Sammons, Yvette Unoarumhi, Darlene Wagner, Matthew Schmerer, Eileen de Feo, Jan Antico, Christine Tran, Matthew Tolentino, Shannon Wickline, Kim Gietzen, Brad Sickler, Jingtao Liu, Eric Allen, Phil Febbo, Nicole L. Washington, Simon White, Geraint Levan, Kelly Schiabor Barrett, Elizabeth Cirulli, Alexandre Bolze, Ary Ascencio, Charlotte Rivera-Garcia, Ryan Cho, Jason Nguyen, Sherry Wang, Jimmy Ramirez, Tyler Cassens, Efren Sandoval, Magnus Isaksson, William Lee, David Becker, Marc Laurent, James Lu, Clinton R. Paden, Duncan MacCannell |
| EPI_ISL_1680308, EPI_ISL_1680309 | UW Virology Lab | UW Virology Lab | Pavitra Roychoudhury, Hong Xie, Lasata Shrestha, Shah Mohamed Bakhash, Michelle Lin, Noah R. Baker, Sean Ellis, Meei-Li Huang, Keith R Jerome, Alexander Greninger |

| | | | |
|---|---|---|---|
| EPI_ISL_1680310 | Helix/Illumina | Centers for Disease Control and Prevention Division of Viral Diseases, Pathogen Discovery | Dakota Howard, Dhwani Batra, Peter W. Cook, Kara Moser, Adrian Paskey, Jason Caravas, Benjamin Rambo-Martin, Shatavia Morrison, Christopher Gulvick, Scott Sammons, Yvette Unoarumhi, Darlene Wagner, Matthew Schmerer, Eileen de Feo, Jan Antico, Christine Tran, Matthew Tolentino, Shannon Wickline, Kim Gietzen, Brad Sickler, Jingtao Liu, Eric Allen, Phil Febbo, Nicole L. Washington, Simon White, Geraint Levan, Kelly Schiabor Barrett, Elizabeth Cirulli, Alexandre Bolze, Ary Ascencio, Charlotte Rivera-Garcia, Ryan Cho, Jason Nguyen, Sherry Wang, Jimmy Ramirez, Tyler Cassens, Efren Sandoval, Magnus Isaksson, William Lee, David Becker, Marc Laurent, James Lu, Clinton R. Paden, Duncan MacCannell |
| EPI_ISL_1680311, EPI_ISL_1680312 | UW Virology Lab | UW Virology Lab | Pavitra Roychoudhury, Hong Xie, Lasata Shrestha, Shah Mohamed Bakhash, Michelle Lin, Noah R. Baker, Sean Ellis, Meei-Li Huang, Keith R Jerome, Alexander Greninger |
| EPI_ISL_1680313 | Helix/Illumina | Centers for Disease Control and Prevention Division of Viral Diseases, Pathogen Discovery | Dakota Howard, Dhwani Batra, Peter W. Cook, Kara Moser, Adrian Paskey, Jason Caravas, Benjamin Rambo-Martin, Shatavia Morrison, Christopher Gulvick, Scott Sammons, Yvette Unoarumhi, Darlene Wagner, Matthew Schmerer, Eileen de Feo, Jan Antico, Christine Tran, Matthew Tolentino, Shannon Wickline, Kim Gietzen, Brad Sickler, Jingtao Liu, Eric Allen, Phil Febbo, Nicole L. Washington, Simon White, Geraint Levan, Kelly Schiabor Barrett, Elizabeth Cirulli, Alexandre Bolze, Ary Ascencio, Charlotte Rivera-Garcia, Ryan Cho, Jason Nguyen, Sherry Wang, Jimmy Ramirez, Tyler Cassens, Efren Sandoval, Magnus Isaksson, William Lee, David Becker, Marc Laurent, James Lu, Clinton R. Paden, Duncan MacCannell |
| EPI_ISL_1680314, EPI_ISL_1680315 | UW Virology Lab | UW Virology Lab | Pavitra Roychoudhury, Hong Xie, Lasata Shrestha, Shah Mohamed Bakhash, Michelle Lin, Noah R. Baker, Sean Ellis, Meei-Li Huang, Keith R Jerome, Alexander Greninger |
| EPI_ISL_1680316 | Helix/Illumina | Centers for Disease Control and Prevention Division of Viral Diseases, Pathogen Discovery | Dakota Howard, Dhwani Batra, Peter W. Cook, Kara Moser, Adrian Paskey, Jason Caravas, Benjamin Rambo-Martin, Shatavia Morrison, Christopher Gulvick, Scott Sammons, Yvette Unoarumhi, Darlene Wagner, Matthew Schmerer, Eileen de Feo, Jan Antico, Christine Tran, Matthew Tolentino, Shannon Wickline, Kim Gietzen, Brad Sickler, Jingtao Liu, Eric Allen, Phil Febbo, Nicole L. Washington, Simon White, Geraint Levan, Kelly Schiabor Barrett, Elizabeth Cirulli, Alexandre Bolze, Ary Ascencio, Charlotte Rivera-Garcia, Ryan Cho, Jason Nguyen, Sherry Wang, Jimmy Ramirez, Tyler Cassens, Efren Sandoval, Magnus Isaksson, William Lee, David Becker, Marc Laurent, James Lu, Clinton R. Paden, Duncan MacCannell |
| EPI_ISL_1680317, EPI_ISL_1680318 | UW Virology Lab | UW Virology Lab | Pavitra Roychoudhury, Hong Xie, Lasata Shrestha, Shah Mohamed Bakhash, Michelle Lin, Noah R. Baker, Sean Ellis, Meei-Li Huang, Keith R Jerome, Alexander Greninger |
| EPI_ISL_1680319 | Helix/Illumina | Centers for Disease Control and Prevention Division of Viral Diseases, Pathogen Discovery | Dakota Howard, Dhwani Batra, Peter W. Cook, Kara Moser, Adrian Paskey, Jason Caravas, Benjamin Rambo-Martin, Shatavia Morrison, Christopher Gulvick, Scott Sammons, Yvette Unoarumhi, Darlene Wagner, Matthew Schmerer, Eileen de Feo, Jan Antico, Christine Tran, Matthew Tolentino, Shannon Wickline, Kim Gietzen, Brad Sickler, Jingtao Liu, Eric Allen, Phil Febbo, Nicole L. Washington, Simon White, Geraint Levan, Kelly Schiabor Barrett, Elizabeth Cirulli, Alexandre Bolze, Ary Ascencio, Charlotte Rivera-Garcia, Ryan Cho, Jason Nguyen, Sherry Wang, Jimmy Ramirez, Tyler Cassens, Efren Sandoval, Magnus Isaksson, William Lee, David Becker, Marc Laurent, James Lu, Clinton R. Paden, Duncan MacCannell |
| EPI_ISL_1680320, EPI_ISL_1680321 | UW Virology Lab | UW Virology Lab | Pavitra Roychoudhury, Hong Xie, Lasata Shrestha, Shah Mohamed Bakhash, Michelle Lin, Noah R. Baker, Sean Ellis, Meei-Li Huang, Keith R Jerome, Alexander Greninger |
| EPI_ISL_1680322 | Helix/Illumina | Centers for Disease Control and Prevention Division of Viral Diseases, Pathogen Discovery | Dakota Howard, Dhwani Batra, Peter W. Cook, Kara Moser, Adrian Paskey, Jason Caravas, Benjamin Rambo-Martin, Shatavia Morrison, Christopher Gulvick, Scott Sammons, Yvette Unoarumhi, Darlene Wagner, Matthew Schmerer, Eileen de Feo, Jan Antico, Christine Tran, Matthew Tolentino, Shannon Wickline, Kim Gietzen, Brad Sickler, Jingtao Liu, Eric Allen, Phil Febbo, Nicole L. Washington, Simon White, Geraint Levan, Kelly Schiabor Barrett, Elizabeth Cirulli, Alexandre Bolze, Ary Ascencio, Charlotte Rivera-Garcia, Ryan Cho, Jason Nguyen, Sherry Wang, Jimmy Ramirez, Tyler Cassens, Efren Sandoval, Magnus Isaksson, William Lee, David Becker, Marc Laurent, James Lu, Clinton R. Paden, Duncan MacCannell |
| EPI_ISL_1680323 | UW Virology Lab | UW Virology Lab | Pavitra Roychoudhury, Hong Xie, Lasata Shrestha, Shah Mohamed Bakhash, Michelle Lin, Noah R. Baker, Sean Ellis, Meei-Li Huang, Keith R Jerome, Alexander Greninger |
| EPI_ISL_1680324, EPI_ISL_1680325 | Helix/Illumina | Centers for Disease Control and Prevention Division of Viral Diseases, Pathogen Discovery | Dakota Howard, Dhwani Batra, Peter W. Cook, Kara Moser, Adrian Paskey, Jason Caravas, Benjamin Rambo-Martin, Shatavia Morrison, Christopher Gulvick, Scott Sammons, Yvette Unoarumhi, Darlene Wagner, Matthew Schmerer, Eileen de Feo, Jan Antico, Christine Tran, Matthew Tolentino, Shannon Wickline, Kim Gietzen, Brad Sickler, Jingtao Liu, Eric Allen, Phil Febbo, Nicole L. Washington, Simon White, Geraint Levan, Kelly Schiabor Barrett, Elizabeth Cirulli, Alexandre Bolze, Ary Ascencio, Charlotte Rivera-Garcia, Ryan Cho, Jason Nguyen, Sherry Wang, Jimmy Ramirez, Tyler Cassens, Efren Sandoval, Magnus Isaksson, William Lee, David Becker, Marc Laurent, James Lu, Clinton R. Paden, Duncan MacCannell |
| EPI_ISL_1680326 | UW Virology Lab | UW Virology Lab | Pavitra Roychoudhury, Hong Xie, Lasata Shrestha, Shah Mohamed Bakhash, Michelle Lin, Noah R. Baker, Sean Ellis, Meei-Li Huang, Keith R Jerome, Alexander Greninger |
| EPI_ISL_1680327, EPI_ISL_1680328, EPI_ISL_1680329 | Helix/Illumina | Centers for Disease Control and Prevention Division of Viral Diseases, Pathogen Discovery | Dakota Howard, Dhwani Batra, Peter W. Cook, Kara Moser, Adrian Paskey, Jason Caravas, Benjamin Rambo-Martin, Shatavia Morrison, Christopher Gulvick, Scott Sammons, Yvette Unoarumhi, Darlene Wagner, Matthew Schmerer, Eileen de Feo, Jan Antico, Christine Tran, Matthew Tolentino, Shannon Wickline, Kim Gietzen, Brad Sickler, Jingtao Liu, Eric Allen, Phil Febbo, Nicole L. Washington, Simon White, Geraint Levan, Kelly Schiabor Barrett, Elizabeth Cirulli, Alexandre Bolze, Ary Ascencio, Charlotte Rivera-Garcia, Ryan Cho, Jason Nguyen, Sherry Wang, Jimmy Ramirez, Tyler Cassens, Efren Sandoval, Magnus Isaksson, William Lee, David Becker, Marc Laurent, James Lu, Clinton R. Paden, Duncan MacCannell |
| EPI_ISL_1680330 | UW Virology Lab | UW Virology Lab | Pavitra Roychoudhury, Hong Xie, Lasata Shrestha, Shah Mohamed Bakhash, Michelle Lin, Noah R. Baker, Sean Ellis, Meei-Li Huang, Keith R Jerome, Alexander Greninger |
| EPI_ISL_1680331 | Helix/Illumina | Centers for Disease Control and Prevention Division of Viral Diseases, Pathogen Discovery | Dakota Howard, Dhwani Batra, Peter W. Cook, Kara Moser, Adrian Paskey, Jason Caravas, Benjamin Rambo-Martin, Shatavia Morrison, Christopher Gulvick, Scott Sammons, Yvette Unoarumhi, Darlene Wagner, Matthew Schmerer, Eileen de Feo, Jan Antico, Christine Tran, Matthew Tolentino, Shannon Wickline, Kim Gietzen, Brad Sickler, Jingtao Liu, Eric Allen, Phil Febbo, Nicole L. Washington, Simon White, Geraint Levan, Kelly Schiabor Barrett, Elizabeth Cirulli, Alexandre Bolze, Ary Ascencio, Charlotte Rivera-Garcia, Ryan Cho, Jason Nguyen, Sherry Wang, Jimmy Ramirez, Tyler Cassens, Efren Sandoval, Magnus Isaksson, William Lee, David Becker, Marc Laurent, James Lu, Clinton R. Paden, Duncan MacCannell |
| EPI_ISL_1680332, EPI_ISL_1680333, EPI_ISL_1680334, EPI_ISL_1680335, EPI_ISL_1680336, EPI_ISL_1680337, EPI_ISL_1680338, EPI_ISL_1680339, EPI_ISL_1680340, EPI_ISL_1680341, EPI_ISL_1680342, EPI_ISL_1680343, EPI_ISL_1680344, EPI_ISL_1680346, EPI_ISL_1680347, EPI_ISL_1680348, EPI_ISL_1680349, EPI_ISL_1680350, EPI_ISL_1680351, EPI_ISL_1680352, EPI_ISL_1680353, EPI_ISL_1680354, EPI_ISL_1680355, EPI_ISL_1680356, EPI_ISL_1680357, EPI_ISL_1680358, EPI_ISL_1680359, EPI_ISL_1680360, EPI_ISL_1680361, EPI_ISL_1680362, EPI_ISL_1680363, EPI_ISL_1680364, EPI_ISL_1680365, EPI_ISL_1680366, EPI_ISL_1680367, EPI_ISL_1680368, EPI_ISL_1680369, EPI_ISL_1680370, EPI_ISL_1680371, EPI_ISL_1680372, EPI_ISL_1680373, EPI_ISL_1680374, EPI_ISL_1680375, EPI_ISL_1680376, EPI_ISL_1680377, EPI_ISL_1680378, EPI_ISL_1680379, EPI_ISL_1680380, EPI_ISL_1680381, EPI_ISL_1680382, EPI_ISL_1680383, EPI_ISL_1680384, EPI_ISL_1680385, EPI_ISL_1680386, EPI_ISL_1680387, EPI_ISL_1680388, EPI_ISL_1680389, EPI_ISL_1680390, EPI_ISL_1680391, EPI_ISL_1680392, EPI_ISL_1680393, EPI_ISL_1680394, EPI_ISL_1680395, EPI_ISL_1680396, EPI_ISL_1680397, EPI_ISL_1680398, EPI_ISL_1680399, EPI_ISL_1680400, EPI_ISL_1680401, EPI_ISL_1680402, EPI_ISL_1680403, EPI_ISL_1680404, EPI_ISL_1680405, EPI_ISL_1680406, EPI_ISL_1680407, EPI_ISL_1680409, EPI_ISL_1680410, EPI_ISL_1680411, EPI_ISL_1680412, EPI_ISL_1680413, EPI_ISL_1680415, EPI_ISL_1680416, EPI_ISL_1680417, EPI_ISL_1680418, EPI_ISL_1680419, EPI_ISL_1680420, EPI_ISL_1680421, EPI_ISL_1680422, EPI_ISL_1680423, EPI_ISL_1680424, EPI_ISL_1680425, EPI_ISL_1680426, EPI_ISL_1680427, EPI_ISL_1680428, EPI_ISL_1680429, EPI_ISL_1680430, EPI_ISL_1680431, EPI_ISL_1680432, EPI_ISL_1680433, EPI_ISL_1680434, EPI_ISL_1680435, EPI_ISL_1680436, EPI_ISL_1680437, EPI_ISL_1680438, EPI_ISL_1680439, EPI_ISL_1680440, EPI_ISL_1680441, EPI_ISL_1680442, EPI_ISL_1680443, EPI_ISL_1680444, EPI_ISL_1680445, EPI_ISL_1680446, EPI_ISL_1680447, EPI_ISL_1680448, EPI_ISL_1680449, EPI_ISL_1680451, EPI_ISL_1680452, EPI_ISL_1680453, EPI_ISL_1680454, EPI_ISL_1680455, EPI_ISL_1680456, EPI_ISL_1680457, EPI_ISL_1680458, EPI_ISL_1680459, EPI_ISL_1680460, EPI_ISL_1680461, EPI_ISL_1680462, EPI_ISL_1680463, EPI_ISL_1680464, EPI_ISL_1680465 | | | |
| see above | UW Virology Lab | UW Virology Lab | Pavitra Roychoudhury, Hong Xie, Lasata Shrestha, Shah Mohamed Bakhash, Michelle Lin, Noah R. Baker, Sean Ellis, Meei-Li Huang, Keith R Jerome, Alexander Greninger |
| EPI_ISL_1680467, EPI_ISL_1680468, EPI_ISL_1680469, EPI_ISL_1680471, EPI_ISL_1680472, EPI_ISL_1680475, EPI_ISL_1680477, EPI_ISL_1680478, EPI_ISL_1680479, EPI_ISL_1680480, EPI_ISL_1680481, EPI_ISL_1680482, EPI_ISL_1680484, EPI_ISL_1680487, EPI_ISL_1680489, EPI_ISL_1680490, EPI_ISL_1680491, EPI_ISL_1680492, EPI_ISL_1680493, EPI_ISL_1680494, EPI_ISL_1680495, EPI_ISL_1680497, EPI_ISL_1680498, EPI_ISL_1680499, EPI_ISL_1680500, EPI_ISL_1680501, EPI_ISL_1680502, EPI_ISL_1680503, EPI_ISL_1680504, EPI_ISL_1680505, EPI_ISL_1680506, EPI_ISL_1680507, EPI_ISL_1680508, EPI_ISL_1680509, EPI_ISL_1680510, EPI_ISL_1680511, EPI_ISL_1680512, EPI_ISL_1680513, EPI_ISL_1680514, EPI_ISL_1680515, EPI_ISL_1680516, EPI_ISL_1680518, EPI_ISL_1680519, EPI_ISL_1680520, EPI_ISL_1680521, EPI_ISL_1680522, EPI_ISL_1680523, EPI_ISL_1680524, EPI_ISL_1680525, EPI_ISL_1680527, EPI_ISL_1680528, EPI_ISL_1680529, EPI_ISL_1680530, EPI_ISL_1680532, EPI_ISL_1680533, EPI_ISL_1680534, EPI_ISL_1680535, EPI_ISL_1680536, EPI_ISL_1680537, EPI_ISL_1680538, EPI_ISL_1680539, EPI_ISL_1680540, EPI_ISL_1680542, EPI_ISL_1680543, EPI_ISL_1680544, EPI_ISL_1680545, EPI_ISL_1680546, EPI_ISL_1680547, EPI_ISL_1680548, EPI_ISL_1680549, EPI_ISL_1680551, EPI_ISL_1680554, EPI_ISL_1680555, EPI_ISL_1680556, EPI_ISL_1680558, EPI_ISL_1680559, EPI_ISL_1680561, EPI_ISL_1680563, EPI_ISL_1680564, EPI_ISL_1680565, EPI_ISL_1680566, EPI_ISL_1680567, EPI_ISL_1680568, EPI_ISL_1680570, EPI_ISL_1680571, EPI_ISL_1680572, EPI_ISL_1680573, EPI_ISL_1680574, EPI_ISL_1680575, EPI_ISL_1680576, EPI_ISL_1680577, EPI_ISL_1680578, EPI_ISL_1680581, EPI_ISL_1680582, EPI_ISL_1680583, EPI_ISL_1680584, EPI_ISL_1680585, EPI_ISL_1680586, EPI_ISL_1680588, EPI_ISL_1680589, EPI_ISL_1680591, EPI_ISL_1680592, EPI_ISL_1680594, EPI_ISL_1680595, EPI_ISL_1680596, EPI_ISL_1680597, EPI_ISL_1680598, EPI_ISL_1680599, EPI_ISL_1680602, EPI_ISL_1680603, EPI_ISL_1680604, EPI_ISL_1680605, EPI_ISL_1680606, EPI_ISL_1680607, EPI_ISL_1680608, EPI_ISL_1680609, EPI_ISL_1680610, EPI_ISL_1680612, EPI_ISL_1680613, EPI_ISL_1680614, EPI_ISL_1680615, EPI_ISL_1680616, EPI_ISL_1680617, EPI_ISL_1680618, EPI_ISL_1680620, EPI_ISL_1680621, EPI_ISL_1680623, EPI_ISL_1680624, EPI_ISL_1680625, EPI_ISL_1680626, EPI_ISL_1680627, EPI_ISL_1680628, EPI_ISL_1680629, EPI_ISL_1680630, EPI_ISL_1680631, EPI_ISL_1680632, EPI_ISL_1680634, EPI_ISL_1680635, EPI_ISL_1680636, EPI_ISL_1680637, EPI_ISL_1680639, EPI_ISL_1680640, EPI_ISL_1680641, EPI_ISL_1680642, EPI_ISL_1680643, EPI_ISL_1680644, EPI_ISL_1680645, EPI_ISL_1680646, EPI_ISL_1680647, EPI_ISL_1680649, EPI_ISL_1680650, EPI_ISL_1680651, EPI_ISL_1680652, EPI_ISL_1680655, EPI_ISL_1680656, EPI_ISL_1680657, EPI_ISL_1680659, EPI_ISL_1680660, EPI_ISL_1680661, EPI_ISL_1680662, EPI_ISL_1680663, EPI_ISL_1680664, EPI_ISL_1680665, EPI_ISL_1680666, EPI_ISL_1680667, EPI_ISL_1680668, EPI_ISL_1680669, EPI_ISL_1680672, EPI_ISL_1680673, EPI_ISL_1680674, EPI_ISL_1680675, EPI_ISL_1680676, EPI_ISL_1680679, EPI_ISL_1680680, EPI_ISL_1680683, EPI_ISL_1680684, EPI_ISL_1680685, EPI_ISL_1680686, EPI_ISL_1680687, EPI_ISL_1680689, EPI_ISL_1680690, EPI_ISL_1680691, EPI_ISL_1680692, EPI_ISL_1680693, EPI_ISL_1680694, EPI_ISL_1680695, EPI_ISL_1680696, EPI_ISL_1680697, EPI_ISL_1680699, EPI_ISL_1680700, EPI_ISL_1680701, EPI_ISL_1680702, EPI_ISL_1680703, EPI_ISL_1680704, EPI_ISL_1680705, EPI_ISL_1680707, EPI_ISL_1680708, EPI_ISL_1680709, EPI_ISL_1680711, EPI_ISL_1680712, EPI_ISL_1680713, EPI_ISL_1680715, EPI_ISL_1680716, EPI_ISL_1680717, EPI_ISL_1680718, EPI_ISL_1680719, EPI_ISL_1680720, EPI_ISL_1680723, EPI_ISL_1680724, EPI_ISL_1680725, EPI_ISL_1680726, EPI_ISL_1680727, EPI_ISL_1680728, EPI_ISL_1680730, EPI_ISL_1680732, EPI_ISL_1680733, EPI_ISL_1680734, EPI_ISL_1680735, EPI_ISL_1680736, EPI_ISL_1680737, EPI_ISL_1680738, EPI_ISL_1680741, EPI_ISL_1680742, EPI_ISL_1680743, EPI_ISL_1680744, EPI_ISL_1680745, EPI_ISL_1680746, EPI_ISL_1680748, EPI_ISL_1680749, EPI_ISL_1680750, EPI_ISL_1680752, EPI_ISL_1680753, EPI_ISL_1680754, EPI_ISL_1680755, EPI_ISL_1680756, EPI_ISL_1680757, EPI_ISL_1680758, EPI_ISL_1680759, EPI_ISL_1680760, EPI_ISL_1680761, EPI_ISL_1680762, EPI_ISL_1680763, EPI_ISL_1680765, EPI_ISL_1680766, EPI_ISL_1680767, EPI_ISL_1680769, EPI_ISL_1680770, EPI_ISL_1680771, EPI_ISL_1680772, EPI_ISL_1680773, EPI_ISL_1680774, EPI_ISL_1680775, EPI_ISL_1680776, EPI_ISL_1680777, EPI_ISL_1680778, EPI_ISL_1680780, EPI_ISL_1680781, EPI_ISL_1680782, EPI_ISL_1680783, EPI_ISL_1680784, EPI_ISL_1680785, EPI_ISL_1680786, EPI_ISL_1680787, EPI_ISL_1680788, EPI_ISL_1680789, EPI_ISL_1680790, EPI_ISL_1680791, EPI_ISL_1680792, EPI_ISL_1680793, EPI_ISL_1680794, EPI_ISL_1680795, EPI_ISL_1680796, EPI_ISL_1680797, EPI_ISL_1680798, EPI_ISL_1680799, EPI_ISL_1680801, EPI_ISL_1680802, EPI_ISL_1680803, EPI_ISL_1680804, EPI_ISL_1680805, EPI_ISL_1680806, EPI_ISL_1680807, EPI_ISL_1680808, EPI_ISL_1680809, EPI_ISL_1680810, EPI_ISL_1680811, EPI_ISL_1680812, EPI_ISL_1680814, EPI_ISL_1680816, EPI_ISL_1680817, EPI_ISL_1680818, EPI_ISL_1680820, EPI_ISL_1680821, EPI_ISL_1680822, EPI_ISL_1680823, EPI_ISL_1680825, EPI_ISL_1680828, EPI_ISL_1680830, EPI_ISL_1680831, EPI_ISL_1680833, EPI_ISL_1680835, EPI_ISL_1680836, EPI_ISL_1680837, EPI_ISL_1680840, EPI_ISL_1680841, EPI_ISL_1680842, EPI_ISL_1680843, EPI_ISL_1680845, EPI_ISL_1680849, EPI_ISL_1680850, EPI_ISL_1680851, EPI_ISL_1680852, EPI_ISL_1680853, EPI_ISL_1680854, EPI_ISL_1680855, EPI_ISL_1680856, EPI_ISL_1680857, EPI_ISL_1680859, EPI_ISL_1680860, EPI_ISL_1680861, EPI_ISL_1680862, EPI_ISL_1680863, EPI_ISL_1680864, EPI_ISL_1680865, EPI_ISL_1680866, EPI_ISL_1680867, EPI_ISL_1680869, EPI_ISL_1680870, EPI_ISL_1680872, EPI_ISL_1680873, EPI_ISL_1680874, EPI_ISL_1680875, EPI_ISL_1680876, EPI_ISL_1680877, EPI_ISL_1680878, EPI_ISL_1680879, EPI_ISL_1680880, EPI_ISL_1680881, EPI_ISL_1680882, EPI_ISL_1680883, EPI_ISL_1680884, EPI_ISL_1680885, EPI_ISL_1680886, EPI_ISL_1680887, EPI_ISL_1680888, EPI_ISL_1680889, EPI_ISL_1680890, EPI_ISL_1680891, EPI_ISL_1680892, EPI_ISL_1680893, EPI_ISL_1680896, EPI_ISL_1680897, EPI_ISL_1680898, EPI_ISL_1680899, EPI_ISL_1680900, EPI_ISL_1680902, EPI_ISL_1680903, EPI_ISL_1680905, EPI_ISL_1680906, EPI_ISL_1680907, EPI_ISL_1680908, EPI_ISL_1680909, EPI_ISL_1680910, EPI_ISL_1680911, EPI_ISL_1680912, EPI_ISL_1680913, EPI_ISL_1680914, EPI_ISL_1680915, EPI_ISL_1680916 | | | |
| see above | Laboratory Corporation of America | Centers for Disease Control and Prevention Division of Viral Diseases, Pathogen Discovery | Dakota Howard, Dhwani Batra, Peter W. Cook, Kara Moser, Adrian Paskey, Jason Caravas, Benjamin Rambo-Martin, Shatavia Morrison, Christopher Gulvick, Scott Sammons, Yvette Unoarumhi, Darlene Wagner, Matthew Schmerer, Minoo Agarwal, Eyad Almasri, Debbie Boles, Ayla Burns, Nuthawin Charoensri, Oren Cohen, Susan Countryman, Mary Ann Cristobal, Bobbi Croy, Suzanne Dale, Hrushikesh Deshmukh, Amanda Douglas, Vincent Drouillon, Marcia Eisenberg, Howard Engler, Rama Ghatti, Prashant Gupta, Susan Hicks, Jake Humphrey, Lax Iyer, Manoj Jain, Mohan Kolli, Brian Krueger, Tim Kuphal, Stanley Letovsky, Michael Levandoski, Craig Lukasik, Jonathan Meltzer, Brian Norvell, Mindy Nye, Scott Parker, Christos Petropoulos, John Pruitt, Steven Ragan, Scott Ryan, Mike Sapeta, Jana Schroth, Suresh Babu Selvaraju, Goran Stevovic, Amanda Suchanek, Andrea Throop, Lyndon Tilson, Thomas Urban, Joe Voshell, Kimberly Wagner, Jonathan Williams, Mary Williamson, Qian Zeng, Tricia Zwiefelhofer, Clinton R. Paden, Duncan MacCannell |
| EPI_ISL_1680917 | Ohio Department of Health Laboratory | Ohio Department of Health Laboratory | Holmes, Jennifer; Eric Brandt, Keoni Omura, Glen McGillivary, Caitlin McDonnell, Jade Mowery, Stephanie Mccracken, Tyler Payne, Kirtana Ramadugu, Erica Leasure, Brent Lee, Kelsey Florek, Heather Blankenship, Allison Black, Quanta Brown, and Tammy Bannerman |
| EPI_ISL_1680918, EPI_ISL_1680919, EPI_ISL_1680920, EPI_ISL_1680921, EPI_ISL_1680922, EPI_ISL_1680923, EPI_ISL_1680924, EPI_ISL_1680925, EPI_ISL_1680926, EPI_ISL_1680927, EPI_ISL_1680928, EPI_ISL_1680929, EPI_ISL_1680930, EPI_ISL_1680932, EPI_ISL_1680934, EPI_ISL_1680935, EPI_ISL_1680936, EPI_ISL_1680937, EPI_ISL_1680939, EPI_ISL_1680940, EPI_ISL_1680941, EPI_ISL_1680942, EPI_ISL_1680943, EPI_ISL_1680944, EPI_ISL_1680945, EPI_ISL_1680946, EPI_ISL_1680947, EPI_ISL_1680948, EPI_ISL_1680949, EPI_ISL_1680950, EPI_ISL_1680951, EPI_ISL_1680952, EPI_ISL_1680953, EPI_ISL_1680954, EPI_ISL_1680955, EPI_ISL_1680956, EPI_ISL_1680957, EPI_ISL_1680958, EPI_ISL_1680959, EPI_ISL_1680960, EPI_ISL_1680961, EPI_ISL_1680962, EPI_ISL_1680963, EPI_ISL_1680964, EPI_ISL_1680965, EPI_ISL_1680966, EPI_ISL_1680967, EPI_ISL_1680968, EPI_ISL_1680969, EPI_ISL_1680970, EPI_ISL_1680971, EPI_ISL_1680972, EPI_ISL_1680973, EPI_ISL_1680974, EPI_ISL_1680975, EPI_ISL_1680976, EPI_ISL_1680977, EPI_ISL_1680978, EPI_ISL_1680979, EPI_ISL_1680980, EPI_ISL_1680981, EPI_ISL_1680982, EPI_ISL_1680983, EPI_ISL_1680984, EPI_ISL_1680985, EPI_ISL_1680987, EPI_ISL_1680988, EPI_ISL_1680989, EPI_ISL_1680991, EPI_ISL_1680992, EPI_ISL_1680993, EPI_ISL_1680994, EPI_ISL_1680995, EPI_ISL_1680997, EPI_ISL_1680998, EPI_ISL_1680999, EPI_ISL_1681000, EPI_ISL_1681001, EPI_ISL_1681002, EPI_ISL_1681003, EPI_ISL_1681004, EPI_ISL_1681005, EPI_ISL_1681006, EPI_ISL_1681007, EPI_ISL_1681008, EPI_ISL_1681010, EPI_ISL_1681011, EPI_ISL_1681012, EPI_ISL_1681013, EPI_ISL_1681014, EPI_ISL_1681015, EPI_ISL_1681016, EPI_ISL_1681017, EPI_ISL_1681019, EPI_ISL_1681020, EPI_ISL_1681021, EPI_ISL_1681023, EPI_ISL_1681024, EPI_ISL_1681025, EPI_ISL_1681026, EPI_ISL_1681027, EPI_ISL_1681028, EPI_ISL_1681029, EPI_ISL_1681030, EPI_ISL_1681031, EPI_ISL_1681032, EPI_ISL_1681033, EPI_ISL_1681034, EPI_ISL_1681035, EPI_ISL_1681036, EPI_ISL_1681037, EPI_ISL_1681039, EPI_ISL_1681040, EPI_ISL_1681041, EPI_ISL_1681042, EPI_ISL_1681044, EPI_ISL_1681045, EPI_ISL_1681047, EPI_ISL_1681048, EPI_ISL_1681049, EPI_ISL_1681050, EPI_ISL_1681051, EPI_ISL_1681052, EPI_ISL_1681053, EPI_ISL_1681054, EPI_ISL_1681055, EPI_ISL_1681056, EPI_ISL_1681058, EPI_ISL_1681060, EPI_ISL_1681061, EPI_ISL_1681062, EPI_ISL_1681063, EPI_ISL_1681064, EPI_ISL_1681065, EPI_ISL_1681066, EPI_ISL_1681067, EPI_ISL_1681068, EPI_ISL_1681070, EPI_ISL_1681071, EPI_ISL_1681072, EPI_ISL_1681073, EPI_ISL_1681074, EPI_ISL_1681075, EPI_ISL_1681076, EPI_ISL_1681077, EPI_ISL_1681078, EPI_ISL_1681079, EPI_ISL_1681081, EPI_ISL_1681082, EPI_ISL_1681083, EPI_ISL_1681084, EPI_ISL_1681085, EPI_ISL_1681086, EPI_ISL_1681087, EPI_ISL_1681088, EPI_ISL_1681089, EPI_ISL_1681090, EPI_ISL_1681091, EPI_ISL_1681092, EPI_ISL_1681093, EPI_ISL_1681094, EPI_ISL_1681095, EPI_ISL_1681096, EPI_ISL_1681097, EPI_ISL_1681098, EPI_ISL_1681099, EPI_ISL_1681100, EPI_ISL_1681101, EPI_ISL_1681103, EPI_ISL_1681104, EPI_ISL_1681105, EPI_ISL_1681107, EPI_ISL_1681108, EPI_ISL_1681109, EPI_ISL_1681110, EPI_ISL_1681111, EPI_ISL_1681112, EPI_ISL_1681113, EPI_ISL_1681114, EPI_ISL_1681115, EPI_ISL_1681116, EPI_ISL_1681117, EPI_ISL_1681118, EPI_ISL_1681119, EPI_ISL_1681121, EPI_ISL_1681122, EPI_ISL_1681123, EPI_ISL_1681124, EPI_ISL_1681125, EPI_ISL_1681126, EPI_ISL_1681127, EPI_ISL_1681128, EPI_ISL_1681129, EPI_ISL_1681130, EPI_ISL_1681131, EPI_ISL_1681132, EPI_ISL_1681133, EPI_ISL_1681135, EPI_ISL_1681137, EPI_ISL_1681138, EPI_ISL_1681139, EPI_ISL_1681140, EPI_ISL_1681142, EPI_ISL_1681145, EPI_ISL_1681146, EPI_ISL_1681148, EPI_ISL_1681149, EPI_ISL_1681150, EPI_ISL_1681151, EPI_ISL_1681152, EPI_ISL_1681153, EPI_ISL_1681154, EPI_ISL_1681155, EPI_ISL_1681156, EPI_ISL_1681157, EPI_ISL_1681158, EPI_ISL_1681159, EPI_ISL_1681160, EPI_ISL_1681161, EPI_ISL_1681162, EPI_ISL_1681163, EPI_ISL_1681164, EPI_ISL_1681165, EPI_ISL_1681167, EPI_ISL_1681168, EPI_ISL_1681170, EPI_ISL_1681171, EPI_ISL_1681172, EPI_ISL_1681173, EPI_ISL_1681174, EPI_ISL_1681176, EPI_ISL_1681177, EPI_ISL_1681178, EPI_ISL_1681179, EPI_ISL_1681181, EPI_ISL_1681182, EPI_ISL_1681183, EPI_ISL_1681185, EPI_ISL_1681188, EPI_ISL_1681189, EPI_ISL_1681190, EPI_ISL_1681192, EPI_ISL_1681193, EPI_ISL_1681194, EPI_ISL_1681195, EPI_ISL_1681196, EPI_ISL_1681197, EPI_ISL_1681198, EPI_ISL_1681199, EPI_ISL_1681200, EPI_ISL_1681201, EPI_ISL_1681202, EPI_ISL_1681204, EPI_ISL_1681205, EPI_ISL_1681206, EPI_ISL_1681207, EPI_ISL_1681208, EPI_ISL_1681209, EPI_ISL_1681210, EPI_ISL_1681212, EPI_ISL_1681213, EPI_ISL_1681215, EPI_ISL_1681216, EPI_ISL_1681217, EPI_ISL_1681218, EPI_ISL_1681219, EPI_ISL_1681221, EPI_ISL_1681222, EPI_ISL_1681224, EPI_ISL_1681225, EPI_ISL_1681226, EPI_ISL_1681227, EPI_ISL_1681228, EPI_ISL_1681229, EPI_ISL_1681230, EPI_ISL_1681231, EPI_ISL_1681232, EPI_ISL_1681233, EPI_ISL_1681234, EPI_ISL_1681235, EPI_ISL_1681236, EPI_ISL_1681237, EPI_ISL_1681238, EPI_ISL_1681239, EPI_ISL_1681240, EPI_ISL_1681241, EPI_ISL_1681242, EPI_ISL_1681243, EPI_ISL_1681244, EPI_ISL_1681245, EPI_ISL_1681247, EPI_ISL_1681249, EPI_ISL_1681251, EPI_ISL_1681252, EPI_ISL_1681253, EPI_ISL_1681254, EPI_ISL_1681255, EPI_ISL_1681256, EPI_ISL_1681258, EPI_ISL_1681259, EPI_ISL_1681260, EPI_ISL_1681261, EPI_ISL_1681262, EPI_ISL_1681263, EPI_ISL_1681265, EPI_ISL_1681266, EPI_ISL_1681267, EPI_ISL_1681268, EPI_ISL_1681270, EPI_ISL_1681272, EPI_ISL_1681273, EPI_ISL_1681275, EPI_ISL_1681276, EPI_ISL_1681277, EPI_ISL_1681278, EPI_ISL_1681279, EPI_ISL_1681280, EPI_ISL_1681281, EPI_ISL_1681282, EPI_ISL_1681283, EPI_ISL_1681285, EPI_ISL_1681286, EPI_ISL_1681287, EPI_ISL_1681290, EPI_ISL_1681292, EPI_ISL_1681293, EPI_ISL_1681296, EPI_ISL_1681297, EPI_ISL_1681299, EPI_ISL_1681300, EPI_ISL_1681301, EPI_ISL_1681303, EPI_ISL_1681304, EPI_ISL_1681305, EPI_ISL_1681306, EPI_ISL_1681307, EPI_ISL_1681308, EPI_ISL_1681309, EPI_ISL_1681311, EPI_ISL_1681313, EPI_ISL_1681314, EPI_ISL_1681315, EPI_ISL_1681316, EPI_ISL_1681317, EPI_ISL_1681318, EPI_ISL_1681319, EPI_ISL_1681320, EPI_ISL_1681321, EPI_ISL_1681323, EPI_ISL_1681324, EPI_ISL_1681325, EPI_ISL_1681326, EPI_ISL_1681328, EPI_ISL_1681329, EPI_ISL_1681331, EPI_ISL_1681332, EPI_ISL_1681333, EPI_ISL_1681334, EPI_ISL_1681335, EPI_ISL_1681336, EPI_ISL_1681337, EPI_ISL_1681338, EPI_ISL_1681339, EPI_ISL_1681340, EPI_ISL_1681341, EPI_ISL_1681343, EPI_ISL_1681344, EPI_ISL_1681345, EPI_ISL_1681346, EPI_ISL_1681347, EPI_ISL_1681348, EPI_ISL_1681349, EPI_ISL_1681350, EPI_ISL_1681351, EPI_ISL_1681352, EPI_ISL_1681354, EPI_ISL_1681355, EPI_ISL_1681356, EPI_ISL_1681358, EPI_ISL_1681359, EPI_ISL_1681360, EPI_ISL_1681361, EPI_ISL_1681362, EPI_ISL_1681363, EPI_ISL_1681366, EPI_ISL_1681367, EPI_ISL_1681368, EPI_ISL_1681369, EPI_ISL_1681370, EPI_ISL_1681371, EPI_ISL_1681372, EPI_ISL_1681373, EPI_ISL_1681374, EPI_ISL_1681375, EPI_ISL_1681376, EPI_ISL_1681377, EPI_ISL_1681379, EPI_ISL_1681380, EPI_ISL_1681381, EPI_ISL_1681382, EPI_ISL_1681383, EPI_ISL_1681384, EPI_ISL_1681385, EPI_ISL_1681386, EPI_ISL_1681387, EPI_ISL_1681389, EPI_ISL_1681390, EPI_ISL_1681391, EPI_ISL_1681392, EPI_ISL_1681394, EPI_ISL_1681395, EPI_ISL_1681396, EPI_ISL_1681398, EPI_ISL_1681399, EPI_ISL_1681400, EPI_ISL_1681401, EPI_ISL_1681402, EPI_ISL_1681405, EPI_ISL_1681406, EPI_ISL_1681407, EPI_ISL_1681408, EPI_ISL_1681410, EPI_ISL_1681411, EPI_ISL_1681413, EPI_ISL_1681414, EPI_ISL_1681415, EPI_ISL_1681417, EPI_ISL_1681418, EPI_ISL_1681420, EPI_ISL_1681421, EPI_ISL_1681422, EPI_ISL_1681423, EPI_ISL_1681424, EPI_ISL_1681425, EPI_ISL_1681426, EPI_ISL_1681427, EPI_ISL_1681428, EPI_ISL_1681429, EPI_ISL_1681430, EPI_ISL_1681431, EPI_ISL_1681432, EPI_ISL_1681433, EPI_ISL_1681434, EPI_ISL_1681435, EPI_ISL_1681437, EPI_ISL_1681438, EPI_ISL_1681439, EPI_ISL_1681440, EPI_ISL_1681441, EPI_ISL_1681442, EPI_ISL_1681444, EPI_ISL_1681445, EPI_ISL_1681447, EPI_ISL_1681448, EPI_ISL_1681449, EPI_ISL_1681450, EPI_ISL_1681451, EPI_ISL_1681453, EPI_ISL_1681454, EPI_ISL_1681455, EPI_ISL_1681457, EPI_ISL_1681459, EPI_ISL_1681460, EPI_ISL_1681461, EPI_ISL_1681463, EPI_ISL_1681464, EPI_ISL_1681465, EPI_ISL_1681466, EPI_ISL_1681467, EPI_ISL_1681468, EPI_ISL_1681469, EPI_ISL_1681470, EPI_ISL_1681472, EPI_ISL_1681473, EPI_ISL_1681474, EPI_ISL_1681475, EPI_ISL_1681476, EPI_ISL_1681477, EPI_ISL_1681478, EPI_ISL_1681479, EPI_ISL_1681480, EPI_ISL_1681481, EPI_ISL_1681482, EPI_ISL_1681483, EPI_ISL_1681485, EPI_ISL_1681486, EPI_ISL_1681488, EPI_ISL_1681489, EPI_ISL_1681490, EPI_ISL_1681491, EPI_ISL_1681492, EPI_ISL_1681494, EPI_ISL_1681495, EPI_ISL_1681496, EPI_ISL_1681497, EPI_ISL_1681498, EPI_ISL_1681499, EPI_ISL_1681500, EPI_ISL_1681502, EPI_ISL_1681503, EPI_ISL_1681504, EPI_ISL_1681505, EPI_ISL_1681506, EPI_ISL_1681510, EPI_ISL_1681511, EPI_ISL_1681512, EPI_ISL_1681513, EPI_ISL_1681514, EPI_ISL_1681516, EPI_ISL_1681518, EPI_ISL_1681519, EPI_ISL_1681521, EPI_ISL_1681522, EPI_ISL_1681523, EPI_ISL_1681524, EPI_ISL_1681525, EPI_ISL_1681527, EPI_ISL_1681528, EPI_ISL_1681530, EPI_ISL_1681531, EPI_ISL_1681532, EPI_ISL_1681533, EPI_ISL_1681534, EPI_ISL_1681535, EPI_ISL_1681537, EPI_ISL_1681538, EPI_ISL_1681539, EPI_ISL_1681540, EPI_ISL_1681542, EPI_ISL_1681545, EPI_ISL_1681546, EPI_ISL_1681548, EPI_ISL_1681549, EPI_ISL_1681551, EPI_ISL_1681552, EPI_ISL_1681553, EPI_ISL_1681555, EPI_ISL_1681556, EPI_ISL_1681558, EPI_ISL_1681559, EPI_ISL_1681560, EPI_ISL_1681561, EPI_ISL_1681562, EPI_ISL_1681563, EPI_ISL_1681567, EPI_ISL_1681569, EPI_ISL_1681570, EPI_ISL_1681571, EPI_ISL_1681572, EPI_ISL_1681573, EPI_ISL_1681574, EPI_ISL_1681575, EPI_ISL_1681576, EPI_ISL_1681577, EPI_ISL_1681578, EPI_ISL_1681579, EPI_ISL_1681580, EPI_ISL_1681581, EPI_ISL_1681582, EPI_ISL_1681583, EPI_ISL_1681584, EPI_ISL_1681585, EPI_ISL_1681586, EPI_ISL_1681587, EPI_ISL_1681588, EPI_ISL_1681590, EPI_ISL_1681591, EPI_ISL_1681592, EPI_ISL_1681593, EPI_ISL_1681594, EPI_ISL_1681595, EPI_ISL_1681598, EPI_ISL_1681599, EPI_ISL_1681601, EPI_ISL_1681607, EPI_ISL_1681608, EPI_ISL_1681610, EPI_ISL_1681612, EPI_ISL_1681613, EPI_ISL_1681614, EPI_ISL_1681615, EPI_ISL_1681616, EPI_ISL_1681617, EPI_ISL_1681618, EPI_ISL_1681619, EPI_ISL_1681620, EPI_ISL_1681621, EPI_ISL_1681622, EPI_ISL_1681623, EPI_ISL_1681624, EPI_ISL_1681625, EPI_ISL_1681626, EPI_ISL_1681627, EPI_ISL_1681628, EPI_ISL_1681629, EPI_ISL_1681630, EPI_ISL_1681631, EPI_ISL_1681632, EPI_ISL_1681633, EPI_ISL_1681634, EPI_ISL_1681635, EPI_ISL_1681636, EPI_ISL_1681638, EPI_ISL_1681639, EPI_ISL_1681640, EPI_ISL_1681641, EPI_ISL_1681642, EPI_ISL_1681643, EPI_ISL_1681646, EPI_ISL_1681647, EPI_ISL_1681650, EPI_ISL_1681651, EPI_ISL_1681652, EPI_ISL_1681653, EPI_ISL_1681654, EPI_ISL_1681655, EPI_ISL_1681656, EPI_ISL_1681657, EPI_ISL_1681658, EPI_ISL_1681659, EPI_ISL_1681660, EPI_ISL_1681661, EPI_ISL_1681662, EPI_ISL_1681663, EPI_ISL_1681664, EPI_ISL_1681665, EPI_ISL_1681666, EPI_ISL_1681667, EPI_ISL_1681669, EPI_ISL_1681670, EPI_ISL_1681671, EPI_ISL_1681672, EPI_ISL_1681673, EPI_ISL_1681674, EPI_ISL_1681675, EPI_ISL_1681676, EPI_ISL_1681677, EPI_ISL_1681678, EPI_ISL_1681679, EPI_ISL_1681681, EPI_ISL_1681682, EPI_ISL_1681684, EPI_ISL_1681685, EPI_ISL_1681686, EPI_ISL_1681687, EPI_ISL_1681688, EPI_ISL_1681689, EPI_ISL_1681690, EPI_ISL_1681691, EPI_ISL_1681692, EPI_ISL_1681694, EPI_ISL_1681695, EPI_ISL_1681696, EPI_ISL_1681697, EPI_ISL_1681698, EPI_ISL_1681699, EPI_ISL_1681700, EPI_ISL_1681701, EPI_ISL_1681702, EPI_ISL_1681705, EPI_ISL_1681709, EPI_ISL_1681710, EPI_ISL_1681711, EPI_ISL_1681715, EPI_ISL_1681717, EPI_ISL_1681718, EPI_ISL_1681719, EPI_ISL_1681720, EPI_ISL_1681721, EPI_ISL_1681723, EPI_ISL_1681725, EPI_ISL_1681726, EPI_ISL_1681727, EPI_ISL_1681728, EPI_ISL_1681729, EPI_ISL_1681733, EPI_ISL_1681734, EPI_ISL_1681736, EPI_ISL_1681737, EPI_ISL_1681738, EPI_ISL_1681739, EPI_ISL_1681741, EPI_ISL_1681742, EPI_ISL_1681743, EPI_ISL_1681744, EPI_ISL_1681745, EPI_ISL_1681746, EPI_ISL_1681748, EPI_ISL_1681750, EPI_ISL_1681751, EPI_ISL_1681752, EPI_ISL_1681753, EPI_ISL_1681754, EPI_ISL_1681755, EPI_ISL_1681756, EPI_ISL_1681757, EPI_ISL_1681758, EPI_ISL_1681759, EPI_ISL_1681760, EPI_ISL_1681761, EPI_ISL_1681763, EPI_ISL_1681764, EPI_ISL_1681765, EPI_ISL_1681766, EPI_ISL_1681767, EPI_ISL_1681768, EPI_ISL_1681769, EPI_ISL_1681771, EPI_ISL_1681772, EPI_ISL_1681774, EPI_ISL_1681775, EPI_ISL_1681776, EPI_ISL_1681777, EPI_ISL_1681778, EPI_ISL_1681779, EPI_ISL_1681780, EPI_ISL_1681781, EPI_ISL_1681782, EPI_ISL_1681783, EPI_ISL_1681785, EPI_ISL_1681786, EPI_ISL_1681788, EPI_ISL_1681789, EPI_ISL_1681790, EPI_ISL_1681791, EPI_ISL_1681792, EPI_ISL_1681793, EPI_ISL_1681794, EPI_ISL_1681795, EPI_ISL_1681796, EPI_ISL_1681798, EPI_ISL_1681799, EPI_ISL_1681802, EPI_ISL_1681803, EPI_ISL_1681804, EPI_ISL_1681805, EPI_ISL_1681807, EPI_ISL_1681808, EPI_ISL_1681809, EPI_ISL_1681810, EPI_ISL_1681811, EPI_ISL_1681812, EPI_ISL_1681813, EPI_ISL_1681814, EPI_ISL_1681815, EPI_ISL_1681817, EPI_ISL_1681818, EPI_ISL_1681819, EPI_ISL_1681820, EPI_ISL_1681821, EPI_ISL_1681822, EPI_ISL_1681823, EPI_ISL_1681826, EPI_ISL_1681827, EPI_ISL_1681828, EPI_ISL_1681829, EPI_ISL_1681830, EPI_ISL_1681831, EPI_ISL_1681832, EPI_ISL_1681833, EPI_ISL_1681834, EPI_ISL_1681835, EPI_ISL_1681836, EPI_ISL_1681837, EPI_ISL_1681838, EPI_ISL_1681839, EPI_ISL_1681840, EPI_ISL_1681841, EPI_ISL_1681842, EPI_ISL_1681844, EPI_ISL_1681845, EPI_ISL_1681846, EPI_ISL_1681847, EPI_ISL_1681848, EPI_ISL_1681849, EPI_ISL_1681851, EPI_ISL_1681852, EPI_ISL_1681853, EPI_ISL_1681855, EPI_ISL_1681857, EPI_ISL_1681858, EPI_ISL_1681859, EPI_ISL_1681860, EPI_ISL_1681862, EPI_ISL_1681863, EPI_ISL_1681864, EPI_ISL_1681866, EPI_ISL_1681868, EPI_ISL_1681870, EPI_ISL_1681871, EPI_ISL_1681872, EPI_ISL_1681873, EPI_ISL_1681874, EPI_ISL_1681875, EPI_ISL_1681877, EPI_ISL_1681878, EPI_ISL_1681879, EPI_ISL_1681880, EPI_ISL_1681881, EPI_ISL_1681882, EPI_ISL_1681884, EPI_ISL_1681885, EPI_ISL_1681886, EPI_ISL_1681887, EPI_ISL_1681888, EPI_ISL_1681889, | | | |

EPI_ISL_1681890, EPI_ISL_1681891, EPI_ISL_1681892, EPI_ISL_1681894, EPI_ISL_1681898, EPI_ISL_1681899, EPI_ISL_1681900, EPI_ISL_1681901, EPI_ISL_1681902, EPI_ISL_1681903, EPI_ISL_1681904, EPI_ISL_1681905, EPI_ISL_1681906, EPI_ISL_1681907, EPI_ISL_1681908, EPI_ISL_1681909, EPI_ISL_1681910, EPI_ISL_1681911, EPI_ISL_1681912, EPI_ISL_1681913, EPI_ISL_1681914, EPI_ISL_1681915, EPI_ISL_1681916, EPI_ISL_1681917, EPI_ISL_1681918, EPI_ISL_1681919, EPI_ISL_1681920, EPI_ISL_1681921, EPI_ISL_1681922, EPI_ISL_1681923, EPI_ISL_1681924, EPI_ISL_1681925, EPI_ISL_1681926, EPI_ISL_1681927, EPI_ISL_1681929, EPI_ISL_1681930, EPI_ISL_1681931, EPI_ISL_1681932, EPI_ISL_1681933, EPI_ISL_1681934, EPI_ISL_1681935, EPI_ISL_1681936, EPI_ISL_1681937, EPI_ISL_1681938

| | | | |
|---|---|---|---|
| see above | Laboratory Corporation of America | Centers for Disease Control and Prevention Division of Viral Diseases, Pathogen Discovery | Dakota Howard, Dhwani Batra, Peter W. Cook, Kara Moser, Adrian Paskey, Jason Caravas, Benjamin Rambo-Martin, Shatavia Morrison, Christopher Gulvick, Scott Sammons, Yvette Unoarumhi, Darlene Wagner, Matthew Schmerer, Minoo Agarwal, Eyad Almasri, Debbie Boles, Ayla Burns, Nuthawin Charoensri, Oren Cohen, Susan Countryman, Mary Ann Cristobal, Bobbi Croy, Suzanne Dale, Hrushikesh Deshmukh, Amanda Douglas, Vincent Drouillon, Marcia Eisenberg, Howard Engler, Rama Ghatti, Prashant Gupta, Susan Hicks, Jake Humphrey, Lax Iyer, Manoj Jain, Mohan Kolli, Brian Krueger, Tim Kuphal, Stanley Letovsky, Michael Levandoski, Craig Lukasik, Jonathan Meltzer, Brian Norvell, Mindy Nye, Scott Parker, Christos Petropoulos, John Pruitt, Steven Ragan, Scott Ryan, Mike Sapeta, Jana Schroth, Suresh Babu Selvaraju, Goran Stevovic, Amanda Suchanek, Andrea Throop, Lyndon Tilson, Thomas Urban, Joe Voshell, Kimberly Wagner, Jonathan Williams, Mary Williamson, Qian Zeng, Tricia Zwiefelhofer, Clinton R. Paden, Duncan MacCannell |
| EPI_ISL_1681940 | Ohio Department of Health Laboratory | Ohio Department of Health Laboratory | Holmes, Jennifer; Eric Brandt, Keoni Omura, Glen McGillivary, Caitlin McDonnell, Jade Mowery, Stephanie Mccracken, Tyler Payne, Kirtana Ramadugu, Erica Leasure, Brent Lee, Kelsey Florek, Heather Blankenship, Allison Black, Quanta Brown, and Tammy Bannerman |

EPI_ISL_1681941, EPI_ISL_1681942, EPI_ISL_1681943, EPI_ISL_1681944, EPI_ISL_1681945, EPI_ISL_1681946, EPI_ISL_1681947, EPI_ISL_1681948, EPI_ISL_1681949, EPI_ISL_1681950, EPI_ISL_1681952, EPI_ISL_1681953, EPI_ISL_1681954, EPI_ISL_1681955, EPI_ISL_1681956, EPI_ISL_1681957, EPI_ISL_1681958, EPI_ISL_1681959, EPI_ISL_1681960, EPI_ISL_1681961, EPI_ISL_1681962, EPI_ISL_1681963, EPI_ISL_1681964, EPI_ISL_1681965, EPI_ISL_1681966, EPI_ISL_1681967, EPI_ISL_1681968, EPI_ISL_1681969, EPI_ISL_1681970, EPI_ISL_1681971, EPI_ISL_1681972, EPI_ISL_1681973, EPI_ISL_1681974, EPI_ISL_1681975, EPI_ISL_1681976, EPI_ISL_1681977, EPI_ISL_1681978, EPI_ISL_1681979, EPI_ISL_1681981, EPI_ISL_1681983, EPI_ISL_1681984, EPI_ISL_1681985, EPI_ISL_1681986, EPI_ISL_1681987, EPI_ISL_1681988, EPI_ISL_1681989, EPI_ISL_1681991, EPI_ISL_1681992, EPI_ISL_1681993, EPI_ISL_1681995, EPI_ISL_1681996, EPI_ISL_1681998, EPI_ISL_1682000, EPI_ISL_1682001, EPI_ISL_1682002, EPI_ISL_1682003, EPI_ISL_1682004, EPI_ISL_1682005, EPI_ISL_1682006, EPI_ISL_1682008, EPI_ISL_1682009, EPI_ISL_1682010, EPI_ISL_1682012, EPI_ISL_1682013, EPI_ISL_1682014, EPI_ISL_1682016, EPI_ISL_1682017, EPI_ISL_1682018, EPI_ISL_1682019, EPI_ISL_1682020, EPI_ISL_1682021, EPI_ISL_1682022, EPI_ISL_1682023, EPI_ISL_1682024, EPI_ISL_1682025, EPI_ISL_1682026, EPI_ISL_1682027, EPI_ISL_1682028, EPI_ISL_1682029, EPI_ISL_1682031, EPI_ISL_1682034, EPI_ISL_1682036, EPI_ISL_1682037, EPI_ISL_1682038, EPI_ISL_1682040, EPI_ISL_1682042, EPI_ISL_1682043, EPI_ISL_1682044, EPI_ISL_1682047, EPI_ISL_1682048, EPI_ISL_1682049, EPI_ISL_1682050, EPI_ISL_1682052, EPI_ISL_1682054, EPI_ISL_1682056, EPI_ISL_1682057, EPI_ISL_1682058, EPI_ISL_1682059, EPI_ISL_1682060, EPI_ISL_1682061, EPI_ISL_1682062, EPI_ISL_1682063, EPI_ISL_1682064, EPI_ISL_1682065, EPI_ISL_1682066, EPI_ISL_1682067, EPI_ISL_1682068, EPI_ISL_1682069, EPI_ISL_1682070, EPI_ISL_1682071, EPI_ISL_1682073, EPI_ISL_1682074, EPI_ISL_1682075, EPI_ISL_1682077, EPI_ISL_1682078, EPI_ISL_1682081, EPI_ISL_1682082, EPI_ISL_1682083, EPI_ISL_1682084, EPI_ISL_1682085, EPI_ISL_1682087, EPI_ISL_1682088, EPI_ISL_1682089, EPI_ISL_1682090, EPI_ISL_1682091, EPI_ISL_1682092, EPI_ISL_1682093, EPI_ISL_1682094, EPI_ISL_1682095, EPI_ISL_1682097, EPI_ISL_1682098, EPI_ISL_1682099, EPI_ISL_1682100, EPI_ISL_1682101, EPI_ISL_1682102, EPI_ISL_1682103, EPI_ISL_1682104, EPI_ISL_1682106, EPI_ISL_1682107, EPI_ISL_1682108, EPI_ISL_1682109, EPI_ISL_1682111, EPI_ISL_1682113, EPI_ISL_1682114, EPI_ISL_1682115, EPI_ISL_1682116, EPI_ISL_1682118, EPI_ISL_1682119, EPI_ISL_1682120, EPI_ISL_1682121, EPI_ISL_1682122, EPI_ISL_1682123, EPI_ISL_1682124, EPI_ISL_1682127, EPI_ISL_1682128, EPI_ISL_1682129, EPI_ISL_1682130, EPI_ISL_1682131, EPI_ISL_1682132, EPI_ISL_1682133, EPI_ISL_1682134, EPI_ISL_1682135, EPI_ISL_1682136, EPI_ISL_1682137, EPI_ISL_1682138, EPI_ISL_1682139, EPI_ISL_1682141, EPI_ISL_1682142, EPI_ISL_1682143, EPI_ISL_1682144, EPI_ISL_1682145, EPI_ISL_1682146, EPI_ISL_1682147, EPI_ISL_1682148, EPI_ISL_1682149, EPI_ISL_1682150, EPI_ISL_1682152, EPI_ISL_1682154, EPI_ISL_1682155, EPI_ISL_1682157, EPI_ISL_1682158, EPI_ISL_1682160, EPI_ISL_1682161, EPI_ISL_1682162, EPI_ISL_1682163, EPI_ISL_1682164, EPI_ISL_1682165, EPI_ISL_1682166, EPI_ISL_1682167, EPI_ISL_1682168, EPI_ISL_1682169, EPI_ISL_1682170, EPI_ISL_1682171, EPI_ISL_1682172, EPI_ISL_1682175, EPI_ISL_1682178, EPI_ISL_1682181, EPI_ISL_1682184, EPI_ISL_1682187, EPI_ISL_1682190, EPI_ISL_1682193, EPI_ISL_1682196, EPI_ISL_1682203, EPI_ISL_1682206, EPI_ISL_1682209, EPI_ISL_1682212, EPI_ISL_1682215, EPI_ISL_1682218, EPI_ISL_1682222, EPI_ISL_1682225, EPI_ISL_1682228, EPI_ISL_1682231, EPI_ISL_1682234, EPI_ISL_1682237, EPI_ISL_1682240, EPI_ISL_1682244, EPI_ISL_1682247, EPI_ISL_1682253, EPI_ISL_1682256, EPI_ISL_1682259, EPI_ISL_1682262, EPI_ISL_1682265, EPI_ISL_1682268, EPI_ISL_1682272, EPI_ISL_1682275, EPI_ISL_1682277, EPI_ISL_1682281, EPI_ISL_1682284, EPI_ISL_1682287, EPI_ISL_1682290, EPI_ISL_1682297, EPI_ISL_1682299, EPI_ISL_1682303, EPI_ISL_1682306, EPI_ISL_1682309, EPI_ISL_1682312, EPI_ISL_1682315, EPI_ISL_1682317, EPI_ISL_1682323, EPI_ISL_1682326, EPI_ISL_1682329, EPI_ISL_1682332, EPI_ISL_1682335, EPI_ISL_1682338, EPI_ISL_1682342, EPI_ISL_1682345, EPI_ISL_1682348, EPI_ISL_1682351, EPI_ISL_1682354, EPI_ISL_1682357, EPI_ISL_1682361, EPI_ISL_1682364, EPI_ISL_1682367, EPI_ISL_1682371, EPI_ISL_1682377, EPI_ISL_1682380, EPI_ISL_1682383, EPI_ISL_1682389, EPI_ISL_1682393, EPI_ISL_1682396, EPI_ISL_1682403, EPI_ISL_1682405, EPI_ISL_1682408, EPI_ISL_1682412, EPI_ISL_1682415, EPI_ISL_1682418, EPI_ISL_1682420, EPI_ISL_1682432, EPI_ISL_1682435, EPI_ISL_1682438, EPI_ISL_1682444, EPI_ISL_1682451, EPI_ISL_1682454, EPI_ISL_1682460, EPI_ISL_1682464, EPI_ISL_1682467, EPI_ISL_1682469, EPI_ISL_1682473, EPI_ISL_1682475, EPI_ISL_1682481, EPI_ISL_1682484, EPI_ISL_1682487, EPI_ISL_1682490, EPI_ISL_1682493, EPI_ISL_1682496, EPI_ISL_1682499, EPI_ISL_1682505, EPI_ISL_1682509, EPI_ISL_1682512, EPI_ISL_1682515, EPI_ISL_1682518, EPI_ISL_1682525, EPI_ISL_1682528, EPI_ISL_1682530, EPI_ISL_1682534, EPI_ISL_1682537, EPI_ISL_1682540, EPI_ISL_1682543, EPI_ISL_1682546, EPI_ISL_1682549, EPI_ISL_1682552, EPI_ISL_1682555, EPI_ISL_1682558, EPI_ISL_1682561, EPI_ISL_1682565, EPI_ISL_1682568, EPI_ISL_1682574, EPI_ISL_1682584, EPI_ISL_1682590, EPI_ISL_1682593, EPI_ISL_1682596, EPI_ISL_1682599, EPI_ISL_1682605, EPI_ISL_1682608, EPI_ISL_1682611, EPI_ISL_1682614, EPI_ISL_1682618, EPI_ISL_1682621, EPI_ISL_1682624, EPI_ISL_1682627, EPI_ISL_1682630, EPI_ISL_1682633, EPI_ISL_1682636, EPI_ISL_1682639, EPI_ISL_1682642, EPI_ISL_1682646, EPI_ISL_1682649, EPI_ISL_1682652, EPI_ISL_1682658, EPI_ISL_1682661, EPI_ISL_1682664, EPI_ISL_1682667, EPI_ISL_1682669, EPI_ISL_1682672, EPI_ISL_1682675, EPI_ISL_1682678, EPI_ISL_1682681, EPI_ISL_1682684, EPI_ISL_1682687, EPI_ISL_1682690, EPI_ISL_1682693, EPI_ISL_1682696, EPI_ISL_1682699, EPI_ISL_1682702, EPI_ISL_1682708, EPI_ISL_1682714, EPI_ISL_1682717, EPI_ISL_1682720, EPI_ISL_1682724, EPI_ISL_1682730, EPI_ISL_1682732, EPI_ISL_1682735, EPI_ISL_1682739, EPI_ISL_1682742, EPI_ISL_1682745, EPI_ISL_1682747, EPI_ISL_1682751, EPI_ISL_1682754, EPI_ISL_1682757, EPI_ISL_1682760, EPI_ISL_1682763, EPI_ISL_1682766, EPI_ISL_1682769, EPI_ISL_1682772, EPI_ISL_1682775, EPI_ISL_1682778, EPI_ISL_1682781, EPI_ISL_1682784, EPI_ISL_1682787, EPI_ISL_1682791, EPI_ISL_1682794, EPI_ISL_1682796, EPI_ISL_1682799, EPI_ISL_1682802, EPI_ISL_1682814, EPI_ISL_1682817, EPI_ISL_1682820, EPI_ISL_1682827, EPI_ISL_1682829, EPI_ISL_1682833, EPI_ISL_1682839, EPI_ISL_1682841, EPI_ISL_1682844, EPI_ISL_1682847, EPI_ISL_1682853, EPI_ISL_1682856, EPI_ISL_1682859, EPI_ISL_1682862, EPI_ISL_1682865, EPI_ISL_1682870, EPI_ISL_1682877, EPI_ISL_1682879, EPI_ISL_1682883, EPI_ISL_1682889, EPI_ISL_1682892, EPI_ISL_1682895, EPI_ISL_1682898, EPI_ISL_1682901, EPI_ISL_1682904, EPI_ISL_1682910, EPI_ISL_1682913, EPI_ISL_1682916, EPI_ISL_1682919, EPI_ISL_1682922, EPI_ISL_1682926, EPI_ISL_1682929, EPI_ISL_1682931, EPI_ISL_1682934, EPI_ISL_1682937, EPI_ISL_1682949, EPI_ISL_1682952, EPI_ISL_1682955, EPI_ISL_1682958, EPI_ISL_1682961, EPI_ISL_1682971, EPI_ISL_1682975, EPI_ISL_1682978, EPI_ISL_1682981, EPI_ISL_1682984, EPI_ISL_1682987, EPI_ISL_1682990, EPI_ISL_1682996, EPI_ISL_1682999, EPI_ISL_1683002, EPI_ISL_1683005, EPI_ISL_1683007, EPI_ISL_1683010, EPI_ISL_1683014, EPI_ISL_1683019, EPI_ISL_1683022, EPI_ISL_1683025, EPI_ISL_1683028, EPI_ISL_1683034, EPI_ISL_1683037, EPI_ISL_1683040, EPI_ISL_1683043, EPI_ISL_1683046, EPI_ISL_1683049, EPI_ISL_1683058, EPI_ISL_1683061, EPI_ISL_1683064, EPI_ISL_1683067, EPI_ISL_1683070, EPI_ISL_1683073, EPI_ISL_1683082, EPI_ISL_1683085, EPI_ISL_1683087, EPI_ISL_1683088, EPI_ISL_1683090, EPI_ISL_1683091, EPI_ISL_1683092, EPI_ISL_1683093, EPI_ISL_1683094, EPI_ISL_1683096, EPI_ISL_1683097, EPI_ISL_1683098, EPI_ISL_1683099, EPI_ISL_1683102, EPI_ISL_1683103, EPI_ISL_1683104, EPI_ISL_1683105, EPI_ISL_1683106, EPI_ISL_1683107, EPI_ISL_1683108, EPI_ISL_1683109, EPI_ISL_1683110, EPI_ISL_1683111, EPI_ISL_1683112, EPI_ISL_1683113, EPI_ISL_1683115, EPI_ISL_1683116, EPI_ISL_1683117

| | | | |
|---|---|---|---|
| see above | Laboratory Corporation of America | Centers for Disease Control and Prevention Division of Viral Diseases, Pathogen Discovery | Dakota Howard, Dhwani Batra, Peter W. Cook, Kara Moser, Adrian Paskey, Jason Caravas, Benjamin Rambo-Martin, Shatavia Morrison, Christopher Gulvick, Scott Sammons, Yvette Unoarumhi, Darlene Wagner, Matthew Schmerer, Minoo Agarwal, Eyad Almasri, Debbie Boles, Ayla Burns, Nuthawin Charoensri, Oren Cohen, Susan Countryman, Mary Ann Cristobal, Bobbi Croy, Suzanne Dale, Hrushikesh Deshmukh, Amanda Douglas, Vincent Drouillon, Marcia Eisenberg, Howard Engler, Rama Ghatti, Prashant Gupta, Susan Hicks, Jake Humphrey, Lax Iyer, Manoj Jain, Mohan Kolli, Brian Krueger, Tim Kuphal, Stanley Letovsky, Michael Levandoski, Craig Lukasik, Jonathan Meltzer, Brian Norvell, Mindy Nye, Scott Parker, Christos Petropoulos, John Pruitt, Steven Ragan, Scott Ryan, Mike Sapeta, Jana Schroth, Suresh Babu Selvaraju, Goran Stevovic, Amanda Suchanek, Andrea Throop, Lyndon Tilson, Thomas Urban, Joe Voshell, Kimberly Wagner, Jonathan Williams, Mary Williamson, Qian Zeng, Tricia Zwiefelhofer, Clinton R. Paden, Duncan MacCannell |
| EPI_ISL_1683118 | Ohio Department of Health Laboratory | Ohio Department of Health Laboratory | Holmes, Jennifer; Eric Brandt, Keoni Omura, Glen McGillivary, Caitlin McDonnell, Jade Mowery, Stephanie Mccracken, Tyler Payne, Kirtana Ramadugu, Erica Leasure, Brent Lee, Kelsey Florek, Heather Blankenship, Allison Black, Quanta Brown, and Tammy Bannerman |

EPI_ISL_1683119, EPI_ISL_1683121, EPI_ISL_1683122, EPI_ISL_1683123, EPI_ISL_1683124, EPI_ISL_1683125, EPI_ISL_1683126, EPI_ISL_1683127, EPI_ISL_1683128, EPI_ISL_1683129, EPI_ISL_1683130, EPI_ISL_1683131, EPI_ISL_1683132, EPI_ISL_1683133, EPI_ISL_1683134, EPI_ISL_1683135, EPI_ISL_1683137, EPI_ISL_1683138, EPI_ISL_1683139, EPI_ISL_1683141, EPI_ISL_1683142, EPI_ISL_1683143, EPI_ISL_1683144, EPI_ISL_1683147, EPI_ISL_1683148, EPI_ISL_1683150, EPI_ISL_1683151, EPI_ISL_1683152, EPI_ISL_1683154, EPI_ISL_1683156, EPI_ISL_1683157, EPI_ISL_1683158, EPI_ISL_1683159, EPI_ISL_1683160, EPI_ISL_1683161, EPI_ISL_1683162, EPI_ISL_1683163, EPI_ISL_1683164, EPI_ISL_1683165, EPI_ISL_1683166, EPI_ISL_1683167, EPI_ISL_1683168, EPI_ISL_1683169, EPI_ISL_1683170, EPI_ISL_1683173, EPI_ISL_1683174, EPI_ISL_1683175, EPI_ISL_1683176, EPI_ISL_1683177, EPI_ISL_1683178, EPI_ISL_1683179, EPI_ISL_1683180, EPI_ISL_1683181, EPI_ISL_1683182, EPI_ISL_1683183, EPI_ISL_1683184, EPI_ISL_1683185, EPI_ISL_1683187, EPI_ISL_1683188, EPI_ISL_1683189, EPI_ISL_1683190, EPI_ISL_1683191, EPI_ISL_1683192, EPI_ISL_1683193, EPI_ISL_1683194, EPI_ISL_1683195, EPI_ISL_1683196, EPI_ISL_1683197, EPI_ISL_1683198, EPI_ISL_1683199, EPI_ISL_1683200, EPI_ISL_1683201, EPI_ISL_1683202, EPI_ISL_1683203, EPI_ISL_1683204, EPI_ISL_1683205, EPI_ISL_1683207, EPI_ISL_1683209, EPI_ISL_1683210, EPI_ISL_1683211, EPI_ISL_1683212, EPI_ISL_1683213, EPI_ISL_1683214, EPI_ISL_1683215, EPI_ISL_1683216, EPI_ISL_1683217, EPI_ISL_1683218, EPI_ISL_1683219, EPI_ISL_1683220, EPI_ISL_1683221, EPI_ISL_1683222, EPI_ISL_1683224, EPI_ISL_1683225, EPI_ISL_1683226, EPI_ISL_1683227, EPI_ISL_1683228, EPI_ISL_1683231, EPI_ISL_1683232, EPI_ISL_1683233, EPI_ISL_1683234, EPI_ISL_1683236, EPI_ISL_1683237, EPI_ISL_1683238, EPI_ISL_1683239, EPI_ISL_1683240, EPI_ISL_1683241, EPI_ISL_1683242, EPI_ISL_1683243, EPI_ISL_1683244, EPI_ISL_1683246, EPI_ISL_1683248, EPI_ISL_1683249, EPI_ISL_1683250, EPI_ISL_1683251, EPI_ISL_1683252, EPI_ISL_1683253, EPI_ISL_1683254, EPI_ISL_1683255, EPI_ISL_1683256, EPI_ISL_1683257, EPI_ISL_1683259, EPI_ISL_1683261, EPI_ISL_1683262, EPI_ISL_1683263, EPI_ISL_1683264, EPI_ISL_1683265, EPI_ISL_1683267, EPI_ISL_1683268, EPI_ISL_1683269, EPI_ISL_1683270, EPI_ISL_1683271, EPI_ISL_1683272, EPI_ISL_1683273, EPI_ISL_1683274, EPI_ISL_1683275, EPI_ISL_1683277, EPI_ISL_1683278, EPI_ISL_1683280, EPI_ISL_1683281, EPI_ISL_1683282, EPI_ISL_1683283, EPI_ISL_1683284, EPI_ISL_1683285, EPI_ISL_1683286, EPI_ISL_1683287, EPI_ISL_1683288, EPI_ISL_1683289, EPI_ISL_1683290, EPI_ISL_1683291, EPI_ISL_1683292, EPI_ISL_1683293, EPI_ISL_1683294, EPI_ISL_1683295, EPI_ISL_1683296, EPI_ISL_1683297, EPI_ISL_1683298, EPI_ISL_1683299, EPI_ISL_1683300, EPI_ISL_1683301, EPI_ISL_1683303, EPI_ISL_1683304, EPI_ISL_1683305, EPI_ISL_1683306, EPI_ISL_1683307, EPI_ISL_1683308, EPI_ISL_1683309, EPI_ISL_1683310, EPI_ISL_1683312, EPI_ISL_1683313, EPI_ISL_1683314, EPI_ISL_1683315, EPI_ISL_1683316, EPI_ISL_1683317, EPI_ISL_1683319, EPI_ISL_1683320, EPI_ISL_1683321, EPI_ISL_1683323, EPI_ISL_1683324, EPI_ISL_1683325, EPI_ISL_1683326, EPI_ISL_1683327, EPI_ISL_1683328, EPI_ISL_1683331, EPI_ISL_1683332, EPI_ISL_1683334, EPI_ISL_1683335, EPI_ISL_1683336, EPI_ISL_1683337, EPI_ISL_1683338, EPI_ISL_1683339, EPI_ISL_1683341, EPI_ISL_1683342, EPI_ISL_1683343, EPI_ISL_1683345, EPI_ISL_1683346, EPI_ISL_1683347, EPI_ISL_1683348, EPI_ISL_1683349, EPI_ISL_1683350, EPI_ISL_1683351, EPI_ISL_1683353, EPI_ISL_1683354, EPI_ISL_1683356, EPI_ISL_1683357, EPI_ISL_1683358, EPI_ISL_1683359, EPI_ISL_1683360, EPI_ISL_1683361, EPI_ISL_1683363, EPI_ISL_1683365, EPI_ISL_1683366, EPI_ISL_1683367, EPI_ISL_1683368, EPI_ISL_1683370, EPI_ISL_1683371, EPI_ISL_1683374, EPI_ISL_1683376, EPI_ISL_1683377, EPI_ISL_1683378, EPI_ISL_1683379, EPI_ISL_1683380, EPI_ISL_1683382, EPI_ISL_1683383, EPI_ISL_1683385, EPI_ISL_1683386, EPI_ISL_1683388, EPI_ISL_1683391, EPI_ISL_1683392, EPI_ISL_1683393, EPI_ISL_1683394, EPI_ISL_1683395, EPI_ISL_1683396, EPI_ISL_1683397, EPI_ISL_1683401, EPI_ISL_1683402, EPI_ISL_1683403, EPI_ISL_1683404, EPI_ISL_1683405, EPI_ISL_1683406, EPI_ISL_1683407, EPI_ISL_1683408, EPI_ISL_1683409, EPI_ISL_1683410, EPI_ISL_1683411, EPI_ISL_1683414, EPI_ISL_1683415, EPI_ISL_1683419, EPI_ISL_1683420, EPI_ISL_1683421, EPI_ISL_1683423, EPI_ISL_1683424, EPI_ISL_1683425, EPI_ISL_1683426, EPI_ISL_1683427, EPI_ISL_1683428, EPI_ISL_1683429, EPI_ISL_1683430, EPI_ISL_1683431, EPI_ISL_1683432, EPI_ISL_1683434, EPI_ISL_1683435, EPI_ISL_1683437, EPI_ISL_1683438, EPI_ISL_1683439, EPI_ISL_1683440, EPI_ISL_1683441, EPI_ISL_1683442, EPI_ISL_1683443, EPI_ISL_1683444, EPI_ISL_1683445, EPI_ISL_1683446, EPI_ISL_1683447, EPI_ISL_1683449, EPI_ISL_1683450, EPI_ISL_1683451, EPI_ISL_1683452, EPI_ISL_1683453, EPI_ISL_1683454, EPI_ISL_1683455, EPI_ISL_1683457, EPI_ISL_1683458, EPI_ISL_1683459, EPI_ISL_1683460, EPI_ISL_1683461, EPI_ISL_1683462, EPI_ISL_1683463, EPI_ISL_1683464, EPI_ISL_1683465, EPI_ISL_1683466, EPI_ISL_1683467, EPI_ISL_1683468, EPI_ISL_1683469, EPI_ISL_1683470, EPI_ISL_1683471, EPI_ISL_1683472, EPI_ISL_1683473, EPI_ISL_1683474, EPI_ISL_1683475, EPI_ISL_1683476, EPI_ISL_1683477, EPI_ISL_1683478, EPI_ISL_1683479, EPI_ISL_1683480, EPI_ISL_1683481, EPI_ISL_1683482, EPI_ISL_1683483, EPI_ISL_1683485, EPI_ISL_1683486, EPI_ISL_1683487, EPI_ISL_1683489, EPI_ISL_1683493, EPI_ISL_1683494, EPI_ISL_1683495, EPI_ISL_1683496, EPI_ISL_1683497, EPI_ISL_1683498, EPI_ISL_1683499, EPI_ISL_1683500, EPI_ISL_1683501, EPI_ISL_1683502, EPI_ISL_1683503, EPI_ISL_1683504, EPI_ISL_1683505, EPI_ISL_1683506, EPI_ISL_1683507, EPI_ISL_1683508, EPI_ISL_1683509, EPI_ISL_1683510, EPI_ISL_1683511, EPI_ISL_1683512, EPI_ISL_1683513, EPI_ISL_1683514, EPI_ISL_1683515, EPI_ISL_1683516, EPI_ISL_1683517, EPI_ISL_1683518, EPI_ISL_1683519, EPI_ISL_1683520, EPI_ISL_1683521, EPI_ISL_1683522, EPI_ISL_1683523, EPI_ISL_1683524, EPI_ISL_1683525, EPI_ISL_1683527, EPI_ISL_1683528, EPI_ISL_1683529, EPI_ISL_1683530, EPI_ISL_1683531, EPI_ISL_1683532, EPI_ISL_1683533, EPI_ISL_1683534, EPI_ISL_1683535, EPI_ISL_1683536, EPI_ISL_1683537, EPI_ISL_1683539, EPI_ISL_1683540, EPI_ISL_1683541, EPI_ISL_1683542, EPI_ISL_1683544, EPI_ISL_1683545, EPI_ISL_1683546, EPI_ISL_1683548, EPI_ISL_1683549, EPI_ISL_1683550, EPI_ISL_1683552, EPI_ISL_1683553, EPI_ISL_1683554, EPI_ISL_1683557, EPI_ISL_1683558, EPI_ISL_1683559, EPI_ISL_1683560, EPI_ISL_1683561, EPI_ISL_1683562, EPI_ISL_1683565, EPI_ISL_1683567, EPI_ISL_1683569, EPI_ISL_1683572, EPI_ISL_1683573, EPI_ISL_1683574, EPI_ISL_1683575, EPI_ISL_1683576, EPI_ISL_1683577, EPI_ISL_1683578, EPI_ISL_1683579, EPI_ISL_1683580, EPI_ISL_1683581, EPI_ISL_1683582, EPI_ISL_1683583, EPI_ISL_1683584, EPI_ISL_1683585, EPI_ISL_1683586, EPI_ISL_1683587, EPI_ISL_1683591, EPI_ISL_1683594, EPI_ISL_1683596, EPI_ISL_1683597, EPI_ISL_1683599, EPI_ISL_1683600, EPI_ISL_1683601, EPI_ISL_1683602, EPI_ISL_1683603, EPI_ISL_1683604, EPI_ISL_1683606, EPI_ISL_1683607, EPI_ISL_1683608, EPI_ISL_1683609, EPI_ISL_1683611, EPI_ISL_1683612, EPI_ISL_1683613, EPI_ISL_1683614, EPI_ISL_1683615, EPI_ISL_1683618, EPI_ISL_1683619, EPI_ISL_1683621, EPI_ISL_1683623, EPI_ISL_1683624, EPI_ISL_1683625, EPI_ISL_1683626, EPI_ISL_1683629, EPI_ISL_1683633, EPI_ISL_1683634, EPI_ISL_1683635, EPI_ISL_1683636, EPI_ISL_1683637, EPI_ISL_1683638, EPI_ISL_1683641, EPI_ISL_1683642, EPI_ISL_1683643, EPI_ISL_1683644, EPI_ISL_1683646, EPI_ISL_1683648, EPI_ISL_1683649, EPI_ISL_1683650, EPI_ISL_1683651, EPI_ISL_1683653, EPI_ISL_1683654, EPI_ISL_1683657, EPI_ISL_1683660, EPI_ISL_1683662, EPI_ISL_1683664, EPI_ISL_1683666, EPI_ISL_1683668, EPI_ISL_1683669, EPI_ISL_1683670, EPI_ISL_1683673, EPI_ISL_1683674, EPI_ISL_1683677, EPI_ISL_1683679, EPI_ISL_1683681, EPI_ISL_1683684, EPI_ISL_1683685, EPI_ISL_1683686, EPI_ISL_1683689, EPI_ISL_1683690, EPI_ISL_1683691, EPI_ISL_1683693, EPI_ISL_1683696, EPI_ISL_1683698, EPI_ISL_1683700, EPI_ISL_1683702, EPI_ISL_1683704, EPI_ISL_1683708, EPI_ISL_1683713, EPI_ISL_1683714, EPI_ISL_1683715, EPI_ISL_1683716, EPI_ISL_1683717, EPI_ISL_1683718, EPI_ISL_1683719, EPI_ISL_1683720, EPI_ISL_1683721, EPI_ISL_1683723, EPI_ISL_1683724, EPI_ISL_1683725, EPI_ISL_1683726, EPI_ISL_1683732, EPI_ISL_1683734, EPI_ISL_1683735, EPI_ISL_1683739, EPI_ISL_1683740, EPI_ISL_1683741, EPI_ISL_1683742, EPI_ISL_1683746, EPI_ISL_1683749, EPI_ISL_1683751, EPI_ISL_1683752, EPI_ISL_1683756, EPI_ISL_1683757, EPI_ISL_1683760, EPI_ISL_1683762, EPI_ISL_1683763, EPI_ISL_1683765, EPI_ISL_1683766, EPI_ISL_1683769, EPI_ISL_1683774, EPI_ISL_1683777, EPI_ISL_1683778, EPI_ISL_1683780, EPI_ISL_1683784, EPI_ISL_1683786, EPI_ISL_1683787, EPI_ISL_1683793, EPI_ISL_1683794, EPI_ISL_1683796, EPI_ISL_1683797, EPI_ISL_1683799, EPI_ISL_1683800, EPI_ISL_1683806, EPI_ISL_1683809, EPI_ISL_1683815, EPI_ISL_1683817, EPI_ISL_1683818, EPI_ISL_1683819, EPI_ISL_1683821, EPI_ISL_1683823, EPI_ISL_1683825, EPI_ISL_1683827, EPI_ISL_1683829, EPI_ISL_1683833, EPI_ISL_1683836, EPI_ISL_1683837, EPI_ISL_1683839, EPI_ISL_1683840, EPI_ISL_1683843, EPI_ISL_1683845, EPI_ISL_1683848, EPI_ISL_1683849, EPI_ISL_1683852, EPI_ISL_1683854, EPI_ISL_1683855, EPI_ISL_1683858, EPI_ISL_1683860, EPI_ISL_1683861, EPI_ISL_1683863, EPI_ISL_1683864, EPI_ISL_1683870, EPI_ISL_1683872, EPI_ISL_1683873, EPI_ISL_1683875, EPI_ISL_1683876, EPI_ISL_1683878, EPI_ISL_1683882, EPI_ISL_1683884, EPI_ISL_1683885, EPI_ISL_1683886, EPI_ISL_1683891, EPI_ISL_1683892, EPI_ISL_1683894, EPI_ISL_1683895, EPI_ISL_1683897, EPI_ISL_1683898, EPI_ISL_1683900, EPI_ISL_1683901, EPI_ISL_1683903, EPI_ISL_1683904, EPI_ISL_1683906, EPI_ISL_1683907, EPI_ISL_1683910, EPI_ISL_1683913, EPI_ISL_1683915, EPI_ISL_1683916, EPI_ISL_1683919, EPI_ISL_1683921, EPI_ISL_1683922, EPI_ISL_1683924, EPI_ISL_1683927, EPI_ISL_1683930, EPI_ISL_1683931, EPI_ISL_1683933, EPI_ISL_1683936, EPI_ISL_1683938, EPI_ISL_1683940, EPI_ISL_1683941, EPI_ISL_1683943, EPI_ISL_1683946, EPI_ISL_1683947, EPI_ISL_1683949, EPI_ISL_1683950, EPI_ISL_1683952, EPI_ISL_1683953, EPI_ISL_1683954, EPI_ISL_1683956, EPI_ISL_1683957, EPI_ISL_1683959, EPI_ISL_1683960, EPI_ISL_1683962, EPI_ISL_1683964, EPI_ISL_1683965, EPI_ISL_1683967, EPI_ISL_1683970, EPI_ISL_1683971, EPI_ISL_1683972, EPI_ISL_1683976, EPI_ISL_1683980, EPI_ISL_1683982, EPI_ISL_1683983, EPI_ISL_1683985, EPI_ISL_1683986, EPI_ISL_1683989, EPI_ISL_1683990, EPI_ISL_1683992, EPI_ISL_1683995, EPI_ISL_1683997, EPI_ISL_1683998, EPI_ISL_1683999, EPI_ISL_1684001, EPI_ISL_1684002, EPI_ISL_1684004, EPI_ISL_1684005, EPI_ISL_1684007, EPI_ISL_1684010, EPI_ISL_1684011, EPI_ISL_1684013, EPI_ISL_1684014, EPI_ISL_1684016, EPI_ISL_1684017, EPI_ISL_1684020, EPI_ISL_1684022, EPI_ISL_1684023, EPI_ISL_1684025, EPI_ISL_1684026, EPI_ISL_1684028, EPI_ISL_1684029, EPI_ISL_1684031, EPI_ISL_1684032, EPI_ISL_1684034, EPI_ISL_1684035, EPI_ISL_1684036, EPI_ISL_1684042, EPI_ISL_1684044, EPI_ISL_1684045, EPI_ISL_1684046, EPI_ISL_1684048, EPI_ISL_1684049, EPI_ISL_1684051, EPI_ISL_1684052, EPI_ISL_1684054, EPI_ISL_1684056, EPI_ISL_1684057, EPI_ISL_1684063, EPI_ISL_1684065, EPI_ISL_1684068, EPI_ISL_1684069, EPI_ISL_1684071, EPI_ISL_1684072, EPI_ISL_1684074, EPI_ISL_1684075, EPI_ISL_1684077, EPI_ISL_1684078, EPI_ISL_1684080, EPI_ISL_1684081, EPI_ISL_1684082, EPI_ISL_1684084, EPI_ISL_1684085, EPI_ISL_1684087, EPI_ISL_1684088, EPI_ISL_1684090, EPI_ISL_1684091, EPI_ISL_1684093, EPI_ISL_1684095, EPI_ISL_1684098, EPI_ISL_1684101, EPI_ISL_1684102, EPI_ISL_1684104, EPI_ISL_1684105, EPI_ISL_1684107, EPI_ISL_1684108, EPI_ISL_1684110, EPI_ISL_1684111, EPI_ISL_1684113, EPI_ISL_1684114, EPI_ISL_1684116, EPI_ISL_1684118, EPI_ISL_1684119, EPI_ISL_1684120, EPI_ISL_1684122, EPI_ISL_1684125, EPI_ISL_1684127, EPI_ISL_1684128, EPI_ISL_1684130, EPI_ISL_1684131, EPI_ISL_1684133, EPI_ISL_1684134, EPI_ISL_1684136, EPI_ISL_1684137, EPI_ISL_1684140, EPI_ISL_1684142, EPI_ISL_1684143, EPI_ISL_1684146, EPI_ISL_1684147, EPI_ISL_1684149, EPI_ISL_1684150, EPI_ISL_1684152, EPI_ISL_1684153, EPI_ISL_1684155, EPI_ISL_1684156, EPI_ISL_1684158, EPI_ISL_1684159, EPI_ISL_1684161, EPI_ISL_1684164, EPI_ISL_1684165, EPI_ISL_1684167, EPI_ISL_1684168, EPI_ISL_1684170, EPI_ISL_1684171, EPI_ISL_1684173, EPI_ISL_1684174, EPI_ISL_1684176, EPI_ISL_1684179, EPI_ISL_1684182, EPI_ISL_1684185, EPI_ISL_1684188, EPI_ISL_1684191, EPI_ISL_1684193, EPI_ISL_1684196, EPI_ISL_1684199, EPI_ISL_1684201, EPI_ISL_1684202, EPI_ISL_1684204, EPI_ISL_1684205, EPI_ISL_1684207, EPI_ISL_1684208, EPI_ISL_1684209, EPI_ISL_1684210, EPI_ISL_1684211, EPI_ISL_1684212, EPI_ISL_1684213, EPI_ISL_1684214, EPI_ISL_1684215, EPI_ISL_1684216, EPI_ISL_1684217, EPI_ISL_1684218, EPI_ISL_1684219, EPI_ISL_1684220, EPI_ISL_1684221, EPI_ISL_1684223, EPI_ISL_1684224, EPI_ISL_1684226, EPI_ISL_1684227, EPI_ISL_1684228, EPI_ISL_1684229, EPI_ISL_1684230, EPI_ISL_1684231, EPI_ISL_1684233, EPI_ISL_1684234, EPI_ISL_1684235, EPI_ISL_1684236, EPI_ISL_1684238, EPI_ISL_1684239, EPI_ISL_1684241, EPI_ISL_1684242, EPI_ISL_1684244, EPI_ISL_1684245, EPI_ISL_1684246, EPI_ISL_1684247, EPI_ISL_1684249, EPI_ISL_1684250, EPI_ISL_1684251, EPI_ISL_1684252, EPI_ISL_1684253, EPI_ISL_1684254, EPI_ISL_1684255, EPI_ISL_1684257, EPI_ISL_1684258, EPI_ISL_1684259, EPI_ISL_1684260, EPI_ISL_1684261, EPI_ISL_1684263, EPI_ISL_1684264, EPI_ISL_1684266, EPI_ISL_1684267, EPI_ISL_1684268, EPI_ISL_1684269, EPI_ISL_1684270, EPI_ISL_1684271, EPI_ISL_1684272, EPI_ISL_1684273, EPI_ISL_1684274, EPI_ISL_1684275, EPI_ISL_1684276, EPI_ISL_1684277, EPI_ISL_1684278, EPI_ISL_1684279, EPI_ISL_1684280, EPI_ISL_1684281, EPI_ISL_1684282, EPI_ISL_1684284, EPI_ISL_1684285, EPI_ISL_1684286, EPI_ISL_1684287, EPI_ISL_1684288, EPI_ISL_1684289, EPI_ISL_1684290, EPI_ISL_1684292, EPI_ISL_1684293, EPI_ISL_1684294, EPI_ISL_1684295, EPI_ISL_1684297, EPI_ISL_1684298, EPI_ISL_1684299, EPI_ISL_1684300, EPI_ISL_1684301, EPI_ISL_1684302, EPI_ISL_1684303, EPI_ISL_1684306, EPI_ISL_1684307, EPI_ISL_1684308, EPI_ISL_1684309, EPI_ISL_1684310, EPI_ISL_1684311, EPI_ISL_1684312, EPI_ISL_1684314, EPI_ISL_1684315, EPI_ISL_1684316, EPI_ISL_1684317, EPI_ISL_1684318, EPI_ISL_1684319, EPI_ISL_1684320, EPI_ISL_1684321, EPI_ISL_1684322, EPI_ISL_1684324, EPI_ISL_1684325, EPI_ISL_1684327, EPI_ISL_1684328, EPI_ISL_1684329, EPI_ISL_1684330, EPI_ISL_1684331, EPI_ISL_1684332, EPI_ISL_1684334, EPI_ISL_1684335, EPI_ISL_1684336, EPI_ISL_1684337, EPI_ISL_1684339, EPI_ISL_1684342, EPI_ISL_1684343, EPI_ISL_1684344, EPI_ISL_1684346, EPI_ISL_1684347, EPI_ISL_1684349, EPI_ISL_1684350, EPI_ISL_1684351, EPI_ISL_1684352, EPI_ISL_1684353, EPI_ISL_1684354, EPI_ISL_1684355, EPI_ISL_1684356, EPI_ISL_1684359, EPI_ISL_1684360, EPI_ISL_1684361, EPI_ISL_1684362, EPI_ISL_1684363, EPI_ISL_1684365, EPI_ISL_1684366, EPI_ISL_1684368, EPI_ISL_1684369, EPI_ISL_1684370, EPI_ISL_1684371, EPI_ISL_1684372, EPI_ISL_1684373, EPI_ISL_1684374, EPI_ISL_1684375, EPI_ISL_1684377, EPI_ISL_1684378, EPI_ISL_1684379, EPI_ISL_1684380, EPI_ISL_1684381, EPI_ISL_1684383, EPI_ISL_1684384, EPI_ISL_1684385, EPI_ISL_1684386, EPI_ISL_1684387, EPI_ISL_1684388, EPI_ISL_1684389, EPI_ISL_1684390, EPI_ISL_1684391, EPI_ISL_1684392, EPI_ISL_1684393, EPI_ISL_1684394, EPI_ISL_1684396, EPI_ISL_1684397, EPI_ISL_1684398, EPI_ISL_1684399, EPI_ISL_1684400, EPI_ISL_1684401, EPI_ISL_1684402, EPI_ISL_1684403, EPI_ISL_1684404, EPI_ISL_1684406, EPI_ISL_1684409, EPI_ISL_1684410, EPI_ISL_1684412, EPI_ISL_1684415, EPI_ISL_1684417, EPI_ISL_1684418, EPI_ISL_1684419, EPI_ISL_1684420, EPI_ISL_1684421, EPI_ISL_1684422, EPI_ISL_1684423, EPI_ISL_1684424, EPI_ISL_1684426, EPI_ISL_1684427, EPI_ISL_1684428, EPI_ISL_1684429, EPI_ISL_1684430, EPI_ISL_1684433, EPI_ISL_1684434, EPI_ISL_1684435, EPI_ISL_1684436, EPI_ISL_1684437, EPI_ISL_1684438, EPI_ISL_1684439, EPI_ISL_1684440, EPI_ISL_1684441, EPI_ISL_1684442, EPI_ISL_1684444, EPI_ISL_1684446, EPI_ISL_1684447, EPI_ISL_1684448, EPI_ISL_1684451, EPI_ISL_1684452, EPI_ISL_1684453, EPI_ISL_1684454, EPI_ISL_1684455, EPI_ISL_1684456, EPI_ISL_1684457, EPI_ISL_1684458, EPI_ISL_1684459, EPI_ISL_1684460, EPI_ISL_1684461, EPI_ISL_1684462, EPI_ISL_1684464, EPI_ISL_1684465, EPI_ISL_1684466, EPI_ISL_1684467, EPI_ISL_1684469, EPI_ISL_1684470, EPI_ISL_1684471, EPI_ISL_1684472, EPI_ISL_1684473, EPI_ISL_1684474, EPI_ISL_1684475, EPI_ISL_1684477, EPI_ISL_1684479, EPI_ISL_1684480, EPI_ISL_1684484, EPI_ISL_1684485, EPI_ISL_1684486, EPI_ISL_1684488, EPI_ISL_1684489, EPI_ISL_1684490, EPI_ISL_1684491, EPI_ISL_1684492, EPI_ISL_1684493, EPI_ISL_1684494, EPI_ISL_1684495, EPI_ISL_1684496, EPI_ISL_1684497, EPI_ISL_1684498, EPI_ISL_1684499, EPI_ISL_1684500, EPI_ISL_1684501, EPI_ISL_1684502, EPI_ISL_1684506, EPI_ISL_1684507, EPI_ISL_1684508, EPI_ISL_1684509, EPI_ISL_1684510, EPI_ISL_1684511, EPI_ISL_1684512, EPI_ISL_1684513, EPI_ISL_1684514, EPI_ISL_1684515, EPI_ISL_1684516, EPI_ISL_1684517, EPI_ISL_1684518, EPI_ISL_1684519, EPI_ISL_1684520, EPI_ISL_1684521, EPI_ISL_1684522, EPI_ISL_1684523, EPI_ISL_1684524, EPI_ISL_1684525, EPI_ISL_1684527, EPI_ISL_1684528, EPI_ISL_1684530, EPI_ISL_1684531, EPI_ISL_1684532, EPI_ISL_1684533, EPI_ISL_1684535, EPI_ISL_1684536, EPI_ISL_1684537, EPI_ISL_1684538, EPI_ISL_1684540, EPI_ISL_1684541, EPI_ISL_1684542, EPI_ISL_1684543, EPI_ISL_1684544, EPI_ISL_1684545, EPI_ISL_1684547, EPI_ISL_1684548, EPI_ISL_1684549, EPI_ISL_1684550, EPI_ISL_1684551, EPI_ISL_1684552, EPI_ISL_1684553, EPI_ISL_1684555, EPI_ISL_1684556, EPI_ISL_1684557, EPI_ISL_1684558, EPI_ISL_1684559, EPI_ISL_1684560, EPI_ISL_1684561, EPI_ISL_1684562, EPI_ISL_1684563, EPI_ISL_1684564, EPI_ISL_1684565, EPI_ISL_1684566, EPI_ISL_1684568, EPI_ISL_1684569, EPI_ISL_1684571, EPI_ISL_1684572, EPI_ISL_1684573, EPI_ISL_1684574, EPI_ISL_1684575, EPI_ISL_1684576, EPI_ISL_1684578, EPI_ISL_1684579, EPI_ISL_1684580, EPI_ISL_1684582, EPI_ISL_1684583, EPI_ISL_1684584, EPI_ISL_1684585, EPI_ISL_1684587, EPI_ISL_1684588, EPI_ISL_1684589, EPI_ISL_1684590, EPI_ISL_1684591, EPI_ISL_1684593, EPI_ISL_1684594, EPI_ISL_1684596, EPI_ISL_1684597, EPI_ISL_1684598, EPI_ISL_1684599, EPI_ISL_1684601, EPI_ISL_1684602, EPI_ISL_1684603, EPI_ISL_1684604, EPI_ISL_1684605, EPI_ISL_1684606, EPI_ISL_1684607, EPI_ISL_1684608, EPI_ISL_1684609, EPI_ISL_1684610, EPI_ISL_1684611, EPI_ISL_1684612, EPI_ISL_1684613, EPI_ISL_1684614, EPI_ISL_1684615, EPI_ISL_1684616, EPI_ISL_1684618, EPI_ISL_1684619, EPI_ISL_1684620, EPI_ISL_1684621, EPI_ISL_1684622, EPI_ISL_1684623, EPI_ISL_1684624, EPI_ISL_1684625, EPI_ISL_1684626, EPI_ISL_1684627, EPI_ISL_1684629, EPI_ISL_1684630, EPI_ISL_1684631, EPI_ISL_1684634, EPI_ISL_1684635, EPI_ISL_1684636, EPI_ISL_1684638, EPI_ISL_1684639, EPI_ISL_1684640, EPI_ISL_1684641, EPI_ISL_1684642, EPI_ISL_1684643, EPI_ISL_1684644, EPI_ISL_1684645, EPI_ISL_1684646, EPI_ISL_1684647, EPI_ISL_1684648, EPI_ISL_1684649, EPI_ISL_1684650, EPI_ISL_1684653, EPI_ISL_1684654, EPI_ISL_1684655, EPI_ISL_1684656, EPI_ISL_1684657, EPI_ISL_1684658, EPI_ISL_1684659, EPI_ISL_1684660, EPI_ISL_1684661, EPI_ISL_1684663, EPI_ISL_1684664, EPI_ISL_1684665, EPI_ISL_1684666, EPI_ISL_1684667, EPI_ISL_1684668, EPI_ISL_1684669, EPI_ISL_1684670, EPI_ISL_1684671, EPI_ISL_1684672, EPI_ISL_1684673, EPI_ISL_1684675, EPI_ISL_1684676, EPI_ISL_1684677, EPI_ISL_1684678, EPI_ISL_1684679, EPI_ISL_1684680, EPI_ISL_1684681, EPI_ISL_1684682, EPI_ISL_1684683, EPI_ISL_1684684, EPI_ISL_1684685, EPI_ISL_1684686, EPI_ISL_1684687, EPI_ISL_1684688, EPI_ISL_1684689, EPI_ISL_1684690, EPI_ISL_1684691, EPI_ISL_1684693, EPI_ISL_1684694, EPI_ISL_1684695, EPI_ISL_1684696, EPI_ISL_1684698, EPI_ISL_1684700, EPI_ISL_1684701, EPI_ISL_1684702, EPI_ISL_1684703, EPI_ISL_1684705, EPI_ISL_1684706, EPI_ISL_1684707, EPI_ISL_1684708, EPI_ISL_1684709, EPI_ISL_1684710, EPI_ISL_1684711, EPI_ISL_1684714, EPI_ISL_1684715, EPI_ISL_1684717, EPI_ISL_1684718, EPI_ISL_1684719, EPI_ISL_1684720, EPI_ISL_1684721, EPI_ISL_1684722, EPI_ISL_1684723, EPI_ISL_1684724, EPI_ISL_1684725, EPI_ISL_1684726, EPI_ISL_1684728, EPI_ISL_1684729, EPI_ISL_1684732, EPI_ISL_1684733, EPI_ISL_1684734, EPI_ISL_1684735, EPI_ISL_1684736, EPI_ISL_1684737, EPI_ISL_1684738, EPI_ISL_1684739, EPI_ISL_1684741, EPI_ISL_1684742, EPI_ISL_1684743, EPI_ISL_1684745, EPI_ISL_1684746, EPI_ISL_1684748, EPI_ISL_1684749, EPI_ISL_1684750, EPI_ISL_1684751, EPI_ISL_1684752, EPI_ISL_1684753, EPI_ISL_1684754, EPI_ISL_1684756, EPI_ISL_1684757, EPI_ISL_1684759, EPI_ISL_1684761, EPI_ISL_1684762, EPI_ISL_1684763, EPI_ISL_1684764, EPI_ISL_1684765, EPI_ISL_1684767, EPI_ISL_1684768, EPI_ISL_1684769, EPI_ISL_1684770, EPI_ISL_1684771, EPI_ISL_1684775, EPI_ISL_1684776, EPI_ISL_1684777, EPI_ISL_1684779, EPI_ISL_1684780

| | | | |
|---|---|---|---|
| see above | Laboratory Corporation of America | Centers for Disease Control and Prevention Division of Viral Diseases, Pathogen Discovery | Dakota Howard, Dhwani Batra, Peter W. Cook, Kara Moser, Adrian Paskey, Jason Caravas, Benjamin Rambo-Martin, Shatavia Morrison, Christopher Gulvick, Scott Sammons, Yvette Unoarumhi, Darlene Wagner, Matthew Schmerer, Minoo Agarwal, Eyad Almasri, Debbie Boles, Ayla Burns, Nuthawin Charoensri, Oren Cohen, Susan Countryman, Mary Ann Cristobal, Bobbi Croy, Suzanne Dale, Hrushikesh Deshmukh, Amanda Douglas, Vincent Drouillon, Marcia Eisenberg, Howard Engler, Rama Ghatti, Prashant Gupta, Susan Hicks, Jake Humphrey, Lax Iyer, Manoj Jain, Mohan Kolli, Brian Krueger, Tim Kuphal, Stanley Letovsky, Michael Levandoski, Craig Lukasik, Jonathan Meltzer, Brian Norvell, Mindy Nye, Scott Parker, Christos Petropoulos, John Pruitt, Steven Ragan, Scott Ryan, Mike Sapeta, Jana Schroth, Suresh Babu Selvaraju, Goran Stevovic, Amanda Suchanek, Andrea Throop, Lyndon Tilson, Thomas Urban, Joe Voshell, Kimberly Wagner, Jonathan Williams, Mary Williamson, Qian Zeng, Tricia Zwiefelhofer, Clinton R. Paden, Duncan MacCannell |
| EPI_ISL_1684781 | Ohio Department of Health Laboratory | Ohio Department of Health Laboratory | Holmes, Jennifer; Eric Brandt, Keoni Omura, Glen McGillivary, Caitlin McDonnell, Jade Mowery, Stephanie Mccracken, Tyler Payne, Kirtana Ramadugu, Erica Leasure, Brent Lee, Kelsey Florek, Heather Blankenship, Allison Black, Quanta Brown, and Tammy Bannerman |

EPI_ISL_1684782, EPI_ISL_1684783, EPI_ISL_1684784, EPI_ISL_1684785, EPI_ISL_1684786, EPI_ISL_1684787, EPI_ISL_1684788, EPI_ISL_1684789, EPI_ISL_1684790, EPI_ISL_1684791, EPI_ISL_1684792, EPI_ISL_1684793, EPI_ISL_1684795, EPI_ISL_1684796, EPI_ISL_1684798, EPI_ISL_1684800, EPI_ISL_1684801, EPI_ISL_1684803, EPI_ISL_1684804, EPI_ISL_1684805, EPI_ISL_1684806, EPI_ISL_1684808, EPI_ISL_1684809, EPI_ISL_1684813, EPI_ISL_1684815, EPI_ISL_1684816, EPI_ISL_1684817, EPI_ISL_1684818, EPI_ISL_1684819, EPI_ISL_1684820, EPI_ISL_1684822, EPI_ISL_1684823, EPI_ISL_1684825, EPI_ISL_1684826, EPI_ISL_1684827, EPI_ISL_1684828, EPI_ISL_1684829, EPI_ISL_1684830, EPI_ISL_1684831, EPI_ISL_1684832, EPI_ISL_1684833, EPI_ISL_1684834, EPI_ISL_1684835, EPI_ISL_1684837, EPI_ISL_1684838, EPI_ISL_1684839, EPI_ISL_1684840, EPI_ISL_1684841, EPI_ISL_1684842, EPI_ISL_1684843, EPI_ISL_1684845, EPI_ISL_1684846, EPI_ISL_1684847, EPI_ISL_1684848, EPI_ISL_1684849, EPI_ISL_1684850, EPI_ISL_1684852, EPI_ISL_1684853, EPI_ISL_1684855, EPI_ISL_1684856, EPI_ISL_1684857, EPI_ISL_1684858, EPI_ISL_1684859, EPI_ISL_1684860, EPI_ISL_1684864, EPI_ISL_1684865, EPI_ISL_1684866, EPI_ISL_1684867, EPI_ISL_1684868, EPI_ISL_1684869, EPI_ISL_1684871, EPI_ISL_1684873, EPI_ISL_1684874, EPI_ISL_1684876, EPI_ISL_1684877, EPI_ISL_1684879, EPI_ISL_1684880, EPI_ISL_1684882, EPI_ISL_1684883, EPI_ISL_1684884, EPI_ISL_1684885, EPI_ISL_1684886, EPI_ISL_1684887, EPI_ISL_1684888, EPI_ISL_1684889, EPI_ISL_1684891, EPI_ISL_1684892, EPI_ISL_1684894, EPI_ISL_1684896, EPI_ISL_1684897, EPI_ISL_1684899, EPI_ISL_1684900, EPI_ISL_1684901, EPI_ISL_1684902, EPI_ISL_1684903, EPI_ISL_1684904, EPI_ISL_1684908, EPI_ISL_1684909, EPI_ISL_1684910, EPI_ISL_1684913, EPI_ISL_1684914, EPI_ISL_1684915, EPI_ISL_1684916, EPI_ISL_1684917, EPI_ISL_1684918, EPI_ISL_1684919, EPI_ISL_1684920, EPI_ISL_1684921, EPI_ISL_1684922, EPI_ISL_1684923, EPI_ISL_1684926, EPI_ISL_1684927, EPI_ISL_1684928, EPI_ISL_1684929, EPI_ISL_1684931, EPI_ISL_1684933, EPI_ISL_1684936, EPI_ISL_1684937, EPI_ISL_1684938, EPI_ISL_1684939, EPI_ISL_1684941, EPI_ISL_1684942, EPI_ISL_1684944, EPI_ISL_1684946, EPI_ISL_1684947, EPI_ISL_1684949, EPI_ISL_1684950, EPI_ISL_1684951, EPI_ISL_1684952, EPI_ISL_1684954, EPI_ISL_1684957, EPI_ISL_1684958, EPI_ISL_1684961, EPI_ISL_1684962, EPI_ISL_1684963, EPI_ISL_1684967, EPI_ISL_1684968, EPI_ISL_1684969, EPI_ISL_1684970, EPI_ISL_1684971, EPI_ISL_1684972, EPI_ISL_1684974, EPI_ISL_1684975, EPI_ISL_1684977, EPI_ISL_1684978, EPI_ISL_1684980, EPI_ISL_1684982, EPI_ISL_1684983, EPI_ISL_1684985, EPI_ISL_1684986, EPI_ISL_1684988, EPI_ISL_1684989, EPI_ISL_1684992, EPI_ISL_1684993, EPI_ISL_1684996, EPI_ISL_1684997, EPI_ISL_1684998, EPI_ISL_1685001, EPI_ISL_1685002, EPI_ISL_1685004, EPI_ISL_1685005, EPI_ISL_1685006, EPI_ISL_1685007, EPI_ISL_1685008, EPI_ISL_1685009, EPI_ISL_1685010, EPI_ISL_1685012, EPI_ISL_1685013, EPI_ISL_1685014, EPI_ISL_1685015, EPI_ISL_1685016, EPI_ISL_1685018, EPI_ISL_1685019, EPI_ISL_1685024, EPI_ISL_1685025, EPI_ISL_1685026, EPI_ISL_1685028, EPI_ISL_1685030, EPI_ISL_1685031, EPI_ISL_1685032, EPI_ISL_1685033, EPI_ISL_1685036, EPI_ISL_1685037, EPI_ISL_1685038, EPI_ISL_1685040, EPI_ISL_1685042, EPI_ISL_1685043, EPI_ISL_1685046, EPI_ISL_1685047, EPI_ISL_1685048, EPI_ISL_1685049, EPI_ISL_1685052, EPI_ISL_1685053, EPI_ISL_1685054, EPI_ISL_1685056, EPI_ISL_1685057, EPI_ISL_1685059, EPI_ISL_1685060, EPI_ISL_1685061, EPI_ISL_1685062, EPI_ISL_1685063, EPI_ISL_1685064, EPI_ISL_1685065, EPI_ISL_1685067, EPI_ISL_1685068, EPI_ISL_1685069, EPI_ISL_1685071, EPI_ISL_1685072, EPI_ISL_1685073, EPI_ISL_1685074, EPI_ISL_1685075, EPI_ISL_1685076, EPI_ISL_1685077, EPI_ISL_1685078, EPI_ISL_1685079, EPI_ISL_1685080, EPI_ISL_1685081, EPI_ISL_1685083, EPI_ISL_1685084, EPI_ISL_1685085, EPI_ISL_1685088, EPI_ISL_1685089, EPI_ISL_1685090, EPI_ISL_1685091, EPI_ISL_1685093, EPI_ISL_1685094, EPI_ISL_1685095, EPI_ISL_1685096, EPI_ISL_1685097, EPI_ISL_1685098, EPI_ISL_1685099, EPI_ISL_1685100, EPI_ISL_1685101, EPI_ISL_1685102, EPI_ISL_1685103, EPI_ISL_1685105, EPI_ISL_1685106, EPI_ISL_1685108, EPI_ISL_1685109, EPI_ISL_1685110, EPI_ISL_1685113, EPI_ISL_1685114, EPI_ISL_1685117, EPI_ISL_1685118, EPI_ISL_1685119, EPI_ISL_1685120, EPI_ISL_1685121, EPI_ISL_1685122, EPI_ISL_1685123, EPI_ISL_1685124, EPI_ISL_1685125, EPI_ISL_1685126, EPI_ISL_1685127, EPI_ISL_1685128, EPI_ISL_1685129, EPI_ISL_1685130, EPI_ISL_1685132, EPI_ISL_1685133, EPI_ISL_1685135, EPI_ISL_1685136, EPI_ISL_1685137, EPI_ISL_1685138, EPI_ISL_1685140, EPI_ISL_1685143,

| | | | |
|---|---|---|---|
| EPI_ISL_1685144, EPI_ISL_1685146, EPI_ISL_1685147, EPI_ISL_1685148, EPI_ISL_1685149, EPI_ISL_1685151, EPI_ISL_1685154, EPI_ISL_1685156, EPI_ISL_1685157, EPI_ISL_1685158, EPI_ISL_1685159, EPI_ISL_1685160, EPI_ISL_1685162, EPI_ISL_1685164, EPI_ISL_1685165, EPI_ISL_1685166, EPI_ISL_1685167, EPI_ISL_1685168, EPI_ISL_1685169, EPI_ISL_1685170, EPI_ISL_1685171, EPI_ISL_1685172, EPI_ISL_1685173, EPI_ISL_1685175, EPI_ISL_1685177, EPI_ISL_1685178, EPI_ISL_1685180, EPI_ISL_1685181, EPI_ISL_1685182, EPI_ISL_1685183, EPI_ISL_1685184, EPI_ISL_1685185, EPI_ISL_1685186, EPI_ISL_1685187, EPI_ISL_1685188, EPI_ISL_1685190, EPI_ISL_1685191, EPI_ISL_1685192, EPI_ISL_1685193, EPI_ISL_1685194, EPI_ISL_1685195, EPI_ISL_1685196, EPI_ISL_1685197, EPI_ISL_1685198, EPI_ISL_1685199, EPI_ISL_1685200, EPI_ISL_1685202, EPI_ISL_1685203, EPI_ISL_1685204, EPI_ISL_1685205, EPI_ISL_1685206, EPI_ISL_1685207, EPI_ISL_1685209, EPI_ISL_1685210, EPI_ISL_1685212, EPI_ISL_1685213, EPI_ISL_1685214, EPI_ISL_1685215, EPI_ISL_1685217, EPI_ISL_1685218, EPI_ISL_1685220, EPI_ISL_1685221, EPI_ISL_1685225, EPI_ISL_1685227, EPI_ISL_1685228 | | | |
| see above | Laboratory Corporation of America | Centers for Disease Control and Prevention Division of Viral Diseases, Pathogen Discovery | Dakota Howard, Dhwani Batra, Peter W. Cook, Kara Moser, Adrian Paskey, Jason Caravas, Benjamin Rambo-Martin, Shatavia Morrison, Christopher Gulvick, Scott Sammons, Yvette Unoarumhi, Darlene Wagner, Matthew Schmerer, Minoo Agarwal, Eyad Almasri, Debbie Boles, Ayla Burns, Nuthawin Charoensri, Oren Cohen, Susan Countryman, Mary Ann Cristobal, Bobbi Croy, Suzanne Dale, Hrushikesh Deshmukh, Amanda Douglas, Vincent Drouillon, Marcia Eisenberg, Howard Engler, Rama Ghatti, Prashant Gupta, Susan Hicks, Jake Humphrey, Lax Iyer, Manoj Jain, Mohan Kolli, Brian Krueger, Tim Kuphal, Stanley Letovsky, Michael Levandoski, Craig Lukasik, Jonathan Meltzer, Brian Norvell, Mindy Nye, Scott Parker, Christos Petropoulos, John Pruitt, Steven Ragan, Scott Ryan, Mike Sapeta, Jana Schroth, Suresh Babu Selvaraju, Goran Stevovic, Amanda Suchanek, Andrea Throop, Lyndon Tilson, Thomas Urban, Joe Voshell, Kimberly Wagner, Jonathan Williams, Mary Williamson, Qian Zeng, Tricia Zwiefelhofer, Clinton R. Paden, Duncan MacCannell |
| EPI_ISL_1685466 | Ohio Department of Health Laboratory | Ohio Department of Health Laboratory | Holmes, Jennifer; Eric Brandt, Keoni Omura, Glen McGillivary, Caitlin McDonnell, Jade Mowery, Stephanie Mccracken, Tyler Payne, Kirtana Ramadugu, Erica Leasure, Brent Lee, Kelsey Florek, Heather Blankenship, Allison Black, Quanta Brown, and Tammy Bannerman |
| EPI_ISL_1685467, EPI_ISL_1685468, EPI_ISL_1685469, EPI_ISL_1685470, EPI_ISL_1685471, EPI_ISL_1685472, EPI_ISL_1685473, EPI_ISL_1685474, EPI_ISL_1685475, EPI_ISL_1685476, EPI_ISL_1685478, EPI_ISL_1685479, EPI_ISL_1685480, EPI_ISL_1685481, EPI_ISL_1685482, EPI_ISL_1685483, EPI_ISL_1685484, EPI_ISL_1685485, EPI_ISL_1685486, EPI_ISL_1685487, EPI_ISL_1685488, EPI_ISL_1685489, EPI_ISL_1685490, EPI_ISL_1685491, EPI_ISL_1685492, EPI_ISL_1685493, EPI_ISL_1685494, EPI_ISL_1685495, EPI_ISL_1685496, EPI_ISL_1685497, EPI_ISL_1685498, EPI_ISL_1685499, EPI_ISL_1685500, EPI_ISL_1685501, EPI_ISL_1685502, EPI_ISL_1685503, EPI_ISL_1685504, EPI_ISL_1685505, EPI_ISL_1685506, EPI_ISL_1685507, EPI_ISL_1685508, EPI_ISL_1685509, EPI_ISL_1685510, EPI_ISL_1685511, EPI_ISL_1685512, EPI_ISL_1685513, EPI_ISL_1685514, EPI_ISL_1685515, EPI_ISL_1685516, EPI_ISL_1685517, EPI_ISL_1685518, EPI_ISL_1685519, EPI_ISL_1685520, EPI_ISL_1685521, EPI_ISL_1685522, EPI_ISL_1685523, EPI_ISL_1685524, EPI_ISL_1685525, EPI_ISL_1685526, EPI_ISL_1685527, EPI_ISL_1685528, EPI_ISL_1685529, EPI_ISL_1685530, EPI_ISL_1685531, EPI_ISL_1685532, EPI_ISL_1685533, EPI_ISL_1685534, EPI_ISL_1685535, EPI_ISL_1685536, EPI_ISL_1685537, EPI_ISL_1685538, EPI_ISL_1685539, EPI_ISL_1685540, EPI_ISL_1685541, EPI_ISL_1685542, EPI_ISL_1685543, EPI_ISL_1685544, EPI_ISL_1685545, EPI_ISL_1685546, EPI_ISL_1685547, EPI_ISL_1685548, EPI_ISL_1685549, EPI_ISL_1685550, EPI_ISL_1685551, EPI_ISL_1685552, EPI_ISL_1685553, EPI_ISL_1685554, EPI_ISL_1685556, EPI_ISL_1685557, EPI_ISL_1685558, EPI_ISL_1685559, EPI_ISL_1685560, EPI_ISL_1685561, EPI_ISL_1685562, EPI_ISL_1685563, EPI_ISL_1685564, EPI_ISL_1685565, EPI_ISL_1685566, EPI_ISL_1685567, EPI_ISL_1685568, EPI_ISL_1685569, EPI_ISL_1685570, EPI_ISL_1685571, EPI_ISL_1685572, EPI_ISL_1685573, EPI_ISL_1685574, EPI_ISL_1685575, EPI_ISL_1685576, EPI_ISL_1685577, EPI_ISL_1685578, EPI_ISL_1685579, EPI_ISL_1685580, EPI_ISL_1685581, EPI_ISL_1685582, EPI_ISL_1685583, EPI_ISL_1685584, EPI_ISL_1685585, EPI_ISL_1685586, EPI_ISL_1685587, EPI_ISL_1685588, EPI_ISL_1685589, EPI_ISL_1685590, EPI_ISL_1685591, EPI_ISL_1685592, EPI_ISL_1685593, EPI_ISL_1685594, EPI_ISL_1685595, EPI_ISL_1685596, EPI_ISL_1685597, EPI_ISL_1685598, EPI_ISL_1685600, EPI_ISL_1685601, EPI_ISL_1685602, EPI_ISL_1685603, EPI_ISL_1685604, EPI_ISL_1685605, EPI_ISL_1685606, EPI_ISL_1685607, EPI_ISL_1685608, EPI_ISL_1685609, EPI_ISL_1685610, EPI_ISL_1685611, EPI_ISL_1685612, EPI_ISL_1685613, EPI_ISL_1685614, EPI_ISL_1685615, EPI_ISL_1685616, EPI_ISL_1685617, EPI_ISL_1685618, EPI_ISL_1685619, EPI_ISL_1685620, EPI_ISL_1685621, EPI_ISL_1685622, EPI_ISL_1685623, EPI_ISL_1685624, EPI_ISL_1685625, EPI_ISL_1685626, EPI_ISL_1685627, EPI_ISL_1685628, EPI_ISL_1685629, EPI_ISL_1685630, EPI_ISL_1685631, EPI_ISL_1685632, EPI_ISL_1685633, EPI_ISL_1685634, EPI_ISL_1685635, EPI_ISL_1685636, EPI_ISL_1685637, EPI_ISL_1685638, EPI_ISL_1685639, EPI_ISL_1685640, EPI_ISL_1685642, EPI_ISL_1685644, EPI_ISL_1685645, EPI_ISL_1685646, EPI_ISL_1685647, EPI_ISL_1685648, EPI_ISL_1685649, EPI_ISL_1685650, EPI_ISL_1685651, EPI_ISL_1685652, EPI_ISL_1685653, EPI_ISL_1685654, EPI_ISL_1685655, EPI_ISL_1685656, EPI_ISL_1685657, EPI_ISL_1685658, EPI_ISL_1685659, EPI_ISL_1685660, EPI_ISL_1685661, EPI_ISL_1685662, EPI_ISL_1685663, EPI_ISL_1685664, EPI_ISL_1685665, EPI_ISL_1685666, EPI_ISL_1685667, EPI_ISL_1685668, EPI_ISL_1685669, EPI_ISL_1685671, EPI_ISL_1685672, EPI_ISL_1685673, EPI_ISL_1685674, EPI_ISL_1685675, EPI_ISL_1685676, EPI_ISL_1685677, EPI_ISL_1685678, EPI_ISL_1685679, EPI_ISL_1685680, EPI_ISL_1685682, EPI_ISL_1685683, EPI_ISL_1685684, EPI_ISL_1685685, EPI_ISL_1685686, EPI_ISL_1685687, EPI_ISL_1685688, EPI_ISL_1685689, EPI_ISL_1685690, EPI_ISL_1685691, EPI_ISL_1685692, EPI_ISL_1685693, EPI_ISL_1685694, EPI_ISL_1685695, EPI_ISL_1685696, EPI_ISL_1685697, EPI_ISL_1685698, EPI_ISL_1685699, EPI_ISL_1685700, EPI_ISL_1685701, EPI_ISL_1685702, EPI_ISL_1685703, EPI_ISL_1685704, EPI_ISL_1685705, EPI_ISL_1685706, EPI_ISL_1685707, EPI_ISL_1685708, EPI_ISL_1685709, EPI_ISL_1685710, EPI_ISL_1685711, EPI_ISL_1685712, EPI_ISL_1685713, EPI_ISL_1685714, EPI_ISL_1685715, EPI_ISL_1685716, EPI_ISL_1685717, EPI_ISL_1685718, EPI_ISL_1685719, EPI_ISL_1685720, EPI_ISL_1685721, EPI_ISL_1685722, EPI_ISL_1685723, EPI_ISL_1685724, EPI_ISL_1685725, EPI_ISL_1685726, EPI_ISL_1685727, EPI_ISL_1685728, EPI_ISL_1685729, EPI_ISL_1685730, EPI_ISL_1685732, EPI_ISL_1685733, EPI_ISL_1685734, EPI_ISL_1685735, EPI_ISL_1685736, EPI_ISL_1685737, EPI_ISL_1685738, EPI_ISL_1685739, EPI_ISL_1685740, EPI_ISL_1685741, EPI_ISL_1685742, EPI_ISL_1685743, EPI_ISL_1685744, EPI_ISL_1685745, EPI_ISL_1685746, EPI_ISL_1685747, EPI_ISL_1685748, EPI_ISL_1685749, EPI_ISL_1685750, EPI_ISL_1685751, EPI_ISL_1685752, EPI_ISL_1685753, EPI_ISL_1685754, EPI_ISL_1685755, EPI_ISL_1685756, EPI_ISL_1685757, EPI_ISL_1685758, EPI_ISL_1685759, EPI_ISL_1685760, EPI_ISL_1685761, EPI_ISL_1685762, EPI_ISL_1685763, EPI_ISL_1685764, EPI_ISL_1685765, EPI_ISL_1685766, EPI_ISL_1685767, EPI_ISL_1685768, EPI_ISL_1685769, EPI_ISL_1685770, EPI_ISL_1685771, EPI_ISL_1685772, EPI_ISL_1685773, EPI_ISL_1685774, EPI_ISL_1685775, EPI_ISL_1685776, EPI_ISL_1685777, EPI_ISL_1685778, EPI_ISL_1685779, EPI_ISL_1685780, EPI_ISL_1685781, EPI_ISL_1685782, EPI_ISL_1685783, EPI_ISL_1685784, EPI_ISL_1685785, EPI_ISL_1685786, EPI_ISL_1685787, EPI_ISL_1685788, EPI_ISL_1685789, EPI_ISL_1685790, EPI_ISL_1685791, EPI_ISL_1685792, EPI_ISL_1685793, EPI_ISL_1685794, EPI_ISL_1685795, EPI_ISL_1685796, EPI_ISL_1685797, EPI_ISL_1685798, EPI_ISL_1685799, EPI_ISL_1685800, EPI_ISL_1685801, EPI_ISL_1685802, EPI_ISL_1685803, EPI_ISL_1685804, EPI_ISL_1685805, EPI_ISL_1685806, EPI_ISL_1685807, EPI_ISL_1685808, EPI_ISL_1685809, EPI_ISL_1685810, EPI_ISL_1685811, EPI_ISL_1685812, EPI_ISL_1685813, EPI_ISL_1685814, EPI_ISL_1685815, EPI_ISL_1685816, EPI_ISL_1685817, EPI_ISL_1685818, EPI_ISL_1685819, EPI_ISL_1685820, EPI_ISL_1685821, EPI_ISL_1685822, EPI_ISL_1685823, EPI_ISL_1685824, EPI_ISL_1685825, EPI_ISL_1685826, EPI_ISL_1685827, EPI_ISL_1685828, EPI_ISL_1685829, EPI_ISL_1685830, EPI_ISL_1685831, EPI_ISL_1685832, EPI_ISL_1685833, EPI_ISL_1685834, EPI_ISL_1685835, EPI_ISL_1685836, EPI_ISL_1685837, EPI_ISL_1685838, EPI_ISL_1685839, EPI_ISL_1685840, EPI_ISL_1685842, EPI_ISL_1685843, EPI_ISL_1685844, EPI_ISL_1685845, EPI_ISL_1685846, EPI_ISL_1685847, EPI_ISL_1685848, EPI_ISL_1685849, EPI_ISL_1685850, EPI_ISL_1685851, EPI_ISL_1685853, EPI_ISL_1685854, EPI_ISL_1685855, EPI_ISL_1685856, EPI_ISL_1685857, EPI_ISL_1685858, EPI_ISL_1685859, EPI_ISL_1685860, EPI_ISL_1685862, EPI_ISL_1685863, EPI_ISL_1685864, EPI_ISL_1685865, EPI_ISL_1685866, EPI_ISL_1685867, EPI_ISL_1685868, EPI_ISL_1685869, EPI_ISL_1685870, EPI_ISL_1685871, EPI_ISL_1685872, EPI_ISL_1685873, EPI_ISL_1685874, EPI_ISL_1685875, EPI_ISL_1685876, EPI_ISL_1685877, EPI_ISL_1685878, EPI_ISL_1685879, EPI_ISL_1685880, EPI_ISL_1685881, EPI_ISL_1685882, EPI_ISL_1685883, EPI_ISL_1685884, EPI_ISL_1685885, EPI_ISL_1685886, EPI_ISL_1685887, EPI_ISL_1685888, EPI_ISL_1685889, EPI_ISL_1685890, EPI_ISL_1685891, EPI_ISL_1685892, EPI_ISL_1685893, EPI_ISL_1685894, EPI_ISL_1685895, EPI_ISL_1685896, EPI_ISL_1685897, EPI_ISL_1685898, EPI_ISL_1685899, EPI_ISL_1685900, EPI_ISL_1685901, EPI_ISL_1685902, EPI_ISL_1685903, EPI_ISL_1685905, EPI_ISL_1685906, EPI_ISL_1685907, EPI_ISL_1685908, EPI_ISL_1685909, EPI_ISL_1685910, EPI_ISL_1685911, EPI_ISL_1685912, EPI_ISL_1685913, EPI_ISL_1685914, EPI_ISL_1685915, EPI_ISL_1685916, EPI_ISL_1685917, EPI_ISL_1685918, EPI_ISL_1685919, EPI_ISL_1685920, EPI_ISL_1685921, EPI_ISL_1685922, EPI_ISL_1685923, EPI_ISL_1685924, EPI_ISL_1685925, EPI_ISL_1685926, EPI_ISL_1685927, EPI_ISL_1685928, EPI_ISL_1685929, EPI_ISL_1685930, EPI_ISL_1685931, EPI_ISL_1685932, EPI_ISL_1685933, EPI_ISL_1685934, EPI_ISL_1685935, EPI_ISL_1685936, EPI_ISL_1685937, EPI_ISL_1685938, EPI_ISL_1685941, EPI_ISL_1685942, EPI_ISL_1685943, EPI_ISL_1685944, EPI_ISL_1685945, EPI_ISL_1685946, EPI_ISL_1685947, EPI_ISL_1685948, EPI_ISL_1685949, EPI_ISL_1685950, EPI_ISL_1685951, EPI_ISL_1685952, EPI_ISL_1685953, EPI_ISL_1685954, EPI_ISL_1685955, EPI_ISL_1685956, EPI_ISL_1685957, EPI_ISL_1685958, EPI_ISL_1685959, EPI_ISL_1685960, EPI_ISL_1685961, EPI_ISL_1685962, EPI_ISL_1685963, EPI_ISL_1685964, EPI_ISL_1685965, EPI_ISL_1685966, EPI_ISL_1685967, EPI_ISL_1685968, EPI_ISL_1685969, EPI_ISL_1685970, EPI_ISL_1685971, EPI_ISL_1685972, EPI_ISL_1685973, EPI_ISL_1685974, EPI_ISL_1685975, EPI_ISL_1685976, EPI_ISL_1685977, EPI_ISL_1685978, EPI_ISL_1685979, EPI_ISL_1685980, EPI_ISL_1685981, EPI_ISL_1685982, EPI_ISL_1685983, EPI_ISL_1685984, EPI_ISL_1685985, EPI_ISL_1685986, EPI_ISL_1685987, EPI_ISL_1685988, EPI_ISL_1685989, EPI_ISL_1685990, EPI_ISL_1685992, EPI_ISL_1685996, EPI_ISL_1685999, EPI_ISL_1686002, EPI_ISL_1686003, EPI_ISL_1686005, EPI_ISL_1686006, EPI_ISL_1686008, EPI_ISL_1686009, EPI_ISL_1686011, EPI_ISL_1686012, EPI_ISL_1686013, EPI_ISL_1686015, EPI_ISL_1686017, EPI_ISL_1686020, EPI_ISL_1686023, EPI_ISL_1686026, EPI_ISL_1686029, EPI_ISL_1686035, EPI_ISL_1686038, EPI_ISL_1686041, EPI_ISL_1686044, EPI_ISL_1686047, EPI_ISL_1686051, EPI_ISL_1686054, EPI_ISL_1686056, EPI_ISL_1686060, EPI_ISL_1686063, EPI_ISL_1686066, EPI_ISL_1686069, EPI_ISL_1686072, EPI_ISL_1686075, EPI_ISL_1686079, EPI_ISL_1686082, EPI_ISL_1686084, EPI_ISL_1686088, EPI_ISL_1686090, EPI_ISL_1686094, EPI_ISL_1686097, EPI_ISL_1686100, EPI_ISL_1686104, EPI_ISL_1686107, EPI_ISL_1686110, EPI_ISL_1686113, EPI_ISL_1686116, EPI_ISL_1686120, EPI_ISL_1686122, EPI_ISL_1686126, EPI_ISL_1686128, EPI_ISL_1686132, EPI_ISL_1686135, EPI_ISL_1686141, EPI_ISL_1686146, EPI_ISL_1686149, EPI_ISL_1686153, EPI_ISL_1686155, EPI_ISL_1686159, EPI_ISL_1686161, EPI_ISL_1686163, EPI_ISL_1686164, EPI_ISL_1686166, EPI_ISL_1686167, EPI_ISL_1686169, EPI_ISL_1686170, EPI_ISL_1686172, EPI_ISL_1686173, EPI_ISL_1686175, EPI_ISL_1686177, EPI_ISL_1686179, EPI_ISL_1686180, EPI_ISL_1686181, EPI_ISL_1686183, EPI_ISL_1686184, EPI_ISL_1686186, EPI_ISL_1686187, EPI_ISL_1686188, EPI_ISL_1686191, EPI_ISL_1686194, EPI_ISL_1686197, EPI_ISL_1686200, EPI_ISL_1686204, EPI_ISL_1686207, EPI_ISL_1686210, EPI_ISL_1686213, EPI_ISL_1686217, EPI_ISL_1686220, EPI_ISL_1686224, EPI_ISL_1686227, EPI_ISL_1686230, EPI_ISL_1686233, EPI_ISL_1686236, EPI_ISL_1686239, EPI_ISL_1686243, EPI_ISL_1686246, EPI_ISL_1686249, EPI_ISL_1686252, EPI_ISL_1686255, EPI_ISL_1686258, EPI_ISL_1686261, EPI_ISL_1686264, EPI_ISL_1686267, EPI_ISL_1686271, EPI_ISL_1686273, EPI_ISL_1686274, EPI_ISL_1686275, EPI_ISL_1686277, EPI_ISL_1686278, EPI_ISL_1686279, EPI_ISL_1686281, EPI_ISL_1686282, EPI_ISL_1686283, EPI_ISL_1686284, EPI_ISL_1686285, EPI_ISL_1686286, EPI_ISL_1686287, EPI_ISL_1686288, EPI_ISL_1686289, EPI_ISL_1686290, EPI_ISL_1686291, EPI_ISL_1686292, EPI_ISL_1686293, EPI_ISL_1686294, EPI_ISL_1686295, EPI_ISL_1686296, EPI_ISL_1686297, EPI_ISL_1686298, EPI_ISL_1686299, EPI_ISL_1686300, EPI_ISL_1686301, EPI_ISL_1686302, EPI_ISL_1686303, EPI_ISL_1686304, EPI_ISL_1686305, EPI_ISL_1686306, EPI_ISL_1686307, EPI_ISL_1686308, EPI_ISL_1686309, EPI_ISL_1686310, EPI_ISL_1686311, EPI_ISL_1686312, EPI_ISL_1686313, EPI_ISL_1686314, EPI_ISL_1686315, EPI_ISL_1686316, EPI_ISL_1686318, EPI_ISL_1686319, EPI_ISL_1686320, EPI_ISL_1686321, EPI_ISL_1686322, EPI_ISL_1686323, EPI_ISL_1686324, EPI_ISL_1686325, EPI_ISL_1686326, EPI_ISL_1686327, EPI_ISL_1686328, EPI_ISL_1686329, EPI_ISL_1686330, EPI_ISL_1686331, EPI_ISL_1686332, EPI_ISL_1686333, EPI_ISL_1686334, EPI_ISL_1686335, EPI_ISL_1686336, EPI_ISL_1686337, EPI_ISL_1686338, EPI_ISL_1686339, EPI_ISL_1686340, EPI_ISL_1686341, EPI_ISL_1686342, EPI_ISL_1686343, EPI_ISL_1686344, EPI_ISL_1686345, EPI_ISL_1686346, EPI_ISL_1686347, EPI_ISL_1686348, EPI_ISL_1686349 | | | |
| see above | Aegis Sciences Corporation | Centers for Disease Control and Prevention Division of Viral Diseases, Pathogen Discovery | Dakota Howard, Dhwani Batra, Peter W. Cook, Kara Moser, Adrian Paskey, Jason Caravas, Benjamin Rambo-Martin, Shatavia Morrison, Christopher Gulvick, Scott Sammons, Yvette Unoarumhi, Darlene Wagner, Matthew Schmerer, Cyndi Clark, Patrick Campbell, Rob Case, Vikramsinha Ghorpade, Holly Houdeshell, Ola Kvalvaag, Dillon Nall, Ethan Sanders, Alec Vest, Shaun Westlund, Matthew Hardison, Clinton R. Paden, Duncan MacCannell |
| EPI_ISL_1686350 | Ohio Department of Health Laboratory | Ohio Department of Health Laboratory | Holmes, Jennifer; Eric Brandt, Keoni Omura, Glen McGillivary, Caitlin McDonnell, Jade Mowery, Stephanie Mccracken, Tyler Payne, Kirtana Ramadugu, Erica Leasure, Brent Lee, Kelsey Florek, Heather Blankenship, Allison Black, Quanta Brown, and Tammy Bannerman |
| EPI_ISL_1686351, EPI_ISL_1686352, EPI_ISL_1686353, EPI_ISL_1686354, EPI_ISL_1686355, EPI_ISL_1686356, EPI_ISL_1686357, EPI_ISL_1686358, EPI_ISL_1686359, EPI_ISL_1686360, EPI_ISL_1686361, EPI_ISL_1686362, EPI_ISL_1686363, EPI_ISL_1686364, EPI_ISL_1686365, EPI_ISL_1686366, EPI_ISL_1686367, EPI_ISL_1686368, EPI_ISL_1686369, EPI_ISL_1686370, EPI_ISL_1686371, EPI_ISL_1686372, EPI_ISL_1686373, EPI_ISL_1686374, EPI_ISL_1686375, EPI_ISL_1686376, EPI_ISL_1686377, EPI_ISL_1686378, EPI_ISL_1686379, EPI_ISL_1686380, EPI_ISL_1686381, EPI_ISL_1686382, EPI_ISL_1686383, EPI_ISL_1686384, EPI_ISL_1686385, EPI_ISL_1686386, EPI_ISL_1686387, EPI_ISL_1686388, EPI_ISL_1686389, EPI_ISL_1686390, EPI_ISL_1686391, EPI_ISL_1686392, EPI_ISL_1686393, EPI_ISL_1686394, EPI_ISL_1686395, EPI_ISL_1686396, EPI_ISL_1686397, EPI_ISL_1686398, EPI_ISL_1686399, EPI_ISL_1686400, EPI_ISL_1686401, EPI_ISL_1686402, EPI_ISL_1686403, EPI_ISL_1686404, EPI_ISL_1686405, EPI_ISL_1686406, EPI_ISL_1686407, EPI_ISL_1686408, EPI_ISL_1686409, EPI_ISL_1686410, EPI_ISL_1686411, EPI_ISL_1686412, EPI_ISL_1686413, EPI_ISL_1686414, EPI_ISL_1686415, EPI_ISL_1686416, EPI_ISL_1686417, EPI_ISL_1686418, EPI_ISL_1686419, EPI_ISL_1686420, EPI_ISL_1686421, EPI_ISL_1686422, EPI_ISL_1686423, EPI_ISL_1686424, EPI_ISL_1686425, EPI_ISL_1686426, EPI_ISL_1686427, EPI_ISL_1686428, EPI_ISL_1686429, EPI_ISL_1686431, EPI_ISL_1686432, EPI_ISL_1686433, EPI_ISL_1686434, EPI_ISL_1686435, EPI_ISL_1686436, EPI_ISL_1686437, EPI_ISL_1686438, EPI_ISL_1686439, EPI_ISL_1686440, EPI_ISL_1686441, EPI_ISL_1686442, EPI_ISL_1686443, EPI_ISL_1686444, EPI_ISL_1686445, EPI_ISL_1686446, EPI_ISL_1686447, EPI_ISL_1686448, EPI_ISL_1686449, EPI_ISL_1686450, EPI_ISL_1686451, EPI_ISL_1686452, EPI_ISL_1686453, EPI_ISL_1686454, EPI_ISL_1686455, EPI_ISL_1686456, EPI_ISL_1686457, EPI_ISL_1686458, EPI_ISL_1686459, EPI_ISL_1686460, EPI_ISL_1686461, EPI_ISL_1686462, EPI_ISL_1686463, EPI_ISL_1686464, EPI_ISL_1686465, EPI_ISL_1686466, EPI_ISL_1686467, EPI_ISL_1686468, EPI_ISL_1686469, EPI_ISL_1686470, EPI_ISL_1686471, EPI_ISL_1686472, EPI_ISL_1686473, EPI_ISL_1686474, EPI_ISL_1686475, EPI_ISL_1686476, EPI_ISL_1686477, EPI_ISL_1686478, EPI_ISL_1686479, EPI_ISL_1686480, EPI_ISL_1686481, EPI_ISL_1686482, EPI_ISL_1686483, EPI_ISL_1686484, EPI_ISL_1686485, EPI_ISL_1686486, EPI_ISL_1686487, EPI_ISL_1686488, EPI_ISL_1686489, EPI_ISL_1686490, EPI_ISL_1686491, EPI_ISL_1686492, EPI_ISL_1686493, EPI_ISL_1686494, EPI_ISL_1686495, EPI_ISL_1686496, EPI_ISL_1686497, EPI_ISL_1686498, EPI_ISL_1686499, EPI_ISL_1686500, EPI_ISL_1686501, EPI_ISL_1686502, EPI_ISL_1686503, EPI_ISL_1686504, EPI_ISL_1686505, EPI_ISL_1686506, EPI_ISL_1686507, EPI_ISL_1686508, EPI_ISL_1686509, EPI_ISL_1686510, EPI_ISL_1686511, EPI_ISL_1686512, EPI_ISL_1686513, EPI_ISL_1686514, EPI_ISL_1686515, EPI_ISL_1686516, EPI_ISL_1686517, EPI_ISL_1686518, EPI_ISL_1686519, EPI_ISL_1686520, EPI_ISL_1686522, EPI_ISL_1686523, EPI_ISL_1686524, EPI_ISL_1686525, EPI_ISL_1686526, EPI_ISL_1686527, EPI_ISL_1686528, EPI_ISL_1686529, EPI_ISL_1686530, EPI_ISL_1686531, EPI_ISL_1686532, EPI_ISL_1686533, EPI_ISL_1686534, EPI_ISL_1686535, EPI_ISL_1686536, EPI_ISL_1686537, EPI_ISL_1686538, EPI_ISL_1686539, EPI_ISL_1686540, EPI_ISL_1686541, EPI_ISL_1686542, EPI_ISL_1686543, EPI_ISL_1686544, EPI_ISL_1686545, EPI_ISL_1686546, EPI_ISL_1686547, EPI_ISL_1686548, EPI_ISL_1686549, EPI_ISL_1686550, EPI_ISL_1686551, EPI_ISL_1686552, EPI_ISL_1686553, EPI_ISL_1686554, EPI_ISL_1686555, EPI_ISL_1686556, EPI_ISL_1686557, EPI_ISL_1686558, EPI_ISL_1686559, EPI_ISL_1686560, EPI_ISL_1686561, EPI_ISL_1686562, EPI_ISL_1686563, EPI_ISL_1686564, EPI_ISL_1686565, EPI_ISL_1686566, EPI_ISL_1686567, EPI_ISL_1686568, EPI_ISL_1686569, EPI_ISL_1686570, EPI_ISL_1686571, EPI_ISL_1686572, EPI_ISL_1686573, EPI_ISL_1686574, EPI_ISL_1686575, EPI_ISL_1686576, EPI_ISL_1686577, EPI_ISL_1686578, EPI_ISL_1686579, EPI_ISL_1686580, EPI_ISL_1686581, EPI_ISL_1686582, EPI_ISL_1686583, EPI_ISL_1686584, EPI_ISL_1686585, EPI_ISL_1686586, EPI_ISL_1686587, EPI_ISL_1686588, EPI_ISL_1686589, EPI_ISL_1686590, EPI_ISL_1686591, EPI_ISL_1686592, EPI_ISL_1686593, EPI_ISL_1686594, EPI_ISL_1686595, EPI_ISL_1686596, EPI_ISL_1686597, EPI_ISL_1686598, EPI_ISL_1686599, EPI_ISL_1686600, EPI_ISL_1686601, EPI_ISL_1686602, EPI_ISL_1686603, EPI_ISL_1686604, EPI_ISL_1686605, EPI_ISL_1686606, EPI_ISL_1686607, EPI_ISL_1686608, EPI_ISL_1686609, EPI_ISL_1686610, EPI_ISL_1686611, EPI_ISL_1686612, EPI_ISL_1686613, EPI_ISL_1686614, EPI_ISL_1686615, EPI_ISL_1686616, EPI_ISL_1686617, EPI_ISL_1686618, EPI_ISL_1686620, EPI_ISL_1686621, EPI_ISL_1686622, EPI_ISL_1686623, EPI_ISL_1686624, EPI_ISL_1686625, EPI_ISL_1686626, EPI_ISL_1686627, EPI_ISL_1686628, EPI_ISL_1686629, EPI_ISL_1686630, EPI_ISL_1686631, EPI_ISL_1686632, EPI_ISL_1686633, EPI_ISL_1686634, EPI_ISL_1686635, EPI_ISL_1686636, EPI_ISL_1686637, EPI_ISL_1686638, EPI_ISL_1686639, EPI_ISL_1686640, EPI_ISL_1686641, EPI_ISL_1686642, EPI_ISL_1686643, EPI_ISL_1686644, EPI_ISL_1686645, EPI_ISL_1686646, EPI_ISL_1686647, EPI_ISL_1686648, EPI_ISL_1686649, EPI_ISL_1686650, EPI_ISL_1686651, EPI_ISL_1686652, EPI_ISL_1686653, EPI_ISL_1686654, EPI_ISL_1686655, EPI_ISL_1686656, EPI_ISL_1686657, EPI_ISL_1686658, EPI_ISL_1686659, EPI_ISL_1686660, EPI_ISL_1686661, EPI_ISL_1686662, EPI_ISL_1686663, EPI_ISL_1686665, EPI_ISL_1686666, EPI_ISL_1686667, EPI_ISL_1686668, EPI_ISL_1686669, EPI_ISL_1686670, EPI_ISL_1686671, EPI_ISL_1686672, EPI_ISL_1686673, EPI_ISL_1686674, EPI_ISL_1686675, EPI_ISL_1686676, EPI_ISL_1686677, EPI_ISL_1686678, EPI_ISL_1686679, EPI_ISL_1686680, EPI_ISL_1686681, EPI_ISL_1686682, EPI_ISL_1686683, EPI_ISL_1686684, EPI_ISL_1686685, EPI_ISL_1686686, EPI_ISL_1686687, EPI_ISL_1686688, EPI_ISL_1686689, EPI_ISL_1686690, EPI_ISL_1686691, EPI_ISL_1686692, EPI_ISL_1686693, EPI_ISL_1686694, EPI_ISL_1686695, EPI_ISL_1686696, EPI_ISL_1686697, EPI_ISL_1686699, EPI_ISL_1686700, EPI_ISL_1686701, EPI_ISL_1686702, EPI_ISL_1686703, EPI_ISL_1686704, EPI_ISL_1686705, EPI_ISL_1686706, EPI_ISL_1686707, EPI_ISL_1686708, EPI_ISL_1686709, EPI_ISL_1686710, EPI_ISL_1686711, EPI_ISL_1686712, EPI_ISL_1686713, EPI_ISL_1686714, EPI_ISL_1686715, EPI_ISL_1686716, EPI_ISL_1686717, EPI_ISL_1686718, EPI_ISL_1686719, EPI_ISL_1686720, EPI_ISL_1686721, EPI_ISL_1686722, EPI_ISL_1686723, EPI_ISL_1686724, EPI_ISL_1686725, EPI_ISL_1686726, EPI_ISL_1686727, EPI_ISL_1686728, EPI_ISL_1686729, EPI_ISL_1686730, EPI_ISL_1686731, EPI_ISL_1686732, EPI_ISL_1686733, EPI_ISL_1686734, EPI_ISL_1686735, EPI_ISL_1686736, EPI_ISL_1686737, EPI_ISL_1686738, EPI_ISL_1686739, EPI_ISL_1686740, EPI_ISL_1686741, EPI_ISL_1686742, EPI_ISL_1686743, EPI_ISL_1686744, EPI_ISL_1686745, EPI_ISL_1686746, EPI_ISL_1686747, EPI_ISL_1686748, EPI_ISL_1686749, EPI_ISL_1686750, EPI_ISL_1686751, EPI_ISL_1686752, EPI_ISL_1686753, EPI_ISL_1686754, EPI_ISL_1686755, EPI_ISL_1686756, EPI_ISL_1686757, EPI_ISL_1686758, EPI_ISL_1686759, EPI_ISL_1686760, EPI_ISL_1686761, EPI_ISL_1686762, EPI_ISL_1686763, EPI_ISL_1686764, EPI_ISL_1686765, EPI_ISL_1686766, EPI_ISL_1686767, EPI_ISL_1686768, EPI_ISL_1686769, EPI_ISL_1686770, EPI_ISL_1686771, EPI_ISL_1686772, EPI_ISL_1686773, EPI_ISL_1686774, EPI_ISL_1686775, EPI_ISL_1686776, EPI_ISL_1686777, EPI_ISL_1686778, EPI_ISL_1686779, EPI_ISL_1686780, EPI_ISL_1686781, EPI_ISL_1686782, EPI_ISL_1686783, EPI_ISL_1686784, EPI_ISL_1686785, EPI_ISL_1686786, EPI_ISL_1686787, EPI_ISL_1686788, EPI_ISL_1686789, EPI_ISL_1686790, EPI_ISL_1686791, EPI_ISL_1686792, EPI_ISL_1686793, EPI_ISL_1686794, EPI_ISL_1686795, EPI_ISL_1686796, EPI_ISL_1686797, EPI_ISL_1686798, EPI_ISL_1686799, EPI_ISL_1686800, EPI_ISL_1686801, EPI_ISL_1686802, EPI_ISL_1686803, EPI_ISL_1686804, EPI_ISL_1686805, EPI_ISL_1686806, EPI_ISL_1686807, EPI_ISL_1686808, EPI_ISL_1686809, EPI_ISL_1686810, EPI_ISL_1686811, EPI_ISL_1686812, EPI_ISL_1686813, EPI_ISL_1686814, EPI_ISL_1686815, EPI_ISL_1686816, EPI_ISL_1686817, EPI_ISL_1686818, EPI_ISL_1686819, EPI_ISL_1686820, EPI_ISL_1686821, EPI_ISL_1686822, EPI_ISL_1686823, EPI_ISL_1686824, EPI_ISL_1686825, EPI_ISL_1686826, EPI_ISL_1686827, EPI_ISL_1686828, EPI_ISL_1686829, EPI_ISL_1686830, EPI_ISL_1686831, EPI_ISL_1686832, EPI_ISL_1686833, EPI_ISL_1686834, EPI_ISL_1686835, EPI_ISL_1686836, EPI_ISL_1686837, EPI_ISL_1686838, EPI_ISL_1686839, EPI_ISL_1686840, EPI_ISL_1686841, EPI_ISL_1686842, EPI_ISL_1686843, EPI_ISL_1686844, EPI_ISL_1686845, EPI_ISL_1686846, EPI_ISL_1686847, EPI_ISL_1686848, EPI_ISL_1686849, EPI_ISL_1686850, EPI_ISL_1686851, EPI_ISL_1686852, EPI_ISL_1686853, EPI_ISL_1686854, EPI_ISL_1686855, EPI_ISL_1686856, EPI_ISL_1686857, EPI_ISL_1686858, EPI_ISL_1686859, EPI_ISL_1686860, EPI_ISL_1686861, EPI_ISL_1686862, EPI_ISL_1686863, EPI_ISL_1686864, EPI_ISL_1686865, EPI_ISL_1686866, EPI_ISL_1686867, EPI_ISL_1686868, EPI_ISL_1686869, EPI_ISL_1686870, EPI_ISL_1686871, EPI_ISL_1686872, EPI_ISL_1686873, EPI_ISL_1686874, EPI_ISL_1686875, EPI_ISL_1686876, EPI_ISL_1686877, EPI_ISL_1686878, EPI_ISL_1686879, EPI_ISL_1686880, EPI_ISL_1686881, EPI_ISL_1686882, EPI_ISL_1686883, EPI_ISL_1686884, EPI_ISL_1686885, EPI_ISL_1686886, EPI_ISL_1686887, EPI_ISL_1686888, EPI_ISL_1686889, EPI_ISL_1686890, EPI_ISL_1686891, EPI_ISL_1686892, EPI_ISL_1686893, EPI_ISL_1686894, EPI_ISL_1686895, EPI_ISL_1686896, EPI_ISL_1686897, EPI_ISL_1686898, EPI_ISL_1686899, EPI_ISL_1686900, EPI_ISL_1686901, EPI_ISL_1686902, EPI_ISL_1686903, EPI_ISL_1686904, EPI_ISL_1686905, EPI_ISL_1686906, EPI_ISL_1686907, EPI_ISL_1686908, EPI_ISL_1686909, EPI_ISL_1686910, EPI_ISL_1686911, EPI_ISL_1686912, EPI_ISL_1686913, EPI_ISL_1686914, EPI_ISL_1686915, EPI_ISL_1686916, EPI_ISL_1686917, EPI_ISL_1686918, EPI_ISL_1686919, EPI_ISL_1686920, EPI_ISL_1686921, EPI_ISL_1686922, EPI_ISL_1686923, EPI_ISL_1686924, EPI_ISL_1686925, EPI_ISL_1686926, EPI_ISL_1686927, EPI_ISL_1686928, EPI_ISL_1686929, EPI_ISL_1686930, EPI_ISL_1686931, EPI_ISL_1686932, EPI_ISL_1686933, EPI_ISL_1686934, EPI_ISL_1686935, EPI_ISL_1686936, EPI_ISL_1686937, EPI_ISL_1686938, EPI_ISL_1686939, EPI_ISL_1686940, EPI_ISL_1686941, EPI_ISL_1686942, EPI_ISL_1686943, EPI_ISL_1686944, EPI_ISL_1686945, EPI_ISL_1686946, EPI_ISL_1686947, EPI_ISL_1686948, EPI_ISL_1686949, EPI_ISL_1686950, EPI_ISL_1686951, EPI_ISL_1686952, EPI_ISL_1686953, EPI_ISL_1686954, EPI_ISL_1686955, EPI_ISL_1686956, EPI_ISL_1686957, EPI_ISL_1686958, EPI_ISL_1686959, EPI_ISL_1686960, EPI_ISL_1686961, EPI_ISL_1686962, EPI_ISL_1686963, EPI_ISL_1686964, EPI_ISL_1686965, EPI_ISL_1686966, EPI_ISL_1686967, EPI_ISL_1686968, EPI_ISL_1686969, EPI_ISL_1686970, EPI_ISL_1686971, EPI_ISL_1686972, EPI_ISL_1686973, EPI_ISL_1686974, EPI_ISL_1686975, EPI_ISL_1686976, EPI_ISL_1686977, EPI_ISL_1686978, EPI_ISL_1686979, EPI_ISL_1686980, EPI_ISL_1686981, EPI_ISL_1686982, EPI_ISL_1686983, EPI_ISL_1686984, EPI_ISL_1686985, EPI_ISL_1686986, EPI_ISL_1686987, EPI_ISL_1686988, EPI_ISL_1686989, EPI_ISL_1686990, EPI_ISL_1686991, EPI_ISL_1686992, EPI_ISL_1686993, EPI_ISL_1686994, EPI_ISL_1686995, EPI_ISL_1686996, EPI_ISL_1686997, EPI_ISL_1686998, EPI_ISL_1686999, EPI_ISL_1687000, EPI_ISL_1687001, EPI_ISL_1687002, EPI_ISL_1687003, EPI_ISL_1687004, EPI_ISL_1687005, EPI_ISL_1687006, EPI_ISL_1687007, EPI_ISL_1687008, EPI_ISL_1687009, EPI_ISL_1687010, EPI_ISL_1687011, EPI_ISL_1687012, EPI_ISL_1687013, EPI_ISL_1687014, EPI_ISL_1687015, EPI_ISL_1687016, EPI_ISL_1687017, EPI_ISL_1687018, EPI_ISL_1687019, EPI_ISL_1687020, EPI_ISL_1687021, EPI_ISL_1687022, EPI_ISL_1687023, EPI_ISL_1687024, EPI_ISL_1687025, EPI_ISL_1687026, EPI_ISL_1687027, EPI_ISL_1687028, EPI_ISL_1687029, EPI_ISL_1687030, EPI_ISL_1687031, EPI_ISL_1687032, EPI_ISL_1687033, EPI_ISL_1687034, EPI_ISL_1687035, EPI_ISL_1687036, EPI_ISL_1687037, EPI_ISL_1687038, EPI_ISL_1687039, EPI_ISL_1687040, EPI_ISL_1687041, EPI_ISL_1687042, EPI_ISL_1687043, EPI_ISL_1687044, EPI_ISL_1687045, EPI_ISL_1687046, EPI_ISL_1687047, EPI_ISL_1687048, EPI_ISL_1687049, EPI_ISL_1687050, EPI_ISL_1687051, EPI_ISL_1687052, EPI_ISL_1687053, EPI_ISL_1687054, EPI_ISL_1687055, EPI_ISL_1687056, EPI_ISL_1687057, EPI_ISL_1687058, EPI_ISL_1687059, EPI_ISL_1687060, EPI_ISL_1687061, EPI_ISL_1687062, EPI_ISL_1687063, EPI_ISL_1687064, EPI_ISL_1687065, EPI_ISL_1687066, EPI_ISL_1687067, EPI_ISL_1687068, EPI_ISL_1687069, EPI_ISL_1687070, EPI_ISL_1687071, EPI_ISL_1687072, EPI_ISL_1687073, EPI_ISL_1687074, EPI_ISL_1687075, EPI_ISL_1687076, EPI_ISL_1687077, EPI_ISL_1687078, EPI_ISL_1687079, EPI_ISL_1687080, EPI_ISL_1687082, EPI_ISL_1687083, EPI_ISL_1687084, EPI_ISL_1687085, EPI_ISL_1687086, EPI_ISL_1687087, EPI_ISL_1687088, EPI_ISL_1687089, EPI_ISL_1687090, EPI_ISL_1687091, EPI_ISL_1687092, EPI_ISL_1687093, EPI_ISL_1687094, EPI_ISL_1687095, EPI_ISL_1687096, EPI_ISL_1687097, EPI_ISL_1687098, EPI_ISL_1687099, EPI_ISL_1687100, EPI_ISL_1687101, EPI_ISL_1687102, EPI_ISL_1687103, EPI_ISL_1687104, EPI_ISL_1687105, EPI_ISL_1687106, EPI_ISL_1687107, EPI_ISL_1687108, EPI_ISL_1687109, EPI_ISL_1687110, EPI_ISL_1687111, EPI_ISL_1687112, EPI_ISL_1687113, EPI_ISL_1687114, EPI_ISL_1687115, EPI_ISL_1687116, EPI_ISL_1687117, EPI_ISL_1687118, EPI_ISL_1687119, EPI_ISL_1687120, EPI_ISL_1687121, EPI_ISL_1687122, EPI_ISL_1687123, EPI_ISL_1687124, EPI_ISL_1687125, EPI_ISL_1687126, EPI_ISL_1687127, EPI_ISL_1687128, EPI_ISL_1687129, EPI_ISL_1687130, EPI_ISL_1687131, EPI_ISL_1687132, EPI_ISL_1687133, EPI_ISL_1687134, EPI_ISL_1687135, EPI_ISL_1687136, EPI_ISL_1687137, EPI_ISL_1687138, EPI_ISL_1687139, EPI_ISL_1687140, EPI_ISL_1687141, EPI_ISL_1687142, EPI_ISL_1687143, EPI_ISL_1687144, EPI_ISL_1687145, EPI_ISL_1687146, EPI_ISL_1687147, EPI_ISL_1687148, EPI_ISL_1687149, EPI_ISL_1687150, EPI_ISL_1687151, EPI_ISL_1687152, EPI_ISL_1687153, EPI_ISL_1687154, EPI_ISL_1687155, EPI_ISL_1687156, EPI_ISL_1687157, EPI_ISL_1687158, EPI_ISL_1687159, EPI_ISL_1687160, EPI_ISL_1687161, EPI_ISL_1687162, EPI_ISL_1687163, EPI_ISL_1687164, EPI_ISL_1687165, EPI_ISL_1687166, EPI_ISL_1687167, EPI_ISL_1687168, EPI_ISL_1687169, EPI_ISL_1687170, EPI_ISL_1687171, EPI_ISL_1687172, EPI_ISL_1687173, EPI_ISL_1687174, EPI_ISL_1687175, EPI_ISL_1687176, EPI_ISL_1687177, EPI_ISL_1687178, EPI_ISL_1687179, EPI_ISL_1687180, EPI_ISL_1687181, EPI_ISL_1687182, EPI_ISL_1687183, EPI_ISL_1687184, EPI_ISL_1687185, EPI_ISL_1687186, EPI_ISL_1687187, EPI_ISL_1687188, EPI_ISL_1687189, EPI_ISL_1687190, EPI_ISL_1687191, EPI_ISL_1687192, EPI_ISL_1687193, EPI_ISL_1687194, EPI_ISL_1687195, EPI_ISL_1687196, EPI_ISL_1687197, EPI_ISL_1687198, EPI_ISL_1687199, EPI_ISL_1687200, EPI_ISL_1687201, EPI_ISL_1687202, EPI_ISL_1687203, EPI_ISL_1687204, EPI_ISL_1687205, EPI_ISL_1687206, EPI_ISL_1687207, EPI_ISL_1687208, EPI_ISL_1687209, EPI_ISL_1687210, EPI_ISL_1687211, EPI_ISL_1687212, EPI_ISL_1687213, EPI_ISL_1687214, EPI_ISL_1687215, EPI_ISL_1687216, EPI_ISL_1687217, EPI_ISL_1687218, EPI_ISL_1687219, EPI_ISL_1687220, EPI_ISL_1687221, EPI_ISL_1687222, EPI_ISL_1687223, EPI_ISL_1687224, EPI_ISL_1687225, EPI_ISL_1687226, EPI_ISL_1687227, EPI_ISL_1687228, EPI_ISL_1687229, EPI_ISL_1687230, EPI_ISL_1687231, EPI_ISL_1687232, EPI_ISL_1687233, EPI_ISL_1687234, EPI_ISL_1687235, EPI_ISL_1687236, EPI_ISL_1687237, EPI_ISL_1687238, EPI_ISL_1687239, EPI_ISL_1687240, EPI_ISL_1687241, EPI_ISL_1687242, EPI_ISL_1687243, EPI_ISL_1687244, EPI_ISL_1687245, EPI_ISL_1687246, EPI_ISL_1687247, EPI_ISL_1687248, EPI_ISL_1687249, EPI_ISL_1687250, EPI_ISL_1687251, EPI_ISL_1687252, EPI_ISL_1687253, EPI_ISL_1687254, EPI_ISL_1687255, EPI_ISL_1687256, EPI_ISL_1687257, EPI_ISL_1687258, EPI_ISL_1687259, EPI_ISL_1687260, EPI_ISL_1687261, EPI_ISL_1687262, EPI_ISL_1687263, EPI_ISL_1687264, EPI_ISL_1687265, EPI_ISL_1687266, EPI_ISL_1687267, EPI_ISL_1687268, EPI_ISL_1687269, EPI_ISL_1687270, EPI_ISL_1687271, EPI_ISL_1687272, EPI_ISL_1687273, EPI_ISL_1687274, EPI_ISL_1687275, EPI_ISL_1687276, EPI_ISL_1687277, EPI_ISL_1687278, EPI_ISL_1687279, EPI_ISL_1687280, EPI_ISL_1687282, EPI_ISL_1687283, EPI_ISL_1687284, EPI_ISL_1687285, EPI_ISL_1687286, EPI_ISL_1687287, EPI_ISL_1687288, EPI_ISL_1687289, EPI_ISL_1687290, EPI_ISL_1687291, EPI_ISL_1687293, EPI_ISL_1687294, EPI_ISL_1687295, EPI_ISL_1687296, EPI_ISL_1687297, EPI_ISL_1687298, EPI_ISL_1687299, EPI_ISL_1687300, EPI_ISL_1687301, EPI_ISL_1687302, EPI_ISL_1687303, EPI_ISL_1687304, EPI_ISL_1687305, EPI_ISL_1687306, EPI_ISL_1687307, EPI_ISL_1687308, EPI_ISL_1687309, EPI_ISL_1687310, EPI_ISL_1687312, EPI_ISL_1687313, EPI_ISL_1687314, EPI_ISL_1687315, EPI_ISL_1687316, EPI_ISL_1687317, EPI_ISL_1687318, EPI_ISL_1687319, EPI_ISL_1687320, EPI_ISL_1687321, EPI_ISL_1687322, EPI_ISL_1687323, EPI_ISL_1687324, EPI_ISL_1687326, EPI_ISL_1687327, EPI_ISL_1687328, EPI_ISL_1687329, EPI_ISL_1687330, EPI_ISL_1687331, EPI_ISL_1687333, EPI_ISL_1687335, EPI_ISL_1687336, EPI_ISL_1687337, EPI_ISL_1687338, EPI_ISL_1687339, EPI_ISL_1687340, EPI_ISL_1687341, EPI_ISL_1687342, EPI_ISL_1687343, EPI_ISL_1687344, EPI_ISL_1687345, EPI_ISL_1687346, EPI_ISL_1687347, EPI_ISL_1687348, EPI_ISL_1687349, EPI_ISL_1687350, EPI_ISL_1687351, EPI_ISL_1687352, EPI_ISL_1687353, EPI_ISL_1687354, EPI_ISL_1687355, EPI_ISL_1687356, EPI_ISL_1687357, EPI_ISL_1687358, EPI_ISL_1687359, EPI_ISL_1687361, EPI_ISL_1687362, EPI_ISL_1687363, EPI_ISL_1687364, EPI_ISL_1687365, EPI_ISL_1687366, EPI_ISL_1687367, EPI_ISL_1687368, EPI_ISL_1687369, EPI_ISL_1687370, EPI_ISL_1687371, EPI_ISL_1687372, EPI_ISL_1687373, EPI_ISL_1687374, EPI_ISL_1687375, EPI_ISL_1687376, EPI_ISL_1687377, EPI_ISL_1687378, EPI_ISL_1687379, EPI_ISL_1687380, EPI_ISL_1687381, EPI_ISL_1687382, EPI_ISL_1687383, EPI_ISL_1687384, EPI_ISL_1687386, EPI_ISL_1687387, EPI_ISL_1687388, EPI_ISL_1687389, EPI_ISL_1687390, EPI_ISL_1687391, EPI_ISL_1687392, EPI_ISL_1687393, EPI_ISL_1687395, EPI_ISL_1687396, EPI_ISL_1687397, EPI_ISL_1687398, EPI_ISL_1687399, EPI_ISL_1687401, EPI_ISL_1687402, EPI_ISL_1687403, EPI_ISL_1687404, EPI_ISL_1687405, EPI_ISL_1687406, EPI_ISL_1687407, EPI_ISL_1687408, EPI_ISL_1687409, EPI_ISL_1687410, EPI_ISL_1687411, EPI_ISL_1687412, EPI_ISL_1687413, EPI_ISL_1687414, EPI_ISL_1687415, EPI_ISL_1687416, EPI_ISL_1687417, EPI_ISL_1687418, EPI_ISL_1687419, EPI_ISL_1687420, EPI_ISL_1687421, EPI_ISL_1687422, EPI_ISL_1687423, EPI_ISL_1687424, EPI_ISL_1687425, EPI_ISL_1687426, EPI_ISL_1687427, EPI_ISL_1687428, EPI_ISL_1687429, EPI_ISL_1687430, EPI_ISL_1687431, EPI_ISL_1687432, EPI_ISL_1687433, EPI_ISL_1687434, EPI_ISL_1687435, EPI_ISL_1687436, EPI_ISL_1687437, EPI_ISL_1687438, EPI_ISL_1687439, EPI_ISL_1687441, EPI_ISL_1687442, EPI_ISL_1687443, EPI_ISL_1687444, EPI_ISL_1687445, EPI_ISL_1687446, EPI_ISL_1687447, EPI_ISL_1687448, EPI_ISL_1687449, EPI_ISL_1687450, EPI_ISL_1687451, EPI_ISL_1687452, EPI_ISL_1687453, EPI_ISL_1687454, EPI_ISL_1687455, EPI_ISL_1687456, EPI_ISL_1687457, EPI_ISL_1687458, EPI_ISL_1687459, EPI_ISL_1687460, EPI_ISL_1687461, EPI_ISL_1687462, EPI_ISL_1687463, EPI_ISL_1687465, EPI_ISL_1687466, EPI_ISL_1687467, EPI_ISL_1687468, EPI_ISL_1687469, EPI_ISL_1687470, EPI_ISL_1687471, EPI_ISL_1687472, EPI_ISL_1687473, EPI_ISL_1687474, EPI_ISL_1687475, EPI_ISL_1687476, EPI_ISL_1687477, EPI_ISL_1687478, EPI_ISL_1687479, EPI_ISL_1687480, EPI_ISL_1687481, EPI_ISL_1687482, EPI_ISL_1687483, EPI_ISL_1687484, EPI_ISL_1687485, EPI_ISL_1687486, EPI_ISL_1687487, EPI_ISL_1687488, EPI_ISL_1687489, EPI_ISL_1687490, EPI_ISL_1687491, EPI_ISL_1687492, EPI_ISL_1687493, EPI_ISL_1687494, EPI_ISL_1687495, EPI_ISL_1687496, EPI_ISL_1687497, EPI_ISL_1687498, EPI_ISL_1687499, EPI_ISL_1687500, EPI_ISL_1687501, EPI_ISL_1687503, EPI_ISL_1687504, EPI_ISL_1687505, EPI_ISL_1687506, EPI_ISL_1687507, EPI_ISL_1687508, EPI_ISL_1687509, EPI_ISL_1687510, EPI_ISL_1687511, EPI_ISL_1687512, EPI_ISL_1687513, EPI_ISL_1687514, EPI_ISL_1687515, EPI_ISL_1687516, EPI_ISL_1687517, EPI_ISL_1687518, EPI_ISL_1687519, EPI_ISL_1687521, EPI_ISL_1687522, EPI_ISL_1687523, EPI_ISL_1687524, EPI_ISL_1687525, EPI_ISL_1687526, EPI_ISL_1687527, EPI_ISL_1687528, EPI_ISL_1687529, EPI_ISL_1687530, EPI_ISL_1687531, EPI_ISL_1687532, EPI_ISL_1687533, EPI_ISL_1687534, EPI_ISL_1687535, EPI_ISL_1687536, EPI_ISL_1687537, EPI_ISL_1687538, EPI_ISL_1687539, EPI_ISL_1687540, EPI_ISL_1687541, EPI_ISL_1687542, EPI_ISL_1687543, EPI_ISL_1687544, EPI_ISL_1687545, EPI_ISL_1687546, EPI_ISL_1687547, EPI_ISL_1687548, EPI_ISL_1687549, EPI_ISL_1687550, EPI_ISL_1687551, EPI_ISL_1687552, EPI_ISL_1687553, EPI_ISL_1687554, EPI_ISL_1687555, EPI_ISL_1687556, EPI_ISL_1687557, EPI_ISL_1687558, EPI_ISL_1687559, EPI_ISL_1687560, EPI_ISL_1687561, EPI_ISL_1687562, EPI_ISL_1687563, EPI_ISL_1687564, EPI_ISL_1687565, EPI_ISL_1687566, EPI_ISL_1687567, EPI_ISL_1687568, EPI_ISL_1687569, EPI_ISL_1687570, EPI_ISL_1687571, EPI_ISL_1687572, EPI_ISL_1687573, EPI_ISL_1687574, EPI_ISL_1687575, EPI_ISL_1687576, EPI_ISL_1687577, EPI_ISL_1687578, EPI_ISL_1687579, EPI_ISL_1687580, EPI_ISL_1687581, EPI_ISL_1687582, EPI_ISL_1687583, EPI_ISL_1687584, EPI_ISL_1687585, EPI_ISL_1687586, EPI_ISL_1687587, EPI_ISL_1687588, EPI_ISL_1687590, EPI_ISL_1687591, EPI_ISL_1687592, EPI_ISL_1687593, EPI_ISL_1687594, EPI_ISL_1687595, EPI_ISL_1687596, EPI_ISL_1687597 | | | |
| see above | Aegis Sciences Corporation | Centers for Disease Control and Prevention Division of Viral Diseases, Pathogen Discovery | Dakota Howard, Dhwani Batra, Peter W. Cook, Kara Moser, Adrian Paskey, Jason Caravas, Benjamin Rambo-Martin, Shatavia Morrison, Christopher Gulvick, Scott Sammons, Yvette Unoarumhi, Darlene Wagner, Matthew Schmerer, Cyndi Clark, Patrick Campbell, Rob Case, Vikramsinha Ghorpade, Holly Houdeshell, Ola Kvalvaag, Dillon Nall, Ethan Sanders, Alec Vest, Shaun Westlund, Matthew Hardison, Clinton R. Paden, Duncan MacCannell |
| EPI_ISL_1687598 | Ohio Department of Health Laboratory | Ohio Department of Health Laboratory | Holmes, Jennifer; Eric Brandt, Keoni Omura, Glen McGillivary, Caitlin McDonnell, Jade Mowery, Stephanie Mccracken, Tyler Payne, Kirtana Ramadugu, Erica Leasure, Brent Lee, Kelsey Florek, Heather Blankenship, Allison Black, Quanta Brown, and Tammy Bannerman |
| EPI_ISL_1687599, EPI_ISL_1687600, EPI_ISL_1687601, EPI_ISL_1687602, EPI_ISL_1687603, EPI_ISL_1687604, EPI_ISL_1687605, EPI_ISL_1687606, EPI_ISL_1687607, EPI_ISL_1687608, EPI_ISL_1687609, EPI_ISL_1687610, EPI_ISL_1687611, EPI_ISL_1687612, EPI_ISL_1687613, EPI_ISL_1687614, EPI_ISL_1687615, EPI_ISL_1687616, EPI_ISL_1687617, EPI_ISL_1687618, EPI_ISL_1687619, EPI_ISL_1687620, EPI_ISL_1687621, EPI_ISL_1687622, EPI_ISL_1687623, EPI_ISL_1687624, EPI_ISL_1687625, EPI_ISL_1687627, EPI_ISL_1687628, EPI_ISL_1687629, EPI_ISL_1687630, EPI_ISL_1687631, EPI_ISL_1687632, EPI_ISL_1687633, EPI_ISL_1687635, EPI_ISL_1687637, EPI_ISL_1687638, EPI_ISL_1687639, EPI_ISL_1687640, EPI_ISL_1687641, EPI_ISL_1687642, EPI_ISL_1687643, EPI_ISL_1687644, EPI_ISL_1687645, | | | |

| | | | |
|---|---|---|---|
| EPI_ISL_1687646, EPI_ISL_1687647, EPI_ISL_1687648, EPI_ISL_1687649, EPI_ISL_1687650, EPI_ISL_1687651, EPI_ISL_1687652, EPI_ISL_1687653, EPI_ISL_1687654, EPI_ISL_1687655, EPI_ISL_1687656, EPI_ISL_1687657, EPI_ISL_1687658, EPI_ISL_1687659, EPI_ISL_1687660, EPI_ISL_1687661, EPI_ISL_1687662, EPI_ISL_1687663, EPI_ISL_1687664, EPI_ISL_1687665, EPI_ISL_1687666, EPI_ISL_1687667, EPI_ISL_1687668, EPI_ISL_1687669, EPI_ISL_1687670, EPI_ISL_1687671, EPI_ISL_1687672, EPI_ISL_1687673, EPI_ISL_1687674, EPI_ISL_1687675, EPI_ISL_1687678, EPI_ISL_1687679, EPI_ISL_1687680, EPI_ISL_1687681, EPI_ISL_1687682, EPI_ISL_1687683, EPI_ISL_1687684, EPI_ISL_1687685, EPI_ISL_1687686, EPI_ISL_1687687, EPI_ISL_1687688, EPI_ISL_1687689, EPI_ISL_1687690, EPI_ISL_1687691, EPI_ISL_1687692, EPI_ISL_1687693, EPI_ISL_1687694, EPI_ISL_1687695, EPI_ISL_1687696, EPI_ISL_1687697, EPI_ISL_1687698, EPI_ISL_1687699, EPI_ISL_1687700, EPI_ISL_1687701, EPI_ISL_1687702, EPI_ISL_1687704, EPI_ISL_1687705, EPI_ISL_1687706, EPI_ISL_1687707, EPI_ISL_1687708, EPI_ISL_1687709, EPI_ISL_1687710, EPI_ISL_1687711, EPI_ISL_1687712, EPI_ISL_1687713, EPI_ISL_1687714, EPI_ISL_1687715, EPI_ISL_1687716, EPI_ISL_1687718, EPI_ISL_1687719, EPI_ISL_1687720, EPI_ISL_1687721, EPI_ISL_1687722, EPI_ISL_1687723, EPI_ISL_1687724, EPI_ISL_1687725, EPI_ISL_1687726, EPI_ISL_1687727, EPI_ISL_1687728, EPI_ISL_1687729, EPI_ISL_1687731, EPI_ISL_1687732, EPI_ISL_1687733, EPI_ISL_1687734, EPI_ISL_1687735, EPI_ISL_1687736, EPI_ISL_1687737, EPI_ISL_1687738, EPI_ISL_1687739, EPI_ISL_1687740, EPI_ISL_1687741, EPI_ISL_1687742, EPI_ISL_1687743, EPI_ISL_1687744, EPI_ISL_1687745, EPI_ISL_1687746, EPI_ISL_1687747, EPI_ISL_1687748, EPI_ISL_1687749, EPI_ISL_1687750, EPI_ISL_1687751, EPI_ISL_1687752, EPI_ISL_1687753, EPI_ISL_1687754, EPI_ISL_1687755, EPI_ISL_1687756, EPI_ISL_1687757, EPI_ISL_1687759, EPI_ISL_1687760, EPI_ISL_1687761, EPI_ISL_1687762, EPI_ISL_1687763, EPI_ISL_1687764, EPI_ISL_1687765, EPI_ISL_1687766, EPI_ISL_1687767, EPI_ISL_1687768, EPI_ISL_1687769, EPI_ISL_1687770, EPI_ISL_1687771, EPI_ISL_1687772, EPI_ISL_1687773, EPI_ISL_1687774, EPI_ISL_1687775, EPI_ISL_1687776, EPI_ISL_1687777, EPI_ISL_1687778, EPI_ISL_1687781, EPI_ISL_1687782, EPI_ISL_1687783, EPI_ISL_1687784, EPI_ISL_1687785, EPI_ISL_1687786, EPI_ISL_1687787, EPI_ISL_1687788, EPI_ISL_1687789, EPI_ISL_1687790, EPI_ISL_1687791, EPI_ISL_1687792, EPI_ISL_1687793, EPI_ISL_1687794, EPI_ISL_1687795, EPI_ISL_1687796, EPI_ISL_1687797, EPI_ISL_1687798, EPI_ISL_1687799, EPI_ISL_1687800, EPI_ISL_1687801, EPI_ISL_1687802, EPI_ISL_1687803, EPI_ISL_1687804, EPI_ISL_1687805, EPI_ISL_1687806, EPI_ISL_1687807, EPI_ISL_1687808, EPI_ISL_1687809, EPI_ISL_1687810, EPI_ISL_1687811, EPI_ISL_1687812, EPI_ISL_1687813, EPI_ISL_1687814, EPI_ISL_1687815, EPI_ISL_1687816, EPI_ISL_1687817, EPI_ISL_1687818, EPI_ISL_1687819, EPI_ISL_1687820, EPI_ISL_1687821, EPI_ISL_1687822, EPI_ISL_1687823, EPI_ISL_1687824, EPI_ISL_1687825, EPI_ISL_1687826, EPI_ISL_1687827, EPI_ISL_1687828, EPI_ISL_1687829, EPI_ISL_1687830, EPI_ISL_1687831, EPI_ISL_1687832, EPI_ISL_1687834, EPI_ISL_1687835, EPI_ISL_1687836, EPI_ISL_1687837, EPI_ISL_1687839, EPI_ISL_1687840, EPI_ISL_1687841, EPI_ISL_1687842, EPI_ISL_1687843, EPI_ISL_1687844, EPI_ISL_1687845, EPI_ISL_1687846, EPI_ISL_1687847, EPI_ISL_1687848, EPI_ISL_1687849, EPI_ISL_1687850, EPI_ISL_1687851, EPI_ISL_1687852, EPI_ISL_1687853, EPI_ISL_1687854, EPI_ISL_1687855, EPI_ISL_1687856, EPI_ISL_1687857, EPI_ISL_1687858, EPI_ISL_1687859, EPI_ISL_1687860, EPI_ISL_1687861, EPI_ISL_1687863, EPI_ISL_1687864, EPI_ISL_1687865, EPI_ISL_1687866, EPI_ISL_1687867, EPI_ISL_1687868, EPI_ISL_1687870, EPI_ISL_1687871, EPI_ISL_1687872, EPI_ISL_1687873, EPI_ISL_1687874, EPI_ISL_1687875, EPI_ISL_1687876, EPI_ISL_1687877, EPI_ISL_1687878, EPI_ISL_1687879, EPI_ISL_1687880, EPI_ISL_1687881, EPI_ISL_1687883, EPI_ISL_1687884, EPI_ISL_1687885, EPI_ISL_1687886, EPI_ISL_1687887, EPI_ISL_1687888, EPI_ISL_1687889, EPI_ISL_1687890, EPI_ISL_1687891, EPI_ISL_1687893, EPI_ISL_1687894, EPI_ISL_1687895, EPI_ISL_1687896, EPI_ISL_1687897, EPI_ISL_1687898, EPI_ISL_1687901, EPI_ISL_1687902, EPI_ISL_1687903, EPI_ISL_1687904, EPI_ISL_1687905, EPI_ISL_1687906, EPI_ISL_1687907, EPI_ISL_1687908, EPI_ISL_1687909, EPI_ISL_1687910, EPI_ISL_1687911, EPI_ISL_1687912, EPI_ISL_1687913, EPI_ISL_1687914, EPI_ISL_1687915, EPI_ISL_1687916, EPI_ISL_1687918, EPI_ISL_1687919, EPI_ISL_1687920, EPI_ISL_1687921, EPI_ISL_1687922, EPI_ISL_1687923, EPI_ISL_1687924, EPI_ISL_1687925, EPI_ISL_1687926, EPI_ISL_1687927, EPI_ISL_1687928, EPI_ISL_1687929, EPI_ISL_1687930, EPI_ISL_1687931, EPI_ISL_1687932, EPI_ISL_1687933, EPI_ISL_1687934, EPI_ISL_1687935, EPI_ISL_1687936, EPI_ISL_1687937, EPI_ISL_1687938, EPI_ISL_1687939, EPI_ISL_1687940, EPI_ISL_1687942, EPI_ISL_1687943, EPI_ISL_1687944, EPI_ISL_1687945, EPI_ISL_1687946, EPI_ISL_1687947, EPI_ISL_1687948, EPI_ISL_1687949, EPI_ISL_1687950, EPI_ISL_1687951, EPI_ISL_1687952, EPI_ISL_1687953, EPI_ISL_1687954, EPI_ISL_1687955, EPI_ISL_1687956, EPI_ISL_1687957, EPI_ISL_1687958, EPI_ISL_1687959, EPI_ISL_1687960, EPI_ISL_1687961, EPI_ISL_1687962, EPI_ISL_1687963, EPI_ISL_1687964, EPI_ISL_1687965, EPI_ISL_1687966, EPI_ISL_1687967, EPI_ISL_1687968, EPI_ISL_1687970, EPI_ISL_1687971, EPI_ISL_1687972, EPI_ISL_1687973, EPI_ISL_1687974, EPI_ISL_1687975, EPI_ISL_1687976, EPI_ISL_1687977, EPI_ISL_1687978, EPI_ISL_1687979, EPI_ISL_1687980, EPI_ISL_1687981, EPI_ISL_1687982, EPI_ISL_1687983, EPI_ISL_1687984, EPI_ISL_1687985, EPI_ISL_1687986, EPI_ISL_1687987, EPI_ISL_1687988, EPI_ISL_1687989, EPI_ISL_1687990, EPI_ISL_1687991, EPI_ISL_1687992, EPI_ISL_1687993, EPI_ISL_1687994, EPI_ISL_1687995, EPI_ISL_1687996, EPI_ISL_1687997, EPI_ISL_1687998, EPI_ISL_1687999, EPI_ISL_1688000, EPI_ISL_1688001, EPI_ISL_1688002, EPI_ISL_1688003, EPI_ISL_1688004, EPI_ISL_1688005, EPI_ISL_1688006, EPI_ISL_1688007, EPI_ISL_1688008, EPI_ISL_1688009, EPI_ISL_1688010, EPI_ISL_1688011, EPI_ISL_1688013, EPI_ISL_1688014, EPI_ISL_1688015, EPI_ISL_1688016, EPI_ISL_1688017, EPI_ISL_1688018, EPI_ISL_1688019, EPI_ISL_1688020, EPI_ISL_1688021, EPI_ISL_1688022, EPI_ISL_1688023, EPI_ISL_1688024, EPI_ISL_1688026, EPI_ISL_1688027, EPI_ISL_1688028, EPI_ISL_1688029, EPI_ISL_1688030, EPI_ISL_1688031, EPI_ISL_1688032, EPI_ISL_1688033, EPI_ISL_1688034, EPI_ISL_1688035, EPI_ISL_1688036, EPI_ISL_1688037, EPI_ISL_1688038, EPI_ISL_1688039, EPI_ISL_1688040, EPI_ISL_1688041, EPI_ISL_1688042, EPI_ISL_1688043, EPI_ISL_1688044, EPI_ISL_1688045, EPI_ISL_1688046, EPI_ISL_1688047, EPI_ISL_1688048, EPI_ISL_1688049, EPI_ISL_1688050, EPI_ISL_1688051, EPI_ISL_1688052, EPI_ISL_1688053, EPI_ISL_1688054, EPI_ISL_1688055, EPI_ISL_1688056, EPI_ISL_1688057, EPI_ISL_1688058, EPI_ISL_1688059, EPI_ISL_1688060, EPI_ISL_1688061, EPI_ISL_1688062, EPI_ISL_1688063, EPI_ISL_1688064, EPI_ISL_1688065, EPI_ISL_1688066, EPI_ISL_1688067, EPI_ISL_1688068, EPI_ISL_1688069, EPI_ISL_1688070, EPI_ISL_1688071, EPI_ISL_1688072, EPI_ISL_1688073, EPI_ISL_1688074, EPI_ISL_1688075, EPI_ISL_1688076, EPI_ISL_1688078, EPI_ISL_1688079, EPI_ISL_1688080, EPI_ISL_1688081, EPI_ISL_1688082, EPI_ISL_1688083, EPI_ISL_1688084, EPI_ISL_1688085, EPI_ISL_1688086, EPI_ISL_1688087, EPI_ISL_1688088, EPI_ISL_1688089, EPI_ISL_1688090, EPI_ISL_1688091, EPI_ISL_1688092, EPI_ISL_1688093, EPI_ISL_1688094, EPI_ISL_1688095, EPI_ISL_1688096, EPI_ISL_1688097, EPI_ISL_1688098, EPI_ISL_1688099, EPI_ISL_1688100, EPI_ISL_1688101, EPI_ISL_1688102, EPI_ISL_1688103, EPI_ISL_1688105, EPI_ISL_1688106, EPI_ISL_1688107, EPI_ISL_1688108, EPI_ISL_1688109, EPI_ISL_1688110, EPI_ISL_1688111, EPI_ISL_1688112, EPI_ISL_1688113, EPI_ISL_1688114, EPI_ISL_1688115, EPI_ISL_1688116, EPI_ISL_1688117, EPI_ISL_1688118, EPI_ISL_1688119, EPI_ISL_1688120, EPI_ISL_1688121, EPI_ISL_1688122, EPI_ISL_1688123, EPI_ISL_1688124, EPI_ISL_1688125, EPI_ISL_1688126, EPI_ISL_1688127, EPI_ISL_1688128, EPI_ISL_1688129, EPI_ISL_1688131, EPI_ISL_1688132, EPI_ISL_1688133, EPI_ISL_1688134, EPI_ISL_1688135, EPI_ISL_1688136, EPI_ISL_1688137, EPI_ISL_1688138, EPI_ISL_1688139, EPI_ISL_1688140, EPI_ISL_1688141, EPI_ISL_1688142, EPI_ISL_1688143, EPI_ISL_1688144, EPI_ISL_1688145, EPI_ISL_1688146, EPI_ISL_1688147, EPI_ISL_1688148, EPI_ISL_1688149, EPI_ISL_1688150, EPI_ISL_1688151, EPI_ISL_1688152, EPI_ISL_1688153, EPI_ISL_1688154, EPI_ISL_1688155, EPI_ISL_1688156, EPI_ISL_1688157, EPI_ISL_1688158, EPI_ISL_1688159, EPI_ISL_1688160, EPI_ISL_1688161, EPI_ISL_1688162, EPI_ISL_1688163, EPI_ISL_1688164, EPI_ISL_1688165, EPI_ISL_1688166, EPI_ISL_1688167, EPI_ISL_1688168, EPI_ISL_1688169, EPI_ISL_1688171, EPI_ISL_1688172, EPI_ISL_1688173, EPI_ISL_1688174, EPI_ISL_1688175, EPI_ISL_1688176, EPI_ISL_1688177, EPI_ISL_1688178, EPI_ISL_1688179, EPI_ISL_1688180, EPI_ISL_1688181, EPI_ISL_1688182, EPI_ISL_1688184, EPI_ISL_1688185, EPI_ISL_1688186, EPI_ISL_1688187, EPI_ISL_1688188, EPI_ISL_1688189, EPI_ISL_1688190, EPI_ISL_1688191, EPI_ISL_1688192, EPI_ISL_1688193, EPI_ISL_1688194, EPI_ISL_1688195, EPI_ISL_1688196, EPI_ISL_1688197, EPI_ISL_1688198, EPI_ISL_1688199, EPI_ISL_1688200, EPI_ISL_1688201, EPI_ISL_1688202, EPI_ISL_1688203, EPI_ISL_1688204, EPI_ISL_1688206, EPI_ISL_1688207, EPI_ISL_1688208, EPI_ISL_1688209, EPI_ISL_1688210, EPI_ISL_1688211, EPI_ISL_1688212, EPI_ISL_1688214, EPI_ISL_1688215, EPI_ISL_1688216, EPI_ISL_1688217, EPI_ISL_1688218, EPI_ISL_1688219, EPI_ISL_1688220, EPI_ISL_1688221, EPI_ISL_1688222, EPI_ISL_1688223, EPI_ISL_1688224, EPI_ISL_1688225, EPI_ISL_1688226, EPI_ISL_1688227, EPI_ISL_1688228, EPI_ISL_1688229, EPI_ISL_1688230, EPI_ISL_1688231, EPI_ISL_1688232, EPI_ISL_1688233, EPI_ISL_1688234, EPI_ISL_1688235, EPI_ISL_1688236, EPI_ISL_1688237, EPI_ISL_1688238, EPI_ISL_1688239, EPI_ISL_1688240, EPI_ISL_1688241, EPI_ISL_1688242, EPI_ISL_1688243, EPI_ISL_1688244, EPI_ISL_1688245, EPI_ISL_1688246, EPI_ISL_1688247, EPI_ISL_1688248, EPI_ISL_1688249, EPI_ISL_1688250, EPI_ISL_1688251, EPI_ISL_1688252, EPI_ISL_1688253, EPI_ISL_1688254, EPI_ISL_1688255, EPI_ISL_1688256, EPI_ISL_1688257, EPI_ISL_1688258, EPI_ISL_1688259, EPI_ISL_1688260, EPI_ISL_1688261, EPI_ISL_1688262, EPI_ISL_1688263, EPI_ISL_1688264, EPI_ISL_1688265, EPI_ISL_1688266, EPI_ISL_1688267, EPI_ISL_1688268, EPI_ISL_1688269, EPI_ISL_1688270, EPI_ISL_1688271, EPI_ISL_1688272, EPI_ISL_1688273, EPI_ISL_1688274, EPI_ISL_1688275, EPI_ISL_1688276, EPI_ISL_1688277, EPI_ISL_1688278, EPI_ISL_1688279, EPI_ISL_1688280, EPI_ISL_1688281, EPI_ISL_1688282, EPI_ISL_1688283, EPI_ISL_1688284, EPI_ISL_1688285, EPI_ISL_1688286, EPI_ISL_1688287, EPI_ISL_1688288, EPI_ISL_1688289, EPI_ISL_1688290, EPI_ISL_1688291, EPI_ISL_1688292, EPI_ISL_1688293, EPI_ISL_1688294, EPI_ISL_1688295, EPI_ISL_1688296, EPI_ISL_1688297, EPI_ISL_1688298, EPI_ISL_1688299, EPI_ISL_1688300, EPI_ISL_1688301, EPI_ISL_1688302, EPI_ISL_1688303, EPI_ISL_1688304, EPI_ISL_1688305, EPI_ISL_1688306, EPI_ISL_1688307, EPI_ISL_1688308, EPI_ISL_1688309, EPI_ISL_1688310, EPI_ISL_1688311, EPI_ISL_1688312, EPI_ISL_1688313, EPI_ISL_1688314, EPI_ISL_1688315, EPI_ISL_1688316, EPI_ISL_1688317, EPI_ISL_1688318, EPI_ISL_1688319, EPI_ISL_1688320, EPI_ISL_1688321, EPI_ISL_1688322, EPI_ISL_1688323, EPI_ISL_1688324, EPI_ISL_1688325, EPI_ISL_1688326, EPI_ISL_1688327, EPI_ISL_1688328, EPI_ISL_1688329, EPI_ISL_1688330, EPI_ISL_1688331, EPI_ISL_1688332, EPI_ISL_1688333, EPI_ISL_1688334, EPI_ISL_1688335, EPI_ISL_1688336, EPI_ISL_1688337, EPI_ISL_1688338, EPI_ISL_1688339, EPI_ISL_1688340, EPI_ISL_1688341, EPI_ISL_1688343, EPI_ISL_1688344, EPI_ISL_1688345, EPI_ISL_1688346, EPI_ISL_1688347, EPI_ISL_1688348, EPI_ISL_1688349, EPI_ISL_1688350, EPI_ISL_1688351, EPI_ISL_1688352, EPI_ISL_1688353, EPI_ISL_1688354, EPI_ISL_1688355, EPI_ISL_1688356, EPI_ISL_1688357, EPI_ISL_1688358, EPI_ISL_1688359, EPI_ISL_1688360, EPI_ISL_1688361, EPI_ISL_1688362, EPI_ISL_1688363, EPI_ISL_1688364, EPI_ISL_1688365, EPI_ISL_1688366, EPI_ISL_1688367, EPI_ISL_1688368, EPI_ISL_1688369, EPI_ISL_1688370, EPI_ISL_1688371, EPI_ISL_1688372, EPI_ISL_1688373, EPI_ISL_1688374, EPI_ISL_1688375, EPI_ISL_1688376, EPI_ISL_1688377, EPI_ISL_1688378, EPI_ISL_1688379, EPI_ISL_1688380, EPI_ISL_1688381, EPI_ISL_1688382, EPI_ISL_1688383, EPI_ISL_1688384, EPI_ISL_1688385, EPI_ISL_1688386, EPI_ISL_1688387, EPI_ISL_1688388, EPI_ISL_1688389, EPI_ISL_1688390, EPI_ISL_1688391, EPI_ISL_1688392, EPI_ISL_1688393, EPI_ISL_1688394, EPI_ISL_1688395, EPI_ISL_1688396, EPI_ISL_1688397, EPI_ISL_1688398, EPI_ISL_1688399, EPI_ISL_1688401, EPI_ISL_1688402, EPI_ISL_1688403, EPI_ISL_1688405, EPI_ISL_1688406, EPI_ISL_1688408, EPI_ISL_1688409, EPI_ISL_1688411, EPI_ISL_1688412, EPI_ISL_1688413, EPI_ISL_1688415, EPI_ISL_1688416, EPI_ISL_1688418, EPI_ISL_1688419, EPI_ISL_1688421, EPI_ISL_1688422, EPI_ISL_1688423, EPI_ISL_1688425, EPI_ISL_1688426, EPI_ISL_1688428, EPI_ISL_1688429, EPI_ISL_1688431, EPI_ISL_1688432, EPI_ISL_1688433, EPI_ISL_1688435, EPI_ISL_1688436, EPI_ISL_1688438, EPI_ISL_1688439, EPI_ISL_1688441, EPI_ISL_1688442, EPI_ISL_1688444, EPI_ISL_1688445, EPI_ISL_1688447, EPI_ISL_1688448, EPI_ISL_1688450, EPI_ISL_1688451, EPI_ISL_1688452, EPI_ISL_1688454, EPI_ISL_1688455, EPI_ISL_1688457, EPI_ISL_1688458, EPI_ISL_1688460, EPI_ISL_1688461, EPI_ISL_1688462, EPI_ISL_1688464, EPI_ISL_1688465, EPI_ISL_1688467, EPI_ISL_1688468, EPI_ISL_1688470, EPI_ISL_1688471, EPI_ISL_1688472, EPI_ISL_1688474, EPI_ISL_1688475, EPI_ISL_1688477, EPI_ISL_1688478, EPI_ISL_1688479, EPI_ISL_1688481, EPI_ISL_1688482, EPI_ISL_1688484, EPI_ISL_1688485, EPI_ISL_1688487, EPI_ISL_1688488, EPI_ISL_1688490, EPI_ISL_1688491, EPI_ISL_1688493, EPI_ISL_1688494, EPI_ISL_1688496, EPI_ISL_1688497, EPI_ISL_1688498, EPI_ISL_1688500, EPI_ISL_1688501, EPI_ISL_1688503, EPI_ISL_1688504, EPI_ISL_1688506, EPI_ISL_1688507, EPI_ISL_1688509, EPI_ISL_1688510, EPI_ISL_1688512, EPI_ISL_1688513, EPI_ISL_1688515, EPI_ISL_1688516, EPI_ISL_1688518, EPI_ISL_1688519, EPI_ISL_1688521, EPI_ISL_1688522, EPI_ISL_1688523, EPI_ISL_1688525, EPI_ISL_1688526, EPI_ISL_1688528, EPI_ISL_1688529, EPI_ISL_1688531, EPI_ISL_1688532, EPI_ISL_1688534, EPI_ISL_1688535, EPI_ISL_1688537, EPI_ISL_1688538, EPI_ISL_1688539, EPI_ISL_1688541, EPI_ISL_1688542, EPI_ISL_1688544, EPI_ISL_1688545, EPI_ISL_1688547, EPI_ISL_1688548, EPI_ISL_1688550, EPI_ISL_1688551, EPI_ISL_1688552, EPI_ISL_1688554, EPI_ISL_1688555, EPI_ISL_1688557, EPI_ISL_1688559, EPI_ISL_1688562, EPI_ISL_1688564, EPI_ISL_1688566, EPI_ISL_1688569, EPI_ISL_1688570, EPI_ISL_1688573, EPI_ISL_1688576, EPI_ISL_1688579, EPI_ISL_1688581, EPI_ISL_1688584, EPI_ISL_1688586, EPI_ISL_1688587, EPI_ISL_1688590, EPI_ISL_1688593, EPI_ISL_1688595, EPI_ISL_1688598, EPI_ISL_1688600, EPI_ISL_1688602, EPI_ISL_1688604, EPI_ISL_1688606, EPI_ISL_1688609, EPI_ISL_1688611, EPI_ISL_1688613, EPI_ISL_1688616, EPI_ISL_1688618, EPI_ISL_1688620, EPI_ISL_1688622, EPI_ISL_1688625, EPI_ISL_1688627, EPI_ISL_1688629, EPI_ISL_1688632, EPI_ISL_1688634, EPI_ISL_1688636, EPI_ISL_1688638, EPI_ISL_1688640, EPI_ISL_1688642, EPI_ISL_1688645, EPI_ISL_1688647 | | | |
| see above | Aegis Sciences Corporation | Centers for Disease Control and Prevention Division of Viral Diseases, Pathogen Discovery | Dakota Howard, Dhwani Batra, Peter W. Cook, Kara Moser, Adrian Paskey, Jason Caravas, Benjamin Rambo-Martin, Shatavia Morrison, Christopher Gulvick, Scott Sammons, Yvette Unoarumhi, Darlene Wagner, Matthew Schmerer, Cyndi Clark, Patrick Campbell, Rob Case, Vikramsinha Ghorpade, Holly Houdeshell, Ola Kvalvaag, Dillon Nall, Ethan Sanders, Alec Vest, Shaun Westlund, Matthew Hardison, Clinton R. Paden, Duncan MacCannell |
| EPI_ISL_1688648 | Sonora Quest Laboratories | TGen North | Jolene Bowers, Heather Centner, Chris French, Hayley Yaglom, Ashlyn Pfeiffer, Darrin Lemmer, Dave Engelthaler, The Arizona COVID Genomics Union (ACGU) |
| EPI_ISL_1688650, EPI_ISL_1688652 | Aegis Sciences Corporation | Centers for Disease Control and Prevention Division of Viral Diseases, Pathogen Discovery | Dakota Howard, Dhwani Batra, Peter W. Cook, Kara Moser, Adrian Paskey, Jason Caravas, Benjamin Rambo-Martin, Shatavia Morrison, Christopher Gulvick, Scott Sammons, Yvette Unoarumhi, Darlene Wagner, Matthew Schmerer, Cyndi Clark, Patrick Campbell, Rob Case, Vikramsinha Ghorpade, Holly Houdeshell, Ola Kvalvaag, Dillon Nall, Ethan Sanders, Alec Vest, Shaun Westlund, Matthew Hardison, Clinton R. Paden, Duncan MacCannell |
| EPI_ISL_1688653 | Sonora Quest Laboratories | TGen North | Jolene Bowers, Heather Centner, Chris French, Hayley Yaglom, Ashlyn Pfeiffer, Darrin Lemmer, Dave Engelthaler, The Arizona COVID Genomics Union (ACGU) |
| EPI_ISL_1688654, EPI_ISL_1688657, EPI_ISL_1688659, EPI_ISL_1688662, EPI_ISL_1688663, EPI_ISL_1688665, EPI_ISL_1688668, EPI_ISL_1688670 | Aegis Sciences Corporation | Centers for Disease Control and Prevention Division of Viral Diseases, Pathogen Discovery | Dakota Howard, Dhwani Batra, Peter W. Cook, Kara Moser, Adrian Paskey, Jason Caravas, Benjamin Rambo-Martin, Shatavia Morrison, Christopher Gulvick, Scott Sammons, Yvette Unoarumhi, Darlene Wagner, Matthew Schmerer, Cyndi Clark, Patrick Campbell, Rob Case, Vikramsinha Ghorpade, Holly Houdeshell, Ola Kvalvaag, Dillon Nall, Ethan Sanders, Alec Vest, Shaun Westlund, Matthew Hardison, Clinton R. Paden, Duncan MacCannell |
| EPI_ISL_1688671 | Sonora Quest Laboratories | TGen North | Jolene Bowers, Heather Centner, Chris French, Hayley Yaglom, Ashlyn Pfeiffer, Darrin Lemmer, Dave Engelthaler, The Arizona COVID Genomics Union (ACGU) |
| EPI_ISL_1688675, EPI_ISL_1688678, EPI_ISL_1688680, EPI_ISL_1688682, EPI_ISL_1688685, EPI_ISL_1688686, EPI_ISL_1688689, EPI_ISL_1688690, EPI_ISL_1688692, EPI_ISL_1688696, EPI_ISL_1688698, EPI_ISL_1688700, EPI_ISL_1688702, EPI_ISL_1688705, EPI_ISL_1688707 | | | |
| see above | Aegis Sciences Corporation | Centers for Disease Control and Prevention Division of Viral Diseases, Pathogen Discovery | Dakota Howard, Dhwani Batra, Peter W. Cook, Kara Moser, Adrian Paskey, Jason Caravas, Benjamin Rambo-Martin, Shatavia Morrison, Christopher Gulvick, Scott Sammons, Yvette Unoarumhi, Darlene Wagner, Matthew Schmerer, Cyndi Clark, Patrick Campbell, Rob Case, Vikramsinha Ghorpade, Holly Houdeshell, Ola Kvalvaag, Dillon Nall, Ethan Sanders, Alec Vest, Shaun Westlund, Matthew Hardison, Clinton R. Paden, Duncan MacCannell |
| EPI_ISL_1688708 | Sonora Quest Laboratories | TGen North | Jolene Bowers, Heather Centner, Chris French, Hayley Yaglom, Ashlyn Pfeiffer, Darrin Lemmer, Dave Engelthaler, The Arizona COVID Genomics Union (ACGU) |
| EPI_ISL_1688709, EPI_ISL_1688712, EPI_ISL_1688714, EPI_ISL_1688717, EPI_ISL_1688719, EPI_ISL_1688721, EPI_ISL_1688723, EPI_ISL_1688725, EPI_ISL_1688728 | Aegis Sciences Corporation | Centers for Disease Control and Prevention Division of Viral Diseases, Pathogen Discovery | Dakota Howard, Dhwani Batra, Peter W. Cook, Kara Moser, Adrian Paskey, Jason Caravas, Benjamin Rambo-Martin, Shatavia Morrison, Christopher Gulvick, Scott Sammons, Yvette Unoarumhi, Darlene Wagner, Matthew Schmerer, Cyndi Clark, Patrick Campbell, Rob Case, Vikramsinha Ghorpade, Holly Houdeshell, Ola Kvalvaag, Dillon Nall, Ethan Sanders, Alec Vest, Shaun Westlund, Matthew Hardison, Clinton R. Paden, Duncan MacCannell |
| EPI_ISL_1688729 | Sonora Quest Laboratories | TGen North | Jolene Bowers, Heather Centner, Chris French, Hayley Yaglom, Ashlyn Pfeiffer, Darrin Lemmer, Dave Engelthaler, The Arizona COVID Genomics Union (ACGU) |
| EPI_ISL_1688730, EPI_ISL_1688733, EPI_ISL_1688735, EPI_ISL_1688738, EPI_ISL_1688739, EPI_ISL_1688742, EPI_ISL_1688744, EPI_ISL_1688747, EPI_ISL_1688749, EPI_ISL_1688752, EPI_ISL_1688754, EPI_ISL_1688756, EPI_ISL_1688759, EPI_ISL_1688761, EPI_ISL_1688764, EPI_ISL_1688766, EPI_ISL_1688768, EPI_ISL_1688770, EPI_ISL_1688772, EPI_ISL_1688775, EPI_ISL_1688777, EPI_ISL_1688779, EPI_ISL_1688781, EPI_ISL_1688784, EPI_ISL_1688786, EPI_ISL_1688788, EPI_ISL_1688791, EPI_ISL_1688793, EPI_ISL_1688796, EPI_ISL_1688798, EPI_ISL_1688801, EPI_ISL_1688803, EPI_ISL_1688806, EPI_ISL_1688807, EPI_ISL_1688809, EPI_ISL_1688812, EPI_ISL_1688818, EPI_ISL_1688820, EPI_ISL_1688822, EPI_ISL_1688825, EPI_ISL_1688827, EPI_ISL_1688829, EPI_ISL_1688831, EPI_ISL_1688835, EPI_ISL_1688837, EPI_ISL_1688839, EPI_ISL_1688841, EPI_ISL_1688843, EPI_ISL_1688846, EPI_ISL_1688848, EPI_ISL_1688850, EPI_ISL_1688852, EPI_ISL_1688855, EPI_ISL_1688857, EPI_ISL_1688859, EPI_ISL_1688862, EPI_ISL_1688864, EPI_ISL_1688866, EPI_ISL_1688868, EPI_ISL_1688871, EPI_ISL_1688872, EPI_ISL_1688875, EPI_ISL_1688877, EPI_ISL_1688882, EPI_ISL_1688884, EPI_ISL_1688887, EPI_ISL_1688889, EPI_ISL_1688891, EPI_ISL_1688894, EPI_ISL_1688896, EPI_ISL_1688899, EPI_ISL_1688900, EPI_ISL_1688903, EPI_ISL_1688904, EPI_ISL_1688907, EPI_ISL_1688909, EPI_ISL_1688912, EPI_ISL_1688914, EPI_ISL_1688917, EPI_ISL_1688920, EPI_ISL_1688924, EPI_ISL_1688926, EPI_ISL_1688928, EPI_ISL_1688930, EPI_ISL_1688932, EPI_ISL_1688936, EPI_ISL_1688937, EPI_ISL_1688940, EPI_ISL_1688942, EPI_ISL_1688944, EPI_ISL_1688947, EPI_ISL_1688949, EPI_ISL_1688952, EPI_ISL_1688954, EPI_ISL_1688957, EPI_ISL_1688958, EPI_ISL_1688961, EPI_ISL_1688963, EPI_ISL_1688966, EPI_ISL_1688968, EPI_ISL_1688970, EPI_ISL_1688974, EPI_ISL_1688977, EPI_ISL_1688979, EPI_ISL_1688982, EPI_ISL_1688984, EPI_ISL_1688987, EPI_ISL_1688989, EPI_ISL_1688992, EPI_ISL_1688994, EPI_ISL_1688996, EPI_ISL_1688998, EPI_ISL_1689001, EPI_ISL_1689003, EPI_ISL_1689006, EPI_ISL_1689008, EPI_ISL_1689011, EPI_ISL_1689013, EPI_ISL_1689016, EPI_ISL_1689018, EPI_ISL_1689021, EPI_ISL_1689023, EPI_ISL_1689026, EPI_ISL_1689028, EPI_ISL_1689031, EPI_ISL_1689033, EPI_ISL_1689035, EPI_ISL_1689037, EPI_ISL_1689040, EPI_ISL_1689042, EPI_ISL_1689043, EPI_ISL_1689046, EPI_ISL_1689048, EPI_ISL_1689051, EPI_ISL_1689053, EPI_ISL_1689056, EPI_ISL_1689058, EPI_ISL_1689061, EPI_ISL_1689063, EPI_ISL_1689065, EPI_ISL_1689068, EPI_ISL_1689070, EPI_ISL_1689073, EPI_ISL_1689075, EPI_ISL_1689077, EPI_ISL_1689079, EPI_ISL_1689082, EPI_ISL_1689086, EPI_ISL_1689089, EPI_ISL_1689093, EPI_ISL_1689097, EPI_ISL_1689099, EPI_ISL_1689101, EPI_ISL_1689104, EPI_ISL_1689105, EPI_ISL_1689109, EPI_ISL_1689111, EPI_ISL_1689112, EPI_ISL_1689115, EPI_ISL_1689117, EPI_ISL_1689120, EPI_ISL_1689122, EPI_ISL_1689124, EPI_ISL_1689127, EPI_ISL_1689129, EPI_ISL_1689132, EPI_ISL_1689136, EPI_ISL_1689139, EPI_ISL_1689141 | | | |
| see above | Aegis Sciences Corporation | Centers for Disease Control and Prevention Division of Viral Diseases, Pathogen Discovery | Dakota Howard, Dhwani Batra, Peter W. Cook, Kara Moser, Adrian Paskey, Jason Caravas, Benjamin Rambo-Martin, Shatavia Morrison, Christopher Gulvick, Scott Sammons, Yvette Unoarumhi, Darlene Wagner, Matthew Schmerer, Cyndi Clark, Patrick Campbell, Rob Case, Vikramsinha Ghorpade, Holly Houdeshell, Ola Kvalvaag, Dillon Nall, Ethan Sanders, Alec Vest, Shaun Westlund, Matthew Hardison, Clinton R. Paden, Duncan MacCannell |
| EPI_ISL_1689143 | Sonora Quest Laboratories | TGen North | Jolene Bowers, Heather Centner, Chris French, Hayley Yaglom, Ashlyn Pfeiffer, Darrin Lemmer, Dave Engelthaler, The Arizona COVID Genomics Union (ACGU) |
| EPI_ISL_1689144, EPI_ISL_1689146, EPI_ISL_1689148, EPI_ISL_1689150, EPI_ISL_1689152, EPI_ISL_1689155, EPI_ISL_1689157, EPI_ISL_1689160, EPI_ISL_1689162, EPI_ISL_1689165, EPI_ISL_1689167, EPI_ISL_1689170, EPI_ISL_1689175, EPI_ISL_1689176, EPI_ISL_1689179, EPI_ISL_1689181, EPI_ISL_1689183, EPI_ISL_1689186 | | | |
| see above | Aegis Sciences Corporation | Centers for Disease Control and Prevention Division of Viral Diseases, Pathogen Discovery | Dakota Howard, Dhwani Batra, Peter W. Cook, Kara Moser, Adrian Paskey, Jason Caravas, Benjamin Rambo-Martin, Shatavia Morrison, Christopher Gulvick, Scott Sammons, Yvette Unoarumhi, Darlene Wagner, Matthew Schmerer, Cyndi Clark, Patrick Campbell, Rob Case, Vikramsinha Ghorpade, Holly Houdeshell, Ola Kvalvaag, Dillon Nall, Ethan Sanders, Alec Vest, Shaun Westlund, Matthew Hardison, Clinton R. Paden, Duncan MacCannell |
| EPI_ISL_1689187 | Sonora Quest Laboratories | TGen North | Jolene Bowers, Heather Centner, Chris French, Hayley Yaglom, Ashlyn Pfeiffer, Darrin Lemmer, Dave Engelthaler, The Arizona COVID Genomics Union (ACGU) |
| EPI_ISL_1689188, EPI_ISL_1689190, EPI_ISL_1689192, EPI_ISL_1689194, EPI_ISL_1689197, EPI_ISL_1689200, EPI_ISL_1689202, EPI_ISL_1689204, EPI_ISL_1689207, EPI_ISL_1689209, EPI_ISL_1689212, EPI_ISL_1689214, EPI_ISL_1689217, EPI_ISL_1689219, EPI_ISL_1689222, EPI_ISL_1689224, EPI_ISL_1689227, EPI_ISL_1689229, EPI_ISL_1689234, EPI_ISL_1689235, EPI_ISL_1689238, EPI_ISL_1689241, EPI_ISL_1689243, EPI_ISL_1689245, EPI_ISL_1689248, EPI_ISL_1689250, EPI_ISL_1689254, EPI_ISL_1689258, EPI_ISL_1689261, EPI_ISL_1689265, EPI_ISL_1689269, EPI_ISL_1689273, EPI_ISL_1689276, EPI_ISL_1689278, EPI_ISL_1689282, EPI_ISL_1689285, EPI_ISL_1689289, EPI_ISL_1689294, EPI_ISL_1689298, EPI_ISL_1689300, EPI_ISL_1689302, EPI_ISL_1689305, EPI_ISL_1689308, EPI_ISL_1689312, EPI_ISL_1689314, EPI_ISL_1689318, EPI_ISL_1689322, EPI_ISL_1689326, EPI_ISL_1689328, EPI_ISL_1689330, EPI_ISL_1689333, EPI_ISL_1689335, EPI_ISL_1689338, EPI_ISL_1689340, EPI_ISL_1689343, EPI_ISL_1689346, EPI_ISL_1689350, EPI_ISL_1689353, EPI_ISL_1689356, EPI_ISL_1689357, EPI_ISL_1689359, EPI_ISL_1689362, EPI_ISL_1689364, EPI_ISL_1689367, EPI_ISL_1689369, EPI_ISL_1689371, EPI_ISL_1689373, EPI_ISL_1689375, EPI_ISL_1689377, EPI_ISL_1689381, EPI_ISL_1689385, EPI_ISL_1689389, EPI_ISL_1689391, EPI_ISL_1689393, EPI_ISL_1689396, EPI_ISL_1689397, EPI_ISL_1689400, EPI_ISL_1689404, EPI_ISL_1689407, EPI_ISL_1689410, EPI_ISL_1689412, EPI_ISL_1689415, EPI_ISL_1689417, EPI_ISL_1689419, EPI_ISL_1689421, EPI_ISL_1689423, EPI_ISL_1689426, EPI_ISL_1689428, EPI_ISL_1689431, EPI_ISL_1689433, EPI_ISL_1689435, EPI_ISL_1689438, EPI_ISL_1689440, EPI_ISL_1689442, EPI_ISL_1689445, EPI_ISL_1689447, EPI_ISL_1689449, EPI_ISL_1689451, EPI_ISL_1689454, EPI_ISL_1689457, EPI_ISL_1689459, EPI_ISL_1689462, EPI_ISL_1689465, EPI_ISL_1689469, EPI_ISL_1689472, EPI_ISL_1689476, EPI_ISL_1689480, EPI_ISL_1689484, EPI_ISL_1689490, EPI_ISL_1689494, EPI_ISL_1689498, EPI_ISL_1689501, EPI_ISL_1689503, EPI_ISL_1689505, EPI_ISL_1689508, EPI_ISL_1689510, EPI_ISL_1689513, EPI_ISL_1689515, EPI_ISL_1689518, EPI_ISL_1689520, EPI_ISL_1689522, EPI_ISL_1689526, EPI_ISL_1689529, EPI_ISL_1689532, EPI_ISL_1689534, EPI_ISL_1689537, EPI_ISL_1689540, EPI_ISL_1689541, EPI_ISL_1689544, EPI_ISL_1689548, EPI_ISL_1689552, EPI_ISL_1689556, EPI_ISL_1689560, EPI_ISL_1689562, EPI_ISL_1689564, EPI_ISL_1689566, EPI_ISL_1689569, EPI_ISL_1689572, EPI_ISL_1689574, EPI_ISL_1689576, EPI_ISL_1689579, EPI_ISL_1689580, EPI_ISL_1689583, EPI_ISL_1689585, EPI_ISL_1689587, EPI_ISL_1689589, EPI_ISL_1689590, EPI_ISL_1689592, EPI_ISL_1689593, EPI_ISL_1689595, EPI_ISL_1689596, EPI_ISL_1689597, EPI_ISL_1689599, EPI_ISL_1689600, EPI_ISL_1689603, EPI_ISL_1689605, EPI_ISL_1689608, EPI_ISL_1689609, EPI_ISL_1689611, EPI_ISL_1689612, EPI_ISL_1689613, EPI_ISL_1689615, EPI_ISL_1689616, EPI_ISL_1689617, EPI_ISL_1689619, EPI_ISL_1689620, EPI_ISL_1689622, EPI_ISL_1689623, EPI_ISL_1689624, EPI_ISL_1689626, EPI_ISL_1689627, EPI_ISL_1689629, EPI_ISL_1689630, EPI_ISL_1689631, EPI_ISL_1689633, EPI_ISL_1689634, EPI_ISL_1689636, EPI_ISL_1689637, EPI_ISL_1689638, EPI_ISL_1689640, EPI_ISL_1689641, EPI_ISL_1689644, EPI_ISL_1689646, EPI_ISL_1689649, EPI_ISL_1689652, EPI_ISL_1689655, EPI_ISL_1689658, EPI_ISL_1689661, EPI_ISL_1689663, EPI_ISL_1689666, EPI_ISL_1689670, EPI_ISL_1689672, EPI_ISL_1689675, EPI_ISL_1689678, EPI_ISL_1689681, EPI_ISL_1689684, EPI_ISL_1689687, EPI_ISL_1689690, EPI_ISL_1689693, EPI_ISL_1689695, EPI_ISL_1689699, EPI_ISL_1689702, EPI_ISL_1689704, EPI_ISL_1689707, EPI_ISL_1689710, EPI_ISL_1689712, EPI_ISL_1689715, EPI_ISL_1689718, EPI_ISL_1689721, EPI_ISL_1689723, EPI_ISL_1689726, EPI_ISL_1689729, EPI_ISL_1689732, EPI_ISL_1689735, EPI_ISL_1689738, EPI_ISL_1689741, EPI_ISL_1689744, EPI_ISL_1689747, EPI_ISL_1689750, EPI_ISL_1689752, EPI_ISL_1689754, EPI_ISL_1689757, EPI_ISL_1689760, EPI_ISL_1689763, EPI_ISL_1689766, EPI_ISL_1689769, EPI_ISL_1689772, EPI_ISL_1689775, EPI_ISL_1689778, EPI_ISL_1689780, EPI_ISL_1689783, EPI_ISL_1689786, EPI_ISL_1689789, EPI_ISL_1689795, EPI_ISL_1689797, EPI_ISL_1689800, EPI_ISL_1689803, EPI_ISL_1689806, EPI_ISL_1689808, EPI_ISL_1689809, EPI_ISL_1689810, EPI_ISL_1689811, EPI_ISL_1689812, EPI_ISL_1689813, EPI_ISL_1689814, EPI_ISL_1689815, EPI_ISL_1689816, EPI_ISL_1689817, EPI_ISL_1689818, EPI_ISL_1689819, EPI_ISL_1689820, EPI_ISL_1689821, EPI_ISL_1689822, EPI_ISL_1689823, EPI_ISL_1689824, EPI_ISL_1689825, EPI_ISL_1689826, EPI_ISL_1689827, EPI_ISL_1689828, EPI_ISL_1689829, EPI_ISL_1689830, EPI_ISL_1689831, EPI_ISL_1689832, EPI_ISL_1689833, EPI_ISL_1689834, EPI_ISL_1689835, EPI_ISL_1689836, EPI_ISL_1689837, EPI_ISL_1689838, EPI_ISL_1689839, EPI_ISL_1689840, EPI_ISL_1689841, EPI_ISL_1689842, EPI_ISL_1689843, EPI_ISL_1689844, EPI_ISL_1689845, EPI_ISL_1689846, EPI_ISL_1689847, EPI_ISL_1689848, EPI_ISL_1689849, EPI_ISL_1689850, EPI_ISL_1689851, EPI_ISL_1689852, EPI_ISL_1689853, EPI_ISL_1689854, EPI_ISL_1689855, | | | |

| | | | |
|---|---|---|---|
| EPI_ISL_1689856, EPI_ISL_1689857, EPI_ISL_1689858, EPI_ISL_1689859, EPI_ISL_1689860, EPI_ISL_1689861, EPI_ISL_1689862, EPI_ISL_1689863, EPI_ISL_1689864, EPI_ISL_1689865, EPI_ISL_1689866, EPI_ISL_1689867, EPI_ISL_1689868, EPI_ISL_1689869, EPI_ISL_1689870, EPI_ISL_1689871, EPI_ISL_1689872, EPI_ISL_1689873, EPI_ISL_1689874, EPI_ISL_1689875, EPI_ISL_1689876, EPI_ISL_1689877, EPI_ISL_1689878, EPI_ISL_1689879, EPI_ISL_1689880, EPI_ISL_1689881, EPI_ISL_1689882, EPI_ISL_1689883, EPI_ISL_1689884, EPI_ISL_1689885, EPI_ISL_1689887, EPI_ISL_1689888, EPI_ISL_1689889, EPI_ISL_1689890, EPI_ISL_1689891, EPI_ISL_1689892, EPI_ISL_1689893, EPI_ISL_1689894, EPI_ISL_1689895, EPI_ISL_1689896, EPI_ISL_1689897, EPI_ISL_1689898, EPI_ISL_1689899, EPI_ISL_1689900, EPI_ISL_1689901, EPI_ISL_1689902, EPI_ISL_1689903, EPI_ISL_1689904, EPI_ISL_1689905, EPI_ISL_1689906, EPI_ISL_1689907, EPI_ISL_1689908, EPI_ISL_1689909, EPI_ISL_1689910, EPI_ISL_1689911, EPI_ISL_1689912, EPI_ISL_1689913, EPI_ISL_1689914, EPI_ISL_1689915, EPI_ISL_1689916, EPI_ISL_1689917, EPI_ISL_1689918, EPI_ISL_1689919, EPI_ISL_1689920, EPI_ISL_1689921, EPI_ISL_1689922, EPI_ISL_1689923, EPI_ISL_1689924, EPI_ISL_1689925, EPI_ISL_1689926, EPI_ISL_1689927, EPI_ISL_1689928, EPI_ISL_1689929, EPI_ISL_1689930, EPI_ISL_1689931, EPI_ISL_1689932, EPI_ISL_1689933, EPI_ISL_1689934, EPI_ISL_1689935, EPI_ISL_1689936, EPI_ISL_1689937, EPI_ISL_1689938, EPI_ISL_1689939, EPI_ISL_1689940, EPI_ISL_1689941, EPI_ISL_1689942, EPI_ISL_1689943, EPI_ISL_1689944, EPI_ISL_1689945, EPI_ISL_1689946, EPI_ISL_1689947, EPI_ISL_1689948, EPI_ISL_1689949, EPI_ISL_1689950, EPI_ISL_1689951, EPI_ISL_1689952, EPI_ISL_1689953, EPI_ISL_1689954, EPI_ISL_1689955, EPI_ISL_1689956, EPI_ISL_1689957, EPI_ISL_1689958, EPI_ISL_1689959, EPI_ISL_1689960, EPI_ISL_1689961, EPI_ISL_1689962, EPI_ISL_1689963, EPI_ISL_1689964, EPI_ISL_1689965, EPI_ISL_1689966, EPI_ISL_1689967, EPI_ISL_1689968, EPI_ISL_1689969, EPI_ISL_1689971, EPI_ISL_1689972, EPI_ISL_1689973, EPI_ISL_1689974, EPI_ISL_1689975, EPI_ISL_1689976, EPI_ISL_1689977, EPI_ISL_1689978, EPI_ISL_1689979, EPI_ISL_1689980, EPI_ISL_1689981, EPI_ISL_1689982 | | | |
| see above | Aegis Sciences Corporation | Centers for Disease Control and Prevention Division of Viral Diseases, Pathogen Discovery | Dakota Howard, Dhwani Batra, Peter W. Cook, Kara Moser, Adrian Paskey, Jason Caravas, Benjamin Rambo-Martin, Shatavia Morrison, Christopher Gulvick, Scott Sammons, Yvette Unoarumhi, Darlene Wagner, Matthew Schmerer, Cyndi Clark, Patrick Campbell, Rob Case, Vikramsinha Ghorpade, Holly Houdeshell, Ola Kvalvaag, Dillon Nall, Ethan Sanders, Alec Vest, Shaun Westlund, Matthew Hardison, Clinton R. Paden, Duncan MacCannell |
| EPI_ISL_1689983 | Ohio Department of Health Laboratory | Ohio Department of Health Laboratory | Holmes, Jennifer; Eric Brandt, Keoni Omura, Glen McGillivary, Caitlin McDonnell, Jade Mowery, Stephanie Mccracken, Tyler Payne, Kirtana Ramadugu, Erica Leasure, Brent Lee, Kelsey Florek, Heather Blankenship, Allison Black, Quanta Brown, and Tammy Bannerman |
| EPI_ISL_1689984, EPI_ISL_1689985, EPI_ISL_1689986, EPI_ISL_1689987, EPI_ISL_1689988, EPI_ISL_1689989, EPI_ISL_1689990, EPI_ISL_1689991, EPI_ISL_1689992, EPI_ISL_1689993, EPI_ISL_1689994, EPI_ISL_1689995, EPI_ISL_1689996, EPI_ISL_1689997, EPI_ISL_1689998, EPI_ISL_1689999, EPI_ISL_1690000, EPI_ISL_1690001, EPI_ISL_1690002, EPI_ISL_1690003, EPI_ISL_1690004, EPI_ISL_1690005, EPI_ISL_1690006, EPI_ISL_1690007, EPI_ISL_1690008, EPI_ISL_1690009, EPI_ISL_1690010, EPI_ISL_1690011, EPI_ISL_1690012, EPI_ISL_1690013, EPI_ISL_1690014, EPI_ISL_1690015, EPI_ISL_1690016, EPI_ISL_1690017, EPI_ISL_1690018, EPI_ISL_1690019, EPI_ISL_1690020, EPI_ISL_1690021, EPI_ISL_1690022, EPI_ISL_1690023, EPI_ISL_1690024, EPI_ISL_1690025, EPI_ISL_1690026, EPI_ISL_1690027, EPI_ISL_1690028, EPI_ISL_1690029, EPI_ISL_1690030, EPI_ISL_1690031, EPI_ISL_1690032, EPI_ISL_1690033, EPI_ISL_1690034, EPI_ISL_1690035, EPI_ISL_1690036, EPI_ISL_1690037, EPI_ISL_1690038, EPI_ISL_1690039, EPI_ISL_1690040, EPI_ISL_1690041, EPI_ISL_1690042, EPI_ISL_1690043, EPI_ISL_1690044, EPI_ISL_1690045, EPI_ISL_1690046, EPI_ISL_1690047, EPI_ISL_1690048, EPI_ISL_1690049, EPI_ISL_1690050, EPI_ISL_1690051, EPI_ISL_1690052, EPI_ISL_1690053, EPI_ISL_1690054, EPI_ISL_1690055, EPI_ISL_1690056, EPI_ISL_1690057, EPI_ISL_1690058, EPI_ISL_1690059, EPI_ISL_1690060, EPI_ISL_1690061, EPI_ISL_1690062, EPI_ISL_1690063, EPI_ISL_1690064, EPI_ISL_1690065, EPI_ISL_1690066, EPI_ISL_1690067, EPI_ISL_1690068, EPI_ISL_1690069, EPI_ISL_1690070, EPI_ISL_1690071, EPI_ISL_1690072, EPI_ISL_1690073, EPI_ISL_1690074, EPI_ISL_1690075, EPI_ISL_1690076, EPI_ISL_1690077, EPI_ISL_1690078, EPI_ISL_1690079, EPI_ISL_1690080, EPI_ISL_1690081, EPI_ISL_1690082, EPI_ISL_1690083, EPI_ISL_1690084, EPI_ISL_1690085, EPI_ISL_1690086, EPI_ISL_1690087, EPI_ISL_1690088, EPI_ISL_1690089, EPI_ISL_1690090, EPI_ISL_1690091, EPI_ISL_1690092, EPI_ISL_1690093, EPI_ISL_1690094, EPI_ISL_1690095, EPI_ISL_1690096, EPI_ISL_1690097, EPI_ISL_1690098, EPI_ISL_1690099, EPI_ISL_1690100, EPI_ISL_1690101, EPI_ISL_1690102, EPI_ISL_1690103, EPI_ISL_1690104, EPI_ISL_1690105, EPI_ISL_1690106, EPI_ISL_1690107, EPI_ISL_1690108, EPI_ISL_1690109, EPI_ISL_1690110, EPI_ISL_1690111, EPI_ISL_1690112, EPI_ISL_1690113, EPI_ISL_1690114, EPI_ISL_1690115, EPI_ISL_1690116, EPI_ISL_1690117, EPI_ISL_1690119, EPI_ISL_1690120, EPI_ISL_1690121, EPI_ISL_1690122, EPI_ISL_1690123, EPI_ISL_1690124, EPI_ISL_1690125, EPI_ISL_1690126, EPI_ISL_1690127, EPI_ISL_1690128, EPI_ISL_1690129, EPI_ISL_1690130, EPI_ISL_1690131, EPI_ISL_1690132, EPI_ISL_1690133, EPI_ISL_1690134, EPI_ISL_1690135, EPI_ISL_1690136, EPI_ISL_1690137, EPI_ISL_1690138, EPI_ISL_1690139, EPI_ISL_1690140, EPI_ISL_1690141, EPI_ISL_1690142, EPI_ISL_1690143, EPI_ISL_1690144, EPI_ISL_1690145, EPI_ISL_1690146, EPI_ISL_1690147, EPI_ISL_1690148, EPI_ISL_1690149, EPI_ISL_1690150, EPI_ISL_1690151, EPI_ISL_1690152, EPI_ISL_1690153, EPI_ISL_1690154, EPI_ISL_1690155, EPI_ISL_1690156, EPI_ISL_1690157, EPI_ISL_1690158, EPI_ISL_1690159, EPI_ISL_1690160, EPI_ISL_1690161, EPI_ISL_1690162, EPI_ISL_1690163, EPI_ISL_1690164, EPI_ISL_1690165, EPI_ISL_1690166, EPI_ISL_1690167, EPI_ISL_1690168, EPI_ISL_1690169, EPI_ISL_1690170, EPI_ISL_1690171, EPI_ISL_1690172, EPI_ISL_1690173, EPI_ISL_1690174, EPI_ISL_1690175, EPI_ISL_1690176, EPI_ISL_1690177, EPI_ISL_1690178, EPI_ISL_1690179, EPI_ISL_1690180, EPI_ISL_1690181, EPI_ISL_1690182, EPI_ISL_1690183, EPI_ISL_1690184, EPI_ISL_1690185, EPI_ISL_1690186, EPI_ISL_1690187, EPI_ISL_1690188, EPI_ISL_1690189, EPI_ISL_1690190, EPI_ISL_1690191, EPI_ISL_1690192, EPI_ISL_1690193, EPI_ISL_1690194, EPI_ISL_1690195, EPI_ISL_1690196, EPI_ISL_1690197, EPI_ISL_1690198, EPI_ISL_1690199, EPI_ISL_1690200, EPI_ISL_1690201, EPI_ISL_1690202, EPI_ISL_1690203, EPI_ISL_1690204, EPI_ISL_1690205, EPI_ISL_1690206, EPI_ISL_1690207, EPI_ISL_1690209, EPI_ISL_1690210, EPI_ISL_1690211, EPI_ISL_1690212, EPI_ISL_1690213, EPI_ISL_1690214, EPI_ISL_1690215, EPI_ISL_1690216, EPI_ISL_1690217, EPI_ISL_1690218, EPI_ISL_1690219, EPI_ISL_1690220, EPI_ISL_1690221, EPI_ISL_1690222, EPI_ISL_1690223, EPI_ISL_1690224, EPI_ISL_1690225, EPI_ISL_1690226, EPI_ISL_1690227, EPI_ISL_1690228, EPI_ISL_1690229, EPI_ISL_1690230, EPI_ISL_1690231, EPI_ISL_1690232, EPI_ISL_1690233, EPI_ISL_1690234, EPI_ISL_1690235, EPI_ISL_1690236, EPI_ISL_1690237, EPI_ISL_1690238, EPI_ISL_1690239, EPI_ISL_1690240, EPI_ISL_1690241, EPI_ISL_1690242, EPI_ISL_1690243, EPI_ISL_1690244, EPI_ISL_1690245, EPI_ISL_1690246, EPI_ISL_1690247, EPI_ISL_1690248, EPI_ISL_1690249, EPI_ISL_1690250, EPI_ISL_1690251, EPI_ISL_1690252, EPI_ISL_1690253, EPI_ISL_1690254, EPI_ISL_1690255, EPI_ISL_1690256, EPI_ISL_1690257, EPI_ISL_1690258, EPI_ISL_1690259, EPI_ISL_1690260, EPI_ISL_1690261, EPI_ISL_1690262, EPI_ISL_1690263, EPI_ISL_1690264, EPI_ISL_1690265, EPI_ISL_1690266, EPI_ISL_1690267, EPI_ISL_1690268, EPI_ISL_1690269, EPI_ISL_1690270, EPI_ISL_1690271, EPI_ISL_1690272, EPI_ISL_1690273, EPI_ISL_1690274, EPI_ISL_1690275, EPI_ISL_1690276, EPI_ISL_1690277 | | | |
| see above | Aegis Sciences Corporation | Centers for Disease Control and Prevention Division of Viral Diseases, Pathogen Discovery | Dakota Howard, Dhwani Batra, Peter W. Cook, Kara Moser, Adrian Paskey, Jason Caravas, Benjamin Rambo-Martin, Shatavia Morrison, Christopher Gulvick, Scott Sammons, Yvette Unoarumhi, Darlene Wagner, Matthew Schmerer, Cyndi Clark, Patrick Campbell, Rob Case, Vikramsinha Ghorpade, Holly Houdeshell, Ola Kvalvaag, Dillon Nall, Ethan Sanders, Alec Vest, Shaun Westlund, Matthew Hardison, Clinton R. Paden, Duncan MacCannell |
| EPI_ISL_1690278, EPI_ISL_1690279, EPI_ISL_1690281, EPI_ISL_1690282, EPI_ISL_1690283, EPI_ISL_1690284, EPI_ISL_1690285, EPI_ISL_1690286, EPI_ISL_1690287, EPI_ISL_1690288, EPI_ISL_1690289, EPI_ISL_1690290, EPI_ISL_1690291, EPI_ISL_1690292, EPI_ISL_1690293, EPI_ISL_1690294, EPI_ISL_1690295, EPI_ISL_1690296, EPI_ISL_1690297, EPI_ISL_1690298, EPI_ISL_1690299, EPI_ISL_1690300, EPI_ISL_1690301, EPI_ISL_1690302, EPI_ISL_1690303, EPI_ISL_1690304, EPI_ISL_1690305, EPI_ISL_1690306, EPI_ISL_1690307, EPI_ISL_1690308, EPI_ISL_1690309, EPI_ISL_1690311, EPI_ISL_1690312, EPI_ISL_1690313, EPI_ISL_1690314, EPI_ISL_1690315, EPI_ISL_1690317, EPI_ISL_1690318, EPI_ISL_1690319, EPI_ISL_1690320, EPI_ISL_1690321, EPI_ISL_1690322, EPI_ISL_1690323, EPI_ISL_1690324, EPI_ISL_1690325, EPI_ISL_1690326, EPI_ISL_1690327, EPI_ISL_1690328, EPI_ISL_1690329, EPI_ISL_1690330, EPI_ISL_1690331, EPI_ISL_1690332, EPI_ISL_1690333, EPI_ISL_1690334, EPI_ISL_1690335, EPI_ISL_1690336, EPI_ISL_1690337, EPI_ISL_1690338, EPI_ISL_1690339 | | | |
| see above | Helix/Illumina | Centers for Disease Control and Prevention Division of Viral Diseases, Pathogen Discovery | Dakota Howard, Dhwani Batra, Peter W. Cook, Kara Moser, Adrian Paskey, Jason Caravas, Benjamin Rambo-Martin, Shatavia Morrison, Christopher Gulvick, Scott Sammons, Yvette Unoarumhi, Darlene Wagner, Matthew Schmerer, Eileen de Feo, Jan Antico, Christine Tran, Matthew Tolentino, Shannon Wickline, Kim Gietzen, Brad Sickler, Jingtao Liu, Eric Allen, Phil Febbo, Nicole L. Washington, Simon White, Geraint Levan, Kelly Schiabor Barrett, Elizabeth Cirulli, Alexandre Bolze, Ary Ascencio, Charlotte Rivera-Garcia, Ryan Cho, Jason Nguyen, Sherry Wang, Jimmy Ramirez, Tyler Cassens, Efren Sandoval, Magnus Isaksson, William Lee, David Becker, Marc Laurent, James Lu, Clinton R. Paden, Duncan MacCannell |
| EPI_ISL_1690340 | Ohio Department of Health Laboratory | Ohio Department of Health Laboratory | Holmes, Jennifer; Eric Brandt, Keoni Omura, Glen McGillivary, Caitlin McDonnell, Jade Mowery, Stephanie Mccracken, Tyler Payne, Kirtana Ramadugu, Erica Leasure, Brent Lee, Kelsey Florek, Heather Blankenship, Allison Black, Quanta Brown, and Tammy Bannerman |
| EPI_ISL_1690341, EPI_ISL_1690342, EPI_ISL_1690343, EPI_ISL_1690344, EPI_ISL_1690345, EPI_ISL_1690346, EPI_ISL_1690347, EPI_ISL_1690348, EPI_ISL_1690349, EPI_ISL_1690350, EPI_ISL_1690351, EPI_ISL_1690352, EPI_ISL_1690353, EPI_ISL_1690354, EPI_ISL_1690355, EPI_ISL_1690356, EPI_ISL_1690357, EPI_ISL_1690358, EPI_ISL_1690359, EPI_ISL_1690360, EPI_ISL_1690361, EPI_ISL_1690362, EPI_ISL_1690363, EPI_ISL_1690364, EPI_ISL_1690365, EPI_ISL_1690366, EPI_ISL_1690367, EPI_ISL_1690368, EPI_ISL_1690369, EPI_ISL_1690370, EPI_ISL_1690371, EPI_ISL_1690372, EPI_ISL_1690373, EPI_ISL_1690374, EPI_ISL_1690375, EPI_ISL_1690376, EPI_ISL_1690377, EPI_ISL_1690378, EPI_ISL_1690379, EPI_ISL_1690380, EPI_ISL_1690381, EPI_ISL_1690384, EPI_ISL_1690387, EPI_ISL_1690390, EPI_ISL_1690392, EPI_ISL_1690393, EPI_ISL_1690394, EPI_ISL_1690395, EPI_ISL_1690396, EPI_ISL_1690397, EPI_ISL_1690398, EPI_ISL_1690399, EPI_ISL_1690400, EPI_ISL_1690401, EPI_ISL_1690402, EPI_ISL_1690403, EPI_ISL_1690404, EPI_ISL_1690405, EPI_ISL_1690407, EPI_ISL_1690408, EPI_ISL_1690409, EPI_ISL_1690410, EPI_ISL_1690411, EPI_ISL_1690412, EPI_ISL_1690413, EPI_ISL_1690414, EPI_ISL_1690415, EPI_ISL_1690417, EPI_ISL_1690418, EPI_ISL_1690420, EPI_ISL_1690421, EPI_ISL_1690422, EPI_ISL_1690423, EPI_ISL_1690424, EPI_ISL_1690425, EPI_ISL_1690426, EPI_ISL_1690427, EPI_ISL_1690428, EPI_ISL_1690429, EPI_ISL_1690430, EPI_ISL_1690431, EPI_ISL_1690433, EPI_ISL_1690434, EPI_ISL_1690435, EPI_ISL_1690436, EPI_ISL_1690437, EPI_ISL_1690438, EPI_ISL_1690439, EPI_ISL_1690440, EPI_ISL_1690441, EPI_ISL_1690442, EPI_ISL_1690443, EPI_ISL_1690444, EPI_ISL_1690445, EPI_ISL_1690446, EPI_ISL_1690447, EPI_ISL_1690448, EPI_ISL_1690449, EPI_ISL_1690450, EPI_ISL_1690451, EPI_ISL_1690452, EPI_ISL_1690453, EPI_ISL_1690454, EPI_ISL_1690455, EPI_ISL_1690456, EPI_ISL_1690457, EPI_ISL_1690458, EPI_ISL_1690459, EPI_ISL_1690460, EPI_ISL_1690461, EPI_ISL_1690462, EPI_ISL_1690463, EPI_ISL_1690464, EPI_ISL_1690465, EPI_ISL_1690466, EPI_ISL_1690467, EPI_ISL_1690468, EPI_ISL_1690469, EPI_ISL_1690470, EPI_ISL_1690471, EPI_ISL_1690472, EPI_ISL_1690473, EPI_ISL_1690474, EPI_ISL_1690475, EPI_ISL_1690476, EPI_ISL_1690477, EPI_ISL_1690478, EPI_ISL_1690479, EPI_ISL_1690480, EPI_ISL_1690481, EPI_ISL_1690482, EPI_ISL_1690483, EPI_ISL_1690484, EPI_ISL_1690485, EPI_ISL_1690486, EPI_ISL_1690487, EPI_ISL_1690488, EPI_ISL_1690489, EPI_ISL_1690490, EPI_ISL_1690491, EPI_ISL_1690492, EPI_ISL_1690493, EPI_ISL_1690494, EPI_ISL_1690495, EPI_ISL_1690496, EPI_ISL_1690497, EPI_ISL_1690498, EPI_ISL_1690499, EPI_ISL_1690500, EPI_ISL_1690501, EPI_ISL_1690502, EPI_ISL_1690503, EPI_ISL_1690504, EPI_ISL_1690505, EPI_ISL_1690506, EPI_ISL_1690507, EPI_ISL_1690508, EPI_ISL_1690509, EPI_ISL_1690510, EPI_ISL_1690511, EPI_ISL_1690512, EPI_ISL_1690513, EPI_ISL_1690514, EPI_ISL_1690515, EPI_ISL_1690516, EPI_ISL_1690517, EPI_ISL_1690518, EPI_ISL_1690519, EPI_ISL_1690520, EPI_ISL_1690521, EPI_ISL_1690522, EPI_ISL_1690523, EPI_ISL_1690524, EPI_ISL_1690525, EPI_ISL_1690526, EPI_ISL_1690527, EPI_ISL_1690528, EPI_ISL_1690529, EPI_ISL_1690530, EPI_ISL_1690531, EPI_ISL_1690532, EPI_ISL_1690533, EPI_ISL_1690534, EPI_ISL_1690535, EPI_ISL_1690536, EPI_ISL_1690537, EPI_ISL_1690538, EPI_ISL_1690539, EPI_ISL_1690540, EPI_ISL_1690541, EPI_ISL_1690542, EPI_ISL_1690543, EPI_ISL_1690544, EPI_ISL_1690545, EPI_ISL_1690546, EPI_ISL_1690547, EPI_ISL_1690548, EPI_ISL_1690549, EPI_ISL_1690550, EPI_ISL_1690552, EPI_ISL_1690553, EPI_ISL_1690554, EPI_ISL_1690555, EPI_ISL_1690556, EPI_ISL_1690557, EPI_ISL_1690558, EPI_ISL_1690559, EPI_ISL_1690560, EPI_ISL_1690561, EPI_ISL_1690562, EPI_ISL_1690563, EPI_ISL_1690564, EPI_ISL_1690565, EPI_ISL_1690566, EPI_ISL_1690567, EPI_ISL_1690568, EPI_ISL_1690569, EPI_ISL_1690570, EPI_ISL_1690571, EPI_ISL_1690572, EPI_ISL_1690573, EPI_ISL_1690574, EPI_ISL_1690575, EPI_ISL_1690576, EPI_ISL_1690577, EPI_ISL_1690578, EPI_ISL_1690579, EPI_ISL_1690580, EPI_ISL_1690581, EPI_ISL_1690582, EPI_ISL_1690583, EPI_ISL_1690584, EPI_ISL_1690585, EPI_ISL_1690586, EPI_ISL_1690587 | | | |
| see above | Helix/Illumina | Centers for Disease Control and Prevention Division of Viral Diseases, Pathogen Discovery | Dakota Howard, Dhwani Batra, Peter W. Cook, Kara Moser, Adrian Paskey, Jason Caravas, Benjamin Rambo-Martin, Shatavia Morrison, Christopher Gulvick, Scott Sammons, Yvette Unoarumhi, Darlene Wagner, Matthew Schmerer, Eileen de Feo, Jan Antico, Christine Tran, Matthew Tolentino, Shannon Wickline, Kim Gietzen, Brad Sickler, Jingtao Liu, Eric Allen, Phil Febbo, Nicole L. Washington, Simon White, Geraint Levan, Kelly Schiabor Barrett, Elizabeth Cirulli, Alexandre Bolze, Ary Ascencio, Charlotte Rivera-Garcia, Ryan Cho, Jason Nguyen, Sherry Wang, Jimmy Ramirez, Tyler Cassens, Efren Sandoval, Magnus Isaksson, William Lee, David Becker, Marc Laurent, James Lu, Clinton R. Paden, Duncan MacCannell |
| EPI_ISL_1690588 | Uniformed Services University Infectious Diseases Clinical Research Program | US Air Force School of Aerospace Medicine | Anthony Fries, Jennifer Meyer, Sarah Purves, Clarise Starr, Elizabeth Macias, Simon Pollett, Brian Agan, Mark Simons, Timothy Burgess |
| EPI_ISL_1690589 | Helix/Illumina | Centers for Disease Control and Prevention Division of Viral Diseases, Pathogen Discovery | Dakota Howard, Dhwani Batra, Peter W. Cook, Kara Moser, Adrian Paskey, Jason Caravas, Benjamin Rambo-Martin, Shatavia Morrison, Christopher Gulvick, Scott Sammons, Yvette Unoarumhi, Darlene Wagner, Matthew Schmerer, Eileen de Feo, Jan Antico, Christine Tran, Matthew Tolentino, Shannon Wickline, Kim Gietzen, Brad Sickler, Jingtao Liu, Eric Allen, Phil Febbo, Nicole L. Washington, Simon White, Geraint Levan, Kelly Schiabor Barrett, Elizabeth Cirulli, Alexandre Bolze, Ary Ascencio, Charlotte Rivera-Garcia, Ryan Cho, Jason Nguyen, Sherry Wang, Jimmy Ramirez, Tyler Cassens, Efren Sandoval, Magnus Isaksson, William Lee, David Becker, Marc Laurent, James Lu, Clinton R. Paden, Duncan MacCannell |
| EPI_ISL_1690590, EPI_ISL_1690591 | Uniformed Services University Infectious Diseases Clinical Research Program | US Air Force School of Aerospace Medicine | Anthony Fries, Jennifer Meyer, Sarah Purves, Clarise Starr, Elizabeth Macias, Simon Pollett, Brian Agan, Mark Simons, Timothy Burgess |
| EPI_ISL_1690592 | Helix/Illumina | Centers for Disease Control and Prevention Division of Viral Diseases, Pathogen Discovery | Dakota Howard, Dhwani Batra, Peter W. Cook, Kara Moser, Adrian Paskey, Jason Caravas, Benjamin Rambo-Martin, Shatavia Morrison, Christopher Gulvick, Scott Sammons, Yvette Unoarumhi, Darlene Wagner, Matthew Schmerer, Eileen de Feo, Jan Antico, Christine Tran, Matthew Tolentino, Shannon Wickline, Kim Gietzen, Brad Sickler, Jingtao Liu, Eric Allen, Phil Febbo, Nicole L. Washington, Simon White, Geraint Levan, Kelly Schiabor Barrett, Elizabeth Cirulli, Alexandre Bolze, Ary Ascencio, Charlotte Rivera-Garcia, Ryan Cho, Jason Nguyen, Sherry Wang, Jimmy Ramirez, Tyler Cassens, Efren Sandoval, Magnus Isaksson, William Lee, David Becker, Marc Laurent, James Lu, Clinton R. Paden, Duncan MacCannell |
| EPI_ISL_1690593 | Uniformed Services University Infectious Diseases Clinical Research Program | US Air Force School of Aerospace Medicine | Anthony Fries, Jennifer Meyer, Sarah Purves, Clarise Starr, Elizabeth Macias, Simon Pollett, Brian Agan, Mark Simons, Timothy Burgess |
| EPI_ISL_1690594, EPI_ISL_1690595, EPI_ISL_1690596, EPI_ISL_1690597, EPI_ISL_1690598, EPI_ISL_1690599, EPI_ISL_1690600, EPI_ISL_1690601, EPI_ISL_1690602, EPI_ISL_1690603, EPI_ISL_1690604, EPI_ISL_1690605, EPI_ISL_1690606, EPI_ISL_1690607, EPI_ISL_1690608, EPI_ISL_1690609, EPI_ISL_1690610, EPI_ISL_1690611, EPI_ISL_1690612, EPI_ISL_1690613, EPI_ISL_1690614, EPI_ISL_1690615, EPI_ISL_1690616, EPI_ISL_1690617, EPI_ISL_1690618, EPI_ISL_1690619, EPI_ISL_1690620, EPI_ISL_1690621, EPI_ISL_1690622, EPI_ISL_1690623, EPI_ISL_1690624, EPI_ISL_1690625, EPI_ISL_1690626, EPI_ISL_1690627, EPI_ISL_1690628, EPI_ISL_1690629, EPI_ISL_1690630, EPI_ISL_1690631, EPI_ISL_1690632, EPI_ISL_1690633, EPI_ISL_1690634, EPI_ISL_1690635, EPI_ISL_1690636, EPI_ISL_1690637, EPI_ISL_1690638, EPI_ISL_1690639, EPI_ISL_1690640, EPI_ISL_1690641, EPI_ISL_1690642, EPI_ISL_1690643, EPI_ISL_1690644, EPI_ISL_1690645, EPI_ISL_1690646, EPI_ISL_1690647, EPI_ISL_1690648, EPI_ISL_1690649, EPI_ISL_1690650, EPI_ISL_1690651, EPI_ISL_1690652, EPI_ISL_1690653, EPI_ISL_1690654, EPI_ISL_1690655, EPI_ISL_1690656, EPI_ISL_1690657, EPI_ISL_1690658, EPI_ISL_1690659, EPI_ISL_1690661, EPI_ISL_1690662, EPI_ISL_1690663, EPI_ISL_1690664, EPI_ISL_1690665, EPI_ISL_1690666, EPI_ISL_1690667, EPI_ISL_1690668, EPI_ISL_1690669, EPI_ISL_1690670, EPI_ISL_1690671, EPI_ISL_1690672, EPI_ISL_1690673, EPI_ISL_1690674, EPI_ISL_1690675, EPI_ISL_1690676, EPI_ISL_1690677, EPI_ISL_1690678, EPI_ISL_1690679, EPI_ISL_1690680, EPI_ISL_1690681, EPI_ISL_1690682, EPI_ISL_1690683, EPI_ISL_1690684, EPI_ISL_1690685, EPI_ISL_1690686, EPI_ISL_1690687, EPI_ISL_1690688, EPI_ISL_1690689, EPI_ISL_1690690, EPI_ISL_1690691, EPI_ISL_1690692, EPI_ISL_1690693, EPI_ISL_1690694, EPI_ISL_1690695, EPI_ISL_1690696, EPI_ISL_1690697, EPI_ISL_1690698, EPI_ISL_1690699, EPI_ISL_1690700, EPI_ISL_1690701, EPI_ISL_1690702, EPI_ISL_1690703, EPI_ISL_1690704, EPI_ISL_1690705, EPI_ISL_1690706, EPI_ISL_1690707, EPI_ISL_1690708, EPI_ISL_1690709, EPI_ISL_1690710, EPI_ISL_1690711, EPI_ISL_1690712, EPI_ISL_1690713, EPI_ISL_1690714, EPI_ISL_1690715, EPI_ISL_1690716, EPI_ISL_1690717, EPI_ISL_1690718, EPI_ISL_1690719, EPI_ISL_1690720, EPI_ISL_1690721, EPI_ISL_1690722, EPI_ISL_1690723, EPI_ISL_1690724, EPI_ISL_1690725, EPI_ISL_1690726, EPI_ISL_1690727, EPI_ISL_1690728, EPI_ISL_1690729, EPI_ISL_1690730, EPI_ISL_1690731, EPI_ISL_1690732, EPI_ISL_1690733, EPI_ISL_1690734, EPI_ISL_1690735, EPI_ISL_1690736, EPI_ISL_1690738, EPI_ISL_1690739, EPI_ISL_1690740, EPI_ISL_1690741, EPI_ISL_1690742, EPI_ISL_1690743, EPI_ISL_1690744, EPI_ISL_1690745, EPI_ISL_1690746, EPI_ISL_1690747, EPI_ISL_1690748, EPI_ISL_1690749, EPI_ISL_1690750, EPI_ISL_1690751, EPI_ISL_1690752, EPI_ISL_1690753, EPI_ISL_1690754, EPI_ISL_1690755, EPI_ISL_1690756, EPI_ISL_1690757, EPI_ISL_1690758, EPI_ISL_1690759, EPI_ISL_1690760, EPI_ISL_1690761, EPI_ISL_1690762, EPI_ISL_1690763, EPI_ISL_1690764, EPI_ISL_1690765, EPI_ISL_1690766, EPI_ISL_1690767, EPI_ISL_1690768, EPI_ISL_1690769, EPI_ISL_1690770, EPI_ISL_1690771, EPI_ISL_1690772, EPI_ISL_1690773, EPI_ISL_1690774, EPI_ISL_1690775, EPI_ISL_1690776, EPI_ISL_1690777, EPI_ISL_1690778, EPI_ISL_1690779, EPI_ISL_1690780, EPI_ISL_1690781, EPI_ISL_1690782, EPI_ISL_1690783, EPI_ISL_1690784, EPI_ISL_1690785, EPI_ISL_1690786, EPI_ISL_1690787, EPI_ISL_1690788, EPI_ISL_1690789, EPI_ISL_1690790, EPI_ISL_1690791, EPI_ISL_1690792, EPI_ISL_1690793, EPI_ISL_1690794, EPI_ISL_1690795, EPI_ISL_1690796, EPI_ISL_1690797, EPI_ISL_1690798, EPI_ISL_1690799, EPI_ISL_1690800, EPI_ISL_1690801, EPI_ISL_1690802, EPI_ISL_1690803, EPI_ISL_1690804, EPI_ISL_1690805, EPI_ISL_1690806, EPI_ISL_1690807, EPI_ISL_1690808, EPI_ISL_1690809, EPI_ISL_1690810, EPI_ISL_1690811, EPI_ISL_1690812, EPI_ISL_1690813, EPI_ISL_1690814, EPI_ISL_1690815, EPI_ISL_1690816, EPI_ISL_1690817, EPI_ISL_1690818, EPI_ISL_1690819, EPI_ISL_1690820, EPI_ISL_1690821, EPI_ISL_1690822, EPI_ISL_1690823, EPI_ISL_1690824, EPI_ISL_1690825, EPI_ISL_1690826, EPI_ISL_1690827, EPI_ISL_1690828, EPI_ISL_1690829, EPI_ISL_1690830, EPI_ISL_1690831, EPI_ISL_1690832, EPI_ISL_1690833, EPI_ISL_1690834, EPI_ISL_1690835, EPI_ISL_1690836, EPI_ISL_1690837, EPI_ISL_1690838, EPI_ISL_1690839, EPI_ISL_1690840, EPI_ISL_1690841, EPI_ISL_1690842, EPI_ISL_1690843, EPI_ISL_1690844, EPI_ISL_1690845, EPI_ISL_1690846, EPI_ISL_1690847, EPI_ISL_1690848, EPI_ISL_1690849, EPI_ISL_1690850, EPI_ISL_1690851, EPI_ISL_1690852, EPI_ISL_1690853, EPI_ISL_1690854, EPI_ISL_1690855, EPI_ISL_1690856, EPI_ISL_1690857, EPI_ISL_1690858, EPI_ISL_1690859, EPI_ISL_1690860, EPI_ISL_1690861, EPI_ISL_1690862, EPI_ISL_1690863, EPI_ISL_1690864, EPI_ISL_1690865, EPI_ISL_1690866, EPI_ISL_1690867, EPI_ISL_1690868, EPI_ISL_1690869, EPI_ISL_1690870, EPI_ISL_1690871, EPI_ISL_1690872, EPI_ISL_1690873, EPI_ISL_1690874, EPI_ISL_1690875, EPI_ISL_1690876, EPI_ISL_1690877, EPI_ISL_1690878, EPI_ISL_1690879, EPI_ISL_1690880, EPI_ISL_1690881, EPI_ISL_1690882, EPI_ISL_1690883, EPI_ISL_1690884, EPI_ISL_1690885, EPI_ISL_1690886, EPI_ISL_1690887, EPI_ISL_1690888, EPI_ISL_1690889, EPI_ISL_1690890, EPI_ISL_1690891, EPI_ISL_1690892, EPI_ISL_1690893, EPI_ISL_1690894, EPI_ISL_1690895, EPI_ISL_1690896, EPI_ISL_1690897, EPI_ISL_1690898, EPI_ISL_1690899, EPI_ISL_1690900, EPI_ISL_1690901, EPI_ISL_1690902, EPI_ISL_1690903, EPI_ISL_1690904, EPI_ISL_1690905, EPI_ISL_1690906, EPI_ISL_1690907, EPI_ISL_1690908, EPI_ISL_1690909, EPI_ISL_1690910, EPI_ISL_1690911, EPI_ISL_1690912, EPI_ISL_1690913, EPI_ISL_1690914, EPI_ISL_1690915, EPI_ISL_1690916, EPI_ISL_1690917, EPI_ISL_1690918, EPI_ISL_1690919, EPI_ISL_1690920, EPI_ISL_1690921, EPI_ISL_1690922, EPI_ISL_1690923, EPI_ISL_1690924, EPI_ISL_1690925, EPI_ISL_1690926, EPI_ISL_1690927, EPI_ISL_1690928, EPI_ISL_1690930, EPI_ISL_1690931, EPI_ISL_1690932, EPI_ISL_1690933, EPI_ISL_1690934, EPI_ISL_1690935, EPI_ISL_1690936, EPI_ISL_1690937, EPI_ISL_1690938, EPI_ISL_1690939, EPI_ISL_1690940, EPI_ISL_1690941, EPI_ISL_1690942, EPI_ISL_1690943, EPI_ISL_1690944, EPI_ISL_1690945, EPI_ISL_1690946, EPI_ISL_1690947, EPI_ISL_1690948, EPI_ISL_1690949, EPI_ISL_1690950, EPI_ISL_1690951, EPI_ISL_1690952, EPI_ISL_1690953, EPI_ISL_1690954, EPI_ISL_1690955, EPI_ISL_1690956, EPI_ISL_1690957, EPI_ISL_1690958, EPI_ISL_1690959, EPI_ISL_1690960, EPI_ISL_1690961, EPI_ISL_1690962, EPI_ISL_1690963, EPI_ISL_1690964, EPI_ISL_1690965, EPI_ISL_1690966, EPI_ISL_1690967, EPI_ISL_1690968, EPI_ISL_1690969, EPI_ISL_1690970, EPI_ISL_1690971, EPI_ISL_1690972, EPI_ISL_1690973, EPI_ISL_1690974, EPI_ISL_1690975, EPI_ISL_1690976, EPI_ISL_1690977, EPI_ISL_1690978, EPI_ISL_1690979, EPI_ISL_1690980, EPI_ISL_1690981, EPI_ISL_1690982, EPI_ISL_1690983, EPI_ISL_1690984, EPI_ISL_1690985, EPI_ISL_1690986, EPI_ISL_1690987, EPI_ISL_1690988, EPI_ISL_1690989, EPI_ISL_1690990, EPI_ISL_1690991, EPI_ISL_1690992, EPI_ISL_1690993, EPI_ISL_1690994, EPI_ISL_1690995, EPI_ISL_1690996, EPI_ISL_1690997, EPI_ISL_1690998, EPI_ISL_1690999, EPI_ISL_1691000, EPI_ISL_1691001, EPI_ISL_1691002, EPI_ISL_1691003, EPI_ISL_1691004, EPI_ISL_1691005, EPI_ISL_1691006, EPI_ISL_1691007, EPI_ISL_1691008, EPI_ISL_1691009, EPI_ISL_1691010, EPI_ISL_1691011, EPI_ISL_1691012, EPI_ISL_1691013, EPI_ISL_1691014, EPI_ISL_1691015, EPI_ISL_1691016, EPI_ISL_1691017, EPI_ISL_1691018, EPI_ISL_1691019, EPI_ISL_1691020, EPI_ISL_1691021, EPI_ISL_1691022, EPI_ISL_1691023, EPI_ISL_1691024, EPI_ISL_1691025, EPI_ISL_1691026, EPI_ISL_1691027, EPI_ISL_1691028, EPI_ISL_1691029, EPI_ISL_1691030, EPI_ISL_1691031, EPI_ISL_1691032, EPI_ISL_1691033, EPI_ISL_1691034, EPI_ISL_1691035, EPI_ISL_1691036, EPI_ISL_1691037, EPI_ISL_1691038, EPI_ISL_1691039, EPI_ISL_1691040, EPI_ISL_1691041, EPI_ISL_1691042, EPI_ISL_1691043, EPI_ISL_1691044, EPI_ISL_1691045, EPI_ISL_1691046, EPI_ISL_1691047, EPI_ISL_1691048, EPI_ISL_1691049, EPI_ISL_1691050, EPI_ISL_1691051, EPI_ISL_1691052, EPI_ISL_1691053, EPI_ISL_1691054, EPI_ISL_1691055, EPI_ISL_1691056, EPI_ISL_1691057, EPI_ISL_1691058, EPI_ISL_1691059, EPI_ISL_1691060, EPI_ISL_1691061, EPI_ISL_1691062, EPI_ISL_1691063, EPI_ISL_1691064, EPI_ISL_1691065, EPI_ISL_1691066, EPI_ISL_1691067, EPI_ISL_1691068, EPI_ISL_1691069, EPI_ISL_1691070, EPI_ISL_1691071, EPI_ISL_1691072, EPI_ISL_1691073, EPI_ISL_1691074, EPI_ISL_1691075, EPI_ISL_1691076, EPI_ISL_1691077, EPI_ISL_1691078, EPI_ISL_1691079, EPI_ISL_1691080, EPI_ISL_1691081, EPI_ISL_1691082, EPI_ISL_1691083, EPI_ISL_1691084, EPI_ISL_1691085, EPI_ISL_1691086, EPI_ISL_1691087, EPI_ISL_1691088, EPI_ISL_1691089, EPI_ISL_1691090, EPI_ISL_1691091, EPI_ISL_1691092, EPI_ISL_1691093, EPI_ISL_1691094, EPI_ISL_1691095, EPI_ISL_1691096, EPI_ISL_1691097, EPI_ISL_1691098, EPI_ISL_1691099, EPI_ISL_1691100, EPI_ISL_1691101, EPI_ISL_1691102, EPI_ISL_1691104, EPI_ISL_1691105, EPI_ISL_1691106, EPI_ISL_1691107, EPI_ISL_1691108, EPI_ISL_1691109, EPI_ISL_1691110, EPI_ISL_1691111, EPI_ISL_1691112, EPI_ISL_1691113, EPI_ISL_1691114, EPI_ISL_1691115, EPI_ISL_1691116, EPI_ISL_1691117, EPI_ISL_1691118, EPI_ISL_1691119, EPI_ISL_1691120, EPI_ISL_1691121, EPI_ISL_1691122, EPI_ISL_1691123, EPI_ISL_1691124, EPI_ISL_1691125, EPI_ISL_1691126, EPI_ISL_1691127, EPI_ISL_1691128, EPI_ISL_1691129, EPI_ISL_1691130, EPI_ISL_1691131, EPI_ISL_1691132, EPI_ISL_1691133, EPI_ISL_1691134, EPI_ISL_1691135, EPI_ISL_1691136, EPI_ISL_1691137, EPI_ISL_1691138, EPI_ISL_1691139, EPI_ISL_1691140, EPI_ISL_1691141, EPI_ISL_1691142, EPI_ISL_1691143, EPI_ISL_1691144, EPI_ISL_1691145, EPI_ISL_1691146, EPI_ISL_1691147, EPI_ISL_1691148, EPI_ISL_1691149, EPI_ISL_1691150, EPI_ISL_1691151, EPI_ISL_1691152, EPI_ISL_1691153, EPI_ISL_1691154, EPI_ISL_1691155, EPI_ISL_1691156, EPI_ISL_1691157, EPI_ISL_1691158, EPI_ISL_1691159, EPI_ISL_1691160, EPI_ISL_1691161, EPI_ISL_1691162, EPI_ISL_1691163, EPI_ISL_1691164, EPI_ISL_1691165, EPI_ISL_1691166, EPI_ISL_1691167, EPI_ISL_1691168, EPI_ISL_1691169, EPI_ISL_1691170, EPI_ISL_1691171, EPI_ISL_1691172, EPI_ISL_1691173, EPI_ISL_1691174, EPI_ISL_1691175, EPI_ISL_1691176, EPI_ISL_1691177, EPI_ISL_1691178, EPI_ISL_1691179, EPI_ISL_1691180, EPI_ISL_1691181, EPI_ISL_1691182, EPI_ISL_1691183, EPI_ISL_1691184, EPI_ISL_1691185, EPI_ISL_1691186, EPI_ISL_1691187, EPI_ISL_1691188, EPI_ISL_1691189, EPI_ISL_1691190, EPI_ISL_1691191, EPI_ISL_1691192, EPI_ISL_1691193, EPI_ISL_1691194, EPI_ISL_1691195, EPI_ISL_1691196, EPI_ISL_1691197, EPI_ISL_1691198, EPI_ISL_1691199, EPI_ISL_1691200, EPI_ISL_1691201, EPI_ISL_1691202, EPI_ISL_1691203, EPI_ISL_1691204, EPI_ISL_1691205, EPI_ISL_1691206, EPI_ISL_1691207, EPI_ISL_1691208, EPI_ISL_1691209, EPI_ISL_1691210, EPI_ISL_1691211, EPI_ISL_1691212, EPI_ISL_1691213, EPI_ISL_1691214, EPI_ISL_1691215, EPI_ISL_1691216, EPI_ISL_1691217, EPI_ISL_1691218, EPI_ISL_1691219, EPI_ISL_1691220, EPI_ISL_1691221, EPI_ISL_1691222, EPI_ISL_1691223, EPI_ISL_1691224, EPI_ISL_1691225, EPI_ISL_1691226, EPI_ISL_1691227, EPI_ISL_1691228, EPI_ISL_1691229, EPI_ISL_1691230, EPI_ISL_1691231, EPI_ISL_1691232, EPI_ISL_1691233, EPI_ISL_1691234, EPI_ISL_1691235, EPI_ISL_1691236, EPI_ISL_1691237, EPI_ISL_1691238, EPI_ISL_1691239, EPI_ISL_1691240, EPI_ISL_1691241, EPI_ISL_1691242, EPI_ISL_1691244, EPI_ISL_1691245, EPI_ISL_1691246, EPI_ISL_1691247, EPI_ISL_1691248, EPI_ISL_1691249, EPI_ISL_1691250, EPI_ISL_1691252, EPI_ISL_1691253, EPI_ISL_1691254, EPI_ISL_1691255, EPI_ISL_1691256, EPI_ISL_1691257, EPI_ISL_1691258, EPI_ISL_1691259, EPI_ISL_1691260, EPI_ISL_1691261, EPI_ISL_1691262, EPI_ISL_1691263, EPI_ISL_1691264, EPI_ISL_1691265, EPI_ISL_1691266, EPI_ISL_1691267, EPI_ISL_1691268, EPI_ISL_1691269, EPI_ISL_1691270, EPI_ISL_1691271, EPI_ISL_1691272, EPI_ISL_1691273, EPI_ISL_1691274, EPI_ISL_1691275, EPI_ISL_1691276, EPI_ISL_1691277, EPI_ISL_1691278, EPI_ISL_1691279, EPI_ISL_1691280, EPI_ISL_1691281, EPI_ISL_1691282, EPI_ISL_1691283, EPI_ISL_1691284, EPI_ISL_1691285, EPI_ISL_1691286, EPI_ISL_1691287, EPI_ISL_1691288, EPI_ISL_1691289, EPI_ISL_1691290, EPI_ISL_1691291, EPI_ISL_1691292, EPI_ISL_1691293, EPI_ISL_1691294, EPI_ISL_1691295, EPI_ISL_1691296, EPI_ISL_1691297, EPI_ISL_1691298, EPI_ISL_1691299, EPI_ISL_1691300, EPI_ISL_1691301, EPI_ISL_1691302, EPI_ISL_1691303, EPI_ISL_1691304, EPI_ISL_1691305, EPI_ISL_1691306, EPI_ISL_1691307, EPI_ISL_1691308, EPI_ISL_1691309, EPI_ISL_1691310, EPI_ISL_1691311, EPI_ISL_1691312, EPI_ISL_1691313, EPI_ISL_1691314, EPI_ISL_1691315, EPI_ISL_1691316, EPI_ISL_1691317, EPI_ISL_1691318, EPI_ISL_1691319, EPI_ISL_1691320, EPI_ISL_1691321, EPI_ISL_1691322, EPI_ISL_1691323, EPI_ISL_1691324, EPI_ISL_1691325, EPI_ISL_1691326, EPI_ISL_1691327, EPI_ISL_1691328, EPI_ISL_1691329, EPI_ISL_1691330, EPI_ISL_1691331, EPI_ISL_1691332, EPI_ISL_1691333, EPI_ISL_1691334, EPI_ISL_1691335, EPI_ISL_1691336, EPI_ISL_1691337, EPI_ISL_1691338, EPI_ISL_1691339, EPI_ISL_1691340, EPI_ISL_1691341, EPI_ISL_1691342, EPI_ISL_1691343, EPI_ISL_1691344, EPI_ISL_1691345, EPI_ISL_1691346, EPI_ISL_1691347, EPI_ISL_1691348, EPI_ISL_1691349, EPI_ISL_1691350, EPI_ISL_1691351, EPI_ISL_1691352, EPI_ISL_1691353, EPI_ISL_1691354, EPI_ISL_1691355, EPI_ISL_1691356, EPI_ISL_1691357, EPI_ISL_1691358, EPI_ISL_1691359, EPI_ISL_1691360, EPI_ISL_1691361, EPI_ISL_1691362, EPI_ISL_1691363, EPI_ISL_1691364, EPI_ISL_1691365, EPI_ISL_1691366, EPI_ISL_1691367, EPI_ISL_1691368, EPI_ISL_1691369, EPI_ISL_1691370, EPI_ISL_1691371, EPI_ISL_1691372, EPI_ISL_1691373, EPI_ISL_1691374, EPI_ISL_1691375, EPI_ISL_1691376, EPI_ISL_1691377, EPI_ISL_1691378, EPI_ISL_1691379, EPI_ISL_1691380, EPI_ISL_1691381, EPI_ISL_1691382, EPI_ISL_1691383, EPI_ISL_1691384, EPI_ISL_1691385, EPI_ISL_1691386, EPI_ISL_1691387, EPI_ISL_1691388, EPI_ISL_1691389, EPI_ISL_1691390, EPI_ISL_1691391, EPI_ISL_1691392, EPI_ISL_1691393, EPI_ISL_1691394 | | | |
| see above | Helix/Illumina | Centers for Disease Control and Prevention Division of Viral Diseases, Pathogen Discovery | Dakota Howard, Dhwani Batra, Peter W. Cook, Kara Moser, Adrian Paskey, Jason Caravas, Benjamin Rambo-Martin, Shatavia Morrison, Christopher Gulvick, Scott Sammons, Yvette Unoarumhi, Darlene Wagner, Matthew Schmerer, Eileen de Feo, Jan Antico, Christine Tran, Matthew Tolentino, Shannon Wickline, Kim Gietzen, Brad Sickler, Jingtao Liu, Eric Allen, Phil Febbo, Nicole L. Washington, Simon White, Geraint Levan, Kelly Schiabor Barrett, Elizabeth Cirulli, Alexandre Bolze, Ary Ascencio, Charlotte Rivera-Garcia, Ryan Cho, Jason Nguyen, Sherry Wang, Jimmy Ramirez, Tyler Cassens, Efren Sandoval, Magnus Isaksson, William Lee, David Becker, Marc Laurent, James Lu, Clinton R. Paden, Duncan MacCannell |

| | | | |
|---|---|---|---|
| EPI_ISL_1691395, EPI_ISL_1691396, EPI_ISL_1691397, EPI_ISL_1691398, EPI_ISL_1691399, EPI_ISL_1691400, EPI_ISL_1691401, EPI_ISL_1691402, EPI_ISL_1691403, EPI_ISL_1691404, EPI_ISL_1691405, EPI_ISL_1691406, EPI_ISL_1691407, EPI_ISL_1691408, EPI_ISL_1691409, EPI_ISL_1691410, EPI_ISL_1691411, EPI_ISL_1691412, EPI_ISL_1691413, EPI_ISL_1691414, EPI_ISL_1691415, EPI_ISL_1691416, EPI_ISL_1691417, EPI_ISL_1691418, EPI_ISL_1691419, EPI_ISL_1691420, EPI_ISL_1691421, EPI_ISL_1691422, EPI_ISL_1691423, EPI_ISL_1691424, EPI_ISL_1691425, EPI_ISL_1691426, EPI_ISL_1691427, EPI_ISL_1691428, EPI_ISL_1691429, EPI_ISL_1691430, EPI_ISL_1691431, EPI_ISL_1691432, EPI_ISL_1691433, EPI_ISL_1691434, EPI_ISL_1691435, EPI_ISL_1691436, EPI_ISL_1691437, EPI_ISL_1691438, EPI_ISL_1691439, EPI_ISL_1691440, EPI_ISL_1691441, EPI_ISL_1691442, EPI_ISL_1691443, EPI_ISL_1691444, EPI_ISL_1691445, EPI_ISL_1691446, EPI_ISL_1691447, EPI_ISL_1691448, EPI_ISL_1691449, EPI_ISL_1691450, EPI_ISL_1691451, EPI_ISL_1691452, EPI_ISL_1691453, EPI_ISL_1691454, EPI_ISL_1691455, EPI_ISL_1691456, EPI_ISL_1691457, EPI_ISL_1691458, EPI_ISL_1691460, EPI_ISL_1691461, EPI_ISL_1691462, EPI_ISL_1691463, EPI_ISL_1691464, EPI_ISL_1691465, EPI_ISL_1691466, EPI_ISL_1691467, EPI_ISL_1691468, EPI_ISL_1691469, EPI_ISL_1691470, EPI_ISL_1691471, EPI_ISL_1691472, EPI_ISL_1691473, EPI_ISL_1691474, EPI_ISL_1691475, EPI_ISL_1691476, EPI_ISL_1691477, EPI_ISL_1691478, EPI_ISL_1691479, EPI_ISL_1691480, EPI_ISL_1691481, EPI_ISL_1691482, EPI_ISL_1691483, EPI_ISL_1691484, EPI_ISL_1691485, EPI_ISL_1691486, EPI_ISL_1691488, EPI_ISL_1691489, EPI_ISL_1691490, EPI_ISL_1691491, EPI_ISL_1691492, EPI_ISL_1691493, EPI_ISL_1691494, EPI_ISL_1691495, EPI_ISL_1691496, EPI_ISL_1691497, EPI_ISL_1691498, EPI_ISL_1691499, EPI_ISL_1691500, EPI_ISL_1691501, EPI_ISL_1691502, EPI_ISL_1691503, EPI_ISL_1691504, EPI_ISL_1691505, EPI_ISL_1691506, EPI_ISL_1691507, EPI_ISL_1691508, EPI_ISL_1691509, EPI_ISL_1691510, EPI_ISL_1691511, EPI_ISL_1691512, EPI_ISL_1691513, EPI_ISL_1691514, EPI_ISL_1691515, EPI_ISL_1691516, EPI_ISL_1691517, EPI_ISL_1691518, EPI_ISL_1691519, EPI_ISL_1691520, EPI_ISL_1691521, EPI_ISL_1691522, EPI_ISL_1691523, EPI_ISL_1691524, EPI_ISL_1691525, EPI_ISL_1691526, EPI_ISL_1691527, EPI_ISL_1691528, EPI_ISL_1691529, EPI_ISL_1691530, EPI_ISL_1691531, EPI_ISL_1691532, EPI_ISL_1691533, EPI_ISL_1691534, EPI_ISL_1691535, EPI_ISL_1691536, EPI_ISL_1691537, EPI_ISL_1691538, EPI_ISL_1691539, EPI_ISL_1691540, EPI_ISL_1691541, EPI_ISL_1691542, EPI_ISL_1691543, EPI_ISL_1691544, EPI_ISL_1691545 | | | |
| see above | Quest Diagnostics Incorporated | Centers for Disease Control and Prevention Division of Viral Diseases, Pathogen Discovery | Dakota Howard, Dhwani Batra, Peter W. Cook, Kara Moser, Adrian Paskey, Jason Caravas, Benjamin Rambo-Martin, Shatavia Morrison, Christopher Gulvick, Scott Sammons, Yvette Unoarumhi, Darlene Wagner, Matthew Schmerer, S. H. Rosenthal, A. Gerasimova, R. M. Kagan, B. Anderson, M. Hua, Y. Liu, L.E. Bernstein, K.E. Livingston, A. Perez, I. A. Shlyakhter, R. V. Rolando, R. Owen, P. Tanpaiboon, F. Lacbawan, Clinton R. Paden, Duncan MacCannell |
| EPI_ISL_1691546 | Ohio Department of Health Laboratory | Ohio Department of Health Laboratory | Holmes, Jennifer; Eric Brandt, Keoni Omura, Glen McGillivary, Caitlin McDonnell, Jade Mowery, Stephanie Mccracken, Tyler Payne, Kirtana Ramadugu, Erica Leasure, Brent Lee, Kelsey Florek, Heather Blankenship, Allison Black, Quanta Brown, and Tammy Bannerman |
| EPI_ISL_1691547, EPI_ISL_1691548, EPI_ISL_1691549, EPI_ISL_1691550, EPI_ISL_1691551, EPI_ISL_1691552, EPI_ISL_1691553, EPI_ISL_1691554, EPI_ISL_1691556, EPI_ISL_1691557, EPI_ISL_1691558, EPI_ISL_1691559, EPI_ISL_1691560, EPI_ISL_1691561, EPI_ISL_1691562, EPI_ISL_1691563, EPI_ISL_1691564, EPI_ISL_1691565, EPI_ISL_1691566, EPI_ISL_1691567, EPI_ISL_1691576, EPI_ISL_1691577, EPI_ISL_1691578, EPI_ISL_1691579, EPI_ISL_1691580, EPI_ISL_1691581, EPI_ISL_1691582, EPI_ISL_1691583, EPI_ISL_1691584, EPI_ISL_1691586, EPI_ISL_1691588, EPI_ISL_1691590, EPI_ISL_1691592, EPI_ISL_1691594, EPI_ISL_1691597, EPI_ISL_1691598, EPI_ISL_1691599, EPI_ISL_1691600, EPI_ISL_1691601, EPI_ISL_1691602, EPI_ISL_1691605, EPI_ISL_1691606, EPI_ISL_1691607, EPI_ISL_1691608, EPI_ISL_1691609, EPI_ISL_1691610, EPI_ISL_1691611, EPI_ISL_1691612, EPI_ISL_1691613, EPI_ISL_1691614, EPI_ISL_1691615, EPI_ISL_1691616, EPI_ISL_1691617, EPI_ISL_1691618, EPI_ISL_1691619, EPI_ISL_1691620, EPI_ISL_1691621, EPI_ISL_1691622, EPI_ISL_1691623, EPI_ISL_1691624, EPI_ISL_1691625, EPI_ISL_1691626, EPI_ISL_1691627, EPI_ISL_1691628, EPI_ISL_1691629, EPI_ISL_1691630, EPI_ISL_1691631, EPI_ISL_1691632, EPI_ISL_1691633, EPI_ISL_1691634, EPI_ISL_1691635, EPI_ISL_1691636, EPI_ISL_1691637, EPI_ISL_1691638, EPI_ISL_1691639, EPI_ISL_1691640, EPI_ISL_1691641, EPI_ISL_1691642, EPI_ISL_1691643, EPI_ISL_1691644, EPI_ISL_1691645, EPI_ISL_1691646, EPI_ISL_1691647, EPI_ISL_1691648, EPI_ISL_1691649, EPI_ISL_1691650, EPI_ISL_1691651, EPI_ISL_1691652, EPI_ISL_1691653, EPI_ISL_1691654, EPI_ISL_1691655, EPI_ISL_1691656, EPI_ISL_1691657, EPI_ISL_1691658, EPI_ISL_1691659, EPI_ISL_1691660, EPI_ISL_1691661, EPI_ISL_1691662, EPI_ISL_1691663, EPI_ISL_1691664, EPI_ISL_1691665, EPI_ISL_1691666, EPI_ISL_1691667, EPI_ISL_1691668, EPI_ISL_1691669, EPI_ISL_1691670, EPI_ISL_1691671, EPI_ISL_1691672, EPI_ISL_1691673, EPI_ISL_1691674, EPI_ISL_1691675, EPI_ISL_1691676, EPI_ISL_1691677, EPI_ISL_1691678, EPI_ISL_1691679, EPI_ISL_1691680, EPI_ISL_1691681, EPI_ISL_1691682, EPI_ISL_1691683, EPI_ISL_1691684, EPI_ISL_1691685, EPI_ISL_1691686, EPI_ISL_1691687, EPI_ISL_1691689, EPI_ISL_1691690, EPI_ISL_1691691, EPI_ISL_1691692, EPI_ISL_1691693, EPI_ISL_1691694, EPI_ISL_1691695, EPI_ISL_1691696, EPI_ISL_1691698, EPI_ISL_1691699, EPI_ISL_1691700, EPI_ISL_1691701, EPI_ISL_1691702, EPI_ISL_1691703, EPI_ISL_1691704, EPI_ISL_1691705, EPI_ISL_1691706, EPI_ISL_1691707, EPI_ISL_1691708, EPI_ISL_1691709, EPI_ISL_1691710, EPI_ISL_1691711, EPI_ISL_1691712, EPI_ISL_1691713, EPI_ISL_1691714, EPI_ISL_1691715, EPI_ISL_1691716, EPI_ISL_1691717, EPI_ISL_1691718, EPI_ISL_1691719, EPI_ISL_1691720, EPI_ISL_1691721, EPI_ISL_1691722, EPI_ISL_1691723, EPI_ISL_1691724, EPI_ISL_1691725, EPI_ISL_1691726, EPI_ISL_1691729, EPI_ISL_1691732, EPI_ISL_1691733, EPI_ISL_1691734, EPI_ISL_1691735, EPI_ISL_1691736, EPI_ISL_1691737, EPI_ISL_1691738, EPI_ISL_1691739, EPI_ISL_1691741, EPI_ISL_1691742, EPI_ISL_1691743, EPI_ISL_1691744, EPI_ISL_1691745, EPI_ISL_1691746, EPI_ISL_1691747, EPI_ISL_1691748, EPI_ISL_1691749, EPI_ISL_1691750, EPI_ISL_1691752, EPI_ISL_1691753, EPI_ISL_1691755, EPI_ISL_1691756, EPI_ISL_1691757, EPI_ISL_1691758, EPI_ISL_1691759, EPI_ISL_1691760, EPI_ISL_1691761, EPI_ISL_1691762, EPI_ISL_1691763, EPI_ISL_1691764, EPI_ISL_1691765, EPI_ISL_1691766, EPI_ISL_1691767, EPI_ISL_1691768, EPI_ISL_1691769, EPI_ISL_1691770, EPI_ISL_1691771, EPI_ISL_1691772, EPI_ISL_1691773, EPI_ISL_1691774, EPI_ISL_1691775, EPI_ISL_1691776, EPI_ISL_1691777, EPI_ISL_1691778, EPI_ISL_1691779, EPI_ISL_1691780, EPI_ISL_1691781, EPI_ISL_1691782, EPI_ISL_1691783, EPI_ISL_1691784, EPI_ISL_1691785, EPI_ISL_1691786, EPI_ISL_1691787, EPI_ISL_1691788, EPI_ISL_1691789, EPI_ISL_1691790, EPI_ISL_1691791, EPI_ISL_1691793, EPI_ISL_1691794, EPI_ISL_1691795, EPI_ISL_1691796, EPI_ISL_1691797, EPI_ISL_1691798, EPI_ISL_1691799, EPI_ISL_1691800, EPI_ISL_1691801, EPI_ISL_1691802, EPI_ISL_1691803, EPI_ISL_1691804, EPI_ISL_1691805, EPI_ISL_1691806, EPI_ISL_1691807, EPI_ISL_1691808, EPI_ISL_1691809, EPI_ISL_1691810, EPI_ISL_1691811, EPI_ISL_1691812, EPI_ISL_1691813, EPI_ISL_1691814, EPI_ISL_1691815, EPI_ISL_1691816, EPI_ISL_1691817, EPI_ISL_1691818, EPI_ISL_1691819, EPI_ISL_1691820, EPI_ISL_1691821, EPI_ISL_1691822, EPI_ISL_1691823, EPI_ISL_1691824, EPI_ISL_1691825, EPI_ISL_1691826, EPI_ISL_1691827, EPI_ISL_1691828, EPI_ISL_1691829, EPI_ISL_1691830, EPI_ISL_1691831, EPI_ISL_1691832, EPI_ISL_1691833, EPI_ISL_1691834, EPI_ISL_1691835, EPI_ISL_1691836, EPI_ISL_1691837, EPI_ISL_1691838, EPI_ISL_1691839, EPI_ISL_1691840, EPI_ISL_1691841, EPI_ISL_1691842, EPI_ISL_1691843, EPI_ISL_1691844, EPI_ISL_1691845, EPI_ISL_1691846, EPI_ISL_1691847, EPI_ISL_1691848, EPI_ISL_1691849, EPI_ISL_1691850, EPI_ISL_1691851, EPI_ISL_1691852, EPI_ISL_1691853, EPI_ISL_1691854, EPI_ISL_1691856, EPI_ISL_1691857, EPI_ISL_1691858, EPI_ISL_1691859, EPI_ISL_1691860, EPI_ISL_1691861, EPI_ISL_1691862, EPI_ISL_1691863, EPI_ISL_1691864, EPI_ISL_1691865, EPI_ISL_1691866, EPI_ISL_1691867, EPI_ISL_1691868, EPI_ISL_1691869, EPI_ISL_1691870, EPI_ISL_1691871, EPI_ISL_1691872, EPI_ISL_1691873, EPI_ISL_1691874, EPI_ISL_1691875, EPI_ISL_1691876, EPI_ISL_1691877, EPI_ISL_1691878, EPI_ISL_1691879, EPI_ISL_1691880, EPI_ISL_1691881, EPI_ISL_1691882, EPI_ISL_1691883, EPI_ISL_1691884, EPI_ISL_1691885, EPI_ISL_1691886, EPI_ISL_1691887, EPI_ISL_1691888, EPI_ISL_1691889, EPI_ISL_1691890, EPI_ISL_1691891, EPI_ISL_1691892, EPI_ISL_1691893, EPI_ISL_1691894, EPI_ISL_1691895, EPI_ISL_1691896, EPI_ISL_1691897, EPI_ISL_1691898, EPI_ISL_1691899, EPI_ISL_1691900, EPI_ISL_1691901, EPI_ISL_1691902, EPI_ISL_1691903, EPI_ISL_1691904, EPI_ISL_1691905, EPI_ISL_1691906, EPI_ISL_1691907, EPI_ISL_1691908, EPI_ISL_1691909, EPI_ISL_1691910, EPI_ISL_1691911, EPI_ISL_1691912, EPI_ISL_1691913, EPI_ISL_1691914, EPI_ISL_1691915, EPI_ISL_1691916, EPI_ISL_1691917, EPI_ISL_1691918, EPI_ISL_1691919, EPI_ISL_1691920, EPI_ISL_1691921, EPI_ISL_1691922, EPI_ISL_1691923, EPI_ISL_1691924, EPI_ISL_1691925, EPI_ISL_1691926, EPI_ISL_1691927, EPI_ISL_1691928, EPI_ISL_1691930, EPI_ISL_1691931, EPI_ISL_1691932, EPI_ISL_1691933, EPI_ISL_1691934, EPI_ISL_1691935, EPI_ISL_1691936, EPI_ISL_1691937, EPI_ISL_1691938, EPI_ISL_1691939, EPI_ISL_1691940, EPI_ISL_1691941, EPI_ISL_1691942, EPI_ISL_1691943, EPI_ISL_1691944, EPI_ISL_1691945, EPI_ISL_1691946, EPI_ISL_1691947, EPI_ISL_1691948, EPI_ISL_1691949, EPI_ISL_1691951, EPI_ISL_1691952, EPI_ISL_1691953, EPI_ISL_1691954, EPI_ISL_1691955, EPI_ISL_1691956, EPI_ISL_1691957, EPI_ISL_1691958, EPI_ISL_1691959, EPI_ISL_1691960, EPI_ISL_1691961, EPI_ISL_1691962, EPI_ISL_1691963, EPI_ISL_1691964, EPI_ISL_1691965, EPI_ISL_1691966, EPI_ISL_1691967, EPI_ISL_1691968, EPI_ISL_1691969, EPI_ISL_1691970, EPI_ISL_1691971, EPI_ISL_1691972, EPI_ISL_1691973, EPI_ISL_1691974, EPI_ISL_1691975, EPI_ISL_1691976, EPI_ISL_1691977, EPI_ISL_1691978, EPI_ISL_1691979, EPI_ISL_1691980, EPI_ISL_1691981, EPI_ISL_1691982, EPI_ISL_1691984, EPI_ISL_1691985, EPI_ISL_1691986, EPI_ISL_1691987, EPI_ISL_1691988, EPI_ISL_1691989, EPI_ISL_1691990, EPI_ISL_1691991, EPI_ISL_1691992, EPI_ISL_1691993, EPI_ISL_1691994, EPI_ISL_1691995, EPI_ISL_1691996, EPI_ISL_1691997, EPI_ISL_1691998, EPI_ISL_1691999, EPI_ISL_1692000, EPI_ISL_1692001, EPI_ISL_1692002, EPI_ISL_1692003, EPI_ISL_1692004, EPI_ISL_1692005, EPI_ISL_1692006, EPI_ISL_1692007, EPI_ISL_1692008, EPI_ISL_1692009, EPI_ISL_1692010, EPI_ISL_1692012, EPI_ISL_1692013, EPI_ISL_1692015, EPI_ISL_1692016, EPI_ISL_1692017, EPI_ISL_1692019, EPI_ISL_1692020, EPI_ISL_1692021, EPI_ISL_1692022, EPI_ISL_1692023, EPI_ISL_1692024, EPI_ISL_1692025, EPI_ISL_1692026, EPI_ISL_1692027, EPI_ISL_1692028, EPI_ISL_1692029, EPI_ISL_1692030, EPI_ISL_1692031, EPI_ISL_1692032, EPI_ISL_1692033, EPI_ISL_1692034, EPI_ISL_1692035, EPI_ISL_1692036, EPI_ISL_1692037, EPI_ISL_1692038, EPI_ISL_1692039, EPI_ISL_1692040, EPI_ISL_1692041, EPI_ISL_1692042, EPI_ISL_1692043, EPI_ISL_1692044, EPI_ISL_1692045, EPI_ISL_1692046, EPI_ISL_1692047, EPI_ISL_1692048, EPI_ISL_1692049, EPI_ISL_1692050, EPI_ISL_1692051, EPI_ISL_1692052, EPI_ISL_1692053, EPI_ISL_1692054, EPI_ISL_1692055, EPI_ISL_1692056, EPI_ISL_1692057, EPI_ISL_1692058, EPI_ISL_1692059, EPI_ISL_1692060, EPI_ISL_1692061, EPI_ISL_1692062, EPI_ISL_1692063, EPI_ISL_1692064, EPI_ISL_1692065, EPI_ISL_1692066, EPI_ISL_1692067, EPI_ISL_1692069, EPI_ISL_1692071, EPI_ISL_1692072, EPI_ISL_1692073, EPI_ISL_1692074, EPI_ISL_1692075, EPI_ISL_1692077, EPI_ISL_1692078, EPI_ISL_1692079, EPI_ISL_1692080, EPI_ISL_1692081, EPI_ISL_1692082, EPI_ISL_1692083, EPI_ISL_1692084, EPI_ISL_1692085, EPI_ISL_1692086, EPI_ISL_1692087, EPI_ISL_1692088, EPI_ISL_1692089, EPI_ISL_1692091, EPI_ISL_1692092, EPI_ISL_1692093, EPI_ISL_1692094, EPI_ISL_1692095, EPI_ISL_1692096, EPI_ISL_1692097, EPI_ISL_1692098, EPI_ISL_1692099, EPI_ISL_1692102, EPI_ISL_1692103, EPI_ISL_1692104, EPI_ISL_1692105, EPI_ISL_1692106, EPI_ISL_1692107, EPI_ISL_1692108, EPI_ISL_1692109, EPI_ISL_1692110, EPI_ISL_1692111, EPI_ISL_1692112, EPI_ISL_1692113, EPI_ISL_1692114, EPI_ISL_1692115, EPI_ISL_1692116, EPI_ISL_1692117, EPI_ISL_1692118, EPI_ISL_1692119, EPI_ISL_1692120, EPI_ISL_1692121, EPI_ISL_1692122, EPI_ISL_1692123, EPI_ISL_1692124, EPI_ISL_1692125, EPI_ISL_1692126, EPI_ISL_1692127, EPI_ISL_1692128, EPI_ISL_1692129, EPI_ISL_1692130, EPI_ISL_1692131, EPI_ISL_1692132, EPI_ISL_1692133, EPI_ISL_1692134, EPI_ISL_1692135, EPI_ISL_1692136, EPI_ISL_1692137, EPI_ISL_1692138, EPI_ISL_1692139, EPI_ISL_1692140, EPI_ISL_1692141, EPI_ISL_1692143, EPI_ISL_1692144, EPI_ISL_1692145, EPI_ISL_1692146, EPI_ISL_1692147, EPI_ISL_1692148, EPI_ISL_1692149, EPI_ISL_1692150, EPI_ISL_1692151, EPI_ISL_1692152, EPI_ISL_1692153, EPI_ISL_1692155, EPI_ISL_1692156, EPI_ISL_1692157, EPI_ISL_1692158, EPI_ISL_1692159, EPI_ISL_1692160, EPI_ISL_1692161, EPI_ISL_1692162, EPI_ISL_1692163, EPI_ISL_1692164, EPI_ISL_1692165, EPI_ISL_1692166, EPI_ISL_1692167, EPI_ISL_1692168, EPI_ISL_1692169, EPI_ISL_1692170, EPI_ISL_1692171, EPI_ISL_1692172, EPI_ISL_1692173, EPI_ISL_1692174, EPI_ISL_1692175, EPI_ISL_1692176, EPI_ISL_1692177, EPI_ISL_1692178, EPI_ISL_1692179, EPI_ISL_1692180, EPI_ISL_1692181, EPI_ISL_1692182, EPI_ISL_1692183, EPI_ISL_1692184, EPI_ISL_1692185, EPI_ISL_1692186, EPI_ISL_1692187, EPI_ISL_1692188, EPI_ISL_1692189, EPI_ISL_1692190, EPI_ISL_1692191, EPI_ISL_1692192, EPI_ISL_1692193, EPI_ISL_1692194, EPI_ISL_1692195, EPI_ISL_1692196, EPI_ISL_1692197, EPI_ISL_1692198, EPI_ISL_1692199, EPI_ISL_1692200, EPI_ISL_1692201, EPI_ISL_1692202, EPI_ISL_1692203, EPI_ISL_1692204, EPI_ISL_1692205, EPI_ISL_1692206, EPI_ISL_1692207, EPI_ISL_1692208, EPI_ISL_1692209, EPI_ISL_1692210, EPI_ISL_1692211, EPI_ISL_1692212, EPI_ISL_1692213, EPI_ISL_1692214, EPI_ISL_1692215, EPI_ISL_1692216, EPI_ISL_1692217, EPI_ISL_1692218, EPI_ISL_1692220, EPI_ISL_1692221, EPI_ISL_1692222, EPI_ISL_1692223, EPI_ISL_1692224, EPI_ISL_1692225, EPI_ISL_1692226, EPI_ISL_1692227, EPI_ISL_1692228, EPI_ISL_1692229, EPI_ISL_1692230, EPI_ISL_1692231, EPI_ISL_1692232, EPI_ISL_1692233, EPI_ISL_1692234, EPI_ISL_1692235, EPI_ISL_1692236, EPI_ISL_1692237, EPI_ISL_1692238, EPI_ISL_1692239, EPI_ISL_1692240, EPI_ISL_1692241, EPI_ISL_1692242, EPI_ISL_1692243, EPI_ISL_1692244, EPI_ISL_1692245, EPI_ISL_1692246, EPI_ISL_1692247, EPI_ISL_1692248, EPI_ISL_1692249, EPI_ISL_1692250, EPI_ISL_1692251, EPI_ISL_1692252, EPI_ISL_1692253, EPI_ISL_1692255, EPI_ISL_1692256, EPI_ISL_1692257, EPI_ISL_1692258, EPI_ISL_1692259, EPI_ISL_1692260, EPI_ISL_1692261, EPI_ISL_1692262, EPI_ISL_1692263, EPI_ISL_1692264, EPI_ISL_1692265, EPI_ISL_1692266, EPI_ISL_1692267, EPI_ISL_1692268, EPI_ISL_1692269, EPI_ISL_1692270, EPI_ISL_1692272, EPI_ISL_1692273, EPI_ISL_1692274, EPI_ISL_1692275, EPI_ISL_1692276, EPI_ISL_1692277, EPI_ISL_1692278, EPI_ISL_1692279, EPI_ISL_1692280, EPI_ISL_1692281, EPI_ISL_1692283, EPI_ISL_1692284, EPI_ISL_1692285, EPI_ISL_1692286, EPI_ISL_1692287, EPI_ISL_1692288, EPI_ISL_1692289, EPI_ISL_1692290, EPI_ISL_1692291, EPI_ISL_1692292, EPI_ISL_1692293, EPI_ISL_1692294, EPI_ISL_1692295, EPI_ISL_1692296, EPI_ISL_1692297, EPI_ISL_1692298, EPI_ISL_1692299, EPI_ISL_1692300, EPI_ISL_1692302, EPI_ISL_1692303, EPI_ISL_1692304, EPI_ISL_1692305, EPI_ISL_1692306, EPI_ISL_1692307, EPI_ISL_1692308, EPI_ISL_1692309, EPI_ISL_1692310, EPI_ISL_1692311, EPI_ISL_1692312, EPI_ISL_1692313, EPI_ISL_1692314, EPI_ISL_1692315, EPI_ISL_1692316, EPI_ISL_1692317, EPI_ISL_1692318, EPI_ISL_1692319, EPI_ISL_1692320, EPI_ISL_1692321, EPI_ISL_1692322, EPI_ISL_1692323, EPI_ISL_1692324, EPI_ISL_1692325, EPI_ISL_1692326, EPI_ISL_1692327, EPI_ISL_1692328, EPI_ISL_1692329, EPI_ISL_1692330, EPI_ISL_1692331, EPI_ISL_1692332, EPI_ISL_1692333, EPI_ISL_1692334, EPI_ISL_1692335, EPI_ISL_1692336, EPI_ISL_1692337, EPI_ISL_1692338, EPI_ISL_1692339, EPI_ISL_1692340, EPI_ISL_1692341, EPI_ISL_1692342, EPI_ISL_1692343, EPI_ISL_1692344, EPI_ISL_1692345, EPI_ISL_1692346, EPI_ISL_1692347, EPI_ISL_1692348, EPI_ISL_1692349, EPI_ISL_1692350, EPI_ISL_1692351, EPI_ISL_1692352, EPI_ISL_1692353, EPI_ISL_1692354, EPI_ISL_1692355, EPI_ISL_1692356, EPI_ISL_1692357, EPI_ISL_1692358, EPI_ISL_1692359, EPI_ISL_1692360, EPI_ISL_1692361, EPI_ISL_1692362, EPI_ISL_1692364, EPI_ISL_1692365, EPI_ISL_1692367, EPI_ISL_1692368, EPI_ISL_1692369, EPI_ISL_1692370, EPI_ISL_1692371, EPI_ISL_1692372, EPI_ISL_1692373, EPI_ISL_1692374, EPI_ISL_1692375, EPI_ISL_1692376, EPI_ISL_1692377, EPI_ISL_1692378, EPI_ISL_1692379, EPI_ISL_1692380, EPI_ISL_1692381, EPI_ISL_1692382, EPI_ISL_1692383, EPI_ISL_1692384, EPI_ISL_1692385, EPI_ISL_1692386, EPI_ISL_1692387, EPI_ISL_1692388, EPI_ISL_1692389, EPI_ISL_1692390, EPI_ISL_1692391, EPI_ISL_1692392, EPI_ISL_1692393, EPI_ISL_1692394, EPI_ISL_1692395, EPI_ISL_1692396, EPI_ISL_1692397, EPI_ISL_1692398, EPI_ISL_1692399, EPI_ISL_1692400, EPI_ISL_1692401, EPI_ISL_1692402, EPI_ISL_1692403, EPI_ISL_1692404, EPI_ISL_1692405, EPI_ISL_1692408, EPI_ISL_1692409, EPI_ISL_1692411, EPI_ISL_1692412, EPI_ISL_1692413, EPI_ISL_1692414, EPI_ISL_1692416, EPI_ISL_1692417, EPI_ISL_1692418, EPI_ISL_1692419, EPI_ISL_1692420, EPI_ISL_1692421, EPI_ISL_1692422, EPI_ISL_1692423, EPI_ISL_1692424, EPI_ISL_1692425, EPI_ISL_1692426, EPI_ISL_1692427, EPI_ISL_1692428, EPI_ISL_1692429, EPI_ISL_1692430, EPI_ISL_1692431, EPI_ISL_1692432, EPI_ISL_1692433, EPI_ISL_1692434, EPI_ISL_1692435, EPI_ISL_1692436, EPI_ISL_1692437, EPI_ISL_1692438, EPI_ISL_1692439, EPI_ISL_1692440, EPI_ISL_1692441, EPI_ISL_1692442, EPI_ISL_1692443, EPI_ISL_1692444, EPI_ISL_1692445, EPI_ISL_1692446, EPI_ISL_1692447, EPI_ISL_1692448, EPI_ISL_1692449, EPI_ISL_1692450, EPI_ISL_1692451, EPI_ISL_1692452, EPI_ISL_1692453, EPI_ISL_1692454, EPI_ISL_1692455, EPI_ISL_1692456, EPI_ISL_1692457, EPI_ISL_1692458, EPI_ISL_1692459, EPI_ISL_1692460, EPI_ISL_1692461, EPI_ISL_1692462, EPI_ISL_1692463, EPI_ISL_1692464, EPI_ISL_1692466, EPI_ISL_1692469, EPI_ISL_1692471, EPI_ISL_1692474, EPI_ISL_1692477, EPI_ISL_1692479, EPI_ISL_1692482, EPI_ISL_1692485, EPI_ISL_1692487, EPI_ISL_1692489, EPI_ISL_1692492, EPI_ISL_1692495, EPI_ISL_1692497, EPI_ISL_1692500, EPI_ISL_1692503, EPI_ISL_1692506, EPI_ISL_1692508, EPI_ISL_1692511, EPI_ISL_1692514, EPI_ISL_1692516, EPI_ISL_1692519, EPI_ISL_1692522, EPI_ISL_1692524, EPI_ISL_1692527, EPI_ISL_1692530, EPI_ISL_1692533, EPI_ISL_1692535, EPI_ISL_1692538, EPI_ISL_1692541, EPI_ISL_1692544, EPI_ISL_1692546, EPI_ISL_1692549, EPI_ISL_1692552, EPI_ISL_1692555, EPI_ISL_1692557, EPI_ISL_1692560, EPI_ISL_1692562, EPI_ISL_1692565, EPI_ISL_1692568, EPI_ISL_1692570, EPI_ISL_1692575, EPI_ISL_1692578, EPI_ISL_1692581, EPI_ISL_1692584, EPI_ISL_1692587, EPI_ISL_1692589, EPI_ISL_1692592, EPI_ISL_1692594, EPI_ISL_1692597, EPI_ISL_1692600, EPI_ISL_1692603, EPI_ISL_1692606, EPI_ISL_1692608, EPI_ISL_1692611, EPI_ISL_1692613, EPI_ISL_1692616, EPI_ISL_1692618, EPI_ISL_1692621, EPI_ISL_1692624, EPI_ISL_1692626, EPI_ISL_1692629, EPI_ISL_1692631, EPI_ISL_1692634, EPI_ISL_1692637, EPI_ISL_1692640, EPI_ISL_1692642, EPI_ISL_1692645, EPI_ISL_1692648, EPI_ISL_1692650, EPI_ISL_1692653, EPI_ISL_1692655, EPI_ISL_1692658, EPI_ISL_1692661, EPI_ISL_1692664, EPI_ISL_1692666, EPI_ISL_1692669, EPI_ISL_1692671, EPI_ISL_1692675, EPI_ISL_1692677, EPI_ISL_1692680, EPI_ISL_1692690, EPI_ISL_1692692, EPI_ISL_1692695, EPI_ISL_1692698, EPI_ISL_1692701, EPI_ISL_1692703, EPI_ISL_1692705, EPI_ISL_1692708, EPI_ISL_1692711, EPI_ISL_1692714, EPI_ISL_1692716, EPI_ISL_1692719, EPI_ISL_1692722, EPI_ISL_1692724, EPI_ISL_1692727, EPI_ISL_1692730, EPI_ISL_1692732, EPI_ISL_1692735, EPI_ISL_1692738, EPI_ISL_1692740, EPI_ISL_1692742, EPI_ISL_1692744, EPI_ISL_1692750, EPI_ISL_1692753, EPI_ISL_1692755, EPI_ISL_1692758, EPI_ISL_1692760, EPI_ISL_1692763, EPI_ISL_1692765, EPI_ISL_1692768, EPI_ISL_1692771, EPI_ISL_1692774, EPI_ISL_1692776, EPI_ISL_1692779, EPI_ISL_1692782, EPI_ISL_1692784, EPI_ISL_1692787, EPI_ISL_1692792, EPI_ISL_1692795, EPI_ISL_1692797, EPI_ISL_1692800, EPI_ISL_1692802, EPI_ISL_1692804, EPI_ISL_1692807, EPI_ISL_1692810, EPI_ISL_1692812, EPI_ISL_1692815, EPI_ISL_1692818, EPI_ISL_1692820, EPI_ISL_1692822, EPI_ISL_1692825, EPI_ISL_1692827, EPI_ISL_1692831, EPI_ISL_1692833, EPI_ISL_1692836, EPI_ISL_1692839, EPI_ISL_1692841, EPI_ISL_1692844, EPI_ISL_1692849, EPI_ISL_1692852, EPI_ISL_1692854, EPI_ISL_1692860, EPI_ISL_1692862, EPI_ISL_1692865, EPI_ISL_1692867, EPI_ISL_1692870, EPI_ISL_1692873, EPI_ISL_1692875, EPI_ISL_1692878, EPI_ISL_1692881, EPI_ISL_1692884, EPI_ISL_1692886, EPI_ISL_1692889, EPI_ISL_1692891, EPI_ISL_1692893, EPI_ISL_1692895, EPI_ISL_1692898, EPI_ISL_1692902, EPI_ISL_1692905, EPI_ISL_1692908, EPI_ISL_1692910, EPI_ISL_1692913, EPI_ISL_1692915, EPI_ISL_1692917, EPI_ISL_1692919, EPI_ISL_1692922, EPI_ISL_1692925, EPI_ISL_1692927, EPI_ISL_1692930, EPI_ISL_1692933, EPI_ISL_1692935, EPI_ISL_1692938, EPI_ISL_1692940, EPI_ISL_1692943, EPI_ISL_1692945, EPI_ISL_1692948, EPI_ISL_1692951, EPI_ISL_1692953, EPI_ISL_1692955, EPI_ISL_1692958, EPI_ISL_1692961, EPI_ISL_1692963, EPI_ISL_1692966, EPI_ISL_1692969, EPI_ISL_1692971, EPI_ISL_1692974, EPI_ISL_1692976, EPI_ISL_1692979, EPI_ISL_1692981, EPI_ISL_1692983, EPI_ISL_1692984, EPI_ISL_1692985, EPI_ISL_1692986, EPI_ISL_1692987, EPI_ISL_1692988, EPI_ISL_1692989, EPI_ISL_1692990, EPI_ISL_1692991, EPI_ISL_1692992, EPI_ISL_1692993, EPI_ISL_1692994, EPI_ISL_1692995, EPI_ISL_1692996, EPI_ISL_1692997, EPI_ISL_1692998, EPI_ISL_1692999, EPI_ISL_1693000, EPI_ISL_1693001, EPI_ISL_1693002, EPI_ISL_1693003, EPI_ISL_1693005, EPI_ISL_1693006, EPI_ISL_1693007, EPI_ISL_1693008, EPI_ISL_1693009, EPI_ISL_1693010, EPI_ISL_1693011, EPI_ISL_1693012, EPI_ISL_1693013, EPI_ISL_1693014, EPI_ISL_1693015, EPI_ISL_1693016, EPI_ISL_1693017, EPI_ISL_1693018, EPI_ISL_1693019, EPI_ISL_1693020, EPI_ISL_1693021, EPI_ISL_1693022, EPI_ISL_1693023, EPI_ISL_1693024, EPI_ISL_1693025, EPI_ISL_1693026, EPI_ISL_1693027, EPI_ISL_1693028, EPI_ISL_1693029, EPI_ISL_1693030, EPI_ISL_1693031, EPI_ISL_1693032, EPI_ISL_1693033, EPI_ISL_1693034, EPI_ISL_1693035, EPI_ISL_1693036, EPI_ISL_1693037, EPI_ISL_1693038, EPI_ISL_1693039, EPI_ISL_1693040, EPI_ISL_1693041, EPI_ISL_1693042, EPI_ISL_1693043, EPI_ISL_1693044, EPI_ISL_1693045, EPI_ISL_1693046, EPI_ISL_1693047, EPI_ISL_1693048, EPI_ISL_1693049, EPI_ISL_1693050, EPI_ISL_1693051, EPI_ISL_1693052, EPI_ISL_1693053, EPI_ISL_1693054, EPI_ISL_1693055, EPI_ISL_1693056, EPI_ISL_1693057, EPI_ISL_1693058, EPI_ISL_1693059, EPI_ISL_1693060, EPI_ISL_1693061, EPI_ISL_1693062, EPI_ISL_1693063, EPI_ISL_1693064, EPI_ISL_1693065, EPI_ISL_1693066, EPI_ISL_1693067, EPI_ISL_1693068, EPI_ISL_1693069, EPI_ISL_1693070, EPI_ISL_1693071, EPI_ISL_1693072, EPI_ISL_1693073, EPI_ISL_1693074, EPI_ISL_1693075, EPI_ISL_1693076, EPI_ISL_1693077, EPI_ISL_1693078, EPI_ISL_1693079, EPI_ISL_1693080, EPI_ISL_1693081, EPI_ISL_1693082, EPI_ISL_1693083, EPI_ISL_1693084, EPI_ISL_1693085, EPI_ISL_1693086, EPI_ISL_1693088, EPI_ISL_1693089, EPI_ISL_1693090, EPI_ISL_1693091, EPI_ISL_1693092, EPI_ISL_1693093, EPI_ISL_1693094, EPI_ISL_1693095, EPI_ISL_1693096, EPI_ISL_1693097, EPI_ISL_1693098, EPI_ISL_1693099, EPI_ISL_1693100, EPI_ISL_1693101, EPI_ISL_1693102, EPI_ISL_1693104, EPI_ISL_1693105, EPI_ISL_1693106, EPI_ISL_1693107, EPI_ISL_1693108, EPI_ISL_1693109, EPI_ISL_1693110, EPI_ISL_1693111, EPI_ISL_1693112, EPI_ISL_1693113, EPI_ISL_1693114, EPI_ISL_1693115, EPI_ISL_1693116, EPI_ISL_1693117, EPI_ISL_1693118, EPI_ISL_1693119, EPI_ISL_1693120, EPI_ISL_1693121, EPI_ISL_1693122, EPI_ISL_1693123, EPI_ISL_1693124, EPI_ISL_1693125, EPI_ISL_1693126, EPI_ISL_1693127, EPI_ISL_1693128, EPI_ISL_1693129, EPI_ISL_1693130, EPI_ISL_1693131, EPI_ISL_1693132, EPI_ISL_1693133, EPI_ISL_1693134, EPI_ISL_1693135, EPI_ISL_1693136, EPI_ISL_1693137, EPI_ISL_1693138, EPI_ISL_1693139, EPI_ISL_1693140, EPI_ISL_1693141, EPI_ISL_1693143, EPI_ISL_1693144, EPI_ISL_1693145, EPI_ISL_1693146, EPI_ISL_1693147, EPI_ISL_1693148, EPI_ISL_1693149, EPI_ISL_1693150, EPI_ISL_1693151, EPI_ISL_1693152, EPI_ISL_1693153, EPI_ISL_1693154, EPI_ISL_1693155, EPI_ISL_1693156, EPI_ISL_1693157, EPI_ISL_1693158, EPI_ISL_1693159, EPI_ISL_1693160, EPI_ISL_1693161, EPI_ISL_1693162, EPI_ISL_1693163, EPI_ISL_1693164, EPI_ISL_1693165, EPI_ISL_1693166, EPI_ISL_1693167, EPI_ISL_1693168, EPI_ISL_1693169, EPI_ISL_1693170, EPI_ISL_1693171, EPI_ISL_1693172, EPI_ISL_1693173, EPI_ISL_1693174, EPI_ISL_1693175, EPI_ISL_1693176, EPI_ISL_1693177, EPI_ISL_1693178, EPI_ISL_1693179, EPI_ISL_1693180, EPI_ISL_1693181, EPI_ISL_1693182, EPI_ISL_1693183, EPI_ISL_1693184, EPI_ISL_1693185, EPI_ISL_1693186, EPI_ISL_1693187, EPI_ISL_1693188, EPI_ISL_1693189, EPI_ISL_1693190, EPI_ISL_1693191, EPI_ISL_1693192, EPI_ISL_1693193, EPI_ISL_1693194, EPI_ISL_1693195, EPI_ISL_1693196, EPI_ISL_1693197, EPI_ISL_1693198, EPI_ISL_1693199, EPI_ISL_1693200, EPI_ISL_1693201, EPI_ISL_1693202, EPI_ISL_1693203, EPI_ISL_1693204, EPI_ISL_1693205, EPI_ISL_1693206, EPI_ISL_1693207, EPI_ISL_1693208, EPI_ISL_1693209, EPI_ISL_1693210, EPI_ISL_1693211, EPI_ISL_1693212, EPI_ISL_1693213, EPI_ISL_1693214, EPI_ISL_1693215, EPI_ISL_1693216, EPI_ISL_1693217, EPI_ISL_1693218, EPI_ISL_1693219, EPI_ISL_1693220, EPI_ISL_1693221, EPI_ISL_1693222, EPI_ISL_1693223, EPI_ISL_1693224, EPI_ISL_1693225, EPI_ISL_1693226, EPI_ISL_1693228, EPI_ISL_1693229, EPI_ISL_1693230, EPI_ISL_1693231, EPI_ISL_1693232, EPI_ISL_1693233, EPI_ISL_1693234, EPI_ISL_1693235, EPI_ISL_1693236, EPI_ISL_1693237, EPI_ISL_1693238, EPI_ISL_1693239, EPI_ISL_1693240, EPI_ISL_1693241, EPI_ISL_1693242, EPI_ISL_1693243, EPI_ISL_1693244, EPI_ISL_1693245, EPI_ISL_1693246, EPI_ISL_1693247, EPI_ISL_1693248, EPI_ISL_1693249, EPI_ISL_1693250, EPI_ISL_1693251, EPI_ISL_1693252, EPI_ISL_1693253, EPI_ISL_1693254, EPI_ISL_1693255, EPI_ISL_1693256, EPI_ISL_1693257, EPI_ISL_1693258, EPI_ISL_1693260, EPI_ISL_1693261, EPI_ISL_1693262, EPI_ISL_1693263, EPI_ISL_1693264, EPI_ISL_1693265, EPI_ISL_1693266, EPI_ISL_1693267, EPI_ISL_1693268, EPI_ISL_1693269, EPI_ISL_1693270, EPI_ISL_1693271, EPI_ISL_1693273, EPI_ISL_1693274, EPI_ISL_1693275, EPI_ISL_1693276, EPI_ISL_1693277, EPI_ISL_1693278, EPI_ISL_1693279, EPI_ISL_1693280, EPI_ISL_1693281, EPI_ISL_1693282, EPI_ISL_1693283, EPI_ISL_1693284, EPI_ISL_1693285, EPI_ISL_1693286, EPI_ISL_1693287, EPI_ISL_1693288, EPI_ISL_1693289, EPI_ISL_1693290, EPI_ISL_1693291, EPI_ISL_1693292, EPI_ISL_1693293, EPI_ISL_1693294, EPI_ISL_1693295, EPI_ISL_1693296, EPI_ISL_1693297, EPI_ISL_1693298, EPI_ISL_1693299, EPI_ISL_1693300, EPI_ISL_1693301, EPI_ISL_1693302, EPI_ISL_1693303, EPI_ISL_1693304, EPI_ISL_1693305, EPI_ISL_1693306, EPI_ISL_1693308, EPI_ISL_1693309, EPI_ISL_1693310, EPI_ISL_1693311, EPI_ISL_1693312, EPI_ISL_1693314, EPI_ISL_1693315, EPI_ISL_1693316, EPI_ISL_1693317, EPI_ISL_1693318, EPI_ISL_1693319, EPI_ISL_1693320, EPI_ISL_1693321, EPI_ISL_1693322, EPI_ISL_1693323, EPI_ISL_1693324, EPI_ISL_1693325, EPI_ISL_1693326, EPI_ISL_1693327, EPI_ISL_1693328, EPI_ISL_1693331, EPI_ISL_1693332, EPI_ISL_1693333, EPI_ISL_1693334, EPI_ISL_1693335, EPI_ISL_1693336, EPI_ISL_1693338, EPI_ISL_1693340, EPI_ISL_1693342, EPI_ISL_1693343, EPI_ISL_1693344, EPI_ISL_1693345, EPI_ISL_1693347, EPI_ISL_1693348, EPI_ISL_1693349, EPI_ISL_1693350, EPI_ISL_1693351, EPI_ISL_1693352, EPI_ISL_1693353, EPI_ISL_1693354, EPI_ISL_1693355, EPI_ISL_1693356, EPI_ISL_1693357, EPI_ISL_1693358, EPI_ISL_1693359, EPI_ISL_1693360, EPI_ISL_1693361, EPI_ISL_1693362, EPI_ISL_1693363, EPI_ISL_1693364, EPI_ISL_1693365, EPI_ISL_1693366, EPI_ISL_1693367, EPI_ISL_1693368, EPI_ISL_1693369, EPI_ISL_1693370, EPI_ISL_1693372, EPI_ISL_1693373, EPI_ISL_1693374, EPI_ISL_1693375, EPI_ISL_1693376, EPI_ISL_1693377, EPI_ISL_1693378, EPI_ISL_1693379, EPI_ISL_1693380, EPI_ISL_1693382, EPI_ISL_1693383, EPI_ISL_1693384, EPI_ISL_1693385, EPI_ISL_1693386, EPI_ISL_1693387, EPI_ISL_1693388, EPI_ISL_1693389, EPI_ISL_1693390, EPI_ISL_1693391, EPI_ISL_1693392, EPI_ISL_1693393, EPI_ISL_1693394, EPI_ISL_1693395, EPI_ISL_1693396, EPI_ISL_1693397, EPI_ISL_1693398, EPI_ISL_1693399, EPI_ISL_1693400, EPI_ISL_1693401, EPI_ISL_1693402, EPI_ISL_1693403, EPI_ISL_1693405, EPI_ISL_1693406, EPI_ISL_1693408, EPI_ISL_1693409, EPI_ISL_1693411, EPI_ISL_1693412, EPI_ISL_1693413, EPI_ISL_1693414, EPI_ISL_1693415, EPI_ISL_1693416, EPI_ISL_1693417, EPI_ISL_1693418, EPI_ISL_1693419, EPI_ISL_1693420, EPI_ISL_1693421, EPI_ISL_1693422, EPI_ISL_1693423, EPI_ISL_1693424, EPI_ISL_1693425, EPI_ISL_1693426, EPI_ISL_1693427, EPI_ISL_1693429, EPI_ISL_1693430, EPI_ISL_1693431, EPI_ISL_1693432, EPI_ISL_1693433, EPI_ISL_1693434, EPI_ISL_1693435, EPI_ISL_1693436, EPI_ISL_1693438, EPI_ISL_1693439, EPI_ISL_1693440, EPI_ISL_1693441, EPI_ISL_1693442, EPI_ISL_1693443, EPI_ISL_1693444, EPI_ISL_1693445, EPI_ISL_1693446, EPI_ISL_1693447, EPI_ISL_1693448, EPI_ISL_1693449, EPI_ISL_1693451, EPI_ISL_1693452, EPI_ISL_1693453, EPI_ISL_1693454, EPI_ISL_1693455, EPI_ISL_1693456, EPI_ISL_1693457, EPI_ISL_1693458, EPI_ISL_1693459, EPI_ISL_1693460, EPI_ISL_1693461, EPI_ISL_1693462, EPI_ISL_1693463, EPI_ISL_1693464, EPI_ISL_1693465, EPI_ISL_1693466, EPI_ISL_1693467, EPI_ISL_1693468, EPI_ISL_1693469, EPI_ISL_1693470, EPI_ISL_1693471, EPI_ISL_1693472, EPI_ISL_1693473, EPI_ISL_1693476, EPI_ISL_1693477, EPI_ISL_1693478, EPI_ISL_1693480, EPI_ISL_1693481, EPI_ISL_1693482, EPI_ISL_1693483, EPI_ISL_1693484, EPI_ISL_1693485, EPI_ISL_1693486, EPI_ISL_1693487, EPI_ISL_1693488, EPI_ISL_1693489, EPI_ISL_1693490, EPI_ISL_1693491, EPI_ISL_1693492, EPI_ISL_1693493, EPI_ISL_1693494, EPI_ISL_1693495, EPI_ISL_1693496, EPI_ISL_1693497, EPI_ISL_1693498, EPI_ISL_1693499, EPI_ISL_1693500, EPI_ISL_1693502, EPI_ISL_1693503, EPI_ISL_1693504, EPI_ISL_1693505, EPI_ISL_1693506, EPI_ISL_1693507, EPI_ISL_1693508, EPI_ISL_1693509, EPI_ISL_1693510, EPI_ISL_1693511, EPI_ISL_1693512, EPI_ISL_1693513, EPI_ISL_1693514, EPI_ISL_1693515, EPI_ISL_1693516, EPI_ISL_1693517, EPI_ISL_1693518, EPI_ISL_1693519, EPI_ISL_1693520, EPI_ISL_1693521, EPI_ISL_1693522, EPI_ISL_1693523, EPI_ISL_1693524, EPI_ISL_1693525, EPI_ISL_1693526, EPI_ISL_1693527, EPI_ISL_1693528, EPI_ISL_1693529, EPI_ISL_1693530, EPI_ISL_1693531, EPI_ISL_1693533, EPI_ISL_1693534, EPI_ISL_1693535, EPI_ISL_1693536, EPI_ISL_1693537, EPI_ISL_1693538, EPI_ISL_1693539, EPI_ISL_1693541, EPI_ISL_1693542, EPI_ISL_1693545, EPI_ISL_1693546, EPI_ISL_1693547, EPI_ISL_1693548, EPI_ISL_1693549, EPI_ISL_1693550, EPI_ISL_1693551, EPI_ISL_1693552, EPI_ISL_1693553, EPI_ISL_1693554, EPI_ISL_1693555, EPI_ISL_1693556, EPI_ISL_1693558, EPI_ISL_1693559, EPI_ISL_1693560, EPI_ISL_1693562, EPI_ISL_1693563, EPI_ISL_1693564, EPI_ISL_1693565, EPI_ISL_1693566, EPI_ISL_1693567, EPI_ISL_1693568, EPI_ISL_1693569, EPI_ISL_1693570, EPI_ISL_1693571, EPI_ISL_1693572, EPI_ISL_1693573, EPI_ISL_1693574, EPI_ISL_1693576, EPI_ISL_1693578, EPI_ISL_1693579, EPI_ISL_1693580, EPI_ISL_1693581, EPI_ISL_1693582, EPI_ISL_1693583, EPI_ISL_1693584, EPI_ISL_1693585, EPI_ISL_1693586, EPI_ISL_1693587, EPI_ISL_1693588, EPI_ISL_1693589, EPI_ISL_1693590, EPI_ISL_1693591, EPI_ISL_1693592, EPI_ISL_1693593, EPI_ISL_1693596, EPI_ISL_1693597, EPI_ISL_1693598, EPI_ISL_1693599, EPI_ISL_1693600, EPI_ISL_1693601, EPI_ISL_1693602, EPI_ISL_1693603, EPI_ISL_1693604, EPI_ISL_1693605, EPI_ISL_1693606, EPI_ISL_1693607, EPI_ISL_1693608, EPI_ISL_1693609, EPI_ISL_1693610, EPI_ISL_1693611, EPI_ISL_1693612, EPI_ISL_1693613, EPI_ISL_1693614, EPI_ISL_1693615, EPI_ISL_1693617, EPI_ISL_1693618, EPI_ISL_1693619, EPI_ISL_1693621, EPI_ISL_1693622, EPI_ISL_1693623, EPI_ISL_1693624, EPI_ISL_1693625, EPI_ISL_1693626, EPI_ISL_1693627, EPI_ISL_1693628, EPI_ISL_1693629, EPI_ISL_1693630, EPI_ISL_1693631, EPI_ISL_1693632, EPI_ISL_1693633, EPI_ISL_1693634, EPI_ISL_1693635, EPI_ISL_1693636, EPI_ISL_1693637, EPI_ISL_1693638, EPI_ISL_1693639, EPI_ISL_1693640, EPI_ISL_1693642, EPI_ISL_1693643, EPI_ISL_1693644, EPI_ISL_1693645, EPI_ISL_1693646, EPI_ISL_1693647, EPI_ISL_1693648, EPI_ISL_1693649, EPI_ISL_1693650, EPI_ISL_1693651, EPI_ISL_1693652, EPI_ISL_1693653, EPI_ISL_1693654, EPI_ISL_1693655, EPI_ISL_1693656, EPI_ISL_1693657, EPI_ISL_1693658, EPI_ISL_1693659, EPI_ISL_1693660, EPI_ISL_1693661, EPI_ISL_1693662, EPI_ISL_1693663, EPI_ISL_1693664, EPI_ISL_1693665, EPI_ISL_1693666, EPI_ISL_1693667, EPI_ISL_1693668, EPI_ISL_1693669, EPI_ISL_1693670, EPI_ISL_1693671, EPI_ISL_1693672, EPI_ISL_1693673, EPI_ISL_1693674, EPI_ISL_1693675, EPI_ISL_1693676, EPI_ISL_1693677, EPI_ISL_1693678, EPI_ISL_1693679, EPI_ISL_1693680, EPI_ISL_1693681, EPI_ISL_1693683, EPI_ISL_1693684, EPI_ISL_1693685, EPI_ISL_1693686, EPI_ISL_1693687, EPI_ISL_1693688, EPI_ISL_1693689, EPI_ISL_1693690, EPI_ISL_1693691, EPI_ISL_1693692, EPI_ISL_1693693, EPI_ISL_1693694, EPI_ISL_1693695, EPI_ISL_1693696, EPI_ISL_1693698, EPI_ISL_1693699, EPI_ISL_1693700, EPI_ISL_1693701, EPI_ISL_1693702, EPI_ISL_1693703, EPI_ISL_1693704, EPI_ISL_1693705, EPI_ISL_1693706, EPI_ISL_1693707, EPI_ISL_1693708, EPI_ISL_1693710, EPI_ISL_1693711, EPI_ISL_1693712, EPI_ISL_1693713, EPI_ISL_1693715, EPI_ISL_1693716, EPI_ISL_1693717, EPI_ISL_1693718, EPI_ISL_1693719, EPI_ISL_1693721, EPI_ISL_1693722, EPI_ISL_1693723, EPI_ISL_1693724, EPI_ISL_1693725, EPI_ISL_1693726, EPI_ISL_1693727, EPI_ISL_1693728, EPI_ISL_1693729, EPI_ISL_1693730, EPI_ISL_1693731, EPI_ISL_1693732, EPI_ISL_1693733, EPI_ISL_1693734, EPI_ISL_1693735, EPI_ISL_1693736, EPI_ISL_1693737, EPI_ISL_1693738, EPI_ISL_1693739, EPI_ISL_1693740, EPI_ISL_1693741, EPI_ISL_1693742, EPI_ISL_1693743, EPI_ISL_1693744, EPI_ISL_1693745, EPI_ISL_1693746, EPI_ISL_1693747, EPI_ISL_1693748, EPI_ISL_1693749, EPI_ISL_1693750, EPI_ISL_1693752, EPI_ISL_1693753, EPI_ISL_1693754, EPI_ISL_1693755, EPI_ISL_1693756, EPI_ISL_1693757, EPI_ISL_1693758, EPI_ISL_1693759, EPI_ISL_1693760, EPI_ISL_1693761, EPI_ISL_1693762, EPI_ISL_1693763, EPI_ISL_1693765, EPI_ISL_1693766, EPI_ISL_1693767, EPI_ISL_1693768, EPI_ISL_1693769, EPI_ISL_1693770, EPI_ISL_1693771, EPI_ISL_1693772, EPI_ISL_1693773, EPI_ISL_1693774, EPI_ISL_1693775, EPI_ISL_1693776, EPI_ISL_1693777, EPI_ISL_1693778, EPI_ISL_1693779, EPI_ISL_1693780, EPI_ISL_1693781, EPI_ISL_1693782, EPI_ISL_1693783, EPI_ISL_1693784, EPI_ISL_1693785, EPI_ISL_1693786, EPI_ISL_1693787, EPI_ISL_1693788, EPI_ISL_1693789, EPI_ISL_1693790, EPI_ISL_1693791, EPI_ISL_1693792, EPI_ISL_1693793, EPI_ISL_1693794, EPI_ISL_1693795, EPI_ISL_1693796, EPI_ISL_1693797, EPI_ISL_1693798, EPI_ISL_1693799, EPI_ISL_1693800, EPI_ISL_1693801, EPI_ISL_1693802, EPI_ISL_1693803, EPI_ISL_1693804, EPI_ISL_1693805, EPI_ISL_1693806, EPI_ISL_1693807, EPI_ISL_1693808, EPI_ISL_1693809, EPI_ISL_1693810, EPI_ISL_1693812, EPI_ISL_1693813, EPI_ISL_1693814, EPI_ISL_1693815, EPI_ISL_1693816, EPI_ISL_1693817, EPI_ISL_1693818, EPI_ISL_1693819, EPI_ISL_1693820, EPI_ISL_1693821, EPI_ISL_1693822, EPI_ISL_1693823, EPI_ISL_1693824, EPI_ISL_1693825, EPI_ISL_1693826, EPI_ISL_1693827, EPI_ISL_1693828, EPI_ISL_1693829, EPI_ISL_1693830, EPI_ISL_1693831, EPI_ISL_1693832, EPI_ISL_1693833, EPI_ISL_1693834, EPI_ISL_1693835, EPI_ISL_1693836, EPI_ISL_1693837, EPI_ISL_1693838, EPI_ISL_1693839, EPI_ISL_1693840, EPI_ISL_1693841, EPI_ISL_1693842, EPI_ISL_1693843, EPI_ISL_1693844, EPI_ISL_1693845, EPI_ISL_1693846, EPI_ISL_1693847, EPI_ISL_1693848, EPI_ISL_1693849, EPI_ISL_1693850, EPI_ISL_1693851, EPI_ISL_1693852, EPI_ISL_1693853, EPI_ISL_1693854, EPI_ISL_1693855, EPI_ISL_1693856, EPI_ISL_1693857, EPI_ISL_1693858, EPI_ISL_1693859, EPI_ISL_1693860, EPI_ISL_1693861, EPI_ISL_1693862, EPI_ISL_1693863, EPI_ISL_1693864, EPI_ISL_1693865, EPI_ISL_1693866, EPI_ISL_1693867, EPI_ISL_1693868, EPI_ISL_1693869, EPI_ISL_1693870, EPI_ISL_1693871, EPI_ISL_1693872, EPI_ISL_1693873, EPI_ISL_1693874, EPI_ISL_1693875, EPI_ISL_1693876, EPI_ISL_1693877, EPI_ISL_1693878, EPI_ISL_1693879, EPI_ISL_1693880, EPI_ISL_1693881, EPI_ISL_1693882, EPI_ISL_1693884, EPI_ISL_1693885, EPI_ISL_1693886, EPI_ISL_1693888, EPI_ISL_1693889, EPI_ISL_1693890, EPI_ISL_1693891, EPI_ISL_1693892, EPI_ISL_1693893, EPI_ISL_1693894, EPI_ISL_1693895, EPI_ISL_1693896, EPI_ISL_1693897, EPI_ISL_1693898, EPI_ISL_1693899, EPI_ISL_1693900, EPI_ISL_1693901, EPI_ISL_1693902, EPI_ISL_1693903, EPI_ISL_1693904, EPI_ISL_1693905, EPI_ISL_1693907, EPI_ISL_1693908, EPI_ISL_1693909, EPI_ISL_1693910, EPI_ISL_1693911, EPI_ISL_1693912, EPI_ISL_1693913, EPI_ISL_1693914, EPI_ISL_1693915, EPI_ISL_1693916, EPI_ISL_1693917, EPI_ISL_1693919, EPI_ISL_1693920, EPI_ISL_1693921, EPI_ISL_1693922, EPI_ISL_1693923, EPI_ISL_1693924, EPI_ISL_1693925, EPI_ISL_1693926, EPI_ISL_1693927, EPI_ISL_1693929, EPI_ISL_1693930, EPI_ISL_1693931, EPI_ISL_1693932, EPI_ISL_1693933, EPI_ISL_1693934, EPI_ISL_1693935, EPI_ISL_1693936, EPI_ISL_1693937, EPI_ISL_1693938, EPI_ISL_1693939, EPI_ISL_1693940, EPI_ISL_1693941, EPI_ISL_1693942, EPI_ISL_1693943, EPI_ISL_1693945, EPI_ISL_1693946, EPI_ISL_1693947, EPI_ISL_1693948, EPI_ISL_1693949, EPI_ISL_1693950, EPI_ISL_1693951, EPI_ISL_1693952, EPI_ISL_1693953, EPI_ISL_1693954, EPI_ISL_1693955, EPI_ISL_1693956, EPI_ISL_1693957, EPI_ISL_1693958, EPI_ISL_1693959, EPI_ISL_1693960, EPI_ISL_1693961, EPI_ISL_1693962, EPI_ISL_1693963, EPI_ISL_1693964, EPI_ISL_1693965, EPI_ISL_1693966, EPI_ISL_1693967, EPI_ISL_1693968, EPI_ISL_1693969, EPI_ISL_1693970, EPI_ISL_1693971, EPI_ISL_1693972, EPI_ISL_1693973, EPI_ISL_1693974, EPI_ISL_1693975, EPI_ISL_1693976, EPI_ISL_1693977, EPI_ISL_1693978, EPI_ISL_1693979, EPI_ISL_1693980, EPI_ISL_1693981, EPI_ISL_1693982, EPI_ISL_1693983, EPI_ISL_1693984, EPI_ISL_1693985, EPI_ISL_1693987, EPI_ISL_1693988, EPI_ISL_1693989, EPI_ISL_1693990, EPI_ISL_1693991, EPI_ISL_1693992, EPI_ISL_1693993, EPI_ISL_1693994, EPI_ISL_1693995, EPI_ISL_1693996, EPI_ISL_1693997, EPI_ISL_1693998, EPI_ISL_1693999, EPI_ISL_1694000, EPI_ISL_1694001, EPI_ISL_1694002, EPI_ISL_1694003, EPI_ISL_1694004, EPI_ISL_1694005, EPI_ISL_1694006, EPI_ISL_1694007, EPI_ISL_1694008, EPI_ISL_1694009, EPI_ISL_1694010, EPI_ISL_1694011, EPI_ISL_1694012, EPI_ISL_1694013, EPI_ISL_1694014, EPI_ISL_1694015, EPI_ISL_1694016, EPI_ISL_1694017, EPI_ISL_1694018, EPI_ISL_1694019, EPI_ISL_1694020, EPI_ISL_1694021, EPI_ISL_1694022, EPI_ISL_1694023, EPI_ISL_1694024, EPI_ISL_1694025, EPI_ISL_1694026, EPI_ISL_1694027, EPI_ISL_1694028, EPI_ISL_1694029, EPI_ISL_1694030, EPI_ISL_1694031, EPI_ISL_1694032, EPI_ISL_1694033, EPI_ISL_1694034, EPI_ISL_1694035, EPI_ISL_1694036, EPI_ISL_1694037, EPI_ISL_1694038, EPI_ISL_1694039, EPI_ISL_1694040, EPI_ISL_1694041, EPI_ISL_1694042, EPI_ISL_1694043, EPI_ISL_1694044, EPI_ISL_1694045, EPI_ISL_1694046, EPI_ISL_1694047, EPI_ISL_1694048, EPI_ISL_1694049, EPI_ISL_1694050, EPI_ISL_1694051, EPI_ISL_1694052, EPI_ISL_1694053, EPI_ISL_1694054, EPI_ISL_1694055, EPI_ISL_1694056, EPI_ISL_1694057, EPI_ISL_1694058, EPI_ISL_1694059, EPI_ISL_1694060, EPI_ISL_1694061, EPI_ISL_1694062, EPI_ISL_1694063, EPI_ISL_1694064, EPI_ISL_1694065, EPI_ISL_1694066, EPI_ISL_1694067, EPI_ISL_1694068, EPI_ISL_1694069, EPI_ISL_1694070, EPI_ISL_1694071, EPI_ISL_1694072, EPI_ISL_1694073, EPI_ISL_1694074, EPI_ISL_1694075, EPI_ISL_1694076, EPI_ISL_1694077, EPI_ISL_1694078, EPI_ISL_1694079, EPI_ISL_1694080, EPI_ISL_1694082, EPI_ISL_1694083, EPI_ISL_1694084, EPI_ISL_1694085, EPI_ISL_1694086, EPI_ISL_1694087, EPI_ISL_1694088, EPI_ISL_1694089, EPI_ISL_1694090, EPI_ISL_1694091, EPI_ISL_1694092, EPI_ISL_1694093, EPI_ISL_1694094, EPI_ISL_1694095, EPI_ISL_1694096, EPI_ISL_1694097, EPI_ISL_1694098, EPI_ISL_1694099, EPI_ISL_1694101, EPI_ISL_1694102, EPI_ISL_1694103, EPI_ISL_1694104, EPI_ISL_1694105, EPI_ISL_1694106, EPI_ISL_1694107, EPI_ISL_1694108, EPI_ISL_1694109, EPI_ISL_1694110, EPI_ISL_1694111, EPI_ISL_1694112, EPI_ISL_1694113, EPI_ISL_1694114, EPI_ISL_1694115, EPI_ISL_1694116, EPI_ISL_1694117, EPI_ISL_1694118, EPI_ISL_1694119, EPI_ISL_1694120, EPI_ISL_1694121, EPI_ISL_1694122, EPI_ISL_1694123, EPI_ISL_1694124, EPI_ISL_1694125, EPI_ISL_1694126, EPI_ISL_1694127, EPI_ISL_1694128, EPI_ISL_1694129, EPI_ISL_1694130, EPI_ISL_1694131, EPI_ISL_1694132, EPI_ISL_1694133, EPI_ISL_1694134, EPI_ISL_1694135, EPI_ISL_1694136, EPI_ISL_1694137, EPI_ISL_1694138, EPI_ISL_1694139, EPI_ISL_1694140, EPI_ISL_1694141, EPI_ISL_1694142, | | | |

| | | | |
|---|---|---|---|
| EPI_ISL_1694143, EPI_ISL_1694145, EPI_ISL_1694146, EPI_ISL_1694147, EPI_ISL_1694148, EPI_ISL_1694149, EPI_ISL_1694150, EPI_ISL_1694151, EPI_ISL_1694152, EPI_ISL_1694153, EPI_ISL_1694154, EPI_ISL_1694155, EPI_ISL_1694156, EPI_ISL_1694157, EPI_ISL_1694158, EPI_ISL_1694159, EPI_ISL_1694160, EPI_ISL_1694161, EPI_ISL_1694162, EPI_ISL_1694163, EPI_ISL_1694165, EPI_ISL_1694166, EPI_ISL_1694167, EPI_ISL_1694168, EPI_ISL_1694169, EPI_ISL_1694170, EPI_ISL_1694171, EPI_ISL_1694172, EPI_ISL_1694173, EPI_ISL_1694174, EPI_ISL_1694175, EPI_ISL_1694176, EPI_ISL_1694177, EPI_ISL_1694178, EPI_ISL_1694179, EPI_ISL_1694180, EPI_ISL_1694181, EPI_ISL_1694182, EPI_ISL_1694183, EPI_ISL_1694184, EPI_ISL_1694185, EPI_ISL_1694186, EPI_ISL_1694187, EPI_ISL_1694188, EPI_ISL_1694189, EPI_ISL_1694190, EPI_ISL_1694191, EPI_ISL_1694192, EPI_ISL_1694193, EPI_ISL_1694194, EPI_ISL_1694195, EPI_ISL_1694196, EPI_ISL_1694197, EPI_ISL_1694198, EPI_ISL_1694199, EPI_ISL_1694200, EPI_ISL_1694201, EPI_ISL_1694202, EPI_ISL_1694203, EPI_ISL_1694204, EPI_ISL_1694205, EPI_ISL_1694206, EPI_ISL_1694207, EPI_ISL_1694208, EPI_ISL_1694209, EPI_ISL_1694210, EPI_ISL_1694211, EPI_ISL_1694212, EPI_ISL_1694213, EPI_ISL_1694214, EPI_ISL_1694215, EPI_ISL_1694217, EPI_ISL_1694218, EPI_ISL_1694220, EPI_ISL_1694221, EPI_ISL_1694222, EPI_ISL_1694223, EPI_ISL_1694224, EPI_ISL_1694225, EPI_ISL_1694226, EPI_ISL_1694227, EPI_ISL_1694228, EPI_ISL_1694229, EPI_ISL_1694230, EPI_ISL_1694231, EPI_ISL_1694232, EPI_ISL_1694233, EPI_ISL_1694234, EPI_ISL_1694235, EPI_ISL_1694236, EPI_ISL_1694237, EPI_ISL_1694238, EPI_ISL_1694239, EPI_ISL_1694240, EPI_ISL_1694241, EPI_ISL_1694242, EPI_ISL_1694243, EPI_ISL_1694244, EPI_ISL_1694245, EPI_ISL_1694246, EPI_ISL_1694247, EPI_ISL_1694248, EPI_ISL_1694249, EPI_ISL_1694251, EPI_ISL_1694252, EPI_ISL_1694253, EPI_ISL_1694254, EPI_ISL_1694255, EPI_ISL_1694256, EPI_ISL_1694257, EPI_ISL_1694258, EPI_ISL_1694259, EPI_ISL_1694260, EPI_ISL_1694261, EPI_ISL_1694262, EPI_ISL_1694263, EPI_ISL_1694264, EPI_ISL_1694265, EPI_ISL_1694266, EPI_ISL_1694267, EPI_ISL_1694268, EPI_ISL_1694269, EPI_ISL_1694270, EPI_ISL_1694271, EPI_ISL_1694272, EPI_ISL_1694273, EPI_ISL_1694274, EPI_ISL_1694275, EPI_ISL_1694276, EPI_ISL_1694277, EPI_ISL_1694278, EPI_ISL_1694279, EPI_ISL_1694280, EPI_ISL_1694281, EPI_ISL_1694282, EPI_ISL_1694283, EPI_ISL_1694284, EPI_ISL_1694285, EPI_ISL_1694286, EPI_ISL_1694287, EPI_ISL_1694288, EPI_ISL_1694289, EPI_ISL_1694290, EPI_ISL_1694291, EPI_ISL_1694292, EPI_ISL_1694293, EPI_ISL_1694294, EPI_ISL_1694295, EPI_ISL_1694296, EPI_ISL_1694297, EPI_ISL_1694298, EPI_ISL_1694299, EPI_ISL_1694300, EPI_ISL_1694301, EPI_ISL_1694302, EPI_ISL_1694303, EPI_ISL_1694304, EPI_ISL_1694305, EPI_ISL_1694306, EPI_ISL_1694307, EPI_ISL_1694308, EPI_ISL_1694309, EPI_ISL_1694310, EPI_ISL_1694311, EPI_ISL_1694312, EPI_ISL_1694313, EPI_ISL_1694314, EPI_ISL_1694315, EPI_ISL_1694316, EPI_ISL_1694317, EPI_ISL_1694318, EPI_ISL_1694319, EPI_ISL_1694320, EPI_ISL_1694321, EPI_ISL_1694322, EPI_ISL_1694323, EPI_ISL_1694324, EPI_ISL_1694325, EPI_ISL_1694326, EPI_ISL_1694327, EPI_ISL_1694328, EPI_ISL_1694329, EPI_ISL_1694330, EPI_ISL_1694331, EPI_ISL_1694332, EPI_ISL_1694333, EPI_ISL_1694334, EPI_ISL_1694335, EPI_ISL_1694336, EPI_ISL_1694337, EPI_ISL_1694338, EPI_ISL_1694339, EPI_ISL_1694340, EPI_ISL_1694341, EPI_ISL_1694342, EPI_ISL_1694343, EPI_ISL_1694344, EPI_ISL_1694345, EPI_ISL_1694346, EPI_ISL_1694347, EPI_ISL_1694348, EPI_ISL_1694349, EPI_ISL_1694350, EPI_ISL_1694351, EPI_ISL_1694353, EPI_ISL_1694354, EPI_ISL_1694355, EPI_ISL_1694356, EPI_ISL_1694357, EPI_ISL_1694358, EPI_ISL_1694360, EPI_ISL_1694361, EPI_ISL_1694362, EPI_ISL_1694363, EPI_ISL_1694364, EPI_ISL_1694365, EPI_ISL_1694366, EPI_ISL_1694367, EPI_ISL_1694368, EPI_ISL_1694369, EPI_ISL_1694370, EPI_ISL_1694371, EPI_ISL_1694372, EPI_ISL_1694373, EPI_ISL_1694374, EPI_ISL_1694375, EPI_ISL_1694376, EPI_ISL_1694377, EPI_ISL_1694378, EPI_ISL_1694379, EPI_ISL_1694380, EPI_ISL_1694381, EPI_ISL_1694382, EPI_ISL_1694383, EPI_ISL_1694384, EPI_ISL_1694385, EPI_ISL_1694386, EPI_ISL_1694387, EPI_ISL_1694388, EPI_ISL_1694389, EPI_ISL_1694390, EPI_ISL_1694391, EPI_ISL_1694392, EPI_ISL_1694393, EPI_ISL_1694394, EPI_ISL_1694395, EPI_ISL_1694397, EPI_ISL_1694398, EPI_ISL_1694399, EPI_ISL_1694400, EPI_ISL_1694401, EPI_ISL_1694402, EPI_ISL_1694403, EPI_ISL_1694404, EPI_ISL_1694405, EPI_ISL_1694406, EPI_ISL_1694407, EPI_ISL_1694408, EPI_ISL_1694409, EPI_ISL_1694410, EPI_ISL_1694411, EPI_ISL_1694412, EPI_ISL_1694413, EPI_ISL_1694414, EPI_ISL_1694415, EPI_ISL_1694416, EPI_ISL_1694417, EPI_ISL_1694418, EPI_ISL_1694419, EPI_ISL_1694420, EPI_ISL_1694421, EPI_ISL_1694422, EPI_ISL_1694423, EPI_ISL_1694424, EPI_ISL_1694425, EPI_ISL_1694426, EPI_ISL_1694427, EPI_ISL_1694428, EPI_ISL_1694429, EPI_ISL_1694430, EPI_ISL_1694431, EPI_ISL_1694432, EPI_ISL_1694433, EPI_ISL_1694434, EPI_ISL_1694435, EPI_ISL_1694436, EPI_ISL_1694437, EPI_ISL_1694438, EPI_ISL_1694439, EPI_ISL_1694440, EPI_ISL_1694441, EPI_ISL_1694442, EPI_ISL_1694443, EPI_ISL_1694444, EPI_ISL_1694445, EPI_ISL_1694446, EPI_ISL_1694447, EPI_ISL_1694448, EPI_ISL_1694449, EPI_ISL_1694450, EPI_ISL_1694451, EPI_ISL_1694452, EPI_ISL_1694453, EPI_ISL_1694454, EPI_ISL_1694455, EPI_ISL_1694456, EPI_ISL_1694457, EPI_ISL_1694458, EPI_ISL_1694459, EPI_ISL_1694460, EPI_ISL_1694461, EPI_ISL_1694462, EPI_ISL_1694463, EPI_ISL_1694464, EPI_ISL_1694465, EPI_ISL_1694466, EPI_ISL_1694467, EPI_ISL_1694468, EPI_ISL_1694469, EPI_ISL_1694470, EPI_ISL_1694471, EPI_ISL_1694472, EPI_ISL_1694473, EPI_ISL_1694474, EPI_ISL_1694475, EPI_ISL_1694476, EPI_ISL_1694477, EPI_ISL_1694478, EPI_ISL_1694479, EPI_ISL_1694480, EPI_ISL_1694481, EPI_ISL_1694482, EPI_ISL_1694483, EPI_ISL_1694484, EPI_ISL_1694485, EPI_ISL_1694486, EPI_ISL_1694487, EPI_ISL_1694488, EPI_ISL_1694489, EPI_ISL_1694490, EPI_ISL_1694491, EPI_ISL_1694492, EPI_ISL_1694493, EPI_ISL_1694494, EPI_ISL_1694495, EPI_ISL_1694496, EPI_ISL_1694497, EPI_ISL_1694498, EPI_ISL_1694499, EPI_ISL_1694500, EPI_ISL_1694501, EPI_ISL_1694502, EPI_ISL_1694503, EPI_ISL_1694504, EPI_ISL_1694505, EPI_ISL_1694506, EPI_ISL_1694507, EPI_ISL_1694508, EPI_ISL_1694509, EPI_ISL_1694510, EPI_ISL_1694511, EPI_ISL_1694512, EPI_ISL_1694513, EPI_ISL_1694514, EPI_ISL_1694515, EPI_ISL_1694516, EPI_ISL_1694517, EPI_ISL_1694518, EPI_ISL_1694519, EPI_ISL_1694520, EPI_ISL_1694521, EPI_ISL_1694523, EPI_ISL_1694524, EPI_ISL_1694525, EPI_ISL_1694526, EPI_ISL_1694527, EPI_ISL_1694528, EPI_ISL_1694529, EPI_ISL_1694530, EPI_ISL_1694531, EPI_ISL_1694532, EPI_ISL_1694533, EPI_ISL_1694534, EPI_ISL_1694535, EPI_ISL_1694536 | | | |
| see above | Infinity Biologix | Centers for Disease Control and Prevention Division of Viral Diseases, Pathogen Discovery | Dakota Howard, Dhwani Batra, Peter W. Cook, Kara Moser, Adrian Paskey, Jason Caravas, Benjamin Rambo-Martin, Shatavia Morrison, Christopher Gulvick, Scott Sammons, Yvette Unoarumhi, Darlene Wagner, Matthew Schmerer, Christian Bixby, Yihe Wang, Jonathan Schultz, Chirayu Goswami, Russ Hager, Robin Grimwood, Clinton R. Paden, Duncan MacCannell |
| EPI_ISL_1694635, EPI_ISL_1694636, EPI_ISL_1694637, EPI_ISL_1694638, EPI_ISL_1694639, EPI_ISL_1694640, EPI_ISL_1694641, EPI_ISL_1694642, EPI_ISL_1694643, EPI_ISL_1694644, EPI_ISL_1694645, EPI_ISL_1694646, EPI_ISL_1694647, EPI_ISL_1694648, EPI_ISL_1694649, EPI_ISL_1694650, EPI_ISL_1694651, EPI_ISL_1694652, EPI_ISL_1694653, EPI_ISL_1694654, EPI_ISL_1694655, EPI_ISL_1694656, EPI_ISL_1694657, EPI_ISL_1694658, EPI_ISL_1694659, EPI_ISL_1694660, EPI_ISL_1694661, EPI_ISL_1694662, EPI_ISL_1694663, EPI_ISL_1694664, EPI_ISL_1694665 | | | |
| see above | Helix/Illumina | Centers for Disease Control and Prevention Division of Viral Diseases, Pathogen Discovery | Dakota Howard, Dhwani Batra, Peter W. Cook, Kara Moser, Adrian Paskey, Jason Caravas, Benjamin Rambo-Martin, Shatavia Morrison, Christopher Gulvick, Scott Sammons, Yvette Unoarumhi, Darlene Wagner, Matthew Schmerer, Eileen de Feo, Jan Antico, Christine Tran, Matthew Tolentino, Shannon Wickline, Kim Gietzen, Brad Sickler, Jingtao Liu, Eric Allen, Phil Febbo, Nicole L. Washington, Simon White, Geraint Levan, Kelly Schiabor Barrett, Elizabeth Cirulli, Alexandre Bolze, Ary Ascencio, Charlotte Rivera-Garcia, Ryan Cho, Jason Nguyen, Sherry Wang, Jimmy Ramirez, Tyler Cassens, Efren Sandoval, Magnus Isaksson, William Lee, David Becker, Marc Laurent, James Lu, Clinton R. Paden, Duncan MacCannell |
| EPI_ISL_1694666, EPI_ISL_1694667, EPI_ISL_1694668, EPI_ISL_1694669, EPI_ISL_1694670, EPI_ISL_1694671, EPI_ISL_1694672, EPI_ISL_1694673 | Quest Diagnostics Incorporated | Centers for Disease Control and Prevention Division of Viral Diseases, Pathogen Discovery | Dakota Howard, Dhwani Batra, Peter W. Cook, Kara Moser, Adrian Paskey, Jason Caravas, Benjamin Rambo-Martin, Shatavia Morrison, Christopher Gulvick, Scott Sammons, Yvette Unoarumhi, Darlene Wagner, Matthew Schmerer, S. H. Rosenthal, A. Gerasimova, R. M. Kagan, B. Anderson, M. Hua, Y. Liu, L.E. Bernstein, K.E. Livingston, A. Perez, I. A. Shlyakhter, R. V. Rolando, R. Owen, P. Tanpaiboon, F. Lacbawan, Clinton R. Paden, Duncan MacCannell |
| EPI_ISL_1694674, EPI_ISL_1694675, EPI_ISL_1694676, EPI_ISL_1694677, EPI_ISL_1694678, EPI_ISL_1694679, EPI_ISL_1694680, EPI_ISL_1694681, EPI_ISL_1694682, EPI_ISL_1694683, EPI_ISL_1694684, EPI_ISL_1694685, EPI_ISL_1694686, EPI_ISL_1694687, EPI_ISL_1694688 | | | |
| see above | Helix/Illumina | Centers for Disease Control and Prevention Division of Viral Diseases, Pathogen Discovery | Dakota Howard, Dhwani Batra, Peter W. Cook, Kara Moser, Adrian Paskey, Jason Caravas, Benjamin Rambo-Martin, Shatavia Morrison, Christopher Gulvick, Scott Sammons, Yvette Unoarumhi, Darlene Wagner, Matthew Schmerer, Eileen de Feo, Jan Antico, Christine Tran, Matthew Tolentino, Shannon Wickline, Kim Gietzen, Brad Sickler, Jingtao Liu, Eric Allen, Phil Febbo, Nicole L. Washington, Simon White, Geraint Levan, Kelly Schiabor Barrett, Elizabeth Cirulli, Alexandre Bolze, Ary Ascencio, Charlotte Rivera-Garcia, Ryan Cho, Jason Nguyen, Sherry Wang, Jimmy Ramirez, Tyler Cassens, Efren Sandoval, Magnus Isaksson, William Lee, David Becker, Marc Laurent, James Lu, Clinton R. Paden, Duncan MacCannell |
| EPI_ISL_1694689, EPI_ISL_1694690, EPI_ISL_1694691, EPI_ISL_1694692, EPI_ISL_1694693, EPI_ISL_1694694, EPI_ISL_1694695 | Quest Diagnostics Incorporated | Centers for Disease Control and Prevention Division of Viral Diseases, Pathogen Discovery | Dakota Howard, Dhwani Batra, Peter W. Cook, Kara Moser, Adrian Paskey, Jason Caravas, Benjamin Rambo-Martin, Shatavia Morrison, Christopher Gulvick, Scott Sammons, Yvette Unoarumhi, Darlene Wagner, Matthew Schmerer, S. H. Rosenthal, A. Gerasimova, R. M. Kagan, B. Anderson, M. Hua, Y. Liu, L.E. Bernstein, K.E. Livingston, A. Perez, I. A. Shlyakhter, R. V. Rolando, R. Owen, P. Tanpaiboon, F. Lacbawan, Clinton R. Paden, Duncan MacCannell |
| EPI_ISL_1694696 | DOHMH Riverside | New York City Public Health Laboratory | Jade Wang, et al. |
| EPI_ISL_1694697, EPI_ISL_1694698 | DOHMH Chelsea | New York City Public Health Laboratory | Jade Wang, et al. |
| EPI_ISL_1694699 | DOHMH Jamaica | New York City Public Health Laboratory | Jade Wang, et al. |
| EPI_ISL_1694700, EPI_ISL_1694701 | DOHMH Riverside | New York City Public Health Laboratory | Jade Wang, et al. |
| EPI_ISL_1694702, EPI_ISL_1694703 | DOHMH Central Harlem | New York City Public Health Laboratory | Jade Wang, et al. |
| EPI_ISL_1694704, EPI_ISL_1694705 | DOHMH Morrisania | New York City Public Health Laboratory | Jade Wang, et al. |
| EPI_ISL_1694706 | DOHMH Chelsea | New York City Public Health Laboratory | Jade Wang, et al. |
| EPI_ISL_1694707 | DOHMH Corona | New York City Public Health Laboratory | Jade Wang, et al. |
| EPI_ISL_1694708 | DOHMH Jamaica | New York City Public Health Laboratory | Jade Wang, et al. |
| EPI_ISL_1694709 | DOHMH Central Harlem | New York City Public Health Laboratory | Jade Wang, et al. |
| EPI_ISL_1694710 | DOHMH Corona | New York City Public Health Laboratory | Jade Wang, et al. |
| EPI_ISL_1694711 | DOHMH Crown Heights | New York City Public Health Laboratory | Jade Wang, et al. |
| EPI_ISL_1694712 | DOHMH Jamaica | New York City Public Health Laboratory | Jade Wang, et al. |
| EPI_ISL_1694713, EPI_ISL_1694714 | DOHMH Corona | New York City Public Health Laboratory | Jade Wang, et al. |
| EPI_ISL_1694715, EPI_ISL_1694716 | DOHMH PHL | New York City Public Health Laboratory | Jade Wang, et al. |
| EPI_ISL_1694717 | DOHMH Morrisania | New York City Public Health Laboratory | Jade Wang, et al. |
| EPI_ISL_1694718 | DOHMH Central Harlem | New York City Public Health Laboratory | Jade Wang, et al. |
| EPI_ISL_1694719 | DOHMH Fort Greene | New York City Public Health Laboratory | Jade Wang, et al. |
| EPI_ISL_1694720 | Poplar Healthcare | New York City Public Health Laboratory | Jade Wang, et al. |
| EPI_ISL_1694721, EPI_ISL_1694722, EPI_ISL_1694723, EPI_ISL_1694724, EPI_ISL_1694725, EPI_ISL_1694726, EPI_ISL_1694727 | OCME Office Of Chief Medical Examiner | New York City Public Health Laboratory | Jade Wang, et al. |
| EPI_ISL_1694728 | DOHMH Crown Heights | New York City Public Health Laboratory | Jade Wang, et al. |
| EPI_ISL_1694729 | DOHMH Morrisania | New York City Public Health Laboratory | Jade Wang, et al. |
| EPI_ISL_1694730 | DOHMH Chelsea | New York City Public Health Laboratory | Jade Wang, et al. |
| EPI_ISL_1694731, EPI_ISL_1694732 | OCME Office Of Chief Medical Examiner | New York City Public Health Laboratory | Jade Wang, et al. |
| EPI_ISL_1694733 | DOHMH Fort Greene | New York City Public Health Laboratory | Jade Wang, et al. |
| EPI_ISL_1694734, EPI_ISL_1694735 | DOHMH Corona | New York City Public Health Laboratory | Jade Wang, et al. |
| EPI_ISL_1694736, EPI_ISL_1694737, EPI_ISL_1694738, EPI_ISL_1694739, EPI_ISL_1694740 | DOHMH Jamaica | New York City Public Health Laboratory | Jade Wang, et al. |
| EPI_ISL_1694741, EPI_ISL_1694742, EPI_ISL_1694743, EPI_ISL_1694744, EPI_ISL_1694745 | DOHMH PHL | New York City Public Health Laboratory | Jade Wang, et al. |

| | | | |
|---|---|---|---|
| EPI_ISL_1694746, EPI_ISL_1694747 | DOHMH Fort Greene | New York City Public Health Laboratory | Jade Wang, et al. |
| EPI_ISL_1694748, EPI_ISL_1694749 | DOHMH PHL | New York City Public Health Laboratory | Jade Wang, et al. |
| EPI_ISL_1694750, EPI_ISL_1694751, EPI_ISL_1694752, EPI_ISL_1694753 | DOHMH Corona | New York City Public Health Laboratory | Jade Wang, et al. |
| EPI_ISL_1694754, EPI_ISL_1694755, EPI_ISL_1694756, EPI_ISL_1694757, EPI_ISL_1694758 | DOHMH Jamaica | New York City Public Health Laboratory | Jade Wang, et al. |
| EPI_ISL_1694759 | DOHMH Central Harlem | New York City Public Health Laboratory | Jade Wang, et al. |
| EPI_ISL_1694760, EPI_ISL_1694761, EPI_ISL_1694762 | Department of Homeless Services | New York City Public Health Laboratory | Jade Wang, et al. |
| EPI_ISL_1694763, EPI_ISL_1694764, EPI_ISL_1694765, EPI_ISL_1694766, EPI_ISL_1694767, EPI_ISL_1694768, EPI_ISL_1694769, EPI_ISL_1694770, EPI_ISL_1694771, EPI_ISL_1694772 | OCME Office Of Chief Medical Examiner | New York City Public Health Laboratory | Jade Wang, et al. |
| EPI_ISL_1694773, EPI_ISL_1694774, EPI_ISL_1694775 | DOHMH Central Harlem | New York City Public Health Laboratory | Jade Wang, et al. |
| EPI_ISL_1694776 | DOHMH Morrisania | New York City Public Health Laboratory | Jade Wang, et al. |
| EPI_ISL_1694946, EPI_ISL_1694947, EPI_ISL_1694948, EPI_ISL_1694949, EPI_ISL_1694950, EPI_ISL_1694951, EPI_ISL_1694952, EPI_ISL_1694953, EPI_ISL_1694954, EPI_ISL_1694955, EPI_ISL_1694956 | | | |
| see above | Quest Diagnostics Incorporated | Centers for Disease Control and Prevention Division of Viral Diseases, Pathogen Discovery | Dakota Howard, Dhwani Batra, Peter W. Cook, Kara Moser, Adrian Paskey, Jason Caravas, Benjamin Rambo-Martin, Shatavia Morrison, Christopher Gulvick, Scott Sammons, Yvette Unoarumhi, Darlene Wagner, Matthew Schmerer, S. H. Rosenthal, A. Gerasimova, R. M. Kagan, B. Anderson, M. Hua, Y. Liu, L.E. Bernstein, K.E. Livingston, A. Perez, I. A. Shlyakhter, R. V. Rolando, R. Owen, P. Tanpaiboon, F. Lacbawan, Clinton R. Paden, Duncan MacCannell |
| EPI_ISL_1694957, EPI_ISL_1694958, EPI_ISL_1694959, EPI_ISL_1694960, EPI_ISL_1694961, EPI_ISL_1694962, EPI_ISL_1694963, EPI_ISL_1694965, EPI_ISL_1694966, EPI_ISL_1694967, EPI_ISL_1694968, EPI_ISL_1694969, EPI_ISL_1694970, EPI_ISL_1694971, EPI_ISL_1694972, EPI_ISL_1694973, EPI_ISL_1694974, EPI_ISL_1694975, EPI_ISL_1694976, EPI_ISL_1694977 | | | |
| see above | Fulgent Genetics | Centers for Disease Control and Prevention Division of Viral Diseases, Pathogen Discovery | Dakota Howard, Dhwani Batra, Peter W. Cook, Kara Moser, Adrian Paskey, Jason Caravas, Benjamin Rambo-Martin, Shatavia Morrison, Christopher Gulvick, Scott Sammons, Yvette Unoarumhi, Darlene Wagner, Matthew Schmerer, Harry Gao, Mickey Li, John Gao, Joseph Fierro, Benafsh Sapra, Becky Tsai, Yan Meng, Doreen Ng, James Xie, Clinton R. Paden, Duncan MacCannell |
| EPI_ISL_1694978, EPI_ISL_1694979, EPI_ISL_1694980, EPI_ISL_1694981, EPI_ISL_1694982, EPI_ISL_1694997, EPI_ISL_1694998, EPI_ISL_1694999, EPI_ISL_1695000, EPI_ISL_1695001, EPI_ISL_1695002, EPI_ISL_1695003, EPI_ISL_1695004, EPI_ISL_1695005, EPI_ISL_1695006, EPI_ISL_1695007, EPI_ISL_1695009, EPI_ISL_1695011, EPI_ISL_1695012, EPI_ISL_1695013, EPI_ISL_1695014, EPI_ISL_1695015, EPI_ISL_1695016, EPI_ISL_1695018, EPI_ISL_1695019, EPI_ISL_1695021, EPI_ISL_1695023, EPI_ISL_1695024, EPI_ISL_1695025, EPI_ISL_1695026, EPI_ISL_1695028, EPI_ISL_1695029 | | | |
| see above | Laboratory Corporation of America | Centers for Disease Control and Prevention Division of Viral Diseases, Pathogen Discovery | Dakota Howard, Dhwani Batra, Peter W. Cook, Kara Moser, Adrian Paskey, Jason Caravas, Benjamin Rambo-Martin, Shatavia Morrison, Christopher Gulvick, Scott Sammons, Yvette Unoarumhi, Darlene Wagner, Matthew Schmerer, Minoo Agarwal, Eyad Almasri, Debbie Boles, Ayla Burns, Nuthawin Charoensri, Oren Cohen, Susan Countryman, Mary Ann Cristobal, Bobbi Croy, Suzanne Dale, Hrushikesh Deshmukh, Amanda Douglas, Vincent Drouillon, Marcia Eisenberg, Howard Engler, Rama Ghatti, Prashant Gupta, Susan Hicks, Jake Humphrey, Lax Iyer, Manoj Jain, Mohan Kolli, Brian Krueger, Tim Kuphal, Stanley Letovsky, Michael Levandoski, Craig Lukasik, Jonathan Meltzer, Brian Norvell, Mindy Nye, Scott Parker, Christos Petropoulos, John Pruitt, Steven Ragan, Scott Ryan, Mike Sapeta, Jana Schroth, Suresh Babu Selvaraju, Goran Stevovic, Amanda Suchanek, Andrea Throop, Lyndon Tilson, Thomas Urban, Joe Voshell, Kimberly Wagner, Jonathan Williams, Mary Williamson, Qian Zeng, Tricia Zwiefelhofer, Clinton R. Paden, Duncan MacCannell |
| EPI_ISL_1695032, EPI_ISL_1695033, EPI_ISL_1695034, EPI_ISL_1695035, EPI_ISL_1695036 | Fulgent Genetics | Centers for Disease Control and Prevention Division of Viral Diseases, Pathogen Discovery | Dakota Howard, Dhwani Batra, Peter W. Cook, Kara Moser, Adrian Paskey, Jason Caravas, Benjamin Rambo-Martin, Shatavia Morrison, Christopher Gulvick, Scott Sammons, Yvette Unoarumhi, Darlene Wagner, Matthew Schmerer, Harry Gao, Mickey Li, John Gao, Joseph Fierro, Benafsh Sapra, Becky Tsai, Yan Meng, Doreen Ng, James Xie, Clinton R. Paden, Duncan MacCannell |
| EPI_ISL_1695037, EPI_ISL_1695038, EPI_ISL_1695039, EPI_ISL_1695040, EPI_ISL_1695041, EPI_ISL_1695042 | Quest Diagnostics Incorporated | Centers for Disease Control and Prevention Division of Viral Diseases, Pathogen Discovery | Dakota Howard, Dhwani Batra, Peter W. Cook, Kara Moser, Adrian Paskey, Jason Caravas, Benjamin Rambo-Martin, Shatavia Morrison, Christopher Gulvick, Scott Sammons, Yvette Unoarumhi, Darlene Wagner, Matthew Schmerer, S. H. Rosenthal, A. Gerasimova, R. M. Kagan, B. Anderson, M. Hua, Y. Liu, L.E. Bernstein, K.E. Livingston, A. Perez, I. A. Shlyakhter, R. V. Rolando, R. Owen, P. Tanpaiboon, F. Lacbawan, Clinton R. Paden, Duncan MacCannell |
| EPI_ISL_1695043, EPI_ISL_1695044, EPI_ISL_1695045, EPI_ISL_1695046, EPI_ISL_1695047, EPI_ISL_1695048, EPI_ISL_1695049 | Helix/Illumina | Centers for Disease Control and Prevention Division of Viral Diseases, Pathogen Discovery | Dakota Howard, Dhwani Batra, Peter W. Cook, Kara Moser, Adrian Paskey, Jason Caravas, Benjamin Rambo-Martin, Shatavia Morrison, Christopher Gulvick, Scott Sammons, Yvette Unoarumhi, Darlene Wagner, Matthew Schmerer, Eileen de Feo, Jan Antico, Christine Tran, Matthew Tolentino, Shannon Wickline, Kim Gietzen, Brad Sickler, Jingtao Liu, Eric Allen, Phil Febbo, Nicole L. Washington, Simon White, Geraint Levan, Kelly Schiabor Barrett, Elizabeth Cirulli, Alexandre Bolze, Ary Ascencio, Charlotte Rivera-Garcia, Ryan Cho, Jason Nguyen, Sherry Wang, Jimmy Ramirez, Tyler Cassens, Efren Sandoval, Magnus Isaksson, William Lee, David Becker, Marc Laurent, James Lu, Clinton R. Paden, Duncan MacCannell |
| EPI_ISL_1695050, EPI_ISL_1695051, EPI_ISL_1695052, EPI_ISL_1695053, EPI_ISL_1695054, EPI_ISL_1695055, EPI_ISL_1695056, EPI_ISL_1695057, EPI_ISL_1695058, EPI_ISL_1695059, EPI_ISL_1695060, EPI_ISL_1695061, EPI_ISL_1695062, EPI_ISL_1695063, EPI_ISL_1695064, EPI_ISL_1695065, EPI_ISL_1695066, EPI_ISL_1695067, EPI_ISL_1695068, EPI_ISL_1695069, EPI_ISL_1695070, EPI_ISL_1695071, EPI_ISL_1695072, EPI_ISL_1695073, EPI_ISL_1695074, EPI_ISL_1695075, EPI_ISL_1695076, EPI_ISL_1695077, EPI_ISL_1695078, EPI_ISL_1695079, EPI_ISL_1695080, EPI_ISL_1695081, EPI_ISL_1695082, EPI_ISL_1695083, EPI_ISL_1695084, EPI_ISL_1695085, EPI_ISL_1695086, EPI_ISL_1695087, EPI_ISL_1695088, EPI_ISL_1695089, EPI_ISL_1695090, EPI_ISL_1695091, EPI_ISL_1695092, EPI_ISL_1695093, EPI_ISL_1695094, EPI_ISL_1695095, EPI_ISL_1695096, EPI_ISL_1695097, EPI_ISL_1695098, EPI_ISL_1695099, EPI_ISL_1695100, EPI_ISL_1695101, EPI_ISL_1695102, EPI_ISL_1695103, EPI_ISL_1695104, EPI_ISL_1695105, EPI_ISL_1695106, EPI_ISL_1695107, EPI_ISL_1695108, EPI_ISL_1695109, EPI_ISL_1695110 | | | |
| see above | Quest Diagnostics Incorporated | Centers for Disease Control and Prevention Division of Viral Diseases, Pathogen Discovery | Dakota Howard, Dhwani Batra, Peter W. Cook, Kara Moser, Adrian Paskey, Jason Caravas, Benjamin Rambo-Martin, Shatavia Morrison, Christopher Gulvick, Scott Sammons, Yvette Unoarumhi, Darlene Wagner, Matthew Schmerer, S. H. Rosenthal, A. Gerasimova, R. M. Kagan, B. Anderson, M. Hua, Y. Liu, L.E. Bernstein, K.E. Livingston, A. Perez, I. A. Shlyakhter, R. V. Rolando, R. Owen, P. Tanpaiboon, F. Lacbawan, Clinton R. Paden, Duncan MacCannell |
| EPI_ISL_1695111, EPI_ISL_1695112, EPI_ISL_1695113, EPI_ISL_1695114, EPI_ISL_1695115, EPI_ISL_1695116, EPI_ISL_1695117, EPI_ISL_1695118, EPI_ISL_1695119, EPI_ISL_1695121, EPI_ISL_1695122, EPI_ISL_1695123, EPI_ISL_1695124, EPI_ISL_1695125, EPI_ISL_1695126, EPI_ISL_1695127, EPI_ISL_1695128, EPI_ISL_1695129, EPI_ISL_1695130, EPI_ISL_1695131, EPI_ISL_1695132, EPI_ISL_1695133, EPI_ISL_1695134, EPI_ISL_1695135, EPI_ISL_1695136, EPI_ISL_1695137, EPI_ISL_1695138, EPI_ISL_1695139, EPI_ISL_1695140, EPI_ISL_1695142, EPI_ISL_1695143, EPI_ISL_1695144, EPI_ISL_1695145, EPI_ISL_1695146, EPI_ISL_1695149, EPI_ISL_1695152, EPI_ISL_1695155, EPI_ISL_1695158, EPI_ISL_1695161, EPI_ISL_1695164, EPI_ISL_1695167, EPI_ISL_1695170, EPI_ISL_1695173, EPI_ISL_1695176, EPI_ISL_1695179, EPI_ISL_1695183, EPI_ISL_1695186, EPI_ISL_1695189, EPI_ISL_1695192, EPI_ISL_1695195, EPI_ISL_1695198, EPI_ISL_1695201, EPI_ISL_1695204, EPI_ISL_1695207, EPI_ISL_1695210, EPI_ISL_1695213, EPI_ISL_1695216, EPI_ISL_1695219, EPI_ISL_1695222, EPI_ISL_1695225, EPI_ISL_1695228, EPI_ISL_1695231, EPI_ISL_1695234, EPI_ISL_1695237, EPI_ISL_1695244, EPI_ISL_1695247, EPI_ISL_1695250, EPI_ISL_1695253, EPI_ISL_1695256, EPI_ISL_1695259, EPI_ISL_1695262, EPI_ISL_1695265, EPI_ISL_1695269, EPI_ISL_1695272, EPI_ISL_1695275, EPI_ISL_1695278, EPI_ISL_1695281, EPI_ISL_1695284, EPI_ISL_1695287, EPI_ISL_1695290, EPI_ISL_1695293, EPI_ISL_1695296, EPI_ISL_1695299, EPI_ISL_1695302, EPI_ISL_1695305, EPI_ISL_1695309, EPI_ISL_1695312, EPI_ISL_1695315, EPI_ISL_1695317, EPI_ISL_1695320, EPI_ISL_1695324, EPI_ISL_1695327, EPI_ISL_1695330, EPI_ISL_1695333, EPI_ISL_1695336, EPI_ISL_1695339, EPI_ISL_1695342, EPI_ISL_1695345, EPI_ISL_1695349, EPI_ISL_1695352, EPI_ISL_1695355, EPI_ISL_1695358, EPI_ISL_1695361, EPI_ISL_1695364, EPI_ISL_1695367, EPI_ISL_1695370, EPI_ISL_1695373, EPI_ISL_1695376, EPI_ISL_1695379, EPI_ISL_1695382, EPI_ISL_1695385, EPI_ISL_1695389, EPI_ISL_1695392, EPI_ISL_1695395, EPI_ISL_1695398, EPI_ISL_1695401, EPI_ISL_1695404, EPI_ISL_1695407, EPI_ISL_1695410, EPI_ISL_1695413, EPI_ISL_1695417, EPI_ISL_1695419, EPI_ISL_1695422, EPI_ISL_1695425, EPI_ISL_1695429, EPI_ISL_1695432, EPI_ISL_1695435, EPI_ISL_1695439, EPI_ISL_1695442, EPI_ISL_1695445, EPI_ISL_1695448, EPI_ISL_1695451, EPI_ISL_1695454, EPI_ISL_1695457, EPI_ISL_1695460, EPI_ISL_1695463, EPI_ISL_1695466, EPI_ISL_1695469, EPI_ISL_1695481, EPI_ISL_1695484, EPI_ISL_1695487, EPI_ISL_1695490, EPI_ISL_1695493, EPI_ISL_1695496, EPI_ISL_1695499, EPI_ISL_1695502, EPI_ISL_1695506, EPI_ISL_1695509, EPI_ISL_1695512, EPI_ISL_1695515, EPI_ISL_1695518, EPI_ISL_1695521, EPI_ISL_1695524, EPI_ISL_1695527, EPI_ISL_1695530, EPI_ISL_1695533, EPI_ISL_1695536, EPI_ISL_1695539, EPI_ISL_1695542, EPI_ISL_1695545, EPI_ISL_1695548, EPI_ISL_1695551, EPI_ISL_1695554, EPI_ISL_1695557, EPI_ISL_1695560, EPI_ISL_1695563, EPI_ISL_1695566, EPI_ISL_1695569, EPI_ISL_1695573, EPI_ISL_1695576, EPI_ISL_1695578, EPI_ISL_1695581, EPI_ISL_1695584, EPI_ISL_1695587, EPI_ISL_1695590, EPI_ISL_1695593, EPI_ISL_1695596, EPI_ISL_1695599, EPI_ISL_1695602, EPI_ISL_1695605, EPI_ISL_1695608 | | | |
| see above | Aegis Sciences Corporation | Centers for Disease Control and Prevention Division of Viral Diseases, Pathogen Discovery | Dakota Howard, Dhwani Batra, Peter W. Cook, Kara Moser, Adrian Paskey, Jason Caravas, Benjamin Rambo-Martin, Shatavia Morrison, Christopher Gulvick, Scott Sammons, Yvette Unoarumhi, Darlene Wagner, Matthew Schmerer, Cyndi Clark, Patrick Campbell, Rob Case, Vikramsinha Ghorpade, Holly Houdeshell, Ola Kvalvaag, Dillon Nall, Ethan Sanders, Alec Vest, Shaun Westlund, Matthew Hardison, Clinton R. Paden, Duncan MacCannell |
| EPI_ISL_1695619, EPI_ISL_1695622, EPI_ISL_1695625, EPI_ISL_1695628, EPI_ISL_1695631, EPI_ISL_1695634, EPI_ISL_1695637, EPI_ISL_1695640, EPI_ISL_1695643, EPI_ISL_1695646, EPI_ISL_1695650, EPI_ISL_1695653, EPI_ISL_1695656, EPI_ISL_1695659, EPI_ISL_1695662, EPI_ISL_1695665, EPI_ISL_1695668, EPI_ISL_1695671, EPI_ISL_1695674, EPI_ISL_1695677, EPI_ISL_1695681, EPI_ISL_1695684, EPI_ISL_1695687, EPI_ISL_1695690, EPI_ISL_1695693, EPI_ISL_1695696, EPI_ISL_1695699, EPI_ISL_1695702, EPI_ISL_1695705, EPI_ISL_1695708, EPI_ISL_1695711, EPI_ISL_1695714, EPI_ISL_1695717, EPI_ISL_1695721, EPI_ISL_1695724, EPI_ISL_1695727, EPI_ISL_1695729, EPI_ISL_1695732, EPI_ISL_1695736, EPI_ISL_1695739, EPI_ISL_1695741, EPI_ISL_1695744, EPI_ISL_1695747, EPI_ISL_1695750, EPI_ISL_1695753, EPI_ISL_1695756, EPI_ISL_1695759, EPI_ISL_1695762, EPI_ISL_1695766, EPI_ISL_1695769 | | | |
| see above | Quest Diagnostics Incorporated | Centers for Disease Control and Prevention Division of Viral Diseases, Pathogen Discovery | Dakota Howard, Dhwani Batra, Peter W. Cook, Kara Moser, Adrian Paskey, Jason Caravas, Benjamin Rambo-Martin, Shatavia Morrison, Christopher Gulvick, Scott Sammons, Yvette Unoarumhi, Darlene Wagner, Matthew Schmerer, S. H. Rosenthal, A. Gerasimova, R. M. Kagan, B. Anderson, M. Hua, Y. Liu, L.E. Bernstein, K.E. Livingston, A. Perez, I. A. Shlyakhter, R. V. Rolando, R. Owen, P. Tanpaiboon, F. Lacbawan, Clinton R. Paden, Duncan MacCannell |
| EPI_ISL_1695781 | NM Dept. Health, Scientific Laboratory Division | Centers for Disease Control and Prevention Division of Viral Diseases, Pathogen Discovery | Mili Sheth, Sarah Nobles, Jasmine Padilla, Mark Burroughs, Shoshona Le, Katie Dillon, Peter Cook, Clinton R. Paden, Dhwani Batra, Krista Queen, Kristen Knipe, Dakota Howard, Yvette Unoarumhi, Darlene Wagner, Matthew Schmerer, Ben L. Rambo-Martin, Kristine Lacek, Sam Shepard, Alison Laufer Halpin, Dave Wentworth, Vivien Dugan, Suxiang Tong, Justin Lee |
| EPI_ISL_1695784, EPI_ISL_1695787 | NJ Public Health and Environmental Laboratories | Centers for Disease Control and Prevention Division of Viral Diseases, Pathogen Discovery | Mili Sheth, Sarah Nobles, Jasmine Padilla, Mark Burroughs, Shoshona Le, Katie Dillon, Peter Cook, Clinton R. Paden, Dhwani Batra, Krista Queen, Kristen Knipe, Dakota Howard, Yvette Unoarumhi, Darlene Wagner, Matthew Schmerer, Ben L. Rambo-Martin, Kristine Lacek, Sam Shepard, Alison Laufer Halpin, Dave Wentworth, Vivien Dugan, Suxiang Tong, Justin Lee |
| EPI_ISL_1695790 | WY Public Health Laboratory | Centers for Disease Control and Prevention Division of Viral Diseases, Pathogen Discovery | Mili Sheth, Sarah Nobles, Jasmine Padilla, Mark Burroughs, Shoshona Le, Katie Dillon, Peter Cook, Clinton R. Paden, Dhwani Batra, Krista Queen, Kristen Knipe, Dakota Howard, Yvette Unoarumhi, Darlene Wagner, Matthew Schmerer, Ben L. Rambo-Martin, Kristine Lacek, Sam Shepard, Alison Laufer Halpin, Dave Wentworth, Vivien Dugan, Suxiang Tong, Justin Lee |
| EPI_ISL_1695793 | NJ Public Health and Environmental | Centers for Disease Control and Prevention | Mili Sheth, Sarah Nobles, Jasmine Padilla, Mark Burroughs, Shoshona Le, Katie Dillon, Peter Cook, Clinton R. Paden, Dhwani Batra, Krista Queen, Kristen Knipe, Dakota Howard, Yvette Unoarumhi, Darlene Wagner, Matthew Schmerer, Ben L. Rambo-Martin, Kristine Lacek, Sam Shepard, |

| | | | |
|---|---|---|---|
| | Laboratories | Division of Viral Diseases, Pathogen Discovery | Alison Laufer Halpin, Dave Wentworth, Vivien Dugan, Suxiang Tong, Justin Lee |
| EPI_ISL_1695796, EPI_ISL_1695799 | NC State Laboratory of Public Health | Centers for Disease Control and Prevention Division of Viral Diseases, Pathogen Discovery | Mili Sheth, Sarah Nobles, Jasmine Padilla, Mark Burroughs, Shoshona Le, Katie Dillon, Peter Cook, Clinton R. Paden, Dhwani Batra, Krista Queen, Kristen Knipe, Dakota Howard, Yvette Unoarumhi, Darlene Wagner, Matthew Schmerer, Ben L. Rambo-Martin, Kristine Lacek, Sam Shepard, Alison Laufer Halpin, Dave Wentworth, Vivien Dugan, Suxiang Tong, Justin Lee |
| EPI_ISL_1695802, EPI_ISL_1695805, EPI_ISL_1695808, EPI_ISL_1695811, EPI_ISL_1695814, EPI_ISL_1695817, EPI_ISL_1695819, EPI_ISL_1695822 | RI State Health Laboratories | Centers for Disease Control and Prevention Division of Viral Diseases, Pathogen Discovery | Mili Sheth, Sarah Nobles, Jasmine Padilla, Mark Burroughs, Shoshona Le, Katie Dillon, Peter Cook, Clinton R. Paden, Dhwani Batra, Krista Queen, Kristen Knipe, Dakota Howard, Yvette Unoarumhi, Darlene Wagner, Matthew Schmerer, Ben L. Rambo-Martin, Kristine Lacek, Sam Shepard, Alison Laufer Halpin, Dave Wentworth, Vivien Dugan, Suxiang Tong, Justin Lee |
| EPI_ISL_1695825 | UT-Unified State Labs: Public Health Utah DOH | Centers for Disease Control and Prevention Division of Viral Diseases, Pathogen Discovery | Mili Sheth, Sarah Nobles, Jasmine Padilla, Mark Burroughs, Shoshona Le, Katie Dillon, Peter Cook, Clinton R. Paden, Dhwani Batra, Krista Queen, Kristen Knipe, Dakota Howard, Yvette Unoarumhi, Darlene Wagner, Matthew Schmerer, Ben L. Rambo-Martin, Kristine Lacek, Sam Shepard, Alison Laufer Halpin, Dave Wentworth, Vivien Dugan, Suxiang Tong, Justin Lee |
| EPI_ISL_1695828 | IL Dept. of Public Health Springfield Laboratory | Centers for Disease Control and Prevention Division of Viral Diseases, Pathogen Discovery | Mili Sheth, Sarah Nobles, Jasmine Padilla, Mark Burroughs, Shoshona Le, Katie Dillon, Peter Cook, Clinton R. Paden, Dhwani Batra, Krista Queen, Kristen Knipe, Dakota Howard, Yvette Unoarumhi, Darlene Wagner, Matthew Schmerer, Ben L. Rambo-Martin, Kristine Lacek, Sam Shepard, Alison Laufer Halpin, Dave Wentworth, Vivien Dugan, Suxiang Tong, Justin Lee |
| EPI_ISL_1695831 | TN Division of Laboratory Services | Centers for Disease Control and Prevention Division of Viral Diseases, Pathogen Discovery | Mili Sheth, Sarah Nobles, Jasmine Padilla, Mark Burroughs, Shoshona Le, Katie Dillon, Peter Cook, Clinton R. Paden, Dhwani Batra, Krista Queen, Kristen Knipe, Dakota Howard, Yvette Unoarumhi, Darlene Wagner, Matthew Schmerer, Ben L. Rambo-Martin, Kristine Lacek, Sam Shepard, Alison Laufer Halpin, Dave Wentworth, Vivien Dugan, Suxiang Tong, Justin Lee |
| EPI_ISL_1695834 | LA Office of Public Health Laboratories | Centers for Disease Control and Prevention Division of Viral Diseases, Pathogen Discovery | Mili Sheth, Sarah Nobles, Jasmine Padilla, Mark Burroughs, Shoshona Le, Katie Dillon, Peter Cook, Clinton R. Paden, Dhwani Batra, Krista Queen, Kristen Knipe, Dakota Howard, Yvette Unoarumhi, Darlene Wagner, Matthew Schmerer, Ben L. Rambo-Martin, Kristine Lacek, Sam Shepard, Alison Laufer Halpin, Dave Wentworth, Vivien Dugan, Suxiang Tong, Justin Lee |
| EPI_ISL_1695837 | PA Department of Health, Bureau of Laboratories | Centers for Disease Control and Prevention Division of Viral Diseases, Pathogen Discovery | Mili Sheth, Sarah Nobles, Jasmine Padilla, Mark Burroughs, Shoshona Le, Katie Dillon, Peter Cook, Clinton R. Paden, Dhwani Batra, Krista Queen, Kristen Knipe, Dakota Howard, Yvette Unoarumhi, Darlene Wagner, Matthew Schmerer, Ben L. Rambo-Martin, Kristine Lacek, Sam Shepard, Alison Laufer Halpin, Dave Wentworth, Vivien Dugan, Suxiang Tong, Justin Lee |
| EPI_ISL_1695840, EPI_ISL_1695843, EPI_ISL_1695846 | DE Public Health Laboratory | Centers for Disease Control and Prevention Division of Viral Diseases, Pathogen Discovery | Mili Sheth, Sarah Nobles, Jasmine Padilla, Mark Burroughs, Shoshona Le, Katie Dillon, Peter Cook, Clinton R. Paden, Dhwani Batra, Krista Queen, Kristen Knipe, Dakota Howard, Yvette Unoarumhi, Darlene Wagner, Matthew Schmerer, Ben L. Rambo-Martin, Kristine Lacek, Sam Shepard, Alison Laufer Halpin, Dave Wentworth, Vivien Dugan, Suxiang Tong, Justin Lee |
| EPI_ISL_1695848, EPI_ISL_1695851, EPI_ISL_1695854, EPI_ISL_1695857, EPI_ISL_1695861, EPI_ISL_1695863, EPI_ISL_1695866, EPI_ISL_1695869, EPI_ISL_1695872, EPI_ISL_1695874, EPI_ISL_1695877, EPI_ISL_1695880, EPI_ISL_1695884 | | | |
| see above | MD DOH Laboratories Administration | Centers for Disease Control and Prevention Division of Viral Diseases, Pathogen Discovery | Mili Sheth, Sarah Nobles, Jasmine Padilla, Mark Burroughs, Shoshona Le, Katie Dillon, Peter Cook, Clinton R. Paden, Dhwani Batra, Krista Queen, Kristen Knipe, Dakota Howard, Yvette Unoarumhi, Darlene Wagner, Matthew Schmerer, Ben L. Rambo-Martin, Kristine Lacek, Sam Shepard, Alison Laufer Halpin, Dave Wentworth, Vivien Dugan, Suxiang Tong, Justin Lee |
| EPI_ISL_1695887, EPI_ISL_1695888 | NM Dept. Health, Scientific Laboratory Division | Centers for Disease Control and Prevention Division of Viral Diseases, Pathogen Discovery | Mili Sheth, Sarah Nobles, Jasmine Padilla, Mark Burroughs, Shoshona Le, Katie Dillon, Peter Cook, Clinton R. Paden, Dhwani Batra, Krista Queen, Kristen Knipe, Dakota Howard, Yvette Unoarumhi, Darlene Wagner, Matthew Schmerer, Ben L. Rambo-Martin, Kristine Lacek, Sam Shepard, Alison Laufer Halpin, Dave Wentworth, Vivien Dugan, Suxiang Tong, Justin Lee |
| EPI_ISL_1695889, EPI_ISL_1695890, EPI_ISL_1695891, EPI_ISL_1695892 | NYSDOH Wadsworth Center, Virology Lab | Centers for Disease Control and Prevention Division of Viral Diseases, Pathogen Discovery | Mili Sheth, Sarah Nobles, Jasmine Padilla, Mark Burroughs, Shoshona Le, Katie Dillon, Peter Cook, Clinton R. Paden, Dhwani Batra, Krista Queen, Kristen Knipe, Dakota Howard, Yvette Unoarumhi, Darlene Wagner, Matthew Schmerer, Ben L. Rambo-Martin, Kristine Lacek, Sam Shepard, Alison Laufer Halpin, Dave Wentworth, Vivien Dugan, Suxiang Tong, Justin Lee |
| EPI_ISL_1695893 | MI - Michigan Department of Health and Human Services - Bureau of Laboratories | Centers for Disease Control and Prevention Division of Viral Diseases, Pathogen Discovery | Mili Sheth, Sarah Nobles, Jasmine Padilla, Mark Burroughs, Shoshona Le, Katie Dillon, Peter Cook, Clinton R. Paden, Dhwani Batra, Krista Queen, Kristen Knipe, Dakota Howard, Yvette Unoarumhi, Darlene Wagner, Matthew Schmerer, Ben L. Rambo-Martin, Kristine Lacek, Sam Shepard, Alison Laufer Halpin, Dave Wentworth, Vivien Dugan, Suxiang Tong, Justin Lee |
| EPI_ISL_1695894 | WA State Department of Health | Centers for Disease Control and Prevention Division of Viral Diseases, Pathogen Discovery | Mili Sheth, Sarah Nobles, Jasmine Padilla, Mark Burroughs, Shoshona Le, Katie Dillon, Peter Cook, Clinton R. Paden, Dhwani Batra, Krista Queen, Kristen Knipe, Dakota Howard, Yvette Unoarumhi, Darlene Wagner, Matthew Schmerer, Ben L. Rambo-Martin, Kristine Lacek, Sam Shepard, Alison Laufer Halpin, Dave Wentworth, Vivien Dugan, Suxiang Tong, Justin Lee |
| EPI_ISL_1695895 | PA Department of Health, Bureau of Laboratories | Centers for Disease Control and Prevention Division of Viral Diseases, Pathogen Discovery | Mili Sheth, Sarah Nobles, Jasmine Padilla, Mark Burroughs, Shoshona Le, Katie Dillon, Peter Cook, Clinton R. Paden, Dhwani Batra, Krista Queen, Kristen Knipe, Dakota Howard, Yvette Unoarumhi, Darlene Wagner, Matthew Schmerer, Ben L. Rambo-Martin, Kristine Lacek, Sam Shepard, Alison Laufer Halpin, Dave Wentworth, Vivien Dugan, Suxiang Tong, Justin Lee |
| EPI_ISL_1695896, EPI_ISL_1695897, EPI_ISL_1695898, EPI_ISL_1695899, EPI_ISL_1695900, EPI_ISL_1695901, EPI_ISL_1695902, EPI_ISL_1695903 | NJ Public Health and Environmental Laboratories | Centers for Disease Control and Prevention Division of Viral Diseases, Pathogen Discovery | Mili Sheth, Sarah Nobles, Jasmine Padilla, Mark Burroughs, Shoshona Le, Katie Dillon, Peter Cook, Clinton R. Paden, Dhwani Batra, Krista Queen, Kristen Knipe, Dakota Howard, Yvette Unoarumhi, Darlene Wagner, Matthew Schmerer, Ben L. Rambo-Martin, Kristine Lacek, Sam Shepard, Alison Laufer Halpin, Dave Wentworth, Vivien Dugan, Suxiang Tong, Justin Lee |
| EPI_ISL_1695904 | WVDHHR - Office of Laboratory Services | Centers for Disease Control and Prevention Division of Viral Diseases, Pathogen Discovery | Mili Sheth, Sarah Nobles, Jasmine Padilla, Mark Burroughs, Shoshona Le, Katie Dillon, Peter Cook, Clinton R. Paden, Dhwani Batra, Krista Queen, Kristen Knipe, Dakota Howard, Yvette Unoarumhi, Darlene Wagner, Matthew Schmerer, Ben L. Rambo-Martin, Kristine Lacek, Sam Shepard, Alison Laufer Halpin, Dave Wentworth, Vivien Dugan, Suxiang Tong, Justin Lee |
| EPI_ISL_1695905 | NJ Public Health and Environmental Laboratories | Centers for Disease Control and Prevention Division of Viral Diseases, Pathogen Discovery | Mili Sheth, Sarah Nobles, Jasmine Padilla, Mark Burroughs, Shoshona Le, Katie Dillon, Peter Cook, Clinton R. Paden, Dhwani Batra, Krista Queen, Kristen Knipe, Dakota Howard, Yvette Unoarumhi, Darlene Wagner, Matthew Schmerer, Ben L. Rambo-Martin, Kristine Lacek, Sam Shepard, Alison Laufer Halpin, Dave Wentworth, Vivien Dugan, Suxiang Tong, Justin Lee |
| EPI_ISL_1695906, EPI_ISL_1695907 | ND Dept. of Health Laboratory Services-Microbiology | Centers for Disease Control and Prevention Division of Viral Diseases, Pathogen Discovery | Mili Sheth, Sarah Nobles, Jasmine Padilla, Mark Burroughs, Shoshona Le, Katie Dillon, Peter Cook, Clinton R. Paden, Dhwani Batra, Krista Queen, Kristen Knipe, Dakota Howard, Yvette Unoarumhi, Darlene Wagner, Matthew Schmerer, Ben L. Rambo-Martin, Kristine Lacek, Sam Shepard, Alison Laufer Halpin, Dave Wentworth, Vivien Dugan, Suxiang Tong, Justin Lee |
| EPI_ISL_1695908, EPI_ISL_1695910 | NJ Public Health and Environmental Laboratories | Centers for Disease Control and Prevention Division of Viral Diseases, Pathogen Discovery | Mili Sheth, Sarah Nobles, Jasmine Padilla, Mark Burroughs, Shoshona Le, Katie Dillon, Peter Cook, Clinton R. Paden, Dhwani Batra, Krista Queen, Kristen Knipe, Dakota Howard, Yvette Unoarumhi, Darlene Wagner, Matthew Schmerer, Ben L. Rambo-Martin, Kristine Lacek, Sam Shepard, Alison Laufer Halpin, Dave Wentworth, Vivien Dugan, Suxiang Tong, Justin Lee |
| EPI_ISL_1695911 | GA Department of Public Health Laboratory | Centers for Disease Control and Prevention Division of Viral Diseases, Pathogen Discovery | Mili Sheth, Sarah Nobles, Jasmine Padilla, Mark Burroughs, Shoshona Le, Katie Dillon, Peter Cook, Clinton R. Paden, Dhwani Batra, Krista Queen, Kristen Knipe, Dakota Howard, Yvette Unoarumhi, Darlene Wagner, Matthew Schmerer, Ben L. Rambo-Martin, Kristine Lacek, Sam Shepard, Alison Laufer Halpin, Dave Wentworth, Vivien Dugan, Suxiang Tong, Justin Lee |
| EPI_ISL_1695912 | MD DOH Laboratories Administration | Centers for Disease Control and Prevention Division of Viral Diseases, Pathogen Discovery | Mili Sheth, Sarah Nobles, Jasmine Padilla, Mark Burroughs, Shoshona Le, Katie Dillon, Peter Cook, Clinton R. Paden, Dhwani Batra, Krista Queen, Kristen Knipe, Dakota Howard, Yvette Unoarumhi, Darlene Wagner, Matthew Schmerer, Ben L. Rambo-Martin, Kristine Lacek, Sam Shepard, Alison Laufer Halpin, Dave Wentworth, Vivien Dugan, Suxiang Tong, Justin Lee |
| EPI_ISL_1695913 | MN PHL Division, Minnesota Department of Health | Centers for Disease Control and Prevention Division of Viral Diseases, Pathogen Discovery | Mili Sheth, Sarah Nobles, Jasmine Padilla, Mark Burroughs, Shoshona Le, Katie Dillon, Peter Cook, Clinton R. Paden, Dhwani Batra, Krista Queen, Kristen Knipe, Dakota Howard, Yvette Unoarumhi, Darlene Wagner, Matthew Schmerer, Ben L. Rambo-Martin, Kristine Lacek, Sam Shepard, Alison Laufer Halpin, Dave Wentworth, Vivien Dugan, Suxiang Tong, Justin Lee |
| EPI_ISL_1695914 | NC State Laboratory of Public Health | Centers for Disease Control and Prevention Division of Viral Diseases, Pathogen Discovery | Mili Sheth, Sarah Nobles, Jasmine Padilla, Mark Burroughs, Shoshona Le, Katie Dillon, Peter Cook, Clinton R. Paden, Dhwani Batra, Krista Queen, Kristen Knipe, Dakota Howard, Yvette Unoarumhi, Darlene Wagner, Matthew Schmerer, Ben L. Rambo-Martin, Kristine Lacek, Sam Shepard, Alison Laufer Halpin, Dave Wentworth, Vivien Dugan, Suxiang Tong, Justin Lee |
| EPI_ISL_1695915 | MT Public Health Laboratory | Centers for Disease Control and Prevention Division of Viral Diseases, Pathogen Discovery | Mili Sheth, Sarah Nobles, Jasmine Padilla, Mark Burroughs, Shoshona Le, Katie Dillon, Peter Cook, Clinton R. Paden, Dhwani Batra, Krista Queen, Kristen Knipe, Dakota Howard, Yvette Unoarumhi, Darlene Wagner, Matthew Schmerer, Ben L. Rambo-Martin, Kristine Lacek, Sam Shepard, Alison Laufer Halpin, Dave Wentworth, Vivien Dugan, Suxiang Tong, Justin Lee |
| EPI_ISL_1695916 | NC State Laboratory of Public Health | Centers for Disease Control and Prevention Division of Viral Diseases, Pathogen Discovery | Mili Sheth, Sarah Nobles, Jasmine Padilla, Mark Burroughs, Shoshona Le, Katie Dillon, Peter Cook, Clinton R. Paden, Dhwani Batra, Krista Queen, Kristen Knipe, Dakota Howard, Yvette Unoarumhi, Darlene Wagner, Matthew Schmerer, Ben L. Rambo-Martin, Kristine Lacek, Sam Shepard, Alison Laufer Halpin, Dave Wentworth, Vivien Dugan, Suxiang Tong, Justin Lee |
| EPI_ISL_1695917 | PA Department of Health, Bureau of Laboratories | Centers for Disease Control and Prevention Division of Viral Diseases, Pathogen Discovery | Mili Sheth, Sarah Nobles, Jasmine Padilla, Mark Burroughs, Shoshona Le, Katie Dillon, Peter Cook, Clinton R. Paden, Dhwani Batra, Krista Queen, Kristen Knipe, Dakota Howard, Yvette Unoarumhi, Darlene Wagner, Matthew Schmerer, Ben L. Rambo-Martin, Kristine Lacek, Sam Shepard, Alison Laufer Halpin, Dave Wentworth, Vivien Dugan, Suxiang Tong, Justin Lee |
| EPI_ISL_1695918 | OK Public Health Laboratory, Oklahoma State DOH | Centers for Disease Control and Prevention Division of Viral Diseases, Pathogen Discovery | Mili Sheth, Sarah Nobles, Jasmine Padilla, Mark Burroughs, Shoshona Le, Katie Dillon, Peter Cook, Clinton R. Paden, Dhwani Batra, Krista Queen, Kristen Knipe, Dakota Howard, Yvette Unoarumhi, Darlene Wagner, Matthew Schmerer, Ben L. Rambo-Martin, Kristine Lacek, Sam Shepard, Alison Laufer Halpin, Dave Wentworth, Vivien Dugan, Suxiang Tong, Justin Lee |
| EPI_ISL_1695919 | RI State Health Laboratories | Centers for Disease Control and Prevention Division of Viral Diseases, Pathogen | Mili Sheth, Sarah Nobles, Jasmine Padilla, Mark Burroughs, Shoshona Le, Katie Dillon, Peter Cook, Clinton R. Paden, Dhwani Batra, Krista Queen, Kristen Knipe, Dakota Howard, Yvette Unoarumhi, Darlene Wagner, Matthew Schmerer, Ben L. Rambo-Martin, Kristine Lacek, Sam Shepard, Alison Laufer Halpin, Dave Wentworth, Vivien Dugan, Suxiang Tong, Justin Lee |

| | | | |
|---|---|---|---|
| | | Discovery | |
| EPI_ISL_1695924 | LA Office of Public Health Laboratories | Centers for Disease Control and Prevention Division of Viral Diseases, Pathogen Discovery | Mili Sheth, Sarah Nobles, Jasmine Padilla, Mark Burroughs, Shoshona Le, Katie Dillon, Peter Cook, Clinton R. Paden, Dhwani Batra, Krista Queen, Kristen Knipe, Dakota Howard, Yvette Unoarumhi, Darlene Wagner, Matthew Schmerer, Ben L. Rambo-Martin, Kristine Lacek, Sam Shepard, Alison Laufer Halpin, Dave Wentworth, Vivien Dugan, Suxiang Tong, Justin Lee |
| EPI_ISL_1695935 | TN Division of Laboratory Services | Centers for Disease Control and Prevention Division of Viral Diseases, Pathogen Discovery | Mili Sheth, Sarah Nobles, Jasmine Padilla, Mark Burroughs, Shoshona Le, Katie Dillon, Peter Cook, Clinton R. Paden, Dhwani Batra, Krista Queen, Kristen Knipe, Dakota Howard, Yvette Unoarumhi, Darlene Wagner, Matthew Schmerer, Ben L. Rambo-Martin, Kristine Lacek, Sam Shepard, Alison Laufer Halpin, Dave Wentworth, Vivien Dugan, Suxiang Tong, Justin Lee |
| EPI_ISL_1695937, EPI_ISL_1695940 | WVDHHR - Office of Laboratory Services | Centers for Disease Control and Prevention Division of Viral Diseases, Pathogen Discovery | Mili Sheth, Sarah Nobles, Jasmine Padilla, Mark Burroughs, Shoshona Le, Katie Dillon, Peter Cook, Clinton R. Paden, Dhwani Batra, Krista Queen, Kristen Knipe, Dakota Howard, Yvette Unoarumhi, Darlene Wagner, Matthew Schmerer, Ben L. Rambo-Martin, Kristine Lacek, Sam Shepard, Alison Laufer Halpin, Dave Wentworth, Vivien Dugan, Suxiang Tong, Justin Lee |
| EPI_ISL_1695942 | RI State Health Laboratories | Centers for Disease Control and Prevention Division of Viral Diseases, Pathogen Discovery | Mili Sheth, Sarah Nobles, Jasmine Padilla, Mark Burroughs, Shoshona Le, Katie Dillon, Peter Cook, Clinton R. Paden, Dhwani Batra, Krista Queen, Kristen Knipe, Dakota Howard, Yvette Unoarumhi, Darlene Wagner, Matthew Schmerer, Ben L. Rambo-Martin, Kristine Lacek, Sam Shepard, Alison Laufer Halpin, Dave Wentworth, Vivien Dugan, Suxiang Tong, Justin Lee |
| EPI_ISL_1695945 | GA Department of Public Health Laboratory | Centers for Disease Control and Prevention Division of Viral Diseases, Pathogen Discovery | Mili Sheth, Sarah Nobles, Jasmine Padilla, Mark Burroughs, Shoshona Le, Katie Dillon, Peter Cook, Clinton R. Paden, Dhwani Batra, Krista Queen, Kristen Knipe, Dakota Howard, Yvette Unoarumhi, Darlene Wagner, Matthew Schmerer, Ben L. Rambo-Martin, Kristine Lacek, Sam Shepard, Alison Laufer Halpin, Dave Wentworth, Vivien Dugan, Suxiang Tong, Justin Lee |
| EPI_ISL_1695955 | OR State PHL-Virology/Immunology Section | Centers for Disease Control and Prevention Division of Viral Diseases, Pathogen Discovery | Mili Sheth, Sarah Nobles, Jasmine Padilla, Mark Burroughs, Shoshona Le, Katie Dillon, Peter Cook, Clinton R. Paden, Dhwani Batra, Krista Queen, Kristen Knipe, Dakota Howard, Yvette Unoarumhi, Darlene Wagner, Matthew Schmerer, Ben L. Rambo-Martin, Kristine Lacek, Sam Shepard, Alison Laufer Halpin, Dave Wentworth, Vivien Dugan, Suxiang Tong, Justin Lee |
| EPI_ISL_1695958 | LA Office of Public Health Laboratories | Centers for Disease Control and Prevention Division of Viral Diseases, Pathogen Discovery | Mili Sheth, Sarah Nobles, Jasmine Padilla, Mark Burroughs, Shoshona Le, Katie Dillon, Peter Cook, Clinton R. Paden, Dhwani Batra, Krista Queen, Kristen Knipe, Dakota Howard, Yvette Unoarumhi, Darlene Wagner, Matthew Schmerer, Ben L. Rambo-Martin, Kristine Lacek, Sam Shepard, Alison Laufer Halpin, Dave Wentworth, Vivien Dugan, Suxiang Tong, Justin Lee |
| EPI_ISL_1695968 | VA-Division of Consolidated Laboratory Services | Centers for Disease Control and Prevention Division of Viral Diseases, Pathogen Discovery | Mili Sheth, Sarah Nobles, Jasmine Padilla, Mark Burroughs, Shoshona Le, Katie Dillon, Peter Cook, Clinton R. Paden, Dhwani Batra, Krista Queen, Kristen Knipe, Dakota Howard, Yvette Unoarumhi, Darlene Wagner, Matthew Schmerer, Ben L. Rambo-Martin, Kristine Lacek, Sam Shepard, Alison Laufer Halpin, Dave Wentworth, Vivien Dugan, Suxiang Tong, Justin Lee |
| EPI_ISL_1695970 | NYC Department of Health and Mental Hygiene | Centers for Disease Control and Prevention Division of Viral Diseases, Pathogen Discovery | Mili Sheth, Sarah Nobles, Jasmine Padilla, Mark Burroughs, Shoshona Le, Katie Dillon, Peter Cook, Clinton R. Paden, Dhwani Batra, Krista Queen, Kristen Knipe, Dakota Howard, Yvette Unoarumhi, Darlene Wagner, Matthew Schmerer, Ben L. Rambo-Martin, Kristine Lacek, Sam Shepard, Alison Laufer Halpin, Dave Wentworth, Vivien Dugan, Suxiang Tong, Justin Lee |
| EPI_ISL_1695973 | DC Public Health Lab/ Dept. of Forensic Sciences | Centers for Disease Control and Prevention Division of Viral Diseases, Pathogen Discovery | Mili Sheth, Sarah Nobles, Jasmine Padilla, Mark Burroughs, Shoshona Le, Katie Dillon, Peter Cook, Clinton R. Paden, Dhwani Batra, Krista Queen, Kristen Knipe, Dakota Howard, Yvette Unoarumhi, Darlene Wagner, Matthew Schmerer, Ben L. Rambo-Martin, Kristine Lacek, Sam Shepard, Alison Laufer Halpin, Dave Wentworth, Vivien Dugan, Suxiang Tong, Justin Lee |
| EPI_ISL_1695976 | MT Public Health Laboratory | Centers for Disease Control and Prevention Division of Viral Diseases, Pathogen Discovery | Mili Sheth, Sarah Nobles, Jasmine Padilla, Mark Burroughs, Shoshona Le, Katie Dillon, Peter Cook, Clinton R. Paden, Dhwani Batra, Krista Queen, Kristen Knipe, Dakota Howard, Yvette Unoarumhi, Darlene Wagner, Matthew Schmerer, Ben L. Rambo-Martin, Kristine Lacek, Sam Shepard, Alison Laufer Halpin, Dave Wentworth, Vivien Dugan, Suxiang Tong, Justin Lee |
| EPI_ISL_1695979 | HI Dept. of Health, State Laboratories Division | Centers for Disease Control and Prevention Division of Viral Diseases, Pathogen Discovery | Mili Sheth, Sarah Nobles, Jasmine Padilla, Mark Burroughs, Shoshona Le, Katie Dillon, Peter Cook, Clinton R. Paden, Dhwani Batra, Krista Queen, Kristen Knipe, Dakota Howard, Yvette Unoarumhi, Darlene Wagner, Matthew Schmerer, Ben L. Rambo-Martin, Kristine Lacek, Sam Shepard, Alison Laufer Halpin, Dave Wentworth, Vivien Dugan, Suxiang Tong, Justin Lee |
| EPI_ISL_1696114, EPI_ISL_1696115, EPI_ISL_1696117, EPI_ISL_1696118, EPI_ISL_1696119, EPI_ISL_1696120, EPI_ISL_1696121, EPI_ISL_1696122, EPI_ISL_1696125, EPI_ISL_1696126, EPI_ISL_1696127, EPI_ISL_1696128, EPI_ISL_1696129, EPI_ISL_1696130, EPI_ISL_1696132, EPI_ISL_1696133, EPI_ISL_1696135, EPI_ISL_1696136, EPI_ISL_1696137, EPI_ISL_1696138, EPI_ISL_1696139 | | | |
| see above | UTGSAF | UTGSAF | Jessica Podnar, Sylvie Beaudenon, Audrey Kelly, Zachary Carver, Anna Battenhouse, Andreas Matouschek |
| EPI_ISL_1696313, EPI_ISL_1696314, EPI_ISL_1696315, EPI_ISL_1696316, EPI_ISL_1696317, EPI_ISL_1696318, EPI_ISL_1696319, EPI_ISL_1696320, EPI_ISL_1696321, EPI_ISL_1696322, EPI_ISL_1696323, EPI_ISL_1696324, EPI_ISL_1696325, EPI_ISL_1696326, EPI_ISL_1696327, EPI_ISL_1696328, EPI_ISL_1696329, EPI_ISL_1696330, EPI_ISL_1696331, EPI_ISL_1696332, EPI_ISL_1696333, EPI_ISL_1696334, EPI_ISL_1696335, EPI_ISL_1696336, EPI_ISL_1696337, EPI_ISL_1696338, EPI_ISL_1696339, EPI_ISL_1696340, EPI_ISL_1696341, EPI_ISL_1696342, EPI_ISL_1696343, EPI_ISL_1696344, EPI_ISL_1696345, EPI_ISL_1696346, EPI_ISL_1696347, EPI_ISL_1696348, EPI_ISL_1696349, EPI_ISL_1696350, EPI_ISL_1696351, EPI_ISL_1696352, EPI_ISL_1696353, EPI_ISL_1696354, EPI_ISL_1696355, EPI_ISL_1696356, EPI_ISL_1696357, EPI_ISL_1696358, EPI_ISL_1696359, EPI_ISL_1696360, EPI_ISL_1696361, EPI_ISL_1696362, EPI_ISL_1696363, EPI_ISL_1696364, EPI_ISL_1696365, EPI_ISL_1696366, EPI_ISL_1696367, EPI_ISL_1696368, EPI_ISL_1696369, EPI_ISL_1696370, EPI_ISL_1696371, EPI_ISL_1696372, EPI_ISL_1696373, EPI_ISL_1696374, EPI_ISL_1696375, EPI_ISL_1696376, EPI_ISL_1696377, EPI_ISL_1696378, EPI_ISL_1696379, EPI_ISL_1696380, EPI_ISL_1696381, EPI_ISL_1696382, EPI_ISL_1696383, EPI_ISL_1696384, EPI_ISL_1696385, EPI_ISL_1696386, EPI_ISL_1696387, EPI_ISL_1696388, EPI_ISL_1696389, EPI_ISL_1696390, EPI_ISL_1696391, EPI_ISL_1696392, EPI_ISL_1696393, EPI_ISL_1696394, EPI_ISL_1696395, EPI_ISL_1696396, EPI_ISL_1696397, EPI_ISL_1696398, EPI_ISL_1696399, EPI_ISL_1696400, EPI_ISL_1696401, EPI_ISL_1696402, EPI_ISL_1696403, EPI_ISL_1696404, EPI_ISL_1696405, EPI_ISL_1696406, EPI_ISL_1696407, EPI_ISL_1696408, EPI_ISL_1696409, EPI_ISL_1696410, EPI_ISL_1696411, EPI_ISL_1696412, EPI_ISL_1696413, EPI_ISL_1696414, EPI_ISL_1696415, EPI_ISL_1696416, EPI_ISL_1696417, EPI_ISL_1696418, EPI_ISL_1696419, EPI_ISL_1696420, EPI_ISL_1696421, EPI_ISL_1696422, EPI_ISL_1696423, EPI_ISL_1696424, EPI_ISL_1696425, EPI_ISL_1696426, EPI_ISL_1696427, EPI_ISL_1696428, EPI_ISL_1696429, EPI_ISL_1696430, EPI_ISL_1696431, EPI_ISL_1696432, EPI_ISL_1696433, EPI_ISL_1696434, EPI_ISL_1696435, EPI_ISL_1696436, EPI_ISL_1696437, EPI_ISL_1696438, EPI_ISL_1696439, EPI_ISL_1696440, EPI_ISL_1696441, EPI_ISL_1696442, EPI_ISL_1696443, EPI_ISL_1696444, EPI_ISL_1696445, EPI_ISL_1696446, EPI_ISL_1696447, EPI_ISL_1696448, EPI_ISL_1696449, EPI_ISL_1696450, EPI_ISL_1696451, EPI_ISL_1696452, EPI_ISL_1696453, EPI_ISL_1696454, EPI_ISL_1696455, EPI_ISL_1696456, EPI_ISL_1696457, EPI_ISL_1696458, EPI_ISL_1696459, EPI_ISL_1696460, EPI_ISL_1696461, EPI_ISL_1696462, EPI_ISL_1696463, EPI_ISL_1696464, EPI_ISL_1696466, EPI_ISL_1696467, EPI_ISL_1696468, EPI_ISL_1696469, EPI_ISL_1696470, EPI_ISL_1696471, EPI_ISL_1696472, EPI_ISL_1696473, EPI_ISL_1696474, EPI_ISL_1696475, EPI_ISL_1696476, EPI_ISL_1696477, EPI_ISL_1696478, EPI_ISL_1696479, EPI_ISL_1696480, EPI_ISL_1696481, EPI_ISL_1696482, EPI_ISL_1696483, EPI_ISL_1696484, EPI_ISL_1696485, EPI_ISL_1696486, EPI_ISL_1696487, EPI_ISL_1696488, EPI_ISL_1696489, EPI_ISL_1696490 | | | |
| see above | Maryland Genomics, Institute for Genome Sciences, University of Maryland School of Medicine | Maryland Genomics, Institute for Genome Sciences, University of Maryland School of Medicine | Tallon, Luke J; Sadzewicz, Lisa D; Humphrys, Mike; Ott, Sandra; Roussey, Holly; Mehta, Aditya; Vavikolanu, Kranthi; Fraser, Claire M; Ravel, Jacques |
| EPI_ISL_1696492, EPI_ISL_1696493, EPI_ISL_1696496 | Idaho Bureau of Laboratories | Idaho Bureau of Laboratories | R. Beukelman, Matthew Charles Burns, Aimee Ceniseros, Robert L. Voermans, Christopher Ball |
| EPI_ISL_1696678, EPI_ISL_1696679, EPI_ISL_1696681, EPI_ISL_1696683, EPI_ISL_1696684, EPI_ISL_1696686, EPI_ISL_1696687, EPI_ISL_1696688, EPI_ISL_1696689, EPI_ISL_1696691, EPI_ISL_1696694, EPI_ISL_1696695, EPI_ISL_1696696, EPI_ISL_1696697, EPI_ISL_1696698, EPI_ISL_1696699, EPI_ISL_1696700, EPI_ISL_1696701, EPI_ISL_1696702, EPI_ISL_1696704, EPI_ISL_1696705, EPI_ISL_1696706, EPI_ISL_1696707, EPI_ISL_1696708, EPI_ISL_1696709, EPI_ISL_1696710, EPI_ISL_1696712, EPI_ISL_1696713, EPI_ISL_1696716, EPI_ISL_1696717, EPI_ISL_1696718, EPI_ISL_1696720, EPI_ISL_1696721, EPI_ISL_1696722, EPI_ISL_1696723, EPI_ISL_1696724, EPI_ISL_1696725, EPI_ISL_1696726, EPI_ISL_1696727, EPI_ISL_1696728, EPI_ISL_1696730, EPI_ISL_1696731, EPI_ISL_1696732, EPI_ISL_1696734, EPI_ISL_1696735, EPI_ISL_1696736, EPI_ISL_1696739, EPI_ISL_1696740, EPI_ISL_1696741, EPI_ISL_1696742, EPI_ISL_1696743, EPI_ISL_1696744, EPI_ISL_1696745 | | | |
| see above | LSUHS Emerging Viral Threat Laboratory | LSUHS Emerging Viral Threat Laboratory | Alexander J. Mijalis, Gregory L. Ware, Rona S. Scott, Jennifer L. Carroll, Jeremy P. Kamil, Maarten Van Diest, Andrew D. Yurochko, John A. Vanchiere, Christopher G. Kevil |
| EPI_ISL_1697375 | Springfield-Greene Co. Health Dept. | Pathogen Discovery, Respiratory Viruses Branch, Division of Viral Diseases, Centers for Disease Control and Prevention | Ying Tao, Yan Li, Anna Kelleher, Jing Zhang, Anna Montmayeur, Brian Lynch, Krista Queen, Anna Uehara, Peter Cook, Han Jia Justin Ng, Rachel Marine, Clinton R. Paden, Haibin Wang, Mark Burroughs, Justin Lee, Adam Retchless, Suxiang Tong |
| EPI_ISL_1697383 | Kansas Health and Environmental Lab | Kansas Health and Environmental Lab | Mike Grose, Jonathan Barnell, Ben Olsen, and Phil Adam |
| EPI_ISL_1697384 | Wichita State University - Molecular Diagnostics Lab | Kansas Health and Environmental Lab | Mike Grose, Jonathan Barnell, Ben Olsen, and Phil Adam |
| EPI_ISL_1697397, EPI_ISL_1697398, EPI_ISL_1697399, EPI_ISL_1697400, EPI_ISL_1697401, EPI_ISL_1697402, EPI_ISL_1697404, EPI_ISL_1697405, EPI_ISL_1697406, EPI_ISL_1697407, EPI_ISL_1697408, EPI_ISL_1697409 | | | |
| see above | Kansas Health and Environmental Lab | Kansas Health and Environmental Lab | Mike Grose, Jonathan Barnell, Ben Olsen, and Phil Adam |
| EPI_ISL_1697413 | Cytocheck Laboratories | Kansas Health and Environmental Lab | Mike Grose, Jonathan Barnell, Ben Olsen, and Phil Adam |
| EPI_ISL_1697414, EPI_ISL_1697415, EPI_ISL_1697416, EPI_ISL_1697417, EPI_ISL_1697418 | Kansas Health and Environmental Lab | Kansas Health and Environmental Lab | Mike Grose, Jonathan Barnell, Ben Olsen, and Phil Adam |
| EPI_ISL_1697419, EPI_ISL_1697421 | Wichita State University - Molecular Diagnostics Lab | Kansas Health and Environmental Lab | Mike Grose, Jonathan Barnell, Ben Olsen, and Phil Adam |
| EPI_ISL_1697422, EPI_ISL_1697423, EPI_ISL_1697424, EPI_ISL_1697425 | Kansas Health and Environmental Lab | Kansas Health and Environmental Lab | Mike Grose, Jonathan Barnell, Ben Olsen, and Phil Adam |
| EPI_ISL_1697426, EPI_ISL_1697427, EPI_ISL_1697428, EPI_ISL_1697429, EPI_ISL_1697430, EPI_ISL_1697431, EPI_ISL_1697432 | Wichita State University - Molecular Diagnostics Lab | Kansas Health and Environmental Lab | Mike Grose, Jonathan Barnell, Ben Olsen, and Phil Adam |
| EPI_ISL_1700669, EPI_ISL_1700670, EPI_ISL_1700671, EPI_ISL_1700672, EPI_ISL_1700673 | NYC Department of Health and Mental Hygiene | Pathogen Discovery, Respiratory Viruses Branch, Division of Viral Diseases, Centers for Disease Control and Prevention | Ying Tao, Yan Li, Anna Kelleher, Jing Zhang, Anna Montmayeur, Brian Lynch, Krista Queen, Anna Uehara, Peter Cook, Han Jia Justin Ng, Rachel Marine, Clinton R. Paden, Haibin Wang, Mark Burroughs, Justin Lee, Adam Retchless, Suxiang Tong |
| EPI_ISL_1700688, EPI_ISL_1700689, EPI_ISL_1700690, EPI_ISL_1700691, EPI_ISL_1700692, EPI_ISL_1700693, EPI_ISL_1700694, EPI_ISL_1700695, EPI_ISL_1700696, EPI_ISL_1700697, EPI_ISL_1700698, EPI_ISL_1700699 | | | |
| see above | Monterey County Public Health Laboratory | Monterey County Public Health Laboratory | Monterey County Public Health Laboratory |
| EPI_ISL_1700700, EPI_ISL_1700701, EPI_ISL_1700702, EPI_ISL_1700703, EPI_ISL_1700704, EPI_ISL_1700705, EPI_ISL_1700706, EPI_ISL_1700707, EPI_ISL_1700708, EPI_ISL_1700709, EPI_ISL_1700710, EPI_ISL_1700711, EPI_ISL_1700712 | | | |

| | | | |
|---|---|---|---|
| see above | NJDOH, Public Health and Environmental Laboratories | NJ_PHEL | Lindsey Bodnar, Shiv K. Verma, Jacquelyn Deverell, Matthew Scarnati, Dana Woell, Byeong Jeong |
| EPI_ISL_1700764 | Clinical Reference Laboratory | Kansas Health and Environmental Lab | Mike Grose, Jonathan Barnell, Ben Olsen, and Phil Adam |
| EPI_ISL_1700765 | Kansas Health and Environmental Lab | Kansas Health and Environmental Lab | Mike Grose, Jonathan Barnell, Ben Olsen, and Phil Adam |
| EPI_ISL_1700766 | Clinical Reference Laboratory | Kansas Health and Environmental Lab | Mike Grose, Jonathan Barnell, Ben Olsen, and Phil Adam |
| EPI_ISL_1700767 | Wichita State University - Molecular Diagnostics Lab | Kansas Health and Environmental Lab | Mike Grose, Jonathan Barnell, Ben Olsen, and Phil Adam |
| EPI_ISL_1700768, EPI_ISL_1700769 | Kansas Health and Environmental Lab | Kansas Health and Environmental Lab | Mike Grose, Jonathan Barnell, Ben Olsen, and Phil Adam |
| EPI_ISL_1700770, EPI_ISL_1700771, EPI_ISL_1700772, EPI_ISL_1700773, EPI_ISL_1700774, EPI_ISL_1700775, EPI_ISL_1700776 | Wichita State University - Molecular Diagnostics Lab | Kansas Health and Environmental Lab | Mike Grose, Jonathan Barnell, Ben Olsen, and Phil Adam |
| EPI_ISL_1700777, EPI_ISL_1700778, EPI_ISL_1700779 | Kansas Health and Environmental Lab | Kansas Health and Environmental Lab | Mike Grose, Jonathan Barnell, Ben Olsen, and Phil Adam |
| EPI_ISL_1700789 | NJDOH, Public Health and Environmental Laboratories | NJ Public Health and Environmental Laboratories | Lindsey Bodnar, Shiv K. Verma, Jacquelyn Deverell, Matthew Scarnati, Dana Woell, Byeong Jeong |
| EPI_ISL_1700795 | LESP Chiapas | Instituto de Diagnostico y Referencia Epidemiologicos (INDRE) | Claudia Wong-Arambula, Abril Rodriguez-Maldonado, Vanessa Rivero-Arredondo, Ariadna Medina-Benitez, Joaquin Quiroz-Mercado, Sergio Rangel-Guerrero, Natividad Cruz-Ortiz, Tatiana Nunez-Garcia, Gisela Barrera-Badillo, Lucia Hernandez-Rivas, Irma Lopez-Martinez, Ernesto Ramirez-Gonzalez. |
| EPI_ISL_1700796, EPI_ISL_1700797, EPI_ISL_1700798, EPI_ISL_1700799 | LESP Nuevo Leon | Instituto de Diagnostico y Referencia Epidemiologicos (INDRE) | Claudia Wong-Arambula, Abril Rodriguez-Maldonado, Vanessa Rivero-Arredondo, Ariadna Medina-Benitez, Joaquin Quiroz-Mercado, Sergio Rangel-Guerrero, Natividad Cruz-Ortiz, Tatiana Nunez-Garcia, Gisela Barrera-Badillo, Lucia Hernandez-Rivas, Irma Lopez-Martinez, Ernesto Ramirez-Gonzalez. |
| EPI_ISL_1700800 | LESP Jalisco | Instituto de Diagnostico y Referencia Epidemiologicos (INDRE) | Claudia Wong-Arambula, Abril Rodriguez-Maldonado, Vanessa Rivero-Arredondo, Ariadna Medina-Benitez, Joaquin Quiroz-Mercado, Sergio Rangel-Guerrero, Natividad Cruz-Ortiz, Tatiana Nunez-Garcia, Gisela Barrera-Badillo, Lucia Hernandez-Rivas, Irma Lopez-Martinez, Ernesto Ramirez-Gonzalez. |
| EPI_ISL_1700801 | LESP Sonora | Instituto de Diagnostico y Referencia Epidemiologicos (INDRE) | Claudia Wong-Arambula, Abril Rodriguez-Maldonado, Vanessa Rivero-Arredondo, Ariadna Medina-Benitez, Joaquin Quiroz-Mercado, Sergio Rangel-Guerrero, Natividad Cruz-Ortiz, Tatiana Nunez-Garcia, Gisela Barrera-Badillo, Lucia Hernandez-Rivas, Irma Lopez-Martinez, Ernesto Ramirez-Gonzalez. |
| EPI_ISL_1700802 | LESP Morelos | Instituto de Diagnostico y Referencia Epidemiologicos (INDRE) | Claudia Wong-Arambula, Abril Rodriguez-Maldonado, Vanessa Rivero-Arredondo, Ariadna Medina-Benitez, Joaquin Quiroz-Mercado, Sergio Rangel-Guerrero, Natividad Cruz-Ortiz, Tatiana Nunez-Garcia, Gisela Barrera-Badillo, Lucia Hernandez-Rivas, Irma Lopez-Martinez, Ernesto Ramirez-Gonzalez. |
| EPI_ISL_1700803, EPI_ISL_1700804, EPI_ISL_1700805, EPI_ISL_1700806 | LESP Queretaro | Instituto de Diagnostico y Referencia Epidemiologicos (INDRE) | Claudia Wong-Arambula, Abril Rodriguez-Maldonado, Vanessa Rivero-Arredondo, Ariadna Medina-Benitez, Joaquin Quiroz-Mercado, Sergio Rangel-Guerrero, Natividad Cruz-Ortiz, Tatiana Nunez-Garcia, Gisela Barrera-Badillo, Lucia Hernandez-Rivas, Irma Lopez-Martinez, Ernesto Ramirez-Gonzalez. |
| EPI_ISL_1700807, EPI_ISL_1700808, EPI_ISL_1700809 | LESP Chihuahua | Instituto de Diagnostico y Referencia Epidemiologicos (INDRE) | Claudia Wong-Arambula, Abril Rodriguez-Maldonado, Vanessa Rivero-Arredondo, Ariadna Medina-Benitez, Joaquin Quiroz-Mercado, Sergio Rangel-Guerrero, Natividad Cruz-Ortiz, Tatiana Nunez-Garcia, Gisela Barrera-Badillo, Lucia Hernandez-Rivas, Irma Lopez-Martinez, Ernesto Ramirez-Gonzalez. |
| EPI_ISL_1700810 | LESP Guerrero | Instituto de Diagnostico y Referencia Epidemiologicos (INDRE) | Claudia Wong-Arambula, Abril Rodriguez-Maldonado, Vanessa Rivero-Arredondo, Ariadna Medina-Benitez, Joaquin Quiroz-Mercado, Sergio Rangel-Guerrero, Natividad Cruz-Ortiz, Tatiana Nunez-Garcia, Gisela Barrera-Badillo, Lucia Hernandez-Rivas, Irma Lopez-Martinez, Ernesto Ramirez-Gonzalez. |
| EPI_ISL_1700811, EPI_ISL_1700812, EPI_ISL_1700813, EPI_ISL_1700814, EPI_ISL_1700815 | LESP Veracruz | Instituto de Diagnostico y Referencia Epidemiologicos (INDRE) | Claudia Wong-Arambula, Abril Rodriguez-Maldonado, Vanessa Rivero-Arredondo, Ariadna Medina-Benitez, Joaquin Quiroz-Mercado, Sergio Rangel-Guerrero, Natividad Cruz-Ortiz, Tatiana Nunez-Garcia, Gisela Barrera-Badillo, Lucia Hernandez-Rivas, Irma Lopez-Martinez, Ernesto Ramirez-Gonzalez. |
| EPI_ISL_1700816 | LESP Campeche | Instituto de Diagnostico y Referencia Epidemiologicos (INDRE) | Claudia Wong-Arambula, Abril Rodriguez-Maldonado, Vanessa Rivero-Arredondo, Ariadna Medina-Benitez, Joaquin Quiroz-Mercado, Sergio Rangel-Guerrero, Natividad Cruz-Ortiz, Tatiana Nunez-Garcia, Gisela Barrera-Badillo, Lucia Hernandez-Rivas, Irma Lopez-Martinez, Ernesto Ramirez-Gonzalez. |
| EPI_ISL_1700817 | LESP Tamaulipas | Instituto de Diagnostico y Referencia Epidemiologicos (INDRE) | Claudia Wong-Arambula, Abril Rodriguez-Maldonado, Vanessa Rivero-Arredondo, Ariadna Medina-Benitez, Joaquin Quiroz-Mercado, Sergio Rangel-Guerrero, Natividad Cruz-Ortiz, Tatiana Nunez-Garcia, Gisela Barrera-Badillo, Lucia Hernandez-Rivas, Irma Lopez-Martinez, Ernesto Ramirez-Gonzalez. |
| EPI_ISL_1700818 | LESP Coahuila | Instituto de Diagnostico y Referencia Epidemiologicos (INDRE) | Claudia Wong-Arambula, Abril Rodriguez-Maldonado, Vanessa Rivero-Arredondo, Ariadna Medina-Benitez, Joaquin Quiroz-Mercado, Sergio Rangel-Guerrero, Natividad Cruz-Ortiz, Tatiana Nunez-Garcia, Gisela Barrera-Badillo, Lucia Hernandez-Rivas, Irma Lopez-Martinez, Ernesto Ramirez-Gonzalez. |
| EPI_ISL_1700819 | LESP Baja California Sur | Instituto de Diagnostico y Referencia Epidemiologicos (INDRE) | Claudia Wong-Arambula, Abril Rodriguez-Maldonado, Vanessa Rivero-Arredondo, Ariadna Medina-Benitez, Joaquin Quiroz-Mercado, Sergio Rangel-Guerrero, Natividad Cruz-Ortiz, Tatiana Nunez-Garcia, Gisela Barrera-Badillo, Lucia Hernandez-Rivas, Irma Lopez-Martinez, Ernesto Ramirez-Gonzalez. |
| EPI_ISL_1700820, EPI_ISL_1700821 | LESP Jalisco | Instituto de Diagnostico y Referencia Epidemiologicos (INDRE) | Claudia Wong-Arambula, Abril Rodriguez-Maldonado, Vanessa Rivero-Arredondo, Ariadna Medina-Benitez, Joaquin Quiroz-Mercado, Sergio Rangel-Guerrero, Natividad Cruz-Ortiz, Tatiana Nunez-Garcia, Gisela Barrera-Badillo, Lucia Hernandez-Rivas, Irma Lopez-Martinez, Ernesto Ramirez-Gonzalez. |
| EPI_ISL_1700822 | LESP Baja California Sur | Instituto de Diagnostico y Referencia Epidemiologicos (INDRE) | Claudia Wong-Arambula, Abril Rodriguez-Maldonado, Vanessa Rivero-Arredondo, Ariadna Medina-Benitez, Joaquin Quiroz-Mercado, Sergio Rangel-Guerrero, Natividad Cruz-Ortiz, Tatiana Nunez-Garcia, Gisela Barrera-Badillo, Lucia Hernandez-Rivas, Irma Lopez-Martinez, Ernesto Ramirez-Gonzalez. |
| EPI_ISL_1700823 | LESP Nayarit | Instituto de Diagnostico y Referencia Epidemiologicos (INDRE) | Claudia Wong-Arambula, Abril Rodriguez-Maldonado, Vanessa Rivero-Arredondo, Ariadna Medina-Benitez, Joaquin Quiroz-Mercado, Sergio Rangel-Guerrero, Natividad Cruz-Ortiz, Tatiana Nunez-Garcia, Gisela Barrera-Badillo, Lucia Hernandez-Rivas, Irma Lopez-Martinez, Ernesto Ramirez-Gonzalez. |
| EPI_ISL_1700824 | LESP Ciudad de Mexico | Instituto de Diagnostico y Referencia Epidemiologicos (INDRE) | Claudia Wong-Arambula, Abril Rodriguez-Maldonado, Vanessa Rivero-Arredondo, Ariadna Medina-Benitez, Joaquin Quiroz-Mercado, Sergio Rangel-Guerrero, Natividad Cruz-Ortiz, Tatiana Nunez-Garcia, Gisela Barrera-Badillo, Lucia Hernandez-Rivas, Irma Lopez-Martinez, Ernesto Ramirez-Gonzalez. |
| EPI_ISL_1700825 | LESP Estado de Mexico | Instituto de Diagnostico y Referencia Epidemiologicos (INDRE) | Claudia Wong-Arambula, Abril Rodriguez-Maldonado, Vanessa Rivero-Arredondo, Ariadna Medina-Benitez, Joaquin Quiroz-Mercado, Sergio Rangel-Guerrero, Natividad Cruz-Ortiz, Tatiana Nunez-Garcia, Gisela Barrera-Badillo, Lucia Hernandez-Rivas, Irma Lopez-Martinez, Ernesto Ramirez-Gonzalez. |
| EPI_ISL_1700826, EPI_ISL_1700827 | LESP Morelos | Instituto de Diagnostico y Referencia Epidemiologicos (INDRE) | Claudia Wong-Arambula, Abril Rodriguez-Maldonado, Vanessa Rivero-Arredondo, Ariadna Medina-Benitez, Joaquin Quiroz-Mercado, Sergio Rangel-Guerrero, Natividad Cruz-Ortiz, Tatiana Nunez-Garcia, Gisela Barrera-Badillo, Lucia Hernandez-Rivas, Irma Lopez-Martinez, Ernesto Ramirez-Gonzalez. |
| EPI_ISL_1700828, EPI_ISL_1700829, EPI_ISL_1700830 | LESP Guanajuato | Instituto de Diagnostico y Referencia Epidemiologicos (INDRE) | Claudia Wong-Arambula, Abril Rodriguez-Maldonado, Vanessa Rivero-Arredondo, Ariadna Medina-Benitez, Joaquin Quiroz-Mercado, Sergio Rangel-Guerrero, Natividad Cruz-Ortiz, Tatiana Nunez-Garcia, Gisela Barrera-Badillo, Lucia Hernandez-Rivas, Irma Lopez-Martinez, Ernesto Ramirez-Gonzalez. |
| EPI_ISL_1700831, EPI_ISL_1700832 | LESP Queretaro | Instituto de Diagnostico y Referencia Epidemiologicos (INDRE) | Claudia Wong-Arambula, Abril Rodriguez-Maldonado, Vanessa Rivero-Arredondo, Ariadna Medina-Benitez, Joaquin Quiroz-Mercado, Sergio Rangel-Guerrero, Natividad Cruz-Ortiz, Tatiana Nunez-Garcia, Gisela Barrera-Badillo, Lucia Hernandez-Rivas, Irma Lopez-Martinez, Ernesto Ramirez-Gonzalez. |
| EPI_ISL_1700833 | LESPZacatecas | Instituto de Diagnostico y Referencia Epidemiologicos (INDRE) | Claudia Wong-Arambula, Abril Rodriguez-Maldonado, Vanessa Rivero-Arredondo, Ariadna Medina-Benitez, Joaquin Quiroz-Mercado, Sergio Rangel-Guerrero, Natividad Cruz-Ortiz, Tatiana Nunez-Garcia, Gisela Barrera-Badillo, Lucia Hernandez-Rivas, Irma Lopez-Martinez, Ernesto Ramirez-Gonzalez. |
| EPI_ISL_1700834, EPI_ISL_1700835 | LESP Morelos | Instituto de Diagnostico y Referencia Epidemiologicos (INDRE) | Claudia Wong-Arambula, Abril Rodriguez-Maldonado, Vanessa Rivero-Arredondo, Ariadna Medina-Benitez, Joaquin Quiroz-Mercado, Sergio Rangel-Guerrero, Natividad Cruz-Ortiz, Tatiana Nunez-Garcia, Gisela Barrera-Badillo, Lucia Hernandez-Rivas, Irma Lopez-Martinez, Ernesto Ramirez-Gonzalez. |
| EPI_ISL_1700836 | LESPZacatecas | Instituto de Diagnostico y Referencia Epidemiologicos (INDRE) | Claudia Wong-Arambula, Abril Rodriguez-Maldonado, Vanessa Rivero-Arredondo, Ariadna Medina-Benitez, Joaquin Quiroz-Mercado, Sergio Rangel-Guerrero, Natividad Cruz-Ortiz, Tatiana Nunez-Garcia, Gisela Barrera-Badillo, Lucia Hernandez-Rivas, Irma Lopez-Martinez, Ernesto Ramirez-Gonzalez. |
| EPI_ISL_1700837, EPI_ISL_1700838 | LESP Baja California | Instituto de Diagnostico y Referencia Epidemiologicos (INDRE) | Claudia Wong-Arambula, Abril Rodriguez-Maldonado, Vanessa Rivero-Arredondo, Ariadna Medina-Benitez, Joaquin Quiroz-Mercado, Sergio Rangel-Guerrero, Natividad Cruz-Ortiz, Tatiana Nunez-Garcia, Gisela Barrera-Badillo, Lucia Hernandez-Rivas, Irma Lopez-Martinez, Ernesto Ramirez-Gonzalez. |
| EPI_ISL_1700839 | LESP Jalisco | Instituto de Diagnostico y Referencia Epidemiologicos (INDRE) | Claudia Wong-Arambula, Abril Rodriguez-Maldonado, Vanessa Rivero-Arredondo, Ariadna Medina-Benitez, Joaquin Quiroz-Mercado, Sergio Rangel-Guerrero, Natividad Cruz-Ortiz, Tatiana Nunez-Garcia, Gisela Barrera-Badillo, Lucia Hernandez-Rivas, Irma Lopez-Martinez, Ernesto Ramirez-Gonzalez. |
| EPI_ISL_1700840 | LESP Morelos | Instituto de Diagnostico y Referencia Epidemiologicos (INDRE) | Claudia Wong-Arambula, Abril Rodriguez-Maldonado, Vanessa Rivero-Arredondo, Ariadna Medina-Benitez, Joaquin Quiroz-Mercado, Sergio Rangel-Guerrero, Natividad Cruz-Ortiz, Tatiana Nunez-Garcia, Gisela Barrera-Badillo, Lucia Hernandez-Rivas, Irma Lopez-Martinez, Ernesto Ramirez-Gonzalez. |
| EPI_ISL_1700841, EPI_ISL_1700842, EPI_ISL_1700843, EPI_ISL_1700844, EPI_ISL_1700845, EPI_ISL_1700846, EPI_ISL_1700847, EPI_ISL_1700848, EPI_ISL_1700849, EPI_ISL_1700850, EPI_ISL_1700851, EPI_ISL_1700852, EPI_ISL_1700853, EPI_ISL_1700854, EPI_ISL_1700855, EPI_ISL_1700856, EPI_ISL_1700857, EPI_ISL_1700858, EPI_ISL_1700859, EPI_ISL_1700860, EPI_ISL_1700861, EPI_ISL_1700862, EPI_ISL_1700863, EPI_ISL_1700864, EPI_ISL_1700865, EPI_ISL_1700866, EPI_ISL_1700867, EPI_ISL_1700868, EPI_ISL_1700869, EPI_ISL_1700870, EPI_ISL_1700871, EPI_ISL_1700872, EPI_ISL_1700873, EPI_ISL_1700874, EPI_ISL_1700875, EPI_ISL_1700876, EPI_ISL_1700877, EPI_ISL_1700878, EPI_ISL_1700879, EPI_ISL_1700880, EPI_ISL_1700881 | | | |
| see above | MD PHL | MD PHL | Maryland Department of Health Laboratories Administration |
| EPI_ISL_1700906, EPI_ISL_1700907, EPI_ISL_1700908, EPI_ISL_1700909, EPI_ISL_1700910, EPI_ISL_1700911, EPI_ISL_1700912, EPI_ISL_1700913, EPI_ISL_1700914, EPI_ISL_1700915, EPI_ISL_1700916, EPI_ISL_1700917, EPI_ISL_1700918, EPI_ISL_1700919, EPI_ISL_1700920, EPI_ISL_1700921, EPI_ISL_1700922, EPI_ISL_1700923, EPI_ISL_1700924, EPI_ISL_1700925, EPI_ISL_1700926, EPI_ISL_1700927, EPI_ISL_1700928, EPI_ISL_1700929, EPI_ISL_1700930, EPI_ISL_1700931, EPI_ISL_1700932, EPI_ISL_1700933, EPI_ISL_1700934, EPI_ISL_1700935, EPI_ISL_1700936, EPI_ISL_1700937, EPI_ISL_1700938, EPI_ISL_1700939, EPI_ISL_1700940, EPI_ISL_1700941, EPI_ISL_1700942, EPI_ISL_1700943, EPI_ISL_1700944, EPI_ISL_1700945, EPI_ISL_1700946, EPI_ISL_1700947, EPI_ISL_1700948, EPI_ISL_1700949, EPI_ISL_1700950, EPI_ISL_1700951, EPI_ISL_1700952, EPI_ISL_1700953, EPI_ISL_1700954, EPI_ISL_1700955, EPI_ISL_1700956, EPI_ISL_1700957, EPI_ISL_1700958, EPI_ISL_1700959, EPI_ISL_1700960, EPI_ISL_1700961, EPI_ISL_1700962, EPI_ISL_1700963, EPI_ISL_1700964, EPI_ISL_1700965, EPI_ISL_1700966, EPI_ISL_1700967, EPI_ISL_1700968, EPI_ISL_1700969, EPI_ISL_1700970, EPI_ISL_1700971, EPI_ISL_1700972, EPI_ISL_1700973, EPI_ISL_1700974, EPI_ISL_1700975, EPI_ISL_1700976, EPI_ISL_1700977, EPI_ISL_1700978, EPI_ISL_1700979, EPI_ISL_1700980, EPI_ISL_1700981, EPI_ISL_1700982, EPI_ISL_1700983, EPI_ISL_1700984, EPI_ISL_1700985, EPI_ISL_1700986, EPI_ISL_1700987, EPI_ISL_1700988, EPI_ISL_1700989, EPI_ISL_1700990, EPI_ISL_1700991, EPI_ISL_1700992, EPI_ISL_1700993, EPI_ISL_1700994, EPI_ISL_1700995, EPI_ISL_1700996, EPI_ISL_1700997, EPI_ISL_1700998, EPI_ISL_1700999, EPI_ISL_1701000, EPI_ISL_1701001, EPI_ISL_1701002, EPI_ISL_1701003, EPI_ISL_1701004, EPI_ISL_1701005, EPI_ISL_1701006, EPI_ISL_1701007, EPI_ISL_1701008, EPI_ISL_1701009, EPI_ISL_1701010, EPI_ISL_1701011, EPI_ISL_1701012, EPI_ISL_1701013, EPI_ISL_1701014, EPI_ISL_1701015, EPI_ISL_1701016, EPI_ISL_1701017, EPI_ISL_1701018, EPI_ISL_1701019, EPI_ISL_1701020, EPI_ISL_1701021, EPI_ISL_1701022, EPI_ISL_1701023, EPI_ISL_1701024, EPI_ISL_1701025, EPI_ISL_1701026, EPI_ISL_1701027, EPI_ISL_1701028, EPI_ISL_1701029, EPI_ISL_1701030, EPI_ISL_1701031, EPI_ISL_1701032, EPI_ISL_1701033, EPI_ISL_1701034, EPI_ISL_1701035, EPI_ISL_1701036, EPI_ISL_1701037, | | | |

EPI_ISL_1701038, EPI_ISL_1701039

| | | | |
|---|---|---|---|
| see above | Utah Public Health Laboratory | Utah Public Health Laboratory | Erin L. Young, Kelly F. Oakeson, Tara Gallagher |
| EPI_ISL_1701040, EPI_ISL_1701041 | San Joaquin County Public Health Lab | Chan-Zuckerberg Biohub | CZB Cliahub Consortium |
| EPI_ISL_1701042, EPI_ISL_1701043, EPI_ISL_1701044, EPI_ISL_1701045 | CA DPH Viral and Rickettsial Disease Laboratory | Chan-Zuckerberg Biohub | CZB Cliahub Consortium |
| EPI_ISL_1701046 | Alameda County Public Health Lab | Chan-Zuckerberg Biohub | CZB Cliahub Consortium |
| EPI_ISL_1701047, EPI_ISL_1701048, EPI_ISL_1701049 | CA DPH Viral and Rickettsial Disease Laboratory | Chan-Zuckerberg Biohub | CZB Cliahub Consortium |
| EPI_ISL_1701050 | San Joaquin County Public Health Lab | Chan-Zuckerberg Biohub | CZB Cliahub Consortium |
| EPI_ISL_1701051 | CA DPH Viral and Rickettsial Disease Laboratory | Chan-Zuckerberg Biohub | CZB Cliahub Consortium |
| EPI_ISL_1701052 | San Joaquin County Public Health Lab | Chan-Zuckerberg Biohub | CZB Cliahub Consortium |
| EPI_ISL_1701053, EPI_ISL_1701054, EPI_ISL_1701055, EPI_ISL_1701056 | CA DPH Viral and Rickettsial Disease Laboratory | Chan-Zuckerberg Biohub | CZB Cliahub Consortium |
| EPI_ISL_1701057 | Alameda County Public Health Lab | Chan-Zuckerberg Biohub | CZB Cliahub Consortium |
| EPI_ISL_1701058, EPI_ISL_1701059, EPI_ISL_1701060 | CA DPH Viral and Rickettsial Disease Laboratory | Chan-Zuckerberg Biohub | CZB Cliahub Consortium |
| EPI_ISL_1701061 | San Joaquin County Public Health Lab | Chan-Zuckerberg Biohub | CZB Cliahub Consortium |
| EPI_ISL_1701062, EPI_ISL_1701063, EPI_ISL_1701064 | CA DPH Viral and Rickettsial Disease Laboratory | Chan-Zuckerberg Biohub | CZB Cliahub Consortium |
| EPI_ISL_1701065 | Alameda County Public Health Lab | Chan-Zuckerberg Biohub | CZB Cliahub Consortium |
| EPI_ISL_1701066 | County of San Luis Obispo Public Health Laboratory | Chan-Zuckerberg Biohub | CZB Cliahub Consortium |
| EPI_ISL_1701067, EPI_ISL_1701068 | CA DPH Viral and Rickettsial Disease Laboratory | Chan-Zuckerberg Biohub | CZB Cliahub Consortium |
| EPI_ISL_1701069 | San Joaquin County Public Health Lab | Chan-Zuckerberg Biohub | CZB Cliahub Consortium |
| EPI_ISL_1701070 | CA DPH Viral and Rickettsial Disease Laboratory | Chan-Zuckerberg Biohub | CZB Cliahub Consortium |
| EPI_ISL_1701071, EPI_ISL_1701072 | Alameda County Public Health Lab | Chan-Zuckerberg Biohub | CZB Cliahub Consortium |
| EPI_ISL_1701073 | County of San Luis Obispo Public Health Laboratory | Chan-Zuckerberg Biohub | CZB Cliahub Consortium |
| EPI_ISL_1701074 | Alameda County Public Health Lab | Chan-Zuckerberg Biohub | CZB Cliahub Consortium |
| EPI_ISL_1701075, EPI_ISL_1701076, EPI_ISL_1701077 | CA DPH Viral and Rickettsial Disease Laboratory | Chan-Zuckerberg Biohub | CZB Cliahub Consortium |
| EPI_ISL_1701078 | Alameda County Public Health Lab | Chan-Zuckerberg Biohub | CZB Cliahub Consortium |
| EPI_ISL_1701079 | San Joaquin County Public Health Lab | Chan-Zuckerberg Biohub | CZB Cliahub Consortium |
| EPI_ISL_1701080, EPI_ISL_1701081, EPI_ISL_1701082, EPI_ISL_1701083, EPI_ISL_1701084 | Alameda County Public Health Lab | Chan-Zuckerberg Biohub | CZB Cliahub Consortium |
| EPI_ISL_1701085 | CA DPH Viral and Rickettsial Disease Laboratory | Chan-Zuckerberg Biohub | CZB Cliahub Consortium |
| EPI_ISL_1701086 | San Joaquin County Public Health Lab | Chan-Zuckerberg Biohub | CZB Cliahub Consortium |
| EPI_ISL_1701087 | Alameda County Public Health Lab | Chan-Zuckerberg Biohub | CZB Cliahub Consortium |
| EPI_ISL_1701088 | County of San Luis Obispo Public Health Laboratory | Chan-Zuckerberg Biohub | CZB Cliahub Consortium |
| EPI_ISL_1701089 | CA DPH Viral and Rickettsial Disease Laboratory | Chan-Zuckerberg Biohub | CZB Cliahub Consortium |
| EPI_ISL_1701090, EPI_ISL_1701091, EPI_ISL_1701092 | Alameda County Public Health Lab | Chan-Zuckerberg Biohub | CZB Cliahub Consortium |
| EPI_ISL_1701093 | County of San Luis Obispo Public Health Laboratory | Chan-Zuckerberg Biohub | CZB Cliahub Consortium |
| EPI_ISL_1701094, EPI_ISL_1701095 | CA DPH Viral and Rickettsial Disease Laboratory | Chan-Zuckerberg Biohub | CZB Cliahub Consortium |
| EPI_ISL_1701096 | San Joaquin County Public Health Lab | Chan-Zuckerberg Biohub | CZB Cliahub Consortium |
| EPI_ISL_1701097, EPI_ISL_1701098 | CA DPH Viral and Rickettsial Disease Laboratory | Chan-Zuckerberg Biohub | CZB Cliahub Consortium |
| EPI_ISL_1701099 | Alameda County Public Health Lab | Chan-Zuckerberg Biohub | CZB Cliahub Consortium |
| EPI_ISL_1701100, EPI_ISL_1701101, EPI_ISL_1701102, EPI_ISL_1701103 | CA DPH Viral and Rickettsial Disease Laboratory | Chan-Zuckerberg Biohub | CZB Cliahub Consortium |
| EPI_ISL_1701104 | Alameda County Public Health Lab | Chan-Zuckerberg Biohub | CZB Cliahub Consortium |
| EPI_ISL_1701105 | CA DPH Viral and Rickettsial Disease Laboratory | Chan-Zuckerberg Biohub | CZB Cliahub Consortium |
| EPI_ISL_1701106 | Alameda County Public Health Lab | Chan-Zuckerberg Biohub | CZB Cliahub Consortium |
| EPI_ISL_1701107, EPI_ISL_1701108, EPI_ISL_1701109, EPI_ISL_1701110, EPI_ISL_1701111, EPI_ISL_1701112, EPI_ISL_1701113, EPI_ISL_1701114 | CA DPH Viral and Rickettsial Disease Laboratory | Chan-Zuckerberg Biohub | CZB Cliahub Consortium |
| EPI_ISL_1701115 | Alameda County Public Health Lab | Chan-Zuckerberg Biohub | CZB Cliahub Consortium |
| EPI_ISL_1701116 | CA DPH Viral and Rickettsial Disease Laboratory | Chan-Zuckerberg Biohub | CZB Cliahub Consortium |
| EPI_ISL_1701117 | San Joaquin County Public Health Lab | Chan-Zuckerberg Biohub | CZB Cliahub Consortium |

| | | | |
|---|---|---|---|
| EPI_ISL_1701118 | CA DPH Viral and Rickettsial Disease Laboratory | Chan-Zuckerberg Biohub | CZB Cliahub Consortium |
| EPI_ISL_1701119, EPI_ISL_1701120 | Alameda County Public Health Lab | Chan-Zuckerberg Biohub | CZB Cliahub Consortium |
| EPI_ISL_1701121, EPI_ISL_1701122 | CA DPH Viral and Rickettsial Disease Laboratory | Chan-Zuckerberg Biohub | CZB Cliahub Consortium |
| EPI_ISL_1701123, EPI_ISL_1701124 | Alameda County Public Health Lab | Chan-Zuckerberg Biohub | CZB Cliahub Consortium |
| EPI_ISL_1701125, EPI_ISL_1701126 | CA DPH Viral and Rickettsial Disease Laboratory | Chan-Zuckerberg Biohub | CZB Cliahub Consortium |
| EPI_ISL_1701127 | Alameda County Public Health Lab | Chan-Zuckerberg Biohub | CZB Cliahub Consortium |
| EPI_ISL_1701128, EPI_ISL_1701129, EPI_ISL_1701130, EPI_ISL_1701131 | CA DPH Viral and Rickettsial Disease Laboratory | Chan-Zuckerberg Biohub | CZB Cliahub Consortium |
| EPI_ISL_1701132 | Alameda County Public Health Lab | Chan-Zuckerberg Biohub | CZB Cliahub Consortium |
| EPI_ISL_1701133, EPI_ISL_1701134 | CA DPH Viral and Rickettsial Disease Laboratory | Chan-Zuckerberg Biohub | CZB Cliahub Consortium |
| EPI_ISL_1701135 | Alameda County Public Health Lab | Chan-Zuckerberg Biohub | CZB Cliahub Consortium |
| EPI_ISL_1701136 | County of San Luis Obispo Public Health Laboratory | Chan-Zuckerberg Biohub | CZB Cliahub Consortium |
| EPI_ISL_1701137 | Alameda County Public Health Lab | Chan-Zuckerberg Biohub | CZB Cliahub Consortium |
| EPI_ISL_1701138, EPI_ISL_1701139, EPI_ISL_1701140, EPI_ISL_1701141 | CA DPH Viral and Rickettsial Disease Laboratory | Chan-Zuckerberg Biohub | CZB Cliahub Consortium |
| EPI_ISL_1701142 | San Joaquin County Public Health Lab | Chan-Zuckerberg Biohub | CZB Cliahub Consortium |
| EPI_ISL_1701143, EPI_ISL_1701144, EPI_ISL_1701145, EPI_ISL_1701146 | CA DPH Viral and Rickettsial Disease Laboratory | Chan-Zuckerberg Biohub | CZB Cliahub Consortium |
| EPI_ISL_1701147, EPI_ISL_1701148 | Alameda County Public Health Lab | Chan-Zuckerberg Biohub | CZB Cliahub Consortium |
| EPI_ISL_1701149 | CA DPH Viral and Rickettsial Disease Laboratory | Chan-Zuckerberg Biohub | CZB Cliahub Consortium |
| EPI_ISL_1701150 | Alameda County Public Health Lab | Chan-Zuckerberg Biohub | CZB Cliahub Consortium |
| EPI_ISL_1701151, EPI_ISL_1701152, EPI_ISL_1701153, EPI_ISL_1701154 | CA DPH Viral and Rickettsial Disease Laboratory | Chan-Zuckerberg Biohub | CZB Cliahub Consortium |
| EPI_ISL_1701155, EPI_ISL_1701156, EPI_ISL_1701157 | Alameda County Public Health Lab | Chan-Zuckerberg Biohub | CZB Cliahub Consortium |
| EPI_ISL_1701158, EPI_ISL_1701159, EPI_ISL_1701160 | CA DPH Viral and Rickettsial Disease Laboratory | Chan-Zuckerberg Biohub | CZB Cliahub Consortium |
| EPI_ISL_1701161, EPI_ISL_1701162 | Alameda County Public Health Lab | Chan-Zuckerberg Biohub | CZB Cliahub Consortium |
| EPI_ISL_1701163, EPI_ISL_1701164, EPI_ISL_1701165, EPI_ISL_1701166 | CA DPH Viral and Rickettsial Disease Laboratory | Chan-Zuckerberg Biohub | CZB Cliahub Consortium |
| EPI_ISL_1701167 | Alameda County Public Health Lab | Chan-Zuckerberg Biohub | CZB Cliahub Consortium |
| EPI_ISL_1701168 | County of San Luis Obispo Public Health Laboratory | Chan-Zuckerberg Biohub | CZB Cliahub Consortium |
| EPI_ISL_1701169, EPI_ISL_1701170 | CA DPH Viral and Rickettsial Disease Laboratory | Chan-Zuckerberg Biohub | CZB Cliahub Consortium |
| EPI_ISL_1701171, EPI_ISL_1701172 | Alameda County Public Health Lab | Chan-Zuckerberg Biohub | CZB Cliahub Consortium |
| EPI_ISL_1701173 | CA DPH Viral and Rickettsial Disease Laboratory | Chan-Zuckerberg Biohub | CZB Cliahub Consortium |
| EPI_ISL_1701174, EPI_ISL_1701175, EPI_ISL_1701176, EPI_ISL_1701177, EPI_ISL_1701178, EPI_ISL_1701179, EPI_ISL_1701180 | Alameda County Public Health Lab | Chan-Zuckerberg Biohub | CZB Cliahub Consortium |
| EPI_ISL_1701181, EPI_ISL_1701182, EPI_ISL_1701183, EPI_ISL_1701184, EPI_ISL_1701185, EPI_ISL_1701186, EPI_ISL_1701187 | CA DPH Viral and Rickettsial Disease Laboratory | Chan-Zuckerberg Biohub | CZB Cliahub Consortium |
| EPI_ISL_1701188 | Alameda County Public Health Lab | Chan-Zuckerberg Biohub | CZB Cliahub Consortium |
| EPI_ISL_1701189 | County of San Luis Obispo Public Health Laboratory | Chan-Zuckerberg Biohub | CZB Cliahub Consortium |
| EPI_ISL_1701190, EPI_ISL_1701191, EPI_ISL_1701192, EPI_ISL_1701193 | CA DPH Viral and Rickettsial Disease Laboratory | Chan-Zuckerberg Biohub | CZB Cliahub Consortium |
| EPI_ISL_1701194 | Alameda County Public Health Lab | Chan-Zuckerberg Biohub | CZB Cliahub Consortium |
| EPI_ISL_1701195, EPI_ISL_1701196, EPI_ISL_1701197, EPI_ISL_1701198, EPI_ISL_1701199, EPI_ISL_1701200 | CA DPH Viral and Rickettsial Disease Laboratory | Chan-Zuckerberg Biohub | CZB Cliahub Consortium |
| EPI_ISL_1701201 | County of San Luis Obispo Public Health Laboratory | Chan-Zuckerberg Biohub | CZB Cliahub Consortium |
| EPI_ISL_1701202, EPI_ISL_1701203 | CA DPH Viral and Rickettsial Disease Laboratory | Chan-Zuckerberg Biohub | CZB Cliahub Consortium |

| | | | |
|---|---|---|---|
| EPI_ISL_1701205 | Alameda County Public Health Lab | Chan-Zuckerberg Biohub | CZB Cliahub Consortium |
| EPI_ISL_1701206, EPI_ISL_1701207 | CA DPH Viral and Rickettsial Disease Laboratory | Chan-Zuckerberg Biohub | CZB Cliahub Consortium |
| EPI_ISL_1701208 | San Joaquin County Public Health Lab | Chan-Zuckerberg Biohub | CZB Cliahub Consortium |
| EPI_ISL_1701209 | CA DPH Viral and Rickettsial Disease Laboratory | Chan-Zuckerberg Biohub | CZB Cliahub Consortium |
| EPI_ISL_1701210 | Alameda County Public Health Lab | Chan-Zuckerberg Biohub | CZB Cliahub Consortium |
| EPI_ISL_1701211 | San Joaquin County Public Health Lab | Chan-Zuckerberg Biohub | CZB Cliahub Consortium |
| EPI_ISL_1701212, EPI_ISL_1701213, EPI_ISL_1701214 | Alameda County Public Health Lab | Chan-Zuckerberg Biohub | CZB Cliahub Consortium |
| EPI_ISL_1701215, EPI_ISL_1701216 | CA DPH Viral and Rickettsial Disease Laboratory | Chan-Zuckerberg Biohub | CZB Cliahub Consortium |
| EPI_ISL_1701217, EPI_ISL_1701218 | Alameda County Public Health Lab | Chan-Zuckerberg Biohub | CZB Cliahub Consortium |
| EPI_ISL_1701219 | County of San Luis Obispo Public Health Laboratory | Chan-Zuckerberg Biohub | CZB Cliahub Consortium |
| EPI_ISL_1701220, EPI_ISL_1701221 | CA DPH Viral and Rickettsial Disease Laboratory | Chan-Zuckerberg Biohub | CZB Cliahub Consortium |
| EPI_ISL_1701222 | Alameda County Public Health Lab | Chan-Zuckerberg Biohub | CZB Cliahub Consortium |
| EPI_ISL_1701223 | CA DPH Viral and Rickettsial Disease Laboratory | Chan-Zuckerberg Biohub | CZB Cliahub Consortium |
| EPI_ISL_1701224 | Alameda County Public Health Lab | Chan-Zuckerberg Biohub | CZB Cliahub Consortium |
| EPI_ISL_1701225, EPI_ISL_1701226, EPI_ISL_1701227, EPI_ISL_1701229 | CA DPH Viral and Rickettsial Disease Laboratory | Chan-Zuckerberg Biohub | CZB Cliahub Consortium |
| EPI_ISL_1701230 | Alameda County Public Health Lab | Chan-Zuckerberg Biohub | CZB Cliahub Consortium |
| EPI_ISL_1701231, EPI_ISL_1701232 | CA DPH Viral and Rickettsial Disease Laboratory | Chan-Zuckerberg Biohub | CZB Cliahub Consortium |
| EPI_ISL_1701233, EPI_ISL_1701234, EPI_ISL_1701235 | Alameda County Public Health Lab | Chan-Zuckerberg Biohub | CZB Cliahub Consortium |
| EPI_ISL_1701236 | San Joaquin County Public Health Lab | Chan-Zuckerberg Biohub | CZB Cliahub Consortium |
| EPI_ISL_1701237, EPI_ISL_1701238, EPI_ISL_1701239, EPI_ISL_1701240, EPI_ISL_1701241, EPI_ISL_1701242, EPI_ISL_1701243, EPI_ISL_1701244 | CA DPH Viral and Rickettsial Disease Laboratory | Chan-Zuckerberg Biohub | CZB Cliahub Consortium |
| EPI_ISL_1701245 | Alameda County Public Health Lab | Chan-Zuckerberg Biohub | CZB Cliahub Consortium |
| EPI_ISL_1701246 | CA DPH Viral and Rickettsial Disease Laboratory | Chan-Zuckerberg Biohub | CZB Cliahub Consortium |
| EPI_ISL_1701247 | Alameda County Public Health Lab | Chan-Zuckerberg Biohub | CZB Cliahub Consortium |
| EPI_ISL_1701248 | CA DPH Viral and Rickettsial Disease Laboratory | Chan-Zuckerberg Biohub | CZB Cliahub Consortium |
| EPI_ISL_1701249, EPI_ISL_1701250 | Alameda County Public Health Lab | Chan-Zuckerberg Biohub | CZB Cliahub Consortium |
| EPI_ISL_1701251 | Orange County Public Health Lab | Chan-Zuckerberg Biohub | CZB Cliahub Consortium |
| EPI_ISL_1701252, EPI_ISL_1701253, EPI_ISL_1701254, EPI_ISL_1701255, EPI_ISL_1701256, EPI_ISL_1701257, EPI_ISL_1701258, EPI_ISL_1701259, EPI_ISL_1701260, EPI_ISL_1701261 | CA DPH Viral and Rickettsial Disease Laboratory | Chan-Zuckerberg Biohub | CZB Cliahub Consortium |
| EPI_ISL_1701262 | San Joaquin County Public Health Lab | Chan-Zuckerberg Biohub | CZB Cliahub Consortium |
| EPI_ISL_1701263 | CA DPH Viral and Rickettsial Disease Laboratory | Chan-Zuckerberg Biohub | CZB Cliahub Consortium |
| EPI_ISL_1701264 | County of San Luis Obispo Public Health Laboratory | Chan-Zuckerberg Biohub | CZB Cliahub Consortium |
| EPI_ISL_1701265 | CA DPH Viral and Rickettsial Disease Laboratory | Chan-Zuckerberg Biohub | CZB Cliahub Consortium |
| EPI_ISL_1701266 | County of San Luis Obispo Public Health Laboratory | Chan-Zuckerberg Biohub | CZB Cliahub Consortium |
| EPI_ISL_1701267 | Alameda County Public Health Lab | Chan-Zuckerberg Biohub | CZB Cliahub Consortium |
| EPI_ISL_1701268 | County of San Luis Obispo Public Health Laboratory | Chan-Zuckerberg Biohub | CZB Cliahub Consortium |
| EPI_ISL_1701269, EPI_ISL_1701270, EPI_ISL_1701271, EPI_ISL_1701272 | CA DPH Viral and Rickettsial Disease Laboratory | Chan-Zuckerberg Biohub | CZB Cliahub Consortium |
| EPI_ISL_1701273, EPI_ISL_1701274 | Alameda County Public Health Lab | Chan-Zuckerberg Biohub | CZB Cliahub Consortium |
| EPI_ISL_1701275, EPI_ISL_1701276 | CA DPH Viral and Rickettsial Disease Laboratory | Chan-Zuckerberg Biohub | CZB Cliahub Consortium |
| EPI_ISL_1701277 | Alameda County Public Health Lab | Chan-Zuckerberg Biohub | CZB Cliahub Consortium |
| EPI_ISL_1701278, EPI_ISL_1701279 | CA DPH Viral and Rickettsial Disease Laboratory | Chan-Zuckerberg Biohub | CZB Cliahub Consortium |
| EPI_ISL_1701280, EPI_ISL_1701281 | San Joaquin County Public Health Lab | Chan-Zuckerberg Biohub | CZB Cliahub Consortium |
| EPI_ISL_1701282, EPI_ISL_1701283, EPI_ISL_1701284 | CA DPH Viral and Rickettsial Disease Laboratory | Chan-Zuckerberg Biohub | CZB Cliahub Consortium |
| EPI_ISL_1701285 | Alameda County Public Health Lab | Chan-Zuckerberg Biohub | CZB Cliahub Consortium |
| EPI_ISL_1701286, | CA DPH Viral and Rickettsial Disease | Chan-Zuckerberg Biohub | CZB Cliahub Consortium |

| | | | |
|---|---|---|---|
| EPI_ISL_1701287, EPI_ISL_1701288 | Laboratory | | |
| EPI_ISL_1701289 | Alameda County Public Health Lab | Chan-Zuckerberg Biohub | CZB Cliahub Consortium |
| EPI_ISL_1701290 | San Joaquin County Public Health Lab | Chan-Zuckerberg Biohub | CZB Cliahub Consortium |
| EPI_ISL_1701291 | Alameda County Public Health Lab | Chan-Zuckerberg Biohub | CZB Cliahub Consortium |
| EPI_ISL_1701292 | CA DPH Viral and Rickettsial Disease Laboratory | Chan-Zuckerberg Biohub | CZB Cliahub Consortium |
| EPI_ISL_1701293 | Alameda County Public Health Lab | Chan-Zuckerberg Biohub | CZB Cliahub Consortium |
| EPI_ISL_1701294 | CA DPH Viral and Rickettsial Disease Laboratory | Chan-Zuckerberg Biohub | CZB Cliahub Consortium |
| EPI_ISL_1701295 | Alameda County Public Health Lab | Chan-Zuckerberg Biohub | CZB Cliahub Consortium |
| EPI_ISL_1701296 | CA DPH Viral and Rickettsial Disease Laboratory | Chan-Zuckerberg Biohub | CZB Cliahub Consortium |
| EPI_ISL_1701297 | County of San Luis Obispo Public Health Laboratory | Chan-Zuckerberg Biohub | CZB Cliahub Consortium |
| EPI_ISL_1701298 | CA DPH Viral and Rickettsial Disease Laboratory | Chan-Zuckerberg Biohub | CZB Cliahub Consortium |
| EPI_ISL_1701299 | Alameda County Public Health Lab | Chan-Zuckerberg Biohub | CZB Cliahub Consortium |
| EPI_ISL_1701300 | CA DPH Viral and Rickettsial Disease Laboratory | Chan-Zuckerberg Biohub | CZB Cliahub Consortium |
| EPI_ISL_1701301, EPI_ISL_1701302, EPI_ISL_1701303 | Alameda County Public Health Lab | Chan-Zuckerberg Biohub | CZB Cliahub Consortium |
| EPI_ISL_1701304, EPI_ISL_1701305 | CA DPH Viral and Rickettsial Disease Laboratory | Chan-Zuckerberg Biohub | CZB Cliahub Consortium |
| EPI_ISL_1701306, EPI_ISL_1701307 | Alameda County Public Health Lab | Chan-Zuckerberg Biohub | CZB Cliahub Consortium |
| EPI_ISL_1701308, EPI_ISL_1701309, EPI_ISL_1701310 | CA DPH Viral and Rickettsial Disease Laboratory | Chan-Zuckerberg Biohub | CZB Cliahub Consortium |
| EPI_ISL_1701396 | Humboldt County Public Health Laboratory | Chan-Zuckerberg Biohub | CZB Cliahub Consortium |
| EPI_ISL_1701397 | Alameda County Public Health Lab | Chan-Zuckerberg Biohub | CZB Cliahub Consortium |
| EPI_ISL_1701398 | Humboldt County Public Health Laboratory | Chan-Zuckerberg Biohub | CZB Cliahub Consortium |
| EPI_ISL_1701399, EPI_ISL_1701400 | Orange County Public Health Lab | Chan-Zuckerberg Biohub | CZB Cliahub Consortium |
| EPI_ISL_1701401 | Santa Clara County Public Health Laboratory | Chan-Zuckerberg Biohub | CZB Cliahub Consortium |
| EPI_ISL_1701402 | Humboldt County Public Health Laboratory | Chan-Zuckerberg Biohub | CZB Cliahub Consortium |
| EPI_ISL_1701403 | Alameda County Public Health Lab | Chan-Zuckerberg Biohub | CZB Cliahub Consortium |
| EPI_ISL_1701404 | Humboldt County Public Health Laboratory | Chan-Zuckerberg Biohub | CZB Cliahub Consortium |
| EPI_ISL_1701405, EPI_ISL_1701406 | Santa Clara County Public Health Laboratory | Chan-Zuckerberg Biohub | CZB Cliahub Consortium |
| EPI_ISL_1701407 | Alameda County Public Health Lab | Chan-Zuckerberg Biohub | CZB Cliahub Consortium |
| EPI_ISL_1701408 | County of San Luis Obispo Public Health Laboratory | Chan-Zuckerberg Biohub | CZB Cliahub Consortium |
| EPI_ISL_1701409 | Alameda County Public Health Lab | Chan-Zuckerberg Biohub | CZB Cliahub Consortium |
| EPI_ISL_1701410 | Santa Clara County Public Health Laboratory | Chan-Zuckerberg Biohub | CZB Cliahub Consortium |
| EPI_ISL_1701411, EPI_ISL_1701412 | Humboldt County Public Health Laboratory | Chan-Zuckerberg Biohub | CZB Cliahub Consortium |
| EPI_ISL_1701413, EPI_ISL_1701414 | Santa Clara County Public Health Laboratory | Chan-Zuckerberg Biohub | CZB Cliahub Consortium |
| EPI_ISL_1701415, EPI_ISL_1701416, EPI_ISL_1701417, EPI_ISL_1701418 | Alameda County Public Health Lab | Chan-Zuckerberg Biohub | CZB Cliahub Consortium |
| EPI_ISL_1701419 | County of San Luis Obispo Public Health Laboratory | Chan-Zuckerberg Biohub | CZB Cliahub Consortium |
| EPI_ISL_1701420 | Humboldt County Public Health Laboratory | Chan-Zuckerberg Biohub | CZB Cliahub Consortium |
| EPI_ISL_1701421 | Alameda County Public Health Lab | Chan-Zuckerberg Biohub | CZB Cliahub Consortium |
| EPI_ISL_1701422 | Humboldt County Public Health Laboratory | Chan-Zuckerberg Biohub | CZB Cliahub Consortium |
| EPI_ISL_1701423 | County of San Luis Obispo Public Health Laboratory | Chan-Zuckerberg Biohub | CZB Cliahub Consortium |
| EPI_ISL_1701424 | Alameda County Public Health Lab | Chan-Zuckerberg Biohub | CZB Cliahub Consortium |
| EPI_ISL_1701426 | Orange County Public Health Lab | Chan-Zuckerberg Biohub | CZB Cliahub Consortium |
| EPI_ISL_1701427 | Alameda County Public Health Lab | Chan-Zuckerberg Biohub | CZB Cliahub Consortium |
| EPI_ISL_1701428, EPI_ISL_1701429 | Humboldt County Public Health Laboratory | Chan-Zuckerberg Biohub | CZB Cliahub Consortium |
| EPI_ISL_1701430, EPI_ISL_1701431 | Orange County Public Health Lab | Chan-Zuckerberg Biohub | CZB Cliahub Consortium |
| EPI_ISL_1701432 | Alameda County Public Health Lab | Chan-Zuckerberg Biohub | CZB Cliahub Consortium |
| EPI_ISL_1701433 | Humboldt County Public Health Laboratory | Chan-Zuckerberg Biohub | CZB Cliahub Consortium |
| EPI_ISL_1701434 | Alameda County Public Health Lab | Chan-Zuckerberg Biohub | CZB Cliahub Consortium |
| EPI_ISL_1701435 | Humboldt County Public Health Laboratory | Chan-Zuckerberg Biohub | CZB Cliahub Consortium |
| EPI_ISL_1701436 | Alameda County Public Health Lab | Chan-Zuckerberg Biohub | CZB Cliahub Consortium |
| EPI_ISL_1701437 | Santa Clara County Public Health Laboratory | Chan-Zuckerberg Biohub | CZB Cliahub Consortium |
| EPI_ISL_1701438 | Orange County Public Health Lab | Chan-Zuckerberg Biohub | CZB Cliahub Consortium |
| EPI_ISL_1701439, EPI_ISL_1701440 | Santa Clara County Public Health Laboratory | Chan-Zuckerberg Biohub | CZB Cliahub Consortium |

| | | | |
|---|---|---|---|
| EPI_ISL_1701441 | Humboldt County Public Health Laboratory | Chan-Zuckerberg Biohub | CZB Cliahub Consortium |
| EPI_ISL_1701442, EPI_ISL_1701443 | Alameda County Public Health Lab | Chan-Zuckerberg Biohub | CZB Cliahub Consortium |
| EPI_ISL_1701444 | Humboldt County Public Health Laboratory | Chan-Zuckerberg Biohub | CZB Cliahub Consortium |
| EPI_ISL_1701445 | Santa Clara County Public Health Laboratory | Chan-Zuckerberg Biohub | CZB Cliahub Consortium |
| EPI_ISL_1701446, EPI_ISL_1701447, EPI_ISL_1701448 | Humboldt County Public Health Laboratory | Chan-Zuckerberg Biohub | CZB Cliahub Consortium |
| EPI_ISL_1701449 | Alameda County Public Health Lab | Chan-Zuckerberg Biohub | CZB Cliahub Consortium |
| EPI_ISL_1701450 | Orange County Public Health Lab | Chan-Zuckerberg Biohub | CZB Cliahub Consortium |
| EPI_ISL_1701451 | Humboldt County Public Health Laboratory | Chan-Zuckerberg Biohub | CZB Cliahub Consortium |
| EPI_ISL_1701452, EPI_ISL_1701453 | Alameda County Public Health Lab | Chan-Zuckerberg Biohub | CZB Cliahub Consortium |
| EPI_ISL_1701454 | Humboldt County Public Health Laboratory | Chan-Zuckerberg Biohub | CZB Cliahub Consortium |
| EPI_ISL_1701455 | Alameda County Public Health Lab | Chan-Zuckerberg Biohub | CZB Cliahub Consortium |
| EPI_ISL_1701456 | California Institute of Technology | Chan-Zuckerberg Biohub | CZB Cliahub Consortium |
| EPI_ISL_1701457 | Santa Clara County Public Health Laboratory | Chan-Zuckerberg Biohub | CZB Cliahub Consortium |
| EPI_ISL_1701458 | Humboldt County Public Health Laboratory | Chan-Zuckerberg Biohub | CZB Cliahub Consortium |
| EPI_ISL_1701459 | Alameda County Public Health Lab | Chan-Zuckerberg Biohub | CZB Cliahub Consortium |
| EPI_ISL_1701460, EPI_ISL_1701461 | County of San Luis Obispo Public Health Laboratory | Chan-Zuckerberg Biohub | CZB Cliahub Consortium |
| EPI_ISL_1701462 | Santa Clara County Public Health Laboratory | Chan-Zuckerberg Biohub | CZB Cliahub Consortium |
| EPI_ISL_1701463 | Alameda County Public Health Lab | Chan-Zuckerberg Biohub | CZB Cliahub Consortium |
| EPI_ISL_1701464 | County of San Luis Obispo Public Health Laboratory | Chan-Zuckerberg Biohub | CZB Cliahub Consortium |
| EPI_ISL_1701465 | Orange County Public Health Lab | Chan-Zuckerberg Biohub | CZB Cliahub Consortium |
| EPI_ISL_1701466 | Alameda County Public Health Lab | Chan-Zuckerberg Biohub | CZB Cliahub Consortium |
| EPI_ISL_1701467 | Santa Clara County Public Health Laboratory | Chan-Zuckerberg Biohub | CZB Cliahub Consortium |
| EPI_ISL_1701468 | County of San Luis Obispo Public Health Laboratory | Chan-Zuckerberg Biohub | CZB Cliahub Consortium |
| EPI_ISL_1701469 | Santa Clara County Public Health Laboratory | Chan-Zuckerberg Biohub | CZB Cliahub Consortium |
| EPI_ISL_1701470 | Alameda County Public Health Lab | Chan-Zuckerberg Biohub | CZB Cliahub Consortium |
| EPI_ISL_1701471 | County of San Luis Obispo Public Health Laboratory | Chan-Zuckerberg Biohub | CZB Cliahub Consortium |
| EPI_ISL_1701472 | Humboldt County Public Health Laboratory | Chan-Zuckerberg Biohub | CZB Cliahub Consortium |
| EPI_ISL_1701473 | California Institute of Technology | Chan-Zuckerberg Biohub | CZB Cliahub Consortium |
| EPI_ISL_1701474 | Humboldt County Public Health Laboratory | Chan-Zuckerberg Biohub | CZB Cliahub Consortium |
| EPI_ISL_1701475 | Alameda County Public Health Lab | Chan-Zuckerberg Biohub | CZB Cliahub Consortium |
| EPI_ISL_1701476 | County of San Luis Obispo Public Health Laboratory | Chan-Zuckerberg Biohub | CZB Cliahub Consortium |
| EPI_ISL_1701477 | Santa Clara County Public Health Laboratory | Chan-Zuckerberg Biohub | CZB Cliahub Consortium |
| EPI_ISL_1701478 | Orange County Public Health Lab | Chan-Zuckerberg Biohub | CZB Cliahub Consortium |
| EPI_ISL_1701479 | Humboldt County Public Health Laboratory | Chan-Zuckerberg Biohub | CZB Cliahub Consortium |
| EPI_ISL_1701480, EPI_ISL_1701481, EPI_ISL_1701482 | Santa Clara County Public Health Laboratory | Chan-Zuckerberg Biohub | CZB Cliahub Consortium |
| EPI_ISL_1701483 | Orange County Public Health Lab | Chan-Zuckerberg Biohub | CZB Cliahub Consortium |
| EPI_ISL_1701484 | Humboldt County Public Health Laboratory | Chan-Zuckerberg Biohub | CZB Cliahub Consortium |
| EPI_ISL_1701485 | Alameda County Public Health Lab | Chan-Zuckerberg Biohub | CZB Cliahub Consortium |
| EPI_ISL_1701486 | Santa Clara County Public Health Laboratory | Chan-Zuckerberg Biohub | CZB Cliahub Consortium |
| EPI_ISL_1701487, EPI_ISL_1701488, EPI_ISL_1701489 | Alameda County Public Health Lab | Chan-Zuckerberg Biohub | CZB Cliahub Consortium |
| EPI_ISL_1701490 | County of San Luis Obispo Public Health Laboratory | Chan-Zuckerberg Biohub | CZB Cliahub Consortium |
| EPI_ISL_1701491 | Orange County Public Health Lab | Chan-Zuckerberg Biohub | CZB Cliahub Consortium |
| EPI_ISL_1701492 | Alameda County Public Health Lab | Chan-Zuckerberg Biohub | CZB Cliahub Consortium |
| EPI_ISL_1701493 | Humboldt County Public Health Laboratory | Chan-Zuckerberg Biohub | CZB Cliahub Consortium |
| EPI_ISL_1701494, EPI_ISL_1701495 | California Institute of Technology | Chan-Zuckerberg Biohub | CZB Cliahub Consortium |
| EPI_ISL_1701496 | Santa Clara County Public Health Laboratory | Chan-Zuckerberg Biohub | CZB Cliahub Consortium |
| EPI_ISL_1701497 | Humboldt County Public Health Laboratory | Chan-Zuckerberg Biohub | CZB Cliahub Consortium |
| EPI_ISL_1701498 | Santa Clara County Public Health Laboratory | Chan-Zuckerberg Biohub | CZB Cliahub Consortium |
| EPI_ISL_1701499 | Alameda County Public Health Lab | Chan-Zuckerberg Biohub | CZB Cliahub Consortium |
| EPI_ISL_1701500 | Humboldt County Public Health Laboratory | Chan-Zuckerberg Biohub | CZB Cliahub Consortium |
| EPI_ISL_1701501, EPI_ISL_1701502 | Santa Clara County Public Health Laboratory | Chan-Zuckerberg Biohub | CZB Cliahub Consortium |

| | | | |
|---|---|---|---|
| EPI_ISL_1701503 | Alameda County Public Health Lab | Chan-Zuckerberg Biohub | CZB Cliahub Consortium |
| EPI_ISL_1701504, EPI_ISL_1701505 | Santa Clara County Public Health Laboratory | Chan-Zuckerberg Biohub | CZB Cliahub Consortium |
| EPI_ISL_1701506 | Alameda County Public Health Lab | Chan-Zuckerberg Biohub | CZB Cliahub Consortium |
| EPI_ISL_1701507 | Humboldt County Public Health Laboratory | Chan-Zuckerberg Biohub | CZB Cliahub Consortium |
| EPI_ISL_1701508 | Alameda County Public Health Lab | Chan-Zuckerberg Biohub | CZB Cliahub Consortium |
| EPI_ISL_1701509 | Humboldt County Public Health Laboratory | Chan-Zuckerberg Biohub | CZB Cliahub Consortium |
| EPI_ISL_1701510 | Santa Clara County Public Health Laboratory | Chan-Zuckerberg Biohub | CZB Cliahub Consortium |
| EPI_ISL_1701511 | Orange County Public Health Lab | Chan-Zuckerberg Biohub | CZB Cliahub Consortium |
| EPI_ISL_1701512 | Alameda County Public Health Lab | Chan-Zuckerberg Biohub | CZB Cliahub Consortium |
| EPI_ISL_1701513 | Orange County Public Health Lab | Chan-Zuckerberg Biohub | CZB Cliahub Consortium |
| EPI_ISL_1701514, EPI_ISL_1701515 | Alameda County Public Health Lab | Chan-Zuckerberg Biohub | CZB Cliahub Consortium |
| EPI_ISL_1701517, EPI_ISL_1701518 | Santa Clara County Public Health Laboratory | Chan-Zuckerberg Biohub | CZB Cliahub Consortium |
| EPI_ISL_1701519 | County of San Luis Obispo Public Health Laboratory | Chan-Zuckerberg Biohub | CZB Cliahub Consortium |
| EPI_ISL_1701520 | Santa Clara County Public Health Laboratory | Chan-Zuckerberg Biohub | CZB Cliahub Consortium |
| EPI_ISL_1701521, EPI_ISL_1701522 | Alameda County Public Health Lab | Chan-Zuckerberg Biohub | CZB Cliahub Consortium |
| EPI_ISL_1701523 | Santa Clara County Public Health Laboratory | Chan-Zuckerberg Biohub | CZB Cliahub Consortium |
| EPI_ISL_1701524 | Alameda County Public Health Lab | Chan-Zuckerberg Biohub | CZB Cliahub Consortium |
| EPI_ISL_1701525, EPI_ISL_1701526 | Santa Clara County Public Health Laboratory | Chan-Zuckerberg Biohub | CZB Cliahub Consortium |
| EPI_ISL_1701527 | Humboldt County Public Health Laboratory | Chan-Zuckerberg Biohub | CZB Cliahub Consortium |
| EPI_ISL_1701528 | Santa Clara County Public Health Laboratory | Chan-Zuckerberg Biohub | CZB Cliahub Consortium |
| EPI_ISL_1701529 | Alameda County Public Health Lab | Chan-Zuckerberg Biohub | CZB Cliahub Consortium |
| EPI_ISL_1701530 | Santa Clara County Public Health Laboratory | Chan-Zuckerberg Biohub | CZB Cliahub Consortium |
| EPI_ISL_1701531 | County of San Luis Obispo Public Health Laboratory | Chan-Zuckerberg Biohub | CZB Cliahub Consortium |
| EPI_ISL_1701532 | Santa Clara County Public Health Laboratory | Chan-Zuckerberg Biohub | CZB Cliahub Consortium |
| EPI_ISL_1701533 | Orange County Public Health Lab | Chan-Zuckerberg Biohub | CZB Cliahub Consortium |
| EPI_ISL_1701534 | Humboldt County Public Health Laboratory | Chan-Zuckerberg Biohub | CZB Cliahub Consortium |
| EPI_ISL_1701535 | Alameda County Public Health Lab | Chan-Zuckerberg Biohub | CZB Cliahub Consortium |
| EPI_ISL_1701536 | Santa Clara County Public Health Laboratory | Chan-Zuckerberg Biohub | CZB Cliahub Consortium |
| EPI_ISL_1701537 | Orange County Public Health Lab | Chan-Zuckerberg Biohub | CZB Cliahub Consortium |
| EPI_ISL_1701538 | Humboldt County Public Health Laboratory | Chan-Zuckerberg Biohub | CZB Cliahub Consortium |
| EPI_ISL_1701539, EPI_ISL_1701540 | County of San Luis Obispo Public Health Laboratory | Chan-Zuckerberg Biohub | CZB Cliahub Consortium |
| EPI_ISL_1701541, EPI_ISL_1701542 | Santa Clara County Public Health Laboratory | Chan-Zuckerberg Biohub | CZB Cliahub Consortium |
| EPI_ISL_1701543 | Orange County Public Health Lab | Chan-Zuckerberg Biohub | CZB Cliahub Consortium |
| EPI_ISL_1701544 | Santa Clara County Public Health Laboratory | Chan-Zuckerberg Biohub | CZB Cliahub Consortium |
| EPI_ISL_1701545 | Orange County Public Health Lab | Chan-Zuckerberg Biohub | CZB Cliahub Consortium |
| EPI_ISL_1701546, EPI_ISL_1701547 | Alameda County Public Health Lab | Chan-Zuckerberg Biohub | CZB Cliahub Consortium |
| EPI_ISL_1701548 | County of San Luis Obispo Public Health Laboratory | Chan-Zuckerberg Biohub | CZB Cliahub Consortium |
| EPI_ISL_1701549 | Humboldt County Public Health Laboratory | Chan-Zuckerberg Biohub | CZB Cliahub Consortium |
| EPI_ISL_1701551, EPI_ISL_1701552, EPI_ISL_1701553 | Alameda County Public Health Lab | Chan-Zuckerberg Biohub | CZB Cliahub Consortium |
| EPI_ISL_1701554 | Santa Clara County Public Health Laboratory | Chan-Zuckerberg Biohub | CZB Cliahub Consortium |
| EPI_ISL_1701555 | Humboldt County Public Health Laboratory | Chan-Zuckerberg Biohub | CZB Cliahub Consortium |
| EPI_ISL_1701556 | Alameda County Public Health Lab | Chan-Zuckerberg Biohub | CZB Cliahub Consortium |
| EPI_ISL_1701557 | County of San Luis Obispo Public Health Laboratory | Chan-Zuckerberg Biohub | CZB Cliahub Consortium |
| EPI_ISL_1701558 | Humboldt County Public Health Laboratory | Chan-Zuckerberg Biohub | CZB Cliahub Consortium |
| EPI_ISL_1701559 | Santa Clara County Public Health Laboratory | Chan-Zuckerberg Biohub | CZB Cliahub Consortium |
| EPI_ISL_1701560 | Humboldt County Public Health Laboratory | Chan-Zuckerberg Biohub | CZB Cliahub Consortium |
| EPI_ISL_1701561 | Alameda County Public Health Lab | Chan-Zuckerberg Biohub | CZB Cliahub Consortium |
| EPI_ISL_1701562 | County of San Luis Obispo Public Health Laboratory | Chan-Zuckerberg Biohub | CZB Cliahub Consortium |
| EPI_ISL_1701563 | Alameda County Public Health Lab | Chan-Zuckerberg Biohub | CZB Cliahub Consortium |
| EPI_ISL_1701564 | Santa Clara County Public Health Laboratory | Chan-Zuckerberg Biohub | CZB Cliahub Consortium |
| EPI_ISL_1701565, EPI_ISL_1701566 | Orange County Public Health Lab | Chan-Zuckerberg Biohub | CZB Cliahub Consortium |

| | | | |
|---|---|---|---|
| EPI_ISL_1701567 | Santa Clara County Public Health Laboratory | Chan-Zuckerberg Biohub | CZB Cliahub Consortium |
| EPI_ISL_1701568 | Humboldt County Public Health Laboratory | Chan-Zuckerberg Biohub | CZB Cliahub Consortium |
| EPI_ISL_1701569 | County of San Luis Obispo Public Health Laboratory | Chan-Zuckerberg Biohub | CZB Cliahub Consortium |
| EPI_ISL_1701570 | Alameda County Public Health Lab | Chan-Zuckerberg Biohub | CZB Cliahub Consortium |
| EPI_ISL_1701571 | Orange County Public Health Lab | Chan-Zuckerberg Biohub | CZB Cliahub Consortium |
| EPI_ISL_1701573, EPI_ISL_1701574, EPI_ISL_1701575 | Santa Clara County Public Health Laboratory | Chan-Zuckerberg Biohub | CZB Cliahub Consortium |
| EPI_ISL_1701576 | Alameda County Public Health Lab | Chan-Zuckerberg Biohub | CZB Cliahub Consortium |
| EPI_ISL_1701577 | Humboldt County Public Health Laboratory | Chan-Zuckerberg Biohub | CZB Cliahub Consortium |
| EPI_ISL_1701578 | Alameda County Public Health Lab | Chan-Zuckerberg Biohub | CZB Cliahub Consortium |
| EPI_ISL_1701579 | Santa Clara County Public Health Laboratory | Chan-Zuckerberg Biohub | CZB Cliahub Consortium |
| EPI_ISL_1701580 | Alameda County Public Health Lab | Chan-Zuckerberg Biohub | CZB Cliahub Consortium |
| EPI_ISL_1701581 | Santa Clara County Public Health Laboratory | Chan-Zuckerberg Biohub | CZB Cliahub Consortium |
| EPI_ISL_1701582 | County of San Luis Obispo Public Health Laboratory | Chan-Zuckerberg Biohub | CZB Cliahub Consortium |
| EPI_ISL_1701583 | Santa Clara County Public Health Laboratory | Chan-Zuckerberg Biohub | CZB Cliahub Consortium |
| EPI_ISL_1701584 | Humboldt County Public Health Laboratory | Chan-Zuckerberg Biohub | CZB Cliahub Consortium |
| EPI_ISL_1701585 | Alameda County Public Health Lab | Chan-Zuckerberg Biohub | CZB Cliahub Consortium |
| EPI_ISL_1701586 | Orange County Public Health Lab | Chan-Zuckerberg Biohub | CZB Cliahub Consortium |
| EPI_ISL_1701587, EPI_ISL_1701588 | Humboldt County Public Health Laboratory | Chan-Zuckerberg Biohub | CZB Cliahub Consortium |
| EPI_ISL_1701589 | Orange County Public Health Lab | Chan-Zuckerberg Biohub | CZB Cliahub Consortium |
| EPI_ISL_1701590 | Humboldt County Public Health Laboratory | Chan-Zuckerberg Biohub | CZB Cliahub Consortium |
| EPI_ISL_1701591 | County of San Luis Obispo Public Health Laboratory | Chan-Zuckerberg Biohub | CZB Cliahub Consortium |
| EPI_ISL_1701592 | Alameda County Public Health Lab | Chan-Zuckerberg Biohub | CZB Cliahub Consortium |
| EPI_ISL_1701593 | Santa Clara County Public Health Laboratory | Chan-Zuckerberg Biohub | CZB Cliahub Consortium |
| EPI_ISL_1701594, EPI_ISL_1701595 | Humboldt County Public Health Laboratory | Chan-Zuckerberg Biohub | CZB Cliahub Consortium |
| EPI_ISL_1701596 | Santa Clara County Public Health Laboratory | Chan-Zuckerberg Biohub | CZB Cliahub Consortium |
| EPI_ISL_1701597 | Humboldt County Public Health Laboratory | Chan-Zuckerberg Biohub | CZB Cliahub Consortium |
| EPI_ISL_1701598 | Santa Clara County Public Health Laboratory | Chan-Zuckerberg Biohub | CZB Cliahub Consortium |
| EPI_ISL_1701599 | Orange County Public Health Lab | Chan-Zuckerberg Biohub | CZB Cliahub Consortium |
| EPI_ISL_1701600, EPI_ISL_1701601 | Humboldt County Public Health Laboratory | Chan-Zuckerberg Biohub | CZB Cliahub Consortium |
| EPI_ISL_1701602 | Santa Clara County Public Health Laboratory | Chan-Zuckerberg Biohub | CZB Cliahub Consortium |
| EPI_ISL_1701603 | Humboldt County Public Health Laboratory | Chan-Zuckerberg Biohub | CZB Cliahub Consortium |
| EPI_ISL_1701604 | County of San Luis Obispo Public Health Laboratory | Chan-Zuckerberg Biohub | CZB Cliahub Consortium |
| EPI_ISL_1701605 | Humboldt County Public Health Laboratory | Chan-Zuckerberg Biohub | CZB Cliahub Consortium |
| EPI_ISL_1701606 | Alameda County Public Health Lab | Chan-Zuckerberg Biohub | CZB Cliahub Consortium |
| EPI_ISL_1701607 | County of San Luis Obispo Public Health Laboratory | Chan-Zuckerberg Biohub | CZB Cliahub Consortium |
| EPI_ISL_1701608 | Humboldt County Public Health Laboratory | Chan-Zuckerberg Biohub | CZB Cliahub Consortium |
| EPI_ISL_1701609 | Santa Clara County Public Health Laboratory | Chan-Zuckerberg Biohub | CZB Cliahub Consortium |
| EPI_ISL_1701610 | Alameda County Public Health Lab | Chan-Zuckerberg Biohub | CZB Cliahub Consortium |
| EPI_ISL_1701611 | Orange County Public Health Lab | Chan-Zuckerberg Biohub | CZB Cliahub Consortium |
| EPI_ISL_1701612 | Santa Clara County Public Health Laboratory | Chan-Zuckerberg Biohub | CZB Cliahub Consortium |
| EPI_ISL_1701613 | Alameda County Public Health Lab | Chan-Zuckerberg Biohub | CZB Cliahub Consortium |
| EPI_ISL_1701614 | Humboldt County Public Health Laboratory | Chan-Zuckerberg Biohub | CZB Cliahub Consortium |
| EPI_ISL_1701616 | Orange County Public Health Lab | Chan-Zuckerberg Biohub | CZB Cliahub Consortium |
| EPI_ISL_1701617, EPI_ISL_1701618 | Humboldt County Public Health Laboratory | Chan-Zuckerberg Biohub | CZB Cliahub Consortium |
| EPI_ISL_1701619, EPI_ISL_1701620 | California Institute of Technology | Chan-Zuckerberg Biohub | CZB Cliahub Consortium |
| EPI_ISL_1701621, EPI_ISL_1701622 | Alameda County Public Health Lab | Chan-Zuckerberg Biohub | CZB Cliahub Consortium |
| EPI_ISL_1701623 | Orange County Public Health Lab | Chan-Zuckerberg Biohub | CZB Cliahub Consortium |
| EPI_ISL_1701624 | Humboldt County Public Health Laboratory | Chan-Zuckerberg Biohub | CZB Cliahub Consortium |
| EPI_ISL_1701625 | Santa Clara County Public Health Laboratory | Chan-Zuckerberg Biohub | CZB Cliahub Consortium |
| EPI_ISL_1701627 | Humboldt County Public Health Laboratory | Chan-Zuckerberg Biohub | CZB Cliahub Consortium |
| EPI_ISL_1701628 | Santa Clara County Public Health Laboratory | Chan-Zuckerberg Biohub | CZB Cliahub Consortium |

| | | | |
|---|---|---|---|
| EPI_ISL_1701629 | Orange County Public Health Lab | Chan-Zuckerberg Biohub | CZB Cliahub Consortium |
| EPI_ISL_1701630, EPI_ISL_1701631 | Santa Clara County Public Health Laboratory | Chan-Zuckerberg Biohub | CZB Cliahub Consortium |
| EPI_ISL_1701632, EPI_ISL_1701633, EPI_ISL_1701635 | Alameda County Public Health Lab | Chan-Zuckerberg Biohub | CZB Cliahub Consortium |
| EPI_ISL_1701636 | Santa Clara County Public Health Laboratory | Chan-Zuckerberg Biohub | CZB Cliahub Consortium |
| EPI_ISL_1701637 | Humboldt County Public Health Laboratory | Chan-Zuckerberg Biohub | CZB Cliahub Consortium |
| EPI_ISL_1701638 | California Institute of Technology | Chan-Zuckerberg Biohub | CZB Cliahub Consortium |
| EPI_ISL_1701639, EPI_ISL_1701640 | Santa Clara County Public Health Laboratory | Chan-Zuckerberg Biohub | CZB Cliahub Consortium |
| EPI_ISL_1701642 | Alameda County Public Health Lab | Chan-Zuckerberg Biohub | CZB Cliahub Consortium |
| EPI_ISL_1701643 | Humboldt County Public Health Laboratory | Chan-Zuckerberg Biohub | CZB Cliahub Consortium |
| EPI_ISL_1701644 | Alameda County Public Health Lab | Chan-Zuckerberg Biohub | CZB Cliahub Consortium |
| EPI_ISL_1701645 | Santa Clara County Public Health Laboratory | Chan-Zuckerberg Biohub | CZB Cliahub Consortium |
| EPI_ISL_1701646 | Orange County Public Health Lab | Chan-Zuckerberg Biohub | CZB Cliahub Consortium |
| EPI_ISL_1701647, EPI_ISL_1701649, EPI_ISL_1701650, EPI_ISL_1701651, EPI_ISL_1701652 | Alameda County Public Health Lab | Chan-Zuckerberg Biohub | CZB Cliahub Consortium |
| EPI_ISL_1701653, EPI_ISL_1701654 | Santa Clara County Public Health Laboratory | Chan-Zuckerberg Biohub | CZB Cliahub Consortium |
| EPI_ISL_1701655 | Alameda County Public Health Lab | Chan-Zuckerberg Biohub | CZB Cliahub Consortium |
| EPI_ISL_1701656 | Santa Clara County Public Health Laboratory | Chan-Zuckerberg Biohub | CZB Cliahub Consortium |
| EPI_ISL_1701657 | Orange County Public Health Lab | Chan-Zuckerberg Biohub | CZB Cliahub Consortium |
| EPI_ISL_1701658 | Humboldt County Public Health Laboratory | Chan-Zuckerberg Biohub | CZB Cliahub Consortium |
| EPI_ISL_1701660 | County of San Luis Obispo Public Health Laboratory | Chan-Zuckerberg Biohub | CZB Cliahub Consortium |
| EPI_ISL_1701680, EPI_ISL_1701681 | Stanford Health Care | Stanford University School of Medicine, Clinical Virology Laboratory | Bernadette Truong, Becky Jiang, Mamdouh Sibai, Daniel Solis, Michelle Verghese, Jacob Miller, Hannah Wang, Fumiko Yamamoto, James Zehnder, Malaya K. Sahoo, and Benjamin A. Pinsky |
| EPI_ISL_1701683, EPI_ISL_1701684, EPI_ISL_1701685, EPI_ISL_1701686, EPI_ISL_1701687, EPI_ISL_1701688, EPI_ISL_1701689, EPI_ISL_1701690, EPI_ISL_1701691, EPI_ISL_1701692, EPI_ISL_1701693, EPI_ISL_1701694, EPI_ISL_1701695, EPI_ISL_1701696, EPI_ISL_1701697, EPI_ISL_1701698, EPI_ISL_1701699, EPI_ISL_1701700, EPI_ISL_1701701, EPI_ISL_1701702, EPI_ISL_1701703, EPI_ISL_1701704, EPI_ISL_1701705, EPI_ISL_1701706, EPI_ISL_1701707, EPI_ISL_1701708, EPI_ISL_1701709, EPI_ISL_1701710, EPI_ISL_1701711, EPI_ISL_1701712, EPI_ISL_1701713, EPI_ISL_1701714, EPI_ISL_1701715, EPI_ISL_1701716 | | | |
| see above | Alaska State Virology Laboratory | Alaska State Virology Laboratory | Stephanie DeRonde, Elva House, Lisa Smith, Ph.D., Jack Chen, Ph.D. |
| EPI_ISL_1701717, EPI_ISL_1701718, EPI_ISL_1701719, EPI_ISL_1701720, EPI_ISL_1701721, EPI_ISL_1701722, EPI_ISL_1701727, EPI_ISL_1701731, EPI_ISL_1701734, EPI_ISL_1701735, EPI_ISL_1701737 | | | |
| see above | Wyoming Public Health Laboratory | Wyoming Public Health Laboratory | Jim Mildenberger, Taylor Fearing, Channing Weber, Ashley Norberg, Chayse Rowley, Marley Goetz, Brian Dominguez, Elliot Thomasson, Brittany Oher, Sam Britz, Cari Sloma, and Rob Christensen |
| EPI_ISL_1701778, EPI_ISL_1701779, EPI_ISL_1701780, EPI_ISL_1701781, EPI_ISL_1701782, EPI_ISL_1701783, EPI_ISL_1701784, EPI_ISL_1701785, EPI_ISL_1701786, EPI_ISL_1701787, EPI_ISL_1701788, EPI_ISL_1701789, EPI_ISL_1701790, EPI_ISL_1701792, EPI_ISL_1701793, EPI_ISL_1701794, EPI_ISL_1701795, EPI_ISL_1701796, EPI_ISL_1701797, EPI_ISL_1701798, EPI_ISL_1701799, EPI_ISL_1701800, EPI_ISL_1701801, EPI_ISL_1701802, EPI_ISL_1701804, EPI_ISL_1701805, EPI_ISL_1701808, EPI_ISL_1701809, EPI_ISL_1701810, EPI_ISL_1701811, EPI_ISL_1701813, EPI_ISL_1701814, EPI_ISL_1701815, EPI_ISL_1701816, EPI_ISL_1701817, EPI_ISL_1701818, EPI_ISL_1701819, EPI_ISL_1701821 | | | |
| see above | OHSU Lab Services Molecular Microbiology Lab | Oregon SARS-CoV-2 Genome Sequencing Center | Brendan L. O'Connell, Sally Grindstaff, Sonia Acharya, Cierra LaBlanc, Kayla Carter, Ruth V. Nichols, Alec J. Hirsch, Donna Hansel, Guang Fan, Xuan Qin, Daniel N. Streblow, William B. Messer, Andrew C. Adey, Benjamin N. Bimber, Brian J. O'Roak |
| EPI_ISL_1701822 | OHSU Lab Services Molecular Microbiology Lab | Oregon SARS-CoV-2 Genome Sequencing Center | Brendan L. O'Connell, Sally Grindstaff, Kayla Carter, Sonia Acharya, Ruth V. Nichols, Alec J. Hirsch, Donna Hansel, Guang Fan, Xuan Qin, Daniel N. Streblow, William B. Messer, Andrew C. Adey, Benjamin N. Bimber, Brian J. O'Roak |
| EPI_ISL_1701823, EPI_ISL_1701824, EPI_ISL_1701825, EPI_ISL_1701827, EPI_ISL_1701829 | OHSU Lab Services Molecular Microbiology Lab | Oregon SARS-CoV-2 Genome Sequencing Center | Brendan L. O'Connell, Sally Grindstaff, Sonia Acharya, Cierra LaBlanc, Kayla Carter, Ruth V. Nichols, Alec J. Hirsch, Donna Hansel, Guang Fan, Xuan Qin, Daniel N. Streblow, William B. Messer, Andrew C. Adey, Benjamin N. Bimber, Brian J. O'Roak |
| EPI_ISL_1701833 | OR State PHL-Virology/Immunology Section | Centers for Disease Control and Prevention Division of Viral Diseases, Pathogen Discovery | Mili Sheth, Sarah Nobles, Jasmine Padilla, Mark Burroughs, Shoshona Le, Katie Dillon, Peter Cook, Clinton R. Paden, Dhwani Batra, Krista Queen, Kristen Knipe, Dakota Howard, Yvette Unoarumhi, Darlene Wagner, Matthew Schmerer, Ben L. Rambo-Martin, Kristine Lacek, Sam Shepard, Alison Laufer Halpin, Dave Wentworth, Vivien Dugan, Suxiang Tong, Justin Lee |
| EPI_ISL_1701834, EPI_ISL_1701835, EPI_ISL_1701836, EPI_ISL_1701837, EPI_ISL_1701838, EPI_ISL_1701839, EPI_ISL_1701840, EPI_ISL_1701841, EPI_ISL_1701842, EPI_ISL_1701843, EPI_ISL_1701844, EPI_ISL_1701845, EPI_ISL_1701846, EPI_ISL_1701847, EPI_ISL_1701848, EPI_ISL_1701849, EPI_ISL_1701850, EPI_ISL_1701851, EPI_ISL_1701852, EPI_ISL_1701853, EPI_ISL_1701854, EPI_ISL_1701855, EPI_ISL_1701856, EPI_ISL_1701857, EPI_ISL_1701858, EPI_ISL_1701859, EPI_ISL_1701860, EPI_ISL_1701861, EPI_ISL_1701862, EPI_ISL_1701863, EPI_ISL_1701864, EPI_ISL_1701865, EPI_ISL_1701866 | | | |
| see above | Helix/Illumina | Centers for Disease Control and Prevention Division of Viral Diseases, Pathogen Discovery | Dakota Howard, Dhwani Batra, Peter W. Cook, Kara Moser, Adrian Paskey, Jason Caravas, Benjamin Rambo-Martin, Shatavia Morrison, Christopher Gulvick, Scott Sammons, Yvette Unoarumhi, Darlene Wagner, Matthew Schmerer, Eileen de Feo, Jan Antico, Christine Tran, Matthew Tolentino, Shannon Wickline, Kim Gietzen, Brad Sickler, Jingtao Liu, Eric Allen, Phil Febbo, Nicole L. Washington, Simon White, Geraint Levan, Kelly Schiabor Barrett, Elizabeth Cirulli, Alexandre Bolze, Ary Ascencio, Charlotte Rivera-Garcia, Ryan Cho, Jason Nguyen, Sherry Wang, Jimmy Ramirez, Tyler Cassens, Efren Sandoval, Magnus Isaksson, William Lee, David Becker, Marc Laurent, James Lu, Clinton R. Paden, Duncan MacCannell |
| EPI_ISL_1701867, EPI_ISL_1701869, EPI_ISL_1701870, EPI_ISL_1701871, EPI_ISL_1701873, EPI_ISL_1701874, EPI_ISL_1701875 | Laboratory Corporation of America | Centers for Disease Control and Prevention Division of Viral Diseases, Pathogen Discovery | Dakota Howard, Dhwani Batra, Peter W. Cook, Kara Moser, Adrian Paskey, Jason Caravas, Benjamin Rambo-Martin, Shatavia Morrison, Christopher Gulvick, Scott Sammons, Yvette Unoarumhi, Darlene Wagner, Matthew Schmerer, Minoo Agarwal, Eyad Almasri, Debbie Boles, Ayla Burns, Nuthawin Charoensri, Oren Cohen, Susan Countryman, Mary Ann Cristobal, Bobbi Croy, Suzanne Dale, Hrushikesh Deshmukh, Amanda Douglas, Vincent Drouillon, Marcia Eisenberg, Howard Engler, Rama Ghatti, Prashant Gupta, Susan Hicks, Jake Humphrey, Lax Iyer, Manoj Jain, Mohan Kolli, Brian Krueger, Tim Kuphal, Stanley Letovsky, Michael Levandoski, Craig Lukasik, Jonathan Meltzer, Brian Norvell, Mindy Nye, Scott Parker, Christos Petropoulos, John Pruitt, Steven Ragan, Scott Ryan, Mike Sapeta, Jana Schroth, Suresh Babu Selvaraju, Goran Stevovic, Amanda Suchanek, Andrea Throop, Lyndon Tilson, Thomas Urban, Joe Voshell, Kimberly Wagner, Jonathan Williams, Mary Williamson, Qian Zeng, Tricia Zwiefelhofer, Clinton R. Paden, Duncan MacCannell |
| EPI_ISL_1701876, EPI_ISL_1701877, EPI_ISL_1701878, EPI_ISL_1701879, EPI_ISL_1701880, EPI_ISL_1701881, EPI_ISL_1701882, EPI_ISL_1701883, EPI_ISL_1701884, EPI_ISL_1701885, EPI_ISL_1701886, EPI_ISL_1701887, EPI_ISL_1701888, EPI_ISL_1701889, EPI_ISL_1701890, EPI_ISL_1701891, EPI_ISL_1701892, EPI_ISL_1701893, EPI_ISL_1701894, EPI_ISL_1701895, EPI_ISL_1701896, EPI_ISL_1701897, EPI_ISL_1701898, EPI_ISL_1701899, EPI_ISL_1701900, EPI_ISL_1701901, EPI_ISL_1701902, EPI_ISL_1701903, EPI_ISL_1701904, EPI_ISL_1701905, EPI_ISL_1701906, EPI_ISL_1701907, EPI_ISL_1701908, EPI_ISL_1701909, EPI_ISL_1701910, EPI_ISL_1701911, EPI_ISL_1701912, EPI_ISL_1701913, EPI_ISL_1701914, EPI_ISL_1701915, EPI_ISL_1701916, EPI_ISL_1701917, EPI_ISL_1701918, EPI_ISL_1701919, EPI_ISL_1701920, EPI_ISL_1701921, EPI_ISL_1701922, EPI_ISL_1701923, EPI_ISL_1701924, EPI_ISL_1701925, EPI_ISL_1701926, EPI_ISL_1701927, EPI_ISL_1701928, EPI_ISL_1701929, EPI_ISL_1701931, EPI_ISL_1701932, EPI_ISL_1701933, EPI_ISL_1701934, EPI_ISL_1701935, EPI_ISL_1701936, EPI_ISL_1701937, EPI_ISL_1701938, EPI_ISL_1701940, EPI_ISL_1701941, EPI_ISL_1701943, EPI_ISL_1701944, EPI_ISL_1701945, EPI_ISL_1701946, EPI_ISL_1701947, EPI_ISL_1701948, EPI_ISL_1701949, EPI_ISL_1701950, EPI_ISL_1701951, EPI_ISL_1701952, EPI_ISL_1701953, EPI_ISL_1701954, EPI_ISL_1701955, EPI_ISL_1701956, EPI_ISL_1701957, EPI_ISL_1701958, EPI_ISL_1701959, EPI_ISL_1701960, EPI_ISL_1701961, EPI_ISL_1701962, EPI_ISL_1701963, EPI_ISL_1701964, EPI_ISL_1701965, EPI_ISL_1701966, EPI_ISL_1701967, EPI_ISL_1701968, EPI_ISL_1701969, EPI_ISL_1701970, EPI_ISL_1701971, EPI_ISL_1701972, EPI_ISL_1701973, EPI_ISL_1701974, EPI_ISL_1701975, EPI_ISL_1701976, EPI_ISL_1701977, EPI_ISL_1701978, EPI_ISL_1701979, EPI_ISL_1701980, EPI_ISL_1701981, EPI_ISL_1701982, EPI_ISL_1701983, EPI_ISL_1701984, EPI_ISL_1701986, EPI_ISL_1701987, EPI_ISL_1701988, EPI_ISL_1701990, EPI_ISL_1701991, EPI_ISL_1701992, EPI_ISL_1701993, EPI_ISL_1701994, EPI_ISL_1701995, EPI_ISL_1701997, EPI_ISL_1701998, EPI_ISL_1701999, EPI_ISL_1702000, EPI_ISL_1702001, EPI_ISL_1702002, EPI_ISL_1702003, EPI_ISL_1702004, EPI_ISL_1702005, EPI_ISL_1702006, EPI_ISL_1702007, EPI_ISL_1702008, EPI_ISL_1702009, EPI_ISL_1702010, EPI_ISL_1702011, EPI_ISL_1702012, EPI_ISL_1702013, EPI_ISL_1702014, EPI_ISL_1702015, EPI_ISL_1702016, EPI_ISL_1702017, EPI_ISL_1702018, EPI_ISL_1702019, EPI_ISL_1702020, EPI_ISL_1702021, EPI_ISL_1702022, EPI_ISL_1702023, EPI_ISL_1702024, EPI_ISL_1702025, EPI_ISL_1702026, EPI_ISL_1702027, EPI_ISL_1702028, EPI_ISL_1702029, EPI_ISL_1702031, EPI_ISL_1702032, EPI_ISL_1702033, EPI_ISL_1702034, EPI_ISL_1702035, EPI_ISL_1702036, EPI_ISL_1702037, EPI_ISL_1702038, EPI_ISL_1702039, EPI_ISL_1702040, EPI_ISL_1702041, EPI_ISL_1702042, EPI_ISL_1702043, EPI_ISL_1702044, EPI_ISL_1702045, EPI_ISL_1702046, EPI_ISL_1702047, EPI_ISL_1702048, EPI_ISL_1702049, EPI_ISL_1702050, EPI_ISL_1702051, EPI_ISL_1702052, EPI_ISL_1702053, EPI_ISL_1702054, EPI_ISL_1702055, EPI_ISL_1702056, EPI_ISL_1702057, EPI_ISL_1702058, EPI_ISL_1702059, EPI_ISL_1702060, EPI_ISL_1702061, EPI_ISL_1702062, EPI_ISL_1702063, EPI_ISL_1702064, EPI_ISL_1702065, EPI_ISL_1702066, EPI_ISL_1702067, EPI_ISL_1702068, EPI_ISL_1702069, EPI_ISL_1702070, EPI_ISL_1702071, EPI_ISL_1702072, EPI_ISL_1702073, EPI_ISL_1702074, EPI_ISL_1702075, EPI_ISL_1702076, EPI_ISL_1702077, EPI_ISL_1702078, EPI_ISL_1702079, EPI_ISL_1702080, EPI_ISL_1702081, EPI_ISL_1702082, EPI_ISL_1702083, EPI_ISL_1702084, EPI_ISL_1702085, EPI_ISL_1702086, EPI_ISL_1702087, EPI_ISL_1702088, EPI_ISL_1702089, EPI_ISL_1702090, EPI_ISL_1702091, EPI_ISL_1702092, EPI_ISL_1702093, EPI_ISL_1702094, EPI_ISL_1702095, EPI_ISL_1702096, EPI_ISL_1702097, EPI_ISL_1702098, EPI_ISL_1702099, EPI_ISL_1702100, EPI_ISL_1702101, EPI_ISL_1702102, EPI_ISL_1702103, EPI_ISL_1702104, EPI_ISL_1702106, EPI_ISL_1702107, EPI_ISL_1702109, EPI_ISL_1702110, EPI_ISL_1702111, EPI_ISL_1702112, EPI_ISL_1702113, EPI_ISL_1702114, EPI_ISL_1702115, EPI_ISL_1702116, EPI_ISL_1702117, EPI_ISL_1702118, EPI_ISL_1702119, EPI_ISL_1702120, EPI_ISL_1702121, EPI_ISL_1702122, EPI_ISL_1702123, EPI_ISL_1702124, EPI_ISL_1702125, EPI_ISL_1702126, EPI_ISL_1702127, EPI_ISL_1702128, EPI_ISL_1702129, EPI_ISL_1702130, EPI_ISL_1702131, EPI_ISL_1702132, EPI_ISL_1702133, EPI_ISL_1702134, EPI_ISL_1702135, EPI_ISL_1702136, EPI_ISL_1702137, EPI_ISL_1702138, EPI_ISL_1702139, EPI_ISL_1702140, EPI_ISL_1702141, EPI_ISL_1702142, EPI_ISL_1702143, EPI_ISL_1702144 | | | |
| see above | Aegis Sciences Corporation | Centers for Disease Control and Prevention Division of Viral Diseases, Pathogen Discovery | Dakota Howard, Dhwani Batra, Peter W. Cook, Kara Moser, Adrian Paskey, Jason Caravas, Benjamin Rambo-Martin, Shatavia Morrison, Christopher Gulvick, Scott Sammons, Yvette Unoarumhi, Darlene Wagner, Matthew Schmerer, Cyndi Clark, Patrick Campbell, Rob Case, Vikramsinha Ghorpade, Holly Houdeshell, Ola Kvalvaag, Dillon Nall, Ethan Sanders, Alec Vest, Shaun Westlund, Matthew Hardison, Clinton R. Paden, Duncan MacCannell |
| EPI_ISL_1702145, EPI_ISL_1702146, EPI_ISL_1702147, EPI_ISL_1702148, EPI_ISL_1702149, EPI_ISL_1702150, EPI_ISL_1702151, EPI_ISL_1702152, EPI_ISL_1702153, EPI_ISL_1702154, EPI_ISL_1702155, EPI_ISL_1702156, EPI_ISL_1702157, EPI_ISL_1702158, EPI_ISL_1702159, EPI_ISL_1702160, EPI_ISL_1702161, EPI_ISL_1702162 | | | |
| see above | Helix/Illumina | Centers for Disease Control and Prevention Division of Viral Diseases, Pathogen Discovery | Dakota Howard, Dhwani Batra, Peter W. Cook, Kara Moser, Adrian Paskey, Jason Caravas, Benjamin Rambo-Martin, Shatavia Morrison, Christopher Gulvick, Scott Sammons, Yvette Unoarumhi, Darlene Wagner, Matthew Schmerer, Eileen de Feo, Jan Antico, Christine Tran, Matthew Tolentino, Shannon Wickline, Kim Gietzen, Brad Sickler, Jingtao Liu, Eric Allen, Phil Febbo, Nicole L. Washington, Simon White, Geraint Levan, Kelly Schiabor Barrett, Elizabeth Cirulli, Alexandre Bolze, Ary Ascencio, Charlotte Rivera-Garcia, Ryan Cho, Jason Nguyen, Sherry Wang, Jimmy Ramirez, Tyler Cassens, Efren Sandoval, Magnus Isaksson, William Lee, David Becker, Marc Laurent, James Lu, Clinton R. Paden, Duncan MacCannell |
| EPI_ISL_1702163, EPI_ISL_1702164, EPI_ISL_1702165, EPI_ISL_1702166, EPI_ISL_1702167, EPI_ISL_1702168, EPI_ISL_1702169, EPI_ISL_1702170, EPI_ISL_1702171, EPI_ISL_1702172, EPI_ISL_1702173, EPI_ISL_1702174, EPI_ISL_1702175, EPI_ISL_1702176, EPI_ISL_1702177, EPI_ISL_1702178 | | | |
| see above | Quest Diagnostics Incorporated | Centers for Disease Control and Prevention Division of Viral Diseases, Pathogen Discovery | Dakota Howard, Dhwani Batra, Peter W. Cook, Kara Moser, Adrian Paskey, Jason Caravas, Benjamin Rambo-Martin, Shatavia Morrison, Christopher Gulvick, Scott Sammons, Yvette Unoarumhi, Darlene Wagner, Matthew Schmerer, S. H. Rosenthal, A. Gerasimova, R. M. Kagan, B. Anderson, M. Hua, Y. Liu, L.E. Bernstein, K.E. Livingston, A. Perez, I. A. Shlyakhter, R. V. Rolando, R. Owen, P. Tanpaiboon, F. Lacbawan, Clinton R. Paden, Duncan MacCannell |
| EPI_ISL_1702179, EPI_ISL_1702180, EPI_ISL_1702181, EPI_ISL_1702182, EPI_ISL_1702183, EPI_ISL_1702184, EPI_ISL_1702185, EPI_ISL_1702186, EPI_ISL_1702187, EPI_ISL_1702188, EPI_ISL_1702189, EPI_ISL_1702190, EPI_ISL_1702192, EPI_ISL_1702193, EPI_ISL_1702194, EPI_ISL_1702195, EPI_ISL_1702196, EPI_ISL_1702197, EPI_ISL_1702198, EPI_ISL_1702199, EPI_ISL_1702200, EPI_ISL_1702201, EPI_ISL_1702202, EPI_ISL_1702203, EPI_ISL_1702204, EPI_ISL_1702205, EPI_ISL_1702206, EPI_ISL_1702207, EPI_ISL_1702208, EPI_ISL_1702209, EPI_ISL_1702210, EPI_ISL_1702211, EPI_ISL_1702212, EPI_ISL_1702213, EPI_ISL_1702214, EPI_ISL_1702215, EPI_ISL_1702216, EPI_ISL_1702217, EPI_ISL_1702218, EPI_ISL_1702219, EPI_ISL_1702220, EPI_ISL_1702221, EPI_ISL_1702222, EPI_ISL_1702223, | | | |

| | | | |
|---|---|---|---|
| EPI_ISL_1702224, EPI_ISL_1702225, EPI_ISL_1702226, EPI_ISL_1702227, EPI_ISL_1702228, EPI_ISL_1702229, EPI_ISL_1702230, EPI_ISL_1702231, EPI_ISL_1702233, EPI_ISL_1702234, EPI_ISL_1702235, EPI_ISL_1702236, EPI_ISL_1702237, EPI_ISL_1702238, EPI_ISL_1702239, EPI_ISL_1702240, EPI_ISL_1702242, EPI_ISL_1702243, EPI_ISL_1702244, EPI_ISL_1702245, EPI_ISL_1702246, EPI_ISL_1702247, EPI_ISL_1702248, EPI_ISL_1702249, EPI_ISL_1702250, EPI_ISL_1702251, EPI_ISL_1702252, EPI_ISL_1702255, EPI_ISL_1702256, EPI_ISL_1702258, EPI_ISL_1702259, EPI_ISL_1702260, EPI_ISL_1702261, EPI_ISL_1702262, EPI_ISL_1702263, EPI_ISL_1702264, EPI_ISL_1702265, EPI_ISL_1702266, EPI_ISL_1702267, EPI_ISL_1702269, EPI_ISL_1702270, EPI_ISL_1702271, EPI_ISL_1702272, EPI_ISL_1702274, EPI_ISL_1702275, EPI_ISL_1702276, EPI_ISL_1702277, EPI_ISL_1702278, EPI_ISL_1702279, EPI_ISL_1702280, EPI_ISL_1702281, EPI_ISL_1702282, EPI_ISL_1702283, EPI_ISL_1702284, EPI_ISL_1702285, EPI_ISL_1702286, EPI_ISL_1702287, EPI_ISL_1702288, EPI_ISL_1702289, EPI_ISL_1702290, EPI_ISL_1702291, EPI_ISL_1702292, EPI_ISL_1702293, EPI_ISL_1702294, EPI_ISL_1702295, EPI_ISL_1702296, EPI_ISL_1702297, EPI_ISL_1702298, EPI_ISL_1702299, EPI_ISL_1702300, EPI_ISL_1702302, EPI_ISL_1702303, EPI_ISL_1702304, EPI_ISL_1702305, EPI_ISL_1702306, EPI_ISL_1702307, EPI_ISL_1702308, EPI_ISL_1702309, EPI_ISL_1702310, EPI_ISL_1702311, EPI_ISL_1702312, EPI_ISL_1702313, EPI_ISL_1702314, EPI_ISL_1702315, EPI_ISL_1702316, EPI_ISL_1702317, EPI_ISL_1702318, EPI_ISL_1702319, EPI_ISL_1702320, EPI_ISL_1702321, EPI_ISL_1702322, EPI_ISL_1702323, EPI_ISL_1702324, EPI_ISL_1702326, EPI_ISL_1702327, EPI_ISL_1702328, EPI_ISL_1702330, EPI_ISL_1702331, EPI_ISL_1702332, EPI_ISL_1702333, EPI_ISL_1702334, EPI_ISL_1702335, EPI_ISL_1702336, EPI_ISL_1702337, EPI_ISL_1702338, EPI_ISL_1702339, EPI_ISL_1702340, EPI_ISL_1702341, EPI_ISL_1702342, EPI_ISL_1702343, EPI_ISL_1702344, EPI_ISL_1702345, EPI_ISL_1702346, EPI_ISL_1702347, EPI_ISL_1702348, EPI_ISL_1702349, EPI_ISL_1702350, EPI_ISL_1702351, EPI_ISL_1702352, EPI_ISL_1702353, EPI_ISL_1702354, EPI_ISL_1702355, EPI_ISL_1702356, EPI_ISL_1702357, EPI_ISL_1702358, EPI_ISL_1702359, EPI_ISL_1702360, EPI_ISL_1702361, EPI_ISL_1702362, EPI_ISL_1702363, EPI_ISL_1702364, EPI_ISL_1702365, EPI_ISL_1702367, EPI_ISL_1702368, EPI_ISL_1702369, EPI_ISL_1702370, EPI_ISL_1702371, EPI_ISL_1702372, EPI_ISL_1702373, EPI_ISL_1702374, EPI_ISL_1702375, EPI_ISL_1702376, EPI_ISL_1702377, EPI_ISL_1702378, EPI_ISL_1702379, EPI_ISL_1702380, EPI_ISL_1702381, EPI_ISL_1702382, EPI_ISL_1702383, EPI_ISL_1702384, EPI_ISL_1702385, EPI_ISL_1702386, EPI_ISL_1702387, EPI_ISL_1702388, EPI_ISL_1702389, EPI_ISL_1702390, EPI_ISL_1702391, EPI_ISL_1702392, EPI_ISL_1702393, EPI_ISL_1702394, EPI_ISL_1702395, EPI_ISL_1702396, EPI_ISL_1702397, EPI_ISL_1702398, EPI_ISL_1702399, EPI_ISL_1702400, EPI_ISL_1702401, EPI_ISL_1702402, EPI_ISL_1702403, EPI_ISL_1702404, EPI_ISL_1702405, EPI_ISL_1702406, EPI_ISL_1702407, EPI_ISL_1702408, EPI_ISL_1702409, EPI_ISL_1702411, EPI_ISL_1702412, EPI_ISL_1702413, EPI_ISL_1702414, EPI_ISL_1702415, EPI_ISL_1702416, EPI_ISL_1702417, EPI_ISL_1702418, EPI_ISL_1702419, EPI_ISL_1702421, EPI_ISL_1702422, EPI_ISL_1702423, EPI_ISL_1702424, EPI_ISL_1702425, EPI_ISL_1702426, EPI_ISL_1702427, EPI_ISL_1702428, EPI_ISL_1702429, EPI_ISL_1702430, EPI_ISL_1702431, EPI_ISL_1702432, EPI_ISL_1702433, EPI_ISL_1702434, EPI_ISL_1702435, EPI_ISL_1702436, EPI_ISL_1702437, EPI_ISL_1702438, EPI_ISL_1702439, EPI_ISL_1702440, EPI_ISL_1702441, EPI_ISL_1702443, EPI_ISL_1702444, EPI_ISL_1702445, EPI_ISL_1702446, EPI_ISL_1702447, EPI_ISL_1702448, EPI_ISL_1702449, EPI_ISL_1702450, EPI_ISL_1702451, EPI_ISL_1702452, EPI_ISL_1702453, EPI_ISL_1702454, EPI_ISL_1702455, EPI_ISL_1702456, EPI_ISL_1702457, EPI_ISL_1702458, EPI_ISL_1702459, EPI_ISL_1702460, EPI_ISL_1702461, EPI_ISL_1702462, EPI_ISL_1702463, EPI_ISL_1702464, EPI_ISL_1702465, EPI_ISL_1702466, EPI_ISL_1702467, EPI_ISL_1702468, EPI_ISL_1702469, EPI_ISL_1702470, EPI_ISL_1702471, EPI_ISL_1702472, EPI_ISL_1702473, EPI_ISL_1702475, EPI_ISL_1702476, EPI_ISL_1702477, EPI_ISL_1702478, EPI_ISL_1702479, EPI_ISL_1702480, EPI_ISL_1702481, EPI_ISL_1702482, EPI_ISL_1702483, EPI_ISL_1702484, EPI_ISL_1702485, EPI_ISL_1702486, EPI_ISL_1702487, EPI_ISL_1702488, EPI_ISL_1702489, EPI_ISL_1702490, EPI_ISL_1702491, EPI_ISL_1702493, EPI_ISL_1702494, EPI_ISL_1702495, EPI_ISL_1702496, EPI_ISL_1702497, EPI_ISL_1702498, EPI_ISL_1702499, EPI_ISL_1702500, EPI_ISL_1702501, EPI_ISL_1702502, EPI_ISL_1702503, EPI_ISL_1702504, EPI_ISL_1702505, EPI_ISL_1702506, EPI_ISL_1702507, EPI_ISL_1702508, EPI_ISL_1702509, EPI_ISL_1702511, EPI_ISL_1702512, EPI_ISL_1702513, EPI_ISL_1702514, EPI_ISL_1702515, EPI_ISL_1702516, EPI_ISL_1702517, EPI_ISL_1702518, EPI_ISL_1702519, EPI_ISL_1702521, EPI_ISL_1702522, EPI_ISL_1702523, EPI_ISL_1702524, EPI_ISL_1702525, EPI_ISL_1702526, EPI_ISL_1702527, EPI_ISL_1702528, EPI_ISL_1702529, EPI_ISL_1702531, EPI_ISL_1702532, EPI_ISL_1702533, EPI_ISL_1702534, EPI_ISL_1702535, EPI_ISL_1702536, EPI_ISL_1702537, EPI_ISL_1702538, EPI_ISL_1702539, EPI_ISL_1702540, EPI_ISL_1702541, EPI_ISL_1702542, EPI_ISL_1702544, EPI_ISL_1702545, EPI_ISL_1702546, EPI_ISL_1702547, EPI_ISL_1702548, EPI_ISL_1702549, EPI_ISL_1702550, EPI_ISL_1702551, EPI_ISL_1702552, EPI_ISL_1702553, EPI_ISL_1702554, EPI_ISL_1702555, EPI_ISL_1702556, EPI_ISL_1702557, EPI_ISL_1702558, EPI_ISL_1702559, EPI_ISL_1702560, EPI_ISL_1702561, EPI_ISL_1702562, EPI_ISL_1702563, EPI_ISL_1702564, EPI_ISL_1702565, EPI_ISL_1702566, EPI_ISL_1702567, EPI_ISL_1702568, EPI_ISL_1702569, EPI_ISL_1702570, EPI_ISL_1702571, EPI_ISL_1702572, EPI_ISL_1702573, EPI_ISL_1702574, EPI_ISL_1702575, EPI_ISL_1702576, EPI_ISL_1702577, EPI_ISL_1702578, EPI_ISL_1702579 | | | |
| see above | Infinity Biologix | Centers for Disease Control and Prevention Division of Viral Diseases, Pathogen Discovery | Dakota Howard, Dhwani Batra, Peter W. Cook, Kara Moser, Adrian Paskey, Jason Caravas, Benjamin Rambo-Martin, Shatavia Morrison, Christopher Gulvick, Scott Sammons, Yvette Unoarumhi, Darlene Wagner, Matthew Schmerer, Christian Bixby, Yihe Wang, Jonathan Schultz, Chirayu Goswami, Russ Hager, Robin Grimwood, Clinton R. Paden, Duncan MacCannell |
| EPI_ISL_1702580, EPI_ISL_1702581, EPI_ISL_1702582, EPI_ISL_1702583, EPI_ISL_1702584, EPI_ISL_1702585, EPI_ISL_1702586, EPI_ISL_1702587, EPI_ISL_1702588, EPI_ISL_1702589, EPI_ISL_1702590, EPI_ISL_1702591, EPI_ISL_1702592, EPI_ISL_1702593, EPI_ISL_1702594, EPI_ISL_1702595, EPI_ISL_1702596, EPI_ISL_1702597, EPI_ISL_1702598, EPI_ISL_1702599, EPI_ISL_1702600, EPI_ISL_1702601, EPI_ISL_1702602, EPI_ISL_1702603, EPI_ISL_1702604, EPI_ISL_1702605, EPI_ISL_1702606, EPI_ISL_1702607, EPI_ISL_1702608, EPI_ISL_1702609, EPI_ISL_1702610, EPI_ISL_1702611, EPI_ISL_1702612, EPI_ISL_1702613, EPI_ISL_1702614, EPI_ISL_1702615, EPI_ISL_1702616, EPI_ISL_1702617, EPI_ISL_1702618, EPI_ISL_1702619, EPI_ISL_1702620, EPI_ISL_1702621, EPI_ISL_1702622, EPI_ISL_1702623, EPI_ISL_1702624, EPI_ISL_1702625, EPI_ISL_1702626, EPI_ISL_1702627, EPI_ISL_1702628, EPI_ISL_1702629, EPI_ISL_1702630, EPI_ISL_1702631, EPI_ISL_1702632, EPI_ISL_1702633, EPI_ISL_1702634, EPI_ISL_1702635, EPI_ISL_1702636, EPI_ISL_1702637, EPI_ISL_1702638, EPI_ISL_1702639, EPI_ISL_1702640, EPI_ISL_1702641, EPI_ISL_1702642, EPI_ISL_1702643, EPI_ISL_1702644, EPI_ISL_1702645, EPI_ISL_1702646, EPI_ISL_1702647, EPI_ISL_1702648, EPI_ISL_1702649, EPI_ISL_1702650, EPI_ISL_1702651, EPI_ISL_1702652, EPI_ISL_1702653, EPI_ISL_1702654, EPI_ISL_1702655, EPI_ISL_1702656, EPI_ISL_1702657, EPI_ISL_1702658, EPI_ISL_1702659, EPI_ISL_1702660, EPI_ISL_1702661, EPI_ISL_1702662, EPI_ISL_1702663, EPI_ISL_1702664, EPI_ISL_1702665, EPI_ISL_1702666, EPI_ISL_1702667, EPI_ISL_1702668, EPI_ISL_1702669, EPI_ISL_1702670, EPI_ISL_1702671, EPI_ISL_1702672, EPI_ISL_1702673, EPI_ISL_1702674, EPI_ISL_1702675, EPI_ISL_1702676, EPI_ISL_1702677, EPI_ISL_1702678, EPI_ISL_1702679, EPI_ISL_1702680, EPI_ISL_1702681, EPI_ISL_1702682, EPI_ISL_1702683, EPI_ISL_1702684, EPI_ISL_1702685, EPI_ISL_1702686, EPI_ISL_1702687, EPI_ISL_1702688, EPI_ISL_1702689, EPI_ISL_1702690, EPI_ISL_1702691, EPI_ISL_1702692, EPI_ISL_1702693, EPI_ISL_1702694, EPI_ISL_1702695, EPI_ISL_1702696, EPI_ISL_1702697, EPI_ISL_1702698, EPI_ISL_1702699, EPI_ISL_1702700, EPI_ISL_1702701, EPI_ISL_1702702, EPI_ISL_1702703, EPI_ISL_1702704, EPI_ISL_1702705, EPI_ISL_1702706, EPI_ISL_1702707, EPI_ISL_1702708, EPI_ISL_1702709, EPI_ISL_1702710, EPI_ISL_1702711, EPI_ISL_1702712, EPI_ISL_1702713, EPI_ISL_1702714, EPI_ISL_1702715, EPI_ISL_1702716, EPI_ISL_1702717, EPI_ISL_1702718 | | | |
| see above | Quest Diagnostics Incorporated | Centers for Disease Control and Prevention Division of Viral Diseases, Pathogen Discovery | Dakota Howard, Dhwani Batra, Peter W. Cook, Kara Moser, Adrian Paskey, Jason Caravas, Benjamin Rambo-Martin, Shatavia Morrison, Christopher Gulvick, Scott Sammons, Yvette Unoarumhi, Darlene Wagner, Matthew Schmerer, S. H. Rosenthal, A. Gerasimova, R. M. Kagan, B. Anderson, M. Hua, Y. Liu, L.E. Bernstein, K.E. Livingston, A. Perez, I. A. Shlyakhter, R. V. Rolando, R. Owen, P. Tanpaiboon, F. Lacbawan, Clinton R. Paden, Duncan MacCannell |
| EPI_ISL_1702719, EPI_ISL_1702720, EPI_ISL_1702721, EPI_ISL_1702722, EPI_ISL_1702723, EPI_ISL_1702724, EPI_ISL_1702725, EPI_ISL_1702726, EPI_ISL_1702727, EPI_ISL_1702728, EPI_ISL_1702729, EPI_ISL_1702730, EPI_ISL_1702731, EPI_ISL_1702732, EPI_ISL_1702733, EPI_ISL_1702734, EPI_ISL_1702735, EPI_ISL_1702736, EPI_ISL_1702737, EPI_ISL_1702738, EPI_ISL_1702739, EPI_ISL_1702740, EPI_ISL_1702741, EPI_ISL_1702742, EPI_ISL_1702743, EPI_ISL_1702744, EPI_ISL_1702745, EPI_ISL_1702746, EPI_ISL_1702747, EPI_ISL_1702748, EPI_ISL_1702749, EPI_ISL_1702750, EPI_ISL_1702751, EPI_ISL_1702752, EPI_ISL_1702753, EPI_ISL_1702754, EPI_ISL_1702755, EPI_ISL_1702756, EPI_ISL_1702757, EPI_ISL_1702758, EPI_ISL_1702759, EPI_ISL_1702760, EPI_ISL_1702761, EPI_ISL_1702762, EPI_ISL_1702763, EPI_ISL_1702764, EPI_ISL_1702765, EPI_ISL_1702766, EPI_ISL_1702767, EPI_ISL_1702768, EPI_ISL_1702769, EPI_ISL_1702770, EPI_ISL_1702771, EPI_ISL_1702772, EPI_ISL_1702773, EPI_ISL_1702774, EPI_ISL_1702775, EPI_ISL_1702776, EPI_ISL_1702777, EPI_ISL_1702778, EPI_ISL_1702779, EPI_ISL_1702780, EPI_ISL_1702781, EPI_ISL_1702782, EPI_ISL_1702783, EPI_ISL_1702784, EPI_ISL_1702785, EPI_ISL_1702786, EPI_ISL_1702787, EPI_ISL_1702788, EPI_ISL_1702789, EPI_ISL_1702790, EPI_ISL_1702791, EPI_ISL_1702792, EPI_ISL_1702793, EPI_ISL_1702794, EPI_ISL_1702795, EPI_ISL_1702797, EPI_ISL_1702798, EPI_ISL_1702799, EPI_ISL_1702800, EPI_ISL_1702801, EPI_ISL_1702802, EPI_ISL_1702803, EPI_ISL_1702804, EPI_ISL_1702805, EPI_ISL_1702806, EPI_ISL_1702807, EPI_ISL_1702808, EPI_ISL_1702809, EPI_ISL_1702810, EPI_ISL_1702811, EPI_ISL_1702812, EPI_ISL_1702813, EPI_ISL_1702814, EPI_ISL_1702815, EPI_ISL_1702816, EPI_ISL_1702817, EPI_ISL_1702818, EPI_ISL_1702819, EPI_ISL_1702820, EPI_ISL_1702821, EPI_ISL_1702822, EPI_ISL_1702823, EPI_ISL_1702824, EPI_ISL_1702825, EPI_ISL_1702826, EPI_ISL_1702827, EPI_ISL_1702828, EPI_ISL_1702829, EPI_ISL_1702830, EPI_ISL_1702831, EPI_ISL_1702832, EPI_ISL_1702833, EPI_ISL_1702834, EPI_ISL_1702836, EPI_ISL_1702837, EPI_ISL_1702838, EPI_ISL_1702839, EPI_ISL_1702840, EPI_ISL_1702841, EPI_ISL_1702842, EPI_ISL_1702843, EPI_ISL_1702844, EPI_ISL_1702845, EPI_ISL_1702846, EPI_ISL_1702847, EPI_ISL_1702848, EPI_ISL_1702849, EPI_ISL_1702850, EPI_ISL_1702851, EPI_ISL_1702852, EPI_ISL_1702853, EPI_ISL_1702854, EPI_ISL_1702855, EPI_ISL_1702856, EPI_ISL_1702857, EPI_ISL_1702858, EPI_ISL_1702860, EPI_ISL_1702861, EPI_ISL_1702862, EPI_ISL_1702863, EPI_ISL_1702864, EPI_ISL_1702865, EPI_ISL_1702866, EPI_ISL_1702867, EPI_ISL_1702868, EPI_ISL_1702869, EPI_ISL_1702870, EPI_ISL_1702871, EPI_ISL_1702872, EPI_ISL_1702873, EPI_ISL_1702874, EPI_ISL_1702875, EPI_ISL_1702876, EPI_ISL_1702877, EPI_ISL_1702878, EPI_ISL_1702879, EPI_ISL_1702880, EPI_ISL_1702881, EPI_ISL_1702882, EPI_ISL_1702883, EPI_ISL_1702884, EPI_ISL_1702885, EPI_ISL_1702886, EPI_ISL_1702887, EPI_ISL_1702888, EPI_ISL_1702889, EPI_ISL_1702890, EPI_ISL_1702891, EPI_ISL_1702892, EPI_ISL_1702893, EPI_ISL_1702894, EPI_ISL_1702895, EPI_ISL_1702896, EPI_ISL_1702897, EPI_ISL_1702898, EPI_ISL_1702899, EPI_ISL_1702900, EPI_ISL_1702901, EPI_ISL_1702902, EPI_ISL_1702903, EPI_ISL_1702904, EPI_ISL_1702905, EPI_ISL_1702906, EPI_ISL_1702907, EPI_ISL_1702908, EPI_ISL_1702909, EPI_ISL_1702910, EPI_ISL_1702911, EPI_ISL_1702912, EPI_ISL_1702913, EPI_ISL_1702914, EPI_ISL_1702915, EPI_ISL_1702916, EPI_ISL_1702917, EPI_ISL_1702918, EPI_ISL_1702919, EPI_ISL_1702920, EPI_ISL_1702921, EPI_ISL_1702922, EPI_ISL_1702923, EPI_ISL_1702924, EPI_ISL_1702925, EPI_ISL_1702926, EPI_ISL_1702927, EPI_ISL_1702928, EPI_ISL_1702929, EPI_ISL_1702930, EPI_ISL_1702931, EPI_ISL_1702932, EPI_ISL_1702933, EPI_ISL_1702934, EPI_ISL_1702935, EPI_ISL_1702936, EPI_ISL_1702937, EPI_ISL_1702938, EPI_ISL_1702939, EPI_ISL_1702940, EPI_ISL_1702941, EPI_ISL_1702942, EPI_ISL_1702943, EPI_ISL_1702944, EPI_ISL_1702945, EPI_ISL_1702946, EPI_ISL_1702947, EPI_ISL_1702948, EPI_ISL_1702949, EPI_ISL_1702950, EPI_ISL_1702951, EPI_ISL_1702952, EPI_ISL_1702953, EPI_ISL_1702954, EPI_ISL_1702955, EPI_ISL_1702956, EPI_ISL_1702957, EPI_ISL_1702958, EPI_ISL_1702959, EPI_ISL_1702960, EPI_ISL_1702961, EPI_ISL_1702963, EPI_ISL_1702964, EPI_ISL_1702965, EPI_ISL_1702966, EPI_ISL_1702967, EPI_ISL_1702968, EPI_ISL_1702969, EPI_ISL_1702970, EPI_ISL_1702971, EPI_ISL_1702972, EPI_ISL_1702973, EPI_ISL_1702974, EPI_ISL_1702975, EPI_ISL_1702976, EPI_ISL_1702977, EPI_ISL_1702978, EPI_ISL_1702979, EPI_ISL_1702980, EPI_ISL_1702981 | | | |
| see above | Aegis Sciences Corporation | Centers for Disease Control and Prevention Division of Viral Diseases, Pathogen Discovery | Dakota Howard, Dhwani Batra, Peter W. Cook, Kara Moser, Adrian Paskey, Jason Caravas, Benjamin Rambo-Martin, Shatavia Morrison, Christopher Gulvick, Scott Sammons, Yvette Unoarumhi, Darlene Wagner, Matthew Schmerer, Cyndi Clark, Patrick Campbell, Rob Case, Vikramsinha Ghorpade, Holly Houdeshell, Ola Kvalvaag, Dillon Nall, Ethan Sanders, Alec Vest, Shaun Westlund, Matthew Hardison, Clinton R. Paden, Duncan MacCannell |
| EPI_ISL_1704679, EPI_ISL_1704680, EPI_ISL_1704681, EPI_ISL_1704682, EPI_ISL_1704683, EPI_ISL_1704684, EPI_ISL_1704685, EPI_ISL_1704686, EPI_ISL_1704687, EPI_ISL_1704688, EPI_ISL_1704689, EPI_ISL_1704690, EPI_ISL_1704691, EPI_ISL_1704692, EPI_ISL_1704693, EPI_ISL_1704694, EPI_ISL_1704695, EPI_ISL_1704696, EPI_ISL_1704697, EPI_ISL_1704698, EPI_ISL_1704699, EPI_ISL_1704700, EPI_ISL_1704701, EPI_ISL_1704702, EPI_ISL_1704703, EPI_ISL_1704704, EPI_ISL_1704705, EPI_ISL_1704706, EPI_ISL_1704707, EPI_ISL_1704708, EPI_ISL_1704709, EPI_ISL_1704710, EPI_ISL_1704711, EPI_ISL_1704712, EPI_ISL_1704713, EPI_ISL_1704714 | | | |
| see above | Northwestern Memorial Hospital | Northwestern University - Ozer Lab | Ramon Lorenzo-Redondo, Lacy M. Simons, Taylor J. Dean, Chad J. Achenbach, Lawrence J. Jennings, Chao Qi, Michael G. Ison, Judd F. Hultquist, Egon A. Ozer |
| EPI_ISL_1704715, EPI_ISL_1704716, EPI_ISL_1704717, EPI_ISL_1704718, EPI_ISL_1704719, EPI_ISL_1704720, EPI_ISL_1704721, EPI_ISL_1704722, EPI_ISL_1704723, EPI_ISL_1704724, EPI_ISL_1704725, EPI_ISL_1704726, EPI_ISL_1704727, EPI_ISL_1704728, EPI_ISL_1704729, EPI_ISL_1704730, EPI_ISL_1704731, EPI_ISL_1704732, EPI_ISL_1704733, EPI_ISL_1704734, EPI_ISL_1704735, EPI_ISL_1704736, EPI_ISL_1704737, EPI_ISL_1704738, EPI_ISL_1704739, EPI_ISL_1704740, EPI_ISL_1704741, EPI_ISL_1704742, EPI_ISL_1704743, EPI_ISL_1704744, EPI_ISL_1704745, EPI_ISL_1704746, EPI_ISL_1704747, EPI_ISL_1704748, EPI_ISL_1704749, EPI_ISL_1704750, EPI_ISL_1704751, EPI_ISL_1704752, EPI_ISL_1704753, EPI_ISL_1704754, EPI_ISL_1704755, EPI_ISL_1704756, EPI_ISL_1704757, EPI_ISL_1704758, EPI_ISL_1704759, EPI_ISL_1704760, EPI_ISL_1704761, EPI_ISL_1704762, EPI_ISL_1704763, EPI_ISL_1704764, EPI_ISL_1704765, EPI_ISL_1704766, EPI_ISL_1704767, EPI_ISL_1704768, EPI_ISL_1704769, EPI_ISL_1704770, EPI_ISL_1704771, EPI_ISL_1704772, EPI_ISL_1704773 | | | |
| see above | Lurie Children's Hospital of Chicago | Northwestern University - Ozer Lab | Ramon Lorenzo-Redondo, Lacy M. Simons, Taylor J. Dean, Michael G. Ison, Xiaotian, Zheng, William J. Muller, Larry K. Kociolek, Judd F. Hultquist, Egon A. Ozer |
| EPI_ISL_1704774, EPI_ISL_1704775, EPI_ISL_1704776, EPI_ISL_1704777, EPI_ISL_1704778, EPI_ISL_1704780, EPI_ISL_1704781, EPI_ISL_1704782, EPI_ISL_1704783, EPI_ISL_1704784, EPI_ISL_1704785 | | | |
| see above | Northwestern Memorial Hospital | Northwestern University - Ozer Lab | Ramon Lorenzo-Redondo, Lacy M. Simons, Taylor J. Dean, Chad J. Achenbach, Lawrence J. Jennings, Chao Qi, Michael G. Ison, Judd F. Hultquist, Egon A. Ozer |
| EPI_ISL_1706094, EPI_ISL_1706095, EPI_ISL_1706096, EPI_ISL_1706097, EPI_ISL_1706098, EPI_ISL_1706099, EPI_ISL_1706100, EPI_ISL_1706101, EPI_ISL_1706102, EPI_ISL_1706103, EPI_ISL_1706104, EPI_ISL_1706105, EPI_ISL_1706106, EPI_ISL_1706107, EPI_ISL_1706108, EPI_ISL_1706109, EPI_ISL_1706110, EPI_ISL_1706111, EPI_ISL_1706112, EPI_ISL_1706113, EPI_ISL_1706114, EPI_ISL_1706115, EPI_ISL_1706116, EPI_ISL_1706117, EPI_ISL_1706118, EPI_ISL_1706119, EPI_ISL_1706120, EPI_ISL_1706121, EPI_ISL_1706122, EPI_ISL_1706123, EPI_ISL_1706124, EPI_ISL_1706125, EPI_ISL_1706126, EPI_ISL_1706127, EPI_ISL_1706128, EPI_ISL_1706129, EPI_ISL_1706130, EPI_ISL_1706131, EPI_ISL_1706132, EPI_ISL_1706133, EPI_ISL_1706134, EPI_ISL_1706135, EPI_ISL_1706136, EPI_ISL_1706137, EPI_ISL_1706138, EPI_ISL_1706139, EPI_ISL_1706140, EPI_ISL_1706141, EPI_ISL_1706142, EPI_ISL_1706143, EPI_ISL_1706144, EPI_ISL_1706145, EPI_ISL_1706146, EPI_ISL_1706147, EPI_ISL_1706148, EPI_ISL_1706149, EPI_ISL_1706150, EPI_ISL_1706151, EPI_ISL_1706152, EPI_ISL_1706153, EPI_ISL_1706154, EPI_ISL_1706155, EPI_ISL_1706156, EPI_ISL_1706157, EPI_ISL_1706158, EPI_ISL_1706159, EPI_ISL_1706160, EPI_ISL_1706161, EPI_ISL_1706162, EPI_ISL_1706163, EPI_ISL_1706164, EPI_ISL_1706165, EPI_ISL_1706166, EPI_ISL_1706167, EPI_ISL_1706168, EPI_ISL_1706169, EPI_ISL_1706170, EPI_ISL_1706171, EPI_ISL_1706172, EPI_ISL_1706173, EPI_ISL_1706174, EPI_ISL_1706175, EPI_ISL_1706176, EPI_ISL_1706177, EPI_ISL_1706178, EPI_ISL_1706179, EPI_ISL_1706180, EPI_ISL_1706181, EPI_ISL_1706182, EPI_ISL_1706183, EPI_ISL_1706184, EPI_ISL_1706185, EPI_ISL_1706186, EPI_ISL_1706187, EPI_ISL_1706188, EPI_ISL_1706189, EPI_ISL_1706190, EPI_ISL_1706191, EPI_ISL_1706192, EPI_ISL_1706193, EPI_ISL_1706194, EPI_ISL_1706195, EPI_ISL_1706196, EPI_ISL_1706197, EPI_ISL_1706198, EPI_ISL_1706199, EPI_ISL_1706200, EPI_ISL_1706201, EPI_ISL_1706202, EPI_ISL_1706203, EPI_ISL_1706204, EPI_ISL_1706205, EPI_ISL_1706206, EPI_ISL_1706207, EPI_ISL_1706208, EPI_ISL_1706209, EPI_ISL_1706210, EPI_ISL_1706211, EPI_ISL_1706212, EPI_ISL_1706213, EPI_ISL_1706214, EPI_ISL_1706215, EPI_ISL_1706216, EPI_ISL_1706217, EPI_ISL_1706218, EPI_ISL_1706219, EPI_ISL_1706220, EPI_ISL_1706221, EPI_ISL_1706222, EPI_ISL_1706223, EPI_ISL_1706224, EPI_ISL_1706225, EPI_ISL_1706226, EPI_ISL_1706227, EPI_ISL_1706228, EPI_ISL_1706229, EPI_ISL_1706230, EPI_ISL_1706231, EPI_ISL_1706232, EPI_ISL_1706233, EPI_ISL_1706234, EPI_ISL_1706235, EPI_ISL_1706236, EPI_ISL_1706237, EPI_ISL_1706238, EPI_ISL_1706239, EPI_ISL_1706240, EPI_ISL_1706241, EPI_ISL_1706242, EPI_ISL_1706243, EPI_ISL_1706244, EPI_ISL_1706245, EPI_ISL_1706246, EPI_ISL_1706247, EPI_ISL_1706248, EPI_ISL_1706249, EPI_ISL_1706250, EPI_ISL_1706251, EPI_ISL_1706252, EPI_ISL_1706253, EPI_ISL_1706254, EPI_ISL_1706255, EPI_ISL_1706256, EPI_ISL_1706257, EPI_ISL_1706258, EPI_ISL_1706259, EPI_ISL_1706260, EPI_ISL_1706261, EPI_ISL_1706262, EPI_ISL_1706263, EPI_ISL_1706264, EPI_ISL_1706265, EPI_ISL_1706266, EPI_ISL_1706267, EPI_ISL_1706268, EPI_ISL_1706269, EPI_ISL_1706270, EPI_ISL_1706271, EPI_ISL_1706272, EPI_ISL_1706273, EPI_ISL_1706274, EPI_ISL_1706275, EPI_ISL_1706276 | | | |
| see above | Maryland Genomics, Institute for Genome Sciences, University of Maryland School of Medicine | Maryland Genomics, Institute for Genome Sciences, University of Maryland School of Medicine | Tallon, Luke J; Sadzewicz, Lisa D; Humphrys, Mike; Ott, Sandra; Roussey, Holly; Mehta, Aditya; Vavikolanu, Kranthi; Fraser, Claire M; Ravel, Jacques |
| EPI_ISL_1706279, EPI_ISL_1706280, EPI_ISL_1706281, EPI_ISL_1706282, EPI_ISL_1706283, EPI_ISL_1706284, EPI_ISL_1706285, EPI_ISL_1706286, EPI_ISL_1706287, EPI_ISL_1706288, EPI_ISL_1706289, EPI_ISL_1706290, EPI_ISL_1706291, EPI_ISL_1706292, EPI_ISL_1706293, EPI_ISL_1706294, EPI_ISL_1706295, EPI_ISL_1706296, EPI_ISL_1706297, EPI_ISL_1706298, EPI_ISL_1706299, EPI_ISL_1706300, EPI_ISL_1706301, EPI_ISL_1706302, EPI_ISL_1706303, EPI_ISL_1706304, EPI_ISL_1706305, EPI_ISL_1706306, EPI_ISL_1706307, EPI_ISL_1706308, EPI_ISL_1706309, EPI_ISL_1706310, EPI_ISL_1706312, EPI_ISL_1706313, EPI_ISL_1706314, EPI_ISL_1706315, EPI_ISL_1706316, EPI_ISL_1706317, EPI_ISL_1706318, EPI_ISL_1706319, EPI_ISL_1706320, EPI_ISL_1706321, EPI_ISL_1706322, EPI_ISL_1706323, EPI_ISL_1706324, EPI_ISL_1706325, EPI_ISL_1706326, EPI_ISL_1706327, EPI_ISL_1706328, EPI_ISL_1706329, EPI_ISL_1706330, EPI_ISL_1706331, EPI_ISL_1706332, EPI_ISL_1706333, EPI_ISL_1706334, EPI_ISL_1706335, EPI_ISL_1706336, EPI_ISL_1706337, EPI_ISL_1706338, EPI_ISL_1706339, EPI_ISL_1706340, EPI_ISL_1706341, EPI_ISL_1706342, EPI_ISL_1706343, EPI_ISL_1706344, EPI_ISL_1706345, EPI_ISL_1706347, EPI_ISL_1706348, EPI_ISL_1706350, EPI_ISL_1706351, EPI_ISL_1706352, EPI_ISL_1706353, EPI_ISL_1706355, EPI_ISL_1706356, EPI_ISL_1706357, EPI_ISL_1706358, EPI_ISL_1706359 | | | |
| see above | Missouri State Public Health Laboratory | Missouri State Public Health Laboratory | Matthew Sinn, Joshua Barry, Ashley New |
| EPI_ISL_1706582, EPI_ISL_1706583, EPI_ISL_1706584 | MD PHL | MD PHL | Maryland Department of Health Laboratories Administration |
| EPI_ISL_1706603, EPI_ISL_1706604 | UCLA Clinical Micro Lab | Los Angeles County Public Health Laboratories | P. Hemarajata et al. |
| EPI_ISL_1706605 | Flow Diagnostics Lab | Los Angeles County Public Health Laboratories | P. Hemarajata et al. |
| EPI_ISL_1706608, EPI_ISL_1706609 | USC Clinical Lab | Los Angeles County Public Health Laboratories | P. Hemarajata et al. |
| EPI_ISL_1707042, EPI_ISL_1707043, EPI_ISL_1707044, EPI_ISL_1707045, EPI_ISL_1707046, EPI_ISL_1707047, EPI_ISL_1707048, EPI_ISL_1707049, EPI_ISL_1707050, EPI_ISL_1707051, EPI_ISL_1707052, EPI_ISL_1707053, EPI_ISL_1707054, EPI_ISL_1707055, EPI_ISL_1707056, EPI_ISL_1707057, EPI_ISL_1707058, EPI_ISL_1707059, EPI_ISL_1707060, EPI_ISL_1707061 | | | |
| see above | UCLA Clinical Micro Lab | Los Angeles County Public Health Laboratories | P. Hemarajata et al. |
| EPI_ISL_1707078, EPI_ISL_1707079, EPI_ISL_1707080, EPI_ISL_1707081, EPI_ISL_1707082, EPI_ISL_1707083, EPI_ISL_1707084, EPI_ISL_1707085, EPI_ISL_1707086, EPI_ISL_1707087, EPI_ISL_1707088, EPI_ISL_1707089, EPI_ISL_1707090, EPI_ISL_1707091, EPI_ISL_1707093, EPI_ISL_1707094, EPI_ISL_1707095, EPI_ISL_1707096, EPI_ISL_1707097, EPI_ISL_1707098 | | | |
| see above | USC Clinical Lab | Los Angeles County Public Health Laboratories | P. Hemarajata et al. |
| EPI_ISL_1707101, EPI_ISL_1707102, EPI_ISL_1707103, EPI_ISL_1707105, EPI_ISL_1707108, EPI_ISL_1707110, EPI_ISL_1707120, EPI_ISL_1707125, EPI_ISL_1707126, EPI_ISL_1707140, EPI_ISL_1707155, EPI_ISL_1707165, EPI_ISL_1707168, EPI_ISL_1707177 | | | |
| see above | Sonora Quest Laboratories | TGen North | "Jolene Bowers, Heather Centner, Chris French, Hayley Yaglom, Ashlyn Pfeiffer, Darrin Lemmer, Dave Engelthaler, The Arizona COVID Genomics Union (ACGU)" |
| EPI_ISL_1707181 | TGen North | TGen North | "Jolene Bowers, Heather Centner, Chris French, Hayley Yaglom, Ashlyn Pfeiffer, Darrin Lemmer, Dave Engelthaler, The Arizona COVID Genomics Union (ACGU)" |
| EPI_ISL_1707191, EPI_ISL_1707201, EPI_ISL_1707226, EPI_ISL_1707267, EPI_ISL_1707304, EPI_ISL_1707327, EPI_ISL_1707348, EPI_ISL_1707349, EPI_ISL_1707350, EPI_ISL_1707352, EPI_ISL_1707354, EPI_ISL_1707355, EPI_ISL_1707356, EPI_ISL_1707360, EPI_ISL_1707368, EPI_ISL_1707369, EPI_ISL_1707374, EPI_ISL_1707377, EPI_ISL_1707382, EPI_ISL_1707383, EPI_ISL_1707385, EPI_ISL_1707389, EPI_ISL_1707391, EPI_ISL_1707393, EPI_ISL_1707395, EPI_ISL_1707396, EPI_ISL_1707399, EPI_ISL_1707400, EPI_ISL_1707403, EPI_ISL_1707404, EPI_ISL_1707405, EPI_ISL_1707409, EPI_ISL_1707410, EPI_ISL_1707412, EPI_ISL_1707413, EPI_ISL_1707417, EPI_ISL_1707418, EPI_ISL_1707419, EPI_ISL_1707421, EPI_ISL_1707422, EPI_ISL_1707426, EPI_ISL_1707428, EPI_ISL_1707431, EPI_ISL_1707433, EPI_ISL_1707435, EPI_ISL_1707439, EPI_ISL_1707440, EPI_ISL_1707446, EPI_ISL_1707449, EPI_ISL_1707456, EPI_ISL_1707457, EPI_ISL_1707460, EPI_ISL_1707461, EPI_ISL_1707462, EPI_ISL_1707464, EPI_ISL_1707466, EPI_ISL_1707468, EPI_ISL_1707470, EPI_ISL_1707477, EPI_ISL_1707478, EPI_ISL_1707482 | | | |
| see above | Sonora Quest Laboratories | TGen North | "Jolene Bowers, Heather Centner, Chris French, Hayley Yaglom, Ashlyn Pfeiffer, Darrin Lemmer, Dave Engelthaler, The Arizona COVID Genomics Union (ACGU)" |
| EPI_ISL_1707675, EPI_ISL_1707676 | Flow Diagnostics Lab | Los Angeles County Public Health Laboratories | P. Hemarajata et al. |
| EPI_ISL_1707677, EPI_ISL_1707678, EPI_ISL_1707679 | Los Angeles County Public Health Laboratories | Los Angeles County Public Health Laboratories | P. Hemarajata et al. |

| | | | |
|---|---|---|---|
| EPI_ISL_1707680, EPI_ISL_1707681, EPI_ISL_1707682, EPI_ISL_1707683, EPI_ISL_1707684, EPI_ISL_1707685 | Fulgent Genetics | Los Angeles County Public Health Laboratories | P. Hemarajata et al. |
| EPI_ISL_1707686, EPI_ISL_1707687 | Los Angeles County Public Health Laboratories | Los Angeles County Public Health Laboratories | P. Hemarajata et al. |
| EPI_ISL_1708232, EPI_ISL_1708233, EPI_ISL_1708234, EPI_ISL_1708235, EPI_ISL_1708237, EPI_ISL_1708238, EPI_ISL_1708239, EPI_ISL_1708240, EPI_ISL_1708241, EPI_ISL_1708242, EPI_ISL_1708243, EPI_ISL_1708244, EPI_ISL_1708245, EPI_ISL_1708246, EPI_ISL_1708247, EPI_ISL_1708248, EPI_ISL_1708249, EPI_ISL_1708250, EPI_ISL_1708252, EPI_ISL_1708253, EPI_ISL_1708254, EPI_ISL_1708255, EPI_ISL_1708256, EPI_ISL_1708257, EPI_ISL_1708258, EPI_ISL_1708259, EPI_ISL_1708260, EPI_ISL_1708261, EPI_ISL_1708262, EPI_ISL_1708263, EPI_ISL_1708264, EPI_ISL_1708265, EPI_ISL_1708266, EPI_ISL_1708267, EPI_ISL_1708268, EPI_ISL_1708269, EPI_ISL_1708270, EPI_ISL_1708271, EPI_ISL_1708272, EPI_ISL_1708273, EPI_ISL_1708274, EPI_ISL_1708275, EPI_ISL_1708276, EPI_ISL_1708277, EPI_ISL_1708278, EPI_ISL_1708279, EPI_ISL_1708280, EPI_ISL_1708281, EPI_ISL_1708282, EPI_ISL_1708283, EPI_ISL_1708284, EPI_ISL_1708285, EPI_ISL_1708286, EPI_ISL_1708287, EPI_ISL_1708289, EPI_ISL_1708291, EPI_ISL_1708292, EPI_ISL_1708293, EPI_ISL_1708294, EPI_ISL_1708295, EPI_ISL_1708296, EPI_ISL_1708297, EPI_ISL_1708298, EPI_ISL_1708299, EPI_ISL_1708300, EPI_ISL_1708301, EPI_ISL_1708302, EPI_ISL_1708303, EPI_ISL_1708304, EPI_ISL_1708305, EPI_ISL_1708306, EPI_ISL_1708307, EPI_ISL_1708308, EPI_ISL_1708309, EPI_ISL_1708310, EPI_ISL_1708311, EPI_ISL_1708312, EPI_ISL_1708313, EPI_ISL_1708314, EPI_ISL_1708315, EPI_ISL_1708316 | | | |
| see above | Illinois Department of Public Health - Springfield Lab | Illinois Department of Public Health - Springfield Lab | Bryan Sim, Gordon McCall |
| EPI_ISL_1708520, EPI_ISL_1708521, EPI_ISL_1708522, EPI_ISL_1708523, EPI_ISL_1708524, EPI_ISL_1708525, EPI_ISL_1708526, EPI_ISL_1708527, EPI_ISL_1708528, EPI_ISL_1708529, EPI_ISL_1708530, EPI_ISL_1708531, EPI_ISL_1708532, EPI_ISL_1708533, EPI_ISL_1708534, EPI_ISL_1708535, EPI_ISL_1708536, EPI_ISL_1708537, EPI_ISL_1708538, EPI_ISL_1708539, EPI_ISL_1708540, EPI_ISL_1708541, EPI_ISL_1708542, EPI_ISL_1708543, EPI_ISL_1708544, EPI_ISL_1708545, EPI_ISL_1708546, EPI_ISL_1708547, EPI_ISL_1708548, EPI_ISL_1708549, EPI_ISL_1708550, EPI_ISL_1708551, EPI_ISL_1708552, EPI_ISL_1708553, EPI_ISL_1708554, EPI_ISL_1708555, EPI_ISL_1708556, EPI_ISL_1708557, EPI_ISL_1708558, EPI_ISL_1708559, EPI_ISL_1708560, EPI_ISL_1708561, EPI_ISL_1708562, EPI_ISL_1708563, EPI_ISL_1708564, EPI_ISL_1708565, EPI_ISL_1708566, EPI_ISL_1708567, EPI_ISL_1708568, EPI_ISL_1708569, EPI_ISL_1708570, EPI_ISL_1708571, EPI_ISL_1708572, EPI_ISL_1708573 | | | |
| see above | Gundersen Clinical Microbiology Laboratory | Kabara Cancer Research Institute | Craig S. Richmond, Paraic A. Kenny |
| EPI_ISL_1708574 | UW Virology Lab | UW Virology Lab | Pavitra Roychoudhury, Hong Xie, Lasata Shrestha, Shah Mohamed Bakhash, Michelle Lin, Noah R. Baker, Sean Ellis, Meei-Li Huang, Keith R Jerome, Alexander Greninger |
| EPI_ISL_1708601, EPI_ISL_1708602, EPI_ISL_1708603, EPI_ISL_1708604 | IN State Department of Health Laboratory Services | IN State Department of Health Laboratory Services | Cassandra Campion, Jamie Yeadon, Brian Pope, Lixia Liu, Kyle Brownlee, Melissa Hindenlang, Ankita Kashikar, Mark Glazier |
| EPI_ISL_1708605 | LESP Coahuila | Instituto de Diagnostico y Referencia Epidemiologicos (INDRE) | Claudia Wong-Arambula, Abril Rodriguez-Maldonado, Vanessa Rivero-Arredondo, Ariadna Medina-Benitez, Joaquin Quiroz-Mercado, Sergio Rangel-Guerrero, Natividad Cruz-Ortiz, Tatiana Nunez-Garcia, Gisela Barrera-Badillo, Lucia Hernandez-Rivas, Irma Lopez-Martinez, Ernesto Ramirez-Gonzalez. |
| EPI_ISL_1708606 | LESP Jalisco | Instituto de Diagnostico y Referencia Epidemiologicos (INDRE) | Claudia Wong-Arambula, Abril Rodriguez-Maldonado, Vanessa Rivero-Arredondo, Ariadna Medina-Benitez, Joaquin Quiroz-Mercado, Sergio Rangel-Guerrero, Natividad Cruz-Ortiz, Tatiana Nunez-Garcia, Gisela Barrera-Badillo, Lucia Hernandez-Rivas, Irma Lopez-Martinez, Ernesto Ramirez-Gonzalez. |
| EPI_ISL_1708607, EPI_ISL_1708608, EPI_ISL_1708609 | LESP Puebla | Instituto de Diagnostico y Referencia Epidemiologicos (INDRE) | Claudia Wong-Arambula, Abril Rodriguez-Maldonado, Vanessa Rivero-Arredondo, Ariadna Medina-Benitez, Joaquin Quiroz-Mercado, Sergio Rangel-Guerrero, Natividad Cruz-Ortiz, Tatiana Nunez-Garcia, Gisela Barrera-Badillo, Lucia Hernandez-Rivas, Irma Lopez-Martinez, Ernesto Ramirez-Gonzalez. |
| EPI_ISL_1708610 | LESP Sinaloa | Instituto de Diagnostico y Referencia Epidemiologicos (INDRE) | Claudia Wong-Arambula, Abril Rodriguez-Maldonado, Vanessa Rivero-Arredondo, Ariadna Medina-Benitez, Joaquin Quiroz-Mercado, Sergio Rangel-Guerrero, Natividad Cruz-Ortiz, Tatiana Nunez-Garcia, Gisela Barrera-Badillo, Lucia Hernandez-Rivas, Irma Lopez-Martinez, Ernesto Ramirez-Gonzalez. |
| EPI_ISL_1708611, EPI_ISL_1708612 | LESP Guanajuato | Instituto de Diagnostico y Referencia Epidemiologicos (INDRE) | Claudia Wong-Arambula, Abril Rodriguez-Maldonado, Vanessa Rivero-Arredondo, Ariadna Medina-Benitez, Joaquin Quiroz-Mercado, Sergio Rangel-Guerrero, Natividad Cruz-Ortiz, Tatiana Nunez-Garcia, Gisela Barrera-Badillo, Lucia Hernandez-Rivas, Irma Lopez-Martinez, Ernesto Ramirez-Gonzalez. |
| EPI_ISL_1708745, EPI_ISL_1708746, EPI_ISL_1708747, EPI_ISL_1708748, EPI_ISL_1708749, EPI_ISL_1708750, EPI_ISL_1708751, EPI_ISL_1708752, EPI_ISL_1708753, EPI_ISL_1708754, EPI_ISL_1708755, EPI_ISL_1708756, EPI_ISL_1708757, EPI_ISL_1708758, EPI_ISL_1708759, EPI_ISL_1708760, EPI_ISL_1708761, EPI_ISL_1708762, EPI_ISL_1708763, EPI_ISL_1708764, EPI_ISL_1708765, EPI_ISL_1708766, EPI_ISL_1708767, EPI_ISL_1708768, EPI_ISL_1708769, EPI_ISL_1708770, EPI_ISL_1708771, EPI_ISL_1708772, EPI_ISL_1708773, EPI_ISL_1708774, EPI_ISL_1708775, EPI_ISL_1708776, EPI_ISL_1708777, EPI_ISL_1708778, EPI_ISL_1708779, EPI_ISL_1708780, EPI_ISL_1708781, EPI_ISL_1708782, EPI_ISL_1708783, EPI_ISL_1708785, EPI_ISL_1708786, EPI_ISL_1708787, EPI_ISL_1708788, EPI_ISL_1708789, EPI_ISL_1708790, EPI_ISL_1708791, EPI_ISL_1708792, EPI_ISL_1708793, EPI_ISL_1708794, EPI_ISL_1708795, EPI_ISL_1708796, EPI_ISL_1708797, EPI_ISL_1708798, EPI_ISL_1708799, EPI_ISL_1708800, EPI_ISL_1708801, EPI_ISL_1708802, EPI_ISL_1708803, EPI_ISL_1708804, EPI_ISL_1708805, EPI_ISL_1708806, EPI_ISL_1708807, EPI_ISL_1708808, EPI_ISL_1708809, EPI_ISL_1708810, EPI_ISL_1708811, EPI_ISL_1708812, EPI_ISL_1708813, EPI_ISL_1708814, EPI_ISL_1708815, EPI_ISL_1708816, EPI_ISL_1708817, EPI_ISL_1708818, EPI_ISL_1708819, EPI_ISL_1708820, EPI_ISL_1708821, EPI_ISL_1708822, EPI_ISL_1708823, EPI_ISL_1708824, EPI_ISL_1708825, EPI_ISL_1708826, EPI_ISL_1708827, EPI_ISL_1708828, EPI_ISL_1708829, EPI_ISL_1708830, EPI_ISL_1708831, EPI_ISL_1708832, EPI_ISL_1708833, EPI_ISL_1708834, EPI_ISL_1708835, EPI_ISL_1708836, EPI_ISL_1708837, EPI_ISL_1708838, EPI_ISL_1708839, EPI_ISL_1708840, EPI_ISL_1708841, EPI_ISL_1708842, EPI_ISL_1708843, EPI_ISL_1708844, EPI_ISL_1708845, EPI_ISL_1708846, EPI_ISL_1708847, EPI_ISL_1708848, EPI_ISL_1708849, EPI_ISL_1708850, EPI_ISL_1708851, EPI_ISL_1708852, EPI_ISL_1708853, EPI_ISL_1708854, EPI_ISL_1708855, EPI_ISL_1708856, EPI_ISL_1708857, EPI_ISL_1708858, EPI_ISL_1708859, EPI_ISL_1708860, EPI_ISL_1708861, EPI_ISL_1708862, EPI_ISL_1708863, EPI_ISL_1708864, EPI_ISL_1708865, EPI_ISL_1708866, EPI_ISL_1708867, EPI_ISL_1708868, EPI_ISL_1708869, EPI_ISL_1708870, EPI_ISL_1708871, EPI_ISL_1708872, EPI_ISL_1708873, EPI_ISL_1708874, EPI_ISL_1708875, EPI_ISL_1708876, EPI_ISL_1708877, EPI_ISL_1708878, EPI_ISL_1708879, EPI_ISL_1708880, EPI_ISL_1708881, EPI_ISL_1708882, EPI_ISL_1708883, EPI_ISL_1708884, EPI_ISL_1708885, EPI_ISL_1708886, EPI_ISL_1708887, EPI_ISL_1708888, EPI_ISL_1708889, EPI_ISL_1708890, EPI_ISL_1708891, EPI_ISL_1708892, EPI_ISL_1708893, EPI_ISL_1708894, EPI_ISL_1708895, EPI_ISL_1708896, EPI_ISL_1708897, EPI_ISL_1708898, EPI_ISL_1708899, EPI_ISL_1708900, EPI_ISL_1708901, EPI_ISL_1708902, EPI_ISL_1708903, EPI_ISL_1708904, EPI_ISL_1708905, EPI_ISL_1708906, EPI_ISL_1708907, EPI_ISL_1708908, EPI_ISL_1708909, EPI_ISL_1708910, EPI_ISL_1708911, EPI_ISL_1708912, EPI_ISL_1708913, EPI_ISL_1708914, EPI_ISL_1708915, EPI_ISL_1708916, EPI_ISL_1708917, EPI_ISL_1708918, EPI_ISL_1708919, EPI_ISL_1708920, EPI_ISL_1708921, EPI_ISL_1708922, EPI_ISL_1708923, EPI_ISL_1708924, EPI_ISL_1708925, EPI_ISL_1708926, EPI_ISL_1708927, EPI_ISL_1708928, EPI_ISL_1708929, EPI_ISL_1708930, EPI_ISL_1708931, EPI_ISL_1708932, EPI_ISL_1708933, EPI_ISL_1708934, EPI_ISL_1708935, EPI_ISL_1708936, EPI_ISL_1708937, EPI_ISL_1708938, EPI_ISL_1708939, EPI_ISL_1708940, EPI_ISL_1708941, EPI_ISL_1708942, EPI_ISL_1708943, EPI_ISL_1708944, EPI_ISL_1708945, EPI_ISL_1708946, EPI_ISL_1708947, EPI_ISL_1708948, EPI_ISL_1708949, EPI_ISL_1708950, EPI_ISL_1708951, EPI_ISL_1708952, EPI_ISL_1708953, EPI_ISL_1708954, EPI_ISL_1708955, EPI_ISL_1708956, EPI_ISL_1708957, EPI_ISL_1708958, EPI_ISL_1708959, EPI_ISL_1708960, EPI_ISL_1708961, EPI_ISL_1708962, EPI_ISL_1708963, EPI_ISL_1708964, EPI_ISL_1708965, EPI_ISL_1708966, EPI_ISL_1708967, EPI_ISL_1708968, EPI_ISL_1708969, EPI_ISL_1708970, EPI_ISL_1708971, EPI_ISL_1708972, EPI_ISL_1708973, EPI_ISL_1708974, EPI_ISL_1708975, EPI_ISL_1708976, EPI_ISL_1708977, EPI_ISL_1708978, EPI_ISL_1708979, EPI_ISL_1708980, EPI_ISL_1708981, EPI_ISL_1708982, EPI_ISL_1708983, EPI_ISL_1708984, EPI_ISL_1708985, EPI_ISL_1708987, EPI_ISL_1708988, EPI_ISL_1708989, EPI_ISL_1708990, EPI_ISL_1708991, EPI_ISL_1708992, EPI_ISL_1708993, EPI_ISL_1708994, EPI_ISL_1708995, EPI_ISL_1708996, EPI_ISL_1708997, EPI_ISL_1708998, EPI_ISL_1708999, EPI_ISL_1709000, EPI_ISL_1709001, EPI_ISL_1709002, EPI_ISL_1709003, EPI_ISL_1709004, EPI_ISL_1709005, EPI_ISL_1709006, EPI_ISL_1709007, EPI_ISL_1709008, EPI_ISL_1709009, EPI_ISL_1709010, EPI_ISL_1709011, EPI_ISL_1709012, EPI_ISL_1709013, EPI_ISL_1709014, EPI_ISL_1709015, EPI_ISL_1709016, EPI_ISL_1709017, EPI_ISL_1709018, EPI_ISL_1709019, EPI_ISL_1709020, EPI_ISL_1709021, EPI_ISL_1709022, EPI_ISL_1709023, EPI_ISL_1709024, EPI_ISL_1709025, EPI_ISL_1709026, EPI_ISL_1709027, EPI_ISL_1709028, EPI_ISL_1709029, EPI_ISL_1709030, EPI_ISL_1709031, EPI_ISL_1709032, EPI_ISL_1709033, EPI_ISL_1709034, EPI_ISL_1709035, EPI_ISL_1709036, EPI_ISL_1709037, EPI_ISL_1709038, EPI_ISL_1709039, EPI_ISL_1709040, EPI_ISL_1709041, EPI_ISL_1709042, EPI_ISL_1709043, EPI_ISL_1709044, EPI_ISL_1709045, EPI_ISL_1709046, EPI_ISL_1709047, EPI_ISL_1709048, EPI_ISL_1709049, EPI_ISL_1709050, EPI_ISL_1709051, EPI_ISL_1709052, EPI_ISL_1709053, EPI_ISL_1709054, EPI_ISL_1709055, EPI_ISL_1709056, EPI_ISL_1709057, EPI_ISL_1709058, EPI_ISL_1709059, EPI_ISL_1709060, EPI_ISL_1709061, EPI_ISL_1709062, EPI_ISL_1709063, EPI_ISL_1709064, EPI_ISL_1709065, EPI_ISL_1709066, EPI_ISL_1709067, EPI_ISL_1709068, EPI_ISL_1709069, EPI_ISL_1709070, EPI_ISL_1709071, EPI_ISL_1709072, EPI_ISL_1709073, EPI_ISL_1709074, EPI_ISL_1709075, EPI_ISL_1709076, EPI_ISL_1709077, EPI_ISL_1709078, EPI_ISL_1709079, EPI_ISL_1709080, EPI_ISL_1709081, EPI_ISL_1709082, EPI_ISL_1709083, EPI_ISL_1709084, EPI_ISL_1709085, EPI_ISL_1709086, EPI_ISL_1709087, EPI_ISL_1709088, EPI_ISL_1709089, EPI_ISL_1709090, EPI_ISL_1709091, EPI_ISL_1709092, EPI_ISL_1709093, EPI_ISL_1709094, EPI_ISL_1709095, EPI_ISL_1709096, EPI_ISL_1709097, EPI_ISL_1709098, EPI_ISL_1709099, EPI_ISL_1709100, EPI_ISL_1709101, EPI_ISL_1709102, EPI_ISL_1709103, EPI_ISL_1709104, EPI_ISL_1709105, EPI_ISL_1709106, EPI_ISL_1709107, EPI_ISL_1709108, EPI_ISL_1709109, EPI_ISL_1709110, EPI_ISL_1709111, EPI_ISL_1709112, EPI_ISL_1709113, EPI_ISL_1709114, EPI_ISL_1709115, EPI_ISL_1709116, EPI_ISL_1709117, EPI_ISL_1709118, EPI_ISL_1709119, EPI_ISL_1709120, EPI_ISL_1709121, EPI_ISL_1709122, EPI_ISL_1709123, EPI_ISL_1709124, EPI_ISL_1709125, EPI_ISL_1709126, EPI_ISL_1709127, EPI_ISL_1709128, EPI_ISL_1709129, EPI_ISL_1709130, EPI_ISL_1709131, EPI_ISL_1709132, EPI_ISL_1709133, EPI_ISL_1709134, EPI_ISL_1709135, EPI_ISL_1709136, EPI_ISL_1709137, EPI_ISL_1709138, EPI_ISL_1709139, EPI_ISL_1709140, EPI_ISL_1709141, EPI_ISL_1709142, EPI_ISL_1709143, EPI_ISL_1709144, EPI_ISL_1709145, EPI_ISL_1709146, EPI_ISL_1709147, EPI_ISL_1709148, EPI_ISL_1709149, EPI_ISL_1709150, EPI_ISL_1709151, EPI_ISL_1709152, EPI_ISL_1709153, EPI_ISL_1709154, EPI_ISL_1709155, EPI_ISL_1709156, EPI_ISL_1709157, EPI_ISL_1709158, EPI_ISL_1709159, EPI_ISL_1709160, EPI_ISL_1709161, EPI_ISL_1709162, EPI_ISL_1709163, EPI_ISL_1709164, EPI_ISL_1709165, EPI_ISL_1709166, EPI_ISL_1709167, EPI_ISL_1709168, EPI_ISL_1709169, EPI_ISL_1709170, EPI_ISL_1709171, EPI_ISL_1709172, EPI_ISL_1709173, EPI_ISL_1709174, EPI_ISL_1709175, EPI_ISL_1709176, EPI_ISL_1709177, EPI_ISL_1709178, EPI_ISL_1709179, EPI_ISL_1709180, EPI_ISL_1709181, EPI_ISL_1709182, EPI_ISL_1709183, EPI_ISL_1709184, EPI_ISL_1709185, EPI_ISL_1709186, EPI_ISL_1709187, EPI_ISL_1709188, EPI_ISL_1709189, EPI_ISL_1709190, EPI_ISL_1709191, EPI_ISL_1709192, EPI_ISL_1709193, EPI_ISL_1709194, EPI_ISL_1709195, EPI_ISL_1709196, EPI_ISL_1709197, EPI_ISL_1709198, EPI_ISL_1709199, EPI_ISL_1709200, EPI_ISL_1709201, EPI_ISL_1709202, EPI_ISL_1709203, EPI_ISL_1709204, EPI_ISL_1709205, EPI_ISL_1709206, EPI_ISL_1709207, EPI_ISL_1709208, EPI_ISL_1709209, EPI_ISL_1709210, EPI_ISL_1709211, EPI_ISL_1709212, EPI_ISL_1709213, EPI_ISL_1709214, EPI_ISL_1709215, EPI_ISL_1709216, EPI_ISL_1709217, EPI_ISL_1709218, EPI_ISL_1709219, EPI_ISL_1709220, EPI_ISL_1709221, EPI_ISL_1709222, EPI_ISL_1709223, EPI_ISL_1709224, EPI_ISL_1709225, EPI_ISL_1709226, EPI_ISL_1709227, EPI_ISL_1709228, EPI_ISL_1709229, EPI_ISL_1709230, EPI_ISL_1709231, EPI_ISL_1709232, EPI_ISL_1709233, EPI_ISL_1709234, EPI_ISL_1709235, EPI_ISL_1709236, EPI_ISL_1709237, EPI_ISL_1709238, EPI_ISL_1709239, EPI_ISL_1709240, EPI_ISL_1709241, EPI_ISL_1709242, EPI_ISL_1709243, EPI_ISL_1709244, EPI_ISL_1709245, EPI_ISL_1709246, EPI_ISL_1709247, EPI_ISL_1709248, EPI_ISL_1709249, EPI_ISL_1709250, EPI_ISL_1709251, EPI_ISL_1709252, EPI_ISL_1709253, EPI_ISL_1709254, EPI_ISL_1709255, EPI_ISL_1709256, EPI_ISL_1709257, EPI_ISL_1709258, EPI_ISL_1709259, EPI_ISL_1709260, EPI_ISL_1709261, EPI_ISL_1709262, EPI_ISL_1709263, EPI_ISL_1709264, EPI_ISL_1709265, EPI_ISL_1709266, EPI_ISL_1709267, EPI_ISL_1709268, EPI_ISL_1709269, EPI_ISL_1709270, EPI_ISL_1709271, EPI_ISL_1709272, EPI_ISL_1709273, EPI_ISL_1709274, EPI_ISL_1709275, EPI_ISL_1709276, EPI_ISL_1709277, EPI_ISL_1709278, EPI_ISL_1709279, EPI_ISL_1709280, EPI_ISL_1709281, EPI_ISL_1709282, EPI_ISL_1709283, EPI_ISL_1709284, EPI_ISL_1709285, EPI_ISL_1709286, EPI_ISL_1709287, EPI_ISL_1709288, EPI_ISL_1709289, EPI_ISL_1709290, EPI_ISL_1709291, EPI_ISL_1709292, EPI_ISL_1709293, EPI_ISL_1709294, EPI_ISL_1709295, EPI_ISL_1709296, EPI_ISL_1709297, EPI_ISL_1709298, EPI_ISL_1709299, EPI_ISL_1709300, EPI_ISL_1709301, EPI_ISL_1709302, EPI_ISL_1709303, EPI_ISL_1709304, EPI_ISL_1709305, EPI_ISL_1709306, EPI_ISL_1709307, EPI_ISL_1709308, EPI_ISL_1709309, EPI_ISL_1709310, EPI_ISL_1709311, EPI_ISL_1709312, EPI_ISL_1709313, EPI_ISL_1709314, EPI_ISL_1709315, EPI_ISL_1709316, EPI_ISL_1709317, EPI_ISL_1709318, EPI_ISL_1709319, EPI_ISL_1709320, EPI_ISL_1709321, EPI_ISL_1709322, EPI_ISL_1709323, EPI_ISL_1709324, EPI_ISL_1709325, EPI_ISL_1709326, EPI_ISL_1709327, EPI_ISL_1709328, EPI_ISL_1709329, EPI_ISL_1709330, EPI_ISL_1709331, EPI_ISL_1709332, EPI_ISL_1709333, EPI_ISL_1709334, EPI_ISL_1709335, EPI_ISL_1709336, EPI_ISL_1709337, EPI_ISL_1709338, EPI_ISL_1709339, EPI_ISL_1709340, EPI_ISL_1709341, EPI_ISL_1709342, EPI_ISL_1709343, EPI_ISL_1709344, EPI_ISL_1709345, EPI_ISL_1709346, EPI_ISL_1709347, EPI_ISL_1709348, EPI_ISL_1709349, EPI_ISL_1709350, EPI_ISL_1709351, EPI_ISL_1709352, EPI_ISL_1709353, EPI_ISL_1709354, EPI_ISL_1709355, EPI_ISL_1709356, EPI_ISL_1709357, EPI_ISL_1709358, EPI_ISL_1709359, EPI_ISL_1709360, EPI_ISL_1709361, EPI_ISL_1709362, EPI_ISL_1709363, EPI_ISL_1709364, EPI_ISL_1709365, EPI_ISL_1709366, EPI_ISL_1709367, EPI_ISL_1709368, EPI_ISL_1709369, EPI_ISL_1709370, EPI_ISL_1709371, EPI_ISL_1709372, EPI_ISL_1709373, EPI_ISL_1709374, EPI_ISL_1709375, EPI_ISL_1709376, EPI_ISL_1709377, EPI_ISL_1709378, EPI_ISL_1709379, EPI_ISL_1709380, EPI_ISL_1709381, EPI_ISL_1709382, EPI_ISL_1709383, EPI_ISL_1709384, EPI_ISL_1709385, EPI_ISL_1709386, EPI_ISL_1709387, EPI_ISL_1709388, EPI_ISL_1709389, EPI_ISL_1709390, EPI_ISL_1709391, EPI_ISL_1709392, EPI_ISL_1709393, EPI_ISL_1709394, EPI_ISL_1709395, EPI_ISL_1709396, EPI_ISL_1709397, EPI_ISL_1709398, EPI_ISL_1709399, EPI_ISL_1709400, EPI_ISL_1709401, EPI_ISL_1709402, EPI_ISL_1709403, EPI_ISL_1709404, EPI_ISL_1709405, EPI_ISL_1709406, EPI_ISL_1709407, EPI_ISL_1709408, EPI_ISL_1709409, EPI_ISL_1709410, EPI_ISL_1709411, EPI_ISL_1709412, EPI_ISL_1709413, EPI_ISL_1709414, EPI_ISL_1709415, EPI_ISL_1709416, EPI_ISL_1709417, EPI_ISL_1709418, EPI_ISL_1709419, EPI_ISL_1709420, EPI_ISL_1709421, EPI_ISL_1709422, EPI_ISL_1709423, EPI_ISL_1709424, EPI_ISL_1709425, EPI_ISL_1709426, EPI_ISL_1709427, EPI_ISL_1709428, EPI_ISL_1709429, EPI_ISL_1709430, EPI_ISL_1709431, EPI_ISL_1709432, EPI_ISL_1709433, EPI_ISL_1709434, EPI_ISL_1709435, EPI_ISL_1709436, EPI_ISL_1709437, EPI_ISL_1709438, EPI_ISL_1709439, EPI_ISL_1709440, EPI_ISL_1709441, EPI_ISL_1709442, EPI_ISL_1709443, EPI_ISL_1709444, EPI_ISL_1709445, EPI_ISL_1709446, EPI_ISL_1709447, EPI_ISL_1709448, EPI_ISL_1709449, EPI_ISL_1709450, EPI_ISL_1709451, EPI_ISL_1709452, EPI_ISL_1709453, EPI_ISL_1709454, EPI_ISL_1709455, EPI_ISL_1709456, EPI_ISL_1709457, EPI_ISL_1709458, EPI_ISL_1709459, EPI_ISL_1709460, EPI_ISL_1709461, EPI_ISL_1709462, EPI_ISL_1709463, EPI_ISL_1709464, EPI_ISL_1709465, EPI_ISL_1709466, EPI_ISL_1709467, EPI_ISL_1709468, EPI_ISL_1709469, EPI_ISL_1709470, EPI_ISL_1709471, EPI_ISL_1709472, EPI_ISL_1709473, EPI_ISL_1709474, EPI_ISL_1709475, EPI_ISL_1709476, EPI_ISL_1709477, EPI_ISL_1709478, EPI_ISL_1709479, EPI_ISL_1709480, EPI_ISL_1709481, EPI_ISL_1709482, EPI_ISL_1709483, EPI_ISL_1709484, EPI_ISL_1709485, EPI_ISL_1709486, EPI_ISL_1709487, EPI_ISL_1709488, EPI_ISL_1709489, EPI_ISL_1709490, EPI_ISL_1709491, EPI_ISL_1709492, EPI_ISL_1709493, EPI_ISL_1709494, EPI_ISL_1709495, EPI_ISL_1709496, EPI_ISL_1709497, EPI_ISL_1709498, EPI_ISL_1709499, EPI_ISL_1709500, EPI_ISL_1709501, EPI_ISL_1709502, EPI_ISL_1709503, EPI_ISL_1709504, EPI_ISL_1709505, EPI_ISL_1709506, EPI_ISL_1709507, EPI_ISL_1709508, EPI_ISL_1709509, EPI_ISL_1709510, EPI_ISL_1709511, EPI_ISL_1709512, EPI_ISL_1709513, EPI_ISL_1709514, EPI_ISL_1709515, EPI_ISL_1709516, EPI_ISL_1709517, EPI_ISL_1709518, EPI_ISL_1709519, EPI_ISL_1709520, EPI_ISL_1709521, EPI_ISL_1709522, EPI_ISL_1709523, EPI_ISL_1709524, EPI_ISL_1709525, EPI_ISL_1709526, EPI_ISL_1709527, EPI_ISL_1709528, EPI_ISL_1709529, EPI_ISL_1709530, EPI_ISL_1709531, EPI_ISL_1709532, EPI_ISL_1709533, EPI_ISL_1709534, EPI_ISL_1709535, EPI_ISL_1709536, EPI_ISL_1709537, EPI_ISL_1709538, EPI_ISL_1709539, EPI_ISL_1709540, EPI_ISL_1709541, EPI_ISL_1709542, EPI_ISL_1709543, EPI_ISL_1709544, EPI_ISL_1709545, EPI_ISL_1709546, EPI_ISL_1709547, EPI_ISL_1709548, EPI_ISL_1709549, EPI_ISL_1709550, EPI_ISL_1709551, EPI_ISL_1709552, EPI_ISL_1709553, EPI_ISL_1709554, EPI_ISL_1709555, EPI_ISL_1709556, EPI_ISL_1709557, EPI_ISL_1709558, EPI_ISL_1709559, EPI_ISL_1709560, EPI_ISL_1709561, EPI_ISL_1709562, EPI_ISL_1709563, EPI_ISL_1709564, EPI_ISL_1709565, EPI_ISL_1709566, EPI_ISL_1709567, EPI_ISL_1709568, EPI_ISL_1709569, EPI_ISL_1709570, EPI_ISL_1709571, EPI_ISL_1709572, EPI_ISL_1709573, EPI_ISL_1709574, EPI_ISL_1709575, EPI_ISL_1709576, EPI_ISL_1709577, EPI_ISL_1709578, EPI_ISL_1709579, EPI_ISL_1709580, EPI_ISL_1709581, EPI_ISL_1709582, EPI_ISL_1709583, EPI_ISL_1709584, EPI_ISL_1709585, EPI_ISL_1709586, EPI_ISL_1709587, EPI_ISL_1709588, EPI_ISL_1709589, EPI_ISL_1709590, EPI_ISL_1709591, EPI_ISL_1709592, EPI_ISL_1709593, EPI_ISL_1709594, EPI_ISL_1709595, EPI_ISL_1709596, EPI_ISL_1709597, EPI_ISL_1709598, EPI_ISL_1709599, EPI_ISL_1709600, EPI_ISL_1709601, EPI_ISL_1709602, EPI_ISL_1709603, EPI_ISL_1709604, EPI_ISL_1709605, EPI_ISL_1709606, EPI_ISL_1709607, EPI_ISL_1709608, EPI_ISL_1709609, EPI_ISL_1709610, EPI_ISL_1709611, EPI_ISL_1709612, EPI_ISL_1709613, EPI_ISL_1709614, EPI_ISL_1709615, EPI_ISL_1709616, EPI_ISL_1709617, EPI_ISL_1709618, EPI_ISL_1709619, EPI_ISL_1709620, EPI_ISL_1709621, EPI_ISL_1709622, EPI_ISL_1709623, EPI_ISL_1709624, EPI_ISL_1709625, EPI_ISL_1709626, EPI_ISL_1709627, EPI_ISL_1709628, EPI_ISL_1709629, EPI_ISL_1709630, EPI_ISL_1709631, EPI_ISL_1709632, EPI_ISL_1709633, EPI_ISL_1709634, EPI_ISL_1709635, EPI_ISL_1709636, EPI_ISL_1709637, EPI_ISL_1709638, EPI_ISL_1709639, EPI_ISL_1709640, EPI_ISL_1709641, EPI_ISL_1709642, EPI_ISL_1709643, EPI_ISL_1709644, EPI_ISL_1709645, EPI_ISL_1709646, EPI_ISL_1709647, EPI_ISL_1709648, EPI_ISL_1709649, EPI_ISL_1709650, EPI_ISL_1709651, EPI_ISL_1709652, EPI_ISL_1709653, EPI_ISL_1709654, EPI_ISL_1709655, EPI_ISL_1709656, EPI_ISL_1709657, EPI_ISL_1709658, EPI_ISL_1709659, EPI_ISL_1709660, EPI_ISL_1709661, EPI_ISL_1709662, EPI_ISL_1709663, EPI_ISL_1709664, EPI_ISL_1709665, EPI_ISL_1709666, EPI_ISL_1709667, EPI_ISL_1709668, EPI_ISL_1709669, EPI_ISL_1709670, EPI_ISL_1709671, EPI_ISL_1709672, EPI_ISL_1709673, EPI_ISL_1709674, EPI_ISL_1709675, EPI_ISL_1709676, EPI_ISL_1709677, EPI_ISL_1709678, EPI_ISL_1709679, EPI_ISL_1709680, EPI_ISL_1709681, EPI_ISL_1709682, EPI_ISL_1709683, EPI_ISL_1709684, EPI_ISL_1709685, EPI_ISL_1709686, EPI_ISL_1709687, EPI_ISL_1709688, EPI_ISL_1709689, EPI_ISL_1709690, EPI_ISL_1709691, EPI_ISL_1709692, EPI_ISL_1709693, EPI_ISL_1709694, EPI_ISL_1709695, EPI_ISL_1709696, EPI_ISL_1709697, EPI_ISL_1709698, EPI_ISL_1709699, EPI_ISL_1709700, EPI_ISL_1709701, EPI_ISL_1709702, EPI_ISL_1709703, EPI_ISL_1709704, EPI_ISL_1709705, EPI_ISL_1709706, EPI_ISL_1709707, EPI_ISL_1709708, EPI_ISL_1709709, EPI_ISL_1709710, EPI_ISL_1709711, EPI_ISL_1709712, EPI_ISL_1709713, EPI_ISL_1709714, EPI_ISL_1709715, EPI_ISL_1709716, EPI_ISL_1709717, EPI_ISL_1709718, EPI_ISL_1709719, EPI_ISL_1709720, EPI_ISL_1709721, EPI_ISL_1709722, EPI_ISL_1709723, EPI_ISL_1709724, EPI_ISL_1709725, EPI_ISL_1709726, EPI_ISL_1709727, EPI_ISL_1709728, EPI_ISL_1709729, EPI_ISL_1709730, EPI_ISL_1709731, EPI_ISL_1709732, EPI_ISL_1709733, EPI_ISL_1709734, EPI_ISL_1709735, EPI_ISL_1709736, EPI_ISL_1709737, EPI_ISL_1709738, EPI_ISL_1709739, EPI_ISL_1709740, EPI_ISL_1709741, EPI_ISL_1709742, EPI_ISL_1709743, EPI_ISL_1709744, EPI_ISL_1709745, EPI_ISL_1709746, EPI_ISL_1709747, EPI_ISL_1709748, EPI_ISL_1709749, EPI_ISL_1709750, EPI_ISL_1709751, EPI_ISL_1709752, EPI_ISL_1709753, EPI_ISL_1709754, EPI_ISL_1709755, EPI_ISL_1709756, EPI_ISL_1709757, EPI_ISL_1709758, EPI_ISL_1709759, EPI_ISL_1709760, EPI_ISL_1709761, EPI_ISL_1709762, EPI_ISL_1709763, EPI_ISL_1709764, EPI_ISL_1709765, EPI_ISL_1709766, EPI_ISL_1709767, EPI_ISL_1709768, EPI_ISL_1709769, EPI_ISL_1709770, EPI_ISL_1709771, EPI_ISL_1709772, EPI_ISL_1709773, EPI_ISL_1709774, EPI_ISL_1709775, EPI_ISL_1709776, EPI_ISL_1709777, EPI_ISL_1709778, EPI_ISL_1709779, EPI_ISL_1709780, EPI_ISL_1709781, EPI_ISL_1709782, EPI_ISL_1709783, EPI_ISL_1709784, EPI_ISL_1709785, EPI_ISL_1709786, EPI_ISL_1709787, EPI_ISL_1709788, EPI_ISL_1709789, EPI_ISL_1709790, EPI_ISL_1709791, EPI_ISL_1709792, EPI_ISL_1709793, EPI_ISL_1709794, EPI_ISL_1709795, EPI_ISL_1709796, EPI_ISL_1709797, EPI_ISL_1709798, EPI_ISL_1709799, EPI_ISL_1709800, EPI_ISL_1709801, EPI_ISL_1709802, EPI_ISL_1709803, EPI_ISL_1709804, EPI_ISL_1709805, EPI_ISL_1709806, EPI_ISL_1709807, EPI_ISL_1709808, EPI_ISL_1709809, EPI_ISL_1709810, EPI_ISL_1709811, EPI_ISL_1709812, EPI_ISL_1709813, EPI_ISL_1709814, EPI_ISL_1709815, EPI_ISL_1709816, EPI_ISL_1709817, EPI_ISL_1709818, EPI_ISL_1709819, EPI_ISL_1709820, EPI_ISL_1709821, EPI_ISL_1709822, EPI_ISL_1709823, EPI_ISL_1709824, EPI_ISL_1709825, EPI_ISL_1709826, EPI_ISL_1709827, EPI_ISL_1709828, EPI_ISL_1709829, EPI_ISL_1709830, EPI_ISL_1709831, EPI_ISL_1709832, EPI_ISL_1709833, EPI_ISL_1709834, EPI_ISL_1709835, EPI_ISL_1709836, EPI_ISL_1709837, EPI_ISL_1709838, EPI_ISL_1709839, EPI_ISL_1709840, EPI_ISL_1709841, EPI_ISL_1709842, EPI_ISL_1709843, EPI_ISL_1709844, EPI_ISL_1709845, EPI_ISL_1709846, EPI_ISL_1709847, EPI_ISL_1709848, EPI_ISL_1709849, EPI_ISL_1709850, EPI_ISL_1709851, EPI_ISL_1709852, EPI_ISL_1709853, EPI_ISL_1709854, EPI_ISL_1709855, EPI_ISL_1709856, EPI_ISL_1709857, EPI_ISL_1709858, EPI_ISL_1709859, EPI_ISL_1709860, EPI_ISL_1709861, EPI_ISL_1709862, EPI_ISL_1709863, EPI_ISL_1709864, EPI_ISL_1709865, EPI_ISL_1709866, EPI_ISL_1709867, EPI_ISL_1709868, EPI_ISL_1709869, EPI_ISL_1709870, EPI_ISL_1709871, EPI_ISL_1709872, EPI_ISL_1709873, EPI_ISL_1709874, EPI_ISL_1709875, EPI_ISL_1709876, EPI_ISL_1709877, EPI_ISL_1709878, EPI_ISL_1709879, EPI_ISL_1709880, EPI_ISL_1709881, EPI_ISL_1709882, EPI_ISL_1709883, EPI_ISL_1709884, EPI_ISL_1709885, EPI_ISL_1709886, EPI_ISL_1709887, EPI_ISL_1709888, EPI_ISL_1709889, EPI_ISL_1709890, EPI_ISL_1709891, EPI_ISL_1709892, EPI_ISL_1709893, EPI_ISL_1709894, EPI_ISL_1709895, EPI_ISL_1709896, EPI_ISL_1709897, EPI_ISL_1709898, EPI_ISL_1709899, EPI_ISL_1709900, EPI_ISL_1709901, EPI_ISL_1709902, EPI_ISL_1709903, EPI_ISL_1709904, EPI_ISL_1709905, EPI_ISL_1709906, EPI_ISL_1709907, EPI_ISL_1709908, EPI_ISL_1709909, EPI_ISL_1709910, EPI_ISL_1709911, EPI_ISL_1709912, EPI_ISL_1709913, EPI_ISL_1709914, EPI_ISL_1709915, EPI_ISL_1709916, EPI_ISL_1709917, EPI_ISL_1709918, EPI_ISL_1709919, EPI_ISL_1709920, EPI_ISL_1709921, EPI_ISL_1709922, EPI_ISL_1709923, EPI_ISL_1709924, EPI_ISL_1709925, EPI_ISL_1709926, EPI_ISL_1709927, EPI_ISL_1709928, EPI_ISL_1709929, EPI_ISL_1709930, EPI_ISL_1709931, EPI_ISL_1709932, EPI_ISL_1709933, EPI_ISL_1709934, EPI_ISL_1709935, EPI_ISL_1709936, EPI_ISL_1709937, EPI_ISL_1709938, EPI_ISL_1709939, EPI_ISL_1709940, EPI_ISL_1709941, EPI_ISL_1709942, EPI_ISL_1709943, EPI_ISL_1709944, EPI_ISL_1709945, EPI_ISL_1709946, EPI_ISL_1709947, EPI_ISL_1709948, EPI_ISL_1709949, EPI_ISL_1709950, EPI_ISL_1709951, EPI_ISL_1709952, EPI_ISL_1709953, EPI_ISL_1709954, EPI_ISL_1709955, EPI_ISL_1709956, EPI_ISL_1709957, EPI_ISL_1709958, EPI_ISL_1709959, EPI_ISL_1709960, EPI_ISL_1709961, EPI_ISL_1709962, EPI_ISL_1709963, EPI_ISL_1709964, EPI_ISL_1709965, EPI_ISL_1709966, EPI_ISL_1709967, EPI_ISL_1709968, EPI_ISL_1709969, EPI_ISL_1709970, EPI_ISL_1709971, EPI_ISL_1709972, EPI_ISL_1709973, EPI_ISL_1709974, EPI_ISL_1709975, EPI_ISL_1709976, EPI_ISL_1709977, EPI_ISL_1709978, EPI_ISL_1709979, EPI_ISL_1709980, EPI_ISL_1709981, EPI_ISL_1709982, EPI_ISL_1709983, EPI_ISL_1709984, EPI_ISL_1709985, EPI_ISL_1709986, EPI_ISL_1709987, EPI_ISL_1709988, EPI_ISL_1709989, EPI_ISL_1709990, EPI_ISL_1709991, EPI_ISL_1709992, EPI_ISL_1709993, EPI_ISL_1709994, EPI_ISL_1709995, EPI_ISL_1709996, EPI_ISL_1709997, EPI_ISL_1709998, EPI_ISL_1709999, EPI_ISL_1710000, EPI_ISL_1710001, EPI_ISL_1710002, EPI_ISL_1710003, EPI_ISL_1710004, EPI_ISL_1710005, EPI_ISL_1710006, EPI_ISL_1710007, EPI_ISL_1710008, EPI_ISL_1710009, EPI_ISL_1710010, EPI_ISL_1710011, EPI_ISL_1710012, EPI_ISL_1710013, EPI_ISL_1710014 | | | |
| see above | MSHS Clinical Microbiology Laboratories | MSHS Pathogen Surveillance Program | Ana S. Gonzalez-Reiche, Hala Alshammary, Mitchell J. Sullivan, Brianne Ciferri, Ajay Obla, Angela Amoako, Mahmoud Awawda, Daniel Floda, Julia Matthews, Ashley Salimbangon, Levy Sominsky, Katherine Beach, Kayla Russo, Charles Gleason, Shelcie Fabre, Giulio Kleiner, Zenab Khan, Bremy Alburquerque, Adriana van de Guchte, Komal Srivastava, Matthew M. Hernandez, Jayeeta Dutta, Denise Jurczyszak, Nancy Francoeur, Betsaida Salom Melo, Irina Oussenko, Gintaras Deikus, Juan Soto, Shwetha Hara Sridhar, Ying-Chih Wang, Kathryn Twyman, Deena R. Altman, Robert Sebra, Adolfo García-Sastre, Marta Luksza, Gopi Patel, Sarah Schaefer, Melissa Gitman, Michael D. Nowak, Alberto Paniz-Mondolfi, Emilia Mia Sordillo, Viviana Simon, Harm van Bakel |
| EPI_ISL_1710048, EPI_ISL_1710049, EPI_ISL_1710050, EPI_ISL_1710051, EPI_ISL_1710052, EPI_ISL_1710053, EPI_ISL_1710054, EPI_ISL_1710055, EPI_ISL_1710056, EPI_ISL_1710057, EPI_ISL_1710058, EPI_ISL_1710059, EPI_ISL_1710060, EPI_ISL_1710061, EPI_ISL_1710062, EPI_ISL_1710063, EPI_ISL_1710064, EPI_ISL_1710065, EPI_ISL_1710066, EPI_ISL_1710067, EPI_ISL_1710068, EPI_ISL_1710069, EPI_ISL_1710070, EPI_ISL_1710071, EPI_ISL_1710072, EPI_ISL_1710073, EPI_ISL_1710074, EPI_ISL_1710075, EPI_ISL_1710076, EPI_ISL_1710077, EPI_ISL_1710078, EPI_ISL_1710079, EPI_ISL_1710080, EPI_ISL_1710081, EPI_ISL_1710082, EPI_ISL_1710083, EPI_ISL_1710084, EPI_ISL_1710085, EPI_ISL_1710086, EPI_ISL_1710087, EPI_ISL_1710088, EPI_ISL_1710089, EPI_ISL_1710090, EPI_ISL_1710091, EPI_ISL_1710092, EPI_ISL_1710093, EPI_ISL_1710094, EPI_ISL_1710095, EPI_ISL_1710096, EPI_ISL_1710097, EPI_ISL_1710098, EPI_ISL_1710099, EPI_ISL_1710100, EPI_ISL_1710101, EPI_ISL_1710102, EPI_ISL_1710103, EPI_ISL_1710105, EPI_ISL_1710106, EPI_ISL_1710107, EPI_ISL_1710108, EPI_ISL_1710109, EPI_ISL_1710110, EPI_ISL_1710111, EPI_ISL_1710112, EPI_ISL_1710113, EPI_ISL_1710114, EPI_ISL_1710115, EPI_ISL_1710116, EPI_ISL_1710117, EPI_ISL_1710118, EPI_ISL_1710119, EPI_ISL_1710120, EPI_ISL_1710121, EPI_ISL_1710122, EPI_ISL_1710123, EPI_ISL_1710124, EPI_ISL_1710125, EPI_ISL_1710126, EPI_ISL_1710127, EPI_ISL_1710128, EPI_ISL_1710129, EPI_ISL_1710130, EPI_ISL_1710131, EPI_ISL_1710132, EPI_ISL_1710133, EPI_ISL_1710134, EPI_ISL_1710135, EPI_ISL_1710136, EPI_ISL_1710137, EPI_ISL_1710138, EPI_ISL_1710139, EPI_ISL_1710140, EPI_ISL_1710141, EPI_ISL_1710142, EPI_ISL_1710143, EPI_ISL_1710144, EPI_ISL_1710145, EPI_ISL_1710146, EPI_ISL_1710147, EPI_ISL_1710149, EPI_ISL_1710150, EPI_ISL_1710151, EPI_ISL_1710152, EPI_ISL_1710153, EPI_ISL_1710154, EPI_ISL_1710155, EPI_ISL_1710156, EPI_ISL_1710157, EPI_ISL_1710159, EPI_ISL_1710160, EPI_ISL_1710161, EPI_ISL_1710162, EPI_ISL_1710163, EPI_ISL_1710166, EPI_ISL_1710167, EPI_ISL_1710168, EPI_ISL_1710169, EPI_ISL_1710170, EPI_ISL_1710171, EPI_ISL_1710172, EPI_ISL_1710173, EPI_ISL_1710174, EPI_ISL_1710176, EPI_ISL_1710177, EPI_ISL_1710178, EPI_ISL_1710179, EPI_ISL_1710180 | | | |
| see above | Broad Institute Clinical Research | Infectious Disease Program, Broad Institute | Siddle,K.J., Adams,G., Pearlman,L., Gladden-Young,A., Vicente,G., Blumenstiel,B., DeFelice,M., Lee,M., McGovern,S., Lagerborg,K., Rudy,M., DeRuff,K., Carter,A., Normandin,E., Bauer,M., Reilly,S., Tomkins-Tinch,C., Loreth,C., Chaluvadi,S., Meldrim,J., Granger,B., Lemieux,J.E., Birren,B.W., |

| | | | |
|---|---|---|---|
| | Sequencing Platform | of Harvard and MIT | Sabeti,P.C., Larkin,K., Dodge,S., Lennon,N., Madoff,L., Brown,C., Gallagher,G., Smole,S., Park,D.J., Gabriel,S., and MacInnis,B.L. |
| EPI_ISL_1710181, EPI_ISL_1710182, EPI_ISL_1710183, EPI_ISL_1710184, EPI_ISL_1710185, EPI_ISL_1710186, EPI_ISL_1710187, EPI_ISL_1710188, EPI_ISL_1710189, EPI_ISL_1710190, EPI_ISL_1710191, EPI_ISL_1710192, EPI_ISL_1710193, EPI_ISL_1710194, EPI_ISL_1710195, EPI_ISL_1710196, EPI_ISL_1710197, EPI_ISL_1710198, EPI_ISL_1710199, EPI_ISL_1710200, EPI_ISL_1710201, EPI_ISL_1710202, EPI_ISL_1710203, EPI_ISL_1710204, EPI_ISL_1710205, EPI_ISL_1710206, EPI_ISL_1710207, EPI_ISL_1710208, EPI_ISL_1710209, EPI_ISL_1710210, EPI_ISL_1710211, EPI_ISL_1710212, EPI_ISL_1710213, EPI_ISL_1710214, EPI_ISL_1710215, EPI_ISL_1710216, EPI_ISL_1710217, EPI_ISL_1710218, EPI_ISL_1710219, EPI_ISL_1710220, EPI_ISL_1710221, EPI_ISL_1710222, EPI_ISL_1710223, EPI_ISL_1710224, EPI_ISL_1710225, EPI_ISL_1710226, EPI_ISL_1710227, EPI_ISL_1710228, EPI_ISL_1710229, EPI_ISL_1710230, EPI_ISL_1710231, EPI_ISL_1710232, EPI_ISL_1710233, EPI_ISL_1710234, EPI_ISL_1710235, EPI_ISL_1710236, EPI_ISL_1710237, EPI_ISL_1710238, EPI_ISL_1710239, EPI_ISL_1710240, EPI_ISL_1710241, EPI_ISL_1710242, EPI_ISL_1710243 | | | |
| see above | Rhode Island Department of Health | Infectious Disease Program, Broad Institute of Harvard and MIT | Siddle,K.J., Azevedo,K., Miller,A., Adams,G., Pearlman,L., Gladden-Young,A., Lagerborg,K., Rudy,M., DeRuff,K., Carter,A., Normandin,E., Bauer,M., Reilly,S., Tomkins-Tinch,C., Loreth,C., Chaluvadi,S., Lemieux,J.E., Birren,B.W., Sabeti,P.C., Huard,R., King,E., Park,D.J., and MacInnis,B.L. |
| EPI_ISL_1710245, EPI_ISL_1710246, EPI_ISL_1710247, EPI_ISL_1710248, EPI_ISL_1710249, EPI_ISL_1710250, EPI_ISL_1710251, EPI_ISL_1710252, EPI_ISL_1710253, EPI_ISL_1710254, EPI_ISL_1710255, EPI_ISL_1710256, EPI_ISL_1710257, EPI_ISL_1710258, EPI_ISL_1710259, EPI_ISL_1710260, EPI_ISL_1710261, EPI_ISL_1710262, EPI_ISL_1710263, EPI_ISL_1710264, EPI_ISL_1710265, EPI_ISL_1710266, EPI_ISL_1710267, EPI_ISL_1710268, EPI_ISL_1710270, EPI_ISL_1710271, EPI_ISL_1710272, EPI_ISL_1710273, EPI_ISL_1710274, EPI_ISL_1710275, EPI_ISL_1710276, EPI_ISL_1710277, EPI_ISL_1710278, EPI_ISL_1710279, EPI_ISL_1710280, EPI_ISL_1710281, EPI_ISL_1710282, EPI_ISL_1710283, EPI_ISL_1710285, EPI_ISL_1710286, EPI_ISL_1710287, EPI_ISL_1710288, EPI_ISL_1710289, EPI_ISL_1710290, EPI_ISL_1710291, EPI_ISL_1710292, EPI_ISL_1710293, EPI_ISL_1710294, EPI_ISL_1710295, EPI_ISL_1710296, EPI_ISL_1710298, EPI_ISL_1710299, EPI_ISL_1710300, EPI_ISL_1710301, EPI_ISL_1710303, EPI_ISL_1710304, EPI_ISL_1710305, EPI_ISL_1710307, EPI_ISL_1710308, EPI_ISL_1710309, EPI_ISL_1710311, EPI_ISL_1710312, EPI_ISL_1710313, EPI_ISL_1710314, EPI_ISL_1710315, EPI_ISL_1710316, EPI_ISL_1710317, EPI_ISL_1710318, EPI_ISL_1710319, EPI_ISL_1710320, EPI_ISL_1710321, EPI_ISL_1710322, EPI_ISL_1710323, EPI_ISL_1710324, EPI_ISL_1710325, EPI_ISL_1710326, EPI_ISL_1710327, EPI_ISL_1710328, EPI_ISL_1710329, EPI_ISL_1710330, EPI_ISL_1710331, EPI_ISL_1710332, EPI_ISL_1710333, EPI_ISL_1710334, EPI_ISL_1710335, EPI_ISL_1710336, EPI_ISL_1710337, EPI_ISL_1710338, EPI_ISL_1710339, EPI_ISL_1710340, EPI_ISL_1710341, EPI_ISL_1710342, EPI_ISL_1710343, EPI_ISL_1710345, EPI_ISL_1710346, EPI_ISL_1710347, EPI_ISL_1710348, EPI_ISL_1710349, EPI_ISL_1710350, EPI_ISL_1710352, EPI_ISL_1710354, EPI_ISL_1710355, EPI_ISL_1710356, EPI_ISL_1710357, EPI_ISL_1710358, EPI_ISL_1710359, EPI_ISL_1710360, EPI_ISL_1710361, EPI_ISL_1710362, EPI_ISL_1710363, EPI_ISL_1710364, EPI_ISL_1710365, EPI_ISL_1710366, EPI_ISL_1710367, EPI_ISL_1710368, EPI_ISL_1710369, EPI_ISL_1710370, EPI_ISL_1710371, EPI_ISL_1710372, EPI_ISL_1710373, EPI_ISL_1710374, EPI_ISL_1710375, EPI_ISL_1710376, EPI_ISL_1710377, EPI_ISL_1710378, EPI_ISL_1710379, EPI_ISL_1710380, EPI_ISL_1710381, EPI_ISL_1710382, EPI_ISL_1710383, EPI_ISL_1710384, EPI_ISL_1710385, EPI_ISL_1710386, EPI_ISL_1710387, EPI_ISL_1710388, EPI_ISL_1710389, EPI_ISL_1710390, EPI_ISL_1710391, EPI_ISL_1710392, EPI_ISL_1710393, EPI_ISL_1710395, EPI_ISL_1710396, EPI_ISL_1710397, EPI_ISL_1710398, EPI_ISL_1710400, EPI_ISL_1710401, EPI_ISL_1710402, EPI_ISL_1710403, EPI_ISL_1710404, EPI_ISL_1710405, EPI_ISL_1710406, EPI_ISL_1710407, EPI_ISL_1710408, EPI_ISL_1710409, EPI_ISL_1710410, EPI_ISL_1710414, EPI_ISL_1710415, EPI_ISL_1710416, EPI_ISL_1710417, EPI_ISL_1710418, EPI_ISL_1710419, EPI_ISL_1710420, EPI_ISL_1710421, EPI_ISL_1710422, EPI_ISL_1710423, EPI_ISL_1710424, EPI_ISL_1710425, EPI_ISL_1710426, EPI_ISL_1710427, EPI_ISL_1710428, EPI_ISL_1710429, EPI_ISL_1710430, EPI_ISL_1710431, EPI_ISL_1710432, EPI_ISL_1710433, EPI_ISL_1710434, EPI_ISL_1710435, EPI_ISL_1710436, EPI_ISL_1710437, EPI_ISL_1710438, EPI_ISL_1710439, EPI_ISL_1710440, EPI_ISL_1710441, EPI_ISL_1710442, EPI_ISL_1710443, EPI_ISL_1710444, EPI_ISL_1710445, EPI_ISL_1710446, EPI_ISL_1710447, EPI_ISL_1710448, EPI_ISL_1710449, EPI_ISL_1710450, EPI_ISL_1710451, EPI_ISL_1710452, EPI_ISL_1710453, EPI_ISL_1710454, EPI_ISL_1710455, EPI_ISL_1710456, EPI_ISL_1710457, EPI_ISL_1710458, EPI_ISL_1710459, EPI_ISL_1710460, EPI_ISL_1710461, EPI_ISL_1710462, EPI_ISL_1710463, EPI_ISL_1710464, EPI_ISL_1710465, EPI_ISL_1710466, EPI_ISL_1710467, EPI_ISL_1710468, EPI_ISL_1710469, EPI_ISL_1710470, EPI_ISL_1710471, EPI_ISL_1710472, EPI_ISL_1710473, EPI_ISL_1710474, EPI_ISL_1710475, EPI_ISL_1710476, EPI_ISL_1710477, EPI_ISL_1710478, EPI_ISL_1710479, EPI_ISL_1710480, EPI_ISL_1710481, EPI_ISL_1710482, EPI_ISL_1710483, EPI_ISL_1710484, EPI_ISL_1710485, EPI_ISL_1710486, EPI_ISL_1710487, EPI_ISL_1710488, EPI_ISL_1710489, EPI_ISL_1710490, EPI_ISL_1710491, EPI_ISL_1710492, EPI_ISL_1710493, EPI_ISL_1710494, EPI_ISL_1710495, EPI_ISL_1710496, EPI_ISL_1710497, EPI_ISL_1710498, EPI_ISL_1710499, EPI_ISL_1710500, EPI_ISL_1710501, EPI_ISL_1710502, EPI_ISL_1710503, EPI_ISL_1710505 | | | |
| see above | Broad Institute Clinical Research Sequencing Platform | Infectious Disease Program, Broad Institute of Harvard and MIT | Siddle,K.J., Adams,G., Pearlman,L., Gladden-Young,A., Vicente,G., Blumenstiel,B., DeFelice,M., Lee,M., McGovern,S., Lagerborg,K., Rudy,M., DeRuff,K., Carter,A., Normandin,E., Bauer,M., Reilly,S., Tomkins-Tinch,C., Loreth,C., Chaluvadi,S., Meldrim,J., Granger,B., Lemieux,J.E., Birren,B.W., Sabeti,P.C., Larkin,K., Dodge,S., Lennon,N., Madoff,L., Brown,C., Gallagher,G., Smole,S., Park,D.J., Gabriel,S., and MacInnis,B.L. |
| EPI_ISL_1710518, EPI_ISL_1710519, EPI_ISL_1710520, EPI_ISL_1710521, EPI_ISL_1710522, EPI_ISL_1710523, EPI_ISL_1710524, EPI_ISL_1710525, EPI_ISL_1710526, EPI_ISL_1710528, EPI_ISL_1710529, EPI_ISL_1710530, EPI_ISL_1710531, EPI_ISL_1710532, EPI_ISL_1710533, EPI_ISL_1710534, EPI_ISL_1710535, EPI_ISL_1710537, EPI_ISL_1710538, EPI_ISL_1710539, EPI_ISL_1710540, EPI_ISL_1710541, EPI_ISL_1710542, EPI_ISL_1710543, EPI_ISL_1710544, EPI_ISL_1710545, EPI_ISL_1710546, EPI_ISL_1710547, EPI_ISL_1710548, EPI_ISL_1710550, EPI_ISL_1710551, EPI_ISL_1710553, EPI_ISL_1710554 | | | |
| see above | CURATIVE | New Mexico Department of Health Scientific Laboratory | Ellie Johnson, D'eldra Malone, Jennifer Benoit, Linda Salazar, Ratheesh Rajan, Anastacia Griego-Fisher |
| EPI_ISL_1710555, EPI_ISL_1710556 | San Juan Regional Medical Center | New Mexico Department of Health Scientific Laboratory | Ellie Johnson, D'eldra Malone, Jennifer Benoit, Linda Salazar, Ratheesh Rajan, Anastacia Griego-Fisher |
| EPI_ISL_1710557, EPI_ISL_1710559, EPI_ISL_1710560 | TriCore Reference Laboratory Inc. | New Mexico Department of Health Scientific Laboratory | Ellie Johnson, D'eldra Malone, Jennifer Benoit, Linda Salazar, Ratheesh Rajan, Anastacia Griego-Fisher |
| EPI_ISL_1710561, EPI_ISL_1710562, EPI_ISL_1710563, EPI_ISL_1710564, EPI_ISL_1710567, EPI_ISL_1710568, EPI_ISL_1710569, EPI_ISL_1710570, EPI_ISL_1710571, EPI_ISL_1710572, EPI_ISL_1710573, EPI_ISL_1710574, EPI_ISL_1710575, EPI_ISL_1710577, EPI_ISL_1710578, EPI_ISL_1710579 | | | |
| see above | New Mexico Department of Health Scientific Laboratory | New Mexico Department of Health Scientific Laboratory | Ellie Johnson, D'eldra Malone, Jennifer Benoit, Linda Salazar, Ratheesh Rajan, Anastacia Griego-Fisher |
| EPI_ISL_1710580, EPI_ISL_1710581, EPI_ISL_1710582, EPI_ISL_1710583 | TriCore Reference Laboratory Inc. | New Mexico Department of Health Scientific Laboratory | Ellie Johnson, D'eldra Malone, Jennifer Benoit, Linda Salazar, Ratheesh Rajan, Anastacia Griego-Fisher |
| EPI_ISL_1710585, EPI_ISL_1710586, EPI_ISL_1710587, EPI_ISL_1710588, EPI_ISL_1710589, EPI_ISL_1710590, EPI_ISL_1710594, EPI_ISL_1710595, EPI_ISL_1710596, EPI_ISL_1710597 | New Mexico Department of Health Scientific Laboratory | New Mexico Department of Health Scientific Laboratory | Ellie Johnson, D'eldra Malone, Jennifer Benoit, Linda Salazar, Ratheesh Rajan, Anastacia Griego-Fisher |
| EPI_ISL_1710599, EPI_ISL_1710600, EPI_ISL_1710601, EPI_ISL_1710602, EPI_ISL_1710603, EPI_ISL_1710604, EPI_ISL_1710605, EPI_ISL_1710606, EPI_ISL_1710607, EPI_ISL_1710608, EPI_ISL_1710609, EPI_ISL_1710610, EPI_ISL_1710611, EPI_ISL_1710612, EPI_ISL_1710613, EPI_ISL_1710614, EPI_ISL_1710615, EPI_ISL_1710616, EPI_ISL_1710617, EPI_ISL_1710618, EPI_ISL_1710619, EPI_ISL_1710620, EPI_ISL_1710621, EPI_ISL_1710622, EPI_ISL_1710623, EPI_ISL_1710624, EPI_ISL_1710625, EPI_ISL_1710626, EPI_ISL_1710627, EPI_ISL_1710628, EPI_ISL_1710629, EPI_ISL_1710630, EPI_ISL_1710631, EPI_ISL_1710632, EPI_ISL_1710633, EPI_ISL_1710634, EPI_ISL_1710635, EPI_ISL_1710636, EPI_ISL_1710637, EPI_ISL_1710638, EPI_ISL_1710639, EPI_ISL_1710640, EPI_ISL_1710641, EPI_ISL_1710642, EPI_ISL_1710643, EPI_ISL_1710644, EPI_ISL_1710645, EPI_ISL_1710646, EPI_ISL_1710647, EPI_ISL_1710653, EPI_ISL_1710654, EPI_ISL_1710655, EPI_ISL_1710656, EPI_ISL_1710657, EPI_ISL_1710658, EPI_ISL_1710659, EPI_ISL_1710660, EPI_ISL_1710661 | | | |
| see above | Massachusetts State Public Health Laboratory | Massachusetts State Public Health Laboratory | Andrew Lang, Timelia Fink, Glen Gallagher, Sandra Smole |
| EPI_ISL_1710699, EPI_ISL_1710700, EPI_ISL_1710702, EPI_ISL_1710703, EPI_ISL_1710704, EPI_ISL_1710705, EPI_ISL_1710706 | School of Pharmacy, Shenandoah University | School of Pharmacy, Shenandoah University | Adams,S.M., Harralson,A.F., Kidd,R.S., Sawyer,G.W. |
| EPI_ISL_1710707, EPI_ISL_1710708, EPI_ISL_1710709, EPI_ISL_1710711, EPI_ISL_1710712, EPI_ISL_1710713, EPI_ISL_1710714, EPI_ISL_1710715, EPI_ISL_1710716, EPI_ISL_1710717, EPI_ISL_1710718, EPI_ISL_1710719, EPI_ISL_1710720, EPI_ISL_1710721, EPI_ISL_1710722, EPI_ISL_1710723, EPI_ISL_1710724, EPI_ISL_1710726, EPI_ISL_1710727, EPI_ISL_1710728, EPI_ISL_1710729, EPI_ISL_1710730, EPI_ISL_1710731, EPI_ISL_1710732, EPI_ISL_1710733, EPI_ISL_1710734, EPI_ISL_1710735, EPI_ISL_1710736, EPI_ISL_1710737, EPI_ISL_1710738, EPI_ISL_1710739, EPI_ISL_1710740, EPI_ISL_1710741, EPI_ISL_1710742, EPI_ISL_1710743, EPI_ISL_1710744, EPI_ISL_1710745, EPI_ISL_1710746, EPI_ISL_1710747, EPI_ISL_1710748, EPI_ISL_1710749, EPI_ISL_1710750, EPI_ISL_1710751, EPI_ISL_1710752, EPI_ISL_1710753, EPI_ISL_1710754, EPI_ISL_1710755, EPI_ISL_1710756, EPI_ISL_1710757, EPI_ISL_1710758, EPI_ISL_1710759, EPI_ISL_1710760, EPI_ISL_1710761, EPI_ISL_1710762, EPI_ISL_1710763, EPI_ISL_1710764, EPI_ISL_1710765, EPI_ISL_1710766, EPI_ISL_1710767, EPI_ISL_1710768, EPI_ISL_1710769, EPI_ISL_1710770, EPI_ISL_1710771, EPI_ISL_1710772, EPI_ISL_1710773, EPI_ISL_1710774, EPI_ISL_1710775, EPI_ISL_1710776, EPI_ISL_1710777, EPI_ISL_1710778, EPI_ISL_1710779, EPI_ISL_1710780, EPI_ISL_1710781, EPI_ISL_1710782, EPI_ISL_1710783, EPI_ISL_1710784, EPI_ISL_1710785, EPI_ISL_1710786, EPI_ISL_1710787, EPI_ISL_1710788, EPI_ISL_1710789 | | | |
| see above | Tampa General Hospital Esoteric Lab | Tampa General Hospital Esoteric Research & Development Lab | Grant Vestal, Deanna Becker, Dominic Uy, Vicki Healer, Amorce Lima, Suzane Silbert |
| EPI_ISL_1710790, EPI_ISL_1710791, EPI_ISL_1710792, EPI_ISL_1710793, EPI_ISL_1710794, EPI_ISL_1710795, EPI_ISL_1710796, EPI_ISL_1710797, EPI_ISL_1710798, EPI_ISL_1710799 | IN State Department of Health Laboratory Services | IN State Department of Health Laboratory Services | Cassandra Campion, Jamie Yeadon, Brian Pope, Lixia Liu, Kyle Brownlee, Melissa Hindenlang, Ankita Kashikar, Mark Glazier |
| EPI_ISL_1710800, EPI_ISL_1710801, EPI_ISL_1710802, EPI_ISL_1710803, EPI_ISL_1710804, EPI_ISL_1710805, EPI_ISL_1710806, EPI_ISL_1710807, EPI_ISL_1710808, EPI_ISL_1710809, EPI_ISL_1710810, EPI_ISL_1710811, EPI_ISL_1710812, EPI_ISL_1710813, EPI_ISL_1710814, EPI_ISL_1710815, EPI_ISL_1710816, EPI_ISL_1710817, EPI_ISL_1710818, EPI_ISL_1710819, EPI_ISL_1710820, EPI_ISL_1710821, EPI_ISL_1710822, EPI_ISL_1710823, EPI_ISL_1710824, EPI_ISL_1710825, EPI_ISL_1710826, EPI_ISL_1710827, EPI_ISL_1710828, EPI_ISL_1710829, EPI_ISL_1710830, EPI_ISL_1710831, EPI_ISL_1710832, EPI_ISL_1710841, EPI_ISL_1710842, EPI_ISL_1710843, EPI_ISL_1710844, EPI_ISL_1710845, EPI_ISL_1710846, EPI_ISL_1710847, EPI_ISL_1710848 | | | |
| see above | UTGSAF | UTGSAF | Jessica Podnar, Sylvie Beaudenon, Audrey Kelly, Zachary Carver, Anna Battenhouse, Andreas Matouschek |
| EPI_ISL_1710867, EPI_ISL_1710868, EPI_ISL_1710869, EPI_ISL_1710870, EPI_ISL_1710871, EPI_ISL_1710872, EPI_ISL_1710873, EPI_ISL_1710874, EPI_ISL_1710875, EPI_ISL_1710876, EPI_ISL_1710877, EPI_ISL_1710878, EPI_ISL_1710879 | | | |
| see above | IN State Department of Health Laboratory Services | IN State Department of Health Laboratory Services | Cassandra Campion, Jamie Yeadon, Brian Pope, Lixia Liu, Kyle Brownlee, Melissa Hindenlang, Ankita Kashikar, Mark Glazier |
| EPI_ISL_1710881, EPI_ISL_1710883, EPI_ISL_1710884, EPI_ISL_1710885, EPI_ISL_1710886, EPI_ISL_1710887, EPI_ISL_1710888, EPI_ISL_1710889, EPI_ISL_1710890, EPI_ISL_1710891, EPI_ISL_1710892, EPI_ISL_1710893, EPI_ISL_1710894, EPI_ISL_1710895, EPI_ISL_1710896, EPI_ISL_1710897, EPI_ISL_1710898, EPI_ISL_1710900, EPI_ISL_1710901, EPI_ISL_1710902, EPI_ISL_1710903, EPI_ISL_1710904, EPI_ISL_1710905, EPI_ISL_1710906, EPI_ISL_1710907, EPI_ISL_1710908, EPI_ISL_1710909, EPI_ISL_1710910, EPI_ISL_1710911, EPI_ISL_1710912, EPI_ISL_1710913, EPI_ISL_1710914, EPI_ISL_1710915, EPI_ISL_1710916, EPI_ISL_1710917, EPI_ISL_1710918, EPI_ISL_1710919, EPI_ISL_1710920, EPI_ISL_1710921, EPI_ISL_1710922, EPI_ISL_1710924, EPI_ISL_1710925, EPI_ISL_1710926, EPI_ISL_1710927, EPI_ISL_1710928, EPI_ISL_1710929, EPI_ISL_1710930, EPI_ISL_1710931, EPI_ISL_1710932, EPI_ISL_1710933, EPI_ISL_1710934, EPI_ISL_1710935, EPI_ISL_1710936, EPI_ISL_1710937, EPI_ISL_1710938, EPI_ISL_1710939, EPI_ISL_1710940, EPI_ISL_1710941, EPI_ISL_1710942, EPI_ISL_1710943, EPI_ISL_1710944, EPI_ISL_1710945, EPI_ISL_1710946, EPI_ISL_1710947, EPI_ISL_1710948, EPI_ISL_1710949, EPI_ISL_1710950, EPI_ISL_1710951, EPI_ISL_1710952, EPI_ISL_1710954, EPI_ISL_1710955, EPI_ISL_1710956, EPI_ISL_1710957, EPI_ISL_1710958, EPI_ISL_1710959, EPI_ISL_1710960, EPI_ISL_1710961, EPI_ISL_1710962 | | | |
| see above | Minnesota Department of Health, Public Health Laboratory | Minnesota Department of Health, Public Health Laboratory | Alexandra Lorentz, Jacob Garfin, Matt Plumb, and Xiong Wang |
| EPI_ISL_1710963 | LESP Quintana Roo | Instituto de Diagnostico y Referencia Epidemiologicos (INDRE) | Claudia Wong-Arambula, Abril Rodriguez-Maldonado, Vanessa Rivero-Arredondo, Ariadna Medina-Benitez, Joaquin Quiroz-Mercado, Sergio Rangel-Guerrero, Natividad Cruz-Ortiz, Tatiana Nunez-Garcia, Gisela Barrera-Badillo, Lucia Hernandez-Rivas, Irma Lopez-Martinez, Ernesto Ramirez-Gonzalez. |
| EPI_ISL_1710964, EPI_ISL_1710965, EPI_ISL_1710966 | LESP Ciudad de Mexico | Instituto de Diagnostico y Referencia Epidemiologicos (INDRE) | Claudia Wong-Arambula, Abril Rodriguez-Maldonado, Vanessa Rivero-Arredondo, Ariadna Medina-Benitez, Joaquin Quiroz-Mercado, Sergio Rangel-Guerrero, Natividad Cruz-Ortiz, Tatiana Nunez-Garcia, Gisela Barrera-Badillo, Lucia Hernandez-Rivas, Irma Lopez-Martinez, Ernesto Ramirez-Gonzalez. |
| EPI_ISL_1710967 | LESP Morelos | Instituto de Diagnostico y Referencia Epidemiologicos (INDRE) | Claudia Wong-Arambula, Abril Rodriguez-Maldonado, Vanessa Rivero-Arredondo, Ariadna Medina-Benitez, Joaquin Quiroz-Mercado, Sergio Rangel-Guerrero, Natividad Cruz-Ortiz, Tatiana Nunez-Garcia, Gisela Barrera-Badillo, Lucia Hernandez-Rivas, Irma Lopez-Martinez, Ernesto Ramirez-Gonzalez. |
| EPI_ISL_1710968, EPI_ISL_1710969 | LESP Guanajuato | Instituto de Diagnostico y Referencia Epidemiologicos (INDRE) | Claudia Wong-Arambula, Abril Rodriguez-Maldonado, Vanessa Rivero-Arredondo, Ariadna Medina-Benitez, Joaquin Quiroz-Mercado, Sergio Rangel-Guerrero, Natividad Cruz-Ortiz, Tatiana Nunez-Garcia, Gisela Barrera-Badillo, Lucia Hernandez-Rivas, Irma Lopez-Martinez, Ernesto Ramirez-Gonzalez. |
| EPI_ISL_1710970 | LESPZacatecas | Instituto de Diagnostico y Referencia Epidemiologicos (INDRE) | Claudia Wong-Arambula, Abril Rodriguez-Maldonado, Vanessa Rivero-Arredondo, Ariadna Medina-Benitez, Joaquin Quiroz-Mercado, Sergio Rangel-Guerrero, Natividad Cruz-Ortiz, Tatiana Nunez-Garcia, Gisela Barrera-Badillo, Lucia Hernandez-Rivas, Irma Lopez-Martinez, Ernesto Ramirez-Gonzalez. |

| | | | |
|---|---|---|---|
| EPI_ISL_1710971, EPI_ISL_1710972 | LESP Baja California | Instituto de Diagnostico y Referencia Epidemiologicos (INDRE) | Claudia Wong-Arambula, Abril Rodriguez-Maldonado, Vanessa Rivero-Arredondo, Ariadna Medina-Benitez, Joaquin Quiroz-Mercado, Sergio Rangel-Guerrero, Natividad Cruz-Ortiz, Tatiana Nunez-Garcia, Gisela Barrera-Badillo, Lucia Hernandez-Rivas, Irma Lopez-Martinez, Ernesto Ramirez-Gonzalez. |
| EPI_ISL_1710973, EPI_ISL_1710974, EPI_ISL_1710975, EPI_ISL_1710976, EPI_ISL_1710977, EPI_ISL_1710978, EPI_ISL_1710979, EPI_ISL_1710980, EPI_ISL_1710981, EPI_ISL_1710982, EPI_ISL_1710983, EPI_ISL_1710984, EPI_ISL_1710985, EPI_ISL_1710986, EPI_ISL_1710987, EPI_ISL_1710988, EPI_ISL_1710989, EPI_ISL_1710990 | | | |
| see above | Minnesota Department of Health, Public Health Laboratory | Minnesota Department of Health, Public Health Laboratory | Alexandra Lorentz, Jacob Garfin, Matt Plumb, and Xiong Wang |
| EPI_ISL_1710991, EPI_ISL_1710992, EPI_ISL_1710993 | M Health Fairview | Minnesota Department of Health, Public Health Laboratory | Alexandra Lorentz, Jacob Garfin, Matt Plumb, and Xiong Wang |
| EPI_ISL_1710994, EPI_ISL_1710995, EPI_ISL_1710996, EPI_ISL_1710997, EPI_ISL_1710998, EPI_ISL_1710999, EPI_ISL_1711000, EPI_ISL_1711001, EPI_ISL_1711002, EPI_ISL_1711003, EPI_ISL_1711004, EPI_ISL_1711005, EPI_ISL_1711006, EPI_ISL_1711007, EPI_ISL_1711008, EPI_ISL_1711009, EPI_ISL_1711010, EPI_ISL_1711011, EPI_ISL_1711012, EPI_ISL_1711013, EPI_ISL_1711014, EPI_ISL_1711015, EPI_ISL_1711016, EPI_ISL_1711017, EPI_ISL_1711018, EPI_ISL_1711019, EPI_ISL_1711020, EPI_ISL_1711021, EPI_ISL_1711022, EPI_ISL_1711023, EPI_ISL_1711024, EPI_ISL_1711025, EPI_ISL_1711026, EPI_ISL_1711027, EPI_ISL_1711028, EPI_ISL_1711029, EPI_ISL_1711030, EPI_ISL_1711031, EPI_ISL_1711032, EPI_ISL_1711038, EPI_ISL_1711039, EPI_ISL_1711040, EPI_ISL_1711041 | | | |
| see above | Minnesota Department of Health, Public Health Laboratory | Minnesota Department of Health, Public Health Laboratory | Alexandra Lorentz, Jacob Garfin, Matt Plumb, and Xiong Wang |
| EPI_ISL_1711042, EPI_ISL_1711044, EPI_ISL_1711045, EPI_ISL_1711046, EPI_ISL_1711047, EPI_ISL_1711048, EPI_ISL_1711049, EPI_ISL_1711051, EPI_ISL_1711052 | IN State Department of Health Laboratory Services | IN State Department of Health Laboratory Services | Cassandra Campion, Jamie Yeadon, Brian Pope, Lixia Liu, Kyle Brownlee, Melissa Hindenlang, Ankita Kashikar, Mark Glazier |
| EPI_ISL_1711053, EPI_ISL_1711054, EPI_ISL_1711055, EPI_ISL_1711056, EPI_ISL_1711057, EPI_ISL_1711058, EPI_ISL_1711059, EPI_ISL_1711060, EPI_ISL_1711061, EPI_ISL_1711063, EPI_ISL_1711065, EPI_ISL_1711066, EPI_ISL_1711067, EPI_ISL_1711070 | | | |
| see above | University of Mississippi Medical Center, Department of Pathology | University of Mississippi Medical Center, Molecular and Genomics Core Facility | Ashley C. Johnson, Ithiel J. Frame, Krishna K. Ayyalasomayajula, Michael R. Garrett, D. Ashley Robinson |
| EPI_ISL_1711071, EPI_ISL_1711072, EPI_ISL_1711073, EPI_ISL_1711074, EPI_ISL_1711075, EPI_ISL_1711076, EPI_ISL_1711077, EPI_ISL_1711078, EPI_ISL_1711079, EPI_ISL_1711080, EPI_ISL_1711081, EPI_ISL_1711082, EPI_ISL_1711083, EPI_ISL_1711084 | | | |
| see above | Vanderbilt University Medical Center | Pathogen Discovery, Respiratory Viruses Branch, Division of Viral Diseases, Centers for Disease Control and Prevention | Ying Tao, Yan Li, Anna Kelleher, Jing Zhang, Anna Montmayeur, Brian Lynch, Krista Queen, Anna Uehara, Peter Cook, Han Jia Justin Ng, Rachel Marine, Clinton R. Paden, Haibin Wang, Mark Burroughs, Justin Lee, Adam Retchless, Suxiang Tong |
| EPI_ISL_1711085, EPI_ISL_1711086, EPI_ISL_1711087, EPI_ISL_1711088, EPI_ISL_1711089, EPI_ISL_1711090, EPI_ISL_1711091, EPI_ISL_1711092, EPI_ISL_1711093, EPI_ISL_1711094, EPI_ISL_1711095, EPI_ISL_1711096, EPI_ISL_1711097, EPI_ISL_1711098, EPI_ISL_1711099, EPI_ISL_1711100, EPI_ISL_1711101, EPI_ISL_1711102, EPI_ISL_1711103, EPI_ISL_1711104, EPI_ISL_1711105, EPI_ISL_1711106, EPI_ISL_1711107, EPI_ISL_1711108, EPI_ISL_1711109, EPI_ISL_1711110, EPI_ISL_1711111, EPI_ISL_1711112, EPI_ISL_1711113, EPI_ISL_1711114, EPI_ISL_1711115, EPI_ISL_1711116, EPI_ISL_1711117, EPI_ISL_1711118, EPI_ISL_1711119, EPI_ISL_1711128, EPI_ISL_1711129, EPI_ISL_1711130, EPI_ISL_1711131, EPI_ISL_1711132, EPI_ISL_1711133, EPI_ISL_1711134, EPI_ISL_1711135, EPI_ISL_1711136, EPI_ISL_1711137, EPI_ISL_1711138, EPI_ISL_1711139, EPI_ISL_1711140, EPI_ISL_1711141, EPI_ISL_1711142, EPI_ISL_1711143, EPI_ISL_1711144, EPI_ISL_1711145, EPI_ISL_1711146, EPI_ISL_1711147, EPI_ISL_1711148, EPI_ISL_1711149, EPI_ISL_1711150, EPI_ISL_1711151, EPI_ISL_1711152, EPI_ISL_1711153, EPI_ISL_1711154, EPI_ISL_1711155, EPI_ISL_1711156, EPI_ISL_1711157, EPI_ISL_1711158, EPI_ISL_1711159, EPI_ISL_1711160, EPI_ISL_1711161, EPI_ISL_1711162, EPI_ISL_1711164, EPI_ISL_1711165, EPI_ISL_1711168, EPI_ISL_1711169, EPI_ISL_1711170, EPI_ISL_1711174, EPI_ISL_1711175, EPI_ISL_1711176, EPI_ISL_1711178, EPI_ISL_1711179, EPI_ISL_1711180, EPI_ISL_1711181, EPI_ISL_1711199, EPI_ISL_1711200, EPI_ISL_1711201, EPI_ISL_1711202, EPI_ISL_1711205, EPI_ISL_1711206, EPI_ISL_1711207, EPI_ISL_1711214, EPI_ISL_1711215, EPI_ISL_1711216, EPI_ISL_1711217, EPI_ISL_1711218, EPI_ISL_1711230, EPI_ISL_1711231, EPI_ISL_1711232, EPI_ISL_1711233, EPI_ISL_1711234, EPI_ISL_1711235, EPI_ISL_1711236, EPI_ISL_1711237, EPI_ISL_1711242, EPI_ISL_1711243, EPI_ISL_1711244, EPI_ISL_1711245, EPI_ISL_1711246, EPI_ISL_1711247, EPI_ISL_1711248, EPI_ISL_1711249, EPI_ISL_1711250, EPI_ISL_1711251, EPI_ISL_1711252, EPI_ISL_1711253, EPI_ISL_1711254, EPI_ISL_1711255 | | | |
| see above | US Air Force School of Aerospace Medicine | US Air Force School of Aerospace Medicine | Anthony Fries, Jennifer Meyer, William Gruner, William Buggele, Amanda Javorina, Sarah Purves, Clarise Starr, Elizabeth Macias |
| EPI_ISL_1711257, EPI_ISL_1711258, EPI_ISL_1711259, EPI_ISL_1711260, EPI_ISL_1711261, EPI_ISL_1711262, EPI_ISL_1711263, EPI_ISL_1711264, EPI_ISL_1711268, EPI_ISL_1711269, EPI_ISL_1711270, EPI_ISL_1711271 | | | |
| see above | Wyoming Public Health Laboratory | Wyoming Public Health Laboratory | Jim Mildenberger, Taylor Fearing, Channing Weber, Ashley Norberg, Chayse Rowley, Marley Goetz, Brian Dominguez, Elliot Thomasson, Brittany Oher, Sam Britz, Cari Sloma, and Rob Christensen |
| EPI_ISL_1711272, EPI_ISL_1711274, EPI_ISL_1711275 | Wyoming Public Health Laboratory | Center for Global Health, University of New Mexico Health Sciences Center | Daryl Domman, Kurt Schwalm, Rob Christensen, Wanda Manley, Cari Sloma, Noah Hull, Valerie Morley, Darrell Dinwiddie |
| EPI_ISL_1711276, EPI_ISL_1711277, EPI_ISL_1711278, EPI_ISL_1711279, EPI_ISL_1711280, EPI_ISL_1711281, EPI_ISL_1711282, EPI_ISL_1711283, EPI_ISL_1711284, EPI_ISL_1711285, EPI_ISL_1711286, EPI_ISL_1711287, EPI_ISL_1711288, EPI_ISL_1711289, EPI_ISL_1711290, EPI_ISL_1711291, EPI_ISL_1711292, EPI_ISL_1711293, EPI_ISL_1711294, EPI_ISL_1711295, EPI_ISL_1711296, EPI_ISL_1711297, EPI_ISL_1711298, EPI_ISL_1711299, EPI_ISL_1711300, EPI_ISL_1711301, EPI_ISL_1711302, EPI_ISL_1711315, EPI_ISL_1711316, EPI_ISL_1711317, EPI_ISL_1711318, EPI_ISL_1711319, EPI_ISL_1711320, EPI_ISL_1711321, EPI_ISL_1711322, EPI_ISL_1711323, EPI_ISL_1711324, EPI_ISL_1711325, EPI_ISL_1711326, EPI_ISL_1711327, EPI_ISL_1711328, EPI_ISL_1711329, EPI_ISL_1711330, EPI_ISL_1711331, EPI_ISL_1711332, EPI_ISL_1711333, EPI_ISL_1711334, EPI_ISL_1711335, EPI_ISL_1711336, EPI_ISL_1711337, EPI_ISL_1711338, EPI_ISL_1711339, EPI_ISL_1711340, EPI_ISL_1711341, EPI_ISL_1711342, EPI_ISL_1711343, EPI_ISL_1711344, EPI_ISL_1711345, EPI_ISL_1711346, EPI_ISL_1711347 | | | |
| see above | Instituto Nacional de Medicina Genomica | Instituto Nacional de Medicina Genomica | Hidalgo-Miranda A, Mendoza-Vargas A, Reyes-Grajeda JP, Cedro-Tanda A, Alcaraz N, Gonzalez-Barrera D, Rangel-DeLeon D, Miranda-Ortiz H, Rosas-Escobar P, Canseco Mendez JC, Munguia-Garza P, Ramirez-Vega O, Garcia-Cardenas FJ, Gonzalez-Woge MA, Herrera-Montalvo LA |
| EPI_ISL_1711348, EPI_ISL_1711349 | Wyoming Public Health Laboratory | Wyoming Public Health Laboratory | Jim Mildenberger, Wanda Manley, Noah Hull, Taylor Fearing, Lynette Gumbleton, Channing Weber, Ashley Norberg, Chayse Rowley, Marley Goetz, Brian Dominguez, Elliot Thomasson, Sam Britz, Cari Sloma, and Rob Christensen |
| EPI_ISL_1711351, EPI_ISL_1711352, EPI_ISL_1711353 | Kansas Health and Environmental Lab | Kansas Health and Environmental Lab | Mike Grose, Jonathan Barnell, Ben Olsen, and Phil Adam |
| EPI_ISL_1711354, EPI_ISL_1711355, EPI_ISL_1711356, EPI_ISL_1711357, EPI_ISL_1711358, EPI_ISL_1711359, EPI_ISL_1711360, EPI_ISL_1711361, EPI_ISL_1711362, EPI_ISL_1711363, EPI_ISL_1711364, EPI_ISL_1711365, EPI_ISL_1711366 | | | |
| see above | Wichita State University - Molecular Diagnostics Lab | Kansas Health and Environmental Lab | Mike Grose, Jonathan Barnell, Ben Olsen, and Phil Adam |
| EPI_ISL_1711367, EPI_ISL_1711368, EPI_ISL_1711369, EPI_ISL_1711370, EPI_ISL_1711371, EPI_ISL_1711372, EPI_ISL_1711373, EPI_ISL_1711374, EPI_ISL_1711375, EPI_ISL_1711376, EPI_ISL_1711377, EPI_ISL_1711378, EPI_ISL_1711379, EPI_ISL_1711380, EPI_ISL_1711381, EPI_ISL_1711382, EPI_ISL_1711383, EPI_ISL_1711384, EPI_ISL_1711385, EPI_ISL_1711386, EPI_ISL_1711387, EPI_ISL_1711388, EPI_ISL_1711389, EPI_ISL_1711390, EPI_ISL_1711391, EPI_ISL_1711392, EPI_ISL_1711393 | | | |
| see above | Wyoming Public Health Laboratory | Center for Global Health, University of New Mexico Health Sciences Center | Daryl Domman, Kurt Schwalm, Rob Christensen, Wanda Manley, Cari Sloma, Noah Hull, Valerie Morley, Darrell Dinwiddie |
| EPI_ISL_1711394, EPI_ISL_1711395, EPI_ISL_1711396, EPI_ISL_1711419 | Wyoming Public Health Laboratory | Wyoming Public Health Laboratory | Jim Mildenberger, Wanda Manley, Noah Hull, Taylor Fearing, Lynette Gumbleton, Channing Weber, Ashley Norberg, Chayse Rowley, Marley Goetz, Brian Dominguez, Elliot Thomasson, Sam Britz, Cari Sloma, and Rob Christensen |
| EPI_ISL_1711420 | FidaLab | Seattle Flu Study | Deborah A. Nickerson, Chris D. Frazar, Jover Lee, Benjamin Pelle, Erica Ryke, Matthew Richardson, Amanda Adler, Elisabeth Brandstetter, Peter D. Han, Kairsten Fay, Misja Ilcisin, Kirsten Lacombe, Thomas R. Sibley, Melissa Truong, Caitlin R. Wolf, Romesh Gautom, Geoff Melly, Brian Hiatt, Philip Dykema, Scott Lindquist, Michael Boeckh, Janet A. Englund, Michael Famulare, Barry R. Lutz, Mark J. Rieder, Lea M. Starita, Matthew Thompson, Helen Y. Chu, Jay Shendure, Trevor Bedford |
| EPI_ISL_1711421, EPI_ISL_1711422, EPI_ISL_1711423 | Atlas Genomics | Seattle Flu Study | Deborah A. Nickerson, Chris D. Frazar, Jover Lee, Benjamin Pelle, Erica Ryke, Matthew Richardson, Amanda Adler, Elisabeth Brandstetter, Peter D. Han, Kairsten Fay, Misja Ilcisin, Kirsten Lacombe, Thomas R. Sibley, Melissa Truong, Caitlin R. Wolf, Romesh Gautom, Geoff Melly, Brian Hiatt, Philip Dykema, Scott Lindquist, Michael Boeckh, Janet A. Englund, Michael Famulare, Barry R. Lutz, Mark J. Rieder, Lea M. Starita, Matthew Thompson, Helen Y. Chu, Jay Shendure, Trevor Bedford |
| EPI_ISL_1711424 | Northwest Laboratories | Seattle Flu Study | Deborah A. Nickerson, Chris D. Frazar, Jover Lee, Benjamin Pelle, Erica Ryke, Matthew Richardson, Amanda Adler, Elisabeth Brandstetter, Peter D. Han, Kairsten Fay, Misja Ilcisin, Kirsten Lacombe, Thomas R. Sibley, Melissa Truong, Caitlin R. Wolf, Romesh Gautom, Geoff Melly, Brian Hiatt, Philip Dykema, Scott Lindquist, Michael Boeckh, Janet A. Englund, Michael Famulare, Barry R. Lutz, Mark J. Rieder, Lea M. Starita, Matthew Thompson, Helen Y. Chu, Jay Shendure, Trevor Bedford |
| EPI_ISL_1711426 | Seattle Flu Study | Seattle Flu Study | Deborah A. Nickerson, Chris D. Frazar, Jover Lee, Benjamin Pelle, Erica Ryke, Matthew Richardson, Amanda Adler, Elisabeth Brandstetter, Peter D. Han, Kairsten Fay, Misja Ilcisin, Kirsten Lacombe, Thomas R. Sibley, Melissa Truong, Caitlin R. Wolf, Karen Cowgill, Stephanie Schrag, Jeff Duchin, Michael Boeckh, Janet A. Englund, Michael Famulare, Barry R. Lutz, Mark J. Rieder, Lea M. Starita, Matthew Thompson, Helen Y. Chu, Trevor Bedford, Jay Shendure |
| EPI_ISL_1711427 | Atlas Genomics | Seattle Flu Study | Deborah A. Nickerson, Chris D. Frazar, Jover Lee, Benjamin Pelle, Erica Ryke, Matthew Richardson, Amanda Adler, Elisabeth Brandstetter, Peter D. Han, Kairsten Fay, Misja Ilcisin, Kirsten Lacombe, Thomas R. Sibley, Melissa Truong, Caitlin R. Wolf, Romesh Gautom, Geoff Melly, Brian Hiatt, Philip Dykema, Scott Lindquist, Michael Boeckh, Janet A. Englund, Michael Famulare, Barry R. Lutz, Mark J. Rieder, Lea M. Starita, Matthew Thompson, Helen Y. Chu, Jay Shendure, Trevor Bedford |
| EPI_ISL_1711428, EPI_ISL_1711429 | Seattle Flu Study | Seattle Flu Study | Deborah A. Nickerson, Chris D. Frazar, Jover Lee, Benjamin Pelle, Erica Ryke, Matthew Richardson, Amanda Adler, Elisabeth Brandstetter, Peter D. Han, Kairsten Fay, Misja Ilcisin, Kirsten Lacombe, Thomas R. Sibley, Melissa Truong, Caitlin R. Wolf, Karen Cowgill, Stephanie Schrag, Jeff Duchin, Michael Boeckh, Janet A. Englund, Michael Famulare, Barry R. Lutz, Mark J. Rieder, Lea M. Starita, Matthew Thompson, Helen Y. Chu, Trevor Bedford, Jay Shendure |
| EPI_ISL_1711430 | Interpath Laboratory | Seattle Flu Study | Deborah A. Nickerson, Chris D. Frazar, Jover Lee, Benjamin Pelle, Erica Ryke, Matthew Richardson, Amanda Adler, Elisabeth Brandstetter, Peter D. Han, Kairsten Fay, Misja Ilcisin, Kirsten Lacombe, Thomas R. Sibley, Melissa Truong, Caitlin R. Wolf, Romesh Gautom, Geoff Melly, Brian Hiatt, Philip Dykema, Scott Lindquist, Michael Boeckh, Janet A. Englund, Michael Famulare, Barry R. Lutz, Mark J. Rieder, Lea M. Starita, Matthew Thompson, Helen Y. Chu, Jay Shendure, Trevor Bedford |
| EPI_ISL_1711432, EPI_ISL_1711433 | Atlas Genomics | Seattle Flu Study | Deborah A. Nickerson, Chris D. Frazar, Jover Lee, Benjamin Pelle, Erica Ryke, Matthew Richardson, Amanda Adler, Elisabeth Brandstetter, Peter D. Han, Kairsten Fay, Misja Ilcisin, Kirsten Lacombe, Thomas R. Sibley, Melissa Truong, Caitlin R. Wolf, Romesh Gautom, Geoff Melly, Brian Hiatt, Philip Dykema, Scott Lindquist, Michael Boeckh, Janet A. Englund, Michael Famulare, Barry R. Lutz, Mark J. Rieder, Lea M. Starita, Matthew Thompson, Helen Y. Chu, Jay Shendure, Trevor Bedford |
| EPI_ISL_1711434 | Northwest Laboratories | Seattle Flu Study | Deborah A. Nickerson, Chris D. Frazar, Jover Lee, Benjamin Pelle, Erica Ryke, Matthew Richardson, Amanda Adler, Elisabeth Brandstetter, Peter D. Han, Kairsten Fay, Misja Ilcisin, Kirsten Lacombe, Thomas R. Sibley, Melissa Truong, Caitlin R. Wolf, Romesh Gautom, Geoff Melly, Brian Hiatt, Philip Dykema, Scott Lindquist, Michael Boeckh, Janet A. Englund, Michael Famulare, Barry R. Lutz, Mark J. Rieder, Lea M. Starita, Matthew Thompson, Helen Y. Chu, Jay Shendure, Trevor Bedford |
| EPI_ISL_1711436 | Atlas Genomics | Seattle Flu Study | Deborah A. Nickerson, Chris D. Frazar, Jover Lee, Benjamin Pelle, Erica Ryke, Matthew Richardson, Amanda Adler, Elisabeth Brandstetter, Peter D. Han, Kairsten Fay, Misja Ilcisin, Kirsten Lacombe, Thomas R. Sibley, Melissa Truong, Caitlin R. Wolf, Romesh Gautom, Geoff Melly, Brian Hiatt, Philip Dykema, Scott Lindquist, Michael Boeckh, Janet A. Englund, Michael Famulare, Barry R. Lutz, Mark J. Rieder, Lea M. Starita, Matthew Thompson, Helen Y. Chu, Jay Shendure, Trevor Bedford |
| EPI_ISL_1711437 | Seattle Flu Study | Seattle Flu Study | Deborah A. Nickerson, Chris D. Frazar, Jover Lee, Benjamin Pelle, Erica Ryke, Matthew Richardson, Amanda Adler, Elisabeth Brandstetter, Peter D. Han, Kairsten Fay, Misja Ilcisin, Kirsten Lacombe, Thomas R. Sibley, Melissa Truong, Caitlin R. Wolf, Michael Boeckh, Janet A. Englund, Michael Famulare, Barry R. Lutz, Mark J. Rieder, Lea M. Starita, Matthew Thompson, Jay Shendure, Trevor Bedford, Helen Y. Chu |
| EPI_ISL_1711438 | Atlas Genomics | Seattle Flu Study | Deborah A. Nickerson, Chris D. Frazar, Jover Lee, Benjamin Pelle, Erica Ryke, Matthew Richardson, Amanda Adler, Elisabeth Brandstetter, Peter D. Han, Kairsten Fay, Misja Ilcisin, Kirsten Lacombe, Thomas R. Sibley, Melissa Truong, Caitlin R. Wolf, Romesh Gautom, Geoff Melly, Brian Hiatt, Philip Dykema, Scott Lindquist, Michael Boeckh, Janet A. Englund, Michael Famulare, Barry R. Lutz, Mark J. Rieder, Lea M. Starita, Matthew Thompson, Helen Y. Chu, Jay Shendure, Trevor Bedford |
| EPI_ISL_1711439 | Interpath Laboratory | Seattle Flu Study | Deborah A. Nickerson, Chris D. Frazar, Jover Lee, Benjamin Pelle, Erica Ryke, Matthew Richardson, Amanda Adler, Elisabeth Brandstetter, Peter D. Han, Kairsten Fay, Misja Ilcisin, Kirsten Lacombe, Thomas R. Sibley, Melissa Truong, Caitlin R. Wolf, Romesh Gautom, Geoff Melly, Brian Hiatt, Philip Dykema, Scott Lindquist, Michael Boeckh, Janet A. Englund, Michael Famulare, Barry R. Lutz, Mark J. Rieder, Lea M. Starita, Matthew Thompson, Helen Y. Chu, Jay Shendure, Trevor Bedford |
| EPI_ISL_1711440 | Seattle Flu Study | Seattle Flu Study | Deborah A. Nickerson, Chris D. Frazar, Jover Lee, Benjamin Pelle, Erica Ryke, Matthew Richardson, Amanda Adler, Elisabeth Brandstetter, Peter D. Han, Kairsten Fay, Misja Ilcisin, Kirsten Lacombe, Thomas R. Sibley, Melissa Truong, Caitlin R. Wolf, Karen Cowgill, Stephanie Schrag, Jeff Duchin, Michael Boeckh, Janet A. Englund, Michael Famulare, Barry R. Lutz, Mark J. Rieder, Lea M. Starita, Matthew Thompson, Helen Y. Chu, Trevor Bedford, Jay Shendure |
| EPI_ISL_1711441 | Seattle Flu Study | Seattle Flu Study | Deborah A. Nickerson, Chris D. Frazar, Jover Lee, Benjamin Pelle, Erica Ryke, Matthew Richardson, Amanda Adler, Elisabeth Brandstetter, Peter D. Han, Kairsten Fay, Misja Ilcisin, Kirsten Lacombe, Thomas R. Sibley, Melissa Truong, Caitlin R. Wolf, Michael Boeckh, Janet A. Englund, |

| | | | |
|---|---|---|---|
| | | | Michael Famulare, Barry R. Lutz, Mark J. Rieder, Lea M. Starita, Matthew Thompson, Jay Shendure, Trevor Bedford, Helen Y. Chu |
| EPI_ISL_1711442 | Interpath Laboratory | Seattle Flu Study | Deborah A. Nickerson, Chris D. Frazar, Jover Lee, Benjamin Pelle, Erica Ryke, Matthew Richardson, Amanda Adler, Elisabeth Brandstetter, Peter D. Han, Kairsten Fay, Misja Ilcisin, Kirsten Lacombe, Thomas R. Sibley, Melissa Truong, Caitlin R. Wolf, Romesh Gautom, Geoff Melly, Brian Hiatt, Philip Dykema, Scott Lindquist, Michael Boeckh, Janet A. Englund, Michael Famulare, Barry R. Lutz, Mark J. Rieder, Lea M. Starita, Matthew Thompson, Helen Y. Chu, Jay Shendure, Trevor Bedford |
| EPI_ISL_1711443 | Atlas Genomics | Seattle Flu Study | Deborah A. Nickerson, Chris D. Frazar, Jover Lee, Benjamin Pelle, Erica Ryke, Matthew Richardson, Amanda Adler, Elisabeth Brandstetter, Peter D. Han, Kairsten Fay, Misja Ilcisin, Kirsten Lacombe, Thomas R. Sibley, Melissa Truong, Caitlin R. Wolf, Romesh Gautom, Geoff Melly, Brian Hiatt, Philip Dykema, Scott Lindquist, Michael Boeckh, Janet A. Englund, Michael Famulare, Barry R. Lutz, Mark J. Rieder, Lea M. Starita, Matthew Thompson, Helen Y. Chu, Jay Shendure, Trevor Bedford |
| EPI_ISL_1711444 | Northwest Laboratories | Seattle Flu Study | Deborah A. Nickerson, Chris D. Frazar, Jover Lee, Benjamin Pelle, Erica Ryke, Matthew Richardson, Amanda Adler, Elisabeth Brandstetter, Peter D. Han, Kairsten Fay, Misja Ilcisin, Kirsten Lacombe, Thomas R. Sibley, Melissa Truong, Caitlin R. Wolf, Romesh Gautom, Geoff Melly, Brian Hiatt, Philip Dykema, Scott Lindquist, Michael Boeckh, Janet A. Englund, Michael Famulare, Barry R. Lutz, Mark J. Rieder, Lea M. Starita, Matthew Thompson, Helen Y. Chu, Jay Shendure, Trevor Bedford |
| EPI_ISL_1711445 | Seattle Flu Study | Seattle Flu Study | Deborah A. Nickerson, Chris D. Frazar, Jover Lee, Benjamin Pelle, Erica Ryke, Matthew Richardson, Amanda Adler, Elisabeth Brandstetter, Peter D. Han, Kairsten Fay, Misja Ilcisin, Kirsten Lacombe, Thomas R. Sibley, Melissa Truong, Caitlin R. Wolf, Michael Boeckh, Janet A. Englund, Michael Famulare, Barry R. Lutz, Mark J. Rieder, Lea M. Starita, Matthew Thompson, Jay Shendure, Trevor Bedford, Helen Y. Chu |
| EPI_ISL_1711446 | Atlas Genomics | Seattle Flu Study | Deborah A. Nickerson, Chris D. Frazar, Jover Lee, Benjamin Pelle, Erica Ryke, Matthew Richardson, Amanda Adler, Elisabeth Brandstetter, Peter D. Han, Kairsten Fay, Misja Ilcisin, Kirsten Lacombe, Thomas R. Sibley, Melissa Truong, Caitlin R. Wolf, Romesh Gautom, Geoff Melly, Brian Hiatt, Philip Dykema, Scott Lindquist, Michael Boeckh, Janet A. Englund, Michael Famulare, Barry R. Lutz, Mark J. Rieder, Lea M. Starita, Matthew Thompson, Helen Y. Chu, Jay Shendure, Trevor Bedford |
| EPI_ISL_1711447 | Seattle Flu Study | Seattle Flu Study | Deborah A. Nickerson, Chris D. Frazar, Jover Lee, Benjamin Pelle, Erica Ryke, Matthew Richardson, Amanda Adler, Elisabeth Brandstetter, Peter D. Han, Kairsten Fay, Misja Ilcisin, Kirsten Lacombe, Thomas R. Sibley, Melissa Truong, Caitlin R. Wolf, Michael Boeckh, Janet A. Englund, Michael Famulare, Barry R. Lutz, Mark J. Rieder, Lea M. Starita, Matthew Thompson, Jay Shendure, Trevor Bedford, Helen Y. Chu |
| EPI_ISL_1711448 | Northwest Laboratories | Seattle Flu Study | Deborah A. Nickerson, Chris D. Frazar, Jover Lee, Benjamin Pelle, Erica Ryke, Matthew Richardson, Amanda Adler, Elisabeth Brandstetter, Peter D. Han, Kairsten Fay, Misja Ilcisin, Kirsten Lacombe, Thomas R. Sibley, Melissa Truong, Caitlin R. Wolf, Romesh Gautom, Geoff Melly, Brian Hiatt, Philip Dykema, Scott Lindquist, Michael Boeckh, Janet A. Englund, Michael Famulare, Barry R. Lutz, Mark J. Rieder, Lea M. Starita, Matthew Thompson, Helen Y. Chu, Jay Shendure, Trevor Bedford |
| EPI_ISL_1711449 | Interpath Laboratory | Seattle Flu Study | Deborah A. Nickerson, Chris D. Frazar, Jover Lee, Benjamin Pelle, Erica Ryke, Matthew Richardson, Amanda Adler, Elisabeth Brandstetter, Peter D. Han, Kairsten Fay, Misja Ilcisin, Kirsten Lacombe, Thomas R. Sibley, Melissa Truong, Caitlin R. Wolf, Romesh Gautom, Geoff Melly, Brian Hiatt, Philip Dykema, Scott Lindquist, Michael Boeckh, Janet A. Englund, Michael Famulare, Barry R. Lutz, Mark J. Rieder, Lea M. Starita, Matthew Thompson, Helen Y. Chu, Jay Shendure, Trevor Bedford |
| EPI_ISL_1711450 | Atlas Genomics | Seattle Flu Study | Deborah A. Nickerson, Chris D. Frazar, Jover Lee, Benjamin Pelle, Erica Ryke, Matthew Richardson, Amanda Adler, Elisabeth Brandstetter, Peter D. Han, Kairsten Fay, Misja Ilcisin, Kirsten Lacombe, Thomas R. Sibley, Melissa Truong, Caitlin R. Wolf, Romesh Gautom, Geoff Melly, Brian Hiatt, Philip Dykema, Scott Lindquist, Michael Boeckh, Janet A. Englund, Michael Famulare, Barry R. Lutz, Mark J. Rieder, Lea M. Starita, Matthew Thompson, Helen Y. Chu, Jay Shendure, Trevor Bedford |
| EPI_ISL_1711451, EPI_ISL_1711452 | Seattle Flu Study | Seattle Flu Study | Deborah A. Nickerson, Chris D. Frazar, Jover Lee, Benjamin Pelle, Erica Ryke, Matthew Richardson, Amanda Adler, Elisabeth Brandstetter, Peter D. Han, Kairsten Fay, Misja Ilcisin, Kirsten Lacombe, Thomas R. Sibley, Melissa Truong, Caitlin R. Wolf, Michael Boeckh, Janet A. Englund, Michael Famulare, Barry R. Lutz, Mark J. Rieder, Lea M. Starita, Matthew Thompson, Jay Shendure, Trevor Bedford, Helen Y. Chu |
| EPI_ISL_1711453 | Interpath Laboratory | Seattle Flu Study | Deborah A. Nickerson, Chris D. Frazar, Jover Lee, Benjamin Pelle, Erica Ryke, Matthew Richardson, Amanda Adler, Elisabeth Brandstetter, Peter D. Han, Kairsten Fay, Misja Ilcisin, Kirsten Lacombe, Thomas R. Sibley, Melissa Truong, Caitlin R. Wolf, Romesh Gautom, Geoff Melly, Brian Hiatt, Philip Dykema, Scott Lindquist, Michael Boeckh, Janet A. Englund, Michael Famulare, Barry R. Lutz, Mark J. Rieder, Lea M. Starita, Matthew Thompson, Helen Y. Chu, Jay Shendure, Trevor Bedford |
| EPI_ISL_1711454 | Atlas Genomics | Seattle Flu Study | Deborah A. Nickerson, Chris D. Frazar, Jover Lee, Benjamin Pelle, Erica Ryke, Matthew Richardson, Amanda Adler, Elisabeth Brandstetter, Peter D. Han, Kairsten Fay, Misja Ilcisin, Kirsten Lacombe, Thomas R. Sibley, Melissa Truong, Caitlin R. Wolf, Romesh Gautom, Geoff Melly, Brian Hiatt, Philip Dykema, Scott Lindquist, Michael Boeckh, Janet A. Englund, Michael Famulare, Barry R. Lutz, Mark J. Rieder, Lea M. Starita, Matthew Thompson, Helen Y. Chu, Jay Shendure, Trevor Bedford |
| EPI_ISL_1711455 | Seattle Flu Study | Seattle Flu Study | Deborah A. Nickerson, Chris D. Frazar, Jover Lee, Benjamin Pelle, Erica Ryke, Matthew Richardson, Amanda Adler, Elisabeth Brandstetter, Peter D. Han, Kairsten Fay, Misja Ilcisin, Kirsten Lacombe, Thomas R. Sibley, Melissa Truong, Caitlin R. Wolf, Michael Boeckh, Janet A. Englund, Michael Famulare, Barry R. Lutz, Mark J. Rieder, Lea M. Starita, Matthew Thompson, Jay Shendure, Trevor Bedford, Helen Y. Chu |
| EPI_ISL_1711456 | Northwest Laboratories | Seattle Flu Study | Deborah A. Nickerson, Chris D. Frazar, Jover Lee, Benjamin Pelle, Erica Ryke, Matthew Richardson, Amanda Adler, Elisabeth Brandstetter, Peter D. Han, Kairsten Fay, Misja Ilcisin, Kirsten Lacombe, Thomas R. Sibley, Melissa Truong, Caitlin R. Wolf, Romesh Gautom, Geoff Melly, Brian Hiatt, Philip Dykema, Scott Lindquist, Michael Boeckh, Janet A. Englund, Michael Famulare, Barry R. Lutz, Mark J. Rieder, Lea M. Starita, Matthew Thompson, Helen Y. Chu, Jay Shendure, Trevor Bedford |
| EPI_ISL_1711457 | Seattle Flu Study | Seattle Flu Study | Deborah A. Nickerson, Chris D. Frazar, Jover Lee, Benjamin Pelle, Erica Ryke, Matthew Richardson, Amanda Adler, Elisabeth Brandstetter, Peter D. Han, Kairsten Fay, Misja Ilcisin, Kirsten Lacombe, Thomas R. Sibley, Melissa Truong, Caitlin R. Wolf, Michael Boeckh, Janet A. Englund, Michael Famulare, Barry R. Lutz, Mark J. Rieder, Lea M. Starita, Matthew Thompson, Jay Shendure, Trevor Bedford, Helen Y. Chu |
| EPI_ISL_1711458 | Northwest Laboratories | Seattle Flu Study | Deborah A. Nickerson, Chris D. Frazar, Jover Lee, Benjamin Pelle, Erica Ryke, Matthew Richardson, Amanda Adler, Elisabeth Brandstetter, Peter D. Han, Kairsten Fay, Misja Ilcisin, Kirsten Lacombe, Thomas R. Sibley, Melissa Truong, Caitlin R. Wolf, Romesh Gautom, Geoff Melly, Brian Hiatt, Philip Dykema, Scott Lindquist, Michael Boeckh, Janet A. Englund, Michael Famulare, Barry R. Lutz, Mark J. Rieder, Lea M. Starita, Matthew Thompson, Helen Y. Chu, Jay Shendure, Trevor Bedford |
| EPI_ISL_1711459 | Atlas Genomics | Seattle Flu Study | Deborah A. Nickerson, Chris D. Frazar, Jover Lee, Benjamin Pelle, Erica Ryke, Matthew Richardson, Amanda Adler, Elisabeth Brandstetter, Peter D. Han, Kairsten Fay, Misja Ilcisin, Kirsten Lacombe, Thomas R. Sibley, Melissa Truong, Caitlin R. Wolf, Romesh Gautom, Geoff Melly, Brian Hiatt, Philip Dykema, Scott Lindquist, Michael Boeckh, Janet A. Englund, Michael Famulare, Barry R. Lutz, Mark J. Rieder, Lea M. Starita, Matthew Thompson, Helen Y. Chu, Jay Shendure, Trevor Bedford |
| EPI_ISL_1711460 | Seattle Flu Study | Seattle Flu Study | Deborah A. Nickerson, Chris D. Frazar, Jover Lee, Benjamin Pelle, Erica Ryke, Matthew Richardson, Amanda Adler, Elisabeth Brandstetter, Peter D. Han, Kairsten Fay, Misja Ilcisin, Kirsten Lacombe, Thomas R. Sibley, Melissa Truong, Caitlin R. Wolf, Michael Boeckh, Janet A. Englund, Michael Famulare, Barry R. Lutz, Mark J. Rieder, Lea M. Starita, Matthew Thompson, Jay Shendure, Trevor Bedford, Helen Y. Chu |
| EPI_ISL_1711461 | Atlas Genomics | Seattle Flu Study | Deborah A. Nickerson, Chris D. Frazar, Jover Lee, Benjamin Pelle, Erica Ryke, Matthew Richardson, Amanda Adler, Elisabeth Brandstetter, Peter D. Han, Kairsten Fay, Misja Ilcisin, Kirsten Lacombe, Thomas R. Sibley, Melissa Truong, Caitlin R. Wolf, Romesh Gautom, Geoff Melly, Brian Hiatt, Philip Dykema, Scott Lindquist, Michael Boeckh, Janet A. Englund, Michael Famulare, Barry R. Lutz, Mark J. Rieder, Lea M. Starita, Matthew Thompson, Helen Y. Chu, Jay Shendure, Trevor Bedford |
| EPI_ISL_1711462 | Northwest Laboratories | Seattle Flu Study | Deborah A. Nickerson, Chris D. Frazar, Jover Lee, Benjamin Pelle, Erica Ryke, Matthew Richardson, Amanda Adler, Elisabeth Brandstetter, Peter D. Han, Kairsten Fay, Misja Ilcisin, Kirsten Lacombe, Thomas R. Sibley, Melissa Truong, Caitlin R. Wolf, Romesh Gautom, Geoff Melly, Brian Hiatt, Philip Dykema, Scott Lindquist, Michael Boeckh, Janet A. Englund, Michael Famulare, Barry R. Lutz, Mark J. Rieder, Lea M. Starita, Matthew Thompson, Helen Y. Chu, Jay Shendure, Trevor Bedford |
| EPI_ISL_1711463 | Evergreen Healthcare | Seattle Flu Study | Deborah A. Nickerson, Chris D. Frazar, Jover Lee, Benjamin Pelle, Erica Ryke, Matthew Richardson, Amanda Adler, Elisabeth Brandstetter, Peter D. Han, Kairsten Fay, Misja Ilcisin, Kirsten Lacombe, Thomas R. Sibley, Melissa Truong, Caitlin R. Wolf, Romesh Gautom, Geoff Melly, Brian Hiatt, Philip Dykema, Scott Lindquist, Michael Boeckh, Janet A. Englund, Michael Famulare, Barry R. Lutz, Mark J. Rieder, Lea M. Starita, Matthew Thompson, Helen Y. Chu, Jay Shendure, Trevor Bedford |
| EPI_ISL_1711464 | Atlas Genomics | Seattle Flu Study | Deborah A. Nickerson, Chris D. Frazar, Jover Lee, Benjamin Pelle, Erica Ryke, Matthew Richardson, Amanda Adler, Elisabeth Brandstetter, Peter D. Han, Kairsten Fay, Misja Ilcisin, Kirsten Lacombe, Thomas R. Sibley, Melissa Truong, Caitlin R. Wolf, Romesh Gautom, Geoff Melly, Brian Hiatt, Philip Dykema, Scott Lindquist, Michael Boeckh, Janet A. Englund, Michael Famulare, Barry R. Lutz, Mark J. Rieder, Lea M. Starita, Matthew Thompson, Helen Y. Chu, Jay Shendure, Trevor Bedford |
| EPI_ISL_1711465 | Seattle Flu Study | Seattle Flu Study | Deborah A. Nickerson, Chris D. Frazar, Jover Lee, Benjamin Pelle, Erica Ryke, Matthew Richardson, Amanda Adler, Elisabeth Brandstetter, Peter D. Han, Kairsten Fay, Misja Ilcisin, Kirsten Lacombe, Thomas R. Sibley, Melissa Truong, Caitlin R. Wolf, Karen Cowgill, Stephanie Schrag, Jeff Duchin, Michael Boeckh, Janet A. Englund, Michael Famulare, Barry R. Lutz, Mark J. Rieder, Lea M. Starita, Matthew Thompson, Helen Y. Chu, Trevor Bedford, Jay Shendure |
| EPI_ISL_1711466 | Atlas Genomics | Seattle Flu Study | Deborah A. Nickerson, Chris D. Frazar, Jover Lee, Benjamin Pelle, Erica Ryke, Matthew Richardson, Amanda Adler, Elisabeth Brandstetter, Peter D. Han, Kairsten Fay, Misja Ilcisin, Kirsten Lacombe, Thomas R. Sibley, Melissa Truong, Caitlin R. Wolf, Romesh Gautom, Geoff Melly, Brian Hiatt, Philip Dykema, Scott Lindquist, Michael Boeckh, Janet A. Englund, Michael Famulare, Barry R. Lutz, Mark J. Rieder, Lea M. Starita, Matthew Thompson, Helen Y. Chu, Jay Shendure, Trevor Bedford |
| EPI_ISL_1711467 | Seattle Flu Study | Seattle Flu Study | Deborah A. Nickerson, Chris D. Frazar, Jover Lee, Benjamin Pelle, Erica Ryke, Matthew Richardson, Amanda Adler, Elisabeth Brandstetter, Peter D. Han, Kairsten Fay, Misja Ilcisin, Kirsten Lacombe, Thomas R. Sibley, Melissa Truong, Caitlin R. Wolf, Michael Boeckh, Janet A. Englund, Michael Famulare, Barry R. Lutz, Mark J. Rieder, Lea M. Starita, Matthew Thompson, Jay Shendure, Trevor Bedford, Helen Y. Chu |
| EPI_ISL_1711468 | Atlas Genomics | Seattle Flu Study | Deborah A. Nickerson, Chris D. Frazar, Jover Lee, Benjamin Pelle, Erica Ryke, Matthew Richardson, Amanda Adler, Elisabeth Brandstetter, Peter D. Han, Kairsten Fay, Misja Ilcisin, Kirsten Lacombe, Thomas R. Sibley, Melissa Truong, Caitlin R. Wolf, Romesh Gautom, Geoff Melly, Brian Hiatt, Philip Dykema, Scott Lindquist, Michael Boeckh, Janet A. Englund, Michael Famulare, Barry R. Lutz, Mark J. Rieder, Lea M. Starita, Matthew Thompson, Helen Y. Chu, Jay Shendure, Trevor Bedford |
| EPI_ISL_1711469 | Interpath Laboratory | Seattle Flu Study | Deborah A. Nickerson, Chris D. Frazar, Jover Lee, Benjamin Pelle, Erica Ryke, Matthew Richardson, Amanda Adler, Elisabeth Brandstetter, Peter D. Han, Kairsten Fay, Misja Ilcisin, Kirsten Lacombe, Thomas R. Sibley, Melissa Truong, Caitlin R. Wolf, Romesh Gautom, Geoff Melly, Brian Hiatt, Philip Dykema, Scott Lindquist, Michael Boeckh, Janet A. Englund, Michael Famulare, Barry R. Lutz, Mark J. Rieder, Lea M. Starita, Matthew Thompson, Helen Y. Chu, Jay Shendure, Trevor Bedford |
| EPI_ISL_1711470, EPI_ISL_1711471, EPI_ISL_1711472 | Atlas Genomics | Seattle Flu Study | Deborah A. Nickerson, Chris D. Frazar, Jover Lee, Benjamin Pelle, Erica Ryke, Matthew Richardson, Amanda Adler, Elisabeth Brandstetter, Peter D. Han, Kairsten Fay, Misja Ilcisin, Kirsten Lacombe, Thomas R. Sibley, Melissa Truong, Caitlin R. Wolf, Romesh Gautom, Geoff Melly, Brian Hiatt, Philip Dykema, Scott Lindquist, Michael Boeckh, Janet A. Englund, Michael Famulare, Barry R. Lutz, Mark J. Rieder, Lea M. Starita, Matthew Thompson, Helen Y. Chu, Jay Shendure, Trevor Bedford |
| EPI_ISL_1711473 | FidaLab | Seattle Flu Study | Deborah A. Nickerson, Chris D. Frazar, Jover Lee, Benjamin Pelle, Erica Ryke, Matthew Richardson, Amanda Adler, Elisabeth Brandstetter, Peter D. Han, Kairsten Fay, Misja Ilcisin, Kirsten Lacombe, Thomas R. Sibley, Melissa Truong, Caitlin R. Wolf, Romesh Gautom, Geoff Melly, Brian Hiatt, Philip Dykema, Scott Lindquist, Michael Boeckh, Janet A. Englund, Michael Famulare, Barry R. Lutz, Mark J. Rieder, Lea M. Starita, Matthew Thompson, Helen Y. Chu, Jay Shendure, Trevor Bedford |
| EPI_ISL_1711474 | Atlas Genomics | Seattle Flu Study | Deborah A. Nickerson, Chris D. Frazar, Jover Lee, Benjamin Pelle, Erica Ryke, Matthew Richardson, Amanda Adler, Elisabeth Brandstetter, Peter D. Han, Kairsten Fay, Misja Ilcisin, Kirsten Lacombe, Thomas R. Sibley, Melissa Truong, Caitlin R. Wolf, Romesh Gautom, Geoff Melly, Brian Hiatt, Philip Dykema, Scott Lindquist, Michael Boeckh, Janet A. Englund, Michael Famulare, Barry R. Lutz, Mark J. Rieder, Lea M. Starita, Matthew Thompson, Helen Y. Chu, Jay Shendure, Trevor Bedford |
| EPI_ISL_1711475 | Northwest Laboratories | Seattle Flu Study | Deborah A. Nickerson, Chris D. Frazar, Jover Lee, Benjamin Pelle, Erica Ryke, Matthew Richardson, Amanda Adler, Elisabeth Brandstetter, Peter D. Han, Kairsten Fay, Misja Ilcisin, Kirsten Lacombe, Thomas R. Sibley, Melissa Truong, Caitlin R. Wolf, Romesh Gautom, Geoff Melly, Brian Hiatt, Philip Dykema, Scott Lindquist, Michael Boeckh, Janet A. Englund, Michael Famulare, Barry R. Lutz, Mark J. Rieder, Lea M. Starita, Matthew Thompson, Helen Y. Chu, Jay Shendure, Trevor Bedford |
| EPI_ISL_1711476 | Atlas Genomics | Seattle Flu Study | Deborah A. Nickerson, Chris D. Frazar, Jover Lee, Benjamin Pelle, Erica Ryke, Matthew Richardson, Amanda Adler, Elisabeth Brandstetter, Peter D. Han, Kairsten Fay, Misja Ilcisin, Kirsten Lacombe, Thomas R. Sibley, Melissa Truong, Caitlin R. Wolf, Romesh Gautom, Geoff Melly, Brian Hiatt, Philip Dykema, Scott Lindquist, Michael Boeckh, Janet A. Englund, Michael Famulare, Barry R. Lutz, Mark J. Rieder, Lea M. Starita, Matthew Thompson, Helen Y. Chu, Jay Shendure, Trevor Bedford |
| EPI_ISL_1711477 | Seattle Flu Study | Seattle Flu Study | Deborah A. Nickerson, Chris D. Frazar, Jover Lee, Benjamin Pelle, Erica Ryke, Matthew Richardson, Amanda Adler, Elisabeth Brandstetter, Peter D. Han, Kairsten Fay, Misja Ilcisin, Kirsten Lacombe, Thomas R. Sibley, Melissa Truong, Caitlin R. Wolf, Karen Cowgill, Stephanie Schrag, Jeff Duchin, Michael Boeckh, Janet A. Englund, Michael Famulare, Barry R. Lutz, Mark J. Rieder, Lea M. Starita, Matthew Thompson, Helen Y. Chu, Trevor Bedford, Jay Shendure |
| EPI_ISL_1711478, EPI_ISL_1711479 | Northwest Laboratories | Seattle Flu Study | Deborah A. Nickerson, Chris D. Frazar, Jover Lee, Benjamin Pelle, Erica Ryke, Matthew Richardson, Amanda Adler, Elisabeth Brandstetter, Peter D. Han, Kairsten Fay, Misja Ilcisin, Kirsten Lacombe, Thomas R. Sibley, Melissa Truong, Caitlin R. Wolf, Romesh Gautom, Geoff Melly, Brian Hiatt, Philip Dykema, Scott Lindquist, Michael Boeckh, Janet A. Englund, Michael Famulare, Barry R. Lutz, Mark J. Rieder, Lea M. Starita, Matthew Thompson, Helen Y. Chu, Jay Shendure, Trevor Bedford |
| EPI_ISL_1711480 | FidaLab | Seattle Flu Study | Deborah A. Nickerson, Chris D. Frazar, Jover Lee, Benjamin Pelle, Erica Ryke, Matthew Richardson, Amanda Adler, Elisabeth Brandstetter, Peter D. Han, Kairsten Fay, Misja Ilcisin, Kirsten Lacombe, Thomas R. Sibley, Melissa Truong, Caitlin R. Wolf, Romesh Gautom, Geoff Melly, Brian Hiatt, Philip Dykema, Scott Lindquist, Michael Boeckh, Janet A. Englund, Michael Famulare, Barry R. Lutz, Mark J. Rieder, Lea M. Starita, Matthew Thompson, Helen Y. Chu, Jay Shendure, Trevor Bedford |
| EPI_ISL_1711481 | Atlas Genomics | Seattle Flu Study | Deborah A. Nickerson, Chris D. Frazar, Jover Lee, Benjamin Pelle, Erica Ryke, Matthew Richardson, Amanda Adler, Elisabeth Brandstetter, Peter D. Han, Kairsten Fay, Misja Ilcisin, Kirsten Lacombe, Thomas R. Sibley, Melissa Truong, Caitlin R. Wolf, Romesh Gautom, Geoff Melly, Brian Hiatt, Philip Dykema, Scott Lindquist, Michael Boeckh, Janet A. Englund, Michael Famulare, Barry R. Lutz, Mark J. Rieder, Lea M. Starita, Matthew Thompson, Helen Y. Chu, Jay Shendure, Trevor Bedford |
| EPI_ISL_1711482 | Seattle Flu Study | Seattle Flu Study | Deborah A. Nickerson, Chris D. Frazar, Jover Lee, Benjamin Pelle, Erica Ryke, Matthew Richardson, Amanda Adler, Elisabeth Brandstetter, Peter D. Han, Kairsten Fay, Misja Ilcisin, Kirsten Lacombe, Thomas R. Sibley, Melissa Truong, Caitlin R. Wolf, Michael Boeckh, Janet A. Englund, Michael Famulare, Barry R. Lutz, Mark J. Rieder, Lea M. Starita, Matthew Thompson, Jay Shendure, Trevor Bedford, Helen Y. Chu |
| EPI_ISL_1711483, EPI_ISL_1711484 | Seattle Flu Study | Seattle Flu Study | Deborah A. Nickerson, Chris D. Frazar, Jover Lee, Benjamin Pelle, Erica Ryke, Matthew Richardson, Amanda Adler, Elisabeth Brandstetter, Peter D. Han, Kairsten Fay, Misja Ilcisin, Kirsten Lacombe, Thomas R. Sibley, Melissa Truong, Caitlin R. Wolf, Karen Cowgill, Stephanie Schrag, Jeff Duchin, Michael Boeckh, Janet A. Englund, Michael Famulare, Barry R. Lutz, Mark J. Rieder, Lea M. Starita, Matthew Thompson, Helen Y. Chu, Trevor Bedford, Jay Shendure |
| EPI_ISL_1711485 | Northwest Laboratories | Seattle Flu Study | Deborah A. Nickerson, Chris D. Frazar, Jover Lee, Benjamin Pelle, Erica Ryke, Matthew Richardson, Amanda Adler, Elisabeth Brandstetter, Peter D. Han, Kairsten Fay, Misja Ilcisin, Kirsten Lacombe, Thomas R. Sibley, Melissa Truong, Caitlin R. Wolf, Romesh Gautom, Geoff Melly, Brian Hiatt, Philip Dykema, Scott Lindquist, Michael Boeckh, Janet A. Englund, Michael Famulare, Barry R. Lutz, Mark J. Rieder, Lea M. Starita, Matthew Thompson, Helen Y. Chu, Jay Shendure, Trevor Bedford |
| EPI_ISL_1711486 | Seattle Flu Study | Seattle Flu Study | Deborah A. Nickerson, Chris D. Frazar, Jover Lee, Benjamin Pelle, Erica Ryke, Matthew Richardson, Amanda Adler, Elisabeth Brandstetter, Peter D. Han, Kairsten Fay, Misja Ilcisin, Kirsten Lacombe, Thomas R. Sibley, Melissa Truong, Caitlin R. Wolf, Karen Cowgill, Stephanie Schrag, Jeff Duchin, Michael Boeckh, Janet A. Englund, Michael Famulare, Barry R. Lutz, Mark J. Rieder, Lea M. Starita, Matthew Thompson, Helen Y. Chu, Trevor Bedford, Jay Shendure |
| EPI_ISL_1711487 | Seattle Flu Study | Seattle Flu Study | Deborah A. Nickerson, Chris D. Frazar, Jover Lee, Benjamin Pelle, Erica Ryke, Matthew Richardson, Amanda Adler, Elisabeth Brandstetter, Peter D. Han, Kairsten Fay, Misja Ilcisin, Kirsten Lacombe, Thomas R. Sibley, Melissa Truong, Caitlin R. Wolf, Michael Boeckh, Janet A. Englund, Michael Famulare, Barry R. Lutz, Mark J. Rieder, Lea M. Starita, Matthew Thompson, Jay Shendure, Trevor Bedford, Helen Y. Chu |
| EPI_ISL_1711488 | YAKAMA INDIAN HEALTH SERVICE | Seattle Flu Study | Deborah A. Nickerson, Chris D. Frazar, Jover Lee, Benjamin Pelle, Erica Ryke, Matthew Richardson, Amanda Adler, Elisabeth Brandstetter, Peter D. Han, Kairsten Fay, Misja Ilcisin, Kirsten Lacombe, Thomas R. Sibley, Melissa Truong, Caitlin R. Wolf, Romesh Gautom, Geoff Melly, Brian Hiatt, Philip Dykema, Scott Lindquist, Michael Boeckh, Janet A. Englund, Michael Famulare, Barry R. Lutz, Mark J. Rieder, Lea M. Starita, Matthew Thompson, Helen Y. Chu, Jay Shendure, Trevor Bedford |
| EPI_ISL_1711489 | Atlas Genomics | Seattle Flu Study | Deborah A. Nickerson, Chris D. Frazar, Jover Lee, Benjamin Pelle, Erica Ryke, Matthew Richardson, Amanda Adler, Elisabeth Brandstetter, Peter D. Han, Kairsten Fay, Misja Ilcisin, Kirsten Lacombe, Thomas R. Sibley, Melissa Truong, Caitlin R. Wolf, Romesh Gautom, Geoff Melly, Brian Hiatt, Philip Dykema, Scott Lindquist, Michael Boeckh, Janet A. Englund, Michael Famulare, Barry R. Lutz, Mark J. Rieder, Lea M. Starita, Matthew Thompson, Helen Y. Chu, Jay Shendure, Trevor Bedford |
| EPI_ISL_1711490 | Interpath Laboratory | Seattle Flu Study | Deborah A. Nickerson, Chris D. Frazar, Jover Lee, Benjamin Pelle, Erica Ryke, Matthew Richardson, Amanda Adler, Elisabeth Brandstetter, Peter D. Han, Kairsten Fay, Misja Ilcisin, Kirsten Lacombe, Thomas R. Sibley, Melissa Truong, Caitlin R. Wolf, Romesh Gautom, Geoff Melly, Brian Hiatt, Philip Dykema, Scott Lindquist, Michael Boeckh, Janet A. Englund, Michael Famulare, Barry R. Lutz, Mark J. Rieder, Lea M. Starita, Matthew Thompson, Helen Y. Chu, Jay Shendure, Trevor Bedford |

| | | | |
|---|---|---|---|
| EPI_ISL_1711491 | Seattle Flu Study | Seattle Flu Study | Deborah A. Nickerson, Chris D. Frazar, Jover Lee, Benjamin Pelle, Erica Ryke, Matthew Richardson, Amanda Adler, Elisabeth Brandstetter, Peter D. Han, Kairsten Fay, Misja Ilcisin, Kirsten Lacombe, Thomas R. Sibley, Melissa Truong, Caitlin R. Wolf, Karen Cowgill, Stephanie Schrag, Jeff Duchin, Michael Boeckh, Janet A. Englund, Michael Famulare, Barry R. Lutz, Mark J. Rieder, Lea M. Starita, Matthew Thompson, Helen Y. Chu, Trevor Bedford, Jay Shendure |
| EPI_ISL_1711492 | Atlas Genomics | Seattle Flu Study | Deborah A. Nickerson, Chris D. Frazar, Jover Lee, Benjamin Pelle, Erica Ryke, Matthew Richardson, Amanda Adler, Elisabeth Brandstetter, Peter D. Han, Kairsten Fay, Misja Ilcisin, Kirsten Lacombe, Thomas R. Sibley, Melissa Truong, Caitlin R. Wolf, Romesh Gautom, Geoff Melly, Brian Hiatt, Philip Dykema, Scott Lindquist, Michael Boeckh, Janet A. Englund, Michael Famulare, Barry R. Lutz, Mark J. Rieder, Lea M. Starita, Matthew Thompson, Helen Y. Chu, Jay Shendure, Trevor Bedford |
| EPI_ISL_1711493 | Interpath Laboratory | Seattle Flu Study | Deborah A. Nickerson, Chris D. Frazar, Jover Lee, Benjamin Pelle, Erica Ryke, Matthew Richardson, Amanda Adler, Elisabeth Brandstetter, Peter D. Han, Kairsten Fay, Misja Ilcisin, Kirsten Lacombe, Thomas R. Sibley, Melissa Truong, Caitlin R. Wolf, Romesh Gautom, Geoff Melly, Brian Hiatt, Philip Dykema, Scott Lindquist, Michael Boeckh, Janet A. Englund, Michael Famulare, Barry R. Lutz, Mark J. Rieder, Lea M. Starita, Matthew Thompson, Helen Y. Chu, Jay Shendure, Trevor Bedford |
| EPI_ISL_1711494, EPI_ISL_1711495 | FidaLab | Seattle Flu Study | Deborah A. Nickerson, Chris D. Frazar, Jover Lee, Benjamin Pelle, Erica Ryke, Matthew Richardson, Amanda Adler, Elisabeth Brandstetter, Peter D. Han, Kairsten Fay, Misja Ilcisin, Kirsten Lacombe, Thomas R. Sibley, Melissa Truong, Caitlin R. Wolf, Romesh Gautom, Geoff Melly, Brian Hiatt, Philip Dykema, Scott Lindquist, Michael Boeckh, Janet A. Englund, Michael Famulare, Barry R. Lutz, Mark J. Rieder, Lea M. Starita, Matthew Thompson, Helen Y. Chu, Jay Shendure, Trevor Bedford |
| EPI_ISL_1711496 | Atlas Genomics | Seattle Flu Study | Deborah A. Nickerson, Chris D. Frazar, Jover Lee, Benjamin Pelle, Erica Ryke, Matthew Richardson, Amanda Adler, Elisabeth Brandstetter, Peter D. Han, Kairsten Fay, Misja Ilcisin, Kirsten Lacombe, Thomas R. Sibley, Melissa Truong, Caitlin R. Wolf, Romesh Gautom, Geoff Melly, Brian Hiatt, Philip Dykema, Scott Lindquist, Michael Boeckh, Janet A. Englund, Michael Famulare, Barry R. Lutz, Mark J. Rieder, Lea M. Starita, Matthew Thompson, Helen Y. Chu, Jay Shendure, Trevor Bedford |
| EPI_ISL_1711497 | Northwest Laboratories | Seattle Flu Study | Deborah A. Nickerson, Chris D. Frazar, Jover Lee, Benjamin Pelle, Erica Ryke, Matthew Richardson, Amanda Adler, Elisabeth Brandstetter, Peter D. Han, Kairsten Fay, Misja Ilcisin, Kirsten Lacombe, Thomas R. Sibley, Melissa Truong, Caitlin R. Wolf, Romesh Gautom, Geoff Melly, Brian Hiatt, Philip Dykema, Scott Lindquist, Michael Boeckh, Janet A. Englund, Michael Famulare, Barry R. Lutz, Mark J. Rieder, Lea M. Starita, Matthew Thompson, Helen Y. Chu, Jay Shendure, Trevor Bedford |
| EPI_ISL_1711498 | Seattle Flu Study | Seattle Flu Study | Deborah A. Nickerson, Chris D. Frazar, Jover Lee, Benjamin Pelle, Erica Ryke, Matthew Richardson, Amanda Adler, Elisabeth Brandstetter, Peter D. Han, Kairsten Fay, Misja Ilcisin, Kirsten Lacombe, Thomas R. Sibley, Melissa Truong, Caitlin R. Wolf, Michael Boeckh, Janet A. Englund, Michael Famulare, Barry R. Lutz, Mark J. Rieder, Lea M. Starita, Matthew Thompson, Jay Shendure, Trevor Bedford, Helen Y. Chu |
| EPI_ISL_1711499 | Atlas Genomics | Seattle Flu Study | Deborah A. Nickerson, Chris D. Frazar, Jover Lee, Benjamin Pelle, Erica Ryke, Matthew Richardson, Amanda Adler, Elisabeth Brandstetter, Peter D. Han, Kairsten Fay, Misja Ilcisin, Kirsten Lacombe, Thomas R. Sibley, Melissa Truong, Caitlin R. Wolf, Romesh Gautom, Geoff Melly, Brian Hiatt, Philip Dykema, Scott Lindquist, Michael Boeckh, Janet A. Englund, Michael Famulare, Barry R. Lutz, Mark J. Rieder, Lea M. Starita, Matthew Thompson, Helen Y. Chu, Jay Shendure, Trevor Bedford |
| EPI_ISL_1711500 | Seattle Flu Study | Seattle Flu Study | Deborah A. Nickerson, Chris D. Frazar, Jover Lee, Benjamin Pelle, Erica Ryke, Matthew Richardson, Amanda Adler, Elisabeth Brandstetter, Peter D. Han, Kairsten Fay, Misja Ilcisin, Kirsten Lacombe, Thomas R. Sibley, Melissa Truong, Caitlin R. Wolf, Karen Cowgill, Stephanie Schrag, Jeff Duchin, Michael Boeckh, Janet A. Englund, Michael Famulare, Barry R. Lutz, Mark J. Rieder, Lea M. Starita, Matthew Thompson, Helen Y. Chu, Trevor Bedford, Jay Shendure |
| EPI_ISL_1711501 | Seattle Flu Study | Seattle Flu Study | Deborah A. Nickerson, Chris D. Frazar, Jover Lee, Benjamin Pelle, Erica Ryke, Matthew Richardson, Amanda Adler, Elisabeth Brandstetter, Peter D. Han, Kairsten Fay, Misja Ilcisin, Kirsten Lacombe, Thomas R. Sibley, Melissa Truong, Caitlin R. Wolf, Michael Boeckh, Janet A. Englund, Michael Famulare, Barry R. Lutz, Mark J. Rieder, Lea M. Starita, Matthew Thompson, Jay Shendure, Trevor Bedford, Helen Y. Chu |
| EPI_ISL_1711502 | Atlas Genomics | Seattle Flu Study | Deborah A. Nickerson, Chris D. Frazar, Jover Lee, Benjamin Pelle, Erica Ryke, Matthew Richardson, Amanda Adler, Elisabeth Brandstetter, Peter D. Han, Kairsten Fay, Misja Ilcisin, Kirsten Lacombe, Thomas R. Sibley, Melissa Truong, Caitlin R. Wolf, Romesh Gautom, Geoff Melly, Brian Hiatt, Philip Dykema, Scott Lindquist, Michael Boeckh, Janet A. Englund, Michael Famulare, Barry R. Lutz, Mark J. Rieder, Lea M. Starita, Matthew Thompson, Helen Y. Chu, Jay Shendure, Trevor Bedford |
| EPI_ISL_1711503 | Seattle Flu Study | Seattle Flu Study | Deborah A. Nickerson, Chris D. Frazar, Jover Lee, Benjamin Pelle, Erica Ryke, Matthew Richardson, Amanda Adler, Elisabeth Brandstetter, Peter D. Han, Kairsten Fay, Misja Ilcisin, Kirsten Lacombe, Thomas R. Sibley, Melissa Truong, Caitlin R. Wolf, Karen Cowgill, Stephanie Schrag, Jeff Duchin, Michael Boeckh, Janet A. Englund, Michael Famulare, Barry R. Lutz, Mark J. Rieder, Lea M. Starita, Matthew Thompson, Helen Y. Chu, Trevor Bedford, Jay Shendure |
| EPI_ISL_1711504 | Atlas Genomics | Seattle Flu Study | Deborah A. Nickerson, Chris D. Frazar, Jover Lee, Benjamin Pelle, Erica Ryke, Matthew Richardson, Amanda Adler, Elisabeth Brandstetter, Peter D. Han, Kairsten Fay, Misja Ilcisin, Kirsten Lacombe, Thomas R. Sibley, Melissa Truong, Caitlin R. Wolf, Romesh Gautom, Geoff Melly, Brian Hiatt, Philip Dykema, Scott Lindquist, Michael Boeckh, Janet A. Englund, Michael Famulare, Barry R. Lutz, Mark J. Rieder, Lea M. Starita, Matthew Thompson, Helen Y. Chu, Jay Shendure, Trevor Bedford |
| EPI_ISL_1711505 | Seattle Flu Study | Seattle Flu Study | Deborah A. Nickerson, Chris D. Frazar, Jover Lee, Benjamin Pelle, Erica Ryke, Matthew Richardson, Amanda Adler, Elisabeth Brandstetter, Peter D. Han, Kairsten Fay, Misja Ilcisin, Kirsten Lacombe, Thomas R. Sibley, Melissa Truong, Caitlin R. Wolf, Karen Cowgill, Stephanie Schrag, Jeff Duchin, Michael Boeckh, Janet A. Englund, Michael Famulare, Barry R. Lutz, Mark J. Rieder, Lea M. Starita, Matthew Thompson, Helen Y. Chu, Trevor Bedford, Jay Shendure |
| EPI_ISL_1711506 | Seattle Flu Study | Seattle Flu Study | Deborah A. Nickerson, Chris D. Frazar, Jover Lee, Benjamin Pelle, Erica Ryke, Matthew Richardson, Amanda Adler, Elisabeth Brandstetter, Peter D. Han, Kairsten Fay, Misja Ilcisin, Kirsten Lacombe, Thomas R. Sibley, Melissa Truong, Caitlin R. Wolf, Michael Boeckh, Janet A. Englund, Michael Famulare, Barry R. Lutz, Mark J. Rieder, Lea M. Starita, Matthew Thompson, Jay Shendure, Trevor Bedford, Helen Y. Chu |
| EPI_ISL_1711507 | Evergreen Healthcare | Seattle Flu Study | Deborah A. Nickerson, Chris D. Frazar, Jover Lee, Benjamin Pelle, Erica Ryke, Matthew Richardson, Amanda Adler, Elisabeth Brandstetter, Peter D. Han, Kairsten Fay, Misja Ilcisin, Kirsten Lacombe, Thomas R. Sibley, Melissa Truong, Caitlin R. Wolf, Romesh Gautom, Geoff Melly, Brian Hiatt, Philip Dykema, Scott Lindquist, Michael Boeckh, Janet A. Englund, Michael Famulare, Barry R. Lutz, Mark J. Rieder, Lea M. Starita, Matthew Thompson, Helen Y. Chu, Jay Shendure, Trevor Bedford |
| EPI_ISL_1711508 | Seattle Flu Study | Seattle Flu Study | Deborah A. Nickerson, Chris D. Frazar, Jover Lee, Benjamin Pelle, Erica Ryke, Matthew Richardson, Amanda Adler, Elisabeth Brandstetter, Peter D. Han, Kairsten Fay, Misja Ilcisin, Kirsten Lacombe, Thomas R. Sibley, Melissa Truong, Caitlin R. Wolf, Michael Boeckh, Janet A. Englund, Michael Famulare, Barry R. Lutz, Mark J. Rieder, Lea M. Starita, Matthew Thompson, Jay Shendure, Trevor Bedford, Helen Y. Chu |
| EPI_ISL_1711509, EPI_ISL_1711510, EPI_ISL_1711511 | Atlas Genomics | Seattle Flu Study | Deborah A. Nickerson, Chris D. Frazar, Jover Lee, Benjamin Pelle, Erica Ryke, Matthew Richardson, Amanda Adler, Elisabeth Brandstetter, Peter D. Han, Kairsten Fay, Misja Ilcisin, Kirsten Lacombe, Thomas R. Sibley, Melissa Truong, Caitlin R. Wolf, Romesh Gautom, Geoff Melly, Brian Hiatt, Philip Dykema, Scott Lindquist, Michael Boeckh, Janet A. Englund, Michael Famulare, Barry R. Lutz, Mark J. Rieder, Lea M. Starita, Matthew Thompson, Helen Y. Chu, Jay Shendure, Trevor Bedford |
| EPI_ISL_1711512 | Seattle Flu Study | Seattle Flu Study | Deborah A. Nickerson, Chris D. Frazar, Jover Lee, Benjamin Pelle, Erica Ryke, Matthew Richardson, Amanda Adler, Elisabeth Brandstetter, Peter D. Han, Kairsten Fay, Misja Ilcisin, Kirsten Lacombe, Thomas R. Sibley, Melissa Truong, Caitlin R. Wolf, Karen Cowgill, Stephanie Schrag, Jeff Duchin, Michael Boeckh, Janet A. Englund, Michael Famulare, Barry R. Lutz, Mark J. Rieder, Lea M. Starita, Matthew Thompson, Helen Y. Chu, Trevor Bedford, Jay Shendure |
| EPI_ISL_1711513 | Seattle Flu Study | Seattle Flu Study | Deborah A. Nickerson, Chris D. Frazar, Jover Lee, Benjamin Pelle, Erica Ryke, Matthew Richardson, Amanda Adler, Elisabeth Brandstetter, Peter D. Han, Kairsten Fay, Misja Ilcisin, Kirsten Lacombe, Thomas R. Sibley, Melissa Truong, Caitlin R. Wolf, Michael Boeckh, Janet A. Englund, Michael Famulare, Barry R. Lutz, Mark J. Rieder, Lea M. Starita, Matthew Thompson, Jay Shendure, Trevor Bedford, Helen Y. Chu |
| EPI_ISL_1711514, EPI_ISL_1711515 | Evergreen Healthcare | Seattle Flu Study | Deborah A. Nickerson, Chris D. Frazar, Jover Lee, Benjamin Pelle, Erica Ryke, Matthew Richardson, Amanda Adler, Elisabeth Brandstetter, Peter D. Han, Kairsten Fay, Misja Ilcisin, Kirsten Lacombe, Thomas R. Sibley, Melissa Truong, Caitlin R. Wolf, Romesh Gautom, Geoff Melly, Brian Hiatt, Philip Dykema, Scott Lindquist, Michael Boeckh, Janet A. Englund, Michael Famulare, Barry R. Lutz, Mark J. Rieder, Lea M. Starita, Matthew Thompson, Helen Y. Chu, Jay Shendure, Trevor Bedford |
| EPI_ISL_1711516 | Atlas Genomics | Seattle Flu Study | Deborah A. Nickerson, Chris D. Frazar, Jover Lee, Benjamin Pelle, Erica Ryke, Matthew Richardson, Amanda Adler, Elisabeth Brandstetter, Peter D. Han, Kairsten Fay, Misja Ilcisin, Kirsten Lacombe, Thomas R. Sibley, Melissa Truong, Caitlin R. Wolf, Romesh Gautom, Geoff Melly, Brian Hiatt, Philip Dykema, Scott Lindquist, Michael Boeckh, Janet A. Englund, Michael Famulare, Barry R. Lutz, Mark J. Rieder, Lea M. Starita, Matthew Thompson, Helen Y. Chu, Jay Shendure, Trevor Bedford |
| EPI_ISL_1711517, EPI_ISL_1711518, EPI_ISL_1711519 | Seattle Flu Study | Seattle Flu Study | Deborah A. Nickerson, Chris D. Frazar, Jover Lee, Benjamin Pelle, Erica Ryke, Matthew Richardson, Amanda Adler, Elisabeth Brandstetter, Peter D. Han, Kairsten Fay, Misja Ilcisin, Kirsten Lacombe, Thomas R. Sibley, Melissa Truong, Caitlin R. Wolf, Karen Cowgill, Stephanie Schrag, Jeff Duchin, Michael Boeckh, Janet A. Englund, Michael Famulare, Barry R. Lutz, Mark J. Rieder, Lea M. Starita, Matthew Thompson, Helen Y. Chu, Trevor Bedford, Jay Shendure |
| EPI_ISL_1711520 | Northwest Laboratories | Seattle Flu Study | Deborah A. Nickerson, Chris D. Frazar, Jover Lee, Benjamin Pelle, Erica Ryke, Matthew Richardson, Amanda Adler, Elisabeth Brandstetter, Peter D. Han, Kairsten Fay, Misja Ilcisin, Kirsten Lacombe, Thomas R. Sibley, Melissa Truong, Caitlin R. Wolf, Romesh Gautom, Geoff Melly, Brian Hiatt, Philip Dykema, Scott Lindquist, Michael Boeckh, Janet A. Englund, Michael Famulare, Barry R. Lutz, Mark J. Rieder, Lea M. Starita, Matthew Thompson, Helen Y. Chu, Jay Shendure, Trevor Bedford |
| EPI_ISL_1711521 | Atlas Genomics | Seattle Flu Study | Deborah A. Nickerson, Chris D. Frazar, Jover Lee, Benjamin Pelle, Erica Ryke, Matthew Richardson, Amanda Adler, Elisabeth Brandstetter, Peter D. Han, Kairsten Fay, Misja Ilcisin, Kirsten Lacombe, Thomas R. Sibley, Melissa Truong, Caitlin R. Wolf, Romesh Gautom, Geoff Melly, Brian Hiatt, Philip Dykema, Scott Lindquist, Michael Boeckh, Janet A. Englund, Michael Famulare, Barry R. Lutz, Mark J. Rieder, Lea M. Starita, Matthew Thompson, Helen Y. Chu, Jay Shendure, Trevor Bedford |
| EPI_ISL_1711522 | Seattle Flu Study | Seattle Flu Study | Deborah A. Nickerson, Chris D. Frazar, Jover Lee, Benjamin Pelle, Erica Ryke, Matthew Richardson, Amanda Adler, Elisabeth Brandstetter, Peter D. Han, Kairsten Fay, Misja Ilcisin, Kirsten Lacombe, Thomas R. Sibley, Melissa Truong, Caitlin R. Wolf, Michael Boeckh, Janet A. Englund, Michael Famulare, Barry R. Lutz, Mark J. Rieder, Lea M. Starita, Matthew Thompson, Jay Shendure, Trevor Bedford, Helen Y. Chu |
| EPI_ISL_1711523 | Seattle Flu Study | Seattle Flu Study | Deborah A. Nickerson, Chris D. Frazar, Jover Lee, Benjamin Pelle, Erica Ryke, Matthew Richardson, Amanda Adler, Elisabeth Brandstetter, Peter D. Han, Kairsten Fay, Misja Ilcisin, Kirsten Lacombe, Thomas R. Sibley, Melissa Truong, Caitlin R. Wolf, Karen Cowgill, Stephanie Schrag, Jeff Duchin, Michael Boeckh, Janet A. Englund, Michael Famulare, Barry R. Lutz, Mark J. Rieder, Lea M. Starita, Matthew Thompson, Helen Y. Chu, Trevor Bedford, Jay Shendure |
| EPI_ISL_1711524, EPI_ISL_1711525 | Northwest Laboratories | Seattle Flu Study | Deborah A. Nickerson, Chris D. Frazar, Jover Lee, Benjamin Pelle, Erica Ryke, Matthew Richardson, Amanda Adler, Elisabeth Brandstetter, Peter D. Han, Kairsten Fay, Misja Ilcisin, Kirsten Lacombe, Thomas R. Sibley, Melissa Truong, Caitlin R. Wolf, Romesh Gautom, Geoff Melly, Brian Hiatt, Philip Dykema, Scott Lindquist, Michael Boeckh, Janet A. Englund, Michael Famulare, Barry R. Lutz, Mark J. Rieder, Lea M. Starita, Matthew Thompson, Helen Y. Chu, Jay Shendure, Trevor Bedford |
| EPI_ISL_1711526, EPI_ISL_1711527 | Evergreen Healthcare | Seattle Flu Study | Deborah A. Nickerson, Chris D. Frazar, Jover Lee, Benjamin Pelle, Erica Ryke, Matthew Richardson, Amanda Adler, Elisabeth Brandstetter, Peter D. Han, Kairsten Fay, Misja Ilcisin, Kirsten Lacombe, Thomas R. Sibley, Melissa Truong, Caitlin R. Wolf, Romesh Gautom, Geoff Melly, Brian Hiatt, Philip Dykema, Scott Lindquist, Michael Boeckh, Janet A. Englund, Michael Famulare, Barry R. Lutz, Mark J. Rieder, Lea M. Starita, Matthew Thompson, Helen Y. Chu, Jay Shendure, Trevor Bedford |
| EPI_ISL_1711528, EPI_ISL_1711529, EPI_ISL_1711530, EPI_ISL_1711531, EPI_ISL_1711623, EPI_ISL_1711624, EPI_ISL_1711625 | Atlas Genomics | Seattle Flu Study | Deborah A. Nickerson, Chris D. Frazar, Jover Lee, Benjamin Pelle, Erica Ryke, Matthew Richardson, Amanda Adler, Elisabeth Brandstetter, Peter D. Han, Kairsten Fay, Misja Ilcisin, Kirsten Lacombe, Thomas R. Sibley, Melissa Truong, Caitlin R. Wolf, Romesh Gautom, Geoff Melly, Brian Hiatt, Philip Dykema, Scott Lindquist, Michael Boeckh, Janet A. Englund, Michael Famulare, Barry R. Lutz, Mark J. Rieder, Lea M. Starita, Matthew Thompson, Helen Y. Chu, Jay Shendure, Trevor Bedford |
| EPI_ISL_1711626, EPI_ISL_1711627 | Seattle Flu Study | Seattle Flu Study | Deborah A. Nickerson, Chris D. Frazar, Jover Lee, Benjamin Pelle, Erica Ryke, Matthew Richardson, Amanda Adler, Elisabeth Brandstetter, Peter D. Han, Kairsten Fay, Misja Ilcisin, Kirsten Lacombe, Thomas R. Sibley, Melissa Truong, Caitlin R. Wolf, Michael Boeckh, Janet A. Englund, Michael Famulare, Barry R. Lutz, Mark J. Rieder, Lea M. Starita, Matthew Thompson, Jay Shendure, Trevor Bedford, Helen Y. Chu |
| EPI_ISL_1711628 | Seattle Flu Study | Seattle Flu Study | Deborah A. Nickerson, Chris D. Frazar, Jover Lee, Benjamin Pelle, Erica Ryke, Matthew Richardson, Amanda Adler, Elisabeth Brandstetter, Peter D. Han, Kairsten Fay, Misja Ilcisin, Kirsten Lacombe, Thomas R. Sibley, Melissa Truong, Caitlin R. Wolf, Karen Cowgill, Stephanie Schrag, Jeff Duchin, Michael Boeckh, Janet A. Englund, Michael Famulare, Barry R. Lutz, Mark J. Rieder, Lea M. Starita, Matthew Thompson, Helen Y. Chu, Trevor Bedford, Jay Shendure |
| EPI_ISL_1711629, EPI_ISL_1711630, EPI_ISL_1711631, EPI_ISL_1711632, EPI_ISL_1711633, EPI_ISL_1711634 | Atlas Genomics | Seattle Flu Study | Deborah A. Nickerson, Chris D. Frazar, Jover Lee, Benjamin Pelle, Erica Ryke, Matthew Richardson, Amanda Adler, Elisabeth Brandstetter, Peter D. Han, Kairsten Fay, Misja Ilcisin, Kirsten Lacombe, Thomas R. Sibley, Melissa Truong, Caitlin R. Wolf, Romesh Gautom, Geoff Melly, Brian Hiatt, Philip Dykema, Scott Lindquist, Michael Boeckh, Janet A. Englund, Michael Famulare, Barry R. Lutz, Mark J. Rieder, Lea M. Starita, Matthew Thompson, Helen Y. Chu, Jay Shendure, Trevor Bedford |
| EPI_ISL_1711636, EPI_ISL_1711637, EPI_ISL_1711638, EPI_ISL_1711639 | Nevada State Public Health Laboratory | Nevada State Public Health Laboratory | Andrew Gorzalski, Mark Pandori |
| EPI_ISL_1711676, EPI_ISL_1711677 | MO State Public Health Laboratory | Centers for Disease Control and Prevention Division of Viral Diseases, Pathogen Discovery | Mili Sheth, Sarah Nobles, Jasmine Padilla, Mark Burroughs, Shoshona Le, Katie Dillon, Peter Cook, Clinton R. Paden, Dhwani Batra, Krista Queen, Kristen Knipe, Dakota Howard, Yvette Unoarumhi, Darlene Wagner, Matthew Schmerer, Ben L. Rambo-Martin, Kristine Lacek, Sam Shepard, Alison Laufer Halpin, Dave Wentworth, Vivien Dugan, Suxiang Tong, Justin Lee |
| EPI_ISL_1711678, EPI_ISL_1711679, EPI_ISL_1711680, EPI_ISL_1711681, EPI_ISL_1711682, EPI_ISL_1711683, EPI_ISL_1711684, EPI_ISL_1711685, EPI_ISL_1711686, EPI_ISL_1711687, EPI_ISL_1711688, EPI_ISL_1711689, EPI_ISL_1711690 | | | |
| see above | PR Public Health Lab | Centers for Disease Control and Prevention Division of Viral Diseases, Pathogen Discovery | Mili Sheth, Sarah Nobles, Jasmine Padilla, Mark Burroughs, Shoshona Le, Katie Dillon, Peter Cook, Clinton R. Paden, Dhwani Batra, Krista Queen, Kristen Knipe, Dakota Howard, Yvette Unoarumhi, Darlene Wagner, Matthew Schmerer, Ben L. Rambo-Martin, Kristine Lacek, Sam Shepard, Alison Laufer Halpin, Dave Wentworth, Vivien Dugan, Suxiang Tong, Justin Lee |
| EPI_ISL_1711691 | SD Public Health Laboratory | Centers for Disease Control and Prevention Division of Viral Diseases, Pathogen Discovery | Mili Sheth, Sarah Nobles, Jasmine Padilla, Mark Burroughs, Shoshona Le, Katie Dillon, Peter Cook, Clinton R. Paden, Dhwani Batra, Krista Queen, Kristen Knipe, Dakota Howard, Yvette Unoarumhi, Darlene Wagner, Matthew Schmerer, Ben L. Rambo-Martin, Kristine Lacek, Sam Shepard, Alison Laufer Halpin, Dave Wentworth, Vivien Dugan, Suxiang Tong, Justin Lee |
| EPI_ISL_1711692, EPI_ISL_1711693, EPI_ISL_1711694, EPI_ISL_1711695, EPI_ISL_1711696, EPI_ISL_1711697, EPI_ISL_1711698, EPI_ISL_1711699, EPI_ISL_1711700, EPI_ISL_1711701, EPI_ISL_1711702, EPI_ISL_1711703 | | | |
| see above | "AR Dept. of Health-PHL, Molecular | Centers for Disease Control and Prevention | Mili Sheth, Sarah Nobles, Jasmine Padilla, Mark Burroughs, Shoshona Le, Katie Dillon, Peter Cook, Clinton R. Paden, Dhwani Batra, Krista Queen, Kristen Knipe, Dakota Howard, Yvette Unoarumhi, Darlene Wagner, Matthew Schmerer, Ben L. Rambo-Martin, Kristine Lacek, Sam Shepard, |

| | | | |
|---|---|---|---|
| | Diagnostics" | Division of Viral Diseases, Pathogen Discovery | Alison Laufer Halpin, Dave Wentworth, Vivien Dugan, Suxiang Tong, Justin Lee |
| EPI_ISL_1711704, EPI_ISL_1711705, EPI_ISL_1711706, EPI_ISL_1711707, EPI_ISL_1711708, EPI_ISL_1711709 | AR Dept. of Health-Public Health Lab | Centers for Disease Control and Prevention Division of Viral Diseases, Pathogen Discovery | Mili Sheth, Sarah Nobles, Jasmine Padilla, Mark Burroughs, Shoshona Le, Katie Dillon, Peter Cook, Clinton R. Paden, Dhwani Batra, Krista Queen, Kristen Knipe, Dakota Howard, Yvette Unoarumhi, Darlene Wagner, Matthew Schmerer, Ben L. Rambo-Martin, Kristine Lacek, Sam Shepard, Alison Laufer Halpin, Dave Wentworth, Vivien Dugan, Suxiang Tong, Justin Lee |
| EPI_ISL_1711710, EPI_ISL_1711711, EPI_ISL_1711712, EPI_ISL_1711713, EPI_ISL_1711714, EPI_ISL_1711715, EPI_ISL_1711716 | CT-Dr. Katherine A. Kelley State Public Health Lab | Centers for Disease Control and Prevention Division of Viral Diseases, Pathogen Discovery | Mili Sheth, Sarah Nobles, Jasmine Padilla, Mark Burroughs, Shoshona Le, Katie Dillon, Peter Cook, Clinton R. Paden, Dhwani Batra, Krista Queen, Kristen Knipe, Dakota Howard, Yvette Unoarumhi, Darlene Wagner, Matthew Schmerer, Ben L. Rambo-Martin, Kristine Lacek, Sam Shepard, Alison Laufer Halpin, Dave Wentworth, Vivien Dugan, Suxiang Tong, Justin Lee |
| EPI_ISL_1711717, EPI_ISL_1711718 | "NYSDOH Wadsworth Center, Virology Lab" | Centers for Disease Control and Prevention Division of Viral Diseases, Pathogen Discovery | Mili Sheth, Sarah Nobles, Jasmine Padilla, Mark Burroughs, Shoshona Le, Katie Dillon, Peter Cook, Clinton R. Paden, Dhwani Batra, Krista Queen, Kristen Knipe, Dakota Howard, Yvette Unoarumhi, Darlene Wagner, Matthew Schmerer, Ben L. Rambo-Martin, Kristine Lacek, Sam Shepard, Alison Laufer Halpin, Dave Wentworth, Vivien Dugan, Suxiang Tong, Justin Lee |
| EPI_ISL_1711719, EPI_ISL_1711720, EPI_ISL_1711721, EPI_ISL_1711722, EPI_ISL_1711723, EPI_ISL_1711724, EPI_ISL_1711725, EPI_ISL_1711726, EPI_ISL_1711727, EPI_ISL_1711728, EPI_ISL_1711729, EPI_ISL_1711730, EPI_ISL_1711731, EPI_ISL_1711732, EPI_ISL_1711733, EPI_ISL_1711734, EPI_ISL_1711735, EPI_ISL_1711736, EPI_ISL_1711737, EPI_ISL_1711738, EPI_ISL_1711739, EPI_ISL_1711740, EPI_ISL_1711741, EPI_ISL_1711742, EPI_ISL_1711743, EPI_ISL_1711744, EPI_ISL_1711745, EPI_ISL_1711746, EPI_ISL_1711747, EPI_ISL_1711748, EPI_ISL_1711749, EPI_ISL_1711750, EPI_ISL_1711751, EPI_ISL_1711752, EPI_ISL_1711753 | | | |
| see above | "NM Dept. Health, Scientific Laboratory Division " | Centers for Disease Control and Prevention Division of Viral Diseases, Pathogen Discovery | Mili Sheth, Sarah Nobles, Jasmine Padilla, Mark Burroughs, Shoshona Le, Katie Dillon, Peter Cook, Clinton R. Paden, Dhwani Batra, Krista Queen, Kristen Knipe, Dakota Howard, Yvette Unoarumhi, Darlene Wagner, Matthew Schmerer, Ben L. Rambo-Martin, Kristine Lacek, Sam Shepard, Alison Laufer Halpin, Dave Wentworth, Vivien Dugan, Suxiang Tong, Justin Lee |
| EPI_ISL_1711754, EPI_ISL_1711755, EPI_ISL_1711756, EPI_ISL_1711757, EPI_ISL_1711758, EPI_ISL_1711759 | "CO Dept. of Public Health and Environment, Lab Services Division" | Centers for Disease Control and Prevention Division of Viral Diseases, Pathogen Discovery | Mili Sheth, Sarah Nobles, Jasmine Padilla, Mark Burroughs, Shoshona Le, Katie Dillon, Peter Cook, Clinton R. Paden, Dhwani Batra, Krista Queen, Kristen Knipe, Dakota Howard, Yvette Unoarumhi, Darlene Wagner, Matthew Schmerer, Ben L. Rambo-Martin, Kristine Lacek, Sam Shepard, Alison Laufer Halpin, Dave Wentworth, Vivien Dugan, Suxiang Tong, Justin Lee |
| EPI_ISL_1711760, EPI_ISL_1711761, EPI_ISL_1711762, EPI_ISL_1711763, EPI_ISL_1711764, EPI_ISL_1711765, EPI_ISL_1711766, EPI_ISL_1711767, EPI_ISL_1711768, EPI_ISL_1711769 | ME Health and Environmental Testing Laboratory | Centers for Disease Control and Prevention Division of Viral Diseases, Pathogen Discovery | Mili Sheth, Sarah Nobles, Jasmine Padilla, Mark Burroughs, Shoshona Le, Katie Dillon, Peter Cook, Clinton R. Paden, Dhwani Batra, Krista Queen, Kristen Knipe, Dakota Howard, Yvette Unoarumhi, Darlene Wagner, Matthew Schmerer, Ben L. Rambo-Martin, Kristine Lacek, Sam Shepard, Alison Laufer Halpin, Dave Wentworth, Vivien Dugan, Suxiang Tong, Justin Lee |
| EPI_ISL_1711770, EPI_ISL_1711771, EPI_ISL_1711772, EPI_ISL_1711773, EPI_ISL_1711774 | NV State Public Health Laboratory | Centers for Disease Control and Prevention Division of Viral Diseases, Pathogen Discovery | Mili Sheth, Sarah Nobles, Jasmine Padilla, Mark Burroughs, Shoshona Le, Katie Dillon, Peter Cook, Clinton R. Paden, Dhwani Batra, Krista Queen, Kristen Knipe, Dakota Howard, Yvette Unoarumhi, Darlene Wagner, Matthew Schmerer, Ben L. Rambo-Martin, Kristine Lacek, Sam Shepard, Alison Laufer Halpin, Dave Wentworth, Vivien Dugan, Suxiang Tong, Justin Lee |
| EPI_ISL_1711775, EPI_ISL_1711776, EPI_ISL_1711777 | OH Department of Health Laboratory | Centers for Disease Control and Prevention Division of Viral Diseases, Pathogen Discovery | Mili Sheth, Sarah Nobles, Jasmine Padilla, Mark Burroughs, Shoshona Le, Katie Dillon, Peter Cook, Clinton R. Paden, Dhwani Batra, Krista Queen, Kristen Knipe, Dakota Howard, Yvette Unoarumhi, Darlene Wagner, Matthew Schmerer, Ben L. Rambo-Martin, Kristine Lacek, Sam Shepard, Alison Laufer Halpin, Dave Wentworth, Vivien Dugan, Suxiang Tong, Justin Lee |
| EPI_ISL_1711778, EPI_ISL_1711779, EPI_ISL_1711780, EPI_ISL_1711781, EPI_ISL_1711782, EPI_ISL_1711783, EPI_ISL_1711784, EPI_ISL_1711785, EPI_ISL_1711786, EPI_ISL_1711787, EPI_ISL_1711788, EPI_ISL_1711789, EPI_ISL_1711790, EPI_ISL_1711791, EPI_ISL_1711792, EPI_ISL_1711793, EPI_ISL_1711794, EPI_ISL_1711795, EPI_ISL_1711796, EPI_ISL_1711797 | | | |
| see above | OR State PHL-Virology/Immunology Section | Centers for Disease Control and Prevention Division of Viral Diseases, Pathogen Discovery | Mili Sheth, Sarah Nobles, Jasmine Padilla, Mark Burroughs, Shoshona Le, Katie Dillon, Peter Cook, Clinton R. Paden, Dhwani Batra, Krista Queen, Kristen Knipe, Dakota Howard, Yvette Unoarumhi, Darlene Wagner, Matthew Schmerer, Ben L. Rambo-Martin, Kristine Lacek, Sam Shepard, Alison Laufer Halpin, Dave Wentworth, Vivien Dugan, Suxiang Tong, Justin Lee |
| EPI_ISL_1711798, EPI_ISL_1711799, EPI_ISL_1711800, EPI_ISL_1711801, EPI_ISL_1711802 | NJ Public Health and Environmental Laboratories | Centers for Disease Control and Prevention Division of Viral Diseases, Pathogen Discovery | Mili Sheth, Sarah Nobles, Jasmine Padilla, Mark Burroughs, Shoshona Le, Katie Dillon, Peter Cook, Clinton R. Paden, Dhwani Batra, Krista Queen, Kristen Knipe, Dakota Howard, Yvette Unoarumhi, Darlene Wagner, Matthew Schmerer, Ben L. Rambo-Martin, Kristine Lacek, Sam Shepard, Alison Laufer Halpin, Dave Wentworth, Vivien Dugan, Suxiang Tong, Justin Lee |
| EPI_ISL_1711803, EPI_ISL_1711804, EPI_ISL_1711805, EPI_ISL_1711806, EPI_ISL_1711807, EPI_ISL_1711808, EPI_ISL_1711809, EPI_ISL_1711810, EPI_ISL_1711811, EPI_ISL_1711812, EPI_ISL_1711813, EPI_ISL_1711814 | | | |
| see above | "OK Public Health Laboratory, Oklahoma State DOH" | Centers for Disease Control and Prevention Division of Viral Diseases, Pathogen Discovery | Mili Sheth, Sarah Nobles, Jasmine Padilla, Mark Burroughs, Shoshona Le, Katie Dillon, Peter Cook, Clinton R. Paden, Dhwani Batra, Krista Queen, Kristen Knipe, Dakota Howard, Yvette Unoarumhi, Darlene Wagner, Matthew Schmerer, Ben L. Rambo-Martin, Kristine Lacek, Sam Shepard, Alison Laufer Halpin, Dave Wentworth, Vivien Dugan, Suxiang Tong, Justin Lee |
| EPI_ISL_1711815, EPI_ISL_1711816, EPI_ISL_1711817, EPI_ISL_1711818, EPI_ISL_1711819 | WVDHHR - Office of Laboratory Services | Centers for Disease Control and Prevention Division of Viral Diseases, Pathogen Discovery | Mili Sheth, Sarah Nobles, Jasmine Padilla, Mark Burroughs, Shoshona Le, Katie Dillon, Peter Cook, Clinton R. Paden, Dhwani Batra, Krista Queen, Kristen Knipe, Dakota Howard, Yvette Unoarumhi, Darlene Wagner, Matthew Schmerer, Ben L. Rambo-Martin, Kristine Lacek, Sam Shepard, Alison Laufer Halpin, Dave Wentworth, Vivien Dugan, Suxiang Tong, Justin Lee |
| EPI_ISL_1711820, EPI_ISL_1711821, EPI_ISL_1711822, EPI_ISL_1711823, EPI_ISL_1711824, EPI_ISL_1711825, EPI_ISL_1711826, EPI_ISL_1711827, EPI_ISL_1711828, EPI_ISL_1711829, EPI_ISL_1711830, EPI_ISL_1711831, EPI_ISL_1711832, EPI_ISL_1711833, EPI_ISL_1711834 | | | |
| see above | MI - Michigan Department of Health and Human Services - Bureau of Laboratories | Centers for Disease Control and Prevention Division of Viral Diseases, Pathogen Discovery | Mili Sheth, Sarah Nobles, Jasmine Padilla, Mark Burroughs, Shoshona Le, Katie Dillon, Peter Cook, Clinton R. Paden, Dhwani Batra, Krista Queen, Kristen Knipe, Dakota Howard, Yvette Unoarumhi, Darlene Wagner, Matthew Schmerer, Ben L. Rambo-Martin, Kristine Lacek, Sam Shepard, Alison Laufer Halpin, Dave Wentworth, Vivien Dugan, Suxiang Tong, Justin Lee |
| EPI_ISL_1711835, EPI_ISL_1711836, EPI_ISL_1711837, EPI_ISL_1711838, EPI_ISL_1711839, EPI_ISL_1711840, EPI_ISL_1711841, EPI_ISL_1711842, EPI_ISL_1711843, EPI_ISL_1711844, EPI_ISL_1711845 | | | |
| see above | LA Office of Public Health Laboratories | Centers for Disease Control and Prevention Division of Viral Diseases, Pathogen Discovery | Mili Sheth, Sarah Nobles, Jasmine Padilla, Mark Burroughs, Shoshona Le, Katie Dillon, Peter Cook, Clinton R. Paden, Dhwani Batra, Krista Queen, Kristen Knipe, Dakota Howard, Yvette Unoarumhi, Darlene Wagner, Matthew Schmerer, Ben L. Rambo-Martin, Kristine Lacek, Sam Shepard, Alison Laufer Halpin, Dave Wentworth, Vivien Dugan, Suxiang Tong, Justin Lee |
| EPI_ISL_1711846, EPI_ISL_1711847, EPI_ISL_1711848, EPI_ISL_1711849, EPI_ISL_1711850, EPI_ISL_1711851, EPI_ISL_1711852, EPI_ISL_1711853, EPI_ISL_1711854, EPI_ISL_1711855, EPI_ISL_1711856, EPI_ISL_1711857, EPI_ISL_1711858, EPI_ISL_1711859, EPI_ISL_1711860, EPI_ISL_1711861 | | | |
| see above | RI State Health Laboratories | Centers for Disease Control and Prevention Division of Viral Diseases, Pathogen Discovery | Mili Sheth, Sarah Nobles, Jasmine Padilla, Mark Burroughs, Shoshona Le, Katie Dillon, Peter Cook, Clinton R. Paden, Dhwani Batra, Krista Queen, Kristen Knipe, Dakota Howard, Yvette Unoarumhi, Darlene Wagner, Matthew Schmerer, Ben L. Rambo-Martin, Kristine Lacek, Sam Shepard, Alison Laufer Halpin, Dave Wentworth, Vivien Dugan, Suxiang Tong, Justin Lee |
| EPI_ISL_1711862, EPI_ISL_1711863, EPI_ISL_1711864, EPI_ISL_1711865, EPI_ISL_1711866, EPI_ISL_1711867, EPI_ISL_1711868, EPI_ISL_1711869, EPI_ISL_1711870, EPI_ISL_1711871, EPI_ISL_1711872, EPI_ISL_1711873, EPI_ISL_1711874 | | | |
| see above | DC Public Health Lab/ Dept. of Forensic Sciences | Centers for Disease Control and Prevention Division of Viral Diseases, Pathogen Discovery | Mili Sheth, Sarah Nobles, Jasmine Padilla, Mark Burroughs, Shoshona Le, Katie Dillon, Peter Cook, Clinton R. Paden, Dhwani Batra, Krista Queen, Kristen Knipe, Dakota Howard, Yvette Unoarumhi, Darlene Wagner, Matthew Schmerer, Ben L. Rambo-Martin, Kristine Lacek, Sam Shepard, Alison Laufer Halpin, Dave Wentworth, Vivien Dugan, Suxiang Tong, Justin Lee |
| EPI_ISL_1711875, EPI_ISL_1711876, EPI_ISL_1711877, EPI_ISL_1711878, EPI_ISL_1711879, EPI_ISL_1711880, EPI_ISL_1711881, EPI_ISL_1711882, EPI_ISL_1711883, EPI_ISL_1711884, EPI_ISL_1711885, EPI_ISL_1711886, EPI_ISL_1711887, EPI_ISL_1711888, EPI_ISL_1711889, EPI_ISL_1711890, EPI_ISL_1711891, EPI_ISL_1711892, EPI_ISL_1711893, EPI_ISL_1711894, EPI_ISL_1711895, EPI_ISL_1711896, EPI_ISL_1711897, EPI_ISL_1711898, EPI_ISL_1711899, EPI_ISL_1711900, EPI_ISL_1711901, EPI_ISL_1711902, EPI_ISL_1711903, EPI_ISL_1711904, EPI_ISL_1711905, EPI_ISL_1711906, EPI_ISL_1711907, EPI_ISL_1711908, EPI_ISL_1711909, EPI_ISL_1711910, EPI_ISL_1711911, EPI_ISL_1711912, EPI_ISL_1711913, EPI_ISL_1711914, EPI_ISL_1711915, EPI_ISL_1711916, EPI_ISL_1711917, EPI_ISL_1711918 | | | |
| see above | "AZ SPHL, Arizona Department of Health Services" | Centers for Disease Control and Prevention Division of Viral Diseases, Pathogen Discovery | Mili Sheth, Sarah Nobles, Jasmine Padilla, Mark Burroughs, Shoshona Le, Katie Dillon, Peter Cook, Clinton R. Paden, Dhwani Batra, Krista Queen, Kristen Knipe, Dakota Howard, Yvette Unoarumhi, Darlene Wagner, Matthew Schmerer, Ben L. Rambo-Martin, Kristine Lacek, Sam Shepard, Alison Laufer Halpin, Dave Wentworth, Vivien Dugan, Suxiang Tong, Justin Lee |
| EPI_ISL_1711919, EPI_ISL_1711920, EPI_ISL_1711921, EPI_ISL_1711922, EPI_ISL_1711923 | VT Dept. of Health Laboratory | Centers for Disease Control and Prevention Division of Viral Diseases, Pathogen Discovery | Mili Sheth, Sarah Nobles, Jasmine Padilla, Mark Burroughs, Shoshona Le, Katie Dillon, Peter Cook, Clinton R. Paden, Dhwani Batra, Krista Queen, Kristen Knipe, Dakota Howard, Yvette Unoarumhi, Darlene Wagner, Matthew Schmerer, Ben L. Rambo-Martin, Kristine Lacek, Sam Shepard, Alison Laufer Halpin, Dave Wentworth, Vivien Dugan, Suxiang Tong, Justin Lee |
| EPI_ISL_1711924, EPI_ISL_1711925, EPI_ISL_1711926, EPI_ISL_1711927, EPI_ISL_1711928, EPI_ISL_1711929, EPI_ISL_1711930, | SC Dept of Health and Env. Control-Bureau of Laboratories | Centers for Disease Control and Prevention Division of Viral Diseases, Pathogen Discovery | Mili Sheth, Sarah Nobles, Jasmine Padilla, Mark Burroughs, Shoshona Le, Katie Dillon, Peter Cook, Clinton R. Paden, Dhwani Batra, Krista Queen, Kristen Knipe, Dakota Howard, Yvette Unoarumhi, Darlene Wagner, Matthew Schmerer, Ben L. Rambo-Martin, Kristine Lacek, Sam Shepard, Alison Laufer Halpin, Dave Wentworth, Vivien Dugan, Suxiang Tong, Justin Lee |

| | | | |
|---|---|---|---|
| EPI_ISL_1711931, EPI_ISL_1711932, EPI_ISL_1711933 | | | |
| EPI_ISL_1711934, EPI_ISL_1711935, EPI_ISL_1711936, EPI_ISL_1711937 | NH Dept. of Health and Human Services Public Health Labs | Centers for Disease Control and Prevention Division of Viral Diseases, Pathogen Discovery | Mili Sheth, Sarah Nobles, Jasmine Padilla, Mark Burroughs, Shoshona Le, Katie Dillon, Peter Cook, Clinton R. Paden, Dhwani Batra, Krista Queen, Kristen Knipe, Dakota Howard, Yvette Unoarumhi, Darlene Wagner, Matthew Schmerer, Ben L. Rambo-Martin, Kristine Lacek, Sam Shepard, Alison Laufer Halpin, Dave Wentworth, Vivien Dugan, Suxiang Tong, Justin Lee |
| EPI_ISL_1711938, EPI_ISL_1711939, EPI_ISL_1711940, EPI_ISL_1711941, EPI_ISL_1711942, EPI_ISL_1711943, EPI_ISL_1711944, EPI_ISL_1711945, EPI_ISL_1711946, EPI_ISL_1711947, EPI_ISL_1711948, EPI_ISL_1711949, EPI_ISL_1711950, EPI_ISL_1711951 | | | |
| see above | NC State Laboratory of Public Health | Centers for Disease Control and Prevention Division of Viral Diseases, Pathogen Discovery | Mili Sheth, Sarah Nobles, Jasmine Padilla, Mark Burroughs, Shoshona Le, Katie Dillon, Peter Cook, Clinton R. Paden, Dhwani Batra, Krista Queen, Kristen Knipe, Dakota Howard, Yvette Unoarumhi, Darlene Wagner, Matthew Schmerer, Ben L. Rambo-Martin, Kristine Lacek, Sam Shepard, Alison Laufer Halpin, Dave Wentworth, Vivien Dugan, Suxiang Tong, Justin Lee |
| EPI_ISL_1711952, EPI_ISL_1711953, EPI_ISL_1711954, EPI_ISL_1711955, EPI_ISL_1711956, EPI_ISL_1711957, EPI_ISL_1711958, EPI_ISL_1711959, EPI_ISL_1711960, EPI_ISL_1711961, EPI_ISL_1711962, EPI_ISL_1711963, EPI_ISL_1711964, EPI_ISL_1711965, EPI_ISL_1711966, EPI_ISL_1711967, EPI_ISL_1711968, EPI_ISL_1711969 | | | |
| see above | GA Department of Public Health Laboratory | Centers for Disease Control and Prevention Division of Viral Diseases, Pathogen Discovery | Mili Sheth, Sarah Nobles, Jasmine Padilla, Mark Burroughs, Shoshona Le, Katie Dillon, Peter Cook, Clinton R. Paden, Dhwani Batra, Krista Queen, Kristen Knipe, Dakota Howard, Yvette Unoarumhi, Darlene Wagner, Matthew Schmerer, Ben L. Rambo-Martin, Kristine Lacek, Sam Shepard, Alison Laufer Halpin, Dave Wentworth, Vivien Dugan, Suxiang Tong, Justin Lee |
| EPI_ISL_1711970, EPI_ISL_1711971, EPI_ISL_1711972, EPI_ISL_1711973 | Quest Diagnostics Incorporated | Centers for Disease Control and Prevention Division of Viral Diseases, Pathogen Discovery | Dakota Howard, Dhwani Batra, Peter W. Cook, Kara Moser, Adrian Paskey, Jason Caravas, Benjamin Rambo-Martin, Shatavia Morrison, Christopher Gulvick, Scott Sammons, Yvette Unoarumhi, Darlene Wagner, Matthew Schmerer, S. H. Rosenthal, A. Gerasimova, R. M. Kagan, B. Anderson, M. Hua, Y. Liu, L.E. Bernstein, K.E. Livingston, A. Perez, I. A. Shlyakhter, R. V. Rolando, R. Owen, P. Tanpaiboon, F. Lacbawan, Clinton R. Paden, Duncan MacCannell |
| EPI_ISL_1711974, EPI_ISL_1711975 | Aegis Sciences Corporation | Centers for Disease Control and Prevention Division of Viral Diseases, Pathogen Discovery | Dakota Howard, Dhwani Batra, Peter W. Cook, Kara Moser, Adrian Paskey, Jason Caravas, Benjamin Rambo-Martin, Shatavia Morrison, Christopher Gulvick, Scott Sammons, Yvette Unoarumhi, Darlene Wagner, Matthew Schmerer, Cyndi Clark, Patrick Campbell, Rob Case, Vikramsinha Ghorpade, Holly Houdeshell, Ola Kvalvaag, Dillon Nall, Ethan Sanders, Alec Vest, Shaun Westlund, Matthew Hardison, Clinton R. Paden, Duncan MacCannell |
| EPI_ISL_1712422, EPI_ISL_1712423, EPI_ISL_1712424, EPI_ISL_1712425, EPI_ISL_1712427, EPI_ISL_1712429, EPI_ISL_1712430, EPI_ISL_1712431, EPI_ISL_1712432, EPI_ISL_1712433, EPI_ISL_1712438, EPI_ISL_1712439, EPI_ISL_1712440, EPI_ISL_1712442, EPI_ISL_1712444, EPI_ISL_1712445, EPI_ISL_1712446, EPI_ISL_1712447, EPI_ISL_1712448, EPI_ISL_1712453, EPI_ISL_1712454, EPI_ISL_1712455, EPI_ISL_1712456, EPI_ISL_1712457, EPI_ISL_1712458, EPI_ISL_1712460, EPI_ISL_1712461, EPI_ISL_1712462, EPI_ISL_1712464, EPI_ISL_1712465, EPI_ISL_1712466, EPI_ISL_1712467, EPI_ISL_1712475, EPI_ISL_1712476, EPI_ISL_1712477, EPI_ISL_1712479, EPI_ISL_1712480, EPI_ISL_1712481, EPI_ISL_1712482, EPI_ISL_1712483, EPI_ISL_1712484, EPI_ISL_1712485, EPI_ISL_1712486, EPI_ISL_1712487, EPI_ISL_1712488, EPI_ISL_1712489, EPI_ISL_1712490, EPI_ISL_1712491, EPI_ISL_1712492, EPI_ISL_1712493, EPI_ISL_1712494, EPI_ISL_1712495, EPI_ISL_1712496, EPI_ISL_1712497, EPI_ISL_1712498, EPI_ISL_1712499, EPI_ISL_1712500, EPI_ISL_1712501, EPI_ISL_1712502, EPI_ISL_1712503, EPI_ISL_1712504, EPI_ISL_1712506, EPI_ISL_1712507, EPI_ISL_1712508, EPI_ISL_1712509, EPI_ISL_1712510, EPI_ISL_1712511, EPI_ISL_1712512, EPI_ISL_1712513, EPI_ISL_1712514, EPI_ISL_1712516, EPI_ISL_1712517, EPI_ISL_1712518, EPI_ISL_1712519, EPI_ISL_1712520, EPI_ISL_1712521, EPI_ISL_1712522, EPI_ISL_1712523, EPI_ISL_1712524, EPI_ISL_1712525, EPI_ISL_1712526, EPI_ISL_1712527, EPI_ISL_1712528, EPI_ISL_1712529, EPI_ISL_1712530, EPI_ISL_1712531, EPI_ISL_1712532, EPI_ISL_1712533, EPI_ISL_1712534, EPI_ISL_1712535, EPI_ISL_1712536, EPI_ISL_1712538, EPI_ISL_1712539, EPI_ISL_1712541, EPI_ISL_1712542, EPI_ISL_1712543, EPI_ISL_1712544, EPI_ISL_1712545, EPI_ISL_1712547, EPI_ISL_1712549, EPI_ISL_1712551, EPI_ISL_1712552, EPI_ISL_1712553, EPI_ISL_1712554, EPI_ISL_1712555, EPI_ISL_1712556, EPI_ISL_1712557, EPI_ISL_1712558, EPI_ISL_1712559, EPI_ISL_1712560, EPI_ISL_1712561, EPI_ISL_1712562 | | | |
| see above | Innovative Genomics Institute, UC Berkeley | Innovative Genomics Institute, UC Berkeley | Stacia Wyman, Phil Frankino, John Boyle, Holly Gildea |
| EPI_ISL_1712654, EPI_ISL_1712655, EPI_ISL_1712656, EPI_ISL_1712657, EPI_ISL_1712658 | Tampa General Hospital Esoteric Lab | Tampa General Hospital Esoteric Research & Development Lab | Grant Vestal, Deanna Becker, Dominic Uy, Vicki Healer, Amorce Lima, Suzane Silbert |
| EPI_ISL_1714756, EPI_ISL_1714757, EPI_ISL_1714758, EPI_ISL_1714759, EPI_ISL_1714760, EPI_ISL_1714761, EPI_ISL_1714762, EPI_ISL_1714763, EPI_ISL_1714764, EPI_ISL_1714765, EPI_ISL_1714766, EPI_ISL_1714767, EPI_ISL_1714768, EPI_ISL_1714769, EPI_ISL_1714770, EPI_ISL_1714771, EPI_ISL_1714772, EPI_ISL_1714773, EPI_ISL_1714774, EPI_ISL_1714775, EPI_ISL_1714776, EPI_ISL_1714777, EPI_ISL_1714778, EPI_ISL_1714779, EPI_ISL_1714780, EPI_ISL_1714781, EPI_ISL_1714782, EPI_ISL_1714783, EPI_ISL_1714784 | | | |
| see above | University Hospitals Translational Laboratory (UHTL), University Hospitals | University Hospitals Translational Laboratory (UHTL), University Hospitals | Sadri,N., Alouani,D., Song,X. |
| EPI_ISL_1714785, EPI_ISL_1714786, EPI_ISL_1714788, EPI_ISL_1714789, EPI_ISL_1714790 | Operational Infectious Diseases, Naval Health Research Center | Operational Infectious Diseases, Naval Health Research Center | Pan,R.W., Underwood,R.S., Balansay-Ames,M.S., Vo,S., Myers,C.A., Fierro,M., Kriner,P., Iniguez-Stevens,E., Moser,K., Phippard,A.E. |
| EPI_ISL_1714791, EPI_ISL_1714792, EPI_ISL_1714793, EPI_ISL_1714794 | Operational Infectious Diseases, Naval Health Research Center | Operational Infectious Diseases, Naval Health Research Center | Pan,R.W., Underwood,R.S., Balansay-Ames,M.S., Myers,C.A. |
| EPI_ISL_1714795 | Operational Infectious Diseases, Naval Health Research Center | Operational Infectious Diseases, Naval Health Research Center | Pan,R.W., Underwood,R.S., Balansay-Ames,M.S., Vo,S., Myers,C.A., Fierro,M., Kriner,P., Iniguez-Stevens,E., Moser,K., Phippard,A.E. |
| EPI_ISL_1715440, EPI_ISL_1715441, EPI_ISL_1715442, EPI_ISL_1715443, EPI_ISL_1715445, EPI_ISL_1715446, EPI_ISL_1715447, EPI_ISL_1715448, EPI_ISL_1715449, EPI_ISL_1715450, EPI_ISL_1715451, EPI_ISL_1715452, EPI_ISL_1715453, EPI_ISL_1715454, EPI_ISL_1715455, EPI_ISL_1715456, EPI_ISL_1715457, EPI_ISL_1715458, EPI_ISL_1715459, EPI_ISL_1715460, EPI_ISL_1715461, EPI_ISL_1715462, EPI_ISL_1715463, EPI_ISL_1715464, EPI_ISL_1715465, EPI_ISL_1715466, EPI_ISL_1715467, EPI_ISL_1715468, EPI_ISL_1715469, EPI_ISL_1715470, EPI_ISL_1715471, EPI_ISL_1715472, EPI_ISL_1715473, EPI_ISL_1715474, EPI_ISL_1715475, EPI_ISL_1715476, EPI_ISL_1715477, EPI_ISL_1715479, EPI_ISL_1715480, EPI_ISL_1715481, EPI_ISL_1715482, EPI_ISL_1715483, EPI_ISL_1715484, EPI_ISL_1715485, EPI_ISL_1715486, EPI_ISL_1715487, EPI_ISL_1715488, EPI_ISL_1715489, EPI_ISL_1715490, EPI_ISL_1715491, EPI_ISL_1715492, EPI_ISL_1715493, EPI_ISL_1715494, EPI_ISL_1715495, EPI_ISL_1715496, EPI_ISL_1715497, EPI_ISL_1715498, EPI_ISL_1715499, EPI_ISL_1715500, EPI_ISL_1715502, EPI_ISL_1715503, EPI_ISL_1715504, EPI_ISL_1715505, EPI_ISL_1715506, EPI_ISL_1715507, EPI_ISL_1715508, EPI_ISL_1715509, EPI_ISL_1715510, EPI_ISL_1715511, EPI_ISL_1715512, EPI_ISL_1715513, EPI_ISL_1715514, EPI_ISL_1715515, EPI_ISL_1715517, EPI_ISL_1715518, EPI_ISL_1715519, EPI_ISL_1715520, EPI_ISL_1715521, EPI_ISL_1715522, EPI_ISL_1715523, EPI_ISL_1715524, EPI_ISL_1715525, EPI_ISL_1715526, EPI_ISL_1715527, EPI_ISL_1715528, EPI_ISL_1715529, EPI_ISL_1715530, EPI_ISL_1715531, EPI_ISL_1715532, EPI_ISL_1715533, EPI_ISL_1715534, EPI_ISL_1715535, EPI_ISL_1715536, EPI_ISL_1715537, EPI_ISL_1715538, EPI_ISL_1715539, EPI_ISL_1715540, EPI_ISL_1715541, EPI_ISL_1715542, EPI_ISL_1715543, EPI_ISL_1715544, EPI_ISL_1715545, EPI_ISL_1715546, EPI_ISL_1715547, EPI_ISL_1715548, EPI_ISL_1715549, EPI_ISL_1715550, EPI_ISL_1715551, EPI_ISL_1715552, EPI_ISL_1715553, EPI_ISL_1715554, EPI_ISL_1715555, EPI_ISL_1715557, EPI_ISL_1715558, EPI_ISL_1715559, EPI_ISL_1715560, EPI_ISL_1715561, EPI_ISL_1715562, EPI_ISL_1715563, EPI_ISL_1715564, EPI_ISL_1715565, EPI_ISL_1715566, EPI_ISL_1715567, EPI_ISL_1715568, EPI_ISL_1715569, EPI_ISL_1715570, EPI_ISL_1715571, EPI_ISL_1715572, EPI_ISL_1715573, EPI_ISL_1715574, EPI_ISL_1715575, EPI_ISL_1715576, EPI_ISL_1715577, EPI_ISL_1715579, EPI_ISL_1715580, EPI_ISL_1715581, EPI_ISL_1715582, EPI_ISL_1715583, EPI_ISL_1715584, EPI_ISL_1715585, EPI_ISL_1715586, EPI_ISL_1715587, EPI_ISL_1715588, EPI_ISL_1715589, EPI_ISL_1715590, EPI_ISL_1715591, EPI_ISL_1715592, EPI_ISL_1715594, EPI_ISL_1715595, EPI_ISL_1715596, EPI_ISL_1715597, EPI_ISL_1715598, EPI_ISL_1715599, EPI_ISL_1715600, EPI_ISL_1715602, EPI_ISL_1715603, EPI_ISL_1715604, EPI_ISL_1715605, EPI_ISL_1715606, EPI_ISL_1715607, EPI_ISL_1715608, EPI_ISL_1715609, EPI_ISL_1715610, EPI_ISL_1715611, EPI_ISL_1715612, EPI_ISL_1715613, EPI_ISL_1715614, EPI_ISL_1715615, EPI_ISL_1715616, EPI_ISL_1715617, EPI_ISL_1715618, EPI_ISL_1715619, EPI_ISL_1715620, EPI_ISL_1715621, EPI_ISL_1715622, EPI_ISL_1715623, EPI_ISL_1715624, EPI_ISL_1715625, EPI_ISL_1715626, EPI_ISL_1715627, EPI_ISL_1715628, EPI_ISL_1715629, EPI_ISL_1715630, EPI_ISL_1715631, EPI_ISL_1715632, EPI_ISL_1715633, EPI_ISL_1715634, EPI_ISL_1715635, EPI_ISL_1715636, EPI_ISL_1715637, EPI_ISL_1715638, EPI_ISL_1715639, EPI_ISL_1715640, EPI_ISL_1715641, EPI_ISL_1715642, EPI_ISL_1715643, EPI_ISL_1715644, EPI_ISL_1715645, EPI_ISL_1715646, EPI_ISL_1715647, EPI_ISL_1715648, EPI_ISL_1715649, EPI_ISL_1715650, EPI_ISL_1715651, EPI_ISL_1715652, EPI_ISL_1715653, EPI_ISL_1715654, EPI_ISL_1715655, EPI_ISL_1715656, EPI_ISL_1715657, EPI_ISL_1715658, EPI_ISL_1715659, EPI_ISL_1715660, EPI_ISL_1715661, EPI_ISL_1715662, EPI_ISL_1715663, EPI_ISL_1715664, EPI_ISL_1715665, EPI_ISL_1715666, EPI_ISL_1715667, EPI_ISL_1715668, EPI_ISL_1715669, EPI_ISL_1715670, EPI_ISL_1715671, EPI_ISL_1715672, EPI_ISL_1715673, EPI_ISL_1715674, EPI_ISL_1715675, EPI_ISL_1715676, EPI_ISL_1715677, EPI_ISL_1715678, EPI_ISL_1715679, EPI_ISL_1715680, EPI_ISL_1715681, EPI_ISL_1715682, EPI_ISL_1715683, EPI_ISL_1715684, EPI_ISL_1715685, EPI_ISL_1715686, EPI_ISL_1715687, EPI_ISL_1715688, EPI_ISL_1715689, EPI_ISL_1715690, EPI_ISL_1715691, EPI_ISL_1715692, EPI_ISL_1715693, EPI_ISL_1715694, EPI_ISL_1715695, EPI_ISL_1715696, EPI_ISL_1715697, EPI_ISL_1715698, EPI_ISL_1715699, EPI_ISL_1715700, EPI_ISL_1715701, EPI_ISL_1715702, EPI_ISL_1715703, EPI_ISL_1715704, EPI_ISL_1715705, EPI_ISL_1715706, EPI_ISL_1715707, EPI_ISL_1715708, EPI_ISL_1715709, EPI_ISL_1715710, EPI_ISL_1715711, EPI_ISL_1715712, EPI_ISL_1715713, EPI_ISL_1715714, EPI_ISL_1715715, EPI_ISL_1715716, EPI_ISL_1715717, EPI_ISL_1715718, EPI_ISL_1715719, EPI_ISL_1715720, EPI_ISL_1715721, EPI_ISL_1715722, EPI_ISL_1715723, EPI_ISL_1715724, EPI_ISL_1715725, EPI_ISL_1715726, EPI_ISL_1715727, EPI_ISL_1715728, EPI_ISL_1715729, EPI_ISL_1715730, EPI_ISL_1715731, EPI_ISL_1715732, EPI_ISL_1715733, EPI_ISL_1715734, EPI_ISL_1715735, EPI_ISL_1715736, EPI_ISL_1715737, EPI_ISL_1715738, EPI_ISL_1715739, EPI_ISL_1715740, EPI_ISL_1715741, EPI_ISL_1715742, EPI_ISL_1715743, EPI_ISL_1715744, EPI_ISL_1715745, EPI_ISL_1715746, EPI_ISL_1715747, EPI_ISL_1715748, EPI_ISL_1715749, EPI_ISL_1715750, EPI_ISL_1715751, EPI_ISL_1715752, EPI_ISL_1715753, EPI_ISL_1715754, EPI_ISL_1715755, EPI_ISL_1715756, EPI_ISL_1715757, EPI_ISL_1715758, EPI_ISL_1715759, EPI_ISL_1715760, EPI_ISL_1715761, EPI_ISL_1715762, EPI_ISL_1715763, EPI_ISL_1715764, EPI_ISL_1715765, EPI_ISL_1715766, EPI_ISL_1715767 | | | |
| see above | UW Virology Lab | UW Virology Lab | Pavitra Roychoudhury, Hong Xie, Lasata Shrestha, Tien V. Nguyen, Shah Mohamed Bakhash, Michelle Lin, Noah R. Baker, Sean Ellis, Meei-Li Huang, Keith R Jerome, Alexander Greninger |
| EPI_ISL_1716536, EPI_ISL_1716538, EPI_ISL_1716540, EPI_ISL_1716542, EPI_ISL_1716543, EPI_ISL_1716545, EPI_ISL_1716546, EPI_ISL_1716547, EPI_ISL_1716548, EPI_ISL_1716549, EPI_ISL_1716550, EPI_ISL_1716551, EPI_ISL_1716552, EPI_ISL_1716553, EPI_ISL_1716554, EPI_ISL_1716555, EPI_ISL_1716556, EPI_ISL_1716557, EPI_ISL_1716558 | | | |
| see above | UTGSAF | UTGSAF | Jessica Podnar, Sylvie Beaudenon, Audrey Kelly, Zachary Carver, Anna Battenhouse, Andreas Matouschek |
| EPI_ISL_1716559, EPI_ISL_1716560, EPI_ISL_1716561, EPI_ISL_1716562, EPI_ISL_1716563 | Arizona State Public Health Laboratory | Arizona State Public Health Laboratory | Trung Huynh, Jessica Escobar, Katherine Fullerton, Nobuko Fukushima, Stacy White, Linda Getsinger, Victor Waddell |
| EPI_ISL_1716828, EPI_ISL_1716829, EPI_ISL_1716830, EPI_ISL_1716831, EPI_ISL_1716832, EPI_ISL_1716833, EPI_ISL_1716834, EPI_ISL_1716835, EPI_ISL_1716836, EPI_ISL_1716837, EPI_ISL_1716838, EPI_ISL_1716839, EPI_ISL_1716840, EPI_ISL_1716841, EPI_ISL_1716842, EPI_ISL_1716843, EPI_ISL_1716844, EPI_ISL_1716845, EPI_ISL_1716846, EPI_ISL_1716847, EPI_ISL_1716848, EPI_ISL_1716849, EPI_ISL_1716850, EPI_ISL_1716851, EPI_ISL_1716852, EPI_ISL_1716853, EPI_ISL_1716854, EPI_ISL_1716855, EPI_ISL_1716856, EPI_ISL_1716857, EPI_ISL_1716858, EPI_ISL_1716859, EPI_ISL_1716860, EPI_ISL_1716861, EPI_ISL_1716862, EPI_ISL_1716863, EPI_ISL_1716864, EPI_ISL_1716865, EPI_ISL_1716866, EPI_ISL_1716867, EPI_ISL_1716868, EPI_ISL_1716869, EPI_ISL_1716870, EPI_ISL_1716871, EPI_ISL_1716872 | | | |
| see above | Tampa General Hospital Esoteric Lab | Tampa General Hospital Esoteric Research & Development Lab | Grant Vestal, Deanna Becker, Dominic Uy, Vicki Healer, Amorce Lima, Suzane Silbert |
| EPI_ISL_1716873, EPI_ISL_1716874, EPI_ISL_1716875, EPI_ISL_1716876 | UCLA Clinical Micro Lab | Los Angeles County Public Health Laboratories | P. Hemarajata et al. |
| EPI_ISL_1716881, EPI_ISL_1716882, EPI_ISL_1716883, EPI_ISL_1716884, EPI_ISL_1716885, EPI_ISL_1716886, EPI_ISL_1716887, EPI_ISL_1716888, EPI_ISL_1716889, EPI_ISL_1716891, EPI_ISL_1716892, EPI_ISL_1716893, EPI_ISL_1716894, EPI_ISL_1716895 | | | |
| see above | USC Clinical Lab | Los Angeles County Public Health Laboratories | P. Hemarajata et al. |
| EPI_ISL_1716896 | Martin Luther King Jr. Community Hospital | Los Angeles County Public Health Laboratories | P. Hemarajata et al. |
| EPI_ISL_1716897, EPI_ISL_1716898 | USC Clinical Lab | Los Angeles County Public Health Laboratories | P. Hemarajata et al. |
| EPI_ISL_1716899 | Martin Luther King Jr. Community Hospital | Los Angeles County Public Health Laboratories | P. Hemarajata et al. |
| EPI_ISL_1716906, EPI_ISL_1716907, EPI_ISL_1716908, EPI_ISL_1716909, EPI_ISL_1716910, EPI_ISL_1716911, EPI_ISL_1716912, EPI_ISL_1716913, EPI_ISL_1716914, EPI_ISL_1716915, EPI_ISL_1716916, EPI_ISL_1716917, EPI_ISL_1716918, EPI_ISL_1716919, EPI_ISL_1716920, EPI_ISL_1716921, EPI_ISL_1716922, EPI_ISL_1716923, EPI_ISL_1716924, EPI_ISL_1716926, EPI_ISL_1716927, EPI_ISL_1716928, EPI_ISL_1716929, EPI_ISL_1716930, EPI_ISL_1716931, EPI_ISL_1716932, EPI_ISL_1716933, EPI_ISL_1716934, EPI_ISL_1716935, EPI_ISL_1716936, EPI_ISL_1716937, EPI_ISL_1716938, EPI_ISL_1716939, EPI_ISL_1716940, EPI_ISL_1716941, EPI_ISL_1716942, EPI_ISL_1716943, EPI_ISL_1716944, EPI_ISL_1716945, EPI_ISL_1716946, EPI_ISL_1716947, EPI_ISL_1716948, EPI_ISL_1716949, EPI_ISL_1716950, EPI_ISL_1716951, EPI_ISL_1716952, EPI_ISL_1716953, EPI_ISL_1716954, EPI_ISL_1716955, EPI_ISL_1716956, EPI_ISL_1716957, EPI_ISL_1716958, EPI_ISL_1716959, EPI_ISL_1716960, EPI_ISL_1716961, EPI_ISL_1716962, EPI_ISL_1716963, EPI_ISL_1716964, EPI_ISL_1716965, EPI_ISL_1716966, EPI_ISL_1716967, EPI_ISL_1716968, EPI_ISL_1716969, EPI_ISL_1716970, EPI_ISL_1716971, EPI_ISL_1716972, EPI_ISL_1716973, EPI_ISL_1716974, EPI_ISL_1716975, EPI_ISL_1716976, EPI_ISL_1716977, EPI_ISL_1716978, EPI_ISL_1716979, EPI_ISL_1716980, EPI_ISL_1716981, EPI_ISL_1716982, EPI_ISL_1716983, EPI_ISL_1716984, EPI_ISL_1716985, EPI_ISL_1716986, EPI_ISL_1716987, EPI_ISL_1716988, EPI_ISL_1716989, EPI_ISL_1716991, EPI_ISL_1716992, EPI_ISL_1716993, EPI_ISL_1716995, EPI_ISL_1716996, EPI_ISL_1716997, EPI_ISL_1716998, EPI_ISL_1716999, EPI_ISL_1717000, EPI_ISL_1717001, EPI_ISL_1717002, EPI_ISL_1717003, EPI_ISL_1717004, EPI_ISL_1717005, EPI_ISL_1717006, EPI_ISL_1717007, EPI_ISL_1717008, EPI_ISL_1717009, EPI_ISL_1717010, EPI_ISL_1717011, EPI_ISL_1717012, EPI_ISL_1717013, EPI_ISL_1717014, EPI_ISL_1717015, EPI_ISL_1717016, EPI_ISL_1717017, EPI_ISL_1717018, EPI_ISL_1717019, EPI_ISL_1717020, EPI_ISL_1717021, EPI_ISL_1717022, EPI_ISL_1717023, EPI_ISL_1717024 | | | |

| | | | |
|---|---|---|---|
| see above | Pathology and Laboratory Medicine Institute, Cleveland Clinic, Ohio, USA | Pathology and Laboratory Medicine Institute, Cleveland Clinic, Ohio, USA | Jessica Spildener, Joy Nakitandwe, Kristen McDonnell, David Plunkett, Zheng Jin Tu, Jay Brock, Yu-Wei Cheng, Gary Procop, Daniel Rhoads, Daniel H. Farkas, David Bosler |
| EPI_ISL_1717107, EPI_ISL_1717108, EPI_ISL_1717109, EPI_ISL_1717110, EPI_ISL_1717111, EPI_ISL_1717112, EPI_ISL_1717113, EPI_ISL_1717114, EPI_ISL_1717115, EPI_ISL_1717117, EPI_ISL_1717118, EPI_ISL_1717119, EPI_ISL_1717120, EPI_ISL_1717121, EPI_ISL_1717122, EPI_ISL_1717123, EPI_ISL_1717124, EPI_ISL_1717125, EPI_ISL_1717126, EPI_ISL_1717127, EPI_ISL_1717128, EPI_ISL_1717130, EPI_ISL_1717131, EPI_ISL_1717132, EPI_ISL_1717133, EPI_ISL_1717134, EPI_ISL_1717135, EPI_ISL_1717136, EPI_ISL_1717137, EPI_ISL_1717138, EPI_ISL_1717139, EPI_ISL_1717140, EPI_ISL_1717141, EPI_ISL_1717142, EPI_ISL_1717143, EPI_ISL_1717144, EPI_ISL_1717145, EPI_ISL_1717146, EPI_ISL_1717147, EPI_ISL_1717148, EPI_ISL_1717149, EPI_ISL_1717150, EPI_ISL_1717151, EPI_ISL_1717153, EPI_ISL_1717154, EPI_ISL_1717156, EPI_ISL_1717157, EPI_ISL_1717159, EPI_ISL_1717160, EPI_ISL_1717161, EPI_ISL_1717162, EPI_ISL_1717164, EPI_ISL_1717165, EPI_ISL_1717166, EPI_ISL_1717167, EPI_ISL_1717168, EPI_ISL_1717170, EPI_ISL_1717171, EPI_ISL_1717172, EPI_ISL_1717173, EPI_ISL_1717174, EPI_ISL_1717175, EPI_ISL_1717176, EPI_ISL_1717177, EPI_ISL_1717178, EPI_ISL_1717179, EPI_ISL_1717180, EPI_ISL_1717181, EPI_ISL_1717182, EPI_ISL_1717183, EPI_ISL_1717184, EPI_ISL_1717185, EPI_ISL_1717186, EPI_ISL_1717187, EPI_ISL_1717188, EPI_ISL_1717189, EPI_ISL_1717190, EPI_ISL_1717191, EPI_ISL_1717192, EPI_ISL_1717193, EPI_ISL_1717194, EPI_ISL_1717195, EPI_ISL_1717196, EPI_ISL_1717197, EPI_ISL_1717198, EPI_ISL_1717199, EPI_ISL_1717200, EPI_ISL_1717201, EPI_ISL_1717202, EPI_ISL_1717203, EPI_ISL_1717204, EPI_ISL_1717205, EPI_ISL_1717206, EPI_ISL_1717207, EPI_ISL_1717208, EPI_ISL_1717209, EPI_ISL_1717210, EPI_ISL_1717211, EPI_ISL_1717213, EPI_ISL_1717215, EPI_ISL_1717216, EPI_ISL_1717217, EPI_ISL_1717218, EPI_ISL_1717219, EPI_ISL_1717220, EPI_ISL_1717221, EPI_ISL_1717223, EPI_ISL_1717224, EPI_ISL_1717225, EPI_ISL_1717226, EPI_ISL_1717228, EPI_ISL_1717229, EPI_ISL_1717230, EPI_ISL_1717231, EPI_ISL_1717232, EPI_ISL_1717233, EPI_ISL_1717234, EPI_ISL_1717236, EPI_ISL_1717237, EPI_ISL_1717238, EPI_ISL_1717239, EPI_ISL_1717240, EPI_ISL_1717241, EPI_ISL_1717242, EPI_ISL_1717243, EPI_ISL_1717244, EPI_ISL_1717245, EPI_ISL_1717246, EPI_ISL_1717247, EPI_ISL_1717248, EPI_ISL_1717249, EPI_ISL_1717250, EPI_ISL_1717251, EPI_ISL_1717252, EPI_ISL_1717254, EPI_ISL_1717255, EPI_ISL_1717257, EPI_ISL_1717258, EPI_ISL_1717259, EPI_ISL_1717260, EPI_ISL_1717261, EPI_ISL_1717262, EPI_ISL_1717263, EPI_ISL_1717264, EPI_ISL_1717265, EPI_ISL_1717266, EPI_ISL_1717267, EPI_ISL_1717268, EPI_ISL_1717269, EPI_ISL_1717271, EPI_ISL_1717272, EPI_ISL_1717273, EPI_ISL_1717274, EPI_ISL_1717276, EPI_ISL_1717277, EPI_ISL_1717278, EPI_ISL_1717279, EPI_ISL_1717281, EPI_ISL_1717282, EPI_ISL_1717283, EPI_ISL_1717284, EPI_ISL_1717285, EPI_ISL_1717286, EPI_ISL_1717287, EPI_ISL_1717288, EPI_ISL_1717289, EPI_ISL_1717290, EPI_ISL_1717291, EPI_ISL_1717292, EPI_ISL_1717294, EPI_ISL_1717295, EPI_ISL_1717296, EPI_ISL_1717297, EPI_ISL_1717298, EPI_ISL_1717299, EPI_ISL_1717300, EPI_ISL_1717301, EPI_ISL_1717302, EPI_ISL_1717304, EPI_ISL_1717305, EPI_ISL_1717306, EPI_ISL_1717308, EPI_ISL_1717309, EPI_ISL_1717310, EPI_ISL_1717311, EPI_ISL_1717312, EPI_ISL_1717313, EPI_ISL_1717314, EPI_ISL_1717315, EPI_ISL_1717317, EPI_ISL_1717318, EPI_ISL_1717319, EPI_ISL_1717320, EPI_ISL_1717321, EPI_ISL_1717322, EPI_ISL_1717323, EPI_ISL_1717325, EPI_ISL_1717326, EPI_ISL_1717327, EPI_ISL_1717328, EPI_ISL_1717329, EPI_ISL_1717330, EPI_ISL_1717331, EPI_ISL_1717332, EPI_ISL_1717333, EPI_ISL_1717334, EPI_ISL_1717335, EPI_ISL_1717336, EPI_ISL_1717337, EPI_ISL_1717338, EPI_ISL_1717339, EPI_ISL_1717341, EPI_ISL_1717342, EPI_ISL_1717343, EPI_ISL_1717345, EPI_ISL_1717346, EPI_ISL_1717347, EPI_ISL_1717348, EPI_ISL_1717349, EPI_ISL_1717350, EPI_ISL_1717351, EPI_ISL_1717352, EPI_ISL_1717353, EPI_ISL_1717354, EPI_ISL_1717356, EPI_ISL_1717357, EPI_ISL_1717358, EPI_ISL_1717359, EPI_ISL_1717360, EPI_ISL_1717361, EPI_ISL_1717362, EPI_ISL_1717363, EPI_ISL_1717364, EPI_ISL_1717365, EPI_ISL_1717366, EPI_ISL_1717367, EPI_ISL_1717368, EPI_ISL_1717369, EPI_ISL_1717370, EPI_ISL_1717371, EPI_ISL_1717372, EPI_ISL_1717373, EPI_ISL_1717374, EPI_ISL_1717375, EPI_ISL_1717376, EPI_ISL_1717379, EPI_ISL_1717380, EPI_ISL_1717381, EPI_ISL_1717382, EPI_ISL_1717383, EPI_ISL_1717384, EPI_ISL_1717385, EPI_ISL_1717387, EPI_ISL_1717388, EPI_ISL_1717389, EPI_ISL_1717390, EPI_ISL_1717391, EPI_ISL_1717392, EPI_ISL_1717394, EPI_ISL_1717395, EPI_ISL_1717396, EPI_ISL_1717397, EPI_ISL_1717398, EPI_ISL_1717399, EPI_ISL_1717400, EPI_ISL_1717401, EPI_ISL_1717403, EPI_ISL_1717404, EPI_ISL_1717405, EPI_ISL_1717406, EPI_ISL_1717407, EPI_ISL_1717408, EPI_ISL_1717410, EPI_ISL_1717411, EPI_ISL_1717412, EPI_ISL_1717413, EPI_ISL_1717414, EPI_ISL_1717415, EPI_ISL_1717417, EPI_ISL_1717418, EPI_ISL_1717419, EPI_ISL_1717420, EPI_ISL_1717421, EPI_ISL_1717422, EPI_ISL_1717424, EPI_ISL_1717425, EPI_ISL_1717426, EPI_ISL_1717427, EPI_ISL_1717428, EPI_ISL_1717429, EPI_ISL_1717430, EPI_ISL_1717431, EPI_ISL_1717432, EPI_ISL_1717433, EPI_ISL_1717435, EPI_ISL_1717436, EPI_ISL_1717437, EPI_ISL_1717438, EPI_ISL_1717439, EPI_ISL_1717440, EPI_ISL_1717441, EPI_ISL_1717442, EPI_ISL_1717444, EPI_ISL_1717445, EPI_ISL_1717446, EPI_ISL_1717447, EPI_ISL_1717448, EPI_ISL_1717449, EPI_ISL_1717450, EPI_ISL_1717451, EPI_ISL_1717452, EPI_ISL_1717453, EPI_ISL_1717454, EPI_ISL_1717455, EPI_ISL_1717457, EPI_ISL_1717459, EPI_ISL_1717461, EPI_ISL_1717462, EPI_ISL_1717464, EPI_ISL_1717465, EPI_ISL_1717466, EPI_ISL_1717468, EPI_ISL_1717469, EPI_ISL_1717470, EPI_ISL_1717471, EPI_ISL_1717473, EPI_ISL_1717474, EPI_ISL_1717475, EPI_ISL_1717476, EPI_ISL_1717477, EPI_ISL_1717478, EPI_ISL_1717479, EPI_ISL_1717480, EPI_ISL_1717481, EPI_ISL_1717483, EPI_ISL_1717484, EPI_ISL_1717485, EPI_ISL_1717486, EPI_ISL_1717487, EPI_ISL_1717489, EPI_ISL_1717490, EPI_ISL_1717491, EPI_ISL_1717495, EPI_ISL_1717496, EPI_ISL_1717497, EPI_ISL_1717498, EPI_ISL_1717499, EPI_ISL_1717500, EPI_ISL_1717501, EPI_ISL_1717502, EPI_ISL_1717503, EPI_ISL_1717505, EPI_ISL_1717506, EPI_ISL_1717507, EPI_ISL_1717508, EPI_ISL_1717509, EPI_ISL_1717510, EPI_ISL_1717511, EPI_ISL_1717512, EPI_ISL_1717513, EPI_ISL_1717514, EPI_ISL_1717515, EPI_ISL_1717516, EPI_ISL_1717518, EPI_ISL_1717519, EPI_ISL_1717520, EPI_ISL_1717521, EPI_ISL_1717522, EPI_ISL_1717523, EPI_ISL_1717524, EPI_ISL_1717525, EPI_ISL_1717527, EPI_ISL_1717528, EPI_ISL_1717529, EPI_ISL_1717530, EPI_ISL_1717531, EPI_ISL_1717533, EPI_ISL_1717535, EPI_ISL_1717536, EPI_ISL_1717537, EPI_ISL_1717538, EPI_ISL_1717539, EPI_ISL_1717540, EPI_ISL_1717541, EPI_ISL_1717542, EPI_ISL_1717543, EPI_ISL_1717544, EPI_ISL_1717545, EPI_ISL_1717546, EPI_ISL_1717547, EPI_ISL_1717548, EPI_ISL_1717550, EPI_ISL_1717551, EPI_ISL_1717552, EPI_ISL_1717553, EPI_ISL_1717554, EPI_ISL_1717555, EPI_ISL_1717557, EPI_ISL_1717558, EPI_ISL_1717559, EPI_ISL_1717560, EPI_ISL_1717561, EPI_ISL_1717562, EPI_ISL_1717563, EPI_ISL_1717564, EPI_ISL_1717566, EPI_ISL_1717567, EPI_ISL_1717568, EPI_ISL_1717569, EPI_ISL_1717570, EPI_ISL_1717571, EPI_ISL_1717572, EPI_ISL_1717573, EPI_ISL_1717574, EPI_ISL_1717575, EPI_ISL_1717576, EPI_ISL_1717577, EPI_ISL_1717578, EPI_ISL_1717579, EPI_ISL_1717580, EPI_ISL_1717581, EPI_ISL_1717582, EPI_ISL_1717583, EPI_ISL_1717584, EPI_ISL_1717586, EPI_ISL_1717587, EPI_ISL_1717588, EPI_ISL_1717590, EPI_ISL_1717591, EPI_ISL_1717592, EPI_ISL_1717593, EPI_ISL_1717594, EPI_ISL_1717596, EPI_ISL_1717597, EPI_ISL_1717598, EPI_ISL_1717599, EPI_ISL_1717600, EPI_ISL_1717601, EPI_ISL_1717602, EPI_ISL_1717603, EPI_ISL_1717604, EPI_ISL_1717605, EPI_ISL_1717606, EPI_ISL_1717607, EPI_ISL_1717608, EPI_ISL_1717609, EPI_ISL_1717610, EPI_ISL_1717612, EPI_ISL_1717613, EPI_ISL_1717614, EPI_ISL_1717615, EPI_ISL_1717616, EPI_ISL_1717617, EPI_ISL_1717618, EPI_ISL_1717619, EPI_ISL_1717621, EPI_ISL_1717622, EPI_ISL_1717623, EPI_ISL_1717624, EPI_ISL_1717626, EPI_ISL_1717627, EPI_ISL_1717628, EPI_ISL_1717629, EPI_ISL_1717630, EPI_ISL_1717631, EPI_ISL_1717632, EPI_ISL_1717634, EPI_ISL_1717635, EPI_ISL_1717638, EPI_ISL_1717639, EPI_ISL_1717640, EPI_ISL_1717641, EPI_ISL_1717642, EPI_ISL_1717643, EPI_ISL_1717646, EPI_ISL_1717647, EPI_ISL_1717648, EPI_ISL_1717649, EPI_ISL_1717650, EPI_ISL_1717651, EPI_ISL_1717655, EPI_ISL_1717656, EPI_ISL_1717657, EPI_ISL_1717658, EPI_ISL_1717659, EPI_ISL_1717660, EPI_ISL_1717661, EPI_ISL_1717662, EPI_ISL_1717663, EPI_ISL_1717664, EPI_ISL_1717665, EPI_ISL_1717666, EPI_ISL_1717667, EPI_ISL_1717668, EPI_ISL_1717670, EPI_ISL_1717671, EPI_ISL_1717672, EPI_ISL_1717673, EPI_ISL_1717674, EPI_ISL_1717675, EPI_ISL_1717676, EPI_ISL_1717677, EPI_ISL_1717678, EPI_ISL_1717679, EPI_ISL_1717680, EPI_ISL_1717681, EPI_ISL_1717682, EPI_ISL_1717683, EPI_ISL_1717684, EPI_ISL_1717685, EPI_ISL_1717686, EPI_ISL_1717687, EPI_ISL_1717688, EPI_ISL_1717689, EPI_ISL_1717690, EPI_ISL_1717691, EPI_ISL_1717692, EPI_ISL_1717693, EPI_ISL_1717694, EPI_ISL_1717696, EPI_ISL_1717697, EPI_ISL_1717698, EPI_ISL_1717699, EPI_ISL_1717700, EPI_ISL_1717701, EPI_ISL_1717702, EPI_ISL_1717705, EPI_ISL_1717706, EPI_ISL_1717707, EPI_ISL_1717708, EPI_ISL_1717709, EPI_ISL_1717710, EPI_ISL_1717712, EPI_ISL_1717713, EPI_ISL_1717715, EPI_ISL_1717716, EPI_ISL_1717717, EPI_ISL_1717720, EPI_ISL_1717721, EPI_ISL_1717722, EPI_ISL_1717723, EPI_ISL_1717724, EPI_ISL_1717725, EPI_ISL_1717726, EPI_ISL_1717727, EPI_ISL_1717728, EPI_ISL_1717729, EPI_ISL_1717730, EPI_ISL_1717731, EPI_ISL_1717732, EPI_ISL_1717733, EPI_ISL_1717734, EPI_ISL_1717735, EPI_ISL_1717737, EPI_ISL_1717738, EPI_ISL_1717739, EPI_ISL_1717740, EPI_ISL_1717743, EPI_ISL_1717744, EPI_ISL_1717745, EPI_ISL_1717746, EPI_ISL_1717747, EPI_ISL_1717748, EPI_ISL_1717749, EPI_ISL_1717750, EPI_ISL_1717751, EPI_ISL_1717752, EPI_ISL_1717753, EPI_ISL_1717754, EPI_ISL_1717755, EPI_ISL_1717756, EPI_ISL_1717757, EPI_ISL_1717758, EPI_ISL_1717759, EPI_ISL_1717761, EPI_ISL_1717762, EPI_ISL_1717763, EPI_ISL_1717764, EPI_ISL_1717765, EPI_ISL_1717766, EPI_ISL_1717767, EPI_ISL_1717768, EPI_ISL_1717769, EPI_ISL_1717770, EPI_ISL_1717771, EPI_ISL_1717772, EPI_ISL_1717773, EPI_ISL_1717774, EPI_ISL_1717775, EPI_ISL_1717776, EPI_ISL_1717778, EPI_ISL_1717780, EPI_ISL_1717781, EPI_ISL_1717782, EPI_ISL_1717783, EPI_ISL_1717785, EPI_ISL_1717786, EPI_ISL_1717787, EPI_ISL_1717788, EPI_ISL_1717789, EPI_ISL_1717790, EPI_ISL_1717791, EPI_ISL_1717792, EPI_ISL_1717793, EPI_ISL_1717794, EPI_ISL_1717795, EPI_ISL_1717798, EPI_ISL_1717799, EPI_ISL_1717800, EPI_ISL_1717801, EPI_ISL_1717802, EPI_ISL_1717803, EPI_ISL_1717805, EPI_ISL_1717806, EPI_ISL_1717807, EPI_ISL_1717808, EPI_ISL_1717809, EPI_ISL_1717810, EPI_ISL_1717811, EPI_ISL_1717812, EPI_ISL_1717813, EPI_ISL_1717814, EPI_ISL_1717815, EPI_ISL_1717816, EPI_ISL_1717817, EPI_ISL_1717818, EPI_ISL_1717819, EPI_ISL_1717821, EPI_ISL_1717822, EPI_ISL_1717823, EPI_ISL_1717824, EPI_ISL_1717825, EPI_ISL_1717826, EPI_ISL_1717827, EPI_ISL_1717828, EPI_ISL_1717829, EPI_ISL_1717830, EPI_ISL_1717831, EPI_ISL_1717832, EPI_ISL_1717833, EPI_ISL_1717834, EPI_ISL_1717836, EPI_ISL_1717837, EPI_ISL_1717838, EPI_ISL_1717839, EPI_ISL_1717840, EPI_ISL_1717841, EPI_ISL_1717842, EPI_ISL_1717843, EPI_ISL_1717844, EPI_ISL_1717845, EPI_ISL_1717846, EPI_ISL_1717847, EPI_ISL_1717848, EPI_ISL_1717849, EPI_ISL_1717850, EPI_ISL_1717851, EPI_ISL_1717852, EPI_ISL_1717853, EPI_ISL_1717854, EPI_ISL_1717855, EPI_ISL_1717856, EPI_ISL_1717857, EPI_ISL_1717858, EPI_ISL_1717859, EPI_ISL_1717860, EPI_ISL_1717861, EPI_ISL_1717862, EPI_ISL_1717864, EPI_ISL_1717865, EPI_ISL_1717866, EPI_ISL_1717867, EPI_ISL_1717869, EPI_ISL_1717870, EPI_ISL_1717871, EPI_ISL_1717872, EPI_ISL_1717873, EPI_ISL_1717874, EPI_ISL_1717875, EPI_ISL_1717876, EPI_ISL_1717877, EPI_ISL_1717878, EPI_ISL_1717879, EPI_ISL_1717881, EPI_ISL_1717882, EPI_ISL_1717883, EPI_ISL_1717884, EPI_ISL_1717885, EPI_ISL_1717886, EPI_ISL_1717887, EPI_ISL_1717888, EPI_ISL_1717889, EPI_ISL_1717890, EPI_ISL_1717892, EPI_ISL_1717893, EPI_ISL_1717894, EPI_ISL_1717895, EPI_ISL_1717896, EPI_ISL_1717897, EPI_ISL_1717898, EPI_ISL_1717899, EPI_ISL_1717900, EPI_ISL_1717901, EPI_ISL_1717902, EPI_ISL_1717904, EPI_ISL_1717905, EPI_ISL_1717906, EPI_ISL_1717907, EPI_ISL_1717908, EPI_ISL_1717909, EPI_ISL_1717910, EPI_ISL_1717911, EPI_ISL_1717912, EPI_ISL_1717913, EPI_ISL_1717914, EPI_ISL_1717915, EPI_ISL_1717916, EPI_ISL_1717917, EPI_ISL_1717918, EPI_ISL_1717920, EPI_ISL_1717921, EPI_ISL_1717922, EPI_ISL_1717923, EPI_ISL_1717924, EPI_ISL_1717925, EPI_ISL_1717926, EPI_ISL_1717927, EPI_ISL_1717928, EPI_ISL_1717929, EPI_ISL_1717930, EPI_ISL_1717931, EPI_ISL_1717936, EPI_ISL_1717938, EPI_ISL_1717939, EPI_ISL_1717940, EPI_ISL_1717941, EPI_ISL_1717942, EPI_ISL_1717943, EPI_ISL_1717944, EPI_ISL_1717945, EPI_ISL_1717946, EPI_ISL_1717948, EPI_ISL_1717950, EPI_ISL_1717951, EPI_ISL_1717952, EPI_ISL_1717953, EPI_ISL_1717954, EPI_ISL_1717956, EPI_ISL_1717957, EPI_ISL_1717959, EPI_ISL_1717960, EPI_ISL_1717962, EPI_ISL_1717963, EPI_ISL_1717964, EPI_ISL_1717965, EPI_ISL_1717966, EPI_ISL_1717967, EPI_ISL_1717968, EPI_ISL_1717969, EPI_ISL_1717972, EPI_ISL_1717973, EPI_ISL_1717974, EPI_ISL_1717976, EPI_ISL_1717977, EPI_ISL_1717979, EPI_ISL_1717980, EPI_ISL_1717981, EPI_ISL_1717982, EPI_ISL_1717983, EPI_ISL_1717984, EPI_ISL_1717985, EPI_ISL_1717986, EPI_ISL_1717987, EPI_ISL_1717988, EPI_ISL_1717990, EPI_ISL_1717991, EPI_ISL_1717992, EPI_ISL_1717993, EPI_ISL_1717994, EPI_ISL_1717996, EPI_ISL_1717997, EPI_ISL_1717998, EPI_ISL_1718000, EPI_ISL_1718002, EPI_ISL_1718003, EPI_ISL_1718005, EPI_ISL_1718006, EPI_ISL_1718008, EPI_ISL_1718009, EPI_ISL_1718012, EPI_ISL_1718013, EPI_ISL_1718015, EPI_ISL_1718017, EPI_ISL_1718019, EPI_ISL_1718020, EPI_ISL_1718022, EPI_ISL_1718023, EPI_ISL_1718025, EPI_ISL_1718027, EPI_ISL_1718028, EPI_ISL_1718030, EPI_ISL_1718031, EPI_ISL_1718032, EPI_ISL_1718033, EPI_ISL_1718034, EPI_ISL_1718035, EPI_ISL_1718036, EPI_ISL_1718038, EPI_ISL_1718039, EPI_ISL_1718040, EPI_ISL_1718041, EPI_ISL_1718042, EPI_ISL_1718044, EPI_ISL_1718046, EPI_ISL_1718048, EPI_ISL_1718060, EPI_ISL_1718062, EPI_ISL_1718063, EPI_ISL_1718067, EPI_ISL_1718069, EPI_ISL_1718070, EPI_ISL_1718071, EPI_ISL_1718072, EPI_ISL_1718074, EPI_ISL_1718075, EPI_ISL_1718076, EPI_ISL_1718077, EPI_ISL_1718080, EPI_ISL_1718082, EPI_ISL_1718083, EPI_ISL_1718084, EPI_ISL_1718085, EPI_ISL_1718087, EPI_ISL_1718088, EPI_ISL_1718089, EPI_ISL_1718093, EPI_ISL_1718094, EPI_ISL_1718095, EPI_ISL_1718097, EPI_ISL_1718098, EPI_ISL_1718100, EPI_ISL_1718101, EPI_ISL_1718102, EPI_ISL_1718103, EPI_ISL_1718104, EPI_ISL_1718106, EPI_ISL_1718107, EPI_ISL_1718110, EPI_ISL_1718111, EPI_ISL_1718112, EPI_ISL_1718113, EPI_ISL_1718114, EPI_ISL_1718117, EPI_ISL_1718118, EPI_ISL_1718119, EPI_ISL_1718121, EPI_ISL_1718124, EPI_ISL_1718125, EPI_ISL_1718126, EPI_ISL_1718127, EPI_ISL_1718128, EPI_ISL_1718131, EPI_ISL_1718132, EPI_ISL_1718133, EPI_ISL_1718134, EPI_ISL_1718135, EPI_ISL_1718137, EPI_ISL_1718138, EPI_ISL_1718139, EPI_ISL_1718140, EPI_ISL_1718141, EPI_ISL_1718142, EPI_ISL_1718143, EPI_ISL_1718144, EPI_ISL_1718145, EPI_ISL_1718146, EPI_ISL_1718147, EPI_ISL_1718149, EPI_ISL_1718150, EPI_ISL_1718151, EPI_ISL_1718152, EPI_ISL_1718153, EPI_ISL_1718155, EPI_ISL_1718156, EPI_ISL_1718157, EPI_ISL_1718160, EPI_ISL_1718161, EPI_ISL_1718162, EPI_ISL_1718164, EPI_ISL_1718166, EPI_ISL_1718168, EPI_ISL_1718169, EPI_ISL_1718171, EPI_ISL_1718172, EPI_ISL_1718173, EPI_ISL_1718174, EPI_ISL_1718175, EPI_ISL_1718176, EPI_ISL_1718177, EPI_ISL_1718179, EPI_ISL_1718180, EPI_ISL_1718181, EPI_ISL_1718184, EPI_ISL_1718185, EPI_ISL_1718187, EPI_ISL_1718188, EPI_ISL_1718189, EPI_ISL_1718190, EPI_ISL_1718191, EPI_ISL_1718193, EPI_ISL_1718194, EPI_ISL_1718195, EPI_ISL_1718196, EPI_ISL_1718197, EPI_ISL_1718199, EPI_ISL_1718201, EPI_ISL_1718202, EPI_ISL_1718203, EPI_ISL_1718205, EPI_ISL_1718206, EPI_ISL_1718208, EPI_ISL_1718209, EPI_ISL_1718210, EPI_ISL_1718211, EPI_ISL_1718212, EPI_ISL_1718213, EPI_ISL_1718214, EPI_ISL_1718215, EPI_ISL_1718216, EPI_ISL_1718217, EPI_ISL_1718218, EPI_ISL_1718219, EPI_ISL_1718220, EPI_ISL_1718222, EPI_ISL_1718224, EPI_ISL_1718225, EPI_ISL_1718226, EPI_ISL_1718227, EPI_ISL_1718229, EPI_ISL_1718231, EPI_ISL_1718232, EPI_ISL_1718235, EPI_ISL_1718239, EPI_ISL_1718240, EPI_ISL_1718241, EPI_ISL_1718242, EPI_ISL_1718243, EPI_ISL_1718246, EPI_ISL_1718247 | | | |
| see above | Pandemic Response Lab - NYC | Pandemic Response Lab, R&D | Henry Lee, Michael Hammerling, Melissa Hopkins, Cybill del Castillo, Shinyoung Clair Kang, William Ward, Pradeep Bugga, Sol Rey, Dylan Law, Katharine Nelson, Haiping Hao, Jon Laurent |
| EPI_ISL_1731407, EPI_ISL_1731408, EPI_ISL_1731409, EPI_ISL_1731410, EPI_ISL_1731411, EPI_ISL_1731412, EPI_ISL_1731413, EPI_ISL_1731414, EPI_ISL_1731415, EPI_ISL_1731416, EPI_ISL_1731417, EPI_ISL_1731418, EPI_ISL_1731419, EPI_ISL_1731420, EPI_ISL_1731421, EPI_ISL_1731422, EPI_ISL_1731423, EPI_ISL_1731424, EPI_ISL_1731425, EPI_ISL_1731426, EPI_ISL_1731427, EPI_ISL_1731428, EPI_ISL_1731429, EPI_ISL_1731430, EPI_ISL_1731431, EPI_ISL_1731432, EPI_ISL_1731433, EPI_ISL_1731434, EPI_ISL_1731435, EPI_ISL_1731436, EPI_ISL_1731437, EPI_ISL_1731438, EPI_ISL_1731439, EPI_ISL_1731440, EPI_ISL_1731441, EPI_ISL_1731442, EPI_ISL_1731443, EPI_ISL_1731444, EPI_ISL_1731445, EPI_ISL_1731446, EPI_ISL_1731447, EPI_ISL_1731448, EPI_ISL_1731449, EPI_ISL_1731450, EPI_ISL_1731451, EPI_ISL_1731452, EPI_ISL_1731453, EPI_ISL_1731454 | | | |
| see above | MD PHL | MD PHL | Maryland Department of Health Laboratories Administration |
| EPI_ISL_1731541 | UVA Clinical Microbiology Lab | Mathers Lab | Hardik I. Parikh, Katie E. Barry, Amy J. Mathers |
| EPI_ISL_1731660, EPI_ISL_1731661, EPI_ISL_1731662, EPI_ISL_1731663, EPI_ISL_1731664, EPI_ISL_1731665, EPI_ISL_1731666, EPI_ISL_1731667, EPI_ISL_1731668, EPI_ISL_1731669, EPI_ISL_1731670, EPI_ISL_1731671, EPI_ISL_1731672, EPI_ISL_1731673, EPI_ISL_1731674, EPI_ISL_1731675, EPI_ISL_1731676, EPI_ISL_1731677, EPI_ISL_1731678, EPI_ISL_1731679, EPI_ISL_1731680, EPI_ISL_1731681, EPI_ISL_1731682, EPI_ISL_1731683, EPI_ISL_1731684, EPI_ISL_1731685, EPI_ISL_1731686, EPI_ISL_1731688, EPI_ISL_1731689, EPI_ISL_1731690, EPI_ISL_1731691, EPI_ISL_1731692, EPI_ISL_1731693, EPI_ISL_1731694, EPI_ISL_1731695, EPI_ISL_1731696, EPI_ISL_1731697, EPI_ISL_1731698, EPI_ISL_1731699, EPI_ISL_1731700, EPI_ISL_1731701, EPI_ISL_1731702, EPI_ISL_1731703 | | | |
| see above | IN State Department of Health Laboratory Services | IN State Department of Health Laboratory Services | Cassandra Campion, Jamie Yeadon, Brian Pope, Lixia Liu, Kyle Brownlee, Melissa Hindenlang, Mark Glazier |
| EPI_ISL_1731763, EPI_ISL_1731764, EPI_ISL_1731765, EPI_ISL_1731766, EPI_ISL_1731767, EPI_ISL_1731769, EPI_ISL_1731770, EPI_ISL_1731771, EPI_ISL_1731772, EPI_ISL_1731773, EPI_ISL_1731774, EPI_ISL_1731776 | | | |
| see above | University of Michigan Clinical Microbiology Laboratory | Lauring Lab, University of Michigan, Department of Microbiology and Immunology | Valesano |
| EPI_ISL_1731778, EPI_ISL_1731779, EPI_ISL_1731780, EPI_ISL_1731781, EPI_ISL_1731782, EPI_ISL_1731783, EPI_ISL_1731784, EPI_ISL_1731785, EPI_ISL_1731786, EPI_ISL_1731788, EPI_ISL_1731789, EPI_ISL_1731790, EPI_ISL_1731791, EPI_ISL_1731792, EPI_ISL_1731793, EPI_ISL_1731795, EPI_ISL_1731796, EPI_ISL_1731797, EPI_ISL_1731798, EPI_ISL_1731799, EPI_ISL_1731800, EPI_ISL_1731801, EPI_ISL_1731802, EPI_ISL_1731803, EPI_ISL_1731805, EPI_ISL_1731806, EPI_ISL_1731807, EPI_ISL_1731809, EPI_ISL_1731810, EPI_ISL_1731811, EPI_ISL_1731812 | | | |
| see above | Wyoming Public Health Laboratory | Wyoming Public Health Laboratory | Jim Mildenberger, Wanda Manley, Noah Hull, Taylor Fearing, Lynette Gumbleton, Channing Weber, Ashley Norberg, Chayse Rowley, Marley Goetz, Brian Dominguez, Elliot Thomasson, Sam Britz, Cari Sloma, and Rob Christensen |
| EPI_ISL_1731814, EPI_ISL_1731816, EPI_ISL_1731817, EPI_ISL_1731819 | Pathology and Laboratory Medicine Institute, Cleveland Clinic, Ohio, USA | Pathology and Laboratory Medicine Institute, Cleveland Clinic, Ohio, USA | Jessica Spildener, Joy Nakitandwe, Kristen McDonnell, David Plunkett, Zheng Jin Tu, Jay Brock, Yu-Wei Cheng, Gary Procop, Daniel Rhoads, Daniel H. Farkas, David Bosler |
| EPI_ISL_1732080, EPI_ISL_1732081, EPI_ISL_1732082, EPI_ISL_1732084, EPI_ISL_1732087, EPI_ISL_1732090, EPI_ISL_1732092, EPI_ISL_1732094, EPI_ISL_1732095, EPI_ISL_1732096, EPI_ISL_1732097, EPI_ISL_1732099, EPI_ISL_1732101, EPI_ISL_1732102, EPI_ISL_1732103, EPI_ISL_1732105, EPI_ISL_1732106, EPI_ISL_1732107, EPI_ISL_1732108, EPI_ISL_1732109, EPI_ISL_1732110, EPI_ISL_1732111, EPI_ISL_1732113, EPI_ISL_1732114, EPI_ISL_1732116, EPI_ISL_1732117, EPI_ISL_1732118, EPI_ISL_1732119, EPI_ISL_1732122, EPI_ISL_1732123, EPI_ISL_1732124, EPI_ISL_1732125, EPI_ISL_1732126, EPI_ISL_1732127, EPI_ISL_1732128, EPI_ISL_1732129, EPI_ISL_1732130, EPI_ISL_1732131, EPI_ISL_1732132, EPI_ISL_1732133, EPI_ISL_1732134, EPI_ISL_1732135, EPI_ISL_1732136, EPI_ISL_1732137, EPI_ISL_1732138, EPI_ISL_1732139, EPI_ISL_1732140 | | | |
| see above | NJDOH, Public Health and Environmental Laboratories | NJ_PHEL | Lindsey Bodnar, Shiv K. Verma, Jacquelyn Deverell, Matthew Scarnati, Dana Woell, Byeong Jeong |
| EPI_ISL_1732142, EPI_ISL_1732143, EPI_ISL_1732144 | Tampa General Hospital Esoteric Lab | Tampa General Hospital Esoteric Research & Development Lab | Grant Vestal, Deanna Becker, Dominic Uy, Vicki Healer, Amorce Lima, Suzane Silbert |
| EPI_ISL_1732285, EPI_ISL_1732286, EPI_ISL_1732287, EPI_ISL_1732288, EPI_ISL_1732289, EPI_ISL_1732290, EPI_ISL_1732291, EPI_ISL_1732292 | Kansas Health and Environmental Lab | Kansas Health and Environmental Lab | Mike Grose, Jonathan Barnell, Ben Olsen, and Phil Adam |
| EPI_ISL_1732293, EPI_ISL_1732294, EPI_ISL_1732295 | Wichita State University - Molecular Diagnostics Lab | Kansas Health and Environmental Lab | Mike Grose, Jonathan Barnell, Ben Olsen, and Phil Adam |
| EPI_ISL_1732345, EPI_ISL_1732346, EPI_ISL_1732347, EPI_ISL_1732348, EPI_ISL_1732349, EPI_ISL_1732350, EPI_ISL_1732351, EPI_ISL_1732352, EPI_ISL_1732353, EPI_ISL_1732354, EPI_ISL_1732355, EPI_ISL_1732356, EPI_ISL_1732357, EPI_ISL_1732358, EPI_ISL_1732359, EPI_ISL_1732360, EPI_ISL_1732361, EPI_ISL_1732362, EPI_ISL_1732363, EPI_ISL_1732364, EPI_ISL_1732365, EPI_ISL_1732366, EPI_ISL_1732367, EPI_ISL_1732368, EPI_ISL_1732369, EPI_ISL_1732370, EPI_ISL_1732371, EPI_ISL_1732372, EPI_ISL_1732373, EPI_ISL_1732374, EPI_ISL_1732375, EPI_ISL_1732376, EPI_ISL_1732377, EPI_ISL_1732378, EPI_ISL_1732379, EPI_ISL_1732380, EPI_ISL_1732381, EPI_ISL_1732382, EPI_ISL_1732383, EPI_ISL_1732384, EPI_ISL_1732385, EPI_ISL_1732386, EPI_ISL_1732387, EPI_ISL_1732388, EPI_ISL_1732389, EPI_ISL_1732390, EPI_ISL_1732391, EPI_ISL_1732392, EPI_ISL_1732393, EPI_ISL_1732394, EPI_ISL_1732395, EPI_ISL_1732396, EPI_ISL_1732397, EPI_ISL_1732398, EPI_ISL_1732399, EPI_ISL_1732400, EPI_ISL_1732401, EPI_ISL_1732402, EPI_ISL_1732403, EPI_ISL_1732404, EPI_ISL_1732405, EPI_ISL_1732406, EPI_ISL_1732407, EPI_ISL_1732408, EPI_ISL_1732409, EPI_ISL_1732410, EPI_ISL_1732411, EPI_ISL_1732412, EPI_ISL_1732413, EPI_ISL_1732414, EPI_ISL_1732415, EPI_ISL_1732416, EPI_ISL_1732417, EPI_ISL_1732418, EPI_ISL_1732419, EPI_ISL_1732420, EPI_ISL_1732421, EPI_ISL_1732422, EPI_ISL_1732423, EPI_ISL_1732424, EPI_ISL_1732425, EPI_ISL_1732426, EPI_ISL_1732427, EPI_ISL_1732428, EPI_ISL_1732429, EPI_ISL_1732430, EPI_ISL_1732431, EPI_ISL_1732432, EPI_ISL_1732433, EPI_ISL_1732434, EPI_ISL_1732435, EPI_ISL_1732436, EPI_ISL_1732437, EPI_ISL_1732438, EPI_ISL_1732439, EPI_ISL_1732440, EPI_ISL_1732441, EPI_ISL_1732442, EPI_ISL_1732443, EPI_ISL_1732444, EPI_ISL_1732445, EPI_ISL_1732446, EPI_ISL_1732447, EPI_ISL_1732448, EPI_ISL_1732449, EPI_ISL_1732450, EPI_ISL_1732451, EPI_ISL_1732452, EPI_ISL_1732453, EPI_ISL_1732454, EPI_ISL_1732455, EPI_ISL_1732456, EPI_ISL_1732457, EPI_ISL_1732458, EPI_ISL_1732459, EPI_ISL_1732460, EPI_ISL_1732461, EPI_ISL_1732462, EPI_ISL_1732463, EPI_ISL_1732464, EPI_ISL_1732465, EPI_ISL_1732466, EPI_ISL_1732467, EPI_ISL_1732468, EPI_ISL_1732469, EPI_ISL_1732470, EPI_ISL_1732471, EPI_ISL_1732472, EPI_ISL_1732473, EPI_ISL_1732474, EPI_ISL_1732475, EPI_ISL_1732476, EPI_ISL_1732477, EPI_ISL_1732478, EPI_ISL_1732479, EPI_ISL_1732480, EPI_ISL_1732481, EPI_ISL_1732482, EPI_ISL_1732483, EPI_ISL_1732484, EPI_ISL_1732485, EPI_ISL_1732486, EPI_ISL_1732487, EPI_ISL_1732488, EPI_ISL_1732489, EPI_ISL_1732490, EPI_ISL_1732491, EPI_ISL_1732492, EPI_ISL_1732493, EPI_ISL_1732494, EPI_ISL_1732495, EPI_ISL_1732496, EPI_ISL_1732497, EPI_ISL_1732498, EPI_ISL_1732499, EPI_ISL_1732500, EPI_ISL_1732501, EPI_ISL_1732502, EPI_ISL_1732503, EPI_ISL_1732504, EPI_ISL_1732505, EPI_ISL_1732506, EPI_ISL_1732507, EPI_ISL_1732508, EPI_ISL_1732509, EPI_ISL_1732510, EPI_ISL_1732511, EPI_ISL_1732512, EPI_ISL_1732513, EPI_ISL_1732514, EPI_ISL_1732515, EPI_ISL_1732516, EPI_ISL_1732517 | | | |
| see above | UPMC Clinical Microbiology Laboratory | Microbial Genome Sequencing Center; Microbial Genomic Epidemiology Laboratory, University of Pittsburgh | Lee H. Harrison, Jane W. Marsh, Marissa P. Griffith, Stephanie L. Mitchell, Vatsala R. Srinivasa, Kady D. Waggle, Daniel J. Snyder, Vaughn S. Cooper |
| EPI_ISL_1732542, EPI_ISL_1732543, EPI_ISL_1732544, EPI_ISL_1732545, EPI_ISL_1732546, EPI_ISL_1732547, EPI_ISL_1732548, EPI_ISL_1732549, EPI_ISL_1732550, EPI_ISL_1732551, EPI_ISL_1732552, EPI_ISL_1732553, EPI_ISL_1732554, EPI_ISL_1732555, EPI_ISL_1732556, EPI_ISL_1732557, EPI_ISL_1732558, EPI_ISL_1732559, EPI_ISL_1732560, EPI_ISL_1732561, EPI_ISL_1732562, EPI_ISL_1732563, EPI_ISL_1732564, EPI_ISL_1732565, EPI_ISL_1732566, EPI_ISL_1732568, EPI_ISL_1732569, EPI_ISL_1732570, EPI_ISL_1732572, EPI_ISL_1732573, EPI_ISL_1732574, EPI_ISL_1732575, EPI_ISL_1732576, EPI_ISL_1732578, EPI_ISL_1732579, EPI_ISL_1732580, EPI_ISL_1732581, EPI_ISL_1732582, EPI_ISL_1732583, EPI_ISL_1732584, EPI_ISL_1732585, EPI_ISL_1732587, EPI_ISL_1732588, EPI_ISL_1732589, EPI_ISL_1732590, EPI_ISL_1732591, EPI_ISL_1732592, EPI_ISL_1732593, EPI_ISL_1732594, EPI_ISL_1732595, EPI_ISL_1732596, EPI_ISL_1732597, EPI_ISL_1732598, EPI_ISL_1732599, EPI_ISL_1732600, EPI_ISL_1732601, EPI_ISL_1732602, EPI_ISL_1732603, EPI_ISL_1732604, EPI_ISL_1732605, EPI_ISL_1732606, EPI_ISL_1732607, EPI_ISL_1732608, EPI_ISL_1732609, EPI_ISL_1732610, EPI_ISL_1732611, EPI_ISL_1732612, EPI_ISL_1732613, EPI_ISL_1732614, EPI_ISL_1732615, EPI_ISL_1732617, EPI_ISL_1732618, EPI_ISL_1732619, EPI_ISL_1732620, EPI_ISL_1732621, EPI_ISL_1732622, EPI_ISL_1732623, EPI_ISL_1732624, EPI_ISL_1732625, EPI_ISL_1732627, EPI_ISL_1732628, EPI_ISL_1732629, EPI_ISL_1732630 | | | |

| | | | |
|---|---|---|---|
| see above | Illinois Department of Public Health - Springfield Lab | Illinois Department of Public Health - Springfield Lab | Bryan Sim, Gordon McCall |
| EPI_ISL_1732633, EPI_ISL_1732634, EPI_ISL_1732635, EPI_ISL_1732636, EPI_ISL_1732637, EPI_ISL_1732638, EPI_ISL_1732639, EPI_ISL_1732640, EPI_ISL_1732641, EPI_ISL_1732642, EPI_ISL_1732643, EPI_ISL_1732644, EPI_ISL_1732645, EPI_ISL_1732646, EPI_ISL_1732647, EPI_ISL_1732649, EPI_ISL_1732650, EPI_ISL_1732651, EPI_ISL_1732652, EPI_ISL_1732653, EPI_ISL_1732654, EPI_ISL_1732655, EPI_ISL_1732656, EPI_ISL_1732657, EPI_ISL_1732658, EPI_ISL_1732659, EPI_ISL_1732660, EPI_ISL_1732661, EPI_ISL_1732662, EPI_ISL_1732663, EPI_ISL_1732664, EPI_ISL_1732665, EPI_ISL_1732666, EPI_ISL_1732667, EPI_ISL_1732668, EPI_ISL_1732669, EPI_ISL_1732670, EPI_ISL_1732671, EPI_ISL_1732672, EPI_ISL_1732673, EPI_ISL_1732674, EPI_ISL_1732675, EPI_ISL_1732676, EPI_ISL_1732677, EPI_ISL_1732678, EPI_ISL_1732679, EPI_ISL_1732680, EPI_ISL_1732681, EPI_ISL_1732682, EPI_ISL_1732683, EPI_ISL_1732684, EPI_ISL_1732685, EPI_ISL_1732686, EPI_ISL_1732687, EPI_ISL_1732688, EPI_ISL_1732690, EPI_ISL_1732691, EPI_ISL_1732692, EPI_ISL_1732693, EPI_ISL_1732694, EPI_ISL_1732695, EPI_ISL_1732696, EPI_ISL_1732697, EPI_ISL_1732698, EPI_ISL_1732699, EPI_ISL_1732700, EPI_ISL_1732701, EPI_ISL_1732702, EPI_ISL_1732703, EPI_ISL_1732704, EPI_ISL_1732705, EPI_ISL_1732706, EPI_ISL_1732707, EPI_ISL_1732708, EPI_ISL_1732709, EPI_ISL_1732710, EPI_ISL_1732711, EPI_ISL_1732712, EPI_ISL_1732713, EPI_ISL_1732714, EPI_ISL_1732715, EPI_ISL_1732716, EPI_ISL_1732717, EPI_ISL_1732718, EPI_ISL_1732719, EPI_ISL_1732720, EPI_ISL_1732721, EPI_ISL_1732722, EPI_ISL_1732723, EPI_ISL_1732724, EPI_ISL_1732725, EPI_ISL_1732726, EPI_ISL_1732727, EPI_ISL_1732728, EPI_ISL_1732729, EPI_ISL_1732730, EPI_ISL_1732731, EPI_ISL_1732732, EPI_ISL_1732733, EPI_ISL_1732734, EPI_ISL_1732735, EPI_ISL_1732736, EPI_ISL_1732737, EPI_ISL_1732738, EPI_ISL_1732739, EPI_ISL_1732740, EPI_ISL_1732741, EPI_ISL_1732743, EPI_ISL_1732744, EPI_ISL_1732745, EPI_ISL_1732746, EPI_ISL_1732747, EPI_ISL_1732748, EPI_ISL_1732749, EPI_ISL_1732750, EPI_ISL_1732751, EPI_ISL_1732752, EPI_ISL_1732753, EPI_ISL_1732754, EPI_ISL_1732755, EPI_ISL_1732756, EPI_ISL_1732757, EPI_ISL_1732758, EPI_ISL_1732759, EPI_ISL_1732760, EPI_ISL_1732762, EPI_ISL_1732763, EPI_ISL_1732764, EPI_ISL_1732765, EPI_ISL_1732766, EPI_ISL_1732767, EPI_ISL_1732768, EPI_ISL_1732769, EPI_ISL_1732770, EPI_ISL_1732771, EPI_ISL_1732772, EPI_ISL_1732773, EPI_ISL_1732774, EPI_ISL_1732775, EPI_ISL_1732776, EPI_ISL_1732777, EPI_ISL_1732778, EPI_ISL_1732779, EPI_ISL_1732780, EPI_ISL_1732781, EPI_ISL_1732782, EPI_ISL_1732783, EPI_ISL_1732784, EPI_ISL_1732785, EPI_ISL_1732786, EPI_ISL_1732787, EPI_ISL_1732788, EPI_ISL_1732789, EPI_ISL_1732790, EPI_ISL_1732791, EPI_ISL_1732792, EPI_ISL_1732793, EPI_ISL_1732794, EPI_ISL_1732795, EPI_ISL_1732796, EPI_ISL_1732797, EPI_ISL_1732798, EPI_ISL_1732799, EPI_ISL_1732800, EPI_ISL_1732801, EPI_ISL_1732802, EPI_ISL_1732803, EPI_ISL_1732804, EPI_ISL_1732805, EPI_ISL_1732806, EPI_ISL_1732807, EPI_ISL_1732808, EPI_ISL_1732809, EPI_ISL_1732810, EPI_ISL_1732811, EPI_ISL_1732812, EPI_ISL_1732813, EPI_ISL_1732814, EPI_ISL_1732815, EPI_ISL_1732816, EPI_ISL_1732817, EPI_ISL_1732818, EPI_ISL_1732819, EPI_ISL_1732820, EPI_ISL_1732821, EPI_ISL_1732822, EPI_ISL_1732823, EPI_ISL_1732824, EPI_ISL_1732825, EPI_ISL_1732826, EPI_ISL_1732827, EPI_ISL_1732828, EPI_ISL_1732829, EPI_ISL_1732831, EPI_ISL_1732832, EPI_ISL_1732833, EPI_ISL_1732834, EPI_ISL_1732835, EPI_ISL_1732836, EPI_ISL_1732837, EPI_ISL_1732838, EPI_ISL_1732839, EPI_ISL_1732840, EPI_ISL_1732841, EPI_ISL_1732842, EPI_ISL_1732843, EPI_ISL_1732844, EPI_ISL_1732845, EPI_ISL_1732846, EPI_ISL_1732847, EPI_ISL_1732848, EPI_ISL_1732849, EPI_ISL_1732850, EPI_ISL_1732852, EPI_ISL_1732853, EPI_ISL_1732854, EPI_ISL_1732855, EPI_ISL_1732856, EPI_ISL_1732857, EPI_ISL_1732858, EPI_ISL_1732859, EPI_ISL_1732860, EPI_ISL_1732861, EPI_ISL_1732862, EPI_ISL_1732863, EPI_ISL_1732864, EPI_ISL_1732865, EPI_ISL_1732866, EPI_ISL_1732867, EPI_ISL_1732868, EPI_ISL_1732869, EPI_ISL_1732870, EPI_ISL_1732871, EPI_ISL_1732872, EPI_ISL_1732873, EPI_ISL_1732874, EPI_ISL_1732875, EPI_ISL_1732876, EPI_ISL_1732877, EPI_ISL_1732878, EPI_ISL_1732879, EPI_ISL_1732880, EPI_ISL_1732881, EPI_ISL_1732882, EPI_ISL_1732883, EPI_ISL_1732884, EPI_ISL_1732886, EPI_ISL_1732887, EPI_ISL_1732888, EPI_ISL_1732889, EPI_ISL_1732890, EPI_ISL_1732891, EPI_ISL_1732892, EPI_ISL_1732893, EPI_ISL_1732894, EPI_ISL_1732895, EPI_ISL_1732896, EPI_ISL_1732897, EPI_ISL_1732898, EPI_ISL_1732899, EPI_ISL_1732900, EPI_ISL_1732901, EPI_ISL_1732903, EPI_ISL_1732904, EPI_ISL_1732905, EPI_ISL_1732906, EPI_ISL_1732907, EPI_ISL_1732908, EPI_ISL_1732909, EPI_ISL_1732911, EPI_ISL_1732912, EPI_ISL_1732914, EPI_ISL_1732920, EPI_ISL_1732924, EPI_ISL_1732925, EPI_ISL_1732926, EPI_ISL_1732927, EPI_ISL_1732928, EPI_ISL_1732929, EPI_ISL_1732930, EPI_ISL_1732931, EPI_ISL_1732932, EPI_ISL_1732933, EPI_ISL_1732934, EPI_ISL_1732935, EPI_ISL_1732936, EPI_ISL_1732937, EPI_ISL_1732939, EPI_ISL_1732940, EPI_ISL_1732941, EPI_ISL_1732942, EPI_ISL_1732943, EPI_ISL_1732944, EPI_ISL_1732945, EPI_ISL_1732946, EPI_ISL_1732947, EPI_ISL_1732948 | | | |
| see above | Infinity Biologix | Centers for Disease Control and Prevention Division of Viral Diseases, Pathogen Discovery | Dakota Howard, Dhwani Batra, Peter W. Cook, Kara Moser, Adrian Paskey, Jason Caravas, Benjamin Rambo-Martin, Shatavia Morrison, Christopher Gulvick, Scott Sammons, Yvette Unoarumhi, Darlene Wagner, Matthew Schmerer, Christian Bixby, Yihe Wang, Jonathan Schultz, Chirayu Goswami, Russ Hager, Robin Grimwood, Clinton R. Paden, Duncan MacCannell |
| EPI_ISL_1732949 | Baylor Scott & White-Temple | Baylor Scott & White-Temple | Ari Rao, Linden Morales, Kimberly Walker, Marcus Volz, Shelby Hendrickson |
| EPI_ISL_1732950 | Infinity Biologix | Centers for Disease Control and Prevention Division of Viral Diseases, Pathogen Discovery | Dakota Howard, Dhwani Batra, Peter W. Cook, Kara Moser, Adrian Paskey, Jason Caravas, Benjamin Rambo-Martin, Shatavia Morrison, Christopher Gulvick, Scott Sammons, Yvette Unoarumhi, Darlene Wagner, Matthew Schmerer, Christian Bixby, Yihe Wang, Jonathan Schultz, Chirayu Goswami, Russ Hager, Robin Grimwood, Clinton R. Paden, Duncan MacCannell |
| EPI_ISL_1732951, EPI_ISL_1732952 | Baylor Scott & White-Temple | Baylor Scott & White-Temple | Ari Rao, Linden Morales, Kimberly Walker, Marcus Volz, Shelby Hendrickson |
| EPI_ISL_1732953 | Infinity Biologix | Centers for Disease Control and Prevention Division of Viral Diseases, Pathogen Discovery | Dakota Howard, Dhwani Batra, Peter W. Cook, Kara Moser, Adrian Paskey, Jason Caravas, Benjamin Rambo-Martin, Shatavia Morrison, Christopher Gulvick, Scott Sammons, Yvette Unoarumhi, Darlene Wagner, Matthew Schmerer, Christian Bixby, Yihe Wang, Jonathan Schultz, Chirayu Goswami, Russ Hager, Robin Grimwood, Clinton R. Paden, Duncan MacCannell |
| EPI_ISL_1732954 | Baylor Scott & White-Temple | Baylor Scott & White-Temple | Ari Rao, Linden Morales, Kimberly Walker, Marcus Volz, Shelby Hendrickson |
| EPI_ISL_1732955 | Infinity Biologix | Centers for Disease Control and Prevention Division of Viral Diseases, Pathogen Discovery | Dakota Howard, Dhwani Batra, Peter W. Cook, Kara Moser, Adrian Paskey, Jason Caravas, Benjamin Rambo-Martin, Shatavia Morrison, Christopher Gulvick, Scott Sammons, Yvette Unoarumhi, Darlene Wagner, Matthew Schmerer, Christian Bixby, Yihe Wang, Jonathan Schultz, Chirayu Goswami, Russ Hager, Robin Grimwood, Clinton R. Paden, Duncan MacCannell |
| EPI_ISL_1732956, EPI_ISL_1732957 | Baylor Scott & White-Temple | Baylor Scott & White-Temple | Ari Rao, Linden Morales, Kimberly Walker, Marcus Volz, Shelby Hendrickson |
| EPI_ISL_1732958 | Infinity Biologix | Centers for Disease Control and Prevention Division of Viral Diseases, Pathogen Discovery | Dakota Howard, Dhwani Batra, Peter W. Cook, Kara Moser, Adrian Paskey, Jason Caravas, Benjamin Rambo-Martin, Shatavia Morrison, Christopher Gulvick, Scott Sammons, Yvette Unoarumhi, Darlene Wagner, Matthew Schmerer, Christian Bixby, Yihe Wang, Jonathan Schultz, Chirayu Goswami, Russ Hager, Robin Grimwood, Clinton R. Paden, Duncan MacCannell |
| EPI_ISL_1732959, EPI_ISL_1732960 | Baylor Scott & White-Temple | Baylor Scott & White-Temple | Ari Rao, Linden Morales, Kimberly Walker, Marcus Volz, Shelby Hendrickson |
| EPI_ISL_1732961 | Infinity Biologix | Centers for Disease Control and Prevention Division of Viral Diseases, Pathogen Discovery | Dakota Howard, Dhwani Batra, Peter W. Cook, Kara Moser, Adrian Paskey, Jason Caravas, Benjamin Rambo-Martin, Shatavia Morrison, Christopher Gulvick, Scott Sammons, Yvette Unoarumhi, Darlene Wagner, Matthew Schmerer, Christian Bixby, Yihe Wang, Jonathan Schultz, Chirayu Goswami, Russ Hager, Robin Grimwood, Clinton R. Paden, Duncan MacCannell |
| EPI_ISL_1732962, EPI_ISL_1732963 | Baylor Scott & White-Temple | Baylor Scott & White-Temple | Ari Rao, Linden Morales, Kimberly Walker, Marcus Volz, Shelby Hendrickson |
| EPI_ISL_1732964 | Infinity Biologix | Centers for Disease Control and Prevention Division of Viral Diseases, Pathogen Discovery | Dakota Howard, Dhwani Batra, Peter W. Cook, Kara Moser, Adrian Paskey, Jason Caravas, Benjamin Rambo-Martin, Shatavia Morrison, Christopher Gulvick, Scott Sammons, Yvette Unoarumhi, Darlene Wagner, Matthew Schmerer, Christian Bixby, Yihe Wang, Jonathan Schultz, Chirayu Goswami, Russ Hager, Robin Grimwood, Clinton R. Paden, Duncan MacCannell |
| EPI_ISL_1732965, EPI_ISL_1732966 | Baylor Scott & White-Temple | Baylor Scott & White-Temple | Ari Rao, Linden Morales, Kimberly Walker, Marcus Volz, Shelby Hendrickson |
| EPI_ISL_1732967 | Infinity Biologix | Centers for Disease Control and Prevention Division of Viral Diseases, Pathogen Discovery | Dakota Howard, Dhwani Batra, Peter W. Cook, Kara Moser, Adrian Paskey, Jason Caravas, Benjamin Rambo-Martin, Shatavia Morrison, Christopher Gulvick, Scott Sammons, Yvette Unoarumhi, Darlene Wagner, Matthew Schmerer, Christian Bixby, Yihe Wang, Jonathan Schultz, Chirayu Goswami, Russ Hager, Robin Grimwood, Clinton R. Paden, Duncan MacCannell |
| EPI_ISL_1732969 | Baylor Scott & White-Temple | Baylor Scott & White-Temple | Ari Rao, Linden Morales, Kimberly Walker, Marcus Volz, Shelby Hendrickson |
| EPI_ISL_1732970 | Infinity Biologix | Centers for Disease Control and Prevention Division of Viral Diseases, Pathogen Discovery | Dakota Howard, Dhwani Batra, Peter W. Cook, Kara Moser, Adrian Paskey, Jason Caravas, Benjamin Rambo-Martin, Shatavia Morrison, Christopher Gulvick, Scott Sammons, Yvette Unoarumhi, Darlene Wagner, Matthew Schmerer, Christian Bixby, Yihe Wang, Jonathan Schultz, Chirayu Goswami, Russ Hager, Robin Grimwood, Clinton R. Paden, Duncan MacCannell |
| EPI_ISL_1732971, EPI_ISL_1732972 | Baylor Scott & White-Temple | Baylor Scott & White-Temple | Ari Rao, Linden Morales, Kimberly Walker, Marcus Volz, Shelby Hendrickson |
| EPI_ISL_1732973 | Infinity Biologix | Centers for Disease Control and Prevention Division of Viral Diseases, Pathogen Discovery | Dakota Howard, Dhwani Batra, Peter W. Cook, Kara Moser, Adrian Paskey, Jason Caravas, Benjamin Rambo-Martin, Shatavia Morrison, Christopher Gulvick, Scott Sammons, Yvette Unoarumhi, Darlene Wagner, Matthew Schmerer, Christian Bixby, Yihe Wang, Jonathan Schultz, Chirayu Goswami, Russ Hager, Robin Grimwood, Clinton R. Paden, Duncan MacCannell |
| EPI_ISL_1732974, EPI_ISL_1732975 | Baylor Scott & White-Temple | Baylor Scott & White-Temple | Ari Rao, Linden Morales, Kimberly Walker, Marcus Volz, Shelby Hendrickson |
| EPI_ISL_1732976 | Infinity Biologix | Centers for Disease Control and Prevention Division of Viral Diseases, Pathogen Discovery | Dakota Howard, Dhwani Batra, Peter W. Cook, Kara Moser, Adrian Paskey, Jason Caravas, Benjamin Rambo-Martin, Shatavia Morrison, Christopher Gulvick, Scott Sammons, Yvette Unoarumhi, Darlene Wagner, Matthew Schmerer, Christian Bixby, Yihe Wang, Jonathan Schultz, Chirayu Goswami, Russ Hager, Robin Grimwood, Clinton R. Paden, Duncan MacCannell |
| EPI_ISL_1732977, EPI_ISL_1732978 | Baylor Scott & White-Temple | Baylor Scott & White-Temple | Ari Rao, Linden Morales, Kimberly Walker, Marcus Volz, Shelby Hendrickson |
| EPI_ISL_1732979 | Infinity Biologix | Centers for Disease Control and Prevention Division of Viral Diseases, Pathogen Discovery | Dakota Howard, Dhwani Batra, Peter W. Cook, Kara Moser, Adrian Paskey, Jason Caravas, Benjamin Rambo-Martin, Shatavia Morrison, Christopher Gulvick, Scott Sammons, Yvette Unoarumhi, Darlene Wagner, Matthew Schmerer, Christian Bixby, Yihe Wang, Jonathan Schultz, Chirayu Goswami, Russ Hager, Robin Grimwood, Clinton R. Paden, Duncan MacCannell |
| EPI_ISL_1732980, EPI_ISL_1732981 | Baylor Scott & White-Temple | Baylor Scott & White-Temple | Ari Rao, Linden Morales, Kimberly Walker, Marcus Volz, Shelby Hendrickson |
| EPI_ISL_1732982 | Infinity Biologix | Centers for Disease Control and Prevention Division of Viral Diseases, Pathogen Discovery | Dakota Howard, Dhwani Batra, Peter W. Cook, Kara Moser, Adrian Paskey, Jason Caravas, Benjamin Rambo-Martin, Shatavia Morrison, Christopher Gulvick, Scott Sammons, Yvette Unoarumhi, Darlene Wagner, Matthew Schmerer, Christian Bixby, Yihe Wang, Jonathan Schultz, Chirayu Goswami, Russ Hager, Robin Grimwood, Clinton R. Paden, Duncan MacCannell |
| EPI_ISL_1732983, EPI_ISL_1732984 | Baylor Scott & White-Temple | Baylor Scott & White-Temple | Ari Rao, Linden Morales, Kimberly Walker, Marcus Volz, Shelby Hendrickson |
| EPI_ISL_1732985 | Infinity Biologix | Centers for Disease Control and Prevention Division of Viral Diseases, Pathogen Discovery | Dakota Howard, Dhwani Batra, Peter W. Cook, Kara Moser, Adrian Paskey, Jason Caravas, Benjamin Rambo-Martin, Shatavia Morrison, Christopher Gulvick, Scott Sammons, Yvette Unoarumhi, Darlene Wagner, Matthew Schmerer, Christian Bixby, Yihe Wang, Jonathan Schultz, Chirayu Goswami, Russ Hager, Robin Grimwood, Clinton R. Paden, Duncan MacCannell |
| EPI_ISL_1732986 | Baylor Scott & White-Temple | Baylor Scott & White-Temple | Ari Rao, Linden Morales, Kimberly Walker, Marcus Volz, Shelby Hendrickson |
| EPI_ISL_1732987 | Infinity Biologix | Centers for Disease Control and Prevention Division of Viral Diseases, Pathogen Discovery | Dakota Howard, Dhwani Batra, Peter W. Cook, Kara Moser, Adrian Paskey, Jason Caravas, Benjamin Rambo-Martin, Shatavia Morrison, Christopher Gulvick, Scott Sammons, Yvette Unoarumhi, Darlene Wagner, Matthew Schmerer, Christian Bixby, Yihe Wang, Jonathan Schultz, Chirayu Goswami, Russ Hager, Robin Grimwood, Clinton R. Paden, Duncan MacCannell |
| EPI_ISL_1732988, EPI_ISL_1732989 | Baylor Scott & White-Temple | Baylor Scott & White-Temple | Ari Rao, Linden Morales, Kimberly Walker, Marcus Volz, Shelby Hendrickson |
| EPI_ISL_1732990 | Infinity Biologix | Centers for Disease Control and Prevention Division of Viral Diseases, Pathogen Discovery | Dakota Howard, Dhwani Batra, Peter W. Cook, Kara Moser, Adrian Paskey, Jason Caravas, Benjamin Rambo-Martin, Shatavia Morrison, Christopher Gulvick, Scott Sammons, Yvette Unoarumhi, Darlene Wagner, Matthew Schmerer, Christian Bixby, Yihe Wang, Jonathan Schultz, Chirayu Goswami, Russ Hager, Robin Grimwood, Clinton R. Paden, Duncan MacCannell |
| EPI_ISL_1732991, EPI_ISL_1732992 | Baylor Scott & White-Temple | Baylor Scott & White-Temple | Ari Rao, Linden Morales, Kimberly Walker, Marcus Volz, Shelby Hendrickson |
| EPI_ISL_1732993 | Infinity Biologix | Centers for Disease Control and Prevention Division of Viral Diseases, Pathogen Discovery | Dakota Howard, Dhwani Batra, Peter W. Cook, Kara Moser, Adrian Paskey, Jason Caravas, Benjamin Rambo-Martin, Shatavia Morrison, Christopher Gulvick, Scott Sammons, Yvette Unoarumhi, Darlene Wagner, Matthew Schmerer, Christian Bixby, Yihe Wang, Jonathan Schultz, Chirayu Goswami, Russ Hager, Robin Grimwood, Clinton R. Paden, Duncan MacCannell |

| | | | |
|---|---|---|---|
| EPI_ISL_1732994, EPI_ISL_1732995 | Baylor Scott & White-Temple | Baylor Scott & White-Temple | Ari Rao, Linden Morales, Kimberly Walker, Marcus Volz, Shelby Hendrickson |
| EPI_ISL_1732996 | Infinity Biologix | Centers for Disease Control and Prevention Division of Viral Diseases, Pathogen Discovery | Dakota Howard, Dhwani Batra, Peter W. Cook, Kara Moser, Adrian Paskey, Jason Caravas, Benjamin Rambo-Martin, Shatavia Morrison, Christopher Gulvick, Scott Sammons, Yvette Unoarumhi, Darlene Wagner, Matthew Schmerer, Christian Bixby, Yihe Wang, Jonathan Schultz, Chirayu Goswami, Russ Hager, Robin Grimwood, Clinton R. Paden, Duncan MacCannell |
| EPI_ISL_1732997, EPI_ISL_1732998 | Baylor Scott & White-Temple | Baylor Scott & White-Temple | Ari Rao, Linden Morales, Kimberly Walker, Marcus Volz, Shelby Hendrickson |
| EPI_ISL_1732999 | Infinity Biologix | Centers for Disease Control and Prevention Division of Viral Diseases, Pathogen Discovery | Dakota Howard, Dhwani Batra, Peter W. Cook, Kara Moser, Adrian Paskey, Jason Caravas, Benjamin Rambo-Martin, Shatavia Morrison, Christopher Gulvick, Scott Sammons, Yvette Unoarumhi, Darlene Wagner, Matthew Schmerer, Christian Bixby, Yihe Wang, Jonathan Schultz, Chirayu Goswami, Russ Hager, Robin Grimwood, Clinton R. Paden, Duncan MacCannell |
| EPI_ISL_1733001 | Baylor Scott & White-Temple | Baylor Scott & White-Temple | Ari Rao, Linden Morales, Kimberly Walker, Marcus Volz, Shelby Hendrickson |
| EPI_ISL_1733002 | Infinity Biologix | Centers for Disease Control and Prevention Division of Viral Diseases, Pathogen Discovery | Dakota Howard, Dhwani Batra, Peter W. Cook, Kara Moser, Adrian Paskey, Jason Caravas, Benjamin Rambo-Martin, Shatavia Morrison, Christopher Gulvick, Scott Sammons, Yvette Unoarumhi, Darlene Wagner, Matthew Schmerer, Christian Bixby, Yihe Wang, Jonathan Schultz, Chirayu Goswami, Russ Hager, Robin Grimwood, Clinton R. Paden, Duncan MacCannell |
| EPI_ISL_1733003 | Baylor Scott & White-Temple | Baylor Scott & White-Temple | Ari Rao, Linden Morales, Kimberly Walker, Marcus Volz, Shelby Hendrickson |
| EPI_ISL_1733004 | Infinity Biologix | Centers for Disease Control and Prevention Division of Viral Diseases, Pathogen Discovery | Dakota Howard, Dhwani Batra, Peter W. Cook, Kara Moser, Adrian Paskey, Jason Caravas, Benjamin Rambo-Martin, Shatavia Morrison, Christopher Gulvick, Scott Sammons, Yvette Unoarumhi, Darlene Wagner, Matthew Schmerer, Christian Bixby, Yihe Wang, Jonathan Schultz, Chirayu Goswami, Russ Hager, Robin Grimwood, Clinton R. Paden, Duncan MacCannell |
| EPI_ISL_1733005 | Baylor Scott & White-Temple | Baylor Scott & White-Temple | Ari Rao, Linden Morales, Kimberly Walker, Marcus Volz, Shelby Hendrickson |
| EPI_ISL_1733006 | Infinity Biologix | Centers for Disease Control and Prevention Division of Viral Diseases, Pathogen Discovery | Dakota Howard, Dhwani Batra, Peter W. Cook, Kara Moser, Adrian Paskey, Jason Caravas, Benjamin Rambo-Martin, Shatavia Morrison, Christopher Gulvick, Scott Sammons, Yvette Unoarumhi, Darlene Wagner, Matthew Schmerer, Christian Bixby, Yihe Wang, Jonathan Schultz, Chirayu Goswami, Russ Hager, Robin Grimwood, Clinton R. Paden, Duncan MacCannell |
| EPI_ISL_1733007, EPI_ISL_1733008 | Baylor Scott & White-Temple | Baylor Scott & White-Temple | Ari Rao, Linden Morales, Kimberly Walker, Marcus Volz, Shelby Hendrickson |
| EPI_ISL_1733009 | Infinity Biologix | Centers for Disease Control and Prevention Division of Viral Diseases, Pathogen Discovery | Dakota Howard, Dhwani Batra, Peter W. Cook, Kara Moser, Adrian Paskey, Jason Caravas, Benjamin Rambo-Martin, Shatavia Morrison, Christopher Gulvick, Scott Sammons, Yvette Unoarumhi, Darlene Wagner, Matthew Schmerer, Christian Bixby, Yihe Wang, Jonathan Schultz, Chirayu Goswami, Russ Hager, Robin Grimwood, Clinton R. Paden, Duncan MacCannell |
| EPI_ISL_1733010, EPI_ISL_1733011 | Baylor Scott & White-Temple | Baylor Scott & White-Temple | Ari Rao, Linden Morales, Kimberly Walker, Marcus Volz, Shelby Hendrickson |
| EPI_ISL_1733012 | Infinity Biologix | Centers for Disease Control and Prevention Division of Viral Diseases, Pathogen Discovery | Dakota Howard, Dhwani Batra, Peter W. Cook, Kara Moser, Adrian Paskey, Jason Caravas, Benjamin Rambo-Martin, Shatavia Morrison, Christopher Gulvick, Scott Sammons, Yvette Unoarumhi, Darlene Wagner, Matthew Schmerer, Christian Bixby, Yihe Wang, Jonathan Schultz, Chirayu Goswami, Russ Hager, Robin Grimwood, Clinton R. Paden, Duncan MacCannell |
| EPI_ISL_1733013, EPI_ISL_1733014 | Baylor Scott & White-Temple | Baylor Scott & White-Temple | Ari Rao, Linden Morales, Kimberly Walker, Marcus Volz, Shelby Hendrickson |
| EPI_ISL_1733015 | Infinity Biologix | Centers for Disease Control and Prevention Division of Viral Diseases, Pathogen Discovery | Dakota Howard, Dhwani Batra, Peter W. Cook, Kara Moser, Adrian Paskey, Jason Caravas, Benjamin Rambo-Martin, Shatavia Morrison, Christopher Gulvick, Scott Sammons, Yvette Unoarumhi, Darlene Wagner, Matthew Schmerer, Christian Bixby, Yihe Wang, Jonathan Schultz, Chirayu Goswami, Russ Hager, Robin Grimwood, Clinton R. Paden, Duncan MacCannell |
| EPI_ISL_1733017 | Baylor Scott & White-Temple | Baylor Scott & White-Temple | Ari Rao, Linden Morales, Kimberly Walker, Marcus Volz, Shelby Hendrickson |
| EPI_ISL_1733018 | Infinity Biologix | Centers for Disease Control and Prevention Division of Viral Diseases, Pathogen Discovery | Dakota Howard, Dhwani Batra, Peter W. Cook, Kara Moser, Adrian Paskey, Jason Caravas, Benjamin Rambo-Martin, Shatavia Morrison, Christopher Gulvick, Scott Sammons, Yvette Unoarumhi, Darlene Wagner, Matthew Schmerer, Christian Bixby, Yihe Wang, Jonathan Schultz, Chirayu Goswami, Russ Hager, Robin Grimwood, Clinton R. Paden, Duncan MacCannell |
| EPI_ISL_1733019, EPI_ISL_1733020 | Baylor Scott & White-Temple | Baylor Scott & White-Temple | Ari Rao, Linden Morales, Kimberly Walker, Marcus Volz, Shelby Hendrickson |
| EPI_ISL_1733021 | Infinity Biologix | Centers for Disease Control and Prevention Division of Viral Diseases, Pathogen Discovery | Dakota Howard, Dhwani Batra, Peter W. Cook, Kara Moser, Adrian Paskey, Jason Caravas, Benjamin Rambo-Martin, Shatavia Morrison, Christopher Gulvick, Scott Sammons, Yvette Unoarumhi, Darlene Wagner, Matthew Schmerer, Christian Bixby, Yihe Wang, Jonathan Schultz, Chirayu Goswami, Russ Hager, Robin Grimwood, Clinton R. Paden, Duncan MacCannell |
| EPI_ISL_1733022, EPI_ISL_1733023 | Baylor Scott & White-Temple | Baylor Scott & White-Temple | Ari Rao, Linden Morales, Kimberly Walker, Marcus Volz, Shelby Hendrickson |
| EPI_ISL_1733024 | Infinity Biologix | Centers for Disease Control and Prevention Division of Viral Diseases, Pathogen Discovery | Dakota Howard, Dhwani Batra, Peter W. Cook, Kara Moser, Adrian Paskey, Jason Caravas, Benjamin Rambo-Martin, Shatavia Morrison, Christopher Gulvick, Scott Sammons, Yvette Unoarumhi, Darlene Wagner, Matthew Schmerer, Christian Bixby, Yihe Wang, Jonathan Schultz, Chirayu Goswami, Russ Hager, Robin Grimwood, Clinton R. Paden, Duncan MacCannell |
| EPI_ISL_1733025, EPI_ISL_1733026 | Baylor Scott & White-Temple | Baylor Scott & White-Temple | Ari Rao, Linden Morales, Kimberly Walker, Marcus Volz, Shelby Hendrickson |
| EPI_ISL_1733027 | Infinity Biologix | Centers for Disease Control and Prevention Division of Viral Diseases, Pathogen Discovery | Dakota Howard, Dhwani Batra, Peter W. Cook, Kara Moser, Adrian Paskey, Jason Caravas, Benjamin Rambo-Martin, Shatavia Morrison, Christopher Gulvick, Scott Sammons, Yvette Unoarumhi, Darlene Wagner, Matthew Schmerer, Christian Bixby, Yihe Wang, Jonathan Schultz, Chirayu Goswami, Russ Hager, Robin Grimwood, Clinton R. Paden, Duncan MacCannell |
| EPI_ISL_1733028, EPI_ISL_1733029 | Baylor Scott & White-Temple | Baylor Scott & White-Temple | Ari Rao, Linden Morales, Kimberly Walker, Marcus Volz, Shelby Hendrickson |
| EPI_ISL_1733030 | Infinity Biologix | Centers for Disease Control and Prevention Division of Viral Diseases, Pathogen Discovery | Dakota Howard, Dhwani Batra, Peter W. Cook, Kara Moser, Adrian Paskey, Jason Caravas, Benjamin Rambo-Martin, Shatavia Morrison, Christopher Gulvick, Scott Sammons, Yvette Unoarumhi, Darlene Wagner, Matthew Schmerer, Christian Bixby, Yihe Wang, Jonathan Schultz, Chirayu Goswami, Russ Hager, Robin Grimwood, Clinton R. Paden, Duncan MacCannell |
| EPI_ISL_1733031 | Baylor Scott & White-Temple | Baylor Scott & White-Temple | Ari Rao, Linden Morales, Kimberly Walker, Marcus Volz, Shelby Hendrickson |
| EPI_ISL_1733032 | Infinity Biologix | Centers for Disease Control and Prevention Division of Viral Diseases, Pathogen Discovery | Dakota Howard, Dhwani Batra, Peter W. Cook, Kara Moser, Adrian Paskey, Jason Caravas, Benjamin Rambo-Martin, Shatavia Morrison, Christopher Gulvick, Scott Sammons, Yvette Unoarumhi, Darlene Wagner, Matthew Schmerer, Christian Bixby, Yihe Wang, Jonathan Schultz, Chirayu Goswami, Russ Hager, Robin Grimwood, Clinton R. Paden, Duncan MacCannell |
| EPI_ISL_1733033, EPI_ISL_1733034 | Baylor Scott & White-Temple | Baylor Scott & White-Temple | Ari Rao, Linden Morales, Kimberly Walker, Marcus Volz, Shelby Hendrickson |
| EPI_ISL_1733035 | Infinity Biologix | Centers for Disease Control and Prevention Division of Viral Diseases, Pathogen Discovery | Dakota Howard, Dhwani Batra, Peter W. Cook, Kara Moser, Adrian Paskey, Jason Caravas, Benjamin Rambo-Martin, Shatavia Morrison, Christopher Gulvick, Scott Sammons, Yvette Unoarumhi, Darlene Wagner, Matthew Schmerer, Christian Bixby, Yihe Wang, Jonathan Schultz, Chirayu Goswami, Russ Hager, Robin Grimwood, Clinton R. Paden, Duncan MacCannell |
| EPI_ISL_1733037 | Baylor Scott & White-Temple | Baylor Scott & White-Temple | Ari Rao, Linden Morales, Kimberly Walker, Marcus Volz, Shelby Hendrickson |
| EPI_ISL_1733038 | Infinity Biologix | Centers for Disease Control and Prevention Division of Viral Diseases, Pathogen Discovery | Dakota Howard, Dhwani Batra, Peter W. Cook, Kara Moser, Adrian Paskey, Jason Caravas, Benjamin Rambo-Martin, Shatavia Morrison, Christopher Gulvick, Scott Sammons, Yvette Unoarumhi, Darlene Wagner, Matthew Schmerer, Christian Bixby, Yihe Wang, Jonathan Schultz, Chirayu Goswami, Russ Hager, Robin Grimwood, Clinton R. Paden, Duncan MacCannell |
| EPI_ISL_1733039, EPI_ISL_1733040 | Baylor Scott & White-Temple | Baylor Scott & White-Temple | Ari Rao, Linden Morales, Kimberly Walker, Marcus Volz, Shelby Hendrickson |
| EPI_ISL_1733041 | Infinity Biologix | Centers for Disease Control and Prevention Division of Viral Diseases, Pathogen Discovery | Dakota Howard, Dhwani Batra, Peter W. Cook, Kara Moser, Adrian Paskey, Jason Caravas, Benjamin Rambo-Martin, Shatavia Morrison, Christopher Gulvick, Scott Sammons, Yvette Unoarumhi, Darlene Wagner, Matthew Schmerer, Christian Bixby, Yihe Wang, Jonathan Schultz, Chirayu Goswami, Russ Hager, Robin Grimwood, Clinton R. Paden, Duncan MacCannell |
| EPI_ISL_1733042 | Baylor Scott & White-Temple | Baylor Scott & White-Temple | Ari Rao, Linden Morales, Kimberly Walker, Marcus Volz, Shelby Hendrickson |
| EPI_ISL_1733043 | Infinity Biologix | Centers for Disease Control and Prevention Division of Viral Diseases, Pathogen Discovery | Dakota Howard, Dhwani Batra, Peter W. Cook, Kara Moser, Adrian Paskey, Jason Caravas, Benjamin Rambo-Martin, Shatavia Morrison, Christopher Gulvick, Scott Sammons, Yvette Unoarumhi, Darlene Wagner, Matthew Schmerer, Christian Bixby, Yihe Wang, Jonathan Schultz, Chirayu Goswami, Russ Hager, Robin Grimwood, Clinton R. Paden, Duncan MacCannell |
| EPI_ISL_1733045 | Baylor Scott & White-Temple | Baylor Scott & White-Temple | Ari Rao, Linden Morales, Kimberly Walker, Marcus Volz, Shelby Hendrickson |
| EPI_ISL_1733046 | Infinity Biologix | Centers for Disease Control and Prevention Division of Viral Diseases, Pathogen Discovery | Dakota Howard, Dhwani Batra, Peter W. Cook, Kara Moser, Adrian Paskey, Jason Caravas, Benjamin Rambo-Martin, Shatavia Morrison, Christopher Gulvick, Scott Sammons, Yvette Unoarumhi, Darlene Wagner, Matthew Schmerer, Christian Bixby, Yihe Wang, Jonathan Schultz, Chirayu Goswami, Russ Hager, Robin Grimwood, Clinton R. Paden, Duncan MacCannell |
| EPI_ISL_1733047, EPI_ISL_1733048 | Baylor Scott & White-Temple | Baylor Scott & White-Temple | Ari Rao, Linden Morales, Kimberly Walker, Marcus Volz, Shelby Hendrickson |
| EPI_ISL_1733049 | Infinity Biologix | Centers for Disease Control and Prevention Division of Viral Diseases, Pathogen Discovery | Dakota Howard, Dhwani Batra, Peter W. Cook, Kara Moser, Adrian Paskey, Jason Caravas, Benjamin Rambo-Martin, Shatavia Morrison, Christopher Gulvick, Scott Sammons, Yvette Unoarumhi, Darlene Wagner, Matthew Schmerer, Christian Bixby, Yihe Wang, Jonathan Schultz, Chirayu Goswami, Russ Hager, Robin Grimwood, Clinton R. Paden, Duncan MacCannell |
| EPI_ISL_1733050, | Baylor Scott & White-Temple | Baylor Scott & White-Temple | Ari Rao, Linden Morales, Kimberly Walker, Marcus Volz, Shelby Hendrickson |

| | | | |
|---|---|---|---|
| EPI_ISL_1733051 | | | |
| EPI_ISL_1733052 | Infinity Biologix | Centers for Disease Control and Prevention Division of Viral Diseases, Pathogen Discovery | Dakota Howard, Dhwani Batra, Peter W. Cook, Kara Moser, Adrian Paskey, Jason Caravas, Benjamin Rambo-Martin, Shatavia Morrison, Christopher Gulvick, Scott Sammons, Yvette Unoarumhi, Darlene Wagner, Matthew Schmerer, Christian Bixby, Yihe Wang, Jonathan Schultz, Chirayu Goswami, Russ Hager, Robin Grimwood, Clinton R. Paden, Duncan MacCannell |
| EPI_ISL_1733053 | Baylor Scott & White-Temple | Baylor Scott & White-Temple | Ari Rao, Linden Morales, Kimberly Walker, Marcus Volz, Shelby Hendrickson |
| EPI_ISL_1733054 | Infinity Biologix | Centers for Disease Control and Prevention Division of Viral Diseases, Pathogen Discovery | Dakota Howard, Dhwani Batra, Peter W. Cook, Kara Moser, Adrian Paskey, Jason Caravas, Benjamin Rambo-Martin, Shatavia Morrison, Christopher Gulvick, Scott Sammons, Yvette Unoarumhi, Darlene Wagner, Matthew Schmerer, Christian Bixby, Yihe Wang, Jonathan Schultz, Chirayu Goswami, Russ Hager, Robin Grimwood, Clinton R. Paden, Duncan MacCannell |
| EPI_ISL_1733055 | Baylor Scott & White-Temple | Baylor Scott & White-Temple | Ari Rao, Linden Morales, Kimberly Walker, Marcus Volz, Shelby Hendrickson |
| EPI_ISL_1733056 | Infinity Biologix | Centers for Disease Control and Prevention Division of Viral Diseases, Pathogen Discovery | Dakota Howard, Dhwani Batra, Peter W. Cook, Kara Moser, Adrian Paskey, Jason Caravas, Benjamin Rambo-Martin, Shatavia Morrison, Christopher Gulvick, Scott Sammons, Yvette Unoarumhi, Darlene Wagner, Matthew Schmerer, Christian Bixby, Yihe Wang, Jonathan Schultz, Chirayu Goswami, Russ Hager, Robin Grimwood, Clinton R. Paden, Duncan MacCannell |
| EPI_ISL_1733057, EPI_ISL_1733058 | Baylor Scott & White-Temple | Baylor Scott & White-Temple | Ari Rao, Linden Morales, Kimberly Walker, Marcus Volz, Shelby Hendrickson |
| EPI_ISL_1733059 | Infinity Biologix | Centers for Disease Control and Prevention Division of Viral Diseases, Pathogen Discovery | Dakota Howard, Dhwani Batra, Peter W. Cook, Kara Moser, Adrian Paskey, Jason Caravas, Benjamin Rambo-Martin, Shatavia Morrison, Christopher Gulvick, Scott Sammons, Yvette Unoarumhi, Darlene Wagner, Matthew Schmerer, Christian Bixby, Yihe Wang, Jonathan Schultz, Chirayu Goswami, Russ Hager, Robin Grimwood, Clinton R. Paden, Duncan MacCannell |
| EPI_ISL_1733060, EPI_ISL_1733061 | Baylor Scott & White-Temple | Baylor Scott & White-Temple | Ari Rao, Linden Morales, Kimberly Walker, Marcus Volz, Shelby Hendrickson |
| EPI_ISL_1733062 | Infinity Biologix | Centers for Disease Control and Prevention Division of Viral Diseases, Pathogen Discovery | Dakota Howard, Dhwani Batra, Peter W. Cook, Kara Moser, Adrian Paskey, Jason Caravas, Benjamin Rambo-Martin, Shatavia Morrison, Christopher Gulvick, Scott Sammons, Yvette Unoarumhi, Darlene Wagner, Matthew Schmerer, Christian Bixby, Yihe Wang, Jonathan Schultz, Chirayu Goswami, Russ Hager, Robin Grimwood, Clinton R. Paden, Duncan MacCannell |
| EPI_ISL_1733063 | Baylor Scott & White-Temple | Baylor Scott & White-Temple | Ari Rao, Linden Morales, Kimberly Walker, Marcus Volz, Shelby Hendrickson |
| EPI_ISL_1733064 | Infinity Biologix | Centers for Disease Control and Prevention Division of Viral Diseases, Pathogen Discovery | Dakota Howard, Dhwani Batra, Peter W. Cook, Kara Moser, Adrian Paskey, Jason Caravas, Benjamin Rambo-Martin, Shatavia Morrison, Christopher Gulvick, Scott Sammons, Yvette Unoarumhi, Darlene Wagner, Matthew Schmerer, Christian Bixby, Yihe Wang, Jonathan Schultz, Chirayu Goswami, Russ Hager, Robin Grimwood, Clinton R. Paden, Duncan MacCannell |
| EPI_ISL_1733065 | Baylor Scott & White-Temple | Baylor Scott & White-Temple | Ari Rao, Linden Morales, Kimberly Walker, Marcus Volz, Shelby Hendrickson |

EPI_ISL_1733066, EPI_ISL_1733068, EPI_ISL_1733069, EPI_ISL_1733070, EPI_ISL_1733071, EPI_ISL_1733072, EPI_ISL_1733073, EPI_ISL_1733074, EPI_ISL_1733075, EPI_ISL_1733076, EPI_ISL_1733077, EPI_ISL_1733078, EPI_ISL_1733079, EPI_ISL_1733080, EPI_ISL_1733081, EPI_ISL_1733082, EPI_ISL_1733083, EPI_ISL_1733084, EPI_ISL_1733085, EPI_ISL_1733086, EPI_ISL_1733087, EPI_ISL_1733088, EPI_ISL_1733089, EPI_ISL_1733090, EPI_ISL_1733091, EPI_ISL_1733092, EPI_ISL_1733093, EPI_ISL_1733094, EPI_ISL_1733095, EPI_ISL_1733096, EPI_ISL_1733097, EPI_ISL_1733098, EPI_ISL_1733099, EPI_ISL_1733100, EPI_ISL_1733101, EPI_ISL_1733102, EPI_ISL_1733103, EPI_ISL_1733104, EPI_ISL_1733105, EPI_ISL_1733106, EPI_ISL_1733107, EPI_ISL_1733108, EPI_ISL_1733109, EPI_ISL_1733110, EPI_ISL_1733111, EPI_ISL_1733112, EPI_ISL_1733113, EPI_ISL_1733114, EPI_ISL_1733115, EPI_ISL_1733116, EPI_ISL_1733117, EPI_ISL_1733118, EPI_ISL_1733120, EPI_ISL_1733121, EPI_ISL_1733122, EPI_ISL_1733123, EPI_ISL_1733124, EPI_ISL_1733125, EPI_ISL_1733126, EPI_ISL_1733127, EPI_ISL_1733128, EPI_ISL_1733129, EPI_ISL_1733130, EPI_ISL_1733131, EPI_ISL_1733133, EPI_ISL_1733134, EPI_ISL_1733135, EPI_ISL_1733136, EPI_ISL_1733137, EPI_ISL_1733138, EPI_ISL_1733139, EPI_ISL_1733140, EPI_ISL_1733141, EPI_ISL_1733142, EPI_ISL_1733143, EPI_ISL_1733144, EPI_ISL_1733145, EPI_ISL_1733146, EPI_ISL_1733147, EPI_ISL_1733148, EPI_ISL_1733149, EPI_ISL_1733150, EPI_ISL_1733151, EPI_ISL_1733153, EPI_ISL_1733154, EPI_ISL_1733155, EPI_ISL_1733156, EPI_ISL_1733157, EPI_ISL_1733158, EPI_ISL_1733159, EPI_ISL_1733160, EPI_ISL_1733161, EPI_ISL_1733162, EPI_ISL_1733163, EPI_ISL_1733164, EPI_ISL_1733165, EPI_ISL_1733166, EPI_ISL_1733167, EPI_ISL_1733168, EPI_ISL_1733169, EPI_ISL_1733170, EPI_ISL_1733171, EPI_ISL_1733172, EPI_ISL_1733173, EPI_ISL_1733174, EPI_ISL_1733175, EPI_ISL_1733176, EPI_ISL_1733177, EPI_ISL_1733178, EPI_ISL_1733179, EPI_ISL_1733180, EPI_ISL_1733181, EPI_ISL_1733182, EPI_ISL_1733183, EPI_ISL_1733184, EPI_ISL_1733185, EPI_ISL_1733186, EPI_ISL_1733187, EPI_ISL_1733188, EPI_ISL_1733189, EPI_ISL_1733190, EPI_ISL_1733191, EPI_ISL_1733192, EPI_ISL_1733193, EPI_ISL_1733194, EPI_ISL_1733195, EPI_ISL_1733196, EPI_ISL_1733197, EPI_ISL_1733198, EPI_ISL_1733199, EPI_ISL_1733200, EPI_ISL_1733201, EPI_ISL_1733202, EPI_ISL_1733203, EPI_ISL_1733204, EPI_ISL_1733205, EPI_ISL_1733206, EPI_ISL_1733207, EPI_ISL_1733208, EPI_ISL_1733209, EPI_ISL_1733210, EPI_ISL_1733211, EPI_ISL_1733212, EPI_ISL_1733214, EPI_ISL_1733215, EPI_ISL_1733216, EPI_ISL_1733217, EPI_ISL_1733218, EPI_ISL_1733219, EPI_ISL_1733220, EPI_ISL_1733221, EPI_ISL_1733222, EPI_ISL_1733223, EPI_ISL_1733224, EPI_ISL_1733225, EPI_ISL_1733226, EPI_ISL_1733227, EPI_ISL_1733228, EPI_ISL_1733229, EPI_ISL_1733230, EPI_ISL_1733231, EPI_ISL_1733232, EPI_ISL_1733233, EPI_ISL_1733234, EPI_ISL_1733235, EPI_ISL_1733236, EPI_ISL_1733237, EPI_ISL_1733238, EPI_ISL_1733239, EPI_ISL_1733240, EPI_ISL_1733241, EPI_ISL_1733242, EPI_ISL_1733243, EPI_ISL_1733244, EPI_ISL_1733245, EPI_ISL_1733246, EPI_ISL_1733247, EPI_ISL_1733248, EPI_ISL_1733249, EPI_ISL_1733250, EPI_ISL_1733251, EPI_ISL_1733252, EPI_ISL_1733253, EPI_ISL_1733254, EPI_ISL_1733255, EPI_ISL_1733257, EPI_ISL_1733258, EPI_ISL_1733259, EPI_ISL_1733260, EPI_ISL_1733261, EPI_ISL_1733262, EPI_ISL_1733263, EPI_ISL_1733264, EPI_ISL_1733265, EPI_ISL_1733266, EPI_ISL_1733267, EPI_ISL_1733268, EPI_ISL_1733269, EPI_ISL_1733270, EPI_ISL_1733271, EPI_ISL_1733272, EPI_ISL_1733273, EPI_ISL_1733274, EPI_ISL_1733275, EPI_ISL_1733276, EPI_ISL_1733277, EPI_ISL_1733278, EPI_ISL_1733280, EPI_ISL_1733281, EPI_ISL_1733282, EPI_ISL_1733283, EPI_ISL_1733284, EPI_ISL_1733285, EPI_ISL_1733286, EPI_ISL_1733287, EPI_ISL_1733288, EPI_ISL_1733289, EPI_ISL_1733290, EPI_ISL_1733291, EPI_ISL_1733293, EPI_ISL_1733294, EPI_ISL_1733295, EPI_ISL_1733296, EPI_ISL_1733297, EPI_ISL_1733298, EPI_ISL_1733299, EPI_ISL_1733300, EPI_ISL_1733301, EPI_ISL_1733302, EPI_ISL_1733303, EPI_ISL_1733304, EPI_ISL_1733305, EPI_ISL_1733306, EPI_ISL_1733307, EPI_ISL_1733308, EPI_ISL_1733309, EPI_ISL_1733310, EPI_ISL_1733311, EPI_ISL_1733312, EPI_ISL_1733313, EPI_ISL_1733314, EPI_ISL_1733315, EPI_ISL_1733316, EPI_ISL_1733317, EPI_ISL_1733318, EPI_ISL_1733319, EPI_ISL_1733320, EPI_ISL_1733321, EPI_ISL_1733322, EPI_ISL_1733323, EPI_ISL_1733324, EPI_ISL_1733325, EPI_ISL_1733326, EPI_ISL_1733327, EPI_ISL_1733328, EPI_ISL_1733329, EPI_ISL_1733330, EPI_ISL_1733331, EPI_ISL_1733332, EPI_ISL_1733333, EPI_ISL_1733334, EPI_ISL_1733335, EPI_ISL_1733336, EPI_ISL_1733337, EPI_ISL_1733338, EPI_ISL_1733339, EPI_ISL_1733340, EPI_ISL_1733341, EPI_ISL_1733342, EPI_ISL_1733343, EPI_ISL_1733344, EPI_ISL_1733345, EPI_ISL_1733346, EPI_ISL_1733348, EPI_ISL_1733349, EPI_ISL_1733350, EPI_ISL_1733352, EPI_ISL_1733353, EPI_ISL_1733354, EPI_ISL_1733355, EPI_ISL_1733356, EPI_ISL_1733357, EPI_ISL_1733358, EPI_ISL_1733359, EPI_ISL_1733360, EPI_ISL_1733361, EPI_ISL_1733362, EPI_ISL_1733363, EPI_ISL_1733364, EPI_ISL_1733365, EPI_ISL_1733366, EPI_ISL_1733367, EPI_ISL_1733368, EPI_ISL_1733369, EPI_ISL_1733370, EPI_ISL_1733371, EPI_ISL_1733372, EPI_ISL_1733374, EPI_ISL_1733375, EPI_ISL_1733376, EPI_ISL_1733377, EPI_ISL_1733378, EPI_ISL_1733379, EPI_ISL_1733380, EPI_ISL_1733381, EPI_ISL_1733382, EPI_ISL_1733383, EPI_ISL_1733384, EPI_ISL_1733385, EPI_ISL_1733386, EPI_ISL_1733387, EPI_ISL_1733388, EPI_ISL_1733389, EPI_ISL_1733390, EPI_ISL_1733391, EPI_ISL_1733392, EPI_ISL_1733393, EPI_ISL_1733394, EPI_ISL_1733395, EPI_ISL_1733396, EPI_ISL_1733397, EPI_ISL_1733398, EPI_ISL_1733399, EPI_ISL_1733400, EPI_ISL_1733401, EPI_ISL_1733402, EPI_ISL_1733403, EPI_ISL_1733404, EPI_ISL_1733405, EPI_ISL_1733406, EPI_ISL_1733407, EPI_ISL_1733408, EPI_ISL_1733409, EPI_ISL_1733410, EPI_ISL_1733411, EPI_ISL_1733412, EPI_ISL_1733414, EPI_ISL_1733415, EPI_ISL_1733416, EPI_ISL_1733417, EPI_ISL_1733418, EPI_ISL_1733419, EPI_ISL_1733420, EPI_ISL_1733421, EPI_ISL_1733422, EPI_ISL_1733423, EPI_ISL_1733424, EPI_ISL_1733425, EPI_ISL_1733426, EPI_ISL_1733427, EPI_ISL_1733428, EPI_ISL_1733429, EPI_ISL_1733430, EPI_ISL_1733431, EPI_ISL_1733432, EPI_ISL_1733433, EPI_ISL_1733434, EPI_ISL_1733435, EPI_ISL_1733436, EPI_ISL_1733437, EPI_ISL_1733438, EPI_ISL_1733439, EPI_ISL_1733440, EPI_ISL_1733441, EPI_ISL_1733442, EPI_ISL_1733443, EPI_ISL_1733444, EPI_ISL_1733445, EPI_ISL_1733446, EPI_ISL_1733447, EPI_ISL_1733448, EPI_ISL_1733449, EPI_ISL_1733450, EPI_ISL_1733451, EPI_ISL_1733452, EPI_ISL_1733453, EPI_ISL_1733454, EPI_ISL_1733455, EPI_ISL_1733456, EPI_ISL_1733457, EPI_ISL_1733458, EPI_ISL_1733459, EPI_ISL_1733460, EPI_ISL_1733461, EPI_ISL_1733462, EPI_ISL_1733463, EPI_ISL_1733464

| | | | |
|---|---|---|---|
| see above | Infinity Biologix | Centers for Disease Control and Prevention Division of Viral Diseases, Pathogen Discovery | Dakota Howard, Dhwani Batra, Peter W. Cook, Kara Moser, Adrian Paskey, Jason Caravas, Benjamin Rambo-Martin, Shatavia Morrison, Christopher Gulvick, Scott Sammons, Yvette Unoarumhi, Darlene Wagner, Matthew Schmerer, Christian Bixby, Yihe Wang, Jonathan Schultz, Chirayu Goswami, Russ Hager, Robin Grimwood, Clinton R. Paden, Duncan MacCannell |
| EPI_ISL_1733465, EPI_ISL_1733466 | Minnesota Department of Health, Public Health Laboratory | Minnesota Department of Health, Public Health Laboratory | Alexandra Lorentz, Jacob Garfin, Matt Plumb, and Xiong Wang |
| EPI_ISL_1733467 | Infinity Biologix | Centers for Disease Control and Prevention Division of Viral Diseases, Pathogen Discovery | Dakota Howard, Dhwani Batra, Peter W. Cook, Kara Moser, Adrian Paskey, Jason Caravas, Benjamin Rambo-Martin, Shatavia Morrison, Christopher Gulvick, Scott Sammons, Yvette Unoarumhi, Darlene Wagner, Matthew Schmerer, Christian Bixby, Yihe Wang, Jonathan Schultz, Chirayu Goswami, Russ Hager, Robin Grimwood, Clinton R. Paden, Duncan MacCannell |
| EPI_ISL_1733468, EPI_ISL_1733470 | Minnesota Department of Health, Public Health Laboratory | Minnesota Department of Health, Public Health Laboratory | Alexandra Lorentz, Jacob Garfin, Matt Plumb, and Xiong Wang |
| EPI_ISL_1733471 | Infinity Biologix | Centers for Disease Control and Prevention Division of Viral Diseases, Pathogen Discovery | Dakota Howard, Dhwani Batra, Peter W. Cook, Kara Moser, Adrian Paskey, Jason Caravas, Benjamin Rambo-Martin, Shatavia Morrison, Christopher Gulvick, Scott Sammons, Yvette Unoarumhi, Darlene Wagner, Matthew Schmerer, Christian Bixby, Yihe Wang, Jonathan Schultz, Chirayu Goswami, Russ Hager, Robin Grimwood, Clinton R. Paden, Duncan MacCannell |
| EPI_ISL_1733472 | Minnesota Department of Health, Public Health Laboratory | Minnesota Department of Health, Public Health Laboratory | Alexandra Lorentz, Jacob Garfin, Matt Plumb, and Xiong Wang |
| EPI_ISL_1733473 | Infinity Biologix | Centers for Disease Control and Prevention Division of Viral Diseases, Pathogen Discovery | Dakota Howard, Dhwani Batra, Peter W. Cook, Kara Moser, Adrian Paskey, Jason Caravas, Benjamin Rambo-Martin, Shatavia Morrison, Christopher Gulvick, Scott Sammons, Yvette Unoarumhi, Darlene Wagner, Matthew Schmerer, Christian Bixby, Yihe Wang, Jonathan Schultz, Chirayu Goswami, Russ Hager, Robin Grimwood, Clinton R. Paden, Duncan MacCannell |
| EPI_ISL_1733474, EPI_ISL_1733475, EPI_ISL_1733477, EPI_ISL_1733478 | Minnesota Department of Health, Public Health Laboratory | Minnesota Department of Health, Public Health Laboratory | Alexandra Lorentz, Jacob Garfin, Matt Plumb, and Xiong Wang |
| EPI_ISL_1733480 | Infinity Biologix | Centers for Disease Control and Prevention Division of Viral Diseases, Pathogen Discovery | Dakota Howard, Dhwani Batra, Peter W. Cook, Kara Moser, Adrian Paskey, Jason Caravas, Benjamin Rambo-Martin, Shatavia Morrison, Christopher Gulvick, Scott Sammons, Yvette Unoarumhi, Darlene Wagner, Matthew Schmerer, Christian Bixby, Yihe Wang, Jonathan Schultz, Chirayu Goswami, Russ Hager, Robin Grimwood, Clinton R. Paden, Duncan MacCannell |
| EPI_ISL_1733481, EPI_ISL_1733482 | Minnesota Department of Health, Public Health Laboratory | Minnesota Department of Health, Public Health Laboratory | Alexandra Lorentz, Jacob Garfin, Matt Plumb, and Xiong Wang |
| EPI_ISL_1733483 | Infinity Biologix | Centers for Disease Control and Prevention Division of Viral Diseases, Pathogen Discovery | Dakota Howard, Dhwani Batra, Peter W. Cook, Kara Moser, Adrian Paskey, Jason Caravas, Benjamin Rambo-Martin, Shatavia Morrison, Christopher Gulvick, Scott Sammons, Yvette Unoarumhi, Darlene Wagner, Matthew Schmerer, Christian Bixby, Yihe Wang, Jonathan Schultz, Chirayu Goswami, Russ Hager, Robin Grimwood, Clinton R. Paden, Duncan MacCannell |
| EPI_ISL_1733484, EPI_ISL_1733485 | Minnesota Department of Health, Public Health Laboratory | Minnesota Department of Health, Public Health Laboratory | Alexandra Lorentz, Jacob Garfin, Matt Plumb, and Xiong Wang |
| EPI_ISL_1733486 | Infinity Biologix | Centers for Disease Control and Prevention Division of Viral Diseases, Pathogen Discovery | Dakota Howard, Dhwani Batra, Peter W. Cook, Kara Moser, Adrian Paskey, Jason Caravas, Benjamin Rambo-Martin, Shatavia Morrison, Christopher Gulvick, Scott Sammons, Yvette Unoarumhi, Darlene Wagner, Matthew Schmerer, Christian Bixby, Yihe Wang, Jonathan Schultz, Chirayu Goswami, Russ Hager, Robin Grimwood, Clinton R. Paden, Duncan MacCannell |
| EPI_ISL_1733487, EPI_ISL_1733488 | Minnesota Department of Health, Public Health Laboratory | Minnesota Department of Health, Public Health Laboratory | Alexandra Lorentz, Jacob Garfin, Matt Plumb, and Xiong Wang |
| EPI_ISL_1733489 | Infinity Biologix | Centers for Disease Control and Prevention Division of Viral Diseases, Pathogen Discovery | Dakota Howard, Dhwani Batra, Peter W. Cook, Kara Moser, Adrian Paskey, Jason Caravas, Benjamin Rambo-Martin, Shatavia Morrison, Christopher Gulvick, Scott Sammons, Yvette Unoarumhi, Darlene Wagner, Matthew Schmerer, Christian Bixby, Yihe Wang, Jonathan Schultz, Chirayu Goswami, Russ Hager, Robin Grimwood, Clinton R. Paden, Duncan MacCannell |
| EPI_ISL_1733490, EPI_ISL_1733491 | Minnesota Department of Health, Public Health Laboratory | Minnesota Department of Health, Public Health Laboratory | Alexandra Lorentz, Jacob Garfin, Matt Plumb, and Xiong Wang |
| EPI_ISL_1733492 | Infinity Biologix | Centers for Disease Control and Prevention Division of Viral Diseases, Pathogen Discovery | Dakota Howard, Dhwani Batra, Peter W. Cook, Kara Moser, Adrian Paskey, Jason Caravas, Benjamin Rambo-Martin, Shatavia Morrison, Christopher Gulvick, Scott Sammons, Yvette Unoarumhi, Darlene Wagner, Matthew Schmerer, Christian Bixby, Yihe Wang, Jonathan Schultz, Chirayu Goswami, Russ Hager, Robin Grimwood, Clinton R. Paden, Duncan MacCannell |
| EPI_ISL_1733493, EPI_ISL_1733494 | Minnesota Department of Health, Public Health Laboratory | Minnesota Department of Health, Public Health Laboratory | Alexandra Lorentz, Jacob Garfin, Matt Plumb, and Xiong Wang |
| EPI_ISL_1733495 | Infinity Biologix | Centers for Disease Control and Prevention Division of Viral Diseases, Pathogen Discovery | Dakota Howard, Dhwani Batra, Peter W. Cook, Kara Moser, Adrian Paskey, Jason Caravas, Benjamin Rambo-Martin, Shatavia Morrison, Christopher Gulvick, Scott Sammons, Yvette Unoarumhi, Darlene Wagner, Matthew Schmerer, Christian Bixby, Yihe Wang, Jonathan Schultz, Chirayu Goswami, Russ Hager, Robin Grimwood, Clinton R. Paden, Duncan MacCannell |
| EPI_ISL_1733496 | Minnesota Department of Health, Public | Minnesota Department of Health, Public | Alexandra Lorentz, Jacob Garfin, Matt Plumb, and Xiong Wang |

| | | | |
|---|---|---|---|
| | Health Laboratory | Health Laboratory | |
| EPI_ISL_1733497 | M Health Fairview | Minnesota Department of Health, Public Health Laboratory | Alexandra Lorentz, Jacob Garfin, Matt Plumb, and Xiong Wang |
| EPI_ISL_1733498 | Infinity Biologix | Centers for Disease Control and Prevention Division of Viral Diseases, Pathogen Discovery | Dakota Howard, Dhwani Batra, Peter W. Cook, Kara Moser, Adrian Paskey, Jason Caravas, Benjamin Rambo-Martin, Shatavia Morrison, Christopher Gulvick, Scott Sammons, Yvette Unoarumhi, Darlene Wagner, Matthew Schmerer, Christian Bixby, Yihe Wang, Jonathan Schultz, Chirayu Goswami, Russ Hager, Robin Grimwood, Clinton R. Paden, Duncan MacCannell |
| EPI_ISL_1733499, EPI_ISL_1733500 | Minnesota Department of Health, Public Health Laboratory | Minnesota Department of Health, Public Health Laboratory | Alexandra Lorentz, Jacob Garfin, Matt Plumb, and Xiong Wang |
| EPI_ISL_1733501 | Infinity Biologix | Centers for Disease Control and Prevention Division of Viral Diseases, Pathogen Discovery | Dakota Howard, Dhwani Batra, Peter W. Cook, Kara Moser, Adrian Paskey, Jason Caravas, Benjamin Rambo-Martin, Shatavia Morrison, Christopher Gulvick, Scott Sammons, Yvette Unoarumhi, Darlene Wagner, Matthew Schmerer, Christian Bixby, Yihe Wang, Jonathan Schultz, Chirayu Goswami, Russ Hager, Robin Grimwood, Clinton R. Paden, Duncan MacCannell |
| EPI_ISL_1733502, EPI_ISL_1733503 | Minnesota Department of Health, Public Health Laboratory | Minnesota Department of Health, Public Health Laboratory | Alexandra Lorentz, Jacob Garfin, Matt Plumb, and Xiong Wang |
| EPI_ISL_1733504 | Infinity Biologix | Centers for Disease Control and Prevention Division of Viral Diseases, Pathogen Discovery | Dakota Howard, Dhwani Batra, Peter W. Cook, Kara Moser, Adrian Paskey, Jason Caravas, Benjamin Rambo-Martin, Shatavia Morrison, Christopher Gulvick, Scott Sammons, Yvette Unoarumhi, Darlene Wagner, Matthew Schmerer, Christian Bixby, Yihe Wang, Jonathan Schultz, Chirayu Goswami, Russ Hager, Robin Grimwood, Clinton R. Paden, Duncan MacCannell |
| EPI_ISL_1733505, EPI_ISL_1733506 | Minnesota Department of Health, Public Health Laboratory | Minnesota Department of Health, Public Health Laboratory | Alexandra Lorentz, Jacob Garfin, Matt Plumb, and Xiong Wang |
| EPI_ISL_1733507 | Infinity Biologix | Centers for Disease Control and Prevention Division of Viral Diseases, Pathogen Discovery | Dakota Howard, Dhwani Batra, Peter W. Cook, Kara Moser, Adrian Paskey, Jason Caravas, Benjamin Rambo-Martin, Shatavia Morrison, Christopher Gulvick, Scott Sammons, Yvette Unoarumhi, Darlene Wagner, Matthew Schmerer, Christian Bixby, Yihe Wang, Jonathan Schultz, Chirayu Goswami, Russ Hager, Robin Grimwood, Clinton R. Paden, Duncan MacCannell |
| EPI_ISL_1733508, EPI_ISL_1733509 | Minnesota Department of Health, Public Health Laboratory | Minnesota Department of Health, Public Health Laboratory | Alexandra Lorentz, Jacob Garfin, Matt Plumb, and Xiong Wang |
| EPI_ISL_1733510 | Infinity Biologix | Centers for Disease Control and Prevention Division of Viral Diseases, Pathogen Discovery | Dakota Howard, Dhwani Batra, Peter W. Cook, Kara Moser, Adrian Paskey, Jason Caravas, Benjamin Rambo-Martin, Shatavia Morrison, Christopher Gulvick, Scott Sammons, Yvette Unoarumhi, Darlene Wagner, Matthew Schmerer, Christian Bixby, Yihe Wang, Jonathan Schultz, Chirayu Goswami, Russ Hager, Robin Grimwood, Clinton R. Paden, Duncan MacCannell |
| EPI_ISL_1733511, EPI_ISL_1733512 | Minnesota Department of Health, Public Health Laboratory | Minnesota Department of Health, Public Health Laboratory | Alexandra Lorentz, Jacob Garfin, Matt Plumb, and Xiong Wang |
| EPI_ISL_1733513 | Infinity Biologix | Centers for Disease Control and Prevention Division of Viral Diseases, Pathogen Discovery | Dakota Howard, Dhwani Batra, Peter W. Cook, Kara Moser, Adrian Paskey, Jason Caravas, Benjamin Rambo-Martin, Shatavia Morrison, Christopher Gulvick, Scott Sammons, Yvette Unoarumhi, Darlene Wagner, Matthew Schmerer, Christian Bixby, Yihe Wang, Jonathan Schultz, Chirayu Goswami, Russ Hager, Robin Grimwood, Clinton R. Paden, Duncan MacCannell |
| EPI_ISL_1733514, EPI_ISL_1733515 | Minnesota Department of Health, Public Health Laboratory | Minnesota Department of Health, Public Health Laboratory | Alexandra Lorentz, Jacob Garfin, Matt Plumb, and Xiong Wang |
| EPI_ISL_1733516 | Infinity Biologix | Centers for Disease Control and Prevention Division of Viral Diseases, Pathogen Discovery | Dakota Howard, Dhwani Batra, Peter W. Cook, Kara Moser, Adrian Paskey, Jason Caravas, Benjamin Rambo-Martin, Shatavia Morrison, Christopher Gulvick, Scott Sammons, Yvette Unoarumhi, Darlene Wagner, Matthew Schmerer, Christian Bixby, Yihe Wang, Jonathan Schultz, Chirayu Goswami, Russ Hager, Robin Grimwood, Clinton R. Paden, Duncan MacCannell |
| EPI_ISL_1733517, EPI_ISL_1733518, EPI_ISL_1733519 | Minnesota Department of Health, Public Health Laboratory | Minnesota Department of Health, Public Health Laboratory | Alexandra Lorentz, Jacob Garfin, Matt Plumb, and Xiong Wang |
| EPI_ISL_1733520 | Infinity Biologix | Centers for Disease Control and Prevention Division of Viral Diseases, Pathogen Discovery | Dakota Howard, Dhwani Batra, Peter W. Cook, Kara Moser, Adrian Paskey, Jason Caravas, Benjamin Rambo-Martin, Shatavia Morrison, Christopher Gulvick, Scott Sammons, Yvette Unoarumhi, Darlene Wagner, Matthew Schmerer, Christian Bixby, Yihe Wang, Jonathan Schultz, Chirayu Goswami, Russ Hager, Robin Grimwood, Clinton R. Paden, Duncan MacCannell |
| EPI_ISL_1733521 | Minnesota Department of Health, Public Health Laboratory | Minnesota Department of Health, Public Health Laboratory | Alexandra Lorentz, Jacob Garfin, Matt Plumb, and Xiong Wang |
| EPI_ISL_1733522 | Infinity Biologix | Centers for Disease Control and Prevention Division of Viral Diseases, Pathogen Discovery | Dakota Howard, Dhwani Batra, Peter W. Cook, Kara Moser, Adrian Paskey, Jason Caravas, Benjamin Rambo-Martin, Shatavia Morrison, Christopher Gulvick, Scott Sammons, Yvette Unoarumhi, Darlene Wagner, Matthew Schmerer, Christian Bixby, Yihe Wang, Jonathan Schultz, Chirayu Goswami, Russ Hager, Robin Grimwood, Clinton R. Paden, Duncan MacCannell |
| EPI_ISL_1733523, EPI_ISL_1733524 | Minnesota Department of Health, Public Health Laboratory | Minnesota Department of Health, Public Health Laboratory | Alexandra Lorentz, Jacob Garfin, Matt Plumb, and Xiong Wang |
| EPI_ISL_1733525 | Infinity Biologix | Centers for Disease Control and Prevention Division of Viral Diseases, Pathogen Discovery | Dakota Howard, Dhwani Batra, Peter W. Cook, Kara Moser, Adrian Paskey, Jason Caravas, Benjamin Rambo-Martin, Shatavia Morrison, Christopher Gulvick, Scott Sammons, Yvette Unoarumhi, Darlene Wagner, Matthew Schmerer, Christian Bixby, Yihe Wang, Jonathan Schultz, Chirayu Goswami, Russ Hager, Robin Grimwood, Clinton R. Paden, Duncan MacCannell |
| EPI_ISL_1733526, EPI_ISL_1733527 | Minnesota Department of Health, Public Health Laboratory | Minnesota Department of Health, Public Health Laboratory | Alexandra Lorentz, Jacob Garfin, Matt Plumb, and Xiong Wang |
| EPI_ISL_1733528 | Infinity Biologix | Centers for Disease Control and Prevention Division of Viral Diseases, Pathogen Discovery | Dakota Howard, Dhwani Batra, Peter W. Cook, Kara Moser, Adrian Paskey, Jason Caravas, Benjamin Rambo-Martin, Shatavia Morrison, Christopher Gulvick, Scott Sammons, Yvette Unoarumhi, Darlene Wagner, Matthew Schmerer, Christian Bixby, Yihe Wang, Jonathan Schultz, Chirayu Goswami, Russ Hager, Robin Grimwood, Clinton R. Paden, Duncan MacCannell |
| EPI_ISL_1733529, EPI_ISL_1733530 | Minnesota Department of Health, Public Health Laboratory | Minnesota Department of Health, Public Health Laboratory | Alexandra Lorentz, Jacob Garfin, Matt Plumb, and Xiong Wang |
| EPI_ISL_1733531 | Infinity Biologix | Centers for Disease Control and Prevention Division of Viral Diseases, Pathogen Discovery | Dakota Howard, Dhwani Batra, Peter W. Cook, Kara Moser, Adrian Paskey, Jason Caravas, Benjamin Rambo-Martin, Shatavia Morrison, Christopher Gulvick, Scott Sammons, Yvette Unoarumhi, Darlene Wagner, Matthew Schmerer, Christian Bixby, Yihe Wang, Jonathan Schultz, Chirayu Goswami, Russ Hager, Robin Grimwood, Clinton R. Paden, Duncan MacCannell |
| EPI_ISL_1733532, EPI_ISL_1733533, EPI_ISL_1733534 | Minnesota Department of Health, Public Health Laboratory | Minnesota Department of Health, Public Health Laboratory | Alexandra Lorentz, Jacob Garfin, Matt Plumb, and Xiong Wang |
| EPI_ISL_1733535 | Infinity Biologix | Centers for Disease Control and Prevention Division of Viral Diseases, Pathogen Discovery | Dakota Howard, Dhwani Batra, Peter W. Cook, Kara Moser, Adrian Paskey, Jason Caravas, Benjamin Rambo-Martin, Shatavia Morrison, Christopher Gulvick, Scott Sammons, Yvette Unoarumhi, Darlene Wagner, Matthew Schmerer, Christian Bixby, Yihe Wang, Jonathan Schultz, Chirayu Goswami, Russ Hager, Robin Grimwood, Clinton R. Paden, Duncan MacCannell |
| EPI_ISL_1733536, EPI_ISL_1733537, EPI_ISL_1733538 | Minnesota Department of Health, Public Health Laboratory | Minnesota Department of Health, Public Health Laboratory | Alexandra Lorentz, Jacob Garfin, Matt Plumb, and Xiong Wang |
| EPI_ISL_1733539 | Infinity Biologix | Centers for Disease Control and Prevention Division of Viral Diseases, Pathogen Discovery | Dakota Howard, Dhwani Batra, Peter W. Cook, Kara Moser, Adrian Paskey, Jason Caravas, Benjamin Rambo-Martin, Shatavia Morrison, Christopher Gulvick, Scott Sammons, Yvette Unoarumhi, Darlene Wagner, Matthew Schmerer, Christian Bixby, Yihe Wang, Jonathan Schultz, Chirayu Goswami, Russ Hager, Robin Grimwood, Clinton R. Paden, Duncan MacCannell |
| EPI_ISL_1733541 | Minnesota Department of Health, Public Health Laboratory | Minnesota Department of Health, Public Health Laboratory | Alexandra Lorentz, Jacob Garfin, Matt Plumb, and Xiong Wang |
| EPI_ISL_1733542 | Infinity Biologix | Centers for Disease Control and Prevention Division of Viral Diseases, Pathogen Discovery | Dakota Howard, Dhwani Batra, Peter W. Cook, Kara Moser, Adrian Paskey, Jason Caravas, Benjamin Rambo-Martin, Shatavia Morrison, Christopher Gulvick, Scott Sammons, Yvette Unoarumhi, Darlene Wagner, Matthew Schmerer, Christian Bixby, Yihe Wang, Jonathan Schultz, Chirayu Goswami, Russ Hager, Robin Grimwood, Clinton R. Paden, Duncan MacCannell |
| EPI_ISL_1733543, EPI_ISL_1733544 | Minnesota Department of Health, Public Health Laboratory | Minnesota Department of Health, Public Health Laboratory | Alexandra Lorentz, Jacob Garfin, Matt Plumb, and Xiong Wang |
| EPI_ISL_1733545 | Infinity Biologix | Centers for Disease Control and Prevention Division of Viral Diseases, Pathogen Discovery | Dakota Howard, Dhwani Batra, Peter W. Cook, Kara Moser, Adrian Paskey, Jason Caravas, Benjamin Rambo-Martin, Shatavia Morrison, Christopher Gulvick, Scott Sammons, Yvette Unoarumhi, Darlene Wagner, Matthew Schmerer, Christian Bixby, Yihe Wang, Jonathan Schultz, Chirayu Goswami, Russ Hager, Robin Grimwood, Clinton R. Paden, Duncan MacCannell |
| EPI_ISL_1733546, EPI_ISL_1733547, EPI_ISL_1733548, EPI_ISL_1733549, EPI_ISL_1733550, EPI_ISL_1733551, EPI_ISL_1733552 | Minnesota Department of Health, Public Health Laboratory | Minnesota Department of Health, Public Health Laboratory | Alexandra Lorentz, Jacob Garfin, Matt Plumb, and Xiong Wang |
| EPI_ISL_1733553 | Infinity Biologix | Centers for Disease Control and Prevention Division of Viral Diseases, Pathogen Discovery | Dakota Howard, Dhwani Batra, Peter W. Cook, Kara Moser, Adrian Paskey, Jason Caravas, Benjamin Rambo-Martin, Shatavia Morrison, Christopher Gulvick, Scott Sammons, Yvette Unoarumhi, Darlene Wagner, Matthew Schmerer, Christian Bixby, Yihe Wang, Jonathan Schultz, Chirayu Goswami, Russ Hager, Robin Grimwood, Clinton R. Paden, Duncan MacCannell |

| | | | |
|---|---|---|---|
| EPI_ISL_1733554, EPI_ISL_1733555 | Minnesota Department of Health, Public Health Laboratory | Minnesota Department of Health, Public Health Laboratory | Alexandra Lorentz, Jacob Garfin, Matt Plumb, and Xiong Wang |
| EPI_ISL_1733556 | Infinity Biologix | Centers for Disease Control and Prevention Division of Viral Diseases, Pathogen Discovery | Dakota Howard, Dhwani Batra, Peter W. Cook, Kara Moser, Adrian Paskey, Jason Caravas, Benjamin Rambo-Martin, Shatavia Morrison, Christopher Gulvick, Scott Sammons, Yvette Unoarumhi, Darlene Wagner, Matthew Schmerer, Christian Bixby, Yihe Wang, Jonathan Schultz, Chirayu Goswami, Russ Hager, Robin Grimwood, Clinton R. Paden, Duncan MacCannell |
| EPI_ISL_1733557, EPI_ISL_1733558 | Minnesota Department of Health, Public Health Laboratory | Minnesota Department of Health, Public Health Laboratory | Alexandra Lorentz, Jacob Garfin, Matt Plumb, and Xiong Wang |
| EPI_ISL_1733559 | Infinity Biologix | Centers for Disease Control and Prevention Division of Viral Diseases, Pathogen Discovery | Dakota Howard, Dhwani Batra, Peter W. Cook, Kara Moser, Adrian Paskey, Jason Caravas, Benjamin Rambo-Martin, Shatavia Morrison, Christopher Gulvick, Scott Sammons, Yvette Unoarumhi, Darlene Wagner, Matthew Schmerer, Christian Bixby, Yihe Wang, Jonathan Schultz, Chirayu Goswami, Russ Hager, Robin Grimwood, Clinton R. Paden, Duncan MacCannell |
| EPI_ISL_1733560, EPI_ISL_1733561 | Minnesota Department of Health, Public Health Laboratory | Minnesota Department of Health, Public Health Laboratory | Alexandra Lorentz, Jacob Garfin, Matt Plumb, and Xiong Wang |
| EPI_ISL_1733562 | Infinity Biologix | Centers for Disease Control and Prevention Division of Viral Diseases, Pathogen Discovery | Dakota Howard, Dhwani Batra, Peter W. Cook, Kara Moser, Adrian Paskey, Jason Caravas, Benjamin Rambo-Martin, Shatavia Morrison, Christopher Gulvick, Scott Sammons, Yvette Unoarumhi, Darlene Wagner, Matthew Schmerer, Christian Bixby, Yihe Wang, Jonathan Schultz, Chirayu Goswami, Russ Hager, Robin Grimwood, Clinton R. Paden, Duncan MacCannell |
| EPI_ISL_1733563, EPI_ISL_1733564 | Minnesota Department of Health, Public Health Laboratory | Minnesota Department of Health, Public Health Laboratory | Alexandra Lorentz, Jacob Garfin, Matt Plumb, and Xiong Wang |
| EPI_ISL_1733565 | Infinity Biologix | Centers for Disease Control and Prevention Division of Viral Diseases, Pathogen Discovery | Dakota Howard, Dhwani Batra, Peter W. Cook, Kara Moser, Adrian Paskey, Jason Caravas, Benjamin Rambo-Martin, Shatavia Morrison, Christopher Gulvick, Scott Sammons, Yvette Unoarumhi, Darlene Wagner, Matthew Schmerer, Christian Bixby, Yihe Wang, Jonathan Schultz, Chirayu Goswami, Russ Hager, Robin Grimwood, Clinton R. Paden, Duncan MacCannell |
| EPI_ISL_1733566, EPI_ISL_1733567 | Minnesota Department of Health, Public Health Laboratory | Minnesota Department of Health, Public Health Laboratory | Alexandra Lorentz, Jacob Garfin, Matt Plumb, and Xiong Wang |
| EPI_ISL_1733568 | Infinity Biologix | Centers for Disease Control and Prevention Division of Viral Diseases, Pathogen Discovery | Dakota Howard, Dhwani Batra, Peter W. Cook, Kara Moser, Adrian Paskey, Jason Caravas, Benjamin Rambo-Martin, Shatavia Morrison, Christopher Gulvick, Scott Sammons, Yvette Unoarumhi, Darlene Wagner, Matthew Schmerer, Christian Bixby, Yihe Wang, Jonathan Schultz, Chirayu Goswami, Russ Hager, Robin Grimwood, Clinton R. Paden, Duncan MacCannell |
| EPI_ISL_1733569, EPI_ISL_1733570 | Minnesota Department of Health, Public Health Laboratory | Minnesota Department of Health, Public Health Laboratory | Alexandra Lorentz, Jacob Garfin, Matt Plumb, and Xiong Wang |
| EPI_ISL_1733571, EPI_ISL_1733572, EPI_ISL_1733573, EPI_ISL_1733574, EPI_ISL_1733575, EPI_ISL_1733576, EPI_ISL_1733577, EPI_ISL_1733578, EPI_ISL_1733579, EPI_ISL_1733580, EPI_ISL_1733581, EPI_ISL_1733582, EPI_ISL_1733583, EPI_ISL_1733584, EPI_ISL_1733585, EPI_ISL_1733586, EPI_ISL_1733587, EPI_ISL_1733588, EPI_ISL_1733590, EPI_ISL_1733591, EPI_ISL_1733592, EPI_ISL_1733593, EPI_ISL_1733594, EPI_ISL_1733595, EPI_ISL_1733596, EPI_ISL_1733597, EPI_ISL_1733598, EPI_ISL_1733599, EPI_ISL_1733600, EPI_ISL_1733601, EPI_ISL_1733602, EPI_ISL_1733604, EPI_ISL_1733605, EPI_ISL_1733606, EPI_ISL_1733608, EPI_ISL_1733609, EPI_ISL_1733610, EPI_ISL_1733611, EPI_ISL_1733612, EPI_ISL_1733613, EPI_ISL_1733615, EPI_ISL_1733616, EPI_ISL_1733617, EPI_ISL_1733618, EPI_ISL_1733619, EPI_ISL_1733620, EPI_ISL_1733622, EPI_ISL_1733623, EPI_ISL_1733624, EPI_ISL_1733625, EPI_ISL_1733626, EPI_ISL_1733627, EPI_ISL_1733628, EPI_ISL_1733630, EPI_ISL_1733631, EPI_ISL_1733632, EPI_ISL_1733633, EPI_ISL_1733634, EPI_ISL_1733635, EPI_ISL_1733636, EPI_ISL_1733637, EPI_ISL_1733638, EPI_ISL_1733639, EPI_ISL_1733640, EPI_ISL_1733641, EPI_ISL_1733642, EPI_ISL_1733643, EPI_ISL_1733644, EPI_ISL_1733645, EPI_ISL_1733646, EPI_ISL_1733647, EPI_ISL_1733648, EPI_ISL_1733649, EPI_ISL_1733650, EPI_ISL_1733651, EPI_ISL_1733652, EPI_ISL_1733653, EPI_ISL_1733654, EPI_ISL_1733655, EPI_ISL_1733656, EPI_ISL_1733657, EPI_ISL_1733658, EPI_ISL_1733659, EPI_ISL_1733660, EPI_ISL_1733661, EPI_ISL_1733662, EPI_ISL_1733663, EPI_ISL_1733665, EPI_ISL_1733666, EPI_ISL_1733667, EPI_ISL_1733668, EPI_ISL_1733669, EPI_ISL_1733670, EPI_ISL_1733671, EPI_ISL_1733672, EPI_ISL_1733673, EPI_ISL_1733674, EPI_ISL_1733676, EPI_ISL_1733677, EPI_ISL_1733678, EPI_ISL_1733679, EPI_ISL_1733680, EPI_ISL_1733681, EPI_ISL_1733682, EPI_ISL_1733683, EPI_ISL_1733685, EPI_ISL_1733686, EPI_ISL_1733687, EPI_ISL_1733688, EPI_ISL_1733689, EPI_ISL_1733690, EPI_ISL_1733691, EPI_ISL_1733692, EPI_ISL_1733693, EPI_ISL_1733694, EPI_ISL_1733695, EPI_ISL_1733696, EPI_ISL_1733697, EPI_ISL_1733699, EPI_ISL_1733700, EPI_ISL_1733701, EPI_ISL_1733702, EPI_ISL_1733703, EPI_ISL_1733704, EPI_ISL_1733705, EPI_ISL_1733706, EPI_ISL_1733707, EPI_ISL_1733708, EPI_ISL_1733709, EPI_ISL_1733710, EPI_ISL_1733711, EPI_ISL_1733712, EPI_ISL_1733713, EPI_ISL_1733714, EPI_ISL_1733715, EPI_ISL_1733716, EPI_ISL_1733717, EPI_ISL_1733718, EPI_ISL_1733719, EPI_ISL_1733721, EPI_ISL_1733722, EPI_ISL_1733723, EPI_ISL_1733724, EPI_ISL_1733725, EPI_ISL_1733726, EPI_ISL_1733727, EPI_ISL_1733728, EPI_ISL_1733729, EPI_ISL_1733730, EPI_ISL_1733731 | | | |
| see above | Infinity Biologix | Centers for Disease Control and Prevention Division of Viral Diseases, Pathogen Discovery | Dakota Howard, Dhwani Batra, Peter W. Cook, Kara Moser, Adrian Paskey, Jason Caravas, Benjamin Rambo-Martin, Shatavia Morrison, Christopher Gulvick, Scott Sammons, Yvette Unoarumhi, Darlene Wagner, Matthew Schmerer, Christian Bixby, Yihe Wang, Jonathan Schultz, Chirayu Goswami, Russ Hager, Robin Grimwood, Clinton R. Paden, Duncan MacCannell |
| EPI_ISL_1733732, EPI_ISL_1733734, EPI_ISL_1733735, EPI_ISL_1733736, EPI_ISL_1733737, EPI_ISL_1733738, EPI_ISL_1733739, EPI_ISL_1733740, EPI_ISL_1733741, EPI_ISL_1733742, EPI_ISL_1733743, EPI_ISL_1733744, EPI_ISL_1733745, EPI_ISL_1733747, EPI_ISL_1733748, EPI_ISL_1733749, EPI_ISL_1733750, EPI_ISL_1733751, EPI_ISL_1733752, EPI_ISL_1733753, EPI_ISL_1733754, EPI_ISL_1733755, EPI_ISL_1733756, EPI_ISL_1733757, EPI_ISL_1733758, EPI_ISL_1733759, EPI_ISL_1733760, EPI_ISL_1733761, EPI_ISL_1733762, EPI_ISL_1733763, EPI_ISL_1733764, EPI_ISL_1733765, EPI_ISL_1733766, EPI_ISL_1733767, EPI_ISL_1733768, EPI_ISL_1733769, EPI_ISL_1733771, EPI_ISL_1733773, EPI_ISL_1733774, EPI_ISL_1733775, EPI_ISL_1733776, EPI_ISL_1733777, EPI_ISL_1733778, EPI_ISL_1733780, EPI_ISL_1733781, EPI_ISL_1733782, EPI_ISL_1733783, EPI_ISL_1733784, EPI_ISL_1733785, EPI_ISL_1733787, EPI_ISL_1733788, EPI_ISL_1733789, EPI_ISL_1733790, EPI_ISL_1733791, EPI_ISL_1733792, EPI_ISL_1733793, EPI_ISL_1733794, EPI_ISL_1733795, EPI_ISL_1733796, EPI_ISL_1733797, EPI_ISL_1733798, EPI_ISL_1733799, EPI_ISL_1733800, EPI_ISL_1733801, EPI_ISL_1733802, EPI_ISL_1733804, EPI_ISL_1733805, EPI_ISL_1733806, EPI_ISL_1733807, EPI_ISL_1733808, EPI_ISL_1733809, EPI_ISL_1733810, EPI_ISL_1733811, EPI_ISL_1733812, EPI_ISL_1733813, EPI_ISL_1733814, EPI_ISL_1733815, EPI_ISL_1733817, EPI_ISL_1733818, EPI_ISL_1733819, EPI_ISL_1733820, EPI_ISL_1733821, EPI_ISL_1733822, EPI_ISL_1733823, EPI_ISL_1733824, EPI_ISL_1733825, EPI_ISL_1733826, EPI_ISL_1733827, EPI_ISL_1733828, EPI_ISL_1733829, EPI_ISL_1733832, EPI_ISL_1733833, EPI_ISL_1733834, EPI_ISL_1733838, EPI_ISL_1733839, EPI_ISL_1733840, EPI_ISL_1733841, EPI_ISL_1733843, EPI_ISL_1733844, EPI_ISL_1733845, EPI_ISL_1733846, EPI_ISL_1733848, EPI_ISL_1733849, EPI_ISL_1733850, EPI_ISL_1733851, EPI_ISL_1733852, EPI_ISL_1733854, EPI_ISL_1733855, EPI_ISL_1733857, EPI_ISL_1733858, EPI_ISL_1733859, EPI_ISL_1733860, EPI_ISL_1733861, EPI_ISL_1733863, EPI_ISL_1733864, EPI_ISL_1733865, EPI_ISL_1733866, EPI_ISL_1733867, EPI_ISL_1733868, EPI_ISL_1733869, EPI_ISL_1733870, EPI_ISL_1733871, EPI_ISL_1733872, EPI_ISL_1733873, EPI_ISL_1733874, EPI_ISL_1733875, EPI_ISL_1733876, EPI_ISL_1733877, EPI_ISL_1733878, EPI_ISL_1733879, EPI_ISL_1733880, EPI_ISL_1733881, EPI_ISL_1733882, EPI_ISL_1733884, EPI_ISL_1733885, EPI_ISL_1733886, EPI_ISL_1733887, EPI_ISL_1733888, EPI_ISL_1733889, EPI_ISL_1733891, EPI_ISL_1733893, EPI_ISL_1733894, EPI_ISL_1733895, EPI_ISL_1733896, EPI_ISL_1733897, EPI_ISL_1733898, EPI_ISL_1733901, EPI_ISL_1733902, EPI_ISL_1733903, EPI_ISL_1733905, EPI_ISL_1733906, EPI_ISL_1733907, EPI_ISL_1733908, EPI_ISL_1733909, EPI_ISL_1733910, EPI_ISL_1733911, EPI_ISL_1733912, EPI_ISL_1733913, EPI_ISL_1733914, EPI_ISL_1733915, EPI_ISL_1733917, EPI_ISL_1733918, EPI_ISL_1733919, EPI_ISL_1733920, EPI_ISL_1733921, EPI_ISL_1733922, EPI_ISL_1733923, EPI_ISL_1733924, EPI_ISL_1733925, EPI_ISL_1733926, EPI_ISL_1733927, EPI_ISL_1733928, EPI_ISL_1733929, EPI_ISL_1733930, EPI_ISL_1733931, EPI_ISL_1733933, EPI_ISL_1733934, EPI_ISL_1733935, EPI_ISL_1733936, EPI_ISL_1733937, EPI_ISL_1733938, EPI_ISL_1733939, EPI_ISL_1733940, EPI_ISL_1733941, EPI_ISL_1733942, EPI_ISL_1733943, EPI_ISL_1733944, EPI_ISL_1733945, EPI_ISL_1733946, EPI_ISL_1733947, EPI_ISL_1733948, EPI_ISL_1733950, EPI_ISL_1733952, EPI_ISL_1733953, EPI_ISL_1733954, EPI_ISL_1733955, EPI_ISL_1733956, EPI_ISL_1733957, EPI_ISL_1733958, EPI_ISL_1733959, EPI_ISL_1733960, EPI_ISL_1733961, EPI_ISL_1733962, EPI_ISL_1733963, EPI_ISL_1733964, EPI_ISL_1733966, EPI_ISL_1733967, EPI_ISL_1733968, EPI_ISL_1733969, EPI_ISL_1733970, EPI_ISL_1733971, EPI_ISL_1733973, EPI_ISL_1733974, EPI_ISL_1733976, EPI_ISL_1733977, EPI_ISL_1733980, EPI_ISL_1733981, EPI_ISL_1733982, EPI_ISL_1733983, EPI_ISL_1733984, EPI_ISL_1733985, EPI_ISL_1733986, EPI_ISL_1733987, EPI_ISL_1733988, EPI_ISL_1733989, EPI_ISL_1733990, EPI_ISL_1733991, EPI_ISL_1733992, EPI_ISL_1733993, EPI_ISL_1733995, EPI_ISL_1733997, EPI_ISL_1733998, EPI_ISL_1733999, EPI_ISL_1734000, EPI_ISL_1734001, EPI_ISL_1734002, EPI_ISL_1734003, EPI_ISL_1734004, EPI_ISL_1734005, EPI_ISL_1734006, EPI_ISL_1734007, EPI_ISL_1734008, EPI_ISL_1734009, EPI_ISL_1734010, EPI_ISL_1734011, EPI_ISL_1734013, EPI_ISL_1734014, EPI_ISL_1734015, EPI_ISL_1734016, EPI_ISL_1734017, EPI_ISL_1734018, EPI_ISL_1734019, EPI_ISL_1734020, EPI_ISL_1734022, EPI_ISL_1734023, EPI_ISL_1734025, EPI_ISL_1734026, EPI_ISL_1734027, EPI_ISL_1734028, EPI_ISL_1734029, EPI_ISL_1734030, EPI_ISL_1734031, EPI_ISL_1734032, EPI_ISL_1734033, EPI_ISL_1734034, EPI_ISL_1734035, EPI_ISL_1734036, EPI_ISL_1734037, EPI_ISL_1734038, EPI_ISL_1734039, EPI_ISL_1734040, EPI_ISL_1734041, EPI_ISL_1734043, EPI_ISL_1734044, EPI_ISL_1734045, EPI_ISL_1734046, EPI_ISL_1734047, EPI_ISL_1734048, EPI_ISL_1734049, EPI_ISL_1734050, EPI_ISL_1734051, EPI_ISL_1734052, EPI_ISL_1734053, EPI_ISL_1734054, EPI_ISL_1734055, EPI_ISL_1734057, EPI_ISL_1734058, EPI_ISL_1734059, EPI_ISL_1734061, EPI_ISL_1734062, EPI_ISL_1734063, EPI_ISL_1734064, EPI_ISL_1734065, EPI_ISL_1734066, EPI_ISL_1734067, EPI_ISL_1734068, EPI_ISL_1734069, EPI_ISL_1734070, EPI_ISL_1734071, EPI_ISL_1734073, EPI_ISL_1734074, EPI_ISL_1734075, EPI_ISL_1734076, EPI_ISL_1734077, EPI_ISL_1734078, EPI_ISL_1734080, EPI_ISL_1734081, EPI_ISL_1734082, EPI_ISL_1734083, EPI_ISL_1734084, EPI_ISL_1734085, EPI_ISL_1734086, EPI_ISL_1734088, EPI_ISL_1734090, EPI_ISL_1734092, EPI_ISL_1734093, EPI_ISL_1734094, EPI_ISL_1734096, EPI_ISL_1734097, EPI_ISL_1734098, EPI_ISL_1734101, EPI_ISL_1734103, EPI_ISL_1734104, EPI_ISL_1734105, EPI_ISL_1734108, EPI_ISL_1734110, EPI_ISL_1734111, EPI_ISL_1734112, EPI_ISL_1734114, EPI_ISL_1734115, EPI_ISL_1734116, EPI_ISL_1734119, EPI_ISL_1734120, EPI_ISL_1734121, EPI_ISL_1734123, EPI_ISL_1734124, EPI_ISL_1734125, EPI_ISL_1734126, EPI_ISL_1734127, EPI_ISL_1734128, EPI_ISL_1734129, EPI_ISL_1734130, EPI_ISL_1734131, EPI_ISL_1734132, EPI_ISL_1734133, EPI_ISL_1734134, EPI_ISL_1734135, EPI_ISL_1734136, EPI_ISL_1734137, EPI_ISL_1734138, EPI_ISL_1734139, EPI_ISL_1734140, EPI_ISL_1734141, EPI_ISL_1734142, EPI_ISL_1734143, EPI_ISL_1734144, EPI_ISL_1734145, EPI_ISL_1734146, EPI_ISL_1734147, EPI_ISL_1734148, EPI_ISL_1734149, EPI_ISL_1734150, EPI_ISL_1734151, EPI_ISL_1734152, EPI_ISL_1734153, EPI_ISL_1734154 | | | |
| see above | Fulgent Genetics | Centers for Disease Control and Prevention Division of Viral Diseases, Pathogen Discovery | Dakota Howard, Dhwani Batra, Peter W. Cook, Kara Moser, Adrian Paskey, Jason Caravas, Benjamin Rambo-Martin, Shatavia Morrison, Christopher Gulvick, Scott Sammons, Yvette Unoarumhi, Darlene Wagner, Matthew Schmerer, Harry Gao, Mickey Li, John Gao, Joseph Fierro, Benafsh Sapra, Becky Tsai, Yan Meng, Doreen Ng, James Xie, Clinton R. Paden, Duncan MacCannell |
| EPI_ISL_1734155, EPI_ISL_1734156 | NCSLPH | NCSLPH | Chase K, Miller MC, Greene S, Glover W |
| EPI_ISL_1734157 | Fulgent Genetics | Centers for Disease Control and Prevention Division of Viral Diseases, Pathogen Discovery | Dakota Howard, Dhwani Batra, Peter W. Cook, Kara Moser, Adrian Paskey, Jason Caravas, Benjamin Rambo-Martin, Shatavia Morrison, Christopher Gulvick, Scott Sammons, Yvette Unoarumhi, Darlene Wagner, Matthew Schmerer, Harry Gao, Mickey Li, John Gao, Joseph Fierro, Benafsh Sapra, Becky Tsai, Yan Meng, Doreen Ng, James Xie, Clinton R. Paden, Duncan MacCannell |
| EPI_ISL_1734158, EPI_ISL_1734159 | NCSLPH | NCSLPH | Chase K, Miller MC, Greene S, Glover W |
| EPI_ISL_1734160 | Fulgent Genetics | Centers for Disease Control and Prevention Division of Viral Diseases, Pathogen Discovery | Dakota Howard, Dhwani Batra, Peter W. Cook, Kara Moser, Adrian Paskey, Jason Caravas, Benjamin Rambo-Martin, Shatavia Morrison, Christopher Gulvick, Scott Sammons, Yvette Unoarumhi, Darlene Wagner, Matthew Schmerer, Harry Gao, Mickey Li, John Gao, Joseph Fierro, Benafsh Sapra, Becky Tsai, Yan Meng, Doreen Ng, James Xie, Clinton R. Paden, Duncan MacCannell |
| EPI_ISL_1734161, EPI_ISL_1734162 | NCSLPH | NCSLPH | Chase K, Miller MC, Greene S, Glover W |
| EPI_ISL_1734163 | Fulgent Genetics | Centers for Disease Control and Prevention Division of Viral Diseases, Pathogen Discovery | Dakota Howard, Dhwani Batra, Peter W. Cook, Kara Moser, Adrian Paskey, Jason Caravas, Benjamin Rambo-Martin, Shatavia Morrison, Christopher Gulvick, Scott Sammons, Yvette Unoarumhi, Darlene Wagner, Matthew Schmerer, Harry Gao, Mickey Li, John Gao, Joseph Fierro, Benafsh Sapra, Becky Tsai, Yan Meng, Doreen Ng, James Xie, Clinton R. Paden, Duncan MacCannell |
| EPI_ISL_1734164, EPI_ISL_1734165 | NCSLPH | NCSLPH | Chase K, Miller MC, Greene S, Glover W |
| EPI_ISL_1734166 | Fulgent Genetics | Centers for Disease Control and Prevention Division of Viral Diseases, Pathogen Discovery | Dakota Howard, Dhwani Batra, Peter W. Cook, Kara Moser, Adrian Paskey, Jason Caravas, Benjamin Rambo-Martin, Shatavia Morrison, Christopher Gulvick, Scott Sammons, Yvette Unoarumhi, Darlene Wagner, Matthew Schmerer, Harry Gao, Mickey Li, John Gao, Joseph Fierro, Benafsh Sapra, Becky Tsai, Yan Meng, Doreen Ng, James Xie, Clinton R. Paden, Duncan MacCannell |
| EPI_ISL_1734167, EPI_ISL_1734168 | NCSLPH | NCSLPH | Chase K, Miller MC, Greene S, Glover W |
| EPI_ISL_1734169 | Fulgent Genetics | Centers for Disease Control and Prevention Division of Viral Diseases, Pathogen Discovery | Dakota Howard, Dhwani Batra, Peter W. Cook, Kara Moser, Adrian Paskey, Jason Caravas, Benjamin Rambo-Martin, Shatavia Morrison, Christopher Gulvick, Scott Sammons, Yvette Unoarumhi, Darlene Wagner, Matthew Schmerer, Harry Gao, Mickey Li, John Gao, Joseph Fierro, Benafsh Sapra, Becky Tsai, Yan Meng, Doreen Ng, James Xie, Clinton R. Paden, Duncan MacCannell |
| EPI_ISL_1734170, EPI_ISL_1734171 | NCSLPH | NCSLPH | Chase K, Miller MC, Greene S, Glover W |
| EPI_ISL_1734172 | Fulgent Genetics | Centers for Disease Control and Prevention Division of Viral Diseases, Pathogen Discovery | Dakota Howard, Dhwani Batra, Peter W. Cook, Kara Moser, Adrian Paskey, Jason Caravas, Benjamin Rambo-Martin, Shatavia Morrison, Christopher Gulvick, Scott Sammons, Yvette Unoarumhi, Darlene Wagner, Matthew Schmerer, Harry Gao, Mickey Li, John Gao, Joseph Fierro, Benafsh Sapra, Becky Tsai, Yan Meng, Doreen Ng, James Xie, Clinton R. Paden, Duncan MacCannell |
| EPI_ISL_1734173, EPI_ISL_1734174 | NCSLPH | NCSLPH | Chase K, Miller MC, Greene S, Glover W |
| EPI_ISL_1734175 | Fulgent Genetics | Centers for Disease Control and Prevention Division of Viral Diseases, Pathogen Discovery | Dakota Howard, Dhwani Batra, Peter W. Cook, Kara Moser, Adrian Paskey, Jason Caravas, Benjamin Rambo-Martin, Shatavia Morrison, Christopher Gulvick, Scott Sammons, Yvette Unoarumhi, Darlene Wagner, Matthew Schmerer, Harry Gao, Mickey Li, John Gao, Joseph Fierro, Benafsh Sapra, Becky Tsai, Yan Meng, Doreen Ng, James Xie, Clinton R. Paden, Duncan MacCannell |
| EPI_ISL_1734176, EPI_ISL_1734177 | NCSLPH | NCSLPH | Chase K, Miller MC, Greene S, Glover W |
| EPI_ISL_1734178 | Fulgent Genetics | Centers for Disease Control and Prevention Division of Viral Diseases, Pathogen | Dakota Howard, Dhwani Batra, Peter W. Cook, Kara Moser, Adrian Paskey, Jason Caravas, Benjamin Rambo-Martin, Shatavia Morrison, Christopher Gulvick, Scott Sammons, Yvette Unoarumhi, Darlene Wagner, Matthew Schmerer, Harry Gao, Mickey Li, John Gao, Joseph Fierro, Benafsh Sapra, Becky Tsai, Yan Meng, Doreen Ng, James Xie, Clinton R. Paden, Duncan MacCannell |

| | | | |
|---|---|---|---|
| | | Discovery | |
| EPI_ISL_1734179, EPI_ISL_1734180 | NCSLPH | NCSLPH | Chase K, Miller MC, Greene S, Glover W |
| EPI_ISL_1734181 | Fulgent Genetics | Centers for Disease Control and Prevention Division of Viral Diseases, Pathogen Discovery | Dakota Howard, Dhwani Batra, Peter W. Cook, Kara Moser, Adrian Paskey, Jason Caravas, Benjamin Rambo-Martin, Shatavia Morrison, Christopher Gulvick, Scott Sammons, Yvette Unoarumhi, Darlene Wagner, Matthew Schmerer, Harry Gao, Mickey Li, John Gao, Joseph Fierro, Benafsh Sapra, Becky Tsai, Yan Meng, Doreen Ng, James Xie, Clinton R. Paden, Duncan MacCannell |
| EPI_ISL_1734182, EPI_ISL_1734183 | NCSLPH | NCSLPH | Chase K, Miller MC, Greene S, Glover W |
| EPI_ISL_1734184 | Fulgent Genetics | Centers for Disease Control and Prevention Division of Viral Diseases, Pathogen Discovery | Dakota Howard, Dhwani Batra, Peter W. Cook, Kara Moser, Adrian Paskey, Jason Caravas, Benjamin Rambo-Martin, Shatavia Morrison, Christopher Gulvick, Scott Sammons, Yvette Unoarumhi, Darlene Wagner, Matthew Schmerer, Harry Gao, Mickey Li, John Gao, Joseph Fierro, Benafsh Sapra, Becky Tsai, Yan Meng, Doreen Ng, James Xie, Clinton R. Paden, Duncan MacCannell |
| EPI_ISL_1734185 | NCSLPH | NCSLPH | Chase K, Miller MC, Greene S, Glover W |
| EPI_ISL_1734186, EPI_ISL_1734187, EPI_ISL_1734188, EPI_ISL_1734189, EPI_ISL_1734190, EPI_ISL_1734191, EPI_ISL_1734192, EPI_ISL_1734193, EPI_ISL_1734194, EPI_ISL_1734195, EPI_ISL_1734196, EPI_ISL_1734197, EPI_ISL_1734198, EPI_ISL_1734199, EPI_ISL_1734200, EPI_ISL_1734201, EPI_ISL_1734202, EPI_ISL_1734203, EPI_ISL_1734204, EPI_ISL_1734205, EPI_ISL_1734206, EPI_ISL_1734207, EPI_ISL_1734208, EPI_ISL_1734209, EPI_ISL_1734210, EPI_ISL_1734211, EPI_ISL_1734212, EPI_ISL_1734213, EPI_ISL_1734214, EPI_ISL_1734215, EPI_ISL_1734216, EPI_ISL_1734218, EPI_ISL_1734219, EPI_ISL_1734220, EPI_ISL_1734221, EPI_ISL_1734223, EPI_ISL_1734224, EPI_ISL_1734225, EPI_ISL_1734226, EPI_ISL_1734227, EPI_ISL_1734228, EPI_ISL_1734229, EPI_ISL_1734231, EPI_ISL_1734233, EPI_ISL_1734234, EPI_ISL_1734235, EPI_ISL_1734236, EPI_ISL_1734237, EPI_ISL_1734238, EPI_ISL_1734239, EPI_ISL_1734240, EPI_ISL_1734241, EPI_ISL_1734242, EPI_ISL_1734243, EPI_ISL_1734244, EPI_ISL_1734245, EPI_ISL_1734246, EPI_ISL_1734247, EPI_ISL_1734248, EPI_ISL_1734249, EPI_ISL_1734250, EPI_ISL_1734251, EPI_ISL_1734252, EPI_ISL_1734253, EPI_ISL_1734254, EPI_ISL_1734256, EPI_ISL_1734257, EPI_ISL_1734258, EPI_ISL_1734259, EPI_ISL_1734260, EPI_ISL_1734261, EPI_ISL_1734262, EPI_ISL_1734263, EPI_ISL_1734264, EPI_ISL_1734265, EPI_ISL_1734266, EPI_ISL_1734267, EPI_ISL_1734268, EPI_ISL_1734269, EPI_ISL_1734270, EPI_ISL_1734271, EPI_ISL_1734272, EPI_ISL_1734273, EPI_ISL_1734274, EPI_ISL_1734275, EPI_ISL_1734276, EPI_ISL_1734277, EPI_ISL_1734278, EPI_ISL_1734279, EPI_ISL_1734280, EPI_ISL_1734281, EPI_ISL_1734282, EPI_ISL_1734283, EPI_ISL_1734284, EPI_ISL_1734286, EPI_ISL_1734287, EPI_ISL_1734289, EPI_ISL_1734290, EPI_ISL_1734291, EPI_ISL_1734292, EPI_ISL_1734293, EPI_ISL_1734294, EPI_ISL_1734295, EPI_ISL_1734296, EPI_ISL_1734297, EPI_ISL_1734299, EPI_ISL_1734300, EPI_ISL_1734301, EPI_ISL_1734302, EPI_ISL_1734303, EPI_ISL_1734304, EPI_ISL_1734305, EPI_ISL_1734306, EPI_ISL_1734307, EPI_ISL_1734309, EPI_ISL_1734311, EPI_ISL_1734312, EPI_ISL_1734313, EPI_ISL_1734314, EPI_ISL_1734315, EPI_ISL_1734316, EPI_ISL_1734317, EPI_ISL_1734318, EPI_ISL_1734319, EPI_ISL_1734320, EPI_ISL_1734321, EPI_ISL_1734323, EPI_ISL_1734324, EPI_ISL_1734325, EPI_ISL_1734326, EPI_ISL_1734327, EPI_ISL_1734328, EPI_ISL_1734329, EPI_ISL_1734330, EPI_ISL_1734331, EPI_ISL_1734333, EPI_ISL_1734334, EPI_ISL_1734335, EPI_ISL_1734336, EPI_ISL_1734337, EPI_ISL_1734338, EPI_ISL_1734339, EPI_ISL_1734340, EPI_ISL_1734341, EPI_ISL_1734342, EPI_ISL_1734343, EPI_ISL_1734344, EPI_ISL_1734345, EPI_ISL_1734346, EPI_ISL_1734347, EPI_ISL_1734348, EPI_ISL_1734349, EPI_ISL_1734350, EPI_ISL_1734351, EPI_ISL_1734352, EPI_ISL_1734353, EPI_ISL_1734354, EPI_ISL_1734355, EPI_ISL_1734356, EPI_ISL_1734358, EPI_ISL_1734359, EPI_ISL_1734360, EPI_ISL_1734361, EPI_ISL_1734362, EPI_ISL_1734363, EPI_ISL_1734364, EPI_ISL_1734365, EPI_ISL_1734366, EPI_ISL_1734367, EPI_ISL_1734368, EPI_ISL_1734369, EPI_ISL_1734370, EPI_ISL_1734371, EPI_ISL_1734372, EPI_ISL_1734373, EPI_ISL_1734374, EPI_ISL_1734375, EPI_ISL_1734376, EPI_ISL_1734377, EPI_ISL_1734378, EPI_ISL_1734379, EPI_ISL_1734380, EPI_ISL_1734381, EPI_ISL_1734382, EPI_ISL_1734383, EPI_ISL_1734384, EPI_ISL_1734386, EPI_ISL_1734388, EPI_ISL_1734389, EPI_ISL_1734390, EPI_ISL_1734391, EPI_ISL_1734392, EPI_ISL_1734393, EPI_ISL_1734394, EPI_ISL_1734395, EPI_ISL_1734396, EPI_ISL_1734398, EPI_ISL_1734399, EPI_ISL_1734400, EPI_ISL_1734401, EPI_ISL_1734402, EPI_ISL_1734403, EPI_ISL_1734404, EPI_ISL_1734405, EPI_ISL_1734406, EPI_ISL_1734407, EPI_ISL_1734408, EPI_ISL_1734410, EPI_ISL_1734411, EPI_ISL_1734412, EPI_ISL_1734413, EPI_ISL_1734414, EPI_ISL_1734415, EPI_ISL_1734416, EPI_ISL_1734417, EPI_ISL_1734418, EPI_ISL_1734419, EPI_ISL_1734420, EPI_ISL_1734421, EPI_ISL_1734423, EPI_ISL_1734424, EPI_ISL_1734425, EPI_ISL_1734427, EPI_ISL_1734428, EPI_ISL_1734430, EPI_ISL_1734431, EPI_ISL_1734432, EPI_ISL_1734433, EPI_ISL_1734434, EPI_ISL_1734435, EPI_ISL_1734436, EPI_ISL_1734437, EPI_ISL_1734438, EPI_ISL_1734439, EPI_ISL_1734440, EPI_ISL_1734441, EPI_ISL_1734442, EPI_ISL_1734443, EPI_ISL_1734444, EPI_ISL_1734446, EPI_ISL_1734447, EPI_ISL_1734448, EPI_ISL_1734449, EPI_ISL_1734450, EPI_ISL_1734451, EPI_ISL_1734452, EPI_ISL_1734453, EPI_ISL_1734454, EPI_ISL_1734455, EPI_ISL_1734456, EPI_ISL_1734457, EPI_ISL_1734458, EPI_ISL_1734459, EPI_ISL_1734460, EPI_ISL_1734461, EPI_ISL_1734462, EPI_ISL_1734464, EPI_ISL_1734465, EPI_ISL_1734466, EPI_ISL_1734467, EPI_ISL_1734469, EPI_ISL_1734470, EPI_ISL_1734471, EPI_ISL_1734472, EPI_ISL_1734473, EPI_ISL_1734474, EPI_ISL_1734475, EPI_ISL_1734476, EPI_ISL_1734477, EPI_ISL_1734478, EPI_ISL_1734479, EPI_ISL_1734480, EPI_ISL_1734481, EPI_ISL_1734482, EPI_ISL_1734483, EPI_ISL_1734484, EPI_ISL_1734485, EPI_ISL_1734486, EPI_ISL_1734487, EPI_ISL_1734488, EPI_ISL_1734489, EPI_ISL_1734490, EPI_ISL_1734491, EPI_ISL_1734492, EPI_ISL_1734493, EPI_ISL_1734494, EPI_ISL_1734495, EPI_ISL_1734496, EPI_ISL_1734497, EPI_ISL_1734498, EPI_ISL_1734499, EPI_ISL_1734500, EPI_ISL_1734501, EPI_ISL_1734502, EPI_ISL_1734503, EPI_ISL_1734504, EPI_ISL_1734506, EPI_ISL_1734508, EPI_ISL_1734509, EPI_ISL_1734510, EPI_ISL_1734512, EPI_ISL_1734513, EPI_ISL_1734514, EPI_ISL_1734515, EPI_ISL_1734517, EPI_ISL_1734518, EPI_ISL_1734519, EPI_ISL_1734520, EPI_ISL_1734521, EPI_ISL_1734523, EPI_ISL_1734525, EPI_ISL_1734526, EPI_ISL_1734527, EPI_ISL_1734528, EPI_ISL_1734529, EPI_ISL_1734530, EPI_ISL_1734532, EPI_ISL_1734533, EPI_ISL_1734534, EPI_ISL_1734535, EPI_ISL_1734536, EPI_ISL_1734537, EPI_ISL_1734538, EPI_ISL_1734539, EPI_ISL_1734540, EPI_ISL_1734542, EPI_ISL_1734543, EPI_ISL_1734544, EPI_ISL_1734545, EPI_ISL_1734546, EPI_ISL_1734547, EPI_ISL_1734549, EPI_ISL_1734550, EPI_ISL_1734551, EPI_ISL_1734552, EPI_ISL_1734553 | | | |
| see above | Fulgent Genetics | Centers for Disease Control and Prevention Division of Viral Diseases, Pathogen Discovery | Dakota Howard, Dhwani Batra, Peter W. Cook, Kara Moser, Adrian Paskey, Jason Caravas, Benjamin Rambo-Martin, Shatavia Morrison, Christopher Gulvick, Scott Sammons, Yvette Unoarumhi, Darlene Wagner, Matthew Schmerer, Harry Gao, Mickey Li, John Gao, Joseph Fierro, Benafsh Sapra, Becky Tsai, Yan Meng, Doreen Ng, James Xie, Clinton R. Paden, Duncan MacCannell |
| EPI_ISL_1734554, EPI_ISL_1734555, EPI_ISL_1734556, EPI_ISL_1734557, EPI_ISL_1734558, EPI_ISL_1734559, EPI_ISL_1734560, EPI_ISL_1734561, EPI_ISL_1734562, EPI_ISL_1734563, EPI_ISL_1734564, EPI_ISL_1734565, EPI_ISL_1734566, EPI_ISL_1734567, EPI_ISL_1734568, EPI_ISL_1734569, EPI_ISL_1734570, EPI_ISL_1734571, EPI_ISL_1734572, EPI_ISL_1734573, EPI_ISL_1734574, EPI_ISL_1734575, EPI_ISL_1734576, EPI_ISL_1734577, EPI_ISL_1734578, EPI_ISL_1734579, EPI_ISL_1734580, EPI_ISL_1734581, EPI_ISL_1734582, EPI_ISL_1734583, EPI_ISL_1734584, EPI_ISL_1734585, EPI_ISL_1734586, EPI_ISL_1734587, EPI_ISL_1734588, EPI_ISL_1734589, EPI_ISL_1734590, EPI_ISL_1734591, EPI_ISL_1734592, EPI_ISL_1734593, EPI_ISL_1734594, EPI_ISL_1734595, EPI_ISL_1734596, EPI_ISL_1734597, EPI_ISL_1734598, EPI_ISL_1734599, EPI_ISL_1734600, EPI_ISL_1734601, EPI_ISL_1734602, EPI_ISL_1734603, EPI_ISL_1734604, EPI_ISL_1734605, EPI_ISL_1734607, EPI_ISL_1734608, EPI_ISL_1734609, EPI_ISL_1734610, EPI_ISL_1734611, EPI_ISL_1734612, EPI_ISL_1734613, EPI_ISL_1734614, EPI_ISL_1734615, EPI_ISL_1734616, EPI_ISL_1734617, EPI_ISL_1734618, EPI_ISL_1734619, EPI_ISL_1734620, EPI_ISL_1734621, EPI_ISL_1734622, EPI_ISL_1734623, EPI_ISL_1734624, EPI_ISL_1734625, EPI_ISL_1734626, EPI_ISL_1734627, EPI_ISL_1734628, EPI_ISL_1734629, EPI_ISL_1734630, EPI_ISL_1734631, EPI_ISL_1734632, EPI_ISL_1734633, EPI_ISL_1734634, EPI_ISL_1734635, EPI_ISL_1734636, EPI_ISL_1734637, EPI_ISL_1734638, EPI_ISL_1734639, EPI_ISL_1734640, EPI_ISL_1734641, EPI_ISL_1734642, EPI_ISL_1734643, EPI_ISL_1734644, EPI_ISL_1734645, EPI_ISL_1734646, EPI_ISL_1734647, EPI_ISL_1734648, EPI_ISL_1734649, EPI_ISL_1734650, EPI_ISL_1734651, EPI_ISL_1734652, EPI_ISL_1734653, EPI_ISL_1734654, EPI_ISL_1734655, EPI_ISL_1734656, EPI_ISL_1734657, EPI_ISL_1734658, EPI_ISL_1734659, EPI_ISL_1734660, EPI_ISL_1734661, EPI_ISL_1734662, EPI_ISL_1734663, EPI_ISL_1734664, EPI_ISL_1734665, EPI_ISL_1734666, EPI_ISL_1734667, EPI_ISL_1734668, EPI_ISL_1734669, EPI_ISL_1734670, EPI_ISL_1734671, EPI_ISL_1734672, EPI_ISL_1734673, EPI_ISL_1734674, EPI_ISL_1734675, EPI_ISL_1734676, EPI_ISL_1734677, EPI_ISL_1734678, EPI_ISL_1734679, EPI_ISL_1734680, EPI_ISL_1734681, EPI_ISL_1734682, EPI_ISL_1734683, EPI_ISL_1734684, EPI_ISL_1734685, EPI_ISL_1734686, EPI_ISL_1734687, EPI_ISL_1734688, EPI_ISL_1734689, EPI_ISL_1734690, EPI_ISL_1734691, EPI_ISL_1734692, EPI_ISL_1734693, EPI_ISL_1734694, EPI_ISL_1734695, EPI_ISL_1734696, EPI_ISL_1734697, EPI_ISL_1734698, EPI_ISL_1734699, EPI_ISL_1734700, EPI_ISL_1734701, EPI_ISL_1734702, EPI_ISL_1734703, EPI_ISL_1734704, EPI_ISL_1734705, EPI_ISL_1734706, EPI_ISL_1734707, EPI_ISL_1734708, EPI_ISL_1734709, EPI_ISL_1734710, EPI_ISL_1734711, EPI_ISL_1734712, EPI_ISL_1734713, EPI_ISL_1734714, EPI_ISL_1734715, EPI_ISL_1734716, EPI_ISL_1734717, EPI_ISL_1734718, EPI_ISL_1734719, EPI_ISL_1734720, EPI_ISL_1734721, EPI_ISL_1734722, EPI_ISL_1734723, EPI_ISL_1734724, EPI_ISL_1734725, EPI_ISL_1734726, EPI_ISL_1734727, EPI_ISL_1734728, EPI_ISL_1734729, EPI_ISL_1734730, EPI_ISL_1734731, EPI_ISL_1734732, EPI_ISL_1734733, EPI_ISL_1734734, EPI_ISL_1734735, EPI_ISL_1734737, EPI_ISL_1734738, EPI_ISL_1734739, EPI_ISL_1734740, EPI_ISL_1734741, EPI_ISL_1734742, EPI_ISL_1734743, EPI_ISL_1734744, EPI_ISL_1734745, EPI_ISL_1734746, EPI_ISL_1734747, EPI_ISL_1734748, EPI_ISL_1734749, EPI_ISL_1734750, EPI_ISL_1734751, EPI_ISL_1734752, EPI_ISL_1734753, EPI_ISL_1734754, EPI_ISL_1734755, EPI_ISL_1734756, EPI_ISL_1734757, EPI_ISL_1734758, EPI_ISL_1734759, EPI_ISL_1734760, EPI_ISL_1734761, EPI_ISL_1734762, EPI_ISL_1734763, EPI_ISL_1734764, EPI_ISL_1734765, EPI_ISL_1734766, EPI_ISL_1734767, EPI_ISL_1734768, EPI_ISL_1734769, EPI_ISL_1734770, EPI_ISL_1734771, EPI_ISL_1734772, EPI_ISL_1734773, EPI_ISL_1734774, EPI_ISL_1734775, EPI_ISL_1734776, EPI_ISL_1734777, EPI_ISL_1734778, EPI_ISL_1734779, EPI_ISL_1734780, EPI_ISL_1734781, EPI_ISL_1734782, EPI_ISL_1734783, EPI_ISL_1734784, EPI_ISL_1734785, EPI_ISL_1734786, EPI_ISL_1734787, EPI_ISL_1734788, EPI_ISL_1734789, EPI_ISL_1734790, EPI_ISL_1734791, EPI_ISL_1734792, EPI_ISL_1734793, EPI_ISL_1734794, EPI_ISL_1734795, EPI_ISL_1734796, EPI_ISL_1734797, EPI_ISL_1734798, EPI_ISL_1734799, EPI_ISL_1734800, EPI_ISL_1734801, EPI_ISL_1734802, EPI_ISL_1734803, EPI_ISL_1734804, EPI_ISL_1734805, EPI_ISL_1734806, EPI_ISL_1734807, EPI_ISL_1734808, EPI_ISL_1734809, EPI_ISL_1734810, EPI_ISL_1734811, EPI_ISL_1734812, EPI_ISL_1734813, EPI_ISL_1734814, EPI_ISL_1734815, EPI_ISL_1734816, EPI_ISL_1734817, EPI_ISL_1734818, EPI_ISL_1734819, EPI_ISL_1734820, EPI_ISL_1734821, EPI_ISL_1734822, EPI_ISL_1734823, EPI_ISL_1734824, EPI_ISL_1734825, EPI_ISL_1734826, EPI_ISL_1734827, EPI_ISL_1734828, EPI_ISL_1734829, EPI_ISL_1734830, EPI_ISL_1734831, EPI_ISL_1734832, EPI_ISL_1734833, EPI_ISL_1734834, EPI_ISL_1734835, EPI_ISL_1734836, EPI_ISL_1734837, EPI_ISL_1734838, EPI_ISL_1734839, EPI_ISL_1734840, EPI_ISL_1734842, EPI_ISL_1734845, EPI_ISL_1734848, EPI_ISL_1734851, EPI_ISL_1734854, EPI_ISL_1734856, EPI_ISL_1734859, EPI_ISL_1734862, EPI_ISL_1734865, EPI_ISL_1734867, EPI_ISL_1734870, EPI_ISL_1734873, EPI_ISL_1734876, EPI_ISL_1734879, EPI_ISL_1734882, EPI_ISL_1734885, EPI_ISL_1734886, EPI_ISL_1734887, EPI_ISL_1734888, EPI_ISL_1734889, EPI_ISL_1734890, EPI_ISL_1734891, EPI_ISL_1734892, EPI_ISL_1734893, EPI_ISL_1734894, EPI_ISL_1734895, EPI_ISL_1734896, EPI_ISL_1734897, EPI_ISL_1734898, EPI_ISL_1734899, EPI_ISL_1734900, EPI_ISL_1734901, EPI_ISL_1734902, EPI_ISL_1734903, EPI_ISL_1734904, EPI_ISL_1734906, EPI_ISL_1734907, EPI_ISL_1734908, EPI_ISL_1734909, EPI_ISL_1734910, EPI_ISL_1734911, EPI_ISL_1734912, EPI_ISL_1734913, EPI_ISL_1734914, EPI_ISL_1734915, EPI_ISL_1734916, EPI_ISL_1734917, EPI_ISL_1734918, EPI_ISL_1734919, EPI_ISL_1734920, EPI_ISL_1734921, EPI_ISL_1734922, EPI_ISL_1734923, EPI_ISL_1734924, EPI_ISL_1734925, EPI_ISL_1734926, EPI_ISL_1734927, EPI_ISL_1734928, EPI_ISL_1734929, EPI_ISL_1734930, EPI_ISL_1734931, EPI_ISL_1734932, EPI_ISL_1734933, EPI_ISL_1734934, EPI_ISL_1734935, EPI_ISL_1734936, EPI_ISL_1734937, EPI_ISL_1734938, EPI_ISL_1734939, EPI_ISL_1734940, EPI_ISL_1734941, EPI_ISL_1734942, EPI_ISL_1734943, EPI_ISL_1734944, EPI_ISL_1734945, EPI_ISL_1734946, EPI_ISL_1734947, EPI_ISL_1734948, EPI_ISL_1734949, EPI_ISL_1734950, EPI_ISL_1734951, EPI_ISL_1734952, EPI_ISL_1734953, EPI_ISL_1734954, EPI_ISL_1734955, EPI_ISL_1734956, EPI_ISL_1734957, EPI_ISL_1734958, EPI_ISL_1734959, EPI_ISL_1734960, EPI_ISL_1734961, EPI_ISL_1734962, EPI_ISL_1734963, EPI_ISL_1734964, EPI_ISL_1734965, EPI_ISL_1734966, EPI_ISL_1734967, EPI_ISL_1734968, EPI_ISL_1734969, EPI_ISL_1734970, EPI_ISL_1734971, EPI_ISL_1734972, EPI_ISL_1734973, EPI_ISL_1734974, EPI_ISL_1734975, EPI_ISL_1734976, EPI_ISL_1734977, EPI_ISL_1734978, EPI_ISL_1734979, EPI_ISL_1734980, EPI_ISL_1734981, EPI_ISL_1734982, EPI_ISL_1734983, EPI_ISL_1734984, EPI_ISL_1734985, EPI_ISL_1734986, EPI_ISL_1734987, EPI_ISL_1734988, EPI_ISL_1734989, EPI_ISL_1734990, EPI_ISL_1734991, EPI_ISL_1734992, EPI_ISL_1734993, EPI_ISL_1734994, EPI_ISL_1734995, EPI_ISL_1734996, EPI_ISL_1734997, EPI_ISL_1734998, EPI_ISL_1734999, EPI_ISL_1735000, EPI_ISL_1735001, EPI_ISL_1735002, EPI_ISL_1735003, EPI_ISL_1735004, EPI_ISL_1735005, EPI_ISL_1735006, EPI_ISL_1735007, EPI_ISL_1735008, EPI_ISL_1735009, EPI_ISL_1735010, EPI_ISL_1735011, EPI_ISL_1735012, EPI_ISL_1735013, EPI_ISL_1735014, EPI_ISL_1735015, EPI_ISL_1735016, EPI_ISL_1735017, EPI_ISL_1735018, EPI_ISL_1735019, EPI_ISL_1735020, EPI_ISL_1735021, EPI_ISL_1735022, EPI_ISL_1735023, EPI_ISL_1735024, EPI_ISL_1735025, EPI_ISL_1735026, EPI_ISL_1735027, EPI_ISL_1735028, EPI_ISL_1735029, EPI_ISL_1735030, EPI_ISL_1735031, EPI_ISL_1735032, EPI_ISL_1735033, EPI_ISL_1735034, EPI_ISL_1735035, EPI_ISL_1735036, EPI_ISL_1735037, EPI_ISL_1735039, EPI_ISL_1735040, EPI_ISL_1735041, EPI_ISL_1735042, EPI_ISL_1735043, EPI_ISL_1735044, EPI_ISL_1735045, EPI_ISL_1735046, EPI_ISL_1735047, EPI_ISL_1735048, EPI_ISL_1735049, EPI_ISL_1735050, EPI_ISL_1735051, EPI_ISL_1735052, EPI_ISL_1735053, EPI_ISL_1735054, EPI_ISL_1735055, EPI_ISL_1735056, EPI_ISL_1735057, EPI_ISL_1735058, EPI_ISL_1735059, EPI_ISL_1735060, EPI_ISL_1735061, EPI_ISL_1735062, EPI_ISL_1735063, EPI_ISL_1735064, EPI_ISL_1735065, EPI_ISL_1735066, EPI_ISL_1735068, EPI_ISL_1735069, EPI_ISL_1735070, EPI_ISL_1735071, EPI_ISL_1735072, EPI_ISL_1735073, EPI_ISL_1735074, EPI_ISL_1735075, EPI_ISL_1735076, EPI_ISL_1735077, EPI_ISL_1735078, EPI_ISL_1735079, EPI_ISL_1735080, EPI_ISL_1735081, EPI_ISL_1735082, EPI_ISL_1735083, EPI_ISL_1735084, EPI_ISL_1735085, EPI_ISL_1735086, EPI_ISL_1735087, EPI_ISL_1735088, EPI_ISL_1735089, EPI_ISL_1735090, EPI_ISL_1735091, EPI_ISL_1735092, EPI_ISL_1735093, EPI_ISL_1735094, EPI_ISL_1735095, EPI_ISL_1735096, EPI_ISL_1735097, EPI_ISL_1735098, EPI_ISL_1735099, EPI_ISL_1735100, EPI_ISL_1735101, EPI_ISL_1735102, EPI_ISL_1735103, EPI_ISL_1735104, EPI_ISL_1735105, EPI_ISL_1735106, EPI_ISL_1735107, EPI_ISL_1735108, EPI_ISL_1735109, EPI_ISL_1735110, EPI_ISL_1735111, EPI_ISL_1735112, EPI_ISL_1735113, EPI_ISL_1735114, EPI_ISL_1735115, EPI_ISL_1735116, EPI_ISL_1735117, EPI_ISL_1735118, EPI_ISL_1735119, EPI_ISL_1735120, EPI_ISL_1735121, EPI_ISL_1735122, EPI_ISL_1735123, EPI_ISL_1735124, EPI_ISL_1735125, EPI_ISL_1735126, EPI_ISL_1735127, EPI_ISL_1735128, EPI_ISL_1735129, EPI_ISL_1735130, EPI_ISL_1735131, EPI_ISL_1735132, EPI_ISL_1735133, EPI_ISL_1735134, EPI_ISL_1735135, EPI_ISL_1735136, EPI_ISL_1735137, EPI_ISL_1735138, EPI_ISL_1735139, EPI_ISL_1735140, EPI_ISL_1735141, EPI_ISL_1735142, EPI_ISL_1735143, EPI_ISL_1735144, EPI_ISL_1735145, EPI_ISL_1735146, EPI_ISL_1735147, EPI_ISL_1735148, EPI_ISL_1735149, EPI_ISL_1735150, EPI_ISL_1735151, EPI_ISL_1735152, EPI_ISL_1735153, EPI_ISL_1735154, EPI_ISL_1735155, EPI_ISL_1735156, EPI_ISL_1735157, EPI_ISL_1735158, EPI_ISL_1735159, EPI_ISL_1735160, EPI_ISL_1735161, EPI_ISL_1735162, EPI_ISL_1735163, EPI_ISL_1735164, EPI_ISL_1735165, EPI_ISL_1735166, EPI_ISL_1735167, EPI_ISL_1735168, EPI_ISL_1735169, EPI_ISL_1735170, EPI_ISL_1735171, EPI_ISL_1735172, EPI_ISL_1735173, EPI_ISL_1735174, EPI_ISL_1735175, EPI_ISL_1735176, EPI_ISL_1735177, EPI_ISL_1735178, EPI_ISL_1735179, EPI_ISL_1735180, EPI_ISL_1735181, EPI_ISL_1735182, EPI_ISL_1735183, EPI_ISL_1735184, EPI_ISL_1735185, EPI_ISL_1735186, EPI_ISL_1735187, EPI_ISL_1735188, EPI_ISL_1735189, EPI_ISL_1735190, EPI_ISL_1735191, EPI_ISL_1735192, EPI_ISL_1735193, EPI_ISL_1735194, EPI_ISL_1735195, EPI_ISL_1735196, EPI_ISL_1735197, EPI_ISL_1735198, EPI_ISL_1735199, EPI_ISL_1735200, EPI_ISL_1735201, EPI_ISL_1735202, EPI_ISL_1735203, EPI_ISL_1735204, EPI_ISL_1735205, EPI_ISL_1735206, EPI_ISL_1735207, EPI_ISL_1735208, EPI_ISL_1735209, EPI_ISL_1735210, EPI_ISL_1735211, EPI_ISL_1735212, EPI_ISL_1735213, EPI_ISL_1735214, EPI_ISL_1735215, EPI_ISL_1735216, EPI_ISL_1735217, EPI_ISL_1735218, EPI_ISL_1735219, EPI_ISL_1735220, EPI_ISL_1735221, EPI_ISL_1735222, EPI_ISL_1735223, EPI_ISL_1735224, EPI_ISL_1735225, EPI_ISL_1735226, EPI_ISL_1735227, EPI_ISL_1735228, EPI_ISL_1735229, EPI_ISL_1735230, EPI_ISL_1735231, EPI_ISL_1735232, EPI_ISL_1735233, EPI_ISL_1735234, EPI_ISL_1735235, EPI_ISL_1735236, EPI_ISL_1735237, EPI_ISL_1735238, EPI_ISL_1735239, EPI_ISL_1735240, EPI_ISL_1735241, EPI_ISL_1735242, EPI_ISL_1735243, EPI_ISL_1735244, EPI_ISL_1735245, EPI_ISL_1735246, EPI_ISL_1735247, EPI_ISL_1735248, EPI_ISL_1735249, EPI_ISL_1735250, EPI_ISL_1735251, EPI_ISL_1735252, EPI_ISL_1735253, EPI_ISL_1735254, EPI_ISL_1735255, EPI_ISL_1735256, EPI_ISL_1735257, EPI_ISL_1735258, EPI_ISL_1735259, EPI_ISL_1735260, EPI_ISL_1735261, EPI_ISL_1735262, EPI_ISL_1735263, EPI_ISL_1735264, EPI_ISL_1735265, EPI_ISL_1735266, EPI_ISL_1735267, EPI_ISL_1735268, EPI_ISL_1735269, EPI_ISL_1735270, EPI_ISL_1735271, EPI_ISL_1735272, EPI_ISL_1735273, EPI_ISL_1735274, EPI_ISL_1735275, EPI_ISL_1735276, EPI_ISL_1735277, EPI_ISL_1735278, EPI_ISL_1735279, EPI_ISL_1735280, EPI_ISL_1735281, EPI_ISL_1735282, EPI_ISL_1735283, EPI_ISL_1735284, EPI_ISL_1735285, EPI_ISL_1735286, EPI_ISL_1735287, EPI_ISL_1735288, EPI_ISL_1735289, EPI_ISL_1735290, EPI_ISL_1735291, EPI_ISL_1735292, EPI_ISL_1735293, EPI_ISL_1735294, EPI_ISL_1735295, EPI_ISL_1735296, EPI_ISL_1735297, EPI_ISL_1735298, EPI_ISL_1735299, EPI_ISL_1735300, EPI_ISL_1735301, EPI_ISL_1735302, EPI_ISL_1735303, EPI_ISL_1735304, EPI_ISL_1735305, EPI_ISL_1735306, EPI_ISL_1735307, EPI_ISL_1735308, EPI_ISL_1735309, EPI_ISL_1735310, EPI_ISL_1735311, EPI_ISL_1735312, EPI_ISL_1735313, EPI_ISL_1735314, EPI_ISL_1735315, EPI_ISL_1735316, EPI_ISL_1735317, EPI_ISL_1735318, EPI_ISL_1735319, EPI_ISL_1735320, EPI_ISL_1735321, EPI_ISL_1735322, EPI_ISL_1735323, EPI_ISL_1735324, EPI_ISL_1735325, EPI_ISL_1735326, EPI_ISL_1735327, EPI_ISL_1735328, EPI_ISL_1735329, EPI_ISL_1735330, EPI_ISL_1735331, EPI_ISL_1735332, EPI_ISL_1735333, EPI_ISL_1735334, EPI_ISL_1735335, EPI_ISL_1735336, EPI_ISL_1735337, EPI_ISL_1735338, EPI_ISL_1735339, EPI_ISL_1735340, EPI_ISL_1735341, EPI_ISL_1735342, EPI_ISL_1735343, EPI_ISL_1735344, EPI_ISL_1735345, EPI_ISL_1735346, EPI_ISL_1735347, EPI_ISL_1735348, EPI_ISL_1735349, EPI_ISL_1735350, EPI_ISL_1735351, EPI_ISL_1735352, EPI_ISL_1735353, EPI_ISL_1735354, EPI_ISL_1735355, EPI_ISL_1735356, EPI_ISL_1735357, EPI_ISL_1735358, EPI_ISL_1735359, EPI_ISL_1735360, EPI_ISL_1735361, EPI_ISL_1735362, EPI_ISL_1735364, EPI_ISL_1735365, EPI_ISL_1735366, EPI_ISL_1735367, EPI_ISL_1735368, EPI_ISL_1735369, EPI_ISL_1735370, EPI_ISL_1735371, EPI_ISL_1735372, EPI_ISL_1735373, EPI_ISL_1735374, EPI_ISL_1735375, EPI_ISL_1735376, EPI_ISL_1735377, EPI_ISL_1735378, EPI_ISL_1735379, EPI_ISL_1735380, EPI_ISL_1735381, EPI_ISL_1735382, EPI_ISL_1735383, EPI_ISL_1735384, EPI_ISL_1735385, EPI_ISL_1735386, EPI_ISL_1735387, EPI_ISL_1735388, EPI_ISL_1735389, EPI_ISL_1735390, EPI_ISL_1735391, EPI_ISL_1735392, EPI_ISL_1735393, EPI_ISL_1735394, EPI_ISL_1735395, EPI_ISL_1735396, EPI_ISL_1735397, EPI_ISL_1735398, EPI_ISL_1735399, EPI_ISL_1735400, EPI_ISL_1735401, EPI_ISL_1735402, EPI_ISL_1735403, EPI_ISL_1735404, EPI_ISL_1735405, EPI_ISL_1735406, EPI_ISL_1735407, EPI_ISL_1735408, EPI_ISL_1735409, EPI_ISL_1735410, EPI_ISL_1735411, EPI_ISL_1735412, EPI_ISL_1735413, EPI_ISL_1735414, EPI_ISL_1735415, EPI_ISL_1735416, EPI_ISL_1735417, EPI_ISL_1735418, EPI_ISL_1735419, EPI_ISL_1735420, EPI_ISL_1735421, EPI_ISL_1735422, EPI_ISL_1735423, EPI_ISL_1735424, EPI_ISL_1735425, EPI_ISL_1735426, EPI_ISL_1735427, EPI_ISL_1735428, EPI_ISL_1735429, EPI_ISL_1735430, EPI_ISL_1735431, EPI_ISL_1735432, EPI_ISL_1735433, EPI_ISL_1735434, EPI_ISL_1735435, EPI_ISL_1735436, EPI_ISL_1735437, EPI_ISL_1735438, EPI_ISL_1735439, EPI_ISL_1735440, EPI_ISL_1735441, EPI_ISL_1735442, EPI_ISL_1735443, EPI_ISL_1735444, EPI_ISL_1735445, EPI_ISL_1735446, EPI_ISL_1735447, EPI_ISL_1735448, EPI_ISL_1735449, EPI_ISL_1735450, EPI_ISL_1735452, EPI_ISL_1735453, EPI_ISL_1735454, EPI_ISL_1735455, EPI_ISL_1735456, EPI_ISL_1735457, EPI_ISL_1735458, EPI_ISL_1735459, EPI_ISL_1735460, EPI_ISL_1735461, EPI_ISL_1735462, EPI_ISL_1735463, EPI_ISL_1735464, EPI_ISL_1735465, EPI_ISL_1735466, EPI_ISL_1735467, EPI_ISL_1735468, EPI_ISL_1735469, EPI_ISL_1735470, EPI_ISL_1735471, EPI_ISL_1735472, EPI_ISL_1735473, EPI_ISL_1735474, EPI_ISL_1735475, EPI_ISL_1735476, EPI_ISL_1735477, EPI_ISL_1735478, EPI_ISL_1735479, EPI_ISL_1735480, EPI_ISL_1735481, EPI_ISL_1735482, EPI_ISL_1735483, EPI_ISL_1735484, EPI_ISL_1735485, EPI_ISL_1735486, EPI_ISL_1735487, EPI_ISL_1735488, EPI_ISL_1735489, EPI_ISL_1735490, EPI_ISL_1735491, EPI_ISL_1735492, EPI_ISL_1735493, EPI_ISL_1735494, EPI_ISL_1735495, EPI_ISL_1735496, EPI_ISL_1735497, EPI_ISL_1735498, EPI_ISL_1735499, EPI_ISL_1735500, EPI_ISL_1735501, EPI_ISL_1735502, EPI_ISL_1735503, EPI_ISL_1735504, EPI_ISL_1735505, EPI_ISL_1735506, EPI_ISL_1735507, EPI_ISL_1735508, EPI_ISL_1735509, EPI_ISL_1735510, EPI_ISL_1735511, EPI_ISL_1735512, EPI_ISL_1735513, EPI_ISL_1735514, EPI_ISL_1735515, EPI_ISL_1735516, EPI_ISL_1735517, EPI_ISL_1735518, EPI_ISL_1735519, EPI_ISL_1735520, EPI_ISL_1735521, EPI_ISL_1735522, EPI_ISL_1735523, EPI_ISL_1735524, EPI_ISL_1735525, EPI_ISL_1735526, EPI_ISL_1735527, EPI_ISL_1735528, EPI_ISL_1735529, EPI_ISL_1735530, EPI_ISL_1735531, EPI_ISL_1735532, EPI_ISL_1735533, EPI_ISL_1735534, EPI_ISL_1735535, EPI_ISL_1735536, EPI_ISL_1735537, EPI_ISL_1735538, EPI_ISL_1735539, EPI_ISL_1735540, EPI_ISL_1735541, EPI_ISL_1735542, EPI_ISL_1735543, EPI_ISL_1735544, EPI_ISL_1735545, EPI_ISL_1735546, EPI_ISL_1735547, EPI_ISL_1735548, EPI_ISL_1735549, EPI_ISL_1735550, EPI_ISL_1735551, EPI_ISL_1735552, EPI_ISL_1735553, EPI_ISL_1735554, EPI_ISL_1735555, EPI_ISL_1735556, EPI_ISL_1735557, EPI_ISL_1735558, EPI_ISL_1735559, EPI_ISL_1735560, EPI_ISL_1735561, EPI_ISL_1735562, EPI_ISL_1735563, EPI_ISL_1735564, EPI_ISL_1735565, EPI_ISL_1735566, EPI_ISL_1735567, EPI_ISL_1735568, EPI_ISL_1735569, EPI_ISL_1735570, EPI_ISL_1735571, EPI_ISL_1735572, EPI_ISL_1735573, EPI_ISL_1735574, EPI_ISL_1735575, EPI_ISL_1735576, EPI_ISL_1735577, EPI_ISL_1735578, EPI_ISL_1735579, EPI_ISL_1735580, EPI_ISL_1735581, EPI_ISL_1735582, EPI_ISL_1735583, EPI_ISL_1735584, EPI_ISL_1735585, EPI_ISL_1735586, EPI_ISL_1735587, EPI_ISL_1735588, EPI_ISL_1735589, EPI_ISL_1735590, EPI_ISL_1735591, EPI_ISL_1735592, EPI_ISL_1735593, EPI_ISL_1735594, EPI_ISL_1735595, EPI_ISL_1735596, EPI_ISL_1735597, EPI_ISL_1735598, EPI_ISL_1735599, EPI_ISL_1735600, EPI_ISL_1735601, EPI_ISL_1735602, EPI_ISL_1735603, EPI_ISL_1735604, EPI_ISL_1735605, EPI_ISL_1735606, EPI_ISL_1735607, EPI_ISL_1735608, EPI_ISL_1735609, EPI_ISL_1735610, EPI_ISL_1735611, EPI_ISL_1735612, EPI_ISL_1735613, EPI_ISL_1735614, EPI_ISL_1735615, EPI_ISL_1735616, EPI_ISL_1735617, EPI_ISL_1735618, EPI_ISL_1735619, EPI_ISL_1735620, EPI_ISL_1735621, EPI_ISL_1735622, EPI_ISL_1735623, EPI_ISL_1735624, EPI_ISL_1735625, EPI_ISL_1735626, EPI_ISL_1735627, EPI_ISL_1735628, EPI_ISL_1735629, EPI_ISL_1735630, EPI_ISL_1735631, EPI_ISL_1735632, EPI_ISL_1735633, EPI_ISL_1735634, EPI_ISL_1735635, EPI_ISL_1735636, EPI_ISL_1735637, EPI_ISL_1735638, EPI_ISL_1735639, EPI_ISL_1735640, EPI_ISL_1735641, EPI_ISL_1735642, EPI_ISL_1735643, EPI_ISL_1735644, EPI_ISL_1735645, EPI_ISL_1735646, EPI_ISL_1735647, EPI_ISL_1735648, EPI_ISL_1735649, EPI_ISL_1735650, EPI_ISL_1735651, EPI_ISL_1735652, EPI_ISL_1735653, EPI_ISL_1735654, EPI_ISL_1735655, EPI_ISL_1735656, EPI_ISL_1735657, EPI_ISL_1735658, EPI_ISL_1735659, EPI_ISL_1735660, EPI_ISL_1735661, EPI_ISL_1735662, EPI_ISL_1735663, EPI_ISL_1735664, EPI_ISL_1735665, EPI_ISL_1735666, EPI_ISL_1735667, EPI_ISL_1735668, EPI_ISL_1735669, EPI_ISL_1735670, EPI_ISL_1735671, EPI_ISL_1735672, EPI_ISL_1735673, EPI_ISL_1735674, EPI_ISL_1735675, EPI_ISL_1735676, EPI_ISL_1735677, EPI_ISL_1735678, EPI_ISL_1735679 | | | |
| see above | Helix/Illumina | Centers for Disease Control and Prevention Division of Viral Diseases, Pathogen Discovery | Dakota Howard, Dhwani Batra, Peter W. Cook, Kara Moser, Adrian Paskey, Jason Caravas, Benjamin Rambo-Martin, Shatavia Morrison, Christopher Gulvick, Scott Sammons, Yvette Unoarumhi, Darlene Wagner, Matthew Schmerer, Eileen de Feo, Jan Antico, Christine Tran, Matthew Tolentino, Shannon Wickline, Kim Gietzen, Brad Sickler, Jingtao Liu, Eric Allen, Phil Febbo, Nicole L. Washington, Simon White, Geraint Levan, Kelly Schiabor Barrett, Elizabeth Cirulli, Alexandre Bolze, Ary Ascencio, Charlotte Rivera-Garcia, Ryan Cho, Jason Nguyen, Sherry Wang, Jimmy Ramirez, Tyler Cassens, Efren Sandoval, Magnus Isaksson, William Lee, David Becker, Marc Laurent, James Lu, Clinton R. Paden, Duncan MacCannell |
| EPI_ISL_1735680, EPI_ISL_1735681, EPI_ISL_1735682, EPI_ISL_1735683, EPI_ISL_1735684, EPI_ISL_1735685, EPI_ISL_1735686, EPI_ISL_1735687, EPI_ISL_1735688, EPI_ISL_1735689, EPI_ISL_1735690, EPI_ISL_1735691, EPI_ISL_1735692, EPI_ISL_1735693, EPI_ISL_1735694, EPI_ISL_1735695, EPI_ISL_1735696, EPI_ISL_1735697, EPI_ISL_1735698, EPI_ISL_1735699, EPI_ISL_1735700, EPI_ISL_1735701, EPI_ISL_1735702, EPI_ISL_1735703, EPI_ISL_1735704, EPI_ISL_1735705, EPI_ISL_1735706, EPI_ISL_1735707, EPI_ISL_1735708, EPI_ISL_1735709, EPI_ISL_1735710, EPI_ISL_1735711, EPI_ISL_1735712, EPI_ISL_1735713, EPI_ISL_1735714, EPI_ISL_1735715, EPI_ISL_1735716, EPI_ISL_1735717, EPI_ISL_1735718, EPI_ISL_1735719, EPI_ISL_1735720, EPI_ISL_1735721, EPI_ISL_1735722, EPI_ISL_1735723, EPI_ISL_1735724, EPI_ISL_1735725, EPI_ISL_1735726, EPI_ISL_1735727, EPI_ISL_1735728, EPI_ISL_1735729, EPI_ISL_1735730, EPI_ISL_1735731, EPI_ISL_1735732, EPI_ISL_1735733, EPI_ISL_1735734, EPI_ISL_1735735, EPI_ISL_1735736, EPI_ISL_1735737, EPI_ISL_1735738, EPI_ISL_1735739, EPI_ISL_1735740, EPI_ISL_1735741, EPI_ISL_1735742, EPI_ISL_1735743, EPI_ISL_1735744, EPI_ISL_1735745, EPI_ISL_1735746, EPI_ISL_1735747, EPI_ISL_1735748, EPI_ISL_1735749, EPI_ISL_1735750, EPI_ISL_1735751, EPI_ISL_1735752, EPI_ISL_1735753, EPI_ISL_1735754, EPI_ISL_1735755, EPI_ISL_1735756, EPI_ISL_1735757, EPI_ISL_1735758, EPI_ISL_1735759, EPI_ISL_1735760, EPI_ISL_1735761, EPI_ISL_1735762, EPI_ISL_1735763, EPI_ISL_1735764, EPI_ISL_1735765, EPI_ISL_1735766, EPI_ISL_1735767, EPI_ISL_1735768, EPI_ISL_1735769, EPI_ISL_1735770, EPI_ISL_1735771, EPI_ISL_1735772, EPI_ISL_1735773, EPI_ISL_1735774, EPI_ISL_1735775, EPI_ISL_1735776, EPI_ISL_1735777, EPI_ISL_1735778, EPI_ISL_1735779, EPI_ISL_1735780, EPI_ISL_1735781, EPI_ISL_1735782, EPI_ISL_1735783, EPI_ISL_1735784, EPI_ISL_1735785, EPI_ISL_1735786, EPI_ISL_1735787, EPI_ISL_1735788, EPI_ISL_1735789, EPI_ISL_1735790, EPI_ISL_1735791, EPI_ISL_1735792, EPI_ISL_1735793, EPI_ISL_1735794, EPI_ISL_1735795, EPI_ISL_1735796, EPI_ISL_1735797, EPI_ISL_1735798, EPI_ISL_1735799, EPI_ISL_1735800, EPI_ISL_1735801, EPI_ISL_1735802, EPI_ISL_1735803, EPI_ISL_1735804, EPI_ISL_1735805, EPI_ISL_1735806, EPI_ISL_1735807, EPI_ISL_1735808, EPI_ISL_1735809, EPI_ISL_1735811, EPI_ISL_1735812, EPI_ISL_1735813, EPI_ISL_1735814, EPI_ISL_1735815, EPI_ISL_1735816, EPI_ISL_1735817, EPI_ISL_1735818, EPI_ISL_1735819, EPI_ISL_1735820, EPI_ISL_1735821, EPI_ISL_1735822, EPI_ISL_1735823, EPI_ISL_1735824, EPI_ISL_1735825, EPI_ISL_1735826, EPI_ISL_1735827, EPI_ISL_1735828, EPI_ISL_1735829, EPI_ISL_1735830, EPI_ISL_1735831, EPI_ISL_1735832, EPI_ISL_1735833, EPI_ISL_1735834, EPI_ISL_1735835, EPI_ISL_1735836, EPI_ISL_1735837, EPI_ISL_1735838, EPI_ISL_1735839, EPI_ISL_1735840, EPI_ISL_1735841, EPI_ISL_1735842, EPI_ISL_1735843, EPI_ISL_1735844, EPI_ISL_1735845, EPI_ISL_1735846, EPI_ISL_1735847, EPI_ISL_1735848, EPI_ISL_1735849, EPI_ISL_1735850, EPI_ISL_1735851, EPI_ISL_1735852, EPI_ISL_1735853, EPI_ISL_1735854, EPI_ISL_1735855, EPI_ISL_1735856, EPI_ISL_1735857, EPI_ISL_1735858, EPI_ISL_1735859, EPI_ISL_1735860, EPI_ISL_1735861, EPI_ISL_1735862, EPI_ISL_1735863, EPI_ISL_1735864, EPI_ISL_1735865, EPI_ISL_1735866, EPI_ISL_1735868, EPI_ISL_1735869, EPI_ISL_1735870, EPI_ISL_1735871, EPI_ISL_1735872, EPI_ISL_1735873, EPI_ISL_1735874, EPI_ISL_1735875, EPI_ISL_1735876, EPI_ISL_1735877, EPI_ISL_1735878, EPI_ISL_1735879, EPI_ISL_1735880, EPI_ISL_1735881, EPI_ISL_1735882, EPI_ISL_1735883, EPI_ISL_1735884, EPI_ISL_1735885, EPI_ISL_1735886, EPI_ISL_1735887, EPI_ISL_1735888, EPI_ISL_1735889, EPI_ISL_1735890, EPI_ISL_1735891, EPI_ISL_1735892, EPI_ISL_1735893, EPI_ISL_1735894, EPI_ISL_1735895, EPI_ISL_1735896, EPI_ISL_1735897, EPI_ISL_1735898, EPI_ISL_1735899, EPI_ISL_1735900, EPI_ISL_1735901, EPI_ISL_1735902, EPI_ISL_1735903, EPI_ISL_1735904, EPI_ISL_1735905, EPI_ISL_1735906, EPI_ISL_1735907, EPI_ISL_1735908, EPI_ISL_1735909, EPI_ISL_1735910, EPI_ISL_1735911, EPI_ISL_1735912, EPI_ISL_1735913, EPI_ISL_1735914, EPI_ISL_1735915, EPI_ISL_1735916, EPI_ISL_1735917, EPI_ISL_1735918, EPI_ISL_1735919, EPI_ISL_1735920, EPI_ISL_1735921, EPI_ISL_1735922, EPI_ISL_1735923, EPI_ISL_1735924, EPI_ISL_1735925, EPI_ISL_1735926, EPI_ISL_1735927, EPI_ISL_1735928, EPI_ISL_1735929, EPI_ISL_1735930, EPI_ISL_1735931, EPI_ISL_1735932, EPI_ISL_1735933, EPI_ISL_1735934, EPI_ISL_1735935, EPI_ISL_1735936, EPI_ISL_1735938, EPI_ISL_1735939, EPI_ISL_1735940, EPI_ISL_1735941, EPI_ISL_1735942, EPI_ISL_1735943, EPI_ISL_1735944, EPI_ISL_1735945, EPI_ISL_1735946, EPI_ISL_1735947, EPI_ISL_1735948, EPI_ISL_1735949, EPI_ISL_1735950, EPI_ISL_1735951, EPI_ISL_1735952, EPI_ISL_1735953, EPI_ISL_1735954, EPI_ISL_1735955, EPI_ISL_1735956, EPI_ISL_1735957, EPI_ISL_1735958, EPI_ISL_1735959, EPI_ISL_1735960, EPI_ISL_1735961, EPI_ISL_1735962, EPI_ISL_1735963, EPI_ISL_1735964, EPI_ISL_1735965, EPI_ISL_1735966, EPI_ISL_1735967, EPI_ISL_1735968, EPI_ISL_1735969, EPI_ISL_1735970, EPI_ISL_1735971, EPI_ISL_1735972, EPI_ISL_1735973, EPI_ISL_1735974, EPI_ISL_1735975, EPI_ISL_1735976, EPI_ISL_1735977, EPI_ISL_1735978, EPI_ISL_1735979, EPI_ISL_1735980, EPI_ISL_1735981, EPI_ISL_1735982, EPI_ISL_1735983, EPI_ISL_1735984, EPI_ISL_1735985, EPI_ISL_1735986, EPI_ISL_1735987, EPI_ISL_1735988, EPI_ISL_1735989, EPI_ISL_1735990, EPI_ISL_1735991, EPI_ISL_1735992, EPI_ISL_1735993, EPI_ISL_1735994, EPI_ISL_1735995, EPI_ISL_1735996, EPI_ISL_1735997, EPI_ISL_1735998, EPI_ISL_1735999, EPI_ISL_1736000, EPI_ISL_1736001, EPI_ISL_1736002, EPI_ISL_1736003, EPI_ISL_1736004, EPI_ISL_1736005, EPI_ISL_1736006, EPI_ISL_1736007, EPI_ISL_1736008, EPI_ISL_1736009, EPI_ISL_1736010, EPI_ISL_1736011, EPI_ISL_1736012, EPI_ISL_1736013, EPI_ISL_1736014, EPI_ISL_1736015, EPI_ISL_1736016, EPI_ISL_1736017, EPI_ISL_1736018, EPI_ISL_1736019, EPI_ISL_1736020, EPI_ISL_1736021, EPI_ISL_1736022, EPI_ISL_1736023, EPI_ISL_1736024, EPI_ISL_1736025, EPI_ISL_1736026, EPI_ISL_1736027, EPI_ISL_1736028, EPI_ISL_1736029, EPI_ISL_1736030, EPI_ISL_1736031, EPI_ISL_1736032, EPI_ISL_1736033, EPI_ISL_1736034, EPI_ISL_1736035, EPI_ISL_1736036, EPI_ISL_1736037, EPI_ISL_1736038, EPI_ISL_1736039, EPI_ISL_1736040, EPI_ISL_1736041, EPI_ISL_1736042, EPI_ISL_1736043, EPI_ISL_1736044, EPI_ISL_1736045, EPI_ISL_1736046, EPI_ISL_1736047, EPI_ISL_1736048, EPI_ISL_1736049, EPI_ISL_1736050, EPI_ISL_1736051, EPI_ISL_1736052, EPI_ISL_1736053, EPI_ISL_1736054, EPI_ISL_1736055, EPI_ISL_1736056, EPI_ISL_1736057, EPI_ISL_1736058, EPI_ISL_1736059, EPI_ISL_1736060, EPI_ISL_1736061, EPI_ISL_1736062, EPI_ISL_1736063, EPI_ISL_1736064, EPI_ISL_1736065, EPI_ISL_1736066, EPI_ISL_1736067, EPI_ISL_1736068, EPI_ISL_1736069, EPI_ISL_1736070, EPI_ISL_1736071, EPI_ISL_1736072, EPI_ISL_1736073, EPI_ISL_1736074, EPI_ISL_1736075, EPI_ISL_1736076, EPI_ISL_1736077, EPI_ISL_1736078, EPI_ISL_1736079, EPI_ISL_1736080, EPI_ISL_1736081, EPI_ISL_1736082, EPI_ISL_1736083, EPI_ISL_1736085, EPI_ISL_1736086, EPI_ISL_1736087, EPI_ISL_1736088, EPI_ISL_1736089, EPI_ISL_1736090, EPI_ISL_1736091, EPI_ISL_1736093, EPI_ISL_1736094, EPI_ISL_1736095, EPI_ISL_1736096, EPI_ISL_1736097, EPI_ISL_1736098, EPI_ISL_1736099, EPI_ISL_1736100, EPI_ISL_1736101, EPI_ISL_1736102, EPI_ISL_1736103, EPI_ISL_1736104, EPI_ISL_1736105, EPI_ISL_1736106, EPI_ISL_1736107, EPI_ISL_1736108, EPI_ISL_1736109, EPI_ISL_1736110, EPI_ISL_1736111, EPI_ISL_1736112, EPI_ISL_1736113, EPI_ISL_1736114, EPI_ISL_1736115, EPI_ISL_1736116, EPI_ISL_1736117, EPI_ISL_1736118, EPI_ISL_1736119, EPI_ISL_1736120, EPI_ISL_1736121, EPI_ISL_1736122, EPI_ISL_1736123, EPI_ISL_1736124, EPI_ISL_1736125, EPI_ISL_1736126, EPI_ISL_1736127, EPI_ISL_1736128, EPI_ISL_1736129, EPI_ISL_1736130, EPI_ISL_1736131, EPI_ISL_1736132, EPI_ISL_1736133, EPI_ISL_1736134, EPI_ISL_1736135, EPI_ISL_1736136, EPI_ISL_1736137, EPI_ISL_1736138, EPI_ISL_1736139, EPI_ISL_1736140, EPI_ISL_1736141, EPI_ISL_1736142, EPI_ISL_1736143, EPI_ISL_1736144, EPI_ISL_1736145, EPI_ISL_1736146, EPI_ISL_1736147, EPI_ISL_1736148, EPI_ISL_1736149, EPI_ISL_1736150, EPI_ISL_1736151, EPI_ISL_1736152, EPI_ISL_1736153, EPI_ISL_1736154, EPI_ISL_1736155 | | | |

| | | | |
|---|---|---|---|
| see above | Aegis Sciences Corporation | Centers for Disease Control and Prevention Division of Viral Diseases, Pathogen Discovery | Dakota Howard, Dhwani Batra, Peter W. Cook, Kara Moser, Adrian Paskey, Jason Caravas, Benjamin Rambo-Martin, Shatavia Morrison, Christopher Gulvick, Scott Sammons, Yvette Unoarumhi, Darlene Wagner, Matthew Schmerer, Cyndi Clark, Patrick Campbell, Rob Case, Vikramsinha Ghorpade, Holly Houdeshell, Ola Kvalvaag, Dillon Nall, Ethan Sanders, Alec Vest, Shaun Westlund, Matthew Hardison, Clinton R. Paden, Duncan MacCannell |
| EPI_ISL_1736156 | Nebraska Public Health Laboratory | NPHL COVID-19 Response Team | NPHL COVID-19 Response Team |
| EPI_ISL_1736157, EPI_ISL_1736158 | Aegis Sciences Corporation | Centers for Disease Control and Prevention Division of Viral Diseases, Pathogen Discovery | Dakota Howard, Dhwani Batra, Peter W. Cook, Kara Moser, Adrian Paskey, Jason Caravas, Benjamin Rambo-Martin, Shatavia Morrison, Christopher Gulvick, Scott Sammons, Yvette Unoarumhi, Darlene Wagner, Matthew Schmerer, Cyndi Clark, Patrick Campbell, Rob Case, Vikramsinha Ghorpade, Holly Houdeshell, Ola Kvalvaag, Dillon Nall, Ethan Sanders, Alec Vest, Shaun Westlund, Matthew Hardison, Clinton R. Paden, Duncan MacCannell |
| EPI_ISL_1736159 | Nebraska Public Health Laboratory | NPHL COVID-19 Response Team | NPHL COVID-19 Response Team |
| EPI_ISL_1736160, EPI_ISL_1736161 | Aegis Sciences Corporation | Centers for Disease Control and Prevention Division of Viral Diseases, Pathogen Discovery | Dakota Howard, Dhwani Batra, Peter W. Cook, Kara Moser, Adrian Paskey, Jason Caravas, Benjamin Rambo-Martin, Shatavia Morrison, Christopher Gulvick, Scott Sammons, Yvette Unoarumhi, Darlene Wagner, Matthew Schmerer, Cyndi Clark, Patrick Campbell, Rob Case, Vikramsinha Ghorpade, Holly Houdeshell, Ola Kvalvaag, Dillon Nall, Ethan Sanders, Alec Vest, Shaun Westlund, Matthew Hardison, Clinton R. Paden, Duncan MacCannell |
| EPI_ISL_1736162 | Nebraska Public Health Laboratory | NPHL COVID-19 Response Team | NPHL COVID-19 Response Team |
| EPI_ISL_1736163, EPI_ISL_1736164 | Aegis Sciences Corporation | Centers for Disease Control and Prevention Division of Viral Diseases, Pathogen Discovery | Dakota Howard, Dhwani Batra, Peter W. Cook, Kara Moser, Adrian Paskey, Jason Caravas, Benjamin Rambo-Martin, Shatavia Morrison, Christopher Gulvick, Scott Sammons, Yvette Unoarumhi, Darlene Wagner, Matthew Schmerer, Cyndi Clark, Patrick Campbell, Rob Case, Vikramsinha Ghorpade, Holly Houdeshell, Ola Kvalvaag, Dillon Nall, Ethan Sanders, Alec Vest, Shaun Westlund, Matthew Hardison, Clinton R. Paden, Duncan MacCannell |
| EPI_ISL_1736165 | Nebraska Public Health Laboratory | NPHL COVID-19 Response Team | NPHL COVID-19 Response Team |
| EPI_ISL_1736166, EPI_ISL_1736167 | Aegis Sciences Corporation | Centers for Disease Control and Prevention Division of Viral Diseases, Pathogen Discovery | Dakota Howard, Dhwani Batra, Peter W. Cook, Kara Moser, Adrian Paskey, Jason Caravas, Benjamin Rambo-Martin, Shatavia Morrison, Christopher Gulvick, Scott Sammons, Yvette Unoarumhi, Darlene Wagner, Matthew Schmerer, Cyndi Clark, Patrick Campbell, Rob Case, Vikramsinha Ghorpade, Holly Houdeshell, Ola Kvalvaag, Dillon Nall, Ethan Sanders, Alec Vest, Shaun Westlund, Matthew Hardison, Clinton R. Paden, Duncan MacCannell |
| EPI_ISL_1736168 | Nebraska Public Health Laboratory | NPHL COVID-19 Response Team | NPHL COVID-19 Response Team |
| EPI_ISL_1736169, EPI_ISL_1736170 | Aegis Sciences Corporation | Centers for Disease Control and Prevention Division of Viral Diseases, Pathogen Discovery | Dakota Howard, Dhwani Batra, Peter W. Cook, Kara Moser, Adrian Paskey, Jason Caravas, Benjamin Rambo-Martin, Shatavia Morrison, Christopher Gulvick, Scott Sammons, Yvette Unoarumhi, Darlene Wagner, Matthew Schmerer, Cyndi Clark, Patrick Campbell, Rob Case, Vikramsinha Ghorpade, Holly Houdeshell, Ola Kvalvaag, Dillon Nall, Ethan Sanders, Alec Vest, Shaun Westlund, Matthew Hardison, Clinton R. Paden, Duncan MacCannell |
| EPI_ISL_1736171 | Nebraska Public Health Laboratory | NPHL COVID-19 Response Team | NPHL COVID-19 Response Team |
| EPI_ISL_1736172, EPI_ISL_1736173 | Aegis Sciences Corporation | Centers for Disease Control and Prevention Division of Viral Diseases, Pathogen Discovery | Dakota Howard, Dhwani Batra, Peter W. Cook, Kara Moser, Adrian Paskey, Jason Caravas, Benjamin Rambo-Martin, Shatavia Morrison, Christopher Gulvick, Scott Sammons, Yvette Unoarumhi, Darlene Wagner, Matthew Schmerer, Cyndi Clark, Patrick Campbell, Rob Case, Vikramsinha Ghorpade, Holly Houdeshell, Ola Kvalvaag, Dillon Nall, Ethan Sanders, Alec Vest, Shaun Westlund, Matthew Hardison, Clinton R. Paden, Duncan MacCannell |
| EPI_ISL_1736174 | Nebraska Public Health Laboratory | NPHL COVID-19 Response Team | NPHL COVID-19 Response Team |
| EPI_ISL_1736175, EPI_ISL_1736176 | Aegis Sciences Corporation | Centers for Disease Control and Prevention Division of Viral Diseases, Pathogen Discovery | Dakota Howard, Dhwani Batra, Peter W. Cook, Kara Moser, Adrian Paskey, Jason Caravas, Benjamin Rambo-Martin, Shatavia Morrison, Christopher Gulvick, Scott Sammons, Yvette Unoarumhi, Darlene Wagner, Matthew Schmerer, Cyndi Clark, Patrick Campbell, Rob Case, Vikramsinha Ghorpade, Holly Houdeshell, Ola Kvalvaag, Dillon Nall, Ethan Sanders, Alec Vest, Shaun Westlund, Matthew Hardison, Clinton R. Paden, Duncan MacCannell |
| EPI_ISL_1736177 | Nebraska Public Health Laboratory | NPHL COVID-19 Response Team | NPHL COVID-19 Response Team |
| EPI_ISL_1736178, EPI_ISL_1736179 | Aegis Sciences Corporation | Centers for Disease Control and Prevention Division of Viral Diseases, Pathogen Discovery | Dakota Howard, Dhwani Batra, Peter W. Cook, Kara Moser, Adrian Paskey, Jason Caravas, Benjamin Rambo-Martin, Shatavia Morrison, Christopher Gulvick, Scott Sammons, Yvette Unoarumhi, Darlene Wagner, Matthew Schmerer, Cyndi Clark, Patrick Campbell, Rob Case, Vikramsinha Ghorpade, Holly Houdeshell, Ola Kvalvaag, Dillon Nall, Ethan Sanders, Alec Vest, Shaun Westlund, Matthew Hardison, Clinton R. Paden, Duncan MacCannell |
| EPI_ISL_1736180 | Nebraska Public Health Laboratory | NPHL COVID-19 Response Team | NPHL COVID-19 Response Team |
| EPI_ISL_1736181, EPI_ISL_1736182 | Aegis Sciences Corporation | Centers for Disease Control and Prevention Division of Viral Diseases, Pathogen Discovery | Dakota Howard, Dhwani Batra, Peter W. Cook, Kara Moser, Adrian Paskey, Jason Caravas, Benjamin Rambo-Martin, Shatavia Morrison, Christopher Gulvick, Scott Sammons, Yvette Unoarumhi, Darlene Wagner, Matthew Schmerer, Cyndi Clark, Patrick Campbell, Rob Case, Vikramsinha Ghorpade, Holly Houdeshell, Ola Kvalvaag, Dillon Nall, Ethan Sanders, Alec Vest, Shaun Westlund, Matthew Hardison, Clinton R. Paden, Duncan MacCannell |
| EPI_ISL_1736183 | Nebraska Public Health Laboratory | NPHL COVID-19 Response Team | NPHL COVID-19 Response Team |
| EPI_ISL_1736184, EPI_ISL_1736185 | Aegis Sciences Corporation | Centers for Disease Control and Prevention Division of Viral Diseases, Pathogen Discovery | Dakota Howard, Dhwani Batra, Peter W. Cook, Kara Moser, Adrian Paskey, Jason Caravas, Benjamin Rambo-Martin, Shatavia Morrison, Christopher Gulvick, Scott Sammons, Yvette Unoarumhi, Darlene Wagner, Matthew Schmerer, Cyndi Clark, Patrick Campbell, Rob Case, Vikramsinha Ghorpade, Holly Houdeshell, Ola Kvalvaag, Dillon Nall, Ethan Sanders, Alec Vest, Shaun Westlund, Matthew Hardison, Clinton R. Paden, Duncan MacCannell |
| EPI_ISL_1736186 | Nebraska Public Health Laboratory | NPHL COVID-19 Response Team | NPHL COVID-19 Response Team |
| EPI_ISL_1736187 | Aegis Sciences Corporation | Centers for Disease Control and Prevention Division of Viral Diseases, Pathogen Discovery | Dakota Howard, Dhwani Batra, Peter W. Cook, Kara Moser, Adrian Paskey, Jason Caravas, Benjamin Rambo-Martin, Shatavia Morrison, Christopher Gulvick, Scott Sammons, Yvette Unoarumhi, Darlene Wagner, Matthew Schmerer, Cyndi Clark, Patrick Campbell, Rob Case, Vikramsinha Ghorpade, Holly Houdeshell, Ola Kvalvaag, Dillon Nall, Ethan Sanders, Alec Vest, Shaun Westlund, Matthew Hardison, Clinton R. Paden, Duncan MacCannell |
| EPI_ISL_1736188 | Nebraska Public Health Laboratory | NPHL COVID-19 Response Team | NPHL COVID-19 Response Team |
| EPI_ISL_1736189, EPI_ISL_1736190 | Aegis Sciences Corporation | Centers for Disease Control and Prevention Division of Viral Diseases, Pathogen Discovery | Dakota Howard, Dhwani Batra, Peter W. Cook, Kara Moser, Adrian Paskey, Jason Caravas, Benjamin Rambo-Martin, Shatavia Morrison, Christopher Gulvick, Scott Sammons, Yvette Unoarumhi, Darlene Wagner, Matthew Schmerer, Cyndi Clark, Patrick Campbell, Rob Case, Vikramsinha Ghorpade, Holly Houdeshell, Ola Kvalvaag, Dillon Nall, Ethan Sanders, Alec Vest, Shaun Westlund, Matthew Hardison, Clinton R. Paden, Duncan MacCannell |
| EPI_ISL_1736191 | Nebraska Public Health Laboratory | NPHL COVID-19 Response Team | NPHL COVID-19 Response Team |
| EPI_ISL_1736192 | Aegis Sciences Corporation | Centers for Disease Control and Prevention Division of Viral Diseases, Pathogen Discovery | Dakota Howard, Dhwani Batra, Peter W. Cook, Kara Moser, Adrian Paskey, Jason Caravas, Benjamin Rambo-Martin, Shatavia Morrison, Christopher Gulvick, Scott Sammons, Yvette Unoarumhi, Darlene Wagner, Matthew Schmerer, Cyndi Clark, Patrick Campbell, Rob Case, Vikramsinha Ghorpade, Holly Houdeshell, Ola Kvalvaag, Dillon Nall, Ethan Sanders, Alec Vest, Shaun Westlund, Matthew Hardison, Clinton R. Paden, Duncan MacCannell |
| EPI_ISL_1736193, EPI_ISL_1736194 | Nebraska Public Health Laboratory | NPHL COVID-19 Response Team | NPHL COVID-19 Response Team |
| EPI_ISL_1736195, EPI_ISL_1736196 | Aegis Sciences Corporation | Centers for Disease Control and Prevention Division of Viral Diseases, Pathogen Discovery | Dakota Howard, Dhwani Batra, Peter W. Cook, Kara Moser, Adrian Paskey, Jason Caravas, Benjamin Rambo-Martin, Shatavia Morrison, Christopher Gulvick, Scott Sammons, Yvette Unoarumhi, Darlene Wagner, Matthew Schmerer, Cyndi Clark, Patrick Campbell, Rob Case, Vikramsinha Ghorpade, Holly Houdeshell, Ola Kvalvaag, Dillon Nall, Ethan Sanders, Alec Vest, Shaun Westlund, Matthew Hardison, Clinton R. Paden, Duncan MacCannell |
| EPI_ISL_1736197 | Nebraska Public Health Laboratory | NPHL COVID-19 Response Team | NPHL COVID-19 Response Team |

EPI_ISL_1736198, EPI_ISL_1736199, EPI_ISL_1736200, EPI_ISL_1736201, EPI_ISL_1736203, EPI_ISL_1736204, EPI_ISL_1736205, EPI_ISL_1736206, EPI_ISL_1736207, EPI_ISL_1736208, EPI_ISL_1736209, EPI_ISL_1736210, EPI_ISL_1736211, EPI_ISL_1736212, EPI_ISL_1736213, EPI_ISL_1736214, EPI_ISL_1736215, EPI_ISL_1736216, EPI_ISL_1736217, EPI_ISL_1736218, EPI_ISL_1736219, EPI_ISL_1736220, EPI_ISL_1736221, EPI_ISL_1736222, EPI_ISL_1736223, EPI_ISL_1736224, EPI_ISL_1736225, EPI_ISL_1736226, EPI_ISL_1736227, EPI_ISL_1736228, EPI_ISL_1736229, EPI_ISL_1736230, EPI_ISL_1736231, EPI_ISL_1736232, EPI_ISL_1736233, EPI_ISL_1736234, EPI_ISL_1736235, EPI_ISL_1736236, EPI_ISL_1736237, EPI_ISL_1736238, EPI_ISL_1736239, EPI_ISL_1736240, EPI_ISL_1736241, EPI_ISL_1736242, EPI_ISL_1736243, EPI_ISL_1736244, EPI_ISL_1736245, EPI_ISL_1736246, EPI_ISL_1736247, EPI_ISL_1736248, EPI_ISL_1736249, EPI_ISL_1736250, EPI_ISL_1736251, EPI_ISL_1736252, EPI_ISL_1736253, EPI_ISL_1736254, EPI_ISL_1736255, EPI_ISL_1736256, EPI_ISL_1736257, EPI_ISL_1736258, EPI_ISL_1736259, EPI_ISL_1736260, EPI_ISL_1736261, EPI_ISL_1736262, EPI_ISL_1736263, EPI_ISL_1736264, EPI_ISL_1736265, EPI_ISL_1736266, EPI_ISL_1736267, EPI_ISL_1736268, EPI_ISL_1736269, EPI_ISL_1736270, EPI_ISL_1736271, EPI_ISL_1736272, EPI_ISL_1736273, EPI_ISL_1736274, EPI_ISL_1736275, EPI_ISL_1736276, EPI_ISL_1736277, EPI_ISL_1736278, EPI_ISL_1736279, EPI_ISL_1736280, EPI_ISL_1736281, EPI_ISL_1736282, EPI_ISL_1736283, EPI_ISL_1736284, EPI_ISL_1736285, EPI_ISL_1736286, EPI_ISL_1736287, EPI_ISL_1736288, EPI_ISL_1736289, EPI_ISL_1736290, EPI_ISL_1736291, EPI_ISL_1736292, EPI_ISL_1736293, EPI_ISL_1736294, EPI_ISL_1736295, EPI_ISL_1736296, EPI_ISL_1736297, EPI_ISL_1736298, EPI_ISL_1736299, EPI_ISL_1736301, EPI_ISL_1736302, EPI_ISL_1736303, EPI_ISL_1736304, EPI_ISL_1736305, EPI_ISL_1736306, EPI_ISL_1736307, EPI_ISL_1736308, EPI_ISL_1736309, EPI_ISL_1736310, EPI_ISL_1736311, EPI_ISL_1736312, EPI_ISL_1736313, EPI_ISL_1736314, EPI_ISL_1736315, EPI_ISL_1736316, EPI_ISL_1736317, EPI_ISL_1736318, EPI_ISL_1736319, EPI_ISL_1736320, EPI_ISL_1736321, EPI_ISL_1736322, EPI_ISL_1736323, EPI_ISL_1736324, EPI_ISL_1736325, EPI_ISL_1736326, EPI_ISL_1736327, EPI_ISL_1736328, EPI_ISL_1736329, EPI_ISL_1736330, EPI_ISL_1736331, EPI_ISL_1736332, EPI_ISL_1736333, EPI_ISL_1736334, EPI_ISL_1736335, EPI_ISL_1736336, EPI_ISL_1736337, EPI_ISL_1736338, EPI_ISL_1736339, EPI_ISL_1736340, EPI_ISL_1736341, EPI_ISL_1736342, EPI_ISL_1736343, EPI_ISL_1736344, EPI_ISL_1736345, EPI_ISL_1736346, EPI_ISL_1736347, EPI_ISL_1736348, EPI_ISL_1736349, EPI_ISL_1736350, EPI_ISL_1736351, EPI_ISL_1736352, EPI_ISL_1736353, EPI_ISL_1736354, EPI_ISL_1736355, EPI_ISL_1736356, EPI_ISL_1736357, EPI_ISL_1736358, EPI_ISL_1736359, EPI_ISL_1736360, EPI_ISL_1736361, EPI_ISL_1736362, EPI_ISL_1736363, EPI_ISL_1736364, EPI_ISL_1736365, EPI_ISL_1736366, EPI_ISL_1736367, EPI_ISL_1736368, EPI_ISL_1736369, EPI_ISL_1736370, EPI_ISL_1736371, EPI_ISL_1736372, EPI_ISL_1736373, EPI_ISL_1736374, EPI_ISL_1736375, EPI_ISL_1736376, EPI_ISL_1736377, EPI_ISL_1736378, EPI_ISL_1736379, EPI_ISL_1736380, EPI_ISL_1736381, EPI_ISL_1736382, EPI_ISL_1736383, EPI_ISL_1736384, EPI_ISL_1736385, EPI_ISL_1736386, EPI_ISL_1736387, EPI_ISL_1736388, EPI_ISL_1736389, EPI_ISL_1736390, EPI_ISL_1736391, EPI_ISL_1736392, EPI_ISL_1736393, EPI_ISL_1736394, EPI_ISL_1736395, EPI_ISL_1736396, EPI_ISL_1736397, EPI_ISL_1736398, EPI_ISL_1736399, EPI_ISL_1736400, EPI_ISL_1736401, EPI_ISL_1736402, EPI_ISL_1736403, EPI_ISL_1736404, EPI_ISL_1736405, EPI_ISL_1736406, EPI_ISL_1736407, EPI_ISL_1736408, EPI_ISL_1736410, EPI_ISL_1736411, EPI_ISL_1736412, EPI_ISL_1736413, EPI_ISL_1736414, EPI_ISL_1736415, EPI_ISL_1736416, EPI_ISL_1736417, EPI_ISL_1736418, EPI_ISL_1736419, EPI_ISL_1736420, EPI_ISL_1736421, EPI_ISL_1736422, EPI_ISL_1736423, EPI_ISL_1736424, EPI_ISL_1736425, EPI_ISL_1736426, EPI_ISL_1736427, EPI_ISL_1736428, EPI_ISL_1736429, EPI_ISL_1736430, EPI_ISL_1736431, EPI_ISL_1736432, EPI_ISL_1736433, EPI_ISL_1736434, EPI_ISL_1736435, EPI_ISL_1736436, EPI_ISL_1736437, EPI_ISL_1736438, EPI_ISL_1736439, EPI_ISL_1736440, EPI_ISL_1736441, EPI_ISL_1736442, EPI_ISL_1736443, EPI_ISL_1736444, EPI_ISL_1736445, EPI_ISL_1736446, EPI_ISL_1736447, EPI_ISL_1736448, EPI_ISL_1736449, EPI_ISL_1736450, EPI_ISL_1736451, EPI_ISL_1736452, EPI_ISL_1736453, EPI_ISL_1736454, EPI_ISL_1736455, EPI_ISL_1736456, EPI_ISL_1736457, EPI_ISL_1736458, EPI_ISL_1736459, EPI_ISL_1736460, EPI_ISL_1736461, EPI_ISL_1736462, EPI_ISL_1736463, EPI_ISL_1736464, EPI_ISL_1736465, EPI_ISL_1736466, EPI_ISL_1736467, EPI_ISL_1736468, EPI_ISL_1736469, EPI_ISL_1736470, EPI_ISL_1736471, EPI_ISL_1736472, EPI_ISL_1736473, EPI_ISL_1736474, EPI_ISL_1736475, EPI_ISL_1736476, EPI_ISL_1736477, EPI_ISL_1736478, EPI_ISL_1736479, EPI_ISL_1736480, EPI_ISL_1736481, EPI_ISL_1736482, EPI_ISL_1736483, EPI_ISL_1736484, EPI_ISL_1736485, EPI_ISL_1736486, EPI_ISL_1736487, EPI_ISL_1736488, EPI_ISL_1736489, EPI_ISL_1736490, EPI_ISL_1736491, EPI_ISL_1736492, EPI_ISL_1736493, EPI_ISL_1736494, EPI_ISL_1736495, EPI_ISL_1736496, EPI_ISL_1736497, EPI_ISL_1736498, EPI_ISL_1736499, EPI_ISL_1736500, EPI_ISL_1736501, EPI_ISL_1736502, EPI_ISL_1736503, EPI_ISL_1736504, EPI_ISL_1736505, EPI_ISL_1736506, EPI_ISL_1736507, EPI_ISL_1736508, EPI_ISL_1736510, EPI_ISL_1736511, EPI_ISL_1736513, EPI_ISL_1736514, EPI_ISL_1736515, EPI_ISL_1736516, EPI_ISL_1736518, EPI_ISL_1736519, EPI_ISL_1736520, EPI_ISL_1736521, EPI_ISL_1736522, EPI_ISL_1736523, EPI_ISL_1736524, EPI_ISL_1736525, EPI_ISL_1736526, EPI_ISL_1736528, EPI_ISL_1736529, EPI_ISL_1736530, EPI_ISL_1736531, EPI_ISL_1736532, EPI_ISL_1736533, EPI_ISL_1736534, EPI_ISL_1736535, EPI_ISL_1736536, EPI_ISL_1736537, EPI_ISL_1736538, EPI_ISL_1736539, EPI_ISL_1736540, EPI_ISL_1736541, EPI_ISL_1736542, EPI_ISL_1736543, EPI_ISL_1736544, EPI_ISL_1736545, EPI_ISL_1736546, EPI_ISL_1736547, EPI_ISL_1736548, EPI_ISL_1736549, EPI_ISL_1736550, EPI_ISL_1736552, EPI_ISL_1736553, EPI_ISL_1736554, EPI_ISL_1736555, EPI_ISL_1736556, EPI_ISL_1736557, EPI_ISL_1736558, EPI_ISL_1736559, EPI_ISL_1736560, EPI_ISL_1736561, EPI_ISL_1736562, EPI_ISL_1736563, EPI_ISL_1736564, EPI_ISL_1736565, EPI_ISL_1736566, EPI_ISL_1736567, EPI_ISL_1736568, EPI_ISL_1736569, EPI_ISL_1736570, EPI_ISL_1736571, EPI_ISL_1736572, EPI_ISL_1736573, EPI_ISL_1736574, EPI_ISL_1736575, EPI_ISL_1736576, EPI_ISL_1736577, EPI_ISL_1736578, EPI_ISL_1736579, EPI_ISL_1736580, EPI_ISL_1736581, EPI_ISL_1736582, EPI_ISL_1736583, EPI_ISL_1736584, EPI_ISL_1736585, EPI_ISL_1736586, EPI_ISL_1736587, EPI_ISL_1736588, EPI_ISL_1736589, EPI_ISL_1736590, EPI_ISL_1736592, EPI_ISL_1736593, EPI_ISL_1736594, EPI_ISL_1736595, EPI_ISL_1736596, EPI_ISL_1736597, EPI_ISL_1736598, EPI_ISL_1736599, EPI_ISL_1736600, EPI_ISL_1736601, EPI_ISL_1736602, EPI_ISL_1736603, EPI_ISL_1736604, EPI_ISL_1736605, EPI_ISL_1736606, EPI_ISL_1736607, EPI_ISL_1736608, EPI_ISL_1736610, EPI_ISL_1736611, EPI_ISL_1736612, EPI_ISL_1736613, EPI_ISL_1736614, EPI_ISL_1736615, EPI_ISL_1736616, EPI_ISL_1736617, EPI_ISL_1736618, EPI_ISL_1736619, EPI_ISL_1736620, EPI_ISL_1736621, EPI_ISL_1736622, EPI_ISL_1736623, EPI_ISL_1736624, EPI_ISL_1736625, EPI_ISL_1736626, EPI_ISL_1736627, EPI_ISL_1736628, EPI_ISL_1736629, EPI_ISL_1736630, EPI_ISL_1736631, EPI_ISL_1736632, EPI_ISL_1736633, EPI_ISL_1736634, EPI_ISL_1736635, EPI_ISL_1736636, EPI_ISL_1736637, EPI_ISL_1736638, EPI_ISL_1736639, EPI_ISL_1736640, EPI_ISL_1736641, EPI_ISL_1736642, EPI_ISL_1736643, EPI_ISL_1736644, EPI_ISL_1736645, EPI_ISL_1736646, EPI_ISL_1736647, EPI_ISL_1736648, EPI_ISL_1736650, EPI_ISL_1736651, EPI_ISL_1736652, EPI_ISL_1736653, EPI_ISL_1736654, EPI_ISL_1736655, EPI_ISL_1736656, EPI_ISL_1736657, EPI_ISL_1736658, EPI_ISL_1736659, EPI_ISL_1736660, EPI_ISL_1736661, EPI_ISL_1736662, EPI_ISL_1736663, EPI_ISL_1736664, EPI_ISL_1736665, EPI_ISL_1736666, EPI_ISL_1736667, EPI_ISL_1736668, EPI_ISL_1736669, EPI_ISL_1736670, EPI_ISL_1736671, EPI_ISL_1736672, EPI_ISL_1736673, EPI_ISL_1736674, EPI_ISL_1736675, EPI_ISL_1736676, EPI_ISL_1736677, EPI_ISL_1736678, EPI_ISL_1736679, EPI_ISL_1736680, EPI_ISL_1736681, EPI_ISL_1736682, EPI_ISL_1736683, EPI_ISL_1736684, EPI_ISL_1736685, EPI_ISL_1736686, EPI_ISL_1736687, EPI_ISL_1736688, EPI_ISL_1736689, EPI_ISL_1736690, EPI_ISL_1736691, EPI_ISL_1736692, EPI_ISL_1736693, EPI_ISL_1736694, EPI_ISL_1736695, EPI_ISL_1736696, EPI_ISL_1736697, EPI_ISL_1736698, EPI_ISL_1736699, EPI_ISL_1736700, EPI_ISL_1736701, EPI_ISL_1736702, EPI_ISL_1736703, EPI_ISL_1736704, EPI_ISL_1736705, EPI_ISL_1736706, EPI_ISL_1736707, EPI_ISL_1736708, EPI_ISL_1736709, EPI_ISL_1736710, EPI_ISL_1736711, EPI_ISL_1736712, EPI_ISL_1736713, EPI_ISL_1736714, EPI_ISL_1736715, EPI_ISL_1736716, EPI_ISL_1736717, EPI_ISL_1736718, EPI_ISL_1736719, EPI_ISL_1736720, EPI_ISL_1736721, EPI_ISL_1736722, EPI_ISL_1736723, EPI_ISL_1736724, EPI_ISL_1736725, EPI_ISL_1736726, EPI_ISL_1736727, EPI_ISL_1736728, EPI_ISL_1736729, EPI_ISL_1736730, EPI_ISL_1736731, EPI_ISL_1736732, EPI_ISL_1736733, EPI_ISL_1736734, EPI_ISL_1736735, EPI_ISL_1736736, EPI_ISL_1736737, EPI_ISL_1736738, EPI_ISL_1736739, EPI_ISL_1736740, EPI_ISL_1736741, EPI_ISL_1736742, EPI_ISL_1736743, EPI_ISL_1736744, EPI_ISL_1736745, EPI_ISL_1736746, EPI_ISL_1736747, EPI_ISL_1736748, EPI_ISL_1736749, EPI_ISL_1736750, EPI_ISL_1736751, EPI_ISL_1736752, EPI_ISL_1736753, EPI_ISL_1736754, EPI_ISL_1736755, EPI_ISL_1736756, EPI_ISL_1736757, EPI_ISL_1736758, EPI_ISL_1736759, EPI_ISL_1736760, EPI_ISL_1736761, EPI_ISL_1736762, EPI_ISL_1736763, EPI_ISL_1736764, EPI_ISL_1736765, EPI_ISL_1736766, EPI_ISL_1736767, EPI_ISL_1736768, EPI_ISL_1736769, EPI_ISL_1736770, EPI_ISL_1736771, EPI_ISL_1736772, EPI_ISL_1736773, EPI_ISL_1736774, EPI_ISL_1736775, EPI_ISL_1736776, EPI_ISL_1736777, EPI_ISL_1736778, EPI_ISL_1736779, EPI_ISL_1736780, EPI_ISL_1736781, EPI_ISL_1736782, EPI_ISL_1736783, EPI_ISL_1736784, EPI_ISL_1736785, EPI_ISL_1736786, EPI_ISL_1736787, EPI_ISL_1736788, EPI_ISL_1736789, EPI_ISL_1736790, EPI_ISL_1736791, EPI_ISL_1736792, EPI_ISL_1736793, EPI_ISL_1736794, EPI_ISL_1736795, EPI_ISL_1736796, EPI_ISL_1736797, EPI_ISL_1736798, EPI_ISL_1736799, EPI_ISL_1736800, EPI_ISL_1736801, EPI_ISL_1736802, EPI_ISL_1736803, EPI_ISL_1736804, EPI_ISL_1736805, EPI_ISL_1736806, EPI_ISL_1736807, EPI_ISL_1736808, EPI_ISL_1736809, EPI_ISL_1736810, EPI_ISL_1736811, EPI_ISL_1736812, EPI_ISL_1736813, EPI_ISL_1736814, EPI_ISL_1736815, EPI_ISL_1736816, EPI_ISL_1736817, EPI_ISL_1736818, EPI_ISL_1736820, EPI_ISL_1736821, EPI_ISL_1736822, EPI_ISL_1736823, EPI_ISL_1736824, EPI_ISL_1736825, EPI_ISL_1736826, EPI_ISL_1736827, EPI_ISL_1736828, EPI_ISL_1736829, EPI_ISL_1736830, EPI_ISL_1736831, EPI_ISL_1736832, EPI_ISL_1736833, EPI_ISL_1736834, EPI_ISL_1736835, EPI_ISL_1736836, EPI_ISL_1736837, EPI_ISL_1736838, EPI_ISL_1736839, EPI_ISL_1736840, EPI_ISL_1736841, EPI_ISL_1736842, EPI_ISL_1736843, EPI_ISL_1736844, EPI_ISL_1736845, EPI_ISL_1736846, EPI_ISL_1736847, EPI_ISL_1736848, EPI_ISL_1736849, EPI_ISL_1736850, EPI_ISL_1736851, EPI_ISL_1736852, EPI_ISL_1736853, EPI_ISL_1736854, EPI_ISL_1736855, EPI_ISL_1736856, EPI_ISL_1736857, EPI_ISL_1736858, EPI_ISL_1736859, EPI_ISL_1736860, EPI_ISL_1736861, EPI_ISL_1736862, EPI_ISL_1736863, EPI_ISL_1736864, EPI_ISL_1736865, EPI_ISL_1736866, EPI_ISL_1736867, EPI_ISL_1736868, EPI_ISL_1736869, EPI_ISL_1736871, EPI_ISL_1736872, EPI_ISL_1736873, EPI_ISL_1736874, EPI_ISL_1736875, EPI_ISL_1736876, EPI_ISL_1736877, EPI_ISL_1736878, EPI_ISL_1736879, EPI_ISL_1736880, EPI_ISL_1736881, EPI_ISL_1736882, EPI_ISL_1736883, EPI_ISL_1736884, EPI_ISL_1736885, EPI_ISL_1736886, EPI_ISL_1736887, EPI_ISL_1736888, EPI_ISL_1736889, EPI_ISL_1736890, EPI_ISL_1736891, EPI_ISL_1736892, EPI_ISL_1736894, EPI_ISL_1736895, EPI_ISL_1736896, EPI_ISL_1736897, EPI_ISL_1736898, EPI_ISL_1736899, EPI_ISL_1736900, EPI_ISL_1736901, EPI_ISL_1736902, EPI_ISL_1736904, EPI_ISL_1736906, EPI_ISL_1736907, EPI_ISL_1736908, EPI_ISL_1736909, EPI_ISL_1736910, EPI_ISL_1736911, EPI_ISL_1736912, EPI_ISL_1736913, EPI_ISL_1736914, EPI_ISL_1736915, EPI_ISL_1736916, EPI_ISL_1736917, EPI_ISL_1736918, EPI_ISL_1736919, EPI_ISL_1736920, EPI_ISL_1736921, EPI_ISL_1736922, EPI_ISL_1736923, EPI_ISL_1736924, EPI_ISL_1736925, EPI_ISL_1736926, EPI_ISL_1736927, EPI_ISL_1736928, EPI_ISL_1736929, EPI_ISL_1736930, EPI_ISL_1736931, EPI_ISL_1736932, EPI_ISL_1736933, EPI_ISL_1736934, EPI_ISL_1736935, EPI_ISL_1736936, EPI_ISL_1736937, EPI_ISL_1736938, EPI_ISL_1736939, EPI_ISL_1736940, EPI_ISL_1736941, EPI_ISL_1736942, EPI_ISL_1736943, EPI_ISL_1736944, EPI_ISL_1736945, EPI_ISL_1736946, EPI_ISL_1736947, EPI_ISL_1736948, EPI_ISL_1736949, EPI_ISL_1736950, EPI_ISL_1736951, EPI_ISL_1736952, EPI_ISL_1736953, EPI_ISL_1736954, EPI_ISL_1736955, EPI_ISL_1736956, EPI_ISL_1736957, EPI_ISL_1736958, EPI_ISL_1736959, EPI_ISL_1736960, EPI_ISL_1736961, EPI_ISL_1736962, EPI_ISL_1736963, EPI_ISL_1736964, EPI_ISL_1736965, EPI_ISL_1736966, EPI_ISL_1736967, EPI_ISL_1736968, EPI_ISL_1736969, EPI_ISL_1736970, EPI_ISL_1736971, EPI_ISL_1736972, EPI_ISL_1736973, EPI_ISL_1736974, EPI_ISL_1736975, EPI_ISL_1736976, EPI_ISL_1736977, EPI_ISL_1736978, EPI_ISL_1736979, EPI_ISL_1736980, EPI_ISL_1736981, EPI_ISL_1736982, EPI_ISL_1736983,

| | | | |
|---|---|---|---|
| EPI_ISL_1736984, EPI_ISL_1736985, EPI_ISL_1736986, EPI_ISL_1736987, EPI_ISL_1736988, EPI_ISL_1736989, EPI_ISL_1736990, EPI_ISL_1736991, EPI_ISL_1736992, EPI_ISL_1736993, EPI_ISL_1736994, EPI_ISL_1736995, EPI_ISL_1736996, EPI_ISL_1736997, EPI_ISL_1736998, EPI_ISL_1736999, EPI_ISL_1737000, EPI_ISL_1737001, EPI_ISL_1737002, EPI_ISL_1737003, EPI_ISL_1737004, EPI_ISL_1737005, EPI_ISL_1737006, EPI_ISL_1737007, EPI_ISL_1737008, EPI_ISL_1737009, EPI_ISL_1737010, EPI_ISL_1737011, EPI_ISL_1737012, EPI_ISL_1737013, EPI_ISL_1737014, EPI_ISL_1737015, EPI_ISL_1737016, EPI_ISL_1737017, EPI_ISL_1737018, EPI_ISL_1737019, EPI_ISL_1737020, EPI_ISL_1737021, EPI_ISL_1737022, EPI_ISL_1737023, EPI_ISL_1737024, EPI_ISL_1737025, EPI_ISL_1737026, EPI_ISL_1737027, EPI_ISL_1737028, EPI_ISL_1737029, EPI_ISL_1737030, EPI_ISL_1737031, EPI_ISL_1737032, EPI_ISL_1737033, EPI_ISL_1737034, EPI_ISL_1737035, EPI_ISL_1737036, EPI_ISL_1737037, EPI_ISL_1737038, EPI_ISL_1737039, EPI_ISL_1737040, EPI_ISL_1737041, EPI_ISL_1737042, EPI_ISL_1737043, EPI_ISL_1737044, EPI_ISL_1737045, EPI_ISL_1737046, EPI_ISL_1737047, EPI_ISL_1737048, EPI_ISL_1737049, EPI_ISL_1737050, EPI_ISL_1737051, EPI_ISL_1737052, EPI_ISL_1737053, EPI_ISL_1737054, EPI_ISL_1737055, EPI_ISL_1737056, EPI_ISL_1737057, EPI_ISL_1737058, EPI_ISL_1737059, EPI_ISL_1737060, EPI_ISL_1737061, EPI_ISL_1737062, EPI_ISL_1737063, EPI_ISL_1737064, EPI_ISL_1737065, EPI_ISL_1737066, EPI_ISL_1737067, EPI_ISL_1737068, EPI_ISL_1737069, EPI_ISL_1737070, EPI_ISL_1737071, EPI_ISL_1737072, EPI_ISL_1737073, EPI_ISL_1737074, EPI_ISL_1737075, EPI_ISL_1737076, EPI_ISL_1737077, EPI_ISL_1737078, EPI_ISL_1737079, EPI_ISL_1737080, EPI_ISL_1737081, EPI_ISL_1737082, EPI_ISL_1737083, EPI_ISL_1737084, EPI_ISL_1737085, EPI_ISL_1737086, EPI_ISL_1737087, EPI_ISL_1737088, EPI_ISL_1737090, EPI_ISL_1737091, EPI_ISL_1737092, EPI_ISL_1737093, EPI_ISL_1737094, EPI_ISL_1737095, EPI_ISL_1737096, EPI_ISL_1737097, EPI_ISL_1737098, EPI_ISL_1737099, EPI_ISL_1737100, EPI_ISL_1737101, EPI_ISL_1737102, EPI_ISL_1737103, EPI_ISL_1737104, EPI_ISL_1737105, EPI_ISL_1737106, EPI_ISL_1737107, EPI_ISL_1737108, EPI_ISL_1737109, EPI_ISL_1737110, EPI_ISL_1737111, EPI_ISL_1737112, EPI_ISL_1737113, EPI_ISL_1737114, EPI_ISL_1737115, EPI_ISL_1737116, EPI_ISL_1737117, EPI_ISL_1737118, EPI_ISL_1737119, EPI_ISL_1737120, EPI_ISL_1737121, EPI_ISL_1737122, EPI_ISL_1737123, EPI_ISL_1737124, EPI_ISL_1737125, EPI_ISL_1737126, EPI_ISL_1737127, EPI_ISL_1737128, EPI_ISL_1737129, EPI_ISL_1737130, EPI_ISL_1737131, EPI_ISL_1737132, EPI_ISL_1737133, EPI_ISL_1737134, EPI_ISL_1737135, EPI_ISL_1737136, EPI_ISL_1737137, EPI_ISL_1737138, EPI_ISL_1737139, EPI_ISL_1737140, EPI_ISL_1737141, EPI_ISL_1737142, EPI_ISL_1737143, EPI_ISL_1737144, EPI_ISL_1737145, EPI_ISL_1737146, EPI_ISL_1737147, EPI_ISL_1737148, EPI_ISL_1737149, EPI_ISL_1737150, EPI_ISL_1737151, EPI_ISL_1737152, EPI_ISL_1737153, EPI_ISL_1737154, EPI_ISL_1737155, EPI_ISL_1737156, EPI_ISL_1737157, EPI_ISL_1737158, EPI_ISL_1737159, EPI_ISL_1737160, EPI_ISL_1737161, EPI_ISL_1737162, EPI_ISL_1737163, EPI_ISL_1737164, EPI_ISL_1737165, EPI_ISL_1737166, EPI_ISL_1737167, EPI_ISL_1737168, EPI_ISL_1737169, EPI_ISL_1737170, EPI_ISL_1737172, EPI_ISL_1737173, EPI_ISL_1737174, EPI_ISL_1737175, EPI_ISL_1737176, EPI_ISL_1737177, EPI_ISL_1737178, EPI_ISL_1737179, EPI_ISL_1737180, EPI_ISL_1737181, EPI_ISL_1737182, EPI_ISL_1737183, EPI_ISL_1737184, EPI_ISL_1737185, EPI_ISL_1737186, EPI_ISL_1737187, EPI_ISL_1737188, EPI_ISL_1737189, EPI_ISL_1737190, EPI_ISL_1737191, EPI_ISL_1737192, EPI_ISL_1737193, EPI_ISL_1737194, EPI_ISL_1737196, EPI_ISL_1737197, EPI_ISL_1737199, EPI_ISL_1737200, EPI_ISL_1737201, EPI_ISL_1737202, EPI_ISL_1737203, EPI_ISL_1737204, EPI_ISL_1737205, EPI_ISL_1737206, EPI_ISL_1737207, EPI_ISL_1737208, EPI_ISL_1737209, EPI_ISL_1737210, EPI_ISL_1737211, EPI_ISL_1737212, EPI_ISL_1737213, EPI_ISL_1737214, EPI_ISL_1737216, EPI_ISL_1737217, EPI_ISL_1737218, EPI_ISL_1737219, EPI_ISL_1737220, EPI_ISL_1737221, EPI_ISL_1737222, EPI_ISL_1737223, EPI_ISL_1737224, EPI_ISL_1737225, EPI_ISL_1737226, EPI_ISL_1737227, EPI_ISL_1737228, EPI_ISL_1737229, EPI_ISL_1737230, EPI_ISL_1737231, EPI_ISL_1737232, EPI_ISL_1737233, EPI_ISL_1737234, EPI_ISL_1737235, EPI_ISL_1737236, EPI_ISL_1737237, EPI_ISL_1737238, EPI_ISL_1737239, EPI_ISL_1737240, EPI_ISL_1737241, EPI_ISL_1737242, EPI_ISL_1737243, EPI_ISL_1737244, EPI_ISL_1737245, EPI_ISL_1737246, EPI_ISL_1737247, EPI_ISL_1737248, EPI_ISL_1737249, EPI_ISL_1737250, EPI_ISL_1737251, EPI_ISL_1737252, EPI_ISL_1737253, EPI_ISL_1737254, EPI_ISL_1737255, EPI_ISL_1737256, EPI_ISL_1737257, EPI_ISL_1737258, EPI_ISL_1737259, EPI_ISL_1737260, EPI_ISL_1737261, EPI_ISL_1737262, EPI_ISL_1737263, EPI_ISL_1737264, EPI_ISL_1737265, EPI_ISL_1737266, EPI_ISL_1737267, EPI_ISL_1737268, EPI_ISL_1737269, EPI_ISL_1737270, EPI_ISL_1737271, EPI_ISL_1737272, EPI_ISL_1737273, EPI_ISL_1737274, EPI_ISL_1737275, EPI_ISL_1737276, EPI_ISL_1737277, EPI_ISL_1737278, EPI_ISL_1737279, EPI_ISL_1737280, EPI_ISL_1737281, EPI_ISL_1737282, EPI_ISL_1737283, EPI_ISL_1737284, EPI_ISL_1737285, EPI_ISL_1737286, EPI_ISL_1737287, EPI_ISL_1737288, EPI_ISL_1737289, EPI_ISL_1737290, EPI_ISL_1737291, EPI_ISL_1737292, EPI_ISL_1737293, EPI_ISL_1737294, EPI_ISL_1737295, EPI_ISL_1737296, EPI_ISL_1737297, EPI_ISL_1737299, EPI_ISL_1737300, EPI_ISL_1737301, EPI_ISL_1737302, EPI_ISL_1737303, EPI_ISL_1737304, EPI_ISL_1737305, EPI_ISL_1737306, EPI_ISL_1737307, EPI_ISL_1737308, EPI_ISL_1737309, EPI_ISL_1737310, EPI_ISL_1737311, EPI_ISL_1737312, EPI_ISL_1737313, EPI_ISL_1737314, EPI_ISL_1737315, EPI_ISL_1737316, EPI_ISL_1737317, EPI_ISL_1737318, EPI_ISL_1737319, EPI_ISL_1737320, EPI_ISL_1737321, EPI_ISL_1737322, EPI_ISL_1737323, EPI_ISL_1737324, EPI_ISL_1737325, EPI_ISL_1737326, EPI_ISL_1737327, EPI_ISL_1737328, EPI_ISL_1737329, EPI_ISL_1737330, EPI_ISL_1737331, EPI_ISL_1737332, EPI_ISL_1737333, EPI_ISL_1737334, EPI_ISL_1737335, EPI_ISL_1737336, EPI_ISL_1737337, EPI_ISL_1737338, EPI_ISL_1737339, EPI_ISL_1737340, EPI_ISL_1737341, EPI_ISL_1737342, EPI_ISL_1737343, EPI_ISL_1737344, EPI_ISL_1737345, EPI_ISL_1737346, EPI_ISL_1737347, EPI_ISL_1737348, EPI_ISL_1737349, EPI_ISL_1737350, EPI_ISL_1737351, EPI_ISL_1737352, EPI_ISL_1737353, EPI_ISL_1737354, EPI_ISL_1737355, EPI_ISL_1737356, EPI_ISL_1737357, EPI_ISL_1737358, EPI_ISL_1737359, EPI_ISL_1737360, EPI_ISL_1737361, EPI_ISL_1737362, EPI_ISL_1737363, EPI_ISL_1737364, EPI_ISL_1737365, EPI_ISL_1737366, EPI_ISL_1737367, EPI_ISL_1737368, EPI_ISL_1737369, EPI_ISL_1737370, EPI_ISL_1737371, EPI_ISL_1737372, EPI_ISL_1737374, EPI_ISL_1737375, EPI_ISL_1737376, EPI_ISL_1737377, EPI_ISL_1737378, EPI_ISL_1737379, EPI_ISL_1737380, EPI_ISL_1737381, EPI_ISL_1737382, EPI_ISL_1737383, EPI_ISL_1737384, EPI_ISL_1737385, EPI_ISL_1737386, EPI_ISL_1737387, EPI_ISL_1737388, EPI_ISL_1737389, EPI_ISL_1737390, EPI_ISL_1737391, EPI_ISL_1737392, EPI_ISL_1737393, EPI_ISL_1737394, EPI_ISL_1737395, EPI_ISL_1737396, EPI_ISL_1737397, EPI_ISL_1737398, EPI_ISL_1737399, EPI_ISL_1737400, EPI_ISL_1737401, EPI_ISL_1737402, EPI_ISL_1737403, EPI_ISL_1737404, EPI_ISL_1737405, EPI_ISL_1737406, EPI_ISL_1737407, EPI_ISL_1737408, EPI_ISL_1737409, EPI_ISL_1737410, EPI_ISL_1737411, EPI_ISL_1737412, EPI_ISL_1737413, EPI_ISL_1737414, EPI_ISL_1737415, EPI_ISL_1737416, EPI_ISL_1737417, EPI_ISL_1737418, EPI_ISL_1737419, EPI_ISL_1737420, EPI_ISL_1737421, EPI_ISL_1737423, EPI_ISL_1737424, EPI_ISL_1737425, EPI_ISL_1737426, EPI_ISL_1737427, EPI_ISL_1737428, EPI_ISL_1737429, EPI_ISL_1737430, EPI_ISL_1737431, EPI_ISL_1737432, EPI_ISL_1737433, EPI_ISL_1737434, EPI_ISL_1737435, EPI_ISL_1737436, EPI_ISL_1737437, EPI_ISL_1737438, EPI_ISL_1737439, EPI_ISL_1737440, EPI_ISL_1737441, EPI_ISL_1737442, EPI_ISL_1737443, EPI_ISL_1737444, EPI_ISL_1737445, EPI_ISL_1737446, EPI_ISL_1737447, EPI_ISL_1737448, EPI_ISL_1737449, EPI_ISL_1737450, EPI_ISL_1737451, EPI_ISL_1737452, EPI_ISL_1737453, EPI_ISL_1737454, EPI_ISL_1737455, EPI_ISL_1737456, EPI_ISL_1737457, EPI_ISL_1737458, EPI_ISL_1737459, EPI_ISL_1737460, EPI_ISL_1737461, EPI_ISL_1737462, EPI_ISL_1737463, EPI_ISL_1737464, EPI_ISL_1737465, EPI_ISL_1737466, EPI_ISL_1737467, EPI_ISL_1737468, EPI_ISL_1737469, EPI_ISL_1737470, EPI_ISL_1737471, EPI_ISL_1737472, EPI_ISL_1737474, EPI_ISL_1737475, EPI_ISL_1737476, EPI_ISL_1737477, EPI_ISL_1737478, EPI_ISL_1737479, EPI_ISL_1737480, EPI_ISL_1737481, EPI_ISL_1737482, EPI_ISL_1737483, EPI_ISL_1737485, EPI_ISL_1737486, EPI_ISL_1737487, EPI_ISL_1737488, EPI_ISL_1737489, EPI_ISL_1737490, EPI_ISL_1737491, EPI_ISL_1737492, EPI_ISL_1737493, EPI_ISL_1737494, EPI_ISL_1737495, EPI_ISL_1737496, EPI_ISL_1737497, EPI_ISL_1737498, EPI_ISL_1737499, EPI_ISL_1737500, EPI_ISL_1737501, EPI_ISL_1737502, EPI_ISL_1737503, EPI_ISL_1737504, EPI_ISL_1737505, EPI_ISL_1737506, EPI_ISL_1737507, EPI_ISL_1737508, EPI_ISL_1737509, EPI_ISL_1737510, EPI_ISL_1737511, EPI_ISL_1737512, EPI_ISL_1737513, EPI_ISL_1737514, EPI_ISL_1737515, EPI_ISL_1737516, EPI_ISL_1737517, EPI_ISL_1737518, EPI_ISL_1737519, EPI_ISL_1737520, EPI_ISL_1737521, EPI_ISL_1737522, EPI_ISL_1737523, EPI_ISL_1737524, EPI_ISL_1737525, EPI_ISL_1737526, EPI_ISL_1737527, EPI_ISL_1737528, EPI_ISL_1737529, EPI_ISL_1737530, EPI_ISL_1737531, EPI_ISL_1737532, EPI_ISL_1737533, EPI_ISL_1737534, EPI_ISL_1737535, EPI_ISL_1737536, EPI_ISL_1737537, EPI_ISL_1737538, EPI_ISL_1737539, EPI_ISL_1737540, EPI_ISL_1737541, EPI_ISL_1737542, EPI_ISL_1737543, EPI_ISL_1737544, EPI_ISL_1737545, EPI_ISL_1737546, EPI_ISL_1737547, EPI_ISL_1737548, EPI_ISL_1737549, EPI_ISL_1737550, EPI_ISL_1737551, EPI_ISL_1737552, EPI_ISL_1737553, EPI_ISL_1737554, EPI_ISL_1737555, EPI_ISL_1737556, EPI_ISL_1737557, EPI_ISL_1737558, EPI_ISL_1737559, EPI_ISL_1737560, EPI_ISL_1737561, EPI_ISL_1737562, EPI_ISL_1737563, EPI_ISL_1737564, EPI_ISL_1737565, EPI_ISL_1737566, EPI_ISL_1737567, EPI_ISL_1737568, EPI_ISL_1737569, EPI_ISL_1737570, EPI_ISL_1737571, EPI_ISL_1737572, EPI_ISL_1737573, EPI_ISL_1737574, EPI_ISL_1737575, EPI_ISL_1737576, EPI_ISL_1737577, EPI_ISL_1737578, EPI_ISL_1737579, EPI_ISL_1737580, EPI_ISL_1737581, EPI_ISL_1737582, EPI_ISL_1737583, EPI_ISL_1737584, EPI_ISL_1737585, EPI_ISL_1737586, EPI_ISL_1737587, EPI_ISL_1737588, EPI_ISL_1737589, EPI_ISL_1737590, EPI_ISL_1737591, EPI_ISL_1737592, EPI_ISL_1737593, EPI_ISL_1737594, EPI_ISL_1737595, EPI_ISL_1737596, EPI_ISL_1737597, EPI_ISL_1737598, EPI_ISL_1737599, EPI_ISL_1737600, EPI_ISL_1737601, EPI_ISL_1737602, EPI_ISL_1737603, EPI_ISL_1737604, EPI_ISL_1737605, EPI_ISL_1737606, EPI_ISL_1737607, EPI_ISL_1737608, EPI_ISL_1737609, EPI_ISL_1737610, EPI_ISL_1737611, EPI_ISL_1737612, EPI_ISL_1737613, EPI_ISL_1737614, EPI_ISL_1737615, EPI_ISL_1737616, EPI_ISL_1737617, EPI_ISL_1737618, EPI_ISL_1737619, EPI_ISL_1737620, EPI_ISL_1737622, EPI_ISL_1737623, EPI_ISL_1737624, EPI_ISL_1737625, EPI_ISL_1737626, EPI_ISL_1737627, EPI_ISL_1737628, EPI_ISL_1737629, EPI_ISL_1737630, EPI_ISL_1737631, EPI_ISL_1737632, EPI_ISL_1737633, EPI_ISL_1737634, EPI_ISL_1737635, EPI_ISL_1737636, EPI_ISL_1737637, EPI_ISL_1737638, EPI_ISL_1737639, EPI_ISL_1737640, EPI_ISL_1737641, EPI_ISL_1737642, EPI_ISL_1737643, EPI_ISL_1737644, EPI_ISL_1737645, EPI_ISL_1737646, EPI_ISL_1737647, EPI_ISL_1737648, EPI_ISL_1737649, EPI_ISL_1737650, EPI_ISL_1737651, EPI_ISL_1737652, EPI_ISL_1737653, EPI_ISL_1737654, EPI_ISL_1737655, EPI_ISL_1737656, EPI_ISL_1737657, EPI_ISL_1737658, EPI_ISL_1737659, EPI_ISL_1737660, EPI_ISL_1737661, EPI_ISL_1737662, EPI_ISL_1737663, EPI_ISL_1737664, EPI_ISL_1737665, EPI_ISL_1737666, EPI_ISL_1737667, EPI_ISL_1737668, EPI_ISL_1737669, EPI_ISL_1737670, EPI_ISL_1737671, EPI_ISL_1737672, EPI_ISL_1737673, EPI_ISL_1737674, EPI_ISL_1737675, EPI_ISL_1737676, EPI_ISL_1737677, EPI_ISL_1737678, EPI_ISL_1737679, EPI_ISL_1737680, EPI_ISL_1737681, EPI_ISL_1737682, EPI_ISL_1737683, EPI_ISL_1737684, EPI_ISL_1737685, EPI_ISL_1737686, EPI_ISL_1737687, EPI_ISL_1737688, EPI_ISL_1737689, EPI_ISL_1737690, EPI_ISL_1737691, EPI_ISL_1737692, EPI_ISL_1737693, EPI_ISL_1737694, EPI_ISL_1737695, EPI_ISL_1737696, EPI_ISL_1737697, EPI_ISL_1737698, EPI_ISL_1737699, EPI_ISL_1737700, EPI_ISL_1737701, EPI_ISL_1737702, EPI_ISL_1737703, EPI_ISL_1737704, EPI_ISL_1737705, EPI_ISL_1737706, EPI_ISL_1737707, EPI_ISL_1737708, EPI_ISL_1737709, EPI_ISL_1737710, EPI_ISL_1737711, EPI_ISL_1737712, EPI_ISL_1737713, EPI_ISL_1737714, EPI_ISL_1737715, EPI_ISL_1737716, EPI_ISL_1737717, EPI_ISL_1737718, EPI_ISL_1737719, EPI_ISL_1737720, EPI_ISL_1737721, EPI_ISL_1737722, EPI_ISL_1737723, EPI_ISL_1737724, EPI_ISL_1737725, EPI_ISL_1737726, EPI_ISL_1737727, EPI_ISL_1737728, EPI_ISL_1737729, EPI_ISL_1737730, EPI_ISL_1737731, EPI_ISL_1737732, EPI_ISL_1737733, EPI_ISL_1737734, EPI_ISL_1737735, EPI_ISL_1737736, EPI_ISL_1737737, EPI_ISL_1737738, EPI_ISL_1737739, EPI_ISL_1737740, EPI_ISL_1737741, EPI_ISL_1737742, EPI_ISL_1737743, EPI_ISL_1737744, EPI_ISL_1737745, EPI_ISL_1737746, EPI_ISL_1737747, EPI_ISL_1737748, EPI_ISL_1737751, EPI_ISL_1737752, EPI_ISL_1737753, EPI_ISL_1737754, EPI_ISL_1737755, EPI_ISL_1737756, EPI_ISL_1737757, EPI_ISL_1737758, EPI_ISL_1737759, EPI_ISL_1737760, EPI_ISL_1737761, EPI_ISL_1737762, EPI_ISL_1737763, EPI_ISL_1737764, EPI_ISL_1737765, EPI_ISL_1737766, EPI_ISL_1737767, EPI_ISL_1737768, EPI_ISL_1737769, EPI_ISL_1737770, EPI_ISL_1737771, EPI_ISL_1737772, EPI_ISL_1737773, EPI_ISL_1737774, EPI_ISL_1737775, EPI_ISL_1737776, EPI_ISL_1737777, EPI_ISL_1737778, EPI_ISL_1737779, EPI_ISL_1737780, EPI_ISL_1737781, EPI_ISL_1737782, EPI_ISL_1737783, EPI_ISL_1737784, EPI_ISL_1737785, EPI_ISL_1737786, EPI_ISL_1737787, EPI_ISL_1737788, EPI_ISL_1737789, EPI_ISL_1737790, EPI_ISL_1737791, EPI_ISL_1737792, EPI_ISL_1737793, EPI_ISL_1737794, EPI_ISL_1737795, EPI_ISL_1737796, EPI_ISL_1737799, EPI_ISL_1737800, EPI_ISL_1737801, EPI_ISL_1737802, EPI_ISL_1737803, EPI_ISL_1737804, EPI_ISL_1737805, EPI_ISL_1737806, EPI_ISL_1737807, EPI_ISL_1737808, EPI_ISL_1737809, EPI_ISL_1737810, EPI_ISL_1737811, EPI_ISL_1737812, EPI_ISL_1737813, EPI_ISL_1737814, EPI_ISL_1737815, EPI_ISL_1737816, EPI_ISL_1737817, EPI_ISL_1737818, EPI_ISL_1737819, EPI_ISL_1737820, EPI_ISL_1737821, EPI_ISL_1737822, EPI_ISL_1737823, EPI_ISL_1737824, EPI_ISL_1737825, EPI_ISL_1737826, EPI_ISL_1737827, EPI_ISL_1737828, EPI_ISL_1737829, EPI_ISL_1737830, EPI_ISL_1737831, EPI_ISL_1737832, EPI_ISL_1737833, EPI_ISL_1737834, EPI_ISL_1737835, EPI_ISL_1737836, EPI_ISL_1737837, EPI_ISL_1737838, EPI_ISL_1737839, EPI_ISL_1737840, EPI_ISL_1737841, EPI_ISL_1737842, EPI_ISL_1737843, EPI_ISL_1737844, EPI_ISL_1737845, EPI_ISL_1737846, EPI_ISL_1737847, EPI_ISL_1737848, EPI_ISL_1737849, EPI_ISL_1737850, EPI_ISL_1737851, EPI_ISL_1737852, EPI_ISL_1737853, EPI_ISL_1737854, EPI_ISL_1737855, EPI_ISL_1737856, EPI_ISL_1737857, EPI_ISL_1737858, EPI_ISL_1737859, EPI_ISL_1737860, EPI_ISL_1737861, EPI_ISL_1737862, EPI_ISL_1737863, EPI_ISL_1737864, EPI_ISL_1737865, EPI_ISL_1737866, EPI_ISL_1737867, EPI_ISL_1737868, EPI_ISL_1737869, EPI_ISL_1737870, EPI_ISL_1737871, EPI_ISL_1737872, EPI_ISL_1737873, EPI_ISL_1737874, EPI_ISL_1737875, EPI_ISL_1737876, EPI_ISL_1737877, EPI_ISL_1737878, EPI_ISL_1737879, EPI_ISL_1737880, EPI_ISL_1737881 | | | |
| see above | Aegis Sciences Corporation | Centers for Disease Control and Prevention Division of Viral Diseases, Pathogen Discovery | Dakota Howard, Dhwani Batra, Peter W. Cook, Kara Moser, Adrian Paskey, Jason Caravas, Benjamin Rambo-Martin, Shatavia Morrison, Christopher Gulvick, Scott Sammons, Yvette Unoarumhi, Darlene Wagner, Matthew Schmerer, Cyndi Clark, Patrick Campbell, Rob Case, Vikramsinha Ghorpade, Holly Houdeshell, Ola Kvalvaag, Dillon Nall, Ethan Sanders, Alec Vest, Shaun Westlund, Matthew Hardison, Clinton R. Paden, Duncan MacCannell |
| EPI_ISL_1737882, EPI_ISL_1737883, EPI_ISL_1737884, EPI_ISL_1737885, EPI_ISL_1737886, EPI_ISL_1737887, EPI_ISL_1737888, EPI_ISL_1737889, EPI_ISL_1737890, EPI_ISL_1737891, EPI_ISL_1737892, EPI_ISL_1737893, EPI_ISL_1737894, EPI_ISL_1737895, EPI_ISL_1737896, EPI_ISL_1737897, EPI_ISL_1737898, EPI_ISL_1737899, EPI_ISL_1737900, EPI_ISL_1737901, EPI_ISL_1737902, EPI_ISL_1737903, EPI_ISL_1737904, EPI_ISL_1737905, EPI_ISL_1737906, EPI_ISL_1737907, EPI_ISL_1737908, EPI_ISL_1737909, EPI_ISL_1737910, EPI_ISL_1737911, EPI_ISL_1737912, EPI_ISL_1737913, EPI_ISL_1737914, EPI_ISL_1737915, EPI_ISL_1737916, EPI_ISL_1737917, EPI_ISL_1737918, EPI_ISL_1737919, EPI_ISL_1737920, EPI_ISL_1737921, EPI_ISL_1737922, EPI_ISL_1737923, EPI_ISL_1737924, EPI_ISL_1737925, EPI_ISL_1737926, EPI_ISL_1737927, EPI_ISL_1737928, EPI_ISL_1737929, EPI_ISL_1737930, EPI_ISL_1737931, EPI_ISL_1737932, EPI_ISL_1737933, EPI_ISL_1737934, EPI_ISL_1737935, EPI_ISL_1737936, EPI_ISL_1737937, EPI_ISL_1737938, EPI_ISL_1737939, EPI_ISL_1737940, EPI_ISL_1737941, EPI_ISL_1737942, EPI_ISL_1737943, EPI_ISL_1737944, EPI_ISL_1737945, EPI_ISL_1737946, EPI_ISL_1737947, EPI_ISL_1737948, EPI_ISL_1737949, EPI_ISL_1737950, EPI_ISL_1737951, EPI_ISL_1737952, EPI_ISL_1737953, EPI_ISL_1737954, EPI_ISL_1737955, EPI_ISL_1737956, EPI_ISL_1737957, EPI_ISL_1737958, EPI_ISL_1737959, EPI_ISL_1737960, EPI_ISL_1737961, EPI_ISL_1737962, EPI_ISL_1737963, EPI_ISL_1737964, EPI_ISL_1737965, EPI_ISL_1737966, EPI_ISL_1737967, EPI_ISL_1737968, EPI_ISL_1737969, EPI_ISL_1737970, EPI_ISL_1737971, EPI_ISL_1737972, EPI_ISL_1737973, EPI_ISL_1737974, EPI_ISL_1737975, EPI_ISL_1737976, EPI_ISL_1737977, EPI_ISL_1737978, EPI_ISL_1737979, EPI_ISL_1737980, EPI_ISL_1737981, EPI_ISL_1737982, EPI_ISL_1737983, EPI_ISL_1737984, EPI_ISL_1737985, EPI_ISL_1737986, EPI_ISL_1737987, EPI_ISL_1737988, EPI_ISL_1737989, EPI_ISL_1737990, EPI_ISL_1737991, EPI_ISL_1737992, EPI_ISL_1737993, EPI_ISL_1737994, EPI_ISL_1737995, EPI_ISL_1737996, EPI_ISL_1737997, EPI_ISL_1737998, EPI_ISL_1737999, EPI_ISL_1738000, EPI_ISL_1738001, EPI_ISL_1738002, EPI_ISL_1738003, EPI_ISL_1738004, EPI_ISL_1738005, EPI_ISL_1738006, EPI_ISL_1738007, EPI_ISL_1738008, EPI_ISL_1738009, EPI_ISL_1738010, EPI_ISL_1738011, EPI_ISL_1738012, EPI_ISL_1738013, EPI_ISL_1738014, EPI_ISL_1738015, EPI_ISL_1738016, EPI_ISL_1738017, EPI_ISL_1738018, EPI_ISL_1738019, EPI_ISL_1738020, EPI_ISL_1738021, EPI_ISL_1738022, EPI_ISL_1738023, EPI_ISL_1738024, EPI_ISL_1738025, EPI_ISL_1738026, EPI_ISL_1738027, EPI_ISL_1738028, EPI_ISL_1738029, EPI_ISL_1738030, EPI_ISL_1738031, EPI_ISL_1738032, EPI_ISL_1738033, EPI_ISL_1738034, EPI_ISL_1738035, EPI_ISL_1738036, EPI_ISL_1738037, EPI_ISL_1738039, EPI_ISL_1738040, EPI_ISL_1738041, EPI_ISL_1738042, EPI_ISL_1738043, EPI_ISL_1738044, EPI_ISL_1738045, EPI_ISL_1738046, EPI_ISL_1738047, EPI_ISL_1738048, EPI_ISL_1738049, EPI_ISL_1738050, EPI_ISL_1738051, EPI_ISL_1738052, EPI_ISL_1738053, EPI_ISL_1738054, EPI_ISL_1738055, EPI_ISL_1738056, EPI_ISL_1738057, EPI_ISL_1738058, EPI_ISL_1738059, EPI_ISL_1738060, EPI_ISL_1738061, EPI_ISL_1738062, EPI_ISL_1738063, EPI_ISL_1738064, EPI_ISL_1738065, EPI_ISL_1738066, EPI_ISL_1738067, EPI_ISL_1738068, EPI_ISL_1738069, EPI_ISL_1738070, EPI_ISL_1738071, EPI_ISL_1738072, EPI_ISL_1738073, EPI_ISL_1738074, EPI_ISL_1738075, EPI_ISL_1738076, EPI_ISL_1738077, EPI_ISL_1738078, EPI_ISL_1738079 | | | |
| see above | Quest Diagnostics Incorporated | Centers for Disease Control and Prevention Division of Viral Diseases, Pathogen Discovery | Dakota Howard, Dhwani Batra, Peter W. Cook, Kara Moser, Adrian Paskey, Jason Caravas, Benjamin Rambo-Martin, Shatavia Morrison, Christopher Gulvick, Scott Sammons, Yvette Unoarumhi, Darlene Wagner, Matthew Schmerer, S. H. Rosenthal, A. Gerasimova, R. M. Kagan, B. Anderson, M. Hua, Y. Liu, L.E. Bernstein, K.E. Livingston, A. Perez, I. A. Shlyakhter, R. V. Rolando, R. Owen, P. Tanpaiboon, F. Lacbawan, Clinton R. Paden, Duncan MacCannell |
| EPI_ISL_1738080 | School of Pharmacy, Shenandoah University | School of Pharmacy, Shenandoah University | Adams,S.M., Harralson,A.F., Kidd,R.S., Sawyer,G.W. |
| EPI_ISL_1738081, EPI_ISL_1738082, EPI_ISL_1738083, EPI_ISL_1738084, EPI_ISL_1738085, EPI_ISL_1738086, EPI_ISL_1738087, EPI_ISL_1738088, EPI_ISL_1738089, EPI_ISL_1738090, EPI_ISL_1738091, EPI_ISL_1738092, EPI_ISL_1738093, EPI_ISL_1738094, EPI_ISL_1738095, EPI_ISL_1738096, EPI_ISL_1738097, EPI_ISL_1738098, EPI_ISL_1738099, EPI_ISL_1738100, EPI_ISL_1738101, EPI_ISL_1738102, EPI_ISL_1738103, EPI_ISL_1738104, EPI_ISL_1738105, EPI_ISL_1738106, EPI_ISL_1738107, EPI_ISL_1738108, EPI_ISL_1738109, EPI_ISL_1738110, EPI_ISL_1738111, EPI_ISL_1738112, EPI_ISL_1738113, EPI_ISL_1738114, EPI_ISL_1738115, EPI_ISL_1738116, EPI_ISL_1738117, EPI_ISL_1738118, EPI_ISL_1738119, EPI_ISL_1738120, EPI_ISL_1738121, EPI_ISL_1738122, EPI_ISL_1738123, EPI_ISL_1738124, EPI_ISL_1738125, EPI_ISL_1738126, EPI_ISL_1738127, EPI_ISL_1738128, EPI_ISL_1738129, EPI_ISL_1738130, EPI_ISL_1738131, EPI_ISL_1738132, EPI_ISL_1738133, EPI_ISL_1738134, EPI_ISL_1738135, EPI_ISL_1738136, EPI_ISL_1738137, EPI_ISL_1738138, EPI_ISL_1738139, EPI_ISL_1738140, EPI_ISL_1738141, EPI_ISL_1738142, EPI_ISL_1738143, EPI_ISL_1738144, EPI_ISL_1738145, EPI_ISL_1738146, EPI_ISL_1738147, EPI_ISL_1738148, EPI_ISL_1738149, EPI_ISL_1738150, EPI_ISL_1738151, EPI_ISL_1738152, EPI_ISL_1738153, EPI_ISL_1738154, EPI_ISL_1738155, EPI_ISL_1738156, EPI_ISL_1738157, EPI_ISL_1738158, EPI_ISL_1738159, EPI_ISL_1738160, EPI_ISL_1738161, EPI_ISL_1738162, EPI_ISL_1738163, EPI_ISL_1738164, EPI_ISL_1738165, EPI_ISL_1738166, EPI_ISL_1738167, EPI_ISL_1738168, EPI_ISL_1738169, EPI_ISL_1738170, EPI_ISL_1738171, EPI_ISL_1738172, EPI_ISL_1738173, EPI_ISL_1738174, EPI_ISL_1738175, EPI_ISL_1738176, EPI_ISL_1738177, EPI_ISL_1738178, EPI_ISL_1738179, EPI_ISL_1738180, EPI_ISL_1738181, EPI_ISL_1738182, EPI_ISL_1738183, EPI_ISL_1738184, EPI_ISL_1738185, EPI_ISL_1738186, EPI_ISL_1738187, EPI_ISL_1738188, EPI_ISL_1738189, EPI_ISL_1738190, EPI_ISL_1738191, EPI_ISL_1738192, EPI_ISL_1738193, EPI_ISL_1738194, EPI_ISL_1738195, EPI_ISL_1738196, EPI_ISL_1738197, EPI_ISL_1738198, EPI_ISL_1738199, EPI_ISL_1738200, EPI_ISL_1738201, EPI_ISL_1738202, EPI_ISL_1738203, EPI_ISL_1738204, EPI_ISL_1738205, EPI_ISL_1738206, EPI_ISL_1738207, EPI_ISL_1738208, EPI_ISL_1738209, EPI_ISL_1738210, EPI_ISL_1738211, EPI_ISL_1738212, EPI_ISL_1738213, EPI_ISL_1738214, EPI_ISL_1738215, EPI_ISL_1738216, EPI_ISL_1738217, EPI_ISL_1738218, EPI_ISL_1738219, EPI_ISL_1738220, EPI_ISL_1738221, EPI_ISL_1738222, EPI_ISL_1738223, EPI_ISL_1738224, EPI_ISL_1738225, EPI_ISL_1738226, EPI_ISL_1738227, EPI_ISL_1738228, EPI_ISL_1738229, EPI_ISL_1738230, EPI_ISL_1738231, EPI_ISL_1738232, EPI_ISL_1738233, EPI_ISL_1738234, EPI_ISL_1738235, EPI_ISL_1738236, EPI_ISL_1738237, EPI_ISL_1738238, EPI_ISL_1738239, EPI_ISL_1738240, EPI_ISL_1738241, EPI_ISL_1738242, EPI_ISL_1738243, EPI_ISL_1738244, EPI_ISL_1738245, EPI_ISL_1738246, EPI_ISL_1738247, EPI_ISL_1738248, EPI_ISL_1738249, EPI_ISL_1738250, EPI_ISL_1738251, EPI_ISL_1738252, EPI_ISL_1738253, EPI_ISL_1738254, EPI_ISL_1738255, EPI_ISL_1738256, EPI_ISL_1738257, EPI_ISL_1738258, EPI_ISL_1738259, EPI_ISL_1738260, EPI_ISL_1738261, EPI_ISL_1738262, EPI_ISL_1738263, EPI_ISL_1738264, EPI_ISL_1738265, EPI_ISL_1738266, EPI_ISL_1738267, EPI_ISL_1738268, EPI_ISL_1738269, EPI_ISL_1738270, EPI_ISL_1738271, EPI_ISL_1738272, EPI_ISL_1738273, EPI_ISL_1738274, EPI_ISL_1738275, EPI_ISL_1738276, EPI_ISL_1738277, EPI_ISL_1738278, EPI_ISL_1738279, EPI_ISL_1738280, EPI_ISL_1738281, EPI_ISL_1738282, EPI_ISL_1738283, EPI_ISL_1738284, EPI_ISL_1738285, EPI_ISL_1738286, EPI_ISL_1738287, EPI_ISL_1738288, EPI_ISL_1738289, EPI_ISL_1738290, EPI_ISL_1738291, EPI_ISL_1738292, EPI_ISL_1738293, EPI_ISL_1738294, EPI_ISL_1738295, EPI_ISL_1738296, EPI_ISL_1738297, EPI_ISL_1738298, EPI_ISL_1738299, EPI_ISL_1738300, EPI_ISL_1738301, EPI_ISL_1738302, EPI_ISL_1738303, EPI_ISL_1738304, EPI_ISL_1738305, EPI_ISL_1738306, EPI_ISL_1738307, EPI_ISL_1738308, EPI_ISL_1738309, EPI_ISL_1738310, EPI_ISL_1738311, EPI_ISL_1738312, EPI_ISL_1738313, EPI_ISL_1738314, EPI_ISL_1738315, EPI_ISL_1738316, EPI_ISL_1738317, EPI_ISL_1738318, EPI_ISL_1738319, EPI_ISL_1738320, EPI_ISL_1738321, EPI_ISL_1738322, EPI_ISL_1738323, EPI_ISL_1738324, EPI_ISL_1738325, EPI_ISL_1738326, EPI_ISL_1738327, EPI_ISL_1738328, EPI_ISL_1738329, EPI_ISL_1738330, EPI_ISL_1738331, EPI_ISL_1738332, EPI_ISL_1738333, EPI_ISL_1738334, EPI_ISL_1738335, EPI_ISL_1738336, EPI_ISL_1738337, EPI_ISL_1738338, EPI_ISL_1738339, EPI_ISL_1738340, EPI_ISL_1738341, EPI_ISL_1738342, EPI_ISL_1738343, EPI_ISL_1738344, EPI_ISL_1738345, EPI_ISL_1738346, EPI_ISL_1738347, EPI_ISL_1738348, EPI_ISL_1738349, EPI_ISL_1738350, EPI_ISL_1738351, EPI_ISL_1738352, EPI_ISL_1738353, EPI_ISL_1738354, EPI_ISL_1738355, EPI_ISL_1738356, EPI_ISL_1738357, EPI_ISL_1738358, EPI_ISL_1738359, EPI_ISL_1738360, EPI_ISL_1738361, EPI_ISL_1738362, EPI_ISL_1738363, EPI_ISL_1738364, EPI_ISL_1738365, EPI_ISL_1738366, EPI_ISL_1738367, EPI_ISL_1738368, EPI_ISL_1738369, EPI_ISL_1738370, EPI_ISL_1738371, EPI_ISL_1738372, EPI_ISL_1738373, EPI_ISL_1738374, EPI_ISL_1738375, EPI_ISL_1738376, EPI_ISL_1738377, EPI_ISL_1738378, EPI_ISL_1738379, EPI_ISL_1738380, EPI_ISL_1738381, EPI_ISL_1738382, EPI_ISL_1738383, EPI_ISL_1738384, EPI_ISL_1738385, EPI_ISL_1738386, EPI_ISL_1738387, EPI_ISL_1738388, EPI_ISL_1738389, EPI_ISL_1738390, EPI_ISL_1738391, EPI_ISL_1738392, EPI_ISL_1738393, EPI_ISL_1738394, EPI_ISL_1738395, EPI_ISL_1738396, EPI_ISL_1738397, EPI_ISL_1738398, EPI_ISL_1738399, EPI_ISL_1738400, EPI_ISL_1738401, EPI_ISL_1738402, EPI_ISL_1738403, EPI_ISL_1738404, EPI_ISL_1738405, EPI_ISL_1738406, EPI_ISL_1738407, EPI_ISL_1738408, EPI_ISL_1738409, EPI_ISL_1738410, EPI_ISL_1738411, EPI_ISL_1738412, EPI_ISL_1738413, EPI_ISL_1738414, EPI_ISL_1738415, EPI_ISL_1738416, EPI_ISL_1738417, EPI_ISL_1738418, EPI_ISL_1738419, EPI_ISL_1738420, EPI_ISL_1738421, EPI_ISL_1738422, EPI_ISL_1738423, EPI_ISL_1738424, EPI_ISL_1738425, EPI_ISL_1738426, EPI_ISL_1738427, EPI_ISL_1738428, EPI_ISL_1738429, EPI_ISL_1738430, EPI_ISL_1738431, EPI_ISL_1738432, EPI_ISL_1738433, EPI_ISL_1738434, EPI_ISL_1738435, EPI_ISL_1738436, EPI_ISL_1738437, EPI_ISL_1738438, EPI_ISL_1738439, EPI_ISL_1738440, EPI_ISL_1738441, EPI_ISL_1738442, EPI_ISL_1738443, EPI_ISL_1738444, EPI_ISL_1738445, EPI_ISL_1738446, EPI_ISL_1738447, EPI_ISL_1738448, EPI_ISL_1738449, EPI_ISL_1738450, EPI_ISL_1738451, EPI_ISL_1738452, EPI_ISL_1738453, EPI_ISL_1738454, EPI_ISL_1738455, EPI_ISL_1738456, EPI_ISL_1738457, EPI_ISL_1738458, EPI_ISL_1738459, EPI_ISL_1738460, EPI_ISL_1738461, EPI_ISL_1738462, EPI_ISL_1738463, EPI_ISL_1738464, EPI_ISL_1738465, EPI_ISL_1738466, EPI_ISL_1738467, EPI_ISL_1738468, EPI_ISL_1738469, EPI_ISL_1738470, EPI_ISL_1738471, EPI_ISL_1738472, EPI_ISL_1738473, EPI_ISL_1738474, EPI_ISL_1738475, EPI_ISL_1738476, EPI_ISL_1738477, EPI_ISL_1738478, EPI_ISL_1738479, EPI_ISL_1738480, EPI_ISL_1738481, EPI_ISL_1738482, EPI_ISL_1738483, EPI_ISL_1738484, EPI_ISL_1738485, EPI_ISL_1738486, EPI_ISL_1738487, EPI_ISL_1738488, EPI_ISL_1738489, EPI_ISL_1738490, EPI_ISL_1738491, EPI_ISL_1738492, EPI_ISL_1738493, EPI_ISL_1738494, EPI_ISL_1738495, EPI_ISL_1738496, EPI_ISL_1738497, EPI_ISL_1738498, EPI_ISL_1738499, EPI_ISL_1738500, EPI_ISL_1738501, EPI_ISL_1738502, EPI_ISL_1738503, EPI_ISL_1738504, EPI_ISL_1738505, EPI_ISL_1738506, EPI_ISL_1738507, EPI_ISL_1738508, EPI_ISL_1738509, EPI_ISL_1738510, EPI_ISL_1738511, EPI_ISL_1738512, EPI_ISL_1738513, EPI_ISL_1738514, EPI_ISL_1738515, EPI_ISL_1738516, EPI_ISL_1738517, EPI_ISL_1738518, EPI_ISL_1738519, EPI_ISL_1738520, EPI_ISL_1738521, EPI_ISL_1738522, EPI_ISL_1738523, EPI_ISL_1738524, EPI_ISL_1738525, EPI_ISL_1738526, EPI_ISL_1738527, EPI_ISL_1738528, EPI_ISL_1738529, EPI_ISL_1738530, EPI_ISL_1738531, EPI_ISL_1738532, EPI_ISL_1738533, EPI_ISL_1738534, EPI_ISL_1738535, EPI_ISL_1738536, EPI_ISL_1738537, EPI_ISL_1738538, EPI_ISL_1738539, EPI_ISL_1738540, EPI_ISL_1738541, EPI_ISL_1738542, EPI_ISL_1738543, EPI_ISL_1738544, EPI_ISL_1738545, EPI_ISL_1738546, EPI_ISL_1738547, EPI_ISL_1738548, EPI_ISL_1738549, EPI_ISL_1738550, EPI_ISL_1738551, EPI_ISL_1738552, EPI_ISL_1738553, EPI_ISL_1738554, EPI_ISL_1738555, EPI_ISL_1738556, EPI_ISL_1738557, EPI_ISL_1738558, EPI_ISL_1738559, EPI_ISL_1738560, EPI_ISL_1738561, EPI_ISL_1738562, EPI_ISL_1738563, EPI_ISL_1738564, EPI_ISL_1738565, EPI_ISL_1738566, EPI_ISL_1738567, EPI_ISL_1738568, EPI_ISL_1738569, EPI_ISL_1738570, EPI_ISL_1738571, EPI_ISL_1738572, EPI_ISL_1738573, EPI_ISL_1738574, EPI_ISL_1738575, EPI_ISL_1738576, EPI_ISL_1738577, EPI_ISL_1738578, EPI_ISL_1738579, EPI_ISL_1738580, EPI_ISL_1738581, EPI_ISL_1738582, EPI_ISL_1738583, EPI_ISL_1738584, EPI_ISL_1738585, EPI_ISL_1738586, EPI_ISL_1738587, EPI_ISL_1738588, EPI_ISL_1738589, EPI_ISL_1738590, EPI_ISL_1738591, EPI_ISL_1738592, EPI_ISL_1738593, EPI_ISL_1738594, EPI_ISL_1738595, EPI_ISL_1738596, EPI_ISL_1738597, EPI_ISL_1738598, EPI_ISL_1738599, EPI_ISL_1738600, EPI_ISL_1738601, EPI_ISL_1738602, EPI_ISL_1738603, EPI_ISL_1738604, EPI_ISL_1738605, EPI_ISL_1738606, EPI_ISL_1738607, EPI_ISL_1738608, EPI_ISL_1738609, EPI_ISL_1738610, EPI_ISL_1738611, EPI_ISL_1738612, EPI_ISL_1738613, EPI_ISL_1738614, EPI_ISL_1738615, EPI_ISL_1738616, EPI_ISL_1738617, EPI_ISL_1738618, EPI_ISL_1738619, EPI_ISL_1738620, EPI_ISL_1738621, EPI_ISL_1738622, EPI_ISL_1738623, EPI_ISL_1738624, EPI_ISL_1738625, EPI_ISL_1738626, EPI_ISL_1738627, EPI_ISL_1738628, EPI_ISL_1738629, EPI_ISL_1738630, EPI_ISL_1738631, EPI_ISL_1738632, EPI_ISL_1738633, EPI_ISL_1738634, EPI_ISL_1738635, EPI_ISL_1738636, EPI_ISL_1738637, EPI_ISL_1738638, EPI_ISL_1738639, EPI_ISL_1738640, EPI_ISL_1738641, EPI_ISL_1738642, EPI_ISL_1738643, EPI_ISL_1738644, EPI_ISL_1738645, EPI_ISL_1738646, EPI_ISL_1738647, EPI_ISL_1738648, EPI_ISL_1738649, EPI_ISL_1738650, EPI_ISL_1738651, EPI_ISL_1738652, EPI_ISL_1738653, EPI_ISL_1738654, EPI_ISL_1738655, EPI_ISL_1738656, EPI_ISL_1738657, EPI_ISL_1738658, EPI_ISL_1738659, EPI_ISL_1738660, EPI_ISL_1738661, EPI_ISL_1738662, EPI_ISL_1738663, EPI_ISL_1738664, EPI_ISL_1738665, EPI_ISL_1738666, EPI_ISL_1738667, EPI_ISL_1738668, EPI_ISL_1738669, EPI_ISL_1738670, EPI_ISL_1738671, EPI_ISL_1738672, EPI_ISL_1738673, EPI_ISL_1738674, EPI_ISL_1738675, EPI_ISL_1738676, EPI_ISL_1738677, EPI_ISL_1738678, EPI_ISL_1738679, EPI_ISL_1738680, EPI_ISL_1738681, EPI_ISL_1738682, EPI_ISL_1738683, EPI_ISL_1738684, EPI_ISL_1738685, EPI_ISL_1738686, EPI_ISL_1738687, EPI_ISL_1738688, EPI_ISL_1738689, EPI_ISL_1738690, EPI_ISL_1738691, EPI_ISL_1738692, EPI_ISL_1738693, EPI_ISL_1738694, EPI_ISL_1738695, EPI_ISL_1738696, EPI_ISL_1738697, EPI_ISL_1738698, EPI_ISL_1738699, EPI_ISL_1738700, EPI_ISL_1738701, EPI_ISL_1738702, EPI_ISL_1738703, EPI_ISL_1738704, EPI_ISL_1738705, EPI_ISL_1738706, EPI_ISL_1738707, EPI_ISL_1738708, EPI_ISL_1738709, EPI_ISL_1738710, EPI_ISL_1738711, EPI_ISL_1738712, EPI_ISL_1738713, EPI_ISL_1738714, EPI_ISL_1738715, EPI_ISL_1738716, EPI_ISL_1738717, EPI_ISL_1738718, EPI_ISL_1738719, EPI_ISL_1738720, EPI_ISL_1738721, EPI_ISL_1738722, EPI_ISL_1738723, EPI_ISL_1738724, EPI_ISL_1738725, EPI_ISL_1738726, EPI_ISL_1738727, EPI_ISL_1738728, EPI_ISL_1738729, EPI_ISL_1738730, EPI_ISL_1738731, EPI_ISL_1738732, EPI_ISL_1738733, EPI_ISL_1738734, EPI_ISL_1738735, EPI_ISL_1738736, EPI_ISL_1738737, EPI_ISL_1738738, EPI_ISL_1738739, EPI_ISL_1738740, EPI_ISL_1738741, EPI_ISL_1738742, EPI_ISL_1738743, EPI_ISL_1738744, EPI_ISL_1738745, EPI_ISL_1738746, EPI_ISL_1738747, EPI_ISL_1738748, EPI_ISL_1738749, EPI_ISL_1738750, EPI_ISL_1738751, EPI_ISL_1738752, EPI_ISL_1738753, EPI_ISL_1738754, EPI_ISL_1738755, EPI_ISL_1738756, EPI_ISL_1738757, EPI_ISL_1738758 | | | |
| see above | Quest Diagnostics Incorporated | Centers for Disease Control and Prevention Division of Viral Diseases, Pathogen Discovery | Dakota Howard, Dhwani Batra, Peter W. Cook, Kara Moser, Adrian Paskey, Jason Caravas, Benjamin Rambo-Martin, Shatavia Morrison, Christopher Gulvick, Scott Sammons, Yvette Unoarumhi, Darlene Wagner, Matthew Schmerer, S. H. Rosenthal, A. Gerasimova, R. M. Kagan, B. Anderson, M. Hua, Y. Liu, L.E. Bernstein, K.E. Livingston, A. Perez, I. A. Shlyakhter, R. V. Rolando, R. Owen, P. Tanpaiboon, F. Lacbawan, Clinton R. Paden, Duncan MacCannell |
| EPI_ISL_1738759, EPI_ISL_1738760, EPI_ISL_1738761, EPI_ISL_1738762, EPI_ISL_1738763, EPI_ISL_1738764, EPI_ISL_1738765, EPI_ISL_1738766, EPI_ISL_1738767, EPI_ISL_1738768, EPI_ISL_1738769, EPI_ISL_1738770, EPI_ISL_1738771, EPI_ISL_1738772, EPI_ISL_1738773, EPI_ISL_1738776, EPI_ISL_1738777, EPI_ISL_1738778, EPI_ISL_1738779, EPI_ISL_1738780 | | | |
| see above | Aegis Sciences Corporation | Centers for Disease Control and Prevention Division of Viral Diseases, Pathogen Discovery | Dakota Howard, Dhwani Batra, Peter W. Cook, Kara Moser, Adrian Paskey, Jason Caravas, Benjamin Rambo-Martin, Shatavia Morrison, Christopher Gulvick, Scott Sammons, Yvette Unoarumhi, Darlene Wagner, Matthew Schmerer, Cyndi Clark, Patrick Campbell, Rob Case, Vikramsinha Ghorpade, Holly Houdeshell, Ola Kvalvaag, Dillon Nall, Ethan Sanders, Alec Vest, Shaun Westlund, Matthew Hardison, Clinton R. Paden, Duncan MacCannell |
| EPI_ISL_1738791, EPI_ISL_1738792, EPI_ISL_1738793, EPI_ISL_1738794, EPI_ISL_1738795, EPI_ISL_1738796, EPI_ISL_1738797, EPI_ISL_1738798, EPI_ISL_1738799, EPI_ISL_1738800, EPI_ISL_1738801, EPI_ISL_1738802, EPI_ISL_1738803, EPI_ISL_1738804 | | | |
| see above | Nebraska Public Health Laboratory | NPHL COVID-19 Response Team | NPHL COVID-19 Response Team |
| EPI_ISL_1739275, EPI_ISL_1739277, EPI_ISL_1739278, EPI_ISL_1739279, EPI_ISL_1739280, EPI_ISL_1739281, EPI_ISL_1739282, EPI_ISL_1739283, EPI_ISL_1739285, EPI_ISL_1739286, EPI_ISL_1739287, EPI_ISL_1739288, EPI_ISL_1739289, EPI_ISL_1739290, EPI_ISL_1739291, EPI_ISL_1739292, EPI_ISL_1739293 | | | |
| see above | Southern Nevada Public Health Laboratory | Southern Nevada Public Health Laboratory | Michael Picker |
| EPI_ISL_1739300, EPI_ISL_1739301, EPI_ISL_1739302, EPI_ISL_1739303, EPI_ISL_1739304, EPI_ISL_1739305, EPI_ISL_1739332, EPI_ISL_1739334, EPI_ISL_1739335, EPI_ISL_1739336, EPI_ISL_1739337, EPI_ISL_1739338 | | | |
| see above | Maryland Genomics, Institute for Genome Sciences, University of | Maryland Genomics, Institute for Genome Sciences, University of Maryland School of | Tallon, Luke J; Sadzewicz, Lisa D; Humphrys, Mike; Ott, Sandra; Roussey, Holly; Mehta, Aditya; Vavikolanu, Kranthi; Fraser, Claire M; Ravel, Jacques |

| | | | |
|---|---|---|---|
| | Maryland School of Medicine | Medicine | |
| EPI_ISL_1739339 | TriCore Reference Laboratory Inc. | New Mexico Department of Health Scientific Laboratory | Ellie Johnson, D'eldra Malone, Jennifer Benoit, Linda Salazar, Ratheesh Rajan, Anastacia Griego-Fisher |
| EPI_ISL_1739426, EPI_ISL_1739427, EPI_ISL_1739428, EPI_ISL_1739429, EPI_ISL_1739430, EPI_ISL_1739431, EPI_ISL_1739432, EPI_ISL_1739433, EPI_ISL_1739434, EPI_ISL_1739436, EPI_ISL_1739437, EPI_ISL_1739438, EPI_ISL_1739439, EPI_ISL_1739440, EPI_ISL_1739441, EPI_ISL_1739442, EPI_ISL_1739443, EPI_ISL_1739444, EPI_ISL_1739445, EPI_ISL_1739446, EPI_ISL_1739447, EPI_ISL_1739449, EPI_ISL_1739450, EPI_ISL_1739451, EPI_ISL_1739452, EPI_ISL_1739453, EPI_ISL_1739454, EPI_ISL_1739455, EPI_ISL_1739456, EPI_ISL_1739457, EPI_ISL_1739458, EPI_ISL_1739459, EPI_ISL_1739460, EPI_ISL_1739461, EPI_ISL_1739462, EPI_ISL_1739463, EPI_ISL_1739464, EPI_ISL_1739465, EPI_ISL_1739466, EPI_ISL_1739467, EPI_ISL_1739468, EPI_ISL_1739470, EPI_ISL_1739471, EPI_ISL_1739472, EPI_ISL_1739473, EPI_ISL_1739474, EPI_ISL_1739475, EPI_ISL_1739476, EPI_ISL_1739477, EPI_ISL_1739478, EPI_ISL_1739479, EPI_ISL_1739480, EPI_ISL_1739481, EPI_ISL_1739482, EPI_ISL_1739483, EPI_ISL_1739484, EPI_ISL_1739485, EPI_ISL_1739486, EPI_ISL_1739487, EPI_ISL_1739488, EPI_ISL_1739489, EPI_ISL_1739490, EPI_ISL_1739491, EPI_ISL_1739492, EPI_ISL_1739493, EPI_ISL_1739494, EPI_ISL_1739495, EPI_ISL_1739496, EPI_ISL_1739497, EPI_ISL_1739498, EPI_ISL_1739499, EPI_ISL_1739501, EPI_ISL_1739503, EPI_ISL_1739504, EPI_ISL_1739505, EPI_ISL_1739506, EPI_ISL_1739507, EPI_ISL_1739508, EPI_ISL_1739509, EPI_ISL_1739510, EPI_ISL_1739511, EPI_ISL_1739512, EPI_ISL_1739513, EPI_ISL_1739514, EPI_ISL_1739515, EPI_ISL_1739516, EPI_ISL_1739517, EPI_ISL_1739518, EPI_ISL_1739519, EPI_ISL_1739520, EPI_ISL_1739521, EPI_ISL_1739522, EPI_ISL_1739523, EPI_ISL_1739524, EPI_ISL_1739525, EPI_ISL_1739526, EPI_ISL_1739527, EPI_ISL_1739528, EPI_ISL_1739529, EPI_ISL_1739530, EPI_ISL_1739531, EPI_ISL_1739532, EPI_ISL_1739533, EPI_ISL_1739534, EPI_ISL_1739535, EPI_ISL_1739536, EPI_ISL_1739537, EPI_ISL_1739538, EPI_ISL_1739539, EPI_ISL_1739540, EPI_ISL_1739541, EPI_ISL_1739542, EPI_ISL_1739543 | | | |
| see above | The Jackson Laboratory | The Jackson Laboratory | Bergeron D, Renzette N, Adams M, Omerza G, Kelly K, Long J, Li L |
| EPI_ISL_1740453, EPI_ISL_1740457, EPI_ISL_1740458, EPI_ISL_1740459, EPI_ISL_1740461, EPI_ISL_1740462, EPI_ISL_1740463, EPI_ISL_1740464, EPI_ISL_1740465, EPI_ISL_1740466, EPI_ISL_1740468, EPI_ISL_1740469, EPI_ISL_1740470, EPI_ISL_1740472, EPI_ISL_1740474, EPI_ISL_1740475, EPI_ISL_1740477, EPI_ISL_1740478, EPI_ISL_1740479, EPI_ISL_1740480, EPI_ISL_1740481, EPI_ISL_1740482, EPI_ISL_1740483, EPI_ISL_1740486, EPI_ISL_1740488, EPI_ISL_1740491 | | | |
| see above | Wyoming Public Health Laboratory | Wyoming Public Health Laboratory | Jim Mildenberger, Wanda Manley, Noah Hull, Taylor Fearing, Lynette Gumbleton, Channing Weber, Ashley Norberg, Chayse Rowley, Marley Goetz, Brian Dominguez, Elliot Thomasson, Sam Britz, Cari Sloma, and Rob Christensen |
| EPI_ISL_1740492, EPI_ISL_1740493, EPI_ISL_1740494, EPI_ISL_1740495, EPI_ISL_1740497, EPI_ISL_1740498 | Wyoming Public Health Laboratory | Wyoming Public Health Laboratory | Jim Mildenberger, Taylor Fearing, Channing Weber, Ashley Norberg, Chayse Rowley, Marley Goetz, Brian Dominguez, Elliot Thomasson, Brittany Oher, Sam Britz, Cari Sloma, and Rob Christensen |
| EPI_ISL_1740502 | MEMORIAL SLOAN KETTERING CANCER CENTER | Wadsworth Center, New York State Department of Health | Kirsten St. George, Daryl M. Lamson, Alexis Russell, Matthew Shudt, Melissa A Leisner, Jonathan Plitnick, Catharine Prussing, Navjot Singh, John Kelly, Erasmus Schneider, Erica Lasek-Nesselquist |
| EPI_ISL_1740503 | MONTEFIORE MEDICAL CENTER LABORATORIES | Wadsworth Center, New York State Department of Health | Kirsten St. George, Daryl M. Lamson, Alexis Russell, Matthew Shudt, Melissa A Leisner, Jonathan Plitnick, Catharine Prussing, Navjot Singh, John Kelly, Erasmus Schneider, Erica Lasek-Nesselquist |
| EPI_ISL_1740504 | NORTHWELL HEALTH LABORATORIES | Wadsworth Center, New York State Department of Health | Kirsten St. George, Daryl M. Lamson, Alexis Russell, Matthew Shudt, Melissa A Leisner, Jonathan Plitnick, Catharine Prussing, Navjot Singh, John Kelly, Erasmus Schneider, Erica Lasek-Nesselquist |
| EPI_ISL_1740505 | MONTEFIORE MEDICAL CENTER LABORATORIES | Wadsworth Center, New York State Department of Health | Kirsten St. George, Daryl M. Lamson, Alexis Russell, Matthew Shudt, Melissa A Leisner, Jonathan Plitnick, Catharine Prussing, Navjot Singh, John Kelly, Erasmus Schneider, Erica Lasek-Nesselquist |
| EPI_ISL_1740506, EPI_ISL_1740507 | TEMPUS LABS INC | Wadsworth Center, New York State Department of Health | Kirsten St. George, Daryl M. Lamson, Alexis Russell, Matthew Shudt, Melissa A Leisner, Jonathan Plitnick, Catharine Prussing, Navjot Singh, John Kelly, Erasmus Schneider, Erica Lasek-Nesselquist |
| EPI_ISL_1740508 | STONY BROOK UNIVERSITY HOSPITAL | Wadsworth Center, New York State Department of Health | Kirsten St. George, Daryl M. Lamson, Alexis Russell, Matthew Shudt, Melissa A Leisner, Jonathan Plitnick, Catharine Prussing, Navjot Singh, John Kelly, Erasmus Schneider, Erica Lasek-Nesselquist |
| EPI_ISL_1740509 | ALBANY MEDICAL CENTER | Wadsworth Center, New York State Department of Health | Kirsten St. George, Daryl M. Lamson, Alexis Russell, Matthew Shudt, Melissa A Leisner, Jonathan Plitnick, Catharine Prussing, Navjot Singh, John Kelly, Erasmus Schneider, Erica Lasek-Nesselquist |
| EPI_ISL_1740510, EPI_ISL_1740511 | BOSTON HEART DIAGNOSTICS CORP | Wadsworth Center, New York State Department of Health | Kirsten St. George, Daryl M. Lamson, Alexis Russell, Matthew Shudt, Melissa A Leisner, Jonathan Plitnick, Catharine Prussing, Navjot Singh, John Kelly, Erasmus Schneider, Erica Lasek-Nesselquist |
| EPI_ISL_1740512, EPI_ISL_1740513 | MONTEFIORE MEDICAL CENTER LABORATORIES | Wadsworth Center, New York State Department of Health | Kirsten St. George, Daryl M. Lamson, Alexis Russell, Matthew Shudt, Melissa A Leisner, Jonathan Plitnick, Catharine Prussing, Navjot Singh, John Kelly, Erasmus Schneider, Erica Lasek-Nesselquist |
| EPI_ISL_1740514, EPI_ISL_1740515 | WESTCHESTER MEDICAL CENTER | Wadsworth Center, New York State Department of Health | Kirsten St. George, Daryl M. Lamson, Alexis Russell, Matthew Shudt, Melissa A Leisner, Jonathan Plitnick, Catharine Prussing, Navjot Singh, John Kelly, Erasmus Schneider, Erica Lasek-Nesselquist |
| EPI_ISL_1740516, EPI_ISL_1740517 | URMC LABS | Wadsworth Center, New York State Department of Health | Kirsten St. George, Daryl M. Lamson, Alexis Russell, Matthew Shudt, Melissa A Leisner, Jonathan Plitnick, Catharine Prussing, Navjot Singh, John Kelly, Erasmus Schneider, Erica Lasek-Nesselquist |
| EPI_ISL_1740518, EPI_ISL_1740519, EPI_ISL_1740520, EPI_ISL_1740521, EPI_ISL_1740522 | NORTHWELL HEALTH LABORATORIES | Wadsworth Center, New York State Department of Health | Kirsten St. George, Daryl M. Lamson, Alexis Russell, Matthew Shudt, Melissa A Leisner, Jonathan Plitnick, Catharine Prussing, Navjot Singh, John Kelly, Erasmus Schneider, Erica Lasek-Nesselquist |
| EPI_ISL_1740530 | WI State Laboratory of Hygiene | Pathogen Discovery, Respiratory Viruses Branch, Division of Viral Diseases, Centers for Disease Control and Prevention | Ying Tao, Yan Li, Anna Kelleher, Jing Zhang, Anna Montmayeur, Brian Lynch, Krista Queen, Anna Uehara, Peter Cook, Han Jia Justin Ng, Rachel Marine, Clinton R. Paden, Haibin Wang, Mark Burroughs, Justin Lee, Adam Retchless, Suxiang Tong |
| EPI_ISL_1741546, EPI_ISL_1741549, EPI_ISL_1741552, EPI_ISL_1741555, EPI_ISL_1741558, EPI_ISL_1741560, EPI_ISL_1741563, EPI_ISL_1741566, EPI_ISL_1741569, EPI_ISL_1741572, EPI_ISL_1741575, EPI_ISL_1741578, EPI_ISL_1741581, EPI_ISL_1741583, EPI_ISL_1741586, EPI_ISL_1741589, EPI_ISL_1741592, EPI_ISL_1741595, EPI_ISL_1741598, EPI_ISL_1741601, EPI_ISL_1741604, EPI_ISL_1741607, EPI_ISL_1741610, EPI_ISL_1742310, EPI_ISL_1742311, EPI_ISL_1742312, EPI_ISL_1742313, EPI_ISL_1742314, EPI_ISL_1742315, EPI_ISL_1742316, EPI_ISL_1742317, EPI_ISL_1742318, EPI_ISL_1742319, EPI_ISL_1742320, EPI_ISL_1742321, EPI_ISL_1742322, EPI_ISL_1742323, EPI_ISL_1742324, EPI_ISL_1742325, EPI_ISL_1742326, EPI_ISL_1742327, EPI_ISL_1742328, EPI_ISL_1742329, EPI_ISL_1742330, EPI_ISL_1742331, EPI_ISL_1742332, EPI_ISL_1742333 | | | |
| see above | Oregon State Public Health Laboratory | Oregon State Public Health Laboratory | Rafia Razzaque, Eugene Yeboah, Vanda Makris, Laura Tsaknaridis, John Fontana and Shane Sevey |
| EPI_ISL_1742334, EPI_ISL_1742336, EPI_ISL_1742337, EPI_ISL_1742338, EPI_ISL_1742339, EPI_ISL_1742340, EPI_ISL_1742341, EPI_ISL_1742342, EPI_ISL_1742343, EPI_ISL_1742344, EPI_ISL_1742345, EPI_ISL_1742346, EPI_ISL_1742347, EPI_ISL_1742348, EPI_ISL_1742349, EPI_ISL_1742350, EPI_ISL_1742351, EPI_ISL_1742352, EPI_ISL_1742353, EPI_ISL_1742354, EPI_ISL_1742355, EPI_ISL_1742356, EPI_ISL_1742357, EPI_ISL_1742358, EPI_ISL_1742359, EPI_ISL_1742360, EPI_ISL_1742361, EPI_ISL_1742362, EPI_ISL_1742363, EPI_ISL_1742365, EPI_ISL_1742366, EPI_ISL_1742367, EPI_ISL_1742368, EPI_ISL_1742369, EPI_ISL_1742370, EPI_ISL_1742371, EPI_ISL_1742372, EPI_ISL_1742373, EPI_ISL_1742374, EPI_ISL_1742375, EPI_ISL_1742376, EPI_ISL_1742377, EPI_ISL_1742378, EPI_ISL_1742379, EPI_ISL_1742380, EPI_ISL_1742381, EPI_ISL_1742382, EPI_ISL_1742383, EPI_ISL_1742384, EPI_ISL_1742385, EPI_ISL_1742386, EPI_ISL_1742387, EPI_ISL_1742388, EPI_ISL_1742389, EPI_ISL_1742390, EPI_ISL_1742391, EPI_ISL_1742392, EPI_ISL_1742393, EPI_ISL_1742394, EPI_ISL_1742395, EPI_ISL_1742396, EPI_ISL_1742397, EPI_ISL_1742398, EPI_ISL_1742399, EPI_ISL_1742400, EPI_ISL_1742401, EPI_ISL_1742402, EPI_ISL_1742403, EPI_ISL_1742404, EPI_ISL_1742405, EPI_ISL_1742406, EPI_ISL_1742407, EPI_ISL_1742408, EPI_ISL_1742409, EPI_ISL_1742410, EPI_ISL_1742411, EPI_ISL_1742412, EPI_ISL_1742413, EPI_ISL_1742414, EPI_ISL_1742415, EPI_ISL_1742416, EPI_ISL_1742417, EPI_ISL_1742418, EPI_ISL_1742419, EPI_ISL_1742420 | | | |
| see above | Illinois Department of Public Health | Illinois Department of Public Health - Chicago Lab | Vineet K. Dhiman, Ira Heimler |
| EPI_ISL_1742421, EPI_ISL_1742422, EPI_ISL_1742423, EPI_ISL_1742424, EPI_ISL_1742425, EPI_ISL_1742426, EPI_ISL_1742427, EPI_ISL_1742428, EPI_ISL_1742429, EPI_ISL_1742430, EPI_ISL_1742431 | | | |
| see above | WVU Rapid Development Lab | WVU and Marshall University Combined Genomics Core Facilities | James Denvir, Peter Stoilov, Peter Perrotta, Wesley Kimble, Ryan Percifield |
| EPI_ISL_1742432, EPI_ISL_1742433, EPI_ISL_1742434, EPI_ISL_1742436, EPI_ISL_1742437, EPI_ISL_1742438, EPI_ISL_1742439, EPI_ISL_1742440, EPI_ISL_1742441, EPI_ISL_1742442, EPI_ISL_1742443, EPI_ISL_1742444, EPI_ISL_1742445, EPI_ISL_1742446, EPI_ISL_1742447, EPI_ISL_1742448, EPI_ISL_1742449, EPI_ISL_1742450, EPI_ISL_1742451, EPI_ISL_1742452, EPI_ISL_1742453, EPI_ISL_1742454, EPI_ISL_1742455, EPI_ISL_1742456, EPI_ISL_1742457, EPI_ISL_1742458, EPI_ISL_1742459, EPI_ISL_1742460 | | | |
| see above | J.W. Ruby Memorial Hospital | WVU and Marshall University Combined Genomics Core Facilities | James Denvir, Peter Stoilov, Peter Perrotta, Wesley Kimble, Ryan Percifield |
| EPI_ISL_1742461, EPI_ISL_1742462, EPI_ISL_1742463, EPI_ISL_1742464, EPI_ISL_1742465, EPI_ISL_1742466, EPI_ISL_1742467, EPI_ISL_1742468, EPI_ISL_1742469, EPI_ISL_1742470, EPI_ISL_1742471, EPI_ISL_1742472, EPI_ISL_1742473, EPI_ISL_1742475, EPI_ISL_1742476, EPI_ISL_1742477, EPI_ISL_1742479, EPI_ISL_1742480, EPI_ISL_1742481, EPI_ISL_1742482, EPI_ISL_1742483, EPI_ISL_1742484, EPI_ISL_1742485, EPI_ISL_1742486, EPI_ISL_1742487, EPI_ISL_1742488, EPI_ISL_1742489, EPI_ISL_1742490, EPI_ISL_1742491, EPI_ISL_1742492, EPI_ISL_1742493, EPI_ISL_1742494, EPI_ISL_1742495, EPI_ISL_1742496, EPI_ISL_1742497, EPI_ISL_1742498, EPI_ISL_1742499, EPI_ISL_1742500, EPI_ISL_1742501, EPI_ISL_1742502, EPI_ISL_1742503, EPI_ISL_1742504, EPI_ISL_1742505, EPI_ISL_1742506, EPI_ISL_1742507, EPI_ISL_1742508, EPI_ISL_1742509, EPI_ISL_1742510, EPI_ISL_1742511, EPI_ISL_1742513, EPI_ISL_1742514, EPI_ISL_1742515, EPI_ISL_1742516, EPI_ISL_1742517, EPI_ISL_1742518, EPI_ISL_1742519, EPI_ISL_1742520, EPI_ISL_1742521, EPI_ISL_1742522 | | | |
| see above | QLabs | WVU and Marshall University Combined Genomics Core Facilities | James Denvir, Peter Stoilov, Peter Perrotta, Wesley Kimble, Ryan Percifield |
| EPI_ISL_1742523, EPI_ISL_1742524, EPI_ISL_1742525, EPI_ISL_1742526, EPI_ISL_1742527, EPI_ISL_1742528, EPI_ISL_1742529, EPI_ISL_1742530, EPI_ISL_1742531, EPI_ISL_1742532, EPI_ISL_1742533, EPI_ISL_1742534, EPI_ISL_1742535, EPI_ISL_1742536, EPI_ISL_1742537, EPI_ISL_1742538, EPI_ISL_1742539, EPI_ISL_1742540, EPI_ISL_1742541, EPI_ISL_1742542, EPI_ISL_1742543, EPI_ISL_1742544 | | | |
| see above | J.W. Ruby Memorial Hospital | WVU and Marshall University Combined Genomics Core Facilities | James Denvir, Peter Stoilov, Peter Perrotta, Wesley Kimble, Ryan Percifield |
| EPI_ISL_1742545 | WVU Rapid Development Lab | WVU and Marshall University Combined Genomics Core Facilities | James Denvir, Peter Stoilov, Peter Perrotta, Wesley Kimble, Ryan Percifield |
| EPI_ISL_1742546, EPI_ISL_1742547, EPI_ISL_1742548, EPI_ISL_1742549, EPI_ISL_1742550, EPI_ISL_1742551, EPI_ISL_1742552, EPI_ISL_1742553, EPI_ISL_1742554, EPI_ISL_1742555, EPI_ISL_1742556, EPI_ISL_1742557, EPI_ISL_1742558, EPI_ISL_1742559, EPI_ISL_1742560, EPI_ISL_1742701, EPI_ISL_1742702, EPI_ISL_1742703, EPI_ISL_1742704, EPI_ISL_1742705, EPI_ISL_1742706, EPI_ISL_1742707, EPI_ISL_1742708, EPI_ISL_1742709, EPI_ISL_1742710, EPI_ISL_1742711 | | | |
| see above | J.W. Ruby Memorial Hospital | WVU and Marshall University Combined Genomics Core Facilities | James Denvir, Peter Stoilov, Peter Perrotta, Wesley Kimble, Ryan Percifield |
| EPI_ISL_1742712, EPI_ISL_1742713 | Berkeley Medical Center | WVU and Marshall University Combined Genomics Core Facilities | James Denvir, Peter Stoilov, Peter Perrotta, Wesley Kimble, Ryan Percifield |
| EPI_ISL_1742714 | Mon Health Medical Center | WVU and Marshall University Combined Genomics Core Facilities | James Denvir, Peter Stoilov, Peter Perrotta, Wesley Kimble, Ryan Percifield |
| EPI_ISL_1742715, EPI_ISL_1742716, EPI_ISL_1742717 | Preston Taylor Community Health Center | WVU and Marshall University Combined Genomics Core Facilities | James Denvir, Peter Stoilov, Peter Perrotta, Wesley Kimble, Ryan Percifield |
| EPI_ISL_1742718, EPI_ISL_1742719, EPI_ISL_1742720, EPI_ISL_1742721, EPI_ISL_1742722, EPI_ISL_1742724, EPI_ISL_1742725, EPI_ISL_1742726, EPI_ISL_1742727, EPI_ISL_1742728, EPI_ISL_1742729, EPI_ISL_1742730, EPI_ISL_1742731, EPI_ISL_1742732, EPI_ISL_1742733, EPI_ISL_1742734, EPI_ISL_1742735, EPI_ISL_1742736, EPI_ISL_1742737, EPI_ISL_1742738, EPI_ISL_1742739, EPI_ISL_1742740, EPI_ISL_1742741, EPI_ISL_1742742, EPI_ISL_1742743, EPI_ISL_1742744, EPI_ISL_1742745, EPI_ISL_1742746, EPI_ISL_1742747, EPI_ISL_1742748, EPI_ISL_1742749 | | | |
| see above | Berkeley Medical Center | WVU and Marshall University Combined Genomics Core Facilities | James Denvir, Peter Stoilov, Peter Perrotta, Wesley Kimble, Ryan Percifield |
| EPI_ISL_1742750, EPI_ISL_1742751, EPI_ISL_1742752, EPI_ISL_1742753, EPI_ISL_1742754, EPI_ISL_1742755, EPI_ISL_1742756, EPI_ISL_1742757, EPI_ISL_1742758, EPI_ISL_1742759, EPI_ISL_1742760, EPI_ISL_1742761, EPI_ISL_1742762, EPI_ISL_1742763, EPI_ISL_1742764, EPI_ISL_1742765, EPI_ISL_1742766, EPI_ISL_1742767, EPI_ISL_1742768, EPI_ISL_1742769, EPI_ISL_1742770, EPI_ISL_1742771, | | | |

| | | | |
|---|---|---|---|
| EPI_ISL_1742772, EPI_ISL_1742773, EPI_ISL_1742774, EPI_ISL_1742775, EPI_ISL_1742776, EPI_ISL_1742777, EPI_ISL_1742778, EPI_ISL_1742779, EPI_ISL_1742780, EPI_ISL_1742781, EPI_ISL_1742782, EPI_ISL_1742783, EPI_ISL_1742784, EPI_ISL_1742785, EPI_ISL_1742786, EPI_ISL_1742787, EPI_ISL_1742788, EPI_ISL_1742789, EPI_ISL_1742790 | | | |
| see above | J.W. Ruby Memorial Hospital | WVU and Marshall University Combined Genomics Core Facilities | James Denvir, Peter Stoilov, Peter Perrotta, Wesley Kimble, Ryan Percifield |
| EPI_ISL_1742791, EPI_ISL_1742792, EPI_ISL_1742793, EPI_ISL_1742794, EPI_ISL_1742795, EPI_ISL_1742796, EPI_ISL_1742797, EPI_ISL_1742798, EPI_ISL_1742799, EPI_ISL_1742800, EPI_ISL_1742801, EPI_ISL_1742802, EPI_ISL_1742803, EPI_ISL_1742804, EPI_ISL_1742805, EPI_ISL_1742806, EPI_ISL_1742807, EPI_ISL_1742808, EPI_ISL_1742809, EPI_ISL_1742810, EPI_ISL_1742811, EPI_ISL_1742812, EPI_ISL_1742813, EPI_ISL_1742814, EPI_ISL_1742815, EPI_ISL_1742816, EPI_ISL_1742817, EPI_ISL_1742818, EPI_ISL_1742819, EPI_ISL_1742820, EPI_ISL_1742821, EPI_ISL_1742822 | | | |
| see above | Berkeley Medical Center | WVU and Marshall University Combined Genomics Core Facilities | James Denvir, Peter Stoilov, Peter Perrotta, Wesley Kimble, Ryan Percifield |
| EPI_ISL_1742823 | Preston Taylor Community Health Center | WVU and Marshall University Combined Genomics Core Facilities | James Denvir, Peter Stoilov, Peter Perrotta, Wesley Kimble, Ryan Percifield |
| EPI_ISL_1742824 | Berkeley Medical Center | WVU and Marshall University Combined Genomics Core Facilities | James Denvir, Peter Stoilov, Peter Perrotta, Wesley Kimble, Ryan Percifield |
| EPI_ISL_1742825 | Mon Health Medical Center | WVU and Marshall University Combined Genomics Core Facilities | James Denvir, Peter Stoilov, Peter Perrotta, Wesley Kimble, Ryan Percifield |
| EPI_ISL_1742826, EPI_ISL_1742827, EPI_ISL_1742828, EPI_ISL_1742830, EPI_ISL_1742831, EPI_ISL_1742832, EPI_ISL_1742833 | QLabs | WVU and Marshall University Combined Genomics Core Facilities | James Denvir, Peter Stoilov, Peter Perrotta, Wesley Kimble, Ryan Percifield |
| EPI_ISL_1742834 | J.W. Ruby Memorial Hospital | WVU and Marshall University Combined Genomics Core Facilities | James Denvir, Peter Stoilov, Peter Perrotta, Wesley Kimble, Ryan Percifield |
| EPI_ISL_1743594, EPI_ISL_1743597, EPI_ISL_1743598, EPI_ISL_1743599, EPI_ISL_1743600, EPI_ISL_1743603, EPI_ISL_1743604, EPI_ISL_1743605, EPI_ISL_1743606, EPI_ISL_1743607, EPI_ISL_1743608, EPI_ISL_1743610, EPI_ISL_1743611, EPI_ISL_1743612, EPI_ISL_1743613, EPI_ISL_1743614, EPI_ISL_1743615, EPI_ISL_1743617, EPI_ISL_1743618, EPI_ISL_1743619, EPI_ISL_1743620, EPI_ISL_1743621, EPI_ISL_1743623, EPI_ISL_1743624, EPI_ISL_1743625, EPI_ISL_1743626, EPI_ISL_1743627, EPI_ISL_1743628, EPI_ISL_1743629, EPI_ISL_1743630, EPI_ISL_1743631, EPI_ISL_1743632, EPI_ISL_1743633, EPI_ISL_1743635, EPI_ISL_1743636, EPI_ISL_1743637, EPI_ISL_1743638, EPI_ISL_1743639, EPI_ISL_1743640, EPI_ISL_1743641, EPI_ISL_1743642, EPI_ISL_1743643, EPI_ISL_1743645, EPI_ISL_1743646, EPI_ISL_1743647, EPI_ISL_1743648, EPI_ISL_1743649, EPI_ISL_1743650, EPI_ISL_1743651, EPI_ISL_1743653, EPI_ISL_1743654, EPI_ISL_1743655, EPI_ISL_1743656, EPI_ISL_1743657, EPI_ISL_1743658, EPI_ISL_1743659, EPI_ISL_1743660, EPI_ISL_1743661, EPI_ISL_1743663, EPI_ISL_1743664, EPI_ISL_1743665, EPI_ISL_1743666, EPI_ISL_1743667, EPI_ISL_1743668, EPI_ISL_1743669, EPI_ISL_1743670, EPI_ISL_1743671, EPI_ISL_1743672, EPI_ISL_1743674, EPI_ISL_1743675, EPI_ISL_1743676, EPI_ISL_1743677, EPI_ISL_1743678, EPI_ISL_1743679, EPI_ISL_1743681, EPI_ISL_1743682, EPI_ISL_1743683, EPI_ISL_1743684, EPI_ISL_1743685, EPI_ISL_1743686 | | | |
| see above | Arizona State University | Arizona State University | Peter T. Skidmore, LaRinda A. Holland, Rabia Maqsood, Nicholas J. Mellor, Joy M. Blain, Valerie Harris, Joshua LaBaer, Vel Murugan, Efrem S. Lim |
| EPI_ISL_1743688, EPI_ISL_1743689, EPI_ISL_1743690, EPI_ISL_1743691, EPI_ISL_1743692, EPI_ISL_1743693, EPI_ISL_1743694, EPI_ISL_1743695, EPI_ISL_1743696, EPI_ISL_1743697, EPI_ISL_1743698, EPI_ISL_1743699, EPI_ISL_1743700, EPI_ISL_1743701, EPI_ISL_1743702, EPI_ISL_1743703, EPI_ISL_1743704, EPI_ISL_1743705, EPI_ISL_1743706, EPI_ISL_1743707, EPI_ISL_1743708, EPI_ISL_1743709, EPI_ISL_1743710, EPI_ISL_1743711, EPI_ISL_1743712, EPI_ISL_1743713, EPI_ISL_1743714 | | | |
| see above | Alaska State Virology Laboratory | Alaska State Virology Laboratory | Stephanie DeRonde, Elva House, Lisa Smith, Ph.D., Jack Chen, Ph.D. |
| EPI_ISL_1743717 | Interpath Laboratory | Seattle Flu Study | Deborah A. Nickerson, Chris D. Frazar, Jover Lee, Benjamin Pelle, Erica Ryke, Matthew Richardson, Amanda Adler, Elisabeth Brandstetter, Peter D. Han, Kairsten Fay, Misja Ilcisin, Kirsten Lacombe, Thomas R. Sibley, Melissa Truong, Caitlin R. Wolf, Romesh Gautom, Geoff Melly, Brian Hiatt, Philip Dykema, Scott Lindquist, Michael Boeckh, Janet A. Englund, Michael Famulare, Barry R. Lutz, Mark J. Rieder, Lea M. Starita, Matthew Thompson, Helen Y. Chu, Jay Shendure, Trevor Bedford |
| EPI_ISL_1743718, EPI_ISL_1743719, EPI_ISL_1743720, EPI_ISL_1743721, EPI_ISL_1743722, EPI_ISL_1743724, EPI_ISL_1743725, EPI_ISL_1743726, EPI_ISL_1743727, EPI_ISL_1743728, EPI_ISL_1743729, EPI_ISL_1743730, EPI_ISL_1743731, EPI_ISL_1743732, EPI_ISL_1743733, EPI_ISL_1743734, EPI_ISL_1743735, EPI_ISL_1743736, EPI_ISL_1743737, EPI_ISL_1743738, EPI_ISL_1743739, EPI_ISL_1743740, EPI_ISL_1743741, EPI_ISL_1743742, EPI_ISL_1743743, EPI_ISL_1743744, EPI_ISL_1743745, EPI_ISL_1743746, EPI_ISL_1743747, EPI_ISL_1743748, EPI_ISL_1743749, EPI_ISL_1743750, EPI_ISL_1743751, EPI_ISL_1743752, EPI_ISL_1743753, EPI_ISL_1743754, EPI_ISL_1743755, EPI_ISL_1743756, EPI_ISL_1743757, EPI_ISL_1743758, EPI_ISL_1743759, EPI_ISL_1743760, EPI_ISL_1743761, EPI_ISL_1743762, EPI_ISL_1743763, EPI_ISL_1743764, EPI_ISL_1743765, EPI_ISL_1743766, EPI_ISL_1743767, EPI_ISL_1743768, EPI_ISL_1743769, EPI_ISL_1743770, EPI_ISL_1743771, EPI_ISL_1743772, EPI_ISL_1743773, EPI_ISL_1743774, EPI_ISL_1743775, EPI_ISL_1743776, EPI_ISL_1743777, EPI_ISL_1743778, EPI_ISL_1743779, EPI_ISL_1743780, EPI_ISL_1743781, EPI_ISL_1743782, EPI_ISL_1743784, EPI_ISL_1743785, EPI_ISL_1743786, EPI_ISL_1743787, EPI_ISL_1743788, EPI_ISL_1743789, EPI_ISL_1743790, EPI_ISL_1743791, EPI_ISL_1743792, EPI_ISL_1743793, EPI_ISL_1743794, EPI_ISL_1743795, EPI_ISL_1743796, EPI_ISL_1743797, EPI_ISL_1743798, EPI_ISL_1743799, EPI_ISL_1743800, EPI_ISL_1743801, EPI_ISL_1743802, EPI_ISL_1743803, EPI_ISL_1743804, EPI_ISL_1743805, EPI_ISL_1743806, EPI_ISL_1743807, EPI_ISL_1743808, EPI_ISL_1743809, EPI_ISL_1743810, EPI_ISL_1743811, EPI_ISL_1743812, EPI_ISL_1743813, EPI_ISL_1743814, EPI_ISL_1743815, EPI_ISL_1743816, EPI_ISL_1743817, EPI_ISL_1743818, EPI_ISL_1743819, EPI_ISL_1743820, EPI_ISL_1743821, EPI_ISL_1743822, EPI_ISL_1743823, EPI_ISL_1743824, EPI_ISL_1743825, EPI_ISL_1743826, EPI_ISL_1743827, EPI_ISL_1743828, EPI_ISL_1743829, EPI_ISL_1743830, EPI_ISL_1743831, EPI_ISL_1743832, EPI_ISL_1743833, EPI_ISL_1743834, EPI_ISL_1743835, EPI_ISL_1743836, EPI_ISL_1743837, EPI_ISL_1743838, EPI_ISL_1743839, EPI_ISL_1743840, EPI_ISL_1743841, EPI_ISL_1743842, EPI_ISL_1743843, EPI_ISL_1743844, EPI_ISL_1743845, EPI_ISL_1743846, EPI_ISL_1743847, EPI_ISL_1743848, EPI_ISL_1743849, EPI_ISL_1743850, EPI_ISL_1743851, EPI_ISL_1743852, EPI_ISL_1743853, EPI_ISL_1743854, EPI_ISL_1743855, EPI_ISL_1743856, EPI_ISL_1743857, EPI_ISL_1743858, EPI_ISL_1743860, EPI_ISL_1743861, EPI_ISL_1743862, EPI_ISL_1743863, EPI_ISL_1743864, EPI_ISL_1743865, EPI_ISL_1743867, EPI_ISL_1743868, EPI_ISL_1743869, EPI_ISL_1743870, EPI_ISL_1743871, EPI_ISL_1743872, EPI_ISL_1743873, EPI_ISL_1743874, EPI_ISL_1743875, EPI_ISL_1743876, EPI_ISL_1743877, EPI_ISL_1743878, EPI_ISL_1743879, EPI_ISL_1743880, EPI_ISL_1743881, EPI_ISL_1743882, EPI_ISL_1743883, EPI_ISL_1743884, EPI_ISL_1743885, EPI_ISL_1743886, EPI_ISL_1743887, EPI_ISL_1743888, EPI_ISL_1743889, EPI_ISL_1743890, EPI_ISL_1743891, EPI_ISL_1743892, EPI_ISL_1743893, EPI_ISL_1743894, EPI_ISL_1743895, EPI_ISL_1743896, EPI_ISL_1743897, EPI_ISL_1743898, EPI_ISL_1743899, EPI_ISL_1743900, EPI_ISL_1743901, EPI_ISL_1743902, EPI_ISL_1743903, EPI_ISL_1743904, EPI_ISL_1743905, EPI_ISL_1743906, EPI_ISL_1743907, EPI_ISL_1743908, EPI_ISL_1743909, EPI_ISL_1743910, EPI_ISL_1743911, EPI_ISL_1743912, EPI_ISL_1743913, EPI_ISL_1743914, EPI_ISL_1743915, EPI_ISL_1743916, EPI_ISL_1743917, EPI_ISL_1743918, EPI_ISL_1743919, EPI_ISL_1743920, EPI_ISL_1743921, EPI_ISL_1743922, EPI_ISL_1743923, EPI_ISL_1743924, EPI_ISL_1743925, EPI_ISL_1743926, EPI_ISL_1743927, EPI_ISL_1743928, EPI_ISL_1743929, EPI_ISL_1743930, EPI_ISL_1743931, EPI_ISL_1743932, EPI_ISL_1743933, EPI_ISL_1743934, EPI_ISL_1743935, EPI_ISL_1743936, EPI_ISL_1743937, EPI_ISL_1743938, EPI_ISL_1743939, EPI_ISL_1743940, EPI_ISL_1743941, EPI_ISL_1743942, EPI_ISL_1743943, EPI_ISL_1743944, EPI_ISL_1743945, EPI_ISL_1743946, EPI_ISL_1743947, EPI_ISL_1743948, EPI_ISL_1743949, EPI_ISL_1743950, EPI_ISL_1743951, EPI_ISL_1743952, EPI_ISL_1743953, EPI_ISL_1743954, EPI_ISL_1743955, EPI_ISL_1743956, EPI_ISL_1743957, EPI_ISL_1743958, EPI_ISL_1743959, EPI_ISL_1743960, EPI_ISL_1743961, EPI_ISL_1743962, EPI_ISL_1743963, EPI_ISL_1743964, EPI_ISL_1743965, EPI_ISL_1743966, EPI_ISL_1743967, EPI_ISL_1743968, EPI_ISL_1743969, EPI_ISL_1743970, EPI_ISL_1743971, EPI_ISL_1743972, EPI_ISL_1743973, EPI_ISL_1743974, EPI_ISL_1743975, EPI_ISL_1743976, EPI_ISL_1743977, EPI_ISL_1743978, EPI_ISL_1743979, EPI_ISL_1743980, EPI_ISL_1743981, EPI_ISL_1743982, EPI_ISL_1743983, EPI_ISL_1743984, EPI_ISL_1743985, EPI_ISL_1743986, EPI_ISL_1743987, EPI_ISL_1743988, EPI_ISL_1743989, EPI_ISL_1743992, EPI_ISL_1743994, EPI_ISL_1743995, EPI_ISL_1743996, EPI_ISL_1743997, EPI_ISL_1743998, EPI_ISL_1743999, EPI_ISL_1744000, EPI_ISL_1744001, EPI_ISL_1744002, EPI_ISL_1744003, EPI_ISL_1744004, EPI_ISL_1744005, EPI_ISL_1744006, EPI_ISL_1744007, EPI_ISL_1744008, EPI_ISL_1744009, EPI_ISL_1744010, EPI_ISL_1744011, EPI_ISL_1744012, EPI_ISL_1744013, EPI_ISL_1744014, EPI_ISL_1744015, EPI_ISL_1744017, EPI_ISL_1744018, EPI_ISL_1744019, EPI_ISL_1744020, EPI_ISL_1744021, EPI_ISL_1744022, EPI_ISL_1744023, EPI_ISL_1744024, EPI_ISL_1744025, EPI_ISL_1744026, EPI_ISL_1744028, EPI_ISL_1744029, EPI_ISL_1744030, EPI_ISL_1744031, EPI_ISL_1744032, EPI_ISL_1744033, EPI_ISL_1744034, EPI_ISL_1744035, EPI_ISL_1744036, EPI_ISL_1744037, EPI_ISL_1744038, EPI_ISL_1744039, EPI_ISL_1744040, EPI_ISL_1744042, EPI_ISL_1744043, EPI_ISL_1744044, EPI_ISL_1744045, EPI_ISL_1744046, EPI_ISL_1744047, EPI_ISL_1744048, EPI_ISL_1744049, EPI_ISL_1744050, EPI_ISL_1744051, EPI_ISL_1744052, EPI_ISL_1744053, EPI_ISL_1744054, EPI_ISL_1744055, EPI_ISL_1744056, EPI_ISL_1744057, EPI_ISL_1744058, EPI_ISL_1744059, EPI_ISL_1744060, EPI_ISL_1744061, EPI_ISL_1744062, EPI_ISL_1744063, EPI_ISL_1744064, EPI_ISL_1744065, EPI_ISL_1744066, EPI_ISL_1744067, EPI_ISL_1744068, EPI_ISL_1744069, EPI_ISL_1744070, EPI_ISL_1744071, EPI_ISL_1744072, EPI_ISL_1744073, EPI_ISL_1744074, EPI_ISL_1744075, EPI_ISL_1744076, EPI_ISL_1744077, EPI_ISL_1744078, EPI_ISL_1744079, EPI_ISL_1744080, EPI_ISL_1744081, EPI_ISL_1744082, EPI_ISL_1744083, EPI_ISL_1744084, EPI_ISL_1744085, EPI_ISL_1744086, EPI_ISL_1744087, EPI_ISL_1744088, EPI_ISL_1744089, EPI_ISL_1744090, EPI_ISL_1744091, EPI_ISL_1744092, EPI_ISL_1744093, EPI_ISL_1744094, EPI_ISL_1744095, EPI_ISL_1744096, EPI_ISL_1744097, EPI_ISL_1744098, EPI_ISL_1744099, EPI_ISL_1744100, EPI_ISL_1744101, EPI_ISL_1744102, EPI_ISL_1744103, EPI_ISL_1744104, EPI_ISL_1744105, EPI_ISL_1744106, EPI_ISL_1744107, EPI_ISL_1744108, EPI_ISL_1744109, EPI_ISL_1744110, EPI_ISL_1744111, EPI_ISL_1744112, EPI_ISL_1744113, EPI_ISL_1744114, EPI_ISL_1744115, EPI_ISL_1744116, EPI_ISL_1744117, EPI_ISL_1744118, EPI_ISL_1744119, EPI_ISL_1744120, EPI_ISL_1744121, EPI_ISL_1744122, EPI_ISL_1744123, EPI_ISL_1744124, EPI_ISL_1744125, EPI_ISL_1744126, EPI_ISL_1744127, EPI_ISL_1744128, EPI_ISL_1744129, EPI_ISL_1744130, EPI_ISL_1744131, EPI_ISL_1744132, EPI_ISL_1744133, EPI_ISL_1744134, EPI_ISL_1744135, EPI_ISL_1744136, EPI_ISL_1744137, EPI_ISL_1744138, EPI_ISL_1744139, EPI_ISL_1744140, EPI_ISL_1744141, EPI_ISL_1744142, EPI_ISL_1744143, EPI_ISL_1744144, EPI_ISL_1744145, EPI_ISL_1744146, EPI_ISL_1744147, EPI_ISL_1744148, EPI_ISL_1744149, EPI_ISL_1744150, EPI_ISL_1744151, EPI_ISL_1744152, EPI_ISL_1744153, EPI_ISL_1744154, EPI_ISL_1744155, EPI_ISL_1744156, EPI_ISL_1744157, EPI_ISL_1744158, EPI_ISL_1744159, EPI_ISL_1744160, EPI_ISL_1744161, EPI_ISL_1744162, EPI_ISL_1744163, EPI_ISL_1744164, EPI_ISL_1744165, EPI_ISL_1744166, EPI_ISL_1744167, EPI_ISL_1744168, EPI_ISL_1744169, EPI_ISL_1744170, EPI_ISL_1744171, EPI_ISL_1744172, EPI_ISL_1744173, EPI_ISL_1744174, EPI_ISL_1744175, EPI_ISL_1744176, EPI_ISL_1744177, EPI_ISL_1744178, EPI_ISL_1744179, EPI_ISL_1744180, EPI_ISL_1744181, EPI_ISL_1744182, EPI_ISL_1744183, EPI_ISL_1744184, EPI_ISL_1744185, EPI_ISL_1744186, EPI_ISL_1744187, EPI_ISL_1744188, EPI_ISL_1744189, EPI_ISL_1744190, EPI_ISL_1744191, EPI_ISL_1744192, EPI_ISL_1744193, EPI_ISL_1744194, EPI_ISL_1744195, EPI_ISL_1744196, EPI_ISL_1744197, EPI_ISL_1744198, EPI_ISL_1744199, EPI_ISL_1744200, EPI_ISL_1744201, EPI_ISL_1744202, EPI_ISL_1744203, EPI_ISL_1744204, EPI_ISL_1744205, EPI_ISL_1744206, EPI_ISL_1744207, EPI_ISL_1744208, EPI_ISL_1744209, EPI_ISL_1744210, EPI_ISL_1744211, EPI_ISL_1744212, EPI_ISL_1744213, EPI_ISL_1744214, EPI_ISL_1744215, EPI_ISL_1744216, EPI_ISL_1744217, EPI_ISL_1744218, EPI_ISL_1744219, EPI_ISL_1744220, EPI_ISL_1744221, EPI_ISL_1744222, EPI_ISL_1744223, EPI_ISL_1744224, EPI_ISL_1744225, EPI_ISL_1744226, EPI_ISL_1744227, EPI_ISL_1744228, EPI_ISL_1744229, EPI_ISL_1744230, EPI_ISL_1744231, EPI_ISL_1744232, EPI_ISL_1744233, EPI_ISL_1744234, EPI_ISL_1744235, EPI_ISL_1744236, EPI_ISL_1744237, EPI_ISL_1744238, EPI_ISL_1744239, EPI_ISL_1744240, EPI_ISL_1744241, EPI_ISL_1744242, EPI_ISL_1744243, EPI_ISL_1744244, EPI_ISL_1744245, EPI_ISL_1744246, EPI_ISL_1744247, EPI_ISL_1744248, EPI_ISL_1744249, EPI_ISL_1744250, EPI_ISL_1744251, EPI_ISL_1744252, EPI_ISL_1744253, EPI_ISL_1744254, EPI_ISL_1744255, EPI_ISL_1744256, EPI_ISL_1744257, EPI_ISL_1744258, EPI_ISL_1744259, EPI_ISL_1744260, EPI_ISL_1744261, EPI_ISL_1744262, EPI_ISL_1744263, EPI_ISL_1744264, EPI_ISL_1744265, EPI_ISL_1744266, EPI_ISL_1744267, EPI_ISL_1744268, EPI_ISL_1744269, EPI_ISL_1744270, EPI_ISL_1744271, EPI_ISL_1744272, EPI_ISL_1744273, EPI_ISL_1744274, EPI_ISL_1744275, EPI_ISL_1744276, EPI_ISL_1744277, EPI_ISL_1744278, EPI_ISL_1744279, EPI_ISL_1744280, EPI_ISL_1744281, EPI_ISL_1744282, EPI_ISL_1744283, EPI_ISL_1744284, EPI_ISL_1744285, EPI_ISL_1744286, EPI_ISL_1744287, EPI_ISL_1744288, EPI_ISL_1744289, EPI_ISL_1744290, EPI_ISL_1744291, EPI_ISL_1744292, EPI_ISL_1744293, EPI_ISL_1744294, EPI_ISL_1744295, EPI_ISL_1744296, EPI_ISL_1744297, EPI_ISL_1744298, EPI_ISL_1744299, EPI_ISL_1744300, EPI_ISL_1744301, EPI_ISL_1744302, EPI_ISL_1744303, EPI_ISL_1744304 | | | |
| see above | Broad Institute Clinical Research Sequencing Platform | Infectious Disease Program, Broad Institute of Harvard and MIT | Siddle,K.J., Adams,G., Pearlman,L., Gladden-Young,A., Vicente,G., Blumenstiel,B., DeFelice,M., Lee,M., McGovern,S., Lagerborg,K., Rudy,M., DeRuff,K., Carter,A., Normandin,E., Bauer,M., Reilly,S., Tomkins-Tinch,C., Loreth,C., Chaluvadi,S., Meldrim,J., Granger,B., Lemieux,J.E., Birren,B.W., Sabeti,P.C., Larkin,K., Dodge,S., Lennon,N., Madoff,L., Brown,C., Gallagher,G., Smole,S., Park,D.J., Gabriel,S., and MacInnis,B.L. |
| EPI_ISL_1744389, EPI_ISL_1744390, EPI_ISL_1744391, EPI_ISL_1744392, EPI_ISL_1744393, EPI_ISL_1744394, EPI_ISL_1744395, EPI_ISL_1744396, EPI_ISL_1744397, EPI_ISL_1744398, EPI_ISL_1744399, EPI_ISL_1744400, EPI_ISL_1744401, EPI_ISL_1744402, EPI_ISL_1744403, EPI_ISL_1744404, EPI_ISL_1744405, EPI_ISL_1744406, EPI_ISL_1744407, EPI_ISL_1744408, EPI_ISL_1744409, EPI_ISL_1744410, EPI_ISL_1744411, EPI_ISL_1744412, EPI_ISL_1744413, EPI_ISL_1744414, EPI_ISL_1744415, EPI_ISL_1744416, EPI_ISL_1744417, EPI_ISL_1744418, EPI_ISL_1744419, EPI_ISL_1744420, EPI_ISL_1744421, EPI_ISL_1744422, EPI_ISL_1744423, EPI_ISL_1744424, EPI_ISL_1744425, EPI_ISL_1744426, EPI_ISL_1744427, EPI_ISL_1744428, EPI_ISL_1744429, EPI_ISL_1744430, EPI_ISL_1744431, EPI_ISL_1744432, EPI_ISL_1744433, EPI_ISL_1744434, EPI_ISL_1744435, EPI_ISL_1744436, EPI_ISL_1744437, EPI_ISL_1744438, EPI_ISL_1744439, EPI_ISL_1744440, EPI_ISL_1744441, EPI_ISL_1744442, EPI_ISL_1744443, EPI_ISL_1744444, EPI_ISL_1744445, EPI_ISL_1744446, EPI_ISL_1744447, EPI_ISL_1744448, EPI_ISL_1744449, EPI_ISL_1744450, EPI_ISL_1744451, EPI_ISL_1744453, EPI_ISL_1744454, EPI_ISL_1744455, EPI_ISL_1744456, EPI_ISL_1744457, EPI_ISL_1744458, EPI_ISL_1744459, EPI_ISL_1744460, EPI_ISL_1744461, EPI_ISL_1744462, EPI_ISL_1744463, EPI_ISL_1744464, EPI_ISL_1744466, EPI_ISL_1744467, EPI_ISL_1744468, EPI_ISL_1744469, EPI_ISL_1744470, EPI_ISL_1744471, EPI_ISL_1744472, EPI_ISL_1744473, EPI_ISL_1744474, EPI_ISL_1744475, EPI_ISL_1744476, EPI_ISL_1744477, EPI_ISL_1744478, EPI_ISL_1744479, EPI_ISL_1744480, EPI_ISL_1744481, EPI_ISL_1744482, EPI_ISL_1744484, EPI_ISL_1744485, EPI_ISL_1744487, EPI_ISL_1744488, EPI_ISL_1744489, EPI_ISL_1744491, EPI_ISL_1744494, EPI_ISL_1744495, EPI_ISL_1744496, EPI_ISL_1744498, EPI_ISL_1744499, EPI_ISL_1744500, EPI_ISL_1744501, EPI_ISL_1744502, EPI_ISL_1744503, EPI_ISL_1744504, EPI_ISL_1744505, EPI_ISL_1744506, EPI_ISL_1744507, EPI_ISL_1744510, EPI_ISL_1744511, EPI_ISL_1744512, EPI_ISL_1744513, EPI_ISL_1744514, EPI_ISL_1744518, EPI_ISL_1744520, EPI_ISL_1744521, EPI_ISL_1744522, EPI_ISL_1744523, EPI_ISL_1744525, EPI_ISL_1744526, EPI_ISL_1744528, EPI_ISL_1744533, EPI_ISL_1744535 | | | |
| see above | University of Michigan Clinical Microbiology Laboratory | Lauring Lab, University of Michigan, Department of Microbiology and Immunology | Valesano |
| EPI_ISL_1745204, EPI_ISL_1745205, EPI_ISL_1745206, EPI_ISL_1745207, EPI_ISL_1745208, EPI_ISL_1745209, EPI_ISL_1745210, EPI_ISL_1745211, EPI_ISL_1745212, EPI_ISL_1745213, EPI_ISL_1745214, EPI_ISL_1745215, EPI_ISL_1745216, EPI_ISL_1745217, EPI_ISL_1745218, EPI_ISL_1745219, EPI_ISL_1745220, EPI_ISL_1745221, EPI_ISL_1745222, EPI_ISL_1745223, EPI_ISL_1745224, EPI_ISL_1745225, EPI_ISL_1745226, EPI_ISL_1745227, EPI_ISL_1745228, EPI_ISL_1745229, EPI_ISL_1745230, EPI_ISL_1745231, EPI_ISL_1745232, EPI_ISL_1745233, EPI_ISL_1745234, EPI_ISL_1745235 | | | |
| see above | Gundersen Clinical Microbiology Laboratory | Kabara Cancer Research Institute | Craig S. Richmond, Paraic A. Kenny |
| EPI_ISL_1747388, EPI_ISL_1747390, EPI_ISL_1747391, EPI_ISL_1747392, EPI_ISL_1747393, EPI_ISL_1747394, EPI_ISL_1747395, EPI_ISL_1747396, EPI_ISL_1747397, EPI_ISL_1747398, EPI_ISL_1747399, EPI_ISL_1747400, EPI_ISL_1747401, EPI_ISL_1747402, EPI_ISL_1747403, EPI_ISL_1747404, EPI_ISL_1747405, EPI_ISL_1747406, EPI_ISL_1747407, EPI_ISL_1747408, EPI_ISL_1747409, EPI_ISL_1747410, EPI_ISL_1747411, EPI_ISL_1747412, EPI_ISL_1747413, EPI_ISL_1747414, EPI_ISL_1747415, EPI_ISL_1747416, EPI_ISL_1747417, EPI_ISL_1747418, EPI_ISL_1747419, EPI_ISL_1747420, EPI_ISL_1747421, EPI_ISL_1747422, EPI_ISL_1747423, EPI_ISL_1747424, EPI_ISL_1747425, EPI_ISL_1747426, EPI_ISL_1747428, EPI_ISL_1747429, EPI_ISL_1747430, EPI_ISL_1747431, EPI_ISL_1747432, EPI_ISL_1747433, EPI_ISL_1747434, EPI_ISL_1747435, EPI_ISL_1747436, EPI_ISL_1747437, EPI_ISL_1747438, EPI_ISL_1747439, EPI_ISL_1747440, EPI_ISL_1747441, EPI_ISL_1747443, EPI_ISL_1747444, EPI_ISL_1747445, EPI_ISL_1747446, EPI_ISL_1747447, EPI_ISL_1747448, EPI_ISL_1747449, EPI_ISL_1747450, EPI_ISL_1747451, EPI_ISL_1747452, EPI_ISL_1747453, EPI_ISL_1747454, EPI_ISL_1747455, EPI_ISL_1747456, EPI_ISL_1747457, EPI_ISL_1747458, EPI_ISL_1747459, EPI_ISL_1747460 | | | |
| see above | Washington State Department of Health Public Health Laboratories | Washington State Department of Health Public Health Laboratories | Drew MacKellar, Philip Dykema, Denny Russell, Joenice Gonzalez, Hannah Gray, Geoff Melly, Vanessa De Los Santos, Darren Lucas, JohnAric Peterson, Avi Singh, Rebecca Cao |
| EPI_ISL_1749378, EPI_ISL_1749379, EPI_ISL_1749380, EPI_ISL_1749381, EPI_ISL_1749382, EPI_ISL_1749383, EPI_ISL_1749384, EPI_ISL_1749385, EPI_ISL_1749386, EPI_ISL_1749388, EPI_ISL_1749390, EPI_ISL_1749391, EPI_ISL_1749392, EPI_ISL_1749393, EPI_ISL_1749394, EPI_ISL_1749396, EPI_ISL_1749397, EPI_ISL_1749399, EPI_ISL_1749401, EPI_ISL_1749403, EPI_ISL_1749404, EPI_ISL_1749406, EPI_ISL_1749408, EPI_ISL_1749409, EPI_ISL_1749410, EPI_ISL_1749411, EPI_ISL_1749412, EPI_ISL_1749413, EPI_ISL_1749414, EPI_ISL_1749415, EPI_ISL_1749416, EPI_ISL_1749417, EPI_ISL_1749418, EPI_ISL_1749419, EPI_ISL_1749421, EPI_ISL_1749422, EPI_ISL_1749423, EPI_ISL_1749424, EPI_ISL_1749425, EPI_ISL_1749426, EPI_ISL_1749427, EPI_ISL_1749428 | | | |
| see above | UTGSAF | UTGSAF | Jessica Podnar, Sylvie Beaudenon, Audrey Kelly, Zachary Carver, Anna Battenhouse, Andreas Matouschek |
| EPI_ISL_1752570, EPI_ISL_1752587, EPI_ISL_1752588, EPI_ISL_1752636, EPI_ISL_1752669, EPI_ISL_1752676, EPI_ISL_1752682 | Ohio Department of Health Laboratory | Ohio Department of Health Laboratory | Holmes, Jennifer; Eric Brandt, Keoni Omura, Glen McGillivary, Caitlin McDonnell, Jade Mowery, Stephanie Mccracken, Tyler Payne, Kirtana Ramadugu, Erica Leasure, Brent Lee, Kelsey Florek, Heather Blankenship, Allison Black, Quanta Brown, and Tammy Bannerman |
| EPI_ISL_1752963, EPI_ISL_1752964, EPI_ISL_1752966, EPI_ISL_1752967, EPI_ISL_1752968, EPI_ISL_1752969, EPI_ISL_1752970, EPI_ISL_1752971, EPI_ISL_1752972, EPI_ISL_1752974, EPI_ISL_1752975, EPI_ISL_1752976, EPI_ISL_1752977, EPI_ISL_1752978, EPI_ISL_1752979, EPI_ISL_1752980, EPI_ISL_1752981, EPI_ISL_1752982, EPI_ISL_1752983, EPI_ISL_1752984, EPI_ISL_1752986, EPI_ISL_1752987, EPI_ISL_1752988, EPI_ISL_1752989, EPI_ISL_1752990, EPI_ISL_1752991, EPI_ISL_1752992, EPI_ISL_1752993, EPI_ISL_1752994, EPI_ISL_1752995 | | | |
| see above | Wyoming Public Health Laboratory | Wyoming Public Health Laboratory | Jim Mildenberger, Wanda Manley, Noah Hull, Taylor Fearing, Lynette Gumbleton, Channing Weber, Ashley Norberg, Chayse Rowley, Marley Goetz, Brian Dominguez, Elliot Thomasson, Sam Britz, Cari Sloma, and Rob Christensen |
| EPI_ISL_1752996, EPI_ISL_1752997, EPI_ISL_1752998, EPI_ISL_1752999, EPI_ISL_1753000, | DC Public Health Lab/ Dept. of Forensic Sciences | DC Public Health Lab/ Dept. of Forensic Sciences | Janis Doss, Scott Nguyen, Elizabeth Zelaya, Sarah Scott, Connie Maza, Monica Mann, Brittany Hamilton, David Payne, Jocelyn Hauser |

| | | | |
|---|---|---|---|
| EPI_ISL_1753001, EPI_ISL_1753002, EPI_ISL_1753003 | | | |
| EPI_ISL_1753006, EPI_ISL_1753007, EPI_ISL_1753008, EPI_ISL_1753009, EPI_ISL_1753011, EPI_ISL_1753012, EPI_ISL_1753013, EPI_ISL_1753014, EPI_ISL_1753015, EPI_ISL_1753016, EPI_ISL_1753017 | | | |
| see above | Wyoming Public Health Laboratory | Wyoming Public Health Laboratory | Jim Mildenberger, Taylor Fearing, Channing Weber, Ashley Norberg, Chayse Rowley, Marley Goetz, Brian Dominguez, Elliot Thomasson, Brittany Oher, Sam Britz, Cari Sloma, and Rob Christensen |
| EPI_ISL_1753019 | GA Department of Public Health Laboratory | Pathogen Discovery, Respiratory Viruses Branch, Division of Viral Diseases, Centers for Disease Control and Prevention | Ying Tao, Yan Li, Anna Kelleher, Jing Zhang, Anna Montmayeur, Brian Lynch, Krista Queen, Anna Uehara, Peter Cook, Han Jia Justin Ng, Rachel Marine, Clinton R. Paden, Haibin Wang, Mark Burroughs, Justin Lee, Adam Retchless, Suxiang Tong |
| EPI_ISL_1753026, EPI_ISL_1753027, EPI_ISL_1753028, EPI_ISL_1753029, EPI_ISL_1753030, EPI_ISL_1753031, EPI_ISL_1753032, EPI_ISL_1753033, EPI_ISL_1753034, EPI_ISL_1753035, EPI_ISL_1753036, EPI_ISL_1753037, EPI_ISL_1753038, EPI_ISL_1753039, EPI_ISL_1753040, EPI_ISL_1753041, EPI_ISL_1753042, EPI_ISL_1753043, EPI_ISL_1753044, EPI_ISL_1753045, EPI_ISL_1753046, EPI_ISL_1753047, EPI_ISL_1753048, EPI_ISL_1753049, EPI_ISL_1753050, EPI_ISL_1753051, EPI_ISL_1753052, EPI_ISL_1753053, EPI_ISL_1753054, EPI_ISL_1753055, EPI_ISL_1753056, EPI_ISL_1753057, EPI_ISL_1753058, EPI_ISL_1753059, EPI_ISL_1753060, EPI_ISL_1753061, EPI_ISL_1753062, EPI_ISL_1753063, EPI_ISL_1753064, EPI_ISL_1753065, EPI_ISL_1753066, EPI_ISL_1753067, EPI_ISL_1753068, EPI_ISL_1753069, EPI_ISL_1753070, EPI_ISL_1753071, EPI_ISL_1753072, EPI_ISL_1753073, EPI_ISL_1753074, EPI_ISL_1753075, EPI_ISL_1753076, EPI_ISL_1753077, EPI_ISL_1753078, EPI_ISL_1753079, EPI_ISL_1753080, EPI_ISL_1753081, EPI_ISL_1753082, EPI_ISL_1753083, EPI_ISL_1753084, EPI_ISL_1753085, EPI_ISL_1753086, EPI_ISL_1753087, EPI_ISL_1753088, EPI_ISL_1753089, EPI_ISL_1753090, EPI_ISL_1753091, EPI_ISL_1753092, EPI_ISL_1753093, EPI_ISL_1753094, EPI_ISL_1753095, EPI_ISL_1753096, EPI_ISL_1753097, EPI_ISL_1753098, EPI_ISL_1753099, EPI_ISL_1753100, EPI_ISL_1753101, EPI_ISL_1753102, EPI_ISL_1753103, EPI_ISL_1753104, EPI_ISL_1753105, EPI_ISL_1753106, EPI_ISL_1753107, EPI_ISL_1753108, EPI_ISL_1753109, EPI_ISL_1753110, EPI_ISL_1753111, EPI_ISL_1753112, EPI_ISL_1753113, EPI_ISL_1753114, EPI_ISL_1753115, EPI_ISL_1753116, EPI_ISL_1753117, EPI_ISL_1753118, EPI_ISL_1753119, EPI_ISL_1753120, EPI_ISL_1753121, EPI_ISL_1753122, EPI_ISL_1753123, EPI_ISL_1753124, EPI_ISL_1753125, EPI_ISL_1753126, EPI_ISL_1753127, EPI_ISL_1753128, EPI_ISL_1753129, EPI_ISL_1753130, EPI_ISL_1753131, EPI_ISL_1753132, EPI_ISL_1753133, EPI_ISL_1753134, EPI_ISL_1753135, EPI_ISL_1753136, EPI_ISL_1753137, EPI_ISL_1753138, EPI_ISL_1753139, EPI_ISL_1753140, EPI_ISL_1753141, EPI_ISL_1753142, EPI_ISL_1753143, EPI_ISL_1753144, EPI_ISL_1753145, EPI_ISL_1753146, EPI_ISL_1753147, EPI_ISL_1753148, EPI_ISL_1753149, EPI_ISL_1753150, EPI_ISL_1753151, EPI_ISL_1753152, EPI_ISL_1753153, EPI_ISL_1753154, EPI_ISL_1753155, EPI_ISL_1753156, EPI_ISL_1753157, EPI_ISL_1753158, EPI_ISL_1753159, EPI_ISL_1753160, EPI_ISL_1753161, EPI_ISL_1753162, EPI_ISL_1753163, EPI_ISL_1753164, EPI_ISL_1753165, EPI_ISL_1753166, EPI_ISL_1753167, EPI_ISL_1753168, EPI_ISL_1753169, EPI_ISL_1753170, EPI_ISL_1753171, EPI_ISL_1753172, EPI_ISL_1753173, EPI_ISL_1753174, EPI_ISL_1753175, EPI_ISL_1753176, EPI_ISL_1753177, EPI_ISL_1753178, EPI_ISL_1753179, EPI_ISL_1753180, EPI_ISL_1753181, EPI_ISL_1753182, EPI_ISL_1753183, EPI_ISL_1753184, EPI_ISL_1753185, EPI_ISL_1753186, EPI_ISL_1753187, EPI_ISL_1753188, EPI_ISL_1753189, EPI_ISL_1753190, EPI_ISL_1753191, EPI_ISL_1753192, EPI_ISL_1753193, EPI_ISL_1753194, EPI_ISL_1753195, EPI_ISL_1753196, EPI_ISL_1753197, EPI_ISL_1753198, EPI_ISL_1753199, EPI_ISL_1753200, EPI_ISL_1753201, EPI_ISL_1753202, EPI_ISL_1753203, EPI_ISL_1753204, EPI_ISL_1753205, EPI_ISL_1753206, EPI_ISL_1753207, EPI_ISL_1753208, EPI_ISL_1753209, EPI_ISL_1753210, EPI_ISL_1753211, EPI_ISL_1753212, EPI_ISL_1753213, EPI_ISL_1753214, EPI_ISL_1753215, EPI_ISL_1753216, EPI_ISL_1753217, EPI_ISL_1753218, EPI_ISL_1753219, EPI_ISL_1753220, EPI_ISL_1753221, EPI_ISL_1753222, EPI_ISL_1753223, EPI_ISL_1753224, EPI_ISL_1753225, EPI_ISL_1753226, EPI_ISL_1753227, EPI_ISL_1753228, EPI_ISL_1753229, EPI_ISL_1753230, EPI_ISL_1753231, EPI_ISL_1753232, EPI_ISL_1753233, EPI_ISL_1753234, EPI_ISL_1753235, EPI_ISL_1753236, EPI_ISL_1753237, EPI_ISL_1753238, EPI_ISL_1753239, EPI_ISL_1753240, EPI_ISL_1753241, EPI_ISL_1753242, EPI_ISL_1753243, EPI_ISL_1753244, EPI_ISL_1753245, EPI_ISL_1753246, EPI_ISL_1753247, EPI_ISL_1753248, EPI_ISL_1753249, EPI_ISL_1753250, EPI_ISL_1753251, EPI_ISL_1753252, EPI_ISL_1753253, EPI_ISL_1753254, EPI_ISL_1753255, EPI_ISL_1753256, EPI_ISL_1753257, EPI_ISL_1753258, EPI_ISL_1753259, EPI_ISL_1753260, EPI_ISL_1753261, EPI_ISL_1753262, EPI_ISL_1753263, EPI_ISL_1753264, EPI_ISL_1753265, EPI_ISL_1753266, EPI_ISL_1753267, EPI_ISL_1753268, EPI_ISL_1753269, EPI_ISL_1753270, EPI_ISL_1753271, EPI_ISL_1753272, EPI_ISL_1753273, EPI_ISL_1753274, EPI_ISL_1753275, EPI_ISL_1753276, EPI_ISL_1753277, EPI_ISL_1753278, EPI_ISL_1753279, EPI_ISL_1753280, EPI_ISL_1753281, EPI_ISL_1753282, EPI_ISL_1753283, EPI_ISL_1753284, EPI_ISL_1753285, EPI_ISL_1753286, EPI_ISL_1753287, EPI_ISL_1753288, EPI_ISL_1753289, EPI_ISL_1753290, EPI_ISL_1753291, EPI_ISL_1753292, EPI_ISL_1753293, EPI_ISL_1753294, EPI_ISL_1753295, EPI_ISL_1753296, EPI_ISL_1753297, EPI_ISL_1753298, EPI_ISL_1753299, EPI_ISL_1753300, EPI_ISL_1753301, EPI_ISL_1753302, EPI_ISL_1753303, EPI_ISL_1753304, EPI_ISL_1753305, EPI_ISL_1753306, EPI_ISL_1753307, EPI_ISL_1753308, EPI_ISL_1753309, EPI_ISL_1753310, EPI_ISL_1753311, EPI_ISL_1753312, EPI_ISL_1753313, EPI_ISL_1753314, EPI_ISL_1753315, EPI_ISL_1753316, EPI_ISL_1753317, EPI_ISL_1753318, EPI_ISL_1753319, EPI_ISL_1753320, EPI_ISL_1753321, EPI_ISL_1753322, EPI_ISL_1753323, EPI_ISL_1753324, EPI_ISL_1753325, EPI_ISL_1753326, EPI_ISL_1753327, EPI_ISL_1753328, EPI_ISL_1753329, EPI_ISL_1753330, EPI_ISL_1753331, EPI_ISL_1753332, EPI_ISL_1753333, EPI_ISL_1753334, EPI_ISL_1753335, EPI_ISL_1753336, EPI_ISL_1753337, EPI_ISL_1753338, EPI_ISL_1753339, EPI_ISL_1753340, EPI_ISL_1753341, EPI_ISL_1753342, EPI_ISL_1753343, EPI_ISL_1753344, EPI_ISL_1753345, EPI_ISL_1753346, EPI_ISL_1753347, EPI_ISL_1753348, EPI_ISL_1753349, EPI_ISL_1753350, EPI_ISL_1753351, EPI_ISL_1753352, EPI_ISL_1753353, EPI_ISL_1753354, EPI_ISL_1753355, EPI_ISL_1753356, EPI_ISL_1753357, EPI_ISL_1753358, EPI_ISL_1753359, EPI_ISL_1753360, EPI_ISL_1753361, EPI_ISL_1753362, EPI_ISL_1753363, EPI_ISL_1753364, EPI_ISL_1753365, EPI_ISL_1753366, EPI_ISL_1753367, EPI_ISL_1753368, EPI_ISL_1753369, EPI_ISL_1753370, EPI_ISL_1753371, EPI_ISL_1753372, EPI_ISL_1753373, EPI_ISL_1753374, EPI_ISL_1753375, EPI_ISL_1753376, EPI_ISL_1753377, EPI_ISL_1753378, EPI_ISL_1753379, EPI_ISL_1753380, EPI_ISL_1753381, EPI_ISL_1753382, EPI_ISL_1753383, EPI_ISL_1753384, EPI_ISL_1753385, EPI_ISL_1753386, EPI_ISL_1753387, EPI_ISL_1753388, EPI_ISL_1753389, EPI_ISL_1753390, EPI_ISL_1753391, EPI_ISL_1753392, EPI_ISL_1753393, EPI_ISL_1753394, EPI_ISL_1753395, EPI_ISL_1753396, EPI_ISL_1753397, EPI_ISL_1753398, EPI_ISL_1753399, EPI_ISL_1753400, EPI_ISL_1753401, EPI_ISL_1753402, EPI_ISL_1753403, EPI_ISL_1753404, EPI_ISL_1753405, EPI_ISL_1753406, EPI_ISL_1753407, EPI_ISL_1753408, EPI_ISL_1753409, EPI_ISL_1753410, EPI_ISL_1753411, EPI_ISL_1753412, EPI_ISL_1753413, EPI_ISL_1753414, EPI_ISL_1753415, EPI_ISL_1753416, EPI_ISL_1753417, EPI_ISL_1753418, EPI_ISL_1753419, EPI_ISL_1753420, EPI_ISL_1753421, EPI_ISL_1753422, EPI_ISL_1753423 | | | |
| see above | Quest Diagnostics Incorporated | Centers for Disease Control and Prevention Division of Viral Diseases, Pathogen Discovery | Dakota Howard, Dhwani Batra, Peter W. Cook, Kara Moser, Adrian Paskey, Jason Caravas, Benjamin Rambo-Martin, Shatavia Morrison, Christopher Gulvick, Scott Sammons, Yvette Unoarumhi, Darlene Wagner, Matthew Schmerer, S. H. Rosenthal, A. Gerasimova, R. M. Kagan, B. Anderson, M. Hua, Y. Liu, L.E. Bernstein, K.E. Livingston, A. Perez, I. A. Shlyakhter, R. V. Rolando, R. Owen, P. Tanpaiboon, F. Lacbawan, Clinton R. Paden, Duncan MacCannell |
| EPI_ISL_1753424, EPI_ISL_1753425, EPI_ISL_1753426, EPI_ISL_1753427, EPI_ISL_1753428, EPI_ISL_1753429, EPI_ISL_1753430, EPI_ISL_1753431, EPI_ISL_1753432, EPI_ISL_1753433, EPI_ISL_1753434, EPI_ISL_1753435, EPI_ISL_1753436, EPI_ISL_1753437, EPI_ISL_1753438, EPI_ISL_1753439, EPI_ISL_1753440, EPI_ISL_1753441, EPI_ISL_1753442, EPI_ISL_1753443, EPI_ISL_1753444, EPI_ISL_1753445, EPI_ISL_1753446, EPI_ISL_1753447, EPI_ISL_1753448, EPI_ISL_1753449, EPI_ISL_1753450 | | | |
| see above | Wyoming Public Health Laboratory | Wyoming Public Health Laboratory | Jim Mildenberger, Wanda Manley, Noah Hull, Taylor Fearing, Lynette Gumbleton, Channing Weber, Ashley Norberg, Chayse Rowley, Marley Goetz, Brian Dominguez, Elliot Thomasson, Sam Britz, Cari Sloma, and Rob Christensen |
| EPI_ISL_1753451, EPI_ISL_1753452, EPI_ISL_1753453, EPI_ISL_1753454, EPI_ISL_1753455, EPI_ISL_1753456, EPI_ISL_1753457, EPI_ISL_1753458, EPI_ISL_1753459, EPI_ISL_1753460, EPI_ISL_1753461, EPI_ISL_1753462, EPI_ISL_1753463, EPI_ISL_1753464, EPI_ISL_1753465, EPI_ISL_1753466, EPI_ISL_1753468, EPI_ISL_1753469, EPI_ISL_1753470, EPI_ISL_1753471, EPI_ISL_1753472, EPI_ISL_1753473, EPI_ISL_1753475, EPI_ISL_1753476, EPI_ISL_1753477, EPI_ISL_1753478, EPI_ISL_1753480, EPI_ISL_1753481, EPI_ISL_1753482, EPI_ISL_1753483, EPI_ISL_1753484, EPI_ISL_1753486, EPI_ISL_1753487, EPI_ISL_1753488, EPI_ISL_1753489, EPI_ISL_1753490, EPI_ISL_1753491, EPI_ISL_1753492, EPI_ISL_1753493 | | | |
| see above | Infinity Biologix | Centers for Disease Control and Prevention Division of Viral Diseases, Pathogen Discovery | Dakota Howard, Dhwani Batra, Peter W. Cook, Kara Moser, Adrian Paskey, Jason Caravas, Benjamin Rambo-Martin, Shatavia Morrison, Christopher Gulvick, Scott Sammons, Yvette Unoarumhi, Darlene Wagner, Matthew Schmerer, Christian Bixby, Yihe Wang, Jonathan Schultz, Chirayu Goswami, Russ Hager, Robin Grimwood, Clinton R. Paden, Duncan MacCannell |
| EPI_ISL_1753494, EPI_ISL_1753495, EPI_ISL_1753496, EPI_ISL_1753497, EPI_ISL_1753498, EPI_ISL_1753499, EPI_ISL_1753500, EPI_ISL_1753501, EPI_ISL_1753502, EPI_ISL_1753503, EPI_ISL_1753504, EPI_ISL_1753505, EPI_ISL_1753506, EPI_ISL_1753507, EPI_ISL_1753508, EPI_ISL_1753509, EPI_ISL_1753510, EPI_ISL_1753511, EPI_ISL_1753512, EPI_ISL_1753513, EPI_ISL_1753514, EPI_ISL_1753515 | | | |
| see above | Helix/Illumina | Centers for Disease Control and Prevention Division of Viral Diseases, Pathogen Discovery | Dakota Howard, Dhwani Batra, Peter W. Cook, Kara Moser, Adrian Paskey, Jason Caravas, Benjamin Rambo-Martin, Shatavia Morrison, Christopher Gulvick, Scott Sammons, Yvette Unoarumhi, Darlene Wagner, Matthew Schmerer, Eileen de Feo, Jan Antico, Christine Tran, Matthew Tolentino, Shannon Wickline, Kim Gietzen, Brad Sickler, Jingtao Liu, Eric Allen, Phil Febbo, Nicole L. Washington, Simon White, Geraint Levan, Kelly Schiabor Barrett, Elizabeth Cirulli, Alexandre Bolze, Ary Ascencio, Charlotte Rivera-Garcia, Ryan Cho, Jason Nguyen, Sherry Wang, Jimmy Ramirez, Tyler Cassens, Efren Sandoval, Magnus Isaksson, William Lee, David Becker, Marc Laurent, James Lu, Clinton R. Paden, Duncan MacCannell |
| EPI_ISL_1753516, EPI_ISL_1753517, EPI_ISL_1753518, EPI_ISL_1753519, EPI_ISL_1753520, EPI_ISL_1753521, EPI_ISL_1753522, EPI_ISL_1753523, EPI_ISL_1753524, EPI_ISL_1753525, EPI_ISL_1753526, EPI_ISL_1753527, EPI_ISL_1753528, EPI_ISL_1753529, EPI_ISL_1753530, EPI_ISL_1753531, EPI_ISL_1753532, EPI_ISL_1753533, EPI_ISL_1753534, EPI_ISL_1753535, EPI_ISL_1753536, EPI_ISL_1753537, EPI_ISL_1753538, EPI_ISL_1753539, EPI_ISL_1753540, EPI_ISL_1753541, EPI_ISL_1753542, EPI_ISL_1753543, EPI_ISL_1753544, EPI_ISL_1753545, EPI_ISL_1753546, EPI_ISL_1753547, EPI_ISL_1753548, EPI_ISL_1753549, EPI_ISL_1753550, EPI_ISL_1753551, EPI_ISL_1753552, EPI_ISL_1753553, EPI_ISL_1753554, EPI_ISL_1753555, EPI_ISL_1753556, EPI_ISL_1753557, EPI_ISL_1753558, EPI_ISL_1753559, EPI_ISL_1753560, EPI_ISL_1753561, EPI_ISL_1753562, EPI_ISL_1753563, EPI_ISL_1753564, EPI_ISL_1753565, EPI_ISL_1753566, EPI_ISL_1753567, EPI_ISL_1753568, EPI_ISL_1753569, EPI_ISL_1753570, EPI_ISL_1753571, EPI_ISL_1753572, EPI_ISL_1753573, EPI_ISL_1753574, EPI_ISL_1753575, EPI_ISL_1753576, EPI_ISL_1753577, EPI_ISL_1753578, EPI_ISL_1753579, EPI_ISL_1753580, EPI_ISL_1753581, EPI_ISL_1753582, EPI_ISL_1753583, EPI_ISL_1753584, EPI_ISL_1753585, EPI_ISL_1753586, EPI_ISL_1753587, EPI_ISL_1753588, EPI_ISL_1753589, EPI_ISL_1753590, EPI_ISL_1753591, EPI_ISL_1753592, EPI_ISL_1753593, EPI_ISL_1753594, EPI_ISL_1753595, EPI_ISL_1753596, EPI_ISL_1753597, EPI_ISL_1753598, EPI_ISL_1753599, EPI_ISL_1753600, EPI_ISL_1753601, EPI_ISL_1753602, EPI_ISL_1753603, EPI_ISL_1753604, EPI_ISL_1753605, EPI_ISL_1753606, EPI_ISL_1753608, EPI_ISL_1753609, EPI_ISL_1753610, EPI_ISL_1753611, EPI_ISL_1753612 | | | |
| see above | Aegis Sciences Corporation | Centers for Disease Control and Prevention Division of Viral Diseases, Pathogen Discovery | Dakota Howard, Dhwani Batra, Peter W. Cook, Kara Moser, Adrian Paskey, Jason Caravas, Benjamin Rambo-Martin, Shatavia Morrison, Christopher Gulvick, Scott Sammons, Yvette Unoarumhi, Darlene Wagner, Matthew Schmerer, Cyndi Clark, Patrick Campbell, Rob Case, Vikramsinha Ghorpade, Holly Houdeshell, Ola Kvalvaag, Dillon Nall, Ethan Sanders, Alec Vest, Shaun Westlund, Matthew Hardison, Clinton R. Paden, Duncan MacCannell |
| EPI_ISL_1753613, EPI_ISL_1753614, EPI_ISL_1753615, EPI_ISL_1753616, EPI_ISL_1753617, EPI_ISL_1753618, EPI_ISL_1753619, EPI_ISL_1753620, EPI_ISL_1753621, EPI_ISL_1753622, EPI_ISL_1753623, EPI_ISL_1753624, EPI_ISL_1753625, EPI_ISL_1753626, EPI_ISL_1753627, EPI_ISL_1753628, EPI_ISL_1753629, EPI_ISL_1753630, EPI_ISL_1753631, EPI_ISL_1753632, EPI_ISL_1753633, EPI_ISL_1753634, EPI_ISL_1753635, EPI_ISL_1753636, EPI_ISL_1753637, EPI_ISL_1753638, EPI_ISL_1753639, EPI_ISL_1753640, EPI_ISL_1753641, EPI_ISL_1753642, EPI_ISL_1753643, EPI_ISL_1753644, EPI_ISL_1753645, EPI_ISL_1753646, EPI_ISL_1753647, EPI_ISL_1753648, EPI_ISL_1753649, EPI_ISL_1753650, EPI_ISL_1753651, EPI_ISL_1753652, EPI_ISL_1753653, EPI_ISL_1753654, EPI_ISL_1753655, EPI_ISL_1753656, EPI_ISL_1753657, EPI_ISL_1753658, EPI_ISL_1753659, EPI_ISL_1753660, EPI_ISL_1753661 | | | |
| see above | Quest Diagnostics Incorporated | Centers for Disease Control and Prevention Division of Viral Diseases, Pathogen Discovery | Dakota Howard, Dhwani Batra, Peter W. Cook, Kara Moser, Adrian Paskey, Jason Caravas, Benjamin Rambo-Martin, Shatavia Morrison, Christopher Gulvick, Scott Sammons, Yvette Unoarumhi, Darlene Wagner, Matthew Schmerer, S. H. Rosenthal, A. Gerasimova, R. M. Kagan, B. Anderson, M. Hua, Y. Liu, L.E. Bernstein, K.E. Livingston, A. Perez, I. A. Shlyakhter, R. V. Rolando, R. Owen, P. Tanpaiboon, F. Lacbawan, Clinton R. Paden, Duncan MacCannell |
| EPI_ISL_1753668, EPI_ISL_1753669, EPI_ISL_1753670, EPI_ISL_1753671, EPI_ISL_1753672, EPI_ISL_1753673, EPI_ISL_1753674 | University of Michigan Clinical Microbiology Laboratory | Lauring Lab, University of Michigan, Department of Microbiology and Immunology | Valesano |
| EPI_ISL_1753844 | Ohio Department of Health Laboratory | Ohio Department of Health Laboratory | Holmes, Jennifer; Eric Brandt, Keoni Omura, Glen McGillivary, Caitlin McDonnell, Jade Mowery, Stephanie Mccracken, Tyler Payne, Kirtana Ramadugu, Erica Leasure, Brent Lee, Kelsey Florek, Heather Blankenship, Allison Black, Quanta Brown, and Tammy Bannerman |
| EPI_ISL_1754139, EPI_ISL_1754140 | LESP Campeche | Instituto de Diagnostico y Referencia Epidemiologicos (INDRE) | Claudia Wong-Arambula, Abril Rodriguez-Maldonado, Vanessa Rivero-Arredondo, Ariadna Medina-Benitez, Joaquin Quiroz-Mercado, Sergio Rangel-Guerrero, Natividad Cruz-Ortiz, Tatiana Nunez-Garcia, Gisela Barrera-Badillo, Lucia Hernandez-Rivas, Irma Lopez-Martinez, Ernesto Ramirez-Gonzalez. |
| EPI_ISL_1754141 | LESP Baja California | Instituto de Diagnostico y Referencia Epidemiologicos (INDRE) | Claudia Wong-Arambula, Abril Rodriguez-Maldonado, Vanessa Rivero-Arredondo, Ariadna Medina-Benitez, Joaquin Quiroz-Mercado, Sergio Rangel-Guerrero, Natividad Cruz-Ortiz, Tatiana Nunez-Garcia, Gisela Barrera-Badillo, Lucia Hernandez-Rivas, Irma Lopez-Martinez, Ernesto Ramirez-Gonzalez. |
| EPI_ISL_1754142, EPI_ISL_1754143, EPI_ISL_1754144, EPI_ISL_1754145, EPI_ISL_1754146, EPI_ISL_1754147, EPI_ISL_1754148, EPI_ISL_1754149, EPI_ISL_1754150, EPI_ISL_1754151, EPI_ISL_1754152, EPI_ISL_1754153, EPI_ISL_1754154, EPI_ISL_1754155, EPI_ISL_1754156, EPI_ISL_1754157, EPI_ISL_1754158, EPI_ISL_1754159, EPI_ISL_1754160, EPI_ISL_1754161, EPI_ISL_1754162, EPI_ISL_1754163, EPI_ISL_1754164, EPI_ISL_1754165, EPI_ISL_1754166, EPI_ISL_1754167, EPI_ISL_1754168, EPI_ISL_1754169, EPI_ISL_1754170, EPI_ISL_1754171, EPI_ISL_1754172, EPI_ISL_1754173, EPI_ISL_1754174, EPI_ISL_1754175, EPI_ISL_1754176, EPI_ISL_1754177, EPI_ISL_1754178, EPI_ISL_1754179, EPI_ISL_1754180, EPI_ISL_1754181, EPI_ISL_1754182, EPI_ISL_1754183, EPI_ISL_1754184 | | | |
| see above | Tampa General Hospital Esoteric Lab | Tampa General Hospital Esoteric Research & Development Lab | Grant Vestal, Deanna Becker, Dominic Uy, Vicki Healer, Amorce Lima, Suzane Silbert |
| EPI_ISL_1754185 | PA Dept of Heath, Bureau of Laboratories | Genomics and Discovery, Respiratory Viruses Branch, Division of Viral Diseases, Centers for Disease Control and Prevention | Ying Tao, Yan Li, Anna Kelleher, Jing Zhang, Anna Montmayeur, Brian Lynch, Krista Queen, Anna Uehara, Peter Cook, Han Jia Justin Ng, Rachel Marine, Clinton R. Paden, Haibin Wang, Mark Burroughs, Justin Lee, Adam Retchless, Suxiang Tong |
| EPI_ISL_1754187, EPI_ISL_1754188 | Ohio Department of Health Laboratory | Ohio Department of Health Laboratory | Holmes, Jennifer; Eric Brandt, Keoni Omura, Glen McGillivary, Caitlin McDonnell, Jade Mowery, Stephanie Mccracken, Tyler Payne, Kirtana Ramadugu, Erica Leasure, Brent Lee, Kelsey Florek, Heather Blankenship, Allison Black, Quanta Brown, and Tammy Bannerman |
| EPI_ISL_1754868, EPI_ISL_1754869, EPI_ISL_1754870, EPI_ISL_1754871, EPI_ISL_1754872, EPI_ISL_1754873, EPI_ISL_1754874, EPI_ISL_1754875, EPI_ISL_1754876, EPI_ISL_1754877, EPI_ISL_1754878, EPI_ISL_1754879, EPI_ISL_1754880, EPI_ISL_1754881, EPI_ISL_1754882, EPI_ISL_1754883, EPI_ISL_1754884, EPI_ISL_1754885, EPI_ISL_1754886, EPI_ISL_1754887, EPI_ISL_1754888, EPI_ISL_1754889, EPI_ISL_1754890, EPI_ISL_1754891, EPI_ISL_1754892, EPI_ISL_1754893, EPI_ISL_1754894, EPI_ISL_1754895, EPI_ISL_1754896 | | | |
| see above | Alaska State Virology Laboratory | Alaska State Virology Laboratory | Stephanie DeRonde, Elva House, Lisa Smith, Ph.D., Jack Chen, Ph.D. |
| EPI_ISL_1755445, EPI_ISL_1755446, EPI_ISL_1755447, EPI_ISL_1755448, EPI_ISL_1755449, EPI_ISL_1755450, EPI_ISL_1755451, EPI_ISL_1755452, EPI_ISL_1755453, EPI_ISL_1755454, EPI_ISL_1755455, EPI_ISL_1755456, EPI_ISL_1755457, EPI_ISL_1755458, EPI_ISL_1755459, EPI_ISL_1755460, EPI_ISL_1755461, EPI_ISL_1755462, EPI_ISL_1755463, EPI_ISL_1755464, EPI_ISL_1755465, EPI_ISL_1755466, EPI_ISL_1755467, EPI_ISL_1755468, EPI_ISL_1755469, EPI_ISL_1755470, EPI_ISL_1755471, EPI_ISL_1755472, EPI_ISL_1755473, EPI_ISL_1755474, EPI_ISL_1755475, EPI_ISL_1755476, EPI_ISL_1755477, EPI_ISL_1755478, EPI_ISL_1755479, EPI_ISL_1755480, EPI_ISL_1755481, EPI_ISL_1755482, EPI_ISL_1755483, EPI_ISL_1755484, EPI_ISL_1755485, EPI_ISL_1755486 | | | |
| see above | NYC Pandemic Response Lab | Wadsworth Center, New York State Department of Health | Kirsten St. George, Daryl M. Lamson, Alexis Russell, Matthew Shudt, Melissa A Leisner, Jonathan Plitnick, Catharine Prussing, Navjot Singh, John Kelly, Erasmus Schneider, Erica Lasek-Nesselquist |
| EPI_ISL_1755487 | SUNY UPSTATE MEDICAL UNIVERSITY | Wadsworth Center, New York State Department of Health | Kirsten St. George, Daryl M. Lamson, Alexis Russell, Matthew Shudt, Melissa A Leisner, Jonathan Plitnick, Catharine Prussing, Navjot Singh, John Kelly, Erasmus Schneider, Erica Lasek-Nesselquist |
| EPI_ISL_1755488 | GLENS FALLS HOSPITAL LABORATORY | Wadsworth Center, New York State Department of Health | Kirsten St. George, Daryl M. Lamson, Alexis Russell, Matthew Shudt, Melissa A Leisner, Jonathan Plitnick, Catharine Prussing, Navjot Singh, John Kelly, Erasmus Schneider, Erica Lasek-Nesselquist |
| EPI_ISL_1755489, EPI_ISL_1755490 | SUNY UPSTATE MEDICAL UNIVERSITY | Wadsworth Center, New York State Department of Health | Kirsten St. George, Daryl M. Lamson, Alexis Russell, Matthew Shudt, Melissa A Leisner, Jonathan Plitnick, Catharine Prussing, Navjot Singh, John Kelly, Erasmus Schneider, Erica Lasek-Nesselquist |
| EPI_ISL_1755491, EPI_ISL_1755492, EPI_ISL_1755493, EPI_ISL_1755494, EPI_ISL_1755495, EPI_ISL_1755496, EPI_ISL_1755497, EPI_ISL_1755498, EPI_ISL_1755499, EPI_ISL_1755500, EPI_ISL_1755501 | | | |
| see above | NYC Pandemic Response Lab | Wadsworth Center, New York State Department of Health | Kirsten St. George, Daryl M. Lamson, Alexis Russell, Matthew Shudt, Melissa A Leisner, Jonathan Plitnick, Catharine Prussing, Navjot Singh, John Kelly, Erasmus Schneider, Erica Lasek-Nesselquist |
| EPI_ISL_1755820, EPI_ISL_1755821, EPI_ISL_1755822, EPI_ISL_1755823, EPI_ISL_1755824, EPI_ISL_1755825, EPI_ISL_1755826, EPI_ISL_1755827, EPI_ISL_1755828, EPI_ISL_1755829, EPI_ISL_1755830, EPI_ISL_1755831, EPI_ISL_1755832, EPI_ISL_1755833, EPI_ISL_1755834 | | | |
| see above | MONTEFIORE MEDICAL CENTER | Wadsworth Center, New York State | Kirsten St. George, Daryl M. Lamson, Alexis Russell, Matthew Shudt, Melissa A Leisner, Jonathan Plitnick, Catharine Prussing, Navjot Singh, John Kelly, Erasmus Schneider, Erica Lasek-Nesselquist |

| | | | |
|---|---|---|---|
| | LABORATORIES | Department of Health | |
| EPI_ISL_1755835 | URMC LABS | Wadsworth Center, New York State Department of Health | Kirsten St. George, Daryl M. Lamson, Alexis Russell, Matthew Shudt, Melissa A Leisner, Jonathan Plitnick, Catharine Prussing, Navjot Singh, John Kelly, Erasmus Schneider, Erica Lasek-Nesselquist |
| EPI_ISL_1755836, EPI_ISL_1755837 | MONTEFIORE MEDICAL CENTER LABORATORIES | Wadsworth Center, New York State Department of Health | Kirsten St. George, Daryl M. Lamson, Alexis Russell, Matthew Shudt, Melissa A Leisner, Jonathan Plitnick, Catharine Prussing, Navjot Singh, John Kelly, Erasmus Schneider, Erica Lasek-Nesselquist |
| EPI_ISL_1755838, EPI_ISL_1755839, EPI_ISL_1755840, EPI_ISL_1755841 | ALBANY MEDICAL CENTER | Wadsworth Center, New York State Department of Health | Kirsten St. George, Daryl M. Lamson, Alexis Russell, Matthew Shudt, Melissa A Leisner, Jonathan Plitnick, Catharine Prussing, Navjot Singh, John Kelly, Erasmus Schneider, Erica Lasek-Nesselquist |
| EPI_ISL_1755842, EPI_ISL_1755843, EPI_ISL_1755844, EPI_ISL_1755845, EPI_ISL_1755846, EPI_ISL_1755847, EPI_ISL_1755848 | SUNY UPSTATE MEDICAL UNIVERSITY | Wadsworth Center, New York State Department of Health | Kirsten St. George, Daryl M. Lamson, Alexis Russell, Matthew Shudt, Melissa A Leisner, Jonathan Plitnick, Catharine Prussing, Navjot Singh, John Kelly, Erasmus Schneider, Erica Lasek-Nesselquist |
| EPI_ISL_1755849, EPI_ISL_1755850 | BOSTON HEART DIAGNOSTICS CORP | Wadsworth Center, New York State Department of Health | Kirsten St. George, Daryl M. Lamson, Alexis Russell, Matthew Shudt, Melissa A Leisner, Jonathan Plitnick, Catharine Prussing, Navjot Singh, John Kelly, Erasmus Schneider, Erica Lasek-Nesselquist |
| EPI_ISL_1755851, EPI_ISL_1755852, EPI_ISL_1755853 | WESTCHESTER MEDICAL CENTER | Wadsworth Center, New York State Department of Health | Kirsten St. George, Daryl M. Lamson, Alexis Russell, Matthew Shudt, Melissa A Leisner, Jonathan Plitnick, Catharine Prussing, Navjot Singh, John Kelly, Erasmus Schneider, Erica Lasek-Nesselquist |
| EPI_ISL_1755854, EPI_ISL_1755855, EPI_ISL_1755856, EPI_ISL_1755857, EPI_ISL_1755858, EPI_ISL_1755859, EPI_ISL_1755860, EPI_ISL_1755861, EPI_ISL_1755862, EPI_ISL_1755863, EPI_ISL_1755864, EPI_ISL_1755865, EPI_ISL_1755866 | | | |
| see above | Columbia University Irving Medical Center | Wadsworth Center, New York State Department of Health | Kirsten St. George, Daryl M. Lamson, Alexis Russell, Matthew Shudt, Melissa A Leisner, Jonathan Plitnick, Catharine Prussing, Navjot Singh, John Kelly, Erasmus Schneider, Erica Lasek-Nesselquist |
| EPI_ISL_1755867, EPI_ISL_1755868 | WESTCHESTER MEDICAL CENTER | Wadsworth Center, New York State Department of Health | Kirsten St. George, Daryl M. Lamson, Alexis Russell, Matthew Shudt, Melissa A Leisner, Jonathan Plitnick, Catharine Prussing, Navjot Singh, John Kelly, Erasmus Schneider, Erica Lasek-Nesselquist |
| EPI_ISL_1755869, EPI_ISL_1755870, EPI_ISL_1755871, EPI_ISL_1755872, EPI_ISL_1755873, EPI_ISL_1755874, EPI_ISL_1755875, EPI_ISL_1755876, EPI_ISL_1755877, EPI_ISL_1755878, EPI_ISL_1755879, EPI_ISL_1755880, EPI_ISL_1755881, EPI_ISL_1755882, EPI_ISL_1755883, EPI_ISL_1755884, EPI_ISL_1755885, EPI_ISL_1755886, EPI_ISL_1755887, EPI_ISL_1755888, EPI_ISL_1755889, EPI_ISL_1755890, EPI_ISL_1755891, EPI_ISL_1755892, EPI_ISL_1755893, EPI_ISL_1755894, EPI_ISL_1755895, EPI_ISL_1755896, EPI_ISL_1755897, EPI_ISL_1755898, EPI_ISL_1755899, EPI_ISL_1755900, EPI_ISL_1755901, EPI_ISL_1755902, EPI_ISL_1755903, EPI_ISL_1755904, EPI_ISL_1755905, EPI_ISL_1755906, EPI_ISL_1755907 | | | |
| see above | URMC LABS | Wadsworth Center, New York State Department of Health | Kirsten St. George, Daryl M. Lamson, Alexis Russell, Matthew Shudt, Melissa A Leisner, Jonathan Plitnick, Catharine Prussing, Navjot Singh, John Kelly, Erasmus Schneider, Erica Lasek-Nesselquist |
| EPI_ISL_1755917, EPI_ISL_1755919, EPI_ISL_1755920, EPI_ISL_1755921, EPI_ISL_1755922, EPI_ISL_1755923, EPI_ISL_1755924, EPI_ISL_1755925, EPI_ISL_1755926, EPI_ISL_1755927, EPI_ISL_1755930, EPI_ISL_1755931, EPI_ISL_1755932, EPI_ISL_1755933, EPI_ISL_1755935, EPI_ISL_1755936, EPI_ISL_1755937, EPI_ISL_1755938 | | | |
| see above | Wyoming Public Health Laboratory | Wyoming Public Health Laboratory | Jim Mildenberger, Taylor Fearing, Channing Weber, Ashley Norberg, Chayse Rowley, Marley Goetz, Brian Dominguez, Elliot Thomasson, Brittany Oher, Sam Britz, Cari Sloma, and Rob Christensen |
| EPI_ISL_1755939 | ALBANY MEDICAL CENTER | Wadsworth Center, New York State Department of Health | Kirsten St. George, Daryl M. Lamson, Alexis Russell, Matthew Shudt, Melissa A Leisner, Jonathan Plitnick, Catharine Prussing, Navjot Singh, John Kelly, Erasmus Schneider, Erica Lasek-Nesselquist |
| EPI_ISL_1755940 | MEMORIAL SLOAN KETTERING CANCER CENTER | Wadsworth Center, New York State Department of Health | Kirsten St. George, Daryl M. Lamson, Alexis Russell, Matthew Shudt, Melissa A Leisner, Jonathan Plitnick, Catharine Prussing, Navjot Singh, John Kelly, Erasmus Schneider, Erica Lasek-Nesselquist |
| EPI_ISL_1755941, EPI_ISL_1755942 | NYC Pandemic Response Lab | Wadsworth Center, New York State Department of Health | Kirsten St. George, Daryl M. Lamson, Alexis Russell, Matthew Shudt, Melissa A Leisner, Jonathan Plitnick, Catharine Prussing, Navjot Singh, John Kelly, Erasmus Schneider, Erica Lasek-Nesselquist |
| EPI_ISL_1755943, EPI_ISL_1755944, EPI_ISL_1755945, EPI_ISL_1755946 | Columbia University Irving Medical Center | Wadsworth Center, New York State Department of Health | Kirsten St. George, Daryl M. Lamson, Alexis Russell, Matthew Shudt, Melissa A Leisner, Jonathan Plitnick, Catharine Prussing, Navjot Singh, John Kelly, Erasmus Schneider, Erica Lasek-Nesselquist |
| EPI_ISL_1755947 | NYC Pandemic Response Lab | Wadsworth Center, New York State Department of Health | Kirsten St. George, Daryl M. Lamson, Alexis Russell, Matthew Shudt, Melissa A Leisner, Jonathan Plitnick, Catharine Prussing, Navjot Singh, John Kelly, Erasmus Schneider, Erica Lasek-Nesselquist |
| EPI_ISL_1755948, EPI_ISL_1755949, EPI_ISL_1755950, EPI_ISL_1755951, EPI_ISL_1755952, EPI_ISL_1755953, EPI_ISL_1755954, EPI_ISL_1755955, EPI_ISL_1755956, EPI_ISL_1755957, EPI_ISL_1755958, EPI_ISL_1755959, EPI_ISL_1755960, EPI_ISL_1755961, EPI_ISL_1755962, EPI_ISL_1755963, EPI_ISL_1755964 | | | |
| see above | MONTEFIORE MEDICAL CENTER LABORATORIES | Wadsworth Center, New York State Department of Health | Kirsten St. George, Daryl M. Lamson, Alexis Russell, Matthew Shudt, Melissa A Leisner, Jonathan Plitnick, Catharine Prussing, Navjot Singh, John Kelly, Erasmus Schneider, Erica Lasek-Nesselquist |
| EPI_ISL_1755965, EPI_ISL_1755966, EPI_ISL_1755967 | WESTCHESTER MEDICAL CENTER | Wadsworth Center, New York State Department of Health | Kirsten St. George, Daryl M. Lamson, Alexis Russell, Matthew Shudt, Melissa A Leisner, Jonathan Plitnick, Catharine Prussing, Navjot Singh, John Kelly, Erasmus Schneider, Erica Lasek-Nesselquist |
| EPI_ISL_1755968, EPI_ISL_1755969, EPI_ISL_1755970, EPI_ISL_1755971, EPI_ISL_1755972, EPI_ISL_1755973, EPI_ISL_1755974, EPI_ISL_1755975, EPI_ISL_1755976, EPI_ISL_1755977, EPI_ISL_1755978, EPI_ISL_1755979, EPI_ISL_1755980, EPI_ISL_1755981, EPI_ISL_1755982, EPI_ISL_1755983, EPI_ISL_1755984, EPI_ISL_1755985, EPI_ISL_1755986, EPI_ISL_1755987, EPI_ISL_1755988, EPI_ISL_1755989, EPI_ISL_1755990, EPI_ISL_1755991 | | | |
| see above | TEMPUS LABS INC | Wadsworth Center, New York State Department of Health | Kirsten St. George, Daryl M. Lamson, Alexis Russell, Matthew Shudt, Melissa A Leisner, Jonathan Plitnick, Catharine Prussing, Navjot Singh, John Kelly, Erasmus Schneider, Erica Lasek-Nesselquist |
| EPI_ISL_1755992, EPI_ISL_1755993, EPI_ISL_1755994, EPI_ISL_1755995, EPI_ISL_1755996, EPI_ISL_1755997, EPI_ISL_1755998, EPI_ISL_1755999 | SARATOGA HOSPITAL LABORATORY | Wadsworth Center, New York State Department of Health | Kirsten St. George, Daryl M. Lamson, Alexis Russell, Matthew Shudt, Melissa A Leisner, Jonathan Plitnick, Catharine Prussing, Navjot Singh, John Kelly, Erasmus Schneider, Erica Lasek-Nesselquist |
| EPI_ISL_1756000 | STONY BROOK UNIVERSITY HOSPITAL | Wadsworth Center, New York State Department of Health | Kirsten St. George, Daryl M. Lamson, Alexis Russell, Matthew Shudt, Melissa A Leisner, Jonathan Plitnick, Catharine Prussing, Navjot Singh, John Kelly, Erasmus Schneider, Erica Lasek-Nesselquist |
| EPI_ISL_1756001, EPI_ISL_1756002, EPI_ISL_1756003, EPI_ISL_1756004, EPI_ISL_1756005, EPI_ISL_1756006, EPI_ISL_1756007, EPI_ISL_1756008, EPI_ISL_1756009, EPI_ISL_1756010, EPI_ISL_1756011, EPI_ISL_1756012, EPI_ISL_1756013, EPI_ISL_1756014, EPI_ISL_1756015, EPI_ISL_1756016, EPI_ISL_1756017, EPI_ISL_1756018, EPI_ISL_1756019, EPI_ISL_1756020 | | | |
| see above | ALBANY MEDICAL CENTER | Wadsworth Center, New York State Department of Health | Kirsten St. George, Daryl M. Lamson, Alexis Russell, Matthew Shudt, Melissa A Leisner, Jonathan Plitnick, Catharine Prussing, Navjot Singh, John Kelly, Erasmus Schneider, Erica Lasek-Nesselquist |
| EPI_ISL_1756326 | Columbia University Irving Medical Center | Wadsworth Center, New York State Department of Health | Kirsten St. George, Daryl M. Lamson, Alexis Russell, Matthew Shudt, Melissa A Leisner, Jonathan Plitnick, Catharine Prussing, Navjot Singh, John Kelly, Erasmus Schneider, Erica Lasek-Nesselquist |
| EPI_ISL_1756327 | NORTHWELL HEALTH LABORATORIES | Wadsworth Center, New York State Department of Health | Kirsten St. George, Daryl M. Lamson, Alexis Russell, Matthew Shudt, Melissa A Leisner, Jonathan Plitnick, Catharine Prussing, Navjot Singh, John Kelly, Erasmus Schneider, Erica Lasek-Nesselquist |
| EPI_ISL_1756328 | Columbia University Irving Medical Center | Wadsworth Center, New York State Department of Health | Kirsten St. George, Daryl M. Lamson, Alexis Russell, Matthew Shudt, Melissa A Leisner, Jonathan Plitnick, Catharine Prussing, Navjot Singh, John Kelly, Erasmus Schneider, Erica Lasek-Nesselquist |
| EPI_ISL_1756329 | ALBANY MEDICAL CENTER | Wadsworth Center, New York State Department of Health | Kirsten St. George, Daryl M. Lamson, Alexis Russell, Matthew Shudt, Melissa A Leisner, Jonathan Plitnick, Catharine Prussing, Navjot Singh, John Kelly, Erasmus Schneider, Erica Lasek-Nesselquist |
| EPI_ISL_1756330 | NORTHWELL HEALTH LABORATORIES | Wadsworth Center, New York State Department of Health | Kirsten St. George, Daryl M. Lamson, Alexis Russell, Matthew Shudt, Melissa A Leisner, Jonathan Plitnick, Catharine Prussing, Navjot Singh, John Kelly, Erasmus Schneider, Erica Lasek-Nesselquist |
| EPI_ISL_1756331, EPI_ISL_1756332, EPI_ISL_1756333, EPI_ISL_1756334, EPI_ISL_1756335, EPI_ISL_1756336, EPI_ISL_1756337 | STONY BROOK UNIVERSITY HOSPITAL | Wadsworth Center, New York State Department of Health | Kirsten St. George, Daryl M. Lamson, Alexis Russell, Matthew Shudt, Melissa A Leisner, Jonathan Plitnick, Catharine Prussing, Navjot Singh, John Kelly, Erasmus Schneider, Erica Lasek-Nesselquist |
| EPI_ISL_1756338, EPI_ISL_1756339, EPI_ISL_1756340, EPI_ISL_1756341 | SUNY UPSTATE MEDICAL UNIVERSITY | Wadsworth Center, New York State Department of Health | Kirsten St. George, Daryl M. Lamson, Alexis Russell, Matthew Shudt, Melissa A Leisner, Jonathan Plitnick, Catharine Prussing, Navjot Singh, John Kelly, Erasmus Schneider, Erica Lasek-Nesselquist |
| EPI_ISL_1756342, EPI_ISL_1756343, EPI_ISL_1756344, EPI_ISL_1756345, EPI_ISL_1756346, EPI_ISL_1756347, EPI_ISL_1756348, EPI_ISL_1756349, EPI_ISL_1756350, EPI_ISL_1756351, EPI_ISL_1756352, EPI_ISL_1756353, EPI_ISL_1756354, EPI_ISL_1756355 | | | |
| see above | MONTEFIORE MEDICAL CENTER LABORATORIES | Wadsworth Center, New York State Department of Health | Kirsten St. George, Daryl M. Lamson, Alexis Russell, Matthew Shudt, Melissa A Leisner, Jonathan Plitnick, Catharine Prussing, Navjot Singh, John Kelly, Erasmus Schneider, Erica Lasek-Nesselquist |
| EPI_ISL_1756356 | BIO-REFERENCE LABORATORIES | Wadsworth Center, New York State Department of Health | Kirsten St. George, Daryl M. Lamson, Alexis Russell, Matthew Shudt, Melissa A Leisner, Jonathan Plitnick, Catharine Prussing, Navjot Singh, John Kelly, Erasmus Schneider, Erica Lasek-Nesselquist |

| | | | |
|---|---|---|---|
| EPI_ISL_1756357, EPI_ISL_1756358, EPI_ISL_1756359, EPI_ISL_1756360, EPI_ISL_1756361, EPI_ISL_1756362, EPI_ISL_1756363, EPI_ISL_1756364, EPI_ISL_1756365, EPI_ISL_1756366, EPI_ISL_1756367 | | | |
| see above | URMC LABS | Wadsworth Center, New York State Department of Health | Kirsten St. George, Daryl M. Lamson, Alexis Russell, Matthew Shudt, Melissa A Leisner, Jonathan Plitnick, Catharine Prussing, Navjot Singh, John Kelly, Erasmus Schneider, Erica Lasek-Nesselquist |
| EPI_ISL_1756368 | ST. FRANCIS HOSPITAL LABORATORY | Wadsworth Center, New York State Department of Health | Kirsten St. George, Daryl M. Lamson, Alexis Russell, Matthew Shudt, Melissa A Leisner, Jonathan Plitnick, Catharine Prussing, Navjot Singh, John Kelly, Erasmus Schneider, Erica Lasek-Nesselquist |
| EPI_ISL_1756369, EPI_ISL_1756370, EPI_ISL_1756371, EPI_ISL_1756372, EPI_ISL_1756373, EPI_ISL_1756374, EPI_ISL_1756375, EPI_ISL_1756376, EPI_ISL_1756377, EPI_ISL_1756378, EPI_ISL_1756379, EPI_ISL_1756380, EPI_ISL_1756381, EPI_ISL_1756382, EPI_ISL_1756383, EPI_ISL_1756384, EPI_ISL_1756385, EPI_ISL_1756386, EPI_ISL_1756387, EPI_ISL_1756388, EPI_ISL_1756389, EPI_ISL_1756390, EPI_ISL_1756391, EPI_ISL_1756392, EPI_ISL_1756393, EPI_ISL_1756394, EPI_ISL_1756395, EPI_ISL_1756396, EPI_ISL_1756397, EPI_ISL_1756398, EPI_ISL_1756399, EPI_ISL_1756400, EPI_ISL_1756401, EPI_ISL_1756402, EPI_ISL_1756403, EPI_ISL_1756404, EPI_ISL_1756405, EPI_ISL_1756406, EPI_ISL_1756407, EPI_ISL_1756408 | | | |
| see above | WESTCHESTER MEDICAL CENTER | Wadsworth Center, New York State Department of Health | Kirsten St. George, Daryl M. Lamson, Alexis Russell, Matthew Shudt, Melissa A Leisner, Jonathan Plitnick, Catharine Prussing, Navjot Singh, John Kelly, Erasmus Schneider, Erica Lasek-Nesselquist |
| EPI_ISL_1756987, EPI_ISL_1756988, EPI_ISL_1756989, EPI_ISL_1756990, EPI_ISL_1756991, EPI_ISL_1756992, EPI_ISL_1756993, EPI_ISL_1756994, EPI_ISL_1756995, EPI_ISL_1756996, EPI_ISL_1756997, EPI_ISL_1756998, EPI_ISL_1756999, EPI_ISL_1757000, EPI_ISL_1757001, EPI_ISL_1757002, EPI_ISL_1757003, EPI_ISL_1757004, EPI_ISL_1757005, EPI_ISL_1757006, EPI_ISL_1757007, EPI_ISL_1757008, EPI_ISL_1757009, EPI_ISL_1757010, EPI_ISL_1757012, EPI_ISL_1757013, EPI_ISL_1757014, EPI_ISL_1757015, EPI_ISL_1757016, EPI_ISL_1757017, EPI_ISL_1757018, EPI_ISL_1757019, EPI_ISL_1757020, EPI_ISL_1757021, EPI_ISL_1757022, EPI_ISL_1757023, EPI_ISL_1757024, EPI_ISL_1757025, EPI_ISL_1757026, EPI_ISL_1757027, EPI_ISL_1757028, EPI_ISL_1757029, EPI_ISL_1757030, EPI_ISL_1757031, EPI_ISL_1757032, EPI_ISL_1757033, EPI_ISL_1757034, EPI_ISL_1757035, EPI_ISL_1757036, EPI_ISL_1757038, EPI_ISL_1757039, EPI_ISL_1757040, EPI_ISL_1757042, EPI_ISL_1757043, EPI_ISL_1757044, EPI_ISL_1757045, EPI_ISL_1757046, EPI_ISL_1757047, EPI_ISL_1757048, EPI_ISL_1757049, EPI_ISL_1757050, EPI_ISL_1757051, EPI_ISL_1757052, EPI_ISL_1757053, EPI_ISL_1757054, EPI_ISL_1757055, EPI_ISL_1757056, EPI_ISL_1757057, EPI_ISL_1757058, EPI_ISL_1757059, EPI_ISL_1757060, EPI_ISL_1757061, EPI_ISL_1757062, EPI_ISL_1757063, EPI_ISL_1757064, EPI_ISL_1757065, EPI_ISL_1757066, EPI_ISL_1757067, EPI_ISL_1757068, EPI_ISL_1757069, EPI_ISL_1757070, EPI_ISL_1757071, EPI_ISL_1757072, EPI_ISL_1757073, EPI_ISL_1757074, EPI_ISL_1757075, EPI_ISL_1757076, EPI_ISL_1757077, EPI_ISL_1757078, EPI_ISL_1757079, EPI_ISL_1757080, EPI_ISL_1757081, EPI_ISL_1757082, EPI_ISL_1757083, EPI_ISL_1757084, EPI_ISL_1757085, EPI_ISL_1757086, EPI_ISL_1757087, EPI_ISL_1757088, EPI_ISL_1757089, EPI_ISL_1757090, EPI_ISL_1757091, EPI_ISL_1757092, EPI_ISL_1757093, EPI_ISL_1757094, EPI_ISL_1757095, EPI_ISL_1757096, EPI_ISL_1757097, EPI_ISL_1757098, EPI_ISL_1757099, EPI_ISL_1757100, EPI_ISL_1757101, EPI_ISL_1757102, EPI_ISL_1757103, EPI_ISL_1757104, EPI_ISL_1757105, EPI_ISL_1757106, EPI_ISL_1757107, EPI_ISL_1757108, EPI_ISL_1757109, EPI_ISL_1757110, EPI_ISL_1757111, EPI_ISL_1757112, EPI_ISL_1757113, EPI_ISL_1757114, EPI_ISL_1757115, EPI_ISL_1757116, EPI_ISL_1757117, EPI_ISL_1757118, EPI_ISL_1757119, EPI_ISL_1757120, EPI_ISL_1757121 | | | |
| see above | Broad Institute Clinical Research Sequencing Platform | Infectious Disease Program, Broad Institute of Harvard and MIT | Siddle,K.J., Adams,G., Pearlman,L., Gladden-Young,A., Vicente,G., Blumenstiel,B., DeFelice,M., Lee,M., McGovern,S., Lagerborg,K., Rudy,M., DeRuff,K., Carter,A., Normandin,E., Bauer,M., Reilly,S., Tomkins-Tinch,C., Loreth,C., Chaluvadi,S., Meldrim,J., Granger,B., Lemieux,J.E., Birren,B.W., Sabeti,P.C., Larkin,K., Dodge,S., Lennon,N., Madoff,L., Brown,C., Gallagher,G., Smole,S., Park,D.J., Gabriel,S., and MacInnis,B.L. |
| EPI_ISL_1757122, EPI_ISL_1757123, EPI_ISL_1757124, EPI_ISL_1757125, EPI_ISL_1757126, EPI_ISL_1757127, EPI_ISL_1757128, EPI_ISL_1757129, EPI_ISL_1757130, EPI_ISL_1757131, EPI_ISL_1757132, EPI_ISL_1757133, EPI_ISL_1757134, EPI_ISL_1757135, EPI_ISL_1757136, EPI_ISL_1757137, EPI_ISL_1757138, EPI_ISL_1757139, EPI_ISL_1757140, EPI_ISL_1757141, EPI_ISL_1757142, EPI_ISL_1757143, EPI_ISL_1757144, EPI_ISL_1757145, EPI_ISL_1757146, EPI_ISL_1757147, EPI_ISL_1757148, EPI_ISL_1757149, EPI_ISL_1757150, EPI_ISL_1757151, EPI_ISL_1757152, EPI_ISL_1757153, EPI_ISL_1757154, EPI_ISL_1757155, EPI_ISL_1757157, EPI_ISL_1757158, EPI_ISL_1757159, EPI_ISL_1757160, EPI_ISL_1757161, EPI_ISL_1757162, EPI_ISL_1757163, EPI_ISL_1757164, EPI_ISL_1757165, EPI_ISL_1757166, EPI_ISL_1757167, EPI_ISL_1757168, EPI_ISL_1757169, EPI_ISL_1757170, EPI_ISL_1757171, EPI_ISL_1757172, EPI_ISL_1757173, EPI_ISL_1757174, EPI_ISL_1757175, EPI_ISL_1757176, EPI_ISL_1757177, EPI_ISL_1757178, EPI_ISL_1757179, EPI_ISL_1757180 | | | |
| see above | Rhode Island Department of Health | Infectious Disease Program, Broad Institute of Harvard and MIT | Siddle,K.J., Azevedo,K., Miller,A., Adams,G., Pearlman,L., Gladden-Young,A., Lagerborg,K., Rudy,M., DeRuff,K., Carter,A., Normandin,E., Bauer,M., Reilly,S., Tomkins-Tinch,C., Loreth,C., Chaluvadi,S., Lemieux,J.E., Birren,B.W., Sabeti,P.C., Huard,R., King,E., Park,D.J., and MacInnis,B.L. |
| EPI_ISL_1757181, EPI_ISL_1757182, EPI_ISL_1757183, EPI_ISL_1757184, EPI_ISL_1757185, EPI_ISL_1757186, EPI_ISL_1757187, EPI_ISL_1757188, EPI_ISL_1757189, EPI_ISL_1757190, EPI_ISL_1757191, EPI_ISL_1757192, EPI_ISL_1757193, EPI_ISL_1757194, EPI_ISL_1757195, EPI_ISL_1757196, EPI_ISL_1757197, EPI_ISL_1757198, EPI_ISL_1757199, EPI_ISL_1757200, EPI_ISL_1757201, EPI_ISL_1757202, EPI_ISL_1757203, EPI_ISL_1757204, EPI_ISL_1757205, EPI_ISL_1757206, EPI_ISL_1757207, EPI_ISL_1757208, EPI_ISL_1757209, EPI_ISL_1757210, EPI_ISL_1757211, EPI_ISL_1757212, EPI_ISL_1757214, EPI_ISL_1757215, EPI_ISL_1757216, EPI_ISL_1757217, EPI_ISL_1757218, EPI_ISL_1757219, EPI_ISL_1757221, EPI_ISL_1757222, EPI_ISL_1757223, EPI_ISL_1757224, EPI_ISL_1757225, EPI_ISL_1757226, EPI_ISL_1757227, EPI_ISL_1757228, EPI_ISL_1757229, EPI_ISL_1757230, EPI_ISL_1757232, EPI_ISL_1757233, EPI_ISL_1757234, EPI_ISL_1757235, EPI_ISL_1757236, EPI_ISL_1757237, EPI_ISL_1757238, EPI_ISL_1757239, EPI_ISL_1757240, EPI_ISL_1757241, EPI_ISL_1757242, EPI_ISL_1757243, EPI_ISL_1757244, EPI_ISL_1757245, EPI_ISL_1757246, EPI_ISL_1757247, EPI_ISL_1757248, EPI_ISL_1757249, EPI_ISL_1757250, EPI_ISL_1757251, EPI_ISL_1757252, EPI_ISL_1757253, EPI_ISL_1757254, EPI_ISL_1757255, EPI_ISL_1757256, EPI_ISL_1757257, EPI_ISL_1757258, EPI_ISL_1757259, EPI_ISL_1757260, EPI_ISL_1757261, EPI_ISL_1757262, EPI_ISL_1757263, EPI_ISL_1757264, EPI_ISL_1757265, EPI_ISL_1757266, EPI_ISL_1757267, EPI_ISL_1757269, EPI_ISL_1757270, EPI_ISL_1757271, EPI_ISL_1757272, EPI_ISL_1757273, EPI_ISL_1757274, EPI_ISL_1757275, EPI_ISL_1757276, EPI_ISL_1757277, EPI_ISL_1757278, EPI_ISL_1757279, EPI_ISL_1757280, EPI_ISL_1757281, EPI_ISL_1757282, EPI_ISL_1757283, EPI_ISL_1757284, EPI_ISL_1757286, EPI_ISL_1757287, EPI_ISL_1757288, EPI_ISL_1757289, EPI_ISL_1757291, EPI_ISL_1757293, EPI_ISL_1757294, EPI_ISL_1757295, EPI_ISL_1757296, EPI_ISL_1757297, EPI_ISL_1757298, EPI_ISL_1757299, EPI_ISL_1757300, EPI_ISL_1757303, EPI_ISL_1757305, EPI_ISL_1757306, EPI_ISL_1757307, EPI_ISL_1757308, EPI_ISL_1757309, EPI_ISL_1757310, EPI_ISL_1757311, EPI_ISL_1757313, EPI_ISL_1757314, EPI_ISL_1757315, EPI_ISL_1757316, EPI_ISL_1757317, EPI_ISL_1757318, EPI_ISL_1757319, EPI_ISL_1757320, EPI_ISL_1757321, EPI_ISL_1757322, EPI_ISL_1757323, EPI_ISL_1757324, EPI_ISL_1757325 | | | |
| see above | Broad Institute Clinical Research Sequencing Platform | Infectious Disease Program, Broad Institute of Harvard and MIT | Siddle,K.J., Adams,G., Pearlman,L., Gladden-Young,A., Vicente,G., Blumenstiel,B., DeFelice,M., Lee,M., McGovern,S., Lagerborg,K., Rudy,M., DeRuff,K., Carter,A., Normandin,E., Bauer,M., Reilly,S., Tomkins-Tinch,C., Loreth,C., Chaluvadi,S., Meldrim,J., Granger,B., Lemieux,J.E., Birren,B.W., Sabeti,P.C., Larkin,K., Dodge,S., Lennon,N., Madoff,L., Brown,C., Gallagher,G., Smole,S., Park,D.J., Gabriel,S., and MacInnis,B.L. |
| EPI_ISL_1757924, EPI_ISL_1757925, EPI_ISL_1757926, EPI_ISL_1757927, EPI_ISL_1757928, EPI_ISL_1757930, EPI_ISL_1757931, EPI_ISL_1757932, EPI_ISL_1757933, EPI_ISL_1757935, EPI_ISL_1757936, EPI_ISL_1757937, EPI_ISL_1757938, EPI_ISL_1757939, EPI_ISL_1757940, EPI_ISL_1757941, EPI_ISL_1757942, EPI_ISL_1757943, EPI_ISL_1757944, EPI_ISL_1757945, EPI_ISL_1757946, EPI_ISL_1757947, EPI_ISL_1757948, EPI_ISL_1757949, EPI_ISL_1757950, EPI_ISL_1757951, EPI_ISL_1757953, EPI_ISL_1757954, EPI_ISL_1757955, EPI_ISL_1757956, EPI_ISL_1757958, EPI_ISL_1757959, EPI_ISL_1757960, EPI_ISL_1757961, EPI_ISL_1757962, EPI_ISL_1757963, EPI_ISL_1757964, EPI_ISL_1757965, EPI_ISL_1757966, EPI_ISL_1757967, EPI_ISL_1757968, EPI_ISL_1757969, EPI_ISL_1757970, EPI_ISL_1757971, EPI_ISL_1757972, EPI_ISL_1757976, EPI_ISL_1757977, EPI_ISL_1757979, EPI_ISL_1757980, EPI_ISL_1757982, EPI_ISL_1757983, EPI_ISL_1757985, EPI_ISL_1757986, EPI_ISL_1757988, EPI_ISL_1757989, EPI_ISL_1757990, EPI_ISL_1757992, EPI_ISL_1757993, EPI_ISL_1757994, EPI_ISL_1757997, EPI_ISL_1757998, EPI_ISL_1757999, EPI_ISL_1758000, EPI_ISL_1758001, EPI_ISL_1758002, EPI_ISL_1758003, EPI_ISL_1758004, EPI_ISL_1758005, EPI_ISL_1758006, EPI_ISL_1758007, EPI_ISL_1758008, EPI_ISL_1758010, EPI_ISL_1758012, EPI_ISL_1758013, EPI_ISL_1758014, EPI_ISL_1758016, EPI_ISL_1758017, EPI_ISL_1758019, EPI_ISL_1758021, EPI_ISL_1758022, EPI_ISL_1758023, EPI_ISL_1758024, EPI_ISL_1758025, EPI_ISL_1758027, EPI_ISL_1758028, EPI_ISL_1758029, EPI_ISL_1758030, EPI_ISL_1758032, EPI_ISL_1758033, EPI_ISL_1758034, EPI_ISL_1758035, EPI_ISL_1758036, EPI_ISL_1758037, EPI_ISL_1758039, EPI_ISL_1758040, EPI_ISL_1758041, EPI_ISL_1758043, EPI_ISL_1758044, EPI_ISL_1758045, EPI_ISL_1758046, EPI_ISL_1758047, EPI_ISL_1758048, EPI_ISL_1758050, EPI_ISL_1758051, EPI_ISL_1758052, EPI_ISL_1758053, EPI_ISL_1758054, EPI_ISL_1758055, EPI_ISL_1758056, EPI_ISL_1758058, EPI_ISL_1758059, EPI_ISL_1758060, EPI_ISL_1758061, EPI_ISL_1758062, EPI_ISL_1758063, EPI_ISL_1758064, EPI_ISL_1758065, EPI_ISL_1758066, EPI_ISL_1758067, EPI_ISL_1758068, EPI_ISL_1758069, EPI_ISL_1758070, EPI_ISL_1758071, EPI_ISL_1758072, EPI_ISL_1758073, EPI_ISL_1758074, EPI_ISL_1758075 | | | |
| see above | Arizona State University | Arizona State University | Rabia Maqsood, LaRinda A. Holland, Peter T. Skidmore, Nicholas J. Mellor, Joy M. Blain, Valerie Harris, Joshua LaBaer, Vel Murugan, Efrem S. Lim |
| EPI_ISL_1758186 | Instituto Nacional de Medicina Genomica | Instituto Nacional de Medicina Genomica | Hidalgo-Miranda A, Mendoza-Vargas A, Reyes-Grajeda JP, Cedro-Tanda A, Alcaraz N, Cisneros-Villanueva M, Gonzalez-Barrera D, Rangel-DeLeon D, Miranda-Ortiz H, Rosas-Escobar P, Canseco Mendez JC, Ramirez-Vega O, Munguia-Garza P, Garcia-Cardenas FJ, Gonzalez-Woge MA, Herrera-Montalvo LA |
| EPI_ISL_1759351, EPI_ISL_1759352, EPI_ISL_1759353, EPI_ISL_1759354, EPI_ISL_1759355, EPI_ISL_1759356, EPI_ISL_1759357, EPI_ISL_1759358, EPI_ISL_1759359, EPI_ISL_1759360, EPI_ISL_1759361, EPI_ISL_1759362, EPI_ISL_1759363, EPI_ISL_1759364, EPI_ISL_1759365, EPI_ISL_1759366, EPI_ISL_1759367, EPI_ISL_1759368, EPI_ISL_1759369, EPI_ISL_1759370, EPI_ISL_1759371, EPI_ISL_1759372, EPI_ISL_1759373 | | | |
| see above | PA Department of Health, Bureau of Laboratories | Pathogen Discovery, Respiratory Viruses Branch, Division of Viral Diseases, Centers for Disease Control and Prevention | Ying Tao, Yan Li, Anna Kelleher, Jing Zhang, Anna Montmayeur, Brian Lynch, Krista Queen, Anna Uehara, Peter Cook, Han Jia Justin Ng, Rachel Marine, Clinton R. Paden, Haibin Wang, Mark Burroughs, Justin Lee, Adam Retchless, Suxiang Tong |
| EPI_ISL_1759374 | OK Public Health Laboratory, Oklahoma State DOH | Pathogen Discovery, Respiratory Viruses Branch, Division of Viral Diseases, Centers for Disease Control and Prevention | Ying Tao, Yan Li, Anna Kelleher, Jing Zhang, Anna Montmayeur, Brian Lynch, Krista Queen, Anna Uehara, Peter Cook, Han Jia Justin Ng, Rachel Marine, Clinton R. Paden, Haibin Wang, Mark Burroughs, Justin Lee, Adam Retchless, Suxiang Tong |
| EPI_ISL_1759375 | LA Office of Public Health Laboratories | Pathogen Discovery, Respiratory Viruses Branch, Division of Viral Diseases, Centers for Disease Control and Prevention | Ying Tao, Yan Li, Anna Kelleher, Jing Zhang, Anna Montmayeur, Brian Lynch, Krista Queen, Anna Uehara, Peter Cook, Han Jia Justin Ng, Rachel Marine, Clinton R. Paden, Haibin Wang, Mark Burroughs, Justin Lee, Adam Retchless, Suxiang Tong |
| EPI_ISL_1759376 | NM Dept. Health, Scientific Laboratory Division | Pathogen Discovery, Respiratory Viruses Branch, Division of Viral Diseases, Centers for Disease Control and Prevention | Ying Tao, Yan Li, Anna Kelleher, Jing Zhang, Anna Montmayeur, Brian Lynch, Krista Queen, Anna Uehara, Peter Cook, Han Jia Justin Ng, Rachel Marine, Clinton R. Paden, Haibin Wang, Mark Burroughs, Justin Lee, Adam Retchless, Suxiang Tong |
| EPI_ISL_1759377, EPI_ISL_1759378 | AR Dept. of Health-PHL, Molecular Diagnostics | Pathogen Discovery, Respiratory Viruses Branch, Division of Viral Diseases, Centers for Disease Control and Prevention | Ying Tao, Yan Li, Anna Kelleher, Jing Zhang, Anna Montmayeur, Brian Lynch, Krista Queen, Anna Uehara, Peter Cook, Han Jia Justin Ng, Rachel Marine, Clinton R. Paden, Haibin Wang, Mark Burroughs, Justin Lee, Adam Retchless, Suxiang Tong |
| EPI_ISL_1759379, EPI_ISL_1759380, EPI_ISL_1759381, EPI_ISL_1759382 | WVDHHR - Office of Laboratory Services | Pathogen Discovery, Respiratory Viruses Branch, Division of Viral Diseases, Centers for Disease Control and Prevention | Ying Tao, Yan Li, Anna Kelleher, Jing Zhang, Anna Montmayeur, Brian Lynch, Krista Queen, Anna Uehara, Peter Cook, Han Jia Justin Ng, Rachel Marine, Clinton R. Paden, Haibin Wang, Mark Burroughs, Justin Lee, Adam Retchless, Suxiang Tong |
| EPI_ISL_1759383, EPI_ISL_1759384, EPI_ISL_1759385 | TN Division of Laboratory Services | Pathogen Discovery, Respiratory Viruses Branch, Division of Viral Diseases, Centers for Disease Control and Prevention | Ying Tao, Yan Li, Anna Kelleher, Jing Zhang, Anna Montmayeur, Brian Lynch, Krista Queen, Anna Uehara, Peter Cook, Han Jia Justin Ng, Rachel Marine, Clinton R. Paden, Haibin Wang, Mark Burroughs, Justin Lee, Adam Retchless, Suxiang Tong |
| EPI_ISL_1759386, EPI_ISL_1759387 | GA Department of Public Health Laboratory | Pathogen Discovery, Respiratory Viruses Branch, Division of Viral Diseases, Centers for Disease Control and Prevention | Ying Tao, Yan Li, Anna Kelleher, Jing Zhang, Anna Montmayeur, Brian Lynch, Krista Queen, Anna Uehara, Peter Cook, Han Jia Justin Ng, Rachel Marine, Clinton R. Paden, Haibin Wang, Mark Burroughs, Justin Lee, Adam Retchless, Suxiang Tong |
| EPI_ISL_1759388, EPI_ISL_1759389 | ME Health and Environmental Testing Laboratory | Pathogen Discovery, Respiratory Viruses Branch, Division of Viral Diseases, Centers for Disease Control and Prevention | Ying Tao, Yan Li, Anna Kelleher, Jing Zhang, Anna Montmayeur, Brian Lynch, Krista Queen, Anna Uehara, Peter Cook, Han Jia Justin Ng, Rachel Marine, Clinton R. Paden, Haibin Wang, Mark Burroughs, Justin Lee, Adam Retchless, Suxiang Tong |
| EPI_ISL_1759390, EPI_ISL_1759391, EPI_ISL_1759392 | NH Dept. of Health and Human Services Public Health Labs | Pathogen Discovery, Respiratory Viruses Branch, Division of Viral Diseases, Centers for Disease Control and Prevention | Ying Tao, Yan Li, Anna Kelleher, Jing Zhang, Anna Montmayeur, Brian Lynch, Krista Queen, Anna Uehara, Peter Cook, Han Jia Justin Ng, Rachel Marine, Clinton R. Paden, Haibin Wang, Mark Burroughs, Justin Lee, Adam Retchless, Suxiang Tong |
| EPI_ISL_1759393 | SD Public Health Laboratory | Pathogen Discovery, Respiratory Viruses Branch, Division of Viral Diseases, Centers for Disease Control and Prevention | Ying Tao, Yan Li, Anna Kelleher, Jing Zhang, Anna Montmayeur, Brian Lynch, Krista Queen, Anna Uehara, Peter Cook, Han Jia Justin Ng, Rachel Marine, Clinton R. Paden, Haibin Wang, Mark Burroughs, Justin Lee, Adam Retchless, Suxiang Tong |
| EPI_ISL_1759394, EPI_ISL_1759395 | ME Health and Environmental Testing Laboratory | Pathogen Discovery, Respiratory Viruses Branch, Division of Viral Diseases, Centers for Disease Control and Prevention | Ying Tao, Yan Li, Anna Kelleher, Jing Zhang, Anna Montmayeur, Brian Lynch, Krista Queen, Anna Uehara, Peter Cook, Han Jia Justin Ng, Rachel Marine, Clinton R. Paden, Haibin Wang, Mark Burroughs, Justin Lee, Adam Retchless, Suxiang Tong |
| EPI_ISL_1759396 | LA Office of Public Health Laboratories | Pathogen Discovery, Respiratory Viruses Branch, Division of Viral Diseases, Centers for Disease Control and Prevention | Ying Tao, Yan Li, Anna Kelleher, Jing Zhang, Anna Montmayeur, Brian Lynch, Krista Queen, Anna Uehara, Peter Cook, Han Jia Justin Ng, Rachel Marine, Clinton R. Paden, Haibin Wang, Mark Burroughs, Justin Lee, Adam Retchless, Suxiang Tong |
| EPI_ISL_1759735, EPI_ISL_1759736, EPI_ISL_1759737 | Washington State Department of Health Public Health Laboratories | Washington State Department of Health Public Health Laboratories | Drew MacKellar, Philip Dykema, Denny Russell, Joenice Gonzalez, Hannah Gray, Geoff Melly, Vanessa De Los Santos, Darren Lucas, JohnAric Peterson, Avi Singh, Rebecca Cao |
| EPI_ISL_1759740, EPI_ISL_1759741, EPI_ISL_1759742, EPI_ISL_1759744, EPI_ISL_1759745, EPI_ISL_1759746 | Seattle Flu Study | Seattle Flu Study | Deborah A. Nickerson, Chris D. Frazar, Jover Lee, Benjamin Pelle, Erica Ryke, Matthew Richardson, Amanda Adler, Elisabeth Brandstetter, Peter D. Han, Kairsten Fay, Misja Ilcisin, Kirsten Lacombe, Thomas R. Sibley, Melissa Truong, Caitlin R. Wolf, Michael Boeckh, Janet A. Englund, Michael Famulare, Barry R. Lutz, Mark J. Rieder, Lea M. Starita, Matthew Thompson, Jay Shendure, Trevor Bedford, Helen Y. Chu |
| EPI_ISL_1759748, EPI_ISL_1759749, EPI_ISL_1759750, EPI_ISL_1759751 | Minnesota Department of Health, Public Health Laboratory | Minnesota Department of Health, Public Health Laboratory | Alexandra Lorentz, Jacob Garfin, Matt Plumb, and Xiong Wang |
| EPI_ISL_1759752, EPI_ISL_1759753, EPI_ISL_1759754, EPI_ISL_1759755, EPI_ISL_1759756, EPI_ISL_1759757, EPI_ISL_1759758, EPI_ISL_1759759, EPI_ISL_1759760, EPI_ISL_1759761, EPI_ISL_1759762, EPI_ISL_1759763, EPI_ISL_1759764, EPI_ISL_1759765, EPI_ISL_1759766 | | | |

| | | | |
|---|---|---|---|
| see above | Mayo Clinic & Mayo Clinic Laboratories | Minnesota Department of Health, Public Health Laboratory | Alexandra Lorentz, Jacob Garfin, Matt Plumb, and Xiong Wang |
| EPI_ISL_1759767, EPI_ISL_1759768, EPI_ISL_1759769, EPI_ISL_1759770, EPI_ISL_1759771, EPI_ISL_1759772, EPI_ISL_1759773, EPI_ISL_1759774, EPI_ISL_1759775, EPI_ISL_1759776, EPI_ISL_1759777, EPI_ISL_1759778, EPI_ISL_1759779, EPI_ISL_1759780, EPI_ISL_1759781, EPI_ISL_1759782, EPI_ISL_1759783, EPI_ISL_1759784, EPI_ISL_1759785, EPI_ISL_1759786, EPI_ISL_1759787, EPI_ISL_1759788, EPI_ISL_1759789, EPI_ISL_1759790, EPI_ISL_1759791, EPI_ISL_1759792, EPI_ISL_1759793, EPI_ISL_1759794, EPI_ISL_1759795, EPI_ISL_1759796, EPI_ISL_1759797, EPI_ISL_1759798, EPI_ISL_1759799, EPI_ISL_1759800, EPI_ISL_1759801, EPI_ISL_1759802, EPI_ISL_1759803, EPI_ISL_1759804 | | | |
| see above | Centracare Laboratory Services | Minnesota Department of Health, Public Health Laboratory | Alexandra Lorentz, Jacob Garfin, Matt Plumb, and Xiong Wang |
| EPI_ISL_1759805, EPI_ISL_1759806, EPI_ISL_1759807, EPI_ISL_1759808, EPI_ISL_1759809, EPI_ISL_1759810, EPI_ISL_1759811 | M Health Fairview | Minnesota Department of Health, Public Health Laboratory | Alexandra Lorentz, Jacob Garfin, Matt Plumb, and Xiong Wang |
| EPI_ISL_1760026, EPI_ISL_1760027, EPI_ISL_1760028, EPI_ISL_1760029, EPI_ISL_1760030, EPI_ISL_1760031, EPI_ISL_1760032, EPI_ISL_1760033, EPI_ISL_1760034, EPI_ISL_1760035, EPI_ISL_1760036, EPI_ISL_1760037, EPI_ISL_1760038, EPI_ISL_1760039, EPI_ISL_1760040, EPI_ISL_1760041, EPI_ISL_1760042, EPI_ISL_1760043, EPI_ISL_1760044, EPI_ISL_1760045, EPI_ISL_1760046, EPI_ISL_1760047, EPI_ISL_1760048, EPI_ISL_1760049, EPI_ISL_1760050, EPI_ISL_1760051, EPI_ISL_1760052, EPI_ISL_1760053, EPI_ISL_1760054, EPI_ISL_1760055, EPI_ISL_1760056, EPI_ISL_1760057, EPI_ISL_1760058, EPI_ISL_1760059, EPI_ISL_1760060, EPI_ISL_1760061, EPI_ISL_1760062, EPI_ISL_1760063, EPI_ISL_1760064, EPI_ISL_1760065, EPI_ISL_1760066, EPI_ISL_1760067, EPI_ISL_1760068, EPI_ISL_1760069, EPI_ISL_1760071, EPI_ISL_1760072, EPI_ISL_1760073, EPI_ISL_1760074, EPI_ISL_1760075, EPI_ISL_1760076, EPI_ISL_1760077, EPI_ISL_1760078, EPI_ISL_1760079, EPI_ISL_1760080, EPI_ISL_1760081, EPI_ISL_1760082, EPI_ISL_1760083, EPI_ISL_1760084, EPI_ISL_1760085, EPI_ISL_1760086, EPI_ISL_1760087, EPI_ISL_1760088, EPI_ISL_1760089, EPI_ISL_1760090, EPI_ISL_1760091, EPI_ISL_1760092, EPI_ISL_1760093, EPI_ISL_1760094, EPI_ISL_1760095, EPI_ISL_1760096, EPI_ISL_1760097, EPI_ISL_1760098, EPI_ISL_1760099, EPI_ISL_1760100, EPI_ISL_1760101, EPI_ISL_1760102, EPI_ISL_1760104, EPI_ISL_1760105, EPI_ISL_1760106, EPI_ISL_1760107, EPI_ISL_1760108, EPI_ISL_1760109, EPI_ISL_1760110, EPI_ISL_1760112, EPI_ISL_1760113, EPI_ISL_1760114 | | | |
| see above | Texas Department of State Health Services (TXDSHS) | Texas Department of State Health Services (TXDSHS) | Rashmi Tuladhar, Bonnie Oh, Jenny Zhang, Maliha Rahman, Mayela Pedrueza, Anita Pokharel, Lorraine Rodriguez, Myong Koag, Chun Wang, Rachel Lee, Grace Kubin |
| EPI_ISL_1760175, EPI_ISL_1760176, EPI_ISL_1760177, EPI_ISL_1760179, EPI_ISL_1760180, EPI_ISL_1760181, EPI_ISL_1760182, EPI_ISL_1760184, EPI_ISL_1760186, EPI_ISL_1760187, EPI_ISL_1760190, EPI_ISL_1760191, EPI_ISL_1760192, EPI_ISL_1760193, EPI_ISL_1760196, EPI_ISL_1760197, EPI_ISL_1760198, EPI_ISL_1760199, EPI_ISL_1760200 | | | |
| see above | Wichita State University - Molecular Diagnostics Lab | Kansas Health and Environmental Lab | Mike Grose, Jonathan Barnell, Ben Olsen, and Phil Adam |
| EPI_ISL_1760201, EPI_ISL_1760202, EPI_ISL_1760204, EPI_ISL_1760205, EPI_ISL_1760206, EPI_ISL_1760207, EPI_ISL_1760208 | Kansas Health and Environmental Lab | Kansas Health and Environmental Lab | Mike Grose, Jonathan Barnell, Ben Olsen, and Phil Adam |
| EPI_ISL_1760210, EPI_ISL_1760211, EPI_ISL_1760212, EPI_ISL_1760213, EPI_ISL_1760214, EPI_ISL_1760215, EPI_ISL_1760216, EPI_ISL_1760217, EPI_ISL_1760218, EPI_ISL_1760219, EPI_ISL_1760220, EPI_ISL_1760221, EPI_ISL_1760222, EPI_ISL_1760223, EPI_ISL_1760224, EPI_ISL_1760225, EPI_ISL_1760226, EPI_ISL_1760227, EPI_ISL_1760228, EPI_ISL_1760229, EPI_ISL_1760230, EPI_ISL_1760231, EPI_ISL_1760232, EPI_ISL_1760233, EPI_ISL_1760234, EPI_ISL_1760235, EPI_ISL_1760236, EPI_ISL_1760237, EPI_ISL_1760238, EPI_ISL_1760239, EPI_ISL_1760240, EPI_ISL_1760241, EPI_ISL_1760242, EPI_ISL_1760243, EPI_ISL_1760244, EPI_ISL_1760245, EPI_ISL_1760246, EPI_ISL_1760247, EPI_ISL_1760248, EPI_ISL_1760249, EPI_ISL_1760250, EPI_ISL_1760251, EPI_ISL_1760252, EPI_ISL_1760253, EPI_ISL_1760254, EPI_ISL_1760255, EPI_ISL_1760256, EPI_ISL_1760257, EPI_ISL_1760258, EPI_ISL_1760259, EPI_ISL_1760260, EPI_ISL_1760261, EPI_ISL_1760262, EPI_ISL_1760263, EPI_ISL_1760264, EPI_ISL_1760265, EPI_ISL_1760266, EPI_ISL_1760267, EPI_ISL_1760268, EPI_ISL_1760269, EPI_ISL_1760270, EPI_ISL_1760271, EPI_ISL_1760272, EPI_ISL_1760273, EPI_ISL_1760274, EPI_ISL_1760275, EPI_ISL_1760276, EPI_ISL_1760277, EPI_ISL_1760278, EPI_ISL_1760279, EPI_ISL_1760280, EPI_ISL_1760281, EPI_ISL_1760282, EPI_ISL_1760283, EPI_ISL_1760284, EPI_ISL_1760285, EPI_ISL_1760286, EPI_ISL_1760287, EPI_ISL_1760288, EPI_ISL_1760289, EPI_ISL_1760290, EPI_ISL_1760291, EPI_ISL_1760292, EPI_ISL_1760293, EPI_ISL_1760294, EPI_ISL_1760295 | | | |
| see above | Centers for Disease Control and Prevention, Dengue Branch | Centers for Disease Control and Prevention, Dengue Branch | Gilberto A. Santiago, Glenda Gonzalez, Betzabel Flores, Keyla Charriez, Gabriela Paz-Bailey, Jorge L. Munoz-Jordan |
| EPI_ISL_1760296, EPI_ISL_1760297, EPI_ISL_1760298, EPI_ISL_1760299 | Puerto Rico Department of Health | Centers for Disease Control and Prevention, Dengue Branch | Gilberto A. Santiago, Glenda Gonzalez, Betzabel Flores, Keyla Charriez, Gabriela Paz-Bailey, Jorge L. Munoz-Jordan |
| EPI_ISL_1760301, EPI_ISL_1760302, EPI_ISL_1760303, EPI_ISL_1760304, EPI_ISL_1760305, EPI_ISL_1760306, EPI_ISL_1760307, EPI_ISL_1760308, EPI_ISL_1760309, EPI_ISL_1760310, EPI_ISL_1760312, EPI_ISL_1760313, EPI_ISL_1760315, EPI_ISL_1760316, EPI_ISL_1760317, EPI_ISL_1760318, EPI_ISL_1760319, EPI_ISL_1760320, EPI_ISL_1760321, EPI_ISL_1760322, EPI_ISL_1760323, EPI_ISL_1760324, EPI_ISL_1760325, EPI_ISL_1760326, EPI_ISL_1760328, EPI_ISL_1760329, EPI_ISL_1760330, EPI_ISL_1760331, EPI_ISL_1760332, EPI_ISL_1760333, EPI_ISL_1760334, EPI_ISL_1760335, EPI_ISL_1760336, EPI_ISL_1760337, EPI_ISL_1760338, EPI_ISL_1760339, EPI_ISL_1760340, EPI_ISL_1760341, EPI_ISL_1760342, EPI_ISL_1760343, EPI_ISL_1760344, EPI_ISL_1760345, EPI_ISL_1760346, EPI_ISL_1760348, EPI_ISL_1760349, EPI_ISL_1760350, EPI_ISL_1760351, EPI_ISL_1760352, EPI_ISL_1760353, EPI_ISL_1760354, EPI_ISL_1760355, EPI_ISL_1760356, EPI_ISL_1760357, EPI_ISL_1760358, EPI_ISL_1760360, EPI_ISL_1760361, EPI_ISL_1760362, EPI_ISL_1760363, EPI_ISL_1760366, EPI_ISL_1760367, EPI_ISL_1760368, EPI_ISL_1760370, EPI_ISL_1760371, EPI_ISL_1760372, EPI_ISL_1760373, EPI_ISL_1760374, EPI_ISL_1760376, EPI_ISL_1760377, EPI_ISL_1760378 | | | |
| see above | Virginia Division of Consolidated Laboratory Services | Virginia Division of Consolidated Laboratory Services | Virginia DCLS |
| EPI_ISL_1760557 | WI State Laboratory of Hygiene | Pathogen Discovery, Respiratory Viruses Branch, Division of Viral Diseases, Centers for Disease Control and Prevention | Ying Tao, Yan Li, Anna Kelleher, Jing Zhang, Anna Montmayeur, Brian Lynch, Krista Queen, Anna Uehara, Peter Cook, Han Jia Justin Ng, Rachel Marine, Clinton R. Paden, Haibin Wang, Mark Burroughs, Justin Lee, Adam Retchless, Suxiang Tong |
| EPI_ISL_1760559, EPI_ISL_1760560, EPI_ISL_1760562, EPI_ISL_1760563, EPI_ISL_1760564, EPI_ISL_1760565, EPI_ISL_1760566, EPI_ISL_1760567, EPI_ISL_1760569, EPI_ISL_1760570, EPI_ISL_1760571, EPI_ISL_1760572, EPI_ISL_1760573, EPI_ISL_1760574, EPI_ISL_1760575, EPI_ISL_1760577, EPI_ISL_1760578, EPI_ISL_1760579, EPI_ISL_1760580, EPI_ISL_1760581, EPI_ISL_1760582, EPI_ISL_1760583, EPI_ISL_1760584, EPI_ISL_1760585, EPI_ISL_1760586, EPI_ISL_1760587, EPI_ISL_1760588, EPI_ISL_1760589, EPI_ISL_1760590, EPI_ISL_1760591, EPI_ISL_1760592, EPI_ISL_1760593, EPI_ISL_1760594, EPI_ISL_1760595, EPI_ISL_1760596, EPI_ISL_1760597, EPI_ISL_1760598, EPI_ISL_1760599, EPI_ISL_1760600, EPI_ISL_1760601, EPI_ISL_1760602, EPI_ISL_1760603, EPI_ISL_1760604, EPI_ISL_1760605, EPI_ISL_1760606, EPI_ISL_1760607, EPI_ISL_1760608, EPI_ISL_1760609, EPI_ISL_1760610, EPI_ISL_1760611, EPI_ISL_1760612, EPI_ISL_1760613 | | | |
| see above | TXDSHS | TXDSHS | Rashmi Tuladhar, Bonnie Oh, Jenny Zhang, Maliha Rahman, Mayela Pedrueza, Anita Pokharel, Lorraine Rodriguez, Myong Koag, Chun Wang, Rachel Lee, Grace Kubin |
| EPI_ISL_1760651, EPI_ISL_1760652, EPI_ISL_1760653, EPI_ISL_1760654, EPI_ISL_1760655, EPI_ISL_1760656, EPI_ISL_1760657, EPI_ISL_1760658, EPI_ISL_1760659, EPI_ISL_1760660, EPI_ISL_1760661, EPI_ISL_1760662, EPI_ISL_1760663, EPI_ISL_1760664, EPI_ISL_1760665, EPI_ISL_1760666, EPI_ISL_1760667, EPI_ISL_1760668, EPI_ISL_1760669, EPI_ISL_1760670, EPI_ISL_1760671, EPI_ISL_1760672, EPI_ISL_1760673, EPI_ISL_1760674, EPI_ISL_1760675, EPI_ISL_1760676, EPI_ISL_1760677, EPI_ISL_1760678, EPI_ISL_1760679, EPI_ISL_1760680, EPI_ISL_1760681, EPI_ISL_1760682, EPI_ISL_1760683, EPI_ISL_1760684, EPI_ISL_1760685, EPI_ISL_1760686, EPI_ISL_1760687, EPI_ISL_1760688, EPI_ISL_1760689, EPI_ISL_1760690, EPI_ISL_1760691, EPI_ISL_1760692, EPI_ISL_1760693, EPI_ISL_1760694, EPI_ISL_1760695, EPI_ISL_1760696, EPI_ISL_1760697, EPI_ISL_1760698 | | | |
| see above | Minnesota Department of Health, Public Health Laboratory | Minnesota Department of Health, Public Health Laboratory | Alexandra Lorentz, Jacob Garfin, Matt Plumb, and Xiong Wang |
| EPI_ISL_1760699, EPI_ISL_1760700, EPI_ISL_1760701, EPI_ISL_1760702, EPI_ISL_1760703, EPI_ISL_1760704, EPI_ISL_1760705, EPI_ISL_1760706, EPI_ISL_1760707, EPI_ISL_1760708, EPI_ISL_1760709, EPI_ISL_1760710, EPI_ISL_1760712, EPI_ISL_1760713, EPI_ISL_1760714 | | | |
| see above | M Health Fairview | Minnesota Department of Health, Public Health Laboratory | Alexandra Lorentz, Jacob Garfin, Matt Plumb, and Xiong Wang |
| EPI_ISL_1760715, EPI_ISL_1760716, EPI_ISL_1760717, EPI_ISL_1760718, EPI_ISL_1760719, EPI_ISL_1760720, EPI_ISL_1760722, EPI_ISL_1760723, EPI_ISL_1760724, EPI_ISL_1760725, EPI_ISL_1760726, EPI_ISL_1760727, EPI_ISL_1760728, EPI_ISL_1760729, EPI_ISL_1760730, EPI_ISL_1760732, EPI_ISL_1760733, EPI_ISL_1760734 | | | |
| see above | Mayo Clinic & Mayo Clinic Laboratories | Minnesota Department of Health, Public Health Laboratory | Alexandra Lorentz, Jacob Garfin, Matt Plumb, and Xiong Wang |
| EPI_ISL_1760735, EPI_ISL_1760737, EPI_ISL_1760738, EPI_ISL_1760739, EPI_ISL_1760740, EPI_ISL_1760741, EPI_ISL_1760742, EPI_ISL_1760743, EPI_ISL_1760744, EPI_ISL_1760745, EPI_ISL_1760746, EPI_ISL_1760747, EPI_ISL_1760748, EPI_ISL_1760750, EPI_ISL_1760751, EPI_ISL_1760752, EPI_ISL_1760753, EPI_ISL_1760754, EPI_ISL_1760755, EPI_ISL_1760756, EPI_ISL_1760757, EPI_ISL_1760758, EPI_ISL_1760759, EPI_ISL_1760760, EPI_ISL_1760761, EPI_ISL_1760762, EPI_ISL_1760763, EPI_ISL_1760764, EPI_ISL_1760765, EPI_ISL_1760766, EPI_ISL_1760767, EPI_ISL_1760768, EPI_ISL_1760769, EPI_ISL_1760770, EPI_ISL_1760771, EPI_ISL_1760772, EPI_ISL_1760773, EPI_ISL_1760774, EPI_ISL_1760775, EPI_ISL_1760776, EPI_ISL_1760777, EPI_ISL_1760778, EPI_ISL_1760779, EPI_ISL_1760780, EPI_ISL_1760781, EPI_ISL_1760782, EPI_ISL_1760783, EPI_ISL_1760784, EPI_ISL_1760785, EPI_ISL_1760786, EPI_ISL_1760787, EPI_ISL_1760788, EPI_ISL_1760789, EPI_ISL_1760790, EPI_ISL_1760791, EPI_ISL_1760792, EPI_ISL_1760793, EPI_ISL_1760794, EPI_ISL_1760795, EPI_ISL_1760796, EPI_ISL_1760797, EPI_ISL_1760798, EPI_ISL_1760799, EPI_ISL_1760800, EPI_ISL_1760801, EPI_ISL_1760802, EPI_ISL_1760804 | | | |
| see above | Minnesota Department of Health, Public Health Laboratory | Minnesota Department of Health, Public Health Laboratory | Alexandra Lorentz, Jacob Garfin, Matt Plumb, and Xiong Wang |
| EPI_ISL_1760805, EPI_ISL_1760806, EPI_ISL_1760807, EPI_ISL_1760808, EPI_ISL_1760809, EPI_ISL_1760811, EPI_ISL_1760812 | Mayo Clinic & Mayo Clinic Laboratories | Minnesota Department of Health, Public Health Laboratory | Alexandra Lorentz, Jacob Garfin, Matt Plumb, and Xiong Wang |
| EPI_ISL_1760818, EPI_ISL_1760819, EPI_ISL_1760820, EPI_ISL_1760846, EPI_ISL_1761037, EPI_ISL_1761133, EPI_ISL_1761134, EPI_ISL_1761135, EPI_ISL_1761212, EPI_ISL_1761305, EPI_ISL_1761306, EPI_ISL_1761307, EPI_ISL_1761370, EPI_ISL_1761371, EPI_ISL_1761395, EPI_ISL_1761396, EPI_ISL_1761397, EPI_ISL_1761398, EPI_ISL_1761399, EPI_ISL_1761450, EPI_ISL_1761451, EPI_ISL_1761452, EPI_ISL_1761453, EPI_ISL_1761454, EPI_ISL_1761455, EPI_ISL_1761456, EPI_ISL_1761457, EPI_ISL_1761458, EPI_ISL_1761462, EPI_ISL_1761552, EPI_ISL_1761553, EPI_ISL_1761581, EPI_ISL_1761582, EPI_ISL_1761583, EPI_ISL_1761584, EPI_ISL_1761585, EPI_ISL_1761610, EPI_ISL_1761611, EPI_ISL_1761612, EPI_ISL_1761640, EPI_ISL_1761641, EPI_ISL_1761642 | | | |
| see above | Montana Public Health Laboratory | Montana Public Health Laboratory | Joy Ritter, Michelle Mozer, Carrie Biskupiak, Deborah Gibson |
| EPI_ISL_1761645, EPI_ISL_1761652, EPI_ISL_1761654, EPI_ISL_1761656, EPI_ISL_1761657, EPI_ISL_1761661, EPI_ISL_1761667, EPI_ISL_1761672, EPI_ISL_1761674, EPI_ISL_1761676, EPI_ISL_1761677, EPI_ISL_1761685, EPI_ISL_1761689, EPI_ISL_1761691, EPI_ISL_1761698, EPI_ISL_1761702, EPI_ISL_1761704, EPI_ISL_1761707, EPI_ISL_1761709, EPI_ISL_1761714, EPI_ISL_1761717, EPI_ISL_1761722, EPI_ISL_1761728, EPI_ISL_1761731, EPI_ISL_1761733, EPI_ISL_1761734 | | | |
| see above | Sonora Quest Laboratories | TGen North | "Jolene Bowers, Heather Centner, Chris French, Hayley Yaglom, Ashlyn Pfeiffer, Darrin Lemmer, Dave Engelthaler, The Arizona COVID Genomics Union (ACGU)" |
| EPI_ISL_1761736 | TGen North | TGen North | "Jolene Bowers, Heather Centner, Chris French, Hayley Yaglom, Ashlyn Pfeiffer, Darrin Lemmer, Dave Engelthaler, The Arizona COVID Genomics Union (ACGU)" |
| EPI_ISL_1761740, EPI_ISL_1761741, EPI_ISL_1761747, EPI_ISL_1761748, EPI_ISL_1761753, EPI_ISL_1761756, EPI_ISL_1761757, EPI_ISL_1761759, EPI_ISL_1761761, EPI_ISL_1761762, EPI_ISL_1761764, EPI_ISL_1761766, EPI_ISL_1761769, EPI_ISL_1761771, EPI_ISL_1761775, EPI_ISL_1761776, EPI_ISL_1761777, EPI_ISL_1761778, EPI_ISL_1761781, EPI_ISL_1761782, EPI_ISL_1761791, EPI_ISL_1761794, EPI_ISL_1761798, EPI_ISL_1761799, EPI_ISL_1761802 | | | |
| see above | Sonora Quest Laboratories | TGen North | "Jolene Bowers, Heather Centner, Chris French, Hayley Yaglom, Ashlyn Pfeiffer, Darrin Lemmer, Dave Engelthaler, The Arizona COVID Genomics Union (ACGU)" |
| EPI_ISL_1761807 | TGen North | TGen North | "Jolene Bowers, Heather Centner, Chris French, Hayley Yaglom, Ashlyn Pfeiffer, Darrin Lemmer, Dave Engelthaler, The Arizona COVID Genomics Union (ACGU)" |
| EPI_ISL_1761811, EPI_ISL_1761812, EPI_ISL_1761813, EPI_ISL_1761827, EPI_ISL_1761828, EPI_ISL_1761830, EPI_ISL_1761833, EPI_ISL_1761839, EPI_ISL_1761842, EPI_ISL_1761843, EPI_ISL_1761845, EPI_ISL_1761848, EPI_ISL_1761854, EPI_ISL_1761857, EPI_ISL_1761858, EPI_ISL_1761859, EPI_ISL_1761860, EPI_ISL_1761862, EPI_ISL_1761865 | | | |
| see above | Sonora Quest Laboratories | TGen North | "Jolene Bowers, Heather Centner, Chris French, Hayley Yaglom, Ashlyn Pfeiffer, Darrin Lemmer, Dave Engelthaler, The Arizona COVID Genomics Union (ACGU)" |
| EPI_ISL_1761870 | Montana Public Health Laboratory | Montana Public Health Laboratory | Joy Ritter, Michelle Mozer, Carrie Biskupiak, Deborah Gibson |
| EPI_ISL_1761871, EPI_ISL_1761872, EPI_ISL_1761873, EPI_ISL_1761874, EPI_ISL_1761875, EPI_ISL_1761876, EPI_ISL_1761877, EPI_ISL_1761878, EPI_ISL_1761879, EPI_ISL_1761880, EPI_ISL_1761881, EPI_ISL_1761882, EPI_ISL_1761883, EPI_ISL_1761884, EPI_ISL_1761885, EPI_ISL_1761886, EPI_ISL_1761887, EPI_ISL_1761888, EPI_ISL_1761889, EPI_ISL_1761890, EPI_ISL_1761891, EPI_ISL_1761892, EPI_ISL_1761893, EPI_ISL_1761894, EPI_ISL_1761895, EPI_ISL_1761896, EPI_ISL_1761897 | | | |
| see above | Quest Diagnostics Incorporated | Centers for Disease Control and Prevention | Dakota Howard, Dhwani Batra, Peter W. Cook, Kara Moser, Adrian Paskey, Jason Caravas, Benjamin Rambo-Martin, Shatavia Morrison, Christopher Gulvick, Scott Sammons, Yvette Unoarumhi, Darlene Wagner, Matthew Schmerer, S. H. Rosenthal, A. Gerasimova, R. M. Kagan, B. |

| | | | |
|---|---|---|---|
| | | Division of Viral Diseases, Pathogen Discovery | Anderson, M. Hua, Y. Liu, L.E. Bernstein, K.E. Livingston, A. Perez, I. A. Shlyakhter, R. V. Rolando, R. Owen, P. Tanpaiboon, F. Lacbawan, Clinton R. Paden, Duncan MacCannell |
| EPI_ISL_1761898 | Montana Public Health Laboratory | Montana Public Health Laboratory | Joy Ritter, Michelle Mozer, Carrie Biskupiak, Deborah Gibson |
| EPI_ISL_1761899, EPI_ISL_1761900, EPI_ISL_1761901, EPI_ISL_1761902, EPI_ISL_1761903, EPI_ISL_1761904, EPI_ISL_1761905, EPI_ISL_1761906, EPI_ISL_1761907, EPI_ISL_1761908, EPI_ISL_1761909, EPI_ISL_1761910, EPI_ISL_1761911, EPI_ISL_1761912, EPI_ISL_1761913, EPI_ISL_1761914, EPI_ISL_1761915, EPI_ISL_1761916, EPI_ISL_1761917, EPI_ISL_1761918, EPI_ISL_1761919, EPI_ISL_1761920, EPI_ISL_1761921, EPI_ISL_1761922, EPI_ISL_1761923, EPI_ISL_1761924, EPI_ISL_1761925, EPI_ISL_1761926, EPI_ISL_1761927, EPI_ISL_1761928, EPI_ISL_1761929, EPI_ISL_1761930, EPI_ISL_1761931, EPI_ISL_1761932, EPI_ISL_1761933, EPI_ISL_1761934, EPI_ISL_1761935, EPI_ISL_1761936, EPI_ISL_1761937, EPI_ISL_1761938, EPI_ISL_1761939, EPI_ISL_1761940, EPI_ISL_1761941, EPI_ISL_1761942, EPI_ISL_1761943, EPI_ISL_1761944, EPI_ISL_1761945, EPI_ISL_1761946, EPI_ISL_1761947, EPI_ISL_1761948, EPI_ISL_1761949, EPI_ISL_1761950, EPI_ISL_1761951, EPI_ISL_1761952, EPI_ISL_1761953, EPI_ISL_1761954, EPI_ISL_1761955, EPI_ISL_1761956, EPI_ISL_1761957, EPI_ISL_1761958, EPI_ISL_1761959, EPI_ISL_1761960, EPI_ISL_1761961, EPI_ISL_1761962, EPI_ISL_1761963, EPI_ISL_1761964, EPI_ISL_1761965, EPI_ISL_1761966, EPI_ISL_1761967, EPI_ISL_1761968, EPI_ISL_1761969, EPI_ISL_1761970, EPI_ISL_1761971, EPI_ISL_1761972, EPI_ISL_1761973, EPI_ISL_1761974, EPI_ISL_1761975, EPI_ISL_1761976, EPI_ISL_1761977, EPI_ISL_1761978, EPI_ISL_1761979, EPI_ISL_1761980, EPI_ISL_1761981, EPI_ISL_1761982, EPI_ISL_1761983, EPI_ISL_1761984, EPI_ISL_1761985, EPI_ISL_1761986, EPI_ISL_1761987, EPI_ISL_1761988, EPI_ISL_1761989, EPI_ISL_1761990, EPI_ISL_1761991, EPI_ISL_1761992, EPI_ISL_1761993, EPI_ISL_1761994, EPI_ISL_1761995, EPI_ISL_1761996, EPI_ISL_1761997, EPI_ISL_1761998, EPI_ISL_1761999, EPI_ISL_1762000, EPI_ISL_1762001, EPI_ISL_1762002, EPI_ISL_1762003, EPI_ISL_1762004, EPI_ISL_1762005, EPI_ISL_1762006, EPI_ISL_1762007, EPI_ISL_1762008, EPI_ISL_1762009, EPI_ISL_1762010, EPI_ISL_1762011, EPI_ISL_1762012, EPI_ISL_1762013, EPI_ISL_1762014, EPI_ISL_1762015, EPI_ISL_1762016, EPI_ISL_1762017, EPI_ISL_1762018, EPI_ISL_1762019, EPI_ISL_1762020, EPI_ISL_1762021, EPI_ISL_1762022, EPI_ISL_1762023, EPI_ISL_1762024, EPI_ISL_1762025, EPI_ISL_1762026, EPI_ISL_1762027, EPI_ISL_1762028, EPI_ISL_1762029, EPI_ISL_1762030, EPI_ISL_1762031, EPI_ISL_1762032, EPI_ISL_1762033, EPI_ISL_1762034, EPI_ISL_1762035, EPI_ISL_1762036, EPI_ISL_1762037, EPI_ISL_1762038, EPI_ISL_1762039, EPI_ISL_1762040, EPI_ISL_1762041, EPI_ISL_1762042, EPI_ISL_1762043, EPI_ISL_1762044, EPI_ISL_1762045, EPI_ISL_1762046, EPI_ISL_1762047, EPI_ISL_1762048, EPI_ISL_1762049, EPI_ISL_1762050, EPI_ISL_1762051 | | | |
| see above | Quest Diagnostics Incorporated | Centers for Disease Control and Prevention Division of Viral Diseases, Pathogen Discovery | Dakota Howard, Dhwani Batra, Peter W. Cook, Kara Moser, Adrian Paskey, Jason Caravas, Benjamin Rambo-Martin, Shatavia Morrison, Christopher Gulvick, Scott Sammons, Yvette Unoarumhi, Darlene Wagner, Matthew Schmerer, S. H. Rosenthal, A. Gerasimova, R. M. Kagan, B. Anderson, M. Hua, Y. Liu, L.E. Bernstein, K.E. Livingston, A. Perez, I. A. Shlyakhter, R. V. Rolando, R. Owen, P. Tanpaiboon, F. Lacbawan, Clinton R. Paden, Duncan MacCannell |
| EPI_ISL_1770503, EPI_ISL_1770507, EPI_ISL_1770509, EPI_ISL_1770513, EPI_ISL_1770518, EPI_ISL_1770522, EPI_ISL_1770526, EPI_ISL_1770530, EPI_ISL_1770534, EPI_ISL_1770539, EPI_ISL_1770543, EPI_ISL_1770548, EPI_ISL_1770552, EPI_ISL_1770557, EPI_ISL_1770561, EPI_ISL_1770565, EPI_ISL_1770569, EPI_ISL_1770572, EPI_ISL_1770577, EPI_ISL_1770581, EPI_ISL_1770585, EPI_ISL_1770589, EPI_ISL_1770594, EPI_ISL_1770598, EPI_ISL_1770603, EPI_ISL_1770607, EPI_ISL_1770611, EPI_ISL_1770615, EPI_ISL_1770620, EPI_ISL_1770624, EPI_ISL_1770629, EPI_ISL_1770633, EPI_ISL_1770637, EPI_ISL_1770641, EPI_ISL_1770646, EPI_ISL_1770651, EPI_ISL_1770655, EPI_ISL_1770659, EPI_ISL_1770662, EPI_ISL_1770666, EPI_ISL_1770669, EPI_ISL_1770673, EPI_ISL_1770678, EPI_ISL_1770682, EPI_ISL_1770686, EPI_ISL_1770690, EPI_ISL_1770695, EPI_ISL_1770699, EPI_ISL_1770703, EPI_ISL_1770707, EPI_ISL_1770710, EPI_ISL_1770714, EPI_ISL_1770717, EPI_ISL_1770721, EPI_ISL_1770726, EPI_ISL_1770730, EPI_ISL_1770735, EPI_ISL_1770739, EPI_ISL_1770743, EPI_ISL_1770747, EPI_ISL_1770752, EPI_ISL_1770755, EPI_ISL_1770759, EPI_ISL_1770764, EPI_ISL_1770768, EPI_ISL_1770773, EPI_ISL_1770777, EPI_ISL_1770782, EPI_ISL_1770786, EPI_ISL_1770790, EPI_ISL_1770794, EPI_ISL_1770799, EPI_ISL_1770803, EPI_ISL_1770807, EPI_ISL_1770811, EPI_ISL_1770816, EPI_ISL_1770820, EPI_ISL_1770824, EPI_ISL_1770826, EPI_ISL_1770829, EPI_ISL_1770833, EPI_ISL_1770838, EPI_ISL_1770842, EPI_ISL_1770846, EPI_ISL_1770851, EPI_ISL_1770855, EPI_ISL_1770859, EPI_ISL_1770862, EPI_ISL_1770866, EPI_ISL_1770870, EPI_ISL_1770874, EPI_ISL_1770878, EPI_ISL_1770882, EPI_ISL_1770886, EPI_ISL_1770890, EPI_ISL_1770894, EPI_ISL_1770898, EPI_ISL_1770902, EPI_ISL_1770905, EPI_ISL_1770910, EPI_ISL_1770914, EPI_ISL_1770918, EPI_ISL_1770922, EPI_ISL_1770925, EPI_ISL_1770929, EPI_ISL_1770933, EPI_ISL_1770937, EPI_ISL_1770941, EPI_ISL_1770945, EPI_ISL_1770949, EPI_ISL_1770953, EPI_ISL_1770957, EPI_ISL_1770961, EPI_ISL_1770965, EPI_ISL_1770970, EPI_ISL_1770974, EPI_ISL_1770979, EPI_ISL_1770983, EPI_ISL_1770987, EPI_ISL_1770992, EPI_ISL_1770996, EPI_ISL_1771000, EPI_ISL_1771005, EPI_ISL_1771009, EPI_ISL_1771014, EPI_ISL_1771018, EPI_ISL_1771023, EPI_ISL_1771027, EPI_ISL_1771032, EPI_ISL_1771036, EPI_ISL_1771045, EPI_ISL_1771050, EPI_ISL_1771054, EPI_ISL_1771059, EPI_ISL_1771063, EPI_ISL_1771065, EPI_ISL_1771070, EPI_ISL_1771073, EPI_ISL_1771077, EPI_ISL_1771081, EPI_ISL_1771086, EPI_ISL_1771090, EPI_ISL_1771095, EPI_ISL_1771099, EPI_ISL_1771103, EPI_ISL_1771108, EPI_ISL_1771111, EPI_ISL_1771115, EPI_ISL_1771119, EPI_ISL_1771123, EPI_ISL_1771127, EPI_ISL_1771132, EPI_ISL_1771136, EPI_ISL_1771141, EPI_ISL_1771145, EPI_ISL_1771150, EPI_ISL_1771154, EPI_ISL_1771158, EPI_ISL_1771162, EPI_ISL_1771166, EPI_ISL_1771170, EPI_ISL_1771175, EPI_ISL_1771178, EPI_ISL_1771183, EPI_ISL_1771187, EPI_ISL_1771191, EPI_ISL_1771195, EPI_ISL_1771199, EPI_ISL_1771203, EPI_ISL_1771207, EPI_ISL_1771211, EPI_ISL_1771215, EPI_ISL_1771219, EPI_ISL_1771222, EPI_ISL_1771226, EPI_ISL_1771230, EPI_ISL_1771235, EPI_ISL_1771239, EPI_ISL_1771241, EPI_ISL_1771245, EPI_ISL_1771248, EPI_ISL_1771252, EPI_ISL_1771255, EPI_ISL_1771259, EPI_ISL_1771264, EPI_ISL_1771268, EPI_ISL_1771273, EPI_ISL_1771277, EPI_ISL_1771281, EPI_ISL_1771284, EPI_ISL_1771287, EPI_ISL_1771292, EPI_ISL_1771296, EPI_ISL_1771300, EPI_ISL_1771305, EPI_ISL_1771309, EPI_ISL_1771312, EPI_ISL_1771316, EPI_ISL_1771320, EPI_ISL_1771327, EPI_ISL_1771331, EPI_ISL_1771335, EPI_ISL_1771340, EPI_ISL_1771344, EPI_ISL_1771349, EPI_ISL_1771353, EPI_ISL_1771357, EPI_ISL_1771361, EPI_ISL_1771365, EPI_ISL_1771369, EPI_ISL_1771373, EPI_ISL_1771377, EPI_ISL_1771380, EPI_ISL_1771383, EPI_ISL_1771388, EPI_ISL_1771392, EPI_ISL_1771397, EPI_ISL_1771401, EPI_ISL_1771405, EPI_ISL_1771410, EPI_ISL_1771414, EPI_ISL_1771418, EPI_ISL_1771422, EPI_ISL_1771427, EPI_ISL_1771431, EPI_ISL_1771435, EPI_ISL_1771439, EPI_ISL_1771444, EPI_ISL_1771448, EPI_ISL_1771452, EPI_ISL_1771457, EPI_ISL_1771461, EPI_ISL_1771466, EPI_ISL_1771470, EPI_ISL_1771474, EPI_ISL_1771478, EPI_ISL_1771482, EPI_ISL_1771486, EPI_ISL_1771491, EPI_ISL_1771495, EPI_ISL_1771499, EPI_ISL_1771502, EPI_ISL_1771507, EPI_ISL_1771511, EPI_ISL_1771515, EPI_ISL_1771519, EPI_ISL_1771524, EPI_ISL_1771527, EPI_ISL_1771531, EPI_ISL_1771536, EPI_ISL_1771540, EPI_ISL_1771544, EPI_ISL_1771548, EPI_ISL_1771553, EPI_ISL_1771557, EPI_ISL_1771562, EPI_ISL_1771566, EPI_ISL_1771571, EPI_ISL_1771575, EPI_ISL_1771578, EPI_ISL_1771582, EPI_ISL_1771586, EPI_ISL_1771589, EPI_ISL_1771593, EPI_ISL_1771598, EPI_ISL_1771602, EPI_ISL_1771606, EPI_ISL_1771610, EPI_ISL_1771614, EPI_ISL_1771619, EPI_ISL_1771623, EPI_ISL_1771627, EPI_ISL_1771632, EPI_ISL_1771636, EPI_ISL_1771641, EPI_ISL_1771646, EPI_ISL_1771650 | | | |
| see above | State Laboratories Division, Hawaii State Department of Health | State Laboratories Division, Hawaii State Department of Health | Pamela O'Brien, Drew Kuwazaki, Ayana Garnet, Razvan Sultana, Edward Desmond |
| EPI_ISL_1785513, EPI_ISL_1785514, EPI_ISL_1785515, EPI_ISL_1785516, EPI_ISL_1785517, EPI_ISL_1785518, EPI_ISL_1785519, EPI_ISL_1785520, EPI_ISL_1785521, EPI_ISL_1785523, EPI_ISL_1785524, EPI_ISL_1785525, EPI_ISL_1785526, EPI_ISL_1785527, EPI_ISL_1785528, EPI_ISL_1785530, EPI_ISL_1785531, EPI_ISL_1785532, EPI_ISL_1785533, EPI_ISL_1785534, EPI_ISL_1785535, EPI_ISL_1785536, EPI_ISL_1785537, EPI_ISL_1785538, EPI_ISL_1785539, EPI_ISL_1785540 | | | |
| see above | University of Michigan Clinical Microbiology Laboratory | Lauring Lab, University of Michigan, Department of Microbiology and Immunology | Valesano |
| EPI_ISL_1785541, EPI_ISL_1785542, EPI_ISL_1785544, EPI_ISL_1785547, EPI_ISL_1785548, EPI_ISL_1785549 | Wyoming Public Health Laboratory | Wyoming Public Health Laboratory | Jim Mildenberger, Wanda Manley, Noah Hull, Taylor Fearing, Lynette Gumbleton, Channing Weber, Ashley Norberg, Chayse Rowley, Marley Goetz, Brian Dominguez, Elliot Thomasson, Sam Britz, Cari Sloma, and Rob Christensen |
| EPI_ISL_1785607, EPI_ISL_1785608 | Vanderbilt University Medical Center | Pathogen Discovery, Respiratory Viruses Branch, Division of Viral Diseases, Centers for Disease Control and Prevention | Ying Tao, Yan Li, Anna Kelleher, Jing Zhang, Anna Montmayeur, Brian Lynch, Krista Queen, Anna Uehara, Peter Cook, Han Jia Justin Ng, Rachel Marine, Clinton R. Paden, Haibin Wang, Mark Burroughs, Justin Lee, Adam Retchless, Suxiang Tong |
| EPI_ISL_1785615, EPI_ISL_1785616, EPI_ISL_1785617, EPI_ISL_1785619, EPI_ISL_1785620, EPI_ISL_1785621, EPI_ISL_1785622, EPI_ISL_1785623, EPI_ISL_1785624, EPI_ISL_1785625, EPI_ISL_1785626, EPI_ISL_1785627, EPI_ISL_1785628, EPI_ISL_1785629, EPI_ISL_1785630, EPI_ISL_1785631, EPI_ISL_1785632, EPI_ISL_1785633, EPI_ISL_1785634, EPI_ISL_1785635, EPI_ISL_1785636, EPI_ISL_1785637, EPI_ISL_1785638, EPI_ISL_1785639, EPI_ISL_1785640, EPI_ISL_1785641, EPI_ISL_1785642, EPI_ISL_1785643, EPI_ISL_1785644, EPI_ISL_1785645, EPI_ISL_1785646, EPI_ISL_1785647, EPI_ISL_1785648, EPI_ISL_1785649, EPI_ISL_1785650, EPI_ISL_1785651, EPI_ISL_1785652, EPI_ISL_1785653, EPI_ISL_1785654, EPI_ISL_1785655, EPI_ISL_1785656, EPI_ISL_1785657, EPI_ISL_1785658, EPI_ISL_1785659, EPI_ISL_1785660, EPI_ISL_1785661, EPI_ISL_1785662, EPI_ISL_1785663, EPI_ISL_1785664, EPI_ISL_1785665, EPI_ISL_1785667, EPI_ISL_1785668, EPI_ISL_1785669, EPI_ISL_1785670, EPI_ISL_1785671, EPI_ISL_1785672, EPI_ISL_1785673, EPI_ISL_1785674, EPI_ISL_1785675, EPI_ISL_1785676, EPI_ISL_1785677, EPI_ISL_1785678, EPI_ISL_1785679, EPI_ISL_1785680, EPI_ISL_1785681, EPI_ISL_1785682, EPI_ISL_1785683, EPI_ISL_1785684, EPI_ISL_1785685, EPI_ISL_1785686, EPI_ISL_1785687, EPI_ISL_1785688, EPI_ISL_1785689, EPI_ISL_1785690, EPI_ISL_1785691, EPI_ISL_1785692, EPI_ISL_1785693, EPI_ISL_1785694, EPI_ISL_1785695, EPI_ISL_1785696, EPI_ISL_1785697, EPI_ISL_1785698, EPI_ISL_1785699, EPI_ISL_1785700, EPI_ISL_1785701, EPI_ISL_1785702, EPI_ISL_1785703, EPI_ISL_1785704, EPI_ISL_1785705, EPI_ISL_1785706, EPI_ISL_1785707, EPI_ISL_1785708, EPI_ISL_1785709, EPI_ISL_1785710, EPI_ISL_1785711, EPI_ISL_1785712, EPI_ISL_1785713, EPI_ISL_1785714, EPI_ISL_1785715, EPI_ISL_1785716, EPI_ISL_1785717, EPI_ISL_1785718, EPI_ISL_1785719, EPI_ISL_1785720, EPI_ISL_1785721, EPI_ISL_1785722, EPI_ISL_1785723, EPI_ISL_1785724, EPI_ISL_1785725, EPI_ISL_1785726, EPI_ISL_1785727, EPI_ISL_1785728, EPI_ISL_1785729, EPI_ISL_1785730, EPI_ISL_1785731, EPI_ISL_1785732, EPI_ISL_1785733, EPI_ISL_1785734, EPI_ISL_1785735, EPI_ISL_1785736, EPI_ISL_1785737, EPI_ISL_1785738, EPI_ISL_1785739, EPI_ISL_1785740, EPI_ISL_1785741, EPI_ISL_1785742, EPI_ISL_1785743, EPI_ISL_1785744, EPI_ISL_1785745, EPI_ISL_1785746, EPI_ISL_1785747, EPI_ISL_1785748, EPI_ISL_1785749, EPI_ISL_1785750, EPI_ISL_1785751, EPI_ISL_1785752, EPI_ISL_1785753, EPI_ISL_1785754, EPI_ISL_1785755, EPI_ISL_1785756, EPI_ISL_1785757, EPI_ISL_1785758, EPI_ISL_1785759, EPI_ISL_1785760, EPI_ISL_1785761, EPI_ISL_1785762, EPI_ISL_1785763, EPI_ISL_1785764, EPI_ISL_1785765, EPI_ISL_1785766, EPI_ISL_1785767, EPI_ISL_1785768, EPI_ISL_1785769, EPI_ISL_1785770, EPI_ISL_1785771, EPI_ISL_1785772, EPI_ISL_1785773, EPI_ISL_1785774, EPI_ISL_1785775, EPI_ISL_1785776, EPI_ISL_1785777, EPI_ISL_1785778, EPI_ISL_1785779, EPI_ISL_1785780, EPI_ISL_1785781, EPI_ISL_1785782, EPI_ISL_1785783, EPI_ISL_1785784, EPI_ISL_1785785, EPI_ISL_1785786, EPI_ISL_1785789, EPI_ISL_1785790, EPI_ISL_1785791, EPI_ISL_1785792, EPI_ISL_1785793, EPI_ISL_1785794, EPI_ISL_1785795, EPI_ISL_1785796, EPI_ISL_1785797, EPI_ISL_1785798, EPI_ISL_1785799, EPI_ISL_1785800, EPI_ISL_1785801, EPI_ISL_1785802, EPI_ISL_1785803, EPI_ISL_1785805, EPI_ISL_1785806, EPI_ISL_1785807, EPI_ISL_1785808, EPI_ISL_1785809, EPI_ISL_1785810, EPI_ISL_1785811, EPI_ISL_1785812, EPI_ISL_1785813, EPI_ISL_1785814, EPI_ISL_1785815, EPI_ISL_1785816, EPI_ISL_1785817, EPI_ISL_1785818, EPI_ISL_1785819, EPI_ISL_1785820, EPI_ISL_1785821, EPI_ISL_1785822, EPI_ISL_1785823, EPI_ISL_1785824, EPI_ISL_1785825, EPI_ISL_1785826, EPI_ISL_1785827, EPI_ISL_1785828, EPI_ISL_1785829, EPI_ISL_1785830, EPI_ISL_1785831, EPI_ISL_1785832, EPI_ISL_1785833, EPI_ISL_1785835, EPI_ISL_1785836, EPI_ISL_1785837, EPI_ISL_1785838, EPI_ISL_1785839, EPI_ISL_1785840, EPI_ISL_1785841, EPI_ISL_1785842, EPI_ISL_1785843, EPI_ISL_1785844, EPI_ISL_1785845, EPI_ISL_1785846, EPI_ISL_1785847, EPI_ISL_1785848, EPI_ISL_1785849, EPI_ISL_1785850, EPI_ISL_1785851, EPI_ISL_1785852, EPI_ISL_1785853, EPI_ISL_1785854, EPI_ISL_1785855, EPI_ISL_1785856, EPI_ISL_1785857, EPI_ISL_1785858, EPI_ISL_1785859, EPI_ISL_1785860, EPI_ISL_1785861, EPI_ISL_1785862, EPI_ISL_1785863, EPI_ISL_1785864, EPI_ISL_1785865, EPI_ISL_1785866, EPI_ISL_1785867, EPI_ISL_1785868, EPI_ISL_1785869, EPI_ISL_1785870, EPI_ISL_1785871, EPI_ISL_1785872, EPI_ISL_1785873, EPI_ISL_1785874, EPI_ISL_1785875, EPI_ISL_1785876, EPI_ISL_1785877, EPI_ISL_1785878, EPI_ISL_1785879, EPI_ISL_1785880, EPI_ISL_1785881, EPI_ISL_1785882, EPI_ISL_1785883, EPI_ISL_1785884, EPI_ISL_1785885, EPI_ISL_1785886, EPI_ISL_1785887, EPI_ISL_1785888, EPI_ISL_1785890, EPI_ISL_1785891, EPI_ISL_1785892, EPI_ISL_1785893, EPI_ISL_1785894, EPI_ISL_1785895, EPI_ISL_1785896, EPI_ISL_1785897, EPI_ISL_1785898, EPI_ISL_1785899, EPI_ISL_1785900, EPI_ISL_1785901, EPI_ISL_1785902, EPI_ISL_1785903, EPI_ISL_1785904, EPI_ISL_1785905, EPI_ISL_1785906, EPI_ISL_1785907, EPI_ISL_1785908, EPI_ISL_1785909, EPI_ISL_1785910, EPI_ISL_1785911, EPI_ISL_1785912, EPI_ISL_1785913, EPI_ISL_1785914, EPI_ISL_1785915, EPI_ISL_1785916, EPI_ISL_1785917, EPI_ISL_1785918, EPI_ISL_1785919, EPI_ISL_1785920, EPI_ISL_1785921, EPI_ISL_1785922, EPI_ISL_1785923, EPI_ISL_1785924, EPI_ISL_1785925, EPI_ISL_1785926, EPI_ISL_1785927, EPI_ISL_1785928, EPI_ISL_1785929, EPI_ISL_1785930, EPI_ISL_1785931, EPI_ISL_1785932, EPI_ISL_1785933, EPI_ISL_1785934, EPI_ISL_1785935, EPI_ISL_1785936, EPI_ISL_1785937, EPI_ISL_1785938, EPI_ISL_1785939, EPI_ISL_1785940, EPI_ISL_1785941, EPI_ISL_1785942, EPI_ISL_1785943, EPI_ISL_1785944, EPI_ISL_1785945, EPI_ISL_1785946, EPI_ISL_1785947, EPI_ISL_1785948, EPI_ISL_1785949, EPI_ISL_1785950, EPI_ISL_1785951, EPI_ISL_1785952, EPI_ISL_1785953, EPI_ISL_1785954, EPI_ISL_1785955, EPI_ISL_1785956, EPI_ISL_1785957, EPI_ISL_1785958, EPI_ISL_1785959, EPI_ISL_1785960, EPI_ISL_1785961, EPI_ISL_1785962, EPI_ISL_1785963, EPI_ISL_1785964, EPI_ISL_1785965, EPI_ISL_1785966, EPI_ISL_1785967, EPI_ISL_1785968 | | | |
| see above | UW Virology Lab | UW Virology Lab | Pavitra Roychoudhury, Hong Xie, Lasata Shrestha, Tien V. Nguyen, Shah Mohamed Bakhash, Michelle Lin, Noah R. Baker, Sean Ellis, Meei-Li Huang, Keith R Jerome, Alexander Greninger |
| EPI_ISL_1785969, EPI_ISL_1785970, EPI_ISL_1785971, EPI_ISL_1785972, EPI_ISL_1785973, EPI_ISL_1785974, EPI_ISL_1785975, EPI_ISL_1785976, EPI_ISL_1785977, EPI_ISL_1785978, EPI_ISL_1785979, EPI_ISL_1785980, EPI_ISL_1785981, EPI_ISL_1785982 | | | |
| see above | DC Public Health Lab/ Dept. of Forensic Sciences | DC Public Health Lab/ Dept. of Forensic Sciences | Janis Doss, Scott Nguyen, Elizabeth Zelaya, Sarah Scott, Connie Maza, Monica Mann, Brittany Hamilton, David Payne, Jocelyn Hauser |
| EPI_ISL_1786504, EPI_ISL_1786505, EPI_ISL_1786506, EPI_ISL_1786507, EPI_ISL_1786509, EPI_ISL_1786510, EPI_ISL_1786511, EPI_ISL_1786512, EPI_ISL_1786513, EPI_ISL_1786515, EPI_ISL_1786516, EPI_ISL_1786517, EPI_ISL_1786518, EPI_ISL_1786519, EPI_ISL_1786520, EPI_ISL_1786521, EPI_ISL_1786522, EPI_ISL_1786523, EPI_ISL_1786524, EPI_ISL_1786525, EPI_ISL_1786526, EPI_ISL_1786527, EPI_ISL_1786528, EPI_ISL_1786529, EPI_ISL_1786530, EPI_ISL_1786531, EPI_ISL_1786532, EPI_ISL_1786533, EPI_ISL_1786534, EPI_ISL_1786535, EPI_ISL_1786536, EPI_ISL_1786537, EPI_ISL_1786538, EPI_ISL_1786539, EPI_ISL_1786542, EPI_ISL_1786543, EPI_ISL_1786544, EPI_ISL_1786545, EPI_ISL_1786546, EPI_ISL_1786547, EPI_ISL_1786548, EPI_ISL_1786549, EPI_ISL_1786550, EPI_ISL_1786551, EPI_ISL_1786552, EPI_ISL_1786554, EPI_ISL_1786556, EPI_ISL_1786557, EPI_ISL_1786558 | | | |
| see above | Wyoming Public Health Laboratory | Wyoming Public Health Laboratory | Jim Mildenberger, Wanda Manley, Noah Hull, Taylor Fearing, Lynette Gumbleton, Channing Weber, Ashley Norberg, Chayse Rowley, Marley Goetz, Brian Dominguez, Elliot Thomasson, Sam Britz, Cari Sloma, and Rob Christensen |
| EPI_ISL_1786571, EPI_ISL_1786572 | PA Department of Health, Bureau of Laboratories | Pathogen Discovery, Respiratory Viruses Branch, Division of Viral Diseases, Centers for Disease Control and Prevention | Ying Tao, Yan Li, Anna Kelleher, Jing Zhang, Anna Montmayeur, Brian Lynch, Krista Queen, Anna Uehara, Peter Cook, Han Jia Justin Ng, Rachel Marine, Clinton R. Paden, Haibin Wang, Mark Burroughs, Justin Lee, Adam Retchless, Suxiang Tong |
| EPI_ISL_1786573 | OK Public Health Laboratory, Oklahoma State DOH | Pathogen Discovery, Respiratory Viruses Branch, Division of Viral Diseases, Centers for Disease Control and Prevention | Ying Tao, Yan Li, Anna Kelleher, Jing Zhang, Anna Montmayeur, Brian Lynch, Krista Queen, Anna Uehara, Peter Cook, Han Jia Justin Ng, Rachel Marine, Clinton R. Paden, Haibin Wang, Mark Burroughs, Justin Lee, Adam Retchless, Suxiang Tong |
| EPI_ISL_1786574 | GA Department of Public Health Laboratory | Pathogen Discovery, Respiratory Viruses Branch, Division of Viral Diseases, Centers for Disease Control and Prevention | Ying Tao, Yan Li, Anna Kelleher, Jing Zhang, Anna Montmayeur, Brian Lynch, Krista Queen, Anna Uehara, Peter Cook, Han Jia Justin Ng, Rachel Marine, Clinton R. Paden, Haibin Wang, Mark Burroughs, Justin Lee, Adam Retchless, Suxiang Tong |
| EPI_ISL_1786575, EPI_ISL_1786576, EPI_ISL_1786577, EPI_ISL_1786578, EPI_ISL_1786579, EPI_ISL_1786580, EPI_ISL_1786581, EPI_ISL_1786582, EPI_ISL_1786583, EPI_ISL_1786584, EPI_ISL_1786585, EPI_ISL_1786586, EPI_ISL_1786587, EPI_ISL_1786588, EPI_ISL_1786589, EPI_ISL_1786590, EPI_ISL_1786591, EPI_ISL_1786592, EPI_ISL_1786593, EPI_ISL_1786594, EPI_ISL_1786595, EPI_ISL_1786596, EPI_ISL_1786600, EPI_ISL_1786601, EPI_ISL_1786602, EPI_ISL_1786603, EPI_ISL_1786604, EPI_ISL_1786605, EPI_ISL_1786606, EPI_ISL_1786607, EPI_ISL_1786608, EPI_ISL_1786609, EPI_ISL_1786610, EPI_ISL_1786611, EPI_ISL_1786612, EPI_ISL_1786613, EPI_ISL_1786614, EPI_ISL_1786615, EPI_ISL_1786616, EPI_ISL_1786617, EPI_ISL_1786618, EPI_ISL_1786619, EPI_ISL_1786620, EPI_ISL_1786622, EPI_ISL_1786623, EPI_ISL_1786624, EPI_ISL_1786625, EPI_ISL_1786626 | | | |
| see above | Wyoming Public Health Laboratory | Wyoming Public Health Laboratory | Jim Mildenberger, Wanda Manley, Noah Hull, Taylor Fearing, Lynette Gumbleton, Channing Weber, Ashley Norberg, Chayse Rowley, Marley Goetz, Brian Dominguez, Elliot Thomasson, Sam Britz, Cari Sloma, and Rob Christensen |
| EPI_ISL_1786627, EPI_ISL_1786628, EPI_ISL_1786716, EPI_ISL_1786717, EPI_ISL_1786718 | University of Wisconsin-Madison AIDS Vaccine Research Laboratories | University of Wisconsin-Madison AIDS Vaccine Research Laboratories | Gage Moreno, Katarina Braun, et al. AIDS Vaccine Research Laboratories |
| EPI_ISL_1786719 | IL Dept. of Public Health Springfield Laboratory | Genomics and Discovery, Respiratory Viruses Branch, Division of Viral Diseases, Centers for Disease Control and Prevention | Ying Tao, Yan Li, Anna Kelleher, Jing Zhang, Anna Montmayeur, Brian Lynch, Krista Queen, Anna Uehara, Peter Cook, Han Jia Justin Ng, Rachel Marine, Clinton R. Paden, Haibin Wang, Mark Burroughs, Justin Lee, Adam Retchless, Suxiang Tong |
| EPI_ISL_1786720, EPI_ISL_1786721, EPI_ISL_1786722, EPI_ISL_1786723, EPI_ISL_1786724, EPI_ISL_1786725, EPI_ISL_1786726, EPI_ISL_1786727, EPI_ISL_1786728, EPI_ISL_1786729, EPI_ISL_1786730, EPI_ISL_1786731, EPI_ISL_1786732, EPI_ISL_1786733, EPI_ISL_1786734, EPI_ISL_1786735, EPI_ISL_1786736, EPI_ISL_1786737, EPI_ISL_1786738, EPI_ISL_1786739, EPI_ISL_1786740, EPI_ISL_1786741, EPI_ISL_1786742, EPI_ISL_1786743, EPI_ISL_1786744, EPI_ISL_1786745, EPI_ISL_1786746, EPI_ISL_1786747, EPI_ISL_1786748, EPI_ISL_1786749, EPI_ISL_1786750, EPI_ISL_1786751, EPI_ISL_1786752, EPI_ISL_1786753, EPI_ISL_1786754, EPI_ISL_1786755, EPI_ISL_1786756, EPI_ISL_1786757, EPI_ISL_1786758, EPI_ISL_1786759, EPI_ISL_1786760, EPI_ISL_1786761, EPI_ISL_1786762, EPI_ISL_1786763, EPI_ISL_1786764, EPI_ISL_1786765, EPI_ISL_1786766, EPI_ISL_1786768, EPI_ISL_1786769, EPI_ISL_1786770, EPI_ISL_1786771, EPI_ISL_1786772, EPI_ISL_1786773, EPI_ISL_1786774, EPI_ISL_1786775, EPI_ISL_1786776, EPI_ISL_1786777, EPI_ISL_1786778, EPI_ISL_1786779, EPI_ISL_1786780, EPI_ISL_1786781, EPI_ISL_1786782, EPI_ISL_1786783, EPI_ISL_1786784, EPI_ISL_1786785, EPI_ISL_1786786, EPI_ISL_1786787, EPI_ISL_1786788, EPI_ISL_1786789, EPI_ISL_1786790, EPI_ISL_1786791, EPI_ISL_1786793, EPI_ISL_1786794, EPI_ISL_1786795, EPI_ISL_1786796, EPI_ISL_1786797, EPI_ISL_1786798, EPI_ISL_1786799, EPI_ISL_1786800, EPI_ISL_1786801, EPI_ISL_1786803, EPI_ISL_1786804, EPI_ISL_1786805, EPI_ISL_1786806, EPI_ISL_1786807, EPI_ISL_1786808, EPI_ISL_1786809, EPI_ISL_1786810, EPI_ISL_1786811, EPI_ISL_1786812, EPI_ISL_1786813, EPI_ISL_1786814, EPI_ISL_1786815, EPI_ISL_1786816, EPI_ISL_1786817, EPI_ISL_1786818, EPI_ISL_1786819, EPI_ISL_1786820, EPI_ISL_1786821, EPI_ISL_1786822, EPI_ISL_1786823, EPI_ISL_1786824, EPI_ISL_1786825, EPI_ISL_1786826, EPI_ISL_1786827, EPI_ISL_1786828, EPI_ISL_1786829, EPI_ISL_1786830, EPI_ISL_1786831, EPI_ISL_1786832, EPI_ISL_1786833, EPI_ISL_1786834, EPI_ISL_1786835, EPI_ISL_1786836, EPI_ISL_1786837, EPI_ISL_1786838, EPI_ISL_1786839, EPI_ISL_1786840, EPI_ISL_1786841, EPI_ISL_1786842, EPI_ISL_1786843, EPI_ISL_1786844, EPI_ISL_1786845, EPI_ISL_1786846, EPI_ISL_1786847, EPI_ISL_1786848, EPI_ISL_1786849, EPI_ISL_1786850, EPI_ISL_1786851, EPI_ISL_1786852, EPI_ISL_1786853, EPI_ISL_1786854, EPI_ISL_1786855, EPI_ISL_1786856, EPI_ISL_1786857, EPI_ISL_1786858, EPI_ISL_1786859, EPI_ISL_1786860, EPI_ISL_1786861, EPI_ISL_1786862, EPI_ISL_1786863, EPI_ISL_1786864, EPI_ISL_1786865, EPI_ISL_1786866, EPI_ISL_1786867, EPI_ISL_1786868, EPI_ISL_1786869, EPI_ISL_1786870, EPI_ISL_1786871, EPI_ISL_1786872, EPI_ISL_1786873, EPI_ISL_1786874, EPI_ISL_1786875, EPI_ISL_1786876, EPI_ISL_1786877, EPI_ISL_1786878, EPI_ISL_1786879, EPI_ISL_1786880, EPI_ISL_1786881, EPI_ISL_1786882, EPI_ISL_1786883, EPI_ISL_1786884, EPI_ISL_1786885, EPI_ISL_1786886, EPI_ISL_1786887, EPI_ISL_1786888, EPI_ISL_1786889, EPI_ISL_1786890, EPI_ISL_1786891, EPI_ISL_1786892, EPI_ISL_1786893, EPI_ISL_1786894, EPI_ISL_1786895, EPI_ISL_1786896, EPI_ISL_1786897, EPI_ISL_1786898, EPI_ISL_1786899, EPI_ISL_1786900, EPI_ISL_1786901, EPI_ISL_1786902, EPI_ISL_1786903, EPI_ISL_1786904, EPI_ISL_1786905, EPI_ISL_1786906, EPI_ISL_1786907, EPI_ISL_1786908, EPI_ISL_1786909, EPI_ISL_1786910, EPI_ISL_1786911, EPI_ISL_1786912, EPI_ISL_1786913, EPI_ISL_1786914, EPI_ISL_1786915, EPI_ISL_1786916, EPI_ISL_1786917, EPI_ISL_1786918, EPI_ISL_1786919, EPI_ISL_1786920, EPI_ISL_1786921, EPI_ISL_1786922, EPI_ISL_1786923, EPI_ISL_1786924, EPI_ISL_1786925, EPI_ISL_1786926, EPI_ISL_1786927, EPI_ISL_1786928, EPI_ISL_1786929, EPI_ISL_1786930, EPI_ISL_1786931, EPI_ISL_1786932, EPI_ISL_1786933, EPI_ISL_1786934, EPI_ISL_1786935, EPI_ISL_1786936, EPI_ISL_1786937, EPI_ISL_1786938, EPI_ISL_1786939, EPI_ISL_1786940, EPI_ISL_1786941, EPI_ISL_1786942, EPI_ISL_1786943, EPI_ISL_1786944, EPI_ISL_1786945, EPI_ISL_1786946, EPI_ISL_1786947, EPI_ISL_1786948, EPI_ISL_1786949, EPI_ISL_1786950, EPI_ISL_1786951, EPI_ISL_1786952, EPI_ISL_1786953, EPI_ISL_1786954, EPI_ISL_1786955, EPI_ISL_1786956, EPI_ISL_1786957, EPI_ISL_1786958, EPI_ISL_1786959, EPI_ISL_1786960, EPI_ISL_1786961, EPI_ISL_1786962, EPI_ISL_1786963, EPI_ISL_1786964, EPI_ISL_1786965, EPI_ISL_1786966, EPI_ISL_1786967, EPI_ISL_1786968, EPI_ISL_1786969, EPI_ISL_1786970, EPI_ISL_1786971, EPI_ISL_1786972, EPI_ISL_1786973, EPI_ISL_1786974, EPI_ISL_1786975, EPI_ISL_1786976, EPI_ISL_1786977, EPI_ISL_1786978, EPI_ISL_1786980, EPI_ISL_1786981, EPI_ISL_1786982, EPI_ISL_1786983, EPI_ISL_1786984, EPI_ISL_1786985, EPI_ISL_1786986, EPI_ISL_1786987, EPI_ISL_1786988, EPI_ISL_1786989, EPI_ISL_1786990, EPI_ISL_1786991, EPI_ISL_1786992, EPI_ISL_1786993, EPI_ISL_1786994, EPI_ISL_1786995, EPI_ISL_1786996, EPI_ISL_1786997, EPI_ISL_1786998, EPI_ISL_1786999, EPI_ISL_1787000, EPI_ISL_1787001, EPI_ISL_1787002, EPI_ISL_1787003, EPI_ISL_1787004, EPI_ISL_1787005, EPI_ISL_1787006, EPI_ISL_1787007, EPI_ISL_1787008, EPI_ISL_1787009, | | | |

| | | | |
|---|---|---|---|
| EPI_ISL_1787010, EPI_ISL_1787011, EPI_ISL_1787012, EPI_ISL_1787013, EPI_ISL_1787014, EPI_ISL_1787015, EPI_ISL_1787016, EPI_ISL_1787017, EPI_ISL_1787018, EPI_ISL_1787019, EPI_ISL_1787020, EPI_ISL_1787021, EPI_ISL_1787022, EPI_ISL_1787023, EPI_ISL_1787024, EPI_ISL_1787025, EPI_ISL_1787026, EPI_ISL_1787027, EPI_ISL_1787028, EPI_ISL_1787029, EPI_ISL_1787030, EPI_ISL_1787031, EPI_ISL_1787032, EPI_ISL_1787033, EPI_ISL_1787034, EPI_ISL_1787035, EPI_ISL_1787036, EPI_ISL_1787037, EPI_ISL_1787038, EPI_ISL_1787039, EPI_ISL_1787040, EPI_ISL_1787041, EPI_ISL_1787042, EPI_ISL_1787043, EPI_ISL_1787044, EPI_ISL_1787045, EPI_ISL_1787046, EPI_ISL_1787047, EPI_ISL_1787048, EPI_ISL_1787049, EPI_ISL_1787050, EPI_ISL_1787051, EPI_ISL_1787052, EPI_ISL_1787053, EPI_ISL_1787054, EPI_ISL_1787055, EPI_ISL_1787056, EPI_ISL_1787057, EPI_ISL_1787058, EPI_ISL_1787059, EPI_ISL_1787060, EPI_ISL_1787061, EPI_ISL_1787062, EPI_ISL_1787063, EPI_ISL_1787064, EPI_ISL_1787065, EPI_ISL_1787066, EPI_ISL_1787067, EPI_ISL_1787068, EPI_ISL_1787069, EPI_ISL_1787070, EPI_ISL_1787071, EPI_ISL_1787072, EPI_ISL_1787073, EPI_ISL_1787074, EPI_ISL_1787075, EPI_ISL_1787076, EPI_ISL_1787077, EPI_ISL_1787078, EPI_ISL_1787079, EPI_ISL_1787080, EPI_ISL_1787081, EPI_ISL_1787082, EPI_ISL_1787083, EPI_ISL_1787084, EPI_ISL_1787085, EPI_ISL_1787086, EPI_ISL_1787087, EPI_ISL_1787088, EPI_ISL_1787089, EPI_ISL_1787090, EPI_ISL_1787091, EPI_ISL_1787092, EPI_ISL_1787093, EPI_ISL_1787094, EPI_ISL_1787095, EPI_ISL_1787096, EPI_ISL_1787097, EPI_ISL_1787098, EPI_ISL_1787099, EPI_ISL_1787100, EPI_ISL_1787101, EPI_ISL_1787102, EPI_ISL_1787103, EPI_ISL_1787104, EPI_ISL_1787105, EPI_ISL_1787106, EPI_ISL_1787107, EPI_ISL_1787108, EPI_ISL_1787109, EPI_ISL_1787110, EPI_ISL_1787111, EPI_ISL_1787112, EPI_ISL_1787113, EPI_ISL_1787114, EPI_ISL_1787115, EPI_ISL_1787116, EPI_ISL_1787117, EPI_ISL_1787118, EPI_ISL_1787119, EPI_ISL_1787120, EPI_ISL_1787121, EPI_ISL_1787122, EPI_ISL_1787123, EPI_ISL_1787124, EPI_ISL_1787125, EPI_ISL_1787126, EPI_ISL_1787127, EPI_ISL_1787128, EPI_ISL_1787129, EPI_ISL_1787130, EPI_ISL_1787131, EPI_ISL_1787132, EPI_ISL_1787134, EPI_ISL_1787135, EPI_ISL_1787136, EPI_ISL_1787137, EPI_ISL_1787138, EPI_ISL_1787139, EPI_ISL_1787140, EPI_ISL_1787141, EPI_ISL_1787142, EPI_ISL_1787143, EPI_ISL_1787144, EPI_ISL_1787145, EPI_ISL_1787146, EPI_ISL_1787147, EPI_ISL_1787148, EPI_ISL_1787149, EPI_ISL_1787150, EPI_ISL_1787151, EPI_ISL_1787152, EPI_ISL_1787153, EPI_ISL_1787154, EPI_ISL_1787155, EPI_ISL_1787156, EPI_ISL_1787157, EPI_ISL_1787158, EPI_ISL_1787159, EPI_ISL_1787160, EPI_ISL_1787161, EPI_ISL_1787162, EPI_ISL_1787163, EPI_ISL_1787164, EPI_ISL_1787165, EPI_ISL_1787166, EPI_ISL_1787167, EPI_ISL_1787168, EPI_ISL_1787169, EPI_ISL_1787170, EPI_ISL_1787171, EPI_ISL_1787172, EPI_ISL_1787173, EPI_ISL_1787174, EPI_ISL_1787175, EPI_ISL_1787176, EPI_ISL_1787177, EPI_ISL_1787178, EPI_ISL_1787179, EPI_ISL_1787180, EPI_ISL_1787181, EPI_ISL_1787182, EPI_ISL_1787183, EPI_ISL_1787184, EPI_ISL_1787185, EPI_ISL_1787186, EPI_ISL_1787187, EPI_ISL_1787188, EPI_ISL_1787189, EPI_ISL_1787190, EPI_ISL_1787191, EPI_ISL_1787192, EPI_ISL_1787193, EPI_ISL_1787194, EPI_ISL_1787195, EPI_ISL_1787196, EPI_ISL_1787197, EPI_ISL_1787198, EPI_ISL_1787199, EPI_ISL_1787200, EPI_ISL_1787201, EPI_ISL_1787202, EPI_ISL_1787203, EPI_ISL_1787204, EPI_ISL_1787205, EPI_ISL_1787206, EPI_ISL_1787207, EPI_ISL_1787208, EPI_ISL_1787209, EPI_ISL_1787210, EPI_ISL_1787211, EPI_ISL_1787212, EPI_ISL_1787213, EPI_ISL_1787214, EPI_ISL_1787215, EPI_ISL_1787216, EPI_ISL_1787217, EPI_ISL_1787218, EPI_ISL_1787219, EPI_ISL_1787220, EPI_ISL_1787221, EPI_ISL_1787222, EPI_ISL_1787223, EPI_ISL_1787224, EPI_ISL_1787225, EPI_ISL_1787226, EPI_ISL_1787227, EPI_ISL_1787228, EPI_ISL_1787229, EPI_ISL_1787230, EPI_ISL_1787231, EPI_ISL_1787232, EPI_ISL_1787233, EPI_ISL_1787234, EPI_ISL_1787235, EPI_ISL_1787236, EPI_ISL_1787237, EPI_ISL_1787238, EPI_ISL_1787239, EPI_ISL_1787240, EPI_ISL_1787241, EPI_ISL_1787242, EPI_ISL_1787243, EPI_ISL_1787244, EPI_ISL_1787245, EPI_ISL_1787246, EPI_ISL_1787247, EPI_ISL_1787248, EPI_ISL_1787249, EPI_ISL_1787250, EPI_ISL_1787251, EPI_ISL_1787252, EPI_ISL_1787253, EPI_ISL_1787255, EPI_ISL_1787256, EPI_ISL_1787257, EPI_ISL_1787258, EPI_ISL_1787259, EPI_ISL_1787260, EPI_ISL_1787261, EPI_ISL_1787262, EPI_ISL_1787263, EPI_ISL_1787264, EPI_ISL_1787265, EPI_ISL_1787266, EPI_ISL_1787267, EPI_ISL_1787268, EPI_ISL_1787269, EPI_ISL_1787270, EPI_ISL_1787271, EPI_ISL_1787272, EPI_ISL_1787273, EPI_ISL_1787274, EPI_ISL_1787275, EPI_ISL_1787276, EPI_ISL_1787277, EPI_ISL_1787278, EPI_ISL_1787279, EPI_ISL_1787280, EPI_ISL_1787281, EPI_ISL_1787282, EPI_ISL_1787283, EPI_ISL_1787284, EPI_ISL_1787285, EPI_ISL_1787286, EPI_ISL_1787287, EPI_ISL_1787288, EPI_ISL_1787289, EPI_ISL_1787290, EPI_ISL_1787291, EPI_ISL_1787292, EPI_ISL_1787293, EPI_ISL_1787294, EPI_ISL_1787295, EPI_ISL_1787296, EPI_ISL_1787297, EPI_ISL_1787298, EPI_ISL_1787299, EPI_ISL_1787300, EPI_ISL_1787301, EPI_ISL_1787302, EPI_ISL_1787303, EPI_ISL_1787304, EPI_ISL_1787305, EPI_ISL_1787306, EPI_ISL_1787307, EPI_ISL_1787308, EPI_ISL_1787309, EPI_ISL_1787310, EPI_ISL_1787311, EPI_ISL_1787312, EPI_ISL_1787313, EPI_ISL_1787314, EPI_ISL_1787315, EPI_ISL_1787316 | | | |
| see above | Broad Institute Clinical Research Sequencing Platform | Infectious Disease Program, Broad Institute of Harvard and MIT | Siddle,K.J., Adams,G., Pearlman,L., Gladden-Young,A., Vicente,G., Blumenstiel,B., DeFelice,M., Lee,M., McGovern,S., Lagerborg,K., Rudy,M., DeRuff,K., Carter,A., Normandin,E., Bauer,M., Reilly,S., Tomkins-Tinch,C., Loreth,C., Chaluvadi,S., Meldrim,J., Granger,B., Lemieux,J.E., Birren,B.W., Sabeti,P.C., Larkin,K., Dodge,S., Lennon,N., Madoff,L., Brown,C., Gallagher,G., Smole,S., Park,D.J., Gabriel,S., and MacInnis,B.L. |
| EPI_ISL_1789120, EPI_ISL_1789122, EPI_ISL_1789123, EPI_ISL_1789124, EPI_ISL_1789125, EPI_ISL_1789126, EPI_ISL_1789127, EPI_ISL_1789128, EPI_ISL_1789129, EPI_ISL_1789130, EPI_ISL_1789131, EPI_ISL_1789133, EPI_ISL_1789134, EPI_ISL_1789135, EPI_ISL_1789136, EPI_ISL_1789137, EPI_ISL_1789138, EPI_ISL_1789139, EPI_ISL_1789140, EPI_ISL_1789141, EPI_ISL_1789142, EPI_ISL_1789143, EPI_ISL_1789144, EPI_ISL_1789145, EPI_ISL_1789147, EPI_ISL_1789148, EPI_ISL_1789152, EPI_ISL_1789153, EPI_ISL_1789154, EPI_ISL_1789155, EPI_ISL_1789156, EPI_ISL_1789157, EPI_ISL_1789158, EPI_ISL_1789159, EPI_ISL_1789160, EPI_ISL_1789161, EPI_ISL_1789162, EPI_ISL_1789163, EPI_ISL_1789164, EPI_ISL_1789165, EPI_ISL_1789166, EPI_ISL_1789167, EPI_ISL_1789168, EPI_ISL_1789169, EPI_ISL_1789170, EPI_ISL_1789171, EPI_ISL_1789173, EPI_ISL_1789174, EPI_ISL_1789175, EPI_ISL_1789176, EPI_ISL_1789177, EPI_ISL_1789178, EPI_ISL_1789180, EPI_ISL_1789181, EPI_ISL_1789182, EPI_ISL_1789184, EPI_ISL_1789185, EPI_ISL_1789186, EPI_ISL_1789187, EPI_ISL_1789188, EPI_ISL_1789189, EPI_ISL_1789190, EPI_ISL_1789191, EPI_ISL_1789192, EPI_ISL_1789193, EPI_ISL_1789194, EPI_ISL_1789195, EPI_ISL_1789196, EPI_ISL_1789197, EPI_ISL_1789198, EPI_ISL_1789199, EPI_ISL_1789200, EPI_ISL_1789201 | | | |
| see above | Illinois Department of Public Health - Springfield Lab | Illinois Department of Public Health - Springfield Lab | Bryan Sim, Gordon McCall |
| EPI_ISL_1789216, EPI_ISL_1789217, EPI_ISL_1789218, EPI_ISL_1789219, EPI_ISL_1789220, EPI_ISL_1789221, EPI_ISL_1789222, EPI_ISL_1789223, EPI_ISL_1789224, EPI_ISL_1789225, EPI_ISL_1789226, EPI_ISL_1789227, EPI_ISL_1789228, EPI_ISL_1789229, EPI_ISL_1789230, EPI_ISL_1789231, EPI_ISL_1789233, EPI_ISL_1789234, EPI_ISL_1789235, EPI_ISL_1789236, EPI_ISL_1789237, EPI_ISL_1789238, EPI_ISL_1789239, EPI_ISL_1789240, EPI_ISL_1789241, EPI_ISL_1789242, EPI_ISL_1789243, EPI_ISL_1789244, EPI_ISL_1789245, EPI_ISL_1789246, EPI_ISL_1789247, EPI_ISL_1789248, EPI_ISL_1789249, EPI_ISL_1789250, EPI_ISL_1789251, EPI_ISL_1789252, EPI_ISL_1789253, EPI_ISL_1789254, EPI_ISL_1789255, EPI_ISL_1789256, EPI_ISL_1789257, EPI_ISL_1789258, EPI_ISL_1789259, EPI_ISL_1789260, EPI_ISL_1789261, EPI_ISL_1789262, EPI_ISL_1789263, EPI_ISL_1789264, EPI_ISL_1789265, EPI_ISL_1789266, EPI_ISL_1789267, EPI_ISL_1789268, EPI_ISL_1789269, EPI_ISL_1789270, EPI_ISL_1789271, EPI_ISL_1789272, EPI_ISL_1789273, EPI_ISL_1789274, EPI_ISL_1789275, EPI_ISL_1789276, EPI_ISL_1789277, EPI_ISL_1789278, EPI_ISL_1789279, EPI_ISL_1789280, EPI_ISL_1789281, EPI_ISL_1789282, EPI_ISL_1789283, EPI_ISL_1789284, EPI_ISL_1789285, EPI_ISL_1789286, EPI_ISL_1789287, EPI_ISL_1789288, EPI_ISL_1789289, EPI_ISL_1789290, EPI_ISL_1789291, EPI_ISL_1789292, EPI_ISL_1789293, EPI_ISL_1789294, EPI_ISL_1789295, EPI_ISL_1789296, EPI_ISL_1789297, EPI_ISL_1789298, EPI_ISL_1789299, EPI_ISL_1789300, EPI_ISL_1789301, EPI_ISL_1789302, EPI_ISL_1789303, EPI_ISL_1789304, EPI_ISL_1789305, EPI_ISL_1789306, EPI_ISL_1789307, EPI_ISL_1789308, EPI_ISL_1789309, EPI_ISL_1789310, EPI_ISL_1789311, EPI_ISL_1789313, EPI_ISL_1789314, EPI_ISL_1789315, EPI_ISL_1789316, EPI_ISL_1789317, EPI_ISL_1789318, EPI_ISL_1789319, EPI_ISL_1789320, EPI_ISL_1789321, EPI_ISL_1789323, EPI_ISL_1789324, EPI_ISL_1789325, EPI_ISL_1789326, EPI_ISL_1789327, EPI_ISL_1789328, EPI_ISL_1789329, EPI_ISL_1789330, EPI_ISL_1789331, EPI_ISL_1789332, EPI_ISL_1789333, EPI_ISL_1789335, EPI_ISL_1789336, EPI_ISL_1789337, EPI_ISL_1789338, EPI_ISL_1789339, EPI_ISL_1789340, EPI_ISL_1789341, EPI_ISL_1789342, EPI_ISL_1789343, EPI_ISL_1789344, EPI_ISL_1789345, EPI_ISL_1789346, EPI_ISL_1789347, EPI_ISL_1789348, EPI_ISL_1789349, EPI_ISL_1789350, EPI_ISL_1789351, EPI_ISL_1789352, EPI_ISL_1789353, EPI_ISL_1789354, EPI_ISL_1789355, EPI_ISL_1789356, EPI_ISL_1789357, EPI_ISL_1789358, EPI_ISL_1789359, EPI_ISL_1789360, EPI_ISL_1789361, EPI_ISL_1789362, EPI_ISL_1789363, EPI_ISL_1789364, EPI_ISL_1789365, EPI_ISL_1789366, EPI_ISL_1789367, EPI_ISL_1789368, EPI_ISL_1789369, EPI_ISL_1789370, EPI_ISL_1789372, EPI_ISL_1789373, EPI_ISL_1789374, EPI_ISL_1789375, EPI_ISL_1789376, EPI_ISL_1789377, EPI_ISL_1789378, EPI_ISL_1789379, EPI_ISL_1789380, EPI_ISL_1789381, EPI_ISL_1789382, EPI_ISL_1789383, EPI_ISL_1789384, EPI_ISL_1789385, EPI_ISL_1789386, EPI_ISL_1789387, EPI_ISL_1789388, EPI_ISL_1789389, EPI_ISL_1789390, EPI_ISL_1789391, EPI_ISL_1789392, EPI_ISL_1789393, EPI_ISL_1789394, EPI_ISL_1789395, EPI_ISL_1789396, EPI_ISL_1789397, EPI_ISL_1789398, EPI_ISL_1789399, EPI_ISL_1789400, EPI_ISL_1789401, EPI_ISL_1789402, EPI_ISL_1789403, EPI_ISL_1789404, EPI_ISL_1789405, EPI_ISL_1789406, EPI_ISL_1789407, EPI_ISL_1789408, EPI_ISL_1789409, EPI_ISL_1789410, EPI_ISL_1789411, EPI_ISL_1789412, EPI_ISL_1789413, EPI_ISL_1789414, EPI_ISL_1789415, EPI_ISL_1789416, EPI_ISL_1789417, EPI_ISL_1789418, EPI_ISL_1789419, EPI_ISL_1789420, EPI_ISL_1789421, EPI_ISL_1789422, EPI_ISL_1789423, EPI_ISL_1789424, EPI_ISL_1789425, EPI_ISL_1789426, EPI_ISL_1789427, EPI_ISL_1789428, EPI_ISL_1789429, EPI_ISL_1789430, EPI_ISL_1789431, EPI_ISL_1789432, EPI_ISL_1789433, EPI_ISL_1789434, EPI_ISL_1789435, EPI_ISL_1789436, EPI_ISL_1789437, EPI_ISL_1789438, EPI_ISL_1789439, EPI_ISL_1789440, EPI_ISL_1789441, EPI_ISL_1789442, EPI_ISL_1789443, EPI_ISL_1789444, EPI_ISL_1789445, EPI_ISL_1789446, EPI_ISL_1789447, EPI_ISL_1789448, EPI_ISL_1789449, EPI_ISL_1789450, EPI_ISL_1789451, EPI_ISL_1789452, EPI_ISL_1789453, EPI_ISL_1789454, EPI_ISL_1789455, EPI_ISL_1789456, EPI_ISL_1789457, EPI_ISL_1789458, EPI_ISL_1789459, EPI_ISL_1789460, EPI_ISL_1789461, EPI_ISL_1789462, EPI_ISL_1789463, EPI_ISL_1789464, EPI_ISL_1789465, EPI_ISL_1789466, EPI_ISL_1789467, EPI_ISL_1789468, EPI_ISL_1789469, EPI_ISL_1789470, EPI_ISL_1789471, EPI_ISL_1789472, EPI_ISL_1789473, EPI_ISL_1789474, EPI_ISL_1789475, EPI_ISL_1789476, EPI_ISL_1789477, EPI_ISL_1789478, EPI_ISL_1789479, EPI_ISL_1789480, EPI_ISL_1789481, EPI_ISL_1789482, EPI_ISL_1789483, EPI_ISL_1789484 | | | |
| see above | UW Virology Lab | UW Virology Lab | Pavitra Roychoudhury, Hong Xie, Lasata Shrestha, Tien V. Nguyen, Shah Mohamed Bakhash, Michelle Lin, Noah R. Baker, Sean Ellis, Meei-Li Huang, Keith R Jerome, Alexander Greninger |
| EPI_ISL_1789545, EPI_ISL_1789546, EPI_ISL_1789547, EPI_ISL_1789548, EPI_ISL_1789549, EPI_ISL_1789550, EPI_ISL_1789551, EPI_ISL_1789552, EPI_ISL_1789553, EPI_ISL_1789554, EPI_ISL_1789555, EPI_ISL_1789556, EPI_ISL_1789557, EPI_ISL_1789558, EPI_ISL_1789559, EPI_ISL_1789560, EPI_ISL_1789561, EPI_ISL_1789562, EPI_ISL_1789563, EPI_ISL_1789564, EPI_ISL_1789565, EPI_ISL_1789566, EPI_ISL_1789567, EPI_ISL_1789568, EPI_ISL_1789569, EPI_ISL_1789570, EPI_ISL_1789571, EPI_ISL_1789573, EPI_ISL_1789574, EPI_ISL_1789575, EPI_ISL_1789576, EPI_ISL_1789577, EPI_ISL_1789578, EPI_ISL_1789580, EPI_ISL_1789581, EPI_ISL_1789582, EPI_ISL_1789583, EPI_ISL_1789584, EPI_ISL_1789585, EPI_ISL_1789586, EPI_ISL_1789588, EPI_ISL_1789589, EPI_ISL_1789590, EPI_ISL_1789591, EPI_ISL_1789592, EPI_ISL_1789593, EPI_ISL_1789594, EPI_ISL_1789595, EPI_ISL_1789596, EPI_ISL_1789597, EPI_ISL_1789598, EPI_ISL_1789599, EPI_ISL_1789601, EPI_ISL_1789602, EPI_ISL_1789603, EPI_ISL_1789604, EPI_ISL_1789605, EPI_ISL_1789606, EPI_ISL_1789607, EPI_ISL_1789609, EPI_ISL_1789610, EPI_ISL_1789611, EPI_ISL_1789612, EPI_ISL_1789613, EPI_ISL_1789614, EPI_ISL_1789615, EPI_ISL_1789616, EPI_ISL_1789617, EPI_ISL_1789618, EPI_ISL_1789619, EPI_ISL_1789620, EPI_ISL_1789621, EPI_ISL_1789622, EPI_ISL_1789623, EPI_ISL_1789624, EPI_ISL_1789625, EPI_ISL_1789626, EPI_ISL_1789627, EPI_ISL_1789628, EPI_ISL_1789629 | | | |
| see above | M Health Fairview | Minnesota Department of Health, Public Health Laboratory | Alexandra Lorentz, Jacob Garfin, Matt Plumb, and Xiong Wang |
| EPI_ISL_1789630, EPI_ISL_1789631, EPI_ISL_1789632, EPI_ISL_1789633, EPI_ISL_1789634, EPI_ISL_1789635, EPI_ISL_1789636, EPI_ISL_1789637, EPI_ISL_1789638, EPI_ISL_1789639, EPI_ISL_1789640, EPI_ISL_1789641, EPI_ISL_1789642, EPI_ISL_1789643, EPI_ISL_1789644, EPI_ISL_1789645, EPI_ISL_1789646, EPI_ISL_1789647, EPI_ISL_1789648, EPI_ISL_1789649, EPI_ISL_1789650, EPI_ISL_1789651, EPI_ISL_1789652, EPI_ISL_1789653, EPI_ISL_1789654, EPI_ISL_1789655, EPI_ISL_1789657, EPI_ISL_1789658, EPI_ISL_1789659, EPI_ISL_1789660, EPI_ISL_1789661, EPI_ISL_1789662, EPI_ISL_1789663, EPI_ISL_1789664, EPI_ISL_1789665, EPI_ISL_1789666, EPI_ISL_1789667, EPI_ISL_1789668, EPI_ISL_1789669, EPI_ISL_1789670 | | | |
| see above | Alaska State Virology Laboratory | Alaska State Virology Laboratory | Stephanie DeRonde, Elva House, Lisa Smith, Ph.D., Jack Chen, Ph.D. |
| EPI_ISL_1789674, EPI_ISL_1789675, EPI_ISL_1789676, EPI_ISL_1789677, EPI_ISL_1789678, EPI_ISL_1789679 | Marshfield Labs | Pathogen Discovery, Respiratory Viruses Branch, Division of Viral Diseases, Centers for Disease Control and Prevention | Ying Tao, Yan Li, Anna Kelleher, Jing Zhang, Anna Montmayeur, Brian Lynch, Krista Queen, Anna Uehara, Peter Cook, Han Jia Justin Ng, Rachel Marine, Clinton R. Paden, Haibin Wang, Mark Burroughs, Justin Lee, Adam Retchless, Suxiang Tong |
| EPI_ISL_1789683, EPI_ISL_1789684, EPI_ISL_1789685, EPI_ISL_1789686, EPI_ISL_1789687, EPI_ISL_1789688, EPI_ISL_1789689, EPI_ISL_1789690, EPI_ISL_1789691, EPI_ISL_1789693, EPI_ISL_1789694 | | | |
| see above | Arizona State University | Arizona State University | Peter T. Skidmore, LaRinda A. Holland, Rabia Maqsood, Nicholas J. Mellor, Joy M. Blain, Valerie Harris, Joshua LaBaer, Vel Murugan, Efrem S. Lim |
| EPI_ISL_1789696, EPI_ISL_1789697, EPI_ISL_1789698, EPI_ISL_1789699 | Laboratorio PGM | Laboratorio de Infectología Molecular, Departamento de Bioquímica y Medicina Molecular,Facultad de Medicina - Universidad Autónoma de Nuevo León | Kame A. Galán-Huerta, María F. Herrera-Saldivar, Natalia Martínez-Acuña, Sonia A. Lozano-Sepúlveda, Daniel Arellanos-Soto, Ana M. Rivas-Estilla, Javier Ramos-Jimenez, Gabriela Elizondo, Eduardo Garza-de-la-Peña |
| EPI_ISL_1789700, EPI_ISL_1789701, EPI_ISL_1789702, EPI_ISL_1789704, EPI_ISL_1789705 | NYC Pandemic Response Lab | Wadsworth Center, New York State Department of Health | Kirsten St. George, Daryl M. Lamson, Alexis Russell, Matthew Shudt, Melissa A Leisner, Jonathan Plitnick, Catharine Prussing, Navjot Singh, John Kelly, Erasmus Schneider, Erica Lasek-Nesselquist |
| EPI_ISL_1789706, EPI_ISL_1789707 | URMC LABS | Wadsworth Center, New York State Department of Health | Kirsten St. George, Daryl M. Lamson, Alexis Russell, Matthew Shudt, Melissa A Leisner, Jonathan Plitnick, Catharine Prussing, Navjot Singh, John Kelly, Erasmus Schneider, Erica Lasek-Nesselquist |
| EPI_ISL_1789708 | NYC Pandemic Response Lab | Wadsworth Center, New York State Department of Health | Kirsten St. George, Daryl M. Lamson, Alexis Russell, Matthew Shudt, Melissa A Leisner, Jonathan Plitnick, Catharine Prussing, Navjot Singh, John Kelly, Erasmus Schneider, Erica Lasek-Nesselquist |
| EPI_ISL_1789709 | SUNY UPSTATE MEDICAL UNIVERSITY | Wadsworth Center, New York State Department of Health | Kirsten St. George, Daryl M. Lamson, Alexis Russell, Matthew Shudt, Melissa A Leisner, Jonathan Plitnick, Catharine Prussing, Navjot Singh, John Kelly, Erasmus Schneider, Erica Lasek-Nesselquist |
| EPI_ISL_1789710, EPI_ISL_1789711 | MONTEFIORE MEDICAL CENTER LABORATORIES | Wadsworth Center, New York State Department of Health | Kirsten St. George, Daryl M. Lamson, Alexis Russell, Matthew Shudt, Melissa A Leisner, Jonathan Plitnick, Catharine Prussing, Navjot Singh, John Kelly, Erasmus Schneider, Erica Lasek-Nesselquist |
| EPI_ISL_1789712 | BOSTON HEART DIAGNOSTICS CORP | Wadsworth Center, New York State Department of Health | Kirsten St. George, Daryl M. Lamson, Alexis Russell, Matthew Shudt, Melissa A Leisner, Jonathan Plitnick, Catharine Prussing, Navjot Singh, John Kelly, Erasmus Schneider, Erica Lasek-Nesselquist |
| EPI_ISL_1789713 | Columbia University Irving Medical Center | Wadsworth Center, New York State Department of Health | Kirsten St. George, Daryl M. Lamson, Alexis Russell, Matthew Shudt, Melissa A Leisner, Jonathan Plitnick, Catharine Prussing, Navjot Singh, John Kelly, Erasmus Schneider, Erica Lasek-Nesselquist |
| EPI_ISL_1789714 | New York Presbyterian Hospital | Wadsworth Center, New York State Department of Health | Kirsten St. George, Daryl M. Lamson, Alexis Russell, Matthew Shudt, Melissa A Leisner, Jonathan Plitnick, Catharine Prussing, Navjot Singh, John Kelly, Erasmus Schneider, Erica Lasek-Nesselquist |
| EPI_ISL_1789715 | MONTEFIORE MEDICAL CENTER LABORATORIES | Wadsworth Center, New York State Department of Health | Kirsten St. George, Daryl M. Lamson, Alexis Russell, Matthew Shudt, Melissa A Leisner, Jonathan Plitnick, Catharine Prussing, Navjot Singh, John Kelly, Erasmus Schneider, Erica Lasek-Nesselquist |
| EPI_ISL_1789716, EPI_ISL_1789717, EPI_ISL_1789718 | ALBANY MEDICAL CENTER | Wadsworth Center, New York State Department of Health | Kirsten St. George, Daryl M. Lamson, Alexis Russell, Matthew Shudt, Melissa A Leisner, Jonathan Plitnick, Catharine Prussing, Navjot Singh, John Kelly, Erasmus Schneider, Erica Lasek-Nesselquist |
| EPI_ISL_1789719 | STONY BROOK UNIVERSITY HOSPITAL | Wadsworth Center, New York State Department of Health | Kirsten St. George, Daryl M. Lamson, Alexis Russell, Matthew Shudt, Melissa A Leisner, Jonathan Plitnick, Catharine Prussing, Navjot Singh, John Kelly, Erasmus Schneider, Erica Lasek-Nesselquist |
| EPI_ISL_1789720, EPI_ISL_1789721 | MONTEFIORE MEDICAL CENTER LABORATORIES | Wadsworth Center, New York State Department of Health | Kirsten St. George, Daryl M. Lamson, Alexis Russell, Matthew Shudt, Melissa A Leisner, Jonathan Plitnick, Catharine Prussing, Navjot Singh, John Kelly, Erasmus Schneider, Erica Lasek-Nesselquist |
| EPI_ISL_1789722, EPI_ISL_1789723 | WESTCHESTER MEDICAL CENTER | Wadsworth Center, New York State Department of Health | Kirsten St. George, Daryl M. Lamson, Alexis Russell, Matthew Shudt, Melissa A Leisner, Jonathan Plitnick, Catharine Prussing, Navjot Singh, John Kelly, Erasmus Schneider, Erica Lasek-Nesselquist |
| EPI_ISL_1789724, EPI_ISL_1789725, EPI_ISL_1789726 | DC Public Health Lab/ Dept. of Forensic Sciences | Pathogen Discovery, Respiratory Viruses Branch, Division of Viral Diseases, Centers for Disease Control and Prevention | Ying Tao, Yan Li, Anna Kelleher, Jing Zhang, Anna Montmayeur, Brian Lynch, Krista Queen, Anna Uehara, Peter Cook, Han Jia Justin Ng, Rachel Marine, Clinton R. Paden, Haibin Wang, Mark Burroughs, Justin Lee, Adam Retchless, Suxiang Tong |
| EPI_ISL_1789778, EPI_ISL_1789779, EPI_ISL_1789780, EPI_ISL_1789781, EPI_ISL_1789782, EPI_ISL_1789783, EPI_ISL_1789784, EPI_ISL_1789785, EPI_ISL_1789786, EPI_ISL_1789787, EPI_ISL_1789788, EPI_ISL_1789789, EPI_ISL_1789790, EPI_ISL_1789791, EPI_ISL_1789792, EPI_ISL_1789793, EPI_ISL_1789794, EPI_ISL_1789795, EPI_ISL_1789796, EPI_ISL_1789797, EPI_ISL_1789798, EPI_ISL_1789799, EPI_ISL_1789800, EPI_ISL_1789801, EPI_ISL_1789802, EPI_ISL_1789803, EPI_ISL_1789804, EPI_ISL_1789805, EPI_ISL_1789806, EPI_ISL_1789807, EPI_ISL_1789808, EPI_ISL_1789809, EPI_ISL_1789810 | | | |
| see above | Houston Health Dept. | Houston Health Dept. | Ryker Penn, Pamela Brown, Adolpho Lara, Yanlai Lai |
| EPI_ISL_1789819, EPI_ISL_1789820, EPI_ISL_1789821, EPI_ISL_1789822, EPI_ISL_1789823, EPI_ISL_1789824, EPI_ISL_1789825, EPI_ISL_1789826, EPI_ISL_1789827, EPI_ISL_1789828, EPI_ISL_1789829, EPI_ISL_1789830, EPI_ISL_1789831, EPI_ISL_1789832, EPI_ISL_1789833, EPI_ISL_1789834, EPI_ISL_1789835, EPI_ISL_1789836, EPI_ISL_1789837, EPI_ISL_1789838, EPI_ISL_1789839 | | | |
| see above | UW Virology Lab | UW Virology Lab | Pavitra Roychoudhury, Hong Xie, Lasata Shrestha, Shah Mohamed Bakhash, Michelle Lin, Noah R. Baker, Sean Ellis, Saraswathi Sathees, Meei-Li Huang, Keith R Jerome, Alexander Greninger |
| EPI_ISL_1789841, EPI_ISL_1789842, EPI_ISL_1789843, EPI_ISL_1789844, EPI_ISL_1789845, EPI_ISL_1789846, EPI_ISL_1789847, EPI_ISL_1789848, EPI_ISL_1789849, EPI_ISL_1789850, EPI_ISL_1789851 | | | |
| see above | Washington University | Pathogen Discovery, Respiratory Viruses | Ying Tao, Yan Li, Anna Kelleher, Jing Zhang, Anna Montmayeur, Brian Lynch, Krista Queen, Anna Uehara, Peter Cook, Han Jia Justin Ng, Rachel Marine, Clinton R. Paden, Haibin Wang, Mark Burroughs, Justin Lee, Adam Retchless, Suxiang Tong |

| | | | |
|---|---|---|---|
| | | Branch, Division of Viral Diseases, Centers for Disease Control and Prevention | |
| EPI_ISL_1789854, EPI_ISL_1789856, EPI_ISL_1789860, EPI_ISL_1789863, EPI_ISL_1789865, EPI_ISL_1789866, EPI_ISL_1789867, EPI_ISL_1789868, EPI_ISL_1789869, EPI_ISL_1789870, EPI_ISL_1789871, EPI_ISL_1789872, EPI_ISL_1789873, EPI_ISL_1789874, EPI_ISL_1789875, EPI_ISL_1789879, EPI_ISL_1789880, EPI_ISL_1789881, EPI_ISL_1789883, EPI_ISL_1789884, EPI_ISL_1789885, EPI_ISL_1789887, EPI_ISL_1789888, EPI_ISL_1789896, EPI_ISL_1789899, EPI_ISL_1789902, EPI_ISL_1789903, EPI_ISL_1789904, EPI_ISL_1789905, EPI_ISL_1789906, EPI_ISL_1789907, EPI_ISL_1789908, EPI_ISL_1789909, EPI_ISL_1789910, EPI_ISL_1789911, EPI_ISL_1789912, EPI_ISL_1789913, EPI_ISL_1789914, EPI_ISL_1789915, EPI_ISL_1789916, EPI_ISL_1789917, EPI_ISL_1789918, EPI_ISL_1789919, EPI_ISL_1789920, EPI_ISL_1789921, EPI_ISL_1789922, EPI_ISL_1789923, EPI_ISL_1789924, EPI_ISL_1789925, EPI_ISL_1789926, EPI_ISL_1789927, EPI_ISL_1789928, EPI_ISL_1789929, EPI_ISL_1789931, EPI_ISL_1789933, EPI_ISL_1789934, EPI_ISL_1789935, EPI_ISL_1789936, EPI_ISL_1789937, EPI_ISL_1789938, EPI_ISL_1789939, EPI_ISL_1789941, EPI_ISL_1789943, EPI_ISL_1789946, EPI_ISL_1789947, EPI_ISL_1789950, EPI_ISL_1789951, EPI_ISL_1789952, EPI_ISL_1789953, EPI_ISL_1789954, EPI_ISL_1789955, EPI_ISL_1789957, EPI_ISL_1789958, EPI_ISL_1789959, EPI_ISL_1789960, EPI_ISL_1789961, EPI_ISL_1789962, EPI_ISL_1789963, EPI_ISL_1789964, EPI_ISL_1789965, EPI_ISL_1789967, EPI_ISL_1789968, EPI_ISL_1789969, EPI_ISL_1789970, EPI_ISL_1789971, EPI_ISL_1789972, EPI_ISL_1789973, EPI_ISL_1789976, EPI_ISL_1789977, EPI_ISL_1789978, EPI_ISL_1789980, EPI_ISL_1789981, EPI_ISL_1789982, EPI_ISL_1789983, EPI_ISL_1789985, EPI_ISL_1789986, EPI_ISL_1789987, EPI_ISL_1789988, EPI_ISL_1789989, EPI_ISL_1789990, EPI_ISL_1789991, EPI_ISL_1789992, EPI_ISL_1789993, EPI_ISL_1789994, EPI_ISL_1789995, EPI_ISL_1789996, EPI_ISL_1789997, EPI_ISL_1789998, EPI_ISL_1789999, EPI_ISL_1790000, EPI_ISL_1790001, EPI_ISL_1790002, EPI_ISL_1790003, EPI_ISL_1790004, EPI_ISL_1790005, EPI_ISL_1790006, EPI_ISL_1790008, EPI_ISL_1790009, EPI_ISL_1790010, EPI_ISL_1790011, EPI_ISL_1790013, EPI_ISL_1790014, EPI_ISL_1790015, EPI_ISL_1790018, EPI_ISL_1790019, EPI_ISL_1790020, EPI_ISL_1790022, EPI_ISL_1790025, EPI_ISL_1790027, EPI_ISL_1790029, EPI_ISL_1790030, EPI_ISL_1790031, EPI_ISL_1790032, EPI_ISL_1790033, EPI_ISL_1790034, EPI_ISL_1790035, EPI_ISL_1790036, EPI_ISL_1790037, EPI_ISL_1790038, EPI_ISL_1790039, EPI_ISL_1790040, EPI_ISL_1790041, EPI_ISL_1790043, EPI_ISL_1790044 | | | |
| see above | NYU Langone Health | Departments of Pathology and Medicine, New York University School of Medicine | Adriana Heguy, Dacia Dimartino, Emily Guzman, Christian Marier, Peter Meyn, Sitharam Ramaswami, Gael Westby, Paul Zappile, Yutong Zhang, Paolo Cotzia, Guiqing Wang |
| EPI_ISL_1791072, EPI_ISL_1791073, EPI_ISL_1791074 | Arizona State University | Arizona State University | Rabia Maqsood, LaRinda A. Holland, Peter T. Skidmore, Nicholas J. Mellor, Joy M. Blain, Valerie Harris, Joshua LaBaer, Vel Murugan, Efrem S. Lim |
| EPI_ISL_1791075, EPI_ISL_1791076, EPI_ISL_1791077, EPI_ISL_1791078, EPI_ISL_1791079, EPI_ISL_1791080, EPI_ISL_1791081, EPI_ISL_1791082, EPI_ISL_1791083, EPI_ISL_1791084, EPI_ISL_1791085, EPI_ISL_1791087, EPI_ISL_1791088, EPI_ISL_1791089, EPI_ISL_1791090, EPI_ISL_1791091, EPI_ISL_1791092, EPI_ISL_1791093, EPI_ISL_1791094, EPI_ISL_1791095, EPI_ISL_1791096, EPI_ISL_1791097, EPI_ISL_1791098, EPI_ISL_1791099, EPI_ISL_1791100, EPI_ISL_1791101, EPI_ISL_1791102, EPI_ISL_1791103, EPI_ISL_1791104, EPI_ISL_1791105, EPI_ISL_1791106, EPI_ISL_1791107, EPI_ISL_1791108, EPI_ISL_1791109, EPI_ISL_1791110, EPI_ISL_1791111, EPI_ISL_1791112, EPI_ISL_1791113, EPI_ISL_1791114, EPI_ISL_1791115, EPI_ISL_1791116, EPI_ISL_1791117, EPI_ISL_1791118, EPI_ISL_1791119, EPI_ISL_1791120, EPI_ISL_1791121, EPI_ISL_1791122, EPI_ISL_1791123, EPI_ISL_1791124, EPI_ISL_1791125, EPI_ISL_1791126, EPI_ISL_1791127, EPI_ISL_1791128, EPI_ISL_1791129, EPI_ISL_1791130, EPI_ISL_1791131, EPI_ISL_1791132, EPI_ISL_1791133, EPI_ISL_1791134, EPI_ISL_1791135, EPI_ISL_1791136, EPI_ISL_1791137, EPI_ISL_1791138, EPI_ISL_1791139, EPI_ISL_1791140 | | | |
| see above | New Mexico Department of Health Scientific Laboratory | Center for Global Health, University of New Mexico Health Sciences Center | Daryl Domman, Kurt Schwalm, Twila Kunde, Joseph Hicks, Anastacia Griego, Michael Edwards, Valerie Morley, Darrell Dinwiddie |
| EPI_ISL_1791317, EPI_ISL_1791318, EPI_ISL_1791319, EPI_ISL_1791320, EPI_ISL_1791321, EPI_ISL_1791322, EPI_ISL_1791323, EPI_ISL_1791324, EPI_ISL_1791325, EPI_ISL_1791326, EPI_ISL_1791327, EPI_ISL_1791328, EPI_ISL_1791329, EPI_ISL_1791330, EPI_ISL_1791331, EPI_ISL_1791332, EPI_ISL_1791333, EPI_ISL_1791334, EPI_ISL_1791335, EPI_ISL_1791336, EPI_ISL_1791337, EPI_ISL_1791338, EPI_ISL_1791339, EPI_ISL_1791340, EPI_ISL_1791341, EPI_ISL_1791342, EPI_ISL_1791343, EPI_ISL_1791344, EPI_ISL_1791345, EPI_ISL_1791346, EPI_ISL_1791347, EPI_ISL_1791348, EPI_ISL_1791349, EPI_ISL_1791350, EPI_ISL_1791351, EPI_ISL_1791352, EPI_ISL_1791353, EPI_ISL_1791354, EPI_ISL_1791355, EPI_ISL_1791356, EPI_ISL_1791357, EPI_ISL_1791358, EPI_ISL_1791359, EPI_ISL_1791360, EPI_ISL_1791361, EPI_ISL_1791362, EPI_ISL_1791363, EPI_ISL_1791364, EPI_ISL_1791365, EPI_ISL_1791366, EPI_ISL_1791367, EPI_ISL_1791368, EPI_ISL_1791369, EPI_ISL_1791370, EPI_ISL_1791371, EPI_ISL_1791372, EPI_ISL_1791373, EPI_ISL_1791374, EPI_ISL_1791375, EPI_ISL_1791376, EPI_ISL_1791377, EPI_ISL_1791378, EPI_ISL_1791379, EPI_ISL_1791380, EPI_ISL_1791381, EPI_ISL_1791382, EPI_ISL_1791383, EPI_ISL_1791384, EPI_ISL_1791385, EPI_ISL_1791386, EPI_ISL_1791387, EPI_ISL_1791388, EPI_ISL_1791389, EPI_ISL_1791390, EPI_ISL_1791391, EPI_ISL_1791392, EPI_ISL_1791393, EPI_ISL_1791394, EPI_ISL_1791395, EPI_ISL_1791396, EPI_ISL_1791397, EPI_ISL_1791398, EPI_ISL_1791399, EPI_ISL_1791400, EPI_ISL_1791401, EPI_ISL_1791402 | | | |
| see above | Missouri State Public Health Laboratory | Missouri State Public Health Laboratory | Matthew Sinn, Joshua Barry, Ashley New |
| EPI_ISL_1791403, EPI_ISL_1791404, EPI_ISL_1791405, EPI_ISL_1791406, EPI_ISL_1791407, EPI_ISL_1791408, EPI_ISL_1791409, EPI_ISL_1791410, EPI_ISL_1791411, EPI_ISL_1791412, EPI_ISL_1791413, EPI_ISL_1791414, EPI_ISL_1791415, EPI_ISL_1791416, EPI_ISL_1791417, EPI_ISL_1791418, EPI_ISL_1791419, EPI_ISL_1791420, EPI_ISL_1791421, EPI_ISL_1791422, EPI_ISL_1791423, EPI_ISL_1791424, EPI_ISL_1791425, EPI_ISL_1791426, EPI_ISL_1791427, EPI_ISL_1791428, EPI_ISL_1791429, EPI_ISL_1791430, EPI_ISL_1791431, EPI_ISL_1791432, EPI_ISL_1791433, EPI_ISL_1791434, EPI_ISL_1791435, EPI_ISL_1791436, EPI_ISL_1791437, EPI_ISL_1791438, EPI_ISL_1791439, EPI_ISL_1791440, EPI_ISL_1791441, EPI_ISL_1791442, EPI_ISL_1791443, EPI_ISL_1791444, EPI_ISL_1791445, EPI_ISL_1791446, EPI_ISL_1791447, EPI_ISL_1791448, EPI_ISL_1791449, EPI_ISL_1791450, EPI_ISL_1791451, EPI_ISL_1791452, EPI_ISL_1791453, EPI_ISL_1791454, EPI_ISL_1791455, EPI_ISL_1791456 | | | |
| see above | San Diego County Public Health Laboratory | Andersen lab at Scripps Research | SEARCH Alliance San Diego with Ashleigh Murphy, Jovan Shephard, Brett Austin |
| EPI_ISL_1791459, EPI_ISL_1791461, EPI_ISL_1791464, EPI_ISL_1791470, EPI_ISL_1791472, EPI_ISL_1791475, EPI_ISL_1791476, EPI_ISL_1791477, EPI_ISL_1791478, EPI_ISL_1791479, EPI_ISL_1791480, EPI_ISL_1791481, EPI_ISL_1791482, EPI_ISL_1791483, EPI_ISL_1791484, EPI_ISL_1791485, EPI_ISL_1791486, EPI_ISL_1791487, EPI_ISL_1791488, EPI_ISL_1791489, EPI_ISL_1791490, EPI_ISL_1791491, EPI_ISL_1791493, EPI_ISL_1791495, EPI_ISL_1791496, EPI_ISL_1791497, EPI_ISL_1791498, EPI_ISL_1791499, EPI_ISL_1791500, EPI_ISL_1791501, EPI_ISL_1791502, EPI_ISL_1791503, EPI_ISL_1791504, EPI_ISL_1791505, EPI_ISL_1791506, EPI_ISL_1791507, EPI_ISL_1791508, EPI_ISL_1791509, EPI_ISL_1791510, EPI_ISL_1791511, EPI_ISL_1791512, EPI_ISL_1791513, EPI_ISL_1791514, EPI_ISL_1791515, EPI_ISL_1791516, EPI_ISL_1791517, EPI_ISL_1791518, EPI_ISL_1791519, EPI_ISL_1791520, EPI_ISL_1791521, EPI_ISL_1791522, EPI_ISL_1791523, EPI_ISL_1791524, EPI_ISL_1791526, EPI_ISL_1791527, EPI_ISL_1791529, EPI_ISL_1791530, EPI_ISL_1791531, EPI_ISL_1791532, EPI_ISL_1791533, EPI_ISL_1791534, EPI_ISL_1791535, EPI_ISL_1791536, EPI_ISL_1791537, EPI_ISL_1791539, EPI_ISL_1791540, EPI_ISL_1791541, EPI_ISL_1791542, EPI_ISL_1791543, EPI_ISL_1791544, EPI_ISL_1791545, EPI_ISL_1791546, EPI_ISL_1791547, EPI_ISL_1791548, EPI_ISL_1791549, EPI_ISL_1791550, EPI_ISL_1791551, EPI_ISL_1791552, EPI_ISL_1791553, EPI_ISL_1791554, EPI_ISL_1791556, EPI_ISL_1791557, EPI_ISL_1791558, EPI_ISL_1791559, EPI_ISL_1791560, EPI_ISL_1791561, EPI_ISL_1791562, EPI_ISL_1791563, EPI_ISL_1791564, EPI_ISL_1791565, EPI_ISL_1791566, EPI_ISL_1791567, EPI_ISL_1791568, EPI_ISL_1791569, EPI_ISL_1791570, EPI_ISL_1791571, EPI_ISL_1791572, EPI_ISL_1791573, EPI_ISL_1791574, EPI_ISL_1791575, EPI_ISL_1791576, EPI_ISL_1791578, EPI_ISL_1791579, EPI_ISL_1791580, EPI_ISL_1791581, EPI_ISL_1791582, EPI_ISL_1791585, EPI_ISL_1791588, EPI_ISL_1791589, EPI_ISL_1791590 | | | |
| see above | University of Oregon COVID-19 MAP Laboratory | University of Oregon Genomics and Cell Characterization Core Facility (GC3F) | Douglas Turnbull, Ariana White, Jeff Bishop, Jason Carriere |
| EPI_ISL_1791813 | MAWD Pathology Group, PA | MAWD Pathology Group, PA | Pradip Manna, PhD, Kelli Fayard, Shelby Herner, London Collins, Kelsi Eastman |
| EPI_ISL_1792927 | Texas Children's Hospital | Texas Children's Microbiome Center | Ruth Ann Luna, Jennifer K. Spinler, Jessica Runge, Sabeen Raza, James Dunn, Ila Singh, James Versalovic, Sridevi Devaraj |
| EPI_ISL_1792928, EPI_ISL_1792929, EPI_ISL_1792930, EPI_ISL_1792931, EPI_ISL_1792932, EPI_ISL_1792933, EPI_ISL_1792934, EPI_ISL_1792935, EPI_ISL_1792936, EPI_ISL_1792937, EPI_ISL_1792938, EPI_ISL_1792939, EPI_ISL_1792940, EPI_ISL_1792941, EPI_ISL_1792942, EPI_ISL_1792943, EPI_ISL_1792944, EPI_ISL_1792945, EPI_ISL_1792946, EPI_ISL_1792947, EPI_ISL_1792948, EPI_ISL_1792949, EPI_ISL_1792950, EPI_ISL_1792951, EPI_ISL_1792952, EPI_ISL_1792953, EPI_ISL_1792954, EPI_ISL_1792955, EPI_ISL_1792956, EPI_ISL_1792957, EPI_ISL_1792958, EPI_ISL_1792959, EPI_ISL_1792960, EPI_ISL_1792961, EPI_ISL_1792962, EPI_ISL_1792963, EPI_ISL_1792964, EPI_ISL_1792965, EPI_ISL_1792966, EPI_ISL_1792967, EPI_ISL_1792968, EPI_ISL_1792969, EPI_ISL_1792970, EPI_ISL_1792971, EPI_ISL_1792972, EPI_ISL_1792973, EPI_ISL_1792974, EPI_ISL_1792975, EPI_ISL_1792976, EPI_ISL_1792977, EPI_ISL_1792978, EPI_ISL_1792979, EPI_ISL_1792980, EPI_ISL_1792981, EPI_ISL_1792982, EPI_ISL_1792983, EPI_ISL_1792984, EPI_ISL_1792985, EPI_ISL_1792986, EPI_ISL_1792987, EPI_ISL_1792988, EPI_ISL_1792989, EPI_ISL_1792990, EPI_ISL_1792991, EPI_ISL_1792992, EPI_ISL_1792993, EPI_ISL_1792994, EPI_ISL_1792995, EPI_ISL_1792996, EPI_ISL_1792997, EPI_ISL_1792998, EPI_ISL_1792999, EPI_ISL_1793000, EPI_ISL_1793001, EPI_ISL_1793002, EPI_ISL_1793003, EPI_ISL_1793004, EPI_ISL_1793005, EPI_ISL_1793006, EPI_ISL_1793007, EPI_ISL_1793008, EPI_ISL_1793009, EPI_ISL_1793010, EPI_ISL_1793011, EPI_ISL_1793012, EPI_ISL_1793013, EPI_ISL_1793015, EPI_ISL_1793016, EPI_ISL_1793017, EPI_ISL_1793018, EPI_ISL_1793019, EPI_ISL_1793020, EPI_ISL_1793021, EPI_ISL_1793022, EPI_ISL_1793023, EPI_ISL_1793024, EPI_ISL_1793025, EPI_ISL_1793026, EPI_ISL_1793027, EPI_ISL_1793028, EPI_ISL_1793029, EPI_ISL_1793030, EPI_ISL_1793031, EPI_ISL_1793032, EPI_ISL_1793033, EPI_ISL_1793034, EPI_ISL_1793035, EPI_ISL_1793036, EPI_ISL_1793037, EPI_ISL_1793038, EPI_ISL_1793039, EPI_ISL_1793040, EPI_ISL_1793041, EPI_ISL_1793042, EPI_ISL_1793043, EPI_ISL_1793044, EPI_ISL_1793045, EPI_ISL_1793046, EPI_ISL_1793047, EPI_ISL_1793048, EPI_ISL_1793049, EPI_ISL_1793050, EPI_ISL_1793051, EPI_ISL_1793052, EPI_ISL_1793053, EPI_ISL_1793054, EPI_ISL_1793055, EPI_ISL_1793056, EPI_ISL_1793057, EPI_ISL_1793058, EPI_ISL_1793059, EPI_ISL_1793060, EPI_ISL_1793061, EPI_ISL_1793062, EPI_ISL_1793063, EPI_ISL_1793064, EPI_ISL_1793065, EPI_ISL_1793066, EPI_ISL_1793067, EPI_ISL_1793068, EPI_ISL_1793069, EPI_ISL_1793070, EPI_ISL_1793071, EPI_ISL_1793072, EPI_ISL_1793073, EPI_ISL_1793074, EPI_ISL_1793075, EPI_ISL_1793076, EPI_ISL_1793077, EPI_ISL_1793078, EPI_ISL_1793079, EPI_ISL_1793080, EPI_ISL_1793081, EPI_ISL_1793082, EPI_ISL_1793083, EPI_ISL_1793084, EPI_ISL_1793085, EPI_ISL_1793086, EPI_ISL_1793087, EPI_ISL_1793088, EPI_ISL_1793089, EPI_ISL_1793090, EPI_ISL_1793091, EPI_ISL_1793092, EPI_ISL_1793093, EPI_ISL_1793094, EPI_ISL_1793095 | | | |
| see above | Yale Clinical Virology Lab | Grubaugh Lab - Yale School of Public Health | Joseph Fauver, Mallery Breban, Isabel Ott, Tara Alpert, Mary Petrone, Anderson Brito, Chantal Vogels, Annie Watkins, Chaney Kalinich, Jessica Rothman, Marie L. Landry, Nathan Grubaugh |
| EPI_ISL_1793385, EPI_ISL_1793386, EPI_ISL_1793387, EPI_ISL_1793388, EPI_ISL_1793389, EPI_ISL_1793390, EPI_ISL_1793391, EPI_ISL_1793392, EPI_ISL_1793393, EPI_ISL_1793394, EPI_ISL_1793395, EPI_ISL_1793396, EPI_ISL_1793397, EPI_ISL_1793398, EPI_ISL_1793399, EPI_ISL_1793400, EPI_ISL_1793401, EPI_ISL_1793402, EPI_ISL_1793403, EPI_ISL_1793404, EPI_ISL_1793405, EPI_ISL_1793406, EPI_ISL_1793407, EPI_ISL_1793408, EPI_ISL_1793409, EPI_ISL_1793410, EPI_ISL_1793411, EPI_ISL_1793412, EPI_ISL_1793413, EPI_ISL_1793414, EPI_ISL_1793415, EPI_ISL_1793416, EPI_ISL_1793417, EPI_ISL_1793418, EPI_ISL_1793419, EPI_ISL_1793420, EPI_ISL_1793421, EPI_ISL_1793422, EPI_ISL_1793423, EPI_ISL_1793424, EPI_ISL_1793425, EPI_ISL_1793426, EPI_ISL_1793427, EPI_ISL_1793428, EPI_ISL_1793429, EPI_ISL_1793430, EPI_ISL_1793431, EPI_ISL_1793432, EPI_ISL_1793433, EPI_ISL_1793434, EPI_ISL_1793435, EPI_ISL_1793436, EPI_ISL_1793437, EPI_ISL_1793438, EPI_ISL_1793439, EPI_ISL_1793440, EPI_ISL_1793441, EPI_ISL_1793442, EPI_ISL_1793443, EPI_ISL_1793444, EPI_ISL_1793445, EPI_ISL_1793446, EPI_ISL_1793447, EPI_ISL_1793448, EPI_ISL_1793449, EPI_ISL_1793450, EPI_ISL_1793451, EPI_ISL_1793452, EPI_ISL_1793453, EPI_ISL_1793454, EPI_ISL_1793455, EPI_ISL_1793456, EPI_ISL_1793457, EPI_ISL_1793458, EPI_ISL_1793459, EPI_ISL_1793460, EPI_ISL_1793461, EPI_ISL_1793462, EPI_ISL_1793463, EPI_ISL_1793464, EPI_ISL_1793465, EPI_ISL_1793466, EPI_ISL_1793467, EPI_ISL_1793468, EPI_ISL_1793469, EPI_ISL_1793470, EPI_ISL_1793471, EPI_ISL_1793472, EPI_ISL_1793473, EPI_ISL_1793474, EPI_ISL_1793475, EPI_ISL_1793477, EPI_ISL_1793478, EPI_ISL_1793479, EPI_ISL_1793480, EPI_ISL_1793481, EPI_ISL_1793482, EPI_ISL_1793483, EPI_ISL_1793484, EPI_ISL_1793485, EPI_ISL_1793486, EPI_ISL_1793488, EPI_ISL_1793489, EPI_ISL_1793490, EPI_ISL_1793491, EPI_ISL_1793492, EPI_ISL_1793493, EPI_ISL_1793495, EPI_ISL_1793497, EPI_ISL_1793498, EPI_ISL_1793499, EPI_ISL_1793500, EPI_ISL_1793501, EPI_ISL_1793502, EPI_ISL_1793503, EPI_ISL_1793504, EPI_ISL_1793505, EPI_ISL_1793506, EPI_ISL_1793507, EPI_ISL_1793508, EPI_ISL_1793509, EPI_ISL_1793510, EPI_ISL_1793511, EPI_ISL_1793512, EPI_ISL_1793513, EPI_ISL_1793514, EPI_ISL_1793515, EPI_ISL_1793516, EPI_ISL_1793517, EPI_ISL_1793519, EPI_ISL_1793520, EPI_ISL_1793521, EPI_ISL_1793522, EPI_ISL_1793523, EPI_ISL_1793524, EPI_ISL_1793525, EPI_ISL_1793526, EPI_ISL_1793527, EPI_ISL_1793528, EPI_ISL_1793529, EPI_ISL_1793530, EPI_ISL_1793531, EPI_ISL_1793532, EPI_ISL_1793533, EPI_ISL_1793534, EPI_ISL_1793535, EPI_ISL_1793536, EPI_ISL_1793537, EPI_ISL_1793538, EPI_ISL_1793539, EPI_ISL_1793540, EPI_ISL_1793541, EPI_ISL_1793542, EPI_ISL_1793543, EPI_ISL_1793544, EPI_ISL_1793545, EPI_ISL_1793546, EPI_ISL_1793547, EPI_ISL_1793548, EPI_ISL_1793549, EPI_ISL_1793550, EPI_ISL_1793551, EPI_ISL_1793552, EPI_ISL_1793553, EPI_ISL_1793554, EPI_ISL_1793555, EPI_ISL_1793556, EPI_ISL_1793557, EPI_ISL_1793559, EPI_ISL_1793560, EPI_ISL_1793561, EPI_ISL_1793562, EPI_ISL_1793563, EPI_ISL_1793564, EPI_ISL_1793565, EPI_ISL_1793566, EPI_ISL_1793567, EPI_ISL_1793568, EPI_ISL_1793569, EPI_ISL_1793570, EPI_ISL_1793571, EPI_ISL_1793572, EPI_ISL_1793573, EPI_ISL_1793574, EPI_ISL_1793575, EPI_ISL_1793576, EPI_ISL_1793577, EPI_ISL_1793578, EPI_ISL_1793579, EPI_ISL_1793580, EPI_ISL_1793581, EPI_ISL_1793582, EPI_ISL_1793583, EPI_ISL_1793584, EPI_ISL_1793586, EPI_ISL_1793588, EPI_ISL_1793589, EPI_ISL_1793590, EPI_ISL_1793591, EPI_ISL_1793592, EPI_ISL_1793593, EPI_ISL_1793594, EPI_ISL_1793595, EPI_ISL_1793596, EPI_ISL_1793597, EPI_ISL_1793598, EPI_ISL_1793599, EPI_ISL_1793600, EPI_ISL_1793601, EPI_ISL_1793602, EPI_ISL_1793603, EPI_ISL_1793604, EPI_ISL_1793605, EPI_ISL_1793607, EPI_ISL_1793608, EPI_ISL_1793609, EPI_ISL_1793610, EPI_ISL_1793611, EPI_ISL_1793612, EPI_ISL_1793613, EPI_ISL_1793614, EPI_ISL_1793615, EPI_ISL_1793616, EPI_ISL_1793617, EPI_ISL_1793618, EPI_ISL_1793619, EPI_ISL_1793620, EPI_ISL_1793621, EPI_ISL_1793623, EPI_ISL_1793624, EPI_ISL_1793625, EPI_ISL_1793626, EPI_ISL_1793627, EPI_ISL_1793628, EPI_ISL_1793630, EPI_ISL_1793631, EPI_ISL_1793632, EPI_ISL_1793633, EPI_ISL_1793634, EPI_ISL_1793636, EPI_ISL_1793637, EPI_ISL_1793638, EPI_ISL_1793639, EPI_ISL_1793640, EPI_ISL_1793641, EPI_ISL_1793642, EPI_ISL_1793644, EPI_ISL_1793645 | | | |
| see above | Yale Clinical Virology Lab | Yale Center for Genomic Analysis | Shrikant Mane, Kaya Bilguvar, Curt Scharfe, Irina Tikhonova, Christopher Castaldi, Brooke Sullivan |
| EPI_ISL_1793843, EPI_ISL_1793844, EPI_ISL_1793845, EPI_ISL_1793846, EPI_ISL_1793847, EPI_ISL_1793848, EPI_ISL_1793849, EPI_ISL_1793850, EPI_ISL_1793851, EPI_ISL_1793852, EPI_ISL_1793853, EPI_ISL_1793854, EPI_ISL_1793855, EPI_ISL_1793856, EPI_ISL_1793857, EPI_ISL_1793858, EPI_ISL_1793859, EPI_ISL_1793860, EPI_ISL_1793861, EPI_ISL_1793862, EPI_ISL_1793863, EPI_ISL_1793864, EPI_ISL_1793865, EPI_ISL_1793866, EPI_ISL_1793867, EPI_ISL_1793868, EPI_ISL_1793869, EPI_ISL_1793870, EPI_ISL_1793871, EPI_ISL_1793872, EPI_ISL_1793873, EPI_ISL_1793874, EPI_ISL_1793875, EPI_ISL_1793876, EPI_ISL_1793877, EPI_ISL_1793878, EPI_ISL_1793879, EPI_ISL_1793880, EPI_ISL_1793881, EPI_ISL_1793882, EPI_ISL_1793883, EPI_ISL_1793884, EPI_ISL_1793885, EPI_ISL_1793886, EPI_ISL_1793887, EPI_ISL_1793888, EPI_ISL_1793889, EPI_ISL_1793890, EPI_ISL_1793891, EPI_ISL_1793892, EPI_ISL_1793893, EPI_ISL_1793894, EPI_ISL_1793895, EPI_ISL_1793896, EPI_ISL_1793897, EPI_ISL_1793898, EPI_ISL_1793899, EPI_ISL_1793900, EPI_ISL_1793901, EPI_ISL_1793902, EPI_ISL_1793903, EPI_ISL_1793904, EPI_ISL_1793905, EPI_ISL_1793906, EPI_ISL_1793907, EPI_ISL_1793908, EPI_ISL_1793909, EPI_ISL_1793910, EPI_ISL_1793911, EPI_ISL_1793912, EPI_ISL_1793913, EPI_ISL_1793914, EPI_ISL_1793915, EPI_ISL_1793916, EPI_ISL_1793917, EPI_ISL_1793918, EPI_ISL_1793919, EPI_ISL_1793920, EPI_ISL_1793921, EPI_ISL_1793922, EPI_ISL_1793923, EPI_ISL_1793924, EPI_ISL_1793925, EPI_ISL_1793926, EPI_ISL_1793927, EPI_ISL_1793928, EPI_ISL_1793929, EPI_ISL_1793930, EPI_ISL_1793931, EPI_ISL_1793932, EPI_ISL_1793933, EPI_ISL_1793934, EPI_ISL_1793935, EPI_ISL_1793936, EPI_ISL_1793937, EPI_ISL_1793938, EPI_ISL_1793939, EPI_ISL_1793940, EPI_ISL_1793941, EPI_ISL_1793942, EPI_ISL_1793943, EPI_ISL_1793944, EPI_ISL_1793945, EPI_ISL_1793946, EPI_ISL_1793947, EPI_ISL_1793948, EPI_ISL_1793949, EPI_ISL_1793950, EPI_ISL_1793951, EPI_ISL_1793952, EPI_ISL_1793953, EPI_ISL_1793954, EPI_ISL_1793955, EPI_ISL_1793956, EPI_ISL_1793957, EPI_ISL_1793958, EPI_ISL_1793959, EPI_ISL_1793960, EPI_ISL_1793961, EPI_ISL_1793962, EPI_ISL_1793963, EPI_ISL_1793964, EPI_ISL_1793965, EPI_ISL_1793966, EPI_ISL_1793967, EPI_ISL_1793968, EPI_ISL_1793969, EPI_ISL_1793970, EPI_ISL_1793971, EPI_ISL_1793972, EPI_ISL_1793973, EPI_ISL_1793974, EPI_ISL_1793975, EPI_ISL_1793976, EPI_ISL_1793977, EPI_ISL_1793978, EPI_ISL_1793979, EPI_ISL_1793980, EPI_ISL_1793981, EPI_ISL_1793982, EPI_ISL_1793983, EPI_ISL_1793984, EPI_ISL_1793985, EPI_ISL_1793986, EPI_ISL_1793987, EPI_ISL_1793988, EPI_ISL_1793989, EPI_ISL_1793990, EPI_ISL_1793991, EPI_ISL_1793992, EPI_ISL_1793993, EPI_ISL_1793994, EPI_ISL_1793995, EPI_ISL_1793996, EPI_ISL_1793997, EPI_ISL_1793998, EPI_ISL_1794000, EPI_ISL_1794001, EPI_ISL_1794002, EPI_ISL_1794003, EPI_ISL_1794004, EPI_ISL_1794005, EPI_ISL_1794007, EPI_ISL_1794008, EPI_ISL_1794009, EPI_ISL_1794010, EPI_ISL_1794011, EPI_ISL_1794012, EPI_ISL_1794013, EPI_ISL_1794014, EPI_ISL_1794015, EPI_ISL_1794016, EPI_ISL_1794017, EPI_ISL_1794018, EPI_ISL_1794019, EPI_ISL_1794020, EPI_ISL_1794021, EPI_ISL_1794022, EPI_ISL_1794023, EPI_ISL_1794024, EPI_ISL_1794025, EPI_ISL_1794026, EPI_ISL_1794027, EPI_ISL_1794028, EPI_ISL_1794029, EPI_ISL_1794030, EPI_ISL_1794032, EPI_ISL_1794035, EPI_ISL_1794036, EPI_ISL_1794037, EPI_ISL_1794038, EPI_ISL_1794040, EPI_ISL_1794041, EPI_ISL_1794043, EPI_ISL_1794045, EPI_ISL_1794046, EPI_ISL_1794047, EPI_ISL_1794051, EPI_ISL_1794052, EPI_ISL_1794053, EPI_ISL_1794054, EPI_ISL_1794055, EPI_ISL_1794057, EPI_ISL_1794058, EPI_ISL_1794059, EPI_ISL_1794060, EPI_ISL_1794061, EPI_ISL_1794062, EPI_ISL_1794064, EPI_ISL_1794065, EPI_ISL_1794067, EPI_ISL_1794068, EPI_ISL_1794069, EPI_ISL_1794070, EPI_ISL_1794071, EPI_ISL_1794072, EPI_ISL_1794073, EPI_ISL_1794075, EPI_ISL_1794077, EPI_ISL_1794078, EPI_ISL_1794079, EPI_ISL_1794080, EPI_ISL_1794081, EPI_ISL_1794082, EPI_ISL_1794083, EPI_ISL_1794084, EPI_ISL_1794085, EPI_ISL_1794086, EPI_ISL_1794087, EPI_ISL_1794088, EPI_ISL_1794089, EPI_ISL_1794090, EPI_ISL_1794091, EPI_ISL_1794093, EPI_ISL_1794094, EPI_ISL_1794095, EPI_ISL_1794097, EPI_ISL_1794098, EPI_ISL_1794099, EPI_ISL_1794101, EPI_ISL_1794102, EPI_ISL_1794103, EPI_ISL_1794104, EPI_ISL_1794105, EPI_ISL_1794106, EPI_ISL_1794107, EPI_ISL_1794108, EPI_ISL_1794109, EPI_ISL_1794110, EPI_ISL_1794111, EPI_ISL_1794112, EPI_ISL_1794113, EPI_ISL_1794114, EPI_ISL_1794115, EPI_ISL_1794116, EPI_ISL_1794118, EPI_ISL_1794119, EPI_ISL_1794120, EPI_ISL_1794121, EPI_ISL_1794122, EPI_ISL_1794123, EPI_ISL_1794124, EPI_ISL_1794125, EPI_ISL_1794126, EPI_ISL_1794127, EPI_ISL_1794128, EPI_ISL_1794129, EPI_ISL_1794130, EPI_ISL_1794131, EPI_ISL_1794132, EPI_ISL_1794134, EPI_ISL_1794135, EPI_ISL_1794136, EPI_ISL_1794137, EPI_ISL_1794138, EPI_ISL_1794140, EPI_ISL_1794141, EPI_ISL_1794142, EPI_ISL_1794143, EPI_ISL_1794144, EPI_ISL_1794145, EPI_ISL_1794148, EPI_ISL_1794149, EPI_ISL_1794150, EPI_ISL_1794151, EPI_ISL_1794153, EPI_ISL_1794154, EPI_ISL_1794155, EPI_ISL_1794157, EPI_ISL_1794158, EPI_ISL_1794159, EPI_ISL_1794161, EPI_ISL_1794162, EPI_ISL_1794163, EPI_ISL_1794164, EPI_ISL_1794165, EPI_ISL_1794166, EPI_ISL_1794167, EPI_ISL_1794168, EPI_ISL_1794169, EPI_ISL_1794170, EPI_ISL_1794171, EPI_ISL_1794172, EPI_ISL_1794173, EPI_ISL_1794174, EPI_ISL_1794175, EPI_ISL_1794176, EPI_ISL_1794177, EPI_ISL_1794178, EPI_ISL_1794179, EPI_ISL_1794180, EPI_ISL_1794181, EPI_ISL_1794182, EPI_ISL_1794183, EPI_ISL_1794184, EPI_ISL_1794185, EPI_ISL_1794186, EPI_ISL_1794187, EPI_ISL_1794188, EPI_ISL_1794189, EPI_ISL_1794190, EPI_ISL_1794191, EPI_ISL_1794192, EPI_ISL_1794193, EPI_ISL_1794194, EPI_ISL_1794195, EPI_ISL_1794196, EPI_ISL_1794197, EPI_ISL_1794198, EPI_ISL_1794199, EPI_ISL_1794200, EPI_ISL_1794201, EPI_ISL_1794202, EPI_ISL_1794203, EPI_ISL_1794204, EPI_ISL_1794205, EPI_ISL_1794206, EPI_ISL_1794207, EPI_ISL_1794208, EPI_ISL_1794209, EPI_ISL_1794210, EPI_ISL_1794211, EPI_ISL_1794212, EPI_ISL_1794213, EPI_ISL_1794214, EPI_ISL_1794215, EPI_ISL_1794216, EPI_ISL_1794217, EPI_ISL_1794218, EPI_ISL_1794219, EPI_ISL_1794220, EPI_ISL_1794221, EPI_ISL_1794222, EPI_ISL_1794223, EPI_ISL_1794224, EPI_ISL_1794225, EPI_ISL_1794226, EPI_ISL_1794227, EPI_ISL_1794228, EPI_ISL_1794229, EPI_ISL_1794230, EPI_ISL_1794231, EPI_ISL_1794232, EPI_ISL_1794233, EPI_ISL_1794234, EPI_ISL_1794235, EPI_ISL_1794236, EPI_ISL_1794237, EPI_ISL_1794238, EPI_ISL_1794239, EPI_ISL_1794240, EPI_ISL_1794241, EPI_ISL_1794242, EPI_ISL_1794243, EPI_ISL_1794244, EPI_ISL_1794245, EPI_ISL_1794246, EPI_ISL_1794247, EPI_ISL_1794248, EPI_ISL_1794249, EPI_ISL_1794250, EPI_ISL_1794251, EPI_ISL_1794252, EPI_ISL_1794253, EPI_ISL_1794254, EPI_ISL_1794255, EPI_ISL_1794256, EPI_ISL_1794257, EPI_ISL_1794258, EPI_ISL_1794259, EPI_ISL_1794260, EPI_ISL_1794261, EPI_ISL_1794262, EPI_ISL_1794263, EPI_ISL_1794264, EPI_ISL_1794265, EPI_ISL_1794266, EPI_ISL_1794267, EPI_ISL_1794268, EPI_ISL_1794269, EPI_ISL_1794270, EPI_ISL_1794271, EPI_ISL_1794272, EPI_ISL_1794273, EPI_ISL_1794274, EPI_ISL_1794275, EPI_ISL_1794276, EPI_ISL_1794277, EPI_ISL_1794278, EPI_ISL_1794279, EPI_ISL_1794280, EPI_ISL_1794281, EPI_ISL_1794282, EPI_ISL_1794283, EPI_ISL_1794284, EPI_ISL_1794285, EPI_ISL_1794286, EPI_ISL_1794287, EPI_ISL_1794288, EPI_ISL_1794289, EPI_ISL_1794292, EPI_ISL_1794293, EPI_ISL_1794294, EPI_ISL_1794295, EPI_ISL_1794296, EPI_ISL_1794297, EPI_ISL_1794298, EPI_ISL_1794300, EPI_ISL_1794301, EPI_ISL_1794302, EPI_ISL_1794303, EPI_ISL_1794304, EPI_ISL_1794305, EPI_ISL_1794306, EPI_ISL_1794307, EPI_ISL_1794308, EPI_ISL_1794309, EPI_ISL_1794310, EPI_ISL_1794311, EPI_ISL_1794312, EPI_ISL_1794313, EPI_ISL_1794314, EPI_ISL_1794315, EPI_ISL_1794316, EPI_ISL_1794317, EPI_ISL_1794318, EPI_ISL_1794319, EPI_ISL_1794320, EPI_ISL_1794321, EPI_ISL_1794322, EPI_ISL_1794323, EPI_ISL_1794324, EPI_ISL_1794325, EPI_ISL_1794326, EPI_ISL_1794327, EPI_ISL_1794328, EPI_ISL_1794329, EPI_ISL_1794330, EPI_ISL_1794331, EPI_ISL_1794332, EPI_ISL_1794333, EPI_ISL_1794334, EPI_ISL_1794335, EPI_ISL_1794337, EPI_ISL_1794338, EPI_ISL_1794339, EPI_ISL_1794340, EPI_ISL_1794341, EPI_ISL_1794342, EPI_ISL_1794343, EPI_ISL_1794344, EPI_ISL_1794345, EPI_ISL_1794346, EPI_ISL_1794347, EPI_ISL_1794348, EPI_ISL_1794349, EPI_ISL_1794350, EPI_ISL_1794351, EPI_ISL_1794352, EPI_ISL_1794353, EPI_ISL_1794354, EPI_ISL_1794356, EPI_ISL_1794357, EPI_ISL_1794358, EPI_ISL_1794359, EPI_ISL_1794360, EPI_ISL_1794361, EPI_ISL_1794362, EPI_ISL_1794363, EPI_ISL_1794364, EPI_ISL_1794365, EPI_ISL_1794366, EPI_ISL_1794367, EPI_ISL_1794368, EPI_ISL_1794369, EPI_ISL_1794370, EPI_ISL_1794371, EPI_ISL_1794372, EPI_ISL_1794373, EPI_ISL_1794374, EPI_ISL_1794375, EPI_ISL_1794376, EPI_ISL_1794377, EPI_ISL_1794378, EPI_ISL_1794379, EPI_ISL_1794380, EPI_ISL_1794381, EPI_ISL_1794382, EPI_ISL_1794383, EPI_ISL_1794384, EPI_ISL_1794385, EPI_ISL_1794386, EPI_ISL_1794387, EPI_ISL_1794388, EPI_ISL_1794389, EPI_ISL_1794390, EPI_ISL_1794391, EPI_ISL_1794392, EPI_ISL_1794393, EPI_ISL_1794394, EPI_ISL_1794395, EPI_ISL_1794396, EPI_ISL_1794397, EPI_ISL_1794398, EPI_ISL_1794399, EPI_ISL_1794400, EPI_ISL_1794401, EPI_ISL_1794402, EPI_ISL_1794403, EPI_ISL_1794404, EPI_ISL_1794405, EPI_ISL_1794406, EPI_ISL_1794407, EPI_ISL_1794408, EPI_ISL_1794409, EPI_ISL_1794410, EPI_ISL_1794411, EPI_ISL_1794412, EPI_ISL_1794413, EPI_ISL_1794414, EPI_ISL_1794415, EPI_ISL_1794417, EPI_ISL_1794421, EPI_ISL_1794423, EPI_ISL_1794425, EPI_ISL_1794427, EPI_ISL_1794428, EPI_ISL_1794430, EPI_ISL_1794437, EPI_ISL_1794442, EPI_ISL_1794444, EPI_ISL_1794445, EPI_ISL_1794446, EPI_ISL_1794447, EPI_ISL_1794448, EPI_ISL_1794449, EPI_ISL_1794451, EPI_ISL_1794454 | | | |
| see above | Wisconsin State Laboratory of Hygiene Communicable Disease Division | Wisconsin State Laboratory of Hygiene Communicable Disease Division | Kelsey R. Florek, Abigail C. Shockey, Alicia J. Mooney, Sara Wagner |
| EPI_ISL_1794455 | Laboratorio PGM | Laboratorio de Infectología Molecular, Departamento de Bioquímica y Medicina Molecular,Facultad de Medicina - Universidad Autónoma de Nuevo León | Kame A. Galán-Huerta, María F. Herrera-Saldivar, Natalia Martínez-Acuña, Sonia A. Lozano-Sepúlveda, Daniel Arellanos-Soto, Ana M. Rivas-Estilla, Javier Ramos-Jimenez, Gabriela Elizondo, Eduardo Garza-de-la-Peña |
| EPI_ISL_1794457, EPI_ISL_1794459, EPI_ISL_1794460, EPI_ISL_1794462, EPI_ISL_1794463, EPI_ISL_1794464, EPI_ISL_1794465, EPI_ISL_1794467, EPI_ISL_1794468, EPI_ISL_1794469, EPI_ISL_1794470, EPI_ISL_1794471, EPI_ISL_1794472, EPI_ISL_1794478, EPI_ISL_1794480, EPI_ISL_1794481, EPI_ISL_1794482, EPI_ISL_1794483, EPI_ISL_1794484, EPI_ISL_1794485, EPI_ISL_1794487, EPI_ISL_1794488, EPI_ISL_1794489, EPI_ISL_1794490, EPI_ISL_1794491, EPI_ISL_1794492, EPI_ISL_1794493, EPI_ISL_1794494, EPI_ISL_1794495, EPI_ISL_1794496, EPI_ISL_1794497, EPI_ISL_1794498, EPI_ISL_1794499, EPI_ISL_1794500, EPI_ISL_1794501, EPI_ISL_1794502, EPI_ISL_1794503, EPI_ISL_1794504, EPI_ISL_1794505, EPI_ISL_1794506, EPI_ISL_1794507, EPI_ISL_1794508, EPI_ISL_1794509, EPI_ISL_1794510, EPI_ISL_1794511, EPI_ISL_1794512, EPI_ISL_1794513, EPI_ISL_1794514, EPI_ISL_1794515, EPI_ISL_1794516, EPI_ISL_1794517, EPI_ISL_1794518, EPI_ISL_1794519, EPI_ISL_1794520, EPI_ISL_1794521, EPI_ISL_1794522, EPI_ISL_1794523, EPI_ISL_1794524, EPI_ISL_1794525, EPI_ISL_1794526, EPI_ISL_1794527, EPI_ISL_1794528, EPI_ISL_1794529, EPI_ISL_1794530, EPI_ISL_1794531, EPI_ISL_1794532, EPI_ISL_1794533, EPI_ISL_1794534, EPI_ISL_1794535, EPI_ISL_1794536, EPI_ISL_1794537, EPI_ISL_1794538, EPI_ISL_1794539, EPI_ISL_1794540, EPI_ISL_1794541, EPI_ISL_1794542, EPI_ISL_1794543, EPI_ISL_1794544, EPI_ISL_1794545, EPI_ISL_1794546, EPI_ISL_1794547, EPI_ISL_1794548, EPI_ISL_1794549, EPI_ISL_1794550, EPI_ISL_1794551, EPI_ISL_1794552, EPI_ISL_1794553, EPI_ISL_1794554, EPI_ISL_1794555, EPI_ISL_1794556, EPI_ISL_1794557, EPI_ISL_1794558, EPI_ISL_1794559, EPI_ISL_1794560, EPI_ISL_1794561, EPI_ISL_1794562, EPI_ISL_1794563, EPI_ISL_1794565, EPI_ISL_1794566, EPI_ISL_1794567, EPI_ISL_1794568, EPI_ISL_1794569, EPI_ISL_1794570, EPI_ISL_1794571, EPI_ISL_1794572, EPI_ISL_1794573, EPI_ISL_1794574, EPI_ISL_1794575 | | | |
| see above | Wisconsin State Laboratory of Hygiene Communicable Disease Division | Wisconsin State Laboratory of Hygiene Communicable Disease Division | Kelsey R. Florek, Abigail C. Shockey, Alicia J. Mooney, Sara Wagner |

| | | | |
|---|---|---|---|
| EPI_ISL_1794576 | Southern Nevada Public Health Laboratory | Southern Nevada Public Health Laboratory | Michael Picker |
| EPI_ISL_1794577, EPI_ISL_1794578, EPI_ISL_1794579, EPI_ISL_1794580, EPI_ISL_1794581, EPI_ISL_1794582, EPI_ISL_1794583, EPI_ISL_1794584, EPI_ISL_1794585, EPI_ISL_1794586, EPI_ISL_1794587, EPI_ISL_1794588, EPI_ISL_1794589, EPI_ISL_1794590, EPI_ISL_1794591, EPI_ISL_1794592, EPI_ISL_1794593, EPI_ISL_1794594, EPI_ISL_1794595, EPI_ISL_1794596, EPI_ISL_1794597, EPI_ISL_1794598, EPI_ISL_1794599, EPI_ISL_1794600, EPI_ISL_1794601, EPI_ISL_1794602, EPI_ISL_1794603, EPI_ISL_1794604, EPI_ISL_1794605, EPI_ISL_1794606, EPI_ISL_1794607, EPI_ISL_1794608, EPI_ISL_1794609, EPI_ISL_1794610, EPI_ISL_1794611, EPI_ISL_1794612, EPI_ISL_1794613, EPI_ISL_1794614, EPI_ISL_1794615 | | | |
| see above | Scripps Medical Laboratory | Andersen lab at Scripps Research | SEARCH Alliance San Diego with Michael Quigley, Ellen Stefanski, Ian Mchardy |
| EPI_ISL_1794616, EPI_ISL_1794617, EPI_ISL_1794618, EPI_ISL_1794619, EPI_ISL_1794620, EPI_ISL_1794621, EPI_ISL_1794622, EPI_ISL_1794623, EPI_ISL_1794624, EPI_ISL_1794625, EPI_ISL_1794626, EPI_ISL_1794627, EPI_ISL_1794628, EPI_ISL_1794629, EPI_ISL_1794630, EPI_ISL_1794631, EPI_ISL_1794632, EPI_ISL_1794633, EPI_ISL_1794634, EPI_ISL_1794635, EPI_ISL_1794636, EPI_ISL_1794637, EPI_ISL_1794638, EPI_ISL_1794639, EPI_ISL_1794640, EPI_ISL_1794641, EPI_ISL_1794642, EPI_ISL_1794643, EPI_ISL_1794644, EPI_ISL_1794645, EPI_ISL_1794646, EPI_ISL_1794647, EPI_ISL_1794648, EPI_ISL_1794649, EPI_ISL_1794650, EPI_ISL_1794651, EPI_ISL_1794652, EPI_ISL_1794653, EPI_ISL_1794654, EPI_ISL_1794655, EPI_ISL_1794656, EPI_ISL_1794657, EPI_ISL_1794658, EPI_ISL_1794659, EPI_ISL_1794660, EPI_ISL_1794661, EPI_ISL_1794662, EPI_ISL_1794663, EPI_ISL_1794664, EPI_ISL_1794665, EPI_ISL_1794666, EPI_ISL_1794667, EPI_ISL_1794668, EPI_ISL_1794669, EPI_ISL_1794670, EPI_ISL_1794671, EPI_ISL_1794672, EPI_ISL_1794673, EPI_ISL_1794674, EPI_ISL_1794675, EPI_ISL_1794676, EPI_ISL_1794677, EPI_ISL_1794678, EPI_ISL_1794679, EPI_ISL_1794680, EPI_ISL_1794681, EPI_ISL_1794682, EPI_ISL_1794683, EPI_ISL_1794684 | | | |
| see above | San Diego County Public Health Laboratory | Andersen lab at Scripps Research | SEARCH Alliance San Diego with Tracy Basler, Jovan Shephard, Brett Austin |
| EPI_ISL_1794685, EPI_ISL_1794686, EPI_ISL_1794687, EPI_ISL_1794688, EPI_ISL_1794689, EPI_ISL_1794690, EPI_ISL_1794691, EPI_ISL_1794692, EPI_ISL_1794693, EPI_ISL_1794694, EPI_ISL_1794695, EPI_ISL_1794696, EPI_ISL_1794697, EPI_ISL_1794698, EPI_ISL_1794699, EPI_ISL_1794700, EPI_ISL_1794701, EPI_ISL_1794702, EPI_ISL_1794703, EPI_ISL_1794704, EPI_ISL_1794705, EPI_ISL_1794706, EPI_ISL_1794707, EPI_ISL_1794708, EPI_ISL_1794709, EPI_ISL_1794710, EPI_ISL_1794711, EPI_ISL_1794712, EPI_ISL_1794713, EPI_ISL_1794714, EPI_ISL_1794715, EPI_ISL_1794716, EPI_ISL_1794717, EPI_ISL_1794718, EPI_ISL_1794719, EPI_ISL_1794720 | | | |
| see above | Sharp HealthCare Laboratory | Andersen lab at Scripps Research | SEARCH Alliance San Diego with Aaron Harding, Jacquelyn Berumen, Cathy Woerle, Liam McGinnis, Art Mendoza, Omid Bakhtar |
| EPI_ISL_1794721, EPI_ISL_1794722, EPI_ISL_1794723, EPI_ISL_1794724, EPI_ISL_1794725, EPI_ISL_1794726, EPI_ISL_1794727, EPI_ISL_1794728, EPI_ISL_1794729, EPI_ISL_1794730, EPI_ISL_1794731, EPI_ISL_1794732, EPI_ISL_1794733, EPI_ISL_1794734, EPI_ISL_1794735, EPI_ISL_1794736, EPI_ISL_1794737, EPI_ISL_1794738, EPI_ISL_1794739, EPI_ISL_1794740, EPI_ISL_1794741 | | | |
| see above | Scripps Medical Laboratory | Andersen lab at Scripps Research | SEARCH Alliance San Diego with Michael Quigley, Ellen Stefanski, Ian Mchardy |
| EPI_ISL_1794742, EPI_ISL_1794743, EPI_ISL_1794744, EPI_ISL_1794745, EPI_ISL_1794746, EPI_ISL_1794747, EPI_ISL_1794748, EPI_ISL_1794749, EPI_ISL_1794750, EPI_ISL_1794751, EPI_ISL_1794752, EPI_ISL_1794753, EPI_ISL_1794754, EPI_ISL_1794755, EPI_ISL_1794756, EPI_ISL_1794757, EPI_ISL_1794758, EPI_ISL_1794759, EPI_ISL_1794760, EPI_ISL_1794761, EPI_ISL_1794762, EPI_ISL_1794763, EPI_ISL_1794764, EPI_ISL_1794765, EPI_ISL_1794766, EPI_ISL_1794767 | | | |
| see above | Sharp HealthCare Laboratory | Andersen lab at Scripps Research | SEARCH Alliance San Diego with Aaron Harding, Jacquelyn Berumen, Cathy Woerle, Liam McGinnis, Art Mendoza, Omid Bakhtar |
| EPI_ISL_1794768, EPI_ISL_1794769, EPI_ISL_1794770, EPI_ISL_1794771, EPI_ISL_1794772, EPI_ISL_1794773, EPI_ISL_1794774, EPI_ISL_1794775, EPI_ISL_1794776, EPI_ISL_1794777, EPI_ISL_1794778, EPI_ISL_1794779, EPI_ISL_1794780, EPI_ISL_1794781 | | | |
| see above | Rady Children's Hospital - San Diego | Andersen lab at Scripps Research | SEARCH Alliance San Diego with Nanda Radamchar, David Dimmock, Linda Luo, Christina Clarke, Kathryn Bouic, Teresa Mueller, Denise Malicki |
| EPI_ISL_1794782, EPI_ISL_1794783, EPI_ISL_1794784, EPI_ISL_1794785, EPI_ISL_1794786 | Sharp HealthCare Laboratory | Andersen lab at Scripps Research | SEARCH Alliance San Diego with Aaron Harding, Jacquelyn Berumen, Cathy Woerle, Liam McGinnis, Art Mendoza, Omid Bakhtar |
| EPI_ISL_1794787, EPI_ISL_1794788, EPI_ISL_1794789, EPI_ISL_1794790, EPI_ISL_1794791, EPI_ISL_1794792, EPI_ISL_1794793, EPI_ISL_1794794, EPI_ISL_1794795, EPI_ISL_1794796, EPI_ISL_1794797, EPI_ISL_1794798, EPI_ISL_1794799, EPI_ISL_1794800, EPI_ISL_1794801, EPI_ISL_1794802, EPI_ISL_1794803, EPI_ISL_1794804, EPI_ISL_1794805, EPI_ISL_1794806, EPI_ISL_1794807, EPI_ISL_1794808, EPI_ISL_1794809, EPI_ISL_1794810, EPI_ISL_1794811, EPI_ISL_1794812, EPI_ISL_1794813, EPI_ISL_1794814, EPI_ISL_1794815, EPI_ISL_1794816, EPI_ISL_1794817, EPI_ISL_1794818, EPI_ISL_1794819, EPI_ISL_1794820, EPI_ISL_1794821, EPI_ISL_1794822, EPI_ISL_1794823, EPI_ISL_1794824, EPI_ISL_1794825, EPI_ISL_1794826, EPI_ISL_1794827, EPI_ISL_1794828, EPI_ISL_1794829, EPI_ISL_1794830, EPI_ISL_1794831, EPI_ISL_1794832, EPI_ISL_1794833, EPI_ISL_1794834, EPI_ISL_1794835, EPI_ISL_1794836, EPI_ISL_1794837, EPI_ISL_1794838, EPI_ISL_1794839, EPI_ISL_1794840, EPI_ISL_1794841, EPI_ISL_1794842, EPI_ISL_1794843, EPI_ISL_1794844, EPI_ISL_1794845, EPI_ISL_1794846, EPI_ISL_1794847, EPI_ISL_1794848, EPI_ISL_1794849, EPI_ISL_1794850, EPI_ISL_1794851, EPI_ISL_1794852, EPI_ISL_1794853 | | | |
| see above | Scripps Medical Laboratory | Andersen lab at Scripps Research | SEARCH Alliance San Diego with Michael Quigley, Ellen Stefanski, Ian Mchardy |
| EPI_ISL_1794854, EPI_ISL_1794855, EPI_ISL_1794856, EPI_ISL_1794857, EPI_ISL_1794858, EPI_ISL_1794859, EPI_ISL_1794860, EPI_ISL_1794861, EPI_ISL_1794862, EPI_ISL_1794863, EPI_ISL_1794864, EPI_ISL_1794865, EPI_ISL_1794866, EPI_ISL_1794867, EPI_ISL_1794868, EPI_ISL_1794869, EPI_ISL_1794870, EPI_ISL_1794871, EPI_ISL_1794872, EPI_ISL_1794873, EPI_ISL_1794874, EPI_ISL_1794875, EPI_ISL_1794876, EPI_ISL_1794877, EPI_ISL_1794878, EPI_ISL_1794879, EPI_ISL_1794880, EPI_ISL_1794881, EPI_ISL_1794882, EPI_ISL_1794883, EPI_ISL_1794884, EPI_ISL_1794885, EPI_ISL_1794886, EPI_ISL_1794887, EPI_ISL_1794888, EPI_ISL_1794889, EPI_ISL_1794890, EPI_ISL_1794891, EPI_ISL_1794892, EPI_ISL_1794893, EPI_ISL_1794894, EPI_ISL_1794895, EPI_ISL_1794896, EPI_ISL_1794897, EPI_ISL_1794898, EPI_ISL_1794899, EPI_ISL_1794900, EPI_ISL_1794901, EPI_ISL_1794902, EPI_ISL_1794903, EPI_ISL_1794904, EPI_ISL_1794905, EPI_ISL_1794906, EPI_ISL_1794907, EPI_ISL_1794908, EPI_ISL_1794909, EPI_ISL_1794910, EPI_ISL_1794911, EPI_ISL_1794912, EPI_ISL_1794913, EPI_ISL_1794914, EPI_ISL_1794915, EPI_ISL_1794916, EPI_ISL_1794917, EPI_ISL_1794918, EPI_ISL_1794919, EPI_ISL_1794920, EPI_ISL_1794921, EPI_ISL_1794922, EPI_ISL_1794923, EPI_ISL_1794924, EPI_ISL_1794925, EPI_ISL_1794926, EPI_ISL_1794927, EPI_ISL_1794928, EPI_ISL_1794929, EPI_ISL_1794930, EPI_ISL_1794931, EPI_ISL_1794932, EPI_ISL_1794933 | | | |
| see above | Sharp HealthCare Laboratory | Andersen lab at Scripps Research | SEARCH Alliance San Diego with Aaron Harding, Jacquelyn Berumen, Cathy Woerle, Liam McGinnis, Art Mendoza, Omid Bakhtar |
| EPI_ISL_1794934, EPI_ISL_1794935, EPI_ISL_1794936 | Scripps Medical Laboratory | Andersen lab at Scripps Research | SEARCH Alliance San Diego with Michael Quigley, Ellen Stefanski, Ian Mchardy |
| EPI_ISL_1794937, EPI_ISL_1794938, EPI_ISL_1794939, EPI_ISL_1794940, EPI_ISL_1794941, EPI_ISL_1794942, EPI_ISL_1794943, EPI_ISL_1794944, EPI_ISL_1794945, EPI_ISL_1794946, EPI_ISL_1794947, EPI_ISL_1794948, EPI_ISL_1794949, EPI_ISL_1794950, EPI_ISL_1794951, EPI_ISL_1794952, EPI_ISL_1794953, EPI_ISL_1794954, EPI_ISL_1794955, EPI_ISL_1794956, EPI_ISL_1794957, EPI_ISL_1794958, EPI_ISL_1794959, EPI_ISL_1794960, EPI_ISL_1794961, EPI_ISL_1794962, EPI_ISL_1794963, EPI_ISL_1794964, EPI_ISL_1794965, EPI_ISL_1794966, EPI_ISL_1794967, EPI_ISL_1794968, EPI_ISL_1794969, EPI_ISL_1794970, EPI_ISL_1794971, EPI_ISL_1794972, EPI_ISL_1794973, EPI_ISL_1794974, EPI_ISL_1794975, EPI_ISL_1794976, EPI_ISL_1794977, EPI_ISL_1794978, EPI_ISL_1794979, EPI_ISL_1794980, EPI_ISL_1794981, EPI_ISL_1794982, EPI_ISL_1794983, EPI_ISL_1794984, EPI_ISL_1794985, EPI_ISL_1794986, EPI_ISL_1794987, EPI_ISL_1794988, EPI_ISL_1794989, EPI_ISL_1794990, EPI_ISL_1794991, EPI_ISL_1794992, EPI_ISL_1794993, EPI_ISL_1794994, EPI_ISL_1794995, EPI_ISL_1794996, EPI_ISL_1794997, EPI_ISL_1794998, EPI_ISL_1794999, EPI_ISL_1795000, EPI_ISL_1795001, EPI_ISL_1795002, EPI_ISL_1795003, EPI_ISL_1795004, EPI_ISL_1795005, EPI_ISL_1795006, EPI_ISL_1795007, EPI_ISL_1795008, EPI_ISL_1795009, EPI_ISL_1795010, EPI_ISL_1795011, EPI_ISL_1795012, EPI_ISL_1795013, EPI_ISL_1795014, EPI_ISL_1795015, EPI_ISL_1795016, EPI_ISL_1795017, EPI_ISL_1795018, EPI_ISL_1795019, EPI_ISL_1795020, EPI_ISL_1795021, EPI_ISL_1795022, EPI_ISL_1795023, EPI_ISL_1795024, EPI_ISL_1795025, EPI_ISL_1795026, EPI_ISL_1795027, EPI_ISL_1795028, EPI_ISL_1795029, EPI_ISL_1795030, EPI_ISL_1795031, EPI_ISL_1795032, EPI_ISL_1795033, EPI_ISL_1795034, EPI_ISL_1795035, EPI_ISL_1795036, EPI_ISL_1795037, EPI_ISL_1795038, EPI_ISL_1795039, EPI_ISL_1795040, EPI_ISL_1795041, EPI_ISL_1795042 | | | |
| see above | San Diego County Public Health Laboratory | Andersen lab at Scripps Research | SEARCH Alliance San Diego with Tracy Basler, Jovan Shephard, Brett Austin |
| EPI_ISL_1795430, EPI_ISL_1795431, EPI_ISL_1795432, EPI_ISL_1795433, EPI_ISL_1795434, EPI_ISL_1795435, EPI_ISL_1795436, EPI_ISL_1795437, EPI_ISL_1795438, EPI_ISL_1795439, EPI_ISL_1795441, EPI_ISL_1795442, EPI_ISL_1795443, EPI_ISL_1795444, EPI_ISL_1795445, EPI_ISL_1795446, EPI_ISL_1795447, EPI_ISL_1795448, EPI_ISL_1795449, EPI_ISL_1795450, EPI_ISL_1795451, EPI_ISL_1795452, EPI_ISL_1795453, EPI_ISL_1795454, EPI_ISL_1795455, EPI_ISL_1795456, EPI_ISL_1795457, EPI_ISL_1795458, EPI_ISL_1795459, EPI_ISL_1795460, EPI_ISL_1795461, EPI_ISL_1795462, EPI_ISL_1795463, EPI_ISL_1795464, EPI_ISL_1795465, EPI_ISL_1795466, EPI_ISL_1795467, EPI_ISL_1795468, EPI_ISL_1795469, EPI_ISL_1795470, EPI_ISL_1795471, EPI_ISL_1795472, EPI_ISL_1795473, EPI_ISL_1795474, EPI_ISL_1795475, EPI_ISL_1795476, EPI_ISL_1795477, EPI_ISL_1795478, EPI_ISL_1795479, EPI_ISL_1795480, EPI_ISL_1795481, EPI_ISL_1795482, EPI_ISL_1795483, EPI_ISL_1795484, EPI_ISL_1795485, EPI_ISL_1795486, EPI_ISL_1795487, EPI_ISL_1795488, EPI_ISL_1795489, EPI_ISL_1795490, EPI_ISL_1795491, EPI_ISL_1795492, EPI_ISL_1795493, EPI_ISL_1795494, EPI_ISL_1795495, EPI_ISL_1795496, EPI_ISL_1795497, EPI_ISL_1795498, EPI_ISL_1795499, EPI_ISL_1795500, EPI_ISL_1795501, EPI_ISL_1795502, EPI_ISL_1795504, EPI_ISL_1795505, EPI_ISL_1795506, EPI_ISL_1795507, EPI_ISL_1795508, EPI_ISL_1795509, EPI_ISL_1795510, EPI_ISL_1795511, EPI_ISL_1795512, EPI_ISL_1795513, EPI_ISL_1795514, EPI_ISL_1795515, EPI_ISL_1795516, EPI_ISL_1795517, EPI_ISL_1795518, EPI_ISL_1795519, EPI_ISL_1795520, EPI_ISL_1795522, EPI_ISL_1795523, EPI_ISL_1795524, EPI_ISL_1795525, EPI_ISL_1795526, EPI_ISL_1795527, EPI_ISL_1795528, EPI_ISL_1795529, EPI_ISL_1795530, EPI_ISL_1795531, EPI_ISL_1795532, EPI_ISL_1795533, EPI_ISL_1795534, EPI_ISL_1795535, EPI_ISL_1795536, EPI_ISL_1795537, EPI_ISL_1795538, EPI_ISL_1795539, EPI_ISL_1795540, EPI_ISL_1795541, EPI_ISL_1795542, EPI_ISL_1795543, EPI_ISL_1795544, EPI_ISL_1795545, EPI_ISL_1795546, EPI_ISL_1795547, EPI_ISL_1795548, EPI_ISL_1795549, EPI_ISL_1795550, EPI_ISL_1795551, EPI_ISL_1795552, EPI_ISL_1795553, EPI_ISL_1795554, EPI_ISL_1795555, EPI_ISL_1795556, EPI_ISL_1795557, EPI_ISL_1795558, EPI_ISL_1795559, EPI_ISL_1795560, EPI_ISL_1795561, EPI_ISL_1795562, EPI_ISL_1795563, EPI_ISL_1795564, EPI_ISL_1795565, EPI_ISL_1795566, EPI_ISL_1795567, EPI_ISL_1795568, EPI_ISL_1795569, EPI_ISL_1795570, EPI_ISL_1795571, EPI_ISL_1795572, EPI_ISL_1795573, EPI_ISL_1795574, EPI_ISL_1795575, EPI_ISL_1795576, EPI_ISL_1795577, EPI_ISL_1795578, EPI_ISL_1795579, EPI_ISL_1795580, EPI_ISL_1795581, EPI_ISL_1795582, EPI_ISL_1795583, EPI_ISL_1795584, EPI_ISL_1795585, EPI_ISL_1795586, EPI_ISL_1795587, EPI_ISL_1795588, EPI_ISL_1795589, EPI_ISL_1795590, EPI_ISL_1795591, EPI_ISL_1795592, EPI_ISL_1795593, EPI_ISL_1795594, EPI_ISL_1795595, EPI_ISL_1795596, EPI_ISL_1795597, EPI_ISL_1795598, EPI_ISL_1795599, EPI_ISL_1795600, EPI_ISL_1795601, EPI_ISL_1795602, EPI_ISL_1795603, EPI_ISL_1795604, EPI_ISL_1795605, EPI_ISL_1795606, EPI_ISL_1795607, EPI_ISL_1795608, EPI_ISL_1795609, EPI_ISL_1795610, EPI_ISL_1795611, EPI_ISL_1795612, EPI_ISL_1795613, EPI_ISL_1795614, EPI_ISL_1795615, EPI_ISL_1795616, EPI_ISL_1795617, EPI_ISL_1795618, EPI_ISL_1795619, EPI_ISL_1795620, EPI_ISL_1795621, EPI_ISL_1795622, EPI_ISL_1795623, EPI_ISL_1795624, EPI_ISL_1795625, EPI_ISL_1795626, EPI_ISL_1795628, EPI_ISL_1795629, EPI_ISL_1795630, EPI_ISL_1795631, EPI_ISL_1795632, EPI_ISL_1795633, EPI_ISL_1795634, EPI_ISL_1795635, EPI_ISL_1795636, EPI_ISL_1795637, EPI_ISL_1795638, EPI_ISL_1795639, EPI_ISL_1795640, EPI_ISL_1795641, EPI_ISL_1795642, EPI_ISL_1795643, EPI_ISL_1795644, EPI_ISL_1795645, EPI_ISL_1795646, EPI_ISL_1795647, EPI_ISL_1795648, EPI_ISL_1795649, EPI_ISL_1795650, EPI_ISL_1795651, EPI_ISL_1795652, EPI_ISL_1795655, EPI_ISL_1795657, EPI_ISL_1795661, EPI_ISL_1795665, EPI_ISL_1795668, EPI_ISL_1795671, EPI_ISL_1795675, EPI_ISL_1795681, EPI_ISL_1795684, EPI_ISL_1795687, EPI_ISL_1795691, EPI_ISL_1795693, EPI_ISL_1795696, EPI_ISL_1795699, EPI_ISL_1795701, EPI_ISL_1795703, EPI_ISL_1795704, EPI_ISL_1795707, EPI_ISL_1795710, EPI_ISL_1795716, EPI_ISL_1795718, EPI_ISL_1795721, EPI_ISL_1795722, EPI_ISL_1795724, EPI_ISL_1795726, EPI_ISL_1795728, EPI_ISL_1795729, EPI_ISL_1795731, EPI_ISL_1795733, EPI_ISL_1795735, EPI_ISL_1795737, EPI_ISL_1795739, EPI_ISL_1795742, EPI_ISL_1795745, EPI_ISL_1795748, EPI_ISL_1795752, EPI_ISL_1795755, EPI_ISL_1795758, EPI_ISL_1795761, EPI_ISL_1795765, EPI_ISL_1795769, EPI_ISL_1795771, EPI_ISL_1795774, EPI_ISL_1795777, EPI_ISL_1795781, EPI_ISL_1795784, EPI_ISL_1795787, EPI_ISL_1795791, EPI_ISL_1795794, EPI_ISL_1795797, EPI_ISL_1795800, EPI_ISL_1795804, EPI_ISL_1795808, EPI_ISL_1795811, EPI_ISL_1795814, EPI_ISL_1795817, EPI_ISL_1795820, EPI_ISL_1795824, EPI_ISL_1795827, EPI_ISL_1795830, EPI_ISL_1795833, EPI_ISL_1795836, EPI_ISL_1795839, EPI_ISL_1795842, EPI_ISL_1795845, EPI_ISL_1795849, EPI_ISL_1795852, EPI_ISL_1795855, EPI_ISL_1795857, EPI_ISL_1795860, EPI_ISL_1795864, EPI_ISL_1795867, EPI_ISL_1795874, EPI_ISL_1795877, EPI_ISL_1795880, EPI_ISL_1795884, EPI_ISL_1795888, EPI_ISL_1795891, EPI_ISL_1795895, EPI_ISL_1795898, EPI_ISL_1795901, EPI_ISL_1795904, EPI_ISL_1795907, EPI_ISL_1795910, EPI_ISL_1795914, EPI_ISL_1795917, EPI_ISL_1795920, EPI_ISL_1795923, EPI_ISL_1795927, EPI_ISL_1795930, EPI_ISL_1795933, EPI_ISL_1795936, EPI_ISL_1795938, EPI_ISL_1795942, EPI_ISL_1795946, EPI_ISL_1795948, EPI_ISL_1795952, EPI_ISL_1795955, EPI_ISL_1795958, EPI_ISL_1795961, EPI_ISL_1795967, EPI_ISL_1795970, EPI_ISL_1795973, EPI_ISL_1795976, EPI_ISL_1795979, EPI_ISL_1795983, EPI_ISL_1795986, EPI_ISL_1795987, EPI_ISL_1795988, EPI_ISL_1795989, EPI_ISL_1795990, EPI_ISL_1795991, EPI_ISL_1795992, EPI_ISL_1795993, EPI_ISL_1795994, EPI_ISL_1795995, EPI_ISL_1795996, EPI_ISL_1795997, EPI_ISL_1795998, EPI_ISL_1795999, EPI_ISL_1796000, EPI_ISL_1796001, EPI_ISL_1796002, EPI_ISL_1796003, EPI_ISL_1796004, EPI_ISL_1796005, EPI_ISL_1796006, EPI_ISL_1796007, EPI_ISL_1796008, EPI_ISL_1796009, EPI_ISL_1796010, EPI_ISL_1796011, EPI_ISL_1796012, EPI_ISL_1796013, EPI_ISL_1796014, EPI_ISL_1796015, EPI_ISL_1796016, EPI_ISL_1796017, EPI_ISL_1796018, EPI_ISL_1796019, EPI_ISL_1796020, EPI_ISL_1796021, EPI_ISL_1796022, EPI_ISL_1796023, EPI_ISL_1796024, EPI_ISL_1796025, EPI_ISL_1796026, EPI_ISL_1796027, EPI_ISL_1796028, EPI_ISL_1796029, EPI_ISL_1796030, EPI_ISL_1796031, EPI_ISL_1796032, EPI_ISL_1796033, EPI_ISL_1796034, EPI_ISL_1796035, EPI_ISL_1796036, EPI_ISL_1796037, EPI_ISL_1796038, EPI_ISL_1796039, EPI_ISL_1796040, EPI_ISL_1796041, EPI_ISL_1796042, EPI_ISL_1796044, EPI_ISL_1796045, EPI_ISL_1796046, EPI_ISL_1796047, EPI_ISL_1796048, EPI_ISL_1796049, EPI_ISL_1796050, EPI_ISL_1796051, EPI_ISL_1796052, EPI_ISL_1796053, EPI_ISL_1796055, EPI_ISL_1796056, EPI_ISL_1796057, EPI_ISL_1796059, EPI_ISL_1796060, EPI_ISL_1796062, EPI_ISL_1796063, EPI_ISL_1796064, EPI_ISL_1796066, EPI_ISL_1796067, EPI_ISL_1796068, EPI_ISL_1796069, EPI_ISL_1796070, EPI_ISL_1796071, EPI_ISL_1796072, EPI_ISL_1796073, EPI_ISL_1796074, EPI_ISL_1796075, EPI_ISL_1796076, EPI_ISL_1796077, EPI_ISL_1796078, EPI_ISL_1796079, EPI_ISL_1796080, EPI_ISL_1796081, EPI_ISL_1796082, EPI_ISL_1796083, EPI_ISL_1796084, EPI_ISL_1796085, EPI_ISL_1796086, EPI_ISL_1796087, EPI_ISL_1796088, EPI_ISL_1796089, EPI_ISL_1796090, EPI_ISL_1796091, EPI_ISL_1796092, EPI_ISL_1796093, EPI_ISL_1796094, EPI_ISL_1796095, EPI_ISL_1796096, EPI_ISL_1796097, EPI_ISL_1796098, EPI_ISL_1796099, EPI_ISL_1796100, EPI_ISL_1796101, EPI_ISL_1796102, EPI_ISL_1796103, EPI_ISL_1796104, EPI_ISL_1796105, EPI_ISL_1796106, EPI_ISL_1796107, EPI_ISL_1796109, EPI_ISL_1796110, EPI_ISL_1796111, EPI_ISL_1796112, EPI_ISL_1796114, EPI_ISL_1796115, EPI_ISL_1796116, EPI_ISL_1796117, EPI_ISL_1796118, EPI_ISL_1796119, EPI_ISL_1796120, EPI_ISL_1796121, EPI_ISL_1796122 | | | |
| see above | Infinity Biologix | Centers for Disease Control and Prevention Division of Viral Diseases, Pathogen Discovery | Dakota Howard, Dhwani Batra, Peter W. Cook, Kara Moser, Adrian Paskey, Jason Caravas, Benjamin Rambo-Martin, Shatavia Morrison, Christopher Gulvick, Scott Sammons, Yvette Unoarumhi, Darlene Wagner, Matthew Schmerer, Christian Bixby, Yihe Wang, Jonathan Schultz, Chirayu Goswami, Russ Hager, Robin Grimwood, Clinton R. Paden, Duncan MacCannell |
| EPI_ISL_1796123, EPI_ISL_1796124, EPI_ISL_1796125, EPI_ISL_1796127, EPI_ISL_1796128, EPI_ISL_1796129, EPI_ISL_1796130, EPI_ISL_1796131, EPI_ISL_1796132, EPI_ISL_1796133, EPI_ISL_1796134, EPI_ISL_1796135, EPI_ISL_1796136, EPI_ISL_1796137, EPI_ISL_1796138, EPI_ISL_1796139, EPI_ISL_1796140, EPI_ISL_1796141, EPI_ISL_1796142, EPI_ISL_1796143, EPI_ISL_1796144, EPI_ISL_1796145, EPI_ISL_1796146, EPI_ISL_1796147, EPI_ISL_1796148, EPI_ISL_1796149, EPI_ISL_1796150, EPI_ISL_1796151, EPI_ISL_1796152, EPI_ISL_1796153, EPI_ISL_1796154, EPI_ISL_1796155, EPI_ISL_1796156, EPI_ISL_1796157, EPI_ISL_1796158, EPI_ISL_1796159, EPI_ISL_1796160, EPI_ISL_1796161, EPI_ISL_1796162, EPI_ISL_1796163, EPI_ISL_1796164, EPI_ISL_1796165, EPI_ISL_1796166, EPI_ISL_1796167, EPI_ISL_1796168, EPI_ISL_1796169, EPI_ISL_1796170, EPI_ISL_1796171, EPI_ISL_1796172, EPI_ISL_1796173, EPI_ISL_1796174, EPI_ISL_1796175, EPI_ISL_1796176, EPI_ISL_1796177, EPI_ISL_1796178, EPI_ISL_1796179, EPI_ISL_1796180, EPI_ISL_1796181, EPI_ISL_1796182, EPI_ISL_1796183, EPI_ISL_1796184, EPI_ISL_1796185, EPI_ISL_1796186, EPI_ISL_1796187, EPI_ISL_1796188, EPI_ISL_1796189, EPI_ISL_1796190, EPI_ISL_1796191, EPI_ISL_1796192, EPI_ISL_1796193, EPI_ISL_1796195, EPI_ISL_1796196, EPI_ISL_1796197, EPI_ISL_1796198, EPI_ISL_1796199, EPI_ISL_1796200, EPI_ISL_1796201, EPI_ISL_1796202, EPI_ISL_1796203, EPI_ISL_1796204, EPI_ISL_1796205, EPI_ISL_1796206, EPI_ISL_1796207, EPI_ISL_1796208, EPI_ISL_1796210, EPI_ISL_1796211, EPI_ISL_1796212, EPI_ISL_1796213, EPI_ISL_1796214, EPI_ISL_1796215, EPI_ISL_1796216, EPI_ISL_1796217, EPI_ISL_1796218, EPI_ISL_1796219, EPI_ISL_1796220, EPI_ISL_1796221, EPI_ISL_1796222, EPI_ISL_1796223, EPI_ISL_1796224, EPI_ISL_1796225, EPI_ISL_1796226, EPI_ISL_1796227, EPI_ISL_1796228, EPI_ISL_1796229, EPI_ISL_1796230, EPI_ISL_1796231, EPI_ISL_1796232, EPI_ISL_1796233, EPI_ISL_1796234, EPI_ISL_1796235, EPI_ISL_1796237, EPI_ISL_1796238, EPI_ISL_1796239, EPI_ISL_1796240, EPI_ISL_1796241, EPI_ISL_1796242, EPI_ISL_1796243, EPI_ISL_1796244, EPI_ISL_1796245, EPI_ISL_1796246, EPI_ISL_1796247, EPI_ISL_1796248, EPI_ISL_1796249, EPI_ISL_1796250, EPI_ISL_1796252, EPI_ISL_1796253, EPI_ISL_1796254, EPI_ISL_1796255, EPI_ISL_1796256, EPI_ISL_1796257, EPI_ISL_1796258, EPI_ISL_1796259, EPI_ISL_1796260, EPI_ISL_1796261, EPI_ISL_1796262, EPI_ISL_1796263, EPI_ISL_1796265, EPI_ISL_1796266, EPI_ISL_1796267, EPI_ISL_1796269, EPI_ISL_1796270, EPI_ISL_1796271, EPI_ISL_1796272, EPI_ISL_1796273, EPI_ISL_1796274, EPI_ISL_1796275, EPI_ISL_1796276, EPI_ISL_1796277, EPI_ISL_1796278, EPI_ISL_1796279, EPI_ISL_1796280, EPI_ISL_1796281, EPI_ISL_1796282, EPI_ISL_1796283, EPI_ISL_1796284, EPI_ISL_1796285, EPI_ISL_1796286, EPI_ISL_1796287, EPI_ISL_1796288, EPI_ISL_1796289, EPI_ISL_1796290, EPI_ISL_1796291, EPI_ISL_1796292, EPI_ISL_1796293, EPI_ISL_1796294, EPI_ISL_1796295, EPI_ISL_1796296, EPI_ISL_1796297, EPI_ISL_1796298, EPI_ISL_1796299, EPI_ISL_1796300, EPI_ISL_1796301, EPI_ISL_1796302, EPI_ISL_1796303, EPI_ISL_1796304, EPI_ISL_1796305, EPI_ISL_1796306, EPI_ISL_1796307, EPI_ISL_1796308, EPI_ISL_1796309, EPI_ISL_1796310, EPI_ISL_1796311, EPI_ISL_1796312, EPI_ISL_1796313, EPI_ISL_1796314, EPI_ISL_1796315, EPI_ISL_1796316, EPI_ISL_1796317, EPI_ISL_1796318, EPI_ISL_1796319, EPI_ISL_1796320, EPI_ISL_1796321, EPI_ISL_1796322, EPI_ISL_1796323, EPI_ISL_1796324, EPI_ISL_1796326, EPI_ISL_1796327, EPI_ISL_1796328, EPI_ISL_1796329, EPI_ISL_1796330, EPI_ISL_1796331, EPI_ISL_1796332, EPI_ISL_1796333, EPI_ISL_1796334, EPI_ISL_1796335, EPI_ISL_1796336, EPI_ISL_1796337, EPI_ISL_1796338, EPI_ISL_1796339, EPI_ISL_1796340, EPI_ISL_1796341, EPI_ISL_1796342, EPI_ISL_1796343, EPI_ISL_1796344, EPI_ISL_1796345, EPI_ISL_1796346, EPI_ISL_1796347, EPI_ISL_1796348, EPI_ISL_1796349, EPI_ISL_1796350, EPI_ISL_1796351, EPI_ISL_1796352, EPI_ISL_1796353, EPI_ISL_1796354, EPI_ISL_1796355, EPI_ISL_1796356, EPI_ISL_1796357, EPI_ISL_1796358, EPI_ISL_1796359, EPI_ISL_1796360, EPI_ISL_1796361, EPI_ISL_1796362, EPI_ISL_1796363, EPI_ISL_1796364, EPI_ISL_1796365, EPI_ISL_1796366, EPI_ISL_1796367, EPI_ISL_1796368, EPI_ISL_1796369, EPI_ISL_1796370, EPI_ISL_1796372, EPI_ISL_1796373, EPI_ISL_1796374, EPI_ISL_1796375, EPI_ISL_1796376, EPI_ISL_1796377, EPI_ISL_1796378, EPI_ISL_1796379, EPI_ISL_1796380, EPI_ISL_1796381, EPI_ISL_1796382, EPI_ISL_1796383, EPI_ISL_1796384, EPI_ISL_1796385, EPI_ISL_1796386, EPI_ISL_1796387, EPI_ISL_1796388, EPI_ISL_1796389, EPI_ISL_1796390, EPI_ISL_1796391, EPI_ISL_1796392, EPI_ISL_1796393, EPI_ISL_1796394, EPI_ISL_1796395, EPI_ISL_1796396, EPI_ISL_1796397, EPI_ISL_1796398, EPI_ISL_1796399, EPI_ISL_1796400, EPI_ISL_1796401, EPI_ISL_1796402, EPI_ISL_1796403, EPI_ISL_1796404, EPI_ISL_1796405, EPI_ISL_1796406, EPI_ISL_1796407, EPI_ISL_1796408, EPI_ISL_1796409, EPI_ISL_1796410, EPI_ISL_1796411, EPI_ISL_1796412, EPI_ISL_1796413, EPI_ISL_1796414, EPI_ISL_1796415, EPI_ISL_1796416, EPI_ISL_1796417, EPI_ISL_1796418, EPI_ISL_1796419, EPI_ISL_1796420, EPI_ISL_1796421, EPI_ISL_1796422, EPI_ISL_1796423, EPI_ISL_1796424, EPI_ISL_1796425, EPI_ISL_1796426, EPI_ISL_1796427, EPI_ISL_1796428, EPI_ISL_1796429, EPI_ISL_1796430, EPI_ISL_1796431, EPI_ISL_1796432, EPI_ISL_1796433, EPI_ISL_1796434, EPI_ISL_1796435, EPI_ISL_1796436, EPI_ISL_1796437, EPI_ISL_1796438, EPI_ISL_1796439, EPI_ISL_1796440, EPI_ISL_1796441, EPI_ISL_1796442, EPI_ISL_1796443, EPI_ISL_1796444, EPI_ISL_1796445, EPI_ISL_1796446, EPI_ISL_1796447, EPI_ISL_1796448, EPI_ISL_1796449, EPI_ISL_1796450, EPI_ISL_1796451, EPI_ISL_1796452, EPI_ISL_1796453, EPI_ISL_1796454, EPI_ISL_1796455, EPI_ISL_1796456, EPI_ISL_1796457, EPI_ISL_1796458, EPI_ISL_1796459, EPI_ISL_1796460, EPI_ISL_1796461, EPI_ISL_1796462, EPI_ISL_1796463, EPI_ISL_1796464, EPI_ISL_1796465, EPI_ISL_1796466, EPI_ISL_1796467, EPI_ISL_1796468, EPI_ISL_1796469, EPI_ISL_1796470, EPI_ISL_1796471, EPI_ISL_1796472, EPI_ISL_1796473, EPI_ISL_1796474, EPI_ISL_1796475, EPI_ISL_1796476, EPI_ISL_1796477, EPI_ISL_1796478, EPI_ISL_1796479, EPI_ISL_1796480, EPI_ISL_1796481, EPI_ISL_1796482, EPI_ISL_1796483, EPI_ISL_1796484, EPI_ISL_1796485, EPI_ISL_1796486, EPI_ISL_1796487, EPI_ISL_1796488, EPI_ISL_1796489, EPI_ISL_1796490, EPI_ISL_1796491, EPI_ISL_1796492, EPI_ISL_1796493, EPI_ISL_1796494, EPI_ISL_1796495, EPI_ISL_1796496, EPI_ISL_1796497, EPI_ISL_1796498, EPI_ISL_1796499, EPI_ISL_1796500, EPI_ISL_1796501, EPI_ISL_1796502, EPI_ISL_1796503, EPI_ISL_1796504, EPI_ISL_1796505, EPI_ISL_1796506, EPI_ISL_1796507, EPI_ISL_1796508, EPI_ISL_1796509, EPI_ISL_1796510, EPI_ISL_1796511, EPI_ISL_1796512, EPI_ISL_1796513, EPI_ISL_1796514, EPI_ISL_1796515, EPI_ISL_1796516, EPI_ISL_1796517, EPI_ISL_1796518, EPI_ISL_1796519, EPI_ISL_1796520, EPI_ISL_1796521, EPI_ISL_1796522, EPI_ISL_1796523, EPI_ISL_1796524, EPI_ISL_1796525, EPI_ISL_1796526, EPI_ISL_1796527, EPI_ISL_1796528, EPI_ISL_1796529, EPI_ISL_1796530, EPI_ISL_1796531, EPI_ISL_1796532, EPI_ISL_1796533, EPI_ISL_1796534, EPI_ISL_1796535, EPI_ISL_1796536, EPI_ISL_1796537, EPI_ISL_1796538, EPI_ISL_1796539, EPI_ISL_1796540, EPI_ISL_1796541, EPI_ISL_1796542, EPI_ISL_1796543, EPI_ISL_1796544, EPI_ISL_1796545, EPI_ISL_1796546, EPI_ISL_1796547, EPI_ISL_1796548, EPI_ISL_1796549, EPI_ISL_1796550, EPI_ISL_1796551, EPI_ISL_1796552, EPI_ISL_1796553, EPI_ISL_1796554, EPI_ISL_1796555, EPI_ISL_1796556, EPI_ISL_1796557, EPI_ISL_1796558, EPI_ISL_1796559, EPI_ISL_1796560, EPI_ISL_1796561, EPI_ISL_1796562, EPI_ISL_1796563, EPI_ISL_1796564, EPI_ISL_1796565, EPI_ISL_1796567, EPI_ISL_1796568, EPI_ISL_1796569, EPI_ISL_1796570, EPI_ISL_1796571, EPI_ISL_1796572, EPI_ISL_1796573, EPI_ISL_1796574, EPI_ISL_1796575, EPI_ISL_1796576, EPI_ISL_1796577, EPI_ISL_1796578, EPI_ISL_1796579, EPI_ISL_1796580, EPI_ISL_1796581, EPI_ISL_1796582, EPI_ISL_1796583, EPI_ISL_1796584, EPI_ISL_1796585, EPI_ISL_1796586, EPI_ISL_1796587, EPI_ISL_1796588, EPI_ISL_1796589, EPI_ISL_1796590, EPI_ISL_1796591, EPI_ISL_1796592, EPI_ISL_1796593, EPI_ISL_1796595, EPI_ISL_1796596, EPI_ISL_1796597, EPI_ISL_1796598, EPI_ISL_1796599, EPI_ISL_1796600, EPI_ISL_1796601, EPI_ISL_1796602, EPI_ISL_1796603, EPI_ISL_1796604, EPI_ISL_1796605, EPI_ISL_1796606, EPI_ISL_1796607, EPI_ISL_1796608, EPI_ISL_1796609, EPI_ISL_1796610, EPI_ISL_1796611, EPI_ISL_1796612, EPI_ISL_1796613, EPI_ISL_1796614, EPI_ISL_1796615, EPI_ISL_1796616, EPI_ISL_1796617, EPI_ISL_1796618, EPI_ISL_1796619, EPI_ISL_1796620, EPI_ISL_1796621, EPI_ISL_1796622, EPI_ISL_1796623, EPI_ISL_1796624, EPI_ISL_1796625, EPI_ISL_1796626, EPI_ISL_1796627, EPI_ISL_1796628, EPI_ISL_1796629, EPI_ISL_1796630, EPI_ISL_1796631, EPI_ISL_1796632, EPI_ISL_1796633, EPI_ISL_1796634, EPI_ISL_1796635, EPI_ISL_1796636, EPI_ISL_1796637, EPI_ISL_1796638, EPI_ISL_1796639, EPI_ISL_1796640, EPI_ISL_1796641, EPI_ISL_1796642, EPI_ISL_1796643, EPI_ISL_1796644, EPI_ISL_1796645, EPI_ISL_1796646, EPI_ISL_1796647, EPI_ISL_1796648, EPI_ISL_1796649, EPI_ISL_1796650, EPI_ISL_1796651, EPI_ISL_1796652, EPI_ISL_1796653, EPI_ISL_1796654, EPI_ISL_1796655, EPI_ISL_1796656, EPI_ISL_1796657, EPI_ISL_1796658, EPI_ISL_1796659, EPI_ISL_1796660, EPI_ISL_1796661, EPI_ISL_1796662, EPI_ISL_1796663, EPI_ISL_1796664, EPI_ISL_1796665, EPI_ISL_1796666, EPI_ISL_1796667, EPI_ISL_1796668, EPI_ISL_1796669, EPI_ISL_1796670, EPI_ISL_1796671, EPI_ISL_1796672, EPI_ISL_1796673, EPI_ISL_1796674, EPI_ISL_1796675, EPI_ISL_1796676, EPI_ISL_1796677, EPI_ISL_1796678, EPI_ISL_1796679, EPI_ISL_1796680, EPI_ISL_1796681, EPI_ISL_1796682, EPI_ISL_1796683, EPI_ISL_1796684, EPI_ISL_1796685, EPI_ISL_1796686, EPI_ISL_1796687, EPI_ISL_1796688, EPI_ISL_1796689, EPI_ISL_1796690, EPI_ISL_1796691, EPI_ISL_1796692, EPI_ISL_1796693, EPI_ISL_1796694, EPI_ISL_1796695, EPI_ISL_1796696, EPI_ISL_1796697, EPI_ISL_1796698, EPI_ISL_1796699, EPI_ISL_1796700, EPI_ISL_1796701, EPI_ISL_1796702, EPI_ISL_1796703, EPI_ISL_1796704, EPI_ISL_1796705, EPI_ISL_1796706, EPI_ISL_1796707, EPI_ISL_1796708, EPI_ISL_1796709, EPI_ISL_1796710, EPI_ISL_1796711, EPI_ISL_1796712, EPI_ISL_1796713, EPI_ISL_1796714, EPI_ISL_1796715, EPI_ISL_1796717, EPI_ISL_1796718, EPI_ISL_1796719, EPI_ISL_1796720, EPI_ISL_1796721, EPI_ISL_1796722, EPI_ISL_1796723, EPI_ISL_1796724, EPI_ISL_1796725, EPI_ISL_1796726, EPI_ISL_1796727, EPI_ISL_1796728, EPI_ISL_1796729, EPI_ISL_1796730, EPI_ISL_1796731, EPI_ISL_1796732, EPI_ISL_1796733, EPI_ISL_1796734, EPI_ISL_1796735, EPI_ISL_1796736, EPI_ISL_1796737, EPI_ISL_1796738, EPI_ISL_1796739, EPI_ISL_1796740, EPI_ISL_1796741, EPI_ISL_1796742, EPI_ISL_1796743, EPI_ISL_1796744, EPI_ISL_1796745, EPI_ISL_1796746, EPI_ISL_1796747, EPI_ISL_1796748, EPI_ISL_1796749, EPI_ISL_1796750, EPI_ISL_1796751, EPI_ISL_1796752, EPI_ISL_1796753, EPI_ISL_1796754, EPI_ISL_1796755, EPI_ISL_1796756, EPI_ISL_1796757, EPI_ISL_1796758, EPI_ISL_1796759, EPI_ISL_1796760, EPI_ISL_1796761, EPI_ISL_1796762, EPI_ISL_1796763, EPI_ISL_1796764, EPI_ISL_1796765, EPI_ISL_1796766, EPI_ISL_1796767, EPI_ISL_1796768, EPI_ISL_1796769, EPI_ISL_1796770, EPI_ISL_1796771, EPI_ISL_1796772, EPI_ISL_1796773, EPI_ISL_1796774, EPI_ISL_1796775, EPI_ISL_1796776, EPI_ISL_1796777, EPI_ISL_1796778, EPI_ISL_1796779, EPI_ISL_1796780, EPI_ISL_1796781, EPI_ISL_1796784, EPI_ISL_1796785, EPI_ISL_1796786, EPI_ISL_1796787, EPI_ISL_1796788, EPI_ISL_1796789, EPI_ISL_1796790, EPI_ISL_1796791, EPI_ISL_1796792, EPI_ISL_1796793, EPI_ISL_1796794, EPI_ISL_1796795, EPI_ISL_1796796, | | | |

| | | | |
|---|---|---|---|
| EPI_ISL_1796797, EPI_ISL_1796798, EPI_ISL_1796799, EPI_ISL_1796800, EPI_ISL_1796801, EPI_ISL_1796802, EPI_ISL_1796803, EPI_ISL_1796804, EPI_ISL_1796805, EPI_ISL_1796806, EPI_ISL_1796807, EPI_ISL_1796808, EPI_ISL_1796809, EPI_ISL_1796810, EPI_ISL_1796811, EPI_ISL_1796812, EPI_ISL_1796813, EPI_ISL_1796814, EPI_ISL_1796815, EPI_ISL_1796816, EPI_ISL_1796817, EPI_ISL_1796818, EPI_ISL_1796819, EPI_ISL_1796820, EPI_ISL_1796821, EPI_ISL_1796822, EPI_ISL_1796823, EPI_ISL_1796824, EPI_ISL_1796825, EPI_ISL_1796826, EPI_ISL_1796827, EPI_ISL_1796828, EPI_ISL_1796829, EPI_ISL_1796830, EPI_ISL_1796831, EPI_ISL_1796832, EPI_ISL_1796833, EPI_ISL_1796834, EPI_ISL_1796835, EPI_ISL_1796836, EPI_ISL_1796837, EPI_ISL_1796838, EPI_ISL_1796839, EPI_ISL_1796840, EPI_ISL_1796841, EPI_ISL_1796842, EPI_ISL_1796843, EPI_ISL_1796844, EPI_ISL_1796845, EPI_ISL_1796846, EPI_ISL_1796847, EPI_ISL_1796848, EPI_ISL_1796849, EPI_ISL_1796850, EPI_ISL_1796851, EPI_ISL_1796852, EPI_ISL_1796853, EPI_ISL_1796854, EPI_ISL_1796855, EPI_ISL_1796856, EPI_ISL_1796857, EPI_ISL_1796858, EPI_ISL_1796859, EPI_ISL_1796860, EPI_ISL_1796861, EPI_ISL_1796862, EPI_ISL_1796863, EPI_ISL_1796864, EPI_ISL_1796865, EPI_ISL_1796866, EPI_ISL_1796867, EPI_ISL_1796868, EPI_ISL_1796869, EPI_ISL_1796870, EPI_ISL_1796871, EPI_ISL_1796872, EPI_ISL_1796873, EPI_ISL_1796874, EPI_ISL_1796875, EPI_ISL_1796876, EPI_ISL_1796877, EPI_ISL_1796878, EPI_ISL_1796879, EPI_ISL_1796880, EPI_ISL_1796881, EPI_ISL_1796882, EPI_ISL_1796883, EPI_ISL_1796884, EPI_ISL_1796885, EPI_ISL_1796886, EPI_ISL_1796887, EPI_ISL_1796888, EPI_ISL_1796889, EPI_ISL_1796890, EPI_ISL_1796891, EPI_ISL_1796892, EPI_ISL_1796893, EPI_ISL_1796894, EPI_ISL_1796895, EPI_ISL_1796896, EPI_ISL_1796897, EPI_ISL_1796898, EPI_ISL_1796899, EPI_ISL_1796900, EPI_ISL_1796901, EPI_ISL_1796902, EPI_ISL_1796903, EPI_ISL_1796904, EPI_ISL_1796905, EPI_ISL_1796906, EPI_ISL_1796907, EPI_ISL_1796908, EPI_ISL_1796909, EPI_ISL_1796910, EPI_ISL_1796911, EPI_ISL_1796912, EPI_ISL_1796913, EPI_ISL_1796914, EPI_ISL_1796915, EPI_ISL_1796916, EPI_ISL_1796917, EPI_ISL_1796918, EPI_ISL_1796919, EPI_ISL_1796920, EPI_ISL_1796921, EPI_ISL_1796922, EPI_ISL_1796923, EPI_ISL_1796924, EPI_ISL_1796925, EPI_ISL_1796926, EPI_ISL_1796927, EPI_ISL_1796928, EPI_ISL_1796929, EPI_ISL_1796930, EPI_ISL_1796931, EPI_ISL_1796932, EPI_ISL_1796933, EPI_ISL_1796934, EPI_ISL_1796935, EPI_ISL_1796936, EPI_ISL_1796937, EPI_ISL_1796938, EPI_ISL_1796939, EPI_ISL_1796940, EPI_ISL_1796941, EPI_ISL_1796942, EPI_ISL_1796943, EPI_ISL_1796944, EPI_ISL_1796945, EPI_ISL_1796946, EPI_ISL_1796947, EPI_ISL_1796948, EPI_ISL_1796949, EPI_ISL_1796950, EPI_ISL_1796951, EPI_ISL_1796952, EPI_ISL_1796953, EPI_ISL_1796954, EPI_ISL_1796955, EPI_ISL_1796956, EPI_ISL_1796957, EPI_ISL_1796958, EPI_ISL_1796959, EPI_ISL_1796960, EPI_ISL_1796961, EPI_ISL_1796962, EPI_ISL_1796963, EPI_ISL_1796964, EPI_ISL_1796965, EPI_ISL_1796966, EPI_ISL_1796967, EPI_ISL_1796968, EPI_ISL_1796969, EPI_ISL_1796970, EPI_ISL_1796971, EPI_ISL_1796972, EPI_ISL_1796973, EPI_ISL_1796974, EPI_ISL_1796975, EPI_ISL_1796976, EPI_ISL_1796977, EPI_ISL_1796978, EPI_ISL_1796979, EPI_ISL_1796980, EPI_ISL_1796981, EPI_ISL_1796982, EPI_ISL_1796983, EPI_ISL_1796984, EPI_ISL_1796985, EPI_ISL_1796986, EPI_ISL_1796987, EPI_ISL_1796988, EPI_ISL_1796989, EPI_ISL_1796990, EPI_ISL_1796991, EPI_ISL_1796992, EPI_ISL_1796993, EPI_ISL_1796994, EPI_ISL_1796995, EPI_ISL_1796996, EPI_ISL_1796997, EPI_ISL_1796998, EPI_ISL_1796999, EPI_ISL_1797000, EPI_ISL_1797001, EPI_ISL_1797002, EPI_ISL_1797003, EPI_ISL_1797004, EPI_ISL_1797005, EPI_ISL_1797006, EPI_ISL_1797007, EPI_ISL_1797008, EPI_ISL_1797009, EPI_ISL_1797010, EPI_ISL_1797011, EPI_ISL_1797012, EPI_ISL_1797013, EPI_ISL_1797014, EPI_ISL_1797015, EPI_ISL_1797016, EPI_ISL_1797017, EPI_ISL_1797018, EPI_ISL_1797019, EPI_ISL_1797020, EPI_ISL_1797021, EPI_ISL_1797022, EPI_ISL_1797023, EPI_ISL_1797024, EPI_ISL_1797025, EPI_ISL_1797026, EPI_ISL_1797027, EPI_ISL_1797028, EPI_ISL_1797029, EPI_ISL_1797030, EPI_ISL_1797031, EPI_ISL_1797032, EPI_ISL_1797033, EPI_ISL_1797034, EPI_ISL_1797035, EPI_ISL_1797036, EPI_ISL_1797037, EPI_ISL_1797038, EPI_ISL_1797039, EPI_ISL_1797040, EPI_ISL_1797041, EPI_ISL_1797042, EPI_ISL_1797043, EPI_ISL_1797044, EPI_ISL_1797045, EPI_ISL_1797046, EPI_ISL_1797047, EPI_ISL_1797048, EPI_ISL_1797049, EPI_ISL_1797050, EPI_ISL_1797051, EPI_ISL_1797052, EPI_ISL_1797053, EPI_ISL_1797054, EPI_ISL_1797055, EPI_ISL_1797056, EPI_ISL_1797057, EPI_ISL_1797058, EPI_ISL_1797059, EPI_ISL_1797060, EPI_ISL_1797061, EPI_ISL_1797062, EPI_ISL_1797063, EPI_ISL_1797064, EPI_ISL_1797065, EPI_ISL_1797066, EPI_ISL_1797067, EPI_ISL_1797068, EPI_ISL_1797069, EPI_ISL_1797070, EPI_ISL_1797071, EPI_ISL_1797072, EPI_ISL_1797073, EPI_ISL_1797074, EPI_ISL_1797075, EPI_ISL_1797076, EPI_ISL_1797077, EPI_ISL_1797078, EPI_ISL_1797079, EPI_ISL_1797080, EPI_ISL_1797081, EPI_ISL_1797082, EPI_ISL_1797083, EPI_ISL_1797084, EPI_ISL_1797085, EPI_ISL_1797086, EPI_ISL_1797087, EPI_ISL_1797088, EPI_ISL_1797089, EPI_ISL_1797090, EPI_ISL_1797091, EPI_ISL_1797092, EPI_ISL_1797093, EPI_ISL_1797094, EPI_ISL_1797095, EPI_ISL_1797096, EPI_ISL_1797097, EPI_ISL_1797098, EPI_ISL_1797099, EPI_ISL_1797100, EPI_ISL_1797101, EPI_ISL_1797102, EPI_ISL_1797103, EPI_ISL_1797104, EPI_ISL_1797105, EPI_ISL_1797106, EPI_ISL_1797107, EPI_ISL_1797108, EPI_ISL_1797109, EPI_ISL_1797110, EPI_ISL_1797111, EPI_ISL_1797112, EPI_ISL_1797113, EPI_ISL_1797114, EPI_ISL_1797115, EPI_ISL_1797116, EPI_ISL_1797117, EPI_ISL_1797118, EPI_ISL_1797119, EPI_ISL_1797120, EPI_ISL_1797121, EPI_ISL_1797122, EPI_ISL_1797123, EPI_ISL_1797124, EPI_ISL_1797125, EPI_ISL_1797126, EPI_ISL_1797127, EPI_ISL_1797128, EPI_ISL_1797129, EPI_ISL_1797130, EPI_ISL_1797131, EPI_ISL_1797132, EPI_ISL_1797133, EPI_ISL_1797134, EPI_ISL_1797135, EPI_ISL_1797136, EPI_ISL_1797137, EPI_ISL_1797138, EPI_ISL_1797139, EPI_ISL_1797140, EPI_ISL_1797141, EPI_ISL_1797142, EPI_ISL_1797143, EPI_ISL_1797144, EPI_ISL_1797145, EPI_ISL_1797146, EPI_ISL_1797147, EPI_ISL_1797148, EPI_ISL_1797149, EPI_ISL_1797150, EPI_ISL_1797151, EPI_ISL_1797152, EPI_ISL_1797153, EPI_ISL_1797154, EPI_ISL_1797155, EPI_ISL_1797156, EPI_ISL_1797157, EPI_ISL_1797158, EPI_ISL_1797159, EPI_ISL_1797160, EPI_ISL_1797161, EPI_ISL_1797162, EPI_ISL_1797163, EPI_ISL_1797164, EPI_ISL_1797165, EPI_ISL_1797166, EPI_ISL_1797167, EPI_ISL_1797168, EPI_ISL_1797169, EPI_ISL_1797170, EPI_ISL_1797171, EPI_ISL_1797172, EPI_ISL_1797173, EPI_ISL_1797174, EPI_ISL_1797175, EPI_ISL_1797176, EPI_ISL_1797177, EPI_ISL_1797178, EPI_ISL_1797179, EPI_ISL_1797180, EPI_ISL_1797181, EPI_ISL_1797182, EPI_ISL_1797183, EPI_ISL_1797184, EPI_ISL_1797185, EPI_ISL_1797186, EPI_ISL_1797187, EPI_ISL_1797188, EPI_ISL_1797189, EPI_ISL_1797190, EPI_ISL_1797191, EPI_ISL_1797192, EPI_ISL_1797193, EPI_ISL_1797194, EPI_ISL_1797195, EPI_ISL_1797196, EPI_ISL_1797197, EPI_ISL_1797198, EPI_ISL_1797199, EPI_ISL_1797200, EPI_ISL_1797201, EPI_ISL_1797202, EPI_ISL_1797203, EPI_ISL_1797204, EPI_ISL_1797205, EPI_ISL_1797206, EPI_ISL_1797207, EPI_ISL_1797208, EPI_ISL_1797209, EPI_ISL_1797210, EPI_ISL_1797211, EPI_ISL_1797212, EPI_ISL_1797213, EPI_ISL_1797214, EPI_ISL_1797215, EPI_ISL_1797216, EPI_ISL_1797217, EPI_ISL_1797218, EPI_ISL_1797219, EPI_ISL_1797220, EPI_ISL_1797221, EPI_ISL_1797222, EPI_ISL_1797223, EPI_ISL_1797224, EPI_ISL_1797225, EPI_ISL_1797226, EPI_ISL_1797227, EPI_ISL_1797228, EPI_ISL_1797229, EPI_ISL_1797230, EPI_ISL_1797231, EPI_ISL_1797232, EPI_ISL_1797233, EPI_ISL_1797234, EPI_ISL_1797235, EPI_ISL_1797236, EPI_ISL_1797237, EPI_ISL_1797238, EPI_ISL_1797239, EPI_ISL_1797240, EPI_ISL_1797241, EPI_ISL_1797242, EPI_ISL_1797243, EPI_ISL_1797244, EPI_ISL_1797245, EPI_ISL_1797246, EPI_ISL_1797247, EPI_ISL_1797248, EPI_ISL_1797249, EPI_ISL_1797250, EPI_ISL_1797251, EPI_ISL_1797252, EPI_ISL_1797253, EPI_ISL_1797254, EPI_ISL_1797255, EPI_ISL_1797256, EPI_ISL_1797257, EPI_ISL_1797258, EPI_ISL_1797259, EPI_ISL_1797260, EPI_ISL_1797261, EPI_ISL_1797262, EPI_ISL_1797263, EPI_ISL_1797264, EPI_ISL_1797265, EPI_ISL_1797266, EPI_ISL_1797267, EPI_ISL_1797268, EPI_ISL_1797269, EPI_ISL_1797270, EPI_ISL_1797271, EPI_ISL_1797272, EPI_ISL_1797273, EPI_ISL_1797274, EPI_ISL_1797275, EPI_ISL_1797276, EPI_ISL_1797277, EPI_ISL_1797278, EPI_ISL_1797279, EPI_ISL_1797280, EPI_ISL_1797281, EPI_ISL_1797282, EPI_ISL_1797283, EPI_ISL_1797284, EPI_ISL_1797285, EPI_ISL_1797286, EPI_ISL_1797287, EPI_ISL_1797288, EPI_ISL_1797289, EPI_ISL_1797290, EPI_ISL_1797291, EPI_ISL_1797292, EPI_ISL_1797293, EPI_ISL_1797294, EPI_ISL_1797295, EPI_ISL_1797296, EPI_ISL_1797297, EPI_ISL_1797298, EPI_ISL_1797299, EPI_ISL_1797300, EPI_ISL_1797301, EPI_ISL_1797302, EPI_ISL_1797303, EPI_ISL_1797304, EPI_ISL_1797305, EPI_ISL_1797306, EPI_ISL_1797307, EPI_ISL_1797308, EPI_ISL_1797309, EPI_ISL_1797310, EPI_ISL_1797311, EPI_ISL_1797312, EPI_ISL_1797313, EPI_ISL_1797314, EPI_ISL_1797315, EPI_ISL_1797316, EPI_ISL_1797317, EPI_ISL_1797318, EPI_ISL_1797319, EPI_ISL_1797320, EPI_ISL_1797321, EPI_ISL_1797322, EPI_ISL_1797323, EPI_ISL_1797324, EPI_ISL_1797325, EPI_ISL_1797326, EPI_ISL_1797327, EPI_ISL_1797328, EPI_ISL_1797329, EPI_ISL_1797330, EPI_ISL_1797331, EPI_ISL_1797332, EPI_ISL_1797333, EPI_ISL_1797334, EPI_ISL_1797335, EPI_ISL_1797336, EPI_ISL_1797337, EPI_ISL_1797338, EPI_ISL_1797339, EPI_ISL_1797340, EPI_ISL_1797341, EPI_ISL_1797342, EPI_ISL_1797343, EPI_ISL_1797347, EPI_ISL_1797351, EPI_ISL_1797352, EPI_ISL_1797353, EPI_ISL_1797354, EPI_ISL_1797355, EPI_ISL_1797356, EPI_ISL_1797357, EPI_ISL_1797358, EPI_ISL_1797359, EPI_ISL_1797360, EPI_ISL_1797361, EPI_ISL_1797362, EPI_ISL_1797363, EPI_ISL_1797364, EPI_ISL_1797365, EPI_ISL_1797366, EPI_ISL_1797367, EPI_ISL_1797368, EPI_ISL_1797369, EPI_ISL_1797370, EPI_ISL_1797371, EPI_ISL_1797372, EPI_ISL_1797373, EPI_ISL_1797374, EPI_ISL_1797375, EPI_ISL_1797376, EPI_ISL_1797377, EPI_ISL_1797378, EPI_ISL_1797379, EPI_ISL_1797380, EPI_ISL_1797381, EPI_ISL_1797382, EPI_ISL_1797383, EPI_ISL_1797384, EPI_ISL_1797385, EPI_ISL_1797386, EPI_ISL_1797387, EPI_ISL_1797388, EPI_ISL_1797389, EPI_ISL_1797390, EPI_ISL_1797391, EPI_ISL_1797392, EPI_ISL_1797393, EPI_ISL_1797394, EPI_ISL_1797395, EPI_ISL_1797396, EPI_ISL_1797397, EPI_ISL_1797398, EPI_ISL_1797399, EPI_ISL_1797400, EPI_ISL_1797401, EPI_ISL_1797402, EPI_ISL_1797403, EPI_ISL_1797404, EPI_ISL_1797405, EPI_ISL_1797406, EPI_ISL_1797407, EPI_ISL_1797408, EPI_ISL_1797409, EPI_ISL_1797410, EPI_ISL_1797411, EPI_ISL_1797412, EPI_ISL_1797413, EPI_ISL_1797414, EPI_ISL_1797415, EPI_ISL_1797416, EPI_ISL_1797417, EPI_ISL_1797418, EPI_ISL_1797419, EPI_ISL_1797420, EPI_ISL_1797421, EPI_ISL_1797422, EPI_ISL_1797423, EPI_ISL_1797424, EPI_ISL_1797425, EPI_ISL_1797426, EPI_ISL_1797427, EPI_ISL_1797428, EPI_ISL_1797429, EPI_ISL_1797430, EPI_ISL_1797431, EPI_ISL_1797432, EPI_ISL_1797433, EPI_ISL_1797434 | | | |
| see above | Helix/Illumina | Centers for Disease Control and Prevention Division of Viral Diseases, Pathogen Discovery | Dakota Howard, Dhwani Batra, Peter W. Cook, Kara Moser, Adrian Paskey, Jason Caravas, Benjamin Rambo-Martin, Shatavia Morrison, Christopher Gulvick, Scott Sammons, Yvette Unoarumhi, Darlene Wagner, Matthew Schmerer, Eileen de Feo, Jan Antico, Christine Tran, Matthew Tolentino, Shannon Wickline, Kim Gietzen, Brad Sickler, Jingtao Liu, Eric Allen, Phil Febbo, Nicole L. Washington, Simon White, Geraint Levan, Kelly Schiabor Barrett, Elizabeth Cirulli, Alexandre Bolze, Ary Ascencio, Charlotte Rivera-Garcia, Ryan Cho, Jason Nguyen, Sherry Wang, Jimmy Ramirez, Tyler Cassens, Efren Sandoval, Magnus Isaksson, William Lee, David Becker, Marc Laurent, James Lu, Clinton R. Paden, Duncan MacCannell |
| EPI_ISL_1797880, EPI_ISL_1797881, EPI_ISL_1797882, EPI_ISL_1797883, EPI_ISL_1797884, EPI_ISL_1797885, EPI_ISL_1797886, EPI_ISL_1797887, EPI_ISL_1797888, EPI_ISL_1797889, EPI_ISL_1797890, EPI_ISL_1797891, EPI_ISL_1797892, EPI_ISL_1797893, EPI_ISL_1797894, EPI_ISL_1797895, EPI_ISL_1797896, EPI_ISL_1797897, EPI_ISL_1797898, EPI_ISL_1797899, EPI_ISL_1797900, EPI_ISL_1797901, EPI_ISL_1797902, EPI_ISL_1797903, EPI_ISL_1797904, EPI_ISL_1797905, EPI_ISL_1797906, EPI_ISL_1797907, EPI_ISL_1797908, EPI_ISL_1797909, EPI_ISL_1797910, EPI_ISL_1797911, EPI_ISL_1797912, EPI_ISL_1797913, EPI_ISL_1797914, EPI_ISL_1797915, EPI_ISL_1797916, EPI_ISL_1797917, EPI_ISL_1797918, EPI_ISL_1797919, EPI_ISL_1797920, EPI_ISL_1797921, EPI_ISL_1797922, EPI_ISL_1797923, EPI_ISL_1797924, EPI_ISL_1797925, EPI_ISL_1797926, EPI_ISL_1797927, EPI_ISL_1797928, EPI_ISL_1797929, EPI_ISL_1797930, EPI_ISL_1797931, EPI_ISL_1797932, EPI_ISL_1797933, EPI_ISL_1797934, EPI_ISL_1797935, EPI_ISL_1797936, EPI_ISL_1797937, EPI_ISL_1797938, EPI_ISL_1797939, EPI_ISL_1797940, EPI_ISL_1797941, EPI_ISL_1797942, EPI_ISL_1797943, EPI_ISL_1797944, EPI_ISL_1797945, EPI_ISL_1797946, EPI_ISL_1797947, EPI_ISL_1797948, EPI_ISL_1797949, EPI_ISL_1797950, EPI_ISL_1797951, EPI_ISL_1797952, EPI_ISL_1797953, EPI_ISL_1797954, EPI_ISL_1797955, EPI_ISL_1797956, EPI_ISL_1797957, EPI_ISL_1797958, EPI_ISL_1797959, EPI_ISL_1797960, EPI_ISL_1797961, EPI_ISL_1797962, EPI_ISL_1797963, EPI_ISL_1797964, EPI_ISL_1797965, EPI_ISL_1797966, EPI_ISL_1797967, EPI_ISL_1797968, EPI_ISL_1797969, EPI_ISL_1797970, EPI_ISL_1797971, EPI_ISL_1797972, EPI_ISL_1797973, EPI_ISL_1797974, EPI_ISL_1797975, EPI_ISL_1797976, EPI_ISL_1797977, EPI_ISL_1797978, EPI_ISL_1797979, EPI_ISL_1797980, EPI_ISL_1797981, EPI_ISL_1797982, EPI_ISL_1797983, EPI_ISL_1797984, EPI_ISL_1797985, EPI_ISL_1797986, EPI_ISL_1797987, EPI_ISL_1797988, EPI_ISL_1797989, EPI_ISL_1797990, EPI_ISL_1797991, EPI_ISL_1797992, EPI_ISL_1797993, EPI_ISL_1797994, EPI_ISL_1797995, EPI_ISL_1797996, EPI_ISL_1797997, EPI_ISL_1797998, EPI_ISL_1797999, EPI_ISL_1798000, EPI_ISL_1798001, EPI_ISL_1798002, EPI_ISL_1798003, EPI_ISL_1798004, EPI_ISL_1798005, EPI_ISL_1798006, EPI_ISL_1798007, EPI_ISL_1798008, EPI_ISL_1798009, EPI_ISL_1798010, EPI_ISL_1798011, EPI_ISL_1798012, EPI_ISL_1798013, EPI_ISL_1798014, EPI_ISL_1798015, EPI_ISL_1798016, EPI_ISL_1798017, EPI_ISL_1798018, EPI_ISL_1798019, EPI_ISL_1798020, EPI_ISL_1798021, EPI_ISL_1798022, EPI_ISL_1798023, EPI_ISL_1798024, EPI_ISL_1798025, EPI_ISL_1798026, EPI_ISL_1798027, EPI_ISL_1798028, EPI_ISL_1798029, EPI_ISL_1798030, EPI_ISL_1798031, EPI_ISL_1798032, EPI_ISL_1798033, EPI_ISL_1798034, EPI_ISL_1798035, EPI_ISL_1798036, EPI_ISL_1798037, EPI_ISL_1798038, EPI_ISL_1798039, EPI_ISL_1798040, EPI_ISL_1798041, EPI_ISL_1798042, EPI_ISL_1798043, EPI_ISL_1798044, EPI_ISL_1798045, EPI_ISL_1798046, EPI_ISL_1798047, EPI_ISL_1798048, EPI_ISL_1798049, EPI_ISL_1798050, EPI_ISL_1798051, EPI_ISL_1798052, EPI_ISL_1798053, EPI_ISL_1798054, EPI_ISL_1798055, EPI_ISL_1798056, EPI_ISL_1798057, EPI_ISL_1798058, EPI_ISL_1798059, EPI_ISL_1798060, EPI_ISL_1798061, EPI_ISL_1798062, EPI_ISL_1798063, EPI_ISL_1798064, EPI_ISL_1798065, EPI_ISL_1798066, EPI_ISL_1798067, EPI_ISL_1798068, EPI_ISL_1798069, EPI_ISL_1798070, EPI_ISL_1798071, EPI_ISL_1798072, EPI_ISL_1798073, EPI_ISL_1798074, EPI_ISL_1798075, EPI_ISL_1798076, EPI_ISL_1798077, EPI_ISL_1798078, EPI_ISL_1798079, EPI_ISL_1798080, EPI_ISL_1798081, EPI_ISL_1798082, EPI_ISL_1798083, EPI_ISL_1798084, EPI_ISL_1798085, EPI_ISL_1798086, EPI_ISL_1798087, EPI_ISL_1798088, EPI_ISL_1798089, EPI_ISL_1798090, EPI_ISL_1798091, EPI_ISL_1798092, EPI_ISL_1798093, EPI_ISL_1798094, EPI_ISL_1798095, EPI_ISL_1798096, EPI_ISL_1798097, EPI_ISL_1798098, EPI_ISL_1798099, EPI_ISL_1798100, EPI_ISL_1798101, EPI_ISL_1798102, EPI_ISL_1798103, EPI_ISL_1798104, EPI_ISL_1798105, EPI_ISL_1798106, EPI_ISL_1798107, EPI_ISL_1798108, EPI_ISL_1798109, EPI_ISL_1798110, EPI_ISL_1798111, EPI_ISL_1798112, EPI_ISL_1798113, EPI_ISL_1798114, EPI_ISL_1798115, EPI_ISL_1798116, EPI_ISL_1798117, EPI_ISL_1798118, EPI_ISL_1798119, EPI_ISL_1798120, EPI_ISL_1798121, EPI_ISL_1798122, EPI_ISL_1798123, EPI_ISL_1798124, EPI_ISL_1798125, EPI_ISL_1798126, EPI_ISL_1798127, EPI_ISL_1798128, EPI_ISL_1798129, EPI_ISL_1798130, EPI_ISL_1798131, EPI_ISL_1798132, EPI_ISL_1798133, EPI_ISL_1798134, EPI_ISL_1798135, EPI_ISL_1798136, EPI_ISL_1798137, EPI_ISL_1798138, EPI_ISL_1798139, EPI_ISL_1798140, EPI_ISL_1798141, EPI_ISL_1798142, EPI_ISL_1798143, EPI_ISL_1798144, EPI_ISL_1798145, EPI_ISL_1798146, EPI_ISL_1798147, EPI_ISL_1798148, EPI_ISL_1798149, EPI_ISL_1798150, EPI_ISL_1798151, EPI_ISL_1798152, EPI_ISL_1798153, EPI_ISL_1798154, EPI_ISL_1798155, EPI_ISL_1798156, EPI_ISL_1798157, EPI_ISL_1798158, EPI_ISL_1798159, EPI_ISL_1798160, EPI_ISL_1798161, EPI_ISL_1798162, EPI_ISL_1798163, EPI_ISL_1798164, EPI_ISL_1798165, EPI_ISL_1798166, EPI_ISL_1798167, EPI_ISL_1798168, EPI_ISL_1798169, EPI_ISL_1798170, EPI_ISL_1798171, EPI_ISL_1798172, EPI_ISL_1798173, EPI_ISL_1798174, EPI_ISL_1798175, EPI_ISL_1798176, EPI_ISL_1798177, EPI_ISL_1798178, EPI_ISL_1798179, EPI_ISL_1798180, EPI_ISL_1798181, EPI_ISL_1798182, EPI_ISL_1798183, EPI_ISL_1798184, EPI_ISL_1798185, EPI_ISL_1798186, EPI_ISL_1798187, EPI_ISL_1798188, EPI_ISL_1798189, EPI_ISL_1798190 | | | |
| see above | Quest Diagnostics Incorporated | Centers for Disease Control and Prevention Division of Viral Diseases, Pathogen Discovery | Dakota Howard, Dhwani Batra, Peter W. Cook, Kara Moser, Adrian Paskey, Jason Caravas, Benjamin Rambo-Martin, Shatavia Morrison, Christopher Gulvick, Scott Sammons, Yvette Unoarumhi, Darlene Wagner, Matthew Schmerer, S. H. Rosenthal, A. Gerasimova, R. M. Kagan, B. Anderson, M. Hua, Y. Liu, L.E. Bernstein, K.E. Livingston, A. Perez, I. A. Shlyakhter, R. V. Rolando, R. Owen, P. Tanpaiboon, F. Lacbawan, Clinton R. Paden, Duncan MacCannell |
| EPI_ISL_1798192, EPI_ISL_1798193, EPI_ISL_1798196, EPI_ISL_1798198, EPI_ISL_1798199, EPI_ISL_1798200, EPI_ISL_1798202, EPI_ISL_1798204, EPI_ISL_1798205, EPI_ISL_1798206, EPI_ISL_1798207, EPI_ISL_1798208, EPI_ISL_1798209, EPI_ISL_1798210, EPI_ISL_1798211, EPI_ISL_1798212, EPI_ISL_1798213, EPI_ISL_1798214, EPI_ISL_1798215, EPI_ISL_1798217, EPI_ISL_1798218, EPI_ISL_1798219, EPI_ISL_1798220, EPI_ISL_1798223, EPI_ISL_1798224, EPI_ISL_1798225, EPI_ISL_1798226, EPI_ISL_1798227, EPI_ISL_1798228, EPI_ISL_1798229, EPI_ISL_1798231, EPI_ISL_1798232, EPI_ISL_1798233, EPI_ISL_1798234, EPI_ISL_1798235, EPI_ISL_1798236, EPI_ISL_1798237, EPI_ISL_1798238, EPI_ISL_1798239, EPI_ISL_1798240, EPI_ISL_1798241, EPI_ISL_1798242, EPI_ISL_1798243, EPI_ISL_1798244, EPI_ISL_1798245, EPI_ISL_1798247, EPI_ISL_1798248, EPI_ISL_1798249, EPI_ISL_1798250, EPI_ISL_1798252, EPI_ISL_1798253, EPI_ISL_1798254, EPI_ISL_1798255, EPI_ISL_1798256, EPI_ISL_1798258, EPI_ISL_1798259, EPI_ISL_1798260, EPI_ISL_1798262, EPI_ISL_1798265, EPI_ISL_1798266, EPI_ISL_1798268, EPI_ISL_1798269, EPI_ISL_1798270, EPI_ISL_1798271, EPI_ISL_1798272, EPI_ISL_1798273, EPI_ISL_1798274, EPI_ISL_1798275, EPI_ISL_1798276, EPI_ISL_1798277, EPI_ISL_1798278, EPI_ISL_1798279, EPI_ISL_1798280, EPI_ISL_1798281, EPI_ISL_1798282, EPI_ISL_1798283, EPI_ISL_1798284, EPI_ISL_1798285, EPI_ISL_1798286, EPI_ISL_1798287, EPI_ISL_1798288, EPI_ISL_1798290, EPI_ISL_1798292, EPI_ISL_1798293, EPI_ISL_1798294, EPI_ISL_1798295, EPI_ISL_1798298, EPI_ISL_1798301, EPI_ISL_1798302, EPI_ISL_1798303, EPI_ISL_1798304, EPI_ISL_1798305, EPI_ISL_1798306, EPI_ISL_1798308, EPI_ISL_1798309, EPI_ISL_1798311, EPI_ISL_1798314, EPI_ISL_1798315, EPI_ISL_1798318 | | | |
| see above | Laboratory Corporation of America | Centers for Disease Control and Prevention Division of Viral Diseases, Pathogen Discovery | Dakota Howard, Dhwani Batra, Peter W. Cook, Kara Moser, Adrian Paskey, Jason Caravas, Benjamin Rambo-Martin, Shatavia Morrison, Christopher Gulvick, Scott Sammons, Yvette Unoarumhi, Darlene Wagner, Matthew Schmerer, Minoo Agarwal, Eyad Almasri, Debbie Boles, Ayla Burns, Nuthawin Charoensri, Oren Cohen, Susan Countryman, Mary Ann Cristobal, Bobbi Croy, Suzanne Dale, Hrushikesh Deshmukh, Amanda Douglas, Vincent Drouillon, Marcia Eisenberg, Howard Engler, Rama Ghatti, Prashant Gupta, Susan Hicks, Jake Humphrey, Lax Iyer, Manoj Jain, Mohan Kolli, Brian Krueger, Tim Kuphal, Stanley Letovsky, Michael Levandoski, Craig Lukasik, Jonathan Meltzer, Brian Norvell, Mindy Nye, Scott Parker, Christos Petropoulos, John Pruitt, Steven Ragan, Scott Ryan, Mike Sapeta, Jana Schroth, Suresh Babu Selvaraju, Goran Stevovic, Amanda Suchanek, Andrea Throop, Lyndon Tilson, Thomas Urban, Joe Voshell, Kimberly Wagner, Jonathan Williams, Mary Williamson, Qian Zeng, Tricia Zwiefelhofer, Clinton R. Paden, Duncan MacCannell |
| EPI_ISL_1798319, EPI_ISL_1798320 | MD DOH Laboratories Administration | Centers for Disease Control and Prevention Division of Viral Diseases, Pathogen Discovery | Mili Sheth, Sarah Nobles, Jasmine Padilla, Mark Burroughs, Shoshona Le, Katie Dillon, Peter Cook, Clinton R. Paden, Dhwani Batra, Krista Queen, Kristen Knipe, Dakota Howard, Yvette Unoarumhi, Darlene Wagner, Matthew Schmerer, Ben L. Rambo-Martin, Kristine Lacek, Sam Shepard, Alison Laufer Halpin, Dave Wentworth, Vivien Dugan, Suxiang Tong, Justin Lee |
| EPI_ISL_1798321 | Laboratory Corporation of America | Centers for Disease Control and Prevention Division of Viral Diseases, Pathogen Discovery | Dakota Howard, Dhwani Batra, Peter W. Cook, Kara Moser, Adrian Paskey, Jason Caravas, Benjamin Rambo-Martin, Shatavia Morrison, Christopher Gulvick, Scott Sammons, Yvette Unoarumhi, Darlene Wagner, Matthew Schmerer, Minoo Agarwal, Eyad Almasri, Debbie Boles, Ayla Burns, Nuthawin Charoensri, Oren Cohen, Susan Countryman, Mary Ann Cristobal, Bobbi Croy, Suzanne Dale, Hrushikesh Deshmukh, Amanda Douglas, Vincent Drouillon, Marcia Eisenberg, Howard Engler, Rama Ghatti, Prashant Gupta, Susan Hicks, Jake Humphrey, Lax Iyer, Manoj Jain, Mohan Kolli, Brian Krueger, Tim Kuphal, Stanley Letovsky, Michael Levandoski, Craig Lukasik, Jonathan Meltzer, Brian Norvell, Mindy Nye, Scott Parker, Christos Petropoulos, John Pruitt, Steven Ragan, Scott Ryan, Mike Sapeta, Jana Schroth, Suresh Babu Selvaraju, Goran Stevovic, Amanda Suchanek, Andrea Throop, Lyndon Tilson, Thomas Urban, Joe Voshell, Kimberly Wagner, Jonathan Williams, Mary Williamson, Qian Zeng, Tricia Zwiefelhofer, Clinton R. Paden, Duncan MacCannell |
| EPI_ISL_1798323, EPI_ISL_1798324 | PR Public Health Lab | Centers for Disease Control and Prevention Division of Viral Diseases, Pathogen Discovery | Mili Sheth, Sarah Nobles, Jasmine Padilla, Mark Burroughs, Shoshona Le, Katie Dillon, Peter Cook, Clinton R. Paden, Dhwani Batra, Krista Queen, Kristen Knipe, Dakota Howard, Yvette Unoarumhi, Darlene Wagner, Matthew Schmerer, Ben L. Rambo-Martin, Kristine Lacek, Sam Shepard, Alison Laufer Halpin, Dave Wentworth, Vivien Dugan, Suxiang Tong, Justin Lee |
| EPI_ISL_1798326 | Laboratory Corporation of America | Centers for Disease Control and Prevention Division of Viral Diseases, Pathogen Discovery | Dakota Howard, Dhwani Batra, Peter W. Cook, Kara Moser, Adrian Paskey, Jason Caravas, Benjamin Rambo-Martin, Shatavia Morrison, Christopher Gulvick, Scott Sammons, Yvette Unoarumhi, Darlene Wagner, Matthew Schmerer, Minoo Agarwal, Eyad Almasri, Debbie Boles, Ayla Burns, Nuthawin Charoensri, Oren Cohen, Susan Countryman, Mary Ann Cristobal, Bobbi Croy, Suzanne Dale, Hrushikesh Deshmukh, Amanda Douglas, Vincent Drouillon, Marcia Eisenberg, Howard Engler, Rama Ghatti, Prashant Gupta, Susan Hicks, Jake Humphrey, Lax Iyer, Manoj Jain, Mohan Kolli, Brian Krueger, Tim Kuphal, Stanley Letovsky, Michael Levandoski, Craig Lukasik, Jonathan Meltzer, Brian Norvell, Mindy Nye, Scott Parker, Christos Petropoulos, John Pruitt, Steven Ragan, Scott Ryan, Mike Sapeta, Jana Schroth, Suresh Babu Selvaraju, Goran Stevovic, Amanda Suchanek, Andrea Throop, Lyndon Tilson, Thomas Urban, Joe Voshell, Kimberly Wagner, Jonathan Williams, Mary Williamson, Qian Zeng, Tricia Zwiefelhofer, Clinton R. Paden, Duncan MacCannell |
| EPI_ISL_1798327 | PR Public Health Lab | Centers for Disease Control and Prevention Division of Viral Diseases, Pathogen Discovery | Mili Sheth, Sarah Nobles, Jasmine Padilla, Mark Burroughs, Shoshona Le, Katie Dillon, Peter Cook, Clinton R. Paden, Dhwani Batra, Krista Queen, Kristen Knipe, Dakota Howard, Yvette Unoarumhi, Darlene Wagner, Matthew Schmerer, Ben L. Rambo-Martin, Kristine Lacek, Sam Shepard, Alison Laufer Halpin, Dave Wentworth, Vivien Dugan, Suxiang Tong, Justin Lee |
| EPI_ISL_1798328 | Laboratory Corporation of America | Centers for Disease Control and Prevention Division of Viral Diseases, Pathogen Discovery | Dakota Howard, Dhwani Batra, Peter W. Cook, Kara Moser, Adrian Paskey, Jason Caravas, Benjamin Rambo-Martin, Shatavia Morrison, Christopher Gulvick, Scott Sammons, Yvette Unoarumhi, Darlene Wagner, Matthew Schmerer, Minoo Agarwal, Eyad Almasri, Debbie Boles, Ayla Burns, Nuthawin Charoensri, Oren Cohen, Susan Countryman, Mary Ann Cristobal, Bobbi Croy, Suzanne Dale, Hrushikesh Deshmukh, Amanda Douglas, Vincent Drouillon, Marcia Eisenberg, Howard Engler, Rama Ghatti, Prashant Gupta, Susan Hicks, Jake Humphrey, Lax Iyer, Manoj Jain, Mohan Kolli, Brian Krueger, Tim Kuphal, Stanley Letovsky, Michael Levandoski, Craig Lukasik, Jonathan Meltzer, Brian Norvell, Mindy Nye, Scott Parker, Christos Petropoulos, John Pruitt, Steven Ragan, Scott Ryan, Mike Sapeta, Jana Schroth, Suresh Babu Selvaraju, Goran Stevovic, Amanda Suchanek, Andrea Throop, Lyndon Tilson, Thomas Urban, Joe Voshell, Kimberly Wagner, Jonathan Williams, Mary Williamson, Qian Zeng, Tricia Zwiefelhofer, Clinton R. Paden, Duncan MacCannell |
| EPI_ISL_1798329 | MD DOH Laboratories Administration | Centers for Disease Control and Prevention Division of Viral Diseases, Pathogen Discovery | Mili Sheth, Sarah Nobles, Jasmine Padilla, Mark Burroughs, Shoshona Le, Katie Dillon, Peter Cook, Clinton R. Paden, Dhwani Batra, Krista Queen, Kristen Knipe, Dakota Howard, Yvette Unoarumhi, Darlene Wagner, Matthew Schmerer, Ben L. Rambo-Martin, Kristine Lacek, Sam Shepard, Alison Laufer Halpin, Dave Wentworth, Vivien Dugan, Suxiang Tong, Justin Lee |
| EPI_ISL_1798330 | Laboratory Corporation of America | Centers for Disease Control and Prevention Division of Viral Diseases, Pathogen Discovery | Dakota Howard, Dhwani Batra, Peter W. Cook, Kara Moser, Adrian Paskey, Jason Caravas, Benjamin Rambo-Martin, Shatavia Morrison, Christopher Gulvick, Scott Sammons, Yvette Unoarumhi, Darlene Wagner, Matthew Schmerer, Minoo Agarwal, Eyad Almasri, Debbie Boles, Ayla Burns, Nuthawin Charoensri, Oren Cohen, Susan Countryman, Mary Ann Cristobal, Bobbi Croy, Suzanne Dale, Hrushikesh Deshmukh, Amanda Douglas, Vincent Drouillon, Marcia Eisenberg, Howard Engler, Rama Ghatti, Prashant Gupta, Susan Hicks, Jake Humphrey, Lax Iyer, Manoj Jain, Mohan Kolli, Brian Krueger, Tim Kuphal, Stanley Letovsky, Michael Levandoski, Craig Lukasik, Jonathan Meltzer, Brian Norvell, Mindy Nye, Scott Parker, Christos Petropoulos, John Pruitt, Steven Ragan, Scott Ryan, Mike Sapeta, Jana Schroth, Suresh Babu Selvaraju, Goran Stevovic, Amanda Suchanek, Andrea Throop, Lyndon Tilson, Thomas Urban, Joe Voshell, Kimberly Wagner, Jonathan Williams, Mary Williamson, Qian Zeng, Tricia Zwiefelhofer, Clinton R. Paden, Duncan MacCannell |
| EPI_ISL_1798331 | MT Public Health Laboratory | Centers for Disease Control and Prevention Division of Viral Diseases, Pathogen Discovery | Mili Sheth, Sarah Nobles, Jasmine Padilla, Mark Burroughs, Shoshona Le, Katie Dillon, Peter Cook, Clinton R. Paden, Dhwani Batra, Krista Queen, Kristen Knipe, Dakota Howard, Yvette Unoarumhi, Darlene Wagner, Matthew Schmerer, Ben L. Rambo-Martin, Kristine Lacek, Sam Shepard, Alison Laufer Halpin, Dave Wentworth, Vivien Dugan, Suxiang Tong, Justin Lee |
| EPI_ISL_1798333 | NJ Public Health and Environmental Laboratories | Centers for Disease Control and Prevention Division of Viral Diseases, Pathogen Discovery | Mili Sheth, Sarah Nobles, Jasmine Padilla, Mark Burroughs, Shoshona Le, Katie Dillon, Peter Cook, Clinton R. Paden, Dhwani Batra, Krista Queen, Kristen Knipe, Dakota Howard, Yvette Unoarumhi, Darlene Wagner, Matthew Schmerer, Ben L. Rambo-Martin, Kristine Lacek, Sam Shepard, Alison Laufer Halpin, Dave Wentworth, Vivien Dugan, Suxiang Tong, Justin Lee |
| EPI_ISL_1798334 | MN PHL Division, Minnesota Department of Health | Centers for Disease Control and Prevention Division of Viral Diseases, Pathogen Discovery | Mili Sheth, Sarah Nobles, Jasmine Padilla, Mark Burroughs, Shoshona Le, Katie Dillon, Peter Cook, Clinton R. Paden, Dhwani Batra, Krista Queen, Kristen Knipe, Dakota Howard, Yvette Unoarumhi, Darlene Wagner, Matthew Schmerer, Ben L. Rambo-Martin, Kristine Lacek, Sam Shepard, Alison Laufer Halpin, Dave Wentworth, Vivien Dugan, Suxiang Tong, Justin Lee |
| EPI_ISL_1798336 | CT-Dr. Katherine A. Kelley State Public Health Lab | Centers for Disease Control and Prevention Division of Viral Diseases, Pathogen Discovery | Mili Sheth, Sarah Nobles, Jasmine Padilla, Mark Burroughs, Shoshona Le, Katie Dillon, Peter Cook, Clinton R. Paden, Dhwani Batra, Krista Queen, Kristen Knipe, Dakota Howard, Yvette Unoarumhi, Darlene Wagner, Matthew Schmerer, Ben L. Rambo-Martin, Kristine Lacek, Sam Shepard, Alison Laufer Halpin, Dave Wentworth, Vivien Dugan, Suxiang Tong, Justin Lee |
| EPI_ISL_1798337 | Laboratory Corporation of America | Centers for Disease Control and Prevention Division of Viral Diseases, Pathogen Discovery | Dakota Howard, Dhwani Batra, Peter W. Cook, Kara Moser, Adrian Paskey, Jason Caravas, Benjamin Rambo-Martin, Shatavia Morrison, Christopher Gulvick, Scott Sammons, Yvette Unoarumhi, Darlene Wagner, Matthew Schmerer, Minoo Agarwal, Eyad Almasri, Debbie Boles, Ayla Burns, Nuthawin Charoensri, Oren Cohen, Susan Countryman, Mary Ann Cristobal, Bobbi Croy, Suzanne Dale, Hrushikesh Deshmukh, Amanda Douglas, Vincent Drouillon, Marcia Eisenberg, Howard Engler, Rama Ghatti, Prashant Gupta, Susan Hicks, Jake Humphrey, Lax Iyer, Manoj Jain, Mohan Kolli, Brian Krueger, Tim Kuphal, Stanley Letovsky, Michael Levandoski, Craig Lukasik, Jonathan Meltzer, Brian Norvell, Mindy Nye, Scott Parker, Christos Petropoulos, John Pruitt, Steven Ragan, Scott Ryan, Mike Sapeta, Jana Schroth, Suresh Babu Selvaraju, Goran Stevovic, Amanda |

| | | | |
|---|---|---|---|
| | | | Suchanek, Andrea Throop, Lyndon Tilson, Thomas Urban, Joe Voshell, Kimberly Wagner, Jonathan Williams, Mary Williamson, Qian Zeng, Tricia Zwiefelhofer, Clinton R. Paden, Duncan MacCannell |
| EPI_ISL_1798339 | MD DOH Laboratories Administration | Centers for Disease Control and Prevention Division of Viral Diseases, Pathogen Discovery | Mili Sheth, Sarah Nobles, Jasmine Padilla, Mark Burroughs, Shoshona Le, Katie Dillon, Peter Cook, Clinton R. Paden, Dhwani Batra, Krista Queen, Kristen Knipe, Dakota Howard, Yvette Unoarumhi, Darlene Wagner, Matthew Schmerer, Ben L. Rambo-Martin, Kristine Lacek, Sam Shepard, Alison Laufer Halpin, Dave Wentworth, Vivien Dugan, Suxiang Tong, Justin Lee |
| EPI_ISL_1798340, EPI_ISL_1798342 | PR Public Health Lab | Centers for Disease Control and Prevention Division of Viral Diseases, Pathogen Discovery | Mili Sheth, Sarah Nobles, Jasmine Padilla, Mark Burroughs, Shoshona Le, Katie Dillon, Peter Cook, Clinton R. Paden, Dhwani Batra, Krista Queen, Kristen Knipe, Dakota Howard, Yvette Unoarumhi, Darlene Wagner, Matthew Schmerer, Ben L. Rambo-Martin, Kristine Lacek, Sam Shepard, Alison Laufer Halpin, Dave Wentworth, Vivien Dugan, Suxiang Tong, Justin Lee |
| EPI_ISL_1798343 | Laboratory Corporation of America | Centers for Disease Control and Prevention Division of Viral Diseases, Pathogen Discovery | Dakota Howard, Dhwani Batra, Peter W. Cook, Kara Moser, Adrian Paskey, Jason Caravas, Benjamin Rambo-Martin, Shatavia Morrison, Christopher Gulvick, Scott Sammons, Yvette Unoarumhi, Darlene Wagner, Matthew Schmerer, Minoo Agarwal, Eyad Almasri, Debbie Boles, Ayla Burns, Nuthawin Charoensri, Oren Cohen, Susan Countryman, Mary Ann Cristobal, Bobbi Croy, Suzanne Dale, Hrushikesh Deshmukh, Amanda Douglas, Vincent Drouillon, Marcia Eisenberg, Howard Engler, Rama Ghatti, Prashant Gupta, Susan Hicks, Jake Humphrey, Lax Iyer, Manoj Jain, Mohan Kolli, Brian Krueger, Tim Kuphal, Stanley Letovsky, Michael Levandoski, Craig Lukasik, Jonathan Meltzer, Brian Norvell, Mindy Nye, Scott Parker, Christos Petropoulos, John Pruitt, Steven Ragan, Scott Ryan, Mike Sapeta, Jana Schroth, Suresh Babu Selvaraju, Goran Stevovic, Amanda Suchanek, Andrea Throop, Lyndon Tilson, Thomas Urban, Joe Voshell, Kimberly Wagner, Jonathan Williams, Mary Williamson, Qian Zeng, Tricia Zwiefelhofer, Clinton R. Paden, Duncan MacCannell |
| EPI_ISL_1798344 | MD DOH Laboratories Administration | Centers for Disease Control and Prevention Division of Viral Diseases, Pathogen Discovery | Mili Sheth, Sarah Nobles, Jasmine Padilla, Mark Burroughs, Shoshona Le, Katie Dillon, Peter Cook, Clinton R. Paden, Dhwani Batra, Krista Queen, Kristen Knipe, Dakota Howard, Yvette Unoarumhi, Darlene Wagner, Matthew Schmerer, Ben L. Rambo-Martin, Kristine Lacek, Sam Shepard, Alison Laufer Halpin, Dave Wentworth, Vivien Dugan, Suxiang Tong, Justin Lee |
| EPI_ISL_1798345 | Laboratory Corporation of America | Centers for Disease Control and Prevention Division of Viral Diseases, Pathogen Discovery | Dakota Howard, Dhwani Batra, Peter W. Cook, Kara Moser, Adrian Paskey, Jason Caravas, Benjamin Rambo-Martin, Shatavia Morrison, Christopher Gulvick, Scott Sammons, Yvette Unoarumhi, Darlene Wagner, Matthew Schmerer, Minoo Agarwal, Eyad Almasri, Debbie Boles, Ayla Burns, Nuthawin Charoensri, Oren Cohen, Susan Countryman, Mary Ann Cristobal, Bobbi Croy, Suzanne Dale, Hrushikesh Deshmukh, Amanda Douglas, Vincent Drouillon, Marcia Eisenberg, Howard Engler, Rama Ghatti, Prashant Gupta, Susan Hicks, Jake Humphrey, Lax Iyer, Manoj Jain, Mohan Kolli, Brian Krueger, Tim Kuphal, Stanley Letovsky, Michael Levandoski, Craig Lukasik, Jonathan Meltzer, Brian Norvell, Mindy Nye, Scott Parker, Christos Petropoulos, John Pruitt, Steven Ragan, Scott Ryan, Mike Sapeta, Jana Schroth, Suresh Babu Selvaraju, Goran Stevovic, Amanda Suchanek, Andrea Throop, Lyndon Tilson, Thomas Urban, Joe Voshell, Kimberly Wagner, Jonathan Williams, Mary Williamson, Qian Zeng, Tricia Zwiefelhofer, Clinton R. Paden, Duncan MacCannell |
| EPI_ISL_1798346 | MD DOH Laboratories Administration | Centers for Disease Control and Prevention Division of Viral Diseases, Pathogen Discovery | Mili Sheth, Sarah Nobles, Jasmine Padilla, Mark Burroughs, Shoshona Le, Katie Dillon, Peter Cook, Clinton R. Paden, Dhwani Batra, Krista Queen, Kristen Knipe, Dakota Howard, Yvette Unoarumhi, Darlene Wagner, Matthew Schmerer, Ben L. Rambo-Martin, Kristine Lacek, Sam Shepard, Alison Laufer Halpin, Dave Wentworth, Vivien Dugan, Suxiang Tong, Justin Lee |
| EPI_ISL_1798347 | Laboratory Corporation of America | Centers for Disease Control and Prevention Division of Viral Diseases, Pathogen Discovery | Dakota Howard, Dhwani Batra, Peter W. Cook, Kara Moser, Adrian Paskey, Jason Caravas, Benjamin Rambo-Martin, Shatavia Morrison, Christopher Gulvick, Scott Sammons, Yvette Unoarumhi, Darlene Wagner, Matthew Schmerer, Minoo Agarwal, Eyad Almasri, Debbie Boles, Ayla Burns, Nuthawin Charoensri, Oren Cohen, Susan Countryman, Mary Ann Cristobal, Bobbi Croy, Suzanne Dale, Hrushikesh Deshmukh, Amanda Douglas, Vincent Drouillon, Marcia Eisenberg, Howard Engler, Rama Ghatti, Prashant Gupta, Susan Hicks, Jake Humphrey, Lax Iyer, Manoj Jain, Mohan Kolli, Brian Krueger, Tim Kuphal, Stanley Letovsky, Michael Levandoski, Craig Lukasik, Jonathan Meltzer, Brian Norvell, Mindy Nye, Scott Parker, Christos Petropoulos, John Pruitt, Steven Ragan, Scott Ryan, Mike Sapeta, Jana Schroth, Suresh Babu Selvaraju, Goran Stevovic, Amanda Suchanek, Andrea Throop, Lyndon Tilson, Thomas Urban, Joe Voshell, Kimberly Wagner, Jonathan Williams, Mary Williamson, Qian Zeng, Tricia Zwiefelhofer, Clinton R. Paden, Duncan MacCannell |
| EPI_ISL_1798348 | MD DOH Laboratories Administration | Centers for Disease Control and Prevention Division of Viral Diseases, Pathogen Discovery | Mili Sheth, Sarah Nobles, Jasmine Padilla, Mark Burroughs, Shoshona Le, Katie Dillon, Peter Cook, Clinton R. Paden, Dhwani Batra, Krista Queen, Kristen Knipe, Dakota Howard, Yvette Unoarumhi, Darlene Wagner, Matthew Schmerer, Ben L. Rambo-Martin, Kristine Lacek, Sam Shepard, Alison Laufer Halpin, Dave Wentworth, Vivien Dugan, Suxiang Tong, Justin Lee |
| EPI_ISL_1798349 | Laboratory Corporation of America | Centers for Disease Control and Prevention Division of Viral Diseases, Pathogen Discovery | Dakota Howard, Dhwani Batra, Peter W. Cook, Kara Moser, Adrian Paskey, Jason Caravas, Benjamin Rambo-Martin, Shatavia Morrison, Christopher Gulvick, Scott Sammons, Yvette Unoarumhi, Darlene Wagner, Matthew Schmerer, Minoo Agarwal, Eyad Almasri, Debbie Boles, Ayla Burns, Nuthawin Charoensri, Oren Cohen, Susan Countryman, Mary Ann Cristobal, Bobbi Croy, Suzanne Dale, Hrushikesh Deshmukh, Amanda Douglas, Vincent Drouillon, Marcia Eisenberg, Howard Engler, Rama Ghatti, Prashant Gupta, Susan Hicks, Jake Humphrey, Lax Iyer, Manoj Jain, Mohan Kolli, Brian Krueger, Tim Kuphal, Stanley Letovsky, Michael Levandoski, Craig Lukasik, Jonathan Meltzer, Brian Norvell, Mindy Nye, Scott Parker, Christos Petropoulos, John Pruitt, Steven Ragan, Scott Ryan, Mike Sapeta, Jana Schroth, Suresh Babu Selvaraju, Goran Stevovic, Amanda Suchanek, Andrea Throop, Lyndon Tilson, Thomas Urban, Joe Voshell, Kimberly Wagner, Jonathan Williams, Mary Williamson, Qian Zeng, Tricia Zwiefelhofer, Clinton R. Paden, Duncan MacCannell |
| EPI_ISL_1798351, EPI_ISL_1798352, EPI_ISL_1798354 | PR Public Health Lab | Centers for Disease Control and Prevention Division of Viral Diseases, Pathogen Discovery | Mili Sheth, Sarah Nobles, Jasmine Padilla, Mark Burroughs, Shoshona Le, Katie Dillon, Peter Cook, Clinton R. Paden, Dhwani Batra, Krista Queen, Kristen Knipe, Dakota Howard, Yvette Unoarumhi, Darlene Wagner, Matthew Schmerer, Ben L. Rambo-Martin, Kristine Lacek, Sam Shepard, Alison Laufer Halpin, Dave Wentworth, Vivien Dugan, Suxiang Tong, Justin Lee |
| EPI_ISL_1798355 | Laboratory Corporation of America | Centers for Disease Control and Prevention Division of Viral Diseases, Pathogen Discovery | Dakota Howard, Dhwani Batra, Peter W. Cook, Kara Moser, Adrian Paskey, Jason Caravas, Benjamin Rambo-Martin, Shatavia Morrison, Christopher Gulvick, Scott Sammons, Yvette Unoarumhi, Darlene Wagner, Matthew Schmerer, Minoo Agarwal, Eyad Almasri, Debbie Boles, Ayla Burns, Nuthawin Charoensri, Oren Cohen, Susan Countryman, Mary Ann Cristobal, Bobbi Croy, Suzanne Dale, Hrushikesh Deshmukh, Amanda Douglas, Vincent Drouillon, Marcia Eisenberg, Howard Engler, Rama Ghatti, Prashant Gupta, Susan Hicks, Jake Humphrey, Lax Iyer, Manoj Jain, Mohan Kolli, Brian Krueger, Tim Kuphal, Stanley Letovsky, Michael Levandoski, Craig Lukasik, Jonathan Meltzer, Brian Norvell, Mindy Nye, Scott Parker, Christos Petropoulos, John Pruitt, Steven Ragan, Scott Ryan, Mike Sapeta, Jana Schroth, Suresh Babu Selvaraju, Goran Stevovic, Amanda Suchanek, Andrea Throop, Lyndon Tilson, Thomas Urban, Joe Voshell, Kimberly Wagner, Jonathan Williams, Mary Williamson, Qian Zeng, Tricia Zwiefelhofer, Clinton R. Paden, Duncan MacCannell |
| EPI_ISL_1798356 | MT Public Health Laboratory | Centers for Disease Control and Prevention Division of Viral Diseases, Pathogen Discovery | Mili Sheth, Sarah Nobles, Jasmine Padilla, Mark Burroughs, Shoshona Le, Katie Dillon, Peter Cook, Clinton R. Paden, Dhwani Batra, Krista Queen, Kristen Knipe, Dakota Howard, Yvette Unoarumhi, Darlene Wagner, Matthew Schmerer, Ben L. Rambo-Martin, Kristine Lacek, Sam Shepard, Alison Laufer Halpin, Dave Wentworth, Vivien Dugan, Suxiang Tong, Justin Lee |
| EPI_ISL_1798357 | Laboratory Corporation of America | Centers for Disease Control and Prevention Division of Viral Diseases, Pathogen Discovery | Dakota Howard, Dhwani Batra, Peter W. Cook, Kara Moser, Adrian Paskey, Jason Caravas, Benjamin Rambo-Martin, Shatavia Morrison, Christopher Gulvick, Scott Sammons, Yvette Unoarumhi, Darlene Wagner, Matthew Schmerer, Minoo Agarwal, Eyad Almasri, Debbie Boles, Ayla Burns, Nuthawin Charoensri, Oren Cohen, Susan Countryman, Mary Ann Cristobal, Bobbi Croy, Suzanne Dale, Hrushikesh Deshmukh, Amanda Douglas, Vincent Drouillon, Marcia Eisenberg, Howard Engler, Rama Ghatti, Prashant Gupta, Susan Hicks, Jake Humphrey, Lax Iyer, Manoj Jain, Mohan Kolli, Brian Krueger, Tim Kuphal, Stanley Letovsky, Michael Levandoski, Craig Lukasik, Jonathan Meltzer, Brian Norvell, Mindy Nye, Scott Parker, Christos Petropoulos, John Pruitt, Steven Ragan, Scott Ryan, Mike Sapeta, Jana Schroth, Suresh Babu Selvaraju, Goran Stevovic, Amanda Suchanek, Andrea Throop, Lyndon Tilson, Thomas Urban, Joe Voshell, Kimberly Wagner, Jonathan Williams, Mary Williamson, Qian Zeng, Tricia Zwiefelhofer, Clinton R. Paden, Duncan MacCannell |
| EPI_ISL_1798358 | MD DOH Laboratories Administration | Centers for Disease Control and Prevention Division of Viral Diseases, Pathogen Discovery | Mili Sheth, Sarah Nobles, Jasmine Padilla, Mark Burroughs, Shoshona Le, Katie Dillon, Peter Cook, Clinton R. Paden, Dhwani Batra, Krista Queen, Kristen Knipe, Dakota Howard, Yvette Unoarumhi, Darlene Wagner, Matthew Schmerer, Ben L. Rambo-Martin, Kristine Lacek, Sam Shepard, Alison Laufer Halpin, Dave Wentworth, Vivien Dugan, Suxiang Tong, Justin Lee |
| EPI_ISL_1798359 | Laboratory Corporation of America | Centers for Disease Control and Prevention Division of Viral Diseases, Pathogen Discovery | Dakota Howard, Dhwani Batra, Peter W. Cook, Kara Moser, Adrian Paskey, Jason Caravas, Benjamin Rambo-Martin, Shatavia Morrison, Christopher Gulvick, Scott Sammons, Yvette Unoarumhi, Darlene Wagner, Matthew Schmerer, Minoo Agarwal, Eyad Almasri, Debbie Boles, Ayla Burns, Nuthawin Charoensri, Oren Cohen, Susan Countryman, Mary Ann Cristobal, Bobbi Croy, Suzanne Dale, Hrushikesh Deshmukh, Amanda Douglas, Vincent Drouillon, Marcia Eisenberg, Howard Engler, Rama Ghatti, Prashant Gupta, Susan Hicks, Jake Humphrey, Lax Iyer, Manoj Jain, Mohan Kolli, Brian Krueger, Tim Kuphal, Stanley Letovsky, Michael Levandoski, Craig Lukasik, Jonathan Meltzer, Brian Norvell, Mindy Nye, Scott Parker, Christos Petropoulos, John Pruitt, Steven Ragan, Scott Ryan, Mike Sapeta, Jana Schroth, Suresh Babu Selvaraju, Goran Stevovic, Amanda Suchanek, Andrea Throop, Lyndon Tilson, Thomas Urban, Joe Voshell, Kimberly Wagner, Jonathan Williams, Mary Williamson, Qian Zeng, Tricia Zwiefelhofer, Clinton R. Paden, Duncan MacCannell |
| EPI_ISL_1798360 | NJ Public Health and Environmental Laboratories | Centers for Disease Control and Prevention Division of Viral Diseases, Pathogen Discovery | Mili Sheth, Sarah Nobles, Jasmine Padilla, Mark Burroughs, Shoshona Le, Katie Dillon, Peter Cook, Clinton R. Paden, Dhwani Batra, Krista Queen, Kristen Knipe, Dakota Howard, Yvette Unoarumhi, Darlene Wagner, Matthew Schmerer, Ben L. Rambo-Martin, Kristine Lacek, Sam Shepard, Alison Laufer Halpin, Dave Wentworth, Vivien Dugan, Suxiang Tong, Justin Lee |
| EPI_ISL_1798362, EPI_ISL_1798363, EPI_ISL_1798365, EPI_ISL_1798366 | Laboratory Corporation of America | Centers for Disease Control and Prevention Division of Viral Diseases, Pathogen Discovery | Dakota Howard, Dhwani Batra, Peter W. Cook, Kara Moser, Adrian Paskey, Jason Caravas, Benjamin Rambo-Martin, Shatavia Morrison, Christopher Gulvick, Scott Sammons, Yvette Unoarumhi, Darlene Wagner, Matthew Schmerer, Minoo Agarwal, Eyad Almasri, Debbie Boles, Ayla Burns, Nuthawin Charoensri, Oren Cohen, Susan Countryman, Mary Ann Cristobal, Bobbi Croy, Suzanne Dale, Hrushikesh Deshmukh, Amanda Douglas, Vincent Drouillon, Marcia Eisenberg, Howard Engler, Rama Ghatti, Prashant Gupta, Susan Hicks, Jake Humphrey, Lax Iyer, Manoj Jain, Mohan Kolli, Brian Krueger, Tim Kuphal, Stanley Letovsky, Michael Levandoski, Craig Lukasik, Jonathan Meltzer, Brian Norvell, Mindy Nye, Scott Parker, Christos Petropoulos, John Pruitt, Steven Ragan, Scott Ryan, Mike Sapeta, Jana Schroth, Suresh Babu Selvaraju, Goran Stevovic, Amanda Suchanek, Andrea Throop, Lyndon Tilson, Thomas Urban, Joe Voshell, Kimberly Wagner, Jonathan Williams, Mary Williamson, Qian Zeng, Tricia Zwiefelhofer, Clinton R. Paden, Duncan MacCannell |
| EPI_ISL_1798367 | MD DOH Laboratories Administration | Centers for Disease Control and Prevention Division of Viral Diseases, Pathogen Discovery | Mili Sheth, Sarah Nobles, Jasmine Padilla, Mark Burroughs, Shoshona Le, Katie Dillon, Peter Cook, Clinton R. Paden, Dhwani Batra, Krista Queen, Kristen Knipe, Dakota Howard, Yvette Unoarumhi, Darlene Wagner, Matthew Schmerer, Ben L. Rambo-Martin, Kristine Lacek, Sam Shepard, Alison Laufer Halpin, Dave Wentworth, Vivien Dugan, Suxiang Tong, Justin Lee |
| EPI_ISL_1798368 | Laboratory Corporation of America | Centers for Disease Control and Prevention Division of Viral Diseases, Pathogen Discovery | Dakota Howard, Dhwani Batra, Peter W. Cook, Kara Moser, Adrian Paskey, Jason Caravas, Benjamin Rambo-Martin, Shatavia Morrison, Christopher Gulvick, Scott Sammons, Yvette Unoarumhi, Darlene Wagner, Matthew Schmerer, Minoo Agarwal, Eyad Almasri, Debbie Boles, Ayla Burns, Nuthawin Charoensri, Oren Cohen, Susan Countryman, Mary Ann Cristobal, Bobbi Croy, Suzanne Dale, Hrushikesh Deshmukh, Amanda Douglas, Vincent Drouillon, Marcia Eisenberg, Howard Engler, Rama Ghatti, Prashant Gupta, Susan Hicks, Jake Humphrey, Lax Iyer, Manoj Jain, Mohan Kolli, Brian Krueger, Tim Kuphal, Stanley Letovsky, Michael Levandoski, Craig Lukasik, Jonathan Meltzer, Brian Norvell, Mindy Nye, Scott Parker, Christos Petropoulos, John Pruitt, Steven Ragan, Scott Ryan, Mike Sapeta, Jana Schroth, Suresh Babu Selvaraju, Goran Stevovic, Amanda Suchanek, Andrea Throop, Lyndon Tilson, Thomas Urban, Joe Voshell, Kimberly Wagner, Jonathan Williams, Mary Williamson, Qian Zeng, Tricia Zwiefelhofer, Clinton R. Paden, Duncan MacCannell |
| EPI_ISL_1798369 | MT Public Health Laboratory | Centers for Disease Control and Prevention Division of Viral Diseases, Pathogen Discovery | Mili Sheth, Sarah Nobles, Jasmine Padilla, Mark Burroughs, Shoshona Le, Katie Dillon, Peter Cook, Clinton R. Paden, Dhwani Batra, Krista Queen, Kristen Knipe, Dakota Howard, Yvette Unoarumhi, Darlene Wagner, Matthew Schmerer, Ben L. Rambo-Martin, Kristine Lacek, Sam Shepard, Alison Laufer Halpin, Dave Wentworth, Vivien Dugan, Suxiang Tong, Justin Lee |
| EPI_ISL_1798370 | Laboratory Corporation of America | Centers for Disease Control and Prevention Division of Viral Diseases, Pathogen Discovery | Dakota Howard, Dhwani Batra, Peter W. Cook, Kara Moser, Adrian Paskey, Jason Caravas, Benjamin Rambo-Martin, Shatavia Morrison, Christopher Gulvick, Scott Sammons, Yvette Unoarumhi, Darlene Wagner, Matthew Schmerer, Minoo Agarwal, Eyad Almasri, Debbie Boles, Ayla Burns, Nuthawin Charoensri, Oren Cohen, Susan Countryman, Mary Ann Cristobal, Bobbi Croy, Suzanne Dale, Hrushikesh Deshmukh, Amanda Douglas, Vincent Drouillon, Marcia Eisenberg, Howard Engler, Rama Ghatti, Prashant Gupta, Susan Hicks, Jake Humphrey, Lax Iyer, Manoj Jain, Mohan Kolli, Brian Krueger, Tim Kuphal, Stanley Letovsky, Michael Levandoski, Craig Lukasik, Jonathan Meltzer, Brian Norvell, Mindy Nye, Scott Parker, Christos Petropoulos, John Pruitt, Steven Ragan, Scott Ryan, Mike Sapeta, Jana Schroth, Suresh Babu Selvaraju, Goran Stevovic, Amanda Suchanek, Andrea Throop, Lyndon Tilson, Thomas Urban, Joe Voshell, Kimberly Wagner, Jonathan Williams, Mary Williamson, Qian Zeng, Tricia Zwiefelhofer, Clinton R. Paden, Duncan MacCannell |
| EPI_ISL_1798371 | MD DOH Laboratories Administration | Centers for Disease Control and Prevention Division of Viral Diseases, Pathogen Discovery | Mili Sheth, Sarah Nobles, Jasmine Padilla, Mark Burroughs, Shoshona Le, Katie Dillon, Peter Cook, Clinton R. Paden, Dhwani Batra, Krista Queen, Kristen Knipe, Dakota Howard, Yvette Unoarumhi, Darlene Wagner, Matthew Schmerer, Ben L. Rambo-Martin, Kristine Lacek, Sam Shepard, Alison Laufer Halpin, Dave Wentworth, Vivien Dugan, Suxiang Tong, Justin Lee |
| EPI_ISL_1798372 | Laboratory Corporation of America | Centers for Disease Control and Prevention Division of Viral Diseases, Pathogen Discovery | Dakota Howard, Dhwani Batra, Peter W. Cook, Kara Moser, Adrian Paskey, Jason Caravas, Benjamin Rambo-Martin, Shatavia Morrison, Christopher Gulvick, Scott Sammons, Yvette Unoarumhi, Darlene Wagner, Matthew Schmerer, Minoo Agarwal, Eyad Almasri, Debbie Boles, Ayla Burns, Nuthawin Charoensri, Oren Cohen, Susan Countryman, Mary Ann Cristobal, Bobbi Croy, Suzanne Dale, Hrushikesh Deshmukh, Amanda Douglas, Vincent Drouillon, Marcia Eisenberg, Howard Engler, Rama Ghatti, Prashant Gupta, Susan Hicks, Jake Humphrey, Lax Iyer, Manoj Jain, Mohan Kolli, Brian Krueger, Tim Kuphal, Stanley Letovsky, Michael Levandoski, Craig Lukasik, Jonathan Meltzer, Brian Norvell, Mindy Nye, Scott Parker, Christos Petropoulos, John Pruitt, Steven Ragan, Scott Ryan, Mike Sapeta, Jana Schroth, Suresh Babu Selvaraju, Goran Stevovic, Amanda Suchanek, Andrea Throop, Lyndon Tilson, Thomas Urban, Joe Voshell, Kimberly Wagner, Jonathan Williams, Mary Williamson, Qian Zeng, Tricia Zwiefelhofer, Clinton R. Paden, Duncan MacCannell |
| EPI_ISL_1798373 | NJ Public Health and Environmental Laboratories | Centers for Disease Control and Prevention Division of Viral Diseases, Pathogen Discovery | Mili Sheth, Sarah Nobles, Jasmine Padilla, Mark Burroughs, Shoshona Le, Katie Dillon, Peter Cook, Clinton R. Paden, Dhwani Batra, Krista Queen, Kristen Knipe, Dakota Howard, Yvette Unoarumhi, Darlene Wagner, Matthew Schmerer, Ben L. Rambo-Martin, Kristine Lacek, Sam Shepard, Alison Laufer Halpin, Dave Wentworth, Vivien Dugan, Suxiang Tong, Justin Lee |
| EPI_ISL_1798374 | Laboratory Corporation of America | Centers for Disease Control and Prevention Division of Viral Diseases, Pathogen Discovery | Dakota Howard, Dhwani Batra, Peter W. Cook, Kara Moser, Adrian Paskey, Jason Caravas, Benjamin Rambo-Martin, Shatavia Morrison, Christopher Gulvick, Scott Sammons, Yvette Unoarumhi, Darlene Wagner, Matthew Schmerer, Minoo Agarwal, Eyad Almasri, Debbie Boles, Ayla Burns, Nuthawin Charoensri, Oren Cohen, Susan Countryman, Mary Ann Cristobal, Bobbi Croy, Suzanne Dale, Hrushikesh Deshmukh, Amanda Douglas, Vincent Drouillon, Marcia Eisenberg, Howard Engler, Rama Ghatti, Prashant Gupta, Susan Hicks, Jake Humphrey, Lax Iyer, Manoj Jain, Mohan Kolli, Brian Krueger, Tim Kuphal, Stanley Letovsky, Michael Levandoski, Craig Lukasik, Jonathan Meltzer, Brian Norvell, Mindy Nye, Scott Parker, Christos Petropoulos, John Pruitt, Steven Ragan, Scott Ryan, Mike Sapeta, Jana Schroth, Suresh Babu Selvaraju, Goran Stevovic, Amanda Suchanek, Andrea Throop, Lyndon Tilson, Thomas Urban, Joe Voshell, Kimberly Wagner, Jonathan Williams, Mary Williamson, Qian Zeng, Tricia Zwiefelhofer, Clinton R. Paden, Duncan MacCannell |
| EPI_ISL_1798375 | MT Public Health Laboratory | Centers for Disease Control and Prevention Division of Viral Diseases, Pathogen Discovery | Mili Sheth, Sarah Nobles, Jasmine Padilla, Mark Burroughs, Shoshona Le, Katie Dillon, Peter Cook, Clinton R. Paden, Dhwani Batra, Krista Queen, Kristen Knipe, Dakota Howard, Yvette Unoarumhi, Darlene Wagner, Matthew Schmerer, Ben L. Rambo-Martin, Kristine Lacek, Sam Shepard, Alison Laufer Halpin, Dave Wentworth, Vivien Dugan, Suxiang Tong, Justin Lee |
| EPI_ISL_1798377 | PR Public Health Lab | Centers for Disease Control and Prevention Division of Viral Diseases, Pathogen Discovery | Mili Sheth, Sarah Nobles, Jasmine Padilla, Mark Burroughs, Shoshona Le, Katie Dillon, Peter Cook, Clinton R. Paden, Dhwani Batra, Krista Queen, Kristen Knipe, Dakota Howard, Yvette Unoarumhi, Darlene Wagner, Matthew Schmerer, Ben L. Rambo-Martin, Kristine Lacek, Sam Shepard, Alison Laufer Halpin, Dave Wentworth, Vivien Dugan, Suxiang Tong, Justin Lee |
| EPI_ISL_1798379 | NJ Public Health and Environmental Laboratories | Centers for Disease Control and Prevention Division of Viral Diseases, Pathogen Discovery | Mili Sheth, Sarah Nobles, Jasmine Padilla, Mark Burroughs, Shoshona Le, Katie Dillon, Peter Cook, Clinton R. Paden, Dhwani Batra, Krista Queen, Kristen Knipe, Dakota Howard, Yvette Unoarumhi, Darlene Wagner, Matthew Schmerer, Ben L. Rambo-Martin, Kristine Lacek, Sam Shepard, Alison Laufer Halpin, Dave Wentworth, Vivien Dugan, Suxiang Tong, Justin Lee |

| | | | |
|---|---|---|---|
| EPI_ISL_1798380 | PR Public Health Lab | Centers for Disease Control and Prevention Division of Viral Diseases, Pathogen Discovery | Mili Sheth, Sarah Nobles, Jasmine Padilla, Mark Burroughs, Shoshona Le, Katie Dillon, Peter Cook, Clinton R. Paden, Dhwani Batra, Krista Queen, Kristen Knipe, Dakota Howard, Yvette Unoarumhi, Darlene Wagner, Matthew Schmerer, Ben L. Rambo-Martin, Kristine Lacek, Sam Shepard, Alison Laufer Halpin, Dave Wentworth, Vivien Dugan, Suxiang Tong, Justin Lee |
| EPI_ISL_1798381 | Laboratory Corporation of America | Centers for Disease Control and Prevention Division of Viral Diseases, Pathogen Discovery | Dakota Howard, Dhwani Batra, Peter W. Cook, Kara Moser, Adrian Paskey, Jason Caravas, Benjamin Rambo-Martin, Shatavia Morrison, Christopher Gulvick, Scott Sammons, Yvette Unoarumhi, Darlene Wagner, Matthew Schmerer, Minoo Agarwal, Eyad Almasri, Debbie Boles, Ayla Burns, Nuthawin Charoensri, Oren Cohen, Susan Countryman, Mary Ann Cristobal, Bobbi Croy, Suzanne Dale, Hrushikesh Deshmukh, Amanda Douglas, Vincent Drouillon, Marcia Eisenberg, Howard Engler, Rama Ghatti, Prashant Gupta, Susan Hicks, Jake Humphrey, Lax Iyer, Manoj Jain, Mohan Kolli, Brian Krueger, Tim Kuphal, Stanley Letovsky, Michael Levandoski, Craig Lukasik, Jonathan Meltzer, Brian Norvell, Mindy Nye, Scott Parker, Christos Petropoulos, John Pruitt, Steven Ragan, Scott Ryan, Mike Sapeta, Jana Schroth, Suresh Babu Selvaraju, Goran Stevovic, Amanda Suchanek, Andrea Throop, Lyndon Tilson, Thomas Urban, Joe Voshell, Kimberly Wagner, Jonathan Williams, Mary Williamson, Qian Zeng, Tricia Zwiefelhofer, Clinton R. Paden, Duncan MacCannell |
| EPI_ISL_1798382 | CT-Dr. Katherine A. Kelley State Public Health Lab | Centers for Disease Control and Prevention Division of Viral Diseases, Pathogen Discovery | Mili Sheth, Sarah Nobles, Jasmine Padilla, Mark Burroughs, Shoshona Le, Katie Dillon, Peter Cook, Clinton R. Paden, Dhwani Batra, Krista Queen, Kristen Knipe, Dakota Howard, Yvette Unoarumhi, Darlene Wagner, Matthew Schmerer, Ben L. Rambo-Martin, Kristine Lacek, Sam Shepard, Alison Laufer Halpin, Dave Wentworth, Vivien Dugan, Suxiang Tong, Justin Lee |
| EPI_ISL_1798384 | MD DOH Laboratories Administration | Centers for Disease Control and Prevention Division of Viral Diseases, Pathogen Discovery | Mili Sheth, Sarah Nobles, Jasmine Padilla, Mark Burroughs, Shoshona Le, Katie Dillon, Peter Cook, Clinton R. Paden, Dhwani Batra, Krista Queen, Kristen Knipe, Dakota Howard, Yvette Unoarumhi, Darlene Wagner, Matthew Schmerer, Ben L. Rambo-Martin, Kristine Lacek, Sam Shepard, Alison Laufer Halpin, Dave Wentworth, Vivien Dugan, Suxiang Tong, Justin Lee |
| EPI_ISL_1798386 | MN PHL Division, Minnesota Department of Health | Centers for Disease Control and Prevention Division of Viral Diseases, Pathogen Discovery | Mili Sheth, Sarah Nobles, Jasmine Padilla, Mark Burroughs, Shoshona Le, Katie Dillon, Peter Cook, Clinton R. Paden, Dhwani Batra, Krista Queen, Kristen Knipe, Dakota Howard, Yvette Unoarumhi, Darlene Wagner, Matthew Schmerer, Ben L. Rambo-Martin, Kristine Lacek, Sam Shepard, Alison Laufer Halpin, Dave Wentworth, Vivien Dugan, Suxiang Tong, Justin Lee |
| EPI_ISL_1798387 | Laboratory Corporation of America | Centers for Disease Control and Prevention Division of Viral Diseases, Pathogen Discovery | Dakota Howard, Dhwani Batra, Peter W. Cook, Kara Moser, Adrian Paskey, Jason Caravas, Benjamin Rambo-Martin, Shatavia Morrison, Christopher Gulvick, Scott Sammons, Yvette Unoarumhi, Darlene Wagner, Matthew Schmerer, Minoo Agarwal, Eyad Almasri, Debbie Boles, Ayla Burns, Nuthawin Charoensri, Oren Cohen, Susan Countryman, Mary Ann Cristobal, Bobbi Croy, Suzanne Dale, Hrushikesh Deshmukh, Amanda Douglas, Vincent Drouillon, Marcia Eisenberg, Howard Engler, Rama Ghatti, Prashant Gupta, Susan Hicks, Jake Humphrey, Lax Iyer, Manoj Jain, Mohan Kolli, Brian Krueger, Tim Kuphal, Stanley Letovsky, Michael Levandoski, Craig Lukasik, Jonathan Meltzer, Brian Norvell, Mindy Nye, Scott Parker, Christos Petropoulos, John Pruitt, Steven Ragan, Scott Ryan, Mike Sapeta, Jana Schroth, Suresh Babu Selvaraju, Goran Stevovic, Amanda Suchanek, Andrea Throop, Lyndon Tilson, Thomas Urban, Joe Voshell, Kimberly Wagner, Jonathan Williams, Mary Williamson, Qian Zeng, Tricia Zwiefelhofer, Clinton R. Paden, Duncan MacCannell |
| EPI_ISL_1798388 | CT-Dr. Katherine A. Kelley State Public Health Lab | Centers for Disease Control and Prevention Division of Viral Diseases, Pathogen Discovery | Mili Sheth, Sarah Nobles, Jasmine Padilla, Mark Burroughs, Shoshona Le, Katie Dillon, Peter Cook, Clinton R. Paden, Dhwani Batra, Krista Queen, Kristen Knipe, Dakota Howard, Yvette Unoarumhi, Darlene Wagner, Matthew Schmerer, Ben L. Rambo-Martin, Kristine Lacek, Sam Shepard, Alison Laufer Halpin, Dave Wentworth, Vivien Dugan, Suxiang Tong, Justin Lee |
| EPI_ISL_1798389 | Laboratory Corporation of America | Centers for Disease Control and Prevention Division of Viral Diseases, Pathogen Discovery | Dakota Howard, Dhwani Batra, Peter W. Cook, Kara Moser, Adrian Paskey, Jason Caravas, Benjamin Rambo-Martin, Shatavia Morrison, Christopher Gulvick, Scott Sammons, Yvette Unoarumhi, Darlene Wagner, Matthew Schmerer, Minoo Agarwal, Eyad Almasri, Debbie Boles, Ayla Burns, Nuthawin Charoensri, Oren Cohen, Susan Countryman, Mary Ann Cristobal, Bobbi Croy, Suzanne Dale, Hrushikesh Deshmukh, Amanda Douglas, Vincent Drouillon, Marcia Eisenberg, Howard Engler, Rama Ghatti, Prashant Gupta, Susan Hicks, Jake Humphrey, Lax Iyer, Manoj Jain, Mohan Kolli, Brian Krueger, Tim Kuphal, Stanley Letovsky, Michael Levandoski, Craig Lukasik, Jonathan Meltzer, Brian Norvell, Mindy Nye, Scott Parker, Christos Petropoulos, John Pruitt, Steven Ragan, Scott Ryan, Mike Sapeta, Jana Schroth, Suresh Babu Selvaraju, Goran Stevovic, Amanda Suchanek, Andrea Throop, Lyndon Tilson, Thomas Urban, Joe Voshell, Kimberly Wagner, Jonathan Williams, Mary Williamson, Qian Zeng, Tricia Zwiefelhofer, Clinton R. Paden, Duncan MacCannell |
| EPI_ISL_1798390 | MN PHL Division, Minnesota Department of Health | Centers for Disease Control and Prevention Division of Viral Diseases, Pathogen Discovery | Mili Sheth, Sarah Nobles, Jasmine Padilla, Mark Burroughs, Shoshona Le, Katie Dillon, Peter Cook, Clinton R. Paden, Dhwani Batra, Krista Queen, Kristen Knipe, Dakota Howard, Yvette Unoarumhi, Darlene Wagner, Matthew Schmerer, Ben L. Rambo-Martin, Kristine Lacek, Sam Shepard, Alison Laufer Halpin, Dave Wentworth, Vivien Dugan, Suxiang Tong, Justin Lee |
| EPI_ISL_1798391 | Laboratory Corporation of America | Centers for Disease Control and Prevention Division of Viral Diseases, Pathogen Discovery | Dakota Howard, Dhwani Batra, Peter W. Cook, Kara Moser, Adrian Paskey, Jason Caravas, Benjamin Rambo-Martin, Shatavia Morrison, Christopher Gulvick, Scott Sammons, Yvette Unoarumhi, Darlene Wagner, Matthew Schmerer, Minoo Agarwal, Eyad Almasri, Debbie Boles, Ayla Burns, Nuthawin Charoensri, Oren Cohen, Susan Countryman, Mary Ann Cristobal, Bobbi Croy, Suzanne Dale, Hrushikesh Deshmukh, Amanda Douglas, Vincent Drouillon, Marcia Eisenberg, Howard Engler, Rama Ghatti, Prashant Gupta, Susan Hicks, Jake Humphrey, Lax Iyer, Manoj Jain, Mohan Kolli, Brian Krueger, Tim Kuphal, Stanley Letovsky, Michael Levandoski, Craig Lukasik, Jonathan Meltzer, Brian Norvell, Mindy Nye, Scott Parker, Christos Petropoulos, John Pruitt, Steven Ragan, Scott Ryan, Mike Sapeta, Jana Schroth, Suresh Babu Selvaraju, Goran Stevovic, Amanda Suchanek, Andrea Throop, Lyndon Tilson, Thomas Urban, Joe Voshell, Kimberly Wagner, Jonathan Williams, Mary Williamson, Qian Zeng, Tricia Zwiefelhofer, Clinton R. Paden, Duncan MacCannell |
| EPI_ISL_1798392 | MN PHL Division, Minnesota Department of Health | Centers for Disease Control and Prevention Division of Viral Diseases, Pathogen Discovery | Mili Sheth, Sarah Nobles, Jasmine Padilla, Mark Burroughs, Shoshona Le, Katie Dillon, Peter Cook, Clinton R. Paden, Dhwani Batra, Krista Queen, Kristen Knipe, Dakota Howard, Yvette Unoarumhi, Darlene Wagner, Matthew Schmerer, Ben L. Rambo-Martin, Kristine Lacek, Sam Shepard, Alison Laufer Halpin, Dave Wentworth, Vivien Dugan, Suxiang Tong, Justin Lee |
| EPI_ISL_1798393 | Laboratory Corporation of America | Centers for Disease Control and Prevention Division of Viral Diseases, Pathogen Discovery | Dakota Howard, Dhwani Batra, Peter W. Cook, Kara Moser, Adrian Paskey, Jason Caravas, Benjamin Rambo-Martin, Shatavia Morrison, Christopher Gulvick, Scott Sammons, Yvette Unoarumhi, Darlene Wagner, Matthew Schmerer, Minoo Agarwal, Eyad Almasri, Debbie Boles, Ayla Burns, Nuthawin Charoensri, Oren Cohen, Susan Countryman, Mary Ann Cristobal, Bobbi Croy, Suzanne Dale, Hrushikesh Deshmukh, Amanda Douglas, Vincent Drouillon, Marcia Eisenberg, Howard Engler, Rama Ghatti, Prashant Gupta, Susan Hicks, Jake Humphrey, Lax Iyer, Manoj Jain, Mohan Kolli, Brian Krueger, Tim Kuphal, Stanley Letovsky, Michael Levandoski, Craig Lukasik, Jonathan Meltzer, Brian Norvell, Mindy Nye, Scott Parker, Christos Petropoulos, John Pruitt, Steven Ragan, Scott Ryan, Mike Sapeta, Jana Schroth, Suresh Babu Selvaraju, Goran Stevovic, Amanda Suchanek, Andrea Throop, Lyndon Tilson, Thomas Urban, Joe Voshell, Kimberly Wagner, Jonathan Williams, Mary Williamson, Qian Zeng, Tricia Zwiefelhofer, Clinton R. Paden, Duncan MacCannell |
| EPI_ISL_1798394 | MD DOH Laboratories Administration | Centers for Disease Control and Prevention Division of Viral Diseases, Pathogen Discovery | Mili Sheth, Sarah Nobles, Jasmine Padilla, Mark Burroughs, Shoshona Le, Katie Dillon, Peter Cook, Clinton R. Paden, Dhwani Batra, Krista Queen, Kristen Knipe, Dakota Howard, Yvette Unoarumhi, Darlene Wagner, Matthew Schmerer, Ben L. Rambo-Martin, Kristine Lacek, Sam Shepard, Alison Laufer Halpin, Dave Wentworth, Vivien Dugan, Suxiang Tong, Justin Lee |
| EPI_ISL_1798395 | Laboratory Corporation of America | Centers for Disease Control and Prevention Division of Viral Diseases, Pathogen Discovery | Dakota Howard, Dhwani Batra, Peter W. Cook, Kara Moser, Adrian Paskey, Jason Caravas, Benjamin Rambo-Martin, Shatavia Morrison, Christopher Gulvick, Scott Sammons, Yvette Unoarumhi, Darlene Wagner, Matthew Schmerer, Minoo Agarwal, Eyad Almasri, Debbie Boles, Ayla Burns, Nuthawin Charoensri, Oren Cohen, Susan Countryman, Mary Ann Cristobal, Bobbi Croy, Suzanne Dale, Hrushikesh Deshmukh, Amanda Douglas, Vincent Drouillon, Marcia Eisenberg, Howard Engler, Rama Ghatti, Prashant Gupta, Susan Hicks, Jake Humphrey, Lax Iyer, Manoj Jain, Mohan Kolli, Brian Krueger, Tim Kuphal, Stanley Letovsky, Michael Levandoski, Craig Lukasik, Jonathan Meltzer, Brian Norvell, Mindy Nye, Scott Parker, Christos Petropoulos, John Pruitt, Steven Ragan, Scott Ryan, Mike Sapeta, Jana Schroth, Suresh Babu Selvaraju, Goran Stevovic, Amanda Suchanek, Andrea Throop, Lyndon Tilson, Thomas Urban, Joe Voshell, Kimberly Wagner, Jonathan Williams, Mary Williamson, Qian Zeng, Tricia Zwiefelhofer, Clinton R. Paden, Duncan MacCannell |
| EPI_ISL_1798396 | CT-Dr. Katherine A. Kelley State Public Health Lab | Centers for Disease Control and Prevention Division of Viral Diseases, Pathogen Discovery | Mili Sheth, Sarah Nobles, Jasmine Padilla, Mark Burroughs, Shoshona Le, Katie Dillon, Peter Cook, Clinton R. Paden, Dhwani Batra, Krista Queen, Kristen Knipe, Dakota Howard, Yvette Unoarumhi, Darlene Wagner, Matthew Schmerer, Ben L. Rambo-Martin, Kristine Lacek, Sam Shepard, Alison Laufer Halpin, Dave Wentworth, Vivien Dugan, Suxiang Tong, Justin Lee |
| EPI_ISL_1798398 | MD DOH Laboratories Administration | Centers for Disease Control and Prevention Division of Viral Diseases, Pathogen Discovery | Mili Sheth, Sarah Nobles, Jasmine Padilla, Mark Burroughs, Shoshona Le, Katie Dillon, Peter Cook, Clinton R. Paden, Dhwani Batra, Krista Queen, Kristen Knipe, Dakota Howard, Yvette Unoarumhi, Darlene Wagner, Matthew Schmerer, Ben L. Rambo-Martin, Kristine Lacek, Sam Shepard, Alison Laufer Halpin, Dave Wentworth, Vivien Dugan, Suxiang Tong, Justin Lee |
| EPI_ISL_1798400 | CT-Dr. Katherine A. Kelley State Public Health Lab | Centers for Disease Control and Prevention Division of Viral Diseases, Pathogen Discovery | Mili Sheth, Sarah Nobles, Jasmine Padilla, Mark Burroughs, Shoshona Le, Katie Dillon, Peter Cook, Clinton R. Paden, Dhwani Batra, Krista Queen, Kristen Knipe, Dakota Howard, Yvette Unoarumhi, Darlene Wagner, Matthew Schmerer, Ben L. Rambo-Martin, Kristine Lacek, Sam Shepard, Alison Laufer Halpin, Dave Wentworth, Vivien Dugan, Suxiang Tong, Justin Lee |
| EPI_ISL_1798401 | Laboratory Corporation of America | Centers for Disease Control and Prevention Division of Viral Diseases, Pathogen Discovery | Dakota Howard, Dhwani Batra, Peter W. Cook, Kara Moser, Adrian Paskey, Jason Caravas, Benjamin Rambo-Martin, Shatavia Morrison, Christopher Gulvick, Scott Sammons, Yvette Unoarumhi, Darlene Wagner, Matthew Schmerer, Minoo Agarwal, Eyad Almasri, Debbie Boles, Ayla Burns, Nuthawin Charoensri, Oren Cohen, Susan Countryman, Mary Ann Cristobal, Bobbi Croy, Suzanne Dale, Hrushikesh Deshmukh, Amanda Douglas, Vincent Drouillon, Marcia Eisenberg, Howard Engler, Rama Ghatti, Prashant Gupta, Susan Hicks, Jake Humphrey, Lax Iyer, Manoj Jain, Mohan Kolli, Brian Krueger, Tim Kuphal, Stanley Letovsky, Michael Levandoski, Craig Lukasik, Jonathan Meltzer, Brian Norvell, Mindy Nye, Scott Parker, Christos Petropoulos, John Pruitt, Steven Ragan, Scott Ryan, Mike Sapeta, Jana Schroth, Suresh Babu Selvaraju, Goran Stevovic, Amanda Suchanek, Andrea Throop, Lyndon Tilson, Thomas Urban, Joe Voshell, Kimberly Wagner, Jonathan Williams, Mary Williamson, Qian Zeng, Tricia Zwiefelhofer, Clinton R. Paden, Duncan MacCannell |
| EPI_ISL_1798402 | MN PHL Division, Minnesota Department of Health | Centers for Disease Control and Prevention Division of Viral Diseases, Pathogen Discovery | Mili Sheth, Sarah Nobles, Jasmine Padilla, Mark Burroughs, Shoshona Le, Katie Dillon, Peter Cook, Clinton R. Paden, Dhwani Batra, Krista Queen, Kristen Knipe, Dakota Howard, Yvette Unoarumhi, Darlene Wagner, Matthew Schmerer, Ben L. Rambo-Martin, Kristine Lacek, Sam Shepard, Alison Laufer Halpin, Dave Wentworth, Vivien Dugan, Suxiang Tong, Justin Lee |
| EPI_ISL_1798403 | Laboratory Corporation of America | Centers for Disease Control and Prevention Division of Viral Diseases, Pathogen Discovery | Dakota Howard, Dhwani Batra, Peter W. Cook, Kara Moser, Adrian Paskey, Jason Caravas, Benjamin Rambo-Martin, Shatavia Morrison, Christopher Gulvick, Scott Sammons, Yvette Unoarumhi, Darlene Wagner, Matthew Schmerer, Minoo Agarwal, Eyad Almasri, Debbie Boles, Ayla Burns, Nuthawin Charoensri, Oren Cohen, Susan Countryman, Mary Ann Cristobal, Bobbi Croy, Suzanne Dale, Hrushikesh Deshmukh, Amanda Douglas, Vincent Drouillon, Marcia Eisenberg, Howard Engler, Rama Ghatti, Prashant Gupta, Susan Hicks, Jake Humphrey, Lax Iyer, Manoj Jain, Mohan Kolli, Brian Krueger, Tim Kuphal, Stanley Letovsky, Michael Levandoski, Craig Lukasik, Jonathan Meltzer, Brian Norvell, Mindy Nye, Scott Parker, Christos Petropoulos, John Pruitt, Steven Ragan, Scott Ryan, Mike Sapeta, Jana Schroth, Suresh Babu Selvaraju, Goran Stevovic, Amanda Suchanek, Andrea Throop, Lyndon Tilson, Thomas Urban, Joe Voshell, Kimberly Wagner, Jonathan Williams, Mary Williamson, Qian Zeng, Tricia Zwiefelhofer, Clinton R. Paden, Duncan MacCannell |
| EPI_ISL_1798404 | MT Public Health Laboratory | Centers for Disease Control and Prevention Division of Viral Diseases, Pathogen Discovery | Mili Sheth, Sarah Nobles, Jasmine Padilla, Mark Burroughs, Shoshona Le, Katie Dillon, Peter Cook, Clinton R. Paden, Dhwani Batra, Krista Queen, Kristen Knipe, Dakota Howard, Yvette Unoarumhi, Darlene Wagner, Matthew Schmerer, Ben L. Rambo-Martin, Kristine Lacek, Sam Shepard, Alison Laufer Halpin, Dave Wentworth, Vivien Dugan, Suxiang Tong, Justin Lee |
| EPI_ISL_1798406 | NJ Public Health and Environmental Laboratories | Centers for Disease Control and Prevention Division of Viral Diseases, Pathogen Discovery | Mili Sheth, Sarah Nobles, Jasmine Padilla, Mark Burroughs, Shoshona Le, Katie Dillon, Peter Cook, Clinton R. Paden, Dhwani Batra, Krista Queen, Kristen Knipe, Dakota Howard, Yvette Unoarumhi, Darlene Wagner, Matthew Schmerer, Ben L. Rambo-Martin, Kristine Lacek, Sam Shepard, Alison Laufer Halpin, Dave Wentworth, Vivien Dugan, Suxiang Tong, Justin Lee |
| EPI_ISL_1798407 | Laboratory Corporation of America | Centers for Disease Control and Prevention Division of Viral Diseases, Pathogen Discovery | Dakota Howard, Dhwani Batra, Peter W. Cook, Kara Moser, Adrian Paskey, Jason Caravas, Benjamin Rambo-Martin, Shatavia Morrison, Christopher Gulvick, Scott Sammons, Yvette Unoarumhi, Darlene Wagner, Matthew Schmerer, Minoo Agarwal, Eyad Almasri, Debbie Boles, Ayla Burns, Nuthawin Charoensri, Oren Cohen, Susan Countryman, Mary Ann Cristobal, Bobbi Croy, Suzanne Dale, Hrushikesh Deshmukh, Amanda Douglas, Vincent Drouillon, Marcia Eisenberg, Howard Engler, Rama Ghatti, Prashant Gupta, Susan Hicks, Jake Humphrey, Lax Iyer, Manoj Jain, Mohan Kolli, Brian Krueger, Tim Kuphal, Stanley Letovsky, Michael Levandoski, Craig Lukasik, Jonathan Meltzer, Brian Norvell, Mindy Nye, Scott Parker, Christos Petropoulos, John Pruitt, Steven Ragan, Scott Ryan, Mike Sapeta, Jana Schroth, Suresh Babu Selvaraju, Goran Stevovic, Amanda Suchanek, Andrea Throop, Lyndon Tilson, Thomas Urban, Joe Voshell, Kimberly Wagner, Jonathan Williams, Mary Williamson, Qian Zeng, Tricia Zwiefelhofer, Clinton R. Paden, Duncan MacCannell |
| EPI_ISL_1798408 | MD DOH Laboratories Administration | Centers for Disease Control and Prevention Division of Viral Diseases, Pathogen Discovery | Mili Sheth, Sarah Nobles, Jasmine Padilla, Mark Burroughs, Shoshona Le, Katie Dillon, Peter Cook, Clinton R. Paden, Dhwani Batra, Krista Queen, Kristen Knipe, Dakota Howard, Yvette Unoarumhi, Darlene Wagner, Matthew Schmerer, Ben L. Rambo-Martin, Kristine Lacek, Sam Shepard, Alison Laufer Halpin, Dave Wentworth, Vivien Dugan, Suxiang Tong, Justin Lee |
| EPI_ISL_1798409 | Laboratory Corporation of America | Centers for Disease Control and Prevention Division of Viral Diseases, Pathogen Discovery | Dakota Howard, Dhwani Batra, Peter W. Cook, Kara Moser, Adrian Paskey, Jason Caravas, Benjamin Rambo-Martin, Shatavia Morrison, Christopher Gulvick, Scott Sammons, Yvette Unoarumhi, Darlene Wagner, Matthew Schmerer, Minoo Agarwal, Eyad Almasri, Debbie Boles, Ayla Burns, Nuthawin Charoensri, Oren Cohen, Susan Countryman, Mary Ann Cristobal, Bobbi Croy, Suzanne Dale, Hrushikesh Deshmukh, Amanda Douglas, Vincent Drouillon, Marcia Eisenberg, Howard Engler, Rama Ghatti, Prashant Gupta, Susan Hicks, Jake Humphrey, Lax Iyer, Manoj Jain, Mohan Kolli, Brian Krueger, Tim Kuphal, Stanley Letovsky, Michael Levandoski, Craig Lukasik, Jonathan Meltzer, Brian Norvell, Mindy Nye, Scott Parker, Christos Petropoulos, John Pruitt, Steven Ragan, Scott Ryan, Mike Sapeta, Jana Schroth, Suresh Babu Selvaraju, Goran Stevovic, Amanda Suchanek, Andrea Throop, Lyndon Tilson, Thomas Urban, Joe Voshell, Kimberly Wagner, Jonathan Williams, Mary Williamson, Qian Zeng, Tricia Zwiefelhofer, Clinton R. Paden, Duncan MacCannell |
| EPI_ISL_1798410 | MT Public Health Laboratory | Centers for Disease Control and Prevention Division of Viral Diseases, Pathogen Discovery | Mili Sheth, Sarah Nobles, Jasmine Padilla, Mark Burroughs, Shoshona Le, Katie Dillon, Peter Cook, Clinton R. Paden, Dhwani Batra, Krista Queen, Kristen Knipe, Dakota Howard, Yvette Unoarumhi, Darlene Wagner, Matthew Schmerer, Ben L. Rambo-Martin, Kristine Lacek, Sam Shepard, Alison Laufer Halpin, Dave Wentworth, Vivien Dugan, Suxiang Tong, Justin Lee |
| EPI_ISL_1798411 | Laboratory Corporation of America | Centers for Disease Control and Prevention Division of Viral Diseases, Pathogen Discovery | Dakota Howard, Dhwani Batra, Peter W. Cook, Kara Moser, Adrian Paskey, Jason Caravas, Benjamin Rambo-Martin, Shatavia Morrison, Christopher Gulvick, Scott Sammons, Yvette Unoarumhi, Darlene Wagner, Matthew Schmerer, Minoo Agarwal, Eyad Almasri, Debbie Boles, Ayla Burns, Nuthawin Charoensri, Oren Cohen, Susan Countryman, Mary Ann Cristobal, Bobbi Croy, Suzanne Dale, Hrushikesh Deshmukh, Amanda Douglas, Vincent Drouillon, Marcia Eisenberg, Howard Engler, Rama Ghatti, Prashant Gupta, Susan Hicks, Jake Humphrey, Lax Iyer, Manoj Jain, Mohan Kolli, Brian Krueger, Tim Kuphal, Stanley Letovsky, Michael Levandoski, Craig Lukasik, Jonathan Meltzer, Brian Norvell, Mindy Nye, Scott Parker, Christos Petropoulos, John Pruitt, Steven Ragan, Scott Ryan, Mike Sapeta, Jana Schroth, Suresh Babu Selvaraju, Goran Stevovic, Amanda Suchanek, Andrea Throop, Lyndon Tilson, Thomas Urban, Joe Voshell, Kimberly Wagner, Jonathan Williams, Mary Williamson, Qian Zeng, Tricia Zwiefelhofer, Clinton R. Paden, Duncan MacCannell |
| EPI_ISL_1798412 | MD DOH Laboratories Administration | Centers for Disease Control and Prevention Division of Viral Diseases, Pathogen Discovery | Mili Sheth, Sarah Nobles, Jasmine Padilla, Mark Burroughs, Shoshona Le, Katie Dillon, Peter Cook, Clinton R. Paden, Dhwani Batra, Krista Queen, Kristen Knipe, Dakota Howard, Yvette Unoarumhi, Darlene Wagner, Matthew Schmerer, Ben L. Rambo-Martin, Kristine Lacek, Sam Shepard, Alison Laufer Halpin, Dave Wentworth, Vivien Dugan, Suxiang Tong, Justin Lee |
| EPI_ISL_1798413, EPI_ISL_1798414 | Laboratory Corporation of America | Centers for Disease Control and Prevention Division of Viral Diseases, Pathogen Discovery | Dakota Howard, Dhwani Batra, Peter W. Cook, Kara Moser, Adrian Paskey, Jason Caravas, Benjamin Rambo-Martin, Shatavia Morrison, Christopher Gulvick, Scott Sammons, Yvette Unoarumhi, Darlene Wagner, Matthew Schmerer, Minoo Agarwal, Eyad Almasri, Debbie Boles, Ayla Burns, Nuthawin Charoensri, Oren Cohen, Susan Countryman, Mary Ann Cristobal, Bobbi Croy, Suzanne Dale, Hrushikesh Deshmukh, Amanda Douglas, Vincent Drouillon, Marcia Eisenberg, Howard Engler, Rama Ghatti, Prashant Gupta, Susan Hicks, Jake Humphrey, Lax Iyer, Manoj Jain, Mohan Kolli, Brian Krueger, Tim Kuphal, Stanley Letovsky, Michael Levandoski, Craig Lukasik, Jonathan Meltzer, Brian Norvell, Mindy Nye, Scott Parker, Christos Petropoulos, John Pruitt, Steven Ragan, Scott Ryan, Mike Sapeta, Jana Schroth, Suresh Babu Selvaraju, Goran Stevovic, Amanda |

| | | | |
|---|---|---|---|
| | | | Suchanek, Andrea Throop, Lyndon Tilson, Thomas Urban, Joe Voshell, Kimberly Wagner, Jonathan Williams, Mary Williamson, Qian Zeng, Tricia Zwiefelhofer, Clinton R. Paden, Duncan MacCannell |
| EPI_ISL_1798415 | MN PHL Division, Minnesota Department of Health | Centers for Disease Control and Prevention Division of Viral Diseases, Pathogen Discovery | Mili Sheth, Sarah Nobles, Jasmine Padilla, Mark Burroughs, Shoshona Le, Katie Dillon, Peter Cook, Clinton R. Paden, Dhwani Batra, Krista Queen, Kristen Knipe, Dakota Howard, Yvette Unoarumhi, Darlene Wagner, Matthew Schmerer, Ben L. Rambo-Martin, Kristine Lacek, Sam Shepard, Alison Laufer Halpin, Dave Wentworth, Vivien Dugan, Suxiang Tong, Justin Lee |
| EPI_ISL_1798417 | CT-Dr. Katherine A. Kelley State Public Health Lab | Centers for Disease Control and Prevention Division of Viral Diseases, Pathogen Discovery | Mili Sheth, Sarah Nobles, Jasmine Padilla, Mark Burroughs, Shoshona Le, Katie Dillon, Peter Cook, Clinton R. Paden, Dhwani Batra, Krista Queen, Kristen Knipe, Dakota Howard, Yvette Unoarumhi, Darlene Wagner, Matthew Schmerer, Ben L. Rambo-Martin, Kristine Lacek, Sam Shepard, Alison Laufer Halpin, Dave Wentworth, Vivien Dugan, Suxiang Tong, Justin Lee |
| EPI_ISL_1798418, EPI_ISL_1798420 | PR Public Health Lab | Centers for Disease Control and Prevention Division of Viral Diseases, Pathogen Discovery | Mili Sheth, Sarah Nobles, Jasmine Padilla, Mark Burroughs, Shoshona Le, Katie Dillon, Peter Cook, Clinton R. Paden, Dhwani Batra, Krista Queen, Kristen Knipe, Dakota Howard, Yvette Unoarumhi, Darlene Wagner, Matthew Schmerer, Ben L. Rambo-Martin, Kristine Lacek, Sam Shepard, Alison Laufer Halpin, Dave Wentworth, Vivien Dugan, Suxiang Tong, Justin Lee |
| EPI_ISL_1798421 | Laboratory Corporation of America | Centers for Disease Control and Prevention Division of Viral Diseases, Pathogen Discovery | Dakota Howard, Dhwani Batra, Peter W. Cook, Kara Moser, Adrian Paskey, Jason Caravas, Benjamin Rambo-Martin, Shatavia Morrison, Christopher Gulvick, Scott Sammons, Yvette Unoarumhi, Darlene Wagner, Matthew Schmerer, Minoo Agarwal, Eyad Almasri, Debbie Boles, Ayla Burns, Nuthawin Charoensri, Oren Cohen, Susan Countryman, Mary Ann Cristobal, Bobbi Croy, Suzanne Dale, Hrushikesh Deshmukh, Amanda Douglas, Vincent Drouillon, Marcia Eisenberg, Howard Engler, Rama Ghatti, Prashant Gupta, Susan Hicks, Jake Humphrey, Lax Iyer, Manoj Jain, Mohan Kolli, Brian Krueger, Tim Kuphal, Stanley Letovsky, Michael Levandoski, Craig Lukasik, Jonathan Meltzer, Brian Norvell, Mindy Nye, Scott Parker, Christos Petropoulos, John Pruitt, Steven Ragan, Scott Ryan, Mike Sapeta, Jana Schroth, Suresh Babu Selvaraju, Goran Stevovic, Amanda Suchanek, Andrea Throop, Lyndon Tilson, Thomas Urban, Joe Voshell, Kimberly Wagner, Jonathan Williams, Mary Williamson, Qian Zeng, Tricia Zwiefelhofer, Clinton R. Paden, Duncan MacCannell |
| EPI_ISL_1798422 | PR Public Health Lab | Centers for Disease Control and Prevention Division of Viral Diseases, Pathogen Discovery | Mili Sheth, Sarah Nobles, Jasmine Padilla, Mark Burroughs, Shoshona Le, Katie Dillon, Peter Cook, Clinton R. Paden, Dhwani Batra, Krista Queen, Kristen Knipe, Dakota Howard, Yvette Unoarumhi, Darlene Wagner, Matthew Schmerer, Ben L. Rambo-Martin, Kristine Lacek, Sam Shepard, Alison Laufer Halpin, Dave Wentworth, Vivien Dugan, Suxiang Tong, Justin Lee |
| EPI_ISL_1798423 | Laboratory Corporation of America | Centers for Disease Control and Prevention Division of Viral Diseases, Pathogen Discovery | Dakota Howard, Dhwani Batra, Peter W. Cook, Kara Moser, Adrian Paskey, Jason Caravas, Benjamin Rambo-Martin, Shatavia Morrison, Christopher Gulvick, Scott Sammons, Yvette Unoarumhi, Darlene Wagner, Matthew Schmerer, Minoo Agarwal, Eyad Almasri, Debbie Boles, Ayla Burns, Nuthawin Charoensri, Oren Cohen, Susan Countryman, Mary Ann Cristobal, Bobbi Croy, Suzanne Dale, Hrushikesh Deshmukh, Amanda Douglas, Vincent Drouillon, Marcia Eisenberg, Howard Engler, Rama Ghatti, Prashant Gupta, Susan Hicks, Jake Humphrey, Lax Iyer, Manoj Jain, Mohan Kolli, Brian Krueger, Tim Kuphal, Stanley Letovsky, Michael Levandoski, Craig Lukasik, Jonathan Meltzer, Brian Norvell, Mindy Nye, Scott Parker, Christos Petropoulos, John Pruitt, Steven Ragan, Scott Ryan, Mike Sapeta, Jana Schroth, Suresh Babu Selvaraju, Goran Stevovic, Amanda Suchanek, Andrea Throop, Lyndon Tilson, Thomas Urban, Joe Voshell, Kimberly Wagner, Jonathan Williams, Mary Williamson, Qian Zeng, Tricia Zwiefelhofer, Clinton R. Paden, Duncan MacCannell |
| EPI_ISL_1798424 | MD DOH Laboratories Administration | Centers for Disease Control and Prevention Division of Viral Diseases, Pathogen Discovery | Mili Sheth, Sarah Nobles, Jasmine Padilla, Mark Burroughs, Shoshona Le, Katie Dillon, Peter Cook, Clinton R. Paden, Dhwani Batra, Krista Queen, Kristen Knipe, Dakota Howard, Yvette Unoarumhi, Darlene Wagner, Matthew Schmerer, Ben L. Rambo-Martin, Kristine Lacek, Sam Shepard, Alison Laufer Halpin, Dave Wentworth, Vivien Dugan, Suxiang Tong, Justin Lee |
| EPI_ISL_1798426 | MT Public Health Laboratory | Centers for Disease Control and Prevention Division of Viral Diseases, Pathogen Discovery | Mili Sheth, Sarah Nobles, Jasmine Padilla, Mark Burroughs, Shoshona Le, Katie Dillon, Peter Cook, Clinton R. Paden, Dhwani Batra, Krista Queen, Kristen Knipe, Dakota Howard, Yvette Unoarumhi, Darlene Wagner, Matthew Schmerer, Ben L. Rambo-Martin, Kristine Lacek, Sam Shepard, Alison Laufer Halpin, Dave Wentworth, Vivien Dugan, Suxiang Tong, Justin Lee |
| EPI_ISL_1798427 | Laboratory Corporation of America | Centers for Disease Control and Prevention Division of Viral Diseases, Pathogen Discovery | Dakota Howard, Dhwani Batra, Peter W. Cook, Kara Moser, Adrian Paskey, Jason Caravas, Benjamin Rambo-Martin, Shatavia Morrison, Christopher Gulvick, Scott Sammons, Yvette Unoarumhi, Darlene Wagner, Matthew Schmerer, Minoo Agarwal, Eyad Almasri, Debbie Boles, Ayla Burns, Nuthawin Charoensri, Oren Cohen, Susan Countryman, Mary Ann Cristobal, Bobbi Croy, Suzanne Dale, Hrushikesh Deshmukh, Amanda Douglas, Vincent Drouillon, Marcia Eisenberg, Howard Engler, Rama Ghatti, Prashant Gupta, Susan Hicks, Jake Humphrey, Lax Iyer, Manoj Jain, Mohan Kolli, Brian Krueger, Tim Kuphal, Stanley Letovsky, Michael Levandoski, Craig Lukasik, Jonathan Meltzer, Brian Norvell, Mindy Nye, Scott Parker, Christos Petropoulos, John Pruitt, Steven Ragan, Scott Ryan, Mike Sapeta, Jana Schroth, Suresh Babu Selvaraju, Goran Stevovic, Amanda Suchanek, Andrea Throop, Lyndon Tilson, Thomas Urban, Joe Voshell, Kimberly Wagner, Jonathan Williams, Mary Williamson, Qian Zeng, Tricia Zwiefelhofer, Clinton R. Paden, Duncan MacCannell |
| EPI_ISL_1798428 | MD DOH Laboratories Administration | Centers for Disease Control and Prevention Division of Viral Diseases, Pathogen Discovery | Mili Sheth, Sarah Nobles, Jasmine Padilla, Mark Burroughs, Shoshona Le, Katie Dillon, Peter Cook, Clinton R. Paden, Dhwani Batra, Krista Queen, Kristen Knipe, Dakota Howard, Yvette Unoarumhi, Darlene Wagner, Matthew Schmerer, Ben L. Rambo-Martin, Kristine Lacek, Sam Shepard, Alison Laufer Halpin, Dave Wentworth, Vivien Dugan, Suxiang Tong, Justin Lee |
| EPI_ISL_1798429 | Laboratory Corporation of America | Centers for Disease Control and Prevention Division of Viral Diseases, Pathogen Discovery | Dakota Howard, Dhwani Batra, Peter W. Cook, Kara Moser, Adrian Paskey, Jason Caravas, Benjamin Rambo-Martin, Shatavia Morrison, Christopher Gulvick, Scott Sammons, Yvette Unoarumhi, Darlene Wagner, Matthew Schmerer, Minoo Agarwal, Eyad Almasri, Debbie Boles, Ayla Burns, Nuthawin Charoensri, Oren Cohen, Susan Countryman, Mary Ann Cristobal, Bobbi Croy, Suzanne Dale, Hrushikesh Deshmukh, Amanda Douglas, Vincent Drouillon, Marcia Eisenberg, Howard Engler, Rama Ghatti, Prashant Gupta, Susan Hicks, Jake Humphrey, Lax Iyer, Manoj Jain, Mohan Kolli, Brian Krueger, Tim Kuphal, Stanley Letovsky, Michael Levandoski, Craig Lukasik, Jonathan Meltzer, Brian Norvell, Mindy Nye, Scott Parker, Christos Petropoulos, John Pruitt, Steven Ragan, Scott Ryan, Mike Sapeta, Jana Schroth, Suresh Babu Selvaraju, Goran Stevovic, Amanda Suchanek, Andrea Throop, Lyndon Tilson, Thomas Urban, Joe Voshell, Kimberly Wagner, Jonathan Williams, Mary Williamson, Qian Zeng, Tricia Zwiefelhofer, Clinton R. Paden, Duncan MacCannell |
| EPI_ISL_1798430 | MD DOH Laboratories Administration | Centers for Disease Control and Prevention Division of Viral Diseases, Pathogen Discovery | Mili Sheth, Sarah Nobles, Jasmine Padilla, Mark Burroughs, Shoshona Le, Katie Dillon, Peter Cook, Clinton R. Paden, Dhwani Batra, Krista Queen, Kristen Knipe, Dakota Howard, Yvette Unoarumhi, Darlene Wagner, Matthew Schmerer, Ben L. Rambo-Martin, Kristine Lacek, Sam Shepard, Alison Laufer Halpin, Dave Wentworth, Vivien Dugan, Suxiang Tong, Justin Lee |
| EPI_ISL_1798431, EPI_ISL_1798432 | Laboratory Corporation of America | Centers for Disease Control and Prevention Division of Viral Diseases, Pathogen Discovery | Dakota Howard, Dhwani Batra, Peter W. Cook, Kara Moser, Adrian Paskey, Jason Caravas, Benjamin Rambo-Martin, Shatavia Morrison, Christopher Gulvick, Scott Sammons, Yvette Unoarumhi, Darlene Wagner, Matthew Schmerer, Minoo Agarwal, Eyad Almasri, Debbie Boles, Ayla Burns, Nuthawin Charoensri, Oren Cohen, Susan Countryman, Mary Ann Cristobal, Bobbi Croy, Suzanne Dale, Hrushikesh Deshmukh, Amanda Douglas, Vincent Drouillon, Marcia Eisenberg, Howard Engler, Rama Ghatti, Prashant Gupta, Susan Hicks, Jake Humphrey, Lax Iyer, Manoj Jain, Mohan Kolli, Brian Krueger, Tim Kuphal, Stanley Letovsky, Michael Levandoski, Craig Lukasik, Jonathan Meltzer, Brian Norvell, Mindy Nye, Scott Parker, Christos Petropoulos, John Pruitt, Steven Ragan, Scott Ryan, Mike Sapeta, Jana Schroth, Suresh Babu Selvaraju, Goran Stevovic, Amanda Suchanek, Andrea Throop, Lyndon Tilson, Thomas Urban, Joe Voshell, Kimberly Wagner, Jonathan Williams, Mary Williamson, Qian Zeng, Tricia Zwiefelhofer, Clinton R. Paden, Duncan MacCannell |
| EPI_ISL_1798433 | PR Public Health Lab | Centers for Disease Control and Prevention Division of Viral Diseases, Pathogen Discovery | Mili Sheth, Sarah Nobles, Jasmine Padilla, Mark Burroughs, Shoshona Le, Katie Dillon, Peter Cook, Clinton R. Paden, Dhwani Batra, Krista Queen, Kristen Knipe, Dakota Howard, Yvette Unoarumhi, Darlene Wagner, Matthew Schmerer, Ben L. Rambo-Martin, Kristine Lacek, Sam Shepard, Alison Laufer Halpin, Dave Wentworth, Vivien Dugan, Suxiang Tong, Justin Lee |
| EPI_ISL_1798434 | NJ Public Health and Environmental Laboratories | Centers for Disease Control and Prevention Division of Viral Diseases, Pathogen Discovery | Mili Sheth, Sarah Nobles, Jasmine Padilla, Mark Burroughs, Shoshona Le, Katie Dillon, Peter Cook, Clinton R. Paden, Dhwani Batra, Krista Queen, Kristen Knipe, Dakota Howard, Yvette Unoarumhi, Darlene Wagner, Matthew Schmerer, Ben L. Rambo-Martin, Kristine Lacek, Sam Shepard, Alison Laufer Halpin, Dave Wentworth, Vivien Dugan, Suxiang Tong, Justin Lee |
| EPI_ISL_1798435, EPI_ISL_1798436 | Laboratory Corporation of America | Centers for Disease Control and Prevention Division of Viral Diseases, Pathogen Discovery | Dakota Howard, Dhwani Batra, Peter W. Cook, Kara Moser, Adrian Paskey, Jason Caravas, Benjamin Rambo-Martin, Shatavia Morrison, Christopher Gulvick, Scott Sammons, Yvette Unoarumhi, Darlene Wagner, Matthew Schmerer, Minoo Agarwal, Eyad Almasri, Debbie Boles, Ayla Burns, Nuthawin Charoensri, Oren Cohen, Susan Countryman, Mary Ann Cristobal, Bobbi Croy, Suzanne Dale, Hrushikesh Deshmukh, Amanda Douglas, Vincent Drouillon, Marcia Eisenberg, Howard Engler, Rama Ghatti, Prashant Gupta, Susan Hicks, Jake Humphrey, Lax Iyer, Manoj Jain, Mohan Kolli, Brian Krueger, Tim Kuphal, Stanley Letovsky, Michael Levandoski, Craig Lukasik, Jonathan Meltzer, Brian Norvell, Mindy Nye, Scott Parker, Christos Petropoulos, John Pruitt, Steven Ragan, Scott Ryan, Mike Sapeta, Jana Schroth, Suresh Babu Selvaraju, Goran Stevovic, Amanda Suchanek, Andrea Throop, Lyndon Tilson, Thomas Urban, Joe Voshell, Kimberly Wagner, Jonathan Williams, Mary Williamson, Qian Zeng, Tricia Zwiefelhofer, Clinton R. Paden, Duncan MacCannell |
| EPI_ISL_1798437 | MT Public Health Laboratory | Centers for Disease Control and Prevention Division of Viral Diseases, Pathogen Discovery | Mili Sheth, Sarah Nobles, Jasmine Padilla, Mark Burroughs, Shoshona Le, Katie Dillon, Peter Cook, Clinton R. Paden, Dhwani Batra, Krista Queen, Kristen Knipe, Dakota Howard, Yvette Unoarumhi, Darlene Wagner, Matthew Schmerer, Ben L. Rambo-Martin, Kristine Lacek, Sam Shepard, Alison Laufer Halpin, Dave Wentworth, Vivien Dugan, Suxiang Tong, Justin Lee |
| EPI_ISL_1798439 | MD DOH Laboratories Administration | Centers for Disease Control and Prevention Division of Viral Diseases, Pathogen Discovery | Mili Sheth, Sarah Nobles, Jasmine Padilla, Mark Burroughs, Shoshona Le, Katie Dillon, Peter Cook, Clinton R. Paden, Dhwani Batra, Krista Queen, Kristen Knipe, Dakota Howard, Yvette Unoarumhi, Darlene Wagner, Matthew Schmerer, Ben L. Rambo-Martin, Kristine Lacek, Sam Shepard, Alison Laufer Halpin, Dave Wentworth, Vivien Dugan, Suxiang Tong, Justin Lee |
| EPI_ISL_1798440, EPI_ISL_1798441 | Laboratory Corporation of America | Centers for Disease Control and Prevention Division of Viral Diseases, Pathogen Discovery | Dakota Howard, Dhwani Batra, Peter W. Cook, Kara Moser, Adrian Paskey, Jason Caravas, Benjamin Rambo-Martin, Shatavia Morrison, Christopher Gulvick, Scott Sammons, Yvette Unoarumhi, Darlene Wagner, Matthew Schmerer, Minoo Agarwal, Eyad Almasri, Debbie Boles, Ayla Burns, Nuthawin Charoensri, Oren Cohen, Susan Countryman, Mary Ann Cristobal, Bobbi Croy, Suzanne Dale, Hrushikesh Deshmukh, Amanda Douglas, Vincent Drouillon, Marcia Eisenberg, Howard Engler, Rama Ghatti, Prashant Gupta, Susan Hicks, Jake Humphrey, Lax Iyer, Manoj Jain, Mohan Kolli, Brian Krueger, Tim Kuphal, Stanley Letovsky, Michael Levandoski, Craig Lukasik, Jonathan Meltzer, Brian Norvell, Mindy Nye, Scott Parker, Christos Petropoulos, John Pruitt, Steven Ragan, Scott Ryan, Mike Sapeta, Jana Schroth, Suresh Babu Selvaraju, Goran Stevovic, Amanda Suchanek, Andrea Throop, Lyndon Tilson, Thomas Urban, Joe Voshell, Kimberly Wagner, Jonathan Williams, Mary Williamson, Qian Zeng, Tricia Zwiefelhofer, Clinton R. Paden, Duncan MacCannell |
| EPI_ISL_1798442 | MD DOH Laboratories Administration | Centers for Disease Control and Prevention Division of Viral Diseases, Pathogen Discovery | Mili Sheth, Sarah Nobles, Jasmine Padilla, Mark Burroughs, Shoshona Le, Katie Dillon, Peter Cook, Clinton R. Paden, Dhwani Batra, Krista Queen, Kristen Knipe, Dakota Howard, Yvette Unoarumhi, Darlene Wagner, Matthew Schmerer, Ben L. Rambo-Martin, Kristine Lacek, Sam Shepard, Alison Laufer Halpin, Dave Wentworth, Vivien Dugan, Suxiang Tong, Justin Lee |
| EPI_ISL_1798443 | Laboratory Corporation of America | Centers for Disease Control and Prevention Division of Viral Diseases, Pathogen Discovery | Dakota Howard, Dhwani Batra, Peter W. Cook, Kara Moser, Adrian Paskey, Jason Caravas, Benjamin Rambo-Martin, Shatavia Morrison, Christopher Gulvick, Scott Sammons, Yvette Unoarumhi, Darlene Wagner, Matthew Schmerer, Minoo Agarwal, Eyad Almasri, Debbie Boles, Ayla Burns, Nuthawin Charoensri, Oren Cohen, Susan Countryman, Mary Ann Cristobal, Bobbi Croy, Suzanne Dale, Hrushikesh Deshmukh, Amanda Douglas, Vincent Drouillon, Marcia Eisenberg, Howard Engler, Rama Ghatti, Prashant Gupta, Susan Hicks, Jake Humphrey, Lax Iyer, Manoj Jain, Mohan Kolli, Brian Krueger, Tim Kuphal, Stanley Letovsky, Michael Levandoski, Craig Lukasik, Jonathan Meltzer, Brian Norvell, Mindy Nye, Scott Parker, Christos Petropoulos, John Pruitt, Steven Ragan, Scott Ryan, Mike Sapeta, Jana Schroth, Suresh Babu Selvaraju, Goran Stevovic, Amanda Suchanek, Andrea Throop, Lyndon Tilson, Thomas Urban, Joe Voshell, Kimberly Wagner, Jonathan Williams, Mary Williamson, Qian Zeng, Tricia Zwiefelhofer, Clinton R. Paden, Duncan MacCannell |
| EPI_ISL_1798444 | MD DOH Laboratories Administration | Centers for Disease Control and Prevention Division of Viral Diseases, Pathogen Discovery | Mili Sheth, Sarah Nobles, Jasmine Padilla, Mark Burroughs, Shoshona Le, Katie Dillon, Peter Cook, Clinton R. Paden, Dhwani Batra, Krista Queen, Kristen Knipe, Dakota Howard, Yvette Unoarumhi, Darlene Wagner, Matthew Schmerer, Ben L. Rambo-Martin, Kristine Lacek, Sam Shepard, Alison Laufer Halpin, Dave Wentworth, Vivien Dugan, Suxiang Tong, Justin Lee |
| EPI_ISL_1798446 | NJ Public Health and Environmental Laboratories | Centers for Disease Control and Prevention Division of Viral Diseases, Pathogen Discovery | Mili Sheth, Sarah Nobles, Jasmine Padilla, Mark Burroughs, Shoshona Le, Katie Dillon, Peter Cook, Clinton R. Paden, Dhwani Batra, Krista Queen, Kristen Knipe, Dakota Howard, Yvette Unoarumhi, Darlene Wagner, Matthew Schmerer, Ben L. Rambo-Martin, Kristine Lacek, Sam Shepard, Alison Laufer Halpin, Dave Wentworth, Vivien Dugan, Suxiang Tong, Justin Lee |
| EPI_ISL_1798447 | Laboratory Corporation of America | Centers for Disease Control and Prevention Division of Viral Diseases, Pathogen Discovery | Dakota Howard, Dhwani Batra, Peter W. Cook, Kara Moser, Adrian Paskey, Jason Caravas, Benjamin Rambo-Martin, Shatavia Morrison, Christopher Gulvick, Scott Sammons, Yvette Unoarumhi, Darlene Wagner, Matthew Schmerer, Minoo Agarwal, Eyad Almasri, Debbie Boles, Ayla Burns, Nuthawin Charoensri, Oren Cohen, Susan Countryman, Mary Ann Cristobal, Bobbi Croy, Suzanne Dale, Hrushikesh Deshmukh, Amanda Douglas, Vincent Drouillon, Marcia Eisenberg, Howard Engler, Rama Ghatti, Prashant Gupta, Susan Hicks, Jake Humphrey, Lax Iyer, Manoj Jain, Mohan Kolli, Brian Krueger, Tim Kuphal, Stanley Letovsky, Michael Levandoski, Craig Lukasik, Jonathan Meltzer, Brian Norvell, Mindy Nye, Scott Parker, Christos Petropoulos, John Pruitt, Steven Ragan, Scott Ryan, Mike Sapeta, Jana Schroth, Suresh Babu Selvaraju, Goran Stevovic, Amanda Suchanek, Andrea Throop, Lyndon Tilson, Thomas Urban, Joe Voshell, Kimberly Wagner, Jonathan Williams, Mary Williamson, Qian Zeng, Tricia Zwiefelhofer, Clinton R. Paden, Duncan MacCannell |
| EPI_ISL_1798448 | MT Public Health Laboratory | Centers for Disease Control and Prevention Division of Viral Diseases, Pathogen Discovery | Mili Sheth, Sarah Nobles, Jasmine Padilla, Mark Burroughs, Shoshona Le, Katie Dillon, Peter Cook, Clinton R. Paden, Dhwani Batra, Krista Queen, Kristen Knipe, Dakota Howard, Yvette Unoarumhi, Darlene Wagner, Matthew Schmerer, Ben L. Rambo-Martin, Kristine Lacek, Sam Shepard, Alison Laufer Halpin, Dave Wentworth, Vivien Dugan, Suxiang Tong, Justin Lee |
| EPI_ISL_1798449 | Laboratory Corporation of America | Centers for Disease Control and Prevention Division of Viral Diseases, Pathogen Discovery | Dakota Howard, Dhwani Batra, Peter W. Cook, Kara Moser, Adrian Paskey, Jason Caravas, Benjamin Rambo-Martin, Shatavia Morrison, Christopher Gulvick, Scott Sammons, Yvette Unoarumhi, Darlene Wagner, Matthew Schmerer, Minoo Agarwal, Eyad Almasri, Debbie Boles, Ayla Burns, Nuthawin Charoensri, Oren Cohen, Susan Countryman, Mary Ann Cristobal, Bobbi Croy, Suzanne Dale, Hrushikesh Deshmukh, Amanda Douglas, Vincent Drouillon, Marcia Eisenberg, Howard Engler, Rama Ghatti, Prashant Gupta, Susan Hicks, Jake Humphrey, Lax Iyer, Manoj Jain, Mohan Kolli, Brian Krueger, Tim Kuphal, Stanley Letovsky, Michael Levandoski, Craig Lukasik, Jonathan Meltzer, Brian Norvell, Mindy Nye, Scott Parker, Christos Petropoulos, John Pruitt, Steven Ragan, Scott Ryan, Mike Sapeta, Jana Schroth, Suresh Babu Selvaraju, Goran Stevovic, Amanda Suchanek, Andrea Throop, Lyndon Tilson, Thomas Urban, Joe Voshell, Kimberly Wagner, Jonathan Williams, Mary Williamson, Qian Zeng, Tricia Zwiefelhofer, Clinton R. Paden, Duncan MacCannell |
| EPI_ISL_1798450 | MD DOH Laboratories Administration | Centers for Disease Control and Prevention Division of Viral Diseases, Pathogen Discovery | Mili Sheth, Sarah Nobles, Jasmine Padilla, Mark Burroughs, Shoshona Le, Katie Dillon, Peter Cook, Clinton R. Paden, Dhwani Batra, Krista Queen, Kristen Knipe, Dakota Howard, Yvette Unoarumhi, Darlene Wagner, Matthew Schmerer, Ben L. Rambo-Martin, Kristine Lacek, Sam Shepard, Alison Laufer Halpin, Dave Wentworth, Vivien Dugan, Suxiang Tong, Justin Lee |
| EPI_ISL_1798451 | Laboratory Corporation of America | Centers for Disease Control and Prevention Division of Viral Diseases, Pathogen Discovery | Dakota Howard, Dhwani Batra, Peter W. Cook, Kara Moser, Adrian Paskey, Jason Caravas, Benjamin Rambo-Martin, Shatavia Morrison, Christopher Gulvick, Scott Sammons, Yvette Unoarumhi, Darlene Wagner, Matthew Schmerer, Minoo Agarwal, Eyad Almasri, Debbie Boles, Ayla Burns, Nuthawin Charoensri, Oren Cohen, Susan Countryman, Mary Ann Cristobal, Bobbi Croy, Suzanne Dale, Hrushikesh Deshmukh, Amanda Douglas, Vincent Drouillon, Marcia Eisenberg, Howard Engler, Rama Ghatti, Prashant Gupta, Susan Hicks, Jake Humphrey, Lax Iyer, Manoj Jain, Mohan Kolli, Brian Krueger, Tim Kuphal, Stanley Letovsky, Michael Levandoski, Craig Lukasik, Jonathan Meltzer, Brian Norvell, Mindy Nye, Scott Parker, Christos Petropoulos, John Pruitt, Steven Ragan, Scott Ryan, Mike Sapeta, Jana Schroth, Suresh Babu Selvaraju, Goran Stevovic, Amanda Suchanek, Andrea Throop, Lyndon Tilson, Thomas Urban, Joe Voshell, Kimberly Wagner, Jonathan Williams, Mary Williamson, Qian Zeng, Tricia Zwiefelhofer, Clinton R. Paden, Duncan MacCannell |

| | | | |
|---|---|---|---|
| EPI_ISL_1798452, EPI_ISL_1798453 | PR Public Health Lab | Centers for Disease Control and Prevention Division of Viral Diseases, Pathogen Discovery | Mili Sheth, Sarah Nobles, Jasmine Padilla, Mark Burroughs, Shoshona Le, Katie Dillon, Peter Cook, Clinton R. Paden, Dhwani Batra, Krista Queen, Kristen Knipe, Dakota Howard, Yvette Unoarumhi, Darlene Wagner, Matthew Schmerer, Ben L. Rambo-Martin, Kristine Lacek, Sam Shepard, Alison Laufer Halpin, Dave Wentworth, Vivien Dugan, Suxiang Tong, Justin Lee |
| EPI_ISL_1798454, EPI_ISL_1798455 | Laboratory Corporation of America | Centers for Disease Control and Prevention Division of Viral Diseases, Pathogen Discovery | Dakota Howard, Dhwani Batra, Peter W. Cook, Kara Moser, Adrian Paskey, Jason Caravas, Benjamin Rambo-Martin, Shatavia Morrison, Christopher Gulvick, Scott Sammons, Yvette Unoarumhi, Darlene Wagner, Matthew Schmerer, Minoo Agarwal, Eyad Almasri, Debbie Boles, Ayla Burns, Nuthawin Charoensri, Oren Cohen, Susan Countryman, Mary Ann Cristobal, Bobbi Croy, Suzanne Dale, Hrushikesh Deshmukh, Amanda Douglas, Vincent Drouillon, Marcia Eisenberg, Howard Engler, Rama Ghatti, Prashant Gupta, Susan Hicks, Jake Humphrey, Lax Iyer, Manoj Jain, Mohan Kolli, Brian Krueger, Tim Kuphal, Stanley Letovsky, Michael Levandoski, Craig Lukasik, Jonathan Meltzer, Brian Norvell, Mindy Nye, Scott Parker, Christos Petropoulos, John Pruitt, Steven Ragan, Scott Ryan, Mike Sapeta, Jana Schroth, Suresh Babu Selvaraju, Goran Stevovic, Amanda Suchanek, Andrea Throop, Lyndon Tilson, Thomas Urban, Joe Voshell, Kimberly Wagner, Jonathan Williams, Mary Williamson, Qian Zeng, Tricia Zwiefelhofer, Clinton R. Paden, Duncan MacCannell |
| EPI_ISL_1798456 | PR Public Health Lab | Centers for Disease Control and Prevention Division of Viral Diseases, Pathogen Discovery | Mili Sheth, Sarah Nobles, Jasmine Padilla, Mark Burroughs, Shoshona Le, Katie Dillon, Peter Cook, Clinton R. Paden, Dhwani Batra, Krista Queen, Kristen Knipe, Dakota Howard, Yvette Unoarumhi, Darlene Wagner, Matthew Schmerer, Ben L. Rambo-Martin, Kristine Lacek, Sam Shepard, Alison Laufer Halpin, Dave Wentworth, Vivien Dugan, Suxiang Tong, Justin Lee |
| EPI_ISL_1798458 | MD DOH Laboratories Administration | Centers for Disease Control and Prevention Division of Viral Diseases, Pathogen Discovery | Mili Sheth, Sarah Nobles, Jasmine Padilla, Mark Burroughs, Shoshona Le, Katie Dillon, Peter Cook, Clinton R. Paden, Dhwani Batra, Krista Queen, Kristen Knipe, Dakota Howard, Yvette Unoarumhi, Darlene Wagner, Matthew Schmerer, Ben L. Rambo-Martin, Kristine Lacek, Sam Shepard, Alison Laufer Halpin, Dave Wentworth, Vivien Dugan, Suxiang Tong, Justin Lee |
| EPI_ISL_1798459 | Laboratory Corporation of America | Centers for Disease Control and Prevention Division of Viral Diseases, Pathogen Discovery | Dakota Howard, Dhwani Batra, Peter W. Cook, Kara Moser, Adrian Paskey, Jason Caravas, Benjamin Rambo-Martin, Shatavia Morrison, Christopher Gulvick, Scott Sammons, Yvette Unoarumhi, Darlene Wagner, Matthew Schmerer, Minoo Agarwal, Eyad Almasri, Debbie Boles, Ayla Burns, Nuthawin Charoensri, Oren Cohen, Susan Countryman, Mary Ann Cristobal, Bobbi Croy, Suzanne Dale, Hrushikesh Deshmukh, Amanda Douglas, Vincent Drouillon, Marcia Eisenberg, Howard Engler, Rama Ghatti, Prashant Gupta, Susan Hicks, Jake Humphrey, Lax Iyer, Manoj Jain, Mohan Kolli, Brian Krueger, Tim Kuphal, Stanley Letovsky, Michael Levandoski, Craig Lukasik, Jonathan Meltzer, Brian Norvell, Mindy Nye, Scott Parker, Christos Petropoulos, John Pruitt, Steven Ragan, Scott Ryan, Mike Sapeta, Jana Schroth, Suresh Babu Selvaraju, Goran Stevovic, Amanda Suchanek, Andrea Throop, Lyndon Tilson, Thomas Urban, Joe Voshell, Kimberly Wagner, Jonathan Williams, Mary Williamson, Qian Zeng, Tricia Zwiefelhofer, Clinton R. Paden, Duncan MacCannell |
| EPI_ISL_1798460 | MT Public Health Laboratory | Centers for Disease Control and Prevention Division of Viral Diseases, Pathogen Discovery | Mili Sheth, Sarah Nobles, Jasmine Padilla, Mark Burroughs, Shoshona Le, Katie Dillon, Peter Cook, Clinton R. Paden, Dhwani Batra, Krista Queen, Kristen Knipe, Dakota Howard, Yvette Unoarumhi, Darlene Wagner, Matthew Schmerer, Ben L. Rambo-Martin, Kristine Lacek, Sam Shepard, Alison Laufer Halpin, Dave Wentworth, Vivien Dugan, Suxiang Tong, Justin Lee |
| EPI_ISL_1798461 | Laboratory Corporation of America | Centers for Disease Control and Prevention Division of Viral Diseases, Pathogen Discovery | Dakota Howard, Dhwani Batra, Peter W. Cook, Kara Moser, Adrian Paskey, Jason Caravas, Benjamin Rambo-Martin, Shatavia Morrison, Christopher Gulvick, Scott Sammons, Yvette Unoarumhi, Darlene Wagner, Matthew Schmerer, Minoo Agarwal, Eyad Almasri, Debbie Boles, Ayla Burns, Nuthawin Charoensri, Oren Cohen, Susan Countryman, Mary Ann Cristobal, Bobbi Croy, Suzanne Dale, Hrushikesh Deshmukh, Amanda Douglas, Vincent Drouillon, Marcia Eisenberg, Howard Engler, Rama Ghatti, Prashant Gupta, Susan Hicks, Jake Humphrey, Lax Iyer, Manoj Jain, Mohan Kolli, Brian Krueger, Tim Kuphal, Stanley Letovsky, Michael Levandoski, Craig Lukasik, Jonathan Meltzer, Brian Norvell, Mindy Nye, Scott Parker, Christos Petropoulos, John Pruitt, Steven Ragan, Scott Ryan, Mike Sapeta, Jana Schroth, Suresh Babu Selvaraju, Goran Stevovic, Amanda Suchanek, Andrea Throop, Lyndon Tilson, Thomas Urban, Joe Voshell, Kimberly Wagner, Jonathan Williams, Mary Williamson, Qian Zeng, Tricia Zwiefelhofer, Clinton R. Paden, Duncan MacCannell |
| EPI_ISL_1798462 | MD DOH Laboratories Administration | Centers for Disease Control and Prevention Division of Viral Diseases, Pathogen Discovery | Mili Sheth, Sarah Nobles, Jasmine Padilla, Mark Burroughs, Shoshona Le, Katie Dillon, Peter Cook, Clinton R. Paden, Dhwani Batra, Krista Queen, Kristen Knipe, Dakota Howard, Yvette Unoarumhi, Darlene Wagner, Matthew Schmerer, Ben L. Rambo-Martin, Kristine Lacek, Sam Shepard, Alison Laufer Halpin, Dave Wentworth, Vivien Dugan, Suxiang Tong, Justin Lee |
| EPI_ISL_1798463 | Laboratory Corporation of America | Centers for Disease Control and Prevention Division of Viral Diseases, Pathogen Discovery | Dakota Howard, Dhwani Batra, Peter W. Cook, Kara Moser, Adrian Paskey, Jason Caravas, Benjamin Rambo-Martin, Shatavia Morrison, Christopher Gulvick, Scott Sammons, Yvette Unoarumhi, Darlene Wagner, Matthew Schmerer, Minoo Agarwal, Eyad Almasri, Debbie Boles, Ayla Burns, Nuthawin Charoensri, Oren Cohen, Susan Countryman, Mary Ann Cristobal, Bobbi Croy, Suzanne Dale, Hrushikesh Deshmukh, Amanda Douglas, Vincent Drouillon, Marcia Eisenberg, Howard Engler, Rama Ghatti, Prashant Gupta, Susan Hicks, Jake Humphrey, Lax Iyer, Manoj Jain, Mohan Kolli, Brian Krueger, Tim Kuphal, Stanley Letovsky, Michael Levandoski, Craig Lukasik, Jonathan Meltzer, Brian Norvell, Mindy Nye, Scott Parker, Christos Petropoulos, John Pruitt, Steven Ragan, Scott Ryan, Mike Sapeta, Jana Schroth, Suresh Babu Selvaraju, Goran Stevovic, Amanda Suchanek, Andrea Throop, Lyndon Tilson, Thomas Urban, Joe Voshell, Kimberly Wagner, Jonathan Williams, Mary Williamson, Qian Zeng, Tricia Zwiefelhofer, Clinton R. Paden, Duncan MacCannell |
| EPI_ISL_1798464 | MN PHL Division, Minnesota Department of Health | Centers for Disease Control and Prevention Division of Viral Diseases, Pathogen Discovery | Mili Sheth, Sarah Nobles, Jasmine Padilla, Mark Burroughs, Shoshona Le, Katie Dillon, Peter Cook, Clinton R. Paden, Dhwani Batra, Krista Queen, Kristen Knipe, Dakota Howard, Yvette Unoarumhi, Darlene Wagner, Matthew Schmerer, Ben L. Rambo-Martin, Kristine Lacek, Sam Shepard, Alison Laufer Halpin, Dave Wentworth, Vivien Dugan, Suxiang Tong, Justin Lee |
| EPI_ISL_1798466, EPI_ISL_1798468 | CT-Dr. Katherine A. Kelley State Public Health Lab | Centers for Disease Control and Prevention Division of Viral Diseases, Pathogen Discovery | Mili Sheth, Sarah Nobles, Jasmine Padilla, Mark Burroughs, Shoshona Le, Katie Dillon, Peter Cook, Clinton R. Paden, Dhwani Batra, Krista Queen, Kristen Knipe, Dakota Howard, Yvette Unoarumhi, Darlene Wagner, Matthew Schmerer, Ben L. Rambo-Martin, Kristine Lacek, Sam Shepard, Alison Laufer Halpin, Dave Wentworth, Vivien Dugan, Suxiang Tong, Justin Lee |
| EPI_ISL_1798469 | Laboratory Corporation of America | Centers for Disease Control and Prevention Division of Viral Diseases, Pathogen Discovery | Dakota Howard, Dhwani Batra, Peter W. Cook, Kara Moser, Adrian Paskey, Jason Caravas, Benjamin Rambo-Martin, Shatavia Morrison, Christopher Gulvick, Scott Sammons, Yvette Unoarumhi, Darlene Wagner, Matthew Schmerer, Minoo Agarwal, Eyad Almasri, Debbie Boles, Ayla Burns, Nuthawin Charoensri, Oren Cohen, Susan Countryman, Mary Ann Cristobal, Bobbi Croy, Suzanne Dale, Hrushikesh Deshmukh, Amanda Douglas, Vincent Drouillon, Marcia Eisenberg, Howard Engler, Rama Ghatti, Prashant Gupta, Susan Hicks, Jake Humphrey, Lax Iyer, Manoj Jain, Mohan Kolli, Brian Krueger, Tim Kuphal, Stanley Letovsky, Michael Levandoski, Craig Lukasik, Jonathan Meltzer, Brian Norvell, Mindy Nye, Scott Parker, Christos Petropoulos, John Pruitt, Steven Ragan, Scott Ryan, Mike Sapeta, Jana Schroth, Suresh Babu Selvaraju, Goran Stevovic, Amanda Suchanek, Andrea Throop, Lyndon Tilson, Thomas Urban, Joe Voshell, Kimberly Wagner, Jonathan Williams, Mary Williamson, Qian Zeng, Tricia Zwiefelhofer, Clinton R. Paden, Duncan MacCannell |
| EPI_ISL_1798470 | OH Department of Health Laboratory | Centers for Disease Control and Prevention Division of Viral Diseases, Pathogen Discovery | Mili Sheth, Sarah Nobles, Jasmine Padilla, Mark Burroughs, Shoshona Le, Katie Dillon, Peter Cook, Clinton R. Paden, Dhwani Batra, Krista Queen, Kristen Knipe, Dakota Howard, Yvette Unoarumhi, Darlene Wagner, Matthew Schmerer, Ben L. Rambo-Martin, Kristine Lacek, Sam Shepard, Alison Laufer Halpin, Dave Wentworth, Vivien Dugan, Suxiang Tong, Justin Lee |
| EPI_ISL_1798471 | Laboratory Corporation of America | Centers for Disease Control and Prevention Division of Viral Diseases, Pathogen Discovery | Dakota Howard, Dhwani Batra, Peter W. Cook, Kara Moser, Adrian Paskey, Jason Caravas, Benjamin Rambo-Martin, Shatavia Morrison, Christopher Gulvick, Scott Sammons, Yvette Unoarumhi, Darlene Wagner, Matthew Schmerer, Minoo Agarwal, Eyad Almasri, Debbie Boles, Ayla Burns, Nuthawin Charoensri, Oren Cohen, Susan Countryman, Mary Ann Cristobal, Bobbi Croy, Suzanne Dale, Hrushikesh Deshmukh, Amanda Douglas, Vincent Drouillon, Marcia Eisenberg, Howard Engler, Rama Ghatti, Prashant Gupta, Susan Hicks, Jake Humphrey, Lax Iyer, Manoj Jain, Mohan Kolli, Brian Krueger, Tim Kuphal, Stanley Letovsky, Michael Levandoski, Craig Lukasik, Jonathan Meltzer, Brian Norvell, Mindy Nye, Scott Parker, Christos Petropoulos, John Pruitt, Steven Ragan, Scott Ryan, Mike Sapeta, Jana Schroth, Suresh Babu Selvaraju, Goran Stevovic, Amanda Suchanek, Andrea Throop, Lyndon Tilson, Thomas Urban, Joe Voshell, Kimberly Wagner, Jonathan Williams, Mary Williamson, Qian Zeng, Tricia Zwiefelhofer, Clinton R. Paden, Duncan MacCannell |
| EPI_ISL_1798472 | OH Department of Health Laboratory | Centers for Disease Control and Prevention Division of Viral Diseases, Pathogen Discovery | Mili Sheth, Sarah Nobles, Jasmine Padilla, Mark Burroughs, Shoshona Le, Katie Dillon, Peter Cook, Clinton R. Paden, Dhwani Batra, Krista Queen, Kristen Knipe, Dakota Howard, Yvette Unoarumhi, Darlene Wagner, Matthew Schmerer, Ben L. Rambo-Martin, Kristine Lacek, Sam Shepard, Alison Laufer Halpin, Dave Wentworth, Vivien Dugan, Suxiang Tong, Justin Lee |
| EPI_ISL_1798473 | WI State Laboratory of Hygiene | Centers for Disease Control and Prevention Division of Viral Diseases, Pathogen Discovery | Mili Sheth, Sarah Nobles, Jasmine Padilla, Mark Burroughs, Shoshona Le, Katie Dillon, Peter Cook, Clinton R. Paden, Dhwani Batra, Krista Queen, Kristen Knipe, Dakota Howard, Yvette Unoarumhi, Darlene Wagner, Matthew Schmerer, Ben L. Rambo-Martin, Kristine Lacek, Sam Shepard, Alison Laufer Halpin, Dave Wentworth, Vivien Dugan, Suxiang Tong, Justin Lee |
| EPI_ISL_1798474, EPI_ISL_1798475 | Laboratory Corporation of America | Centers for Disease Control and Prevention Division of Viral Diseases, Pathogen Discovery | Dakota Howard, Dhwani Batra, Peter W. Cook, Kara Moser, Adrian Paskey, Jason Caravas, Benjamin Rambo-Martin, Shatavia Morrison, Christopher Gulvick, Scott Sammons, Yvette Unoarumhi, Darlene Wagner, Matthew Schmerer, Minoo Agarwal, Eyad Almasri, Debbie Boles, Ayla Burns, Nuthawin Charoensri, Oren Cohen, Susan Countryman, Mary Ann Cristobal, Bobbi Croy, Suzanne Dale, Hrushikesh Deshmukh, Amanda Douglas, Vincent Drouillon, Marcia Eisenberg, Howard Engler, Rama Ghatti, Prashant Gupta, Susan Hicks, Jake Humphrey, Lax Iyer, Manoj Jain, Mohan Kolli, Brian Krueger, Tim Kuphal, Stanley Letovsky, Michael Levandoski, Craig Lukasik, Jonathan Meltzer, Brian Norvell, Mindy Nye, Scott Parker, Christos Petropoulos, John Pruitt, Steven Ragan, Scott Ryan, Mike Sapeta, Jana Schroth, Suresh Babu Selvaraju, Goran Stevovic, Amanda Suchanek, Andrea Throop, Lyndon Tilson, Thomas Urban, Joe Voshell, Kimberly Wagner, Jonathan Williams, Mary Williamson, Qian Zeng, Tricia Zwiefelhofer, Clinton R. Paden, Duncan MacCannell |
| EPI_ISL_1798476 | IL Dept. of Public Health Springfield Laboratory | Centers for Disease Control and Prevention Division of Viral Diseases, Pathogen Discovery | Mili Sheth, Sarah Nobles, Jasmine Padilla, Mark Burroughs, Shoshona Le, Katie Dillon, Peter Cook, Clinton R. Paden, Dhwani Batra, Krista Queen, Kristen Knipe, Dakota Howard, Yvette Unoarumhi, Darlene Wagner, Matthew Schmerer, Ben L. Rambo-Martin, Kristine Lacek, Sam Shepard, Alison Laufer Halpin, Dave Wentworth, Vivien Dugan, Suxiang Tong, Justin Lee |
| EPI_ISL_1798477 | Laboratory Corporation of America | Centers for Disease Control and Prevention Division of Viral Diseases, Pathogen Discovery | Dakota Howard, Dhwani Batra, Peter W. Cook, Kara Moser, Adrian Paskey, Jason Caravas, Benjamin Rambo-Martin, Shatavia Morrison, Christopher Gulvick, Scott Sammons, Yvette Unoarumhi, Darlene Wagner, Matthew Schmerer, Minoo Agarwal, Eyad Almasri, Debbie Boles, Ayla Burns, Nuthawin Charoensri, Oren Cohen, Susan Countryman, Mary Ann Cristobal, Bobbi Croy, Suzanne Dale, Hrushikesh Deshmukh, Amanda Douglas, Vincent Drouillon, Marcia Eisenberg, Howard Engler, Rama Ghatti, Prashant Gupta, Susan Hicks, Jake Humphrey, Lax Iyer, Manoj Jain, Mohan Kolli, Brian Krueger, Tim Kuphal, Stanley Letovsky, Michael Levandoski, Craig Lukasik, Jonathan Meltzer, Brian Norvell, Mindy Nye, Scott Parker, Christos Petropoulos, John Pruitt, Steven Ragan, Scott Ryan, Mike Sapeta, Jana Schroth, Suresh Babu Selvaraju, Goran Stevovic, Amanda Suchanek, Andrea Throop, Lyndon Tilson, Thomas Urban, Joe Voshell, Kimberly Wagner, Jonathan Williams, Mary Williamson, Qian Zeng, Tricia Zwiefelhofer, Clinton R. Paden, Duncan MacCannell |
| EPI_ISL_1798478 | OH Department of Health Laboratory | Centers for Disease Control and Prevention Division of Viral Diseases, Pathogen Discovery | Mili Sheth, Sarah Nobles, Jasmine Padilla, Mark Burroughs, Shoshona Le, Katie Dillon, Peter Cook, Clinton R. Paden, Dhwani Batra, Krista Queen, Kristen Knipe, Dakota Howard, Yvette Unoarumhi, Darlene Wagner, Matthew Schmerer, Ben L. Rambo-Martin, Kristine Lacek, Sam Shepard, Alison Laufer Halpin, Dave Wentworth, Vivien Dugan, Suxiang Tong, Justin Lee |
| EPI_ISL_1798479 | Laboratory Corporation of America | Centers for Disease Control and Prevention Division of Viral Diseases, Pathogen Discovery | Dakota Howard, Dhwani Batra, Peter W. Cook, Kara Moser, Adrian Paskey, Jason Caravas, Benjamin Rambo-Martin, Shatavia Morrison, Christopher Gulvick, Scott Sammons, Yvette Unoarumhi, Darlene Wagner, Matthew Schmerer, Minoo Agarwal, Eyad Almasri, Debbie Boles, Ayla Burns, Nuthawin Charoensri, Oren Cohen, Susan Countryman, Mary Ann Cristobal, Bobbi Croy, Suzanne Dale, Hrushikesh Deshmukh, Amanda Douglas, Vincent Drouillon, Marcia Eisenberg, Howard Engler, Rama Ghatti, Prashant Gupta, Susan Hicks, Jake Humphrey, Lax Iyer, Manoj Jain, Mohan Kolli, Brian Krueger, Tim Kuphal, Stanley Letovsky, Michael Levandoski, Craig Lukasik, Jonathan Meltzer, Brian Norvell, Mindy Nye, Scott Parker, Christos Petropoulos, John Pruitt, Steven Ragan, Scott Ryan, Mike Sapeta, Jana Schroth, Suresh Babu Selvaraju, Goran Stevovic, Amanda Suchanek, Andrea Throop, Lyndon Tilson, Thomas Urban, Joe Voshell, Kimberly Wagner, Jonathan Williams, Mary Williamson, Qian Zeng, Tricia Zwiefelhofer, Clinton R. Paden, Duncan MacCannell |
| EPI_ISL_1798480 | IA State Hygienic Laboratory | Centers for Disease Control and Prevention Division of Viral Diseases, Pathogen Discovery | Mili Sheth, Sarah Nobles, Jasmine Padilla, Mark Burroughs, Shoshona Le, Katie Dillon, Peter Cook, Clinton R. Paden, Dhwani Batra, Krista Queen, Kristen Knipe, Dakota Howard, Yvette Unoarumhi, Darlene Wagner, Matthew Schmerer, Ben L. Rambo-Martin, Kristine Lacek, Sam Shepard, Alison Laufer Halpin, Dave Wentworth, Vivien Dugan, Suxiang Tong, Justin Lee |
| EPI_ISL_1798481 | Laboratory Corporation of America | Centers for Disease Control and Prevention Division of Viral Diseases, Pathogen Discovery | Dakota Howard, Dhwani Batra, Peter W. Cook, Kara Moser, Adrian Paskey, Jason Caravas, Benjamin Rambo-Martin, Shatavia Morrison, Christopher Gulvick, Scott Sammons, Yvette Unoarumhi, Darlene Wagner, Matthew Schmerer, Minoo Agarwal, Eyad Almasri, Debbie Boles, Ayla Burns, Nuthawin Charoensri, Oren Cohen, Susan Countryman, Mary Ann Cristobal, Bobbi Croy, Suzanne Dale, Hrushikesh Deshmukh, Amanda Douglas, Vincent Drouillon, Marcia Eisenberg, Howard Engler, Rama Ghatti, Prashant Gupta, Susan Hicks, Jake Humphrey, Lax Iyer, Manoj Jain, Mohan Kolli, Brian Krueger, Tim Kuphal, Stanley Letovsky, Michael Levandoski, Craig Lukasik, Jonathan Meltzer, Brian Norvell, Mindy Nye, Scott Parker, Christos Petropoulos, John Pruitt, Steven Ragan, Scott Ryan, Mike Sapeta, Jana Schroth, Suresh Babu Selvaraju, Goran Stevovic, Amanda Suchanek, Andrea Throop, Lyndon Tilson, Thomas Urban, Joe Voshell, Kimberly Wagner, Jonathan Williams, Mary Williamson, Qian Zeng, Tricia Zwiefelhofer, Clinton R. Paden, Duncan MacCannell |
| EPI_ISL_1798482 | IA State Hygienic Laboratory | Centers for Disease Control and Prevention Division of Viral Diseases, Pathogen Discovery | Mili Sheth, Sarah Nobles, Jasmine Padilla, Mark Burroughs, Shoshona Le, Katie Dillon, Peter Cook, Clinton R. Paden, Dhwani Batra, Krista Queen, Kristen Knipe, Dakota Howard, Yvette Unoarumhi, Darlene Wagner, Matthew Schmerer, Ben L. Rambo-Martin, Kristine Lacek, Sam Shepard, Alison Laufer Halpin, Dave Wentworth, Vivien Dugan, Suxiang Tong, Justin Lee |
| EPI_ISL_1798484 | UT-Unified State Labs: Public Health Utah DOH | Centers for Disease Control and Prevention Division of Viral Diseases, Pathogen Discovery | Mili Sheth, Sarah Nobles, Jasmine Padilla, Mark Burroughs, Shoshona Le, Katie Dillon, Peter Cook, Clinton R. Paden, Dhwani Batra, Krista Queen, Kristen Knipe, Dakota Howard, Yvette Unoarumhi, Darlene Wagner, Matthew Schmerer, Ben L. Rambo-Martin, Kristine Lacek, Sam Shepard, Alison Laufer Halpin, Dave Wentworth, Vivien Dugan, Suxiang Tong, Justin Lee |
| EPI_ISL_1798486, EPI_ISL_1798488, EPI_ISL_1798490 | OH Department of Health Laboratory | Centers for Disease Control and Prevention Division of Viral Diseases, Pathogen Discovery | Mili Sheth, Sarah Nobles, Jasmine Padilla, Mark Burroughs, Shoshona Le, Katie Dillon, Peter Cook, Clinton R. Paden, Dhwani Batra, Krista Queen, Kristen Knipe, Dakota Howard, Yvette Unoarumhi, Darlene Wagner, Matthew Schmerer, Ben L. Rambo-Martin, Kristine Lacek, Sam Shepard, Alison Laufer Halpin, Dave Wentworth, Vivien Dugan, Suxiang Tong, Justin Lee |
| EPI_ISL_1798491 | Laboratory Corporation of America | Centers for Disease Control and Prevention Division of Viral Diseases, Pathogen Discovery | Dakota Howard, Dhwani Batra, Peter W. Cook, Kara Moser, Adrian Paskey, Jason Caravas, Benjamin Rambo-Martin, Shatavia Morrison, Christopher Gulvick, Scott Sammons, Yvette Unoarumhi, Darlene Wagner, Matthew Schmerer, Minoo Agarwal, Eyad Almasri, Debbie Boles, Ayla Burns, Nuthawin Charoensri, Oren Cohen, Susan Countryman, Mary Ann Cristobal, Bobbi Croy, Suzanne Dale, Hrushikesh Deshmukh, Amanda Douglas, Vincent Drouillon, Marcia Eisenberg, Howard Engler, Rama Ghatti, Prashant Gupta, Susan Hicks, Jake Humphrey, Lax Iyer, Manoj Jain, Mohan Kolli, Brian Krueger, Tim Kuphal, Stanley Letovsky, Michael Levandoski, Craig Lukasik, Jonathan Meltzer, Brian Norvell, Mindy Nye, Scott Parker, Christos Petropoulos, John Pruitt, Steven Ragan, Scott Ryan, Mike Sapeta, Jana Schroth, Suresh Babu Selvaraju, Goran Stevovic, Amanda Suchanek, Andrea Throop, Lyndon Tilson, Thomas Urban, Joe Voshell, Kimberly Wagner, Jonathan Williams, Mary Williamson, Qian Zeng, Tricia Zwiefelhofer, Clinton R. Paden, Duncan MacCannell |
| EPI_ISL_1798492, EPI_ISL_1798494 | WI State Laboratory of Hygiene | Centers for Disease Control and Prevention Division of Viral Diseases, Pathogen Discovery | Mili Sheth, Sarah Nobles, Jasmine Padilla, Mark Burroughs, Shoshona Le, Katie Dillon, Peter Cook, Clinton R. Paden, Dhwani Batra, Krista Queen, Kristen Knipe, Dakota Howard, Yvette Unoarumhi, Darlene Wagner, Matthew Schmerer, Ben L. Rambo-Martin, Kristine Lacek, Sam Shepard, Alison Laufer Halpin, Dave Wentworth, Vivien Dugan, Suxiang Tong, Justin Lee |
| EPI_ISL_1798496 | IL Dept. of Public Health Springfield Laboratory | Centers for Disease Control and Prevention Division of Viral Diseases, Pathogen | Mili Sheth, Sarah Nobles, Jasmine Padilla, Mark Burroughs, Shoshona Le, Katie Dillon, Peter Cook, Clinton R. Paden, Dhwani Batra, Krista Queen, Kristen Knipe, Dakota Howard, Yvette Unoarumhi, Darlene Wagner, Matthew Schmerer, Ben L. Rambo-Martin, Kristine Lacek, Sam Shepard, Alison Laufer Halpin, Dave Wentworth, Vivien Dugan, Suxiang Tong, Justin Lee |

| | | | |
|---|---|---|---|
| | | Discovery | |
| EPI_ISL_1798497 | Laboratory Corporation of America | Centers for Disease Control and Prevention Division of Viral Diseases, Pathogen Discovery | Dakota Howard, Dhwani Batra, Peter W. Cook, Kara Moser, Adrian Paskey, Jason Caravas, Benjamin Rambo-Martin, Shatavia Morrison, Christopher Gulvick, Scott Sammons, Yvette Unoarumhi, Darlene Wagner, Matthew Schmerer, Minoo Agarwal, Eyad Almasri, Debbie Boles, Ayla Burns, Nuthawin Charoensri, Oren Cohen, Susan Countryman, Mary Ann Cristobal, Bobbi Croy, Suzanne Dale, Hrushikesh Deshmukh, Amanda Douglas, Vincent Drouillon, Marcia Eisenberg, Howard Engler, Rama Ghatti, Prashant Gupta, Susan Hicks, Jake Humphrey, Lax Iyer, Manoj Jain, Mohan Kolli, Brian Krueger, Tim Kuphal, Stanley Letovsky, Michael Levandoski, Craig Lukasik, Jonathan Meltzer, Brian Norvell, Mindy Nye, Scott Parker, Christos Petropoulos, John Pruitt, Steven Ragan, Scott Ryan, Mike Sapeta, Jana Schroth, Suresh Babu Selvaraju, Goran Stevovic, Amanda Suchanek, Andrea Throop, Lyndon Tilson, Thomas Urban, Joe Voshell, Kimberly Wagner, Jonathan Williams, Mary Williamson, Qian Zeng, Tricia Zwiefelhofer, Clinton R. Paden, Duncan MacCannell |
| EPI_ISL_1798498 | IL Dept. of Public Health Springfield Laboratory | Centers for Disease Control and Prevention Division of Viral Diseases, Pathogen Discovery | Mili Sheth, Sarah Nobles, Jasmine Padilla, Mark Burroughs, Shoshona Le, Katie Dillon, Peter Cook, Clinton R. Paden, Dhwani Batra, Krista Queen, Kristen Knipe, Dakota Howard, Yvette Unoarumhi, Darlene Wagner, Matthew Schmerer, Ben L. Rambo-Martin, Kristine Lacek, Sam Shepard, Alison Laufer Halpin, Dave Wentworth, Vivien Dugan, Suxiang Tong, Justin Lee |
| EPI_ISL_1798499 | Laboratory Corporation of America | Centers for Disease Control and Prevention Division of Viral Diseases, Pathogen Discovery | Dakota Howard, Dhwani Batra, Peter W. Cook, Kara Moser, Adrian Paskey, Jason Caravas, Benjamin Rambo-Martin, Shatavia Morrison, Christopher Gulvick, Scott Sammons, Yvette Unoarumhi, Darlene Wagner, Matthew Schmerer, Minoo Agarwal, Eyad Almasri, Debbie Boles, Ayla Burns, Nuthawin Charoensri, Oren Cohen, Susan Countryman, Mary Ann Cristobal, Bobbi Croy, Suzanne Dale, Hrushikesh Deshmukh, Amanda Douglas, Vincent Drouillon, Marcia Eisenberg, Howard Engler, Rama Ghatti, Prashant Gupta, Susan Hicks, Jake Humphrey, Lax Iyer, Manoj Jain, Mohan Kolli, Brian Krueger, Tim Kuphal, Stanley Letovsky, Michael Levandoski, Craig Lukasik, Jonathan Meltzer, Brian Norvell, Mindy Nye, Scott Parker, Christos Petropoulos, John Pruitt, Steven Ragan, Scott Ryan, Mike Sapeta, Jana Schroth, Suresh Babu Selvaraju, Goran Stevovic, Amanda Suchanek, Andrea Throop, Lyndon Tilson, Thomas Urban, Joe Voshell, Kimberly Wagner, Jonathan Williams, Mary Williamson, Qian Zeng, Tricia Zwiefelhofer, Clinton R. Paden, Duncan MacCannell |
| EPI_ISL_1798500 | OH Department of Health Laboratory | Centers for Disease Control and Prevention Division of Viral Diseases, Pathogen Discovery | Mili Sheth, Sarah Nobles, Jasmine Padilla, Mark Burroughs, Shoshona Le, Katie Dillon, Peter Cook, Clinton R. Paden, Dhwani Batra, Krista Queen, Kristen Knipe, Dakota Howard, Yvette Unoarumhi, Darlene Wagner, Matthew Schmerer, Ben L. Rambo-Martin, Kristine Lacek, Sam Shepard, Alison Laufer Halpin, Dave Wentworth, Vivien Dugan, Suxiang Tong, Justin Lee |
| EPI_ISL_1798501 | Laboratory Corporation of America | Centers for Disease Control and Prevention Division of Viral Diseases, Pathogen Discovery | Dakota Howard, Dhwani Batra, Peter W. Cook, Kara Moser, Adrian Paskey, Jason Caravas, Benjamin Rambo-Martin, Shatavia Morrison, Christopher Gulvick, Scott Sammons, Yvette Unoarumhi, Darlene Wagner, Matthew Schmerer, Minoo Agarwal, Eyad Almasri, Debbie Boles, Ayla Burns, Nuthawin Charoensri, Oren Cohen, Susan Countryman, Mary Ann Cristobal, Bobbi Croy, Suzanne Dale, Hrushikesh Deshmukh, Amanda Douglas, Vincent Drouillon, Marcia Eisenberg, Howard Engler, Rama Ghatti, Prashant Gupta, Susan Hicks, Jake Humphrey, Lax Iyer, Manoj Jain, Mohan Kolli, Brian Krueger, Tim Kuphal, Stanley Letovsky, Michael Levandoski, Craig Lukasik, Jonathan Meltzer, Brian Norvell, Mindy Nye, Scott Parker, Christos Petropoulos, John Pruitt, Steven Ragan, Scott Ryan, Mike Sapeta, Jana Schroth, Suresh Babu Selvaraju, Goran Stevovic, Amanda Suchanek, Andrea Throop, Lyndon Tilson, Thomas Urban, Joe Voshell, Kimberly Wagner, Jonathan Williams, Mary Williamson, Qian Zeng, Tricia Zwiefelhofer, Clinton R. Paden, Duncan MacCannell |
| EPI_ISL_1798502 | OH Department of Health Laboratory | Centers for Disease Control and Prevention Division of Viral Diseases, Pathogen Discovery | Mili Sheth, Sarah Nobles, Jasmine Padilla, Mark Burroughs, Shoshona Le, Katie Dillon, Peter Cook, Clinton R. Paden, Dhwani Batra, Krista Queen, Kristen Knipe, Dakota Howard, Yvette Unoarumhi, Darlene Wagner, Matthew Schmerer, Ben L. Rambo-Martin, Kristine Lacek, Sam Shepard, Alison Laufer Halpin, Dave Wentworth, Vivien Dugan, Suxiang Tong, Justin Lee |
| EPI_ISL_1798503 | Laboratory Corporation of America | Centers for Disease Control and Prevention Division of Viral Diseases, Pathogen Discovery | Dakota Howard, Dhwani Batra, Peter W. Cook, Kara Moser, Adrian Paskey, Jason Caravas, Benjamin Rambo-Martin, Shatavia Morrison, Christopher Gulvick, Scott Sammons, Yvette Unoarumhi, Darlene Wagner, Matthew Schmerer, Minoo Agarwal, Eyad Almasri, Debbie Boles, Ayla Burns, Nuthawin Charoensri, Oren Cohen, Susan Countryman, Mary Ann Cristobal, Bobbi Croy, Suzanne Dale, Hrushikesh Deshmukh, Amanda Douglas, Vincent Drouillon, Marcia Eisenberg, Howard Engler, Rama Ghatti, Prashant Gupta, Susan Hicks, Jake Humphrey, Lax Iyer, Manoj Jain, Mohan Kolli, Brian Krueger, Tim Kuphal, Stanley Letovsky, Michael Levandoski, Craig Lukasik, Jonathan Meltzer, Brian Norvell, Mindy Nye, Scott Parker, Christos Petropoulos, John Pruitt, Steven Ragan, Scott Ryan, Mike Sapeta, Jana Schroth, Suresh Babu Selvaraju, Goran Stevovic, Amanda Suchanek, Andrea Throop, Lyndon Tilson, Thomas Urban, Joe Voshell, Kimberly Wagner, Jonathan Williams, Mary Williamson, Qian Zeng, Tricia Zwiefelhofer, Clinton R. Paden, Duncan MacCannell |
| EPI_ISL_1798504 | OH Department of Health Laboratory | Centers for Disease Control and Prevention Division of Viral Diseases, Pathogen Discovery | Mili Sheth, Sarah Nobles, Jasmine Padilla, Mark Burroughs, Shoshona Le, Katie Dillon, Peter Cook, Clinton R. Paden, Dhwani Batra, Krista Queen, Kristen Knipe, Dakota Howard, Yvette Unoarumhi, Darlene Wagner, Matthew Schmerer, Ben L. Rambo-Martin, Kristine Lacek, Sam Shepard, Alison Laufer Halpin, Dave Wentworth, Vivien Dugan, Suxiang Tong, Justin Lee |
| EPI_ISL_1798505 | Laboratory Corporation of America | Centers for Disease Control and Prevention Division of Viral Diseases, Pathogen Discovery | Dakota Howard, Dhwani Batra, Peter W. Cook, Kara Moser, Adrian Paskey, Jason Caravas, Benjamin Rambo-Martin, Shatavia Morrison, Christopher Gulvick, Scott Sammons, Yvette Unoarumhi, Darlene Wagner, Matthew Schmerer, Minoo Agarwal, Eyad Almasri, Debbie Boles, Ayla Burns, Nuthawin Charoensri, Oren Cohen, Susan Countryman, Mary Ann Cristobal, Bobbi Croy, Suzanne Dale, Hrushikesh Deshmukh, Amanda Douglas, Vincent Drouillon, Marcia Eisenberg, Howard Engler, Rama Ghatti, Prashant Gupta, Susan Hicks, Jake Humphrey, Lax Iyer, Manoj Jain, Mohan Kolli, Brian Krueger, Tim Kuphal, Stanley Letovsky, Michael Levandoski, Craig Lukasik, Jonathan Meltzer, Brian Norvell, Mindy Nye, Scott Parker, Christos Petropoulos, John Pruitt, Steven Ragan, Scott Ryan, Mike Sapeta, Jana Schroth, Suresh Babu Selvaraju, Goran Stevovic, Amanda Suchanek, Andrea Throop, Lyndon Tilson, Thomas Urban, Joe Voshell, Kimberly Wagner, Jonathan Williams, Mary Williamson, Qian Zeng, Tricia Zwiefelhofer, Clinton R. Paden, Duncan MacCannell |
| EPI_ISL_1798506 | OH Department of Health Laboratory | Centers for Disease Control and Prevention Division of Viral Diseases, Pathogen Discovery | Mili Sheth, Sarah Nobles, Jasmine Padilla, Mark Burroughs, Shoshona Le, Katie Dillon, Peter Cook, Clinton R. Paden, Dhwani Batra, Krista Queen, Kristen Knipe, Dakota Howard, Yvette Unoarumhi, Darlene Wagner, Matthew Schmerer, Ben L. Rambo-Martin, Kristine Lacek, Sam Shepard, Alison Laufer Halpin, Dave Wentworth, Vivien Dugan, Suxiang Tong, Justin Lee |
| EPI_ISL_1798507 | Laboratory Corporation of America | Centers for Disease Control and Prevention Division of Viral Diseases, Pathogen Discovery | Dakota Howard, Dhwani Batra, Peter W. Cook, Kara Moser, Adrian Paskey, Jason Caravas, Benjamin Rambo-Martin, Shatavia Morrison, Christopher Gulvick, Scott Sammons, Yvette Unoarumhi, Darlene Wagner, Matthew Schmerer, Minoo Agarwal, Eyad Almasri, Debbie Boles, Ayla Burns, Nuthawin Charoensri, Oren Cohen, Susan Countryman, Mary Ann Cristobal, Bobbi Croy, Suzanne Dale, Hrushikesh Deshmukh, Amanda Douglas, Vincent Drouillon, Marcia Eisenberg, Howard Engler, Rama Ghatti, Prashant Gupta, Susan Hicks, Jake Humphrey, Lax Iyer, Manoj Jain, Mohan Kolli, Brian Krueger, Tim Kuphal, Stanley Letovsky, Michael Levandoski, Craig Lukasik, Jonathan Meltzer, Brian Norvell, Mindy Nye, Scott Parker, Christos Petropoulos, John Pruitt, Steven Ragan, Scott Ryan, Mike Sapeta, Jana Schroth, Suresh Babu Selvaraju, Goran Stevovic, Amanda Suchanek, Andrea Throop, Lyndon Tilson, Thomas Urban, Joe Voshell, Kimberly Wagner, Jonathan Williams, Mary Williamson, Qian Zeng, Tricia Zwiefelhofer, Clinton R. Paden, Duncan MacCannell |
| EPI_ISL_1798508, EPI_ISL_1798510 | IL Dept. of Public Health Springfield Laboratory | Centers for Disease Control and Prevention Division of Viral Diseases, Pathogen Discovery | Mili Sheth, Sarah Nobles, Jasmine Padilla, Mark Burroughs, Shoshona Le, Katie Dillon, Peter Cook, Clinton R. Paden, Dhwani Batra, Krista Queen, Kristen Knipe, Dakota Howard, Yvette Unoarumhi, Darlene Wagner, Matthew Schmerer, Ben L. Rambo-Martin, Kristine Lacek, Sam Shepard, Alison Laufer Halpin, Dave Wentworth, Vivien Dugan, Suxiang Tong, Justin Lee |
| EPI_ISL_1798511 | Laboratory Corporation of America | Centers for Disease Control and Prevention Division of Viral Diseases, Pathogen Discovery | Dakota Howard, Dhwani Batra, Peter W. Cook, Kara Moser, Adrian Paskey, Jason Caravas, Benjamin Rambo-Martin, Shatavia Morrison, Christopher Gulvick, Scott Sammons, Yvette Unoarumhi, Darlene Wagner, Matthew Schmerer, Minoo Agarwal, Eyad Almasri, Debbie Boles, Ayla Burns, Nuthawin Charoensri, Oren Cohen, Susan Countryman, Mary Ann Cristobal, Bobbi Croy, Suzanne Dale, Hrushikesh Deshmukh, Amanda Douglas, Vincent Drouillon, Marcia Eisenberg, Howard Engler, Rama Ghatti, Prashant Gupta, Susan Hicks, Jake Humphrey, Lax Iyer, Manoj Jain, Mohan Kolli, Brian Krueger, Tim Kuphal, Stanley Letovsky, Michael Levandoski, Craig Lukasik, Jonathan Meltzer, Brian Norvell, Mindy Nye, Scott Parker, Christos Petropoulos, John Pruitt, Steven Ragan, Scott Ryan, Mike Sapeta, Jana Schroth, Suresh Babu Selvaraju, Goran Stevovic, Amanda Suchanek, Andrea Throop, Lyndon Tilson, Thomas Urban, Joe Voshell, Kimberly Wagner, Jonathan Williams, Mary Williamson, Qian Zeng, Tricia Zwiefelhofer, Clinton R. Paden, Duncan MacCannell |
| EPI_ISL_1798512 | IA State Hygienic Laboratory | Centers for Disease Control and Prevention Division of Viral Diseases, Pathogen Discovery | Mili Sheth, Sarah Nobles, Jasmine Padilla, Mark Burroughs, Shoshona Le, Katie Dillon, Peter Cook, Clinton R. Paden, Dhwani Batra, Krista Queen, Kristen Knipe, Dakota Howard, Yvette Unoarumhi, Darlene Wagner, Matthew Schmerer, Ben L. Rambo-Martin, Kristine Lacek, Sam Shepard, Alison Laufer Halpin, Dave Wentworth, Vivien Dugan, Suxiang Tong, Justin Lee |
| EPI_ISL_1798513, EPI_ISL_1798514 | Laboratory Corporation of America | Centers for Disease Control and Prevention Division of Viral Diseases, Pathogen Discovery | Dakota Howard, Dhwani Batra, Peter W. Cook, Kara Moser, Adrian Paskey, Jason Caravas, Benjamin Rambo-Martin, Shatavia Morrison, Christopher Gulvick, Scott Sammons, Yvette Unoarumhi, Darlene Wagner, Matthew Schmerer, Minoo Agarwal, Eyad Almasri, Debbie Boles, Ayla Burns, Nuthawin Charoensri, Oren Cohen, Susan Countryman, Mary Ann Cristobal, Bobbi Croy, Suzanne Dale, Hrushikesh Deshmukh, Amanda Douglas, Vincent Drouillon, Marcia Eisenberg, Howard Engler, Rama Ghatti, Prashant Gupta, Susan Hicks, Jake Humphrey, Lax Iyer, Manoj Jain, Mohan Kolli, Brian Krueger, Tim Kuphal, Stanley Letovsky, Michael Levandoski, Craig Lukasik, Jonathan Meltzer, Brian Norvell, Mindy Nye, Scott Parker, Christos Petropoulos, John Pruitt, Steven Ragan, Scott Ryan, Mike Sapeta, Jana Schroth, Suresh Babu Selvaraju, Goran Stevovic, Amanda Suchanek, Andrea Throop, Lyndon Tilson, Thomas Urban, Joe Voshell, Kimberly Wagner, Jonathan Williams, Mary Williamson, Qian Zeng, Tricia Zwiefelhofer, Clinton R. Paden, Duncan MacCannell |
| EPI_ISL_1798515 | OH Department of Health Laboratory | Centers for Disease Control and Prevention Division of Viral Diseases, Pathogen Discovery | Mili Sheth, Sarah Nobles, Jasmine Padilla, Mark Burroughs, Shoshona Le, Katie Dillon, Peter Cook, Clinton R. Paden, Dhwani Batra, Krista Queen, Kristen Knipe, Dakota Howard, Yvette Unoarumhi, Darlene Wagner, Matthew Schmerer, Ben L. Rambo-Martin, Kristine Lacek, Sam Shepard, Alison Laufer Halpin, Dave Wentworth, Vivien Dugan, Suxiang Tong, Justin Lee |
| EPI_ISL_1798516 | Laboratory Corporation of America | Centers for Disease Control and Prevention Division of Viral Diseases, Pathogen Discovery | Dakota Howard, Dhwani Batra, Peter W. Cook, Kara Moser, Adrian Paskey, Jason Caravas, Benjamin Rambo-Martin, Shatavia Morrison, Christopher Gulvick, Scott Sammons, Yvette Unoarumhi, Darlene Wagner, Matthew Schmerer, Minoo Agarwal, Eyad Almasri, Debbie Boles, Ayla Burns, Nuthawin Charoensri, Oren Cohen, Susan Countryman, Mary Ann Cristobal, Bobbi Croy, Suzanne Dale, Hrushikesh Deshmukh, Amanda Douglas, Vincent Drouillon, Marcia Eisenberg, Howard Engler, Rama Ghatti, Prashant Gupta, Susan Hicks, Jake Humphrey, Lax Iyer, Manoj Jain, Mohan Kolli, Brian Krueger, Tim Kuphal, Stanley Letovsky, Michael Levandoski, Craig Lukasik, Jonathan Meltzer, Brian Norvell, Mindy Nye, Scott Parker, Christos Petropoulos, John Pruitt, Steven Ragan, Scott Ryan, Mike Sapeta, Jana Schroth, Suresh Babu Selvaraju, Goran Stevovic, Amanda Suchanek, Andrea Throop, Lyndon Tilson, Thomas Urban, Joe Voshell, Kimberly Wagner, Jonathan Williams, Mary Williamson, Qian Zeng, Tricia Zwiefelhofer, Clinton R. Paden, Duncan MacCannell |
| EPI_ISL_1798517 | OH Department of Health Laboratory | Centers for Disease Control and Prevention Division of Viral Diseases, Pathogen Discovery | Mili Sheth, Sarah Nobles, Jasmine Padilla, Mark Burroughs, Shoshona Le, Katie Dillon, Peter Cook, Clinton R. Paden, Dhwani Batra, Krista Queen, Kristen Knipe, Dakota Howard, Yvette Unoarumhi, Darlene Wagner, Matthew Schmerer, Ben L. Rambo-Martin, Kristine Lacek, Sam Shepard, Alison Laufer Halpin, Dave Wentworth, Vivien Dugan, Suxiang Tong, Justin Lee |
| EPI_ISL_1798518 | Laboratory Corporation of America | Centers for Disease Control and Prevention Division of Viral Diseases, Pathogen Discovery | Dakota Howard, Dhwani Batra, Peter W. Cook, Kara Moser, Adrian Paskey, Jason Caravas, Benjamin Rambo-Martin, Shatavia Morrison, Christopher Gulvick, Scott Sammons, Yvette Unoarumhi, Darlene Wagner, Matthew Schmerer, Minoo Agarwal, Eyad Almasri, Debbie Boles, Ayla Burns, Nuthawin Charoensri, Oren Cohen, Susan Countryman, Mary Ann Cristobal, Bobbi Croy, Suzanne Dale, Hrushikesh Deshmukh, Amanda Douglas, Vincent Drouillon, Marcia Eisenberg, Howard Engler, Rama Ghatti, Prashant Gupta, Susan Hicks, Jake Humphrey, Lax Iyer, Manoj Jain, Mohan Kolli, Brian Krueger, Tim Kuphal, Stanley Letovsky, Michael Levandoski, Craig Lukasik, Jonathan Meltzer, Brian Norvell, Mindy Nye, Scott Parker, Christos Petropoulos, John Pruitt, Steven Ragan, Scott Ryan, Mike Sapeta, Jana Schroth, Suresh Babu Selvaraju, Goran Stevovic, Amanda Suchanek, Andrea Throop, Lyndon Tilson, Thomas Urban, Joe Voshell, Kimberly Wagner, Jonathan Williams, Mary Williamson, Qian Zeng, Tricia Zwiefelhofer, Clinton R. Paden, Duncan MacCannell |
| EPI_ISL_1798519 | OH Department of Health Laboratory | Centers for Disease Control and Prevention Division of Viral Diseases, Pathogen Discovery | Mili Sheth, Sarah Nobles, Jasmine Padilla, Mark Burroughs, Shoshona Le, Katie Dillon, Peter Cook, Clinton R. Paden, Dhwani Batra, Krista Queen, Kristen Knipe, Dakota Howard, Yvette Unoarumhi, Darlene Wagner, Matthew Schmerer, Ben L. Rambo-Martin, Kristine Lacek, Sam Shepard, Alison Laufer Halpin, Dave Wentworth, Vivien Dugan, Suxiang Tong, Justin Lee |
| EPI_ISL_1798521 | AZ SPHL, Arizona Department of Health Services | Centers for Disease Control and Prevention Division of Viral Diseases, Pathogen Discovery | Mili Sheth, Sarah Nobles, Jasmine Padilla, Mark Burroughs, Shoshona Le, Katie Dillon, Peter Cook, Clinton R. Paden, Dhwani Batra, Krista Queen, Kristen Knipe, Dakota Howard, Yvette Unoarumhi, Darlene Wagner, Matthew Schmerer, Ben L. Rambo-Martin, Kristine Lacek, Sam Shepard, Alison Laufer Halpin, Dave Wentworth, Vivien Dugan, Suxiang Tong, Justin Lee |
| EPI_ISL_1798522 | Laboratory Corporation of America | Centers for Disease Control and Prevention Division of Viral Diseases, Pathogen Discovery | Dakota Howard, Dhwani Batra, Peter W. Cook, Kara Moser, Adrian Paskey, Jason Caravas, Benjamin Rambo-Martin, Shatavia Morrison, Christopher Gulvick, Scott Sammons, Yvette Unoarumhi, Darlene Wagner, Matthew Schmerer, Minoo Agarwal, Eyad Almasri, Debbie Boles, Ayla Burns, Nuthawin Charoensri, Oren Cohen, Susan Countryman, Mary Ann Cristobal, Bobbi Croy, Suzanne Dale, Hrushikesh Deshmukh, Amanda Douglas, Vincent Drouillon, Marcia Eisenberg, Howard Engler, Rama Ghatti, Prashant Gupta, Susan Hicks, Jake Humphrey, Lax Iyer, Manoj Jain, Mohan Kolli, Brian Krueger, Tim Kuphal, Stanley Letovsky, Michael Levandoski, Craig Lukasik, Jonathan Meltzer, Brian Norvell, Mindy Nye, Scott Parker, Christos Petropoulos, John Pruitt, Steven Ragan, Scott Ryan, Mike Sapeta, Jana Schroth, Suresh Babu Selvaraju, Goran Stevovic, Amanda Suchanek, Andrea Throop, Lyndon Tilson, Thomas Urban, Joe Voshell, Kimberly Wagner, Jonathan Williams, Mary Williamson, Qian Zeng, Tricia Zwiefelhofer, Clinton R. Paden, Duncan MacCannell |
| EPI_ISL_1798523 | WI State Laboratory of Hygiene | Centers for Disease Control and Prevention Division of Viral Diseases, Pathogen Discovery | Mili Sheth, Sarah Nobles, Jasmine Padilla, Mark Burroughs, Shoshona Le, Katie Dillon, Peter Cook, Clinton R. Paden, Dhwani Batra, Krista Queen, Kristen Knipe, Dakota Howard, Yvette Unoarumhi, Darlene Wagner, Matthew Schmerer, Ben L. Rambo-Martin, Kristine Lacek, Sam Shepard, Alison Laufer Halpin, Dave Wentworth, Vivien Dugan, Suxiang Tong, Justin Lee |
| EPI_ISL_1798525 | IA State Hygienic Laboratory | Centers for Disease Control and Prevention Division of Viral Diseases, Pathogen Discovery | Mili Sheth, Sarah Nobles, Jasmine Padilla, Mark Burroughs, Shoshona Le, Katie Dillon, Peter Cook, Clinton R. Paden, Dhwani Batra, Krista Queen, Kristen Knipe, Dakota Howard, Yvette Unoarumhi, Darlene Wagner, Matthew Schmerer, Ben L. Rambo-Martin, Kristine Lacek, Sam Shepard, Alison Laufer Halpin, Dave Wentworth, Vivien Dugan, Suxiang Tong, Justin Lee |
| EPI_ISL_1798527 | IL Dept. of Public Health Springfield Laboratory | Centers for Disease Control and Prevention Division of Viral Diseases, Pathogen Discovery | Mili Sheth, Sarah Nobles, Jasmine Padilla, Mark Burroughs, Shoshona Le, Katie Dillon, Peter Cook, Clinton R. Paden, Dhwani Batra, Krista Queen, Kristen Knipe, Dakota Howard, Yvette Unoarumhi, Darlene Wagner, Matthew Schmerer, Ben L. Rambo-Martin, Kristine Lacek, Sam Shepard, Alison Laufer Halpin, Dave Wentworth, Vivien Dugan, Suxiang Tong, Justin Lee |
| EPI_ISL_1798528 | WI State Laboratory of Hygiene | Centers for Disease Control and Prevention Division of Viral Diseases, Pathogen Discovery | Mili Sheth, Sarah Nobles, Jasmine Padilla, Mark Burroughs, Shoshona Le, Katie Dillon, Peter Cook, Clinton R. Paden, Dhwani Batra, Krista Queen, Kristen Knipe, Dakota Howard, Yvette Unoarumhi, Darlene Wagner, Matthew Schmerer, Ben L. Rambo-Martin, Kristine Lacek, Sam Shepard, Alison Laufer Halpin, Dave Wentworth, Vivien Dugan, Suxiang Tong, Justin Lee |
| EPI_ISL_1798529 | Laboratory Corporation of America | Centers for Disease Control and Prevention Division of Viral Diseases, Pathogen Discovery | Dakota Howard, Dhwani Batra, Peter W. Cook, Kara Moser, Adrian Paskey, Jason Caravas, Benjamin Rambo-Martin, Shatavia Morrison, Christopher Gulvick, Scott Sammons, Yvette Unoarumhi, Darlene Wagner, Matthew Schmerer, Minoo Agarwal, Eyad Almasri, Debbie Boles, Ayla Burns, Nuthawin Charoensri, Oren Cohen, Susan Countryman, Mary Ann Cristobal, Bobbi Croy, Suzanne Dale, Hrushikesh Deshmukh, Amanda Douglas, Vincent Drouillon, Marcia Eisenberg, Howard Engler, Rama Ghatti, Prashant Gupta, Susan Hicks, Jake Humphrey, Lax Iyer, Manoj Jain, Mohan Kolli, Brian Krueger, Tim Kuphal, Stanley Letovsky, Michael Levandoski, Craig Lukasik, Jonathan Meltzer, Brian Norvell, Mindy Nye, Scott Parker, Christos Petropoulos, John Pruitt, Steven Ragan, Scott Ryan, Mike Sapeta, Jana Schroth, Suresh Babu Selvaraju, Goran Stevovic, Amanda Suchanek, Andrea Throop, Lyndon Tilson, Thomas Urban, Joe Voshell, Kimberly Wagner, Jonathan Williams, Mary Williamson, Qian Zeng, Tricia Zwiefelhofer, Clinton R. Paden, Duncan MacCannell |
| EPI_ISL_1798530 | IL Dept. of Public Health Springfield Laboratory | Centers for Disease Control and Prevention Division of Viral Diseases, Pathogen Discovery | Mili Sheth, Sarah Nobles, Jasmine Padilla, Mark Burroughs, Shoshona Le, Katie Dillon, Peter Cook, Clinton R. Paden, Dhwani Batra, Krista Queen, Kristen Knipe, Dakota Howard, Yvette Unoarumhi, Darlene Wagner, Matthew Schmerer, Ben L. Rambo-Martin, Kristine Lacek, Sam Shepard, Alison Laufer Halpin, Dave Wentworth, Vivien Dugan, Suxiang Tong, Justin Lee |

| | | | |
|---|---|---|---|
| EPI_ISL_1798531 | Laboratory Corporation of America | Centers for Disease Control and Prevention Division of Viral Diseases, Pathogen Discovery | Dakota Howard, Dhwani Batra, Peter W. Cook, Kara Moser, Adrian Paskey, Jason Caravas, Benjamin Rambo-Martin, Shatavia Morrison, Christopher Gulvick, Scott Sammons, Yvette Unoarumhi, Darlene Wagner, Matthew Schmerer, Minoo Agarwal, Eyad Almasri, Debbie Boles, Ayla Burns, Nuthawin Charoensri, Oren Cohen, Susan Countryman, Mary Ann Cristobal, Bobbi Croy, Suzanne Dale, Hrushikesh Deshmukh, Amanda Douglas, Vincent Drouillon, Marcia Eisenberg, Howard Engler, Rama Ghatti, Prashant Gupta, Susan Hicks, Jake Humphrey, Lax Iyer, Manoj Jain, Mohan Kolli, Brian Krueger, Tim Kuphal, Stanley Letovsky, Michael Levandoski, Craig Lukasik, Jonathan Meltzer, Brian Norvell, Mindy Nye, Scott Parker, Christos Petropoulos, John Pruitt, Steven Ragan, Scott Ryan, Mike Sapeta, Jana Schroth, Suresh Babu Selvaraju, Goran Stevovic, Amanda Suchanek, Andrea Throop, Lyndon Tilson, Thomas Urban, Joe Voshell, Kimberly Wagner, Jonathan Williams, Mary Williamson, Qian Zeng, Tricia Zwiefelhofer, Clinton R. Paden, Duncan MacCannell |
| EPI_ISL_1798532, EPI_ISL_1798534 | OH Department of Health Laboratory | Centers for Disease Control and Prevention Division of Viral Diseases, Pathogen Discovery | Mili Sheth, Sarah Nobles, Jasmine Padilla, Mark Burroughs, Shoshona Le, Katie Dillon, Peter Cook, Clinton R. Paden, Dhwani Batra, Krista Queen, Kristen Knipe, Dakota Howard, Yvette Unoarumhi, Darlene Wagner, Matthew Schmerer, Ben L. Rambo-Martin, Kristine Lacek, Sam Shepard, Alison Laufer Halpin, Dave Wentworth, Vivien Dugan, Suxiang Tong, Justin Lee |
| EPI_ISL_1798535 | Laboratory Corporation of America | Centers for Disease Control and Prevention Division of Viral Diseases, Pathogen Discovery | Dakota Howard, Dhwani Batra, Peter W. Cook, Kara Moser, Adrian Paskey, Jason Caravas, Benjamin Rambo-Martin, Shatavia Morrison, Christopher Gulvick, Scott Sammons, Yvette Unoarumhi, Darlene Wagner, Matthew Schmerer, Minoo Agarwal, Eyad Almasri, Debbie Boles, Ayla Burns, Nuthawin Charoensri, Oren Cohen, Susan Countryman, Mary Ann Cristobal, Bobbi Croy, Suzanne Dale, Hrushikesh Deshmukh, Amanda Douglas, Vincent Drouillon, Marcia Eisenberg, Howard Engler, Rama Ghatti, Prashant Gupta, Susan Hicks, Jake Humphrey, Lax Iyer, Manoj Jain, Mohan Kolli, Brian Krueger, Tim Kuphal, Stanley Letovsky, Michael Levandoski, Craig Lukasik, Jonathan Meltzer, Brian Norvell, Mindy Nye, Scott Parker, Christos Petropoulos, John Pruitt, Steven Ragan, Scott Ryan, Mike Sapeta, Jana Schroth, Suresh Babu Selvaraju, Goran Stevovic, Amanda Suchanek, Andrea Throop, Lyndon Tilson, Thomas Urban, Joe Voshell, Kimberly Wagner, Jonathan Williams, Mary Williamson, Qian Zeng, Tricia Zwiefelhofer, Clinton R. Paden, Duncan MacCannell |
| EPI_ISL_1798536 | NE Public Health Laboratory | Centers for Disease Control and Prevention Division of Viral Diseases, Pathogen Discovery | Mili Sheth, Sarah Nobles, Jasmine Padilla, Mark Burroughs, Shoshona Le, Katie Dillon, Peter Cook, Clinton R. Paden, Dhwani Batra, Krista Queen, Kristen Knipe, Dakota Howard, Yvette Unoarumhi, Darlene Wagner, Matthew Schmerer, Ben L. Rambo-Martin, Kristine Lacek, Sam Shepard, Alison Laufer Halpin, Dave Wentworth, Vivien Dugan, Suxiang Tong, Justin Lee |
| EPI_ISL_1798537 | Laboratory Corporation of America | Centers for Disease Control and Prevention Division of Viral Diseases, Pathogen Discovery | Dakota Howard, Dhwani Batra, Peter W. Cook, Kara Moser, Adrian Paskey, Jason Caravas, Benjamin Rambo-Martin, Shatavia Morrison, Christopher Gulvick, Scott Sammons, Yvette Unoarumhi, Darlene Wagner, Matthew Schmerer, Minoo Agarwal, Eyad Almasri, Debbie Boles, Ayla Burns, Nuthawin Charoensri, Oren Cohen, Susan Countryman, Mary Ann Cristobal, Bobbi Croy, Suzanne Dale, Hrushikesh Deshmukh, Amanda Douglas, Vincent Drouillon, Marcia Eisenberg, Howard Engler, Rama Ghatti, Prashant Gupta, Susan Hicks, Jake Humphrey, Lax Iyer, Manoj Jain, Mohan Kolli, Brian Krueger, Tim Kuphal, Stanley Letovsky, Michael Levandoski, Craig Lukasik, Jonathan Meltzer, Brian Norvell, Mindy Nye, Scott Parker, Christos Petropoulos, John Pruitt, Steven Ragan, Scott Ryan, Mike Sapeta, Jana Schroth, Suresh Babu Selvaraju, Goran Stevovic, Amanda Suchanek, Andrea Throop, Lyndon Tilson, Thomas Urban, Joe Voshell, Kimberly Wagner, Jonathan Williams, Mary Williamson, Qian Zeng, Tricia Zwiefelhofer, Clinton R. Paden, Duncan MacCannell |
| EPI_ISL_1798538 | OH Department of Health Laboratory | Centers for Disease Control and Prevention Division of Viral Diseases, Pathogen Discovery | Mili Sheth, Sarah Nobles, Jasmine Padilla, Mark Burroughs, Shoshona Le, Katie Dillon, Peter Cook, Clinton R. Paden, Dhwani Batra, Krista Queen, Kristen Knipe, Dakota Howard, Yvette Unoarumhi, Darlene Wagner, Matthew Schmerer, Ben L. Rambo-Martin, Kristine Lacek, Sam Shepard, Alison Laufer Halpin, Dave Wentworth, Vivien Dugan, Suxiang Tong, Justin Lee |
| EPI_ISL_1798540 | UT-Unified State Labs: Public Health Utah DOH | Centers for Disease Control and Prevention Division of Viral Diseases, Pathogen Discovery | Mili Sheth, Sarah Nobles, Jasmine Padilla, Mark Burroughs, Shoshona Le, Katie Dillon, Peter Cook, Clinton R. Paden, Dhwani Batra, Krista Queen, Kristen Knipe, Dakota Howard, Yvette Unoarumhi, Darlene Wagner, Matthew Schmerer, Ben L. Rambo-Martin, Kristine Lacek, Sam Shepard, Alison Laufer Halpin, Dave Wentworth, Vivien Dugan, Suxiang Tong, Justin Lee |
| EPI_ISL_1798541 | Laboratory Corporation of America | Centers for Disease Control and Prevention Division of Viral Diseases, Pathogen Discovery | Dakota Howard, Dhwani Batra, Peter W. Cook, Kara Moser, Adrian Paskey, Jason Caravas, Benjamin Rambo-Martin, Shatavia Morrison, Christopher Gulvick, Scott Sammons, Yvette Unoarumhi, Darlene Wagner, Matthew Schmerer, Minoo Agarwal, Eyad Almasri, Debbie Boles, Ayla Burns, Nuthawin Charoensri, Oren Cohen, Susan Countryman, Mary Ann Cristobal, Bobbi Croy, Suzanne Dale, Hrushikesh Deshmukh, Amanda Douglas, Vincent Drouillon, Marcia Eisenberg, Howard Engler, Rama Ghatti, Prashant Gupta, Susan Hicks, Jake Humphrey, Lax Iyer, Manoj Jain, Mohan Kolli, Brian Krueger, Tim Kuphal, Stanley Letovsky, Michael Levandoski, Craig Lukasik, Jonathan Meltzer, Brian Norvell, Mindy Nye, Scott Parker, Christos Petropoulos, John Pruitt, Steven Ragan, Scott Ryan, Mike Sapeta, Jana Schroth, Suresh Babu Selvaraju, Goran Stevovic, Amanda Suchanek, Andrea Throop, Lyndon Tilson, Thomas Urban, Joe Voshell, Kimberly Wagner, Jonathan Williams, Mary Williamson, Qian Zeng, Tricia Zwiefelhofer, Clinton R. Paden, Duncan MacCannell |
| EPI_ISL_1798542 | NE Public Health Laboratory | Centers for Disease Control and Prevention Division of Viral Diseases, Pathogen Discovery | Mili Sheth, Sarah Nobles, Jasmine Padilla, Mark Burroughs, Shoshona Le, Katie Dillon, Peter Cook, Clinton R. Paden, Dhwani Batra, Krista Queen, Kristen Knipe, Dakota Howard, Yvette Unoarumhi, Darlene Wagner, Matthew Schmerer, Ben L. Rambo-Martin, Kristine Lacek, Sam Shepard, Alison Laufer Halpin, Dave Wentworth, Vivien Dugan, Suxiang Tong, Justin Lee |
| EPI_ISL_1798544 | Laboratory Corporation of America | Centers for Disease Control and Prevention Division of Viral Diseases, Pathogen Discovery | Dakota Howard, Dhwani Batra, Peter W. Cook, Kara Moser, Adrian Paskey, Jason Caravas, Benjamin Rambo-Martin, Shatavia Morrison, Christopher Gulvick, Scott Sammons, Yvette Unoarumhi, Darlene Wagner, Matthew Schmerer, Minoo Agarwal, Eyad Almasri, Debbie Boles, Ayla Burns, Nuthawin Charoensri, Oren Cohen, Susan Countryman, Mary Ann Cristobal, Bobbi Croy, Suzanne Dale, Hrushikesh Deshmukh, Amanda Douglas, Vincent Drouillon, Marcia Eisenberg, Howard Engler, Rama Ghatti, Prashant Gupta, Susan Hicks, Jake Humphrey, Lax Iyer, Manoj Jain, Mohan Kolli, Brian Krueger, Tim Kuphal, Stanley Letovsky, Michael Levandoski, Craig Lukasik, Jonathan Meltzer, Brian Norvell, Mindy Nye, Scott Parker, Christos Petropoulos, John Pruitt, Steven Ragan, Scott Ryan, Mike Sapeta, Jana Schroth, Suresh Babu Selvaraju, Goran Stevovic, Amanda Suchanek, Andrea Throop, Lyndon Tilson, Thomas Urban, Joe Voshell, Kimberly Wagner, Jonathan Williams, Mary Williamson, Qian Zeng, Tricia Zwiefelhofer, Clinton R. Paden, Duncan MacCannell |
| EPI_ISL_1798545 | OH Department of Health Laboratory | Centers for Disease Control and Prevention Division of Viral Diseases, Pathogen Discovery | Mili Sheth, Sarah Nobles, Jasmine Padilla, Mark Burroughs, Shoshona Le, Katie Dillon, Peter Cook, Clinton R. Paden, Dhwani Batra, Krista Queen, Kristen Knipe, Dakota Howard, Yvette Unoarumhi, Darlene Wagner, Matthew Schmerer, Ben L. Rambo-Martin, Kristine Lacek, Sam Shepard, Alison Laufer Halpin, Dave Wentworth, Vivien Dugan, Suxiang Tong, Justin Lee |
| EPI_ISL_1798546 | Laboratory Corporation of America | Centers for Disease Control and Prevention Division of Viral Diseases, Pathogen Discovery | Dakota Howard, Dhwani Batra, Peter W. Cook, Kara Moser, Adrian Paskey, Jason Caravas, Benjamin Rambo-Martin, Shatavia Morrison, Christopher Gulvick, Scott Sammons, Yvette Unoarumhi, Darlene Wagner, Matthew Schmerer, Minoo Agarwal, Eyad Almasri, Debbie Boles, Ayla Burns, Nuthawin Charoensri, Oren Cohen, Susan Countryman, Mary Ann Cristobal, Bobbi Croy, Suzanne Dale, Hrushikesh Deshmukh, Amanda Douglas, Vincent Drouillon, Marcia Eisenberg, Howard Engler, Rama Ghatti, Prashant Gupta, Susan Hicks, Jake Humphrey, Lax Iyer, Manoj Jain, Mohan Kolli, Brian Krueger, Tim Kuphal, Stanley Letovsky, Michael Levandoski, Craig Lukasik, Jonathan Meltzer, Brian Norvell, Mindy Nye, Scott Parker, Christos Petropoulos, John Pruitt, Steven Ragan, Scott Ryan, Mike Sapeta, Jana Schroth, Suresh Babu Selvaraju, Goran Stevovic, Amanda Suchanek, Andrea Throop, Lyndon Tilson, Thomas Urban, Joe Voshell, Kimberly Wagner, Jonathan Williams, Mary Williamson, Qian Zeng, Tricia Zwiefelhofer, Clinton R. Paden, Duncan MacCannell |
| EPI_ISL_1798547 | OH Department of Health Laboratory | Centers for Disease Control and Prevention Division of Viral Diseases, Pathogen Discovery | Mili Sheth, Sarah Nobles, Jasmine Padilla, Mark Burroughs, Shoshona Le, Katie Dillon, Peter Cook, Clinton R. Paden, Dhwani Batra, Krista Queen, Kristen Knipe, Dakota Howard, Yvette Unoarumhi, Darlene Wagner, Matthew Schmerer, Ben L. Rambo-Martin, Kristine Lacek, Sam Shepard, Alison Laufer Halpin, Dave Wentworth, Vivien Dugan, Suxiang Tong, Justin Lee |
| EPI_ISL_1798548 | Laboratory Corporation of America | Centers for Disease Control and Prevention Division of Viral Diseases, Pathogen Discovery | Dakota Howard, Dhwani Batra, Peter W. Cook, Kara Moser, Adrian Paskey, Jason Caravas, Benjamin Rambo-Martin, Shatavia Morrison, Christopher Gulvick, Scott Sammons, Yvette Unoarumhi, Darlene Wagner, Matthew Schmerer, Minoo Agarwal, Eyad Almasri, Debbie Boles, Ayla Burns, Nuthawin Charoensri, Oren Cohen, Susan Countryman, Mary Ann Cristobal, Bobbi Croy, Suzanne Dale, Hrushikesh Deshmukh, Amanda Douglas, Vincent Drouillon, Marcia Eisenberg, Howard Engler, Rama Ghatti, Prashant Gupta, Susan Hicks, Jake Humphrey, Lax Iyer, Manoj Jain, Mohan Kolli, Brian Krueger, Tim Kuphal, Stanley Letovsky, Michael Levandoski, Craig Lukasik, Jonathan Meltzer, Brian Norvell, Mindy Nye, Scott Parker, Christos Petropoulos, John Pruitt, Steven Ragan, Scott Ryan, Mike Sapeta, Jana Schroth, Suresh Babu Selvaraju, Goran Stevovic, Amanda Suchanek, Andrea Throop, Lyndon Tilson, Thomas Urban, Joe Voshell, Kimberly Wagner, Jonathan Williams, Mary Williamson, Qian Zeng, Tricia Zwiefelhofer, Clinton R. Paden, Duncan MacCannell |
| EPI_ISL_1798549 | OH Department of Health Laboratory | Centers for Disease Control and Prevention Division of Viral Diseases, Pathogen Discovery | Mili Sheth, Sarah Nobles, Jasmine Padilla, Mark Burroughs, Shoshona Le, Katie Dillon, Peter Cook, Clinton R. Paden, Dhwani Batra, Krista Queen, Kristen Knipe, Dakota Howard, Yvette Unoarumhi, Darlene Wagner, Matthew Schmerer, Ben L. Rambo-Martin, Kristine Lacek, Sam Shepard, Alison Laufer Halpin, Dave Wentworth, Vivien Dugan, Suxiang Tong, Justin Lee |
| EPI_ISL_1798551 | NE Public Health Laboratory | Centers for Disease Control and Prevention Division of Viral Diseases, Pathogen Discovery | Mili Sheth, Sarah Nobles, Jasmine Padilla, Mark Burroughs, Shoshona Le, Katie Dillon, Peter Cook, Clinton R. Paden, Dhwani Batra, Krista Queen, Kristen Knipe, Dakota Howard, Yvette Unoarumhi, Darlene Wagner, Matthew Schmerer, Ben L. Rambo-Martin, Kristine Lacek, Sam Shepard, Alison Laufer Halpin, Dave Wentworth, Vivien Dugan, Suxiang Tong, Justin Lee |
| EPI_ISL_1798552 | Laboratory Corporation of America | Centers for Disease Control and Prevention Division of Viral Diseases, Pathogen Discovery | Dakota Howard, Dhwani Batra, Peter W. Cook, Kara Moser, Adrian Paskey, Jason Caravas, Benjamin Rambo-Martin, Shatavia Morrison, Christopher Gulvick, Scott Sammons, Yvette Unoarumhi, Darlene Wagner, Matthew Schmerer, Minoo Agarwal, Eyad Almasri, Debbie Boles, Ayla Burns, Nuthawin Charoensri, Oren Cohen, Susan Countryman, Mary Ann Cristobal, Bobbi Croy, Suzanne Dale, Hrushikesh Deshmukh, Amanda Douglas, Vincent Drouillon, Marcia Eisenberg, Howard Engler, Rama Ghatti, Prashant Gupta, Susan Hicks, Jake Humphrey, Lax Iyer, Manoj Jain, Mohan Kolli, Brian Krueger, Tim Kuphal, Stanley Letovsky, Michael Levandoski, Craig Lukasik, Jonathan Meltzer, Brian Norvell, Mindy Nye, Scott Parker, Christos Petropoulos, John Pruitt, Steven Ragan, Scott Ryan, Mike Sapeta, Jana Schroth, Suresh Babu Selvaraju, Goran Stevovic, Amanda Suchanek, Andrea Throop, Lyndon Tilson, Thomas Urban, Joe Voshell, Kimberly Wagner, Jonathan Williams, Mary Williamson, Qian Zeng, Tricia Zwiefelhofer, Clinton R. Paden, Duncan MacCannell |
| EPI_ISL_1798553 | IA State Hygienic Laboratory | Centers for Disease Control and Prevention Division of Viral Diseases, Pathogen Discovery | Mili Sheth, Sarah Nobles, Jasmine Padilla, Mark Burroughs, Shoshona Le, Katie Dillon, Peter Cook, Clinton R. Paden, Dhwani Batra, Krista Queen, Kristen Knipe, Dakota Howard, Yvette Unoarumhi, Darlene Wagner, Matthew Schmerer, Ben L. Rambo-Martin, Kristine Lacek, Sam Shepard, Alison Laufer Halpin, Dave Wentworth, Vivien Dugan, Suxiang Tong, Justin Lee |
| EPI_ISL_1798554 | OH Department of Health Laboratory | Centers for Disease Control and Prevention Division of Viral Diseases, Pathogen Discovery | Mili Sheth, Sarah Nobles, Jasmine Padilla, Mark Burroughs, Shoshona Le, Katie Dillon, Peter Cook, Clinton R. Paden, Dhwani Batra, Krista Queen, Kristen Knipe, Dakota Howard, Yvette Unoarumhi, Darlene Wagner, Matthew Schmerer, Ben L. Rambo-Martin, Kristine Lacek, Sam Shepard, Alison Laufer Halpin, Dave Wentworth, Vivien Dugan, Suxiang Tong, Justin Lee |
| EPI_ISL_1798555 | AZ SPHL, Arizona Department of Health Services | Centers for Disease Control and Prevention Division of Viral Diseases, Pathogen Discovery | Mili Sheth, Sarah Nobles, Jasmine Padilla, Mark Burroughs, Shoshona Le, Katie Dillon, Peter Cook, Clinton R. Paden, Dhwani Batra, Krista Queen, Kristen Knipe, Dakota Howard, Yvette Unoarumhi, Darlene Wagner, Matthew Schmerer, Ben L. Rambo-Martin, Kristine Lacek, Sam Shepard, Alison Laufer Halpin, Dave Wentworth, Vivien Dugan, Suxiang Tong, Justin Lee |
| EPI_ISL_1798556 | Laboratory Corporation of America | Centers for Disease Control and Prevention Division of Viral Diseases, Pathogen Discovery | Dakota Howard, Dhwani Batra, Peter W. Cook, Kara Moser, Adrian Paskey, Jason Caravas, Benjamin Rambo-Martin, Shatavia Morrison, Christopher Gulvick, Scott Sammons, Yvette Unoarumhi, Darlene Wagner, Matthew Schmerer, Minoo Agarwal, Eyad Almasri, Debbie Boles, Ayla Burns, Nuthawin Charoensri, Oren Cohen, Susan Countryman, Mary Ann Cristobal, Bobbi Croy, Suzanne Dale, Hrushikesh Deshmukh, Amanda Douglas, Vincent Drouillon, Marcia Eisenberg, Howard Engler, Rama Ghatti, Prashant Gupta, Susan Hicks, Jake Humphrey, Lax Iyer, Manoj Jain, Mohan Kolli, Brian Krueger, Tim Kuphal, Stanley Letovsky, Michael Levandoski, Craig Lukasik, Jonathan Meltzer, Brian Norvell, Mindy Nye, Scott Parker, Christos Petropoulos, John Pruitt, Steven Ragan, Scott Ryan, Mike Sapeta, Jana Schroth, Suresh Babu Selvaraju, Goran Stevovic, Amanda Suchanek, Andrea Throop, Lyndon Tilson, Thomas Urban, Joe Voshell, Kimberly Wagner, Jonathan Williams, Mary Williamson, Qian Zeng, Tricia Zwiefelhofer, Clinton R. Paden, Duncan MacCannell |
| EPI_ISL_1798557 | IL Dept. of Public Health Springfield Laboratory | Centers for Disease Control and Prevention Division of Viral Diseases, Pathogen Discovery | Mili Sheth, Sarah Nobles, Jasmine Padilla, Mark Burroughs, Shoshona Le, Katie Dillon, Peter Cook, Clinton R. Paden, Dhwani Batra, Krista Queen, Kristen Knipe, Dakota Howard, Yvette Unoarumhi, Darlene Wagner, Matthew Schmerer, Ben L. Rambo-Martin, Kristine Lacek, Sam Shepard, Alison Laufer Halpin, Dave Wentworth, Vivien Dugan, Suxiang Tong, Justin Lee |
| EPI_ISL_1798558 | Laboratory Corporation of America | Centers for Disease Control and Prevention Division of Viral Diseases, Pathogen Discovery | Dakota Howard, Dhwani Batra, Peter W. Cook, Kara Moser, Adrian Paskey, Jason Caravas, Benjamin Rambo-Martin, Shatavia Morrison, Christopher Gulvick, Scott Sammons, Yvette Unoarumhi, Darlene Wagner, Matthew Schmerer, Minoo Agarwal, Eyad Almasri, Debbie Boles, Ayla Burns, Nuthawin Charoensri, Oren Cohen, Susan Countryman, Mary Ann Cristobal, Bobbi Croy, Suzanne Dale, Hrushikesh Deshmukh, Amanda Douglas, Vincent Drouillon, Marcia Eisenberg, Howard Engler, Rama Ghatti, Prashant Gupta, Susan Hicks, Jake Humphrey, Lax Iyer, Manoj Jain, Mohan Kolli, Brian Krueger, Tim Kuphal, Stanley Letovsky, Michael Levandoski, Craig Lukasik, Jonathan Meltzer, Brian Norvell, Mindy Nye, Scott Parker, Christos Petropoulos, John Pruitt, Steven Ragan, Scott Ryan, Mike Sapeta, Jana Schroth, Suresh Babu Selvaraju, Goran Stevovic, Amanda Suchanek, Andrea Throop, Lyndon Tilson, Thomas Urban, Joe Voshell, Kimberly Wagner, Jonathan Williams, Mary Williamson, Qian Zeng, Tricia Zwiefelhofer, Clinton R. Paden, Duncan MacCannell |
| EPI_ISL_1798559 | WI State Laboratory of Hygiene | Centers for Disease Control and Prevention Division of Viral Diseases, Pathogen Discovery | Mili Sheth, Sarah Nobles, Jasmine Padilla, Mark Burroughs, Shoshona Le, Katie Dillon, Peter Cook, Clinton R. Paden, Dhwani Batra, Krista Queen, Kristen Knipe, Dakota Howard, Yvette Unoarumhi, Darlene Wagner, Matthew Schmerer, Ben L. Rambo-Martin, Kristine Lacek, Sam Shepard, Alison Laufer Halpin, Dave Wentworth, Vivien Dugan, Suxiang Tong, Justin Lee |
| EPI_ISL_1798560 | Laboratory Corporation of America | Centers for Disease Control and Prevention Division of Viral Diseases, Pathogen Discovery | Dakota Howard, Dhwani Batra, Peter W. Cook, Kara Moser, Adrian Paskey, Jason Caravas, Benjamin Rambo-Martin, Shatavia Morrison, Christopher Gulvick, Scott Sammons, Yvette Unoarumhi, Darlene Wagner, Matthew Schmerer, Minoo Agarwal, Eyad Almasri, Debbie Boles, Ayla Burns, Nuthawin Charoensri, Oren Cohen, Susan Countryman, Mary Ann Cristobal, Bobbi Croy, Suzanne Dale, Hrushikesh Deshmukh, Amanda Douglas, Vincent Drouillon, Marcia Eisenberg, Howard Engler, Rama Ghatti, Prashant Gupta, Susan Hicks, Jake Humphrey, Lax Iyer, Manoj Jain, Mohan Kolli, Brian Krueger, Tim Kuphal, Stanley Letovsky, Michael Levandoski, Craig Lukasik, Jonathan Meltzer, Brian Norvell, Mindy Nye, Scott Parker, Christos Petropoulos, John Pruitt, Steven Ragan, Scott Ryan, Mike Sapeta, Jana Schroth, Suresh Babu Selvaraju, Goran Stevovic, Amanda Suchanek, Andrea Throop, Lyndon Tilson, Thomas Urban, Joe Voshell, Kimberly Wagner, Jonathan Williams, Mary Williamson, Qian Zeng, Tricia Zwiefelhofer, Clinton R. Paden, Duncan MacCannell |
| EPI_ISL_1798561 | IL Dept. of Public Health Springfield Laboratory | Centers for Disease Control and Prevention Division of Viral Diseases, Pathogen Discovery | Mili Sheth, Sarah Nobles, Jasmine Padilla, Mark Burroughs, Shoshona Le, Katie Dillon, Peter Cook, Clinton R. Paden, Dhwani Batra, Krista Queen, Kristen Knipe, Dakota Howard, Yvette Unoarumhi, Darlene Wagner, Matthew Schmerer, Ben L. Rambo-Martin, Kristine Lacek, Sam Shepard, Alison Laufer Halpin, Dave Wentworth, Vivien Dugan, Suxiang Tong, Justin Lee |
| EPI_ISL_1798562 | Laboratory Corporation of America | Centers for Disease Control and Prevention Division of Viral Diseases, Pathogen Discovery | Dakota Howard, Dhwani Batra, Peter W. Cook, Kara Moser, Adrian Paskey, Jason Caravas, Benjamin Rambo-Martin, Shatavia Morrison, Christopher Gulvick, Scott Sammons, Yvette Unoarumhi, Darlene Wagner, Matthew Schmerer, Minoo Agarwal, Eyad Almasri, Debbie Boles, Ayla Burns, Nuthawin Charoensri, Oren Cohen, Susan Countryman, Mary Ann Cristobal, Bobbi Croy, Suzanne Dale, Hrushikesh Deshmukh, Amanda Douglas, Vincent Drouillon, Marcia Eisenberg, Howard Engler, Rama Ghatti, Prashant Gupta, Susan Hicks, Jake Humphrey, Lax Iyer, Manoj Jain, Mohan Kolli, Brian Krueger, Tim Kuphal, Stanley Letovsky, Michael Levandoski, Craig Lukasik, Jonathan Meltzer, Brian Norvell, Mindy Nye, Scott Parker, Christos Petropoulos, John Pruitt, Steven Ragan, Scott Ryan, Mike Sapeta, Jana Schroth, Suresh Babu Selvaraju, Goran Stevovic, Amanda Suchanek, Andrea Throop, Lyndon Tilson, Thomas Urban, Joe Voshell, Kimberly Wagner, Jonathan Williams, Mary Williamson, Qian Zeng, Tricia Zwiefelhofer, Clinton R. Paden, Duncan MacCannell |
| EPI_ISL_1798563 | OH Department of Health Laboratory | Centers for Disease Control and Prevention Division of Viral Diseases, Pathogen Discovery | Mili Sheth, Sarah Nobles, Jasmine Padilla, Mark Burroughs, Shoshona Le, Katie Dillon, Peter Cook, Clinton R. Paden, Dhwani Batra, Krista Queen, Kristen Knipe, Dakota Howard, Yvette Unoarumhi, Darlene Wagner, Matthew Schmerer, Ben L. Rambo-Martin, Kristine Lacek, Sam Shepard, Alison Laufer Halpin, Dave Wentworth, Vivien Dugan, Suxiang Tong, Justin Lee |
| EPI_ISL_1798564 | Laboratory Corporation of America | Centers for Disease Control and Prevention Division of Viral Diseases, Pathogen | Dakota Howard, Dhwani Batra, Peter W. Cook, Kara Moser, Adrian Paskey, Jason Caravas, Benjamin Rambo-Martin, Shatavia Morrison, Christopher Gulvick, Scott Sammons, Yvette Unoarumhi, Darlene Wagner, Matthew Schmerer, Minoo Agarwal, Eyad Almasri, Debbie Boles, Ayla Burns, Nuthawin Charoensri, Oren Cohen, Susan Countryman, Mary Ann Cristobal, Bobbi Croy, Suzanne Dale, Hrushikesh Deshmukh, Amanda Douglas, Vincent Drouillon, Marcia Eisenberg, Howard Engler, Rama Ghatti, Prashant Gupta, Susan Hicks, Jake Humphrey, Lax Iyer, Manoj Jain, Mohan |

| | | | |
|---|---|---|---|
| | | Discovery | Kolli, Brian Krueger, Tim Kuphal, Stanley Letovsky, Michael Levandoski, Craig Lukasik, Jonathan Meltzer, Brian Norvell, Mindy Nye, Scott Parker, Christos Petropoulos, John Pruitt, Steven Ragan, Scott Ryan, Mike Sapeta, Jana Schroth, Suresh Babu Selvaraju, Goran Stevovic, Amanda Suchanek, Andrea Throop, Lyndon Tilson, Thomas Urban, Joe Voshell, Kimberly Wagner, Jonathan Williams, Mary Williamson, Qian Zeng, Tricia Zwiefelhofer, Clinton R. Paden, Duncan MacCannell |
| EPI_ISL_1798565 | UT-Unified State Labs: Public Health Utah DOH | Centers for Disease Control and Prevention Division of Viral Diseases, Pathogen Discovery | Mili Sheth, Sarah Nobles, Jasmine Padilla, Mark Burroughs, Shoshona Le, Katie Dillon, Peter Cook, Clinton R. Paden, Dhwani Batra, Krista Queen, Kristen Knipe, Dakota Howard, Yvette Unoarumhi, Darlene Wagner, Matthew Schmerer, Ben L. Rambo-Martin, Kristine Lacek, Sam Shepard, Alison Laufer Halpin, Dave Wentworth, Vivien Dugan, Suxiang Tong, Justin Lee |
| EPI_ISL_1798566 | OH Department of Health Laboratory | Centers for Disease Control and Prevention Division of Viral Diseases, Pathogen Discovery | Mili Sheth, Sarah Nobles, Jasmine Padilla, Mark Burroughs, Shoshona Le, Katie Dillon, Peter Cook, Clinton R. Paden, Dhwani Batra, Krista Queen, Kristen Knipe, Dakota Howard, Yvette Unoarumhi, Darlene Wagner, Matthew Schmerer, Ben L. Rambo-Martin, Kristine Lacek, Sam Shepard, Alison Laufer Halpin, Dave Wentworth, Vivien Dugan, Suxiang Tong, Justin Lee |
| EPI_ISL_1798567 | Laboratory Corporation of America | Centers for Disease Control and Prevention Division of Viral Diseases, Pathogen Discovery | Dakota Howard, Dhwani Batra, Peter W. Cook, Kara Moser, Adrian Paskey, Jason Caravas, Benjamin Rambo-Martin, Shatavia Morrison, Christopher Gulvick, Scott Sammons, Yvette Unoarumhi, Darlene Wagner, Matthew Schmerer, Minoo Agarwal, Eyad Almasri, Debbie Boles, Ayla Burns, Nuthawin Charoensri, Oren Cohen, Susan Countryman, Mary Ann Cristobal, Bobbi Croy, Suzanne Dale, Hrushikesh Deshmukh, Amanda Douglas, Vincent Drouillon, Marcia Eisenberg, Howard Engler, Rama Ghatti, Prashant Gupta, Susan Hicks, Jake Humphrey, Lax Iyer, Manoj Jain, Mohan Kolli, Brian Krueger, Tim Kuphal, Stanley Letovsky, Michael Levandoski, Craig Lukasik, Jonathan Meltzer, Brian Norvell, Mindy Nye, Scott Parker, Christos Petropoulos, John Pruitt, Steven Ragan, Scott Ryan, Mike Sapeta, Jana Schroth, Suresh Babu Selvaraju, Goran Stevovic, Amanda Suchanek, Andrea Throop, Lyndon Tilson, Thomas Urban, Joe Voshell, Kimberly Wagner, Jonathan Williams, Mary Williamson, Qian Zeng, Tricia Zwiefelhofer, Clinton R. Paden, Duncan MacCannell |
| EPI_ISL_1798568 | NE Public Health Laboratory | Centers for Disease Control and Prevention Division of Viral Diseases, Pathogen Discovery | Mili Sheth, Sarah Nobles, Jasmine Padilla, Mark Burroughs, Shoshona Le, Katie Dillon, Peter Cook, Clinton R. Paden, Dhwani Batra, Krista Queen, Kristen Knipe, Dakota Howard, Yvette Unoarumhi, Darlene Wagner, Matthew Schmerer, Ben L. Rambo-Martin, Kristine Lacek, Sam Shepard, Alison Laufer Halpin, Dave Wentworth, Vivien Dugan, Suxiang Tong, Justin Lee |
| EPI_ISL_1798569, EPI_ISL_1798570 | Laboratory Corporation of America | Centers for Disease Control and Prevention Division of Viral Diseases, Pathogen Discovery | Dakota Howard, Dhwani Batra, Peter W. Cook, Kara Moser, Adrian Paskey, Jason Caravas, Benjamin Rambo-Martin, Shatavia Morrison, Christopher Gulvick, Scott Sammons, Yvette Unoarumhi, Darlene Wagner, Matthew Schmerer, Minoo Agarwal, Eyad Almasri, Debbie Boles, Ayla Burns, Nuthawin Charoensri, Oren Cohen, Susan Countryman, Mary Ann Cristobal, Bobbi Croy, Suzanne Dale, Hrushikesh Deshmukh, Amanda Douglas, Vincent Drouillon, Marcia Eisenberg, Howard Engler, Rama Ghatti, Prashant Gupta, Susan Hicks, Jake Humphrey, Lax Iyer, Manoj Jain, Mohan Kolli, Brian Krueger, Tim Kuphal, Stanley Letovsky, Michael Levandoski, Craig Lukasik, Jonathan Meltzer, Brian Norvell, Mindy Nye, Scott Parker, Christos Petropoulos, John Pruitt, Steven Ragan, Scott Ryan, Mike Sapeta, Jana Schroth, Suresh Babu Selvaraju, Goran Stevovic, Amanda Suchanek, Andrea Throop, Lyndon Tilson, Thomas Urban, Joe Voshell, Kimberly Wagner, Jonathan Williams, Mary Williamson, Qian Zeng, Tricia Zwiefelhofer, Clinton R. Paden, Duncan MacCannell |
| EPI_ISL_1798571 | IL Dept. of Public Health Springfield Laboratory | Centers for Disease Control and Prevention Division of Viral Diseases, Pathogen Discovery | Mili Sheth, Sarah Nobles, Jasmine Padilla, Mark Burroughs, Shoshona Le, Katie Dillon, Peter Cook, Clinton R. Paden, Dhwani Batra, Krista Queen, Kristen Knipe, Dakota Howard, Yvette Unoarumhi, Darlene Wagner, Matthew Schmerer, Ben L. Rambo-Martin, Kristine Lacek, Sam Shepard, Alison Laufer Halpin, Dave Wentworth, Vivien Dugan, Suxiang Tong, Justin Lee |
| EPI_ISL_1798572 | Laboratory Corporation of America | Centers for Disease Control and Prevention Division of Viral Diseases, Pathogen Discovery | Dakota Howard, Dhwani Batra, Peter W. Cook, Kara Moser, Adrian Paskey, Jason Caravas, Benjamin Rambo-Martin, Shatavia Morrison, Christopher Gulvick, Scott Sammons, Yvette Unoarumhi, Darlene Wagner, Matthew Schmerer, Minoo Agarwal, Eyad Almasri, Debbie Boles, Ayla Burns, Nuthawin Charoensri, Oren Cohen, Susan Countryman, Mary Ann Cristobal, Bobbi Croy, Suzanne Dale, Hrushikesh Deshmukh, Amanda Douglas, Vincent Drouillon, Marcia Eisenberg, Howard Engler, Rama Ghatti, Prashant Gupta, Susan Hicks, Jake Humphrey, Lax Iyer, Manoj Jain, Mohan Kolli, Brian Krueger, Tim Kuphal, Stanley Letovsky, Michael Levandoski, Craig Lukasik, Jonathan Meltzer, Brian Norvell, Mindy Nye, Scott Parker, Christos Petropoulos, John Pruitt, Steven Ragan, Scott Ryan, Mike Sapeta, Jana Schroth, Suresh Babu Selvaraju, Goran Stevovic, Amanda Suchanek, Andrea Throop, Lyndon Tilson, Thomas Urban, Joe Voshell, Kimberly Wagner, Jonathan Williams, Mary Williamson, Qian Zeng, Tricia Zwiefelhofer, Clinton R. Paden, Duncan MacCannell |
| EPI_ISL_1798573 | WI State Laboratory of Hygiene | Centers for Disease Control and Prevention Division of Viral Diseases, Pathogen Discovery | Mili Sheth, Sarah Nobles, Jasmine Padilla, Mark Burroughs, Shoshona Le, Katie Dillon, Peter Cook, Clinton R. Paden, Dhwani Batra, Krista Queen, Kristen Knipe, Dakota Howard, Yvette Unoarumhi, Darlene Wagner, Matthew Schmerer, Ben L. Rambo-Martin, Kristine Lacek, Sam Shepard, Alison Laufer Halpin, Dave Wentworth, Vivien Dugan, Suxiang Tong, Justin Lee |
| EPI_ISL_1798574 | Laboratory Corporation of America | Centers for Disease Control and Prevention Division of Viral Diseases, Pathogen Discovery | Dakota Howard, Dhwani Batra, Peter W. Cook, Kara Moser, Adrian Paskey, Jason Caravas, Benjamin Rambo-Martin, Shatavia Morrison, Christopher Gulvick, Scott Sammons, Yvette Unoarumhi, Darlene Wagner, Matthew Schmerer, Minoo Agarwal, Eyad Almasri, Debbie Boles, Ayla Burns, Nuthawin Charoensri, Oren Cohen, Susan Countryman, Mary Ann Cristobal, Bobbi Croy, Suzanne Dale, Hrushikesh Deshmukh, Amanda Douglas, Vincent Drouillon, Marcia Eisenberg, Howard Engler, Rama Ghatti, Prashant Gupta, Susan Hicks, Jake Humphrey, Lax Iyer, Manoj Jain, Mohan Kolli, Brian Krueger, Tim Kuphal, Stanley Letovsky, Michael Levandoski, Craig Lukasik, Jonathan Meltzer, Brian Norvell, Mindy Nye, Scott Parker, Christos Petropoulos, John Pruitt, Steven Ragan, Scott Ryan, Mike Sapeta, Jana Schroth, Suresh Babu Selvaraju, Goran Stevovic, Amanda Suchanek, Andrea Throop, Lyndon Tilson, Thomas Urban, Joe Voshell, Kimberly Wagner, Jonathan Williams, Mary Williamson, Qian Zeng, Tricia Zwiefelhofer, Clinton R. Paden, Duncan MacCannell |
| EPI_ISL_1798575 | IL Dept. of Public Health Springfield Laboratory | Centers for Disease Control and Prevention Division of Viral Diseases, Pathogen Discovery | Mili Sheth, Sarah Nobles, Jasmine Padilla, Mark Burroughs, Shoshona Le, Katie Dillon, Peter Cook, Clinton R. Paden, Dhwani Batra, Krista Queen, Kristen Knipe, Dakota Howard, Yvette Unoarumhi, Darlene Wagner, Matthew Schmerer, Ben L. Rambo-Martin, Kristine Lacek, Sam Shepard, Alison Laufer Halpin, Dave Wentworth, Vivien Dugan, Suxiang Tong, Justin Lee |
| EPI_ISL_1798576 | Laboratory Corporation of America | Centers for Disease Control and Prevention Division of Viral Diseases, Pathogen Discovery | Dakota Howard, Dhwani Batra, Peter W. Cook, Kara Moser, Adrian Paskey, Jason Caravas, Benjamin Rambo-Martin, Shatavia Morrison, Christopher Gulvick, Scott Sammons, Yvette Unoarumhi, Darlene Wagner, Matthew Schmerer, Minoo Agarwal, Eyad Almasri, Debbie Boles, Ayla Burns, Nuthawin Charoensri, Oren Cohen, Susan Countryman, Mary Ann Cristobal, Bobbi Croy, Suzanne Dale, Hrushikesh Deshmukh, Amanda Douglas, Vincent Drouillon, Marcia Eisenberg, Howard Engler, Rama Ghatti, Prashant Gupta, Susan Hicks, Jake Humphrey, Lax Iyer, Manoj Jain, Mohan Kolli, Brian Krueger, Tim Kuphal, Stanley Letovsky, Michael Levandoski, Craig Lukasik, Jonathan Meltzer, Brian Norvell, Mindy Nye, Scott Parker, Christos Petropoulos, John Pruitt, Steven Ragan, Scott Ryan, Mike Sapeta, Jana Schroth, Suresh Babu Selvaraju, Goran Stevovic, Amanda Suchanek, Andrea Throop, Lyndon Tilson, Thomas Urban, Joe Voshell, Kimberly Wagner, Jonathan Williams, Mary Williamson, Qian Zeng, Tricia Zwiefelhofer, Clinton R. Paden, Duncan MacCannell |
| EPI_ISL_1798577 | IA State Hygienic Laboratory | Centers for Disease Control and Prevention Division of Viral Diseases, Pathogen Discovery | Mili Sheth, Sarah Nobles, Jasmine Padilla, Mark Burroughs, Shoshona Le, Katie Dillon, Peter Cook, Clinton R. Paden, Dhwani Batra, Krista Queen, Kristen Knipe, Dakota Howard, Yvette Unoarumhi, Darlene Wagner, Matthew Schmerer, Ben L. Rambo-Martin, Kristine Lacek, Sam Shepard, Alison Laufer Halpin, Dave Wentworth, Vivien Dugan, Suxiang Tong, Justin Lee |
| EPI_ISL_1798578 | Laboratory Corporation of America | Centers for Disease Control and Prevention Division of Viral Diseases, Pathogen Discovery | Dakota Howard, Dhwani Batra, Peter W. Cook, Kara Moser, Adrian Paskey, Jason Caravas, Benjamin Rambo-Martin, Shatavia Morrison, Christopher Gulvick, Scott Sammons, Yvette Unoarumhi, Darlene Wagner, Matthew Schmerer, Minoo Agarwal, Eyad Almasri, Debbie Boles, Ayla Burns, Nuthawin Charoensri, Oren Cohen, Susan Countryman, Mary Ann Cristobal, Bobbi Croy, Suzanne Dale, Hrushikesh Deshmukh, Amanda Douglas, Vincent Drouillon, Marcia Eisenberg, Howard Engler, Rama Ghatti, Prashant Gupta, Susan Hicks, Jake Humphrey, Lax Iyer, Manoj Jain, Mohan Kolli, Brian Krueger, Tim Kuphal, Stanley Letovsky, Michael Levandoski, Craig Lukasik, Jonathan Meltzer, Brian Norvell, Mindy Nye, Scott Parker, Christos Petropoulos, John Pruitt, Steven Ragan, Scott Ryan, Mike Sapeta, Jana Schroth, Suresh Babu Selvaraju, Goran Stevovic, Amanda Suchanek, Andrea Throop, Lyndon Tilson, Thomas Urban, Joe Voshell, Kimberly Wagner, Jonathan Williams, Mary Williamson, Qian Zeng, Tricia Zwiefelhofer, Clinton R. Paden, Duncan MacCannell |
| EPI_ISL_1798579 | OH Department of Health Laboratory | Centers for Disease Control and Prevention Division of Viral Diseases, Pathogen Discovery | Mili Sheth, Sarah Nobles, Jasmine Padilla, Mark Burroughs, Shoshona Le, Katie Dillon, Peter Cook, Clinton R. Paden, Dhwani Batra, Krista Queen, Kristen Knipe, Dakota Howard, Yvette Unoarumhi, Darlene Wagner, Matthew Schmerer, Ben L. Rambo-Martin, Kristine Lacek, Sam Shepard, Alison Laufer Halpin, Dave Wentworth, Vivien Dugan, Suxiang Tong, Justin Lee |
| EPI_ISL_1798580 | Laboratory Corporation of America | Centers for Disease Control and Prevention Division of Viral Diseases, Pathogen Discovery | Dakota Howard, Dhwani Batra, Peter W. Cook, Kara Moser, Adrian Paskey, Jason Caravas, Benjamin Rambo-Martin, Shatavia Morrison, Christopher Gulvick, Scott Sammons, Yvette Unoarumhi, Darlene Wagner, Matthew Schmerer, Minoo Agarwal, Eyad Almasri, Debbie Boles, Ayla Burns, Nuthawin Charoensri, Oren Cohen, Susan Countryman, Mary Ann Cristobal, Bobbi Croy, Suzanne Dale, Hrushikesh Deshmukh, Amanda Douglas, Vincent Drouillon, Marcia Eisenberg, Howard Engler, Rama Ghatti, Prashant Gupta, Susan Hicks, Jake Humphrey, Lax Iyer, Manoj Jain, Mohan Kolli, Brian Krueger, Tim Kuphal, Stanley Letovsky, Michael Levandoski, Craig Lukasik, Jonathan Meltzer, Brian Norvell, Mindy Nye, Scott Parker, Christos Petropoulos, John Pruitt, Steven Ragan, Scott Ryan, Mike Sapeta, Jana Schroth, Suresh Babu Selvaraju, Goran Stevovic, Amanda Suchanek, Andrea Throop, Lyndon Tilson, Thomas Urban, Joe Voshell, Kimberly Wagner, Jonathan Williams, Mary Williamson, Qian Zeng, Tricia Zwiefelhofer, Clinton R. Paden, Duncan MacCannell |
| EPI_ISL_1798581, EPI_ISL_1798583 | IL Dept. of Public Health Springfield Laboratory | Centers for Disease Control and Prevention Division of Viral Diseases, Pathogen Discovery | Mili Sheth, Sarah Nobles, Jasmine Padilla, Mark Burroughs, Shoshona Le, Katie Dillon, Peter Cook, Clinton R. Paden, Dhwani Batra, Krista Queen, Kristen Knipe, Dakota Howard, Yvette Unoarumhi, Darlene Wagner, Matthew Schmerer, Ben L. Rambo-Martin, Kristine Lacek, Sam Shepard, Alison Laufer Halpin, Dave Wentworth, Vivien Dugan, Suxiang Tong, Justin Lee |
| EPI_ISL_1798584 | Laboratory Corporation of America | Centers for Disease Control and Prevention Division of Viral Diseases, Pathogen Discovery | Dakota Howard, Dhwani Batra, Peter W. Cook, Kara Moser, Adrian Paskey, Jason Caravas, Benjamin Rambo-Martin, Shatavia Morrison, Christopher Gulvick, Scott Sammons, Yvette Unoarumhi, Darlene Wagner, Matthew Schmerer, Minoo Agarwal, Eyad Almasri, Debbie Boles, Ayla Burns, Nuthawin Charoensri, Oren Cohen, Susan Countryman, Mary Ann Cristobal, Bobbi Croy, Suzanne Dale, Hrushikesh Deshmukh, Amanda Douglas, Vincent Drouillon, Marcia Eisenberg, Howard Engler, Rama Ghatti, Prashant Gupta, Susan Hicks, Jake Humphrey, Lax Iyer, Manoj Jain, Mohan Kolli, Brian Krueger, Tim Kuphal, Stanley Letovsky, Michael Levandoski, Craig Lukasik, Jonathan Meltzer, Brian Norvell, Mindy Nye, Scott Parker, Christos Petropoulos, John Pruitt, Steven Ragan, Scott Ryan, Mike Sapeta, Jana Schroth, Suresh Babu Selvaraju, Goran Stevovic, Amanda Suchanek, Andrea Throop, Lyndon Tilson, Thomas Urban, Joe Voshell, Kimberly Wagner, Jonathan Williams, Mary Williamson, Qian Zeng, Tricia Zwiefelhofer, Clinton R. Paden, Duncan MacCannell |
| EPI_ISL_1798585 | WI State Laboratory of Hygiene | Centers for Disease Control and Prevention Division of Viral Diseases, Pathogen Discovery | Mili Sheth, Sarah Nobles, Jasmine Padilla, Mark Burroughs, Shoshona Le, Katie Dillon, Peter Cook, Clinton R. Paden, Dhwani Batra, Krista Queen, Kristen Knipe, Dakota Howard, Yvette Unoarumhi, Darlene Wagner, Matthew Schmerer, Ben L. Rambo-Martin, Kristine Lacek, Sam Shepard, Alison Laufer Halpin, Dave Wentworth, Vivien Dugan, Suxiang Tong, Justin Lee |
| EPI_ISL_1798586 | IA State Hygienic Laboratory | Centers for Disease Control and Prevention Division of Viral Diseases, Pathogen Discovery | Mili Sheth, Sarah Nobles, Jasmine Padilla, Mark Burroughs, Shoshona Le, Katie Dillon, Peter Cook, Clinton R. Paden, Dhwani Batra, Krista Queen, Kristen Knipe, Dakota Howard, Yvette Unoarumhi, Darlene Wagner, Matthew Schmerer, Ben L. Rambo-Martin, Kristine Lacek, Sam Shepard, Alison Laufer Halpin, Dave Wentworth, Vivien Dugan, Suxiang Tong, Justin Lee |
| EPI_ISL_1798587 | Laboratory Corporation of America | Centers for Disease Control and Prevention Division of Viral Diseases, Pathogen Discovery | Dakota Howard, Dhwani Batra, Peter W. Cook, Kara Moser, Adrian Paskey, Jason Caravas, Benjamin Rambo-Martin, Shatavia Morrison, Christopher Gulvick, Scott Sammons, Yvette Unoarumhi, Darlene Wagner, Matthew Schmerer, Minoo Agarwal, Eyad Almasri, Debbie Boles, Ayla Burns, Nuthawin Charoensri, Oren Cohen, Susan Countryman, Mary Ann Cristobal, Bobbi Croy, Suzanne Dale, Hrushikesh Deshmukh, Amanda Douglas, Vincent Drouillon, Marcia Eisenberg, Howard Engler, Rama Ghatti, Prashant Gupta, Susan Hicks, Jake Humphrey, Lax Iyer, Manoj Jain, Mohan Kolli, Brian Krueger, Tim Kuphal, Stanley Letovsky, Michael Levandoski, Craig Lukasik, Jonathan Meltzer, Brian Norvell, Mindy Nye, Scott Parker, Christos Petropoulos, John Pruitt, Steven Ragan, Scott Ryan, Mike Sapeta, Jana Schroth, Suresh Babu Selvaraju, Goran Stevovic, Amanda Suchanek, Andrea Throop, Lyndon Tilson, Thomas Urban, Joe Voshell, Kimberly Wagner, Jonathan Williams, Mary Williamson, Qian Zeng, Tricia Zwiefelhofer, Clinton R. Paden, Duncan MacCannell |
| EPI_ISL_1798589 | OH Department of Health Laboratory | Centers for Disease Control and Prevention Division of Viral Diseases, Pathogen Discovery | Mili Sheth, Sarah Nobles, Jasmine Padilla, Mark Burroughs, Shoshona Le, Katie Dillon, Peter Cook, Clinton R. Paden, Dhwani Batra, Krista Queen, Kristen Knipe, Dakota Howard, Yvette Unoarumhi, Darlene Wagner, Matthew Schmerer, Ben L. Rambo-Martin, Kristine Lacek, Sam Shepard, Alison Laufer Halpin, Dave Wentworth, Vivien Dugan, Suxiang Tong, Justin Lee |
| EPI_ISL_1798590 | Laboratory Corporation of America | Centers for Disease Control and Prevention Division of Viral Diseases, Pathogen Discovery | Dakota Howard, Dhwani Batra, Peter W. Cook, Kara Moser, Adrian Paskey, Jason Caravas, Benjamin Rambo-Martin, Shatavia Morrison, Christopher Gulvick, Scott Sammons, Yvette Unoarumhi, Darlene Wagner, Matthew Schmerer, Minoo Agarwal, Eyad Almasri, Debbie Boles, Ayla Burns, Nuthawin Charoensri, Oren Cohen, Susan Countryman, Mary Ann Cristobal, Bobbi Croy, Suzanne Dale, Hrushikesh Deshmukh, Amanda Douglas, Vincent Drouillon, Marcia Eisenberg, Howard Engler, Rama Ghatti, Prashant Gupta, Susan Hicks, Jake Humphrey, Lax Iyer, Manoj Jain, Mohan Kolli, Brian Krueger, Tim Kuphal, Stanley Letovsky, Michael Levandoski, Craig Lukasik, Jonathan Meltzer, Brian Norvell, Mindy Nye, Scott Parker, Christos Petropoulos, John Pruitt, Steven Ragan, Scott Ryan, Mike Sapeta, Jana Schroth, Suresh Babu Selvaraju, Goran Stevovic, Amanda Suchanek, Andrea Throop, Lyndon Tilson, Thomas Urban, Joe Voshell, Kimberly Wagner, Jonathan Williams, Mary Williamson, Qian Zeng, Tricia Zwiefelhofer, Clinton R. Paden, Duncan MacCannell |
| EPI_ISL_1798591 | OH Department of Health Laboratory | Centers for Disease Control and Prevention Division of Viral Diseases, Pathogen Discovery | Mili Sheth, Sarah Nobles, Jasmine Padilla, Mark Burroughs, Shoshona Le, Katie Dillon, Peter Cook, Clinton R. Paden, Dhwani Batra, Krista Queen, Kristen Knipe, Dakota Howard, Yvette Unoarumhi, Darlene Wagner, Matthew Schmerer, Ben L. Rambo-Martin, Kristine Lacek, Sam Shepard, Alison Laufer Halpin, Dave Wentworth, Vivien Dugan, Suxiang Tong, Justin Lee |
| EPI_ISL_1798592 | Laboratory Corporation of America | Centers for Disease Control and Prevention Division of Viral Diseases, Pathogen Discovery | Dakota Howard, Dhwani Batra, Peter W. Cook, Kara Moser, Adrian Paskey, Jason Caravas, Benjamin Rambo-Martin, Shatavia Morrison, Christopher Gulvick, Scott Sammons, Yvette Unoarumhi, Darlene Wagner, Matthew Schmerer, Minoo Agarwal, Eyad Almasri, Debbie Boles, Ayla Burns, Nuthawin Charoensri, Oren Cohen, Susan Countryman, Mary Ann Cristobal, Bobbi Croy, Suzanne Dale, Hrushikesh Deshmukh, Amanda Douglas, Vincent Drouillon, Marcia Eisenberg, Howard Engler, Rama Ghatti, Prashant Gupta, Susan Hicks, Jake Humphrey, Lax Iyer, Manoj Jain, Mohan Kolli, Brian Krueger, Tim Kuphal, Stanley Letovsky, Michael Levandoski, Craig Lukasik, Jonathan Meltzer, Brian Norvell, Mindy Nye, Scott Parker, Christos Petropoulos, John Pruitt, Steven Ragan, Scott Ryan, Mike Sapeta, Jana Schroth, Suresh Babu Selvaraju, Goran Stevovic, Amanda Suchanek, Andrea Throop, Lyndon Tilson, Thomas Urban, Joe Voshell, Kimberly Wagner, Jonathan Williams, Mary Williamson, Qian Zeng, Tricia Zwiefelhofer, Clinton R. Paden, Duncan MacCannell |
| EPI_ISL_1798593, EPI_ISL_1798595 | IA State Hygienic Laboratory | Centers for Disease Control and Prevention Division of Viral Diseases, Pathogen Discovery | Mili Sheth, Sarah Nobles, Jasmine Padilla, Mark Burroughs, Shoshona Le, Katie Dillon, Peter Cook, Clinton R. Paden, Dhwani Batra, Krista Queen, Kristen Knipe, Dakota Howard, Yvette Unoarumhi, Darlene Wagner, Matthew Schmerer, Ben L. Rambo-Martin, Kristine Lacek, Sam Shepard, Alison Laufer Halpin, Dave Wentworth, Vivien Dugan, Suxiang Tong, Justin Lee |
| EPI_ISL_1798596 | Laboratory Corporation of America | Centers for Disease Control and Prevention Division of Viral Diseases, Pathogen Discovery | Dakota Howard, Dhwani Batra, Peter W. Cook, Kara Moser, Adrian Paskey, Jason Caravas, Benjamin Rambo-Martin, Shatavia Morrison, Christopher Gulvick, Scott Sammons, Yvette Unoarumhi, Darlene Wagner, Matthew Schmerer, Minoo Agarwal, Eyad Almasri, Debbie Boles, Ayla Burns, Nuthawin Charoensri, Oren Cohen, Susan Countryman, Mary Ann Cristobal, Bobbi Croy, Suzanne Dale, Hrushikesh Deshmukh, Amanda Douglas, Vincent Drouillon, Marcia Eisenberg, Howard Engler, Rama Ghatti, Prashant Gupta, Susan Hicks, Jake Humphrey, Lax Iyer, Manoj Jain, Mohan Kolli, Brian Krueger, Tim Kuphal, Stanley Letovsky, Michael Levandoski, Craig Lukasik, Jonathan Meltzer, Brian Norvell, Mindy Nye, Scott Parker, Christos Petropoulos, John Pruitt, Steven Ragan, Scott Ryan, Mike Sapeta, Jana Schroth, Suresh Babu Selvaraju, Goran Stevovic, Amanda Suchanek, Andrea Throop, Lyndon Tilson, Thomas Urban, Joe Voshell, Kimberly Wagner, Jonathan Williams, Mary Williamson, Qian Zeng, Tricia Zwiefelhofer, Clinton R. Paden, Duncan MacCannell |
| EPI_ISL_1798597 | OH Department of Health Laboratory | Centers for Disease Control and Prevention Division of Viral Diseases, Pathogen Discovery | Mili Sheth, Sarah Nobles, Jasmine Padilla, Mark Burroughs, Shoshona Le, Katie Dillon, Peter Cook, Clinton R. Paden, Dhwani Batra, Krista Queen, Kristen Knipe, Dakota Howard, Yvette Unoarumhi, Darlene Wagner, Matthew Schmerer, Ben L. Rambo-Martin, Kristine Lacek, Sam Shepard, Alison Laufer Halpin, Dave Wentworth, Vivien Dugan, Suxiang Tong, Justin Lee |
| EPI_ISL_1798599, EPI_ISL_1798600 | IL Dept. of Public Health Springfield Laboratory | Centers for Disease Control and Prevention Division of Viral Diseases, Pathogen Discovery | Mili Sheth, Sarah Nobles, Jasmine Padilla, Mark Burroughs, Shoshona Le, Katie Dillon, Peter Cook, Clinton R. Paden, Dhwani Batra, Krista Queen, Kristen Knipe, Dakota Howard, Yvette Unoarumhi, Darlene Wagner, Matthew Schmerer, Ben L. Rambo-Martin, Kristine Lacek, Sam Shepard, Alison Laufer Halpin, Dave Wentworth, Vivien Dugan, Suxiang Tong, Justin Lee |

| | | | |
|---|---|---|---|
| EPI_ISL_1798601 | Laboratory Corporation of America | Centers for Disease Control and Prevention Division of Viral Diseases, Pathogen Discovery | Dakota Howard, Dhwani Batra, Peter W. Cook, Kara Moser, Adrian Paskey, Jason Caravas, Benjamin Rambo-Martin, Shatavia Morrison, Christopher Gulvick, Scott Sammons, Yvette Unoarumhi, Darlene Wagner, Matthew Schmerer, Minoo Agarwal, Eyad Almasri, Debbie Boles, Ayla Burns, Nuthawin Charoensri, Oren Cohen, Susan Countryman, Mary Ann Cristobal, Bobbi Croy, Suzanne Dale, Hrushikesh Deshmukh, Amanda Douglas, Vincent Drouillon, Marcia Eisenberg, Howard Engler, Rama Ghatti, Prashant Gupta, Susan Hicks, Jake Humphrey, Lax Iyer, Manoj Jain, Mohan Kolli, Brian Krueger, Tim Kuphal, Stanley Letovsky, Michael Levandoski, Craig Lukasik, Jonathan Meltzer, Brian Norvell, Mindy Nye, Scott Parker, Christos Petropoulos, John Pruitt, Steven Ragan, Scott Ryan, Mike Sapeta, Jana Schroth, Suresh Babu Selvaraju, Goran Stevovic, Amanda Suchanek, Andrea Throop, Lyndon Tilson, Thomas Urban, Joe Voshell, Kimberly Wagner, Jonathan Williams, Mary Williamson, Qian Zeng, Tricia Zwiefelhofer, Clinton R. Paden, Duncan MacCannell |
| EPI_ISL_1798603 | WI State Laboratory of Hygiene | Centers for Disease Control and Prevention Division of Viral Diseases, Pathogen Discovery | Mili Sheth, Sarah Nobles, Jasmine Padilla, Mark Burroughs, Shoshona Le, Katie Dillon, Peter Cook, Clinton R. Paden, Dhwani Batra, Krista Queen, Kristen Knipe, Dakota Howard, Yvette Unoarumhi, Darlene Wagner, Matthew Schmerer, Ben L. Rambo-Martin, Kristine Lacek, Sam Shepard, Alison Laufer Halpin, Dave Wentworth, Vivien Dugan, Suxiang Tong, Justin Lee |
| EPI_ISL_1798605 | IA State Hygienic Laboratory | Centers for Disease Control and Prevention Division of Viral Diseases, Pathogen Discovery | Mili Sheth, Sarah Nobles, Jasmine Padilla, Mark Burroughs, Shoshona Le, Katie Dillon, Peter Cook, Clinton R. Paden, Dhwani Batra, Krista Queen, Kristen Knipe, Dakota Howard, Yvette Unoarumhi, Darlene Wagner, Matthew Schmerer, Ben L. Rambo-Martin, Kristine Lacek, Sam Shepard, Alison Laufer Halpin, Dave Wentworth, Vivien Dugan, Suxiang Tong, Justin Lee |
| EPI_ISL_1798606 | Laboratory Corporation of America | Centers for Disease Control and Prevention Division of Viral Diseases, Pathogen Discovery | Dakota Howard, Dhwani Batra, Peter W. Cook, Kara Moser, Adrian Paskey, Jason Caravas, Benjamin Rambo-Martin, Shatavia Morrison, Christopher Gulvick, Scott Sammons, Yvette Unoarumhi, Darlene Wagner, Matthew Schmerer, Minoo Agarwal, Eyad Almasri, Debbie Boles, Ayla Burns, Nuthawin Charoensri, Oren Cohen, Susan Countryman, Mary Ann Cristobal, Bobbi Croy, Suzanne Dale, Hrushikesh Deshmukh, Amanda Douglas, Vincent Drouillon, Marcia Eisenberg, Howard Engler, Rama Ghatti, Prashant Gupta, Susan Hicks, Jake Humphrey, Lax Iyer, Manoj Jain, Mohan Kolli, Brian Krueger, Tim Kuphal, Stanley Letovsky, Michael Levandoski, Craig Lukasik, Jonathan Meltzer, Brian Norvell, Mindy Nye, Scott Parker, Christos Petropoulos, John Pruitt, Steven Ragan, Scott Ryan, Mike Sapeta, Jana Schroth, Suresh Babu Selvaraju, Goran Stevovic, Amanda Suchanek, Andrea Throop, Lyndon Tilson, Thomas Urban, Joe Voshell, Kimberly Wagner, Jonathan Williams, Mary Williamson, Qian Zeng, Tricia Zwiefelhofer, Clinton R. Paden, Duncan MacCannell |
| EPI_ISL_1798607 | NE Public Health Laboratory | Centers for Disease Control and Prevention Division of Viral Diseases, Pathogen Discovery | Mili Sheth, Sarah Nobles, Jasmine Padilla, Mark Burroughs, Shoshona Le, Katie Dillon, Peter Cook, Clinton R. Paden, Dhwani Batra, Krista Queen, Kristen Knipe, Dakota Howard, Yvette Unoarumhi, Darlene Wagner, Matthew Schmerer, Ben L. Rambo-Martin, Kristine Lacek, Sam Shepard, Alison Laufer Halpin, Dave Wentworth, Vivien Dugan, Suxiang Tong, Justin Lee |
| EPI_ISL_1798608 | Laboratory Corporation of America | Centers for Disease Control and Prevention Division of Viral Diseases, Pathogen Discovery | Dakota Howard, Dhwani Batra, Peter W. Cook, Kara Moser, Adrian Paskey, Jason Caravas, Benjamin Rambo-Martin, Shatavia Morrison, Christopher Gulvick, Scott Sammons, Yvette Unoarumhi, Darlene Wagner, Matthew Schmerer, Minoo Agarwal, Eyad Almasri, Debbie Boles, Ayla Burns, Nuthawin Charoensri, Oren Cohen, Susan Countryman, Mary Ann Cristobal, Bobbi Croy, Suzanne Dale, Hrushikesh Deshmukh, Amanda Douglas, Vincent Drouillon, Marcia Eisenberg, Howard Engler, Rama Ghatti, Prashant Gupta, Susan Hicks, Jake Humphrey, Lax Iyer, Manoj Jain, Mohan Kolli, Brian Krueger, Tim Kuphal, Stanley Letovsky, Michael Levandoski, Craig Lukasik, Jonathan Meltzer, Brian Norvell, Mindy Nye, Scott Parker, Christos Petropoulos, John Pruitt, Steven Ragan, Scott Ryan, Mike Sapeta, Jana Schroth, Suresh Babu Selvaraju, Goran Stevovic, Amanda Suchanek, Andrea Throop, Lyndon Tilson, Thomas Urban, Joe Voshell, Kimberly Wagner, Jonathan Williams, Mary Williamson, Qian Zeng, Tricia Zwiefelhofer, Clinton R. Paden, Duncan MacCannell |
| EPI_ISL_1798609 | OH Department of Health Laboratory | Centers for Disease Control and Prevention Division of Viral Diseases, Pathogen Discovery | Mili Sheth, Sarah Nobles, Jasmine Padilla, Mark Burroughs, Shoshona Le, Katie Dillon, Peter Cook, Clinton R. Paden, Dhwani Batra, Krista Queen, Kristen Knipe, Dakota Howard, Yvette Unoarumhi, Darlene Wagner, Matthew Schmerer, Ben L. Rambo-Martin, Kristine Lacek, Sam Shepard, Alison Laufer Halpin, Dave Wentworth, Vivien Dugan, Suxiang Tong, Justin Lee |
| EPI_ISL_1798610 | NE Public Health Laboratory | Centers for Disease Control and Prevention Division of Viral Diseases, Pathogen Discovery | Mili Sheth, Sarah Nobles, Jasmine Padilla, Mark Burroughs, Shoshona Le, Katie Dillon, Peter Cook, Clinton R. Paden, Dhwani Batra, Krista Queen, Kristen Knipe, Dakota Howard, Yvette Unoarumhi, Darlene Wagner, Matthew Schmerer, Ben L. Rambo-Martin, Kristine Lacek, Sam Shepard, Alison Laufer Halpin, Dave Wentworth, Vivien Dugan, Suxiang Tong, Justin Lee |
| EPI_ISL_1798611 | Laboratory Corporation of America | Centers for Disease Control and Prevention Division of Viral Diseases, Pathogen Discovery | Dakota Howard, Dhwani Batra, Peter W. Cook, Kara Moser, Adrian Paskey, Jason Caravas, Benjamin Rambo-Martin, Shatavia Morrison, Christopher Gulvick, Scott Sammons, Yvette Unoarumhi, Darlene Wagner, Matthew Schmerer, Minoo Agarwal, Eyad Almasri, Debbie Boles, Ayla Burns, Nuthawin Charoensri, Oren Cohen, Susan Countryman, Mary Ann Cristobal, Bobbi Croy, Suzanne Dale, Hrushikesh Deshmukh, Amanda Douglas, Vincent Drouillon, Marcia Eisenberg, Howard Engler, Rama Ghatti, Prashant Gupta, Susan Hicks, Jake Humphrey, Lax Iyer, Manoj Jain, Mohan Kolli, Brian Krueger, Tim Kuphal, Stanley Letovsky, Michael Levandoski, Craig Lukasik, Jonathan Meltzer, Brian Norvell, Mindy Nye, Scott Parker, Christos Petropoulos, John Pruitt, Steven Ragan, Scott Ryan, Mike Sapeta, Jana Schroth, Suresh Babu Selvaraju, Goran Stevovic, Amanda Suchanek, Andrea Throop, Lyndon Tilson, Thomas Urban, Joe Voshell, Kimberly Wagner, Jonathan Williams, Mary Williamson, Qian Zeng, Tricia Zwiefelhofer, Clinton R. Paden, Duncan MacCannell |
| EPI_ISL_1798612 | OH Department of Health Laboratory | Centers for Disease Control and Prevention Division of Viral Diseases, Pathogen Discovery | Mili Sheth, Sarah Nobles, Jasmine Padilla, Mark Burroughs, Shoshona Le, Katie Dillon, Peter Cook, Clinton R. Paden, Dhwani Batra, Krista Queen, Kristen Knipe, Dakota Howard, Yvette Unoarumhi, Darlene Wagner, Matthew Schmerer, Ben L. Rambo-Martin, Kristine Lacek, Sam Shepard, Alison Laufer Halpin, Dave Wentworth, Vivien Dugan, Suxiang Tong, Justin Lee |
| EPI_ISL_1798613 | Laboratory Corporation of America | Centers for Disease Control and Prevention Division of Viral Diseases, Pathogen Discovery | Dakota Howard, Dhwani Batra, Peter W. Cook, Kara Moser, Adrian Paskey, Jason Caravas, Benjamin Rambo-Martin, Shatavia Morrison, Christopher Gulvick, Scott Sammons, Yvette Unoarumhi, Darlene Wagner, Matthew Schmerer, Minoo Agarwal, Eyad Almasri, Debbie Boles, Ayla Burns, Nuthawin Charoensri, Oren Cohen, Susan Countryman, Mary Ann Cristobal, Bobbi Croy, Suzanne Dale, Hrushikesh Deshmukh, Amanda Douglas, Vincent Drouillon, Marcia Eisenberg, Howard Engler, Rama Ghatti, Prashant Gupta, Susan Hicks, Jake Humphrey, Lax Iyer, Manoj Jain, Mohan Kolli, Brian Krueger, Tim Kuphal, Stanley Letovsky, Michael Levandoski, Craig Lukasik, Jonathan Meltzer, Brian Norvell, Mindy Nye, Scott Parker, Christos Petropoulos, John Pruitt, Steven Ragan, Scott Ryan, Mike Sapeta, Jana Schroth, Suresh Babu Selvaraju, Goran Stevovic, Amanda Suchanek, Andrea Throop, Lyndon Tilson, Thomas Urban, Joe Voshell, Kimberly Wagner, Jonathan Williams, Mary Williamson, Qian Zeng, Tricia Zwiefelhofer, Clinton R. Paden, Duncan MacCannell |
| EPI_ISL_1798614 | UT-Unified State Labs: Public Health Utah DOH | Centers for Disease Control and Prevention Division of Viral Diseases, Pathogen Discovery | Mili Sheth, Sarah Nobles, Jasmine Padilla, Mark Burroughs, Shoshona Le, Katie Dillon, Peter Cook, Clinton R. Paden, Dhwani Batra, Krista Queen, Kristen Knipe, Dakota Howard, Yvette Unoarumhi, Darlene Wagner, Matthew Schmerer, Ben L. Rambo-Martin, Kristine Lacek, Sam Shepard, Alison Laufer Halpin, Dave Wentworth, Vivien Dugan, Suxiang Tong, Justin Lee |
| EPI_ISL_1798615 | Laboratory Corporation of America | Centers for Disease Control and Prevention Division of Viral Diseases, Pathogen Discovery | Dakota Howard, Dhwani Batra, Peter W. Cook, Kara Moser, Adrian Paskey, Jason Caravas, Benjamin Rambo-Martin, Shatavia Morrison, Christopher Gulvick, Scott Sammons, Yvette Unoarumhi, Darlene Wagner, Matthew Schmerer, Minoo Agarwal, Eyad Almasri, Debbie Boles, Ayla Burns, Nuthawin Charoensri, Oren Cohen, Susan Countryman, Mary Ann Cristobal, Bobbi Croy, Suzanne Dale, Hrushikesh Deshmukh, Amanda Douglas, Vincent Drouillon, Marcia Eisenberg, Howard Engler, Rama Ghatti, Prashant Gupta, Susan Hicks, Jake Humphrey, Lax Iyer, Manoj Jain, Mohan Kolli, Brian Krueger, Tim Kuphal, Stanley Letovsky, Michael Levandoski, Craig Lukasik, Jonathan Meltzer, Brian Norvell, Mindy Nye, Scott Parker, Christos Petropoulos, John Pruitt, Steven Ragan, Scott Ryan, Mike Sapeta, Jana Schroth, Suresh Babu Selvaraju, Goran Stevovic, Amanda Suchanek, Andrea Throop, Lyndon Tilson, Thomas Urban, Joe Voshell, Kimberly Wagner, Jonathan Williams, Mary Williamson, Qian Zeng, Tricia Zwiefelhofer, Clinton R. Paden, Duncan MacCannell |
| EPI_ISL_1798616 | CO Dept. of Public Health and Environment, Lab Services Division | Centers for Disease Control and Prevention Division of Viral Diseases, Pathogen Discovery | Mili Sheth, Sarah Nobles, Jasmine Padilla, Mark Burroughs, Shoshona Le, Katie Dillon, Peter Cook, Clinton R. Paden, Dhwani Batra, Krista Queen, Kristen Knipe, Dakota Howard, Yvette Unoarumhi, Darlene Wagner, Matthew Schmerer, Ben L. Rambo-Martin, Kristine Lacek, Sam Shepard, Alison Laufer Halpin, Dave Wentworth, Vivien Dugan, Suxiang Tong, Justin Lee |
| EPI_ISL_1798617 | Laboratory Corporation of America | Centers for Disease Control and Prevention Division of Viral Diseases, Pathogen Discovery | Dakota Howard, Dhwani Batra, Peter W. Cook, Kara Moser, Adrian Paskey, Jason Caravas, Benjamin Rambo-Martin, Shatavia Morrison, Christopher Gulvick, Scott Sammons, Yvette Unoarumhi, Darlene Wagner, Matthew Schmerer, Minoo Agarwal, Eyad Almasri, Debbie Boles, Ayla Burns, Nuthawin Charoensri, Oren Cohen, Susan Countryman, Mary Ann Cristobal, Bobbi Croy, Suzanne Dale, Hrushikesh Deshmukh, Amanda Douglas, Vincent Drouillon, Marcia Eisenberg, Howard Engler, Rama Ghatti, Prashant Gupta, Susan Hicks, Jake Humphrey, Lax Iyer, Manoj Jain, Mohan Kolli, Brian Krueger, Tim Kuphal, Stanley Letovsky, Michael Levandoski, Craig Lukasik, Jonathan Meltzer, Brian Norvell, Mindy Nye, Scott Parker, Christos Petropoulos, John Pruitt, Steven Ragan, Scott Ryan, Mike Sapeta, Jana Schroth, Suresh Babu Selvaraju, Goran Stevovic, Amanda Suchanek, Andrea Throop, Lyndon Tilson, Thomas Urban, Joe Voshell, Kimberly Wagner, Jonathan Williams, Mary Williamson, Qian Zeng, Tricia Zwiefelhofer, Clinton R. Paden, Duncan MacCannell |
| EPI_ISL_1798618, EPI_ISL_1798620 | CO Dept. of Public Health and Environment, Lab Services Division | Centers for Disease Control and Prevention Division of Viral Diseases, Pathogen Discovery | Mili Sheth, Sarah Nobles, Jasmine Padilla, Mark Burroughs, Shoshona Le, Katie Dillon, Peter Cook, Clinton R. Paden, Dhwani Batra, Krista Queen, Kristen Knipe, Dakota Howard, Yvette Unoarumhi, Darlene Wagner, Matthew Schmerer, Ben L. Rambo-Martin, Kristine Lacek, Sam Shepard, Alison Laufer Halpin, Dave Wentworth, Vivien Dugan, Suxiang Tong, Justin Lee |
| EPI_ISL_1798621 | Laboratory Corporation of America | Centers for Disease Control and Prevention Division of Viral Diseases, Pathogen Discovery | Dakota Howard, Dhwani Batra, Peter W. Cook, Kara Moser, Adrian Paskey, Jason Caravas, Benjamin Rambo-Martin, Shatavia Morrison, Christopher Gulvick, Scott Sammons, Yvette Unoarumhi, Darlene Wagner, Matthew Schmerer, Minoo Agarwal, Eyad Almasri, Debbie Boles, Ayla Burns, Nuthawin Charoensri, Oren Cohen, Susan Countryman, Mary Ann Cristobal, Bobbi Croy, Suzanne Dale, Hrushikesh Deshmukh, Amanda Douglas, Vincent Drouillon, Marcia Eisenberg, Howard Engler, Rama Ghatti, Prashant Gupta, Susan Hicks, Jake Humphrey, Lax Iyer, Manoj Jain, Mohan Kolli, Brian Krueger, Tim Kuphal, Stanley Letovsky, Michael Levandoski, Craig Lukasik, Jonathan Meltzer, Brian Norvell, Mindy Nye, Scott Parker, Christos Petropoulos, John Pruitt, Steven Ragan, Scott Ryan, Mike Sapeta, Jana Schroth, Suresh Babu Selvaraju, Goran Stevovic, Amanda Suchanek, Andrea Throop, Lyndon Tilson, Thomas Urban, Joe Voshell, Kimberly Wagner, Jonathan Williams, Mary Williamson, Qian Zeng, Tricia Zwiefelhofer, Clinton R. Paden, Duncan MacCannell |
| EPI_ISL_1798622 | CO Dept. of Public Health and Environment, Lab Services Division | Centers for Disease Control and Prevention Division of Viral Diseases, Pathogen Discovery | Mili Sheth, Sarah Nobles, Jasmine Padilla, Mark Burroughs, Shoshona Le, Katie Dillon, Peter Cook, Clinton R. Paden, Dhwani Batra, Krista Queen, Kristen Knipe, Dakota Howard, Yvette Unoarumhi, Darlene Wagner, Matthew Schmerer, Ben L. Rambo-Martin, Kristine Lacek, Sam Shepard, Alison Laufer Halpin, Dave Wentworth, Vivien Dugan, Suxiang Tong, Justin Lee |
| EPI_ISL_1798623 | Laboratory Corporation of America | Centers for Disease Control and Prevention Division of Viral Diseases, Pathogen Discovery | Dakota Howard, Dhwani Batra, Peter W. Cook, Kara Moser, Adrian Paskey, Jason Caravas, Benjamin Rambo-Martin, Shatavia Morrison, Christopher Gulvick, Scott Sammons, Yvette Unoarumhi, Darlene Wagner, Matthew Schmerer, Minoo Agarwal, Eyad Almasri, Debbie Boles, Ayla Burns, Nuthawin Charoensri, Oren Cohen, Susan Countryman, Mary Ann Cristobal, Bobbi Croy, Suzanne Dale, Hrushikesh Deshmukh, Amanda Douglas, Vincent Drouillon, Marcia Eisenberg, Howard Engler, Rama Ghatti, Prashant Gupta, Susan Hicks, Jake Humphrey, Lax Iyer, Manoj Jain, Mohan Kolli, Brian Krueger, Tim Kuphal, Stanley Letovsky, Michael Levandoski, Craig Lukasik, Jonathan Meltzer, Brian Norvell, Mindy Nye, Scott Parker, Christos Petropoulos, John Pruitt, Steven Ragan, Scott Ryan, Mike Sapeta, Jana Schroth, Suresh Babu Selvaraju, Goran Stevovic, Amanda Suchanek, Andrea Throop, Lyndon Tilson, Thomas Urban, Joe Voshell, Kimberly Wagner, Jonathan Williams, Mary Williamson, Qian Zeng, Tricia Zwiefelhofer, Clinton R. Paden, Duncan MacCannell |
| EPI_ISL_1798624 | CO Dept. of Public Health and Environment, Lab Services Division | Centers for Disease Control and Prevention Division of Viral Diseases, Pathogen Discovery | Mili Sheth, Sarah Nobles, Jasmine Padilla, Mark Burroughs, Shoshona Le, Katie Dillon, Peter Cook, Clinton R. Paden, Dhwani Batra, Krista Queen, Kristen Knipe, Dakota Howard, Yvette Unoarumhi, Darlene Wagner, Matthew Schmerer, Ben L. Rambo-Martin, Kristine Lacek, Sam Shepard, Alison Laufer Halpin, Dave Wentworth, Vivien Dugan, Suxiang Tong, Justin Lee |
| EPI_ISL_1798625 | Laboratory Corporation of America | Centers for Disease Control and Prevention Division of Viral Diseases, Pathogen Discovery | Dakota Howard, Dhwani Batra, Peter W. Cook, Kara Moser, Adrian Paskey, Jason Caravas, Benjamin Rambo-Martin, Shatavia Morrison, Christopher Gulvick, Scott Sammons, Yvette Unoarumhi, Darlene Wagner, Matthew Schmerer, Minoo Agarwal, Eyad Almasri, Debbie Boles, Ayla Burns, Nuthawin Charoensri, Oren Cohen, Susan Countryman, Mary Ann Cristobal, Bobbi Croy, Suzanne Dale, Hrushikesh Deshmukh, Amanda Douglas, Vincent Drouillon, Marcia Eisenberg, Howard Engler, Rama Ghatti, Prashant Gupta, Susan Hicks, Jake Humphrey, Lax Iyer, Manoj Jain, Mohan Kolli, Brian Krueger, Tim Kuphal, Stanley Letovsky, Michael Levandoski, Craig Lukasik, Jonathan Meltzer, Brian Norvell, Mindy Nye, Scott Parker, Christos Petropoulos, John Pruitt, Steven Ragan, Scott Ryan, Mike Sapeta, Jana Schroth, Suresh Babu Selvaraju, Goran Stevovic, Amanda Suchanek, Andrea Throop, Lyndon Tilson, Thomas Urban, Joe Voshell, Kimberly Wagner, Jonathan Williams, Mary Williamson, Qian Zeng, Tricia Zwiefelhofer, Clinton R. Paden, Duncan MacCannell |
| EPI_ISL_1798626 | CO Dept. of Public Health and Environment, Lab Services Division | Centers for Disease Control and Prevention Division of Viral Diseases, Pathogen Discovery | Mili Sheth, Sarah Nobles, Jasmine Padilla, Mark Burroughs, Shoshona Le, Katie Dillon, Peter Cook, Clinton R. Paden, Dhwani Batra, Krista Queen, Kristen Knipe, Dakota Howard, Yvette Unoarumhi, Darlene Wagner, Matthew Schmerer, Ben L. Rambo-Martin, Kristine Lacek, Sam Shepard, Alison Laufer Halpin, Dave Wentworth, Vivien Dugan, Suxiang Tong, Justin Lee |
| EPI_ISL_1798627 | Laboratory Corporation of America | Centers for Disease Control and Prevention Division of Viral Diseases, Pathogen Discovery | Dakota Howard, Dhwani Batra, Peter W. Cook, Kara Moser, Adrian Paskey, Jason Caravas, Benjamin Rambo-Martin, Shatavia Morrison, Christopher Gulvick, Scott Sammons, Yvette Unoarumhi, Darlene Wagner, Matthew Schmerer, Minoo Agarwal, Eyad Almasri, Debbie Boles, Ayla Burns, Nuthawin Charoensri, Oren Cohen, Susan Countryman, Mary Ann Cristobal, Bobbi Croy, Suzanne Dale, Hrushikesh Deshmukh, Amanda Douglas, Vincent Drouillon, Marcia Eisenberg, Howard Engler, Rama Ghatti, Prashant Gupta, Susan Hicks, Jake Humphrey, Lax Iyer, Manoj Jain, Mohan Kolli, Brian Krueger, Tim Kuphal, Stanley Letovsky, Michael Levandoski, Craig Lukasik, Jonathan Meltzer, Brian Norvell, Mindy Nye, Scott Parker, Christos Petropoulos, John Pruitt, Steven Ragan, Scott Ryan, Mike Sapeta, Jana Schroth, Suresh Babu Selvaraju, Goran Stevovic, Amanda Suchanek, Andrea Throop, Lyndon Tilson, Thomas Urban, Joe Voshell, Kimberly Wagner, Jonathan Williams, Mary Williamson, Qian Zeng, Tricia Zwiefelhofer, Clinton R. Paden, Duncan MacCannell |
| EPI_ISL_1798628 | CO Dept. of Public Health and Environment, Lab Services Division | Centers for Disease Control and Prevention Division of Viral Diseases, Pathogen Discovery | Mili Sheth, Sarah Nobles, Jasmine Padilla, Mark Burroughs, Shoshona Le, Katie Dillon, Peter Cook, Clinton R. Paden, Dhwani Batra, Krista Queen, Kristen Knipe, Dakota Howard, Yvette Unoarumhi, Darlene Wagner, Matthew Schmerer, Ben L. Rambo-Martin, Kristine Lacek, Sam Shepard, Alison Laufer Halpin, Dave Wentworth, Vivien Dugan, Suxiang Tong, Justin Lee |
| EPI_ISL_1798629 | Laboratory Corporation of America | Centers for Disease Control and Prevention Division of Viral Diseases, Pathogen Discovery | Dakota Howard, Dhwani Batra, Peter W. Cook, Kara Moser, Adrian Paskey, Jason Caravas, Benjamin Rambo-Martin, Shatavia Morrison, Christopher Gulvick, Scott Sammons, Yvette Unoarumhi, Darlene Wagner, Matthew Schmerer, Minoo Agarwal, Eyad Almasri, Debbie Boles, Ayla Burns, Nuthawin Charoensri, Oren Cohen, Susan Countryman, Mary Ann Cristobal, Bobbi Croy, Suzanne Dale, Hrushikesh Deshmukh, Amanda Douglas, Vincent Drouillon, Marcia Eisenberg, Howard Engler, Rama Ghatti, Prashant Gupta, Susan Hicks, Jake Humphrey, Lax Iyer, Manoj Jain, Mohan Kolli, Brian Krueger, Tim Kuphal, Stanley Letovsky, Michael Levandoski, Craig Lukasik, Jonathan Meltzer, Brian Norvell, Mindy Nye, Scott Parker, Christos Petropoulos, John Pruitt, Steven Ragan, Scott Ryan, Mike Sapeta, Jana Schroth, Suresh Babu Selvaraju, Goran Stevovic, Amanda Suchanek, Andrea Throop, Lyndon Tilson, Thomas Urban, Joe Voshell, Kimberly Wagner, Jonathan Williams, Mary Williamson, Qian Zeng, Tricia Zwiefelhofer, Clinton R. Paden, Duncan MacCannell |
| EPI_ISL_1798631, EPI_ISL_1798633 | CO Dept. of Public Health and Environment, Lab Services Division | Centers for Disease Control and Prevention Division of Viral Diseases, Pathogen Discovery | Mili Sheth, Sarah Nobles, Jasmine Padilla, Mark Burroughs, Shoshona Le, Katie Dillon, Peter Cook, Clinton R. Paden, Dhwani Batra, Krista Queen, Kristen Knipe, Dakota Howard, Yvette Unoarumhi, Darlene Wagner, Matthew Schmerer, Ben L. Rambo-Martin, Kristine Lacek, Sam Shepard, Alison Laufer Halpin, Dave Wentworth, Vivien Dugan, Suxiang Tong, Justin Lee |
| EPI_ISL_1798634 | Laboratory Corporation of America | Centers for Disease Control and Prevention Division of Viral Diseases, Pathogen Discovery | Dakota Howard, Dhwani Batra, Peter W. Cook, Kara Moser, Adrian Paskey, Jason Caravas, Benjamin Rambo-Martin, Shatavia Morrison, Christopher Gulvick, Scott Sammons, Yvette Unoarumhi, Darlene Wagner, Matthew Schmerer, Minoo Agarwal, Eyad Almasri, Debbie Boles, Ayla Burns, Nuthawin Charoensri, Oren Cohen, Susan Countryman, Mary Ann Cristobal, Bobbi Croy, Suzanne Dale, Hrushikesh Deshmukh, Amanda Douglas, Vincent Drouillon, Marcia Eisenberg, Howard Engler, Rama Ghatti, Prashant Gupta, Susan Hicks, Jake Humphrey, Lax Iyer, Manoj Jain, Mohan Kolli, Brian Krueger, Tim Kuphal, Stanley Letovsky, Michael Levandoski, Craig Lukasik, Jonathan Meltzer, Brian Norvell, Mindy Nye, Scott Parker, Christos Petropoulos, John Pruitt, Steven Ragan, Scott Ryan, Mike Sapeta, Jana Schroth, Suresh Babu Selvaraju, Goran Stevovic, Amanda Suchanek, Andrea Throop, Lyndon Tilson, Thomas Urban, Joe Voshell, Kimberly Wagner, Jonathan Williams, Mary Williamson, Qian Zeng, Tricia Zwiefelhofer, Clinton R. Paden, Duncan MacCannell |
| EPI_ISL_1798635, EPI_ISL_1798636 | CO Dept. of Public Health and Environment, Lab Services Division | Centers for Disease Control and Prevention Division of Viral Diseases, Pathogen Discovery | Mili Sheth, Sarah Nobles, Jasmine Padilla, Mark Burroughs, Shoshona Le, Katie Dillon, Peter Cook, Clinton R. Paden, Dhwani Batra, Krista Queen, Kristen Knipe, Dakota Howard, Yvette Unoarumhi, Darlene Wagner, Matthew Schmerer, Ben L. Rambo-Martin, Kristine Lacek, Sam Shepard, Alison Laufer Halpin, Dave Wentworth, Vivien Dugan, Suxiang Tong, Justin Lee |

| | | | |
|---|---|---|---|
| EPI_ISL_1798637 | Laboratory Corporation of America | Centers for Disease Control and Prevention Division of Viral Diseases, Pathogen Discovery | Dakota Howard, Dhwani Batra, Peter W. Cook, Kara Moser, Adrian Paskey, Jason Caravas, Benjamin Rambo-Martin, Shatavia Morrison, Christopher Gulvick, Scott Sammons, Yvette Unoarumhi, Darlene Wagner, Matthew Schmerer, Minoo Agarwal, Eyad Almasri, Debbie Boles, Ayla Burns, Nuthawin Charoensri, Oren Cohen, Susan Countryman, Mary Ann Cristobal, Bobbi Croy, Suzanne Dale, Hrushikesh Deshmukh, Amanda Douglas, Vincent Drouillon, Marcia Eisenberg, Howard Engler, Rama Ghatti, Prashant Gupta, Susan Hicks, Jake Humphrey, Lax Iyer, Manoj Jain, Mohan Kolli, Brian Krueger, Tim Kuphal, Stanley Letovsky, Michael Levandoski, Craig Lukasik, Jonathan Meltzer, Brian Norvell, Mindy Nye, Scott Parker, Christos Petropoulos, John Pruitt, Steven Ragan, Scott Ryan, Mike Sapeta, Jana Schroth, Suresh Babu Selvaraju, Goran Stevovic, Amanda Suchanek, Andrea Throop, Lyndon Tilson, Thomas Urban, Joe Voshell, Kimberly Wagner, Jonathan Williams, Mary Williamson, Qian Zeng, Tricia Zwiefelhofer, Clinton R. Paden, Duncan MacCannell |
| EPI_ISL_1798638 | CO Dept. of Public Health and Environment, Lab Services Division | Centers for Disease Control and Prevention Division of Viral Diseases, Pathogen Discovery | Mili Sheth, Sarah Nobles, Jasmine Padilla, Mark Burroughs, Shoshona Le, Katie Dillon, Peter Cook, Clinton R. Paden, Dhwani Batra, Krista Queen, Kristen Knipe, Dakota Howard, Yvette Unoarumhi, Darlene Wagner, Matthew Schmerer, Ben L. Rambo-Martin, Kristine Lacek, Sam Shepard, Alison Laufer Halpin, Dave Wentworth, Vivien Dugan, Suxiang Tong, Justin Lee |
| EPI_ISL_1798639 | Laboratory Corporation of America | Centers for Disease Control and Prevention Division of Viral Diseases, Pathogen Discovery | Dakota Howard, Dhwani Batra, Peter W. Cook, Kara Moser, Adrian Paskey, Jason Caravas, Benjamin Rambo-Martin, Shatavia Morrison, Christopher Gulvick, Scott Sammons, Yvette Unoarumhi, Darlene Wagner, Matthew Schmerer, Minoo Agarwal, Eyad Almasri, Debbie Boles, Ayla Burns, Nuthawin Charoensri, Oren Cohen, Susan Countryman, Mary Ann Cristobal, Bobbi Croy, Suzanne Dale, Hrushikesh Deshmukh, Amanda Douglas, Vincent Drouillon, Marcia Eisenberg, Howard Engler, Rama Ghatti, Prashant Gupta, Susan Hicks, Jake Humphrey, Lax Iyer, Manoj Jain, Mohan Kolli, Brian Krueger, Tim Kuphal, Stanley Letovsky, Michael Levandoski, Craig Lukasik, Jonathan Meltzer, Brian Norvell, Mindy Nye, Scott Parker, Christos Petropoulos, John Pruitt, Steven Ragan, Scott Ryan, Mike Sapeta, Jana Schroth, Suresh Babu Selvaraju, Goran Stevovic, Amanda Suchanek, Andrea Throop, Lyndon Tilson, Thomas Urban, Joe Voshell, Kimberly Wagner, Jonathan Williams, Mary Williamson, Qian Zeng, Tricia Zwiefelhofer, Clinton R. Paden, Duncan MacCannell |
| EPI_ISL_1798640, EPI_ISL_1798642 | CO Dept. of Public Health and Environment, Lab Services Division | Centers for Disease Control and Prevention Division of Viral Diseases, Pathogen Discovery | Mili Sheth, Sarah Nobles, Jasmine Padilla, Mark Burroughs, Shoshona Le, Katie Dillon, Peter Cook, Clinton R. Paden, Dhwani Batra, Krista Queen, Kristen Knipe, Dakota Howard, Yvette Unoarumhi, Darlene Wagner, Matthew Schmerer, Ben L. Rambo-Martin, Kristine Lacek, Sam Shepard, Alison Laufer Halpin, Dave Wentworth, Vivien Dugan, Suxiang Tong, Justin Lee |
| EPI_ISL_1798643, EPI_ISL_1798644 | Laboratory Corporation of America | Centers for Disease Control and Prevention Division of Viral Diseases, Pathogen Discovery | Dakota Howard, Dhwani Batra, Peter W. Cook, Kara Moser, Adrian Paskey, Jason Caravas, Benjamin Rambo-Martin, Shatavia Morrison, Christopher Gulvick, Scott Sammons, Yvette Unoarumhi, Darlene Wagner, Matthew Schmerer, Minoo Agarwal, Eyad Almasri, Debbie Boles, Ayla Burns, Nuthawin Charoensri, Oren Cohen, Susan Countryman, Mary Ann Cristobal, Bobbi Croy, Suzanne Dale, Hrushikesh Deshmukh, Amanda Douglas, Vincent Drouillon, Marcia Eisenberg, Howard Engler, Rama Ghatti, Prashant Gupta, Susan Hicks, Jake Humphrey, Lax Iyer, Manoj Jain, Mohan Kolli, Brian Krueger, Tim Kuphal, Stanley Letovsky, Michael Levandoski, Craig Lukasik, Jonathan Meltzer, Brian Norvell, Mindy Nye, Scott Parker, Christos Petropoulos, John Pruitt, Steven Ragan, Scott Ryan, Mike Sapeta, Jana Schroth, Suresh Babu Selvaraju, Goran Stevovic, Amanda Suchanek, Andrea Throop, Lyndon Tilson, Thomas Urban, Joe Voshell, Kimberly Wagner, Jonathan Williams, Mary Williamson, Qian Zeng, Tricia Zwiefelhofer, Clinton R. Paden, Duncan MacCannell |
| EPI_ISL_1798645, EPI_ISL_1798646, EPI_ISL_1798648 | CO Dept. of Public Health and Environment, Lab Services Division | Centers for Disease Control and Prevention Division of Viral Diseases, Pathogen Discovery | Mili Sheth, Sarah Nobles, Jasmine Padilla, Mark Burroughs, Shoshona Le, Katie Dillon, Peter Cook, Clinton R. Paden, Dhwani Batra, Krista Queen, Kristen Knipe, Dakota Howard, Yvette Unoarumhi, Darlene Wagner, Matthew Schmerer, Ben L. Rambo-Martin, Kristine Lacek, Sam Shepard, Alison Laufer Halpin, Dave Wentworth, Vivien Dugan, Suxiang Tong, Justin Lee |
| EPI_ISL_1798649 | Laboratory Corporation of America | Centers for Disease Control and Prevention Division of Viral Diseases, Pathogen Discovery | Dakota Howard, Dhwani Batra, Peter W. Cook, Kara Moser, Adrian Paskey, Jason Caravas, Benjamin Rambo-Martin, Shatavia Morrison, Christopher Gulvick, Scott Sammons, Yvette Unoarumhi, Darlene Wagner, Matthew Schmerer, Minoo Agarwal, Eyad Almasri, Debbie Boles, Ayla Burns, Nuthawin Charoensri, Oren Cohen, Susan Countryman, Mary Ann Cristobal, Bobbi Croy, Suzanne Dale, Hrushikesh Deshmukh, Amanda Douglas, Vincent Drouillon, Marcia Eisenberg, Howard Engler, Rama Ghatti, Prashant Gupta, Susan Hicks, Jake Humphrey, Lax Iyer, Manoj Jain, Mohan Kolli, Brian Krueger, Tim Kuphal, Stanley Letovsky, Michael Levandoski, Craig Lukasik, Jonathan Meltzer, Brian Norvell, Mindy Nye, Scott Parker, Christos Petropoulos, John Pruitt, Steven Ragan, Scott Ryan, Mike Sapeta, Jana Schroth, Suresh Babu Selvaraju, Goran Stevovic, Amanda Suchanek, Andrea Throop, Lyndon Tilson, Thomas Urban, Joe Voshell, Kimberly Wagner, Jonathan Williams, Mary Williamson, Qian Zeng, Tricia Zwiefelhofer, Clinton R. Paden, Duncan MacCannell |
| EPI_ISL_1798650 | CO Dept. of Public Health and Environment, Lab Services Division | Centers for Disease Control and Prevention Division of Viral Diseases, Pathogen Discovery | Mili Sheth, Sarah Nobles, Jasmine Padilla, Mark Burroughs, Shoshona Le, Katie Dillon, Peter Cook, Clinton R. Paden, Dhwani Batra, Krista Queen, Kristen Knipe, Dakota Howard, Yvette Unoarumhi, Darlene Wagner, Matthew Schmerer, Ben L. Rambo-Martin, Kristine Lacek, Sam Shepard, Alison Laufer Halpin, Dave Wentworth, Vivien Dugan, Suxiang Tong, Justin Lee |
| EPI_ISL_1798651 | Laboratory Corporation of America | Centers for Disease Control and Prevention Division of Viral Diseases, Pathogen Discovery | Dakota Howard, Dhwani Batra, Peter W. Cook, Kara Moser, Adrian Paskey, Jason Caravas, Benjamin Rambo-Martin, Shatavia Morrison, Christopher Gulvick, Scott Sammons, Yvette Unoarumhi, Darlene Wagner, Matthew Schmerer, Minoo Agarwal, Eyad Almasri, Debbie Boles, Ayla Burns, Nuthawin Charoensri, Oren Cohen, Susan Countryman, Mary Ann Cristobal, Bobbi Croy, Suzanne Dale, Hrushikesh Deshmukh, Amanda Douglas, Vincent Drouillon, Marcia Eisenberg, Howard Engler, Rama Ghatti, Prashant Gupta, Susan Hicks, Jake Humphrey, Lax Iyer, Manoj Jain, Mohan Kolli, Brian Krueger, Tim Kuphal, Stanley Letovsky, Michael Levandoski, Craig Lukasik, Jonathan Meltzer, Brian Norvell, Mindy Nye, Scott Parker, Christos Petropoulos, John Pruitt, Steven Ragan, Scott Ryan, Mike Sapeta, Jana Schroth, Suresh Babu Selvaraju, Goran Stevovic, Amanda Suchanek, Andrea Throop, Lyndon Tilson, Thomas Urban, Joe Voshell, Kimberly Wagner, Jonathan Williams, Mary Williamson, Qian Zeng, Tricia Zwiefelhofer, Clinton R. Paden, Duncan MacCannell |
| EPI_ISL_1798652 | CO Dept. of Public Health and Environment, Lab Services Division | Centers for Disease Control and Prevention Division of Viral Diseases, Pathogen Discovery | Mili Sheth, Sarah Nobles, Jasmine Padilla, Mark Burroughs, Shoshona Le, Katie Dillon, Peter Cook, Clinton R. Paden, Dhwani Batra, Krista Queen, Kristen Knipe, Dakota Howard, Yvette Unoarumhi, Darlene Wagner, Matthew Schmerer, Ben L. Rambo-Martin, Kristine Lacek, Sam Shepard, Alison Laufer Halpin, Dave Wentworth, Vivien Dugan, Suxiang Tong, Justin Lee |
| EPI_ISL_1798653 | Laboratory Corporation of America | Centers for Disease Control and Prevention Division of Viral Diseases, Pathogen Discovery | Dakota Howard, Dhwani Batra, Peter W. Cook, Kara Moser, Adrian Paskey, Jason Caravas, Benjamin Rambo-Martin, Shatavia Morrison, Christopher Gulvick, Scott Sammons, Yvette Unoarumhi, Darlene Wagner, Matthew Schmerer, Minoo Agarwal, Eyad Almasri, Debbie Boles, Ayla Burns, Nuthawin Charoensri, Oren Cohen, Susan Countryman, Mary Ann Cristobal, Bobbi Croy, Suzanne Dale, Hrushikesh Deshmukh, Amanda Douglas, Vincent Drouillon, Marcia Eisenberg, Howard Engler, Rama Ghatti, Prashant Gupta, Susan Hicks, Jake Humphrey, Lax Iyer, Manoj Jain, Mohan Kolli, Brian Krueger, Tim Kuphal, Stanley Letovsky, Michael Levandoski, Craig Lukasik, Jonathan Meltzer, Brian Norvell, Mindy Nye, Scott Parker, Christos Petropoulos, John Pruitt, Steven Ragan, Scott Ryan, Mike Sapeta, Jana Schroth, Suresh Babu Selvaraju, Goran Stevovic, Amanda Suchanek, Andrea Throop, Lyndon Tilson, Thomas Urban, Joe Voshell, Kimberly Wagner, Jonathan Williams, Mary Williamson, Qian Zeng, Tricia Zwiefelhofer, Clinton R. Paden, Duncan MacCannell |
| EPI_ISL_1798654 | CO Dept. of Public Health and Environment, Lab Services Division | Centers for Disease Control and Prevention Division of Viral Diseases, Pathogen Discovery | Mili Sheth, Sarah Nobles, Jasmine Padilla, Mark Burroughs, Shoshona Le, Katie Dillon, Peter Cook, Clinton R. Paden, Dhwani Batra, Krista Queen, Kristen Knipe, Dakota Howard, Yvette Unoarumhi, Darlene Wagner, Matthew Schmerer, Ben L. Rambo-Martin, Kristine Lacek, Sam Shepard, Alison Laufer Halpin, Dave Wentworth, Vivien Dugan, Suxiang Tong, Justin Lee |
| EPI_ISL_1798655 | Laboratory Corporation of America | Centers for Disease Control and Prevention Division of Viral Diseases, Pathogen Discovery | Dakota Howard, Dhwani Batra, Peter W. Cook, Kara Moser, Adrian Paskey, Jason Caravas, Benjamin Rambo-Martin, Shatavia Morrison, Christopher Gulvick, Scott Sammons, Yvette Unoarumhi, Darlene Wagner, Matthew Schmerer, Minoo Agarwal, Eyad Almasri, Debbie Boles, Ayla Burns, Nuthawin Charoensri, Oren Cohen, Susan Countryman, Mary Ann Cristobal, Bobbi Croy, Suzanne Dale, Hrushikesh Deshmukh, Amanda Douglas, Vincent Drouillon, Marcia Eisenberg, Howard Engler, Rama Ghatti, Prashant Gupta, Susan Hicks, Jake Humphrey, Lax Iyer, Manoj Jain, Mohan Kolli, Brian Krueger, Tim Kuphal, Stanley Letovsky, Michael Levandoski, Craig Lukasik, Jonathan Meltzer, Brian Norvell, Mindy Nye, Scott Parker, Christos Petropoulos, John Pruitt, Steven Ragan, Scott Ryan, Mike Sapeta, Jana Schroth, Suresh Babu Selvaraju, Goran Stevovic, Amanda Suchanek, Andrea Throop, Lyndon Tilson, Thomas Urban, Joe Voshell, Kimberly Wagner, Jonathan Williams, Mary Williamson, Qian Zeng, Tricia Zwiefelhofer, Clinton R. Paden, Duncan MacCannell |
| EPI_ISL_1798656 | CO Dept. of Public Health and Environment, Lab Services Division | Centers for Disease Control and Prevention Division of Viral Diseases, Pathogen Discovery | Mili Sheth, Sarah Nobles, Jasmine Padilla, Mark Burroughs, Shoshona Le, Katie Dillon, Peter Cook, Clinton R. Paden, Dhwani Batra, Krista Queen, Kristen Knipe, Dakota Howard, Yvette Unoarumhi, Darlene Wagner, Matthew Schmerer, Ben L. Rambo-Martin, Kristine Lacek, Sam Shepard, Alison Laufer Halpin, Dave Wentworth, Vivien Dugan, Suxiang Tong, Justin Lee |
| EPI_ISL_1798657 | Laboratory Corporation of America | Centers for Disease Control and Prevention Division of Viral Diseases, Pathogen Discovery | Dakota Howard, Dhwani Batra, Peter W. Cook, Kara Moser, Adrian Paskey, Jason Caravas, Benjamin Rambo-Martin, Shatavia Morrison, Christopher Gulvick, Scott Sammons, Yvette Unoarumhi, Darlene Wagner, Matthew Schmerer, Minoo Agarwal, Eyad Almasri, Debbie Boles, Ayla Burns, Nuthawin Charoensri, Oren Cohen, Susan Countryman, Mary Ann Cristobal, Bobbi Croy, Suzanne Dale, Hrushikesh Deshmukh, Amanda Douglas, Vincent Drouillon, Marcia Eisenberg, Howard Engler, Rama Ghatti, Prashant Gupta, Susan Hicks, Jake Humphrey, Lax Iyer, Manoj Jain, Mohan Kolli, Brian Krueger, Tim Kuphal, Stanley Letovsky, Michael Levandoski, Craig Lukasik, Jonathan Meltzer, Brian Norvell, Mindy Nye, Scott Parker, Christos Petropoulos, John Pruitt, Steven Ragan, Scott Ryan, Mike Sapeta, Jana Schroth, Suresh Babu Selvaraju, Goran Stevovic, Amanda Suchanek, Andrea Throop, Lyndon Tilson, Thomas Urban, Joe Voshell, Kimberly Wagner, Jonathan Williams, Mary Williamson, Qian Zeng, Tricia Zwiefelhofer, Clinton R. Paden, Duncan MacCannell |
| EPI_ISL_1798658 | CO Dept. of Public Health and Environment, Lab Services Division | Centers for Disease Control and Prevention Division of Viral Diseases, Pathogen Discovery | Mili Sheth, Sarah Nobles, Jasmine Padilla, Mark Burroughs, Shoshona Le, Katie Dillon, Peter Cook, Clinton R. Paden, Dhwani Batra, Krista Queen, Kristen Knipe, Dakota Howard, Yvette Unoarumhi, Darlene Wagner, Matthew Schmerer, Ben L. Rambo-Martin, Kristine Lacek, Sam Shepard, Alison Laufer Halpin, Dave Wentworth, Vivien Dugan, Suxiang Tong, Justin Lee |
| EPI_ISL_1798659 | Laboratory Corporation of America | Centers for Disease Control and Prevention Division of Viral Diseases, Pathogen Discovery | Dakota Howard, Dhwani Batra, Peter W. Cook, Kara Moser, Adrian Paskey, Jason Caravas, Benjamin Rambo-Martin, Shatavia Morrison, Christopher Gulvick, Scott Sammons, Yvette Unoarumhi, Darlene Wagner, Matthew Schmerer, Minoo Agarwal, Eyad Almasri, Debbie Boles, Ayla Burns, Nuthawin Charoensri, Oren Cohen, Susan Countryman, Mary Ann Cristobal, Bobbi Croy, Suzanne Dale, Hrushikesh Deshmukh, Amanda Douglas, Vincent Drouillon, Marcia Eisenberg, Howard Engler, Rama Ghatti, Prashant Gupta, Susan Hicks, Jake Humphrey, Lax Iyer, Manoj Jain, Mohan Kolli, Brian Krueger, Tim Kuphal, Stanley Letovsky, Michael Levandoski, Craig Lukasik, Jonathan Meltzer, Brian Norvell, Mindy Nye, Scott Parker, Christos Petropoulos, John Pruitt, Steven Ragan, Scott Ryan, Mike Sapeta, Jana Schroth, Suresh Babu Selvaraju, Goran Stevovic, Amanda Suchanek, Andrea Throop, Lyndon Tilson, Thomas Urban, Joe Voshell, Kimberly Wagner, Jonathan Williams, Mary Williamson, Qian Zeng, Tricia Zwiefelhofer, Clinton R. Paden, Duncan MacCannell |
| EPI_ISL_1798660, EPI_ISL_1798662 | CO Dept. of Public Health and Environment, Lab Services Division | Centers for Disease Control and Prevention Division of Viral Diseases, Pathogen Discovery | Mili Sheth, Sarah Nobles, Jasmine Padilla, Mark Burroughs, Shoshona Le, Katie Dillon, Peter Cook, Clinton R. Paden, Dhwani Batra, Krista Queen, Kristen Knipe, Dakota Howard, Yvette Unoarumhi, Darlene Wagner, Matthew Schmerer, Ben L. Rambo-Martin, Kristine Lacek, Sam Shepard, Alison Laufer Halpin, Dave Wentworth, Vivien Dugan, Suxiang Tong, Justin Lee |
| EPI_ISL_1798663 | Laboratory Corporation of America | Centers for Disease Control and Prevention Division of Viral Diseases, Pathogen Discovery | Dakota Howard, Dhwani Batra, Peter W. Cook, Kara Moser, Adrian Paskey, Jason Caravas, Benjamin Rambo-Martin, Shatavia Morrison, Christopher Gulvick, Scott Sammons, Yvette Unoarumhi, Darlene Wagner, Matthew Schmerer, Minoo Agarwal, Eyad Almasri, Debbie Boles, Ayla Burns, Nuthawin Charoensri, Oren Cohen, Susan Countryman, Mary Ann Cristobal, Bobbi Croy, Suzanne Dale, Hrushikesh Deshmukh, Amanda Douglas, Vincent Drouillon, Marcia Eisenberg, Howard Engler, Rama Ghatti, Prashant Gupta, Susan Hicks, Jake Humphrey, Lax Iyer, Manoj Jain, Mohan Kolli, Brian Krueger, Tim Kuphal, Stanley Letovsky, Michael Levandoski, Craig Lukasik, Jonathan Meltzer, Brian Norvell, Mindy Nye, Scott Parker, Christos Petropoulos, John Pruitt, Steven Ragan, Scott Ryan, Mike Sapeta, Jana Schroth, Suresh Babu Selvaraju, Goran Stevovic, Amanda Suchanek, Andrea Throop, Lyndon Tilson, Thomas Urban, Joe Voshell, Kimberly Wagner, Jonathan Williams, Mary Williamson, Qian Zeng, Tricia Zwiefelhofer, Clinton R. Paden, Duncan MacCannell |
| EPI_ISL_1798664, EPI_ISL_1798666 | TN Division of Laboratory Services | Centers for Disease Control and Prevention Division of Viral Diseases, Pathogen Discovery | Mili Sheth, Sarah Nobles, Jasmine Padilla, Mark Burroughs, Shoshona Le, Katie Dillon, Peter Cook, Clinton R. Paden, Dhwani Batra, Krista Queen, Kristen Knipe, Dakota Howard, Yvette Unoarumhi, Darlene Wagner, Matthew Schmerer, Ben L. Rambo-Martin, Kristine Lacek, Sam Shepard, Alison Laufer Halpin, Dave Wentworth, Vivien Dugan, Suxiang Tong, Justin Lee |
| EPI_ISL_1798667, EPI_ISL_1798668 | Laboratory Corporation of America | Centers for Disease Control and Prevention Division of Viral Diseases, Pathogen Discovery | Dakota Howard, Dhwani Batra, Peter W. Cook, Kara Moser, Adrian Paskey, Jason Caravas, Benjamin Rambo-Martin, Shatavia Morrison, Christopher Gulvick, Scott Sammons, Yvette Unoarumhi, Darlene Wagner, Matthew Schmerer, Minoo Agarwal, Eyad Almasri, Debbie Boles, Ayla Burns, Nuthawin Charoensri, Oren Cohen, Susan Countryman, Mary Ann Cristobal, Bobbi Croy, Suzanne Dale, Hrushikesh Deshmukh, Amanda Douglas, Vincent Drouillon, Marcia Eisenberg, Howard Engler, Rama Ghatti, Prashant Gupta, Susan Hicks, Jake Humphrey, Lax Iyer, Manoj Jain, Mohan Kolli, Brian Krueger, Tim Kuphal, Stanley Letovsky, Michael Levandoski, Craig Lukasik, Jonathan Meltzer, Brian Norvell, Mindy Nye, Scott Parker, Christos Petropoulos, John Pruitt, Steven Ragan, Scott Ryan, Mike Sapeta, Jana Schroth, Suresh Babu Selvaraju, Goran Stevovic, Amanda Suchanek, Andrea Throop, Lyndon Tilson, Thomas Urban, Joe Voshell, Kimberly Wagner, Jonathan Williams, Mary Williamson, Qian Zeng, Tricia Zwiefelhofer, Clinton R. Paden, Duncan MacCannell |
| EPI_ISL_1798669, EPI_ISL_1798670, EPI_ISL_1798672 | TN Division of Laboratory Services | Centers for Disease Control and Prevention Division of Viral Diseases, Pathogen Discovery | Mili Sheth, Sarah Nobles, Jasmine Padilla, Mark Burroughs, Shoshona Le, Katie Dillon, Peter Cook, Clinton R. Paden, Dhwani Batra, Krista Queen, Kristen Knipe, Dakota Howard, Yvette Unoarumhi, Darlene Wagner, Matthew Schmerer, Ben L. Rambo-Martin, Kristine Lacek, Sam Shepard, Alison Laufer Halpin, Dave Wentworth, Vivien Dugan, Suxiang Tong, Justin Lee |
| EPI_ISL_1798673 | Laboratory Corporation of America | Centers for Disease Control and Prevention Division of Viral Diseases, Pathogen Discovery | Dakota Howard, Dhwani Batra, Peter W. Cook, Kara Moser, Adrian Paskey, Jason Caravas, Benjamin Rambo-Martin, Shatavia Morrison, Christopher Gulvick, Scott Sammons, Yvette Unoarumhi, Darlene Wagner, Matthew Schmerer, Minoo Agarwal, Eyad Almasri, Debbie Boles, Ayla Burns, Nuthawin Charoensri, Oren Cohen, Susan Countryman, Mary Ann Cristobal, Bobbi Croy, Suzanne Dale, Hrushikesh Deshmukh, Amanda Douglas, Vincent Drouillon, Marcia Eisenberg, Howard Engler, Rama Ghatti, Prashant Gupta, Susan Hicks, Jake Humphrey, Lax Iyer, Manoj Jain, Mohan Kolli, Brian Krueger, Tim Kuphal, Stanley Letovsky, Michael Levandoski, Craig Lukasik, Jonathan Meltzer, Brian Norvell, Mindy Nye, Scott Parker, Christos Petropoulos, John Pruitt, Steven Ragan, Scott Ryan, Mike Sapeta, Jana Schroth, Suresh Babu Selvaraju, Goran Stevovic, Amanda Suchanek, Andrea Throop, Lyndon Tilson, Thomas Urban, Joe Voshell, Kimberly Wagner, Jonathan Williams, Mary Williamson, Qian Zeng, Tricia Zwiefelhofer, Clinton R. Paden, Duncan MacCannell |
| EPI_ISL_1798674 | TN Division of Laboratory Services | Centers for Disease Control and Prevention Division of Viral Diseases, Pathogen Discovery | Mili Sheth, Sarah Nobles, Jasmine Padilla, Mark Burroughs, Shoshona Le, Katie Dillon, Peter Cook, Clinton R. Paden, Dhwani Batra, Krista Queen, Kristen Knipe, Dakota Howard, Yvette Unoarumhi, Darlene Wagner, Matthew Schmerer, Ben L. Rambo-Martin, Kristine Lacek, Sam Shepard, Alison Laufer Halpin, Dave Wentworth, Vivien Dugan, Suxiang Tong, Justin Lee |
| EPI_ISL_1798675 | Laboratory Corporation of America | Centers for Disease Control and Prevention Division of Viral Diseases, Pathogen Discovery | Dakota Howard, Dhwani Batra, Peter W. Cook, Kara Moser, Adrian Paskey, Jason Caravas, Benjamin Rambo-Martin, Shatavia Morrison, Christopher Gulvick, Scott Sammons, Yvette Unoarumhi, Darlene Wagner, Matthew Schmerer, Minoo Agarwal, Eyad Almasri, Debbie Boles, Ayla Burns, Nuthawin Charoensri, Oren Cohen, Susan Countryman, Mary Ann Cristobal, Bobbi Croy, Suzanne Dale, Hrushikesh Deshmukh, Amanda Douglas, Vincent Drouillon, Marcia Eisenberg, Howard Engler, Rama Ghatti, Prashant Gupta, Susan Hicks, Jake Humphrey, Lax Iyer, Manoj Jain, Mohan Kolli, Brian Krueger, Tim Kuphal, Stanley Letovsky, Michael Levandoski, Craig Lukasik, Jonathan Meltzer, Brian Norvell, Mindy Nye, Scott Parker, Christos Petropoulos, John Pruitt, Steven Ragan, Scott Ryan, Mike Sapeta, Jana Schroth, Suresh Babu Selvaraju, Goran Stevovic, Amanda Suchanek, Andrea Throop, Lyndon Tilson, Thomas Urban, Joe Voshell, Kimberly Wagner, Jonathan Williams, Mary Williamson, Qian Zeng, Tricia Zwiefelhofer, Clinton R. Paden, Duncan MacCannell |
| EPI_ISL_1798676 | TN Division of Laboratory Services | Centers for Disease Control and Prevention Division of Viral Diseases, Pathogen Discovery | Mili Sheth, Sarah Nobles, Jasmine Padilla, Mark Burroughs, Shoshona Le, Katie Dillon, Peter Cook, Clinton R. Paden, Dhwani Batra, Krista Queen, Kristen Knipe, Dakota Howard, Yvette Unoarumhi, Darlene Wagner, Matthew Schmerer, Ben L. Rambo-Martin, Kristine Lacek, Sam Shepard, Alison Laufer Halpin, Dave Wentworth, Vivien Dugan, Suxiang Tong, Justin Lee |
| EPI_ISL_1798677 | Laboratory Corporation of America | Centers for Disease Control and Prevention Division of Viral Diseases, Pathogen Discovery | Dakota Howard, Dhwani Batra, Peter W. Cook, Kara Moser, Adrian Paskey, Jason Caravas, Benjamin Rambo-Martin, Shatavia Morrison, Christopher Gulvick, Scott Sammons, Yvette Unoarumhi, Darlene Wagner, Matthew Schmerer, Minoo Agarwal, Eyad Almasri, Debbie Boles, Ayla Burns, Nuthawin Charoensri, Oren Cohen, Susan Countryman, Mary Ann Cristobal, Bobbi Croy, Suzanne Dale, Hrushikesh Deshmukh, Amanda Douglas, Vincent Drouillon, Marcia Eisenberg, Howard Engler, Rama Ghatti, Prashant Gupta, Susan Hicks, Jake Humphrey, Lax Iyer, Manoj Jain, Mohan Kolli, Brian Krueger, Tim Kuphal, Stanley Letovsky, Michael Levandoski, Craig Lukasik, Jonathan Meltzer, Brian Norvell, Mindy Nye, Scott Parker, Christos Petropoulos, John Pruitt, Steven Ragan, Scott Ryan, Mike Sapeta, Jana Schroth, Suresh Babu Selvaraju, Goran Stevovic, Amanda Suchanek, Andrea Throop, Lyndon Tilson, Thomas Urban, Joe Voshell, Kimberly Wagner, Jonathan Williams, Mary Williamson, Qian Zeng, Tricia Zwiefelhofer, Clinton R. Paden, Duncan MacCannell |
| EPI_ISL_1798678, EPI_ISL_1798680 | TN Division of Laboratory Services | Centers for Disease Control and Prevention Division of Viral Diseases, Pathogen Discovery | Mili Sheth, Sarah Nobles, Jasmine Padilla, Mark Burroughs, Shoshona Le, Katie Dillon, Peter Cook, Clinton R. Paden, Dhwani Batra, Krista Queen, Kristen Knipe, Dakota Howard, Yvette Unoarumhi, Darlene Wagner, Matthew Schmerer, Ben L. Rambo-Martin, Kristine Lacek, Sam Shepard, Alison Laufer Halpin, Dave Wentworth, Vivien Dugan, Suxiang Tong, Justin Lee |

| | | | |
|---|---|---|---|
| EPI_ISL_1798681 | Laboratory Corporation of America | Centers for Disease Control and Prevention Division of Viral Diseases, Pathogen Discovery | Dakota Howard, Dhwani Batra, Peter W. Cook, Kara Moser, Adrian Paskey, Jason Caravas, Benjamin Rambo-Martin, Shatavia Morrison, Christopher Gulvick, Scott Sammons, Yvette Unoarumhi, Darlene Wagner, Matthew Schmerer, Minoo Agarwal, Eyad Almasri, Debbie Boles, Ayla Burns, Nuthawin Charoensri, Oren Cohen, Susan Countryman, Mary Ann Cristobal, Bobbi Croy, Suzanne Dale, Hrushikesh Deshmukh, Amanda Douglas, Vincent Drouillon, Marcia Eisenberg, Howard Engler, Rama Ghatti, Prashant Gupta, Susan Hicks, Jake Humphrey, Lax Iyer, Manoj Jain, Mohan Kolli, Brian Krueger, Tim Kuphal, Stanley Letovsky, Michael Levandoski, Craig Lukasik, Jonathan Meltzer, Brian Norvell, Mindy Nye, Scott Parker, Christos Petropoulos, John Pruitt, Steven Ragan, Scott Ryan, Mike Sapeta, Jana Schroth, Suresh Babu Selvaraju, Goran Stevovic, Amanda Suchanek, Andrea Throop, Lyndon Tilson, Thomas Urban, Joe Voshell, Kimberly Wagner, Jonathan Williams, Mary Williamson, Qian Zeng, Tricia Zwiefelhofer, Clinton R. Paden, Duncan MacCannell |
| EPI_ISL_1798682, EPI_ISL_1798684 | TN Division of Laboratory Services | Centers for Disease Control and Prevention Division of Viral Diseases, Pathogen Discovery | Mili Sheth, Sarah Nobles, Jasmine Padilla, Mark Burroughs, Shoshona Le, Katie Dillon, Peter Cook, Clinton R. Paden, Dhwani Batra, Krista Queen, Kristen Knipe, Dakota Howard, Yvette Unoarumhi, Darlene Wagner, Matthew Schmerer, Ben L. Rambo-Martin, Kristine Lacek, Sam Shepard, Alison Laufer Halpin, Dave Wentworth, Vivien Dugan, Suxiang Tong, Justin Lee |
| EPI_ISL_1798685, EPI_ISL_1798686 | Laboratory Corporation of America | Centers for Disease Control and Prevention Division of Viral Diseases, Pathogen Discovery | Dakota Howard, Dhwani Batra, Peter W. Cook, Kara Moser, Adrian Paskey, Jason Caravas, Benjamin Rambo-Martin, Shatavia Morrison, Christopher Gulvick, Scott Sammons, Yvette Unoarumhi, Darlene Wagner, Matthew Schmerer, Minoo Agarwal, Eyad Almasri, Debbie Boles, Ayla Burns, Nuthawin Charoensri, Oren Cohen, Susan Countryman, Mary Ann Cristobal, Bobbi Croy, Suzanne Dale, Hrushikesh Deshmukh, Amanda Douglas, Vincent Drouillon, Marcia Eisenberg, Howard Engler, Rama Ghatti, Prashant Gupta, Susan Hicks, Jake Humphrey, Lax Iyer, Manoj Jain, Mohan Kolli, Brian Krueger, Tim Kuphal, Stanley Letovsky, Michael Levandoski, Craig Lukasik, Jonathan Meltzer, Brian Norvell, Mindy Nye, Scott Parker, Christos Petropoulos, John Pruitt, Steven Ragan, Scott Ryan, Mike Sapeta, Jana Schroth, Suresh Babu Selvaraju, Goran Stevovic, Amanda Suchanek, Andrea Throop, Lyndon Tilson, Thomas Urban, Joe Voshell, Kimberly Wagner, Jonathan Williams, Mary Williamson, Qian Zeng, Tricia Zwiefelhofer, Clinton R. Paden, Duncan MacCannell |
| EPI_ISL_1798687 | TN Division of Laboratory Services | Centers for Disease Control and Prevention Division of Viral Diseases, Pathogen Discovery | Mili Sheth, Sarah Nobles, Jasmine Padilla, Mark Burroughs, Shoshona Le, Katie Dillon, Peter Cook, Clinton R. Paden, Dhwani Batra, Krista Queen, Kristen Knipe, Dakota Howard, Yvette Unoarumhi, Darlene Wagner, Matthew Schmerer, Ben L. Rambo-Martin, Kristine Lacek, Sam Shepard, Alison Laufer Halpin, Dave Wentworth, Vivien Dugan, Suxiang Tong, Justin Lee |
| EPI_ISL_1798688 | Laboratory Corporation of America | Centers for Disease Control and Prevention Division of Viral Diseases, Pathogen Discovery | Dakota Howard, Dhwani Batra, Peter W. Cook, Kara Moser, Adrian Paskey, Jason Caravas, Benjamin Rambo-Martin, Shatavia Morrison, Christopher Gulvick, Scott Sammons, Yvette Unoarumhi, Darlene Wagner, Matthew Schmerer, Minoo Agarwal, Eyad Almasri, Debbie Boles, Ayla Burns, Nuthawin Charoensri, Oren Cohen, Susan Countryman, Mary Ann Cristobal, Bobbi Croy, Suzanne Dale, Hrushikesh Deshmukh, Amanda Douglas, Vincent Drouillon, Marcia Eisenberg, Howard Engler, Rama Ghatti, Prashant Gupta, Susan Hicks, Jake Humphrey, Lax Iyer, Manoj Jain, Mohan Kolli, Brian Krueger, Tim Kuphal, Stanley Letovsky, Michael Levandoski, Craig Lukasik, Jonathan Meltzer, Brian Norvell, Mindy Nye, Scott Parker, Christos Petropoulos, John Pruitt, Steven Ragan, Scott Ryan, Mike Sapeta, Jana Schroth, Suresh Babu Selvaraju, Goran Stevovic, Amanda Suchanek, Andrea Throop, Lyndon Tilson, Thomas Urban, Joe Voshell, Kimberly Wagner, Jonathan Williams, Mary Williamson, Qian Zeng, Tricia Zwiefelhofer, Clinton R. Paden, Duncan MacCannell |
| EPI_ISL_1798689, EPI_ISL_1798691 | TN Division of Laboratory Services | Centers for Disease Control and Prevention Division of Viral Diseases, Pathogen Discovery | Mili Sheth, Sarah Nobles, Jasmine Padilla, Mark Burroughs, Shoshona Le, Katie Dillon, Peter Cook, Clinton R. Paden, Dhwani Batra, Krista Queen, Kristen Knipe, Dakota Howard, Yvette Unoarumhi, Darlene Wagner, Matthew Schmerer, Ben L. Rambo-Martin, Kristine Lacek, Sam Shepard, Alison Laufer Halpin, Dave Wentworth, Vivien Dugan, Suxiang Tong, Justin Lee |
| EPI_ISL_1798692 | Laboratory Corporation of America | Centers for Disease Control and Prevention Division of Viral Diseases, Pathogen Discovery | Dakota Howard, Dhwani Batra, Peter W. Cook, Kara Moser, Adrian Paskey, Jason Caravas, Benjamin Rambo-Martin, Shatavia Morrison, Christopher Gulvick, Scott Sammons, Yvette Unoarumhi, Darlene Wagner, Matthew Schmerer, Minoo Agarwal, Eyad Almasri, Debbie Boles, Ayla Burns, Nuthawin Charoensri, Oren Cohen, Susan Countryman, Mary Ann Cristobal, Bobbi Croy, Suzanne Dale, Hrushikesh Deshmukh, Amanda Douglas, Vincent Drouillon, Marcia Eisenberg, Howard Engler, Rama Ghatti, Prashant Gupta, Susan Hicks, Jake Humphrey, Lax Iyer, Manoj Jain, Mohan Kolli, Brian Krueger, Tim Kuphal, Stanley Letovsky, Michael Levandoski, Craig Lukasik, Jonathan Meltzer, Brian Norvell, Mindy Nye, Scott Parker, Christos Petropoulos, John Pruitt, Steven Ragan, Scott Ryan, Mike Sapeta, Jana Schroth, Suresh Babu Selvaraju, Goran Stevovic, Amanda Suchanek, Andrea Throop, Lyndon Tilson, Thomas Urban, Joe Voshell, Kimberly Wagner, Jonathan Williams, Mary Williamson, Qian Zeng, Tricia Zwiefelhofer, Clinton R. Paden, Duncan MacCannell |
| EPI_ISL_1798693 | TN Division of Laboratory Services | Centers for Disease Control and Prevention Division of Viral Diseases, Pathogen Discovery | Mili Sheth, Sarah Nobles, Jasmine Padilla, Mark Burroughs, Shoshona Le, Katie Dillon, Peter Cook, Clinton R. Paden, Dhwani Batra, Krista Queen, Kristen Knipe, Dakota Howard, Yvette Unoarumhi, Darlene Wagner, Matthew Schmerer, Ben L. Rambo-Martin, Kristine Lacek, Sam Shepard, Alison Laufer Halpin, Dave Wentworth, Vivien Dugan, Suxiang Tong, Justin Lee |
| EPI_ISL_1798695, EPI_ISL_1798696, EPI_ISL_1798698 | RI State Health Laboratories | Centers for Disease Control and Prevention Division of Viral Diseases, Pathogen Discovery | Mili Sheth, Sarah Nobles, Jasmine Padilla, Mark Burroughs, Shoshona Le, Katie Dillon, Peter Cook, Clinton R. Paden, Dhwani Batra, Krista Queen, Kristen Knipe, Dakota Howard, Yvette Unoarumhi, Darlene Wagner, Matthew Schmerer, Ben L. Rambo-Martin, Kristine Lacek, Sam Shepard, Alison Laufer Halpin, Dave Wentworth, Vivien Dugan, Suxiang Tong, Justin Lee |
| EPI_ISL_1798699 | Laboratory Corporation of America | Centers for Disease Control and Prevention Division of Viral Diseases, Pathogen Discovery | Dakota Howard, Dhwani Batra, Peter W. Cook, Kara Moser, Adrian Paskey, Jason Caravas, Benjamin Rambo-Martin, Shatavia Morrison, Christopher Gulvick, Scott Sammons, Yvette Unoarumhi, Darlene Wagner, Matthew Schmerer, Minoo Agarwal, Eyad Almasri, Debbie Boles, Ayla Burns, Nuthawin Charoensri, Oren Cohen, Susan Countryman, Mary Ann Cristobal, Bobbi Croy, Suzanne Dale, Hrushikesh Deshmukh, Amanda Douglas, Vincent Drouillon, Marcia Eisenberg, Howard Engler, Rama Ghatti, Prashant Gupta, Susan Hicks, Jake Humphrey, Lax Iyer, Manoj Jain, Mohan Kolli, Brian Krueger, Tim Kuphal, Stanley Letovsky, Michael Levandoski, Craig Lukasik, Jonathan Meltzer, Brian Norvell, Mindy Nye, Scott Parker, Christos Petropoulos, John Pruitt, Steven Ragan, Scott Ryan, Mike Sapeta, Jana Schroth, Suresh Babu Selvaraju, Goran Stevovic, Amanda Suchanek, Andrea Throop, Lyndon Tilson, Thomas Urban, Joe Voshell, Kimberly Wagner, Jonathan Williams, Mary Williamson, Qian Zeng, Tricia Zwiefelhofer, Clinton R. Paden, Duncan MacCannell |
| EPI_ISL_1798700 | RI State Health Laboratories | Centers for Disease Control and Prevention Division of Viral Diseases, Pathogen Discovery | Mili Sheth, Sarah Nobles, Jasmine Padilla, Mark Burroughs, Shoshona Le, Katie Dillon, Peter Cook, Clinton R. Paden, Dhwani Batra, Krista Queen, Kristen Knipe, Dakota Howard, Yvette Unoarumhi, Darlene Wagner, Matthew Schmerer, Ben L. Rambo-Martin, Kristine Lacek, Sam Shepard, Alison Laufer Halpin, Dave Wentworth, Vivien Dugan, Suxiang Tong, Justin Lee |
| EPI_ISL_1798702 | Laboratory Corporation of America | Centers for Disease Control and Prevention Division of Viral Diseases, Pathogen Discovery | Dakota Howard, Dhwani Batra, Peter W. Cook, Kara Moser, Adrian Paskey, Jason Caravas, Benjamin Rambo-Martin, Shatavia Morrison, Christopher Gulvick, Scott Sammons, Yvette Unoarumhi, Darlene Wagner, Matthew Schmerer, Minoo Agarwal, Eyad Almasri, Debbie Boles, Ayla Burns, Nuthawin Charoensri, Oren Cohen, Susan Countryman, Mary Ann Cristobal, Bobbi Croy, Suzanne Dale, Hrushikesh Deshmukh, Amanda Douglas, Vincent Drouillon, Marcia Eisenberg, Howard Engler, Rama Ghatti, Prashant Gupta, Susan Hicks, Jake Humphrey, Lax Iyer, Manoj Jain, Mohan Kolli, Brian Krueger, Tim Kuphal, Stanley Letovsky, Michael Levandoski, Craig Lukasik, Jonathan Meltzer, Brian Norvell, Mindy Nye, Scott Parker, Christos Petropoulos, John Pruitt, Steven Ragan, Scott Ryan, Mike Sapeta, Jana Schroth, Suresh Babu Selvaraju, Goran Stevovic, Amanda Suchanek, Andrea Throop, Lyndon Tilson, Thomas Urban, Joe Voshell, Kimberly Wagner, Jonathan Williams, Mary Williamson, Qian Zeng, Tricia Zwiefelhofer, Clinton R. Paden, Duncan MacCannell |
| EPI_ISL_1798703 | RI State Health Laboratories | Centers for Disease Control and Prevention Division of Viral Diseases, Pathogen Discovery | Mili Sheth, Sarah Nobles, Jasmine Padilla, Mark Burroughs, Shoshona Le, Katie Dillon, Peter Cook, Clinton R. Paden, Dhwani Batra, Krista Queen, Kristen Knipe, Dakota Howard, Yvette Unoarumhi, Darlene Wagner, Matthew Schmerer, Ben L. Rambo-Martin, Kristine Lacek, Sam Shepard, Alison Laufer Halpin, Dave Wentworth, Vivien Dugan, Suxiang Tong, Justin Lee |
| EPI_ISL_1798704 | Laboratory Corporation of America | Centers for Disease Control and Prevention Division of Viral Diseases, Pathogen Discovery | Dakota Howard, Dhwani Batra, Peter W. Cook, Kara Moser, Adrian Paskey, Jason Caravas, Benjamin Rambo-Martin, Shatavia Morrison, Christopher Gulvick, Scott Sammons, Yvette Unoarumhi, Darlene Wagner, Matthew Schmerer, Minoo Agarwal, Eyad Almasri, Debbie Boles, Ayla Burns, Nuthawin Charoensri, Oren Cohen, Susan Countryman, Mary Ann Cristobal, Bobbi Croy, Suzanne Dale, Hrushikesh Deshmukh, Amanda Douglas, Vincent Drouillon, Marcia Eisenberg, Howard Engler, Rama Ghatti, Prashant Gupta, Susan Hicks, Jake Humphrey, Lax Iyer, Manoj Jain, Mohan Kolli, Brian Krueger, Tim Kuphal, Stanley Letovsky, Michael Levandoski, Craig Lukasik, Jonathan Meltzer, Brian Norvell, Mindy Nye, Scott Parker, Christos Petropoulos, John Pruitt, Steven Ragan, Scott Ryan, Mike Sapeta, Jana Schroth, Suresh Babu Selvaraju, Goran Stevovic, Amanda Suchanek, Andrea Throop, Lyndon Tilson, Thomas Urban, Joe Voshell, Kimberly Wagner, Jonathan Williams, Mary Williamson, Qian Zeng, Tricia Zwiefelhofer, Clinton R. Paden, Duncan MacCannell |
| EPI_ISL_1798705 | RI State Health Laboratories | Centers for Disease Control and Prevention Division of Viral Diseases, Pathogen Discovery | Mili Sheth, Sarah Nobles, Jasmine Padilla, Mark Burroughs, Shoshona Le, Katie Dillon, Peter Cook, Clinton R. Paden, Dhwani Batra, Krista Queen, Kristen Knipe, Dakota Howard, Yvette Unoarumhi, Darlene Wagner, Matthew Schmerer, Ben L. Rambo-Martin, Kristine Lacek, Sam Shepard, Alison Laufer Halpin, Dave Wentworth, Vivien Dugan, Suxiang Tong, Justin Lee |
| EPI_ISL_1798706 | Laboratory Corporation of America | Centers for Disease Control and Prevention Division of Viral Diseases, Pathogen Discovery | Dakota Howard, Dhwani Batra, Peter W. Cook, Kara Moser, Adrian Paskey, Jason Caravas, Benjamin Rambo-Martin, Shatavia Morrison, Christopher Gulvick, Scott Sammons, Yvette Unoarumhi, Darlene Wagner, Matthew Schmerer, Minoo Agarwal, Eyad Almasri, Debbie Boles, Ayla Burns, Nuthawin Charoensri, Oren Cohen, Susan Countryman, Mary Ann Cristobal, Bobbi Croy, Suzanne Dale, Hrushikesh Deshmukh, Amanda Douglas, Vincent Drouillon, Marcia Eisenberg, Howard Engler, Rama Ghatti, Prashant Gupta, Susan Hicks, Jake Humphrey, Lax Iyer, Manoj Jain, Mohan Kolli, Brian Krueger, Tim Kuphal, Stanley Letovsky, Michael Levandoski, Craig Lukasik, Jonathan Meltzer, Brian Norvell, Mindy Nye, Scott Parker, Christos Petropoulos, John Pruitt, Steven Ragan, Scott Ryan, Mike Sapeta, Jana Schroth, Suresh Babu Selvaraju, Goran Stevovic, Amanda Suchanek, Andrea Throop, Lyndon Tilson, Thomas Urban, Joe Voshell, Kimberly Wagner, Jonathan Williams, Mary Williamson, Qian Zeng, Tricia Zwiefelhofer, Clinton R. Paden, Duncan MacCannell |
| EPI_ISL_1798707, EPI_ISL_1798708, EPI_ISL_1798710 | RI State Health Laboratories | Centers for Disease Control and Prevention Division of Viral Diseases, Pathogen Discovery | Mili Sheth, Sarah Nobles, Jasmine Padilla, Mark Burroughs, Shoshona Le, Katie Dillon, Peter Cook, Clinton R. Paden, Dhwani Batra, Krista Queen, Kristen Knipe, Dakota Howard, Yvette Unoarumhi, Darlene Wagner, Matthew Schmerer, Ben L. Rambo-Martin, Kristine Lacek, Sam Shepard, Alison Laufer Halpin, Dave Wentworth, Vivien Dugan, Suxiang Tong, Justin Lee |
| EPI_ISL_1798711 | Laboratory Corporation of America | Centers for Disease Control and Prevention Division of Viral Diseases, Pathogen Discovery | Dakota Howard, Dhwani Batra, Peter W. Cook, Kara Moser, Adrian Paskey, Jason Caravas, Benjamin Rambo-Martin, Shatavia Morrison, Christopher Gulvick, Scott Sammons, Yvette Unoarumhi, Darlene Wagner, Matthew Schmerer, Minoo Agarwal, Eyad Almasri, Debbie Boles, Ayla Burns, Nuthawin Charoensri, Oren Cohen, Susan Countryman, Mary Ann Cristobal, Bobbi Croy, Suzanne Dale, Hrushikesh Deshmukh, Amanda Douglas, Vincent Drouillon, Marcia Eisenberg, Howard Engler, Rama Ghatti, Prashant Gupta, Susan Hicks, Jake Humphrey, Lax Iyer, Manoj Jain, Mohan Kolli, Brian Krueger, Tim Kuphal, Stanley Letovsky, Michael Levandoski, Craig Lukasik, Jonathan Meltzer, Brian Norvell, Mindy Nye, Scott Parker, Christos Petropoulos, John Pruitt, Steven Ragan, Scott Ryan, Mike Sapeta, Jana Schroth, Suresh Babu Selvaraju, Goran Stevovic, Amanda Suchanek, Andrea Throop, Lyndon Tilson, Thomas Urban, Joe Voshell, Kimberly Wagner, Jonathan Williams, Mary Williamson, Qian Zeng, Tricia Zwiefelhofer, Clinton R. Paden, Duncan MacCannell |
| EPI_ISL_1798713 | RI State Health Laboratories | Centers for Disease Control and Prevention Division of Viral Diseases, Pathogen Discovery | Mili Sheth, Sarah Nobles, Jasmine Padilla, Mark Burroughs, Shoshona Le, Katie Dillon, Peter Cook, Clinton R. Paden, Dhwani Batra, Krista Queen, Kristen Knipe, Dakota Howard, Yvette Unoarumhi, Darlene Wagner, Matthew Schmerer, Ben L. Rambo-Martin, Kristine Lacek, Sam Shepard, Alison Laufer Halpin, Dave Wentworth, Vivien Dugan, Suxiang Tong, Justin Lee |
| EPI_ISL_1798714 | Laboratory Corporation of America | Centers for Disease Control and Prevention Division of Viral Diseases, Pathogen Discovery | Dakota Howard, Dhwani Batra, Peter W. Cook, Kara Moser, Adrian Paskey, Jason Caravas, Benjamin Rambo-Martin, Shatavia Morrison, Christopher Gulvick, Scott Sammons, Yvette Unoarumhi, Darlene Wagner, Matthew Schmerer, Minoo Agarwal, Eyad Almasri, Debbie Boles, Ayla Burns, Nuthawin Charoensri, Oren Cohen, Susan Countryman, Mary Ann Cristobal, Bobbi Croy, Suzanne Dale, Hrushikesh Deshmukh, Amanda Douglas, Vincent Drouillon, Marcia Eisenberg, Howard Engler, Rama Ghatti, Prashant Gupta, Susan Hicks, Jake Humphrey, Lax Iyer, Manoj Jain, Mohan Kolli, Brian Krueger, Tim Kuphal, Stanley Letovsky, Michael Levandoski, Craig Lukasik, Jonathan Meltzer, Brian Norvell, Mindy Nye, Scott Parker, Christos Petropoulos, John Pruitt, Steven Ragan, Scott Ryan, Mike Sapeta, Jana Schroth, Suresh Babu Selvaraju, Goran Stevovic, Amanda Suchanek, Andrea Throop, Lyndon Tilson, Thomas Urban, Joe Voshell, Kimberly Wagner, Jonathan Williams, Mary Williamson, Qian Zeng, Tricia Zwiefelhofer, Clinton R. Paden, Duncan MacCannell |
| EPI_ISL_1798715, EPI_ISL_1798716 | RI State Health Laboratories | Centers for Disease Control and Prevention Division of Viral Diseases, Pathogen Discovery | Mili Sheth, Sarah Nobles, Jasmine Padilla, Mark Burroughs, Shoshona Le, Katie Dillon, Peter Cook, Clinton R. Paden, Dhwani Batra, Krista Queen, Kristen Knipe, Dakota Howard, Yvette Unoarumhi, Darlene Wagner, Matthew Schmerer, Ben L. Rambo-Martin, Kristine Lacek, Sam Shepard, Alison Laufer Halpin, Dave Wentworth, Vivien Dugan, Suxiang Tong, Justin Lee |
| EPI_ISL_1798717 | Laboratory Corporation of America | Centers for Disease Control and Prevention Division of Viral Diseases, Pathogen Discovery | Dakota Howard, Dhwani Batra, Peter W. Cook, Kara Moser, Adrian Paskey, Jason Caravas, Benjamin Rambo-Martin, Shatavia Morrison, Christopher Gulvick, Scott Sammons, Yvette Unoarumhi, Darlene Wagner, Matthew Schmerer, Minoo Agarwal, Eyad Almasri, Debbie Boles, Ayla Burns, Nuthawin Charoensri, Oren Cohen, Susan Countryman, Mary Ann Cristobal, Bobbi Croy, Suzanne Dale, Hrushikesh Deshmukh, Amanda Douglas, Vincent Drouillon, Marcia Eisenberg, Howard Engler, Rama Ghatti, Prashant Gupta, Susan Hicks, Jake Humphrey, Lax Iyer, Manoj Jain, Mohan Kolli, Brian Krueger, Tim Kuphal, Stanley Letovsky, Michael Levandoski, Craig Lukasik, Jonathan Meltzer, Brian Norvell, Mindy Nye, Scott Parker, Christos Petropoulos, John Pruitt, Steven Ragan, Scott Ryan, Mike Sapeta, Jana Schroth, Suresh Babu Selvaraju, Goran Stevovic, Amanda Suchanek, Andrea Throop, Lyndon Tilson, Thomas Urban, Joe Voshell, Kimberly Wagner, Jonathan Williams, Mary Williamson, Qian Zeng, Tricia Zwiefelhofer, Clinton R. Paden, Duncan MacCannell |
| EPI_ISL_1798719, EPI_ISL_1798720, EPI_ISL_1798722 | RI State Health Laboratories | Centers for Disease Control and Prevention Division of Viral Diseases, Pathogen Discovery | Mili Sheth, Sarah Nobles, Jasmine Padilla, Mark Burroughs, Shoshona Le, Katie Dillon, Peter Cook, Clinton R. Paden, Dhwani Batra, Krista Queen, Kristen Knipe, Dakota Howard, Yvette Unoarumhi, Darlene Wagner, Matthew Schmerer, Ben L. Rambo-Martin, Kristine Lacek, Sam Shepard, Alison Laufer Halpin, Dave Wentworth, Vivien Dugan, Suxiang Tong, Justin Lee |
| EPI_ISL_1798723 | Laboratory Corporation of America | Centers for Disease Control and Prevention Division of Viral Diseases, Pathogen Discovery | Dakota Howard, Dhwani Batra, Peter W. Cook, Kara Moser, Adrian Paskey, Jason Caravas, Benjamin Rambo-Martin, Shatavia Morrison, Christopher Gulvick, Scott Sammons, Yvette Unoarumhi, Darlene Wagner, Matthew Schmerer, Minoo Agarwal, Eyad Almasri, Debbie Boles, Ayla Burns, Nuthawin Charoensri, Oren Cohen, Susan Countryman, Mary Ann Cristobal, Bobbi Croy, Suzanne Dale, Hrushikesh Deshmukh, Amanda Douglas, Vincent Drouillon, Marcia Eisenberg, Howard Engler, Rama Ghatti, Prashant Gupta, Susan Hicks, Jake Humphrey, Lax Iyer, Manoj Jain, Mohan Kolli, Brian Krueger, Tim Kuphal, Stanley Letovsky, Michael Levandoski, Craig Lukasik, Jonathan Meltzer, Brian Norvell, Mindy Nye, Scott Parker, Christos Petropoulos, John Pruitt, Steven Ragan, Scott Ryan, Mike Sapeta, Jana Schroth, Suresh Babu Selvaraju, Goran Stevovic, Amanda Suchanek, Andrea Throop, Lyndon Tilson, Thomas Urban, Joe Voshell, Kimberly Wagner, Jonathan Williams, Mary Williamson, Qian Zeng, Tricia Zwiefelhofer, Clinton R. Paden, Duncan MacCannell |
| EPI_ISL_1798724, EPI_ISL_1798726, EPI_ISL_1798728 | RI State Health Laboratories | Centers for Disease Control and Prevention Division of Viral Diseases, Pathogen Discovery | Mili Sheth, Sarah Nobles, Jasmine Padilla, Mark Burroughs, Shoshona Le, Katie Dillon, Peter Cook, Clinton R. Paden, Dhwani Batra, Krista Queen, Kristen Knipe, Dakota Howard, Yvette Unoarumhi, Darlene Wagner, Matthew Schmerer, Ben L. Rambo-Martin, Kristine Lacek, Sam Shepard, Alison Laufer Halpin, Dave Wentworth, Vivien Dugan, Suxiang Tong, Justin Lee |
| EPI_ISL_1798729 | Laboratory Corporation of America | Centers for Disease Control and Prevention Division of Viral Diseases, Pathogen Discovery | Dakota Howard, Dhwani Batra, Peter W. Cook, Kara Moser, Adrian Paskey, Jason Caravas, Benjamin Rambo-Martin, Shatavia Morrison, Christopher Gulvick, Scott Sammons, Yvette Unoarumhi, Darlene Wagner, Matthew Schmerer, Minoo Agarwal, Eyad Almasri, Debbie Boles, Ayla Burns, Nuthawin Charoensri, Oren Cohen, Susan Countryman, Mary Ann Cristobal, Bobbi Croy, Suzanne Dale, Hrushikesh Deshmukh, Amanda Douglas, Vincent Drouillon, Marcia Eisenberg, Howard Engler, Rama Ghatti, Prashant Gupta, Susan Hicks, Jake Humphrey, Lax Iyer, Manoj Jain, Mohan Kolli, Brian Krueger, Tim Kuphal, Stanley Letovsky, Michael Levandoski, Craig Lukasik, Jonathan Meltzer, Brian Norvell, Mindy Nye, Scott Parker, Christos Petropoulos, John Pruitt, Steven Ragan, Scott Ryan, Mike Sapeta, Jana Schroth, Suresh Babu Selvaraju, Goran Stevovic, Amanda Suchanek, Andrea Throop, Lyndon Tilson, Thomas Urban, Joe Voshell, Kimberly Wagner, Jonathan Williams, Mary Williamson, Qian Zeng, Tricia Zwiefelhofer, Clinton R. Paden, Duncan MacCannell |
| EPI_ISL_1798730 | RI State Health Laboratories | Centers for Disease Control and Prevention Division of Viral Diseases, Pathogen Discovery | Mili Sheth, Sarah Nobles, Jasmine Padilla, Mark Burroughs, Shoshona Le, Katie Dillon, Peter Cook, Clinton R. Paden, Dhwani Batra, Krista Queen, Kristen Knipe, Dakota Howard, Yvette Unoarumhi, Darlene Wagner, Matthew Schmerer, Ben L. Rambo-Martin, Kristine Lacek, Sam Shepard, Alison Laufer Halpin, Dave Wentworth, Vivien Dugan, Suxiang Tong, Justin Lee |
| EPI_ISL_1798731 | Laboratory Corporation of America | Centers for Disease Control and Prevention Division of Viral Diseases, Pathogen Discovery | Dakota Howard, Dhwani Batra, Peter W. Cook, Kara Moser, Adrian Paskey, Jason Caravas, Benjamin Rambo-Martin, Shatavia Morrison, Christopher Gulvick, Scott Sammons, Yvette Unoarumhi, Darlene Wagner, Matthew Schmerer, Minoo Agarwal, Eyad Almasri, Debbie Boles, Ayla Burns, Nuthawin Charoensri, Oren Cohen, Susan Countryman, Mary Ann Cristobal, Bobbi Croy, Suzanne Dale, Hrushikesh Deshmukh, Amanda Douglas, Vincent Drouillon, Marcia Eisenberg, Howard Engler, Rama Ghatti, Prashant Gupta, Susan Hicks, Jake Humphrey, Lax Iyer, Manoj Jain, Mohan Kolli, Brian Krueger, Tim Kuphal, Stanley Letovsky, Michael Levandoski, Craig Lukasik, Jonathan Meltzer, Brian Norvell, Mindy Nye, Scott Parker, Christos Petropoulos, John Pruitt, Steven Ragan, Scott Ryan, Mike Sapeta, Jana Schroth, Suresh Babu Selvaraju, Goran Stevovic, Amanda Suchanek, Andrea Throop, Lyndon Tilson, Thomas Urban, Joe Voshell, Kimberly Wagner, Jonathan Williams, Mary Williamson, Qian Zeng, Tricia Zwiefelhofer, Clinton R. Paden, Duncan MacCannell |

| | | | |
|---|---|---|---|
| EPI_ISL_1798732, EPI_ISL_1798734 | RI State Health Laboratories | Centers for Disease Control and Prevention Division of Viral Diseases, Pathogen Discovery | Mili Sheth, Sarah Nobles, Jasmine Padilla, Mark Burroughs, Shoshona Le, Katie Dillon, Peter Cook, Clinton R. Paden, Dhwani Batra, Krista Queen, Kristen Knipe, Dakota Howard, Yvette Unoarumhi, Darlene Wagner, Matthew Schmerer, Ben L. Rambo-Martin, Kristine Lacek, Sam Shepard, Alison Laufer Halpin, Dave Wentworth, Vivien Dugan, Suxiang Tong, Justin Lee |
| EPI_ISL_1798735 | Laboratory Corporation of America | Centers for Disease Control and Prevention Division of Viral Diseases, Pathogen Discovery | Dakota Howard, Dhwani Batra, Peter W. Cook, Kara Moser, Adrian Paskey, Jason Caravas, Benjamin Rambo-Martin, Shatavia Morrison, Christopher Gulvick, Scott Sammons, Yvette Unoarumhi, Darlene Wagner, Matthew Schmerer, Minoo Agarwal, Eyad Almasri, Debbie Boles, Ayla Burns, Nuthawin Charoensri, Oren Cohen, Susan Countryman, Mary Ann Cristobal, Bobbi Croy, Suzanne Dale, Hrushikesh Deshmukh, Amanda Douglas, Vincent Drouillon, Marcia Eisenberg, Howard Engler, Rama Ghatti, Prashant Gupta, Susan Hicks, Jake Humphrey, Lax Iyer, Manoj Jain, Mohan Kolli, Brian Krueger, Tim Kuphal, Stanley Letovsky, Michael Levandoski, Craig Lukasik, Jonathan Meltzer, Brian Norvell, Mindy Nye, Scott Parker, Christos Petropoulos, John Pruitt, Steven Ragan, Scott Ryan, Mike Sapeta, Jana Schroth, Suresh Babu Selvaraju, Goran Stevovic, Amanda Suchanek, Andrea Throop, Lyndon Tilson, Thomas Urban, Joe Voshell, Kimberly Wagner, Jonathan Williams, Mary Williamson, Qian Zeng, Tricia Zwiefelhofer, Clinton R. Paden, Duncan MacCannell |
| EPI_ISL_1798736 | RI State Health Laboratories | Centers for Disease Control and Prevention Division of Viral Diseases, Pathogen Discovery | Mili Sheth, Sarah Nobles, Jasmine Padilla, Mark Burroughs, Shoshona Le, Katie Dillon, Peter Cook, Clinton R. Paden, Dhwani Batra, Krista Queen, Kristen Knipe, Dakota Howard, Yvette Unoarumhi, Darlene Wagner, Matthew Schmerer, Ben L. Rambo-Martin, Kristine Lacek, Sam Shepard, Alison Laufer Halpin, Dave Wentworth, Vivien Dugan, Suxiang Tong, Justin Lee |
| EPI_ISL_1798737, EPI_ISL_1798738 | Laboratory Corporation of America | Centers for Disease Control and Prevention Division of Viral Diseases, Pathogen Discovery | Dakota Howard, Dhwani Batra, Peter W. Cook, Kara Moser, Adrian Paskey, Jason Caravas, Benjamin Rambo-Martin, Shatavia Morrison, Christopher Gulvick, Scott Sammons, Yvette Unoarumhi, Darlene Wagner, Matthew Schmerer, Minoo Agarwal, Eyad Almasri, Debbie Boles, Ayla Burns, Nuthawin Charoensri, Oren Cohen, Susan Countryman, Mary Ann Cristobal, Bobbi Croy, Suzanne Dale, Hrushikesh Deshmukh, Amanda Douglas, Vincent Drouillon, Marcia Eisenberg, Howard Engler, Rama Ghatti, Prashant Gupta, Susan Hicks, Jake Humphrey, Lax Iyer, Manoj Jain, Mohan Kolli, Brian Krueger, Tim Kuphal, Stanley Letovsky, Michael Levandoski, Craig Lukasik, Jonathan Meltzer, Brian Norvell, Mindy Nye, Scott Parker, Christos Petropoulos, John Pruitt, Steven Ragan, Scott Ryan, Mike Sapeta, Jana Schroth, Suresh Babu Selvaraju, Goran Stevovic, Amanda Suchanek, Andrea Throop, Lyndon Tilson, Thomas Urban, Joe Voshell, Kimberly Wagner, Jonathan Williams, Mary Williamson, Qian Zeng, Tricia Zwiefelhofer, Clinton R. Paden, Duncan MacCannell |
| EPI_ISL_1798739 | RI State Health Laboratories | Centers for Disease Control and Prevention Division of Viral Diseases, Pathogen Discovery | Mili Sheth, Sarah Nobles, Jasmine Padilla, Mark Burroughs, Shoshona Le, Katie Dillon, Peter Cook, Clinton R. Paden, Dhwani Batra, Krista Queen, Kristen Knipe, Dakota Howard, Yvette Unoarumhi, Darlene Wagner, Matthew Schmerer, Ben L. Rambo-Martin, Kristine Lacek, Sam Shepard, Alison Laufer Halpin, Dave Wentworth, Vivien Dugan, Suxiang Tong, Justin Lee |
| EPI_ISL_1798740 | Laboratory Corporation of America | Centers for Disease Control and Prevention Division of Viral Diseases, Pathogen Discovery | Dakota Howard, Dhwani Batra, Peter W. Cook, Kara Moser, Adrian Paskey, Jason Caravas, Benjamin Rambo-Martin, Shatavia Morrison, Christopher Gulvick, Scott Sammons, Yvette Unoarumhi, Darlene Wagner, Matthew Schmerer, Minoo Agarwal, Eyad Almasri, Debbie Boles, Ayla Burns, Nuthawin Charoensri, Oren Cohen, Susan Countryman, Mary Ann Cristobal, Bobbi Croy, Suzanne Dale, Hrushikesh Deshmukh, Amanda Douglas, Vincent Drouillon, Marcia Eisenberg, Howard Engler, Rama Ghatti, Prashant Gupta, Susan Hicks, Jake Humphrey, Lax Iyer, Manoj Jain, Mohan Kolli, Brian Krueger, Tim Kuphal, Stanley Letovsky, Michael Levandoski, Craig Lukasik, Jonathan Meltzer, Brian Norvell, Mindy Nye, Scott Parker, Christos Petropoulos, John Pruitt, Steven Ragan, Scott Ryan, Mike Sapeta, Jana Schroth, Suresh Babu Selvaraju, Goran Stevovic, Amanda Suchanek, Andrea Throop, Lyndon Tilson, Thomas Urban, Joe Voshell, Kimberly Wagner, Jonathan Williams, Mary Williamson, Qian Zeng, Tricia Zwiefelhofer, Clinton R. Paden, Duncan MacCannell |
| EPI_ISL_1798741 | RI State Health Laboratories | Centers for Disease Control and Prevention Division of Viral Diseases, Pathogen Discovery | Mili Sheth, Sarah Nobles, Jasmine Padilla, Mark Burroughs, Shoshona Le, Katie Dillon, Peter Cook, Clinton R. Paden, Dhwani Batra, Krista Queen, Kristen Knipe, Dakota Howard, Yvette Unoarumhi, Darlene Wagner, Matthew Schmerer, Ben L. Rambo-Martin, Kristine Lacek, Sam Shepard, Alison Laufer Halpin, Dave Wentworth, Vivien Dugan, Suxiang Tong, Justin Lee |
| EPI_ISL_1798742 | DE Public Health Laboratory | Centers for Disease Control and Prevention Division of Viral Diseases, Pathogen Discovery | Mili Sheth, Sarah Nobles, Jasmine Padilla, Mark Burroughs, Shoshona Le, Katie Dillon, Peter Cook, Clinton R. Paden, Dhwani Batra, Krista Queen, Kristen Knipe, Dakota Howard, Yvette Unoarumhi, Darlene Wagner, Matthew Schmerer, Ben L. Rambo-Martin, Kristine Lacek, Sam Shepard, Alison Laufer Halpin, Dave Wentworth, Vivien Dugan, Suxiang Tong, Justin Lee |
| EPI_ISL_1798743 | Laboratory Corporation of America | Centers for Disease Control and Prevention Division of Viral Diseases, Pathogen Discovery | Dakota Howard, Dhwani Batra, Peter W. Cook, Kara Moser, Adrian Paskey, Jason Caravas, Benjamin Rambo-Martin, Shatavia Morrison, Christopher Gulvick, Scott Sammons, Yvette Unoarumhi, Darlene Wagner, Matthew Schmerer, Minoo Agarwal, Eyad Almasri, Debbie Boles, Ayla Burns, Nuthawin Charoensri, Oren Cohen, Susan Countryman, Mary Ann Cristobal, Bobbi Croy, Suzanne Dale, Hrushikesh Deshmukh, Amanda Douglas, Vincent Drouillon, Marcia Eisenberg, Howard Engler, Rama Ghatti, Prashant Gupta, Susan Hicks, Jake Humphrey, Lax Iyer, Manoj Jain, Mohan Kolli, Brian Krueger, Tim Kuphal, Stanley Letovsky, Michael Levandoski, Craig Lukasik, Jonathan Meltzer, Brian Norvell, Mindy Nye, Scott Parker, Christos Petropoulos, John Pruitt, Steven Ragan, Scott Ryan, Mike Sapeta, Jana Schroth, Suresh Babu Selvaraju, Goran Stevovic, Amanda Suchanek, Andrea Throop, Lyndon Tilson, Thomas Urban, Joe Voshell, Kimberly Wagner, Jonathan Williams, Mary Williamson, Qian Zeng, Tricia Zwiefelhofer, Clinton R. Paden, Duncan MacCannell |
| EPI_ISL_1798744 | DE Public Health Laboratory | Centers for Disease Control and Prevention Division of Viral Diseases, Pathogen Discovery | Mili Sheth, Sarah Nobles, Jasmine Padilla, Mark Burroughs, Shoshona Le, Katie Dillon, Peter Cook, Clinton R. Paden, Dhwani Batra, Krista Queen, Kristen Knipe, Dakota Howard, Yvette Unoarumhi, Darlene Wagner, Matthew Schmerer, Ben L. Rambo-Martin, Kristine Lacek, Sam Shepard, Alison Laufer Halpin, Dave Wentworth, Vivien Dugan, Suxiang Tong, Justin Lee |
| EPI_ISL_1798745, EPI_ISL_1798746 | Laboratory Corporation of America | Centers for Disease Control and Prevention Division of Viral Diseases, Pathogen Discovery | Dakota Howard, Dhwani Batra, Peter W. Cook, Kara Moser, Adrian Paskey, Jason Caravas, Benjamin Rambo-Martin, Shatavia Morrison, Christopher Gulvick, Scott Sammons, Yvette Unoarumhi, Darlene Wagner, Matthew Schmerer, Minoo Agarwal, Eyad Almasri, Debbie Boles, Ayla Burns, Nuthawin Charoensri, Oren Cohen, Susan Countryman, Mary Ann Cristobal, Bobbi Croy, Suzanne Dale, Hrushikesh Deshmukh, Amanda Douglas, Vincent Drouillon, Marcia Eisenberg, Howard Engler, Rama Ghatti, Prashant Gupta, Susan Hicks, Jake Humphrey, Lax Iyer, Manoj Jain, Mohan Kolli, Brian Krueger, Tim Kuphal, Stanley Letovsky, Michael Levandoski, Craig Lukasik, Jonathan Meltzer, Brian Norvell, Mindy Nye, Scott Parker, Christos Petropoulos, John Pruitt, Steven Ragan, Scott Ryan, Mike Sapeta, Jana Schroth, Suresh Babu Selvaraju, Goran Stevovic, Amanda Suchanek, Andrea Throop, Lyndon Tilson, Thomas Urban, Joe Voshell, Kimberly Wagner, Jonathan Williams, Mary Williamson, Qian Zeng, Tricia Zwiefelhofer, Clinton R. Paden, Duncan MacCannell |
| EPI_ISL_1798747 | DE Public Health Laboratory | Centers for Disease Control and Prevention Division of Viral Diseases, Pathogen Discovery | Mili Sheth, Sarah Nobles, Jasmine Padilla, Mark Burroughs, Shoshona Le, Katie Dillon, Peter Cook, Clinton R. Paden, Dhwani Batra, Krista Queen, Kristen Knipe, Dakota Howard, Yvette Unoarumhi, Darlene Wagner, Matthew Schmerer, Ben L. Rambo-Martin, Kristine Lacek, Sam Shepard, Alison Laufer Halpin, Dave Wentworth, Vivien Dugan, Suxiang Tong, Justin Lee |
| EPI_ISL_1798748 | Laboratory Corporation of America | Centers for Disease Control and Prevention Division of Viral Diseases, Pathogen Discovery | Dakota Howard, Dhwani Batra, Peter W. Cook, Kara Moser, Adrian Paskey, Jason Caravas, Benjamin Rambo-Martin, Shatavia Morrison, Christopher Gulvick, Scott Sammons, Yvette Unoarumhi, Darlene Wagner, Matthew Schmerer, Minoo Agarwal, Eyad Almasri, Debbie Boles, Ayla Burns, Nuthawin Charoensri, Oren Cohen, Susan Countryman, Mary Ann Cristobal, Bobbi Croy, Suzanne Dale, Hrushikesh Deshmukh, Amanda Douglas, Vincent Drouillon, Marcia Eisenberg, Howard Engler, Rama Ghatti, Prashant Gupta, Susan Hicks, Jake Humphrey, Lax Iyer, Manoj Jain, Mohan Kolli, Brian Krueger, Tim Kuphal, Stanley Letovsky, Michael Levandoski, Craig Lukasik, Jonathan Meltzer, Brian Norvell, Mindy Nye, Scott Parker, Christos Petropoulos, John Pruitt, Steven Ragan, Scott Ryan, Mike Sapeta, Jana Schroth, Suresh Babu Selvaraju, Goran Stevovic, Amanda Suchanek, Andrea Throop, Lyndon Tilson, Thomas Urban, Joe Voshell, Kimberly Wagner, Jonathan Williams, Mary Williamson, Qian Zeng, Tricia Zwiefelhofer, Clinton R. Paden, Duncan MacCannell |
| EPI_ISL_1798749 | DE Public Health Laboratory | Centers for Disease Control and Prevention Division of Viral Diseases, Pathogen Discovery | Mili Sheth, Sarah Nobles, Jasmine Padilla, Mark Burroughs, Shoshona Le, Katie Dillon, Peter Cook, Clinton R. Paden, Dhwani Batra, Krista Queen, Kristen Knipe, Dakota Howard, Yvette Unoarumhi, Darlene Wagner, Matthew Schmerer, Ben L. Rambo-Martin, Kristine Lacek, Sam Shepard, Alison Laufer Halpin, Dave Wentworth, Vivien Dugan, Suxiang Tong, Justin Lee |
| EPI_ISL_1798750 | Laboratory Corporation of America | Centers for Disease Control and Prevention Division of Viral Diseases, Pathogen Discovery | Dakota Howard, Dhwani Batra, Peter W. Cook, Kara Moser, Adrian Paskey, Jason Caravas, Benjamin Rambo-Martin, Shatavia Morrison, Christopher Gulvick, Scott Sammons, Yvette Unoarumhi, Darlene Wagner, Matthew Schmerer, Minoo Agarwal, Eyad Almasri, Debbie Boles, Ayla Burns, Nuthawin Charoensri, Oren Cohen, Susan Countryman, Mary Ann Cristobal, Bobbi Croy, Suzanne Dale, Hrushikesh Deshmukh, Amanda Douglas, Vincent Drouillon, Marcia Eisenberg, Howard Engler, Rama Ghatti, Prashant Gupta, Susan Hicks, Jake Humphrey, Lax Iyer, Manoj Jain, Mohan Kolli, Brian Krueger, Tim Kuphal, Stanley Letovsky, Michael Levandoski, Craig Lukasik, Jonathan Meltzer, Brian Norvell, Mindy Nye, Scott Parker, Christos Petropoulos, John Pruitt, Steven Ragan, Scott Ryan, Mike Sapeta, Jana Schroth, Suresh Babu Selvaraju, Goran Stevovic, Amanda Suchanek, Andrea Throop, Lyndon Tilson, Thomas Urban, Joe Voshell, Kimberly Wagner, Jonathan Williams, Mary Williamson, Qian Zeng, Tricia Zwiefelhofer, Clinton R. Paden, Duncan MacCannell |
| EPI_ISL_1798751, EPI_ISL_1798754, EPI_ISL_1798756, EPI_ISL_1798757 | DE Public Health Laboratory | Centers for Disease Control and Prevention Division of Viral Diseases, Pathogen Discovery | Mili Sheth, Sarah Nobles, Jasmine Padilla, Mark Burroughs, Shoshona Le, Katie Dillon, Peter Cook, Clinton R. Paden, Dhwani Batra, Krista Queen, Kristen Knipe, Dakota Howard, Yvette Unoarumhi, Darlene Wagner, Matthew Schmerer, Ben L. Rambo-Martin, Kristine Lacek, Sam Shepard, Alison Laufer Halpin, Dave Wentworth, Vivien Dugan, Suxiang Tong, Justin Lee |
| EPI_ISL_1798758 | Laboratory Corporation of America | Centers for Disease Control and Prevention Division of Viral Diseases, Pathogen Discovery | Dakota Howard, Dhwani Batra, Peter W. Cook, Kara Moser, Adrian Paskey, Jason Caravas, Benjamin Rambo-Martin, Shatavia Morrison, Christopher Gulvick, Scott Sammons, Yvette Unoarumhi, Darlene Wagner, Matthew Schmerer, Minoo Agarwal, Eyad Almasri, Debbie Boles, Ayla Burns, Nuthawin Charoensri, Oren Cohen, Susan Countryman, Mary Ann Cristobal, Bobbi Croy, Suzanne Dale, Hrushikesh Deshmukh, Amanda Douglas, Vincent Drouillon, Marcia Eisenberg, Howard Engler, Rama Ghatti, Prashant Gupta, Susan Hicks, Jake Humphrey, Lax Iyer, Manoj Jain, Mohan Kolli, Brian Krueger, Tim Kuphal, Stanley Letovsky, Michael Levandoski, Craig Lukasik, Jonathan Meltzer, Brian Norvell, Mindy Nye, Scott Parker, Christos Petropoulos, John Pruitt, Steven Ragan, Scott Ryan, Mike Sapeta, Jana Schroth, Suresh Babu Selvaraju, Goran Stevovic, Amanda Suchanek, Andrea Throop, Lyndon Tilson, Thomas Urban, Joe Voshell, Kimberly Wagner, Jonathan Williams, Mary Williamson, Qian Zeng, Tricia Zwiefelhofer, Clinton R. Paden, Duncan MacCannell |
| EPI_ISL_1798759 | DE Public Health Laboratory | Centers for Disease Control and Prevention Division of Viral Diseases, Pathogen Discovery | Mili Sheth, Sarah Nobles, Jasmine Padilla, Mark Burroughs, Shoshona Le, Katie Dillon, Peter Cook, Clinton R. Paden, Dhwani Batra, Krista Queen, Kristen Knipe, Dakota Howard, Yvette Unoarumhi, Darlene Wagner, Matthew Schmerer, Ben L. Rambo-Martin, Kristine Lacek, Sam Shepard, Alison Laufer Halpin, Dave Wentworth, Vivien Dugan, Suxiang Tong, Justin Lee |
| EPI_ISL_1798761 | Laboratory Corporation of America | Centers for Disease Control and Prevention Division of Viral Diseases, Pathogen Discovery | Dakota Howard, Dhwani Batra, Peter W. Cook, Kara Moser, Adrian Paskey, Jason Caravas, Benjamin Rambo-Martin, Shatavia Morrison, Christopher Gulvick, Scott Sammons, Yvette Unoarumhi, Darlene Wagner, Matthew Schmerer, Minoo Agarwal, Eyad Almasri, Debbie Boles, Ayla Burns, Nuthawin Charoensri, Oren Cohen, Susan Countryman, Mary Ann Cristobal, Bobbi Croy, Suzanne Dale, Hrushikesh Deshmukh, Amanda Douglas, Vincent Drouillon, Marcia Eisenberg, Howard Engler, Rama Ghatti, Prashant Gupta, Susan Hicks, Jake Humphrey, Lax Iyer, Manoj Jain, Mohan Kolli, Brian Krueger, Tim Kuphal, Stanley Letovsky, Michael Levandoski, Craig Lukasik, Jonathan Meltzer, Brian Norvell, Mindy Nye, Scott Parker, Christos Petropoulos, John Pruitt, Steven Ragan, Scott Ryan, Mike Sapeta, Jana Schroth, Suresh Babu Selvaraju, Goran Stevovic, Amanda Suchanek, Andrea Throop, Lyndon Tilson, Thomas Urban, Joe Voshell, Kimberly Wagner, Jonathan Williams, Mary Williamson, Qian Zeng, Tricia Zwiefelhofer, Clinton R. Paden, Duncan MacCannell |
| EPI_ISL_1798762, EPI_ISL_1798764 | DE Public Health Laboratory | Centers for Disease Control and Prevention Division of Viral Diseases, Pathogen Discovery | Mili Sheth, Sarah Nobles, Jasmine Padilla, Mark Burroughs, Shoshona Le, Katie Dillon, Peter Cook, Clinton R. Paden, Dhwani Batra, Krista Queen, Kristen Knipe, Dakota Howard, Yvette Unoarumhi, Darlene Wagner, Matthew Schmerer, Ben L. Rambo-Martin, Kristine Lacek, Sam Shepard, Alison Laufer Halpin, Dave Wentworth, Vivien Dugan, Suxiang Tong, Justin Lee |
| EPI_ISL_1798765 | Laboratory Corporation of America | Centers for Disease Control and Prevention Division of Viral Diseases, Pathogen Discovery | Dakota Howard, Dhwani Batra, Peter W. Cook, Kara Moser, Adrian Paskey, Jason Caravas, Benjamin Rambo-Martin, Shatavia Morrison, Christopher Gulvick, Scott Sammons, Yvette Unoarumhi, Darlene Wagner, Matthew Schmerer, Minoo Agarwal, Eyad Almasri, Debbie Boles, Ayla Burns, Nuthawin Charoensri, Oren Cohen, Susan Countryman, Mary Ann Cristobal, Bobbi Croy, Suzanne Dale, Hrushikesh Deshmukh, Amanda Douglas, Vincent Drouillon, Marcia Eisenberg, Howard Engler, Rama Ghatti, Prashant Gupta, Susan Hicks, Jake Humphrey, Lax Iyer, Manoj Jain, Mohan Kolli, Brian Krueger, Tim Kuphal, Stanley Letovsky, Michael Levandoski, Craig Lukasik, Jonathan Meltzer, Brian Norvell, Mindy Nye, Scott Parker, Christos Petropoulos, John Pruitt, Steven Ragan, Scott Ryan, Mike Sapeta, Jana Schroth, Suresh Babu Selvaraju, Goran Stevovic, Amanda Suchanek, Andrea Throop, Lyndon Tilson, Thomas Urban, Joe Voshell, Kimberly Wagner, Jonathan Williams, Mary Williamson, Qian Zeng, Tricia Zwiefelhofer, Clinton R. Paden, Duncan MacCannell |
| EPI_ISL_1798766, EPI_ISL_1798768 | CDPH, Viral and Rickettsial Disease Laboratory | Centers for Disease Control and Prevention Division of Viral Diseases, Pathogen Discovery | Mili Sheth, Sarah Nobles, Jasmine Padilla, Mark Burroughs, Shoshona Le, Katie Dillon, Peter Cook, Clinton R. Paden, Dhwani Batra, Krista Queen, Kristen Knipe, Dakota Howard, Yvette Unoarumhi, Darlene Wagner, Matthew Schmerer, Ben L. Rambo-Martin, Kristine Lacek, Sam Shepard, Alison Laufer Halpin, Dave Wentworth, Vivien Dugan, Suxiang Tong, Justin Lee |
| EPI_ISL_1798769 | Laboratory Corporation of America | Centers for Disease Control and Prevention Division of Viral Diseases, Pathogen Discovery | Dakota Howard, Dhwani Batra, Peter W. Cook, Kara Moser, Adrian Paskey, Jason Caravas, Benjamin Rambo-Martin, Shatavia Morrison, Christopher Gulvick, Scott Sammons, Yvette Unoarumhi, Darlene Wagner, Matthew Schmerer, Minoo Agarwal, Eyad Almasri, Debbie Boles, Ayla Burns, Nuthawin Charoensri, Oren Cohen, Susan Countryman, Mary Ann Cristobal, Bobbi Croy, Suzanne Dale, Hrushikesh Deshmukh, Amanda Douglas, Vincent Drouillon, Marcia Eisenberg, Howard Engler, Rama Ghatti, Prashant Gupta, Susan Hicks, Jake Humphrey, Lax Iyer, Manoj Jain, Mohan Kolli, Brian Krueger, Tim Kuphal, Stanley Letovsky, Michael Levandoski, Craig Lukasik, Jonathan Meltzer, Brian Norvell, Mindy Nye, Scott Parker, Christos Petropoulos, John Pruitt, Steven Ragan, Scott Ryan, Mike Sapeta, Jana Schroth, Suresh Babu Selvaraju, Goran Stevovic, Amanda Suchanek, Andrea Throop, Lyndon Tilson, Thomas Urban, Joe Voshell, Kimberly Wagner, Jonathan Williams, Mary Williamson, Qian Zeng, Tricia Zwiefelhofer, Clinton R. Paden, Duncan MacCannell |
| EPI_ISL_1798770, EPI_ISL_1798772, EPI_ISL_1798774 | CDPH, Viral and Rickettsial Disease Laboratory | Centers for Disease Control and Prevention Division of Viral Diseases, Pathogen Discovery | Mili Sheth, Sarah Nobles, Jasmine Padilla, Mark Burroughs, Shoshona Le, Katie Dillon, Peter Cook, Clinton R. Paden, Dhwani Batra, Krista Queen, Kristen Knipe, Dakota Howard, Yvette Unoarumhi, Darlene Wagner, Matthew Schmerer, Ben L. Rambo-Martin, Kristine Lacek, Sam Shepard, Alison Laufer Halpin, Dave Wentworth, Vivien Dugan, Suxiang Tong, Justin Lee |
| EPI_ISL_1798775, EPI_ISL_1798776 | Laboratory Corporation of America | Centers for Disease Control and Prevention Division of Viral Diseases, Pathogen Discovery | Dakota Howard, Dhwani Batra, Peter W. Cook, Kara Moser, Adrian Paskey, Jason Caravas, Benjamin Rambo-Martin, Shatavia Morrison, Christopher Gulvick, Scott Sammons, Yvette Unoarumhi, Darlene Wagner, Matthew Schmerer, Minoo Agarwal, Eyad Almasri, Debbie Boles, Ayla Burns, Nuthawin Charoensri, Oren Cohen, Susan Countryman, Mary Ann Cristobal, Bobbi Croy, Suzanne Dale, Hrushikesh Deshmukh, Amanda Douglas, Vincent Drouillon, Marcia Eisenberg, Howard Engler, Rama Ghatti, Prashant Gupta, Susan Hicks, Jake Humphrey, Lax Iyer, Manoj Jain, Mohan Kolli, Brian Krueger, Tim Kuphal, Stanley Letovsky, Michael Levandoski, Craig Lukasik, Jonathan Meltzer, Brian Norvell, Mindy Nye, Scott Parker, Christos Petropoulos, John Pruitt, Steven Ragan, Scott Ryan, Mike Sapeta, Jana Schroth, Suresh Babu Selvaraju, Goran Stevovic, Amanda Suchanek, Andrea Throop, Lyndon Tilson, Thomas Urban, Joe Voshell, Kimberly Wagner, Jonathan Williams, Mary Williamson, Qian Zeng, Tricia Zwiefelhofer, Clinton R. Paden, Duncan MacCannell |
| EPI_ISL_1798777 | CDPH, Viral and Rickettsial Disease Laboratory | Centers for Disease Control and Prevention Division of Viral Diseases, Pathogen Discovery | Mili Sheth, Sarah Nobles, Jasmine Padilla, Mark Burroughs, Shoshona Le, Katie Dillon, Peter Cook, Clinton R. Paden, Dhwani Batra, Krista Queen, Kristen Knipe, Dakota Howard, Yvette Unoarumhi, Darlene Wagner, Matthew Schmerer, Ben L. Rambo-Martin, Kristine Lacek, Sam Shepard, Alison Laufer Halpin, Dave Wentworth, Vivien Dugan, Suxiang Tong, Justin Lee |
| EPI_ISL_1798778 | Laboratory Corporation of America | Centers for Disease Control and Prevention Division of Viral Diseases, Pathogen Discovery | Dakota Howard, Dhwani Batra, Peter W. Cook, Kara Moser, Adrian Paskey, Jason Caravas, Benjamin Rambo-Martin, Shatavia Morrison, Christopher Gulvick, Scott Sammons, Yvette Unoarumhi, Darlene Wagner, Matthew Schmerer, Minoo Agarwal, Eyad Almasri, Debbie Boles, Ayla Burns, Nuthawin Charoensri, Oren Cohen, Susan Countryman, Mary Ann Cristobal, Bobbi Croy, Suzanne Dale, Hrushikesh Deshmukh, Amanda Douglas, Vincent Drouillon, Marcia Eisenberg, Howard Engler, Rama Ghatti, Prashant Gupta, Susan Hicks, Jake Humphrey, Lax Iyer, Manoj Jain, Mohan Kolli, Brian Krueger, Tim Kuphal, Stanley Letovsky, Michael Levandoski, Craig Lukasik, Jonathan Meltzer, Brian Norvell, Mindy Nye, Scott Parker, Christos Petropoulos, John Pruitt, Steven Ragan, Scott Ryan, Mike Sapeta, Jana Schroth, Suresh Babu Selvaraju, Goran Stevovic, Amanda Suchanek, Andrea Throop, Lyndon Tilson, Thomas Urban, Joe Voshell, Kimberly Wagner, Jonathan Williams, Mary Williamson, Qian Zeng, Tricia Zwiefelhofer, Clinton R. Paden, Duncan MacCannell |
| EPI_ISL_1798779, EPI_ISL_1798781 | CDPH, Viral and Rickettsial Disease Laboratory | Centers for Disease Control and Prevention Division of Viral Diseases, Pathogen Discovery | Mili Sheth, Sarah Nobles, Jasmine Padilla, Mark Burroughs, Shoshona Le, Katie Dillon, Peter Cook, Clinton R. Paden, Dhwani Batra, Krista Queen, Kristen Knipe, Dakota Howard, Yvette Unoarumhi, Darlene Wagner, Matthew Schmerer, Ben L. Rambo-Martin, Kristine Lacek, Sam Shepard, Alison Laufer Halpin, Dave Wentworth, Vivien Dugan, Suxiang Tong, Justin Lee |

| | | | |
|---|---|---|---|
| EPI_ISL_1798782 | Laboratory Corporation of America | Centers for Disease Control and Prevention Division of Viral Diseases, Pathogen Discovery | Dakota Howard, Dhwani Batra, Peter W. Cook, Kara Moser, Adrian Paskey, Jason Caravas, Benjamin Rambo-Martin, Shatavia Morrison, Christopher Gulvick, Scott Sammons, Yvette Unoarumhi, Darlene Wagner, Matthew Schmerer, Minoo Agarwal, Eyad Almasri, Debbie Boles, Ayla Burns, Nuthawin Charoensri, Oren Cohen, Susan Countryman, Mary Ann Cristobal, Bobbi Croy, Suzanne Dale, Hrushikesh Deshmukh, Amanda Douglas, Vincent Drouillon, Marcia Eisenberg, Howard Engler, Rama Ghatti, Prashant Gupta, Susan Hicks, Jake Humphrey, Lax Iyer, Manoj Jain, Mohan Kolli, Brian Krueger, Tim Kuphal, Stanley Letovsky, Michael Levandoski, Craig Lukasik, Jonathan Meltzer, Brian Norvell, Mindy Nye, Scott Parker, Christos Petropoulos, John Pruitt, Steven Ragan, Scott Ryan, Mike Sapeta, Jana Schroth, Suresh Babu Selvaraju, Goran Stevovic, Amanda Suchanek, Andrea Throop, Lyndon Tilson, Thomas Urban, Joe Voshell, Kimberly Wagner, Jonathan Williams, Mary Williamson, Qian Zeng, Tricia Zwiefelhofer, Clinton R. Paden, Duncan MacCannell |
| EPI_ISL_1798783 | CDPH, Viral and Rickettsial Disease Laboratory | Centers for Disease Control and Prevention Division of Viral Diseases, Pathogen Discovery | Mili Sheth, Sarah Nobles, Jasmine Padilla, Mark Burroughs, Shoshona Le, Katie Dillon, Peter Cook, Clinton R. Paden, Dhwani Batra, Krista Queen, Kristen Knipe, Dakota Howard, Yvette Unoarumhi, Darlene Wagner, Matthew Schmerer, Ben L. Rambo-Martin, Kristine Lacek, Sam Shepard, Alison Laufer Halpin, Dave Wentworth, Vivien Dugan, Suxiang Tong, Justin Lee |
| EPI_ISL_1798784 | Laboratory Corporation of America | Centers for Disease Control and Prevention Division of Viral Diseases, Pathogen Discovery | Dakota Howard, Dhwani Batra, Peter W. Cook, Kara Moser, Adrian Paskey, Jason Caravas, Benjamin Rambo-Martin, Shatavia Morrison, Christopher Gulvick, Scott Sammons, Yvette Unoarumhi, Darlene Wagner, Matthew Schmerer, Minoo Agarwal, Eyad Almasri, Debbie Boles, Ayla Burns, Nuthawin Charoensri, Oren Cohen, Susan Countryman, Mary Ann Cristobal, Bobbi Croy, Suzanne Dale, Hrushikesh Deshmukh, Amanda Douglas, Vincent Drouillon, Marcia Eisenberg, Howard Engler, Rama Ghatti, Prashant Gupta, Susan Hicks, Jake Humphrey, Lax Iyer, Manoj Jain, Mohan Kolli, Brian Krueger, Tim Kuphal, Stanley Letovsky, Michael Levandoski, Craig Lukasik, Jonathan Meltzer, Brian Norvell, Mindy Nye, Scott Parker, Christos Petropoulos, John Pruitt, Steven Ragan, Scott Ryan, Mike Sapeta, Jana Schroth, Suresh Babu Selvaraju, Goran Stevovic, Amanda Suchanek, Andrea Throop, Lyndon Tilson, Thomas Urban, Joe Voshell, Kimberly Wagner, Jonathan Williams, Mary Williamson, Qian Zeng, Tricia Zwiefelhofer, Clinton R. Paden, Duncan MacCannell |
| EPI_ISL_1798785 | CDPH, Viral and Rickettsial Disease Laboratory | Centers for Disease Control and Prevention Division of Viral Diseases, Pathogen Discovery | Mili Sheth, Sarah Nobles, Jasmine Padilla, Mark Burroughs, Shoshona Le, Katie Dillon, Peter Cook, Clinton R. Paden, Dhwani Batra, Krista Queen, Kristen Knipe, Dakota Howard, Yvette Unoarumhi, Darlene Wagner, Matthew Schmerer, Ben L. Rambo-Martin, Kristine Lacek, Sam Shepard, Alison Laufer Halpin, Dave Wentworth, Vivien Dugan, Suxiang Tong, Justin Lee |
| EPI_ISL_1798786 | Laboratory Corporation of America | Centers for Disease Control and Prevention Division of Viral Diseases, Pathogen Discovery | Dakota Howard, Dhwani Batra, Peter W. Cook, Kara Moser, Adrian Paskey, Jason Caravas, Benjamin Rambo-Martin, Shatavia Morrison, Christopher Gulvick, Scott Sammons, Yvette Unoarumhi, Darlene Wagner, Matthew Schmerer, Minoo Agarwal, Eyad Almasri, Debbie Boles, Ayla Burns, Nuthawin Charoensri, Oren Cohen, Susan Countryman, Mary Ann Cristobal, Bobbi Croy, Suzanne Dale, Hrushikesh Deshmukh, Amanda Douglas, Vincent Drouillon, Marcia Eisenberg, Howard Engler, Rama Ghatti, Prashant Gupta, Susan Hicks, Jake Humphrey, Lax Iyer, Manoj Jain, Mohan Kolli, Brian Krueger, Tim Kuphal, Stanley Letovsky, Michael Levandoski, Craig Lukasik, Jonathan Meltzer, Brian Norvell, Mindy Nye, Scott Parker, Christos Petropoulos, John Pruitt, Steven Ragan, Scott Ryan, Mike Sapeta, Jana Schroth, Suresh Babu Selvaraju, Goran Stevovic, Amanda Suchanek, Andrea Throop, Lyndon Tilson, Thomas Urban, Joe Voshell, Kimberly Wagner, Jonathan Williams, Mary Williamson, Qian Zeng, Tricia Zwiefelhofer, Clinton R. Paden, Duncan MacCannell |
| EPI_ISL_1798787, EPI_ISL_1798789 | CDPH, Viral and Rickettsial Disease Laboratory | Centers for Disease Control and Prevention Division of Viral Diseases, Pathogen Discovery | Mili Sheth, Sarah Nobles, Jasmine Padilla, Mark Burroughs, Shoshona Le, Katie Dillon, Peter Cook, Clinton R. Paden, Dhwani Batra, Krista Queen, Kristen Knipe, Dakota Howard, Yvette Unoarumhi, Darlene Wagner, Matthew Schmerer, Ben L. Rambo-Martin, Kristine Lacek, Sam Shepard, Alison Laufer Halpin, Dave Wentworth, Vivien Dugan, Suxiang Tong, Justin Lee |
| EPI_ISL_1798790 | Laboratory Corporation of America | Centers for Disease Control and Prevention Division of Viral Diseases, Pathogen Discovery | Dakota Howard, Dhwani Batra, Peter W. Cook, Kara Moser, Adrian Paskey, Jason Caravas, Benjamin Rambo-Martin, Shatavia Morrison, Christopher Gulvick, Scott Sammons, Yvette Unoarumhi, Darlene Wagner, Matthew Schmerer, Minoo Agarwal, Eyad Almasri, Debbie Boles, Ayla Burns, Nuthawin Charoensri, Oren Cohen, Susan Countryman, Mary Ann Cristobal, Bobbi Croy, Suzanne Dale, Hrushikesh Deshmukh, Amanda Douglas, Vincent Drouillon, Marcia Eisenberg, Howard Engler, Rama Ghatti, Prashant Gupta, Susan Hicks, Jake Humphrey, Lax Iyer, Manoj Jain, Mohan Kolli, Brian Krueger, Tim Kuphal, Stanley Letovsky, Michael Levandoski, Craig Lukasik, Jonathan Meltzer, Brian Norvell, Mindy Nye, Scott Parker, Christos Petropoulos, John Pruitt, Steven Ragan, Scott Ryan, Mike Sapeta, Jana Schroth, Suresh Babu Selvaraju, Goran Stevovic, Amanda Suchanek, Andrea Throop, Lyndon Tilson, Thomas Urban, Joe Voshell, Kimberly Wagner, Jonathan Williams, Mary Williamson, Qian Zeng, Tricia Zwiefelhofer, Clinton R. Paden, Duncan MacCannell |
| EPI_ISL_1798791 | CDPH, Viral and Rickettsial Disease Laboratory | Centers for Disease Control and Prevention Division of Viral Diseases, Pathogen Discovery | Mili Sheth, Sarah Nobles, Jasmine Padilla, Mark Burroughs, Shoshona Le, Katie Dillon, Peter Cook, Clinton R. Paden, Dhwani Batra, Krista Queen, Kristen Knipe, Dakota Howard, Yvette Unoarumhi, Darlene Wagner, Matthew Schmerer, Ben L. Rambo-Martin, Kristine Lacek, Sam Shepard, Alison Laufer Halpin, Dave Wentworth, Vivien Dugan, Suxiang Tong, Justin Lee |
| EPI_ISL_1798792 | Laboratory Corporation of America | Centers for Disease Control and Prevention Division of Viral Diseases, Pathogen Discovery | Dakota Howard, Dhwani Batra, Peter W. Cook, Kara Moser, Adrian Paskey, Jason Caravas, Benjamin Rambo-Martin, Shatavia Morrison, Christopher Gulvick, Scott Sammons, Yvette Unoarumhi, Darlene Wagner, Matthew Schmerer, Minoo Agarwal, Eyad Almasri, Debbie Boles, Ayla Burns, Nuthawin Charoensri, Oren Cohen, Susan Countryman, Mary Ann Cristobal, Bobbi Croy, Suzanne Dale, Hrushikesh Deshmukh, Amanda Douglas, Vincent Drouillon, Marcia Eisenberg, Howard Engler, Rama Ghatti, Prashant Gupta, Susan Hicks, Jake Humphrey, Lax Iyer, Manoj Jain, Mohan Kolli, Brian Krueger, Tim Kuphal, Stanley Letovsky, Michael Levandoski, Craig Lukasik, Jonathan Meltzer, Brian Norvell, Mindy Nye, Scott Parker, Christos Petropoulos, John Pruitt, Steven Ragan, Scott Ryan, Mike Sapeta, Jana Schroth, Suresh Babu Selvaraju, Goran Stevovic, Amanda Suchanek, Andrea Throop, Lyndon Tilson, Thomas Urban, Joe Voshell, Kimberly Wagner, Jonathan Williams, Mary Williamson, Qian Zeng, Tricia Zwiefelhofer, Clinton R. Paden, Duncan MacCannell |
| EPI_ISL_1798793 | CDPH, Viral and Rickettsial Disease Laboratory | Centers for Disease Control and Prevention Division of Viral Diseases, Pathogen Discovery | Mili Sheth, Sarah Nobles, Jasmine Padilla, Mark Burroughs, Shoshona Le, Katie Dillon, Peter Cook, Clinton R. Paden, Dhwani Batra, Krista Queen, Kristen Knipe, Dakota Howard, Yvette Unoarumhi, Darlene Wagner, Matthew Schmerer, Ben L. Rambo-Martin, Kristine Lacek, Sam Shepard, Alison Laufer Halpin, Dave Wentworth, Vivien Dugan, Suxiang Tong, Justin Lee |
| EPI_ISL_1798794 | Laboratory Corporation of America | Centers for Disease Control and Prevention Division of Viral Diseases, Pathogen Discovery | Dakota Howard, Dhwani Batra, Peter W. Cook, Kara Moser, Adrian Paskey, Jason Caravas, Benjamin Rambo-Martin, Shatavia Morrison, Christopher Gulvick, Scott Sammons, Yvette Unoarumhi, Darlene Wagner, Matthew Schmerer, Minoo Agarwal, Eyad Almasri, Debbie Boles, Ayla Burns, Nuthawin Charoensri, Oren Cohen, Susan Countryman, Mary Ann Cristobal, Bobbi Croy, Suzanne Dale, Hrushikesh Deshmukh, Amanda Douglas, Vincent Drouillon, Marcia Eisenberg, Howard Engler, Rama Ghatti, Prashant Gupta, Susan Hicks, Jake Humphrey, Lax Iyer, Manoj Jain, Mohan Kolli, Brian Krueger, Tim Kuphal, Stanley Letovsky, Michael Levandoski, Craig Lukasik, Jonathan Meltzer, Brian Norvell, Mindy Nye, Scott Parker, Christos Petropoulos, John Pruitt, Steven Ragan, Scott Ryan, Mike Sapeta, Jana Schroth, Suresh Babu Selvaraju, Goran Stevovic, Amanda Suchanek, Andrea Throop, Lyndon Tilson, Thomas Urban, Joe Voshell, Kimberly Wagner, Jonathan Williams, Mary Williamson, Qian Zeng, Tricia Zwiefelhofer, Clinton R. Paden, Duncan MacCannell |
| EPI_ISL_1798795 | CDPH, Viral and Rickettsial Disease Laboratory | Centers for Disease Control and Prevention Division of Viral Diseases, Pathogen Discovery | Mili Sheth, Sarah Nobles, Jasmine Padilla, Mark Burroughs, Shoshona Le, Katie Dillon, Peter Cook, Clinton R. Paden, Dhwani Batra, Krista Queen, Kristen Knipe, Dakota Howard, Yvette Unoarumhi, Darlene Wagner, Matthew Schmerer, Ben L. Rambo-Martin, Kristine Lacek, Sam Shepard, Alison Laufer Halpin, Dave Wentworth, Vivien Dugan, Suxiang Tong, Justin Lee |
| EPI_ISL_1798796 | Laboratory Corporation of America | Centers for Disease Control and Prevention Division of Viral Diseases, Pathogen Discovery | Dakota Howard, Dhwani Batra, Peter W. Cook, Kara Moser, Adrian Paskey, Jason Caravas, Benjamin Rambo-Martin, Shatavia Morrison, Christopher Gulvick, Scott Sammons, Yvette Unoarumhi, Darlene Wagner, Matthew Schmerer, Minoo Agarwal, Eyad Almasri, Debbie Boles, Ayla Burns, Nuthawin Charoensri, Oren Cohen, Susan Countryman, Mary Ann Cristobal, Bobbi Croy, Suzanne Dale, Hrushikesh Deshmukh, Amanda Douglas, Vincent Drouillon, Marcia Eisenberg, Howard Engler, Rama Ghatti, Prashant Gupta, Susan Hicks, Jake Humphrey, Lax Iyer, Manoj Jain, Mohan Kolli, Brian Krueger, Tim Kuphal, Stanley Letovsky, Michael Levandoski, Craig Lukasik, Jonathan Meltzer, Brian Norvell, Mindy Nye, Scott Parker, Christos Petropoulos, John Pruitt, Steven Ragan, Scott Ryan, Mike Sapeta, Jana Schroth, Suresh Babu Selvaraju, Goran Stevovic, Amanda Suchanek, Andrea Throop, Lyndon Tilson, Thomas Urban, Joe Voshell, Kimberly Wagner, Jonathan Williams, Mary Williamson, Qian Zeng, Tricia Zwiefelhofer, Clinton R. Paden, Duncan MacCannell |
| EPI_ISL_1798797 | CDPH, Viral and Rickettsial Disease Laboratory | Centers for Disease Control and Prevention Division of Viral Diseases, Pathogen Discovery | Mili Sheth, Sarah Nobles, Jasmine Padilla, Mark Burroughs, Shoshona Le, Katie Dillon, Peter Cook, Clinton R. Paden, Dhwani Batra, Krista Queen, Kristen Knipe, Dakota Howard, Yvette Unoarumhi, Darlene Wagner, Matthew Schmerer, Ben L. Rambo-Martin, Kristine Lacek, Sam Shepard, Alison Laufer Halpin, Dave Wentworth, Vivien Dugan, Suxiang Tong, Justin Lee |
| EPI_ISL_1798798 | Laboratory Corporation of America | Centers for Disease Control and Prevention Division of Viral Diseases, Pathogen Discovery | Dakota Howard, Dhwani Batra, Peter W. Cook, Kara Moser, Adrian Paskey, Jason Caravas, Benjamin Rambo-Martin, Shatavia Morrison, Christopher Gulvick, Scott Sammons, Yvette Unoarumhi, Darlene Wagner, Matthew Schmerer, Minoo Agarwal, Eyad Almasri, Debbie Boles, Ayla Burns, Nuthawin Charoensri, Oren Cohen, Susan Countryman, Mary Ann Cristobal, Bobbi Croy, Suzanne Dale, Hrushikesh Deshmukh, Amanda Douglas, Vincent Drouillon, Marcia Eisenberg, Howard Engler, Rama Ghatti, Prashant Gupta, Susan Hicks, Jake Humphrey, Lax Iyer, Manoj Jain, Mohan Kolli, Brian Krueger, Tim Kuphal, Stanley Letovsky, Michael Levandoski, Craig Lukasik, Jonathan Meltzer, Brian Norvell, Mindy Nye, Scott Parker, Christos Petropoulos, John Pruitt, Steven Ragan, Scott Ryan, Mike Sapeta, Jana Schroth, Suresh Babu Selvaraju, Goran Stevovic, Amanda Suchanek, Andrea Throop, Lyndon Tilson, Thomas Urban, Joe Voshell, Kimberly Wagner, Jonathan Williams, Mary Williamson, Qian Zeng, Tricia Zwiefelhofer, Clinton R. Paden, Duncan MacCannell |
| EPI_ISL_1798799 | CDPH, Viral and Rickettsial Disease Laboratory | Centers for Disease Control and Prevention Division of Viral Diseases, Pathogen Discovery | Mili Sheth, Sarah Nobles, Jasmine Padilla, Mark Burroughs, Shoshona Le, Katie Dillon, Peter Cook, Clinton R. Paden, Dhwani Batra, Krista Queen, Kristen Knipe, Dakota Howard, Yvette Unoarumhi, Darlene Wagner, Matthew Schmerer, Ben L. Rambo-Martin, Kristine Lacek, Sam Shepard, Alison Laufer Halpin, Dave Wentworth, Vivien Dugan, Suxiang Tong, Justin Lee |
| EPI_ISL_1798800 | Laboratory Corporation of America | Centers for Disease Control and Prevention Division of Viral Diseases, Pathogen Discovery | Dakota Howard, Dhwani Batra, Peter W. Cook, Kara Moser, Adrian Paskey, Jason Caravas, Benjamin Rambo-Martin, Shatavia Morrison, Christopher Gulvick, Scott Sammons, Yvette Unoarumhi, Darlene Wagner, Matthew Schmerer, Minoo Agarwal, Eyad Almasri, Debbie Boles, Ayla Burns, Nuthawin Charoensri, Oren Cohen, Susan Countryman, Mary Ann Cristobal, Bobbi Croy, Suzanne Dale, Hrushikesh Deshmukh, Amanda Douglas, Vincent Drouillon, Marcia Eisenberg, Howard Engler, Rama Ghatti, Prashant Gupta, Susan Hicks, Jake Humphrey, Lax Iyer, Manoj Jain, Mohan Kolli, Brian Krueger, Tim Kuphal, Stanley Letovsky, Michael Levandoski, Craig Lukasik, Jonathan Meltzer, Brian Norvell, Mindy Nye, Scott Parker, Christos Petropoulos, John Pruitt, Steven Ragan, Scott Ryan, Mike Sapeta, Jana Schroth, Suresh Babu Selvaraju, Goran Stevovic, Amanda Suchanek, Andrea Throop, Lyndon Tilson, Thomas Urban, Joe Voshell, Kimberly Wagner, Jonathan Williams, Mary Williamson, Qian Zeng, Tricia Zwiefelhofer, Clinton R. Paden, Duncan MacCannell |
| EPI_ISL_1798801 | CDPH, Viral and Rickettsial Disease Laboratory | Centers for Disease Control and Prevention Division of Viral Diseases, Pathogen Discovery | Mili Sheth, Sarah Nobles, Jasmine Padilla, Mark Burroughs, Shoshona Le, Katie Dillon, Peter Cook, Clinton R. Paden, Dhwani Batra, Krista Queen, Kristen Knipe, Dakota Howard, Yvette Unoarumhi, Darlene Wagner, Matthew Schmerer, Ben L. Rambo-Martin, Kristine Lacek, Sam Shepard, Alison Laufer Halpin, Dave Wentworth, Vivien Dugan, Suxiang Tong, Justin Lee |
| EPI_ISL_1798802 | PA Department of Health, Bureau of Laboratories | Centers for Disease Control and Prevention Division of Viral Diseases, Pathogen Discovery | Mili Sheth, Sarah Nobles, Jasmine Padilla, Mark Burroughs, Shoshona Le, Katie Dillon, Peter Cook, Clinton R. Paden, Dhwani Batra, Krista Queen, Kristen Knipe, Dakota Howard, Yvette Unoarumhi, Darlene Wagner, Matthew Schmerer, Ben L. Rambo-Martin, Kristine Lacek, Sam Shepard, Alison Laufer Halpin, Dave Wentworth, Vivien Dugan, Suxiang Tong, Justin Lee |
| EPI_ISL_1798803 | Laboratory Corporation of America | Centers for Disease Control and Prevention Division of Viral Diseases, Pathogen Discovery | Dakota Howard, Dhwani Batra, Peter W. Cook, Kara Moser, Adrian Paskey, Jason Caravas, Benjamin Rambo-Martin, Shatavia Morrison, Christopher Gulvick, Scott Sammons, Yvette Unoarumhi, Darlene Wagner, Matthew Schmerer, Minoo Agarwal, Eyad Almasri, Debbie Boles, Ayla Burns, Nuthawin Charoensri, Oren Cohen, Susan Countryman, Mary Ann Cristobal, Bobbi Croy, Suzanne Dale, Hrushikesh Deshmukh, Amanda Douglas, Vincent Drouillon, Marcia Eisenberg, Howard Engler, Rama Ghatti, Prashant Gupta, Susan Hicks, Jake Humphrey, Lax Iyer, Manoj Jain, Mohan Kolli, Brian Krueger, Tim Kuphal, Stanley Letovsky, Michael Levandoski, Craig Lukasik, Jonathan Meltzer, Brian Norvell, Mindy Nye, Scott Parker, Christos Petropoulos, John Pruitt, Steven Ragan, Scott Ryan, Mike Sapeta, Jana Schroth, Suresh Babu Selvaraju, Goran Stevovic, Amanda Suchanek, Andrea Throop, Lyndon Tilson, Thomas Urban, Joe Voshell, Kimberly Wagner, Jonathan Williams, Mary Williamson, Qian Zeng, Tricia Zwiefelhofer, Clinton R. Paden, Duncan MacCannell |
| EPI_ISL_1798804 | PA Department of Health, Bureau of Laboratories | Centers for Disease Control and Prevention Division of Viral Diseases, Pathogen Discovery | Mili Sheth, Sarah Nobles, Jasmine Padilla, Mark Burroughs, Shoshona Le, Katie Dillon, Peter Cook, Clinton R. Paden, Dhwani Batra, Krista Queen, Kristen Knipe, Dakota Howard, Yvette Unoarumhi, Darlene Wagner, Matthew Schmerer, Ben L. Rambo-Martin, Kristine Lacek, Sam Shepard, Alison Laufer Halpin, Dave Wentworth, Vivien Dugan, Suxiang Tong, Justin Lee |
| EPI_ISL_1798805 | Laboratory Corporation of America | Centers for Disease Control and Prevention Division of Viral Diseases, Pathogen Discovery | Dakota Howard, Dhwani Batra, Peter W. Cook, Kara Moser, Adrian Paskey, Jason Caravas, Benjamin Rambo-Martin, Shatavia Morrison, Christopher Gulvick, Scott Sammons, Yvette Unoarumhi, Darlene Wagner, Matthew Schmerer, Minoo Agarwal, Eyad Almasri, Debbie Boles, Ayla Burns, Nuthawin Charoensri, Oren Cohen, Susan Countryman, Mary Ann Cristobal, Bobbi Croy, Suzanne Dale, Hrushikesh Deshmukh, Amanda Douglas, Vincent Drouillon, Marcia Eisenberg, Howard Engler, Rama Ghatti, Prashant Gupta, Susan Hicks, Jake Humphrey, Lax Iyer, Manoj Jain, Mohan Kolli, Brian Krueger, Tim Kuphal, Stanley Letovsky, Michael Levandoski, Craig Lukasik, Jonathan Meltzer, Brian Norvell, Mindy Nye, Scott Parker, Christos Petropoulos, John Pruitt, Steven Ragan, Scott Ryan, Mike Sapeta, Jana Schroth, Suresh Babu Selvaraju, Goran Stevovic, Amanda Suchanek, Andrea Throop, Lyndon Tilson, Thomas Urban, Joe Voshell, Kimberly Wagner, Jonathan Williams, Mary Williamson, Qian Zeng, Tricia Zwiefelhofer, Clinton R. Paden, Duncan MacCannell |
| EPI_ISL_1798806 | PA Department of Health, Bureau of Laboratories | Centers for Disease Control and Prevention Division of Viral Diseases, Pathogen Discovery | Mili Sheth, Sarah Nobles, Jasmine Padilla, Mark Burroughs, Shoshona Le, Katie Dillon, Peter Cook, Clinton R. Paden, Dhwani Batra, Krista Queen, Kristen Knipe, Dakota Howard, Yvette Unoarumhi, Darlene Wagner, Matthew Schmerer, Ben L. Rambo-Martin, Kristine Lacek, Sam Shepard, Alison Laufer Halpin, Dave Wentworth, Vivien Dugan, Suxiang Tong, Justin Lee |
| EPI_ISL_1798807 | Laboratory Corporation of America | Centers for Disease Control and Prevention Division of Viral Diseases, Pathogen Discovery | Dakota Howard, Dhwani Batra, Peter W. Cook, Kara Moser, Adrian Paskey, Jason Caravas, Benjamin Rambo-Martin, Shatavia Morrison, Christopher Gulvick, Scott Sammons, Yvette Unoarumhi, Darlene Wagner, Matthew Schmerer, Minoo Agarwal, Eyad Almasri, Debbie Boles, Ayla Burns, Nuthawin Charoensri, Oren Cohen, Susan Countryman, Mary Ann Cristobal, Bobbi Croy, Suzanne Dale, Hrushikesh Deshmukh, Amanda Douglas, Vincent Drouillon, Marcia Eisenberg, Howard Engler, Rama Ghatti, Prashant Gupta, Susan Hicks, Jake Humphrey, Lax Iyer, Manoj Jain, Mohan Kolli, Brian Krueger, Tim Kuphal, Stanley Letovsky, Michael Levandoski, Craig Lukasik, Jonathan Meltzer, Brian Norvell, Mindy Nye, Scott Parker, Christos Petropoulos, John Pruitt, Steven Ragan, Scott Ryan, Mike Sapeta, Jana Schroth, Suresh Babu Selvaraju, Goran Stevovic, Amanda Suchanek, Andrea Throop, Lyndon Tilson, Thomas Urban, Joe Voshell, Kimberly Wagner, Jonathan Williams, Mary Williamson, Qian Zeng, Tricia Zwiefelhofer, Clinton R. Paden, Duncan MacCannell |
| EPI_ISL_1798808 | PA Department of Health, Bureau of Laboratories | Centers for Disease Control and Prevention Division of Viral Diseases, Pathogen Discovery | Mili Sheth, Sarah Nobles, Jasmine Padilla, Mark Burroughs, Shoshona Le, Katie Dillon, Peter Cook, Clinton R. Paden, Dhwani Batra, Krista Queen, Kristen Knipe, Dakota Howard, Yvette Unoarumhi, Darlene Wagner, Matthew Schmerer, Ben L. Rambo-Martin, Kristine Lacek, Sam Shepard, Alison Laufer Halpin, Dave Wentworth, Vivien Dugan, Suxiang Tong, Justin Lee |
| EPI_ISL_1798809 | Laboratory Corporation of America | Centers for Disease Control and Prevention Division of Viral Diseases, Pathogen Discovery | Dakota Howard, Dhwani Batra, Peter W. Cook, Kara Moser, Adrian Paskey, Jason Caravas, Benjamin Rambo-Martin, Shatavia Morrison, Christopher Gulvick, Scott Sammons, Yvette Unoarumhi, Darlene Wagner, Matthew Schmerer, Minoo Agarwal, Eyad Almasri, Debbie Boles, Ayla Burns, Nuthawin Charoensri, Oren Cohen, Susan Countryman, Mary Ann Cristobal, Bobbi Croy, Suzanne Dale, Hrushikesh Deshmukh, Amanda Douglas, Vincent Drouillon, Marcia Eisenberg, Howard Engler, Rama Ghatti, Prashant Gupta, Susan Hicks, Jake Humphrey, Lax Iyer, Manoj Jain, Mohan Kolli, Brian Krueger, Tim Kuphal, Stanley Letovsky, Michael Levandoski, Craig Lukasik, Jonathan Meltzer, Brian Norvell, Mindy Nye, Scott Parker, Christos Petropoulos, John Pruitt, Steven Ragan, Scott Ryan, Mike Sapeta, Jana Schroth, Suresh Babu Selvaraju, Goran Stevovic, Amanda Suchanek, Andrea Throop, Lyndon Tilson, Thomas Urban, Joe Voshell, Kimberly Wagner, Jonathan Williams, Mary Williamson, Qian Zeng, Tricia Zwiefelhofer, Clinton R. Paden, Duncan MacCannell |
| EPI_ISL_1798810 | PA Department of Health, Bureau of Laboratories | Centers for Disease Control and Prevention Division of Viral Diseases, Pathogen Discovery | Mili Sheth, Sarah Nobles, Jasmine Padilla, Mark Burroughs, Shoshona Le, Katie Dillon, Peter Cook, Clinton R. Paden, Dhwani Batra, Krista Queen, Kristen Knipe, Dakota Howard, Yvette Unoarumhi, Darlene Wagner, Matthew Schmerer, Ben L. Rambo-Martin, Kristine Lacek, Sam Shepard, Alison Laufer Halpin, Dave Wentworth, Vivien Dugan, Suxiang Tong, Justin Lee |
| EPI_ISL_1798811 | Laboratory Corporation of America | Centers for Disease Control and Prevention Division of Viral Diseases, Pathogen Discovery | Dakota Howard, Dhwani Batra, Peter W. Cook, Kara Moser, Adrian Paskey, Jason Caravas, Benjamin Rambo-Martin, Shatavia Morrison, Christopher Gulvick, Scott Sammons, Yvette Unoarumhi, Darlene Wagner, Matthew Schmerer, Minoo Agarwal, Eyad Almasri, Debbie Boles, Ayla Burns, Nuthawin Charoensri, Oren Cohen, Susan Countryman, Mary Ann Cristobal, Bobbi Croy, Suzanne Dale, Hrushikesh Deshmukh, Amanda Douglas, Vincent Drouillon, Marcia Eisenberg, Howard Engler, Rama Ghatti, Prashant Gupta, Susan Hicks, Jake Humphrey, Lax Iyer, Manoj Jain, Mohan Kolli, Brian Krueger, Tim Kuphal, Stanley Letovsky, Michael Levandoski, Craig Lukasik, Jonathan Meltzer, Brian Norvell, Mindy Nye, Scott Parker, Christos Petropoulos, John Pruitt, Steven Ragan, Scott Ryan, Mike Sapeta, Jana Schroth, Suresh Babu Selvaraju, Goran Stevovic, Amanda |

| | | | |
|---|---|---|---|
| | | | Suchanek, Andrea Throop, Lyndon Tilson, Thomas Urban, Joe Voshell, Kimberly Wagner, Jonathan Williams, Mary Williamson, Qian Zeng, Tricia Zwiefelhofer, Clinton R. Paden, Duncan MacCannell |
| EPI_ISL_1798812, EPI_ISL_1798814 | PA Department of Health, Bureau of Laboratories | Centers for Disease Control and Prevention Division of Viral Diseases, Pathogen Discovery | Mili Sheth, Sarah Nobles, Jasmine Padilla, Mark Burroughs, Shoshona Le, Katie Dillon, Peter Cook, Clinton R. Paden, Dhwani Batra, Krista Queen, Kristen Knipe, Dakota Howard, Yvette Unoarumhi, Darlene Wagner, Matthew Schmerer, Ben L. Rambo-Martin, Kristine Lacek, Sam Shepard, Alison Laufer Halpin, Dave Wentworth, Vivien Dugan, Suxiang Tong, Justin Lee |
| EPI_ISL_1798815 | Laboratory Corporation of America | Centers for Disease Control and Prevention Division of Viral Diseases, Pathogen Discovery | Dakota Howard, Dhwani Batra, Peter W. Cook, Kara Moser, Adrian Paskey, Jason Caravas, Benjamin Rambo-Martin, Shatavia Morrison, Christopher Gulvick, Scott Sammons, Yvette Unoarumhi, Darlene Wagner, Matthew Schmerer, Minoo Agarwal, Eyad Almasri, Debbie Boles, Ayla Burns, Nuthawin Charoensri, Oren Cohen, Susan Countryman, Mary Ann Cristobal, Bobbi Croy, Suzanne Dale, Hrushikesh Deshmukh, Amanda Douglas, Vincent Drouillon, Marcia Eisenberg, Howard Engler, Rama Ghatti, Prashant Gupta, Susan Hicks, Jake Humphrey, Lax Iyer, Manoj Jain, Mohan Kolli, Brian Krueger, Tim Kuphal, Stanley Letovsky, Michael Levandoski, Craig Lukasik, Jonathan Meltzer, Brian Norvell, Mindy Nye, Scott Parker, Christos Petropoulos, John Pruitt, Steven Ragan, Scott Ryan, Mike Sapeta, Jana Schroth, Suresh Babu Selvaraju, Goran Stevovic, Amanda Suchanek, Andrea Throop, Lyndon Tilson, Thomas Urban, Joe Voshell, Kimberly Wagner, Jonathan Williams, Mary Williamson, Qian Zeng, Tricia Zwiefelhofer, Clinton R. Paden, Duncan MacCannell |
| EPI_ISL_1798816 | PA Department of Health, Bureau of Laboratories | Centers for Disease Control and Prevention Division of Viral Diseases, Pathogen Discovery | Mili Sheth, Sarah Nobles, Jasmine Padilla, Mark Burroughs, Shoshona Le, Katie Dillon, Peter Cook, Clinton R. Paden, Dhwani Batra, Krista Queen, Kristen Knipe, Dakota Howard, Yvette Unoarumhi, Darlene Wagner, Matthew Schmerer, Ben L. Rambo-Martin, Kristine Lacek, Sam Shepard, Alison Laufer Halpin, Dave Wentworth, Vivien Dugan, Suxiang Tong, Justin Lee |
| EPI_ISL_1798817 | Laboratory Corporation of America | Centers for Disease Control and Prevention Division of Viral Diseases, Pathogen Discovery | Dakota Howard, Dhwani Batra, Peter W. Cook, Kara Moser, Adrian Paskey, Jason Caravas, Benjamin Rambo-Martin, Shatavia Morrison, Christopher Gulvick, Scott Sammons, Yvette Unoarumhi, Darlene Wagner, Matthew Schmerer, Minoo Agarwal, Eyad Almasri, Debbie Boles, Ayla Burns, Nuthawin Charoensri, Oren Cohen, Susan Countryman, Mary Ann Cristobal, Bobbi Croy, Suzanne Dale, Hrushikesh Deshmukh, Amanda Douglas, Vincent Drouillon, Marcia Eisenberg, Howard Engler, Rama Ghatti, Prashant Gupta, Susan Hicks, Jake Humphrey, Lax Iyer, Manoj Jain, Mohan Kolli, Brian Krueger, Tim Kuphal, Stanley Letovsky, Michael Levandoski, Craig Lukasik, Jonathan Meltzer, Brian Norvell, Mindy Nye, Scott Parker, Christos Petropoulos, John Pruitt, Steven Ragan, Scott Ryan, Mike Sapeta, Jana Schroth, Suresh Babu Selvaraju, Goran Stevovic, Amanda Suchanek, Andrea Throop, Lyndon Tilson, Thomas Urban, Joe Voshell, Kimberly Wagner, Jonathan Williams, Mary Williamson, Qian Zeng, Tricia Zwiefelhofer, Clinton R. Paden, Duncan MacCannell |
| EPI_ISL_1798818 | PA Department of Health, Bureau of Laboratories | Centers for Disease Control and Prevention Division of Viral Diseases, Pathogen Discovery | Mili Sheth, Sarah Nobles, Jasmine Padilla, Mark Burroughs, Shoshona Le, Katie Dillon, Peter Cook, Clinton R. Paden, Dhwani Batra, Krista Queen, Kristen Knipe, Dakota Howard, Yvette Unoarumhi, Darlene Wagner, Matthew Schmerer, Ben L. Rambo-Martin, Kristine Lacek, Sam Shepard, Alison Laufer Halpin, Dave Wentworth, Vivien Dugan, Suxiang Tong, Justin Lee |
| EPI_ISL_1798819 | Laboratory Corporation of America | Centers for Disease Control and Prevention Division of Viral Diseases, Pathogen Discovery | Dakota Howard, Dhwani Batra, Peter W. Cook, Kara Moser, Adrian Paskey, Jason Caravas, Benjamin Rambo-Martin, Shatavia Morrison, Christopher Gulvick, Scott Sammons, Yvette Unoarumhi, Darlene Wagner, Matthew Schmerer, Minoo Agarwal, Eyad Almasri, Debbie Boles, Ayla Burns, Nuthawin Charoensri, Oren Cohen, Susan Countryman, Mary Ann Cristobal, Bobbi Croy, Suzanne Dale, Hrushikesh Deshmukh, Amanda Douglas, Vincent Drouillon, Marcia Eisenberg, Howard Engler, Rama Ghatti, Prashant Gupta, Susan Hicks, Jake Humphrey, Lax Iyer, Manoj Jain, Mohan Kolli, Brian Krueger, Tim Kuphal, Stanley Letovsky, Michael Levandoski, Craig Lukasik, Jonathan Meltzer, Brian Norvell, Mindy Nye, Scott Parker, Christos Petropoulos, John Pruitt, Steven Ragan, Scott Ryan, Mike Sapeta, Jana Schroth, Suresh Babu Selvaraju, Goran Stevovic, Amanda Suchanek, Andrea Throop, Lyndon Tilson, Thomas Urban, Joe Voshell, Kimberly Wagner, Jonathan Williams, Mary Williamson, Qian Zeng, Tricia Zwiefelhofer, Clinton R. Paden, Duncan MacCannell |
| EPI_ISL_1798820 | PA Department of Health, Bureau of Laboratories | Centers for Disease Control and Prevention Division of Viral Diseases, Pathogen Discovery | Mili Sheth, Sarah Nobles, Jasmine Padilla, Mark Burroughs, Shoshona Le, Katie Dillon, Peter Cook, Clinton R. Paden, Dhwani Batra, Krista Queen, Kristen Knipe, Dakota Howard, Yvette Unoarumhi, Darlene Wagner, Matthew Schmerer, Ben L. Rambo-Martin, Kristine Lacek, Sam Shepard, Alison Laufer Halpin, Dave Wentworth, Vivien Dugan, Suxiang Tong, Justin Lee |
| EPI_ISL_1798821 | Laboratory Corporation of America | Centers for Disease Control and Prevention Division of Viral Diseases, Pathogen Discovery | Dakota Howard, Dhwani Batra, Peter W. Cook, Kara Moser, Adrian Paskey, Jason Caravas, Benjamin Rambo-Martin, Shatavia Morrison, Christopher Gulvick, Scott Sammons, Yvette Unoarumhi, Darlene Wagner, Matthew Schmerer, Minoo Agarwal, Eyad Almasri, Debbie Boles, Ayla Burns, Nuthawin Charoensri, Oren Cohen, Susan Countryman, Mary Ann Cristobal, Bobbi Croy, Suzanne Dale, Hrushikesh Deshmukh, Amanda Douglas, Vincent Drouillon, Marcia Eisenberg, Howard Engler, Rama Ghatti, Prashant Gupta, Susan Hicks, Jake Humphrey, Lax Iyer, Manoj Jain, Mohan Kolli, Brian Krueger, Tim Kuphal, Stanley Letovsky, Michael Levandoski, Craig Lukasik, Jonathan Meltzer, Brian Norvell, Mindy Nye, Scott Parker, Christos Petropoulos, John Pruitt, Steven Ragan, Scott Ryan, Mike Sapeta, Jana Schroth, Suresh Babu Selvaraju, Goran Stevovic, Amanda Suchanek, Andrea Throop, Lyndon Tilson, Thomas Urban, Joe Voshell, Kimberly Wagner, Jonathan Williams, Mary Williamson, Qian Zeng, Tricia Zwiefelhofer, Clinton R. Paden, Duncan MacCannell |
| EPI_ISL_1798822 | PA Department of Health, Bureau of Laboratories | Centers for Disease Control and Prevention Division of Viral Diseases, Pathogen Discovery | Mili Sheth, Sarah Nobles, Jasmine Padilla, Mark Burroughs, Shoshona Le, Katie Dillon, Peter Cook, Clinton R. Paden, Dhwani Batra, Krista Queen, Kristen Knipe, Dakota Howard, Yvette Unoarumhi, Darlene Wagner, Matthew Schmerer, Ben L. Rambo-Martin, Kristine Lacek, Sam Shepard, Alison Laufer Halpin, Dave Wentworth, Vivien Dugan, Suxiang Tong, Justin Lee |
| EPI_ISL_1798823 | Laboratory Corporation of America | Centers for Disease Control and Prevention Division of Viral Diseases, Pathogen Discovery | Dakota Howard, Dhwani Batra, Peter W. Cook, Kara Moser, Adrian Paskey, Jason Caravas, Benjamin Rambo-Martin, Shatavia Morrison, Christopher Gulvick, Scott Sammons, Yvette Unoarumhi, Darlene Wagner, Matthew Schmerer, Minoo Agarwal, Eyad Almasri, Debbie Boles, Ayla Burns, Nuthawin Charoensri, Oren Cohen, Susan Countryman, Mary Ann Cristobal, Bobbi Croy, Suzanne Dale, Hrushikesh Deshmukh, Amanda Douglas, Vincent Drouillon, Marcia Eisenberg, Howard Engler, Rama Ghatti, Prashant Gupta, Susan Hicks, Jake Humphrey, Lax Iyer, Manoj Jain, Mohan Kolli, Brian Krueger, Tim Kuphal, Stanley Letovsky, Michael Levandoski, Craig Lukasik, Jonathan Meltzer, Brian Norvell, Mindy Nye, Scott Parker, Christos Petropoulos, John Pruitt, Steven Ragan, Scott Ryan, Mike Sapeta, Jana Schroth, Suresh Babu Selvaraju, Goran Stevovic, Amanda Suchanek, Andrea Throop, Lyndon Tilson, Thomas Urban, Joe Voshell, Kimberly Wagner, Jonathan Williams, Mary Williamson, Qian Zeng, Tricia Zwiefelhofer, Clinton R. Paden, Duncan MacCannell |
| EPI_ISL_1798824, EPI_ISL_1798826, EPI_ISL_1798828 | PA Department of Health, Bureau of Laboratories | Centers for Disease Control and Prevention Division of Viral Diseases, Pathogen Discovery | Mili Sheth, Sarah Nobles, Jasmine Padilla, Mark Burroughs, Shoshona Le, Katie Dillon, Peter Cook, Clinton R. Paden, Dhwani Batra, Krista Queen, Kristen Knipe, Dakota Howard, Yvette Unoarumhi, Darlene Wagner, Matthew Schmerer, Ben L. Rambo-Martin, Kristine Lacek, Sam Shepard, Alison Laufer Halpin, Dave Wentworth, Vivien Dugan, Suxiang Tong, Justin Lee |
| EPI_ISL_1798829 | Laboratory Corporation of America | Centers for Disease Control and Prevention Division of Viral Diseases, Pathogen Discovery | Dakota Howard, Dhwani Batra, Peter W. Cook, Kara Moser, Adrian Paskey, Jason Caravas, Benjamin Rambo-Martin, Shatavia Morrison, Christopher Gulvick, Scott Sammons, Yvette Unoarumhi, Darlene Wagner, Matthew Schmerer, Minoo Agarwal, Eyad Almasri, Debbie Boles, Ayla Burns, Nuthawin Charoensri, Oren Cohen, Susan Countryman, Mary Ann Cristobal, Bobbi Croy, Suzanne Dale, Hrushikesh Deshmukh, Amanda Douglas, Vincent Drouillon, Marcia Eisenberg, Howard Engler, Rama Ghatti, Prashant Gupta, Susan Hicks, Jake Humphrey, Lax Iyer, Manoj Jain, Mohan Kolli, Brian Krueger, Tim Kuphal, Stanley Letovsky, Michael Levandoski, Craig Lukasik, Jonathan Meltzer, Brian Norvell, Mindy Nye, Scott Parker, Christos Petropoulos, John Pruitt, Steven Ragan, Scott Ryan, Mike Sapeta, Jana Schroth, Suresh Babu Selvaraju, Goran Stevovic, Amanda Suchanek, Andrea Throop, Lyndon Tilson, Thomas Urban, Joe Voshell, Kimberly Wagner, Jonathan Williams, Mary Williamson, Qian Zeng, Tricia Zwiefelhofer, Clinton R. Paden, Duncan MacCannell |
| EPI_ISL_1798830 | CA-Los Angeles County Public Health Laboratory | Centers for Disease Control and Prevention Division of Viral Diseases, Pathogen Discovery | Mili Sheth, Sarah Nobles, Jasmine Padilla, Mark Burroughs, Shoshona Le, Katie Dillon, Peter Cook, Clinton R. Paden, Dhwani Batra, Krista Queen, Kristen Knipe, Dakota Howard, Yvette Unoarumhi, Darlene Wagner, Matthew Schmerer, Ben L. Rambo-Martin, Kristine Lacek, Sam Shepard, Alison Laufer Halpin, Dave Wentworth, Vivien Dugan, Suxiang Tong, Justin Lee |
| EPI_ISL_1798831 | Laboratory Corporation of America | Centers for Disease Control and Prevention Division of Viral Diseases, Pathogen Discovery | Dakota Howard, Dhwani Batra, Peter W. Cook, Kara Moser, Adrian Paskey, Jason Caravas, Benjamin Rambo-Martin, Shatavia Morrison, Christopher Gulvick, Scott Sammons, Yvette Unoarumhi, Darlene Wagner, Matthew Schmerer, Minoo Agarwal, Eyad Almasri, Debbie Boles, Ayla Burns, Nuthawin Charoensri, Oren Cohen, Susan Countryman, Mary Ann Cristobal, Bobbi Croy, Suzanne Dale, Hrushikesh Deshmukh, Amanda Douglas, Vincent Drouillon, Marcia Eisenberg, Howard Engler, Rama Ghatti, Prashant Gupta, Susan Hicks, Jake Humphrey, Lax Iyer, Manoj Jain, Mohan Kolli, Brian Krueger, Tim Kuphal, Stanley Letovsky, Michael Levandoski, Craig Lukasik, Jonathan Meltzer, Brian Norvell, Mindy Nye, Scott Parker, Christos Petropoulos, John Pruitt, Steven Ragan, Scott Ryan, Mike Sapeta, Jana Schroth, Suresh Babu Selvaraju, Goran Stevovic, Amanda Suchanek, Andrea Throop, Lyndon Tilson, Thomas Urban, Joe Voshell, Kimberly Wagner, Jonathan Williams, Mary Williamson, Qian Zeng, Tricia Zwiefelhofer, Clinton R. Paden, Duncan MacCannell |
| EPI_ISL_1798832 | CA-Los Angeles County Public Health Laboratory | Centers for Disease Control and Prevention Division of Viral Diseases, Pathogen Discovery | Mili Sheth, Sarah Nobles, Jasmine Padilla, Mark Burroughs, Shoshona Le, Katie Dillon, Peter Cook, Clinton R. Paden, Dhwani Batra, Krista Queen, Kristen Knipe, Dakota Howard, Yvette Unoarumhi, Darlene Wagner, Matthew Schmerer, Ben L. Rambo-Martin, Kristine Lacek, Sam Shepard, Alison Laufer Halpin, Dave Wentworth, Vivien Dugan, Suxiang Tong, Justin Lee |
| EPI_ISL_1798833 | Laboratory Corporation of America | Centers for Disease Control and Prevention Division of Viral Diseases, Pathogen Discovery | Dakota Howard, Dhwani Batra, Peter W. Cook, Kara Moser, Adrian Paskey, Jason Caravas, Benjamin Rambo-Martin, Shatavia Morrison, Christopher Gulvick, Scott Sammons, Yvette Unoarumhi, Darlene Wagner, Matthew Schmerer, Minoo Agarwal, Eyad Almasri, Debbie Boles, Ayla Burns, Nuthawin Charoensri, Oren Cohen, Susan Countryman, Mary Ann Cristobal, Bobbi Croy, Suzanne Dale, Hrushikesh Deshmukh, Amanda Douglas, Vincent Drouillon, Marcia Eisenberg, Howard Engler, Rama Ghatti, Prashant Gupta, Susan Hicks, Jake Humphrey, Lax Iyer, Manoj Jain, Mohan Kolli, Brian Krueger, Tim Kuphal, Stanley Letovsky, Michael Levandoski, Craig Lukasik, Jonathan Meltzer, Brian Norvell, Mindy Nye, Scott Parker, Christos Petropoulos, John Pruitt, Steven Ragan, Scott Ryan, Mike Sapeta, Jana Schroth, Suresh Babu Selvaraju, Goran Stevovic, Amanda Suchanek, Andrea Throop, Lyndon Tilson, Thomas Urban, Joe Voshell, Kimberly Wagner, Jonathan Williams, Mary Williamson, Qian Zeng, Tricia Zwiefelhofer, Clinton R. Paden, Duncan MacCannell |
| EPI_ISL_1798834 | CA-Los Angeles County Public Health Laboratory | Centers for Disease Control and Prevention Division of Viral Diseases, Pathogen Discovery | Mili Sheth, Sarah Nobles, Jasmine Padilla, Mark Burroughs, Shoshona Le, Katie Dillon, Peter Cook, Clinton R. Paden, Dhwani Batra, Krista Queen, Kristen Knipe, Dakota Howard, Yvette Unoarumhi, Darlene Wagner, Matthew Schmerer, Ben L. Rambo-Martin, Kristine Lacek, Sam Shepard, Alison Laufer Halpin, Dave Wentworth, Vivien Dugan, Suxiang Tong, Justin Lee |
| EPI_ISL_1798835 | Laboratory Corporation of America | Centers for Disease Control and Prevention Division of Viral Diseases, Pathogen Discovery | Dakota Howard, Dhwani Batra, Peter W. Cook, Kara Moser, Adrian Paskey, Jason Caravas, Benjamin Rambo-Martin, Shatavia Morrison, Christopher Gulvick, Scott Sammons, Yvette Unoarumhi, Darlene Wagner, Matthew Schmerer, Minoo Agarwal, Eyad Almasri, Debbie Boles, Ayla Burns, Nuthawin Charoensri, Oren Cohen, Susan Countryman, Mary Ann Cristobal, Bobbi Croy, Suzanne Dale, Hrushikesh Deshmukh, Amanda Douglas, Vincent Drouillon, Marcia Eisenberg, Howard Engler, Rama Ghatti, Prashant Gupta, Susan Hicks, Jake Humphrey, Lax Iyer, Manoj Jain, Mohan Kolli, Brian Krueger, Tim Kuphal, Stanley Letovsky, Michael Levandoski, Craig Lukasik, Jonathan Meltzer, Brian Norvell, Mindy Nye, Scott Parker, Christos Petropoulos, John Pruitt, Steven Ragan, Scott Ryan, Mike Sapeta, Jana Schroth, Suresh Babu Selvaraju, Goran Stevovic, Amanda Suchanek, Andrea Throop, Lyndon Tilson, Thomas Urban, Joe Voshell, Kimberly Wagner, Jonathan Williams, Mary Williamson, Qian Zeng, Tricia Zwiefelhofer, Clinton R. Paden, Duncan MacCannell |
| EPI_ISL_1798836 | CA-Los Angeles County Public Health Laboratory | Centers for Disease Control and Prevention Division of Viral Diseases, Pathogen Discovery | Mili Sheth, Sarah Nobles, Jasmine Padilla, Mark Burroughs, Shoshona Le, Katie Dillon, Peter Cook, Clinton R. Paden, Dhwani Batra, Krista Queen, Kristen Knipe, Dakota Howard, Yvette Unoarumhi, Darlene Wagner, Matthew Schmerer, Ben L. Rambo-Martin, Kristine Lacek, Sam Shepard, Alison Laufer Halpin, Dave Wentworth, Vivien Dugan, Suxiang Tong, Justin Lee |
| EPI_ISL_1798837, EPI_ISL_1798838 | Laboratory Corporation of America | Centers for Disease Control and Prevention Division of Viral Diseases, Pathogen Discovery | Dakota Howard, Dhwani Batra, Peter W. Cook, Kara Moser, Adrian Paskey, Jason Caravas, Benjamin Rambo-Martin, Shatavia Morrison, Christopher Gulvick, Scott Sammons, Yvette Unoarumhi, Darlene Wagner, Matthew Schmerer, Minoo Agarwal, Eyad Almasri, Debbie Boles, Ayla Burns, Nuthawin Charoensri, Oren Cohen, Susan Countryman, Mary Ann Cristobal, Bobbi Croy, Suzanne Dale, Hrushikesh Deshmukh, Amanda Douglas, Vincent Drouillon, Marcia Eisenberg, Howard Engler, Rama Ghatti, Prashant Gupta, Susan Hicks, Jake Humphrey, Lax Iyer, Manoj Jain, Mohan Kolli, Brian Krueger, Tim Kuphal, Stanley Letovsky, Michael Levandoski, Craig Lukasik, Jonathan Meltzer, Brian Norvell, Mindy Nye, Scott Parker, Christos Petropoulos, John Pruitt, Steven Ragan, Scott Ryan, Mike Sapeta, Jana Schroth, Suresh Babu Selvaraju, Goran Stevovic, Amanda Suchanek, Andrea Throop, Lyndon Tilson, Thomas Urban, Joe Voshell, Kimberly Wagner, Jonathan Williams, Mary Williamson, Qian Zeng, Tricia Zwiefelhofer, Clinton R. Paden, Duncan MacCannell |
| EPI_ISL_1798839 | CA-Los Angeles County Public Health Laboratory | Centers for Disease Control and Prevention Division of Viral Diseases, Pathogen Discovery | Mili Sheth, Sarah Nobles, Jasmine Padilla, Mark Burroughs, Shoshona Le, Katie Dillon, Peter Cook, Clinton R. Paden, Dhwani Batra, Krista Queen, Kristen Knipe, Dakota Howard, Yvette Unoarumhi, Darlene Wagner, Matthew Schmerer, Ben L. Rambo-Martin, Kristine Lacek, Sam Shepard, Alison Laufer Halpin, Dave Wentworth, Vivien Dugan, Suxiang Tong, Justin Lee |
| EPI_ISL_1798840 | Laboratory Corporation of America | Centers for Disease Control and Prevention Division of Viral Diseases, Pathogen Discovery | Dakota Howard, Dhwani Batra, Peter W. Cook, Kara Moser, Adrian Paskey, Jason Caravas, Benjamin Rambo-Martin, Shatavia Morrison, Christopher Gulvick, Scott Sammons, Yvette Unoarumhi, Darlene Wagner, Matthew Schmerer, Minoo Agarwal, Eyad Almasri, Debbie Boles, Ayla Burns, Nuthawin Charoensri, Oren Cohen, Susan Countryman, Mary Ann Cristobal, Bobbi Croy, Suzanne Dale, Hrushikesh Deshmukh, Amanda Douglas, Vincent Drouillon, Marcia Eisenberg, Howard Engler, Rama Ghatti, Prashant Gupta, Susan Hicks, Jake Humphrey, Lax Iyer, Manoj Jain, Mohan Kolli, Brian Krueger, Tim Kuphal, Stanley Letovsky, Michael Levandoski, Craig Lukasik, Jonathan Meltzer, Brian Norvell, Mindy Nye, Scott Parker, Christos Petropoulos, John Pruitt, Steven Ragan, Scott Ryan, Mike Sapeta, Jana Schroth, Suresh Babu Selvaraju, Goran Stevovic, Amanda Suchanek, Andrea Throop, Lyndon Tilson, Thomas Urban, Joe Voshell, Kimberly Wagner, Jonathan Williams, Mary Williamson, Qian Zeng, Tricia Zwiefelhofer, Clinton R. Paden, Duncan MacCannell |
| EPI_ISL_1798841, EPI_ISL_1798843, EPI_ISL_1798844 | CA-Los Angeles County Public Health Laboratory | Centers for Disease Control and Prevention Division of Viral Diseases, Pathogen Discovery | Mili Sheth, Sarah Nobles, Jasmine Padilla, Mark Burroughs, Shoshona Le, Katie Dillon, Peter Cook, Clinton R. Paden, Dhwani Batra, Krista Queen, Kristen Knipe, Dakota Howard, Yvette Unoarumhi, Darlene Wagner, Matthew Schmerer, Ben L. Rambo-Martin, Kristine Lacek, Sam Shepard, Alison Laufer Halpin, Dave Wentworth, Vivien Dugan, Suxiang Tong, Justin Lee |
| EPI_ISL_1798845 | Laboratory Corporation of America | Centers for Disease Control and Prevention Division of Viral Diseases, Pathogen Discovery | Dakota Howard, Dhwani Batra, Peter W. Cook, Kara Moser, Adrian Paskey, Jason Caravas, Benjamin Rambo-Martin, Shatavia Morrison, Christopher Gulvick, Scott Sammons, Yvette Unoarumhi, Darlene Wagner, Matthew Schmerer, Minoo Agarwal, Eyad Almasri, Debbie Boles, Ayla Burns, Nuthawin Charoensri, Oren Cohen, Susan Countryman, Mary Ann Cristobal, Bobbi Croy, Suzanne Dale, Hrushikesh Deshmukh, Amanda Douglas, Vincent Drouillon, Marcia Eisenberg, Howard Engler, Rama Ghatti, Prashant Gupta, Susan Hicks, Jake Humphrey, Lax Iyer, Manoj Jain, Mohan Kolli, Brian Krueger, Tim Kuphal, Stanley Letovsky, Michael Levandoski, Craig Lukasik, Jonathan Meltzer, Brian Norvell, Mindy Nye, Scott Parker, Christos Petropoulos, John Pruitt, Steven Ragan, Scott Ryan, Mike Sapeta, Jana Schroth, Suresh Babu Selvaraju, Goran Stevovic, Amanda Suchanek, Andrea Throop, Lyndon Tilson, Thomas Urban, Joe Voshell, Kimberly Wagner, Jonathan Williams, Mary Williamson, Qian Zeng, Tricia Zwiefelhofer, Clinton R. Paden, Duncan MacCannell |
| EPI_ISL_1798846 | CA-Los Angeles County Public Health Laboratory | Centers for Disease Control and Prevention Division of Viral Diseases, Pathogen Discovery | Mili Sheth, Sarah Nobles, Jasmine Padilla, Mark Burroughs, Shoshona Le, Katie Dillon, Peter Cook, Clinton R. Paden, Dhwani Batra, Krista Queen, Kristen Knipe, Dakota Howard, Yvette Unoarumhi, Darlene Wagner, Matthew Schmerer, Ben L. Rambo-Martin, Kristine Lacek, Sam Shepard, Alison Laufer Halpin, Dave Wentworth, Vivien Dugan, Suxiang Tong, Justin Lee |
| EPI_ISL_1798847 | Laboratory Corporation of America | Centers for Disease Control and Prevention Division of Viral Diseases, Pathogen Discovery | Dakota Howard, Dhwani Batra, Peter W. Cook, Kara Moser, Adrian Paskey, Jason Caravas, Benjamin Rambo-Martin, Shatavia Morrison, Christopher Gulvick, Scott Sammons, Yvette Unoarumhi, Darlene Wagner, Matthew Schmerer, Minoo Agarwal, Eyad Almasri, Debbie Boles, Ayla Burns, Nuthawin Charoensri, Oren Cohen, Susan Countryman, Mary Ann Cristobal, Bobbi Croy, Suzanne Dale, Hrushikesh Deshmukh, Amanda Douglas, Vincent Drouillon, Marcia Eisenberg, Howard Engler, Rama Ghatti, Prashant Gupta, Susan Hicks, Jake Humphrey, Lax Iyer, Manoj Jain, Mohan Kolli, Brian Krueger, Tim Kuphal, Stanley Letovsky, Michael Levandoski, Craig Lukasik, Jonathan Meltzer, Brian Norvell, Mindy Nye, Scott Parker, Christos Petropoulos, John Pruitt, Steven Ragan, Scott Ryan, Mike Sapeta, Jana Schroth, Suresh Babu Selvaraju, Goran Stevovic, Amanda Suchanek, Andrea Throop, Lyndon Tilson, Thomas Urban, Joe Voshell, Kimberly Wagner, Jonathan Williams, Mary Williamson, Qian Zeng, Tricia Zwiefelhofer, Clinton R. Paden, Duncan MacCannell |
| EPI_ISL_1798848 | CA-Los Angeles County Public Health Laboratory | Centers for Disease Control and Prevention Division of Viral Diseases, Pathogen Discovery | Mili Sheth, Sarah Nobles, Jasmine Padilla, Mark Burroughs, Shoshona Le, Katie Dillon, Peter Cook, Clinton R. Paden, Dhwani Batra, Krista Queen, Kristen Knipe, Dakota Howard, Yvette Unoarumhi, Darlene Wagner, Matthew Schmerer, Ben L. Rambo-Martin, Kristine Lacek, Sam Shepard, Alison Laufer Halpin, Dave Wentworth, Vivien Dugan, Suxiang Tong, Justin Lee |
| EPI_ISL_1798849 | Laboratory Corporation of America | Centers for Disease Control and Prevention Division of Viral Diseases, Pathogen | Dakota Howard, Dhwani Batra, Peter W. Cook, Kara Moser, Adrian Paskey, Jason Caravas, Benjamin Rambo-Martin, Shatavia Morrison, Christopher Gulvick, Scott Sammons, Yvette Unoarumhi, Darlene Wagner, Matthew Schmerer, Minoo Agarwal, Eyad Almasri, Debbie Boles, Ayla Burns, Nuthawin Charoensri, Oren Cohen, Susan Countryman, Mary Ann Cristobal, Bobbi Croy, Suzanne Dale, Hrushikesh Deshmukh, Amanda Douglas, Vincent Drouillon, Marcia Eisenberg, Howard Engler, Rama Ghatti, Prashant Gupta, Susan Hicks, Jake Humphrey, Lax Iyer, Manoj Jain, Mohan |

| | | | |
|---|---|---|---|
| | | Discovery | Kolli, Brian Krueger, Tim Kuphal, Stanley Letovsky, Michael Levandoski, Craig Lukasik, Jonathan Meltzer, Brian Norvell, Mindy Nye, Scott Parker, Christos Petropoulos, John Pruitt, Steven Ragan, Scott Ryan, Mike Sapeta, Jana Schroth, Suresh Babu Selvaraju, Goran Stevovic, Amanda Suchanek, Andrea Throop, Lyndon Tilson, Thomas Urban, Joe Voshell, Kimberly Wagner, Jonathan Williams, Mary Williamson, Qian Zeng, Tricia Zwiefelhofer, Clinton R. Paden, Duncan MacCannell |
| EPI_ISL_1798850 | CA-Los Angeles County Public Health Laboratory | Centers for Disease Control and Prevention Division of Viral Diseases, Pathogen Discovery | Mili Sheth, Sarah Nobles, Jasmine Padilla, Mark Burroughs, Shoshona Le, Katie Dillon, Peter Cook, Clinton R. Paden, Dhwani Batra, Krista Queen, Kristen Knipe, Dakota Howard, Yvette Unoarumhi, Darlene Wagner, Matthew Schmerer, Ben L. Rambo-Martin, Kristine Lacek, Sam Shepard, Alison Laufer Halpin, Dave Wentworth, Vivien Dugan, Suxiang Tong, Justin Lee |
| EPI_ISL_1798851, EPI_ISL_1798852 | Laboratory Corporation of America | Centers for Disease Control and Prevention Division of Viral Diseases, Pathogen Discovery | Dakota Howard, Dhwani Batra, Peter W. Cook, Kara Moser, Adrian Paskey, Jason Caravas, Benjamin Rambo-Martin, Shatavia Morrison, Christopher Gulvick, Scott Sammons, Yvette Unoarumhi, Darlene Wagner, Matthew Schmerer, Minoo Agarwal, Eyad Almasri, Debbie Boles, Ayla Burns, Nuthawin Charoensri, Oren Cohen, Susan Countryman, Mary Ann Cristobal, Bobbi Croy, Suzanne Dale, Hrushikesh Deshmukh, Amanda Douglas, Vincent Drouillon, Marcia Eisenberg, Howard Engler, Rama Ghatti, Prashant Gupta, Susan Hicks, Jake Humphrey, Lax Iyer, Manoj Jain, Mohan Kolli, Brian Krueger, Tim Kuphal, Stanley Letovsky, Michael Levandoski, Craig Lukasik, Jonathan Meltzer, Brian Norvell, Mindy Nye, Scott Parker, Christos Petropoulos, John Pruitt, Steven Ragan, Scott Ryan, Mike Sapeta, Jana Schroth, Suresh Babu Selvaraju, Goran Stevovic, Amanda Suchanek, Andrea Throop, Lyndon Tilson, Thomas Urban, Joe Voshell, Kimberly Wagner, Jonathan Williams, Mary Williamson, Qian Zeng, Tricia Zwiefelhofer, Clinton R. Paden, Duncan MacCannell |
| EPI_ISL_1798853 | CA-Los Angeles County Public Health Laboratory | Centers for Disease Control and Prevention Division of Viral Diseases, Pathogen Discovery | Mili Sheth, Sarah Nobles, Jasmine Padilla, Mark Burroughs, Shoshona Le, Katie Dillon, Peter Cook, Clinton R. Paden, Dhwani Batra, Krista Queen, Kristen Knipe, Dakota Howard, Yvette Unoarumhi, Darlene Wagner, Matthew Schmerer, Ben L. Rambo-Martin, Kristine Lacek, Sam Shepard, Alison Laufer Halpin, Dave Wentworth, Vivien Dugan, Suxiang Tong, Justin Lee |
| EPI_ISL_1798854 | Laboratory Corporation of America | Centers for Disease Control and Prevention Division of Viral Diseases, Pathogen Discovery | Dakota Howard, Dhwani Batra, Peter W. Cook, Kara Moser, Adrian Paskey, Jason Caravas, Benjamin Rambo-Martin, Shatavia Morrison, Christopher Gulvick, Scott Sammons, Yvette Unoarumhi, Darlene Wagner, Matthew Schmerer, Minoo Agarwal, Eyad Almasri, Debbie Boles, Ayla Burns, Nuthawin Charoensri, Oren Cohen, Susan Countryman, Mary Ann Cristobal, Bobbi Croy, Suzanne Dale, Hrushikesh Deshmukh, Amanda Douglas, Vincent Drouillon, Marcia Eisenberg, Howard Engler, Rama Ghatti, Prashant Gupta, Susan Hicks, Jake Humphrey, Lax Iyer, Manoj Jain, Mohan Kolli, Brian Krueger, Tim Kuphal, Stanley Letovsky, Michael Levandoski, Craig Lukasik, Jonathan Meltzer, Brian Norvell, Mindy Nye, Scott Parker, Christos Petropoulos, John Pruitt, Steven Ragan, Scott Ryan, Mike Sapeta, Jana Schroth, Suresh Babu Selvaraju, Goran Stevovic, Amanda Suchanek, Andrea Throop, Lyndon Tilson, Thomas Urban, Joe Voshell, Kimberly Wagner, Jonathan Williams, Mary Williamson, Qian Zeng, Tricia Zwiefelhofer, Clinton R. Paden, Duncan MacCannell |
| EPI_ISL_1798855, EPI_ISL_1798857, EPI_ISL_1798858 | CA-Los Angeles County Public Health Laboratory | Centers for Disease Control and Prevention Division of Viral Diseases, Pathogen Discovery | Mili Sheth, Sarah Nobles, Jasmine Padilla, Mark Burroughs, Shoshona Le, Katie Dillon, Peter Cook, Clinton R. Paden, Dhwani Batra, Krista Queen, Kristen Knipe, Dakota Howard, Yvette Unoarumhi, Darlene Wagner, Matthew Schmerer, Ben L. Rambo-Martin, Kristine Lacek, Sam Shepard, Alison Laufer Halpin, Dave Wentworth, Vivien Dugan, Suxiang Tong, Justin Lee |
| EPI_ISL_1798859 | Laboratory Corporation of America | Centers for Disease Control and Prevention Division of Viral Diseases, Pathogen Discovery | Dakota Howard, Dhwani Batra, Peter W. Cook, Kara Moser, Adrian Paskey, Jason Caravas, Benjamin Rambo-Martin, Shatavia Morrison, Christopher Gulvick, Scott Sammons, Yvette Unoarumhi, Darlene Wagner, Matthew Schmerer, Minoo Agarwal, Eyad Almasri, Debbie Boles, Ayla Burns, Nuthawin Charoensri, Oren Cohen, Susan Countryman, Mary Ann Cristobal, Bobbi Croy, Suzanne Dale, Hrushikesh Deshmukh, Amanda Douglas, Vincent Drouillon, Marcia Eisenberg, Howard Engler, Rama Ghatti, Prashant Gupta, Susan Hicks, Jake Humphrey, Lax Iyer, Manoj Jain, Mohan Kolli, Brian Krueger, Tim Kuphal, Stanley Letovsky, Michael Levandoski, Craig Lukasik, Jonathan Meltzer, Brian Norvell, Mindy Nye, Scott Parker, Christos Petropoulos, John Pruitt, Steven Ragan, Scott Ryan, Mike Sapeta, Jana Schroth, Suresh Babu Selvaraju, Goran Stevovic, Amanda Suchanek, Andrea Throop, Lyndon Tilson, Thomas Urban, Joe Voshell, Kimberly Wagner, Jonathan Williams, Mary Williamson, Qian Zeng, Tricia Zwiefelhofer, Clinton R. Paden, Duncan MacCannell |
| EPI_ISL_1798860 | CA-Los Angeles County Public Health Laboratory | Centers for Disease Control and Prevention Division of Viral Diseases, Pathogen Discovery | Mili Sheth, Sarah Nobles, Jasmine Padilla, Mark Burroughs, Shoshona Le, Katie Dillon, Peter Cook, Clinton R. Paden, Dhwani Batra, Krista Queen, Kristen Knipe, Dakota Howard, Yvette Unoarumhi, Darlene Wagner, Matthew Schmerer, Ben L. Rambo-Martin, Kristine Lacek, Sam Shepard, Alison Laufer Halpin, Dave Wentworth, Vivien Dugan, Suxiang Tong, Justin Lee |
| EPI_ISL_1798861 | Laboratory Corporation of America | Centers for Disease Control and Prevention Division of Viral Diseases, Pathogen Discovery | Dakota Howard, Dhwani Batra, Peter W. Cook, Kara Moser, Adrian Paskey, Jason Caravas, Benjamin Rambo-Martin, Shatavia Morrison, Christopher Gulvick, Scott Sammons, Yvette Unoarumhi, Darlene Wagner, Matthew Schmerer, Minoo Agarwal, Eyad Almasri, Debbie Boles, Ayla Burns, Nuthawin Charoensri, Oren Cohen, Susan Countryman, Mary Ann Cristobal, Bobbi Croy, Suzanne Dale, Hrushikesh Deshmukh, Amanda Douglas, Vincent Drouillon, Marcia Eisenberg, Howard Engler, Rama Ghatti, Prashant Gupta, Susan Hicks, Jake Humphrey, Lax Iyer, Manoj Jain, Mohan Kolli, Brian Krueger, Tim Kuphal, Stanley Letovsky, Michael Levandoski, Craig Lukasik, Jonathan Meltzer, Brian Norvell, Mindy Nye, Scott Parker, Christos Petropoulos, John Pruitt, Steven Ragan, Scott Ryan, Mike Sapeta, Jana Schroth, Suresh Babu Selvaraju, Goran Stevovic, Amanda Suchanek, Andrea Throop, Lyndon Tilson, Thomas Urban, Joe Voshell, Kimberly Wagner, Jonathan Williams, Mary Williamson, Qian Zeng, Tricia Zwiefelhofer, Clinton R. Paden, Duncan MacCannell |
| EPI_ISL_1798862 | CA-Los Angeles County Public Health Laboratory | Centers for Disease Control and Prevention Division of Viral Diseases, Pathogen Discovery | Mili Sheth, Sarah Nobles, Jasmine Padilla, Mark Burroughs, Shoshona Le, Katie Dillon, Peter Cook, Clinton R. Paden, Dhwani Batra, Krista Queen, Kristen Knipe, Dakota Howard, Yvette Unoarumhi, Darlene Wagner, Matthew Schmerer, Ben L. Rambo-Martin, Kristine Lacek, Sam Shepard, Alison Laufer Halpin, Dave Wentworth, Vivien Dugan, Suxiang Tong, Justin Lee |
| EPI_ISL_1798863 | Laboratory Corporation of America | Centers for Disease Control and Prevention Division of Viral Diseases, Pathogen Discovery | Dakota Howard, Dhwani Batra, Peter W. Cook, Kara Moser, Adrian Paskey, Jason Caravas, Benjamin Rambo-Martin, Shatavia Morrison, Christopher Gulvick, Scott Sammons, Yvette Unoarumhi, Darlene Wagner, Matthew Schmerer, Minoo Agarwal, Eyad Almasri, Debbie Boles, Ayla Burns, Nuthawin Charoensri, Oren Cohen, Susan Countryman, Mary Ann Cristobal, Bobbi Croy, Suzanne Dale, Hrushikesh Deshmukh, Amanda Douglas, Vincent Drouillon, Marcia Eisenberg, Howard Engler, Rama Ghatti, Prashant Gupta, Susan Hicks, Jake Humphrey, Lax Iyer, Manoj Jain, Mohan Kolli, Brian Krueger, Tim Kuphal, Stanley Letovsky, Michael Levandoski, Craig Lukasik, Jonathan Meltzer, Brian Norvell, Mindy Nye, Scott Parker, Christos Petropoulos, John Pruitt, Steven Ragan, Scott Ryan, Mike Sapeta, Jana Schroth, Suresh Babu Selvaraju, Goran Stevovic, Amanda Suchanek, Andrea Throop, Lyndon Tilson, Thomas Urban, Joe Voshell, Kimberly Wagner, Jonathan Williams, Mary Williamson, Qian Zeng, Tricia Zwiefelhofer, Clinton R. Paden, Duncan MacCannell |
| EPI_ISL_1798864, EPI_ISL_1798866, EPI_ISL_1798868 | CA-Los Angeles County Public Health Laboratory | Centers for Disease Control and Prevention Division of Viral Diseases, Pathogen Discovery | Mili Sheth, Sarah Nobles, Jasmine Padilla, Mark Burroughs, Shoshona Le, Katie Dillon, Peter Cook, Clinton R. Paden, Dhwani Batra, Krista Queen, Kristen Knipe, Dakota Howard, Yvette Unoarumhi, Darlene Wagner, Matthew Schmerer, Ben L. Rambo-Martin, Kristine Lacek, Sam Shepard, Alison Laufer Halpin, Dave Wentworth, Vivien Dugan, Suxiang Tong, Justin Lee |
| EPI_ISL_1798869 | Laboratory Corporation of America | Centers for Disease Control and Prevention Division of Viral Diseases, Pathogen Discovery | Dakota Howard, Dhwani Batra, Peter W. Cook, Kara Moser, Adrian Paskey, Jason Caravas, Benjamin Rambo-Martin, Shatavia Morrison, Christopher Gulvick, Scott Sammons, Yvette Unoarumhi, Darlene Wagner, Matthew Schmerer, Minoo Agarwal, Eyad Almasri, Debbie Boles, Ayla Burns, Nuthawin Charoensri, Oren Cohen, Susan Countryman, Mary Ann Cristobal, Bobbi Croy, Suzanne Dale, Hrushikesh Deshmukh, Amanda Douglas, Vincent Drouillon, Marcia Eisenberg, Howard Engler, Rama Ghatti, Prashant Gupta, Susan Hicks, Jake Humphrey, Lax Iyer, Manoj Jain, Mohan Kolli, Brian Krueger, Tim Kuphal, Stanley Letovsky, Michael Levandoski, Craig Lukasik, Jonathan Meltzer, Brian Norvell, Mindy Nye, Scott Parker, Christos Petropoulos, John Pruitt, Steven Ragan, Scott Ryan, Mike Sapeta, Jana Schroth, Suresh Babu Selvaraju, Goran Stevovic, Amanda Suchanek, Andrea Throop, Lyndon Tilson, Thomas Urban, Joe Voshell, Kimberly Wagner, Jonathan Williams, Mary Williamson, Qian Zeng, Tricia Zwiefelhofer, Clinton R. Paden, Duncan MacCannell |
| EPI_ISL_1798870 | DC Public Health Lab/ Dept. of Forensic Sciences | Centers for Disease Control and Prevention Division of Viral Diseases, Pathogen Discovery | Mili Sheth, Sarah Nobles, Jasmine Padilla, Mark Burroughs, Shoshona Le, Katie Dillon, Peter Cook, Clinton R. Paden, Dhwani Batra, Krista Queen, Kristen Knipe, Dakota Howard, Yvette Unoarumhi, Darlene Wagner, Matthew Schmerer, Ben L. Rambo-Martin, Kristine Lacek, Sam Shepard, Alison Laufer Halpin, Dave Wentworth, Vivien Dugan, Suxiang Tong, Justin Lee |
| EPI_ISL_1798871 | Laboratory Corporation of America | Centers for Disease Control and Prevention Division of Viral Diseases, Pathogen Discovery | Dakota Howard, Dhwani Batra, Peter W. Cook, Kara Moser, Adrian Paskey, Jason Caravas, Benjamin Rambo-Martin, Shatavia Morrison, Christopher Gulvick, Scott Sammons, Yvette Unoarumhi, Darlene Wagner, Matthew Schmerer, Minoo Agarwal, Eyad Almasri, Debbie Boles, Ayla Burns, Nuthawin Charoensri, Oren Cohen, Susan Countryman, Mary Ann Cristobal, Bobbi Croy, Suzanne Dale, Hrushikesh Deshmukh, Amanda Douglas, Vincent Drouillon, Marcia Eisenberg, Howard Engler, Rama Ghatti, Prashant Gupta, Susan Hicks, Jake Humphrey, Lax Iyer, Manoj Jain, Mohan Kolli, Brian Krueger, Tim Kuphal, Stanley Letovsky, Michael Levandoski, Craig Lukasik, Jonathan Meltzer, Brian Norvell, Mindy Nye, Scott Parker, Christos Petropoulos, John Pruitt, Steven Ragan, Scott Ryan, Mike Sapeta, Jana Schroth, Suresh Babu Selvaraju, Goran Stevovic, Amanda Suchanek, Andrea Throop, Lyndon Tilson, Thomas Urban, Joe Voshell, Kimberly Wagner, Jonathan Williams, Mary Williamson, Qian Zeng, Tricia Zwiefelhofer, Clinton R. Paden, Duncan MacCannell |
| EPI_ISL_1798873 | DC Public Health Lab/ Dept. of Forensic Sciences | Centers for Disease Control and Prevention Division of Viral Diseases, Pathogen Discovery | Mili Sheth, Sarah Nobles, Jasmine Padilla, Mark Burroughs, Shoshona Le, Katie Dillon, Peter Cook, Clinton R. Paden, Dhwani Batra, Krista Queen, Kristen Knipe, Dakota Howard, Yvette Unoarumhi, Darlene Wagner, Matthew Schmerer, Ben L. Rambo-Martin, Kristine Lacek, Sam Shepard, Alison Laufer Halpin, Dave Wentworth, Vivien Dugan, Suxiang Tong, Justin Lee |
| EPI_ISL_1798874 | Laboratory Corporation of America | Centers for Disease Control and Prevention Division of Viral Diseases, Pathogen Discovery | Dakota Howard, Dhwani Batra, Peter W. Cook, Kara Moser, Adrian Paskey, Jason Caravas, Benjamin Rambo-Martin, Shatavia Morrison, Christopher Gulvick, Scott Sammons, Yvette Unoarumhi, Darlene Wagner, Matthew Schmerer, Minoo Agarwal, Eyad Almasri, Debbie Boles, Ayla Burns, Nuthawin Charoensri, Oren Cohen, Susan Countryman, Mary Ann Cristobal, Bobbi Croy, Suzanne Dale, Hrushikesh Deshmukh, Amanda Douglas, Vincent Drouillon, Marcia Eisenberg, Howard Engler, Rama Ghatti, Prashant Gupta, Susan Hicks, Jake Humphrey, Lax Iyer, Manoj Jain, Mohan Kolli, Brian Krueger, Tim Kuphal, Stanley Letovsky, Michael Levandoski, Craig Lukasik, Jonathan Meltzer, Brian Norvell, Mindy Nye, Scott Parker, Christos Petropoulos, John Pruitt, Steven Ragan, Scott Ryan, Mike Sapeta, Jana Schroth, Suresh Babu Selvaraju, Goran Stevovic, Amanda Suchanek, Andrea Throop, Lyndon Tilson, Thomas Urban, Joe Voshell, Kimberly Wagner, Jonathan Williams, Mary Williamson, Qian Zeng, Tricia Zwiefelhofer, Clinton R. Paden, Duncan MacCannell |
| EPI_ISL_1798875, EPI_ISL_1798876 | DC Public Health Lab/ Dept. of Forensic Sciences | Centers for Disease Control and Prevention Division of Viral Diseases, Pathogen Discovery | Mili Sheth, Sarah Nobles, Jasmine Padilla, Mark Burroughs, Shoshona Le, Katie Dillon, Peter Cook, Clinton R. Paden, Dhwani Batra, Krista Queen, Kristen Knipe, Dakota Howard, Yvette Unoarumhi, Darlene Wagner, Matthew Schmerer, Ben L. Rambo-Martin, Kristine Lacek, Sam Shepard, Alison Laufer Halpin, Dave Wentworth, Vivien Dugan, Suxiang Tong, Justin Lee |
| EPI_ISL_1798878 | Laboratory Corporation of America | Centers for Disease Control and Prevention Division of Viral Diseases, Pathogen Discovery | Dakota Howard, Dhwani Batra, Peter W. Cook, Kara Moser, Adrian Paskey, Jason Caravas, Benjamin Rambo-Martin, Shatavia Morrison, Christopher Gulvick, Scott Sammons, Yvette Unoarumhi, Darlene Wagner, Matthew Schmerer, Minoo Agarwal, Eyad Almasri, Debbie Boles, Ayla Burns, Nuthawin Charoensri, Oren Cohen, Susan Countryman, Mary Ann Cristobal, Bobbi Croy, Suzanne Dale, Hrushikesh Deshmukh, Amanda Douglas, Vincent Drouillon, Marcia Eisenberg, Howard Engler, Rama Ghatti, Prashant Gupta, Susan Hicks, Jake Humphrey, Lax Iyer, Manoj Jain, Mohan Kolli, Brian Krueger, Tim Kuphal, Stanley Letovsky, Michael Levandoski, Craig Lukasik, Jonathan Meltzer, Brian Norvell, Mindy Nye, Scott Parker, Christos Petropoulos, John Pruitt, Steven Ragan, Scott Ryan, Mike Sapeta, Jana Schroth, Suresh Babu Selvaraju, Goran Stevovic, Amanda Suchanek, Andrea Throop, Lyndon Tilson, Thomas Urban, Joe Voshell, Kimberly Wagner, Jonathan Williams, Mary Williamson, Qian Zeng, Tricia Zwiefelhofer, Clinton R. Paden, Duncan MacCannell |
| EPI_ISL_1798879 | DC Public Health Lab/ Dept. of Forensic Sciences | Centers for Disease Control and Prevention Division of Viral Diseases, Pathogen Discovery | Mili Sheth, Sarah Nobles, Jasmine Padilla, Mark Burroughs, Shoshona Le, Katie Dillon, Peter Cook, Clinton R. Paden, Dhwani Batra, Krista Queen, Kristen Knipe, Dakota Howard, Yvette Unoarumhi, Darlene Wagner, Matthew Schmerer, Ben L. Rambo-Martin, Kristine Lacek, Sam Shepard, Alison Laufer Halpin, Dave Wentworth, Vivien Dugan, Suxiang Tong, Justin Lee |
| EPI_ISL_1798880 | Laboratory Corporation of America | Centers for Disease Control and Prevention Division of Viral Diseases, Pathogen Discovery | Dakota Howard, Dhwani Batra, Peter W. Cook, Kara Moser, Adrian Paskey, Jason Caravas, Benjamin Rambo-Martin, Shatavia Morrison, Christopher Gulvick, Scott Sammons, Yvette Unoarumhi, Darlene Wagner, Matthew Schmerer, Minoo Agarwal, Eyad Almasri, Debbie Boles, Ayla Burns, Nuthawin Charoensri, Oren Cohen, Susan Countryman, Mary Ann Cristobal, Bobbi Croy, Suzanne Dale, Hrushikesh Deshmukh, Amanda Douglas, Vincent Drouillon, Marcia Eisenberg, Howard Engler, Rama Ghatti, Prashant Gupta, Susan Hicks, Jake Humphrey, Lax Iyer, Manoj Jain, Mohan Kolli, Brian Krueger, Tim Kuphal, Stanley Letovsky, Michael Levandoski, Craig Lukasik, Jonathan Meltzer, Brian Norvell, Mindy Nye, Scott Parker, Christos Petropoulos, John Pruitt, Steven Ragan, Scott Ryan, Mike Sapeta, Jana Schroth, Suresh Babu Selvaraju, Goran Stevovic, Amanda Suchanek, Andrea Throop, Lyndon Tilson, Thomas Urban, Joe Voshell, Kimberly Wagner, Jonathan Williams, Mary Williamson, Qian Zeng, Tricia Zwiefelhofer, Clinton R. Paden, Duncan MacCannell |
| EPI_ISL_1798881 | DC Public Health Lab/ Dept. of Forensic Sciences | Centers for Disease Control and Prevention Division of Viral Diseases, Pathogen Discovery | Mili Sheth, Sarah Nobles, Jasmine Padilla, Mark Burroughs, Shoshona Le, Katie Dillon, Peter Cook, Clinton R. Paden, Dhwani Batra, Krista Queen, Kristen Knipe, Dakota Howard, Yvette Unoarumhi, Darlene Wagner, Matthew Schmerer, Ben L. Rambo-Martin, Kristine Lacek, Sam Shepard, Alison Laufer Halpin, Dave Wentworth, Vivien Dugan, Suxiang Tong, Justin Lee |
| EPI_ISL_1798882 | Laboratory Corporation of America | Centers for Disease Control and Prevention Division of Viral Diseases, Pathogen Discovery | Dakota Howard, Dhwani Batra, Peter W. Cook, Kara Moser, Adrian Paskey, Jason Caravas, Benjamin Rambo-Martin, Shatavia Morrison, Christopher Gulvick, Scott Sammons, Yvette Unoarumhi, Darlene Wagner, Matthew Schmerer, Minoo Agarwal, Eyad Almasri, Debbie Boles, Ayla Burns, Nuthawin Charoensri, Oren Cohen, Susan Countryman, Mary Ann Cristobal, Bobbi Croy, Suzanne Dale, Hrushikesh Deshmukh, Amanda Douglas, Vincent Drouillon, Marcia Eisenberg, Howard Engler, Rama Ghatti, Prashant Gupta, Susan Hicks, Jake Humphrey, Lax Iyer, Manoj Jain, Mohan Kolli, Brian Krueger, Tim Kuphal, Stanley Letovsky, Michael Levandoski, Craig Lukasik, Jonathan Meltzer, Brian Norvell, Mindy Nye, Scott Parker, Christos Petropoulos, John Pruitt, Steven Ragan, Scott Ryan, Mike Sapeta, Jana Schroth, Suresh Babu Selvaraju, Goran Stevovic, Amanda Suchanek, Andrea Throop, Lyndon Tilson, Thomas Urban, Joe Voshell, Kimberly Wagner, Jonathan Williams, Mary Williamson, Qian Zeng, Tricia Zwiefelhofer, Clinton R. Paden, Duncan MacCannell |
| EPI_ISL_1798883 | DC Public Health Lab/ Dept. of Forensic Sciences | Centers for Disease Control and Prevention Division of Viral Diseases, Pathogen Discovery | Mili Sheth, Sarah Nobles, Jasmine Padilla, Mark Burroughs, Shoshona Le, Katie Dillon, Peter Cook, Clinton R. Paden, Dhwani Batra, Krista Queen, Kristen Knipe, Dakota Howard, Yvette Unoarumhi, Darlene Wagner, Matthew Schmerer, Ben L. Rambo-Martin, Kristine Lacek, Sam Shepard, Alison Laufer Halpin, Dave Wentworth, Vivien Dugan, Suxiang Tong, Justin Lee |
| EPI_ISL_1798884 | Laboratory Corporation of America | Centers for Disease Control and Prevention Division of Viral Diseases, Pathogen Discovery | Dakota Howard, Dhwani Batra, Peter W. Cook, Kara Moser, Adrian Paskey, Jason Caravas, Benjamin Rambo-Martin, Shatavia Morrison, Christopher Gulvick, Scott Sammons, Yvette Unoarumhi, Darlene Wagner, Matthew Schmerer, Minoo Agarwal, Eyad Almasri, Debbie Boles, Ayla Burns, Nuthawin Charoensri, Oren Cohen, Susan Countryman, Mary Ann Cristobal, Bobbi Croy, Suzanne Dale, Hrushikesh Deshmukh, Amanda Douglas, Vincent Drouillon, Marcia Eisenberg, Howard Engler, Rama Ghatti, Prashant Gupta, Susan Hicks, Jake Humphrey, Lax Iyer, Manoj Jain, Mohan Kolli, Brian Krueger, Tim Kuphal, Stanley Letovsky, Michael Levandoski, Craig Lukasik, Jonathan Meltzer, Brian Norvell, Mindy Nye, Scott Parker, Christos Petropoulos, John Pruitt, Steven Ragan, Scott Ryan, Mike Sapeta, Jana Schroth, Suresh Babu Selvaraju, Goran Stevovic, Amanda Suchanek, Andrea Throop, Lyndon Tilson, Thomas Urban, Joe Voshell, Kimberly Wagner, Jonathan Williams, Mary Williamson, Qian Zeng, Tricia Zwiefelhofer, Clinton R. Paden, Duncan MacCannell |
| EPI_ISL_1798885 | DC Public Health Lab/ Dept. of Forensic Sciences | Centers for Disease Control and Prevention Division of Viral Diseases, Pathogen Discovery | Mili Sheth, Sarah Nobles, Jasmine Padilla, Mark Burroughs, Shoshona Le, Katie Dillon, Peter Cook, Clinton R. Paden, Dhwani Batra, Krista Queen, Kristen Knipe, Dakota Howard, Yvette Unoarumhi, Darlene Wagner, Matthew Schmerer, Ben L. Rambo-Martin, Kristine Lacek, Sam Shepard, Alison Laufer Halpin, Dave Wentworth, Vivien Dugan, Suxiang Tong, Justin Lee |
| EPI_ISL_1798886, EPI_ISL_1798887 | Laboratory Corporation of America | Centers for Disease Control and Prevention Division of Viral Diseases, Pathogen Discovery | Dakota Howard, Dhwani Batra, Peter W. Cook, Kara Moser, Adrian Paskey, Jason Caravas, Benjamin Rambo-Martin, Shatavia Morrison, Christopher Gulvick, Scott Sammons, Yvette Unoarumhi, Darlene Wagner, Matthew Schmerer, Minoo Agarwal, Eyad Almasri, Debbie Boles, Ayla Burns, Nuthawin Charoensri, Oren Cohen, Susan Countryman, Mary Ann Cristobal, Bobbi Croy, Suzanne Dale, Hrushikesh Deshmukh, Amanda Douglas, Vincent Drouillon, Marcia Eisenberg, Howard Engler, Rama Ghatti, Prashant Gupta, Susan Hicks, Jake Humphrey, Lax Iyer, Manoj Jain, Mohan Kolli, Brian Krueger, Tim Kuphal, Stanley Letovsky, Michael Levandoski, Craig Lukasik, Jonathan Meltzer, Brian Norvell, Mindy Nye, Scott Parker, Christos Petropoulos, John Pruitt, Steven Ragan, Scott Ryan, Mike Sapeta, Jana Schroth, Suresh Babu Selvaraju, Goran Stevovic, Amanda Suchanek, Andrea Throop, Lyndon Tilson, Thomas Urban, Joe Voshell, Kimberly Wagner, Jonathan Williams, Mary Williamson, Qian Zeng, Tricia Zwiefelhofer, Clinton R. Paden, Duncan MacCannell |
| EPI_ISL_1798888 | DC Public Health Lab/ Dept. of Forensic Sciences | Centers for Disease Control and Prevention Division of Viral Diseases, Pathogen Discovery | Mili Sheth, Sarah Nobles, Jasmine Padilla, Mark Burroughs, Shoshona Le, Katie Dillon, Peter Cook, Clinton R. Paden, Dhwani Batra, Krista Queen, Kristen Knipe, Dakota Howard, Yvette Unoarumhi, Darlene Wagner, Matthew Schmerer, Ben L. Rambo-Martin, Kristine Lacek, Sam Shepard, Alison Laufer Halpin, Dave Wentworth, Vivien Dugan, Suxiang Tong, Justin Lee |
| EPI_ISL_1798889 | Laboratory Corporation of America | Centers for Disease Control and Prevention Division of Viral Diseases, Pathogen | Dakota Howard, Dhwani Batra, Peter W. Cook, Kara Moser, Adrian Paskey, Jason Caravas, Benjamin Rambo-Martin, Shatavia Morrison, Christopher Gulvick, Scott Sammons, Yvette Unoarumhi, Darlene Wagner, Matthew Schmerer, Minoo Agarwal, Eyad Almasri, Debbie Boles, Ayla Burns, Nuthawin Charoensri, Oren Cohen, Susan Countryman, Mary Ann Cristobal, Bobbi Croy, Suzanne Dale, Hrushikesh Deshmukh, Amanda Douglas, Vincent Drouillon, Marcia Eisenberg, Howard Engler, Rama Ghatti, Prashant Gupta, Susan Hicks, Jake Humphrey, Lax Iyer, Manoj Jain, Mohan |

| | | | |
|---|---|---|---|
| | | Discovery | Kolli, Brian Krueger, Tim Kuphal, Stanley Letovsky, Michael Levandoski, Craig Lukasik, Jonathan Meltzer, Brian Norvell, Mindy Nye, Scott Parker, Christos Petropoulos, John Pruitt, Steven Ragan, Scott Ryan, Mike Sapeta, Jana Schroth, Suresh Babu Selvaraju, Goran Stevovic, Amanda Suchanek, Andrea Throop, Lyndon Tilson, Thomas Urban, Joe Voshell, Kimberly Wagner, Jonathan Williams, Mary Williamson, Qian Zeng, Tricia Zwiefelhofer, Clinton R. Paden, Duncan MacCannell |
| EPI_ISL_1798890 | GA Department of Public Health Laboratory | Centers for Disease Control and Prevention Division of Viral Diseases, Pathogen Discovery | Mili Sheth, Sarah Nobles, Jasmine Padilla, Mark Burroughs, Shoshona Le, Katie Dillon, Peter Cook, Clinton R. Paden, Dhwani Batra, Krista Queen, Kristen Knipe, Dakota Howard, Yvette Unoarumhi, Darlene Wagner, Matthew Schmerer, Ben L. Rambo-Martin, Kristine Lacek, Sam Shepard, Alison Laufer Halpin, Dave Wentworth, Vivien Dugan, Suxiang Tong, Justin Lee |
| EPI_ISL_1798891 | Laboratory Corporation of America | Centers for Disease Control and Prevention Division of Viral Diseases, Pathogen Discovery | Dakota Howard, Dhwani Batra, Peter W. Cook, Kara Moser, Adrian Paskey, Jason Caravas, Benjamin Rambo-Martin, Shatavia Morrison, Christopher Gulvick, Scott Sammons, Yvette Unoarumhi, Darlene Wagner, Matthew Schmerer, Minoo Agarwal, Eyad Almasri, Debbie Boles, Ayla Burns, Nuthawin Charoensri, Oren Cohen, Susan Countryman, Mary Ann Cristobal, Bobbi Croy, Suzanne Dale, Hrushikesh Deshmukh, Amanda Douglas, Vincent Drouillon, Marcia Eisenberg, Howard Engler, Rama Ghatti, Prashant Gupta, Susan Hicks, Jake Humphrey, Lax Iyer, Manoj Jain, Mohan Kolli, Brian Krueger, Tim Kuphal, Stanley Letovsky, Michael Levandoski, Craig Lukasik, Jonathan Meltzer, Brian Norvell, Mindy Nye, Scott Parker, Christos Petropoulos, John Pruitt, Steven Ragan, Scott Ryan, Mike Sapeta, Jana Schroth, Suresh Babu Selvaraju, Goran Stevovic, Amanda Suchanek, Andrea Throop, Lyndon Tilson, Thomas Urban, Joe Voshell, Kimberly Wagner, Jonathan Williams, Mary Williamson, Qian Zeng, Tricia Zwiefelhofer, Clinton R. Paden, Duncan MacCannell |
| EPI_ISL_1798892 | GA Department of Public Health Laboratory | Centers for Disease Control and Prevention Division of Viral Diseases, Pathogen Discovery | Mili Sheth, Sarah Nobles, Jasmine Padilla, Mark Burroughs, Shoshona Le, Katie Dillon, Peter Cook, Clinton R. Paden, Dhwani Batra, Krista Queen, Kristen Knipe, Dakota Howard, Yvette Unoarumhi, Darlene Wagner, Matthew Schmerer, Ben L. Rambo-Martin, Kristine Lacek, Sam Shepard, Alison Laufer Halpin, Dave Wentworth, Vivien Dugan, Suxiang Tong, Justin Lee |
| EPI_ISL_1798893 | Laboratory Corporation of America | Centers for Disease Control and Prevention Division of Viral Diseases, Pathogen Discovery | Dakota Howard, Dhwani Batra, Peter W. Cook, Kara Moser, Adrian Paskey, Jason Caravas, Benjamin Rambo-Martin, Shatavia Morrison, Christopher Gulvick, Scott Sammons, Yvette Unoarumhi, Darlene Wagner, Matthew Schmerer, Minoo Agarwal, Eyad Almasri, Debbie Boles, Ayla Burns, Nuthawin Charoensri, Oren Cohen, Susan Countryman, Mary Ann Cristobal, Bobbi Croy, Suzanne Dale, Hrushikesh Deshmukh, Amanda Douglas, Vincent Drouillon, Marcia Eisenberg, Howard Engler, Rama Ghatti, Prashant Gupta, Susan Hicks, Jake Humphrey, Lax Iyer, Manoj Jain, Mohan Kolli, Brian Krueger, Tim Kuphal, Stanley Letovsky, Michael Levandoski, Craig Lukasik, Jonathan Meltzer, Brian Norvell, Mindy Nye, Scott Parker, Christos Petropoulos, John Pruitt, Steven Ragan, Scott Ryan, Mike Sapeta, Jana Schroth, Suresh Babu Selvaraju, Goran Stevovic, Amanda Suchanek, Andrea Throop, Lyndon Tilson, Thomas Urban, Joe Voshell, Kimberly Wagner, Jonathan Williams, Mary Williamson, Qian Zeng, Tricia Zwiefelhofer, Clinton R. Paden, Duncan MacCannell |
| EPI_ISL_1798894 | GA Department of Public Health Laboratory | Centers for Disease Control and Prevention Division of Viral Diseases, Pathogen Discovery | Mili Sheth, Sarah Nobles, Jasmine Padilla, Mark Burroughs, Shoshona Le, Katie Dillon, Peter Cook, Clinton R. Paden, Dhwani Batra, Krista Queen, Kristen Knipe, Dakota Howard, Yvette Unoarumhi, Darlene Wagner, Matthew Schmerer, Ben L. Rambo-Martin, Kristine Lacek, Sam Shepard, Alison Laufer Halpin, Dave Wentworth, Vivien Dugan, Suxiang Tong, Justin Lee |
| EPI_ISL_1798895 | Laboratory Corporation of America | Centers for Disease Control and Prevention Division of Viral Diseases, Pathogen Discovery | Dakota Howard, Dhwani Batra, Peter W. Cook, Kara Moser, Adrian Paskey, Jason Caravas, Benjamin Rambo-Martin, Shatavia Morrison, Christopher Gulvick, Scott Sammons, Yvette Unoarumhi, Darlene Wagner, Matthew Schmerer, Minoo Agarwal, Eyad Almasri, Debbie Boles, Ayla Burns, Nuthawin Charoensri, Oren Cohen, Susan Countryman, Mary Ann Cristobal, Bobbi Croy, Suzanne Dale, Hrushikesh Deshmukh, Amanda Douglas, Vincent Drouillon, Marcia Eisenberg, Howard Engler, Rama Ghatti, Prashant Gupta, Susan Hicks, Jake Humphrey, Lax Iyer, Manoj Jain, Mohan Kolli, Brian Krueger, Tim Kuphal, Stanley Letovsky, Michael Levandoski, Craig Lukasik, Jonathan Meltzer, Brian Norvell, Mindy Nye, Scott Parker, Christos Petropoulos, John Pruitt, Steven Ragan, Scott Ryan, Mike Sapeta, Jana Schroth, Suresh Babu Selvaraju, Goran Stevovic, Amanda Suchanek, Andrea Throop, Lyndon Tilson, Thomas Urban, Joe Voshell, Kimberly Wagner, Jonathan Williams, Mary Williamson, Qian Zeng, Tricia Zwiefelhofer, Clinton R. Paden, Duncan MacCannell |
| EPI_ISL_1798896 | GA Department of Public Health Laboratory | Centers for Disease Control and Prevention Division of Viral Diseases, Pathogen Discovery | Mili Sheth, Sarah Nobles, Jasmine Padilla, Mark Burroughs, Shoshona Le, Katie Dillon, Peter Cook, Clinton R. Paden, Dhwani Batra, Krista Queen, Kristen Knipe, Dakota Howard, Yvette Unoarumhi, Darlene Wagner, Matthew Schmerer, Ben L. Rambo-Martin, Kristine Lacek, Sam Shepard, Alison Laufer Halpin, Dave Wentworth, Vivien Dugan, Suxiang Tong, Justin Lee |
| EPI_ISL_1798897 | Laboratory Corporation of America | Centers for Disease Control and Prevention Division of Viral Diseases, Pathogen Discovery | Dakota Howard, Dhwani Batra, Peter W. Cook, Kara Moser, Adrian Paskey, Jason Caravas, Benjamin Rambo-Martin, Shatavia Morrison, Christopher Gulvick, Scott Sammons, Yvette Unoarumhi, Darlene Wagner, Matthew Schmerer, Minoo Agarwal, Eyad Almasri, Debbie Boles, Ayla Burns, Nuthawin Charoensri, Oren Cohen, Susan Countryman, Mary Ann Cristobal, Bobbi Croy, Suzanne Dale, Hrushikesh Deshmukh, Amanda Douglas, Vincent Drouillon, Marcia Eisenberg, Howard Engler, Rama Ghatti, Prashant Gupta, Susan Hicks, Jake Humphrey, Lax Iyer, Manoj Jain, Mohan Kolli, Brian Krueger, Tim Kuphal, Stanley Letovsky, Michael Levandoski, Craig Lukasik, Jonathan Meltzer, Brian Norvell, Mindy Nye, Scott Parker, Christos Petropoulos, John Pruitt, Steven Ragan, Scott Ryan, Mike Sapeta, Jana Schroth, Suresh Babu Selvaraju, Goran Stevovic, Amanda Suchanek, Andrea Throop, Lyndon Tilson, Thomas Urban, Joe Voshell, Kimberly Wagner, Jonathan Williams, Mary Williamson, Qian Zeng, Tricia Zwiefelhofer, Clinton R. Paden, Duncan MacCannell |
| EPI_ISL_1798899 | GA Department of Public Health Laboratory | Centers for Disease Control and Prevention Division of Viral Diseases, Pathogen Discovery | Mili Sheth, Sarah Nobles, Jasmine Padilla, Mark Burroughs, Shoshona Le, Katie Dillon, Peter Cook, Clinton R. Paden, Dhwani Batra, Krista Queen, Kristen Knipe, Dakota Howard, Yvette Unoarumhi, Darlene Wagner, Matthew Schmerer, Ben L. Rambo-Martin, Kristine Lacek, Sam Shepard, Alison Laufer Halpin, Dave Wentworth, Vivien Dugan, Suxiang Tong, Justin Lee |
| EPI_ISL_1798900 | Laboratory Corporation of America | Centers for Disease Control and Prevention Division of Viral Diseases, Pathogen Discovery | Dakota Howard, Dhwani Batra, Peter W. Cook, Kara Moser, Adrian Paskey, Jason Caravas, Benjamin Rambo-Martin, Shatavia Morrison, Christopher Gulvick, Scott Sammons, Yvette Unoarumhi, Darlene Wagner, Matthew Schmerer, Minoo Agarwal, Eyad Almasri, Debbie Boles, Ayla Burns, Nuthawin Charoensri, Oren Cohen, Susan Countryman, Mary Ann Cristobal, Bobbi Croy, Suzanne Dale, Hrushikesh Deshmukh, Amanda Douglas, Vincent Drouillon, Marcia Eisenberg, Howard Engler, Rama Ghatti, Prashant Gupta, Susan Hicks, Jake Humphrey, Lax Iyer, Manoj Jain, Mohan Kolli, Brian Krueger, Tim Kuphal, Stanley Letovsky, Michael Levandoski, Craig Lukasik, Jonathan Meltzer, Brian Norvell, Mindy Nye, Scott Parker, Christos Petropoulos, John Pruitt, Steven Ragan, Scott Ryan, Mike Sapeta, Jana Schroth, Suresh Babu Selvaraju, Goran Stevovic, Amanda Suchanek, Andrea Throop, Lyndon Tilson, Thomas Urban, Joe Voshell, Kimberly Wagner, Jonathan Williams, Mary Williamson, Qian Zeng, Tricia Zwiefelhofer, Clinton R. Paden, Duncan MacCannell |
| EPI_ISL_1798901 | GA Department of Public Health Laboratory | Centers for Disease Control and Prevention Division of Viral Diseases, Pathogen Discovery | Mili Sheth, Sarah Nobles, Jasmine Padilla, Mark Burroughs, Shoshona Le, Katie Dillon, Peter Cook, Clinton R. Paden, Dhwani Batra, Krista Queen, Kristen Knipe, Dakota Howard, Yvette Unoarumhi, Darlene Wagner, Matthew Schmerer, Ben L. Rambo-Martin, Kristine Lacek, Sam Shepard, Alison Laufer Halpin, Dave Wentworth, Vivien Dugan, Suxiang Tong, Justin Lee |
| EPI_ISL_1798902 | Guam Public Health Laboratory | Centers for Disease Control and Prevention Division of Viral Diseases, Pathogen Discovery | Mili Sheth, Sarah Nobles, Jasmine Padilla, Mark Burroughs, Shoshona Le, Katie Dillon, Peter Cook, Clinton R. Paden, Dhwani Batra, Krista Queen, Kristen Knipe, Dakota Howard, Yvette Unoarumhi, Darlene Wagner, Matthew Schmerer, Ben L. Rambo-Martin, Kristine Lacek, Sam Shepard, Alison Laufer Halpin, Dave Wentworth, Vivien Dugan, Suxiang Tong, Justin Lee |
| EPI_ISL_1798903 | Laboratory Corporation of America | Centers for Disease Control and Prevention Division of Viral Diseases, Pathogen Discovery | Dakota Howard, Dhwani Batra, Peter W. Cook, Kara Moser, Adrian Paskey, Jason Caravas, Benjamin Rambo-Martin, Shatavia Morrison, Christopher Gulvick, Scott Sammons, Yvette Unoarumhi, Darlene Wagner, Matthew Schmerer, Minoo Agarwal, Eyad Almasri, Debbie Boles, Ayla Burns, Nuthawin Charoensri, Oren Cohen, Susan Countryman, Mary Ann Cristobal, Bobbi Croy, Suzanne Dale, Hrushikesh Deshmukh, Amanda Douglas, Vincent Drouillon, Marcia Eisenberg, Howard Engler, Rama Ghatti, Prashant Gupta, Susan Hicks, Jake Humphrey, Lax Iyer, Manoj Jain, Mohan Kolli, Brian Krueger, Tim Kuphal, Stanley Letovsky, Michael Levandoski, Craig Lukasik, Jonathan Meltzer, Brian Norvell, Mindy Nye, Scott Parker, Christos Petropoulos, John Pruitt, Steven Ragan, Scott Ryan, Mike Sapeta, Jana Schroth, Suresh Babu Selvaraju, Goran Stevovic, Amanda Suchanek, Andrea Throop, Lyndon Tilson, Thomas Urban, Joe Voshell, Kimberly Wagner, Jonathan Williams, Mary Williamson, Qian Zeng, Tricia Zwiefelhofer, Clinton R. Paden, Duncan MacCannell |
| EPI_ISL_1798904, EPI_ISL_1798906 | Guam Public Health Laboratory | Centers for Disease Control and Prevention Division of Viral Diseases, Pathogen Discovery | Mili Sheth, Sarah Nobles, Jasmine Padilla, Mark Burroughs, Shoshona Le, Katie Dillon, Peter Cook, Clinton R. Paden, Dhwani Batra, Krista Queen, Kristen Knipe, Dakota Howard, Yvette Unoarumhi, Darlene Wagner, Matthew Schmerer, Ben L. Rambo-Martin, Kristine Lacek, Sam Shepard, Alison Laufer Halpin, Dave Wentworth, Vivien Dugan, Suxiang Tong, Justin Lee |

EPI_ISL_1798908, EPI_ISL_1798909, EPI_ISL_1798910, EPI_ISL_1798911, EPI_ISL_1798912, EPI_ISL_1798913, EPI_ISL_1798914, EPI_ISL_1798915, EPI_ISL_1798916, EPI_ISL_1798917, EPI_ISL_1798918, EPI_ISL_1798919, EPI_ISL_1798920, EPI_ISL_1798921, EPI_ISL_1798922, EPI_ISL_1798923, EPI_ISL_1798924, EPI_ISL_1798925, EPI_ISL_1798926, EPI_ISL_1798928, EPI_ISL_1798929, EPI_ISL_1798930,
EPI_ISL_1798931, EPI_ISL_1798932, EPI_ISL_1798933, EPI_ISL_1798934, EPI_ISL_1798935, EPI_ISL_1798937, EPI_ISL_1798938, EPI_ISL_1798939, EPI_ISL_1798940, EPI_ISL_1798941, EPI_ISL_1798942, EPI_ISL_1798943, EPI_ISL_1798944, EPI_ISL_1798945, EPI_ISL_1798946, EPI_ISL_1798947, EPI_ISL_1798948, EPI_ISL_1798950, EPI_ISL_1798952, EPI_ISL_1798953, EPI_ISL_1798954, EPI_ISL_1798955,
EPI_ISL_1798956, EPI_ISL_1798957, EPI_ISL_1798958, EPI_ISL_1798959, EPI_ISL_1798961, EPI_ISL_1798962, EPI_ISL_1798963, EPI_ISL_1798964, EPI_ISL_1798965, EPI_ISL_1798966, EPI_ISL_1798967, EPI_ISL_1798969, EPI_ISL_1798970, EPI_ISL_1798971, EPI_ISL_1798972, EPI_ISL_1798973, EPI_ISL_1798974, EPI_ISL_1798976, EPI_ISL_1798978, EPI_ISL_1798980, EPI_ISL_1798982, EPI_ISL_1798983,
EPI_ISL_1798984, EPI_ISL_1798985, EPI_ISL_1798987, EPI_ISL_1798988, EPI_ISL_1798989, EPI_ISL_1798991, EPI_ISL_1798992, EPI_ISL_1798993, EPI_ISL_1798994, EPI_ISL_1798995, EPI_ISL_1798996, EPI_ISL_1798997, EPI_ISL_1799000, EPI_ISL_1799001, EPI_ISL_1799002, EPI_ISL_1799003, EPI_ISL_1799004, EPI_ISL_1799005, EPI_ISL_1799006, EPI_ISL_1799007, EPI_ISL_1799008, EPI_ISL_1799009,
EPI_ISL_1799010, EPI_ISL_1799011, EPI_ISL_1799015, EPI_ISL_1799017, EPI_ISL_1799018, EPI_ISL_1799019, EPI_ISL_1799020, EPI_ISL_1799021, EPI_ISL_1799025, EPI_ISL_1799027, EPI_ISL_1799028, EPI_ISL_1799029, EPI_ISL_1799030, EPI_ISL_1799031, EPI_ISL_1799032, EPI_ISL_1799034, EPI_ISL_1799036, EPI_ISL_1799037, EPI_ISL_1799039, EPI_ISL_1799040, EPI_ISL_1799041, EPI_ISL_1799042,
EPI_ISL_1799043, EPI_ISL_1799044, EPI_ISL_1799045, EPI_ISL_1799046, EPI_ISL_1799047, EPI_ISL_1799048, EPI_ISL_1799049, EPI_ISL_1799050, EPI_ISL_1799051, EPI_ISL_1799052, EPI_ISL_1799053, EPI_ISL_1799054, EPI_ISL_1799055, EPI_ISL_1799056, EPI_ISL_1799057, EPI_ISL_1799058, EPI_ISL_1799059, EPI_ISL_1799060, EPI_ISL_1799061, EPI_ISL_1799062, EPI_ISL_1799063, EPI_ISL_1799064,
EPI_ISL_1799066, EPI_ISL_1799067, EPI_ISL_1799068, EPI_ISL_1799069, EPI_ISL_1799070, EPI_ISL_1799072, EPI_ISL_1799073, EPI_ISL_1799076, EPI_ISL_1799077, EPI_ISL_1799080, EPI_ISL_1799081, EPI_ISL_1799082, EPI_ISL_1799084, EPI_ISL_1799085, EPI_ISL_1799088, EPI_ISL_1799090, EPI_ISL_1799091, EPI_ISL_1799092, EPI_ISL_1799093, EPI_ISL_1799094, EPI_ISL_1799095, EPI_ISL_1799097,
EPI_ISL_1799098, EPI_ISL_1799099, EPI_ISL_1799100, EPI_ISL_1799101, EPI_ISL_1799102, EPI_ISL_1799103, EPI_ISL_1799104, EPI_ISL_1799106, EPI_ISL_1799107, EPI_ISL_1799108, EPI_ISL_1799110, EPI_ISL_1799111, EPI_ISL_1799112, EPI_ISL_1799114, EPI_ISL_1799115, EPI_ISL_1799116, EPI_ISL_1799117, EPI_ISL_1799118, EPI_ISL_1799119, EPI_ISL_1799120, EPI_ISL_1799121, EPI_ISL_1799122,
EPI_ISL_1799123, EPI_ISL_1799125, EPI_ISL_1799126, EPI_ISL_1799127, EPI_ISL_1799128, EPI_ISL_1799129, EPI_ISL_1799130, EPI_ISL_1799131, EPI_ISL_1799132, EPI_ISL_1799134, EPI_ISL_1799135, EPI_ISL_1799136, EPI_ISL_1799137, EPI_ISL_1799138, EPI_ISL_1799141, EPI_ISL_1799143, EPI_ISL_1799144, EPI_ISL_1799145, EPI_ISL_1799146, EPI_ISL_1799147, EPI_ISL_1799149, EPI_ISL_1799150,
EPI_ISL_1799151, EPI_ISL_1799152, EPI_ISL_1799154, EPI_ISL_1799155, EPI_ISL_1799156, EPI_ISL_1799158, EPI_ISL_1799160, EPI_ISL_1799162, EPI_ISL_1799163, EPI_ISL_1799164, EPI_ISL_1799165, EPI_ISL_1799166, EPI_ISL_1799167, EPI_ISL_1799168, EPI_ISL_1799170, EPI_ISL_1799171, EPI_ISL_1799172, EPI_ISL_1799173, EPI_ISL_1799174, EPI_ISL_1799176, EPI_ISL_1799177, EPI_ISL_1799179,
EPI_ISL_1799180, EPI_ISL_1799181, EPI_ISL_1799182, EPI_ISL_1799183, EPI_ISL_1799184, EPI_ISL_1799185, EPI_ISL_1799186, EPI_ISL_1799187, EPI_ISL_1799189, EPI_ISL_1799190, EPI_ISL_1799192, EPI_ISL_1799193, EPI_ISL_1799195, EPI_ISL_1799196, EPI_ISL_1799197, EPI_ISL_1799198, EPI_ISL_1799199, EPI_ISL_1799200, EPI_ISL_1799201, EPI_ISL_1799202, EPI_ISL_1799203, EPI_ISL_1799204,
EPI_ISL_1799206, EPI_ISL_1799208, EPI_ISL_1799209, EPI_ISL_1799210, EPI_ISL_1799211, EPI_ISL_1799212, EPI_ISL_1799213, EPI_ISL_1799214, EPI_ISL_1799216, EPI_ISL_1799217, EPI_ISL_1799218, EPI_ISL_1799219, EPI_ISL_1799220, EPI_ISL_1799221, EPI_ISL_1799222, EPI_ISL_1799223, EPI_ISL_1799224, EPI_ISL_1799225, EPI_ISL_1799228, EPI_ISL_1799229, EPI_ISL_1799230, EPI_ISL_1799231,
EPI_ISL_1799233, EPI_ISL_1799234, EPI_ISL_1799235, EPI_ISL_1799236, EPI_ISL_1799237, EPI_ISL_1799238, EPI_ISL_1799239, EPI_ISL_1799240, EPI_ISL_1799242, EPI_ISL_1799243, EPI_ISL_1799244, EPI_ISL_1799245, EPI_ISL_1799246, EPI_ISL_1799247, EPI_ISL_1799248, EPI_ISL_1799249, EPI_ISL_1799251, EPI_ISL_1799252, EPI_ISL_1799253, EPI_ISL_1799254, EPI_ISL_1799256, EPI_ISL_1799257,
EPI_ISL_1799259, EPI_ISL_1799260, EPI_ISL_1799262, EPI_ISL_1799263, EPI_ISL_1799264, EPI_ISL_1799265, EPI_ISL_1799267, EPI_ISL_1799268, EPI_ISL_1799270, EPI_ISL_1799273, EPI_ISL_1799274, EPI_ISL_1799275, EPI_ISL_1799276, EPI_ISL_1799277, EPI_ISL_1799278, EPI_ISL_1799280, EPI_ISL_1799281, EPI_ISL_1799283, EPI_ISL_1799286, EPI_ISL_1799290, EPI_ISL_1799291, EPI_ISL_1799292,
EPI_ISL_1799294, EPI_ISL_1799296, EPI_ISL_1799297, EPI_ISL_1799298, EPI_ISL_1799299, EPI_ISL_1799300, EPI_ISL_1799301, EPI_ISL_1799302, EPI_ISL_1799303, EPI_ISL_1799304, EPI_ISL_1799305, EPI_ISL_1799307, EPI_ISL_1799309, EPI_ISL_1799310, EPI_ISL_1799311, EPI_ISL_1799312, EPI_ISL_1799313, EPI_ISL_1799314, EPI_ISL_1799316, EPI_ISL_1799317, EPI_ISL_1799318, EPI_ISL_1799319,
EPI_ISL_1799320, EPI_ISL_1799321, EPI_ISL_1799322, EPI_ISL_1799323, EPI_ISL_1799324, EPI_ISL_1799325, EPI_ISL_1799326, EPI_ISL_1799327, EPI_ISL_1799329, EPI_ISL_1799330, EPI_ISL_1799332, EPI_ISL_1799333, EPI_ISL_1799334, EPI_ISL_1799335, EPI_ISL_1799336, EPI_ISL_1799337, EPI_ISL_1799338, EPI_ISL_1799339, EPI_ISL_1799340, EPI_ISL_1799341, EPI_ISL_1799342, EPI_ISL_1799343,
EPI_ISL_1799344, EPI_ISL_1799345, EPI_ISL_1799347, EPI_ISL_1799348, EPI_ISL_1799349, EPI_ISL_1799350, EPI_ISL_1799351, EPI_ISL_1799352, EPI_ISL_1799353, EPI_ISL_1799354, EPI_ISL_1799355, EPI_ISL_1799356, EPI_ISL_1799357, EPI_ISL_1799359, EPI_ISL_1799360, EPI_ISL_1799361, EPI_ISL_1799362, EPI_ISL_1799363, EPI_ISL_1799364, EPI_ISL_1799366, EPI_ISL_1799367, EPI_ISL_1799368,
EPI_ISL_1799369, EPI_ISL_1799372, EPI_ISL_1799373, EPI_ISL_1799375, EPI_ISL_1799376, EPI_ISL_1799377, EPI_ISL_1799378, EPI_ISL_1799379, EPI_ISL_1799380, EPI_ISL_1799381, EPI_ISL_1799382, EPI_ISL_1799383, EPI_ISL_1799384, EPI_ISL_1799385, EPI_ISL_1799386, EPI_ISL_1799388, EPI_ISL_1799389, EPI_ISL_1799392, EPI_ISL_1799396, EPI_ISL_1799397, EPI_ISL_1799398, EPI_ISL_1799399,
EPI_ISL_1799401, EPI_ISL_1799402, EPI_ISL_1799403, EPI_ISL_1799405, EPI_ISL_1799406, EPI_ISL_1799407, EPI_ISL_1799408, EPI_ISL_1799409, EPI_ISL_1799410, EPI_ISL_1799411, EPI_ISL_1799412, EPI_ISL_1799414, EPI_ISL_1799415, EPI_ISL_1799416, EPI_ISL_1799417, EPI_ISL_1799418, EPI_ISL_1799419, EPI_ISL_1799421, EPI_ISL_1799422, EPI_ISL_1799423, EPI_ISL_1799424, EPI_ISL_1799425,
EPI_ISL_1799429, EPI_ISL_1799430, EPI_ISL_1799431, EPI_ISL_1799432, EPI_ISL_1799434, EPI_ISL_1799435, EPI_ISL_1799436, EPI_ISL_1799437, EPI_ISL_1799440, EPI_ISL_1799441, EPI_ISL_1799446, EPI_ISL_1799447, EPI_ISL_1799448, EPI_ISL_1799450, EPI_ISL_1799451, EPI_ISL_1799453, EPI_ISL_1799454, EPI_ISL_1799455, EPI_ISL_1799456, EPI_ISL_1799457, EPI_ISL_1799458, EPI_ISL_1799460,
EPI_ISL_1799461, EPI_ISL_1799462, EPI_ISL_1799463, EPI_ISL_1799464, EPI_ISL_1799465, EPI_ISL_1799466, EPI_ISL_1799467, EPI_ISL_1799470, EPI_ISL_1799471, EPI_ISL_1799472, EPI_ISL_1799473, EPI_ISL_1799474, EPI_ISL_1799475, EPI_ISL_1799476, EPI_ISL_1799478, EPI_ISL_1799480, EPI_ISL_1799482, EPI_ISL_1799485, EPI_ISL_1799486, EPI_ISL_1799488, EPI_ISL_1799490, EPI_ISL_1799491,
EPI_ISL_1799493, EPI_ISL_1799495, EPI_ISL_1799496, EPI_ISL_1799497, EPI_ISL_1799500, EPI_ISL_1799503, EPI_ISL_1799506, EPI_ISL_1799510, EPI_ISL_1799513, EPI_ISL_1799514, EPI_ISL_1799515, EPI_ISL_1799516, EPI_ISL_1799517, EPI_ISL_1799519, EPI_ISL_1799520, EPI_ISL_1799521, EPI_ISL_1799522, EPI_ISL_1799523, EPI_ISL_1799524, EPI_ISL_1799526, EPI_ISL_1799527, EPI_ISL_1799528,
EPI_ISL_1799529, EPI_ISL_1799530, EPI_ISL_1799531, EPI_ISL_1799532, EPI_ISL_1799533, EPI_ISL_1799534, EPI_ISL_1799535, EPI_ISL_1799538, EPI_ISL_1799539, EPI_ISL_1799540, EPI_ISL_1799542, EPI_ISL_1799543, EPI_ISL_1799544, EPI_ISL_1799545, EPI_ISL_1799546, EPI_ISL_1799549, EPI_ISL_1799550, EPI_ISL_1799551, EPI_ISL_1799553, EPI_ISL_1799554, EPI_ISL_1799555, EPI_ISL_1799557,
EPI_ISL_1799558, EPI_ISL_1799559, EPI_ISL_1799560, EPI_ISL_1799561, EPI_ISL_1799562, EPI_ISL_1799563, EPI_ISL_1799564, EPI_ISL_1799566, EPI_ISL_1799567, EPI_ISL_1799569, EPI_ISL_1799572, EPI_ISL_1799573, EPI_ISL_1799575, EPI_ISL_1799577, EPI_ISL_1799578, EPI_ISL_1799579, EPI_ISL_1799580, EPI_ISL_1799581, EPI_ISL_1799582, EPI_ISL_1799583, EPI_ISL_1799585, EPI_ISL_1799587,
EPI_ISL_1799588, EPI_ISL_1799589, EPI_ISL_1799593, EPI_ISL_1799594, EPI_ISL_1799595, EPI_ISL_1799596, EPI_ISL_1799597, EPI_ISL_1799598, EPI_ISL_1799599, EPI_ISL_1799600, EPI_ISL_1799601, EPI_ISL_1799602, EPI_ISL_1799603, EPI_ISL_1799604, EPI_ISL_1799607, EPI_ISL_1799611, EPI_ISL_1799612, EPI_ISL_1799613, EPI_ISL_1799614, EPI_ISL_1799615, EPI_ISL_1799617, EPI_ISL_1799618,
EPI_ISL_1799619, EPI_ISL_1799620, EPI_ISL_1799621, EPI_ISL_1799622, EPI_ISL_1799623, EPI_ISL_1799624, EPI_ISL_1799625, EPI_ISL_1799626, EPI_ISL_1799627, EPI_ISL_1799628, EPI_ISL_1799629, EPI_ISL_1799630, EPI_ISL_1799631, EPI_ISL_1799632, EPI_ISL_1799633, EPI_ISL_1799634, EPI_ISL_1799635, EPI_ISL_1799636, EPI_ISL_1799637, EPI_ISL_1799638, EPI_ISL_1799640, EPI_ISL_1799641,
EPI_ISL_1799642, EPI_ISL_1799643, EPI_ISL_1799645, EPI_ISL_1799646, EPI_ISL_1799647, EPI_ISL_1799648, EPI_ISL_1799651, EPI_ISL_1799652, EPI_ISL_1799654, EPI_ISL_1799655, EPI_ISL_1799657, EPI_ISL_1799658, EPI_ISL_1799659, EPI_ISL_1799660, EPI_ISL_1799661, EPI_ISL_1799662, EPI_ISL_1799663, EPI_ISL_1799664, EPI_ISL_1799665, EPI_ISL_1799667, EPI_ISL_1799668, EPI_ISL_1799669,
EPI_ISL_1799670, EPI_ISL_1799671, EPI_ISL_1799673, EPI_ISL_1799674, EPI_ISL_1799675, EPI_ISL_1799676, EPI_ISL_1799677, EPI_ISL_1799678, EPI_ISL_1799679, EPI_ISL_1799680, EPI_ISL_1799681, EPI_ISL_1799682, EPI_ISL_1799683, EPI_ISL_1799685, EPI_ISL_1799686, EPI_ISL_1799687, EPI_ISL_1799688, EPI_ISL_1799689, EPI_ISL_1799690, EPI_ISL_1799691, EPI_ISL_1799694, EPI_ISL_1799695,
EPI_ISL_1799696, EPI_ISL_1799697, EPI_ISL_1799698, EPI_ISL_1799699, EPI_ISL_1799700, EPI_ISL_1799701, EPI_ISL_1799703, EPI_ISL_1799705, EPI_ISL_1799706, EPI_ISL_1799707, EPI_ISL_1799708, EPI_ISL_1799709, EPI_ISL_1799711, EPI_ISL_1799712, EPI_ISL_1799713, EPI_ISL_1799714, EPI_ISL_1799715, EPI_ISL_1799716, EPI_ISL_1799718, EPI_ISL_1799719, EPI_ISL_1799720, EPI_ISL_1799721,
EPI_ISL_1799722, EPI_ISL_1799724, EPI_ISL_1799726, EPI_ISL_1799727, EPI_ISL_1799730, EPI_ISL_1799731, EPI_ISL_1799732, EPI_ISL_1799733, EPI_ISL_1799734, EPI_ISL_1799736, EPI_ISL_1799737, EPI_ISL_1799738, EPI_ISL_1799739, EPI_ISL_1799740, EPI_ISL_1799741, EPI_ISL_1799742, EPI_ISL_1799743, EPI_ISL_1799744, EPI_ISL_1799747, EPI_ISL_1799748, EPI_ISL_1799750, EPI_ISL_1799751,
EPI_ISL_1799753, EPI_ISL_1799755, EPI_ISL_1799756, EPI_ISL_1799757, EPI_ISL_1799758, EPI_ISL_1799760, EPI_ISL_1799761, EPI_ISL_1799762, EPI_ISL_1799763, EPI_ISL_1799765, EPI_ISL_1799767, EPI_ISL_1799769, EPI_ISL_1799770, EPI_ISL_1799772, EPI_ISL_1799774, EPI_ISL_1799775, EPI_ISL_1799776, EPI_ISL_1799779, EPI_ISL_1799780, EPI_ISL_1799781, EPI_ISL_1799783, EPI_ISL_1799784,
EPI_ISL_1799785, EPI_ISL_1799786, EPI_ISL_1799787, EPI_ISL_1799790, EPI_ISL_1799792, EPI_ISL_1799793, EPI_ISL_1799794, EPI_ISL_1799795, EPI_ISL_1799796, EPI_ISL_1799797, EPI_ISL_1799798, EPI_ISL_1799799, EPI_ISL_1799800, EPI_ISL_1799801, EPI_ISL_1799802, EPI_ISL_1799803, EPI_ISL_1799804, EPI_ISL_1799805, EPI_ISL_1799806, EPI_ISL_1799810, EPI_ISL_1799811, EPI_ISL_1799812,
EPI_ISL_1799814, EPI_ISL_1799817, EPI_ISL_1799818, EPI_ISL_1799819, EPI_ISL_1799820, EPI_ISL_1799822, EPI_ISL_1799823, EPI_ISL_1799824, EPI_ISL_1799826, EPI_ISL_1799828, EPI_ISL_1799829, EPI_ISL_1799830, EPI_ISL_1799831, EPI_ISL_1799832, EPI_ISL_1799833, EPI_ISL_1799834, EPI_ISL_1799836, EPI_ISL_1799837, EPI_ISL_1799838, EPI_ISL_1799843, EPI_ISL_1799844, EPI_ISL_1799845,
EPI_ISL_1799846, EPI_ISL_1799848, EPI_ISL_1799849, EPI_ISL_1799850, EPI_ISL_1799851, EPI_ISL_1799852, EPI_ISL_1799853, EPI_ISL_1799854, EPI_ISL_1799855, EPI_ISL_1799857, EPI_ISL_1799858, EPI_ISL_1799860, EPI_ISL_1799861, EPI_ISL_1799862, EPI_ISL_1799863, EPI_ISL_1799864, EPI_ISL_1799866, EPI_ISL_1799868, EPI_ISL_1799869, EPI_ISL_1799870, EPI_ISL_1799871, EPI_ISL_1799872,
EPI_ISL_1799873, EPI_ISL_1799875, EPI_ISL_1799876, EPI_ISL_1799877, EPI_ISL_1799878, EPI_ISL_1799879, EPI_ISL_1799880, EPI_ISL_1799881, EPI_ISL_1799882, EPI_ISL_1799883, EPI_ISL_1799884, EPI_ISL_1799885, EPI_ISL_1799886, EPI_ISL_1799888, EPI_ISL_1799889, EPI_ISL_1799890, EPI_ISL_1799891, EPI_ISL_1799892, EPI_ISL_1799893, EPI_ISL_1799894, EPI_ISL_1799895, EPI_ISL_1799896,
EPI_ISL_1799898, EPI_ISL_1799900, EPI_ISL_1799901, EPI_ISL_1799903, EPI_ISL_1799905, EPI_ISL_1799906, EPI_ISL_1799907, EPI_ISL_1799909, EPI_ISL_1799910, EPI_ISL_1799911, EPI_ISL_1799912, EPI_ISL_1799913, EPI_ISL_1799914, EPI_ISL_1799915, EPI_ISL_1799916, EPI_ISL_1799917, EPI_ISL_1799918, EPI_ISL_1799920, EPI_ISL_1799921, EPI_ISL_1799922, EPI_ISL_1799923, EPI_ISL_1799925,
EPI_ISL_1799926, EPI_ISL_1799927, EPI_ISL_1799928, EPI_ISL_1799929, EPI_ISL_1799930, EPI_ISL_1799931, EPI_ISL_1799932, EPI_ISL_1799933, EPI_ISL_1799934, EPI_ISL_1799935, EPI_ISL_1799936, EPI_ISL_1799937, EPI_ISL_1799938, EPI_ISL_1799940, EPI_ISL_1799941, EPI_ISL_1799942, EPI_ISL_1799943, EPI_ISL_1799944, EPI_ISL_1799945, EPI_ISL_1799946, EPI_ISL_1799947, EPI_ISL_1799948,
EPI_ISL_1799950, EPI_ISL_1799951, EPI_ISL_1799952, EPI_ISL_1799953, EPI_ISL_1799954, EPI_ISL_1799955, EPI_ISL_1799956, EPI_ISL_1799957, EPI_ISL_1799958, EPI_ISL_1799959, EPI_ISL_1799960, EPI_ISL_1799961, EPI_ISL_1799962, EPI_ISL_1799963, EPI_ISL_1799964, EPI_ISL_1799966, EPI_ISL_1799967, EPI_ISL_1799968, EPI_ISL_1799969, EPI_ISL_1799971, EPI_ISL_1799972, EPI_ISL_1799974,
EPI_ISL_1799978, EPI_ISL_1799979, EPI_ISL_1799982, EPI_ISL_1799984, EPI_ISL_1799985, EPI_ISL_1799986, EPI_ISL_1799988, EPI_ISL_1799989, EPI_ISL_1799991, EPI_ISL_1799992, EPI_ISL_1799993, EPI_ISL_1799994, EPI_ISL_1799995, EPI_ISL_1799996, EPI_ISL_1799997, EPI_ISL_1799998, EPI_ISL_1799999, EPI_ISL_1800000, EPI_ISL_1800001, EPI_ISL_1800002, EPI_ISL_1800003, EPI_ISL_1800004,
EPI_ISL_1800005, EPI_ISL_1800006, EPI_ISL_1800007, EPI_ISL_1800008, EPI_ISL_1800009, EPI_ISL_1800012, EPI_ISL_1800013, EPI_ISL_1800015, EPI_ISL_1800016, EPI_ISL_1800018, EPI_ISL_1800019, EPI_ISL_1800020, EPI_ISL_1800022, EPI_ISL_1800023, EPI_ISL_1800024, EPI_ISL_1800025, EPI_ISL_1800026, EPI_ISL_1800028, EPI_ISL_1800031, EPI_ISL_1800032, EPI_ISL_1800033, EPI_ISL_1800034,
EPI_ISL_1800035, EPI_ISL_1800037, EPI_ISL_1800039, EPI_ISL_1800040, EPI_ISL_1800041, EPI_ISL_1800044, EPI_ISL_1800045, EPI_ISL_1800046, EPI_ISL_1800047, EPI_ISL_1800050, EPI_ISL_1800051, EPI_ISL_1800052, EPI_ISL_1800053, EPI_ISL_1800054, EPI_ISL_1800055, EPI_ISL_1800056, EPI_ISL_1800057, EPI_ISL_1800058, EPI_ISL_1800059, EPI_ISL_1800062, EPI_ISL_1800063, EPI_ISL_1800064,
EPI_ISL_1800066, EPI_ISL_1800068, EPI_ISL_1800069, EPI_ISL_1800070, EPI_ISL_1800072, EPI_ISL_1800073, EPI_ISL_1800074, EPI_ISL_1800075, EPI_ISL_1800076, EPI_ISL_1800077, EPI_ISL_1800078, EPI_ISL_1800079, EPI_ISL_1800081, EPI_ISL_1800082, EPI_ISL_1800084, EPI_ISL_1800085, EPI_ISL_1800086, EPI_ISL_1800087, EPI_ISL_1800089, EPI_ISL_1800090, EPI_ISL_1800091, EPI_ISL_1800092,
EPI_ISL_1800093, EPI_ISL_1800094, EPI_ISL_1800095, EPI_ISL_1800096, EPI_ISL_1800099, EPI_ISL_1800100, EPI_ISL_1800101, EPI_ISL_1800102, EPI_ISL_1800103, EPI_ISL_1800104, EPI_ISL_1800105, EPI_ISL_1800106, EPI_ISL_1800107, EPI_ISL_1800108, EPI_ISL_1800109, EPI_ISL_1800110, EPI_ISL_1800111, EPI_ISL_1800112, EPI_ISL_1800113, EPI_ISL_1800114, EPI_ISL_1800115, EPI_ISL_1800116,
EPI_ISL_1800118, EPI_ISL_1800119, EPI_ISL_1800120, EPI_ISL_1800121, EPI_ISL_1800122, EPI_ISL_1800123, EPI_ISL_1800124, EPI_ISL_1800125, EPI_ISL_1800126, EPI_ISL_1800127, EPI_ISL_1800128, EPI_ISL_1800129, EPI_ISL_1800131, EPI_ISL_1800132, EPI_ISL_1800133, EPI_ISL_1800134, EPI_ISL_1800135, EPI_ISL_1800136, EPI_ISL_1800137, EPI_ISL_1800138, EPI_ISL_1800139, EPI_ISL_1800140,
EPI_ISL_1800141, EPI_ISL_1800142, EPI_ISL_1800143, EPI_ISL_1800145, EPI_ISL_1800147, EPI_ISL_1800149, EPI_ISL_1800150, EPI_ISL_1800151, EPI_ISL_1800152, EPI_ISL_1800153, EPI_ISL_1800154, EPI_ISL_1800155, EPI_ISL_1800156, EPI_ISL_1800158, EPI_ISL_1800159, EPI_ISL_1800160, EPI_ISL_1800161, EPI_ISL_1800162, EPI_ISL_1800163, EPI_ISL_1800164, EPI_ISL_1800165, EPI_ISL_1800167,
EPI_ISL_1800169, EPI_ISL_1800170, EPI_ISL_1800171, EPI_ISL_1800172, EPI_ISL_1800173, EPI_ISL_1800174, EPI_ISL_1800175, EPI_ISL_1800176, EPI_ISL_1800177, EPI_ISL_1800178, EPI_ISL_1800179, EPI_ISL_1800180, EPI_ISL_1800181, EPI_ISL_1800182, EPI_ISL_1800185, EPI_ISL_1800186, EPI_ISL_1800187, EPI_ISL_1800188, EPI_ISL_1800189, EPI_ISL_1800190, EPI_ISL_1800191, EPI_ISL_1800192,
EPI_ISL_1800195, EPI_ISL_1800198, EPI_ISL_1800199, EPI_ISL_1800200, EPI_ISL_1800201, EPI_ISL_1800202, EPI_ISL_1800203, EPI_ISL_1800204, EPI_ISL_1800206, EPI_ISL_1800207, EPI_ISL_1800209, EPI_ISL_1800211, EPI_ISL_1800212, EPI_ISL_1800213, EPI_ISL_1800216, EPI_ISL_1800217, EPI_ISL_1800218, EPI_ISL_1800219, EPI_ISL_1800220, EPI_ISL_1800221, EPI_ISL_1800222, EPI_ISL_1800224,
EPI_ISL_1800225, EPI_ISL_1800226, EPI_ISL_1800227, EPI_ISL_1800228, EPI_ISL_1800229, EPI_ISL_1800230, EPI_ISL_1800231, EPI_ISL_1800232, EPI_ISL_1800233, EPI_ISL_1800234, EPI_ISL_1800235, EPI_ISL_1800236, EPI_ISL_1800237, EPI_ISL_1800238, EPI_ISL_1800239, EPI_ISL_1800240, EPI_ISL_1800243, EPI_ISL_1800245, EPI_ISL_1800246, EPI_ISL_1800247, EPI_ISL_1800248, EPI_ISL_1800249,
EPI_ISL_1800250, EPI_ISL_1800251, EPI_ISL_1800252, EPI_ISL_1800254, EPI_ISL_1800255, EPI_ISL_1800256, EPI_ISL_1800257, EPI_ISL_1800258, EPI_ISL_1800259, EPI_ISL_1800260, EPI_ISL_1800261, EPI_ISL_1800263, EPI_ISL_1800264, EPI_ISL_1800267, EPI_ISL_1800269, EPI_ISL_1800270, EPI_ISL_1800271, EPI_ISL_1800272, EPI_ISL_1800273, EPI_ISL_1800274, EPI_ISL_1800277, EPI_ISL_1800278,
EPI_ISL_1800280, EPI_ISL_1800281, EPI_ISL_1800283, EPI_ISL_1800285, EPI_ISL_1800286, EPI_ISL_1800287, EPI_ISL_1800288, EPI_ISL_1800289, EPI_ISL_1800290, EPI_ISL_1800291, EPI_ISL_1800292, EPI_ISL_1800293, EPI_ISL_1800294, EPI_ISL_1800295, EPI_ISL_1800296, EPI_ISL_1800297, EPI_ISL_1800298, EPI_ISL_1800299, EPI_ISL_1800300, EPI_ISL_1800301, EPI_ISL_1800303, EPI_ISL_1800304,
EPI_ISL_1800305, EPI_ISL_1800306, EPI_ISL_1800307, EPI_ISL_1800308, EPI_ISL_1800309, EPI_ISL_1800310, EPI_ISL_1800311, EPI_ISL_1800312, EPI_ISL_1800316, EPI_ISL_1800317, EPI_ISL_1800319, EPI_ISL_1800320, EPI_ISL_1800321, EPI_ISL_1800322, EPI_ISL_1800324, EPI_ISL_1800325, EPI_ISL_1800326, EPI_ISL_1800327, EPI_ISL_1800328, EPI_ISL_1800329, EPI_ISL_1800330, EPI_ISL_1800331,
EPI_ISL_1800332, EPI_ISL_1800334, EPI_ISL_1800335, EPI_ISL_1800336, EPI_ISL_1800337, EPI_ISL_1800338, EPI_ISL_1800339, EPI_ISL_1800340, EPI_ISL_1800341, EPI_ISL_1800342, EPI_ISL_1800343, EPI_ISL_1800344, EPI_ISL_1800346, EPI_ISL_1800347, EPI_ISL_1800348, EPI_ISL_1800349, EPI_ISL_1800350, EPI_ISL_1800351, EPI_ISL_1800352, EPI_ISL_1800353, EPI_ISL_1800354, EPI_ISL_1800355,
EPI_ISL_1800356, EPI_ISL_1800357, EPI_ISL_1800358, EPI_ISL_1800359, EPI_ISL_1800360, EPI_ISL_1800361, EPI_ISL_1800362, EPI_ISL_1800363, EPI_ISL_1800364, EPI_ISL_1800365, EPI_ISL_1800366, EPI_ISL_1800367, EPI_ISL_1800368, EPI_ISL_1800369, EPI_ISL_1800370, EPI_ISL_1800371, EPI_ISL_1800372, EPI_ISL_1800373, EPI_ISL_1800374, EPI_ISL_1800375, EPI_ISL_1800376, EPI_ISL_1800378,
EPI_ISL_1800379, EPI_ISL_1800380, EPI_ISL_1800383, EPI_ISL_1800384, EPI_ISL_1800385, EPI_ISL_1800386, EPI_ISL_1800387, EPI_ISL_1800388, EPI_ISL_1800389, EPI_ISL_1800390, EPI_ISL_1800392, EPI_ISL_1800393, EPI_ISL_1800394, EPI_ISL_1800395, EPI_ISL_1800396, EPI_ISL_1800397, EPI_ISL_1800399, EPI_ISL_1800400, EPI_ISL_1800401, EPI_ISL_1800403, EPI_ISL_1800404, EPI_ISL_1800405,

EPI_ISL_1800407, EPI_ISL_1800408, EPI_ISL_1800409, EPI_ISL_1800410, EPI_ISL_1800411, EPI_ISL_1800412, EPI_ISL_1800413, EPI_ISL_1800414, EPI_ISL_1800415, EPI_ISL_1800416, EPI_ISL_1800417, EPI_ISL_1800419, EPI_ISL_1800420, EPI_ISL_1800421, EPI_ISL_1800422, EPI_ISL_1800423, EPI_ISL_1800424, EPI_ISL_1800425, EPI_ISL_1800426, EPI_ISL_1800427, EPI_ISL_1800428, EPI_ISL_1800429, EPI_ISL_1800431, EPI_ISL_1800432, EPI_ISL_1800433, EPI_ISL_1800434, EPI_ISL_1800438, EPI_ISL_1800439, EPI_ISL_1800440, EPI_ISL_1800441, EPI_ISL_1800442, EPI_ISL_1800443, EPI_ISL_1800445, EPI_ISL_1800447, EPI_ISL_1800448, EPI_ISL_1800449, EPI_ISL_1800450, EPI_ISL_1800451, EPI_ISL_1800452, EPI_ISL_1800453, EPI_ISL_1800454, EPI_ISL_1800455, EPI_ISL_1800457, EPI_ISL_1800458, EPI_ISL_1800459, EPI_ISL_1800461, EPI_ISL_1800462, EPI_ISL_1800463, EPI_ISL_1800464, EPI_ISL_1800465, EPI_ISL_1800466, EPI_ISL_1800467, EPI_ISL_1800468, EPI_ISL_1800469, EPI_ISL_1800470, EPI_ISL_1800471, EPI_ISL_1800472, EPI_ISL_1800473, EPI_ISL_1800474, EPI_ISL_1800475, EPI_ISL_1800476, EPI_ISL_1800477, EPI_ISL_1800478, EPI_ISL_1800479, EPI_ISL_1800480, EPI_ISL_1800483, EPI_ISL_1800484, EPI_ISL_1800485, EPI_ISL_1800486, EPI_ISL_1800487, EPI_ISL_1800488, EPI_ISL_1800489, EPI_ISL_1800490, EPI_ISL_1800491, EPI_ISL_1800492, EPI_ISL_1800493, EPI_ISL_1800494, EPI_ISL_1800496, EPI_ISL_1800497, EPI_ISL_1800498, EPI_ISL_1800500, EPI_ISL_1800501, EPI_ISL_1800502, EPI_ISL_1800503, EPI_ISL_1800504, EPI_ISL_1800505, EPI_ISL_1800506, EPI_ISL_1800507, EPI_ISL_1800508, EPI_ISL_1800509, EPI_ISL_1800510, EPI_ISL_1800511, EPI_ISL_1800512, EPI_ISL_1800513, EPI_ISL_1800514, EPI_ISL_1800515, EPI_ISL_1800516, EPI_ISL_1800517, EPI_ISL_1800519, EPI_ISL_1800520, EPI_ISL_1800521, EPI_ISL_1800522, EPI_ISL_1800523, EPI_ISL_1800524, EPI_ISL_1800525, EPI_ISL_1800527, EPI_ISL_1800528, EPI_ISL_1800530, EPI_ISL_1800531, EPI_ISL_1800532, EPI_ISL_1800533, EPI_ISL_1800534, EPI_ISL_1800536, EPI_ISL_1800537, EPI_ISL_1800538, EPI_ISL_1800539, EPI_ISL_1800540, EPI_ISL_1800541, EPI_ISL_1800542, EPI_ISL_1800543, EPI_ISL_1800544, EPI_ISL_1800545, EPI_ISL_1800547, EPI_ISL_1800548, EPI_ISL_1800549, EPI_ISL_1800550, EPI_ISL_1800552, EPI_ISL_1800555, EPI_ISL_1800556, EPI_ISL_1800557, EPI_ISL_1800559, EPI_ISL_1800560, EPI_ISL_1800562, EPI_ISL_1800563, EPI_ISL_1800564, EPI_ISL_1800565, EPI_ISL_1800568, EPI_ISL_1800569, EPI_ISL_1800571, EPI_ISL_1800572, EPI_ISL_1800573, EPI_ISL_1800574, EPI_ISL_1800575, EPI_ISL_1800576, EPI_ISL_1800577, EPI_ISL_1800578, EPI_ISL_1800579, EPI_ISL_1800584, EPI_ISL_1800585, EPI_ISL_1800586, EPI_ISL_1800587, EPI_ISL_1800588, EPI_ISL_1800589, EPI_ISL_1800590, EPI_ISL_1800591, EPI_ISL_1800592, EPI_ISL_1800593, EPI_ISL_1800594, EPI_ISL_1800596, EPI_ISL_1800597, EPI_ISL_1800598, EPI_ISL_1800599, EPI_ISL_1800600, EPI_ISL_1800601, EPI_ISL_1800602, EPI_ISL_1800604, EPI_ISL_1800606, EPI_ISL_1800607, EPI_ISL_1800608, EPI_ISL_1800609, EPI_ISL_1800611, EPI_ISL_1800612, EPI_ISL_1800613, EPI_ISL_1800614, EPI_ISL_1800615, EPI_ISL_1800616, EPI_ISL_1800617, EPI_ISL_1800618, EPI_ISL_1800619, EPI_ISL_1800620, EPI_ISL_1800621, EPI_ISL_1800622, EPI_ISL_1800623, EPI_ISL_1800624, EPI_ISL_1800625, EPI_ISL_1800626, EPI_ISL_1800627, EPI_ISL_1800628, EPI_ISL_1800629, EPI_ISL_1800630, EPI_ISL_1800631, EPI_ISL_1800632, EPI_ISL_1800633, EPI_ISL_1800634, EPI_ISL_1800635, EPI_ISL_1800636, EPI_ISL_1800637, EPI_ISL_1800638, EPI_ISL_1800639, EPI_ISL_1800641, EPI_ISL_1800643, EPI_ISL_1800645, EPI_ISL_1800646, EPI_ISL_1800647, EPI_ISL_1800648, EPI_ISL_1800650, EPI_ISL_1800651, EPI_ISL_1800652, EPI_ISL_1800653, EPI_ISL_1800654, EPI_ISL_1800655, EPI_ISL_1800656, EPI_ISL_1800657, EPI_ISL_1800658, EPI_ISL_1800660, EPI_ISL_1800661, EPI_ISL_1800662, EPI_ISL_1800663, EPI_ISL_1800664, EPI_ISL_1800665, EPI_ISL_1800666, EPI_ISL_1800667, EPI_ISL_1800668, EPI_ISL_1800670, EPI_ISL_1800671, EPI_ISL_1800673, EPI_ISL_1800675, EPI_ISL_1800676, EPI_ISL_1800680, EPI_ISL_1800681, EPI_ISL_1800682, EPI_ISL_1800683, EPI_ISL_1800684, EPI_ISL_1800685, EPI_ISL_1800687, EPI_ISL_1800688, EPI_ISL_1800689, EPI_ISL_1800690, EPI_ISL_1800691, EPI_ISL_1800693, EPI_ISL_1800694, EPI_ISL_1800695, EPI_ISL_1800697, EPI_ISL_1800698, EPI_ISL_1800699, EPI_ISL_1800700, EPI_ISL_1800701, EPI_ISL_1800702, EPI_ISL_1800703, EPI_ISL_1800704, EPI_ISL_1800706, EPI_ISL_1800710, EPI_ISL_1800712, EPI_ISL_1800713, EPI_ISL_1800714, EPI_ISL_1800717, EPI_ISL_1800718, EPI_ISL_1800719, EPI_ISL_1800720, EPI_ISL_1800721, EPI_ISL_1800723, EPI_ISL_1800724, EPI_ISL_1800725, EPI_ISL_1800727, EPI_ISL_1800728, EPI_ISL_1800729, EPI_ISL_1800730, EPI_ISL_1800731, EPI_ISL_1800733, EPI_ISL_1800735, EPI_ISL_1800736, EPI_ISL_1800737, EPI_ISL_1800738, EPI_ISL_1800740, EPI_ISL_1800741, EPI_ISL_1800742, EPI_ISL_1800744, EPI_ISL_1800745, EPI_ISL_1800746, EPI_ISL_1800747, EPI_ISL_1800748, EPI_ISL_1800749, EPI_ISL_1800750, EPI_ISL_1800752, EPI_ISL_1800753, EPI_ISL_1800754, EPI_ISL_1800756, EPI_ISL_1800757, EPI_ISL_1800758, EPI_ISL_1800759, EPI_ISL_1800761, EPI_ISL_1800762, EPI_ISL_1800763, EPI_ISL_1800764, EPI_ISL_1800765, EPI_ISL_1800766, EPI_ISL_1800769, EPI_ISL_1800771, EPI_ISL_1800772, EPI_ISL_1800773, EPI_ISL_1800775, EPI_ISL_1800776, EPI_ISL_1800777, EPI_ISL_1800778, EPI_ISL_1800779, EPI_ISL_1800780, EPI_ISL_1800781, EPI_ISL_1800782, EPI_ISL_1800783, EPI_ISL_1800784, EPI_ISL_1800786, EPI_ISL_1800789, EPI_ISL_1800790, EPI_ISL_1800791, EPI_ISL_1800792, EPI_ISL_1800793, EPI_ISL_1800794, EPI_ISL_1800795, EPI_ISL_1800796, EPI_ISL_1800797, EPI_ISL_1800798, EPI_ISL_1800799, EPI_ISL_1800800, EPI_ISL_1800802, EPI_ISL_1800803, EPI_ISL_1800805, EPI_ISL_1800806, EPI_ISL_1800808, EPI_ISL_1800810, EPI_ISL_1800811, EPI_ISL_1800812, EPI_ISL_1800813, EPI_ISL_1800814, EPI_ISL_1800819, EPI_ISL_1800820, EPI_ISL_1800821, EPI_ISL_1800822, EPI_ISL_1800823, EPI_ISL_1800824, EPI_ISL_1800825, EPI_ISL_1800826, EPI_ISL_1800827, EPI_ISL_1800828, EPI_ISL_1800829, EPI_ISL_1800831, EPI_ISL_1800832, EPI_ISL_1800833, EPI_ISL_1800834, EPI_ISL_1800835, EPI_ISL_1800836, EPI_ISL_1800837, EPI_ISL_1800839, EPI_ISL_1800840, EPI_ISL_1800841, EPI_ISL_1800842, EPI_ISL_1800843, EPI_ISL_1800844, EPI_ISL_1800845, EPI_ISL_1800846, EPI_ISL_1800847, EPI_ISL_1800849, EPI_ISL_1800850, EPI_ISL_1800851, EPI_ISL_1800852, EPI_ISL_1800853, EPI_ISL_1800854, EPI_ISL_1800855, EPI_ISL_1800856, EPI_ISL_1800858, EPI_ISL_1800860, EPI_ISL_1800861, EPI_ISL_1800862, EPI_ISL_1800863, EPI_ISL_1800865, EPI_ISL_1800866, EPI_ISL_1800867, EPI_ISL_1800868, EPI_ISL_1800869, EPI_ISL_1800871, EPI_ISL_1800873, EPI_ISL_1800875, EPI_ISL_1800876, EPI_ISL_1800877, EPI_ISL_1800879, EPI_ISL_1800880, EPI_ISL_1800881, EPI_ISL_1800886, EPI_ISL_1800887, EPI_ISL_1800889, EPI_ISL_1800890, EPI_ISL_1800892, EPI_ISL_1800893, EPI_ISL_1800894, EPI_ISL_1800895, EPI_ISL_1800896, EPI_ISL_1800897, EPI_ISL_1800898, EPI_ISL_1800899, EPI_ISL_1800902, EPI_ISL_1800903, EPI_ISL_1800905, EPI_ISL_1800906, EPI_ISL_1800908, EPI_ISL_1800913, EPI_ISL_1800914, EPI_ISL_1800916, EPI_ISL_1800917, EPI_ISL_1800918, EPI_ISL_1800919, EPI_ISL_1800920, EPI_ISL_1800921, EPI_ISL_1800922, EPI_ISL_1800923, EPI_ISL_1800924, EPI_ISL_1800925, EPI_ISL_1800926, EPI_ISL_1800927, EPI_ISL_1800928, EPI_ISL_1800930, EPI_ISL_1800931, EPI_ISL_1800932, EPI_ISL_1800936, EPI_ISL_1800937, EPI_ISL_1800938, EPI_ISL_1800940, EPI_ISL_1800942, EPI_ISL_1800943, EPI_ISL_1800944, EPI_ISL_1800945, EPI_ISL_1800946, EPI_ISL_1800947, EPI_ISL_1800948, EPI_ISL_1800949, EPI_ISL_1800950, EPI_ISL_1800951, EPI_ISL_1800952, EPI_ISL_1800953, EPI_ISL_1800954, EPI_ISL_1800955, EPI_ISL_1800956, EPI_ISL_1800957, EPI_ISL_1800958, EPI_ISL_1800959, EPI_ISL_1800960, EPI_ISL_1800961, EPI_ISL_1800963, EPI_ISL_1800964, EPI_ISL_1800966, EPI_ISL_1800967, EPI_ISL_1800968, EPI_ISL_1800969, EPI_ISL_1800971, EPI_ISL_1800972, EPI_ISL_1800973, EPI_ISL_1800974, EPI_ISL_1800975, EPI_ISL_1800977, EPI_ISL_1800978, EPI_ISL_1800979, EPI_ISL_1800980, EPI_ISL_1800981, EPI_ISL_1800982, EPI_ISL_1800983, EPI_ISL_1800984, EPI_ISL_1800985, EPI_ISL_1800986, EPI_ISL_1800987, EPI_ISL_1800988, EPI_ISL_1800989, EPI_ISL_1800990, EPI_ISL_1800991, EPI_ISL_1800994, EPI_ISL_1800995, EPI_ISL_1800996, EPI_ISL_1800997, EPI_ISL_1800998, EPI_ISL_1800999, EPI_ISL_1801002, EPI_ISL_1801003, EPI_ISL_1801004, EPI_ISL_1801005, EPI_ISL_1801006, EPI_ISL_1801007, EPI_ISL_1801009, EPI_ISL_1801010, EPI_ISL_1801011, EPI_ISL_1801012, EPI_ISL_1801014, EPI_ISL_1801016, EPI_ISL_1801017, EPI_ISL_1801018, EPI_ISL_1801019, EPI_ISL_1801020, EPI_ISL_1801021, EPI_ISL_1801022, EPI_ISL_1801023, EPI_ISL_1801024, EPI_ISL_1801025, EPI_ISL_1801026, EPI_ISL_1801027, EPI_ISL_1801028, EPI_ISL_1801029, EPI_ISL_1801030, EPI_ISL_1801031, EPI_ISL_1801032, EPI_ISL_1801034, EPI_ISL_1801035, EPI_ISL_1801036, EPI_ISL_1801040, EPI_ISL_1801041, EPI_ISL_1801042, EPI_ISL_1801043, EPI_ISL_1801044, EPI_ISL_1801045, EPI_ISL_1801046, EPI_ISL_1801047, EPI_ISL_1801048, EPI_ISL_1801049, EPI_ISL_1801050, EPI_ISL_1801051, EPI_ISL_1801056, EPI_ISL_1801057, EPI_ISL_1801059, EPI_ISL_1801060, EPI_ISL_1801061, EPI_ISL_1801062, EPI_ISL_1801064, EPI_ISL_1801065, EPI_ISL_1801066, EPI_ISL_1801067, EPI_ISL_1801071, EPI_ISL_1801074, EPI_ISL_1801075, EPI_ISL_1801076, EPI_ISL_1801077, EPI_ISL_1801078, EPI_ISL_1801079, EPI_ISL_1801081, EPI_ISL_1801082, EPI_ISL_1801083, EPI_ISL_1801084, EPI_ISL_1801085, EPI_ISL_1801086, EPI_ISL_1801087, EPI_ISL_1801088, EPI_ISL_1801089, EPI_ISL_1801090, EPI_ISL_1801091, EPI_ISL_1801092, EPI_ISL_1801095, EPI_ISL_1801096, EPI_ISL_1801097, EPI_ISL_1801098, EPI_ISL_1801099, EPI_ISL_1801100, EPI_ISL_1801101, EPI_ISL_1801102, EPI_ISL_1801104, EPI_ISL_1801105, EPI_ISL_1801106, EPI_ISL_1801107, EPI_ISL_1801108, EPI_ISL_1801109, EPI_ISL_1801110, EPI_ISL_1801111, EPI_ISL_1801112, EPI_ISL_1801113, EPI_ISL_1801114, EPI_ISL_1801115, EPI_ISL_1801117, EPI_ISL_1801118, EPI_ISL_1801121, EPI_ISL_1801122, EPI_ISL_1801123, EPI_ISL_1801124, EPI_ISL_1801126, EPI_ISL_1801127, EPI_ISL_1801128, EPI_ISL_1801129, EPI_ISL_1801130, EPI_ISL_1801131, EPI_ISL_1801132, EPI_ISL_1801133, EPI_ISL_1801135, EPI_ISL_1801136, EPI_ISL_1801137, EPI_ISL_1801138, EPI_ISL_1801139, EPI_ISL_1801140, EPI_ISL_1801141, EPI_ISL_1801142, EPI_ISL_1801143, EPI_ISL_1801144, EPI_ISL_1801145, EPI_ISL_1801146, EPI_ISL_1801147

see above | Laboratory Corporation of America | Centers for Disease Control and Prevention Division of Viral Diseases, Pathogen Discovery | Dakota Howard, Dhwani Batra, Peter W. Cook, Kara Moser, Adrian Paskey, Jason Caravas, Benjamin Rambo-Martin, Shatavia Morrison, Christopher Gulvick, Scott Sammons, Yvette Unoarumhi, Darlene Wagner, Matthew Schmerer, Minoo Agarwal, Eyad Almasri, Debbie Boles, Ayla Burns, Nuthawin Charoensri, Oren Cohen, Susan Countryman, Mary Ann Cristobal, Bobbi Croy, Suzanne Dale, Hrushikesh Deshmukh, Amanda Douglas, Vincent Drouillon, Marcia Eisenberg, Howard Engler, Rama Ghatti, Prashant Gupta, Susan Hicks, Jake Humphrey, Lax Iyer, Manoj Jain, Mohan Kolli, Brian Krueger, Tim Kuphal, Stanley Letovsky, Michael Levandoski, Craig Lukasik, Jonathan Meltzer, Brian Norvell, Mindy Nye, Scott Parker, Christos Petropoulos, John Pruitt, Steven Ragan, Scott Ryan, Mike Sapeta, Jana Schroth, Suresh Babu Selvaraju, Goran Stevovic, Amanda Suchanek, Andrea Throop, Lyndon Tilson, Thomas Urban, Joe Voshell, Kimberly Wagner, Jonathan Williams, Mary Williamson, Qian Zeng, Tricia Zwiefelhofer, Clinton R. Paden, Duncan MacCannell

EPI_ISL_1801149 | Ohio Department of Health Laboratory | Ohio Department of Health Laboratory | Holmes, Jennifer; Eric Brandt, Keoni Omura, Glen McGillivary, Caitlin McDonnell, Jade Mowery, Stephanie Mccracken, Tyler Payne, Kirtana Ramadugu, Erica Leasure, Brent Lee, Kelsey Florek, Heather Blankenship, Allison Black, Quanta Brown, and Tammy Bannerman

EPI_ISL_1801150, EPI_ISL_1801151, EPI_ISL_1801152, EPI_ISL_1801153, EPI_ISL_1801154, EPI_ISL_1801155, EPI_ISL_1801156, EPI_ISL_1801157, EPI_ISL_1801158, EPI_ISL_1801159, EPI_ISL_1801160, EPI_ISL_1801161, EPI_ISL_1801162, EPI_ISL_1801163, EPI_ISL_1801164, EPI_ISL_1801165, EPI_ISL_1801166, EPI_ISL_1801167, EPI_ISL_1801168, EPI_ISL_1801170, EPI_ISL_1801171, EPI_ISL_1801172, EPI_ISL_1801173, EPI_ISL_1801174, EPI_ISL_1801175, EPI_ISL_1801176, EPI_ISL_1801177, EPI_ISL_1801178, EPI_ISL_1801179, EPI_ISL_1801180, EPI_ISL_1801181, EPI_ISL_1801183, EPI_ISL_1801184, EPI_ISL_1801185, EPI_ISL_1801186, EPI_ISL_1801187, EPI_ISL_1801188, EPI_ISL_1801189, EPI_ISL_1801190, EPI_ISL_1801192, EPI_ISL_1801193, EPI_ISL_1801194, EPI_ISL_1801195, EPI_ISL_1801196, EPI_ISL_1801197, EPI_ISL_1801198, EPI_ISL_1801200, EPI_ISL_1801201, EPI_ISL_1801204, EPI_ISL_1801205, EPI_ISL_1801206, EPI_ISL_1801207, EPI_ISL_1801208, EPI_ISL_1801209, EPI_ISL_1801210, EPI_ISL_1801211, EPI_ISL_1801212, EPI_ISL_1801213, EPI_ISL_1801214, EPI_ISL_1801215, EPI_ISL_1801216, EPI_ISL_1801217, EPI_ISL_1801218, EPI_ISL_1801219, EPI_ISL_1801220, EPI_ISL_1801221, EPI_ISL_1801222, EPI_ISL_1801223, EPI_ISL_1801224, EPI_ISL_1801225, EPI_ISL_1801226, EPI_ISL_1801227, EPI_ISL_1801228, EPI_ISL_1801229, EPI_ISL_1801230, EPI_ISL_1801231, EPI_ISL_1801232, EPI_ISL_1801234, EPI_ISL_1801235, EPI_ISL_1801237, EPI_ISL_1801239, EPI_ISL_1801240, EPI_ISL_1801241, EPI_ISL_1801242, EPI_ISL_1801243, EPI_ISL_1801246, EPI_ISL_1801247, EPI_ISL_1801248, EPI_ISL_1801249, EPI_ISL_1801251, EPI_ISL_1801252, EPI_ISL_1801253, EPI_ISL_1801254, EPI_ISL_1801255, EPI_ISL_1801256, EPI_ISL_1801257, EPI_ISL_1801258, EPI_ISL_1801259, EPI_ISL_1801260, EPI_ISL_1801261, EPI_ISL_1801262, EPI_ISL_1801264, EPI_ISL_1801265, EPI_ISL_1801266, EPI_ISL_1801267, EPI_ISL_1801270, EPI_ISL_1801271, EPI_ISL_1801272, EPI_ISL_1801273, EPI_ISL_1801274, EPI_ISL_1801275, EPI_ISL_1801276, EPI_ISL_1801277, EPI_ISL_1801278, EPI_ISL_1801279, EPI_ISL_1801280, EPI_ISL_1801281, EPI_ISL_1801282, EPI_ISL_1801283, EPI_ISL_1801285, EPI_ISL_1801286, EPI_ISL_1801287, EPI_ISL_1801288, EPI_ISL_1801289, EPI_ISL_1801290, EPI_ISL_1801292, EPI_ISL_1801293, EPI_ISL_1801294, EPI_ISL_1801295, EPI_ISL_1801297, EPI_ISL_1801298, EPI_ISL_1801299, EPI_ISL_1801301, EPI_ISL_1801302, EPI_ISL_1801303, EPI_ISL_1801304, EPI_ISL_1801305, EPI_ISL_1801306, EPI_ISL_1801307, EPI_ISL_1801308, EPI_ISL_1801309, EPI_ISL_1801311, EPI_ISL_1801312, EPI_ISL_1801313, EPI_ISL_1801314, EPI_ISL_1801315, EPI_ISL_1801316, EPI_ISL_1801317, EPI_ISL_1801318, EPI_ISL_1801319, EPI_ISL_1801320, EPI_ISL_1801321, EPI_ISL_1801322, EPI_ISL_1801323, EPI_ISL_1801325, EPI_ISL_1801326, EPI_ISL_1801328, EPI_ISL_1801329, EPI_ISL_1801330, EPI_ISL_1801331, EPI_ISL_1801333, EPI_ISL_1801334, EPI_ISL_1801335, EPI_ISL_1801336, EPI_ISL_1801337, EPI_ISL_1801339, EPI_ISL_1801340, EPI_ISL_1801341, EPI_ISL_1801342, EPI_ISL_1801343, EPI_ISL_1801344, EPI_ISL_1801348, EPI_ISL_1801349, EPI_ISL_1801350, EPI_ISL_1801351, EPI_ISL_1801352, EPI_ISL_1801353, EPI_ISL_1801354, EPI_ISL_1801355, EPI_ISL_1801356, EPI_ISL_1801357, EPI_ISL_1801358, EPI_ISL_1801359, EPI_ISL_1801360, EPI_ISL_1801362, EPI_ISL_1801364, EPI_ISL_1801365, EPI_ISL_1801368, EPI_ISL_1801369, EPI_ISL_1801370, EPI_ISL_1801371, EPI_ISL_1801372, EPI_ISL_1801373, EPI_ISL_1801375, EPI_ISL_1801376, EPI_ISL_1801377, EPI_ISL_1801378, EPI_ISL_1801379, EPI_ISL_1801380, EPI_ISL_1801381, EPI_ISL_1801382, EPI_ISL_1801383, EPI_ISL_1801384, EPI_ISL_1801385, EPI_ISL_1801386, EPI_ISL_1801387, EPI_ISL_1801390, EPI_ISL_1801391, EPI_ISL_1801393, EPI_ISL_1801395, EPI_ISL_1801396, EPI_ISL_1801397, EPI_ISL_1801398, EPI_ISL_1801400, EPI_ISL_1801401, EPI_ISL_1801403, EPI_ISL_1801404, EPI_ISL_1801407, EPI_ISL_1801408, EPI_ISL_1801409, EPI_ISL_1801410, EPI_ISL_1801411, EPI_ISL_1801414, EPI_ISL_1801415, EPI_ISL_1801416, EPI_ISL_1801417, EPI_ISL_1801418, EPI_ISL_1801419, EPI_ISL_1801420, EPI_ISL_1801421, EPI_ISL_1801422, EPI_ISL_1801424, EPI_ISL_1801425

see above | Laboratory Corporation of America | Centers for Disease Control and Prevention Division of Viral Diseases, Pathogen Discovery | Dakota Howard, Dhwani Batra, Peter W. Cook, Kara Moser, Adrian Paskey, Jason Caravas, Benjamin Rambo-Martin, Shatavia Morrison, Christopher Gulvick, Scott Sammons, Yvette Unoarumhi, Darlene Wagner, Matthew Schmerer, Minoo Agarwal, Eyad Almasri, Debbie Boles, Ayla Burns, Nuthawin Charoensri, Oren Cohen, Susan Countryman, Mary Ann Cristobal, Bobbi Croy, Suzanne Dale, Hrushikesh Deshmukh, Amanda Douglas, Vincent Drouillon, Marcia Eisenberg, Howard Engler, Rama Ghatti, Prashant Gupta, Susan Hicks, Jake Humphrey, Lax Iyer, Manoj Jain, Mohan Kolli, Brian Krueger, Tim Kuphal, Stanley Letovsky, Michael Levandoski, Craig Lukasik, Jonathan Meltzer, Brian Norvell, Mindy Nye, Scott Parker, Christos Petropoulos, John Pruitt, Steven Ragan, Scott Ryan, Mike Sapeta, Jana Schroth, Suresh Babu Selvaraju, Goran Stevovic, Amanda Suchanek, Andrea Throop, Lyndon Tilson, Thomas Urban, Joe Voshell, Kimberly Wagner, Jonathan Williams, Mary Williamson, Qian Zeng, Tricia Zwiefelhofer, Clinton R. Paden, Duncan MacCannell

EPI_ISL_1801426 | Ohio Department of Health Laboratory | Ohio Department of Health Laboratory | Holmes, Jennifer; Eric Brandt, Keoni Omura, Glen McGillivary, Caitlin McDonnell, Jade Mowery, Stephanie Mccracken, Tyler Payne, Kirtana Ramadugu, Erica Leasure, Brent Lee, Kelsey Florek, Heather Blankenship, Allison Black, Quanta Brown, and Tammy Bannerman

EPI_ISL_1801427, EPI_ISL_1801428, EPI_ISL_1801429, EPI_ISL_1801430, EPI_ISL_1801431, EPI_ISL_1801432, EPI_ISL_1801433, EPI_ISL_1801434, EPI_ISL_1801435, EPI_ISL_1801437, EPI_ISL_1801438, EPI_ISL_1801439, EPI_ISL_1801440, EPI_ISL_1801441, EPI_ISL_1801442, EPI_ISL_1801443, EPI_ISL_1801444, EPI_ISL_1801450, EPI_ISL_1801451, EPI_ISL_1801453, EPI_ISL_1801454, EPI_ISL_1801455, EPI_ISL_1801456, EPI_ISL_1801457, EPI_ISL_1801458, EPI_ISL_1801459, EPI_ISL_1801460, EPI_ISL_1801461, EPI_ISL_1801462, EPI_ISL_1801463, EPI_ISL_1801464, EPI_ISL_1801465, EPI_ISL_1801466, EPI_ISL_1801467, EPI_ISL_1801468, EPI_ISL_1801469, EPI_ISL_1801470, EPI_ISL_1801471, EPI_ISL_1801472, EPI_ISL_1801473, EPI_ISL_1801474, EPI_ISL_1801475, EPI_ISL_1801476, EPI_ISL_1801477, EPI_ISL_1801478, EPI_ISL_1801480, EPI_ISL_1801481, EPI_ISL_1801483, EPI_ISL_1801484, EPI_ISL_1801486, EPI_ISL_1801487, EPI_ISL_1801489, EPI_ISL_1801490, EPI_ISL_1801491, EPI_ISL_1801493, EPI_ISL_1801494, EPI_ISL_1801495, EPI_ISL_1801496, EPI_ISL_1801497, EPI_ISL_1801498, EPI_ISL_1801499, EPI_ISL_1801500, EPI_ISL_1801501, EPI_ISL_1801502, EPI_ISL_1801503, EPI_ISL_1801504, EPI_ISL_1801505, EPI_ISL_1801506, EPI_ISL_1801507, EPI_ISL_1801508, EPI_ISL_1801509, EPI_ISL_1801510, EPI_ISL_1801511, EPI_ISL_1801512, EPI_ISL_1801513, EPI_ISL_1801514, EPI_ISL_1801517, EPI_ISL_1801518, EPI_ISL_1801519, EPI_ISL_1801520, EPI_ISL_1801521, EPI_ISL_1801522, EPI_ISL_1801523, EPI_ISL_1801524, EPI_ISL_1801526, EPI_ISL_1801527, EPI_ISL_1801528, EPI_ISL_1801529, EPI_ISL_1801532, EPI_ISL_1801533, EPI_ISL_1801534, EPI_ISL_1801535, EPI_ISL_1801536, EPI_ISL_1801537, EPI_ISL_1801538, EPI_ISL_1801539, EPI_ISL_1801540, EPI_ISL_1801542, EPI_ISL_1801543, EPI_ISL_1801544, EPI_ISL_1801545, EPI_ISL_1801546, EPI_ISL_1801547, EPI_ISL_1801549, EPI_ISL_1801550, EPI_ISL_1801551, EPI_ISL_1801552, EPI_ISL_1801553, EPI_ISL_1801555, EPI_ISL_1801556, EPI_ISL_1801557, EPI_ISL_1801559, EPI_ISL_1801560, EPI_ISL_1801561, EPI_ISL_1801562, EPI_ISL_1801563, EPI_ISL_1801565, EPI_ISL_1801566, EPI_ISL_1801567, EPI_ISL_1801568, EPI_ISL_1801569, EPI_ISL_1801570, EPI_ISL_1801572, EPI_ISL_1801573, EPI_ISL_1801574, EPI_ISL_1801575, EPI_ISL_1801576, EPI_ISL_1801577, EPI_ISL_1801578, EPI_ISL_1801579, EPI_ISL_1801580, EPI_ISL_1801581, EPI_ISL_1801582, EPI_ISL_1801583, EPI_ISL_1801585, EPI_ISL_1801586, EPI_ISL_1801587, EPI_ISL_1801588, EPI_ISL_1801589, EPI_ISL_1801590, EPI_ISL_1801591, EPI_ISL_1801592, EPI_ISL_1801593, EPI_ISL_1801594, EPI_ISL_1801595, EPI_ISL_1801596, EPI_ISL_1801599, EPI_ISL_1801601, EPI_ISL_1801602, EPI_ISL_1801603, EPI_ISL_1801604, EPI_ISL_1801605, EPI_ISL_1801606, EPI_ISL_1801607, EPI_ISL_1801609, EPI_ISL_1801610, EPI_ISL_1801612, EPI_ISL_1801613, EPI_ISL_1801614, EPI_ISL_1801615, EPI_ISL_1801616, EPI_ISL_1801617, EPI_ISL_1801619, EPI_ISL_1801620, EPI_ISL_1801621, EPI_ISL_1801622, EPI_ISL_1801624, EPI_ISL_1801626, EPI_ISL_1801627, EPI_ISL_1801628, EPI_ISL_1801629, EPI_ISL_1801630, EPI_ISL_1801631, EPI_ISL_1801632, EPI_ISL_1801633, EPI_ISL_1801634, EPI_ISL_1801635, EPI_ISL_1801636, EPI_ISL_1801637, EPI_ISL_1801638, EPI_ISL_1801639, EPI_ISL_1801640, EPI_ISL_1801641, EPI_ISL_1801642, EPI_ISL_1801643, EPI_ISL_1801645, EPI_ISL_1801646, EPI_ISL_1801647, EPI_ISL_1801648, EPI_ISL_1801649, EPI_ISL_1801650, EPI_ISL_1801651, EPI_ISL_1801652, EPI_ISL_1801653, EPI_ISL_1801654, EPI_ISL_1801655, EPI_ISL_1801657, EPI_ISL_1801658, EPI_ISL_1801659, EPI_ISL_1801660, EPI_ISL_1801661

see above | Laboratory Corporation of America | Centers for Disease Control and Prevention Division of Viral Diseases, Pathogen Discovery | Dakota Howard, Dhwani Batra, Peter W. Cook, Kara Moser, Adrian Paskey, Jason Caravas, Benjamin Rambo-Martin, Shatavia Morrison, Christopher Gulvick, Scott Sammons, Yvette Unoarumhi, Darlene Wagner, Matthew Schmerer, Minoo Agarwal, Eyad Almasri, Debbie Boles, Ayla Burns, Nuthawin Charoensri, Oren Cohen, Susan Countryman, Mary Ann Cristobal, Bobbi Croy, Suzanne Dale, Hrushikesh Deshmukh, Amanda Douglas, Vincent Drouillon, Marcia Eisenberg, Howard Engler, Rama Ghatti, Prashant Gupta, Susan Hicks, Jake Humphrey, Lax Iyer, Manoj Jain, Mohan Kolli, Brian Krueger, Tim Kuphal, Stanley Letovsky, Michael Levandoski, Craig Lukasik, Jonathan Meltzer, Brian Norvell, Mindy Nye, Scott Parker, Christos Petropoulos, John Pruitt, Steven Ragan, Scott Ryan, Mike Sapeta, Jana Schroth, Suresh Babu Selvaraju, Goran Stevovic, Amanda Suchanek, Andrea Throop, Lyndon Tilson, Thomas Urban, Joe Voshell, Kimberly Wagner, Jonathan Williams, Mary Williamson, Qian Zeng, Tricia Zwiefelhofer, Clinton R. Paden, Duncan MacCannell

EPI_ISL_1801662 | Ohio Department of Health Laboratory | Ohio Department of Health Laboratory | Holmes, Jennifer; Eric Brandt, Keoni Omura, Glen McGillivary, Caitlin McDonnell, Jade Mowery, Stephanie Mccracken, Tyler Payne, Kirtana Ramadugu, Erica Leasure, Brent Lee, Kelsey Florek, Heather Blankenship, Allison Black, Quanta Brown, and Tammy Bannerman

EPI_ISL_1801663, EPI_ISL_1801664, EPI_ISL_1801665, EPI_ISL_1801667, EPI_ISL_1801668, EPI_ISL_1801669, EPI_ISL_1801670, EPI_ISL_1801671, EPI_ISL_1801672, EPI_ISL_1801674, EPI_ISL_1801675, EPI_ISL_1801676, EPI_ISL_1801677, EPI_ISL_1801678, EPI_ISL_1801679, EPI_ISL_1801680, EPI_ISL_1801681, EPI_ISL_1801682, EPI_ISL_1801683, EPI_ISL_1801684, EPI_ISL_1801685, EPI_ISL_1801686, EPI_ISL_1801687, EPI_ISL_1801688, EPI_ISL_1801689, EPI_ISL_1801690, EPI_ISL_1801691, EPI_ISL_1801692, EPI_ISL_1801693, EPI_ISL_1801694, EPI_ISL_1801695, EPI_ISL_1801696, EPI_ISL_1801697, EPI_ISL_1801698, EPI_ISL_1801699, EPI_ISL_1801700, EPI_ISL_1801701, EPI_ISL_1801702, EPI_ISL_1801703, EPI_ISL_1801704, EPI_ISL_1801706, EPI_ISL_1801707, EPI_ISL_1801708, EPI_ISL_1801709, EPI_ISL_1801710, EPI_ISL_1801711, EPI_ISL_1801712, EPI_ISL_1801713, EPI_ISL_1801714, EPI_ISL_1801715, EPI_ISL_1801716, EPI_ISL_1801717, EPI_ISL_1801718, EPI_ISL_1801719, EPI_ISL_1801720, EPI_ISL_1801721, EPI_ISL_1801722, EPI_ISL_1801723, EPI_ISL_1801724, EPI_ISL_1801725, EPI_ISL_1801726, EPI_ISL_1801727, EPI_ISL_1801728, EPI_ISL_1801729, EPI_ISL_1801732, EPI_ISL_1801739, EPI_ISL_1801743, EPI_ISL_1801746, EPI_ISL_1801750, EPI_ISL_1801752, EPI_ISL_1801754, EPI_ISL_1801755, EPI_ISL_1801757, EPI_ISL_1801759, EPI_ISL_1801761, EPI_ISL_1801762, EPI_ISL_1801764, EPI_ISL_1801766, EPI_ISL_1801768, EPI_ISL_1801769, EPI_ISL_1801773, EPI_ISL_1801775, EPI_ISL_1801776, EPI_ISL_1801780, EPI_ISL_1801781, EPI_ISL_1801783, EPI_ISL_1801785, EPI_ISL_1801786, EPI_ISL_1801788, EPI_ISL_1801790, EPI_ISL_1801792, EPI_ISL_1801794, EPI_ISL_1801796, EPI_ISL_1801798, EPI_ISL_1801799, EPI_ISL_1801800, EPI_ISL_1801801, EPI_ISL_1801802, EPI_ISL_1801803, EPI_ISL_1801804, EPI_ISL_1801805, EPI_ISL_1801806, EPI_ISL_1801807, EPI_ISL_1801808, EPI_ISL_1801809, EPI_ISL_1801810, EPI_ISL_1801811, EPI_ISL_1801812, EPI_ISL_1801814, EPI_ISL_1801815, EPI_ISL_1801816, EPI_ISL_1801817, EPI_ISL_1801818, EPI_ISL_1801819, EPI_ISL_1801820, EPI_ISL_1801821, EPI_ISL_1801822, EPI_ISL_1801825, EPI_ISL_1801827, EPI_ISL_1801828, EPI_ISL_1801829, EPI_ISL_1801830, EPI_ISL_1801831, EPI_ISL_1801832, EPI_ISL_1801833, EPI_ISL_1801834, EPI_ISL_1801835, EPI_ISL_1801837, EPI_ISL_1801838, EPI_ISL_1801839, EPI_ISL_1801840, EPI_ISL_1801841, EPI_ISL_1801842, EPI_ISL_1801845, EPI_ISL_1801846, EPI_ISL_1801847, EPI_ISL_1801849, EPI_ISL_1801853, EPI_ISL_1801854, EPI_ISL_1801855, EPI_ISL_1801856, EPI_ISL_1801857, EPI_ISL_1801858, EPI_ISL_1801859, EPI_ISL_1801860, EPI_ISL_1801861, EPI_ISL_1801862, EPI_ISL_1801863, EPI_ISL_1801864, EPI_ISL_1801865, EPI_ISL_1801866, EPI_ISL_1801867, EPI_ISL_1801868, EPI_ISL_1801869, EPI_ISL_1801870, EPI_ISL_1801871, EPI_ISL_1801872, EPI_ISL_1801873, EPI_ISL_1801874, EPI_ISL_1801875, EPI_ISL_1801876, EPI_ISL_1801877, EPI_ISL_1801878, EPI_ISL_1801880, EPI_ISL_1801882, EPI_ISL_1801883, EPI_ISL_1801884, EPI_ISL_1801885, EPI_ISL_1801886, EPI_ISL_1801887, EPI_ISL_1801888, EPI_ISL_1801889, EPI_ISL_1801890, EPI_ISL_1801891, EPI_ISL_1801892, EPI_ISL_1801893, EPI_ISL_1801894, EPI_ISL_1801895, EPI_ISL_1801896, EPI_ISL_1801897, EPI_ISL_1801898, EPI_ISL_1801900, EPI_ISL_1801901, EPI_ISL_1801904, EPI_ISL_1801905, EPI_ISL_1801907, EPI_ISL_1801908, EPI_ISL_1801911, EPI_ISL_1801912, EPI_ISL_1801913, EPI_ISL_1801914

see above | Laboratory Corporation of America | Centers for Disease Control and Prevention Division of Viral Diseases, Pathogen Discovery | Dakota Howard, Dhwani Batra, Peter W. Cook, Kara Moser, Adrian Paskey, Jason Caravas, Benjamin Rambo-Martin, Shatavia Morrison, Christopher Gulvick, Scott Sammons, Yvette Unoarumhi, Darlene Wagner, Matthew Schmerer, Minoo Agarwal, Eyad Almasri, Debbie Boles, Ayla Burns, Nuthawin Charoensri, Oren Cohen, Susan Countryman, Mary Ann Cristobal, Bobbi Croy, Suzanne Dale, Hrushikesh Deshmukh, Amanda Douglas, Vincent Drouillon, Marcia Eisenberg, Howard Engler, Rama Ghatti, Prashant Gupta, Susan Hicks, Jake Humphrey, Lax Iyer, Manoj Jain, Mohan Kolli, Brian Krueger, Tim Kuphal, Stanley Letovsky, Michael Levandoski, Craig Lukasik, Jonathan Meltzer, Brian Norvell, Mindy Nye, Scott Parker, Christos Petropoulos, John Pruitt, Steven Ragan, Scott Ryan, Mike Sapeta, Jana Schroth, Suresh Babu Selvaraju, Goran Stevovic, Amanda Suchanek, Andrea Throop, Lyndon Tilson, Thomas Urban, Joe Voshell, Kimberly Wagner, Jonathan Williams, Mary Williamson, Qian Zeng, Tricia Zwiefelhofer, Clinton R. Paden, Duncan MacCannell

EPI_ISL_1801915 | Ohio Department of Health Laboratory | Ohio Department of Health Laboratory | Holmes, Jennifer; Eric Brandt, Keoni Omura, Glen McGillivary, Caitlin McDonnell, Jade Mowery, Stephanie Mccracken, Tyler Payne, Kirtana Ramadugu, Erica Leasure, Brent Lee, Kelsey Florek, Heather Blankenship, Allison Black, Quanta Brown, and Tammy Bannerman

EPI_ISL_1801916, EPI_ISL_1801917, EPI_ISL_1801918, EPI_ISL_1801919, EPI_ISL_1801920, EPI_ISL_1801921, EPI_ISL_1801922, EPI_ISL_1801923, EPI_ISL_1801924, EPI_ISL_1801925, EPI_ISL_1801926, EPI_ISL_1801927, EPI_ISL_1801928, EPI_ISL_1801929, EPI_ISL_1801930, EPI_ISL_1801931, EPI_ISL_1801932, EPI_ISL_1801933, EPI_ISL_1801935, EPI_ISL_1801936, EPI_ISL_1801937, EPI_ISL_1801938, EPI_ISL_1801939, EPI_ISL_1801940, EPI_ISL_1801941, EPI_ISL_1801942, EPI_ISL_1801943, EPI_ISL_1801944, EPI_ISL_1801946, EPI_ISL_1801947, EPI_ISL_1801948, EPI_ISL_1801950, EPI_ISL_1801951, EPI_ISL_1801952, EPI_ISL_1801953, EPI_ISL_1801954, EPI_ISL_1801955, EPI_ISL_1801956, EPI_ISL_1801957, EPI_ISL_1801958, EPI_ISL_1801959, EPI_ISL_1801960, EPI_ISL_1801961, EPI_ISL_1801962, EPI_ISL_1801963, EPI_ISL_1801964, EPI_ISL_1801965, EPI_ISL_1801966, EPI_ISL_1801967, EPI_ISL_1801968, EPI_ISL_1801970, EPI_ISL_1801971, EPI_ISL_1801972, EPI_ISL_1801973, EPI_ISL_1801974, EPI_ISL_1801975, EPI_ISL_1801976, EPI_ISL_1801977, EPI_ISL_1801978, EPI_ISL_1801979, EPI_ISL_1801980, EPI_ISL_1801981, EPI_ISL_1801982, EPI_ISL_1801984, EPI_ISL_1801985, EPI_ISL_1801986, EPI_ISL_1801987, EPI_ISL_1801988, EPI_ISL_1801989, EPI_ISL_1801990, EPI_ISL_1801992, EPI_ISL_1801993, EPI_ISL_1801994, EPI_ISL_1801995, EPI_ISL_1801996, EPI_ISL_1801997, EPI_ISL_1801998, EPI_ISL_1802001, EPI_ISL_1802003, EPI_ISL_1802004, EPI_ISL_1802005, EPI_ISL_1802006, EPI_ISL_1802007, EPI_ISL_1802008, EPI_ISL_1802009, EPI_ISL_1802010, EPI_ISL_1802011, EPI_ISL_1802013, EPI_ISL_1802015, EPI_ISL_1802016, EPI_ISL_1802017, EPI_ISL_1802019, EPI_ISL_1802020, EPI_ISL_1802021, EPI_ISL_1802022, EPI_ISL_1802023, EPI_ISL_1802024, EPI_ISL_1802025, EPI_ISL_1802026, EPI_ISL_1802027, EPI_ISL_1802028, EPI_ISL_1802029, EPI_ISL_1802030, EPI_ISL_1802031, EPI_ISL_1802032, EPI_ISL_1802034, EPI_ISL_1802036, EPI_ISL_1802037, EPI_ISL_1802038, EPI_ISL_1802039, EPI_ISL_1802041, EPI_ISL_1802043, EPI_ISL_1802044, EPI_ISL_1802045, EPI_ISL_1802047, EPI_ISL_1802048, EPI_ISL_1802049, EPI_ISL_1802050, EPI_ISL_1802051, EPI_ISL_1802052, EPI_ISL_1802053, EPI_ISL_1802054, EPI_ISL_1802055, EPI_ISL_1802056, EPI_ISL_1802057, EPI_ISL_1802058, EPI_ISL_1802059, EPI_ISL_1802060, EPI_ISL_1802062, EPI_ISL_1802063, EPI_ISL_1802064, EPI_ISL_1802065, EPI_ISL_1802066, EPI_ISL_1802067, EPI_ISL_1802068, EPI_ISL_1802069, EPI_ISL_1802071, EPI_ISL_1802072, EPI_ISL_1802073, EPI_ISL_1802074

see above | Laboratory Corporation of America | Centers for Disease Control and Prevention Division of Viral Diseases, Pathogen Discovery | Dakota Howard, Dhwani Batra, Peter W. Cook, Kara Moser, Adrian Paskey, Jason Caravas, Benjamin Rambo-Martin, Shatavia Morrison, Christopher Gulvick, Scott Sammons, Yvette Unoarumhi, Darlene Wagner, Matthew Schmerer, Minoo Agarwal, Eyad Almasri, Debbie Boles, Ayla Burns, Nuthawin Charoensri, Oren Cohen, Susan Countryman, Mary Ann Cristobal, Bobbi Croy, Suzanne Dale, Hrushikesh Deshmukh, Amanda Douglas, Vincent Drouillon, Marcia Eisenberg, Howard Engler, Rama Ghatti, Prashant Gupta, Susan Hicks, Jake Humphrey, Lax Iyer, Manoj Jain, Mohan Kolli, Brian Krueger, Tim Kuphal, Stanley Letovsky, Michael Levandoski, Craig Lukasik, Jonathan Meltzer, Brian Norvell, Mindy Nye, Scott Parker, Christos Petropoulos, John Pruitt, Steven Ragan, Scott Ryan, Mike Sapeta, Jana Schroth, Suresh Babu Selvaraju, Goran Stevovic, Amanda Suchanek, Andrea Throop, Lyndon Tilson, Thomas Urban, Joe Voshell, Kimberly Wagner, Jonathan Williams, Mary Williamson, Qian Zeng, Tricia Zwiefelhofer, Clinton R. Paden, Duncan MacCannell

EPI_ISL_1802075 | MAWD Pathology Group, PA | MAWD Pathology Group, PA | Pradip Manna, PhD, Kelli Fayard, Shelby Herner, London Collins, Kelsi Eastman

EPI_ISL_1802077, EPI_ISL_1802078, EPI_ISL_1802079, EPI_ISL_1802080, EPI_ISL_1802081, EPI_ISL_1802082, EPI_ISL_1802083, EPI_ISL_1802084, EPI_ISL_1802085, EPI_ISL_1802087, EPI_ISL_1802088, EPI_ISL_1802089, EPI_ISL_1802090, EPI_ISL_1802091, EPI_ISL_1802093, EPI_ISL_1802094, EPI_ISL_1802095, EPI_ISL_1802096, EPI_ISL_1802097, EPI_ISL_1802098, EPI_ISL_1802099, EPI_ISL_1802100, EPI_ISL_1802101, EPI_ISL_1802104, EPI_ISL_1802105, EPI_ISL_1802106, EPI_ISL_1802107, EPI_ISL_1802108, EPI_ISL_1802109, EPI_ISL_1802110, EPI_ISL_1802111, EPI_ISL_1802112, EPI_ISL_1802113, EPI_ISL_1802114, EPI_ISL_1802115, EPI_ISL_1802116, EPI_ISL_1802117, EPI_ISL_1802118, EPI_ISL_1802119, EPI_ISL_1802120, EPI_ISL_1802121, EPI_ISL_1802122, EPI_ISL_1802123, EPI_ISL_1802124, EPI_ISL_1802125, EPI_ISL_1802126, EPI_ISL_1802127, EPI_ISL_1802128, EPI_ISL_1802129, EPI_ISL_1802130, EPI_ISL_1802131, EPI_ISL_1802132, EPI_ISL_1802133, EPI_ISL_1802134, EPI_ISL_1802135, EPI_ISL_1802137, EPI_ISL_1802138, EPI_ISL_1802139, EPI_ISL_1802140, EPI_ISL_1802142, EPI_ISL_1802143, EPI_ISL_1802144, EPI_ISL_1802145, EPI_ISL_1802146, EPI_ISL_1802147, EPI_ISL_1802148, EPI_ISL_1802149, EPI_ISL_1802150, EPI_ISL_1802151, EPI_ISL_1802152, EPI_ISL_1802153, EPI_ISL_1802154, EPI_ISL_1802155, EPI_ISL_1802156, EPI_ISL_1802157, EPI_ISL_1802158, EPI_ISL_1802159, EPI_ISL_1802160, EPI_ISL_1802161, EPI_ISL_1802162, EPI_ISL_1802163, EPI_ISL_1802164, EPI_ISL_1802167, EPI_ISL_1802168, EPI_ISL_1802171, EPI_ISL_1802172, EPI_ISL_1802174, EPI_ISL_1802176, EPI_ISL_1802177, EPI_ISL_1802178, EPI_ISL_1802179, EPI_ISL_1802180, EPI_ISL_1802181, EPI_ISL_1802182, EPI_ISL_1802184, EPI_ISL_1802185, EPI_ISL_1802186, EPI_ISL_1802187, EPI_ISL_1802189, EPI_ISL_1802190, EPI_ISL_1802191, EPI_ISL_1802193, EPI_ISL_1802194, EPI_ISL_1802195, EPI_ISL_1802196, EPI_ISL_1802197, EPI_ISL_1802198, EPI_ISL_1802199, EPI_ISL_1802200, EPI_ISL_1802201, EPI_ISL_1802202, EPI_ISL_1802203, EPI_ISL_1802204, EPI_ISL_1802205, EPI_ISL_1802206, EPI_ISL_1802207, EPI_ISL_1802208, EPI_ISL_1802209, EPI_ISL_1802210, EPI_ISL_1802211, EPI_ISL_1802213, EPI_ISL_1802214, EPI_ISL_1802215, EPI_ISL_1802216, EPI_ISL_1802217, EPI_ISL_1802218, EPI_ISL_1802219, EPI_ISL_1802220, EPI_ISL_1802222, EPI_ISL_1802223, EPI_ISL_1802224, EPI_ISL_1802225, EPI_ISL_1802226, EPI_ISL_1802229, EPI_ISL_1802231, EPI_ISL_1802232, EPI_ISL_1802233, EPI_ISL_1802234, EPI_ISL_1802235, EPI_ISL_1802237, EPI_ISL_1802238, EPI_ISL_1802239, EPI_ISL_1802240, EPI_ISL_1802241, EPI_ISL_1802242, EPI_ISL_1802243, EPI_ISL_1802244, EPI_ISL_1802245, EPI_ISL_1802246, EPI_ISL_1802247, EPI_ISL_1802248, EPI_ISL_1802249, EPI_ISL_1802250, EPI_ISL_1802251, EPI_ISL_1802252, EPI_ISL_1802253, EPI_ISL_1802254, EPI_ISL_1802255, EPI_ISL_1802256, EPI_ISL_1802257, EPI_ISL_1802258, EPI_ISL_1802259, EPI_ISL_1802260, EPI_ISL_1802262, EPI_ISL_1802263, EPI_ISL_1802268, EPI_ISL_1802269, EPI_ISL_1802270, EPI_ISL_1802271, EPI_ISL_1802272, EPI_ISL_1802273, EPI_ISL_1802274, EPI_ISL_1802275, EPI_ISL_1802276, EPI_ISL_1802278, EPI_ISL_1802283, EPI_ISL_1802284, EPI_ISL_1802285, EPI_ISL_1802286, EPI_ISL_1802287, EPI_ISL_1802289, EPI_ISL_1802290, EPI_ISL_1802291, EPI_ISL_1802292, EPI_ISL_1802293, EPI_ISL_1802294, EPI_ISL_1802295, EPI_ISL_1802296, EPI_ISL_1802297, EPI_ISL_1802298, EPI_ISL_1802299, EPI_ISL_1802300, EPI_ISL_1802301, EPI_ISL_1802302, EPI_ISL_1802303, EPI_ISL_1802304, EPI_ISL_1802305, EPI_ISL_1802306, EPI_ISL_1802307, EPI_ISL_1802308, EPI_ISL_1802309, EPI_ISL_1802311, EPI_ISL_1802312, EPI_ISL_1802313, EPI_ISL_1802314, EPI_ISL_1802316, EPI_ISL_1802318, EPI_ISL_1802319, EPI_ISL_1802320, EPI_ISL_1802321, EPI_ISL_1802322, EPI_ISL_1802324, EPI_ISL_1802325, EPI_ISL_1802326, EPI_ISL_1802327, EPI_ISL_1802328, EPI_ISL_1802329, EPI_ISL_1802330, EPI_ISL_1802331, EPI_ISL_1802332, EPI_ISL_1802333, EPI_ISL_1802334, EPI_ISL_1802335, EPI_ISL_1802336, EPI_ISL_1802337, EPI_ISL_1802340, EPI_ISL_1802341, EPI_ISL_1802342, EPI_ISL_1802343, EPI_ISL_1802344, EPI_ISL_1802345, EPI_ISL_1802346, EPI_ISL_1802347, EPI_ISL_1802349, EPI_ISL_1802350, EPI_ISL_1802351, EPI_ISL_1802352, EPI_ISL_1802353, EPI_ISL_1802354, EPI_ISL_1802355, EPI_ISL_1802357, EPI_ISL_1802358, EPI_ISL_1802360, EPI_ISL_1802361, EPI_ISL_1802362, EPI_ISL_1802363, EPI_ISL_1802364, EPI_ISL_1802365, EPI_ISL_1802367, EPI_ISL_1802368, EPI_ISL_1802370, EPI_ISL_1802371, EPI_ISL_1802372, EPI_ISL_1802374, EPI_ISL_1802376, EPI_ISL_1802377, EPI_ISL_1802378, EPI_ISL_1802379, EPI_ISL_1802380, EPI_ISL_1802381, EPI_ISL_1802382, EPI_ISL_1802383, EPI_ISL_1802384, EPI_ISL_1802385, EPI_ISL_1802386, EPI_ISL_1802387, EPI_ISL_1802388, EPI_ISL_1802390, EPI_ISL_1802392, EPI_ISL_1802393, EPI_ISL_1802394, EPI_ISL_1802395, EPI_ISL_1802396, EPI_ISL_1802397, EPI_ISL_1802398, EPI_ISL_1802399, EPI_ISL_1802400, EPI_ISL_1802401, EPI_ISL_1802402, EPI_ISL_1802403, EPI_ISL_1802404, EPI_ISL_1802405, EPI_ISL_1802406, EPI_ISL_1802407, EPI_ISL_1802408, EPI_ISL_1802409,

| | | | |
|---|---|---|---|
| EPI_ISL_1802410, EPI_ISL_1802411, EPI_ISL_1802412, EPI_ISL_1802415, EPI_ISL_1802416, EPI_ISL_1802417, EPI_ISL_1802418, EPI_ISL_1802421, EPI_ISL_1802422, EPI_ISL_1802423, EPI_ISL_1802424, EPI_ISL_1802425, EPI_ISL_1802427, EPI_ISL_1802428, EPI_ISL_1802429, EPI_ISL_1802430, EPI_ISL_1802431, EPI_ISL_1802433, EPI_ISL_1802434, EPI_ISL_1802435, EPI_ISL_1802436, EPI_ISL_1802437, EPI_ISL_1802438, EPI_ISL_1802439, EPI_ISL_1802440, EPI_ISL_1802441, EPI_ISL_1802442, EPI_ISL_1802443, EPI_ISL_1802445, EPI_ISL_1802447, EPI_ISL_1802448, EPI_ISL_1802449, EPI_ISL_1802450, EPI_ISL_1802451, EPI_ISL_1802452, EPI_ISL_1802453, EPI_ISL_1802454, EPI_ISL_1802455, EPI_ISL_1802456, EPI_ISL_1802458, EPI_ISL_1802459, EPI_ISL_1802460, EPI_ISL_1802461, EPI_ISL_1802462, EPI_ISL_1802463, EPI_ISL_1802464, EPI_ISL_1802466, EPI_ISL_1802467, EPI_ISL_1802468, EPI_ISL_1802469, EPI_ISL_1802470, EPI_ISL_1802471, EPI_ISL_1802472, EPI_ISL_1802473, EPI_ISL_1802474, EPI_ISL_1802475, EPI_ISL_1802476, EPI_ISL_1802477, EPI_ISL_1802478, EPI_ISL_1802479, EPI_ISL_1802480, EPI_ISL_1802481, EPI_ISL_1802482, EPI_ISL_1802483, EPI_ISL_1802484, EPI_ISL_1802486, EPI_ISL_1802487, EPI_ISL_1802488, EPI_ISL_1802489, EPI_ISL_1802490, EPI_ISL_1802491, EPI_ISL_1802492, EPI_ISL_1802493, EPI_ISL_1802494, EPI_ISL_1802496, EPI_ISL_1802497, EPI_ISL_1802498, EPI_ISL_1802499, EPI_ISL_1802502, EPI_ISL_1802503, EPI_ISL_1802504, EPI_ISL_1802505, EPI_ISL_1802507, EPI_ISL_1802510, EPI_ISL_1802513, EPI_ISL_1802514, EPI_ISL_1802516, EPI_ISL_1802517, EPI_ISL_1802518, EPI_ISL_1802519, EPI_ISL_1802520, EPI_ISL_1802521, EPI_ISL_1802522, EPI_ISL_1802523, EPI_ISL_1802524, EPI_ISL_1802525, EPI_ISL_1802526, EPI_ISL_1802527, EPI_ISL_1802528, EPI_ISL_1802529, EPI_ISL_1802530, EPI_ISL_1802531, EPI_ISL_1802532, EPI_ISL_1802533, EPI_ISL_1802534, EPI_ISL_1802535, EPI_ISL_1802536, EPI_ISL_1802537, EPI_ISL_1802538, EPI_ISL_1802539, EPI_ISL_1802540, EPI_ISL_1802541, EPI_ISL_1802542, EPI_ISL_1802544, EPI_ISL_1802545, EPI_ISL_1802546, EPI_ISL_1802547, EPI_ISL_1802548, EPI_ISL_1802549, EPI_ISL_1802550, EPI_ISL_1802551, EPI_ISL_1802553, EPI_ISL_1802554, EPI_ISL_1802555, EPI_ISL_1802556, EPI_ISL_1802557, EPI_ISL_1802558, EPI_ISL_1802559, EPI_ISL_1802560, EPI_ISL_1802561, EPI_ISL_1802563, EPI_ISL_1802564, EPI_ISL_1802565, EPI_ISL_1802566, EPI_ISL_1802567, EPI_ISL_1802568, EPI_ISL_1802569, EPI_ISL_1802571, EPI_ISL_1802573, EPI_ISL_1802574, EPI_ISL_1802575, EPI_ISL_1802576, EPI_ISL_1802577, EPI_ISL_1802578, EPI_ISL_1802579, EPI_ISL_1802580, EPI_ISL_1802581, EPI_ISL_1802582, EPI_ISL_1802583, EPI_ISL_1802584, EPI_ISL_1802585, EPI_ISL_1802586, EPI_ISL_1802587, EPI_ISL_1802589, EPI_ISL_1802590, EPI_ISL_1802591, EPI_ISL_1802592, EPI_ISL_1802593, EPI_ISL_1802594, EPI_ISL_1802595, EPI_ISL_1802596, EPI_ISL_1802597, EPI_ISL_1802598, EPI_ISL_1802599, EPI_ISL_1802600, EPI_ISL_1802601, EPI_ISL_1802602, EPI_ISL_1802603, EPI_ISL_1802604, EPI_ISL_1802605, EPI_ISL_1802606, EPI_ISL_1802607, EPI_ISL_1802608, EPI_ISL_1802609, EPI_ISL_1802610, EPI_ISL_1802611, EPI_ISL_1802612, EPI_ISL_1802613, EPI_ISL_1802614, EPI_ISL_1802615, EPI_ISL_1802616, EPI_ISL_1802617, EPI_ISL_1802619, EPI_ISL_1802620, EPI_ISL_1802621, EPI_ISL_1802622, EPI_ISL_1802623, EPI_ISL_1802624, EPI_ISL_1802625, EPI_ISL_1802626, EPI_ISL_1802627, EPI_ISL_1802628, EPI_ISL_1802629, EPI_ISL_1802630, EPI_ISL_1802631, EPI_ISL_1802632, EPI_ISL_1802633, EPI_ISL_1802634, EPI_ISL_1802635, EPI_ISL_1802636, EPI_ISL_1802637, EPI_ISL_1802638, EPI_ISL_1802639, EPI_ISL_1802640, EPI_ISL_1802641, EPI_ISL_1802642, EPI_ISL_1802643, EPI_ISL_1802644, EPI_ISL_1802645, EPI_ISL_1802646, EPI_ISL_1802647, EPI_ISL_1802648, EPI_ISL_1802649, EPI_ISL_1802650, EPI_ISL_1802651, EPI_ISL_1802652, EPI_ISL_1802653, EPI_ISL_1802654, EPI_ISL_1802655, EPI_ISL_1802656, EPI_ISL_1802658, EPI_ISL_1802659, EPI_ISL_1802660, EPI_ISL_1802661, EPI_ISL_1802662, EPI_ISL_1802664 | | | |
| see above | Laboratory Corporation of America | Centers for Disease Control and Prevention Division of Viral Diseases, Pathogen Discovery | Dakota Howard, Dhwani Batra, Peter W. Cook, Kara Moser, Adrian Paskey, Jason Caravas, Benjamin Rambo-Martin, Shatavia Morrison, Christopher Gulvick, Scott Sammons, Yvette Unoarumhi, Darlene Wagner, Matthew Schmerer, Minoo Agarwal, Eyad Almasri, Debbie Boles, Ayla Burns, Nuthawin Charoensri, Oren Cohen, Susan Countryman, Mary Ann Cristobal, Bobbi Croy, Suzanne Dale, Hrushikesh Deshmukh, Amanda Douglas, Vincent Drouillon, Marcia Eisenberg, Howard Engler, Rama Ghatti, Prashant Gupta, Susan Hicks, Jake Humphrey, Lax Iyer, Manoj Jain, Mohan Kolli, Brian Krueger, Tim Kuphal, Stanley Letovsky, Michael Levandoski, Craig Lukasik, Jonathan Meltzer, Brian Norvell, Mindy Nye, Scott Parker, Christos Petropoulos, John Pruitt, Steven Ragan, Scott Ryan, Mike Sapeta, Jana Schroth, Suresh Babu Selvaraju, Goran Stevovic, Amanda Suchanek, Andrea Throop, Lyndon Tilson, Thomas Urban, Joe Voshell, Kimberly Wagner, Jonathan Williams, Mary Williamson, Qian Zeng, Tricia Zwiefelhofer, Clinton R. Paden, Duncan MacCannell |
| EPI_ISL_1802665, EPI_ISL_1802666, EPI_ISL_1802667, EPI_ISL_1802668, EPI_ISL_1802670, EPI_ISL_1802671, EPI_ISL_1802672, EPI_ISL_1802673, EPI_ISL_1802674, EPI_ISL_1802675, EPI_ISL_1802676, EPI_ISL_1802677, EPI_ISL_1802678, EPI_ISL_1802680, EPI_ISL_1802681, EPI_ISL_1802682, EPI_ISL_1802683, EPI_ISL_1802684, EPI_ISL_1802685, EPI_ISL_1802687, EPI_ISL_1802688, EPI_ISL_1802689, EPI_ISL_1802690, EPI_ISL_1802691, EPI_ISL_1802693, EPI_ISL_1802694, EPI_ISL_1802695, EPI_ISL_1802696, EPI_ISL_1802697, EPI_ISL_1802698, EPI_ISL_1802699 | | | |
| see above | Fulgent Genetics | Centers for Disease Control and Prevention Division of Viral Diseases, Pathogen Discovery | Dakota Howard, Dhwani Batra, Peter W. Cook, Kara Moser, Adrian Paskey, Jason Caravas, Benjamin Rambo-Martin, Shatavia Morrison, Christopher Gulvick, Scott Sammons, Yvette Unoarumhi, Darlene Wagner, Matthew Schmerer, Harry Gao, Mickey Li, John Gao, Joseph Fierro, Benafsh Sapra, Becky Tsai, Yan Meng, Doreen Ng, James Xie, Clinton R. Paden, Duncan MacCannell |
| EPI_ISL_1802700, EPI_ISL_1802701, EPI_ISL_1802702, EPI_ISL_1802703, EPI_ISL_1802704, EPI_ISL_1802705, EPI_ISL_1802706, EPI_ISL_1802707, EPI_ISL_1802708, EPI_ISL_1802709, EPI_ISL_1802710, EPI_ISL_1802711, EPI_ISL_1802712, EPI_ISL_1802713, EPI_ISL_1802714, EPI_ISL_1802715, EPI_ISL_1802716, EPI_ISL_1802717, EPI_ISL_1802718, EPI_ISL_1802719, EPI_ISL_1802720, EPI_ISL_1802721, EPI_ISL_1802722, EPI_ISL_1802723, EPI_ISL_1802724, EPI_ISL_1802725, EPI_ISL_1802726, EPI_ISL_1802727, EPI_ISL_1802728, EPI_ISL_1802729, EPI_ISL_1802730, EPI_ISL_1802731, EPI_ISL_1802732, EPI_ISL_1802733, EPI_ISL_1802734, EPI_ISL_1802735, EPI_ISL_1802736, EPI_ISL_1802737, EPI_ISL_1802738, EPI_ISL_1802739, EPI_ISL_1802740, EPI_ISL_1802741, EPI_ISL_1802742, EPI_ISL_1802743, EPI_ISL_1802744, EPI_ISL_1802745, EPI_ISL_1802746, EPI_ISL_1802747, EPI_ISL_1802748, EPI_ISL_1802749, EPI_ISL_1802750, EPI_ISL_1802751, EPI_ISL_1802752, EPI_ISL_1802753, EPI_ISL_1802754, EPI_ISL_1802755, EPI_ISL_1802756, EPI_ISL_1802757, EPI_ISL_1802758, EPI_ISL_1802759, EPI_ISL_1802760, EPI_ISL_1802761, EPI_ISL_1802762, EPI_ISL_1802763, EPI_ISL_1802764, EPI_ISL_1802765, EPI_ISL_1802766, EPI_ISL_1802767, EPI_ISL_1802768, EPI_ISL_1802769, EPI_ISL_1802770, EPI_ISL_1802771, EPI_ISL_1802772, EPI_ISL_1802773, EPI_ISL_1802774, EPI_ISL_1802775, EPI_ISL_1802776, EPI_ISL_1802777, EPI_ISL_1802778, EPI_ISL_1802779, EPI_ISL_1802780, EPI_ISL_1802781, EPI_ISL_1802782, EPI_ISL_1802783, EPI_ISL_1802784, EPI_ISL_1802785, EPI_ISL_1802786, EPI_ISL_1802787, EPI_ISL_1802788, EPI_ISL_1802789, EPI_ISL_1802790, EPI_ISL_1802791, EPI_ISL_1802792, EPI_ISL_1802793, EPI_ISL_1802794, EPI_ISL_1802795, EPI_ISL_1802796, EPI_ISL_1802797, EPI_ISL_1802798, EPI_ISL_1802799, EPI_ISL_1802800, EPI_ISL_1802801, EPI_ISL_1802802, EPI_ISL_1802803, EPI_ISL_1802804, EPI_ISL_1802805, EPI_ISL_1802806, EPI_ISL_1802807, EPI_ISL_1802808, EPI_ISL_1802809, EPI_ISL_1802810, EPI_ISL_1802811, EPI_ISL_1802812, EPI_ISL_1802813, EPI_ISL_1802814, EPI_ISL_1802815, EPI_ISL_1802816, EPI_ISL_1802817, EPI_ISL_1802818, EPI_ISL_1802819, EPI_ISL_1802820, EPI_ISL_1802821, EPI_ISL_1802822, EPI_ISL_1802823, EPI_ISL_1802824, EPI_ISL_1802825, EPI_ISL_1802826, EPI_ISL_1802827, EPI_ISL_1802828, EPI_ISL_1802829, EPI_ISL_1802830, EPI_ISL_1802831, EPI_ISL_1802832, EPI_ISL_1802833, EPI_ISL_1802834, EPI_ISL_1802835, EPI_ISL_1802836, EPI_ISL_1802837, EPI_ISL_1802838, EPI_ISL_1802839, EPI_ISL_1802840, EPI_ISL_1802841, EPI_ISL_1802842, EPI_ISL_1802843, EPI_ISL_1802844, EPI_ISL_1802845, EPI_ISL_1802846, EPI_ISL_1802848, EPI_ISL_1802849, EPI_ISL_1802850, EPI_ISL_1802851, EPI_ISL_1802852, EPI_ISL_1802853, EPI_ISL_1802854, EPI_ISL_1802855, EPI_ISL_1802856, EPI_ISL_1802857, EPI_ISL_1802858, EPI_ISL_1802859, EPI_ISL_1802860, EPI_ISL_1802861, EPI_ISL_1802862, EPI_ISL_1802863, EPI_ISL_1802864, EPI_ISL_1802865, EPI_ISL_1802866, EPI_ISL_1802867, EPI_ISL_1802868, EPI_ISL_1802869, EPI_ISL_1802870, EPI_ISL_1802871, EPI_ISL_1802873, EPI_ISL_1802874, EPI_ISL_1802875, EPI_ISL_1802876, EPI_ISL_1802877, EPI_ISL_1802878, EPI_ISL_1802879, EPI_ISL_1802880, EPI_ISL_1802881, EPI_ISL_1802882, EPI_ISL_1802883, EPI_ISL_1802884, EPI_ISL_1802885, EPI_ISL_1802886, EPI_ISL_1802887, EPI_ISL_1802888, EPI_ISL_1802889, EPI_ISL_1802890, EPI_ISL_1802891, EPI_ISL_1802892, EPI_ISL_1802893, EPI_ISL_1802894, EPI_ISL_1802895, EPI_ISL_1802896, EPI_ISL_1802897, EPI_ISL_1802898, EPI_ISL_1802899, EPI_ISL_1802900, EPI_ISL_1802901, EPI_ISL_1802902, EPI_ISL_1802903, EPI_ISL_1802904, EPI_ISL_1802905, EPI_ISL_1802906, EPI_ISL_1802907, EPI_ISL_1802908, EPI_ISL_1802909, EPI_ISL_1802910, EPI_ISL_1802911, EPI_ISL_1802912, EPI_ISL_1802913, EPI_ISL_1802914, EPI_ISL_1802915, EPI_ISL_1802916, EPI_ISL_1802917, EPI_ISL_1802918, EPI_ISL_1802919, EPI_ISL_1802920, EPI_ISL_1802921, EPI_ISL_1802922, EPI_ISL_1802923, EPI_ISL_1802924, EPI_ISL_1802925, EPI_ISL_1802926, EPI_ISL_1802927, EPI_ISL_1802928, EPI_ISL_1802929, EPI_ISL_1802930, EPI_ISL_1802931, EPI_ISL_1802932, EPI_ISL_1802933, EPI_ISL_1802934, EPI_ISL_1802935, EPI_ISL_1802936, EPI_ISL_1802937, EPI_ISL_1802938, EPI_ISL_1802939, EPI_ISL_1802940, EPI_ISL_1802941, EPI_ISL_1802942, EPI_ISL_1802943, EPI_ISL_1802944, EPI_ISL_1802945, EPI_ISL_1802946, EPI_ISL_1802947, EPI_ISL_1802948, EPI_ISL_1802949, EPI_ISL_1802950, EPI_ISL_1802951, EPI_ISL_1802952, EPI_ISL_1802953, EPI_ISL_1802954, EPI_ISL_1802955, EPI_ISL_1802956, EPI_ISL_1802957, EPI_ISL_1802958, EPI_ISL_1802959, EPI_ISL_1802960, EPI_ISL_1802961, EPI_ISL_1802962, EPI_ISL_1802963, EPI_ISL_1802964, EPI_ISL_1802965, EPI_ISL_1802966, EPI_ISL_1802967, EPI_ISL_1802968, EPI_ISL_1802969, EPI_ISL_1802970, EPI_ISL_1802971, EPI_ISL_1802972, EPI_ISL_1802974, EPI_ISL_1802975, EPI_ISL_1802976, EPI_ISL_1802977, EPI_ISL_1802978, EPI_ISL_1802979, EPI_ISL_1802980, EPI_ISL_1802981, EPI_ISL_1802982, EPI_ISL_1802983, EPI_ISL_1802984, EPI_ISL_1802985, EPI_ISL_1802986, EPI_ISL_1802987, EPI_ISL_1802988, EPI_ISL_1802989, EPI_ISL_1802990, EPI_ISL_1802991, EPI_ISL_1802992, EPI_ISL_1802993, EPI_ISL_1802995, EPI_ISL_1802996, EPI_ISL_1802997, EPI_ISL_1802998, EPI_ISL_1802999, EPI_ISL_1803000, EPI_ISL_1803001, EPI_ISL_1803002, EPI_ISL_1803003, EPI_ISL_1803004, EPI_ISL_1803005, EPI_ISL_1803006, EPI_ISL_1803007, EPI_ISL_1803008, EPI_ISL_1803009, EPI_ISL_1803010, EPI_ISL_1803011, EPI_ISL_1803012, EPI_ISL_1803013, EPI_ISL_1803014, EPI_ISL_1803015, EPI_ISL_1803016, EPI_ISL_1803017, EPI_ISL_1803018, EPI_ISL_1803019, EPI_ISL_1803020, EPI_ISL_1803021, EPI_ISL_1803022, EPI_ISL_1803023, EPI_ISL_1803024, EPI_ISL_1803025, EPI_ISL_1803026, EPI_ISL_1803027, EPI_ISL_1803028, EPI_ISL_1803029, EPI_ISL_1803030, EPI_ISL_1803032, EPI_ISL_1803033, EPI_ISL_1803034, EPI_ISL_1803035, EPI_ISL_1803037, EPI_ISL_1803038, EPI_ISL_1803039, EPI_ISL_1803040, EPI_ISL_1803042, EPI_ISL_1803043, EPI_ISL_1803044, EPI_ISL_1803045, EPI_ISL_1803046, EPI_ISL_1803047, EPI_ISL_1803048, EPI_ISL_1803049, EPI_ISL_1803050, EPI_ISL_1803051, EPI_ISL_1803052, EPI_ISL_1803053, EPI_ISL_1803054, EPI_ISL_1803055, EPI_ISL_1803056, EPI_ISL_1803057, EPI_ISL_1803058, EPI_ISL_1803059, EPI_ISL_1803060, EPI_ISL_1803061, EPI_ISL_1803062, EPI_ISL_1803063, EPI_ISL_1803065, EPI_ISL_1803066, EPI_ISL_1803067, EPI_ISL_1803068, EPI_ISL_1803069, EPI_ISL_1803070, EPI_ISL_1803071, EPI_ISL_1803072, EPI_ISL_1803073, EPI_ISL_1803075, EPI_ISL_1803076, EPI_ISL_1803077, EPI_ISL_1803078, EPI_ISL_1803079, EPI_ISL_1803080, EPI_ISL_1803081, EPI_ISL_1803083, EPI_ISL_1803084, EPI_ISL_1803085, EPI_ISL_1803086, EPI_ISL_1803087, EPI_ISL_1803088, EPI_ISL_1803089, EPI_ISL_1803090, EPI_ISL_1803091, EPI_ISL_1803092, EPI_ISL_1803093, EPI_ISL_1803095, EPI_ISL_1803096, EPI_ISL_1803097, EPI_ISL_1803098, EPI_ISL_1803099, EPI_ISL_1803100, EPI_ISL_1803101, EPI_ISL_1803102, EPI_ISL_1803103, EPI_ISL_1803104, EPI_ISL_1803105, EPI_ISL_1803106, EPI_ISL_1803107, EPI_ISL_1803108, EPI_ISL_1803109, EPI_ISL_1803110, EPI_ISL_1803111, EPI_ISL_1803112, EPI_ISL_1803113, EPI_ISL_1803114, EPI_ISL_1803115, EPI_ISL_1803116, EPI_ISL_1803118, EPI_ISL_1803119, EPI_ISL_1803120, EPI_ISL_1803121, EPI_ISL_1803122, EPI_ISL_1803123, EPI_ISL_1803124, EPI_ISL_1803125, EPI_ISL_1803126, EPI_ISL_1803127, EPI_ISL_1803128, EPI_ISL_1803129, EPI_ISL_1803130, EPI_ISL_1803131, EPI_ISL_1803132, EPI_ISL_1803133, EPI_ISL_1803134, EPI_ISL_1803135, EPI_ISL_1803136, EPI_ISL_1803137, EPI_ISL_1803138, EPI_ISL_1803139, EPI_ISL_1803140, EPI_ISL_1803141, EPI_ISL_1803142, EPI_ISL_1803143, EPI_ISL_1803144, EPI_ISL_1803145, EPI_ISL_1803146, EPI_ISL_1803147, EPI_ISL_1803148, EPI_ISL_1803149, EPI_ISL_1803150, EPI_ISL_1803151, EPI_ISL_1803152, EPI_ISL_1803153, EPI_ISL_1803154, EPI_ISL_1803155, EPI_ISL_1803156, EPI_ISL_1803157, EPI_ISL_1803158, EPI_ISL_1803159, EPI_ISL_1803160, EPI_ISL_1803161, EPI_ISL_1803162, EPI_ISL_1803163, EPI_ISL_1803164, EPI_ISL_1803165, EPI_ISL_1803166, EPI_ISL_1803167, EPI_ISL_1803168, EPI_ISL_1803169, EPI_ISL_1803170, EPI_ISL_1803171, EPI_ISL_1803172, EPI_ISL_1803173, EPI_ISL_1803174, EPI_ISL_1803175, EPI_ISL_1803176, EPI_ISL_1803177, EPI_ISL_1803178, EPI_ISL_1803179, EPI_ISL_1803180, EPI_ISL_1803181, EPI_ISL_1803182, EPI_ISL_1803183, EPI_ISL_1803184, EPI_ISL_1803185, EPI_ISL_1803186, EPI_ISL_1803187, EPI_ISL_1803188, EPI_ISL_1803189, EPI_ISL_1803190, EPI_ISL_1803191, EPI_ISL_1803192, EPI_ISL_1803193, EPI_ISL_1803194, EPI_ISL_1803195, EPI_ISL_1803196, EPI_ISL_1803197, EPI_ISL_1803198, EPI_ISL_1803199, EPI_ISL_1803200, EPI_ISL_1803201, EPI_ISL_1803202, EPI_ISL_1803203, EPI_ISL_1803204, EPI_ISL_1803205, EPI_ISL_1803206, EPI_ISL_1803207, EPI_ISL_1803208, EPI_ISL_1803209, EPI_ISL_1803210, EPI_ISL_1803211, EPI_ISL_1803212, EPI_ISL_1803213, EPI_ISL_1803214, EPI_ISL_1803215, EPI_ISL_1803216, EPI_ISL_1803217, EPI_ISL_1803218, EPI_ISL_1803219, EPI_ISL_1803220, EPI_ISL_1803221, EPI_ISL_1803222, EPI_ISL_1803223, EPI_ISL_1803224, EPI_ISL_1803225, EPI_ISL_1803226, EPI_ISL_1803227, EPI_ISL_1803228, EPI_ISL_1803229, EPI_ISL_1803230, EPI_ISL_1803234, EPI_ISL_1803237, EPI_ISL_1803240, EPI_ISL_1803243, EPI_ISL_1803247, EPI_ISL_1803250, EPI_ISL_1803253, EPI_ISL_1803256, EPI_ISL_1803260, EPI_ISL_1803263, EPI_ISL_1803268, EPI_ISL_1803270, EPI_ISL_1803277, EPI_ISL_1803279, EPI_ISL_1803281, EPI_ISL_1803283, EPI_ISL_1803285, EPI_ISL_1803287, EPI_ISL_1803289, EPI_ISL_1803290, EPI_ISL_1803292, EPI_ISL_1803293, EPI_ISL_1803295, EPI_ISL_1803296, EPI_ISL_1803297, EPI_ISL_1803298, EPI_ISL_1803299, EPI_ISL_1803300, EPI_ISL_1803301, EPI_ISL_1803302, EPI_ISL_1803303, EPI_ISL_1803304, EPI_ISL_1803305, EPI_ISL_1803306, EPI_ISL_1803307, EPI_ISL_1803308, EPI_ISL_1803309, EPI_ISL_1803310, EPI_ISL_1803311, EPI_ISL_1803312, EPI_ISL_1803313, EPI_ISL_1803314, EPI_ISL_1803315, EPI_ISL_1803316, EPI_ISL_1803317, EPI_ISL_1803318, EPI_ISL_1803319, EPI_ISL_1803320, EPI_ISL_1803321, EPI_ISL_1803322, EPI_ISL_1803323, EPI_ISL_1803324, EPI_ISL_1803325, EPI_ISL_1803326, EPI_ISL_1803327, EPI_ISL_1803328, EPI_ISL_1803329, EPI_ISL_1803330, EPI_ISL_1803331, EPI_ISL_1803332, EPI_ISL_1803333, EPI_ISL_1803334, EPI_ISL_1803335, EPI_ISL_1803336, EPI_ISL_1803337, EPI_ISL_1803338, EPI_ISL_1803339, EPI_ISL_1803340, EPI_ISL_1803341, EPI_ISL_1803342, EPI_ISL_1803343, EPI_ISL_1803344, EPI_ISL_1803345, EPI_ISL_1803346, EPI_ISL_1803347, EPI_ISL_1803348, EPI_ISL_1803349, EPI_ISL_1803350, EPI_ISL_1803351, EPI_ISL_1803352, EPI_ISL_1803353, EPI_ISL_1803354, EPI_ISL_1803355, EPI_ISL_1803356, EPI_ISL_1803357, EPI_ISL_1803358, EPI_ISL_1803359, EPI_ISL_1803360, EPI_ISL_1803361, EPI_ISL_1803362, EPI_ISL_1803363, EPI_ISL_1803364, EPI_ISL_1803365, EPI_ISL_1803366, EPI_ISL_1803367, EPI_ISL_1803368, EPI_ISL_1803369, EPI_ISL_1803370, EPI_ISL_1803372, EPI_ISL_1803373, EPI_ISL_1803374, EPI_ISL_1803375, EPI_ISL_1803376, EPI_ISL_1803377, EPI_ISL_1803378, EPI_ISL_1803379, EPI_ISL_1803380, EPI_ISL_1803381, EPI_ISL_1803382, EPI_ISL_1803383, EPI_ISL_1803384, EPI_ISL_1803385, EPI_ISL_1803386, EPI_ISL_1803387, EPI_ISL_1803388, EPI_ISL_1803389, EPI_ISL_1803390, EPI_ISL_1803391, EPI_ISL_1803392, EPI_ISL_1803393, EPI_ISL_1803394, EPI_ISL_1803395, EPI_ISL_1803396, EPI_ISL_1803397, EPI_ISL_1803398, EPI_ISL_1803399, EPI_ISL_1803400, EPI_ISL_1803401, EPI_ISL_1803402, EPI_ISL_1803403, EPI_ISL_1803404, EPI_ISL_1803405, EPI_ISL_1803406, EPI_ISL_1803407, EPI_ISL_1803408, EPI_ISL_1803409, EPI_ISL_1803410, EPI_ISL_1803411, EPI_ISL_1803412, EPI_ISL_1803413, EPI_ISL_1803414, EPI_ISL_1803415, EPI_ISL_1803416, EPI_ISL_1803417, EPI_ISL_1803418, EPI_ISL_1803419, EPI_ISL_1803420, EPI_ISL_1803421, EPI_ISL_1803422, EPI_ISL_1803423, EPI_ISL_1803425, EPI_ISL_1803426, EPI_ISL_1803427, EPI_ISL_1803428, EPI_ISL_1803429, EPI_ISL_1803430, EPI_ISL_1803431, EPI_ISL_1803432, EPI_ISL_1803433, EPI_ISL_1803434, EPI_ISL_1803435, EPI_ISL_1803436, EPI_ISL_1803437, EPI_ISL_1803438, EPI_ISL_1803439, EPI_ISL_1803440, EPI_ISL_1803441, EPI_ISL_1803442, EPI_ISL_1803443, EPI_ISL_1803444, EPI_ISL_1803445, EPI_ISL_1803446, EPI_ISL_1803447, EPI_ISL_1803448, EPI_ISL_1803449, EPI_ISL_1803450, EPI_ISL_1803451, EPI_ISL_1803452, EPI_ISL_1803453, EPI_ISL_1803454, EPI_ISL_1803455, EPI_ISL_1803456, EPI_ISL_1803457, EPI_ISL_1803458, EPI_ISL_1803459, EPI_ISL_1803460, EPI_ISL_1803461, EPI_ISL_1803462, EPI_ISL_1803463, EPI_ISL_1803464, EPI_ISL_1803465, EPI_ISL_1803466, EPI_ISL_1803467, EPI_ISL_1803468, EPI_ISL_1803469, EPI_ISL_1803470, EPI_ISL_1803471, EPI_ISL_1803472, EPI_ISL_1803473, EPI_ISL_1803474, EPI_ISL_1803475, EPI_ISL_1803476, EPI_ISL_1803477, EPI_ISL_1803478, EPI_ISL_1803479, EPI_ISL_1803480, EPI_ISL_1803481, EPI_ISL_1803482, EPI_ISL_1803483, EPI_ISL_1803484, EPI_ISL_1803485, EPI_ISL_1803486, EPI_ISL_1803487, EPI_ISL_1803488, EPI_ISL_1803489, EPI_ISL_1803490, EPI_ISL_1803491, EPI_ISL_1803492, EPI_ISL_1803493, EPI_ISL_1803494, EPI_ISL_1803495, EPI_ISL_1803496, EPI_ISL_1803497, EPI_ISL_1803498, EPI_ISL_1803499, EPI_ISL_1803500, EPI_ISL_1803501 | | | |
| see above | Infinity Biologix | Centers for Disease Control and Prevention Division of Viral Diseases, Pathogen Discovery | Dakota Howard, Dhwani Batra, Peter W. Cook, Kara Moser, Adrian Paskey, Jason Caravas, Benjamin Rambo-Martin, Shatavia Morrison, Christopher Gulvick, Scott Sammons, Yvette Unoarumhi, Darlene Wagner, Matthew Schmerer, Christian Bixby, Yihe Wang, Jonathan Schultz, Chirayu Goswami, Russ Hager, Robin Grimwood, Clinton R. Paden, Duncan MacCannell |
| EPI_ISL_1803502, EPI_ISL_1803503, EPI_ISL_1803504, EPI_ISL_1803505, EPI_ISL_1803506, EPI_ISL_1803507, EPI_ISL_1803508, EPI_ISL_1803509, EPI_ISL_1803510, EPI_ISL_1803512, EPI_ISL_1803513, EPI_ISL_1803514, EPI_ISL_1803515, EPI_ISL_1803516, EPI_ISL_1803517, EPI_ISL_1803518, EPI_ISL_1803519, EPI_ISL_1803520, EPI_ISL_1803521, EPI_ISL_1803522, EPI_ISL_1803523, EPI_ISL_1803524, EPI_ISL_1803525, EPI_ISL_1803526, EPI_ISL_1803527, EPI_ISL_1803528, EPI_ISL_1803529, EPI_ISL_1803530, EPI_ISL_1803531, EPI_ISL_1803532, EPI_ISL_1803533, EPI_ISL_1803534, EPI_ISL_1803535, EPI_ISL_1803536, EPI_ISL_1803537, EPI_ISL_1803538, EPI_ISL_1803539, EPI_ISL_1803540, EPI_ISL_1803541, EPI_ISL_1803542, EPI_ISL_1803543, EPI_ISL_1803544, EPI_ISL_1803545, EPI_ISL_1803546, EPI_ISL_1803547, EPI_ISL_1803548, EPI_ISL_1803549, EPI_ISL_1803550, EPI_ISL_1803551, EPI_ISL_1803552, EPI_ISL_1803553, EPI_ISL_1803554, EPI_ISL_1803555, EPI_ISL_1803556, EPI_ISL_1803557, EPI_ISL_1803558, EPI_ISL_1803559, EPI_ISL_1803560, EPI_ISL_1803561, EPI_ISL_1803562, EPI_ISL_1803563, EPI_ISL_1803564, EPI_ISL_1803565, EPI_ISL_1803566, EPI_ISL_1803567, EPI_ISL_1803568, EPI_ISL_1803569, EPI_ISL_1803570, EPI_ISL_1803571, EPI_ISL_1803572, EPI_ISL_1803573, EPI_ISL_1803574, EPI_ISL_1803575, EPI_ISL_1803576, EPI_ISL_1803577, EPI_ISL_1803578, EPI_ISL_1803579, EPI_ISL_1803580, EPI_ISL_1803581, EPI_ISL_1803582, EPI_ISL_1803583, EPI_ISL_1803584, EPI_ISL_1803585, EPI_ISL_1803586, EPI_ISL_1803587, EPI_ISL_1803588, EPI_ISL_1803589, EPI_ISL_1803590, EPI_ISL_1803591, EPI_ISL_1803592, EPI_ISL_1803593, EPI_ISL_1803594, EPI_ISL_1803595, EPI_ISL_1803596, EPI_ISL_1803597, EPI_ISL_1803598, EPI_ISL_1803599, EPI_ISL_1803600, EPI_ISL_1803601, EPI_ISL_1803602, EPI_ISL_1803603, EPI_ISL_1803604, EPI_ISL_1803605, EPI_ISL_1803606, EPI_ISL_1803607, EPI_ISL_1803608, EPI_ISL_1803609, EPI_ISL_1803610, EPI_ISL_1803611, EPI_ISL_1803612, EPI_ISL_1803613, EPI_ISL_1803614, EPI_ISL_1803615, EPI_ISL_1803616, EPI_ISL_1803617, EPI_ISL_1803618, EPI_ISL_1803619, EPI_ISL_1803620, EPI_ISL_1803621, EPI_ISL_1803622, EPI_ISL_1803623, EPI_ISL_1803624, EPI_ISL_1803625, EPI_ISL_1803626, EPI_ISL_1803627, EPI_ISL_1803628, EPI_ISL_1803629, EPI_ISL_1803630, EPI_ISL_1803631, EPI_ISL_1803632, EPI_ISL_1803633, EPI_ISL_1803634, EPI_ISL_1803635, EPI_ISL_1803636, EPI_ISL_1803637, EPI_ISL_1803638, EPI_ISL_1803639, EPI_ISL_1803640, EPI_ISL_1803641, EPI_ISL_1803642, EPI_ISL_1803643, EPI_ISL_1803644, EPI_ISL_1803645, EPI_ISL_1803646, EPI_ISL_1803647, EPI_ISL_1803648, EPI_ISL_1803649, EPI_ISL_1803650, EPI_ISL_1803651, EPI_ISL_1803652, EPI_ISL_1803653, EPI_ISL_1803654, EPI_ISL_1803655, EPI_ISL_1803656, EPI_ISL_1803657, EPI_ISL_1803658, EPI_ISL_1803659, EPI_ISL_1803660, EPI_ISL_1803661, EPI_ISL_1803662, EPI_ISL_1803663, EPI_ISL_1803664, EPI_ISL_1803665, EPI_ISL_1803666, EPI_ISL_1803667, EPI_ISL_1803668, EPI_ISL_1803669, EPI_ISL_1803670, EPI_ISL_1803671, EPI_ISL_1803672, EPI_ISL_1803673, EPI_ISL_1803674, EPI_ISL_1803675, EPI_ISL_1803676, EPI_ISL_1803677, EPI_ISL_1803678, EPI_ISL_1803679, EPI_ISL_1803680, EPI_ISL_1803681, EPI_ISL_1803682, EPI_ISL_1803683, EPI_ISL_1803684, EPI_ISL_1803685, EPI_ISL_1803686, EPI_ISL_1803687, EPI_ISL_1803688, EPI_ISL_1803689, EPI_ISL_1803690, EPI_ISL_1803691, EPI_ISL_1803692, EPI_ISL_1803693, EPI_ISL_1803694, EPI_ISL_1803695, EPI_ISL_1803696, EPI_ISL_1803697, EPI_ISL_1803698, EPI_ISL_1803699, EPI_ISL_1803700, EPI_ISL_1803701, EPI_ISL_1803702, EPI_ISL_1803703, EPI_ISL_1803704, EPI_ISL_1803705, EPI_ISL_1803706, EPI_ISL_1803707, EPI_ISL_1803708, EPI_ISL_1803709, EPI_ISL_1803710, EPI_ISL_1803712, EPI_ISL_1803713, EPI_ISL_1803714, EPI_ISL_1803715, EPI_ISL_1803716, EPI_ISL_1803717, EPI_ISL_1803718, EPI_ISL_1803719, EPI_ISL_1803720, EPI_ISL_1803721, EPI_ISL_1803722, EPI_ISL_1803723, EPI_ISL_1803724, EPI_ISL_1803725, EPI_ISL_1803726, EPI_ISL_1803727, EPI_ISL_1803728, EPI_ISL_1803729, EPI_ISL_1803730, EPI_ISL_1803731, EPI_ISL_1803732, EPI_ISL_1803733, EPI_ISL_1803734, EPI_ISL_1803735, EPI_ISL_1803736, EPI_ISL_1803737, EPI_ISL_1803738, EPI_ISL_1803739, EPI_ISL_1803740, EPI_ISL_1803741, EPI_ISL_1803742, EPI_ISL_1803743, EPI_ISL_1803744, EPI_ISL_1803745, EPI_ISL_1803746, EPI_ISL_1803747, EPI_ISL_1803748, EPI_ISL_1803749, EPI_ISL_1803750, EPI_ISL_1803751, EPI_ISL_1803752, EPI_ISL_1803753, EPI_ISL_1803754, EPI_ISL_1803755, EPI_ISL_1803756, EPI_ISL_1803757, EPI_ISL_1803758, EPI_ISL_1803759, EPI_ISL_1803760, EPI_ISL_1803761, EPI_ISL_1803762, EPI_ISL_1803763, EPI_ISL_1803764, EPI_ISL_1803765, EPI_ISL_1803766, EPI_ISL_1803767, EPI_ISL_1803768, EPI_ISL_1803769, EPI_ISL_1803770, EPI_ISL_1803771, EPI_ISL_1803772, EPI_ISL_1803773, EPI_ISL_1803774, EPI_ISL_1803775, EPI_ISL_1803776, EPI_ISL_1803777, EPI_ISL_1803778, EPI_ISL_1803779, EPI_ISL_1803780, EPI_ISL_1803781, EPI_ISL_1803782, EPI_ISL_1803783, EPI_ISL_1803784, EPI_ISL_1803785, EPI_ISL_1803786, EPI_ISL_1803787, EPI_ISL_1803788, EPI_ISL_1803789, EPI_ISL_1803790, EPI_ISL_1803791, EPI_ISL_1803792, EPI_ISL_1803793, EPI_ISL_1803795 | | | |
| see above | Helix/Illumina | Centers for Disease Control and Prevention Division of Viral Diseases, Pathogen Discovery | Dakota Howard, Dhwani Batra, Peter W. Cook, Kara Moser, Adrian Paskey, Jason Caravas, Benjamin Rambo-Martin, Shatavia Morrison, Christopher Gulvick, Scott Sammons, Yvette Unoarumhi, Darlene Wagner, Matthew Schmerer, Eileen de Feo, Jan Antico, Christine Tran, Matthew Tolentino, Shannon Wickline, Kim Gietzen, Brad Sickler, Jingtao Liu, Eric Allen, Phil Febbo, Nicole L. Washington, Simon White, Geraint Levan, Kelly Schiabor Barrett, Elizabeth Cirulli, Alexandre Bolze, Ary Ascencio, Charlotte Rivera-Garcia, Ryan Cho, Jason Nguyen, Sherry Wang, Jimmy Ramirez, Tyler Cassens, Efren Sandoval, Magnus Isaksson, William Lee, David Becker, Marc Laurent, James Lu, Clinton R. Paden, Duncan MacCannell |
| EPI_ISL_1803796, EPI_ISL_1803797 | University of Michigan Clinical Microbiology Laboratory | Lauring Lab, University of Michigan, Department of Microbiology and Immunology | Valesano |
| EPI_ISL_1803798, EPI_ISL_1803799, EPI_ISL_1803800, EPI_ISL_1803801, EPI_ISL_1803802, EPI_ISL_1803803, EPI_ISL_1803804, EPI_ISL_1803805, EPI_ISL_1803806, EPI_ISL_1803807, EPI_ISL_1803808, EPI_ISL_1803810, EPI_ISL_1803811, EPI_ISL_1803812, EPI_ISL_1803813, EPI_ISL_1803814, EPI_ISL_1803815, EPI_ISL_1803816, EPI_ISL_1803817, EPI_ISL_1803818, EPI_ISL_1803819, EPI_ISL_1803820, EPI_ISL_1803821, EPI_ISL_1803822, EPI_ISL_1803823, EPI_ISL_1803824, EPI_ISL_1803825, EPI_ISL_1803826, EPI_ISL_1803827, EPI_ISL_1803828, EPI_ISL_1803829, EPI_ISL_1803830, EPI_ISL_1803831, EPI_ISL_1803832, EPI_ISL_1803833, EPI_ISL_1803834, EPI_ISL_1803835, EPI_ISL_1803836, EPI_ISL_1803837, EPI_ISL_1803838, EPI_ISL_1803839, EPI_ISL_1803840, EPI_ISL_1803841, EPI_ISL_1803842, EPI_ISL_1803843, EPI_ISL_1803844, EPI_ISL_1803845, EPI_ISL_1803846, EPI_ISL_1803847, EPI_ISL_1803848, EPI_ISL_1803849, EPI_ISL_1803850, EPI_ISL_1803851, EPI_ISL_1803852, EPI_ISL_1803853, EPI_ISL_1803854, EPI_ISL_1803855, EPI_ISL_1803856, EPI_ISL_1803857, EPI_ISL_1803858, EPI_ISL_1803859, EPI_ISL_1803860, EPI_ISL_1803861, EPI_ISL_1803862, EPI_ISL_1803863, EPI_ISL_1803864, EPI_ISL_1803865, EPI_ISL_1803866, EPI_ISL_1803867, EPI_ISL_1803868, EPI_ISL_1803869, EPI_ISL_1803871, EPI_ISL_1803872, EPI_ISL_1803873, EPI_ISL_1803874, EPI_ISL_1803875, EPI_ISL_1803876, EPI_ISL_1803877, EPI_ISL_1803878, EPI_ISL_1803879, EPI_ISL_1803880, EPI_ISL_1803881, EPI_ISL_1803882, EPI_ISL_1803883, EPI_ISL_1803884, EPI_ISL_1803885, EPI_ISL_1803886, EPI_ISL_1803887, EPI_ISL_1803888, EPI_ISL_1803889, EPI_ISL_1803890, EPI_ISL_1803891, EPI_ISL_1803892, EPI_ISL_1803893, EPI_ISL_1803894, EPI_ISL_1803895, EPI_ISL_1803896, EPI_ISL_1803897, EPI_ISL_1803898, EPI_ISL_1803899, EPI_ISL_1803900, EPI_ISL_1803901, EPI_ISL_1803902, EPI_ISL_1803903, EPI_ISL_1803904, EPI_ISL_1803905, EPI_ISL_1803906, EPI_ISL_1803907, EPI_ISL_1803908, EPI_ISL_1803909, EPI_ISL_1803910, EPI_ISL_1803911, EPI_ISL_1803912, EPI_ISL_1803913, EPI_ISL_1803914, EPI_ISL_1803915, EPI_ISL_1803916, EPI_ISL_1803917, EPI_ISL_1803918, EPI_ISL_1803919, EPI_ISL_1803920, EPI_ISL_1803921, EPI_ISL_1803922, EPI_ISL_1803923, EPI_ISL_1803924, EPI_ISL_1803925, EPI_ISL_1803926, EPI_ISL_1803927, EPI_ISL_1803928, EPI_ISL_1803929, EPI_ISL_1803930, EPI_ISL_1803931, EPI_ISL_1803932, EPI_ISL_1803933, EPI_ISL_1803934, EPI_ISL_1803935, EPI_ISL_1803936, EPI_ISL_1803937, EPI_ISL_1803938, EPI_ISL_1803939, EPI_ISL_1803940, EPI_ISL_1803941, EPI_ISL_1803942, EPI_ISL_1803943, EPI_ISL_1803944, EPI_ISL_1803945, EPI_ISL_1803946, EPI_ISL_1803947, EPI_ISL_1803948, EPI_ISL_1803949, EPI_ISL_1803950, EPI_ISL_1803951, EPI_ISL_1803952, EPI_ISL_1803953, EPI_ISL_1803954, EPI_ISL_1803955, EPI_ISL_1803956, EPI_ISL_1803957, EPI_ISL_1803958, EPI_ISL_1803959, EPI_ISL_1803960, EPI_ISL_1803961, EPI_ISL_1803962, EPI_ISL_1803963, EPI_ISL_1803964, EPI_ISL_1803965, EPI_ISL_1803966, EPI_ISL_1803967, EPI_ISL_1803968, EPI_ISL_1803969, EPI_ISL_1803970, EPI_ISL_1803971, EPI_ISL_1803972, EPI_ISL_1803973, EPI_ISL_1803974, EPI_ISL_1803975, EPI_ISL_1803976, EPI_ISL_1803977, EPI_ISL_1803978, EPI_ISL_1803979, EPI_ISL_1803980, EPI_ISL_1803981, EPI_ISL_1803982, EPI_ISL_1803983, EPI_ISL_1803984, EPI_ISL_1803985, EPI_ISL_1803986, EPI_ISL_1803987, EPI_ISL_1803988, EPI_ISL_1803989, EPI_ISL_1803990, EPI_ISL_1803991, EPI_ISL_1803992, EPI_ISL_1803993, EPI_ISL_1803994, EPI_ISL_1803995, EPI_ISL_1803996, EPI_ISL_1803997, EPI_ISL_1803998, EPI_ISL_1803999, EPI_ISL_1804000, EPI_ISL_1804001, EPI_ISL_1804002, EPI_ISL_1804003, EPI_ISL_1804004, EPI_ISL_1804005, EPI_ISL_1804006, EPI_ISL_1804007, EPI_ISL_1804008, EPI_ISL_1804009, EPI_ISL_1804010, EPI_ISL_1804011, EPI_ISL_1804012, EPI_ISL_1804013, EPI_ISL_1804014, EPI_ISL_1804015, EPI_ISL_1804016, EPI_ISL_1804017, EPI_ISL_1804018, EPI_ISL_1804019, EPI_ISL_1804020, EPI_ISL_1804021, EPI_ISL_1804022, EPI_ISL_1804023, EPI_ISL_1804024, EPI_ISL_1804025, EPI_ISL_1804026, EPI_ISL_1804027, EPI_ISL_1804028, EPI_ISL_1804029, EPI_ISL_1804030, EPI_ISL_1804031, EPI_ISL_1804032, EPI_ISL_1804033, EPI_ISL_1804034, EPI_ISL_1804035, EPI_ISL_1804036, EPI_ISL_1804037, EPI_ISL_1804038, EPI_ISL_1804039, EPI_ISL_1804040, EPI_ISL_1804041, EPI_ISL_1804042, EPI_ISL_1804043, EPI_ISL_1804044, EPI_ISL_1804045, EPI_ISL_1804046, EPI_ISL_1804047, EPI_ISL_1804048, EPI_ISL_1804049, EPI_ISL_1804050, EPI_ISL_1804051, EPI_ISL_1804052, EPI_ISL_1804053, EPI_ISL_1804054, EPI_ISL_1804055, EPI_ISL_1804056, EPI_ISL_1804057, EPI_ISL_1804058, EPI_ISL_1804059, EPI_ISL_1804060, EPI_ISL_1804061, EPI_ISL_1804062, EPI_ISL_1804063, EPI_ISL_1804064, EPI_ISL_1804065, EPI_ISL_1804066, EPI_ISL_1804067, EPI_ISL_1804068, EPI_ISL_1804069, EPI_ISL_1804070, EPI_ISL_1804071, EPI_ISL_1804072, EPI_ISL_1804073, EPI_ISL_1804074, EPI_ISL_1804075, EPI_ISL_1804076, EPI_ISL_1804077, EPI_ISL_1804078, EPI_ISL_1804079, EPI_ISL_1804080, EPI_ISL_1804081, EPI_ISL_1804082, EPI_ISL_1804084, EPI_ISL_1804085, EPI_ISL_1804086, EPI_ISL_1804087, EPI_ISL_1804088, EPI_ISL_1804089, EPI_ISL_1804090, EPI_ISL_1804091, EPI_ISL_1804092, EPI_ISL_1804093, EPI_ISL_1804095, EPI_ISL_1804096, EPI_ISL_1804097, EPI_ISL_1804098, EPI_ISL_1804099, EPI_ISL_1804100, EPI_ISL_1804101, EPI_ISL_1804102, EPI_ISL_1804103, EPI_ISL_1804104, EPI_ISL_1804105, EPI_ISL_1804106, EPI_ISL_1804107, EPI_ISL_1804108, EPI_ISL_1804109, EPI_ISL_1804110, EPI_ISL_1804111, EPI_ISL_1804112, EPI_ISL_1804113, EPI_ISL_1804114, EPI_ISL_1804115, EPI_ISL_1804116, EPI_ISL_1804117, EPI_ISL_1804118, EPI_ISL_1804119, EPI_ISL_1804120, EPI_ISL_1804121, EPI_ISL_1804122, EPI_ISL_1804123, EPI_ISL_1804124, EPI_ISL_1804125, EPI_ISL_1804126, EPI_ISL_1804127, EPI_ISL_1804128, EPI_ISL_1804129, EPI_ISL_1804130, EPI_ISL_1804131, EPI_ISL_1804132, EPI_ISL_1804133, EPI_ISL_1804134, EPI_ISL_1804135, EPI_ISL_1804136, EPI_ISL_1804137, EPI_ISL_1804138, EPI_ISL_1804139, EPI_ISL_1804141, EPI_ISL_1804142, EPI_ISL_1804143, EPI_ISL_1804144, EPI_ISL_1804145, EPI_ISL_1804146, EPI_ISL_1804147, EPI_ISL_1804148, EPI_ISL_1804149, EPI_ISL_1804150, EPI_ISL_1804151, EPI_ISL_1804152, EPI_ISL_1804153, EPI_ISL_1804154, EPI_ISL_1804155, EPI_ISL_1804156, EPI_ISL_1804157, EPI_ISL_1804158, EPI_ISL_1804159, EPI_ISL_1804160, EPI_ISL_1804161, EPI_ISL_1804162, EPI_ISL_1804163, EPI_ISL_1804164, EPI_ISL_1804165, EPI_ISL_1804166, EPI_ISL_1804167, EPI_ISL_1804168, EPI_ISL_1804169, EPI_ISL_1804170, EPI_ISL_1804171, EPI_ISL_1804172, EPI_ISL_1804173, EPI_ISL_1804174, EPI_ISL_1804175, EPI_ISL_1804176, EPI_ISL_1804177, EPI_ISL_1804178, EPI_ISL_1804179, EPI_ISL_1804180, EPI_ISL_1804181, EPI_ISL_1804182, EPI_ISL_1804183, EPI_ISL_1804184, EPI_ISL_1804185, EPI_ISL_1804186, EPI_ISL_1804187, EPI_ISL_1804188, EPI_ISL_1804189, EPI_ISL_1804190, EPI_ISL_1804191, EPI_ISL_1804192, EPI_ISL_1804193, EPI_ISL_1804194, EPI_ISL_1804195, EPI_ISL_1804196, EPI_ISL_1804197, EPI_ISL_1804198, EPI_ISL_1804199, EPI_ISL_1804200, EPI_ISL_1804201, EPI_ISL_1804202, EPI_ISL_1804203, EPI_ISL_1804204, EPI_ISL_1804205, EPI_ISL_1804206, EPI_ISL_1804207, EPI_ISL_1804208, EPI_ISL_1804209, EPI_ISL_1804210, EPI_ISL_1804211, EPI_ISL_1804212, EPI_ISL_1804213, EPI_ISL_1804214, EPI_ISL_1804215, EPI_ISL_1804216, EPI_ISL_1804217, EPI_ISL_1804218, EPI_ISL_1804219 | | | |
| see above | Helix/Illumina | Centers for Disease Control and Prevention Division of Viral Diseases, Pathogen Discovery | Dakota Howard, Dhwani Batra, Peter W. Cook, Kara Moser, Adrian Paskey, Jason Caravas, Benjamin Rambo-Martin, Shatavia Morrison, Christopher Gulvick, Scott Sammons, Yvette Unoarumhi, Darlene Wagner, Matthew Schmerer, Eileen de Feo, Jan Antico, Christine Tran, Matthew Tolentino, Shannon Wickline, Kim Gietzen, Brad Sickler, Jingtao Liu, Eric Allen, Phil Febbo, Nicole L. Washington, Simon White, Geraint Levan, Kelly Schiabor Barrett, Elizabeth Cirulli, Alexandre Bolze, Ary Ascencio, Charlotte Rivera-Garcia, Ryan Cho, Jason Nguyen, Sherry Wang, Jimmy Ramirez, Tyler Cassens, Efren Sandoval, Magnus Isaksson, William Lee, David Becker, Marc Laurent, James Lu, Clinton R. Paden, Duncan MacCannell |
| EPI_ISL_1804220 | MAWD Pathology Group, PA | MAWD Pathology Group, PA | Pradip Manna, Ph D, Kelli Fayard, Shelby Herner, London Collins, Kelsi Eastman |
| EPI_ISL_1804221, EPI_ISL_1804222, EPI_ISL_1804223, EPI_ISL_1804224, EPI_ISL_1804225, EPI_ISL_1804226, EPI_ISL_1804227, EPI_ISL_1804228, EPI_ISL_1804229, EPI_ISL_1804230, EPI_ISL_1804231, EPI_ISL_1804232, EPI_ISL_1804233, EPI_ISL_1804234, EPI_ISL_1804235, EPI_ISL_1804236, EPI_ISL_1804237, EPI_ISL_1804238, EPI_ISL_1804239, EPI_ISL_1804240, EPI_ISL_1804241, EPI_ISL_1804242, EPI_ISL_1804243, EPI_ISL_1804244, EPI_ISL_1804245, EPI_ISL_1804246, EPI_ISL_1804247, EPI_ISL_1804248, EPI_ISL_1804249, EPI_ISL_1804250, EPI_ISL_1804251, EPI_ISL_1804252, EPI_ISL_1804253, EPI_ISL_1804254, EPI_ISL_1804255, EPI_ISL_1804256, EPI_ISL_1804257, EPI_ISL_1804258, EPI_ISL_1804259, EPI_ISL_1804260, EPI_ISL_1804261, EPI_ISL_1804262, EPI_ISL_1804263, EPI_ISL_1804264, EPI_ISL_1804265, EPI_ISL_1804266, EPI_ISL_1804267, EPI_ISL_1804268, EPI_ISL_1804269, EPI_ISL_1804270, EPI_ISL_1804271, EPI_ISL_1804272, EPI_ISL_1804273, EPI_ISL_1804274, EPI_ISL_1804275, EPI_ISL_1804276, EPI_ISL_1804277, EPI_ISL_1804278, EPI_ISL_1804279, EPI_ISL_1804280, EPI_ISL_1804281, EPI_ISL_1804282, EPI_ISL_1804283, EPI_ISL_1804284, EPI_ISL_1804285, EPI_ISL_1804286, EPI_ISL_1804287, EPI_ISL_1804288, EPI_ISL_1804289, EPI_ISL_1804290, EPI_ISL_1804291, EPI_ISL_1804292, EPI_ISL_1804293, EPI_ISL_1804294, EPI_ISL_1804295, EPI_ISL_1804296, EPI_ISL_1804297, EPI_ISL_1804298, EPI_ISL_1804299, EPI_ISL_1804300, EPI_ISL_1804301, EPI_ISL_1804302, EPI_ISL_1804303, EPI_ISL_1804304, EPI_ISL_1804305, EPI_ISL_1804306, EPI_ISL_1804307, EPI_ISL_1804308, EPI_ISL_1804309, EPI_ISL_1804310, EPI_ISL_1804311, EPI_ISL_1804312, EPI_ISL_1804313, EPI_ISL_1804314, EPI_ISL_1804315, EPI_ISL_1804316, EPI_ISL_1804317, EPI_ISL_1804318, EPI_ISL_1804319, EPI_ISL_1804320, EPI_ISL_1804321, EPI_ISL_1804322, EPI_ISL_1804323, EPI_ISL_1804324, EPI_ISL_1804325, EPI_ISL_1804326, EPI_ISL_1804327, EPI_ISL_1804328, EPI_ISL_1804329, EPI_ISL_1804330, EPI_ISL_1804331, | | | |

| | | | |
|---|---|---|---|
| EPI_ISL_1804332, EPI_ISL_1804333, EPI_ISL_1804334, EPI_ISL_1804335, EPI_ISL_1804336, EPI_ISL_1804337, EPI_ISL_1804338, EPI_ISL_1804339, EPI_ISL_1804340, EPI_ISL_1804341, EPI_ISL_1804342, EPI_ISL_1804343, EPI_ISL_1804344, EPI_ISL_1804345, EPI_ISL_1804346, EPI_ISL_1804347, EPI_ISL_1804348, EPI_ISL_1804349, EPI_ISL_1804350, EPI_ISL_1804351, EPI_ISL_1804352, EPI_ISL_1804353, EPI_ISL_1804354, EPI_ISL_1804355, EPI_ISL_1804356, EPI_ISL_1804357, EPI_ISL_1804358, EPI_ISL_1804359, EPI_ISL_1804360, EPI_ISL_1804361, EPI_ISL_1804362, EPI_ISL_1804363, EPI_ISL_1804364, EPI_ISL_1804365, EPI_ISL_1804366, EPI_ISL_1804367, EPI_ISL_1804368, EPI_ISL_1804369, EPI_ISL_1804370, EPI_ISL_1804371, EPI_ISL_1804372, EPI_ISL_1804373, EPI_ISL_1804374, EPI_ISL_1804375, EPI_ISL_1804376, EPI_ISL_1804377, EPI_ISL_1804378, EPI_ISL_1804379, EPI_ISL_1804380, EPI_ISL_1804381, EPI_ISL_1804382, EPI_ISL_1804383, EPI_ISL_1804384, EPI_ISL_1804385, EPI_ISL_1804386, EPI_ISL_1804387, EPI_ISL_1804388, EPI_ISL_1804389, EPI_ISL_1804390, EPI_ISL_1804391, EPI_ISL_1804392, EPI_ISL_1804393, EPI_ISL_1804394, EPI_ISL_1804395, EPI_ISL_1804396, EPI_ISL_1804397, EPI_ISL_1804398, EPI_ISL_1804399, EPI_ISL_1804400, EPI_ISL_1804401, EPI_ISL_1804402, EPI_ISL_1804403, EPI_ISL_1804404, EPI_ISL_1804405, EPI_ISL_1804406, EPI_ISL_1804407, EPI_ISL_1804408, EPI_ISL_1804409, EPI_ISL_1804410 | | | |
| see above | Helix/Illumina | Centers for Disease Control and Prevention Division of Viral Diseases, Pathogen Discovery | Dakota Howard, Dhwani Batra, Peter W. Cook, Kara Moser, Adrian Paskey, Jason Caravas, Benjamin Rambo-Martin, Shatavia Morrison, Christopher Gulvick, Scott Sammons, Yvette Unoarumhi, Darlene Wagner, Matthew Schmerer, Eileen de Feo, Jan Antico, Christine Tran, Matthew Tolentino, Shannon Wickline, Kim Gietzen, Brad Sickler, Jingtao Liu, Eric Allen, Phil Febbo, Nicole L. Washington, Simon White, Geraint Levan, Kelly Schiabor Barrett, Elizabeth Cirulli, Alexandre Bolze, Ary Ascencio, Charlotte Rivera-Garcia, Ryan Cho, Jason Nguyen, Sherry Wang, Jimmy Ramirez, Tyler Cassens, Efren Sandoval, Magnus Isaksson, William Lee, David Becker, Marc Laurent, James Lu, Clinton R. Paden, Duncan MacCannell |
| EPI_ISL_1804411 | MAWD Pathology Group, PA | MAWD Pathology Group, PA | Pradip Manna, Ph D, Kelli Fayard, Shelby Herner, London Collins, Kelsi Eastman |
| EPI_ISL_1804412, EPI_ISL_1804413, EPI_ISL_1804414, EPI_ISL_1804415, EPI_ISL_1804416, EPI_ISL_1804417, EPI_ISL_1804418, EPI_ISL_1804419, EPI_ISL_1804420, EPI_ISL_1804421, EPI_ISL_1804422, EPI_ISL_1804423, EPI_ISL_1804425, EPI_ISL_1804426, EPI_ISL_1804427, EPI_ISL_1804428, EPI_ISL_1804429, EPI_ISL_1804430, EPI_ISL_1804431, EPI_ISL_1804432, EPI_ISL_1804433, EPI_ISL_1804434, EPI_ISL_1804435, EPI_ISL_1804436, EPI_ISL_1804437, EPI_ISL_1804438, EPI_ISL_1804439, EPI_ISL_1804440, EPI_ISL_1804441, EPI_ISL_1804442, EPI_ISL_1804443, EPI_ISL_1804444, EPI_ISL_1804445, EPI_ISL_1804447, EPI_ISL_1804448, EPI_ISL_1804449, EPI_ISL_1804450, EPI_ISL_1804451, EPI_ISL_1804452, EPI_ISL_1804453, EPI_ISL_1804454, EPI_ISL_1804455, EPI_ISL_1804456, EPI_ISL_1804457, EPI_ISL_1804458, EPI_ISL_1804459, EPI_ISL_1804460, EPI_ISL_1804461, EPI_ISL_1804462, EPI_ISL_1804463, EPI_ISL_1804464, EPI_ISL_1804465, EPI_ISL_1804466, EPI_ISL_1804467, EPI_ISL_1804468, EPI_ISL_1804469, EPI_ISL_1804470, EPI_ISL_1804471, EPI_ISL_1804472, EPI_ISL_1804473, EPI_ISL_1804474, EPI_ISL_1804475, EPI_ISL_1804476, EPI_ISL_1804477, EPI_ISL_1804478, EPI_ISL_1804479, EPI_ISL_1804480, EPI_ISL_1804481, EPI_ISL_1804482, EPI_ISL_1804483, EPI_ISL_1804484, EPI_ISL_1804485, EPI_ISL_1804486, EPI_ISL_1804487, EPI_ISL_1804488, EPI_ISL_1804489, EPI_ISL_1804490, EPI_ISL_1804491, EPI_ISL_1804492, EPI_ISL_1804493, EPI_ISL_1804494, EPI_ISL_1804495, EPI_ISL_1804496, EPI_ISL_1804497, EPI_ISL_1804498, EPI_ISL_1804499, EPI_ISL_1804500, EPI_ISL_1804501, EPI_ISL_1804502, EPI_ISL_1804503, EPI_ISL_1804504, EPI_ISL_1804505, EPI_ISL_1804506, EPI_ISL_1804507, EPI_ISL_1804508, EPI_ISL_1804509, EPI_ISL_1804510, EPI_ISL_1804511, EPI_ISL_1804512, EPI_ISL_1804513, EPI_ISL_1804514, EPI_ISL_1804515, EPI_ISL_1804516, EPI_ISL_1804517, EPI_ISL_1804518, EPI_ISL_1804519, EPI_ISL_1804520, EPI_ISL_1804521, EPI_ISL_1804522, EPI_ISL_1804523, EPI_ISL_1804524, EPI_ISL_1804525, EPI_ISL_1804526, EPI_ISL_1804527, EPI_ISL_1804528, EPI_ISL_1804529, EPI_ISL_1804530, EPI_ISL_1804531, EPI_ISL_1804532, EPI_ISL_1804533, EPI_ISL_1804534, EPI_ISL_1804535, EPI_ISL_1804536, EPI_ISL_1804537, EPI_ISL_1804538, EPI_ISL_1804539, EPI_ISL_1804540, EPI_ISL_1804541, EPI_ISL_1804542, EPI_ISL_1804543, EPI_ISL_1804544, EPI_ISL_1804545, EPI_ISL_1804546, EPI_ISL_1804547, EPI_ISL_1804548, EPI_ISL_1804549, EPI_ISL_1804550, EPI_ISL_1804551, EPI_ISL_1804552, EPI_ISL_1804553, EPI_ISL_1804554, EPI_ISL_1804555, EPI_ISL_1804556, EPI_ISL_1804557, EPI_ISL_1804558, EPI_ISL_1804559, EPI_ISL_1804560, EPI_ISL_1804561, EPI_ISL_1804562, EPI_ISL_1804563, EPI_ISL_1804564, EPI_ISL_1804565, EPI_ISL_1804566, EPI_ISL_1804567, EPI_ISL_1804568, EPI_ISL_1804569, EPI_ISL_1804570, EPI_ISL_1804571, EPI_ISL_1804572, EPI_ISL_1804573, EPI_ISL_1804574, EPI_ISL_1804575, EPI_ISL_1804576, EPI_ISL_1804577, EPI_ISL_1804578, EPI_ISL_1804579, EPI_ISL_1804580, EPI_ISL_1804581, EPI_ISL_1804582, EPI_ISL_1804583, EPI_ISL_1804584, EPI_ISL_1804585, EPI_ISL_1804586, EPI_ISL_1804587, EPI_ISL_1804588, EPI_ISL_1804589, EPI_ISL_1804590, EPI_ISL_1804591, EPI_ISL_1804592, EPI_ISL_1804593, EPI_ISL_1804594, EPI_ISL_1804595, EPI_ISL_1804596, EPI_ISL_1804597, EPI_ISL_1804598, EPI_ISL_1804599, EPI_ISL_1804600, EPI_ISL_1804601, EPI_ISL_1804602, EPI_ISL_1804603, EPI_ISL_1804604, EPI_ISL_1804605, EPI_ISL_1804606, EPI_ISL_1804607, EPI_ISL_1804608, EPI_ISL_1804609, EPI_ISL_1804610, EPI_ISL_1804611, EPI_ISL_1804612, EPI_ISL_1804613, EPI_ISL_1804615, EPI_ISL_1804616, EPI_ISL_1804617, EPI_ISL_1804618, EPI_ISL_1804619, EPI_ISL_1804620, EPI_ISL_1804621, EPI_ISL_1804622, EPI_ISL_1804623, EPI_ISL_1804624, EPI_ISL_1804625, EPI_ISL_1804626, EPI_ISL_1804627, EPI_ISL_1804628, EPI_ISL_1804629, EPI_ISL_1804630, EPI_ISL_1804631, EPI_ISL_1804632, EPI_ISL_1804633, EPI_ISL_1804634, EPI_ISL_1804635, EPI_ISL_1804636, EPI_ISL_1804637, EPI_ISL_1804638, EPI_ISL_1804639, EPI_ISL_1804640, EPI_ISL_1804641, EPI_ISL_1804642, EPI_ISL_1804643, EPI_ISL_1804644, EPI_ISL_1804645, EPI_ISL_1804646, EPI_ISL_1804647, EPI_ISL_1804648, EPI_ISL_1804649, EPI_ISL_1804650, EPI_ISL_1804651, EPI_ISL_1804652, EPI_ISL_1804653, EPI_ISL_1804654, EPI_ISL_1804655, EPI_ISL_1804656, EPI_ISL_1804657, EPI_ISL_1804658, EPI_ISL_1804659, EPI_ISL_1804660, EPI_ISL_1804661, EPI_ISL_1804662, EPI_ISL_1804663, EPI_ISL_1804664, EPI_ISL_1804665, EPI_ISL_1804666, EPI_ISL_1804667, EPI_ISL_1804668, EPI_ISL_1804669, EPI_ISL_1804670, EPI_ISL_1804671, EPI_ISL_1804672, EPI_ISL_1804673, EPI_ISL_1804674, EPI_ISL_1804675, EPI_ISL_1804676, EPI_ISL_1804678, EPI_ISL_1804679, EPI_ISL_1804680, EPI_ISL_1804682, EPI_ISL_1804683, EPI_ISL_1804684, EPI_ISL_1804685, EPI_ISL_1804686, EPI_ISL_1804687, EPI_ISL_1804688, EPI_ISL_1804689, EPI_ISL_1804690, EPI_ISL_1804691, EPI_ISL_1804692, EPI_ISL_1804693, EPI_ISL_1804694, EPI_ISL_1804695, EPI_ISL_1804696, EPI_ISL_1804697, EPI_ISL_1804698, EPI_ISL_1804699, EPI_ISL_1804700, EPI_ISL_1804701, EPI_ISL_1804702, EPI_ISL_1804703, EPI_ISL_1804704, EPI_ISL_1804705, EPI_ISL_1804706, EPI_ISL_1804707, EPI_ISL_1804708, EPI_ISL_1804709, EPI_ISL_1804710, EPI_ISL_1804711, EPI_ISL_1804712, EPI_ISL_1804713, EPI_ISL_1804714, EPI_ISL_1804715, EPI_ISL_1804716, EPI_ISL_1804717, EPI_ISL_1804718, EPI_ISL_1804719, EPI_ISL_1804720, EPI_ISL_1804721, EPI_ISL_1804722, EPI_ISL_1804723, EPI_ISL_1804724, EPI_ISL_1804725, EPI_ISL_1804726, EPI_ISL_1804727, EPI_ISL_1804728, EPI_ISL_1804729, EPI_ISL_1804730, EPI_ISL_1804731, EPI_ISL_1804732, EPI_ISL_1804733, EPI_ISL_1804734, EPI_ISL_1804735, EPI_ISL_1804736, EPI_ISL_1804737, EPI_ISL_1804738, EPI_ISL_1804739, EPI_ISL_1804740, EPI_ISL_1804741, EPI_ISL_1804742, EPI_ISL_1804743, EPI_ISL_1804744, EPI_ISL_1804745, EPI_ISL_1804746, EPI_ISL_1804747, EPI_ISL_1804748, EPI_ISL_1804749, EPI_ISL_1804750, EPI_ISL_1804751, EPI_ISL_1804752, EPI_ISL_1804753, EPI_ISL_1804754, EPI_ISL_1804755, EPI_ISL_1804756, EPI_ISL_1804757, EPI_ISL_1804758, EPI_ISL_1804759, EPI_ISL_1804760, EPI_ISL_1804761, EPI_ISL_1804762, EPI_ISL_1804763, EPI_ISL_1804764, EPI_ISL_1804765, EPI_ISL_1804766, EPI_ISL_1804767, EPI_ISL_1804768, EPI_ISL_1804769, EPI_ISL_1804770, EPI_ISL_1804771, EPI_ISL_1804772, EPI_ISL_1804773, EPI_ISL_1804774, EPI_ISL_1804775, EPI_ISL_1804776, EPI_ISL_1804777, EPI_ISL_1804778, EPI_ISL_1804779, EPI_ISL_1804780, EPI_ISL_1804781, EPI_ISL_1804782, EPI_ISL_1804783, EPI_ISL_1804784, EPI_ISL_1804785, EPI_ISL_1804786, EPI_ISL_1804787, EPI_ISL_1804788, EPI_ISL_1804789, EPI_ISL_1804790, EPI_ISL_1804791, EPI_ISL_1804792, EPI_ISL_1804793, EPI_ISL_1804794, EPI_ISL_1804795, EPI_ISL_1804796, EPI_ISL_1804797, EPI_ISL_1804798, EPI_ISL_1804799, EPI_ISL_1804800, EPI_ISL_1804801, EPI_ISL_1804802, EPI_ISL_1804803, EPI_ISL_1804804, EPI_ISL_1804805, EPI_ISL_1804806, EPI_ISL_1804807, EPI_ISL_1804808, EPI_ISL_1804809, EPI_ISL_1804810, EPI_ISL_1804811 | | | |
| see above | Helix/Illumina | Centers for Disease Control and Prevention Division of Viral Diseases, Pathogen Discovery | Dakota Howard, Dhwani Batra, Peter W. Cook, Kara Moser, Adrian Paskey, Jason Caravas, Benjamin Rambo-Martin, Shatavia Morrison, Christopher Gulvick, Scott Sammons, Yvette Unoarumhi, Darlene Wagner, Matthew Schmerer, Eileen de Feo, Jan Antico, Christine Tran, Matthew Tolentino, Shannon Wickline, Kim Gietzen, Brad Sickler, Jingtao Liu, Eric Allen, Phil Febbo, Nicole L. Washington, Simon White, Geraint Levan, Kelly Schiabor Barrett, Elizabeth Cirulli, Alexandre Bolze, Ary Ascencio, Charlotte Rivera-Garcia, Ryan Cho, Jason Nguyen, Sherry Wang, Jimmy Ramirez, Tyler Cassens, Efren Sandoval, Magnus Isaksson, William Lee, David Becker, Marc Laurent, James Lu, Clinton R. Paden, Duncan MacCannell |
| EPI_ISL_1804812, EPI_ISL_1804813, EPI_ISL_1804814, EPI_ISL_1804815, EPI_ISL_1804816, EPI_ISL_1804817, EPI_ISL_1804818, EPI_ISL_1804819, EPI_ISL_1804820 | Quest Diagnostics Incorporated | Centers for Disease Control and Prevention Division of Viral Diseases, Pathogen Discovery | Dakota Howard, Dhwani Batra, Peter W. Cook, Kara Moser, Adrian Paskey, Jason Caravas, Benjamin Rambo-Martin, Shatavia Morrison, Christopher Gulvick, Scott Sammons, Yvette Unoarumhi, Darlene Wagner, Matthew Schmerer, S. H. Rosenthal, A. Gerasimova, R. M. Kagan, B. Anderson, M. Hua, Y. Liu, L.E. Bernstein, K.E. Livingston, A. Perez, I. A. Shlyakhter, R. V. Rolando, R. Owen, P. Tanpaiboon, F. Lacbawan, Clinton R. Paden, Duncan MacCannell |
| EPI_ISL_1804941 | Department of Homeless Services | New York City Public Health Laboratory | Jade Wang, et al. |
| EPI_ISL_1804942 | DOHMH Jamaica | New York City Public Health Laboratory | Jade Wang, et al. |
| EPI_ISL_1804943 | DOHMH Corona | New York City Public Health Laboratory | Jade Wang, et al. |
| EPI_ISL_1804944 | DOHMH PHL | New York City Public Health Laboratory | Jade Wang, et al. |
| EPI_ISL_1804945 | DOHMH Central Harlem | New York City Public Health Laboratory | Jade Wang, et al. |
| EPI_ISL_1804946 | DOHMH Chelsea | New York City Public Health Laboratory | Jade Wang, et al. |
| EPI_ISL_1804947 | Department of Homeless Services | New York City Public Health Laboratory | Jade Wang, et al. |
| EPI_ISL_1804948, EPI_ISL_1804949 | OCME Office Of Chief Medical Examiner | New York City Public Health Laboratory | Jade Wang, et al. |
| EPI_ISL_1804950 | DOHMH Chelsea | New York City Public Health Laboratory | Jade Wang, et al. |
| EPI_ISL_1804951 | DOHMH PHL | New York City Public Health Laboratory | Jade Wang, et al. |
| EPI_ISL_1804952, EPI_ISL_1804953 | DOHMH Crown Heights | New York City Public Health Laboratory | Jade Wang, et al. |
| EPI_ISL_1804954, EPI_ISL_1804955 | DOHMH Central Harlem | New York City Public Health Laboratory | Jade Wang, et al. |
| EPI_ISL_1804956 | DOHMH Morrisania | New York City Public Health Laboratory | Jade Wang, et al. |
| EPI_ISL_1804957 | DOHMH Corona | New York City Public Health Laboratory | Jade Wang, et al. |
| EPI_ISL_1804958, EPI_ISL_1804959 | DOHMH Jamaica | New York City Public Health Laboratory | Jade Wang, et al. |
| EPI_ISL_1804960 | DOHMH Corona | New York City Public Health Laboratory | Jade Wang, et al. |
| EPI_ISL_1804961 | OCME Office Of Chief Medical Examiner | New York City Public Health Laboratory | Jade Wang, et al. |
| EPI_ISL_1805057, EPI_ISL_1805058, EPI_ISL_1805059, EPI_ISL_1805060, EPI_ISL_1805061, EPI_ISL_1805062, EPI_ISL_1805063, EPI_ISL_1805064, EPI_ISL_1805065, EPI_ISL_1805066, EPI_ISL_1805067, EPI_ISL_1805068, EPI_ISL_1805069, EPI_ISL_1805070, EPI_ISL_1805071, EPI_ISL_1805072, EPI_ISL_1805073, EPI_ISL_1805074, EPI_ISL_1805075, EPI_ISL_1805076, EPI_ISL_1805077, EPI_ISL_1805078, EPI_ISL_1805079, EPI_ISL_1805080, EPI_ISL_1805081, EPI_ISL_1805082, EPI_ISL_1805083, EPI_ISL_1805084, EPI_ISL_1805085, EPI_ISL_1805086, EPI_ISL_1805087, EPI_ISL_1805088, EPI_ISL_1805089, EPI_ISL_1805090, EPI_ISL_1805091, EPI_ISL_1805092, EPI_ISL_1805093, EPI_ISL_1805094, EPI_ISL_1805095, EPI_ISL_1805096, EPI_ISL_1805097, EPI_ISL_1805098, EPI_ISL_1805099, EPI_ISL_1805100, EPI_ISL_1805101, EPI_ISL_1805102, EPI_ISL_1805103, EPI_ISL_1805104, EPI_ISL_1805105, EPI_ISL_1805106, EPI_ISL_1805107, EPI_ISL_1805108, EPI_ISL_1805109, EPI_ISL_1805110, EPI_ISL_1805111, EPI_ISL_1805112, EPI_ISL_1805113, EPI_ISL_1805114, EPI_ISL_1805115, EPI_ISL_1805116, EPI_ISL_1805117, EPI_ISL_1805118, EPI_ISL_1805119, EPI_ISL_1805120, EPI_ISL_1805121, EPI_ISL_1805122, EPI_ISL_1805123, EPI_ISL_1805124, EPI_ISL_1805125, EPI_ISL_1805126, EPI_ISL_1805127, EPI_ISL_1805128, EPI_ISL_1805129, EPI_ISL_1805130, EPI_ISL_1805132, EPI_ISL_1805133, EPI_ISL_1805134, EPI_ISL_1805135, EPI_ISL_1805136, EPI_ISL_1805137, EPI_ISL_1805138, EPI_ISL_1805139, EPI_ISL_1805140, EPI_ISL_1805141, EPI_ISL_1805142, EPI_ISL_1805143, EPI_ISL_1805144, EPI_ISL_1805145, EPI_ISL_1805146, EPI_ISL_1805147, EPI_ISL_1805148, EPI_ISL_1805149, EPI_ISL_1805150, EPI_ISL_1805151, EPI_ISL_1805152, EPI_ISL_1805153, EPI_ISL_1805154, EPI_ISL_1805155, EPI_ISL_1805156, EPI_ISL_1805157, EPI_ISL_1805158, EPI_ISL_1805159, EPI_ISL_1805160, EPI_ISL_1805161, EPI_ISL_1805162, EPI_ISL_1805163, EPI_ISL_1805164, EPI_ISL_1805165, EPI_ISL_1805166, EPI_ISL_1805167, EPI_ISL_1805168, EPI_ISL_1805169, EPI_ISL_1805170, EPI_ISL_1805171, EPI_ISL_1805172, EPI_ISL_1805173, EPI_ISL_1805174, EPI_ISL_1805175, EPI_ISL_1805176, EPI_ISL_1805177, EPI_ISL_1805178, EPI_ISL_1805179, EPI_ISL_1805180, EPI_ISL_1805181, EPI_ISL_1805182, EPI_ISL_1805183, EPI_ISL_1805184, EPI_ISL_1805186, EPI_ISL_1805187, EPI_ISL_1805188, EPI_ISL_1805189, EPI_ISL_1805190, EPI_ISL_1805191, EPI_ISL_1805192, EPI_ISL_1805193, EPI_ISL_1805194, EPI_ISL_1805195, EPI_ISL_1805196, EPI_ISL_1805197, EPI_ISL_1805198, EPI_ISL_1805199, EPI_ISL_1805200, EPI_ISL_1805201, EPI_ISL_1805202, EPI_ISL_1805203, EPI_ISL_1805204, EPI_ISL_1805205, EPI_ISL_1805206, EPI_ISL_1805207, EPI_ISL_1805208, EPI_ISL_1805209, EPI_ISL_1805210, EPI_ISL_1805211, EPI_ISL_1805212, EPI_ISL_1805213, EPI_ISL_1805214, EPI_ISL_1805215, EPI_ISL_1805216, EPI_ISL_1805217, EPI_ISL_1805218, EPI_ISL_1805219, EPI_ISL_1805220, EPI_ISL_1805221, EPI_ISL_1805222, EPI_ISL_1805223, EPI_ISL_1805224, EPI_ISL_1805225, EPI_ISL_1805226, EPI_ISL_1805227, EPI_ISL_1805228, EPI_ISL_1805229, EPI_ISL_1805230, EPI_ISL_1805231, EPI_ISL_1805232, EPI_ISL_1805233, EPI_ISL_1805234, EPI_ISL_1805235, EPI_ISL_1805236, EPI_ISL_1805237, EPI_ISL_1805238, EPI_ISL_1805239, EPI_ISL_1805240, EPI_ISL_1805241, EPI_ISL_1805242, EPI_ISL_1805243, EPI_ISL_1805244 | | | |
| see above | Maryland Genomics, Institute for Genome Sciences, University of Maryland School of Medicine | Maryland Genomics, Institute for Genome Sciences, University of Maryland School of Medicine | Tallon, Luke J; Sadzewicz, Lisa D; Humphrys, Mike; Ott, Sandra; Roussey, Holly; Mehta, Aditya; Vavikolanu, Kranthi; Fraser, Claire M; Ravel, Jacques |
| EPI_ISL_1805245, EPI_ISL_1805246, EPI_ISL_1805247, EPI_ISL_1805248, EPI_ISL_1805249, EPI_ISL_1805250, EPI_ISL_1805251, EPI_ISL_1805252, EPI_ISL_1805253, EPI_ISL_1805254, EPI_ISL_1805255, EPI_ISL_1805256, EPI_ISL_1805257, EPI_ISL_1805258 | | | |
| see above | GA Department of Public Health | GA Department of Public Health | Stacy Reeves, Jonathan Edwards, Cynthia Dixey, Tonia Parrott, Aliyah Fields, Taylor Smith |
| EPI_ISL_1805259, EPI_ISL_1805260, EPI_ISL_1805261, EPI_ISL_1805262, EPI_ISL_1805263, EPI_ISL_1805264, EPI_ISL_1805265, EPI_ISL_1805266, EPI_ISL_1805267, EPI_ISL_1805268, EPI_ISL_1805269, EPI_ISL_1805270, EPI_ISL_1805271, EPI_ISL_1805272, EPI_ISL_1805273, EPI_ISL_1805274, EPI_ISL_1805275, EPI_ISL_1805276, EPI_ISL_1805277, EPI_ISL_1805278, EPI_ISL_1805279, EPI_ISL_1805280, EPI_ISL_1805281, EPI_ISL_1805282, EPI_ISL_1805283, EPI_ISL_1805284, EPI_ISL_1805285, EPI_ISL_1805286, EPI_ISL_1805287, EPI_ISL_1805288, EPI_ISL_1805289, EPI_ISL_1805290, EPI_ISL_1805291, EPI_ISL_1805292, EPI_ISL_1805293, EPI_ISL_1805294, EPI_ISL_1805295, EPI_ISL_1805296, EPI_ISL_1805297, EPI_ISL_1805298, EPI_ISL_1805299, EPI_ISL_1805300, EPI_ISL_1805301, EPI_ISL_1805302, EPI_ISL_1805303, EPI_ISL_1805304, EPI_ISL_1805306, EPI_ISL_1805307, EPI_ISL_1805308, EPI_ISL_1805309, EPI_ISL_1805310, EPI_ISL_1805311, EPI_ISL_1805312, EPI_ISL_1805313, EPI_ISL_1805314, EPI_ISL_1805315, EPI_ISL_1805316, EPI_ISL_1805317, EPI_ISL_1805318, EPI_ISL_1805319 | | | |
| see above | M Health Fairview | Minnesota Department of Health, Public Health Laboratory | Alexandra Lorentz, Jacob Garfin, Matt Plumb, and Xiong Wang |
| EPI_ISL_1805320, EPI_ISL_1805321 | Mayo Clinic & Mayo Clinic Laboratories | Minnesota Department of Health, Public Health Laboratory | Alexandra Lorentz, Jacob Garfin, Matt Plumb, and Xiong Wang |
| EPI_ISL_1805322, EPI_ISL_1805324, EPI_ISL_1805325, EPI_ISL_1805326 | M Health Fairview | Minnesota Department of Health, Public Health Laboratory | Alexandra Lorentz, Jacob Garfin, Matt Plumb, and Xiong Wang |
| EPI_ISL_1805327, EPI_ISL_1805328, EPI_ISL_1805329, EPI_ISL_1805330, EPI_ISL_1805331, EPI_ISL_1805332, EPI_ISL_1805333, EPI_ISL_1805334, EPI_ISL_1805335, EPI_ISL_1805336, EPI_ISL_1805337, EPI_ISL_1805338, EPI_ISL_1805339, EPI_ISL_1805341, EPI_ISL_1805342, EPI_ISL_1805343, EPI_ISL_1805344, EPI_ISL_1805345, EPI_ISL_1805346, EPI_ISL_1805347, EPI_ISL_1805348, EPI_ISL_1805349, EPI_ISL_1805351, EPI_ISL_1805353, EPI_ISL_1805354, EPI_ISL_1805355, EPI_ISL_1805357, EPI_ISL_1805360, EPI_ISL_1805361, EPI_ISL_1805362, EPI_ISL_1805363, EPI_ISL_1805364, EPI_ISL_1805365 | | | |
| see above | Curative | New Mexico Department of Health Scientific Laboratory | Ellie Johnson, D'eldra Malone, Jennifer Benoit, Ratheesh Rajan, Linda Salazar, Anastacia Griego-Fisher |
| EPI_ISL_1805368, EPI_ISL_1805369, EPI_ISL_1805370, EPI_ISL_1805371, EPI_ISL_1805372, EPI_ISL_1805373, EPI_ISL_1805374, EPI_ISL_1805375, EPI_ISL_1805376, EPI_ISL_1805377, EPI_ISL_1805378, EPI_ISL_1805379, EPI_ISL_1805380, EPI_ISL_1805381, EPI_ISL_1805382, EPI_ISL_1805383, EPI_ISL_1805384, EPI_ISL_1805385, EPI_ISL_1805386, EPI_ISL_1805387, EPI_ISL_1805388, EPI_ISL_1805389, EPI_ISL_1805390, EPI_ISL_1805391 | | | |
| see above | New Mexico Department of Health Scientific Laboratory | New Mexico Department of Health Scientific Laboratory | Ellie Johnson, D'eldra Malone, Jennifer Benoit, Ratheesh Rajan, Linda Salazar, Anastacia Griego-Fisher |
| EPI_ISL_1805392 | PHS/IHS Hospital-Jicarilla Service Unit | New Mexico Department of Health Scientific Laboratory | Ellie Johnson, D'eldra Malone, Jennifer Benoit, Ratheesh Rajan, Linda Salazar, Anastacia Griego-Fisher |

| | | | |
|---|---|---|---|
| EPI_ISL_1805393, EPI_ISL_1805394 | New Mexico Department of Health Scientific Laboratory | New Mexico Department of Health Scientific Laboratory | Ellie Johnson, D'eldra Malone, Jennifer Benoit, Ratheesh Rajan, Linda Salazar, Anastacia Griego-Fisher |
| EPI_ISL_1805395 | Holy Cross Hospital | New Mexico Department of Health Scientific Laboratory | Ellie Johnson, D'eldra Malone, Jennifer Benoit, Ratheesh Rajan, Linda Salazar, Anastacia Griego-Fisher |
| EPI_ISL_1805396, EPI_ISL_1805397, EPI_ISL_1805398 | New Mexico Department of Health Scientific Laboratory | New Mexico Department of Health Scientific Laboratory | Ellie Johnson, D'eldra Malone, Jennifer Benoit, Ratheesh Rajan, Linda Salazar, Anastacia Griego-Fisher |
| EPI_ISL_1805399, EPI_ISL_1805400 | Sierra Vista Hospital | New Mexico Department of Health Scientific Laboratory | Ellie Johnson, D'eldra Malone, Jennifer Benoit, Ratheesh Rajan, Linda Salazar, Anastacia Griego-Fisher |
| EPI_ISL_1805401, EPI_ISL_1805402, EPI_ISL_1805403, EPI_ISL_1805404, EPI_ISL_1805405, EPI_ISL_1805406, EPI_ISL_1805407, EPI_ISL_1805408 | San Juan Regional Medical Center | New Mexico Department of Health Scientific Laboratory | Ellie Johnson, D'eldra Malone, Jennifer Benoit, Ratheesh Rajan, Linda Salazar, Anastacia Griego-Fisher |
| EPI_ISL_1805409 | PHS/IHS Hospital-Jicarilla Service Unit | New Mexico Department of Health Scientific Laboratory | Ellie Johnson, D'eldra Malone, Jennifer Benoit, Ratheesh Rajan, Linda Salazar, Anastacia Griego-Fisher |
| EPI_ISL_1805410, EPI_ISL_1805411, EPI_ISL_1805412 | M Health Fairview | Minnesota Department of Health, Public Health Laboratory | Alexandra Lorentz, Jacob Garfin, Matt Plumb, and Xiong Wang |
| EPI_ISL_1805431, EPI_ISL_1805432, EPI_ISL_1805433, EPI_ISL_1805434, EPI_ISL_1805435, EPI_ISL_1805436, EPI_ISL_1805437, EPI_ISL_1805438, EPI_ISL_1805439, EPI_ISL_1805440, EPI_ISL_1805441 | | | |
| see above | Kansas Health and Environmental Lab | Kansas Health and Environmental Lab | Mike Grose, Jonathan Barnell, Ben Olsen, and Phil Adam |
| EPI_ISL_1805442, EPI_ISL_1805443, EPI_ISL_1805444, EPI_ISL_1805445, EPI_ISL_1805446 | Wichita State University - Molecular Diagnostics Lab | Kansas Health and Environmental Lab | Mike Grose, Jonathan Barnell, Ben Olsen, and Phil Adam |
| EPI_ISL_1805448 | Kansas Health and Environmental Lab | Kansas Health and Environmental Lab | Mike Grose, Jonathan Barnell, Ben Olsen, and Phil Adam |
| EPI_ISL_1805449, EPI_ISL_1805450, EPI_ISL_1805451 | Wichita State University - Molecular Diagnostics Lab | Kansas Health and Environmental Lab | Mike Grose, Jonathan Barnell, Ben Olsen, and Phil Adam |
| EPI_ISL_1805452 | Cytocheck Laboratories | Kansas Health and Environmental Lab | Mike Grose, Jonathan Barnell, Ben Olsen, and Phil Adam |
| EPI_ISL_1805454, EPI_ISL_1805457, EPI_ISL_1805458, EPI_ISL_1805459, EPI_ISL_1805460 | Wichita State University - Molecular Diagnostics Lab | Kansas Health and Environmental Lab | Mike Grose, Jonathan Barnell, Ben Olsen, and Phil Adam |
| EPI_ISL_1805461, EPI_ISL_1805462, EPI_ISL_1805463, EPI_ISL_1805464 | Kansas Health and Environmental Lab | Kansas Health and Environmental Lab | Mike Grose, Jonathan Barnell, Ben Olsen, and Phil Adam |
| EPI_ISL_1805465 | LESP Queretaro | Instituto de Diagnostico y Referencia Epidemiologicos (INDRE) | Claudia Wong-Arambula, Abril Rodriguez-Maldonado, Vanessa Rivero-Arredondo, Ariadna Medina-Benitez, Joaquin Quiroz-Mercado, Sergio Rangel-Guerrero, Natividad Cruz-Ortiz, Tatiana Nunez-Garcia, Gisela Barrera-Badillo, Lucia Hernandez-Rivas, Irma Lopez-Martinez, Ernesto Ramirez-Gonzalez. |
| EPI_ISL_1805466, EPI_ISL_1805467, EPI_ISL_1805468, EPI_ISL_1805469 | LESP Chihuahua | Instituto de Diagnostico y Referencia Epidemiologicos (INDRE) | Claudia Wong-Arambula, Abril Rodriguez-Maldonado, Vanessa Rivero-Arredondo, Ariadna Medina-Benitez, Joaquin Quiroz-Mercado, Sergio Rangel-Guerrero, Natividad Cruz-Ortiz, Tatiana Nunez-Garcia, Gisela Barrera-Badillo, Lucia Hernandez-Rivas, Irma Lopez-Martinez, Ernesto Ramirez-Gonzalez. |
| EPI_ISL_1805470 | LESP Quintana Roo | Instituto de Diagnostico y Referencia Epidemiologicos (INDRE) | Claudia Wong-Arambula, Abril Rodriguez-Maldonado, Vanessa Rivero-Arredondo, Ariadna Medina-Benitez, Joaquin Quiroz-Mercado, Sergio Rangel-Guerrero, Natividad Cruz-Ortiz, Tatiana Nunez-Garcia, Gisela Barrera-Badillo, Lucia Hernandez-Rivas, Irma Lopez-Martinez, Ernesto Ramirez-Gonzalez. |
| EPI_ISL_1805471 | LESP Coahuila | Instituto de Diagnostico y Referencia Epidemiologicos (INDRE) | Claudia Wong-Arambula, Abril Rodriguez-Maldonado, Vanessa Rivero-Arredondo, Ariadna Medina-Benitez, Joaquin Quiroz-Mercado, Sergio Rangel-Guerrero, Natividad Cruz-Ortiz, Tatiana Nunez-Garcia, Gisela Barrera-Badillo, Lucia Hernandez-Rivas, Irma Lopez-Martinez, Ernesto Ramirez-Gonzalez. |
| EPI_ISL_1805472 | LESP Sonora | Instituto de Diagnostico y Referencia Epidemiologicos (INDRE) | Claudia Wong-Arambula, Abril Rodriguez-Maldonado, Vanessa Rivero-Arredondo, Ariadna Medina-Benitez, Joaquin Quiroz-Mercado, Sergio Rangel-Guerrero, Natividad Cruz-Ortiz, Tatiana Nunez-Garcia, Gisela Barrera-Badillo, Lucia Hernandez-Rivas, Irma Lopez-Martinez, Ernesto Ramirez-Gonzalez. |
| EPI_ISL_1805473 | LESP Nuevo Leon | Instituto de Diagnostico y Referencia Epidemiologicos (INDRE) | Claudia Wong-Arambula, Abril Rodriguez-Maldonado, Vanessa Rivero-Arredondo, Ariadna Medina-Benitez, Joaquin Quiroz-Mercado, Sergio Rangel-Guerrero, Natividad Cruz-Ortiz, Tatiana Nunez-Garcia, Gisela Barrera-Badillo, Lucia Hernandez-Rivas, Irma Lopez-Martinez, Ernesto Ramirez-Gonzalez. |
| EPI_ISL_1805475, EPI_ISL_1805476, EPI_ISL_1805477, EPI_ISL_1805479 | LESP Chihuahua | Instituto de Diagnostico y Referencia Epidemiologicos (INDRE) | Claudia Wong-Arambula, Abril Rodriguez-Maldonado, Vanessa Rivero-Arredondo, Ariadna Medina-Benitez, Joaquin Quiroz-Mercado, Sergio Rangel-Guerrero, Natividad Cruz-Ortiz, Tatiana Nunez-Garcia, Gisela Barrera-Badillo, Lucia Hernandez-Rivas, Irma Lopez-Martinez, Ernesto Ramirez-Gonzalez. |
| EPI_ISL_1805480 | LESP Ciudad de Mexico | Instituto de Diagnostico y Referencia Epidemiologicos (INDRE) | Claudia Wong-Arambula, Abril Rodriguez-Maldonado, Vanessa Rivero-Arredondo, Ariadna Medina-Benitez, Joaquin Quiroz-Mercado, Sergio Rangel-Guerrero, Natividad Cruz-Ortiz, Tatiana Nunez-Garcia, Gisela Barrera-Badillo, Lucia Hernandez-Rivas, Irma Lopez-Martinez, Ernesto Ramirez-Gonzalez. |
| EPI_ISL_1805481 | LESP Quintana Roo | Instituto de Diagnostico y Referencia Epidemiologicos (INDRE) | Claudia Wong-Arambula, Abril Rodriguez-Maldonado, Vanessa Rivero-Arredondo, Ariadna Medina-Benitez, Joaquin Quiroz-Mercado, Sergio Rangel-Guerrero, Natividad Cruz-Ortiz, Tatiana Nunez-Garcia, Gisela Barrera-Badillo, Lucia Hernandez-Rivas, Irma Lopez-Martinez, Ernesto Ramirez-Gonzalez. |
| EPI_ISL_1805482 | LESP Oaxaca | Instituto de Diagnostico y Referencia Epidemiologicos (INDRE) | Claudia Wong-Arambula, Abril Rodriguez-Maldonado, Vanessa Rivero-Arredondo, Ariadna Medina-Benitez, Joaquin Quiroz-Mercado, Sergio Rangel-Guerrero, Natividad Cruz-Ortiz, Tatiana Nunez-Garcia, Gisela Barrera-Badillo, Lucia Hernandez-Rivas, Irma Lopez-Martinez, Ernesto Ramirez-Gonzalez. |
| EPI_ISL_1805483 | LESP Hidalgo | Instituto de Diagnostico y Referencia Epidemiologicos (INDRE) | Claudia Wong-Arambula, Abril Rodriguez-Maldonado, Vanessa Rivero-Arredondo, Ariadna Medina-Benitez, Joaquin Quiroz-Mercado, Sergio Rangel-Guerrero, Natividad Cruz-Ortiz, Tatiana Nunez-Garcia, Gisela Barrera-Badillo, Lucia Hernandez-Rivas, Irma Lopez-Martinez, Ernesto Ramirez-Gonzalez. |
| EPI_ISL_1805484 | LESP Nuevo Leon | Instituto de Diagnostico y Referencia Epidemiologicos (INDRE) | Claudia Wong-Arambula, Abril Rodriguez-Maldonado, Vanessa Rivero-Arredondo, Ariadna Medina-Benitez, Joaquin Quiroz-Mercado, Sergio Rangel-Guerrero, Natividad Cruz-Ortiz, Tatiana Nunez-Garcia, Gisela Barrera-Badillo, Lucia Hernandez-Rivas, Irma Lopez-Martinez, Ernesto Ramirez-Gonzalez. |
| EPI_ISL_1805485 | LESP Oaxaca | Instituto de Diagnostico y Referencia Epidemiologicos (INDRE) | Claudia Wong-Arambula, Abril Rodriguez-Maldonado, Vanessa Rivero-Arredondo, Ariadna Medina-Benitez, Joaquin Quiroz-Mercado, Sergio Rangel-Guerrero, Natividad Cruz-Ortiz, Tatiana Nunez-Garcia, Gisela Barrera-Badillo, Lucia Hernandez-Rivas, Irma Lopez-Martinez, Ernesto Ramirez-Gonzalez. |
| EPI_ISL_1805486 | LESP Ciudad de Mexico | Instituto de Diagnostico y Referencia Epidemiologicos (INDRE) | Claudia Wong-Arambula, Abril Rodriguez-Maldonado, Vanessa Rivero-Arredondo, Ariadna Medina-Benitez, Joaquin Quiroz-Mercado, Sergio Rangel-Guerrero, Natividad Cruz-Ortiz, Tatiana Nunez-Garcia, Gisela Barrera-Badillo, Lucia Hernandez-Rivas, Irma Lopez-Martinez, Ernesto Ramirez-Gonzalez. |
| EPI_ISL_1805487 | LESP Baja California Sur | Instituto de Diagnostico y Referencia Epidemiologicos (INDRE) | Claudia Wong-Arambula, Abril Rodriguez-Maldonado, Vanessa Rivero-Arredondo, Ariadna Medina-Benitez, Joaquin Quiroz-Mercado, Sergio Rangel-Guerrero, Natividad Cruz-Ortiz, Tatiana Nunez-Garcia, Gisela Barrera-Badillo, Lucia Hernandez-Rivas, Irma Lopez-Martinez, Ernesto Ramirez-Gonzalez. |
| EPI_ISL_1805488, EPI_ISL_1805489 | LESP Jalisco | Instituto de Diagnostico y Referencia Epidemiologicos (INDRE) | Claudia Wong-Arambula, Abril Rodriguez-Maldonado, Vanessa Rivero-Arredondo, Ariadna Medina-Benitez, Joaquin Quiroz-Mercado, Sergio Rangel-Guerrero, Natividad Cruz-Ortiz, Tatiana Nunez-Garcia, Gisela Barrera-Badillo, Lucia Hernandez-Rivas, Irma Lopez-Martinez, Ernesto Ramirez-Gonzalez. |
| EPI_ISL_1805490 | LESP Nuevo Leon | Instituto de Diagnostico y Referencia Epidemiologicos (INDRE) | Claudia Wong-Arambula, Abril Rodriguez-Maldonado, Vanessa Rivero-Arredondo, Ariadna Medina-Benitez, Joaquin Quiroz-Mercado, Sergio Rangel-Guerrero, Natividad Cruz-Ortiz, Tatiana Nunez-Garcia, Gisela Barrera-Badillo, Lucia Hernandez-Rivas, Irma Lopez-Martinez, Ernesto Ramirez-Gonzalez. |
| EPI_ISL_1805491 | LESP Jalisco | Instituto de Diagnostico y Referencia Epidemiologicos (INDRE) | Claudia Wong-Arambula, Abril Rodriguez-Maldonado, Vanessa Rivero-Arredondo, Ariadna Medina-Benitez, Joaquin Quiroz-Mercado, Sergio Rangel-Guerrero, Natividad Cruz-Ortiz, Tatiana Nunez-Garcia, Gisela Barrera-Badillo, Lucia Hernandez-Rivas, Irma Lopez-Martinez, Ernesto Ramirez-Gonzalez. |
| EPI_ISL_1805492 | LESP Sonora | Instituto de Diagnostico y Referencia Epidemiologicos (INDRE) | Claudia Wong-Arambula, Abril Rodriguez-Maldonado, Vanessa Rivero-Arredondo, Ariadna Medina-Benitez, Joaquin Quiroz-Mercado, Sergio Rangel-Guerrero, Natividad Cruz-Ortiz, Tatiana Nunez-Garcia, Gisela Barrera-Badillo, Lucia Hernandez-Rivas, Irma Lopez-Martinez, Ernesto Ramirez-Gonzalez. |
| EPI_ISL_1805493 | LESP Baja California | Instituto de Diagnostico y Referencia Epidemiologicos (INDRE) | Claudia Wong-Arambula, Abril Rodriguez-Maldonado, Vanessa Rivero-Arredondo, Ariadna Medina-Benitez, Joaquin Quiroz-Mercado, Sergio Rangel-Guerrero, Natividad Cruz-Ortiz, Tatiana Nunez-Garcia, Gisela Barrera-Badillo, Lucia Hernandez-Rivas, Irma Lopez-Martinez, Ernesto Ramirez-Gonzalez. |
| EPI_ISL_1805494 | LESPZacatecas | Instituto de Diagnostico y Referencia Epidemiologicos (INDRE) | Claudia Wong-Arambula, Abril Rodriguez-Maldonado, Vanessa Rivero-Arredondo, Ariadna Medina-Benitez, Joaquin Quiroz-Mercado, Sergio Rangel-Guerrero, Natividad Cruz-Ortiz, Tatiana Nunez-Garcia, Gisela Barrera-Badillo, Lucia Hernandez-Rivas, Irma Lopez-Martinez, Ernesto Ramirez-Gonzalez. |
| EPI_ISL_1805495, EPI_ISL_1805496 | LESP Jalisco/Unidad de Patologia | Instituto de Diagnostico y Referencia Epidemiologicos (INDRE) | Claudia Wong-Arambula, Abril Rodriguez-Maldonado, Vanessa Rivero-Arredondo, Ariadna Medina-Benitez, Joaquin Quiroz-Mercado, Sergio Rangel-Guerrero, Natividad Cruz-Ortiz, Tatiana Nunez-Garcia, Gisela Barrera-Badillo, Lucia Hernandez-Rivas, Irma Lopez-Martinez, Ernesto Ramirez-Gonzalez. |
| EPI_ISL_1805497 | LESP Jalisco | Instituto de Diagnostico y Referencia Epidemiologicos (INDRE) | Claudia Wong-Arambula, Abril Rodriguez-Maldonado, Vanessa Rivero-Arredondo, Ariadna Medina-Benitez, Joaquin Quiroz-Mercado, Sergio Rangel-Guerrero, Natividad Cruz-Ortiz, Tatiana Nunez-Garcia, Gisela Barrera-Badillo, Lucia Hernandez-Rivas, Irma Lopez-Martinez, Ernesto Ramirez-Gonzalez. |

| | | | |
|---|---|---|---|
| EPI_ISL_1805498 | LESP Guanajuato | Instituto de Diagnostico y Referencia Epidemiologicos (INDRE) | Claudia Wong-Arambula, Abril Rodriguez-Maldonado, Vanessa Rivero-Arredondo, Ariadna Medina-Benitez, Joaquin Quiroz-Mercado, Sergio Rangel-Guerrero, Natividad Cruz-Ortiz, Tatiana Nunez-Garcia, Gisela Barrera-Badillo, Lucia Hernandez-Rivas, Irma Lopez-Martinez, Ernesto Ramirez-Gonzalez. |
| EPI_ISL_1805499, EPI_ISL_1805500 | LESP Nuevo Leon | Instituto de Diagnostico y Referencia Epidemiologicos (INDRE) | Claudia Wong-Arambula, Abril Rodriguez-Maldonado, Vanessa Rivero-Arredondo, Ariadna Medina-Benitez, Joaquin Quiroz-Mercado, Sergio Rangel-Guerrero, Natividad Cruz-Ortiz, Tatiana Nunez-Garcia, Gisela Barrera-Badillo, Lucia Hernandez-Rivas, Irma Lopez-Martinez, Ernesto Ramirez-Gonzalez. |
| EPI_ISL_1805501, EPI_ISL_1805502 | LESP Yucatan | Instituto de Diagnostico y Referencia Epidemiologicos (INDRE) | Claudia Wong-Arambula, Abril Rodriguez-Maldonado, Vanessa Rivero-Arredondo, Ariadna Medina-Benitez, Joaquin Quiroz-Mercado, Sergio Rangel-Guerrero, Natividad Cruz-Ortiz, Tatiana Nunez-Garcia, Gisela Barrera-Badillo, Lucia Hernandez-Rivas, Irma Lopez-Martinez, Ernesto Ramirez-Gonzalez. |
| EPI_ISL_1805503, EPI_ISL_1805504 | LESP Nuevo Leon | Instituto de Diagnostico y Referencia Epidemiologicos (INDRE) | Claudia Wong-Arambula, Abril Rodriguez-Maldonado, Vanessa Rivero-Arredondo, Ariadna Medina-Benitez, Joaquin Quiroz-Mercado, Sergio Rangel-Guerrero, Natividad Cruz-Ortiz, Tatiana Nunez-Garcia, Gisela Barrera-Badillo, Lucia Hernandez-Rivas, Irma Lopez-Martinez, Ernesto Ramirez-Gonzalez. |
| EPI_ISL_1805506, EPI_ISL_1805507 | Wyoming Public Health Laboratory | Wyoming Public Health Laboratory | Jim Mildenberger, Wanda Manley, Noah Hull, Taylor Fearing, Lynette Gumbleton, Channing Weber, Ashley Norberg, Chayse Rowley, Marley Goetz, Brian Dominguez, Elliot Thomasson, Sam Britz, Cari Sloma, and Rob Christensen |
| EPI_ISL_1805510, EPI_ISL_1805511, EPI_ISL_1805512, EPI_ISL_1805513, EPI_ISL_1805514, EPI_ISL_1805515, EPI_ISL_1805516, EPI_ISL_1805517, EPI_ISL_1805518, EPI_ISL_1805519, EPI_ISL_1805520, EPI_ISL_1805521, EPI_ISL_1805522, EPI_ISL_1805523, EPI_ISL_1805524, EPI_ISL_1805525, EPI_ISL_1805526 | | | |
| see above | UW Virology Lab | UW Virology Lab | Pavitra Roychoudhury, Hong Xie, Lasata Shrestha, Tien V. Nguyen, Shah Mohamed Bakhash, Michelle Lin, Noah R. Baker, Ricardo Perez, Sean Ellis, Nathan Breit, Robert J. Livingston, Meei-Li Huang, Keith R Jerome, Patrick Mathias, Alexander Greninger |
| EPI_ISL_1805527, EPI_ISL_1805528, EPI_ISL_1805529, EPI_ISL_1805530, EPI_ISL_1805531, EPI_ISL_1805532, EPI_ISL_1805533, EPI_ISL_1805534, EPI_ISL_1805535, EPI_ISL_1805536, EPI_ISL_1805537, EPI_ISL_1805538, EPI_ISL_1805539, EPI_ISL_1805540, EPI_ISL_1805541, EPI_ISL_1805542, EPI_ISL_1805543, EPI_ISL_1805544, EPI_ISL_1805545, EPI_ISL_1805546, EPI_ISL_1805547, EPI_ISL_1805548, EPI_ISL_1805549, EPI_ISL_1805550, EPI_ISL_1805551, EPI_ISL_1805552, EPI_ISL_1805553, EPI_ISL_1805554, EPI_ISL_1805555, EPI_ISL_1805556, EPI_ISL_1805557, EPI_ISL_1805558, EPI_ISL_1805559, EPI_ISL_1805560, EPI_ISL_1805561, EPI_ISL_1805562, EPI_ISL_1805563, EPI_ISL_1805564, EPI_ISL_1805565, EPI_ISL_1805566, EPI_ISL_1805567, EPI_ISL_1805568, EPI_ISL_1805569, EPI_ISL_1805570, EPI_ISL_1805571 | | | |
| see above | Virginia Division of Consolidated Laboratory Services | Virginia Division of Consolidated Laboratory Services | Virginia DCLS |
| EPI_ISL_1805573 | DC Public Health Lab/ Dept. of Forensic Sciences | DC Public Health Lab/ Dept. of Forensic Sciences | Janis Doss, Scott Nguyen, Elizabeth Zelaya, Sarah Scott, Connie Maza, Monica Mann, Brittany Hamilton, David Payne, Jocelyn Hauser |
| EPI_ISL_1805574, EPI_ISL_1805575, EPI_ISL_1805576, EPI_ISL_1805577, EPI_ISL_1805578, EPI_ISL_1805579, EPI_ISL_1805580, EPI_ISL_1805582, EPI_ISL_1805583, EPI_ISL_1805584, EPI_ISL_1805585, EPI_ISL_1805586, EPI_ISL_1805587, EPI_ISL_1805588, EPI_ISL_1805589, EPI_ISL_1805590, EPI_ISL_1805591, EPI_ISL_1805592, EPI_ISL_1805593, EPI_ISL_1805594, EPI_ISL_1805595, EPI_ISL_1805596, EPI_ISL_1805597, EPI_ISL_1805598, EPI_ISL_1805599, EPI_ISL_1805600, EPI_ISL_1805601, EPI_ISL_1805602, EPI_ISL_1805605, EPI_ISL_1805606, EPI_ISL_1805607, EPI_ISL_1805608, EPI_ISL_1805609, EPI_ISL_1805613, EPI_ISL_1805614, EPI_ISL_1805615, EPI_ISL_1805616, EPI_ISL_1805617, EPI_ISL_1805618, EPI_ISL_1805619, EPI_ISL_1805620, EPI_ISL_1805621, EPI_ISL_1805622, EPI_ISL_1805623, EPI_ISL_1805624, EPI_ISL_1805625, EPI_ISL_1805626, EPI_ISL_1805627, EPI_ISL_1805628, EPI_ISL_1805629, EPI_ISL_1805630, EPI_ISL_1805662, EPI_ISL_1805663, EPI_ISL_1805664, EPI_ISL_1805665, EPI_ISL_1805666, EPI_ISL_1805667, EPI_ISL_1805668, EPI_ISL_1805669, EPI_ISL_1805670, EPI_ISL_1805671, EPI_ISL_1805672, EPI_ISL_1805673, EPI_ISL_1805674, EPI_ISL_1805675, EPI_ISL_1805676, EPI_ISL_1805677, EPI_ISL_1805678, EPI_ISL_1805679, EPI_ISL_1805680, EPI_ISL_1805681, EPI_ISL_1805682, EPI_ISL_1805684, EPI_ISL_1805685, EPI_ISL_1805686, EPI_ISL_1805687, EPI_ISL_1805688, EPI_ISL_1805689, EPI_ISL_1805691, EPI_ISL_1805692, EPI_ISL_1805693, EPI_ISL_1805694, EPI_ISL_1805695 | | | |
| see above | Virginia Division of Consolidated Laboratory Services | Virginia Division of Consolidated Laboratory Services | Virginia DCLS |
| EPI_ISL_1805719, EPI_ISL_1805720, EPI_ISL_1805721, EPI_ISL_1805722, EPI_ISL_1805723, EPI_ISL_1805724, EPI_ISL_1805725, EPI_ISL_1805726, EPI_ISL_1805727, EPI_ISL_1805728, EPI_ISL_1805729, EPI_ISL_1805730, EPI_ISL_1805731, EPI_ISL_1805732, EPI_ISL_1805733, EPI_ISL_1805734, EPI_ISL_1805735, EPI_ISL_1805736, EPI_ISL_1805737, EPI_ISL_1805738, EPI_ISL_1805739 | | | |
| see above | County of San Diego Health and Human Services Agency | Pathogen Discovery, Respiratory Viruses Branch, Division of Viral Diseases, Centers for Disease Control and Prevention | Ying Tao, Yan Li, Anna Kelleher, Jing Zhang, Anna Montmayeur, Brian Lynch, Krista Queen, Anna Uehara, Peter Cook, Han Jia Justin Ng, Rachel Marine, Clinton R. Paden, Haibin Wang, Mark Burroughs, Justin Lee, Adam Retchless, Suxiang Tong |
| EPI_ISL_1805748, EPI_ISL_1805750, EPI_ISL_1805754, EPI_ISL_1805755, EPI_ISL_1805756, EPI_ISL_1805757, EPI_ISL_1805758, EPI_ISL_1805759, EPI_ISL_1805760, EPI_ISL_1805766, EPI_ISL_1805767, EPI_ISL_1805768, EPI_ISL_1805769, EPI_ISL_1805770, EPI_ISL_1805771, EPI_ISL_1805772, EPI_ISL_1805773 | | | |
| see above | University of Texas at Austin Genomic Sequencing and Analysis Facility (UTGSAF) | University of Texas at Austin Genomic Sequencing and Analysis Facility (UTGSAF) | Jessica Podnar, Sylvie Beaudenon, Audrey Kelly, Zachary Carver, Anna Battenhouse, Andreas Matouschek |
| EPI_ISL_1805774, EPI_ISL_1805775, EPI_ISL_1805776 | Kansas Health and Environmental Lab | Kansas Health and Environmental Lab | Mike Grose, Jonathan Barnell, Ben Olsen, and Phil Adam |
| EPI_ISL_1805777, EPI_ISL_1805778, EPI_ISL_1805779, EPI_ISL_1805780 | Wichita State University - Molecular Diagnostics Lab | Kansas Health and Environmental Lab | Mike Grose, Jonathan Barnell, Ben Olsen, and Phil Adam |
| EPI_ISL_1805782, EPI_ISL_1805783, EPI_ISL_1805785 | Kansas Health and Environmental Lab | Kansas Health and Environmental Lab | Mike Grose, Jonathan Barnell, Ben Olsen, and Phil Adam |
| EPI_ISL_1805786 | Clinical Reference Laboratory | Kansas Health and Environmental Lab | Mike Grose, Jonathan Barnell, Ben Olsen, and Phil Adam |
| EPI_ISL_1805791, EPI_ISL_1805792, EPI_ISL_1805793, EPI_ISL_1805794, EPI_ISL_1805795, EPI_ISL_1805796, EPI_ISL_1805797, EPI_ISL_1805798, EPI_ISL_1805799, EPI_ISL_1805800, EPI_ISL_1805801, EPI_ISL_1805802, EPI_ISL_1805803, EPI_ISL_1805804, EPI_ISL_1805805, EPI_ISL_1805806, EPI_ISL_1805807, EPI_ISL_1805808, EPI_ISL_1805809, EPI_ISL_1805810, EPI_ISL_1805811, EPI_ISL_1805813, EPI_ISL_1805814, EPI_ISL_1805815, EPI_ISL_1805816, EPI_ISL_1805817, EPI_ISL_1805818, EPI_ISL_1805819, EPI_ISL_1805820, EPI_ISL_1805821, EPI_ISL_1805822, EPI_ISL_1805823, EPI_ISL_1805824, EPI_ISL_1805825, EPI_ISL_1805826, EPI_ISL_1805827, EPI_ISL_1805828, EPI_ISL_1805829, EPI_ISL_1805830, EPI_ISL_1805831, EPI_ISL_1805832, EPI_ISL_1805833, EPI_ISL_1805834, EPI_ISL_1805835, EPI_ISL_1805836, EPI_ISL_1805837, EPI_ISL_1805838, EPI_ISL_1805839, EPI_ISL_1805840, EPI_ISL_1805841, EPI_ISL_1805842, EPI_ISL_1805843, EPI_ISL_1805844, EPI_ISL_1805845, EPI_ISL_1805846, EPI_ISL_1805847, EPI_ISL_1805848, EPI_ISL_1805849, EPI_ISL_1805850, EPI_ISL_1805851, EPI_ISL_1805852, EPI_ISL_1805853, EPI_ISL_1805854, EPI_ISL_1805855, EPI_ISL_1805856, EPI_ISL_1805857, EPI_ISL_1805858, EPI_ISL_1805859, EPI_ISL_1805860, EPI_ISL_1805861, EPI_ISL_1805862, EPI_ISL_1805863, EPI_ISL_1805864, EPI_ISL_1805865, EPI_ISL_1805866, EPI_ISL_1805867, EPI_ISL_1805868, EPI_ISL_1805869, EPI_ISL_1805870, EPI_ISL_1805871, EPI_ISL_1805872, EPI_ISL_1805873, EPI_ISL_1805874, EPI_ISL_1805875, EPI_ISL_1805876, EPI_ISL_1805877, EPI_ISL_1805878, EPI_ISL_1805879, EPI_ISL_1805880, EPI_ISL_1805881, EPI_ISL_1805883, EPI_ISL_1805884, EPI_ISL_1805885, EPI_ISL_1805886, EPI_ISL_1805887, EPI_ISL_1805888, EPI_ISL_1805889, EPI_ISL_1805890, EPI_ISL_1805892, EPI_ISL_1805894, EPI_ISL_1805895, EPI_ISL_1805896, EPI_ISL_1805897, EPI_ISL_1805898, EPI_ISL_1805899, EPI_ISL_1805901, EPI_ISL_1805902, EPI_ISL_1805904, EPI_ISL_1805905, EPI_ISL_1805906, EPI_ISL_1805907, EPI_ISL_1805908, EPI_ISL_1805909, EPI_ISL_1805910, EPI_ISL_1805911, EPI_ISL_1805912, EPI_ISL_1805913, EPI_ISL_1805914, EPI_ISL_1805915, EPI_ISL_1805916, EPI_ISL_1805917, EPI_ISL_1805918, EPI_ISL_1805919, EPI_ISL_1805920, EPI_ISL_1805921, EPI_ISL_1805922, EPI_ISL_1805923, EPI_ISL_1805924, EPI_ISL_1805925, EPI_ISL_1805926, EPI_ISL_1805927, EPI_ISL_1805928, EPI_ISL_1805929, EPI_ISL_1805930, EPI_ISL_1805931 | | | |
| see above | Johns Hopkins Hospital Department of Pathology | Johns Hopkins Hospital Department of Pathology | C. Paul Morris, Chun Huai Luo, Adannaya Amadi, Matthew Schwartz, Heba H. Mostafa |
| EPI_ISL_1805935, EPI_ISL_1805937, EPI_ISL_1805939, EPI_ISL_1805941, EPI_ISL_1805943, EPI_ISL_1805945, EPI_ISL_1805946, EPI_ISL_1805947, EPI_ISL_1805948, EPI_ISL_1805950, EPI_ISL_1805951, EPI_ISL_1805953 | | | |
| see above | Wyoming Public Health Laboratory | Wyoming Public Health Laboratory | Jim Mildenberger, Taylor Fearing, Channing Weber, Ashley Norberg, Chayse Rowley, Marley Goetz, Brian Dominguez, Elliot Thomasson, Brittany Oher, Sam Britz, Cari Sloma, and Rob Christensen |
| EPI_ISL_1806183, EPI_ISL_1806186, EPI_ISL_1806188, EPI_ISL_1806191, EPI_ISL_1806194, EPI_ISL_1806197, EPI_ISL_1806200, EPI_ISL_1806203, EPI_ISL_1806206, EPI_ISL_1806209, EPI_ISL_1806212, EPI_ISL_1806215, EPI_ISL_1806218, EPI_ISL_1806220, EPI_ISL_1806223, EPI_ISL_1806226, EPI_ISL_1806229, EPI_ISL_1806232, EPI_ISL_1806235, EPI_ISL_1806238, EPI_ISL_1806241, EPI_ISL_1806244, EPI_ISL_1806247, EPI_ISL_1806250, EPI_ISL_1806253, EPI_ISL_1806256, EPI_ISL_1806259, EPI_ISL_1806262, EPI_ISL_1806265, EPI_ISL_1806268, EPI_ISL_1806270, EPI_ISL_1806274, EPI_ISL_1806277, EPI_ISL_1806280, EPI_ISL_1806283, EPI_ISL_1806286, EPI_ISL_1806289, EPI_ISL_1806292, EPI_ISL_1806296, EPI_ISL_1806298, EPI_ISL_1806301, EPI_ISL_1806305, EPI_ISL_1806307, EPI_ISL_1806310, EPI_ISL_1806313, EPI_ISL_1806317, EPI_ISL_1806320, EPI_ISL_1806323, EPI_ISL_1806326, EPI_ISL_1806329, EPI_ISL_1806332, EPI_ISL_1806335, EPI_ISL_1806338, EPI_ISL_1806341, EPI_ISL_1806345, EPI_ISL_1806347 | | | |
| see above | Idaho Bureau of Laboratories | Center for Global Health, University of New Mexico Health Sciences Center | Daryl Domman, Kurt Schwalm, Valerie Morley, Matthew Burns, Robert Voermans, Christopher Ball, Darrell Dinwiddie |
| EPI_ISL_1807187 | New Mexico Department of Health Scientific Laboratory | Center for Global Health, University of New Mexico Health Sciences Center | Daryl Domman, Kurt Schwalm, Twila Kunde, Joseph Hicks, Anastacia Griego, Michael Edwards, Valerie Morley, Darrell Dinwiddie |
| EPI_ISL_1807190, EPI_ISL_1807191, EPI_ISL_1807192, EPI_ISL_1807193, EPI_ISL_1807194, EPI_ISL_1807195, EPI_ISL_1807196, EPI_ISL_1807197, EPI_ISL_1807198, EPI_ISL_1807199, EPI_ISL_1807200, EPI_ISL_1807201, EPI_ISL_1807202, EPI_ISL_1807203, EPI_ISL_1807204, EPI_ISL_1807205 | | | |
| see above | Wichita State University - Molecular Diagnostics Lab | Kansas Health and Environmental Lab | Mike Grose, Jonathan Barnell, Ben Olsen, and Phil Adam |
| EPI_ISL_1807209, EPI_ISL_1807210, EPI_ISL_1807211, EPI_ISL_1807212, EPI_ISL_1807213, EPI_ISL_1807214, EPI_ISL_1807215, EPI_ISL_1807216, EPI_ISL_1807217 | GA Department of Public Health | GA Department of Public Health | Stacy Reeves, Jonathan Edwards, Cynthia Dixey, Tonia Parrott, Aliyah Fields, Taylor Smith |
| EPI_ISL_1807218, EPI_ISL_1807219, EPI_ISL_1807220, EPI_ISL_1807221, EPI_ISL_1807222, EPI_ISL_1807223, EPI_ISL_1807224, EPI_ISL_1807225, EPI_ISL_1807226, EPI_ISL_1807228, EPI_ISL_1807229, EPI_ISL_1807230, EPI_ISL_1807231, EPI_ISL_1807232, EPI_ISL_1807233, EPI_ISL_1807234, EPI_ISL_1807235, EPI_ISL_1807236, EPI_ISL_1807237, EPI_ISL_1807238, EPI_ISL_1807239, EPI_ISL_1807240, EPI_ISL_1807241, EPI_ISL_1807242, EPI_ISL_1807243, EPI_ISL_1807244, EPI_ISL_1807245, EPI_ISL_1807246, EPI_ISL_1807247, EPI_ISL_1807249, EPI_ISL_1807251, EPI_ISL_1807252, EPI_ISL_1807253, EPI_ISL_1807254, EPI_ISL_1807255, EPI_ISL_1807256, EPI_ISL_1807257, EPI_ISL_1807258, EPI_ISL_1807259, EPI_ISL_1807260, EPI_ISL_1807261, EPI_ISL_1807262, EPI_ISL_1807263, EPI_ISL_1807264, EPI_ISL_1807265, EPI_ISL_1807266, EPI_ISL_1807267, EPI_ISL_1807268, EPI_ISL_1807269, EPI_ISL_1807270, EPI_ISL_1807271, EPI_ISL_1807272, EPI_ISL_1807273, EPI_ISL_1807274, EPI_ISL_1807276, EPI_ISL_1807277, EPI_ISL_1807278, EPI_ISL_1807279, EPI_ISL_1807280, EPI_ISL_1807281, EPI_ISL_1807282, EPI_ISL_1807283, EPI_ISL_1807284, EPI_ISL_1807285, EPI_ISL_1807286, EPI_ISL_1807287, EPI_ISL_1807288, EPI_ISL_1807290, EPI_ISL_1807291, EPI_ISL_1807292, EPI_ISL_1807293, EPI_ISL_1807294, EPI_ISL_1807295, EPI_ISL_1807296, EPI_ISL_1807297, EPI_ISL_1807298, EPI_ISL_1807299, EPI_ISL_1807300 | | | |
| see above | Texas Department of State Health Services (TXDSHS) | Texas Department of State Health Services (TXDSHS) | Rashmi Tuladhar, Bonnie Oh, Jenny Zhang, Maliha Rahman, Mayela Pedrueza, Anita Pokharel, Lorraine Rodriguez, Myong Koag, Chun Wang, Rachel Lee, Grace Kubin |
| EPI_ISL_1807301, EPI_ISL_1807302, EPI_ISL_1807303, EPI_ISL_1807304, EPI_ISL_1807305, EPI_ISL_1807306, EPI_ISL_1807307, EPI_ISL_1807308, EPI_ISL_1807309 | GA Department of Public Health | GA Department of Public Health | Stacy Reeves, Jonathan Edwards, Cynthia Dixey, Tonia Parrott, Aliyah Fields, Taylor Smith |
| EPI_ISL_1807319, EPI_ISL_1807323, EPI_ISL_1807324, EPI_ISL_1807325, EPI_ISL_1807326, EPI_ISL_1807327, EPI_ISL_1807329, EPI_ISL_1807330, EPI_ISL_1807331, EPI_ISL_1807332, EPI_ISL_1807333, EPI_ISL_1807336, EPI_ISL_1807337, EPI_ISL_1807338, EPI_ISL_1807339, EPI_ISL_1807340, EPI_ISL_1807341, EPI_ISL_1807342, EPI_ISL_1807343, EPI_ISL_1807344, EPI_ISL_1807345, EPI_ISL_1807347, EPI_ISL_1807348, EPI_ISL_1807349, EPI_ISL_1807350, EPI_ISL_1807351, EPI_ISL_1807353, EPI_ISL_1807354, EPI_ISL_1807355, EPI_ISL_1807356, EPI_ISL_1807361, EPI_ISL_1807363, EPI_ISL_1807365, EPI_ISL_1807366, EPI_ISL_1807373 | | | |
| see above | Instituto Nacional de Medicina Genomica | Instituto Nacional de Medicina Genomica | Hidalgo-Miranda A, Mendoza-Vargas A, Reyes-Grajeda JP, Cedro-Tanda A, Alcaraz N, Gonzalez-Barrera D, Rangel-DeLeon D, Munguia-Garza P, Ramirez-Vega O, Garcia-Cardenas FJ, Gonzalez-Woge MA, Herrera-Montalvo LA |
| EPI_ISL_1809447, EPI_ISL_1809449, EPI_ISL_1809451, EPI_ISL_1809453, EPI_ISL_1809455, EPI_ISL_1809457, EPI_ISL_1809459, EPI_ISL_1809461, EPI_ISL_1809463, EPI_ISL_1809465, EPI_ISL_1809467, EPI_ISL_1809469, EPI_ISL_1809471, EPI_ISL_1809472, EPI_ISL_1809474, EPI_ISL_1809476, EPI_ISL_1809478, EPI_ISL_1809480, EPI_ISL_1809482, EPI_ISL_1809484, EPI_ISL_1809486, EPI_ISL_1809488, | | | |

| | | | |
|---|---|---|---|
| EPI_ISL_1809490, EPI_ISL_1809492, EPI_ISL_1809493, EPI_ISL_1809495, EPI_ISL_1809499, EPI_ISL_1809501, EPI_ISL_1809503, EPI_ISL_1809507, EPI_ISL_1809509, EPI_ISL_1809513, EPI_ISL_1809515, EPI_ISL_1809521, EPI_ISL_1809523, EPI_ISL_1809524, EPI_ISL_1809526, EPI_ISL_1809528, EPI_ISL_1809530, EPI_ISL_1809532, EPI_ISL_1809536, EPI_ISL_1809540, EPI_ISL_1809542, EPI_ISL_1809544, EPI_ISL_1809545, EPI_ISL_1809549, EPI_ISL_1809553, EPI_ISL_1809555, EPI_ISL_1809559, EPI_ISL_1809561, EPI_ISL_1809566, EPI_ISL_1809568, EPI_ISL_1809570, EPI_ISL_1809572, EPI_ISL_1809574, EPI_ISL_1809576, EPI_ISL_1809578, EPI_ISL_1809580, EPI_ISL_1809581, EPI_ISL_1809583, EPI_ISL_1809585, EPI_ISL_1809587, EPI_ISL_1809589, EPI_ISL_1809591, EPI_ISL_1809593, EPI_ISL_1809595, EPI_ISL_1809597, EPI_ISL_1809599, EPI_ISL_1809600, EPI_ISL_1809602, EPI_ISL_1809606, EPI_ISL_1809607, EPI_ISL_1809609, EPI_ISL_1809613, EPI_ISL_1809615, EPI_ISL_1809619, EPI_ISL_1809623, EPI_ISL_1809625, EPI_ISL_1809629, EPI_ISL_1809635, EPI_ISL_1809637, EPI_ISL_1809639, EPI_ISL_1809644, EPI_ISL_1809646, EPI_ISL_1809648, EPI_ISL_1809650, EPI_ISL_1809652, EPI_ISL_1809654, EPI_ISL_1809656, EPI_ISL_1809658, EPI_ISL_1809660, EPI_ISL_1809663, EPI_ISL_1809672, EPI_ISL_1809674, EPI_ISL_1809676, EPI_ISL_1809677, EPI_ISL_1809679, EPI_ISL_1809681, EPI_ISL_1809683, EPI_ISL_1809685, EPI_ISL_1809687, EPI_ISL_1809689, EPI_ISL_1809691, EPI_ISL_1809695, EPI_ISL_1809697, EPI_ISL_1809698, EPI_ISL_1809701, EPI_ISL_1809702, EPI_ISL_1809704, EPI_ISL_1809706, EPI_ISL_1809708, EPI_ISL_1809710, EPI_ISL_1809712, EPI_ISL_1809714, EPI_ISL_1809716, EPI_ISL_1809718, EPI_ISL_1809720, EPI_ISL_1809722, EPI_ISL_1809724, EPI_ISL_1809726, EPI_ISL_1809728, EPI_ISL_1809732 | | | |
| see above | Illinois Department of Public Health | Illinois Department of Public Health - Chicago Lab | Vineet K. Dhiman, Ira Heimler, Joel Price |
| EPI_ISL_1810831, EPI_ISL_1810832, EPI_ISL_1810833, EPI_ISL_1810834, EPI_ISL_1810835, EPI_ISL_1810837, EPI_ISL_1810838, EPI_ISL_1810839, EPI_ISL_1810840, EPI_ISL_1810841, EPI_ISL_1810842, EPI_ISL_1810843, EPI_ISL_1810844, EPI_ISL_1810845, EPI_ISL_1810846, EPI_ISL_1810847, EPI_ISL_1810848, EPI_ISL_1810849, EPI_ISL_1810850, EPI_ISL_1810852, EPI_ISL_1810853, EPI_ISL_1810855, EPI_ISL_1810856, EPI_ISL_1810858, EPI_ISL_1810859, EPI_ISL_1810862, EPI_ISL_1810863, EPI_ISL_1810864, EPI_ISL_1810865, EPI_ISL_1810866, EPI_ISL_1810867, EPI_ISL_1810868, EPI_ISL_1810869, EPI_ISL_1810870, EPI_ISL_1810871, EPI_ISL_1810872, EPI_ISL_1810873, EPI_ISL_1810874, EPI_ISL_1810875, EPI_ISL_1810876, EPI_ISL_1810877, EPI_ISL_1810878, EPI_ISL_1810879, EPI_ISL_1810881, EPI_ISL_1810882, EPI_ISL_1810883, EPI_ISL_1810884, EPI_ISL_1810885, EPI_ISL_1810886, EPI_ISL_1810887, EPI_ISL_1810888, EPI_ISL_1810889, EPI_ISL_1810890, EPI_ISL_1810891, EPI_ISL_1810892, EPI_ISL_1810893, EPI_ISL_1810894, EPI_ISL_1810895, EPI_ISL_1810896, EPI_ISL_1810898, EPI_ISL_1810899, EPI_ISL_1810900, EPI_ISL_1810901, EPI_ISL_1810902, EPI_ISL_1810903, EPI_ISL_1810904, EPI_ISL_1810905, EPI_ISL_1810906, EPI_ISL_1810907, EPI_ISL_1810908, EPI_ISL_1810909, EPI_ISL_1810910, EPI_ISL_1810911, EPI_ISL_1810912, EPI_ISL_1810916, EPI_ISL_1810917, EPI_ISL_1810918, EPI_ISL_1810919, EPI_ISL_1810920, EPI_ISL_1810921, EPI_ISL_1810922, EPI_ISL_1810923 | | | |
| see above | InDRE | Instituto Nacional de Medicina Genomica | Hidalgo-Miranda A, Mendoza-Vargas A, Reyes-Grajeda JP, Cedro-Tanda A, Gisela Barrera-Badillo, Irma Lopez-Martinez, Jose Ernesto Ramirez González, Gonzalez-Barrera D, Rangel-DeLeon D, Munguia-Garza P, Garcia-Cardenas FJ, Gonzalez-Woge MA, Herrera-Montalvo LA. |
| EPI_ISL_1810946 | Moderna Inc. | Moderna Inc. | Yamuna D. Paila; Rolando Pajon |
| EPI_ISL_1811368, EPI_ISL_1811370, EPI_ISL_1811371, EPI_ISL_1811372, EPI_ISL_1811373, EPI_ISL_1811374, EPI_ISL_1811375, EPI_ISL_1811376, EPI_ISL_1811377, EPI_ISL_1811378 | IN State Department of Health Laboratory Services | IN State Department of Health Laboratory Services | Cassandra Campion, Jamie Yeadon, Brian Pope, Lixia Liu, Kyle Brownlee, Melissa Hindenlang, Mark Glazier |
| EPI_ISL_1811379, EPI_ISL_1811380, EPI_ISL_1811381, EPI_ISL_1811382, EPI_ISL_1811383, EPI_ISL_1811384, EPI_ISL_1811385, EPI_ISL_1811386, EPI_ISL_1811387, EPI_ISL_1811388, EPI_ISL_1811389, EPI_ISL_1811390, EPI_ISL_1811391, EPI_ISL_1811392, EPI_ISL_1811393, EPI_ISL_1811394, EPI_ISL_1811395, EPI_ISL_1811396, EPI_ISL_1811397, EPI_ISL_1811398, EPI_ISL_1811399, EPI_ISL_1811400, EPI_ISL_1811401, EPI_ISL_1811402, EPI_ISL_1811403, EPI_ISL_1811404, EPI_ISL_1811405, EPI_ISL_1811406, EPI_ISL_1811407, EPI_ISL_1811408, EPI_ISL_1811409, EPI_ISL_1811410, EPI_ISL_1811411, EPI_ISL_1811412, EPI_ISL_1811413, EPI_ISL_1811414, EPI_ISL_1811415, EPI_ISL_1811416, EPI_ISL_1811417, EPI_ISL_1811418, EPI_ISL_1811419, EPI_ISL_1811420, EPI_ISL_1811421, EPI_ISL_1811422, EPI_ISL_1811423, EPI_ISL_1811424, EPI_ISL_1811425, EPI_ISL_1811426, EPI_ISL_1811427, EPI_ISL_1811428, EPI_ISL_1811429, EPI_ISL_1811430, EPI_ISL_1811431, EPI_ISL_1811432, EPI_ISL_1811433, EPI_ISL_1811434, EPI_ISL_1811435, EPI_ISL_1811436, EPI_ISL_1811437, EPI_ISL_1811438, EPI_ISL_1811439, EPI_ISL_1811440, EPI_ISL_1811441, EPI_ISL_1811442, EPI_ISL_1811443, EPI_ISL_1811444, EPI_ISL_1811445, EPI_ISL_1811446, EPI_ISL_1811447, EPI_ISL_1811448, EPI_ISL_1811449, EPI_ISL_1811450, EPI_ISL_1811451, EPI_ISL_1811452, EPI_ISL_1811453, EPI_ISL_1811454, EPI_ISL_1811455, EPI_ISL_1811456, EPI_ISL_1811457, EPI_ISL_1811458, EPI_ISL_1811459, EPI_ISL_1811460, EPI_ISL_1811461, EPI_ISL_1811462, EPI_ISL_1811463, EPI_ISL_1811464, EPI_ISL_1811465, EPI_ISL_1811466, EPI_ISL_1811467, EPI_ISL_1811468, EPI_ISL_1811469, EPI_ISL_1811470, EPI_ISL_1811471, EPI_ISL_1811472, EPI_ISL_1811473, EPI_ISL_1811474, EPI_ISL_1811475, EPI_ISL_1811476, EPI_ISL_1811477, EPI_ISL_1811478, EPI_ISL_1811479, EPI_ISL_1811480, EPI_ISL_1811481, EPI_ISL_1811482, EPI_ISL_1811483, EPI_ISL_1811484, EPI_ISL_1811485, EPI_ISL_1811486, EPI_ISL_1811487, EPI_ISL_1811488, EPI_ISL_1811489, EPI_ISL_1811490, EPI_ISL_1811491, EPI_ISL_1811492, EPI_ISL_1811493, EPI_ISL_1811494, EPI_ISL_1811495, EPI_ISL_1811496, EPI_ISL_1811497, EPI_ISL_1811498, EPI_ISL_1811499, EPI_ISL_1811500, EPI_ISL_1811501, EPI_ISL_1811502, EPI_ISL_1811503, EPI_ISL_1811504, EPI_ISL_1811505, EPI_ISL_1811506, EPI_ISL_1811507, EPI_ISL_1811508, EPI_ISL_1811509, EPI_ISL_1811510, EPI_ISL_1811511, EPI_ISL_1811512, EPI_ISL_1811513, EPI_ISL_1811514, EPI_ISL_1811515, EPI_ISL_1811516, EPI_ISL_1811517, EPI_ISL_1811519, EPI_ISL_1811520, EPI_ISL_1811521, EPI_ISL_1811522, EPI_ISL_1811523, EPI_ISL_1811524, EPI_ISL_1811525, EPI_ISL_1811526, EPI_ISL_1811527, EPI_ISL_1811528, EPI_ISL_1811529, EPI_ISL_1811530, EPI_ISL_1811531, EPI_ISL_1811532, EPI_ISL_1811533, EPI_ISL_1811534, EPI_ISL_1811535, EPI_ISL_1811536, EPI_ISL_1811537, EPI_ISL_1811538, EPI_ISL_1811539, EPI_ISL_1811540, EPI_ISL_1811541, EPI_ISL_1811542, EPI_ISL_1811543, EPI_ISL_1811544, EPI_ISL_1811545 | | | |
| see above | Laboratorio Central de Epidemiología (LCE) | Instituto de Biotecnología de la UNAM | Julio Elias Alvarado-Yaah, Carlos F. Arias, Santiago Ávila-Ríos, Víctor Hugo Borja-Aburto, Celia Boukadida, Juan Bautista Chale-Dzul, Célida Duque-Molina, José Antonio Enciso-Moreno, Gloria Elena Espinosa-Ayala, Fernando Fontove-Herrera, Victor Eduardo Garcia-Arias, Concepción Grajales-Muñiz, Ricardo Grande, Alfredo Herrera-Estrella, Carla Ivón Herrera-Najera, Pavel Isa, Brenda Irasema Maldonado-Meza, Bernardo Martínez-Miguel, Margarita Matías-Florentino, María Guadalupe de Jesús Mireles-Rivera, Gloria María Molina-Salinas, Hector Montoya-Fuentes, José Esteban Muñoz-Medina, José de Jesús Nuñez-Contreras, Alicia Ocaña-Mondragón, Luis Alberto Ochoa-Carrera, Hector Esteban Paz-Juárez, Francisco Pulido, Helen Haydee Fernanda Ramírez-Plascencia, Angel Gustavo Salas-Lais, Alejandro Sanchez-Flores, Clara Esperanza Santacruz-Tinoco, María Guadalupe Santiago-Mauricio, Nelly Sélem-Mojica, Blanca Taboada, Gloria Vazquez |
| EPI_ISL_1811546, EPI_ISL_1811547, EPI_ISL_1811548, EPI_ISL_1811549, EPI_ISL_1811550, EPI_ISL_1811551, EPI_ISL_1811552, EPI_ISL_1811553, EPI_ISL_1811554, EPI_ISL_1811555, EPI_ISL_1811556, EPI_ISL_1811557, EPI_ISL_1811558, EPI_ISL_1811559, EPI_ISL_1811560, EPI_ISL_1811561, EPI_ISL_1811562, EPI_ISL_1811563, EPI_ISL_1811564, EPI_ISL_1811565, EPI_ISL_1811566, EPI_ISL_1811567, EPI_ISL_1811568, EPI_ISL_1811569, EPI_ISL_1811570, EPI_ISL_1811571, EPI_ISL_1811572, EPI_ISL_1811573, EPI_ISL_1811574, EPI_ISL_1811575, EPI_ISL_1811576, EPI_ISL_1811577, EPI_ISL_1811578, EPI_ISL_1811579, EPI_ISL_1811580, EPI_ISL_1811581, EPI_ISL_1811583, EPI_ISL_1811584, EPI_ISL_1811585, EPI_ISL_1811586, EPI_ISL_1811587, EPI_ISL_1811588, EPI_ISL_1811589, EPI_ISL_1811590, EPI_ISL_1811591, EPI_ISL_1811592, EPI_ISL_1811593, EPI_ISL_1811594, EPI_ISL_1811595, EPI_ISL_1811596, EPI_ISL_1811597, EPI_ISL_1811598, EPI_ISL_1811599, EPI_ISL_1811600, EPI_ISL_1811601, EPI_ISL_1811602, EPI_ISL_1811603, EPI_ISL_1811604, EPI_ISL_1811605, EPI_ISL_1811606, EPI_ISL_1811607, EPI_ISL_1811608, EPI_ISL_1811609, EPI_ISL_1811610, EPI_ISL_1811611, EPI_ISL_1811612, EPI_ISL_1811613, EPI_ISL_1811614, EPI_ISL_1811615, EPI_ISL_1811616, EPI_ISL_1811617, EPI_ISL_1811618, EPI_ISL_1811619, EPI_ISL_1811620, EPI_ISL_1811621, EPI_ISL_1811622, EPI_ISL_1811623, EPI_ISL_1811624, EPI_ISL_1811625, EPI_ISL_1811626, EPI_ISL_1811627, EPI_ISL_1811628, EPI_ISL_1811629, EPI_ISL_1811630, EPI_ISL_1811631, EPI_ISL_1811632, EPI_ISL_1811633, EPI_ISL_1811634, EPI_ISL_1811635, EPI_ISL_1811636, EPI_ISL_1811637, EPI_ISL_1811638, EPI_ISL_1811639, EPI_ISL_1811640, EPI_ISL_1811641 | | | |
| see above | MD PHL | MD PHL | Maryland Department of Health Laboratories Administration |
| EPI_ISL_1811642, EPI_ISL_1811643 | EXCITE Lab | Andersen lab at Scripps Research | Nicole L Washington, Simon White, Geraint Levan, Kelly Schiabor Barrett, Elizabeth Cirulli, Alexandre Bolze, Charlotte Rivera-Garcia, Ryan Cho, Jason Nguyen, Sherry Wang, Jimmy Ramirez, Celena Andrade, Alice Summerfield, Tyler Cassens, Efren Sandoval, Francisco Tanudjaja, Magnus Isaksson, William Lee, David Becker, Marc Laurent, James Lu + SEARCH |
| EPI_ISL_1811644, EPI_ISL_1811645, EPI_ISL_1811646, EPI_ISL_1811647, EPI_ISL_1811648 | EXCITE Lab | Andersen lab at Scripps Research | David Pride, Sharon Reed, Chip Schooley, Angela Scioscia, Natasha Martin Cheryl Anderson, Sawyer Farmer, Abigail Schnapper, Helena Tubb, Tommy Valles + SEARCH |
| EPI_ISL_1811649 | EXCITE Lab | Andersen lab at Scripps Research | Nicole L Washington, Simon White, Geraint Levan, Kelly Schiabor Barrett, Elizabeth Cirulli, Alexandre Bolze, Charlotte Rivera-Garcia, Ryan Cho, Jason Nguyen, Sherry Wang, Jimmy Ramirez, Celena Andrade, Alice Summerfield, Tyler Cassens, Efren Sandoval, Francisco Tanudjaja, Magnus Isaksson, William Lee, David Becker, Marc Laurent, James Lu + SEARCH |
| EPI_ISL_1811650 | EXCITE Lab | Andersen lab at Scripps Research | David Pride, Sharon Reed, Chip Schooley, Angela Scioscia, Natasha Martin Cheryl Anderson, Sawyer Farmer, Abigail Schnapper, Helena Tubb, Tommy Valles + SEARCH |
| EPI_ISL_1811651, EPI_ISL_1811652, EPI_ISL_1811653, EPI_ISL_1811654 | EXCITE Lab | Andersen lab at Scripps Research | Nicole L Washington, Simon White, Geraint Levan, Kelly Schiabor Barrett, Elizabeth Cirulli, Alexandre Bolze, Charlotte Rivera-Garcia, Ryan Cho, Jason Nguyen, Sherry Wang, Jimmy Ramirez, Celena Andrade, Alice Summerfield, Tyler Cassens, Efren Sandoval, Francisco Tanudjaja, Magnus Isaksson, William Lee, David Becker, Marc Laurent, James Lu + SEARCH |
| EPI_ISL_1811655, EPI_ISL_1811656, EPI_ISL_1811657 | EXCITE Lab | Andersen lab at Scripps Research | David Pride, Sharon Reed, Chip Schooley, Angela Scioscia, Natasha Martin Cheryl Anderson, Sawyer Farmer, Abigail Schnapper, Helena Tubb, Tommy Valles + SEARCH |
| EPI_ISL_1811658, EPI_ISL_1811659 | EXCITE Lab | Andersen lab at Scripps Research | Nicole L Washington, Simon White, Geraint Levan, Kelly Schiabor Barrett, Elizabeth Cirulli, Alexandre Bolze, Charlotte Rivera-Garcia, Ryan Cho, Jason Nguyen, Sherry Wang, Jimmy Ramirez, Celena Andrade, Alice Summerfield, Tyler Cassens, Efren Sandoval, Francisco Tanudjaja, Magnus Isaksson, William Lee, David Becker, Marc Laurent, James Lu + SEARCH |
| EPI_ISL_1811660, EPI_ISL_1811661 | EXCITE Lab | Andersen lab at Scripps Research | David Pride, Sharon Reed, Chip Schooley, Angela Scioscia, Natasha Martin Cheryl Anderson, Sawyer Farmer, Abigail Schnapper, Helena Tubb, Tommy Valles + SEARCH |
| EPI_ISL_1811662 | EXCITE Lab | Andersen lab at Scripps Research | Nicole L Washington, Simon White, Geraint Levan, Kelly Schiabor Barrett, Elizabeth Cirulli, Alexandre Bolze, Charlotte Rivera-Garcia, Ryan Cho, Jason Nguyen, Sherry Wang, Jimmy Ramirez, Celena Andrade, Alice Summerfield, Tyler Cassens, Efren Sandoval, Francisco Tanudjaja, Magnus Isaksson, William Lee, David Becker, Marc Laurent, James Lu + SEARCH |
| EPI_ISL_1811663, EPI_ISL_1811664, EPI_ISL_1811665, EPI_ISL_1811666 | EXCITE Lab | Andersen lab at Scripps Research | David Pride, Sharon Reed, Chip Schooley, Angela Scioscia, Natasha Martin Cheryl Anderson, Sawyer Farmer, Abigail Schnapper, Helena Tubb, Tommy Valles + SEARCH |
| EPI_ISL_1811667 | EXCITE Lab | Andersen lab at Scripps Research | Sydney Morgan + Marni Jacobs + Celestine Magallanes + Holly Valentine + Louise C. Laurent + SEARCH |
| EPI_ISL_1811668, EPI_ISL_1811669 | EXCITE Lab | Andersen lab at Scripps Research | David Pride, Sharon Reed, Chip Schooley, Angela Scioscia, Natasha Martin Cheryl Anderson, Sawyer Farmer, Abigail Schnapper, Helena Tubb, Tommy Valles + SEARCH |
| EPI_ISL_1811670, EPI_ISL_1811671 | EXCITE Lab | Andersen lab at Scripps Research | Nicole L Washington, Simon White, Geraint Levan, Kelly Schiabor Barrett, Elizabeth Cirulli, Alexandre Bolze, Charlotte Rivera-Garcia, Ryan Cho, Jason Nguyen, Sherry Wang, Jimmy Ramirez, Celena Andrade, Alice Summerfield, Tyler Cassens, Efren Sandoval, Francisco Tanudjaja, Magnus Isaksson, William Lee, David Becker, Marc Laurent, James Lu + SEARCH |
| EPI_ISL_1811672, EPI_ISL_1811673 | EXCITE Lab | Andersen lab at Scripps Research | David Pride, Sharon Reed, Chip Schooley, Angela Scioscia, Natasha Martin Cheryl Anderson, Sawyer Farmer, Abigail Schnapper, Helena Tubb, Tommy Valles + SEARCH |
| EPI_ISL_1811674, EPI_ISL_1811675 | EXCITE Lab | Andersen lab at Scripps Research | Nicole L Washington, Simon White, Geraint Levan, Kelly Schiabor Barrett, Elizabeth Cirulli, Alexandre Bolze, Charlotte Rivera-Garcia, Ryan Cho, Jason Nguyen, Sherry Wang, Jimmy Ramirez, Celena Andrade, Alice Summerfield, Tyler Cassens, Efren Sandoval, Francisco Tanudjaja, Magnus Isaksson, William Lee, David Becker, Marc Laurent, James Lu + SEARCH |
| EPI_ISL_1811676 | EXCITE Lab | Andersen lab at Scripps Research | Chip Schooley, Natasha Martin, Cheryl Anderson, Angela Scioscia, Smruthi Karthikeyan, Greg Humphrey, Sawyer Farmer, Abigail Schnapper, Helena Tubb, Tommy Valles + SEARCH |
| EPI_ISL_1811677 | EXCITE Lab | Andersen lab at Scripps Research | Sydney Morgan + Marni Jacobs + Celestine Magallanes + Holly Valentine + Louise C. Laurent + SEARCH |
| EPI_ISL_1811678, EPI_ISL_1811679 | EXCITE Lab | Andersen lab at Scripps Research | David Pride, Sharon Reed, Chip Schooley, Angela Scioscia, Natasha Martin Cheryl Anderson, Sawyer Farmer, Abigail Schnapper, Helena Tubb, Tommy Valles + SEARCH |
| EPI_ISL_1811680 | EXCITE Lab | Andersen lab at Scripps Research | Sydney Morgan + Marni Jacobs + Celestine Magallanes + Holly Valentine + Louise C. Laurent + SEARCH |
| EPI_ISL_1811681, EPI_ISL_1811682, EPI_ISL_1811683 | EXCITE Lab | Andersen lab at Scripps Research | Nicole L Washington, Simon White, Geraint Levan, Kelly Schiabor Barrett, Elizabeth Cirulli, Alexandre Bolze, Charlotte Rivera-Garcia, Ryan Cho, Jason Nguyen, Sherry Wang, Jimmy Ramirez, Celena Andrade, Alice Summerfield, Tyler Cassens, Efren Sandoval, Francisco Tanudjaja, Magnus Isaksson, William Lee, David Becker, Marc Laurent, James Lu + SEARCH |
| EPI_ISL_1811684, EPI_ISL_1811685, EPI_ISL_1811686 | EXCITE Lab | Andersen lab at Scripps Research | David Pride, Sharon Reed, Chip Schooley, Angela Scioscia, Natasha Martin Cheryl Anderson, Sawyer Farmer, Abigail Schnapper, Helena Tubb, Tommy Valles + SEARCH |
| EPI_ISL_1811687 | EXCITE Lab | Andersen lab at Scripps Research | Rebecca Fielding-Miller, Laura Kohn, Patricia Gonzalez-Zuniga, Fitri Carrissa Wijaya, Vinton Omaleki, Marlene Flores, Araz Majnoonian, Megan Nguyen, Ashkan Hassani, Smruthi Karthikeyan, Rodolfo Salido, Victor Cantu, Greg Humphrey, Sawyer Farmer, Abigail Schnapper, Helena Tubb, Tommy Valles + SEARCH |
| EPI_ISL_1811688, EPI_ISL_1811689, EPI_ISL_1811690, EPI_ISL_1811691, EPI_ISL_1811692, EPI_ISL_1811693, EPI_ISL_1811694, EPI_ISL_1811695, EPI_ISL_1811696, EPI_ISL_1811697, EPI_ISL_1811698 | | | |
| see above | EXCITE Lab | Andersen lab at Scripps Research | Nicole L Washington, Simon White, Geraint Levan, Kelly Schiabor Barrett, Elizabeth Cirulli, Alexandre Bolze, Charlotte Rivera-Garcia, Ryan Cho, Jason Nguyen, Sherry Wang, Jimmy Ramirez, Celena Andrade, Alice Summerfield, Tyler Cassens, Efren Sandoval, Francisco Tanudjaja, Magnus Isaksson, William Lee, David Becker, Marc Laurent, James Lu + SEARCH |
| EPI_ISL_1811699 | EXCITE Lab | Andersen lab at Scripps Research | Chip Schooley, Natasha Martin, Cheryl Anderson, Angela Scioscia, Smruthi Karthikeyan, Greg Humphrey, Sawyer Farmer, Abigail Schnapper, Helena Tubb, Tommy Valles + SEARCH |

| | | | |
|---|---|---|---|
| EPI_ISL_1811700, EPI_ISL_1811701, EPI_ISL_1811702, EPI_ISL_1811703, EPI_ISL_1811704, EPI_ISL_1811705, EPI_ISL_1811706, EPI_ISL_1811707, EPI_ISL_1811708 | EXCITE Lab | Andersen lab at Scripps Research | Nicole L Washington, Simon White, Geraint Levan, Kelly Schiabor Barrett, Elizabeth Cirulli, Alexandre Bolze, Charlotte Rivera-Garcia, Ryan Cho, Jason Nguyen, Sherry Wang, Jimmy Ramirez, Celena Andrade, Alice Summerfield, Tyler Cassens, Efren Sandoval, Francisco Tanudjaja, Magnus Isaksson, William Lee, David Becker, Marc Laurent, James Lu + SEARCH |
| EPI_ISL_1811709 | EXCITE Lab | Andersen lab at Scripps Research | David Pride, Sharon Reed, Chip Schooley, Angela Scioscia, Natasha Martin Cheryl Anderson, Sawyer Farmer, Abigail Schnapper, Helena Tubb, Tommy Valles + SEARCH |
| EPI_ISL_1811710, EPI_ISL_1811711, EPI_ISL_1811712 | EXCITE Lab | Andersen lab at Scripps Research | Nicole L Washington, Simon White, Geraint Levan, Kelly Schiabor Barrett, Elizabeth Cirulli, Alexandre Bolze, Charlotte Rivera-Garcia, Ryan Cho, Jason Nguyen, Sherry Wang, Jimmy Ramirez, Celena Andrade, Alice Summerfield, Tyler Cassens, Efren Sandoval, Francisco Tanudjaja, Magnus Isaksson, William Lee, David Becker, Marc Laurent, James Lu + SEARCH |
| EPI_ISL_1811713 | EXCITE Lab | Andersen lab at Scripps Research | David Pride, Sharon Reed, Chip Schooley, Angela Scioscia, Natasha Martin Cheryl Anderson, Sawyer Farmer, Abigail Schnapper, Helena Tubb, Tommy Valles + SEARCH |
| EPI_ISL_1811714, EPI_ISL_1811715 | EXCITE Lab | Andersen lab at Scripps Research | Nicole L Washington, Simon White, Geraint Levan, Kelly Schiabor Barrett, Elizabeth Cirulli, Alexandre Bolze, Charlotte Rivera-Garcia, Ryan Cho, Jason Nguyen, Sherry Wang, Jimmy Ramirez, Celena Andrade, Alice Summerfield, Tyler Cassens, Efren Sandoval, Francisco Tanudjaja, Magnus Isaksson, William Lee, David Becker, Marc Laurent, James Lu + SEARCH |
| EPI_ISL_1811716 | EXCITE Lab | Andersen lab at Scripps Research | Sydney Morgan + Marni Jacobs + Celestine Magallanes + Holly Valentine + Louise C. Laurent + SEARCH |
| EPI_ISL_1811717, EPI_ISL_1811718, EPI_ISL_1811719, EPI_ISL_1811720, EPI_ISL_1811721 | EXCITE Lab | Andersen lab at Scripps Research | Nicole L Washington, Simon White, Geraint Levan, Kelly Schiabor Barrett, Elizabeth Cirulli, Alexandre Bolze, Charlotte Rivera-Garcia, Ryan Cho, Jason Nguyen, Sherry Wang, Jimmy Ramirez, Celena Andrade, Alice Summerfield, Tyler Cassens, Efren Sandoval, Francisco Tanudjaja, Magnus Isaksson, William Lee, David Becker, Marc Laurent, James Lu + SEARCH |
| EPI_ISL_1811722, EPI_ISL_1811723 | EXCITE Lab | Andersen lab at Scripps Research | David Pride, Sharon Reed, Chip Schooley, Angela Scioscia, Natasha Martin Cheryl Anderson, Sawyer Farmer, Abigail Schnapper, Helena Tubb, Tommy Valles + SEARCH |
| EPI_ISL_1811724, EPI_ISL_1811725, EPI_ISL_1811726, EPI_ISL_1811727 | EXCITE Lab | Andersen lab at Scripps Research | Nicole L Washington, Simon White, Geraint Levan, Kelly Schiabor Barrett, Elizabeth Cirulli, Alexandre Bolze, Charlotte Rivera-Garcia, Ryan Cho, Jason Nguyen, Sherry Wang, Jimmy Ramirez, Celena Andrade, Alice Summerfield, Tyler Cassens, Efren Sandoval, Francisco Tanudjaja, Magnus Isaksson, William Lee, David Becker, Marc Laurent, James Lu + SEARCH |
| EPI_ISL_1811728 | EXCITE Lab | Andersen lab at Scripps Research | Chip Schooley, Natasha Martin, Cheryl Anderson, Angela Scioscia, Smruthi Karthikeyan, Greg Humphrey, Sawyer Farmer, Abigail Schnapper, Helena Tubb, Tommy Valles + SEARCH |
| EPI_ISL_1811729, EPI_ISL_1811730, EPI_ISL_1811731, EPI_ISL_1811732, EPI_ISL_1811733 | EXCITE Lab | Andersen lab at Scripps Research | Nicole L Washington, Simon White, Geraint Levan, Kelly Schiabor Barrett, Elizabeth Cirulli, Alexandre Bolze, Charlotte Rivera-Garcia, Ryan Cho, Jason Nguyen, Sherry Wang, Jimmy Ramirez, Celena Andrade, Alice Summerfield, Tyler Cassens, Efren Sandoval, Francisco Tanudjaja, Magnus Isaksson, William Lee, David Becker, Marc Laurent, James Lu + SEARCH |
| EPI_ISL_1811734 | EXCITE Lab | Andersen lab at Scripps Research | David Pride, Sharon Reed, Chip Schooley, Angela Scioscia, Natasha Martin Cheryl Anderson, Sawyer Farmer, Abigail Schnapper, Helena Tubb, Tommy Valles + SEARCH |
| EPI_ISL_1811735, EPI_ISL_1811736, EPI_ISL_1811737, EPI_ISL_1811738, EPI_ISL_1811739, EPI_ISL_1811740, EPI_ISL_1811741, EPI_ISL_1811742, EPI_ISL_1811743, EPI_ISL_1811744, EPI_ISL_1811745, EPI_ISL_1811746, EPI_ISL_1811747, EPI_ISL_1811748, EPI_ISL_1811749, EPI_ISL_1811750 | | | |
| see above | EXCITE Lab | Andersen lab at Scripps Research | Nicole L Washington, Simon White, Geraint Levan, Kelly Schiabor Barrett, Elizabeth Cirulli, Alexandre Bolze, Charlotte Rivera-Garcia, Ryan Cho, Jason Nguyen, Sherry Wang, Jimmy Ramirez, Celena Andrade, Alice Summerfield, Tyler Cassens, Efren Sandoval, Francisco Tanudjaja, Magnus Isaksson, William Lee, David Becker, Marc Laurent, James Lu + SEARCH |
| EPI_ISL_1811751 | EXCITE Lab | Andersen lab at Scripps Research | David Pride, Sharon Reed, Chip Schooley, Angela Scioscia, Natasha Martin Cheryl Anderson, Sawyer Farmer, Abigail Schnapper, Helena Tubb, Tommy Valles + SEARCH |
| EPI_ISL_1811752, EPI_ISL_1811753, EPI_ISL_1811754, EPI_ISL_1811755, EPI_ISL_1811756, EPI_ISL_1811757, EPI_ISL_1811758, EPI_ISL_1811759, EPI_ISL_1811760, EPI_ISL_1811761, EPI_ISL_1811762, EPI_ISL_1811763, EPI_ISL_1811764 | | | |
| see above | EXCITE Lab | Andersen lab at Scripps Research | Nicole L Washington, Simon White, Geraint Levan, Kelly Schiabor Barrett, Elizabeth Cirulli, Alexandre Bolze, Charlotte Rivera-Garcia, Ryan Cho, Jason Nguyen, Sherry Wang, Jimmy Ramirez, Celena Andrade, Alice Summerfield, Tyler Cassens, Efren Sandoval, Francisco Tanudjaja, Magnus Isaksson, William Lee, David Becker, Marc Laurent, James Lu + SEARCH |
| EPI_ISL_1811765 | EXCITE Lab | Andersen lab at Scripps Research | Chip Schooley, Natasha Martin, Cheryl Anderson, Angela Scioscia, Smruthi Karthikeyan, Greg Humphrey, Sawyer Farmer, Abigail Schnapper, Helena Tubb, Tommy Valles + SEARCH |
| EPI_ISL_1811766 | EXCITE Lab | Andersen lab at Scripps Research | Nicole L Washington, Simon White, Geraint Levan, Kelly Schiabor Barrett, Elizabeth Cirulli, Alexandre Bolze, Charlotte Rivera-Garcia, Ryan Cho, Jason Nguyen, Sherry Wang, Jimmy Ramirez, Celena Andrade, Alice Summerfield, Tyler Cassens, Efren Sandoval, Francisco Tanudjaja, Magnus Isaksson, William Lee, David Becker, Marc Laurent, James Lu + SEARCH |
| EPI_ISL_1811767 | EXCITE Lab | Andersen lab at Scripps Research | Fred Wu, Christine Mitchell + SEARCH |
| EPI_ISL_1811768, EPI_ISL_1811769, EPI_ISL_1811770, EPI_ISL_1811771, EPI_ISL_1811772, EPI_ISL_1811773, EPI_ISL_1811774, EPI_ISL_1811775, EPI_ISL_1811776, EPI_ISL_1811777, EPI_ISL_1811778, EPI_ISL_1811779, EPI_ISL_1811780, EPI_ISL_1811781, EPI_ISL_1811782, EPI_ISL_1811783, EPI_ISL_1811784, EPI_ISL_1811785, EPI_ISL_1811786, EPI_ISL_1811787, EPI_ISL_1811788, EPI_ISL_1811789 | | | |
| see above | EXCITE Lab | Andersen lab at Scripps Research | Nicole L Washington, Simon White, Geraint Levan, Kelly Schiabor Barrett, Elizabeth Cirulli, Alexandre Bolze, Charlotte Rivera-Garcia, Ryan Cho, Jason Nguyen, Sherry Wang, Jimmy Ramirez, Celena Andrade, Alice Summerfield, Tyler Cassens, Efren Sandoval, Francisco Tanudjaja, Magnus Isaksson, William Lee, David Becker, Marc Laurent, James Lu + SEARCH |
| EPI_ISL_1811790, EPI_ISL_1811791 | EXCITE Lab | Andersen lab at Scripps Research | David Pride, Sharon Reed, Chip Schooley, Angela Scioscia, Natasha Martin Cheryl Anderson, Sawyer Farmer, Abigail Schnapper, Helena Tubb, Tommy Valles + SEARCH |
| EPI_ISL_1811792, EPI_ISL_1811793, EPI_ISL_1811794 | EXCITE Lab | Andersen lab at Scripps Research | Sydney Morgan + Marni Jacobs + Celestine Magallanes + Holly Valentine + Louise C. Laurent + SEARCH |
| EPI_ISL_1811795, EPI_ISL_1811796, EPI_ISL_1811797, EPI_ISL_1811798, EPI_ISL_1811799, EPI_ISL_1811800, EPI_ISL_1811801, EPI_ISL_1811802, EPI_ISL_1811803, EPI_ISL_1811804, EPI_ISL_1811805, EPI_ISL_1811806, EPI_ISL_1811807 | | | |
| see above | EXCITE Lab | Andersen lab at Scripps Research | Nicole L Washington, Simon White, Geraint Levan, Kelly Schiabor Barrett, Elizabeth Cirulli, Alexandre Bolze, Charlotte Rivera-Garcia, Ryan Cho, Jason Nguyen, Sherry Wang, Jimmy Ramirez, Celena Andrade, Alice Summerfield, Tyler Cassens, Efren Sandoval, Francisco Tanudjaja, Magnus Isaksson, William Lee, David Becker, Marc Laurent, James Lu + SEARCH |
| EPI_ISL_1811808 | EXCITE Lab | Andersen lab at Scripps Research | Chip Schooley, Natasha Martin, Cheryl Anderson, Angela Scioscia, Smruthi Karthikeyan, Greg Humphrey, Sawyer Farmer, Abigail Schnapper, Helena Tubb, Tommy Valles + SEARCH |
| EPI_ISL_1811809, EPI_ISL_1811810, EPI_ISL_1811811 | EXCITE Lab | Andersen lab at Scripps Research | Nicole L Washington, Simon White, Geraint Levan, Kelly Schiabor Barrett, Elizabeth Cirulli, Alexandre Bolze, Charlotte Rivera-Garcia, Ryan Cho, Jason Nguyen, Sherry Wang, Jimmy Ramirez, Celena Andrade, Alice Summerfield, Tyler Cassens, Efren Sandoval, Francisco Tanudjaja, Magnus Isaksson, William Lee, David Becker, Marc Laurent, James Lu + SEARCH |
| EPI_ISL_1811812 | EXCITE Lab | Andersen lab at Scripps Research | David Pride, Sharon Reed, Chip Schooley, Angela Scioscia, Natasha Martin Cheryl Anderson, Sawyer Farmer, Abigail Schnapper, Helena Tubb, Tommy Valles + SEARCH |
| EPI_ISL_1811813, EPI_ISL_1811814, EPI_ISL_1811815, EPI_ISL_1811816, EPI_ISL_1811817, EPI_ISL_1811818, EPI_ISL_1811819, EPI_ISL_1811820 | EXCITE Lab | Andersen lab at Scripps Research | Nicole L Washington, Simon White, Geraint Levan, Kelly Schiabor Barrett, Elizabeth Cirulli, Alexandre Bolze, Charlotte Rivera-Garcia, Ryan Cho, Jason Nguyen, Sherry Wang, Jimmy Ramirez, Celena Andrade, Alice Summerfield, Tyler Cassens, Efren Sandoval, Francisco Tanudjaja, Magnus Isaksson, William Lee, David Becker, Marc Laurent, James Lu + SEARCH |
| EPI_ISL_1811821 | EXCITE Lab | Andersen lab at Scripps Research | David Pride, Sharon Reed, Chip Schooley, Angela Scioscia, Natasha Martin Cheryl Anderson, Sawyer Farmer, Abigail Schnapper, Helena Tubb, Tommy Valles + SEARCH |
| EPI_ISL_1811822, EPI_ISL_1811823 | EXCITE Lab | Andersen lab at Scripps Research | Nicole L Washington, Simon White, Geraint Levan, Kelly Schiabor Barrett, Elizabeth Cirulli, Alexandre Bolze, Charlotte Rivera-Garcia, Ryan Cho, Jason Nguyen, Sherry Wang, Jimmy Ramirez, Celena Andrade, Alice Summerfield, Tyler Cassens, Efren Sandoval, Francisco Tanudjaja, Magnus Isaksson, William Lee, David Becker, Marc Laurent, James Lu + SEARCH |
| EPI_ISL_1811824 | EXCITE Lab | Andersen lab at Scripps Research | David Pride, Sharon Reed, Chip Schooley, Angela Scioscia, Natasha Martin Cheryl Anderson, Sawyer Farmer, Abigail Schnapper, Helena Tubb, Tommy Valles + SEARCH |
| EPI_ISL_1811825, EPI_ISL_1811826, EPI_ISL_1811827, EPI_ISL_1811828, EPI_ISL_1811829, EPI_ISL_1811830, EPI_ISL_1811831, EPI_ISL_1811832, EPI_ISL_1811833 | EXCITE Lab | Andersen lab at Scripps Research | Nicole L Washington, Simon White, Geraint Levan, Kelly Schiabor Barrett, Elizabeth Cirulli, Alexandre Bolze, Charlotte Rivera-Garcia, Ryan Cho, Jason Nguyen, Sherry Wang, Jimmy Ramirez, Celena Andrade, Alice Summerfield, Tyler Cassens, Efren Sandoval, Francisco Tanudjaja, Magnus Isaksson, William Lee, David Becker, Marc Laurent, James Lu + SEARCH |
| EPI_ISL_1811834 | EXCITE Lab | Andersen lab at Scripps Research | Chip Schooley, Natasha Martin, Cheryl Anderson, Angela Scioscia, Smruthi Karthikeyan, Greg Humphrey, Sawyer Farmer, Abigail Schnapper, Helena Tubb, Tommy Valles + SEARCH |
| EPI_ISL_1811835 | EXCITE Lab | Andersen lab at Scripps Research | David Pride, Sharon Reed, Chip Schooley, Angela Scioscia, Natasha Martin Cheryl Anderson, Sawyer Farmer, Abigail Schnapper, Helena Tubb, Tommy Valles + SEARCH |
| EPI_ISL_1811836 | EXCITE Lab | Andersen lab at Scripps Research | Nicole L Washington, Simon White, Geraint Levan, Kelly Schiabor Barrett, Elizabeth Cirulli, Alexandre Bolze, Charlotte Rivera-Garcia, Ryan Cho, Jason Nguyen, Sherry Wang, Jimmy Ramirez, Celena Andrade, Alice Summerfield, Tyler Cassens, Efren Sandoval, Francisco Tanudjaja, Magnus Isaksson, William Lee, David Becker, Marc Laurent, James Lu + SEARCH |
| EPI_ISL_1811837, EPI_ISL_1811838 | EXCITE Lab | Andersen lab at Scripps Research | David Pride, Sharon Reed, Chip Schooley, Angela Scioscia, Natasha Martin Cheryl Anderson, Sawyer Farmer, Abigail Schnapper, Helena Tubb, Tommy Valles + SEARCH |
| EPI_ISL_1811839, EPI_ISL_1811840 | EXCITE Lab | Andersen lab at Scripps Research | Nicole L Washington, Simon White, Geraint Levan, Kelly Schiabor Barrett, Elizabeth Cirulli, Alexandre Bolze, Charlotte Rivera-Garcia, Ryan Cho, Jason Nguyen, Sherry Wang, Jimmy Ramirez, Celena Andrade, Alice Summerfield, Tyler Cassens, Efren Sandoval, Francisco Tanudjaja, Magnus Isaksson, William Lee, David Becker, Marc Laurent, James Lu + SEARCH |
| EPI_ISL_1811841, | EXCITE Lab | Andersen lab at Scripps Research | David Pride, Sharon Reed, Chip Schooley, Angela Scioscia, Natasha Martin Cheryl Anderson, Sawyer Farmer, Abigail Schnapper, Helena Tubb, Tommy Valles + SEARCH |

| | | | |
|---|---|---|---|
| EPI_ISL_1811842 | | | |
| EPI_ISL_1811843, EPI_ISL_1811844, EPI_ISL_1811845, EPI_ISL_1811846 | EXCITE Lab | Andersen lab at Scripps Research | Nicole L Washington, Simon White, Geraint Levan, Kelly Schiabor Barrett, Elizabeth Cirulli, Alexandre Bolze, Charlotte Rivera-Garcia, Ryan Cho, Jason Nguyen, Sherry Wang, Jimmy Ramirez, Celena Andrade, Alice Summerfield, Tyler Cassens, Efren Sandoval, Francisco Tanudjaja, Magnus Isaksson, William Lee, David Becker, Marc Laurent, James Lu + SEARCH |
| EPI_ISL_1811847 | EXCITE Lab | Andersen lab at Scripps Research | David Pride, Sharon Reed, Chip Schooley, Angela Scioscia, Natasha Martin Cheryl Anderson, Sawyer Farmer, Abigail Schnapper, Helena Tubb, Tommy Valles + SEARCH |
| EPI_ISL_1811848 | EXCITE Lab | Andersen lab at Scripps Research | Chip Schooley, Natasha Martin, Cheryl Anderson, Angela Scioscia, Smruthi Karthikeyan, Greg Humphrey, Sawyer Farmer, Abigail Schnapper, Helena Tubb, Tommy Valles + SEARCH |
| EPI_ISL_1811849 | EXCITE Lab | Andersen lab at Scripps Research | David Pride, Sharon Reed, Chip Schooley, Angela Scioscia, Natasha Martin Cheryl Anderson, Sawyer Farmer, Abigail Schnapper, Helena Tubb, Tommy Valles + SEARCH |
| EPI_ISL_1811850 | EXCITE Lab | Andersen lab at Scripps Research | Nicole L Washington, Simon White, Geraint Levan, Kelly Schiabor Barrett, Elizabeth Cirulli, Alexandre Bolze, Charlotte Rivera-Garcia, Ryan Cho, Jason Nguyen, Sherry Wang, Jimmy Ramirez, Celena Andrade, Alice Summerfield, Tyler Cassens, Efren Sandoval, Francisco Tanudjaja, Magnus Isaksson, William Lee, David Becker, Marc Laurent, James Lu + SEARCH |
| EPI_ISL_1811851 | EXCITE Lab | Andersen lab at Scripps Research | Sydney Morgan + Marni Jacobs + Celestine Magallanes + Holly Valentine + Louise C. Laurent + SEARCH |
| EPI_ISL_1811852, EPI_ISL_1811853, EPI_ISL_1811854, EPI_ISL_1811855, EPI_ISL_1811856 | EXCITE Lab | Andersen lab at Scripps Research | Nicole L Washington, Simon White, Geraint Levan, Kelly Schiabor Barrett, Elizabeth Cirulli, Alexandre Bolze, Charlotte Rivera-Garcia, Ryan Cho, Jason Nguyen, Sherry Wang, Jimmy Ramirez, Celena Andrade, Alice Summerfield, Tyler Cassens, Efren Sandoval, Francisco Tanudjaja, Magnus Isaksson, William Lee, David Becker, Marc Laurent, James Lu + SEARCH |
| EPI_ISL_1811857 | EXCITE Lab | Andersen lab at Scripps Research | David Pride, Sharon Reed, Chip Schooley, Angela Scioscia, Natasha Martin Cheryl Anderson, Sawyer Farmer, Abigail Schnapper, Helena Tubb, Tommy Valles + SEARCH |
| EPI_ISL_1811858, EPI_ISL_1811859, EPI_ISL_1811860 | EXCITE Lab | Andersen lab at Scripps Research | Nicole L Washington, Simon White, Geraint Levan, Kelly Schiabor Barrett, Elizabeth Cirulli, Alexandre Bolze, Charlotte Rivera-Garcia, Ryan Cho, Jason Nguyen, Sherry Wang, Jimmy Ramirez, Celena Andrade, Alice Summerfield, Tyler Cassens, Efren Sandoval, Francisco Tanudjaja, Magnus Isaksson, William Lee, David Becker, Marc Laurent, James Lu + SEARCH |
| EPI_ISL_1811861 | EXCITE Lab | Andersen lab at Scripps Research | David Pride, Sharon Reed, Chip Schooley, Angela Scioscia, Natasha Martin Cheryl Anderson, Sawyer Farmer, Abigail Schnapper, Helena Tubb, Tommy Valles + SEARCH |
| EPI_ISL_1811862 | EXCITE Lab | Andersen lab at Scripps Research | Sydney Morgan + Marni Jacobs + Celestine Magallanes + Holly Valentine + Louise C. Laurent + SEARCH |
| EPI_ISL_1811863, EPI_ISL_1811864, EPI_ISL_1811865 | EXCITE Lab | Andersen lab at Scripps Research | Nicole L Washington, Simon White, Geraint Levan, Kelly Schiabor Barrett, Elizabeth Cirulli, Alexandre Bolze, Charlotte Rivera-Garcia, Ryan Cho, Jason Nguyen, Sherry Wang, Jimmy Ramirez, Celena Andrade, Alice Summerfield, Tyler Cassens, Efren Sandoval, Francisco Tanudjaja, Magnus Isaksson, William Lee, David Becker, Marc Laurent, James Lu + SEARCH |
| EPI_ISL_1811866, EPI_ISL_1811867 | EXCITE Lab | Andersen lab at Scripps Research | David Pride, Sharon Reed, Chip Schooley, Angela Scioscia, Natasha Martin Cheryl Anderson, Sawyer Farmer, Abigail Schnapper, Helena Tubb, Tommy Valles + SEARCH |
| EPI_ISL_1811868 | EXCITE Lab | Andersen lab at Scripps Research | Nicole L Washington, Simon White, Geraint Levan, Kelly Schiabor Barrett, Elizabeth Cirulli, Alexandre Bolze, Charlotte Rivera-Garcia, Ryan Cho, Jason Nguyen, Sherry Wang, Jimmy Ramirez, Celena Andrade, Alice Summerfield, Tyler Cassens, Efren Sandoval, Francisco Tanudjaja, Magnus Isaksson, William Lee, David Becker, Marc Laurent, James Lu + SEARCH |
| EPI_ISL_1811869 | EXCITE Lab | Andersen lab at Scripps Research | Sydney Morgan + Marni Jacobs + Celestine Magallanes + Holly Valentine + Louise C. Laurent + SEARCH |
| EPI_ISL_1811870, EPI_ISL_1811871, EPI_ISL_1811872, EPI_ISL_1811873, EPI_ISL_1811874, EPI_ISL_1811875, EPI_ISL_1811876 | EXCITE Lab | Andersen lab at Scripps Research | Nicole L Washington, Simon White, Geraint Levan, Kelly Schiabor Barrett, Elizabeth Cirulli, Alexandre Bolze, Charlotte Rivera-Garcia, Ryan Cho, Jason Nguyen, Sherry Wang, Jimmy Ramirez, Celena Andrade, Alice Summerfield, Tyler Cassens, Efren Sandoval, Francisco Tanudjaja, Magnus Isaksson, William Lee, David Becker, Marc Laurent, James Lu + SEARCH |
| EPI_ISL_1811877 | EXCITE Lab | Andersen lab at Scripps Research | David Pride, Sharon Reed, Chip Schooley, Angela Scioscia, Natasha Martin Cheryl Anderson, Sawyer Farmer, Abigail Schnapper, Helena Tubb, Tommy Valles + SEARCH |
| EPI_ISL_1811878, EPI_ISL_1811879, EPI_ISL_1811880, EPI_ISL_1811881, EPI_ISL_1811882 | EXCITE Lab | Andersen lab at Scripps Research | Nicole L Washington, Simon White, Geraint Levan, Kelly Schiabor Barrett, Elizabeth Cirulli, Alexandre Bolze, Charlotte Rivera-Garcia, Ryan Cho, Jason Nguyen, Sherry Wang, Jimmy Ramirez, Celena Andrade, Alice Summerfield, Tyler Cassens, Efren Sandoval, Francisco Tanudjaja, Magnus Isaksson, William Lee, David Becker, Marc Laurent, James Lu + SEARCH |
| EPI_ISL_1811883 | EXCITE Lab | Andersen lab at Scripps Research | David Pride, Sharon Reed, Chip Schooley, Angela Scioscia, Natasha Martin Cheryl Anderson, Sawyer Farmer, Abigail Schnapper, Helena Tubb, Tommy Valles + SEARCH |
| EPI_ISL_1811884, EPI_ISL_1811885 | EXCITE Lab | Andersen lab at Scripps Research | Nicole L Washington, Simon White, Geraint Levan, Kelly Schiabor Barrett, Elizabeth Cirulli, Alexandre Bolze, Charlotte Rivera-Garcia, Ryan Cho, Jason Nguyen, Sherry Wang, Jimmy Ramirez, Celena Andrade, Alice Summerfield, Tyler Cassens, Efren Sandoval, Francisco Tanudjaja, Magnus Isaksson, William Lee, David Becker, Marc Laurent, James Lu + SEARCH |
| EPI_ISL_1811886 | EXCITE Lab | Andersen lab at Scripps Research | David Pride, Sharon Reed, Chip Schooley, Angela Scioscia, Natasha Martin Cheryl Anderson, Sawyer Farmer, Abigail Schnapper, Helena Tubb, Tommy Valles + SEARCH |
| EPI_ISL_1811887, EPI_ISL_1811888 | EXCITE Lab | Andersen lab at Scripps Research | Nicole L Washington, Simon White, Geraint Levan, Kelly Schiabor Barrett, Elizabeth Cirulli, Alexandre Bolze, Charlotte Rivera-Garcia, Ryan Cho, Jason Nguyen, Sherry Wang, Jimmy Ramirez, Celena Andrade, Alice Summerfield, Tyler Cassens, Efren Sandoval, Francisco Tanudjaja, Magnus Isaksson, William Lee, David Becker, Marc Laurent, James Lu + SEARCH |
| EPI_ISL_1811889 | EXCITE Lab | Andersen lab at Scripps Research | David Pride, Sharon Reed, Chip Schooley, Angela Scioscia, Natasha Martin Cheryl Anderson, Sawyer Farmer, Abigail Schnapper, Helena Tubb, Tommy Valles + SEARCH |
| EPI_ISL_1811890 | EXCITE Lab | Andersen lab at Scripps Research | Nicole L Washington, Simon White, Geraint Levan, Kelly Schiabor Barrett, Elizabeth Cirulli, Alexandre Bolze, Charlotte Rivera-Garcia, Ryan Cho, Jason Nguyen, Sherry Wang, Jimmy Ramirez, Celena Andrade, Alice Summerfield, Tyler Cassens, Efren Sandoval, Francisco Tanudjaja, Magnus Isaksson, William Lee, David Becker, Marc Laurent, James Lu + SEARCH |
| EPI_ISL_1811891, EPI_ISL_1811892, EPI_ISL_1811893 | EXCITE Lab | Andersen lab at Scripps Research | David Pride, Sharon Reed, Chip Schooley, Angela Scioscia, Natasha Martin Cheryl Anderson, Sawyer Farmer, Abigail Schnapper, Helena Tubb, Tommy Valles + SEARCH |
| EPI_ISL_1811894 | EXCITE Lab | Andersen lab at Scripps Research | Nicole L Washington, Simon White, Geraint Levan, Kelly Schiabor Barrett, Elizabeth Cirulli, Alexandre Bolze, Charlotte Rivera-Garcia, Ryan Cho, Jason Nguyen, Sherry Wang, Jimmy Ramirez, Celena Andrade, Alice Summerfield, Tyler Cassens, Efren Sandoval, Francisco Tanudjaja, Magnus Isaksson, William Lee, David Becker, Marc Laurent, James Lu + SEARCH |
| EPI_ISL_1811895 | EXCITE Lab | Andersen lab at Scripps Research | David Pride, Sharon Reed, Chip Schooley, Angela Scioscia, Natasha Martin Cheryl Anderson, Sawyer Farmer, Abigail Schnapper, Helena Tubb, Tommy Valles + SEARCH |
| EPI_ISL_1811896, EPI_ISL_1811897, EPI_ISL_1811898, EPI_ISL_1811899, EPI_ISL_1811900, EPI_ISL_1811901, EPI_ISL_1811902, EPI_ISL_1811903, EPI_ISL_1811904 | EXCITE Lab | Andersen lab at Scripps Research | Nicole L Washington, Simon White, Geraint Levan, Kelly Schiabor Barrett, Elizabeth Cirulli, Alexandre Bolze, Charlotte Rivera-Garcia, Ryan Cho, Jason Nguyen, Sherry Wang, Jimmy Ramirez, Celena Andrade, Alice Summerfield, Tyler Cassens, Efren Sandoval, Francisco Tanudjaja, Magnus Isaksson, William Lee, David Becker, Marc Laurent, James Lu + SEARCH |
| EPI_ISL_1811905 | EXCITE Lab | Andersen lab at Scripps Research | David Pride, Sharon Reed, Chip Schooley, Angela Scioscia, Natasha Martin Cheryl Anderson, Sawyer Farmer, Abigail Schnapper, Helena Tubb, Tommy Valles + SEARCH |
| EPI_ISL_1811906, EPI_ISL_1811907, EPI_ISL_1811908, EPI_ISL_1811909, EPI_ISL_1811910, EPI_ISL_1811911, EPI_ISL_1811912, EPI_ISL_1811913, EPI_ISL_1811914, EPI_ISL_1811915, EPI_ISL_1811916, EPI_ISL_1811917, EPI_ISL_1811918, EPI_ISL_1811919, EPI_ISL_1811920, EPI_ISL_1811921, EPI_ISL_1811922, EPI_ISL_1811923 | | | |
| see above | EXCITE Lab | Andersen lab at Scripps Research | Nicole L Washington, Simon White, Geraint Levan, Kelly Schiabor Barrett, Elizabeth Cirulli, Alexandre Bolze, Charlotte Rivera-Garcia, Ryan Cho, Jason Nguyen, Sherry Wang, Jimmy Ramirez, Celena Andrade, Alice Summerfield, Tyler Cassens, Efren Sandoval, Francisco Tanudjaja, Magnus Isaksson, William Lee, David Becker, Marc Laurent, James Lu + SEARCH |
| EPI_ISL_1811924 | EXCITE Lab | Andersen lab at Scripps Research | David Pride, Sharon Reed, Chip Schooley, Angela Scioscia, Natasha Martin Cheryl Anderson, Sawyer Farmer, Abigail Schnapper, Helena Tubb, Tommy Valles + SEARCH |
| EPI_ISL_1811925, EPI_ISL_1811926, EPI_ISL_1811927, EPI_ISL_1811928, EPI_ISL_1811929, EPI_ISL_1811930, EPI_ISL_1811931 | EXCITE Lab | Andersen lab at Scripps Research | Nicole L Washington, Simon White, Geraint Levan, Kelly Schiabor Barrett, Elizabeth Cirulli, Alexandre Bolze, Charlotte Rivera-Garcia, Ryan Cho, Jason Nguyen, Sherry Wang, Jimmy Ramirez, Celena Andrade, Alice Summerfield, Tyler Cassens, Efren Sandoval, Francisco Tanudjaja, Magnus Isaksson, William Lee, David Becker, Marc Laurent, James Lu + SEARCH |
| EPI_ISL_1811932 | EXCITE Lab | Andersen lab at Scripps Research | David Pride, Sharon Reed, Chip Schooley, Angela Scioscia, Natasha Martin Cheryl Anderson, Sawyer Farmer, Abigail Schnapper, Helena Tubb, Tommy Valles + SEARCH |
| EPI_ISL_1811933, EPI_ISL_1811934, EPI_ISL_1811935, EPI_ISL_1811936, EPI_ISL_1811937, EPI_ISL_1811938 | EXCITE Lab | Andersen lab at Scripps Research | Nicole L Washington, Simon White, Geraint Levan, Kelly Schiabor Barrett, Elizabeth Cirulli, Alexandre Bolze, Charlotte Rivera-Garcia, Ryan Cho, Jason Nguyen, Sherry Wang, Jimmy Ramirez, Celena Andrade, Alice Summerfield, Tyler Cassens, Efren Sandoval, Francisco Tanudjaja, Magnus Isaksson, William Lee, David Becker, Marc Laurent, James Lu + SEARCH |
| EPI_ISL_1811939 | EXCITE Lab | Andersen lab at Scripps Research | David Pride, Sharon Reed, Chip Schooley, Angela Scioscia, Natasha Martin Cheryl Anderson, Sawyer Farmer, Abigail Schnapper, Helena Tubb, Tommy Valles + SEARCH |
| EPI_ISL_1811940 | EXCITE Lab | Andersen lab at Scripps Research | Sydney Morgan + Marni Jacobs + Celestine Magallanes + Holly Valentine + Louise C. Laurent + SEARCH |
| EPI_ISL_1811941 | EXCITE Lab | Andersen lab at Scripps Research | Nicole L Washington, Simon White, Geraint Levan, Kelly Schiabor Barrett, Elizabeth Cirulli, Alexandre Bolze, Charlotte Rivera-Garcia, Ryan Cho, Jason Nguyen, Sherry Wang, Jimmy Ramirez, Celena Andrade, Alice Summerfield, Tyler Cassens, Efren Sandoval, Francisco Tanudjaja, Magnus Isaksson, William Lee, David Becker, Marc Laurent, James Lu + SEARCH |
| EPI_ISL_1811942 | EXCITE Lab | Andersen lab at Scripps Research | David Pride, Sharon Reed, Chip Schooley, Angela Scioscia, Natasha Martin Cheryl Anderson, Sawyer Farmer, Abigail Schnapper, Helena Tubb, Tommy Valles + SEARCH |
| EPI_ISL_1811943, | EXCITE Lab | Andersen lab at Scripps Research | Nicole L Washington, Simon White, Geraint Levan, Kelly Schiabor Barrett, Elizabeth Cirulli, Alexandre Bolze, Charlotte Rivera-Garcia, Ryan Cho, Jason Nguyen, Sherry Wang, Jimmy Ramirez, Celena Andrade, Alice Summerfield, Tyler Cassens, Efren Sandoval, Francisco Tanudjaja, Magnus |

| | | | |
|---|---|---|---|
| EPI_ISL_1811944 | | | Isaksson, William Lee, David Becker, Marc Laurent, James Lu + SEARCH |
| EPI_ISL_1811945, EPI_ISL_1811946 | EXCITE Lab | Andersen lab at Scripps Research | David Pride, Sharon Reed, Chip Schooley, Angela Scioscia, Natasha Martin Cheryl Anderson, Sawyer Farmer, Abigail Schnapper, Helena Tubb, Tommy Valles + SEARCH |
| EPI_ISL_1811947, EPI_ISL_1811948, EPI_ISL_1811949, EPI_ISL_1811950, EPI_ISL_1811951, EPI_ISL_1811952, EPI_ISL_1811953, EPI_ISL_1811954, EPI_ISL_1811955, EPI_ISL_1811956, EPI_ISL_1811957, EPI_ISL_1811958, EPI_ISL_1811959, EPI_ISL_1811960, EPI_ISL_1811961, EPI_ISL_1811962, EPI_ISL_1811963, EPI_ISL_1811964, EPI_ISL_1811965 | | | |
| see above | EXCITE Lab | Andersen lab at Scripps Research | Nicole L Washington, Simon White, Geraint Levan, Kelly Schiabor Barrett, Elizabeth Cirulli, Alexandre Bolze, Charlotte Rivera-Garcia, Ryan Cho, Jason Nguyen, Sherry Wang, Jimmy Ramirez, Celena Andrade, Alice Summerfield, Tyler Cassens, Efren Sandoval, Francisco Tanudjaja, Magnus Isaksson, William Lee, David Becker, Marc Laurent, James Lu + SEARCH |
| EPI_ISL_1811966 | EXCITE Lab | Andersen lab at Scripps Research | David Pride, Sharon Reed, Chip Schooley, Angela Scioscia, Natasha Martin Cheryl Anderson, Sawyer Farmer, Abigail Schnapper, Helena Tubb, Tommy Valles + SEARCH |
| EPI_ISL_1811967, EPI_ISL_1811968, EPI_ISL_1811969 | EXCITE Lab | Andersen lab at Scripps Research | Nicole L Washington, Simon White, Geraint Levan, Kelly Schiabor Barrett, Elizabeth Cirulli, Alexandre Bolze, Charlotte Rivera-Garcia, Ryan Cho, Jason Nguyen, Sherry Wang, Jimmy Ramirez, Celena Andrade, Alice Summerfield, Tyler Cassens, Efren Sandoval, Francisco Tanudjaja, Magnus Isaksson, William Lee, David Becker, Marc Laurent, James Lu + SEARCH |
| EPI_ISL_1811970 | EXCITE Lab | Andersen lab at Scripps Research | David Pride, Sharon Reed, Chip Schooley, Angela Scioscia, Natasha Martin Cheryl Anderson, Sawyer Farmer, Abigail Schnapper, Helena Tubb, Tommy Valles + SEARCH |
| EPI_ISL_1811971, EPI_ISL_1811972, EPI_ISL_1811973, EPI_ISL_1811974, EPI_ISL_1811975, EPI_ISL_1811976 | EXCITE Lab | Andersen lab at Scripps Research | Nicole L Washington, Simon White, Geraint Levan, Kelly Schiabor Barrett, Elizabeth Cirulli, Alexandre Bolze, Charlotte Rivera-Garcia, Ryan Cho, Jason Nguyen, Sherry Wang, Jimmy Ramirez, Celena Andrade, Alice Summerfield, Tyler Cassens, Efren Sandoval, Francisco Tanudjaja, Magnus Isaksson, William Lee, David Becker, Marc Laurent, James Lu + SEARCH |
| EPI_ISL_1811977 | EXCITE Lab | Andersen lab at Scripps Research | David Pride, Sharon Reed, Chip Schooley, Angela Scioscia, Natasha Martin Cheryl Anderson, Sawyer Farmer, Abigail Schnapper, Helena Tubb, Tommy Valles + SEARCH |
| EPI_ISL_1811978, EPI_ISL_1811979, EPI_ISL_1811980, EPI_ISL_1811981, EPI_ISL_1811982 | EXCITE Lab | Andersen lab at Scripps Research | Nicole L Washington, Simon White, Geraint Levan, Kelly Schiabor Barrett, Elizabeth Cirulli, Alexandre Bolze, Charlotte Rivera-Garcia, Ryan Cho, Jason Nguyen, Sherry Wang, Jimmy Ramirez, Celena Andrade, Alice Summerfield, Tyler Cassens, Efren Sandoval, Francisco Tanudjaja, Magnus Isaksson, William Lee, David Becker, Marc Laurent, James Lu + SEARCH |
| EPI_ISL_1811983 | EXCITE Lab | Andersen lab at Scripps Research | David Pride, Sharon Reed, Chip Schooley, Angela Scioscia, Natasha Martin Cheryl Anderson, Sawyer Farmer, Abigail Schnapper, Helena Tubb, Tommy Valles + SEARCH |
| EPI_ISL_1811984, EPI_ISL_1811985 | EXCITE Lab | Andersen lab at Scripps Research | Nicole L Washington, Simon White, Geraint Levan, Kelly Schiabor Barrett, Elizabeth Cirulli, Alexandre Bolze, Charlotte Rivera-Garcia, Ryan Cho, Jason Nguyen, Sherry Wang, Jimmy Ramirez, Celena Andrade, Alice Summerfield, Tyler Cassens, Efren Sandoval, Francisco Tanudjaja, Magnus Isaksson, William Lee, David Becker, Marc Laurent, James Lu + SEARCH |
| EPI_ISL_1811986 | EXCITE Lab | Andersen lab at Scripps Research | David Pride, Sharon Reed, Chip Schooley, Angela Scioscia, Natasha Martin Cheryl Anderson, Sawyer Farmer, Abigail Schnapper, Helena Tubb, Tommy Valles + SEARCH |
| EPI_ISL_1811987, EPI_ISL_1811988, EPI_ISL_1811989, EPI_ISL_1811990, EPI_ISL_1811991, EPI_ISL_1811992, EPI_ISL_1811993, EPI_ISL_1811994, EPI_ISL_1811995, EPI_ISL_1811996, EPI_ISL_1811997, EPI_ISL_1811998, EPI_ISL_1811999, EPI_ISL_1812000, EPI_ISL_1812001, EPI_ISL_1812002, EPI_ISL_1812003, EPI_ISL_1812004, EPI_ISL_1812005, EPI_ISL_1812006 | | | |
| see above | EXCITE Lab | Andersen lab at Scripps Research | Nicole L Washington, Simon White, Geraint Levan, Kelly Schiabor Barrett, Elizabeth Cirulli, Alexandre Bolze, Charlotte Rivera-Garcia, Ryan Cho, Jason Nguyen, Sherry Wang, Jimmy Ramirez, Celena Andrade, Alice Summerfield, Tyler Cassens, Efren Sandoval, Francisco Tanudjaja, Magnus Isaksson, William Lee, David Becker, Marc Laurent, James Lu + SEARCH |
| EPI_ISL_1812007 | EXCITE Lab | Andersen lab at Scripps Research | David Pride, Sharon Reed, Chip Schooley, Angela Scioscia, Natasha Martin Cheryl Anderson, Sawyer Farmer, Abigail Schnapper, Helena Tubb, Tommy Valles + SEARCH |
| EPI_ISL_1812008, EPI_ISL_1812009, EPI_ISL_1812010, EPI_ISL_1812011, EPI_ISL_1812012 | EXCITE Lab | Andersen lab at Scripps Research | Nicole L Washington, Simon White, Geraint Levan, Kelly Schiabor Barrett, Elizabeth Cirulli, Alexandre Bolze, Charlotte Rivera-Garcia, Ryan Cho, Jason Nguyen, Sherry Wang, Jimmy Ramirez, Celena Andrade, Alice Summerfield, Tyler Cassens, Efren Sandoval, Francisco Tanudjaja, Magnus Isaksson, William Lee, David Becker, Marc Laurent, James Lu + SEARCH |
| EPI_ISL_1812013 | EXCITE Lab | Andersen lab at Scripps Research | David Pride, Sharon Reed, Chip Schooley, Angela Scioscia, Natasha Martin Cheryl Anderson, Sawyer Farmer, Abigail Schnapper, Helena Tubb, Tommy Valles + SEARCH |
| EPI_ISL_1812014, EPI_ISL_1812015, EPI_ISL_1812016, EPI_ISL_1812017, EPI_ISL_1812018, EPI_ISL_1812019, EPI_ISL_1812020, EPI_ISL_1812021 | EXCITE Lab | Andersen lab at Scripps Research | Nicole L Washington, Simon White, Geraint Levan, Kelly Schiabor Barrett, Elizabeth Cirulli, Alexandre Bolze, Charlotte Rivera-Garcia, Ryan Cho, Jason Nguyen, Sherry Wang, Jimmy Ramirez, Celena Andrade, Alice Summerfield, Tyler Cassens, Efren Sandoval, Francisco Tanudjaja, Magnus Isaksson, William Lee, David Becker, Marc Laurent, James Lu + SEARCH |
| EPI_ISL_1812022 | EXCITE Lab | Andersen lab at Scripps Research | David Pride, Sharon Reed, Chip Schooley, Angela Scioscia, Natasha Martin Cheryl Anderson, Sawyer Farmer, Abigail Schnapper, Helena Tubb, Tommy Valles + SEARCH |
| EPI_ISL_1812023, EPI_ISL_1812024, EPI_ISL_1812025, EPI_ISL_1812026, EPI_ISL_1812027, EPI_ISL_1812028, EPI_ISL_1812029, EPI_ISL_1812030, EPI_ISL_1812031, EPI_ISL_1812032 | EXCITE Lab | Andersen lab at Scripps Research | Nicole L Washington, Simon White, Geraint Levan, Kelly Schiabor Barrett, Elizabeth Cirulli, Alexandre Bolze, Charlotte Rivera-Garcia, Ryan Cho, Jason Nguyen, Sherry Wang, Jimmy Ramirez, Celena Andrade, Alice Summerfield, Tyler Cassens, Efren Sandoval, Francisco Tanudjaja, Magnus Isaksson, William Lee, David Becker, Marc Laurent, James Lu + SEARCH |
| EPI_ISL_1812033 | EXCITE Lab | Andersen lab at Scripps Research | David Pride, Sharon Reed, Chip Schooley, Angela Scioscia, Natasha Martin Cheryl Anderson, Sawyer Farmer, Abigail Schnapper, Helena Tubb, Tommy Valles + SEARCH |
| EPI_ISL_1812034, EPI_ISL_1812035, EPI_ISL_1812036, EPI_ISL_1812037 | EXCITE Lab | Andersen lab at Scripps Research | Nicole L Washington, Simon White, Geraint Levan, Kelly Schiabor Barrett, Elizabeth Cirulli, Alexandre Bolze, Charlotte Rivera-Garcia, Ryan Cho, Jason Nguyen, Sherry Wang, Jimmy Ramirez, Celena Andrade, Alice Summerfield, Tyler Cassens, Efren Sandoval, Francisco Tanudjaja, Magnus Isaksson, William Lee, David Becker, Marc Laurent, James Lu + SEARCH |
| EPI_ISL_1812038 | EXCITE Lab | Andersen lab at Scripps Research | David Pride, Sharon Reed, Chip Schooley, Angela Scioscia, Natasha Martin Cheryl Anderson, Sawyer Farmer, Abigail Schnapper, Helena Tubb, Tommy Valles + SEARCH |
| EPI_ISL_1812039 | EXCITE Lab | Andersen lab at Scripps Research | Sydney Morgan + Marni Jacobs + Celestine Magallanes + Holly Valentine + Louise C. Laurent + SEARCH |
| EPI_ISL_1812040, EPI_ISL_1812041, EPI_ISL_1812042 | EXCITE Lab | Andersen lab at Scripps Research | Nicole L Washington, Simon White, Geraint Levan, Kelly Schiabor Barrett, Elizabeth Cirulli, Alexandre Bolze, Charlotte Rivera-Garcia, Ryan Cho, Jason Nguyen, Sherry Wang, Jimmy Ramirez, Celena Andrade, Alice Summerfield, Tyler Cassens, Efren Sandoval, Francisco Tanudjaja, Magnus Isaksson, William Lee, David Becker, Marc Laurent, James Lu + SEARCH |
| EPI_ISL_1812043 | EXCITE Lab | Andersen lab at Scripps Research | David Pride, Sharon Reed, Chip Schooley, Angela Scioscia, Natasha Martin Cheryl Anderson, Sawyer Farmer, Abigail Schnapper, Helena Tubb, Tommy Valles + SEARCH |
| EPI_ISL_1812044, EPI_ISL_1812045, EPI_ISL_1812046 | EXCITE Lab | Andersen lab at Scripps Research | Nicole L Washington, Simon White, Geraint Levan, Kelly Schiabor Barrett, Elizabeth Cirulli, Alexandre Bolze, Charlotte Rivera-Garcia, Ryan Cho, Jason Nguyen, Sherry Wang, Jimmy Ramirez, Celena Andrade, Alice Summerfield, Tyler Cassens, Efren Sandoval, Francisco Tanudjaja, Magnus Isaksson, William Lee, David Becker, Marc Laurent, James Lu + SEARCH |
| EPI_ISL_1812047 | EXCITE Lab | Andersen lab at Scripps Research | David Pride, Sharon Reed, Chip Schooley, Angela Scioscia, Natasha Martin Cheryl Anderson, Sawyer Farmer, Abigail Schnapper, Helena Tubb, Tommy Valles + SEARCH |
| EPI_ISL_1812048, EPI_ISL_1812049, EPI_ISL_1812050, EPI_ISL_1812051, EPI_ISL_1812052, EPI_ISL_1812053, EPI_ISL_1812054, EPI_ISL_1812055, EPI_ISL_1812056, EPI_ISL_1812057, EPI_ISL_1812058, EPI_ISL_1812059, EPI_ISL_1812060, EPI_ISL_1812061, EPI_ISL_1812062, EPI_ISL_1812063, EPI_ISL_1812064, EPI_ISL_1812065, EPI_ISL_1812066, EPI_ISL_1812067, EPI_ISL_1812068, EPI_ISL_1812069, EPI_ISL_1812070, EPI_ISL_1812071, EPI_ISL_1812072 | | | |
| see above | EXCITE Lab | Andersen lab at Scripps Research | Nicole L Washington, Simon White, Geraint Levan, Kelly Schiabor Barrett, Elizabeth Cirulli, Alexandre Bolze, Charlotte Rivera-Garcia, Ryan Cho, Jason Nguyen, Sherry Wang, Jimmy Ramirez, Celena Andrade, Alice Summerfield, Tyler Cassens, Efren Sandoval, Francisco Tanudjaja, Magnus Isaksson, William Lee, David Becker, Marc Laurent, James Lu + SEARCH |
| EPI_ISL_1812073 | EXCITE Lab | Andersen lab at Scripps Research | David Pride, Sharon Reed, Chip Schooley, Angela Scioscia, Natasha Martin Cheryl Anderson, Sawyer Farmer, Abigail Schnapper, Helena Tubb, Tommy Valles + SEARCH |
| EPI_ISL_1812074, EPI_ISL_1812075, EPI_ISL_1812076, EPI_ISL_1812077, EPI_ISL_1812078, EPI_ISL_1812079, EPI_ISL_1812080, EPI_ISL_1812081, EPI_ISL_1812082, EPI_ISL_1812083, EPI_ISL_1812084, EPI_ISL_1812085, EPI_ISL_1812086, EPI_ISL_1812087, EPI_ISL_1812088, EPI_ISL_1812089, EPI_ISL_1812090, EPI_ISL_1812091, EPI_ISL_1812092, EPI_ISL_1812093, EPI_ISL_1812094, EPI_ISL_1812095, EPI_ISL_1812096, EPI_ISL_1812097, EPI_ISL_1812098, EPI_ISL_1812099, EPI_ISL_1812100, EPI_ISL_1812101, EPI_ISL_1812102, EPI_ISL_1812103, EPI_ISL_1812104, EPI_ISL_1812105 | | | |
| see above | EXCITE Lab | Andersen lab at Scripps Research | Nicole L Washington, Simon White, Geraint Levan, Kelly Schiabor Barrett, Elizabeth Cirulli, Alexandre Bolze, Charlotte Rivera-Garcia, Ryan Cho, Jason Nguyen, Sherry Wang, Jimmy Ramirez, Celena Andrade, Alice Summerfield, Tyler Cassens, Efren Sandoval, Francisco Tanudjaja, Magnus Isaksson, William Lee, David Becker, Marc Laurent, James Lu + SEARCH |
| EPI_ISL_1812106 | EXCITE Lab | Andersen lab at Scripps Research | David Pride, Sharon Reed, Chip Schooley, Angela Scioscia, Natasha Martin Cheryl Anderson, Sawyer Farmer, Abigail Schnapper, Helena Tubb, Tommy Valles + SEARCH |
| EPI_ISL_1812107, EPI_ISL_1812108, EPI_ISL_1812109, EPI_ISL_1812110, EPI_ISL_1812111, EPI_ISL_1812112, EPI_ISL_1812113, EPI_ISL_1812114, EPI_ISL_1812115, EPI_ISL_1812116 | EXCITE Lab | Andersen lab at Scripps Research | Nicole L Washington, Simon White, Geraint Levan, Kelly Schiabor Barrett, Elizabeth Cirulli, Alexandre Bolze, Charlotte Rivera-Garcia, Ryan Cho, Jason Nguyen, Sherry Wang, Jimmy Ramirez, Celena Andrade, Alice Summerfield, Tyler Cassens, Efren Sandoval, Francisco Tanudjaja, Magnus Isaksson, William Lee, David Becker, Marc Laurent, James Lu + SEARCH |
| EPI_ISL_1812117 | EXCITE Lab | Andersen lab at Scripps Research | David Pride, Sharon Reed, Chip Schooley, Angela Scioscia, Natasha Martin Cheryl Anderson, Sawyer Farmer, Abigail Schnapper, Helena Tubb, Tommy Valles + SEARCH |
| EPI_ISL_1812118 | EXCITE Lab | Andersen lab at Scripps Research | Nicole L Washington, Simon White, Geraint Levan, Kelly Schiabor Barrett, Elizabeth Cirulli, Alexandre Bolze, Charlotte Rivera-Garcia, Ryan Cho, Jason Nguyen, Sherry Wang, Jimmy Ramirez, Celena Andrade, Alice Summerfield, Tyler Cassens, Efren Sandoval, Francisco Tanudjaja, Magnus |

| | | | |
|---|---|---|---|
| | | | Isaksson, William Lee, David Becker, Marc Laurent, James Lu + SEARCH |
| EPI_ISL_1812119 | EXCITE Lab | Andersen lab at Scripps Research | Chip Schooley, Natasha Martin, Cheryl Anderson, Angela Scioscia, Smruthi Karthikeyan, Greg Humphrey, Sawyer Farmer, Abigail Schnapper, Helena Tubb, Tommy Valles + SEARCH |
| EPI_ISL_1812120, EPI_ISL_1812121, EPI_ISL_1812122, EPI_ISL_1812123, EPI_ISL_1812124, EPI_ISL_1812125, EPI_ISL_1812126, EPI_ISL_1812127, EPI_ISL_1812128, EPI_ISL_1812129, EPI_ISL_1812130, EPI_ISL_1812131, EPI_ISL_1812132, EPI_ISL_1812133, EPI_ISL_1812134, EPI_ISL_1812135, EPI_ISL_1812136, EPI_ISL_1812137, EPI_ISL_1812138 | | | |
| see above | EXCITE Lab | Andersen lab at Scripps Research | Nicole L Washington, Simon White, Geraint Levan, Kelly Schiabor Barrett, Elizabeth Cirulli, Alexandre Bolze, Charlotte Rivera-Garcia, Ryan Cho, Jason Nguyen, Sherry Wang, Jimmy Ramirez, Celena Andrade, Alice Summerfield, Tyler Cassens, Efren Sandoval, Francisco Tanudjaja, Magnus Isaksson, William Lee, David Becker, Marc Laurent, James Lu + SEARCH |
| EPI_ISL_1812139, EPI_ISL_1812140 | EXCITE Lab | Andersen lab at Scripps Research | David Pride, Sharon Reed, Chip Schooley, Angela Scioscia, Natasha Martin Cheryl Anderson, Sawyer Farmer, Abigail Schnapper, Helena Tubb, Tommy Valles + SEARCH |
| EPI_ISL_1812141, EPI_ISL_1812142 | EXCITE Lab | Andersen lab at Scripps Research | Nicole L Washington, Simon White, Geraint Levan, Kelly Schiabor Barrett, Elizabeth Cirulli, Alexandre Bolze, Charlotte Rivera-Garcia, Ryan Cho, Jason Nguyen, Sherry Wang, Jimmy Ramirez, Celena Andrade, Alice Summerfield, Tyler Cassens, Efren Sandoval, Francisco Tanudjaja, Magnus Isaksson, William Lee, David Becker, Marc Laurent, James Lu + SEARCH |
| EPI_ISL_1812143 | EXCITE Lab | Andersen lab at Scripps Research | David Pride, Sharon Reed, Chip Schooley, Angela Scioscia, Natasha Martin Cheryl Anderson, Sawyer Farmer, Abigail Schnapper, Helena Tubb, Tommy Valles + SEARCH |
| EPI_ISL_1812144, EPI_ISL_1812145, EPI_ISL_1812146, EPI_ISL_1812147 | EXCITE Lab | Andersen lab at Scripps Research | Nicole L Washington, Simon White, Geraint Levan, Kelly Schiabor Barrett, Elizabeth Cirulli, Alexandre Bolze, Charlotte Rivera-Garcia, Ryan Cho, Jason Nguyen, Sherry Wang, Jimmy Ramirez, Celena Andrade, Alice Summerfield, Tyler Cassens, Efren Sandoval, Francisco Tanudjaja, Magnus Isaksson, William Lee, David Becker, Marc Laurent, James Lu + SEARCH |
| EPI_ISL_1812148 | EXCITE Lab | Andersen lab at Scripps Research | David Pride, Sharon Reed, Chip Schooley, Angela Scioscia, Natasha Martin Cheryl Anderson, Sawyer Farmer, Abigail Schnapper, Helena Tubb, Tommy Valles + SEARCH |
| EPI_ISL_1812149, EPI_ISL_1812150, EPI_ISL_1812151, EPI_ISL_1812152 | EXCITE Lab | Andersen lab at Scripps Research | Nicole L Washington, Simon White, Geraint Levan, Kelly Schiabor Barrett, Elizabeth Cirulli, Alexandre Bolze, Charlotte Rivera-Garcia, Ryan Cho, Jason Nguyen, Sherry Wang, Jimmy Ramirez, Celena Andrade, Alice Summerfield, Tyler Cassens, Efren Sandoval, Francisco Tanudjaja, Magnus Isaksson, William Lee, David Becker, Marc Laurent, James Lu + SEARCH |
| EPI_ISL_1812153 | EXCITE Lab | Andersen lab at Scripps Research | David Pride, Sharon Reed, Chip Schooley, Angela Scioscia, Natasha Martin Cheryl Anderson, Sawyer Farmer, Abigail Schnapper, Helena Tubb, Tommy Valles + SEARCH |
| EPI_ISL_1812154, EPI_ISL_1812155, EPI_ISL_1812156, EPI_ISL_1812157, EPI_ISL_1812158 | EXCITE Lab | Andersen lab at Scripps Research | Nicole L Washington, Simon White, Geraint Levan, Kelly Schiabor Barrett, Elizabeth Cirulli, Alexandre Bolze, Charlotte Rivera-Garcia, Ryan Cho, Jason Nguyen, Sherry Wang, Jimmy Ramirez, Celena Andrade, Alice Summerfield, Tyler Cassens, Efren Sandoval, Francisco Tanudjaja, Magnus Isaksson, William Lee, David Becker, Marc Laurent, James Lu + SEARCH |
| EPI_ISL_1812159 | EXCITE Lab | Andersen lab at Scripps Research | David Pride, Sharon Reed, Chip Schooley, Angela Scioscia, Natasha Martin Cheryl Anderson, Sawyer Farmer, Abigail Schnapper, Helena Tubb, Tommy Valles + SEARCH |
| EPI_ISL_1812160, EPI_ISL_1812161, EPI_ISL_1812162, EPI_ISL_1812163, EPI_ISL_1812164, EPI_ISL_1812165, EPI_ISL_1812166, EPI_ISL_1812167, EPI_ISL_1812168, EPI_ISL_1812169, EPI_ISL_1812170, EPI_ISL_1812171, EPI_ISL_1812172 | | | |
| see above | EXCITE Lab | Andersen lab at Scripps Research | Nicole L Washington, Simon White, Geraint Levan, Kelly Schiabor Barrett, Elizabeth Cirulli, Alexandre Bolze, Charlotte Rivera-Garcia, Ryan Cho, Jason Nguyen, Sherry Wang, Jimmy Ramirez, Celena Andrade, Alice Summerfield, Tyler Cassens, Efren Sandoval, Francisco Tanudjaja, Magnus Isaksson, William Lee, David Becker, Marc Laurent, James Lu + SEARCH |
| EPI_ISL_1812173 | EXCITE Lab | Andersen lab at Scripps Research | David Pride, Sharon Reed, Chip Schooley, Angela Scioscia, Natasha Martin Cheryl Anderson, Sawyer Farmer, Abigail Schnapper, Helena Tubb, Tommy Valles + SEARCH |
| EPI_ISL_1812174, EPI_ISL_1812175, EPI_ISL_1812176, EPI_ISL_1812177 | EXCITE Lab | Andersen lab at Scripps Research | Nicole L Washington, Simon White, Geraint Levan, Kelly Schiabor Barrett, Elizabeth Cirulli, Alexandre Bolze, Charlotte Rivera-Garcia, Ryan Cho, Jason Nguyen, Sherry Wang, Jimmy Ramirez, Celena Andrade, Alice Summerfield, Tyler Cassens, Efren Sandoval, Francisco Tanudjaja, Magnus Isaksson, William Lee, David Becker, Marc Laurent, James Lu + SEARCH |
| EPI_ISL_1812178 | EXCITE Lab | Andersen lab at Scripps Research | David Pride, Sharon Reed, Chip Schooley, Angela Scioscia, Natasha Martin Cheryl Anderson, Sawyer Farmer, Abigail Schnapper, Helena Tubb, Tommy Valles + SEARCH |
| EPI_ISL_1812179, EPI_ISL_1812180, EPI_ISL_1812181, EPI_ISL_1812182, EPI_ISL_1812183, EPI_ISL_1812184, EPI_ISL_1812185, EPI_ISL_1812186, EPI_ISL_1812187, EPI_ISL_1812188, EPI_ISL_1812189, EPI_ISL_1812190, EPI_ISL_1812191, EPI_ISL_1812192, EPI_ISL_1812193, EPI_ISL_1812194, EPI_ISL_1812195, EPI_ISL_1812196, EPI_ISL_1812197, EPI_ISL_1812198 | | | |
| see above | EXCITE Lab | Andersen lab at Scripps Research | Nicole L Washington, Simon White, Geraint Levan, Kelly Schiabor Barrett, Elizabeth Cirulli, Alexandre Bolze, Charlotte Rivera-Garcia, Ryan Cho, Jason Nguyen, Sherry Wang, Jimmy Ramirez, Celena Andrade, Alice Summerfield, Tyler Cassens, Efren Sandoval, Francisco Tanudjaja, Magnus Isaksson, William Lee, David Becker, Marc Laurent, James Lu + SEARCH |
| EPI_ISL_1812199 | EXCITE Lab | Andersen lab at Scripps Research | David Pride, Sharon Reed, Chip Schooley, Angela Scioscia, Natasha Martin Cheryl Anderson, Sawyer Farmer, Abigail Schnapper, Helena Tubb, Tommy Valles + SEARCH |
| EPI_ISL_1812200, EPI_ISL_1812201, EPI_ISL_1812202, EPI_ISL_1812203, EPI_ISL_1812204, EPI_ISL_1812205, EPI_ISL_1812206, EPI_ISL_1812207, EPI_ISL_1812208, EPI_ISL_1812209, EPI_ISL_1812210, EPI_ISL_1812211, EPI_ISL_1812212, EPI_ISL_1812213, EPI_ISL_1812214, EPI_ISL_1812215, EPI_ISL_1812216 | | | |
| see above | EXCITE Lab | Andersen lab at Scripps Research | Nicole L Washington, Simon White, Geraint Levan, Kelly Schiabor Barrett, Elizabeth Cirulli, Alexandre Bolze, Charlotte Rivera-Garcia, Ryan Cho, Jason Nguyen, Sherry Wang, Jimmy Ramirez, Celena Andrade, Alice Summerfield, Tyler Cassens, Efren Sandoval, Francisco Tanudjaja, Magnus Isaksson, William Lee, David Becker, Marc Laurent, James Lu + SEARCH |
| EPI_ISL_1812217 | EXCITE Lab | Andersen lab at Scripps Research | David Pride, Sharon Reed, Chip Schooley, Angela Scioscia, Natasha Martin Cheryl Anderson, Sawyer Farmer, Abigail Schnapper, Helena Tubb, Tommy Valles + SEARCH |
| EPI_ISL_1812218, EPI_ISL_1812219, EPI_ISL_1812220, EPI_ISL_1812221, EPI_ISL_1812222, EPI_ISL_1812223, EPI_ISL_1812224, EPI_ISL_1812225, EPI_ISL_1812226, EPI_ISL_1812227, EPI_ISL_1812228, EPI_ISL_1812229 | | | |
| see above | EXCITE Lab | Andersen lab at Scripps Research | Nicole L Washington, Simon White, Geraint Levan, Kelly Schiabor Barrett, Elizabeth Cirulli, Alexandre Bolze, Charlotte Rivera-Garcia, Ryan Cho, Jason Nguyen, Sherry Wang, Jimmy Ramirez, Celena Andrade, Alice Summerfield, Tyler Cassens, Efren Sandoval, Francisco Tanudjaja, Magnus Isaksson, William Lee, David Becker, Marc Laurent, James Lu + SEARCH |
| EPI_ISL_1812230 | EXCITE Lab | Andersen lab at Scripps Research | David Pride, Sharon Reed, Chip Schooley, Angela Scioscia, Natasha Martin Cheryl Anderson, Sawyer Farmer, Abigail Schnapper, Helena Tubb, Tommy Valles + SEARCH |
| EPI_ISL_1812231, EPI_ISL_1812232, EPI_ISL_1812233, EPI_ISL_1812234, EPI_ISL_1812235, EPI_ISL_1812236, EPI_ISL_1812237, EPI_ISL_1812238, EPI_ISL_1812239 | EXCITE Lab | Andersen lab at Scripps Research | Nicole L Washington, Simon White, Geraint Levan, Kelly Schiabor Barrett, Elizabeth Cirulli, Alexandre Bolze, Charlotte Rivera-Garcia, Ryan Cho, Jason Nguyen, Sherry Wang, Jimmy Ramirez, Celena Andrade, Alice Summerfield, Tyler Cassens, Efren Sandoval, Francisco Tanudjaja, Magnus Isaksson, William Lee, David Becker, Marc Laurent, James Lu + SEARCH |
| EPI_ISL_1812240 | EXCITE Lab | Andersen lab at Scripps Research | David Pride, Sharon Reed, Chip Schooley, Angela Scioscia, Natasha Martin Cheryl Anderson, Sawyer Farmer, Abigail Schnapper, Helena Tubb, Tommy Valles + SEARCH |
| EPI_ISL_1812241, EPI_ISL_1812242, EPI_ISL_1812243, EPI_ISL_1812244, EPI_ISL_1812245, EPI_ISL_1812246, EPI_ISL_1812247, EPI_ISL_1812248, EPI_ISL_1812249, EPI_ISL_1812250, EPI_ISL_1812251, EPI_ISL_1812252, EPI_ISL_1812253, EPI_ISL_1812254, EPI_ISL_1812255, EPI_ISL_1812256, EPI_ISL_1812257, EPI_ISL_1812258, EPI_ISL_1812259, EPI_ISL_1812260 | | | |
| see above | EXCITE Lab | Andersen lab at Scripps Research | Nicole L Washington, Simon White, Geraint Levan, Kelly Schiabor Barrett, Elizabeth Cirulli, Alexandre Bolze, Charlotte Rivera-Garcia, Ryan Cho, Jason Nguyen, Sherry Wang, Jimmy Ramirez, Celena Andrade, Alice Summerfield, Tyler Cassens, Efren Sandoval, Francisco Tanudjaja, Magnus Isaksson, William Lee, David Becker, Marc Laurent, James Lu + SEARCH |
| EPI_ISL_1812261 | EXCITE Lab | Andersen lab at Scripps Research | David Pride, Sharon Reed, Chip Schooley, Angela Scioscia, Natasha Martin Cheryl Anderson, Sawyer Farmer, Abigail Schnapper, Helena Tubb, Tommy Valles + SEARCH |
| EPI_ISL_1812262, EPI_ISL_1812263, EPI_ISL_1812264, EPI_ISL_1812265, EPI_ISL_1812266 | EXCITE Lab | Andersen lab at Scripps Research | Nicole L Washington, Simon White, Geraint Levan, Kelly Schiabor Barrett, Elizabeth Cirulli, Alexandre Bolze, Charlotte Rivera-Garcia, Ryan Cho, Jason Nguyen, Sherry Wang, Jimmy Ramirez, Celena Andrade, Alice Summerfield, Tyler Cassens, Efren Sandoval, Francisco Tanudjaja, Magnus Isaksson, William Lee, David Becker, Marc Laurent, James Lu + SEARCH |
| EPI_ISL_1812267 | EXCITE Lab | Andersen lab at Scripps Research | David Pride, Sharon Reed, Chip Schooley, Angela Scioscia, Natasha Martin Cheryl Anderson, Sawyer Farmer, Abigail Schnapper, Helena Tubb, Tommy Valles + SEARCH |
| EPI_ISL_1812268, EPI_ISL_1812269, EPI_ISL_1812270 | EXCITE Lab | Andersen lab at Scripps Research | Nicole L Washington, Simon White, Geraint Levan, Kelly Schiabor Barrett, Elizabeth Cirulli, Alexandre Bolze, Charlotte Rivera-Garcia, Ryan Cho, Jason Nguyen, Sherry Wang, Jimmy Ramirez, Celena Andrade, Alice Summerfield, Tyler Cassens, Efren Sandoval, Francisco Tanudjaja, Magnus Isaksson, William Lee, David Becker, Marc Laurent, James Lu + SEARCH |
| EPI_ISL_1812271 | EXCITE Lab | Andersen lab at Scripps Research | David Pride, Sharon Reed, Chip Schooley, Angela Scioscia, Natasha Martin Cheryl Anderson, Sawyer Farmer, Abigail Schnapper, Helena Tubb, Tommy Valles + SEARCH |
| EPI_ISL_1812272, EPI_ISL_1812273, EPI_ISL_1812274, EPI_ISL_1812275, EPI_ISL_1812276, EPI_ISL_1812277, EPI_ISL_1812278, EPI_ISL_1812279, EPI_ISL_1812280 | EXCITE Lab | Andersen lab at Scripps Research | Nicole L Washington, Simon White, Geraint Levan, Kelly Schiabor Barrett, Elizabeth Cirulli, Alexandre Bolze, Charlotte Rivera-Garcia, Ryan Cho, Jason Nguyen, Sherry Wang, Jimmy Ramirez, Celena Andrade, Alice Summerfield, Tyler Cassens, Efren Sandoval, Francisco Tanudjaja, Magnus Isaksson, William Lee, David Becker, Marc Laurent, James Lu + SEARCH |
| EPI_ISL_1812281, EPI_ISL_1812282, EPI_ISL_1812283 | EXCITE Lab | Andersen lab at Scripps Research | David Pride, Sharon Reed, Chip Schooley, Angela Scioscia, Natasha Martin Cheryl Anderson, Sawyer Farmer, Abigail Schnapper, Helena Tubb, Tommy Valles + SEARCH |
| EPI_ISL_1812284, EPI_ISL_1812285, EPI_ISL_1812286, EPI_ISL_1812287, EPI_ISL_1812288, EPI_ISL_1812289, EPI_ISL_1812290 | EXCITE Lab | Andersen lab at Scripps Research | Nicole L Washington, Simon White, Geraint Levan, Kelly Schiabor Barrett, Elizabeth Cirulli, Alexandre Bolze, Charlotte Rivera-Garcia, Ryan Cho, Jason Nguyen, Sherry Wang, Jimmy Ramirez, Celena Andrade, Alice Summerfield, Tyler Cassens, Efren Sandoval, Francisco Tanudjaja, Magnus Isaksson, William Lee, David Becker, Marc Laurent, James Lu + SEARCH |
| EPI_ISL_1812291 | EXCITE Lab | Andersen lab at Scripps Research | Sydney Morgan + Marni Jacobs + Celestine Magallanes + Holly Valentine + Louise C. Laurent + SEARCH |
| EPI_ISL_1812292, EPI_ISL_1812293, EPI_ISL_1812294, EPI_ISL_1812295, EPI_ISL_1812296, EPI_ISL_1812297, EPI_ISL_1812298, EPI_ISL_1812299, EPI_ISL_1812300, EPI_ISL_1812301, EPI_ISL_1812302, EPI_ISL_1812303, EPI_ISL_1812304, EPI_ISL_1812305, EPI_ISL_1812306, EPI_ISL_1812307, EPI_ISL_1812308, EPI_ISL_1812309, EPI_ISL_1812310, EPI_ISL_1812311, EPI_ISL_1812312, EPI_ISL_1812313, EPI_ISL_1812314, EPI_ISL_1812315, EPI_ISL_1812316, EPI_ISL_1812317, EPI_ISL_1812318, EPI_ISL_1812319 | | | |

| | | | |
|---|---|---|---|
| see above | EXCITE Lab | Andersen lab at Scripps Research | Nicole L Washington, Simon White, Geraint Levan, Kelly Schiabor Barrett, Elizabeth Cirulli, Alexandre Bolze, Charlotte Rivera-Garcia, Ryan Cho, Jason Nguyen, Sherry Wang, Jimmy Ramirez, Celena Andrade, Alice Summerfield, Tyler Cassens, Efren Sandoval, Francisco Tanudjaja, Magnus Isaksson, William Lee, David Becker, Marc Laurent, James Lu + SEARCH |
| EPI_ISL_1812320 | EXCITE Lab | Andersen lab at Scripps Research | David Pride, Sharon Reed, Chip Schooley, Angela Scioscia, Natasha Martin Cheryl Anderson, Sawyer Farmer, Abigail Schnapper, Helena Tubb, Tommy Valles + SEARCH |
| EPI_ISL_1812321 | EXCITE Lab | Andersen lab at Scripps Research | Nicole L Washington, Simon White, Geraint Levan, Kelly Schiabor Barrett, Elizabeth Cirulli, Alexandre Bolze, Charlotte Rivera-Garcia, Ryan Cho, Jason Nguyen, Sherry Wang, Jimmy Ramirez, Celena Andrade, Alice Summerfield, Tyler Cassens, Efren Sandoval, Francisco Tanudjaja, Magnus Isaksson, William Lee, David Becker, Marc Laurent, James Lu + SEARCH |
| EPI_ISL_1812322 | EXCITE Lab | Andersen lab at Scripps Research | David Pride, Sharon Reed, Chip Schooley, Angela Scioscia, Natasha Martin Cheryl Anderson, Sawyer Farmer, Abigail Schnapper, Helena Tubb, Tommy Valles + SEARCH |
| EPI_ISL_1812323, EPI_ISL_1812324, EPI_ISL_1812325, EPI_ISL_1812326, EPI_ISL_1812327, EPI_ISL_1812328, EPI_ISL_1812329 | EXCITE Lab | Andersen lab at Scripps Research | Nicole L Washington, Simon White, Geraint Levan, Kelly Schiabor Barrett, Elizabeth Cirulli, Alexandre Bolze, Charlotte Rivera-Garcia, Ryan Cho, Jason Nguyen, Sherry Wang, Jimmy Ramirez, Celena Andrade, Alice Summerfield, Tyler Cassens, Efren Sandoval, Francisco Tanudjaja, Magnus Isaksson, William Lee, David Becker, Marc Laurent, James Lu + SEARCH |
| EPI_ISL_1812330 | EXCITE Lab | Andersen lab at Scripps Research | David Pride, Sharon Reed, Chip Schooley, Angela Scioscia, Natasha Martin Cheryl Anderson, Sawyer Farmer, Abigail Schnapper, Helena Tubb, Tommy Valles + SEARCH |
| EPI_ISL_1812331, EPI_ISL_1812332, EPI_ISL_1812333, EPI_ISL_1812334, EPI_ISL_1812335, EPI_ISL_1812336, EPI_ISL_1812337, EPI_ISL_1812338, EPI_ISL_1812339, EPI_ISL_1812340, EPI_ISL_1812341, EPI_ISL_1812342, EPI_ISL_1812343, EPI_ISL_1812344, EPI_ISL_1812345, EPI_ISL_1812346 | | | |
| see above | EXCITE Lab | Andersen lab at Scripps Research | Nicole L Washington, Simon White, Geraint Levan, Kelly Schiabor Barrett, Elizabeth Cirulli, Alexandre Bolze, Charlotte Rivera-Garcia, Ryan Cho, Jason Nguyen, Sherry Wang, Jimmy Ramirez, Celena Andrade, Alice Summerfield, Tyler Cassens, Efren Sandoval, Francisco Tanudjaja, Magnus Isaksson, William Lee, David Becker, Marc Laurent, James Lu + SEARCH |
| EPI_ISL_1812347 | EXCITE Lab | Andersen lab at Scripps Research | David Pride, Sharon Reed, Chip Schooley, Angela Scioscia, Natasha Martin Cheryl Anderson, Sawyer Farmer, Abigail Schnapper, Helena Tubb, Tommy Valles + SEARCH |
| EPI_ISL_1812348, EPI_ISL_1812349, EPI_ISL_1812350, EPI_ISL_1812351, EPI_ISL_1812352, EPI_ISL_1812353, EPI_ISL_1812354, EPI_ISL_1812355, EPI_ISL_1812356, EPI_ISL_1812357, EPI_ISL_1812358, EPI_ISL_1812359, EPI_ISL_1812360 | | | |
| see above | EXCITE Lab | Andersen lab at Scripps Research | Nicole L Washington, Simon White, Geraint Levan, Kelly Schiabor Barrett, Elizabeth Cirulli, Alexandre Bolze, Charlotte Rivera-Garcia, Ryan Cho, Jason Nguyen, Sherry Wang, Jimmy Ramirez, Celena Andrade, Alice Summerfield, Tyler Cassens, Efren Sandoval, Francisco Tanudjaja, Magnus Isaksson, William Lee, David Becker, Marc Laurent, James Lu + SEARCH |
| EPI_ISL_1812361 | EXCITE Lab | Andersen lab at Scripps Research | David Pride, Sharon Reed, Chip Schooley, Angela Scioscia, Natasha Martin Cheryl Anderson, Sawyer Farmer, Abigail Schnapper, Helena Tubb, Tommy Valles + SEARCH |
| EPI_ISL_1812362 | EXCITE Lab | Andersen lab at Scripps Research | Nicole L Washington, Simon White, Geraint Levan, Kelly Schiabor Barrett, Elizabeth Cirulli, Alexandre Bolze, Charlotte Rivera-Garcia, Ryan Cho, Jason Nguyen, Sherry Wang, Jimmy Ramirez, Celena Andrade, Alice Summerfield, Tyler Cassens, Efren Sandoval, Francisco Tanudjaja, Magnus Isaksson, William Lee, David Becker, Marc Laurent, James Lu + SEARCH |
| EPI_ISL_1812363 | EXCITE Lab | Andersen lab at Scripps Research | David Pride, Sharon Reed, Chip Schooley, Angela Scioscia, Natasha Martin Cheryl Anderson, Sawyer Farmer, Abigail Schnapper, Helena Tubb, Tommy Valles + SEARCH |
| EPI_ISL_1812364, EPI_ISL_1812365, EPI_ISL_1812366 | EXCITE Lab | Andersen lab at Scripps Research | Nicole L Washington, Simon White, Geraint Levan, Kelly Schiabor Barrett, Elizabeth Cirulli, Alexandre Bolze, Charlotte Rivera-Garcia, Ryan Cho, Jason Nguyen, Sherry Wang, Jimmy Ramirez, Celena Andrade, Alice Summerfield, Tyler Cassens, Efren Sandoval, Francisco Tanudjaja, Magnus Isaksson, William Lee, David Becker, Marc Laurent, James Lu + SEARCH |
| EPI_ISL_1812367 | EXCITE Lab | Andersen lab at Scripps Research | Sydney Morgan + Marni Jacobs + Celestine Magallanes + Holly Valentine + Louise C. Laurent + SEARCH |
| EPI_ISL_1812368, EPI_ISL_1812369, EPI_ISL_1812370 | EXCITE Lab | Andersen lab at Scripps Research | Nicole L Washington, Simon White, Geraint Levan, Kelly Schiabor Barrett, Elizabeth Cirulli, Alexandre Bolze, Charlotte Rivera-Garcia, Ryan Cho, Jason Nguyen, Sherry Wang, Jimmy Ramirez, Celena Andrade, Alice Summerfield, Tyler Cassens, Efren Sandoval, Francisco Tanudjaja, Magnus Isaksson, William Lee, David Becker, Marc Laurent, James Lu + SEARCH |
| EPI_ISL_1812371 | EXCITE Lab | Andersen lab at Scripps Research | David Pride, Sharon Reed, Chip Schooley, Angela Scioscia, Natasha Martin Cheryl Anderson, Sawyer Farmer, Abigail Schnapper, Helena Tubb, Tommy Valles + SEARCH |
| EPI_ISL_1812372, EPI_ISL_1812373, EPI_ISL_1812374, EPI_ISL_1812375, EPI_ISL_1812376, EPI_ISL_1812377, EPI_ISL_1812378, EPI_ISL_1812379, EPI_ISL_1812380 | EXCITE Lab | Andersen lab at Scripps Research | Nicole L Washington, Simon White, Geraint Levan, Kelly Schiabor Barrett, Elizabeth Cirulli, Alexandre Bolze, Charlotte Rivera-Garcia, Ryan Cho, Jason Nguyen, Sherry Wang, Jimmy Ramirez, Celena Andrade, Alice Summerfield, Tyler Cassens, Efren Sandoval, Francisco Tanudjaja, Magnus Isaksson, William Lee, David Becker, Marc Laurent, James Lu + SEARCH |
| EPI_ISL_1812381 | EXCITE Lab | Andersen lab at Scripps Research | David Pride, Sharon Reed, Chip Schooley, Angela Scioscia, Natasha Martin Cheryl Anderson, Sawyer Farmer, Abigail Schnapper, Helena Tubb, Tommy Valles + SEARCH |
| EPI_ISL_1812382 | EXCITE Lab | Andersen lab at Scripps Research | Nicole L Washington, Simon White, Geraint Levan, Kelly Schiabor Barrett, Elizabeth Cirulli, Alexandre Bolze, Charlotte Rivera-Garcia, Ryan Cho, Jason Nguyen, Sherry Wang, Jimmy Ramirez, Celena Andrade, Alice Summerfield, Tyler Cassens, Efren Sandoval, Francisco Tanudjaja, Magnus Isaksson, William Lee, David Becker, Marc Laurent, James Lu + SEARCH |
| EPI_ISL_1812383 | EXCITE Lab | Andersen lab at Scripps Research | David Pride, Sharon Reed, Chip Schooley, Angela Scioscia, Natasha Martin Cheryl Anderson, Sawyer Farmer, Abigail Schnapper, Helena Tubb, Tommy Valles + SEARCH |
| EPI_ISL_1812384, EPI_ISL_1812385, EPI_ISL_1812386, EPI_ISL_1812387 | EXCITE Lab | Andersen lab at Scripps Research | Nicole L Washington, Simon White, Geraint Levan, Kelly Schiabor Barrett, Elizabeth Cirulli, Alexandre Bolze, Charlotte Rivera-Garcia, Ryan Cho, Jason Nguyen, Sherry Wang, Jimmy Ramirez, Celena Andrade, Alice Summerfield, Tyler Cassens, Efren Sandoval, Francisco Tanudjaja, Magnus Isaksson, William Lee, David Becker, Marc Laurent, James Lu + SEARCH |
| EPI_ISL_1812388 | EXCITE Lab | Andersen lab at Scripps Research | David Pride, Sharon Reed, Chip Schooley, Angela Scioscia, Natasha Martin Cheryl Anderson, Sawyer Farmer, Abigail Schnapper, Helena Tubb, Tommy Valles + SEARCH |
| EPI_ISL_1812389, EPI_ISL_1812390 | EXCITE Lab | Andersen lab at Scripps Research | Nicole L Washington, Simon White, Geraint Levan, Kelly Schiabor Barrett, Elizabeth Cirulli, Alexandre Bolze, Charlotte Rivera-Garcia, Ryan Cho, Jason Nguyen, Sherry Wang, Jimmy Ramirez, Celena Andrade, Alice Summerfield, Tyler Cassens, Efren Sandoval, Francisco Tanudjaja, Magnus Isaksson, William Lee, David Becker, Marc Laurent, James Lu + SEARCH |
| EPI_ISL_1812391 | EXCITE Lab | Andersen lab at Scripps Research | David Pride, Sharon Reed, Chip Schooley, Angela Scioscia, Natasha Martin Cheryl Anderson, Sawyer Farmer, Abigail Schnapper, Helena Tubb, Tommy Valles + SEARCH |
| EPI_ISL_1812392, EPI_ISL_1812393, EPI_ISL_1812394 | EXCITE Lab | Andersen lab at Scripps Research | Nicole L Washington, Simon White, Geraint Levan, Kelly Schiabor Barrett, Elizabeth Cirulli, Alexandre Bolze, Charlotte Rivera-Garcia, Ryan Cho, Jason Nguyen, Sherry Wang, Jimmy Ramirez, Celena Andrade, Alice Summerfield, Tyler Cassens, Efren Sandoval, Francisco Tanudjaja, Magnus Isaksson, William Lee, David Becker, Marc Laurent, James Lu + SEARCH |
| EPI_ISL_1812395 | EXCITE Lab | Andersen lab at Scripps Research | David Pride, Sharon Reed, Chip Schooley, Angela Scioscia, Natasha Martin Cheryl Anderson, Sawyer Farmer, Abigail Schnapper, Helena Tubb, Tommy Valles + SEARCH |
| EPI_ISL_1812396, EPI_ISL_1812397, EPI_ISL_1812398, EPI_ISL_1812399, EPI_ISL_1812400 | EXCITE Lab | Andersen lab at Scripps Research | Nicole L Washington, Simon White, Geraint Levan, Kelly Schiabor Barrett, Elizabeth Cirulli, Alexandre Bolze, Charlotte Rivera-Garcia, Ryan Cho, Jason Nguyen, Sherry Wang, Jimmy Ramirez, Celena Andrade, Alice Summerfield, Tyler Cassens, Efren Sandoval, Francisco Tanudjaja, Magnus Isaksson, William Lee, David Becker, Marc Laurent, James Lu + SEARCH |
| EPI_ISL_1812401 | EXCITE Lab | Andersen lab at Scripps Research | David Pride, Sharon Reed, Chip Schooley, Angela Scioscia, Natasha Martin Cheryl Anderson, Sawyer Farmer, Abigail Schnapper, Helena Tubb, Tommy Valles + SEARCH |
| EPI_ISL_1812402, EPI_ISL_1812403, EPI_ISL_1812404, EPI_ISL_1812405, EPI_ISL_1812406, EPI_ISL_1812407, EPI_ISL_1812408, EPI_ISL_1812409, EPI_ISL_1812410, EPI_ISL_1812411 | EXCITE Lab | Andersen lab at Scripps Research | Nicole L Washington, Simon White, Geraint Levan, Kelly Schiabor Barrett, Elizabeth Cirulli, Alexandre Bolze, Charlotte Rivera-Garcia, Ryan Cho, Jason Nguyen, Sherry Wang, Jimmy Ramirez, Celena Andrade, Alice Summerfield, Tyler Cassens, Efren Sandoval, Francisco Tanudjaja, Magnus Isaksson, William Lee, David Becker, Marc Laurent, James Lu + SEARCH |
| EPI_ISL_1812412 | EXCITE Lab | Andersen lab at Scripps Research | David Pride, Sharon Reed, Chip Schooley, Angela Scioscia, Natasha Martin Cheryl Anderson, Sawyer Farmer, Abigail Schnapper, Helena Tubb, Tommy Valles + SEARCH |
| EPI_ISL_1812413, EPI_ISL_1812414, EPI_ISL_1812415, EPI_ISL_1812416 | EXCITE Lab | Andersen lab at Scripps Research | Nicole L Washington, Simon White, Geraint Levan, Kelly Schiabor Barrett, Elizabeth Cirulli, Alexandre Bolze, Charlotte Rivera-Garcia, Ryan Cho, Jason Nguyen, Sherry Wang, Jimmy Ramirez, Celena Andrade, Alice Summerfield, Tyler Cassens, Efren Sandoval, Francisco Tanudjaja, Magnus Isaksson, William Lee, David Becker, Marc Laurent, James Lu + SEARCH |
| EPI_ISL_1812417 | EXCITE Lab | Andersen lab at Scripps Research | Fred Wu, Christine Mitchell + SEARCH |
| EPI_ISL_1812418, EPI_ISL_1812419, EPI_ISL_1812420, EPI_ISL_1812421, EPI_ISL_1812422, EPI_ISL_1812423, EPI_ISL_1812424, EPI_ISL_1812425, EPI_ISL_1812426, EPI_ISL_1812427, EPI_ISL_1812428, EPI_ISL_1812429, EPI_ISL_1812430, EPI_ISL_1812431 | | | |
| see above | EXCITE Lab | Andersen lab at Scripps Research | Nicole L Washington, Simon White, Geraint Levan, Kelly Schiabor Barrett, Elizabeth Cirulli, Alexandre Bolze, Charlotte Rivera-Garcia, Ryan Cho, Jason Nguyen, Sherry Wang, Jimmy Ramirez, Celena Andrade, Alice Summerfield, Tyler Cassens, Efren Sandoval, Francisco Tanudjaja, Magnus Isaksson, William Lee, David Becker, Marc Laurent, James Lu + SEARCH |
| EPI_ISL_1812432 | EXCITE Lab | Andersen lab at Scripps Research | David Pride, Sharon Reed, Chip Schooley, Angela Scioscia, Natasha Martin Cheryl Anderson, Sawyer Farmer, Abigail Schnapper, Helena Tubb, Tommy Valles + SEARCH |
| EPI_ISL_1812433, EPI_ISL_1812434, EPI_ISL_1812435, EPI_ISL_1812436 | EXCITE Lab | Andersen lab at Scripps Research | Nicole L Washington, Simon White, Geraint Levan, Kelly Schiabor Barrett, Elizabeth Cirulli, Alexandre Bolze, Charlotte Rivera-Garcia, Ryan Cho, Jason Nguyen, Sherry Wang, Jimmy Ramirez, Celena Andrade, Alice Summerfield, Tyler Cassens, Efren Sandoval, Francisco Tanudjaja, Magnus Isaksson, William Lee, David Becker, Marc Laurent, James Lu + SEARCH |
| EPI_ISL_1812437 | EXCITE Lab | Andersen lab at Scripps Research | David Pride, Sharon Reed, Chip Schooley, Angela Scioscia, Natasha Martin Cheryl Anderson, Sawyer Farmer, Abigail Schnapper, Helena Tubb, Tommy Valles + SEARCH |
| EPI_ISL_1812438, EPI_ISL_1812439, EPI_ISL_1812440, | EXCITE Lab | Andersen lab at Scripps Research | Nicole L Washington, Simon White, Geraint Levan, Kelly Schiabor Barrett, Elizabeth Cirulli, Alexandre Bolze, Charlotte Rivera-Garcia, Ryan Cho, Jason Nguyen, Sherry Wang, Jimmy Ramirez, Celena Andrade, Alice Summerfield, Tyler Cassens, Efren Sandoval, Francisco Tanudjaja, Magnus Isaksson, William Lee, David Becker, Marc Laurent, James Lu + SEARCH |

| | | | |
|---|---|---|---|
| EPI_ISL_1812441, EPI_ISL_1812442, EPI_ISL_1812443, EPI_ISL_1812444 | | | |
| EPI_ISL_1812445 | EXCITE Lab | Andersen lab at Scripps Research | David Pride, Sharon Reed, Chip Schooley, Angela Scioscia, Natasha Martin Cheryl Anderson, Sawyer Farmer, Abigail Schnapper, Helena Tubb, Tommy Valles + SEARCH |
| EPI_ISL_1812446, EPI_ISL_1812447 | EXCITE Lab | Andersen lab at Scripps Research | Nicole L Washington, Simon White, Geraint Levan, Kelly Schiabor Barrett, Elizabeth Cirulli, Alexandre Bolze, Charlotte Rivera-Garcia, Ryan Cho, Jason Nguyen, Sherry Wang, Jimmy Ramirez, Celena Andrade, Alice Summerfield, Tyler Cassens, Efren Sandoval, Francisco Tanudjaja, Magnus Isaksson, William Lee, David Becker, Marc Laurent, James Lu + SEARCH |
| EPI_ISL_1812448, EPI_ISL_1812449 | EXCITE Lab | Andersen lab at Scripps Research | David Pride, Sharon Reed, Chip Schooley, Angela Scioscia, Natasha Martin Cheryl Anderson, Sawyer Farmer, Abigail Schnapper, Helena Tubb, Tommy Valles + SEARCH |
| EPI_ISL_1812450, EPI_ISL_1812451, EPI_ISL_1812452, EPI_ISL_1812453, EPI_ISL_1812454, EPI_ISL_1812455, EPI_ISL_1812456, EPI_ISL_1812457, EPI_ISL_1812458, EPI_ISL_1812459, EPI_ISL_1812460, EPI_ISL_1812461, EPI_ISL_1812462, EPI_ISL_1812463, EPI_ISL_1812464 | | | |
| see above | EXCITE Lab | Andersen lab at Scripps Research | Nicole L Washington, Simon White, Geraint Levan, Kelly Schiabor Barrett, Elizabeth Cirulli, Alexandre Bolze, Charlotte Rivera-Garcia, Ryan Cho, Jason Nguyen, Sherry Wang, Jimmy Ramirez, Celena Andrade, Alice Summerfield, Tyler Cassens, Efren Sandoval, Francisco Tanudjaja, Magnus Isaksson, William Lee, David Becker, Marc Laurent, James Lu + SEARCH |
| EPI_ISL_1812465 | EXCITE Lab | Andersen lab at Scripps Research | David Pride, Sharon Reed, Chip Schooley, Angela Scioscia, Natasha Martin Cheryl Anderson, Sawyer Farmer, Abigail Schnapper, Helena Tubb, Tommy Valles + SEARCH |
| EPI_ISL_1812466, EPI_ISL_1812467, EPI_ISL_1812468, EPI_ISL_1812469, EPI_ISL_1812470, EPI_ISL_1812471, EPI_ISL_1812472 | EXCITE Lab | Andersen lab at Scripps Research | Nicole L Washington, Simon White, Geraint Levan, Kelly Schiabor Barrett, Elizabeth Cirulli, Alexandre Bolze, Charlotte Rivera-Garcia, Ryan Cho, Jason Nguyen, Sherry Wang, Jimmy Ramirez, Celena Andrade, Alice Summerfield, Tyler Cassens, Efren Sandoval, Francisco Tanudjaja, Magnus Isaksson, William Lee, David Becker, Marc Laurent, James Lu + SEARCH |
| EPI_ISL_1812473 | EXCITE Lab | Andersen lab at Scripps Research | David Pride, Sharon Reed, Chip Schooley, Angela Scioscia, Natasha Martin Cheryl Anderson, Sawyer Farmer, Abigail Schnapper, Helena Tubb, Tommy Valles + SEARCH |
| EPI_ISL_1812474, EPI_ISL_1812475, EPI_ISL_1812476, EPI_ISL_1812477, EPI_ISL_1812478 | EXCITE Lab | Andersen lab at Scripps Research | Nicole L Washington, Simon White, Geraint Levan, Kelly Schiabor Barrett, Elizabeth Cirulli, Alexandre Bolze, Charlotte Rivera-Garcia, Ryan Cho, Jason Nguyen, Sherry Wang, Jimmy Ramirez, Celena Andrade, Alice Summerfield, Tyler Cassens, Efren Sandoval, Francisco Tanudjaja, Magnus Isaksson, William Lee, David Becker, Marc Laurent, James Lu + SEARCH |
| EPI_ISL_1812479 | EXCITE Lab | Andersen lab at Scripps Research | David Pride, Sharon Reed, Chip Schooley, Angela Scioscia, Natasha Martin Cheryl Anderson, Sawyer Farmer, Abigail Schnapper, Helena Tubb, Tommy Valles + SEARCH |
| EPI_ISL_1812480, EPI_ISL_1812481, EPI_ISL_1812482, EPI_ISL_1812483, EPI_ISL_1812484, EPI_ISL_1812485, EPI_ISL_1812486, EPI_ISL_1812487, EPI_ISL_1812488, EPI_ISL_1812489, EPI_ISL_1812490, EPI_ISL_1812491 | | | |
| see above | EXCITE Lab | Andersen lab at Scripps Research | Nicole L Washington, Simon White, Geraint Levan, Kelly Schiabor Barrett, Elizabeth Cirulli, Alexandre Bolze, Charlotte Rivera-Garcia, Ryan Cho, Jason Nguyen, Sherry Wang, Jimmy Ramirez, Celena Andrade, Alice Summerfield, Tyler Cassens, Efren Sandoval, Francisco Tanudjaja, Magnus Isaksson, William Lee, David Becker, Marc Laurent, James Lu + SEARCH |
| EPI_ISL_1812492 | EXCITE Lab | Andersen lab at Scripps Research | Sydney Morgan + Marni Jacobs + Celestine Magallanes + Holly Valentine + Louise C. Laurent + SEARCH |
| EPI_ISL_1812493, EPI_ISL_1812494 | EXCITE Lab | Andersen lab at Scripps Research | Nicole L Washington, Simon White, Geraint Levan, Kelly Schiabor Barrett, Elizabeth Cirulli, Alexandre Bolze, Charlotte Rivera-Garcia, Ryan Cho, Jason Nguyen, Sherry Wang, Jimmy Ramirez, Celena Andrade, Alice Summerfield, Tyler Cassens, Efren Sandoval, Francisco Tanudjaja, Magnus Isaksson, William Lee, David Becker, Marc Laurent, James Lu + SEARCH |
| EPI_ISL_1812495 | EXCITE Lab | Andersen lab at Scripps Research | David Pride, Sharon Reed, Chip Schooley, Angela Scioscia, Natasha Martin Cheryl Anderson, Sawyer Farmer, Abigail Schnapper, Helena Tubb, Tommy Valles + SEARCH |
| EPI_ISL_1812496, EPI_ISL_1812497, EPI_ISL_1812498 | EXCITE Lab | Andersen lab at Scripps Research | Nicole L Washington, Simon White, Geraint Levan, Kelly Schiabor Barrett, Elizabeth Cirulli, Alexandre Bolze, Charlotte Rivera-Garcia, Ryan Cho, Jason Nguyen, Sherry Wang, Jimmy Ramirez, Celena Andrade, Alice Summerfield, Tyler Cassens, Efren Sandoval, Francisco Tanudjaja, Magnus Isaksson, William Lee, David Becker, Marc Laurent, James Lu + SEARCH |
| EPI_ISL_1812499 | EXCITE Lab | Andersen lab at Scripps Research | David Pride, Sharon Reed, Chip Schooley, Angela Scioscia, Natasha Martin Cheryl Anderson, Sawyer Farmer, Abigail Schnapper, Helena Tubb, Tommy Valles + SEARCH |
| EPI_ISL_1812500, EPI_ISL_1812501, EPI_ISL_1812502, EPI_ISL_1812503, EPI_ISL_1812504, EPI_ISL_1812505, EPI_ISL_1812506, EPI_ISL_1812507 | EXCITE Lab | Andersen lab at Scripps Research | Nicole L Washington, Simon White, Geraint Levan, Kelly Schiabor Barrett, Elizabeth Cirulli, Alexandre Bolze, Charlotte Rivera-Garcia, Ryan Cho, Jason Nguyen, Sherry Wang, Jimmy Ramirez, Celena Andrade, Alice Summerfield, Tyler Cassens, Efren Sandoval, Francisco Tanudjaja, Magnus Isaksson, William Lee, David Becker, Marc Laurent, James Lu + SEARCH |
| EPI_ISL_1812508 | EXCITE Lab | Andersen lab at Scripps Research | Chip Schooley, Natasha Martin, Cheryl Anderson, Angela Scioscia, Smruthi Karthikeyan, Greg Humphrey, Sawyer Farmer, Abigail Schnapper, Helena Tubb, Tommy Valles + SEARCH |
| EPI_ISL_1812509 | EXCITE Lab | Andersen lab at Scripps Research | David Pride, Sharon Reed, Chip Schooley, Angela Scioscia, Natasha Martin Cheryl Anderson, Sawyer Farmer, Abigail Schnapper, Helena Tubb, Tommy Valles + SEARCH |
| EPI_ISL_1812510, EPI_ISL_1812511 | EXCITE Lab | Andersen lab at Scripps Research | Nicole L Washington, Simon White, Geraint Levan, Kelly Schiabor Barrett, Elizabeth Cirulli, Alexandre Bolze, Charlotte Rivera-Garcia, Ryan Cho, Jason Nguyen, Sherry Wang, Jimmy Ramirez, Celena Andrade, Alice Summerfield, Tyler Cassens, Efren Sandoval, Francisco Tanudjaja, Magnus Isaksson, William Lee, David Becker, Marc Laurent, James Lu + SEARCH |
| EPI_ISL_1812512 | EXCITE Lab | Andersen lab at Scripps Research | Chip Schooley, Natasha Martin, Cheryl Anderson, Angela Scioscia, Smruthi Karthikeyan, Greg Humphrey, Sawyer Farmer, Abigail Schnapper, Helena Tubb, Tommy Valles + SEARCH |
| EPI_ISL_1812513, EPI_ISL_1812514, EPI_ISL_1812515, EPI_ISL_1812516, EPI_ISL_1812517 | EXCITE Lab | Andersen lab at Scripps Research | Nicole L Washington, Simon White, Geraint Levan, Kelly Schiabor Barrett, Elizabeth Cirulli, Alexandre Bolze, Charlotte Rivera-Garcia, Ryan Cho, Jason Nguyen, Sherry Wang, Jimmy Ramirez, Celena Andrade, Alice Summerfield, Tyler Cassens, Efren Sandoval, Francisco Tanudjaja, Magnus Isaksson, William Lee, David Becker, Marc Laurent, James Lu + SEARCH |
| EPI_ISL_1812518 | EXCITE Lab | Andersen lab at Scripps Research | David Pride, Sharon Reed, Chip Schooley, Angela Scioscia, Natasha Martin Cheryl Anderson, Sawyer Farmer, Abigail Schnapper, Helena Tubb, Tommy Valles + SEARCH |
| EPI_ISL_1812519 | EXCITE Lab | Andersen lab at Scripps Research | Nicole L Washington, Simon White, Geraint Levan, Kelly Schiabor Barrett, Elizabeth Cirulli, Alexandre Bolze, Charlotte Rivera-Garcia, Ryan Cho, Jason Nguyen, Sherry Wang, Jimmy Ramirez, Celena Andrade, Alice Summerfield, Tyler Cassens, Efren Sandoval, Francisco Tanudjaja, Magnus Isaksson, William Lee, David Becker, Marc Laurent, James Lu + SEARCH |
| EPI_ISL_1812520 | EXCITE Lab | Andersen lab at Scripps Research | Sydney Morgan + Marni Jacobs + Celestine Magallanes + Holly Valentine + Louise C. Laurent + SEARCH |
| EPI_ISL_1812521, EPI_ISL_1812522, EPI_ISL_1812523, EPI_ISL_1812524, EPI_ISL_1812525, EPI_ISL_1812526, EPI_ISL_1812527, EPI_ISL_1812528, EPI_ISL_1812529, EPI_ISL_1812530, EPI_ISL_1812531, EPI_ISL_1812532, EPI_ISL_1812533, EPI_ISL_1812534, EPI_ISL_1812535 | | | |
| see above | EXCITE Lab | Andersen lab at Scripps Research | Nicole L Washington, Simon White, Geraint Levan, Kelly Schiabor Barrett, Elizabeth Cirulli, Alexandre Bolze, Charlotte Rivera-Garcia, Ryan Cho, Jason Nguyen, Sherry Wang, Jimmy Ramirez, Celena Andrade, Alice Summerfield, Tyler Cassens, Efren Sandoval, Francisco Tanudjaja, Magnus Isaksson, William Lee, David Becker, Marc Laurent, James Lu + SEARCH |
| EPI_ISL_1812536 | EXCITE Lab | Andersen lab at Scripps Research | David Pride, Sharon Reed, Chip Schooley, Angela Scioscia, Natasha Martin Cheryl Anderson, Sawyer Farmer, Abigail Schnapper, Helena Tubb, Tommy Valles + SEARCH |
| EPI_ISL_1812537, EPI_ISL_1812538, EPI_ISL_1812539, EPI_ISL_1812540, EPI_ISL_1812541, EPI_ISL_1812542, EPI_ISL_1812543, EPI_ISL_1812544, EPI_ISL_1812545, EPI_ISL_1812546, EPI_ISL_1812547 | | | |
| see above | EXCITE Lab | Andersen lab at Scripps Research | Nicole L Washington, Simon White, Geraint Levan, Kelly Schiabor Barrett, Elizabeth Cirulli, Alexandre Bolze, Charlotte Rivera-Garcia, Ryan Cho, Jason Nguyen, Sherry Wang, Jimmy Ramirez, Celena Andrade, Alice Summerfield, Tyler Cassens, Efren Sandoval, Francisco Tanudjaja, Magnus Isaksson, William Lee, David Becker, Marc Laurent, James Lu + SEARCH |
| EPI_ISL_1812548 | EXCITE Lab | Andersen lab at Scripps Research | David Pride, Sharon Reed, Chip Schooley, Angela Scioscia, Natasha Martin Cheryl Anderson, Sawyer Farmer, Abigail Schnapper, Helena Tubb, Tommy Valles + SEARCH |
| EPI_ISL_1812549, EPI_ISL_1812550, EPI_ISL_1812551, EPI_ISL_1812552 | EXCITE Lab | Andersen lab at Scripps Research | Nicole L Washington, Simon White, Geraint Levan, Kelly Schiabor Barrett, Elizabeth Cirulli, Alexandre Bolze, Charlotte Rivera-Garcia, Ryan Cho, Jason Nguyen, Sherry Wang, Jimmy Ramirez, Celena Andrade, Alice Summerfield, Tyler Cassens, Efren Sandoval, Francisco Tanudjaja, Magnus Isaksson, William Lee, David Becker, Marc Laurent, James Lu + SEARCH |
| EPI_ISL_1812553 | EXCITE Lab | Andersen lab at Scripps Research | Sydney Morgan + Marni Jacobs + Celestine Magallanes + Holly Valentine + Louise C. Laurent + SEARCH |
| EPI_ISL_1812554, EPI_ISL_1812555, EPI_ISL_1812556, EPI_ISL_1812557 | EXCITE Lab | Andersen lab at Scripps Research | Nicole L Washington, Simon White, Geraint Levan, Kelly Schiabor Barrett, Elizabeth Cirulli, Alexandre Bolze, Charlotte Rivera-Garcia, Ryan Cho, Jason Nguyen, Sherry Wang, Jimmy Ramirez, Celena Andrade, Alice Summerfield, Tyler Cassens, Efren Sandoval, Francisco Tanudjaja, Magnus Isaksson, William Lee, David Becker, Marc Laurent, James Lu + SEARCH |
| EPI_ISL_1812558 | EXCITE Lab | Andersen lab at Scripps Research | David Pride, Sharon Reed, Chip Schooley, Angela Scioscia, Natasha Martin Cheryl Anderson, Sawyer Farmer, Abigail Schnapper, Helena Tubb, Tommy Valles + SEARCH |
| EPI_ISL_1812559, EPI_ISL_1812560, EPI_ISL_1812561, EPI_ISL_1812562 | EXCITE Lab | Andersen lab at Scripps Research | Nicole L Washington, Simon White, Geraint Levan, Kelly Schiabor Barrett, Elizabeth Cirulli, Alexandre Bolze, Charlotte Rivera-Garcia, Ryan Cho, Jason Nguyen, Sherry Wang, Jimmy Ramirez, Celena Andrade, Alice Summerfield, Tyler Cassens, Efren Sandoval, Francisco Tanudjaja, Magnus Isaksson, William Lee, David Becker, Marc Laurent, James Lu + SEARCH |
| EPI_ISL_1812563 | EXCITE Lab | Andersen lab at Scripps Research | David Pride, Sharon Reed, Chip Schooley, Angela Scioscia, Natasha Martin Cheryl Anderson, Sawyer Farmer, Abigail Schnapper, Helena Tubb, Tommy Valles + SEARCH |
| EPI_ISL_1812564, EPI_ISL_1812565 | EXCITE Lab | Andersen lab at Scripps Research | Nicole L Washington, Simon White, Geraint Levan, Kelly Schiabor Barrett, Elizabeth Cirulli, Alexandre Bolze, Charlotte Rivera-Garcia, Ryan Cho, Jason Nguyen, Sherry Wang, Jimmy Ramirez, Celena Andrade, Alice Summerfield, Tyler Cassens, Efren Sandoval, Francisco Tanudjaja, Magnus Isaksson, William Lee, David Becker, Marc Laurent, James Lu + SEARCH |
| EPI_ISL_1812566 | EXCITE Lab | Andersen lab at Scripps Research | David Pride, Sharon Reed, Chip Schooley, Angela Scioscia, Natasha Martin Cheryl Anderson, Sawyer Farmer, Abigail Schnapper, Helena Tubb, Tommy Valles + SEARCH |
| EPI_ISL_1812567, EPI_ISL_1812568, EPI_ISL_1812569, EPI_ISL_1812570, EPI_ISL_1812571, EPI_ISL_1812572, EPI_ISL_1812573, EPI_ISL_1812574, EPI_ISL_1812575, EPI_ISL_1812576, EPI_ISL_1812577, EPI_ISL_1812578, EPI_ISL_1812579, EPI_ISL_1812580, EPI_ISL_1812581, EPI_ISL_1812582, EPI_ISL_1812583, EPI_ISL_1812584, EPI_ISL_1812585 | | | |
| see above | EXCITE Lab | Andersen lab at Scripps Research | Nicole L Washington, Simon White, Geraint Levan, Kelly Schiabor Barrett, Elizabeth Cirulli, Alexandre Bolze, Charlotte Rivera-Garcia, Ryan Cho, Jason Nguyen, Sherry Wang, Jimmy Ramirez, Celena Andrade, Alice Summerfield, Tyler Cassens, Efren Sandoval, Francisco Tanudjaja, Magnus Isaksson, William Lee, David Becker, Marc Laurent, James Lu + SEARCH |
| EPI_ISL_1812586 | EXCITE Lab | Andersen lab at Scripps Research | David Pride, Sharon Reed, Chip Schooley, Angela Scioscia, Natasha Martin Cheryl Anderson, Sawyer Farmer, Abigail Schnapper, Helena Tubb, Tommy Valles + SEARCH |
| EPI_ISL_1812587 | EXCITE Lab | Andersen lab at Scripps Research | Nicole L Washington, Simon White, Geraint Levan, Kelly Schiabor Barrett, Elizabeth Cirulli, Alexandre Bolze, Charlotte Rivera-Garcia, Ryan Cho, Jason Nguyen, Sherry Wang, Jimmy Ramirez, Celena Andrade, Alice Summerfield, Tyler Cassens, Efren Sandoval, Francisco Tanudjaja, Magnus |

| | | | |
|---|---|---|---|
| | | | Isaksson, William Lee, David Becker, Marc Laurent, James Lu + SEARCH |
| EPI_ISL_1812588 | EXCITE Lab | Andersen lab at Scripps Research | David Pride, Sharon Reed, Chip Schooley, Angela Scioscia, Natasha Martin Cheryl Anderson, Sawyer Farmer, Abigail Schnapper, Helena Tubb, Tommy Valles + SEARCH |
| EPI_ISL_1812589 | EXCITE Lab | Andersen lab at Scripps Research | Nicole L Washington, Simon White, Geraint Levan, Kelly Schiabor Barrett, Elizabeth Cirulli, Alexandre Bolze, Charlotte Rivera-Garcia, Ryan Cho, Jason Nguyen, Sherry Wang, Jimmy Ramirez, Celena Andrade, Alice Summerfield, Tyler Cassens, Efren Sandoval, Francisco Tanudjaja, Magnus Isaksson, William Lee, David Becker, Marc Laurent, James Lu + SEARCH |
| EPI_ISL_1812590 | EXCITE Lab | Andersen lab at Scripps Research | Sydney Morgan + Marni Jacobs + Celestine Magallanes + Holly Valentine + Louise C. Laurent + SEARCH |
| EPI_ISL_1812591, EPI_ISL_1812592, EPI_ISL_1812593, EPI_ISL_1812594 | EXCITE Lab | Andersen lab at Scripps Research | Nicole L Washington, Simon White, Geraint Levan, Kelly Schiabor Barrett, Elizabeth Cirulli, Alexandre Bolze, Charlotte Rivera-Garcia, Ryan Cho, Jason Nguyen, Sherry Wang, Jimmy Ramirez, Celena Andrade, Alice Summerfield, Tyler Cassens, Efren Sandoval, Francisco Tanudjaja, Magnus Isaksson, William Lee, David Becker, Marc Laurent, James Lu + SEARCH |
| EPI_ISL_1812595 | EXCITE Lab | Andersen lab at Scripps Research | David Pride, Sharon Reed, Chip Schooley, Angela Scioscia, Natasha Martin Cheryl Anderson, Sawyer Farmer, Abigail Schnapper, Helena Tubb, Tommy Valles + SEARCH |
| EPI_ISL_1812596, EPI_ISL_1812597, EPI_ISL_1812598, EPI_ISL_1812599, EPI_ISL_1812600, EPI_ISL_1812601, EPI_ISL_1812602, EPI_ISL_1812603, EPI_ISL_1812604, EPI_ISL_1812605, EPI_ISL_1812606, EPI_ISL_1812607, EPI_ISL_1812608 | | | |
| see above | EXCITE Lab | Andersen lab at Scripps Research | Nicole L Washington, Simon White, Geraint Levan, Kelly Schiabor Barrett, Elizabeth Cirulli, Alexandre Bolze, Charlotte Rivera-Garcia, Ryan Cho, Jason Nguyen, Sherry Wang, Jimmy Ramirez, Celena Andrade, Alice Summerfield, Tyler Cassens, Efren Sandoval, Francisco Tanudjaja, Magnus Isaksson, William Lee, David Becker, Marc Laurent, James Lu + SEARCH |
| EPI_ISL_1812609 | EXCITE Lab | Andersen lab at Scripps Research | David Pride, Sharon Reed, Chip Schooley, Angela Scioscia, Natasha Martin Cheryl Anderson, Sawyer Farmer, Abigail Schnapper, Helena Tubb, Tommy Valles + SEARCH |
| EPI_ISL_1812610, EPI_ISL_1812611, EPI_ISL_1812612, EPI_ISL_1812613, EPI_ISL_1812614, EPI_ISL_1812615, EPI_ISL_1812616, EPI_ISL_1812617, EPI_ISL_1812618, EPI_ISL_1812619, EPI_ISL_1812620, EPI_ISL_1812621, EPI_ISL_1812622 | | | |
| see above | EXCITE Lab | Andersen lab at Scripps Research | Nicole L Washington, Simon White, Geraint Levan, Kelly Schiabor Barrett, Elizabeth Cirulli, Alexandre Bolze, Charlotte Rivera-Garcia, Ryan Cho, Jason Nguyen, Sherry Wang, Jimmy Ramirez, Celena Andrade, Alice Summerfield, Tyler Cassens, Efren Sandoval, Francisco Tanudjaja, Magnus Isaksson, William Lee, David Becker, Marc Laurent, James Lu + SEARCH |
| EPI_ISL_1812623 | EXCITE Lab | Andersen lab at Scripps Research | David Pride, Sharon Reed, Chip Schooley, Angela Scioscia, Natasha Martin Cheryl Anderson, Sawyer Farmer, Abigail Schnapper, Helena Tubb, Tommy Valles + SEARCH |
| EPI_ISL_1812624, EPI_ISL_1812625, EPI_ISL_1812626, EPI_ISL_1812627, EPI_ISL_1812628, EPI_ISL_1812629 | EXCITE Lab | Andersen lab at Scripps Research | Nicole L Washington, Simon White, Geraint Levan, Kelly Schiabor Barrett, Elizabeth Cirulli, Alexandre Bolze, Charlotte Rivera-Garcia, Ryan Cho, Jason Nguyen, Sherry Wang, Jimmy Ramirez, Celena Andrade, Alice Summerfield, Tyler Cassens, Efren Sandoval, Francisco Tanudjaja, Magnus Isaksson, William Lee, David Becker, Marc Laurent, James Lu + SEARCH |
| EPI_ISL_1812630 | EXCITE Lab | Andersen lab at Scripps Research | David Pride, Sharon Reed, Chip Schooley, Angela Scioscia, Natasha Martin Cheryl Anderson, Sawyer Farmer, Abigail Schnapper, Helena Tubb, Tommy Valles + SEARCH |
| EPI_ISL_1812631, EPI_ISL_1812632, EPI_ISL_1812633, EPI_ISL_1812634, EPI_ISL_1812635, EPI_ISL_1812636, EPI_ISL_1812637 | EXCITE Lab | Andersen lab at Scripps Research | Nicole L Washington, Simon White, Geraint Levan, Kelly Schiabor Barrett, Elizabeth Cirulli, Alexandre Bolze, Charlotte Rivera-Garcia, Ryan Cho, Jason Nguyen, Sherry Wang, Jimmy Ramirez, Celena Andrade, Alice Summerfield, Tyler Cassens, Efren Sandoval, Francisco Tanudjaja, Magnus Isaksson, William Lee, David Becker, Marc Laurent, James Lu + SEARCH |
| EPI_ISL_1812638 | EXCITE Lab | Andersen lab at Scripps Research | Sydney Morgan + Marni Jacobs + Celestine Magallanes + Holly Valentine + Louise C. Laurent + SEARCH |
| EPI_ISL_1812639, EPI_ISL_1812640, EPI_ISL_1812641, EPI_ISL_1812642, EPI_ISL_1812643, EPI_ISL_1812644, EPI_ISL_1812645 | EXCITE Lab | Andersen lab at Scripps Research | Nicole L Washington, Simon White, Geraint Levan, Kelly Schiabor Barrett, Elizabeth Cirulli, Alexandre Bolze, Charlotte Rivera-Garcia, Ryan Cho, Jason Nguyen, Sherry Wang, Jimmy Ramirez, Celena Andrade, Alice Summerfield, Tyler Cassens, Efren Sandoval, Francisco Tanudjaja, Magnus Isaksson, William Lee, David Becker, Marc Laurent, James Lu + SEARCH |
| EPI_ISL_1812646 | EXCITE Lab | Andersen lab at Scripps Research | David Pride, Sharon Reed, Chip Schooley, Angela Scioscia, Natasha Martin Cheryl Anderson, Sawyer Farmer, Abigail Schnapper, Helena Tubb, Tommy Valles + SEARCH |
| EPI_ISL_1812647, EPI_ISL_1812648, EPI_ISL_1812649 | EXCITE Lab | Andersen lab at Scripps Research | Nicole L Washington, Simon White, Geraint Levan, Kelly Schiabor Barrett, Elizabeth Cirulli, Alexandre Bolze, Charlotte Rivera-Garcia, Ryan Cho, Jason Nguyen, Sherry Wang, Jimmy Ramirez, Celena Andrade, Alice Summerfield, Tyler Cassens, Efren Sandoval, Francisco Tanudjaja, Magnus Isaksson, William Lee, David Becker, Marc Laurent, James Lu + SEARCH |
| EPI_ISL_1812650 | EXCITE Lab | Andersen lab at Scripps Research | David Pride, Sharon Reed, Chip Schooley, Angela Scioscia, Natasha Martin Cheryl Anderson, Sawyer Farmer, Abigail Schnapper, Helena Tubb, Tommy Valles + SEARCH |
| EPI_ISL_1812651, EPI_ISL_1812652, EPI_ISL_1812653, EPI_ISL_1812654, EPI_ISL_1812655, EPI_ISL_1812656 | EXCITE Lab | Andersen lab at Scripps Research | Nicole L Washington, Simon White, Geraint Levan, Kelly Schiabor Barrett, Elizabeth Cirulli, Alexandre Bolze, Charlotte Rivera-Garcia, Ryan Cho, Jason Nguyen, Sherry Wang, Jimmy Ramirez, Celena Andrade, Alice Summerfield, Tyler Cassens, Efren Sandoval, Francisco Tanudjaja, Magnus Isaksson, William Lee, David Becker, Marc Laurent, James Lu + SEARCH |
| EPI_ISL_1812657 | EXCITE Lab | Andersen lab at Scripps Research | David Pride, Sharon Reed, Chip Schooley, Angela Scioscia, Natasha Martin Cheryl Anderson, Sawyer Farmer, Abigail Schnapper, Helena Tubb, Tommy Valles + SEARCH |
| EPI_ISL_1812658, EPI_ISL_1812659, EPI_ISL_1812660, EPI_ISL_1812661, EPI_ISL_1812662, EPI_ISL_1812663, EPI_ISL_1812664, EPI_ISL_1812665 | EXCITE Lab | Andersen lab at Scripps Research | Nicole L Washington, Simon White, Geraint Levan, Kelly Schiabor Barrett, Elizabeth Cirulli, Alexandre Bolze, Charlotte Rivera-Garcia, Ryan Cho, Jason Nguyen, Sherry Wang, Jimmy Ramirez, Celena Andrade, Alice Summerfield, Tyler Cassens, Efren Sandoval, Francisco Tanudjaja, Magnus Isaksson, William Lee, David Becker, Marc Laurent, James Lu + SEARCH |
| EPI_ISL_1812666 | EXCITE Lab | Andersen lab at Scripps Research | David Pride, Sharon Reed, Chip Schooley, Angela Scioscia, Natasha Martin Cheryl Anderson, Sawyer Farmer, Abigail Schnapper, Helena Tubb, Tommy Valles + SEARCH |
| EPI_ISL_1812667 | EXCITE Lab | Andersen lab at Scripps Research | Nicole L Washington, Simon White, Geraint Levan, Kelly Schiabor Barrett, Elizabeth Cirulli, Alexandre Bolze, Charlotte Rivera-Garcia, Ryan Cho, Jason Nguyen, Sherry Wang, Jimmy Ramirez, Celena Andrade, Alice Summerfield, Tyler Cassens, Efren Sandoval, Francisco Tanudjaja, Magnus Isaksson, William Lee, David Becker, Marc Laurent, James Lu + SEARCH |
| EPI_ISL_1812668 | EXCITE Lab | Andersen lab at Scripps Research | David Pride, Sharon Reed, Chip Schooley, Angela Scioscia, Natasha Martin Cheryl Anderson, Sawyer Farmer, Abigail Schnapper, Helena Tubb, Tommy Valles + SEARCH |
| EPI_ISL_1812669, EPI_ISL_1812670 | EXCITE Lab | Andersen lab at Scripps Research | Nicole L Washington, Simon White, Geraint Levan, Kelly Schiabor Barrett, Elizabeth Cirulli, Alexandre Bolze, Charlotte Rivera-Garcia, Ryan Cho, Jason Nguyen, Sherry Wang, Jimmy Ramirez, Celena Andrade, Alice Summerfield, Tyler Cassens, Efren Sandoval, Francisco Tanudjaja, Magnus Isaksson, William Lee, David Becker, Marc Laurent, James Lu + SEARCH |
| EPI_ISL_1812671 | EXCITE Lab | Andersen lab at Scripps Research | Sydney Morgan + Marni Jacobs + Celestine Magallanes + Holly Valentine + Louise C. Laurent + SEARCH |
| EPI_ISL_1812672 | EXCITE Lab | Andersen lab at Scripps Research | Nicole L Washington, Simon White, Geraint Levan, Kelly Schiabor Barrett, Elizabeth Cirulli, Alexandre Bolze, Charlotte Rivera-Garcia, Ryan Cho, Jason Nguyen, Sherry Wang, Jimmy Ramirez, Celena Andrade, Alice Summerfield, Tyler Cassens, Efren Sandoval, Francisco Tanudjaja, Magnus Isaksson, William Lee, David Becker, Marc Laurent, James Lu + SEARCH |
| EPI_ISL_1812673 | EXCITE Lab | Andersen lab at Scripps Research | David Pride, Sharon Reed, Chip Schooley, Angela Scioscia, Natasha Martin Cheryl Anderson, Sawyer Farmer, Abigail Schnapper, Helena Tubb, Tommy Valles + SEARCH |
| EPI_ISL_1812674, EPI_ISL_1812675, EPI_ISL_1812676, EPI_ISL_1812677, EPI_ISL_1812678, EPI_ISL_1812679 | EXCITE Lab | Andersen lab at Scripps Research | Nicole L Washington, Simon White, Geraint Levan, Kelly Schiabor Barrett, Elizabeth Cirulli, Alexandre Bolze, Charlotte Rivera-Garcia, Ryan Cho, Jason Nguyen, Sherry Wang, Jimmy Ramirez, Celena Andrade, Alice Summerfield, Tyler Cassens, Efren Sandoval, Francisco Tanudjaja, Magnus Isaksson, William Lee, David Becker, Marc Laurent, James Lu + SEARCH |
| EPI_ISL_1812680 | EXCITE Lab | Andersen lab at Scripps Research | David Pride, Sharon Reed, Chip Schooley, Angela Scioscia, Natasha Martin Cheryl Anderson, Sawyer Farmer, Abigail Schnapper, Helena Tubb, Tommy Valles + SEARCH |
| EPI_ISL_1812681, EPI_ISL_1812682 | EXCITE Lab | Andersen lab at Scripps Research | Nicole L Washington, Simon White, Geraint Levan, Kelly Schiabor Barrett, Elizabeth Cirulli, Alexandre Bolze, Charlotte Rivera-Garcia, Ryan Cho, Jason Nguyen, Sherry Wang, Jimmy Ramirez, Celena Andrade, Alice Summerfield, Tyler Cassens, Efren Sandoval, Francisco Tanudjaja, Magnus Isaksson, William Lee, David Becker, Marc Laurent, James Lu + SEARCH |
| EPI_ISL_1812683 | EXCITE Lab | Andersen lab at Scripps Research | Sydney Morgan + Marni Jacobs + Celestine Magallanes + Holly Valentine + Louise C. Laurent + SEARCH |
| EPI_ISL_1812684, EPI_ISL_1812685, EPI_ISL_1812686, EPI_ISL_1812687, EPI_ISL_1812688, EPI_ISL_1812689, EPI_ISL_1812690, EPI_ISL_1812691, EPI_ISL_1812692, EPI_ISL_1812693, EPI_ISL_1812694, EPI_ISL_1812695, EPI_ISL_1812696, EPI_ISL_1812697, EPI_ISL_1812698, EPI_ISL_1812699, EPI_ISL_1812700, EPI_ISL_1812701 | | | |
| see above | EXCITE Lab | Andersen lab at Scripps Research | Nicole L Washington, Simon White, Geraint Levan, Kelly Schiabor Barrett, Elizabeth Cirulli, Alexandre Bolze, Charlotte Rivera-Garcia, Ryan Cho, Jason Nguyen, Sherry Wang, Jimmy Ramirez, Celena Andrade, Alice Summerfield, Tyler Cassens, Efren Sandoval, Francisco Tanudjaja, Magnus Isaksson, William Lee, David Becker, Marc Laurent, James Lu + SEARCH |
| EPI_ISL_1812702 | EXCITE Lab | Andersen lab at Scripps Research | David Pride, Sharon Reed, Chip Schooley, Angela Scioscia, Natasha Martin Cheryl Anderson, Sawyer Farmer, Abigail Schnapper, Helena Tubb, Tommy Valles + SEARCH |
| EPI_ISL_1812703, EPI_ISL_1812704, EPI_ISL_1812705, EPI_ISL_1812706, EPI_ISL_1812707, EPI_ISL_1812708, EPI_ISL_1812709, EPI_ISL_1812710, EPI_ISL_1812711, EPI_ISL_1812712, EPI_ISL_1812713, EPI_ISL_1812714, EPI_ISL_1812715, EPI_ISL_1812716 | | | |
| see above | EXCITE Lab | Andersen lab at Scripps Research | Nicole L Washington, Simon White, Geraint Levan, Kelly Schiabor Barrett, Elizabeth Cirulli, Alexandre Bolze, Charlotte Rivera-Garcia, Ryan Cho, Jason Nguyen, Sherry Wang, Jimmy Ramirez, Celena Andrade, Alice Summerfield, Tyler Cassens, Efren Sandoval, Francisco Tanudjaja, Magnus Isaksson, William Lee, David Becker, Marc Laurent, James Lu + SEARCH |
| EPI_ISL_1812717 | EXCITE Lab | Andersen lab at Scripps Research | David Pride, Sharon Reed, Chip Schooley, Angela Scioscia, Natasha Martin Cheryl Anderson, Sawyer Farmer, Abigail Schnapper, Helena Tubb, Tommy Valles + SEARCH |
| EPI_ISL_1812718, EPI_ISL_1812719, EPI_ISL_1812720, EPI_ISL_1812721, EPI_ISL_1812722, EPI_ISL_1812723, EPI_ISL_1812724, EPI_ISL_1812725, EPI_ISL_1812726, EPI_ISL_1812727, EPI_ISL_1812728, EPI_ISL_1812729, EPI_ISL_1812730, EPI_ISL_1812731, EPI_ISL_1812732, EPI_ISL_1812733, EPI_ISL_1812734, EPI_ISL_1812735, EPI_ISL_1812736, EPI_ISL_1812737, EPI_ISL_1812738, EPI_ISL_1812739, EPI_ISL_1812740 | | | |
| see above | EXCITE Lab | Andersen lab at Scripps Research | Nicole L Washington, Simon White, Geraint Levan, Kelly Schiabor Barrett, Elizabeth Cirulli, Alexandre Bolze, Charlotte Rivera-Garcia, Ryan Cho, Jason Nguyen, Sherry Wang, Jimmy Ramirez, Celena Andrade, Alice Summerfield, Tyler Cassens, Efren Sandoval, Francisco Tanudjaja, Magnus |

| | | | |
|---|---|---|---|
| | | | Isaksson, William Lee, David Becker, Marc Laurent, James Lu + SEARCH |
| EPI_ISL_1812741 | EXCITE Lab | Andersen lab at Scripps Research | Chip Schooley, Natasha Martin, Cheryl Anderson, Angela Scioscia, Smruthi Karthikeyan, Greg Humphrey, Sawyer Farmer, Abigail Schnapper, Helena Tubb, Tommy Valles + SEARCH |
| EPI_ISL_1812742 | EXCITE Lab | Andersen lab at Scripps Research | Nicole L Washington, Simon White, Geraint Levan, Kelly Schiabor Barrett, Elizabeth Cirulli, Alexandre Bolze, Charlotte Rivera-Garcia, Ryan Cho, Jason Nguyen, Sherry Wang, Jimmy Ramirez, Celena Andrade, Alice Summerfield, Tyler Cassens, Efren Sandoval, Francisco Tanudjaja, Magnus Isaksson, William Lee, David Becker, Marc Laurent, James Lu + SEARCH |
| EPI_ISL_1812743 | EXCITE Lab | Andersen lab at Scripps Research | David Pride, Sharon Reed, Chip Schooley, Angela Scioscia, Natasha Martin Cheryl Anderson, Sawyer Farmer, Abigail Schnapper, Helena Tubb, Tommy Valles + SEARCH |
| EPI_ISL_1812744, EPI_ISL_1812745, EPI_ISL_1812746, EPI_ISL_1812747, EPI_ISL_1812748, EPI_ISL_1812749, EPI_ISL_1812750, EPI_ISL_1812751, EPI_ISL_1812752, EPI_ISL_1812753, EPI_ISL_1812754, EPI_ISL_1812755, EPI_ISL_1812756 | | | |
| see above | EXCITE Lab | Andersen lab at Scripps Research | Nicole L Washington, Simon White, Geraint Levan, Kelly Schiabor Barrett, Elizabeth Cirulli, Alexandre Bolze, Charlotte Rivera-Garcia, Ryan Cho, Jason Nguyen, Sherry Wang, Jimmy Ramirez, Celena Andrade, Alice Summerfield, Tyler Cassens, Efren Sandoval, Francisco Tanudjaja, Magnus Isaksson, William Lee, David Becker, Marc Laurent, James Lu + SEARCH |
| EPI_ISL_1812757, EPI_ISL_1812758 | EXCITE Lab | Andersen lab at Scripps Research | David Pride, Sharon Reed, Chip Schooley, Angela Scioscia, Natasha Martin Cheryl Anderson, Sawyer Farmer, Abigail Schnapper, Helena Tubb, Tommy Valles + SEARCH |
| EPI_ISL_1812759, EPI_ISL_1812760, EPI_ISL_1812761, EPI_ISL_1812762 | EXCITE Lab | Andersen lab at Scripps Research | Nicole L Washington, Simon White, Geraint Levan, Kelly Schiabor Barrett, Elizabeth Cirulli, Alexandre Bolze, Charlotte Rivera-Garcia, Ryan Cho, Jason Nguyen, Sherry Wang, Jimmy Ramirez, Celena Andrade, Alice Summerfield, Tyler Cassens, Efren Sandoval, Francisco Tanudjaja, Magnus Isaksson, William Lee, David Becker, Marc Laurent, James Lu + SEARCH |
| EPI_ISL_1812763 | EXCITE Lab | Andersen lab at Scripps Research | Chip Schooley, Natasha Martin, Cheryl Anderson, Angela Scioscia, Smruthi Karthikeyan, Greg Humphrey, Sawyer Farmer, Abigail Schnapper, Helena Tubb, Tommy Valles + SEARCH |
| EPI_ISL_1812764, EPI_ISL_1812765 | EXCITE Lab | Andersen lab at Scripps Research | Nicole L Washington, Simon White, Geraint Levan, Kelly Schiabor Barrett, Elizabeth Cirulli, Alexandre Bolze, Charlotte Rivera-Garcia, Ryan Cho, Jason Nguyen, Sherry Wang, Jimmy Ramirez, Celena Andrade, Alice Summerfield, Tyler Cassens, Efren Sandoval, Francisco Tanudjaja, Magnus Isaksson, William Lee, David Becker, Marc Laurent, James Lu + SEARCH |
| EPI_ISL_1812766 | EXCITE Lab | Andersen lab at Scripps Research | David Pride, Sharon Reed, Chip Schooley, Angela Scioscia, Natasha Martin Cheryl Anderson, Sawyer Farmer, Abigail Schnapper, Helena Tubb, Tommy Valles + SEARCH |
| EPI_ISL_1812767, EPI_ISL_1812768 | EXCITE Lab | Andersen lab at Scripps Research | Nicole L Washington, Simon White, Geraint Levan, Kelly Schiabor Barrett, Elizabeth Cirulli, Alexandre Bolze, Charlotte Rivera-Garcia, Ryan Cho, Jason Nguyen, Sherry Wang, Jimmy Ramirez, Celena Andrade, Alice Summerfield, Tyler Cassens, Efren Sandoval, Francisco Tanudjaja, Magnus Isaksson, William Lee, David Becker, Marc Laurent, James Lu + SEARCH |
| EPI_ISL_1812769 | EXCITE Lab | Andersen lab at Scripps Research | David Pride, Sharon Reed, Chip Schooley, Angela Scioscia, Natasha Martin Cheryl Anderson, Sawyer Farmer, Abigail Schnapper, Helena Tubb, Tommy Valles + SEARCH |
| EPI_ISL_1812770, EPI_ISL_1812771, EPI_ISL_1812772, EPI_ISL_1812773, EPI_ISL_1812774, EPI_ISL_1812775, EPI_ISL_1812776 | EXCITE Lab | Andersen lab at Scripps Research | Nicole L Washington, Simon White, Geraint Levan, Kelly Schiabor Barrett, Elizabeth Cirulli, Alexandre Bolze, Charlotte Rivera-Garcia, Ryan Cho, Jason Nguyen, Sherry Wang, Jimmy Ramirez, Celena Andrade, Alice Summerfield, Tyler Cassens, Efren Sandoval, Francisco Tanudjaja, Magnus Isaksson, William Lee, David Becker, Marc Laurent, James Lu + SEARCH |
| EPI_ISL_1812777 | EXCITE Lab | Andersen lab at Scripps Research | Sydney Morgan + Marni Jacobs + Celestine Magallanes + Holly Valentine + Louise C. Laurent + SEARCH |
| EPI_ISL_1812778, EPI_ISL_1812779 | EXCITE Lab | Andersen lab at Scripps Research | Nicole L Washington, Simon White, Geraint Levan, Kelly Schiabor Barrett, Elizabeth Cirulli, Alexandre Bolze, Charlotte Rivera-Garcia, Ryan Cho, Jason Nguyen, Sherry Wang, Jimmy Ramirez, Celena Andrade, Alice Summerfield, Tyler Cassens, Efren Sandoval, Francisco Tanudjaja, Magnus Isaksson, William Lee, David Becker, Marc Laurent, James Lu + SEARCH |
| EPI_ISL_1812780 | EXCITE Lab | Andersen lab at Scripps Research | David Pride, Sharon Reed, Chip Schooley, Angela Scioscia, Natasha Martin Cheryl Anderson, Sawyer Farmer, Abigail Schnapper, Helena Tubb, Tommy Valles + SEARCH |
| EPI_ISL_1812781, EPI_ISL_1812782 | EXCITE Lab | Andersen lab at Scripps Research | Nicole L Washington, Simon White, Geraint Levan, Kelly Schiabor Barrett, Elizabeth Cirulli, Alexandre Bolze, Charlotte Rivera-Garcia, Ryan Cho, Jason Nguyen, Sherry Wang, Jimmy Ramirez, Celena Andrade, Alice Summerfield, Tyler Cassens, Efren Sandoval, Francisco Tanudjaja, Magnus Isaksson, William Lee, David Becker, Marc Laurent, James Lu + SEARCH |
| EPI_ISL_1812783, EPI_ISL_1812784 | EXCITE Lab | Andersen lab at Scripps Research | David Pride, Sharon Reed, Chip Schooley, Angela Scioscia, Natasha Martin Cheryl Anderson, Sawyer Farmer, Abigail Schnapper, Helena Tubb, Tommy Valles + SEARCH |
| EPI_ISL_1812785, EPI_ISL_1812786, EPI_ISL_1812787, EPI_ISL_1812788, EPI_ISL_1812789, EPI_ISL_1812790, EPI_ISL_1812791, EPI_ISL_1812792, EPI_ISL_1812793, EPI_ISL_1812794, EPI_ISL_1812795, EPI_ISL_1812796, EPI_ISL_1812797 | | | |
| see above | EXCITE Lab | Andersen lab at Scripps Research | Nicole L Washington, Simon White, Geraint Levan, Kelly Schiabor Barrett, Elizabeth Cirulli, Alexandre Bolze, Charlotte Rivera-Garcia, Ryan Cho, Jason Nguyen, Sherry Wang, Jimmy Ramirez, Celena Andrade, Alice Summerfield, Tyler Cassens, Efren Sandoval, Francisco Tanudjaja, Magnus Isaksson, William Lee, David Becker, Marc Laurent, James Lu + SEARCH |
| EPI_ISL_1812798 | EXCITE Lab | Andersen lab at Scripps Research | David Pride, Sharon Reed, Chip Schooley, Angela Scioscia, Natasha Martin Cheryl Anderson, Sawyer Farmer, Abigail Schnapper, Helena Tubb, Tommy Valles + SEARCH |
| EPI_ISL_1812799 | EXCITE Lab | Andersen lab at Scripps Research | Sydney Morgan + Marni Jacobs + Celestine Magallanes + Holly Valentine + Louise C. Laurent + SEARCH |
| EPI_ISL_1812800, EPI_ISL_1812801, EPI_ISL_1812802, EPI_ISL_1812803, EPI_ISL_1812804, EPI_ISL_1812805, EPI_ISL_1812806, EPI_ISL_1812807, EPI_ISL_1812808, EPI_ISL_1812809 | EXCITE Lab | Andersen lab at Scripps Research | Nicole L Washington, Simon White, Geraint Levan, Kelly Schiabor Barrett, Elizabeth Cirulli, Alexandre Bolze, Charlotte Rivera-Garcia, Ryan Cho, Jason Nguyen, Sherry Wang, Jimmy Ramirez, Celena Andrade, Alice Summerfield, Tyler Cassens, Efren Sandoval, Francisco Tanudjaja, Magnus Isaksson, William Lee, David Becker, Marc Laurent, James Lu + SEARCH |
| EPI_ISL_1812810 | EXCITE Lab | Andersen lab at Scripps Research | David Pride, Sharon Reed, Chip Schooley, Angela Scioscia, Natasha Martin Cheryl Anderson, Sawyer Farmer, Abigail Schnapper, Helena Tubb, Tommy Valles + SEARCH |
| EPI_ISL_1812811, EPI_ISL_1812812 | EXCITE Lab | Andersen lab at Scripps Research | Nicole L Washington, Simon White, Geraint Levan, Kelly Schiabor Barrett, Elizabeth Cirulli, Alexandre Bolze, Charlotte Rivera-Garcia, Ryan Cho, Jason Nguyen, Sherry Wang, Jimmy Ramirez, Celena Andrade, Alice Summerfield, Tyler Cassens, Efren Sandoval, Francisco Tanudjaja, Magnus Isaksson, William Lee, David Becker, Marc Laurent, James Lu + SEARCH |
| EPI_ISL_1812813 | EXCITE Lab | Andersen lab at Scripps Research | David Pride, Sharon Reed, Chip Schooley, Angela Scioscia, Natasha Martin Cheryl Anderson, Sawyer Farmer, Abigail Schnapper, Helena Tubb, Tommy Valles + SEARCH |
| EPI_ISL_1812814, EPI_ISL_1812815, EPI_ISL_1812816, EPI_ISL_1812817, EPI_ISL_1812818 | EXCITE Lab | Andersen lab at Scripps Research | Nicole L Washington, Simon White, Geraint Levan, Kelly Schiabor Barrett, Elizabeth Cirulli, Alexandre Bolze, Charlotte Rivera-Garcia, Ryan Cho, Jason Nguyen, Sherry Wang, Jimmy Ramirez, Celena Andrade, Alice Summerfield, Tyler Cassens, Efren Sandoval, Francisco Tanudjaja, Magnus Isaksson, William Lee, David Becker, Marc Laurent, James Lu + SEARCH |
| EPI_ISL_1812819 | EXCITE Lab | Andersen lab at Scripps Research | David Pride, Sharon Reed, Chip Schooley, Angela Scioscia, Natasha Martin Cheryl Anderson, Sawyer Farmer, Abigail Schnapper, Helena Tubb, Tommy Valles + SEARCH |
| EPI_ISL_1812820, EPI_ISL_1812821, EPI_ISL_1812822, EPI_ISL_1812823, EPI_ISL_1812824, EPI_ISL_1812825 | EXCITE Lab | Andersen lab at Scripps Research | Nicole L Washington, Simon White, Geraint Levan, Kelly Schiabor Barrett, Elizabeth Cirulli, Alexandre Bolze, Charlotte Rivera-Garcia, Ryan Cho, Jason Nguyen, Sherry Wang, Jimmy Ramirez, Celena Andrade, Alice Summerfield, Tyler Cassens, Efren Sandoval, Francisco Tanudjaja, Magnus Isaksson, William Lee, David Becker, Marc Laurent, James Lu + SEARCH |
| EPI_ISL_1812826 | EXCITE Lab | Andersen lab at Scripps Research | David Pride, Sharon Reed, Chip Schooley, Angela Scioscia, Natasha Martin Cheryl Anderson, Sawyer Farmer, Abigail Schnapper, Helena Tubb, Tommy Valles + SEARCH |
| EPI_ISL_1812827 | EXCITE Lab | Andersen lab at Scripps Research | Chip Schooley, Natasha Martin, Cheryl Anderson, Angela Scioscia, Smruthi Karthikeyan, Greg Humphrey, Sawyer Farmer, Abigail Schnapper, Helena Tubb, Tommy Valles + SEARCH |
| EPI_ISL_1812828, EPI_ISL_1812829, EPI_ISL_1812830, EPI_ISL_1812831, EPI_ISL_1812832, EPI_ISL_1812833, EPI_ISL_1812834 | EXCITE Lab | Andersen lab at Scripps Research | Nicole L Washington, Simon White, Geraint Levan, Kelly Schiabor Barrett, Elizabeth Cirulli, Alexandre Bolze, Charlotte Rivera-Garcia, Ryan Cho, Jason Nguyen, Sherry Wang, Jimmy Ramirez, Celena Andrade, Alice Summerfield, Tyler Cassens, Efren Sandoval, Francisco Tanudjaja, Magnus Isaksson, William Lee, David Becker, Marc Laurent, James Lu + SEARCH |
| EPI_ISL_1812835 | EXCITE Lab | Andersen lab at Scripps Research | Chip Schooley, Natasha Martin, Cheryl Anderson, Angela Scioscia, Smruthi Karthikeyan, Greg Humphrey, Sawyer Farmer, Abigail Schnapper, Helena Tubb, Tommy Valles + SEARCH |
| EPI_ISL_1812836, EPI_ISL_1812837, EPI_ISL_1812838, EPI_ISL_1812839, EPI_ISL_1812840, EPI_ISL_1812841, EPI_ISL_1812842, EPI_ISL_1812843, EPI_ISL_1812844 | EXCITE Lab | Andersen lab at Scripps Research | Nicole L Washington, Simon White, Geraint Levan, Kelly Schiabor Barrett, Elizabeth Cirulli, Alexandre Bolze, Charlotte Rivera-Garcia, Ryan Cho, Jason Nguyen, Sherry Wang, Jimmy Ramirez, Celena Andrade, Alice Summerfield, Tyler Cassens, Efren Sandoval, Francisco Tanudjaja, Magnus Isaksson, William Lee, David Becker, Marc Laurent, James Lu + SEARCH |
| EPI_ISL_1812845 | EXCITE Lab | Andersen lab at Scripps Research | David Pride, Sharon Reed, Chip Schooley, Angela Scioscia, Natasha Martin Cheryl Anderson, Sawyer Farmer, Abigail Schnapper, Helena Tubb, Tommy Valles + SEARCH |
| EPI_ISL_1812846, EPI_ISL_1812847, EPI_ISL_1812848, EPI_ISL_1812849 | EXCITE Lab | Andersen lab at Scripps Research | Nicole L Washington, Simon White, Geraint Levan, Kelly Schiabor Barrett, Elizabeth Cirulli, Alexandre Bolze, Charlotte Rivera-Garcia, Ryan Cho, Jason Nguyen, Sherry Wang, Jimmy Ramirez, Celena Andrade, Alice Summerfield, Tyler Cassens, Efren Sandoval, Francisco Tanudjaja, Magnus Isaksson, William Lee, David Becker, Marc Laurent, James Lu + SEARCH |
| EPI_ISL_1812850 | EXCITE Lab | Andersen lab at Scripps Research | David Pride, Sharon Reed, Chip Schooley, Angela Scioscia, Natasha Martin Cheryl Anderson, Sawyer Farmer, Abigail Schnapper, Helena Tubb, Tommy Valles + SEARCH |
| EPI_ISL_1812851 | EXCITE Lab | Andersen lab at Scripps Research | Fred Wu, Christine Mitchell + SEARCH |

| | | | |
|---|---|---|---|
| EPI_ISL_1812852, EPI_ISL_1812853 | EXCITE Lab | Andersen lab at Scripps Research | Nicole L Washington, Simon White, Geraint Levan, Kelly Schiabor Barrett, Elizabeth Cirulli, Alexandre Bolze, Charlotte Rivera-Garcia, Ryan Cho, Jason Nguyen, Sherry Wang, Jimmy Ramirez, Celena Andrade, Alice Summerfield, Tyler Cassens, Efren Sandoval, Francisco Tanudjaja, Magnus Isaksson, William Lee, David Becker, Marc Laurent, James Lu + SEARCH |
| EPI_ISL_1812854 | EXCITE Lab | Andersen lab at Scripps Research | David Pride, Sharon Reed, Chip Schooley, Angela Scioscia, Natasha Martin Cheryl Anderson, Sawyer Farmer, Abigail Schnapper, Helena Tubb, Tommy Valles + SEARCH |
| EPI_ISL_1812855, EPI_ISL_1812856, EPI_ISL_1812857, EPI_ISL_1812858 | EXCITE Lab | Andersen lab at Scripps Research | Nicole L Washington, Simon White, Geraint Levan, Kelly Schiabor Barrett, Elizabeth Cirulli, Alexandre Bolze, Charlotte Rivera-Garcia, Ryan Cho, Jason Nguyen, Sherry Wang, Jimmy Ramirez, Celena Andrade, Alice Summerfield, Tyler Cassens, Efren Sandoval, Francisco Tanudjaja, Magnus Isaksson, William Lee, David Becker, Marc Laurent, James Lu + SEARCH |
| EPI_ISL_1812859 | EXCITE Lab | Andersen lab at Scripps Research | David Pride, Sharon Reed, Chip Schooley, Angela Scioscia, Natasha Martin Cheryl Anderson, Sawyer Farmer, Abigail Schnapper, Helena Tubb, Tommy Valles + SEARCH |
| EPI_ISL_1812860, EPI_ISL_1812861, EPI_ISL_1812862, EPI_ISL_1812863, EPI_ISL_1812864, EPI_ISL_1812865, EPI_ISL_1812866, EPI_ISL_1812867 | EXCITE Lab | Andersen lab at Scripps Research | Nicole L Washington, Simon White, Geraint Levan, Kelly Schiabor Barrett, Elizabeth Cirulli, Alexandre Bolze, Charlotte Rivera-Garcia, Ryan Cho, Jason Nguyen, Sherry Wang, Jimmy Ramirez, Celena Andrade, Alice Summerfield, Tyler Cassens, Efren Sandoval, Francisco Tanudjaja, Magnus Isaksson, William Lee, David Becker, Marc Laurent, James Lu + SEARCH |
| EPI_ISL_1812868 | EXCITE Lab | Andersen lab at Scripps Research | David Pride, Sharon Reed, Chip Schooley, Angela Scioscia, Natasha Martin Cheryl Anderson, Sawyer Farmer, Abigail Schnapper, Helena Tubb, Tommy Valles + SEARCH |
| EPI_ISL_1812869 | EXCITE Lab | Andersen lab at Scripps Research | Nicole L Washington, Simon White, Geraint Levan, Kelly Schiabor Barrett, Elizabeth Cirulli, Alexandre Bolze, Charlotte Rivera-Garcia, Ryan Cho, Jason Nguyen, Sherry Wang, Jimmy Ramirez, Celena Andrade, Alice Summerfield, Tyler Cassens, Efren Sandoval, Francisco Tanudjaja, Magnus Isaksson, William Lee, David Becker, Marc Laurent, James Lu + SEARCH |
| EPI_ISL_1812870 | EXCITE Lab | Andersen lab at Scripps Research | David Pride, Sharon Reed, Chip Schooley, Angela Scioscia, Natasha Martin Cheryl Anderson, Sawyer Farmer, Abigail Schnapper, Helena Tubb, Tommy Valles + SEARCH |
| EPI_ISL_1812871, EPI_ISL_1812872, EPI_ISL_1812873, EPI_ISL_1812874, EPI_ISL_1812875, EPI_ISL_1812876, EPI_ISL_1812877, EPI_ISL_1812878, EPI_ISL_1812879, EPI_ISL_1812880, EPI_ISL_1812881, EPI_ISL_1812882, EPI_ISL_1812883, EPI_ISL_1812884, EPI_ISL_1812885, EPI_ISL_1812886, EPI_ISL_1812887, EPI_ISL_1812888, EPI_ISL_1812889, EPI_ISL_1812890, EPI_ISL_1812891, EPI_ISL_1812892, EPI_ISL_1812893, EPI_ISL_1812894, EPI_ISL_1812895, EPI_ISL_1812896, EPI_ISL_1812897, EPI_ISL_1812898, EPI_ISL_1812899 | | | |
| see above | EXCITE Lab | Andersen lab at Scripps Research | Nicole L Washington, Simon White, Geraint Levan, Kelly Schiabor Barrett, Elizabeth Cirulli, Alexandre Bolze, Charlotte Rivera-Garcia, Ryan Cho, Jason Nguyen, Sherry Wang, Jimmy Ramirez, Celena Andrade, Alice Summerfield, Tyler Cassens, Efren Sandoval, Francisco Tanudjaja, Magnus Isaksson, William Lee, David Becker, Marc Laurent, James Lu + SEARCH |
| EPI_ISL_1812900 | EXCITE Lab | Andersen lab at Scripps Research | David Pride, Sharon Reed, Chip Schooley, Angela Scioscia, Natasha Martin Cheryl Anderson, Sawyer Farmer, Abigail Schnapper, Helena Tubb, Tommy Valles + SEARCH |
| EPI_ISL_1812901, EPI_ISL_1812902 | EXCITE Lab | Andersen lab at Scripps Research | Nicole L Washington, Simon White, Geraint Levan, Kelly Schiabor Barrett, Elizabeth Cirulli, Alexandre Bolze, Charlotte Rivera-Garcia, Ryan Cho, Jason Nguyen, Sherry Wang, Jimmy Ramirez, Celena Andrade, Alice Summerfield, Tyler Cassens, Efren Sandoval, Francisco Tanudjaja, Magnus Isaksson, William Lee, David Becker, Marc Laurent, James Lu + SEARCH |
| EPI_ISL_1812903 | EXCITE Lab | Andersen lab at Scripps Research | David Pride, Sharon Reed, Chip Schooley, Angela Scioscia, Natasha Martin Cheryl Anderson, Sawyer Farmer, Abigail Schnapper, Helena Tubb, Tommy Valles + SEARCH |
| EPI_ISL_1812904 | EXCITE Lab | Andersen lab at Scripps Research | Nicole L Washington, Simon White, Geraint Levan, Kelly Schiabor Barrett, Elizabeth Cirulli, Alexandre Bolze, Charlotte Rivera-Garcia, Ryan Cho, Jason Nguyen, Sherry Wang, Jimmy Ramirez, Celena Andrade, Alice Summerfield, Tyler Cassens, Efren Sandoval, Francisco Tanudjaja, Magnus Isaksson, William Lee, David Becker, Marc Laurent, James Lu + SEARCH |
| EPI_ISL_1812905 | EXCITE Lab | Andersen lab at Scripps Research | Sydney Morgan + Marni Jacobs + Celestine Magallanes + Holly Valentine + Louise C. Laurent + SEARCH |
| EPI_ISL_1812906, EPI_ISL_1812907, EPI_ISL_1812908, EPI_ISL_1812909 | EXCITE Lab | Andersen lab at Scripps Research | Nicole L Washington, Simon White, Geraint Levan, Kelly Schiabor Barrett, Elizabeth Cirulli, Alexandre Bolze, Charlotte Rivera-Garcia, Ryan Cho, Jason Nguyen, Sherry Wang, Jimmy Ramirez, Celena Andrade, Alice Summerfield, Tyler Cassens, Efren Sandoval, Francisco Tanudjaja, Magnus Isaksson, William Lee, David Becker, Marc Laurent, James Lu + SEARCH |
| EPI_ISL_1812910 | EXCITE Lab | Andersen lab at Scripps Research | David Pride, Sharon Reed, Chip Schooley, Angela Scioscia, Natasha Martin Cheryl Anderson, Sawyer Farmer, Abigail Schnapper, Helena Tubb, Tommy Valles + SEARCH |
| EPI_ISL_1812911, EPI_ISL_1812912 | EXCITE Lab | Andersen lab at Scripps Research | Nicole L Washington, Simon White, Geraint Levan, Kelly Schiabor Barrett, Elizabeth Cirulli, Alexandre Bolze, Charlotte Rivera-Garcia, Ryan Cho, Jason Nguyen, Sherry Wang, Jimmy Ramirez, Celena Andrade, Alice Summerfield, Tyler Cassens, Efren Sandoval, Francisco Tanudjaja, Magnus Isaksson, William Lee, David Becker, Marc Laurent, James Lu + SEARCH |
| EPI_ISL_1812913 | EXCITE Lab | Andersen lab at Scripps Research | David Pride, Sharon Reed, Chip Schooley, Angela Scioscia, Natasha Martin Cheryl Anderson, Sawyer Farmer, Abigail Schnapper, Helena Tubb, Tommy Valles + SEARCH |
| EPI_ISL_1812914, EPI_ISL_1812915, EPI_ISL_1812916 | EXCITE Lab | Andersen lab at Scripps Research | Nicole L Washington, Simon White, Geraint Levan, Kelly Schiabor Barrett, Elizabeth Cirulli, Alexandre Bolze, Charlotte Rivera-Garcia, Ryan Cho, Jason Nguyen, Sherry Wang, Jimmy Ramirez, Celena Andrade, Alice Summerfield, Tyler Cassens, Efren Sandoval, Francisco Tanudjaja, Magnus Isaksson, William Lee, David Becker, Marc Laurent, James Lu + SEARCH |
| EPI_ISL_1812917 | EXCITE Lab | Andersen lab at Scripps Research | Sydney Morgan + Marni Jacobs + Celestine Magallanes + Holly Valentine + Louise C. Laurent + SEARCH |
| EPI_ISL_1812918, EPI_ISL_1812919, EPI_ISL_1812920, EPI_ISL_1812921, EPI_ISL_1812922, EPI_ISL_1812923 | EXCITE Lab | Andersen lab at Scripps Research | Nicole L Washington, Simon White, Geraint Levan, Kelly Schiabor Barrett, Elizabeth Cirulli, Alexandre Bolze, Charlotte Rivera-Garcia, Ryan Cho, Jason Nguyen, Sherry Wang, Jimmy Ramirez, Celena Andrade, Alice Summerfield, Tyler Cassens, Efren Sandoval, Francisco Tanudjaja, Magnus Isaksson, William Lee, David Becker, Marc Laurent, James Lu + SEARCH |
| EPI_ISL_1812924 | EXCITE Lab | Andersen lab at Scripps Research | David Pride, Sharon Reed, Chip Schooley, Angela Scioscia, Natasha Martin Cheryl Anderson, Sawyer Farmer, Abigail Schnapper, Helena Tubb, Tommy Valles + SEARCH |
| EPI_ISL_1812925 | EXCITE Lab | Andersen lab at Scripps Research | Sydney Morgan + Marni Jacobs + Celestine Magallanes + Holly Valentine + Louise C. Laurent + SEARCH |
| EPI_ISL_1812926, EPI_ISL_1812927 | EXCITE Lab | Andersen lab at Scripps Research | Nicole L Washington, Simon White, Geraint Levan, Kelly Schiabor Barrett, Elizabeth Cirulli, Alexandre Bolze, Charlotte Rivera-Garcia, Ryan Cho, Jason Nguyen, Sherry Wang, Jimmy Ramirez, Celena Andrade, Alice Summerfield, Tyler Cassens, Efren Sandoval, Francisco Tanudjaja, Magnus Isaksson, William Lee, David Becker, Marc Laurent, James Lu + SEARCH |
| EPI_ISL_1812928 | EXCITE Lab | Andersen lab at Scripps Research | Chip Schooley, Natasha Martin, Cheryl Anderson, Angela Scioscia, Smruthi Karthikeyan, Greg Humphrey, Sawyer Farmer, Abigail Schnapper, Helena Tubb, Tommy Valles + SEARCH |
| EPI_ISL_1812929 | EXCITE Lab | Andersen lab at Scripps Research | David Pride, Sharon Reed, Chip Schooley, Angela Scioscia, Natasha Martin Cheryl Anderson, Sawyer Farmer, Abigail Schnapper, Helena Tubb, Tommy Valles + SEARCH |
| EPI_ISL_1812930, EPI_ISL_1812931, EPI_ISL_1812932, EPI_ISL_1812933, EPI_ISL_1812934, EPI_ISL_1812935, EPI_ISL_1812936, EPI_ISL_1812937, EPI_ISL_1812938, EPI_ISL_1812939, EPI_ISL_1812940, EPI_ISL_1812941, EPI_ISL_1812942 | | | |
| see above | EXCITE Lab | Andersen lab at Scripps Research | Nicole L Washington, Simon White, Geraint Levan, Kelly Schiabor Barrett, Elizabeth Cirulli, Alexandre Bolze, Charlotte Rivera-Garcia, Ryan Cho, Jason Nguyen, Sherry Wang, Jimmy Ramirez, Celena Andrade, Alice Summerfield, Tyler Cassens, Efren Sandoval, Francisco Tanudjaja, Magnus Isaksson, William Lee, David Becker, Marc Laurent, James Lu + SEARCH |
| EPI_ISL_1812943 | EXCITE Lab | Andersen lab at Scripps Research | Chip Schooley, Natasha Martin, Cheryl Anderson, Angela Scioscia, Smruthi Karthikeyan, Greg Humphrey, Sawyer Farmer, Abigail Schnapper, Helena Tubb, Tommy Valles + SEARCH |
| EPI_ISL_1812944 | EXCITE Lab | Andersen lab at Scripps Research | Nicole L Washington, Simon White, Geraint Levan, Kelly Schiabor Barrett, Elizabeth Cirulli, Alexandre Bolze, Charlotte Rivera-Garcia, Ryan Cho, Jason Nguyen, Sherry Wang, Jimmy Ramirez, Celena Andrade, Alice Summerfield, Tyler Cassens, Efren Sandoval, Francisco Tanudjaja, Magnus Isaksson, William Lee, David Becker, Marc Laurent, James Lu + SEARCH |
| EPI_ISL_1812945 | EXCITE Lab | Andersen lab at Scripps Research | Chip Schooley, Natasha Martin, Cheryl Anderson, Angela Scioscia, Smruthi Karthikeyan, Greg Humphrey, Sawyer Farmer, Abigail Schnapper, Helena Tubb, Tommy Valles + SEARCH |
| EPI_ISL_1812946, EPI_ISL_1812947, EPI_ISL_1812948, EPI_ISL_1812949, EPI_ISL_1812950, EPI_ISL_1812951 | EXCITE Lab | Andersen lab at Scripps Research | Nicole L Washington, Simon White, Geraint Levan, Kelly Schiabor Barrett, Elizabeth Cirulli, Alexandre Bolze, Charlotte Rivera-Garcia, Ryan Cho, Jason Nguyen, Sherry Wang, Jimmy Ramirez, Celena Andrade, Alice Summerfield, Tyler Cassens, Efren Sandoval, Francisco Tanudjaja, Magnus Isaksson, William Lee, David Becker, Marc Laurent, James Lu + SEARCH |
| EPI_ISL_1812952 | EXCITE Lab | Andersen lab at Scripps Research | David Pride, Sharon Reed, Chip Schooley, Angela Scioscia, Natasha Martin Cheryl Anderson, Sawyer Farmer, Abigail Schnapper, Helena Tubb, Tommy Valles + SEARCH |
| EPI_ISL_1812953, EPI_ISL_1812954 | EXCITE Lab | Andersen lab at Scripps Research | Nicole L Washington, Simon White, Geraint Levan, Kelly Schiabor Barrett, Elizabeth Cirulli, Alexandre Bolze, Charlotte Rivera-Garcia, Ryan Cho, Jason Nguyen, Sherry Wang, Jimmy Ramirez, Celena Andrade, Alice Summerfield, Tyler Cassens, Efren Sandoval, Francisco Tanudjaja, Magnus Isaksson, William Lee, David Becker, Marc Laurent, James Lu + SEARCH |
| EPI_ISL_1812955 | EXCITE Lab | Andersen lab at Scripps Research | David Pride, Sharon Reed, Chip Schooley, Angela Scioscia, Natasha Martin Cheryl Anderson, Sawyer Farmer, Abigail Schnapper, Helena Tubb, Tommy Valles + SEARCH |
| EPI_ISL_1812956 | EXCITE Lab | Andersen lab at Scripps Research | Nicole L Washington, Simon White, Geraint Levan, Kelly Schiabor Barrett, Elizabeth Cirulli, Alexandre Bolze, Charlotte Rivera-Garcia, Ryan Cho, Jason Nguyen, Sherry Wang, Jimmy Ramirez, Celena Andrade, Alice Summerfield, Tyler Cassens, Efren Sandoval, Francisco Tanudjaja, Magnus Isaksson, William Lee, David Becker, Marc Laurent, James Lu + SEARCH |
| EPI_ISL_1812957 | EXCITE Lab | Andersen lab at Scripps Research | David Pride, Sharon Reed, Chip Schooley, Angela Scioscia, Natasha Martin Cheryl Anderson, Sawyer Farmer, Abigail Schnapper, Helena Tubb, Tommy Valles + SEARCH |
| EPI_ISL_1812958 | EXCITE Lab | Andersen lab at Scripps Research | Nicole L Washington, Simon White, Geraint Levan, Kelly Schiabor Barrett, Elizabeth Cirulli, Alexandre Bolze, Charlotte Rivera-Garcia, Ryan Cho, Jason Nguyen, Sherry Wang, Jimmy Ramirez, Celena Andrade, Alice Summerfield, Tyler Cassens, Efren Sandoval, Francisco Tanudjaja, Magnus Isaksson, William Lee, David Becker, Marc Laurent, James Lu + SEARCH |
| EPI_ISL_1812959 | EXCITE Lab | Andersen lab at Scripps Research | David Pride, Sharon Reed, Chip Schooley, Angela Scioscia, Natasha Martin Cheryl Anderson, Sawyer Farmer, Abigail Schnapper, Helena Tubb, Tommy Valles + SEARCH |
| EPI_ISL_1812960, EPI_ISL_1812961, EPI_ISL_1812962 | EXCITE Lab | Andersen lab at Scripps Research | Nicole L Washington, Simon White, Geraint Levan, Kelly Schiabor Barrett, Elizabeth Cirulli, Alexandre Bolze, Charlotte Rivera-Garcia, Ryan Cho, Jason Nguyen, Sherry Wang, Jimmy Ramirez, Celena Andrade, Alice Summerfield, Tyler Cassens, Efren Sandoval, Francisco Tanudjaja, Magnus Isaksson, William Lee, David Becker, Marc Laurent, James Lu + SEARCH |
| EPI_ISL_1812963 | EXCITE Lab | Andersen lab at Scripps Research | Sydney Morgan + Marni Jacobs + Celestine Magallanes + Holly Valentine + Louise C. Laurent + SEARCH |
| EPI_ISL_1812964, EPI_ISL_1812965, EPI_ISL_1812966, EPI_ISL_1812967, EPI_ISL_1812968, EPI_ISL_1812969, EPI_ISL_1812970, EPI_ISL_1812971, EPI_ISL_1812972, EPI_ISL_1812973, EPI_ISL_1812974, EPI_ISL_1812975, EPI_ISL_1812976, EPI_ISL_1812977, EPI_ISL_1812978, EPI_ISL_1812979, EPI_ISL_1812980 | | | |
| see above | EXCITE Lab | Andersen lab at Scripps Research | Nicole L Washington, Simon White, Geraint Levan, Kelly Schiabor Barrett, Elizabeth Cirulli, Alexandre Bolze, Charlotte Rivera-Garcia, Ryan Cho, Jason Nguyen, Sherry Wang, Jimmy Ramirez, Celena Andrade, Alice Summerfield, Tyler Cassens, Efren Sandoval, Francisco Tanudjaja, Magnus Isaksson, William Lee, David Becker, Marc Laurent, James Lu + SEARCH |
| EPI_ISL_1812981 | EXCITE Lab | Andersen lab at Scripps Research | David Pride, Sharon Reed, Chip Schooley, Angela Scioscia, Natasha Martin Cheryl Anderson, Sawyer Farmer, Abigail Schnapper, Helena Tubb, Tommy Valles + SEARCH |
| EPI_ISL_1812982, EPI_ISL_1812983, EPI_ISL_1812984, EPI_ISL_1812985 | EXCITE Lab | Andersen lab at Scripps Research | Nicole L Washington, Simon White, Geraint Levan, Kelly Schiabor Barrett, Elizabeth Cirulli, Alexandre Bolze, Charlotte Rivera-Garcia, Ryan Cho, Jason Nguyen, Sherry Wang, Jimmy Ramirez, Celena Andrade, Alice Summerfield, Tyler Cassens, Efren Sandoval, Francisco Tanudjaja, Magnus Isaksson, William Lee, David Becker, Marc Laurent, James Lu + SEARCH |

| | | | |
|---|---|---|---|
| EPI_ISL_1812986, EPI_ISL_1812987 | EXCITE Lab | Andersen lab at Scripps Research | David Pride, Sharon Reed, Chip Schooley, Angela Scioscia, Natasha Martin Cheryl Anderson, Sawyer Farmer, Abigail Schnapper, Helena Tubb, Tommy Valles + SEARCH |
| EPI_ISL_1812988 | EXCITE Lab | Andersen lab at Scripps Research | Fred Wu, Christine Mitchell + SEARCH |
| EPI_ISL_1812989, EPI_ISL_1812990, EPI_ISL_1812991, EPI_ISL_1812992, EPI_ISL_1812993 | EXCITE Lab | Andersen lab at Scripps Research | Nicole L Washington, Simon White, Geraint Levan, Kelly Schiabor Barrett, Elizabeth Cirulli, Alexandre Bolze, Charlotte Rivera-Garcia, Ryan Cho, Jason Nguyen, Sherry Wang, Jimmy Ramirez, Celena Andrade, Alice Summerfield, Tyler Cassens, Efren Sandoval, Francisco Tanudjaja, Magnus Isaksson, William Lee, David Becker, Marc Laurent, James Lu + SEARCH |
| EPI_ISL_1812994 | EXCITE Lab | Andersen lab at Scripps Research | David Pride, Sharon Reed, Chip Schooley, Angela Scioscia, Natasha Martin Cheryl Anderson, Sawyer Farmer, Abigail Schnapper, Helena Tubb, Tommy Valles + SEARCH |
| EPI_ISL_1812995, EPI_ISL_1812996, EPI_ISL_1812997, EPI_ISL_1812998, EPI_ISL_1812999, EPI_ISL_1813000, EPI_ISL_1813001 | EXCITE Lab | Andersen lab at Scripps Research | Nicole L Washington, Simon White, Geraint Levan, Kelly Schiabor Barrett, Elizabeth Cirulli, Alexandre Bolze, Charlotte Rivera-Garcia, Ryan Cho, Jason Nguyen, Sherry Wang, Jimmy Ramirez, Celena Andrade, Alice Summerfield, Tyler Cassens, Efren Sandoval, Francisco Tanudjaja, Magnus Isaksson, William Lee, David Becker, Marc Laurent, James Lu + SEARCH |
| EPI_ISL_1813002 | EXCITE Lab | Andersen lab at Scripps Research | David Pride, Sharon Reed, Chip Schooley, Angela Scioscia, Natasha Martin Cheryl Anderson, Sawyer Farmer, Abigail Schnapper, Helena Tubb, Tommy Valles + SEARCH |
| EPI_ISL_1813003, EPI_ISL_1813004, EPI_ISL_1813005 | EXCITE Lab | Andersen lab at Scripps Research | Nicole L Washington, Simon White, Geraint Levan, Kelly Schiabor Barrett, Elizabeth Cirulli, Alexandre Bolze, Charlotte Rivera-Garcia, Ryan Cho, Jason Nguyen, Sherry Wang, Jimmy Ramirez, Celena Andrade, Alice Summerfield, Tyler Cassens, Efren Sandoval, Francisco Tanudjaja, Magnus Isaksson, William Lee, David Becker, Marc Laurent, James Lu + SEARCH |
| EPI_ISL_1813006 | EXCITE Lab | Andersen lab at Scripps Research | David Pride, Sharon Reed, Chip Schooley, Angela Scioscia, Natasha Martin Cheryl Anderson, Sawyer Farmer, Abigail Schnapper, Helena Tubb, Tommy Valles + SEARCH |
| EPI_ISL_1813007, EPI_ISL_1813008, EPI_ISL_1813009, EPI_ISL_1813010, EPI_ISL_1813011 | EXCITE Lab | Andersen lab at Scripps Research | Nicole L Washington, Simon White, Geraint Levan, Kelly Schiabor Barrett, Elizabeth Cirulli, Alexandre Bolze, Charlotte Rivera-Garcia, Ryan Cho, Jason Nguyen, Sherry Wang, Jimmy Ramirez, Celena Andrade, Alice Summerfield, Tyler Cassens, Efren Sandoval, Francisco Tanudjaja, Magnus Isaksson, William Lee, David Becker, Marc Laurent, James Lu + SEARCH |
| EPI_ISL_1813012 | EXCITE Lab | Andersen lab at Scripps Research | David Pride, Sharon Reed, Chip Schooley, Angela Scioscia, Natasha Martin Cheryl Anderson, Sawyer Farmer, Abigail Schnapper, Helena Tubb, Tommy Valles + SEARCH |
| EPI_ISL_1813013, EPI_ISL_1813014, EPI_ISL_1813015, EPI_ISL_1813016, EPI_ISL_1813017 | EXCITE Lab | Andersen lab at Scripps Research | Nicole L Washington, Simon White, Geraint Levan, Kelly Schiabor Barrett, Elizabeth Cirulli, Alexandre Bolze, Charlotte Rivera-Garcia, Ryan Cho, Jason Nguyen, Sherry Wang, Jimmy Ramirez, Celena Andrade, Alice Summerfield, Tyler Cassens, Efren Sandoval, Francisco Tanudjaja, Magnus Isaksson, William Lee, David Becker, Marc Laurent, James Lu + SEARCH |
| EPI_ISL_1813018 | EXCITE Lab | Andersen lab at Scripps Research | Chip Schooley, Natasha Martin, Cheryl Anderson, Angela Scioscia, Smruthi Karthikeyan, Greg Humphrey, Sawyer Farmer, Abigail Schnapper, Helena Tubb, Tommy Valles + SEARCH |
| EPI_ISL_1813019, EPI_ISL_1813020, EPI_ISL_1813021, EPI_ISL_1813022, EPI_ISL_1813023, EPI_ISL_1813024, EPI_ISL_1813025, EPI_ISL_1813026, EPI_ISL_1813027 | EXCITE Lab | Andersen lab at Scripps Research | Nicole L Washington, Simon White, Geraint Levan, Kelly Schiabor Barrett, Elizabeth Cirulli, Alexandre Bolze, Charlotte Rivera-Garcia, Ryan Cho, Jason Nguyen, Sherry Wang, Jimmy Ramirez, Celena Andrade, Alice Summerfield, Tyler Cassens, Efren Sandoval, Francisco Tanudjaja, Magnus Isaksson, William Lee, David Becker, Marc Laurent, James Lu + SEARCH |
| EPI_ISL_1813028 | EXCITE Lab | Andersen lab at Scripps Research | David Pride, Sharon Reed, Chip Schooley, Angela Scioscia, Natasha Martin Cheryl Anderson, Sawyer Farmer, Abigail Schnapper, Helena Tubb, Tommy Valles + SEARCH |
| EPI_ISL_1813029, EPI_ISL_1813030, EPI_ISL_1813031, EPI_ISL_1813032, EPI_ISL_1813033, EPI_ISL_1813034, EPI_ISL_1813035, EPI_ISL_1813036, EPI_ISL_1813037, EPI_ISL_1813038, EPI_ISL_1813039 | | | |
| see above | EXCITE Lab | Andersen lab at Scripps Research | Nicole L Washington, Simon White, Geraint Levan, Kelly Schiabor Barrett, Elizabeth Cirulli, Alexandre Bolze, Charlotte Rivera-Garcia, Ryan Cho, Jason Nguyen, Sherry Wang, Jimmy Ramirez, Celena Andrade, Alice Summerfield, Tyler Cassens, Efren Sandoval, Francisco Tanudjaja, Magnus Isaksson, William Lee, David Becker, Marc Laurent, James Lu + SEARCH |
| EPI_ISL_1813040 | EXCITE Lab | Andersen lab at Scripps Research | David Pride, Sharon Reed, Chip Schooley, Angela Scioscia, Natasha Martin Cheryl Anderson, Sawyer Farmer, Abigail Schnapper, Helena Tubb, Tommy Valles + SEARCH |
| EPI_ISL_1813041 | EXCITE Lab | Andersen lab at Scripps Research | Nicole L Washington, Simon White, Geraint Levan, Kelly Schiabor Barrett, Elizabeth Cirulli, Alexandre Bolze, Charlotte Rivera-Garcia, Ryan Cho, Jason Nguyen, Sherry Wang, Jimmy Ramirez, Celena Andrade, Alice Summerfield, Tyler Cassens, Efren Sandoval, Francisco Tanudjaja, Magnus Isaksson, William Lee, David Becker, Marc Laurent, James Lu + SEARCH |
| EPI_ISL_1813042 | EXCITE Lab | Andersen lab at Scripps Research | David Pride, Sharon Reed, Chip Schooley, Angela Scioscia, Natasha Martin Cheryl Anderson, Sawyer Farmer, Abigail Schnapper, Helena Tubb, Tommy Valles + SEARCH |
| EPI_ISL_1813043, EPI_ISL_1813044, EPI_ISL_1813045 | EXCITE Lab | Andersen lab at Scripps Research | Nicole L Washington, Simon White, Geraint Levan, Kelly Schiabor Barrett, Elizabeth Cirulli, Alexandre Bolze, Charlotte Rivera-Garcia, Ryan Cho, Jason Nguyen, Sherry Wang, Jimmy Ramirez, Celena Andrade, Alice Summerfield, Tyler Cassens, Efren Sandoval, Francisco Tanudjaja, Magnus Isaksson, William Lee, David Becker, Marc Laurent, James Lu + SEARCH |
| EPI_ISL_1813046 | EXCITE Lab | Andersen lab at Scripps Research | David Pride, Sharon Reed, Chip Schooley, Angela Scioscia, Natasha Martin Cheryl Anderson, Sawyer Farmer, Abigail Schnapper, Helena Tubb, Tommy Valles + SEARCH |
| EPI_ISL_1813047, EPI_ISL_1813048, EPI_ISL_1813049, EPI_ISL_1813050, EPI_ISL_1813051, EPI_ISL_1813052 | EXCITE Lab | Andersen lab at Scripps Research | Nicole L Washington, Simon White, Geraint Levan, Kelly Schiabor Barrett, Elizabeth Cirulli, Alexandre Bolze, Charlotte Rivera-Garcia, Ryan Cho, Jason Nguyen, Sherry Wang, Jimmy Ramirez, Celena Andrade, Alice Summerfield, Tyler Cassens, Efren Sandoval, Francisco Tanudjaja, Magnus Isaksson, William Lee, David Becker, Marc Laurent, James Lu + SEARCH |
| EPI_ISL_1813053 | EXCITE Lab | Andersen lab at Scripps Research | David Pride, Sharon Reed, Chip Schooley, Angela Scioscia, Natasha Martin Cheryl Anderson, Sawyer Farmer, Abigail Schnapper, Helena Tubb, Tommy Valles + SEARCH |
| EPI_ISL_1813054 | EXCITE Lab | Andersen lab at Scripps Research | Sydney Morgan + Marni Jacobs + Celestine Magallanes + Holly Valentine + Louise C. Laurent + SEARCH |
| EPI_ISL_1813055 | EXCITE Lab | Andersen lab at Scripps Research | Nicole L Washington, Simon White, Geraint Levan, Kelly Schiabor Barrett, Elizabeth Cirulli, Alexandre Bolze, Charlotte Rivera-Garcia, Ryan Cho, Jason Nguyen, Sherry Wang, Jimmy Ramirez, Celena Andrade, Alice Summerfield, Tyler Cassens, Efren Sandoval, Francisco Tanudjaja, Magnus Isaksson, William Lee, David Becker, Marc Laurent, James Lu + SEARCH |
| EPI_ISL_1813056 | EXCITE Lab | Andersen lab at Scripps Research | David Pride, Sharon Reed, Chip Schooley, Angela Scioscia, Natasha Martin Cheryl Anderson, Sawyer Farmer, Abigail Schnapper, Helena Tubb, Tommy Valles + SEARCH |
| EPI_ISL_1813057, EPI_ISL_1813058 | EXCITE Lab | Andersen lab at Scripps Research | Nicole L Washington, Simon White, Geraint Levan, Kelly Schiabor Barrett, Elizabeth Cirulli, Alexandre Bolze, Charlotte Rivera-Garcia, Ryan Cho, Jason Nguyen, Sherry Wang, Jimmy Ramirez, Celena Andrade, Alice Summerfield, Tyler Cassens, Efren Sandoval, Francisco Tanudjaja, Magnus Isaksson, William Lee, David Becker, Marc Laurent, James Lu + SEARCH |
| EPI_ISL_1813059 | EXCITE Lab | Andersen lab at Scripps Research | David Pride, Sharon Reed, Chip Schooley, Angela Scioscia, Natasha Martin Cheryl Anderson, Sawyer Farmer, Abigail Schnapper, Helena Tubb, Tommy Valles + SEARCH |
| EPI_ISL_1813060, EPI_ISL_1813061, EPI_ISL_1813062 | EXCITE Lab | Andersen lab at Scripps Research | Nicole L Washington, Simon White, Geraint Levan, Kelly Schiabor Barrett, Elizabeth Cirulli, Alexandre Bolze, Charlotte Rivera-Garcia, Ryan Cho, Jason Nguyen, Sherry Wang, Jimmy Ramirez, Celena Andrade, Alice Summerfield, Tyler Cassens, Efren Sandoval, Francisco Tanudjaja, Magnus Isaksson, William Lee, David Becker, Marc Laurent, James Lu + SEARCH |
| EPI_ISL_1813063 | EXCITE Lab | Andersen lab at Scripps Research | David Pride, Sharon Reed, Chip Schooley, Angela Scioscia, Natasha Martin Cheryl Anderson, Sawyer Farmer, Abigail Schnapper, Helena Tubb, Tommy Valles + SEARCH |
| EPI_ISL_1813064 | EXCITE Lab | Andersen lab at Scripps Research | Nicole L Washington, Simon White, Geraint Levan, Kelly Schiabor Barrett, Elizabeth Cirulli, Alexandre Bolze, Charlotte Rivera-Garcia, Ryan Cho, Jason Nguyen, Sherry Wang, Jimmy Ramirez, Celena Andrade, Alice Summerfield, Tyler Cassens, Efren Sandoval, Francisco Tanudjaja, Magnus Isaksson, William Lee, David Becker, Marc Laurent, James Lu + SEARCH |
| EPI_ISL_1813065, EPI_ISL_1813066 | EXCITE Lab | Andersen lab at Scripps Research | David Pride, Sharon Reed, Chip Schooley, Angela Scioscia, Natasha Martin Cheryl Anderson, Sawyer Farmer, Abigail Schnapper, Helena Tubb, Tommy Valles + SEARCH |
| EPI_ISL_1813067, EPI_ISL_1813068, EPI_ISL_1813069, EPI_ISL_1813070, EPI_ISL_1813071 | EXCITE Lab | Andersen lab at Scripps Research | Nicole L Washington, Simon White, Geraint Levan, Kelly Schiabor Barrett, Elizabeth Cirulli, Alexandre Bolze, Charlotte Rivera-Garcia, Ryan Cho, Jason Nguyen, Sherry Wang, Jimmy Ramirez, Celena Andrade, Alice Summerfield, Tyler Cassens, Efren Sandoval, Francisco Tanudjaja, Magnus Isaksson, William Lee, David Becker, Marc Laurent, James Lu + SEARCH |
| EPI_ISL_1813072 | EXCITE Lab | Andersen lab at Scripps Research | David Pride, Sharon Reed, Chip Schooley, Angela Scioscia, Natasha Martin Cheryl Anderson, Sawyer Farmer, Abigail Schnapper, Helena Tubb, Tommy Valles + SEARCH |
| EPI_ISL_1813073, EPI_ISL_1813074, EPI_ISL_1813075, EPI_ISL_1813076, EPI_ISL_1813077, EPI_ISL_1813078 | EXCITE Lab | Andersen lab at Scripps Research | Nicole L Washington, Simon White, Geraint Levan, Kelly Schiabor Barrett, Elizabeth Cirulli, Alexandre Bolze, Charlotte Rivera-Garcia, Ryan Cho, Jason Nguyen, Sherry Wang, Jimmy Ramirez, Celena Andrade, Alice Summerfield, Tyler Cassens, Efren Sandoval, Francisco Tanudjaja, Magnus Isaksson, William Lee, David Becker, Marc Laurent, James Lu + SEARCH |
| EPI_ISL_1813079 | EXCITE Lab | Andersen lab at Scripps Research | David Pride, Sharon Reed, Chip Schooley, Angela Scioscia, Natasha Martin Cheryl Anderson, Sawyer Farmer, Abigail Schnapper, Helena Tubb, Tommy Valles + SEARCH |
| EPI_ISL_1813080, EPI_ISL_1813081, EPI_ISL_1813082, EPI_ISL_1813083, EPI_ISL_1813084, EPI_ISL_1813085, EPI_ISL_1813086, EPI_ISL_1813087, EPI_ISL_1813088, EPI_ISL_1813089, EPI_ISL_1813090, EPI_ISL_1813091, EPI_ISL_1813092 | | | |
| see above | EXCITE Lab | Andersen lab at Scripps Research | Nicole L Washington, Simon White, Geraint Levan, Kelly Schiabor Barrett, Elizabeth Cirulli, Alexandre Bolze, Charlotte Rivera-Garcia, Ryan Cho, Jason Nguyen, Sherry Wang, Jimmy Ramirez, Celena Andrade, Alice Summerfield, Tyler Cassens, Efren Sandoval, Francisco Tanudjaja, Magnus Isaksson, William Lee, David Becker, Marc Laurent, James Lu + SEARCH |
| EPI_ISL_1813093 | EXCITE Lab | Andersen lab at Scripps Research | Sydney Morgan + Marni Jacobs + Celestine Magallanes + Holly Valentine + Louise C. Laurent + SEARCH |

| | | | |
|---|---|---|---|
| EPI_ISL_1813094, EPI_ISL_1813095, EPI_ISL_1813096 | EXCITE Lab | Andersen lab at Scripps Research | Nicole L Washington, Simon White, Geraint Levan, Kelly Schiabor Barrett, Elizabeth Cirulli, Alexandre Bolze, Charlotte Rivera-Garcia, Ryan Cho, Jason Nguyen, Sherry Wang, Jimmy Ramirez, Celena Andrade, Alice Summerfield, Tyler Cassens, Efren Sandoval, Francisco Tanudjaja, Magnus Isaksson, William Lee, David Becker, Marc Laurent, James Lu + SEARCH |
| EPI_ISL_1813097 | EXCITE Lab | Andersen lab at Scripps Research | Sydney Morgan + Marni Jacobs + Celestine Magallanes + Holly Valentine + Louise C. Laurent + SEARCH |
| EPI_ISL_1813098 | EXCITE Lab | Andersen lab at Scripps Research | Nicole L Washington, Simon White, Geraint Levan, Kelly Schiabor Barrett, Elizabeth Cirulli, Alexandre Bolze, Charlotte Rivera-Garcia, Ryan Cho, Jason Nguyen, Sherry Wang, Jimmy Ramirez, Celena Andrade, Alice Summerfield, Tyler Cassens, Efren Sandoval, Francisco Tanudjaja, Magnus Isaksson, William Lee, David Becker, Marc Laurent, James Lu + SEARCH |
| EPI_ISL_1813099 | EXCITE Lab | Andersen lab at Scripps Research | David Pride, Sharon Reed, Chip Schooley, Angela Scioscia, Natasha Martin Cheryl Anderson, Sawyer Farmer, Abigail Schnapper, Helena Tubb, Tommy Valles + SEARCH |
| EPI_ISL_1813100, EPI_ISL_1813101, EPI_ISL_1813102, EPI_ISL_1813103, EPI_ISL_1813104 | EXCITE Lab | Andersen lab at Scripps Research | Nicole L Washington, Simon White, Geraint Levan, Kelly Schiabor Barrett, Elizabeth Cirulli, Alexandre Bolze, Charlotte Rivera-Garcia, Ryan Cho, Jason Nguyen, Sherry Wang, Jimmy Ramirez, Celena Andrade, Alice Summerfield, Tyler Cassens, Efren Sandoval, Francisco Tanudjaja, Magnus Isaksson, William Lee, David Becker, Marc Laurent, James Lu + SEARCH |
| EPI_ISL_1813105 | EXCITE Lab | Andersen lab at Scripps Research | David Pride, Sharon Reed, Chip Schooley, Angela Scioscia, Natasha Martin Cheryl Anderson, Sawyer Farmer, Abigail Schnapper, Helena Tubb, Tommy Valles + SEARCH |
| EPI_ISL_1813106, EPI_ISL_1813107, EPI_ISL_1813108, EPI_ISL_1813109, EPI_ISL_1813110 | EXCITE Lab | Andersen lab at Scripps Research | Nicole L Washington, Simon White, Geraint Levan, Kelly Schiabor Barrett, Elizabeth Cirulli, Alexandre Bolze, Charlotte Rivera-Garcia, Ryan Cho, Jason Nguyen, Sherry Wang, Jimmy Ramirez, Celena Andrade, Alice Summerfield, Tyler Cassens, Efren Sandoval, Francisco Tanudjaja, Magnus Isaksson, William Lee, David Becker, Marc Laurent, James Lu + SEARCH |
| EPI_ISL_1813111 | EXCITE Lab | Andersen lab at Scripps Research | David Pride, Sharon Reed, Chip Schooley, Angela Scioscia, Natasha Martin Cheryl Anderson, Sawyer Farmer, Abigail Schnapper, Helena Tubb, Tommy Valles + SEARCH |
| EPI_ISL_1813112 | EXCITE Lab | Andersen lab at Scripps Research | Nicole L Washington, Simon White, Geraint Levan, Kelly Schiabor Barrett, Elizabeth Cirulli, Alexandre Bolze, Charlotte Rivera-Garcia, Ryan Cho, Jason Nguyen, Sherry Wang, Jimmy Ramirez, Celena Andrade, Alice Summerfield, Tyler Cassens, Efren Sandoval, Francisco Tanudjaja, Magnus Isaksson, William Lee, David Becker, Marc Laurent, James Lu + SEARCH |
| EPI_ISL_1813113 | EXCITE Lab | Andersen lab at Scripps Research | David Pride, Sharon Reed, Chip Schooley, Angela Scioscia, Natasha Martin Cheryl Anderson, Sawyer Farmer, Abigail Schnapper, Helena Tubb, Tommy Valles + SEARCH |
| EPI_ISL_1813114, EPI_ISL_1813115, EPI_ISL_1813116, EPI_ISL_1813117, EPI_ISL_1813118, EPI_ISL_1813119, EPI_ISL_1813120, EPI_ISL_1813121, EPI_ISL_1813122, EPI_ISL_1813123, EPI_ISL_1813124, EPI_ISL_1813125, EPI_ISL_1813126, EPI_ISL_1813127, EPI_ISL_1813128, EPI_ISL_1813129, EPI_ISL_1813130, EPI_ISL_1813131, EPI_ISL_1813132, EPI_ISL_1813133 | | | |
| see above | EXCITE Lab | Andersen lab at Scripps Research | Nicole L Washington, Simon White, Geraint Levan, Kelly Schiabor Barrett, Elizabeth Cirulli, Alexandre Bolze, Charlotte Rivera-Garcia, Ryan Cho, Jason Nguyen, Sherry Wang, Jimmy Ramirez, Celena Andrade, Alice Summerfield, Tyler Cassens, Efren Sandoval, Francisco Tanudjaja, Magnus Isaksson, William Lee, David Becker, Marc Laurent, James Lu + SEARCH |
| EPI_ISL_1813134 | EXCITE Lab | Andersen lab at Scripps Research | Sydney Morgan + Marni Jacobs + Celestine Magallanes + Holly Valentine + Louise C. Laurent + SEARCH |
| EPI_ISL_1813135, EPI_ISL_1813136, EPI_ISL_1813137, EPI_ISL_1813138 | EXCITE Lab | Andersen lab at Scripps Research | Nicole L Washington, Simon White, Geraint Levan, Kelly Schiabor Barrett, Elizabeth Cirulli, Alexandre Bolze, Charlotte Rivera-Garcia, Ryan Cho, Jason Nguyen, Sherry Wang, Jimmy Ramirez, Celena Andrade, Alice Summerfield, Tyler Cassens, Efren Sandoval, Francisco Tanudjaja, Magnus Isaksson, William Lee, David Becker, Marc Laurent, James Lu + SEARCH |
| EPI_ISL_1813139 | EXCITE Lab | Andersen lab at Scripps Research | David Pride, Sharon Reed, Chip Schooley, Angela Scioscia, Natasha Martin Cheryl Anderson, Sawyer Farmer, Abigail Schnapper, Helena Tubb, Tommy Valles + SEARCH |
| EPI_ISL_1813140, EPI_ISL_1813141, EPI_ISL_1813142, EPI_ISL_1813143, EPI_ISL_1813144 | EXCITE Lab | Andersen lab at Scripps Research | Nicole L Washington, Simon White, Geraint Levan, Kelly Schiabor Barrett, Elizabeth Cirulli, Alexandre Bolze, Charlotte Rivera-Garcia, Ryan Cho, Jason Nguyen, Sherry Wang, Jimmy Ramirez, Celena Andrade, Alice Summerfield, Tyler Cassens, Efren Sandoval, Francisco Tanudjaja, Magnus Isaksson, William Lee, David Becker, Marc Laurent, James Lu + SEARCH |
| EPI_ISL_1813145 | EXCITE Lab | Andersen lab at Scripps Research | David Pride, Sharon Reed, Chip Schooley, Angela Scioscia, Natasha Martin Cheryl Anderson, Sawyer Farmer, Abigail Schnapper, Helena Tubb, Tommy Valles + SEARCH |
| EPI_ISL_1813146, EPI_ISL_1813147, EPI_ISL_1813148, EPI_ISL_1813149, EPI_ISL_1813150, EPI_ISL_1813151, EPI_ISL_1813152, EPI_ISL_1813153, EPI_ISL_1813154, EPI_ISL_1813155, EPI_ISL_1813156, EPI_ISL_1813157, EPI_ISL_1813158, EPI_ISL_1813159, EPI_ISL_1813160, EPI_ISL_1813161, EPI_ISL_1813162, EPI_ISL_1813163, EPI_ISL_1813164, EPI_ISL_1813165, EPI_ISL_1813166, EPI_ISL_1813167, EPI_ISL_1813168 | | | |
| see above | EXCITE Lab | Andersen lab at Scripps Research | Nicole L Washington, Simon White, Geraint Levan, Kelly Schiabor Barrett, Elizabeth Cirulli, Alexandre Bolze, Charlotte Rivera-Garcia, Ryan Cho, Jason Nguyen, Sherry Wang, Jimmy Ramirez, Celena Andrade, Alice Summerfield, Tyler Cassens, Efren Sandoval, Francisco Tanudjaja, Magnus Isaksson, William Lee, David Becker, Marc Laurent, James Lu + SEARCH |
| EPI_ISL_1813169 | EXCITE Lab | Andersen lab at Scripps Research | David Pride, Sharon Reed, Chip Schooley, Angela Scioscia, Natasha Martin Cheryl Anderson, Sawyer Farmer, Abigail Schnapper, Helena Tubb, Tommy Valles + SEARCH |
| EPI_ISL_1813170, EPI_ISL_1813171, EPI_ISL_1813172, EPI_ISL_1813173, EPI_ISL_1813174, EPI_ISL_1813175 | EXCITE Lab | Andersen lab at Scripps Research | Nicole L Washington, Simon White, Geraint Levan, Kelly Schiabor Barrett, Elizabeth Cirulli, Alexandre Bolze, Charlotte Rivera-Garcia, Ryan Cho, Jason Nguyen, Sherry Wang, Jimmy Ramirez, Celena Andrade, Alice Summerfield, Tyler Cassens, Efren Sandoval, Francisco Tanudjaja, Magnus Isaksson, William Lee, David Becker, Marc Laurent, James Lu + SEARCH |
| EPI_ISL_1813176, EPI_ISL_1813177 | EXCITE Lab | Andersen lab at Scripps Research | David Pride, Sharon Reed, Chip Schooley, Angela Scioscia, Natasha Martin Cheryl Anderson, Sawyer Farmer, Abigail Schnapper, Helena Tubb, Tommy Valles + SEARCH |
| EPI_ISL_1813178, EPI_ISL_1813179, EPI_ISL_1813180, EPI_ISL_1813181, EPI_ISL_1813182, EPI_ISL_1813183, EPI_ISL_1813184, EPI_ISL_1813185 | EXCITE Lab | Andersen lab at Scripps Research | Nicole L Washington, Simon White, Geraint Levan, Kelly Schiabor Barrett, Elizabeth Cirulli, Alexandre Bolze, Charlotte Rivera-Garcia, Ryan Cho, Jason Nguyen, Sherry Wang, Jimmy Ramirez, Celena Andrade, Alice Summerfield, Tyler Cassens, Efren Sandoval, Francisco Tanudjaja, Magnus Isaksson, William Lee, David Becker, Marc Laurent, James Lu + SEARCH |
| EPI_ISL_1813186, EPI_ISL_1813187 | EXCITE Lab | Andersen lab at Scripps Research | Chip Schooley, Natasha Martin, Cheryl Anderson, Angela Scioscia, Smruthi Karthikeyan, Greg Humphrey, Sawyer Farmer, Abigail Schnapper, Helena Tubb, Tommy Valles + SEARCH |
| EPI_ISL_1813188, EPI_ISL_1813189, EPI_ISL_1813190, EPI_ISL_1813191, EPI_ISL_1813192, EPI_ISL_1813193, EPI_ISL_1813194, EPI_ISL_1813195, EPI_ISL_1813196, EPI_ISL_1813197, EPI_ISL_1813198, EPI_ISL_1813199, EPI_ISL_1813200, EPI_ISL_1813201, EPI_ISL_1813202, EPI_ISL_1813203, EPI_ISL_1813204, EPI_ISL_1813205, EPI_ISL_1813206, EPI_ISL_1813207, EPI_ISL_1813208, EPI_ISL_1813209, EPI_ISL_1813210, EPI_ISL_1813211, EPI_ISL_1813212, EPI_ISL_1813213, EPI_ISL_1813214 | | | |
| see above | EXCITE Lab | Andersen lab at Scripps Research | Nicole L Washington, Simon White, Geraint Levan, Kelly Schiabor Barrett, Elizabeth Cirulli, Alexandre Bolze, Charlotte Rivera-Garcia, Ryan Cho, Jason Nguyen, Sherry Wang, Jimmy Ramirez, Celena Andrade, Alice Summerfield, Tyler Cassens, Efren Sandoval, Francisco Tanudjaja, Magnus Isaksson, William Lee, David Becker, Marc Laurent, James Lu + SEARCH |
| EPI_ISL_1813215 | EXCITE Lab | Andersen lab at Scripps Research | Chip Schooley, Natasha Martin, Cheryl Anderson, Angela Scioscia, Smruthi Karthikeyan, Greg Humphrey, Sawyer Farmer, Abigail Schnapper, Helena Tubb, Tommy Valles + SEARCH |
| EPI_ISL_1813216, EPI_ISL_1813217, EPI_ISL_1813218, EPI_ISL_1813219, EPI_ISL_1813220, EPI_ISL_1813221, EPI_ISL_1813222, EPI_ISL_1813223, EPI_ISL_1813224, EPI_ISL_1813225, EPI_ISL_1813226, EPI_ISL_1813227, EPI_ISL_1813228, EPI_ISL_1813229, EPI_ISL_1813230, EPI_ISL_1813231, EPI_ISL_1813232, EPI_ISL_1813233, EPI_ISL_1813234, EPI_ISL_1813235, EPI_ISL_1813236, EPI_ISL_1813237, EPI_ISL_1813238 | | | |
| see above | EXCITE Lab | Andersen lab at Scripps Research | Nicole L Washington, Simon White, Geraint Levan, Kelly Schiabor Barrett, Elizabeth Cirulli, Alexandre Bolze, Charlotte Rivera-Garcia, Ryan Cho, Jason Nguyen, Sherry Wang, Jimmy Ramirez, Celena Andrade, Alice Summerfield, Tyler Cassens, Efren Sandoval, Francisco Tanudjaja, Magnus Isaksson, William Lee, David Becker, Marc Laurent, James Lu + SEARCH |
| EPI_ISL_1813239 | EXCITE Lab | Andersen lab at Scripps Research | Chip Schooley, Natasha Martin, Cheryl Anderson, Angela Scioscia, Smruthi Karthikeyan, Greg Humphrey, Sawyer Farmer, Abigail Schnapper, Helena Tubb, Tommy Valles + SEARCH |
| EPI_ISL_1813240, EPI_ISL_1813241, EPI_ISL_1813242, EPI_ISL_1813243, EPI_ISL_1813244, EPI_ISL_1813245, EPI_ISL_1813246, EPI_ISL_1813247, EPI_ISL_1813248, EPI_ISL_1813249, EPI_ISL_1813250, EPI_ISL_1813251, EPI_ISL_1813252, EPI_ISL_1813253, EPI_ISL_1813254, EPI_ISL_1813255, EPI_ISL_1813256, EPI_ISL_1813257, EPI_ISL_1813258, EPI_ISL_1813259 | | | |
| see above | EXCITE Lab | Andersen lab at Scripps Research | Nicole L Washington, Simon White, Geraint Levan, Kelly Schiabor Barrett, Elizabeth Cirulli, Alexandre Bolze, Charlotte Rivera-Garcia, Ryan Cho, Jason Nguyen, Sherry Wang, Jimmy Ramirez, Celena Andrade, Alice Summerfield, Tyler Cassens, Efren Sandoval, Francisco Tanudjaja, Magnus Isaksson, William Lee, David Becker, Marc Laurent, James Lu + SEARCH |
| EPI_ISL_1813260, EPI_ISL_1813261 | EXCITE Lab | Andersen lab at Scripps Research | David Pride, Sharon Reed, Chip Schooley, Angela Scioscia, Natasha Martin Cheryl Anderson, Sawyer Farmer, Abigail Schnapper, Helena Tubb, Tommy Valles + SEARCH |
| EPI_ISL_1813262 | EXCITE Lab | Andersen lab at Scripps Research | Nicole L Washington, Simon White, Geraint Levan, Kelly Schiabor Barrett, Elizabeth Cirulli, Alexandre Bolze, Charlotte Rivera-Garcia, Ryan Cho, Jason Nguyen, Sherry Wang, Jimmy Ramirez, Celena Andrade, Alice Summerfield, Tyler Cassens, Efren Sandoval, Francisco Tanudjaja, Magnus Isaksson, William Lee, David Becker, Marc Laurent, James Lu + SEARCH |
| EPI_ISL_1813263 | EXCITE Lab | Andersen lab at Scripps Research | David Pride, Sharon Reed, Chip Schooley, Angela Scioscia, Natasha Martin Cheryl Anderson, Sawyer Farmer, Abigail Schnapper, Helena Tubb, Tommy Valles + SEARCH |
| EPI_ISL_1813264 | EXCITE Lab | Andersen lab at Scripps Research | Nicole L Washington, Simon White, Geraint Levan, Kelly Schiabor Barrett, Elizabeth Cirulli, Alexandre Bolze, Charlotte Rivera-Garcia, Ryan Cho, Jason Nguyen, Sherry Wang, Jimmy Ramirez, Celena Andrade, Alice Summerfield, Tyler Cassens, Efren Sandoval, Francisco Tanudjaja, Magnus Isaksson, William Lee, David Becker, Marc Laurent, James Lu + SEARCH |
| EPI_ISL_1813265 | EXCITE Lab | Andersen lab at Scripps Research | David Pride, Sharon Reed, Chip Schooley, Angela Scioscia, Natasha Martin Cheryl Anderson, Sawyer Farmer, Abigail Schnapper, Helena Tubb, Tommy Valles + SEARCH |
| EPI_ISL_1813266, EPI_ISL_1813267, EPI_ISL_1813268 | EXCITE Lab | Andersen lab at Scripps Research | Nicole L Washington, Simon White, Geraint Levan, Kelly Schiabor Barrett, Elizabeth Cirulli, Alexandre Bolze, Charlotte Rivera-Garcia, Ryan Cho, Jason Nguyen, Sherry Wang, Jimmy Ramirez, Celena Andrade, Alice Summerfield, Tyler Cassens, Efren Sandoval, Francisco Tanudjaja, Magnus Isaksson, William Lee, David Becker, Marc Laurent, James Lu + SEARCH |
| EPI_ISL_1813269 | EXCITE Lab | Andersen lab at Scripps Research | Chip Schooley, Natasha Martin, Cheryl Anderson, Angela Scioscia, Smruthi Karthikeyan, Greg Humphrey, Sawyer Farmer, Abigail Schnapper, Helena Tubb, Tommy Valles + SEARCH |
| EPI_ISL_1813270, EPI_ISL_1813271, EPI_ISL_1813272, EPI_ISL_1813273, EPI_ISL_1813274, EPI_ISL_1813275, EPI_ISL_1813276, EPI_ISL_1813277, | EXCITE Lab | Andersen lab at Scripps Research | Nicole L Washington, Simon White, Geraint Levan, Kelly Schiabor Barrett, Elizabeth Cirulli, Alexandre Bolze, Charlotte Rivera-Garcia, Ryan Cho, Jason Nguyen, Sherry Wang, Jimmy Ramirez, Celena Andrade, Alice Summerfield, Tyler Cassens, Efren Sandoval, Francisco Tanudjaja, Magnus Isaksson, William Lee, David Becker, Marc Laurent, James Lu + SEARCH |

| | | | |
|---|---|---|---|
| EPI_ISL_1813278 | | | |
| EPI_ISL_1813279 | EXCITE Lab | Andersen lab at Scripps Research | David Pride, Sharon Reed, Chip Schooley, Angela Scioscia, Natasha Martin Cheryl Anderson, Sawyer Farmer, Abigail Schnapper, Helena Tubb, Tommy Valles + SEARCH |
| EPI_ISL_1813280, EPI_ISL_1813281, EPI_ISL_1813282 | EXCITE Lab | Andersen lab at Scripps Research | Nicole L Washington, Simon White, Geraint Levan, Kelly Schiabor Barrett, Elizabeth Cirulli, Alexandre Bolze, Charlotte Rivera-Garcia, Ryan Cho, Jason Nguyen, Sherry Wang, Jimmy Ramirez, Celena Andrade, Alice Summerfield, Tyler Cassens, Efren Sandoval, Francisco Tanudjaja, Magnus Isaksson, William Lee, David Becker, Marc Laurent, James Lu + SEARCH |
| EPI_ISL_1813283 | EXCITE Lab | Andersen lab at Scripps Research | Chip Schooley, Natasha Martin, Cheryl Anderson, Angela Scioscia, Smruthi Karthikeyan, Greg Humphrey, Sawyer Farmer, Abigail Schnapper, Helena Tubb, Tommy Valles + SEARCH |
| EPI_ISL_1813284, EPI_ISL_1813285, EPI_ISL_1813286, EPI_ISL_1813287, EPI_ISL_1813288, EPI_ISL_1813289, EPI_ISL_1813290, EPI_ISL_1813291, EPI_ISL_1813292, EPI_ISL_1813293, EPI_ISL_1813294, EPI_ISL_1813295, EPI_ISL_1813296, EPI_ISL_1813297, EPI_ISL_1813298, EPI_ISL_1813299, EPI_ISL_1813300, EPI_ISL_1813301 | | | |
| see above | EXCITE Lab | Andersen lab at Scripps Research | Nicole L Washington, Simon White, Geraint Levan, Kelly Schiabor Barrett, Elizabeth Cirulli, Alexandre Bolze, Charlotte Rivera-Garcia, Ryan Cho, Jason Nguyen, Sherry Wang, Jimmy Ramirez, Celena Andrade, Alice Summerfield, Tyler Cassens, Efren Sandoval, Francisco Tanudjaja, Magnus Isaksson, William Lee, David Becker, Marc Laurent, James Lu + SEARCH |
| EPI_ISL_1813302 | EXCITE Lab | Andersen lab at Scripps Research | David Pride, Sharon Reed, Chip Schooley, Angela Scioscia, Natasha Martin Cheryl Anderson, Sawyer Farmer, Abigail Schnapper, Helena Tubb, Tommy Valles + SEARCH |
| EPI_ISL_1813303 | EXCITE Lab | Andersen lab at Scripps Research | Nicole L Washington, Simon White, Geraint Levan, Kelly Schiabor Barrett, Elizabeth Cirulli, Alexandre Bolze, Charlotte Rivera-Garcia, Ryan Cho, Jason Nguyen, Sherry Wang, Jimmy Ramirez, Celena Andrade, Alice Summerfield, Tyler Cassens, Efren Sandoval, Francisco Tanudjaja, Magnus Isaksson, William Lee, David Becker, Marc Laurent, James Lu + SEARCH |
| EPI_ISL_1813304 | EXCITE Lab | Andersen lab at Scripps Research | Sydney Morgan + Marni Jacobs + Celestine Magallanes + Holly Valentine + Louise C. Laurent + SEARCH |
| EPI_ISL_1813305 | EXCITE Lab | Andersen lab at Scripps Research | Nicole L Washington, Simon White, Geraint Levan, Kelly Schiabor Barrett, Elizabeth Cirulli, Alexandre Bolze, Charlotte Rivera-Garcia, Ryan Cho, Jason Nguyen, Sherry Wang, Jimmy Ramirez, Celena Andrade, Alice Summerfield, Tyler Cassens, Efren Sandoval, Francisco Tanudjaja, Magnus Isaksson, William Lee, David Becker, Marc Laurent, James Lu + SEARCH |
| EPI_ISL_1813306 | EXCITE Lab | Andersen lab at Scripps Research | David Pride, Sharon Reed, Chip Schooley, Angela Scioscia, Natasha Martin Cheryl Anderson, Sawyer Farmer, Abigail Schnapper, Helena Tubb, Tommy Valles + SEARCH |
| EPI_ISL_1813307, EPI_ISL_1813308, EPI_ISL_1813309, EPI_ISL_1813310, EPI_ISL_1813311, EPI_ISL_1813312, EPI_ISL_1813313, EPI_ISL_1813314, EPI_ISL_1813315, EPI_ISL_1813316, EPI_ISL_1813317, EPI_ISL_1813318, EPI_ISL_1813319, EPI_ISL_1813320, EPI_ISL_1813321, EPI_ISL_1813322, EPI_ISL_1813323, EPI_ISL_1813324, EPI_ISL_1813325, EPI_ISL_1813326, EPI_ISL_1813327, EPI_ISL_1813328, EPI_ISL_1813329, EPI_ISL_1813330, EPI_ISL_1813331, EPI_ISL_1813332, EPI_ISL_1813333, EPI_ISL_1813334 | | | |
| see above | EXCITE Lab | Andersen lab at Scripps Research | Nicole L Washington, Simon White, Geraint Levan, Kelly Schiabor Barrett, Elizabeth Cirulli, Alexandre Bolze, Charlotte Rivera-Garcia, Ryan Cho, Jason Nguyen, Sherry Wang, Jimmy Ramirez, Celena Andrade, Alice Summerfield, Tyler Cassens, Efren Sandoval, Francisco Tanudjaja, Magnus Isaksson, William Lee, David Becker, Marc Laurent, James Lu + SEARCH |
| EPI_ISL_1813335 | EXCITE Lab | Andersen lab at Scripps Research | Chip Schooley, Natasha Martin, Cheryl Anderson, Angela Scioscia, Smruthi Karthikeyan, Greg Humphrey, Sawyer Farmer, Abigail Schnapper, Helena Tubb, Tommy Valles + SEARCH |
| EPI_ISL_1813336, EPI_ISL_1813337, EPI_ISL_1813338, EPI_ISL_1813339 | EXCITE Lab | Andersen lab at Scripps Research | Nicole L Washington, Simon White, Geraint Levan, Kelly Schiabor Barrett, Elizabeth Cirulli, Alexandre Bolze, Charlotte Rivera-Garcia, Ryan Cho, Jason Nguyen, Sherry Wang, Jimmy Ramirez, Celena Andrade, Alice Summerfield, Tyler Cassens, Efren Sandoval, Francisco Tanudjaja, Magnus Isaksson, William Lee, David Becker, Marc Laurent, James Lu + SEARCH |
| EPI_ISL_1813340 | EXCITE Lab | Andersen lab at Scripps Research | David Pride, Sharon Reed, Chip Schooley, Angela Scioscia, Natasha Martin Cheryl Anderson, Sawyer Farmer, Abigail Schnapper, Helena Tubb, Tommy Valles + SEARCH |
| EPI_ISL_1813341 | EXCITE Lab | Andersen lab at Scripps Research | Nicole L Washington, Simon White, Geraint Levan, Kelly Schiabor Barrett, Elizabeth Cirulli, Alexandre Bolze, Charlotte Rivera-Garcia, Ryan Cho, Jason Nguyen, Sherry Wang, Jimmy Ramirez, Celena Andrade, Alice Summerfield, Tyler Cassens, Efren Sandoval, Francisco Tanudjaja, Magnus Isaksson, William Lee, David Becker, Marc Laurent, James Lu + SEARCH |
| EPI_ISL_1813342 | EXCITE Lab | Andersen lab at Scripps Research | David Pride, Sharon Reed, Chip Schooley, Angela Scioscia, Natasha Martin Cheryl Anderson, Sawyer Farmer, Abigail Schnapper, Helena Tubb, Tommy Valles + SEARCH |
| EPI_ISL_1813343, EPI_ISL_1813344, EPI_ISL_1813345, EPI_ISL_1813346, EPI_ISL_1813347, EPI_ISL_1813348, EPI_ISL_1813349, EPI_ISL_1813350, EPI_ISL_1813351, EPI_ISL_1813352, EPI_ISL_1813353, EPI_ISL_1813354, EPI_ISL_1813355, EPI_ISL_1813356, EPI_ISL_1813357, EPI_ISL_1813358, EPI_ISL_1813359 | | | |
| see above | EXCITE Lab | Andersen lab at Scripps Research | Nicole L Washington, Simon White, Geraint Levan, Kelly Schiabor Barrett, Elizabeth Cirulli, Alexandre Bolze, Charlotte Rivera-Garcia, Ryan Cho, Jason Nguyen, Sherry Wang, Jimmy Ramirez, Celena Andrade, Alice Summerfield, Tyler Cassens, Efren Sandoval, Francisco Tanudjaja, Magnus Isaksson, William Lee, David Becker, Marc Laurent, James Lu + SEARCH |
| EPI_ISL_1813360 | EXCITE Lab | Andersen lab at Scripps Research | David Pride, Sharon Reed, Chip Schooley, Angela Scioscia, Natasha Martin Cheryl Anderson, Sawyer Farmer, Abigail Schnapper, Helena Tubb, Tommy Valles + SEARCH |
| EPI_ISL_1813361, EPI_ISL_1813362, EPI_ISL_1813363, EPI_ISL_1813364, EPI_ISL_1813365, EPI_ISL_1813366, EPI_ISL_1813367, EPI_ISL_1813368, EPI_ISL_1813369, EPI_ISL_1813370, EPI_ISL_1813371, EPI_ISL_1813372, EPI_ISL_1813373, EPI_ISL_1813374, EPI_ISL_1813375, EPI_ISL_1813376 | | | |
| see above | EXCITE Lab | Andersen lab at Scripps Research | Nicole L Washington, Simon White, Geraint Levan, Kelly Schiabor Barrett, Elizabeth Cirulli, Alexandre Bolze, Charlotte Rivera-Garcia, Ryan Cho, Jason Nguyen, Sherry Wang, Jimmy Ramirez, Celena Andrade, Alice Summerfield, Tyler Cassens, Efren Sandoval, Francisco Tanudjaja, Magnus Isaksson, William Lee, David Becker, Marc Laurent, James Lu + SEARCH |
| EPI_ISL_1813377 | EXCITE Lab | Andersen lab at Scripps Research | David Pride, Sharon Reed, Chip Schooley, Angela Scioscia, Natasha Martin Cheryl Anderson, Sawyer Farmer, Abigail Schnapper, Helena Tubb, Tommy Valles + SEARCH |
| EPI_ISL_1813378, EPI_ISL_1813379, EPI_ISL_1813380, EPI_ISL_1813381, EPI_ISL_1813382, EPI_ISL_1813383, EPI_ISL_1813384, EPI_ISL_1813385 | EXCITE Lab | Andersen lab at Scripps Research | Nicole L Washington, Simon White, Geraint Levan, Kelly Schiabor Barrett, Elizabeth Cirulli, Alexandre Bolze, Charlotte Rivera-Garcia, Ryan Cho, Jason Nguyen, Sherry Wang, Jimmy Ramirez, Celena Andrade, Alice Summerfield, Tyler Cassens, Efren Sandoval, Francisco Tanudjaja, Magnus Isaksson, William Lee, David Becker, Marc Laurent, James Lu + SEARCH |
| EPI_ISL_1813386 | EXCITE Lab | Andersen lab at Scripps Research | David Pride, Sharon Reed, Chip Schooley, Angela Scioscia, Natasha Martin Cheryl Anderson, Sawyer Farmer, Abigail Schnapper, Helena Tubb, Tommy Valles + SEARCH |
| EPI_ISL_1813387, EPI_ISL_1813388, EPI_ISL_1813389, EPI_ISL_1813390, EPI_ISL_1813391, EPI_ISL_1813392, EPI_ISL_1813393, EPI_ISL_1813394, EPI_ISL_1813395, EPI_ISL_1813396, EPI_ISL_1813397, EPI_ISL_1813398 | | | |
| see above | EXCITE Lab | Andersen lab at Scripps Research | Nicole L Washington, Simon White, Geraint Levan, Kelly Schiabor Barrett, Elizabeth Cirulli, Alexandre Bolze, Charlotte Rivera-Garcia, Ryan Cho, Jason Nguyen, Sherry Wang, Jimmy Ramirez, Celena Andrade, Alice Summerfield, Tyler Cassens, Efren Sandoval, Francisco Tanudjaja, Magnus Isaksson, William Lee, David Becker, Marc Laurent, James Lu + SEARCH |
| EPI_ISL_1813399 | EXCITE Lab | Andersen lab at Scripps Research | David Pride, Sharon Reed, Chip Schooley, Angela Scioscia, Natasha Martin Cheryl Anderson, Sawyer Farmer, Abigail Schnapper, Helena Tubb, Tommy Valles + SEARCH |
| EPI_ISL_1813400, EPI_ISL_1813401, EPI_ISL_1813402, EPI_ISL_1813403, EPI_ISL_1813404 | EXCITE Lab | Andersen lab at Scripps Research | Nicole L Washington, Simon White, Geraint Levan, Kelly Schiabor Barrett, Elizabeth Cirulli, Alexandre Bolze, Charlotte Rivera-Garcia, Ryan Cho, Jason Nguyen, Sherry Wang, Jimmy Ramirez, Celena Andrade, Alice Summerfield, Tyler Cassens, Efren Sandoval, Francisco Tanudjaja, Magnus Isaksson, William Lee, David Becker, Marc Laurent, James Lu + SEARCH |
| EPI_ISL_1813405 | EXCITE Lab | Andersen lab at Scripps Research | David Pride, Sharon Reed, Chip Schooley, Angela Scioscia, Natasha Martin Cheryl Anderson, Sawyer Farmer, Abigail Schnapper, Helena Tubb, Tommy Valles + SEARCH |
| EPI_ISL_1813406, EPI_ISL_1813407, EPI_ISL_1813408 | EXCITE Lab | Andersen lab at Scripps Research | Nicole L Washington, Simon White, Geraint Levan, Kelly Schiabor Barrett, Elizabeth Cirulli, Alexandre Bolze, Charlotte Rivera-Garcia, Ryan Cho, Jason Nguyen, Sherry Wang, Jimmy Ramirez, Celena Andrade, Alice Summerfield, Tyler Cassens, Efren Sandoval, Francisco Tanudjaja, Magnus Isaksson, William Lee, David Becker, Marc Laurent, James Lu + SEARCH |
| EPI_ISL_1813409 | EXCITE Lab | Andersen lab at Scripps Research | David Pride, Sharon Reed, Chip Schooley, Angela Scioscia, Natasha Martin Cheryl Anderson, Sawyer Farmer, Abigail Schnapper, Helena Tubb, Tommy Valles + SEARCH |
| EPI_ISL_1813410, EPI_ISL_1813411 | EXCITE Lab | Andersen lab at Scripps Research | Nicole L Washington, Simon White, Geraint Levan, Kelly Schiabor Barrett, Elizabeth Cirulli, Alexandre Bolze, Charlotte Rivera-Garcia, Ryan Cho, Jason Nguyen, Sherry Wang, Jimmy Ramirez, Celena Andrade, Alice Summerfield, Tyler Cassens, Efren Sandoval, Francisco Tanudjaja, Magnus Isaksson, William Lee, David Becker, Marc Laurent, James Lu + SEARCH |
| EPI_ISL_1813412 | EXCITE Lab | Andersen lab at Scripps Research | Chip Schooley, Natasha Martin, Cheryl Anderson, Angela Scioscia, Smruthi Karthikeyan, Greg Humphrey, Sawyer Farmer, Abigail Schnapper, Helena Tubb, Tommy Valles + SEARCH |
| EPI_ISL_1813413, EPI_ISL_1813414, EPI_ISL_1813415, EPI_ISL_1813416, EPI_ISL_1813417, EPI_ISL_1813418, EPI_ISL_1813419, EPI_ISL_1813420 | EXCITE Lab | Andersen lab at Scripps Research | Nicole L Washington, Simon White, Geraint Levan, Kelly Schiabor Barrett, Elizabeth Cirulli, Alexandre Bolze, Charlotte Rivera-Garcia, Ryan Cho, Jason Nguyen, Sherry Wang, Jimmy Ramirez, Celena Andrade, Alice Summerfield, Tyler Cassens, Efren Sandoval, Francisco Tanudjaja, Magnus Isaksson, William Lee, David Becker, Marc Laurent, James Lu + SEARCH |
| EPI_ISL_1813421 | EXCITE Lab | Andersen lab at Scripps Research | David Pride, Sharon Reed, Chip Schooley, Angela Scioscia, Natasha Martin Cheryl Anderson, Sawyer Farmer, Abigail Schnapper, Helena Tubb, Tommy Valles + SEARCH |
| EPI_ISL_1813422, EPI_ISL_1813423, EPI_ISL_1813424 | EXCITE Lab | Andersen lab at Scripps Research | Nicole L Washington, Simon White, Geraint Levan, Kelly Schiabor Barrett, Elizabeth Cirulli, Alexandre Bolze, Charlotte Rivera-Garcia, Ryan Cho, Jason Nguyen, Sherry Wang, Jimmy Ramirez, Celena Andrade, Alice Summerfield, Tyler Cassens, Efren Sandoval, Francisco Tanudjaja, Magnus Isaksson, William Lee, David Becker, Marc Laurent, James Lu + SEARCH |
| EPI_ISL_1813425 | EXCITE Lab | Andersen lab at Scripps Research | David Pride, Sharon Reed, Chip Schooley, Angela Scioscia, Natasha Martin Cheryl Anderson, Sawyer Farmer, Abigail Schnapper, Helena Tubb, Tommy Valles + SEARCH |
| EPI_ISL_1813426, EPI_ISL_1813427, EPI_ISL_1813428, EPI_ISL_1813429 | EXCITE Lab | Andersen lab at Scripps Research | Nicole L Washington, Simon White, Geraint Levan, Kelly Schiabor Barrett, Elizabeth Cirulli, Alexandre Bolze, Charlotte Rivera-Garcia, Ryan Cho, Jason Nguyen, Sherry Wang, Jimmy Ramirez, Celena Andrade, Alice Summerfield, Tyler Cassens, Efren Sandoval, Francisco Tanudjaja, Magnus Isaksson, William Lee, David Becker, Marc Laurent, James Lu + SEARCH |
| EPI_ISL_1813430 | EXCITE Lab | Andersen lab at Scripps Research | David Pride, Sharon Reed, Chip Schooley, Angela Scioscia, Natasha Martin Cheryl Anderson, Sawyer Farmer, Abigail Schnapper, Helena Tubb, Tommy Valles + SEARCH |
| EPI_ISL_1813431, EPI_ISL_1813432, EPI_ISL_1813433, EPI_ISL_1813434, EPI_ISL_1813435, EPI_ISL_1813436, EPI_ISL_1813437, EPI_ISL_1813438, EPI_ISL_1813439, EPI_ISL_1813440, EPI_ISL_1813441, EPI_ISL_1813442, EPI_ISL_1813443, EPI_ISL_1813444, EPI_ISL_1813445, EPI_ISL_1813446 | | | |
| see above | EXCITE Lab | Andersen lab at Scripps Research | Nicole L Washington, Simon White, Geraint Levan, Kelly Schiabor Barrett, Elizabeth Cirulli, Alexandre Bolze, Charlotte Rivera-Garcia, Ryan Cho, Jason Nguyen, Sherry Wang, Jimmy Ramirez, Celena Andrade, Alice Summerfield, Tyler Cassens, Efren Sandoval, Francisco Tanudjaja, Magnus Isaksson, William Lee, David Becker, Marc Laurent, James Lu + SEARCH |
| EPI_ISL_1813447, EPI_ISL_1813448 | EXCITE Lab | Andersen lab at Scripps Research | Chip Schooley, Natasha Martin, Cheryl Anderson, Angela Scioscia, Smruthi Karthikeyan, Greg Humphrey, Sawyer Farmer, Abigail Schnapper, Helena Tubb, Tommy Valles + SEARCH |
| EPI_ISL_1813449, EPI_ISL_1813450, EPI_ISL_1813451, EPI_ISL_1813452, EPI_ISL_1813453, EPI_ISL_1813454, EPI_ISL_1813455, EPI_ISL_1813456, EPI_ISL_1813457, EPI_ISL_1813458, EPI_ISL_1813459, EPI_ISL_1813460, EPI_ISL_1813461 | | | |
| see above | EXCITE Lab | Andersen lab at Scripps Research | Nicole L Washington, Simon White, Geraint Levan, Kelly Schiabor Barrett, Elizabeth Cirulli, Alexandre Bolze, Charlotte Rivera-Garcia, Ryan Cho, Jason Nguyen, Sherry Wang, Jimmy Ramirez, Celena Andrade, Alice Summerfield, Tyler Cassens, Efren Sandoval, Francisco Tanudjaja, Magnus Isaksson, William Lee, David Becker, Marc Laurent, James Lu + SEARCH |
| EPI_ISL_1813462 | EXCITE Lab | Andersen lab at Scripps Research | Chip Schooley, Natasha Martin, Cheryl Anderson, Angela Scioscia, Smruthi Karthikeyan, Greg Humphrey, Sawyer Farmer, Abigail Schnapper, Helena Tubb, Tommy Valles + SEARCH |

| | | | |
|---|---|---|---|
| EPI_ISL_1813463 | EXCITE Lab | Andersen lab at Scripps Research | David Pride, Sharon Reed, Chip Schooley, Angela Scioscia, Natasha Martin Cheryl Anderson, Sawyer Farmer, Abigail Schnapper, Helena Tubb, Tommy Valles + SEARCH |
| EPI_ISL_1813464, EPI_ISL_1813465, EPI_ISL_1813466, EPI_ISL_1813467, EPI_ISL_1813468, EPI_ISL_1813469, EPI_ISL_1813470, EPI_ISL_1813471, EPI_ISL_1813472, EPI_ISL_1813473, EPI_ISL_1813474 | | | |
| see above | EXCITE Lab | Andersen lab at Scripps Research | Nicole L Washington, Simon White, Geraint Levan, Kelly Schiabor Barrett, Elizabeth Cirulli, Alexandre Bolze, Charlotte Rivera-Garcia, Ryan Cho, Jason Nguyen, Sherry Wang, Jimmy Ramirez, Celena Andrade, Alice Summerfield, Tyler Cassens, Efren Sandoval, Francisco Tanudjaja, Magnus Isaksson, William Lee, David Becker, Marc Laurent, James Lu + SEARCH |
| EPI_ISL_1813475 | EXCITE Lab | Andersen lab at Scripps Research | David Pride, Sharon Reed, Chip Schooley, Angela Scioscia, Natasha Martin Cheryl Anderson, Sawyer Farmer, Abigail Schnapper, Helena Tubb, Tommy Valles + SEARCH |
| EPI_ISL_1813476, EPI_ISL_1813477, EPI_ISL_1813478, EPI_ISL_1813479, EPI_ISL_1813480, EPI_ISL_1813481, EPI_ISL_1813482, EPI_ISL_1813483, EPI_ISL_1813484, EPI_ISL_1813485, EPI_ISL_1813486, EPI_ISL_1813487, EPI_ISL_1813488, EPI_ISL_1813489, EPI_ISL_1813490, EPI_ISL_1813491, EPI_ISL_1813492 | | | |
| see above | EXCITE Lab | Andersen lab at Scripps Research | Nicole L Washington, Simon White, Geraint Levan, Kelly Schiabor Barrett, Elizabeth Cirulli, Alexandre Bolze, Charlotte Rivera-Garcia, Ryan Cho, Jason Nguyen, Sherry Wang, Jimmy Ramirez, Celena Andrade, Alice Summerfield, Tyler Cassens, Efren Sandoval, Francisco Tanudjaja, Magnus Isaksson, William Lee, David Becker, Marc Laurent, James Lu + SEARCH |
| EPI_ISL_1813493 | EXCITE Lab | Andersen lab at Scripps Research | Fred Wu, Christine Mitchell + SEARCH |
| EPI_ISL_1813494, EPI_ISL_1813495, EPI_ISL_1813496 | EXCITE Lab | Andersen lab at Scripps Research | Nicole L Washington, Simon White, Geraint Levan, Kelly Schiabor Barrett, Elizabeth Cirulli, Alexandre Bolze, Charlotte Rivera-Garcia, Ryan Cho, Jason Nguyen, Sherry Wang, Jimmy Ramirez, Celena Andrade, Alice Summerfield, Tyler Cassens, Efren Sandoval, Francisco Tanudjaja, Magnus Isaksson, William Lee, David Becker, Marc Laurent, James Lu + SEARCH |
| EPI_ISL_1813497 | EXCITE Lab | Andersen lab at Scripps Research | David Pride, Sharon Reed, Chip Schooley, Angela Scioscia, Natasha Martin Cheryl Anderson, Sawyer Farmer, Abigail Schnapper, Helena Tubb, Tommy Valles + SEARCH |
| EPI_ISL_1813498, EPI_ISL_1813499 | EXCITE Lab | Andersen lab at Scripps Research | Nicole L Washington, Simon White, Geraint Levan, Kelly Schiabor Barrett, Elizabeth Cirulli, Alexandre Bolze, Charlotte Rivera-Garcia, Ryan Cho, Jason Nguyen, Sherry Wang, Jimmy Ramirez, Celena Andrade, Alice Summerfield, Tyler Cassens, Efren Sandoval, Francisco Tanudjaja, Magnus Isaksson, William Lee, David Becker, Marc Laurent, James Lu + SEARCH |
| EPI_ISL_1813500 | EXCITE Lab | Andersen lab at Scripps Research | David Pride, Sharon Reed, Chip Schooley, Angela Scioscia, Natasha Martin Cheryl Anderson, Sawyer Farmer, Abigail Schnapper, Helena Tubb, Tommy Valles + SEARCH |
| EPI_ISL_1813501, EPI_ISL_1813502, EPI_ISL_1813503, EPI_ISL_1813504, EPI_ISL_1813505, EPI_ISL_1813506, EPI_ISL_1813507 | EXCITE Lab | Andersen lab at Scripps Research | Nicole L Washington, Simon White, Geraint Levan, Kelly Schiabor Barrett, Elizabeth Cirulli, Alexandre Bolze, Charlotte Rivera-Garcia, Ryan Cho, Jason Nguyen, Sherry Wang, Jimmy Ramirez, Celena Andrade, Alice Summerfield, Tyler Cassens, Efren Sandoval, Francisco Tanudjaja, Magnus Isaksson, William Lee, David Becker, Marc Laurent, James Lu + SEARCH |
| EPI_ISL_1813508 | EXCITE Lab | Andersen lab at Scripps Research | David Pride, Sharon Reed, Chip Schooley, Angela Scioscia, Natasha Martin Cheryl Anderson, Sawyer Farmer, Abigail Schnapper, Helena Tubb, Tommy Valles + SEARCH |
| EPI_ISL_1813509, EPI_ISL_1813510, EPI_ISL_1813511 | EXCITE Lab | Andersen lab at Scripps Research | Nicole L Washington, Simon White, Geraint Levan, Kelly Schiabor Barrett, Elizabeth Cirulli, Alexandre Bolze, Charlotte Rivera-Garcia, Ryan Cho, Jason Nguyen, Sherry Wang, Jimmy Ramirez, Celena Andrade, Alice Summerfield, Tyler Cassens, Efren Sandoval, Francisco Tanudjaja, Magnus Isaksson, William Lee, David Becker, Marc Laurent, James Lu + SEARCH |
| EPI_ISL_1813512 | EXCITE Lab | Andersen lab at Scripps Research | David Pride, Sharon Reed, Chip Schooley, Angela Scioscia, Natasha Martin Cheryl Anderson, Sawyer Farmer, Abigail Schnapper, Helena Tubb, Tommy Valles + SEARCH |
| EPI_ISL_1813513, EPI_ISL_1813514, EPI_ISL_1813515 | EXCITE Lab | Andersen lab at Scripps Research | Nicole L Washington, Simon White, Geraint Levan, Kelly Schiabor Barrett, Elizabeth Cirulli, Alexandre Bolze, Charlotte Rivera-Garcia, Ryan Cho, Jason Nguyen, Sherry Wang, Jimmy Ramirez, Celena Andrade, Alice Summerfield, Tyler Cassens, Efren Sandoval, Francisco Tanudjaja, Magnus Isaksson, William Lee, David Becker, Marc Laurent, James Lu + SEARCH |
| EPI_ISL_1813516 | EXCITE Lab | Andersen lab at Scripps Research | David Pride, Sharon Reed, Chip Schooley, Angela Scioscia, Natasha Martin Cheryl Anderson, Sawyer Farmer, Abigail Schnapper, Helena Tubb, Tommy Valles + SEARCH |
| EPI_ISL_1813517, EPI_ISL_1813518, EPI_ISL_1813519, EPI_ISL_1813520, EPI_ISL_1813521, EPI_ISL_1813522, EPI_ISL_1813523, EPI_ISL_1813524, EPI_ISL_1813525, EPI_ISL_1813526, EPI_ISL_1813527, EPI_ISL_1813528, EPI_ISL_1813529, EPI_ISL_1813530, EPI_ISL_1813531, EPI_ISL_1813532, EPI_ISL_1813533, EPI_ISL_1813534, EPI_ISL_1813535, EPI_ISL_1813536, EPI_ISL_1813537, EPI_ISL_1813538, EPI_ISL_1813539 | | | |
| see above | EXCITE Lab | Andersen lab at Scripps Research | Nicole L Washington, Simon White, Geraint Levan, Kelly Schiabor Barrett, Elizabeth Cirulli, Alexandre Bolze, Charlotte Rivera-Garcia, Ryan Cho, Jason Nguyen, Sherry Wang, Jimmy Ramirez, Celena Andrade, Alice Summerfield, Tyler Cassens, Efren Sandoval, Francisco Tanudjaja, Magnus Isaksson, William Lee, David Becker, Marc Laurent, James Lu + SEARCH |
| EPI_ISL_1813540 | EXCITE Lab | Andersen lab at Scripps Research | Chip Schooley, Natasha Martin, Cheryl Anderson, Angela Scioscia, Smruthi Karthikeyan, Greg Humphrey, Sawyer Farmer, Abigail Schnapper, Helena Tubb, Tommy Valles + SEARCH |
| EPI_ISL_1813541, EPI_ISL_1813542, EPI_ISL_1813543, EPI_ISL_1813544, EPI_ISL_1813545, EPI_ISL_1813546, EPI_ISL_1813547, EPI_ISL_1813548, EPI_ISL_1813549, EPI_ISL_1813550, EPI_ISL_1813551, EPI_ISL_1813552, EPI_ISL_1813553 | | | |
| see above | EXCITE Lab | Andersen lab at Scripps Research | Nicole L Washington, Simon White, Geraint Levan, Kelly Schiabor Barrett, Elizabeth Cirulli, Alexandre Bolze, Charlotte Rivera-Garcia, Ryan Cho, Jason Nguyen, Sherry Wang, Jimmy Ramirez, Celena Andrade, Alice Summerfield, Tyler Cassens, Efren Sandoval, Francisco Tanudjaja, Magnus Isaksson, William Lee, David Becker, Marc Laurent, James Lu + SEARCH |
| EPI_ISL_1813554 | EXCITE Lab | Andersen lab at Scripps Research | David Pride, Sharon Reed, Chip Schooley, Angela Scioscia, Natasha Martin Cheryl Anderson, Sawyer Farmer, Abigail Schnapper, Helena Tubb, Tommy Valles + SEARCH |
| EPI_ISL_1813555, EPI_ISL_1813556, EPI_ISL_1813557, EPI_ISL_1813558 | EXCITE Lab | Andersen lab at Scripps Research | Nicole L Washington, Simon White, Geraint Levan, Kelly Schiabor Barrett, Elizabeth Cirulli, Alexandre Bolze, Charlotte Rivera-Garcia, Ryan Cho, Jason Nguyen, Sherry Wang, Jimmy Ramirez, Celena Andrade, Alice Summerfield, Tyler Cassens, Efren Sandoval, Francisco Tanudjaja, Magnus Isaksson, William Lee, David Becker, Marc Laurent, James Lu + SEARCH |
| EPI_ISL_1813559 | EXCITE Lab | Andersen lab at Scripps Research | David Pride, Sharon Reed, Chip Schooley, Angela Scioscia, Natasha Martin Cheryl Anderson, Sawyer Farmer, Abigail Schnapper, Helena Tubb, Tommy Valles + SEARCH |
| EPI_ISL_1813560, EPI_ISL_1813561, EPI_ISL_1813562, EPI_ISL_1813563, EPI_ISL_1813564, EPI_ISL_1813565, EPI_ISL_1813566, EPI_ISL_1813567, EPI_ISL_1813568, EPI_ISL_1813569, EPI_ISL_1813570, EPI_ISL_1813571, EPI_ISL_1813572, EPI_ISL_1813573, EPI_ISL_1813574, EPI_ISL_1813575, EPI_ISL_1813576, EPI_ISL_1813577, EPI_ISL_1813578, EPI_ISL_1813579, EPI_ISL_1813580, EPI_ISL_1813581 | | | |
| see above | EXCITE Lab | Andersen lab at Scripps Research | Nicole L Washington, Simon White, Geraint Levan, Kelly Schiabor Barrett, Elizabeth Cirulli, Alexandre Bolze, Charlotte Rivera-Garcia, Ryan Cho, Jason Nguyen, Sherry Wang, Jimmy Ramirez, Celena Andrade, Alice Summerfield, Tyler Cassens, Efren Sandoval, Francisco Tanudjaja, Magnus Isaksson, William Lee, David Becker, Marc Laurent, James Lu + SEARCH |
| EPI_ISL_1813582 | EXCITE Lab | Andersen lab at Scripps Research | Chip Schooley, Natasha Martin, Cheryl Anderson, Angela Scioscia, Smruthi Karthikeyan, Greg Humphrey, Sawyer Farmer, Abigail Schnapper, Helena Tubb, Tommy Valles + SEARCH |
| EPI_ISL_1813583, EPI_ISL_1813584, EPI_ISL_1813585, EPI_ISL_1813586, EPI_ISL_1813587, EPI_ISL_1813588, EPI_ISL_1813589 | EXCITE Lab | Andersen lab at Scripps Research | Nicole L Washington, Simon White, Geraint Levan, Kelly Schiabor Barrett, Elizabeth Cirulli, Alexandre Bolze, Charlotte Rivera-Garcia, Ryan Cho, Jason Nguyen, Sherry Wang, Jimmy Ramirez, Celena Andrade, Alice Summerfield, Tyler Cassens, Efren Sandoval, Francisco Tanudjaja, Magnus Isaksson, William Lee, David Becker, Marc Laurent, James Lu + SEARCH |
| EPI_ISL_1813590 | EXCITE Lab | Andersen lab at Scripps Research | J. Joelle Donofrio-Odmann, Brent Brainard+ SEARCH |
| EPI_ISL_1813591, EPI_ISL_1813592, EPI_ISL_1813593, EPI_ISL_1813594, EPI_ISL_1813595, EPI_ISL_1813596, EPI_ISL_1813597, EPI_ISL_1813598, EPI_ISL_1813599 | EXCITE Lab | Andersen lab at Scripps Research | Nicole L Washington, Simon White, Geraint Levan, Kelly Schiabor Barrett, Elizabeth Cirulli, Alexandre Bolze, Charlotte Rivera-Garcia, Ryan Cho, Jason Nguyen, Sherry Wang, Jimmy Ramirez, Celena Andrade, Alice Summerfield, Tyler Cassens, Efren Sandoval, Francisco Tanudjaja, Magnus Isaksson, William Lee, David Becker, Marc Laurent, James Lu + SEARCH |
| EPI_ISL_1813600 | EXCITE Lab | Andersen lab at Scripps Research | David Pride, Sharon Reed, Chip Schooley, Angela Scioscia, Natasha Martin Cheryl Anderson, Sawyer Farmer, Abigail Schnapper, Helena Tubb, Tommy Valles + SEARCH |
| EPI_ISL_1813601, EPI_ISL_1813602, EPI_ISL_1813603, EPI_ISL_1813604, EPI_ISL_1813605, EPI_ISL_1813606, EPI_ISL_1813607, EPI_ISL_1813608, EPI_ISL_1813609, EPI_ISL_1813610, EPI_ISL_1813611, EPI_ISL_1813612, EPI_ISL_1813613, EPI_ISL_1813614, EPI_ISL_1813615, EPI_ISL_1813616, EPI_ISL_1813617, EPI_ISL_1813618, EPI_ISL_1813619, EPI_ISL_1813620, EPI_ISL_1813621, EPI_ISL_1813622, EPI_ISL_1813623, EPI_ISL_1813624, EPI_ISL_1813625 | | | |
| see above | EXCITE Lab | Andersen lab at Scripps Research | Nicole L Washington, Simon White, Geraint Levan, Kelly Schiabor Barrett, Elizabeth Cirulli, Alexandre Bolze, Charlotte Rivera-Garcia, Ryan Cho, Jason Nguyen, Sherry Wang, Jimmy Ramirez, Celena Andrade, Alice Summerfield, Tyler Cassens, Efren Sandoval, Francisco Tanudjaja, Magnus Isaksson, William Lee, David Becker, Marc Laurent, James Lu + SEARCH |
| EPI_ISL_1813626 | EXCITE Lab | Andersen lab at Scripps Research | David Pride, Sharon Reed, Chip Schooley, Angela Scioscia, Natasha Martin Cheryl Anderson, Sawyer Farmer, Abigail Schnapper, Helena Tubb, Tommy Valles + SEARCH |
| EPI_ISL_1813627, EPI_ISL_1813628, EPI_ISL_1813629 | EXCITE Lab | Andersen lab at Scripps Research | Nicole L Washington, Simon White, Geraint Levan, Kelly Schiabor Barrett, Elizabeth Cirulli, Alexandre Bolze, Charlotte Rivera-Garcia, Ryan Cho, Jason Nguyen, Sherry Wang, Jimmy Ramirez, Celena Andrade, Alice Summerfield, Tyler Cassens, Efren Sandoval, Francisco Tanudjaja, Magnus Isaksson, William Lee, David Becker, Marc Laurent, James Lu + SEARCH |
| EPI_ISL_1813630 | EXCITE Lab | Andersen lab at Scripps Research | David Pride, Sharon Reed, Chip Schooley, Angela Scioscia, Natasha Martin Cheryl Anderson, Sawyer Farmer, Abigail Schnapper, Helena Tubb, Tommy Valles + SEARCH |
| EPI_ISL_1813631, EPI_ISL_1813632, EPI_ISL_1813633, EPI_ISL_1813634, EPI_ISL_1813635, EPI_ISL_1813636, EPI_ISL_1813637, EPI_ISL_1813638, EPI_ISL_1813639, EPI_ISL_1813640, EPI_ISL_1813641, EPI_ISL_1813642, EPI_ISL_1813643, EPI_ISL_1813644, EPI_ISL_1813645, EPI_ISL_1813646, EPI_ISL_1813647, EPI_ISL_1813648, EPI_ISL_1813649, EPI_ISL_1813650, EPI_ISL_1813651, EPI_ISL_1813652 | | | |
| see above | EXCITE Lab | Andersen lab at Scripps Research | Nicole L Washington, Simon White, Geraint Levan, Kelly Schiabor Barrett, Elizabeth Cirulli, Alexandre Bolze, Charlotte Rivera-Garcia, Ryan Cho, Jason Nguyen, Sherry Wang, Jimmy Ramirez, Celena Andrade, Alice Summerfield, Tyler Cassens, Efren Sandoval, Francisco Tanudjaja, Magnus Isaksson, William Lee, David Becker, Marc Laurent, James Lu + SEARCH |
| EPI_ISL_1813653 | EXCITE Lab | Andersen lab at Scripps Research | Chip Schooley, Natasha Martin, Cheryl Anderson, Angela Scioscia, Smruthi Karthikeyan, Greg Humphrey, Sawyer Farmer, Abigail Schnapper, Helena Tubb, Tommy Valles + SEARCH |
| EPI_ISL_1813654, EPI_ISL_1813655, EPI_ISL_1813656, EPI_ISL_1813657, EPI_ISL_1813658, EPI_ISL_1813659, EPI_ISL_1813660, EPI_ISL_1813661, EPI_ISL_1813662, EPI_ISL_1813663, EPI_ISL_1813664, EPI_ISL_1813665 | | | |
| see above | EXCITE Lab | Andersen lab at Scripps Research | Nicole L Washington, Simon White, Geraint Levan, Kelly Schiabor Barrett, Elizabeth Cirulli, Alexandre Bolze, Charlotte Rivera-Garcia, Ryan Cho, Jason Nguyen, Sherry Wang, Jimmy Ramirez, Celena Andrade, Alice Summerfield, Tyler Cassens, Efren Sandoval, Francisco Tanudjaja, Magnus Isaksson, William Lee, David Becker, Marc Laurent, James Lu + SEARCH |
| EPI_ISL_1813666 | EXCITE Lab | Andersen lab at Scripps Research | David Pride, Sharon Reed, Chip Schooley, Angela Scioscia, Natasha Martin Cheryl Anderson, Sawyer Farmer, Abigail Schnapper, Helena Tubb, Tommy Valles + SEARCH |
| EPI_ISL_1813667, EPI_ISL_1813668 | EXCITE Lab | Andersen lab at Scripps Research | Nicole L Washington, Simon White, Geraint Levan, Kelly Schiabor Barrett, Elizabeth Cirulli, Alexandre Bolze, Charlotte Rivera-Garcia, Ryan Cho, Jason Nguyen, Sherry Wang, Jimmy Ramirez, Celena Andrade, Alice Summerfield, Tyler Cassens, Efren Sandoval, Francisco Tanudjaja, Magnus Isaksson, William Lee, David Becker, Marc Laurent, James Lu + SEARCH |
| EPI_ISL_1813669 | EXCITE Lab | Andersen lab at Scripps Research | Chip Schooley, Natasha Martin, Cheryl Anderson, Angela Scioscia, Smruthi Karthikeyan, Greg Humphrey, Sawyer Farmer, Abigail Schnapper, Helena Tubb, Tommy Valles + SEARCH |
| EPI_ISL_1813670, | EXCITE Lab | Andersen lab at Scripps Research | Nicole L Washington, Simon White, Geraint Levan, Kelly Schiabor Barrett, Elizabeth Cirulli, Alexandre Bolze, Charlotte Rivera-Garcia, Ryan Cho, Jason Nguyen, Sherry Wang, Jimmy Ramirez, Celena Andrade, Alice Summerfield, Tyler Cassens, Efren Sandoval, Francisco Tanudjaja, Magnus |

| | | | |
|---|---|---|---|
| EPI_ISL_1813671, EPI_ISL_1813672, EPI_ISL_1813673, EPI_ISL_1813674, EPI_ISL_1813675, EPI_ISL_1813676 | | | Isaksson, William Lee, David Becker, Marc Laurent, James Lu + SEARCH |
| EPI_ISL_1813677 | EXCITE Lab | Andersen lab at Scripps Research | David Pride, Sharon Reed, Chip Schooley, Angela Scioscia, Natasha Martin Cheryl Anderson, Sawyer Farmer, Abigail Schnapper, Helena Tubb, Tommy Valles + SEARCH |
| EPI_ISL_1813678, EPI_ISL_1813679, EPI_ISL_1813680, EPI_ISL_1813681, EPI_ISL_1813682, EPI_ISL_1813683, EPI_ISL_1813684, EPI_ISL_1813685, EPI_ISL_1813686, EPI_ISL_1813687 | EXCITE Lab | Andersen lab at Scripps Research | Nicole L Washington, Simon White, Geraint Levan, Kelly Schiabor Barrett, Elizabeth Cirulli, Alexandre Bolze, Charlotte Rivera-Garcia, Ryan Cho, Jason Nguyen, Sherry Wang, Jimmy Ramirez, Celena Andrade, Alice Summerfield, Tyler Cassens, Efren Sandoval, Francisco Tanudjaja, Magnus Isaksson, William Lee, David Becker, Marc Laurent, James Lu + SEARCH |
| EPI_ISL_1813688 | EXCITE Lab | Andersen lab at Scripps Research | David Pride, Sharon Reed, Chip Schooley, Angela Scioscia, Natasha Martin Cheryl Anderson, Sawyer Farmer, Abigail Schnapper, Helena Tubb, Tommy Valles + SEARCH |
| EPI_ISL_1813689, EPI_ISL_1813690, EPI_ISL_1813691, EPI_ISL_1813692, EPI_ISL_1813693, EPI_ISL_1813694, EPI_ISL_1813695, EPI_ISL_1813696, EPI_ISL_1813697, EPI_ISL_1813698, EPI_ISL_1813699, EPI_ISL_1813700 | | | |
| see above | EXCITE Lab | Andersen lab at Scripps Research | Nicole L Washington, Simon White, Geraint Levan, Kelly Schiabor Barrett, Elizabeth Cirulli, Alexandre Bolze, Charlotte Rivera-Garcia, Ryan Cho, Jason Nguyen, Sherry Wang, Jimmy Ramirez, Celena Andrade, Alice Summerfield, Tyler Cassens, Efren Sandoval, Francisco Tanudjaja, Magnus Isaksson, William Lee, David Becker, Marc Laurent, James Lu + SEARCH |
| EPI_ISL_1813701 | EXCITE Lab | Andersen lab at Scripps Research | Fred Wu, Christine Mitchell + SEARCH |
| EPI_ISL_1813702 | EXCITE Lab | Andersen lab at Scripps Research | David Pride, Sharon Reed, Chip Schooley, Angela Scioscia, Natasha Martin Cheryl Anderson, Sawyer Farmer, Abigail Schnapper, Helena Tubb, Tommy Valles + SEARCH |
| EPI_ISL_1813703, EPI_ISL_1813704, EPI_ISL_1813705, EPI_ISL_1813706, EPI_ISL_1813707, EPI_ISL_1813708, EPI_ISL_1813709, EPI_ISL_1813710 | EXCITE Lab | Andersen lab at Scripps Research | Nicole L Washington, Simon White, Geraint Levan, Kelly Schiabor Barrett, Elizabeth Cirulli, Alexandre Bolze, Charlotte Rivera-Garcia, Ryan Cho, Jason Nguyen, Sherry Wang, Jimmy Ramirez, Celena Andrade, Alice Summerfield, Tyler Cassens, Efren Sandoval, Francisco Tanudjaja, Magnus Isaksson, William Lee, David Becker, Marc Laurent, James Lu + SEARCH |
| EPI_ISL_1813711 | EXCITE Lab | Andersen lab at Scripps Research | David Pride, Sharon Reed, Chip Schooley, Angela Scioscia, Natasha Martin Cheryl Anderson, Sawyer Farmer, Abigail Schnapper, Helena Tubb, Tommy Valles + SEARCH |
| EPI_ISL_1813712, EPI_ISL_1813713, EPI_ISL_1813714, EPI_ISL_1813715, EPI_ISL_1813716, EPI_ISL_1813717, EPI_ISL_1813718, EPI_ISL_1813719, EPI_ISL_1813720, EPI_ISL_1813721, EPI_ISL_1813722, EPI_ISL_1813723, EPI_ISL_1813724, EPI_ISL_1813725, EPI_ISL_1813726, EPI_ISL_1813727, EPI_ISL_1813728, EPI_ISL_1813729, EPI_ISL_1813730, EPI_ISL_1813731, EPI_ISL_1813732, EPI_ISL_1813733, EPI_ISL_1813734, EPI_ISL_1813735, EPI_ISL_1813736, EPI_ISL_1813737, EPI_ISL_1813738, EPI_ISL_1813739, EPI_ISL_1813740, EPI_ISL_1813741, EPI_ISL_1813742, EPI_ISL_1813743 | | | |
| see above | EXCITE Lab | Andersen lab at Scripps Research | Nicole L Washington, Simon White, Geraint Levan, Kelly Schiabor Barrett, Elizabeth Cirulli, Alexandre Bolze, Charlotte Rivera-Garcia, Ryan Cho, Jason Nguyen, Sherry Wang, Jimmy Ramirez, Celena Andrade, Alice Summerfield, Tyler Cassens, Efren Sandoval, Francisco Tanudjaja, Magnus Isaksson, William Lee, David Becker, Marc Laurent, James Lu + SEARCH |
| EPI_ISL_1813744 | EXCITE Lab | Andersen lab at Scripps Research | Chip Schooley, Natasha Martin, Cheryl Anderson, Angela Scioscia, Smruthi Karthikeyan, Greg Humphrey, Sawyer Farmer, Abigail Schnapper, Helena Tubb, Tommy Valles + SEARCH |
| EPI_ISL_1813745, EPI_ISL_1813746, EPI_ISL_1813747, EPI_ISL_1813748, EPI_ISL_1813749, EPI_ISL_1813750, EPI_ISL_1813751, EPI_ISL_1813752, EPI_ISL_1813753, EPI_ISL_1813754, EPI_ISL_1813755, EPI_ISL_1813756, EPI_ISL_1813757, EPI_ISL_1813758, EPI_ISL_1813759, EPI_ISL_1813760, EPI_ISL_1813761, EPI_ISL_1813762, EPI_ISL_1813763, EPI_ISL_1813764, EPI_ISL_1813765 | | | |
| see above | EXCITE Lab | Andersen lab at Scripps Research | Nicole L Washington, Simon White, Geraint Levan, Kelly Schiabor Barrett, Elizabeth Cirulli, Alexandre Bolze, Charlotte Rivera-Garcia, Ryan Cho, Jason Nguyen, Sherry Wang, Jimmy Ramirez, Celena Andrade, Alice Summerfield, Tyler Cassens, Efren Sandoval, Francisco Tanudjaja, Magnus Isaksson, William Lee, David Becker, Marc Laurent, James Lu + SEARCH |
| EPI_ISL_1813766 | EXCITE Lab | Andersen lab at Scripps Research | Chip Schooley, Natasha Martin, Cheryl Anderson, Angela Scioscia, Smruthi Karthikeyan, Greg Humphrey, Sawyer Farmer, Abigail Schnapper, Helena Tubb, Tommy Valles + SEARCH |
| EPI_ISL_1813767, EPI_ISL_1813768, EPI_ISL_1813769 | EXCITE Lab | Andersen lab at Scripps Research | Nicole L Washington, Simon White, Geraint Levan, Kelly Schiabor Barrett, Elizabeth Cirulli, Alexandre Bolze, Charlotte Rivera-Garcia, Ryan Cho, Jason Nguyen, Sherry Wang, Jimmy Ramirez, Celena Andrade, Alice Summerfield, Tyler Cassens, Efren Sandoval, Francisco Tanudjaja, Magnus Isaksson, William Lee, David Becker, Marc Laurent, James Lu + SEARCH |
| EPI_ISL_1813770 | EXCITE Lab | Andersen lab at Scripps Research | David Pride, Sharon Reed, Chip Schooley, Angela Scioscia, Natasha Martin Cheryl Anderson, Sawyer Farmer, Abigail Schnapper, Helena Tubb, Tommy Valles + SEARCH |
| EPI_ISL_1813771, EPI_ISL_1813772 | EXCITE Lab | Andersen lab at Scripps Research | Nicole L Washington, Simon White, Geraint Levan, Kelly Schiabor Barrett, Elizabeth Cirulli, Alexandre Bolze, Charlotte Rivera-Garcia, Ryan Cho, Jason Nguyen, Sherry Wang, Jimmy Ramirez, Celena Andrade, Alice Summerfield, Tyler Cassens, Efren Sandoval, Francisco Tanudjaja, Magnus Isaksson, William Lee, David Becker, Marc Laurent, James Lu + SEARCH |
| EPI_ISL_1813773 | EXCITE Lab | Andersen lab at Scripps Research | J. Joelle Donofrio-Odmann, Brent Brainard+ SEARCH |
| EPI_ISL_1813774, EPI_ISL_1813775, EPI_ISL_1813776, EPI_ISL_1813777, EPI_ISL_1813778, EPI_ISL_1813779, EPI_ISL_1813780, EPI_ISL_1813781, EPI_ISL_1813782 | EXCITE Lab | Andersen lab at Scripps Research | Nicole L Washington, Simon White, Geraint Levan, Kelly Schiabor Barrett, Elizabeth Cirulli, Alexandre Bolze, Charlotte Rivera-Garcia, Ryan Cho, Jason Nguyen, Sherry Wang, Jimmy Ramirez, Celena Andrade, Alice Summerfield, Tyler Cassens, Efren Sandoval, Francisco Tanudjaja, Magnus Isaksson, William Lee, David Becker, Marc Laurent, James Lu + SEARCH |
| EPI_ISL_1813783 | EXCITE Lab | Andersen lab at Scripps Research | Sydney Morgan + Marni Jacobs + Celestine Magallanes + Holly Valentine + Louise C. Laurent + SEARCH |
| EPI_ISL_1813784, EPI_ISL_1813785, EPI_ISL_1813786, EPI_ISL_1813787, EPI_ISL_1813788, EPI_ISL_1813789, EPI_ISL_1813790 | EXCITE Lab | Andersen lab at Scripps Research | Nicole L Washington, Simon White, Geraint Levan, Kelly Schiabor Barrett, Elizabeth Cirulli, Alexandre Bolze, Charlotte Rivera-Garcia, Ryan Cho, Jason Nguyen, Sherry Wang, Jimmy Ramirez, Celena Andrade, Alice Summerfield, Tyler Cassens, Efren Sandoval, Francisco Tanudjaja, Magnus Isaksson, William Lee, David Becker, Marc Laurent, James Lu + SEARCH |
| EPI_ISL_1813791 | EXCITE Lab | Andersen lab at Scripps Research | David Pride, Sharon Reed, Chip Schooley, Angela Scioscia, Natasha Martin Cheryl Anderson, Sawyer Farmer, Abigail Schnapper, Helena Tubb, Tommy Valles + SEARCH |
| EPI_ISL_1813792, EPI_ISL_1813793, EPI_ISL_1813794, EPI_ISL_1813795, EPI_ISL_1813796, EPI_ISL_1813797, EPI_ISL_1813798, EPI_ISL_1813799, EPI_ISL_1813800, EPI_ISL_1813801, EPI_ISL_1813802, EPI_ISL_1813803, EPI_ISL_1813804, EPI_ISL_1813805 | | | |
| see above | EXCITE Lab | Andersen lab at Scripps Research | Nicole L Washington, Simon White, Geraint Levan, Kelly Schiabor Barrett, Elizabeth Cirulli, Alexandre Bolze, Charlotte Rivera-Garcia, Ryan Cho, Jason Nguyen, Sherry Wang, Jimmy Ramirez, Celena Andrade, Alice Summerfield, Tyler Cassens, Efren Sandoval, Francisco Tanudjaja, Magnus Isaksson, William Lee, David Becker, Marc Laurent, James Lu + SEARCH |
| EPI_ISL_1813806 | EXCITE Lab | Andersen lab at Scripps Research | David Pride, Sharon Reed, Chip Schooley, Angela Scioscia, Natasha Martin Cheryl Anderson, Sawyer Farmer, Abigail Schnapper, Helena Tubb, Tommy Valles + SEARCH |
| EPI_ISL_1813807 | EXCITE Lab | Andersen lab at Scripps Research | Chip Schooley, Natasha Martin, Cheryl Anderson, Angela Scioscia, Smruthi Karthikeyan, Greg Humphrey, Sawyer Farmer, Abigail Schnapper, Helena Tubb, Tommy Valles + SEARCH |
| EPI_ISL_1813808, EPI_ISL_1813809, EPI_ISL_1813810 | EXCITE Lab | Andersen lab at Scripps Research | Nicole L Washington, Simon White, Geraint Levan, Kelly Schiabor Barrett, Elizabeth Cirulli, Alexandre Bolze, Charlotte Rivera-Garcia, Ryan Cho, Jason Nguyen, Sherry Wang, Jimmy Ramirez, Celena Andrade, Alice Summerfield, Tyler Cassens, Efren Sandoval, Francisco Tanudjaja, Magnus Isaksson, William Lee, David Becker, Marc Laurent, James Lu + SEARCH |
| EPI_ISL_1813811 | EXCITE Lab | Andersen lab at Scripps Research | Chip Schooley, Natasha Martin, Cheryl Anderson, Angela Scioscia, Smruthi Karthikeyan, Greg Humphrey, Sawyer Farmer, Abigail Schnapper, Helena Tubb, Tommy Valles + SEARCH |
| EPI_ISL_1813812, EPI_ISL_1813813, EPI_ISL_1813814, EPI_ISL_1813815, EPI_ISL_1813816, EPI_ISL_1813817, EPI_ISL_1813818, EPI_ISL_1813819 | EXCITE Lab | Andersen lab at Scripps Research | Nicole L Washington, Simon White, Geraint Levan, Kelly Schiabor Barrett, Elizabeth Cirulli, Alexandre Bolze, Charlotte Rivera-Garcia, Ryan Cho, Jason Nguyen, Sherry Wang, Jimmy Ramirez, Celena Andrade, Alice Summerfield, Tyler Cassens, Efren Sandoval, Francisco Tanudjaja, Magnus Isaksson, William Lee, David Becker, Marc Laurent, James Lu + SEARCH |
| EPI_ISL_1813820 | EXCITE Lab | Andersen lab at Scripps Research | David Pride, Sharon Reed, Chip Schooley, Angela Scioscia, Natasha Martin Cheryl Anderson, Sawyer Farmer, Abigail Schnapper, Helena Tubb, Tommy Valles + SEARCH |
| EPI_ISL_1813821, EPI_ISL_1813822 | EXCITE Lab | Andersen lab at Scripps Research | Nicole L Washington, Simon White, Geraint Levan, Kelly Schiabor Barrett, Elizabeth Cirulli, Alexandre Bolze, Charlotte Rivera-Garcia, Ryan Cho, Jason Nguyen, Sherry Wang, Jimmy Ramirez, Celena Andrade, Alice Summerfield, Tyler Cassens, Efren Sandoval, Francisco Tanudjaja, Magnus Isaksson, William Lee, David Becker, Marc Laurent, James Lu + SEARCH |
| EPI_ISL_1813823 | EXCITE Lab | Andersen lab at Scripps Research | Chip Schooley, Natasha Martin, Cheryl Anderson, Angela Scioscia, Smruthi Karthikeyan, Greg Humphrey, Sawyer Farmer, Abigail Schnapper, Helena Tubb, Tommy Valles + SEARCH |
| EPI_ISL_1813824, EPI_ISL_1813825, EPI_ISL_1813826, EPI_ISL_1813827, EPI_ISL_1813828, EPI_ISL_1813829, EPI_ISL_1813830, EPI_ISL_1813831, EPI_ISL_1813832, EPI_ISL_1813833, EPI_ISL_1813834, EPI_ISL_1813835, EPI_ISL_1813836, EPI_ISL_1813837, EPI_ISL_1813838, EPI_ISL_1813839, EPI_ISL_1813840, EPI_ISL_1813841, EPI_ISL_1813842, EPI_ISL_1813843, EPI_ISL_1813844, EPI_ISL_1813845, EPI_ISL_1813846, EPI_ISL_1813847, EPI_ISL_1813848, EPI_ISL_1813849, EPI_ISL_1813850 | | | |
| see above | EXCITE Lab | Andersen lab at Scripps Research | Nicole L Washington, Simon White, Geraint Levan, Kelly Schiabor Barrett, Elizabeth Cirulli, Alexandre Bolze, Charlotte Rivera-Garcia, Ryan Cho, Jason Nguyen, Sherry Wang, Jimmy Ramirez, Celena Andrade, Alice Summerfield, Tyler Cassens, Efren Sandoval, Francisco Tanudjaja, Magnus Isaksson, William Lee, David Becker, Marc Laurent, James Lu + SEARCH |
| EPI_ISL_1813851 | EXCITE Lab | Andersen lab at Scripps Research | David Pride, Sharon Reed, Chip Schooley, Angela Scioscia, Natasha Martin Cheryl Anderson, Sawyer Farmer, Abigail Schnapper, Helena Tubb, Tommy Valles + SEARCH |
| EPI_ISL_1813852, | EXCITE Lab | Andersen lab at Scripps Research | Nicole L Washington, Simon White, Geraint Levan, Kelly Schiabor Barrett, Elizabeth Cirulli, Alexandre Bolze, Charlotte Rivera-Garcia, Ryan Cho, Jason Nguyen, Sherry Wang, Jimmy Ramirez, Celena Andrade, Alice Summerfield, Tyler Cassens, Efren Sandoval, Francisco Tanudjaja, Magnus |

| | | | |
|---|---|---|---|
| EPI_ISL_1813853, EPI_ISL_1813854, EPI_ISL_1813855, EPI_ISL_1813856, EPI_ISL_1813857, EPI_ISL_1813858, EPI_ISL_1813859 | | | Isaksson, William Lee, David Becker, Marc Laurent, James Lu + SEARCH |
| EPI_ISL_1813860 | EXCITE Lab | Andersen lab at Scripps Research | Chip Schooley, Natasha Martin, Cheryl Anderson, Angela Scioscia, Smruthi Karthikeyan, Greg Humphrey, Sawyer Farmer, Abigail Schnapper, Helena Tubb, Tommy Valles + SEARCH |
| EPI_ISL_1813861, EPI_ISL_1813862, EPI_ISL_1813863, EPI_ISL_1813864, EPI_ISL_1813865, EPI_ISL_1813866, EPI_ISL_1813867 | EXCITE Lab | Andersen lab at Scripps Research | Nicole L Washington, Simon White, Geraint Levan, Kelly Schiabor Barrett, Elizabeth Cirulli, Alexandre Bolze, Charlotte Rivera-Garcia, Ryan Cho, Jason Nguyen, Sherry Wang, Jimmy Ramirez, Celena Andrade, Alice Summerfield, Tyler Cassens, Efren Sandoval, Francisco Tanudjaja, Magnus Isaksson, William Lee, David Becker, Marc Laurent, James Lu + SEARCH |
| EPI_ISL_1813868 | EXCITE Lab | Andersen lab at Scripps Research | David Pride, Sharon Reed, Chip Schooley, Angela Scioscia, Natasha Martin Cheryl Anderson, Sawyer Farmer, Abigail Schnapper, Helena Tubb, Tommy Valles + SEARCH |
| EPI_ISL_1813869, EPI_ISL_1813870, EPI_ISL_1813871, EPI_ISL_1813872, EPI_ISL_1813873, EPI_ISL_1813874, EPI_ISL_1813875, EPI_ISL_1813876, EPI_ISL_1813877, EPI_ISL_1813878, EPI_ISL_1813879, EPI_ISL_1813880, EPI_ISL_1813881 | | | |
| see above | EXCITE Lab | Andersen lab at Scripps Research | Nicole L Washington, Simon White, Geraint Levan, Kelly Schiabor Barrett, Elizabeth Cirulli, Alexandre Bolze, Charlotte Rivera-Garcia, Ryan Cho, Jason Nguyen, Sherry Wang, Jimmy Ramirez, Celena Andrade, Alice Summerfield, Tyler Cassens, Efren Sandoval, Francisco Tanudjaja, Magnus Isaksson, William Lee, David Becker, Marc Laurent, James Lu + SEARCH |
| EPI_ISL_1813882 | EXCITE Lab | Andersen lab at Scripps Research | Chip Schooley, Natasha Martin, Cheryl Anderson, Angela Scioscia, Smruthi Karthikeyan, Greg Humphrey, Sawyer Farmer, Abigail Schnapper, Helena Tubb, Tommy Valles + SEARCH |
| EPI_ISL_1813883, EPI_ISL_1813884 | EXCITE Lab | Andersen lab at Scripps Research | Nicole L Washington, Simon White, Geraint Levan, Kelly Schiabor Barrett, Elizabeth Cirulli, Alexandre Bolze, Charlotte Rivera-Garcia, Ryan Cho, Jason Nguyen, Sherry Wang, Jimmy Ramirez, Celena Andrade, Alice Summerfield, Tyler Cassens, Efren Sandoval, Francisco Tanudjaja, Magnus Isaksson, William Lee, David Becker, Marc Laurent, James Lu + SEARCH |
| EPI_ISL_1813885 | EXCITE Lab | Andersen lab at Scripps Research | Chip Schooley, Natasha Martin, Cheryl Anderson, Angela Scioscia, Smruthi Karthikeyan, Greg Humphrey, Sawyer Farmer, Abigail Schnapper, Helena Tubb, Tommy Valles + SEARCH |
| EPI_ISL_1813886, EPI_ISL_1813887 | EXCITE Lab | Andersen lab at Scripps Research | Nicole L Washington, Simon White, Geraint Levan, Kelly Schiabor Barrett, Elizabeth Cirulli, Alexandre Bolze, Charlotte Rivera-Garcia, Ryan Cho, Jason Nguyen, Sherry Wang, Jimmy Ramirez, Celena Andrade, Alice Summerfield, Tyler Cassens, Efren Sandoval, Francisco Tanudjaja, Magnus Isaksson, William Lee, David Becker, Marc Laurent, James Lu + SEARCH |
| EPI_ISL_1813888 | EXCITE Lab | Andersen lab at Scripps Research | David Pride, Sharon Reed, Chip Schooley, Angela Scioscia, Natasha Martin Cheryl Anderson, Sawyer Farmer, Abigail Schnapper, Helena Tubb, Tommy Valles + SEARCH |
| EPI_ISL_1813889, EPI_ISL_1813890, EPI_ISL_1813891, EPI_ISL_1813892 | EXCITE Lab | Andersen lab at Scripps Research | Nicole L Washington, Simon White, Geraint Levan, Kelly Schiabor Barrett, Elizabeth Cirulli, Alexandre Bolze, Charlotte Rivera-Garcia, Ryan Cho, Jason Nguyen, Sherry Wang, Jimmy Ramirez, Celena Andrade, Alice Summerfield, Tyler Cassens, Efren Sandoval, Francisco Tanudjaja, Magnus Isaksson, William Lee, David Becker, Marc Laurent, James Lu + SEARCH |
| EPI_ISL_1813893 | EXCITE Lab | Andersen lab at Scripps Research | Chip Schooley, Natasha Martin, Cheryl Anderson, Angela Scioscia, Smruthi Karthikeyan, Greg Humphrey, Sawyer Farmer, Abigail Schnapper, Helena Tubb, Tommy Valles + SEARCH |
| EPI_ISL_1813894, EPI_ISL_1813895, EPI_ISL_1813896, EPI_ISL_1813897, EPI_ISL_1813898 | EXCITE Lab | Andersen lab at Scripps Research | Nicole L Washington, Simon White, Geraint Levan, Kelly Schiabor Barrett, Elizabeth Cirulli, Alexandre Bolze, Charlotte Rivera-Garcia, Ryan Cho, Jason Nguyen, Sherry Wang, Jimmy Ramirez, Celena Andrade, Alice Summerfield, Tyler Cassens, Efren Sandoval, Francisco Tanudjaja, Magnus Isaksson, William Lee, David Becker, Marc Laurent, James Lu + SEARCH |
| EPI_ISL_1813899 | EXCITE Lab | Andersen lab at Scripps Research | David Pride, Sharon Reed, Chip Schooley, Angela Scioscia, Natasha Martin Cheryl Anderson, Sawyer Farmer, Abigail Schnapper, Helena Tubb, Tommy Valles + SEARCH |
| EPI_ISL_1813900, EPI_ISL_1813901, EPI_ISL_1813902, EPI_ISL_1813903, EPI_ISL_1813904, EPI_ISL_1813905, EPI_ISL_1813906, EPI_ISL_1813907, EPI_ISL_1813908, EPI_ISL_1813909, EPI_ISL_1813910, EPI_ISL_1813911, EPI_ISL_1813912, EPI_ISL_1813913, EPI_ISL_1813914, EPI_ISL_1813915, EPI_ISL_1813916, EPI_ISL_1813917 | | | |
| see above | EXCITE Lab | Andersen lab at Scripps Research | Nicole L Washington, Simon White, Geraint Levan, Kelly Schiabor Barrett, Elizabeth Cirulli, Alexandre Bolze, Charlotte Rivera-Garcia, Ryan Cho, Jason Nguyen, Sherry Wang, Jimmy Ramirez, Celena Andrade, Alice Summerfield, Tyler Cassens, Efren Sandoval, Francisco Tanudjaja, Magnus Isaksson, William Lee, David Becker, Marc Laurent, James Lu + SEARCH |
| EPI_ISL_1813918 | EXCITE Lab | Andersen lab at Scripps Research | David Pride, Sharon Reed, Chip Schooley, Angela Scioscia, Natasha Martin Cheryl Anderson, Sawyer Farmer, Abigail Schnapper, Helena Tubb, Tommy Valles + SEARCH |
| EPI_ISL_1813919, EPI_ISL_1813920, EPI_ISL_1813921, EPI_ISL_1813922, EPI_ISL_1813923, EPI_ISL_1813924 | EXCITE Lab | Andersen lab at Scripps Research | Nicole L Washington, Simon White, Geraint Levan, Kelly Schiabor Barrett, Elizabeth Cirulli, Alexandre Bolze, Charlotte Rivera-Garcia, Ryan Cho, Jason Nguyen, Sherry Wang, Jimmy Ramirez, Celena Andrade, Alice Summerfield, Tyler Cassens, Efren Sandoval, Francisco Tanudjaja, Magnus Isaksson, William Lee, David Becker, Marc Laurent, James Lu + SEARCH |
| EPI_ISL_1813925 | EXCITE Lab | Andersen lab at Scripps Research | David Pride, Sharon Reed, Chip Schooley, Angela Scioscia, Natasha Martin Cheryl Anderson, Sawyer Farmer, Abigail Schnapper, Helena Tubb, Tommy Valles + SEARCH |
| EPI_ISL_1813926, EPI_ISL_1813927, EPI_ISL_1813928, EPI_ISL_1813929, EPI_ISL_1813930, EPI_ISL_1813931, EPI_ISL_1813932, EPI_ISL_1813933, EPI_ISL_1813934, EPI_ISL_1813935, EPI_ISL_1813936, EPI_ISL_1813937, EPI_ISL_1813938, EPI_ISL_1813939, EPI_ISL_1813940, EPI_ISL_1813941, EPI_ISL_1813942, EPI_ISL_1813943, EPI_ISL_1813944, EPI_ISL_1813945, EPI_ISL_1813946, EPI_ISL_1813947, EPI_ISL_1813948, EPI_ISL_1813949, EPI_ISL_1813950, EPI_ISL_1813951, EPI_ISL_1813952, EPI_ISL_1813953 | | | |
| see above | EXCITE Lab | Andersen lab at Scripps Research | Nicole L Washington, Simon White, Geraint Levan, Kelly Schiabor Barrett, Elizabeth Cirulli, Alexandre Bolze, Charlotte Rivera-Garcia, Ryan Cho, Jason Nguyen, Sherry Wang, Jimmy Ramirez, Celena Andrade, Alice Summerfield, Tyler Cassens, Efren Sandoval, Francisco Tanudjaja, Magnus Isaksson, William Lee, David Becker, Marc Laurent, James Lu + SEARCH |
| EPI_ISL_1813954 | EXCITE Lab | Andersen lab at Scripps Research | David Pride, Sharon Reed, Chip Schooley, Angela Scioscia, Natasha Martin Cheryl Anderson, Sawyer Farmer, Abigail Schnapper, Helena Tubb, Tommy Valles + SEARCH |
| EPI_ISL_1813955, EPI_ISL_1813956, EPI_ISL_1813957 | EXCITE Lab | Andersen lab at Scripps Research | Nicole L Washington, Simon White, Geraint Levan, Kelly Schiabor Barrett, Elizabeth Cirulli, Alexandre Bolze, Charlotte Rivera-Garcia, Ryan Cho, Jason Nguyen, Sherry Wang, Jimmy Ramirez, Celena Andrade, Alice Summerfield, Tyler Cassens, Efren Sandoval, Francisco Tanudjaja, Magnus Isaksson, William Lee, David Becker, Marc Laurent, James Lu + SEARCH |
| EPI_ISL_1813958 | EXCITE Lab | Andersen lab at Scripps Research | Chip Schooley, Natasha Martin, Cheryl Anderson, Angela Scioscia, Smruthi Karthikeyan, Greg Humphrey, Sawyer Farmer, Abigail Schnapper, Helena Tubb, Tommy Valles + SEARCH |
| EPI_ISL_1813959, EPI_ISL_1813960, EPI_ISL_1813961, EPI_ISL_1813962, EPI_ISL_1813963, EPI_ISL_1813964, EPI_ISL_1813965, EPI_ISL_1813966, EPI_ISL_1813967, EPI_ISL_1813968 | EXCITE Lab | Andersen lab at Scripps Research | Nicole L Washington, Simon White, Geraint Levan, Kelly Schiabor Barrett, Elizabeth Cirulli, Alexandre Bolze, Charlotte Rivera-Garcia, Ryan Cho, Jason Nguyen, Sherry Wang, Jimmy Ramirez, Celena Andrade, Alice Summerfield, Tyler Cassens, Efren Sandoval, Francisco Tanudjaja, Magnus Isaksson, William Lee, David Becker, Marc Laurent, James Lu + SEARCH |
| EPI_ISL_1813969 | EXCITE Lab | Andersen lab at Scripps Research | Rebecca Fielding-Miller, Laura Kohn, Patricia Gonzalez-Zuniga, Fitri Carrissa Wijaya, Vinton Omaleki, Marlene Flores, Araz Majnoonian, Megan Nguyen, Ashkan Hassani, Smruthi Karthikeyan, Rodolfo Salido, Victor Cantu, Greg Humphrey, Sawyer Farmer, Abigail Schnapper, Helena Tubb, Tommy Valles + SEARCH |
| EPI_ISL_1813970, EPI_ISL_1813971 | EXCITE Lab | Andersen lab at Scripps Research | Nicole L Washington, Simon White, Geraint Levan, Kelly Schiabor Barrett, Elizabeth Cirulli, Alexandre Bolze, Charlotte Rivera-Garcia, Ryan Cho, Jason Nguyen, Sherry Wang, Jimmy Ramirez, Celena Andrade, Alice Summerfield, Tyler Cassens, Efren Sandoval, Francisco Tanudjaja, Magnus Isaksson, William Lee, David Becker, Marc Laurent, James Lu + SEARCH |
| EPI_ISL_1813972 | EXCITE Lab | Andersen lab at Scripps Research | J. Joelle Donofrio-Odmann, Brent Brainard+ SEARCH |
| EPI_ISL_1813973, EPI_ISL_1813974 | EXCITE Lab | Andersen lab at Scripps Research | Nicole L Washington, Simon White, Geraint Levan, Kelly Schiabor Barrett, Elizabeth Cirulli, Alexandre Bolze, Charlotte Rivera-Garcia, Ryan Cho, Jason Nguyen, Sherry Wang, Jimmy Ramirez, Celena Andrade, Alice Summerfield, Tyler Cassens, Efren Sandoval, Francisco Tanudjaja, Magnus Isaksson, William Lee, David Becker, Marc Laurent, James Lu + SEARCH |
| EPI_ISL_1813975 | EXCITE Lab | Andersen lab at Scripps Research | David Pride, Sharon Reed, Chip Schooley, Angela Scioscia, Natasha Martin Cheryl Anderson, Sawyer Farmer, Abigail Schnapper, Helena Tubb, Tommy Valles + SEARCH |
| EPI_ISL_1813976, EPI_ISL_1813977, EPI_ISL_1813978, EPI_ISL_1813979, EPI_ISL_1813980, EPI_ISL_1813981, EPI_ISL_1813982, EPI_ISL_1813983, EPI_ISL_1813984, EPI_ISL_1813985, EPI_ISL_1813986, EPI_ISL_1813987, EPI_ISL_1813988, EPI_ISL_1813989, EPI_ISL_1813990, EPI_ISL_1813991, EPI_ISL_1813992, EPI_ISL_1813993, EPI_ISL_1813994, EPI_ISL_1813995, EPI_ISL_1813996, EPI_ISL_1813997 | | | |
| see above | EXCITE Lab | Andersen lab at Scripps Research | Nicole L Washington, Simon White, Geraint Levan, Kelly Schiabor Barrett, Elizabeth Cirulli, Alexandre Bolze, Charlotte Rivera-Garcia, Ryan Cho, Jason Nguyen, Sherry Wang, Jimmy Ramirez, Celena Andrade, Alice Summerfield, Tyler Cassens, Efren Sandoval, Francisco Tanudjaja, Magnus Isaksson, William Lee, David Becker, Marc Laurent, James Lu + SEARCH |
| EPI_ISL_1813998 | EXCITE Lab | Andersen lab at Scripps Research | Chip Schooley, Natasha Martin, Cheryl Anderson, Angela Scioscia, Smruthi Karthikeyan, Greg Humphrey, Sawyer Farmer, Abigail Schnapper, Helena Tubb, Tommy Valles + SEARCH |
| EPI_ISL_1813999, EPI_ISL_1814000, EPI_ISL_1814001, EPI_ISL_1814002, EPI_ISL_1814003, EPI_ISL_1814004, EPI_ISL_1814005, EPI_ISL_1814006, EPI_ISL_1814007, EPI_ISL_1814008, EPI_ISL_1814009, EPI_ISL_1814010, EPI_ISL_1814011 | | | |
| see above | EXCITE Lab | Andersen lab at Scripps Research | Nicole L Washington, Simon White, Geraint Levan, Kelly Schiabor Barrett, Elizabeth Cirulli, Alexandre Bolze, Charlotte Rivera-Garcia, Ryan Cho, Jason Nguyen, Sherry Wang, Jimmy Ramirez, Celena Andrade, Alice Summerfield, Tyler Cassens, Efren Sandoval, Francisco Tanudjaja, Magnus Isaksson, William Lee, David Becker, Marc Laurent, James Lu + SEARCH |
| EPI_ISL_1814012 | EXCITE Lab | Andersen lab at Scripps Research | Chip Schooley, Natasha Martin, Cheryl Anderson, Angela Scioscia, Smruthi Karthikeyan, Greg Humphrey, Sawyer Farmer, Abigail Schnapper, Helena Tubb, Tommy Valles + SEARCH |
| EPI_ISL_1814013, EPI_ISL_1814014, EPI_ISL_1814015, EPI_ISL_1814016, EPI_ISL_1814017, EPI_ISL_1814018, EPI_ISL_1814019, EPI_ISL_1814020, EPI_ISL_1814021, EPI_ISL_1814022, EPI_ISL_1814023, EPI_ISL_1814024, EPI_ISL_1814025 | | | |
| see above | EXCITE Lab | Andersen lab at Scripps Research | Nicole L Washington, Simon White, Geraint Levan, Kelly Schiabor Barrett, Elizabeth Cirulli, Alexandre Bolze, Charlotte Rivera-Garcia, Ryan Cho, Jason Nguyen, Sherry Wang, Jimmy Ramirez, Celena Andrade, Alice Summerfield, Tyler Cassens, Efren Sandoval, Francisco Tanudjaja, Magnus Isaksson, William Lee, David Becker, Marc Laurent, James Lu + SEARCH |
| EPI_ISL_1814026 | EXCITE Lab | Andersen lab at Scripps Research | David Pride, Sharon Reed, Chip Schooley, Angela Scioscia, Natasha Martin Cheryl Anderson, Sawyer Farmer, Abigail Schnapper, Helena Tubb, Tommy Valles + SEARCH |
| EPI_ISL_1814027, EPI_ISL_1814028, EPI_ISL_1814029, EPI_ISL_1814030, EPI_ISL_1814031, EPI_ISL_1814032, EPI_ISL_1814033, EPI_ISL_1814034, EPI_ISL_1814035, EPI_ISL_1814036, EPI_ISL_1814037, EPI_ISL_1814038, EPI_ISL_1814039, EPI_ISL_1814040, EPI_ISL_1814041, EPI_ISL_1814042, EPI_ISL_1814043, EPI_ISL_1814044, EPI_ISL_1814045, EPI_ISL_1814046, EPI_ISL_1814047, EPI_ISL_1814048 | | | |
| see above | EXCITE Lab | Andersen lab at Scripps Research | Nicole L Washington, Simon White, Geraint Levan, Kelly Schiabor Barrett, Elizabeth Cirulli, Alexandre Bolze, Charlotte Rivera-Garcia, Ryan Cho, Jason Nguyen, Sherry Wang, Jimmy Ramirez, Celena Andrade, Alice Summerfield, Tyler Cassens, Efren Sandoval, Francisco Tanudjaja, Magnus Isaksson, William Lee, David Becker, Marc Laurent, James Lu + SEARCH |
| EPI_ISL_1814049 | EXCITE Lab | Andersen lab at Scripps Research | David Pride, Sharon Reed, Chip Schooley, Angela Scioscia, Natasha Martin Cheryl Anderson, Sawyer Farmer, Abigail Schnapper, Helena Tubb, Tommy Valles + SEARCH |

| | | | |
|---|---|---|---|
| EPI_ISL_1814050, EPI_ISL_1814051, EPI_ISL_1814052, EPI_ISL_1814053, EPI_ISL_1814054 | EXCITE Lab | Andersen lab at Scripps Research | Nicole L Washington, Simon White, Geraint Levan, Kelly Schiabor Barrett, Elizabeth Cirulli, Alexandre Bolze, Charlotte Rivera-Garcia, Ryan Cho, Jason Nguyen, Sherry Wang, Jimmy Ramirez, Celena Andrade, Alice Summerfield, Tyler Cassens, Efren Sandoval, Francisco Tanudjaja, Magnus Isaksson, William Lee, David Becker, Marc Laurent, James Lu + SEARCH |
| EPI_ISL_1814055 | EXCITE Lab | Andersen lab at Scripps Research | David Pride, Sharon Reed, Chip Schooley, Angela Scioscia, Natasha Martin Cheryl Anderson, Sawyer Farmer, Abigail Schnapper, Helena Tubb, Tommy Valles + SEARCH |
| EPI_ISL_1814056, EPI_ISL_1814057, EPI_ISL_1814058, EPI_ISL_1814059, EPI_ISL_1814060, EPI_ISL_1814061, EPI_ISL_1814062, EPI_ISL_1814063, EPI_ISL_1814064, EPI_ISL_1814065 | EXCITE Lab | Andersen lab at Scripps Research | Nicole L Washington, Simon White, Geraint Levan, Kelly Schiabor Barrett, Elizabeth Cirulli, Alexandre Bolze, Charlotte Rivera-Garcia, Ryan Cho, Jason Nguyen, Sherry Wang, Jimmy Ramirez, Celena Andrade, Alice Summerfield, Tyler Cassens, Efren Sandoval, Francisco Tanudjaja, Magnus Isaksson, William Lee, David Becker, Marc Laurent, James Lu + SEARCH |
| EPI_ISL_1814066 | EXCITE Lab | Andersen lab at Scripps Research | David Pride, Sharon Reed, Chip Schooley, Angela Scioscia, Natasha Martin Cheryl Anderson, Sawyer Farmer, Abigail Schnapper, Helena Tubb, Tommy Valles + SEARCH |
| EPI_ISL_1814067, EPI_ISL_1814068, EPI_ISL_1814069, EPI_ISL_1814070, EPI_ISL_1814071, EPI_ISL_1814072 | EXCITE Lab | Andersen lab at Scripps Research | Nicole L Washington, Simon White, Geraint Levan, Kelly Schiabor Barrett, Elizabeth Cirulli, Alexandre Bolze, Charlotte Rivera-Garcia, Ryan Cho, Jason Nguyen, Sherry Wang, Jimmy Ramirez, Celena Andrade, Alice Summerfield, Tyler Cassens, Efren Sandoval, Francisco Tanudjaja, Magnus Isaksson, William Lee, David Becker, Marc Laurent, James Lu + SEARCH |
| EPI_ISL_1814073 | EXCITE Lab | Andersen lab at Scripps Research | Chip Schooley, Natasha Martin, Cheryl Anderson, Angela Scioscia, Smruthi Karthikeyan, Greg Humphrey, Sawyer Farmer, Abigail Schnapper, Helena Tubb, Tommy Valles + SEARCH |
| EPI_ISL_1814074, EPI_ISL_1814075, EPI_ISL_1814076, EPI_ISL_1814077, EPI_ISL_1814078, EPI_ISL_1814079, EPI_ISL_1814080, EPI_ISL_1814081, EPI_ISL_1814082, EPI_ISL_1814083, EPI_ISL_1814084, EPI_ISL_1814085, EPI_ISL_1814086, EPI_ISL_1814087, EPI_ISL_1814088, EPI_ISL_1814089, EPI_ISL_1814090, EPI_ISL_1814091, EPI_ISL_1814092, EPI_ISL_1814093, EPI_ISL_1814094, EPI_ISL_1814095, EPI_ISL_1814096 | | | |
| see above | EXCITE Lab | Andersen lab at Scripps Research | Nicole L Washington, Simon White, Geraint Levan, Kelly Schiabor Barrett, Elizabeth Cirulli, Alexandre Bolze, Charlotte Rivera-Garcia, Ryan Cho, Jason Nguyen, Sherry Wang, Jimmy Ramirez, Celena Andrade, Alice Summerfield, Tyler Cassens, Efren Sandoval, Francisco Tanudjaja, Magnus Isaksson, William Lee, David Becker, Marc Laurent, James Lu + SEARCH |
| EPI_ISL_1814097 | EXCITE Lab | Andersen lab at Scripps Research | Rebecca Fielding-Miller, Laura Kohn, Patricia Gonzalez-Zuniga, Fitri Carrissa Wijaya, Vinton Omaleki, Marlene Flores, Araz Majnoonian, Megan Nguyen, Ashkan Hassani, Smruthi Karthikeyan, Rodolfo Salido, Victor Cantu, Greg Humphrey, Sawyer Farmer, Abigail Schnapper, Helena Tubb, Tommy Valles + SEARCH |
| EPI_ISL_1814098, EPI_ISL_1814099, EPI_ISL_1814100, EPI_ISL_1814101, EPI_ISL_1814102, EPI_ISL_1814103, EPI_ISL_1814104, EPI_ISL_1814105, EPI_ISL_1814106, EPI_ISL_1814107 | EXCITE Lab | Andersen lab at Scripps Research | Nicole L Washington, Simon White, Geraint Levan, Kelly Schiabor Barrett, Elizabeth Cirulli, Alexandre Bolze, Charlotte Rivera-Garcia, Ryan Cho, Jason Nguyen, Sherry Wang, Jimmy Ramirez, Celena Andrade, Alice Summerfield, Tyler Cassens, Efren Sandoval, Francisco Tanudjaja, Magnus Isaksson, William Lee, David Becker, Marc Laurent, James Lu + SEARCH |
| EPI_ISL_1814108 | EXCITE Lab | Andersen lab at Scripps Research | J. Joelle Donofrio-Odmann, Brent Brainard+ SEARCH |
| EPI_ISL_1814109, EPI_ISL_1814110, EPI_ISL_1814111, EPI_ISL_1814112, EPI_ISL_1814113 | EXCITE Lab | Andersen lab at Scripps Research | Nicole L Washington, Simon White, Geraint Levan, Kelly Schiabor Barrett, Elizabeth Cirulli, Alexandre Bolze, Charlotte Rivera-Garcia, Ryan Cho, Jason Nguyen, Sherry Wang, Jimmy Ramirez, Celena Andrade, Alice Summerfield, Tyler Cassens, Efren Sandoval, Francisco Tanudjaja, Magnus Isaksson, William Lee, David Becker, Marc Laurent, James Lu + SEARCH |
| EPI_ISL_1814114 | EXCITE Lab | Andersen lab at Scripps Research | David Pride, Sharon Reed, Chip Schooley, Angela Scioscia, Natasha Martin Cheryl Anderson, Sawyer Farmer, Abigail Schnapper, Helena Tubb, Tommy Valles + SEARCH |
| EPI_ISL_1814115, EPI_ISL_1814116, EPI_ISL_1814117, EPI_ISL_1814118, EPI_ISL_1814119, EPI_ISL_1814120, EPI_ISL_1814121, EPI_ISL_1814122, EPI_ISL_1814123, EPI_ISL_1814124, EPI_ISL_1814125, EPI_ISL_1814126, EPI_ISL_1814127, EPI_ISL_1814128, EPI_ISL_1814129, EPI_ISL_1814130, EPI_ISL_1814131, EPI_ISL_1814132, EPI_ISL_1814133, EPI_ISL_1814134, EPI_ISL_1814135, EPI_ISL_1814136, EPI_ISL_1814137, EPI_ISL_1814138, EPI_ISL_1814139, EPI_ISL_1814140, EPI_ISL_1814141, EPI_ISL_1814142, EPI_ISL_1814143, EPI_ISL_1814144, EPI_ISL_1814145, EPI_ISL_1814146, EPI_ISL_1814147, EPI_ISL_1814148, EPI_ISL_1814149, EPI_ISL_1814150, EPI_ISL_1814151 | | | |
| see above | EXCITE Lab | Andersen lab at Scripps Research | Nicole L Washington, Simon White, Geraint Levan, Kelly Schiabor Barrett, Elizabeth Cirulli, Alexandre Bolze, Charlotte Rivera-Garcia, Ryan Cho, Jason Nguyen, Sherry Wang, Jimmy Ramirez, Celena Andrade, Alice Summerfield, Tyler Cassens, Efren Sandoval, Francisco Tanudjaja, Magnus Isaksson, William Lee, David Becker, Marc Laurent, James Lu + SEARCH |
| EPI_ISL_1814152 | EXCITE Lab | Andersen lab at Scripps Research | Chip Schooley, Natasha Martin, Cheryl Anderson, Angela Scioscia, Smruthi Karthikeyan, Greg Humphrey, Sawyer Farmer, Abigail Schnapper, Helena Tubb, Tommy Valles + SEARCH |
| EPI_ISL_1814153, EPI_ISL_1814154, EPI_ISL_1814155, EPI_ISL_1814156, EPI_ISL_1814157, EPI_ISL_1814158, EPI_ISL_1814159, EPI_ISL_1814160, EPI_ISL_1814161, EPI_ISL_1814162, EPI_ISL_1814163, EPI_ISL_1814164, EPI_ISL_1814165, EPI_ISL_1814166, EPI_ISL_1814167, EPI_ISL_1814168, EPI_ISL_1814169, EPI_ISL_1814170, EPI_ISL_1814171, EPI_ISL_1814172, EPI_ISL_1814173, EPI_ISL_1814174, EPI_ISL_1814175, EPI_ISL_1814176, EPI_ISL_1814177, EPI_ISL_1814178 | | | |
| see above | EXCITE Lab | Andersen lab at Scripps Research | Nicole L Washington, Simon White, Geraint Levan, Kelly Schiabor Barrett, Elizabeth Cirulli, Alexandre Bolze, Charlotte Rivera-Garcia, Ryan Cho, Jason Nguyen, Sherry Wang, Jimmy Ramirez, Celena Andrade, Alice Summerfield, Tyler Cassens, Efren Sandoval, Francisco Tanudjaja, Magnus Isaksson, William Lee, David Becker, Marc Laurent, James Lu + SEARCH |
| EPI_ISL_1814179 | EXCITE Lab | Andersen lab at Scripps Research | Rebecca Fielding-Miller, Laura Kohn, Patricia Gonzalez-Zuniga, Fitri Carrissa Wijaya, Vinton Omaleki, Marlene Flores, Araz Majnoonian, Megan Nguyen, Ashkan Hassani, Smruthi Karthikeyan, Rodolfo Salido, Victor Cantu, Greg Humphrey, Sawyer Farmer, Abigail Schnapper, Helena Tubb, Tommy Valles + SEARCH |
| EPI_ISL_1814180, EPI_ISL_1814181, EPI_ISL_1814182, EPI_ISL_1814183, EPI_ISL_1814184, EPI_ISL_1814185, EPI_ISL_1814186, EPI_ISL_1814187, EPI_ISL_1814188 | EXCITE Lab | Andersen lab at Scripps Research | Nicole L Washington, Simon White, Geraint Levan, Kelly Schiabor Barrett, Elizabeth Cirulli, Alexandre Bolze, Charlotte Rivera-Garcia, Ryan Cho, Jason Nguyen, Sherry Wang, Jimmy Ramirez, Celena Andrade, Alice Summerfield, Tyler Cassens, Efren Sandoval, Francisco Tanudjaja, Magnus Isaksson, William Lee, David Becker, Marc Laurent, James Lu + SEARCH |
| EPI_ISL_1814189 | EXCITE Lab | Andersen lab at Scripps Research | Chip Schooley, Natasha Martin, Cheryl Anderson, Angela Scioscia, Smruthi Karthikeyan, Greg Humphrey, Sawyer Farmer, Abigail Schnapper, Helena Tubb, Tommy Valles + SEARCH |
| EPI_ISL_1814190, EPI_ISL_1814191, EPI_ISL_1814192 | EXCITE Lab | Andersen lab at Scripps Research | Nicole L Washington, Simon White, Geraint Levan, Kelly Schiabor Barrett, Elizabeth Cirulli, Alexandre Bolze, Charlotte Rivera-Garcia, Ryan Cho, Jason Nguyen, Sherry Wang, Jimmy Ramirez, Celena Andrade, Alice Summerfield, Tyler Cassens, Efren Sandoval, Francisco Tanudjaja, Magnus Isaksson, William Lee, David Becker, Marc Laurent, James Lu + SEARCH |
| EPI_ISL_1814193 | EXCITE Lab | Andersen lab at Scripps Research | Chip Schooley, Natasha Martin, Cheryl Anderson, Angela Scioscia, Smruthi Karthikeyan, Greg Humphrey, Sawyer Farmer, Abigail Schnapper, Helena Tubb, Tommy Valles + SEARCH |
| EPI_ISL_1814194 | EXCITE Lab | Andersen lab at Scripps Research | Nicole L Washington, Simon White, Geraint Levan, Kelly Schiabor Barrett, Elizabeth Cirulli, Alexandre Bolze, Charlotte Rivera-Garcia, Ryan Cho, Jason Nguyen, Sherry Wang, Jimmy Ramirez, Celena Andrade, Alice Summerfield, Tyler Cassens, Efren Sandoval, Francisco Tanudjaja, Magnus Isaksson, William Lee, David Becker, Marc Laurent, James Lu + SEARCH |
| EPI_ISL_1814195 | EXCITE Lab | Andersen lab at Scripps Research | Chip Schooley, Natasha Martin, Cheryl Anderson, Angela Scioscia, Smruthi Karthikeyan, Greg Humphrey, Sawyer Farmer, Abigail Schnapper, Helena Tubb, Tommy Valles + SEARCH |
| EPI_ISL_1814196, EPI_ISL_1814197, EPI_ISL_1814198, EPI_ISL_1814199, EPI_ISL_1814200, EPI_ISL_1814201, EPI_ISL_1814202, EPI_ISL_1814203, EPI_ISL_1814204, EPI_ISL_1814205, EPI_ISL_1814206, EPI_ISL_1814207, EPI_ISL_1814208 | | | |
| see above | EXCITE Lab | Andersen lab at Scripps Research | Nicole L Washington, Simon White, Geraint Levan, Kelly Schiabor Barrett, Elizabeth Cirulli, Alexandre Bolze, Charlotte Rivera-Garcia, Ryan Cho, Jason Nguyen, Sherry Wang, Jimmy Ramirez, Celena Andrade, Alice Summerfield, Tyler Cassens, Efren Sandoval, Francisco Tanudjaja, Magnus Isaksson, William Lee, David Becker, Marc Laurent, James Lu + SEARCH |
| EPI_ISL_1814209 | EXCITE Lab | Andersen lab at Scripps Research | Chip Schooley, Natasha Martin, Cheryl Anderson, Angela Scioscia, Smruthi Karthikeyan, Greg Humphrey, Sawyer Farmer, Abigail Schnapper, Helena Tubb, Tommy Valles + SEARCH |
| EPI_ISL_1814210, EPI_ISL_1814211, EPI_ISL_1814212, EPI_ISL_1814213, EPI_ISL_1814214, EPI_ISL_1814215, EPI_ISL_1814216, EPI_ISL_1814217, EPI_ISL_1814218, EPI_ISL_1814219, EPI_ISL_1814220, EPI_ISL_1814221, EPI_ISL_1814222, EPI_ISL_1814223, EPI_ISL_1814224, EPI_ISL_1814225, EPI_ISL_1814226, EPI_ISL_1814227, EPI_ISL_1814228, EPI_ISL_1814229, EPI_ISL_1814230, EPI_ISL_1814231, EPI_ISL_1814232, EPI_ISL_1814233, EPI_ISL_1814234, EPI_ISL_1814235, EPI_ISL_1814236 | | | |
| see above | EXCITE Lab | Andersen lab at Scripps Research | Nicole L Washington, Simon White, Geraint Levan, Kelly Schiabor Barrett, Elizabeth Cirulli, Alexandre Bolze, Charlotte Rivera-Garcia, Ryan Cho, Jason Nguyen, Sherry Wang, Jimmy Ramirez, Celena Andrade, Alice Summerfield, Tyler Cassens, Efren Sandoval, Francisco Tanudjaja, Magnus Isaksson, William Lee, David Becker, Marc Laurent, James Lu + SEARCH |
| EPI_ISL_1814237 | EXCITE Lab | Andersen lab at Scripps Research | David Pride, Sharon Reed, Chip Schooley, Angela Scioscia, Natasha Martin Cheryl Anderson, Sawyer Farmer, Abigail Schnapper, Helena Tubb, Tommy Valles + SEARCH |
| EPI_ISL_1814238 | EXCITE Lab | Andersen lab at Scripps Research | Chip Schooley, Natasha Martin, Cheryl Anderson, Angela Scioscia, Smruthi Karthikeyan, Greg Humphrey, Sawyer Farmer, Abigail Schnapper, Helena Tubb, Tommy Valles + SEARCH |
| EPI_ISL_1814239, EPI_ISL_1814240, EPI_ISL_1814241 | EXCITE Lab | Andersen lab at Scripps Research | Nicole L Washington, Simon White, Geraint Levan, Kelly Schiabor Barrett, Elizabeth Cirulli, Alexandre Bolze, Charlotte Rivera-Garcia, Ryan Cho, Jason Nguyen, Sherry Wang, Jimmy Ramirez, Celena Andrade, Alice Summerfield, Tyler Cassens, Efren Sandoval, Francisco Tanudjaja, Magnus Isaksson, William Lee, David Becker, Marc Laurent, James Lu + SEARCH |
| EPI_ISL_1814242 | EXCITE Lab | Andersen lab at Scripps Research | David Pride, Sharon Reed, Chip Schooley, Angela Scioscia, Natasha Martin Cheryl Anderson, Sawyer Farmer, Abigail Schnapper, Helena Tubb, Tommy Valles + SEARCH |
| EPI_ISL_1814243, EPI_ISL_1814244, EPI_ISL_1814245, EPI_ISL_1814246, EPI_ISL_1814247, EPI_ISL_1814248, EPI_ISL_1814249, EPI_ISL_1814250, EPI_ISL_1814251, EPI_ISL_1814252, EPI_ISL_1814253, EPI_ISL_1814254, EPI_ISL_1814255, EPI_ISL_1814256, EPI_ISL_1814257, EPI_ISL_1814258, EPI_ISL_1814259, EPI_ISL_1814260, EPI_ISL_1814261, EPI_ISL_1814262, EPI_ISL_1814263, EPI_ISL_1814264, EPI_ISL_1814265, EPI_ISL_1814266, EPI_ISL_1814267, EPI_ISL_1814268, EPI_ISL_1814269, EPI_ISL_1814270, EPI_ISL_1814271 | | | |
| see above | EXCITE Lab | Andersen lab at Scripps Research | Nicole L Washington, Simon White, Geraint Levan, Kelly Schiabor Barrett, Elizabeth Cirulli, Alexandre Bolze, Charlotte Rivera-Garcia, Ryan Cho, Jason Nguyen, Sherry Wang, Jimmy Ramirez, Celena Andrade, Alice Summerfield, Tyler Cassens, Efren Sandoval, Francisco Tanudjaja, Magnus Isaksson, William Lee, David Becker, Marc Laurent, James Lu + SEARCH |
| EPI_ISL_1814272 | EXCITE Lab | Andersen lab at Scripps Research | David Pride, Sharon Reed, Chip Schooley, Angela Scioscia, Natasha Martin Cheryl Anderson, Sawyer Farmer, Abigail Schnapper, Helena Tubb, Tommy Valles + SEARCH |

| | | | |
|---|---|---|---|
| EPI_ISL_1814273, EPI_ISL_1814274, EPI_ISL_1814275, EPI_ISL_1814276, EPI_ISL_1814277, EPI_ISL_1814278, EPI_ISL_1814279 | EXCITE Lab | Andersen lab at Scripps Research | Nicole L Washington, Simon White, Geraint Levan, Kelly Schiabor Barrett, Elizabeth Cirulli, Alexandre Bolze, Charlotte Rivera-Garcia, Ryan Cho, Jason Nguyen, Sherry Wang, Jimmy Ramirez, Celena Andrade, Alice Summerfield, Tyler Cassens, Efren Sandoval, Francisco Tanudjaja, Magnus Isaksson, William Lee, David Becker, Marc Laurent, James Lu + SEARCH |
| EPI_ISL_1814280 | EXCITE Lab | Andersen lab at Scripps Research | David Pride, Sharon Reed, Chip Schooley, Angela Scioscia, Natasha Martin Cheryl Anderson, Sawyer Farmer, Abigail Schnapper, Helena Tubb, Tommy Valles + SEARCH |
| EPI_ISL_1814281, EPI_ISL_1814282, EPI_ISL_1814283, EPI_ISL_1814284 | EXCITE Lab | Andersen lab at Scripps Research | Nicole L Washington, Simon White, Geraint Levan, Kelly Schiabor Barrett, Elizabeth Cirulli, Alexandre Bolze, Charlotte Rivera-Garcia, Ryan Cho, Jason Nguyen, Sherry Wang, Jimmy Ramirez, Celena Andrade, Alice Summerfield, Tyler Cassens, Efren Sandoval, Francisco Tanudjaja, Magnus Isaksson, William Lee, David Becker, Marc Laurent, James Lu + SEARCH |
| EPI_ISL_1814285, EPI_ISL_1814286 | EXCITE Lab | Andersen lab at Scripps Research | Chip Schooley, Natasha Martin, Cheryl Anderson, Angela Scioscia, Smruthi Karthikeyan, Greg Humphrey, Sawyer Farmer, Abigail Schnapper, Helena Tubb, Tommy Valles + SEARCH |
| EPI_ISL_1814287, EPI_ISL_1814288, EPI_ISL_1814289, EPI_ISL_1814290, EPI_ISL_1814291 | EXCITE Lab | Andersen lab at Scripps Research | Nicole L Washington, Simon White, Geraint Levan, Kelly Schiabor Barrett, Elizabeth Cirulli, Alexandre Bolze, Charlotte Rivera-Garcia, Ryan Cho, Jason Nguyen, Sherry Wang, Jimmy Ramirez, Celena Andrade, Alice Summerfield, Tyler Cassens, Efren Sandoval, Francisco Tanudjaja, Magnus Isaksson, William Lee, David Becker, Marc Laurent, James Lu + SEARCH |
| EPI_ISL_1814292 | EXCITE Lab | Andersen lab at Scripps Research | Chip Schooley, Natasha Martin, Cheryl Anderson, Angela Scioscia, Smruthi Karthikeyan, Greg Humphrey, Sawyer Farmer, Abigail Schnapper, Helena Tubb, Tommy Valles + SEARCH |
| EPI_ISL_1814293, EPI_ISL_1814294, EPI_ISL_1814295, EPI_ISL_1814296, EPI_ISL_1814297, EPI_ISL_1814298, EPI_ISL_1814299, EPI_ISL_1814300, EPI_ISL_1814301, EPI_ISL_1814302, EPI_ISL_1814303 | | | |
| see above | EXCITE Lab | Andersen lab at Scripps Research | Nicole L Washington, Simon White, Geraint Levan, Kelly Schiabor Barrett, Elizabeth Cirulli, Alexandre Bolze, Charlotte Rivera-Garcia, Ryan Cho, Jason Nguyen, Sherry Wang, Jimmy Ramirez, Celena Andrade, Alice Summerfield, Tyler Cassens, Efren Sandoval, Francisco Tanudjaja, Magnus Isaksson, William Lee, David Becker, Marc Laurent, James Lu + SEARCH |
| EPI_ISL_1814304 | EXCITE Lab | Andersen lab at Scripps Research | Sydney Morgan + Marni Jacobs + Celestine Magallanes + Holly Valentine + Louise C. Laurent + SEARCH |
| EPI_ISL_1814305, EPI_ISL_1814306, EPI_ISL_1814307, EPI_ISL_1814308, EPI_ISL_1814309, EPI_ISL_1814310, EPI_ISL_1814311, EPI_ISL_1814312, EPI_ISL_1814313, EPI_ISL_1814314, EPI_ISL_1814315, EPI_ISL_1814316, EPI_ISL_1814317, EPI_ISL_1814318, EPI_ISL_1814319, EPI_ISL_1814320, EPI_ISL_1814321, EPI_ISL_1814322, EPI_ISL_1814323, EPI_ISL_1814324 | | | |
| see above | EXCITE Lab | Andersen lab at Scripps Research | Nicole L Washington, Simon White, Geraint Levan, Kelly Schiabor Barrett, Elizabeth Cirulli, Alexandre Bolze, Charlotte Rivera-Garcia, Ryan Cho, Jason Nguyen, Sherry Wang, Jimmy Ramirez, Celena Andrade, Alice Summerfield, Tyler Cassens, Efren Sandoval, Francisco Tanudjaja, Magnus Isaksson, William Lee, David Becker, Marc Laurent, James Lu + SEARCH |
| EPI_ISL_1814325 | EXCITE Lab | Andersen lab at Scripps Research | David Pride, Sharon Reed, Chip Schooley, Angela Scioscia, Natasha Martin Cheryl Anderson, Sawyer Farmer, Abigail Schnapper, Helena Tubb, Tommy Valles + SEARCH |
| EPI_ISL_1814326, EPI_ISL_1814327, EPI_ISL_1814328, EPI_ISL_1814329, EPI_ISL_1814330, EPI_ISL_1814331, EPI_ISL_1814332, EPI_ISL_1814333, EPI_ISL_1814334, EPI_ISL_1814335, EPI_ISL_1814336, EPI_ISL_1814337, EPI_ISL_1814338, EPI_ISL_1814339, EPI_ISL_1814340, EPI_ISL_1814341, EPI_ISL_1814342, EPI_ISL_1814343, EPI_ISL_1814344, EPI_ISL_1814345, EPI_ISL_1814346 | | | |
| see above | EXCITE Lab | Andersen lab at Scripps Research | Nicole L Washington, Simon White, Geraint Levan, Kelly Schiabor Barrett, Elizabeth Cirulli, Alexandre Bolze, Charlotte Rivera-Garcia, Ryan Cho, Jason Nguyen, Sherry Wang, Jimmy Ramirez, Celena Andrade, Alice Summerfield, Tyler Cassens, Efren Sandoval, Francisco Tanudjaja, Magnus Isaksson, William Lee, David Becker, Marc Laurent, James Lu + SEARCH |
| EPI_ISL_1814347 | EXCITE Lab | Andersen lab at Scripps Research | David Pride, Sharon Reed, Chip Schooley, Angela Scioscia, Natasha Martin Cheryl Anderson, Sawyer Farmer, Abigail Schnapper, Helena Tubb, Tommy Valles + SEARCH |
| EPI_ISL_1814348 | EXCITE Lab | Andersen lab at Scripps Research | Sydney Morgan + Marni Jacobs + Celestine Magallanes + Holly Valentine + Louise C. Laurent + SEARCH |
| EPI_ISL_1814349, EPI_ISL_1814350, EPI_ISL_1814351, EPI_ISL_1814352, EPI_ISL_1814353, EPI_ISL_1814354, EPI_ISL_1814355, EPI_ISL_1814356, EPI_ISL_1814357, EPI_ISL_1814358, EPI_ISL_1814359, EPI_ISL_1814360, EPI_ISL_1814361, EPI_ISL_1814362, EPI_ISL_1814363, EPI_ISL_1814364, EPI_ISL_1814365, EPI_ISL_1814366 | | | |
| see above | EXCITE Lab | Andersen lab at Scripps Research | Nicole L Washington, Simon White, Geraint Levan, Kelly Schiabor Barrett, Elizabeth Cirulli, Alexandre Bolze, Charlotte Rivera-Garcia, Ryan Cho, Jason Nguyen, Sherry Wang, Jimmy Ramirez, Celena Andrade, Alice Summerfield, Tyler Cassens, Efren Sandoval, Francisco Tanudjaja, Magnus Isaksson, William Lee, David Becker, Marc Laurent, James Lu + SEARCH |
| EPI_ISL_1814367 | EXCITE Lab | Andersen lab at Scripps Research | Chip Schooley, Natasha Martin, Cheryl Anderson, Angela Scioscia, Smruthi Karthikeyan, Greg Humphrey, Sawyer Farmer, Abigail Schnapper, Helena Tubb, Tommy Valles + SEARCH |
| EPI_ISL_1814368, EPI_ISL_1814369, EPI_ISL_1814370, EPI_ISL_1814371, EPI_ISL_1814372, EPI_ISL_1814373, EPI_ISL_1814374, EPI_ISL_1814375, EPI_ISL_1814376, EPI_ISL_1814377, EPI_ISL_1814378, EPI_ISL_1814379, EPI_ISL_1814380, EPI_ISL_1814381, EPI_ISL_1814382, EPI_ISL_1814383, EPI_ISL_1814384, EPI_ISL_1814385, EPI_ISL_1814386, EPI_ISL_1814387, EPI_ISL_1814388, EPI_ISL_1814389, EPI_ISL_1814390, EPI_ISL_1814391, EPI_ISL_1814392, EPI_ISL_1814393, EPI_ISL_1814394, EPI_ISL_1814395 | | | |
| see above | EXCITE Lab | Andersen lab at Scripps Research | Nicole L Washington, Simon White, Geraint Levan, Kelly Schiabor Barrett, Elizabeth Cirulli, Alexandre Bolze, Charlotte Rivera-Garcia, Ryan Cho, Jason Nguyen, Sherry Wang, Jimmy Ramirez, Celena Andrade, Alice Summerfield, Tyler Cassens, Efren Sandoval, Francisco Tanudjaja, Magnus Isaksson, William Lee, David Becker, Marc Laurent, James Lu + SEARCH |
| EPI_ISL_1814396 | EXCITE Lab | Andersen lab at Scripps Research | Chip Schooley, Natasha Martin, Cheryl Anderson, Angela Scioscia, Smruthi Karthikeyan, Greg Humphrey, Sawyer Farmer, Abigail Schnapper, Helena Tubb, Tommy Valles + SEARCH |
| EPI_ISL_1814397, EPI_ISL_1814398, EPI_ISL_1814399, EPI_ISL_1814400, EPI_ISL_1814401, EPI_ISL_1814402 | EXCITE Lab | Andersen lab at Scripps Research | Nicole L Washington, Simon White, Geraint Levan, Kelly Schiabor Barrett, Elizabeth Cirulli, Alexandre Bolze, Charlotte Rivera-Garcia, Ryan Cho, Jason Nguyen, Sherry Wang, Jimmy Ramirez, Celena Andrade, Alice Summerfield, Tyler Cassens, Efren Sandoval, Francisco Tanudjaja, Magnus Isaksson, William Lee, David Becker, Marc Laurent, James Lu + SEARCH |
| EPI_ISL_1814403 | EXCITE Lab | Andersen lab at Scripps Research | Chip Schooley, Natasha Martin, Cheryl Anderson, Angela Scioscia, Smruthi Karthikeyan, Greg Humphrey, Sawyer Farmer, Abigail Schnapper, Helena Tubb, Tommy Valles + SEARCH |
| EPI_ISL_1814404, EPI_ISL_1814405, EPI_ISL_1814406, EPI_ISL_1814407, EPI_ISL_1814408, EPI_ISL_1814409, EPI_ISL_1814410, EPI_ISL_1814411 | EXCITE Lab | Andersen lab at Scripps Research | Nicole L Washington, Simon White, Geraint Levan, Kelly Schiabor Barrett, Elizabeth Cirulli, Alexandre Bolze, Charlotte Rivera-Garcia, Ryan Cho, Jason Nguyen, Sherry Wang, Jimmy Ramirez, Celena Andrade, Alice Summerfield, Tyler Cassens, Efren Sandoval, Francisco Tanudjaja, Magnus Isaksson, William Lee, David Becker, Marc Laurent, James Lu + SEARCH |
| EPI_ISL_1814412 | EXCITE Lab | Andersen lab at Scripps Research | Chip Schooley, Natasha Martin, Cheryl Anderson, Angela Scioscia, Smruthi Karthikeyan, Greg Humphrey, Sawyer Farmer, Abigail Schnapper, Helena Tubb, Tommy Valles + SEARCH |
| EPI_ISL_1814413, EPI_ISL_1814414, EPI_ISL_1814415, EPI_ISL_1814416 | EXCITE Lab | Andersen lab at Scripps Research | Nicole L Washington, Simon White, Geraint Levan, Kelly Schiabor Barrett, Elizabeth Cirulli, Alexandre Bolze, Charlotte Rivera-Garcia, Ryan Cho, Jason Nguyen, Sherry Wang, Jimmy Ramirez, Celena Andrade, Alice Summerfield, Tyler Cassens, Efren Sandoval, Francisco Tanudjaja, Magnus Isaksson, William Lee, David Becker, Marc Laurent, James Lu + SEARCH |
| EPI_ISL_1814417 | EXCITE Lab | Andersen lab at Scripps Research | Chip Schooley, Natasha Martin, Cheryl Anderson, Angela Scioscia, Smruthi Karthikeyan, Greg Humphrey, Sawyer Farmer, Abigail Schnapper, Helena Tubb, Tommy Valles + SEARCH |
| EPI_ISL_1814418, EPI_ISL_1814419, EPI_ISL_1814420, EPI_ISL_1814421, EPI_ISL_1814422, EPI_ISL_1814423, EPI_ISL_1814424, EPI_ISL_1814425, EPI_ISL_1814426, EPI_ISL_1814427, EPI_ISL_1814428, EPI_ISL_1814429, EPI_ISL_1814430, EPI_ISL_1814431, EPI_ISL_1814432, EPI_ISL_1814433, EPI_ISL_1814434, EPI_ISL_1814435, EPI_ISL_1814436, EPI_ISL_1814437, EPI_ISL_1814438, EPI_ISL_1814439, EPI_ISL_1814440, EPI_ISL_1814441, EPI_ISL_1814442, EPI_ISL_1814443, EPI_ISL_1814444, EPI_ISL_1814445, EPI_ISL_1814446, EPI_ISL_1814447, EPI_ISL_1814448, EPI_ISL_1814449, EPI_ISL_1814450, EPI_ISL_1814451 | | | |
| see above | EXCITE Lab | Andersen lab at Scripps Research | Nicole L Washington, Simon White, Geraint Levan, Kelly Schiabor Barrett, Elizabeth Cirulli, Alexandre Bolze, Charlotte Rivera-Garcia, Ryan Cho, Jason Nguyen, Sherry Wang, Jimmy Ramirez, Celena Andrade, Alice Summerfield, Tyler Cassens, Efren Sandoval, Francisco Tanudjaja, Magnus Isaksson, William Lee, David Becker, Marc Laurent, James Lu + SEARCH |
| EPI_ISL_1814452 | EXCITE Lab | Andersen lab at Scripps Research | Chip Schooley, Natasha Martin, Cheryl Anderson, Angela Scioscia, Smruthi Karthikeyan, Greg Humphrey, Sawyer Farmer, Abigail Schnapper, Helena Tubb, Tommy Valles + SEARCH |
| EPI_ISL_1814453, EPI_ISL_1814454, EPI_ISL_1814455, EPI_ISL_1814456, EPI_ISL_1814457, EPI_ISL_1814458, EPI_ISL_1814459 | EXCITE Lab | Andersen lab at Scripps Research | Nicole L Washington, Simon White, Geraint Levan, Kelly Schiabor Barrett, Elizabeth Cirulli, Alexandre Bolze, Charlotte Rivera-Garcia, Ryan Cho, Jason Nguyen, Sherry Wang, Jimmy Ramirez, Celena Andrade, Alice Summerfield, Tyler Cassens, Efren Sandoval, Francisco Tanudjaja, Magnus Isaksson, William Lee, David Becker, Marc Laurent, James Lu + SEARCH |
| EPI_ISL_1814460 | EXCITE Lab | Andersen lab at Scripps Research | David Pride, Sharon Reed, Chip Schooley, Angela Scioscia, Natasha Martin Cheryl Anderson, Sawyer Farmer, Abigail Schnapper, Helena Tubb, Tommy Valles + SEARCH |
| EPI_ISL_1814461, EPI_ISL_1814462, EPI_ISL_1814463 | EXCITE Lab | Andersen lab at Scripps Research | Nicole L Washington, Simon White, Geraint Levan, Kelly Schiabor Barrett, Elizabeth Cirulli, Alexandre Bolze, Charlotte Rivera-Garcia, Ryan Cho, Jason Nguyen, Sherry Wang, Jimmy Ramirez, Celena Andrade, Alice Summerfield, Tyler Cassens, Efren Sandoval, Francisco Tanudjaja, Magnus Isaksson, William Lee, David Becker, Marc Laurent, James Lu + SEARCH |
| EPI_ISL_1814464 | EXCITE Lab | Andersen lab at Scripps Research | Chip Schooley, Natasha Martin, Cheryl Anderson, Angela Scioscia, Smruthi Karthikeyan, Greg Humphrey, Sawyer Farmer, Abigail Schnapper, Helena Tubb, Tommy Valles + SEARCH |
| EPI_ISL_1814465, EPI_ISL_1814466, EPI_ISL_1814467, EPI_ISL_1814468, EPI_ISL_1814469, EPI_ISL_1814470, EPI_ISL_1814471, EPI_ISL_1814472, EPI_ISL_1814473, EPI_ISL_1814474, EPI_ISL_1814475, EPI_ISL_1814476, EPI_ISL_1814477, EPI_ISL_1814478, EPI_ISL_1814479, EPI_ISL_1814480, EPI_ISL_1814481, EPI_ISL_1814482, EPI_ISL_1814483, EPI_ISL_1814484, EPI_ISL_1814485 | | | |
| see above | EXCITE Lab | Andersen lab at Scripps Research | Nicole L Washington, Simon White, Geraint Levan, Kelly Schiabor Barrett, Elizabeth Cirulli, Alexandre Bolze, Charlotte Rivera-Garcia, Ryan Cho, Jason Nguyen, Sherry Wang, Jimmy Ramirez, Celena Andrade, Alice Summerfield, Tyler Cassens, Efren Sandoval, Francisco Tanudjaja, Magnus Isaksson, William Lee, David Becker, Marc Laurent, James Lu + SEARCH |
| EPI_ISL_1814486 | EXCITE Lab | Andersen lab at Scripps Research | Chip Schooley, Natasha Martin, Cheryl Anderson, Angela Scioscia, Smruthi Karthikeyan, Greg Humphrey, Sawyer Farmer, Abigail Schnapper, Helena Tubb, Tommy Valles + SEARCH |
| EPI_ISL_1814487, EPI_ISL_1814488 | EXCITE Lab | Andersen lab at Scripps Research | Nicole L Washington, Simon White, Geraint Levan, Kelly Schiabor Barrett, Elizabeth Cirulli, Alexandre Bolze, Charlotte Rivera-Garcia, Ryan Cho, Jason Nguyen, Sherry Wang, Jimmy Ramirez, Celena Andrade, Alice Summerfield, Tyler Cassens, Efren Sandoval, Francisco Tanudjaja, Magnus Isaksson, William Lee, David Becker, Marc Laurent, James Lu + SEARCH |
| EPI_ISL_1814489 | EXCITE Lab | Andersen lab at Scripps Research | Rebecca Fielding-Miller, Laura Kohn, Patricia Gonzalez-Zuniga, Fitri Carrissa Wijaya, Vinton Omaleki, Marlene Flores, Araz Majnoonian, Megan Nguyen, Ashkan Hassani, Smruthi Karthikeyan, Rodolfo Salido, Victor Cantu, Greg Humphrey, Sawyer Farmer, Abigail Schnapper, Helena Tubb, Tommy Valles + SEARCH |
| EPI_ISL_1814490, EPI_ISL_1814491, EPI_ISL_1814492, EPI_ISL_1814493, | EXCITE Lab | Andersen lab at Scripps Research | Nicole L Washington, Simon White, Geraint Levan, Kelly Schiabor Barrett, Elizabeth Cirulli, Alexandre Bolze, Charlotte Rivera-Garcia, Ryan Cho, Jason Nguyen, Sherry Wang, Jimmy Ramirez, Celena Andrade, Alice Summerfield, Tyler Cassens, Efren Sandoval, Francisco Tanudjaja, Magnus Isaksson, William Lee, David Becker, Marc Laurent, James Lu + SEARCH |

| | | | |
|---|---|---|---|
| EPI_ISL_1814494, EPI_ISL_1814495 | | | |
| EPI_ISL_1814496 | EXCITE Lab | Andersen lab at Scripps Research | David Pride, Sharon Reed, Chip Schooley, Angela Scioscia, Natasha Martin Cheryl Anderson, Sawyer Farmer, Abigail Schnapper, Helena Tubb, Tommy Valles + SEARCH |
| EPI_ISL_1814497, EPI_ISL_1814498, EPI_ISL_1814499, EPI_ISL_1814500 | EXCITE Lab | Andersen lab at Scripps Research | Nicole L Washington, Simon White, Geraint Levan, Kelly Schiabor Barrett, Elizabeth Cirulli, Alexandre Bolze, Charlotte Rivera-Garcia, Ryan Cho, Jason Nguyen, Sherry Wang, Jimmy Ramirez, Celena Andrade, Alice Summerfield, Tyler Cassens, Efren Sandoval, Francisco Tanudjaja, Magnus Isaksson, William Lee, David Becker, Marc Laurent, James Lu + SEARCH |
| EPI_ISL_1814501 | EXCITE Lab | Andersen lab at Scripps Research | Chip Schooley, Natasha Martin, Cheryl Anderson, Angela Scioscia, Smruthi Karthikeyan, Greg Humphrey, Sawyer Farmer, Abigail Schnapper, Helena Tubb, Tommy Valles + SEARCH |
| EPI_ISL_1814502 | EXCITE Lab | Andersen lab at Scripps Research | Nicole L Washington, Simon White, Geraint Levan, Kelly Schiabor Barrett, Elizabeth Cirulli, Alexandre Bolze, Charlotte Rivera-Garcia, Ryan Cho, Jason Nguyen, Sherry Wang, Jimmy Ramirez, Celena Andrade, Alice Summerfield, Tyler Cassens, Efren Sandoval, Francisco Tanudjaja, Magnus Isaksson, William Lee, David Becker, Marc Laurent, James Lu + SEARCH |
| EPI_ISL_1814503 | EXCITE Lab | Andersen lab at Scripps Research | Chip Schooley, Natasha Martin, Cheryl Anderson, Angela Scioscia, Smruthi Karthikeyan, Greg Humphrey, Sawyer Farmer, Abigail Schnapper, Helena Tubb, Tommy Valles + SEARCH |
| EPI_ISL_1814504, EPI_ISL_1814505, EPI_ISL_1814506, EPI_ISL_1814507 | EXCITE Lab | Andersen lab at Scripps Research | Nicole L Washington, Simon White, Geraint Levan, Kelly Schiabor Barrett, Elizabeth Cirulli, Alexandre Bolze, Charlotte Rivera-Garcia, Ryan Cho, Jason Nguyen, Sherry Wang, Jimmy Ramirez, Celena Andrade, Alice Summerfield, Tyler Cassens, Efren Sandoval, Francisco Tanudjaja, Magnus Isaksson, William Lee, David Becker, Marc Laurent, James Lu + SEARCH |
| EPI_ISL_1814508 | EXCITE Lab | Andersen lab at Scripps Research | Fred Wu, Christine Mitchell + SEARCH |
| EPI_ISL_1814509, EPI_ISL_1814510, EPI_ISL_1814511, EPI_ISL_1814512 | EXCITE Lab | Andersen lab at Scripps Research | Nicole L Washington, Simon White, Geraint Levan, Kelly Schiabor Barrett, Elizabeth Cirulli, Alexandre Bolze, Charlotte Rivera-Garcia, Ryan Cho, Jason Nguyen, Sherry Wang, Jimmy Ramirez, Celena Andrade, Alice Summerfield, Tyler Cassens, Efren Sandoval, Francisco Tanudjaja, Magnus Isaksson, William Lee, David Becker, Marc Laurent, James Lu + SEARCH |
| EPI_ISL_1814513 | EXCITE Lab | Andersen lab at Scripps Research | Chip Schooley, Natasha Martin, Cheryl Anderson, Angela Scioscia, Smruthi Karthikeyan, Greg Humphrey, Sawyer Farmer, Abigail Schnapper, Helena Tubb, Tommy Valles + SEARCH |
| EPI_ISL_1814514, EPI_ISL_1814515, EPI_ISL_1814516, EPI_ISL_1814517, EPI_ISL_1814518, EPI_ISL_1814519, EPI_ISL_1814520, EPI_ISL_1814521, EPI_ISL_1814522, EPI_ISL_1814523, EPI_ISL_1814524, EPI_ISL_1814525, EPI_ISL_1814526, EPI_ISL_1814527 | | | |
| see above | EXCITE Lab | Andersen lab at Scripps Research | Nicole L Washington, Simon White, Geraint Levan, Kelly Schiabor Barrett, Elizabeth Cirulli, Alexandre Bolze, Charlotte Rivera-Garcia, Ryan Cho, Jason Nguyen, Sherry Wang, Jimmy Ramirez, Celena Andrade, Alice Summerfield, Tyler Cassens, Efren Sandoval, Francisco Tanudjaja, Magnus Isaksson, William Lee, David Becker, Marc Laurent, James Lu + SEARCH |
| EPI_ISL_1814528, EPI_ISL_1814529 | EXCITE Lab | Andersen lab at Scripps Research | Chip Schooley, Natasha Martin, Cheryl Anderson, Angela Scioscia, Smruthi Karthikeyan, Greg Humphrey, Sawyer Farmer, Abigail Schnapper, Helena Tubb, Tommy Valles + SEARCH |
| EPI_ISL_1814530, EPI_ISL_1814531, EPI_ISL_1814532, EPI_ISL_1814533, EPI_ISL_1814534, EPI_ISL_1814535, EPI_ISL_1814536, EPI_ISL_1814537, EPI_ISL_1814538 | EXCITE Lab | Andersen lab at Scripps Research | Nicole L Washington, Simon White, Geraint Levan, Kelly Schiabor Barrett, Elizabeth Cirulli, Alexandre Bolze, Charlotte Rivera-Garcia, Ryan Cho, Jason Nguyen, Sherry Wang, Jimmy Ramirez, Celena Andrade, Alice Summerfield, Tyler Cassens, Efren Sandoval, Francisco Tanudjaja, Magnus Isaksson, William Lee, David Becker, Marc Laurent, James Lu + SEARCH |
| EPI_ISL_1814539 | EXCITE Lab | Andersen lab at Scripps Research | David Pride, Sharon Reed, Chip Schooley, Angela Scioscia, Natasha Martin Cheryl Anderson, Sawyer Farmer, Abigail Schnapper, Helena Tubb, Tommy Valles + SEARCH |
| EPI_ISL_1814540 | EXCITE Lab | Andersen lab at Scripps Research | Nicole L Washington, Simon White, Geraint Levan, Kelly Schiabor Barrett, Elizabeth Cirulli, Alexandre Bolze, Charlotte Rivera-Garcia, Ryan Cho, Jason Nguyen, Sherry Wang, Jimmy Ramirez, Celena Andrade, Alice Summerfield, Tyler Cassens, Efren Sandoval, Francisco Tanudjaja, Magnus Isaksson, William Lee, David Becker, Marc Laurent, James Lu + SEARCH |
| EPI_ISL_1814541 | EXCITE Lab | Andersen lab at Scripps Research | Fred Wu, Christine Mitchell + SEARCH |
| EPI_ISL_1814542 | EXCITE Lab | Andersen lab at Scripps Research | Chip Schooley, Natasha Martin, Cheryl Anderson, Angela Scioscia, Smruthi Karthikeyan, Greg Humphrey, Sawyer Farmer, Abigail Schnapper, Helena Tubb, Tommy Valles + SEARCH |
| EPI_ISL_1814543, EPI_ISL_1814544, EPI_ISL_1814545, EPI_ISL_1814546, EPI_ISL_1814547, EPI_ISL_1814548, EPI_ISL_1814549 | EXCITE Lab | Andersen lab at Scripps Research | Nicole L Washington, Simon White, Geraint Levan, Kelly Schiabor Barrett, Elizabeth Cirulli, Alexandre Bolze, Charlotte Rivera-Garcia, Ryan Cho, Jason Nguyen, Sherry Wang, Jimmy Ramirez, Celena Andrade, Alice Summerfield, Tyler Cassens, Efren Sandoval, Francisco Tanudjaja, Magnus Isaksson, William Lee, David Becker, Marc Laurent, James Lu + SEARCH |
| EPI_ISL_1814550 | EXCITE Lab | Andersen lab at Scripps Research | David Pride, Sharon Reed, Chip Schooley, Angela Scioscia, Natasha Martin Cheryl Anderson, Sawyer Farmer, Abigail Schnapper, Helena Tubb, Tommy Valles + SEARCH |
| EPI_ISL_1814551, EPI_ISL_1814552, EPI_ISL_1814553 | EXCITE Lab | Andersen lab at Scripps Research | Nicole L Washington, Simon White, Geraint Levan, Kelly Schiabor Barrett, Elizabeth Cirulli, Alexandre Bolze, Charlotte Rivera-Garcia, Ryan Cho, Jason Nguyen, Sherry Wang, Jimmy Ramirez, Celena Andrade, Alice Summerfield, Tyler Cassens, Efren Sandoval, Francisco Tanudjaja, Magnus Isaksson, William Lee, David Becker, Marc Laurent, James Lu + SEARCH |
| EPI_ISL_1814554 | EXCITE Lab | Andersen lab at Scripps Research | J. Joelle Donofrio-Odmann, Brent Brainard+ SEARCH |
| EPI_ISL_1814555, EPI_ISL_1814556, EPI_ISL_1814557, EPI_ISL_1814558, EPI_ISL_1814559, EPI_ISL_1814560 | EXCITE Lab | Andersen lab at Scripps Research | Nicole L Washington, Simon White, Geraint Levan, Kelly Schiabor Barrett, Elizabeth Cirulli, Alexandre Bolze, Charlotte Rivera-Garcia, Ryan Cho, Jason Nguyen, Sherry Wang, Jimmy Ramirez, Celena Andrade, Alice Summerfield, Tyler Cassens, Efren Sandoval, Francisco Tanudjaja, Magnus Isaksson, William Lee, David Becker, Marc Laurent, James Lu + SEARCH |
| EPI_ISL_1814561 | EXCITE Lab | Andersen lab at Scripps Research | Chip Schooley, Natasha Martin, Cheryl Anderson, Angela Scioscia, Smruthi Karthikeyan, Greg Humphrey, Sawyer Farmer, Abigail Schnapper, Helena Tubb, Tommy Valles + SEARCH |
| EPI_ISL_1814562, EPI_ISL_1814563, EPI_ISL_1814564 | EXCITE Lab | Andersen lab at Scripps Research | Nicole L Washington, Simon White, Geraint Levan, Kelly Schiabor Barrett, Elizabeth Cirulli, Alexandre Bolze, Charlotte Rivera-Garcia, Ryan Cho, Jason Nguyen, Sherry Wang, Jimmy Ramirez, Celena Andrade, Alice Summerfield, Tyler Cassens, Efren Sandoval, Francisco Tanudjaja, Magnus Isaksson, William Lee, David Becker, Marc Laurent, James Lu + SEARCH |
| EPI_ISL_1814565 | EXCITE Lab | Andersen lab at Scripps Research | David Pride, Sharon Reed, Chip Schooley, Angela Scioscia, Natasha Martin Cheryl Anderson, Sawyer Farmer, Abigail Schnapper, Helena Tubb, Tommy Valles + SEARCH |
| EPI_ISL_1814566 | EXCITE Lab | Andersen lab at Scripps Research | Nicole L Washington, Simon White, Geraint Levan, Kelly Schiabor Barrett, Elizabeth Cirulli, Alexandre Bolze, Charlotte Rivera-Garcia, Ryan Cho, Jason Nguyen, Sherry Wang, Jimmy Ramirez, Celena Andrade, Alice Summerfield, Tyler Cassens, Efren Sandoval, Francisco Tanudjaja, Magnus Isaksson, William Lee, David Becker, Marc Laurent, James Lu + SEARCH |
| EPI_ISL_1814567 | EXCITE Lab | Andersen lab at Scripps Research | Chip Schooley, Natasha Martin, Cheryl Anderson, Angela Scioscia, Smruthi Karthikeyan, Greg Humphrey, Sawyer Farmer, Abigail Schnapper, Helena Tubb, Tommy Valles + SEARCH |
| EPI_ISL_1814568, EPI_ISL_1814569, EPI_ISL_1814570, EPI_ISL_1814571, EPI_ISL_1814572, EPI_ISL_1814573, EPI_ISL_1814574, EPI_ISL_1814575, EPI_ISL_1814576, EPI_ISL_1814577, EPI_ISL_1814578, EPI_ISL_1814579, EPI_ISL_1814580, EPI_ISL_1814581, EPI_ISL_1814582, EPI_ISL_1814583, EPI_ISL_1814584, EPI_ISL_1814585, EPI_ISL_1814586, EPI_ISL_1814587, EPI_ISL_1814588, EPI_ISL_1814589, EPI_ISL_1814590, EPI_ISL_1814591, EPI_ISL_1814592 | | | |
| see above | EXCITE Lab | Andersen lab at Scripps Research | Nicole L Washington, Simon White, Geraint Levan, Kelly Schiabor Barrett, Elizabeth Cirulli, Alexandre Bolze, Charlotte Rivera-Garcia, Ryan Cho, Jason Nguyen, Sherry Wang, Jimmy Ramirez, Celena Andrade, Alice Summerfield, Tyler Cassens, Efren Sandoval, Francisco Tanudjaja, Magnus Isaksson, William Lee, David Becker, Marc Laurent, James Lu + SEARCH |
| EPI_ISL_1814593, EPI_ISL_1814594 | EXCITE Lab | Andersen lab at Scripps Research | Chip Schooley, Natasha Martin, Cheryl Anderson, Angela Scioscia, Smruthi Karthikeyan, Greg Humphrey, Sawyer Farmer, Abigail Schnapper, Helena Tubb, Tommy Valles + SEARCH |
| EPI_ISL_1814595 | EXCITE Lab | Andersen lab at Scripps Research | Nicole L Washington, Simon White, Geraint Levan, Kelly Schiabor Barrett, Elizabeth Cirulli, Alexandre Bolze, Charlotte Rivera-Garcia, Ryan Cho, Jason Nguyen, Sherry Wang, Jimmy Ramirez, Celena Andrade, Alice Summerfield, Tyler Cassens, Efren Sandoval, Francisco Tanudjaja, Magnus Isaksson, William Lee, David Becker, Marc Laurent, James Lu + SEARCH |
| EPI_ISL_1814596 | EXCITE Lab | Andersen lab at Scripps Research | Rebecca Fielding-Miller, Laura Kohn, Patricia Gonzalez-Zuniga, Fitri Carrissa Wijaya, Vinton Omaleki, Marlene Flores, Araz Majnoonian, Megan Nguyen, Ashkan Hassani, Smruthi Karthikeyan, Rodolfo Salido, Victor Cantu, Greg Humphrey, Sawyer Farmer, Abigail Schnapper, Helena Tubb, Tommy Valles + SEARCH |
| EPI_ISL_1814597, EPI_ISL_1814598, EPI_ISL_1814599, EPI_ISL_1814600 | EXCITE Lab | Andersen lab at Scripps Research | Nicole L Washington, Simon White, Geraint Levan, Kelly Schiabor Barrett, Elizabeth Cirulli, Alexandre Bolze, Charlotte Rivera-Garcia, Ryan Cho, Jason Nguyen, Sherry Wang, Jimmy Ramirez, Celena Andrade, Alice Summerfield, Tyler Cassens, Efren Sandoval, Francisco Tanudjaja, Magnus Isaksson, William Lee, David Becker, Marc Laurent, James Lu + SEARCH |
| EPI_ISL_1814601 | EXCITE Lab | Andersen lab at Scripps Research | David Pride, Sharon Reed, Chip Schooley, Angela Scioscia, Natasha Martin Cheryl Anderson, Sawyer Farmer, Abigail Schnapper, Helena Tubb, Tommy Valles + SEARCH |
| EPI_ISL_1814602 | EXCITE Lab | Andersen lab at Scripps Research | Nicole L Washington, Simon White, Geraint Levan, Kelly Schiabor Barrett, Elizabeth Cirulli, Alexandre Bolze, Charlotte Rivera-Garcia, Ryan Cho, Jason Nguyen, Sherry Wang, Jimmy Ramirez, Celena Andrade, Alice Summerfield, Tyler Cassens, Efren Sandoval, Francisco Tanudjaja, Magnus Isaksson, William Lee, David Becker, Marc Laurent, James Lu + SEARCH |
| EPI_ISL_1814603 | EXCITE Lab | Andersen lab at Scripps Research | David Pride, Sharon Reed, Chip Schooley, Angela Scioscia, Natasha Martin Cheryl Anderson, Sawyer Farmer, Abigail Schnapper, Helena Tubb, Tommy Valles + SEARCH |
| EPI_ISL_1814604, EPI_ISL_1814605, EPI_ISL_1814606, EPI_ISL_1814607 | EXCITE Lab | Andersen lab at Scripps Research | Nicole L Washington, Simon White, Geraint Levan, Kelly Schiabor Barrett, Elizabeth Cirulli, Alexandre Bolze, Charlotte Rivera-Garcia, Ryan Cho, Jason Nguyen, Sherry Wang, Jimmy Ramirez, Celena Andrade, Alice Summerfield, Tyler Cassens, Efren Sandoval, Francisco Tanudjaja, Magnus Isaksson, William Lee, David Becker, Marc Laurent, James Lu + SEARCH |
| EPI_ISL_1814608 | EXCITE Lab | Andersen lab at Scripps Research | Chip Schooley, Natasha Martin, Cheryl Anderson, Angela Scioscia, Smruthi Karthikeyan, Greg Humphrey, Sawyer Farmer, Abigail Schnapper, Helena Tubb, Tommy Valles + SEARCH |
| EPI_ISL_1814609, EPI_ISL_1814610, EPI_ISL_1814611, EPI_ISL_1814612, | EXCITE Lab | Andersen lab at Scripps Research | Nicole L Washington, Simon White, Geraint Levan, Kelly Schiabor Barrett, Elizabeth Cirulli, Alexandre Bolze, Charlotte Rivera-Garcia, Ryan Cho, Jason Nguyen, Sherry Wang, Jimmy Ramirez, Celena Andrade, Alice Summerfield, Tyler Cassens, Efren Sandoval, Francisco Tanudjaja, Magnus Isaksson, William Lee, David Becker, Marc Laurent, James Lu + SEARCH |

| | | | |
|---|---|---|---|
| EPI_ISL_1814613, EPI_ISL_1814614 | | | |
| EPI_ISL_1814615 | EXCITE Lab | Andersen lab at Scripps Research | J. Joelle Donofrio-Odmann, Brent Brainard+ SEARCH |
| EPI_ISL_1814616, EPI_ISL_1814617, EPI_ISL_1814618, EPI_ISL_1814619, EPI_ISL_1814620, EPI_ISL_1814621, EPI_ISL_1814622, EPI_ISL_1814623, EPI_ISL_1814624, EPI_ISL_1814625, EPI_ISL_1814626, EPI_ISL_1814627, EPI_ISL_1814628, EPI_ISL_1814629, EPI_ISL_1814630, EPI_ISL_1814631, EPI_ISL_1814632, EPI_ISL_1814633, EPI_ISL_1814634, EPI_ISL_1814635, EPI_ISL_1814636 | | | |
| see above | EXCITE Lab | Andersen lab at Scripps Research | Nicole L Washington, Simon White, Geraint Levan, Kelly Schiabor Barrett, Elizabeth Cirulli, Alexandre Bolze, Charlotte Rivera-Garcia, Ryan Cho, Jason Nguyen, Sherry Wang, Jimmy Ramirez, Celena Andrade, Alice Summerfield, Tyler Cassens, Efren Sandoval, Francisco Tanudjaja, Magnus Isaksson, William Lee, David Becker, Marc Laurent, James Lu + SEARCH |
| EPI_ISL_1814637 | EXCITE Lab | Andersen lab at Scripps Research | David Pride, Sharon Reed, Chip Schooley, Angela Scioscia, Natasha Martin Cheryl Anderson, Sawyer Farmer, Abigail Schnapper, Helena Tubb, Tommy Valles + SEARCH |
| EPI_ISL_1814638, EPI_ISL_1814639 | EXCITE Lab | Andersen lab at Scripps Research | Nicole L Washington, Simon White, Geraint Levan, Kelly Schiabor Barrett, Elizabeth Cirulli, Alexandre Bolze, Charlotte Rivera-Garcia, Ryan Cho, Jason Nguyen, Sherry Wang, Jimmy Ramirez, Celena Andrade, Alice Summerfield, Tyler Cassens, Efren Sandoval, Francisco Tanudjaja, Magnus Isaksson, William Lee, David Becker, Marc Laurent, James Lu + SEARCH |
| EPI_ISL_1814640 | EXCITE Lab | Andersen lab at Scripps Research | David Pride, Sharon Reed, Chip Schooley, Angela Scioscia, Natasha Martin Cheryl Anderson, Sawyer Farmer, Abigail Schnapper, Helena Tubb, Tommy Valles + SEARCH |
| EPI_ISL_1814641, EPI_ISL_1814642, EPI_ISL_1814643, EPI_ISL_1814644, EPI_ISL_1814645, EPI_ISL_1814646, EPI_ISL_1814647, EPI_ISL_1814648 | EXCITE Lab | Andersen lab at Scripps Research | Nicole L Washington, Simon White, Geraint Levan, Kelly Schiabor Barrett, Elizabeth Cirulli, Alexandre Bolze, Charlotte Rivera-Garcia, Ryan Cho, Jason Nguyen, Sherry Wang, Jimmy Ramirez, Celena Andrade, Alice Summerfield, Tyler Cassens, Efren Sandoval, Francisco Tanudjaja, Magnus Isaksson, William Lee, David Becker, Marc Laurent, James Lu + SEARCH |
| EPI_ISL_1814649 | EXCITE Lab | Andersen lab at Scripps Research | David Pride, Sharon Reed, Chip Schooley, Angela Scioscia, Natasha Martin Cheryl Anderson, Sawyer Farmer, Abigail Schnapper, Helena Tubb, Tommy Valles + SEARCH |
| EPI_ISL_1814650, EPI_ISL_1814651, EPI_ISL_1814652, EPI_ISL_1814653, EPI_ISL_1814654, EPI_ISL_1814655, EPI_ISL_1814656, EPI_ISL_1814657, EPI_ISL_1814658, EPI_ISL_1814659 | EXCITE Lab | Andersen lab at Scripps Research | Nicole L Washington, Simon White, Geraint Levan, Kelly Schiabor Barrett, Elizabeth Cirulli, Alexandre Bolze, Charlotte Rivera-Garcia, Ryan Cho, Jason Nguyen, Sherry Wang, Jimmy Ramirez, Celena Andrade, Alice Summerfield, Tyler Cassens, Efren Sandoval, Francisco Tanudjaja, Magnus Isaksson, William Lee, David Becker, Marc Laurent, James Lu + SEARCH |
| EPI_ISL_1814660 | EXCITE Lab | Andersen lab at Scripps Research | Rebecca Fielding-Miller, Laura Kohn, Patricia Gonzalez-Zuniga, Fitri Carrissa Wijaya, Vinton Omaleki, Marlene Flores, Araz Majnoonian, Megan Nguyen, Ashkan Hassani, Smruthi Karthikeyan, Rodolfo Salido, Victor Cantu, Greg Humphrey, Sawyer Farmer, Abigail Schnapper, Helena Tubb, Tommy Valles + SEARCH |
| EPI_ISL_1814661, EPI_ISL_1814662, EPI_ISL_1814663 | EXCITE Lab | Andersen lab at Scripps Research | Nicole L Washington, Simon White, Geraint Levan, Kelly Schiabor Barrett, Elizabeth Cirulli, Alexandre Bolze, Charlotte Rivera-Garcia, Ryan Cho, Jason Nguyen, Sherry Wang, Jimmy Ramirez, Celena Andrade, Alice Summerfield, Tyler Cassens, Efren Sandoval, Francisco Tanudjaja, Magnus Isaksson, William Lee, David Becker, Marc Laurent, James Lu + SEARCH |
| EPI_ISL_1814664, EPI_ISL_1814665 | EXCITE Lab | Andersen lab at Scripps Research | David Pride, Sharon Reed, Chip Schooley, Angela Scioscia, Natasha Martin Cheryl Anderson, Sawyer Farmer, Abigail Schnapper, Helena Tubb, Tommy Valles + SEARCH |
| EPI_ISL_1814666, EPI_ISL_1814667, EPI_ISL_1814668, EPI_ISL_1814669, EPI_ISL_1814670, EPI_ISL_1814671, EPI_ISL_1814672, EPI_ISL_1814673, EPI_ISL_1814674, EPI_ISL_1814675, EPI_ISL_1814676, EPI_ISL_1814677, EPI_ISL_1814678 | | | |
| see above | EXCITE Lab | Andersen lab at Scripps Research | Nicole L Washington, Simon White, Geraint Levan, Kelly Schiabor Barrett, Elizabeth Cirulli, Alexandre Bolze, Charlotte Rivera-Garcia, Ryan Cho, Jason Nguyen, Sherry Wang, Jimmy Ramirez, Celena Andrade, Alice Summerfield, Tyler Cassens, Efren Sandoval, Francisco Tanudjaja, Magnus Isaksson, William Lee, David Becker, Marc Laurent, James Lu + SEARCH |
| EPI_ISL_1814679 | EXCITE Lab | Andersen lab at Scripps Research | J. Joelle Donofrio-Odmann, Brent Brainard+ SEARCH |
| EPI_ISL_1814680 | EXCITE Lab | Andersen lab at Scripps Research | Nicole L Washington, Simon White, Geraint Levan, Kelly Schiabor Barrett, Elizabeth Cirulli, Alexandre Bolze, Charlotte Rivera-Garcia, Ryan Cho, Jason Nguyen, Sherry Wang, Jimmy Ramirez, Celena Andrade, Alice Summerfield, Tyler Cassens, Efren Sandoval, Francisco Tanudjaja, Magnus Isaksson, William Lee, David Becker, Marc Laurent, James Lu + SEARCH |
| EPI_ISL_1814681 | EXCITE Lab | Andersen lab at Scripps Research | David Pride, Sharon Reed, Chip Schooley, Angela Scioscia, Natasha Martin Cheryl Anderson, Sawyer Farmer, Abigail Schnapper, Helena Tubb, Tommy Valles + SEARCH |
| EPI_ISL_1814682, EPI_ISL_1814683, EPI_ISL_1814684, EPI_ISL_1814685, EPI_ISL_1814686, EPI_ISL_1814687, EPI_ISL_1814688, EPI_ISL_1814689, EPI_ISL_1814690, EPI_ISL_1814691, EPI_ISL_1814692 | | | |
| see above | EXCITE Lab | Andersen lab at Scripps Research | Nicole L Washington, Simon White, Geraint Levan, Kelly Schiabor Barrett, Elizabeth Cirulli, Alexandre Bolze, Charlotte Rivera-Garcia, Ryan Cho, Jason Nguyen, Sherry Wang, Jimmy Ramirez, Celena Andrade, Alice Summerfield, Tyler Cassens, Efren Sandoval, Francisco Tanudjaja, Magnus Isaksson, William Lee, David Becker, Marc Laurent, James Lu + SEARCH |
| EPI_ISL_1814693, EPI_ISL_1814694 | EXCITE Lab | Andersen lab at Scripps Research | David Pride, Sharon Reed, Chip Schooley, Angela Scioscia, Natasha Martin Cheryl Anderson, Sawyer Farmer, Abigail Schnapper, Helena Tubb, Tommy Valles + SEARCH |
| EPI_ISL_1814695 | EXCITE Lab | Andersen lab at Scripps Research | Nicole L Washington, Simon White, Geraint Levan, Kelly Schiabor Barrett, Elizabeth Cirulli, Alexandre Bolze, Charlotte Rivera-Garcia, Ryan Cho, Jason Nguyen, Sherry Wang, Jimmy Ramirez, Celena Andrade, Alice Summerfield, Tyler Cassens, Efren Sandoval, Francisco Tanudjaja, Magnus Isaksson, William Lee, David Becker, Marc Laurent, James Lu + SEARCH |
| EPI_ISL_1814696 | EXCITE Lab | Andersen lab at Scripps Research | David Pride, Sharon Reed, Chip Schooley, Angela Scioscia, Natasha Martin Cheryl Anderson, Sawyer Farmer, Abigail Schnapper, Helena Tubb, Tommy Valles + SEARCH |
| EPI_ISL_1814697, EPI_ISL_1814698, EPI_ISL_1814699, EPI_ISL_1814700, EPI_ISL_1814701, EPI_ISL_1814702, EPI_ISL_1814703, EPI_ISL_1814704, EPI_ISL_1814705, EPI_ISL_1814706, EPI_ISL_1814707, EPI_ISL_1814708, EPI_ISL_1814709, EPI_ISL_1814710, EPI_ISL_1814711, EPI_ISL_1814712, EPI_ISL_1814713, EPI_ISL_1814714, EPI_ISL_1814715, EPI_ISL_1814716 | | | |
| see above | EXCITE Lab | Andersen lab at Scripps Research | Nicole L Washington, Simon White, Geraint Levan, Kelly Schiabor Barrett, Elizabeth Cirulli, Alexandre Bolze, Charlotte Rivera-Garcia, Ryan Cho, Jason Nguyen, Sherry Wang, Jimmy Ramirez, Celena Andrade, Alice Summerfield, Tyler Cassens, Efren Sandoval, Francisco Tanudjaja, Magnus Isaksson, William Lee, David Becker, Marc Laurent, James Lu + SEARCH |
| EPI_ISL_1814717 | EXCITE Lab | Andersen lab at Scripps Research | David Pride, Sharon Reed, Chip Schooley, Angela Scioscia, Natasha Martin Cheryl Anderson, Sawyer Farmer, Abigail Schnapper, Helena Tubb, Tommy Valles + SEARCH |
| EPI_ISL_1814718, EPI_ISL_1814719, EPI_ISL_1814720, EPI_ISL_1814721, EPI_ISL_1814722, EPI_ISL_1814723 | EXCITE Lab | Andersen lab at Scripps Research | Nicole L Washington, Simon White, Geraint Levan, Kelly Schiabor Barrett, Elizabeth Cirulli, Alexandre Bolze, Charlotte Rivera-Garcia, Ryan Cho, Jason Nguyen, Sherry Wang, Jimmy Ramirez, Celena Andrade, Alice Summerfield, Tyler Cassens, Efren Sandoval, Francisco Tanudjaja, Magnus Isaksson, William Lee, David Becker, Marc Laurent, James Lu + SEARCH |
| EPI_ISL_1814724 | EXCITE Lab | Andersen lab at Scripps Research | David Pride, Sharon Reed, Chip Schooley, Angela Scioscia, Natasha Martin Cheryl Anderson, Sawyer Farmer, Abigail Schnapper, Helena Tubb, Tommy Valles + SEARCH |
| EPI_ISL_1814725, EPI_ISL_1814726, EPI_ISL_1814727, EPI_ISL_1814728, EPI_ISL_1814729, EPI_ISL_1814730, EPI_ISL_1814731, EPI_ISL_1814732, EPI_ISL_1814733, EPI_ISL_1814734, EPI_ISL_1814735, EPI_ISL_1814736, EPI_ISL_1814737, EPI_ISL_1814738, EPI_ISL_1814739, EPI_ISL_1814740, EPI_ISL_1814741, EPI_ISL_1814742, EPI_ISL_1814743 | | | |
| see above | EXCITE Lab | Andersen lab at Scripps Research | Nicole L Washington, Simon White, Geraint Levan, Kelly Schiabor Barrett, Elizabeth Cirulli, Alexandre Bolze, Charlotte Rivera-Garcia, Ryan Cho, Jason Nguyen, Sherry Wang, Jimmy Ramirez, Celena Andrade, Alice Summerfield, Tyler Cassens, Efren Sandoval, Francisco Tanudjaja, Magnus Isaksson, William Lee, David Becker, Marc Laurent, James Lu + SEARCH |
| EPI_ISL_1814744 | EXCITE Lab | Andersen lab at Scripps Research | David Pride, Sharon Reed, Chip Schooley, Angela Scioscia, Natasha Martin Cheryl Anderson, Sawyer Farmer, Abigail Schnapper, Helena Tubb, Tommy Valles + SEARCH |
| EPI_ISL_1814745, EPI_ISL_1814746, EPI_ISL_1814747, EPI_ISL_1814748, EPI_ISL_1814749 | EXCITE Lab | Andersen lab at Scripps Research | Nicole L Washington, Simon White, Geraint Levan, Kelly Schiabor Barrett, Elizabeth Cirulli, Alexandre Bolze, Charlotte Rivera-Garcia, Ryan Cho, Jason Nguyen, Sherry Wang, Jimmy Ramirez, Celena Andrade, Alice Summerfield, Tyler Cassens, Efren Sandoval, Francisco Tanudjaja, Magnus Isaksson, William Lee, David Becker, Marc Laurent, James Lu + SEARCH |
| EPI_ISL_1814750 | EXCITE Lab | Andersen lab at Scripps Research | Chip Schooley, Natasha Martin, Cheryl Anderson, Angela Scioscia, Smruthi Karthikeyan, Greg Humphrey, Sawyer Farmer, Abigail Schnapper, Helena Tubb, Tommy Valles + SEARCH |
| EPI_ISL_1814751, EPI_ISL_1814752, EPI_ISL_1814753, EPI_ISL_1814754, EPI_ISL_1814755, EPI_ISL_1814756, EPI_ISL_1814757, EPI_ISL_1814758, EPI_ISL_1814759 | EXCITE Lab | Andersen lab at Scripps Research | Nicole L Washington, Simon White, Geraint Levan, Kelly Schiabor Barrett, Elizabeth Cirulli, Alexandre Bolze, Charlotte Rivera-Garcia, Ryan Cho, Jason Nguyen, Sherry Wang, Jimmy Ramirez, Celena Andrade, Alice Summerfield, Tyler Cassens, Efren Sandoval, Francisco Tanudjaja, Magnus Isaksson, William Lee, David Becker, Marc Laurent, James Lu + SEARCH |
| EPI_ISL_1814760 | EXCITE Lab | Andersen lab at Scripps Research | Rebecca Fielding-Miller, Laura Kohn, Patricia Gonzalez-Zuniga, Fitri Carrissa Wijaya, Vinton Omaleki, Marlene Flores, Araz Majnoonian, Megan Nguyen, Ashkan Hassani, Smruthi Karthikeyan, Rodolfo Salido, Victor Cantu, Greg Humphrey, Sawyer Farmer, Abigail Schnapper, Helena Tubb, Tommy Valles + SEARCH |
| EPI_ISL_1814761, EPI_ISL_1814762, EPI_ISL_1814763, EPI_ISL_1814764, EPI_ISL_1814765 | EXCITE Lab | Andersen lab at Scripps Research | Nicole L Washington, Simon White, Geraint Levan, Kelly Schiabor Barrett, Elizabeth Cirulli, Alexandre Bolze, Charlotte Rivera-Garcia, Ryan Cho, Jason Nguyen, Sherry Wang, Jimmy Ramirez, Celena Andrade, Alice Summerfield, Tyler Cassens, Efren Sandoval, Francisco Tanudjaja, Magnus Isaksson, William Lee, David Becker, Marc Laurent, James Lu + SEARCH |
| EPI_ISL_1814766 | EXCITE Lab | Andersen lab at Scripps Research | David Pride, Sharon Reed, Chip Schooley, Angela Scioscia, Natasha Martin Cheryl Anderson, Sawyer Farmer, Abigail Schnapper, Helena Tubb, Tommy Valles + SEARCH |
| EPI_ISL_1814767, EPI_ISL_1814768, EPI_ISL_1814769, EPI_ISL_1814770, EPI_ISL_1814771, | EXCITE Lab | Andersen lab at Scripps Research | Nicole L Washington, Simon White, Geraint Levan, Kelly Schiabor Barrett, Elizabeth Cirulli, Alexandre Bolze, Charlotte Rivera-Garcia, Ryan Cho, Jason Nguyen, Sherry Wang, Jimmy Ramirez, Celena Andrade, Alice Summerfield, Tyler Cassens, Efren Sandoval, Francisco Tanudjaja, Magnus Isaksson, William Lee, David Becker, Marc Laurent, James Lu + SEARCH |

| | | | |
|---|---|---|---|
| EPI_ISL_1814772, EPI_ISL_1814773, EPI_ISL_1814774, EPI_ISL_1814775 | | | |
| EPI_ISL_1814776, EPI_ISL_1814777 | EXCITE Lab | Andersen lab at Scripps Research | Chip Schooley, Natasha Martin, Cheryl Anderson, Angela Scioscia, Smruthi Karthikeyan, Greg Humphrey, Sawyer Farmer, Abigail Schnapper, Helena Tubb, Tommy Valles + SEARCH |
| EPI_ISL_1814778, EPI_ISL_1814779, EPI_ISL_1814780, EPI_ISL_1814781, EPI_ISL_1814782, EPI_ISL_1814783, EPI_ISL_1814784, EPI_ISL_1814785, EPI_ISL_1814786, EPI_ISL_1814787, EPI_ISL_1814788, EPI_ISL_1814789, EPI_ISL_1814790, EPI_ISL_1814791, EPI_ISL_1814792, EPI_ISL_1814793, EPI_ISL_1814794, EPI_ISL_1814795, EPI_ISL_1814796, EPI_ISL_1814797, EPI_ISL_1814798, EPI_ISL_1814799, EPI_ISL_1814800, EPI_ISL_1814801, EPI_ISL_1814802 | | | |
| see above | EXCITE Lab | Andersen lab at Scripps Research | Nicole L Washington, Simon White, Geraint Levan, Kelly Schiabor Barrett, Elizabeth Cirulli, Alexandre Bolze, Charlotte Rivera-Garcia, Ryan Cho, Jason Nguyen, Sherry Wang, Jimmy Ramirez, Celena Andrade, Alice Summerfield, Tyler Cassens, Efren Sandoval, Francisco Tanudjaja, Magnus Isaksson, William Lee, David Becker, Marc Laurent, James Lu + SEARCH |
| EPI_ISL_1814803 | EXCITE Lab | Andersen lab at Scripps Research | Chip Schooley, Natasha Martin, Cheryl Anderson, Angela Scioscia, Smruthi Karthikeyan, Greg Humphrey, Sawyer Farmer, Abigail Schnapper, Helena Tubb, Tommy Valles + SEARCH |
| EPI_ISL_1814804, EPI_ISL_1814805, EPI_ISL_1814806, EPI_ISL_1814807, EPI_ISL_1814808, EPI_ISL_1814809, EPI_ISL_1814810, EPI_ISL_1814811, EPI_ISL_1814812 | EXCITE Lab | Andersen lab at Scripps Research | Nicole L Washington, Simon White, Geraint Levan, Kelly Schiabor Barrett, Elizabeth Cirulli, Alexandre Bolze, Charlotte Rivera-Garcia, Ryan Cho, Jason Nguyen, Sherry Wang, Jimmy Ramirez, Celena Andrade, Alice Summerfield, Tyler Cassens, Efren Sandoval, Francisco Tanudjaja, Magnus Isaksson, William Lee, David Becker, Marc Laurent, James Lu + SEARCH |
| EPI_ISL_1814813 | EXCITE Lab | Andersen lab at Scripps Research | Chip Schooley, Natasha Martin, Cheryl Anderson, Angela Scioscia, Smruthi Karthikeyan, Greg Humphrey, Sawyer Farmer, Abigail Schnapper, Helena Tubb, Tommy Valles + SEARCH |
| EPI_ISL_1814814 | EXCITE Lab | Andersen lab at Scripps Research | Nicole L Washington, Simon White, Geraint Levan, Kelly Schiabor Barrett, Elizabeth Cirulli, Alexandre Bolze, Charlotte Rivera-Garcia, Ryan Cho, Jason Nguyen, Sherry Wang, Jimmy Ramirez, Celena Andrade, Alice Summerfield, Tyler Cassens, Efren Sandoval, Francisco Tanudjaja, Magnus Isaksson, William Lee, David Becker, Marc Laurent, James Lu + SEARCH |
| EPI_ISL_1814815 | EXCITE Lab | Andersen lab at Scripps Research | David Pride, Sharon Reed, Chip Schooley, Angela Scioscia, Natasha Martin Cheryl Anderson, Sawyer Farmer, Abigail Schnapper, Helena Tubb, Tommy Valles + SEARCH |
| EPI_ISL_1814816, EPI_ISL_1814817, EPI_ISL_1814818 | EXCITE Lab | Andersen lab at Scripps Research | Nicole L Washington, Simon White, Geraint Levan, Kelly Schiabor Barrett, Elizabeth Cirulli, Alexandre Bolze, Charlotte Rivera-Garcia, Ryan Cho, Jason Nguyen, Sherry Wang, Jimmy Ramirez, Celena Andrade, Alice Summerfield, Tyler Cassens, Efren Sandoval, Francisco Tanudjaja, Magnus Isaksson, William Lee, David Becker, Marc Laurent, James Lu + SEARCH |
| EPI_ISL_1814819 | EXCITE Lab | Andersen lab at Scripps Research | Chip Schooley, Natasha Martin, Cheryl Anderson, Angela Scioscia, Smruthi Karthikeyan, Greg Humphrey, Sawyer Farmer, Abigail Schnapper, Helena Tubb, Tommy Valles + SEARCH |
| EPI_ISL_1814820, EPI_ISL_1814821, EPI_ISL_1814822, EPI_ISL_1814823, EPI_ISL_1814824, EPI_ISL_1814825, EPI_ISL_1814826, EPI_ISL_1814827, EPI_ISL_1814828, EPI_ISL_1814829, EPI_ISL_1814830, EPI_ISL_1814831, EPI_ISL_1814832, EPI_ISL_1814833, EPI_ISL_1814834, EPI_ISL_1814835, EPI_ISL_1814836, EPI_ISL_1814837 | | | |
| see above | EXCITE Lab | Andersen lab at Scripps Research | Nicole L Washington, Simon White, Geraint Levan, Kelly Schiabor Barrett, Elizabeth Cirulli, Alexandre Bolze, Charlotte Rivera-Garcia, Ryan Cho, Jason Nguyen, Sherry Wang, Jimmy Ramirez, Celena Andrade, Alice Summerfield, Tyler Cassens, Efren Sandoval, Francisco Tanudjaja, Magnus Isaksson, William Lee, David Becker, Marc Laurent, James Lu + SEARCH |
| EPI_ISL_1814838 | EXCITE Lab | Andersen lab at Scripps Research | Chip Schooley, Natasha Martin, Cheryl Anderson, Angela Scioscia, Smruthi Karthikeyan, Greg Humphrey, Sawyer Farmer, Abigail Schnapper, Helena Tubb, Tommy Valles + SEARCH |
| EPI_ISL_1814839, EPI_ISL_1814840, EPI_ISL_1814841 | EXCITE Lab | Andersen lab at Scripps Research | Nicole L Washington, Simon White, Geraint Levan, Kelly Schiabor Barrett, Elizabeth Cirulli, Alexandre Bolze, Charlotte Rivera-Garcia, Ryan Cho, Jason Nguyen, Sherry Wang, Jimmy Ramirez, Celena Andrade, Alice Summerfield, Tyler Cassens, Efren Sandoval, Francisco Tanudjaja, Magnus Isaksson, William Lee, David Becker, Marc Laurent, James Lu + SEARCH |
| EPI_ISL_1814842 | EXCITE Lab | Andersen lab at Scripps Research | Fred Wu, Christine Mitchell + SEARCH |
| EPI_ISL_1814843, EPI_ISL_1814844, EPI_ISL_1814845, EPI_ISL_1814846, EPI_ISL_1814847, EPI_ISL_1814848, EPI_ISL_1814849, EPI_ISL_1814850, EPI_ISL_1814851, EPI_ISL_1814852 | EXCITE Lab | Andersen lab at Scripps Research | Nicole L Washington, Simon White, Geraint Levan, Kelly Schiabor Barrett, Elizabeth Cirulli, Alexandre Bolze, Charlotte Rivera-Garcia, Ryan Cho, Jason Nguyen, Sherry Wang, Jimmy Ramirez, Celena Andrade, Alice Summerfield, Tyler Cassens, Efren Sandoval, Francisco Tanudjaja, Magnus Isaksson, William Lee, David Becker, Marc Laurent, James Lu + SEARCH |
| EPI_ISL_1814853 | EXCITE Lab | Andersen lab at Scripps Research | J. Joelle Donofrio-Odmann, Brent Brainard+ SEARCH |
| EPI_ISL_1814854 | EXCITE Lab | Andersen lab at Scripps Research | David Pride, Sharon Reed, Chip Schooley, Angela Scioscia, Natasha Martin Cheryl Anderson, Sawyer Farmer, Abigail Schnapper, Helena Tubb, Tommy Valles + SEARCH |
| EPI_ISL_1814855, EPI_ISL_1814856, EPI_ISL_1814857, EPI_ISL_1814858, EPI_ISL_1814859, EPI_ISL_1814860, EPI_ISL_1814861, EPI_ISL_1814862, EPI_ISL_1814863, EPI_ISL_1814864, EPI_ISL_1814865, EPI_ISL_1814866 | | | |
| see above | EXCITE Lab | Andersen lab at Scripps Research | Nicole L Washington, Simon White, Geraint Levan, Kelly Schiabor Barrett, Elizabeth Cirulli, Alexandre Bolze, Charlotte Rivera-Garcia, Ryan Cho, Jason Nguyen, Sherry Wang, Jimmy Ramirez, Celena Andrade, Alice Summerfield, Tyler Cassens, Efren Sandoval, Francisco Tanudjaja, Magnus Isaksson, William Lee, David Becker, Marc Laurent, James Lu + SEARCH |
| EPI_ISL_1814867 | EXCITE Lab | Andersen lab at Scripps Research | David Pride, Sharon Reed, Chip Schooley, Angela Scioscia, Natasha Martin Cheryl Anderson, Sawyer Farmer, Abigail Schnapper, Helena Tubb, Tommy Valles + SEARCH |
| EPI_ISL_1814868, EPI_ISL_1814869 | EXCITE Lab | Andersen lab at Scripps Research | Nicole L Washington, Simon White, Geraint Levan, Kelly Schiabor Barrett, Elizabeth Cirulli, Alexandre Bolze, Charlotte Rivera-Garcia, Ryan Cho, Jason Nguyen, Sherry Wang, Jimmy Ramirez, Celena Andrade, Alice Summerfield, Tyler Cassens, Efren Sandoval, Francisco Tanudjaja, Magnus Isaksson, William Lee, David Becker, Marc Laurent, James Lu + SEARCH |
| EPI_ISL_1814870 | EXCITE Lab | Andersen lab at Scripps Research | Chip Schooley, Natasha Martin, Cheryl Anderson, Angela Scioscia, Smruthi Karthikeyan, Greg Humphrey, Sawyer Farmer, Abigail Schnapper, Helena Tubb, Tommy Valles + SEARCH |
| EPI_ISL_1814871, EPI_ISL_1814872 | EXCITE Lab | Andersen lab at Scripps Research | Nicole L Washington, Simon White, Geraint Levan, Kelly Schiabor Barrett, Elizabeth Cirulli, Alexandre Bolze, Charlotte Rivera-Garcia, Ryan Cho, Jason Nguyen, Sherry Wang, Jimmy Ramirez, Celena Andrade, Alice Summerfield, Tyler Cassens, Efren Sandoval, Francisco Tanudjaja, Magnus Isaksson, William Lee, David Becker, Marc Laurent, James Lu + SEARCH |
| EPI_ISL_1814873 | EXCITE Lab | Andersen lab at Scripps Research | Chip Schooley, Natasha Martin, Cheryl Anderson, Angela Scioscia, Smruthi Karthikeyan, Greg Humphrey, Sawyer Farmer, Abigail Schnapper, Helena Tubb, Tommy Valles + SEARCH |
| EPI_ISL_1814874, EPI_ISL_1814875, EPI_ISL_1814876, EPI_ISL_1814877, EPI_ISL_1814878, EPI_ISL_1814879, EPI_ISL_1814880, EPI_ISL_1814881, EPI_ISL_1814882, EPI_ISL_1814883 | EXCITE Lab | Andersen lab at Scripps Research | Nicole L Washington, Simon White, Geraint Levan, Kelly Schiabor Barrett, Elizabeth Cirulli, Alexandre Bolze, Charlotte Rivera-Garcia, Ryan Cho, Jason Nguyen, Sherry Wang, Jimmy Ramirez, Celena Andrade, Alice Summerfield, Tyler Cassens, Efren Sandoval, Francisco Tanudjaja, Magnus Isaksson, William Lee, David Becker, Marc Laurent, James Lu + SEARCH |
| EPI_ISL_1814884 | EXCITE Lab | Andersen lab at Scripps Research | David Pride, Sharon Reed, Chip Schooley, Angela Scioscia, Natasha Martin Cheryl Anderson, Sawyer Farmer, Abigail Schnapper, Helena Tubb, Tommy Valles + SEARCH |
| EPI_ISL_1814885, EPI_ISL_1814886, EPI_ISL_1814887, EPI_ISL_1814888, EPI_ISL_1814889, EPI_ISL_1814890, EPI_ISL_1814891, EPI_ISL_1814892, EPI_ISL_1814893, EPI_ISL_1814894, EPI_ISL_1814895, EPI_ISL_1814896, EPI_ISL_1814897, EPI_ISL_1814898, EPI_ISL_1814899, EPI_ISL_1814900, EPI_ISL_1814901, EPI_ISL_1814902, EPI_ISL_1814903, EPI_ISL_1814904, EPI_ISL_1814905, EPI_ISL_1814906, EPI_ISL_1814907, EPI_ISL_1814908, EPI_ISL_1814909, EPI_ISL_1814910 | | | |
| see above | EXCITE Lab | Andersen lab at Scripps Research | Nicole L Washington, Simon White, Geraint Levan, Kelly Schiabor Barrett, Elizabeth Cirulli, Alexandre Bolze, Charlotte Rivera-Garcia, Ryan Cho, Jason Nguyen, Sherry Wang, Jimmy Ramirez, Celena Andrade, Alice Summerfield, Tyler Cassens, Efren Sandoval, Francisco Tanudjaja, Magnus Isaksson, William Lee, David Becker, Marc Laurent, James Lu + SEARCH |
| EPI_ISL_1814911 | EXCITE Lab | Andersen lab at Scripps Research | Chip Schooley, Natasha Martin, Cheryl Anderson, Angela Scioscia, Smruthi Karthikeyan, Greg Humphrey, Sawyer Farmer, Abigail Schnapper, Helena Tubb, Tommy Valles + SEARCH |
| EPI_ISL_1814912, EPI_ISL_1814913, EPI_ISL_1814914, EPI_ISL_1814915, EPI_ISL_1814916, EPI_ISL_1814917, EPI_ISL_1814918 | EXCITE Lab | Andersen lab at Scripps Research | Nicole L Washington, Simon White, Geraint Levan, Kelly Schiabor Barrett, Elizabeth Cirulli, Alexandre Bolze, Charlotte Rivera-Garcia, Ryan Cho, Jason Nguyen, Sherry Wang, Jimmy Ramirez, Celena Andrade, Alice Summerfield, Tyler Cassens, Efren Sandoval, Francisco Tanudjaja, Magnus Isaksson, William Lee, David Becker, Marc Laurent, James Lu + SEARCH |
| EPI_ISL_1814919 | EXCITE Lab | Andersen lab at Scripps Research | Chip Schooley, Natasha Martin, Cheryl Anderson, Angela Scioscia, Smruthi Karthikeyan, Greg Humphrey, Sawyer Farmer, Abigail Schnapper, Helena Tubb, Tommy Valles + SEARCH |
| EPI_ISL_1814920, EPI_ISL_1814921, EPI_ISL_1814922, EPI_ISL_1814923, EPI_ISL_1814924, EPI_ISL_1814925, EPI_ISL_1814926, EPI_ISL_1814927, EPI_ISL_1814928, EPI_ISL_1814929, EPI_ISL_1814930, EPI_ISL_1814931, EPI_ISL_1814932, EPI_ISL_1814933, EPI_ISL_1814934 | | | |
| see above | EXCITE Lab | Andersen lab at Scripps Research | Nicole L Washington, Simon White, Geraint Levan, Kelly Schiabor Barrett, Elizabeth Cirulli, Alexandre Bolze, Charlotte Rivera-Garcia, Ryan Cho, Jason Nguyen, Sherry Wang, Jimmy Ramirez, Celena Andrade, Alice Summerfield, Tyler Cassens, Efren Sandoval, Francisco Tanudjaja, Magnus Isaksson, William Lee, David Becker, Marc Laurent, James Lu + SEARCH |
| EPI_ISL_1814935 | EXCITE Lab | Andersen lab at Scripps Research | David Pride, Sharon Reed, Chip Schooley, Angela Scioscia, Natasha Martin Cheryl Anderson, Sawyer Farmer, Abigail Schnapper, Helena Tubb, Tommy Valles + SEARCH |
| EPI_ISL_1814936, EPI_ISL_1814937, EPI_ISL_1814938, EPI_ISL_1814939, EPI_ISL_1814940, EPI_ISL_1814941, EPI_ISL_1814942, EPI_ISL_1814943, | EXCITE Lab | Andersen lab at Scripps Research | Nicole L Washington, Simon White, Geraint Levan, Kelly Schiabor Barrett, Elizabeth Cirulli, Alexandre Bolze, Charlotte Rivera-Garcia, Ryan Cho, Jason Nguyen, Sherry Wang, Jimmy Ramirez, Celena Andrade, Alice Summerfield, Tyler Cassens, Efren Sandoval, Francisco Tanudjaja, Magnus Isaksson, William Lee, David Becker, Marc Laurent, James Lu + SEARCH |

| | | | |
|---|---|---|---|
| EPI_ISL_1814944 | | | |
| EPI_ISL_1814945 | EXCITE Lab | Andersen lab at Scripps Research | David Pride, Sharon Reed, Chip Schooley, Angela Scioscia, Natasha Martin Cheryl Anderson, Sawyer Farmer, Abigail Schnapper, Helena Tubb, Tommy Valles + SEARCH |
| EPI_ISL_1814946, EPI_ISL_1814947, EPI_ISL_1814948, EPI_ISL_1814949, EPI_ISL_1814950, EPI_ISL_1814951, EPI_ISL_1814952, EPI_ISL_1814953, EPI_ISL_1814954, EPI_ISL_1814955, EPI_ISL_1814956, EPI_ISL_1814957, EPI_ISL_1814958, EPI_ISL_1814959, EPI_ISL_1814960, EPI_ISL_1814961, EPI_ISL_1814962 | | | |
| see above | EXCITE Lab | Andersen lab at Scripps Research | Nicole L Washington, Simon White, Geraint Levan, Kelly Schiabor Barrett, Elizabeth Cirulli, Alexandre Bolze, Charlotte Rivera-Garcia, Ryan Cho, Jason Nguyen, Sherry Wang, Jimmy Ramirez, Celena Andrade, Alice Summerfield, Tyler Cassens, Efren Sandoval, Francisco Tanudjaja, Magnus Isaksson, William Lee, David Becker, Marc Laurent, James Lu + SEARCH |
| EPI_ISL_1814963 | EXCITE Lab | Andersen lab at Scripps Research | David Pride, Sharon Reed, Chip Schooley, Angela Scioscia, Natasha Martin Cheryl Anderson, Sawyer Farmer, Abigail Schnapper, Helena Tubb, Tommy Valles + SEARCH |
| EPI_ISL_1814964, EPI_ISL_1814965, EPI_ISL_1814966 | EXCITE Lab | Andersen lab at Scripps Research | Nicole L Washington, Simon White, Geraint Levan, Kelly Schiabor Barrett, Elizabeth Cirulli, Alexandre Bolze, Charlotte Rivera-Garcia, Ryan Cho, Jason Nguyen, Sherry Wang, Jimmy Ramirez, Celena Andrade, Alice Summerfield, Tyler Cassens, Efren Sandoval, Francisco Tanudjaja, Magnus Isaksson, William Lee, David Becker, Marc Laurent, James Lu + SEARCH |
| EPI_ISL_1814967 | EXCITE Lab | Andersen lab at Scripps Research | Chip Schooley, Natasha Martin, Cheryl Anderson, Angela Scioscia, Smruthi Karthikeyan, Greg Humphrey, Sawyer Farmer, Abigail Schnapper, Helena Tubb, Tommy Valles + SEARCH |
| EPI_ISL_1814968 | EXCITE Lab | Andersen lab at Scripps Research | David Pride, Sharon Reed, Chip Schooley, Angela Scioscia, Natasha Martin Cheryl Anderson, Sawyer Farmer, Abigail Schnapper, Helena Tubb, Tommy Valles + SEARCH |
| EPI_ISL_1814969, EPI_ISL_1814970, EPI_ISL_1814971, EPI_ISL_1814972, EPI_ISL_1814973 | EXCITE Lab | Andersen lab at Scripps Research | Nicole L Washington, Simon White, Geraint Levan, Kelly Schiabor Barrett, Elizabeth Cirulli, Alexandre Bolze, Charlotte Rivera-Garcia, Ryan Cho, Jason Nguyen, Sherry Wang, Jimmy Ramirez, Celena Andrade, Alice Summerfield, Tyler Cassens, Efren Sandoval, Francisco Tanudjaja, Magnus Isaksson, William Lee, David Becker, Marc Laurent, James Lu + SEARCH |
| EPI_ISL_1814974 | EXCITE Lab | Andersen lab at Scripps Research | David Pride, Sharon Reed, Chip Schooley, Angela Scioscia, Natasha Martin Cheryl Anderson, Sawyer Farmer, Abigail Schnapper, Helena Tubb, Tommy Valles + SEARCH |
| EPI_ISL_1814975, EPI_ISL_1814976, EPI_ISL_1814977, EPI_ISL_1814978, EPI_ISL_1814979, EPI_ISL_1814980, EPI_ISL_1814981, EPI_ISL_1814982, EPI_ISL_1814983, EPI_ISL_1814984, EPI_ISL_1814985, EPI_ISL_1814986, EPI_ISL_1814987, EPI_ISL_1814988, EPI_ISL_1814989, EPI_ISL_1814990 | | | |
| see above | EXCITE Lab | Andersen lab at Scripps Research | Nicole L Washington, Simon White, Geraint Levan, Kelly Schiabor Barrett, Elizabeth Cirulli, Alexandre Bolze, Charlotte Rivera-Garcia, Ryan Cho, Jason Nguyen, Sherry Wang, Jimmy Ramirez, Celena Andrade, Alice Summerfield, Tyler Cassens, Efren Sandoval, Francisco Tanudjaja, Magnus Isaksson, William Lee, David Becker, Marc Laurent, James Lu + SEARCH |
| EPI_ISL_1814991 | EXCITE Lab | Andersen lab at Scripps Research | Chip Schooley, Natasha Martin, Cheryl Anderson, Angela Scioscia, Smruthi Karthikeyan, Greg Humphrey, Sawyer Farmer, Abigail Schnapper, Helena Tubb, Tommy Valles + SEARCH |
| EPI_ISL_1814992, EPI_ISL_1814993, EPI_ISL_1814994, EPI_ISL_1814995, EPI_ISL_1814996, EPI_ISL_1814997, EPI_ISL_1814998, EPI_ISL_1814999, EPI_ISL_1815000, EPI_ISL_1815001, EPI_ISL_1815002, EPI_ISL_1815003, EPI_ISL_1815004, EPI_ISL_1815005, EPI_ISL_1815006, EPI_ISL_1815007, EPI_ISL_1815008 | | | |
| see above | EXCITE Lab | Andersen lab at Scripps Research | Nicole L Washington, Simon White, Geraint Levan, Kelly Schiabor Barrett, Elizabeth Cirulli, Alexandre Bolze, Charlotte Rivera-Garcia, Ryan Cho, Jason Nguyen, Sherry Wang, Jimmy Ramirez, Celena Andrade, Alice Summerfield, Tyler Cassens, Efren Sandoval, Francisco Tanudjaja, Magnus Isaksson, William Lee, David Becker, Marc Laurent, James Lu + SEARCH |
| EPI_ISL_1815009 | EXCITE Lab | Andersen lab at Scripps Research | Chip Schooley, Natasha Martin, Cheryl Anderson, Angela Scioscia, Smruthi Karthikeyan, Greg Humphrey, Sawyer Farmer, Abigail Schnapper, Helena Tubb, Tommy Valles + SEARCH |
| EPI_ISL_1815010 | EXCITE Lab | Andersen lab at Scripps Research | Sydney Morgan + Marni Jacobs + Celestine Magallanes + Holly Valentine + Louise C. Laurent + SEARCH |
| EPI_ISL_1815011 | EXCITE Lab | Andersen lab at Scripps Research | Nicole L Washington, Simon White, Geraint Levan, Kelly Schiabor Barrett, Elizabeth Cirulli, Alexandre Bolze, Charlotte Rivera-Garcia, Ryan Cho, Jason Nguyen, Sherry Wang, Jimmy Ramirez, Celena Andrade, Alice Summerfield, Tyler Cassens, Efren Sandoval, Francisco Tanudjaja, Magnus Isaksson, William Lee, David Becker, Marc Laurent, James Lu + SEARCH |
| EPI_ISL_1815012 | EXCITE Lab | Andersen lab at Scripps Research | Chip Schooley, Natasha Martin, Cheryl Anderson, Angela Scioscia, Smruthi Karthikeyan, Greg Humphrey, Sawyer Farmer, Abigail Schnapper, Helena Tubb, Tommy Valles + SEARCH |
| EPI_ISL_1815013, EPI_ISL_1815014, EPI_ISL_1815015, EPI_ISL_1815016, EPI_ISL_1815017, EPI_ISL_1815018, EPI_ISL_1815019, EPI_ISL_1815020, EPI_ISL_1815021 | EXCITE Lab | Andersen lab at Scripps Research | Nicole L Washington, Simon White, Geraint Levan, Kelly Schiabor Barrett, Elizabeth Cirulli, Alexandre Bolze, Charlotte Rivera-Garcia, Ryan Cho, Jason Nguyen, Sherry Wang, Jimmy Ramirez, Celena Andrade, Alice Summerfield, Tyler Cassens, Efren Sandoval, Francisco Tanudjaja, Magnus Isaksson, William Lee, David Becker, Marc Laurent, James Lu + SEARCH |
| EPI_ISL_1815022 | EXCITE Lab | Andersen lab at Scripps Research | David Pride, Sharon Reed, Chip Schooley, Angela Scioscia, Natasha Martin Cheryl Anderson, Sawyer Farmer, Abigail Schnapper, Helena Tubb, Tommy Valles + SEARCH |
| EPI_ISL_1815023, EPI_ISL_1815024, EPI_ISL_1815025, EPI_ISL_1815026, EPI_ISL_1815027, EPI_ISL_1815028, EPI_ISL_1815029 | EXCITE Lab | Andersen lab at Scripps Research | Nicole L Washington, Simon White, Geraint Levan, Kelly Schiabor Barrett, Elizabeth Cirulli, Alexandre Bolze, Charlotte Rivera-Garcia, Ryan Cho, Jason Nguyen, Sherry Wang, Jimmy Ramirez, Celena Andrade, Alice Summerfield, Tyler Cassens, Efren Sandoval, Francisco Tanudjaja, Magnus Isaksson, William Lee, David Becker, Marc Laurent, James Lu + SEARCH |
| EPI_ISL_1815030 | EXCITE Lab | Andersen lab at Scripps Research | Rebecca Fielding-Miller, Laura Kohn, Patricia Gonzalez-Zuniga, Fitri Carrissa Wijaya, Vinton Omaleki, Marlene Flores, Araz Majnoonian, Megan Nguyen, Ashkan Hassani, Smruthi Karthikeyan, Rodolfo Salido, Victor Cantu, Greg Humphrey, Sawyer Farmer, Abigail Schnapper, Helena Tubb, Tommy Valles + SEARCH |
| EPI_ISL_1815031, EPI_ISL_1815032, EPI_ISL_1815033, EPI_ISL_1815034, EPI_ISL_1815035, EPI_ISL_1815036, EPI_ISL_1815037, EPI_ISL_1815038 | EXCITE Lab | Andersen lab at Scripps Research | Nicole L Washington, Simon White, Geraint Levan, Kelly Schiabor Barrett, Elizabeth Cirulli, Alexandre Bolze, Charlotte Rivera-Garcia, Ryan Cho, Jason Nguyen, Sherry Wang, Jimmy Ramirez, Celena Andrade, Alice Summerfield, Tyler Cassens, Efren Sandoval, Francisco Tanudjaja, Magnus Isaksson, William Lee, David Becker, Marc Laurent, James Lu + SEARCH |
| EPI_ISL_1815039 | EXCITE Lab | Andersen lab at Scripps Research | David Pride, Sharon Reed, Chip Schooley, Angela Scioscia, Natasha Martin Cheryl Anderson, Sawyer Farmer, Abigail Schnapper, Helena Tubb, Tommy Valles + SEARCH |
| EPI_ISL_1815040, EPI_ISL_1815041, EPI_ISL_1815042, EPI_ISL_1815043, EPI_ISL_1815044, EPI_ISL_1815045, EPI_ISL_1815046 | EXCITE Lab | Andersen lab at Scripps Research | Nicole L Washington, Simon White, Geraint Levan, Kelly Schiabor Barrett, Elizabeth Cirulli, Alexandre Bolze, Charlotte Rivera-Garcia, Ryan Cho, Jason Nguyen, Sherry Wang, Jimmy Ramirez, Celena Andrade, Alice Summerfield, Tyler Cassens, Efren Sandoval, Francisco Tanudjaja, Magnus Isaksson, William Lee, David Becker, Marc Laurent, James Lu + SEARCH |
| EPI_ISL_1815047 | EXCITE Lab | Andersen lab at Scripps Research | Chip Schooley, Natasha Martin, Cheryl Anderson, Angela Scioscia, Smruthi Karthikeyan, Greg Humphrey, Sawyer Farmer, Abigail Schnapper, Helena Tubb, Tommy Valles + SEARCH |
| EPI_ISL_1815048, EPI_ISL_1815049, EPI_ISL_1815050, EPI_ISL_1815051, EPI_ISL_1815052, EPI_ISL_1815053, EPI_ISL_1815054, EPI_ISL_1815055, EPI_ISL_1815056, EPI_ISL_1815057, EPI_ISL_1815058 | | | |
| see above | EXCITE Lab | Andersen lab at Scripps Research | Nicole L Washington, Simon White, Geraint Levan, Kelly Schiabor Barrett, Elizabeth Cirulli, Alexandre Bolze, Charlotte Rivera-Garcia, Ryan Cho, Jason Nguyen, Sherry Wang, Jimmy Ramirez, Celena Andrade, Alice Summerfield, Tyler Cassens, Efren Sandoval, Francisco Tanudjaja, Magnus Isaksson, William Lee, David Becker, Marc Laurent, James Lu + SEARCH |
| EPI_ISL_1815059 | EXCITE Lab | Andersen lab at Scripps Research | Sydney Morgan + Marni Jacobs + Celestine Magallanes + Holly Valentine + Louise C. Laurent + SEARCH |
| EPI_ISL_1815060, EPI_ISL_1815061, EPI_ISL_1815062, EPI_ISL_1815063, EPI_ISL_1815064, EPI_ISL_1815065, EPI_ISL_1815066 | EXCITE Lab | Andersen lab at Scripps Research | Nicole L Washington, Simon White, Geraint Levan, Kelly Schiabor Barrett, Elizabeth Cirulli, Alexandre Bolze, Charlotte Rivera-Garcia, Ryan Cho, Jason Nguyen, Sherry Wang, Jimmy Ramirez, Celena Andrade, Alice Summerfield, Tyler Cassens, Efren Sandoval, Francisco Tanudjaja, Magnus Isaksson, William Lee, David Becker, Marc Laurent, James Lu + SEARCH |
| EPI_ISL_1815067 | EXCITE Lab | Andersen lab at Scripps Research | Chip Schooley, Natasha Martin, Cheryl Anderson, Angela Scioscia, Smruthi Karthikeyan, Greg Humphrey, Sawyer Farmer, Abigail Schnapper, Helena Tubb, Tommy Valles + SEARCH |
| EPI_ISL_1815068, EPI_ISL_1815069, EPI_ISL_1815070, EPI_ISL_1815071, EPI_ISL_1815072, EPI_ISL_1815073, EPI_ISL_1815074, EPI_ISL_1815075, EPI_ISL_1815076, EPI_ISL_1815077, EPI_ISL_1815078, EPI_ISL_1815079, EPI_ISL_1815080, EPI_ISL_1815081, EPI_ISL_1815082, EPI_ISL_1815083, EPI_ISL_1815084, EPI_ISL_1815085, EPI_ISL_1815086, EPI_ISL_1815087, EPI_ISL_1815088, EPI_ISL_1815089, EPI_ISL_1815090, EPI_ISL_1815091 | | | |
| see above | EXCITE Lab | Andersen lab at Scripps Research | Nicole L Washington, Simon White, Geraint Levan, Kelly Schiabor Barrett, Elizabeth Cirulli, Alexandre Bolze, Charlotte Rivera-Garcia, Ryan Cho, Jason Nguyen, Sherry Wang, Jimmy Ramirez, Celena Andrade, Alice Summerfield, Tyler Cassens, Efren Sandoval, Francisco Tanudjaja, Magnus Isaksson, William Lee, David Becker, Marc Laurent, James Lu + SEARCH |
| EPI_ISL_1815092 | EXCITE Lab | Andersen lab at Scripps Research | David Pride, Sharon Reed, Chip Schooley, Angela Scioscia, Natasha Martin Cheryl Anderson, Sawyer Farmer, Abigail Schnapper, Helena Tubb, Tommy Valles + SEARCH |
| EPI_ISL_1815093, EPI_ISL_1815094, EPI_ISL_1815095, EPI_ISL_1815096, EPI_ISL_1815097, EPI_ISL_1815098, EPI_ISL_1815099 | EXCITE Lab | Andersen lab at Scripps Research | Nicole L Washington, Simon White, Geraint Levan, Kelly Schiabor Barrett, Elizabeth Cirulli, Alexandre Bolze, Charlotte Rivera-Garcia, Ryan Cho, Jason Nguyen, Sherry Wang, Jimmy Ramirez, Celena Andrade, Alice Summerfield, Tyler Cassens, Efren Sandoval, Francisco Tanudjaja, Magnus Isaksson, William Lee, David Becker, Marc Laurent, James Lu + SEARCH |
| EPI_ISL_1815100, EPI_ISL_1815101 | EXCITE Lab | Andersen lab at Scripps Research | Chip Schooley, Natasha Martin, Cheryl Anderson, Angela Scioscia, Smruthi Karthikeyan, Greg Humphrey, Sawyer Farmer, Abigail Schnapper, Helena Tubb, Tommy Valles + SEARCH |
| EPI_ISL_1815102, EPI_ISL_1815103, EPI_ISL_1815104, EPI_ISL_1815105, EPI_ISL_1815106, EPI_ISL_1815107, EPI_ISL_1815108, EPI_ISL_1815109, EPI_ISL_1815110, EPI_ISL_1815111, EPI_ISL_1815112, EPI_ISL_1815113, EPI_ISL_1815114, EPI_ISL_1815115, EPI_ISL_1815116, EPI_ISL_1815117, EPI_ISL_1815118, EPI_ISL_1815119, EPI_ISL_1815120, EPI_ISL_1815121 | | | |
| see above | EXCITE Lab | Andersen lab at Scripps Research | Nicole L Washington, Simon White, Geraint Levan, Kelly Schiabor Barrett, Elizabeth Cirulli, Alexandre Bolze, Charlotte Rivera-Garcia, Ryan Cho, Jason Nguyen, Sherry Wang, Jimmy Ramirez, Celena Andrade, Alice Summerfield, Tyler Cassens, Efren Sandoval, Francisco Tanudjaja, Magnus |

| | | | |
|---|---|---|---|
| | | | Isaksson, William Lee, David Becker, Marc Laurent, James Lu + SEARCH |
| EPI_ISL_1815122 | EXCITE Lab | Andersen lab at Scripps Research | Chip Schooley, Natasha Martin, Cheryl Anderson, Angela Scioscia, Smruthi Karthikeyan, Greg Humphrey, Sawyer Farmer, Abigail Schnapper, Helena Tubb, Tommy Valles + SEARCH |
| EPI_ISL_1815123, EPI_ISL_1815124, EPI_ISL_1815125, EPI_ISL_1815126 | EXCITE Lab | Andersen lab at Scripps Research | Nicole L Washington, Simon White, Geraint Levan, Kelly Schiabor Barrett, Elizabeth Cirulli, Alexandre Bolze, Charlotte Rivera-Garcia, Ryan Cho, Jason Nguyen, Sherry Wang, Jimmy Ramirez, Celena Andrade, Alice Summerfield, Tyler Cassens, Efren Sandoval, Francisco Tanudjaja, Magnus Isaksson, William Lee, David Becker, Marc Laurent, James Lu + SEARCH |
| EPI_ISL_1815127 | EXCITE Lab | Andersen lab at Scripps Research | David Pride, Sharon Reed, Chip Schooley, Angela Scioscia, Natasha Martin Cheryl Anderson, Sawyer Farmer, Abigail Schnapper, Helena Tubb, Tommy Valles + SEARCH |
| EPI_ISL_1815128, EPI_ISL_1815129, EPI_ISL_1815130, EPI_ISL_1815131, EPI_ISL_1815132, EPI_ISL_1815133, EPI_ISL_1815134, EPI_ISL_1815135, EPI_ISL_1815136 | EXCITE Lab | Andersen lab at Scripps Research | Nicole L Washington, Simon White, Geraint Levan, Kelly Schiabor Barrett, Elizabeth Cirulli, Alexandre Bolze, Charlotte Rivera-Garcia, Ryan Cho, Jason Nguyen, Sherry Wang, Jimmy Ramirez, Celena Andrade, Alice Summerfield, Tyler Cassens, Efren Sandoval, Francisco Tanudjaja, Magnus Isaksson, William Lee, David Becker, Marc Laurent, James Lu + SEARCH |
| EPI_ISL_1815137 | EXCITE Lab | Andersen lab at Scripps Research | David Pride, Sharon Reed, Chip Schooley, Angela Scioscia, Natasha Martin Cheryl Anderson, Sawyer Farmer, Abigail Schnapper, Helena Tubb, Tommy Valles + SEARCH |
| EPI_ISL_1815138, EPI_ISL_1815139, EPI_ISL_1815140, EPI_ISL_1815141 | EXCITE Lab | Andersen lab at Scripps Research | Nicole L Washington, Simon White, Geraint Levan, Kelly Schiabor Barrett, Elizabeth Cirulli, Alexandre Bolze, Charlotte Rivera-Garcia, Ryan Cho, Jason Nguyen, Sherry Wang, Jimmy Ramirez, Celena Andrade, Alice Summerfield, Tyler Cassens, Efren Sandoval, Francisco Tanudjaja, Magnus Isaksson, William Lee, David Becker, Marc Laurent, James Lu + SEARCH |
| EPI_ISL_1815142 | EXCITE Lab | Andersen lab at Scripps Research | David Pride, Sharon Reed, Chip Schooley, Angela Scioscia, Natasha Martin Cheryl Anderson, Sawyer Farmer, Abigail Schnapper, Helena Tubb, Tommy Valles + SEARCH |
| EPI_ISL_1815143, EPI_ISL_1815144, EPI_ISL_1815145, EPI_ISL_1815146, EPI_ISL_1815147, EPI_ISL_1815148, EPI_ISL_1815149, EPI_ISL_1815150, EPI_ISL_1815151, EPI_ISL_1815152, EPI_ISL_1815153, EPI_ISL_1815154, EPI_ISL_1815155, EPI_ISL_1815156, EPI_ISL_1815157, EPI_ISL_1815158, EPI_ISL_1815159, EPI_ISL_1815160, EPI_ISL_1815161, EPI_ISL_1815162, EPI_ISL_1815163, EPI_ISL_1815164, EPI_ISL_1815165, EPI_ISL_1815166, EPI_ISL_1815167, EPI_ISL_1815168, EPI_ISL_1815169, EPI_ISL_1815170, EPI_ISL_1815171, EPI_ISL_1815172, EPI_ISL_1815173, EPI_ISL_1815174, EPI_ISL_1815175 | | | |
| see above | EXCITE Lab | Andersen lab at Scripps Research | Nicole L Washington, Simon White, Geraint Levan, Kelly Schiabor Barrett, Elizabeth Cirulli, Alexandre Bolze, Charlotte Rivera-Garcia, Ryan Cho, Jason Nguyen, Sherry Wang, Jimmy Ramirez, Celena Andrade, Alice Summerfield, Tyler Cassens, Efren Sandoval, Francisco Tanudjaja, Magnus Isaksson, William Lee, David Becker, Marc Laurent, James Lu + SEARCH |
| EPI_ISL_1815176 | EXCITE Lab | Andersen lab at Scripps Research | Chip Schooley, Natasha Martin, Cheryl Anderson, Angela Scioscia, Smruthi Karthikeyan, Greg Humphrey, Sawyer Farmer, Abigail Schnapper, Helena Tubb, Tommy Valles + SEARCH |
| EPI_ISL_1815177 | EXCITE Lab | Andersen lab at Scripps Research | Nicole L Washington, Simon White, Geraint Levan, Kelly Schiabor Barrett, Elizabeth Cirulli, Alexandre Bolze, Charlotte Rivera-Garcia, Ryan Cho, Jason Nguyen, Sherry Wang, Jimmy Ramirez, Celena Andrade, Alice Summerfield, Tyler Cassens, Efren Sandoval, Francisco Tanudjaja, Magnus Isaksson, William Lee, David Becker, Marc Laurent, James Lu + SEARCH |
| EPI_ISL_1815178 | EXCITE Lab | Andersen lab at Scripps Research | David Pride, Sharon Reed, Chip Schooley, Angela Scioscia, Natasha Martin Cheryl Anderson, Sawyer Farmer, Abigail Schnapper, Helena Tubb, Tommy Valles + SEARCH |
| EPI_ISL_1815179, EPI_ISL_1815180, EPI_ISL_1815181, EPI_ISL_1815182, EPI_ISL_1815183, EPI_ISL_1815184, EPI_ISL_1815185, EPI_ISL_1815186, EPI_ISL_1815187, EPI_ISL_1815188, EPI_ISL_1815189, EPI_ISL_1815190, EPI_ISL_1815191, EPI_ISL_1815192, EPI_ISL_1815193, EPI_ISL_1815194, EPI_ISL_1815195, EPI_ISL_1815196, EPI_ISL_1815197, EPI_ISL_1815198, EPI_ISL_1815199, EPI_ISL_1815200, EPI_ISL_1815201 | | | |
| see above | EXCITE Lab | Andersen lab at Scripps Research | Nicole L Washington, Simon White, Geraint Levan, Kelly Schiabor Barrett, Elizabeth Cirulli, Alexandre Bolze, Charlotte Rivera-Garcia, Ryan Cho, Jason Nguyen, Sherry Wang, Jimmy Ramirez, Celena Andrade, Alice Summerfield, Tyler Cassens, Efren Sandoval, Francisco Tanudjaja, Magnus Isaksson, William Lee, David Becker, Marc Laurent, James Lu + SEARCH |
| EPI_ISL_1815202 | EXCITE Lab | Andersen lab at Scripps Research | Chip Schooley, Natasha Martin, Cheryl Anderson, Angela Scioscia, Smruthi Karthikeyan, Greg Humphrey, Sawyer Farmer, Abigail Schnapper, Helena Tubb, Tommy Valles + SEARCH |
| EPI_ISL_1815203, EPI_ISL_1815204, EPI_ISL_1815205, EPI_ISL_1815206 | EXCITE Lab | Andersen lab at Scripps Research | Nicole L Washington, Simon White, Geraint Levan, Kelly Schiabor Barrett, Elizabeth Cirulli, Alexandre Bolze, Charlotte Rivera-Garcia, Ryan Cho, Jason Nguyen, Sherry Wang, Jimmy Ramirez, Celena Andrade, Alice Summerfield, Tyler Cassens, Efren Sandoval, Francisco Tanudjaja, Magnus Isaksson, William Lee, David Becker, Marc Laurent, James Lu + SEARCH |
| EPI_ISL_1815207 | EXCITE Lab | Andersen lab at Scripps Research | David Pride, Sharon Reed, Chip Schooley, Angela Scioscia, Natasha Martin Cheryl Anderson, Sawyer Farmer, Abigail Schnapper, Helena Tubb, Tommy Valles + SEARCH |
| EPI_ISL_1815208 | EXCITE Lab | Andersen lab at Scripps Research | Chip Schooley, Natasha Martin, Cheryl Anderson, Angela Scioscia, Smruthi Karthikeyan, Greg Humphrey, Sawyer Farmer, Abigail Schnapper, Helena Tubb, Tommy Valles + SEARCH |
| EPI_ISL_1815209 | EXCITE Lab | Andersen lab at Scripps Research | Sydney Morgan + Marni Jacobs + Celestine Magallanes + Holly Valentine + Louise C. Laurent + SEARCH |
| EPI_ISL_1815210, EPI_ISL_1815211, EPI_ISL_1815212, EPI_ISL_1815213, EPI_ISL_1815214, EPI_ISL_1815215, EPI_ISL_1815216, EPI_ISL_1815217, EPI_ISL_1815218, EPI_ISL_1815219, EPI_ISL_1815220, EPI_ISL_1815221, EPI_ISL_1815222, EPI_ISL_1815223 | | | |
| see above | EXCITE Lab | Andersen lab at Scripps Research | Nicole L Washington, Simon White, Geraint Levan, Kelly Schiabor Barrett, Elizabeth Cirulli, Alexandre Bolze, Charlotte Rivera-Garcia, Ryan Cho, Jason Nguyen, Sherry Wang, Jimmy Ramirez, Celena Andrade, Alice Summerfield, Tyler Cassens, Efren Sandoval, Francisco Tanudjaja, Magnus Isaksson, William Lee, David Becker, Marc Laurent, James Lu + SEARCH |
| EPI_ISL_1815224, EPI_ISL_1815225 | EXCITE Lab | Andersen lab at Scripps Research | Chip Schooley, Natasha Martin, Cheryl Anderson, Angela Scioscia, Smruthi Karthikeyan, Greg Humphrey, Sawyer Farmer, Abigail Schnapper, Helena Tubb, Tommy Valles + SEARCH |
| EPI_ISL_1815226, EPI_ISL_1815227, EPI_ISL_1815228, EPI_ISL_1815229, EPI_ISL_1815230, EPI_ISL_1815231, EPI_ISL_1815232, EPI_ISL_1815233, EPI_ISL_1815234, EPI_ISL_1815235, EPI_ISL_1815236, EPI_ISL_1815237, EPI_ISL_1815238 | | | |
| see above | EXCITE Lab | Andersen lab at Scripps Research | Nicole L Washington, Simon White, Geraint Levan, Kelly Schiabor Barrett, Elizabeth Cirulli, Alexandre Bolze, Charlotte Rivera-Garcia, Ryan Cho, Jason Nguyen, Sherry Wang, Jimmy Ramirez, Celena Andrade, Alice Summerfield, Tyler Cassens, Efren Sandoval, Francisco Tanudjaja, Magnus Isaksson, William Lee, David Becker, Marc Laurent, James Lu + SEARCH |
| EPI_ISL_1815239 | EXCITE Lab | Andersen lab at Scripps Research | Chip Schooley, Natasha Martin, Cheryl Anderson, Angela Scioscia, Smruthi Karthikeyan, Greg Humphrey, Sawyer Farmer, Abigail Schnapper, Helena Tubb, Tommy Valles + SEARCH |
| EPI_ISL_1815240 | EXCITE Lab | Andersen lab at Scripps Research | Fred Wu, Christine Mitchell + SEARCH |
| EPI_ISL_1815241, EPI_ISL_1815242, EPI_ISL_1815243, EPI_ISL_1815244, EPI_ISL_1815245 | EXCITE Lab | Andersen lab at Scripps Research | Nicole L Washington, Simon White, Geraint Levan, Kelly Schiabor Barrett, Elizabeth Cirulli, Alexandre Bolze, Charlotte Rivera-Garcia, Ryan Cho, Jason Nguyen, Sherry Wang, Jimmy Ramirez, Celena Andrade, Alice Summerfield, Tyler Cassens, Efren Sandoval, Francisco Tanudjaja, Magnus Isaksson, William Lee, David Becker, Marc Laurent, James Lu + SEARCH |
| EPI_ISL_1815246 | EXCITE Lab | Andersen lab at Scripps Research | Chip Schooley, Natasha Martin, Cheryl Anderson, Angela Scioscia, Smruthi Karthikeyan, Greg Humphrey, Sawyer Farmer, Abigail Schnapper, Helena Tubb, Tommy Valles + SEARCH |
| EPI_ISL_1815247 | EXCITE Lab | Andersen lab at Scripps Research | Nicole L Washington, Simon White, Geraint Levan, Kelly Schiabor Barrett, Elizabeth Cirulli, Alexandre Bolze, Charlotte Rivera-Garcia, Ryan Cho, Jason Nguyen, Sherry Wang, Jimmy Ramirez, Celena Andrade, Alice Summerfield, Tyler Cassens, Efren Sandoval, Francisco Tanudjaja, Magnus Isaksson, William Lee, David Becker, Marc Laurent, James Lu + SEARCH |
| EPI_ISL_1815248 | EXCITE Lab | Andersen lab at Scripps Research | David Pride, Sharon Reed, Chip Schooley, Angela Scioscia, Natasha Martin Cheryl Anderson, Sawyer Farmer, Abigail Schnapper, Helena Tubb, Tommy Valles + SEARCH |
| EPI_ISL_1815249, EPI_ISL_1815250, EPI_ISL_1815251, EPI_ISL_1815252, EPI_ISL_1815253, EPI_ISL_1815254, EPI_ISL_1815255, EPI_ISL_1815256, EPI_ISL_1815257, EPI_ISL_1815258, EPI_ISL_1815259, EPI_ISL_1815260, EPI_ISL_1815261, EPI_ISL_1815262, EPI_ISL_1815263, EPI_ISL_1815264, EPI_ISL_1815265, EPI_ISL_1815266, EPI_ISL_1815267, EPI_ISL_1815268, EPI_ISL_1815269, EPI_ISL_1815270, EPI_ISL_1815271, EPI_ISL_1815272, EPI_ISL_1815273, EPI_ISL_1815274, EPI_ISL_1815275, EPI_ISL_1815276, EPI_ISL_1815277, EPI_ISL_1815278, EPI_ISL_1815279, EPI_ISL_1815280, EPI_ISL_1815281, EPI_ISL_1815282, EPI_ISL_1815283, EPI_ISL_1815284 | | | |
| see above | EXCITE Lab | Andersen lab at Scripps Research | Nicole L Washington, Simon White, Geraint Levan, Kelly Schiabor Barrett, Elizabeth Cirulli, Alexandre Bolze, Charlotte Rivera-Garcia, Ryan Cho, Jason Nguyen, Sherry Wang, Jimmy Ramirez, Celena Andrade, Alice Summerfield, Tyler Cassens, Efren Sandoval, Francisco Tanudjaja, Magnus Isaksson, William Lee, David Becker, Marc Laurent, James Lu + SEARCH |
| EPI_ISL_1815285 | EXCITE Lab | Andersen lab at Scripps Research | Chip Schooley, Natasha Martin, Cheryl Anderson, Angela Scioscia, Smruthi Karthikeyan, Greg Humphrey, Sawyer Farmer, Abigail Schnapper, Helena Tubb, Tommy Valles + SEARCH |
| EPI_ISL_1815286, EPI_ISL_1815287, EPI_ISL_1815288, EPI_ISL_1815289, EPI_ISL_1815290, EPI_ISL_1815291, EPI_ISL_1815292, EPI_ISL_1815293, EPI_ISL_1815294, EPI_ISL_1815295, EPI_ISL_1815296, EPI_ISL_1815297, EPI_ISL_1815298 | | | |
| see above | EXCITE Lab | Andersen lab at Scripps Research | Nicole L Washington, Simon White, Geraint Levan, Kelly Schiabor Barrett, Elizabeth Cirulli, Alexandre Bolze, Charlotte Rivera-Garcia, Ryan Cho, Jason Nguyen, Sherry Wang, Jimmy Ramirez, Celena Andrade, Alice Summerfield, Tyler Cassens, Efren Sandoval, Francisco Tanudjaja, Magnus Isaksson, William Lee, David Becker, Marc Laurent, James Lu + SEARCH |
| EPI_ISL_1815299 | EXCITE Lab | Andersen lab at Scripps Research | J. Joelle Donofrio-Odmann, Brent Brainard+ SEARCH |
| EPI_ISL_1815300, EPI_ISL_1815301, EPI_ISL_1815302 | EXCITE Lab | Andersen lab at Scripps Research | Nicole L Washington, Simon White, Geraint Levan, Kelly Schiabor Barrett, Elizabeth Cirulli, Alexandre Bolze, Charlotte Rivera-Garcia, Ryan Cho, Jason Nguyen, Sherry Wang, Jimmy Ramirez, Celena Andrade, Alice Summerfield, Tyler Cassens, Efren Sandoval, Francisco Tanudjaja, Magnus Isaksson, William Lee, David Becker, Marc Laurent, James Lu + SEARCH |
| EPI_ISL_1815303 | EXCITE Lab | Andersen lab at Scripps Research | David Pride, Sharon Reed, Chip Schooley, Angela Scioscia, Natasha Martin Cheryl Anderson, Sawyer Farmer, Abigail Schnapper, Helena Tubb, Tommy Valles + SEARCH |
| EPI_ISL_1815304, EPI_ISL_1815305, EPI_ISL_1815306, EPI_ISL_1815307, EPI_ISL_1815308, EPI_ISL_1815309, EPI_ISL_1815310, EPI_ISL_1815311, EPI_ISL_1815312, EPI_ISL_1815313, EPI_ISL_1815314, EPI_ISL_1815315, EPI_ISL_1815316, EPI_ISL_1815317, EPI_ISL_1815318, EPI_ISL_1815319, EPI_ISL_1815320, EPI_ISL_1815321, EPI_ISL_1815322, EPI_ISL_1815323 | | | |
| see above | EXCITE Lab | Andersen lab at Scripps Research | Nicole L Washington, Simon White, Geraint Levan, Kelly Schiabor Barrett, Elizabeth Cirulli, Alexandre Bolze, Charlotte Rivera-Garcia, Ryan Cho, Jason Nguyen, Sherry Wang, Jimmy Ramirez, Celena Andrade, Alice Summerfield, Tyler Cassens, Efren Sandoval, Francisco Tanudjaja, Magnus Isaksson, William Lee, David Becker, Marc Laurent, James Lu + SEARCH |
| EPI_ISL_1815324 | EXCITE Lab | Andersen lab at Scripps Research | Chip Schooley, Natasha Martin, Cheryl Anderson, Angela Scioscia, Smruthi Karthikeyan, Greg Humphrey, Sawyer Farmer, Abigail Schnapper, Helena Tubb, Tommy Valles + SEARCH |
| EPI_ISL_1815325, EPI_ISL_1815326, EPI_ISL_1815327, EPI_ISL_1815328, EPI_ISL_1815329, EPI_ISL_1815330, EPI_ISL_1815331, EPI_ISL_1815332, EPI_ISL_1815333, EPI_ISL_1815334, EPI_ISL_1815335, EPI_ISL_1815336, EPI_ISL_1815337, EPI_ISL_1815338, EPI_ISL_1815339, EPI_ISL_1815340, EPI_ISL_1815341 | | | |
| see above | EXCITE Lab | Andersen lab at Scripps Research | Nicole L Washington, Simon White, Geraint Levan, Kelly Schiabor Barrett, Elizabeth Cirulli, Alexandre Bolze, Charlotte Rivera-Garcia, Ryan Cho, Jason Nguyen, Sherry Wang, Jimmy Ramirez, Celena Andrade, Alice Summerfield, Tyler Cassens, Efren Sandoval, Francisco Tanudjaja, Magnus Isaksson, William Lee, David Becker, Marc Laurent, James Lu + SEARCH |
| EPI_ISL_1815342 | EXCITE Lab | Andersen lab at Scripps Research | David Pride, Sharon Reed, Chip Schooley, Angela Scioscia, Natasha Martin Cheryl Anderson, Sawyer Farmer, Abigail Schnapper, Helena Tubb, Tommy Valles + SEARCH |
| EPI_ISL_1815343, EPI_ISL_1815344, EPI_ISL_1815345, EPI_ISL_1815346, EPI_ISL_1815347, EPI_ISL_1815348, EPI_ISL_1815349, EPI_ISL_1815350 | EXCITE Lab | Andersen lab at Scripps Research | Nicole L Washington, Simon White, Geraint Levan, Kelly Schiabor Barrett, Elizabeth Cirulli, Alexandre Bolze, Charlotte Rivera-Garcia, Ryan Cho, Jason Nguyen, Sherry Wang, Jimmy Ramirez, Celena Andrade, Alice Summerfield, Tyler Cassens, Efren Sandoval, Francisco Tanudjaja, Magnus Isaksson, William Lee, David Becker, Marc Laurent, James Lu + SEARCH |

| | | | |
|---|---|---|---|
| EPI_ISL_1815351 | EXCITE Lab | Andersen lab at Scripps Research | David Pride, Sharon Reed, Chip Schooley, Angela Scioscia, Natasha Martin Cheryl Anderson, Sawyer Farmer, Abigail Schnapper, Helena Tubb, Tommy Valles + SEARCH |
| EPI_ISL_1815352, EPI_ISL_1815353, EPI_ISL_1815354 | EXCITE Lab | Andersen lab at Scripps Research | Nicole L Washington, Simon White, Geraint Levan, Kelly Schiabor Barrett, Elizabeth Cirulli, Alexandre Bolze, Charlotte Rivera-Garcia, Ryan Cho, Jason Nguyen, Sherry Wang, Jimmy Ramirez, Celena Andrade, Alice Summerfield, Tyler Cassens, Efren Sandoval, Francisco Tanudjaja, Magnus Isaksson, William Lee, David Becker, Marc Laurent, James Lu + SEARCH |
| EPI_ISL_1815355 | EXCITE Lab | Andersen lab at Scripps Research | Chip Schooley, Natasha Martin, Cheryl Anderson, Angela Scioscia, Smruthi Karthikeyan, Greg Humphrey, Sawyer Farmer, Abigail Schnapper, Helena Tubb, Tommy Valles + SEARCH |
| EPI_ISL_1815356, EPI_ISL_1815357, EPI_ISL_1815358, EPI_ISL_1815359, EPI_ISL_1815360, EPI_ISL_1815361, EPI_ISL_1815362, EPI_ISL_1815363, EPI_ISL_1815364, EPI_ISL_1815365, EPI_ISL_1815366, EPI_ISL_1815367, EPI_ISL_1815368, EPI_ISL_1815369, EPI_ISL_1815370, EPI_ISL_1815371 | | | |
| see above | EXCITE Lab | Andersen lab at Scripps Research | Nicole L Washington, Simon White, Geraint Levan, Kelly Schiabor Barrett, Elizabeth Cirulli, Alexandre Bolze, Charlotte Rivera-Garcia, Ryan Cho, Jason Nguyen, Sherry Wang, Jimmy Ramirez, Celena Andrade, Alice Summerfield, Tyler Cassens, Efren Sandoval, Francisco Tanudjaja, Magnus Isaksson, William Lee, David Becker, Marc Laurent, James Lu + SEARCH |
| EPI_ISL_1815372 | EXCITE Lab | Andersen lab at Scripps Research | Rebecca Fielding-Miller, Laura Kohn, Patricia Gonzalez-Zuniga, Fitri Carrissa Wijaya, Vinton Omaleki, Marlene Flores, Araz Majnoonian, Megan Nguyen, Ashkan Hassani, Smruthi Karthikeyan, Rodolfo Salido, Victor Cantu, Greg Humphrey, Sawyer Farmer, Abigail Schnapper, Helena Tubb, Tommy Valles + SEARCH |
| EPI_ISL_1815373 | EXCITE Lab | Andersen lab at Scripps Research | Fred Wu, Christine Mitchell + SEARCH |
| EPI_ISL_1815374, EPI_ISL_1815375, EPI_ISL_1815376 | EXCITE Lab | Andersen lab at Scripps Research | Nicole L Washington, Simon White, Geraint Levan, Kelly Schiabor Barrett, Elizabeth Cirulli, Alexandre Bolze, Charlotte Rivera-Garcia, Ryan Cho, Jason Nguyen, Sherry Wang, Jimmy Ramirez, Celena Andrade, Alice Summerfield, Tyler Cassens, Efren Sandoval, Francisco Tanudjaja, Magnus Isaksson, William Lee, David Becker, Marc Laurent, James Lu + SEARCH |
| EPI_ISL_1815377 | EXCITE Lab | Andersen lab at Scripps Research | Sydney Morgan + Marni Jacobs + Celestine Magallanes + Holly Valentine + Louise C. Laurent + SEARCH |
| EPI_ISL_1815378, EPI_ISL_1815379 | EXCITE Lab | Andersen lab at Scripps Research | Nicole L Washington, Simon White, Geraint Levan, Kelly Schiabor Barrett, Elizabeth Cirulli, Alexandre Bolze, Charlotte Rivera-Garcia, Ryan Cho, Jason Nguyen, Sherry Wang, Jimmy Ramirez, Celena Andrade, Alice Summerfield, Tyler Cassens, Efren Sandoval, Francisco Tanudjaja, Magnus Isaksson, William Lee, David Becker, Marc Laurent, James Lu + SEARCH |
| EPI_ISL_1815380 | EXCITE Lab | Andersen lab at Scripps Research | David Pride, Sharon Reed, Chip Schooley, Angela Scioscia, Natasha Martin Cheryl Anderson, Sawyer Farmer, Abigail Schnapper, Helena Tubb, Tommy Valles + SEARCH |
| EPI_ISL_1815381, EPI_ISL_1815382, EPI_ISL_1815383, EPI_ISL_1815384, EPI_ISL_1815385, EPI_ISL_1815386, EPI_ISL_1815387, EPI_ISL_1815388, EPI_ISL_1815389, EPI_ISL_1815390, EPI_ISL_1815391, EPI_ISL_1815392, EPI_ISL_1815393, EPI_ISL_1815394, EPI_ISL_1815395, EPI_ISL_1815396, EPI_ISL_1815397, EPI_ISL_1815398, EPI_ISL_1815399, EPI_ISL_1815400 | | | |
| see above | EXCITE Lab | Andersen lab at Scripps Research | Nicole L Washington, Simon White, Geraint Levan, Kelly Schiabor Barrett, Elizabeth Cirulli, Alexandre Bolze, Charlotte Rivera-Garcia, Ryan Cho, Jason Nguyen, Sherry Wang, Jimmy Ramirez, Celena Andrade, Alice Summerfield, Tyler Cassens, Efren Sandoval, Francisco Tanudjaja, Magnus Isaksson, William Lee, David Becker, Marc Laurent, James Lu + SEARCH |
| EPI_ISL_1815401 | EXCITE Lab | Andersen lab at Scripps Research | Rebecca Fielding-Miller, Laura Kohn, Patricia Gonzalez-Zuniga, Fitri Carrissa Wijaya, Vinton Omaleki, Marlene Flores, Araz Majnoonian, Megan Nguyen, Ashkan Hassani, Smruthi Karthikeyan, Rodolfo Salido, Victor Cantu, Greg Humphrey, Sawyer Farmer, Abigail Schnapper, Helena Tubb, Tommy Valles + SEARCH |
| EPI_ISL_1815402, EPI_ISL_1815403, EPI_ISL_1815404, EPI_ISL_1815405, EPI_ISL_1815406, EPI_ISL_1815407, EPI_ISL_1815408, EPI_ISL_1815409, EPI_ISL_1815410, EPI_ISL_1815411, EPI_ISL_1815412, EPI_ISL_1815413, EPI_ISL_1815414, EPI_ISL_1815415, EPI_ISL_1815416, EPI_ISL_1815417, EPI_ISL_1815418, EPI_ISL_1815419, EPI_ISL_1815420, EPI_ISL_1815421, EPI_ISL_1815422, EPI_ISL_1815423, EPI_ISL_1815424, EPI_ISL_1815425 | | | |
| see above | EXCITE Lab | Andersen lab at Scripps Research | Nicole L Washington, Simon White, Geraint Levan, Kelly Schiabor Barrett, Elizabeth Cirulli, Alexandre Bolze, Charlotte Rivera-Garcia, Ryan Cho, Jason Nguyen, Sherry Wang, Jimmy Ramirez, Celena Andrade, Alice Summerfield, Tyler Cassens, Efren Sandoval, Francisco Tanudjaja, Magnus Isaksson, William Lee, David Becker, Marc Laurent, James Lu + SEARCH |
| EPI_ISL_1815426 | EXCITE Lab | Andersen lab at Scripps Research | Chip Schooley, Natasha Martin, Cheryl Anderson, Angela Scioscia, Smruthi Karthikeyan, Greg Humphrey, Sawyer Farmer, Abigail Schnapper, Helena Tubb, Tommy Valles + SEARCH |
| EPI_ISL_1815427, EPI_ISL_1815428, EPI_ISL_1815429, EPI_ISL_1815430, EPI_ISL_1815431, EPI_ISL_1815432, EPI_ISL_1815433, EPI_ISL_1815434, EPI_ISL_1815435 | EXCITE Lab | Andersen lab at Scripps Research | Nicole L Washington, Simon White, Geraint Levan, Kelly Schiabor Barrett, Elizabeth Cirulli, Alexandre Bolze, Charlotte Rivera-Garcia, Ryan Cho, Jason Nguyen, Sherry Wang, Jimmy Ramirez, Celena Andrade, Alice Summerfield, Tyler Cassens, Efren Sandoval, Francisco Tanudjaja, Magnus Isaksson, William Lee, David Becker, Marc Laurent, James Lu + SEARCH |
| EPI_ISL_1815436 | EXCITE Lab | Andersen lab at Scripps Research | Chip Schooley, Natasha Martin, Cheryl Anderson, Angela Scioscia, Smruthi Karthikeyan, Greg Humphrey, Sawyer Farmer, Abigail Schnapper, Helena Tubb, Tommy Valles + SEARCH |
| EPI_ISL_1815437, EPI_ISL_1815438, EPI_ISL_1815439, EPI_ISL_1815440, EPI_ISL_1815441, EPI_ISL_1815442, EPI_ISL_1815443, EPI_ISL_1815444, EPI_ISL_1815445, EPI_ISL_1815446, EPI_ISL_1815447, EPI_ISL_1815448, EPI_ISL_1815449, EPI_ISL_1815450, EPI_ISL_1815451, EPI_ISL_1815452, EPI_ISL_1815453, EPI_ISL_1815454, EPI_ISL_1815455, EPI_ISL_1815456, EPI_ISL_1815457, EPI_ISL_1815458, EPI_ISL_1815459, EPI_ISL_1815460 | | | |
| see above | EXCITE Lab | Andersen lab at Scripps Research | Nicole L Washington, Simon White, Geraint Levan, Kelly Schiabor Barrett, Elizabeth Cirulli, Alexandre Bolze, Charlotte Rivera-Garcia, Ryan Cho, Jason Nguyen, Sherry Wang, Jimmy Ramirez, Celena Andrade, Alice Summerfield, Tyler Cassens, Efren Sandoval, Francisco Tanudjaja, Magnus Isaksson, William Lee, David Becker, Marc Laurent, James Lu + SEARCH |
| EPI_ISL_1815461 | EXCITE Lab | Andersen lab at Scripps Research | Chip Schooley, Natasha Martin, Cheryl Anderson, Angela Scioscia, Smruthi Karthikeyan, Greg Humphrey, Sawyer Farmer, Abigail Schnapper, Helena Tubb, Tommy Valles + SEARCH |
| EPI_ISL_1815462, EPI_ISL_1815463, EPI_ISL_1815464, EPI_ISL_1815465, EPI_ISL_1815466, EPI_ISL_1815467, EPI_ISL_1815468, EPI_ISL_1815469, EPI_ISL_1815470, EPI_ISL_1815471, EPI_ISL_1815472, EPI_ISL_1815473 | | | |
| see above | EXCITE Lab | Andersen lab at Scripps Research | Nicole L Washington, Simon White, Geraint Levan, Kelly Schiabor Barrett, Elizabeth Cirulli, Alexandre Bolze, Charlotte Rivera-Garcia, Ryan Cho, Jason Nguyen, Sherry Wang, Jimmy Ramirez, Celena Andrade, Alice Summerfield, Tyler Cassens, Efren Sandoval, Francisco Tanudjaja, Magnus Isaksson, William Lee, David Becker, Marc Laurent, James Lu + SEARCH |
| EPI_ISL_1815474 | EXCITE Lab | Andersen lab at Scripps Research | Fred Wu, Christine Mitchell + SEARCH |
| EPI_ISL_1815475, EPI_ISL_1815476, EPI_ISL_1815477, EPI_ISL_1815478, EPI_ISL_1815479, EPI_ISL_1815480, EPI_ISL_1815481, EPI_ISL_1815482, EPI_ISL_1815483, EPI_ISL_1815484, EPI_ISL_1815485, EPI_ISL_1815486, EPI_ISL_1815487, EPI_ISL_1815488, EPI_ISL_1815489, EPI_ISL_1815490, EPI_ISL_1815491, EPI_ISL_1815492 | | | |
| see above | EXCITE Lab | Andersen lab at Scripps Research | Nicole L Washington, Simon White, Geraint Levan, Kelly Schiabor Barrett, Elizabeth Cirulli, Alexandre Bolze, Charlotte Rivera-Garcia, Ryan Cho, Jason Nguyen, Sherry Wang, Jimmy Ramirez, Celena Andrade, Alice Summerfield, Tyler Cassens, Efren Sandoval, Francisco Tanudjaja, Magnus Isaksson, William Lee, David Becker, Marc Laurent, James Lu + SEARCH |
| EPI_ISL_1815493 | EXCITE Lab | Andersen lab at Scripps Research | Chip Schooley, Natasha Martin, Cheryl Anderson, Angela Scioscia, Smruthi Karthikeyan, Greg Humphrey, Sawyer Farmer, Abigail Schnapper, Helena Tubb, Tommy Valles + SEARCH |
| EPI_ISL_1815494 | EXCITE Lab | Andersen lab at Scripps Research | Nicole L Washington, Simon White, Geraint Levan, Kelly Schiabor Barrett, Elizabeth Cirulli, Alexandre Bolze, Charlotte Rivera-Garcia, Ryan Cho, Jason Nguyen, Sherry Wang, Jimmy Ramirez, Celena Andrade, Alice Summerfield, Tyler Cassens, Efren Sandoval, Francisco Tanudjaja, Magnus Isaksson, William Lee, David Becker, Marc Laurent, James Lu + SEARCH |
| EPI_ISL_1815495 | EXCITE Lab | Andersen lab at Scripps Research | David Pride, Sharon Reed, Chip Schooley, Angela Scioscia, Natasha Martin Cheryl Anderson, Sawyer Farmer, Abigail Schnapper, Helena Tubb, Tommy Valles + SEARCH |
| EPI_ISL_1815496, EPI_ISL_1815497, EPI_ISL_1815498, EPI_ISL_1815499, EPI_ISL_1815500, EPI_ISL_1815501, EPI_ISL_1815502, EPI_ISL_1815503, EPI_ISL_1815504, EPI_ISL_1815505, EPI_ISL_1815506, EPI_ISL_1815507, EPI_ISL_1815508, EPI_ISL_1815509 | | | |
| see above | EXCITE Lab | Andersen lab at Scripps Research | Nicole L Washington, Simon White, Geraint Levan, Kelly Schiabor Barrett, Elizabeth Cirulli, Alexandre Bolze, Charlotte Rivera-Garcia, Ryan Cho, Jason Nguyen, Sherry Wang, Jimmy Ramirez, Celena Andrade, Alice Summerfield, Tyler Cassens, Efren Sandoval, Francisco Tanudjaja, Magnus Isaksson, William Lee, David Becker, Marc Laurent, James Lu + SEARCH |
| EPI_ISL_1815510 | EXCITE Lab | Andersen lab at Scripps Research | Chip Schooley, Natasha Martin, Cheryl Anderson, Angela Scioscia, Smruthi Karthikeyan, Greg Humphrey, Sawyer Farmer, Abigail Schnapper, Helena Tubb, Tommy Valles + SEARCH |
| EPI_ISL_1815511 | EXCITE Lab | Andersen lab at Scripps Research | Nicole L Washington, Simon White, Geraint Levan, Kelly Schiabor Barrett, Elizabeth Cirulli, Alexandre Bolze, Charlotte Rivera-Garcia, Ryan Cho, Jason Nguyen, Sherry Wang, Jimmy Ramirez, Celena Andrade, Alice Summerfield, Tyler Cassens, Efren Sandoval, Francisco Tanudjaja, Magnus Isaksson, William Lee, David Becker, Marc Laurent, James Lu + SEARCH |
| EPI_ISL_1815512 | EXCITE Lab | Andersen lab at Scripps Research | Sydney Morgan + Marni Jacobs + Celestine Magallanes + Holly Valentine + Louise C. Laurent + SEARCH |
| EPI_ISL_1815513, EPI_ISL_1815514 | EXCITE Lab | Andersen lab at Scripps Research | Nicole L Washington, Simon White, Geraint Levan, Kelly Schiabor Barrett, Elizabeth Cirulli, Alexandre Bolze, Charlotte Rivera-Garcia, Ryan Cho, Jason Nguyen, Sherry Wang, Jimmy Ramirez, Celena Andrade, Alice Summerfield, Tyler Cassens, Efren Sandoval, Francisco Tanudjaja, Magnus Isaksson, William Lee, David Becker, Marc Laurent, James Lu + SEARCH |
| EPI_ISL_1815515 | EXCITE Lab | Andersen lab at Scripps Research | David Pride, Sharon Reed, Chip Schooley, Angela Scioscia, Natasha Martin Cheryl Anderson, Sawyer Farmer, Abigail Schnapper, Helena Tubb, Tommy Valles + SEARCH |
| EPI_ISL_1815516, EPI_ISL_1815517, EPI_ISL_1815518 | EXCITE Lab | Andersen lab at Scripps Research | Nicole L Washington, Simon White, Geraint Levan, Kelly Schiabor Barrett, Elizabeth Cirulli, Alexandre Bolze, Charlotte Rivera-Garcia, Ryan Cho, Jason Nguyen, Sherry Wang, Jimmy Ramirez, Celena Andrade, Alice Summerfield, Tyler Cassens, Efren Sandoval, Francisco Tanudjaja, Magnus Isaksson, William Lee, David Becker, Marc Laurent, James Lu + SEARCH |
| EPI_ISL_1815519 | EXCITE Lab | Andersen lab at Scripps Research | David Pride, Sharon Reed, Chip Schooley, Angela Scioscia, Natasha Martin Cheryl Anderson, Sawyer Farmer, Abigail Schnapper, Helena Tubb, Tommy Valles + SEARCH |
| EPI_ISL_1815520, EPI_ISL_1815521, EPI_ISL_1815522, EPI_ISL_1815523, EPI_ISL_1815524 | EXCITE Lab | Andersen lab at Scripps Research | Nicole L Washington, Simon White, Geraint Levan, Kelly Schiabor Barrett, Elizabeth Cirulli, Alexandre Bolze, Charlotte Rivera-Garcia, Ryan Cho, Jason Nguyen, Sherry Wang, Jimmy Ramirez, Celena Andrade, Alice Summerfield, Tyler Cassens, Efren Sandoval, Francisco Tanudjaja, Magnus Isaksson, William Lee, David Becker, Marc Laurent, James Lu + SEARCH |
| EPI_ISL_1815525 | EXCITE Lab | Andersen lab at Scripps Research | Chip Schooley, Natasha Martin, Cheryl Anderson, Angela Scioscia, Smruthi Karthikeyan, Greg Humphrey, Sawyer Farmer, Abigail Schnapper, Helena Tubb, Tommy Valles + SEARCH |
| EPI_ISL_1815526, EPI_ISL_1815527, EPI_ISL_1815528, EPI_ISL_1815529 | EXCITE Lab | Andersen lab at Scripps Research | Nicole L Washington, Simon White, Geraint Levan, Kelly Schiabor Barrett, Elizabeth Cirulli, Alexandre Bolze, Charlotte Rivera-Garcia, Ryan Cho, Jason Nguyen, Sherry Wang, Jimmy Ramirez, Celena Andrade, Alice Summerfield, Tyler Cassens, Efren Sandoval, Francisco Tanudjaja, Magnus Isaksson, William Lee, David Becker, Marc Laurent, James Lu + SEARCH |
| EPI_ISL_1815530 | EXCITE Lab | Andersen lab at Scripps Research | Rebecca Fielding-Miller, Laura Kohn, Patricia Gonzalez-Zuniga, Fitri Carrissa Wijaya, Vinton Omaleki, Marlene Flores, Araz Majnoonian, Megan Nguyen, Ashkan Hassani, Smruthi Karthikeyan, Rodolfo Salido, Victor Cantu, Greg Humphrey, Sawyer Farmer, Abigail Schnapper, Helena Tubb, Tommy Valles + SEARCH |
| EPI_ISL_1815531, EPI_ISL_1815532, EPI_ISL_1815533, EPI_ISL_1815534, EPI_ISL_1815535 | EXCITE Lab | Andersen lab at Scripps Research | Nicole L Washington, Simon White, Geraint Levan, Kelly Schiabor Barrett, Elizabeth Cirulli, Alexandre Bolze, Charlotte Rivera-Garcia, Ryan Cho, Jason Nguyen, Sherry Wang, Jimmy Ramirez, Celena Andrade, Alice Summerfield, Tyler Cassens, Efren Sandoval, Francisco Tanudjaja, Magnus Isaksson, William Lee, David Becker, Marc Laurent, James Lu + SEARCH |
| EPI_ISL_1815536 | EXCITE Lab | Andersen lab at Scripps Research | Chip Schooley, Natasha Martin, Cheryl Anderson, Angela Scioscia, Smruthi Karthikeyan, Greg Humphrey, Sawyer Farmer, Abigail Schnapper, Helena Tubb, Tommy Valles + SEARCH |
| EPI_ISL_1815537, EPI_ISL_1815538, EPI_ISL_1815539, EPI_ISL_1815540, EPI_ISL_1815541, EPI_ISL_1815542, EPI_ISL_1815543, EPI_ISL_1815544, EPI_ISL_1815545, EPI_ISL_1815546, EPI_ISL_1815547, EPI_ISL_1815548, EPI_ISL_1815549, EPI_ISL_1815550, EPI_ISL_1815551, EPI_ISL_1815552, EPI_ISL_1815553 | | | |
| see above | EXCITE Lab | Andersen lab at Scripps Research | Nicole L Washington, Simon White, Geraint Levan, Kelly Schiabor Barrett, Elizabeth Cirulli, Alexandre Bolze, Charlotte Rivera-Garcia, Ryan Cho, Jason Nguyen, Sherry Wang, Jimmy Ramirez, Celena Andrade, Alice Summerfield, Tyler Cassens, Efren Sandoval, Francisco Tanudjaja, Magnus Isaksson, William Lee, David Becker, Marc Laurent, James Lu + SEARCH |

| | | | |
|---|---|---|---|
| EPI_ISL_1815554 | EXCITE Lab | Andersen lab at Scripps Research | David Pride, Sharon Reed, Chip Schooley, Angela Scioscia, Natasha Martin Cheryl Anderson, Sawyer Farmer, Abigail Schnapper, Helena Tubb, Tommy Valles + SEARCH |
| EPI_ISL_1815555, EPI_ISL_1815556, EPI_ISL_1815557, EPI_ISL_1815558, EPI_ISL_1815559 | EXCITE Lab | Andersen lab at Scripps Research | Nicole L Washington, Simon White, Geraint Levan, Kelly Schiabor Barrett, Elizabeth Cirulli, Alexandre Bolze, Charlotte Rivera-Garcia, Ryan Cho, Jason Nguyen, Sherry Wang, Jimmy Ramirez, Celena Andrade, Alice Summerfield, Tyler Cassens, Efren Sandoval, Francisco Tanudjaja, Magnus Isaksson, William Lee, David Becker, Marc Laurent, James Lu + SEARCH |
| EPI_ISL_1815560 | EXCITE Lab | Andersen lab at Scripps Research | Chip Schooley, Natasha Martin, Cheryl Anderson, Angela Scioscia, Smruthi Karthikeyan, Greg Humphrey, Sawyer Farmer, Abigail Schnapper, Helena Tubb, Tommy Valles + SEARCH |
| EPI_ISL_1815561, EPI_ISL_1815562, EPI_ISL_1815563, EPI_ISL_1815564, EPI_ISL_1815565 | EXCITE Lab | Andersen lab at Scripps Research | Nicole L Washington, Simon White, Geraint Levan, Kelly Schiabor Barrett, Elizabeth Cirulli, Alexandre Bolze, Charlotte Rivera-Garcia, Ryan Cho, Jason Nguyen, Sherry Wang, Jimmy Ramirez, Celena Andrade, Alice Summerfield, Tyler Cassens, Efren Sandoval, Francisco Tanudjaja, Magnus Isaksson, William Lee, David Becker, Marc Laurent, James Lu + SEARCH |
| EPI_ISL_1815566 | EXCITE Lab | Andersen lab at Scripps Research | Sydney Morgan + Marni Jacobs + Celestine Magallanes + Holly Valentine + Louise C. Laurent + SEARCH |
| EPI_ISL_1815567 | EXCITE Lab | Andersen lab at Scripps Research | Nicole L Washington, Simon White, Geraint Levan, Kelly Schiabor Barrett, Elizabeth Cirulli, Alexandre Bolze, Charlotte Rivera-Garcia, Ryan Cho, Jason Nguyen, Sherry Wang, Jimmy Ramirez, Celena Andrade, Alice Summerfield, Tyler Cassens, Efren Sandoval, Francisco Tanudjaja, Magnus Isaksson, William Lee, David Becker, Marc Laurent, James Lu + SEARCH |
| EPI_ISL_1815568 | EXCITE Lab | Andersen lab at Scripps Research | Chip Schooley, Natasha Martin, Cheryl Anderson, Angela Scioscia, Smruthi Karthikeyan, Greg Humphrey, Sawyer Farmer, Abigail Schnapper, Helena Tubb, Tommy Valles + SEARCH |
| EPI_ISL_1815569, EPI_ISL_1815570, EPI_ISL_1815571, EPI_ISL_1815572, EPI_ISL_1815573, EPI_ISL_1815574, EPI_ISL_1815575, EPI_ISL_1815576, EPI_ISL_1815577, EPI_ISL_1815578, EPI_ISL_1815579, EPI_ISL_1815580, EPI_ISL_1815581, EPI_ISL_1815582, EPI_ISL_1815583, EPI_ISL_1815584, EPI_ISL_1815585 | | | |
| see above | EXCITE Lab | Andersen lab at Scripps Research | Nicole L Washington, Simon White, Geraint Levan, Kelly Schiabor Barrett, Elizabeth Cirulli, Alexandre Bolze, Charlotte Rivera-Garcia, Ryan Cho, Jason Nguyen, Sherry Wang, Jimmy Ramirez, Celena Andrade, Alice Summerfield, Tyler Cassens, Efren Sandoval, Francisco Tanudjaja, Magnus Isaksson, William Lee, David Becker, Marc Laurent, James Lu + SEARCH |
| EPI_ISL_1815586 | EXCITE Lab | Andersen lab at Scripps Research | David Pride, Sharon Reed, Chip Schooley, Angela Scioscia, Natasha Martin Cheryl Anderson, Sawyer Farmer, Abigail Schnapper, Helena Tubb, Tommy Valles + SEARCH |
| EPI_ISL_1815587, EPI_ISL_1815588, EPI_ISL_1815589, EPI_ISL_1815590, EPI_ISL_1815591, EPI_ISL_1815592, EPI_ISL_1815593 | EXCITE Lab | Andersen lab at Scripps Research | Nicole L Washington, Simon White, Geraint Levan, Kelly Schiabor Barrett, Elizabeth Cirulli, Alexandre Bolze, Charlotte Rivera-Garcia, Ryan Cho, Jason Nguyen, Sherry Wang, Jimmy Ramirez, Celena Andrade, Alice Summerfield, Tyler Cassens, Efren Sandoval, Francisco Tanudjaja, Magnus Isaksson, William Lee, David Becker, Marc Laurent, James Lu + SEARCH |
| EPI_ISL_1815594 | EXCITE Lab | Andersen lab at Scripps Research | Sydney Morgan + Marni Jacobs + Celestine Magallanes + Holly Valentine + Louise C. Laurent + SEARCH |
| EPI_ISL_1815595, EPI_ISL_1815596, EPI_ISL_1815597, EPI_ISL_1815598, EPI_ISL_1815599, EPI_ISL_1815600, EPI_ISL_1815601, EPI_ISL_1815602 | EXCITE Lab | Andersen lab at Scripps Research | Nicole L Washington, Simon White, Geraint Levan, Kelly Schiabor Barrett, Elizabeth Cirulli, Alexandre Bolze, Charlotte Rivera-Garcia, Ryan Cho, Jason Nguyen, Sherry Wang, Jimmy Ramirez, Celena Andrade, Alice Summerfield, Tyler Cassens, Efren Sandoval, Francisco Tanudjaja, Magnus Isaksson, William Lee, David Becker, Marc Laurent, James Lu + SEARCH |
| EPI_ISL_1815603 | EXCITE Lab | Andersen lab at Scripps Research | David Pride, Sharon Reed, Chip Schooley, Angela Scioscia, Natasha Martin Cheryl Anderson, Sawyer Farmer, Abigail Schnapper, Helena Tubb, Tommy Valles + SEARCH |
| EPI_ISL_1815604, EPI_ISL_1815605, EPI_ISL_1815606, EPI_ISL_1815607, EPI_ISL_1815608, EPI_ISL_1815609, EPI_ISL_1815610 | EXCITE Lab | Andersen lab at Scripps Research | Nicole L Washington, Simon White, Geraint Levan, Kelly Schiabor Barrett, Elizabeth Cirulli, Alexandre Bolze, Charlotte Rivera-Garcia, Ryan Cho, Jason Nguyen, Sherry Wang, Jimmy Ramirez, Celena Andrade, Alice Summerfield, Tyler Cassens, Efren Sandoval, Francisco Tanudjaja, Magnus Isaksson, William Lee, David Becker, Marc Laurent, James Lu + SEARCH |
| EPI_ISL_1815611 | EXCITE Lab | Andersen lab at Scripps Research | Sydney Morgan + Marni Jacobs + Celestine Magallanes + Holly Valentine + Louise C. Laurent + SEARCH |
| EPI_ISL_1815612, EPI_ISL_1815613 | EXCITE Lab | Andersen lab at Scripps Research | Nicole L Washington, Simon White, Geraint Levan, Kelly Schiabor Barrett, Elizabeth Cirulli, Alexandre Bolze, Charlotte Rivera-Garcia, Ryan Cho, Jason Nguyen, Sherry Wang, Jimmy Ramirez, Celena Andrade, Alice Summerfield, Tyler Cassens, Efren Sandoval, Francisco Tanudjaja, Magnus Isaksson, William Lee, David Becker, Marc Laurent, James Lu + SEARCH |
| EPI_ISL_1815614 | EXCITE Lab | Andersen lab at Scripps Research | Chip Schooley, Natasha Martin, Cheryl Anderson, Angela Scioscia, Smruthi Karthikeyan, Greg Humphrey, Sawyer Farmer, Abigail Schnapper, Helena Tubb, Tommy Valles + SEARCH |
| EPI_ISL_1815615, EPI_ISL_1815616, EPI_ISL_1815617, EPI_ISL_1815618, EPI_ISL_1815619, EPI_ISL_1815620 | EXCITE Lab | Andersen lab at Scripps Research | Nicole L Washington, Simon White, Geraint Levan, Kelly Schiabor Barrett, Elizabeth Cirulli, Alexandre Bolze, Charlotte Rivera-Garcia, Ryan Cho, Jason Nguyen, Sherry Wang, Jimmy Ramirez, Celena Andrade, Alice Summerfield, Tyler Cassens, Efren Sandoval, Francisco Tanudjaja, Magnus Isaksson, William Lee, David Becker, Marc Laurent, James Lu + SEARCH |
| EPI_ISL_1815621 | EXCITE Lab | Andersen lab at Scripps Research | Chip Schooley, Natasha Martin, Cheryl Anderson, Angela Scioscia, Smruthi Karthikeyan, Greg Humphrey, Sawyer Farmer, Abigail Schnapper, Helena Tubb, Tommy Valles + SEARCH |
| EPI_ISL_1815622, EPI_ISL_1815623, EPI_ISL_1815624, EPI_ISL_1815625, EPI_ISL_1815626, EPI_ISL_1815627 | EXCITE Lab | Andersen lab at Scripps Research | Nicole L Washington, Simon White, Geraint Levan, Kelly Schiabor Barrett, Elizabeth Cirulli, Alexandre Bolze, Charlotte Rivera-Garcia, Ryan Cho, Jason Nguyen, Sherry Wang, Jimmy Ramirez, Celena Andrade, Alice Summerfield, Tyler Cassens, Efren Sandoval, Francisco Tanudjaja, Magnus Isaksson, William Lee, David Becker, Marc Laurent, James Lu + SEARCH |
| EPI_ISL_1815628 | EXCITE Lab | Andersen lab at Scripps Research | Chip Schooley, Natasha Martin, Cheryl Anderson, Angela Scioscia, Smruthi Karthikeyan, Greg Humphrey, Sawyer Farmer, Abigail Schnapper, Helena Tubb, Tommy Valles + SEARCH |
| EPI_ISL_1815629, EPI_ISL_1815630, EPI_ISL_1815631, EPI_ISL_1815632, EPI_ISL_1815633, EPI_ISL_1815634 | EXCITE Lab | Andersen lab at Scripps Research | Nicole L Washington, Simon White, Geraint Levan, Kelly Schiabor Barrett, Elizabeth Cirulli, Alexandre Bolze, Charlotte Rivera-Garcia, Ryan Cho, Jason Nguyen, Sherry Wang, Jimmy Ramirez, Celena Andrade, Alice Summerfield, Tyler Cassens, Efren Sandoval, Francisco Tanudjaja, Magnus Isaksson, William Lee, David Becker, Marc Laurent, James Lu + SEARCH |
| EPI_ISL_1815635 | EXCITE Lab | Andersen lab at Scripps Research | Fred Wu, Christine Mitchell + SEARCH |
| EPI_ISL_1815636, EPI_ISL_1815637, EPI_ISL_1815638, EPI_ISL_1815639, EPI_ISL_1815640, EPI_ISL_1815641, EPI_ISL_1815642, EPI_ISL_1815643, EPI_ISL_1815644, EPI_ISL_1815645, EPI_ISL_1815646, EPI_ISL_1815647, EPI_ISL_1815648, EPI_ISL_1815649, EPI_ISL_1815650, EPI_ISL_1815651 | | | |
| see above | EXCITE Lab | Andersen lab at Scripps Research | Nicole L Washington, Simon White, Geraint Levan, Kelly Schiabor Barrett, Elizabeth Cirulli, Alexandre Bolze, Charlotte Rivera-Garcia, Ryan Cho, Jason Nguyen, Sherry Wang, Jimmy Ramirez, Celena Andrade, Alice Summerfield, Tyler Cassens, Efren Sandoval, Francisco Tanudjaja, Magnus Isaksson, William Lee, David Becker, Marc Laurent, James Lu + SEARCH |
| EPI_ISL_1815652 | EXCITE Lab | Andersen lab at Scripps Research | Sydney Morgan + Marni Jacobs + Celestine Magallanes + Holly Valentine + Louise C. Laurent + SEARCH |
| EPI_ISL_1815653, EPI_ISL_1815654, EPI_ISL_1815655, EPI_ISL_1815656, EPI_ISL_1815657, EPI_ISL_1815658 | EXCITE Lab | Andersen lab at Scripps Research | Nicole L Washington, Simon White, Geraint Levan, Kelly Schiabor Barrett, Elizabeth Cirulli, Alexandre Bolze, Charlotte Rivera-Garcia, Ryan Cho, Jason Nguyen, Sherry Wang, Jimmy Ramirez, Celena Andrade, Alice Summerfield, Tyler Cassens, Efren Sandoval, Francisco Tanudjaja, Magnus Isaksson, William Lee, David Becker, Marc Laurent, James Lu + SEARCH |
| EPI_ISL_1815659 | EXCITE Lab | Andersen lab at Scripps Research | Chip Schooley, Natasha Martin, Cheryl Anderson, Angela Scioscia, Smruthi Karthikeyan, Greg Humphrey, Sawyer Farmer, Abigail Schnapper, Helena Tubb, Tommy Valles + SEARCH |
| EPI_ISL_1815660, EPI_ISL_1815661, EPI_ISL_1815662 | EXCITE Lab | Andersen lab at Scripps Research | Nicole L Washington, Simon White, Geraint Levan, Kelly Schiabor Barrett, Elizabeth Cirulli, Alexandre Bolze, Charlotte Rivera-Garcia, Ryan Cho, Jason Nguyen, Sherry Wang, Jimmy Ramirez, Celena Andrade, Alice Summerfield, Tyler Cassens, Efren Sandoval, Francisco Tanudjaja, Magnus Isaksson, William Lee, David Becker, Marc Laurent, James Lu + SEARCH |
| EPI_ISL_1815663 | EXCITE Lab | Andersen lab at Scripps Research | Fred Wu, Christine Mitchell + SEARCH |
| EPI_ISL_1815664, EPI_ISL_1815665, EPI_ISL_1815666, EPI_ISL_1815667, EPI_ISL_1815668, EPI_ISL_1815669, EPI_ISL_1815670, EPI_ISL_1815671, EPI_ISL_1815672, EPI_ISL_1815673, EPI_ISL_1815674, EPI_ISL_1815675, EPI_ISL_1815676 | | | |
| see above | EXCITE Lab | Andersen lab at Scripps Research | Nicole L Washington, Simon White, Geraint Levan, Kelly Schiabor Barrett, Elizabeth Cirulli, Alexandre Bolze, Charlotte Rivera-Garcia, Ryan Cho, Jason Nguyen, Sherry Wang, Jimmy Ramirez, Celena Andrade, Alice Summerfield, Tyler Cassens, Efren Sandoval, Francisco Tanudjaja, Magnus Isaksson, William Lee, David Becker, Marc Laurent, James Lu + SEARCH |
| EPI_ISL_1815677 | EXCITE Lab | Andersen lab at Scripps Research | Sydney Morgan + Marni Jacobs + Celestine Magallanes + Holly Valentine + Louise C. Laurent + SEARCH |
| EPI_ISL_1815678, EPI_ISL_1815679, EPI_ISL_1815680, EPI_ISL_1815681, EPI_ISL_1815682, EPI_ISL_1815683, EPI_ISL_1815684, EPI_ISL_1815685, EPI_ISL_1815686, EPI_ISL_1815687, EPI_ISL_1815688, EPI_ISL_1815689, EPI_ISL_1815690, EPI_ISL_1815691, EPI_ISL_1815692, EPI_ISL_1815693, EPI_ISL_1815694, EPI_ISL_1815695, EPI_ISL_1815696, EPI_ISL_1815697, EPI_ISL_1815698, EPI_ISL_1815699, EPI_ISL_1815700, EPI_ISL_1815701, EPI_ISL_1815702 | | | |
| see above | EXCITE Lab | Andersen lab at Scripps Research | Nicole L Washington, Simon White, Geraint Levan, Kelly Schiabor Barrett, Elizabeth Cirulli, Alexandre Bolze, Charlotte Rivera-Garcia, Ryan Cho, Jason Nguyen, Sherry Wang, Jimmy Ramirez, Celena Andrade, Alice Summerfield, Tyler Cassens, Efren Sandoval, Francisco Tanudjaja, Magnus Isaksson, William Lee, David Becker, Marc Laurent, James Lu + SEARCH |
| EPI_ISL_1815703 | EXCITE Lab | Andersen lab at Scripps Research | Chip Schooley, Natasha Martin, Cheryl Anderson, Angela Scioscia, Smruthi Karthikeyan, Greg Humphrey, Sawyer Farmer, Abigail Schnapper, Helena Tubb, Tommy Valles + SEARCH |
| EPI_ISL_1815704, EPI_ISL_1815705, EPI_ISL_1815706, EPI_ISL_1815707, EPI_ISL_1815708, EPI_ISL_1815709, EPI_ISL_1815710, EPI_ISL_1815711, EPI_ISL_1815712, EPI_ISL_1815713, EPI_ISL_1815714, EPI_ISL_1815715 | | | |

| | | | |
|---|---|---|---|
| see above | EXCITE Lab | Andersen lab at Scripps Research | Nicole L Washington, Simon White, Geraint Levan, Kelly Schiabor Barrett, Elizabeth Cirulli, Alexandre Bolze, Charlotte Rivera-Garcia, Ryan Cho, Jason Nguyen, Sherry Wang, Jimmy Ramirez, Celena Andrade, Alice Summerfield, Tyler Cassens, Efren Sandoval, Francisco Tanudjaja, Magnus Isaksson, William Lee, David Becker, Marc Laurent, James Lu + SEARCH |
| EPI_ISL_1815716 | EXCITE Lab | Andersen lab at Scripps Research | Chip Schooley, Natasha Martin, Cheryl Anderson, Angela Scioscia, Smruthi Karthikeyan, Greg Humphrey, Sawyer Farmer, Abigail Schnapper, Helena Tubb, Tommy Valles + SEARCH |
| EPI_ISL_1815717, EPI_ISL_1815718 | EXCITE Lab | Andersen lab at Scripps Research | Nicole L Washington, Simon White, Geraint Levan, Kelly Schiabor Barrett, Elizabeth Cirulli, Alexandre Bolze, Charlotte Rivera-Garcia, Ryan Cho, Jason Nguyen, Sherry Wang, Jimmy Ramirez, Celena Andrade, Alice Summerfield, Tyler Cassens, Efren Sandoval, Francisco Tanudjaja, Magnus Isaksson, William Lee, David Becker, Marc Laurent, James Lu + SEARCH |
| EPI_ISL_1815719 | EXCITE Lab | Andersen lab at Scripps Research | Chip Schooley, Natasha Martin, Cheryl Anderson, Angela Scioscia, Smruthi Karthikeyan, Greg Humphrey, Sawyer Farmer, Abigail Schnapper, Helena Tubb, Tommy Valles + SEARCH |
| EPI_ISL_1815720, EPI_ISL_1815721, EPI_ISL_1815722, EPI_ISL_1815723, EPI_ISL_1815724, EPI_ISL_1815725, EPI_ISL_1815726, EPI_ISL_1815727, EPI_ISL_1815728, EPI_ISL_1815729 | EXCITE Lab | Andersen lab at Scripps Research | Nicole L Washington, Simon White, Geraint Levan, Kelly Schiabor Barrett, Elizabeth Cirulli, Alexandre Bolze, Charlotte Rivera-Garcia, Ryan Cho, Jason Nguyen, Sherry Wang, Jimmy Ramirez, Celena Andrade, Alice Summerfield, Tyler Cassens, Efren Sandoval, Francisco Tanudjaja, Magnus Isaksson, William Lee, David Becker, Marc Laurent, James Lu + SEARCH |
| EPI_ISL_1815730 | EXCITE Lab | Andersen lab at Scripps Research | David Pride, Sharon Reed, Chip Schooley, Angela Scioscia, Natasha Martin Cheryl Anderson, Sawyer Farmer, Abigail Schnapper, Helena Tubb, Tommy Valles + SEARCH |
| EPI_ISL_1815731, EPI_ISL_1815732, EPI_ISL_1815733 | EXCITE Lab | Andersen lab at Scripps Research | Nicole L Washington, Simon White, Geraint Levan, Kelly Schiabor Barrett, Elizabeth Cirulli, Alexandre Bolze, Charlotte Rivera-Garcia, Ryan Cho, Jason Nguyen, Sherry Wang, Jimmy Ramirez, Celena Andrade, Alice Summerfield, Tyler Cassens, Efren Sandoval, Francisco Tanudjaja, Magnus Isaksson, William Lee, David Becker, Marc Laurent, James Lu + SEARCH |
| EPI_ISL_1815734 | EXCITE Lab | Andersen lab at Scripps Research | David Pride, Sharon Reed, Chip Schooley, Angela Scioscia, Natasha Martin Cheryl Anderson, Sawyer Farmer, Abigail Schnapper, Helena Tubb, Tommy Valles + SEARCH |
| EPI_ISL_1815735, EPI_ISL_1815736, EPI_ISL_1815737, EPI_ISL_1815738, EPI_ISL_1815739, EPI_ISL_1815740, EPI_ISL_1815741, EPI_ISL_1815742, EPI_ISL_1815743, EPI_ISL_1815744, EPI_ISL_1815745, EPI_ISL_1815746, EPI_ISL_1815747 | | | |
| see above | EXCITE Lab | Andersen lab at Scripps Research | Nicole L Washington, Simon White, Geraint Levan, Kelly Schiabor Barrett, Elizabeth Cirulli, Alexandre Bolze, Charlotte Rivera-Garcia, Ryan Cho, Jason Nguyen, Sherry Wang, Jimmy Ramirez, Celena Andrade, Alice Summerfield, Tyler Cassens, Efren Sandoval, Francisco Tanudjaja, Magnus Isaksson, William Lee, David Becker, Marc Laurent, James Lu + SEARCH |
| EPI_ISL_1815748 | EXCITE Lab | Andersen lab at Scripps Research | David Pride, Sharon Reed, Chip Schooley, Angela Scioscia, Natasha Martin Cheryl Anderson, Sawyer Farmer, Abigail Schnapper, Helena Tubb, Tommy Valles + SEARCH |
| EPI_ISL_1815749, EPI_ISL_1815750, EPI_ISL_1815751, EPI_ISL_1815752, EPI_ISL_1815753, EPI_ISL_1815754, EPI_ISL_1815755, EPI_ISL_1815756, EPI_ISL_1815757, EPI_ISL_1815758, EPI_ISL_1815759, EPI_ISL_1815760, EPI_ISL_1815761, EPI_ISL_1815762, EPI_ISL_1815763 | | | |
| see above | EXCITE Lab | Andersen lab at Scripps Research | Nicole L Washington, Simon White, Geraint Levan, Kelly Schiabor Barrett, Elizabeth Cirulli, Alexandre Bolze, Charlotte Rivera-Garcia, Ryan Cho, Jason Nguyen, Sherry Wang, Jimmy Ramirez, Celena Andrade, Alice Summerfield, Tyler Cassens, Efren Sandoval, Francisco Tanudjaja, Magnus Isaksson, William Lee, David Becker, Marc Laurent, James Lu + SEARCH |
| EPI_ISL_1815764 | EXCITE Lab | Andersen lab at Scripps Research | Sydney Morgan + Marni Jacobs + Celestine Magallanes + Holly Valentine + Louise C. Laurent + SEARCH |
| EPI_ISL_1815765, EPI_ISL_1815766, EPI_ISL_1815767, EPI_ISL_1815768, EPI_ISL_1815769, EPI_ISL_1815770, EPI_ISL_1815771, EPI_ISL_1815772, EPI_ISL_1815773, EPI_ISL_1815774, EPI_ISL_1815775, EPI_ISL_1815776, EPI_ISL_1815777, EPI_ISL_1815778, EPI_ISL_1815779, EPI_ISL_1815780, EPI_ISL_1815781, EPI_ISL_1815782, EPI_ISL_1815783 | | | |
| see above | EXCITE Lab | Andersen lab at Scripps Research | Nicole L Washington, Simon White, Geraint Levan, Kelly Schiabor Barrett, Elizabeth Cirulli, Alexandre Bolze, Charlotte Rivera-Garcia, Ryan Cho, Jason Nguyen, Sherry Wang, Jimmy Ramirez, Celena Andrade, Alice Summerfield, Tyler Cassens, Efren Sandoval, Francisco Tanudjaja, Magnus Isaksson, William Lee, David Becker, Marc Laurent, James Lu + SEARCH |
| EPI_ISL_1815784 | EXCITE Lab | Andersen lab at Scripps Research | David Pride, Sharon Reed, Chip Schooley, Angela Scioscia, Natasha Martin Cheryl Anderson, Sawyer Farmer, Abigail Schnapper, Helena Tubb, Tommy Valles + SEARCH |
| EPI_ISL_1815785, EPI_ISL_1815786, EPI_ISL_1815787, EPI_ISL_1815788 | EXCITE Lab | Andersen lab at Scripps Research | Nicole L Washington, Simon White, Geraint Levan, Kelly Schiabor Barrett, Elizabeth Cirulli, Alexandre Bolze, Charlotte Rivera-Garcia, Ryan Cho, Jason Nguyen, Sherry Wang, Jimmy Ramirez, Celena Andrade, Alice Summerfield, Tyler Cassens, Efren Sandoval, Francisco Tanudjaja, Magnus Isaksson, William Lee, David Becker, Marc Laurent, James Lu + SEARCH |
| EPI_ISL_1815789 | EXCITE Lab | Andersen lab at Scripps Research | David Pride, Sharon Reed, Chip Schooley, Angela Scioscia, Natasha Martin Cheryl Anderson, Sawyer Farmer, Abigail Schnapper, Helena Tubb, Tommy Valles + SEARCH |
| EPI_ISL_1815790 | EXCITE Lab | Andersen lab at Scripps Research | Chip Schooley, Natasha Martin, Cheryl Anderson, Angela Scioscia, Smruthi Karthikeyan, Greg Humphrey, Sawyer Farmer, Abigail Schnapper, Helena Tubb, Tommy Valles + SEARCH |
| EPI_ISL_1815791, EPI_ISL_1815792, EPI_ISL_1815793, EPI_ISL_1815794 | EXCITE Lab | Andersen lab at Scripps Research | Nicole L Washington, Simon White, Geraint Levan, Kelly Schiabor Barrett, Elizabeth Cirulli, Alexandre Bolze, Charlotte Rivera-Garcia, Ryan Cho, Jason Nguyen, Sherry Wang, Jimmy Ramirez, Celena Andrade, Alice Summerfield, Tyler Cassens, Efren Sandoval, Francisco Tanudjaja, Magnus Isaksson, William Lee, David Becker, Marc Laurent, James Lu + SEARCH |
| EPI_ISL_1815795 | EXCITE Lab | Andersen lab at Scripps Research | Chip Schooley, Natasha Martin, Cheryl Anderson, Angela Scioscia, Smruthi Karthikeyan, Greg Humphrey, Sawyer Farmer, Abigail Schnapper, Helena Tubb, Tommy Valles + SEARCH |
| EPI_ISL_1815796, EPI_ISL_1815797, EPI_ISL_1815798, EPI_ISL_1815799 | EXCITE Lab | Andersen lab at Scripps Research | Nicole L Washington, Simon White, Geraint Levan, Kelly Schiabor Barrett, Elizabeth Cirulli, Alexandre Bolze, Charlotte Rivera-Garcia, Ryan Cho, Jason Nguyen, Sherry Wang, Jimmy Ramirez, Celena Andrade, Alice Summerfield, Tyler Cassens, Efren Sandoval, Francisco Tanudjaja, Magnus Isaksson, William Lee, David Becker, Marc Laurent, James Lu + SEARCH |
| EPI_ISL_1815800, EPI_ISL_1815801 | EXCITE Lab | Andersen lab at Scripps Research | Chip Schooley, Natasha Martin, Cheryl Anderson, Angela Scioscia, Smruthi Karthikeyan, Greg Humphrey, Sawyer Farmer, Abigail Schnapper, Helena Tubb, Tommy Valles + SEARCH |
| EPI_ISL_1815802 | EXCITE Lab | Andersen lab at Scripps Research | Nicole L Washington, Simon White, Geraint Levan, Kelly Schiabor Barrett, Elizabeth Cirulli, Alexandre Bolze, Charlotte Rivera-Garcia, Ryan Cho, Jason Nguyen, Sherry Wang, Jimmy Ramirez, Celena Andrade, Alice Summerfield, Tyler Cassens, Efren Sandoval, Francisco Tanudjaja, Magnus Isaksson, William Lee, David Becker, Marc Laurent, James Lu + SEARCH |
| EPI_ISL_1815803 | EXCITE Lab | Andersen lab at Scripps Research | Chip Schooley, Natasha Martin, Cheryl Anderson, Angela Scioscia, Smruthi Karthikeyan, Greg Humphrey, Sawyer Farmer, Abigail Schnapper, Helena Tubb, Tommy Valles + SEARCH |
| EPI_ISL_1815804 | EXCITE Lab | Andersen lab at Scripps Research | Sydney Morgan + Marni Jacobs + Celestine Magallanes + Holly Valentine + Louise C. Laurent + SEARCH |
| EPI_ISL_1815805, EPI_ISL_1815806, EPI_ISL_1815807, EPI_ISL_1815808, EPI_ISL_1815809, EPI_ISL_1815810, EPI_ISL_1815811, EPI_ISL_1815812 | EXCITE Lab | Andersen lab at Scripps Research | Nicole L Washington, Simon White, Geraint Levan, Kelly Schiabor Barrett, Elizabeth Cirulli, Alexandre Bolze, Charlotte Rivera-Garcia, Ryan Cho, Jason Nguyen, Sherry Wang, Jimmy Ramirez, Celena Andrade, Alice Summerfield, Tyler Cassens, Efren Sandoval, Francisco Tanudjaja, Magnus Isaksson, William Lee, David Becker, Marc Laurent, James Lu + SEARCH |
| EPI_ISL_1815813 | EXCITE Lab | Andersen lab at Scripps Research | Chip Schooley, Natasha Martin, Cheryl Anderson, Angela Scioscia, Smruthi Karthikeyan, Greg Humphrey, Sawyer Farmer, Abigail Schnapper, Helena Tubb, Tommy Valles + SEARCH |
| EPI_ISL_1815814 | EXCITE Lab | Andersen lab at Scripps Research | David Pride, Sharon Reed, Chip Schooley, Angela Scioscia, Natasha Martin Cheryl Anderson, Sawyer Farmer, Abigail Schnapper, Helena Tubb, Tommy Valles + SEARCH |
| EPI_ISL_1815815, EPI_ISL_1815816, EPI_ISL_1815817, EPI_ISL_1815818, EPI_ISL_1815819, EPI_ISL_1815820, EPI_ISL_1815821, EPI_ISL_1815822, EPI_ISL_1815823 | EXCITE Lab | Andersen lab at Scripps Research | Nicole L Washington, Simon White, Geraint Levan, Kelly Schiabor Barrett, Elizabeth Cirulli, Alexandre Bolze, Charlotte Rivera-Garcia, Ryan Cho, Jason Nguyen, Sherry Wang, Jimmy Ramirez, Celena Andrade, Alice Summerfield, Tyler Cassens, Efren Sandoval, Francisco Tanudjaja, Magnus Isaksson, William Lee, David Becker, Marc Laurent, James Lu + SEARCH |
| EPI_ISL_1815824 | EXCITE Lab | Andersen lab at Scripps Research | Chip Schooley, Natasha Martin, Cheryl Anderson, Angela Scioscia, Smruthi Karthikeyan, Greg Humphrey, Sawyer Farmer, Abigail Schnapper, Helena Tubb, Tommy Valles + SEARCH |
| EPI_ISL_1815825 | EXCITE Lab | Andersen lab at Scripps Research | Nicole L Washington, Simon White, Geraint Levan, Kelly Schiabor Barrett, Elizabeth Cirulli, Alexandre Bolze, Charlotte Rivera-Garcia, Ryan Cho, Jason Nguyen, Sherry Wang, Jimmy Ramirez, Celena Andrade, Alice Summerfield, Tyler Cassens, Efren Sandoval, Francisco Tanudjaja, Magnus Isaksson, William Lee, David Becker, Marc Laurent, James Lu + SEARCH |
| EPI_ISL_1815826 | EXCITE Lab | Andersen lab at Scripps Research | Fred Wu, Christine Mitchell + SEARCH |
| EPI_ISL_1815827, EPI_ISL_1815828, EPI_ISL_1815829, EPI_ISL_1815830, EPI_ISL_1815831, EPI_ISL_1815832, EPI_ISL_1815833, EPI_ISL_1815834, EPI_ISL_1815835, EPI_ISL_1815836, EPI_ISL_1815837, EPI_ISL_1815838, EPI_ISL_1815839, EPI_ISL_1815840, EPI_ISL_1815841, EPI_ISL_1815842, EPI_ISL_1815843 | | | |
| see above | EXCITE Lab | Andersen lab at Scripps Research | Nicole L Washington, Simon White, Geraint Levan, Kelly Schiabor Barrett, Elizabeth Cirulli, Alexandre Bolze, Charlotte Rivera-Garcia, Ryan Cho, Jason Nguyen, Sherry Wang, Jimmy Ramirez, Celena Andrade, Alice Summerfield, Tyler Cassens, Efren Sandoval, Francisco Tanudjaja, Magnus Isaksson, William Lee, David Becker, Marc Laurent, James Lu + SEARCH |
| EPI_ISL_1815844 | EXCITE Lab | Andersen lab at Scripps Research | Chip Schooley, Natasha Martin, Cheryl Anderson, Angela Scioscia, Smruthi Karthikeyan, Greg Humphrey, Sawyer Farmer, Abigail Schnapper, Helena Tubb, Tommy Valles + SEARCH |
| EPI_ISL_1815845, EPI_ISL_1815846, EPI_ISL_1815847, EPI_ISL_1815848, EPI_ISL_1815849, EPI_ISL_1815850, EPI_ISL_1815851, EPI_ISL_1815852, EPI_ISL_1815853, EPI_ISL_1815854, EPI_ISL_1815855, EPI_ISL_1815856 | | | |
| see above | EXCITE Lab | Andersen lab at Scripps Research | Nicole L Washington, Simon White, Geraint Levan, Kelly Schiabor Barrett, Elizabeth Cirulli, Alexandre Bolze, Charlotte Rivera-Garcia, Ryan Cho, Jason Nguyen, Sherry Wang, Jimmy Ramirez, Celena Andrade, Alice Summerfield, Tyler Cassens, Efren Sandoval, Francisco Tanudjaja, Magnus Isaksson, William Lee, David Becker, Marc Laurent, James Lu + SEARCH |
| EPI_ISL_1815857 | EXCITE Lab | Andersen lab at Scripps Research | David Pride, Sharon Reed, Chip Schooley, Angela Scioscia, Natasha Martin Cheryl Anderson, Sawyer Farmer, Abigail Schnapper, Helena Tubb, Tommy Valles + SEARCH |
| EPI_ISL_1815858 | EXCITE Lab | Andersen lab at Scripps Research | Chip Schooley, Natasha Martin, Cheryl Anderson, Angela Scioscia, Smruthi Karthikeyan, Greg Humphrey, Sawyer Farmer, Abigail Schnapper, Helena Tubb, Tommy Valles + SEARCH |
| EPI_ISL_1815859, EPI_ISL_1815860, EPI_ISL_1815861, EPI_ISL_1815862, EPI_ISL_1815863, | EXCITE Lab | Andersen lab at Scripps Research | Nicole L Washington, Simon White, Geraint Levan, Kelly Schiabor Barrett, Elizabeth Cirulli, Alexandre Bolze, Charlotte Rivera-Garcia, Ryan Cho, Jason Nguyen, Sherry Wang, Jimmy Ramirez, Celena Andrade, Alice Summerfield, Tyler Cassens, Efren Sandoval, Francisco Tanudjaja, Magnus Isaksson, William Lee, David Becker, Marc Laurent, James Lu + SEARCH |

| | | | |
|---|---|---|---|
| EPI_ISL_1815864, EPI_ISL_1815865, EPI_ISL_1815866 | | | |
| EPI_ISL_1815867 | EXCITE Lab | Andersen lab at Scripps Research | Chip Schooley, Natasha Martin, Cheryl Anderson, Angela Scioscia, Smruthi Karthikeyan, Greg Humphrey, Sawyer Farmer, Abigail Schnapper, Helena Tubb, Tommy Valles + SEARCH |
| EPI_ISL_1815868, EPI_ISL_1815869, EPI_ISL_1815870 | EXCITE Lab | Andersen lab at Scripps Research | Nicole L Washington, Simon White, Geraint Levan, Kelly Schiabor Barrett, Elizabeth Cirulli, Alexandre Bolze, Charlotte Rivera-Garcia, Ryan Cho, Jason Nguyen, Sherry Wang, Jimmy Ramirez, Celena Andrade, Alice Summerfield, Tyler Cassens, Efren Sandoval, Francisco Tanudjaja, Magnus Isaksson, William Lee, David Becker, Marc Laurent, James Lu + SEARCH |
| EPI_ISL_1815871 | EXCITE Lab | Andersen lab at Scripps Research | Chip Schooley, Natasha Martin, Cheryl Anderson, Angela Scioscia, Smruthi Karthikeyan, Greg Humphrey, Sawyer Farmer, Abigail Schnapper, Helena Tubb, Tommy Valles + SEARCH |
| EPI_ISL_1815872, EPI_ISL_1815873, EPI_ISL_1815874, EPI_ISL_1815875, EPI_ISL_1815876, EPI_ISL_1815877, EPI_ISL_1815878, EPI_ISL_1815879, EPI_ISL_1815880, EPI_ISL_1815881, EPI_ISL_1815882, EPI_ISL_1815883, EPI_ISL_1815884, EPI_ISL_1815885, EPI_ISL_1815886, EPI_ISL_1815887, EPI_ISL_1815888 | | | |
| see above | EXCITE Lab | Andersen lab at Scripps Research | Nicole L Washington, Simon White, Geraint Levan, Kelly Schiabor Barrett, Elizabeth Cirulli, Alexandre Bolze, Charlotte Rivera-Garcia, Ryan Cho, Jason Nguyen, Sherry Wang, Jimmy Ramirez, Celena Andrade, Alice Summerfield, Tyler Cassens, Efren Sandoval, Francisco Tanudjaja, Magnus Isaksson, William Lee, David Becker, Marc Laurent, James Lu + SEARCH |
| EPI_ISL_1815889 | EXCITE Lab | Andersen lab at Scripps Research | Fred Wu, Christine Mitchell + SEARCH |
| EPI_ISL_1815890, EPI_ISL_1815891, EPI_ISL_1815892, EPI_ISL_1815893, EPI_ISL_1815894, EPI_ISL_1815895 | EXCITE Lab | Andersen lab at Scripps Research | Nicole L Washington, Simon White, Geraint Levan, Kelly Schiabor Barrett, Elizabeth Cirulli, Alexandre Bolze, Charlotte Rivera-Garcia, Ryan Cho, Jason Nguyen, Sherry Wang, Jimmy Ramirez, Celena Andrade, Alice Summerfield, Tyler Cassens, Efren Sandoval, Francisco Tanudjaja, Magnus Isaksson, William Lee, David Becker, Marc Laurent, James Lu + SEARCH |
| EPI_ISL_1815896 | EXCITE Lab | Andersen lab at Scripps Research | Chip Schooley, Natasha Martin, Cheryl Anderson, Angela Scioscia, Smruthi Karthikeyan, Greg Humphrey, Sawyer Farmer, Abigail Schnapper, Helena Tubb, Tommy Valles + SEARCH |
| EPI_ISL_1815897, EPI_ISL_1815898, EPI_ISL_1815899, EPI_ISL_1815900, EPI_ISL_1815901, EPI_ISL_1815902 | EXCITE Lab | Andersen lab at Scripps Research | Nicole L Washington, Simon White, Geraint Levan, Kelly Schiabor Barrett, Elizabeth Cirulli, Alexandre Bolze, Charlotte Rivera-Garcia, Ryan Cho, Jason Nguyen, Sherry Wang, Jimmy Ramirez, Celena Andrade, Alice Summerfield, Tyler Cassens, Efren Sandoval, Francisco Tanudjaja, Magnus Isaksson, William Lee, David Becker, Marc Laurent, James Lu + SEARCH |
| EPI_ISL_1815903 | EXCITE Lab | Andersen lab at Scripps Research | Sydney Morgan + Marni Jacobs + Celestine Magallanes + Holly Valentine + Louise C. Laurent + SEARCH |
| EPI_ISL_1815904, EPI_ISL_1815905 | EXCITE Lab | Andersen lab at Scripps Research | Nicole L Washington, Simon White, Geraint Levan, Kelly Schiabor Barrett, Elizabeth Cirulli, Alexandre Bolze, Charlotte Rivera-Garcia, Ryan Cho, Jason Nguyen, Sherry Wang, Jimmy Ramirez, Celena Andrade, Alice Summerfield, Tyler Cassens, Efren Sandoval, Francisco Tanudjaja, Magnus Isaksson, William Lee, David Becker, Marc Laurent, James Lu + SEARCH |
| EPI_ISL_1815906, EPI_ISL_1815907 | EXCITE Lab | Andersen lab at Scripps Research | Chip Schooley, Natasha Martin, Cheryl Anderson, Angela Scioscia, Smruthi Karthikeyan, Greg Humphrey, Sawyer Farmer, Abigail Schnapper, Helena Tubb, Tommy Valles + SEARCH |
| EPI_ISL_1815908, EPI_ISL_1815909, EPI_ISL_1815910, EPI_ISL_1815911, EPI_ISL_1815912, EPI_ISL_1815913, EPI_ISL_1815914 | EXCITE Lab | Andersen lab at Scripps Research | Nicole L Washington, Simon White, Geraint Levan, Kelly Schiabor Barrett, Elizabeth Cirulli, Alexandre Bolze, Charlotte Rivera-Garcia, Ryan Cho, Jason Nguyen, Sherry Wang, Jimmy Ramirez, Celena Andrade, Alice Summerfield, Tyler Cassens, Efren Sandoval, Francisco Tanudjaja, Magnus Isaksson, William Lee, David Becker, Marc Laurent, James Lu + SEARCH |
| EPI_ISL_1815915 | EXCITE Lab | Andersen lab at Scripps Research | David Pride, Sharon Reed, Chip Schooley, Angela Scioscia, Natasha Martin Cheryl Anderson, Sawyer Farmer, Abigail Schnapper, Helena Tubb, Tommy Valles + SEARCH |
| EPI_ISL_1815916, EPI_ISL_1815917, EPI_ISL_1815918, EPI_ISL_1815919, EPI_ISL_1815920, EPI_ISL_1815921, EPI_ISL_1815922, EPI_ISL_1815923, EPI_ISL_1815924, EPI_ISL_1815925, EPI_ISL_1815926, EPI_ISL_1815927, EPI_ISL_1815928, EPI_ISL_1815929 | | | |
| see above | EXCITE Lab | Andersen lab at Scripps Research | Nicole L Washington, Simon White, Geraint Levan, Kelly Schiabor Barrett, Elizabeth Cirulli, Alexandre Bolze, Charlotte Rivera-Garcia, Ryan Cho, Jason Nguyen, Sherry Wang, Jimmy Ramirez, Celena Andrade, Alice Summerfield, Tyler Cassens, Efren Sandoval, Francisco Tanudjaja, Magnus Isaksson, William Lee, David Becker, Marc Laurent, James Lu + SEARCH |
| EPI_ISL_1815930 | EXCITE Lab | Andersen lab at Scripps Research | Chip Schooley, Natasha Martin, Cheryl Anderson, Angela Scioscia, Smruthi Karthikeyan, Greg Humphrey, Sawyer Farmer, Abigail Schnapper, Helena Tubb, Tommy Valles + SEARCH |
| EPI_ISL_1815931, EPI_ISL_1815932, EPI_ISL_1815933, EPI_ISL_1815934, EPI_ISL_1815935, EPI_ISL_1815936, EPI_ISL_1815937, EPI_ISL_1815938, EPI_ISL_1815939, EPI_ISL_1815940 | EXCITE Lab | Andersen lab at Scripps Research | Nicole L Washington, Simon White, Geraint Levan, Kelly Schiabor Barrett, Elizabeth Cirulli, Alexandre Bolze, Charlotte Rivera-Garcia, Ryan Cho, Jason Nguyen, Sherry Wang, Jimmy Ramirez, Celena Andrade, Alice Summerfield, Tyler Cassens, Efren Sandoval, Francisco Tanudjaja, Magnus Isaksson, William Lee, David Becker, Marc Laurent, James Lu + SEARCH |
| EPI_ISL_1815941 | EXCITE Lab | Andersen lab at Scripps Research | Chip Schooley, Natasha Martin, Cheryl Anderson, Angela Scioscia, Smruthi Karthikeyan, Greg Humphrey, Sawyer Farmer, Abigail Schnapper, Helena Tubb, Tommy Valles + SEARCH |
| EPI_ISL_1815942 | EXCITE Lab | Andersen lab at Scripps Research | Fred Wu, Christine Mitchell + SEARCH |
| EPI_ISL_1815943 | EXCITE Lab | Andersen lab at Scripps Research | Nicole L Washington, Simon White, Geraint Levan, Kelly Schiabor Barrett, Elizabeth Cirulli, Alexandre Bolze, Charlotte Rivera-Garcia, Ryan Cho, Jason Nguyen, Sherry Wang, Jimmy Ramirez, Celena Andrade, Alice Summerfield, Tyler Cassens, Efren Sandoval, Francisco Tanudjaja, Magnus Isaksson, William Lee, David Becker, Marc Laurent, James Lu + SEARCH |
| EPI_ISL_1815944 | EXCITE Lab | Andersen lab at Scripps Research | Chip Schooley, Natasha Martin, Cheryl Anderson, Angela Scioscia, Smruthi Karthikeyan, Greg Humphrey, Sawyer Farmer, Abigail Schnapper, Helena Tubb, Tommy Valles + SEARCH |
| EPI_ISL_1815945, EPI_ISL_1815946, EPI_ISL_1815947, EPI_ISL_1815948, EPI_ISL_1815949, EPI_ISL_1815950, EPI_ISL_1815951, EPI_ISL_1815952, EPI_ISL_1815953, EPI_ISL_1815954, EPI_ISL_1815955 | | | |
| see above | EXCITE Lab | Andersen lab at Scripps Research | Nicole L Washington, Simon White, Geraint Levan, Kelly Schiabor Barrett, Elizabeth Cirulli, Alexandre Bolze, Charlotte Rivera-Garcia, Ryan Cho, Jason Nguyen, Sherry Wang, Jimmy Ramirez, Celena Andrade, Alice Summerfield, Tyler Cassens, Efren Sandoval, Francisco Tanudjaja, Magnus Isaksson, William Lee, David Becker, Marc Laurent, James Lu + SEARCH |
| EPI_ISL_1815956 | EXCITE Lab | Andersen lab at Scripps Research | Fred Wu, Christine Mitchell + SEARCH |
| EPI_ISL_1815957, EPI_ISL_1815958 | EXCITE Lab | Andersen lab at Scripps Research | Nicole L Washington, Simon White, Geraint Levan, Kelly Schiabor Barrett, Elizabeth Cirulli, Alexandre Bolze, Charlotte Rivera-Garcia, Ryan Cho, Jason Nguyen, Sherry Wang, Jimmy Ramirez, Celena Andrade, Alice Summerfield, Tyler Cassens, Efren Sandoval, Francisco Tanudjaja, Magnus Isaksson, William Lee, David Becker, Marc Laurent, James Lu + SEARCH |
| EPI_ISL_1815959 | EXCITE Lab | Andersen lab at Scripps Research | David Pride, Sharon Reed, Chip Schooley, Angela Scioscia, Natasha Martin Cheryl Anderson, Sawyer Farmer, Abigail Schnapper, Helena Tubb, Tommy Valles + SEARCH |
| EPI_ISL_1815960, EPI_ISL_1815961, EPI_ISL_1815962 | EXCITE Lab | Andersen lab at Scripps Research | Nicole L Washington, Simon White, Geraint Levan, Kelly Schiabor Barrett, Elizabeth Cirulli, Alexandre Bolze, Charlotte Rivera-Garcia, Ryan Cho, Jason Nguyen, Sherry Wang, Jimmy Ramirez, Celena Andrade, Alice Summerfield, Tyler Cassens, Efren Sandoval, Francisco Tanudjaja, Magnus Isaksson, William Lee, David Becker, Marc Laurent, James Lu + SEARCH |
| EPI_ISL_1815963 | EXCITE Lab | Andersen lab at Scripps Research | David Pride, Sharon Reed, Chip Schooley, Angela Scioscia, Natasha Martin Cheryl Anderson, Sawyer Farmer, Abigail Schnapper, Helena Tubb, Tommy Valles + SEARCH |
| EPI_ISL_1815964, EPI_ISL_1815965, EPI_ISL_1815966, EPI_ISL_1815967, EPI_ISL_1815968, EPI_ISL_1815969, EPI_ISL_1815970, EPI_ISL_1815971, EPI_ISL_1815972, EPI_ISL_1815973, EPI_ISL_1815974 | | | |
| see above | EXCITE Lab | Andersen lab at Scripps Research | Nicole L Washington, Simon White, Geraint Levan, Kelly Schiabor Barrett, Elizabeth Cirulli, Alexandre Bolze, Charlotte Rivera-Garcia, Ryan Cho, Jason Nguyen, Sherry Wang, Jimmy Ramirez, Celena Andrade, Alice Summerfield, Tyler Cassens, Efren Sandoval, Francisco Tanudjaja, Magnus Isaksson, William Lee, David Becker, Marc Laurent, James Lu + SEARCH |
| EPI_ISL_1815975 | EXCITE Lab | Andersen lab at Scripps Research | Fred Wu, Christine Mitchell + SEARCH |
| EPI_ISL_1815976, EPI_ISL_1815977, EPI_ISL_1815978, EPI_ISL_1815979, EPI_ISL_1815980, EPI_ISL_1815981, EPI_ISL_1815982 | EXCITE Lab | Andersen lab at Scripps Research | Nicole L Washington, Simon White, Geraint Levan, Kelly Schiabor Barrett, Elizabeth Cirulli, Alexandre Bolze, Charlotte Rivera-Garcia, Ryan Cho, Jason Nguyen, Sherry Wang, Jimmy Ramirez, Celena Andrade, Alice Summerfield, Tyler Cassens, Efren Sandoval, Francisco Tanudjaja, Magnus Isaksson, William Lee, David Becker, Marc Laurent, James Lu + SEARCH |
| EPI_ISL_1815983 | EXCITE Lab | Andersen lab at Scripps Research | David Pride, Sharon Reed, Chip Schooley, Angela Scioscia, Natasha Martin Cheryl Anderson, Sawyer Farmer, Abigail Schnapper, Helena Tubb, Tommy Valles + SEARCH |
| EPI_ISL_1815984 | EXCITE Lab | Andersen lab at Scripps Research | Sydney Morgan + Marni Jacobs + Celestine Magallanes + Holly Valentine + Louise C. Laurent + SEARCH |
| EPI_ISL_1815985, EPI_ISL_1815986, EPI_ISL_1815987, EPI_ISL_1815988, EPI_ISL_1815989, EPI_ISL_1815990 | EXCITE Lab | Andersen lab at Scripps Research | Nicole L Washington, Simon White, Geraint Levan, Kelly Schiabor Barrett, Elizabeth Cirulli, Alexandre Bolze, Charlotte Rivera-Garcia, Ryan Cho, Jason Nguyen, Sherry Wang, Jimmy Ramirez, Celena Andrade, Alice Summerfield, Tyler Cassens, Efren Sandoval, Francisco Tanudjaja, Magnus Isaksson, William Lee, David Becker, Marc Laurent, James Lu + SEARCH |
| EPI_ISL_1815991 | EXCITE Lab | Andersen lab at Scripps Research | David Pride, Sharon Reed, Chip Schooley, Angela Scioscia, Natasha Martin Cheryl Anderson, Sawyer Farmer, Abigail Schnapper, Helena Tubb, Tommy Valles + SEARCH |
| EPI_ISL_1815992, EPI_ISL_1815993, EPI_ISL_1815994, EPI_ISL_1815995, EPI_ISL_1815996, EPI_ISL_1815997, EPI_ISL_1815998, EPI_ISL_1815999, EPI_ISL_1816000, EPI_ISL_1816001, EPI_ISL_1816002, EPI_ISL_1816003, EPI_ISL_1816004, EPI_ISL_1816005 | | | |
| see above | EXCITE Lab | Andersen lab at Scripps Research | Nicole L Washington, Simon White, Geraint Levan, Kelly Schiabor Barrett, Elizabeth Cirulli, Alexandre Bolze, Charlotte Rivera-Garcia, Ryan Cho, Jason Nguyen, Sherry Wang, Jimmy Ramirez, Celena Andrade, Alice Summerfield, Tyler Cassens, Efren Sandoval, Francisco Tanudjaja, Magnus Isaksson, William Lee, David Becker, Marc Laurent, James Lu + SEARCH |
| EPI_ISL_1816006 | EXCITE Lab | Andersen lab at Scripps Research | Chip Schooley, Natasha Martin, Cheryl Anderson, Angela Scioscia, Smruthi Karthikeyan, Greg Humphrey, Sawyer Farmer, Abigail Schnapper, Helena Tubb, Tommy Valles + SEARCH |

| | | | |
|---|---|---|---|
| EPI_ISL_1816007, EPI_ISL_1816008, EPI_ISL_1816009, EPI_ISL_1816010, EPI_ISL_1816011, EPI_ISL_1816012, EPI_ISL_1816013, EPI_ISL_1816014 | EXCITE Lab | Andersen lab at Scripps Research | Nicole L Washington, Simon White, Geraint Levan, Kelly Schiabor Barrett, Elizabeth Cirulli, Alexandre Bolze, Charlotte Rivera-Garcia, Ryan Cho, Jason Nguyen, Sherry Wang, Jimmy Ramirez, Celena Andrade, Alice Summerfield, Tyler Cassens, Efren Sandoval, Francisco Tanudjaja, Magnus Isaksson, William Lee, David Becker, Marc Laurent, James Lu + SEARCH |
| EPI_ISL_1816015 | EXCITE Lab | Andersen lab at Scripps Research | Chip Schooley, Natasha Martin, Cheryl Anderson, Angela Scioscia, Smruthi Karthikeyan, Greg Humphrey, Sawyer Farmer, Abigail Schnapper, Helena Tubb, Tommy Valles + SEARCH |
| EPI_ISL_1816016, EPI_ISL_1816017, EPI_ISL_1816018 | EXCITE Lab | Andersen lab at Scripps Research | Nicole L Washington, Simon White, Geraint Levan, Kelly Schiabor Barrett, Elizabeth Cirulli, Alexandre Bolze, Charlotte Rivera-Garcia, Ryan Cho, Jason Nguyen, Sherry Wang, Jimmy Ramirez, Celena Andrade, Alice Summerfield, Tyler Cassens, Efren Sandoval, Francisco Tanudjaja, Magnus Isaksson, William Lee, David Becker, Marc Laurent, James Lu + SEARCH |
| EPI_ISL_1816019 | EXCITE Lab | Andersen lab at Scripps Research | David Pride, Sharon Reed, Chip Schooley, Angela Scioscia, Natasha Martin Cheryl Anderson, Sawyer Farmer, Abigail Schnapper, Helena Tubb, Tommy Valles + SEARCH |
| EPI_ISL_1816020 | EXCITE Lab | Andersen lab at Scripps Research | Nicole L Washington, Simon White, Geraint Levan, Kelly Schiabor Barrett, Elizabeth Cirulli, Alexandre Bolze, Charlotte Rivera-Garcia, Ryan Cho, Jason Nguyen, Sherry Wang, Jimmy Ramirez, Celena Andrade, Alice Summerfield, Tyler Cassens, Efren Sandoval, Francisco Tanudjaja, Magnus Isaksson, William Lee, David Becker, Marc Laurent, James Lu + SEARCH |
| EPI_ISL_1816021 | EXCITE Lab | Andersen lab at Scripps Research | Chip Schooley, Natasha Martin, Cheryl Anderson, Angela Scioscia, Smruthi Karthikeyan, Greg Humphrey, Sawyer Farmer, Abigail Schnapper, Helena Tubb, Tommy Valles + SEARCH |
| EPI_ISL_1816022, EPI_ISL_1816023, EPI_ISL_1816024, EPI_ISL_1816025, EPI_ISL_1816026, EPI_ISL_1816027, EPI_ISL_1816028, EPI_ISL_1816029, EPI_ISL_1816030, EPI_ISL_1816031, EPI_ISL_1816032 | | | |
| see above | EXCITE Lab | Andersen lab at Scripps Research | Nicole L Washington, Simon White, Geraint Levan, Kelly Schiabor Barrett, Elizabeth Cirulli, Alexandre Bolze, Charlotte Rivera-Garcia, Ryan Cho, Jason Nguyen, Sherry Wang, Jimmy Ramirez, Celena Andrade, Alice Summerfield, Tyler Cassens, Efren Sandoval, Francisco Tanudjaja, Magnus Isaksson, William Lee, David Becker, Marc Laurent, James Lu + SEARCH |
| EPI_ISL_1816033 | EXCITE Lab | Andersen lab at Scripps Research | Chip Schooley, Natasha Martin, Cheryl Anderson, Angela Scioscia, Smruthi Karthikeyan, Greg Humphrey, Sawyer Farmer, Abigail Schnapper, Helena Tubb, Tommy Valles + SEARCH |
| EPI_ISL_1816034, EPI_ISL_1816035, EPI_ISL_1816036 | EXCITE Lab | Andersen lab at Scripps Research | Nicole L Washington, Simon White, Geraint Levan, Kelly Schiabor Barrett, Elizabeth Cirulli, Alexandre Bolze, Charlotte Rivera-Garcia, Ryan Cho, Jason Nguyen, Sherry Wang, Jimmy Ramirez, Celena Andrade, Alice Summerfield, Tyler Cassens, Efren Sandoval, Francisco Tanudjaja, Magnus Isaksson, William Lee, David Becker, Marc Laurent, James Lu + SEARCH |
| EPI_ISL_1816037 | EXCITE Lab | Andersen lab at Scripps Research | David Pride, Sharon Reed, Chip Schooley, Angela Scioscia, Natasha Martin Cheryl Anderson, Sawyer Farmer, Abigail Schnapper, Helena Tubb, Tommy Valles + SEARCH |
| EPI_ISL_1816038, EPI_ISL_1816039 | EXCITE Lab | Andersen lab at Scripps Research | Nicole L Washington, Simon White, Geraint Levan, Kelly Schiabor Barrett, Elizabeth Cirulli, Alexandre Bolze, Charlotte Rivera-Garcia, Ryan Cho, Jason Nguyen, Sherry Wang, Jimmy Ramirez, Celena Andrade, Alice Summerfield, Tyler Cassens, Efren Sandoval, Francisco Tanudjaja, Magnus Isaksson, William Lee, David Becker, Marc Laurent, James Lu + SEARCH |
| EPI_ISL_1816040 | EXCITE Lab | Andersen lab at Scripps Research | David Pride, Sharon Reed, Chip Schooley, Angela Scioscia, Natasha Martin Cheryl Anderson, Sawyer Farmer, Abigail Schnapper, Helena Tubb, Tommy Valles + SEARCH |
| EPI_ISL_1816041, EPI_ISL_1816042, EPI_ISL_1816043, EPI_ISL_1816044 | EXCITE Lab | Andersen lab at Scripps Research | Nicole L Washington, Simon White, Geraint Levan, Kelly Schiabor Barrett, Elizabeth Cirulli, Alexandre Bolze, Charlotte Rivera-Garcia, Ryan Cho, Jason Nguyen, Sherry Wang, Jimmy Ramirez, Celena Andrade, Alice Summerfield, Tyler Cassens, Efren Sandoval, Francisco Tanudjaja, Magnus Isaksson, William Lee, David Becker, Marc Laurent, James Lu + SEARCH |
| EPI_ISL_1816045 | EXCITE Lab | Andersen lab at Scripps Research | Chip Schooley, Natasha Martin, Cheryl Anderson, Angela Scioscia, Smruthi Karthikeyan, Greg Humphrey, Sawyer Farmer, Abigail Schnapper, Helena Tubb, Tommy Valles + SEARCH |
| EPI_ISL_1816046, EPI_ISL_1816047 | EXCITE Lab | Andersen lab at Scripps Research | Nicole L Washington, Simon White, Geraint Levan, Kelly Schiabor Barrett, Elizabeth Cirulli, Alexandre Bolze, Charlotte Rivera-Garcia, Ryan Cho, Jason Nguyen, Sherry Wang, Jimmy Ramirez, Celena Andrade, Alice Summerfield, Tyler Cassens, Efren Sandoval, Francisco Tanudjaja, Magnus Isaksson, William Lee, David Becker, Marc Laurent, James Lu + SEARCH |
| EPI_ISL_1816048 | EXCITE Lab | Andersen lab at Scripps Research | Sydney Morgan + Marni Jacobs + Celestine Magallanes + Holly Valentine + Louise C. Laurent + SEARCH |
| EPI_ISL_1816049, EPI_ISL_1816050, EPI_ISL_1816051, EPI_ISL_1816052, EPI_ISL_1816053, EPI_ISL_1816054, EPI_ISL_1816055, EPI_ISL_1816056, EPI_ISL_1816057, EPI_ISL_1816058, EPI_ISL_1816059, EPI_ISL_1816060, EPI_ISL_1816061 | | | |
| see above | EXCITE Lab | Andersen lab at Scripps Research | Nicole L Washington, Simon White, Geraint Levan, Kelly Schiabor Barrett, Elizabeth Cirulli, Alexandre Bolze, Charlotte Rivera-Garcia, Ryan Cho, Jason Nguyen, Sherry Wang, Jimmy Ramirez, Celena Andrade, Alice Summerfield, Tyler Cassens, Efren Sandoval, Francisco Tanudjaja, Magnus Isaksson, William Lee, David Becker, Marc Laurent, James Lu + SEARCH |
| EPI_ISL_1816062 | EXCITE Lab | Andersen lab at Scripps Research | David Pride, Sharon Reed, Chip Schooley, Angela Scioscia, Natasha Martin Cheryl Anderson, Sawyer Farmer, Abigail Schnapper, Helena Tubb, Tommy Valles + SEARCH |
| EPI_ISL_1816063 | EXCITE Lab | Andersen lab at Scripps Research | Rebecca Fielding-Miller, Laura Kohn, Patricia Gonzalez-Zuniga, Fitri Carrissa Wijaya, Vinton Omaleki, Marlene Flores, Araz Majnoonian, Megan Nguyen, Ashkan Hassani, Smruthi Karthikeyan, Rodolfo Salido, Victor Cantu, Greg Humphrey, Sawyer Farmer, Abigail Schnapper, Helena Tubb, Tommy Valles + SEARCH |
| EPI_ISL_1816064, EPI_ISL_1816065 | EXCITE Lab | Andersen lab at Scripps Research | Nicole L Washington, Simon White, Geraint Levan, Kelly Schiabor Barrett, Elizabeth Cirulli, Alexandre Bolze, Charlotte Rivera-Garcia, Ryan Cho, Jason Nguyen, Sherry Wang, Jimmy Ramirez, Celena Andrade, Alice Summerfield, Tyler Cassens, Efren Sandoval, Francisco Tanudjaja, Magnus Isaksson, William Lee, David Becker, Marc Laurent, James Lu + SEARCH |
| EPI_ISL_1816066 | EXCITE Lab | Andersen lab at Scripps Research | David Pride, Sharon Reed, Chip Schooley, Angela Scioscia, Natasha Martin Cheryl Anderson, Sawyer Farmer, Abigail Schnapper, Helena Tubb, Tommy Valles + SEARCH |
| EPI_ISL_1816067 | EXCITE Lab | Andersen lab at Scripps Research | Nicole L Washington, Simon White, Geraint Levan, Kelly Schiabor Barrett, Elizabeth Cirulli, Alexandre Bolze, Charlotte Rivera-Garcia, Ryan Cho, Jason Nguyen, Sherry Wang, Jimmy Ramirez, Celena Andrade, Alice Summerfield, Tyler Cassens, Efren Sandoval, Francisco Tanudjaja, Magnus Isaksson, William Lee, David Becker, Marc Laurent, James Lu + SEARCH |
| EPI_ISL_1816068 | EXCITE Lab | Andersen lab at Scripps Research | Chip Schooley, Natasha Martin, Cheryl Anderson, Angela Scioscia, Smruthi Karthikeyan, Greg Humphrey, Sawyer Farmer, Abigail Schnapper, Helena Tubb, Tommy Valles + SEARCH |
| EPI_ISL_1816069, EPI_ISL_1816070, EPI_ISL_1816071, EPI_ISL_1816072 | EXCITE Lab | Andersen lab at Scripps Research | Nicole L Washington, Simon White, Geraint Levan, Kelly Schiabor Barrett, Elizabeth Cirulli, Alexandre Bolze, Charlotte Rivera-Garcia, Ryan Cho, Jason Nguyen, Sherry Wang, Jimmy Ramirez, Celena Andrade, Alice Summerfield, Tyler Cassens, Efren Sandoval, Francisco Tanudjaja, Magnus Isaksson, William Lee, David Becker, Marc Laurent, James Lu + SEARCH |
| EPI_ISL_1816073 | EXCITE Lab | Andersen lab at Scripps Research | David Pride, Sharon Reed, Chip Schooley, Angela Scioscia, Natasha Martin Cheryl Anderson, Sawyer Farmer, Abigail Schnapper, Helena Tubb, Tommy Valles + SEARCH |
| EPI_ISL_1816074, EPI_ISL_1816075, EPI_ISL_1816076, EPI_ISL_1816077, EPI_ISL_1816078 | EXCITE Lab | Andersen lab at Scripps Research | Nicole L Washington, Simon White, Geraint Levan, Kelly Schiabor Barrett, Elizabeth Cirulli, Alexandre Bolze, Charlotte Rivera-Garcia, Ryan Cho, Jason Nguyen, Sherry Wang, Jimmy Ramirez, Celena Andrade, Alice Summerfield, Tyler Cassens, Efren Sandoval, Francisco Tanudjaja, Magnus Isaksson, William Lee, David Becker, Marc Laurent, James Lu + SEARCH |
| EPI_ISL_1816079 | EXCITE Lab | Andersen lab at Scripps Research | David Pride, Sharon Reed, Chip Schooley, Angela Scioscia, Natasha Martin Cheryl Anderson, Sawyer Farmer, Abigail Schnapper, Helena Tubb, Tommy Valles + SEARCH |
| EPI_ISL_1816080 | EXCITE Lab | Andersen lab at Scripps Research | Sydney Morgan + Marni Jacobs + Celestine Magallanes + Holly Valentine + Louise C. Laurent + SEARCH |
| EPI_ISL_1816081 | EXCITE Lab | Andersen lab at Scripps Research | David Pride, Sharon Reed, Chip Schooley, Angela Scioscia, Natasha Martin Cheryl Anderson, Sawyer Farmer, Abigail Schnapper, Helena Tubb, Tommy Valles + SEARCH |
| EPI_ISL_1816082, EPI_ISL_1816083, EPI_ISL_1816084, EPI_ISL_1816085 | EXCITE Lab | Andersen lab at Scripps Research | Nicole L Washington, Simon White, Geraint Levan, Kelly Schiabor Barrett, Elizabeth Cirulli, Alexandre Bolze, Charlotte Rivera-Garcia, Ryan Cho, Jason Nguyen, Sherry Wang, Jimmy Ramirez, Celena Andrade, Alice Summerfield, Tyler Cassens, Efren Sandoval, Francisco Tanudjaja, Magnus Isaksson, William Lee, David Becker, Marc Laurent, James Lu + SEARCH |
| EPI_ISL_1816086 | EXCITE Lab | Andersen lab at Scripps Research | Chip Schooley, Natasha Martin, Cheryl Anderson, Angela Scioscia, Smruthi Karthikeyan, Greg Humphrey, Sawyer Farmer, Abigail Schnapper, Helena Tubb, Tommy Valles + SEARCH |
| EPI_ISL_1816087 | EXCITE Lab | Andersen lab at Scripps Research | David Pride, Sharon Reed, Chip Schooley, Angela Scioscia, Natasha Martin Cheryl Anderson, Sawyer Farmer, Abigail Schnapper, Helena Tubb, Tommy Valles + SEARCH |
| EPI_ISL_1816088, EPI_ISL_1816089, EPI_ISL_1816090, EPI_ISL_1816091, EPI_ISL_1816092, EPI_ISL_1816093 | EXCITE Lab | Andersen lab at Scripps Research | Nicole L Washington, Simon White, Geraint Levan, Kelly Schiabor Barrett, Elizabeth Cirulli, Alexandre Bolze, Charlotte Rivera-Garcia, Ryan Cho, Jason Nguyen, Sherry Wang, Jimmy Ramirez, Celena Andrade, Alice Summerfield, Tyler Cassens, Efren Sandoval, Francisco Tanudjaja, Magnus Isaksson, William Lee, David Becker, Marc Laurent, James Lu + SEARCH |
| EPI_ISL_1816094 | EXCITE Lab | Andersen lab at Scripps Research | Chip Schooley, Natasha Martin, Cheryl Anderson, Angela Scioscia, Smruthi Karthikeyan, Greg Humphrey, Sawyer Farmer, Abigail Schnapper, Helena Tubb, Tommy Valles + SEARCH |
| EPI_ISL_1816095, EPI_ISL_1816096, EPI_ISL_1816097, EPI_ISL_1816098, EPI_ISL_1816099, EPI_ISL_1816100, EPI_ISL_1816101, EPI_ISL_1816102, EPI_ISL_1816103, EPI_ISL_1816104, EPI_ISL_1816105, EPI_ISL_1816106, EPI_ISL_1816107, EPI_ISL_1816108, EPI_ISL_1816109, EPI_ISL_1816110, EPI_ISL_1816111, EPI_ISL_1816112, EPI_ISL_1816113, EPI_ISL_1816114, EPI_ISL_1816115, EPI_ISL_1816116 | | | |
| see above | EXCITE Lab | Andersen lab at Scripps Research | Nicole L Washington, Simon White, Geraint Levan, Kelly Schiabor Barrett, Elizabeth Cirulli, Alexandre Bolze, Charlotte Rivera-Garcia, Ryan Cho, Jason Nguyen, Sherry Wang, Jimmy Ramirez, Celena Andrade, Alice Summerfield, Tyler Cassens, Efren Sandoval, Francisco Tanudjaja, Magnus Isaksson, William Lee, David Becker, Marc Laurent, James Lu + SEARCH |
| EPI_ISL_1816117 | EXCITE Lab | Andersen lab at Scripps Research | David Pride, Sharon Reed, Chip Schooley, Angela Scioscia, Natasha Martin Cheryl Anderson, Sawyer Farmer, Abigail Schnapper, Helena Tubb, Tommy Valles + SEARCH |
| EPI_ISL_1816118, EPI_ISL_1816119, EPI_ISL_1816120, EPI_ISL_1816121, EPI_ISL_1816122 | EXCITE Lab | Andersen lab at Scripps Research | Nicole L Washington, Simon White, Geraint Levan, Kelly Schiabor Barrett, Elizabeth Cirulli, Alexandre Bolze, Charlotte Rivera-Garcia, Ryan Cho, Jason Nguyen, Sherry Wang, Jimmy Ramirez, Celena Andrade, Alice Summerfield, Tyler Cassens, Efren Sandoval, Francisco Tanudjaja, Magnus Isaksson, William Lee, David Becker, Marc Laurent, James Lu + SEARCH |
| EPI_ISL_1816123 | EXCITE Lab | Andersen lab at Scripps Research | Sydney Morgan + Marni Jacobs + Celestine Magallanes + Holly Valentine + Louise C. Laurent + SEARCH |
| EPI_ISL_1816124, EPI_ISL_1816125, EPI_ISL_1816126 | EXCITE Lab | Andersen lab at Scripps Research | Nicole L Washington, Simon White, Geraint Levan, Kelly Schiabor Barrett, Elizabeth Cirulli, Alexandre Bolze, Charlotte Rivera-Garcia, Ryan Cho, Jason Nguyen, Sherry Wang, Jimmy Ramirez, Celena Andrade, Alice Summerfield, Tyler Cassens, Efren Sandoval, Francisco Tanudjaja, Magnus Isaksson, William Lee, David Becker, Marc Laurent, James Lu + SEARCH |
| EPI_ISL_1816127 | EXCITE Lab | Andersen lab at Scripps Research | David Pride, Sharon Reed, Chip Schooley, Angela Scioscia, Natasha Martin Cheryl Anderson, Sawyer Farmer, Abigail Schnapper, Helena Tubb, Tommy Valles + SEARCH |
| EPI_ISL_1816128 | EXCITE Lab | Andersen lab at Scripps Research | Nicole L Washington, Simon White, Geraint Levan, Kelly Schiabor Barrett, Elizabeth Cirulli, Alexandre Bolze, Charlotte Rivera-Garcia, Ryan Cho, Jason Nguyen, Sherry Wang, Jimmy Ramirez, Celena Andrade, Alice Summerfield, Tyler Cassens, Efren Sandoval, Francisco Tanudjaja, Magnus Isaksson, William Lee, David Becker, Marc Laurent, James Lu + SEARCH |
| EPI_ISL_1816129 | EXCITE Lab | Andersen lab at Scripps Research | David Pride, Sharon Reed, Chip Schooley, Angela Scioscia, Natasha Martin Cheryl Anderson, Sawyer Farmer, Abigail Schnapper, Helena Tubb, Tommy Valles + SEARCH |

| | | | |
|---|---|---|---|
| EPI_ISL_1816130, EPI_ISL_1816131, EPI_ISL_1816132, EPI_ISL_1816133, EPI_ISL_1816134 | EXCITE Lab | Andersen lab at Scripps Research | Nicole L Washington, Simon White, Geraint Levan, Kelly Schiabor Barrett, Elizabeth Cirulli, Alexandre Bolze, Charlotte Rivera-Garcia, Ryan Cho, Jason Nguyen, Sherry Wang, Jimmy Ramirez, Celena Andrade, Alice Summerfield, Tyler Cassens, Efren Sandoval, Francisco Tanudjaja, Magnus Isaksson, William Lee, David Becker, Marc Laurent, James Lu + SEARCH |
| EPI_ISL_1816135 | EXCITE Lab | Andersen lab at Scripps Research | Chip Schooley, Natasha Martin, Cheryl Anderson, Angela Scioscia, Smruthi Karthikeyan, Greg Humphrey, Sawyer Farmer, Abigail Schnapper, Helena Tubb, Tommy Valles + SEARCH |
| EPI_ISL_1816136, EPI_ISL_1816137, EPI_ISL_1816138, EPI_ISL_1816139, EPI_ISL_1816140, EPI_ISL_1816141, EPI_ISL_1816142, EPI_ISL_1816143, EPI_ISL_1816144, EPI_ISL_1816145, EPI_ISL_1816146, EPI_ISL_1816147, EPI_ISL_1816148, EPI_ISL_1816149, EPI_ISL_1816150, EPI_ISL_1816151, EPI_ISL_1816152, EPI_ISL_1816153, EPI_ISL_1816154, EPI_ISL_1816155, EPI_ISL_1816156, EPI_ISL_1816157, EPI_ISL_1816158, EPI_ISL_1816159, EPI_ISL_1816160, EPI_ISL_1816161, EPI_ISL_1816162, EPI_ISL_1816163, EPI_ISL_1816164, EPI_ISL_1816165, EPI_ISL_1816166, EPI_ISL_1816167, EPI_ISL_1816168, EPI_ISL_1816169, EPI_ISL_1816170 | | | |
| see above | EXCITE Lab | Andersen lab at Scripps Research | Nicole L Washington, Simon White, Geraint Levan, Kelly Schiabor Barrett, Elizabeth Cirulli, Alexandre Bolze, Charlotte Rivera-Garcia, Ryan Cho, Jason Nguyen, Sherry Wang, Jimmy Ramirez, Celena Andrade, Alice Summerfield, Tyler Cassens, Efren Sandoval, Francisco Tanudjaja, Magnus Isaksson, William Lee, David Becker, Marc Laurent, James Lu + SEARCH |
| EPI_ISL_1816171 | EXCITE Lab | Andersen lab at Scripps Research | David Pride, Sharon Reed, Chip Schooley, Angela Scioscia, Natasha Martin Cheryl Anderson, Sawyer Farmer, Abigail Schnapper, Helena Tubb, Tommy Valles + SEARCH |
| EPI_ISL_1816172, EPI_ISL_1816173, EPI_ISL_1816174, EPI_ISL_1816175, EPI_ISL_1816176 | EXCITE Lab | Andersen lab at Scripps Research | Nicole L Washington, Simon White, Geraint Levan, Kelly Schiabor Barrett, Elizabeth Cirulli, Alexandre Bolze, Charlotte Rivera-Garcia, Ryan Cho, Jason Nguyen, Sherry Wang, Jimmy Ramirez, Celena Andrade, Alice Summerfield, Tyler Cassens, Efren Sandoval, Francisco Tanudjaja, Magnus Isaksson, William Lee, David Becker, Marc Laurent, James Lu + SEARCH |
| EPI_ISL_1816177 | EXCITE Lab | Andersen lab at Scripps Research | David Pride, Sharon Reed, Chip Schooley, Angela Scioscia, Natasha Martin Cheryl Anderson, Sawyer Farmer, Abigail Schnapper, Helena Tubb, Tommy Valles + SEARCH |
| EPI_ISL_1816178 | EXCITE Lab | Andersen lab at Scripps Research | Sydney Morgan + Marni Jacobs + Celestine Magallanes + Holly Valentine + Louise C. Laurent + SEARCH |
| EPI_ISL_1816179, EPI_ISL_1816180, EPI_ISL_1816181, EPI_ISL_1816182, EPI_ISL_1816183, EPI_ISL_1816184 | EXCITE Lab | Andersen lab at Scripps Research | Nicole L Washington, Simon White, Geraint Levan, Kelly Schiabor Barrett, Elizabeth Cirulli, Alexandre Bolze, Charlotte Rivera-Garcia, Ryan Cho, Jason Nguyen, Sherry Wang, Jimmy Ramirez, Celena Andrade, Alice Summerfield, Tyler Cassens, Efren Sandoval, Francisco Tanudjaja, Magnus Isaksson, William Lee, David Becker, Marc Laurent, James Lu + SEARCH |
| EPI_ISL_1816185 | EXCITE Lab | Andersen lab at Scripps Research | Chip Schooley, Natasha Martin, Cheryl Anderson, Angela Scioscia, Smruthi Karthikeyan, Greg Humphrey, Sawyer Farmer, Abigail Schnapper, Helena Tubb, Tommy Valles + SEARCH |
| EPI_ISL_1816186, EPI_ISL_1816187, EPI_ISL_1816188, EPI_ISL_1816189, EPI_ISL_1816190, EPI_ISL_1816191, EPI_ISL_1816192, EPI_ISL_1816193, EPI_ISL_1816194, EPI_ISL_1816195, EPI_ISL_1816196 | | | |
| see above | EXCITE Lab | Andersen lab at Scripps Research | Nicole L Washington, Simon White, Geraint Levan, Kelly Schiabor Barrett, Elizabeth Cirulli, Alexandre Bolze, Charlotte Rivera-Garcia, Ryan Cho, Jason Nguyen, Sherry Wang, Jimmy Ramirez, Celena Andrade, Alice Summerfield, Tyler Cassens, Efren Sandoval, Francisco Tanudjaja, Magnus Isaksson, William Lee, David Becker, Marc Laurent, James Lu + SEARCH |
| EPI_ISL_1816197 | EXCITE Lab | Andersen lab at Scripps Research | David Pride, Sharon Reed, Chip Schooley, Angela Scioscia, Natasha Martin Cheryl Anderson, Sawyer Farmer, Abigail Schnapper, Helena Tubb, Tommy Valles + SEARCH |
| EPI_ISL_1816198, EPI_ISL_1816199, EPI_ISL_1816200, EPI_ISL_1816201, EPI_ISL_1816202, EPI_ISL_1816203, EPI_ISL_1816204, EPI_ISL_1816205 | EXCITE Lab | Andersen lab at Scripps Research | Nicole L Washington, Simon White, Geraint Levan, Kelly Schiabor Barrett, Elizabeth Cirulli, Alexandre Bolze, Charlotte Rivera-Garcia, Ryan Cho, Jason Nguyen, Sherry Wang, Jimmy Ramirez, Celena Andrade, Alice Summerfield, Tyler Cassens, Efren Sandoval, Francisco Tanudjaja, Magnus Isaksson, William Lee, David Becker, Marc Laurent, James Lu + SEARCH |
| EPI_ISL_1816206 | EXCITE Lab | Andersen lab at Scripps Research | David Pride, Sharon Reed, Chip Schooley, Angela Scioscia, Natasha Martin Cheryl Anderson, Sawyer Farmer, Abigail Schnapper, Helena Tubb, Tommy Valles + SEARCH |
| EPI_ISL_1816207, EPI_ISL_1816208, EPI_ISL_1816209, EPI_ISL_1816210, EPI_ISL_1816211, EPI_ISL_1816212, EPI_ISL_1816213, EPI_ISL_1816214, EPI_ISL_1816215, EPI_ISL_1816216, EPI_ISL_1816217, EPI_ISL_1816218, EPI_ISL_1816219 | | | |
| see above | EXCITE Lab | Andersen lab at Scripps Research | Nicole L Washington, Simon White, Geraint Levan, Kelly Schiabor Barrett, Elizabeth Cirulli, Alexandre Bolze, Charlotte Rivera-Garcia, Ryan Cho, Jason Nguyen, Sherry Wang, Jimmy Ramirez, Celena Andrade, Alice Summerfield, Tyler Cassens, Efren Sandoval, Francisco Tanudjaja, Magnus Isaksson, William Lee, David Becker, Marc Laurent, James Lu + SEARCH |
| EPI_ISL_1816220 | EXCITE Lab | Andersen lab at Scripps Research | Sydney Morgan + Marni Jacobs + Celestine Magallanes + Holly Valentine + Louise C. Laurent + SEARCH |
| EPI_ISL_1816221, EPI_ISL_1816222, EPI_ISL_1816223, EPI_ISL_1816224, EPI_ISL_1816225, EPI_ISL_1816226, EPI_ISL_1816227, EPI_ISL_1816228, EPI_ISL_1816229, EPI_ISL_1816230, EPI_ISL_1816231 | | | |
| see above | EXCITE Lab | Andersen lab at Scripps Research | Nicole L Washington, Simon White, Geraint Levan, Kelly Schiabor Barrett, Elizabeth Cirulli, Alexandre Bolze, Charlotte Rivera-Garcia, Ryan Cho, Jason Nguyen, Sherry Wang, Jimmy Ramirez, Celena Andrade, Alice Summerfield, Tyler Cassens, Efren Sandoval, Francisco Tanudjaja, Magnus Isaksson, William Lee, David Becker, Marc Laurent, James Lu + SEARCH |
| EPI_ISL_1816232 | EXCITE Lab | Andersen lab at Scripps Research | Chip Schooley, Natasha Martin, Cheryl Anderson, Angela Scioscia, Smruthi Karthikeyan, Greg Humphrey, Sawyer Farmer, Abigail Schnapper, Helena Tubb, Tommy Valles + SEARCH |
| EPI_ISL_1816233, EPI_ISL_1816234, EPI_ISL_1816235 | EXCITE Lab | Andersen lab at Scripps Research | Nicole L Washington, Simon White, Geraint Levan, Kelly Schiabor Barrett, Elizabeth Cirulli, Alexandre Bolze, Charlotte Rivera-Garcia, Ryan Cho, Jason Nguyen, Sherry Wang, Jimmy Ramirez, Celena Andrade, Alice Summerfield, Tyler Cassens, Efren Sandoval, Francisco Tanudjaja, Magnus Isaksson, William Lee, David Becker, Marc Laurent, James Lu + SEARCH |
| EPI_ISL_1816236 | EXCITE Lab | Andersen lab at Scripps Research | David Pride, Sharon Reed, Chip Schooley, Angela Scioscia, Natasha Martin Cheryl Anderson, Sawyer Farmer, Abigail Schnapper, Helena Tubb, Tommy Valles + SEARCH |
| EPI_ISL_1816237, EPI_ISL_1816238, EPI_ISL_1816239, EPI_ISL_1816240 | EXCITE Lab | Andersen lab at Scripps Research | Nicole L Washington, Simon White, Geraint Levan, Kelly Schiabor Barrett, Elizabeth Cirulli, Alexandre Bolze, Charlotte Rivera-Garcia, Ryan Cho, Jason Nguyen, Sherry Wang, Jimmy Ramirez, Celena Andrade, Alice Summerfield, Tyler Cassens, Efren Sandoval, Francisco Tanudjaja, Magnus Isaksson, William Lee, David Becker, Marc Laurent, James Lu + SEARCH |
| EPI_ISL_1816241 | EXCITE Lab | Andersen lab at Scripps Research | David Pride, Sharon Reed, Chip Schooley, Angela Scioscia, Natasha Martin Cheryl Anderson, Sawyer Farmer, Abigail Schnapper, Helena Tubb, Tommy Valles + SEARCH |
| EPI_ISL_1816242, EPI_ISL_1816243, EPI_ISL_1816244, EPI_ISL_1816245 | EXCITE Lab | Andersen lab at Scripps Research | Nicole L Washington, Simon White, Geraint Levan, Kelly Schiabor Barrett, Elizabeth Cirulli, Alexandre Bolze, Charlotte Rivera-Garcia, Ryan Cho, Jason Nguyen, Sherry Wang, Jimmy Ramirez, Celena Andrade, Alice Summerfield, Tyler Cassens, Efren Sandoval, Francisco Tanudjaja, Magnus Isaksson, William Lee, David Becker, Marc Laurent, James Lu + SEARCH |
| EPI_ISL_1816246 | EXCITE Lab | Andersen lab at Scripps Research | David Pride, Sharon Reed, Chip Schooley, Angela Scioscia, Natasha Martin Cheryl Anderson, Sawyer Farmer, Abigail Schnapper, Helena Tubb, Tommy Valles + SEARCH |
| EPI_ISL_1816247, EPI_ISL_1816248, EPI_ISL_1816249, EPI_ISL_1816250 | EXCITE Lab | Andersen lab at Scripps Research | Nicole L Washington, Simon White, Geraint Levan, Kelly Schiabor Barrett, Elizabeth Cirulli, Alexandre Bolze, Charlotte Rivera-Garcia, Ryan Cho, Jason Nguyen, Sherry Wang, Jimmy Ramirez, Celena Andrade, Alice Summerfield, Tyler Cassens, Efren Sandoval, Francisco Tanudjaja, Magnus Isaksson, William Lee, David Becker, Marc Laurent, James Lu + SEARCH |
| EPI_ISL_1816251 | EXCITE Lab | Andersen lab at Scripps Research | Sydney Morgan + Marni Jacobs + Celestine Magallanes + Holly Valentine + Louise C. Laurent + SEARCH |
| EPI_ISL_1816252 | EXCITE Lab | Andersen lab at Scripps Research | J. Joelle Donofrio-Odmann, Brent Brainard+ SEARCH |
| EPI_ISL_1816253, EPI_ISL_1816254, EPI_ISL_1816255, EPI_ISL_1816256, EPI_ISL_1816257, EPI_ISL_1816258, EPI_ISL_1816259, EPI_ISL_1816260, EPI_ISL_1816261 | EXCITE Lab | Andersen lab at Scripps Research | Nicole L Washington, Simon White, Geraint Levan, Kelly Schiabor Barrett, Elizabeth Cirulli, Alexandre Bolze, Charlotte Rivera-Garcia, Ryan Cho, Jason Nguyen, Sherry Wang, Jimmy Ramirez, Celena Andrade, Alice Summerfield, Tyler Cassens, Efren Sandoval, Francisco Tanudjaja, Magnus Isaksson, William Lee, David Becker, Marc Laurent, James Lu + SEARCH |
| EPI_ISL_1816262, EPI_ISL_1816263, EPI_ISL_1816264, EPI_ISL_1816265 | EXCITE Lab | Andersen lab at Scripps Research | David Pride, Sharon Reed, Chip Schooley, Angela Scioscia, Natasha Martin Cheryl Anderson, Sawyer Farmer, Abigail Schnapper, Helena Tubb, Tommy Valles + SEARCH |
| EPI_ISL_1816266, EPI_ISL_1816267 | EXCITE Lab | Andersen lab at Scripps Research | Nicole L Washington, Simon White, Geraint Levan, Kelly Schiabor Barrett, Elizabeth Cirulli, Alexandre Bolze, Charlotte Rivera-Garcia, Ryan Cho, Jason Nguyen, Sherry Wang, Jimmy Ramirez, Celena Andrade, Alice Summerfield, Tyler Cassens, Efren Sandoval, Francisco Tanudjaja, Magnus Isaksson, William Lee, David Becker, Marc Laurent, James Lu + SEARCH |
| EPI_ISL_1816268 | EXCITE Lab | Andersen lab at Scripps Research | David Pride, Sharon Reed, Chip Schooley, Angela Scioscia, Natasha Martin Cheryl Anderson, Sawyer Farmer, Abigail Schnapper, Helena Tubb, Tommy Valles + SEARCH |
| EPI_ISL_1816269 | EXCITE Lab | Andersen lab at Scripps Research | Nicole L Washington, Simon White, Geraint Levan, Kelly Schiabor Barrett, Elizabeth Cirulli, Alexandre Bolze, Charlotte Rivera-Garcia, Ryan Cho, Jason Nguyen, Sherry Wang, Jimmy Ramirez, Celena Andrade, Alice Summerfield, Tyler Cassens, Efren Sandoval, Francisco Tanudjaja, Magnus Isaksson, William Lee, David Becker, Marc Laurent, James Lu + SEARCH |
| EPI_ISL_1816270 | EXCITE Lab | Andersen lab at Scripps Research | David Pride, Sharon Reed, Chip Schooley, Angela Scioscia, Natasha Martin Cheryl Anderson, Sawyer Farmer, Abigail Schnapper, Helena Tubb, Tommy Valles + SEARCH |
| EPI_ISL_1816271, EPI_ISL_1816272, EPI_ISL_1816273, EPI_ISL_1816274, EPI_ISL_1816275, EPI_ISL_1816276 | EXCITE Lab | Andersen lab at Scripps Research | Nicole L Washington, Simon White, Geraint Levan, Kelly Schiabor Barrett, Elizabeth Cirulli, Alexandre Bolze, Charlotte Rivera-Garcia, Ryan Cho, Jason Nguyen, Sherry Wang, Jimmy Ramirez, Celena Andrade, Alice Summerfield, Tyler Cassens, Efren Sandoval, Francisco Tanudjaja, Magnus Isaksson, William Lee, David Becker, Marc Laurent, James Lu + SEARCH |
| EPI_ISL_1816277 | EXCITE Lab | Andersen lab at Scripps Research | David Pride, Sharon Reed, Chip Schooley, Angela Scioscia, Natasha Martin Cheryl Anderson, Sawyer Farmer, Abigail Schnapper, Helena Tubb, Tommy Valles + SEARCH |
| EPI_ISL_1816278, | EXCITE Lab | Andersen lab at Scripps Research | Nicole L Washington, Simon White, Geraint Levan, Kelly Schiabor Barrett, Elizabeth Cirulli, Alexandre Bolze, Charlotte Rivera-Garcia, Ryan Cho, Jason Nguyen, Sherry Wang, Jimmy Ramirez, Celena Andrade, Alice Summerfield, Tyler Cassens, Efren Sandoval, Francisco Tanudjaja, Magnus |

| | | | |
|---|---|---|---|
| EPI_ISL_1816279, EPI_ISL_1816280, EPI_ISL_1816281, EPI_ISL_1816282, EPI_ISL_1816283, EPI_ISL_1816284, EPI_ISL_1816285 | | | Isaksson, William Lee, David Becker, Marc Laurent, James Lu + SEARCH |
| EPI_ISL_1816286 | EXCITE Lab | Andersen lab at Scripps Research | Chip Schooley, Natasha Martin, Cheryl Anderson, Angela Scioscia, Smruthi Karthikeyan, Greg Humphrey, Sawyer Farmer, Abigail Schnapper, Helena Tubb, Tommy Valles + SEARCH |
| EPI_ISL_1816287, EPI_ISL_1816288, EPI_ISL_1816289 | EXCITE Lab | Andersen lab at Scripps Research | Nicole L Washington, Simon White, Geraint Levan, Kelly Schiabor Barrett, Elizabeth Cirulli, Alexandre Bolze, Charlotte Rivera-Garcia, Ryan Cho, Jason Nguyen, Sherry Wang, Jimmy Ramirez, Celena Andrade, Alice Summerfield, Tyler Cassens, Efren Sandoval, Francisco Tanudjaja, Magnus Isaksson, William Lee, David Becker, Marc Laurent, James Lu + SEARCH |
| EPI_ISL_1816290 | EXCITE Lab | Andersen lab at Scripps Research | David Pride, Sharon Reed, Chip Schooley, Angela Scioscia, Natasha Martin Cheryl Anderson, Sawyer Farmer, Abigail Schnapper, Helena Tubb, Tommy Valles + SEARCH |
| EPI_ISL_1816291 | EXCITE Lab | Andersen lab at Scripps Research | Chip Schooley, Natasha Martin, Cheryl Anderson, Angela Scioscia, Smruthi Karthikeyan, Greg Humphrey, Sawyer Farmer, Abigail Schnapper, Helena Tubb, Tommy Valles + SEARCH |
| EPI_ISL_1816292, EPI_ISL_1816293, EPI_ISL_1816294, EPI_ISL_1816295, EPI_ISL_1816296 | EXCITE Lab | Andersen lab at Scripps Research | Nicole L Washington, Simon White, Geraint Levan, Kelly Schiabor Barrett, Elizabeth Cirulli, Alexandre Bolze, Charlotte Rivera-Garcia, Ryan Cho, Jason Nguyen, Sherry Wang, Jimmy Ramirez, Celena Andrade, Alice Summerfield, Tyler Cassens, Efren Sandoval, Francisco Tanudjaja, Magnus Isaksson, William Lee, David Becker, Marc Laurent, James Lu + SEARCH |
| EPI_ISL_1816297 | EXCITE Lab | Andersen lab at Scripps Research | David Pride, Sharon Reed, Chip Schooley, Angela Scioscia, Natasha Martin Cheryl Anderson, Sawyer Farmer, Abigail Schnapper, Helena Tubb, Tommy Valles + SEARCH |
| EPI_ISL_1816298, EPI_ISL_1816299, EPI_ISL_1816300, EPI_ISL_1816301, EPI_ISL_1816302, EPI_ISL_1816303, EPI_ISL_1816304, EPI_ISL_1816305, EPI_ISL_1816306, EPI_ISL_1816307, EPI_ISL_1816308 | | | |
| see above | EXCITE Lab | Andersen lab at Scripps Research | Nicole L Washington, Simon White, Geraint Levan, Kelly Schiabor Barrett, Elizabeth Cirulli, Alexandre Bolze, Charlotte Rivera-Garcia, Ryan Cho, Jason Nguyen, Sherry Wang, Jimmy Ramirez, Celena Andrade, Alice Summerfield, Tyler Cassens, Efren Sandoval, Francisco Tanudjaja, Magnus Isaksson, William Lee, David Becker, Marc Laurent, James Lu + SEARCH |
| EPI_ISL_1816309 | EXCITE Lab | Andersen lab at Scripps Research | Chip Schooley, Natasha Martin, Cheryl Anderson, Angela Scioscia, Smruthi Karthikeyan, Greg Humphrey, Sawyer Farmer, Abigail Schnapper, Helena Tubb, Tommy Valles + SEARCH |
| EPI_ISL_1816310, EPI_ISL_1816311, EPI_ISL_1816312, EPI_ISL_1816313, EPI_ISL_1816314, EPI_ISL_1816315, EPI_ISL_1816316, EPI_ISL_1816317 | EXCITE Lab | Andersen lab at Scripps Research | Nicole L Washington, Simon White, Geraint Levan, Kelly Schiabor Barrett, Elizabeth Cirulli, Alexandre Bolze, Charlotte Rivera-Garcia, Ryan Cho, Jason Nguyen, Sherry Wang, Jimmy Ramirez, Celena Andrade, Alice Summerfield, Tyler Cassens, Efren Sandoval, Francisco Tanudjaja, Magnus Isaksson, William Lee, David Becker, Marc Laurent, James Lu + SEARCH |
| EPI_ISL_1816318 | EXCITE Lab | Andersen lab at Scripps Research | Chip Schooley, Natasha Martin, Cheryl Anderson, Angela Scioscia, Smruthi Karthikeyan, Greg Humphrey, Sawyer Farmer, Abigail Schnapper, Helena Tubb, Tommy Valles + SEARCH |
| EPI_ISL_1816319 | EXCITE Lab | Andersen lab at Scripps Research | Rebecca Fielding-Miller, Laura Kohn, Patricia Gonzalez-Zuniga, Fitri Carrissa Wijaya, Vinton Omaleki, Marlene Flores, Araz Majnoonian, Megan Nguyen, Ashkan Hassani, Smruthi Karthikeyan, Rodolfo Salido, Victor Cantu, Greg Humphrey, Sawyer Farmer, Abigail Schnapper, Helena Tubb, Tommy Valles + SEARCH |
| EPI_ISL_1816320, EPI_ISL_1816321, EPI_ISL_1816322 | EXCITE Lab | Andersen lab at Scripps Research | Nicole L Washington, Simon White, Geraint Levan, Kelly Schiabor Barrett, Elizabeth Cirulli, Alexandre Bolze, Charlotte Rivera-Garcia, Ryan Cho, Jason Nguyen, Sherry Wang, Jimmy Ramirez, Celena Andrade, Alice Summerfield, Tyler Cassens, Efren Sandoval, Francisco Tanudjaja, Magnus Isaksson, William Lee, David Becker, Marc Laurent, James Lu + SEARCH |
| EPI_ISL_1816323 | EXCITE Lab | Andersen lab at Scripps Research | Chip Schooley, Natasha Martin, Cheryl Anderson, Angela Scioscia, Smruthi Karthikeyan, Greg Humphrey, Sawyer Farmer, Abigail Schnapper, Helena Tubb, Tommy Valles + SEARCH |
| EPI_ISL_1816324, EPI_ISL_1816325, EPI_ISL_1816326, EPI_ISL_1816327, EPI_ISL_1816328, EPI_ISL_1816329, EPI_ISL_1816330, EPI_ISL_1816331 | EXCITE Lab | Andersen lab at Scripps Research | Nicole L Washington, Simon White, Geraint Levan, Kelly Schiabor Barrett, Elizabeth Cirulli, Alexandre Bolze, Charlotte Rivera-Garcia, Ryan Cho, Jason Nguyen, Sherry Wang, Jimmy Ramirez, Celena Andrade, Alice Summerfield, Tyler Cassens, Efren Sandoval, Francisco Tanudjaja, Magnus Isaksson, William Lee, David Becker, Marc Laurent, James Lu + SEARCH |
| EPI_ISL_1816332 | EXCITE Lab | Andersen lab at Scripps Research | Fred Wu, Christine Mitchell + SEARCH |
| EPI_ISL_1816333, EPI_ISL_1816334, EPI_ISL_1816335 | EXCITE Lab | Andersen lab at Scripps Research | Nicole L Washington, Simon White, Geraint Levan, Kelly Schiabor Barrett, Elizabeth Cirulli, Alexandre Bolze, Charlotte Rivera-Garcia, Ryan Cho, Jason Nguyen, Sherry Wang, Jimmy Ramirez, Celena Andrade, Alice Summerfield, Tyler Cassens, Efren Sandoval, Francisco Tanudjaja, Magnus Isaksson, William Lee, David Becker, Marc Laurent, James Lu + SEARCH |
| EPI_ISL_1816336 | EXCITE Lab | Andersen lab at Scripps Research | Chip Schooley, Natasha Martin, Cheryl Anderson, Angela Scioscia, Smruthi Karthikeyan, Greg Humphrey, Sawyer Farmer, Abigail Schnapper, Helena Tubb, Tommy Valles + SEARCH |
| EPI_ISL_1816337, EPI_ISL_1816338, EPI_ISL_1816339, EPI_ISL_1816340, EPI_ISL_1816341, EPI_ISL_1816342, EPI_ISL_1816343, EPI_ISL_1816344, EPI_ISL_1816345, EPI_ISL_1816346, EPI_ISL_1816347, EPI_ISL_1816348, EPI_ISL_1816349 | | | |
| see above | EXCITE Lab | Andersen lab at Scripps Research | Nicole L Washington, Simon White, Geraint Levan, Kelly Schiabor Barrett, Elizabeth Cirulli, Alexandre Bolze, Charlotte Rivera-Garcia, Ryan Cho, Jason Nguyen, Sherry Wang, Jimmy Ramirez, Celena Andrade, Alice Summerfield, Tyler Cassens, Efren Sandoval, Francisco Tanudjaja, Magnus Isaksson, William Lee, David Becker, Marc Laurent, James Lu + SEARCH |
| EPI_ISL_1816350 | EXCITE Lab | Andersen lab at Scripps Research | David Pride, Sharon Reed, Chip Schooley, Angela Scioscia, Natasha Martin Cheryl Anderson, Sawyer Farmer, Abigail Schnapper, Helena Tubb, Tommy Valles + SEARCH |
| EPI_ISL_1816351, EPI_ISL_1816352, EPI_ISL_1816353, EPI_ISL_1816354, EPI_ISL_1816355, EPI_ISL_1816356, EPI_ISL_1816357, EPI_ISL_1816358, EPI_ISL_1816359, EPI_ISL_1816360 | EXCITE Lab | Andersen lab at Scripps Research | Nicole L Washington, Simon White, Geraint Levan, Kelly Schiabor Barrett, Elizabeth Cirulli, Alexandre Bolze, Charlotte Rivera-Garcia, Ryan Cho, Jason Nguyen, Sherry Wang, Jimmy Ramirez, Celena Andrade, Alice Summerfield, Tyler Cassens, Efren Sandoval, Francisco Tanudjaja, Magnus Isaksson, William Lee, David Becker, Marc Laurent, James Lu + SEARCH |
| EPI_ISL_1816361 | EXCITE Lab | Andersen lab at Scripps Research | David Pride, Sharon Reed, Chip Schooley, Angela Scioscia, Natasha Martin Cheryl Anderson, Sawyer Farmer, Abigail Schnapper, Helena Tubb, Tommy Valles + SEARCH |
| EPI_ISL_1816362, EPI_ISL_1816363, EPI_ISL_1816364, EPI_ISL_1816365, EPI_ISL_1816366, EPI_ISL_1816367, EPI_ISL_1816368, EPI_ISL_1816369, EPI_ISL_1816370, EPI_ISL_1816371, EPI_ISL_1816372, EPI_ISL_1816373, EPI_ISL_1816374, EPI_ISL_1816375, EPI_ISL_1816376, EPI_ISL_1816377 | | | |
| see above | EXCITE Lab | Andersen lab at Scripps Research | Nicole L Washington, Simon White, Geraint Levan, Kelly Schiabor Barrett, Elizabeth Cirulli, Alexandre Bolze, Charlotte Rivera-Garcia, Ryan Cho, Jason Nguyen, Sherry Wang, Jimmy Ramirez, Celena Andrade, Alice Summerfield, Tyler Cassens, Efren Sandoval, Francisco Tanudjaja, Magnus Isaksson, William Lee, David Becker, Marc Laurent, James Lu + SEARCH |
| EPI_ISL_1816378 | EXCITE Lab | Andersen lab at Scripps Research | Chip Schooley, Natasha Martin, Cheryl Anderson, Angela Scioscia, Smruthi Karthikeyan, Greg Humphrey, Sawyer Farmer, Abigail Schnapper, Helena Tubb, Tommy Valles + SEARCH |
| EPI_ISL_1816379, EPI_ISL_1816380, EPI_ISL_1816381, EPI_ISL_1816382, EPI_ISL_1816383, EPI_ISL_1816384, EPI_ISL_1816385, EPI_ISL_1816386, EPI_ISL_1816387, EPI_ISL_1816388, EPI_ISL_1816389, EPI_ISL_1816390 | | | |
| see above | EXCITE Lab | Andersen lab at Scripps Research | Nicole L Washington, Simon White, Geraint Levan, Kelly Schiabor Barrett, Elizabeth Cirulli, Alexandre Bolze, Charlotte Rivera-Garcia, Ryan Cho, Jason Nguyen, Sherry Wang, Jimmy Ramirez, Celena Andrade, Alice Summerfield, Tyler Cassens, Efren Sandoval, Francisco Tanudjaja, Magnus Isaksson, William Lee, David Becker, Marc Laurent, James Lu + SEARCH |
| EPI_ISL_1816391 | EXCITE Lab | Andersen lab at Scripps Research | Chip Schooley, Natasha Martin, Cheryl Anderson, Angela Scioscia, Smruthi Karthikeyan, Greg Humphrey, Sawyer Farmer, Abigail Schnapper, Helena Tubb, Tommy Valles + SEARCH |
| EPI_ISL_1816392, EPI_ISL_1816393 | EXCITE Lab | Andersen lab at Scripps Research | Nicole L Washington, Simon White, Geraint Levan, Kelly Schiabor Barrett, Elizabeth Cirulli, Alexandre Bolze, Charlotte Rivera-Garcia, Ryan Cho, Jason Nguyen, Sherry Wang, Jimmy Ramirez, Celena Andrade, Alice Summerfield, Tyler Cassens, Efren Sandoval, Francisco Tanudjaja, Magnus Isaksson, William Lee, David Becker, Marc Laurent, James Lu + SEARCH |
| EPI_ISL_1816394 | EXCITE Lab | Andersen lab at Scripps Research | David Pride, Sharon Reed, Chip Schooley, Angela Scioscia, Natasha Martin Cheryl Anderson, Sawyer Farmer, Abigail Schnapper, Helena Tubb, Tommy Valles + SEARCH |
| EPI_ISL_1816395, EPI_ISL_1816396, EPI_ISL_1816397, EPI_ISL_1816398, EPI_ISL_1816399, EPI_ISL_1816400, EPI_ISL_1816401, EPI_ISL_1816402, EPI_ISL_1816403, EPI_ISL_1816404 | EXCITE Lab | Andersen lab at Scripps Research | Nicole L Washington, Simon White, Geraint Levan, Kelly Schiabor Barrett, Elizabeth Cirulli, Alexandre Bolze, Charlotte Rivera-Garcia, Ryan Cho, Jason Nguyen, Sherry Wang, Jimmy Ramirez, Celena Andrade, Alice Summerfield, Tyler Cassens, Efren Sandoval, Francisco Tanudjaja, Magnus Isaksson, William Lee, David Becker, Marc Laurent, James Lu + SEARCH |
| EPI_ISL_1816405 | EXCITE Lab | Andersen lab at Scripps Research | David Pride, Sharon Reed, Chip Schooley, Angela Scioscia, Natasha Martin Cheryl Anderson, Sawyer Farmer, Abigail Schnapper, Helena Tubb, Tommy Valles + SEARCH |
| EPI_ISL_1816406, EPI_ISL_1816407 | EXCITE Lab | Andersen lab at Scripps Research | Nicole L Washington, Simon White, Geraint Levan, Kelly Schiabor Barrett, Elizabeth Cirulli, Alexandre Bolze, Charlotte Rivera-Garcia, Ryan Cho, Jason Nguyen, Sherry Wang, Jimmy Ramirez, Celena Andrade, Alice Summerfield, Tyler Cassens, Efren Sandoval, Francisco Tanudjaja, Magnus Isaksson, William Lee, David Becker, Marc Laurent, James Lu + SEARCH |
| EPI_ISL_1816408 | EXCITE Lab | Andersen lab at Scripps Research | Fred Wu, Christine Mitchell + SEARCH |
| EPI_ISL_1816409, EPI_ISL_1816410, EPI_ISL_1816411, | EXCITE Lab | Andersen lab at Scripps Research | Nicole L Washington, Simon White, Geraint Levan, Kelly Schiabor Barrett, Elizabeth Cirulli, Alexandre Bolze, Charlotte Rivera-Garcia, Ryan Cho, Jason Nguyen, Sherry Wang, Jimmy Ramirez, Celena Andrade, Alice Summerfield, Tyler Cassens, Efren Sandoval, Francisco Tanudjaja, Magnus Isaksson, William Lee, David Becker, Marc Laurent, James Lu + SEARCH |

| | | | |
|---|---|---|---|
| EPI_ISL_1816412, EPI_ISL_1816413, EPI_ISL_1816414, EPI_ISL_1816415, EPI_ISL_1816416, EPI_ISL_1816417, EPI_ISL_1816418 | | | |
| EPI_ISL_1816419 | EXCITE Lab | Andersen lab at Scripps Research | Chip Schooley, Natasha Martin, Cheryl Anderson, Angela Scioscia, Smruthi Karthikeyan, Greg Humphrey, Sawyer Farmer, Abigail Schnapper, Helena Tubb, Tommy Valles + SEARCH |
| EPI_ISL_1816420 | EXCITE Lab | Andersen lab at Scripps Research | Nicole L Washington, Simon White, Geraint Levan, Kelly Schiabor Barrett, Elizabeth Cirulli, Alexandre Bolze, Charlotte Rivera-Garcia, Ryan Cho, Jason Nguyen, Sherry Wang, Jimmy Ramirez, Celena Andrade, Alice Summerfield, Tyler Cassens, Efren Sandoval, Francisco Tanudjaja, Magnus Isaksson, William Lee, David Becker, Marc Laurent, James Lu + SEARCH |
| EPI_ISL_1816421 | EXCITE Lab | Andersen lab at Scripps Research | David Pride, Sharon Reed, Chip Schooley, Angela Scioscia, Natasha Martin Cheryl Anderson, Sawyer Farmer, Abigail Schnapper, Helena Tubb, Tommy Valles + SEARCH |
| EPI_ISL_1816422, EPI_ISL_1816423, EPI_ISL_1816424, EPI_ISL_1816425, EPI_ISL_1816426, EPI_ISL_1816427, EPI_ISL_1816428, EPI_ISL_1816429, EPI_ISL_1816430, EPI_ISL_1816431, EPI_ISL_1816432, EPI_ISL_1816433, EPI_ISL_1816434, EPI_ISL_1816435, EPI_ISL_1816436, EPI_ISL_1816437, EPI_ISL_1816438, EPI_ISL_1816439, EPI_ISL_1816440, EPI_ISL_1816441, EPI_ISL_1816442, EPI_ISL_1816443, EPI_ISL_1816444, EPI_ISL_1816445, EPI_ISL_1816446, EPI_ISL_1816447 | | | |
| see above | EXCITE Lab | Andersen lab at Scripps Research | Nicole L Washington, Simon White, Geraint Levan, Kelly Schiabor Barrett, Elizabeth Cirulli, Alexandre Bolze, Charlotte Rivera-Garcia, Ryan Cho, Jason Nguyen, Sherry Wang, Jimmy Ramirez, Celena Andrade, Alice Summerfield, Tyler Cassens, Efren Sandoval, Francisco Tanudjaja, Magnus Isaksson, William Lee, David Becker, Marc Laurent, James Lu + SEARCH |
| EPI_ISL_1816448 | EXCITE Lab | Andersen lab at Scripps Research | David Pride, Sharon Reed, Chip Schooley, Angela Scioscia, Natasha Martin Cheryl Anderson, Sawyer Farmer, Abigail Schnapper, Helena Tubb, Tommy Valles + SEARCH |
| EPI_ISL_1816449, EPI_ISL_1816450, EPI_ISL_1816451, EPI_ISL_1816452, EPI_ISL_1816453, EPI_ISL_1816454, EPI_ISL_1816455, EPI_ISL_1816456, EPI_ISL_1816457, EPI_ISL_1816458, EPI_ISL_1816459, EPI_ISL_1816460, EPI_ISL_1816461, EPI_ISL_1816462, EPI_ISL_1816463, EPI_ISL_1816464, EPI_ISL_1816465, EPI_ISL_1816466, EPI_ISL_1816467, EPI_ISL_1816468, EPI_ISL_1816469, EPI_ISL_1816470, EPI_ISL_1816471, EPI_ISL_1816472, EPI_ISL_1816473 | | | |
| see above | EXCITE Lab | Andersen lab at Scripps Research | Nicole L Washington, Simon White, Geraint Levan, Kelly Schiabor Barrett, Elizabeth Cirulli, Alexandre Bolze, Charlotte Rivera-Garcia, Ryan Cho, Jason Nguyen, Sherry Wang, Jimmy Ramirez, Celena Andrade, Alice Summerfield, Tyler Cassens, Efren Sandoval, Francisco Tanudjaja, Magnus Isaksson, William Lee, David Becker, Marc Laurent, James Lu + SEARCH |
| EPI_ISL_1816474 | EXCITE Lab | Andersen lab at Scripps Research | Chip Schooley, Natasha Martin, Cheryl Anderson, Angela Scioscia, Smruthi Karthikeyan, Greg Humphrey, Sawyer Farmer, Abigail Schnapper, Helena Tubb, Tommy Valles + SEARCH |
| EPI_ISL_1816475, EPI_ISL_1816476 | EXCITE Lab | Andersen lab at Scripps Research | Nicole L Washington, Simon White, Geraint Levan, Kelly Schiabor Barrett, Elizabeth Cirulli, Alexandre Bolze, Charlotte Rivera-Garcia, Ryan Cho, Jason Nguyen, Sherry Wang, Jimmy Ramirez, Celena Andrade, Alice Summerfield, Tyler Cassens, Efren Sandoval, Francisco Tanudjaja, Magnus Isaksson, William Lee, David Becker, Marc Laurent, James Lu + SEARCH |
| EPI_ISL_1816477 | EXCITE Lab | Andersen lab at Scripps Research | Sydney Morgan + Marni Jacobs + Celestine Magallanes + Holly Valentine + Louise C. Laurent + SEARCH |
| EPI_ISL_1816478, EPI_ISL_1816479, EPI_ISL_1816480, EPI_ISL_1816481, EPI_ISL_1816482, EPI_ISL_1816483, EPI_ISL_1816484, EPI_ISL_1816485, EPI_ISL_1816486, EPI_ISL_1816487, EPI_ISL_1816488 | | | |
| see above | EXCITE Lab | Andersen lab at Scripps Research | Nicole L Washington, Simon White, Geraint Levan, Kelly Schiabor Barrett, Elizabeth Cirulli, Alexandre Bolze, Charlotte Rivera-Garcia, Ryan Cho, Jason Nguyen, Sherry Wang, Jimmy Ramirez, Celena Andrade, Alice Summerfield, Tyler Cassens, Efren Sandoval, Francisco Tanudjaja, Magnus Isaksson, William Lee, David Becker, Marc Laurent, James Lu + SEARCH |
| EPI_ISL_1816489 | EXCITE Lab | Andersen lab at Scripps Research | Sydney Morgan + Marni Jacobs + Celestine Magallanes + Holly Valentine + Louise C. Laurent + SEARCH |
| EPI_ISL_1816490, EPI_ISL_1816491 | EXCITE Lab | Andersen lab at Scripps Research | Nicole L Washington, Simon White, Geraint Levan, Kelly Schiabor Barrett, Elizabeth Cirulli, Alexandre Bolze, Charlotte Rivera-Garcia, Ryan Cho, Jason Nguyen, Sherry Wang, Jimmy Ramirez, Celena Andrade, Alice Summerfield, Tyler Cassens, Efren Sandoval, Francisco Tanudjaja, Magnus Isaksson, William Lee, David Becker, Marc Laurent, James Lu + SEARCH |
| EPI_ISL_1816492 | EXCITE Lab | Andersen lab at Scripps Research | J. Joelle Donofrio-Odmann, Brent Brainard+ SEARCH |
| EPI_ISL_1816493, EPI_ISL_1816494, EPI_ISL_1816495, EPI_ISL_1816496 | EXCITE Lab | Andersen lab at Scripps Research | Nicole L Washington, Simon White, Geraint Levan, Kelly Schiabor Barrett, Elizabeth Cirulli, Alexandre Bolze, Charlotte Rivera-Garcia, Ryan Cho, Jason Nguyen, Sherry Wang, Jimmy Ramirez, Celena Andrade, Alice Summerfield, Tyler Cassens, Efren Sandoval, Francisco Tanudjaja, Magnus Isaksson, William Lee, David Becker, Marc Laurent, James Lu + SEARCH |
| EPI_ISL_1816497 | EXCITE Lab | Andersen lab at Scripps Research | David Pride, Sharon Reed, Chip Schooley, Angela Scioscia, Natasha Martin Cheryl Anderson, Sawyer Farmer, Abigail Schnapper, Helena Tubb, Tommy Valles + SEARCH |
| EPI_ISL_1816498, EPI_ISL_1816499 | EXCITE Lab | Andersen lab at Scripps Research | Nicole L Washington, Simon White, Geraint Levan, Kelly Schiabor Barrett, Elizabeth Cirulli, Alexandre Bolze, Charlotte Rivera-Garcia, Ryan Cho, Jason Nguyen, Sherry Wang, Jimmy Ramirez, Celena Andrade, Alice Summerfield, Tyler Cassens, Efren Sandoval, Francisco Tanudjaja, Magnus Isaksson, William Lee, David Becker, Marc Laurent, James Lu + SEARCH |
| EPI_ISL_1816500 | EXCITE Lab | Andersen lab at Scripps Research | David Pride, Sharon Reed, Chip Schooley, Angela Scioscia, Natasha Martin Cheryl Anderson, Sawyer Farmer, Abigail Schnapper, Helena Tubb, Tommy Valles + SEARCH |
| EPI_ISL_1816501 | EXCITE Lab | Andersen lab at Scripps Research | J. Joelle Donofrio-Odmann, Brent Brainard+ SEARCH |
| EPI_ISL_1816502, EPI_ISL_1816503 | EXCITE Lab | Andersen lab at Scripps Research | Nicole L Washington, Simon White, Geraint Levan, Kelly Schiabor Barrett, Elizabeth Cirulli, Alexandre Bolze, Charlotte Rivera-Garcia, Ryan Cho, Jason Nguyen, Sherry Wang, Jimmy Ramirez, Celena Andrade, Alice Summerfield, Tyler Cassens, Efren Sandoval, Francisco Tanudjaja, Magnus Isaksson, William Lee, David Becker, Marc Laurent, James Lu + SEARCH |
| EPI_ISL_1816504 | EXCITE Lab | Andersen lab at Scripps Research | Chip Schooley, Natasha Martin, Cheryl Anderson, Angela Scioscia, Smruthi Karthikeyan, Greg Humphrey, Sawyer Farmer, Abigail Schnapper, Helena Tubb, Tommy Valles + SEARCH |
| EPI_ISL_1816505, EPI_ISL_1816506, EPI_ISL_1816507, EPI_ISL_1816508, EPI_ISL_1816509, EPI_ISL_1816510, EPI_ISL_1816511, EPI_ISL_1816512, EPI_ISL_1816513, EPI_ISL_1816514, EPI_ISL_1816515, EPI_ISL_1816516, EPI_ISL_1816517 | | | |
| see above | EXCITE Lab | Andersen lab at Scripps Research | Nicole L Washington, Simon White, Geraint Levan, Kelly Schiabor Barrett, Elizabeth Cirulli, Alexandre Bolze, Charlotte Rivera-Garcia, Ryan Cho, Jason Nguyen, Sherry Wang, Jimmy Ramirez, Celena Andrade, Alice Summerfield, Tyler Cassens, Efren Sandoval, Francisco Tanudjaja, Magnus Isaksson, William Lee, David Becker, Marc Laurent, James Lu + SEARCH |
| EPI_ISL_1816518 | EXCITE Lab | Andersen lab at Scripps Research | Sydney Morgan + Marni Jacobs + Celestine Magallanes + Holly Valentine + Louise C. Laurent + SEARCH |
| EPI_ISL_1816519, EPI_ISL_1816520, EPI_ISL_1816521, EPI_ISL_1816522, EPI_ISL_1816523, EPI_ISL_1816524 | EXCITE Lab | Andersen lab at Scripps Research | Nicole L Washington, Simon White, Geraint Levan, Kelly Schiabor Barrett, Elizabeth Cirulli, Alexandre Bolze, Charlotte Rivera-Garcia, Ryan Cho, Jason Nguyen, Sherry Wang, Jimmy Ramirez, Celena Andrade, Alice Summerfield, Tyler Cassens, Efren Sandoval, Francisco Tanudjaja, Magnus Isaksson, William Lee, David Becker, Marc Laurent, James Lu + SEARCH |
| EPI_ISL_1816525 | EXCITE Lab | Andersen lab at Scripps Research | David Pride, Sharon Reed, Chip Schooley, Angela Scioscia, Natasha Martin Cheryl Anderson, Sawyer Farmer, Abigail Schnapper, Helena Tubb, Tommy Valles + SEARCH |
| EPI_ISL_1816526, EPI_ISL_1816527, EPI_ISL_1816528, EPI_ISL_1816529, EPI_ISL_1816530, EPI_ISL_1816531, EPI_ISL_1816532, EPI_ISL_1816533, EPI_ISL_1816534, EPI_ISL_1816535, EPI_ISL_1816536, EPI_ISL_1816537, EPI_ISL_1816538 | | | |
| see above | EXCITE Lab | Andersen lab at Scripps Research | Nicole L Washington, Simon White, Geraint Levan, Kelly Schiabor Barrett, Elizabeth Cirulli, Alexandre Bolze, Charlotte Rivera-Garcia, Ryan Cho, Jason Nguyen, Sherry Wang, Jimmy Ramirez, Celena Andrade, Alice Summerfield, Tyler Cassens, Efren Sandoval, Francisco Tanudjaja, Magnus Isaksson, William Lee, David Becker, Marc Laurent, James Lu + SEARCH |
| EPI_ISL_1816539 | EXCITE Lab | Andersen lab at Scripps Research | Sydney Morgan + Marni Jacobs + Celestine Magallanes + Holly Valentine + Louise C. Laurent + SEARCH |
| EPI_ISL_1816540, EPI_ISL_1816541, EPI_ISL_1816542, EPI_ISL_1816543, EPI_ISL_1816544 | EXCITE Lab | Andersen lab at Scripps Research | Nicole L Washington, Simon White, Geraint Levan, Kelly Schiabor Barrett, Elizabeth Cirulli, Alexandre Bolze, Charlotte Rivera-Garcia, Ryan Cho, Jason Nguyen, Sherry Wang, Jimmy Ramirez, Celena Andrade, Alice Summerfield, Tyler Cassens, Efren Sandoval, Francisco Tanudjaja, Magnus Isaksson, William Lee, David Becker, Marc Laurent, James Lu + SEARCH |
| EPI_ISL_1816545 | EXCITE Lab | Andersen lab at Scripps Research | David Pride, Sharon Reed, Chip Schooley, Angela Scioscia, Natasha Martin Cheryl Anderson, Sawyer Farmer, Abigail Schnapper, Helena Tubb, Tommy Valles + SEARCH |
| EPI_ISL_1816546, EPI_ISL_1816547, EPI_ISL_1816548, EPI_ISL_1816549, EPI_ISL_1816550, EPI_ISL_1816551, EPI_ISL_1816552, EPI_ISL_1816553, EPI_ISL_1816554 | EXCITE Lab | Andersen lab at Scripps Research | Nicole L Washington, Simon White, Geraint Levan, Kelly Schiabor Barrett, Elizabeth Cirulli, Alexandre Bolze, Charlotte Rivera-Garcia, Ryan Cho, Jason Nguyen, Sherry Wang, Jimmy Ramirez, Celena Andrade, Alice Summerfield, Tyler Cassens, Efren Sandoval, Francisco Tanudjaja, Magnus Isaksson, William Lee, David Becker, Marc Laurent, James Lu + SEARCH |
| EPI_ISL_1816555 | EXCITE Lab | Andersen lab at Scripps Research | Sydney Morgan + Marni Jacobs + Celestine Magallanes + Holly Valentine + Louise C. Laurent + SEARCH |
| EPI_ISL_1816556 | EXCITE Lab | Andersen lab at Scripps Research | David Pride, Sharon Reed, Chip Schooley, Angela Scioscia, Natasha Martin Cheryl Anderson, Sawyer Farmer, Abigail Schnapper, Helena Tubb, Tommy Valles + SEARCH |
| EPI_ISL_1816557 | EXCITE Lab | Andersen lab at Scripps Research | Sydney Morgan + Marni Jacobs + Celestine Magallanes + Holly Valentine + Louise C. Laurent + SEARCH |
| EPI_ISL_1816558, EPI_ISL_1816559, EPI_ISL_1816560, EPI_ISL_1816561 | EXCITE Lab | Andersen lab at Scripps Research | Nicole L Washington, Simon White, Geraint Levan, Kelly Schiabor Barrett, Elizabeth Cirulli, Alexandre Bolze, Charlotte Rivera-Garcia, Ryan Cho, Jason Nguyen, Sherry Wang, Jimmy Ramirez, Celena Andrade, Alice Summerfield, Tyler Cassens, Efren Sandoval, Francisco Tanudjaja, Magnus Isaksson, William Lee, David Becker, Marc Laurent, James Lu + SEARCH |
| EPI_ISL_1816562 | EXCITE Lab | Andersen lab at Scripps Research | Rebecca Fielding-Miller, Laura Kohn, Patricia Gonzalez-Zuniga, Fitri Carrissa Wijaya, Vinton Omaleki, Marlene Flores, Araz Majnoonian, Megan Nguyen, Ashkan Hassani, Smruthi Karthikeyan, Rodolfo Salido, Victor Cantu, Greg Humphrey, Sawyer Farmer, Abigail Schnapper, Helena Tubb, Tommy Valles + SEARCH |
| EPI_ISL_1816563, EPI_ISL_1816564, EPI_ISL_1816565, EPI_ISL_1816566, EPI_ISL_1816567, EPI_ISL_1816568, EPI_ISL_1816569, EPI_ISL_1816570, EPI_ISL_1816571, EPI_ISL_1816572, EPI_ISL_1816573, EPI_ISL_1816574 | | | |
| see above | EXCITE Lab | Andersen lab at Scripps Research | Nicole L Washington, Simon White, Geraint Levan, Kelly Schiabor Barrett, Elizabeth Cirulli, Alexandre Bolze, Charlotte Rivera-Garcia, Ryan Cho, Jason Nguyen, Sherry Wang, Jimmy Ramirez, Celena Andrade, Alice Summerfield, Tyler Cassens, Efren Sandoval, Francisco Tanudjaja, Magnus Isaksson, William Lee, David Becker, Marc Laurent, James Lu + SEARCH |
| EPI_ISL_1816575 | EXCITE Lab | Andersen lab at Scripps Research | Sydney Morgan + Marni Jacobs + Celestine Magallanes + Holly Valentine + Louise C. Laurent + SEARCH |
| EPI_ISL_1816576, EPI_ISL_1816577, EPI_ISL_1816578, EPI_ISL_1816579, | EXCITE Lab | Andersen lab at Scripps Research | Nicole L Washington, Simon White, Geraint Levan, Kelly Schiabor Barrett, Elizabeth Cirulli, Alexandre Bolze, Charlotte Rivera-Garcia, Ryan Cho, Jason Nguyen, Sherry Wang, Jimmy Ramirez, Celena Andrade, Alice Summerfield, Tyler Cassens, Efren Sandoval, Francisco Tanudjaja, Magnus Isaksson, William Lee, David Becker, Marc Laurent, James Lu + SEARCH |

| | | | |
|---|---|---|---|
| EPI_ISL_1816580, EPI_ISL_1816581, EPI_ISL_1816582 | | | |
| EPI_ISL_1816583 | EXCITE Lab | Andersen lab at Scripps Research | David Pride, Sharon Reed, Chip Schooley, Angela Scioscia, Natasha Martin Cheryl Anderson, Sawyer Farmer, Abigail Schnapper, Helena Tubb, Tommy Valles + SEARCH |
| EPI_ISL_1816584 | EXCITE Lab | Andersen lab at Scripps Research | Nicole L Washington, Simon White, Geraint Levan, Kelly Schiabor Barrett, Elizabeth Cirulli, Alexandre Bolze, Charlotte Rivera-Garcia, Ryan Cho, Jason Nguyen, Sherry Wang, Jimmy Ramirez, Celena Andrade, Alice Summerfield, Tyler Cassens, Efren Sandoval, Francisco Tanudjaja, Magnus Isaksson, William Lee, David Becker, Marc Laurent, James Lu + SEARCH |
| EPI_ISL_1816585 | EXCITE Lab | Andersen lab at Scripps Research | David Pride, Sharon Reed, Chip Schooley, Angela Scioscia, Natasha Martin Cheryl Anderson, Sawyer Farmer, Abigail Schnapper, Helena Tubb, Tommy Valles + SEARCH |
| EPI_ISL_1816586, EPI_ISL_1816587 | EXCITE Lab | Andersen lab at Scripps Research | Nicole L Washington, Simon White, Geraint Levan, Kelly Schiabor Barrett, Elizabeth Cirulli, Alexandre Bolze, Charlotte Rivera-Garcia, Ryan Cho, Jason Nguyen, Sherry Wang, Jimmy Ramirez, Celena Andrade, Alice Summerfield, Tyler Cassens, Efren Sandoval, Francisco Tanudjaja, Magnus Isaksson, William Lee, David Becker, Marc Laurent, James Lu + SEARCH |
| EPI_ISL_1816588, EPI_ISL_1816589 | EXCITE Lab | Andersen lab at Scripps Research | David Pride, Sharon Reed, Chip Schooley, Angela Scioscia, Natasha Martin Cheryl Anderson, Sawyer Farmer, Abigail Schnapper, Helena Tubb, Tommy Valles + SEARCH |
| EPI_ISL_1816590, EPI_ISL_1816591, EPI_ISL_1816592, EPI_ISL_1816593, EPI_ISL_1816594, EPI_ISL_1816595, EPI_ISL_1816596 | EXCITE Lab | Andersen lab at Scripps Research | Nicole L Washington, Simon White, Geraint Levan, Kelly Schiabor Barrett, Elizabeth Cirulli, Alexandre Bolze, Charlotte Rivera-Garcia, Ryan Cho, Jason Nguyen, Sherry Wang, Jimmy Ramirez, Celena Andrade, Alice Summerfield, Tyler Cassens, Efren Sandoval, Francisco Tanudjaja, Magnus Isaksson, William Lee, David Becker, Marc Laurent, James Lu + SEARCH |
| EPI_ISL_1816597 | EXCITE Lab | Andersen lab at Scripps Research | Sydney Morgan + Marni Jacobs + Celestine Magallanes + Holly Valentine + Louise C. Laurent + SEARCH |
| EPI_ISL_1816598, EPI_ISL_1816599, EPI_ISL_1816600, EPI_ISL_1816601, EPI_ISL_1816602, EPI_ISL_1816603, EPI_ISL_1816604, EPI_ISL_1816605, EPI_ISL_1816606, EPI_ISL_1816607, EPI_ISL_1816608, EPI_ISL_1816609 | | | |
| see above | EXCITE Lab | Andersen lab at Scripps Research | Nicole L Washington, Simon White, Geraint Levan, Kelly Schiabor Barrett, Elizabeth Cirulli, Alexandre Bolze, Charlotte Rivera-Garcia, Ryan Cho, Jason Nguyen, Sherry Wang, Jimmy Ramirez, Celena Andrade, Alice Summerfield, Tyler Cassens, Efren Sandoval, Francisco Tanudjaja, Magnus Isaksson, William Lee, David Becker, Marc Laurent, James Lu + SEARCH |
| EPI_ISL_1816610 | EXCITE Lab | Andersen lab at Scripps Research | Chip Schooley, Natasha Martin, Cheryl Anderson, Angela Scioscia, Smruthi Karthikeyan, Greg Humphrey, Sawyer Farmer, Abigail Schnapper, Helena Tubb, Tommy Valles + SEARCH |
| EPI_ISL_1816611, EPI_ISL_1816612, EPI_ISL_1816613, EPI_ISL_1816614, EPI_ISL_1816615, EPI_ISL_1816616, EPI_ISL_1816617, EPI_ISL_1816618, EPI_ISL_1816619, EPI_ISL_1816620, EPI_ISL_1816621 | | | |
| see above | EXCITE Lab | Andersen lab at Scripps Research | Nicole L Washington, Simon White, Geraint Levan, Kelly Schiabor Barrett, Elizabeth Cirulli, Alexandre Bolze, Charlotte Rivera-Garcia, Ryan Cho, Jason Nguyen, Sherry Wang, Jimmy Ramirez, Celena Andrade, Alice Summerfield, Tyler Cassens, Efren Sandoval, Francisco Tanudjaja, Magnus Isaksson, William Lee, David Becker, Marc Laurent, James Lu + SEARCH |
| EPI_ISL_1816622 | EXCITE Lab | Andersen lab at Scripps Research | David Pride, Sharon Reed, Chip Schooley, Angela Scioscia, Natasha Martin Cheryl Anderson, Sawyer Farmer, Abigail Schnapper, Helena Tubb, Tommy Valles + SEARCH |
| EPI_ISL_1816623, EPI_ISL_1816624, EPI_ISL_1816625, EPI_ISL_1816626, EPI_ISL_1816627, EPI_ISL_1816628, EPI_ISL_1816629, EPI_ISL_1816630 | EXCITE Lab | Andersen lab at Scripps Research | Nicole L Washington, Simon White, Geraint Levan, Kelly Schiabor Barrett, Elizabeth Cirulli, Alexandre Bolze, Charlotte Rivera-Garcia, Ryan Cho, Jason Nguyen, Sherry Wang, Jimmy Ramirez, Celena Andrade, Alice Summerfield, Tyler Cassens, Efren Sandoval, Francisco Tanudjaja, Magnus Isaksson, William Lee, David Becker, Marc Laurent, James Lu + SEARCH |
| EPI_ISL_1816631 | EXCITE Lab | Andersen lab at Scripps Research | David Pride, Sharon Reed, Chip Schooley, Angela Scioscia, Natasha Martin Cheryl Anderson, Sawyer Farmer, Abigail Schnapper, Helena Tubb, Tommy Valles + SEARCH |
| EPI_ISL_1816632 | EXCITE Lab | Andersen lab at Scripps Research | Nicole L Washington, Simon White, Geraint Levan, Kelly Schiabor Barrett, Elizabeth Cirulli, Alexandre Bolze, Charlotte Rivera-Garcia, Ryan Cho, Jason Nguyen, Sherry Wang, Jimmy Ramirez, Celena Andrade, Alice Summerfield, Tyler Cassens, Efren Sandoval, Francisco Tanudjaja, Magnus Isaksson, William Lee, David Becker, Marc Laurent, James Lu + SEARCH |
| EPI_ISL_1816633 | EXCITE Lab | Andersen lab at Scripps Research | Sydney Morgan + Marni Jacobs + Celestine Magallanes + Holly Valentine + Louise C. Laurent + SEARCH |
| EPI_ISL_1816634, EPI_ISL_1816635, EPI_ISL_1816636, EPI_ISL_1816637, EPI_ISL_1816638, EPI_ISL_1816639, EPI_ISL_1816640, EPI_ISL_1816641, EPI_ISL_1816642, EPI_ISL_1816643, EPI_ISL_1816644, EPI_ISL_1816645 | | | |
| see above | EXCITE Lab | Andersen lab at Scripps Research | Nicole L Washington, Simon White, Geraint Levan, Kelly Schiabor Barrett, Elizabeth Cirulli, Alexandre Bolze, Charlotte Rivera-Garcia, Ryan Cho, Jason Nguyen, Sherry Wang, Jimmy Ramirez, Celena Andrade, Alice Summerfield, Tyler Cassens, Efren Sandoval, Francisco Tanudjaja, Magnus Isaksson, William Lee, David Becker, Marc Laurent, James Lu + SEARCH |
| EPI_ISL_1816646 | EXCITE Lab | Andersen lab at Scripps Research | David Pride, Sharon Reed, Chip Schooley, Angela Scioscia, Natasha Martin Cheryl Anderson, Sawyer Farmer, Abigail Schnapper, Helena Tubb, Tommy Valles + SEARCH |
| EPI_ISL_1816647, EPI_ISL_1816648, EPI_ISL_1816649 | EXCITE Lab | Andersen lab at Scripps Research | Nicole L Washington, Simon White, Geraint Levan, Kelly Schiabor Barrett, Elizabeth Cirulli, Alexandre Bolze, Charlotte Rivera-Garcia, Ryan Cho, Jason Nguyen, Sherry Wang, Jimmy Ramirez, Celena Andrade, Alice Summerfield, Tyler Cassens, Efren Sandoval, Francisco Tanudjaja, Magnus Isaksson, William Lee, David Becker, Marc Laurent, James Lu + SEARCH |
| EPI_ISL_1816650 | EXCITE Lab | Andersen lab at Scripps Research | David Pride, Sharon Reed, Chip Schooley, Angela Scioscia, Natasha Martin Cheryl Anderson, Sawyer Farmer, Abigail Schnapper, Helena Tubb, Tommy Valles + SEARCH |
| EPI_ISL_1816651, EPI_ISL_1816652, EPI_ISL_1816653, EPI_ISL_1816654, EPI_ISL_1816655, EPI_ISL_1816656, EPI_ISL_1816657, EPI_ISL_1816658 | EXCITE Lab | Andersen lab at Scripps Research | Nicole L Washington, Simon White, Geraint Levan, Kelly Schiabor Barrett, Elizabeth Cirulli, Alexandre Bolze, Charlotte Rivera-Garcia, Ryan Cho, Jason Nguyen, Sherry Wang, Jimmy Ramirez, Celena Andrade, Alice Summerfield, Tyler Cassens, Efren Sandoval, Francisco Tanudjaja, Magnus Isaksson, William Lee, David Becker, Marc Laurent, James Lu + SEARCH |
| EPI_ISL_1816659 | EXCITE Lab | Andersen lab at Scripps Research | David Pride, Sharon Reed, Chip Schooley, Angela Scioscia, Natasha Martin Cheryl Anderson, Sawyer Farmer, Abigail Schnapper, Helena Tubb, Tommy Valles + SEARCH |
| EPI_ISL_1816660, EPI_ISL_1816661, EPI_ISL_1816662, EPI_ISL_1816663, EPI_ISL_1816664, EPI_ISL_1816665, EPI_ISL_1816666, EPI_ISL_1816667, EPI_ISL_1816668, EPI_ISL_1816669, EPI_ISL_1816670, EPI_ISL_1816671, EPI_ISL_1816672, EPI_ISL_1816673 | | | |
| see above | EXCITE Lab | Andersen lab at Scripps Research | Nicole L Washington, Simon White, Geraint Levan, Kelly Schiabor Barrett, Elizabeth Cirulli, Alexandre Bolze, Charlotte Rivera-Garcia, Ryan Cho, Jason Nguyen, Sherry Wang, Jimmy Ramirez, Celena Andrade, Alice Summerfield, Tyler Cassens, Efren Sandoval, Francisco Tanudjaja, Magnus Isaksson, William Lee, David Becker, Marc Laurent, James Lu + SEARCH |
| EPI_ISL_1816674 | EXCITE Lab | Andersen lab at Scripps Research | David Pride, Sharon Reed, Chip Schooley, Angela Scioscia, Natasha Martin Cheryl Anderson, Sawyer Farmer, Abigail Schnapper, Helena Tubb, Tommy Valles + SEARCH |
| EPI_ISL_1816675, EPI_ISL_1816676, EPI_ISL_1816677, EPI_ISL_1816678, EPI_ISL_1816679, EPI_ISL_1816680, EPI_ISL_1816681, EPI_ISL_1816682, EPI_ISL_1816683, EPI_ISL_1816684, EPI_ISL_1816685, EPI_ISL_1816686, EPI_ISL_1816687, EPI_ISL_1816688, EPI_ISL_1816689 | | | |
| see above | EXCITE Lab | Andersen lab at Scripps Research | Nicole L Washington, Simon White, Geraint Levan, Kelly Schiabor Barrett, Elizabeth Cirulli, Alexandre Bolze, Charlotte Rivera-Garcia, Ryan Cho, Jason Nguyen, Sherry Wang, Jimmy Ramirez, Celena Andrade, Alice Summerfield, Tyler Cassens, Efren Sandoval, Francisco Tanudjaja, Magnus Isaksson, William Lee, David Becker, Marc Laurent, James Lu + SEARCH |
| EPI_ISL_1816690 | EXCITE Lab | Andersen lab at Scripps Research | David Pride, Sharon Reed, Chip Schooley, Angela Scioscia, Natasha Martin Cheryl Anderson, Sawyer Farmer, Abigail Schnapper, Helena Tubb, Tommy Valles + SEARCH |
| EPI_ISL_1816691, EPI_ISL_1816692, EPI_ISL_1816693, EPI_ISL_1816694, EPI_ISL_1816695, EPI_ISL_1816696 | EXCITE Lab | Andersen lab at Scripps Research | Nicole L Washington, Simon White, Geraint Levan, Kelly Schiabor Barrett, Elizabeth Cirulli, Alexandre Bolze, Charlotte Rivera-Garcia, Ryan Cho, Jason Nguyen, Sherry Wang, Jimmy Ramirez, Celena Andrade, Alice Summerfield, Tyler Cassens, Efren Sandoval, Francisco Tanudjaja, Magnus Isaksson, William Lee, David Becker, Marc Laurent, James Lu + SEARCH |
| EPI_ISL_1816697 | EXCITE Lab | Andersen lab at Scripps Research | Sydney Morgan + Marni Jacobs + Celestine Magallanes + Holly Valentine + Louise C. Laurent + SEARCH |
| EPI_ISL_1816698, EPI_ISL_1816699, EPI_ISL_1816700, EPI_ISL_1816701, EPI_ISL_1816702, EPI_ISL_1816703, EPI_ISL_1816704, EPI_ISL_1816705, EPI_ISL_1816706, EPI_ISL_1816707, EPI_ISL_1816708, EPI_ISL_1816709 | | | |
| see above | EXCITE Lab | Andersen lab at Scripps Research | Nicole L Washington, Simon White, Geraint Levan, Kelly Schiabor Barrett, Elizabeth Cirulli, Alexandre Bolze, Charlotte Rivera-Garcia, Ryan Cho, Jason Nguyen, Sherry Wang, Jimmy Ramirez, Celena Andrade, Alice Summerfield, Tyler Cassens, Efren Sandoval, Francisco Tanudjaja, Magnus Isaksson, William Lee, David Becker, Marc Laurent, James Lu + SEARCH |
| EPI_ISL_1816710 | EXCITE Lab | Andersen lab at Scripps Research | Sydney Morgan + Marni Jacobs + Celestine Magallanes + Holly Valentine + Louise C. Laurent + SEARCH |
| EPI_ISL_1816711, EPI_ISL_1816712, EPI_ISL_1816713, EPI_ISL_1816714, EPI_ISL_1816715, EPI_ISL_1816716 | EXCITE Lab | Andersen lab at Scripps Research | Nicole L Washington, Simon White, Geraint Levan, Kelly Schiabor Barrett, Elizabeth Cirulli, Alexandre Bolze, Charlotte Rivera-Garcia, Ryan Cho, Jason Nguyen, Sherry Wang, Jimmy Ramirez, Celena Andrade, Alice Summerfield, Tyler Cassens, Efren Sandoval, Francisco Tanudjaja, Magnus Isaksson, William Lee, David Becker, Marc Laurent, James Lu + SEARCH |
| EPI_ISL_1816717, EPI_ISL_1816718 | EXCITE Lab | Andersen lab at Scripps Research | David Pride, Sharon Reed, Chip Schooley, Angela Scioscia, Natasha Martin Cheryl Anderson, Sawyer Farmer, Abigail Schnapper, Helena Tubb, Tommy Valles + SEARCH |
| EPI_ISL_1816719, EPI_ISL_1816720, EPI_ISL_1816721, EPI_ISL_1816722, EPI_ISL_1816723, EPI_ISL_1816724, EPI_ISL_1816725 | EXCITE Lab | Andersen lab at Scripps Research | Nicole L Washington, Simon White, Geraint Levan, Kelly Schiabor Barrett, Elizabeth Cirulli, Alexandre Bolze, Charlotte Rivera-Garcia, Ryan Cho, Jason Nguyen, Sherry Wang, Jimmy Ramirez, Celena Andrade, Alice Summerfield, Tyler Cassens, Efren Sandoval, Francisco Tanudjaja, Magnus Isaksson, William Lee, David Becker, Marc Laurent, James Lu + SEARCH |
| EPI_ISL_1816726 | EXCITE Lab | Andersen lab at Scripps Research | Sydney Morgan + Marni Jacobs + Celestine Magallanes + Holly Valentine + Louise C. Laurent + SEARCH |
| EPI_ISL_1816727, | EXCITE Lab | Andersen lab at Scripps Research | Nicole L Washington, Simon White, Geraint Levan, Kelly Schiabor Barrett, Elizabeth Cirulli, Alexandre Bolze, Charlotte Rivera-Garcia, Ryan Cho, Jason Nguyen, Sherry Wang, Jimmy Ramirez, Celena Andrade, Alice Summerfield, Tyler Cassens, Efren Sandoval, Francisco Tanudjaja, Magnus |

| | | | |
|---|---|---|---|
| EPI_ISL_1816728, EPI_ISL_1816729 | | | Isaksson, William Lee, David Becker, Marc Laurent, James Lu + SEARCH |
| EPI_ISL_1816730 | EXCITE Lab | Andersen lab at Scripps Research | David Pride, Sharon Reed, Chip Schooley, Angela Scioscia, Natasha Martin Cheryl Anderson, Sawyer Farmer, Abigail Schnapper, Helena Tubb, Tommy Valles + SEARCH |
| EPI_ISL_1816731, EPI_ISL_1816732, EPI_ISL_1816733, EPI_ISL_1816734, EPI_ISL_1816735, EPI_ISL_1816736, EPI_ISL_1816737, EPI_ISL_1816738, EPI_ISL_1816739, EPI_ISL_1816740, EPI_ISL_1816741 | | | |
| see above | EXCITE Lab | Andersen lab at Scripps Research | Nicole L Washington, Simon White, Geraint Levan, Kelly Schiabor Barrett, Elizabeth Cirulli, Alexandre Bolze, Charlotte Rivera-Garcia, Ryan Cho, Jason Nguyen, Sherry Wang, Jimmy Ramirez, Celena Andrade, Alice Summerfield, Tyler Cassens, Efren Sandoval, Francisco Tanudjaja, Magnus Isaksson, William Lee, David Becker, Marc Laurent, James Lu + SEARCH |
| EPI_ISL_1816742 | EXCITE Lab | Andersen lab at Scripps Research | David Pride, Sharon Reed, Chip Schooley, Angela Scioscia, Natasha Martin Cheryl Anderson, Sawyer Farmer, Abigail Schnapper, Helena Tubb, Tommy Valles + SEARCH |
| EPI_ISL_1816743 | EXCITE Lab | Andersen lab at Scripps Research | Nicole L Washington, Simon White, Geraint Levan, Kelly Schiabor Barrett, Elizabeth Cirulli, Alexandre Bolze, Charlotte Rivera-Garcia, Ryan Cho, Jason Nguyen, Sherry Wang, Jimmy Ramirez, Celena Andrade, Alice Summerfield, Tyler Cassens, Efren Sandoval, Francisco Tanudjaja, Magnus Isaksson, William Lee, David Becker, Marc Laurent, James Lu + SEARCH |
| EPI_ISL_1816744 | EXCITE Lab | Andersen lab at Scripps Research | David Pride, Sharon Reed, Chip Schooley, Angela Scioscia, Natasha Martin Cheryl Anderson, Sawyer Farmer, Abigail Schnapper, Helena Tubb, Tommy Valles + SEARCH |
| EPI_ISL_1816745, EPI_ISL_1816746, EPI_ISL_1816747, EPI_ISL_1816748, EPI_ISL_1816749 | EXCITE Lab | Andersen lab at Scripps Research | Nicole L Washington, Simon White, Geraint Levan, Kelly Schiabor Barrett, Elizabeth Cirulli, Alexandre Bolze, Charlotte Rivera-Garcia, Ryan Cho, Jason Nguyen, Sherry Wang, Jimmy Ramirez, Celena Andrade, Alice Summerfield, Tyler Cassens, Efren Sandoval, Francisco Tanudjaja, Magnus Isaksson, William Lee, David Becker, Marc Laurent, James Lu + SEARCH |
| EPI_ISL_1816750 | EXCITE Lab | Andersen lab at Scripps Research | David Pride, Sharon Reed, Chip Schooley, Angela Scioscia, Natasha Martin Cheryl Anderson, Sawyer Farmer, Abigail Schnapper, Helena Tubb, Tommy Valles + SEARCH |
| EPI_ISL_1816751, EPI_ISL_1816752 | EXCITE Lab | Andersen lab at Scripps Research | Nicole L Washington, Simon White, Geraint Levan, Kelly Schiabor Barrett, Elizabeth Cirulli, Alexandre Bolze, Charlotte Rivera-Garcia, Ryan Cho, Jason Nguyen, Sherry Wang, Jimmy Ramirez, Celena Andrade, Alice Summerfield, Tyler Cassens, Efren Sandoval, Francisco Tanudjaja, Magnus Isaksson, William Lee, David Becker, Marc Laurent, James Lu + SEARCH |
| EPI_ISL_1816753 | EXCITE Lab | Andersen lab at Scripps Research | Sydney Morgan + Marni Jacobs + Celestine Magallanes + Holly Valentine + Louise C. Laurent + SEARCH |
| EPI_ISL_1816754, EPI_ISL_1816755, EPI_ISL_1816756, EPI_ISL_1816757, EPI_ISL_1816758, EPI_ISL_1816759 | EXCITE Lab | Andersen lab at Scripps Research | Nicole L Washington, Simon White, Geraint Levan, Kelly Schiabor Barrett, Elizabeth Cirulli, Alexandre Bolze, Charlotte Rivera-Garcia, Ryan Cho, Jason Nguyen, Sherry Wang, Jimmy Ramirez, Celena Andrade, Alice Summerfield, Tyler Cassens, Efren Sandoval, Francisco Tanudjaja, Magnus Isaksson, William Lee, David Becker, Marc Laurent, James Lu + SEARCH |
| EPI_ISL_1816760, EPI_ISL_1816761, EPI_ISL_1816762 | EXCITE Lab | Andersen lab at Scripps Research | David Pride, Sharon Reed, Chip Schooley, Angela Scioscia, Natasha Martin Cheryl Anderson, Sawyer Farmer, Abigail Schnapper, Helena Tubb, Tommy Valles + SEARCH |
| EPI_ISL_1816763 | EXCITE Lab | Andersen lab at Scripps Research | Nicole L Washington, Simon White, Geraint Levan, Kelly Schiabor Barrett, Elizabeth Cirulli, Alexandre Bolze, Charlotte Rivera-Garcia, Ryan Cho, Jason Nguyen, Sherry Wang, Jimmy Ramirez, Celena Andrade, Alice Summerfield, Tyler Cassens, Efren Sandoval, Francisco Tanudjaja, Magnus Isaksson, William Lee, David Becker, Marc Laurent, James Lu + SEARCH |
| EPI_ISL_1816764 | EXCITE Lab | Andersen lab at Scripps Research | David Pride, Sharon Reed, Chip Schooley, Angela Scioscia, Natasha Martin Cheryl Anderson, Sawyer Farmer, Abigail Schnapper, Helena Tubb, Tommy Valles + SEARCH |
| EPI_ISL_1816765, EPI_ISL_1816766, EPI_ISL_1816767, EPI_ISL_1816768, EPI_ISL_1816769, EPI_ISL_1816770 | EXCITE Lab | Andersen lab at Scripps Research | Nicole L Washington, Simon White, Geraint Levan, Kelly Schiabor Barrett, Elizabeth Cirulli, Alexandre Bolze, Charlotte Rivera-Garcia, Ryan Cho, Jason Nguyen, Sherry Wang, Jimmy Ramirez, Celena Andrade, Alice Summerfield, Tyler Cassens, Efren Sandoval, Francisco Tanudjaja, Magnus Isaksson, William Lee, David Becker, Marc Laurent, James Lu + SEARCH |
| EPI_ISL_1816771 | EXCITE Lab | Andersen lab at Scripps Research | David Pride, Sharon Reed, Chip Schooley, Angela Scioscia, Natasha Martin Cheryl Anderson, Sawyer Farmer, Abigail Schnapper, Helena Tubb, Tommy Valles + SEARCH |
| EPI_ISL_1816772, EPI_ISL_1816773, EPI_ISL_1816774 | EXCITE Lab | Andersen lab at Scripps Research | Nicole L Washington, Simon White, Geraint Levan, Kelly Schiabor Barrett, Elizabeth Cirulli, Alexandre Bolze, Charlotte Rivera-Garcia, Ryan Cho, Jason Nguyen, Sherry Wang, Jimmy Ramirez, Celena Andrade, Alice Summerfield, Tyler Cassens, Efren Sandoval, Francisco Tanudjaja, Magnus Isaksson, William Lee, David Becker, Marc Laurent, James Lu + SEARCH |
| EPI_ISL_1816775 | EXCITE Lab | Andersen lab at Scripps Research | Sydney Morgan + Marni Jacobs + Celestine Magallanes + Holly Valentine + Louise C. Laurent + SEARCH |
| EPI_ISL_1816776 | EXCITE Lab | Andersen lab at Scripps Research | Nicole L Washington, Simon White, Geraint Levan, Kelly Schiabor Barrett, Elizabeth Cirulli, Alexandre Bolze, Charlotte Rivera-Garcia, Ryan Cho, Jason Nguyen, Sherry Wang, Jimmy Ramirez, Celena Andrade, Alice Summerfield, Tyler Cassens, Efren Sandoval, Francisco Tanudjaja, Magnus Isaksson, William Lee, David Becker, Marc Laurent, James Lu + SEARCH |
| EPI_ISL_1816777 | EXCITE Lab | Andersen lab at Scripps Research | David Pride, Sharon Reed, Chip Schooley, Angela Scioscia, Natasha Martin Cheryl Anderson, Sawyer Farmer, Abigail Schnapper, Helena Tubb, Tommy Valles + SEARCH |
| EPI_ISL_1816778, EPI_ISL_1816779, EPI_ISL_1816780, EPI_ISL_1816781 | EXCITE Lab | Andersen lab at Scripps Research | Nicole L Washington, Simon White, Geraint Levan, Kelly Schiabor Barrett, Elizabeth Cirulli, Alexandre Bolze, Charlotte Rivera-Garcia, Ryan Cho, Jason Nguyen, Sherry Wang, Jimmy Ramirez, Celena Andrade, Alice Summerfield, Tyler Cassens, Efren Sandoval, Francisco Tanudjaja, Magnus Isaksson, William Lee, David Becker, Marc Laurent, James Lu + SEARCH |
| EPI_ISL_1816782 | EXCITE Lab | Andersen lab at Scripps Research | David Pride, Sharon Reed, Chip Schooley, Angela Scioscia, Natasha Martin Cheryl Anderson, Sawyer Farmer, Abigail Schnapper, Helena Tubb, Tommy Valles + SEARCH |
| EPI_ISL_1816783 | EXCITE Lab | Andersen lab at Scripps Research | Nicole L Washington, Simon White, Geraint Levan, Kelly Schiabor Barrett, Elizabeth Cirulli, Alexandre Bolze, Charlotte Rivera-Garcia, Ryan Cho, Jason Nguyen, Sherry Wang, Jimmy Ramirez, Celena Andrade, Alice Summerfield, Tyler Cassens, Efren Sandoval, Francisco Tanudjaja, Magnus Isaksson, William Lee, David Becker, Marc Laurent, James Lu + SEARCH |
| EPI_ISL_1816784 | EXCITE Lab | Andersen lab at Scripps Research | Sydney Morgan + Marni Jacobs + Celestine Magallanes + Holly Valentine + Louise C. Laurent + SEARCH |
| EPI_ISL_1816785, EPI_ISL_1816786, EPI_ISL_1816787 | EXCITE Lab | Andersen lab at Scripps Research | Nicole L Washington, Simon White, Geraint Levan, Kelly Schiabor Barrett, Elizabeth Cirulli, Alexandre Bolze, Charlotte Rivera-Garcia, Ryan Cho, Jason Nguyen, Sherry Wang, Jimmy Ramirez, Celena Andrade, Alice Summerfield, Tyler Cassens, Efren Sandoval, Francisco Tanudjaja, Magnus Isaksson, William Lee, David Becker, Marc Laurent, James Lu + SEARCH |
| EPI_ISL_1816788, EPI_ISL_1816789 | EXCITE Lab | Andersen lab at Scripps Research | Sydney Morgan + Marni Jacobs + Celestine Magallanes + Holly Valentine + Louise C. Laurent + SEARCH |
| EPI_ISL_1816790, EPI_ISL_1816791 | EXCITE Lab | Andersen lab at Scripps Research | Nicole L Washington, Simon White, Geraint Levan, Kelly Schiabor Barrett, Elizabeth Cirulli, Alexandre Bolze, Charlotte Rivera-Garcia, Ryan Cho, Jason Nguyen, Sherry Wang, Jimmy Ramirez, Celena Andrade, Alice Summerfield, Tyler Cassens, Efren Sandoval, Francisco Tanudjaja, Magnus Isaksson, William Lee, David Becker, Marc Laurent, James Lu + SEARCH |
| EPI_ISL_1816792 | EXCITE Lab | Andersen lab at Scripps Research | David Pride, Sharon Reed, Chip Schooley, Angela Scioscia, Natasha Martin Cheryl Anderson, Sawyer Farmer, Abigail Schnapper, Helena Tubb, Tommy Valles + SEARCH |
| EPI_ISL_1816793, EPI_ISL_1816794, EPI_ISL_1816795, EPI_ISL_1816796, EPI_ISL_1816797, EPI_ISL_1816798, EPI_ISL_1816799 | EXCITE Lab | Andersen lab at Scripps Research | Nicole L Washington, Simon White, Geraint Levan, Kelly Schiabor Barrett, Elizabeth Cirulli, Alexandre Bolze, Charlotte Rivera-Garcia, Ryan Cho, Jason Nguyen, Sherry Wang, Jimmy Ramirez, Celena Andrade, Alice Summerfield, Tyler Cassens, Efren Sandoval, Francisco Tanudjaja, Magnus Isaksson, William Lee, David Becker, Marc Laurent, James Lu + SEARCH |
| EPI_ISL_1816800 | EXCITE Lab | Andersen lab at Scripps Research | David Pride, Sharon Reed, Chip Schooley, Angela Scioscia, Natasha Martin Cheryl Anderson, Sawyer Farmer, Abigail Schnapper, Helena Tubb, Tommy Valles + SEARCH |
| EPI_ISL_1816801, EPI_ISL_1816802, EPI_ISL_1816803, EPI_ISL_1816804, EPI_ISL_1816805, EPI_ISL_1816806, EPI_ISL_1816807 | EXCITE Lab | Andersen lab at Scripps Research | Nicole L Washington, Simon White, Geraint Levan, Kelly Schiabor Barrett, Elizabeth Cirulli, Alexandre Bolze, Charlotte Rivera-Garcia, Ryan Cho, Jason Nguyen, Sherry Wang, Jimmy Ramirez, Celena Andrade, Alice Summerfield, Tyler Cassens, Efren Sandoval, Francisco Tanudjaja, Magnus Isaksson, William Lee, David Becker, Marc Laurent, James Lu + SEARCH |
| EPI_ISL_1816808 | EXCITE Lab | Andersen lab at Scripps Research | Sydney Morgan + Marni Jacobs + Celestine Magallanes + Holly Valentine + Louise C. Laurent + SEARCH |
| EPI_ISL_1816809 | EXCITE Lab | Andersen lab at Scripps Research | David Pride, Sharon Reed, Chip Schooley, Angela Scioscia, Natasha Martin Cheryl Anderson, Sawyer Farmer, Abigail Schnapper, Helena Tubb, Tommy Valles + SEARCH |
| EPI_ISL_1816810 | EXCITE Lab | Andersen lab at Scripps Research | Sydney Morgan + Marni Jacobs + Celestine Magallanes + Holly Valentine + Louise C. Laurent + SEARCH |
| EPI_ISL_1816811 | EXCITE Lab | Andersen lab at Scripps Research | Chip Schooley, Natasha Martin, Cheryl Anderson, Angela Scioscia, Smruthi Karthikeyan, Greg Humphrey, Sawyer Farmer, Abigail Schnapper, Helena Tubb, Tommy Valles + SEARCH |
| EPI_ISL_1816812, EPI_ISL_1816813, EPI_ISL_1816814 | EXCITE Lab | Andersen lab at Scripps Research | Nicole L Washington, Simon White, Geraint Levan, Kelly Schiabor Barrett, Elizabeth Cirulli, Alexandre Bolze, Charlotte Rivera-Garcia, Ryan Cho, Jason Nguyen, Sherry Wang, Jimmy Ramirez, Celena Andrade, Alice Summerfield, Tyler Cassens, Efren Sandoval, Francisco Tanudjaja, Magnus Isaksson, William Lee, David Becker, Marc Laurent, James Lu + SEARCH |
| EPI_ISL_1816815 | EXCITE Lab | Andersen lab at Scripps Research | David Pride, Sharon Reed, Chip Schooley, Angela Scioscia, Natasha Martin Cheryl Anderson, Sawyer Farmer, Abigail Schnapper, Helena Tubb, Tommy Valles + SEARCH |
| EPI_ISL_1816816 | EXCITE Lab | Andersen lab at Scripps Research | Nicole L Washington, Simon White, Geraint Levan, Kelly Schiabor Barrett, Elizabeth Cirulli, Alexandre Bolze, Charlotte Rivera-Garcia, Ryan Cho, Jason Nguyen, Sherry Wang, Jimmy Ramirez, Celena Andrade, Alice Summerfield, Tyler Cassens, Efren Sandoval, Francisco Tanudjaja, Magnus Isaksson, William Lee, David Becker, Marc Laurent, James Lu + SEARCH |
| EPI_ISL_1816817 | EXCITE Lab | Andersen lab at Scripps Research | David Pride, Sharon Reed, Chip Schooley, Angela Scioscia, Natasha Martin Cheryl Anderson, Sawyer Farmer, Abigail Schnapper, Helena Tubb, Tommy Valles + SEARCH |
| EPI_ISL_1816818 | EXCITE Lab | Andersen lab at Scripps Research | Fred Wu, Christine Mitchell + SEARCH |
| EPI_ISL_1816819, EPI_ISL_1816820 | EXCITE Lab | Andersen lab at Scripps Research | Nicole L Washington, Simon White, Geraint Levan, Kelly Schiabor Barrett, Elizabeth Cirulli, Alexandre Bolze, Charlotte Rivera-Garcia, Ryan Cho, Jason Nguyen, Sherry Wang, Jimmy Ramirez, Celena Andrade, Alice Summerfield, Tyler Cassens, Efren Sandoval, Francisco Tanudjaja, Magnus Isaksson, William Lee, David Becker, Marc Laurent, James Lu + SEARCH |
| EPI_ISL_1816821 | EXCITE Lab | Andersen lab at Scripps Research | David Pride, Sharon Reed, Chip Schooley, Angela Scioscia, Natasha Martin Cheryl Anderson, Sawyer Farmer, Abigail Schnapper, Helena Tubb, Tommy Valles + SEARCH |

| | | | |
|---|---|---|---|
| EPI_ISL_1816822, EPI_ISL_1816823 | EXCITE Lab | Andersen lab at Scripps Research | Nicole L Washington, Simon White, Geraint Levan, Kelly Schiabor Barrett, Elizabeth Cirulli, Alexandre Bolze, Charlotte Rivera-Garcia, Ryan Cho, Jason Nguyen, Sherry Wang, Jimmy Ramirez, Celena Andrade, Alice Summerfield, Tyler Cassens, Efren Sandoval, Francisco Tanudjaja, Magnus Isaksson, William Lee, David Becker, Marc Laurent, James Lu + SEARCH |
| EPI_ISL_1816824 | EXCITE Lab | Andersen lab at Scripps Research | Fred Wu, Christine Mitchell + SEARCH |
| EPI_ISL_1816825 | EXCITE Lab | Andersen lab at Scripps Research | David Pride, Sharon Reed, Chip Schooley, Angela Scioscia, Natasha Martin Cheryl Anderson, Sawyer Farmer, Abigail Schnapper, Helena Tubb, Tommy Valles + SEARCH |
| EPI_ISL_1816826, EPI_ISL_1816827 | EXCITE Lab | Andersen lab at Scripps Research | Nicole L Washington, Simon White, Geraint Levan, Kelly Schiabor Barrett, Elizabeth Cirulli, Alexandre Bolze, Charlotte Rivera-Garcia, Ryan Cho, Jason Nguyen, Sherry Wang, Jimmy Ramirez, Celena Andrade, Alice Summerfield, Tyler Cassens, Efren Sandoval, Francisco Tanudjaja, Magnus Isaksson, William Lee, David Becker, Marc Laurent, James Lu + SEARCH |
| EPI_ISL_1816828 | EXCITE Lab | Andersen lab at Scripps Research | Sydney Morgan + Marni Jacobs + Celestine Magallanes + Holly Valentine + Louise C. Laurent + SEARCH |
| EPI_ISL_1816829 | EXCITE Lab | Andersen lab at Scripps Research | Chip Schooley, Natasha Martin, Cheryl Anderson, Angela Scioscia, Smruthi Karthikeyan, Greg Humphrey, Sawyer Farmer, Abigail Schnapper, Helena Tubb, Tommy Valles + SEARCH |
| EPI_ISL_1816830 | EXCITE Lab | Andersen lab at Scripps Research | David Pride, Sharon Reed, Chip Schooley, Angela Scioscia, Natasha Martin Cheryl Anderson, Sawyer Farmer, Abigail Schnapper, Helena Tubb, Tommy Valles + SEARCH |
| EPI_ISL_1816831 | EXCITE Lab | Andersen lab at Scripps Research | Sydney Morgan + Marni Jacobs + Celestine Magallanes + Holly Valentine + Louise C. Laurent + SEARCH |
| EPI_ISL_1816832, EPI_ISL_1816833, EPI_ISL_1816834 | EXCITE Lab | Andersen lab at Scripps Research | David Pride, Sharon Reed, Chip Schooley, Angela Scioscia, Natasha Martin Cheryl Anderson, Sawyer Farmer, Abigail Schnapper, Helena Tubb, Tommy Valles + SEARCH |
| EPI_ISL_1816835 | EXCITE Lab | Andersen lab at Scripps Research | Nicole L Washington, Simon White, Geraint Levan, Kelly Schiabor Barrett, Elizabeth Cirulli, Alexandre Bolze, Charlotte Rivera-Garcia, Ryan Cho, Jason Nguyen, Sherry Wang, Jimmy Ramirez, Celena Andrade, Alice Summerfield, Tyler Cassens, Efren Sandoval, Francisco Tanudjaja, Magnus Isaksson, William Lee, David Becker, Marc Laurent, James Lu + SEARCH |
| EPI_ISL_1816836 | EXCITE Lab | Andersen lab at Scripps Research | Sydney Morgan + Marni Jacobs + Celestine Magallanes + Holly Valentine + Louise C. Laurent + SEARCH |
| EPI_ISL_1816837 | EXCITE Lab | Andersen lab at Scripps Research | Nicole L Washington, Simon White, Geraint Levan, Kelly Schiabor Barrett, Elizabeth Cirulli, Alexandre Bolze, Charlotte Rivera-Garcia, Ryan Cho, Jason Nguyen, Sherry Wang, Jimmy Ramirez, Celena Andrade, Alice Summerfield, Tyler Cassens, Efren Sandoval, Francisco Tanudjaja, Magnus Isaksson, William Lee, David Becker, Marc Laurent, James Lu + SEARCH |
| EPI_ISL_1816838 | EXCITE Lab | Andersen lab at Scripps Research | David Pride, Sharon Reed, Chip Schooley, Angela Scioscia, Natasha Martin Cheryl Anderson, Sawyer Farmer, Abigail Schnapper, Helena Tubb, Tommy Valles + SEARCH |
| EPI_ISL_1816839, EPI_ISL_1816840, EPI_ISL_1816841, EPI_ISL_1816842, EPI_ISL_1816843, EPI_ISL_1816844 | EXCITE Lab | Andersen lab at Scripps Research | Nicole L Washington, Simon White, Geraint Levan, Kelly Schiabor Barrett, Elizabeth Cirulli, Alexandre Bolze, Charlotte Rivera-Garcia, Ryan Cho, Jason Nguyen, Sherry Wang, Jimmy Ramirez, Celena Andrade, Alice Summerfield, Tyler Cassens, Efren Sandoval, Francisco Tanudjaja, Magnus Isaksson, William Lee, David Becker, Marc Laurent, James Lu + SEARCH |
| EPI_ISL_1816845, EPI_ISL_1816846 | EXCITE Lab | Andersen lab at Scripps Research | David Pride, Sharon Reed, Chip Schooley, Angela Scioscia, Natasha Martin Cheryl Anderson, Sawyer Farmer, Abigail Schnapper, Helena Tubb, Tommy Valles + SEARCH |
| EPI_ISL_1816847, EPI_ISL_1816848, EPI_ISL_1816849 | EXCITE Lab | Andersen lab at Scripps Research | Nicole L Washington, Simon White, Geraint Levan, Kelly Schiabor Barrett, Elizabeth Cirulli, Alexandre Bolze, Charlotte Rivera-Garcia, Ryan Cho, Jason Nguyen, Sherry Wang, Jimmy Ramirez, Celena Andrade, Alice Summerfield, Tyler Cassens, Efren Sandoval, Francisco Tanudjaja, Magnus Isaksson, William Lee, David Becker, Marc Laurent, James Lu + SEARCH |
| EPI_ISL_1816850 | EXCITE Lab | Andersen lab at Scripps Research | David Pride, Sharon Reed, Chip Schooley, Angela Scioscia, Natasha Martin Cheryl Anderson, Sawyer Farmer, Abigail Schnapper, Helena Tubb, Tommy Valles + SEARCH |
| EPI_ISL_1816851 | EXCITE Lab | Andersen lab at Scripps Research | Nicole L Washington, Simon White, Geraint Levan, Kelly Schiabor Barrett, Elizabeth Cirulli, Alexandre Bolze, Charlotte Rivera-Garcia, Ryan Cho, Jason Nguyen, Sherry Wang, Jimmy Ramirez, Celena Andrade, Alice Summerfield, Tyler Cassens, Efren Sandoval, Francisco Tanudjaja, Magnus Isaksson, William Lee, David Becker, Marc Laurent, James Lu + SEARCH |
| EPI_ISL_1816852, EPI_ISL_1816853, EPI_ISL_1816854, EPI_ISL_1816855, EPI_ISL_1816856 | EXCITE Lab | Andersen lab at Scripps Research | David Pride, Sharon Reed, Chip Schooley, Angela Scioscia, Natasha Martin Cheryl Anderson, Sawyer Farmer, Abigail Schnapper, Helena Tubb, Tommy Valles + SEARCH |
| EPI_ISL_1816857 | EXCITE Lab | Andersen lab at Scripps Research | Nicole L Washington, Simon White, Geraint Levan, Kelly Schiabor Barrett, Elizabeth Cirulli, Alexandre Bolze, Charlotte Rivera-Garcia, Ryan Cho, Jason Nguyen, Sherry Wang, Jimmy Ramirez, Celena Andrade, Alice Summerfield, Tyler Cassens, Efren Sandoval, Francisco Tanudjaja, Magnus Isaksson, William Lee, David Becker, Marc Laurent, James Lu + SEARCH |
| EPI_ISL_1816858, EPI_ISL_1816859, EPI_ISL_1816860 | EXCITE Lab | Andersen lab at Scripps Research | David Pride, Sharon Reed, Chip Schooley, Angela Scioscia, Natasha Martin Cheryl Anderson, Sawyer Farmer, Abigail Schnapper, Helena Tubb, Tommy Valles + SEARCH |
| EPI_ISL_1816861, EPI_ISL_1816862 | EXCITE Lab | Andersen lab at Scripps Research | Nicole L Washington, Simon White, Geraint Levan, Kelly Schiabor Barrett, Elizabeth Cirulli, Alexandre Bolze, Charlotte Rivera-Garcia, Ryan Cho, Jason Nguyen, Sherry Wang, Jimmy Ramirez, Celena Andrade, Alice Summerfield, Tyler Cassens, Efren Sandoval, Francisco Tanudjaja, Magnus Isaksson, William Lee, David Becker, Marc Laurent, James Lu + SEARCH |
| EPI_ISL_1816863 | EXCITE Lab | Andersen lab at Scripps Research | Sydney Morgan + Marni Jacobs + Celestine Magallanes + Holly Valentine + Louise C. Laurent + SEARCH |
| EPI_ISL_1816864 | EXCITE Lab | Andersen lab at Scripps Research | Nicole L Washington, Simon White, Geraint Levan, Kelly Schiabor Barrett, Elizabeth Cirulli, Alexandre Bolze, Charlotte Rivera-Garcia, Ryan Cho, Jason Nguyen, Sherry Wang, Jimmy Ramirez, Celena Andrade, Alice Summerfield, Tyler Cassens, Efren Sandoval, Francisco Tanudjaja, Magnus Isaksson, William Lee, David Becker, Marc Laurent, James Lu + SEARCH |
| EPI_ISL_1816865 | EXCITE Lab | Andersen lab at Scripps Research | David Pride, Sharon Reed, Chip Schooley, Angela Scioscia, Natasha Martin Cheryl Anderson, Sawyer Farmer, Abigail Schnapper, Helena Tubb, Tommy Valles + SEARCH |
| EPI_ISL_1816866, EPI_ISL_1816867 | EXCITE Lab | Andersen lab at Scripps Research | Nicole L Washington, Simon White, Geraint Levan, Kelly Schiabor Barrett, Elizabeth Cirulli, Alexandre Bolze, Charlotte Rivera-Garcia, Ryan Cho, Jason Nguyen, Sherry Wang, Jimmy Ramirez, Celena Andrade, Alice Summerfield, Tyler Cassens, Efren Sandoval, Francisco Tanudjaja, Magnus Isaksson, William Lee, David Becker, Marc Laurent, James Lu + SEARCH |
| EPI_ISL_1816868 | EXCITE Lab | Andersen lab at Scripps Research | Sydney Morgan + Marni Jacobs + Celestine Magallanes + Holly Valentine + Louise C. Laurent + SEARCH |
| EPI_ISL_1816869 | EXCITE Lab | Andersen lab at Scripps Research | David Pride, Sharon Reed, Chip Schooley, Angela Scioscia, Natasha Martin Cheryl Anderson, Sawyer Farmer, Abigail Schnapper, Helena Tubb, Tommy Valles + SEARCH |
| EPI_ISL_1816870 | EXCITE Lab | Andersen lab at Scripps Research | Sydney Morgan + Marni Jacobs + Celestine Magallanes + Holly Valentine + Louise C. Laurent + SEARCH |
| EPI_ISL_1816871, EPI_ISL_1816872, EPI_ISL_1816873, EPI_ISL_1816874 | EXCITE Lab | Andersen lab at Scripps Research | David Pride, Sharon Reed, Chip Schooley, Angela Scioscia, Natasha Martin Cheryl Anderson, Sawyer Farmer, Abigail Schnapper, Helena Tubb, Tommy Valles + SEARCH |
| EPI_ISL_1816875 | EXCITE Lab | Andersen lab at Scripps Research | Nicole L Washington, Simon White, Geraint Levan, Kelly Schiabor Barrett, Elizabeth Cirulli, Alexandre Bolze, Charlotte Rivera-Garcia, Ryan Cho, Jason Nguyen, Sherry Wang, Jimmy Ramirez, Celena Andrade, Alice Summerfield, Tyler Cassens, Efren Sandoval, Francisco Tanudjaja, Magnus Isaksson, William Lee, David Becker, Marc Laurent, James Lu + SEARCH |
| EPI_ISL_1816876 | EXCITE Lab | Andersen lab at Scripps Research | David Pride, Sharon Reed, Chip Schooley, Angela Scioscia, Natasha Martin Cheryl Anderson, Sawyer Farmer, Abigail Schnapper, Helena Tubb, Tommy Valles + SEARCH |
| EPI_ISL_1816877 | EXCITE Lab | Andersen lab at Scripps Research | Sydney Morgan + Marni Jacobs + Celestine Magallanes + Holly Valentine + Louise C. Laurent + SEARCH |
| EPI_ISL_1816878 | EXCITE Lab | Andersen lab at Scripps Research | David Pride, Sharon Reed, Chip Schooley, Angela Scioscia, Natasha Martin Cheryl Anderson, Sawyer Farmer, Abigail Schnapper, Helena Tubb, Tommy Valles + SEARCH |
| EPI_ISL_1816879 | EXCITE Lab | Andersen lab at Scripps Research | Rebecca Fielding-Miller, Laura Kohn, Patricia Gonzalez-Zuniga, Fitri Carrissa Wijaya, Vinton Omaleki, Marlene Flores, Araz Majnoonian, Megan Nguyen, Ashkan Hassani, Smruthi Karthikeyan, Rodolfo Salido, Victor Cantu, Greg Humphrey, Sawyer Farmer, Abigail Schnapper, Helena Tubb, Tommy Valles + SEARCH |
| EPI_ISL_1816880 | EXCITE Lab | Andersen lab at Scripps Research | David Pride, Sharon Reed, Chip Schooley, Angela Scioscia, Natasha Martin Cheryl Anderson, Sawyer Farmer, Abigail Schnapper, Helena Tubb, Tommy Valles + SEARCH |
| EPI_ISL_1816881 | EXCITE Lab | Andersen lab at Scripps Research | Nicole L Washington, Simon White, Geraint Levan, Kelly Schiabor Barrett, Elizabeth Cirulli, Alexandre Bolze, Charlotte Rivera-Garcia, Ryan Cho, Jason Nguyen, Sherry Wang, Jimmy Ramirez, Celena Andrade, Alice Summerfield, Tyler Cassens, Efren Sandoval, Francisco Tanudjaja, Magnus Isaksson, William Lee, David Becker, Marc Laurent, James Lu + SEARCH |
| EPI_ISL_1816882, EPI_ISL_1816883 | EXCITE Lab | Andersen lab at Scripps Research | David Pride, Sharon Reed, Chip Schooley, Angela Scioscia, Natasha Martin Cheryl Anderson, Sawyer Farmer, Abigail Schnapper, Helena Tubb, Tommy Valles + SEARCH |
| EPI_ISL_1816884 | EXCITE Lab | Andersen lab at Scripps Research | Nicole L Washington, Simon White, Geraint Levan, Kelly Schiabor Barrett, Elizabeth Cirulli, Alexandre Bolze, Charlotte Rivera-Garcia, Ryan Cho, Jason Nguyen, Sherry Wang, Jimmy Ramirez, Celena Andrade, Alice Summerfield, Tyler Cassens, Efren Sandoval, Francisco Tanudjaja, Magnus Isaksson, William Lee, David Becker, Marc Laurent, James Lu + SEARCH |
| EPI_ISL_1816885, EPI_ISL_1816886 | EXCITE Lab | Andersen lab at Scripps Research | David Pride, Sharon Reed, Chip Schooley, Angela Scioscia, Natasha Martin Cheryl Anderson, Sawyer Farmer, Abigail Schnapper, Helena Tubb, Tommy Valles + SEARCH |
| EPI_ISL_1816887 | EXCITE Lab | Andersen lab at Scripps Research | Nicole L Washington, Simon White, Geraint Levan, Kelly Schiabor Barrett, Elizabeth Cirulli, Alexandre Bolze, Charlotte Rivera-Garcia, Ryan Cho, Jason Nguyen, Sherry Wang, Jimmy Ramirez, Celena Andrade, Alice Summerfield, Tyler Cassens, Efren Sandoval, Francisco Tanudjaja, Magnus Isaksson, William Lee, David Becker, Marc Laurent, James Lu + SEARCH |
| EPI_ISL_1816888 | EXCITE Lab | Andersen lab at Scripps Research | Sydney Morgan + Marni Jacobs + Celestine Magallanes + Holly Valentine + Louise C. Laurent + SEARCH |
| EPI_ISL_1816889 | EXCITE Lab | Andersen lab at Scripps Research | David Pride, Sharon Reed, Chip Schooley, Angela Scioscia, Natasha Martin Cheryl Anderson, Sawyer Farmer, Abigail Schnapper, Helena Tubb, Tommy Valles + SEARCH |
| EPI_ISL_1816890, EPI_ISL_1816891, EPI_ISL_1816892, EPI_ISL_1816893 | EXCITE Lab | Andersen lab at Scripps Research | Nicole L Washington, Simon White, Geraint Levan, Kelly Schiabor Barrett, Elizabeth Cirulli, Alexandre Bolze, Charlotte Rivera-Garcia, Ryan Cho, Jason Nguyen, Sherry Wang, Jimmy Ramirez, Celena Andrade, Alice Summerfield, Tyler Cassens, Efren Sandoval, Francisco Tanudjaja, Magnus Isaksson, William Lee, David Becker, Marc Laurent, James Lu + SEARCH |
| EPI_ISL_1816894 | EXCITE Lab | Andersen lab at Scripps Research | Fred Wu, Christine Mitchell + SEARCH |
| EPI_ISL_1816895 | EXCITE Lab | Andersen lab at Scripps Research | David Pride, Sharon Reed, Chip Schooley, Angela Scioscia, Natasha Martin Cheryl Anderson, Sawyer Farmer, Abigail Schnapper, Helena Tubb, Tommy Valles + SEARCH |
| EPI_ISL_1816896 | EXCITE Lab | Andersen lab at Scripps Research | Sydney Morgan + Marni Jacobs + Celestine Magallanes + Holly Valentine + Louise C. Laurent + SEARCH |
| EPI_ISL_1816897 | EXCITE Lab | Andersen lab at Scripps Research | David Pride, Sharon Reed, Chip Schooley, Angela Scioscia, Natasha Martin Cheryl Anderson, Sawyer Farmer, Abigail Schnapper, Helena Tubb, Tommy Valles + SEARCH |
| EPI_ISL_1816898 | EXCITE Lab | Andersen lab at Scripps Research | Sydney Morgan + Marni Jacobs + Celestine Magallanes + Holly Valentine + Louise C. Laurent + SEARCH |
| EPI_ISL_1816899, | EXCITE Lab | Andersen lab at Scripps Research | David Pride, Sharon Reed, Chip Schooley, Angela Scioscia, Natasha Martin Cheryl Anderson, Sawyer Farmer, Abigail Schnapper, Helena Tubb, Tommy Valles + SEARCH |

| | | | |
|---|---|---|---|
| EPI_ISL_1816900, EPI_ISL_1816901 | | | |
| EPI_ISL_1816902 | EXCITE Lab | Andersen lab at Scripps Research | Rebecca Fielding-Miller, Laura Kohn, Patricia Gonzalez-Zuniga, Fitri Carrissa Wijaya, Vinton Omaleki, Marlene Flores, Araz Majnoonian, Megan Nguyen, Ashkan Hassani, Smruthi Karthikeyan, Rodolfo Salido, Victor Cantu, Greg Humphrey, Sawyer Farmer, Abigail Schnapper, Helena Tubb, Tommy Valles + SEARCH |
| EPI_ISL_1816903, EPI_ISL_1816904, EPI_ISL_1816905 | EXCITE Lab | Andersen lab at Scripps Research | Nicole L Washington, Simon White, Geraint Levan, Kelly Schiabor Barrett, Elizabeth Cirulli, Alexandre Bolze, Charlotte Rivera-Garcia, Ryan Cho, Jason Nguyen, Sherry Wang, Jimmy Ramirez, Celena Andrade, Alice Summerfield, Tyler Cassens, Efren Sandoval, Francisco Tanudjaja, Magnus Isaksson, William Lee, David Becker, Marc Laurent, James Lu + SEARCH |
| EPI_ISL_1816906 | EXCITE Lab | Andersen lab at Scripps Research | Sydney Morgan + Marni Jacobs + Celestine Magallanes + Holly Valentine + Louise C. Laurent + SEARCH |
| EPI_ISL_1816907 | EXCITE Lab | Andersen lab at Scripps Research | Nicole L Washington, Simon White, Geraint Levan, Kelly Schiabor Barrett, Elizabeth Cirulli, Alexandre Bolze, Charlotte Rivera-Garcia, Ryan Cho, Jason Nguyen, Sherry Wang, Jimmy Ramirez, Celena Andrade, Alice Summerfield, Tyler Cassens, Efren Sandoval, Francisco Tanudjaja, Magnus Isaksson, William Lee, David Becker, Marc Laurent, James Lu + SEARCH |
| EPI_ISL_1816908 | EXCITE Lab | Andersen lab at Scripps Research | David Pride, Sharon Reed, Chip Schooley, Angela Scioscia, Natasha Martin Cheryl Anderson, Sawyer Farmer, Abigail Schnapper, Helena Tubb, Tommy Valles + SEARCH |
| EPI_ISL_1816909 | EXCITE Lab | Andersen lab at Scripps Research | Nicole L Washington, Simon White, Geraint Levan, Kelly Schiabor Barrett, Elizabeth Cirulli, Alexandre Bolze, Charlotte Rivera-Garcia, Ryan Cho, Jason Nguyen, Sherry Wang, Jimmy Ramirez, Celena Andrade, Alice Summerfield, Tyler Cassens, Efren Sandoval, Francisco Tanudjaja, Magnus Isaksson, William Lee, David Becker, Marc Laurent, James Lu + SEARCH |
| EPI_ISL_1816910 | EXCITE Lab | Andersen lab at Scripps Research | David Pride, Sharon Reed, Chip Schooley, Angela Scioscia, Natasha Martin Cheryl Anderson, Sawyer Farmer, Abigail Schnapper, Helena Tubb, Tommy Valles + SEARCH |
| EPI_ISL_1816911 | EXCITE Lab | Andersen lab at Scripps Research | Nicole L Washington, Simon White, Geraint Levan, Kelly Schiabor Barrett, Elizabeth Cirulli, Alexandre Bolze, Charlotte Rivera-Garcia, Ryan Cho, Jason Nguyen, Sherry Wang, Jimmy Ramirez, Celena Andrade, Alice Summerfield, Tyler Cassens, Efren Sandoval, Francisco Tanudjaja, Magnus Isaksson, William Lee, David Becker, Marc Laurent, James Lu + SEARCH |
| EPI_ISL_1816912 | EXCITE Lab | Andersen lab at Scripps Research | David Pride, Sharon Reed, Chip Schooley, Angela Scioscia, Natasha Martin Cheryl Anderson, Sawyer Farmer, Abigail Schnapper, Helena Tubb, Tommy Valles + SEARCH |
| EPI_ISL_1816913 | EXCITE Lab | Andersen lab at Scripps Research | Nicole L Washington, Simon White, Geraint Levan, Kelly Schiabor Barrett, Elizabeth Cirulli, Alexandre Bolze, Charlotte Rivera-Garcia, Ryan Cho, Jason Nguyen, Sherry Wang, Jimmy Ramirez, Celena Andrade, Alice Summerfield, Tyler Cassens, Efren Sandoval, Francisco Tanudjaja, Magnus Isaksson, William Lee, David Becker, Marc Laurent, James Lu + SEARCH |
| EPI_ISL_1816914 | EXCITE Lab | Andersen lab at Scripps Research | David Pride, Sharon Reed, Chip Schooley, Angela Scioscia, Natasha Martin Cheryl Anderson, Sawyer Farmer, Abigail Schnapper, Helena Tubb, Tommy Valles + SEARCH |
| EPI_ISL_1816915 | EXCITE Lab | Andersen lab at Scripps Research | Nicole L Washington, Simon White, Geraint Levan, Kelly Schiabor Barrett, Elizabeth Cirulli, Alexandre Bolze, Charlotte Rivera-Garcia, Ryan Cho, Jason Nguyen, Sherry Wang, Jimmy Ramirez, Celena Andrade, Alice Summerfield, Tyler Cassens, Efren Sandoval, Francisco Tanudjaja, Magnus Isaksson, William Lee, David Becker, Marc Laurent, James Lu + SEARCH |
| EPI_ISL_1816916 | The Ohio State University Applied Microbiology Services Laboratory | The Ohio State University Applied Microbiology Services Laboratory | Seth A. Faith PhD |
| EPI_ISL_1817555, EPI_ISL_1817865, EPI_ISL_1817928 | MAWD Pathology Group, PA | MAWD Pathology Group, PA | Pradip Manna, Ph D, Kelli Fayard, Shelby Herner, London Collins, Kelsi Eastman |
| EPI_ISL_1817989, EPI_ISL_1817990, EPI_ISL_1817991, EPI_ISL_1817992, EPI_ISL_1817993, EPI_ISL_1817994, EPI_ISL_1817995, EPI_ISL_1817997, EPI_ISL_1817998 | Infinity Biologix | Centers for Disease Control and Prevention Division of Viral Diseases, Pathogen Discovery | Dakota Howard, Dhwani Batra, Peter W. Cook, Kara Moser, Adrian Paskey, Jason Caravas, Benjamin Rambo-Martin, Shatavia Morrison, Christopher Gulvick, Scott Sammons, Yvette Unoarumhi, Darlene Wagner, Matthew Schmerer, Christian Bixby, Yihe Wang, Jonathan Schultz, Chirayu Goswami, Russ Hager, Robin Grimwood, Clinton R. Paden, Duncan MacCannell |
| EPI_ISL_1817999, EPI_ISL_1818000, EPI_ISL_1818001, EPI_ISL_1818002, EPI_ISL_1818003, EPI_ISL_1818004, EPI_ISL_1818005, EPI_ISL_1818006, EPI_ISL_1818007, EPI_ISL_1818008, EPI_ISL_1818010, EPI_ISL_1818011, EPI_ISL_1818012, EPI_ISL_1818013, EPI_ISL_1818014, EPI_ISL_1818015, EPI_ISL_1818016, EPI_ISL_1818017, EPI_ISL_1818018, EPI_ISL_1818019, EPI_ISL_1818020, EPI_ISL_1818021, EPI_ISL_1818022, EPI_ISL_1818023, EPI_ISL_1818024, EPI_ISL_1818025, EPI_ISL_1818026, EPI_ISL_1818027, EPI_ISL_1818028, EPI_ISL_1818029, EPI_ISL_1818030, EPI_ISL_1818031, EPI_ISL_1818032, EPI_ISL_1818033, EPI_ISL_1818034, EPI_ISL_1818035, EPI_ISL_1818036, EPI_ISL_1818037, EPI_ISL_1818038, EPI_ISL_1818039, EPI_ISL_1818040, EPI_ISL_1818041, EPI_ISL_1818042, EPI_ISL_1818043, EPI_ISL_1818044, EPI_ISL_1818045, EPI_ISL_1818046, EPI_ISL_1818047, EPI_ISL_1818048, EPI_ISL_1818049, EPI_ISL_1818050 | | | |
| see above | Helix/Illumina | Centers for Disease Control and Prevention Division of Viral Diseases, Pathogen Discovery | Dakota Howard, Dhwani Batra, Peter W. Cook, Kara Moser, Adrian Paskey, Jason Caravas, Benjamin Rambo-Martin, Shatavia Morrison, Christopher Gulvick, Scott Sammons, Yvette Unoarumhi, Darlene Wagner, Matthew Schmerer, Eileen de Feo, Jan Antico, Christine Tran, Matthew Tolentino, Shannon Wickline, Kim Gietzen, Brad Sickler, Jingtao Liu, Eric Allen, Phil Febbo, Nicole L. Washington, Simon White, Geraint Levan, Kelly Schiabor Barrett, Elizabeth Cirulli, Alexandre Bolze, Ary Ascencio, Charlotte Rivera-Garcia, Ryan Cho, Jason Nguyen, Sherry Wang, Jimmy Ramirez, Tyler Cassens, Efren Sandoval, Magnus Isaksson, William Lee, David Becker, Marc Laurent, James Lu, Clinton R. Paden, Duncan MacCannell |
| EPI_ISL_1818051, EPI_ISL_1818052, EPI_ISL_1818053, EPI_ISL_1818054, EPI_ISL_1818055, EPI_ISL_1818056, EPI_ISL_1818057, EPI_ISL_1818058, EPI_ISL_1818059, EPI_ISL_1818060, EPI_ISL_1818061, EPI_ISL_1818062, EPI_ISL_1818063, EPI_ISL_1818064, EPI_ISL_1818065, EPI_ISL_1818066, EPI_ISL_1818067, EPI_ISL_1818068, EPI_ISL_1818069, EPI_ISL_1818070, EPI_ISL_1818071 | | | |
| see above | Quest Diagnostics Incorporated | Centers for Disease Control and Prevention Division of Viral Diseases, Pathogen Discovery | Dakota Howard, Dhwani Batra, Peter W. Cook, Kara Moser, Adrian Paskey, Jason Caravas, Benjamin Rambo-Martin, Shatavia Morrison, Christopher Gulvick, Scott Sammons, Yvette Unoarumhi, Darlene Wagner, Matthew Schmerer, S. H. Rosenthal, A. Gerasimova, R. M. Kagan, B. Anderson, M. Hua, Y. Liu, L.E. Bernstein, K.E. Livingston, A. Perez, I. A. Shlyakhter, R. V. Rolando, R. Owen, P. Tanpaiboon, F. Lacbawan, Clinton R. Paden, Duncan MacCannell |
| EPI_ISL_1818072 | UT-Unified State Labs: Public Health Utah DOH | Centers for Disease Control and Prevention Division of Viral Diseases, Pathogen Discovery | Mili Sheth, Sarah Nobles, Jasmine Padilla, Mark Burroughs, Shoshona Le, Katie Dillon, Peter Cook, Clinton R. Paden, Dhwani Batra, Krista Queen, Kristen Knipe, Dakota Howard, Yvette Unoarumhi, Darlene Wagner, Matthew Schmerer, Ben L. Rambo-Martin, Kristine Lacek, Sam Shepard, Alison Laufer Halpin, Dave Wentworth, Vivien Dugan, Suxiang Tong, Justin Lee |
| EPI_ISL_1818073 | Guam Public Health Laboratory | Centers for Disease Control and Prevention Division of Viral Diseases, Pathogen Discovery | Mili Sheth, Sarah Nobles, Jasmine Padilla, Mark Burroughs, Shoshona Le, Katie Dillon, Peter Cook, Clinton R. Paden, Dhwani Batra, Krista Queen, Kristen Knipe, Dakota Howard, Yvette Unoarumhi, Darlene Wagner, Matthew Schmerer, Ben L. Rambo-Martin, Kristine Lacek, Sam Shepard, Alison Laufer Halpin, Dave Wentworth, Vivien Dugan, Suxiang Tong, Justin Lee |
| EPI_ISL_1818074, EPI_ISL_1818075, EPI_ISL_1818077, EPI_ISL_1818078 | Laboratory Corporation of America | Centers for Disease Control and Prevention Division of Viral Diseases, Pathogen Discovery | Dakota Howard, Dhwani Batra, Peter W. Cook, Kara Moser, Adrian Paskey, Jason Caravas, Benjamin Rambo-Martin, Shatavia Morrison, Christopher Gulvick, Scott Sammons, Yvette Unoarumhi, Darlene Wagner, Matthew Schmerer, Minoo Agarwal, Eyad Almasri, Debbie Boles, Ayla Burns, Nuthawin Charoensri, Oren Cohen, Susan Countryman, Mary Ann Cristobal, Bobbi Croy, Suzanne Dale, Hrushikesh Deshmukh, Amanda Douglas, Vincent Drouillon, Marcia Eisenberg, Howard Engler, Rama Ghatti, Prashant Gupta, Susan Hicks, Jake Humphrey, Lax Iyer, Manoj Jain, Mohan Kolli, Brian Krueger, Tim Kuphal, Stanley Letovsky, Michael Levandoski, Craig Lukasik, Jonathan Meltzer, Brian Norvell, Mindy Nye, Scott Parker, Christos Petropoulos, John Pruitt, Steven Ragan, Scott Ryan, Mike Sapeta, Jana Schroth, Suresh Babu Selvaraju, Goran Stevovic, Amanda Suchanek, Andrea Throop, Lyndon Tilson, Thomas Urban, Joe Voshell, Kimberly Wagner, Jonathan Williams, Mary Williamson, Qian Zeng, Tricia Zwiefelhofer, Clinton R. Paden, Duncan MacCannell |
| EPI_ISL_1818079, EPI_ISL_1818080, EPI_ISL_1818081, EPI_ISL_1818082, EPI_ISL_1818083, EPI_ISL_1818084, EPI_ISL_1818085, EPI_ISL_1818086, EPI_ISL_1818087, EPI_ISL_1818088, EPI_ISL_1818089, EPI_ISL_1818090, EPI_ISL_1818091, EPI_ISL_1818092, EPI_ISL_1818093, EPI_ISL_1818094, EPI_ISL_1818096, EPI_ISL_1818097, EPI_ISL_1818098, EPI_ISL_1818099, EPI_ISL_1818100, EPI_ISL_1818101, EPI_ISL_1818103, EPI_ISL_1818104, EPI_ISL_1818105, EPI_ISL_1818106, EPI_ISL_1818107, EPI_ISL_1818108, EPI_ISL_1818109, EPI_ISL_1818110, EPI_ISL_1818111, EPI_ISL_1818112, EPI_ISL_1818113, EPI_ISL_1818114, EPI_ISL_1818115, EPI_ISL_1818116, EPI_ISL_1818117, EPI_ISL_1818118, EPI_ISL_1818119, EPI_ISL_1818120, EPI_ISL_1818121, EPI_ISL_1818122, EPI_ISL_1818123, EPI_ISL_1818124, EPI_ISL_1818125, EPI_ISL_1818126, EPI_ISL_1818127, EPI_ISL_1818129, EPI_ISL_1818130, EPI_ISL_1818131, EPI_ISL_1818132, EPI_ISL_1818133, EPI_ISL_1818134, EPI_ISL_1818135, EPI_ISL_1818136, EPI_ISL_1818137, EPI_ISL_1818138, EPI_ISL_1818139 | | | |
| see above | Infinity Biologix | Centers for Disease Control and Prevention Division of Viral Diseases, Pathogen Discovery | Dakota Howard, Dhwani Batra, Peter W. Cook, Kara Moser, Adrian Paskey, Jason Caravas, Benjamin Rambo-Martin, Shatavia Morrison, Christopher Gulvick, Scott Sammons, Yvette Unoarumhi, Darlene Wagner, Matthew Schmerer, Christian Bixby, Yihe Wang, Jonathan Schultz, Chirayu Goswami, Russ Hager, Robin Grimwood, Clinton R. Paden, Duncan MacCannell |
| EPI_ISL_1818140, EPI_ISL_1818141, EPI_ISL_1818142, EPI_ISL_1818143, EPI_ISL_1818144, EPI_ISL_1818145, EPI_ISL_1818146, EPI_ISL_1818147, EPI_ISL_1818148, EPI_ISL_1818149, EPI_ISL_1818150, EPI_ISL_1818151, EPI_ISL_1818152, EPI_ISL_1818153, EPI_ISL_1818154, EPI_ISL_1818155, EPI_ISL_1818156, EPI_ISL_1818157, EPI_ISL_1818158, EPI_ISL_1818159, EPI_ISL_1818160, EPI_ISL_1818161, EPI_ISL_1818162, EPI_ISL_1818163, EPI_ISL_1818164, EPI_ISL_1818165, EPI_ISL_1818166, EPI_ISL_1818167, EPI_ISL_1818168, EPI_ISL_1818169, EPI_ISL_1818170, EPI_ISL_1818171, EPI_ISL_1818172, EPI_ISL_1818173, EPI_ISL_1818174, EPI_ISL_1818175, EPI_ISL_1818176, EPI_ISL_1818177, EPI_ISL_1818178, EPI_ISL_1818179, EPI_ISL_1818180, EPI_ISL_1818181, EPI_ISL_1818182, EPI_ISL_1818183, EPI_ISL_1818184, EPI_ISL_1818185, EPI_ISL_1818186, EPI_ISL_1818187, EPI_ISL_1818188, EPI_ISL_1818189, EPI_ISL_1818190, EPI_ISL_1818191, EPI_ISL_1818192, EPI_ISL_1818193, EPI_ISL_1818194, EPI_ISL_1818195, EPI_ISL_1818196, EPI_ISL_1818197, EPI_ISL_1818198, EPI_ISL_1818199, EPI_ISL_1818200, EPI_ISL_1818201, EPI_ISL_1818202, EPI_ISL_1818203, EPI_ISL_1818204, EPI_ISL_1818205, EPI_ISL_1818206, EPI_ISL_1818207, EPI_ISL_1818208, EPI_ISL_1818209, EPI_ISL_1818210, EPI_ISL_1818211, EPI_ISL_1818212, EPI_ISL_1818213, EPI_ISL_1818214, EPI_ISL_1818215, EPI_ISL_1818216, EPI_ISL_1818217, EPI_ISL_1818218, EPI_ISL_1818219, EPI_ISL_1818220, EPI_ISL_1818221, EPI_ISL_1818222, EPI_ISL_1818223, EPI_ISL_1818224, EPI_ISL_1818225, EPI_ISL_1818226, EPI_ISL_1818227, EPI_ISL_1818228, EPI_ISL_1818229, EPI_ISL_1818230, EPI_ISL_1818231, EPI_ISL_1818232, EPI_ISL_1818233, EPI_ISL_1818234, EPI_ISL_1818235, EPI_ISL_1818236, EPI_ISL_1818237, EPI_ISL_1818238, EPI_ISL_1818239, EPI_ISL_1818240, EPI_ISL_1818241, EPI_ISL_1818242, EPI_ISL_1818243, EPI_ISL_1818244, EPI_ISL_1818245, EPI_ISL_1818246, EPI_ISL_1818247, EPI_ISL_1818248, EPI_ISL_1818249, EPI_ISL_1818250, EPI_ISL_1818251, EPI_ISL_1818252, EPI_ISL_1818253, EPI_ISL_1818254, EPI_ISL_1818255, EPI_ISL_1818256, EPI_ISL_1818257, EPI_ISL_1818258, EPI_ISL_1818259, EPI_ISL_1818260, EPI_ISL_1818261, EPI_ISL_1818262, EPI_ISL_1818263, EPI_ISL_1818264, EPI_ISL_1818265, EPI_ISL_1818266, EPI_ISL_1818267, EPI_ISL_1818268, EPI_ISL_1818269, EPI_ISL_1818270, EPI_ISL_1818271, EPI_ISL_1818272, EPI_ISL_1818273, EPI_ISL_1818274, EPI_ISL_1818275, EPI_ISL_1818276, EPI_ISL_1818277, EPI_ISL_1818278, EPI_ISL_1818279, EPI_ISL_1818280, EPI_ISL_1818281, EPI_ISL_1818282, EPI_ISL_1818283, EPI_ISL_1818284, EPI_ISL_1818285, EPI_ISL_1818286, EPI_ISL_1818287, EPI_ISL_1818288, EPI_ISL_1818289, EPI_ISL_1818290, EPI_ISL_1818291, EPI_ISL_1818292, EPI_ISL_1818293, EPI_ISL_1818294, EPI_ISL_1818295, EPI_ISL_1818296, EPI_ISL_1818297, EPI_ISL_1818298, EPI_ISL_1818299, EPI_ISL_1818300, EPI_ISL_1818301, EPI_ISL_1818302, EPI_ISL_1818303, EPI_ISL_1818304, EPI_ISL_1818305, EPI_ISL_1818306, EPI_ISL_1818307, EPI_ISL_1818308, EPI_ISL_1818309, EPI_ISL_1818310, EPI_ISL_1818311, EPI_ISL_1818312, EPI_ISL_1818313, EPI_ISL_1818314, EPI_ISL_1818315, EPI_ISL_1818316, EPI_ISL_1818317, EPI_ISL_1818318, EPI_ISL_1818319, EPI_ISL_1818320, EPI_ISL_1818321, EPI_ISL_1818322, EPI_ISL_1818323, EPI_ISL_1818324, EPI_ISL_1818325, EPI_ISL_1818326, EPI_ISL_1818327, EPI_ISL_1818328, EPI_ISL_1818329, EPI_ISL_1818330, EPI_ISL_1818331, EPI_ISL_1818332, EPI_ISL_1818333, EPI_ISL_1818334, EPI_ISL_1818335, EPI_ISL_1818336, EPI_ISL_1818337, EPI_ISL_1818338, EPI_ISL_1818339, EPI_ISL_1818340, EPI_ISL_1818341, EPI_ISL_1818342, EPI_ISL_1818343, EPI_ISL_1818344, EPI_ISL_1818345, EPI_ISL_1818346, EPI_ISL_1818347, EPI_ISL_1818348, EPI_ISL_1818349, EPI_ISL_1818350, EPI_ISL_1818351, EPI_ISL_1818352, EPI_ISL_1818353, EPI_ISL_1818354, EPI_ISL_1818355, EPI_ISL_1818356, EPI_ISL_1818357, EPI_ISL_1818358, EPI_ISL_1818359, EPI_ISL_1818360, EPI_ISL_1818361, EPI_ISL_1818362, EPI_ISL_1818363, EPI_ISL_1818364, EPI_ISL_1818365, EPI_ISL_1818366, EPI_ISL_1818367, EPI_ISL_1818368, EPI_ISL_1818369, EPI_ISL_1818370, EPI_ISL_1818371, EPI_ISL_1818372, EPI_ISL_1818373, EPI_ISL_1818374, EPI_ISL_1818375, EPI_ISL_1818376, EPI_ISL_1818377, EPI_ISL_1818378, EPI_ISL_1818379, EPI_ISL_1818380, EPI_ISL_1818381, EPI_ISL_1818382, EPI_ISL_1818383, EPI_ISL_1818384, EPI_ISL_1818385, EPI_ISL_1818386, EPI_ISL_1818387, EPI_ISL_1818388, EPI_ISL_1818389, EPI_ISL_1818390, EPI_ISL_1818391, EPI_ISL_1818392, EPI_ISL_1818393, EPI_ISL_1818394, EPI_ISL_1818395, EPI_ISL_1818396, EPI_ISL_1818397, EPI_ISL_1818398, EPI_ISL_1818399, EPI_ISL_1818400, EPI_ISL_1818401, EPI_ISL_1818402, EPI_ISL_1818403, EPI_ISL_1818404, EPI_ISL_1818405, EPI_ISL_1818406, EPI_ISL_1818407, EPI_ISL_1818408, EPI_ISL_1818409, EPI_ISL_1818410, EPI_ISL_1818411, EPI_ISL_1818412, EPI_ISL_1818413, EPI_ISL_1818414, EPI_ISL_1818415, EPI_ISL_1818416, EPI_ISL_1818417, EPI_ISL_1818418, EPI_ISL_1818419, EPI_ISL_1818420, EPI_ISL_1818421, EPI_ISL_1818422, EPI_ISL_1818423, EPI_ISL_1818424, EPI_ISL_1818425, EPI_ISL_1818426, EPI_ISL_1818427, EPI_ISL_1818428, EPI_ISL_1818429, EPI_ISL_1818430, EPI_ISL_1818431, EPI_ISL_1818432, EPI_ISL_1818433, EPI_ISL_1818434, EPI_ISL_1818435, EPI_ISL_1818436, EPI_ISL_1818437, EPI_ISL_1818438, EPI_ISL_1818439, EPI_ISL_1818440, EPI_ISL_1818441, EPI_ISL_1818442, EPI_ISL_1818443, EPI_ISL_1818444, EPI_ISL_1818445, EPI_ISL_1818446, EPI_ISL_1818447, EPI_ISL_1818448, EPI_ISL_1818449, EPI_ISL_1818450, EPI_ISL_1818451, EPI_ISL_1818452, EPI_ISL_1818453, EPI_ISL_1818454, EPI_ISL_1818455, EPI_ISL_1818456, EPI_ISL_1818457, EPI_ISL_1818458, EPI_ISL_1818459, EPI_ISL_1818460, EPI_ISL_1818461, EPI_ISL_1818462, EPI_ISL_1818463, EPI_ISL_1818464, EPI_ISL_1818465, EPI_ISL_1818466, EPI_ISL_1818467, EPI_ISL_1818468, EPI_ISL_1818469, EPI_ISL_1818470, EPI_ISL_1818471, EPI_ISL_1818472, EPI_ISL_1818473, EPI_ISL_1818474, EPI_ISL_1818475, EPI_ISL_1818476, EPI_ISL_1818477, EPI_ISL_1818478, EPI_ISL_1818479, EPI_ISL_1818480, EPI_ISL_1818481, EPI_ISL_1818482, EPI_ISL_1818483, EPI_ISL_1818484, EPI_ISL_1818485, EPI_ISL_1818486, EPI_ISL_1818487, EPI_ISL_1818488, EPI_ISL_1818489, EPI_ISL_1818490, EPI_ISL_1818491, EPI_ISL_1818492, EPI_ISL_1818493, EPI_ISL_1818494, EPI_ISL_1818495, EPI_ISL_1818496, EPI_ISL_1818497, EPI_ISL_1818498, EPI_ISL_1818499, EPI_ISL_1818500, EPI_ISL_1818501, EPI_ISL_1818502, EPI_ISL_1818503, EPI_ISL_1818504, EPI_ISL_1818505, EPI_ISL_1818506, EPI_ISL_1818507, EPI_ISL_1818508, EPI_ISL_1818509, EPI_ISL_1818510, EPI_ISL_1818511, EPI_ISL_1818512, EPI_ISL_1818513, EPI_ISL_1818514, EPI_ISL_1818515, EPI_ISL_1818516, EPI_ISL_1818517, EPI_ISL_1818518, EPI_ISL_1818519, EPI_ISL_1818520, EPI_ISL_1818521, EPI_ISL_1818522, EPI_ISL_1818523, EPI_ISL_1818524, EPI_ISL_1818525, EPI_ISL_1818526, EPI_ISL_1818527, EPI_ISL_1818528, EPI_ISL_1818529, EPI_ISL_1818530, EPI_ISL_1818531, EPI_ISL_1818532, EPI_ISL_1818533, EPI_ISL_1818534, EPI_ISL_1818535, EPI_ISL_1818536, EPI_ISL_1818537, EPI_ISL_1818538, EPI_ISL_1818539, EPI_ISL_1818540, EPI_ISL_1818541, EPI_ISL_1818542, EPI_ISL_1818543, EPI_ISL_1818544, EPI_ISL_1818545, EPI_ISL_1818546, EPI_ISL_1818547, EPI_ISL_1818548, EPI_ISL_1818549, EPI_ISL_1818550, EPI_ISL_1818551, EPI_ISL_1818552, EPI_ISL_1818553, EPI_ISL_1818554, EPI_ISL_1818555, EPI_ISL_1818556, EPI_ISL_1818557, EPI_ISL_1818558, EPI_ISL_1818559, EPI_ISL_1818560, EPI_ISL_1818561, EPI_ISL_1818562, EPI_ISL_1818563, EPI_ISL_1818564, EPI_ISL_1818565, EPI_ISL_1818566, EPI_ISL_1818567, EPI_ISL_1818568, EPI_ISL_1818569, EPI_ISL_1818570, EPI_ISL_1818571, EPI_ISL_1818572, EPI_ISL_1818573 | | | |
| see above | Quest Diagnostics Incorporated | Centers for Disease Control and Prevention Division of Viral Diseases, Pathogen Discovery | Dakota Howard, Dhwani Batra, Peter W. Cook, Kara Moser, Adrian Paskey, Jason Caravas, Benjamin Rambo-Martin, Shatavia Morrison, Christopher Gulvick, Scott Sammons, Yvette Unoarumhi, Darlene Wagner, Matthew Schmerer, S. H. Rosenthal, A. Gerasimova, R. M. Kagan, B. Anderson, M. Hua, Y. Liu, L.E. Bernstein, K.E. Livingston, A. Perez, I. A. Shlyakhter, R. V. Rolando, R. Owen, P. Tanpaiboon, F. Lacbawan, Clinton R. Paden, Duncan MacCannell |
| EPI_ISL_1818574, EPI_ISL_1818575, EPI_ISL_1818576, EPI_ISL_1818577, EPI_ISL_1818578, EPI_ISL_1818579, EPI_ISL_1818580, EPI_ISL_1818581, EPI_ISL_1818582 | Infinity Biologix | Centers for Disease Control and Prevention Division of Viral Diseases, Pathogen Discovery | Dakota Howard, Dhwani Batra, Peter W. Cook, Kara Moser, Adrian Paskey, Jason Caravas, Benjamin Rambo-Martin, Shatavia Morrison, Christopher Gulvick, Scott Sammons, Yvette Unoarumhi, Darlene Wagner, Matthew Schmerer, Christian Bixby, Yihe Wang, Jonathan Schultz, Chirayu Goswami, Russ Hager, Robin Grimwood, Clinton R. Paden, Duncan MacCannell |
| EPI_ISL_1818654, EPI_ISL_1818655, EPI_ISL_1818656, EPI_ISL_1818657, EPI_ISL_1818658, EPI_ISL_1818659, EPI_ISL_1818660, EPI_ISL_1818661, EPI_ISL_1818662, EPI_ISL_1818663, EPI_ISL_1818664, EPI_ISL_1818665, EPI_ISL_1818666, EPI_ISL_1818667, EPI_ISL_1818668, EPI_ISL_1818669, EPI_ISL_1818672, EPI_ISL_1818673, EPI_ISL_1818674, EPI_ISL_1818768, EPI_ISL_1818769, EPI_ISL_1818770, EPI_ISL_1818771, EPI_ISL_1818773, EPI_ISL_1818775, EPI_ISL_1818776, EPI_ISL_1818777, EPI_ISL_1818778, EPI_ISL_1818779 | | | |
| see above | IN State Department of Health | IN State Department of Health Laboratory | Cassandra Campion, Jamie Yeadon, Brian Pope, Lixia Liu, Kyle Brownlee, Melissa Hindenlang, Mark Glazier |

| | | | |
|---|---|---|---|
| | Laboratory Services | Services | |
| EPI_ISL_1819078, EPI_ISL_1819079, EPI_ISL_1819080, EPI_ISL_1819081 | Moderna Inc. | Moderna Inc. | Yamuna Paila; Rolando Pajon |
| EPI_ISL_1819263, EPI_ISL_1819265, EPI_ISL_1819266, EPI_ISL_1819267, EPI_ISL_1819268, EPI_ISL_1819269, EPI_ISL_1819270, EPI_ISL_1819271, EPI_ISL_1819272, EPI_ISL_1819273, EPI_ISL_1819274, EPI_ISL_1819275, EPI_ISL_1819276, EPI_ISL_1819277, EPI_ISL_1819278, EPI_ISL_1819279, EPI_ISL_1819280 | | | |
| see above | DC Public Health Lab/ Dept. of Forensic Sciences | DC Public Health Lab/ Dept. of Forensic Sciences | Janis Doss, Scott Nguyen, Elizabeth Zelaya, Sarah Scott, Connie Maza, Monica Mann, Brittany Hamilton, David Payne, Jocelyn Hauser |
| EPI_ISL_1819282, EPI_ISL_1819283, EPI_ISL_1819284, EPI_ISL_1819285, EPI_ISL_1819286, EPI_ISL_1819287, EPI_ISL_1819288, EPI_ISL_1819289, EPI_ISL_1819290, EPI_ISL_1819291, EPI_ISL_1819292, EPI_ISL_1819293, EPI_ISL_1819295, EPI_ISL_1819296 | | | |
| see above | Wichita State University - Molecular Diagnostics Lab | Kansas Health and Environmental Lab | Mike Grose, Jonathan Barnell, Ben Olsen, and Phil Adam |
| EPI_ISL_1819297, EPI_ISL_1819298 | PA Department of Health, Bureau of Laboratories | Pathogen Discovery, Respiratory Viruses Branch, Division of Viral Diseases, Centers for Disease Control and Prevention | Ying Tao, Yan Li, Anna Kelleher, Jing Zhang, Anna Montmayeur, Brian Lynch, Krista Queen, Anna Uehara, Peter Cook, Han Jia Justin Ng, Rachel Marine, Clinton R. Paden, Haibin Wang, Mark Burroughs, Justin Lee, Adam Retchless, Suxiang Tong |
| EPI_ISL_1819299, EPI_ISL_1819300 | GA Department of Public Health Laboratory | Pathogen Discovery, Respiratory Viruses Branch, Division of Viral Diseases, Centers for Disease Control and Prevention | Ying Tao, Yan Li, Anna Kelleher, Jing Zhang, Anna Montmayeur, Brian Lynch, Krista Queen, Anna Uehara, Peter Cook, Han Jia Justin Ng, Rachel Marine, Clinton R. Paden, Haibin Wang, Mark Burroughs, Justin Lee, Adam Retchless, Suxiang Tong |
| EPI_ISL_1820910, EPI_ISL_1820911, EPI_ISL_1820915, EPI_ISL_1820916, EPI_ISL_1820917, EPI_ISL_1820919, EPI_ISL_1820920, EPI_ISL_1820921, EPI_ISL_1820922, EPI_ISL_1820923, EPI_ISL_1820924 | | | |
| see above | Wyoming Public Health Laboratory | Wyoming Public Health Laboratory | Jim Mildenberger, Taylor Fearing, Channing Weber, Ashley Norberg, Chayse Rowley, Marley Goetz, Brian Dominguez, Elliot Thomasson, Brittany Oher, Sam Britz, Cari Sloma, and Rob Christensen |
| EPI_ISL_1820969 | MAWD Pathology Group, PA | MAWD Pathology Group, PA | Pradip Manna, Ph D, Kelli Fayard, Shelby Herner, London Collins, Kelsi Eastman |
| EPI_ISL_1820970, EPI_ISL_1820971, EPI_ISL_1820972, EPI_ISL_1820973, EPI_ISL_1820974, EPI_ISL_1820975, EPI_ISL_1820976, EPI_ISL_1820977, EPI_ISL_1820978, EPI_ISL_1820979, EPI_ISL_1820980, EPI_ISL_1820981, EPI_ISL_1820982, EPI_ISL_1820983, EPI_ISL_1820984, EPI_ISL_1820985, EPI_ISL_1820986, EPI_ISL_1820987, EPI_ISL_1820988, EPI_ISL_1820989, EPI_ISL_1820990, EPI_ISL_1820991, EPI_ISL_1820992, EPI_ISL_1820993, EPI_ISL_1820994, EPI_ISL_1820995, EPI_ISL_1820996, EPI_ISL_1820997, EPI_ISL_1820998, EPI_ISL_1820999, EPI_ISL_1821000, EPI_ISL_1821001, EPI_ISL_1821002, EPI_ISL_1821003, EPI_ISL_1821004, EPI_ISL_1821005, EPI_ISL_1821006, EPI_ISL_1821007, EPI_ISL_1821008, EPI_ISL_1821009, EPI_ISL_1821010, EPI_ISL_1821011, EPI_ISL_1821012, EPI_ISL_1821013, EPI_ISL_1821014, EPI_ISL_1821015, EPI_ISL_1821016, EPI_ISL_1821017, EPI_ISL_1821018, EPI_ISL_1821019, EPI_ISL_1821020, EPI_ISL_1821021, EPI_ISL_1821022, EPI_ISL_1821023, EPI_ISL_1821024, EPI_ISL_1821030, EPI_ISL_1821031, EPI_ISL_1821032, EPI_ISL_1821033, EPI_ISL_1821034, EPI_ISL_1821035, EPI_ISL_1821036, EPI_ISL_1821037, EPI_ISL_1821038, EPI_ISL_1821039, EPI_ISL_1821040, EPI_ISL_1821041, EPI_ISL_1821042, EPI_ISL_1821043, EPI_ISL_1821044 | | | |
| see above | Minnesota Department of Health, Public Health Laboratory | Minnesota Department of Health, Public Health Laboratory | Alexandra Lorentz, Jacob Garfin, Matt Plumb, and Xiong Wang |
| EPI_ISL_1821045 | MAWD Pathology Group, PA | MAWD Pathology Group, PA | Pradip Manna, Ph D, Kelli Fayard, Shelby Herner, London Collins, Kelsi Eastman |
| EPI_ISL_1821046, EPI_ISL_1821047, EPI_ISL_1821048, EPI_ISL_1821049, EPI_ISL_1821050, EPI_ISL_1821051, EPI_ISL_1821052, EPI_ISL_1821053, EPI_ISL_1821054, EPI_ISL_1821055, EPI_ISL_1821056, EPI_ISL_1821057, EPI_ISL_1821058, EPI_ISL_1821060, EPI_ISL_1821061 | | | |
| see above | Maine HETL | Health and Environmental Testing Laboratory | Matluk,N., Dewey,H., Martha,J., Barter,M., Lynch,R., Munger,H. and Tewhey,R. |
| EPI_ISL_1821078, EPI_ISL_1821079, EPI_ISL_1821080, EPI_ISL_1821081, EPI_ISL_1821082 | MAWD Pathology Group, PA | MAWD Pathology Group, PA | Pradip Manna, Ph D, Kelli Fayard, Shelby Herner, London Collins, Kelsi Eastman |
| EPI_ISL_1821083, EPI_ISL_1821084, EPI_ISL_1821085, EPI_ISL_1821087, EPI_ISL_1821088, EPI_ISL_1821089, EPI_ISL_1821090, EPI_ISL_1821091, EPI_ISL_1821092, EPI_ISL_1821093, EPI_ISL_1821094, EPI_ISL_1821095, EPI_ISL_1821096, EPI_ISL_1821097 | | | |
| see above | Nebraska Public Health Laboratory | NPHL COVID-19 Response Team | NPHL COVID-19 Response Team |
| EPI_ISL_1821098, EPI_ISL_1821099 | MAWD Pathology Group, PA | MAWD Pathology Group, PA | Pradip Manna, Ph D, Kelli Fayard, Shelby Herner, London Collins, Kelsi Eastman |
| EPI_ISL_1821100 | TN Division of Laboratory Services | Pathogen Discovery, Respiratory Viruses Branch, Division of Viral Diseases, Centers for Disease Control and Prevention | Ying Tao, Yan Li, Anna Kelleher, Jing Zhang, Anna Montmayeur, Brian Lynch, Krista Queen, Anna Uehara, Peter Cook, Han Jia Justin Ng, Rachel Marine, Clinton R. Paden, Haibin Wang, Mark Burroughs, Justin Lee, Adam Retchless, Suxiang Tong |
| EPI_ISL_1821101 | MAWD Pathology Group, PA | MAWD Pathology Group, PA | Pradip Manna, Ph D, Kelli Fayard, Shelby Herner, London Collins, Kelsi Eastman |
| EPI_ISL_1821102 | LESP Queretaro | Instituto de Diagnostico y Referencia Epidemiologicos (INDRE) | Claudia Wong-Arambula, Abril Rodriguez-Maldonado, Vanessa Rivero-Arredondo, Ariadna Medina-Benitez, Joaquin Quiroz-Mercado, Sergio Rangel-Guerrero, Natividad Cruz-Ortiz, Tatiana Nunez-Garcia, Gisela Barrera-Badillo, Lucia Hernandez-Rivas, Irma Lopez-Martinez, Ernesto Ramirez-Gonzalez. |
| EPI_ISL_1821103 | LESP Chihuahua | Instituto de Diagnostico y Referencia Epidemiologicos (INDRE) | Claudia Wong-Arambula, Abril Rodriguez-Maldonado, Vanessa Rivero-Arredondo, Ariadna Medina-Benitez, Joaquin Quiroz-Mercado, Sergio Rangel-Guerrero, Natividad Cruz-Ortiz, Tatiana Nunez-Garcia, Gisela Barrera-Badillo, Lucia Hernandez-Rivas, Irma Lopez-Martinez, Ernesto Ramirez-Gonzalez. |
| EPI_ISL_1821104, EPI_ISL_1821105 | LESP Baja California Sur | Instituto de Diagnostico y Referencia Epidemiologicos (INDRE) | Claudia Wong-Arambula, Abril Rodriguez-Maldonado, Vanessa Rivero-Arredondo, Ariadna Medina-Benitez, Joaquin Quiroz-Mercado, Sergio Rangel-Guerrero, Natividad Cruz-Ortiz, Tatiana Nunez-Garcia, Gisela Barrera-Badillo, Lucia Hernandez-Rivas, Irma Lopez-Martinez, Ernesto Ramirez-Gonzalez. |
| EPI_ISL_1821106 | LESP Ciudad de Mexico | Instituto de Diagnostico y Referencia Epidemiologicos (INDRE) | Claudia Wong-Arambula, Abril Rodriguez-Maldonado, Vanessa Rivero-Arredondo, Ariadna Medina-Benitez, Joaquin Quiroz-Mercado, Sergio Rangel-Guerrero, Natividad Cruz-Ortiz, Tatiana Nunez-Garcia, Gisela Barrera-Badillo, Lucia Hernandez-Rivas, Irma Lopez-Martinez, Ernesto Ramirez-Gonzalez. |
| EPI_ISL_1821107 | LESP Colima | Instituto de Diagnostico y Referencia Epidemiologicos (INDRE) | Claudia Wong-Arambula, Abril Rodriguez-Maldonado, Vanessa Rivero-Arredondo, Ariadna Medina-Benitez, Joaquin Quiroz-Mercado, Sergio Rangel-Guerrero, Natividad Cruz-Ortiz, Tatiana Nunez-Garcia, Gisela Barrera-Badillo, Lucia Hernandez-Rivas, Irma Lopez-Martinez, Ernesto Ramirez-Gonzalez. |
| EPI_ISL_1821108, EPI_ISL_1821109, EPI_ISL_1821110, EPI_ISL_1821111 | LESP Guanajuato | Instituto de Diagnostico y Referencia Epidemiologicos (INDRE) | Claudia Wong-Arambula, Abril Rodriguez-Maldonado, Vanessa Rivero-Arredondo, Ariadna Medina-Benitez, Joaquin Quiroz-Mercado, Sergio Rangel-Guerrero, Natividad Cruz-Ortiz, Tatiana Nunez-Garcia, Gisela Barrera-Badillo, Lucia Hernandez-Rivas, Irma Lopez-Martinez, Ernesto Ramirez-Gonzalez. |
| EPI_ISL_1821112, EPI_ISL_1821113, EPI_ISL_1821114, EPI_ISL_1821115, EPI_ISL_1821116, EPI_ISL_1821117, EPI_ISL_1821118 | LESP Nuevo Leon | Instituto de Diagnostico y Referencia Epidemiologicos (INDRE) | Claudia Wong-Arambula, Abril Rodriguez-Maldonado, Vanessa Rivero-Arredondo, Ariadna Medina-Benitez, Joaquin Quiroz-Mercado, Sergio Rangel-Guerrero, Natividad Cruz-Ortiz, Tatiana Nunez-Garcia, Gisela Barrera-Badillo, Lucia Hernandez-Rivas, Irma Lopez-Martinez, Ernesto Ramirez-Gonzalez. |
| EPI_ISL_1821119, EPI_ISL_1821120, EPI_ISL_1821121, EPI_ISL_1821122 | LESP Puebla | Instituto de Diagnostico y Referencia Epidemiologicos (INDRE) | Claudia Wong-Arambula, Abril Rodriguez-Maldonado, Vanessa Rivero-Arredondo, Ariadna Medina-Benitez, Joaquin Quiroz-Mercado, Sergio Rangel-Guerrero, Natividad Cruz-Ortiz, Tatiana Nunez-Garcia, Gisela Barrera-Badillo, Lucia Hernandez-Rivas, Irma Lopez-Martinez, Ernesto Ramirez-Gonzalez. |
| EPI_ISL_1821123 | LESP Guerrero | Instituto de Diagnostico y Referencia Epidemiologicos (INDRE) | Claudia Wong-Arambula, Abril Rodriguez-Maldonado, Vanessa Rivero-Arredondo, Ariadna Medina-Benitez, Joaquin Quiroz-Mercado, Sergio Rangel-Guerrero, Natividad Cruz-Ortiz, Tatiana Nunez-Garcia, Gisela Barrera-Badillo, Lucia Hernandez-Rivas, Irma Lopez-Martinez, Ernesto Ramirez-Gonzalez. |
| EPI_ISL_1821124 | LESP Puebla | Instituto de Diagnostico y Referencia Epidemiologicos (INDRE) | Claudia Wong-Arambula, Abril Rodriguez-Maldonado, Vanessa Rivero-Arredondo, Ariadna Medina-Benitez, Joaquin Quiroz-Mercado, Sergio Rangel-Guerrero, Natividad Cruz-Ortiz, Tatiana Nunez-Garcia, Gisela Barrera-Badillo, Lucia Hernandez-Rivas, Irma Lopez-Martinez, Ernesto Ramirez-Gonzalez. |
| EPI_ISL_1821125 | LESP Chihuahua | Instituto de Diagnostico y Referencia Epidemiologicos (INDRE) | Claudia Wong-Arambula, Abril Rodriguez-Maldonado, Vanessa Rivero-Arredondo, Ariadna Medina-Benitez, Joaquin Quiroz-Mercado, Sergio Rangel-Guerrero, Natividad Cruz-Ortiz, Tatiana Nunez-Garcia, Gisela Barrera-Badillo, Lucia Hernandez-Rivas, Irma Lopez-Martinez, Ernesto Ramirez-Gonzalez. |
| EPI_ISL_1821126 | LESP Sonora | Instituto de Diagnostico y Referencia Epidemiologicos (INDRE) | Claudia Wong-Arambula, Abril Rodriguez-Maldonado, Vanessa Rivero-Arredondo, Ariadna Medina-Benitez, Joaquin Quiroz-Mercado, Sergio Rangel-Guerrero, Natividad Cruz-Ortiz, Tatiana Nunez-Garcia, Gisela Barrera-Badillo, Lucia Hernandez-Rivas, Irma Lopez-Martinez, Ernesto Ramirez-Gonzalez. |
| EPI_ISL_1821127 | LESP Chihuahua | Instituto de Diagnostico y Referencia Epidemiologicos (INDRE) | Claudia Wong-Arambula, Abril Rodriguez-Maldonado, Vanessa Rivero-Arredondo, Ariadna Medina-Benitez, Joaquin Quiroz-Mercado, Sergio Rangel-Guerrero, Natividad Cruz-Ortiz, Tatiana Nunez-Garcia, Gisela Barrera-Badillo, Lucia Hernandez-Rivas, Irma Lopez-Martinez, Ernesto Ramirez-Gonzalez. |
| EPI_ISL_1821128 | LESP Sonora | Instituto de Diagnostico y Referencia Epidemiologicos (INDRE) | Claudia Wong-Arambula, Abril Rodriguez-Maldonado, Vanessa Rivero-Arredondo, Ariadna Medina-Benitez, Joaquin Quiroz-Mercado, Sergio Rangel-Guerrero, Natividad Cruz-Ortiz, Tatiana Nunez-Garcia, Gisela Barrera-Badillo, Lucia Hernandez-Rivas, Irma Lopez-Martinez, Ernesto Ramirez-Gonzalez. |
| EPI_ISL_1821129, EPI_ISL_1821130, EPI_ISL_1821131 | LESP Jalisco | Instituto de Diagnostico y Referencia Epidemiologicos (INDRE) | Claudia Wong-Arambula, Abril Rodriguez-Maldonado, Vanessa Rivero-Arredondo, Ariadna Medina-Benitez, Joaquin Quiroz-Mercado, Sergio Rangel-Guerrero, Natividad Cruz-Ortiz, Tatiana Nunez-Garcia, Gisela Barrera-Badillo, Lucia Hernandez-Rivas, Irma Lopez-Martinez, Ernesto Ramirez-Gonzalez. |
| EPI_ISL_1821132, EPI_ISL_1821133, EPI_ISL_1821134, EPI_ISL_1821135, EPI_ISL_1821136 | LESP Queretaro | Instituto de Diagnostico y Referencia Epidemiologicos (INDRE) | Claudia Wong-Arambula, Abril Rodriguez-Maldonado, Vanessa Rivero-Arredondo, Ariadna Medina-Benitez, Joaquin Quiroz-Mercado, Sergio Rangel-Guerrero, Natividad Cruz-Ortiz, Tatiana Nunez-Garcia, Gisela Barrera-Badillo, Lucia Hernandez-Rivas, Irma Lopez-Martinez, Ernesto Ramirez-Gonzalez. |
| EPI_ISL_1821137, EPI_ISL_1821138, EPI_ISL_1821139, EPI_ISL_1821140, EPI_ISL_1821141, EPI_ISL_1821142, EPI_ISL_1821143, EPI_ISL_1821144, EPI_ISL_1821145, EPI_ISL_1821147, EPI_ISL_1821148 | | | |
| see above | LESP Morelos | Instituto de Diagnostico y Referencia Epidemiologicos (INDRE) | Claudia Wong-Arambula, Abril Rodriguez-Maldonado, Vanessa Rivero-Arredondo, Ariadna Medina-Benitez, Joaquin Quiroz-Mercado, Sergio Rangel-Guerrero, Natividad Cruz-Ortiz, Tatiana Nunez-Garcia, Gisela Barrera-Badillo, Lucia Hernandez-Rivas, Irma Lopez-Martinez, Ernesto Ramirez-Gonzalez. |
| EPI_ISL_1821149, EPI_ISL_1821150, | LESP Guerrero | Instituto de Diagnostico y Referencia Epidemiologicos (INDRE) | Claudia Wong-Arambula, Abril Rodriguez-Maldonado, Vanessa Rivero-Arredondo, Ariadna Medina-Benitez, Joaquin Quiroz-Mercado, Sergio Rangel-Guerrero, Natividad Cruz-Ortiz, Tatiana Nunez-Garcia, Gisela Barrera-Badillo, Lucia Hernandez-Rivas, Irma Lopez-Martinez, Ernesto Ramirez-Gonzalez. |

| | | | |
|---|---|---|---|
| EPI_ISL_1821151 | | | |
| EPI_ISL_1821152, EPI_ISL_1821153, EPI_ISL_1821154, EPI_ISL_1821155 | LESP Yucatan | Instituto de Diagnostico y Referencia Epidemiologicos (INDRE) | Claudia Wong-Arambula, Abril Rodriguez-Maldonado, Vanessa Rivero-Arredondo, Ariadna Medina-Benitez, Joaquin Quiroz-Mercado, Sergio Rangel-Guerrero, Natividad Cruz-Ortiz, Tatiana Nunez-Garcia, Gisela Barrera-Badillo, Lucia Hernandez-Rivas, Irma Lopez-Martinez, Ernesto Ramirez-Gonzalez. |
| EPI_ISL_1821156, EPI_ISL_1821157 | LESP Sinaloa | Instituto de Diagnostico y Referencia Epidemiologicos (INDRE) | Claudia Wong-Arambula, Abril Rodriguez-Maldonado, Vanessa Rivero-Arredondo, Ariadna Medina-Benitez, Joaquin Quiroz-Mercado, Sergio Rangel-Guerrero, Natividad Cruz-Ortiz, Tatiana Nunez-Garcia, Gisela Barrera-Badillo, Lucia Hernandez-Rivas, Irma Lopez-Martinez, Ernesto Ramirez-Gonzalez. |
| EPI_ISL_1821158, EPI_ISL_1821159, EPI_ISL_1821160 | LESP Tamaulipas | Instituto de Diagnostico y Referencia Epidemiologicos (INDRE) | Claudia Wong-Arambula, Abril Rodriguez-Maldonado, Vanessa Rivero-Arredondo, Ariadna Medina-Benitez, Joaquin Quiroz-Mercado, Sergio Rangel-Guerrero, Natividad Cruz-Ortiz, Tatiana Nunez-Garcia, Gisela Barrera-Badillo, Lucia Hernandez-Rivas, Irma Lopez-Martinez, Ernesto Ramirez-Gonzalez. |
| EPI_ISL_1821161 | LESP Nayarit | Instituto de Diagnostico y Referencia Epidemiologicos (INDRE) | Claudia Wong-Arambula, Abril Rodriguez-Maldonado, Vanessa Rivero-Arredondo, Ariadna Medina-Benitez, Joaquin Quiroz-Mercado, Sergio Rangel-Guerrero, Natividad Cruz-Ortiz, Tatiana Nunez-Garcia, Gisela Barrera-Badillo, Lucia Hernandez-Rivas, Irma Lopez-Martinez, Ernesto Ramirez-Gonzalez. |
| EPI_ISL_1821162 | LESP Oaxaca | Instituto de Diagnostico y Referencia Epidemiologicos (INDRE) | Claudia Wong-Arambula, Abril Rodriguez-Maldonado, Vanessa Rivero-Arredondo, Ariadna Medina-Benitez, Joaquin Quiroz-Mercado, Sergio Rangel-Guerrero, Natividad Cruz-Ortiz, Tatiana Nunez-Garcia, Gisela Barrera-Badillo, Lucia Hernandez-Rivas, Irma Lopez-Martinez, Ernesto Ramirez-Gonzalez. |
| EPI_ISL_1821163, EPI_ISL_1821164, EPI_ISL_1821165 | LESP Veracruz | Instituto de Diagnostico y Referencia Epidemiologicos (INDRE) | Claudia Wong-Arambula, Abril Rodriguez-Maldonado, Vanessa Rivero-Arredondo, Ariadna Medina-Benitez, Joaquin Quiroz-Mercado, Sergio Rangel-Guerrero, Natividad Cruz-Ortiz, Tatiana Nunez-Garcia, Gisela Barrera-Badillo, Lucia Hernandez-Rivas, Irma Lopez-Martinez, Ernesto Ramirez-Gonzalez. |
| EPI_ISL_1821166 | LESP Tlaxcala | Instituto de Diagnostico y Referencia Epidemiologicos (INDRE) | Claudia Wong-Arambula, Abril Rodriguez-Maldonado, Vanessa Rivero-Arredondo, Ariadna Medina-Benitez, Joaquin Quiroz-Mercado, Sergio Rangel-Guerrero, Natividad Cruz-Ortiz, Tatiana Nunez-Garcia, Gisela Barrera-Badillo, Lucia Hernandez-Rivas, Irma Lopez-Martinez, Ernesto Ramirez-Gonzalez. |
| EPI_ISL_1821167, EPI_ISL_1821168 | LESP Chihuahua | Instituto de Diagnostico y Referencia Epidemiologicos (INDRE) | Claudia Wong-Arambula, Abril Rodriguez-Maldonado, Vanessa Rivero-Arredondo, Ariadna Medina-Benitez, Joaquin Quiroz-Mercado, Sergio Rangel-Guerrero, Natividad Cruz-Ortiz, Tatiana Nunez-Garcia, Gisela Barrera-Badillo, Lucia Hernandez-Rivas, Irma Lopez-Martinez, Ernesto Ramirez-Gonzalez. |
| EPI_ISL_1821169, EPI_ISL_1821170 | LESP Ciudad de Mexico | Instituto de Diagnostico y Referencia Epidemiologicos (INDRE) | Claudia Wong-Arambula, Abril Rodriguez-Maldonado, Vanessa Rivero-Arredondo, Ariadna Medina-Benitez, Joaquin Quiroz-Mercado, Sergio Rangel-Guerrero, Natividad Cruz-Ortiz, Tatiana Nunez-Garcia, Gisela Barrera-Badillo, Lucia Hernandez-Rivas, Irma Lopez-Martinez, Ernesto Ramirez-Gonzalez. |
| EPI_ISL_1821171, EPI_ISL_1821172 | LESP Quintana Roo | Instituto de Diagnostico y Referencia Epidemiologicos (INDRE) | Claudia Wong-Arambula, Abril Rodriguez-Maldonado, Vanessa Rivero-Arredondo, Ariadna Medina-Benitez, Joaquin Quiroz-Mercado, Sergio Rangel-Guerrero, Natividad Cruz-Ortiz, Tatiana Nunez-Garcia, Gisela Barrera-Badillo, Lucia Hernandez-Rivas, Irma Lopez-Martinez, Ernesto Ramirez-Gonzalez. |
| EPI_ISL_1821173, EPI_ISL_1821174, EPI_ISL_1821175, EPI_ISL_1821176, EPI_ISL_1821177, EPI_ISL_1821178, EPI_ISL_1821179 | LESP Zacatecas | Instituto de Diagnostico y Referencia Epidemiologicos (INDRE) | Claudia Wong-Arambula, Abril Rodriguez-Maldonado, Vanessa Rivero-Arredondo, Ariadna Medina-Benitez, Joaquin Quiroz-Mercado, Sergio Rangel-Guerrero, Natividad Cruz-Ortiz, Tatiana Nunez-Garcia, Gisela Barrera-Badillo, Lucia Hernandez-Rivas, Irma Lopez-Martinez, Ernesto Ramirez-Gonzalez. |
| EPI_ISL_1821180, EPI_ISL_1821181, EPI_ISL_1821182, EPI_ISL_1821183, EPI_ISL_1821184 | LESP Baja California Sur | Instituto de Diagnostico y Referencia Epidemiologicos (INDRE) | Claudia Wong-Arambula, Abril Rodriguez-Maldonado, Vanessa Rivero-Arredondo, Ariadna Medina-Benitez, Joaquin Quiroz-Mercado, Sergio Rangel-Guerrero, Natividad Cruz-Ortiz, Tatiana Nunez-Garcia, Gisela Barrera-Badillo, Lucia Hernandez-Rivas, Irma Lopez-Martinez, Ernesto Ramirez-Gonzalez. |
| EPI_ISL_1821185, EPI_ISL_1821186 | LESP Jalisco | Instituto de Diagnostico y Referencia Epidemiologicos (INDRE) | Claudia Wong-Arambula, Abril Rodriguez-Maldonado, Vanessa Rivero-Arredondo, Ariadna Medina-Benitez, Joaquin Quiroz-Mercado, Sergio Rangel-Guerrero, Natividad Cruz-Ortiz, Tatiana Nunez-Garcia, Gisela Barrera-Badillo, Lucia Hernandez-Rivas, Irma Lopez-Martinez, Ernesto Ramirez-Gonzalez. |
| EPI_ISL_1821187, EPI_ISL_1821188, EPI_ISL_1821189, EPI_ISL_1821190 | LESP Ciudad de Mexico | Instituto de Diagnostico y Referencia Epidemiologicos (INDRE) | Claudia Wong-Arambula, Abril Rodriguez-Maldonado, Vanessa Rivero-Arredondo, Ariadna Medina-Benitez, Joaquin Quiroz-Mercado, Sergio Rangel-Guerrero, Natividad Cruz-Ortiz, Tatiana Nunez-Garcia, Gisela Barrera-Badillo, Lucia Hernandez-Rivas, Irma Lopez-Martinez, Ernesto Ramirez-Gonzalez. |
| EPI_ISL_1821191, EPI_ISL_1821192 | LESP Queretaro | Instituto de Diagnostico y Referencia Epidemiologicos (INDRE) | Claudia Wong-Arambula, Abril Rodriguez-Maldonado, Vanessa Rivero-Arredondo, Ariadna Medina-Benitez, Joaquin Quiroz-Mercado, Sergio Rangel-Guerrero, Natividad Cruz-Ortiz, Tatiana Nunez-Garcia, Gisela Barrera-Badillo, Lucia Hernandez-Rivas, Irma Lopez-Martinez, Ernesto Ramirez-Gonzalez. |
| EPI_ISL_1821193 | LESP Tabasco | Instituto de Diagnostico y Referencia Epidemiologicos (INDRE) | Claudia Wong-Arambula, Abril Rodriguez-Maldonado, Vanessa Rivero-Arredondo, Ariadna Medina-Benitez, Joaquin Quiroz-Mercado, Sergio Rangel-Guerrero, Natividad Cruz-Ortiz, Tatiana Nunez-Garcia, Gisela Barrera-Badillo, Lucia Hernandez-Rivas, Irma Lopez-Martinez, Ernesto Ramirez-Gonzalez. |
| EPI_ISL_1821194, EPI_ISL_1821195, EPI_ISL_1821196 | LESP Hidalgo | Instituto de Diagnostico y Referencia Epidemiologicos (INDRE) | Claudia Wong-Arambula, Abril Rodriguez-Maldonado, Vanessa Rivero-Arredondo, Ariadna Medina-Benitez, Joaquin Quiroz-Mercado, Sergio Rangel-Guerrero, Natividad Cruz-Ortiz, Tatiana Nunez-Garcia, Gisela Barrera-Badillo, Lucia Hernandez-Rivas, Irma Lopez-Martinez, Ernesto Ramirez-Gonzalez. |
| EPI_ISL_1821197, EPI_ISL_1821198 | LESP Jalisco | Instituto de Diagnostico y Referencia Epidemiologicos (INDRE) | Claudia Wong-Arambula, Abril Rodriguez-Maldonado, Vanessa Rivero-Arredondo, Ariadna Medina-Benitez, Joaquin Quiroz-Mercado, Sergio Rangel-Guerrero, Natividad Cruz-Ortiz, Tatiana Nunez-Garcia, Gisela Barrera-Badillo, Lucia Hernandez-Rivas, Irma Lopez-Martinez, Ernesto Ramirez-Gonzalez. |
| EPI_ISL_1821199 | LESP Sinaloa | Instituto de Diagnostico y Referencia Epidemiologicos (INDRE) | Claudia Wong-Arambula, Abril Rodriguez-Maldonado, Vanessa Rivero-Arredondo, Ariadna Medina-Benitez, Joaquin Quiroz-Mercado, Sergio Rangel-Guerrero, Natividad Cruz-Ortiz, Tatiana Nunez-Garcia, Gisela Barrera-Badillo, Lucia Hernandez-Rivas, Irma Lopez-Martinez, Ernesto Ramirez-Gonzalez. |
| EPI_ISL_1821200 | LESP Jalisco | Instituto de Diagnostico y Referencia Epidemiologicos (INDRE) | Claudia Wong-Arambula, Abril Rodriguez-Maldonado, Vanessa Rivero-Arredondo, Ariadna Medina-Benitez, Joaquin Quiroz-Mercado, Sergio Rangel-Guerrero, Natividad Cruz-Ortiz, Tatiana Nunez-Garcia, Gisela Barrera-Badillo, Lucia Hernandez-Rivas, Irma Lopez-Martinez, Ernesto Ramirez-Gonzalez. |
| EPI_ISL_1821201, EPI_ISL_1821202, EPI_ISL_1821203 | LESP Baja California | Instituto de Diagnostico y Referencia Epidemiologicos (INDRE) | Claudia Wong-Arambula, Abril Rodriguez-Maldonado, Vanessa Rivero-Arredondo, Ariadna Medina-Benitez, Joaquin Quiroz-Mercado, Sergio Rangel-Guerrero, Natividad Cruz-Ortiz, Tatiana Nunez-Garcia, Gisela Barrera-Badillo, Lucia Hernandez-Rivas, Irma Lopez-Martinez, Ernesto Ramirez-Gonzalez. |
| EPI_ISL_1821274, EPI_ISL_1821275, EPI_ISL_1821276, EPI_ISL_1821277, EPI_ISL_1821278, EPI_ISL_1821279, EPI_ISL_1821280, EPI_ISL_1821281, EPI_ISL_1821282, EPI_ISL_1821283, EPI_ISL_1821284, EPI_ISL_1821285, EPI_ISL_1821286, EPI_ISL_1821287, EPI_ISL_1821288, EPI_ISL_1821289, EPI_ISL_1821290, EPI_ISL_1821291, EPI_ISL_1821292, EPI_ISL_1821293, EPI_ISL_1821294 | | | |
| see above | Nebraska Public Health Laboratory | NPHL COVID-19 Response Team | NPHL COVID-19 Response Team |
| EPI_ISL_1821295, EPI_ISL_1821296, EPI_ISL_1821297, EPI_ISL_1821298, EPI_ISL_1821299, EPI_ISL_1821300, EPI_ISL_1821301, EPI_ISL_1821302, EPI_ISL_1821303, EPI_ISL_1821304, EPI_ISL_1821305, EPI_ISL_1821306, EPI_ISL_1821307, EPI_ISL_1821308, EPI_ISL_1821309, EPI_ISL_1821310, EPI_ISL_1821311, EPI_ISL_1821314 | | | |
| see above | University of Wisconsin-Madison AIDS Vaccine Research Laboratories | University of Wisconsin-Madison AIDS Vaccine Research Laboratories | Gage Moreno, Katarina Braun, et al. AIDS Vaccine Research Laboratories |
| EPI_ISL_1821315, EPI_ISL_1821316 | CA DPH Viral and Rickettsial Disease Laboratory | Chan-Zuckerberg Biohub | CZB Cliahub Consortium |
| EPI_ISL_1821317 | Santa Clara County Public Health Laboratory | Chan-Zuckerberg Biohub | CZB Cliahub Consortium |
| EPI_ISL_1821318 | CA DPH Viral and Rickettsial Disease Laboratory | Chan-Zuckerberg Biohub | CZB Cliahub Consortium |
| EPI_ISL_1821319, EPI_ISL_1821320 | Santa Clara County Public Health Laboratory | Chan-Zuckerberg Biohub | CZB Cliahub Consortium |
| EPI_ISL_1821321, EPI_ISL_1821322 | CA DPH Viral and Rickettsial Disease Laboratory | Chan-Zuckerberg Biohub | CZB Cliahub Consortium |
| EPI_ISL_1821323 | County of San Luis Obispo Public Health Laboratory | Chan-Zuckerberg Biohub | CZB Cliahub Consortium |
| EPI_ISL_1821324 | Santa Clara County Public Health Laboratory | Chan-Zuckerberg Biohub | CZB Cliahub Consortium |
| EPI_ISL_1821325 | CA DPH Viral and Rickettsial Disease Laboratory | Chan-Zuckerberg Biohub | CZB Cliahub Consortium |
| EPI_ISL_1821326 | County of San Luis Obispo Public Health Laboratory | Chan-Zuckerberg Biohub | CZB Cliahub Consortium |
| EPI_ISL_1821327, EPI_ISL_1821328, EPI_ISL_1821329, EPI_ISL_1821330, EPI_ISL_1821331 | Santa Clara County Public Health Laboratory | Chan-Zuckerberg Biohub | CZB Cliahub Consortium |
| EPI_ISL_1821332 | County of San Luis Obispo Public Health Laboratory | Chan-Zuckerberg Biohub | CZB Cliahub Consortium |
| EPI_ISL_1821333 | Santa Clara County Public Health Laboratory | Chan-Zuckerberg Biohub | CZB Cliahub Consortium |
| EPI_ISL_1821334, EPI_ISL_1821335, | CA DPH Viral and Rickettsial Disease Laboratory | Chan-Zuckerberg Biohub | CZB Cliahub Consortium |

| | | | |
|---|---|---|---|
| EPI_ISL_1821336 | | | |
| EPI_ISL_1821337, EPI_ISL_1821338, EPI_ISL_1821339 | Santa Clara County Public Health Laboratory | Chan-Zuckerberg Biohub | CZB Cliahub Consortium |
| EPI_ISL_1821340 | CA DPH Viral and Rickettsial Disease Laboratory | Chan-Zuckerberg Biohub | CZB Cliahub Consortium |
| EPI_ISL_1821341, EPI_ISL_1821342 | Santa Clara County Public Health Laboratory | Chan-Zuckerberg Biohub | CZB Cliahub Consortium |
| EPI_ISL_1821343 | CA DPH Viral and Rickettsial Disease Laboratory | Chan-Zuckerberg Biohub | CZB Cliahub Consortium |
| EPI_ISL_1821344 | Santa Clara County Public Health Laboratory | Chan-Zuckerberg Biohub | CZB Cliahub Consortium |
| EPI_ISL_1821345 | CA DPH Viral and Rickettsial Disease Laboratory | Chan-Zuckerberg Biohub | CZB Cliahub Consortium |
| EPI_ISL_1821346 | Santa Clara County Public Health Laboratory | Chan-Zuckerberg Biohub | CZB Cliahub Consortium |
| EPI_ISL_1821347, EPI_ISL_1821348, EPI_ISL_1821349 | CA DPH Viral and Rickettsial Disease Laboratory | Chan-Zuckerberg Biohub | CZB Cliahub Consortium |
| EPI_ISL_1821350 | Santa Clara County Public Health Laboratory | Chan-Zuckerberg Biohub | CZB Cliahub Consortium |
| EPI_ISL_1821351, EPI_ISL_1821352, EPI_ISL_1821353 | CA DPH Viral and Rickettsial Disease Laboratory | Chan-Zuckerberg Biohub | CZB Cliahub Consortium |
| EPI_ISL_1821354 | County of San Luis Obispo Public Health Laboratory | Chan-Zuckerberg Biohub | CZB Cliahub Consortium |
| EPI_ISL_1821355 | Santa Clara County Public Health Laboratory | Chan-Zuckerberg Biohub | CZB Cliahub Consortium |
| EPI_ISL_1821356, EPI_ISL_1821357 | CA DPH Viral and Rickettsial Disease Laboratory | Chan-Zuckerberg Biohub | CZB Cliahub Consortium |
| EPI_ISL_1821358, EPI_ISL_1821359 | County of San Luis Obispo Public Health Laboratory | Chan-Zuckerberg Biohub | CZB Cliahub Consortium |
| EPI_ISL_1821360, EPI_ISL_1821361 | CA DPH Viral and Rickettsial Disease Laboratory | Chan-Zuckerberg Biohub | CZB Cliahub Consortium |
| EPI_ISL_1821362, EPI_ISL_1821363 | County of San Luis Obispo Public Health Laboratory | Chan-Zuckerberg Biohub | CZB Cliahub Consortium |
| EPI_ISL_1821364 | CA DPH Viral and Rickettsial Disease Laboratory | Chan-Zuckerberg Biohub | CZB Cliahub Consortium |
| EPI_ISL_1821365, EPI_ISL_1821366, EPI_ISL_1821367, EPI_ISL_1821368 | County of San Luis Obispo Public Health Laboratory | Chan-Zuckerberg Biohub | CZB Cliahub Consortium |
| EPI_ISL_1821369, EPI_ISL_1821370 | CA DPH Viral and Rickettsial Disease Laboratory | Chan-Zuckerberg Biohub | CZB Cliahub Consortium |
| EPI_ISL_1821371, EPI_ISL_1821372, EPI_ISL_1821373, EPI_ISL_1821374, EPI_ISL_1821375, EPI_ISL_1821376, EPI_ISL_1821377, EPI_ISL_1821378 | County of San Luis Obispo Public Health Laboratory | Chan-Zuckerberg Biohub | CZB Cliahub Consortium |
| EPI_ISL_1821379 | CA DPH Viral and Rickettsial Disease Laboratory | Chan-Zuckerberg Biohub | CZB Cliahub Consortium |
| EPI_ISL_1821380, EPI_ISL_1821381 | County of San Luis Obispo Public Health Laboratory | Chan-Zuckerberg Biohub | CZB Cliahub Consortium |
| EPI_ISL_1821382 | CA DPH Viral and Rickettsial Disease Laboratory | Chan-Zuckerberg Biohub | CZB Cliahub Consortium |
| EPI_ISL_1821383, EPI_ISL_1821384, EPI_ISL_1821385, EPI_ISL_1821386, EPI_ISL_1821387, EPI_ISL_1821388, EPI_ISL_1821389, EPI_ISL_1821390, EPI_ISL_1821391 | County of San Luis Obispo Public Health Laboratory | Chan-Zuckerberg Biohub | CZB Cliahub Consortium |
| EPI_ISL_1821392 | Santa Clara County Public Health Laboratory | Chan-Zuckerberg Biohub | CZB Cliahub Consortium |
| EPI_ISL_1821393 | CA DPH Viral and Rickettsial Disease Laboratory | Chan-Zuckerberg Biohub | CZB Cliahub Consortium |
| EPI_ISL_1821394 | County of San Luis Obispo Public Health Laboratory | Chan-Zuckerberg Biohub | CZB Cliahub Consortium |
| EPI_ISL_1821395 | CA DPH Viral and Rickettsial Disease Laboratory | Chan-Zuckerberg Biohub | CZB Cliahub Consortium |
| EPI_ISL_1821398 | Santa Clara County Public Health Laboratory | Chan-Zuckerberg Biohub | CZB Cliahub Consortium |
| EPI_ISL_1821399 | County of San Luis Obispo Public Health Laboratory | Chan-Zuckerberg Biohub | CZB Cliahub Consortium |
| EPI_ISL_1821400, EPI_ISL_1821401 | CA DPH Viral and Rickettsial Disease Laboratory | Chan-Zuckerberg Biohub | CZB Cliahub Consortium |
| EPI_ISL_1821402 | Santa Clara County Public Health Laboratory | Chan-Zuckerberg Biohub | CZB Cliahub Consortium |
| EPI_ISL_1821403 | CA DPH Viral and Rickettsial Disease Laboratory | Chan-Zuckerberg Biohub | CZB Cliahub Consortium |
| EPI_ISL_1821404 | County of San Luis Obispo Public Health Laboratory | Chan-Zuckerberg Biohub | CZB Cliahub Consortium |
| EPI_ISL_1821405 | Santa Clara County Public Health Laboratory | Chan-Zuckerberg Biohub | CZB Cliahub Consortium |
| EPI_ISL_1821406, EPI_ISL_1821407 | CA DPH Viral and Rickettsial Disease Laboratory | Chan-Zuckerberg Biohub | CZB Cliahub Consortium |
| EPI_ISL_1821408 | County of San Luis Obispo Public Health Laboratory | Chan-Zuckerberg Biohub | CZB Cliahub Consortium |

| | | | |
|---|---|---|---|
| EPI_ISL_1821409, EPI_ISL_1821410, EPI_ISL_1821411, EPI_ISL_1821412 | CA DPH Viral and Rickettsial Disease Laboratory | Chan-Zuckerberg Biohub | CZB Cliahub Consortium |
| EPI_ISL_1821413, EPI_ISL_1821414 | Santa Clara County Public Health Laboratory | Chan-Zuckerberg Biohub | CZB Cliahub Consortium |
| EPI_ISL_1821415, EPI_ISL_1821416, EPI_ISL_1821417, EPI_ISL_1821418 | CA DPH Viral and Rickettsial Disease Laboratory | Chan-Zuckerberg Biohub | CZB Cliahub Consortium |
| EPI_ISL_1821419 | Santa Clara County Public Health Laboratory | Chan-Zuckerberg Biohub | CZB Cliahub Consortium |
| EPI_ISL_1821420, EPI_ISL_1821421, EPI_ISL_1821422, EPI_ISL_1821423, EPI_ISL_1821424, EPI_ISL_1821425 | CA DPH Viral and Rickettsial Disease Laboratory | Chan-Zuckerberg Biohub | CZB Cliahub Consortium |
| EPI_ISL_1821426, EPI_ISL_1821427, EPI_ISL_1821428 | Santa Clara County Public Health Laboratory | Chan-Zuckerberg Biohub | CZB Cliahub Consortium |
| EPI_ISL_1821429, EPI_ISL_1821430, EPI_ISL_1821431, EPI_ISL_1821432, EPI_ISL_1821433 | CA DPH Viral and Rickettsial Disease Laboratory | Chan-Zuckerberg Biohub | CZB Cliahub Consortium |
| EPI_ISL_1821434 | County of San Luis Obispo Public Health Laboratory | Chan-Zuckerberg Biohub | CZB Cliahub Consortium |
| EPI_ISL_1821435, EPI_ISL_1821436 | CA DPH Viral and Rickettsial Disease Laboratory | Chan-Zuckerberg Biohub | CZB Cliahub Consortium |
| EPI_ISL_1821437 | Santa Clara County Public Health Laboratory | Chan-Zuckerberg Biohub | CZB Cliahub Consortium |
| EPI_ISL_1821438, EPI_ISL_1821439, EPI_ISL_1821440, EPI_ISL_1821441, EPI_ISL_1821442 | CA DPH Viral and Rickettsial Disease Laboratory | Chan-Zuckerberg Biohub | CZB Cliahub Consortium |
| EPI_ISL_1821443 | Santa Clara County Public Health Laboratory | Chan-Zuckerberg Biohub | CZB Cliahub Consortium |
| EPI_ISL_1821444 | CA DPH Viral and Rickettsial Disease Laboratory | Chan-Zuckerberg Biohub | CZB Cliahub Consortium |
| EPI_ISL_1821445 | Santa Clara County Public Health Laboratory | Chan-Zuckerberg Biohub | CZB Cliahub Consortium |
| EPI_ISL_1821446 | CA DPH Viral and Rickettsial Disease Laboratory | Chan-Zuckerberg Biohub | CZB Cliahub Consortium |
| EPI_ISL_1821447 | County of San Luis Obispo Public Health Laboratory | Chan-Zuckerberg Biohub | CZB Cliahub Consortium |
| EPI_ISL_1821448, EPI_ISL_1821449, EPI_ISL_1821450, EPI_ISL_1821451, EPI_ISL_1821452, EPI_ISL_1821453 | CA DPH Viral and Rickettsial Disease Laboratory | Chan-Zuckerberg Biohub | CZB Cliahub Consortium |
| EPI_ISL_1821454 | County of San Luis Obispo Public Health Laboratory | Chan-Zuckerberg Biohub | CZB Cliahub Consortium |
| EPI_ISL_1821455 | Santa Clara County Public Health Laboratory | Chan-Zuckerberg Biohub | CZB Cliahub Consortium |
| EPI_ISL_1821456, EPI_ISL_1821457 | CA DPH Viral and Rickettsial Disease Laboratory | Chan-Zuckerberg Biohub | CZB Cliahub Consortium |
| EPI_ISL_1821458 | Santa Clara County Public Health Laboratory | Chan-Zuckerberg Biohub | CZB Cliahub Consortium |
| EPI_ISL_1821459 | CA DPH Viral and Rickettsial Disease Laboratory | Chan-Zuckerberg Biohub | CZB Cliahub Consortium |
| EPI_ISL_1821460, EPI_ISL_1821461 | Santa Clara County Public Health Laboratory | Chan-Zuckerberg Biohub | CZB Cliahub Consortium |
| EPI_ISL_1821462 | CA DPH Viral and Rickettsial Disease Laboratory | Chan-Zuckerberg Biohub | CZB Cliahub Consortium |
| EPI_ISL_1821463 | County of San Luis Obispo Public Health Laboratory | Chan-Zuckerberg Biohub | CZB Cliahub Consortium |
| EPI_ISL_1821464, EPI_ISL_1821465, EPI_ISL_1821466, EPI_ISL_1821467 | CA DPH Viral and Rickettsial Disease Laboratory | Chan-Zuckerberg Biohub | CZB Cliahub Consortium |
| EPI_ISL_1821468 | County of San Luis Obispo Public Health Laboratory | Chan-Zuckerberg Biohub | CZB Cliahub Consortium |
| EPI_ISL_1821469 | CA DPH Viral and Rickettsial Disease Laboratory | Chan-Zuckerberg Biohub | CZB Cliahub Consortium |
| EPI_ISL_1821470 | Santa Clara County Public Health Laboratory | Chan-Zuckerberg Biohub | CZB Cliahub Consortium |
| EPI_ISL_1821471, EPI_ISL_1821472 | CA DPH Viral and Rickettsial Disease Laboratory | Chan-Zuckerberg Biohub | CZB Cliahub Consortium |
| EPI_ISL_1821660, EPI_ISL_1821661, EPI_ISL_1821662, EPI_ISL_1821663, EPI_ISL_1821664, EPI_ISL_1821665, EPI_ISL_1821666, EPI_ISL_1821667, EPI_ISL_1821668, EPI_ISL_1821669, EPI_ISL_1821670, EPI_ISL_1821671, EPI_ISL_1821672, EPI_ISL_1821673, EPI_ISL_1821674, EPI_ISL_1821675, EPI_ISL_1821676, EPI_ISL_1821677, EPI_ISL_1821678, EPI_ISL_1821679, EPI_ISL_1821680, EPI_ISL_1821681, EPI_ISL_1821682, EPI_ISL_1821683, EPI_ISL_1821684, EPI_ISL_1821685, EPI_ISL_1821686, EPI_ISL_1821687, EPI_ISL_1821688, EPI_ISL_1821689, EPI_ISL_1821690, EPI_ISL_1821691, EPI_ISL_1821692, EPI_ISL_1821693, EPI_ISL_1821694, EPI_ISL_1821695, EPI_ISL_1821696, EPI_ISL_1821703, EPI_ISL_1821704, EPI_ISL_1821705, EPI_ISL_1821706, EPI_ISL_1821707, EPI_ISL_1821708, EPI_ISL_1821709, EPI_ISL_1821710, EPI_ISL_1821711, EPI_ISL_1821712, EPI_ISL_1821713, EPI_ISL_1821714, EPI_ISL_1821715, EPI_ISL_1821716, EPI_ISL_1821717, EPI_ISL_1821718, EPI_ISL_1821719, EPI_ISL_1821720, EPI_ISL_1821721 | | | |
| see above | UTMG Pathology LLC | UTMG Pathology LLC | Parasakthy Kumaravelu, Timothy Hodge, Kelsey Matande, Tiara Hymon, Vickie Baselski, Abdallah Azouz |
| EPI_ISL_1821777, EPI_ISL_1821778, EPI_ISL_1821779, EPI_ISL_1821780, EPI_ISL_1821781, EPI_ISL_1821782, EPI_ISL_1821783, EPI_ISL_1821784, EPI_ISL_1821785, EPI_ISL_1821786, EPI_ISL_1821787, EPI_ISL_1821788, EPI_ISL_1821790, EPI_ISL_1821791, EPI_ISL_1821792, EPI_ISL_1821793 | | | |
| see above | Wyoming Public Health Laboratory | Wyoming Public Health Laboratory | Jim Mildenberger, Wanda Manley, Noah Hull, Taylor Fearing, Lynette Gumbleton, Channing Weber, Ashley Norberg, Chayse Rowley, Marley Goetz, Brian Dominguez, Elliot Thomasson, Sam Britz, Cari Sloma, and Rob Christensen |
| EPI_ISL_1821886, EPI_ISL_1821887, EPI_ISL_1821888, EPI_ISL_1821889, EPI_ISL_1821890, EPI_ISL_1821891, EPI_ISL_1821892, EPI_ISL_1821893, EPI_ISL_1821894, EPI_ISL_1821895, EPI_ISL_1821896, EPI_ISL_1821897, EPI_ISL_1821898, EPI_ISL_1821899, EPI_ISL_1821900, EPI_ISL_1821901, EPI_ISL_1821902, EPI_ISL_1821903, EPI_ISL_1821904, EPI_ISL_1821905, EPI_ISL_1821906, EPI_ISL_1821907, EPI_ISL_1821908, EPI_ISL_1821909, EPI_ISL_1821910, EPI_ISL_1821911, EPI_ISL_1821912, EPI_ISL_1821913, EPI_ISL_1821914, EPI_ISL_1821915, EPI_ISL_1821916, EPI_ISL_1821917, EPI_ISL_1821918 | | | |
| see above | Alaska State Virology Laboratory | Alaska State Virology Laboratory | Stephanie DeRonde, Elva House, Jacob Zidek, Lisa Smith, Ph.D., Jack Chen, Ph.D. |
| EPI_ISL_1821919, EPI_ISL_1821921, EPI_ISL_1821925, EPI_ISL_1821927, EPI_ISL_1821929, EPI_ISL_1821931, EPI_ISL_1821932, EPI_ISL_1821934, EPI_ISL_1821935, EPI_ISL_1821936, EPI_ISL_1821938, EPI_ISL_1821939, EPI_ISL_1821940, EPI_ISL_1821941, EPI_ISL_1821942, EPI_ISL_1821943, EPI_ISL_1821944, EPI_ISL_1821950, EPI_ISL_1821952, EPI_ISL_1821953, EPI_ISL_1821958, EPI_ISL_1821959, EPI_ISL_1821960, EPI_ISL_1821961, EPI_ISL_1821962, EPI_ISL_1821963, EPI_ISL_1821964, EPI_ISL_1821965, EPI_ISL_1821966, EPI_ISL_1821967, EPI_ISL_1821968, EPI_ISL_1821969, EPI_ISL_1821970, EPI_ISL_1821971, EPI_ISL_1821972, EPI_ISL_1821973, EPI_ISL_1821974, EPI_ISL_1821975, EPI_ISL_1821976, EPI_ISL_1821977, EPI_ISL_1821978, EPI_ISL_1821979, EPI_ISL_1821980 | | | |
| see above | Texas Children's Hospital | Texas Children's Microbiome Center | Ruth Ann Luna, Jennifer K. Spinler, James Dunn, James Versalovic, Ila Singh |

| | | | |
|---|---|---|---|
| EPI_ISL_1821981, EPI_ISL_1821982, EPI_ISL_1821983, EPI_ISL_1821984, EPI_ISL_1821985, EPI_ISL_1821986, EPI_ISL_1821988, EPI_ISL_1821989, EPI_ISL_1821990, EPI_ISL_1821991, EPI_ISL_1821992, EPI_ISL_1821993, EPI_ISL_1821994, EPI_ISL_1821995, EPI_ISL_1821996, EPI_ISL_1821997, EPI_ISL_1821998, EPI_ISL_1821999, EPI_ISL_1822000, EPI_ISL_1822001, EPI_ISL_1822002, EPI_ISL_1822003, EPI_ISL_1822004, EPI_ISL_1822005, EPI_ISL_1822006, EPI_ISL_1822007, EPI_ISL_1822008, EPI_ISL_1822009, EPI_ISL_1822010, EPI_ISL_1822011, EPI_ISL_1822012, EPI_ISL_1822013, EPI_ISL_1822014, EPI_ISL_1822015, EPI_ISL_1822016, EPI_ISL_1822018, EPI_ISL_1822020, EPI_ISL_1822022, EPI_ISL_1822023, EPI_ISL_1822024, EPI_ISL_1822025, EPI_ISL_1822026, EPI_ISL_1822027, EPI_ISL_1822028, EPI_ISL_1822030, EPI_ISL_1822031, EPI_ISL_1822032, EPI_ISL_1822033, EPI_ISL_1822034, EPI_ISL_1822035, EPI_ISL_1822036, EPI_ISL_1822037, EPI_ISL_1822038, EPI_ISL_1822039, EPI_ISL_1822040, EPI_ISL_1822041, EPI_ISL_1822042, EPI_ISL_1822043, EPI_ISL_1822044, EPI_ISL_1822045, EPI_ISL_1822046, EPI_ISL_1822047, EPI_ISL_1822049, EPI_ISL_1822050, EPI_ISL_1822052, EPI_ISL_1822053, EPI_ISL_1822054, EPI_ISL_1822056, EPI_ISL_1822057, EPI_ISL_1822058, EPI_ISL_1822059, EPI_ISL_1822060, EPI_ISL_1822061, EPI_ISL_1822062, EPI_ISL_1822063, EPI_ISL_1822064, EPI_ISL_1822065, EPI_ISL_1822072, EPI_ISL_1822074, EPI_ISL_1822075, EPI_ISL_1822076, EPI_ISL_1822077, EPI_ISL_1822078, EPI_ISL_1822079, EPI_ISL_1822080, EPI_ISL_1822081, EPI_ISL_1822082, EPI_ISL_1822083, EPI_ISL_1822084 | | | |
| see above | University of Wisconsin-Madison AIDS Vaccine Research Laboratories | University of Wisconsin-Madison AIDS Vaccine Research Laboratories | Gage Moreno, Katarina Braun, et al. AIDS Vaccine Research Laboratories |
| EPI_ISL_1822085, EPI_ISL_1822086, EPI_ISL_1822087, EPI_ISL_1822088, EPI_ISL_1822089, EPI_ISL_1822090, EPI_ISL_1822091 | North Carolina State Laboratory of Public Health (NCSLPH) | North Carolina State Laboratory of Public Health (NCSLPH) | Chase K, Miller MC, Greene S, Glover W |
| EPI_ISL_1822092, EPI_ISL_1822093, EPI_ISL_1822094, EPI_ISL_1822095, EPI_ISL_1822096, EPI_ISL_1822097, EPI_ISL_1822098, EPI_ISL_1822099, EPI_ISL_1822100, EPI_ISL_1822101, EPI_ISL_1822102, EPI_ISL_1822103, EPI_ISL_1822104, EPI_ISL_1822105, EPI_ISL_1822106, EPI_ISL_1822107, EPI_ISL_1822108, EPI_ISL_1822109, EPI_ISL_1822110, EPI_ISL_1822111, EPI_ISL_1822112, EPI_ISL_1822113, EPI_ISL_1822114, EPI_ISL_1822115, EPI_ISL_1822116, EPI_ISL_1822117, EPI_ISL_1822118, EPI_ISL_1822119, EPI_ISL_1822120, EPI_ISL_1822121, EPI_ISL_1822122, EPI_ISL_1822123, EPI_ISL_1822124, EPI_ISL_1822125, EPI_ISL_1822126, EPI_ISL_1822127, EPI_ISL_1822128, EPI_ISL_1822129, EPI_ISL_1822130, EPI_ISL_1822131 | | | |
| see above | Virginia Division of Consolidated Laboratory Services | Virginia Division of Consolidated Laboratory Services | Virginia DCLS |
| EPI_ISL_1822600 | Baylor Scott & White-Irving | Baylor Scott & White - Temple | Ari Rao, Linden Morales, Kimberly Walker, Marcus Volz, Shelby Hendrickson |
| EPI_ISL_1822612, EPI_ISL_1822613, EPI_ISL_1822614, EPI_ISL_1822615, EPI_ISL_1822618, EPI_ISL_1822620, EPI_ISL_1822621, EPI_ISL_1822622, EPI_ISL_1822723, EPI_ISL_1822728, EPI_ISL_1822729, EPI_ISL_1822735, EPI_ISL_1822736 | | | |
| see above | Nevada State Public Health Laboratory | Nevada State Public Health Laboratory | Andrew Gorzalski, Mark Pandori |
| EPI_ISL_1822785, EPI_ISL_1822786 | Northwest Laboratories | Seattle Flu Study | Deborah A. Nickerson, Chris D. Frazar, Jover Lee, Benjamin Pelle, Erica Ryke, Matthew Richardson, Amanda Adler, Elisabeth Brandstetter, Peter D. Han, Kairsten Fay, Misja Ilcisin, Kirsten Lacombe, Thomas R. Sibley, Melissa Truong, Caitlin R. Wolf, Romesh Gautom, Geoff Melly, Brian Hiatt, Philip Dykema, Scott Lindquist, Michael Boeckh, Janet A. Englund, Michael Famulare, Barry R. Lutz, Mark J. Rieder, Lea M. Starita, Matthew Thompson, Helen Y. Chu, Jay Shendure, Trevor Bedford |
| EPI_ISL_1822787 | Atlas Genomics | Seattle Flu Study | Deborah A. Nickerson, Chris D. Frazar, Jover Lee, Benjamin Pelle, Erica Ryke, Matthew Richardson, Amanda Adler, Elisabeth Brandstetter, Peter D. Han, Kairsten Fay, Misja Ilcisin, Kirsten Lacombe, Thomas R. Sibley, Melissa Truong, Caitlin R. Wolf, Romesh Gautom, Geoff Melly, Brian Hiatt, Philip Dykema, Scott Lindquist, Michael Boeckh, Janet A. Englund, Michael Famulare, Barry R. Lutz, Mark J. Rieder, Lea M. Starita, Matthew Thompson, Helen Y. Chu, Jay Shendure, Trevor Bedford |
| EPI_ISL_1822788 | Northwest Laboratories | Seattle Flu Study | Deborah A. Nickerson, Chris D. Frazar, Jover Lee, Benjamin Pelle, Erica Ryke, Matthew Richardson, Amanda Adler, Elisabeth Brandstetter, Peter D. Han, Kairsten Fay, Misja Ilcisin, Kirsten Lacombe, Thomas R. Sibley, Melissa Truong, Caitlin R. Wolf, Romesh Gautom, Geoff Melly, Brian Hiatt, Philip Dykema, Scott Lindquist, Michael Boeckh, Janet A. Englund, Michael Famulare, Barry R. Lutz, Mark J. Rieder, Lea M. Starita, Matthew Thompson, Helen Y. Chu, Jay Shendure, Trevor Bedford |
| EPI_ISL_1822789 | Atlas Genomics | Seattle Flu Study | Deborah A. Nickerson, Chris D. Frazar, Jover Lee, Benjamin Pelle, Erica Ryke, Matthew Richardson, Amanda Adler, Elisabeth Brandstetter, Peter D. Han, Kairsten Fay, Misja Ilcisin, Kirsten Lacombe, Thomas R. Sibley, Melissa Truong, Caitlin R. Wolf, Romesh Gautom, Geoff Melly, Brian Hiatt, Philip Dykema, Scott Lindquist, Michael Boeckh, Janet A. Englund, Michael Famulare, Barry R. Lutz, Mark J. Rieder, Lea M. Starita, Matthew Thompson, Helen Y. Chu, Jay Shendure, Trevor Bedford |
| EPI_ISL_1822790 | Northwest Laboratories | Seattle Flu Study | Deborah A. Nickerson, Chris D. Frazar, Jover Lee, Benjamin Pelle, Erica Ryke, Matthew Richardson, Amanda Adler, Elisabeth Brandstetter, Peter D. Han, Kairsten Fay, Misja Ilcisin, Kirsten Lacombe, Thomas R. Sibley, Melissa Truong, Caitlin R. Wolf, Romesh Gautom, Geoff Melly, Brian Hiatt, Philip Dykema, Scott Lindquist, Michael Boeckh, Janet A. Englund, Michael Famulare, Barry R. Lutz, Mark J. Rieder, Lea M. Starita, Matthew Thompson, Helen Y. Chu, Jay Shendure, Trevor Bedford |
| EPI_ISL_1822791 | Incyte Diagnostics Spokane | Seattle Flu Study | Deborah A. Nickerson, Chris D. Frazar, Jover Lee, Benjamin Pelle, Erica Ryke, Matthew Richardson, Amanda Adler, Elisabeth Brandstetter, Peter D. Han, Kairsten Fay, Misja Ilcisin, Kirsten Lacombe, Thomas R. Sibley, Melissa Truong, Caitlin R. Wolf, Romesh Gautom, Geoff Melly, Brian Hiatt, Philip Dykema, Scott Lindquist, Michael Boeckh, Janet A. Englund, Michael Famulare, Barry R. Lutz, Mark J. Rieder, Lea M. Starita, Matthew Thompson, Helen Y. Chu, Jay Shendure, Trevor Bedford |
| EPI_ISL_1822792 | Altius Institute | Seattle Flu Study | Deborah A. Nickerson, Chris D. Frazar, Jover Lee, Benjamin Pelle, Erica Ryke, Matthew Richardson, Amanda Adler, Elisabeth Brandstetter, Peter D. Han, Kairsten Fay, Misja Ilcisin, Kirsten Lacombe, Thomas R. Sibley, Melissa Truong, Caitlin R. Wolf, Ryan Alexander, Daniel Bates, Rebecca Bruders, Stephanie DeBaun, Clem Green, Muhammad Halimun, Jessica Halow, Kneshay Harper, Matt Hartman, Andrew Meuser, Alex Nguyen, Truong Nguyen, Sofia Olsson, Sadie Patraw, Hannah Petersen, Tobias Ragoczy, Joshua Richards, Jacob Rodriguez, John Stamatoyannopoulos, Julia Wald, Olivia Waltner, Michael Boeckh, Janet A. Englund, Michael Famulare, Barry R. Lutz, Mark J. Rieder, Lea M. Starita, Matthew Thompson, Helen Y. Chu, Jay Shendure, Trevor Bedford |
| EPI_ISL_1822793 | Northwest Laboratories | Seattle Flu Study | Deborah A. Nickerson, Chris D. Frazar, Jover Lee, Benjamin Pelle, Erica Ryke, Matthew Richardson, Amanda Adler, Elisabeth Brandstetter, Peter D. Han, Kairsten Fay, Misja Ilcisin, Kirsten Lacombe, Thomas R. Sibley, Melissa Truong, Caitlin R. Wolf, Romesh Gautom, Geoff Melly, Brian Hiatt, Philip Dykema, Scott Lindquist, Michael Boeckh, Janet A. Englund, Michael Famulare, Barry R. Lutz, Mark J. Rieder, Lea M. Starita, Matthew Thompson, Helen Y. Chu, Jay Shendure, Trevor Bedford |
| EPI_ISL_1822794 | Atlas Genomics | Seattle Flu Study | Deborah A. Nickerson, Chris D. Frazar, Jover Lee, Benjamin Pelle, Erica Ryke, Matthew Richardson, Amanda Adler, Elisabeth Brandstetter, Peter D. Han, Kairsten Fay, Misja Ilcisin, Kirsten Lacombe, Thomas R. Sibley, Melissa Truong, Caitlin R. Wolf, Romesh Gautom, Geoff Melly, Brian Hiatt, Philip Dykema, Scott Lindquist, Michael Boeckh, Janet A. Englund, Michael Famulare, Barry R. Lutz, Mark J. Rieder, Lea M. Starita, Matthew Thompson, Helen Y. Chu, Jay Shendure, Trevor Bedford |
| EPI_ISL_1822795 | Northwest Laboratories | Seattle Flu Study | Deborah A. Nickerson, Chris D. Frazar, Jover Lee, Benjamin Pelle, Erica Ryke, Matthew Richardson, Amanda Adler, Elisabeth Brandstetter, Peter D. Han, Kairsten Fay, Misja Ilcisin, Kirsten Lacombe, Thomas R. Sibley, Melissa Truong, Caitlin R. Wolf, Romesh Gautom, Geoff Melly, Brian Hiatt, Philip Dykema, Scott Lindquist, Michael Boeckh, Janet A. Englund, Michael Famulare, Barry R. Lutz, Mark J. Rieder, Lea M. Starita, Matthew Thompson, Helen Y. Chu, Jay Shendure, Trevor Bedford |
| EPI_ISL_1822796, EPI_ISL_1822797, EPI_ISL_1822798, EPI_ISL_1822799 | Altius Institute | Seattle Flu Study | Deborah A. Nickerson, Chris D. Frazar, Jover Lee, Benjamin Pelle, Erica Ryke, Matthew Richardson, Amanda Adler, Elisabeth Brandstetter, Peter D. Han, Kairsten Fay, Misja Ilcisin, Kirsten Lacombe, Thomas R. Sibley, Melissa Truong, Caitlin R. Wolf, Ryan Alexander, Daniel Bates, Rebecca Bruders, Stephanie DeBaun, Clem Green, Muhammad Halimun, Jessica Halow, Kneshay Harper, Matt Hartman, Andrew Meuser, Alex Nguyen, Truong Nguyen, Sofia Olsson, Sadie Patraw, Hannah Petersen, Tobias Ragoczy, Joshua Richards, Jacob Rodriguez, John Stamatoyannopoulos, Julia Wald, Olivia Waltner, Michael Boeckh, Janet A. Englund, Michael Famulare, Barry R. Lutz, Mark J. Rieder, Lea M. Starita, Matthew Thompson, Helen Y. Chu, Jay Shendure, Trevor Bedford |
| EPI_ISL_1822800 | Atlas Genomics | Seattle Flu Study | Deborah A. Nickerson, Chris D. Frazar, Jover Lee, Benjamin Pelle, Erica Ryke, Matthew Richardson, Amanda Adler, Elisabeth Brandstetter, Peter D. Han, Kairsten Fay, Misja Ilcisin, Kirsten Lacombe, Thomas R. Sibley, Melissa Truong, Caitlin R. Wolf, Romesh Gautom, Geoff Melly, Brian Hiatt, Philip Dykema, Scott Lindquist, Michael Boeckh, Janet A. Englund, Michael Famulare, Barry R. Lutz, Mark J. Rieder, Lea M. Starita, Matthew Thompson, Helen Y. Chu, Jay Shendure, Trevor Bedford |
| EPI_ISL_1822801 | Northwest Laboratories | Seattle Flu Study | Deborah A. Nickerson, Chris D. Frazar, Jover Lee, Benjamin Pelle, Erica Ryke, Matthew Richardson, Amanda Adler, Elisabeth Brandstetter, Peter D. Han, Kairsten Fay, Misja Ilcisin, Kirsten Lacombe, Thomas R. Sibley, Melissa Truong, Caitlin R. Wolf, Romesh Gautom, Geoff Melly, Brian Hiatt, Philip Dykema, Scott Lindquist, Michael Boeckh, Janet A. Englund, Michael Famulare, Barry R. Lutz, Mark J. Rieder, Lea M. Starita, Matthew Thompson, Helen Y. Chu, Jay Shendure, Trevor Bedford |
| EPI_ISL_1822802, EPI_ISL_1822803, EPI_ISL_1822804 | Atlas Genomics | Seattle Flu Study | Deborah A. Nickerson, Chris D. Frazar, Jover Lee, Benjamin Pelle, Erica Ryke, Matthew Richardson, Amanda Adler, Elisabeth Brandstetter, Peter D. Han, Kairsten Fay, Misja Ilcisin, Kirsten Lacombe, Thomas R. Sibley, Melissa Truong, Caitlin R. Wolf, Romesh Gautom, Geoff Melly, Brian Hiatt, Philip Dykema, Scott Lindquist, Michael Boeckh, Janet A. Englund, Michael Famulare, Barry R. Lutz, Mark J. Rieder, Lea M. Starita, Matthew Thompson, Helen Y. Chu, Jay Shendure, Trevor Bedford |
| EPI_ISL_1822805, EPI_ISL_1822806 | Altius Institute | Seattle Flu Study | Deborah A. Nickerson, Chris D. Frazar, Jover Lee, Benjamin Pelle, Erica Ryke, Matthew Richardson, Amanda Adler, Elisabeth Brandstetter, Peter D. Han, Kairsten Fay, Misja Ilcisin, Kirsten Lacombe, Thomas R. Sibley, Melissa Truong, Caitlin R. Wolf, Ryan Alexander, Daniel Bates, Rebecca Bruders, Stephanie DeBaun, Clem Green, Muhammad Halimun, Jessica Halow, Kneshay Harper, Matt Hartman, Andrew Meuser, Alex Nguyen, Truong Nguyen, Sofia Olsson, Sadie Patraw, Hannah Petersen, Tobias Ragoczy, Joshua Richards, Jacob Rodriguez, John Stamatoyannopoulos, Julia Wald, Olivia Waltner, Michael Boeckh, Janet A. Englund, Michael Famulare, Barry R. Lutz, Mark J. Rieder, Lea M. Starita, Matthew Thompson, Helen Y. Chu, Jay Shendure, Trevor Bedford |
| EPI_ISL_1822807 | Northwest Laboratories | Seattle Flu Study | Deborah A. Nickerson, Chris D. Frazar, Jover Lee, Benjamin Pelle, Erica Ryke, Matthew Richardson, Amanda Adler, Elisabeth Brandstetter, Peter D. Han, Kairsten Fay, Misja Ilcisin, Kirsten Lacombe, Thomas R. Sibley, Melissa Truong, Caitlin R. Wolf, Romesh Gautom, Geoff Melly, Brian Hiatt, Philip Dykema, Scott Lindquist, Michael Boeckh, Janet A. Englund, Michael Famulare, Barry R. Lutz, Mark J. Rieder, Lea M. Starita, Matthew Thompson, Helen Y. Chu, Jay Shendure, Trevor Bedford |
| EPI_ISL_1822808, EPI_ISL_1822809, EPI_ISL_1822810, EPI_ISL_1822811, EPI_ISL_1822812, EPI_ISL_1822813, EPI_ISL_1822814, EPI_ISL_1822815 | Altius Institute | Seattle Flu Study | Deborah A. Nickerson, Chris D. Frazar, Jover Lee, Benjamin Pelle, Erica Ryke, Matthew Richardson, Amanda Adler, Elisabeth Brandstetter, Peter D. Han, Kairsten Fay, Misja Ilcisin, Kirsten Lacombe, Thomas R. Sibley, Melissa Truong, Caitlin R. Wolf, Ryan Alexander, Daniel Bates, Rebecca Bruders, Stephanie DeBaun, Clem Green, Muhammad Halimun, Jessica Halow, Kneshay Harper, Matt Hartman, Andrew Meuser, Alex Nguyen, Truong Nguyen, Sofia Olsson, Sadie Patraw, Hannah Petersen, Tobias Ragoczy, Joshua Richards, Jacob Rodriguez, John Stamatoyannopoulos, Julia Wald, Olivia Waltner, Michael Boeckh, Janet A. Englund, Michael Famulare, Barry R. Lutz, Mark J. Rieder, Lea M. Starita, Matthew Thompson, Helen Y. Chu, Jay Shendure, Trevor Bedford |
| EPI_ISL_1822816, EPI_ISL_1822817 | Atlas Genomics | Seattle Flu Study | Deborah A. Nickerson, Chris D. Frazar, Jover Lee, Benjamin Pelle, Erica Ryke, Matthew Richardson, Amanda Adler, Elisabeth Brandstetter, Peter D. Han, Kairsten Fay, Misja Ilcisin, Kirsten Lacombe, Thomas R. Sibley, Melissa Truong, Caitlin R. Wolf, Romesh Gautom, Geoff Melly, Brian Hiatt, Philip Dykema, Scott Lindquist, Michael Boeckh, Janet A. Englund, Michael Famulare, Barry R. Lutz, Mark J. Rieder, Lea M. Starita, Matthew Thompson, Helen Y. Chu, Jay Shendure, Trevor Bedford |
| EPI_ISL_1822818, EPI_ISL_1822819 | Altius Institute | Seattle Flu Study | Deborah A. Nickerson, Chris D. Frazar, Jover Lee, Benjamin Pelle, Erica Ryke, Matthew Richardson, Amanda Adler, Elisabeth Brandstetter, Peter D. Han, Kairsten Fay, Misja Ilcisin, Kirsten Lacombe, Thomas R. Sibley, Melissa Truong, Caitlin R. Wolf, Ryan Alexander, Daniel Bates, Rebecca Bruders, Stephanie DeBaun, Clem Green, Muhammad Halimun, Jessica Halow, Kneshay Harper, Matt Hartman, Andrew Meuser, Alex Nguyen, Truong Nguyen, Sofia Olsson, Sadie Patraw, Hannah Petersen, Tobias Ragoczy, Joshua Richards, Jacob Rodriguez, John Stamatoyannopoulos, Julia Wald, Olivia Waltner, Michael Boeckh, Janet A. Englund, Michael Famulare, Barry R. Lutz, Mark J. Rieder, Lea M. Starita, Matthew Thompson, Helen Y. Chu, Jay Shendure, Trevor Bedford |
| EPI_ISL_1822820 | Atlas Genomics | Seattle Flu Study | Deborah A. Nickerson, Chris D. Frazar, Jover Lee, Benjamin Pelle, Erica Ryke, Matthew Richardson, Amanda Adler, Elisabeth Brandstetter, Peter D. Han, Kairsten Fay, Misja Ilcisin, Kirsten Lacombe, Thomas R. Sibley, Melissa Truong, Caitlin R. Wolf, Romesh Gautom, Geoff Melly, Brian Hiatt, Philip Dykema, Scott Lindquist, Michael Boeckh, Janet A. Englund, Michael Famulare, Barry R. Lutz, Mark J. Rieder, Lea M. Starita, Matthew Thompson, Helen Y. Chu, Jay Shendure, Trevor Bedford |
| EPI_ISL_1822821, EPI_ISL_1822822, EPI_ISL_1822823 | Altius Institute | Seattle Flu Study | Deborah A. Nickerson, Chris D. Frazar, Jover Lee, Benjamin Pelle, Erica Ryke, Matthew Richardson, Amanda Adler, Elisabeth Brandstetter, Peter D. Han, Kairsten Fay, Misja Ilcisin, Kirsten Lacombe, Thomas R. Sibley, Melissa Truong, Caitlin R. Wolf, Ryan Alexander, Daniel Bates, Rebecca Bruders, Stephanie DeBaun, Clem Green, Muhammad Halimun, Jessica Halow, Kneshay Harper, Matt Hartman, Andrew Meuser, Alex Nguyen, Truong Nguyen, Sofia Olsson, Sadie Patraw, Hannah Petersen, Tobias Ragoczy, Joshua Richards, Jacob Rodriguez, John Stamatoyannopoulos, Julia Wald, Olivia Waltner, Michael Boeckh, Janet A. Englund, Michael Famulare, Barry R. Lutz, Mark J. Rieder, Lea M. Starita, Matthew Thompson, Helen Y. Chu, Jay Shendure, Trevor Bedford |
| EPI_ISL_1822824 | Northwest Laboratories | Seattle Flu Study | Deborah A. Nickerson, Chris D. Frazar, Jover Lee, Benjamin Pelle, Erica Ryke, Matthew Richardson, Amanda Adler, Elisabeth Brandstetter, Peter D. Han, Kairsten Fay, Misja Ilcisin, Kirsten Lacombe, Thomas R. Sibley, Melissa Truong, Caitlin R. Wolf, Romesh Gautom, Geoff Melly, Brian Hiatt, Philip Dykema, Scott Lindquist, Michael Boeckh, Janet A. Englund, Michael Famulare, Barry R. Lutz, Mark J. Rieder, Lea M. Starita, Matthew Thompson, Helen Y. Chu, Jay Shendure, Trevor Bedford |
| EPI_ISL_1822825 | Altius Institute | Seattle Flu Study | Deborah A. Nickerson, Chris D. Frazar, Jover Lee, Benjamin Pelle, Erica Ryke, Matthew Richardson, Amanda Adler, Elisabeth Brandstetter, Peter D. Han, Kairsten Fay, Misja Ilcisin, Kirsten Lacombe, Thomas R. Sibley, Melissa Truong, Caitlin R. Wolf, Ryan Alexander, Daniel Bates, Rebecca Bruders, Stephanie DeBaun, Clem Green, Muhammad Halimun, Jessica Halow, Kneshay Harper, Matt Hartman, Andrew Meuser, Alex Nguyen, Truong Nguyen, Sofia Olsson, Sadie Patraw, Hannah Petersen, Tobias Ragoczy, Joshua Richards, Jacob Rodriguez, John Stamatoyannopoulos, Julia Wald, Olivia Waltner, Michael Boeckh, Janet A. Englund, Michael Famulare, Barry R. Lutz, Mark J. Rieder, Lea M. Starita, Matthew Thompson, Helen Y. Chu, Jay Shendure, Trevor Bedford |
| EPI_ISL_1822826 | Northwest Laboratories | Seattle Flu Study | Deborah A. Nickerson, Chris D. Frazar, Jover Lee, Benjamin Pelle, Erica Ryke, Matthew Richardson, Amanda Adler, Elisabeth Brandstetter, Peter D. Han, Kairsten Fay, Misja Ilcisin, Kirsten Lacombe, Thomas R. Sibley, Melissa Truong, Caitlin R. Wolf, Romesh Gautom, Geoff Melly, Brian Hiatt, Philip Dykema, Scott Lindquist, Michael Boeckh, Janet A. Englund, Michael Famulare, Barry R. Lutz, Mark J. Rieder, Lea M. Starita, Matthew Thompson, Helen Y. Chu, Jay Shendure, Trevor Bedford |
| EPI_ISL_1822827, EPI_ISL_1822828, EPI_ISL_1822829, EPI_ISL_1822830, EPI_ISL_1822831, EPI_ISL_1822832 | Altius Institute | Seattle Flu Study | Deborah A. Nickerson, Chris D. Frazar, Jover Lee, Benjamin Pelle, Erica Ryke, Matthew Richardson, Amanda Adler, Elisabeth Brandstetter, Peter D. Han, Kairsten Fay, Misja Ilcisin, Kirsten Lacombe, Thomas R. Sibley, Melissa Truong, Caitlin R. Wolf, Ryan Alexander, Daniel Bates, Rebecca Bruders, Stephanie DeBaun, Clem Green, Muhammad Halimun, Jessica Halow, Kneshay Harper, Matt Hartman, Andrew Meuser, Alex Nguyen, Truong Nguyen, Sofia Olsson, Sadie Patraw, Hannah Petersen, Tobias Ragoczy, Joshua Richards, Jacob Rodriguez, John Stamatoyannopoulos, Julia Wald, Olivia Waltner, Michael Boeckh, Janet A. Englund, Michael Famulare, Barry R. Lutz, Mark J. Rieder, Lea M. Starita, Matthew Thompson, Helen Y. Chu, Jay Shendure, Trevor Bedford |
| EPI_ISL_1822833, EPI_ISL_1822834 | Northwest Laboratories | Seattle Flu Study | Deborah A. Nickerson, Chris D. Frazar, Jover Lee, Benjamin Pelle, Erica Ryke, Matthew Richardson, Amanda Adler, Elisabeth Brandstetter, Peter D. Han, Kairsten Fay, Misja Ilcisin, Kirsten Lacombe, Thomas R. Sibley, Melissa Truong, Caitlin R. Wolf, Romesh Gautom, Geoff Melly, Brian Hiatt, Philip Dykema, Scott Lindquist, Michael Boeckh, Janet A. Englund, Michael Famulare, Barry R. Lutz, Mark J. Rieder, Lea M. Starita, Matthew Thompson, Helen Y. Chu, Jay Shendure, Trevor Bedford |
| EPI_ISL_1822835 | Altius Institute | Seattle Flu Study | Deborah A. Nickerson, Chris D. Frazar, Jover Lee, Benjamin Pelle, Erica Ryke, Matthew Richardson, Amanda Adler, Elisabeth Brandstetter, Peter D. Han, Kairsten Fay, Misja Ilcisin, Kirsten Lacombe, Thomas R. Sibley, Melissa Truong, Caitlin R. Wolf, Ryan Alexander, Daniel Bates, Rebecca Bruders, Stephanie DeBaun, Clem Green, Muhammad Halimun, Jessica Halow, Kneshay Harper, Matt Hartman, Andrew Meuser, Alex Nguyen, Truong Nguyen, Sofia Olsson, Sadie Patraw, Hannah Petersen, Tobias Ragoczy, Joshua Richards, Jacob Rodriguez, John Stamatoyannopoulos, Julia Wald, Olivia Waltner, Michael Boeckh, Janet A. Englund, Michael Famulare, Barry R. Lutz, Mark J. Rieder, Lea M. Starita, Matthew Thompson, Helen Y. Chu, Jay Shendure, Trevor Bedford |
| EPI_ISL_1822836 | Northwest Laboratories | Seattle Flu Study | Deborah A. Nickerson, Chris D. Frazar, Jover Lee, Benjamin Pelle, Erica Ryke, Matthew Richardson, Amanda Adler, Elisabeth Brandstetter, Peter D. Han, Kairsten Fay, Misja Ilcisin, Kirsten Lacombe, Thomas R. Sibley, Melissa Truong, Caitlin R. Wolf, Romesh Gautom, Geoff Melly, Brian |

| | | | |
|---|---|---|---|
| | | | Hiatt, Philip Dykema, Scott Lindquist, Michael Boeckh, Janet A. Englund, Michael Famulare, Barry R. Lutz, Mark J. Rieder, Lea M. Starita, Matthew Thompson, Helen Y. Chu, Jay Shendure, Trevor Bedford |
| EPI_ISL_1822837 | Altius Institute | Seattle Flu Study | Deborah A. Nickerson, Chris D. Frazar, Jover Lee, Benjamin Pelle, Erica Ryke, Matthew Richardson, Amanda Adler, Elisabeth Brandstetter, Peter D. Han, Kairsten Fay, Misja Ilcisin, Kirsten Lacombe, Thomas R. Sibley, Melissa Truong, Caitlin R. Wolf, Ryan Alexander, Daniel Bates, Rebecca Bruders, Stephanie DeBaun, Clem Green, Muhammad Halimun, Jessica Halow, Kneshay Harper, Matt Hartman, Andrew Meuser, Alex Nguyen, Truong Nguyen, Sofia Olsson, Sadie Patraw, Hannah Petersen, Tobias Ragoczy, Joshua Richards, Jacob Rodriguez, John Stamatoyannopoulos, Julia Wald, Olivia Waltner, Michael Boeckh, Janet A. Englund, Michael Famulare, Barry R. Lutz, Mark J. Rieder, Lea M. Starita, Matthew Thompson, Helen Y. Chu, Jay Shendure, Trevor Bedford |
| EPI_ISL_1822838 | Northwest Laboratories | Seattle Flu Study | Deborah A. Nickerson, Chris D. Frazar, Jover Lee, Benjamin Pelle, Erica Ryke, Matthew Richardson, Amanda Adler, Elisabeth Brandstetter, Peter D. Han, Kairsten Fay, Misja Ilcisin, Kirsten Lacombe, Thomas R. Sibley, Melissa Truong, Caitlin R. Wolf, Romesh Gautom, Geoff Melly, Brian Hiatt, Philip Dykema, Scott Lindquist, Michael Boeckh, Janet A. Englund, Michael Famulare, Barry R. Lutz, Mark J. Rieder, Lea M. Starita, Matthew Thompson, Helen Y. Chu, Jay Shendure, Trevor Bedford |
| EPI_ISL_1822839, EPI_ISL_1822840 | Altius Institute | Seattle Flu Study | Deborah A. Nickerson, Chris D. Frazar, Jover Lee, Benjamin Pelle, Erica Ryke, Matthew Richardson, Amanda Adler, Elisabeth Brandstetter, Peter D. Han, Kairsten Fay, Misja Ilcisin, Kirsten Lacombe, Thomas R. Sibley, Melissa Truong, Caitlin R. Wolf, Ryan Alexander, Daniel Bates, Rebecca Bruders, Stephanie DeBaun, Clem Green, Muhammad Halimun, Jessica Halow, Kneshay Harper, Matt Hartman, Andrew Meuser, Alex Nguyen, Truong Nguyen, Sofia Olsson, Sadie Patraw, Hannah Petersen, Tobias Ragoczy, Joshua Richards, Jacob Rodriguez, John Stamatoyannopoulos, Julia Wald, Olivia Waltner, Michael Boeckh, Janet A. Englund, Michael Famulare, Barry R. Lutz, Mark J. Rieder, Lea M. Starita, Matthew Thompson, Helen Y. Chu, Jay Shendure, Trevor Bedford |
| EPI_ISL_1822841, EPI_ISL_1822842, EPI_ISL_1822843 | Atlas Genomics | Seattle Flu Study | Deborah A. Nickerson, Chris D. Frazar, Jover Lee, Benjamin Pelle, Erica Ryke, Matthew Richardson, Amanda Adler, Elisabeth Brandstetter, Peter D. Han, Kairsten Fay, Misja Ilcisin, Kirsten Lacombe, Thomas R. Sibley, Melissa Truong, Caitlin R. Wolf, Romesh Gautom, Geoff Melly, Brian Hiatt, Philip Dykema, Scott Lindquist, Michael Boeckh, Janet A. Englund, Michael Famulare, Barry R. Lutz, Mark J. Rieder, Lea M. Starita, Matthew Thompson, Helen Y. Chu, Jay Shendure, Trevor Bedford |
| EPI_ISL_1822844 | Altius Institute | Seattle Flu Study | Deborah A. Nickerson, Chris D. Frazar, Jover Lee, Benjamin Pelle, Erica Ryke, Matthew Richardson, Amanda Adler, Elisabeth Brandstetter, Peter D. Han, Kairsten Fay, Misja Ilcisin, Kirsten Lacombe, Thomas R. Sibley, Melissa Truong, Caitlin R. Wolf, Ryan Alexander, Daniel Bates, Rebecca Bruders, Stephanie DeBaun, Clem Green, Muhammad Halimun, Jessica Halow, Kneshay Harper, Matt Hartman, Andrew Meuser, Alex Nguyen, Truong Nguyen, Sofia Olsson, Sadie Patraw, Hannah Petersen, Tobias Ragoczy, Joshua Richards, Jacob Rodriguez, John Stamatoyannopoulos, Julia Wald, Olivia Waltner, Michael Boeckh, Janet A. Englund, Michael Famulare, Barry R. Lutz, Mark J. Rieder, Lea M. Starita, Matthew Thompson, Helen Y. Chu, Jay Shendure, Trevor Bedford |
| EPI_ISL_1822845 | Atlas Genomics | Seattle Flu Study | Deborah A. Nickerson, Chris D. Frazar, Jover Lee, Benjamin Pelle, Erica Ryke, Matthew Richardson, Amanda Adler, Elisabeth Brandstetter, Peter D. Han, Kairsten Fay, Misja Ilcisin, Kirsten Lacombe, Thomas R. Sibley, Melissa Truong, Caitlin R. Wolf, Romesh Gautom, Geoff Melly, Brian Hiatt, Philip Dykema, Scott Lindquist, Michael Boeckh, Janet A. Englund, Michael Famulare, Barry R. Lutz, Mark J. Rieder, Lea M. Starita, Matthew Thompson, Helen Y. Chu, Jay Shendure, Trevor Bedford |
| EPI_ISL_1822846 | Altius Institute | Seattle Flu Study | Deborah A. Nickerson, Chris D. Frazar, Jover Lee, Benjamin Pelle, Erica Ryke, Matthew Richardson, Amanda Adler, Elisabeth Brandstetter, Peter D. Han, Kairsten Fay, Misja Ilcisin, Kirsten Lacombe, Thomas R. Sibley, Melissa Truong, Caitlin R. Wolf, Ryan Alexander, Daniel Bates, Rebecca Bruders, Stephanie DeBaun, Clem Green, Muhammad Halimun, Jessica Halow, Kneshay Harper, Matt Hartman, Andrew Meuser, Alex Nguyen, Truong Nguyen, Sofia Olsson, Sadie Patraw, Hannah Petersen, Tobias Ragoczy, Joshua Richards, Jacob Rodriguez, John Stamatoyannopoulos, Julia Wald, Olivia Waltner, Michael Boeckh, Janet A. Englund, Michael Famulare, Barry R. Lutz, Mark J. Rieder, Lea M. Starita, Matthew Thompson, Helen Y. Chu, Jay Shendure, Trevor Bedford |
| EPI_ISL_1822847 | Atlas Genomics | Seattle Flu Study | Deborah A. Nickerson, Chris D. Frazar, Jover Lee, Benjamin Pelle, Erica Ryke, Matthew Richardson, Amanda Adler, Elisabeth Brandstetter, Peter D. Han, Kairsten Fay, Misja Ilcisin, Kirsten Lacombe, Thomas R. Sibley, Melissa Truong, Caitlin R. Wolf, Romesh Gautom, Geoff Melly, Brian Hiatt, Philip Dykema, Scott Lindquist, Michael Boeckh, Janet A. Englund, Michael Famulare, Barry R. Lutz, Mark J. Rieder, Lea M. Starita, Matthew Thompson, Helen Y. Chu, Jay Shendure, Trevor Bedford |
| EPI_ISL_1822848 | Altius Institute | Seattle Flu Study | Deborah A. Nickerson, Chris D. Frazar, Jover Lee, Benjamin Pelle, Erica Ryke, Matthew Richardson, Amanda Adler, Elisabeth Brandstetter, Peter D. Han, Kairsten Fay, Misja Ilcisin, Kirsten Lacombe, Thomas R. Sibley, Melissa Truong, Caitlin R. Wolf, Ryan Alexander, Daniel Bates, Rebecca Bruders, Stephanie DeBaun, Clem Green, Muhammad Halimun, Jessica Halow, Kneshay Harper, Matt Hartman, Andrew Meuser, Alex Nguyen, Truong Nguyen, Sofia Olsson, Sadie Patraw, Hannah Petersen, Tobias Ragoczy, Joshua Richards, Jacob Rodriguez, John Stamatoyannopoulos, Julia Wald, Olivia Waltner, Michael Boeckh, Janet A. Englund, Michael Famulare, Barry R. Lutz, Mark J. Rieder, Lea M. Starita, Matthew Thompson, Helen Y. Chu, Jay Shendure, Trevor Bedford |
| EPI_ISL_1822849 | Atlas Genomics | Seattle Flu Study | Deborah A. Nickerson, Chris D. Frazar, Jover Lee, Benjamin Pelle, Erica Ryke, Matthew Richardson, Amanda Adler, Elisabeth Brandstetter, Peter D. Han, Kairsten Fay, Misja Ilcisin, Kirsten Lacombe, Thomas R. Sibley, Melissa Truong, Caitlin R. Wolf, Romesh Gautom, Geoff Melly, Brian Hiatt, Philip Dykema, Scott Lindquist, Michael Boeckh, Janet A. Englund, Michael Famulare, Barry R. Lutz, Mark J. Rieder, Lea M. Starita, Matthew Thompson, Helen Y. Chu, Jay Shendure, Trevor Bedford |
| EPI_ISL_1822850, EPI_ISL_1822851 | Altius Institute | Seattle Flu Study | Deborah A. Nickerson, Chris D. Frazar, Jover Lee, Benjamin Pelle, Erica Ryke, Matthew Richardson, Amanda Adler, Elisabeth Brandstetter, Peter D. Han, Kairsten Fay, Misja Ilcisin, Kirsten Lacombe, Thomas R. Sibley, Melissa Truong, Caitlin R. Wolf, Ryan Alexander, Daniel Bates, Rebecca Bruders, Stephanie DeBaun, Clem Green, Muhammad Halimun, Jessica Halow, Kneshay Harper, Matt Hartman, Andrew Meuser, Alex Nguyen, Truong Nguyen, Sofia Olsson, Sadie Patraw, Hannah Petersen, Tobias Ragoczy, Joshua Richards, Jacob Rodriguez, John Stamatoyannopoulos, Julia Wald, Olivia Waltner, Michael Boeckh, Janet A. Englund, Michael Famulare, Barry R. Lutz, Mark J. Rieder, Lea M. Starita, Matthew Thompson, Helen Y. Chu, Jay Shendure, Trevor Bedford |
| EPI_ISL_1822852, EPI_ISL_1822853 | Atlas Genomics | Seattle Flu Study | Deborah A. Nickerson, Chris D. Frazar, Jover Lee, Benjamin Pelle, Erica Ryke, Matthew Richardson, Amanda Adler, Elisabeth Brandstetter, Peter D. Han, Kairsten Fay, Misja Ilcisin, Kirsten Lacombe, Thomas R. Sibley, Melissa Truong, Caitlin R. Wolf, Romesh Gautom, Geoff Melly, Brian Hiatt, Philip Dykema, Scott Lindquist, Michael Boeckh, Janet A. Englund, Michael Famulare, Barry R. Lutz, Mark J. Rieder, Lea M. Starita, Matthew Thompson, Helen Y. Chu, Jay Shendure, Trevor Bedford |
| EPI_ISL_1822854, EPI_ISL_1822855 | Altius Institute | Seattle Flu Study | Deborah A. Nickerson, Chris D. Frazar, Jover Lee, Benjamin Pelle, Erica Ryke, Matthew Richardson, Amanda Adler, Elisabeth Brandstetter, Peter D. Han, Kairsten Fay, Misja Ilcisin, Kirsten Lacombe, Thomas R. Sibley, Melissa Truong, Caitlin R. Wolf, Ryan Alexander, Daniel Bates, Rebecca Bruders, Stephanie DeBaun, Clem Green, Muhammad Halimun, Jessica Halow, Kneshay Harper, Matt Hartman, Andrew Meuser, Alex Nguyen, Truong Nguyen, Sofia Olsson, Sadie Patraw, Hannah Petersen, Tobias Ragoczy, Joshua Richards, Jacob Rodriguez, John Stamatoyannopoulos, Julia Wald, Olivia Waltner, Michael Boeckh, Janet A. Englund, Michael Famulare, Barry R. Lutz, Mark J. Rieder, Lea M. Starita, Matthew Thompson, Helen Y. Chu, Jay Shendure, Trevor Bedford |
| EPI_ISL_1822856, EPI_ISL_1822857 | Atlas Genomics | Seattle Flu Study | Deborah A. Nickerson, Chris D. Frazar, Jover Lee, Benjamin Pelle, Erica Ryke, Matthew Richardson, Amanda Adler, Elisabeth Brandstetter, Peter D. Han, Kairsten Fay, Misja Ilcisin, Kirsten Lacombe, Thomas R. Sibley, Melissa Truong, Caitlin R. Wolf, Romesh Gautom, Geoff Melly, Brian Hiatt, Philip Dykema, Scott Lindquist, Michael Boeckh, Janet A. Englund, Michael Famulare, Barry R. Lutz, Mark J. Rieder, Lea M. Starita, Matthew Thompson, Helen Y. Chu, Jay Shendure, Trevor Bedford |
| EPI_ISL_1822858, EPI_ISL_1822859, EPI_ISL_1822860 | Altius Institute | Seattle Flu Study | Deborah A. Nickerson, Chris D. Frazar, Jover Lee, Benjamin Pelle, Erica Ryke, Matthew Richardson, Amanda Adler, Elisabeth Brandstetter, Peter D. Han, Kairsten Fay, Misja Ilcisin, Kirsten Lacombe, Thomas R. Sibley, Melissa Truong, Caitlin R. Wolf, Ryan Alexander, Daniel Bates, Rebecca Bruders, Stephanie DeBaun, Clem Green, Muhammad Halimun, Jessica Halow, Kneshay Harper, Matt Hartman, Andrew Meuser, Alex Nguyen, Truong Nguyen, Sofia Olsson, Sadie Patraw, Hannah Petersen, Tobias Ragoczy, Joshua Richards, Jacob Rodriguez, John Stamatoyannopoulos, Julia Wald, Olivia Waltner, Michael Boeckh, Janet A. Englund, Michael Famulare, Barry R. Lutz, Mark J. Rieder, Lea M. Starita, Matthew Thompson, Helen Y. Chu, Jay Shendure, Trevor Bedford |
| EPI_ISL_1822861 | Atlas Genomics | Seattle Flu Study | Deborah A. Nickerson, Chris D. Frazar, Jover Lee, Benjamin Pelle, Erica Ryke, Matthew Richardson, Amanda Adler, Elisabeth Brandstetter, Peter D. Han, Kairsten Fay, Misja Ilcisin, Kirsten Lacombe, Thomas R. Sibley, Melissa Truong, Caitlin R. Wolf, Romesh Gautom, Geoff Melly, Brian Hiatt, Philip Dykema, Scott Lindquist, Michael Boeckh, Janet A. Englund, Michael Famulare, Barry R. Lutz, Mark J. Rieder, Lea M. Starita, Matthew Thompson, Helen Y. Chu, Jay Shendure, Trevor Bedford |
| EPI_ISL_1822862 | Altius Institute | Seattle Flu Study | Deborah A. Nickerson, Chris D. Frazar, Jover Lee, Benjamin Pelle, Erica Ryke, Matthew Richardson, Amanda Adler, Elisabeth Brandstetter, Peter D. Han, Kairsten Fay, Misja Ilcisin, Kirsten Lacombe, Thomas R. Sibley, Melissa Truong, Caitlin R. Wolf, Ryan Alexander, Daniel Bates, Rebecca Bruders, Stephanie DeBaun, Clem Green, Muhammad Halimun, Jessica Halow, Kneshay Harper, Matt Hartman, Andrew Meuser, Alex Nguyen, Truong Nguyen, Sofia Olsson, Sadie Patraw, Hannah Petersen, Tobias Ragoczy, Joshua Richards, Jacob Rodriguez, John Stamatoyannopoulos, Julia Wald, Olivia Waltner, Michael Boeckh, Janet A. Englund, Michael Famulare, Barry R. Lutz, Mark J. Rieder, Lea M. Starita, Matthew Thompson, Helen Y. Chu, Jay Shendure, Trevor Bedford |
| EPI_ISL_1822863, EPI_ISL_1822864 | Northwest Laboratories | Seattle Flu Study | Deborah A. Nickerson, Chris D. Frazar, Jover Lee, Benjamin Pelle, Erica Ryke, Matthew Richardson, Amanda Adler, Elisabeth Brandstetter, Peter D. Han, Kairsten Fay, Misja Ilcisin, Kirsten Lacombe, Thomas R. Sibley, Melissa Truong, Caitlin R. Wolf, Romesh Gautom, Geoff Melly, Brian Hiatt, Philip Dykema, Scott Lindquist, Michael Boeckh, Janet A. Englund, Michael Famulare, Barry R. Lutz, Mark J. Rieder, Lea M. Starita, Matthew Thompson, Helen Y. Chu, Jay Shendure, Trevor Bedford |
| EPI_ISL_1822865, EPI_ISL_1822866 | Altius Institute | Seattle Flu Study | Deborah A. Nickerson, Chris D. Frazar, Jover Lee, Benjamin Pelle, Erica Ryke, Matthew Richardson, Amanda Adler, Elisabeth Brandstetter, Peter D. Han, Kairsten Fay, Misja Ilcisin, Kirsten Lacombe, Thomas R. Sibley, Melissa Truong, Caitlin R. Wolf, Ryan Alexander, Daniel Bates, Rebecca Bruders, Stephanie DeBaun, Clem Green, Muhammad Halimun, Jessica Halow, Kneshay Harper, Matt Hartman, Andrew Meuser, Alex Nguyen, Truong Nguyen, Sofia Olsson, Sadie Patraw, Hannah Petersen, Tobias Ragoczy, Joshua Richards, Jacob Rodriguez, John Stamatoyannopoulos, Julia Wald, Olivia Waltner, Michael Boeckh, Janet A. Englund, Michael Famulare, Barry R. Lutz, Mark J. Rieder, Lea M. Starita, Matthew Thompson, Helen Y. Chu, Jay Shendure, Trevor Bedford |
| EPI_ISL_1822867 | Northwest Laboratories | Seattle Flu Study | Deborah A. Nickerson, Chris D. Frazar, Jover Lee, Benjamin Pelle, Erica Ryke, Matthew Richardson, Amanda Adler, Elisabeth Brandstetter, Peter D. Han, Kairsten Fay, Misja Ilcisin, Kirsten Lacombe, Thomas R. Sibley, Melissa Truong, Caitlin R. Wolf, Romesh Gautom, Geoff Melly, Brian Hiatt, Philip Dykema, Scott Lindquist, Michael Boeckh, Janet A. Englund, Michael Famulare, Barry R. Lutz, Mark J. Rieder, Lea M. Starita, Matthew Thompson, Helen Y. Chu, Jay Shendure, Trevor Bedford |
| EPI_ISL_1822868, EPI_ISL_1822869 | Altius Institute | Seattle Flu Study | Deborah A. Nickerson, Chris D. Frazar, Jover Lee, Benjamin Pelle, Erica Ryke, Matthew Richardson, Amanda Adler, Elisabeth Brandstetter, Peter D. Han, Kairsten Fay, Misja Ilcisin, Kirsten Lacombe, Thomas R. Sibley, Melissa Truong, Caitlin R. Wolf, Ryan Alexander, Daniel Bates, Rebecca Bruders, Stephanie DeBaun, Clem Green, Muhammad Halimun, Jessica Halow, Kneshay Harper, Matt Hartman, Andrew Meuser, Alex Nguyen, Truong Nguyen, Sofia Olsson, Sadie Patraw, Hannah Petersen, Tobias Ragoczy, Joshua Richards, Jacob Rodriguez, John Stamatoyannopoulos, Julia Wald, Olivia Waltner, Michael Boeckh, Janet A. Englund, Michael Famulare, Barry R. Lutz, Mark J. Rieder, Lea M. Starita, Matthew Thompson, Helen Y. Chu, Jay Shendure, Trevor Bedford |
| EPI_ISL_1822870 | Northwest Laboratories | Seattle Flu Study | Deborah A. Nickerson, Chris D. Frazar, Jover Lee, Benjamin Pelle, Erica Ryke, Matthew Richardson, Amanda Adler, Elisabeth Brandstetter, Peter D. Han, Kairsten Fay, Misja Ilcisin, Kirsten Lacombe, Thomas R. Sibley, Melissa Truong, Caitlin R. Wolf, Romesh Gautom, Geoff Melly, Brian Hiatt, Philip Dykema, Scott Lindquist, Michael Boeckh, Janet A. Englund, Michael Famulare, Barry R. Lutz, Mark J. Rieder, Lea M. Starita, Matthew Thompson, Helen Y. Chu, Jay Shendure, Trevor Bedford |
| EPI_ISL_1822871 | Altius Institute | Seattle Flu Study | Deborah A. Nickerson, Chris D. Frazar, Jover Lee, Benjamin Pelle, Erica Ryke, Matthew Richardson, Amanda Adler, Elisabeth Brandstetter, Peter D. Han, Kairsten Fay, Misja Ilcisin, Kirsten Lacombe, Thomas R. Sibley, Melissa Truong, Caitlin R. Wolf, Ryan Alexander, Daniel Bates, Rebecca Bruders, Stephanie DeBaun, Clem Green, Muhammad Halimun, Jessica Halow, Kneshay Harper, Matt Hartman, Andrew Meuser, Alex Nguyen, Truong Nguyen, Sofia Olsson, Sadie Patraw, Hannah Petersen, Tobias Ragoczy, Joshua Richards, Jacob Rodriguez, John Stamatoyannopoulos, Julia Wald, Olivia Waltner, Michael Boeckh, Janet A. Englund, Michael Famulare, Barry R. Lutz, Mark J. Rieder, Lea M. Starita, Matthew Thompson, Helen Y. Chu, Jay Shendure, Trevor Bedford |
| EPI_ISL_1822872 | Atlas Genomics | Seattle Flu Study | Deborah A. Nickerson, Chris D. Frazar, Jover Lee, Benjamin Pelle, Erica Ryke, Matthew Richardson, Amanda Adler, Elisabeth Brandstetter, Peter D. Han, Kairsten Fay, Misja Ilcisin, Kirsten Lacombe, Thomas R. Sibley, Melissa Truong, Caitlin R. Wolf, Romesh Gautom, Geoff Melly, Brian Hiatt, Philip Dykema, Scott Lindquist, Michael Boeckh, Janet A. Englund, Michael Famulare, Barry R. Lutz, Mark J. Rieder, Lea M. Starita, Matthew Thompson, Helen Y. Chu, Jay Shendure, Trevor Bedford |
| EPI_ISL_1822873, EPI_ISL_1822874 | Northwest Laboratories | Seattle Flu Study | Deborah A. Nickerson, Chris D. Frazar, Jover Lee, Benjamin Pelle, Erica Ryke, Matthew Richardson, Amanda Adler, Elisabeth Brandstetter, Peter D. Han, Kairsten Fay, Misja Ilcisin, Kirsten Lacombe, Thomas R. Sibley, Melissa Truong, Caitlin R. Wolf, Romesh Gautom, Geoff Melly, Brian Hiatt, Philip Dykema, Scott Lindquist, Michael Boeckh, Janet A. Englund, Michael Famulare, Barry R. Lutz, Mark J. Rieder, Lea M. Starita, Matthew Thompson, Helen Y. Chu, Jay Shendure, Trevor Bedford |
| EPI_ISL_1822875 | Altius Institute | Seattle Flu Study | Deborah A. Nickerson, Chris D. Frazar, Jover Lee, Benjamin Pelle, Erica Ryke, Matthew Richardson, Amanda Adler, Elisabeth Brandstetter, Peter D. Han, Kairsten Fay, Misja Ilcisin, Kirsten Lacombe, Thomas R. Sibley, Melissa Truong, Caitlin R. Wolf, Ryan Alexander, Daniel Bates, Rebecca Bruders, Stephanie DeBaun, Clem Green, Muhammad Halimun, Jessica Halow, Kneshay Harper, Matt Hartman, Andrew Meuser, Alex Nguyen, Truong Nguyen, Sofia Olsson, Sadie Patraw, Hannah Petersen, Tobias Ragoczy, Joshua Richards, Jacob Rodriguez, John Stamatoyannopoulos, Julia Wald, Olivia Waltner, Michael Boeckh, Janet A. Englund, Michael Famulare, Barry R. Lutz, Mark J. Rieder, Lea M. Starita, Matthew Thompson, Helen Y. Chu, Jay Shendure, Trevor Bedford |
| EPI_ISL_1822876 | Atlas Genomics | Seattle Flu Study | Deborah A. Nickerson, Chris D. Frazar, Jover Lee, Benjamin Pelle, Erica Ryke, Matthew Richardson, Amanda Adler, Elisabeth Brandstetter, Peter D. Han, Kairsten Fay, Misja Ilcisin, Kirsten Lacombe, Thomas R. Sibley, Melissa Truong, Caitlin R. Wolf, Romesh Gautom, Geoff Melly, Brian Hiatt, Philip Dykema, Scott Lindquist, Michael Boeckh, Janet A. Englund, Michael Famulare, Barry R. Lutz, Mark J. Rieder, Lea M. Starita, Matthew Thompson, Helen Y. Chu, Jay Shendure, Trevor Bedford |
| EPI_ISL_1822877, EPI_ISL_1822878 | Altius Institute | Seattle Flu Study | Deborah A. Nickerson, Chris D. Frazar, Jover Lee, Benjamin Pelle, Erica Ryke, Matthew Richardson, Amanda Adler, Elisabeth Brandstetter, Peter D. Han, Kairsten Fay, Misja Ilcisin, Kirsten Lacombe, Thomas R. Sibley, Melissa Truong, Caitlin R. Wolf, Ryan Alexander, Daniel Bates, Rebecca Bruders, Stephanie DeBaun, Clem Green, Muhammad Halimun, Jessica Halow, Kneshay Harper, Matt Hartman, Andrew Meuser, Alex Nguyen, Truong Nguyen, Sofia Olsson, Sadie Patraw, Hannah Petersen, Tobias Ragoczy, Joshua Richards, Jacob Rodriguez, John Stamatoyannopoulos, Julia Wald, Olivia Waltner, Michael Boeckh, Janet A. Englund, Michael Famulare, Barry R. Lutz, Mark J. Rieder, Lea M. Starita, Matthew Thompson, Helen Y. Chu, Jay Shendure, Trevor Bedford |
| EPI_ISL_1822879 | Atlas Genomics | Seattle Flu Study | Deborah A. Nickerson, Chris D. Frazar, Jover Lee, Benjamin Pelle, Erica Ryke, Matthew Richardson, Amanda Adler, Elisabeth Brandstetter, Peter D. Han, Kairsten Fay, Misja Ilcisin, Kirsten Lacombe, Thomas R. Sibley, Melissa Truong, Caitlin R. Wolf, Romesh Gautom, Geoff Melly, Brian Hiatt, Philip Dykema, Scott Lindquist, Michael Boeckh, Janet A. Englund, Michael Famulare, Barry R. Lutz, Mark J. Rieder, Lea M. Starita, Matthew Thompson, Helen Y. Chu, Jay Shendure, Trevor Bedford |
| EPI_ISL_1822880, EPI_ISL_1822881 | Altius Institute | Seattle Flu Study | Deborah A. Nickerson, Chris D. Frazar, Jover Lee, Benjamin Pelle, Erica Ryke, Matthew Richardson, Amanda Adler, Elisabeth Brandstetter, Peter D. Han, Kairsten Fay, Misja Ilcisin, Kirsten Lacombe, Thomas R. Sibley, Melissa Truong, Caitlin R. Wolf, Ryan Alexander, Daniel Bates, Rebecca Bruders, Stephanie DeBaun, Clem Green, Muhammad Halimun, Jessica Halow, Kneshay Harper, Matt Hartman, Andrew Meuser, Alex Nguyen, Truong Nguyen, Sofia Olsson, Sadie Patraw, Hannah Petersen, Tobias Ragoczy, Joshua Richards, Jacob Rodriguez, John Stamatoyannopoulos, Julia Wald, Olivia Waltner, Michael Boeckh, Janet A. Englund, Michael Famulare, Barry R. Lutz, Mark J. Rieder, Lea M. Starita, Matthew Thompson, Helen Y. Chu, Jay Shendure, Trevor Bedford |
| EPI_ISL_1822882, EPI_ISL_1822883 | Atlas Genomics | Seattle Flu Study | Deborah A. Nickerson, Chris D. Frazar, Jover Lee, Benjamin Pelle, Erica Ryke, Matthew Richardson, Amanda Adler, Elisabeth Brandstetter, Peter D. Han, Kairsten Fay, Misja Ilcisin, Kirsten Lacombe, Thomas R. Sibley, Melissa Truong, Caitlin R. Wolf, Romesh Gautom, Geoff Melly, Brian Hiatt, Philip Dykema, Scott Lindquist, Michael Boeckh, Janet A. Englund, Michael Famulare, Barry R. Lutz, Mark J. Rieder, Lea M. Starita, Matthew Thompson, Helen Y. Chu, Jay Shendure, Trevor Bedford |
| EPI_ISL_1822884, EPI_ISL_1822885, EPI_ISL_1822886 | Altius Institute | Seattle Flu Study | Deborah A. Nickerson, Chris D. Frazar, Jover Lee, Benjamin Pelle, Erica Ryke, Matthew Richardson, Amanda Adler, Elisabeth Brandstetter, Peter D. Han, Kairsten Fay, Misja Ilcisin, Kirsten Lacombe, Thomas R. Sibley, Melissa Truong, Caitlin R. Wolf, Ryan Alexander, Daniel Bates, Rebecca Bruders, Stephanie DeBaun, Clem Green, Muhammad Halimun, Jessica Halow, Kneshay Harper, Matt Hartman, Andrew Meuser, Alex Nguyen, Truong Nguyen, Sofia Olsson, Sadie Patraw, Hannah Petersen, Tobias Ragoczy, Joshua Richards, Jacob Rodriguez, John Stamatoyannopoulos, Julia Wald, Olivia Waltner, Michael Boeckh, Janet A. Englund, Michael Famulare, Barry R. Lutz, Mark J. Rieder, Lea M. Starita, Matthew Thompson, Helen Y. Chu, Jay Shendure, Trevor Bedford |
| EPI_ISL_1822887, EPI_ISL_1822888 | Atlas Genomics | Seattle Flu Study | Deborah A. Nickerson, Chris D. Frazar, Jover Lee, Benjamin Pelle, Erica Ryke, Matthew Richardson, Amanda Adler, Elisabeth Brandstetter, Peter D. Han, Kairsten Fay, Misja Ilcisin, Kirsten Lacombe, Thomas R. Sibley, Melissa Truong, Caitlin R. Wolf, Romesh Gautom, Geoff Melly, Brian Hiatt, Philip Dykema, Scott Lindquist, Michael Boeckh, Janet A. Englund, Michael Famulare, Barry R. Lutz, Mark J. Rieder, Lea M. Starita, Matthew Thompson, Helen Y. Chu, Jay Shendure, Trevor Bedford |
| EPI_ISL_1822889 | Altius Institute | Seattle Flu Study | Deborah A. Nickerson, Chris D. Frazar, Jover Lee, Benjamin Pelle, Erica Ryke, Matthew Richardson, Amanda Adler, Elisabeth Brandstetter, Peter D. Han, Kairsten Fay, Misja Ilcisin, Kirsten Lacombe, Thomas R. Sibley, Melissa Truong, Caitlin R. Wolf, Ryan Alexander, Daniel Bates, Rebecca Bruders, Stephanie DeBaun, Clem Green, Muhammad Halimun, Jessica Halow, Kneshay Harper, Matt Hartman, Andrew Meuser, Alex Nguyen, Truong Nguyen, Sofia Olsson, Sadie Patraw, Hannah Petersen, Tobias Ragoczy, Joshua Richards, Jacob Rodriguez, John Stamatoyannopoulos, Julia Wald, Olivia Waltner, Michael Boeckh, Janet A. Englund, Michael Famulare, Barry R. Lutz, Mark J. Rieder, Lea M. Starita, Matthew Thompson, Helen Y. Chu, Jay Shendure, Trevor Bedford |
| EPI_ISL_1822890 | Atlas Genomics | Seattle Flu Study | Deborah A. Nickerson, Chris D. Frazar, Jover Lee, Benjamin Pelle, Erica Ryke, Matthew Richardson, Amanda Adler, Elisabeth Brandstetter, Peter D. Han, Kairsten Fay, Misja Ilcisin, Kirsten Lacombe, Thomas R. Sibley, Melissa Truong, Caitlin R. Wolf, Romesh Gautom, Geoff Melly, Brian Hiatt, Philip Dykema, Scott Lindquist, Michael Boeckh, Janet A. Englund, Michael Famulare, Barry R. Lutz, Mark J. Rieder, Lea M. Starita, Matthew Thompson, Helen Y. Chu, Jay Shendure, Trevor Bedford |
| EPI_ISL_1822891 | Northwest Laboratories | Seattle Flu Study | Deborah A. Nickerson, Chris D. Frazar, Jover Lee, Benjamin Pelle, Erica Ryke, Matthew Richardson, Amanda Adler, Elisabeth Brandstetter, Peter D. Han, Kairsten Fay, Misja Ilcisin, Kirsten Lacombe, Thomas R. Sibley, Melissa Truong, Caitlin R. Wolf, Romesh Gautom, Geoff Melly, Brian Hiatt, Philip Dykema, Scott Lindquist, Michael Boeckh, Janet A. Englund, Michael Famulare, Barry R. Lutz, Mark J. Rieder, Lea M. Starita, Matthew Thompson, Helen Y. Chu, Jay Shendure, Trevor Bedford |
| EPI_ISL_1822892, EPI_ISL_1822893 | Atlas Genomics | Seattle Flu Study | Deborah A. Nickerson, Chris D. Frazar, Jover Lee, Benjamin Pelle, Erica Ryke, Matthew Richardson, Amanda Adler, Elisabeth Brandstetter, Peter D. Han, Kairsten Fay, Misja Ilcisin, Kirsten Lacombe, Thomas R. Sibley, Melissa Truong, Caitlin R. Wolf, Romesh Gautom, Geoff Melly, Brian Hiatt, Philip Dykema, Scott Lindquist, Michael Boeckh, Janet A. Englund, Michael Famulare, Barry R. Lutz, Mark J. Rieder, Lea M. Starita, Matthew Thompson, Helen Y. Chu, Jay Shendure, Trevor Bedford |

| | | | |
|---|---|---|---|
| EPI_ISL_1822894 | Northwest Laboratories | Seattle Flu Study | Deborah A. Nickerson, Chris D. Frazar, Jover Lee, Benjamin Pelle, Erica Ryke, Matthew Richardson, Amanda Adler, Elisabeth Brandstetter, Peter D. Han, Kairsten Fay, Misja Ilcisin, Kirsten Lacombe, Thomas R. Sibley, Melissa Truong, Caitlin R. Wolf, Romesh Gautom, Geoff Melly, Brian Hiatt, Philip Dykema, Scott Lindquist, Michael Boeckh, Janet A. Englund, Michael Famulare, Barry R. Lutz, Mark J. Rieder, Lea M. Starita, Matthew Thompson, Helen Y. Chu, Jay Shendure, Trevor Bedford |
| EPI_ISL_1822895, EPI_ISL_1822896, EPI_ISL_1822897, EPI_ISL_1822898 | Altius Institute | Seattle Flu Study | Deborah A. Nickerson, Chris D. Frazar, Jover Lee, Benjamin Pelle, Erica Ryke, Matthew Richardson, Amanda Adler, Elisabeth Brandstetter, Peter D. Han, Kairsten Fay, Misja Ilcisin, Kirsten Lacombe, Thomas R. Sibley, Melissa Truong, Caitlin R. Wolf, Ryan Alexander, Daniel Bates, Rebecca Bruders, Stephanie DeBaun, Clem Green, Muhammad Halimun, Jessica Halow, Kneshay Harper, Matt Hartman, Andrew Meuser, Alex Nguyen, Truong Nguyen, Sofia Olsson, Sadie Patraw, Hannah Petersen, Tobias Ragoczy, Joshua Richards, Jacob Rodriguez, John Stamatoyannopoulos, Julia Wald, Olivia Waltner, Michael Boeckh, Janet A. Englund, Michael Famulare, Barry R. Lutz, Mark J. Rieder, Lea M. Starita, Matthew Thompson, Helen Y. Chu, Jay Shendure, Trevor Bedford |
| EPI_ISL_1822899, EPI_ISL_1822900 | Northwest Laboratories | Seattle Flu Study | Deborah A. Nickerson, Chris D. Frazar, Jover Lee, Benjamin Pelle, Erica Ryke, Matthew Richardson, Amanda Adler, Elisabeth Brandstetter, Peter D. Han, Kairsten Fay, Misja Ilcisin, Kirsten Lacombe, Thomas R. Sibley, Melissa Truong, Caitlin R. Wolf, Romesh Gautom, Geoff Melly, Brian Hiatt, Philip Dykema, Scott Lindquist, Michael Boeckh, Janet A. Englund, Michael Famulare, Barry R. Lutz, Mark J. Rieder, Lea M. Starita, Matthew Thompson, Helen Y. Chu, Jay Shendure, Trevor Bedford |
| EPI_ISL_1822901 | Altius Institute | Seattle Flu Study | Deborah A. Nickerson, Chris D. Frazar, Jover Lee, Benjamin Pelle, Erica Ryke, Matthew Richardson, Amanda Adler, Elisabeth Brandstetter, Peter D. Han, Kairsten Fay, Misja Ilcisin, Kirsten Lacombe, Thomas R. Sibley, Melissa Truong, Caitlin R. Wolf, Ryan Alexander, Daniel Bates, Rebecca Bruders, Stephanie DeBaun, Clem Green, Muhammad Halimun, Jessica Halow, Kneshay Harper, Matt Hartman, Andrew Meuser, Alex Nguyen, Truong Nguyen, Sofia Olsson, Sadie Patraw, Hannah Petersen, Tobias Ragoczy, Joshua Richards, Jacob Rodriguez, John Stamatoyannopoulos, Julia Wald, Olivia Waltner, Michael Boeckh, Janet A. Englund, Michael Famulare, Barry R. Lutz, Mark J. Rieder, Lea M. Starita, Matthew Thompson, Helen Y. Chu, Jay Shendure, Trevor Bedford |
| EPI_ISL_1822902 | Atlas Genomics | Seattle Flu Study | Deborah A. Nickerson, Chris D. Frazar, Jover Lee, Benjamin Pelle, Erica Ryke, Matthew Richardson, Amanda Adler, Elisabeth Brandstetter, Peter D. Han, Kairsten Fay, Misja Ilcisin, Kirsten Lacombe, Thomas R. Sibley, Melissa Truong, Caitlin R. Wolf, Romesh Gautom, Geoff Melly, Brian Hiatt, Philip Dykema, Scott Lindquist, Michael Boeckh, Janet A. Englund, Michael Famulare, Barry R. Lutz, Mark J. Rieder, Lea M. Starita, Matthew Thompson, Helen Y. Chu, Jay Shendure, Trevor Bedford |
| EPI_ISL_1822903 | Altius Institute | Seattle Flu Study | Deborah A. Nickerson, Chris D. Frazar, Jover Lee, Benjamin Pelle, Erica Ryke, Matthew Richardson, Amanda Adler, Elisabeth Brandstetter, Peter D. Han, Kairsten Fay, Misja Ilcisin, Kirsten Lacombe, Thomas R. Sibley, Melissa Truong, Caitlin R. Wolf, Ryan Alexander, Daniel Bates, Rebecca Bruders, Stephanie DeBaun, Clem Green, Muhammad Halimun, Jessica Halow, Kneshay Harper, Matt Hartman, Andrew Meuser, Alex Nguyen, Truong Nguyen, Sofia Olsson, Sadie Patraw, Hannah Petersen, Tobias Ragoczy, Joshua Richards, Jacob Rodriguez, John Stamatoyannopoulos, Julia Wald, Olivia Waltner, Michael Boeckh, Janet A. Englund, Michael Famulare, Barry R. Lutz, Mark J. Rieder, Lea M. Starita, Matthew Thompson, Helen Y. Chu, Jay Shendure, Trevor Bedford |
| EPI_ISL_1822904 | Northwest Laboratories | Seattle Flu Study | Deborah A. Nickerson, Chris D. Frazar, Jover Lee, Benjamin Pelle, Erica Ryke, Matthew Richardson, Amanda Adler, Elisabeth Brandstetter, Peter D. Han, Kairsten Fay, Misja Ilcisin, Kirsten Lacombe, Thomas R. Sibley, Melissa Truong, Caitlin R. Wolf, Romesh Gautom, Geoff Melly, Brian Hiatt, Philip Dykema, Scott Lindquist, Michael Boeckh, Janet A. Englund, Michael Famulare, Barry R. Lutz, Mark J. Rieder, Lea M. Starita, Matthew Thompson, Helen Y. Chu, Jay Shendure, Trevor Bedford |
| EPI_ISL_1822905, EPI_ISL_1822906, EPI_ISL_1822907, EPI_ISL_1822908, EPI_ISL_1822909 | Altius Institute | Seattle Flu Study | Deborah A. Nickerson, Chris D. Frazar, Jover Lee, Benjamin Pelle, Erica Ryke, Matthew Richardson, Amanda Adler, Elisabeth Brandstetter, Peter D. Han, Kairsten Fay, Misja Ilcisin, Kirsten Lacombe, Thomas R. Sibley, Melissa Truong, Caitlin R. Wolf, Ryan Alexander, Daniel Bates, Rebecca Bruders, Stephanie DeBaun, Clem Green, Muhammad Halimun, Jessica Halow, Kneshay Harper, Matt Hartman, Andrew Meuser, Alex Nguyen, Truong Nguyen, Sofia Olsson, Sadie Patraw, Hannah Petersen, Tobias Ragoczy, Joshua Richards, Jacob Rodriguez, John Stamatoyannopoulos, Julia Wald, Olivia Waltner, Michael Boeckh, Janet A. Englund, Michael Famulare, Barry R. Lutz, Mark J. Rieder, Lea M. Starita, Matthew Thompson, Helen Y. Chu, Jay Shendure, Trevor Bedford |
| EPI_ISL_1822910 | Northwest Laboratories | Seattle Flu Study | Deborah A. Nickerson, Chris D. Frazar, Jover Lee, Benjamin Pelle, Erica Ryke, Matthew Richardson, Amanda Adler, Elisabeth Brandstetter, Peter D. Han, Kairsten Fay, Misja Ilcisin, Kirsten Lacombe, Thomas R. Sibley, Melissa Truong, Caitlin R. Wolf, Romesh Gautom, Geoff Melly, Brian Hiatt, Philip Dykema, Scott Lindquist, Michael Boeckh, Janet A. Englund, Michael Famulare, Barry R. Lutz, Mark J. Rieder, Lea M. Starita, Matthew Thompson, Helen Y. Chu, Jay Shendure, Trevor Bedford |
| EPI_ISL_1822911, EPI_ISL_1822912 | Atlas Genomics | Seattle Flu Study | Deborah A. Nickerson, Chris D. Frazar, Jover Lee, Benjamin Pelle, Erica Ryke, Matthew Richardson, Amanda Adler, Elisabeth Brandstetter, Peter D. Han, Kairsten Fay, Misja Ilcisin, Kirsten Lacombe, Thomas R. Sibley, Melissa Truong, Caitlin R. Wolf, Romesh Gautom, Geoff Melly, Brian Hiatt, Philip Dykema, Scott Lindquist, Michael Boeckh, Janet A. Englund, Michael Famulare, Barry R. Lutz, Mark J. Rieder, Lea M. Starita, Matthew Thompson, Helen Y. Chu, Jay Shendure, Trevor Bedford |
| EPI_ISL_1822913 | Incyte Diagnostics Spokane | Seattle Flu Study | Deborah A. Nickerson, Chris D. Frazar, Jover Lee, Benjamin Pelle, Erica Ryke, Matthew Richardson, Amanda Adler, Elisabeth Brandstetter, Peter D. Han, Kairsten Fay, Misja Ilcisin, Kirsten Lacombe, Thomas R. Sibley, Melissa Truong, Caitlin R. Wolf, Romesh Gautom, Geoff Melly, Brian Hiatt, Philip Dykema, Scott Lindquist, Michael Boeckh, Janet A. Englund, Michael Famulare, Barry R. Lutz, Mark J. Rieder, Lea M. Starita, Matthew Thompson, Helen Y. Chu, Jay Shendure, Trevor Bedford |
| EPI_ISL_1822914 | Atlas Genomics | Seattle Flu Study | Deborah A. Nickerson, Chris D. Frazar, Jover Lee, Benjamin Pelle, Erica Ryke, Matthew Richardson, Amanda Adler, Elisabeth Brandstetter, Peter D. Han, Kairsten Fay, Misja Ilcisin, Kirsten Lacombe, Thomas R. Sibley, Melissa Truong, Caitlin R. Wolf, Romesh Gautom, Geoff Melly, Brian Hiatt, Philip Dykema, Scott Lindquist, Michael Boeckh, Janet A. Englund, Michael Famulare, Barry R. Lutz, Mark J. Rieder, Lea M. Starita, Matthew Thompson, Helen Y. Chu, Jay Shendure, Trevor Bedford |
| EPI_ISL_1822915 | Altius Institute | Seattle Flu Study | Deborah A. Nickerson, Chris D. Frazar, Jover Lee, Benjamin Pelle, Erica Ryke, Matthew Richardson, Amanda Adler, Elisabeth Brandstetter, Peter D. Han, Kairsten Fay, Misja Ilcisin, Kirsten Lacombe, Thomas R. Sibley, Melissa Truong, Caitlin R. Wolf, Ryan Alexander, Daniel Bates, Rebecca Bruders, Stephanie DeBaun, Clem Green, Muhammad Halimun, Jessica Halow, Kneshay Harper, Matt Hartman, Andrew Meuser, Alex Nguyen, Truong Nguyen, Sofia Olsson, Sadie Patraw, Hannah Petersen, Tobias Ragoczy, Joshua Richards, Jacob Rodriguez, John Stamatoyannopoulos, Julia Wald, Olivia Waltner, Michael Boeckh, Janet A. Englund, Michael Famulare, Barry R. Lutz, Mark J. Rieder, Lea M. Starita, Matthew Thompson, Helen Y. Chu, Jay Shendure, Trevor Bedford |
| EPI_ISL_1822916 | Northwest Laboratories | Seattle Flu Study | Deborah A. Nickerson, Chris D. Frazar, Jover Lee, Benjamin Pelle, Erica Ryke, Matthew Richardson, Amanda Adler, Elisabeth Brandstetter, Peter D. Han, Kairsten Fay, Misja Ilcisin, Kirsten Lacombe, Thomas R. Sibley, Melissa Truong, Caitlin R. Wolf, Romesh Gautom, Geoff Melly, Brian Hiatt, Philip Dykema, Scott Lindquist, Michael Boeckh, Janet A. Englund, Michael Famulare, Barry R. Lutz, Mark J. Rieder, Lea M. Starita, Matthew Thompson, Helen Y. Chu, Jay Shendure, Trevor Bedford |
| EPI_ISL_1822917 | Atlas Genomics | Seattle Flu Study | Deborah A. Nickerson, Chris D. Frazar, Jover Lee, Benjamin Pelle, Erica Ryke, Matthew Richardson, Amanda Adler, Elisabeth Brandstetter, Peter D. Han, Kairsten Fay, Misja Ilcisin, Kirsten Lacombe, Thomas R. Sibley, Melissa Truong, Caitlin R. Wolf, Romesh Gautom, Geoff Melly, Brian Hiatt, Philip Dykema, Scott Lindquist, Michael Boeckh, Janet A. Englund, Michael Famulare, Barry R. Lutz, Mark J. Rieder, Lea M. Starita, Matthew Thompson, Helen Y. Chu, Jay Shendure, Trevor Bedford |
| EPI_ISL_1822918 | Altius Institute | Seattle Flu Study | Deborah A. Nickerson, Chris D. Frazar, Jover Lee, Benjamin Pelle, Erica Ryke, Matthew Richardson, Amanda Adler, Elisabeth Brandstetter, Peter D. Han, Kairsten Fay, Misja Ilcisin, Kirsten Lacombe, Thomas R. Sibley, Melissa Truong, Caitlin R. Wolf, Ryan Alexander, Daniel Bates, Rebecca Bruders, Stephanie DeBaun, Clem Green, Muhammad Halimun, Jessica Halow, Kneshay Harper, Matt Hartman, Andrew Meuser, Alex Nguyen, Truong Nguyen, Sofia Olsson, Sadie Patraw, Hannah Petersen, Tobias Ragoczy, Joshua Richards, Jacob Rodriguez, John Stamatoyannopoulos, Julia Wald, Olivia Waltner, Michael Boeckh, Janet A. Englund, Michael Famulare, Barry R. Lutz, Mark J. Rieder, Lea M. Starita, Matthew Thompson, Helen Y. Chu, Jay Shendure, Trevor Bedford |
| EPI_ISL_1822919, EPI_ISL_1822920 | Atlas Genomics | Seattle Flu Study | Deborah A. Nickerson, Chris D. Frazar, Jover Lee, Benjamin Pelle, Erica Ryke, Matthew Richardson, Amanda Adler, Elisabeth Brandstetter, Peter D. Han, Kairsten Fay, Misja Ilcisin, Kirsten Lacombe, Thomas R. Sibley, Melissa Truong, Caitlin R. Wolf, Romesh Gautom, Geoff Melly, Brian Hiatt, Philip Dykema, Scott Lindquist, Michael Boeckh, Janet A. Englund, Michael Famulare, Barry R. Lutz, Mark J. Rieder, Lea M. Starita, Matthew Thompson, Helen Y. Chu, Jay Shendure, Trevor Bedford |
| EPI_ISL_1822921 | Altius Institute | Seattle Flu Study | Deborah A. Nickerson, Chris D. Frazar, Jover Lee, Benjamin Pelle, Erica Ryke, Matthew Richardson, Amanda Adler, Elisabeth Brandstetter, Peter D. Han, Kairsten Fay, Misja Ilcisin, Kirsten Lacombe, Thomas R. Sibley, Melissa Truong, Caitlin R. Wolf, Ryan Alexander, Daniel Bates, Rebecca Bruders, Stephanie DeBaun, Clem Green, Muhammad Halimun, Jessica Halow, Kneshay Harper, Matt Hartman, Andrew Meuser, Alex Nguyen, Truong Nguyen, Sofia Olsson, Sadie Patraw, Hannah Petersen, Tobias Ragoczy, Joshua Richards, Jacob Rodriguez, John Stamatoyannopoulos, Julia Wald, Olivia Waltner, Michael Boeckh, Janet A. Englund, Michael Famulare, Barry R. Lutz, Mark J. Rieder, Lea M. Starita, Matthew Thompson, Helen Y. Chu, Jay Shendure, Trevor Bedford |
| EPI_ISL_1822922 | Northwest Laboratories | Seattle Flu Study | Deborah A. Nickerson, Chris D. Frazar, Jover Lee, Benjamin Pelle, Erica Ryke, Matthew Richardson, Amanda Adler, Elisabeth Brandstetter, Peter D. Han, Kairsten Fay, Misja Ilcisin, Kirsten Lacombe, Thomas R. Sibley, Melissa Truong, Caitlin R. Wolf, Romesh Gautom, Geoff Melly, Brian Hiatt, Philip Dykema, Scott Lindquist, Michael Boeckh, Janet A. Englund, Michael Famulare, Barry R. Lutz, Mark J. Rieder, Lea M. Starita, Matthew Thompson, Helen Y. Chu, Jay Shendure, Trevor Bedford |
| EPI_ISL_1822923 | Atlas Genomics | Seattle Flu Study | Deborah A. Nickerson, Chris D. Frazar, Jover Lee, Benjamin Pelle, Erica Ryke, Matthew Richardson, Amanda Adler, Elisabeth Brandstetter, Peter D. Han, Kairsten Fay, Misja Ilcisin, Kirsten Lacombe, Thomas R. Sibley, Melissa Truong, Caitlin R. Wolf, Romesh Gautom, Geoff Melly, Brian Hiatt, Philip Dykema, Scott Lindquist, Michael Boeckh, Janet A. Englund, Michael Famulare, Barry R. Lutz, Mark J. Rieder, Lea M. Starita, Matthew Thompson, Helen Y. Chu, Jay Shendure, Trevor Bedford |
| EPI_ISL_1822924 | Altius Institute | Seattle Flu Study | Deborah A. Nickerson, Chris D. Frazar, Jover Lee, Benjamin Pelle, Erica Ryke, Matthew Richardson, Amanda Adler, Elisabeth Brandstetter, Peter D. Han, Kairsten Fay, Misja Ilcisin, Kirsten Lacombe, Thomas R. Sibley, Melissa Truong, Caitlin R. Wolf, Ryan Alexander, Daniel Bates, Rebecca Bruders, Stephanie DeBaun, Clem Green, Muhammad Halimun, Jessica Halow, Kneshay Harper, Matt Hartman, Andrew Meuser, Alex Nguyen, Truong Nguyen, Sofia Olsson, Sadie Patraw, Hannah Petersen, Tobias Ragoczy, Joshua Richards, Jacob Rodriguez, John Stamatoyannopoulos, Julia Wald, Olivia Waltner, Michael Boeckh, Janet A. Englund, Michael Famulare, Barry R. Lutz, Mark J. Rieder, Lea M. Starita, Matthew Thompson, Helen Y. Chu, Jay Shendure, Trevor Bedford |
| EPI_ISL_1822925 | Northwest Laboratories | Seattle Flu Study | Deborah A. Nickerson, Chris D. Frazar, Jover Lee, Benjamin Pelle, Erica Ryke, Matthew Richardson, Amanda Adler, Elisabeth Brandstetter, Peter D. Han, Kairsten Fay, Misja Ilcisin, Kirsten Lacombe, Thomas R. Sibley, Melissa Truong, Caitlin R. Wolf, Romesh Gautom, Geoff Melly, Brian Hiatt, Philip Dykema, Scott Lindquist, Michael Boeckh, Janet A. Englund, Michael Famulare, Barry R. Lutz, Mark J. Rieder, Lea M. Starita, Matthew Thompson, Helen Y. Chu, Jay Shendure, Trevor Bedford |
| EPI_ISL_1822926 | Altius Institute | Seattle Flu Study | Deborah A. Nickerson, Chris D. Frazar, Jover Lee, Benjamin Pelle, Erica Ryke, Matthew Richardson, Amanda Adler, Elisabeth Brandstetter, Peter D. Han, Kairsten Fay, Misja Ilcisin, Kirsten Lacombe, Thomas R. Sibley, Melissa Truong, Caitlin R. Wolf, Ryan Alexander, Daniel Bates, Rebecca Bruders, Stephanie DeBaun, Clem Green, Muhammad Halimun, Jessica Halow, Kneshay Harper, Matt Hartman, Andrew Meuser, Alex Nguyen, Truong Nguyen, Sofia Olsson, Sadie Patraw, Hannah Petersen, Tobias Ragoczy, Joshua Richards, Jacob Rodriguez, John Stamatoyannopoulos, Julia Wald, Olivia Waltner, Michael Boeckh, Janet A. Englund, Michael Famulare, Barry R. Lutz, Mark J. Rieder, Lea M. Starita, Matthew Thompson, Helen Y. Chu, Jay Shendure, Trevor Bedford |
| EPI_ISL_1822927 | Northwest Laboratories | Seattle Flu Study | Deborah A. Nickerson, Chris D. Frazar, Jover Lee, Benjamin Pelle, Erica Ryke, Matthew Richardson, Amanda Adler, Elisabeth Brandstetter, Peter D. Han, Kairsten Fay, Misja Ilcisin, Kirsten Lacombe, Thomas R. Sibley, Melissa Truong, Caitlin R. Wolf, Romesh Gautom, Geoff Melly, Brian Hiatt, Philip Dykema, Scott Lindquist, Michael Boeckh, Janet A. Englund, Michael Famulare, Barry R. Lutz, Mark J. Rieder, Lea M. Starita, Matthew Thompson, Helen Y. Chu, Jay Shendure, Trevor Bedford |
| EPI_ISL_1822928 | Altius Institute | Seattle Flu Study | Deborah A. Nickerson, Chris D. Frazar, Jover Lee, Benjamin Pelle, Erica Ryke, Matthew Richardson, Amanda Adler, Elisabeth Brandstetter, Peter D. Han, Kairsten Fay, Misja Ilcisin, Kirsten Lacombe, Thomas R. Sibley, Melissa Truong, Caitlin R. Wolf, Ryan Alexander, Daniel Bates, Rebecca Bruders, Stephanie DeBaun, Clem Green, Muhammad Halimun, Jessica Halow, Kneshay Harper, Matt Hartman, Andrew Meuser, Alex Nguyen, Truong Nguyen, Sofia Olsson, Sadie Patraw, Hannah Petersen, Tobias Ragoczy, Joshua Richards, Jacob Rodriguez, John Stamatoyannopoulos, Julia Wald, Olivia Waltner, Michael Boeckh, Janet A. Englund, Michael Famulare, Barry R. Lutz, Mark J. Rieder, Lea M. Starita, Matthew Thompson, Helen Y. Chu, Jay Shendure, Trevor Bedford |
| EPI_ISL_1822929 | Incyte Diagnostics Spokane | Seattle Flu Study | Deborah A. Nickerson, Chris D. Frazar, Jover Lee, Benjamin Pelle, Erica Ryke, Matthew Richardson, Amanda Adler, Elisabeth Brandstetter, Peter D. Han, Kairsten Fay, Misja Ilcisin, Kirsten Lacombe, Thomas R. Sibley, Melissa Truong, Caitlin R. Wolf, Romesh Gautom, Geoff Melly, Brian Hiatt, Philip Dykema, Scott Lindquist, Michael Boeckh, Janet A. Englund, Michael Famulare, Barry R. Lutz, Mark J. Rieder, Lea M. Starita, Matthew Thompson, Helen Y. Chu, Jay Shendure, Trevor Bedford |
| EPI_ISL_1822930, EPI_ISL_1822931 | Altius Institute | Seattle Flu Study | Deborah A. Nickerson, Chris D. Frazar, Jover Lee, Benjamin Pelle, Erica Ryke, Matthew Richardson, Amanda Adler, Elisabeth Brandstetter, Peter D. Han, Kairsten Fay, Misja Ilcisin, Kirsten Lacombe, Thomas R. Sibley, Melissa Truong, Caitlin R. Wolf, Ryan Alexander, Daniel Bates, Rebecca Bruders, Stephanie DeBaun, Clem Green, Muhammad Halimun, Jessica Halow, Kneshay Harper, Matt Hartman, Andrew Meuser, Alex Nguyen, Truong Nguyen, Sofia Olsson, Sadie Patraw, Hannah Petersen, Tobias Ragoczy, Joshua Richards, Jacob Rodriguez, John Stamatoyannopoulos, Julia Wald, Olivia Waltner, Michael Boeckh, Janet A. Englund, Michael Famulare, Barry R. Lutz, Mark J. Rieder, Lea M. Starita, Matthew Thompson, Helen Y. Chu, Jay Shendure, Trevor Bedford |
| EPI_ISL_1822932 | Incyte Diagnostics Spokane | Seattle Flu Study | Deborah A. Nickerson, Chris D. Frazar, Jover Lee, Benjamin Pelle, Erica Ryke, Matthew Richardson, Amanda Adler, Elisabeth Brandstetter, Peter D. Han, Kairsten Fay, Misja Ilcisin, Kirsten Lacombe, Thomas R. Sibley, Melissa Truong, Caitlin R. Wolf, Romesh Gautom, Geoff Melly, Brian Hiatt, Philip Dykema, Scott Lindquist, Michael Boeckh, Janet A. Englund, Michael Famulare, Barry R. Lutz, Mark J. Rieder, Lea M. Starita, Matthew Thompson, Helen Y. Chu, Jay Shendure, Trevor Bedford |
| EPI_ISL_1822933 | Altius Institute | Seattle Flu Study | Deborah A. Nickerson, Chris D. Frazar, Jover Lee, Benjamin Pelle, Erica Ryke, Matthew Richardson, Amanda Adler, Elisabeth Brandstetter, Peter D. Han, Kairsten Fay, Misja Ilcisin, Kirsten Lacombe, Thomas R. Sibley, Melissa Truong, Caitlin R. Wolf, Ryan Alexander, Daniel Bates, Rebecca Bruders, Stephanie DeBaun, Clem Green, Muhammad Halimun, Jessica Halow, Kneshay Harper, Matt Hartman, Andrew Meuser, Alex Nguyen, Truong Nguyen, Sofia Olsson, Sadie Patraw, Hannah Petersen, Tobias Ragoczy, Joshua Richards, Jacob Rodriguez, John Stamatoyannopoulos, Julia Wald, Olivia Waltner, Michael Boeckh, Janet A. Englund, Michael Famulare, Barry R. Lutz, Mark J. Rieder, Lea M. Starita, Matthew Thompson, Helen Y. Chu, Jay Shendure, Trevor Bedford |
| EPI_ISL_1822934, EPI_ISL_1822935, EPI_ISL_1822936 | Northwest Laboratories | Seattle Flu Study | Deborah A. Nickerson, Chris D. Frazar, Jover Lee, Benjamin Pelle, Erica Ryke, Matthew Richardson, Amanda Adler, Elisabeth Brandstetter, Peter D. Han, Kairsten Fay, Misja Ilcisin, Kirsten Lacombe, Thomas R. Sibley, Melissa Truong, Caitlin R. Wolf, Romesh Gautom, Geoff Melly, Brian Hiatt, Philip Dykema, Scott Lindquist, Michael Boeckh, Janet A. Englund, Michael Famulare, Barry R. Lutz, Mark J. Rieder, Lea M. Starita, Matthew Thompson, Helen Y. Chu, Jay Shendure, Trevor Bedford |
| EPI_ISL_1822937 | Altius Institute | Seattle Flu Study | Deborah A. Nickerson, Chris D. Frazar, Jover Lee, Benjamin Pelle, Erica Ryke, Matthew Richardson, Amanda Adler, Elisabeth Brandstetter, Peter D. Han, Kairsten Fay, Misja Ilcisin, Kirsten Lacombe, Thomas R. Sibley, Melissa Truong, Caitlin R. Wolf, Ryan Alexander, Daniel Bates, Rebecca Bruders, Stephanie DeBaun, Clem Green, Muhammad Halimun, Jessica Halow, Kneshay Harper, Matt Hartman, Andrew Meuser, Alex Nguyen, Truong Nguyen, Sofia Olsson, Sadie Patraw, Hannah Petersen, Tobias Ragoczy, Joshua Richards, Jacob Rodriguez, John Stamatoyannopoulos, Julia Wald, Olivia Waltner, Michael Boeckh, Janet A. Englund, Michael Famulare, Barry R. Lutz, Mark J. Rieder, Lea M. Starita, Matthew Thompson, Helen Y. Chu, Jay Shendure, Trevor Bedford |
| EPI_ISL_1822938, EPI_ISL_1822939, EPI_ISL_1822940, EPI_ISL_1822941 | Northwest Laboratories | Seattle Flu Study | Deborah A. Nickerson, Chris D. Frazar, Jover Lee, Benjamin Pelle, Erica Ryke, Matthew Richardson, Amanda Adler, Elisabeth Brandstetter, Peter D. Han, Kairsten Fay, Misja Ilcisin, Kirsten Lacombe, Thomas R. Sibley, Melissa Truong, Caitlin R. Wolf, Romesh Gautom, Geoff Melly, Brian Hiatt, Philip Dykema, Scott Lindquist, Michael Boeckh, Janet A. Englund, Michael Famulare, Barry R. Lutz, Mark J. Rieder, Lea M. Starita, Matthew Thompson, Helen Y. Chu, Jay Shendure, Trevor Bedford |
| EPI_ISL_1822942 | Altius Institute | Seattle Flu Study | Deborah A. Nickerson, Chris D. Frazar, Jover Lee, Benjamin Pelle, Erica Ryke, Matthew Richardson, Amanda Adler, Elisabeth Brandstetter, Peter D. Han, Kairsten Fay, Misja Ilcisin, Kirsten Lacombe, Thomas R. Sibley, Melissa Truong, Caitlin R. Wolf, Ryan Alexander, Daniel Bates, Rebecca Bruders, Stephanie DeBaun, Clem Green, Muhammad Halimun, Jessica Halow, Kneshay Harper, Matt Hartman, Andrew Meuser, Alex Nguyen, Truong Nguyen, Sofia Olsson, Sadie Patraw, Hannah Petersen, Tobias Ragoczy, Joshua Richards, Jacob Rodriguez, John Stamatoyannopoulos, Julia Wald, Olivia Waltner, Michael Boeckh, Janet A. Englund, Michael Famulare, Barry R. Lutz, Mark J. Rieder, Lea M. Starita, Matthew Thompson, Helen Y. Chu, Jay Shendure, Trevor Bedford |
| EPI_ISL_1822943 | Northwest Laboratories | Seattle Flu Study | Deborah A. Nickerson, Chris D. Frazar, Jover Lee, Benjamin Pelle, Erica Ryke, Matthew Richardson, Amanda Adler, Elisabeth Brandstetter, Peter D. Han, Kairsten Fay, Misja Ilcisin, Kirsten Lacombe, Thomas R. Sibley, Melissa Truong, Caitlin R. Wolf, Romesh Gautom, Geoff Melly, Brian Hiatt, Philip Dykema, Scott Lindquist, Michael Boeckh, Janet A. Englund, Michael Famulare, Barry R. Lutz, Mark J. Rieder, Lea M. Starita, Matthew Thompson, Helen Y. Chu, Jay Shendure, Trevor Bedford |
| EPI_ISL_1822944, EPI_ISL_1822945, EPI_ISL_1822946 | Altius Institute | Seattle Flu Study | Deborah A. Nickerson, Chris D. Frazar, Jover Lee, Benjamin Pelle, Erica Ryke, Matthew Richardson, Amanda Adler, Elisabeth Brandstetter, Peter D. Han, Kairsten Fay, Misja Ilcisin, Kirsten Lacombe, Thomas R. Sibley, Melissa Truong, Caitlin R. Wolf, Ryan Alexander, Daniel Bates, Rebecca Bruders, Stephanie DeBaun, Clem Green, Muhammad Halimun, Jessica Halow, Kneshay Harper, Matt Hartman, Andrew Meuser, Alex Nguyen, Truong Nguyen, Sofia Olsson, Sadie Patraw, Hannah Petersen, Tobias Ragoczy, Joshua Richards, Jacob Rodriguez, John Stamatoyannopoulos, Julia Wald, Olivia Waltner, Michael Boeckh, Janet A. Englund, Michael Famulare, Barry R. Lutz, Mark J. Rieder, Lea M. Starita, Matthew Thompson, Helen Y. Chu, Jay Shendure, Trevor Bedford |
| EPI_ISL_1822947, EPI_ISL_1822948, EPI_ISL_1822949, EPI_ISL_1822950, | Atlas Genomics | Seattle Flu Study | Deborah A. Nickerson, Chris D. Frazar, Jover Lee, Benjamin Pelle, Erica Ryke, Matthew Richardson, Amanda Adler, Elisabeth Brandstetter, Peter D. Han, Kairsten Fay, Misja Ilcisin, Kirsten Lacombe, Thomas R. Sibley, Melissa Truong, Caitlin R. Wolf, Romesh Gautom, Geoff Melly, Brian Hiatt, Philip Dykema, Scott Lindquist, Michael Boeckh, Janet A. Englund, Michael Famulare, Barry R. Lutz, Mark J. Rieder, Lea M. Starita, Matthew Thompson, Helen Y. Chu, Jay Shendure, Trevor Bedford |

| | | | |
|---|---|---|---|
| EPI_ISL_1822951 | | | |
| EPI_ISL_1822952, EPI_ISL_1822953, EPI_ISL_1822954, EPI_ISL_1822955 | Northwest Laboratories | Seattle Flu Study | Deborah A. Nickerson, Chris D. Frazar, Jover Lee, Benjamin Pelle, Erica Ryke, Matthew Richardson, Amanda Adler, Elisabeth Brandstetter, Peter D. Han, Kairsten Fay, Misja Ilcisin, Kirsten Lacombe, Thomas R. Sibley, Melissa Truong, Caitlin R. Wolf, Romesh Gautom, Geoff Melly, Brian Hiatt, Philip Dykema, Scott Lindquist, Michael Boeckh, Janet A. Englund, Michael Famulare, Barry R. Lutz, Mark J. Rieder, Lea M. Starita, Matthew Thompson, Helen Y. Chu, Jay Shendure, Trevor Bedford |
| EPI_ISL_1822956, EPI_ISL_1822957 | Altius Institute | Seattle Flu Study | Deborah A. Nickerson, Chris D. Frazar, Jover Lee, Benjamin Pelle, Erica Ryke, Matthew Richardson, Amanda Adler, Elisabeth Brandstetter, Peter D. Han, Kairsten Fay, Misja Ilcisin, Kirsten Lacombe, Thomas R. Sibley, Melissa Truong, Caitlin R. Wolf, Ryan Alexander, Daniel Bates, Rebecca Bruders, Stephanie DeBaun, Clem Green, Muhammad Halimun, Jessica Halow, Kneshay Harper, Matt Hartman, Andrew Meuser, Alex Nguyen, Truong Nguyen, Sofia Olsson, Sadie Patraw, Hannah Petersen, Tobias Ragoczy, Joshua Richards, Jacob Rodriguez, John Stamatoyannopoulos, Julia Wald, Olivia Waltner, Michael Boeckh, Janet A. Englund, Michael Famulare, Barry R. Lutz, Mark J. Rieder, Lea M. Starita, Matthew Thompson, Helen Y. Chu, Jay Shendure, Trevor Bedford |
| EPI_ISL_1822958 | Northwest Laboratories | Seattle Flu Study | Deborah A. Nickerson, Chris D. Frazar, Jover Lee, Benjamin Pelle, Erica Ryke, Matthew Richardson, Amanda Adler, Elisabeth Brandstetter, Peter D. Han, Kairsten Fay, Misja Ilcisin, Kirsten Lacombe, Thomas R. Sibley, Melissa Truong, Caitlin R. Wolf, Romesh Gautom, Geoff Melly, Brian Hiatt, Philip Dykema, Scott Lindquist, Michael Boeckh, Janet A. Englund, Michael Famulare, Barry R. Lutz, Mark J. Rieder, Lea M. Starita, Matthew Thompson, Helen Y. Chu, Jay Shendure, Trevor Bedford |
| EPI_ISL_1823095 | Altius Institute | Seattle Flu Study | Deborah A. Nickerson, Chris D. Frazar, Jover Lee, Benjamin Pelle, Erica Ryke, Matthew Richardson, Amanda Adler, Elisabeth Brandstetter, Peter D. Han, Kairsten Fay, Misja Ilcisin, Kirsten Lacombe, Thomas R. Sibley, Melissa Truong, Caitlin R. Wolf, Ryan Alexander, Daniel Bates, Rebecca Bruders, Stephanie DeBaun, Clem Green, Muhammad Halimun, Jessica Halow, Kneshay Harper, Matt Hartman, Andrew Meuser, Alex Nguyen, Truong Nguyen, Sofia Olsson, Sadie Patraw, Hannah Petersen, Tobias Ragoczy, Joshua Richards, Jacob Rodriguez, John Stamatoyannopoulos, Julia Wald, Olivia Waltner, Michael Boeckh, Janet A. Englund, Michael Famulare, Barry R. Lutz, Mark J. Rieder, Lea M. Starita, Matthew Thompson, Helen Y. Chu, Jay Shendure, Trevor Bedford |
| EPI_ISL_1823096 | Northwest Laboratories | Seattle Flu Study | Deborah A. Nickerson, Chris D. Frazar, Jover Lee, Benjamin Pelle, Erica Ryke, Matthew Richardson, Amanda Adler, Elisabeth Brandstetter, Peter D. Han, Kairsten Fay, Misja Ilcisin, Kirsten Lacombe, Thomas R. Sibley, Melissa Truong, Caitlin R. Wolf, Romesh Gautom, Geoff Melly, Brian Hiatt, Philip Dykema, Scott Lindquist, Michael Boeckh, Janet A. Englund, Michael Famulare, Barry R. Lutz, Mark J. Rieder, Lea M. Starita, Matthew Thompson, Helen Y. Chu, Jay Shendure, Trevor Bedford |
| EPI_ISL_1823097, EPI_ISL_1823098 | Altius Institute | Seattle Flu Study | Deborah A. Nickerson, Chris D. Frazar, Jover Lee, Benjamin Pelle, Erica Ryke, Matthew Richardson, Amanda Adler, Elisabeth Brandstetter, Peter D. Han, Kairsten Fay, Misja Ilcisin, Kirsten Lacombe, Thomas R. Sibley, Melissa Truong, Caitlin R. Wolf, Ryan Alexander, Daniel Bates, Rebecca Bruders, Stephanie DeBaun, Clem Green, Muhammad Halimun, Jessica Halow, Kneshay Harper, Matt Hartman, Andrew Meuser, Alex Nguyen, Truong Nguyen, Sofia Olsson, Sadie Patraw, Hannah Petersen, Tobias Ragoczy, Joshua Richards, Jacob Rodriguez, John Stamatoyannopoulos, Julia Wald, Olivia Waltner, Michael Boeckh, Janet A. Englund, Michael Famulare, Barry R. Lutz, Mark J. Rieder, Lea M. Starita, Matthew Thompson, Helen Y. Chu, Jay Shendure, Trevor Bedford |
| EPI_ISL_1823099 | Atlas Genomics | Seattle Flu Study | Deborah A. Nickerson, Chris D. Frazar, Jover Lee, Benjamin Pelle, Erica Ryke, Matthew Richardson, Amanda Adler, Elisabeth Brandstetter, Peter D. Han, Kairsten Fay, Misja Ilcisin, Kirsten Lacombe, Thomas R. Sibley, Melissa Truong, Caitlin R. Wolf, Romesh Gautom, Geoff Melly, Brian Hiatt, Philip Dykema, Scott Lindquist, Michael Boeckh, Janet A. Englund, Michael Famulare, Barry R. Lutz, Mark J. Rieder, Lea M. Starita, Matthew Thompson, Helen Y. Chu, Jay Shendure, Trevor Bedford |
| EPI_ISL_1823100 | Incyte Diagnostics Spokane | Seattle Flu Study | Deborah A. Nickerson, Chris D. Frazar, Jover Lee, Benjamin Pelle, Erica Ryke, Matthew Richardson, Amanda Adler, Elisabeth Brandstetter, Peter D. Han, Kairsten Fay, Misja Ilcisin, Kirsten Lacombe, Thomas R. Sibley, Melissa Truong, Caitlin R. Wolf, Romesh Gautom, Geoff Melly, Brian Hiatt, Philip Dykema, Scott Lindquist, Michael Boeckh, Janet A. Englund, Michael Famulare, Barry R. Lutz, Mark J. Rieder, Lea M. Starita, Matthew Thompson, Helen Y. Chu, Jay Shendure, Trevor Bedford |
| EPI_ISL_1823101, EPI_ISL_1823102, EPI_ISL_1823103, EPI_ISL_1823104 | Northwest Laboratories | Seattle Flu Study | Deborah A. Nickerson, Chris D. Frazar, Jover Lee, Benjamin Pelle, Erica Ryke, Matthew Richardson, Amanda Adler, Elisabeth Brandstetter, Peter D. Han, Kairsten Fay, Misja Ilcisin, Kirsten Lacombe, Thomas R. Sibley, Melissa Truong, Caitlin R. Wolf, Romesh Gautom, Geoff Melly, Brian Hiatt, Philip Dykema, Scott Lindquist, Michael Boeckh, Janet A. Englund, Michael Famulare, Barry R. Lutz, Mark J. Rieder, Lea M. Starita, Matthew Thompson, Helen Y. Chu, Jay Shendure, Trevor Bedford |
| EPI_ISL_1823105, EPI_ISL_1823106, EPI_ISL_1823107 | Altius Institute | Seattle Flu Study | Deborah A. Nickerson, Chris D. Frazar, Jover Lee, Benjamin Pelle, Erica Ryke, Matthew Richardson, Amanda Adler, Elisabeth Brandstetter, Peter D. Han, Kairsten Fay, Misja Ilcisin, Kirsten Lacombe, Thomas R. Sibley, Melissa Truong, Caitlin R. Wolf, Ryan Alexander, Daniel Bates, Rebecca Bruders, Stephanie DeBaun, Clem Green, Muhammad Halimun, Jessica Halow, Kneshay Harper, Matt Hartman, Andrew Meuser, Alex Nguyen, Truong Nguyen, Sofia Olsson, Sadie Patraw, Hannah Petersen, Tobias Ragoczy, Joshua Richards, Jacob Rodriguez, John Stamatoyannopoulos, Julia Wald, Olivia Waltner, Michael Boeckh, Janet A. Englund, Michael Famulare, Barry R. Lutz, Mark J. Rieder, Lea M. Starita, Matthew Thompson, Helen Y. Chu, Jay Shendure, Trevor Bedford |
| EPI_ISL_1823109, EPI_ISL_1823110, EPI_ISL_1823111, EPI_ISL_1823112, EPI_ISL_1823113, EPI_ISL_1823114, EPI_ISL_1823115, EPI_ISL_1823116, EPI_ISL_1823117, EPI_ISL_1823119, EPI_ISL_1823121, EPI_ISL_1823123, EPI_ISL_1823124 | | | |
| see above | Wichita State University - Molecular Diagnostics Lab | Kansas Health and Environmental Lab | Mike Grose, Jonathan Barnell, Ben Olsen, and Phil Adam |
| EPI_ISL_1823126, EPI_ISL_1823127, EPI_ISL_1823128, EPI_ISL_1823130, EPI_ISL_1823131, EPI_ISL_1823132, EPI_ISL_1823133, EPI_ISL_1823134, EPI_ISL_1823135 | Kansas Health and Environmental Lab | Kansas Health and Environmental Lab | Mike Grose, Jonathan Barnell, Ben Olsen, and Phil Adam |
| EPI_ISL_1823136, EPI_ISL_1823137, EPI_ISL_1823138, EPI_ISL_1823139, EPI_ISL_1823140, EPI_ISL_1823141, EPI_ISL_1823142, EPI_ISL_1823143, EPI_ISL_1823144, EPI_ISL_1823145, EPI_ISL_1823146, EPI_ISL_1823147, EPI_ISL_1823148, EPI_ISL_1823149, EPI_ISL_1823150, EPI_ISL_1823151, EPI_ISL_1823152, EPI_ISL_1823153, EPI_ISL_1823154, EPI_ISL_1823155, EPI_ISL_1823156, EPI_ISL_1823157, EPI_ISL_1823160, EPI_ISL_1823161, EPI_ISL_1823162, EPI_ISL_1823163, EPI_ISL_1823164 | | | |
| see above | South Dakota Public Health Laboratory | South Dakota Public Health Laboratory | Jacob Garfin and Chris Carlson |
| EPI_ISL_1823165, EPI_ISL_1823166, EPI_ISL_1823167, EPI_ISL_1823168, EPI_ISL_1823169, EPI_ISL_1823170, EPI_ISL_1823171, EPI_ISL_1823172, EPI_ISL_1823173, EPI_ISL_1823174, EPI_ISL_1823176 | | | |
| see above | Wyoming Public Health Laboratory | Wyoming Public Health Laboratory | Jim Mildenberger, Wanda Manley, Noah Hull, Taylor Fearing, Lynette Gumbleton, Channing Weber, Ashley Norberg, Chayse Rowley, Marley Goetz, Brian Dominguez, Elliot Thomasson, Sam Britz, Cari Sloma, and Rob Christensen |
| EPI_ISL_1823404, EPI_ISL_1823405, EPI_ISL_1823406, EPI_ISL_1823407, EPI_ISL_1823408, EPI_ISL_1823409, EPI_ISL_1823410, EPI_ISL_1823411, EPI_ISL_1823412, EPI_ISL_1823413, EPI_ISL_1823414, EPI_ISL_1823415, EPI_ISL_1823416, EPI_ISL_1823417, EPI_ISL_1823418, EPI_ISL_1823419, EPI_ISL_1823420, EPI_ISL_1823421, EPI_ISL_1823422, EPI_ISL_1823423, EPI_ISL_1823424, EPI_ISL_1823425, EPI_ISL_1823426, EPI_ISL_1823427, EPI_ISL_1823428, EPI_ISL_1823429, EPI_ISL_1823430, EPI_ISL_1823431, EPI_ISL_1823432, EPI_ISL_1823433, EPI_ISL_1823434, EPI_ISL_1823435, EPI_ISL_1823436, EPI_ISL_1823437, EPI_ISL_1823438, EPI_ISL_1823439, EPI_ISL_1823440, EPI_ISL_1823441, EPI_ISL_1823442, EPI_ISL_1823443, EPI_ISL_1823444, EPI_ISL_1823445, EPI_ISL_1823446, EPI_ISL_1823447, EPI_ISL_1823448, EPI_ISL_1823449, EPI_ISL_1823450, EPI_ISL_1823451, EPI_ISL_1823452, EPI_ISL_1823453, EPI_ISL_1823454, EPI_ISL_1823455, EPI_ISL_1823456, EPI_ISL_1823457, EPI_ISL_1823458, EPI_ISL_1823459, EPI_ISL_1823460, EPI_ISL_1823461, EPI_ISL_1823462, EPI_ISL_1823463, EPI_ISL_1823464, EPI_ISL_1823465, EPI_ISL_1823466, EPI_ISL_1823467, EPI_ISL_1823468, EPI_ISL_1823469, EPI_ISL_1823470, EPI_ISL_1823471, EPI_ISL_1823472, EPI_ISL_1823473, EPI_ISL_1823474, EPI_ISL_1823475, EPI_ISL_1823476, EPI_ISL_1823477, EPI_ISL_1823478, EPI_ISL_1823479, EPI_ISL_1823480, EPI_ISL_1823481, EPI_ISL_1823482, EPI_ISL_1823483, EPI_ISL_1823484, EPI_ISL_1823485, EPI_ISL_1823486, EPI_ISL_1823487, EPI_ISL_1823488, EPI_ISL_1823489, EPI_ISL_1823490, EPI_ISL_1823491, EPI_ISL_1823492, EPI_ISL_1823493, EPI_ISL_1823494, EPI_ISL_1823495, EPI_ISL_1823496, EPI_ISL_1823497, EPI_ISL_1823498, EPI_ISL_1823499, EPI_ISL_1823501, EPI_ISL_1823502, EPI_ISL_1823503, EPI_ISL_1823504, EPI_ISL_1823505, EPI_ISL_1823506, EPI_ISL_1823507, EPI_ISL_1823508, EPI_ISL_1823509, EPI_ISL_1823510, EPI_ISL_1823511, EPI_ISL_1823512, EPI_ISL_1823513, EPI_ISL_1823514, EPI_ISL_1823515, EPI_ISL_1823516, EPI_ISL_1823517, EPI_ISL_1823518, EPI_ISL_1823519, EPI_ISL_1823520, EPI_ISL_1823521, EPI_ISL_1823522, EPI_ISL_1823523, EPI_ISL_1823524, EPI_ISL_1823525, EPI_ISL_1823526, EPI_ISL_1823527, EPI_ISL_1823528, EPI_ISL_1823529, EPI_ISL_1823530, EPI_ISL_1823531, EPI_ISL_1823532, EPI_ISL_1823533, EPI_ISL_1823534, EPI_ISL_1823535, EPI_ISL_1823536, EPI_ISL_1823537, EPI_ISL_1823538, EPI_ISL_1823539, EPI_ISL_1823540, EPI_ISL_1823541, EPI_ISL_1823542, EPI_ISL_1823543, EPI_ISL_1823544, EPI_ISL_1823545, EPI_ISL_1823546, EPI_ISL_1823547, EPI_ISL_1823548, EPI_ISL_1823549, EPI_ISL_1823550, EPI_ISL_1823551, EPI_ISL_1823552, EPI_ISL_1823553, EPI_ISL_1823554, EPI_ISL_1823555, EPI_ISL_1823556, EPI_ISL_1823557, EPI_ISL_1823558, EPI_ISL_1823559, EPI_ISL_1823560, EPI_ISL_1823561, EPI_ISL_1823562, EPI_ISL_1823563, EPI_ISL_1823564, EPI_ISL_1823565, EPI_ISL_1823566, EPI_ISL_1823567, EPI_ISL_1823568 | | | |
| see above | Indiana Animal Disease Diagnostic Laboratory | Carpi Laboratory - Purdue University | Jack Dorman, Ilinca I Ciubotariu, Nicole M Perry, Jobin J Kattoor, Lev Gorenstein, Abebe A Fola, G Kenitra Hendrix, Rebecca P Wilkes, Giovanna Carpi |
| EPI_ISL_1823569, EPI_ISL_1823570, EPI_ISL_1823571 | FL Bureau of Public Health Laboratories-Miami | Centers for Disease Control and Prevention Division of Viral Diseases, Pathogen Discovery | Mili Sheth, Sarah Nobles, Jasmine Padilla, Mark Burroughs, Shoshona Le, Katie Dillon, Peter Cook, Clinton R. Paden, Dhwani Batra, Krista Queen, Kristen Knipe, Dakota Howard, Yvette Unoarumhi, Darlene Wagner, Matthew Schmerer, Ben L. Rambo-Martin, Kristine Lacek, Sam Shepard, Alison Laufer Halpin, Dave Wentworth, Vivien Dugan, Suxiang Tong, Justin Lee |
| EPI_ISL_1823572, EPI_ISL_1823573, EPI_ISL_1823574, EPI_ISL_1823575, EPI_ISL_1823576, EPI_ISL_1823577, EPI_ISL_1823578, EPI_ISL_1823579, EPI_ISL_1823580, EPI_ISL_1823581, EPI_ISL_1823582 | | | |
| see above | SD Public Health Laboratory | Centers for Disease Control and Prevention Division of Viral Diseases, Pathogen Discovery | Mili Sheth, Sarah Nobles, Jasmine Padilla, Mark Burroughs, Shoshona Le, Katie Dillon, Peter Cook, Clinton R. Paden, Dhwani Batra, Krista Queen, Kristen Knipe, Dakota Howard, Yvette Unoarumhi, Darlene Wagner, Matthew Schmerer, Ben L. Rambo-Martin, Kristine Lacek, Sam Shepard, Alison Laufer Halpin, Dave Wentworth, Vivien Dugan, Suxiang Tong, Justin Lee |
| EPI_ISL_1823583, EPI_ISL_1823584, EPI_ISL_1823585, EPI_ISL_1823586, EPI_ISL_1823587, EPI_ISL_1823588, EPI_ISL_1823589 | Guam Public Health Laboratory | Centers for Disease Control and Prevention Division of Viral Diseases, Pathogen Discovery | Mili Sheth, Sarah Nobles, Jasmine Padilla, Mark Burroughs, Shoshona Le, Katie Dillon, Peter Cook, Clinton R. Paden, Dhwani Batra, Krista Queen, Kristen Knipe, Dakota Howard, Yvette Unoarumhi, Darlene Wagner, Matthew Schmerer, Ben L. Rambo-Martin, Kristine Lacek, Sam Shepard, Alison Laufer Halpin, Dave Wentworth, Vivien Dugan, Suxiang Tong, Justin Lee |
| EPI_ISL_1823590, EPI_ISL_1823591 | SC Dept of Health and Env. Control-Bureau of Laboratories | Centers for Disease Control and Prevention Division of Viral Diseases, Pathogen Discovery | Mili Sheth, Sarah Nobles, Jasmine Padilla, Mark Burroughs, Shoshona Le, Katie Dillon, Peter Cook, Clinton R. Paden, Dhwani Batra, Krista Queen, Kristen Knipe, Dakota Howard, Yvette Unoarumhi, Darlene Wagner, Matthew Schmerer, Ben L. Rambo-Martin, Kristine Lacek, Sam Shepard, Alison Laufer Halpin, Dave Wentworth, Vivien Dugan, Suxiang Tong, Justin Lee |
| EPI_ISL_1823592, EPI_ISL_1823593, EPI_ISL_1823594, EPI_ISL_1823595, EPI_ISL_1823596, EPI_ISL_1823597, EPI_ISL_1823598, EPI_ISL_1823599, EPI_ISL_1823600, EPI_ISL_1823601, EPI_ISL_1823602, EPI_ISL_1823603, EPI_ISL_1823604, EPI_ISL_1823605, EPI_ISL_1823606, EPI_ISL_1823607, EPI_ISL_1823608, EPI_ISL_1823609, EPI_ISL_1823610 | | | |
| see above | PR Public Health Lab | Centers for Disease Control and Prevention Division of Viral Diseases, Pathogen Discovery | Mili Sheth, Sarah Nobles, Jasmine Padilla, Mark Burroughs, Shoshona Le, Katie Dillon, Peter Cook, Clinton R. Paden, Dhwani Batra, Krista Queen, Kristen Knipe, Dakota Howard, Yvette Unoarumhi, Darlene Wagner, Matthew Schmerer, Ben L. Rambo-Martin, Kristine Lacek, Sam Shepard, Alison Laufer Halpin, Dave Wentworth, Vivien Dugan, Suxiang Tong, Justin Lee |
| EPI_ISL_1823611, EPI_ISL_1823612, EPI_ISL_1823613, EPI_ISL_1823614, EPI_ISL_1823615 | Philadelphia Department of Public Health | Centers for Disease Control and Prevention Division of Viral Diseases, Pathogen Discovery | Mili Sheth, Sarah Nobles, Jasmine Padilla, Mark Burroughs, Shoshona Le, Katie Dillon, Peter Cook, Clinton R. Paden, Dhwani Batra, Krista Queen, Kristen Knipe, Dakota Howard, Yvette Unoarumhi, Darlene Wagner, Matthew Schmerer, Ben L. Rambo-Martin, Kristine Lacek, Sam Shepard, Alison Laufer Halpin, Dave Wentworth, Vivien Dugan, Suxiang Tong, Justin Lee |
| EPI_ISL_1823616, EPI_ISL_1823617, EPI_ISL_1823618, EPI_ISL_1823619, EPI_ISL_1823620, EPI_ISL_1823621, EPI_ISL_1823622, EPI_ISL_1823623, EPI_ISL_1823624, EPI_ISL_1823625, EPI_ISL_1823626, EPI_ISL_1823627 | | | |
| see above | KY State Public Health Lab | Centers for Disease Control and Prevention Division of Viral Diseases, Pathogen Discovery | Mili Sheth, Sarah Nobles, Jasmine Padilla, Mark Burroughs, Shoshona Le, Katie Dillon, Peter Cook, Clinton R. Paden, Dhwani Batra, Krista Queen, Kristen Knipe, Dakota Howard, Yvette Unoarumhi, Darlene Wagner, Matthew Schmerer, Ben L. Rambo-Martin, Kristine Lacek, Sam Shepard, Alison Laufer Halpin, Dave Wentworth, Vivien Dugan, Suxiang Tong, Justin Lee |
| EPI_ISL_1823628, EPI_ISL_1823629, EPI_ISL_1823630, EPI_ISL_1823631, EPI_ISL_1823632, EPI_ISL_1823633, EPI_ISL_1823634 | IA State Hygienic Laboratory | Centers for Disease Control and Prevention Division of Viral Diseases, Pathogen Discovery | Mili Sheth, Sarah Nobles, Jasmine Padilla, Mark Burroughs, Shoshona Le, Katie Dillon, Peter Cook, Clinton R. Paden, Dhwani Batra, Krista Queen, Kristen Knipe, Dakota Howard, Yvette Unoarumhi, Darlene Wagner, Matthew Schmerer, Ben L. Rambo-Martin, Kristine Lacek, Sam Shepard, Alison Laufer Halpin, Dave Wentworth, Vivien Dugan, Suxiang Tong, Justin Lee |
| EPI_ISL_1823635, EPI_ISL_1823636, EPI_ISL_1823637, | ND Dept. of Health Laboratory Services-Microbiology | Centers for Disease Control and Prevention Division of Viral Diseases, Pathogen Discovery | Mili Sheth, Sarah Nobles, Jasmine Padilla, Mark Burroughs, Shoshona Le, Katie Dillon, Peter Cook, Clinton R. Paden, Dhwani Batra, Krista Queen, Kristen Knipe, Dakota Howard, Yvette Unoarumhi, Darlene Wagner, Matthew Schmerer, Ben L. Rambo-Martin, Kristine Lacek, Sam Shepard, Alison Laufer Halpin, Dave Wentworth, Vivien Dugan, Suxiang Tong, Justin Lee |

| | | | |
|---|---|---|---|
| EPI_ISL_1823638, EPI_ISL_1823639, EPI_ISL_1823640, EPI_ISL_1823641, EPI_ISL_1823642 | | | |
| EPI_ISL_1823643, EPI_ISL_1823644, EPI_ISL_1823645, EPI_ISL_1823646, EPI_ISL_1823647, EPI_ISL_1823648 | "MN PHL Division, Minnesota Department of Health" | Centers for Disease Control and Prevention Division of Viral Diseases, Pathogen Discovery | Mili Sheth, Sarah Nobles, Jasmine Padilla, Mark Burroughs, Shoshona Le, Katie Dillon, Peter Cook, Clinton R. Paden, Dhwani Batra, Krista Queen, Kristen Knipe, Dakota Howard, Yvette Unoarumhi, Darlene Wagner, Matthew Schmerer, Ben L. Rambo-Martin, Kristine Lacek, Sam Shepard, Alison Laufer Halpin, Dave Wentworth, Vivien Dugan, Suxiang Tong, Justin Lee |
| EPI_ISL_1823649, EPI_ISL_1823650, EPI_ISL_1823651, EPI_ISL_1823652, EPI_ISL_1823653, EPI_ISL_1823654, EPI_ISL_1823655, EPI_ISL_1823656, EPI_ISL_1823657 | OR State PHL-Virology/Immunology Section | Centers for Disease Control and Prevention Division of Viral Diseases, Pathogen Discovery | Mili Sheth, Sarah Nobles, Jasmine Padilla, Mark Burroughs, Shoshona Le, Katie Dillon, Peter Cook, Clinton R. Paden, Dhwani Batra, Krista Queen, Kristen Knipe, Dakota Howard, Yvette Unoarumhi, Darlene Wagner, Matthew Schmerer, Ben L. Rambo-Martin, Kristine Lacek, Sam Shepard, Alison Laufer Halpin, Dave Wentworth, Vivien Dugan, Suxiang Tong, Justin Lee |
| EPI_ISL_1823658, EPI_ISL_1823659, EPI_ISL_1823660, EPI_ISL_1823661, EPI_ISL_1823662, EPI_ISL_1823663, EPI_ISL_1823664, EPI_ISL_1823665, EPI_ISL_1823666, EPI_ISL_1823667, EPI_ISL_1823668, EPI_ISL_1823669, EPI_ISL_1823670, EPI_ISL_1823671, EPI_ISL_1823672, EPI_ISL_1823673, EPI_ISL_1823674, EPI_ISL_1823675 | | | |
| see above | KS Health and Environmental Laboratories | Centers for Disease Control and Prevention Division of Viral Diseases, Pathogen Discovery | Mili Sheth, Sarah Nobles, Jasmine Padilla, Mark Burroughs, Shoshona Le, Katie Dillon, Peter Cook, Clinton R. Paden, Dhwani Batra, Krista Queen, Kristen Knipe, Dakota Howard, Yvette Unoarumhi, Darlene Wagner, Matthew Schmerer, Ben L. Rambo-Martin, Kristine Lacek, Sam Shepard, Alison Laufer Halpin, Dave Wentworth, Vivien Dugan, Suxiang Tong, Justin Lee |
| EPI_ISL_1823676, EPI_ISL_1823677, EPI_ISL_1823678, EPI_ISL_1823679, EPI_ISL_1823680, EPI_ISL_1823681, EPI_ISL_1823682, EPI_ISL_1823683, EPI_ISL_1823684, EPI_ISL_1823685, EPI_ISL_1823686 | | | |
| see above | FL Bureau of Public Health Laboratories-Tampa | Centers for Disease Control and Prevention Division of Viral Diseases, Pathogen Discovery | Mili Sheth, Sarah Nobles, Jasmine Padilla, Mark Burroughs, Shoshona Le, Katie Dillon, Peter Cook, Clinton R. Paden, Dhwani Batra, Krista Queen, Kristen Knipe, Dakota Howard, Yvette Unoarumhi, Darlene Wagner, Matthew Schmerer, Ben L. Rambo-Martin, Kristine Lacek, Sam Shepard, Alison Laufer Halpin, Dave Wentworth, Vivien Dugan, Suxiang Tong, Justin Lee |
| EPI_ISL_1823687 | FL Bur. of Public Health Laboratories-Jacksonville | Centers for Disease Control and Prevention Division of Viral Diseases, Pathogen Discovery | Mili Sheth, Sarah Nobles, Jasmine Padilla, Mark Burroughs, Shoshona Le, Katie Dillon, Peter Cook, Clinton R. Paden, Dhwani Batra, Krista Queen, Kristen Knipe, Dakota Howard, Yvette Unoarumhi, Darlene Wagner, Matthew Schmerer, Ben L. Rambo-Martin, Kristine Lacek, Sam Shepard, Alison Laufer Halpin, Dave Wentworth, Vivien Dugan, Suxiang Tong, Justin Lee |
| EPI_ISL_1823688, EPI_ISL_1823689, EPI_ISL_1823690, EPI_ISL_1823691, EPI_ISL_1823692, EPI_ISL_1823693, EPI_ISL_1823694, EPI_ISL_1823695, EPI_ISL_1823696, EPI_ISL_1823697, EPI_ISL_1823698, EPI_ISL_1823699, EPI_ISL_1823700 | | | |
| see above | ID Bureau of Laboratories | Centers for Disease Control and Prevention Division of Viral Diseases, Pathogen Discovery | Mili Sheth, Sarah Nobles, Jasmine Padilla, Mark Burroughs, Shoshona Le, Katie Dillon, Peter Cook, Clinton R. Paden, Dhwani Batra, Krista Queen, Kristen Knipe, Dakota Howard, Yvette Unoarumhi, Darlene Wagner, Matthew Schmerer, Ben L. Rambo-Martin, Kristine Lacek, Sam Shepard, Alison Laufer Halpin, Dave Wentworth, Vivien Dugan, Suxiang Tong, Justin Lee |
| EPI_ISL_1823701, EPI_ISL_1823702, EPI_ISL_1823703, EPI_ISL_1823704, EPI_ISL_1823705, EPI_ISL_1823706, EPI_ISL_1823707, EPI_ISL_1823708, EPI_ISL_1823709, EPI_ISL_1823710, EPI_ISL_1823711, EPI_ISL_1823712 | | | |
| see above | TN Division of Laboratory Services | Centers for Disease Control and Prevention Division of Viral Diseases, Pathogen Discovery | Mili Sheth, Sarah Nobles, Jasmine Padilla, Mark Burroughs, Shoshona Le, Katie Dillon, Peter Cook, Clinton R. Paden, Dhwani Batra, Krista Queen, Kristen Knipe, Dakota Howard, Yvette Unoarumhi, Darlene Wagner, Matthew Schmerer, Ben L. Rambo-Martin, Kristine Lacek, Sam Shepard, Alison Laufer Halpin, Dave Wentworth, Vivien Dugan, Suxiang Tong, Justin Lee |
| EPI_ISL_1823713, EPI_ISL_1823714, EPI_ISL_1823715, EPI_ISL_1823716, EPI_ISL_1823717, EPI_ISL_1823718, EPI_ISL_1823719, EPI_ISL_1823720, EPI_ISL_1823721, EPI_ISL_1823722, EPI_ISL_1823723, EPI_ISL_1823724, EPI_ISL_1823725, EPI_ISL_1823726, EPI_ISL_1823727, EPI_ISL_1823728, EPI_ISL_1823729, EPI_ISL_1823730, EPI_ISL_1823731, EPI_ISL_1823732, EPI_ISL_1823733, EPI_ISL_1823734, EPI_ISL_1823735, EPI_ISL_1823736, EPI_ISL_1823737, EPI_ISL_1823738, EPI_ISL_1823739, EPI_ISL_1823740, EPI_ISL_1823741, EPI_ISL_1823742, EPI_ISL_1823743, EPI_ISL_1823744, EPI_ISL_1823745, EPI_ISL_1823746, EPI_ISL_1823747 | | | |
| see above | CA-Los Angeles County Public Health Laboratory | Centers for Disease Control and Prevention Division of Viral Diseases, Pathogen Discovery | Mili Sheth, Sarah Nobles, Jasmine Padilla, Mark Burroughs, Shoshona Le, Katie Dillon, Peter Cook, Clinton R. Paden, Dhwani Batra, Krista Queen, Kristen Knipe, Dakota Howard, Yvette Unoarumhi, Darlene Wagner, Matthew Schmerer, Ben L. Rambo-Martin, Kristine Lacek, Sam Shepard, Alison Laufer Halpin, Dave Wentworth, Vivien Dugan, Suxiang Tong, Justin Lee |
| EPI_ISL_1823748, EPI_ISL_1823749, EPI_ISL_1823750, EPI_ISL_1823751, EPI_ISL_1823752, EPI_ISL_1823753, EPI_ISL_1823754, EPI_ISL_1823755, EPI_ISL_1823756, EPI_ISL_1823757, EPI_ISL_1823758, EPI_ISL_1823759, EPI_ISL_1823760, EPI_ISL_1823761, EPI_ISL_1823762, EPI_ISL_1823763, EPI_ISL_1823764, EPI_ISL_1823765, EPI_ISL_1823766, EPI_ISL_1823767, EPI_ISL_1823768, EPI_ISL_1823769, EPI_ISL_1823770, EPI_ISL_1823771, EPI_ISL_1823772, EPI_ISL_1823773 | | | |
| see above | RI State Health Laboratories | Centers for Disease Control and Prevention Division of Viral Diseases, Pathogen Discovery | Mili Sheth, Sarah Nobles, Jasmine Padilla, Mark Burroughs, Shoshona Le, Katie Dillon, Peter Cook, Clinton R. Paden, Dhwani Batra, Krista Queen, Kristen Knipe, Dakota Howard, Yvette Unoarumhi, Darlene Wagner, Matthew Schmerer, Ben L. Rambo-Martin, Kristine Lacek, Sam Shepard, Alison Laufer Halpin, Dave Wentworth, Vivien Dugan, Suxiang Tong, Justin Lee |
| EPI_ISL_1823774, EPI_ISL_1823775, EPI_ISL_1823776, EPI_ISL_1823777, EPI_ISL_1823778, EPI_ISL_1823779, EPI_ISL_1823780 | "NM Dept. Health, Scientific Laboratory Division " | Centers for Disease Control and Prevention Division of Viral Diseases, Pathogen Discovery | Mili Sheth, Sarah Nobles, Jasmine Padilla, Mark Burroughs, Shoshona Le, Katie Dillon, Peter Cook, Clinton R. Paden, Dhwani Batra, Krista Queen, Kristen Knipe, Dakota Howard, Yvette Unoarumhi, Darlene Wagner, Matthew Schmerer, Ben L. Rambo-Martin, Kristine Lacek, Sam Shepard, Alison Laufer Halpin, Dave Wentworth, Vivien Dugan, Suxiang Tong, Justin Lee |
| EPI_ISL_1823781, EPI_ISL_1823782, EPI_ISL_1823783, EPI_ISL_1823784, EPI_ISL_1823785, EPI_ISL_1823786, EPI_ISL_1823787, EPI_ISL_1823788 | IL Department of Public Health Chicago Laboratory | Centers for Disease Control and Prevention Division of Viral Diseases, Pathogen Discovery | Mili Sheth, Sarah Nobles, Jasmine Padilla, Mark Burroughs, Shoshona Le, Katie Dillon, Peter Cook, Clinton R. Paden, Dhwani Batra, Krista Queen, Kristen Knipe, Dakota Howard, Yvette Unoarumhi, Darlene Wagner, Matthew Schmerer, Ben L. Rambo-Martin, Kristine Lacek, Sam Shepard, Alison Laufer Halpin, Dave Wentworth, Vivien Dugan, Suxiang Tong, Justin Lee |
| EPI_ISL_1823789, EPI_ISL_1823790, EPI_ISL_1823791, EPI_ISL_1823792 | ME Health and Environmental Testing Laboratory | Centers for Disease Control and Prevention Division of Viral Diseases, Pathogen Discovery | Mili Sheth, Sarah Nobles, Jasmine Padilla, Mark Burroughs, Shoshona Le, Katie Dillon, Peter Cook, Clinton R. Paden, Dhwani Batra, Krista Queen, Kristen Knipe, Dakota Howard, Yvette Unoarumhi, Darlene Wagner, Matthew Schmerer, Ben L. Rambo-Martin, Kristine Lacek, Sam Shepard, Alison Laufer Halpin, Dave Wentworth, Vivien Dugan, Suxiang Tong, Justin Lee |
| EPI_ISL_1823793, EPI_ISL_1823794, EPI_ISL_1823795, EPI_ISL_1823796 | WVDHHR - Office of Laboratory Services | Centers for Disease Control and Prevention Division of Viral Diseases, Pathogen Discovery | Mili Sheth, Sarah Nobles, Jasmine Padilla, Mark Burroughs, Shoshona Le, Katie Dillon, Peter Cook, Clinton R. Paden, Dhwani Batra, Krista Queen, Kristen Knipe, Dakota Howard, Yvette Unoarumhi, Darlene Wagner, Matthew Schmerer, Ben L. Rambo-Martin, Kristine Lacek, Sam Shepard, Alison Laufer Halpin, Dave Wentworth, Vivien Dugan, Suxiang Tong, Justin Lee |
| EPI_ISL_1823797, EPI_ISL_1823798, EPI_ISL_1823799, EPI_ISL_1823800 | "AR Dept. of Health-PHL, Molecular Diagnostics" | Centers for Disease Control and Prevention Division of Viral Diseases, Pathogen Discovery | Mili Sheth, Sarah Nobles, Jasmine Padilla, Mark Burroughs, Shoshona Le, Katie Dillon, Peter Cook, Clinton R. Paden, Dhwani Batra, Krista Queen, Kristen Knipe, Dakota Howard, Yvette Unoarumhi, Darlene Wagner, Matthew Schmerer, Ben L. Rambo-Martin, Kristine Lacek, Sam Shepard, Alison Laufer Halpin, Dave Wentworth, Vivien Dugan, Suxiang Tong, Justin Lee |
| EPI_ISL_1823801, EPI_ISL_1823802, EPI_ISL_1823803, EPI_ISL_1823804, EPI_ISL_1823805, EPI_ISL_1823806, EPI_ISL_1823807, EPI_ISL_1823808, EPI_ISL_1823809, EPI_ISL_1823810, EPI_ISL_1823811, EPI_ISL_1823812, EPI_ISL_1823813, EPI_ISL_1823814 | | | |
| see above | LA Office of Public Health Laboratories | Centers for Disease Control and Prevention Division of Viral Diseases, Pathogen Discovery | Mili Sheth, Sarah Nobles, Jasmine Padilla, Mark Burroughs, Shoshona Le, Katie Dillon, Peter Cook, Clinton R. Paden, Dhwani Batra, Krista Queen, Kristen Knipe, Dakota Howard, Yvette Unoarumhi, Darlene Wagner, Matthew Schmerer, Ben L. Rambo-Martin, Kristine Lacek, Sam Shepard, Alison Laufer Halpin, Dave Wentworth, Vivien Dugan, Suxiang Tong, Justin Lee |
| EPI_ISL_1823815, EPI_ISL_1823816, EPI_ISL_1823817, EPI_ISL_1823818, EPI_ISL_1823819, EPI_ISL_1823820, EPI_ISL_1823821, EPI_ISL_1823822, EPI_ISL_1823823, EPI_ISL_1823824, EPI_ISL_1823825, EPI_ISL_1823826, EPI_ISL_1823827, EPI_ISL_1823828, EPI_ISL_1823829, EPI_ISL_1823830, EPI_ISL_1823831, EPI_ISL_1823832, EPI_ISL_1823833 | | | |
| see above | WA State Department of Health | Centers for Disease Control and Prevention Division of Viral Diseases, Pathogen Discovery | Mili Sheth, Sarah Nobles, Jasmine Padilla, Mark Burroughs, Shoshona Le, Katie Dillon, Peter Cook, Clinton R. Paden, Dhwani Batra, Krista Queen, Kristen Knipe, Dakota Howard, Yvette Unoarumhi, Darlene Wagner, Matthew Schmerer, Ben L. Rambo-Martin, Kristine Lacek, Sam Shepard, Alison Laufer Halpin, Dave Wentworth, Vivien Dugan, Suxiang Tong, Justin Lee |
| EPI_ISL_1823834 | "AZ SPHL, Arizona Department of Health Services" | Centers for Disease Control and Prevention Division of Viral Diseases, Pathogen Discovery | Mili Sheth, Sarah Nobles, Jasmine Padilla, Mark Burroughs, Shoshona Le, Katie Dillon, Peter Cook, Clinton R. Paden, Dhwani Batra, Krista Queen, Kristen Knipe, Dakota Howard, Yvette Unoarumhi, Darlene Wagner, Matthew Schmerer, Ben L. Rambo-Martin, Kristine Lacek, Sam Shepard, Alison Laufer Halpin, Dave Wentworth, Vivien Dugan, Suxiang Tong, Justin Lee |
| EPI_ISL_1823835, EPI_ISL_1823836, EPI_ISL_1823837, EPI_ISL_1823838, EPI_ISL_1823839, EPI_ISL_1823840 | UT-Unified State Labs: Public Health Utah DOH | Centers for Disease Control and Prevention Division of Viral Diseases, Pathogen Discovery | Mili Sheth, Sarah Nobles, Jasmine Padilla, Mark Burroughs, Shoshona Le, Katie Dillon, Peter Cook, Clinton R. Paden, Dhwani Batra, Krista Queen, Kristen Knipe, Dakota Howard, Yvette Unoarumhi, Darlene Wagner, Matthew Schmerer, Ben L. Rambo-Martin, Kristine Lacek, Sam Shepard, Alison Laufer Halpin, Dave Wentworth, Vivien Dugan, Suxiang Tong, Justin Lee |
| EPI_ISL_1823841, EPI_ISL_1823842, EPI_ISL_1823843, EPI_ISL_1823844, EPI_ISL_1823845, | NE Public Health Laboratory | Centers for Disease Control and Prevention Division of Viral Diseases, Pathogen Discovery | Mili Sheth, Sarah Nobles, Jasmine Padilla, Mark Burroughs, Shoshona Le, Katie Dillon, Peter Cook, Clinton R. Paden, Dhwani Batra, Krista Queen, Kristen Knipe, Dakota Howard, Yvette Unoarumhi, Darlene Wagner, Matthew Schmerer, Ben L. Rambo-Martin, Kristine Lacek, Sam Shepard, Alison Laufer Halpin, Dave Wentworth, Vivien Dugan, Suxiang Tong, Justin Lee |

| | | | |
|---|---|---|---|
| EPI_ISL_1823846, EPI_ISL_1823847, EPI_ISL_1823848, EPI_ISL_1823849, EPI_ISL_1823850 | | | |
| EPI_ISL_1823851, EPI_ISL_1823852, EPI_ISL_1823853, EPI_ISL_1823854, EPI_ISL_1823855, EPI_ISL_1823856, EPI_ISL_1823857, EPI_ISL_1823858, EPI_ISL_1823859, EPI_ISL_1823860 | OH Department of Health Laboratory | Centers for Disease Control and Prevention Division of Viral Diseases, Pathogen Discovery | Mili Sheth, Sarah Nobles, Jasmine Padilla, Mark Burroughs, Shoshona Le, Katie Dillon, Peter Cook, Clinton R. Paden, Dhwani Batra, Krista Queen, Kristen Knipe, Dakota Howard, Yvette Unoarumhi, Darlene Wagner, Matthew Schmerer, Ben L. Rambo-Martin, Kristine Lacek, Sam Shepard, Alison Laufer Halpin, Dave Wentworth, Vivien Dugan, Suxiang Tong, Justin Lee |
| EPI_ISL_1823861, EPI_ISL_1823862, EPI_ISL_1823863, EPI_ISL_1823864, EPI_ISL_1823865 | "PA Department of Health, Bureau of Laboratories" | Centers for Disease Control and Prevention Division of Viral Diseases, Pathogen Discovery | Mili Sheth, Sarah Nobles, Jasmine Padilla, Mark Burroughs, Shoshona Le, Katie Dillon, Peter Cook, Clinton R. Paden, Dhwani Batra, Krista Queen, Kristen Knipe, Dakota Howard, Yvette Unoarumhi, Darlene Wagner, Matthew Schmerer, Ben L. Rambo-Martin, Kristine Lacek, Sam Shepard, Alison Laufer Halpin, Dave Wentworth, Vivien Dugan, Suxiang Tong, Justin Lee |
| EPI_ISL_1823866, EPI_ISL_1823867, EPI_ISL_1823868, EPI_ISL_1823869 | "OK Public Health Laboratory, Oklahoma State DOH" | Centers for Disease Control and Prevention Division of Viral Diseases, Pathogen Discovery | Mili Sheth, Sarah Nobles, Jasmine Padilla, Mark Burroughs, Shoshona Le, Katie Dillon, Peter Cook, Clinton R. Paden, Dhwani Batra, Krista Queen, Kristen Knipe, Dakota Howard, Yvette Unoarumhi, Darlene Wagner, Matthew Schmerer, Ben L. Rambo-Martin, Kristine Lacek, Sam Shepard, Alison Laufer Halpin, Dave Wentworth, Vivien Dugan, Suxiang Tong, Justin Lee |
| EPI_ISL_1823870, EPI_ISL_1823871, EPI_ISL_1823872, EPI_ISL_1823873, EPI_ISL_1823874, EPI_ISL_1823875, EPI_ISL_1823876, EPI_ISL_1823877, EPI_ISL_1823878, EPI_ISL_1823879, EPI_ISL_1823880, EPI_ISL_1823881, EPI_ISL_1823882, EPI_ISL_1823883, EPI_ISL_1823884, EPI_ISL_1823885, EPI_ISL_1823886, EPI_ISL_1823887, EPI_ISL_1823888, EPI_ISL_1823889, EPI_ISL_1823890, EPI_ISL_1823891, EPI_ISL_1823892, EPI_ISL_1823893, EPI_ISL_1823894, EPI_ISL_1823895, EPI_ISL_1823896, EPI_ISL_1823897, EPI_ISL_1823898 | | | |
| see above | VA-Division of Consolidated Laboratory Services | Centers for Disease Control and Prevention Division of Viral Diseases, Pathogen Discovery | Mili Sheth, Sarah Nobles, Jasmine Padilla, Mark Burroughs, Shoshona Le, Katie Dillon, Peter Cook, Clinton R. Paden, Dhwani Batra, Krista Queen, Kristen Knipe, Dakota Howard, Yvette Unoarumhi, Darlene Wagner, Matthew Schmerer, Ben L. Rambo-Martin, Kristine Lacek, Sam Shepard, Alison Laufer Halpin, Dave Wentworth, Vivien Dugan, Suxiang Tong, Justin Lee |
| EPI_ISL_1823899, EPI_ISL_1823900, EPI_ISL_1823901, EPI_ISL_1823902, EPI_ISL_1823903, EPI_ISL_1823904, EPI_ISL_1823905, EPI_ISL_1823906, EPI_ISL_1823907, EPI_ISL_1823908, EPI_ISL_1823909, EPI_ISL_1823910 | | | |
| see above | NC State Laboratory of Public Health | Centers for Disease Control and Prevention Division of Viral Diseases, Pathogen Discovery | Mili Sheth, Sarah Nobles, Jasmine Padilla, Mark Burroughs, Shoshona Le, Katie Dillon, Peter Cook, Clinton R. Paden, Dhwani Batra, Krista Queen, Kristen Knipe, Dakota Howard, Yvette Unoarumhi, Darlene Wagner, Matthew Schmerer, Ben L. Rambo-Martin, Kristine Lacek, Sam Shepard, Alison Laufer Halpin, Dave Wentworth, Vivien Dugan, Suxiang Tong, Justin Lee |
| EPI_ISL_1823911, EPI_ISL_1823912, EPI_ISL_1823913, EPI_ISL_1823914, EPI_ISL_1823915, EPI_ISL_1823916, EPI_ISL_1823917, EPI_ISL_1823918, EPI_ISL_1823919, EPI_ISL_1823920, EPI_ISL_1823921, EPI_ISL_1823922, EPI_ISL_1823923, EPI_ISL_1823924, EPI_ISL_1823925, EPI_ISL_1823926, EPI_ISL_1823927, EPI_ISL_1823928 | | | |
| see above | MI - Michigan Department of Health and Human Services - Bureau of Laboratories | Centers for Disease Control and Prevention Division of Viral Diseases, Pathogen Discovery | Mili Sheth, Sarah Nobles, Jasmine Padilla, Mark Burroughs, Shoshona Le, Katie Dillon, Peter Cook, Clinton R. Paden, Dhwani Batra, Krista Queen, Kristen Knipe, Dakota Howard, Yvette Unoarumhi, Darlene Wagner, Matthew Schmerer, Ben L. Rambo-Martin, Kristine Lacek, Sam Shepard, Alison Laufer Halpin, Dave Wentworth, Vivien Dugan, Suxiang Tong, Justin Lee |
| EPI_ISL_1823929, EPI_ISL_1823930, EPI_ISL_1823931, EPI_ISL_1823932, EPI_ISL_1823933, EPI_ISL_1823934, EPI_ISL_1823935, EPI_ISL_1823936, EPI_ISL_1823937, EPI_ISL_1823938, EPI_ISL_1823939, EPI_ISL_1823940, EPI_ISL_1823941, EPI_ISL_1823942, EPI_ISL_1823943 | | | |
| see above | "AK State Public Health Lab, State Health Department" | Centers for Disease Control and Prevention Division of Viral Diseases, Pathogen Discovery | Mili Sheth, Sarah Nobles, Jasmine Padilla, Mark Burroughs, Shoshona Le, Katie Dillon, Peter Cook, Clinton R. Paden, Dhwani Batra, Krista Queen, Kristen Knipe, Dakota Howard, Yvette Unoarumhi, Darlene Wagner, Matthew Schmerer, Ben L. Rambo-Martin, Kristine Lacek, Sam Shepard, Alison Laufer Halpin, Dave Wentworth, Vivien Dugan, Suxiang Tong, Justin Lee |
| EPI_ISL_1823944, EPI_ISL_1823945, EPI_ISL_1823946, EPI_ISL_1823947, EPI_ISL_1823948, EPI_ISL_1823949, EPI_ISL_1823950, EPI_ISL_1823951, EPI_ISL_1823952, EPI_ISL_1823953, EPI_ISL_1823954, EPI_ISL_1823955, EPI_ISL_1823956, EPI_ISL_1823957, EPI_ISL_1823958, EPI_ISL_1823959, EPI_ISL_1823960, EPI_ISL_1823961, EPI_ISL_1823962, EPI_ISL_1823963, EPI_ISL_1823964, EPI_ISL_1823965, EPI_ISL_1823966, EPI_ISL_1823967, EPI_ISL_1823968, EPI_ISL_1823969, EPI_ISL_1823970, EPI_ISL_1823971, EPI_ISL_1823972, EPI_ISL_1823973 | | | |
| see above | NYC Department of Health and Mental Hygiene | Centers for Disease Control and Prevention Division of Viral Diseases, Pathogen Discovery | Mili Sheth, Sarah Nobles, Jasmine Padilla, Mark Burroughs, Shoshona Le, Katie Dillon, Peter Cook, Clinton R. Paden, Dhwani Batra, Krista Queen, Kristen Knipe, Dakota Howard, Yvette Unoarumhi, Darlene Wagner, Matthew Schmerer, Ben L. Rambo-Martin, Kristine Lacek, Sam Shepard, Alison Laufer Halpin, Dave Wentworth, Vivien Dugan, Suxiang Tong, Justin Lee |
| EPI_ISL_1823974, EPI_ISL_1823975, EPI_ISL_1823976, EPI_ISL_1823977, EPI_ISL_1823978, EPI_ISL_1823979, EPI_ISL_1823980, EPI_ISL_1823981, EPI_ISL_1823982, EPI_ISL_1823983, EPI_ISL_1823984, EPI_ISL_1823985, EPI_ISL_1823986, EPI_ISL_1823987, EPI_ISL_1823988, EPI_ISL_1823989, EPI_ISL_1823990, EPI_ISL_1823991, EPI_ISL_1823992, EPI_ISL_1823993, EPI_ISL_1823994, EPI_ISL_1823995, EPI_ISL_1823996 | | | |
| see above | NJ Public Health and Environmental Laboratories | Centers for Disease Control and Prevention Division of Viral Diseases, Pathogen Discovery | Mili Sheth, Sarah Nobles, Jasmine Padilla, Mark Burroughs, Shoshona Le, Katie Dillon, Peter Cook, Clinton R. Paden, Dhwani Batra, Krista Queen, Kristen Knipe, Dakota Howard, Yvette Unoarumhi, Darlene Wagner, Matthew Schmerer, Ben L. Rambo-Martin, Kristine Lacek, Sam Shepard, Alison Laufer Halpin, Dave Wentworth, Vivien Dugan, Suxiang Tong, Justin Lee |
| EPI_ISL_1823997, EPI_ISL_1823998, EPI_ISL_1823999, EPI_ISL_1824000, EPI_ISL_1824001, EPI_ISL_1824002, EPI_ISL_1824003, EPI_ISL_1824004, EPI_ISL_1824005, EPI_ISL_1824006, EPI_ISL_1824007 | | | |
| see above | NH Dept. of Health and Human Services Public Health Labs | Centers for Disease Control and Prevention Division of Viral Diseases, Pathogen Discovery | Mili Sheth, Sarah Nobles, Jasmine Padilla, Mark Burroughs, Shoshona Le, Katie Dillon, Peter Cook, Clinton R. Paden, Dhwani Batra, Krista Queen, Kristen Knipe, Dakota Howard, Yvette Unoarumhi, Darlene Wagner, Matthew Schmerer, Ben L. Rambo-Martin, Kristine Lacek, Sam Shepard, Alison Laufer Halpin, Dave Wentworth, Vivien Dugan, Suxiang Tong, Justin Lee |
| EPI_ISL_1824008, EPI_ISL_1824009, EPI_ISL_1824010, EPI_ISL_1824011, EPI_ISL_1824012, EPI_ISL_1824013, EPI_ISL_1824014, EPI_ISL_1824015, EPI_ISL_1824016, EPI_ISL_1824017, EPI_ISL_1824018, EPI_ISL_1824019, EPI_ISL_1824020, EPI_ISL_1824021, EPI_ISL_1824022, EPI_ISL_1824023, EPI_ISL_1824024, EPI_ISL_1824025, EPI_ISL_1824026, EPI_ISL_1824027, EPI_ISL_1824028, EPI_ISL_1824029, EPI_ISL_1824030, EPI_ISL_1824031, EPI_ISL_1824032 | | | |
| see above | WY Public Health Laboratory | Centers for Disease Control and Prevention Division of Viral Diseases, Pathogen Discovery | Mili Sheth, Sarah Nobles, Jasmine Padilla, Mark Burroughs, Shoshona Le, Katie Dillon, Peter Cook, Clinton R. Paden, Dhwani Batra, Krista Queen, Kristen Knipe, Dakota Howard, Yvette Unoarumhi, Darlene Wagner, Matthew Schmerer, Ben L. Rambo-Martin, Kristine Lacek, Sam Shepard, Alison Laufer Halpin, Dave Wentworth, Vivien Dugan, Suxiang Tong, Justin Lee |
| EPI_ISL_1824033, EPI_ISL_1824034, EPI_ISL_1824035 | "NYSDOH Wadsworth Center, Virology Lab" | Centers for Disease Control and Prevention Division of Viral Diseases, Pathogen Discovery | Mili Sheth, Sarah Nobles, Jasmine Padilla, Mark Burroughs, Shoshona Le, Katie Dillon, Peter Cook, Clinton R. Paden, Dhwani Batra, Krista Queen, Kristen Knipe, Dakota Howard, Yvette Unoarumhi, Darlene Wagner, Matthew Schmerer, Ben L. Rambo-Martin, Kristine Lacek, Sam Shepard, Alison Laufer Halpin, Dave Wentworth, Vivien Dugan, Suxiang Tong, Justin Lee |
| EPI_ISL_1824036, EPI_ISL_1824037, EPI_ISL_1824038, EPI_ISL_1824039, EPI_ISL_1824040, EPI_ISL_1824041, EPI_ISL_1824042, EPI_ISL_1824043, EPI_ISL_1824044, EPI_ISL_1824045, EPI_ISL_1824046, EPI_ISL_1824047, EPI_ISL_1824048, EPI_ISL_1824049, EPI_ISL_1824050, EPI_ISL_1824051, EPI_ISL_1824052, EPI_ISL_1824053, EPI_ISL_1824054, EPI_ISL_1824055, EPI_ISL_1824056, EPI_ISL_1824057 | | | |
| see above | IN State Department of Health Laboratory Services | Centers for Disease Control and Prevention Division of Viral Diseases, Pathogen Discovery | Mili Sheth, Sarah Nobles, Jasmine Padilla, Mark Burroughs, Shoshona Le, Katie Dillon, Peter Cook, Clinton R. Paden, Dhwani Batra, Krista Queen, Kristen Knipe, Dakota Howard, Yvette Unoarumhi, Darlene Wagner, Matthew Schmerer, Ben L. Rambo-Martin, Kristine Lacek, Sam Shepard, Alison Laufer Halpin, Dave Wentworth, Vivien Dugan, Suxiang Tong, Justin Lee |
| EPI_ISL_1824085, EPI_ISL_1824086, EPI_ISL_1824087, EPI_ISL_1824088, EPI_ISL_1824089, EPI_ISL_1824090, EPI_ISL_1824091, EPI_ISL_1824092, EPI_ISL_1824094, EPI_ISL_1824095, EPI_ISL_1824098, EPI_ISL_1824101, EPI_ISL_1824102, EPI_ISL_1824104, EPI_ISL_1824106, EPI_ISL_1824107, EPI_ISL_1824109, EPI_ISL_1824110 | | | |
| see above | Santa Clara Valley Medical Center | Santa Clara County public Health Laboratory | Santa Clara County Public Health Department |
| EPI_ISL_1824111 | Kaiser San Jose | Santa Clara County public Health Laboratory | Santa Clara County Public Health Department |
| EPI_ISL_1824120 | Kaiser Santa Clara | Santa Clara County public Health Laboratory | Santa Clara County Public Health Department |
| EPI_ISL_1824148, EPI_ISL_1824149, EPI_ISL_1824150 | The Ohio State University Applied Microbiology Services Laboratory | The Ohio State University Applied Microbiology Services Laboratory | Seth A. Faith PhD |
| EPI_ISL_1824151, EPI_ISL_1824152, EPI_ISL_1824153, EPI_ISL_1824154, EPI_ISL_1824155, EPI_ISL_1824156, EPI_ISL_1824157, EPI_ISL_1824158, EPI_ISL_1824159, EPI_ISL_1824160, EPI_ISL_1824161, EPI_ISL_1824162, EPI_ISL_1824163, EPI_ISL_1824164, EPI_ISL_1824165, EPI_ISL_1824166, EPI_ISL_1824167, EPI_ISL_1824168, EPI_ISL_1824169, EPI_ISL_1824170, EPI_ISL_1824171, EPI_ISL_1824172, EPI_ISL_1824173, EPI_ISL_1824174, EPI_ISL_1824175, EPI_ISL_1824176, EPI_ISL_1824177, EPI_ISL_1824178 | | | |
| see above | Gundersen Clinical Microbiology Laboratory | Kabara Cancer Research Institute | Craig S. Richmond, Paraic A. Kenny |
| EPI_ISL_1824189, EPI_ISL_1824190, EPI_ISL_1824191, EPI_ISL_1824192, EPI_ISL_1824193, EPI_ISL_1824195, EPI_ISL_1824196, EPI_ISL_1824197, EPI_ISL_1824199, EPI_ISL_1824200, EPI_ISL_1824201, EPI_ISL_1824202, EPI_ISL_1824203, EPI_ISL_1824204, EPI_ISL_1824205, EPI_ISL_1824206, EPI_ISL_1824207, EPI_ISL_1824208, EPI_ISL_1824209, EPI_ISL_1824210, EPI_ISL_1824211, EPI_ISL_1824212, EPI_ISL_1824213, EPI_ISL_1824214, EPI_ISL_1824216, EPI_ISL_1824217, EPI_ISL_1824218, EPI_ISL_1824219, EPI_ISL_1824220, EPI_ISL_1824221, EPI_ISL_1824222, EPI_ISL_1824223, EPI_ISL_1824224, EPI_ISL_1824225, EPI_ISL_1824226, EPI_ISL_1824227, EPI_ISL_1824229, EPI_ISL_1824231, EPI_ISL_1824233, EPI_ISL_1824234, EPI_ISL_1824235, EPI_ISL_1824236, EPI_ISL_1824237, EPI_ISL_1824238, EPI_ISL_1824239, EPI_ISL_1824240, EPI_ISL_1824241, EPI_ISL_1824242, EPI_ISL_1824243, EPI_ISL_1824244, EPI_ISL_1824245, EPI_ISL_1824246, EPI_ISL_1824247, EPI_ISL_1824248, EPI_ISL_1824249, EPI_ISL_1824250, EPI_ISL_1824251, EPI_ISL_1824252, EPI_ISL_1824253, EPI_ISL_1824254, EPI_ISL_1824255, EPI_ISL_1824256, EPI_ISL_1824257, EPI_ISL_1824258, EPI_ISL_1824259, EPI_ISL_1824260, EPI_ISL_1824261, EPI_ISL_1824262, EPI_ISL_1824263, EPI_ISL_1824264, EPI_ISL_1824265, EPI_ISL_1824266, EPI_ISL_1824267, EPI_ISL_1824268, EPI_ISL_1824269, EPI_ISL_1824270, EPI_ISL_1824271, EPI_ISL_1824272, EPI_ISL_1824273, EPI_ISL_1824274, EPI_ISL_1824275, EPI_ISL_1824276, EPI_ISL_1824277, EPI_ISL_1824278, EPI_ISL_1824279, EPI_ISL_1824280, EPI_ISL_1824281, EPI_ISL_1824282, EPI_ISL_1824283, EPI_ISL_1824284, EPI_ISL_1824285, EPI_ISL_1824286, EPI_ISL_1824287, EPI_ISL_1824288, EPI_ISL_1824289, EPI_ISL_1824290, EPI_ISL_1824291, EPI_ISL_1824292, EPI_ISL_1824293, EPI_ISL_1824294, EPI_ISL_1824295 | | | |
| see above | Pathology and Laboratory Medicine Institute, Cleveland Clinic, Ohio, USA | Pathology and Laboratory Medicine Institute, Cleveland Clinic, Ohio, USA | Jessica Spildener, Joy Nakitandwe, Kristen McDonnell, David Plunkett, Zheng Jin Tu, Jay Brock, Yu-Wei Cheng, Gary Procop, Daniel Rhoads, Daniel H. Farkas, David Bosler |
| EPI_ISL_1824296, EPI_ISL_1824297, EPI_ISL_1824298, EPI_ISL_1824299, EPI_ISL_1824301, EPI_ISL_1824303, EPI_ISL_1824304, EPI_ISL_1824306, EPI_ISL_1824307, EPI_ISL_1824309, EPI_ISL_1824310, EPI_ISL_1824313, EPI_ISL_1824317, EPI_ISL_1824321, EPI_ISL_1824323, EPI_ISL_1824325, EPI_ISL_1824328, EPI_ISL_1824335, EPI_ISL_1824341, EPI_ISL_1824342, EPI_ISL_1824353, EPI_ISL_1824354, EPI_ISL_1824355, EPI_ISL_1824358, EPI_ISL_1824361, EPI_ISL_1824362, EPI_ISL_1824363, EPI_ISL_1824364, EPI_ISL_1824366, EPI_ISL_1824368, EPI_ISL_1824370, EPI_ISL_1824371, EPI_ISL_1824372, EPI_ISL_1824374, EPI_ISL_1824375, EPI_ISL_1824376, EPI_ISL_1824377, EPI_ISL_1824383, EPI_ISL_1824387, EPI_ISL_1824388, EPI_ISL_1824389, EPI_ISL_1824391, EPI_ISL_1824394, EPI_ISL_1824395, EPI_ISL_1824397, EPI_ISL_1824399, EPI_ISL_1824400, EPI_ISL_1824401, EPI_ISL_1824402, EPI_ISL_1824403, EPI_ISL_1824405, EPI_ISL_1824407, EPI_ISL_1824410, EPI_ISL_1824413, EPI_ISL_1824414, EPI_ISL_1824415, EPI_ISL_1824416 | | | |
| see above | Chiu Laboratory, University of California, San Francisco | Chiu Laboratory, University of California, San Francisco | Charles Chiu, Venice Servellita, Candace Wang, Alicia Sotomayor-Gonzalez, Yueyuan Zhang, Daniel Wong |
| EPI_ISL_1824419, EPI_ISL_1824420, EPI_ISL_1824421, EPI_ISL_1824422, EPI_ISL_1824423, EPI_ISL_1824424, EPI_ISL_1824425, EPI_ISL_1824426, EPI_ISL_1824427, EPI_ISL_1824428, EPI_ISL_1824429, EPI_ISL_1824430, EPI_ISL_1824431, EPI_ISL_1824432, EPI_ISL_1824433, EPI_ISL_1824434, EPI_ISL_1824435, EPI_ISL_1824436, EPI_ISL_1824437, EPI_ISL_1824438, EPI_ISL_1824439, EPI_ISL_1824440, EPI_ISL_1824441, EPI_ISL_1824442, EPI_ISL_1824443, EPI_ISL_1824444, EPI_ISL_1824445, EPI_ISL_1824446, EPI_ISL_1824447, EPI_ISL_1824448, EPI_ISL_1824449, EPI_ISL_1824450, EPI_ISL_1824451, EPI_ISL_1824452, EPI_ISL_1824453, EPI_ISL_1824454, EPI_ISL_1824455, EPI_ISL_1824456, EPI_ISL_1824457, EPI_ISL_1824458, EPI_ISL_1824459, EPI_ISL_1824460, EPI_ISL_1824461, EPI_ISL_1824462, EPI_ISL_1824463, EPI_ISL_1824464, EPI_ISL_1824465, EPI_ISL_1824466, EPI_ISL_1824467, EPI_ISL_1824468, EPI_ISL_1824469, EPI_ISL_1824470, EPI_ISL_1824471, EPI_ISL_1824472, EPI_ISL_1824473, EPI_ISL_1824474, EPI_ISL_1824475, EPI_ISL_1824476, EPI_ISL_1824477, EPI_ISL_1824478, EPI_ISL_1824479, EPI_ISL_1824480, EPI_ISL_1824481, EPI_ISL_1824482, EPI_ISL_1824483, EPI_ISL_1824484, EPI_ISL_1824485, EPI_ISL_1824486, EPI_ISL_1824487, EPI_ISL_1824488, EPI_ISL_1824489, EPI_ISL_1824490, EPI_ISL_1824491, EPI_ISL_1824492, EPI_ISL_1824493, EPI_ISL_1824494, EPI_ISL_1824495, EPI_ISL_1824496, EPI_ISL_1824497, EPI_ISL_1824498 | | | |
| see above | Instituto Nacional de Medicina | Centro de Investigación en Enfermedades | Matias-Florentino M, Perez-Garcia M, Hidalgo-Miranda A, Mendoza-Vargas A, Reyes-Grajeda JP, Cedro-Tanda A, Arriaga-Canon C, Herrera-Montalvo LA, Reyes-Teran G, Avila-Rios S, Boukadida C |

| | | | |
|---|---|---|---|
| | Genomica | Infecciosas (CIENI), Instituto Nacional de Enfermedades Respiratorias (INER) | |
| EPI_ISL_1824566 | Gundersen Clinical Microbiology Laboratory | Kabara Cancer Research Institute | Craig S. Richmond, Paraic A. Kenny |
| EPI_ISL_1824686, EPI_ISL_1824687, EPI_ISL_1824688, EPI_ISL_1824690, EPI_ISL_1824691, EPI_ISL_1824693, EPI_ISL_1824694, EPI_ISL_1824695, EPI_ISL_1824696 | Oregon State University TRACE Project | OSU Center for Genome Research and Biocomputing | Oregon State University TRACE Project |
| EPI_ISL_1824720, EPI_ISL_1824721, EPI_ISL_1824723, EPI_ISL_1824724, EPI_ISL_1824726, EPI_ISL_1824727, EPI_ISL_1824729, EPI_ISL_1824731, EPI_ISL_1824732, EPI_ISL_1824733, EPI_ISL_1824734, EPI_ISL_1824738, EPI_ISL_1824743, EPI_ISL_1824744, EPI_ISL_1824747, EPI_ISL_1824749, EPI_ISL_1824750, EPI_ISL_1824751, EPI_ISL_1824752, EPI_ISL_1824755, EPI_ISL_1824756, EPI_ISL_1824757, EPI_ISL_1824759, EPI_ISL_1824760, EPI_ISL_1824761, EPI_ISL_1824763, EPI_ISL_1824765, EPI_ISL_1824766, EPI_ISL_1824767, EPI_ISL_1824768, EPI_ISL_1824769, EPI_ISL_1824771, EPI_ISL_1824772, EPI_ISL_1824775, EPI_ISL_1824777, EPI_ISL_1824778, EPI_ISL_1824780, EPI_ISL_1824782, EPI_ISL_1824784, EPI_ISL_1824786, EPI_ISL_1824787, EPI_ISL_1824788, EPI_ISL_1824789, EPI_ISL_1824791, EPI_ISL_1824793, EPI_ISL_1824794, EPI_ISL_1824795, EPI_ISL_1824796, EPI_ISL_1824797, EPI_ISL_1824800, EPI_ISL_1824801, EPI_ISL_1824802, EPI_ISL_1824803, EPI_ISL_1824805, EPI_ISL_1824806, EPI_ISL_1824807, EPI_ISL_1824808, EPI_ISL_1824810, EPI_ISL_1824812, EPI_ISL_1824813, EPI_ISL_1824815, EPI_ISL_1824816, EPI_ISL_1824821, EPI_ISL_1824822, EPI_ISL_1824824, EPI_ISL_1824825, EPI_ISL_1824827, EPI_ISL_1824829, EPI_ISL_1824830, EPI_ISL_1824831, EPI_ISL_1824832, EPI_ISL_1824833, EPI_ISL_1824836, EPI_ISL_1824838, EPI_ISL_1824839, EPI_ISL_1824842, EPI_ISL_1824843, EPI_ISL_1824844, EPI_ISL_1824846, EPI_ISL_1824847, EPI_ISL_1824848, EPI_ISL_1824851, EPI_ISL_1824852, EPI_ISL_1824853, EPI_ISL_1824854, EPI_ISL_1824855, EPI_ISL_1824856, EPI_ISL_1824857, EPI_ISL_1824858, EPI_ISL_1824859, EPI_ISL_1824860, EPI_ISL_1824861, EPI_ISL_1824862, EPI_ISL_1824864, EPI_ISL_1824865, EPI_ISL_1824866, EPI_ISL_1824867, EPI_ISL_1824868, EPI_ISL_1824869, EPI_ISL_1824870, EPI_ISL_1824871, EPI_ISL_1824872, EPI_ISL_1824873, EPI_ISL_1824874, EPI_ISL_1824875, EPI_ISL_1824876, EPI_ISL_1824877, EPI_ISL_1824878, EPI_ISL_1824880, EPI_ISL_1824881, EPI_ISL_1824882, EPI_ISL_1824883, EPI_ISL_1824885, EPI_ISL_1824886, EPI_ISL_1824887, EPI_ISL_1824888, EPI_ISL_1824889, EPI_ISL_1824893, EPI_ISL_1824894, EPI_ISL_1824895, EPI_ISL_1824896, EPI_ISL_1824897, EPI_ISL_1824900, EPI_ISL_1824901, EPI_ISL_1824902, EPI_ISL_1824903, EPI_ISL_1824904, EPI_ISL_1824905, EPI_ISL_1824907, EPI_ISL_1824908, EPI_ISL_1824910, EPI_ISL_1824913, EPI_ISL_1824914, EPI_ISL_1824915, EPI_ISL_1824916, EPI_ISL_1824919, EPI_ISL_1824924, EPI_ISL_1824926, EPI_ISL_1824928, EPI_ISL_1824931, EPI_ISL_1824932, EPI_ISL_1824933, EPI_ISL_1824934, EPI_ISL_1824938, EPI_ISL_1824943, EPI_ISL_1824944, EPI_ISL_1824945, EPI_ISL_1824946, EPI_ISL_1824947, EPI_ISL_1824952, EPI_ISL_1824954, EPI_ISL_1824955, EPI_ISL_1824959, EPI_ISL_1824960, EPI_ISL_1824961, EPI_ISL_1824962, EPI_ISL_1824963, EPI_ISL_1824964, EPI_ISL_1824965, EPI_ISL_1824966, EPI_ISL_1824967, EPI_ISL_1824969, EPI_ISL_1824970, EPI_ISL_1824971, EPI_ISL_1824974, EPI_ISL_1824975, EPI_ISL_1824976, EPI_ISL_1824977, EPI_ISL_1824978, EPI_ISL_1824981, EPI_ISL_1824983, EPI_ISL_1824984, EPI_ISL_1824985, EPI_ISL_1824987, EPI_ISL_1824988, EPI_ISL_1824989, EPI_ISL_1824991, EPI_ISL_1824993, EPI_ISL_1824994, EPI_ISL_1824995, EPI_ISL_1824996, EPI_ISL_1824998, EPI_ISL_1825000, EPI_ISL_1825002, EPI_ISL_1825003, EPI_ISL_1825004, EPI_ISL_1825005, EPI_ISL_1825006, EPI_ISL_1825007, EPI_ISL_1825009, EPI_ISL_1825010, EPI_ISL_1825011, EPI_ISL_1825012, EPI_ISL_1825013, EPI_ISL_1825015, EPI_ISL_1825016, EPI_ISL_1825017, EPI_ISL_1825020, EPI_ISL_1825021, EPI_ISL_1825022, EPI_ISL_1825024, EPI_ISL_1825025, EPI_ISL_1825026, EPI_ISL_1825028, EPI_ISL_1825029, EPI_ISL_1825030, EPI_ISL_1825031, EPI_ISL_1825033, EPI_ISL_1825034, EPI_ISL_1825035, EPI_ISL_1825036, EPI_ISL_1825037, EPI_ISL_1825038, EPI_ISL_1825039, EPI_ISL_1825040, EPI_ISL_1825041, EPI_ISL_1825042, EPI_ISL_1825043, EPI_ISL_1825044, EPI_ISL_1825045, EPI_ISL_1825046, EPI_ISL_1825047, EPI_ISL_1825048, EPI_ISL_1825049, EPI_ISL_1825050, EPI_ISL_1825051, EPI_ISL_1825052, EPI_ISL_1825053, EPI_ISL_1825054, EPI_ISL_1825055, EPI_ISL_1825056, EPI_ISL_1825057, EPI_ISL_1825058, EPI_ISL_1825059, EPI_ISL_1825060, EPI_ISL_1825061, EPI_ISL_1825062, EPI_ISL_1825063, EPI_ISL_1825064, EPI_ISL_1825065, EPI_ISL_1825066, EPI_ISL_1825067, EPI_ISL_1825068, EPI_ISL_1825069, EPI_ISL_1825070, EPI_ISL_1825071, EPI_ISL_1825072, EPI_ISL_1825073, EPI_ISL_1825075, EPI_ISL_1825080, EPI_ISL_1825081, EPI_ISL_1825082 | | | |
| see above | Color Genomics | Chiu Laboratory, University of California, San Francisco | Charles Chiu, Venice Servellita, Candace Wang, Alicia Sotomayor-Gonzalez, Yueyuan Zhang, Daniel Wong, Scott Topper, Madyson Wanner, Erika Torres, Alicia Zhou |
| EPI_ISL_1825118 | Curative | New Mexico Department of Health Scientific Laboratory | Ellie Johnson, D'eldra Malone, Jennifer Benoit, Ratheesh Rajan, Linda Salazar, Anastacia Griego-Fisher |
| EPI_ISL_1825119, EPI_ISL_1825120, EPI_ISL_1825121, EPI_ISL_1825122, EPI_ISL_1825123, EPI_ISL_1825124, EPI_ISL_1825125, EPI_ISL_1825126, EPI_ISL_1825129, EPI_ISL_1825130, EPI_ISL_1825131, EPI_ISL_1825132, EPI_ISL_1825133, EPI_ISL_1825134, EPI_ISL_1825135, EPI_ISL_1825137, EPI_ISL_1825138, EPI_ISL_1825139 | | | |
| see above | Idaho Bureau of Laboratories | Idaho Bureau of Laboratories | R. Beukelman, Matthew Charles Burns, Aimee Ceniseros, Robert L. Voermans, Christopher Ball |
| EPI_ISL_1825141, EPI_ISL_1825142, EPI_ISL_1825143, EPI_ISL_1825144, EPI_ISL_1825145, EPI_ISL_1825146, EPI_ISL_1825147, EPI_ISL_1825148, EPI_ISL_1825149, EPI_ISL_1825150, EPI_ISL_1825151, EPI_ISL_1825152, EPI_ISL_1825153, EPI_ISL_1825154, EPI_ISL_1825155, EPI_ISL_1825156, EPI_ISL_1825157, EPI_ISL_1825158, EPI_ISL_1825159, EPI_ISL_1825160, EPI_ISL_1825161, EPI_ISL_1825163, EPI_ISL_1825164, EPI_ISL_1825165, EPI_ISL_1825166, EPI_ISL_1825167, EPI_ISL_1825168, EPI_ISL_1825170, EPI_ISL_1825171, EPI_ISL_1825172, EPI_ISL_1825173, EPI_ISL_1825174, EPI_ISL_1825175, EPI_ISL_1825176, EPI_ISL_1825177, EPI_ISL_1825178, EPI_ISL_1825179, EPI_ISL_1825180, EPI_ISL_1825181, EPI_ISL_1825182, EPI_ISL_1825183, EPI_ISL_1825184, EPI_ISL_1825185, EPI_ISL_1825186, EPI_ISL_1825187, EPI_ISL_1825188, EPI_ISL_1825189, EPI_ISL_1825191, EPI_ISL_1825192, EPI_ISL_1825193, EPI_ISL_1825194, EPI_ISL_1825195, EPI_ISL_1825196, EPI_ISL_1825197, EPI_ISL_1825198, EPI_ISL_1825199, EPI_ISL_1825200, EPI_ISL_1825201, EPI_ISL_1825202, EPI_ISL_1825203, EPI_ISL_1825204, EPI_ISL_1825205, EPI_ISL_1825206, EPI_ISL_1825207, EPI_ISL_1825208, EPI_ISL_1825209, EPI_ISL_1825210, EPI_ISL_1825211, EPI_ISL_1825212, EPI_ISL_1825213, EPI_ISL_1825214, EPI_ISL_1825215, EPI_ISL_1825216, EPI_ISL_1825217, EPI_ISL_1825218, EPI_ISL_1825219, EPI_ISL_1825220, EPI_ISL_1825221, EPI_ISL_1825222, EPI_ISL_1825223, EPI_ISL_1825224, EPI_ISL_1825225, EPI_ISL_1825226, EPI_ISL_1825227, EPI_ISL_1825228, EPI_ISL_1825229, EPI_ISL_1825230, EPI_ISL_1825231, EPI_ISL_1825232, EPI_ISL_1825233, EPI_ISL_1825234, EPI_ISL_1825235, EPI_ISL_1825236, EPI_ISL_1825237, EPI_ISL_1825238, EPI_ISL_1825239, EPI_ISL_1825240, EPI_ISL_1825241, EPI_ISL_1825242, EPI_ISL_1825243, EPI_ISL_1825244, EPI_ISL_1825245, EPI_ISL_1825246, EPI_ISL_1825247, EPI_ISL_1825248, EPI_ISL_1825249, EPI_ISL_1825250, EPI_ISL_1825251, EPI_ISL_1825252, EPI_ISL_1825253, EPI_ISL_1825254, EPI_ISL_1825255, EPI_ISL_1825256, EPI_ISL_1825257, EPI_ISL_1825258, EPI_ISL_1825259, EPI_ISL_1825260, EPI_ISL_1825261, EPI_ISL_1825262, EPI_ISL_1825263, EPI_ISL_1825264, EPI_ISL_1825265, EPI_ISL_1825266, EPI_ISL_1825267, EPI_ISL_1825268, EPI_ISL_1825269, EPI_ISL_1825270, EPI_ISL_1825271, EPI_ISL_1825272, EPI_ISL_1825274, EPI_ISL_1825275, EPI_ISL_1825276, EPI_ISL_1825277, EPI_ISL_1825279, EPI_ISL_1825280, EPI_ISL_1825281, EPI_ISL_1825282, EPI_ISL_1825283, EPI_ISL_1825284, EPI_ISL_1825285, EPI_ISL_1825286, EPI_ISL_1825287, EPI_ISL_1825288, EPI_ISL_1825289, EPI_ISL_1825291, EPI_ISL_1825292, EPI_ISL_1825293, EPI_ISL_1825294, EPI_ISL_1825295, EPI_ISL_1825296, EPI_ISL_1825297, EPI_ISL_1825298, EPI_ISL_1825299, EPI_ISL_1825300, EPI_ISL_1825301, EPI_ISL_1825302, EPI_ISL_1825303, EPI_ISL_1825304, EPI_ISL_1825305, EPI_ISL_1825306, EPI_ISL_1825307, EPI_ISL_1825308, EPI_ISL_1825309, EPI_ISL_1825310, EPI_ISL_1825311, EPI_ISL_1825312, EPI_ISL_1825313, EPI_ISL_1825314, EPI_ISL_1825315, EPI_ISL_1825317, EPI_ISL_1825319, EPI_ISL_1825320, EPI_ISL_1825321, EPI_ISL_1825322, EPI_ISL_1825323, EPI_ISL_1825324, EPI_ISL_1825325, EPI_ISL_1825326, EPI_ISL_1825327, EPI_ISL_1825329, EPI_ISL_1825330, EPI_ISL_1825331, EPI_ISL_1825332, EPI_ISL_1825333, EPI_ISL_1825334, EPI_ISL_1825335, EPI_ISL_1825336, EPI_ISL_1825337, EPI_ISL_1825338, EPI_ISL_1825339, EPI_ISL_1825340, EPI_ISL_1825341, EPI_ISL_1825342, EPI_ISL_1825343, EPI_ISL_1825344, EPI_ISL_1825345, EPI_ISL_1825346, EPI_ISL_1825347, EPI_ISL_1825348, EPI_ISL_1825349, EPI_ISL_1825350, EPI_ISL_1825351, EPI_ISL_1825352, EPI_ISL_1825353, EPI_ISL_1825354, EPI_ISL_1825355, EPI_ISL_1825356, EPI_ISL_1825357, EPI_ISL_1825358, EPI_ISL_1825359, EPI_ISL_1825360, EPI_ISL_1825361, EPI_ISL_1825362, EPI_ISL_1825363, EPI_ISL_1825364, EPI_ISL_1825365, EPI_ISL_1825366, EPI_ISL_1825367, EPI_ISL_1825368, EPI_ISL_1825369, EPI_ISL_1825370, EPI_ISL_1825371, EPI_ISL_1825372, EPI_ISL_1825373, EPI_ISL_1825374, EPI_ISL_1825375, EPI_ISL_1825376, EPI_ISL_1825377, EPI_ISL_1825378, EPI_ISL_1825379, EPI_ISL_1825380, EPI_ISL_1825381, EPI_ISL_1825382, EPI_ISL_1825383, EPI_ISL_1825384, EPI_ISL_1825385, EPI_ISL_1825386, EPI_ISL_1825387, EPI_ISL_1825388, EPI_ISL_1825389, EPI_ISL_1825390, EPI_ISL_1825391, EPI_ISL_1825392, EPI_ISL_1825393, EPI_ISL_1825394, EPI_ISL_1825395, EPI_ISL_1825396, EPI_ISL_1825397, EPI_ISL_1825398, EPI_ISL_1825399, EPI_ISL_1825400, EPI_ISL_1825401, EPI_ISL_1825402, EPI_ISL_1825403, EPI_ISL_1825404, EPI_ISL_1825405, EPI_ISL_1825406, EPI_ISL_1825407, EPI_ISL_1825408, EPI_ISL_1825409, EPI_ISL_1825410, EPI_ISL_1825411, EPI_ISL_1825412, EPI_ISL_1825413, EPI_ISL_1825414, EPI_ISL_1825415, EPI_ISL_1825416, EPI_ISL_1825417, EPI_ISL_1825418, EPI_ISL_1825419, EPI_ISL_1825421, EPI_ISL_1825422, EPI_ISL_1825423, EPI_ISL_1825424, EPI_ISL_1825425, EPI_ISL_1825426, EPI_ISL_1825427, EPI_ISL_1825428, EPI_ISL_1825429, EPI_ISL_1825430, EPI_ISL_1825431, EPI_ISL_1825432, EPI_ISL_1825433, EPI_ISL_1825434, EPI_ISL_1825435, EPI_ISL_1825436, EPI_ISL_1825437, EPI_ISL_1825439, EPI_ISL_1825440, EPI_ISL_1825441, EPI_ISL_1825442, EPI_ISL_1825443, EPI_ISL_1825444, EPI_ISL_1825445, EPI_ISL_1825446, EPI_ISL_1825447, EPI_ISL_1825448, EPI_ISL_1825449, EPI_ISL_1825450, EPI_ISL_1825451, EPI_ISL_1825452, EPI_ISL_1825453, EPI_ISL_1825454, EPI_ISL_1825455, EPI_ISL_1825456, EPI_ISL_1825457, EPI_ISL_1825458, EPI_ISL_1825459, EPI_ISL_1825460, EPI_ISL_1825461, EPI_ISL_1825462, EPI_ISL_1825463, EPI_ISL_1825464, EPI_ISL_1825465, EPI_ISL_1825466, EPI_ISL_1825467, EPI_ISL_1825468, EPI_ISL_1825469, EPI_ISL_1825470, EPI_ISL_1825471, EPI_ISL_1825472, EPI_ISL_1825473, EPI_ISL_1825474, EPI_ISL_1825475, EPI_ISL_1825476, EPI_ISL_1825477, EPI_ISL_1825478, EPI_ISL_1825479, EPI_ISL_1825480, EPI_ISL_1825481, EPI_ISL_1825482, EPI_ISL_1825483, EPI_ISL_1825484, EPI_ISL_1825485, EPI_ISL_1825486, EPI_ISL_1825487, EPI_ISL_1825488, EPI_ISL_1825489, EPI_ISL_1825490, EPI_ISL_1825491, EPI_ISL_1825492, EPI_ISL_1825493, EPI_ISL_1825494, EPI_ISL_1825495, EPI_ISL_1825496, EPI_ISL_1825497, EPI_ISL_1825498, EPI_ISL_1825499, EPI_ISL_1825500, EPI_ISL_1825501, EPI_ISL_1825502, EPI_ISL_1825503, EPI_ISL_1825504, EPI_ISL_1825505, EPI_ISL_1825506, EPI_ISL_1825507, EPI_ISL_1825508, EPI_ISL_1825509, EPI_ISL_1825510, EPI_ISL_1825511, EPI_ISL_1825512, EPI_ISL_1825513, EPI_ISL_1825514, EPI_ISL_1825515, EPI_ISL_1825516, EPI_ISL_1825517, EPI_ISL_1825518, EPI_ISL_1825519, EPI_ISL_1825520, EPI_ISL_1825521, EPI_ISL_1825522, EPI_ISL_1825523, EPI_ISL_1825524, EPI_ISL_1825525, EPI_ISL_1825526, EPI_ISL_1825527, EPI_ISL_1825528, EPI_ISL_1825529, EPI_ISL_1825530, EPI_ISL_1825531, EPI_ISL_1825532, EPI_ISL_1825533, EPI_ISL_1825534, EPI_ISL_1825535, EPI_ISL_1825536, EPI_ISL_1825537, EPI_ISL_1825538, EPI_ISL_1825539, EPI_ISL_1825540, EPI_ISL_1825541, EPI_ISL_1825542, EPI_ISL_1825543, EPI_ISL_1825544, EPI_ISL_1825545, EPI_ISL_1825546, EPI_ISL_1825547, EPI_ISL_1825548, EPI_ISL_1825549, EPI_ISL_1825550, EPI_ISL_1825551, EPI_ISL_1825552, EPI_ISL_1825553, EPI_ISL_1825554, EPI_ISL_1825555, EPI_ISL_1825556, EPI_ISL_1825557, EPI_ISL_1825558, EPI_ISL_1825559, EPI_ISL_1825560, EPI_ISL_1825561, EPI_ISL_1825562, EPI_ISL_1825563, EPI_ISL_1825564, EPI_ISL_1825565, EPI_ISL_1825566, EPI_ISL_1825567, EPI_ISL_1825568, EPI_ISL_1825569, EPI_ISL_1825570, EPI_ISL_1825571, EPI_ISL_1825572, EPI_ISL_1825573, EPI_ISL_1825574, EPI_ISL_1825575, EPI_ISL_1825576, EPI_ISL_1825577, EPI_ISL_1825578, EPI_ISL_1825579, EPI_ISL_1825580, EPI_ISL_1825581, EPI_ISL_1825582, EPI_ISL_1825583, EPI_ISL_1825584, EPI_ISL_1825585, EPI_ISL_1825586, EPI_ISL_1825587, EPI_ISL_1825588, EPI_ISL_1825589, EPI_ISL_1825590, EPI_ISL_1825591, EPI_ISL_1825592, EPI_ISL_1825593, EPI_ISL_1825594, EPI_ISL_1825595, EPI_ISL_1825596, EPI_ISL_1825597, EPI_ISL_1825598, EPI_ISL_1825599, EPI_ISL_1825600, EPI_ISL_1825601, EPI_ISL_1825602, EPI_ISL_1825603, EPI_ISL_1825604, EPI_ISL_1825605, EPI_ISL_1825606, EPI_ISL_1825607, EPI_ISL_1825608, EPI_ISL_1825609, EPI_ISL_1825610, EPI_ISL_1825611, EPI_ISL_1825612, EPI_ISL_1825613, EPI_ISL_1825614, EPI_ISL_1825615, EPI_ISL_1825616, EPI_ISL_1825617, EPI_ISL_1825618, EPI_ISL_1825619, EPI_ISL_1825620, EPI_ISL_1825621, EPI_ISL_1825622, EPI_ISL_1825623, EPI_ISL_1825624, EPI_ISL_1825625, EPI_ISL_1825626, EPI_ISL_1825627, EPI_ISL_1825628, EPI_ISL_1825629, EPI_ISL_1825630, EPI_ISL_1825631, EPI_ISL_1825632, EPI_ISL_1825633, EPI_ISL_1825634, EPI_ISL_1825635, EPI_ISL_1825637, EPI_ISL_1825638, EPI_ISL_1825639, EPI_ISL_1825640, EPI_ISL_1825641, EPI_ISL_1825642, EPI_ISL_1825643, EPI_ISL_1825644, EPI_ISL_1825645, EPI_ISL_1825646, EPI_ISL_1825647, EPI_ISL_1825648, EPI_ISL_1825649, EPI_ISL_1825650, EPI_ISL_1825651, EPI_ISL_1825652, EPI_ISL_1825653, EPI_ISL_1825654, EPI_ISL_1825655, EPI_ISL_1825656, EPI_ISL_1825657, EPI_ISL_1825658, EPI_ISL_1825659, EPI_ISL_1825660, EPI_ISL_1825661, EPI_ISL_1825663, EPI_ISL_1825664, EPI_ISL_1825665, EPI_ISL_1825666, EPI_ISL_1825667, EPI_ISL_1825668, EPI_ISL_1825669, EPI_ISL_1825670, EPI_ISL_1825671, EPI_ISL_1825672, EPI_ISL_1825673, EPI_ISL_1825674, EPI_ISL_1825675 | | | |
| see above | Broad Institute Clinical Research Sequencing Platform | Infectious Disease Program, Broad Institute of Harvard and MIT | Siddle,K.J., Adams,G., Pearlman,L., Gladden-Young,A., Vicente,G., Blumenstiel,B., DeFelice,M., Lee,M., McGovern,S., Lagerborg,K., Rudy,M., DeRuff,K., Carter,A., Normandin,E., Bauer,M., Reilly,S., Tomkins-Tinch,C., Loreth,C., Chaluvadi,S., Meldrim,J., Granger,B., Lemieux,J.E., Birren,B.W., Sabeti,P.C., Larkin,K., Dodge,S., Lennon,N., Madoff,L., Brown,C., Gallagher,G., Smole,S., Park,D.J., Gabriel,S., and MacInnis,B.L. |
| EPI_ISL_1825694, EPI_ISL_1825695, EPI_ISL_1825696, EPI_ISL_1825697, EPI_ISL_1825698, EPI_ISL_1825699, EPI_ISL_1825700, EPI_ISL_1825701, EPI_ISL_1825702, EPI_ISL_1825703, EPI_ISL_1825704, EPI_ISL_1825705, EPI_ISL_1825706, EPI_ISL_1825707, EPI_ISL_1825708, EPI_ISL_1825709, EPI_ISL_1825710, EPI_ISL_1825711, EPI_ISL_1825712, EPI_ISL_1825713, EPI_ISL_1825714, EPI_ISL_1825715, EPI_ISL_1825716, EPI_ISL_1825717, EPI_ISL_1825718, EPI_ISL_1825719, EPI_ISL_1825720, EPI_ISL_1825721, EPI_ISL_1825723, EPI_ISL_1825724, EPI_ISL_1825725, EPI_ISL_1825726, EPI_ISL_1825728, EPI_ISL_1825729, EPI_ISL_1825731, EPI_ISL_1825732, EPI_ISL_1825733, EPI_ISL_1825734, EPI_ISL_1825735, EPI_ISL_1825736, EPI_ISL_1825740, EPI_ISL_1825741, EPI_ISL_1825742, EPI_ISL_1825743, EPI_ISL_1825744, EPI_ISL_1825745, EPI_ISL_1825747, EPI_ISL_1825748, EPI_ISL_1825749, EPI_ISL_1825750, EPI_ISL_1825751, EPI_ISL_1825753, EPI_ISL_1825754, EPI_ISL_1825755, EPI_ISL_1825756, EPI_ISL_1825757, EPI_ISL_1825758, EPI_ISL_1825759, EPI_ISL_1825760, EPI_ISL_1825761, EPI_ISL_1825762, EPI_ISL_1825763, EPI_ISL_1825764, EPI_ISL_1825765, EPI_ISL_1825766, EPI_ISL_1825767, EPI_ISL_1825768, EPI_ISL_1825770, EPI_ISL_1825771, EPI_ISL_1825772, EPI_ISL_1825773, EPI_ISL_1825774, EPI_ISL_1825775, EPI_ISL_1825776, EPI_ISL_1825777, EPI_ISL_1825778, EPI_ISL_1825779, EPI_ISL_1825780, EPI_ISL_1825781, EPI_ISL_1825782, EPI_ISL_1825783, EPI_ISL_1825784, EPI_ISL_1825785, EPI_ISL_1825786, EPI_ISL_1825787, EPI_ISL_1825788, EPI_ISL_1825789, EPI_ISL_1825790, EPI_ISL_1825791, EPI_ISL_1825792, EPI_ISL_1825793, EPI_ISL_1825794, EPI_ISL_1825795, EPI_ISL_1825796, EPI_ISL_1825797, EPI_ISL_1825798, EPI_ISL_1825799, EPI_ISL_1825800, EPI_ISL_1825801, EPI_ISL_1825802, EPI_ISL_1825803, EPI_ISL_1825804, EPI_ISL_1825805, EPI_ISL_1825806, EPI_ISL_1825807, EPI_ISL_1825808, EPI_ISL_1825809, EPI_ISL_1825810, EPI_ISL_1825811, EPI_ISL_1825812, EPI_ISL_1825813, EPI_ISL_1825814, EPI_ISL_1825815, EPI_ISL_1825816, EPI_ISL_1825817, EPI_ISL_1825818, EPI_ISL_1825819, EPI_ISL_1825820, EPI_ISL_1825821, EPI_ISL_1825822, EPI_ISL_1825823, EPI_ISL_1825824, EPI_ISL_1825825, EPI_ISL_1825826, EPI_ISL_1825827, EPI_ISL_1825828, EPI_ISL_1825829, EPI_ISL_1825830, EPI_ISL_1825831, EPI_ISL_1825832, EPI_ISL_1825833, EPI_ISL_1825834, EPI_ISL_1825835, EPI_ISL_1825836, EPI_ISL_1825837, EPI_ISL_1825838, EPI_ISL_1825839, EPI_ISL_1825840, EPI_ISL_1825841, EPI_ISL_1825842, EPI_ISL_1825843, EPI_ISL_1825844, EPI_ISL_1825845, EPI_ISL_1825846, EPI_ISL_1825847, EPI_ISL_1825848, EPI_ISL_1825849, EPI_ISL_1825850, EPI_ISL_1825851, EPI_ISL_1825852, EPI_ISL_1825853, EPI_ISL_1825854, EPI_ISL_1825855, EPI_ISL_1825856, EPI_ISL_1825857, EPI_ISL_1825858, EPI_ISL_1825859, EPI_ISL_1825860, EPI_ISL_1825861, EPI_ISL_1825862, EPI_ISL_1825863, EPI_ISL_1825864, EPI_ISL_1825865, EPI_ISL_1825866, EPI_ISL_1825867, EPI_ISL_1825868, EPI_ISL_1825869, EPI_ISL_1825870, EPI_ISL_1825871, EPI_ISL_1825872, EPI_ISL_1825873, EPI_ISL_1825874, EPI_ISL_1825875, EPI_ISL_1825876 | | | |
| see above | Illinois Department of Public Health - Springfield Lab | Illinois Department of Public Health - Springfield Lab | Bryan Sim, Gordon McCall |
| EPI_ISL_1825880, EPI_ISL_1825882, EPI_ISL_1825883, EPI_ISL_1825884, EPI_ISL_1825888, EPI_ISL_1825889, EPI_ISL_1825890, EPI_ISL_1825891, EPI_ISL_1825894, EPI_ISL_1825895, EPI_ISL_1825897, EPI_ISL_1825902, EPI_ISL_1825903, EPI_ISL_1825904, EPI_ISL_1825905, EPI_ISL_1825906, EPI_ISL_1825909, EPI_ISL_1825910, EPI_ISL_1825911, EPI_ISL_1825913, EPI_ISL_1825917, EPI_ISL_1825918, EPI_ISL_1825919, EPI_ISL_1825924, EPI_ISL_1825925, EPI_ISL_1825928, EPI_ISL_1825930, EPI_ISL_1825931, EPI_ISL_1825932, EPI_ISL_1825933, EPI_ISL_1825939, EPI_ISL_1825940, EPI_ISL_1825941, EPI_ISL_1825942, EPI_ISL_1825943, EPI_ISL_1825944, EPI_ISL_1825945, EPI_ISL_1825946, EPI_ISL_1825947, EPI_ISL_1825948, EPI_ISL_1825949, EPI_ISL_1825950, EPI_ISL_1825951, EPI_ISL_1825952, EPI_ISL_1825953, EPI_ISL_1825954, EPI_ISL_1825955, EPI_ISL_1825956, EPI_ISL_1825957, EPI_ISL_1825958, EPI_ISL_1825959, EPI_ISL_1825960, EPI_ISL_1825961, EPI_ISL_1825962, EPI_ISL_1825963, EPI_ISL_1825964, EPI_ISL_1825965, EPI_ISL_1825966, EPI_ISL_1825967, EPI_ISL_1825968, EPI_ISL_1825969, EPI_ISL_1825970, EPI_ISL_1825971, EPI_ISL_1825972, EPI_ISL_1825973, EPI_ISL_1825974, EPI_ISL_1825975, EPI_ISL_1825976, EPI_ISL_1825977, EPI_ISL_1825978, EPI_ISL_1825979, EPI_ISL_1825980, EPI_ISL_1825981, EPI_ISL_1825982, EPI_ISL_1825983, EPI_ISL_1825984, EPI_ISL_1825985, EPI_ISL_1825986, EPI_ISL_1825987, EPI_ISL_1825988, EPI_ISL_1825989, EPI_ISL_1825990, EPI_ISL_1825991, EPI_ISL_1825992, EPI_ISL_1825993, EPI_ISL_1825994, EPI_ISL_1825995, EPI_ISL_1825996, EPI_ISL_1825997, EPI_ISL_1825998, EPI_ISL_1825999, EPI_ISL_1826000, EPI_ISL_1826001, EPI_ISL_1826002, EPI_ISL_1826003, EPI_ISL_1826004, EPI_ISL_1826005, EPI_ISL_1826006, EPI_ISL_1826007, EPI_ISL_1826008, EPI_ISL_1826009, EPI_ISL_1826010, EPI_ISL_1826011, EPI_ISL_1826012, EPI_ISL_1826013, EPI_ISL_1826014, EPI_ISL_1826015, EPI_ISL_1826016, EPI_ISL_1826017, EPI_ISL_1826018, EPI_ISL_1826019, EPI_ISL_1826020, EPI_ISL_1826021, EPI_ISL_1826022, EPI_ISL_1826023, EPI_ISL_1826024, EPI_ISL_1826025, EPI_ISL_1826026, EPI_ISL_1826027, EPI_ISL_1826028, EPI_ISL_1826029, EPI_ISL_1826030, EPI_ISL_1826031, EPI_ISL_1826032, EPI_ISL_1826033, EPI_ISL_1826034, EPI_ISL_1826035, EPI_ISL_1826036, EPI_ISL_1826037, EPI_ISL_1826038, EPI_ISL_1826039, EPI_ISL_1826040, EPI_ISL_1826041, EPI_ISL_1826042, EPI_ISL_1826043, EPI_ISL_1826044, EPI_ISL_1826045, EPI_ISL_1826046, EPI_ISL_1826047, EPI_ISL_1826048, EPI_ISL_1826049, EPI_ISL_1826050, EPI_ISL_1826051, EPI_ISL_1826052, EPI_ISL_1826053, EPI_ISL_1826054, EPI_ISL_1826055, EPI_ISL_1826056, EPI_ISL_1826057, EPI_ISL_1826058, EPI_ISL_1826059, EPI_ISL_1826060, EPI_ISL_1826061, EPI_ISL_1826062, EPI_ISL_1826063, EPI_ISL_1826064, EPI_ISL_1826065, EPI_ISL_1826066, EPI_ISL_1826067, EPI_ISL_1826068, EPI_ISL_1826069, EPI_ISL_1826070, EPI_ISL_1826071, EPI_ISL_1826072, EPI_ISL_1826073, EPI_ISL_1826074, EPI_ISL_1826075, EPI_ISL_1826076, EPI_ISL_1826077, EPI_ISL_1826078, EPI_ISL_1826079, EPI_ISL_1826080, EPI_ISL_1826081, EPI_ISL_1826086, EPI_ISL_1826087, EPI_ISL_1826088, EPI_ISL_1826089, EPI_ISL_1826090, EPI_ISL_1826091, EPI_ISL_1826092, EPI_ISL_1826093, EPI_ISL_1826094, EPI_ISL_1826095, EPI_ISL_1826096, EPI_ISL_1826097, EPI_ISL_1826098, EPI_ISL_1826099, EPI_ISL_1826100, EPI_ISL_1826101, EPI_ISL_1826102, EPI_ISL_1826103, EPI_ISL_1826104, EPI_ISL_1826105, EPI_ISL_1826106, EPI_ISL_1826107, EPI_ISL_1826108, EPI_ISL_1826109, EPI_ISL_1826110, EPI_ISL_1826111, EPI_ISL_1826112, EPI_ISL_1826113, EPI_ISL_1826114, EPI_ISL_1826115, EPI_ISL_1826116, EPI_ISL_1826117, EPI_ISL_1826118, EPI_ISL_1826119, EPI_ISL_1826120, EPI_ISL_1826121, EPI_ISL_1826122, EPI_ISL_1826123, EPI_ISL_1826124, EPI_ISL_1826125, EPI_ISL_1826126, EPI_ISL_1826127, EPI_ISL_1826128, EPI_ISL_1826129, EPI_ISL_1826130, EPI_ISL_1826131, EPI_ISL_1826132, EPI_ISL_1826133, EPI_ISL_1826134, EPI_ISL_1826135, EPI_ISL_1826136, EPI_ISL_1826137, EPI_ISL_1826138, EPI_ISL_1826139, EPI_ISL_1826140, EPI_ISL_1826141, EPI_ISL_1826142, EPI_ISL_1826143, EPI_ISL_1826144, EPI_ISL_1826145, EPI_ISL_1826146, EPI_ISL_1826147, EPI_ISL_1826148, EPI_ISL_1826149, EPI_ISL_1826150, EPI_ISL_1826151, EPI_ISL_1826152, EPI_ISL_1826153, EPI_ISL_1826154, EPI_ISL_1826155, EPI_ISL_1826156, EPI_ISL_1826157, EPI_ISL_1826158, EPI_ISL_1826159, EPI_ISL_1826160, EPI_ISL_1826161, EPI_ISL_1826162, EPI_ISL_1826163, EPI_ISL_1826164, EPI_ISL_1826165, EPI_ISL_1826166, EPI_ISL_1826167, EPI_ISL_1826168, EPI_ISL_1826169, EPI_ISL_1826170, EPI_ISL_1826171, EPI_ISL_1826172, EPI_ISL_1826173, EPI_ISL_1826174, EPI_ISL_1826175, EPI_ISL_1826176, EPI_ISL_1826177, EPI_ISL_1826178, EPI_ISL_1826179, EPI_ISL_1826180, EPI_ISL_1826181, EPI_ISL_1826182, EPI_ISL_1826183, EPI_ISL_1826184, EPI_ISL_1826185, EPI_ISL_1826186, EPI_ISL_1826187, EPI_ISL_1826188, EPI_ISL_1826189, EPI_ISL_1826190, EPI_ISL_1826191, EPI_ISL_1826192, EPI_ISL_1826193, EPI_ISL_1826194, EPI_ISL_1826195, EPI_ISL_1826196, EPI_ISL_1826197, EPI_ISL_1826198, EPI_ISL_1826199, EPI_ISL_1826200, EPI_ISL_1826201, EPI_ISL_1826202, EPI_ISL_1826203, EPI_ISL_1826204, EPI_ISL_1826205, EPI_ISL_1826206, EPI_ISL_1826207, EPI_ISL_1826208, EPI_ISL_1826209, EPI_ISL_1826210, EPI_ISL_1826211, EPI_ISL_1826212, EPI_ISL_1826213, EPI_ISL_1826214, EPI_ISL_1826215, EPI_ISL_1826216, EPI_ISL_1826217, EPI_ISL_1826218, EPI_ISL_1826219, EPI_ISL_1826220, EPI_ISL_1826221, EPI_ISL_1826222, EPI_ISL_1826223, EPI_ISL_1826224, EPI_ISL_1826225, EPI_ISL_1826226, EPI_ISL_1826227, EPI_ISL_1826228, EPI_ISL_1826229, EPI_ISL_1826230, EPI_ISL_1826231, EPI_ISL_1826232, EPI_ISL_1826233, EPI_ISL_1826234, EPI_ISL_1826235, EPI_ISL_1826236, EPI_ISL_1826237, EPI_ISL_1826238, EPI_ISL_1826239, EPI_ISL_1826240 | | | |
| see above | UW Virology Lab | UW Virology Lab | Pavitra Roychoudhury, Hong Xie, Lasata Shrestha, Tien V. Nguyen, Shah Mohamed Bakhash, Michelle Lin, Noah R. Baker, Sean Ellis, Meei-Li Huang, Keith R Jerome, Alexander Greninger |
| EPI_ISL_1826241, EPI_ISL_1826242, EPI_ISL_1826243, EPI_ISL_1826244, EPI_ISL_1826245, EPI_ISL_1826246, EPI_ISL_1826247, EPI_ISL_1826248, EPI_ISL_1826249, EPI_ISL_1826250, EPI_ISL_1826251, EPI_ISL_1826252, EPI_ISL_1826253, EPI_ISL_1826254, EPI_ISL_1826255, EPI_ISL_1826256, EPI_ISL_1826257, EPI_ISL_1826258, EPI_ISL_1826259, EPI_ISL_1826260, EPI_ISL_1826261, EPI_ISL_1826262, EPI_ISL_1826263, EPI_ISL_1826264, EPI_ISL_1826265, EPI_ISL_1826266, EPI_ISL_1826267, EPI_ISL_1826268, EPI_ISL_1826269, EPI_ISL_1826270, EPI_ISL_1826271, EPI_ISL_1826272, EPI_ISL_1826273, EPI_ISL_1826274, EPI_ISL_1826275, EPI_ISL_1826276, EPI_ISL_1826277, EPI_ISL_1826278, EPI_ISL_1826279, EPI_ISL_1826280, EPI_ISL_1826281, EPI_ISL_1826282, EPI_ISL_1826283, EPI_ISL_1826284, EPI_ISL_1826285, EPI_ISL_1826286, EPI_ISL_1826287, EPI_ISL_1826288, EPI_ISL_1826289, EPI_ISL_1826290, EPI_ISL_1826291, EPI_ISL_1826292, EPI_ISL_1826293, EPI_ISL_1826294, EPI_ISL_1826295, EPI_ISL_1826296, EPI_ISL_1826297, EPI_ISL_1826298, EPI_ISL_1826299, EPI_ISL_1826300, EPI_ISL_1826301, EPI_ISL_1826302, EPI_ISL_1826303, EPI_ISL_1826304, EPI_ISL_1826305, EPI_ISL_1826306, EPI_ISL_1826307, EPI_ISL_1826308, EPI_ISL_1826309, EPI_ISL_1826310, EPI_ISL_1826311, EPI_ISL_1826312, EPI_ISL_1826313, EPI_ISL_1826314, EPI_ISL_1826315, EPI_ISL_1826316, EPI_ISL_1826324, EPI_ISL_1826325, EPI_ISL_1826326, EPI_ISL_1826327, EPI_ISL_1826328, EPI_ISL_1826329, EPI_ISL_1826330, EPI_ISL_1826331, EPI_ISL_1826332, EPI_ISL_1826333, EPI_ISL_1826334, EPI_ISL_1826335, EPI_ISL_1826336, EPI_ISL_1826337, EPI_ISL_1826338, EPI_ISL_1826339, EPI_ISL_1826340, EPI_ISL_1826341, EPI_ISL_1826342, EPI_ISL_1826343, EPI_ISL_1826344, EPI_ISL_1826345, EPI_ISL_1826346, EPI_ISL_1826347, EPI_ISL_1826348, EPI_ISL_1826349, EPI_ISL_1826350, EPI_ISL_1826351, EPI_ISL_1826352, EPI_ISL_1826353, EPI_ISL_1826354, EPI_ISL_1826355, EPI_ISL_1826356, EPI_ISL_1826357, EPI_ISL_1826358, EPI_ISL_1826359, EPI_ISL_1826360, EPI_ISL_1826361, EPI_ISL_1826362, EPI_ISL_1826363, EPI_ISL_1826364, EPI_ISL_1826365, EPI_ISL_1826366, EPI_ISL_1826367, EPI_ISL_1826368, EPI_ISL_1826369, EPI_ISL_1826370, EPI_ISL_1826371, EPI_ISL_1826372, EPI_ISL_1826373, EPI_ISL_1826374, EPI_ISL_1826375, EPI_ISL_1826376, EPI_ISL_1826377, EPI_ISL_1826378, EPI_ISL_1826379, EPI_ISL_1826380, EPI_ISL_1826381, EPI_ISL_1826382, EPI_ISL_1826383, EPI_ISL_1826384, EPI_ISL_1826385, EPI_ISL_1826386, EPI_ISL_1826387, EPI_ISL_1826388, EPI_ISL_1826389, EPI_ISL_1826390, EPI_ISL_1826391, EPI_ISL_1826392, EPI_ISL_1826393, EPI_ISL_1826394, EPI_ISL_1826395, EPI_ISL_1826396, EPI_ISL_1826397, EPI_ISL_1826398, EPI_ISL_1826399, EPI_ISL_1826400, EPI_ISL_1826401, EPI_ISL_1826402, EPI_ISL_1826403, EPI_ISL_1826404, EPI_ISL_1826405, EPI_ISL_1826406, EPI_ISL_1826407, EPI_ISL_1826408, EPI_ISL_1826409, EPI_ISL_1826410, EPI_ISL_1826411, EPI_ISL_1826412, EPI_ISL_1826413, EPI_ISL_1826414, EPI_ISL_1826415, EPI_ISL_1826416, EPI_ISL_1826417, EPI_ISL_1826418, EPI_ISL_1826419, EPI_ISL_1826420, EPI_ISL_1826421, EPI_ISL_1826422, EPI_ISL_1826423, EPI_ISL_1826424, EPI_ISL_1826425, EPI_ISL_1826426, EPI_ISL_1826427, EPI_ISL_1826428, EPI_ISL_1826429, EPI_ISL_1826430, EPI_ISL_1826431, EPI_ISL_1826432, EPI_ISL_1826433, EPI_ISL_1826434, EPI_ISL_1826435, EPI_ISL_1826436, EPI_ISL_1826437, EPI_ISL_1826438, EPI_ISL_1826439, EPI_ISL_1826440, EPI_ISL_1826441, EPI_ISL_1826442, EPI_ISL_1826443, EPI_ISL_1826444, EPI_ISL_1826445, EPI_ISL_1826446, EPI_ISL_1826447, EPI_ISL_1826448, EPI_ISL_1826449, EPI_ISL_1826450, EPI_ISL_1826451, EPI_ISL_1826452, EPI_ISL_1826453, EPI_ISL_1826454, EPI_ISL_1826455, EPI_ISL_1826456, EPI_ISL_1826457, EPI_ISL_1826458, EPI_ISL_1826459, EPI_ISL_1826460, EPI_ISL_1826461, EPI_ISL_1826462, EPI_ISL_1826463, EPI_ISL_1826464, EPI_ISL_1826465, EPI_ISL_1826466, EPI_ISL_1826467, EPI_ISL_1826468, EPI_ISL_1826469, EPI_ISL_1826470, EPI_ISL_1826471, EPI_ISL_1826472 | | | |
| see above | Maine Health and Environmental Testing Laboratory | Tewhey Lab, The Jackson Laboratory | Matluk,N., Dewey,H., Iosue,F., Barter,M., Lynch,R., Munger,H. and Tewhey,R. |
| EPI_ISL_1826473, EPI_ISL_1826474, EPI_ISL_1826475, EPI_ISL_1826476, EPI_ISL_1826477, EPI_ISL_1826478, EPI_ISL_1826479, EPI_ISL_1826480, EPI_ISL_1826481, EPI_ISL_1826482, EPI_ISL_1826483, EPI_ISL_1826484, EPI_ISL_1826485, EPI_ISL_1826486, EPI_ISL_1826487, EPI_ISL_1826488, EPI_ISL_1826489, EPI_ISL_1826490, EPI_ISL_1826491, EPI_ISL_1826492, EPI_ISL_1826493, EPI_ISL_1826494, EPI_ISL_1826495, EPI_ISL_1826496, EPI_ISL_1826497, EPI_ISL_1826498, EPI_ISL_1826499, EPI_ISL_1826500, EPI_ISL_1826501, EPI_ISL_1826502, EPI_ISL_1826503, EPI_ISL_1826504, EPI_ISL_1826505, EPI_ISL_1826507, EPI_ISL_1826508, EPI_ISL_1826509, EPI_ISL_1826510, EPI_ISL_1826511, EPI_ISL_1826512, EPI_ISL_1826513, EPI_ISL_1826514, EPI_ISL_1826515, EPI_ISL_1826516, EPI_ISL_1826517, EPI_ISL_1826518, EPI_ISL_1826519, EPI_ISL_1826520, EPI_ISL_1826521, EPI_ISL_1826522, EPI_ISL_1826523, EPI_ISL_1826524, EPI_ISL_1826525, EPI_ISL_1826526, EPI_ISL_1826527, EPI_ISL_1826528, EPI_ISL_1826529, EPI_ISL_1826530, EPI_ISL_1826531, EPI_ISL_1826532, EPI_ISL_1826533, EPI_ISL_1826534, EPI_ISL_1826535, EPI_ISL_1826536, EPI_ISL_1826537, EPI_ISL_1826538, EPI_ISL_1826539, EPI_ISL_1826540, EPI_ISL_1826541, EPI_ISL_1826542, EPI_ISL_1826543, EPI_ISL_1826544, EPI_ISL_1826545, EPI_ISL_1826546, EPI_ISL_1826547, EPI_ISL_1826548, EPI_ISL_1826549, EPI_ISL_1826550, EPI_ISL_1826551, EPI_ISL_1826552, EPI_ISL_1826553, EPI_ISL_1826554, EPI_ISL_1826555, EPI_ISL_1826556, EPI_ISL_1826557 | | | |
| see above | The Jackson Laboratory | The Jackson Laboratory | Bergeron D, Renzette N, Adams M, Omerza G, Kelly K, Long J, Li L |
| EPI_ISL_1826991, | Arizona State Public Health Laboratory | Arizona State Public Health Laboratory | Trung Huynh, Jessica Escobar, Katherine Fullerton, Nobuko Fukushima, Stacy White, Linda Getsinger, Victor Waddell |

| | | | |
|---|---|---|---|
| EPI_ISL_1826993, EPI_ISL_1826994, EPI_ISL_1826995, EPI_ISL_1826996, EPI_ISL_1826997 | | | |
| EPI_ISL_1827548, EPI_ISL_1827549, EPI_ISL_1827550, EPI_ISL_1827551, EPI_ISL_1827552, EPI_ISL_1827554 | University of Wisconsin-Madison AIDS Vaccine Research Laboratories | University of Wisconsin-Madison AIDS Vaccine Research Laboratories | Gage Moreno, Katarina Braun, et al. AIDS Vaccine Research Laboratories |
| EPI_ISL_1827711, EPI_ISL_1827712, EPI_ISL_1827713, EPI_ISL_1827714, EPI_ISL_1827715, EPI_ISL_1827716, EPI_ISL_1827717, EPI_ISL_1827718, EPI_ISL_1827720, EPI_ISL_1827721, EPI_ISL_1827722, EPI_ISL_1827723, EPI_ISL_1827724, EPI_ISL_1827725, EPI_ISL_1827726 | | | |
| see above | UW Virology Lab | UW Virology Lab | Pavitra Roychoudhury, Hong Xie, Lasata Shrestha, Tien V. Nguyen, Shah Mohamed Bakhash, Michelle Lin, Noah R. Baker, Ricardo Perez, Sean Ellis, Nathan Breit, Robert J. Livingston, Meei-Li Huang, Keith R Jerome, Patrick Mathias, Alexander Greninger |
| EPI_ISL_1827954, EPI_ISL_1827955, EPI_ISL_1827956, EPI_ISL_1827957, EPI_ISL_1827958, EPI_ISL_1827959, EPI_ISL_1827960, EPI_ISL_1827961, EPI_ISL_1827962, EPI_ISL_1827963, EPI_ISL_1827965, EPI_ISL_1827966, EPI_ISL_1827968, EPI_ISL_1827969, EPI_ISL_1827972, EPI_ISL_1827973, EPI_ISL_1827974, EPI_ISL_1827975, EPI_ISL_1827976, EPI_ISL_1827977, EPI_ISL_1827978, EPI_ISL_1827980, EPI_ISL_1827981, EPI_ISL_1827982, EPI_ISL_1827983, EPI_ISL_1827984, EPI_ISL_1827985, EPI_ISL_1827986, EPI_ISL_1827987, EPI_ISL_1827990, EPI_ISL_1827991, EPI_ISL_1827992, EPI_ISL_1827993, EPI_ISL_1827995, EPI_ISL_1827996, EPI_ISL_1827997, EPI_ISL_1827999, EPI_ISL_1828001, EPI_ISL_1828002, EPI_ISL_1828004, EPI_ISL_1828006, EPI_ISL_1828008, EPI_ISL_1828009, EPI_ISL_1828010, EPI_ISL_1828014, EPI_ISL_1828015, EPI_ISL_1828016, EPI_ISL_1828018, EPI_ISL_1828019, EPI_ISL_1828020, EPI_ISL_1828021, EPI_ISL_1828022, EPI_ISL_1828023, EPI_ISL_1828024, EPI_ISL_1828025, EPI_ISL_1828026, EPI_ISL_1828027, EPI_ISL_1828028, EPI_ISL_1828031, EPI_ISL_1828032, EPI_ISL_1828033, EPI_ISL_1828034, EPI_ISL_1828035, EPI_ISL_1828036, EPI_ISL_1828037, EPI_ISL_1828038, EPI_ISL_1828039, EPI_ISL_1828040, EPI_ISL_1828042, EPI_ISL_1828043, EPI_ISL_1828045, EPI_ISL_1828046, EPI_ISL_1828047, EPI_ISL_1828048, EPI_ISL_1828050, EPI_ISL_1828051, EPI_ISL_1828052, EPI_ISL_1828053, EPI_ISL_1828054, EPI_ISL_1828055, EPI_ISL_1828056, EPI_ISL_1828058, EPI_ISL_1828060, EPI_ISL_1828062, EPI_ISL_1828063, EPI_ISL_1828065, EPI_ISL_1828067, EPI_ISL_1828068, EPI_ISL_1828069, EPI_ISL_1828071, EPI_ISL_1828072, EPI_ISL_1828073, EPI_ISL_1828074, EPI_ISL_1828075, EPI_ISL_1828076, EPI_ISL_1828077, EPI_ISL_1828079, EPI_ISL_1828080, EPI_ISL_1828081, EPI_ISL_1828082, EPI_ISL_1828083, EPI_ISL_1828085, EPI_ISL_1828086, EPI_ISL_1828087, EPI_ISL_1828089, EPI_ISL_1828090, EPI_ISL_1828093, EPI_ISL_1828095, EPI_ISL_1828096, EPI_ISL_1828098, EPI_ISL_1828099, EPI_ISL_1828101, EPI_ISL_1828102, EPI_ISL_1828103, EPI_ISL_1828104, EPI_ISL_1828106, EPI_ISL_1828110, EPI_ISL_1828111, EPI_ISL_1828113, EPI_ISL_1828114, EPI_ISL_1828115, EPI_ISL_1828117, EPI_ISL_1828118, EPI_ISL_1828119, EPI_ISL_1828120, EPI_ISL_1828121, EPI_ISL_1828124, EPI_ISL_1828125, EPI_ISL_1828127, EPI_ISL_1828129, EPI_ISL_1828130, EPI_ISL_1828131, EPI_ISL_1828132, EPI_ISL_1828134, EPI_ISL_1828135, EPI_ISL_1828136, EPI_ISL_1828137, EPI_ISL_1828138, EPI_ISL_1828139, EPI_ISL_1828141, EPI_ISL_1828142, EPI_ISL_1828143, EPI_ISL_1828144, EPI_ISL_1828145, EPI_ISL_1828146, EPI_ISL_1828147, EPI_ISL_1828149, EPI_ISL_1828150, EPI_ISL_1828151, EPI_ISL_1828152, EPI_ISL_1828153, EPI_ISL_1828155, EPI_ISL_1828156, EPI_ISL_1828158, EPI_ISL_1828159, EPI_ISL_1828161, EPI_ISL_1828162, EPI_ISL_1828163, EPI_ISL_1828164, EPI_ISL_1828165, EPI_ISL_1828167, EPI_ISL_1828168, EPI_ISL_1828169, EPI_ISL_1828170, EPI_ISL_1828172, EPI_ISL_1828174, EPI_ISL_1828175, EPI_ISL_1828176, EPI_ISL_1828177, EPI_ISL_1828179, EPI_ISL_1828181, EPI_ISL_1828182, EPI_ISL_1828183, EPI_ISL_1828184, EPI_ISL_1828185, EPI_ISL_1828186, EPI_ISL_1828187, EPI_ISL_1828188, EPI_ISL_1828189, EPI_ISL_1828190, EPI_ISL_1828193, EPI_ISL_1828194, EPI_ISL_1828196, EPI_ISL_1828197, EPI_ISL_1828199, EPI_ISL_1828200, EPI_ISL_1828202, EPI_ISL_1828203, EPI_ISL_1828204, EPI_ISL_1828205, EPI_ISL_1828206, EPI_ISL_1828207, EPI_ISL_1828209, EPI_ISL_1828210, EPI_ISL_1828211, EPI_ISL_1828213, EPI_ISL_1828215, EPI_ISL_1828216, EPI_ISL_1828217, EPI_ISL_1828218, EPI_ISL_1828219, EPI_ISL_1828220, EPI_ISL_1828221, EPI_ISL_1828222, EPI_ISL_1828223, EPI_ISL_1828224, EPI_ISL_1828225, EPI_ISL_1828227, EPI_ISL_1828228, EPI_ISL_1828229, EPI_ISL_1828230, EPI_ISL_1828232, EPI_ISL_1828233, EPI_ISL_1828234, EPI_ISL_1828235, EPI_ISL_1828238, EPI_ISL_1828239, EPI_ISL_1828240, EPI_ISL_1828242, EPI_ISL_1828243, EPI_ISL_1828244, EPI_ISL_1828245, EPI_ISL_1828246, EPI_ISL_1828247, EPI_ISL_1828248, EPI_ISL_1828250, EPI_ISL_1828251, EPI_ISL_1828252, EPI_ISL_1828253, EPI_ISL_1828254, EPI_ISL_1828255, EPI_ISL_1828257, EPI_ISL_1828258, EPI_ISL_1828259, EPI_ISL_1828260, EPI_ISL_1828261, EPI_ISL_1828262, EPI_ISL_1828263, EPI_ISL_1828264, EPI_ISL_1828265, EPI_ISL_1828266, EPI_ISL_1828267, EPI_ISL_1828268, EPI_ISL_1828270, EPI_ISL_1828271, EPI_ISL_1828272, EPI_ISL_1828273, EPI_ISL_1828274, EPI_ISL_1828276, EPI_ISL_1828278, EPI_ISL_1828279, EPI_ISL_1828280, EPI_ISL_1828281, EPI_ISL_1828282, EPI_ISL_1828284, EPI_ISL_1828285, EPI_ISL_1828287, EPI_ISL_1828288, EPI_ISL_1828290, EPI_ISL_1828291, EPI_ISL_1828292, EPI_ISL_1828295, EPI_ISL_1828296, EPI_ISL_1828298, EPI_ISL_1828299, EPI_ISL_1828300, EPI_ISL_1828301, EPI_ISL_1828302, EPI_ISL_1828303, EPI_ISL_1828304, EPI_ISL_1828305, EPI_ISL_1828306, EPI_ISL_1828307, EPI_ISL_1828309, EPI_ISL_1828310, EPI_ISL_1828311, EPI_ISL_1828313, EPI_ISL_1828314, EPI_ISL_1828315, EPI_ISL_1828316, EPI_ISL_1828317, EPI_ISL_1828318, EPI_ISL_1828319, EPI_ISL_1828320, EPI_ISL_1828322, EPI_ISL_1828323, EPI_ISL_1828324, EPI_ISL_1828325, EPI_ISL_1828326, EPI_ISL_1828327, EPI_ISL_1828328, EPI_ISL_1828329, EPI_ISL_1828330, EPI_ISL_1828331, EPI_ISL_1828332, EPI_ISL_1828333, EPI_ISL_1828335, EPI_ISL_1828336, EPI_ISL_1828338, EPI_ISL_1828339, EPI_ISL_1828340, EPI_ISL_1828341, EPI_ISL_1828342, EPI_ISL_1828343, EPI_ISL_1828344, EPI_ISL_1828346, EPI_ISL_1828348, EPI_ISL_1828349, EPI_ISL_1828350, EPI_ISL_1828351, EPI_ISL_1828352, EPI_ISL_1828353, EPI_ISL_1828354, EPI_ISL_1828355, EPI_ISL_1828356, EPI_ISL_1828357, EPI_ISL_1828358, EPI_ISL_1828359, EPI_ISL_1828360, EPI_ISL_1828361, EPI_ISL_1828362, EPI_ISL_1828363, EPI_ISL_1828364, EPI_ISL_1828365, EPI_ISL_1828366, EPI_ISL_1828367, EPI_ISL_1828368, EPI_ISL_1828369, EPI_ISL_1828370, EPI_ISL_1828371, EPI_ISL_1828372, EPI_ISL_1828373, EPI_ISL_1828374, EPI_ISL_1828376, EPI_ISL_1828378, EPI_ISL_1828380, EPI_ISL_1828381, EPI_ISL_1828382, EPI_ISL_1828384, EPI_ISL_1828385, EPI_ISL_1828386, EPI_ISL_1828387, EPI_ISL_1828388, EPI_ISL_1828389, EPI_ISL_1828391, EPI_ISL_1828392, EPI_ISL_1828394, EPI_ISL_1828395, EPI_ISL_1828396, EPI_ISL_1828397, EPI_ISL_1828398, EPI_ISL_1828399, EPI_ISL_1828400, EPI_ISL_1828401, EPI_ISL_1828402, EPI_ISL_1828403, EPI_ISL_1828404, EPI_ISL_1828405, EPI_ISL_1828406, EPI_ISL_1828407, EPI_ISL_1828408, EPI_ISL_1828409, EPI_ISL_1828410, EPI_ISL_1828411, EPI_ISL_1828412, EPI_ISL_1828413, EPI_ISL_1828414, EPI_ISL_1828415, EPI_ISL_1828416, EPI_ISL_1828417, EPI_ISL_1828418, EPI_ISL_1828419, EPI_ISL_1828420, EPI_ISL_1828421, EPI_ISL_1828422, EPI_ISL_1828423, EPI_ISL_1828424, EPI_ISL_1828425, EPI_ISL_1828426, EPI_ISL_1828428, EPI_ISL_1828429, EPI_ISL_1828430, EPI_ISL_1828431, EPI_ISL_1828432, EPI_ISL_1828433, EPI_ISL_1828434, EPI_ISL_1828435, EPI_ISL_1828436, EPI_ISL_1828437, EPI_ISL_1828438, EPI_ISL_1828439, EPI_ISL_1828440, EPI_ISL_1828441, EPI_ISL_1828442, EPI_ISL_1828443, EPI_ISL_1828444, EPI_ISL_1828445, EPI_ISL_1828446, EPI_ISL_1828447, EPI_ISL_1828448, EPI_ISL_1828449, EPI_ISL_1828450, EPI_ISL_1828451, EPI_ISL_1828452, EPI_ISL_1828453, EPI_ISL_1828454, EPI_ISL_1828455, EPI_ISL_1828456, EPI_ISL_1828457, EPI_ISL_1828458, EPI_ISL_1828459, EPI_ISL_1828460, EPI_ISL_1828461, EPI_ISL_1828462, EPI_ISL_1828463, EPI_ISL_1828464, EPI_ISL_1828465, EPI_ISL_1828466, EPI_ISL_1828467, EPI_ISL_1828468, EPI_ISL_1828469, EPI_ISL_1828470, EPI_ISL_1828471, EPI_ISL_1828472, EPI_ISL_1828473, EPI_ISL_1828474, EPI_ISL_1828475, EPI_ISL_1828476, EPI_ISL_1828477, EPI_ISL_1828478, EPI_ISL_1828479, EPI_ISL_1828480, EPI_ISL_1828481, EPI_ISL_1828482, EPI_ISL_1828483, EPI_ISL_1828484, EPI_ISL_1828485, EPI_ISL_1828486, EPI_ISL_1828487, EPI_ISL_1828488, EPI_ISL_1828489, EPI_ISL_1828490, EPI_ISL_1828491, EPI_ISL_1828492, EPI_ISL_1828493, EPI_ISL_1828494, EPI_ISL_1828495, EPI_ISL_1828496, EPI_ISL_1828497, EPI_ISL_1828498, EPI_ISL_1828499, EPI_ISL_1828500, EPI_ISL_1828502, EPI_ISL_1828503, EPI_ISL_1828504, EPI_ISL_1828505, EPI_ISL_1828506, EPI_ISL_1828507, EPI_ISL_1828508, EPI_ISL_1828509, EPI_ISL_1828510, EPI_ISL_1828511, EPI_ISL_1828512, EPI_ISL_1828513, EPI_ISL_1828514, EPI_ISL_1828515, EPI_ISL_1828516, EPI_ISL_1828517, EPI_ISL_1828519, EPI_ISL_1828520, EPI_ISL_1828521, EPI_ISL_1828522, EPI_ISL_1828523, EPI_ISL_1828524, EPI_ISL_1828525, EPI_ISL_1828526, EPI_ISL_1828527, EPI_ISL_1828528, EPI_ISL_1828530, EPI_ISL_1828531, EPI_ISL_1828532, EPI_ISL_1828534, EPI_ISL_1828535, EPI_ISL_1828536, EPI_ISL_1828537, EPI_ISL_1828538, EPI_ISL_1828539, EPI_ISL_1828540, EPI_ISL_1828541, EPI_ISL_1828542, EPI_ISL_1828543, EPI_ISL_1828544, EPI_ISL_1828545, EPI_ISL_1828546, EPI_ISL_1828547, EPI_ISL_1828548, EPI_ISL_1828550, EPI_ISL_1828551, EPI_ISL_1828552, EPI_ISL_1828553, EPI_ISL_1828554, EPI_ISL_1828555, EPI_ISL_1828556, EPI_ISL_1828558, EPI_ISL_1828559, EPI_ISL_1828560, EPI_ISL_1828562, EPI_ISL_1828563, EPI_ISL_1828564, EPI_ISL_1828565, EPI_ISL_1828566, EPI_ISL_1828567, EPI_ISL_1828568, EPI_ISL_1828569, EPI_ISL_1828570, EPI_ISL_1828571, EPI_ISL_1828572, EPI_ISL_1828573, EPI_ISL_1828574, EPI_ISL_1828575, EPI_ISL_1828576, EPI_ISL_1828577, EPI_ISL_1828578, EPI_ISL_1828579, EPI_ISL_1828580, EPI_ISL_1828581, EPI_ISL_1828582, EPI_ISL_1828583, EPI_ISL_1828584, EPI_ISL_1828585, EPI_ISL_1828586, EPI_ISL_1828587, EPI_ISL_1828588, EPI_ISL_1828590, EPI_ISL_1828591, EPI_ISL_1828592, EPI_ISL_1828593, EPI_ISL_1828594, EPI_ISL_1828595, EPI_ISL_1828596, EPI_ISL_1828597, EPI_ISL_1828598, EPI_ISL_1828599, EPI_ISL_1828600, EPI_ISL_1828601, EPI_ISL_1828602, EPI_ISL_1828603, EPI_ISL_1828604, EPI_ISL_1828605, EPI_ISL_1828606, EPI_ISL_1828607, EPI_ISL_1828608, EPI_ISL_1828609, EPI_ISL_1828610, EPI_ISL_1828611, EPI_ISL_1828612, EPI_ISL_1828613, EPI_ISL_1828614, EPI_ISL_1828615, EPI_ISL_1828616, EPI_ISL_1828617, EPI_ISL_1828618, EPI_ISL_1828619, EPI_ISL_1828620, EPI_ISL_1828621, EPI_ISL_1828622, EPI_ISL_1828623, EPI_ISL_1828624, EPI_ISL_1828625, EPI_ISL_1828626, EPI_ISL_1828627, EPI_ISL_1828628, EPI_ISL_1828629, EPI_ISL_1828630, EPI_ISL_1828631, EPI_ISL_1828632, EPI_ISL_1828633, EPI_ISL_1828634, EPI_ISL_1828635, EPI_ISL_1828636, EPI_ISL_1828637, EPI_ISL_1828638, EPI_ISL_1828639, EPI_ISL_1828640, EPI_ISL_1828641, EPI_ISL_1828642, EPI_ISL_1828643, EPI_ISL_1828644, EPI_ISL_1828645, EPI_ISL_1828646, EPI_ISL_1828647, EPI_ISL_1828648, EPI_ISL_1828649, EPI_ISL_1828650, EPI_ISL_1828651, EPI_ISL_1828652, EPI_ISL_1828653, EPI_ISL_1828654, EPI_ISL_1828655, EPI_ISL_1828657, EPI_ISL_1828658, EPI_ISL_1828659, EPI_ISL_1828661, EPI_ISL_1828662, EPI_ISL_1828663, EPI_ISL_1828664, EPI_ISL_1828665, EPI_ISL_1828666, EPI_ISL_1828667, EPI_ISL_1828668, EPI_ISL_1828669, EPI_ISL_1828670, EPI_ISL_1828671, EPI_ISL_1828672, EPI_ISL_1828673, EPI_ISL_1828674, EPI_ISL_1828675, EPI_ISL_1828676, EPI_ISL_1828677, EPI_ISL_1828678, EPI_ISL_1828679, EPI_ISL_1828681, EPI_ISL_1828682, EPI_ISL_1828683, EPI_ISL_1828684, EPI_ISL_1828685, EPI_ISL_1828686, EPI_ISL_1828687, EPI_ISL_1828688, EPI_ISL_1828689, EPI_ISL_1828690, EPI_ISL_1828691, EPI_ISL_1828692, EPI_ISL_1828693, EPI_ISL_1828694, EPI_ISL_1828695, EPI_ISL_1828697 | | | |
| see above | Pandemic Response Lab - NYC | Pandemic Response Lab, R&D | Henry Lee, Michael Hammerling, Melissa Hopkins, Cybill del Castillo, Shinyoung Clair Kang, William Ward, Pradeep Bugga, Sol Rey, Dylan Law, Katharine Nelson, Haiping Hao, Jon Laurent |
| EPI_ISL_1834186, EPI_ISL_1834187, EPI_ISL_1834188, EPI_ISL_1834189, EPI_ISL_1834190, EPI_ISL_1834191, EPI_ISL_1834192, EPI_ISL_1834193, EPI_ISL_1834194, EPI_ISL_1834195, EPI_ISL_1834196, EPI_ISL_1834197, EPI_ISL_1834198, EPI_ISL_1834199, EPI_ISL_1834200, EPI_ISL_1834201, EPI_ISL_1834202, EPI_ISL_1834203, EPI_ISL_1834204, EPI_ISL_1834205, EPI_ISL_1834206, EPI_ISL_1834207, EPI_ISL_1834208, EPI_ISL_1834209, EPI_ISL_1834210, EPI_ISL_1834211, EPI_ISL_1834212, EPI_ISL_1834213, EPI_ISL_1834214, EPI_ISL_1834215, EPI_ISL_1834216, EPI_ISL_1834217, EPI_ISL_1834218, EPI_ISL_1834219, EPI_ISL_1834220, EPI_ISL_1834221, EPI_ISL_1834222, EPI_ISL_1834223, EPI_ISL_1834224, EPI_ISL_1834225, EPI_ISL_1834226, EPI_ISL_1834227, EPI_ISL_1834228, EPI_ISL_1834229, EPI_ISL_1834230, EPI_ISL_1834231, EPI_ISL_1834232, EPI_ISL_1834233, EPI_ISL_1834234, EPI_ISL_1834235, EPI_ISL_1834236, EPI_ISL_1834237, EPI_ISL_1834238, EPI_ISL_1834239, EPI_ISL_1834240, EPI_ISL_1834241, EPI_ISL_1834242, EPI_ISL_1834243, EPI_ISL_1834244, EPI_ISL_1834245, EPI_ISL_1834246, EPI_ISL_1834247, EPI_ISL_1834248, EPI_ISL_1834249, EPI_ISL_1834250, EPI_ISL_1834251, EPI_ISL_1834252, EPI_ISL_1834253, EPI_ISL_1834254, EPI_ISL_1834255, EPI_ISL_1834256, EPI_ISL_1834257, EPI_ISL_1834258, EPI_ISL_1834259, EPI_ISL_1834260, EPI_ISL_1834261, EPI_ISL_1834262, EPI_ISL_1834263, EPI_ISL_1834264, EPI_ISL_1834265, EPI_ISL_1834266, EPI_ISL_1834267, EPI_ISL_1834268, EPI_ISL_1834269, EPI_ISL_1834270, EPI_ISL_1834271, EPI_ISL_1834272, EPI_ISL_1834273, EPI_ISL_1834274, EPI_ISL_1834275, EPI_ISL_1834276, EPI_ISL_1834277, EPI_ISL_1834278, EPI_ISL_1834279, EPI_ISL_1834280, EPI_ISL_1834281, EPI_ISL_1834282, EPI_ISL_1834283, EPI_ISL_1834284, EPI_ISL_1834285, EPI_ISL_1834286, EPI_ISL_1834287, EPI_ISL_1834288, EPI_ISL_1834289, EPI_ISL_1834290, EPI_ISL_1834291, EPI_ISL_1834292, EPI_ISL_1834293, EPI_ISL_1834294, EPI_ISL_1834295, EPI_ISL_1834296, EPI_ISL_1834297, EPI_ISL_1834298, EPI_ISL_1834299, EPI_ISL_1834300, EPI_ISL_1834301, EPI_ISL_1834302, EPI_ISL_1834303, EPI_ISL_1834304, EPI_ISL_1834305, EPI_ISL_1834306, EPI_ISL_1834307, EPI_ISL_1834308, EPI_ISL_1834309, EPI_ISL_1834310, EPI_ISL_1834311, EPI_ISL_1834312, EPI_ISL_1834313, EPI_ISL_1834314, EPI_ISL_1834315, EPI_ISL_1834316, EPI_ISL_1834317, EPI_ISL_1834318, EPI_ISL_1834319, EPI_ISL_1834320, EPI_ISL_1834321, EPI_ISL_1834322, EPI_ISL_1834323, EPI_ISL_1834324, EPI_ISL_1834325, EPI_ISL_1834326, EPI_ISL_1834327, EPI_ISL_1834328, EPI_ISL_1834329, EPI_ISL_1834330, EPI_ISL_1834331, EPI_ISL_1834332, EPI_ISL_1834333, EPI_ISL_1834334, EPI_ISL_1834335, EPI_ISL_1834336, EPI_ISL_1834337, EPI_ISL_1834338, EPI_ISL_1834339, EPI_ISL_1834340, EPI_ISL_1834341, EPI_ISL_1834342, EPI_ISL_1834343, EPI_ISL_1834344, EPI_ISL_1834345, EPI_ISL_1834346, EPI_ISL_1834347, EPI_ISL_1834348, EPI_ISL_1834349, EPI_ISL_1834350, EPI_ISL_1834351, EPI_ISL_1834352, EPI_ISL_1834353, EPI_ISL_1834354, EPI_ISL_1834355, EPI_ISL_1834356, EPI_ISL_1834357, EPI_ISL_1834358, EPI_ISL_1834359, EPI_ISL_1834360, EPI_ISL_1834361, EPI_ISL_1834362, EPI_ISL_1834363, EPI_ISL_1834364, EPI_ISL_1834365, EPI_ISL_1834366, EPI_ISL_1834367, EPI_ISL_1834368, EPI_ISL_1834369, EPI_ISL_1834370, EPI_ISL_1834371, EPI_ISL_1834372, EPI_ISL_1834373, EPI_ISL_1834374, EPI_ISL_1834375, EPI_ISL_1834376, EPI_ISL_1834377, EPI_ISL_1834378, EPI_ISL_1834379, EPI_ISL_1834380, EPI_ISL_1834381, EPI_ISL_1834382, EPI_ISL_1834383, EPI_ISL_1834384, EPI_ISL_1834385, EPI_ISL_1834386, EPI_ISL_1834387, EPI_ISL_1834388, EPI_ISL_1834389, EPI_ISL_1834390, EPI_ISL_1834391, EPI_ISL_1834392, EPI_ISL_1834393, EPI_ISL_1834394, EPI_ISL_1834395, EPI_ISL_1834396, EPI_ISL_1834397, EPI_ISL_1834398, EPI_ISL_1834399, EPI_ISL_1834400, EPI_ISL_1834401, EPI_ISL_1834402, EPI_ISL_1834403, EPI_ISL_1834404, EPI_ISL_1834405, EPI_ISL_1834406, EPI_ISL_1834407, EPI_ISL_1834408, EPI_ISL_1834409, EPI_ISL_1834410, EPI_ISL_1834411, EPI_ISL_1834412, EPI_ISL_1834413, EPI_ISL_1834414, EPI_ISL_1834415, EPI_ISL_1834416, EPI_ISL_1834417, EPI_ISL_1834418, EPI_ISL_1834419, EPI_ISL_1834420, EPI_ISL_1834421, EPI_ISL_1834422, EPI_ISL_1834423, EPI_ISL_1834424, EPI_ISL_1834425, EPI_ISL_1834426, EPI_ISL_1834427, EPI_ISL_1834428, EPI_ISL_1834429, EPI_ISL_1834430, EPI_ISL_1834431, EPI_ISL_1834432, EPI_ISL_1834433, EPI_ISL_1834434, EPI_ISL_1834435, EPI_ISL_1834436, EPI_ISL_1834437, EPI_ISL_1834438, EPI_ISL_1834439, EPI_ISL_1834440, EPI_ISL_1834441, EPI_ISL_1834442 | | | |
| see above | Quest Diagnostics Incorporated | Centers for Disease Control and Prevention Division of Viral Diseases, Pathogen Discovery | Dakota Howard, Dhwani Batra, Peter W. Cook, Kara Moser, Adrian Paskey, Jason Caravas, Benjamin Rambo-Martin, Shatavia Morrison, Christopher Gulvick, Scott Sammons, Yvette Unoarumhi, Darlene Wagner, Matthew Schmerer, S. H. Rosenthal, A. Gerasimova, R. M. Kagan, B. Anderson, M. Hua, Y. Liu, L.E. Bernstein, K.E. Livingston, A. Perez, I. A. Shlyakhter, R. V. Rolando, R. Owen, P. Tanpaiboon, F. Lacbawan, Clinton R. Paden, Duncan MacCannell |
| EPI_ISL_1834443, EPI_ISL_1834444, EPI_ISL_1834445, EPI_ISL_1834446, EPI_ISL_1834448, EPI_ISL_1834449, EPI_ISL_1834450, EPI_ISL_1834451, EPI_ISL_1834452, EPI_ISL_1834453, EPI_ISL_1834454, EPI_ISL_1834455, EPI_ISL_1834456, EPI_ISL_1834457, EPI_ISL_1834458, EPI_ISL_1834459, EPI_ISL_1834460, EPI_ISL_1834461, EPI_ISL_1834462, EPI_ISL_1834463, EPI_ISL_1834464, EPI_ISL_1834465, EPI_ISL_1834466, EPI_ISL_1834467, EPI_ISL_1834468, EPI_ISL_1834469, EPI_ISL_1834470, EPI_ISL_1834471, EPI_ISL_1834472, EPI_ISL_1834473, EPI_ISL_1834474, EPI_ISL_1834475, EPI_ISL_1834476, EPI_ISL_1834477, EPI_ISL_1834478, EPI_ISL_1834479, EPI_ISL_1834480, EPI_ISL_1834481, EPI_ISL_1834482, EPI_ISL_1834483, EPI_ISL_1834484, EPI_ISL_1834485, EPI_ISL_1834486, EPI_ISL_1834487, EPI_ISL_1834488, EPI_ISL_1834489, EPI_ISL_1834490, EPI_ISL_1834491, EPI_ISL_1834492, EPI_ISL_1834493, EPI_ISL_1834494, EPI_ISL_1834495, EPI_ISL_1834496, EPI_ISL_1834497, EPI_ISL_1834498, EPI_ISL_1834499, EPI_ISL_1834500, EPI_ISL_1834501, EPI_ISL_1834502, EPI_ISL_1834503, EPI_ISL_1834504, EPI_ISL_1834505, EPI_ISL_1834506, EPI_ISL_1834507, EPI_ISL_1834508, EPI_ISL_1834509, EPI_ISL_1834510, EPI_ISL_1834511, EPI_ISL_1834512, EPI_ISL_1834513, EPI_ISL_1834514, EPI_ISL_1834515, EPI_ISL_1834516, EPI_ISL_1834517, EPI_ISL_1834518, EPI_ISL_1834519, EPI_ISL_1834520, EPI_ISL_1834521, EPI_ISL_1834522, EPI_ISL_1834523, EPI_ISL_1834524, EPI_ISL_1834525, EPI_ISL_1834526, EPI_ISL_1834527, EPI_ISL_1834528, EPI_ISL_1834529, EPI_ISL_1834531, EPI_ISL_1834532, EPI_ISL_1834533, EPI_ISL_1834534, EPI_ISL_1834535, EPI_ISL_1834536, EPI_ISL_1834537, EPI_ISL_1834538, EPI_ISL_1834539, EPI_ISL_1834540, EPI_ISL_1834541, EPI_ISL_1834542, EPI_ISL_1834543, EPI_ISL_1834544, EPI_ISL_1834545, EPI_ISL_1834546, EPI_ISL_1834547, EPI_ISL_1834548, EPI_ISL_1834549, EPI_ISL_1834550, EPI_ISL_1834551, EPI_ISL_1834552, EPI_ISL_1834553, EPI_ISL_1834554, EPI_ISL_1834555, EPI_ISL_1834556, EPI_ISL_1834557, EPI_ISL_1834558, EPI_ISL_1834559, EPI_ISL_1834560, EPI_ISL_1834561, EPI_ISL_1834562, EPI_ISL_1834563, EPI_ISL_1834564, EPI_ISL_1834565, EPI_ISL_1834566, EPI_ISL_1834567, EPI_ISL_1834568, EPI_ISL_1834569, EPI_ISL_1834570, EPI_ISL_1834571, EPI_ISL_1834572, EPI_ISL_1834573, EPI_ISL_1834574, EPI_ISL_1834575, EPI_ISL_1834576, EPI_ISL_1834577, EPI_ISL_1834578, EPI_ISL_1834579, EPI_ISL_1834580, EPI_ISL_1834581, EPI_ISL_1834582, EPI_ISL_1834583, EPI_ISL_1834584, EPI_ISL_1834585, EPI_ISL_1834586, EPI_ISL_1834587, EPI_ISL_1834588, EPI_ISL_1834589, EPI_ISL_1834590, EPI_ISL_1834591, EPI_ISL_1834592, EPI_ISL_1834593, EPI_ISL_1834594, EPI_ISL_1834595, EPI_ISL_1834596, EPI_ISL_1834597, EPI_ISL_1834598, EPI_ISL_1834599, EPI_ISL_1834600, EPI_ISL_1834601, EPI_ISL_1834602, EPI_ISL_1834603, EPI_ISL_1834604, EPI_ISL_1834605, EPI_ISL_1834606, EPI_ISL_1834607, EPI_ISL_1834608, EPI_ISL_1834609, EPI_ISL_1834610, EPI_ISL_1834611, EPI_ISL_1834612, EPI_ISL_1834613, EPI_ISL_1834615, EPI_ISL_1834616, EPI_ISL_1834617, EPI_ISL_1834618, EPI_ISL_1834619, EPI_ISL_1834620, EPI_ISL_1834621, EPI_ISL_1834622, EPI_ISL_1834623, EPI_ISL_1834624, EPI_ISL_1834625, EPI_ISL_1834626, EPI_ISL_1834627, EPI_ISL_1834628, EPI_ISL_1834629, EPI_ISL_1834630, EPI_ISL_1834631, EPI_ISL_1834632, EPI_ISL_1834633, EPI_ISL_1834634, EPI_ISL_1834635, EPI_ISL_1834636, EPI_ISL_1834637, EPI_ISL_1834638, EPI_ISL_1834639, EPI_ISL_1834640, EPI_ISL_1834641, EPI_ISL_1834642, EPI_ISL_1834643, EPI_ISL_1834644, EPI_ISL_1834645, EPI_ISL_1834646, EPI_ISL_1834647, EPI_ISL_1834648, EPI_ISL_1834649, EPI_ISL_1834650, EPI_ISL_1834651, EPI_ISL_1834652, EPI_ISL_1834653, EPI_ISL_1834654, EPI_ISL_1834655, EPI_ISL_1834656, EPI_ISL_1834657, EPI_ISL_1834658, EPI_ISL_1834659, EPI_ISL_1834660, EPI_ISL_1834661, EPI_ISL_1834662, EPI_ISL_1834663, EPI_ISL_1834664, EPI_ISL_1834665, EPI_ISL_1834666, EPI_ISL_1834667, EPI_ISL_1834668, EPI_ISL_1834669, EPI_ISL_1834670, EPI_ISL_1834671, EPI_ISL_1834673, EPI_ISL_1834674, EPI_ISL_1834675, EPI_ISL_1834676, EPI_ISL_1834677, EPI_ISL_1834678, EPI_ISL_1834679, EPI_ISL_1834680, EPI_ISL_1834681, EPI_ISL_1834682, EPI_ISL_1834683, EPI_ISL_1834684, EPI_ISL_1834685, EPI_ISL_1834686, EPI_ISL_1834687, EPI_ISL_1834688, EPI_ISL_1834689, EPI_ISL_1834690, EPI_ISL_1834691, EPI_ISL_1834692, EPI_ISL_1834693, EPI_ISL_1834694, EPI_ISL_1834695, EPI_ISL_1834696, EPI_ISL_1834697, EPI_ISL_1834698, EPI_ISL_1834699, EPI_ISL_1834700, EPI_ISL_1834701, EPI_ISL_1834702, EPI_ISL_1834703, EPI_ISL_1834704, EPI_ISL_1834705, EPI_ISL_1834706, EPI_ISL_1834707, EPI_ISL_1834708, EPI_ISL_1834709, EPI_ISL_1834710, EPI_ISL_1834711, EPI_ISL_1834712, EPI_ISL_1834713, EPI_ISL_1834714, EPI_ISL_1834715, EPI_ISL_1834716, EPI_ISL_1834717, EPI_ISL_1834718, EPI_ISL_1834719, EPI_ISL_1834720, EPI_ISL_1834721, EPI_ISL_1834722, EPI_ISL_1834723, EPI_ISL_1834724, EPI_ISL_1834725, EPI_ISL_1834726, EPI_ISL_1834727, EPI_ISL_1834728, EPI_ISL_1834729, EPI_ISL_1834730, EPI_ISL_1834731, EPI_ISL_1834732, EPI_ISL_1834733, EPI_ISL_1834734, EPI_ISL_1834735, EPI_ISL_1834736, EPI_ISL_1834737, EPI_ISL_1834738, EPI_ISL_1834739, EPI_ISL_1834740, EPI_ISL_1834741, EPI_ISL_1834742, EPI_ISL_1834743, EPI_ISL_1834744, EPI_ISL_1834745, EPI_ISL_1834747, EPI_ISL_1834748, EPI_ISL_1834749, EPI_ISL_1834750, EPI_ISL_1834751, EPI_ISL_1834752, EPI_ISL_1834753, EPI_ISL_1834754, EPI_ISL_1834755, EPI_ISL_1834756, EPI_ISL_1834757, EPI_ISL_1834758, EPI_ISL_1834759, EPI_ISL_1834760, EPI_ISL_1834761, EPI_ISL_1834762, EPI_ISL_1834763, EPI_ISL_1834764, EPI_ISL_1834765, EPI_ISL_1834766, EPI_ISL_1834767, EPI_ISL_1834768, EPI_ISL_1834769, EPI_ISL_1834770, EPI_ISL_1834771, EPI_ISL_1834772, EPI_ISL_1834773, EPI_ISL_1834774, EPI_ISL_1834775, EPI_ISL_1834776, EPI_ISL_1834777, EPI_ISL_1834778, EPI_ISL_1834779, EPI_ISL_1834780, EPI_ISL_1834781, EPI_ISL_1834782, EPI_ISL_1834783, EPI_ISL_1834784, EPI_ISL_1834785, EPI_ISL_1834786, EPI_ISL_1834787, EPI_ISL_1834788, EPI_ISL_1834789, EPI_ISL_1834790, EPI_ISL_1834791, EPI_ISL_1834792, EPI_ISL_1834793, EPI_ISL_1834794, EPI_ISL_1834795, EPI_ISL_1834796, EPI_ISL_1834797, EPI_ISL_1834798, EPI_ISL_1834799, EPI_ISL_1834800, EPI_ISL_1834801, EPI_ISL_1834802, EPI_ISL_1834803, EPI_ISL_1834804, EPI_ISL_1834805, EPI_ISL_1834806, EPI_ISL_1834807, EPI_ISL_1834808, EPI_ISL_1834809, EPI_ISL_1834810, EPI_ISL_1834811, EPI_ISL_1834812, EPI_ISL_1834813, EPI_ISL_1834814, EPI_ISL_1834815, EPI_ISL_1834816, EPI_ISL_1834817, EPI_ISL_1834818, EPI_ISL_1834819, EPI_ISL_1834820, EPI_ISL_1834821, EPI_ISL_1834822, EPI_ISL_1834823, EPI_ISL_1834824, EPI_ISL_1834825, EPI_ISL_1834826, EPI_ISL_1834827, EPI_ISL_1834828, EPI_ISL_1834829, EPI_ISL_1834831, EPI_ISL_1834832, EPI_ISL_1834833, EPI_ISL_1834834, EPI_ISL_1834835, EPI_ISL_1834836, EPI_ISL_1834837, EPI_ISL_1834838, EPI_ISL_1834839, EPI_ISL_1834840, EPI_ISL_1834841, EPI_ISL_1834842, EPI_ISL_1834843, EPI_ISL_1834844, EPI_ISL_1834845, EPI_ISL_1834846, EPI_ISL_1834847, EPI_ISL_1834848, EPI_ISL_1834849, EPI_ISL_1834850, EPI_ISL_1834851, EPI_ISL_1834852, EPI_ISL_1834853, EPI_ISL_1834854, EPI_ISL_1834855, EPI_ISL_1834856, EPI_ISL_1834857, EPI_ISL_1834858, EPI_ISL_1834859, EPI_ISL_1834860, EPI_ISL_1834861, EPI_ISL_1834862, EPI_ISL_1834863, EPI_ISL_1834864, EPI_ISL_1834865, EPI_ISL_1834866, EPI_ISL_1834867, EPI_ISL_1834868, EPI_ISL_1834869, EPI_ISL_1834870, EPI_ISL_1834871, EPI_ISL_1834872, EPI_ISL_1834873, EPI_ISL_1834874, EPI_ISL_1834875, EPI_ISL_1834876, EPI_ISL_1834877, EPI_ISL_1834878, EPI_ISL_1834879, EPI_ISL_1834880, EPI_ISL_1834881, EPI_ISL_1834882, EPI_ISL_1834883, EPI_ISL_1834884, EPI_ISL_1834885, EPI_ISL_1834886, EPI_ISL_1834887, EPI_ISL_1834888, EPI_ISL_1834889, EPI_ISL_1834890, EPI_ISL_1834891, EPI_ISL_1834892, EPI_ISL_1834893, EPI_ISL_1834894, EPI_ISL_1834895, EPI_ISL_1834896, EPI_ISL_1834897, EPI_ISL_1834898, EPI_ISL_1834899, EPI_ISL_1834900, EPI_ISL_1834901, EPI_ISL_1834902, EPI_ISL_1834903, EPI_ISL_1834904, EPI_ISL_1834905, EPI_ISL_1834906, EPI_ISL_1834907, EPI_ISL_1834908, EPI_ISL_1834909, EPI_ISL_1834910, EPI_ISL_1834911, EPI_ISL_1834912, EPI_ISL_1834913, EPI_ISL_1834914, EPI_ISL_1834915, EPI_ISL_1834916, EPI_ISL_1834917, EPI_ISL_1834918, EPI_ISL_1834919, EPI_ISL_1834920, EPI_ISL_1834921, EPI_ISL_1834922, EPI_ISL_1834923, EPI_ISL_1834925, EPI_ISL_1834926, EPI_ISL_1834927, EPI_ISL_1834928, EPI_ISL_1834929, EPI_ISL_1834930, EPI_ISL_1834931, EPI_ISL_1834932, EPI_ISL_1834933, EPI_ISL_1834934, EPI_ISL_1834935, EPI_ISL_1834936, EPI_ISL_1834937, EPI_ISL_1834938, EPI_ISL_1834939, EPI_ISL_1834940, EPI_ISL_1834941, EPI_ISL_1834942, EPI_ISL_1834943, EPI_ISL_1834944, EPI_ISL_1834945, EPI_ISL_1834946, EPI_ISL_1834947, EPI_ISL_1834948, EPI_ISL_1834949, EPI_ISL_1834950, EPI_ISL_1834951, EPI_ISL_1834952, EPI_ISL_1834953, EPI_ISL_1834954, EPI_ISL_1834955, EPI_ISL_1834956, EPI_ISL_1834957, EPI_ISL_1834958, EPI_ISL_1834959, EPI_ISL_1834960, EPI_ISL_1834961, EPI_ISL_1834962, EPI_ISL_1834963, EPI_ISL_1834964, EPI_ISL_1834965, EPI_ISL_1834966, EPI_ISL_1834967, EPI_ISL_1834968, EPI_ISL_1834969, EPI_ISL_1834970, EPI_ISL_1834971, EPI_ISL_1834972, EPI_ISL_1834973, EPI_ISL_1834974, EPI_ISL_1834975, EPI_ISL_1834976, EPI_ISL_1834977, EPI_ISL_1834978, EPI_ISL_1834979, EPI_ISL_1834980, EPI_ISL_1834981, EPI_ISL_1834982, EPI_ISL_1834983, EPI_ISL_1834984, EPI_ISL_1834985, EPI_ISL_1834986, EPI_ISL_1834987, EPI_ISL_1834988, EPI_ISL_1834989, EPI_ISL_1834990, EPI_ISL_1834991, EPI_ISL_1834992, EPI_ISL_1834993, EPI_ISL_1834994, EPI_ISL_1834995, EPI_ISL_1834996, EPI_ISL_1834997, EPI_ISL_1834998, EPI_ISL_1834999, EPI_ISL_1835000, EPI_ISL_1835001, EPI_ISL_1835002, EPI_ISL_1835003, EPI_ISL_1835004, EPI_ISL_1835005, EPI_ISL_1835006, EPI_ISL_1835007, EPI_ISL_1835008, EPI_ISL_1835009, EPI_ISL_1835010, EPI_ISL_1835011, EPI_ISL_1835012, EPI_ISL_1835013, EPI_ISL_1835014, EPI_ISL_1835015, EPI_ISL_1835016, EPI_ISL_1835017, EPI_ISL_1835018, EPI_ISL_1835019, EPI_ISL_1835020, EPI_ISL_1835021, EPI_ISL_1835022, EPI_ISL_1835023, EPI_ISL_1835024, EPI_ISL_1835025, EPI_ISL_1835026, EPI_ISL_1835027, EPI_ISL_1835028, EPI_ISL_1835029, EPI_ISL_1835030, EPI_ISL_1835031, EPI_ISL_1835032, EPI_ISL_1835033, EPI_ISL_1835034, EPI_ISL_1835035, EPI_ISL_1835036, EPI_ISL_1835037, EPI_ISL_1835038, EPI_ISL_1835039, EPI_ISL_1835040, EPI_ISL_1835041, EPI_ISL_1835042, EPI_ISL_1835043, EPI_ISL_1835044, EPI_ISL_1835045, EPI_ISL_1835046, EPI_ISL_1835047, EPI_ISL_1835048, EPI_ISL_1835049, EPI_ISL_1835050, EPI_ISL_1835051, EPI_ISL_1835052, EPI_ISL_1835053, EPI_ISL_1835054, EPI_ISL_1835055, EPI_ISL_1835056, EPI_ISL_1835057, EPI_ISL_1835058, EPI_ISL_1835059, EPI_ISL_1835060, EPI_ISL_1835062, EPI_ISL_1835063, EPI_ISL_1835064, EPI_ISL_1835065, EPI_ISL_1835066, EPI_ISL_1835067, EPI_ISL_1835068, EPI_ISL_1835069, EPI_ISL_1835070, EPI_ISL_1835071, EPI_ISL_1835072, EPI_ISL_1835073, EPI_ISL_1835074, EPI_ISL_1835075, EPI_ISL_1835076, EPI_ISL_1835077, EPI_ISL_1835078, EPI_ISL_1835079, EPI_ISL_1835080, EPI_ISL_1835081, EPI_ISL_1835082, EPI_ISL_1835083, EPI_ISL_1835084, EPI_ISL_1835085, EPI_ISL_1835086, EPI_ISL_1835087, EPI_ISL_1835088, EPI_ISL_1835089, EPI_ISL_1835090, EPI_ISL_1835091, EPI_ISL_1835092, EPI_ISL_1835093, EPI_ISL_1835094, EPI_ISL_1835095, EPI_ISL_1835096, EPI_ISL_1835097, EPI_ISL_1835098, EPI_ISL_1835099, EPI_ISL_1835100, EPI_ISL_1835101, EPI_ISL_1835102, EPI_ISL_1835103, EPI_ISL_1835104, EPI_ISL_1835105, EPI_ISL_1835106, EPI_ISL_1835107, EPI_ISL_1835108, EPI_ISL_1835109, EPI_ISL_1835110, EPI_ISL_1835112, EPI_ISL_1835113, EPI_ISL_1835114, EPI_ISL_1835115, EPI_ISL_1835116, EPI_ISL_1835117, EPI_ISL_1835118, EPI_ISL_1835119, EPI_ISL_1835120, EPI_ISL_1835121, EPI_ISL_1835122, EPI_ISL_1835123, EPI_ISL_1835124, EPI_ISL_1835125, EPI_ISL_1835126, EPI_ISL_1835127, EPI_ISL_1835128, EPI_ISL_1835129, EPI_ISL_1835130, EPI_ISL_1835131, EPI_ISL_1835132, EPI_ISL_1835133, EPI_ISL_1835134, EPI_ISL_1835135, EPI_ISL_1835136, EPI_ISL_1835137, EPI_ISL_1835138, EPI_ISL_1835139, EPI_ISL_1835140, EPI_ISL_1835141, EPI_ISL_1835142, EPI_ISL_1835143, EPI_ISL_1835144, EPI_ISL_1835145, EPI_ISL_1835146, EPI_ISL_1835147, EPI_ISL_1835148, EPI_ISL_1835149, EPI_ISL_1835150, EPI_ISL_1835151, EPI_ISL_1835152, EPI_ISL_1835153, EPI_ISL_1835154, EPI_ISL_1835156, EPI_ISL_1835157, EPI_ISL_1835158, EPI_ISL_1835159, EPI_ISL_1835160, EPI_ISL_1835161, EPI_ISL_1835162, EPI_ISL_1835163, EPI_ISL_1835164, EPI_ISL_1835165, EPI_ISL_1835166, EPI_ISL_1835167, EPI_ISL_1835168, EPI_ISL_1835169, EPI_ISL_1835170, EPI_ISL_1835171, EPI_ISL_1835172, EPI_ISL_1835173, EPI_ISL_1835174, EPI_ISL_1835175, EPI_ISL_1835176, EPI_ISL_1835177, EPI_ISL_1835178, EPI_ISL_1835179, EPI_ISL_1835180, EPI_ISL_1835181, EPI_ISL_1835182, EPI_ISL_1835183, EPI_ISL_1835184, EPI_ISL_1835185, EPI_ISL_1835187, EPI_ISL_1835188, EPI_ISL_1835189, EPI_ISL_1835190, EPI_ISL_1835191, EPI_ISL_1835192, EPI_ISL_1835193, EPI_ISL_1835194, EPI_ISL_1835195, EPI_ISL_1835196, EPI_ISL_1835197, EPI_ISL_1835198, EPI_ISL_1835199, EPI_ISL_1835200, EPI_ISL_1835201, EPI_ISL_1835202, EPI_ISL_1835203, EPI_ISL_1835204, EPI_ISL_1835205, EPI_ISL_1835206, EPI_ISL_1835207, EPI_ISL_1835208, EPI_ISL_1835209, EPI_ISL_1835210, EPI_ISL_1835211, EPI_ISL_1835212, EPI_ISL_1835213, EPI_ISL_1835214, EPI_ISL_1835215, EPI_ISL_1835216, EPI_ISL_1835217, EPI_ISL_1835218, EPI_ISL_1835219, EPI_ISL_1835220, EPI_ISL_1835221, EPI_ISL_1835222, EPI_ISL_1835223, EPI_ISL_1835224, EPI_ISL_1835225, EPI_ISL_1835226, EPI_ISL_1835227, EPI_ISL_1835228, EPI_ISL_1835229, EPI_ISL_1835230, EPI_ISL_1835231, EPI_ISL_1835232, EPI_ISL_1835233, EPI_ISL_1835234, EPI_ISL_1835235, EPI_ISL_1835236, EPI_ISL_1835237, EPI_ISL_1835238, EPI_ISL_1835239, EPI_ISL_1835240, EPI_ISL_1835241, EPI_ISL_1835242, EPI_ISL_1835243, EPI_ISL_1835244, EPI_ISL_1835245, EPI_ISL_1835246, EPI_ISL_1835247, EPI_ISL_1835248, EPI_ISL_1835249, EPI_ISL_1835250, EPI_ISL_1835251, EPI_ISL_1835252, EPI_ISL_1835253, EPI_ISL_1835254, EPI_ISL_1835255, EPI_ISL_1835256, EPI_ISL_1835257, EPI_ISL_1835258, EPI_ISL_1835259, EPI_ISL_1835260, EPI_ISL_1835261, EPI_ISL_1835262, EPI_ISL_1835263, EPI_ISL_1835264, EPI_ISL_1835265, EPI_ISL_1835266, EPI_ISL_1835267, EPI_ISL_1835268, EPI_ISL_1835269, EPI_ISL_1835270, EPI_ISL_1835271, EPI_ISL_1835272, EPI_ISL_1835273, EPI_ISL_1835274, EPI_ISL_1835275, EPI_ISL_1835276, EPI_ISL_1835277, EPI_ISL_1835278, EPI_ISL_1835279, EPI_ISL_1835280, EPI_ISL_1835281, EPI_ISL_1835282, EPI_ISL_1835283, EPI_ISL_1835284, EPI_ISL_1835285, EPI_ISL_1835286, EPI_ISL_1835287, EPI_ISL_1835288, EPI_ISL_1835289, EPI_ISL_1835290, EPI_ISL_1835291, EPI_ISL_1835292, EPI_ISL_1835293, EPI_ISL_1835294, EPI_ISL_1835295, EPI_ISL_1835296, EPI_ISL_1835297, EPI_ISL_1835298, EPI_ISL_1835299, EPI_ISL_1835300, EPI_ISL_1835301, EPI_ISL_1835302, EPI_ISL_1835303, EPI_ISL_1835304, EPI_ISL_1835305, EPI_ISL_1835306, EPI_ISL_1835307, EPI_ISL_1835309, EPI_ISL_1835310, EPI_ISL_1835311, EPI_ISL_1835312, EPI_ISL_1835313, EPI_ISL_1835314, EPI_ISL_1835315, EPI_ISL_1835316, EPI_ISL_1835317, EPI_ISL_1835318, EPI_ISL_1835319, EPI_ISL_1835320, EPI_ISL_1835321, EPI_ISL_1835322, EPI_ISL_1835323, EPI_ISL_1835324, EPI_ISL_1835325, EPI_ISL_1835326, EPI_ISL_1835327, EPI_ISL_1835328, EPI_ISL_1835329, EPI_ISL_1835330, EPI_ISL_1835331, EPI_ISL_1835332, EPI_ISL_1835333, EPI_ISL_1835334, EPI_ISL_1835335, EPI_ISL_1835336, EPI_ISL_1835337, EPI_ISL_1835338, EPI_ISL_1835339, EPI_ISL_1835340, EPI_ISL_1835341, EPI_ISL_1835342, EPI_ISL_1835343, EPI_ISL_1835344, EPI_ISL_1835345, EPI_ISL_1835346, EPI_ISL_1835348, EPI_ISL_1835349, EPI_ISL_1835350, EPI_ISL_1835351, EPI_ISL_1835352, EPI_ISL_1835353, EPI_ISL_1835354, EPI_ISL_1835355, EPI_ISL_1835356, EPI_ISL_1835357, EPI_ISL_1835358, EPI_ISL_1835359, EPI_ISL_1835360, EPI_ISL_1835361, EPI_ISL_1835362, EPI_ISL_1835363, EPI_ISL_1835364, EPI_ISL_1835365, EPI_ISL_1835366, EPI_ISL_1835367, EPI_ISL_1835368, EPI_ISL_1835369, EPI_ISL_1835370, EPI_ISL_1835371, EPI_ISL_1835372, EPI_ISL_1835373, EPI_ISL_1835374, EPI_ISL_1835375, EPI_ISL_1835376, EPI_ISL_1835377, EPI_ISL_1835378, EPI_ISL_1835379, EPI_ISL_1835380, EPI_ISL_1835381, EPI_ISL_1835382, EPI_ISL_1835383, EPI_ISL_1835384, EPI_ISL_1835385, EPI_ISL_1835386, EPI_ISL_1835387, EPI_ISL_1835388, EPI_ISL_1835389, EPI_ISL_1835390, EPI_ISL_1835391, EPI_ISL_1835392, EPI_ISL_1835393, EPI_ISL_1835394, EPI_ISL_1835395, EPI_ISL_1835396, EPI_ISL_1835397, EPI_ISL_1835398, EPI_ISL_1835399, EPI_ISL_1835400, EPI_ISL_1835401, EPI_ISL_1835402, EPI_ISL_1835403, EPI_ISL_1835404, EPI_ISL_1835405, EPI_ISL_1835406, EPI_ISL_1835407, EPI_ISL_1835408, EPI_ISL_1835409, EPI_ISL_1835410, EPI_ISL_1835411, EPI_ISL_1835412, EPI_ISL_1835413, EPI_ISL_1835414, EPI_ISL_1835415, EPI_ISL_1835416, EPI_ISL_1835417, EPI_ISL_1835418, EPI_ISL_1835419, EPI_ISL_1835420, EPI_ISL_1835421, EPI_ISL_1835422, EPI_ISL_1835423, EPI_ISL_1835424, EPI_ISL_1835425, EPI_ISL_1835426, EPI_ISL_1835427, EPI_ISL_1835428, EPI_ISL_1835429, EPI_ISL_1835430, EPI_ISL_1835431, EPI_ISL_1835432, EPI_ISL_1835433, EPI_ISL_1835434, EPI_ISL_1835435, EPI_ISL_1835436, EPI_ISL_1835437, EPI_ISL_1835438, EPI_ISL_1835439, EPI_ISL_1835440, EPI_ISL_1835441, EPI_ISL_1835442, EPI_ISL_1835443, EPI_ISL_1835444, EPI_ISL_1835445, EPI_ISL_1835446, EPI_ISL_1835447, EPI_ISL_1835448, EPI_ISL_1835449, EPI_ISL_1835450, EPI_ISL_1835451, EPI_ISL_1835452, EPI_ISL_1835453, EPI_ISL_1835454, EPI_ISL_1835455, EPI_ISL_1835456, EPI_ISL_1835457, EPI_ISL_1835458, EPI_ISL_1835459, EPI_ISL_1835460, EPI_ISL_1835461, EPI_ISL_1835462, EPI_ISL_1835463, EPI_ISL_1835464, EPI_ISL_1835465, EPI_ISL_1835466, EPI_ISL_1835467, EPI_ISL_1835468, EPI_ISL_1835469, EPI_ISL_1835470, EPI_ISL_1835471, EPI_ISL_1835472, EPI_ISL_1835473, EPI_ISL_1835474, EPI_ISL_1835475, EPI_ISL_1835476, EPI_ISL_1835477, EPI_ISL_1835478, EPI_ISL_1835479, EPI_ISL_1835480, EPI_ISL_1835481, EPI_ISL_1835482, EPI_ISL_1835483, EPI_ISL_1835484, EPI_ISL_1835485, EPI_ISL_1835486, EPI_ISL_1835487, EPI_ISL_1835488, EPI_ISL_1835489, EPI_ISL_1835490, EPI_ISL_1835491, EPI_ISL_1835492, EPI_ISL_1835493, EPI_ISL_1835494, EPI_ISL_1835495, EPI_ISL_1835497, EPI_ISL_1835498, EPI_ISL_1835499, EPI_ISL_1835500, EPI_ISL_1835501, EPI_ISL_1835502, EPI_ISL_1835503, EPI_ISL_1835504, EPI_ISL_1835505, EPI_ISL_1835506, EPI_ISL_1835507, EPI_ISL_1835508, EPI_ISL_1835509, EPI_ISL_1835510, EPI_ISL_1835511, EPI_ISL_1835512, EPI_ISL_1835513, EPI_ISL_1835514, EPI_ISL_1835515, EPI_ISL_1835517, EPI_ISL_1835518, EPI_ISL_1835519, EPI_ISL_1835520, EPI_ISL_1835521, EPI_ISL_1835522, EPI_ISL_1835523, EPI_ISL_1835524, EPI_ISL_1835525, EPI_ISL_1835526, EPI_ISL_1835527, EPI_ISL_1835528, EPI_ISL_1835529, EPI_ISL_1835530, EPI_ISL_1835531, EPI_ISL_1835532, EPI_ISL_1835533, EPI_ISL_1835534, EPI_ISL_1835535, | | | |

EPI_ISL_1835536, EPI_ISL_1835537, EPI_ISL_1835538, EPI_ISL_1835539, EPI_ISL_1835540, EPI_ISL_1835541, EPI_ISL_1835542, EPI_ISL_1835543, EPI_ISL_1835544, EPI_ISL_1835545, EPI_ISL_1835546, EPI_ISL_1835547, EPI_ISL_1835548, EPI_ISL_1835549, EPI_ISL_1835550, EPI_ISL_1835551, EPI_ISL_1835552, EPI_ISL_1835553, EPI_ISL_1835554, EPI_ISL_1835555, EPI_ISL_1835556, EPI_ISL_1835557,
EPI_ISL_1835558, EPI_ISL_1835559, EPI_ISL_1835560, EPI_ISL_1835561, EPI_ISL_1835562, EPI_ISL_1835563, EPI_ISL_1835564, EPI_ISL_1835565, EPI_ISL_1835566, EPI_ISL_1835567, EPI_ISL_1835568, EPI_ISL_1835569, EPI_ISL_1835570, EPI_ISL_1835571, EPI_ISL_1835572, EPI_ISL_1835573, EPI_ISL_1835574, EPI_ISL_1835575, EPI_ISL_1835576, EPI_ISL_1835577, EPI_ISL_1835578, EPI_ISL_1835579,
EPI_ISL_1835580, EPI_ISL_1835582, EPI_ISL_1835583, EPI_ISL_1835584, EPI_ISL_1835585, EPI_ISL_1835586, EPI_ISL_1835587, EPI_ISL_1835588, EPI_ISL_1835589, EPI_ISL_1835590, EPI_ISL_1835591, EPI_ISL_1835593, EPI_ISL_1835595, EPI_ISL_1835596, EPI_ISL_1835597, EPI_ISL_1835598, EPI_ISL_1835599, EPI_ISL_1835600, EPI_ISL_1835601, EPI_ISL_1835602, EPI_ISL_1835603, EPI_ISL_1835604,
EPI_ISL_1835605, EPI_ISL_1835606, EPI_ISL_1835607, EPI_ISL_1835608, EPI_ISL_1835609, EPI_ISL_1835610, EPI_ISL_1835611, EPI_ISL_1835612, EPI_ISL_1835613, EPI_ISL_1835614, EPI_ISL_1835615, EPI_ISL_1835616, EPI_ISL_1835617, EPI_ISL_1835618, EPI_ISL_1835619, EPI_ISL_1835620, EPI_ISL_1835621, EPI_ISL_1835622, EPI_ISL_1835623, EPI_ISL_1835624, EPI_ISL_1835625, EPI_ISL_1835626,
EPI_ISL_1835627, EPI_ISL_1835628, EPI_ISL_1835629, EPI_ISL_1835630, EPI_ISL_1835631, EPI_ISL_1835632, EPI_ISL_1835633, EPI_ISL_1835634, EPI_ISL_1835635, EPI_ISL_1835636, EPI_ISL_1835637, EPI_ISL_1835638, EPI_ISL_1835639, EPI_ISL_1835640, EPI_ISL_1835641, EPI_ISL_1835642, EPI_ISL_1835643, EPI_ISL_1835644, EPI_ISL_1835645, EPI_ISL_1835646, EPI_ISL_1835647, EPI_ISL_1835648,
EPI_ISL_1835649, EPI_ISL_1835650, EPI_ISL_1835651, EPI_ISL_1835652, EPI_ISL_1835653, EPI_ISL_1835654, EPI_ISL_1835655, EPI_ISL_1835656, EPI_ISL_1835657, EPI_ISL_1835658, EPI_ISL_1835659, EPI_ISL_1835660, EPI_ISL_1835661, EPI_ISL_1835662, EPI_ISL_1835663, EPI_ISL_1835664, EPI_ISL_1835665, EPI_ISL_1835666, EPI_ISL_1835667, EPI_ISL_1835668, EPI_ISL_1835669, EPI_ISL_1835670,
EPI_ISL_1835671, EPI_ISL_1835672, EPI_ISL_1835673, EPI_ISL_1835674, EPI_ISL_1835675, EPI_ISL_1835676, EPI_ISL_1835677, EPI_ISL_1835678, EPI_ISL_1835679, EPI_ISL_1835680, EPI_ISL_1835681, EPI_ISL_1835682, EPI_ISL_1835683, EPI_ISL_1835684, EPI_ISL_1835685, EPI_ISL_1835686, EPI_ISL_1835687, EPI_ISL_1835688, EPI_ISL_1835689, EPI_ISL_1835690, EPI_ISL_1835691, EPI_ISL_1835692,
EPI_ISL_1835693, EPI_ISL_1835694, EPI_ISL_1835695, EPI_ISL_1835696, EPI_ISL_1835697, EPI_ISL_1835698, EPI_ISL_1835699, EPI_ISL_1835700, EPI_ISL_1835701, EPI_ISL_1835702, EPI_ISL_1835703, EPI_ISL_1835704, EPI_ISL_1835705, EPI_ISL_1835706, EPI_ISL_1835707, EPI_ISL_1835708, EPI_ISL_1835709, EPI_ISL_1835710, EPI_ISL_1835711, EPI_ISL_1835712, EPI_ISL_1835713, EPI_ISL_1835714,
EPI_ISL_1835715, EPI_ISL_1835716, EPI_ISL_1835717, EPI_ISL_1835718, EPI_ISL_1835719, EPI_ISL_1835720, EPI_ISL_1835721, EPI_ISL_1835722, EPI_ISL_1835723, EPI_ISL_1835724, EPI_ISL_1835725, EPI_ISL_1835726, EPI_ISL_1835727, EPI_ISL_1835728, EPI_ISL_1835729, EPI_ISL_1835730, EPI_ISL_1835731, EPI_ISL_1835732, EPI_ISL_1835733, EPI_ISL_1835734, EPI_ISL_1835735, EPI_ISL_1835736,
EPI_ISL_1835737, EPI_ISL_1835738, EPI_ISL_1835739, EPI_ISL_1835740, EPI_ISL_1835741, EPI_ISL_1835742, EPI_ISL_1835743, EPI_ISL_1835744, EPI_ISL_1835745, EPI_ISL_1835746, EPI_ISL_1835747, EPI_ISL_1835748, EPI_ISL_1835749, EPI_ISL_1835750, EPI_ISL_1835751, EPI_ISL_1835752, EPI_ISL_1835753, EPI_ISL_1835754, EPI_ISL_1835755, EPI_ISL_1835756, EPI_ISL_1835757, EPI_ISL_1835758,
EPI_ISL_1835759, EPI_ISL_1835760, EPI_ISL_1835761, EPI_ISL_1835762, EPI_ISL_1835763, EPI_ISL_1835764, EPI_ISL_1835765, EPI_ISL_1835766, EPI_ISL_1835767, EPI_ISL_1835768, EPI_ISL_1835769, EPI_ISL_1835770, EPI_ISL_1835771, EPI_ISL_1835772, EPI_ISL_1835773, EPI_ISL_1835774, EPI_ISL_1835775, EPI_ISL_1835776, EPI_ISL_1835777, EPI_ISL_1835778, EPI_ISL_1835779, EPI_ISL_1835780,
EPI_ISL_1835781, EPI_ISL_1835782, EPI_ISL_1835783, EPI_ISL_1835784, EPI_ISL_1835785, EPI_ISL_1835786, EPI_ISL_1835787, EPI_ISL_1835788, EPI_ISL_1835789, EPI_ISL_1835790, EPI_ISL_1835791, EPI_ISL_1835792, EPI_ISL_1835793, EPI_ISL_1835794, EPI_ISL_1835795, EPI_ISL_1835796, EPI_ISL_1835797, EPI_ISL_1835798, EPI_ISL_1835799, EPI_ISL_1835800, EPI_ISL_1835801, EPI_ISL_1835802,
EPI_ISL_1835803, EPI_ISL_1835804, EPI_ISL_1835805, EPI_ISL_1835806, EPI_ISL_1835807, EPI_ISL_1835808, EPI_ISL_1835809, EPI_ISL_1835810, EPI_ISL_1835811, EPI_ISL_1835812, EPI_ISL_1835813, EPI_ISL_1835814, EPI_ISL_1835817, EPI_ISL_1835818, EPI_ISL_1835819, EPI_ISL_1835820, EPI_ISL_1835821, EPI_ISL_1835822, EPI_ISL_1835823, EPI_ISL_1835824, EPI_ISL_1835825, EPI_ISL_1835826,
EPI_ISL_1835827, EPI_ISL_1835828, EPI_ISL_1835829, EPI_ISL_1835830, EPI_ISL_1835831, EPI_ISL_1835832, EPI_ISL_1835833, EPI_ISL_1835834, EPI_ISL_1835835, EPI_ISL_1835836, EPI_ISL_1835837, EPI_ISL_1835838, EPI_ISL_1835839, EPI_ISL_1835840, EPI_ISL_1835841, EPI_ISL_1835842, EPI_ISL_1835843, EPI_ISL_1835844, EPI_ISL_1835845, EPI_ISL_1835846, EPI_ISL_1835847, EPI_ISL_1835848,
EPI_ISL_1835849, EPI_ISL_1835850, EPI_ISL_1835852, EPI_ISL_1835853, EPI_ISL_1835854, EPI_ISL_1835855, EPI_ISL_1835856, EPI_ISL_1835857, EPI_ISL_1835858, EPI_ISL_1835859, EPI_ISL_1835860, EPI_ISL_1835861, EPI_ISL_1835862, EPI_ISL_1835863, EPI_ISL_1835864, EPI_ISL_1835865, EPI_ISL_1835866, EPI_ISL_1835867, EPI_ISL_1835868, EPI_ISL_1835869, EPI_ISL_1835870, EPI_ISL_1835871,
EPI_ISL_1835872, EPI_ISL_1835873, EPI_ISL_1835874, EPI_ISL_1835875, EPI_ISL_1835876, EPI_ISL_1835877, EPI_ISL_1835878, EPI_ISL_1835879, EPI_ISL_1835880, EPI_ISL_1835881, EPI_ISL_1835882, EPI_ISL_1835883, EPI_ISL_1835884, EPI_ISL_1835885, EPI_ISL_1835886, EPI_ISL_1835887, EPI_ISL_1835888, EPI_ISL_1835889, EPI_ISL_1835890, EPI_ISL_1835891, EPI_ISL_1835892, EPI_ISL_1835893,
EPI_ISL_1835894, EPI_ISL_1835895, EPI_ISL_1835896, EPI_ISL_1835897, EPI_ISL_1835898, EPI_ISL_1835899, EPI_ISL_1835900, EPI_ISL_1835901, EPI_ISL_1835902, EPI_ISL_1835903, EPI_ISL_1835904, EPI_ISL_1835905, EPI_ISL_1835906, EPI_ISL_1835907, EPI_ISL_1835908, EPI_ISL_1835909, EPI_ISL_1835910, EPI_ISL_1835911, EPI_ISL_1835912, EPI_ISL_1835913, EPI_ISL_1835914, EPI_ISL_1835915,
EPI_ISL_1835916, EPI_ISL_1835917, EPI_ISL_1835918, EPI_ISL_1835919, EPI_ISL_1835920, EPI_ISL_1835921, EPI_ISL_1835922, EPI_ISL_1835923, EPI_ISL_1835924, EPI_ISL_1835925, EPI_ISL_1835926, EPI_ISL_1835927, EPI_ISL_1835928, EPI_ISL_1835929, EPI_ISL_1835930, EPI_ISL_1835931, EPI_ISL_1835932, EPI_ISL_1835933, EPI_ISL_1835934, EPI_ISL_1835935, EPI_ISL_1835936, EPI_ISL_1835938,
EPI_ISL_1835939, EPI_ISL_1835940, EPI_ISL_1835941, EPI_ISL_1835942, EPI_ISL_1835943, EPI_ISL_1835944, EPI_ISL_1835945, EPI_ISL_1835946, EPI_ISL_1835947, EPI_ISL_1835948, EPI_ISL_1835949, EPI_ISL_1835950, EPI_ISL_1835951, EPI_ISL_1835952, EPI_ISL_1835953, EPI_ISL_1835954, EPI_ISL_1835955, EPI_ISL_1835956, EPI_ISL_1835957, EPI_ISL_1835958, EPI_ISL_1835959, EPI_ISL_1835960,
EPI_ISL_1835961, EPI_ISL_1835962, EPI_ISL_1835963, EPI_ISL_1835964, EPI_ISL_1835965, EPI_ISL_1835966, EPI_ISL_1835967, EPI_ISL_1835968, EPI_ISL_1835969, EPI_ISL_1835970, EPI_ISL_1835971, EPI_ISL_1835972, EPI_ISL_1835973, EPI_ISL_1835974, EPI_ISL_1835975, EPI_ISL_1835976, EPI_ISL_1835977, EPI_ISL_1835978, EPI_ISL_1835979, EPI_ISL_1835980, EPI_ISL_1835981, EPI_ISL_1835982,
EPI_ISL_1835983, EPI_ISL_1835984, EPI_ISL_1835985, EPI_ISL_1835986, EPI_ISL_1835987, EPI_ISL_1835988, EPI_ISL_1835989, EPI_ISL_1835990, EPI_ISL_1835991, EPI_ISL_1835992, EPI_ISL_1835993, EPI_ISL_1835994, EPI_ISL_1835995, EPI_ISL_1835996, EPI_ISL_1835997, EPI_ISL_1835998, EPI_ISL_1835999, EPI_ISL_1836000, EPI_ISL_1836001, EPI_ISL_1836002, EPI_ISL_1836003, EPI_ISL_1836004,
EPI_ISL_1836005, EPI_ISL_1836006, EPI_ISL_1836007, EPI_ISL_1836008, EPI_ISL_1836009, EPI_ISL_1836010, EPI_ISL_1836011, EPI_ISL_1836012, EPI_ISL_1836013, EPI_ISL_1836014, EPI_ISL_1836016, EPI_ISL_1836017, EPI_ISL_1836018, EPI_ISL_1836019, EPI_ISL_1836020, EPI_ISL_1836021, EPI_ISL_1836022, EPI_ISL_1836023, EPI_ISL_1836024, EPI_ISL_1836025, EPI_ISL_1836026, EPI_ISL_1836027,
EPI_ISL_1836028, EPI_ISL_1836030, EPI_ISL_1836031, EPI_ISL_1836032, EPI_ISL_1836033, EPI_ISL_1836034, EPI_ISL_1836035, EPI_ISL_1836036, EPI_ISL_1836037, EPI_ISL_1836038, EPI_ISL_1836039, EPI_ISL_1836040, EPI_ISL_1836041, EPI_ISL_1836042, EPI_ISL_1836043, EPI_ISL_1836044, EPI_ISL_1836045, EPI_ISL_1836046, EPI_ISL_1836047, EPI_ISL_1836048, EPI_ISL_1836049, EPI_ISL_1836050,
EPI_ISL_1836051, EPI_ISL_1836052, EPI_ISL_1836053, EPI_ISL_1836054, EPI_ISL_1836055, EPI_ISL_1836056, EPI_ISL_1836057, EPI_ISL_1836058, EPI_ISL_1836059, EPI_ISL_1836060, EPI_ISL_1836061, EPI_ISL_1836062, EPI_ISL_1836063, EPI_ISL_1836064, EPI_ISL_1836065, EPI_ISL_1836066, EPI_ISL_1836067, EPI_ISL_1836068, EPI_ISL_1836069, EPI_ISL_1836070, EPI_ISL_1836071, EPI_ISL_1836072,
EPI_ISL_1836073, EPI_ISL_1836074, EPI_ISL_1836075, EPI_ISL_1836076, EPI_ISL_1836077, EPI_ISL_1836078, EPI_ISL_1836079, EPI_ISL_1836080, EPI_ISL_1836081, EPI_ISL_1836082, EPI_ISL_1836083, EPI_ISL_1836084, EPI_ISL_1836085, EPI_ISL_1836086, EPI_ISL_1836087, EPI_ISL_1836088, EPI_ISL_1836089, EPI_ISL_1836090, EPI_ISL_1836091, EPI_ISL_1836092, EPI_ISL_1836093, EPI_ISL_1836094,
EPI_ISL_1836095, EPI_ISL_1836096, EPI_ISL_1836097, EPI_ISL_1836099, EPI_ISL_1836100, EPI_ISL_1836101, EPI_ISL_1836102, EPI_ISL_1836103, EPI_ISL_1836104, EPI_ISL_1836105, EPI_ISL_1836106, EPI_ISL_1836107, EPI_ISL_1836108, EPI_ISL_1836109, EPI_ISL_1836110, EPI_ISL_1836111, EPI_ISL_1836112, EPI_ISL_1836113, EPI_ISL_1836114, EPI_ISL_1836115, EPI_ISL_1836116, EPI_ISL_1836117,
EPI_ISL_1836118, EPI_ISL_1836119, EPI_ISL_1836120, EPI_ISL_1836121, EPI_ISL_1836122, EPI_ISL_1836123, EPI_ISL_1836124, EPI_ISL_1836125, EPI_ISL_1836126, EPI_ISL_1836127, EPI_ISL_1836128, EPI_ISL_1836129, EPI_ISL_1836130, EPI_ISL_1836131, EPI_ISL_1836132, EPI_ISL_1836133, EPI_ISL_1836134, EPI_ISL_1836135, EPI_ISL_1836136, EPI_ISL_1836137, EPI_ISL_1836138, EPI_ISL_1836139,
EPI_ISL_1836140, EPI_ISL_1836141, EPI_ISL_1836142, EPI_ISL_1836143, EPI_ISL_1836144, EPI_ISL_1836145, EPI_ISL_1836146, EPI_ISL_1836147, EPI_ISL_1836148, EPI_ISL_1836149, EPI_ISL_1836150, EPI_ISL_1836151, EPI_ISL_1836153, EPI_ISL_1836154, EPI_ISL_1836155, EPI_ISL_1836156, EPI_ISL_1836157, EPI_ISL_1836158, EPI_ISL_1836159, EPI_ISL_1836160, EPI_ISL_1836161, EPI_ISL_1836162,
EPI_ISL_1836163, EPI_ISL_1836164, EPI_ISL_1836165, EPI_ISL_1836166, EPI_ISL_1836167, EPI_ISL_1836168, EPI_ISL_1836169, EPI_ISL_1836170, EPI_ISL_1836171, EPI_ISL_1836172, EPI_ISL_1836173, EPI_ISL_1836174, EPI_ISL_1836175, EPI_ISL_1836176, EPI_ISL_1836177, EPI_ISL_1836178, EPI_ISL_1836179, EPI_ISL_1836180, EPI_ISL_1836181, EPI_ISL_1836182, EPI_ISL_1836183, EPI_ISL_1836184,
EPI_ISL_1836185, EPI_ISL_1836186, EPI_ISL_1836188, EPI_ISL_1836189, EPI_ISL_1836190, EPI_ISL_1836191, EPI_ISL_1836192, EPI_ISL_1836193, EPI_ISL_1836194, EPI_ISL_1836195, EPI_ISL_1836196, EPI_ISL_1836197, EPI_ISL_1836198, EPI_ISL_1836199, EPI_ISL_1836200, EPI_ISL_1836201, EPI_ISL_1836202, EPI_ISL_1836203, EPI_ISL_1836204, EPI_ISL_1836205, EPI_ISL_1836206, EPI_ISL_1836207,
EPI_ISL_1836208, EPI_ISL_1836209, EPI_ISL_1836210, EPI_ISL_1836211, EPI_ISL_1836212, EPI_ISL_1836213, EPI_ISL_1836214, EPI_ISL_1836215, EPI_ISL_1836216, EPI_ISL_1836217, EPI_ISL_1836218, EPI_ISL_1836219, EPI_ISL_1836220, EPI_ISL_1836221, EPI_ISL_1836222, EPI_ISL_1836223, EPI_ISL_1836224, EPI_ISL_1836225, EPI_ISL_1836226, EPI_ISL_1836227, EPI_ISL_1836228, EPI_ISL_1836229,
EPI_ISL_1836230, EPI_ISL_1836231, EPI_ISL_1836232, EPI_ISL_1836233, EPI_ISL_1836234, EPI_ISL_1836235, EPI_ISL_1836236, EPI_ISL_1836237, EPI_ISL_1836238, EPI_ISL_1836240, EPI_ISL_1836241, EPI_ISL_1836242, EPI_ISL_1836243, EPI_ISL_1836244, EPI_ISL_1836245, EPI_ISL_1836246, EPI_ISL_1836247, EPI_ISL_1836248, EPI_ISL_1836249, EPI_ISL_1836250, EPI_ISL_1836251, EPI_ISL_1836252,
EPI_ISL_1836253, EPI_ISL_1836254, EPI_ISL_1836255, EPI_ISL_1836256, EPI_ISL_1836257, EPI_ISL_1836258, EPI_ISL_1836259, EPI_ISL_1836260, EPI_ISL_1836261, EPI_ISL_1836262, EPI_ISL_1836264, EPI_ISL_1836265, EPI_ISL_1836266, EPI_ISL_1836267, EPI_ISL_1836268, EPI_ISL_1836269, EPI_ISL_1836270, EPI_ISL_1836271, EPI_ISL_1836272, EPI_ISL_1836273, EPI_ISL_1836274, EPI_ISL_1836275,
EPI_ISL_1836276, EPI_ISL_1836277, EPI_ISL_1836278, EPI_ISL_1836279, EPI_ISL_1836280, EPI_ISL_1836281, EPI_ISL_1836282, EPI_ISL_1836283, EPI_ISL_1836284, EPI_ISL_1836285, EPI_ISL_1836286, EPI_ISL_1836287, EPI_ISL_1836288, EPI_ISL_1836289, EPI_ISL_1836290, EPI_ISL_1836291, EPI_ISL_1836292, EPI_ISL_1836293, EPI_ISL_1836294, EPI_ISL_1836295, EPI_ISL_1836296, EPI_ISL_1836297,
EPI_ISL_1836298, EPI_ISL_1836299, EPI_ISL_1836300, EPI_ISL_1836301, EPI_ISL_1836302, EPI_ISL_1836303, EPI_ISL_1836304, EPI_ISL_1836305, EPI_ISL_1836306, EPI_ISL_1836307, EPI_ISL_1836308, EPI_ISL_1836309, EPI_ISL_1836310, EPI_ISL_1836311, EPI_ISL_1836312, EPI_ISL_1836313, EPI_ISL_1836314, EPI_ISL_1836315, EPI_ISL_1836316, EPI_ISL_1836317, EPI_ISL_1836318, EPI_ISL_1836319,
EPI_ISL_1836320, EPI_ISL_1836321, EPI_ISL_1836322, EPI_ISL_1836323, EPI_ISL_1836324, EPI_ISL_1836325, EPI_ISL_1836326, EPI_ISL_1836327, EPI_ISL_1836328, EPI_ISL_1836329, EPI_ISL_1836330, EPI_ISL_1836331, EPI_ISL_1836332, EPI_ISL_1836333, EPI_ISL_1836334, EPI_ISL_1836335, EPI_ISL_1836336, EPI_ISL_1836337, EPI_ISL_1836338, EPI_ISL_1836339, EPI_ISL_1836340, EPI_ISL_1836341,
EPI_ISL_1836342, EPI_ISL_1836343, EPI_ISL_1836344, EPI_ISL_1836345, EPI_ISL_1836346, EPI_ISL_1836347, EPI_ISL_1836348, EPI_ISL_1836349, EPI_ISL_1836350, EPI_ISL_1836351, EPI_ISL_1836352, EPI_ISL_1836353, EPI_ISL_1836354, EPI_ISL_1836355, EPI_ISL_1836356, EPI_ISL_1836357, EPI_ISL_1836358, EPI_ISL_1836359, EPI_ISL_1836360, EPI_ISL_1836362, EPI_ISL_1836363, EPI_ISL_1836364,
EPI_ISL_1836365, EPI_ISL_1836366, EPI_ISL_1836367, EPI_ISL_1836368, EPI_ISL_1836369, EPI_ISL_1836370, EPI_ISL_1836371, EPI_ISL_1836372, EPI_ISL_1836373, EPI_ISL_1836374, EPI_ISL_1836375, EPI_ISL_1836376, EPI_ISL_1836377, EPI_ISL_1836378, EPI_ISL_1836379, EPI_ISL_1836380, EPI_ISL_1836381, EPI_ISL_1836382, EPI_ISL_1836383, EPI_ISL_1836384, EPI_ISL_1836385, EPI_ISL_1836386,
EPI_ISL_1836387, EPI_ISL_1836388, EPI_ISL_1836389, EPI_ISL_1836391, EPI_ISL_1836392, EPI_ISL_1836393, EPI_ISL_1836394, EPI_ISL_1836395, EPI_ISL_1836396, EPI_ISL_1836397, EPI_ISL_1836398, EPI_ISL_1836399, EPI_ISL_1836400, EPI_ISL_1836401, EPI_ISL_1836402, EPI_ISL_1836403, EPI_ISL_1836404, EPI_ISL_1836405, EPI_ISL_1836406, EPI_ISL_1836407, EPI_ISL_1836408, EPI_ISL_1836409,
EPI_ISL_1836410, EPI_ISL_1836411, EPI_ISL_1836412, EPI_ISL_1836413, EPI_ISL_1836414, EPI_ISL_1836415, EPI_ISL_1836416, EPI_ISL_1836417, EPI_ISL_1836418, EPI_ISL_1836419, EPI_ISL_1836420, EPI_ISL_1836421, EPI_ISL_1836422, EPI_ISL_1836423, EPI_ISL_1836424, EPI_ISL_1836425, EPI_ISL_1836426, EPI_ISL_1836427, EPI_ISL_1836428, EPI_ISL_1836429, EPI_ISL_1836430, EPI_ISL_1836431,
EPI_ISL_1836432, EPI_ISL_1836433, EPI_ISL_1836434, EPI_ISL_1836435, EPI_ISL_1836436, EPI_ISL_1836437, EPI_ISL_1836438, EPI_ISL_1836439, EPI_ISL_1836440, EPI_ISL_1836441, EPI_ISL_1836442, EPI_ISL_1836443, EPI_ISL_1836444, EPI_ISL_1836445, EPI_ISL_1836446, EPI_ISL_1836448, EPI_ISL_1836449, EPI_ISL_1836450, EPI_ISL_1836451, EPI_ISL_1836452, EPI_ISL_1836453, EPI_ISL_1836454,
EPI_ISL_1836455, EPI_ISL_1836456, EPI_ISL_1836457, EPI_ISL_1836458, EPI_ISL_1836459, EPI_ISL_1836460, EPI_ISL_1836461, EPI_ISL_1836462, EPI_ISL_1836463, EPI_ISL_1836464, EPI_ISL_1836465, EPI_ISL_1836466, EPI_ISL_1836467, EPI_ISL_1836468, EPI_ISL_1836469, EPI_ISL_1836470, EPI_ISL_1836471, EPI_ISL_1836472, EPI_ISL_1836473, EPI_ISL_1836474, EPI_ISL_1836475, EPI_ISL_1836476,
EPI_ISL_1836477, EPI_ISL_1836478, EPI_ISL_1836479, EPI_ISL_1836480, EPI_ISL_1836481, EPI_ISL_1836482, EPI_ISL_1836483, EPI_ISL_1836484, EPI_ISL_1836485, EPI_ISL_1836486, EPI_ISL_1836487, EPI_ISL_1836488, EPI_ISL_1836489, EPI_ISL_1836490, EPI_ISL_1836491, EPI_ISL_1836492, EPI_ISL_1836493, EPI_ISL_1836494, EPI_ISL_1836495, EPI_ISL_1836496, EPI_ISL_1836497, EPI_ISL_1836498,
EPI_ISL_1836499, EPI_ISL_1836500, EPI_ISL_1836501, EPI_ISL_1836502, EPI_ISL_1836503, EPI_ISL_1836504, EPI_ISL_1836505, EPI_ISL_1836506, EPI_ISL_1836507, EPI_ISL_1836508, EPI_ISL_1836509, EPI_ISL_1836510, EPI_ISL_1836511, EPI_ISL_1836512, EPI_ISL_1836513, EPI_ISL_1836514, EPI_ISL_1836515, EPI_ISL_1836516, EPI_ISL_1836517, EPI_ISL_1836518, EPI_ISL_1836519, EPI_ISL_1836520,
EPI_ISL_1836521, EPI_ISL_1836522, EPI_ISL_1836523, EPI_ISL_1836524, EPI_ISL_1836525, EPI_ISL_1836526, EPI_ISL_1836527, EPI_ISL_1836528, EPI_ISL_1836529, EPI_ISL_1836530, EPI_ISL_1836531, EPI_ISL_1836532, EPI_ISL_1836533, EPI_ISL_1836534, EPI_ISL_1836535, EPI_ISL_1836536, EPI_ISL_1836537, EPI_ISL_1836538, EPI_ISL_1836539, EPI_ISL_1836540, EPI_ISL_1836541, EPI_ISL_1836542,
EPI_ISL_1836543, EPI_ISL_1836544, EPI_ISL_1836545, EPI_ISL_1836546, EPI_ISL_1836547, EPI_ISL_1836548, EPI_ISL_1836549, EPI_ISL_1836550, EPI_ISL_1836551, EPI_ISL_1836552, EPI_ISL_1836553, EPI_ISL_1836554, EPI_ISL_1836555, EPI_ISL_1836556, EPI_ISL_1836557, EPI_ISL_1836558, EPI_ISL_1836559, EPI_ISL_1836560, EPI_ISL_1836561, EPI_ISL_1836562, EPI_ISL_1836563, EPI_ISL_1836564,
EPI_ISL_1836565, EPI_ISL_1836566, EPI_ISL_1836567, EPI_ISL_1836568, EPI_ISL_1836569, EPI_ISL_1836570, EPI_ISL_1836571, EPI_ISL_1836572, EPI_ISL_1836573, EPI_ISL_1836574, EPI_ISL_1836575, EPI_ISL_1836576, EPI_ISL_1836577, EPI_ISL_1836578, EPI_ISL_1836579, EPI_ISL_1836580, EPI_ISL_1836581, EPI_ISL_1836582, EPI_ISL_1836583, EPI_ISL_1836584, EPI_ISL_1836585, EPI_ISL_1836586,
EPI_ISL_1836587, EPI_ISL_1836588, EPI_ISL_1836589, EPI_ISL_1836590, EPI_ISL_1836591, EPI_ISL_1836592, EPI_ISL_1836593, EPI_ISL_1836594, EPI_ISL_1836595, EPI_ISL_1836596, EPI_ISL_1836597, EPI_ISL_1836598, EPI_ISL_1836599, EPI_ISL_1836600, EPI_ISL_1836601, EPI_ISL_1836602, EPI_ISL_1836603, EPI_ISL_1836604, EPI_ISL_1836605, EPI_ISL_1836606, EPI_ISL_1836607, EPI_ISL_1836608,
EPI_ISL_1836609, EPI_ISL_1836610, EPI_ISL_1836611, EPI_ISL_1836612, EPI_ISL_1836613, EPI_ISL_1836614, EPI_ISL_1836615, EPI_ISL_1836616, EPI_ISL_1836617, EPI_ISL_1836618, EPI_ISL_1836619, EPI_ISL_1836620, EPI_ISL_1836621, EPI_ISL_1836622, EPI_ISL_1836623, EPI_ISL_1836624, EPI_ISL_1836625, EPI_ISL_1836626, EPI_ISL_1836627, EPI_ISL_1836628, EPI_ISL_1836629, EPI_ISL_1836630,
EPI_ISL_1836631, EPI_ISL_1836632, EPI_ISL_1836633, EPI_ISL_1836634, EPI_ISL_1836635, EPI_ISL_1836636, EPI_ISL_1836637, EPI_ISL_1836638, EPI_ISL_1836639, EPI_ISL_1836640, EPI_ISL_1836641, EPI_ISL_1836642, EPI_ISL_1836643, EPI_ISL_1836644, EPI_ISL_1836645, EPI_ISL_1836646, EPI_ISL_1836647, EPI_ISL_1836648, EPI_ISL_1836649, EPI_ISL_1836650, EPI_ISL_1836651, EPI_ISL_1836652,
EPI_ISL_1836653, EPI_ISL_1836654, EPI_ISL_1836655, EPI_ISL_1836656, EPI_ISL_1836657, EPI_ISL_1836658, EPI_ISL_1836659, EPI_ISL_1836660, EPI_ISL_1836661, EPI_ISL_1836662, EPI_ISL_1836663, EPI_ISL_1836664, EPI_ISL_1836665, EPI_ISL_1836666, EPI_ISL_1836667, EPI_ISL_1836668, EPI_ISL_1836669, EPI_ISL_1836670, EPI_ISL_1836671, EPI_ISL_1836672, EPI_ISL_1836673, EPI_ISL_1836674,
EPI_ISL_1836675, EPI_ISL_1836676, EPI_ISL_1836677, EPI_ISL_1836678, EPI_ISL_1836679, EPI_ISL_1836680, EPI_ISL_1836681, EPI_ISL_1836682, EPI_ISL_1836683, EPI_ISL_1836684, EPI_ISL_1836685, EPI_ISL_1836686, EPI_ISL_1836687, EPI_ISL_1836688, EPI_ISL_1836689, EPI_ISL_1836690, EPI_ISL_1836691, EPI_ISL_1836692, EPI_ISL_1836693, EPI_ISL_1836694, EPI_ISL_1836695, EPI_ISL_1836696,
EPI_ISL_1836697, EPI_ISL_1836698, EPI_ISL_1836699, EPI_ISL_1836700, EPI_ISL_1836701, EPI_ISL_1836702, EPI_ISL_1836703, EPI_ISL_1836704, EPI_ISL_1836705, EPI_ISL_1836706, EPI_ISL_1836707, EPI_ISL_1836708, EPI_ISL_1836710, EPI_ISL_1836711, EPI_ISL_1836712, EPI_ISL_1836713, EPI_ISL_1836714, EPI_ISL_1836715, EPI_ISL_1836716, EPI_ISL_1836717, EPI_ISL_1836718, EPI_ISL_1836719,
EPI_ISL_1836720, EPI_ISL_1836721, EPI_ISL_1836722, EPI_ISL_1836723, EPI_ISL_1836724, EPI_ISL_1836725, EPI_ISL_1836726, EPI_ISL_1836727, EPI_ISL_1836728, EPI_ISL_1836729, EPI_ISL_1836730, EPI_ISL_1836731, EPI_ISL_1836732, EPI_ISL_1836733, EPI_ISL_1836734, EPI_ISL_1836735, EPI_ISL_1836736, EPI_ISL_1836737, EPI_ISL_1836738, EPI_ISL_1836739, EPI_ISL_1836740, EPI_ISL_1836741,
EPI_ISL_1836742, EPI_ISL_1836743, EPI_ISL_1836744, EPI_ISL_1836745, EPI_ISL_1836746, EPI_ISL_1836747, EPI_ISL_1836748, EPI_ISL_1836749, EPI_ISL_1836750, EPI_ISL_1836751, EPI_ISL_1836753, EPI_ISL_1836754, EPI_ISL_1836755, EPI_ISL_1836756, EPI_ISL_1836757, EPI_ISL_1836758, EPI_ISL_1836759, EPI_ISL_1836760, EPI_ISL_1836761, EPI_ISL_1836762, EPI_ISL_1836763, EPI_ISL_1836764,
EPI_ISL_1836765, EPI_ISL_1836766, EPI_ISL_1836767, EPI_ISL_1836768, EPI_ISL_1836769, EPI_ISL_1836770, EPI_ISL_1836771, EPI_ISL_1836772, EPI_ISL_1836773, EPI_ISL_1836774, EPI_ISL_1836775, EPI_ISL_1836776, EPI_ISL_1836777, EPI_ISL_1836778, EPI_ISL_1836779, EPI_ISL_1836780, EPI_ISL_1836781, EPI_ISL_1836782, EPI_ISL_1836783, EPI_ISL_1836784, EPI_ISL_1836785, EPI_ISL_1836786,
EPI_ISL_1836787, EPI_ISL_1836788, EPI_ISL_1836789, EPI_ISL_1836790, EPI_ISL_1836791, EPI_ISL_1836792, EPI_ISL_1836793, EPI_ISL_1836794, EPI_ISL_1836795, EPI_ISL_1836796, EPI_ISL_1836797, EPI_ISL_1836798, EPI_ISL_1836799, EPI_ISL_1836800, EPI_ISL_1836801, EPI_ISL_1836802, EPI_ISL_1836803, EPI_ISL_1836804, EPI_ISL_1836805, EPI_ISL_1836806, EPI_ISL_1836807, EPI_ISL_1836808,
EPI_ISL_1836809, EPI_ISL_1836810, EPI_ISL_1836811, EPI_ISL_1836812, EPI_ISL_1836813, EPI_ISL_1836814, EPI_ISL_1836815, EPI_ISL_1836816, EPI_ISL_1836817, EPI_ISL_1836818, EPI_ISL_1836819, EPI_ISL_1836820, EPI_ISL_1836821, EPI_ISL_1836822, EPI_ISL_1836823, EPI_ISL_1836824, EPI_ISL_1836825, EPI_ISL_1836826, EPI_ISL_1836827, EPI_ISL_1836828, EPI_ISL_1836829, EPI_ISL_1836830,
EPI_ISL_1836831, EPI_ISL_1836832, EPI_ISL_1836833, EPI_ISL_1836834, EPI_ISL_1836835, EPI_ISL_1836836, EPI_ISL_1836837, EPI_ISL_1836838, EPI_ISL_1836839, EPI_ISL_1836840, EPI_ISL_1836841, EPI_ISL_1836842, EPI_ISL_1836843, EPI_ISL_1836844, EPI_ISL_1836845, EPI_ISL_1836846, EPI_ISL_1836847, EPI_ISL_1836848, EPI_ISL_1836849, EPI_ISL_1836850, EPI_ISL_1836851, EPI_ISL_1836852,
EPI_ISL_1836853, EPI_ISL_1836854, EPI_ISL_1836855, EPI_ISL_1836856, EPI_ISL_1836857, EPI_ISL_1836858, EPI_ISL_1836859, EPI_ISL_1836860, EPI_ISL_1836861, EPI_ISL_1836862, EPI_ISL_1836863, EPI_ISL_1836864, EPI_ISL_1836865, EPI_ISL_1836866, EPI_ISL_1836867, EPI_ISL_1836868, EPI_ISL_1836869, EPI_ISL_1836870, EPI_ISL_1836871, EPI_ISL_1836872, EPI_ISL_1836873, EPI_ISL_1836874,
EPI_ISL_1836875, EPI_ISL_1836876, EPI_ISL_1836877, EPI_ISL_1836878, EPI_ISL_1836879, EPI_ISL_1836880, EPI_ISL_1836881, EPI_ISL_1836882, EPI_ISL_1836883, EPI_ISL_1836884, EPI_ISL_1836885, EPI_ISL_1836886, EPI_ISL_1836887, EPI_ISL_1836888, EPI_ISL_1836889, EPI_ISL_1836890, EPI_ISL_1836891, EPI_ISL_1836892, EPI_ISL_1836893, EPI_ISL_1836894, EPI_ISL_1836895, EPI_ISL_1836896,
EPI_ISL_1836897, EPI_ISL_1836898, EPI_ISL_1836899, EPI_ISL_1836900, EPI_ISL_1836901, EPI_ISL_1836902, EPI_ISL_1836903, EPI_ISL_1836904, EPI_ISL_1836905, EPI_ISL_1836906, EPI_ISL_1836907, EPI_ISL_1836908, EPI_ISL_1836909, EPI_ISL_1836910, EPI_ISL_1836911, EPI_ISL_1836912, EPI_ISL_1836913, EPI_ISL_1836914, EPI_ISL_1836915, EPI_ISL_1836916, EPI_ISL_1836917, EPI_ISL_1836918,
EPI_ISL_1836919, EPI_ISL_1836920, EPI_ISL_1836921, EPI_ISL_1836922, EPI_ISL_1836923, EPI_ISL_1836924, EPI_ISL_1836925, EPI_ISL_1836926, EPI_ISL_1836927, EPI_ISL_1836928, EPI_ISL_1836929, EPI_ISL_1836930, EPI_ISL_1836931, EPI_ISL_1836932, EPI_ISL_1836933, EPI_ISL_1836934, EPI_ISL_1836935, EPI_ISL_1836936, EPI_ISL_1836937, EPI_ISL_1836938, EPI_ISL_1836939, EPI_ISL_1836940,
EPI_ISL_1836941, EPI_ISL_1836942, EPI_ISL_1836943, EPI_ISL_1836944, EPI_ISL_1836945, EPI_ISL_1836946, EPI_ISL_1836947, EPI_ISL_1836948, EPI_ISL_1836949, EPI_ISL_1836950, EPI_ISL_1836951, EPI_ISL_1836952, EPI_ISL_1836953, EPI_ISL_1836954, EPI_ISL_1836955, EPI_ISL_1836956, EPI_ISL_1836957, EPI_ISL_1836958, EPI_ISL_1836959, EPI_ISL_1836960, EPI_ISL_1836961, EPI_ISL_1836962,
EPI_ISL_1836963, EPI_ISL_1836964, EPI_ISL_1836965, EPI_ISL_1836967, EPI_ISL_1836968, EPI_ISL_1836969, EPI_ISL_1836972, EPI_ISL_1836973, EPI_ISL_1836974, EPI_ISL_1836975, EPI_ISL_1836976, EPI_ISL_1836977, EPI_ISL_1836979, EPI_ISL_1836981, EPI_ISL_1836982, EPI_ISL_1836983, EPI_ISL_1836985, EPI_ISL_1836986, EPI_ISL_1836987, EPI_ISL_1836988, EPI_ISL_1836989, EPI_ISL_1836990,
EPI_ISL_1836991, EPI_ISL_1836992, EPI_ISL_1836993, EPI_ISL_1836994, EPI_ISL_1836995, EPI_ISL_1836996, EPI_ISL_1836997, EPI_ISL_1836998, EPI_ISL_1836999, EPI_ISL_1837000, EPI_ISL_1837001, EPI_ISL_1837002, EPI_ISL_1837003, EPI_ISL_1837004, EPI_ISL_1837005, EPI_ISL_1837006, EPI_ISL_1837008, EPI_ISL_1837009, EPI_ISL_1837010, EPI_ISL_1837011, EPI_ISL_1837012, EPI_ISL_1837013,
EPI_ISL_1837014, EPI_ISL_1837015, EPI_ISL_1837016, EPI_ISL_1837017, EPI_ISL_1837018, EPI_ISL_1837019, EPI_ISL_1837020, EPI_ISL_1837021, EPI_ISL_1837022, EPI_ISL_1837023, EPI_ISL_1837024, EPI_ISL_1837025, EPI_ISL_1837027, EPI_ISL_1837028, EPI_ISL_1837029, EPI_ISL_1837030, EPI_ISL_1837031, EPI_ISL_1837032, EPI_ISL_1837033, EPI_ISL_1837034, EPI_ISL_1837035, EPI_ISL_1837036,
EPI_ISL_1837037, EPI_ISL_1837038, EPI_ISL_1837039, EPI_ISL_1837040, EPI_ISL_1837041, EPI_ISL_1837042, EPI_ISL_1837043, EPI_ISL_1837044, EPI_ISL_1837045, EPI_ISL_1837046, EPI_ISL_1837047, EPI_ISL_1837048, EPI_ISL_1837049, EPI_ISL_1837050, EPI_ISL_1837051, EPI_ISL_1837052, EPI_ISL_1837053, EPI_ISL_1837054, EPI_ISL_1837055, EPI_ISL_1837056, EPI_ISL_1837057, EPI_ISL_1837058,
EPI_ISL_1837059, EPI_ISL_1837060, EPI_ISL_1837061, EPI_ISL_1837062, EPI_ISL_1837063, EPI_ISL_1837064, EPI_ISL_1837065, EPI_ISL_1837067, EPI_ISL_1837068, EPI_ISL_1837069, EPI_ISL_1837070, EPI_ISL_1837071, EPI_ISL_1837072, EPI_ISL_1837073, EPI_ISL_1837074, EPI_ISL_1837075, EPI_ISL_1837076, EPI_ISL_1837077, EPI_ISL_1837078, EPI_ISL_1837079, EPI_ISL_1837080, EPI_ISL_1837081,
EPI_ISL_1837082, EPI_ISL_1837083, EPI_ISL_1837084, EPI_ISL_1837085, EPI_ISL_1837086, EPI_ISL_1837087, EPI_ISL_1837088, EPI_ISL_1837089, EPI_ISL_1837090, EPI_ISL_1837091, EPI_ISL_1837092, EPI_ISL_1837093, EPI_ISL_1837094, EPI_ISL_1837095, EPI_ISL_1837096, EPI_ISL_1837097, EPI_ISL_1837098, EPI_ISL_1837099, EPI_ISL_1837100, EPI_ISL_1837101, EPI_ISL_1837102, EPI_ISL_1837103,
EPI_ISL_1837104, EPI_ISL_1837105, EPI_ISL_1837106, EPI_ISL_1837107, EPI_ISL_1837108, EPI_ISL_1837109, EPI_ISL_1837110, EPI_ISL_1837111, EPI_ISL_1837112, EPI_ISL_1837113, EPI_ISL_1837114, EPI_ISL_1837115, EPI_ISL_1837116, EPI_ISL_1837117, EPI_ISL_1837118, EPI_ISL_1837119, EPI_ISL_1837120, EPI_ISL_1837121, EPI_ISL_1837122, EPI_ISL_1837123, EPI_ISL_1837124, EPI_ISL_1837125,
EPI_ISL_1837126, EPI_ISL_1837127, EPI_ISL_1837128, EPI_ISL_1837129, EPI_ISL_1837130, EPI_ISL_1837131, EPI_ISL_1837132, EPI_ISL_1837133, EPI_ISL_1837134, EPI_ISL_1837135, EPI_ISL_1837136, EPI_ISL_1837137, EPI_ISL_1837138, EPI_ISL_1837139, EPI_ISL_1837140, EPI_ISL_1837141, EPI_ISL_1837142, EPI_ISL_1837143, EPI_ISL_1837144, EPI_ISL_1837145, EPI_ISL_1837146, EPI_ISL_1837147,
EPI_ISL_1837148, EPI_ISL_1837149, EPI_ISL_1837150, EPI_ISL_1837151, EPI_ISL_1837152, EPI_ISL_1837153, EPI_ISL_1837154, EPI_ISL_1837155, EPI_ISL_1837156, EPI_ISL_1837157, EPI_ISL_1837158, EPI_ISL_1837159, EPI_ISL_1837160, EPI_ISL_1837161, EPI_ISL_1837162, EPI_ISL_1837163, EPI_ISL_1837164, EPI_ISL_1837165, EPI_ISL_1837166, EPI_ISL_1837167, EPI_ISL_1837168, EPI_ISL_1837169,
EPI_ISL_1837170, EPI_ISL_1837171, EPI_ISL_1837172, EPI_ISL_1837173, EPI_ISL_1837174, EPI_ISL_1837175, EPI_ISL_1837176, EPI_ISL_1837177, EPI_ISL_1837178, EPI_ISL_1837179, EPI_ISL_1837180, EPI_ISL_1837181, EPI_ISL_1837183, EPI_ISL_1837184, EPI_ISL_1837185, EPI_ISL_1837186, EPI_ISL_1837187, EPI_ISL_1837188, EPI_ISL_1837189, EPI_ISL_1837190, EPI_ISL_1837191, EPI_ISL_1837192,
EPI_ISL_1837193, EPI_ISL_1837194, EPI_ISL_1837195, EPI_ISL_1837196, EPI_ISL_1837198, EPI_ISL_1837199, EPI_ISL_1837200, EPI_ISL_1837201, EPI_ISL_1837202, EPI_ISL_1837203, EPI_ISL_1837204, EPI_ISL_1837205, EPI_ISL_1837206, EPI_ISL_1837207, EPI_ISL_1837208, EPI_ISL_1837209, EPI_ISL_1837210, EPI_ISL_1837211, EPI_ISL_1837212, EPI_ISL_1837213, EPI_ISL_1837214, EPI_ISL_1837215,
EPI_ISL_1837216, EPI_ISL_1837217, EPI_ISL_1837218, EPI_ISL_1837219, EPI_ISL_1837220, EPI_ISL_1837221, EPI_ISL_1837222, EPI_ISL_1837223, EPI_ISL_1837224, EPI_ISL_1837225, EPI_ISL_1837226, EPI_ISL_1837227, EPI_ISL_1837228, EPI_ISL_1837229, EPI_ISL_1837230, EPI_ISL_1837231, EPI_ISL_1837232, EPI_ISL_1837233, EPI_ISL_1837234, EPI_ISL_1837235, EPI_ISL_1837236, EPI_ISL_1837237,
EPI_ISL_1837238, EPI_ISL_1837239, EPI_ISL_1837240, EPI_ISL_1837241, EPI_ISL_1837242, EPI_ISL_1837243, EPI_ISL_1837244, EPI_ISL_1837245, EPI_ISL_1837247, EPI_ISL_1837248, EPI_ISL_1837249, EPI_ISL_1837250, EPI_ISL_1837251, EPI_ISL_1837252, EPI_ISL_1837253, EPI_ISL_1837254, EPI_ISL_1837255, EPI_ISL_1837256, EPI_ISL_1837257, EPI_ISL_1837258, EPI_ISL_1837259, EPI_ISL_1837260,
EPI_ISL_1837261, EPI_ISL_1837262, EPI_ISL_1837263, EPI_ISL_1837264, EPI_ISL_1837265, EPI_ISL_1837266, EPI_ISL_1837267, EPI_ISL_1837268, EPI_ISL_1837269, EPI_ISL_1837270, EPI_ISL_1837271, EPI_ISL_1837272, EPI_ISL_1837273, EPI_ISL_1837274, EPI_ISL_1837275, EPI_ISL_1837276, EPI_ISL_1837277, EPI_ISL_1837278, EPI_ISL_1837279, EPI_ISL_1837280, EPI_ISL_1837281, EPI_ISL_1837282,
EPI_ISL_1837283, EPI_ISL_1837284, EPI_ISL_1837285, EPI_ISL_1837286, EPI_ISL_1837287, EPI_ISL_1837288, EPI_ISL_1837289, EPI_ISL_1837290, EPI_ISL_1837291, EPI_ISL_1837292, EPI_ISL_1837293, EPI_ISL_1837294, EPI_ISL_1837295, EPI_ISL_1837296, EPI_ISL_1837297, EPI_ISL_1837298, EPI_ISL_1837299, EPI_ISL_1837300, EPI_ISL_1837301, EPI_ISL_1837302, EPI_ISL_1837303, EPI_ISL_1837304,
EPI_ISL_1837305, EPI_ISL_1837306, EPI_ISL_1837307, EPI_ISL_1837308, EPI_ISL_1837309, EPI_ISL_1837310, EPI_ISL_1837312, EPI_ISL_1837313, EPI_ISL_1837314, EPI_ISL_1837315, EPI_ISL_1837316, EPI_ISL_1837317, EPI_ISL_1837318, EPI_ISL_1837319, EPI_ISL_1837320, EPI_ISL_1837321, EPI_ISL_1837322, EPI_ISL_1837323, EPI_ISL_1837324, EPI_ISL_1837325, EPI_ISL_1837326, EPI_ISL_1837327,
EPI_ISL_1837328, EPI_ISL_1837329, EPI_ISL_1837330, EPI_ISL_1837331, EPI_ISL_1837332, EPI_ISL_1837333, EPI_ISL_1837334, EPI_ISL_1837336, EPI_ISL_1837337, EPI_ISL_1837338, EPI_ISL_1837339, EPI_ISL_1837340, EPI_ISL_1837341, EPI_ISL_1837342, EPI_ISL_1837343, EPI_ISL_1837344, EPI_ISL_1837345, EPI_ISL_1837346, EPI_ISL_1837347, EPI_ISL_1837348, EPI_ISL_1837349, EPI_ISL_1837350,
EPI_ISL_1837351, EPI_ISL_1837352, EPI_ISL_1837353, EPI_ISL_1837354, EPI_ISL_1837355, EPI_ISL_1837356, EPI_ISL_1837357, EPI_ISL_1837358, EPI_ISL_1837359, EPI_ISL_1837360, EPI_ISL_1837361, EPI_ISL_1837362, EPI_ISL_1837363, EPI_ISL_1837364, EPI_ISL_1837365, EPI_ISL_1837366, EPI_ISL_1837368, EPI_ISL_1837369, EPI_ISL_1837370, EPI_ISL_1837371, EPI_ISL_1837372, EPI_ISL_1837373,
EPI_ISL_1837374, EPI_ISL_1837375, EPI_ISL_1837376, EPI_ISL_1837377, EPI_ISL_1837378, EPI_ISL_1837379, EPI_ISL_1837380, EPI_ISL_1837381, EPI_ISL_1837382, EPI_ISL_1837383, EPI_ISL_1837384, EPI_ISL_1837385, EPI_ISL_1837386, EPI_ISL_1837387, EPI_ISL_1837388, EPI_ISL_1837389, EPI_ISL_1837390, EPI_ISL_1837391, EPI_ISL_1837392, EPI_ISL_1837393, EPI_ISL_1837394, EPI_ISL_1837395,
EPI_ISL_1837396, EPI_ISL_1837397, EPI_ISL_1837398, EPI_ISL_1837399, EPI_ISL_1837400, EPI_ISL_1837401, EPI_ISL_1837402, EPI_ISL_1837403, EPI_ISL_1837404, EPI_ISL_1837405, EPI_ISL_1837406, EPI_ISL_1837407, EPI_ISL_1837408, EPI_ISL_1837409, EPI_ISL_1837410, EPI_ISL_1837411, EPI_ISL_1837412, EPI_ISL_1837413, EPI_ISL_1837414, EPI_ISL_1837416, EPI_ISL_1837417, EPI_ISL_1837418,
EPI_ISL_1837419, EPI_ISL_1837420, EPI_ISL_1837421, EPI_ISL_1837422, EPI_ISL_1837423, EPI_ISL_1837424, EPI_ISL_1837425, EPI_ISL_1837426, EPI_ISL_1837427, EPI_ISL_1837428, EPI_ISL_1837429, EPI_ISL_1837430, EPI_ISL_1837431, EPI_ISL_1837432, EPI_ISL_1837433, EPI_ISL_1837434, EPI_ISL_1837435, EPI_ISL_1837436, EPI_ISL_1837437, EPI_ISL_1837438, EPI_ISL_1837439, EPI_ISL_1837441,
EPI_ISL_1837442, EPI_ISL_1837443, EPI_ISL_1837444, EPI_ISL_1837445, EPI_ISL_1837446, EPI_ISL_1837447, EPI_ISL_1837448, EPI_ISL_1837449, EPI_ISL_1837450, EPI_ISL_1837451, EPI_ISL_1837452, EPI_ISL_1837453, EPI_ISL_1837454, EPI_ISL_1837455, EPI_ISL_1837456, EPI_ISL_1837457, EPI_ISL_1837458, EPI_ISL_1837459, EPI_ISL_1837460, EPI_ISL_1837461, EPI_ISL_1837462, EPI_ISL_1837463,
EPI_ISL_1837464, EPI_ISL_1837465, EPI_ISL_1837466, EPI_ISL_1837467, EPI_ISL_1837468, EPI_ISL_1837469, EPI_ISL_1837470, EPI_ISL_1837471, EPI_ISL_1837472, EPI_ISL_1837473, EPI_ISL_1837474, EPI_ISL_1837475, EPI_ISL_1837476, EPI_ISL_1837477, EPI_ISL_1837478, EPI_ISL_1837479, EPI_ISL_1837480, EPI_ISL_1837481, EPI_ISL_1837482, EPI_ISL_1837483, EPI_ISL_1837484, EPI_ISL_1837485,
EPI_ISL_1837486, EPI_ISL_1837487, EPI_ISL_1837488, EPI_ISL_1837489, EPI_ISL_1837490, EPI_ISL_1837491, EPI_ISL_1837492, EPI_ISL_1837493, EPI_ISL_1837494, EPI_ISL_1837495, EPI_ISL_1837496, EPI_ISL_1837497, EPI_ISL_1837498, EPI_ISL_1837499, EPI_ISL_1837500, EPI_ISL_1837501, EPI_ISL_1837502, EPI_ISL_1837503, EPI_ISL_1837504, EPI_ISL_1837505, EPI_ISL_1837506, EPI_ISL_1837507,
EPI_ISL_1837508, EPI_ISL_1837509, EPI_ISL_1837510, EPI_ISL_1837511, EPI_ISL_1837512, EPI_ISL_1837513, EPI_ISL_1837514, EPI_ISL_1837515, EPI_ISL_1837516, EPI_ISL_1837517, EPI_ISL_1837518, EPI_ISL_1837519, EPI_ISL_1837520, EPI_ISL_1837521, EPI_ISL_1837522, EPI_ISL_1837523, EPI_ISL_1837524, EPI_ISL_1837525, EPI_ISL_1837526, EPI_ISL_1837527, EPI_ISL_1837528, EPI_ISL_1837530,
EPI_ISL_1837531, EPI_ISL_1837532, EPI_ISL_1837533, EPI_ISL_1837534, EPI_ISL_1837535, EPI_ISL_1837536, EPI_ISL_1837537, EPI_ISL_1837538, EPI_ISL_1837539, EPI_ISL_1837540, EPI_ISL_1837541, EPI_ISL_1837542, EPI_ISL_1837543, EPI_ISL_1837544, EPI_ISL_1837545, EPI_ISL_1837546, EPI_ISL_1837547, EPI_ISL_1837548, EPI_ISL_1837549, EPI_ISL_1837550, EPI_ISL_1837551, EPI_ISL_1837552,
EPI_ISL_1837553, EPI_ISL_1837554, EPI_ISL_1837555, EPI_ISL_1837556, EPI_ISL_1837557, EPI_ISL_1837558, EPI_ISL_1837559, EPI_ISL_1837560, EPI_ISL_1837561, EPI_ISL_1837562, EPI_ISL_1837563, EPI_ISL_1837564, EPI_ISL_1837566, EPI_ISL_1837567, EPI_ISL_1837568, EPI_ISL_1837569, EPI_ISL_1837570, EPI_ISL_1837572, EPI_ISL_1837573, EPI_ISL_1837574, EPI_ISL_1837575, EPI_ISL_1837576,
EPI_ISL_1837577, EPI_ISL_1837578, EPI_ISL_1837579, EPI_ISL_1837580, EPI_ISL_1837581, EPI_ISL_1837582, EPI_ISL_1837583, EPI_ISL_1837584, EPI_ISL_1837585, EPI_ISL_1837586, EPI_ISL_1837587, EPI_ISL_1837588, EPI_ISL_1837589, EPI_ISL_1837590, EPI_ISL_1837591, EPI_ISL_1837592, EPI_ISL_1837593, EPI_ISL_1837594, EPI_ISL_1837595, EPI_ISL_1837596, EPI_ISL_1837597, EPI_ISL_1837598,
EPI_ISL_1837599, EPI_ISL_1837600, EPI_ISL_1837601, EPI_ISL_1837602, EPI_ISL_1837603, EPI_ISL_1837604, EPI_ISL_1837605, EPI_ISL_1837606, EPI_ISL_1837608, EPI_ISL_1837609, EPI_ISL_1837610, EPI_ISL_1837611, EPI_ISL_1837612, EPI_ISL_1837613, EPI_ISL_1837614, EPI_ISL_1837615, EPI_ISL_1837616, EPI_ISL_1837617, EPI_ISL_1837618, EPI_ISL_1837619, EPI_ISL_1837621, EPI_ISL_1837622,
EPI_ISL_1837623, EPI_ISL_1837624, EPI_ISL_1837625, EPI_ISL_1837626, EPI_ISL_1837627, EPI_ISL_1837628, EPI_ISL_1837629, EPI_ISL_1837630, EPI_ISL_1837631, EPI_ISL_1837632, EPI_ISL_1837633, EPI_ISL_1837634, EPI_ISL_1837635, EPI_ISL_1837636, EPI_ISL_1837637, EPI_ISL_1837638, EPI_ISL_1837639, EPI_ISL_1837640, EPI_ISL_1837641, EPI_ISL_1837642, EPI_ISL_1837643, EPI_ISL_1837645,
EPI_ISL_1837646, EPI_ISL_1837647, EPI_ISL_1837648, EPI_ISL_1837649, EPI_ISL_1837650, EPI_ISL_1837651, EPI_ISL_1837652, EPI_ISL_1837653, EPI_ISL_1837654, EPI_ISL_1837655, EPI_ISL_1837656, EPI_ISL_1837657, EPI_ISL_1837658, EPI_ISL_1837659, EPI_ISL_1837660, EPI_ISL_1837661, EPI_ISL_1837662, EPI_ISL_1837663, EPI_ISL_1837664, EPI_ISL_1837665, EPI_ISL_1837666, EPI_ISL_1837667,
EPI_ISL_1837668, EPI_ISL_1837669, EPI_ISL_1837670, EPI_ISL_1837671, EPI_ISL_1837672, EPI_ISL_1837673, EPI_ISL_1837674, EPI_ISL_1837675, EPI_ISL_1837676, EPI_ISL_1837677, EPI_ISL_1837678, EPI_ISL_1837679, EPI_ISL_1837680, EPI_ISL_1837681, EPI_ISL_1837682, EPI_ISL_1837683, EPI_ISL_1837684, EPI_ISL_1837685, EPI_ISL_1837686, EPI_ISL_1837687, EPI_ISL_1837688, EPI_ISL_1837689,
EPI_ISL_1837690, EPI_ISL_1837691, EPI_ISL_1837692, EPI_ISL_1837693, EPI_ISL_1837694, EPI_ISL_1837695, EPI_ISL_1837696, EPI_ISL_1837697, EPI_ISL_1837698, EPI_ISL_1837699, EPI_ISL_1837700, EPI_ISL_1837701, EPI_ISL_1837702, EPI_ISL_1837703, EPI_ISL_1837704, EPI_ISL_1837705, EPI_ISL_1837706, EPI_ISL_1837707, EPI_ISL_1837708, EPI_ISL_1837709, EPI_ISL_1837710, EPI_ISL_1837711,
EPI_ISL_1837712, EPI_ISL_1837713, EPI_ISL_1837714, EPI_ISL_1837715, EPI_ISL_1837716, EPI_ISL_1837717, EPI_ISL_1837718, EPI_ISL_1837719, EPI_ISL_1837720, EPI_ISL_1837721, EPI_ISL_1837722, EPI_ISL_1837723, EPI_ISL_1837724, EPI_ISL_1837725, EPI_ISL_1837726, EPI_ISL_1837727, EPI_ISL_1837728, EPI_ISL_1837729, EPI_ISL_1837730, EPI_ISL_1837731, EPI_ISL_1837732, EPI_ISL_1837733,
EPI_ISL_1837734, EPI_ISL_1837735, EPI_ISL_1837736, EPI_ISL_1837737, EPI_ISL_1837738, EPI_ISL_1837739, EPI_ISL_1837740, EPI_ISL_1837741, EPI_ISL_1837742, EPI_ISL_1837743, EPI_ISL_1837744, EPI_ISL_1837745, EPI_ISL_1837746, EPI_ISL_1837747, EPI_ISL_1837748, EPI_ISL_1837749, EPI_ISL_1837750, EPI_ISL_1837751, EPI_ISL_1837752, EPI_ISL_1837753, EPI_ISL_1837754, EPI_ISL_1837755,
EPI_ISL_1837756, EPI_ISL_1837757, EPI_ISL_1837758, EPI_ISL_1837759, EPI_ISL_1837760, EPI_ISL_1837761, EPI_ISL_1837762, EPI_ISL_1837763, EPI_ISL_1837764, EPI_ISL_1837765, EPI_ISL_1837766, EPI_ISL_1837767, EPI_ISL_1837768, EPI_ISL_1837769, EPI_ISL_1837770, EPI_ISL_1837771, EPI_ISL_1837772, EPI_ISL_1837773, EPI_ISL_1837774, EPI_ISL_1837775, EPI_ISL_1837776, EPI_ISL_1837777,
EPI_ISL_1837778, EPI_ISL_1837779, EPI_ISL_1837780, EPI_ISL_1837781, EPI_ISL_1837782, EPI_ISL_1837783, EPI_ISL_1837784, EPI_ISL_1837785, EPI_ISL_1837786, EPI_ISL_1837787, EPI_ISL_1837788, EPI_ISL_1837789, EPI_ISL_1837790, EPI_ISL_1837791, EPI_ISL_1837792, EPI_ISL_1837793, EPI_ISL_1837794, EPI_ISL_1837795, EPI_ISL_1837796, EPI_ISL_1837797, EPI_ISL_1837798, EPI_ISL_1837799,
EPI_ISL_1837800, EPI_ISL_1837801, EPI_ISL_1837802, EPI_ISL_1837803, EPI_ISL_1837804, EPI_ISL_1837805, EPI_ISL_1837806, EPI_ISL_1837807, EPI_ISL_1837808, EPI_ISL_1837812, EPI_ISL_1837813, EPI_ISL_1837814, EPI_ISL_1837815, EPI_ISL_1837816, EPI_ISL_1837817, EPI_ISL_1837818, EPI_ISL_1837819, EPI_ISL_1837820, EPI_ISL_1837821, EPI_ISL_1837822, EPI_ISL_1837823, EPI_ISL_1837824,
EPI_ISL_1837825, EPI_ISL_1837826, EPI_ISL_1837827, EPI_ISL_1837828, EPI_ISL_1837829, EPI_ISL_1837830, EPI_ISL_1837831, EPI_ISL_1837832, EPI_ISL_1837833, EPI_ISL_1837834, EPI_ISL_1837835, EPI_ISL_1837836, EPI_ISL_1837837, EPI_ISL_1837838, EPI_ISL_1837839, EPI_ISL_1837840, EPI_ISL_1837841, EPI_ISL_1837842, EPI_ISL_1837843, EPI_ISL_1837844, EPI_ISL_1837845, EPI_ISL_1837846,
EPI_ISL_1837847, EPI_ISL_1837848, EPI_ISL_1837849, EPI_ISL_1837850, EPI_ISL_1837851, EPI_ISL_1837852, EPI_ISL_1837853, EPI_ISL_1837854, EPI_ISL_1837855, EPI_ISL_1837856, EPI_ISL_1837857, EPI_ISL_1837858, EPI_ISL_1837859, EPI_ISL_1837860, EPI_ISL_1837861, EPI_ISL_1837862, EPI_ISL_1837863, EPI_ISL_1837864, EPI_ISL_1837865, EPI_ISL_1837866, EPI_ISL_1837867, EPI_ISL_1837868,
EPI_ISL_1837869, EPI_ISL_1837870, EPI_ISL_1837871, EPI_ISL_1837872, EPI_ISL_1837873, EPI_ISL_1837874, EPI_ISL_1837875, EPI_ISL_1837876, EPI_ISL_1837877, EPI_ISL_1837878, EPI_ISL_1837879, EPI_ISL_1837880, EPI_ISL_1837881, EPI_ISL_1837882, EPI_ISL_1837883, EPI_ISL_1837884, EPI_ISL_1837885, EPI_ISL_1837886, EPI_ISL_1837887, EPI_ISL_1837888, EPI_ISL_1837889, EPI_ISL_1837890,
EPI_ISL_1837891, EPI_ISL_1837892, EPI_ISL_1837893, EPI_ISL_1837894, EPI_ISL_1837895, EPI_ISL_1837896, EPI_ISL_1837897, EPI_ISL_1837898, EPI_ISL_1837899, EPI_ISL_1837900, EPI_ISL_1837901, EPI_ISL_1837902, EPI_ISL_1837903, EPI_ISL_1837904, EPI_ISL_1837905, EPI_ISL_1837906, EPI_ISL_1837907, EPI_ISL_1837908, EPI_ISL_1837909, EPI_ISL_1837910, EPI_ISL_1837911, EPI_ISL_1837912,
EPI_ISL_1837913, EPI_ISL_1837914, EPI_ISL_1837915, EPI_ISL_1837916, EPI_ISL_1837917, EPI_ISL_1837918, EPI_ISL_1837919, EPI_ISL_1837920, EPI_ISL_1837921, EPI_ISL_1837922, EPI_ISL_1837923, EPI_ISL_1837924, EPI_ISL_1837925, EPI_ISL_1837926, EPI_ISL_1837927, EPI_ISL_1837928, EPI_ISL_1837929, EPI_ISL_1837930, EPI_ISL_1837931, EPI_ISL_1837932, EPI_ISL_1837933, EPI_ISL_1837934,
EPI_ISL_1837935, EPI_ISL_1837936, EPI_ISL_1837937, EPI_ISL_1837938, EPI_ISL_1837939, EPI_ISL_1837940, EPI_ISL_1837941, EPI_ISL_1837942, EPI_ISL_1837943, EPI_ISL_1837944, EPI_ISL_1837945, EPI_ISL_1837946, EPI_ISL_1837947, EPI_ISL_1837948, EPI_ISL_1837949, EPI_ISL_1837950, EPI_ISL_1837951, EPI_ISL_1837952, EPI_ISL_1837953, EPI_ISL_1837954, EPI_ISL_1837955, EPI_ISL_1837956,
EPI_ISL_1837957, EPI_ISL_1837958, EPI_ISL_1837959, EPI_ISL_1837960, EPI_ISL_1837961, EPI_ISL_1837962, EPI_ISL_1837963, EPI_ISL_1837964, EPI_ISL_1837965, EPI_ISL_1837966, EPI_ISL_1837967, EPI_ISL_1837968, EPI_ISL_1837969, EPI_ISL_1837970, EPI_ISL_1837971, EPI_ISL_1837972, EPI_ISL_1837973, EPI_ISL_1837974, EPI_ISL_1837975, EPI_ISL_1837976, EPI_ISL_1837977, EPI_ISL_1837978,
EPI_ISL_1837979, EPI_ISL_1837980, EPI_ISL_1837981, EPI_ISL_1837982, EPI_ISL_1837983, EPI_ISL_1837984, EPI_ISL_1837985, EPI_ISL_1837986, EPI_ISL_1837987, EPI_ISL_1837989, EPI_ISL_1837990, EPI_ISL_1837992, EPI_ISL_1837994, EPI_ISL_1837995, EPI_ISL_1837997, EPI_ISL_1837999, EPI_ISL_1838000, EPI_ISL_1838002, EPI_ISL_1838003, EPI_ISL_1838005, EPI_ISL_1838006, EPI_ISL_1838008,

| | | | |
|---|---|---|---|
| EPI_ISL_1838009, EPI_ISL_1838011, EPI_ISL_1838013, EPI_ISL_1838014, EPI_ISL_1838016, EPI_ISL_1838017, EPI_ISL_1838019, EPI_ISL_1838020, EPI_ISL_1838022, EPI_ISL_1838024, EPI_ISL_1838025, EPI_ISL_1838027, EPI_ISL_1838028, EPI_ISL_1838030, EPI_ISL_1838031, EPI_ISL_1838033, EPI_ISL_1838034, EPI_ISL_1838036, EPI_ISL_1838038, EPI_ISL_1838039, EPI_ISL_1838041, EPI_ISL_1838043, EPI_ISL_1838044, EPI_ISL_1838046, EPI_ISL_1838048, EPI_ISL_1838049, EPI_ISL_1838051, EPI_ISL_1838053, EPI_ISL_1838054, EPI_ISL_1838056, EPI_ISL_1838058, EPI_ISL_1838059, EPI_ISL_1838061, EPI_ISL_1838062, EPI_ISL_1838064, EPI_ISL_1838066, EPI_ISL_1838067, EPI_ISL_1838069, EPI_ISL_1838071, EPI_ISL_1838072, EPI_ISL_1838074, EPI_ISL_1838076, EPI_ISL_1838077, EPI_ISL_1838079, EPI_ISL_1838082, EPI_ISL_1838084, EPI_ISL_1838085, EPI_ISL_1838087, EPI_ISL_1838089, EPI_ISL_1838090, EPI_ISL_1838092, EPI_ISL_1838093, EPI_ISL_1838096, EPI_ISL_1838098, EPI_ISL_1838099, EPI_ISL_1838101, EPI_ISL_1838103, EPI_ISL_1838104, EPI_ISL_1838105, EPI_ISL_1838107, EPI_ISL_1838108, EPI_ISL_1838110, EPI_ISL_1838112, EPI_ISL_1838113, EPI_ISL_1838115, EPI_ISL_1838116, EPI_ISL_1838118, EPI_ISL_1838120, EPI_ISL_1838121, EPI_ISL_1838124, EPI_ISL_1838127, EPI_ISL_1838130, EPI_ISL_1838133, EPI_ISL_1838135, EPI_ISL_1838137, EPI_ISL_1838138, EPI_ISL_1838141, EPI_ISL_1838144, EPI_ISL_1838146, EPI_ISL_1838147, EPI_ISL_1838150, EPI_ISL_1838154, EPI_ISL_1838157, EPI_ISL_1838160, EPI_ISL_1838163, EPI_ISL_1838167, EPI_ISL_1838170, EPI_ISL_1838174, EPI_ISL_1838177, EPI_ISL_1838180, EPI_ISL_1838181, EPI_ISL_1838183, EPI_ISL_1838184, EPI_ISL_1838187, EPI_ISL_1838190, EPI_ISL_1838192, EPI_ISL_1838193, EPI_ISL_1838197, EPI_ISL_1838199, EPI_ISL_1838202, EPI_ISL_1838204, EPI_ISL_1838206, EPI_ISL_1838208, EPI_ISL_1838211, EPI_ISL_1838213, EPI_ISL_1838216, EPI_ISL_1838218, EPI_ISL_1838221, EPI_ISL_1838223, EPI_ISL_1838226, EPI_ISL_1838228, EPI_ISL_1838231, EPI_ISL_1838233, EPI_ISL_1838236, EPI_ISL_1838238, EPI_ISL_1838240, EPI_ISL_1838243, EPI_ISL_1838247, EPI_ISL_1838249, EPI_ISL_1838252, EPI_ISL_1838255, EPI_ISL_1838258, EPI_ISL_1838261, EPI_ISL_1838263, EPI_ISL_1838266, EPI_ISL_1838268, EPI_ISL_1838270, EPI_ISL_1838273, EPI_ISL_1838276, EPI_ISL_1838279, EPI_ISL_1838281, EPI_ISL_1838283, EPI_ISL_1838286, EPI_ISL_1838289, EPI_ISL_1838291, EPI_ISL_1838294, EPI_ISL_1838297, EPI_ISL_1838300, EPI_ISL_1838302, EPI_ISL_1838305, EPI_ISL_1838308, EPI_ISL_1838310, EPI_ISL_1838313, EPI_ISL_1838316, EPI_ISL_1838319, EPI_ISL_1838321, EPI_ISL_1838324, EPI_ISL_1838326, EPI_ISL_1838329, EPI_ISL_1838331, EPI_ISL_1838334, EPI_ISL_1838336, EPI_ISL_1838339, EPI_ISL_1838341, EPI_ISL_1838344, EPI_ISL_1838347, EPI_ISL_1838349, EPI_ISL_1838351, EPI_ISL_1838353, EPI_ISL_1838354, EPI_ISL_1838355, EPI_ISL_1838357, EPI_ISL_1838358, EPI_ISL_1838360, EPI_ISL_1838363, EPI_ISL_1838365, EPI_ISL_1838368, EPI_ISL_1838371, EPI_ISL_1838373, EPI_ISL_1838375, EPI_ISL_1838376, EPI_ISL_1838378, EPI_ISL_1838379, EPI_ISL_1838381, EPI_ISL_1838382, EPI_ISL_1838383, EPI_ISL_1838386, EPI_ISL_1838388, EPI_ISL_1838391, EPI_ISL_1838394, EPI_ISL_1838396, EPI_ISL_1838399, EPI_ISL_1838401, EPI_ISL_1838404, EPI_ISL_1838407, EPI_ISL_1838409, EPI_ISL_1838412, EPI_ISL_1838415, EPI_ISL_1838416, EPI_ISL_1838417, EPI_ISL_1838419, EPI_ISL_1838420, EPI_ISL_1838421, EPI_ISL_1838423, EPI_ISL_1838424, EPI_ISL_1838425, EPI_ISL_1838427, EPI_ISL_1838428, EPI_ISL_1838429, EPI_ISL_1838431, EPI_ISL_1838432, EPI_ISL_1838433, EPI_ISL_1838434, EPI_ISL_1838436, EPI_ISL_1838437, EPI_ISL_1838438, EPI_ISL_1838440, EPI_ISL_1838441, EPI_ISL_1838442, EPI_ISL_1838444, EPI_ISL_1838447, EPI_ISL_1838449, EPI_ISL_1838451 | | | |
| see above | Aegis Sciences Corporation | Centers for Disease Control and Prevention Division of Viral Diseases, Pathogen Discovery | Dakota Howard, Dhwani Batra, Peter W. Cook, Kara Moser, Adrian Paskey, Jason Caravas, Benjamin Rambo-Martin, Shatavia Morrison, Christopher Gulvick, Scott Sammons, Yvette Unoarumhi, Darlene Wagner, Matthew Schmerer, Cyndi Clark, Patrick Campbell, Rob Case, Vikramsinha Ghorpade, Holly Houdeshell, Ola Kvalvaag, Dillon Nall, Ethan Sanders, Alec Vest, Shaun Westlund, Matthew Hardison, Clinton R. Paden, Duncan MacCannell |
| EPI_ISL_1838921, EPI_ISL_1838922, EPI_ISL_1838923, EPI_ISL_1838924, EPI_ISL_1838925, EPI_ISL_1838926, EPI_ISL_1838927, EPI_ISL_1838928, EPI_ISL_1838929, EPI_ISL_1838930, EPI_ISL_1838931, EPI_ISL_1838932, EPI_ISL_1838933, EPI_ISL_1838934, EPI_ISL_1838935, EPI_ISL_1838936, EPI_ISL_1838937, EPI_ISL_1838938, EPI_ISL_1838939, EPI_ISL_1838940, EPI_ISL_1838941, EPI_ISL_1838942, EPI_ISL_1838943, EPI_ISL_1838944, EPI_ISL_1838945, EPI_ISL_1838946, EPI_ISL_1838947, EPI_ISL_1838948, EPI_ISL_1838949, EPI_ISL_1838950, EPI_ISL_1838951, EPI_ISL_1838952, EPI_ISL_1838953, EPI_ISL_1838954, EPI_ISL_1838955, EPI_ISL_1838956, EPI_ISL_1838957, EPI_ISL_1838958, EPI_ISL_1838959, EPI_ISL_1838960, EPI_ISL_1838961, EPI_ISL_1838962, EPI_ISL_1838963, EPI_ISL_1838964, EPI_ISL_1838965, EPI_ISL_1838966, EPI_ISL_1838967, EPI_ISL_1838968, EPI_ISL_1838969, EPI_ISL_1838970, EPI_ISL_1838971, EPI_ISL_1838972, EPI_ISL_1838973, EPI_ISL_1838974, EPI_ISL_1838975, EPI_ISL_1838976, EPI_ISL_1838977, EPI_ISL_1838978, EPI_ISL_1838979, EPI_ISL_1838980, EPI_ISL_1838981, EPI_ISL_1838982, EPI_ISL_1838983, EPI_ISL_1838984, EPI_ISL_1838985, EPI_ISL_1838986, EPI_ISL_1838987, EPI_ISL_1838988, EPI_ISL_1838989, EPI_ISL_1838990, EPI_ISL_1838991, EPI_ISL_1838992, EPI_ISL_1838993, EPI_ISL_1838994, EPI_ISL_1838995, EPI_ISL_1838996, EPI_ISL_1838997, EPI_ISL_1838998, EPI_ISL_1838999, EPI_ISL_1839000, EPI_ISL_1839001, EPI_ISL_1839002, EPI_ISL_1839003, EPI_ISL_1839004, EPI_ISL_1839005, EPI_ISL_1839006, EPI_ISL_1839007, EPI_ISL_1839008, EPI_ISL_1839009, EPI_ISL_1839010, EPI_ISL_1839011, EPI_ISL_1839012, EPI_ISL_1839013, EPI_ISL_1839014, EPI_ISL_1839015, EPI_ISL_1839016, EPI_ISL_1839017, EPI_ISL_1839018, EPI_ISL_1839019, EPI_ISL_1839020, EPI_ISL_1839021, EPI_ISL_1839022, EPI_ISL_1839023, EPI_ISL_1839024, EPI_ISL_1839025, EPI_ISL_1839026, EPI_ISL_1839027, EPI_ISL_1839028, EPI_ISL_1839029, EPI_ISL_1839030, EPI_ISL_1839031, EPI_ISL_1839032, EPI_ISL_1839033, EPI_ISL_1839034, EPI_ISL_1839035, EPI_ISL_1839036, EPI_ISL_1839037, EPI_ISL_1839038, EPI_ISL_1839039, EPI_ISL_1839040, EPI_ISL_1839041, EPI_ISL_1839042, EPI_ISL_1839043, EPI_ISL_1839044, EPI_ISL_1839045, EPI_ISL_1839046, EPI_ISL_1839047, EPI_ISL_1839048, EPI_ISL_1839049, EPI_ISL_1839050, EPI_ISL_1839051, EPI_ISL_1839052, EPI_ISL_1839053, EPI_ISL_1839054, EPI_ISL_1839055, EPI_ISL_1839056 | | | |
| see above | Quest Diagnostics Incorporated | Centers for Disease Control and Prevention Division of Viral Diseases, Pathogen Discovery | Dakota Howard, Dhwani Batra, Peter W. Cook, Kara Moser, Adrian Paskey, Jason Caravas, Benjamin Rambo-Martin, Shatavia Morrison, Christopher Gulvick, Scott Sammons, Yvette Unoarumhi, Darlene Wagner, Matthew Schmerer, S. H. Rosenthal, A. Gerasimova, R. M. Kagan, B. Anderson, M. Hua, Y. Liu, L.E. Bernstein, K.E. Livingston, A. Perez, I. A. Shlyakhter, R. V. Rolando, R. Owen, P. Tanpaiboon, F. Lacbawan, Clinton R. Paden, Duncan MacCannell |
| EPI_ISL_1839057, EPI_ISL_1839058, EPI_ISL_1839059, EPI_ISL_1839060, EPI_ISL_1839061, EPI_ISL_1839063, EPI_ISL_1839064, EPI_ISL_1839066, EPI_ISL_1839067, EPI_ISL_1839068, EPI_ISL_1839069, EPI_ISL_1839070, EPI_ISL_1839071, EPI_ISL_1839073, EPI_ISL_1839074, EPI_ISL_1839075, EPI_ISL_1839076, EPI_ISL_1839077, EPI_ISL_1839078, EPI_ISL_1839079, EPI_ISL_1839080, EPI_ISL_1839081, EPI_ISL_1839082, EPI_ISL_1839083, EPI_ISL_1839084, EPI_ISL_1839085, EPI_ISL_1839086, EPI_ISL_1839089, EPI_ISL_1839090, EPI_ISL_1839091, EPI_ISL_1839092, EPI_ISL_1839093, EPI_ISL_1839094, EPI_ISL_1839095, EPI_ISL_1839096, EPI_ISL_1839097, EPI_ISL_1839098, EPI_ISL_1839099, EPI_ISL_1839100, EPI_ISL_1839102, EPI_ISL_1839104, EPI_ISL_1839105, EPI_ISL_1839106, EPI_ISL_1839107, EPI_ISL_1839108, EPI_ISL_1839109, EPI_ISL_1839110, EPI_ISL_1839111, EPI_ISL_1839112, EPI_ISL_1839113, EPI_ISL_1839115, EPI_ISL_1839116, EPI_ISL_1839117, EPI_ISL_1839118, EPI_ISL_1839119, EPI_ISL_1839120, EPI_ISL_1839122, EPI_ISL_1839123, EPI_ISL_1839125, EPI_ISL_1839126, EPI_ISL_1839127, EPI_ISL_1839128, EPI_ISL_1839129, EPI_ISL_1839130, EPI_ISL_1839131, EPI_ISL_1839132, EPI_ISL_1839133, EPI_ISL_1839134, EPI_ISL_1839135, EPI_ISL_1839137, EPI_ISL_1839140, EPI_ISL_1839141, EPI_ISL_1839142, EPI_ISL_1839143, EPI_ISL_1839144, EPI_ISL_1839145, EPI_ISL_1839146, EPI_ISL_1839147, EPI_ISL_1839148, EPI_ISL_1839149, EPI_ISL_1839150, EPI_ISL_1839151, EPI_ISL_1839152, EPI_ISL_1839153, EPI_ISL_1839154, EPI_ISL_1839156, EPI_ISL_1839157, EPI_ISL_1839158, EPI_ISL_1839159, EPI_ISL_1839161, EPI_ISL_1839162, EPI_ISL_1839163, EPI_ISL_1839164, EPI_ISL_1839165, EPI_ISL_1839166, EPI_ISL_1839167, EPI_ISL_1839168, EPI_ISL_1839169, EPI_ISL_1839170, EPI_ISL_1839173, EPI_ISL_1839174, EPI_ISL_1839175, EPI_ISL_1839177, EPI_ISL_1839178, EPI_ISL_1839179, EPI_ISL_1839180, EPI_ISL_1839181, EPI_ISL_1839182, EPI_ISL_1839183, EPI_ISL_1839186, EPI_ISL_1839187, EPI_ISL_1839188, EPI_ISL_1839189, EPI_ISL_1839190, EPI_ISL_1839191, EPI_ISL_1839192, EPI_ISL_1839193, EPI_ISL_1839194, EPI_ISL_1839195, EPI_ISL_1839196, EPI_ISL_1839198, EPI_ISL_1839199, EPI_ISL_1839200, EPI_ISL_1839201, EPI_ISL_1839202, EPI_ISL_1839203, EPI_ISL_1839204, EPI_ISL_1839205, EPI_ISL_1839206, EPI_ISL_1839207, EPI_ISL_1839208, EPI_ISL_1839209, EPI_ISL_1839210, EPI_ISL_1839211, EPI_ISL_1839212, EPI_ISL_1839213, EPI_ISL_1839215, EPI_ISL_1839216, EPI_ISL_1839217, EPI_ISL_1839219, EPI_ISL_1839220, EPI_ISL_1839221, EPI_ISL_1839222, EPI_ISL_1839224, EPI_ISL_1839225, EPI_ISL_1839226, EPI_ISL_1839227, EPI_ISL_1839228, EPI_ISL_1839229, EPI_ISL_1839230, EPI_ISL_1839231, EPI_ISL_1839232, EPI_ISL_1839233, EPI_ISL_1839234, EPI_ISL_1839235, EPI_ISL_1839236, EPI_ISL_1839237, EPI_ISL_1839239, EPI_ISL_1839240, EPI_ISL_1839241, EPI_ISL_1839242, EPI_ISL_1839243, EPI_ISL_1839244, EPI_ISL_1839245, EPI_ISL_1839246, EPI_ISL_1839247, EPI_ISL_1839248, EPI_ISL_1839249, EPI_ISL_1839250, EPI_ISL_1839252, EPI_ISL_1839253, EPI_ISL_1839254, EPI_ISL_1839255, EPI_ISL_1839256, EPI_ISL_1839257, EPI_ISL_1839258, EPI_ISL_1839259, EPI_ISL_1839260, EPI_ISL_1839261, EPI_ISL_1839262, EPI_ISL_1839263, EPI_ISL_1839264, EPI_ISL_1839265, EPI_ISL_1839266, EPI_ISL_1839267, EPI_ISL_1839268, EPI_ISL_1839269, EPI_ISL_1839270, EPI_ISL_1839271, EPI_ISL_1839272, EPI_ISL_1839273, EPI_ISL_1839274, EPI_ISL_1839275, EPI_ISL_1839276, EPI_ISL_1839277, EPI_ISL_1839278, EPI_ISL_1839279, EPI_ISL_1839280, EPI_ISL_1839282, EPI_ISL_1839283, EPI_ISL_1839284, EPI_ISL_1839286, EPI_ISL_1839287, EPI_ISL_1839288, EPI_ISL_1839289, EPI_ISL_1839290, EPI_ISL_1839291, EPI_ISL_1839292, EPI_ISL_1839293, EPI_ISL_1839294, EPI_ISL_1839295, EPI_ISL_1839296, EPI_ISL_1839297, EPI_ISL_1839298, EPI_ISL_1839299, EPI_ISL_1839300, EPI_ISL_1839301, EPI_ISL_1839302, EPI_ISL_1839303, EPI_ISL_1839304, EPI_ISL_1839305, EPI_ISL_1839306, EPI_ISL_1839307, EPI_ISL_1839308, EPI_ISL_1839309, EPI_ISL_1839310, EPI_ISL_1839311, EPI_ISL_1839312, EPI_ISL_1839313, EPI_ISL_1839314, EPI_ISL_1839315, EPI_ISL_1839316, EPI_ISL_1839317, EPI_ISL_1839318, EPI_ISL_1839319, EPI_ISL_1839320, EPI_ISL_1839321, EPI_ISL_1839322, EPI_ISL_1839323, EPI_ISL_1839324, EPI_ISL_1839325, EPI_ISL_1839326, EPI_ISL_1839327, EPI_ISL_1839328, EPI_ISL_1839329, EPI_ISL_1839330, EPI_ISL_1839331, EPI_ISL_1839332, EPI_ISL_1839333, EPI_ISL_1839334, EPI_ISL_1839335, EPI_ISL_1839336, EPI_ISL_1839337, EPI_ISL_1839338, EPI_ISL_1839339, EPI_ISL_1839340, EPI_ISL_1839341, EPI_ISL_1839342, EPI_ISL_1839343, EPI_ISL_1839344, EPI_ISL_1839345, EPI_ISL_1839347, EPI_ISL_1839348, EPI_ISL_1839349, EPI_ISL_1839350, EPI_ISL_1839351, EPI_ISL_1839352, EPI_ISL_1839353, EPI_ISL_1839354, EPI_ISL_1839355, EPI_ISL_1839356, EPI_ISL_1839358, EPI_ISL_1839359, EPI_ISL_1839361, EPI_ISL_1839363, EPI_ISL_1839364, EPI_ISL_1839365, EPI_ISL_1839366, EPI_ISL_1839367, EPI_ISL_1839368, EPI_ISL_1839371, EPI_ISL_1839373, EPI_ISL_1839374, EPI_ISL_1839375, EPI_ISL_1839376, EPI_ISL_1839377, EPI_ISL_1839378, EPI_ISL_1839379, EPI_ISL_1839380, EPI_ISL_1839381, EPI_ISL_1839382, EPI_ISL_1839384, EPI_ISL_1839386, EPI_ISL_1839387, EPI_ISL_1839388, EPI_ISL_1839389, EPI_ISL_1839390, EPI_ISL_1839391, EPI_ISL_1839392, EPI_ISL_1839393, EPI_ISL_1839394, EPI_ISL_1839395, EPI_ISL_1839396, EPI_ISL_1839397, EPI_ISL_1839398, EPI_ISL_1839399, EPI_ISL_1839400, EPI_ISL_1839402, EPI_ISL_1839404, EPI_ISL_1839405, EPI_ISL_1839406, EPI_ISL_1839407, EPI_ISL_1839411, EPI_ISL_1839412, EPI_ISL_1839413, EPI_ISL_1839414, EPI_ISL_1839415, EPI_ISL_1839416, EPI_ISL_1839417, EPI_ISL_1839418, EPI_ISL_1839420, EPI_ISL_1839421, EPI_ISL_1839422, EPI_ISL_1839424, EPI_ISL_1839425, EPI_ISL_1839426, EPI_ISL_1839427, EPI_ISL_1839428, EPI_ISL_1839429, EPI_ISL_1839430, EPI_ISL_1839432, EPI_ISL_1839433, EPI_ISL_1839434, EPI_ISL_1839435, EPI_ISL_1839436, EPI_ISL_1839437, EPI_ISL_1839439, EPI_ISL_1839440, EPI_ISL_1839441, EPI_ISL_1839442, EPI_ISL_1839443, EPI_ISL_1839444, EPI_ISL_1839445, EPI_ISL_1839446, EPI_ISL_1839447, EPI_ISL_1839448, EPI_ISL_1839449, EPI_ISL_1839451, EPI_ISL_1839452, EPI_ISL_1839453, EPI_ISL_1839454, EPI_ISL_1839455, EPI_ISL_1839457, EPI_ISL_1839458, EPI_ISL_1839459, EPI_ISL_1839460, EPI_ISL_1839461, EPI_ISL_1839462, EPI_ISL_1839463, EPI_ISL_1839464, EPI_ISL_1839466, EPI_ISL_1839468, EPI_ISL_1839469, EPI_ISL_1839470, EPI_ISL_1839471, EPI_ISL_1839473, EPI_ISL_1839474, EPI_ISL_1839475, EPI_ISL_1839476, EPI_ISL_1839477, EPI_ISL_1839481, EPI_ISL_1839487, EPI_ISL_1839489, EPI_ISL_1839493, EPI_ISL_1839498, EPI_ISL_1839499, EPI_ISL_1839500, EPI_ISL_1839502, EPI_ISL_1839503, EPI_ISL_1839505, EPI_ISL_1839506, EPI_ISL_1839508, EPI_ISL_1839512, EPI_ISL_1839513, EPI_ISL_1839514, EPI_ISL_1839516, EPI_ISL_1839517, EPI_ISL_1839518, EPI_ISL_1839519, EPI_ISL_1839521, EPI_ISL_1839522, EPI_ISL_1839523, EPI_ISL_1839524, EPI_ISL_1839525, EPI_ISL_1839527, EPI_ISL_1839528, EPI_ISL_1839529, EPI_ISL_1839530, EPI_ISL_1839531, EPI_ISL_1839532, EPI_ISL_1839533, EPI_ISL_1839534, EPI_ISL_1839535, EPI_ISL_1839536, EPI_ISL_1839537, EPI_ISL_1839538, EPI_ISL_1839539, EPI_ISL_1839540, EPI_ISL_1839541, EPI_ISL_1839542, EPI_ISL_1839543, EPI_ISL_1839544, EPI_ISL_1839545, EPI_ISL_1839546, EPI_ISL_1839547, EPI_ISL_1839548, EPI_ISL_1839549, EPI_ISL_1839550, EPI_ISL_1839552, EPI_ISL_1839553, EPI_ISL_1839554, EPI_ISL_1839555, EPI_ISL_1839556, EPI_ISL_1839557, EPI_ISL_1839558, EPI_ISL_1839559, EPI_ISL_1839560, EPI_ISL_1839561, EPI_ISL_1839562, EPI_ISL_1839563, EPI_ISL_1839564, EPI_ISL_1839565, EPI_ISL_1839566, EPI_ISL_1839567, EPI_ISL_1839569, EPI_ISL_1839570, EPI_ISL_1839571, EPI_ISL_1839572, EPI_ISL_1839573, EPI_ISL_1839574, EPI_ISL_1839575, EPI_ISL_1839576, EPI_ISL_1839577, EPI_ISL_1839578, EPI_ISL_1839579, EPI_ISL_1839580, EPI_ISL_1839581, EPI_ISL_1839582, EPI_ISL_1839583, EPI_ISL_1839584, EPI_ISL_1839585, EPI_ISL_1839586, EPI_ISL_1839587, EPI_ISL_1839588, EPI_ISL_1839589, EPI_ISL_1839590, EPI_ISL_1839591, EPI_ISL_1839592, EPI_ISL_1839593, EPI_ISL_1839594, EPI_ISL_1839595, EPI_ISL_1839596, EPI_ISL_1839597, EPI_ISL_1839598, EPI_ISL_1839599, EPI_ISL_1839600, EPI_ISL_1839601, EPI_ISL_1839602, EPI_ISL_1839603, EPI_ISL_1839604, EPI_ISL_1839605, EPI_ISL_1839606, EPI_ISL_1839607, EPI_ISL_1839608, EPI_ISL_1839609, EPI_ISL_1839610, EPI_ISL_1839611, EPI_ISL_1839613, EPI_ISL_1839614, EPI_ISL_1839615, EPI_ISL_1839616, EPI_ISL_1839617, EPI_ISL_1839618, EPI_ISL_1839619, EPI_ISL_1839620, EPI_ISL_1839621, EPI_ISL_1839623, EPI_ISL_1839624, EPI_ISL_1839625, EPI_ISL_1839626, EPI_ISL_1839627, EPI_ISL_1839628, EPI_ISL_1839629, EPI_ISL_1839630, EPI_ISL_1839632, EPI_ISL_1839633, EPI_ISL_1839634, EPI_ISL_1839635, EPI_ISL_1839636, EPI_ISL_1839637, EPI_ISL_1839638, EPI_ISL_1839639, EPI_ISL_1839641, EPI_ISL_1839642, EPI_ISL_1839644, EPI_ISL_1839645, EPI_ISL_1839646, EPI_ISL_1839647, EPI_ISL_1839648, EPI_ISL_1839649, EPI_ISL_1839650, EPI_ISL_1839651, EPI_ISL_1839652, EPI_ISL_1839653, EPI_ISL_1839654, EPI_ISL_1839655, EPI_ISL_1839656, EPI_ISL_1839658, EPI_ISL_1839659, EPI_ISL_1839660, EPI_ISL_1839662, EPI_ISL_1839663, EPI_ISL_1839664, EPI_ISL_1839665, EPI_ISL_1839666, EPI_ISL_1839667, EPI_ISL_1839668, EPI_ISL_1839669, EPI_ISL_1839670, EPI_ISL_1839672, EPI_ISL_1839674, EPI_ISL_1839675, EPI_ISL_1839676, EPI_ISL_1839677, EPI_ISL_1839679, EPI_ISL_1839680, EPI_ISL_1839681, EPI_ISL_1839682, EPI_ISL_1839683, EPI_ISL_1839684, EPI_ISL_1839685, EPI_ISL_1839686, EPI_ISL_1839687, EPI_ISL_1839688, EPI_ISL_1839689, EPI_ISL_1839690, EPI_ISL_1839691, EPI_ISL_1839692, EPI_ISL_1839693, EPI_ISL_1839694, EPI_ISL_1839695, EPI_ISL_1839696, EPI_ISL_1839697, EPI_ISL_1839699, EPI_ISL_1839700, EPI_ISL_1839702, EPI_ISL_1839703, EPI_ISL_1839705, EPI_ISL_1839706, EPI_ISL_1839707, EPI_ISL_1839708, EPI_ISL_1839709, EPI_ISL_1839710, EPI_ISL_1839711, EPI_ISL_1839712, EPI_ISL_1839713, EPI_ISL_1839714, EPI_ISL_1839715, EPI_ISL_1839717, EPI_ISL_1839718, EPI_ISL_1839719, EPI_ISL_1839720, EPI_ISL_1839721, EPI_ISL_1839722, EPI_ISL_1839723, EPI_ISL_1839724, EPI_ISL_1839725, EPI_ISL_1839726, EPI_ISL_1839727, EPI_ISL_1839728, EPI_ISL_1839729, EPI_ISL_1839730, EPI_ISL_1839731, EPI_ISL_1839732, EPI_ISL_1839733, EPI_ISL_1839734, EPI_ISL_1839735, EPI_ISL_1839736, EPI_ISL_1839737, EPI_ISL_1839738, EPI_ISL_1839739, EPI_ISL_1839740, EPI_ISL_1839741, EPI_ISL_1839742, EPI_ISL_1839743, EPI_ISL_1839744, EPI_ISL_1839745, EPI_ISL_1839746, EPI_ISL_1839747, EPI_ISL_1839748, EPI_ISL_1839749, EPI_ISL_1839750, EPI_ISL_1839751, EPI_ISL_1839752, EPI_ISL_1839754, EPI_ISL_1839755, EPI_ISL_1839757, EPI_ISL_1839758, EPI_ISL_1839759, EPI_ISL_1839760, EPI_ISL_1839761, EPI_ISL_1839762, EPI_ISL_1839763, EPI_ISL_1839764, EPI_ISL_1839766, EPI_ISL_1839767, EPI_ISL_1839768, EPI_ISL_1839769, EPI_ISL_1839771, EPI_ISL_1839772, EPI_ISL_1839773, EPI_ISL_1839774, EPI_ISL_1839775, EPI_ISL_1839776, EPI_ISL_1839777, EPI_ISL_1839778, EPI_ISL_1839779, EPI_ISL_1839780, EPI_ISL_1839781, EPI_ISL_1839782, EPI_ISL_1839783, EPI_ISL_1839784, EPI_ISL_1839785, EPI_ISL_1839787, EPI_ISL_1839788, EPI_ISL_1839789, EPI_ISL_1839790, EPI_ISL_1839792, EPI_ISL_1839793, EPI_ISL_1839794, EPI_ISL_1839795, EPI_ISL_1839796, EPI_ISL_1839797, EPI_ISL_1839798, EPI_ISL_1839799, EPI_ISL_1839800, EPI_ISL_1839801, EPI_ISL_1839802, EPI_ISL_1839804, EPI_ISL_1839805, EPI_ISL_1839806, EPI_ISL_1839807, EPI_ISL_1839808, EPI_ISL_1839810, EPI_ISL_1839811, EPI_ISL_1839812, EPI_ISL_1839813, EPI_ISL_1839814, EPI_ISL_1839815, EPI_ISL_1839816, EPI_ISL_1839817, EPI_ISL_1839818, EPI_ISL_1839819, EPI_ISL_1839820, EPI_ISL_1839821, EPI_ISL_1839822, EPI_ISL_1839823, EPI_ISL_1839824, EPI_ISL_1839825, EPI_ISL_1839826, EPI_ISL_1839828, EPI_ISL_1839829, EPI_ISL_1839830, EPI_ISL_1839831, EPI_ISL_1839832, EPI_ISL_1839833, EPI_ISL_1839834, EPI_ISL_1839835, EPI_ISL_1839836, EPI_ISL_1839837, EPI_ISL_1839838, EPI_ISL_1839839, EPI_ISL_1839840, EPI_ISL_1839841, EPI_ISL_1839842, EPI_ISL_1839843, EPI_ISL_1839844, EPI_ISL_1839845, EPI_ISL_1839846, EPI_ISL_1839847, EPI_ISL_1839848, EPI_ISL_1839849, EPI_ISL_1839850, EPI_ISL_1839851, EPI_ISL_1839852, EPI_ISL_1839853, EPI_ISL_1839855, EPI_ISL_1839856, EPI_ISL_1839857, EPI_ISL_1839858, EPI_ISL_1839859, EPI_ISL_1839860, EPI_ISL_1839861, EPI_ISL_1839862, EPI_ISL_1839863, EPI_ISL_1839864, EPI_ISL_1839866, EPI_ISL_1839867, EPI_ISL_1839868, EPI_ISL_1839869, EPI_ISL_1839870, EPI_ISL_1839871, EPI_ISL_1839872, EPI_ISL_1839873, EPI_ISL_1839874, EPI_ISL_1839875, EPI_ISL_1839876, EPI_ISL_1839877, EPI_ISL_1839878, EPI_ISL_1839879, EPI_ISL_1839880, EPI_ISL_1839881, EPI_ISL_1839882, EPI_ISL_1839883, EPI_ISL_1839884, EPI_ISL_1839885, EPI_ISL_1839886, EPI_ISL_1839888, EPI_ISL_1839889, EPI_ISL_1839890, EPI_ISL_1839891, EPI_ISL_1839892, EPI_ISL_1839893, EPI_ISL_1839894, EPI_ISL_1839895, EPI_ISL_1839896, EPI_ISL_1839897, EPI_ISL_1839898, EPI_ISL_1839899, EPI_ISL_1839900, EPI_ISL_1839901, EPI_ISL_1839902, EPI_ISL_1839903, EPI_ISL_1839904, EPI_ISL_1839905, EPI_ISL_1839906, EPI_ISL_1839907, EPI_ISL_1839908, EPI_ISL_1839909, EPI_ISL_1839910, EPI_ISL_1839911, EPI_ISL_1839912, EPI_ISL_1839913, EPI_ISL_1839914, EPI_ISL_1839915, EPI_ISL_1839916, EPI_ISL_1839917, EPI_ISL_1839918, EPI_ISL_1839919, EPI_ISL_1839920, EPI_ISL_1839921, EPI_ISL_1839923, EPI_ISL_1839924, EPI_ISL_1839925, EPI_ISL_1839926, EPI_ISL_1839927, EPI_ISL_1839928, EPI_ISL_1839930, EPI_ISL_1839931, EPI_ISL_1839932, EPI_ISL_1839936, EPI_ISL_1839937, EPI_ISL_1839938, EPI_ISL_1839939, EPI_ISL_1839940, EPI_ISL_1839941, EPI_ISL_1839942, EPI_ISL_1839943, EPI_ISL_1839944, EPI_ISL_1839945, EPI_ISL_1839946, EPI_ISL_1839947, EPI_ISL_1839948, EPI_ISL_1839949, EPI_ISL_1839951, EPI_ISL_1839952, EPI_ISL_1839953, EPI_ISL_1839954, EPI_ISL_1839955, EPI_ISL_1839956, EPI_ISL_1839957, EPI_ISL_1839958, EPI_ISL_1839959, EPI_ISL_1839961, EPI_ISL_1839962, EPI_ISL_1839963, EPI_ISL_1839964, EPI_ISL_1839965, EPI_ISL_1839966, EPI_ISL_1839967, EPI_ISL_1839968, EPI_ISL_1839969, EPI_ISL_1839970, EPI_ISL_1839971, EPI_ISL_1839972, EPI_ISL_1839973, EPI_ISL_1839974, EPI_ISL_1839975, EPI_ISL_1839977, EPI_ISL_1839978, EPI_ISL_1839979, EPI_ISL_1839980, EPI_ISL_1839981, EPI_ISL_1839983, EPI_ISL_1839984, EPI_ISL_1839985, EPI_ISL_1839986, EPI_ISL_1839987, EPI_ISL_1839989, EPI_ISL_1839990, EPI_ISL_1839991, EPI_ISL_1839992, EPI_ISL_1839993, EPI_ISL_1839994, EPI_ISL_1839996, EPI_ISL_1839997, EPI_ISL_1839998, EPI_ISL_1839999, EPI_ISL_1840000, EPI_ISL_1840001, EPI_ISL_1840002, EPI_ISL_1840003, EPI_ISL_1840004, EPI_ISL_1840005, EPI_ISL_1840006, EPI_ISL_1840007, EPI_ISL_1840008, EPI_ISL_1840009, EPI_ISL_1840010, EPI_ISL_1840011, EPI_ISL_1840012, EPI_ISL_1840013, EPI_ISL_1840014, EPI_ISL_1840016, EPI_ISL_1840017, EPI_ISL_1840019, EPI_ISL_1840021, EPI_ISL_1840022, EPI_ISL_1840023, EPI_ISL_1840024, EPI_ISL_1840025, EPI_ISL_1840027, EPI_ISL_1840028, EPI_ISL_1840029, EPI_ISL_1840030, EPI_ISL_1840031, EPI_ISL_1840032, EPI_ISL_1840033, EPI_ISL_1840034, EPI_ISL_1840035, EPI_ISL_1840036, EPI_ISL_1840037, EPI_ISL_1840038, EPI_ISL_1840039, EPI_ISL_1840040, EPI_ISL_1840041, EPI_ISL_1840042, EPI_ISL_1840043, EPI_ISL_1840044, EPI_ISL_1840045, EPI_ISL_1840046, EPI_ISL_1840048, EPI_ISL_1840049, EPI_ISL_1840050, EPI_ISL_1840051, EPI_ISL_1840052, EPI_ISL_1840054, EPI_ISL_1840056, EPI_ISL_1840057, EPI_ISL_1840058, EPI_ISL_1840059, EPI_ISL_1840060, EPI_ISL_1840061, EPI_ISL_1840062, EPI_ISL_1840063, EPI_ISL_1840064, EPI_ISL_1840065, EPI_ISL_1840066, EPI_ISL_1840067, EPI_ISL_1840068, EPI_ISL_1840069, EPI_ISL_1840071, EPI_ISL_1840072, EPI_ISL_1840073, EPI_ISL_1840074, EPI_ISL_1840075, EPI_ISL_1840076, EPI_ISL_1840078, EPI_ISL_1840079, EPI_ISL_1840080, EPI_ISL_1840081, EPI_ISL_1840082, EPI_ISL_1840083, EPI_ISL_1840084, EPI_ISL_1840085, EPI_ISL_1840087, EPI_ISL_1840088, EPI_ISL_1840089, EPI_ISL_1840090, EPI_ISL_1840091, EPI_ISL_1840092, EPI_ISL_1840093, EPI_ISL_1840094, EPI_ISL_1840095, EPI_ISL_1840096, EPI_ISL_1840097, EPI_ISL_1840098, EPI_ISL_1840100, EPI_ISL_1840101, EPI_ISL_1840102, EPI_ISL_1840103, EPI_ISL_1840104, EPI_ISL_1840105, EPI_ISL_1840106, EPI_ISL_1840107, EPI_ISL_1840109, EPI_ISL_1840110, EPI_ISL_1840111, EPI_ISL_1840112, EPI_ISL_1840113, EPI_ISL_1840114, EPI_ISL_1840115, EPI_ISL_1840116, EPI_ISL_1840117, EPI_ISL_1840118, EPI_ISL_1840119, EPI_ISL_1840120, EPI_ISL_1840121, EPI_ISL_1840122, EPI_ISL_1840123, EPI_ISL_1840125, EPI_ISL_1840126, EPI_ISL_1840128, EPI_ISL_1840130, EPI_ISL_1840131, EPI_ISL_1840133, EPI_ISL_1840135, EPI_ISL_1840136, EPI_ISL_1840138, EPI_ISL_1840139, EPI_ISL_1840141, EPI_ISL_1840144, EPI_ISL_1840145, EPI_ISL_1840148, EPI_ISL_1840150, EPI_ISL_1840152, EPI_ISL_1840153, EPI_ISL_1840155, EPI_ISL_1840156, EPI_ISL_1840157, EPI_ISL_1840160, EPI_ISL_1840161, EPI_ISL_1840164, EPI_ISL_1840165, EPI_ISL_1840167 | | | |
| see above | Fulgent Genetics | Centers for Disease Control and Prevention Division of Viral Diseases, Pathogen Discovery | Dakota Howard, Dhwani Batra, Peter W. Cook, Kara Moser, Adrian Paskey, Jason Caravas, Benjamin Rambo-Martin, Shatavia Morrison, Christopher Gulvick, Scott Sammons, Yvette Unoarumhi, Darlene Wagner, Matthew Schmerer, Harry Gao, Mickey Li, John Gao, Joseph Fierro, Benafsh Sapra, Becky Tsai, Yan Meng, Doreen Ng, James Xie, Clinton R. Paden, Duncan MacCannell |
| EPI_ISL_1840252, EPI_ISL_1840253, EPI_ISL_1840254, EPI_ISL_1840255, EPI_ISL_1840256, EPI_ISL_1840257, EPI_ISL_1840258, EPI_ISL_1840259, EPI_ISL_1840260, EPI_ISL_1840261, EPI_ISL_1840262, EPI_ISL_1840263, EPI_ISL_1840264, EPI_ISL_1840265, EPI_ISL_1840266, EPI_ISL_1840267, EPI_ISL_1840268, EPI_ISL_1840269, EPI_ISL_1840270, EPI_ISL_1840271, EPI_ISL_1840272, EPI_ISL_1840273, EPI_ISL_1840274, EPI_ISL_1840275, EPI_ISL_1840276, EPI_ISL_1840277, EPI_ISL_1840278, EPI_ISL_1840279, EPI_ISL_1840280, EPI_ISL_1840281, EPI_ISL_1840282, EPI_ISL_1840283, EPI_ISL_1840284, EPI_ISL_1840285, EPI_ISL_1840286, EPI_ISL_1840287, EPI_ISL_1840288, EPI_ISL_1840289, EPI_ISL_1840290, EPI_ISL_1840291, EPI_ISL_1840292, EPI_ISL_1840293, EPI_ISL_1840294, EPI_ISL_1840295, EPI_ISL_1840296, EPI_ISL_1840297, EPI_ISL_1840298, EPI_ISL_1840299, EPI_ISL_1840300, EPI_ISL_1840301, EPI_ISL_1840302, EPI_ISL_1840303, EPI_ISL_1840304, EPI_ISL_1840305, EPI_ISL_1840306, EPI_ISL_1840307, EPI_ISL_1840308, EPI_ISL_1840309, EPI_ISL_1840310, EPI_ISL_1840311, EPI_ISL_1840312, EPI_ISL_1840313, EPI_ISL_1840314, EPI_ISL_1840315, EPI_ISL_1840316, EPI_ISL_1840317 | | | |
| see above | Quest Diagnostics Incorporated | Centers for Disease Control and Prevention Division of Viral Diseases, Pathogen Discovery | Dakota Howard, Dhwani Batra, Peter W. Cook, Kara Moser, Adrian Paskey, Jason Caravas, Benjamin Rambo-Martin, Shatavia Morrison, Christopher Gulvick, Scott Sammons, Yvette Unoarumhi, Darlene Wagner, Matthew Schmerer, S. H. Rosenthal, A. Gerasimova, R. M. Kagan, B. Anderson, M. Hua, Y. Liu, L.E. Bernstein, K.E. Livingston, A. Perez, I. A. Shlyakhter, R. V. Rolando, R. Owen, P. Tanpaiboon, F. Lacbawan, Clinton R. Paden, Duncan MacCannell |
| EPI_ISL_1840318 | MAWD Pathology Group, PA | MAWD Pathology Group, PA | Pradip Manna, Ph D, Kelli Fayard, Shelby Herner, London Collins, Kelsi Eastman |
| EPI_ISL_1840319, EPI_ISL_1840320, EPI_ISL_1840321, EPI_ISL_1840322, EPI_ISL_1840323, EPI_ISL_1840324, EPI_ISL_1840325, EPI_ISL_1840326, EPI_ISL_1840327, EPI_ISL_1840328, EPI_ISL_1840329, EPI_ISL_1840330, EPI_ISL_1840331, EPI_ISL_1840332, EPI_ISL_1840333, EPI_ISL_1840334, EPI_ISL_1840335, EPI_ISL_1840336, EPI_ISL_1840337, EPI_ISL_1840338, EPI_ISL_1840339, EPI_ISL_1840340, EPI_ISL_1840341, EPI_ISL_1840342, EPI_ISL_1840343, EPI_ISL_1840344, EPI_ISL_1840345, EPI_ISL_1840346, EPI_ISL_1840347, EPI_ISL_1840348, EPI_ISL_1840349, EPI_ISL_1840350, EPI_ISL_1840351, EPI_ISL_1840352, EPI_ISL_1840353, EPI_ISL_1840354, EPI_ISL_1840355, EPI_ISL_1840356, EPI_ISL_1840357, EPI_ISL_1840358, EPI_ISL_1840359, EPI_ISL_1840360, EPI_ISL_1840361, EPI_ISL_1840362, EPI_ISL_1840363, EPI_ISL_1840364, EPI_ISL_1840365, EPI_ISL_1840366, EPI_ISL_1840367, EPI_ISL_1840368, EPI_ISL_1840369, EPI_ISL_1840370, EPI_ISL_1840371, EPI_ISL_1840372, EPI_ISL_1840373, EPI_ISL_1840374, EPI_ISL_1840375, EPI_ISL_1840376, EPI_ISL_1840377, EPI_ISL_1840378, EPI_ISL_1840379, EPI_ISL_1840380, EPI_ISL_1840381, EPI_ISL_1840382, EPI_ISL_1840383, EPI_ISL_1840384, EPI_ISL_1840385, EPI_ISL_1840386, EPI_ISL_1840387, EPI_ISL_1840388, EPI_ISL_1840389, EPI_ISL_1840390, EPI_ISL_1840391, EPI_ISL_1840392, EPI_ISL_1840393, EPI_ISL_1840394, EPI_ISL_1840395, EPI_ISL_1840396, EPI_ISL_1840397, EPI_ISL_1840398, EPI_ISL_1840399, EPI_ISL_1840400, EPI_ISL_1840401, EPI_ISL_1840402, EPI_ISL_1840403, EPI_ISL_1840404, EPI_ISL_1840405, EPI_ISL_1840406, EPI_ISL_1840407, EPI_ISL_1840408, EPI_ISL_1840409, EPI_ISL_1840410, EPI_ISL_1840411, EPI_ISL_1840412, EPI_ISL_1840413, EPI_ISL_1840414, EPI_ISL_1840415, EPI_ISL_1840416, EPI_ISL_1840417, EPI_ISL_1840418, EPI_ISL_1840419, EPI_ISL_1840420, EPI_ISL_1840421, EPI_ISL_1840422, EPI_ISL_1840423, EPI_ISL_1840424, EPI_ISL_1840425, EPI_ISL_1840426, EPI_ISL_1840427, EPI_ISL_1840428, EPI_ISL_1840429, EPI_ISL_1840430, EPI_ISL_1840431, EPI_ISL_1840432, EPI_ISL_1840433, EPI_ISL_1840434, EPI_ISL_1840435, EPI_ISL_1840436, EPI_ISL_1840437, EPI_ISL_1840438, EPI_ISL_1840439, EPI_ISL_1840440, EPI_ISL_1840441, EPI_ISL_1840442, EPI_ISL_1840443, EPI_ISL_1840444, EPI_ISL_1840445, EPI_ISL_1840446, EPI_ISL_1840447, EPI_ISL_1840448, EPI_ISL_1840449, EPI_ISL_1840450, EPI_ISL_1840451, EPI_ISL_1840452, EPI_ISL_1840453, EPI_ISL_1840454, EPI_ISL_1840455, EPI_ISL_1840456, EPI_ISL_1840457, EPI_ISL_1840458, EPI_ISL_1840459, EPI_ISL_1840460, EPI_ISL_1840461, EPI_ISL_1840462, EPI_ISL_1840463, EPI_ISL_1840464, EPI_ISL_1840465, EPI_ISL_1840466, EPI_ISL_1840467, EPI_ISL_1840468, EPI_ISL_1840469, EPI_ISL_1840470, EPI_ISL_1840471, EPI_ISL_1840472, EPI_ISL_1840473, EPI_ISL_1840474, EPI_ISL_1840475, EPI_ISL_1840476, EPI_ISL_1840477, EPI_ISL_1840478, EPI_ISL_1840479, EPI_ISL_1840480, EPI_ISL_1840481, EPI_ISL_1840482, EPI_ISL_1840483, EPI_ISL_1840484, EPI_ISL_1840486, EPI_ISL_1840488, EPI_ISL_1840489, EPI_ISL_1840491, EPI_ISL_1840493, EPI_ISL_1840494, EPI_ISL_1840496, EPI_ISL_1840498, EPI_ISL_1840500, EPI_ISL_1840502, EPI_ISL_1840503, EPI_ISL_1840505, EPI_ISL_1840506, EPI_ISL_1840508, EPI_ISL_1840510, EPI_ISL_1840511, EPI_ISL_1840513, EPI_ISL_1840515, EPI_ISL_1840518, EPI_ISL_1840520, EPI_ISL_1840522, EPI_ISL_1840525, EPI_ISL_1840526, EPI_ISL_1840528, EPI_ISL_1840531, EPI_ISL_1840532, EPI_ISL_1840534, EPI_ISL_1840537, EPI_ISL_1840538, EPI_ISL_1840540, EPI_ISL_1840543, EPI_ISL_1840544, EPI_ISL_1840546, EPI_ISL_1840548, EPI_ISL_1840549, EPI_ISL_1840551, EPI_ISL_1840554, EPI_ISL_1840555, EPI_ISL_1840558, EPI_ISL_1840559, EPI_ISL_1840561, EPI_ISL_1840563, EPI_ISL_1840564, EPI_ISL_1840566, EPI_ISL_1840567, EPI_ISL_1840569, EPI_ISL_1840570, EPI_ISL_1840573, EPI_ISL_1840575, EPI_ISL_1840577, EPI_ISL_1840578, EPI_ISL_1840581, EPI_ISL_1840582, EPI_ISL_1840584, EPI_ISL_1840586, EPI_ISL_1840589, EPI_ISL_1840590, EPI_ISL_1840592, EPI_ISL_1840594, EPI_ISL_1840596, EPI_ISL_1840598, EPI_ISL_1840600, EPI_ISL_1840602, EPI_ISL_1840604, EPI_ISL_1840606, EPI_ISL_1840607, EPI_ISL_1840609, EPI_ISL_1840611, EPI_ISL_1840614, EPI_ISL_1840616, EPI_ISL_1840618, EPI_ISL_1840619, EPI_ISL_1840621, EPI_ISL_1840623, EPI_ISL_1840625, EPI_ISL_1840626, EPI_ISL_1840628 | | | |
| see above | Quest Diagnostics Incorporated | Centers for Disease Control and Prevention Division of Viral Diseases, Pathogen Discovery | Dakota Howard, Dhwani Batra, Peter W. Cook, Kara Moser, Adrian Paskey, Jason Caravas, Benjamin Rambo-Martin, Shatavia Morrison, Christopher Gulvick, Scott Sammons, Yvette Unoarumhi, Darlene Wagner, Matthew Schmerer, S. H. Rosenthal, A. Gerasimova, R. M. Kagan, B. Anderson, M. Hua, Y. Liu, L.E. Bernstein, K.E. Livingston, A. Perez, I. A. Shlyakhter, R. V. Rolando, R. Owen, P. Tanpaiboon, F. Lacbawan, Clinton R. Paden, Duncan MacCannell |
| EPI_ISL_1840656, EPI_ISL_1840669 | MAWD Pathology Group, PA | MAWD Pathology Group, PA | Pradip Manna, Ph D, Kelli Fayard, Shelby Herner, London Collins, Kelsi Eastman |
| EPI_ISL_1840670, EPI_ISL_1840671, EPI_ISL_1840672, EPI_ISL_1840673, EPI_ISL_1840674, EPI_ISL_1840675, EPI_ISL_1840676, EPI_ISL_1840677, EPI_ISL_1840678, EPI_ISL_1840679, EPI_ISL_1840680, EPI_ISL_1840681, EPI_ISL_1840682, EPI_ISL_1840683, EPI_ISL_1840684, EPI_ISL_1840685, EPI_ISL_1840686, EPI_ISL_1840687, EPI_ISL_1840688, EPI_ISL_1840689, EPI_ISL_1840690, EPI_ISL_1840691, EPI_ISL_1840692 | | | |
| see above | Helix/Illumina | Centers for Disease Control and Prevention Division of Viral Diseases, Pathogen Discovery | Dakota Howard, Dhwani Batra, Peter W. Cook, Kara Moser, Adrian Paskey, Jason Caravas, Benjamin Rambo-Martin, Shatavia Morrison, Christopher Gulvick, Scott Sammons, Yvette Unoarumhi, Darlene Wagner, Matthew Schmerer, Eileen de Feo, Jan Antico, Christine Tran, Matthew Tolentino, Shannon Wickline, Kim Gietzen, Brad Sickler, Jingtao Liu, Eric Allen, Phil Febbo, Nicole L. Washington, Simon White, Geraint Levan, Kelly Schiabor Barrett, Elizabeth Cirulli, Alexandre Bolze, Ary Ascencio, Charlotte Rivera-Garcia, Ryan Cho, Jason Nguyen, Sherry Wang, Jimmy Ramirez, Tyler Cassens, Efren Sandoval, Magnus Isaksson, William Lee, David Becker, Marc Laurent, James Lu, Clinton R. Paden, Duncan MacCannell |
| EPI_ISL_1840693 | Baylor Scott & White-Temple | Baylor Scott & White-Temple | Ari Rao, Linden Morales, Kimberly Walker, Marcus Volz, Shelby Hendrickson |
| EPI_ISL_1840860, EPI_ISL_1840861 | MAWD Pathology Group, PA | MAWD Pathology Group, PA | Pradip Manna, Ph D, Kelli Fayard, Shelby Herner, London Collins, Kelsi Eastman |
| EPI_ISL_1841013, EPI_ISL_1841014, EPI_ISL_1841015, EPI_ISL_1841017, EPI_ISL_1841018, EPI_ISL_1841019, EPI_ISL_1841021, EPI_ISL_1841022, EPI_ISL_1841023, EPI_ISL_1841024, EPI_ISL_1841026, EPI_ISL_1841027, EPI_ISL_1841028, EPI_ISL_1841029, EPI_ISL_1841030, EPI_ISL_1841031, EPI_ISL_1841032, EPI_ISL_1841033, EPI_ISL_1841034, EPI_ISL_1841035, EPI_ISL_1841036, EPI_ISL_1841037, | | | |

| | | | |
|---|---|---|---|
| EPI_ISL_1841040, EPI_ISL_1841041, EPI_ISL_1841042, EPI_ISL_1841043, EPI_ISL_1841044, EPI_ISL_1841045, EPI_ISL_1841046, EPI_ISL_1841047, EPI_ISL_1841048, EPI_ISL_1841049, EPI_ISL_1841050, EPI_ISL_1841051, EPI_ISL_1841052 | | | |
| see above | IN State Department of Health Laboratory Services | IN State Department of Health Laboratory Services | Cassandra Campion, Jamie Yeadon, Brian Pope, Lixia Liu, Kyle Brownlee, Melissa Hindenlang, Mark Glazier |
| EPI_ISL_1841228, EPI_ISL_1841229 | Baylor Scott & White-Temple | Baylor Scott & White-Temple | Ari Rao, Linden Morales, Kimberly Walker, Marcus Volz, Shelby Hendrickson |
| EPI_ISL_1841392 | County of San Diego Health and Human Services Agency | Pathogen Discovery, Respiratory Viruses Branch, Division of Viral Diseases, Centers for Disease Control and Prevention | Ying Tao, Yan Li, Anna Kelleher, Jing Zhang, Anna Montmayeur, Brian Lynch, Krista Queen, Anna Uehara, Peter Cook, Han Jia Justin Ng, Rachel Marine, Clinton R. Paden, Haibin Wang, Mark Burroughs, Justin Lee, Adam Retchless, Suxiang Tong |
| EPI_ISL_1841393 | TN Division of Laboratory Services | Genomics and Discovery, Respiratory Viruses Branch, Division of Viral Diseases, Centers for Disease Control and Prevention | Ying Tao, Yan Li, Anna Kelleher, Jing Zhang, Anna Montmayeur, Brian Lynch, Krista Queen, Anna Uehara, Peter Cook, Han Jia Justin Ng, Rachel Marine, Clinton R. Paden, Haibin Wang, Mark Burroughs, Justin Lee, Adam Retchless, Suxiang Tong |
| EPI_ISL_1842777 | MAWD Pathology Group, PA | MAWD Pathology Group, PA | Pradip Manna, Ph D, Kelli Fayard, Shelby Herner, London Collins, Kelsi Eastman |
| EPI_ISL_1848351 | Oregon State Public Health Laboratory | Oregon State Public Health Laboratory | Rafia Razzaque, Eugene Yeboah, Vanda Makris, Laura Tsaknaridis, John Fontana and Shane Sevey |
| EPI_ISL_1849024, EPI_ISL_1849029, EPI_ISL_1849034, EPI_ISL_1849039, EPI_ISL_1849043, EPI_ISL_1849045, EPI_ISL_1849047, EPI_ISL_1849049, EPI_ISL_1849051, EPI_ISL_1849053, EPI_ISL_1849058, EPI_ISL_1849063, EPI_ISL_1849068, EPI_ISL_1849074, EPI_ISL_1849079, EPI_ISL_1849083, EPI_ISL_1849088, EPI_ISL_1849092, EPI_ISL_1849097, EPI_ISL_1849102, EPI_ISL_1849107, EPI_ISL_1849112, EPI_ISL_1849117, EPI_ISL_1849122, EPI_ISL_1849127, EPI_ISL_1849132, EPI_ISL_1849137, EPI_ISL_1849142, EPI_ISL_1849146, EPI_ISL_1849150, EPI_ISL_1849155, EPI_ISL_1849161, EPI_ISL_1849166, EPI_ISL_1849171, EPI_ISL_1849176, EPI_ISL_1849181, EPI_ISL_1849186, EPI_ISL_1849197, EPI_ISL_1849202, EPI_ISL_1849207, EPI_ISL_1849212, EPI_ISL_1849217, EPI_ISL_1849222, EPI_ISL_1849226, EPI_ISL_1849231, EPI_ISL_1849237, EPI_ISL_1849242, EPI_ISL_1849247, EPI_ISL_1849252, EPI_ISL_1849257, EPI_ISL_1849262, EPI_ISL_1849267, EPI_ISL_1849272, EPI_ISL_1849277, EPI_ISL_1849282, EPI_ISL_1849288, EPI_ISL_1849293, EPI_ISL_1849298, EPI_ISL_1849304, EPI_ISL_1849309, EPI_ISL_1849315, EPI_ISL_1849319, EPI_ISL_1849324, EPI_ISL_1849329, EPI_ISL_1849333, EPI_ISL_1849339, EPI_ISL_1849342, EPI_ISL_1849348, EPI_ISL_1849353, EPI_ISL_1849358, EPI_ISL_1849363, EPI_ISL_1849369, EPI_ISL_1849374, EPI_ISL_1849384, EPI_ISL_1849389, EPI_ISL_1849393, EPI_ISL_1849398, EPI_ISL_1849403, EPI_ISL_1849408, EPI_ISL_1849413, EPI_ISL_1849416, EPI_ISL_1849420, EPI_ISL_1849424, EPI_ISL_1849429, EPI_ISL_1849434, EPI_ISL_1849438, EPI_ISL_1849443, EPI_ISL_1849449, EPI_ISL_1849453, EPI_ISL_1849458, EPI_ISL_1849463, EPI_ISL_1849468, EPI_ISL_1849473, EPI_ISL_1849478, EPI_ISL_1849484, EPI_ISL_1849489, EPI_ISL_1849494, EPI_ISL_1849498, EPI_ISL_1849500, EPI_ISL_1849502, EPI_ISL_1849504, EPI_ISL_1849506, EPI_ISL_1849511, EPI_ISL_1849516, EPI_ISL_1849521, EPI_ISL_1849526, EPI_ISL_1849530, EPI_ISL_1849536, EPI_ISL_1849541, EPI_ISL_1849546, EPI_ISL_1849553, EPI_ISL_1849555, EPI_ISL_1849557, EPI_ISL_1849559, EPI_ISL_1849564, EPI_ISL_1849569, EPI_ISL_1849571, EPI_ISL_1849573, EPI_ISL_1849575, EPI_ISL_1849577, EPI_ISL_1849582, EPI_ISL_1849586, EPI_ISL_1849591, EPI_ISL_1849595, EPI_ISL_1849600, EPI_ISL_1849605, EPI_ISL_1849610, EPI_ISL_1849615, EPI_ISL_1849620, EPI_ISL_1849625, EPI_ISL_1849630, EPI_ISL_1849635, EPI_ISL_1849640, EPI_ISL_1849645, EPI_ISL_1849649, EPI_ISL_1849654, EPI_ISL_1849656, EPI_ISL_1849657, EPI_ISL_1849659, EPI_ISL_1849661, EPI_ISL_1849666, EPI_ISL_1849671, EPI_ISL_1849676, EPI_ISL_1849680, EPI_ISL_1849685, EPI_ISL_1849690, EPI_ISL_1849694, EPI_ISL_1849699, EPI_ISL_1849704, EPI_ISL_1849708, EPI_ISL_1849713, EPI_ISL_1849717, EPI_ISL_1849723, EPI_ISL_1849727, EPI_ISL_1849733, EPI_ISL_1849737, EPI_ISL_1849739, EPI_ISL_1849741, EPI_ISL_1849743, EPI_ISL_1849744, EPI_ISL_1849746, EPI_ISL_1849751, EPI_ISL_1849756, EPI_ISL_1849757, EPI_ISL_1849759, EPI_ISL_1849761, EPI_ISL_1849763, EPI_ISL_1849768, EPI_ISL_1849773, EPI_ISL_1849775, EPI_ISL_1849777, EPI_ISL_1849779, EPI_ISL_1849781, EPI_ISL_1849786, EPI_ISL_1849792, EPI_ISL_1849796, EPI_ISL_1849801, EPI_ISL_1849805, EPI_ISL_1849810, EPI_ISL_1849815, EPI_ISL_1849820, EPI_ISL_1849825, EPI_ISL_1849827, EPI_ISL_1849829, EPI_ISL_1849831, EPI_ISL_1849833, EPI_ISL_1849838, EPI_ISL_1849843, EPI_ISL_1849847, EPI_ISL_1849851, EPI_ISL_1849857, EPI_ISL_1849861, EPI_ISL_1849866, EPI_ISL_1849871, EPI_ISL_1849875, EPI_ISL_1849880, EPI_ISL_1849885, EPI_ISL_1849889, EPI_ISL_1849894, EPI_ISL_1849898, EPI_ISL_1849902, EPI_ISL_1849904, EPI_ISL_1849906, EPI_ISL_1849908, EPI_ISL_1849913, EPI_ISL_1849918, EPI_ISL_1849923, EPI_ISL_1849927, EPI_ISL_1849932, EPI_ISL_1849937, EPI_ISL_1849943, EPI_ISL_1849951, EPI_ISL_1849956, EPI_ISL_1849961, EPI_ISL_1849967, EPI_ISL_1849972, EPI_ISL_1849977, EPI_ISL_1849983, EPI_ISL_1849988, EPI_ISL_1849993, EPI_ISL_1849999, EPI_ISL_1850003, EPI_ISL_1850008, EPI_ISL_1850013, EPI_ISL_1850018, EPI_ISL_1850024, EPI_ISL_1850029, EPI_ISL_1850034, EPI_ISL_1850040, EPI_ISL_1850044, EPI_ISL_1850049, EPI_ISL_1850054, EPI_ISL_1850059, EPI_ISL_1850064, EPI_ISL_1850070, EPI_ISL_1850072, EPI_ISL_1850076, EPI_ISL_1850081, EPI_ISL_1850086, EPI_ISL_1850090, EPI_ISL_1850095, EPI_ISL_1850100, EPI_ISL_1850105, EPI_ISL_1850110, EPI_ISL_1850115, EPI_ISL_1850121, EPI_ISL_1850126, EPI_ISL_1850130, EPI_ISL_1850135, EPI_ISL_1850140, EPI_ISL_1850145, EPI_ISL_1850149, EPI_ISL_1850155, EPI_ISL_1850161, EPI_ISL_1850166, EPI_ISL_1850171, EPI_ISL_1850176, EPI_ISL_1850181, EPI_ISL_1850185, EPI_ISL_1850191, EPI_ISL_1850196, EPI_ISL_1850201, EPI_ISL_1850206, EPI_ISL_1850211, EPI_ISL_1850216, EPI_ISL_1850221, EPI_ISL_1850226, EPI_ISL_1850231, EPI_ISL_1850236, EPI_ISL_1850241, EPI_ISL_1850246, EPI_ISL_1850251, EPI_ISL_1850256, EPI_ISL_1850260, EPI_ISL_1850265, EPI_ISL_1850271, EPI_ISL_1850276, EPI_ISL_1850281, EPI_ISL_1850287, EPI_ISL_1850292, EPI_ISL_1850296, EPI_ISL_1850298, EPI_ISL_1850303, EPI_ISL_1850308, EPI_ISL_1850313, EPI_ISL_1850318, EPI_ISL_1850323, EPI_ISL_1850328, EPI_ISL_1850333, EPI_ISL_1850335, EPI_ISL_1850337, EPI_ISL_1850339, EPI_ISL_1850344, EPI_ISL_1850349, EPI_ISL_1850353, EPI_ISL_1850358, EPI_ISL_1850362, EPI_ISL_1850367, EPI_ISL_1850378, EPI_ISL_1850383, EPI_ISL_1850387, EPI_ISL_1850392, EPI_ISL_1850398, EPI_ISL_1850403, EPI_ISL_1850408, EPI_ISL_1850413, EPI_ISL_1850418, EPI_ISL_1850424, EPI_ISL_1850429, EPI_ISL_1850434, EPI_ISL_1850439, EPI_ISL_1850444, EPI_ISL_1850449, EPI_ISL_1850455, EPI_ISL_1850459, EPI_ISL_1850464, EPI_ISL_1850469, EPI_ISL_1850475, EPI_ISL_1850480, EPI_ISL_1850485, EPI_ISL_1850491, EPI_ISL_1850496, EPI_ISL_1850502, EPI_ISL_1850507, EPI_ISL_1850513, EPI_ISL_1850518, EPI_ISL_1850522, EPI_ISL_1850527, EPI_ISL_1850530, EPI_ISL_1850535 | | | |
| see above | UW Virology Lab | UW Virology Lab | Pavitra Roychoudhury, Hong Xie, Lasata Shrestha, Tien V. Nguyen, Shah Mohamed Bakhash, Michelle Lin, Noah R. Baker, Sean Ellis, Meei-Li Huang, Keith R Jerome, Alexander Greninger |
| EPI_ISL_1853447, EPI_ISL_1853448, EPI_ISL_1853451, EPI_ISL_1853452 | Wyoming Public Health Laboratory | Wyoming Public Health Laboratory | Jim Mildenberger, Wanda Manley, Noah Hull, Taylor Fearing, Lynette Gumbleton, Channing Weber, Ashley Norberg, Chayse Rowley, Marley Goetz, Brian Dominguez, Elliot Thomasson, Sam Britz, Cari Sloma, and Rob Christensen |
| EPI_ISL_1853453 | Avellino Labs | Avellino Labs | Milos Pjanic |
| EPI_ISL_1853455, EPI_ISL_1853457, EPI_ISL_1853459, EPI_ISL_1853460, EPI_ISL_1853461, EPI_ISL_1853462, EPI_ISL_1853463, EPI_ISL_1853464, EPI_ISL_1853465, EPI_ISL_1853466, EPI_ISL_1853468 | | | |
| see above | Wyoming Public Health Laboratory | Wyoming Public Health Laboratory | Jim Mildenberger, Wanda Manley, Noah Hull, Taylor Fearing, Lynette Gumbleton, Channing Weber, Ashley Norberg, Chayse Rowley, Marley Goetz, Brian Dominguez, Elliot Thomasson, Sam Britz, Cari Sloma, and Rob Christensen |
| EPI_ISL_1853470, EPI_ISL_1853565 | Avellino Labs | Avellino Labs | Milos Pjanic |
| EPI_ISL_1854475, EPI_ISL_1854476, EPI_ISL_1854477, EPI_ISL_1854478, EPI_ISL_1854479, EPI_ISL_1854480, EPI_ISL_1854483, EPI_ISL_1854484, EPI_ISL_1854485, EPI_ISL_1854486, EPI_ISL_1854487, EPI_ISL_1854488, EPI_ISL_1854489, EPI_ISL_1854490, EPI_ISL_1854491, EPI_ISL_1854492, EPI_ISL_1854493, EPI_ISL_1854494, EPI_ISL_1854495, EPI_ISL_1854496, EPI_ISL_1854497, EPI_ISL_1854498, EPI_ISL_1854499, EPI_ISL_1854500, EPI_ISL_1854501, EPI_ISL_1854503, EPI_ISL_1854504, EPI_ISL_1854505, EPI_ISL_1854506, EPI_ISL_1854507, EPI_ISL_1854508, EPI_ISL_1854509, EPI_ISL_1854510, EPI_ISL_1854511, EPI_ISL_1854512, EPI_ISL_1854513 | | | |
| see above | Wyoming Public Health Laboratory | Wyoming Public Health Laboratory | Jim Mildenberger, Wanda Manley, Noah Hull, Taylor Fearing, Lynette Gumbleton, Channing Weber, Ashley Norberg, Chayse Rowley, Marley Goetz, Brian Dominguez, Elliot Thomasson, Sam Britz, Cari Sloma, and Rob Christensen |
| EPI_ISL_1854691 | SUNY UPSTATE MEDICAL UNIVERSITY | Wadsworth Center, New York State Department of Health | Kirsten St. George, Daryl M. Lamson, Alexis Russell, Matthew Shudt, Melissa A Leisner, Jonathan Plitnick, Catharine Prussing, Navjot Singh, John Kelly, Erasmus Schneider, Erica Lasek-Nesselquist |
| EPI_ISL_1854692 | ALBANY MEDICAL CENTER | Wadsworth Center, New York State Department of Health | Kirsten St. George, Daryl M. Lamson, Alexis Russell, Matthew Shudt, Melissa A Leisner, Jonathan Plitnick, Catharine Prussing, Navjot Singh, John Kelly, Erasmus Schneider, Erica Lasek-Nesselquist |
| EPI_ISL_1854693, EPI_ISL_1854694, EPI_ISL_1854695 | URMC LABS | Wadsworth Center, New York State Department of Health | Kirsten St. George, Daryl M. Lamson, Alexis Russell, Matthew Shudt, Melissa A Leisner, Jonathan Plitnick, Catharine Prussing, Navjot Singh, John Kelly, Erasmus Schneider, Erica Lasek-Nesselquist |
| EPI_ISL_1854696, EPI_ISL_1854697, EPI_ISL_1854698, EPI_ISL_1854699, EPI_ISL_1854700, EPI_ISL_1854701, EPI_ISL_1854702, EPI_ISL_1854703, EPI_ISL_1854704, EPI_ISL_1854705, EPI_ISL_1854706, EPI_ISL_1854707, EPI_ISL_1854708, EPI_ISL_1854709, EPI_ISL_1854710, EPI_ISL_1854711, EPI_ISL_1854712, EPI_ISL_1854713, EPI_ISL_1854714, EPI_ISL_1854715, EPI_ISL_1854716, EPI_ISL_1854717, EPI_ISL_1854718, EPI_ISL_1854719, EPI_ISL_1854720, EPI_ISL_1854721, EPI_ISL_1854722, EPI_ISL_1854723, EPI_ISL_1854724, EPI_ISL_1854725, EPI_ISL_1854726, EPI_ISL_1854727, EPI_ISL_1854728, EPI_ISL_1854729, EPI_ISL_1854730, EPI_ISL_1854731, EPI_ISL_1854732, EPI_ISL_1854733, EPI_ISL_1854734, EPI_ISL_1854735, EPI_ISL_1854736, EPI_ISL_1854737, EPI_ISL_1854738, EPI_ISL_1854739 | | | |
| see above | NYC Pandemic Response Lab | Wadsworth Center, New York State Department of Health | Kirsten St. George, Daryl M. Lamson, Alexis Russell, Matthew Shudt, Melissa A Leisner, Jonathan Plitnick, Catharine Prussing, Navjot Singh, John Kelly, Erasmus Schneider, Erica Lasek-Nesselquist |
| EPI_ISL_1854740, EPI_ISL_1854741, EPI_ISL_1854742, EPI_ISL_1854743, EPI_ISL_1854744, EPI_ISL_1854745, EPI_ISL_1854746, EPI_ISL_1854747, EPI_ISL_1854748, EPI_ISL_1854749, EPI_ISL_1854750, EPI_ISL_1854751, EPI_ISL_1854752, EPI_ISL_1854753, EPI_ISL_1854754 | | | |
| see above | Wichita State University - Molecular Diagnostics Lab | Kansas Health and Environmental Lab | Mike Grose, Jonathan Barnell, Ben Olsen, and Phil Adam |
| EPI_ISL_1854795, EPI_ISL_1854796, EPI_ISL_1854797 | UNC-CH COVID Surveillance Lab | Jeremy Wang | Jeremy Wang, Alexander Rubinsteyn, Melissa Miller, Corbin Jones, Amy James Loftis, Amir Barzin, Susan Fiscus |
| EPI_ISL_1854798, EPI_ISL_1854799, EPI_ISL_1854800, EPI_ISL_1854801, EPI_ISL_1854802, EPI_ISL_1854803, EPI_ISL_1854804, EPI_ISL_1854805, EPI_ISL_1854806, EPI_ISL_1854807, EPI_ISL_1854808, EPI_ISL_1854809, EPI_ISL_1854810, EPI_ISL_1854811, EPI_ISL_1854812, EPI_ISL_1854813, EPI_ISL_1854814, EPI_ISL_1854815, EPI_ISL_1854816, EPI_ISL_1854817, EPI_ISL_1854818, EPI_ISL_1854819, EPI_ISL_1854820, EPI_ISL_1854821, EPI_ISL_1854822, EPI_ISL_1854823, EPI_ISL_1854824, EPI_ISL_1854825, EPI_ISL_1854826, EPI_ISL_1854827, EPI_ISL_1854828, EPI_ISL_1854829, EPI_ISL_1854830, EPI_ISL_1854831, EPI_ISL_1854832, EPI_ISL_1854833, EPI_ISL_1854834, EPI_ISL_1854835, EPI_ISL_1854836, EPI_ISL_1854837, EPI_ISL_1854838, EPI_ISL_1854839, EPI_ISL_1854840, EPI_ISL_1854841, EPI_ISL_1854842, EPI_ISL_1854843, EPI_ISL_1854844, EPI_ISL_1854845, EPI_ISL_1854846, EPI_ISL_1854847, EPI_ISL_1854848, EPI_ISL_1854849, EPI_ISL_1854850, EPI_ISL_1854851, EPI_ISL_1854852, EPI_ISL_1854853, EPI_ISL_1854854, EPI_ISL_1854855, EPI_ISL_1854856, EPI_ISL_1854857, EPI_ISL_1854858, EPI_ISL_1854859, EPI_ISL_1854860, EPI_ISL_1854861, EPI_ISL_1854862, EPI_ISL_1854863, EPI_ISL_1854864, EPI_ISL_1854865, EPI_ISL_1854866, EPI_ISL_1854867, EPI_ISL_1854868, EPI_ISL_1854869 | | | |
| see above | Clinical Molecular Microbiology Laboratory, UNC Hospitals | Jeremy Wang | Jeremy Wang, Alexander Rubinsteyn, Colleen Rice, Jason Smedberg, Shawn Hawken, Melissa Miller, Corbin Jones, Robert Hagan |
| EPI_ISL_1854870, EPI_ISL_1854871, EPI_ISL_1854872, EPI_ISL_1854873, EPI_ISL_1854874, EPI_ISL_1854875, EPI_ISL_1854876, EPI_ISL_1854877, EPI_ISL_1854878, EPI_ISL_1854879, EPI_ISL_1854880, EPI_ISL_1854881, EPI_ISL_1854882, EPI_ISL_1854883, EPI_ISL_1854884, EPI_ISL_1854885, EPI_ISL_1854888, EPI_ISL_1854889, EPI_ISL_1854890, EPI_ISL_1854892, EPI_ISL_1854894, EPI_ISL_1854896, EPI_ISL_1854897, EPI_ISL_1854898, EPI_ISL_1854899, EPI_ISL_1854900, EPI_ISL_1854901, EPI_ISL_1854902, EPI_ISL_1854903, EPI_ISL_1854904, EPI_ISL_1854905, EPI_ISL_1854906, EPI_ISL_1854907 | | | |
| see above | UNC-CH COVID Surveillance Lab | Jeremy Wang | Jeremy Wang, Alexander Rubinsteyn, Melissa Miller, Corbin Jones, Amy James Loftis, Amir Barzin, Susan Fiscus |
| EPI_ISL_1854909, EPI_ISL_1854910, EPI_ISL_1854912, EPI_ISL_1854913, EPI_ISL_1854915, EPI_ISL_1854917, EPI_ISL_1854918, EPI_ISL_1854919, EPI_ISL_1854922, EPI_ISL_1854923, EPI_ISL_1854925, EPI_ISL_1854929, EPI_ISL_1854930, EPI_ISL_1854931, EPI_ISL_1854932, EPI_ISL_1854933, EPI_ISL_1854934, EPI_ISL_1854936, EPI_ISL_1854937, EPI_ISL_1854938, EPI_ISL_1854940, EPI_ISL_1854941, EPI_ISL_1854944, EPI_ISL_1854946, EPI_ISL_1854947, EPI_ISL_1854949, EPI_ISL_1854950, EPI_ISL_1854955, EPI_ISL_1854956, EPI_ISL_1854957, EPI_ISL_1854959, EPI_ISL_1854962, EPI_ISL_1854963, EPI_ISL_1854966, EPI_ISL_1854967, EPI_ISL_1854968, EPI_ISL_1854970, EPI_ISL_1854974, EPI_ISL_1854975, EPI_ISL_1854976, EPI_ISL_1854977, EPI_ISL_1854978, EPI_ISL_1854980 | | | |
| see above | Clinical Molecular Microbiology Laboratory, UNC Hospitals | Jeremy Wang | Jeremy Wang, Alexander Rubinsteyn, Colleen Rice, Jason Smedberg, Shawn Hawken, Melissa Miller, Corbin Jones, Robert Hagan |
| EPI_ISL_1855103 | MONTEFIORE MEDICAL CENTER LABORATORIES | Wadsworth Center, New York State Department of Health | Kirsten St. George, Daryl M. Lamson, Alexis Russell, Matthew Shudt, Melissa A Leisner, Jonathan Plitnick, Catharine Prussing, Navjot Singh, John Kelly, Erasmus Schneider, Erica Lasek-Nesselquist |
| EPI_ISL_1855104 | TEMPUS LABS INC | Wadsworth Center, New York State Department of Health | Kirsten St. George, Daryl M. Lamson, Alexis Russell, Matthew Shudt, Melissa A Leisner, Jonathan Plitnick, Catharine Prussing, Navjot Singh, John Kelly, Erasmus Schneider, Erica Lasek-Nesselquist |
| EPI_ISL_1855105, EPI_ISL_1855106, EPI_ISL_1855107, EPI_ISL_1855108, EPI_ISL_1855109, EPI_ISL_1855110, EPI_ISL_1855111, EPI_ISL_1855112, EPI_ISL_1855113, EPI_ISL_1855114, EPI_ISL_1855115 | | | |
| see above | SUNY UPSTATE MEDICAL UNIVERSITY | Wadsworth Center, New York State Department of Health | Kirsten St. George, Daryl M. Lamson, Alexis Russell, Matthew Shudt, Melissa A Leisner, Jonathan Plitnick, Catharine Prussing, Navjot Singh, John Kelly, Erasmus Schneider, Erica Lasek-Nesselquist |
| EPI_ISL_1855116, EPI_ISL_1855117, EPI_ISL_1855118 | ALBANY MEDICAL CENTER | Wadsworth Center, New York State Department of Health | Kirsten St. George, Daryl M. Lamson, Alexis Russell, Matthew Shudt, Melissa A Leisner, Jonathan Plitnick, Catharine Prussing, Navjot Singh, John Kelly, Erasmus Schneider, Erica Lasek-Nesselquist |
| EPI_ISL_1855119, EPI_ISL_1855120, EPI_ISL_1855121, EPI_ISL_1855122, EPI_ISL_1855123, EPI_ISL_1855124, EPI_ISL_1855125, EPI_ISL_1855126, EPI_ISL_1855127, EPI_ISL_1855128, EPI_ISL_1855129, EPI_ISL_1855130, EPI_ISL_1855131, EPI_ISL_1855132, EPI_ISL_1855133, EPI_ISL_1855134, EPI_ISL_1855135, EPI_ISL_1855136, EPI_ISL_1855137, EPI_ISL_1855138, EPI_ISL_1855139, EPI_ISL_1855140, EPI_ISL_1855141, EPI_ISL_1855142, EPI_ISL_1855143 | | | |
| see above | URMC LABS | Wadsworth Center, New York State Department of Health | Kirsten St. George, Daryl M. Lamson, Alexis Russell, Matthew Shudt, Melissa A Leisner, Jonathan Plitnick, Catharine Prussing, Navjot Singh, John Kelly, Erasmus Schneider, Erica Lasek-Nesselquist |
| EPI_ISL_1855144, EPI_ISL_1855145, EPI_ISL_1855146, EPI_ISL_1855147, EPI_ISL_1855148, EPI_ISL_1855149 | GLENS FALLS HOSPITAL LABORATORY | Wadsworth Center, New York State Department of Health | Kirsten St. George, Daryl M. Lamson, Alexis Russell, Matthew Shudt, Melissa A Leisner, Jonathan Plitnick, Catharine Prussing, Navjot Singh, John Kelly, Erasmus Schneider, Erica Lasek-Nesselquist |
| EPI_ISL_1855150, EPI_ISL_1855151, EPI_ISL_1855152, EPI_ISL_1855153, EPI_ISL_1855154, EPI_ISL_1855155, EPI_ISL_1855156, EPI_ISL_1855157, EPI_ISL_1855158, EPI_ISL_1855159, EPI_ISL_1855160, EPI_ISL_1855161, EPI_ISL_1855162, EPI_ISL_1855163, EPI_ISL_1855164, EPI_ISL_1855165, EPI_ISL_1855166, EPI_ISL_1855167 | | | |
| see above | TEMPUS LABS INC | Wadsworth Center, New York State | Kirsten St. George, Daryl M. Lamson, Alexis Russell, Matthew Shudt, Melissa A Leisner, Jonathan Plitnick, Catharine Prussing, Navjot Singh, John Kelly, Erasmus Schneider, Erica Lasek-Nesselquist |

| | | | |
|---|---|---|---|
| | | Department of Health | |
| EPI_ISL_1855168, EPI_ISL_1855169, EPI_ISL_1855170, EPI_ISL_1855171, EPI_ISL_1855172, EPI_ISL_1855173, EPI_ISL_1855174, EPI_ISL_1855175, EPI_ISL_1855176, EPI_ISL_1855177, EPI_ISL_1855178, EPI_ISL_1855179, EPI_ISL_1855180, EPI_ISL_1855181, EPI_ISL_1855182, EPI_ISL_1855183, EPI_ISL_1855184, EPI_ISL_1855185 | | | |
| see above | URMC LABS | Wadsworth Center, New York State Department of Health | Kirsten St. George, Daryl M. Lamson, Alexis Russell, Matthew Shudt, Melissa A Leisner, Jonathan Plitnick, Catharine Prussing, Navjot Singh, John Kelly, Erasmus Schneider, Erica Lasek-Nesselquist |
| EPI_ISL_1855186, EPI_ISL_1855187 | NYC Pandemic Response Lab | Wadsworth Center, New York State Department of Health | Kirsten St. George, Daryl M. Lamson, Alexis Russell, Matthew Shudt, Melissa A Leisner, Jonathan Plitnick, Catharine Prussing, Navjot Singh, John Kelly, Erasmus Schneider, Erica Lasek-Nesselquist |
| EPI_ISL_1855188, EPI_ISL_1855189, EPI_ISL_1855190, EPI_ISL_1855191, EPI_ISL_1855192, EPI_ISL_1855193, EPI_ISL_1855194, EPI_ISL_1855195, EPI_ISL_1855196 | MONTEFIORE MEDICAL CENTER LABORATORIES | Wadsworth Center, New York State Department of Health | Kirsten St. George, Daryl M. Lamson, Alexis Russell, Matthew Shudt, Melissa A Leisner, Jonathan Plitnick, Catharine Prussing, Navjot Singh, John Kelly, Erasmus Schneider, Erica Lasek-Nesselquist |
| EPI_ISL_1855197, EPI_ISL_1855198, EPI_ISL_1855199, EPI_ISL_1855200, EPI_ISL_1855201 | NYC Pandemic Response Lab | Wadsworth Center, New York State Department of Health | Kirsten St. George, Daryl M. Lamson, Alexis Russell, Matthew Shudt, Melissa A Leisner, Jonathan Plitnick, Catharine Prussing, Navjot Singh, John Kelly, Erasmus Schneider, Erica Lasek-Nesselquist |
| EPI_ISL_1855202, EPI_ISL_1855203, EPI_ISL_1855204, EPI_ISL_1855205, EPI_ISL_1855206, EPI_ISL_1855207, EPI_ISL_1855208, EPI_ISL_1855209 | MONTEFIORE MEDICAL CENTER LABORATORIES | Wadsworth Center, New York State Department of Health | Kirsten St. George, Daryl M. Lamson, Alexis Russell, Matthew Shudt, Melissa A Leisner, Jonathan Plitnick, Catharine Prussing, Navjot Singh, John Kelly, Erasmus Schneider, Erica Lasek-Nesselquist |
| EPI_ISL_1855210 | MEMORIAL SLOAN KETTERING CANCER CENTER | Wadsworth Center, New York State Department of Health | Kirsten St. George, Daryl M. Lamson, Alexis Russell, Matthew Shudt, Melissa A Leisner, Jonathan Plitnick, Catharine Prussing, Navjot Singh, John Kelly, Erasmus Schneider, Erica Lasek-Nesselquist |
| EPI_ISL_1855211, EPI_ISL_1855212, EPI_ISL_1855213, EPI_ISL_1855214 | New York Presbyterian Hospital | Wadsworth Center, New York State Department of Health | Kirsten St. George, Daryl M. Lamson, Alexis Russell, Matthew Shudt, Melissa A Leisner, Jonathan Plitnick, Catharine Prussing, Navjot Singh, John Kelly, Erasmus Schneider, Erica Lasek-Nesselquist |
| EPI_ISL_1855215, EPI_ISL_1855216, EPI_ISL_1855217, EPI_ISL_1855218, EPI_ISL_1855219 | MONTEFIORE MEDICAL CENTER LABORATORIES | Wadsworth Center, New York State Department of Health | Kirsten St. George, Daryl M. Lamson, Alexis Russell, Matthew Shudt, Melissa A Leisner, Jonathan Plitnick, Catharine Prussing, Navjot Singh, John Kelly, Erasmus Schneider, Erica Lasek-Nesselquist |
| EPI_ISL_1855220 | SARATOGA HOSPITAL LABORATORY | Wadsworth Center, New York State Department of Health | Kirsten St. George, Daryl M. Lamson, Alexis Russell, Matthew Shudt, Melissa A Leisner, Jonathan Plitnick, Catharine Prussing, Navjot Singh, John Kelly, Erasmus Schneider, Erica Lasek-Nesselquist |
| EPI_ISL_1855221, EPI_ISL_1855222, EPI_ISL_1855223, EPI_ISL_1855224, EPI_ISL_1855225 | SUNY UPSTATE MEDICAL UNIVERSITY | Wadsworth Center, New York State Department of Health | Kirsten St. George, Daryl M. Lamson, Alexis Russell, Matthew Shudt, Melissa A Leisner, Jonathan Plitnick, Catharine Prussing, Navjot Singh, John Kelly, Erasmus Schneider, Erica Lasek-Nesselquist |
| EPI_ISL_1855226, EPI_ISL_1855227, EPI_ISL_1855228, EPI_ISL_1855229, EPI_ISL_1855230, EPI_ISL_1855231, EPI_ISL_1855232, EPI_ISL_1855233, EPI_ISL_1855234, EPI_ISL_1855235, EPI_ISL_1855236, EPI_ISL_1855237, EPI_ISL_1855238, EPI_ISL_1855239, EPI_ISL_1855240 | | | |
| see above | URMC LABS | Wadsworth Center, New York State Department of Health | Kirsten St. George, Daryl M. Lamson, Alexis Russell, Matthew Shudt, Melissa A Leisner, Jonathan Plitnick, Catharine Prussing, Navjot Singh, John Kelly, Erasmus Schneider, Erica Lasek-Nesselquist |
| EPI_ISL_1855241, EPI_ISL_1855242, EPI_ISL_1855243 | GLENS FALLS HOSPITAL LABORATORY | Wadsworth Center, New York State Department of Health | Kirsten St. George, Daryl M. Lamson, Alexis Russell, Matthew Shudt, Melissa A Leisner, Jonathan Plitnick, Catharine Prussing, Navjot Singh, John Kelly, Erasmus Schneider, Erica Lasek-Nesselquist |
| EPI_ISL_1855244, EPI_ISL_1855245, EPI_ISL_1855246, EPI_ISL_1855247, EPI_ISL_1855248 | TEMPUS LABS INC | Wadsworth Center, New York State Department of Health | Kirsten St. George, Daryl M. Lamson, Alexis Russell, Matthew Shudt, Melissa A Leisner, Jonathan Plitnick, Catharine Prussing, Navjot Singh, John Kelly, Erasmus Schneider, Erica Lasek-Nesselquist |
| EPI_ISL_1855249, EPI_ISL_1855250, EPI_ISL_1855251, EPI_ISL_1855252, EPI_ISL_1855253, EPI_ISL_1855254, EPI_ISL_1855255, EPI_ISL_1855256, EPI_ISL_1855257, EPI_ISL_1855258, EPI_ISL_1855259, EPI_ISL_1855260, EPI_ISL_1855261, EPI_ISL_1855262, EPI_ISL_1855263, EPI_ISL_1855264, EPI_ISL_1855265, EPI_ISL_1855266, EPI_ISL_1855267, EPI_ISL_1855268, EPI_ISL_1855269, EPI_ISL_1855270, EPI_ISL_1855271, EPI_ISL_1855272 | | | |
| see above | URMC LABS | Wadsworth Center, New York State Department of Health | Kirsten St. George, Daryl M. Lamson, Alexis Russell, Matthew Shudt, Melissa A Leisner, Jonathan Plitnick, Catharine Prussing, Navjot Singh, John Kelly, Erasmus Schneider, Erica Lasek-Nesselquist |
| EPI_ISL_1855273, EPI_ISL_1855274, EPI_ISL_1855275 | SARATOGA HOSPITAL LABORATORY | Wadsworth Center, New York State Department of Health | Kirsten St. George, Daryl M. Lamson, Alexis Russell, Matthew Shudt, Melissa A Leisner, Jonathan Plitnick, Catharine Prussing, Navjot Singh, John Kelly, Erasmus Schneider, Erica Lasek-Nesselquist |
| EPI_ISL_1855276, EPI_ISL_1855277, EPI_ISL_1855278, EPI_ISL_1855279, EPI_ISL_1855280, EPI_ISL_1855281, EPI_ISL_1855282, EPI_ISL_1855283, EPI_ISL_1855284, EPI_ISL_1855285 | URMC LABS | Wadsworth Center, New York State Department of Health | Kirsten St. George, Daryl M. Lamson, Alexis Russell, Matthew Shudt, Melissa A Leisner, Jonathan Plitnick, Catharine Prussing, Navjot Singh, John Kelly, Erasmus Schneider, Erica Lasek-Nesselquist |
| EPI_ISL_1855286, EPI_ISL_1855287, EPI_ISL_1855288, EPI_ISL_1855289, EPI_ISL_1855290, EPI_ISL_1855291, EPI_ISL_1855292, EPI_ISL_1855293 | Columbia University Irving Medical Center | Wadsworth Center, New York State Department of Health | Kirsten St. George, Daryl M. Lamson, Alexis Russell, Matthew Shudt, Melissa A Leisner, Jonathan Plitnick, Catharine Prussing, Navjot Singh, John Kelly, Erasmus Schneider, Erica Lasek-Nesselquist |
| EPI_ISL_1855294, EPI_ISL_1855295, EPI_ISL_1855296, EPI_ISL_1855297, EPI_ISL_1855298, EPI_ISL_1855299, EPI_ISL_1855300, EPI_ISL_1855301, EPI_ISL_1855302, EPI_ISL_1855303, EPI_ISL_1855304, EPI_ISL_1855305, EPI_ISL_1855306, EPI_ISL_1855307, EPI_ISL_1855308, EPI_ISL_1855309 | | | |
| see above | URMC LABS | Wadsworth Center, New York State Department of Health | Kirsten St. George, Daryl M. Lamson, Alexis Russell, Matthew Shudt, Melissa A Leisner, Jonathan Plitnick, Catharine Prussing, Navjot Singh, John Kelly, Erasmus Schneider, Erica Lasek-Nesselquist |
| EPI_ISL_1855310, EPI_ISL_1855311, EPI_ISL_1855312, EPI_ISL_1855313, EPI_ISL_1855314, EPI_ISL_1855315, EPI_ISL_1855316, EPI_ISL_1855317, EPI_ISL_1855318, EPI_ISL_1855319, EPI_ISL_1855320, EPI_ISL_1855321, EPI_ISL_1855322, EPI_ISL_1855323 | | | |
| see above | BOSTON HEART DIAGNOSTICS CORP | Wadsworth Center, New York State Department of Health | Kirsten St. George, Daryl M. Lamson, Alexis Russell, Matthew Shudt, Melissa A Leisner, Jonathan Plitnick, Catharine Prussing, Navjot Singh, John Kelly, Erasmus Schneider, Erica Lasek-Nesselquist |
| EPI_ISL_1855324, EPI_ISL_1855325, EPI_ISL_1855326, EPI_ISL_1855327, EPI_ISL_1855328, EPI_ISL_1855329, EPI_ISL_1855330, EPI_ISL_1855331, EPI_ISL_1855332, EPI_ISL_1855333 | MONTEFIORE MEDICAL CENTER LABORATORIES | Wadsworth Center, New York State Department of Health | Kirsten St. George, Daryl M. Lamson, Alexis Russell, Matthew Shudt, Melissa A Leisner, Jonathan Plitnick, Catharine Prussing, Navjot Singh, John Kelly, Erasmus Schneider, Erica Lasek-Nesselquist |
| EPI_ISL_1855334, EPI_ISL_1855335, EPI_ISL_1855336, EPI_ISL_1855337, EPI_ISL_1855338, EPI_ISL_1855339, EPI_ISL_1855340, EPI_ISL_1855341, EPI_ISL_1855342, EPI_ISL_1855343, EPI_ISL_1855344, EPI_ISL_1855345, EPI_ISL_1855346, EPI_ISL_1855347 | | | |

| | | | |
|---|---|---|---|
| see above | WESTCHESTER MEDICAL CENTER | Wadsworth Center, New York State Department of Health | Kirsten St. George, Daryl M. Lamson, Alexis Russell, Matthew Shudt, Melissa A Leisner, Jonathan Plitnick, Catharine Prussing, Navjot Singh, John Kelly, Erasmus Schneider, Erica Lasek-Nesselquist |
| EPI_ISL_1855348, EPI_ISL_1855349, EPI_ISL_1855350, EPI_ISL_1855351, EPI_ISL_1855352, EPI_ISL_1855353 | NORTH SHORE UNIVERSITY HOSPITAL | Wadsworth Center, New York State Department of Health | Kirsten St. George, Daryl M. Lamson, Alexis Russell, Matthew Shudt, Melissa A Leisner, Jonathan Plitnick, Catharine Prussing, Navjot Singh, John Kelly, Erasmus Schneider, Erica Lasek-Nesselquist |
| EPI_ISL_1855354, EPI_ISL_1855355, EPI_ISL_1855356, EPI_ISL_1855357, EPI_ISL_1855358, EPI_ISL_1855359, EPI_ISL_1855360 | SUNY UPSTATE MEDICAL UNIVERSITY | Wadsworth Center, New York State Department of Health | Kirsten St. George, Daryl M. Lamson, Alexis Russell, Matthew Shudt, Melissa A Leisner, Jonathan Plitnick, Catharine Prussing, Navjot Singh, John Kelly, Erasmus Schneider, Erica Lasek-Nesselquist |
| EPI_ISL_1855361, EPI_ISL_1855362, EPI_ISL_1855363, EPI_ISL_1855364, EPI_ISL_1855365, EPI_ISL_1855366, EPI_ISL_1855367, EPI_ISL_1855368, EPI_ISL_1855369, EPI_ISL_1855370, EPI_ISL_1855371, EPI_ISL_1855372, EPI_ISL_1855373, EPI_ISL_1855374, EPI_ISL_1855375, EPI_ISL_1855376, EPI_ISL_1855377, EPI_ISL_1855378, EPI_ISL_1855379, EPI_ISL_1855380, EPI_ISL_1855381, EPI_ISL_1855382, EPI_ISL_1855383 | | | |
| see above | University of Wisconsin-Madison AIDS Vaccine Research Laboratories | University of Wisconsin-Madison AIDS Vaccine Research Laboratories | Gage Moreno, Katarina Braun, et al. AIDS Vaccine Research Laboratories |
| EPI_ISL_1855386, EPI_ISL_1855387, EPI_ISL_1855389 | Pathology and Laboratory Medicine Institute, Cleveland Clinic, Ohio, USA | Pathology and Laboratory Medicine Institute, Cleveland Clinic, Ohio, USA | Jessica Spildener, Joy Nakitandwe, Kristen McDonnell, David Plunkett, Zheng Jin Tu, Jay Brock, Yu-Wei Cheng, Gary Procop, Daniel Rhoads, Daniel H. Farkas, David Bosler |
| EPI_ISL_1855850, EPI_ISL_1855851, EPI_ISL_1855852, EPI_ISL_1855853, EPI_ISL_1855854, EPI_ISL_1855855, EPI_ISL_1855856, EPI_ISL_1855857, EPI_ISL_1855858, EPI_ISL_1855859, EPI_ISL_1855860, EPI_ISL_1855861, EPI_ISL_1855862, EPI_ISL_1855863, EPI_ISL_1855864, EPI_ISL_1855865, EPI_ISL_1855866, EPI_ISL_1855867, EPI_ISL_1855868, EPI_ISL_1855869, EPI_ISL_1855870, EPI_ISL_1855871, EPI_ISL_1855872, EPI_ISL_1855873, EPI_ISL_1855874, EPI_ISL_1855875, EPI_ISL_1855876, EPI_ISL_1855877, EPI_ISL_1855878, EPI_ISL_1855879, EPI_ISL_1855880, EPI_ISL_1855881, EPI_ISL_1855882, EPI_ISL_1855883, EPI_ISL_1855884, EPI_ISL_1855885, EPI_ISL_1855886, EPI_ISL_1855887, EPI_ISL_1855888, EPI_ISL_1855889, EPI_ISL_1855890, EPI_ISL_1855891, EPI_ISL_1855892, EPI_ISL_1855893, EPI_ISL_1855894, EPI_ISL_1855895, EPI_ISL_1855896, EPI_ISL_1855897, EPI_ISL_1855898, EPI_ISL_1855899, EPI_ISL_1855900, EPI_ISL_1855901, EPI_ISL_1855902, EPI_ISL_1855903, EPI_ISL_1855904, EPI_ISL_1855905, EPI_ISL_1855906, EPI_ISL_1855907, EPI_ISL_1855908, EPI_ISL_1855909, EPI_ISL_1855910 | | | |
| see above | Baylor Scott & White-Temple | Baylor Scott & White-Temple | Ari Rao, Linden Morales, Kimberly Walker, Marcus Volz, Shelby Hendrickson |
| EPI_ISL_1855911, EPI_ISL_1855912, EPI_ISL_1855913, EPI_ISL_1855914, EPI_ISL_1855915, EPI_ISL_1855916, EPI_ISL_1855917, EPI_ISL_1855918, EPI_ISL_1855919, EPI_ISL_1855920, EPI_ISL_1855921, EPI_ISL_1855922, EPI_ISL_1855923, EPI_ISL_1855924, EPI_ISL_1855925, EPI_ISL_1855926, EPI_ISL_1855927, EPI_ISL_1855928, EPI_ISL_1855929, EPI_ISL_1855930, EPI_ISL_1855931, EPI_ISL_1855932, EPI_ISL_1855933, EPI_ISL_1855934, EPI_ISL_1855935, EPI_ISL_1855936, EPI_ISL_1855937, EPI_ISL_1855938 | | | |
| see above | University of Michigan Clinical Microbiology Laboratory | Lauring Lab, University of Michigan, Department of Microbiology and Immunology | Valesano |
| EPI_ISL_1855942, EPI_ISL_1855943, EPI_ISL_1855944, EPI_ISL_1855946, EPI_ISL_1855947, EPI_ISL_1855948, EPI_ISL_1855949, EPI_ISL_1855950, EPI_ISL_1855951, EPI_ISL_1855952, EPI_ISL_1855953, EPI_ISL_1855954, EPI_ISL_1855956, EPI_ISL_1855957, EPI_ISL_1855958, EPI_ISL_1855961, EPI_ISL_1855962, EPI_ISL_1855965, EPI_ISL_1855966, EPI_ISL_1855967, EPI_ISL_1855968, EPI_ISL_1855969, EPI_ISL_1855970, EPI_ISL_1855971, EPI_ISL_1855972, EPI_ISL_1855973, EPI_ISL_1855974, EPI_ISL_1855975, EPI_ISL_1855976, EPI_ISL_1855977, EPI_ISL_1855979, EPI_ISL_1855980, EPI_ISL_1855981, EPI_ISL_1855983, EPI_ISL_1855984, EPI_ISL_1855985, EPI_ISL_1855986, EPI_ISL_1855987, EPI_ISL_1855988, EPI_ISL_1855989, EPI_ISL_1855990, EPI_ISL_1855991, EPI_ISL_1855992, EPI_ISL_1855993, EPI_ISL_1855994, EPI_ISL_1855995, EPI_ISL_1855996, EPI_ISL_1855997, EPI_ISL_1855998, EPI_ISL_1855999, EPI_ISL_1856000, EPI_ISL_1856001, EPI_ISL_1856002, EPI_ISL_1856003, EPI_ISL_1856004, EPI_ISL_1856005, EPI_ISL_1856006, EPI_ISL_1856007, EPI_ISL_1856008, EPI_ISL_1856009, EPI_ISL_1856010, EPI_ISL_1856011, EPI_ISL_1856012, EPI_ISL_1856013, EPI_ISL_1856014, EPI_ISL_1856015, EPI_ISL_1856016, EPI_ISL_1856017, EPI_ISL_1856018, EPI_ISL_1856020, EPI_ISL_1856021, EPI_ISL_1856022, EPI_ISL_1856023, EPI_ISL_1856024, EPI_ISL_1856025, EPI_ISL_1856026, EPI_ISL_1856027, EPI_ISL_1856028, EPI_ISL_1856029, EPI_ISL_1856030, EPI_ISL_1856031, EPI_ISL_1856032, EPI_ISL_1856033, EPI_ISL_1856034, EPI_ISL_1856035, EPI_ISL_1856036, EPI_ISL_1856037, EPI_ISL_1856038, EPI_ISL_1856040, EPI_ISL_1856041, EPI_ISL_1856042, EPI_ISL_1856044, EPI_ISL_1856045, EPI_ISL_1856046, EPI_ISL_1856047, EPI_ISL_1856048, EPI_ISL_1856049, EPI_ISL_1856050, EPI_ISL_1856051, EPI_ISL_1856052, EPI_ISL_1856053, EPI_ISL_1856054, EPI_ISL_1856055, EPI_ISL_1856056, EPI_ISL_1856057, EPI_ISL_1856058, EPI_ISL_1856059, EPI_ISL_1856060, EPI_ISL_1856061, EPI_ISL_1856062, EPI_ISL_1856063, EPI_ISL_1856064, EPI_ISL_1856065, EPI_ISL_1856066, EPI_ISL_1856067, EPI_ISL_1856068, EPI_ISL_1856069, EPI_ISL_1856070, EPI_ISL_1856071, EPI_ISL_1856072, EPI_ISL_1856073, EPI_ISL_1856074, EPI_ISL_1856075, EPI_ISL_1856076, EPI_ISL_1856077, EPI_ISL_1856079, EPI_ISL_1856080, EPI_ISL_1856081, EPI_ISL_1856082, EPI_ISL_1856083, EPI_ISL_1856084, EPI_ISL_1856085, EPI_ISL_1856086, EPI_ISL_1856087, EPI_ISL_1856088, EPI_ISL_1856089, EPI_ISL_1856090, EPI_ISL_1856091, EPI_ISL_1856092, EPI_ISL_1856093, EPI_ISL_1856094, EPI_ISL_1856095, EPI_ISL_1856096, EPI_ISL_1856097, EPI_ISL_1856098, EPI_ISL_1856099, EPI_ISL_1856100, EPI_ISL_1856101, EPI_ISL_1856102, EPI_ISL_1856103, EPI_ISL_1856104, EPI_ISL_1856105, EPI_ISL_1856106, EPI_ISL_1856107, EPI_ISL_1856108, EPI_ISL_1856109, EPI_ISL_1856110, EPI_ISL_1856111, EPI_ISL_1856112, EPI_ISL_1856113, EPI_ISL_1856114, EPI_ISL_1856115, EPI_ISL_1856116, EPI_ISL_1856117, EPI_ISL_1856118, EPI_ISL_1856119, EPI_ISL_1856120, EPI_ISL_1856121, EPI_ISL_1856122, EPI_ISL_1856123, EPI_ISL_1856124, EPI_ISL_1856125, EPI_ISL_1856126, EPI_ISL_1856127, EPI_ISL_1856128, EPI_ISL_1856130, EPI_ISL_1856131, EPI_ISL_1856133, EPI_ISL_1856134, EPI_ISL_1856135, EPI_ISL_1856136, EPI_ISL_1856137, EPI_ISL_1856138, EPI_ISL_1856140, EPI_ISL_1856141, EPI_ISL_1856142, EPI_ISL_1856143, EPI_ISL_1856144, EPI_ISL_1856145, EPI_ISL_1856147, EPI_ISL_1856149, EPI_ISL_1856151, EPI_ISL_1856152, EPI_ISL_1856153, EPI_ISL_1856154, EPI_ISL_1856155, EPI_ISL_1856156, EPI_ISL_1856157, EPI_ISL_1856159, EPI_ISL_1856160, EPI_ISL_1856161, EPI_ISL_1856162, EPI_ISL_1856163, EPI_ISL_1856165, EPI_ISL_1856166, EPI_ISL_1856168, EPI_ISL_1856169, EPI_ISL_1856170, EPI_ISL_1856171, EPI_ISL_1856172, EPI_ISL_1856173, EPI_ISL_1856176, EPI_ISL_1856179, EPI_ISL_1856184, EPI_ISL_1856185, EPI_ISL_1856187, EPI_ISL_1856188, EPI_ISL_1856189, EPI_ISL_1856191, EPI_ISL_1856192, EPI_ISL_1856193, EPI_ISL_1856194, EPI_ISL_1856195, EPI_ISL_1856196, EPI_ISL_1856197, EPI_ISL_1856200, EPI_ISL_1856201, EPI_ISL_1856202, EPI_ISL_1856203, EPI_ISL_1856204, EPI_ISL_1856205, EPI_ISL_1856206, EPI_ISL_1856207, EPI_ISL_1856208, EPI_ISL_1856209, EPI_ISL_1856210, EPI_ISL_1856211, EPI_ISL_1856212, EPI_ISL_1856213, EPI_ISL_1856214, EPI_ISL_1856215, EPI_ISL_1856216, EPI_ISL_1856217, EPI_ISL_1856218, EPI_ISL_1856219, EPI_ISL_1856220, EPI_ISL_1856221, EPI_ISL_1856222, EPI_ISL_1856223, EPI_ISL_1856224, EPI_ISL_1856225, EPI_ISL_1856226, EPI_ISL_1856227, EPI_ISL_1856229, EPI_ISL_1856230, EPI_ISL_1856231, EPI_ISL_1856232, EPI_ISL_1856233, EPI_ISL_1856234, EPI_ISL_1856235, EPI_ISL_1856236, EPI_ISL_1856237, EPI_ISL_1856238, EPI_ISL_1856239, EPI_ISL_1856240, EPI_ISL_1856241, EPI_ISL_1856242, EPI_ISL_1856243, EPI_ISL_1856244, EPI_ISL_1856245, EPI_ISL_1856246, EPI_ISL_1856247, EPI_ISL_1856248, EPI_ISL_1856249, EPI_ISL_1856250, EPI_ISL_1856251, EPI_ISL_1856252, EPI_ISL_1856253, EPI_ISL_1856254, EPI_ISL_1856255, EPI_ISL_1856256, EPI_ISL_1856257, EPI_ISL_1856258, EPI_ISL_1856259, EPI_ISL_1856260, EPI_ISL_1856261, EPI_ISL_1856262, EPI_ISL_1856263, EPI_ISL_1856264, EPI_ISL_1856265, EPI_ISL_1856266, EPI_ISL_1856267, EPI_ISL_1856268, EPI_ISL_1856269, EPI_ISL_1856270, EPI_ISL_1856271, EPI_ISL_1856272, EPI_ISL_1856274, EPI_ISL_1856275, EPI_ISL_1856276, EPI_ISL_1856277, EPI_ISL_1856278, EPI_ISL_1856279, EPI_ISL_1856280, EPI_ISL_1856281, EPI_ISL_1856282, EPI_ISL_1856283, EPI_ISL_1856284, EPI_ISL_1856285, EPI_ISL_1856286, EPI_ISL_1856287, EPI_ISL_1856288, EPI_ISL_1856289, EPI_ISL_1856290, EPI_ISL_1856291, EPI_ISL_1856292, EPI_ISL_1856293, EPI_ISL_1856295, EPI_ISL_1856296, EPI_ISL_1856297, EPI_ISL_1856298, EPI_ISL_1856299, EPI_ISL_1856300, EPI_ISL_1856301, EPI_ISL_1856302, EPI_ISL_1856303, EPI_ISL_1856304, EPI_ISL_1856307, EPI_ISL_1856308, EPI_ISL_1856309, EPI_ISL_1856310, EPI_ISL_1856311, EPI_ISL_1856314, EPI_ISL_1856315, EPI_ISL_1856316, EPI_ISL_1856317, EPI_ISL_1856318, EPI_ISL_1856319, EPI_ISL_1856320, EPI_ISL_1856321, EPI_ISL_1856323, EPI_ISL_1856325, EPI_ISL_1856326, EPI_ISL_1856328, EPI_ISL_1856329, EPI_ISL_1856330, EPI_ISL_1856332, EPI_ISL_1856333, EPI_ISL_1856334, EPI_ISL_1856335, EPI_ISL_1856336, EPI_ISL_1856337, EPI_ISL_1856338, EPI_ISL_1856339, EPI_ISL_1856340, EPI_ISL_1856341, EPI_ISL_1856343, EPI_ISL_1856344, EPI_ISL_1856345, EPI_ISL_1856346, EPI_ISL_1856347, EPI_ISL_1856348, EPI_ISL_1856349, EPI_ISL_1856351, EPI_ISL_1856352, EPI_ISL_1856353, EPI_ISL_1856354, EPI_ISL_1856355, EPI_ISL_1856356, EPI_ISL_1856358, EPI_ISL_1856360, EPI_ISL_1856361, EPI_ISL_1856362, EPI_ISL_1856363, EPI_ISL_1856364, EPI_ISL_1856367, EPI_ISL_1856368, EPI_ISL_1856369, EPI_ISL_1856371, EPI_ISL_1856372, EPI_ISL_1856373, EPI_ISL_1856374, EPI_ISL_1856375, EPI_ISL_1856376, EPI_ISL_1856377, EPI_ISL_1856379, EPI_ISL_1856381, EPI_ISL_1856382, EPI_ISL_1856383, EPI_ISL_1856384, EPI_ISL_1856385, EPI_ISL_1856386, EPI_ISL_1856387, EPI_ISL_1856388, EPI_ISL_1856389, EPI_ISL_1856391, EPI_ISL_1856394, EPI_ISL_1856395, EPI_ISL_1856398, EPI_ISL_1856399, EPI_ISL_1856400, EPI_ISL_1856401, EPI_ISL_1856403, EPI_ISL_1856404, EPI_ISL_1856405, EPI_ISL_1856406, EPI_ISL_1856408, EPI_ISL_1856409, EPI_ISL_1856410, EPI_ISL_1856411, EPI_ISL_1856412, EPI_ISL_1856413, EPI_ISL_1856414, EPI_ISL_1856415, EPI_ISL_1856416, EPI_ISL_1856417, EPI_ISL_1856418, EPI_ISL_1856420, EPI_ISL_1856423, EPI_ISL_1856424, EPI_ISL_1856425, EPI_ISL_1856427, EPI_ISL_1856428, EPI_ISL_1856430, EPI_ISL_1856431, EPI_ISL_1856432, EPI_ISL_1856436, EPI_ISL_1856437, EPI_ISL_1856438, EPI_ISL_1856439, EPI_ISL_1856440, EPI_ISL_1856441, EPI_ISL_1856442, EPI_ISL_1856443, EPI_ISL_1856444, EPI_ISL_1856445, EPI_ISL_1856446, EPI_ISL_1856447, EPI_ISL_1856448, EPI_ISL_1856449, EPI_ISL_1856450, EPI_ISL_1856451, EPI_ISL_1856452, EPI_ISL_1856453, EPI_ISL_1856454, EPI_ISL_1856455, EPI_ISL_1856456, EPI_ISL_1856457, EPI_ISL_1856458, EPI_ISL_1856459, EPI_ISL_1856460, EPI_ISL_1856461, EPI_ISL_1856462, EPI_ISL_1856463, EPI_ISL_1856464, EPI_ISL_1856465, EPI_ISL_1856466, EPI_ISL_1856467, EPI_ISL_1856468, EPI_ISL_1856469, EPI_ISL_1856470, EPI_ISL_1856471, EPI_ISL_1856472, EPI_ISL_1856473, EPI_ISL_1856474, EPI_ISL_1856476, EPI_ISL_1856477, EPI_ISL_1856478, EPI_ISL_1856479, EPI_ISL_1856480, EPI_ISL_1856481, EPI_ISL_1856482, EPI_ISL_1856483, EPI_ISL_1856484, EPI_ISL_1856485, EPI_ISL_1856486, EPI_ISL_1856487, EPI_ISL_1856488, EPI_ISL_1856489, EPI_ISL_1856490, EPI_ISL_1856491, EPI_ISL_1856492, EPI_ISL_1856493, EPI_ISL_1856494, EPI_ISL_1856495, EPI_ISL_1856496, EPI_ISL_1856497, EPI_ISL_1856499, EPI_ISL_1856500, EPI_ISL_1856501, EPI_ISL_1856502, EPI_ISL_1856503, EPI_ISL_1856504, EPI_ISL_1856505, EPI_ISL_1856507, EPI_ISL_1856508, EPI_ISL_1856509, EPI_ISL_1856510, EPI_ISL_1856511, EPI_ISL_1856512, EPI_ISL_1856513, EPI_ISL_1856514, EPI_ISL_1856515, EPI_ISL_1856516, EPI_ISL_1856518, EPI_ISL_1856519, EPI_ISL_1856520, EPI_ISL_1856522, EPI_ISL_1856523, EPI_ISL_1856524, EPI_ISL_1856525, EPI_ISL_1856526, EPI_ISL_1856527, EPI_ISL_1856528, EPI_ISL_1856529, EPI_ISL_1856530, EPI_ISL_1856531, EPI_ISL_1856532, EPI_ISL_1856533, EPI_ISL_1856534, EPI_ISL_1856536, EPI_ISL_1856538, EPI_ISL_1856539, EPI_ISL_1856540, EPI_ISL_1856541, EPI_ISL_1856543, EPI_ISL_1856544, EPI_ISL_1856545, EPI_ISL_1856546, EPI_ISL_1856547, EPI_ISL_1856548, EPI_ISL_1856549, EPI_ISL_1856550, EPI_ISL_1856551, EPI_ISL_1856552, EPI_ISL_1856553, EPI_ISL_1856554, EPI_ISL_1856555, EPI_ISL_1856556, EPI_ISL_1856557, EPI_ISL_1856558, EPI_ISL_1856560, EPI_ISL_1856562, EPI_ISL_1856563, EPI_ISL_1856564, EPI_ISL_1856565, EPI_ISL_1856566, EPI_ISL_1856567, EPI_ISL_1856568, EPI_ISL_1856569, EPI_ISL_1856570, EPI_ISL_1856571, EPI_ISL_1856572, EPI_ISL_1856573, EPI_ISL_1856574, EPI_ISL_1856575, EPI_ISL_1856576, EPI_ISL_1856577, EPI_ISL_1856578, EPI_ISL_1856579, EPI_ISL_1856580, EPI_ISL_1856581, EPI_ISL_1856582, EPI_ISL_1856583, EPI_ISL_1856584, EPI_ISL_1856586, EPI_ISL_1856587, EPI_ISL_1856588, EPI_ISL_1856590, EPI_ISL_1856591, EPI_ISL_1856592, EPI_ISL_1856593, EPI_ISL_1856594, EPI_ISL_1856595, EPI_ISL_1856596, EPI_ISL_1856597, EPI_ISL_1856598, EPI_ISL_1856599, EPI_ISL_1856600, EPI_ISL_1856601, EPI_ISL_1856602, EPI_ISL_1856603, EPI_ISL_1856604, EPI_ISL_1856605, EPI_ISL_1856606, EPI_ISL_1856607, EPI_ISL_1856608, EPI_ISL_1856609, EPI_ISL_1856610, EPI_ISL_1856611, EPI_ISL_1856612, EPI_ISL_1856613, EPI_ISL_1856614, EPI_ISL_1856615, EPI_ISL_1856616, EPI_ISL_1856617, EPI_ISL_1856618, EPI_ISL_1856619, EPI_ISL_1856620, EPI_ISL_1856621, EPI_ISL_1856622, EPI_ISL_1856623, EPI_ISL_1856624, EPI_ISL_1856625, EPI_ISL_1856626, EPI_ISL_1856627, EPI_ISL_1856628, EPI_ISL_1856629, EPI_ISL_1856630, EPI_ISL_1856631, EPI_ISL_1856632, EPI_ISL_1856633, EPI_ISL_1856634, EPI_ISL_1856636, EPI_ISL_1856637, EPI_ISL_1856638, EPI_ISL_1856639, EPI_ISL_1856640, EPI_ISL_1856641, EPI_ISL_1856642, EPI_ISL_1856643, EPI_ISL_1856644, EPI_ISL_1856645, EPI_ISL_1856646, EPI_ISL_1856647, EPI_ISL_1856648, EPI_ISL_1856649, EPI_ISL_1856650, EPI_ISL_1856651, EPI_ISL_1856652, EPI_ISL_1856653, EPI_ISL_1856654, EPI_ISL_1856655, EPI_ISL_1856656, EPI_ISL_1856657, EPI_ISL_1856658, EPI_ISL_1856659, EPI_ISL_1856660, EPI_ISL_1856661, EPI_ISL_1856662, EPI_ISL_1856664, EPI_ISL_1856666, EPI_ISL_1856667, EPI_ISL_1856668, EPI_ISL_1856670, EPI_ISL_1856672, EPI_ISL_1856674, EPI_ISL_1856675, EPI_ISL_1856676, EPI_ISL_1856677, EPI_ISL_1856678, EPI_ISL_1856679, EPI_ISL_1856680, EPI_ISL_1856681, EPI_ISL_1856682, EPI_ISL_1856683, EPI_ISL_1856684, EPI_ISL_1856685, EPI_ISL_1856687, EPI_ISL_1856688, EPI_ISL_1856689, EPI_ISL_1856690, EPI_ISL_1856691, EPI_ISL_1856692, EPI_ISL_1856693, EPI_ISL_1856695, EPI_ISL_1856697, EPI_ISL_1856698, EPI_ISL_1856701, EPI_ISL_1856703, EPI_ISL_1856704, EPI_ISL_1856705, EPI_ISL_1856706, EPI_ISL_1856708, EPI_ISL_1856709, EPI_ISL_1856711, EPI_ISL_1856714, EPI_ISL_1856715, EPI_ISL_1856716, EPI_ISL_1856717, EPI_ISL_1856719, EPI_ISL_1856724, EPI_ISL_1856725, EPI_ISL_1856727, EPI_ISL_1856728, EPI_ISL_1856730, EPI_ISL_1856733, EPI_ISL_1856735, EPI_ISL_1856740, EPI_ISL_1856741, EPI_ISL_1856742, EPI_ISL_1856743, EPI_ISL_1856744, EPI_ISL_1856745, EPI_ISL_1856746, EPI_ISL_1856747, EPI_ISL_1856748, EPI_ISL_1856749, EPI_ISL_1856750, EPI_ISL_1856751, EPI_ISL_1856752, EPI_ISL_1856753, EPI_ISL_1856754, EPI_ISL_1856755, EPI_ISL_1856756, EPI_ISL_1856757, EPI_ISL_1856758, EPI_ISL_1856759, EPI_ISL_1856760, EPI_ISL_1856761, EPI_ISL_1856762, EPI_ISL_1856763, EPI_ISL_1856764, EPI_ISL_1856765, EPI_ISL_1856766, EPI_ISL_1856767, EPI_ISL_1856768, EPI_ISL_1856769, EPI_ISL_1856771, EPI_ISL_1856772, EPI_ISL_1856773, EPI_ISL_1856774, EPI_ISL_1856775, EPI_ISL_1856776, EPI_ISL_1856777, EPI_ISL_1856779, EPI_ISL_1856780, EPI_ISL_1856781, EPI_ISL_1856782, EPI_ISL_1856783, EPI_ISL_1856784, EPI_ISL_1856785, EPI_ISL_1856786, EPI_ISL_1856787, EPI_ISL_1856788, EPI_ISL_1856789, EPI_ISL_1856790, EPI_ISL_1856791, EPI_ISL_1856792, EPI_ISL_1856793, EPI_ISL_1856794, EPI_ISL_1856795, EPI_ISL_1856796, EPI_ISL_1856797, EPI_ISL_1856798, EPI_ISL_1856799, EPI_ISL_1856800, EPI_ISL_1856801, EPI_ISL_1856802, EPI_ISL_1856803, EPI_ISL_1856804, EPI_ISL_1856805, EPI_ISL_1856806, EPI_ISL_1856807, EPI_ISL_1856808, EPI_ISL_1856809, EPI_ISL_1856810, EPI_ISL_1856811, EPI_ISL_1856812, EPI_ISL_1856813, EPI_ISL_1856814, EPI_ISL_1856815, EPI_ISL_1856816, EPI_ISL_1856817, EPI_ISL_1856818, EPI_ISL_1856819, EPI_ISL_1856820, EPI_ISL_1856821, EPI_ISL_1856822, EPI_ISL_1856823, EPI_ISL_1856824, EPI_ISL_1856825, EPI_ISL_1856826, EPI_ISL_1856827, EPI_ISL_1856828, EPI_ISL_1856829, EPI_ISL_1856830, EPI_ISL_1856831, EPI_ISL_1856832, EPI_ISL_1856833, EPI_ISL_1856836, EPI_ISL_1856839, EPI_ISL_1856840, EPI_ISL_1856841, EPI_ISL_1856842, EPI_ISL_1856843, EPI_ISL_1856844, EPI_ISL_1856845, EPI_ISL_1856847, EPI_ISL_1856848, EPI_ISL_1856849, EPI_ISL_1856850, EPI_ISL_1856851, EPI_ISL_1856853, EPI_ISL_1856854, EPI_ISL_1856855, EPI_ISL_1856856, EPI_ISL_1856858, EPI_ISL_1856860, EPI_ISL_1856861, EPI_ISL_1856863, EPI_ISL_1856865, EPI_ISL_1856866, EPI_ISL_1856868, EPI_ISL_1856870, EPI_ISL_1856871, EPI_ISL_1856872, EPI_ISL_1856873, EPI_ISL_1856874, EPI_ISL_1856875, EPI_ISL_1856876, EPI_ISL_1856877, EPI_ISL_1856878, EPI_ISL_1856879, EPI_ISL_1856880, EPI_ISL_1856881, EPI_ISL_1856882, EPI_ISL_1856883, EPI_ISL_1856884, EPI_ISL_1856885, EPI_ISL_1856886, EPI_ISL_1856887, EPI_ISL_1856888, EPI_ISL_1856889, EPI_ISL_1856890, EPI_ISL_1856891, EPI_ISL_1856892, EPI_ISL_1856893, EPI_ISL_1856894, EPI_ISL_1856895, EPI_ISL_1856896, EPI_ISL_1856897, EPI_ISL_1856898, EPI_ISL_1856899, EPI_ISL_1856900 | | | |
| see above | UVA Clinical Microbiology Lab | Mathers Lab | Hardik I. Parikh, Katie E. Barry, Amy J. Mathers |
| EPI_ISL_1856901, EPI_ISL_1856902, EPI_ISL_1856903, EPI_ISL_1856904, EPI_ISL_1856905, EPI_ISL_1856906, EPI_ISL_1856907, EPI_ISL_1856908, EPI_ISL_1856909, EPI_ISL_1856910, EPI_ISL_1856911, EPI_ISL_1856912, EPI_ISL_1856913, EPI_ISL_1856914, EPI_ISL_1856915, EPI_ISL_1856916, EPI_ISL_1856917, EPI_ISL_1856918, EPI_ISL_1856919, EPI_ISL_1856920, EPI_ISL_1856921, EPI_ISL_1856922, EPI_ISL_1856923, EPI_ISL_1856924, EPI_ISL_1856925, EPI_ISL_1856926, EPI_ISL_1856927, EPI_ISL_1856928, EPI_ISL_1856929, EPI_ISL_1856930, EPI_ISL_1856931, EPI_ISL_1856933, EPI_ISL_1856934, EPI_ISL_1856935, EPI_ISL_1856936, EPI_ISL_1856937, EPI_ISL_1856938, EPI_ISL_1856939, EPI_ISL_1856940, EPI_ISL_1856941, EPI_ISL_1856942, EPI_ISL_1856943, EPI_ISL_1856944, EPI_ISL_1856945, EPI_ISL_1856946, EPI_ISL_1856947, EPI_ISL_1856948, EPI_ISL_1856949, EPI_ISL_1856950, EPI_ISL_1856951, EPI_ISL_1856952, EPI_ISL_1856953, EPI_ISL_1856954, EPI_ISL_1856955, EPI_ISL_1856956, EPI_ISL_1856957, EPI_ISL_1856958, EPI_ISL_1856959, EPI_ISL_1856960, EPI_ISL_1856961, EPI_ISL_1856963, EPI_ISL_1856964, EPI_ISL_1856965, EPI_ISL_1856966, EPI_ISL_1856967, EPI_ISL_1856968, EPI_ISL_1856972, EPI_ISL_1856973, EPI_ISL_1856974, EPI_ISL_1856976, EPI_ISL_1856977, EPI_ISL_1856978, EPI_ISL_1856979, EPI_ISL_1856980, EPI_ISL_1856981, EPI_ISL_1856982 | | | |
| see above | Texas Department of State Health Services (TXDSHS) | Texas Department of State Health Services (TXDSHS) | Rashmi Tuladhar, Bonnie Oh, Jenny Zhang, Maliha Rahman, Mayela Pedrueza, Anita Pokharel, Lorraine Rodriguez, Myong Koag, Chun Wang, Rachel Lee, Grace Kubin |
| EPI_ISL_1857035, EPI_ISL_1857036, EPI_ISL_1857037, EPI_ISL_1857038, EPI_ISL_1857039, EPI_ISL_1857040, EPI_ISL_1857041, EPI_ISL_1857042, EPI_ISL_1857043, EPI_ISL_1857044, EPI_ISL_1857045, EPI_ISL_1857046, EPI_ISL_1857047, EPI_ISL_1857048, EPI_ISL_1857049, EPI_ISL_1857050, EPI_ISL_1857051, EPI_ISL_1857052, EPI_ISL_1857053, EPI_ISL_1857054, EPI_ISL_1857055, EPI_ISL_1857056, EPI_ISL_1857058, EPI_ISL_1857059, EPI_ISL_1857060, EPI_ISL_1857061, EPI_ISL_1857062, EPI_ISL_1857063, EPI_ISL_1857065, EPI_ISL_1857066, EPI_ISL_1857067, EPI_ISL_1857069, EPI_ISL_1857070 | | | |
| see above | Wichita State University - Molecular Diagnostics Lab | Kansas Health and Environmental Lab | Mike Grose, Jonathan Barnell, Ben Olsen, and Phil Adam |
| EPI_ISL_1857071, EPI_ISL_1857072, EPI_ISL_1857073, EPI_ISL_1857074, EPI_ISL_1857075, EPI_ISL_1857076, EPI_ISL_1857077, EPI_ISL_1857079, EPI_ISL_1857080, EPI_ISL_1857082, EPI_ISL_1857083, EPI_ISL_1857084 | | | |
| see above | Kansas Health and Environmental Lab | Kansas Health and Environmental Lab | Mike Grose, Jonathan Barnell, Ben Olsen, and Phil Adam |
| EPI_ISL_1857085, EPI_ISL_1857086, EPI_ISL_1857087, EPI_ISL_1857088, EPI_ISL_1857089, EPI_ISL_1857090, EPI_ISL_1857091, EPI_ISL_1857092, EPI_ISL_1857093 | Wichita State University - Molecular Diagnostics Lab | Kansas Health and Environmental Lab | Mike Grose, Jonathan Barnell, Ben Olsen, and Phil Adam |
| EPI_ISL_1857099 | LESP Nuevo Leon | Instituto de Diagnostico y Referencia Epidemiologicos (INDRE) | Claudia Wong-Arambula, Abril Rodriguez-Maldonado, Vanessa Rivero-Arredondo, Ariadna Medina-Benitez, Joaquin Quiroz-Mercado, Sergio Rangel-Guerrero, Natividad Cruz-Ortiz, Tatiana Nunez-Garcia, Gisela Barrera-Badillo, Lucia Hernandez-Rivas, Irma Lopez-Martinez, Ernesto Ramirez-Gonzalez. |
| EPI_ISL_1857100 | LESP Puebla | Instituto de Diagnostico y Referencia Epidemiologicos (INDRE) | Claudia Wong-Arambula, Abril Rodriguez-Maldonado, Vanessa Rivero-Arredondo, Ariadna Medina-Benitez, Joaquin Quiroz-Mercado, Sergio Rangel-Guerrero, Natividad Cruz-Ortiz, Tatiana Nunez-Garcia, Gisela Barrera-Badillo, Lucia Hernandez-Rivas, Irma Lopez-Martinez, Ernesto Ramirez-Gonzalez. |
| EPI_ISL_1857101 | LESP Guanajuato | Instituto de Diagnostico y Referencia Epidemiologicos (INDRE) | Claudia Wong-Arambula, Abril Rodriguez-Maldonado, Vanessa Rivero-Arredondo, Ariadna Medina-Benitez, Joaquin Quiroz-Mercado, Sergio Rangel-Guerrero, Natividad Cruz-Ortiz, Tatiana Nunez-Garcia, Gisela Barrera-Badillo, Lucia Hernandez-Rivas, Irma Lopez-Martinez, Ernesto Ramirez-Gonzalez. |
| EPI_ISL_1857102, EPI_ISL_1857103, EPI_ISL_1857104, EPI_ISL_1857105 | LESP Ciudad de Mexico | Instituto de Diagnostico y Referencia Epidemiologicos (INDRE) | Claudia Wong-Arambula, Abril Rodriguez-Maldonado, Vanessa Rivero-Arredondo, Ariadna Medina-Benitez, Joaquin Quiroz-Mercado, Sergio Rangel-Guerrero, Natividad Cruz-Ortiz, Tatiana Nunez-Garcia, Gisela Barrera-Badillo, Lucia Hernandez-Rivas, Irma Lopez-Martinez, Ernesto Ramirez-Gonzalez. |

| | | | |
|---|---|---|---|
| EPI_ISL_1857106, EPI_ISL_1857107, EPI_ISL_1857108, EPI_ISL_1857109, EPI_ISL_1857110, EPI_ISL_1857111, EPI_ISL_1857112, EPI_ISL_1857113, EPI_ISL_1857114, EPI_ISL_1857115, EPI_ISL_1857116, EPI_ISL_1857117, EPI_ISL_1857118, EPI_ISL_1857119, EPI_ISL_1857120, EPI_ISL_1857121, EPI_ISL_1857122, EPI_ISL_1857123, EPI_ISL_1857124, EPI_ISL_1857125, EPI_ISL_1857126, EPI_ISL_1857127, EPI_ISL_1857128, EPI_ISL_1857129, EPI_ISL_1857130, EPI_ISL_1857131, EPI_ISL_1857132, EPI_ISL_1857133, EPI_ISL_1857134, EPI_ISL_1857135, EPI_ISL_1857136, EPI_ISL_1857137, EPI_ISL_1857138, EPI_ISL_1857139, EPI_ISL_1857140, EPI_ISL_1857141, EPI_ISL_1857142, EPI_ISL_1857143, EPI_ISL_1857144, EPI_ISL_1857145, EPI_ISL_1857146, EPI_ISL_1857147, EPI_ISL_1857148, EPI_ISL_1857149, EPI_ISL_1857150, EPI_ISL_1857151, EPI_ISL_1857152, EPI_ISL_1857153, EPI_ISL_1857154, EPI_ISL_1857155, EPI_ISL_1857156, EPI_ISL_1857157, EPI_ISL_1857158, EPI_ISL_1857159, EPI_ISL_1857160, EPI_ISL_1857161, EPI_ISL_1857162, EPI_ISL_1857163, EPI_ISL_1857164, EPI_ISL_1857165, EPI_ISL_1857166, EPI_ISL_1857167, EPI_ISL_1857168, EPI_ISL_1857169, EPI_ISL_1857170, EPI_ISL_1857171, EPI_ISL_1857172, EPI_ISL_1857173, EPI_ISL_1857174, EPI_ISL_1857175, EPI_ISL_1857176, EPI_ISL_1857177, EPI_ISL_1857178, EPI_ISL_1857179, EPI_ISL_1857180, EPI_ISL_1857181, EPI_ISL_1857182, EPI_ISL_1857183, EPI_ISL_1857184, EPI_ISL_1857185, EPI_ISL_1857186, EPI_ISL_1857187, EPI_ISL_1857188, EPI_ISL_1857189, EPI_ISL_1857190, EPI_ISL_1857191, EPI_ISL_1857192, EPI_ISL_1857193, EPI_ISL_1857194, EPI_ISL_1857195, EPI_ISL_1857196, EPI_ISL_1857197, EPI_ISL_1857198, EPI_ISL_1857199 | | | |
| see above | M Health Fairview | Minnesota Department of Health, Public Health Laboratory | Alexandra Lorentz, Jacob Garfin, Matt Plumb, and Xiong Wang |
| EPI_ISL_1857200 | LESP Tamaulipas | Instituto de Diagnostico y Referencia Epidemiologicos (INDRE) | Claudia Wong-Arambula, Abril Rodriguez-Maldonado, Vanessa Rivero-Arredondo, Ariadna Medina-Benitez, Joaquin Quiroz-Mercado, Sergio Rangel-Guerrero, Natividad Cruz-Ortiz, Tatiana Nunez-Garcia, Gisela Barrera-Badillo, Lucia Hernandez-Rivas, Irma Lopez-Martinez, Ernesto Ramirez-Gonzalez. |
| EPI_ISL_1857201 | LESP Guerrero | Instituto de Diagnostico y Referencia Epidemiologicos (INDRE) | Claudia Wong-Arambula, Abril Rodriguez-Maldonado, Vanessa Rivero-Arredondo, Ariadna Medina-Benitez, Joaquin Quiroz-Mercado, Sergio Rangel-Guerrero, Natividad Cruz-Ortiz, Tatiana Nunez-Garcia, Gisela Barrera-Badillo, Lucia Hernandez-Rivas, Irma Lopez-Martinez, Ernesto Ramirez-Gonzalez. |
| EPI_ISL_1857202, EPI_ISL_1857204, EPI_ISL_1857205, EPI_ISL_1857206 | LESP Yucatan | Instituto de Diagnostico y Referencia Epidemiologicos (INDRE) | Claudia Wong-Arambula, Abril Rodriguez-Maldonado, Vanessa Rivero-Arredondo, Ariadna Medina-Benitez, Joaquin Quiroz-Mercado, Sergio Rangel-Guerrero, Natividad Cruz-Ortiz, Tatiana Nunez-Garcia, Gisela Barrera-Badillo, Lucia Hernandez-Rivas, Irma Lopez-Martinez, Ernesto Ramirez-Gonzalez. |
| EPI_ISL_1857207 | LESP Chiapas | Instituto de Diagnostico y Referencia Epidemiologicos (INDRE) | Claudia Wong-Arambula, Abril Rodriguez-Maldonado, Vanessa Rivero-Arredondo, Ariadna Medina-Benitez, Joaquin Quiroz-Mercado, Sergio Rangel-Guerrero, Natividad Cruz-Ortiz, Tatiana Nunez-Garcia, Gisela Barrera-Badillo, Lucia Hernandez-Rivas, Irma Lopez-Martinez, Ernesto Ramirez-Gonzalez. |
| EPI_ISL_1857208, EPI_ISL_1857209, EPI_ISL_1857210, EPI_ISL_1857211, EPI_ISL_1857212, EPI_ISL_1857214, EPI_ISL_1857215, EPI_ISL_1857216 | LESP Queretaro | Instituto de Diagnostico y Referencia Epidemiologicos (INDRE) | Claudia Wong-Arambula, Abril Rodriguez-Maldonado, Vanessa Rivero-Arredondo, Ariadna Medina-Benitez, Joaquin Quiroz-Mercado, Sergio Rangel-Guerrero, Natividad Cruz-Ortiz, Tatiana Nunez-Garcia, Gisela Barrera-Badillo, Lucia Hernandez-Rivas, Irma Lopez-Martinez, Ernesto Ramirez-Gonzalez. |
| EPI_ISL_1857217 | LESP Ciudad de Mexico | Instituto de Diagnostico y Referencia Epidemiologicos (INDRE) | Claudia Wong-Arambula, Abril Rodriguez-Maldonado, Vanessa Rivero-Arredondo, Ariadna Medina-Benitez, Joaquin Quiroz-Mercado, Sergio Rangel-Guerrero, Natividad Cruz-Ortiz, Tatiana Nunez-Garcia, Gisela Barrera-Badillo, Lucia Hernandez-Rivas, Irma Lopez-Martinez, Ernesto Ramirez-Gonzalez. |
| EPI_ISL_1857218 | LESP Queretaro | Instituto de Diagnostico y Referencia Epidemiologicos (INDRE) | Claudia Wong-Arambula, Abril Rodriguez-Maldonado, Vanessa Rivero-Arredondo, Ariadna Medina-Benitez, Joaquin Quiroz-Mercado, Sergio Rangel-Guerrero, Natividad Cruz-Ortiz, Tatiana Nunez-Garcia, Gisela Barrera-Badillo, Lucia Hernandez-Rivas, Irma Lopez-Martinez, Ernesto Ramirez-Gonzalez. |
| EPI_ISL_1857219, EPI_ISL_1857220, EPI_ISL_1857221, EPI_ISL_1857222, EPI_ISL_1857223 | LESP Chihuahua | Instituto de Diagnostico y Referencia Epidemiologicos (INDRE) | Claudia Wong-Arambula, Abril Rodriguez-Maldonado, Vanessa Rivero-Arredondo, Ariadna Medina-Benitez, Joaquin Quiroz-Mercado, Sergio Rangel-Guerrero, Natividad Cruz-Ortiz, Tatiana Nunez-Garcia, Gisela Barrera-Badillo, Lucia Hernandez-Rivas, Irma Lopez-Martinez, Ernesto Ramirez-Gonzalez. |
| EPI_ISL_1857224, EPI_ISL_1857225, EPI_ISL_1857226, EPI_ISL_1857227 | LESP Baja California Sur | Instituto de Diagnostico y Referencia Epidemiologicos (INDRE) | Claudia Wong-Arambula, Abril Rodriguez-Maldonado, Vanessa Rivero-Arredondo, Ariadna Medina-Benitez, Joaquin Quiroz-Mercado, Sergio Rangel-Guerrero, Natividad Cruz-Ortiz, Tatiana Nunez-Garcia, Gisela Barrera-Badillo, Lucia Hernandez-Rivas, Irma Lopez-Martinez, Ernesto Ramirez-Gonzalez. |
| EPI_ISL_1857228, EPI_ISL_1857229 | LESP Quintana Roo | Instituto de Diagnostico y Referencia Epidemiologicos (INDRE) | Claudia Wong-Arambula, Abril Rodriguez-Maldonado, Vanessa Rivero-Arredondo, Ariadna Medina-Benitez, Joaquin Quiroz-Mercado, Sergio Rangel-Guerrero, Natividad Cruz-Ortiz, Tatiana Nunez-Garcia, Gisela Barrera-Badillo, Lucia Hernandez-Rivas, Irma Lopez-Martinez, Ernesto Ramirez-Gonzalez. |
| EPI_ISL_1857230 | LESP Jalisco | Instituto de Diagnostico y Referencia Epidemiologicos (INDRE) | Claudia Wong-Arambula, Abril Rodriguez-Maldonado, Vanessa Rivero-Arredondo, Ariadna Medina-Benitez, Joaquin Quiroz-Mercado, Sergio Rangel-Guerrero, Natividad Cruz-Ortiz, Tatiana Nunez-Garcia, Gisela Barrera-Badillo, Lucia Hernandez-Rivas, Irma Lopez-Martinez, Ernesto Ramirez-Gonzalez. |
| EPI_ISL_1857232 | LESP Coahuila | Instituto de Diagnostico y Referencia Epidemiologicos (INDRE) | Claudia Wong-Arambula, Abril Rodriguez-Maldonado, Vanessa Rivero-Arredondo, Ariadna Medina-Benitez, Joaquin Quiroz-Mercado, Sergio Rangel-Guerrero, Natividad Cruz-Ortiz, Tatiana Nunez-Garcia, Gisela Barrera-Badillo, Lucia Hernandez-Rivas, Irma Lopez-Martinez, Ernesto Ramirez-Gonzalez. |
| EPI_ISL_1857233, EPI_ISL_1857234 | LESP Hidalgo | Instituto de Diagnostico y Referencia Epidemiologicos (INDRE) | Claudia Wong-Arambula, Abril Rodriguez-Maldonado, Vanessa Rivero-Arredondo, Ariadna Medina-Benitez, Joaquin Quiroz-Mercado, Sergio Rangel-Guerrero, Natividad Cruz-Ortiz, Tatiana Nunez-Garcia, Gisela Barrera-Badillo, Lucia Hernandez-Rivas, Irma Lopez-Martinez, Ernesto Ramirez-Gonzalez. |
| EPI_ISL_1857235 | LESP Baja California Sur | Instituto de Diagnostico y Referencia Epidemiologicos (INDRE) | Claudia Wong-Arambula, Abril Rodriguez-Maldonado, Vanessa Rivero-Arredondo, Ariadna Medina-Benitez, Joaquin Quiroz-Mercado, Sergio Rangel-Guerrero, Natividad Cruz-Ortiz, Tatiana Nunez-Garcia, Gisela Barrera-Badillo, Lucia Hernandez-Rivas, Irma Lopez-Martinez, Ernesto Ramirez-Gonzalez. |
| EPI_ISL_1857236, EPI_ISL_1857237, EPI_ISL_1857238, EPI_ISL_1857239 | LESP Guanajuato | Instituto de Diagnostico y Referencia Epidemiologicos (INDRE) | Claudia Wong-Arambula, Abril Rodriguez-Maldonado, Vanessa Rivero-Arredondo, Ariadna Medina-Benitez, Joaquin Quiroz-Mercado, Sergio Rangel-Guerrero, Natividad Cruz-Ortiz, Tatiana Nunez-Garcia, Gisela Barrera-Badillo, Lucia Hernandez-Rivas, Irma Lopez-Martinez, Ernesto Ramirez-Gonzalez. |
| EPI_ISL_1857241, EPI_ISL_1857242, EPI_ISL_1857243 | LESP Puebla | Instituto de Diagnostico y Referencia Epidemiologicos (INDRE) | Claudia Wong-Arambula, Abril Rodriguez-Maldonado, Vanessa Rivero-Arredondo, Ariadna Medina-Benitez, Joaquin Quiroz-Mercado, Sergio Rangel-Guerrero, Natividad Cruz-Ortiz, Tatiana Nunez-Garcia, Gisela Barrera-Badillo, Lucia Hernandez-Rivas, Irma Lopez-Martinez, Ernesto Ramirez-Gonzalez. |
| EPI_ISL_1857244 | LESP Jalisco | Instituto de Diagnostico y Referencia Epidemiologicos (INDRE) | Claudia Wong-Arambula, Abril Rodriguez-Maldonado, Vanessa Rivero-Arredondo, Ariadna Medina-Benitez, Joaquin Quiroz-Mercado, Sergio Rangel-Guerrero, Natividad Cruz-Ortiz, Tatiana Nunez-Garcia, Gisela Barrera-Badillo, Lucia Hernandez-Rivas, Irma Lopez-Martinez, Ernesto Ramirez-Gonzalez. |
| EPI_ISL_1857245, EPI_ISL_1857246 | LESP Queretaro | Instituto de Diagnostico y Referencia Epidemiologicos (INDRE) | Claudia Wong-Arambula, Abril Rodriguez-Maldonado, Vanessa Rivero-Arredondo, Ariadna Medina-Benitez, Joaquin Quiroz-Mercado, Sergio Rangel-Guerrero, Natividad Cruz-Ortiz, Tatiana Nunez-Garcia, Gisela Barrera-Badillo, Lucia Hernandez-Rivas, Irma Lopez-Martinez, Ernesto Ramirez-Gonzalez. |
| EPI_ISL_1857248, EPI_ISL_1857249, EPI_ISL_1857250, EPI_ISL_1857251 | LESP Morelos | Instituto de Diagnostico y Referencia Epidemiologicos (INDRE) | Claudia Wong-Arambula, Abril Rodriguez-Maldonado, Vanessa Rivero-Arredondo, Ariadna Medina-Benitez, Joaquin Quiroz-Mercado, Sergio Rangel-Guerrero, Natividad Cruz-Ortiz, Tatiana Nunez-Garcia, Gisela Barrera-Badillo, Lucia Hernandez-Rivas, Irma Lopez-Martinez, Ernesto Ramirez-Gonzalez. |
| EPI_ISL_1857252 | LESP Guerrero | Instituto de Diagnostico y Referencia Epidemiologicos (INDRE) | Claudia Wong-Arambula, Abril Rodriguez-Maldonado, Vanessa Rivero-Arredondo, Ariadna Medina-Benitez, Joaquin Quiroz-Mercado, Sergio Rangel-Guerrero, Natividad Cruz-Ortiz, Tatiana Nunez-Garcia, Gisela Barrera-Badillo, Lucia Hernandez-Rivas, Irma Lopez-Martinez, Ernesto Ramirez-Gonzalez. |
| EPI_ISL_1857253 | LESP Yucatan | Instituto de Diagnostico y Referencia Epidemiologicos (INDRE) | Claudia Wong-Arambula, Abril Rodriguez-Maldonado, Vanessa Rivero-Arredondo, Ariadna Medina-Benitez, Joaquin Quiroz-Mercado, Sergio Rangel-Guerrero, Natividad Cruz-Ortiz, Tatiana Nunez-Garcia, Gisela Barrera-Badillo, Lucia Hernandez-Rivas, Irma Lopez-Martinez, Ernesto Ramirez-Gonzalez. |
| EPI_ISL_1857254 | LESP Tamaulipas | Instituto de Diagnostico y Referencia Epidemiologicos (INDRE) | Claudia Wong-Arambula, Abril Rodriguez-Maldonado, Vanessa Rivero-Arredondo, Ariadna Medina-Benitez, Joaquin Quiroz-Mercado, Sergio Rangel-Guerrero, Natividad Cruz-Ortiz, Tatiana Nunez-Garcia, Gisela Barrera-Badillo, Lucia Hernandez-Rivas, Irma Lopez-Martinez, Ernesto Ramirez-Gonzalez. |
| EPI_ISL_1857255, EPI_ISL_1857256 | LESP Oaxaca | Instituto de Diagnostico y Referencia Epidemiologicos (INDRE) | Claudia Wong-Arambula, Abril Rodriguez-Maldonado, Vanessa Rivero-Arredondo, Ariadna Medina-Benitez, Joaquin Quiroz-Mercado, Sergio Rangel-Guerrero, Natividad Cruz-Ortiz, Tatiana Nunez-Garcia, Gisela Barrera-Badillo, Lucia Hernandez-Rivas, Irma Lopez-Martinez, Ernesto Ramirez-Gonzalez. |
| EPI_ISL_1857257 | LESP Tlaxcala | Instituto de Diagnostico y Referencia Epidemiologicos (INDRE) | Claudia Wong-Arambula, Abril Rodriguez-Maldonado, Vanessa Rivero-Arredondo, Ariadna Medina-Benitez, Joaquin Quiroz-Mercado, Sergio Rangel-Guerrero, Natividad Cruz-Ortiz, Tatiana Nunez-Garcia, Gisela Barrera-Badillo, Lucia Hernandez-Rivas, Irma Lopez-Martinez, Ernesto Ramirez-Gonzalez. |
| EPI_ISL_1857258, EPI_ISL_1857259, EPI_ISL_1857260, EPI_ISL_1857262 | LESP Chihuahua | Instituto de Diagnostico y Referencia Epidemiologicos (INDRE) | Claudia Wong-Arambula, Abril Rodriguez-Maldonado, Vanessa Rivero-Arredondo, Ariadna Medina-Benitez, Joaquin Quiroz-Mercado, Sergio Rangel-Guerrero, Natividad Cruz-Ortiz, Tatiana Nunez-Garcia, Gisela Barrera-Badillo, Lucia Hernandez-Rivas, Irma Lopez-Martinez, Ernesto Ramirez-Gonzalez. |
| EPI_ISL_1857263 | LESP Coahuila | Instituto de Diagnostico y Referencia Epidemiologicos (INDRE) | Claudia Wong-Arambula, Abril Rodriguez-Maldonado, Vanessa Rivero-Arredondo, Ariadna Medina-Benitez, Joaquin Quiroz-Mercado, Sergio Rangel-Guerrero, Natividad Cruz-Ortiz, Tatiana Nunez-Garcia, Gisela Barrera-Badillo, Lucia Hernandez-Rivas, Irma Lopez-Martinez, Ernesto Ramirez-Gonzalez. |
| EPI_ISL_1857264, EPI_ISL_1857265 | LESP Zacatecas | Instituto de Diagnostico y Referencia Epidemiologicos (INDRE) | Claudia Wong-Arambula, Abril Rodriguez-Maldonado, Vanessa Rivero-Arredondo, Ariadna Medina-Benitez, Joaquin Quiroz-Mercado, Sergio Rangel-Guerrero, Natividad Cruz-Ortiz, Tatiana Nunez-Garcia, Gisela Barrera-Badillo, Lucia Hernandez-Rivas, Irma Lopez-Martinez, Ernesto Ramirez-Gonzalez. |
| EPI_ISL_1857266, EPI_ISL_1857267 | LESP Baja California Sur | Instituto de Diagnostico y Referencia Epidemiologicos (INDRE) | Claudia Wong-Arambula, Abril Rodriguez-Maldonado, Vanessa Rivero-Arredondo, Ariadna Medina-Benitez, Joaquin Quiroz-Mercado, Sergio Rangel-Guerrero, Natividad Cruz-Ortiz, Tatiana Nunez-Garcia, Gisela Barrera-Badillo, Lucia Hernandez-Rivas, Irma Lopez-Martinez, Ernesto Ramirez-Gonzalez. |
| EPI_ISL_1857268, EPI_ISL_1857269, EPI_ISL_1857272, EPI_ISL_1857273, EPI_ISL_1857274, EPI_ISL_1857275 | LESP Ciudad de Mexico | Instituto de Diagnostico y Referencia Epidemiologicos (INDRE) | Claudia Wong-Arambula, Abril Rodriguez-Maldonado, Vanessa Rivero-Arredondo, Ariadna Medina-Benitez, Joaquin Quiroz-Mercado, Sergio Rangel-Guerrero, Natividad Cruz-Ortiz, Tatiana Nunez-Garcia, Gisela Barrera-Badillo, Lucia Hernandez-Rivas, Irma Lopez-Martinez, Ernesto Ramirez-Gonzalez. |
| EPI_ISL_1857276 | LESP Colima | Instituto de Diagnostico y Referencia Epidemiologicos (INDRE) | Claudia Wong-Arambula, Abril Rodriguez-Maldonado, Vanessa Rivero-Arredondo, Ariadna Medina-Benitez, Joaquin Quiroz-Mercado, Sergio Rangel-Guerrero, Natividad Cruz-Ortiz, Tatiana Nunez-Garcia, Gisela Barrera-Badillo, Lucia Hernandez-Rivas, Irma Lopez-Martinez, Ernesto Ramirez-Gonzalez. |
| EPI_ISL_1857295 | Instituto Nacional de Medicina Genomica | Instituto Nacional de Medicina Genomica | Hidalgo-Miranda A, Mendoza-Vargas A, Reyes-Grajeda JP, Cedro-Tanda A, Alcaraz N, Cisneros-Villanueva M, Orjeda-Rodriguez M, Gonzalez-Barrera D, Rangel-DeLeon D, Miranda-Ortiz H, Rosas-Escobar P, Canseco Mendez JC, Munguia-Garza P, Ramirez-Vega O, Garcia-Cardenas FJ, Gonzalez-Woge MA, Herrera-Montalvo LA |
| EPI_ISL_1857296, EPI_ISL_1857297, EPI_ISL_1857298, EPI_ISL_1857299, EPI_ISL_1857300, EPI_ISL_1857301, EPI_ISL_1857302, EPI_ISL_1857303, EPI_ISL_1857304, EPI_ISL_1857305, EPI_ISL_1857306, EPI_ISL_1857307, EPI_ISL_1857308, EPI_ISL_1857309, EPI_ISL_1857310, EPI_ISL_1857311, EPI_ISL_1857312, EPI_ISL_1857313, EPI_ISL_1857314, EPI_ISL_1857315, EPI_ISL_1857316, EPI_ISL_1857317, EPI_ISL_1857318, EPI_ISL_1857319, EPI_ISL_1857320, EPI_ISL_1857321, EPI_ISL_1857322, EPI_ISL_1857323, EPI_ISL_1857324, EPI_ISL_1857325, EPI_ISL_1857326, EPI_ISL_1857327, EPI_ISL_1857328, EPI_ISL_1857329, EPI_ISL_1857330, EPI_ISL_1857331, EPI_ISL_1857332, EPI_ISL_1857333, EPI_ISL_1857334, EPI_ISL_1857335, EPI_ISL_1857336, EPI_ISL_1857337, EPI_ISL_1857338, EPI_ISL_1857339, EPI_ISL_1857340, EPI_ISL_1857341, EPI_ISL_1857342, EPI_ISL_1857343, EPI_ISL_1857344, EPI_ISL_1857345, EPI_ISL_1857346, EPI_ISL_1857347, EPI_ISL_1857348, EPI_ISL_1857349, EPI_ISL_1857350, EPI_ISL_1857351, EPI_ISL_1857352, EPI_ISL_1857353, EPI_ISL_1857354, EPI_ISL_1857355, EPI_ISL_1857356, EPI_ISL_1857357, EPI_ISL_1857358, EPI_ISL_1857359, EPI_ISL_1857360, EPI_ISL_1857361, | | | |

| | | | |
|---|---|---|---|
| EPI_ISL_1857362, EPI_ISL_1857363, EPI_ISL_1857364, EPI_ISL_1857365, EPI_ISL_1857366, EPI_ISL_1857367, EPI_ISL_1857368, EPI_ISL_1857369, EPI_ISL_1857371, EPI_ISL_1857372, EPI_ISL_1857373, EPI_ISL_1857374, EPI_ISL_1857375, EPI_ISL_1857376, EPI_ISL_1857377, EPI_ISL_1857378, EPI_ISL_1857379, EPI_ISL_1857380, EPI_ISL_1857381, EPI_ISL_1857382, EPI_ISL_1857383, EPI_ISL_1857384, EPI_ISL_1857385, EPI_ISL_1857386, EPI_ISL_1857387, EPI_ISL_1857388, EPI_ISL_1857389, EPI_ISL_1857390, EPI_ISL_1857391, EPI_ISL_1857392, EPI_ISL_1857393, EPI_ISL_1857394, EPI_ISL_1857395, EPI_ISL_1857396, EPI_ISL_1857397, EPI_ISL_1857398, EPI_ISL_1857399, EPI_ISL_1857400, EPI_ISL_1857401, EPI_ISL_1857402, EPI_ISL_1857403, EPI_ISL_1857404, EPI_ISL_1857405, EPI_ISL_1857406, EPI_ISL_1857408, EPI_ISL_1857409, EPI_ISL_1857411, EPI_ISL_1857412, EPI_ISL_1857413, EPI_ISL_1857414, EPI_ISL_1857415, EPI_ISL_1857416, EPI_ISL_1857417, EPI_ISL_1857418, EPI_ISL_1857419, EPI_ISL_1857420, EPI_ISL_1857421, EPI_ISL_1857422, EPI_ISL_1857423, EPI_ISL_1857424, EPI_ISL_1857425, EPI_ISL_1857426, EPI_ISL_1857427, EPI_ISL_1857429, EPI_ISL_1857431, EPI_ISL_1857432, EPI_ISL_1857433, EPI_ISL_1857434, EPI_ISL_1857435, EPI_ISL_1857436, EPI_ISL_1857437, EPI_ISL_1857438, EPI_ISL_1857439, EPI_ISL_1857440, EPI_ISL_1857441, EPI_ISL_1857442, EPI_ISL_1857443, EPI_ISL_1857445, EPI_ISL_1857446, EPI_ISL_1857447, EPI_ISL_1857448, EPI_ISL_1857449, EPI_ISL_1857450, EPI_ISL_1857451, EPI_ISL_1857452, EPI_ISL_1857453, EPI_ISL_1857455, EPI_ISL_1857456, EPI_ISL_1857457, EPI_ISL_1857458, EPI_ISL_1857461, EPI_ISL_1857462, EPI_ISL_1857463, EPI_ISL_1857464, EPI_ISL_1857465, EPI_ISL_1857466 | | | |
| see above | Wyoming Public Health Laboratory | Wyoming Public Health Laboratory | Jim Mildenberger, Taylor Fearing, Channing Weber, Ashley Norberg, Chayse Rowley, Marley Goetz, Brian Dominguez, Elliot Thomasson, Brittany Oher, Sam Britz, Cari Sloma, and Rob Christensen |
| EPI_ISL_1857468, EPI_ISL_1857469, EPI_ISL_1857470, EPI_ISL_1857471, EPI_ISL_1857472, EPI_ISL_1857474, EPI_ISL_1857475, EPI_ISL_1857476, EPI_ISL_1857477, EPI_ISL_1857479, EPI_ISL_1857480, EPI_ISL_1857481, EPI_ISL_1857482, EPI_ISL_1857483, EPI_ISL_1857484, EPI_ISL_1857485, EPI_ISL_1857486, EPI_ISL_1857487 | | | |
| see above | Southern Nevada Public Health Laboratory | Southern Nevada Public Health Laboratory | Michael Picker |
| EPI_ISL_1857489, EPI_ISL_1857490, EPI_ISL_1857491, EPI_ISL_1857492, EPI_ISL_1857493, EPI_ISL_1857494, EPI_ISL_1857495, EPI_ISL_1857496, EPI_ISL_1857497, EPI_ISL_1857498, EPI_ISL_1857499, EPI_ISL_1857500, EPI_ISL_1857501, EPI_ISL_1857502, EPI_ISL_1857503, EPI_ISL_1857504, EPI_ISL_1857505, EPI_ISL_1857506, EPI_ISL_1857507, EPI_ISL_1857508, EPI_ISL_1857509, EPI_ISL_1857510, EPI_ISL_1857511, EPI_ISL_1857512, EPI_ISL_1857513, EPI_ISL_1857514, EPI_ISL_1857515, EPI_ISL_1857516, EPI_ISL_1857517, EPI_ISL_1857518, EPI_ISL_1857519, EPI_ISL_1857520, EPI_ISL_1857521, EPI_ISL_1857522, EPI_ISL_1857523, EPI_ISL_1857524, EPI_ISL_1857525, EPI_ISL_1857526, EPI_ISL_1857527, EPI_ISL_1857528, EPI_ISL_1857529, EPI_ISL_1857530, EPI_ISL_1857531, EPI_ISL_1857532, EPI_ISL_1857533, EPI_ISL_1857534, EPI_ISL_1857535, EPI_ISL_1857536, EPI_ISL_1857537, EPI_ISL_1857538, EPI_ISL_1857539, EPI_ISL_1857540, EPI_ISL_1857541, EPI_ISL_1857542, EPI_ISL_1857543, EPI_ISL_1857544, EPI_ISL_1857545, EPI_ISL_1857546, EPI_ISL_1857547, EPI_ISL_1857548, EPI_ISL_1857549, EPI_ISL_1857550, EPI_ISL_1857551, EPI_ISL_1857552, EPI_ISL_1857553, EPI_ISL_1857554, EPI_ISL_1857555, EPI_ISL_1857556, EPI_ISL_1857557, EPI_ISL_1857558, EPI_ISL_1857559, EPI_ISL_1857560, EPI_ISL_1857561, EPI_ISL_1857562, EPI_ISL_1857563, EPI_ISL_1857564, EPI_ISL_1857565, EPI_ISL_1857566, EPI_ISL_1857567, EPI_ISL_1857568, EPI_ISL_1857569, EPI_ISL_1857570, EPI_ISL_1857571, EPI_ISL_1857572, EPI_ISL_1857573, EPI_ISL_1857574, EPI_ISL_1857575, EPI_ISL_1857576, EPI_ISL_1857577 | | | |
| see above | Minnesota Department of Health, Public Health Laboratory | Minnesota Department of Health, Public Health Laboratory | Alexandra Lorentz, Jacob Garfin, Matt Plumb, and Xiong Wang |
| EPI_ISL_1860086, EPI_ISL_1860088, EPI_ISL_1860090, EPI_ISL_1860094, EPI_ISL_1860095, EPI_ISL_1860100, EPI_ISL_1860104, EPI_ISL_1860107, EPI_ISL_1860112, EPI_ISL_1860116, EPI_ISL_1860124, EPI_ISL_1860128, EPI_ISL_1860133, EPI_ISL_1860136, EPI_ISL_1860150 | | | |
| see above | Wyoming Public Health Laboratory | Wyoming Public Health Laboratory | Jim Mildenberger, Taylor Fearing, Channing Weber, Ashley Norberg, Chayse Rowley, Marley Goetz, Brian Dominguez, Elliot Thomasson, Brittany Oher, Sam Britz, Cari Sloma, and Rob Christensen |
| EPI_ISL_1866418, EPI_ISL_1866423 | WVT Laboratory, Corvallis, Oregon | OSU Center for Genome Research and Biocomputing | Oregon State University TRACE Project |
| EPI_ISL_1867240, EPI_ISL_1867251, EPI_ISL_1867256, EPI_ISL_1867262, EPI_ISL_1867274, EPI_ISL_1867276, EPI_ISL_1867278, EPI_ISL_1867280, EPI_ISL_1867282, EPI_ISL_1867284, EPI_ISL_1867286, EPI_ISL_1867288, EPI_ISL_1867290, EPI_ISL_1867292, EPI_ISL_1867294, EPI_ISL_1867296, EPI_ISL_1867304, EPI_ISL_1867311, EPI_ISL_1867318, EPI_ISL_1867320, EPI_ISL_1867322, EPI_ISL_1867324, EPI_ISL_1867326, EPI_ISL_1867329, EPI_ISL_1867335, EPI_ISL_1867341, EPI_ISL_1867346, EPI_ISL_1867350, EPI_ISL_1867352, EPI_ISL_1867354, EPI_ISL_1867356, EPI_ISL_1867365, EPI_ISL_1867370, EPI_ISL_1867376, EPI_ISL_1867378, EPI_ISL_1867380, EPI_ISL_1867382, EPI_ISL_1867384, EPI_ISL_1867386, EPI_ISL_1867394, EPI_ISL_1867401, EPI_ISL_1867407, EPI_ISL_1867409, EPI_ISL_1867413, EPI_ISL_1867415, EPI_ISL_1867417, EPI_ISL_1867424, EPI_ISL_1867430, EPI_ISL_1867436, EPI_ISL_1867443, EPI_ISL_1867444, EPI_ISL_1867446, EPI_ISL_1867450, EPI_ISL_1867452, EPI_ISL_1867454, EPI_ISL_1867456, EPI_ISL_1867458, EPI_ISL_1867460, EPI_ISL_1867462, EPI_ISL_1867465, EPI_ISL_1867467, EPI_ISL_1867469, EPI_ISL_1867470 | | | |
| see above | Arizona State University | Arizona State University | Rabia Maqsood, LaRinda A. Holland, Matthew F. Smith, Peter T. Skidmore, Nicholas J. Mellor, Joy M. Blain, Valerie Harris, Joshua LaBaer, Vel Murugan, Efrem S. Lim |
| EPI_ISL_1892486, EPI_ISL_1892488, EPI_ISL_1892490, EPI_ISL_1892494, EPI_ISL_1892496, EPI_ISL_1892498, EPI_ISL_1892500, EPI_ISL_1892502, EPI_ISL_1892504, EPI_ISL_1892506, EPI_ISL_1892507, EPI_ISL_1892509, EPI_ISL_1892511, EPI_ISL_1892513, EPI_ISL_1892515, EPI_ISL_1892517, EPI_ISL_1892519, EPI_ISL_1892521, EPI_ISL_1892523, EPI_ISL_1892524, EPI_ISL_1892526, EPI_ISL_1892528, EPI_ISL_1892530, EPI_ISL_1892533, EPI_ISL_1892535, EPI_ISL_1892537, EPI_ISL_1892539, EPI_ISL_1892540, EPI_ISL_1892542, EPI_ISL_1892545, EPI_ISL_1892547, EPI_ISL_1892549, EPI_ISL_1892551, EPI_ISL_1892553, EPI_ISL_1892555, EPI_ISL_1892557, EPI_ISL_1892559, EPI_ISL_1892564, EPI_ISL_1892569, EPI_ISL_1892571, EPI_ISL_1892573, EPI_ISL_1892575, EPI_ISL_1892577, EPI_ISL_1892583, EPI_ISL_1892585, EPI_ISL_1892586, EPI_ISL_1892588, EPI_ISL_1892590, EPI_ISL_1892592, EPI_ISL_1892596, EPI_ISL_1892599, EPI_ISL_1892604, EPI_ISL_1892609, EPI_ISL_1892613, EPI_ISL_1892615, EPI_ISL_1892617 | | | |
| see above | NJDOH, Public Health and Environmental Laboratories | NJ_PHEL | Lindsey Bodnar, Shiv K. Verma, Jacquelyn Deverell, Matthew Scarnati, Dana Woell, Byeong Jeong |
| EPI_ISL_1893796, EPI_ISL_1893798, EPI_ISL_1893800, EPI_ISL_1893802, EPI_ISL_1893805, EPI_ISL_1893811, EPI_ISL_1893813, EPI_ISL_1893815, EPI_ISL_1893818, EPI_ISL_1893820, EPI_ISL_1893822, EPI_ISL_1893829, EPI_ISL_1893836, EPI_ISL_1893843, EPI_ISL_1893849, EPI_ISL_1893856, EPI_ISL_1893861, EPI_ISL_1893867, EPI_ISL_1893869, EPI_ISL_1893872, EPI_ISL_1893875, EPI_ISL_1893877, EPI_ISL_1893879, EPI_ISL_1893881, EPI_ISL_1893883, EPI_ISL_1893886, EPI_ISL_1893888, EPI_ISL_1893890, EPI_ISL_1893892, EPI_ISL_1893894, EPI_ISL_1893897, EPI_ISL_1893904, EPI_ISL_1893906, EPI_ISL_1893908, EPI_ISL_1893910, EPI_ISL_1895852, EPI_ISL_1895857, EPI_ISL_1895862, EPI_ISL_1895868, EPI_ISL_1895873, EPI_ISL_1895878, EPI_ISL_1895884, EPI_ISL_1895889, EPI_ISL_1895893, EPI_ISL_1895899, EPI_ISL_1895904, EPI_ISL_1895909, EPI_ISL_1895915, EPI_ISL_1895921, EPI_ISL_1895926, EPI_ISL_1895932, EPI_ISL_1895938, EPI_ISL_1895943, EPI_ISL_1895949, EPI_ISL_1895954, EPI_ISL_1895959, EPI_ISL_1895964, EPI_ISL_1895969, EPI_ISL_1895974, EPI_ISL_1895980, EPI_ISL_1895984, EPI_ISL_1895990, EPI_ISL_1895995, EPI_ISL_1896000, EPI_ISL_1896005, EPI_ISL_1896010, EPI_ISL_1896015, EPI_ISL_1896020, EPI_ISL_1896025, EPI_ISL_1896030, EPI_ISL_1896035, EPI_ISL_1896041 | | | |
| see above | Alaska State Virology Laboratory | Alaska State Virology Laboratory | Stephanie DeRonde, Elva House, Jacob Zidek, Lisa Smith, Ph.D., Jack Chen, Ph.D. |
| EPI_ISL_1897258, EPI_ISL_1897259, EPI_ISL_1897260, EPI_ISL_1897261, EPI_ISL_1897262, EPI_ISL_1897263, EPI_ISL_1897264, EPI_ISL_1897265, EPI_ISL_1897266, EPI_ISL_1897268, EPI_ISL_1897269, EPI_ISL_1897270, EPI_ISL_1897271, EPI_ISL_1897272, EPI_ISL_1897273, EPI_ISL_1897274, EPI_ISL_1897275, EPI_ISL_1897277, EPI_ISL_1897278, EPI_ISL_1897279, EPI_ISL_1897280, EPI_ISL_1897281, EPI_ISL_1897282, EPI_ISL_1897283, EPI_ISL_1897284, EPI_ISL_1897286, EPI_ISL_1897287, EPI_ISL_1897288, EPI_ISL_1897289, EPI_ISL_1897290, EPI_ISL_1897291, EPI_ISL_1897293, EPI_ISL_1897294, EPI_ISL_1897296, EPI_ISL_1897297, EPI_ISL_1897298, EPI_ISL_1897299, EPI_ISL_1897300 | | | |
| see above | WVT Laboratory, Corvallis, Oregon | OSU Center for Genome Research and Biocomputing | Oregon State University TRACE Project |
| EPI_ISL_1897351, EPI_ISL_1897352, EPI_ISL_1897353, EPI_ISL_1897354 | Altius Institute | Seattle Flu Study | Deborah A. Nickerson, Chris D. Frazar, Jover Lee, Benjamin Pelle, Erica Ryke, Matthew Richardson, Amanda Adler, Elisabeth Brandstetter, Peter D. Han, Kairsten Fay, Misja Ilcisin, Kirsten Lacombe, Thomas R. Sibley, Melissa Truong, Caitlin R. Wolf, Ryan Alexander, Daniel Bates, Rebecca Bruders, Stephanie DeBaun, Clem Green, Muhammad Halimun, Jessica Halow, Kneshay Harper, Matt Hartman, Andrew Meuser, Alex Nguyen, Truong Nguyen, Sofia Olsson, Sadie Patraw, Hannah Petersen, Tobias Ragoczy, Joshua Richards, Jacob Rodriguez, John Stamatoyannopoulos, Julia Wald, Olivia Waltner, Michael Boeckh, Janet A. Englund, Michael Famulare, Barry R. Lutz, Mark J. Rieder, Lea M. Starita, Matthew Thompson, Helen Y. Chu, Jay Shendure, Trevor Bedford |
| EPI_ISL_1897355, EPI_ISL_1897356, EPI_ISL_1897357, EPI_ISL_1897358, EPI_ISL_1897359, EPI_ISL_1897360, EPI_ISL_1897361, EPI_ISL_1897362, EPI_ISL_1897363, EPI_ISL_1897364, EPI_ISL_1897365, EPI_ISL_1897366, EPI_ISL_1897367, EPI_ISL_1897368, EPI_ISL_1897369, EPI_ISL_1897370, EPI_ISL_1897371, EPI_ISL_1897372, EPI_ISL_1897373, EPI_ISL_1897374, EPI_ISL_1897375, EPI_ISL_1897376, EPI_ISL_1897377, EPI_ISL_1897378, EPI_ISL_1897379, EPI_ISL_1897380, EPI_ISL_1897381, EPI_ISL_1897382, EPI_ISL_1897383, EPI_ISL_1897384, EPI_ISL_1897385, EPI_ISL_1897386, EPI_ISL_1897387, EPI_ISL_1897388, EPI_ISL_1897389, EPI_ISL_1897390, EPI_ISL_1897391, EPI_ISL_1897392, EPI_ISL_1897393, EPI_ISL_1897394, EPI_ISL_1897395, EPI_ISL_1897396, EPI_ISL_1897397, EPI_ISL_1897398, EPI_ISL_1897399, EPI_ISL_1897400, EPI_ISL_1897401, EPI_ISL_1897402, EPI_ISL_1897403, EPI_ISL_1897404, EPI_ISL_1897405, EPI_ISL_1897406, EPI_ISL_1897407, EPI_ISL_1897408, EPI_ISL_1897409, EPI_ISL_1897410, EPI_ISL_1897411, EPI_ISL_1897412, EPI_ISL_1897413, EPI_ISL_1897414, EPI_ISL_1897415, EPI_ISL_1897416, EPI_ISL_1897417, EPI_ISL_1897418, EPI_ISL_1897419, EPI_ISL_1897420, EPI_ISL_1897421, EPI_ISL_1897422, EPI_ISL_1897423, EPI_ISL_1897424, EPI_ISL_1897425, EPI_ISL_1897426, EPI_ISL_1897427, EPI_ISL_1897428, EPI_ISL_1897429, EPI_ISL_1897430, EPI_ISL_1897431, EPI_ISL_1897432, EPI_ISL_1897433 | | | |
| see above | Washington State Department of Health Public Health Laboratories | Washington State Department of Health Public Health Laboratories | Drew MacKellar, Philip Dykema, Denny Russell, Joenice Gonzalez, Hannah Gray, Geoff Melly, Vanessa De Los Santos, Darren Lucas, JohnAric Peterson, Avi Singh, Rebecca Cao |
| EPI_ISL_1897434, EPI_ISL_1897435, EPI_ISL_1897436, EPI_ISL_1897437, EPI_ISL_1897438, EPI_ISL_1897439, EPI_ISL_1897440, EPI_ISL_1897441, EPI_ISL_1897442, EPI_ISL_1897443, EPI_ISL_1897444, EPI_ISL_1897445, EPI_ISL_1897446, EPI_ISL_1897447, EPI_ISL_1897448, EPI_ISL_1897449, EPI_ISL_1897450, EPI_ISL_1897451, EPI_ISL_1897452, EPI_ISL_1897453, EPI_ISL_1897454, EPI_ISL_1897455, EPI_ISL_1897456, EPI_ISL_1897457, EPI_ISL_1897458, EPI_ISL_1897459, EPI_ISL_1897460, EPI_ISL_1897461, EPI_ISL_1897462, EPI_ISL_1897463, EPI_ISL_1897464, EPI_ISL_1897465, EPI_ISL_1897466, EPI_ISL_1897467, EPI_ISL_1897468, EPI_ISL_1897469, EPI_ISL_1897470, EPI_ISL_1897471, EPI_ISL_1897472, EPI_ISL_1897473, EPI_ISL_1897474, EPI_ISL_1897475, EPI_ISL_1897476, EPI_ISL_1897477, EPI_ISL_1897478, EPI_ISL_1897479, EPI_ISL_1897480, EPI_ISL_1897481, EPI_ISL_1897482, EPI_ISL_1897483, EPI_ISL_1897484 | | | |
| see above | Oregon State Public Health Laboratory | Oregon State Public Health Laboratory | Rafia Razzaque, Eugene Yeboah, Vanda Makris, Laura Tsaknaridis, John Fontana and Shane Sevey |
| EPI_ISL_1897485, EPI_ISL_1897486, EPI_ISL_1897487, EPI_ISL_1897488, EPI_ISL_1897489, EPI_ISL_1897490, EPI_ISL_1897491, EPI_ISL_1897492, EPI_ISL_1897493, EPI_ISL_1897494, EPI_ISL_1897495, EPI_ISL_1897496, EPI_ISL_1897497, EPI_ISL_1897498, EPI_ISL_1897499, EPI_ISL_1897500, EPI_ISL_1897501, EPI_ISL_1897502, EPI_ISL_1897503, EPI_ISL_1897504, EPI_ISL_1897505, EPI_ISL_1897506, EPI_ISL_1897507, EPI_ISL_1897508, EPI_ISL_1897509, EPI_ISL_1897510, EPI_ISL_1897511, EPI_ISL_1897512, EPI_ISL_1897513, EPI_ISL_1897514, EPI_ISL_1897515, EPI_ISL_1897516, EPI_ISL_1897517 | | | |
| see above | WVT Laboratory, Corvallis, Oregon | OSU Center for Genome Research and Biocomputing | Oregon State University TRACE Project |
| EPI_ISL_1897645, EPI_ISL_1897647, EPI_ISL_1897919, EPI_ISL_1899161 | MAWD Pathology Group, PA | MAWD Pathology Group, PA | Pradip Manna, Ph D, Kelli Fayard, Shelby Herner, London Collins, Kelsi Eastman |
| EPI_ISL_1900040, EPI_ISL_1900043, EPI_ISL_1900046, EPI_ISL_1900048, EPI_ISL_1900051, EPI_ISL_1900054, EPI_ISL_1900057, EPI_ISL_1900059, EPI_ISL_1900062, EPI_ISL_1900064, EPI_ISL_1900067, EPI_ISL_1900070, EPI_ISL_1900074, EPI_ISL_1900077, EPI_ISL_1900079, EPI_ISL_1900082, EPI_ISL_1900083, EPI_ISL_1900084, EPI_ISL_1900086, EPI_ISL_1900089, EPI_ISL_1900092, EPI_ISL_1900094, EPI_ISL_1900097, EPI_ISL_1900100, EPI_ISL_1900102, EPI_ISL_1900105 | | | |
| see above | Quest Diagnostics Incorporated | Centers for Disease Control and Prevention Division of Viral Diseases, Pathogen Discovery | Dakota Howard, Dhwani Batra, Peter W. Cook, Kara Moser, Adrian Paskey, Jason Caravas, Benjamin Rambo-Martin, Shatavia Morrison, Christopher Gulvick, Scott Sammons, Yvette Unoarumhi, Darlene Wagner, Matthew Schmerer, S. H. Rosenthal, A. Gerasimova, R. M. Kagan, B. Anderson, M. Hua, Y. Liu, L.E. Bernstein, K.E. Livingston, A. Perez, I. A. Shlyakhter, R. V. Rolando, R. Owen, P. Tanpaiboon, F. Lacbawan, Clinton R. Paden, Duncan MacCannell |
| EPI_ISL_1900114 | VA-Division of Consolidated Laboratory Services | Centers for Disease Control and Prevention Division of Viral Diseases, Pathogen Discovery | Mili Sheth, Sarah Nobles, Jasmine Padilla, Mark Burroughs, Shoshona Le, Katie Dillon, Peter Cook, Clinton R. Paden, Dhwani Batra, Krista Queen, Kristen Knipe, Dakota Howard, Yvette Unoarumhi, Darlene Wagner, Matthew Schmerer, Ben L. Rambo-Martin, Kristine Lacek, Sam Shepard, Alison Laufer Halpin, Dave Wentworth, Vivien Dugan, Suxiang Tong, Justin Lee |
| EPI_ISL_1900121, EPI_ISL_1900124, EPI_ISL_1900128, EPI_ISL_1900132, EPI_ISL_1900134, EPI_ISL_1900137, EPI_ISL_1900139, EPI_ISL_1900142, EPI_ISL_1900144, EPI_ISL_1900147, EPI_ISL_1900149, EPI_ISL_1900152, EPI_ISL_1900155 | | | |
| see above | Quest Diagnostics Incorporated | Centers for Disease Control and Prevention Division of Viral Diseases, Pathogen Discovery | Dakota Howard, Dhwani Batra, Peter W. Cook, Kara Moser, Adrian Paskey, Jason Caravas, Benjamin Rambo-Martin, Shatavia Morrison, Christopher Gulvick, Scott Sammons, Yvette Unoarumhi, Darlene Wagner, Matthew Schmerer, S. H. Rosenthal, A. Gerasimova, R. M. Kagan, B. Anderson, M. Hua, Y. Liu, L.E. Bernstein, K.E. Livingston, A. Perez, I. A. Shlyakhter, R. V. Rolando, R. Owen, P. Tanpaiboon, F. Lacbawan, Clinton R. Paden, Duncan MacCannell |
| EPI_ISL_1900164, EPI_ISL_1900166, EPI_ISL_1900168, EPI_ISL_1900171, EPI_ISL_1900174, EPI_ISL_1900178, EPI_ISL_1900181, EPI_ISL_1900184, EPI_ISL_1900187, EPI_ISL_1900191, EPI_ISL_1900193, EPI_ISL_1900196, EPI_ISL_1900199, EPI_ISL_1900201, EPI_ISL_1900203, EPI_ISL_1900207, EPI_ISL_1900208, EPI_ISL_1900210, EPI_ISL_1900212, EPI_ISL_1900215, EPI_ISL_1900217, EPI_ISL_1900220, EPI_ISL_1900222, EPI_ISL_1900225, EPI_ISL_1900229, EPI_ISL_1900231, EPI_ISL_1900233, EPI_ISL_1900236, EPI_ISL_1900239, EPI_ISL_1900241, EPI_ISL_1900244, EPI_ISL_1900246, EPI_ISL_1900249, EPI_ISL_1900252, EPI_ISL_1900255, EPI_ISL_1900259, EPI_ISL_1900263, EPI_ISL_1900268, EPI_ISL_1900270, EPI_ISL_1900273, EPI_ISL_1900275, EPI_ISL_1900278, EPI_ISL_1900280, EPI_ISL_1900283, EPI_ISL_1900286, EPI_ISL_1900289, EPI_ISL_1900292, EPI_ISL_1900295, EPI_ISL_1900298, EPI_ISL_1900300, EPI_ISL_1900303 | | | |
| see above | Aegis Sciences Corporation | Centers for Disease Control and Prevention Division of Viral Diseases, Pathogen Discovery | Dakota Howard, Dhwani Batra, Peter W. Cook, Kara Moser, Adrian Paskey, Jason Caravas, Benjamin Rambo-Martin, Shatavia Morrison, Christopher Gulvick, Scott Sammons, Yvette Unoarumhi, Darlene Wagner, Matthew Schmerer, Cyndi Clark, Patrick Campbell, Rob Case, Vikramsinha Ghorpade, Holly Houdeshell, Ola Kvalvaag, Dillon Nall, Ethan Sanders, Alec Vest, Shaun Westlund, Matthew Hardison, Clinton R. Paden, Duncan MacCannell |
| EPI_ISL_1900304 | MAWD Pathology Group, PA | MAWD Pathology Group, PA | Pradip Manna, Ph D, Kelli Fayard, Shelby Herner, London Collins, Kelsi Eastman |
| EPI_ISL_1900306, EPI_ISL_1900309, EPI_ISL_1900311, EPI_ISL_1900314, EPI_ISL_1900317, EPI_ISL_1900320, EPI_ISL_1900322, EPI_ISL_1900325, EPI_ISL_1900328, EPI_ISL_1900330, EPI_ISL_1900333, EPI_ISL_1900334, EPI_ISL_1900335, EPI_ISL_1900336, EPI_ISL_1900337, EPI_ISL_1900338, EPI_ISL_1900339, EPI_ISL_1900340, EPI_ISL_1900341, EPI_ISL_1900342, EPI_ISL_1900343, EPI_ISL_1900344, EPI_ISL_1900345, EPI_ISL_1900346, EPI_ISL_1900347, EPI_ISL_1900348, EPI_ISL_1900349, EPI_ISL_1900350, EPI_ISL_1900351, EPI_ISL_1900352, EPI_ISL_1900353, EPI_ISL_1900354, EPI_ISL_1900355, EPI_ISL_1900356, EPI_ISL_1900357, EPI_ISL_1900358, EPI_ISL_1900359, EPI_ISL_1900360, EPI_ISL_1900361, EPI_ISL_1900363, EPI_ISL_1900364, EPI_ISL_1900365, EPI_ISL_1900366, EPI_ISL_1900367, EPI_ISL_1900368, EPI_ISL_1900369, EPI_ISL_1900370, EPI_ISL_1900371, EPI_ISL_1900372, EPI_ISL_1900373, EPI_ISL_1900374, EPI_ISL_1900375, EPI_ISL_1900376, EPI_ISL_1900377, EPI_ISL_1900378, EPI_ISL_1900379, EPI_ISL_1900380, EPI_ISL_1900381, EPI_ISL_1900382, EPI_ISL_1900383, EPI_ISL_1900384, EPI_ISL_1900385, EPI_ISL_1900386, EPI_ISL_1900387, EPI_ISL_1900388, EPI_ISL_1900389, EPI_ISL_1900390, EPI_ISL_1900391, EPI_ISL_1900392, EPI_ISL_1900393, EPI_ISL_1900394, EPI_ISL_1900396, EPI_ISL_1900397, EPI_ISL_1900398, EPI_ISL_1900399, EPI_ISL_1900400, EPI_ISL_1900401, EPI_ISL_1900402, EPI_ISL_1900403, EPI_ISL_1900404, EPI_ISL_1900405, EPI_ISL_1900406, EPI_ISL_1900407, EPI_ISL_1900408, EPI_ISL_1900409, EPI_ISL_1900410, EPI_ISL_1900411, EPI_ISL_1900412, EPI_ISL_1900413, EPI_ISL_1900414, EPI_ISL_1900415, EPI_ISL_1900416, EPI_ISL_1900417, EPI_ISL_1900418, EPI_ISL_1900419, EPI_ISL_1900420, EPI_ISL_1900421, EPI_ISL_1900422, EPI_ISL_1900423, EPI_ISL_1900424 | | | |
| see above | Aegis Sciences Corporation | Centers for Disease Control and Prevention Division of Viral Diseases, Pathogen Discovery | Dakota Howard, Dhwani Batra, Peter W. Cook, Kara Moser, Adrian Paskey, Jason Caravas, Benjamin Rambo-Martin, Shatavia Morrison, Christopher Gulvick, Scott Sammons, Yvette Unoarumhi, Darlene Wagner, Matthew Schmerer, Cyndi Clark, Patrick Campbell, Rob Case, Vikramsinha Ghorpade, Holly Houdeshell, Ola Kvalvaag, Dillon Nall, Ethan Sanders, Alec Vest, Shaun Westlund, Matthew Hardison, Clinton R. Paden, Duncan MacCannell |
| EPI_ISL_1900425, EPI_ISL_1900426, EPI_ISL_1900427, EPI_ISL_1900428, EPI_ISL_1900429, EPI_ISL_1900430, EPI_ISL_1900431, EPI_ISL_1900432, EPI_ISL_1900433, EPI_ISL_1900434, EPI_ISL_1900435 | | | |
| see above | Quest Diagnostics Incorporated | Centers for Disease Control and Prevention Division of Viral Diseases, Pathogen Discovery | Dakota Howard, Dhwani Batra, Peter W. Cook, Kara Moser, Adrian Paskey, Jason Caravas, Benjamin Rambo-Martin, Shatavia Morrison, Christopher Gulvick, Scott Sammons, Yvette Unoarumhi, Darlene Wagner, Matthew Schmerer, S. H. Rosenthal, A. Gerasimova, R. M. Kagan, B. Anderson, M. Hua, Y. Liu, L.E. Bernstein, K.E. Livingston, A. Perez, I. A. Shlyakhter, R. V. Rolando, R. Owen, P. Tanpaiboon, F. Lacbawan, Clinton R. Paden, Duncan MacCannell |
| EPI_ISL_1900436 | Fulgent Genetics | Centers for Disease Control and Prevention Division of Viral Diseases, Pathogen | Dakota Howard, Dhwani Batra, Peter W. Cook, Kara Moser, Adrian Paskey, Jason Caravas, Benjamin Rambo-Martin, Shatavia Morrison, Christopher Gulvick, Scott Sammons, Yvette Unoarumhi, Darlene Wagner, Matthew Schmerer, Harry Gao, Mickey Li, John Gao, Joseph Fierro, Benafsh Sapra, Becky Tsai, Yan Meng, Doreen Ng, James Xie, Clinton R. Paden, Duncan MacCannell |

| | | | |
|---|---|---|---|
| | | Discovery | |
| EPI_ISL_1900437, EPI_ISL_1900438, EPI_ISL_1900439, EPI_ISL_1900440, EPI_ISL_1900441, EPI_ISL_1900442, EPI_ISL_1900443, EPI_ISL_1900444, EPI_ISL_1900445, EPI_ISL_1900446, EPI_ISL_1900447, EPI_ISL_1900448, EPI_ISL_1900449, EPI_ISL_1900450, EPI_ISL_1900451, EPI_ISL_1900452, EPI_ISL_1900453, EPI_ISL_1900454, EPI_ISL_1900455, EPI_ISL_1900456, EPI_ISL_1900457, EPI_ISL_1900458, EPI_ISL_1900459, EPI_ISL_1900460, EPI_ISL_1900461, EPI_ISL_1900462, EPI_ISL_1900463, EPI_ISL_1900464, EPI_ISL_1900465 | | | |
| see above | Quest Diagnostics Incorporated | Centers for Disease Control and Prevention Division of Viral Diseases, Pathogen Discovery | Dakota Howard, Dhwani Batra, Peter W. Cook, Kara Moser, Adrian Paskey, Jason Caravas, Benjamin Rambo-Martin, Shatavia Morrison, Christopher Gulvick, Scott Sammons, Yvette Unoarumhi, Darlene Wagner, Matthew Schmerer, S. H. Rosenthal, A. Gerasimova, R. M. Kagan, B. Anderson, M. Hua, Y. Liu, L.E. Bernstein, K.E. Livingston, A. Perez, I. A. Shlyakhter, R. V. Rolando, R. Owen, P. Tanpaiboon, F. Lacbawan, Clinton R. Paden, Duncan MacCannell |
| EPI_ISL_1900466, EPI_ISL_1901105 | MAWD Pathology Group, PA | MAWD Pathology Group, PA | Pradip Manna, Ph D, Kelli Fayard, Shelby Herner, London Collins, Kelsi Eastman |
| EPI_ISL_1902544, EPI_ISL_1902996, EPI_ISL_1903598, EPI_ISL_1904562 | Ohio Department of Health Laboratory | Ohio Department of Health Laboratory | Holmes, Jennifer; Eric Brandt, Keoni Omura, Glen McGillivary, Caitlin McDonnell, Jade Mowery, Stephanie Mccracken, Tyler Payne, Kirtana Ramadugu, Erica Leasure, Brent Lee, Kelsey Florek, Heather Blankenship, Allison Black, Quanta Brown, and Tammy Bannerman |
| EPI_ISL_1905184, EPI_ISL_1905185, EPI_ISL_1905187, EPI_ISL_1905188, EPI_ISL_1905189, EPI_ISL_1905191, EPI_ISL_1905194, EPI_ISL_1905196, EPI_ISL_1905199, EPI_ISL_1905202, EPI_ISL_1905204, EPI_ISL_1905206, EPI_ISL_1905209, EPI_ISL_1905213, EPI_ISL_1905215, EPI_ISL_1905217, EPI_ISL_1905218, EPI_ISL_1905219, EPI_ISL_1905220, EPI_ISL_1905221, EPI_ISL_1905222, EPI_ISL_1905223, EPI_ISL_1905224, EPI_ISL_1905225, EPI_ISL_1905226, EPI_ISL_1905227, EPI_ISL_1905228, EPI_ISL_1905229, EPI_ISL_1905230, EPI_ISL_1905231, EPI_ISL_1905232, EPI_ISL_1905233, EPI_ISL_1905234, EPI_ISL_1905235, EPI_ISL_1905236, EPI_ISL_1905237, EPI_ISL_1905238, EPI_ISL_1905239, EPI_ISL_1905240, EPI_ISL_1905241, EPI_ISL_1905242, EPI_ISL_1905243, EPI_ISL_1905244, EPI_ISL_1905245, EPI_ISL_1905246, EPI_ISL_1905247, EPI_ISL_1905249, EPI_ISL_1905250, EPI_ISL_1905251, EPI_ISL_1905252, EPI_ISL_1905270, EPI_ISL_1905273, EPI_ISL_1905275, EPI_ISL_1905437, EPI_ISL_1905438, EPI_ISL_1905439, EPI_ISL_1905440, EPI_ISL_1905441, EPI_ISL_1905443, EPI_ISL_1905445, EPI_ISL_1905446, EPI_ISL_1905447, EPI_ISL_1905448, EPI_ISL_1905449, EPI_ISL_1905450, EPI_ISL_1905451, EPI_ISL_1905452, EPI_ISL_1905453, EPI_ISL_1905454, EPI_ISL_1905455, EPI_ISL_1905457, EPI_ISL_1905458, EPI_ISL_1905459, EPI_ISL_1905461, EPI_ISL_1905462, EPI_ISL_1905463, EPI_ISL_1905464, EPI_ISL_1905465, EPI_ISL_1905466, EPI_ISL_1905467, EPI_ISL_1905468, EPI_ISL_1905469, EPI_ISL_1905470, EPI_ISL_1905471, EPI_ISL_1905472, EPI_ISL_1905474, EPI_ISL_1905475, EPI_ISL_1905476, EPI_ISL_1905478, EPI_ISL_1905479, EPI_ISL_1905480, EPI_ISL_1905481, EPI_ISL_1905482, EPI_ISL_1905483, EPI_ISL_1905484, EPI_ISL_1905486, EPI_ISL_1905487, EPI_ISL_1905488, EPI_ISL_1905489, EPI_ISL_1905491, EPI_ISL_1905492, EPI_ISL_1905493, EPI_ISL_1905494, EPI_ISL_1905495, EPI_ISL_1905496, EPI_ISL_1905498, EPI_ISL_1905499, EPI_ISL_1905500, EPI_ISL_1905501, EPI_ISL_1905502, EPI_ISL_1905503, EPI_ISL_1905504, EPI_ISL_1905505, EPI_ISL_1905506, EPI_ISL_1905507, EPI_ISL_1905508, EPI_ISL_1905509, EPI_ISL_1905510, EPI_ISL_1905511, EPI_ISL_1905512, EPI_ISL_1905513, EPI_ISL_1905514, EPI_ISL_1905515, EPI_ISL_1905516, EPI_ISL_1905517, EPI_ISL_1905518, EPI_ISL_1905520, EPI_ISL_1905521, EPI_ISL_1905522, EPI_ISL_1905523, EPI_ISL_1905524, EPI_ISL_1905525, EPI_ISL_1905661, EPI_ISL_1905662, EPI_ISL_1905663, EPI_ISL_1905664, EPI_ISL_1905665, EPI_ISL_1905666, EPI_ISL_1905667, EPI_ISL_1905668, EPI_ISL_1905669, EPI_ISL_1905670, EPI_ISL_1905671, EPI_ISL_1905672, EPI_ISL_1905673, EPI_ISL_1905674, EPI_ISL_1905675, EPI_ISL_1905676, EPI_ISL_1905677, EPI_ISL_1905678, EPI_ISL_1905679, EPI_ISL_1905680, EPI_ISL_1905681, EPI_ISL_1905682, EPI_ISL_1905683, EPI_ISL_1905684, EPI_ISL_1905685, EPI_ISL_1905686, EPI_ISL_1905687, EPI_ISL_1905688, EPI_ISL_1905689, EPI_ISL_1905690, EPI_ISL_1905691, EPI_ISL_1905693, EPI_ISL_1905694, EPI_ISL_1905695, EPI_ISL_1905696, EPI_ISL_1905697, EPI_ISL_1905698, EPI_ISL_1905699, EPI_ISL_1905700, EPI_ISL_1905702, EPI_ISL_1905703, EPI_ISL_1905704, EPI_ISL_1905705, EPI_ISL_1905706, EPI_ISL_1905707, EPI_ISL_1905708, EPI_ISL_1905710, EPI_ISL_1905711, EPI_ISL_1905712, EPI_ISL_1905713, EPI_ISL_1905714, EPI_ISL_1905715, EPI_ISL_1905716, EPI_ISL_1905717, EPI_ISL_1905718, EPI_ISL_1905719, EPI_ISL_1905720, EPI_ISL_1905721, EPI_ISL_1905722, EPI_ISL_1905724, EPI_ISL_1905725, EPI_ISL_1905726, EPI_ISL_1905727, EPI_ISL_1905730, EPI_ISL_1905731, EPI_ISL_1905732, EPI_ISL_1905733, EPI_ISL_1905734, EPI_ISL_1905735, EPI_ISL_1905736, EPI_ISL_1905737, EPI_ISL_1905738, EPI_ISL_1905740, EPI_ISL_1905741, EPI_ISL_1905742, EPI_ISL_1905744, EPI_ISL_1905746, EPI_ISL_1905747, EPI_ISL_1905748, EPI_ISL_1905749, EPI_ISL_1905753, EPI_ISL_1905754, EPI_ISL_1905758, EPI_ISL_1905759, EPI_ISL_1905760, EPI_ISL_1905761, EPI_ISL_1905762, EPI_ISL_1905763, EPI_ISL_1905765, EPI_ISL_1905766, EPI_ISL_1905767, EPI_ISL_1905768, EPI_ISL_1905770, EPI_ISL_1905773, EPI_ISL_1905774, EPI_ISL_1905775, EPI_ISL_1905776, EPI_ISL_1905778, EPI_ISL_1905779, EPI_ISL_1905780, EPI_ISL_1905781, EPI_ISL_1905782, EPI_ISL_1905783, EPI_ISL_1905784, EPI_ISL_1905785, EPI_ISL_1905786, EPI_ISL_1905788, EPI_ISL_1905789, EPI_ISL_1905790, EPI_ISL_1905792, EPI_ISL_1905794, EPI_ISL_1905795, EPI_ISL_1905796, EPI_ISL_1905797, EPI_ISL_1905798, EPI_ISL_1905799, EPI_ISL_1905801, EPI_ISL_1905804, EPI_ISL_1905805, EPI_ISL_1905806, EPI_ISL_1905807, EPI_ISL_1905809, EPI_ISL_1905812, EPI_ISL_1905816, EPI_ISL_1905817, EPI_ISL_1905818, EPI_ISL_1905820, EPI_ISL_1905821, EPI_ISL_1905822, EPI_ISL_1905823, EPI_ISL_1905824, EPI_ISL_1905825, EPI_ISL_1905826, EPI_ISL_1905827, EPI_ISL_1905828, EPI_ISL_1905829, EPI_ISL_1905830, EPI_ISL_1905831, EPI_ISL_1905832, EPI_ISL_1905833, EPI_ISL_1905837, EPI_ISL_1905839, EPI_ISL_1905840 | | | |
| see above | Michigan Department of Health and Human Services, Bureau of Laboratories | Michigan Department of Health and Human Services, Bureau of Laboratories | Blankenship HM, Riner D, Soehnlen MK |
| EPI_ISL_1905841 | Quest Diagnostics Incorporated | Centers for Disease Control and Prevention Division of Viral Diseases, Pathogen Discovery | Dakota Howard, Dhwani Batra, Peter W. Cook, Kara Moser, Adrian Paskey, Jason Caravas, Benjamin Rambo-Martin, Shatavia Morrison, Christopher Gulvick, Scott Sammons, Yvette Unoarumhi, Darlene Wagner, Matthew Schmerer, S. H. Rosenthal, A. Gerasimova, R. M. Kagan, B. Anderson, M. Hua, Y. Liu, L.E. Bernstein, K.E. Livingston, A. Perez, I. A. Shlyakhter, R. V. Rolando, R. Owen, P. Tanpaiboon, F. Lacbawan, Clinton R. Paden, Duncan MacCannell |
| EPI_ISL_1905842 | Michigan Department of Health and Human Services, Bureau of Laboratories | Michigan Department of Health and Human Services, Bureau of Laboratories | Blankenship HM, Riner D, Soehnlen MK |
| EPI_ISL_1905843 | Quest Diagnostics Incorporated | Centers for Disease Control and Prevention Division of Viral Diseases, Pathogen Discovery | Dakota Howard, Dhwani Batra, Peter W. Cook, Kara Moser, Adrian Paskey, Jason Caravas, Benjamin Rambo-Martin, Shatavia Morrison, Christopher Gulvick, Scott Sammons, Yvette Unoarumhi, Darlene Wagner, Matthew Schmerer, S. H. Rosenthal, A. Gerasimova, R. M. Kagan, B. Anderson, M. Hua, Y. Liu, L.E. Bernstein, K.E. Livingston, A. Perez, I. A. Shlyakhter, R. V. Rolando, R. Owen, P. Tanpaiboon, F. Lacbawan, Clinton R. Paden, Duncan MacCannell |
| EPI_ISL_1905844 | Michigan Department of Health and Human Services, Bureau of Laboratories | Michigan Department of Health and Human Services, Bureau of Laboratories | Blankenship HM, Riner D, Soehnlen MK |
| EPI_ISL_1905845 | Quest Diagnostics Incorporated | Centers for Disease Control and Prevention Division of Viral Diseases, Pathogen Discovery | Dakota Howard, Dhwani Batra, Peter W. Cook, Kara Moser, Adrian Paskey, Jason Caravas, Benjamin Rambo-Martin, Shatavia Morrison, Christopher Gulvick, Scott Sammons, Yvette Unoarumhi, Darlene Wagner, Matthew Schmerer, S. H. Rosenthal, A. Gerasimova, R. M. Kagan, B. Anderson, M. Hua, Y. Liu, L.E. Bernstein, K.E. Livingston, A. Perez, I. A. Shlyakhter, R. V. Rolando, R. Owen, P. Tanpaiboon, F. Lacbawan, Clinton R. Paden, Duncan MacCannell |
| EPI_ISL_1905846, EPI_ISL_1905847 | Michigan Department of Health and Human Services, Bureau of Laboratories | Michigan Department of Health and Human Services, Bureau of Laboratories | Blankenship HM, Riner D, Soehnlen MK |
| EPI_ISL_1905848 | Quest Diagnostics Incorporated | Centers for Disease Control and Prevention Division of Viral Diseases, Pathogen Discovery | Dakota Howard, Dhwani Batra, Peter W. Cook, Kara Moser, Adrian Paskey, Jason Caravas, Benjamin Rambo-Martin, Shatavia Morrison, Christopher Gulvick, Scott Sammons, Yvette Unoarumhi, Darlene Wagner, Matthew Schmerer, S. H. Rosenthal, A. Gerasimova, R. M. Kagan, B. Anderson, M. Hua, Y. Liu, L.E. Bernstein, K.E. Livingston, A. Perez, I. A. Shlyakhter, R. V. Rolando, R. Owen, P. Tanpaiboon, F. Lacbawan, Clinton R. Paden, Duncan MacCannell |
| EPI_ISL_1905849 | Michigan Department of Health and Human Services, Bureau of Laboratories | Michigan Department of Health and Human Services, Bureau of Laboratories | Blankenship HM, Riner D, Soehnlen MK |
| EPI_ISL_1905850, EPI_ISL_1905852 | Quest Diagnostics Incorporated | Centers for Disease Control and Prevention Division of Viral Diseases, Pathogen Discovery | Dakota Howard, Dhwani Batra, Peter W. Cook, Kara Moser, Adrian Paskey, Jason Caravas, Benjamin Rambo-Martin, Shatavia Morrison, Christopher Gulvick, Scott Sammons, Yvette Unoarumhi, Darlene Wagner, Matthew Schmerer, S. H. Rosenthal, A. Gerasimova, R. M. Kagan, B. Anderson, M. Hua, Y. Liu, L.E. Bernstein, K.E. Livingston, A. Perez, I. A. Shlyakhter, R. V. Rolando, R. Owen, P. Tanpaiboon, F. Lacbawan, Clinton R. Paden, Duncan MacCannell |
| EPI_ISL_1905853, EPI_ISL_1905854 | Michigan Department of Health and Human Services, Bureau of Laboratories | Michigan Department of Health and Human Services, Bureau of Laboratories | Blankenship HM, Riner D, Soehnlen MK |
| EPI_ISL_1905857 | Quest Diagnostics Incorporated | Centers for Disease Control and Prevention Division of Viral Diseases, Pathogen Discovery | Dakota Howard, Dhwani Batra, Peter W. Cook, Kara Moser, Adrian Paskey, Jason Caravas, Benjamin Rambo-Martin, Shatavia Morrison, Christopher Gulvick, Scott Sammons, Yvette Unoarumhi, Darlene Wagner, Matthew Schmerer, S. H. Rosenthal, A. Gerasimova, R. M. Kagan, B. Anderson, M. Hua, Y. Liu, L.E. Bernstein, K.E. Livingston, A. Perez, I. A. Shlyakhter, R. V. Rolando, R. Owen, P. Tanpaiboon, F. Lacbawan, Clinton R. Paden, Duncan MacCannell |
| EPI_ISL_1905858 | Michigan Department of Health and Human Services, Bureau of Laboratories | Michigan Department of Health and Human Services, Bureau of Laboratories | Blankenship HM, Riner D, Soehnlen MK |
| EPI_ISL_1905859 | Quest Diagnostics Incorporated | Centers for Disease Control and Prevention Division of Viral Diseases, Pathogen Discovery | Dakota Howard, Dhwani Batra, Peter W. Cook, Kara Moser, Adrian Paskey, Jason Caravas, Benjamin Rambo-Martin, Shatavia Morrison, Christopher Gulvick, Scott Sammons, Yvette Unoarumhi, Darlene Wagner, Matthew Schmerer, S. H. Rosenthal, A. Gerasimova, R. M. Kagan, B. Anderson, M. Hua, Y. Liu, L.E. Bernstein, K.E. Livingston, A. Perez, I. A. Shlyakhter, R. V. Rolando, R. Owen, P. Tanpaiboon, F. Lacbawan, Clinton R. Paden, Duncan MacCannell |
| EPI_ISL_1905860, EPI_ISL_1905861, EPI_ISL_1905862 | Michigan Department of Health and Human Services, Bureau of Laboratories | Michigan Department of Health and Human Services, Bureau of Laboratories | Blankenship HM, Riner D, Soehnlen MK |
| EPI_ISL_1905863, EPI_ISL_1905866 | Quest Diagnostics Incorporated | Centers for Disease Control and Prevention Division of Viral Diseases, Pathogen Discovery | Dakota Howard, Dhwani Batra, Peter W. Cook, Kara Moser, Adrian Paskey, Jason Caravas, Benjamin Rambo-Martin, Shatavia Morrison, Christopher Gulvick, Scott Sammons, Yvette Unoarumhi, Darlene Wagner, Matthew Schmerer, S. H. Rosenthal, A. Gerasimova, R. M. Kagan, B. Anderson, M. Hua, Y. Liu, L.E. Bernstein, K.E. Livingston, A. Perez, I. A. Shlyakhter, R. V. Rolando, R. Owen, P. Tanpaiboon, F. Lacbawan, Clinton R. Paden, Duncan MacCannell |
| EPI_ISL_1905867, EPI_ISL_1905868 | Michigan Department of Health and Human Services, Bureau of Laboratories | Michigan Department of Health and Human Services, Bureau of Laboratories | Blankenship HM, Riner D, Soehnlen MK |
| EPI_ISL_1905869 | Quest Diagnostics Incorporated | Centers for Disease Control and Prevention Division of Viral Diseases, Pathogen Discovery | Dakota Howard, Dhwani Batra, Peter W. Cook, Kara Moser, Adrian Paskey, Jason Caravas, Benjamin Rambo-Martin, Shatavia Morrison, Christopher Gulvick, Scott Sammons, Yvette Unoarumhi, Darlene Wagner, Matthew Schmerer, S. H. Rosenthal, A. Gerasimova, R. M. Kagan, B. Anderson, M. Hua, Y. Liu, L.E. Bernstein, K.E. Livingston, A. Perez, I. A. Shlyakhter, R. V. Rolando, R. Owen, P. Tanpaiboon, F. Lacbawan, Clinton R. Paden, Duncan MacCannell |
| EPI_ISL_1905870 | Michigan Department of Health and Human Services, Bureau of Laboratories | Michigan Department of Health and Human Services, Bureau of Laboratories | Blankenship HM, Riner D, Soehnlen MK |
| EPI_ISL_1905871 | Quest Diagnostics Incorporated | Centers for Disease Control and Prevention Division of Viral Diseases, Pathogen Discovery | Dakota Howard, Dhwani Batra, Peter W. Cook, Kara Moser, Adrian Paskey, Jason Caravas, Benjamin Rambo-Martin, Shatavia Morrison, Christopher Gulvick, Scott Sammons, Yvette Unoarumhi, Darlene Wagner, Matthew Schmerer, S. H. Rosenthal, A. Gerasimova, R. M. Kagan, B. Anderson, M. Hua, Y. Liu, L.E. Bernstein, K.E. Livingston, A. Perez, I. A. Shlyakhter, R. V. Rolando, R. Owen, P. Tanpaiboon, F. Lacbawan, Clinton R. Paden, Duncan MacCannell |
| EPI_ISL_1905872 | Michigan Department of Health and Human Services, Bureau of Laboratories | Michigan Department of Health and Human Services, Bureau of Laboratories | Blankenship HM, Riner D, Soehnlen MK |
| EPI_ISL_1905873 | Quest Diagnostics Incorporated | Centers for Disease Control and Prevention Division of Viral Diseases, Pathogen Discovery | Dakota Howard, Dhwani Batra, Peter W. Cook, Kara Moser, Adrian Paskey, Jason Caravas, Benjamin Rambo-Martin, Shatavia Morrison, Christopher Gulvick, Scott Sammons, Yvette Unoarumhi, Darlene Wagner, Matthew Schmerer, S. H. Rosenthal, A. Gerasimova, R. M. Kagan, B. Anderson, M. Hua, Y. Liu, L.E. Bernstein, K.E. Livingston, A. Perez, I. A. Shlyakhter, R. V. Rolando, R. Owen, P. Tanpaiboon, F. Lacbawan, Clinton R. Paden, Duncan MacCannell |
| EPI_ISL_1905874 | Michigan Department of Health and Human Services, Bureau of Laboratories | Michigan Department of Health and Human Services, Bureau of Laboratories | Blankenship HM, Riner D, Soehnlen MK |
| EPI_ISL_1905875 | Quest Diagnostics Incorporated | Centers for Disease Control and Prevention Division of Viral Diseases, Pathogen Discovery | Dakota Howard, Dhwani Batra, Peter W. Cook, Kara Moser, Adrian Paskey, Jason Caravas, Benjamin Rambo-Martin, Shatavia Morrison, Christopher Gulvick, Scott Sammons, Yvette Unoarumhi, Darlene Wagner, Matthew Schmerer, S. H. Rosenthal, A. Gerasimova, R. M. Kagan, B. Anderson, M. Hua, Y. Liu, L.E. Bernstein, K.E. Livingston, A. Perez, I. A. Shlyakhter, R. V. Rolando, R. Owen, P. Tanpaiboon, F. Lacbawan, Clinton R. Paden, Duncan MacCannell |
| EPI_ISL_1905876 | Michigan Department of Health and Human Services, Bureau of Laboratories | Michigan Department of Health and Human Services, Bureau of Laboratories | Blankenship HM, Riner D, Soehnlen MK |
| EPI_ISL_1905877 | Quest Diagnostics Incorporated | Centers for Disease Control and Prevention Division of Viral Diseases, Pathogen Discovery | Dakota Howard, Dhwani Batra, Peter W. Cook, Kara Moser, Adrian Paskey, Jason Caravas, Benjamin Rambo-Martin, Shatavia Morrison, Christopher Gulvick, Scott Sammons, Yvette Unoarumhi, Darlene Wagner, Matthew Schmerer, S. H. Rosenthal, A. Gerasimova, R. M. Kagan, B. Anderson, M. Hua, Y. Liu, L.E. Bernstein, K.E. Livingston, A. Perez, I. A. Shlyakhter, R. V. Rolando, R. Owen, P. Tanpaiboon, F. Lacbawan, Clinton R. Paden, Duncan MacCannell |
| EPI_ISL_1905879 | Michigan Department of Health and Human Services, Bureau of Laboratories | Michigan Department of Health and Human Services, Bureau of Laboratories | Blankenship HM, Riner D, Soehnlen MK |
| EPI_ISL_1905880, EPI_ISL_1905881, EPI_ISL_1905883 | Quest Diagnostics Incorporated | Centers for Disease Control and Prevention Division of Viral Diseases, Pathogen Discovery | Dakota Howard, Dhwani Batra, Peter W. Cook, Kara Moser, Adrian Paskey, Jason Caravas, Benjamin Rambo-Martin, Shatavia Morrison, Christopher Gulvick, Scott Sammons, Yvette Unoarumhi, Darlene Wagner, Matthew Schmerer, S. H. Rosenthal, A. Gerasimova, R. M. Kagan, B. Anderson, M. Hua, Y. Liu, L.E. Bernstein, K.E. Livingston, A. Perez, I. A. Shlyakhter, R. V. Rolando, R. Owen, P. Tanpaiboon, F. Lacbawan, Clinton R. Paden, Duncan MacCannell |

| | | | |
|---|---|---|---|
| EPI_ISL_1905884 | Michigan Department of Health and Human Services, Bureau of Laboratories | Michigan Department of Health and Human Services, Bureau of Laboratories | Blankenship HM, Riner D, Soehnlen MK |
| EPI_ISL_1905885 | Quest Diagnostics Incorporated | Centers for Disease Control and Prevention Division of Viral Diseases, Pathogen Discovery | Dakota Howard, Dhwani Batra, Peter W. Cook, Kara Moser, Adrian Paskey, Jason Caravas, Benjamin Rambo-Martin, Shatavia Morrison, Christopher Gulvick, Scott Sammons, Yvette Unoarumhi, Darlene Wagner, Matthew Schmerer, S. H. Rosenthal, A. Gerasimova, R. M. Kagan, B. Anderson, M. Hua, Y. Liu, L.E. Bernstein, K.E. Livingston, A. Perez, I. A. Shlyakhter, R. V. Rolando, R. Owen, P. Tanpaiboon, F. Lacbawan, Clinton R. Paden, Duncan MacCannell |
| EPI_ISL_1905886, EPI_ISL_1905887 | Michigan Department of Health and Human Services, Bureau of Laboratories | Michigan Department of Health and Human Services, Bureau of Laboratories | Blankenship HM, Riner D, Soehnlen MK |
| EPI_ISL_1905888 | Quest Diagnostics Incorporated | Centers for Disease Control and Prevention Division of Viral Diseases, Pathogen Discovery | Dakota Howard, Dhwani Batra, Peter W. Cook, Kara Moser, Adrian Paskey, Jason Caravas, Benjamin Rambo-Martin, Shatavia Morrison, Christopher Gulvick, Scott Sammons, Yvette Unoarumhi, Darlene Wagner, Matthew Schmerer, S. H. Rosenthal, A. Gerasimova, R. M. Kagan, B. Anderson, M. Hua, Y. Liu, L.E. Bernstein, K.E. Livingston, A. Perez, I. A. Shlyakhter, R. V. Rolando, R. Owen, P. Tanpaiboon, F. Lacbawan, Clinton R. Paden, Duncan MacCannell |
| EPI_ISL_1905889, EPI_ISL_1905890, EPI_ISL_1905891 | Michigan Department of Health and Human Services, Bureau of Laboratories | Michigan Department of Health and Human Services, Bureau of Laboratories | Blankenship HM, Riner D, Soehnlen MK |
| EPI_ISL_1905892 | Quest Diagnostics Incorporated | Centers for Disease Control and Prevention Division of Viral Diseases, Pathogen Discovery | Dakota Howard, Dhwani Batra, Peter W. Cook, Kara Moser, Adrian Paskey, Jason Caravas, Benjamin Rambo-Martin, Shatavia Morrison, Christopher Gulvick, Scott Sammons, Yvette Unoarumhi, Darlene Wagner, Matthew Schmerer, S. H. Rosenthal, A. Gerasimova, R. M. Kagan, B. Anderson, M. Hua, Y. Liu, L.E. Bernstein, K.E. Livingston, A. Perez, I. A. Shlyakhter, R. V. Rolando, R. Owen, P. Tanpaiboon, F. Lacbawan, Clinton R. Paden, Duncan MacCannell |
| EPI_ISL_1905893 | Michigan Department of Health and Human Services, Bureau of Laboratories | Michigan Department of Health and Human Services, Bureau of Laboratories | Blankenship HM, Riner D, Soehnlen MK |
| EPI_ISL_1905894 | Quest Diagnostics Incorporated | Centers for Disease Control and Prevention Division of Viral Diseases, Pathogen Discovery | Dakota Howard, Dhwani Batra, Peter W. Cook, Kara Moser, Adrian Paskey, Jason Caravas, Benjamin Rambo-Martin, Shatavia Morrison, Christopher Gulvick, Scott Sammons, Yvette Unoarumhi, Darlene Wagner, Matthew Schmerer, S. H. Rosenthal, A. Gerasimova, R. M. Kagan, B. Anderson, M. Hua, Y. Liu, L.E. Bernstein, K.E. Livingston, A. Perez, I. A. Shlyakhter, R. V. Rolando, R. Owen, P. Tanpaiboon, F. Lacbawan, Clinton R. Paden, Duncan MacCannell |
| EPI_ISL_1905896 | Michigan Department of Health and Human Services, Bureau of Laboratories | Michigan Department of Health and Human Services, Bureau of Laboratories | Blankenship HM, Riner D, Soehnlen MK |
| EPI_ISL_1905897 | Quest Diagnostics Incorporated | Centers for Disease Control and Prevention Division of Viral Diseases, Pathogen Discovery | Dakota Howard, Dhwani Batra, Peter W. Cook, Kara Moser, Adrian Paskey, Jason Caravas, Benjamin Rambo-Martin, Shatavia Morrison, Christopher Gulvick, Scott Sammons, Yvette Unoarumhi, Darlene Wagner, Matthew Schmerer, S. H. Rosenthal, A. Gerasimova, R. M. Kagan, B. Anderson, M. Hua, Y. Liu, L.E. Bernstein, K.E. Livingston, A. Perez, I. A. Shlyakhter, R. V. Rolando, R. Owen, P. Tanpaiboon, F. Lacbawan, Clinton R. Paden, Duncan MacCannell |
| EPI_ISL_1905898 | Michigan Department of Health and Human Services, Bureau of Laboratories | Michigan Department of Health and Human Services, Bureau of Laboratories | Blankenship HM, Riner D, Soehnlen MK |
| EPI_ISL_1905899 | Quest Diagnostics Incorporated | Centers for Disease Control and Prevention Division of Viral Diseases, Pathogen Discovery | Dakota Howard, Dhwani Batra, Peter W. Cook, Kara Moser, Adrian Paskey, Jason Caravas, Benjamin Rambo-Martin, Shatavia Morrison, Christopher Gulvick, Scott Sammons, Yvette Unoarumhi, Darlene Wagner, Matthew Schmerer, S. H. Rosenthal, A. Gerasimova, R. M. Kagan, B. Anderson, M. Hua, Y. Liu, L.E. Bernstein, K.E. Livingston, A. Perez, I. A. Shlyakhter, R. V. Rolando, R. Owen, P. Tanpaiboon, F. Lacbawan, Clinton R. Paden, Duncan MacCannell |
| EPI_ISL_1905900 | Michigan Department of Health and Human Services, Bureau of Laboratories | Michigan Department of Health and Human Services, Bureau of Laboratories | Blankenship HM, Riner D, Soehnlen MK |
| EPI_ISL_1905901, EPI_ISL_1905903 | Quest Diagnostics Incorporated | Centers for Disease Control and Prevention Division of Viral Diseases, Pathogen Discovery | Dakota Howard, Dhwani Batra, Peter W. Cook, Kara Moser, Adrian Paskey, Jason Caravas, Benjamin Rambo-Martin, Shatavia Morrison, Christopher Gulvick, Scott Sammons, Yvette Unoarumhi, Darlene Wagner, Matthew Schmerer, S. H. Rosenthal, A. Gerasimova, R. M. Kagan, B. Anderson, M. Hua, Y. Liu, L.E. Bernstein, K.E. Livingston, A. Perez, I. A. Shlyakhter, R. V. Rolando, R. Owen, P. Tanpaiboon, F. Lacbawan, Clinton R. Paden, Duncan MacCannell |
| EPI_ISL_1905904 | Michigan Department of Health and Human Services, Bureau of Laboratories | Michigan Department of Health and Human Services, Bureau of Laboratories | Blankenship HM, Riner D, Soehnlen MK |
| EPI_ISL_1905905 | Quest Diagnostics Incorporated | Centers for Disease Control and Prevention Division of Viral Diseases, Pathogen Discovery | Dakota Howard, Dhwani Batra, Peter W. Cook, Kara Moser, Adrian Paskey, Jason Caravas, Benjamin Rambo-Martin, Shatavia Morrison, Christopher Gulvick, Scott Sammons, Yvette Unoarumhi, Darlene Wagner, Matthew Schmerer, S. H. Rosenthal, A. Gerasimova, R. M. Kagan, B. Anderson, M. Hua, Y. Liu, L.E. Bernstein, K.E. Livingston, A. Perez, I. A. Shlyakhter, R. V. Rolando, R. Owen, P. Tanpaiboon, F. Lacbawan, Clinton R. Paden, Duncan MacCannell |
| EPI_ISL_1905906 | Michigan Department of Health and Human Services, Bureau of Laboratories | Michigan Department of Health and Human Services, Bureau of Laboratories | Blankenship HM, Riner D, Soehnlen MK |
| EPI_ISL_1905907 | Quest Diagnostics Incorporated | Centers for Disease Control and Prevention Division of Viral Diseases, Pathogen Discovery | Dakota Howard, Dhwani Batra, Peter W. Cook, Kara Moser, Adrian Paskey, Jason Caravas, Benjamin Rambo-Martin, Shatavia Morrison, Christopher Gulvick, Scott Sammons, Yvette Unoarumhi, Darlene Wagner, Matthew Schmerer, S. H. Rosenthal, A. Gerasimova, R. M. Kagan, B. Anderson, M. Hua, Y. Liu, L.E. Bernstein, K.E. Livingston, A. Perez, I. A. Shlyakhter, R. V. Rolando, R. Owen, P. Tanpaiboon, F. Lacbawan, Clinton R. Paden, Duncan MacCannell |
| EPI_ISL_1905908 | Michigan Department of Health and Human Services, Bureau of Laboratories | Michigan Department of Health and Human Services, Bureau of Laboratories | Blankenship HM, Riner D, Soehnlen MK |
| EPI_ISL_1905909 | Quest Diagnostics Incorporated | Centers for Disease Control and Prevention Division of Viral Diseases, Pathogen Discovery | Dakota Howard, Dhwani Batra, Peter W. Cook, Kara Moser, Adrian Paskey, Jason Caravas, Benjamin Rambo-Martin, Shatavia Morrison, Christopher Gulvick, Scott Sammons, Yvette Unoarumhi, Darlene Wagner, Matthew Schmerer, S. H. Rosenthal, A. Gerasimova, R. M. Kagan, B. Anderson, M. Hua, Y. Liu, L.E. Bernstein, K.E. Livingston, A. Perez, I. A. Shlyakhter, R. V. Rolando, R. Owen, P. Tanpaiboon, F. Lacbawan, Clinton R. Paden, Duncan MacCannell |
| EPI_ISL_1905910 | Michigan Department of Health and Human Services, Bureau of Laboratories | Michigan Department of Health and Human Services, Bureau of Laboratories | Blankenship HM, Riner D, Soehnlen MK |
| EPI_ISL_1905911 | Quest Diagnostics Incorporated | Centers for Disease Control and Prevention Division of Viral Diseases, Pathogen Discovery | Dakota Howard, Dhwani Batra, Peter W. Cook, Kara Moser, Adrian Paskey, Jason Caravas, Benjamin Rambo-Martin, Shatavia Morrison, Christopher Gulvick, Scott Sammons, Yvette Unoarumhi, Darlene Wagner, Matthew Schmerer, S. H. Rosenthal, A. Gerasimova, R. M. Kagan, B. Anderson, M. Hua, Y. Liu, L.E. Bernstein, K.E. Livingston, A. Perez, I. A. Shlyakhter, R. V. Rolando, R. Owen, P. Tanpaiboon, F. Lacbawan, Clinton R. Paden, Duncan MacCannell |
| EPI_ISL_1905912, EPI_ISL_1905913 | Michigan Department of Health and Human Services, Bureau of Laboratories | Michigan Department of Health and Human Services, Bureau of Laboratories | Blankenship HM, Riner D, Soehnlen MK |
| EPI_ISL_1905914 | Quest Diagnostics Incorporated | Centers for Disease Control and Prevention Division of Viral Diseases, Pathogen Discovery | Dakota Howard, Dhwani Batra, Peter W. Cook, Kara Moser, Adrian Paskey, Jason Caravas, Benjamin Rambo-Martin, Shatavia Morrison, Christopher Gulvick, Scott Sammons, Yvette Unoarumhi, Darlene Wagner, Matthew Schmerer, S. H. Rosenthal, A. Gerasimova, R. M. Kagan, B. Anderson, M. Hua, Y. Liu, L.E. Bernstein, K.E. Livingston, A. Perez, I. A. Shlyakhter, R. V. Rolando, R. Owen, P. Tanpaiboon, F. Lacbawan, Clinton R. Paden, Duncan MacCannell |
| EPI_ISL_1905915, EPI_ISL_1905916, EPI_ISL_1905917 | Michigan Department of Health and Human Services, Bureau of Laboratories | Michigan Department of Health and Human Services, Bureau of Laboratories | Blankenship HM, Riner D, Soehnlen MK |
| EPI_ISL_1905918 | Quest Diagnostics Incorporated | Centers for Disease Control and Prevention Division of Viral Diseases, Pathogen Discovery | Dakota Howard, Dhwani Batra, Peter W. Cook, Kara Moser, Adrian Paskey, Jason Caravas, Benjamin Rambo-Martin, Shatavia Morrison, Christopher Gulvick, Scott Sammons, Yvette Unoarumhi, Darlene Wagner, Matthew Schmerer, S. H. Rosenthal, A. Gerasimova, R. M. Kagan, B. Anderson, M. Hua, Y. Liu, L.E. Bernstein, K.E. Livingston, A. Perez, I. A. Shlyakhter, R. V. Rolando, R. Owen, P. Tanpaiboon, F. Lacbawan, Clinton R. Paden, Duncan MacCannell |
| EPI_ISL_1905919, EPI_ISL_1905921 | Michigan Department of Health and Human Services, Bureau of Laboratories | Michigan Department of Health and Human Services, Bureau of Laboratories | Blankenship HM, Riner D, Soehnlen MK |
| EPI_ISL_1905922 | Quest Diagnostics Incorporated | Centers for Disease Control and Prevention Division of Viral Diseases, Pathogen Discovery | Dakota Howard, Dhwani Batra, Peter W. Cook, Kara Moser, Adrian Paskey, Jason Caravas, Benjamin Rambo-Martin, Shatavia Morrison, Christopher Gulvick, Scott Sammons, Yvette Unoarumhi, Darlene Wagner, Matthew Schmerer, S. H. Rosenthal, A. Gerasimova, R. M. Kagan, B. Anderson, M. Hua, Y. Liu, L.E. Bernstein, K.E. Livingston, A. Perez, I. A. Shlyakhter, R. V. Rolando, R. Owen, P. Tanpaiboon, F. Lacbawan, Clinton R. Paden, Duncan MacCannell |
| EPI_ISL_1905923 | Michigan Department of Health and Human Services, Bureau of Laboratories | Michigan Department of Health and Human Services, Bureau of Laboratories | Blankenship HM, Riner D, Soehnlen MK |
| EPI_ISL_1905925 | Quest Diagnostics Incorporated | Centers for Disease Control and Prevention Division of Viral Diseases, Pathogen Discovery | Dakota Howard, Dhwani Batra, Peter W. Cook, Kara Moser, Adrian Paskey, Jason Caravas, Benjamin Rambo-Martin, Shatavia Morrison, Christopher Gulvick, Scott Sammons, Yvette Unoarumhi, Darlene Wagner, Matthew Schmerer, S. H. Rosenthal, A. Gerasimova, R. M. Kagan, B. Anderson, M. Hua, Y. Liu, L.E. Bernstein, K.E. Livingston, A. Perez, I. A. Shlyakhter, R. V. Rolando, R. Owen, P. Tanpaiboon, F. Lacbawan, Clinton R. Paden, Duncan MacCannell |
| EPI_ISL_1905926 | Michigan Department of Health and Human Services, Bureau of Laboratories | Michigan Department of Health and Human Services, Bureau of Laboratories | Blankenship HM, Riner D, Soehnlen MK |
| EPI_ISL_1905927 | Quest Diagnostics Incorporated | Centers for Disease Control and Prevention Division of Viral Diseases, Pathogen Discovery | Dakota Howard, Dhwani Batra, Peter W. Cook, Kara Moser, Adrian Paskey, Jason Caravas, Benjamin Rambo-Martin, Shatavia Morrison, Christopher Gulvick, Scott Sammons, Yvette Unoarumhi, Darlene Wagner, Matthew Schmerer, S. H. Rosenthal, A. Gerasimova, R. M. Kagan, B. Anderson, M. Hua, Y. Liu, L.E. Bernstein, K.E. Livingston, A. Perez, I. A. Shlyakhter, R. V. Rolando, R. Owen, P. Tanpaiboon, F. Lacbawan, Clinton R. Paden, Duncan MacCannell |
| EPI_ISL_1905928 | Michigan Department of Health and Human Services, Bureau of Laboratories | Michigan Department of Health and Human Services, Bureau of Laboratories | Blankenship HM, Riner D, Soehnlen MK |
| EPI_ISL_1905929 | Quest Diagnostics Incorporated | Centers for Disease Control and Prevention Division of Viral Diseases, Pathogen Discovery | Dakota Howard, Dhwani Batra, Peter W. Cook, Kara Moser, Adrian Paskey, Jason Caravas, Benjamin Rambo-Martin, Shatavia Morrison, Christopher Gulvick, Scott Sammons, Yvette Unoarumhi, Darlene Wagner, Matthew Schmerer, S. H. Rosenthal, A. Gerasimova, R. M. Kagan, B. Anderson, M. Hua, Y. Liu, L.E. Bernstein, K.E. Livingston, A. Perez, I. A. Shlyakhter, R. V. Rolando, R. Owen, P. Tanpaiboon, F. Lacbawan, Clinton R. Paden, Duncan MacCannell |
| EPI_ISL_1905930 | Michigan Department of Health and Human Services, Bureau of Laboratories | Michigan Department of Health and Human Services, Bureau of Laboratories | Blankenship HM, Riner D, Soehnlen MK |
| EPI_ISL_1905931 | Quest Diagnostics Incorporated | Centers for Disease Control and Prevention Division of Viral Diseases, Pathogen Discovery | Dakota Howard, Dhwani Batra, Peter W. Cook, Kara Moser, Adrian Paskey, Jason Caravas, Benjamin Rambo-Martin, Shatavia Morrison, Christopher Gulvick, Scott Sammons, Yvette Unoarumhi, Darlene Wagner, Matthew Schmerer, S. H. Rosenthal, A. Gerasimova, R. M. Kagan, B. Anderson, M. Hua, Y. Liu, L.E. Bernstein, K.E. Livingston, A. Perez, I. A. Shlyakhter, R. V. Rolando, R. Owen, P. Tanpaiboon, F. Lacbawan, Clinton R. Paden, Duncan MacCannell |
| EPI_ISL_1905932, EPI_ISL_1905933 | Michigan Department of Health and Human Services, Bureau of Laboratories | Michigan Department of Health and Human Services, Bureau of Laboratories | Blankenship HM, Riner D, Soehnlen MK |

| | | | |
|---|---|---|---|
| EPI_ISL_1905934 | Quest Diagnostics Incorporated | Centers for Disease Control and Prevention Division of Viral Diseases, Pathogen Discovery | Dakota Howard, Dhwani Batra, Peter W. Cook, Kara Moser, Adrian Paskey, Jason Caravas, Benjamin Rambo-Martin, Shatavia Morrison, Christopher Gulvick, Scott Sammons, Yvette Unoarumhi, Darlene Wagner, Matthew Schmerer, S. H. Rosenthal, A. Gerasimova, R. M. Kagan, B. Anderson, M. Hua, Y. Liu, L.E. Bernstein, K.E. Livingston, A. Perez, I. A. Shlyakhter, R. V. Rolando, R. Owen, P. Tanpaiboon, F. Lacbawan, Clinton R. Paden, Duncan MacCannell |
| EPI_ISL_1905935, EPI_ISL_1905936, EPI_ISL_1905937 | Michigan Department of Health and Human Services, Bureau of Laboratories | Michigan Department of Health and Human Services, Bureau of Laboratories | Blankenship HM, Riner D, Soehnlen MK |
| EPI_ISL_1905938 | Quest Diagnostics Incorporated | Centers for Disease Control and Prevention Division of Viral Diseases, Pathogen Discovery | Dakota Howard, Dhwani Batra, Peter W. Cook, Kara Moser, Adrian Paskey, Jason Caravas, Benjamin Rambo-Martin, Shatavia Morrison, Christopher Gulvick, Scott Sammons, Yvette Unoarumhi, Darlene Wagner, Matthew Schmerer, S. H. Rosenthal, A. Gerasimova, R. M. Kagan, B. Anderson, M. Hua, Y. Liu, L.E. Bernstein, K.E. Livingston, A. Perez, I. A. Shlyakhter, R. V. Rolando, R. Owen, P. Tanpaiboon, F. Lacbawan, Clinton R. Paden, Duncan MacCannell |
| EPI_ISL_1905939 | Michigan Department of Health and Human Services, Bureau of Laboratories | Michigan Department of Health and Human Services, Bureau of Laboratories | Blankenship HM, Riner D, Soehnlen MK |
| EPI_ISL_1905940 | Quest Diagnostics Incorporated | Centers for Disease Control and Prevention Division of Viral Diseases, Pathogen Discovery | Dakota Howard, Dhwani Batra, Peter W. Cook, Kara Moser, Adrian Paskey, Jason Caravas, Benjamin Rambo-Martin, Shatavia Morrison, Christopher Gulvick, Scott Sammons, Yvette Unoarumhi, Darlene Wagner, Matthew Schmerer, S. H. Rosenthal, A. Gerasimova, R. M. Kagan, B. Anderson, M. Hua, Y. Liu, L.E. Bernstein, K.E. Livingston, A. Perez, I. A. Shlyakhter, R. V. Rolando, R. Owen, P. Tanpaiboon, F. Lacbawan, Clinton R. Paden, Duncan MacCannell |
| EPI_ISL_1905941 | Michigan Department of Health and Human Services, Bureau of Laboratories | Michigan Department of Health and Human Services, Bureau of Laboratories | Blankenship HM, Riner D, Soehnlen MK |
| EPI_ISL_1905942 | Quest Diagnostics Incorporated | Centers for Disease Control and Prevention Division of Viral Diseases, Pathogen Discovery | Dakota Howard, Dhwani Batra, Peter W. Cook, Kara Moser, Adrian Paskey, Jason Caravas, Benjamin Rambo-Martin, Shatavia Morrison, Christopher Gulvick, Scott Sammons, Yvette Unoarumhi, Darlene Wagner, Matthew Schmerer, S. H. Rosenthal, A. Gerasimova, R. M. Kagan, B. Anderson, M. Hua, Y. Liu, L.E. Bernstein, K.E. Livingston, A. Perez, I. A. Shlyakhter, R. V. Rolando, R. Owen, P. Tanpaiboon, F. Lacbawan, Clinton R. Paden, Duncan MacCannell |
| EPI_ISL_1905943, EPI_ISL_1905944 | Michigan Department of Health and Human Services, Bureau of Laboratories | Michigan Department of Health and Human Services, Bureau of Laboratories | Blankenship HM, Riner D, Soehnlen MK |
| EPI_ISL_1905945, EPI_ISL_1905946 | Quest Diagnostics Incorporated | Centers for Disease Control and Prevention Division of Viral Diseases, Pathogen Discovery | Dakota Howard, Dhwani Batra, Peter W. Cook, Kara Moser, Adrian Paskey, Jason Caravas, Benjamin Rambo-Martin, Shatavia Morrison, Christopher Gulvick, Scott Sammons, Yvette Unoarumhi, Darlene Wagner, Matthew Schmerer, S. H. Rosenthal, A. Gerasimova, R. M. Kagan, B. Anderson, M. Hua, Y. Liu, L.E. Bernstein, K.E. Livingston, A. Perez, I. A. Shlyakhter, R. V. Rolando, R. Owen, P. Tanpaiboon, F. Lacbawan, Clinton R. Paden, Duncan MacCannell |
| EPI_ISL_1905947, EPI_ISL_1905948, EPI_ISL_1905949 | Michigan Department of Health and Human Services, Bureau of Laboratories | Michigan Department of Health and Human Services, Bureau of Laboratories | Blankenship HM, Riner D, Soehnlen MK |
| EPI_ISL_1905950 | Quest Diagnostics Incorporated | Centers for Disease Control and Prevention Division of Viral Diseases, Pathogen Discovery | Dakota Howard, Dhwani Batra, Peter W. Cook, Kara Moser, Adrian Paskey, Jason Caravas, Benjamin Rambo-Martin, Shatavia Morrison, Christopher Gulvick, Scott Sammons, Yvette Unoarumhi, Darlene Wagner, Matthew Schmerer, S. H. Rosenthal, A. Gerasimova, R. M. Kagan, B. Anderson, M. Hua, Y. Liu, L.E. Bernstein, K.E. Livingston, A. Perez, I. A. Shlyakhter, R. V. Rolando, R. Owen, P. Tanpaiboon, F. Lacbawan, Clinton R. Paden, Duncan MacCannell |
| EPI_ISL_1905951 | Michigan Department of Health and Human Services, Bureau of Laboratories | Michigan Department of Health and Human Services, Bureau of Laboratories | Blankenship HM, Riner D, Soehnlen MK |
| EPI_ISL_1905952 | Quest Diagnostics Incorporated | Centers for Disease Control and Prevention Division of Viral Diseases, Pathogen Discovery | Dakota Howard, Dhwani Batra, Peter W. Cook, Kara Moser, Adrian Paskey, Jason Caravas, Benjamin Rambo-Martin, Shatavia Morrison, Christopher Gulvick, Scott Sammons, Yvette Unoarumhi, Darlene Wagner, Matthew Schmerer, S. H. Rosenthal, A. Gerasimova, R. M. Kagan, B. Anderson, M. Hua, Y. Liu, L.E. Bernstein, K.E. Livingston, A. Perez, I. A. Shlyakhter, R. V. Rolando, R. Owen, P. Tanpaiboon, F. Lacbawan, Clinton R. Paden, Duncan MacCannell |
| EPI_ISL_1905953 | Michigan Department of Health and Human Services, Bureau of Laboratories | Michigan Department of Health and Human Services, Bureau of Laboratories | Blankenship HM, Riner D, Soehnlen MK |
| EPI_ISL_1905954 | Quest Diagnostics Incorporated | Centers for Disease Control and Prevention Division of Viral Diseases, Pathogen Discovery | Dakota Howard, Dhwani Batra, Peter W. Cook, Kara Moser, Adrian Paskey, Jason Caravas, Benjamin Rambo-Martin, Shatavia Morrison, Christopher Gulvick, Scott Sammons, Yvette Unoarumhi, Darlene Wagner, Matthew Schmerer, S. H. Rosenthal, A. Gerasimova, R. M. Kagan, B. Anderson, M. Hua, Y. Liu, L.E. Bernstein, K.E. Livingston, A. Perez, I. A. Shlyakhter, R. V. Rolando, R. Owen, P. Tanpaiboon, F. Lacbawan, Clinton R. Paden, Duncan MacCannell |
| EPI_ISL_1905955 | Michigan Department of Health and Human Services, Bureau of Laboratories | Michigan Department of Health and Human Services, Bureau of Laboratories | Blankenship HM, Riner D, Soehnlen MK |
| EPI_ISL_1905956 | Quest Diagnostics Incorporated | Centers for Disease Control and Prevention Division of Viral Diseases, Pathogen Discovery | Dakota Howard, Dhwani Batra, Peter W. Cook, Kara Moser, Adrian Paskey, Jason Caravas, Benjamin Rambo-Martin, Shatavia Morrison, Christopher Gulvick, Scott Sammons, Yvette Unoarumhi, Darlene Wagner, Matthew Schmerer, S. H. Rosenthal, A. Gerasimova, R. M. Kagan, B. Anderson, M. Hua, Y. Liu, L.E. Bernstein, K.E. Livingston, A. Perez, I. A. Shlyakhter, R. V. Rolando, R. Owen, P. Tanpaiboon, F. Lacbawan, Clinton R. Paden, Duncan MacCannell |
| EPI_ISL_1905957 | Michigan Department of Health and Human Services, Bureau of Laboratories | Michigan Department of Health and Human Services, Bureau of Laboratories | Blankenship HM, Riner D, Soehnlen MK |
| EPI_ISL_1905958 | Quest Diagnostics Incorporated | Centers for Disease Control and Prevention Division of Viral Diseases, Pathogen Discovery | Dakota Howard, Dhwani Batra, Peter W. Cook, Kara Moser, Adrian Paskey, Jason Caravas, Benjamin Rambo-Martin, Shatavia Morrison, Christopher Gulvick, Scott Sammons, Yvette Unoarumhi, Darlene Wagner, Matthew Schmerer, S. H. Rosenthal, A. Gerasimova, R. M. Kagan, B. Anderson, M. Hua, Y. Liu, L.E. Bernstein, K.E. Livingston, A. Perez, I. A. Shlyakhter, R. V. Rolando, R. Owen, P. Tanpaiboon, F. Lacbawan, Clinton R. Paden, Duncan MacCannell |
| EPI_ISL_1905959 | Michigan Department of Health and Human Services, Bureau of Laboratories | Michigan Department of Health and Human Services, Bureau of Laboratories | Blankenship HM, Riner D, Soehnlen MK |
| EPI_ISL_1905960 | Quest Diagnostics Incorporated | Centers for Disease Control and Prevention Division of Viral Diseases, Pathogen Discovery | Dakota Howard, Dhwani Batra, Peter W. Cook, Kara Moser, Adrian Paskey, Jason Caravas, Benjamin Rambo-Martin, Shatavia Morrison, Christopher Gulvick, Scott Sammons, Yvette Unoarumhi, Darlene Wagner, Matthew Schmerer, S. H. Rosenthal, A. Gerasimova, R. M. Kagan, B. Anderson, M. Hua, Y. Liu, L.E. Bernstein, K.E. Livingston, A. Perez, I. A. Shlyakhter, R. V. Rolando, R. Owen, P. Tanpaiboon, F. Lacbawan, Clinton R. Paden, Duncan MacCannell |
| EPI_ISL_1905961, EPI_ISL_1905962, EPI_ISL_1905963 | Michigan Department of Health and Human Services, Bureau of Laboratories | Michigan Department of Health and Human Services, Bureau of Laboratories | Blankenship HM, Riner D, Soehnlen MK |
| EPI_ISL_1905964 | Quest Diagnostics Incorporated | Centers for Disease Control and Prevention Division of Viral Diseases, Pathogen Discovery | Dakota Howard, Dhwani Batra, Peter W. Cook, Kara Moser, Adrian Paskey, Jason Caravas, Benjamin Rambo-Martin, Shatavia Morrison, Christopher Gulvick, Scott Sammons, Yvette Unoarumhi, Darlene Wagner, Matthew Schmerer, S. H. Rosenthal, A. Gerasimova, R. M. Kagan, B. Anderson, M. Hua, Y. Liu, L.E. Bernstein, K.E. Livingston, A. Perez, I. A. Shlyakhter, R. V. Rolando, R. Owen, P. Tanpaiboon, F. Lacbawan, Clinton R. Paden, Duncan MacCannell |
| EPI_ISL_1905965, EPI_ISL_1905966 | Michigan Department of Health and Human Services, Bureau of Laboratories | Michigan Department of Health and Human Services, Bureau of Laboratories | Blankenship HM, Riner D, Soehnlen MK |
| EPI_ISL_1905967 | Quest Diagnostics Incorporated | Centers for Disease Control and Prevention Division of Viral Diseases, Pathogen Discovery | Dakota Howard, Dhwani Batra, Peter W. Cook, Kara Moser, Adrian Paskey, Jason Caravas, Benjamin Rambo-Martin, Shatavia Morrison, Christopher Gulvick, Scott Sammons, Yvette Unoarumhi, Darlene Wagner, Matthew Schmerer, S. H. Rosenthal, A. Gerasimova, R. M. Kagan, B. Anderson, M. Hua, Y. Liu, L.E. Bernstein, K.E. Livingston, A. Perez, I. A. Shlyakhter, R. V. Rolando, R. Owen, P. Tanpaiboon, F. Lacbawan, Clinton R. Paden, Duncan MacCannell |
| EPI_ISL_1905968 | Michigan Department of Health and Human Services, Bureau of Laboratories | Michigan Department of Health and Human Services, Bureau of Laboratories | Blankenship HM, Riner D, Soehnlen MK |
| EPI_ISL_1905969 | Quest Diagnostics Incorporated | Centers for Disease Control and Prevention Division of Viral Diseases, Pathogen Discovery | Dakota Howard, Dhwani Batra, Peter W. Cook, Kara Moser, Adrian Paskey, Jason Caravas, Benjamin Rambo-Martin, Shatavia Morrison, Christopher Gulvick, Scott Sammons, Yvette Unoarumhi, Darlene Wagner, Matthew Schmerer, S. H. Rosenthal, A. Gerasimova, R. M. Kagan, B. Anderson, M. Hua, Y. Liu, L.E. Bernstein, K.E. Livingston, A. Perez, I. A. Shlyakhter, R. V. Rolando, R. Owen, P. Tanpaiboon, F. Lacbawan, Clinton R. Paden, Duncan MacCannell |
| EPI_ISL_1905970 | Michigan Department of Health and Human Services, Bureau of Laboratories | Michigan Department of Health and Human Services, Bureau of Laboratories | Blankenship HM, Riner D, Soehnlen MK |
| EPI_ISL_1905971 | Quest Diagnostics Incorporated | Centers for Disease Control and Prevention Division of Viral Diseases, Pathogen Discovery | Dakota Howard, Dhwani Batra, Peter W. Cook, Kara Moser, Adrian Paskey, Jason Caravas, Benjamin Rambo-Martin, Shatavia Morrison, Christopher Gulvick, Scott Sammons, Yvette Unoarumhi, Darlene Wagner, Matthew Schmerer, S. H. Rosenthal, A. Gerasimova, R. M. Kagan, B. Anderson, M. Hua, Y. Liu, L.E. Bernstein, K.E. Livingston, A. Perez, I. A. Shlyakhter, R. V. Rolando, R. Owen, P. Tanpaiboon, F. Lacbawan, Clinton R. Paden, Duncan MacCannell |
| EPI_ISL_1905972 | Michigan Department of Health and Human Services, Bureau of Laboratories | Michigan Department of Health and Human Services, Bureau of Laboratories | Blankenship HM, Riner D, Soehnlen MK |
| EPI_ISL_1905973 | Quest Diagnostics Incorporated | Centers for Disease Control and Prevention Division of Viral Diseases, Pathogen Discovery | Dakota Howard, Dhwani Batra, Peter W. Cook, Kara Moser, Adrian Paskey, Jason Caravas, Benjamin Rambo-Martin, Shatavia Morrison, Christopher Gulvick, Scott Sammons, Yvette Unoarumhi, Darlene Wagner, Matthew Schmerer, S. H. Rosenthal, A. Gerasimova, R. M. Kagan, B. Anderson, M. Hua, Y. Liu, L.E. Bernstein, K.E. Livingston, A. Perez, I. A. Shlyakhter, R. V. Rolando, R. Owen, P. Tanpaiboon, F. Lacbawan, Clinton R. Paden, Duncan MacCannell |
| EPI_ISL_1905974, EPI_ISL_1905975 | Michigan Department of Health and Human Services, Bureau of Laboratories | Michigan Department of Health and Human Services, Bureau of Laboratories | Blankenship HM, Riner D, Soehnlen MK |
| EPI_ISL_1905976 | Quest Diagnostics Incorporated | Centers for Disease Control and Prevention Division of Viral Diseases, Pathogen Discovery | Dakota Howard, Dhwani Batra, Peter W. Cook, Kara Moser, Adrian Paskey, Jason Caravas, Benjamin Rambo-Martin, Shatavia Morrison, Christopher Gulvick, Scott Sammons, Yvette Unoarumhi, Darlene Wagner, Matthew Schmerer, S. H. Rosenthal, A. Gerasimova, R. M. Kagan, B. Anderson, M. Hua, Y. Liu, L.E. Bernstein, K.E. Livingston, A. Perez, I. A. Shlyakhter, R. V. Rolando, R. Owen, P. Tanpaiboon, F. Lacbawan, Clinton R. Paden, Duncan MacCannell |
| EPI_ISL_1905977 | Michigan Department of Health and Human Services, Bureau of Laboratories | Michigan Department of Health and Human Services, Bureau of Laboratories | Blankenship HM, Riner D, Soehnlen MK |
| EPI_ISL_1905978 | Quest Diagnostics Incorporated | Centers for Disease Control and Prevention Division of Viral Diseases, Pathogen Discovery | Dakota Howard, Dhwani Batra, Peter W. Cook, Kara Moser, Adrian Paskey, Jason Caravas, Benjamin Rambo-Martin, Shatavia Morrison, Christopher Gulvick, Scott Sammons, Yvette Unoarumhi, Darlene Wagner, Matthew Schmerer, S. H. Rosenthal, A. Gerasimova, R. M. Kagan, B. Anderson, M. Hua, Y. Liu, L.E. Bernstein, K.E. Livingston, A. Perez, I. A. Shlyakhter, R. V. Rolando, R. Owen, P. Tanpaiboon, F. Lacbawan, Clinton R. Paden, Duncan MacCannell |
| EPI_ISL_1905979 | Michigan Department of Health and Human Services, Bureau of Laboratories | Michigan Department of Health and Human Services, Bureau of Laboratories | Blankenship HM, Riner D, Soehnlen MK |
| EPI_ISL_1905980 | Quest Diagnostics Incorporated | Centers for Disease Control and Prevention | Dakota Howard, Dhwani Batra, Peter W. Cook, Kara Moser, Adrian Paskey, Jason Caravas, Benjamin Rambo-Martin, Shatavia Morrison, Christopher Gulvick, Scott Sammons, Yvette Unoarumhi, Darlene Wagner, Matthew Schmerer, S. H. Rosenthal, A. Gerasimova, R. M. Kagan, B. |

| | | | |
|---|---|---|---|
| | | Division of Viral Diseases, Pathogen Discovery | Anderson, M. Hua, Y. Liu, L.E. Bernstein, K.E. Livingston, A. Perez, I. A. Shlyakhter, R. V. Rolando, R. Owen, P. Tanpaiboon, F. Lacbawan, Clinton R. Paden, Duncan MacCannell |
| EPI_ISL_1905981 | Michigan Department of Health and Human Services, Bureau of Laboratories | Michigan Department of Health and Human Services, Bureau of Laboratories | Blankenship HM, Riner D, Soehnlen MK |
| EPI_ISL_1905982, EPI_ISL_1905984 | Quest Diagnostics Incorporated | Centers for Disease Control and Prevention Division of Viral Diseases, Pathogen Discovery | Dakota Howard, Dhwani Batra, Peter W. Cook, Kara Moser, Adrian Paskey, Jason Caravas, Benjamin Rambo-Martin, Shatavia Morrison, Christopher Gulvick, Scott Sammons, Yvette Unoarumhi, Darlene Wagner, Matthew Schmerer, S. H. Rosenthal, A. Gerasimova, R. M. Kagan, B. Anderson, M. Hua, Y. Liu, L.E. Bernstein, K.E. Livingston, A. Perez, I. A. Shlyakhter, R. V. Rolando, R. Owen, P. Tanpaiboon, F. Lacbawan, Clinton R. Paden, Duncan MacCannell |
| EPI_ISL_1905985, EPI_ISL_1905986 | Michigan Department of Health and Human Services, Bureau of Laboratories | Michigan Department of Health and Human Services, Bureau of Laboratories | Blankenship HM, Riner D, Soehnlen MK |
| EPI_ISL_1905987 | Quest Diagnostics Incorporated | Centers for Disease Control and Prevention Division of Viral Diseases, Pathogen Discovery | Dakota Howard, Dhwani Batra, Peter W. Cook, Kara Moser, Adrian Paskey, Jason Caravas, Benjamin Rambo-Martin, Shatavia Morrison, Christopher Gulvick, Scott Sammons, Yvette Unoarumhi, Darlene Wagner, Matthew Schmerer, S. H. Rosenthal, A. Gerasimova, R. M. Kagan, B. Anderson, M. Hua, Y. Liu, L.E. Bernstein, K.E. Livingston, A. Perez, I. A. Shlyakhter, R. V. Rolando, R. Owen, P. Tanpaiboon, F. Lacbawan, Clinton R. Paden, Duncan MacCannell |
| EPI_ISL_1905988 | Michigan Department of Health and Human Services, Bureau of Laboratories | Michigan Department of Health and Human Services, Bureau of Laboratories | Blankenship HM, Riner D, Soehnlen MK |
| EPI_ISL_1905989 | Quest Diagnostics Incorporated | Centers for Disease Control and Prevention Division of Viral Diseases, Pathogen Discovery | Dakota Howard, Dhwani Batra, Peter W. Cook, Kara Moser, Adrian Paskey, Jason Caravas, Benjamin Rambo-Martin, Shatavia Morrison, Christopher Gulvick, Scott Sammons, Yvette Unoarumhi, Darlene Wagner, Matthew Schmerer, S. H. Rosenthal, A. Gerasimova, R. M. Kagan, B. Anderson, M. Hua, Y. Liu, L.E. Bernstein, K.E. Livingston, A. Perez, I. A. Shlyakhter, R. V. Rolando, R. Owen, P. Tanpaiboon, F. Lacbawan, Clinton R. Paden, Duncan MacCannell |
| EPI_ISL_1905990, EPI_ISL_1905991 | Michigan Department of Health and Human Services, Bureau of Laboratories | Michigan Department of Health and Human Services, Bureau of Laboratories | Blankenship HM, Riner D, Soehnlen MK |
| EPI_ISL_1905992 | Quest Diagnostics Incorporated | Centers for Disease Control and Prevention Division of Viral Diseases, Pathogen Discovery | Dakota Howard, Dhwani Batra, Peter W. Cook, Kara Moser, Adrian Paskey, Jason Caravas, Benjamin Rambo-Martin, Shatavia Morrison, Christopher Gulvick, Scott Sammons, Yvette Unoarumhi, Darlene Wagner, Matthew Schmerer, S. H. Rosenthal, A. Gerasimova, R. M. Kagan, B. Anderson, M. Hua, Y. Liu, L.E. Bernstein, K.E. Livingston, A. Perez, I. A. Shlyakhter, R. V. Rolando, R. Owen, P. Tanpaiboon, F. Lacbawan, Clinton R. Paden, Duncan MacCannell |
| EPI_ISL_1905993 | Michigan Department of Health and Human Services, Bureau of Laboratories | Michigan Department of Health and Human Services, Bureau of Laboratories | Blankenship HM, Riner D, Soehnlen MK |
| EPI_ISL_1905994 | Quest Diagnostics Incorporated | Centers for Disease Control and Prevention Division of Viral Diseases, Pathogen Discovery | Dakota Howard, Dhwani Batra, Peter W. Cook, Kara Moser, Adrian Paskey, Jason Caravas, Benjamin Rambo-Martin, Shatavia Morrison, Christopher Gulvick, Scott Sammons, Yvette Unoarumhi, Darlene Wagner, Matthew Schmerer, S. H. Rosenthal, A. Gerasimova, R. M. Kagan, B. Anderson, M. Hua, Y. Liu, L.E. Bernstein, K.E. Livingston, A. Perez, I. A. Shlyakhter, R. V. Rolando, R. Owen, P. Tanpaiboon, F. Lacbawan, Clinton R. Paden, Duncan MacCannell |
| EPI_ISL_1905995 | Michigan Department of Health and Human Services, Bureau of Laboratories | Michigan Department of Health and Human Services, Bureau of Laboratories | Blankenship HM, Riner D, Soehnlen MK |
| EPI_ISL_1905996 | Quest Diagnostics Incorporated | Centers for Disease Control and Prevention Division of Viral Diseases, Pathogen Discovery | Dakota Howard, Dhwani Batra, Peter W. Cook, Kara Moser, Adrian Paskey, Jason Caravas, Benjamin Rambo-Martin, Shatavia Morrison, Christopher Gulvick, Scott Sammons, Yvette Unoarumhi, Darlene Wagner, Matthew Schmerer, S. H. Rosenthal, A. Gerasimova, R. M. Kagan, B. Anderson, M. Hua, Y. Liu, L.E. Bernstein, K.E. Livingston, A. Perez, I. A. Shlyakhter, R. V. Rolando, R. Owen, P. Tanpaiboon, F. Lacbawan, Clinton R. Paden, Duncan MacCannell |
| EPI_ISL_1905997, EPI_ISL_1905998 | Michigan Department of Health and Human Services, Bureau of Laboratories | Michigan Department of Health and Human Services, Bureau of Laboratories | Blankenship HM, Riner D, Soehnlen MK |
| EPI_ISL_1905999 | Quest Diagnostics Incorporated | Centers for Disease Control and Prevention Division of Viral Diseases, Pathogen Discovery | Dakota Howard, Dhwani Batra, Peter W. Cook, Kara Moser, Adrian Paskey, Jason Caravas, Benjamin Rambo-Martin, Shatavia Morrison, Christopher Gulvick, Scott Sammons, Yvette Unoarumhi, Darlene Wagner, Matthew Schmerer, S. H. Rosenthal, A. Gerasimova, R. M. Kagan, B. Anderson, M. Hua, Y. Liu, L.E. Bernstein, K.E. Livingston, A. Perez, I. A. Shlyakhter, R. V. Rolando, R. Owen, P. Tanpaiboon, F. Lacbawan, Clinton R. Paden, Duncan MacCannell |
| EPI_ISL_1906000 | Michigan Department of Health and Human Services, Bureau of Laboratories | Michigan Department of Health and Human Services, Bureau of Laboratories | Blankenship HM, Riner D, Soehnlen MK |
| EPI_ISL_1906001 | Quest Diagnostics Incorporated | Centers for Disease Control and Prevention Division of Viral Diseases, Pathogen Discovery | Dakota Howard, Dhwani Batra, Peter W. Cook, Kara Moser, Adrian Paskey, Jason Caravas, Benjamin Rambo-Martin, Shatavia Morrison, Christopher Gulvick, Scott Sammons, Yvette Unoarumhi, Darlene Wagner, Matthew Schmerer, S. H. Rosenthal, A. Gerasimova, R. M. Kagan, B. Anderson, M. Hua, Y. Liu, L.E. Bernstein, K.E. Livingston, A. Perez, I. A. Shlyakhter, R. V. Rolando, R. Owen, P. Tanpaiboon, F. Lacbawan, Clinton R. Paden, Duncan MacCannell |
| EPI_ISL_1906002 | Michigan Department of Health and Human Services, Bureau of Laboratories | Michigan Department of Health and Human Services, Bureau of Laboratories | Blankenship HM, Riner D, Soehnlen MK |
| EPI_ISL_1906003 | Quest Diagnostics Incorporated | Centers for Disease Control and Prevention Division of Viral Diseases, Pathogen Discovery | Dakota Howard, Dhwani Batra, Peter W. Cook, Kara Moser, Adrian Paskey, Jason Caravas, Benjamin Rambo-Martin, Shatavia Morrison, Christopher Gulvick, Scott Sammons, Yvette Unoarumhi, Darlene Wagner, Matthew Schmerer, S. H. Rosenthal, A. Gerasimova, R. M. Kagan, B. Anderson, M. Hua, Y. Liu, L.E. Bernstein, K.E. Livingston, A. Perez, I. A. Shlyakhter, R. V. Rolando, R. Owen, P. Tanpaiboon, F. Lacbawan, Clinton R. Paden, Duncan MacCannell |
| EPI_ISL_1906004, EPI_ISL_1906005, EPI_ISL_1906006 | Michigan Department of Health and Human Services, Bureau of Laboratories | Michigan Department of Health and Human Services, Bureau of Laboratories | Blankenship HM, Riner D, Soehnlen MK |
| EPI_ISL_1906007 | Quest Diagnostics Incorporated | Centers for Disease Control and Prevention Division of Viral Diseases, Pathogen Discovery | Dakota Howard, Dhwani Batra, Peter W. Cook, Kara Moser, Adrian Paskey, Jason Caravas, Benjamin Rambo-Martin, Shatavia Morrison, Christopher Gulvick, Scott Sammons, Yvette Unoarumhi, Darlene Wagner, Matthew Schmerer, S. H. Rosenthal, A. Gerasimova, R. M. Kagan, B. Anderson, M. Hua, Y. Liu, L.E. Bernstein, K.E. Livingston, A. Perez, I. A. Shlyakhter, R. V. Rolando, R. Owen, P. Tanpaiboon, F. Lacbawan, Clinton R. Paden, Duncan MacCannell |
| EPI_ISL_1906008 | Michigan Department of Health and Human Services, Bureau of Laboratories | Michigan Department of Health and Human Services, Bureau of Laboratories | Blankenship HM, Riner D, Soehnlen MK |
| EPI_ISL_1906009 | Quest Diagnostics Incorporated | Centers for Disease Control and Prevention Division of Viral Diseases, Pathogen Discovery | Dakota Howard, Dhwani Batra, Peter W. Cook, Kara Moser, Adrian Paskey, Jason Caravas, Benjamin Rambo-Martin, Shatavia Morrison, Christopher Gulvick, Scott Sammons, Yvette Unoarumhi, Darlene Wagner, Matthew Schmerer, S. H. Rosenthal, A. Gerasimova, R. M. Kagan, B. Anderson, M. Hua, Y. Liu, L.E. Bernstein, K.E. Livingston, A. Perez, I. A. Shlyakhter, R. V. Rolando, R. Owen, P. Tanpaiboon, F. Lacbawan, Clinton R. Paden, Duncan MacCannell |
| EPI_ISL_1906010 | Michigan Department of Health and Human Services, Bureau of Laboratories | Michigan Department of Health and Human Services, Bureau of Laboratories | Blankenship HM, Riner D, Soehnlen MK |
| EPI_ISL_1906011 | Quest Diagnostics Incorporated | Centers for Disease Control and Prevention Division of Viral Diseases, Pathogen Discovery | Dakota Howard, Dhwani Batra, Peter W. Cook, Kara Moser, Adrian Paskey, Jason Caravas, Benjamin Rambo-Martin, Shatavia Morrison, Christopher Gulvick, Scott Sammons, Yvette Unoarumhi, Darlene Wagner, Matthew Schmerer, S. H. Rosenthal, A. Gerasimova, R. M. Kagan, B. Anderson, M. Hua, Y. Liu, L.E. Bernstein, K.E. Livingston, A. Perez, I. A. Shlyakhter, R. V. Rolando, R. Owen, P. Tanpaiboon, F. Lacbawan, Clinton R. Paden, Duncan MacCannell |
| EPI_ISL_1906012 | Michigan Department of Health and Human Services, Bureau of Laboratories | Michigan Department of Health and Human Services, Bureau of Laboratories | Blankenship HM, Riner D, Soehnlen MK |
| EPI_ISL_1906013 | Quest Diagnostics Incorporated | Centers for Disease Control and Prevention Division of Viral Diseases, Pathogen Discovery | Dakota Howard, Dhwani Batra, Peter W. Cook, Kara Moser, Adrian Paskey, Jason Caravas, Benjamin Rambo-Martin, Shatavia Morrison, Christopher Gulvick, Scott Sammons, Yvette Unoarumhi, Darlene Wagner, Matthew Schmerer, S. H. Rosenthal, A. Gerasimova, R. M. Kagan, B. Anderson, M. Hua, Y. Liu, L.E. Bernstein, K.E. Livingston, A. Perez, I. A. Shlyakhter, R. V. Rolando, R. Owen, P. Tanpaiboon, F. Lacbawan, Clinton R. Paden, Duncan MacCannell |
| EPI_ISL_1906014, EPI_ISL_1906015 | Michigan Department of Health and Human Services, Bureau of Laboratories | Michigan Department of Health and Human Services, Bureau of Laboratories | Blankenship HM, Riner D, Soehnlen MK |
| EPI_ISL_1906016 | Quest Diagnostics Incorporated | Centers for Disease Control and Prevention Division of Viral Diseases, Pathogen Discovery | Dakota Howard, Dhwani Batra, Peter W. Cook, Kara Moser, Adrian Paskey, Jason Caravas, Benjamin Rambo-Martin, Shatavia Morrison, Christopher Gulvick, Scott Sammons, Yvette Unoarumhi, Darlene Wagner, Matthew Schmerer, S. H. Rosenthal, A. Gerasimova, R. M. Kagan, B. Anderson, M. Hua, Y. Liu, L.E. Bernstein, K.E. Livingston, A. Perez, I. A. Shlyakhter, R. V. Rolando, R. Owen, P. Tanpaiboon, F. Lacbawan, Clinton R. Paden, Duncan MacCannell |
| EPI_ISL_1906017, EPI_ISL_1906018, EPI_ISL_1906019 | Michigan Department of Health and Human Services, Bureau of Laboratories | Michigan Department of Health and Human Services, Bureau of Laboratories | Blankenship HM, Riner D, Soehnlen MK |
| EPI_ISL_1906020 | Quest Diagnostics Incorporated | Centers for Disease Control and Prevention Division of Viral Diseases, Pathogen Discovery | Dakota Howard, Dhwani Batra, Peter W. Cook, Kara Moser, Adrian Paskey, Jason Caravas, Benjamin Rambo-Martin, Shatavia Morrison, Christopher Gulvick, Scott Sammons, Yvette Unoarumhi, Darlene Wagner, Matthew Schmerer, S. H. Rosenthal, A. Gerasimova, R. M. Kagan, B. Anderson, M. Hua, Y. Liu, L.E. Bernstein, K.E. Livingston, A. Perez, I. A. Shlyakhter, R. V. Rolando, R. Owen, P. Tanpaiboon, F. Lacbawan, Clinton R. Paden, Duncan MacCannell |
| EPI_ISL_1906021 | Michigan Department of Health and Human Services, Bureau of Laboratories | Michigan Department of Health and Human Services, Bureau of Laboratories | Blankenship HM, Riner D, Soehnlen MK |
| EPI_ISL_1906022 | Quest Diagnostics Incorporated | Centers for Disease Control and Prevention Division of Viral Diseases, Pathogen Discovery | Dakota Howard, Dhwani Batra, Peter W. Cook, Kara Moser, Adrian Paskey, Jason Caravas, Benjamin Rambo-Martin, Shatavia Morrison, Christopher Gulvick, Scott Sammons, Yvette Unoarumhi, Darlene Wagner, Matthew Schmerer, S. H. Rosenthal, A. Gerasimova, R. M. Kagan, B. Anderson, M. Hua, Y. Liu, L.E. Bernstein, K.E. Livingston, A. Perez, I. A. Shlyakhter, R. V. Rolando, R. Owen, P. Tanpaiboon, F. Lacbawan, Clinton R. Paden, Duncan MacCannell |
| EPI_ISL_1906023 | Michigan Department of Health and Human Services, Bureau of Laboratories | Michigan Department of Health and Human Services, Bureau of Laboratories | Blankenship HM, Riner D, Soehnlen MK |
| EPI_ISL_1906024 | Quest Diagnostics Incorporated | Centers for Disease Control and Prevention Division of Viral Diseases, Pathogen Discovery | Dakota Howard, Dhwani Batra, Peter W. Cook, Kara Moser, Adrian Paskey, Jason Caravas, Benjamin Rambo-Martin, Shatavia Morrison, Christopher Gulvick, Scott Sammons, Yvette Unoarumhi, Darlene Wagner, Matthew Schmerer, S. H. Rosenthal, A. Gerasimova, R. M. Kagan, B. Anderson, M. Hua, Y. Liu, L.E. Bernstein, K.E. Livingston, A. Perez, I. A. Shlyakhter, R. V. Rolando, R. Owen, P. Tanpaiboon, F. Lacbawan, Clinton R. Paden, Duncan MacCannell |
| EPI_ISL_1906025 | Michigan Department of Health and Human Services, Bureau of Laboratories | Michigan Department of Health and Human Services, Bureau of Laboratories | Blankenship HM, Riner D, Soehnlen MK |
| EPI_ISL_1906026 | Quest Diagnostics Incorporated | Centers for Disease Control and Prevention Division of Viral Diseases, Pathogen Discovery | Dakota Howard, Dhwani Batra, Peter W. Cook, Kara Moser, Adrian Paskey, Jason Caravas, Benjamin Rambo-Martin, Shatavia Morrison, Christopher Gulvick, Scott Sammons, Yvette Unoarumhi, Darlene Wagner, Matthew Schmerer, S. H. Rosenthal, A. Gerasimova, R. M. Kagan, B. Anderson, M. Hua, Y. Liu, L.E. Bernstein, K.E. Livingston, A. Perez, I. A. Shlyakhter, R. V. Rolando, R. Owen, P. Tanpaiboon, F. Lacbawan, Clinton R. Paden, Duncan MacCannell |

| | | | |
|---|---|---|---|
| EPI_ISL_1906027, EPI_ISL_1906029 | Michigan Department of Health and Human Services, Bureau of Laboratories | Michigan Department of Health and Human Services, Bureau of Laboratories | Blankenship HM, Riner D, Soehnlen MK |
| EPI_ISL_1906030 | Quest Diagnostics Incorporated | Centers for Disease Control and Prevention Division of Viral Diseases, Pathogen Discovery | Dakota Howard, Dhwani Batra, Peter W. Cook, Kara Moser, Adrian Paskey, Jason Caravas, Benjamin Rambo-Martin, Shatavia Morrison, Christopher Gulvick, Scott Sammons, Yvette Unoarumhi, Darlene Wagner, Matthew Schmerer, S. H. Rosenthal, A. Gerasimova, R. M. Kagan, B. Anderson, M. Hua, Y. Liu, L.E. Bernstein, K.E. Livingston, A. Perez, I. A. Shlyakhter, R. V. Rolando, R. Owen, P. Tanpaiboon, F. Lacbawan, Clinton R. Paden, Duncan MacCannell |
| EPI_ISL_1906031 | Michigan Department of Health and Human Services, Bureau of Laboratories | Michigan Department of Health and Human Services, Bureau of Laboratories | Blankenship HM, Riner D, Soehnlen MK |
| EPI_ISL_1906032 | Quest Diagnostics Incorporated | Centers for Disease Control and Prevention Division of Viral Diseases, Pathogen Discovery | Dakota Howard, Dhwani Batra, Peter W. Cook, Kara Moser, Adrian Paskey, Jason Caravas, Benjamin Rambo-Martin, Shatavia Morrison, Christopher Gulvick, Scott Sammons, Yvette Unoarumhi, Darlene Wagner, Matthew Schmerer, S. H. Rosenthal, A. Gerasimova, R. M. Kagan, B. Anderson, M. Hua, Y. Liu, L.E. Bernstein, K.E. Livingston, A. Perez, I. A. Shlyakhter, R. V. Rolando, R. Owen, P. Tanpaiboon, F. Lacbawan, Clinton R. Paden, Duncan MacCannell |
| EPI_ISL_1906033, EPI_ISL_1906034, EPI_ISL_1906035 | Michigan Department of Health and Human Services, Bureau of Laboratories | Michigan Department of Health and Human Services, Bureau of Laboratories | Blankenship HM, Riner D, Soehnlen MK |
| EPI_ISL_1906036 | Quest Diagnostics Incorporated | Centers for Disease Control and Prevention Division of Viral Diseases, Pathogen Discovery | Dakota Howard, Dhwani Batra, Peter W. Cook, Kara Moser, Adrian Paskey, Jason Caravas, Benjamin Rambo-Martin, Shatavia Morrison, Christopher Gulvick, Scott Sammons, Yvette Unoarumhi, Darlene Wagner, Matthew Schmerer, S. H. Rosenthal, A. Gerasimova, R. M. Kagan, B. Anderson, M. Hua, Y. Liu, L.E. Bernstein, K.E. Livingston, A. Perez, I. A. Shlyakhter, R. V. Rolando, R. Owen, P. Tanpaiboon, F. Lacbawan, Clinton R. Paden, Duncan MacCannell |
| EPI_ISL_1906037 | Michigan Department of Health and Human Services, Bureau of Laboratories | Michigan Department of Health and Human Services, Bureau of Laboratories | Blankenship HM, Riner D, Soehnlen MK |
| EPI_ISL_1906039 | Quest Diagnostics Incorporated | Centers for Disease Control and Prevention Division of Viral Diseases, Pathogen Discovery | Dakota Howard, Dhwani Batra, Peter W. Cook, Kara Moser, Adrian Paskey, Jason Caravas, Benjamin Rambo-Martin, Shatavia Morrison, Christopher Gulvick, Scott Sammons, Yvette Unoarumhi, Darlene Wagner, Matthew Schmerer, S. H. Rosenthal, A. Gerasimova, R. M. Kagan, B. Anderson, M. Hua, Y. Liu, L.E. Bernstein, K.E. Livingston, A. Perez, I. A. Shlyakhter, R. V. Rolando, R. Owen, P. Tanpaiboon, F. Lacbawan, Clinton R. Paden, Duncan MacCannell |
| EPI_ISL_1906040 | Michigan Department of Health and Human Services, Bureau of Laboratories | Michigan Department of Health and Human Services, Bureau of Laboratories | Blankenship HM, Riner D, Soehnlen MK |
| EPI_ISL_1906041 | Quest Diagnostics Incorporated | Centers for Disease Control and Prevention Division of Viral Diseases, Pathogen Discovery | Dakota Howard, Dhwani Batra, Peter W. Cook, Kara Moser, Adrian Paskey, Jason Caravas, Benjamin Rambo-Martin, Shatavia Morrison, Christopher Gulvick, Scott Sammons, Yvette Unoarumhi, Darlene Wagner, Matthew Schmerer, S. H. Rosenthal, A. Gerasimova, R. M. Kagan, B. Anderson, M. Hua, Y. Liu, L.E. Bernstein, K.E. Livingston, A. Perez, I. A. Shlyakhter, R. V. Rolando, R. Owen, P. Tanpaiboon, F. Lacbawan, Clinton R. Paden, Duncan MacCannell |
| EPI_ISL_1906042 | Michigan Department of Health and Human Services, Bureau of Laboratories | Michigan Department of Health and Human Services, Bureau of Laboratories | Blankenship HM, Riner D, Soehnlen MK |
| EPI_ISL_1906043 | Quest Diagnostics Incorporated | Centers for Disease Control and Prevention Division of Viral Diseases, Pathogen Discovery | Dakota Howard, Dhwani Batra, Peter W. Cook, Kara Moser, Adrian Paskey, Jason Caravas, Benjamin Rambo-Martin, Shatavia Morrison, Christopher Gulvick, Scott Sammons, Yvette Unoarumhi, Darlene Wagner, Matthew Schmerer, S. H. Rosenthal, A. Gerasimova, R. M. Kagan, B. Anderson, M. Hua, Y. Liu, L.E. Bernstein, K.E. Livingston, A. Perez, I. A. Shlyakhter, R. V. Rolando, R. Owen, P. Tanpaiboon, F. Lacbawan, Clinton R. Paden, Duncan MacCannell |
| EPI_ISL_1906044 | Michigan Department of Health and Human Services, Bureau of Laboratories | Michigan Department of Health and Human Services, Bureau of Laboratories | Blankenship HM, Riner D, Soehnlen MK |
| EPI_ISL_1906045 | Quest Diagnostics Incorporated | Centers for Disease Control and Prevention Division of Viral Diseases, Pathogen Discovery | Dakota Howard, Dhwani Batra, Peter W. Cook, Kara Moser, Adrian Paskey, Jason Caravas, Benjamin Rambo-Martin, Shatavia Morrison, Christopher Gulvick, Scott Sammons, Yvette Unoarumhi, Darlene Wagner, Matthew Schmerer, S. H. Rosenthal, A. Gerasimova, R. M. Kagan, B. Anderson, M. Hua, Y. Liu, L.E. Bernstein, K.E. Livingston, A. Perez, I. A. Shlyakhter, R. V. Rolando, R. Owen, P. Tanpaiboon, F. Lacbawan, Clinton R. Paden, Duncan MacCannell |
| EPI_ISL_1906046, EPI_ISL_1906047 | Michigan Department of Health and Human Services, Bureau of Laboratories | Michigan Department of Health and Human Services, Bureau of Laboratories | Blankenship HM, Riner D, Soehnlen MK |
| EPI_ISL_1906048 | Quest Diagnostics Incorporated | Centers for Disease Control and Prevention Division of Viral Diseases, Pathogen Discovery | Dakota Howard, Dhwani Batra, Peter W. Cook, Kara Moser, Adrian Paskey, Jason Caravas, Benjamin Rambo-Martin, Shatavia Morrison, Christopher Gulvick, Scott Sammons, Yvette Unoarumhi, Darlene Wagner, Matthew Schmerer, S. H. Rosenthal, A. Gerasimova, R. M. Kagan, B. Anderson, M. Hua, Y. Liu, L.E. Bernstein, K.E. Livingston, A. Perez, I. A. Shlyakhter, R. V. Rolando, R. Owen, P. Tanpaiboon, F. Lacbawan, Clinton R. Paden, Duncan MacCannell |
| EPI_ISL_1906049 | Michigan Department of Health and Human Services, Bureau of Laboratories | Michigan Department of Health and Human Services, Bureau of Laboratories | Blankenship HM, Riner D, Soehnlen MK |
| EPI_ISL_1906050 | Quest Diagnostics Incorporated | Centers for Disease Control and Prevention Division of Viral Diseases, Pathogen Discovery | Dakota Howard, Dhwani Batra, Peter W. Cook, Kara Moser, Adrian Paskey, Jason Caravas, Benjamin Rambo-Martin, Shatavia Morrison, Christopher Gulvick, Scott Sammons, Yvette Unoarumhi, Darlene Wagner, Matthew Schmerer, S. H. Rosenthal, A. Gerasimova, R. M. Kagan, B. Anderson, M. Hua, Y. Liu, L.E. Bernstein, K.E. Livingston, A. Perez, I. A. Shlyakhter, R. V. Rolando, R. Owen, P. Tanpaiboon, F. Lacbawan, Clinton R. Paden, Duncan MacCannell |
| EPI_ISL_1906051, EPI_ISL_1906052, EPI_ISL_1906053 | Michigan Department of Health and Human Services, Bureau of Laboratories | Michigan Department of Health and Human Services, Bureau of Laboratories | Blankenship HM, Riner D, Soehnlen MK |
| EPI_ISL_1906054 | Quest Diagnostics Incorporated | Centers for Disease Control and Prevention Division of Viral Diseases, Pathogen Discovery | Dakota Howard, Dhwani Batra, Peter W. Cook, Kara Moser, Adrian Paskey, Jason Caravas, Benjamin Rambo-Martin, Shatavia Morrison, Christopher Gulvick, Scott Sammons, Yvette Unoarumhi, Darlene Wagner, Matthew Schmerer, S. H. Rosenthal, A. Gerasimova, R. M. Kagan, B. Anderson, M. Hua, Y. Liu, L.E. Bernstein, K.E. Livingston, A. Perez, I. A. Shlyakhter, R. V. Rolando, R. Owen, P. Tanpaiboon, F. Lacbawan, Clinton R. Paden, Duncan MacCannell |
| EPI_ISL_1906055 | Michigan Department of Health and Human Services, Bureau of Laboratories | Michigan Department of Health and Human Services, Bureau of Laboratories | Blankenship HM, Riner D, Soehnlen MK |
| EPI_ISL_1906056 | Quest Diagnostics Incorporated | Centers for Disease Control and Prevention Division of Viral Diseases, Pathogen Discovery | Dakota Howard, Dhwani Batra, Peter W. Cook, Kara Moser, Adrian Paskey, Jason Caravas, Benjamin Rambo-Martin, Shatavia Morrison, Christopher Gulvick, Scott Sammons, Yvette Unoarumhi, Darlene Wagner, Matthew Schmerer, S. H. Rosenthal, A. Gerasimova, R. M. Kagan, B. Anderson, M. Hua, Y. Liu, L.E. Bernstein, K.E. Livingston, A. Perez, I. A. Shlyakhter, R. V. Rolando, R. Owen, P. Tanpaiboon, F. Lacbawan, Clinton R. Paden, Duncan MacCannell |
| EPI_ISL_1906057, EPI_ISL_1906058, EPI_ISL_1906059, EPI_ISL_1906060 | Michigan Department of Health and Human Services, Bureau of Laboratories | Michigan Department of Health and Human Services, Bureau of Laboratories | Blankenship HM, Riner D, Soehnlen MK |
| EPI_ISL_1906061 | Quest Diagnostics Incorporated | Centers for Disease Control and Prevention Division of Viral Diseases, Pathogen Discovery | Dakota Howard, Dhwani Batra, Peter W. Cook, Kara Moser, Adrian Paskey, Jason Caravas, Benjamin Rambo-Martin, Shatavia Morrison, Christopher Gulvick, Scott Sammons, Yvette Unoarumhi, Darlene Wagner, Matthew Schmerer, S. H. Rosenthal, A. Gerasimova, R. M. Kagan, B. Anderson, M. Hua, Y. Liu, L.E. Bernstein, K.E. Livingston, A. Perez, I. A. Shlyakhter, R. V. Rolando, R. Owen, P. Tanpaiboon, F. Lacbawan, Clinton R. Paden, Duncan MacCannell |
| EPI_ISL_1906062 | Michigan Department of Health and Human Services, Bureau of Laboratories | Michigan Department of Health and Human Services, Bureau of Laboratories | Blankenship HM, Riner D, Soehnlen MK |
| EPI_ISL_1906063 | Quest Diagnostics Incorporated | Centers for Disease Control and Prevention Division of Viral Diseases, Pathogen Discovery | Dakota Howard, Dhwani Batra, Peter W. Cook, Kara Moser, Adrian Paskey, Jason Caravas, Benjamin Rambo-Martin, Shatavia Morrison, Christopher Gulvick, Scott Sammons, Yvette Unoarumhi, Darlene Wagner, Matthew Schmerer, S. H. Rosenthal, A. Gerasimova, R. M. Kagan, B. Anderson, M. Hua, Y. Liu, L.E. Bernstein, K.E. Livingston, A. Perez, I. A. Shlyakhter, R. V. Rolando, R. Owen, P. Tanpaiboon, F. Lacbawan, Clinton R. Paden, Duncan MacCannell |
| EPI_ISL_1906064 | Michigan Department of Health and Human Services, Bureau of Laboratories | Michigan Department of Health and Human Services, Bureau of Laboratories | Blankenship HM, Riner D, Soehnlen MK |
| EPI_ISL_1906065 | Quest Diagnostics Incorporated | Centers for Disease Control and Prevention Division of Viral Diseases, Pathogen Discovery | Dakota Howard, Dhwani Batra, Peter W. Cook, Kara Moser, Adrian Paskey, Jason Caravas, Benjamin Rambo-Martin, Shatavia Morrison, Christopher Gulvick, Scott Sammons, Yvette Unoarumhi, Darlene Wagner, Matthew Schmerer, S. H. Rosenthal, A. Gerasimova, R. M. Kagan, B. Anderson, M. Hua, Y. Liu, L.E. Bernstein, K.E. Livingston, A. Perez, I. A. Shlyakhter, R. V. Rolando, R. Owen, P. Tanpaiboon, F. Lacbawan, Clinton R. Paden, Duncan MacCannell |
| EPI_ISL_1906066 | Michigan Department of Health and Human Services, Bureau of Laboratories | Michigan Department of Health and Human Services, Bureau of Laboratories | Blankenship HM, Riner D, Soehnlen MK |
| EPI_ISL_1906068, EPI_ISL_1906070 | Quest Diagnostics Incorporated | Centers for Disease Control and Prevention Division of Viral Diseases, Pathogen Discovery | Dakota Howard, Dhwani Batra, Peter W. Cook, Kara Moser, Adrian Paskey, Jason Caravas, Benjamin Rambo-Martin, Shatavia Morrison, Christopher Gulvick, Scott Sammons, Yvette Unoarumhi, Darlene Wagner, Matthew Schmerer, S. H. Rosenthal, A. Gerasimova, R. M. Kagan, B. Anderson, M. Hua, Y. Liu, L.E. Bernstein, K.E. Livingston, A. Perez, I. A. Shlyakhter, R. V. Rolando, R. Owen, P. Tanpaiboon, F. Lacbawan, Clinton R. Paden, Duncan MacCannell |
| EPI_ISL_1906071 | Michigan Department of Health and Human Services, Bureau of Laboratories | Michigan Department of Health and Human Services, Bureau of Laboratories | Blankenship HM, Riner D, Soehnlen MK |
| EPI_ISL_1906072, EPI_ISL_1906074, EPI_ISL_1906076, EPI_ISL_1906079 | Quest Diagnostics Incorporated | Centers for Disease Control and Prevention Division of Viral Diseases, Pathogen Discovery | Dakota Howard, Dhwani Batra, Peter W. Cook, Kara Moser, Adrian Paskey, Jason Caravas, Benjamin Rambo-Martin, Shatavia Morrison, Christopher Gulvick, Scott Sammons, Yvette Unoarumhi, Darlene Wagner, Matthew Schmerer, S. H. Rosenthal, A. Gerasimova, R. M. Kagan, B. Anderson, M. Hua, Y. Liu, L.E. Bernstein, K.E. Livingston, A. Perez, I. A. Shlyakhter, R. V. Rolando, R. Owen, P. Tanpaiboon, F. Lacbawan, Clinton R. Paden, Duncan MacCannell |
| EPI_ISL_1906080 | Michigan Department of Health and Human Services, Bureau of Laboratories | Michigan Department of Health and Human Services, Bureau of Laboratories | Blankenship HM, Riner D, Soehnlen MK |
| EPI_ISL_1906081, EPI_ISL_1906083, EPI_ISL_1906084 | Quest Diagnostics Incorporated | Centers for Disease Control and Prevention Division of Viral Diseases, Pathogen Discovery | Dakota Howard, Dhwani Batra, Peter W. Cook, Kara Moser, Adrian Paskey, Jason Caravas, Benjamin Rambo-Martin, Shatavia Morrison, Christopher Gulvick, Scott Sammons, Yvette Unoarumhi, Darlene Wagner, Matthew Schmerer, S. H. Rosenthal, A. Gerasimova, R. M. Kagan, B. Anderson, M. Hua, Y. Liu, L.E. Bernstein, K.E. Livingston, A. Perez, I. A. Shlyakhter, R. V. Rolando, R. Owen, P. Tanpaiboon, F. Lacbawan, Clinton R. Paden, Duncan MacCannell |
| EPI_ISL_1906085, EPI_ISL_1906086, EPI_ISL_1906087 | Michigan Department of Health and Human Services, Bureau of Laboratories | Michigan Department of Health and Human Services, Bureau of Laboratories | Blankenship HM, Riner D, Soehnlen MK |

| | | | |
|---|---|---|---|
| EPI_ISL_1906088 | Quest Diagnostics Incorporated | Centers for Disease Control and Prevention Division of Viral Diseases, Pathogen Discovery | Dakota Howard, Dhwani Batra, Peter W. Cook, Kara Moser, Adrian Paskey, Jason Caravas, Benjamin Rambo-Martin, Shatavia Morrison, Christopher Gulvick, Scott Sammons, Yvette Unoarumhi, Darlene Wagner, Matthew Schmerer, S. H. Rosenthal, A. Gerasimova, R. M. Kagan, B. Anderson, M. Hua, Y. Liu, L.E. Bernstein, K.E. Livingston, A. Perez, I. A. Shlyakhter, R. V. Rolando, R. Owen, P. Tanpaiboon, F. Lacbawan, Clinton R. Paden, Duncan MacCannell |
| EPI_ISL_1906090 | Michigan Department of Health and Human Services, Bureau of Laboratories | Michigan Department of Health and Human Services, Bureau of Laboratories | Blankenship HM, Riner D, Soehnlen MK |
| EPI_ISL_1906091, EPI_ISL_1906095 | Quest Diagnostics Incorporated | Centers for Disease Control and Prevention Division of Viral Diseases, Pathogen Discovery | Dakota Howard, Dhwani Batra, Peter W. Cook, Kara Moser, Adrian Paskey, Jason Caravas, Benjamin Rambo-Martin, Shatavia Morrison, Christopher Gulvick, Scott Sammons, Yvette Unoarumhi, Darlene Wagner, Matthew Schmerer, S. H. Rosenthal, A. Gerasimova, R. M. Kagan, B. Anderson, M. Hua, Y. Liu, L.E. Bernstein, K.E. Livingston, A. Perez, I. A. Shlyakhter, R. V. Rolando, R. Owen, P. Tanpaiboon, F. Lacbawan, Clinton R. Paden, Duncan MacCannell |
| EPI_ISL_1906096 | Michigan Department of Health and Human Services, Bureau of Laboratories | Michigan Department of Health and Human Services, Bureau of Laboratories | Blankenship HM, Riner D, Soehnlen MK |
| EPI_ISL_1906098 | Quest Diagnostics Incorporated | Centers for Disease Control and Prevention Division of Viral Diseases, Pathogen Discovery | Dakota Howard, Dhwani Batra, Peter W. Cook, Kara Moser, Adrian Paskey, Jason Caravas, Benjamin Rambo-Martin, Shatavia Morrison, Christopher Gulvick, Scott Sammons, Yvette Unoarumhi, Darlene Wagner, Matthew Schmerer, S. H. Rosenthal, A. Gerasimova, R. M. Kagan, B. Anderson, M. Hua, Y. Liu, L.E. Bernstein, K.E. Livingston, A. Perez, I. A. Shlyakhter, R. V. Rolando, R. Owen, P. Tanpaiboon, F. Lacbawan, Clinton R. Paden, Duncan MacCannell |
| EPI_ISL_1906099 | Michigan Department of Health and Human Services, Bureau of Laboratories | Michigan Department of Health and Human Services, Bureau of Laboratories | Blankenship HM, Riner D, Soehnlen MK |
| EPI_ISL_1906100, EPI_ISL_1906102, EPI_ISL_1906105, EPI_ISL_1906107 | Quest Diagnostics Incorporated | Centers for Disease Control and Prevention Division of Viral Diseases, Pathogen Discovery | Dakota Howard, Dhwani Batra, Peter W. Cook, Kara Moser, Adrian Paskey, Jason Caravas, Benjamin Rambo-Martin, Shatavia Morrison, Christopher Gulvick, Scott Sammons, Yvette Unoarumhi, Darlene Wagner, Matthew Schmerer, S. H. Rosenthal, A. Gerasimova, R. M. Kagan, B. Anderson, M. Hua, Y. Liu, L.E. Bernstein, K.E. Livingston, A. Perez, I. A. Shlyakhter, R. V. Rolando, R. Owen, P. Tanpaiboon, F. Lacbawan, Clinton R. Paden, Duncan MacCannell |
| EPI_ISL_1906108 | Michigan Department of Health and Human Services, Bureau of Laboratories | Michigan Department of Health and Human Services, Bureau of Laboratories | Blankenship HM, Riner D, Soehnlen MK |
| EPI_ISL_1906109 | Quest Diagnostics Incorporated | Centers for Disease Control and Prevention Division of Viral Diseases, Pathogen Discovery | Dakota Howard, Dhwani Batra, Peter W. Cook, Kara Moser, Adrian Paskey, Jason Caravas, Benjamin Rambo-Martin, Shatavia Morrison, Christopher Gulvick, Scott Sammons, Yvette Unoarumhi, Darlene Wagner, Matthew Schmerer, S. H. Rosenthal, A. Gerasimova, R. M. Kagan, B. Anderson, M. Hua, Y. Liu, L.E. Bernstein, K.E. Livingston, A. Perez, I. A. Shlyakhter, R. V. Rolando, R. Owen, P. Tanpaiboon, F. Lacbawan, Clinton R. Paden, Duncan MacCannell |
| EPI_ISL_1906110 | Michigan Department of Health and Human Services, Bureau of Laboratories | Michigan Department of Health and Human Services, Bureau of Laboratories | Blankenship HM, Riner D, Soehnlen MK |
| EPI_ISL_1906111 | Quest Diagnostics Incorporated | Centers for Disease Control and Prevention Division of Viral Diseases, Pathogen Discovery | Dakota Howard, Dhwani Batra, Peter W. Cook, Kara Moser, Adrian Paskey, Jason Caravas, Benjamin Rambo-Martin, Shatavia Morrison, Christopher Gulvick, Scott Sammons, Yvette Unoarumhi, Darlene Wagner, Matthew Schmerer, S. H. Rosenthal, A. Gerasimova, R. M. Kagan, B. Anderson, M. Hua, Y. Liu, L.E. Bernstein, K.E. Livingston, A. Perez, I. A. Shlyakhter, R. V. Rolando, R. Owen, P. Tanpaiboon, F. Lacbawan, Clinton R. Paden, Duncan MacCannell |
| EPI_ISL_1906113, EPI_ISL_1906114 | Michigan Department of Health and Human Services, Bureau of Laboratories | Michigan Department of Health and Human Services, Bureau of Laboratories | Blankenship HM, Riner D, Soehnlen MK |
| EPI_ISL_1906115, EPI_ISL_1906117 | Quest Diagnostics Incorporated | Centers for Disease Control and Prevention Division of Viral Diseases, Pathogen Discovery | Dakota Howard, Dhwani Batra, Peter W. Cook, Kara Moser, Adrian Paskey, Jason Caravas, Benjamin Rambo-Martin, Shatavia Morrison, Christopher Gulvick, Scott Sammons, Yvette Unoarumhi, Darlene Wagner, Matthew Schmerer, S. H. Rosenthal, A. Gerasimova, R. M. Kagan, B. Anderson, M. Hua, Y. Liu, L.E. Bernstein, K.E. Livingston, A. Perez, I. A. Shlyakhter, R. V. Rolando, R. Owen, P. Tanpaiboon, F. Lacbawan, Clinton R. Paden, Duncan MacCannell |
| EPI_ISL_1906118 | Michigan Department of Health and Human Services, Bureau of Laboratories | Michigan Department of Health and Human Services, Bureau of Laboratories | Blankenship HM, Riner D, Soehnlen MK |
| EPI_ISL_1906119, EPI_ISL_1906121 | Quest Diagnostics Incorporated | Centers for Disease Control and Prevention Division of Viral Diseases, Pathogen Discovery | Dakota Howard, Dhwani Batra, Peter W. Cook, Kara Moser, Adrian Paskey, Jason Caravas, Benjamin Rambo-Martin, Shatavia Morrison, Christopher Gulvick, Scott Sammons, Yvette Unoarumhi, Darlene Wagner, Matthew Schmerer, S. H. Rosenthal, A. Gerasimova, R. M. Kagan, B. Anderson, M. Hua, Y. Liu, L.E. Bernstein, K.E. Livingston, A. Perez, I. A. Shlyakhter, R. V. Rolando, R. Owen, P. Tanpaiboon, F. Lacbawan, Clinton R. Paden, Duncan MacCannell |
| EPI_ISL_1906122 | Michigan Department of Health and Human Services, Bureau of Laboratories | Michigan Department of Health and Human Services, Bureau of Laboratories | Blankenship HM, Riner D, Soehnlen MK |
| EPI_ISL_1906123, EPI_ISL_1906126 | Quest Diagnostics Incorporated | Centers for Disease Control and Prevention Division of Viral Diseases, Pathogen Discovery | Dakota Howard, Dhwani Batra, Peter W. Cook, Kara Moser, Adrian Paskey, Jason Caravas, Benjamin Rambo-Martin, Shatavia Morrison, Christopher Gulvick, Scott Sammons, Yvette Unoarumhi, Darlene Wagner, Matthew Schmerer, S. H. Rosenthal, A. Gerasimova, R. M. Kagan, B. Anderson, M. Hua, Y. Liu, L.E. Bernstein, K.E. Livingston, A. Perez, I. A. Shlyakhter, R. V. Rolando, R. Owen, P. Tanpaiboon, F. Lacbawan, Clinton R. Paden, Duncan MacCannell |
| EPI_ISL_1906127 | Michigan Department of Health and Human Services, Bureau of Laboratories | Michigan Department of Health and Human Services, Bureau of Laboratories | Blankenship HM, Riner D, Soehnlen MK |
| EPI_ISL_1906128 | Quest Diagnostics Incorporated | Centers for Disease Control and Prevention Division of Viral Diseases, Pathogen Discovery | Dakota Howard, Dhwani Batra, Peter W. Cook, Kara Moser, Adrian Paskey, Jason Caravas, Benjamin Rambo-Martin, Shatavia Morrison, Christopher Gulvick, Scott Sammons, Yvette Unoarumhi, Darlene Wagner, Matthew Schmerer, S. H. Rosenthal, A. Gerasimova, R. M. Kagan, B. Anderson, M. Hua, Y. Liu, L.E. Bernstein, K.E. Livingston, A. Perez, I. A. Shlyakhter, R. V. Rolando, R. Owen, P. Tanpaiboon, F. Lacbawan, Clinton R. Paden, Duncan MacCannell |
| EPI_ISL_1906129 | Michigan Department of Health and Human Services, Bureau of Laboratories | Michigan Department of Health and Human Services, Bureau of Laboratories | Blankenship HM, Riner D, Soehnlen MK |
| EPI_ISL_1906130 | Quest Diagnostics Incorporated | Centers for Disease Control and Prevention Division of Viral Diseases, Pathogen Discovery | Dakota Howard, Dhwani Batra, Peter W. Cook, Kara Moser, Adrian Paskey, Jason Caravas, Benjamin Rambo-Martin, Shatavia Morrison, Christopher Gulvick, Scott Sammons, Yvette Unoarumhi, Darlene Wagner, Matthew Schmerer, S. H. Rosenthal, A. Gerasimova, R. M. Kagan, B. Anderson, M. Hua, Y. Liu, L.E. Bernstein, K.E. Livingston, A. Perez, I. A. Shlyakhter, R. V. Rolando, R. Owen, P. Tanpaiboon, F. Lacbawan, Clinton R. Paden, Duncan MacCannell |
| EPI_ISL_1906132 | Michigan Department of Health and Human Services, Bureau of Laboratories | Michigan Department of Health and Human Services, Bureau of Laboratories | Blankenship HM, Riner D, Soehnlen MK |
| EPI_ISL_1906134, EPI_ISL_1906136 | Quest Diagnostics Incorporated | Centers for Disease Control and Prevention Division of Viral Diseases, Pathogen Discovery | Dakota Howard, Dhwani Batra, Peter W. Cook, Kara Moser, Adrian Paskey, Jason Caravas, Benjamin Rambo-Martin, Shatavia Morrison, Christopher Gulvick, Scott Sammons, Yvette Unoarumhi, Darlene Wagner, Matthew Schmerer, S. H. Rosenthal, A. Gerasimova, R. M. Kagan, B. Anderson, M. Hua, Y. Liu, L.E. Bernstein, K.E. Livingston, A. Perez, I. A. Shlyakhter, R. V. Rolando, R. Owen, P. Tanpaiboon, F. Lacbawan, Clinton R. Paden, Duncan MacCannell |
| EPI_ISL_1906138 | Michigan Department of Health and Human Services, Bureau of Laboratories | Michigan Department of Health and Human Services, Bureau of Laboratories | Blankenship HM, Riner D, Soehnlen MK |
| EPI_ISL_1906139 | Quest Diagnostics Incorporated | Centers for Disease Control and Prevention Division of Viral Diseases, Pathogen Discovery | Dakota Howard, Dhwani Batra, Peter W. Cook, Kara Moser, Adrian Paskey, Jason Caravas, Benjamin Rambo-Martin, Shatavia Morrison, Christopher Gulvick, Scott Sammons, Yvette Unoarumhi, Darlene Wagner, Matthew Schmerer, S. H. Rosenthal, A. Gerasimova, R. M. Kagan, B. Anderson, M. Hua, Y. Liu, L.E. Bernstein, K.E. Livingston, A. Perez, I. A. Shlyakhter, R. V. Rolando, R. Owen, P. Tanpaiboon, F. Lacbawan, Clinton R. Paden, Duncan MacCannell |
| EPI_ISL_1906140 | Michigan Department of Health and Human Services, Bureau of Laboratories | Michigan Department of Health and Human Services, Bureau of Laboratories | Blankenship HM, Riner D, Soehnlen MK |
| EPI_ISL_1906141, EPI_ISL_1906143, EPI_ISL_1906145 | Quest Diagnostics Incorporated | Centers for Disease Control and Prevention Division of Viral Diseases, Pathogen Discovery | Dakota Howard, Dhwani Batra, Peter W. Cook, Kara Moser, Adrian Paskey, Jason Caravas, Benjamin Rambo-Martin, Shatavia Morrison, Christopher Gulvick, Scott Sammons, Yvette Unoarumhi, Darlene Wagner, Matthew Schmerer, S. H. Rosenthal, A. Gerasimova, R. M. Kagan, B. Anderson, M. Hua, Y. Liu, L.E. Bernstein, K.E. Livingston, A. Perez, I. A. Shlyakhter, R. V. Rolando, R. Owen, P. Tanpaiboon, F. Lacbawan, Clinton R. Paden, Duncan MacCannell |
| EPI_ISL_1906146 | Michigan Department of Health and Human Services, Bureau of Laboratories | Michigan Department of Health and Human Services, Bureau of Laboratories | Blankenship HM, Riner D, Soehnlen MK |
| EPI_ISL_1906147, EPI_ISL_1906151 | Quest Diagnostics Incorporated | Centers for Disease Control and Prevention Division of Viral Diseases, Pathogen Discovery | Dakota Howard, Dhwani Batra, Peter W. Cook, Kara Moser, Adrian Paskey, Jason Caravas, Benjamin Rambo-Martin, Shatavia Morrison, Christopher Gulvick, Scott Sammons, Yvette Unoarumhi, Darlene Wagner, Matthew Schmerer, S. H. Rosenthal, A. Gerasimova, R. M. Kagan, B. Anderson, M. Hua, Y. Liu, L.E. Bernstein, K.E. Livingston, A. Perez, I. A. Shlyakhter, R. V. Rolando, R. Owen, P. Tanpaiboon, F. Lacbawan, Clinton R. Paden, Duncan MacCannell |
| EPI_ISL_1906152, EPI_ISL_1906153 | Michigan Department of Health and Human Services, Bureau of Laboratories | Michigan Department of Health and Human Services, Bureau of Laboratories | Blankenship HM, Riner D, Soehnlen MK |
| EPI_ISL_1906154 | Quest Diagnostics Incorporated | Centers for Disease Control and Prevention Division of Viral Diseases, Pathogen Discovery | Dakota Howard, Dhwani Batra, Peter W. Cook, Kara Moser, Adrian Paskey, Jason Caravas, Benjamin Rambo-Martin, Shatavia Morrison, Christopher Gulvick, Scott Sammons, Yvette Unoarumhi, Darlene Wagner, Matthew Schmerer, S. H. Rosenthal, A. Gerasimova, R. M. Kagan, B. Anderson, M. Hua, Y. Liu, L.E. Bernstein, K.E. Livingston, A. Perez, I. A. Shlyakhter, R. V. Rolando, R. Owen, P. Tanpaiboon, F. Lacbawan, Clinton R. Paden, Duncan MacCannell |
| EPI_ISL_1906155 | Michigan Department of Health and Human Services, Bureau of Laboratories | Michigan Department of Health and Human Services, Bureau of Laboratories | Blankenship HM, Riner D, Soehnlen MK |
| EPI_ISL_1906156 | Quest Diagnostics Incorporated | Centers for Disease Control and Prevention Division of Viral Diseases, Pathogen Discovery | Dakota Howard, Dhwani Batra, Peter W. Cook, Kara Moser, Adrian Paskey, Jason Caravas, Benjamin Rambo-Martin, Shatavia Morrison, Christopher Gulvick, Scott Sammons, Yvette Unoarumhi, Darlene Wagner, Matthew Schmerer, S. H. Rosenthal, A. Gerasimova, R. M. Kagan, B. Anderson, M. Hua, Y. Liu, L.E. Bernstein, K.E. Livingston, A. Perez, I. A. Shlyakhter, R. V. Rolando, R. Owen, P. Tanpaiboon, F. Lacbawan, Clinton R. Paden, Duncan MacCannell |
| EPI_ISL_1906157 | Michigan Department of Health and Human Services, Bureau of Laboratories | Michigan Department of Health and Human Services, Bureau of Laboratories | Blankenship HM, Riner D, Soehnlen MK |
| EPI_ISL_1906158 | Quest Diagnostics Incorporated | Centers for Disease Control and Prevention Division of Viral Diseases, Pathogen Discovery | Dakota Howard, Dhwani Batra, Peter W. Cook, Kara Moser, Adrian Paskey, Jason Caravas, Benjamin Rambo-Martin, Shatavia Morrison, Christopher Gulvick, Scott Sammons, Yvette Unoarumhi, Darlene Wagner, Matthew Schmerer, S. H. Rosenthal, A. Gerasimova, R. M. Kagan, B. Anderson, M. Hua, Y. Liu, L.E. Bernstein, K.E. Livingston, A. Perez, I. A. Shlyakhter, R. V. Rolando, R. Owen, P. Tanpaiboon, F. Lacbawan, Clinton R. Paden, Duncan MacCannell |
| EPI_ISL_1906159 | Michigan Department of Health and Human Services, Bureau of Laboratories | Michigan Department of Health and Human Services, Bureau of Laboratories | Blankenship HM, Riner D, Soehnlen MK |
| EPI_ISL_1906160, EPI_ISL_1906162 | Quest Diagnostics Incorporated | Centers for Disease Control and Prevention Division of Viral Diseases, Pathogen Discovery | Dakota Howard, Dhwani Batra, Peter W. Cook, Kara Moser, Adrian Paskey, Jason Caravas, Benjamin Rambo-Martin, Shatavia Morrison, Christopher Gulvick, Scott Sammons, Yvette Unoarumhi, Darlene Wagner, Matthew Schmerer, S. H. Rosenthal, A. Gerasimova, R. M. Kagan, B. Anderson, M. Hua, Y. Liu, L.E. Bernstein, K.E. Livingston, A. Perez, I. A. Shlyakhter, R. V. Rolando, R. Owen, P. Tanpaiboon, F. Lacbawan, Clinton R. Paden, Duncan MacCannell |

| | | | |
|---|---|---|---|
| EPI_ISL_1906163 | Michigan Department of Health and Human Services, Bureau of Laboratories | Michigan Department of Health and Human Services, Bureau of Laboratories | Blankenship HM, Riner D, Soehnlen MK |
| EPI_ISL_1906164 | Quest Diagnostics Incorporated | Centers for Disease Control and Prevention Division of Viral Diseases, Pathogen Discovery | Dakota Howard, Dhwani Batra, Peter W. Cook, Kara Moser, Adrian Paskey, Jason Caravas, Benjamin Rambo-Martin, Shatavia Morrison, Christopher Gulvick, Scott Sammons, Yvette Unoarumhi, Darlene Wagner, Matthew Schmerer, S. H. Rosenthal, A. Gerasimova, R. M. Kagan, B. Anderson, M. Hua, Y. Liu, L.E. Bernstein, K.E. Livingston, A. Perez, I. A. Shlyakhter, R. V. Rolando, R. Owen, P. Tanpaiboon, F. Lacbawan, Clinton R. Paden, Duncan MacCannell |
| EPI_ISL_1906165 | Michigan Department of Health and Human Services, Bureau of Laboratories | Michigan Department of Health and Human Services, Bureau of Laboratories | Blankenship HM, Riner D, Soehnlen MK |
| EPI_ISL_1906166 | Quest Diagnostics Incorporated | Centers for Disease Control and Prevention Division of Viral Diseases, Pathogen Discovery | Dakota Howard, Dhwani Batra, Peter W. Cook, Kara Moser, Adrian Paskey, Jason Caravas, Benjamin Rambo-Martin, Shatavia Morrison, Christopher Gulvick, Scott Sammons, Yvette Unoarumhi, Darlene Wagner, Matthew Schmerer, S. H. Rosenthal, A. Gerasimova, R. M. Kagan, B. Anderson, M. Hua, Y. Liu, L.E. Bernstein, K.E. Livingston, A. Perez, I. A. Shlyakhter, R. V. Rolando, R. Owen, P. Tanpaiboon, F. Lacbawan, Clinton R. Paden, Duncan MacCannell |
| EPI_ISL_1906167 | Michigan Department of Health and Human Services, Bureau of Laboratories | Michigan Department of Health and Human Services, Bureau of Laboratories | Blankenship HM, Riner D, Soehnlen MK |
| EPI_ISL_1906168, EPI_ISL_1906170 | Quest Diagnostics Incorporated | Centers for Disease Control and Prevention Division of Viral Diseases, Pathogen Discovery | Dakota Howard, Dhwani Batra, Peter W. Cook, Kara Moser, Adrian Paskey, Jason Caravas, Benjamin Rambo-Martin, Shatavia Morrison, Christopher Gulvick, Scott Sammons, Yvette Unoarumhi, Darlene Wagner, Matthew Schmerer, S. H. Rosenthal, A. Gerasimova, R. M. Kagan, B. Anderson, M. Hua, Y. Liu, L.E. Bernstein, K.E. Livingston, A. Perez, I. A. Shlyakhter, R. V. Rolando, R. Owen, P. Tanpaiboon, F. Lacbawan, Clinton R. Paden, Duncan MacCannell |
| EPI_ISL_1906171 | Michigan Department of Health and Human Services, Bureau of Laboratories | Michigan Department of Health and Human Services, Bureau of Laboratories | Blankenship HM, Riner D, Soehnlen MK |
| EPI_ISL_1906172 | Quest Diagnostics Incorporated | Centers for Disease Control and Prevention Division of Viral Diseases, Pathogen Discovery | Dakota Howard, Dhwani Batra, Peter W. Cook, Kara Moser, Adrian Paskey, Jason Caravas, Benjamin Rambo-Martin, Shatavia Morrison, Christopher Gulvick, Scott Sammons, Yvette Unoarumhi, Darlene Wagner, Matthew Schmerer, S. H. Rosenthal, A. Gerasimova, R. M. Kagan, B. Anderson, M. Hua, Y. Liu, L.E. Bernstein, K.E. Livingston, A. Perez, I. A. Shlyakhter, R. V. Rolando, R. Owen, P. Tanpaiboon, F. Lacbawan, Clinton R. Paden, Duncan MacCannell |
| EPI_ISL_1906173, EPI_ISL_1906174 | Michigan Department of Health and Human Services, Bureau of Laboratories | Michigan Department of Health and Human Services, Bureau of Laboratories | Blankenship HM, Riner D, Soehnlen MK |
| EPI_ISL_1906175 | Quest Diagnostics Incorporated | Centers for Disease Control and Prevention Division of Viral Diseases, Pathogen Discovery | Dakota Howard, Dhwani Batra, Peter W. Cook, Kara Moser, Adrian Paskey, Jason Caravas, Benjamin Rambo-Martin, Shatavia Morrison, Christopher Gulvick, Scott Sammons, Yvette Unoarumhi, Darlene Wagner, Matthew Schmerer, S. H. Rosenthal, A. Gerasimova, R. M. Kagan, B. Anderson, M. Hua, Y. Liu, L.E. Bernstein, K.E. Livingston, A. Perez, I. A. Shlyakhter, R. V. Rolando, R. Owen, P. Tanpaiboon, F. Lacbawan, Clinton R. Paden, Duncan MacCannell |
| EPI_ISL_1906176 | Michigan Department of Health and Human Services, Bureau of Laboratories | Michigan Department of Health and Human Services, Bureau of Laboratories | Blankenship HM, Riner D, Soehnlen MK |
| EPI_ISL_1906177 | Quest Diagnostics Incorporated | Centers for Disease Control and Prevention Division of Viral Diseases, Pathogen Discovery | Dakota Howard, Dhwani Batra, Peter W. Cook, Kara Moser, Adrian Paskey, Jason Caravas, Benjamin Rambo-Martin, Shatavia Morrison, Christopher Gulvick, Scott Sammons, Yvette Unoarumhi, Darlene Wagner, Matthew Schmerer, S. H. Rosenthal, A. Gerasimova, R. M. Kagan, B. Anderson, M. Hua, Y. Liu, L.E. Bernstein, K.E. Livingston, A. Perez, I. A. Shlyakhter, R. V. Rolando, R. Owen, P. Tanpaiboon, F. Lacbawan, Clinton R. Paden, Duncan MacCannell |
| EPI_ISL_1906178 | Michigan Department of Health and Human Services, Bureau of Laboratories | Michigan Department of Health and Human Services, Bureau of Laboratories | Blankenship HM, Riner D, Soehnlen MK |
| EPI_ISL_1906179, EPI_ISL_1906181, EPI_ISL_1906183 | Quest Diagnostics Incorporated | Centers for Disease Control and Prevention Division of Viral Diseases, Pathogen Discovery | Dakota Howard, Dhwani Batra, Peter W. Cook, Kara Moser, Adrian Paskey, Jason Caravas, Benjamin Rambo-Martin, Shatavia Morrison, Christopher Gulvick, Scott Sammons, Yvette Unoarumhi, Darlene Wagner, Matthew Schmerer, S. H. Rosenthal, A. Gerasimova, R. M. Kagan, B. Anderson, M. Hua, Y. Liu, L.E. Bernstein, K.E. Livingston, A. Perez, I. A. Shlyakhter, R. V. Rolando, R. Owen, P. Tanpaiboon, F. Lacbawan, Clinton R. Paden, Duncan MacCannell |
| EPI_ISL_1906185, EPI_ISL_1906186 | Michigan Department of Health and Human Services, Bureau of Laboratories | Michigan Department of Health and Human Services, Bureau of Laboratories | Blankenship HM, Riner D, Soehnlen MK |
| EPI_ISL_1906187 | Quest Diagnostics Incorporated | Centers for Disease Control and Prevention Division of Viral Diseases, Pathogen Discovery | Dakota Howard, Dhwani Batra, Peter W. Cook, Kara Moser, Adrian Paskey, Jason Caravas, Benjamin Rambo-Martin, Shatavia Morrison, Christopher Gulvick, Scott Sammons, Yvette Unoarumhi, Darlene Wagner, Matthew Schmerer, S. H. Rosenthal, A. Gerasimova, R. M. Kagan, B. Anderson, M. Hua, Y. Liu, L.E. Bernstein, K.E. Livingston, A. Perez, I. A. Shlyakhter, R. V. Rolando, R. Owen, P. Tanpaiboon, F. Lacbawan, Clinton R. Paden, Duncan MacCannell |
| EPI_ISL_1906188, EPI_ISL_1906189 | Michigan Department of Health and Human Services, Bureau of Laboratories | Michigan Department of Health and Human Services, Bureau of Laboratories | Blankenship HM, Riner D, Soehnlen MK |
| EPI_ISL_1906190 | Quest Diagnostics Incorporated | Centers for Disease Control and Prevention Division of Viral Diseases, Pathogen Discovery | Dakota Howard, Dhwani Batra, Peter W. Cook, Kara Moser, Adrian Paskey, Jason Caravas, Benjamin Rambo-Martin, Shatavia Morrison, Christopher Gulvick, Scott Sammons, Yvette Unoarumhi, Darlene Wagner, Matthew Schmerer, S. H. Rosenthal, A. Gerasimova, R. M. Kagan, B. Anderson, M. Hua, Y. Liu, L.E. Bernstein, K.E. Livingston, A. Perez, I. A. Shlyakhter, R. V. Rolando, R. Owen, P. Tanpaiboon, F. Lacbawan, Clinton R. Paden, Duncan MacCannell |
| EPI_ISL_1906191 | Michigan Department of Health and Human Services, Bureau of Laboratories | Michigan Department of Health and Human Services, Bureau of Laboratories | Blankenship HM, Riner D, Soehnlen MK |
| EPI_ISL_1906192 | Quest Diagnostics Incorporated | Centers for Disease Control and Prevention Division of Viral Diseases, Pathogen Discovery | Dakota Howard, Dhwani Batra, Peter W. Cook, Kara Moser, Adrian Paskey, Jason Caravas, Benjamin Rambo-Martin, Shatavia Morrison, Christopher Gulvick, Scott Sammons, Yvette Unoarumhi, Darlene Wagner, Matthew Schmerer, S. H. Rosenthal, A. Gerasimova, R. M. Kagan, B. Anderson, M. Hua, Y. Liu, L.E. Bernstein, K.E. Livingston, A. Perez, I. A. Shlyakhter, R. V. Rolando, R. Owen, P. Tanpaiboon, F. Lacbawan, Clinton R. Paden, Duncan MacCannell |
| EPI_ISL_1906193, EPI_ISL_1906194 | Michigan Department of Health and Human Services, Bureau of Laboratories | Michigan Department of Health and Human Services, Bureau of Laboratories | Blankenship HM, Riner D, Soehnlen MK |
| EPI_ISL_1906195 | Quest Diagnostics Incorporated | Centers for Disease Control and Prevention Division of Viral Diseases, Pathogen Discovery | Dakota Howard, Dhwani Batra, Peter W. Cook, Kara Moser, Adrian Paskey, Jason Caravas, Benjamin Rambo-Martin, Shatavia Morrison, Christopher Gulvick, Scott Sammons, Yvette Unoarumhi, Darlene Wagner, Matthew Schmerer, S. H. Rosenthal, A. Gerasimova, R. M. Kagan, B. Anderson, M. Hua, Y. Liu, L.E. Bernstein, K.E. Livingston, A. Perez, I. A. Shlyakhter, R. V. Rolando, R. Owen, P. Tanpaiboon, F. Lacbawan, Clinton R. Paden, Duncan MacCannell |
| EPI_ISL_1906196 | Michigan Department of Health and Human Services, Bureau of Laboratories | Michigan Department of Health and Human Services, Bureau of Laboratories | Blankenship HM, Riner D, Soehnlen MK |
| EPI_ISL_1906197, EPI_ISL_1906199 | Quest Diagnostics Incorporated | Centers for Disease Control and Prevention Division of Viral Diseases, Pathogen Discovery | Dakota Howard, Dhwani Batra, Peter W. Cook, Kara Moser, Adrian Paskey, Jason Caravas, Benjamin Rambo-Martin, Shatavia Morrison, Christopher Gulvick, Scott Sammons, Yvette Unoarumhi, Darlene Wagner, Matthew Schmerer, S. H. Rosenthal, A. Gerasimova, R. M. Kagan, B. Anderson, M. Hua, Y. Liu, L.E. Bernstein, K.E. Livingston, A. Perez, I. A. Shlyakhter, R. V. Rolando, R. Owen, P. Tanpaiboon, F. Lacbawan, Clinton R. Paden, Duncan MacCannell |
| EPI_ISL_1906200 | Michigan Department of Health and Human Services, Bureau of Laboratories | Michigan Department of Health and Human Services, Bureau of Laboratories | Blankenship HM, Riner D, Soehnlen MK |
| EPI_ISL_1906202, EPI_ISL_1906204 | Quest Diagnostics Incorporated | Centers for Disease Control and Prevention Division of Viral Diseases, Pathogen Discovery | Dakota Howard, Dhwani Batra, Peter W. Cook, Kara Moser, Adrian Paskey, Jason Caravas, Benjamin Rambo-Martin, Shatavia Morrison, Christopher Gulvick, Scott Sammons, Yvette Unoarumhi, Darlene Wagner, Matthew Schmerer, S. H. Rosenthal, A. Gerasimova, R. M. Kagan, B. Anderson, M. Hua, Y. Liu, L.E. Bernstein, K.E. Livingston, A. Perez, I. A. Shlyakhter, R. V. Rolando, R. Owen, P. Tanpaiboon, F. Lacbawan, Clinton R. Paden, Duncan MacCannell |
| EPI_ISL_1906205 | Michigan Department of Health and Human Services, Bureau of Laboratories | Michigan Department of Health and Human Services, Bureau of Laboratories | Blankenship HM, Riner D, Soehnlen MK |
| EPI_ISL_1906206 | Quest Diagnostics Incorporated | Centers for Disease Control and Prevention Division of Viral Diseases, Pathogen Discovery | Dakota Howard, Dhwani Batra, Peter W. Cook, Kara Moser, Adrian Paskey, Jason Caravas, Benjamin Rambo-Martin, Shatavia Morrison, Christopher Gulvick, Scott Sammons, Yvette Unoarumhi, Darlene Wagner, Matthew Schmerer, S. H. Rosenthal, A. Gerasimova, R. M. Kagan, B. Anderson, M. Hua, Y. Liu, L.E. Bernstein, K.E. Livingston, A. Perez, I. A. Shlyakhter, R. V. Rolando, R. Owen, P. Tanpaiboon, F. Lacbawan, Clinton R. Paden, Duncan MacCannell |
| EPI_ISL_1906207 | Michigan Department of Health and Human Services, Bureau of Laboratories | Michigan Department of Health and Human Services, Bureau of Laboratories | Blankenship HM, Riner D, Soehnlen MK |
| EPI_ISL_1906208, EPI_ISL_1906210 | Quest Diagnostics Incorporated | Centers for Disease Control and Prevention Division of Viral Diseases, Pathogen Discovery | Dakota Howard, Dhwani Batra, Peter W. Cook, Kara Moser, Adrian Paskey, Jason Caravas, Benjamin Rambo-Martin, Shatavia Morrison, Christopher Gulvick, Scott Sammons, Yvette Unoarumhi, Darlene Wagner, Matthew Schmerer, S. H. Rosenthal, A. Gerasimova, R. M. Kagan, B. Anderson, M. Hua, Y. Liu, L.E. Bernstein, K.E. Livingston, A. Perez, I. A. Shlyakhter, R. V. Rolando, R. Owen, P. Tanpaiboon, F. Lacbawan, Clinton R. Paden, Duncan MacCannell |
| EPI_ISL_1906211 | Michigan Department of Health and Human Services, Bureau of Laboratories | Michigan Department of Health and Human Services, Bureau of Laboratories | Blankenship HM, Riner D, Soehnlen MK |
| EPI_ISL_1906213 | Quest Diagnostics Incorporated | Centers for Disease Control and Prevention Division of Viral Diseases, Pathogen Discovery | Dakota Howard, Dhwani Batra, Peter W. Cook, Kara Moser, Adrian Paskey, Jason Caravas, Benjamin Rambo-Martin, Shatavia Morrison, Christopher Gulvick, Scott Sammons, Yvette Unoarumhi, Darlene Wagner, Matthew Schmerer, S. H. Rosenthal, A. Gerasimova, R. M. Kagan, B. Anderson, M. Hua, Y. Liu, L.E. Bernstein, K.E. Livingston, A. Perez, I. A. Shlyakhter, R. V. Rolando, R. Owen, P. Tanpaiboon, F. Lacbawan, Clinton R. Paden, Duncan MacCannell |
| EPI_ISL_1906214 | Michigan Department of Health and Human Services, Bureau of Laboratories | Michigan Department of Health and Human Services, Bureau of Laboratories | Blankenship HM, Riner D, Soehnlen MK |
| EPI_ISL_1906215 | Quest Diagnostics Incorporated | Centers for Disease Control and Prevention Division of Viral Diseases, Pathogen Discovery | Dakota Howard, Dhwani Batra, Peter W. Cook, Kara Moser, Adrian Paskey, Jason Caravas, Benjamin Rambo-Martin, Shatavia Morrison, Christopher Gulvick, Scott Sammons, Yvette Unoarumhi, Darlene Wagner, Matthew Schmerer, S. H. Rosenthal, A. Gerasimova, R. M. Kagan, B. Anderson, M. Hua, Y. Liu, L.E. Bernstein, K.E. Livingston, A. Perez, I. A. Shlyakhter, R. V. Rolando, R. Owen, P. Tanpaiboon, F. Lacbawan, Clinton R. Paden, Duncan MacCannell |
| EPI_ISL_1906216 | Michigan Department of Health and Human Services, Bureau of Laboratories | Michigan Department of Health and Human Services, Bureau of Laboratories | Blankenship HM, Riner D, Soehnlen MK |
| EPI_ISL_1906217 | Quest Diagnostics Incorporated | Centers for Disease Control and Prevention Division of Viral Diseases, Pathogen Discovery | Dakota Howard, Dhwani Batra, Peter W. Cook, Kara Moser, Adrian Paskey, Jason Caravas, Benjamin Rambo-Martin, Shatavia Morrison, Christopher Gulvick, Scott Sammons, Yvette Unoarumhi, Darlene Wagner, Matthew Schmerer, S. H. Rosenthal, A. Gerasimova, R. M. Kagan, B. Anderson, M. Hua, Y. Liu, L.E. Bernstein, K.E. Livingston, A. Perez, I. A. Shlyakhter, R. V. Rolando, R. Owen, P. Tanpaiboon, F. Lacbawan, Clinton R. Paden, Duncan MacCannell |
| EPI_ISL_1906218 | Michigan Department of Health and Human Services, Bureau of Laboratories | Michigan Department of Health and Human Services, Bureau of Laboratories | Blankenship HM, Riner D, Soehnlen MK |
| EPI_ISL_1906219 | Quest Diagnostics Incorporated | Centers for Disease Control and Prevention | Dakota Howard, Dhwani Batra, Peter W. Cook, Kara Moser, Adrian Paskey, Jason Caravas, Benjamin Rambo-Martin, Shatavia Morrison, Christopher Gulvick, Scott Sammons, Yvette Unoarumhi, Darlene Wagner, Matthew Schmerer, S. H. Rosenthal, A. Gerasimova, R. M. Kagan, B. |

| | | | |
|---|---|---|---|
| | | Division of Viral Diseases, Pathogen Discovery | Anderson, M. Hua, Y. Liu, L.E. Bernstein, K.E. Livingston, A. Perez, I. A. Shlyakhter, R. V. Rolando, R. Owen, P. Tanpaiboon, F. Lacbawan, Clinton R. Paden, Duncan MacCannell |
| EPI_ISL_1906220 | Michigan Department of Health and Human Services, Bureau of Laboratories | Michigan Department of Health and Human Services, Bureau of Laboratories | Blankenship HM, Riner D, Soehnlen MK |
| EPI_ISL_1906221 | Quest Diagnostics Incorporated | Centers for Disease Control and Prevention Division of Viral Diseases, Pathogen Discovery | Dakota Howard, Dhwani Batra, Peter W. Cook, Kara Moser, Adrian Paskey, Jason Caravas, Benjamin Rambo-Martin, Shatavia Morrison, Christopher Gulvick, Scott Sammons, Yvette Unoarumhi, Darlene Wagner, Matthew Schmerer, S. H. Rosenthal, A. Gerasimova, R. M. Kagan, B. Anderson, M. Hua, Y. Liu, L.E. Bernstein, K.E. Livingston, A. Perez, I. A. Shlyakhter, R. V. Rolando, R. Owen, P. Tanpaiboon, F. Lacbawan, Clinton R. Paden, Duncan MacCannell |
| EPI_ISL_1906222 | Michigan Department of Health and Human Services, Bureau of Laboratories | Michigan Department of Health and Human Services, Bureau of Laboratories | Blankenship HM, Riner D, Soehnlen MK |
| EPI_ISL_1906223 | Quest Diagnostics Incorporated | Centers for Disease Control and Prevention Division of Viral Diseases, Pathogen Discovery | Dakota Howard, Dhwani Batra, Peter W. Cook, Kara Moser, Adrian Paskey, Jason Caravas, Benjamin Rambo-Martin, Shatavia Morrison, Christopher Gulvick, Scott Sammons, Yvette Unoarumhi, Darlene Wagner, Matthew Schmerer, S. H. Rosenthal, A. Gerasimova, R. M. Kagan, B. Anderson, M. Hua, Y. Liu, L.E. Bernstein, K.E. Livingston, A. Perez, I. A. Shlyakhter, R. V. Rolando, R. Owen, P. Tanpaiboon, F. Lacbawan, Clinton R. Paden, Duncan MacCannell |
| EPI_ISL_1906224 | Michigan Department of Health and Human Services, Bureau of Laboratories | Michigan Department of Health and Human Services, Bureau of Laboratories | Blankenship HM, Riner D, Soehnlen MK |
| EPI_ISL_1906225, EPI_ISL_1906227, EPI_ISL_1906229 | Quest Diagnostics Incorporated | Centers for Disease Control and Prevention Division of Viral Diseases, Pathogen Discovery | Dakota Howard, Dhwani Batra, Peter W. Cook, Kara Moser, Adrian Paskey, Jason Caravas, Benjamin Rambo-Martin, Shatavia Morrison, Christopher Gulvick, Scott Sammons, Yvette Unoarumhi, Darlene Wagner, Matthew Schmerer, S. H. Rosenthal, A. Gerasimova, R. M. Kagan, B. Anderson, M. Hua, Y. Liu, L.E. Bernstein, K.E. Livingston, A. Perez, I. A. Shlyakhter, R. V. Rolando, R. Owen, P. Tanpaiboon, F. Lacbawan, Clinton R. Paden, Duncan MacCannell |
| EPI_ISL_1906230, EPI_ISL_1906231 | Michigan Department of Health and Human Services, Bureau of Laboratories | Michigan Department of Health and Human Services, Bureau of Laboratories | Blankenship HM, Riner D, Soehnlen MK |
| EPI_ISL_1906232, EPI_ISL_1906233 | Quest Diagnostics Incorporated | Centers for Disease Control and Prevention Division of Viral Diseases, Pathogen Discovery | Dakota Howard, Dhwani Batra, Peter W. Cook, Kara Moser, Adrian Paskey, Jason Caravas, Benjamin Rambo-Martin, Shatavia Morrison, Christopher Gulvick, Scott Sammons, Yvette Unoarumhi, Darlene Wagner, Matthew Schmerer, S. H. Rosenthal, A. Gerasimova, R. M. Kagan, B. Anderson, M. Hua, Y. Liu, L.E. Bernstein, K.E. Livingston, A. Perez, I. A. Shlyakhter, R. V. Rolando, R. Owen, P. Tanpaiboon, F. Lacbawan, Clinton R. Paden, Duncan MacCannell |
| EPI_ISL_1906235 | Michigan Department of Health and Human Services, Bureau of Laboratories | Michigan Department of Health and Human Services, Bureau of Laboratories | Blankenship HM, Riner D, Soehnlen MK |
| EPI_ISL_1906236 | Quest Diagnostics Incorporated | Centers for Disease Control and Prevention Division of Viral Diseases, Pathogen Discovery | Dakota Howard, Dhwani Batra, Peter W. Cook, Kara Moser, Adrian Paskey, Jason Caravas, Benjamin Rambo-Martin, Shatavia Morrison, Christopher Gulvick, Scott Sammons, Yvette Unoarumhi, Darlene Wagner, Matthew Schmerer, S. H. Rosenthal, A. Gerasimova, R. M. Kagan, B. Anderson, M. Hua, Y. Liu, L.E. Bernstein, K.E. Livingston, A. Perez, I. A. Shlyakhter, R. V. Rolando, R. Owen, P. Tanpaiboon, F. Lacbawan, Clinton R. Paden, Duncan MacCannell |
| EPI_ISL_1906237, EPI_ISL_1906238 | Michigan Department of Health and Human Services, Bureau of Laboratories | Michigan Department of Health and Human Services, Bureau of Laboratories | Blankenship HM, Riner D, Soehnlen MK |
| EPI_ISL_1906239 | Quest Diagnostics Incorporated | Centers for Disease Control and Prevention Division of Viral Diseases, Pathogen Discovery | Dakota Howard, Dhwani Batra, Peter W. Cook, Kara Moser, Adrian Paskey, Jason Caravas, Benjamin Rambo-Martin, Shatavia Morrison, Christopher Gulvick, Scott Sammons, Yvette Unoarumhi, Darlene Wagner, Matthew Schmerer, S. H. Rosenthal, A. Gerasimova, R. M. Kagan, B. Anderson, M. Hua, Y. Liu, L.E. Bernstein, K.E. Livingston, A. Perez, I. A. Shlyakhter, R. V. Rolando, R. Owen, P. Tanpaiboon, F. Lacbawan, Clinton R. Paden, Duncan MacCannell |
| EPI_ISL_1906240 | Michigan Department of Health and Human Services, Bureau of Laboratories | Michigan Department of Health and Human Services, Bureau of Laboratories | Blankenship HM, Riner D, Soehnlen MK |
| EPI_ISL_1906241, EPI_ISL_1906242 | Quest Diagnostics Incorporated | Centers for Disease Control and Prevention Division of Viral Diseases, Pathogen Discovery | Dakota Howard, Dhwani Batra, Peter W. Cook, Kara Moser, Adrian Paskey, Jason Caravas, Benjamin Rambo-Martin, Shatavia Morrison, Christopher Gulvick, Scott Sammons, Yvette Unoarumhi, Darlene Wagner, Matthew Schmerer, S. H. Rosenthal, A. Gerasimova, R. M. Kagan, B. Anderson, M. Hua, Y. Liu, L.E. Bernstein, K.E. Livingston, A. Perez, I. A. Shlyakhter, R. V. Rolando, R. Owen, P. Tanpaiboon, F. Lacbawan, Clinton R. Paden, Duncan MacCannell |
| EPI_ISL_1906243 | Michigan Department of Health and Human Services, Bureau of Laboratories | Michigan Department of Health and Human Services, Bureau of Laboratories | Blankenship HM, Riner D, Soehnlen MK |
| EPI_ISL_1906244 | Quest Diagnostics Incorporated | Centers for Disease Control and Prevention Division of Viral Diseases, Pathogen Discovery | Dakota Howard, Dhwani Batra, Peter W. Cook, Kara Moser, Adrian Paskey, Jason Caravas, Benjamin Rambo-Martin, Shatavia Morrison, Christopher Gulvick, Scott Sammons, Yvette Unoarumhi, Darlene Wagner, Matthew Schmerer, S. H. Rosenthal, A. Gerasimova, R. M. Kagan, B. Anderson, M. Hua, Y. Liu, L.E. Bernstein, K.E. Livingston, A. Perez, I. A. Shlyakhter, R. V. Rolando, R. Owen, P. Tanpaiboon, F. Lacbawan, Clinton R. Paden, Duncan MacCannell |
| EPI_ISL_1906245, EPI_ISL_1906246 | Michigan Department of Health and Human Services, Bureau of Laboratories | Michigan Department of Health and Human Services, Bureau of Laboratories | Blankenship HM, Riner D, Soehnlen MK |
| EPI_ISL_1906248 | Quest Diagnostics Incorporated | Centers for Disease Control and Prevention Division of Viral Diseases, Pathogen Discovery | Dakota Howard, Dhwani Batra, Peter W. Cook, Kara Moser, Adrian Paskey, Jason Caravas, Benjamin Rambo-Martin, Shatavia Morrison, Christopher Gulvick, Scott Sammons, Yvette Unoarumhi, Darlene Wagner, Matthew Schmerer, S. H. Rosenthal, A. Gerasimova, R. M. Kagan, B. Anderson, M. Hua, Y. Liu, L.E. Bernstein, K.E. Livingston, A. Perez, I. A. Shlyakhter, R. V. Rolando, R. Owen, P. Tanpaiboon, F. Lacbawan, Clinton R. Paden, Duncan MacCannell |
| EPI_ISL_1906249 | Michigan Department of Health and Human Services, Bureau of Laboratories | Michigan Department of Health and Human Services, Bureau of Laboratories | Blankenship HM, Riner D, Soehnlen MK |
| EPI_ISL_1906250 | Quest Diagnostics Incorporated | Centers for Disease Control and Prevention Division of Viral Diseases, Pathogen Discovery | Dakota Howard, Dhwani Batra, Peter W. Cook, Kara Moser, Adrian Paskey, Jason Caravas, Benjamin Rambo-Martin, Shatavia Morrison, Christopher Gulvick, Scott Sammons, Yvette Unoarumhi, Darlene Wagner, Matthew Schmerer, S. H. Rosenthal, A. Gerasimova, R. M. Kagan, B. Anderson, M. Hua, Y. Liu, L.E. Bernstein, K.E. Livingston, A. Perez, I. A. Shlyakhter, R. V. Rolando, R. Owen, P. Tanpaiboon, F. Lacbawan, Clinton R. Paden, Duncan MacCannell |
| EPI_ISL_1906251, EPI_ISL_1906252 | Michigan Department of Health and Human Services, Bureau of Laboratories | Michigan Department of Health and Human Services, Bureau of Laboratories | Blankenship HM, Riner D, Soehnlen MK |
| EPI_ISL_1906253 | Quest Diagnostics Incorporated | Centers for Disease Control and Prevention Division of Viral Diseases, Pathogen Discovery | Dakota Howard, Dhwani Batra, Peter W. Cook, Kara Moser, Adrian Paskey, Jason Caravas, Benjamin Rambo-Martin, Shatavia Morrison, Christopher Gulvick, Scott Sammons, Yvette Unoarumhi, Darlene Wagner, Matthew Schmerer, S. H. Rosenthal, A. Gerasimova, R. M. Kagan, B. Anderson, M. Hua, Y. Liu, L.E. Bernstein, K.E. Livingston, A. Perez, I. A. Shlyakhter, R. V. Rolando, R. Owen, P. Tanpaiboon, F. Lacbawan, Clinton R. Paden, Duncan MacCannell |
| EPI_ISL_1906254 | Michigan Department of Health and Human Services, Bureau of Laboratories | Michigan Department of Health and Human Services, Bureau of Laboratories | Blankenship HM, Riner D, Soehnlen MK |
| EPI_ISL_1906255 | Quest Diagnostics Incorporated | Centers for Disease Control and Prevention Division of Viral Diseases, Pathogen Discovery | Dakota Howard, Dhwani Batra, Peter W. Cook, Kara Moser, Adrian Paskey, Jason Caravas, Benjamin Rambo-Martin, Shatavia Morrison, Christopher Gulvick, Scott Sammons, Yvette Unoarumhi, Darlene Wagner, Matthew Schmerer, S. H. Rosenthal, A. Gerasimova, R. M. Kagan, B. Anderson, M. Hua, Y. Liu, L.E. Bernstein, K.E. Livingston, A. Perez, I. A. Shlyakhter, R. V. Rolando, R. Owen, P. Tanpaiboon, F. Lacbawan, Clinton R. Paden, Duncan MacCannell |
| EPI_ISL_1906256, EPI_ISL_1906257 | Michigan Department of Health and Human Services, Bureau of Laboratories | Michigan Department of Health and Human Services, Bureau of Laboratories | Blankenship HM, Riner D, Soehnlen MK |
| EPI_ISL_1906258 | Quest Diagnostics Incorporated | Centers for Disease Control and Prevention Division of Viral Diseases, Pathogen Discovery | Dakota Howard, Dhwani Batra, Peter W. Cook, Kara Moser, Adrian Paskey, Jason Caravas, Benjamin Rambo-Martin, Shatavia Morrison, Christopher Gulvick, Scott Sammons, Yvette Unoarumhi, Darlene Wagner, Matthew Schmerer, S. H. Rosenthal, A. Gerasimova, R. M. Kagan, B. Anderson, M. Hua, Y. Liu, L.E. Bernstein, K.E. Livingston, A. Perez, I. A. Shlyakhter, R. V. Rolando, R. Owen, P. Tanpaiboon, F. Lacbawan, Clinton R. Paden, Duncan MacCannell |
| EPI_ISL_1906259 | Michigan Department of Health and Human Services, Bureau of Laboratories | Michigan Department of Health and Human Services, Bureau of Laboratories | Blankenship HM, Riner D, Soehnlen MK |
| EPI_ISL_1906260 | Quest Diagnostics Incorporated | Centers for Disease Control and Prevention Division of Viral Diseases, Pathogen Discovery | Dakota Howard, Dhwani Batra, Peter W. Cook, Kara Moser, Adrian Paskey, Jason Caravas, Benjamin Rambo-Martin, Shatavia Morrison, Christopher Gulvick, Scott Sammons, Yvette Unoarumhi, Darlene Wagner, Matthew Schmerer, S. H. Rosenthal, A. Gerasimova, R. M. Kagan, B. Anderson, M. Hua, Y. Liu, L.E. Bernstein, K.E. Livingston, A. Perez, I. A. Shlyakhter, R. V. Rolando, R. Owen, P. Tanpaiboon, F. Lacbawan, Clinton R. Paden, Duncan MacCannell |
| EPI_ISL_1906261 | Michigan Department of Health and Human Services, Bureau of Laboratories | Michigan Department of Health and Human Services, Bureau of Laboratories | Blankenship HM, Riner D, Soehnlen MK |
| EPI_ISL_1906262 | Quest Diagnostics Incorporated | Centers for Disease Control and Prevention Division of Viral Diseases, Pathogen Discovery | Dakota Howard, Dhwani Batra, Peter W. Cook, Kara Moser, Adrian Paskey, Jason Caravas, Benjamin Rambo-Martin, Shatavia Morrison, Christopher Gulvick, Scott Sammons, Yvette Unoarumhi, Darlene Wagner, Matthew Schmerer, S. H. Rosenthal, A. Gerasimova, R. M. Kagan, B. Anderson, M. Hua, Y. Liu, L.E. Bernstein, K.E. Livingston, A. Perez, I. A. Shlyakhter, R. V. Rolando, R. Owen, P. Tanpaiboon, F. Lacbawan, Clinton R. Paden, Duncan MacCannell |
| EPI_ISL_1906263 | Michigan Department of Health and Human Services, Bureau of Laboratories | Michigan Department of Health and Human Services, Bureau of Laboratories | Blankenship HM, Riner D, Soehnlen MK |
| EPI_ISL_1906264 | Quest Diagnostics Incorporated | Centers for Disease Control and Prevention Division of Viral Diseases, Pathogen Discovery | Dakota Howard, Dhwani Batra, Peter W. Cook, Kara Moser, Adrian Paskey, Jason Caravas, Benjamin Rambo-Martin, Shatavia Morrison, Christopher Gulvick, Scott Sammons, Yvette Unoarumhi, Darlene Wagner, Matthew Schmerer, S. H. Rosenthal, A. Gerasimova, R. M. Kagan, B. Anderson, M. Hua, Y. Liu, L.E. Bernstein, K.E. Livingston, A. Perez, I. A. Shlyakhter, R. V. Rolando, R. Owen, P. Tanpaiboon, F. Lacbawan, Clinton R. Paden, Duncan MacCannell |
| EPI_ISL_1906265 | Michigan Department of Health and Human Services, Bureau of Laboratories | Michigan Department of Health and Human Services, Bureau of Laboratories | Blankenship HM, Riner D, Soehnlen MK |
| EPI_ISL_1906266 | Quest Diagnostics Incorporated | Centers for Disease Control and Prevention Division of Viral Diseases, Pathogen Discovery | Dakota Howard, Dhwani Batra, Peter W. Cook, Kara Moser, Adrian Paskey, Jason Caravas, Benjamin Rambo-Martin, Shatavia Morrison, Christopher Gulvick, Scott Sammons, Yvette Unoarumhi, Darlene Wagner, Matthew Schmerer, S. H. Rosenthal, A. Gerasimova, R. M. Kagan, B. Anderson, M. Hua, Y. Liu, L.E. Bernstein, K.E. Livingston, A. Perez, I. A. Shlyakhter, R. V. Rolando, R. Owen, P. Tanpaiboon, F. Lacbawan, Clinton R. Paden, Duncan MacCannell |
| EPI_ISL_1906267 | Michigan Department of Health and Human Services, Bureau of Laboratories | Michigan Department of Health and Human Services, Bureau of Laboratories | Blankenship HM, Riner D, Soehnlen MK |
| EPI_ISL_1906268 | Quest Diagnostics Incorporated | Centers for Disease Control and Prevention Division of Viral Diseases, Pathogen Discovery | Dakota Howard, Dhwani Batra, Peter W. Cook, Kara Moser, Adrian Paskey, Jason Caravas, Benjamin Rambo-Martin, Shatavia Morrison, Christopher Gulvick, Scott Sammons, Yvette Unoarumhi, Darlene Wagner, Matthew Schmerer, S. H. Rosenthal, A. Gerasimova, R. M. Kagan, B. Anderson, M. Hua, Y. Liu, L.E. Bernstein, K.E. Livingston, A. Perez, I. A. Shlyakhter, R. V. Rolando, R. Owen, P. Tanpaiboon, F. Lacbawan, Clinton R. Paden, Duncan MacCannell |
| EPI_ISL_1906269 | Michigan Department of Health and Human Services, Bureau of Laboratories | Michigan Department of Health and Human Services, Bureau of Laboratories | Blankenship HM, Riner D, Soehnlen MK |

| | | | |
|---|---|---|---|
| EPI_ISL_1906270, EPI_ISL_1906272 | Quest Diagnostics Incorporated | Centers for Disease Control and Prevention Division of Viral Diseases, Pathogen Discovery | Dakota Howard, Dhwani Batra, Peter W. Cook, Kara Moser, Adrian Paskey, Jason Caravas, Benjamin Rambo-Martin, Shatavia Morrison, Christopher Gulvick, Scott Sammons, Yvette Unoarumhi, Darlene Wagner, Matthew Schmerer, S. H. Rosenthal, A. Gerasimova, R. M. Kagan, B. Anderson, M. Hua, Y. Liu, L.E. Bernstein, K.E. Livingston, A. Perez, I. A. Shlyakhter, R. V. Rolando, R. Owen, P. Tanpaiboon, F. Lacbawan, Clinton R. Paden, Duncan MacCannell |
| EPI_ISL_1906273 | Michigan Department of Health and Human Services, Bureau of Laboratories | Michigan Department of Health and Human Services, Bureau of Laboratories | Blankenship HM, Riner D, Soehnlen MK |
| EPI_ISL_1906274 | Quest Diagnostics Incorporated | Centers for Disease Control and Prevention Division of Viral Diseases, Pathogen Discovery | Dakota Howard, Dhwani Batra, Peter W. Cook, Kara Moser, Adrian Paskey, Jason Caravas, Benjamin Rambo-Martin, Shatavia Morrison, Christopher Gulvick, Scott Sammons, Yvette Unoarumhi, Darlene Wagner, Matthew Schmerer, S. H. Rosenthal, A. Gerasimova, R. M. Kagan, B. Anderson, M. Hua, Y. Liu, L.E. Bernstein, K.E. Livingston, A. Perez, I. A. Shlyakhter, R. V. Rolando, R. Owen, P. Tanpaiboon, F. Lacbawan, Clinton R. Paden, Duncan MacCannell |
| EPI_ISL_1906275 | Michigan Department of Health and Human Services, Bureau of Laboratories | Michigan Department of Health and Human Services, Bureau of Laboratories | Blankenship HM, Riner D, Soehnlen MK |
| EPI_ISL_1906276 | Quest Diagnostics Incorporated | Centers for Disease Control and Prevention Division of Viral Diseases, Pathogen Discovery | Dakota Howard, Dhwani Batra, Peter W. Cook, Kara Moser, Adrian Paskey, Jason Caravas, Benjamin Rambo-Martin, Shatavia Morrison, Christopher Gulvick, Scott Sammons, Yvette Unoarumhi, Darlene Wagner, Matthew Schmerer, S. H. Rosenthal, A. Gerasimova, R. M. Kagan, B. Anderson, M. Hua, Y. Liu, L.E. Bernstein, K.E. Livingston, A. Perez, I. A. Shlyakhter, R. V. Rolando, R. Owen, P. Tanpaiboon, F. Lacbawan, Clinton R. Paden, Duncan MacCannell |
| EPI_ISL_1906277, EPI_ISL_1906278 | Michigan Department of Health and Human Services, Bureau of Laboratories | Michigan Department of Health and Human Services, Bureau of Laboratories | Blankenship HM, Riner D, Soehnlen MK |
| EPI_ISL_1906279, EPI_ISL_1906281 | Quest Diagnostics Incorporated | Centers for Disease Control and Prevention Division of Viral Diseases, Pathogen Discovery | Dakota Howard, Dhwani Batra, Peter W. Cook, Kara Moser, Adrian Paskey, Jason Caravas, Benjamin Rambo-Martin, Shatavia Morrison, Christopher Gulvick, Scott Sammons, Yvette Unoarumhi, Darlene Wagner, Matthew Schmerer, S. H. Rosenthal, A. Gerasimova, R. M. Kagan, B. Anderson, M. Hua, Y. Liu, L.E. Bernstein, K.E. Livingston, A. Perez, I. A. Shlyakhter, R. V. Rolando, R. Owen, P. Tanpaiboon, F. Lacbawan, Clinton R. Paden, Duncan MacCannell |
| EPI_ISL_1906283 | Michigan Department of Health and Human Services, Bureau of Laboratories | Michigan Department of Health and Human Services, Bureau of Laboratories | Blankenship HM, Riner D, Soehnlen MK |
| EPI_ISL_1906284 | Quest Diagnostics Incorporated | Centers for Disease Control and Prevention Division of Viral Diseases, Pathogen Discovery | Dakota Howard, Dhwani Batra, Peter W. Cook, Kara Moser, Adrian Paskey, Jason Caravas, Benjamin Rambo-Martin, Shatavia Morrison, Christopher Gulvick, Scott Sammons, Yvette Unoarumhi, Darlene Wagner, Matthew Schmerer, S. H. Rosenthal, A. Gerasimova, R. M. Kagan, B. Anderson, M. Hua, Y. Liu, L.E. Bernstein, K.E. Livingston, A. Perez, I. A. Shlyakhter, R. V. Rolando, R. Owen, P. Tanpaiboon, F. Lacbawan, Clinton R. Paden, Duncan MacCannell |
| EPI_ISL_1906285 | Michigan Department of Health and Human Services, Bureau of Laboratories | Michigan Department of Health and Human Services, Bureau of Laboratories | Blankenship HM, Riner D, Soehnlen MK |
| EPI_ISL_1906286, EPI_ISL_1906288 | Quest Diagnostics Incorporated | Centers for Disease Control and Prevention Division of Viral Diseases, Pathogen Discovery | Dakota Howard, Dhwani Batra, Peter W. Cook, Kara Moser, Adrian Paskey, Jason Caravas, Benjamin Rambo-Martin, Shatavia Morrison, Christopher Gulvick, Scott Sammons, Yvette Unoarumhi, Darlene Wagner, Matthew Schmerer, S. H. Rosenthal, A. Gerasimova, R. M. Kagan, B. Anderson, M. Hua, Y. Liu, L.E. Bernstein, K.E. Livingston, A. Perez, I. A. Shlyakhter, R. V. Rolando, R. Owen, P. Tanpaiboon, F. Lacbawan, Clinton R. Paden, Duncan MacCannell |
| EPI_ISL_1906289 | Michigan Department of Health and Human Services, Bureau of Laboratories | Michigan Department of Health and Human Services, Bureau of Laboratories | Blankenship HM, Riner D, Soehnlen MK |
| EPI_ISL_1906290 | Quest Diagnostics Incorporated | Centers for Disease Control and Prevention Division of Viral Diseases, Pathogen Discovery | Dakota Howard, Dhwani Batra, Peter W. Cook, Kara Moser, Adrian Paskey, Jason Caravas, Benjamin Rambo-Martin, Shatavia Morrison, Christopher Gulvick, Scott Sammons, Yvette Unoarumhi, Darlene Wagner, Matthew Schmerer, S. H. Rosenthal, A. Gerasimova, R. M. Kagan, B. Anderson, M. Hua, Y. Liu, L.E. Bernstein, K.E. Livingston, A. Perez, I. A. Shlyakhter, R. V. Rolando, R. Owen, P. Tanpaiboon, F. Lacbawan, Clinton R. Paden, Duncan MacCannell |
| EPI_ISL_1906291 | Michigan Department of Health and Human Services, Bureau of Laboratories | Michigan Department of Health and Human Services, Bureau of Laboratories | Blankenship HM, Riner D, Soehnlen MK |
| EPI_ISL_1906292 | Quest Diagnostics Incorporated | Centers for Disease Control and Prevention Division of Viral Diseases, Pathogen Discovery | Dakota Howard, Dhwani Batra, Peter W. Cook, Kara Moser, Adrian Paskey, Jason Caravas, Benjamin Rambo-Martin, Shatavia Morrison, Christopher Gulvick, Scott Sammons, Yvette Unoarumhi, Darlene Wagner, Matthew Schmerer, S. H. Rosenthal, A. Gerasimova, R. M. Kagan, B. Anderson, M. Hua, Y. Liu, L.E. Bernstein, K.E. Livingston, A. Perez, I. A. Shlyakhter, R. V. Rolando, R. Owen, P. Tanpaiboon, F. Lacbawan, Clinton R. Paden, Duncan MacCannell |
| EPI_ISL_1906293, EPI_ISL_1906294 | Michigan Department of Health and Human Services, Bureau of Laboratories | Michigan Department of Health and Human Services, Bureau of Laboratories | Blankenship HM, Riner D, Soehnlen MK |
| EPI_ISL_1906295 | Quest Diagnostics Incorporated | Centers for Disease Control and Prevention Division of Viral Diseases, Pathogen Discovery | Dakota Howard, Dhwani Batra, Peter W. Cook, Kara Moser, Adrian Paskey, Jason Caravas, Benjamin Rambo-Martin, Shatavia Morrison, Christopher Gulvick, Scott Sammons, Yvette Unoarumhi, Darlene Wagner, Matthew Schmerer, S. H. Rosenthal, A. Gerasimova, R. M. Kagan, B. Anderson, M. Hua, Y. Liu, L.E. Bernstein, K.E. Livingston, A. Perez, I. A. Shlyakhter, R. V. Rolando, R. Owen, P. Tanpaiboon, F. Lacbawan, Clinton R. Paden, Duncan MacCannell |
| EPI_ISL_1906296 | Michigan Department of Health and Human Services, Bureau of Laboratories | Michigan Department of Health and Human Services, Bureau of Laboratories | Blankenship HM, Riner D, Soehnlen MK |
| EPI_ISL_1906297 | Quest Diagnostics Incorporated | Centers for Disease Control and Prevention Division of Viral Diseases, Pathogen Discovery | Dakota Howard, Dhwani Batra, Peter W. Cook, Kara Moser, Adrian Paskey, Jason Caravas, Benjamin Rambo-Martin, Shatavia Morrison, Christopher Gulvick, Scott Sammons, Yvette Unoarumhi, Darlene Wagner, Matthew Schmerer, S. H. Rosenthal, A. Gerasimova, R. M. Kagan, B. Anderson, M. Hua, Y. Liu, L.E. Bernstein, K.E. Livingston, A. Perez, I. A. Shlyakhter, R. V. Rolando, R. Owen, P. Tanpaiboon, F. Lacbawan, Clinton R. Paden, Duncan MacCannell |
| EPI_ISL_1906298 | Michigan Department of Health and Human Services, Bureau of Laboratories | Michigan Department of Health and Human Services, Bureau of Laboratories | Blankenship HM, Riner D, Soehnlen MK |
| EPI_ISL_1906299 | Quest Diagnostics Incorporated | Centers for Disease Control and Prevention Division of Viral Diseases, Pathogen Discovery | Dakota Howard, Dhwani Batra, Peter W. Cook, Kara Moser, Adrian Paskey, Jason Caravas, Benjamin Rambo-Martin, Shatavia Morrison, Christopher Gulvick, Scott Sammons, Yvette Unoarumhi, Darlene Wagner, Matthew Schmerer, S. H. Rosenthal, A. Gerasimova, R. M. Kagan, B. Anderson, M. Hua, Y. Liu, L.E. Bernstein, K.E. Livingston, A. Perez, I. A. Shlyakhter, R. V. Rolando, R. Owen, P. Tanpaiboon, F. Lacbawan, Clinton R. Paden, Duncan MacCannell |
| EPI_ISL_1906300 | Michigan Department of Health and Human Services, Bureau of Laboratories | Michigan Department of Health and Human Services, Bureau of Laboratories | Blankenship HM, Riner D, Soehnlen MK |
| EPI_ISL_1906301 | Quest Diagnostics Incorporated | Centers for Disease Control and Prevention Division of Viral Diseases, Pathogen Discovery | Dakota Howard, Dhwani Batra, Peter W. Cook, Kara Moser, Adrian Paskey, Jason Caravas, Benjamin Rambo-Martin, Shatavia Morrison, Christopher Gulvick, Scott Sammons, Yvette Unoarumhi, Darlene Wagner, Matthew Schmerer, S. H. Rosenthal, A. Gerasimova, R. M. Kagan, B. Anderson, M. Hua, Y. Liu, L.E. Bernstein, K.E. Livingston, A. Perez, I. A. Shlyakhter, R. V. Rolando, R. Owen, P. Tanpaiboon, F. Lacbawan, Clinton R. Paden, Duncan MacCannell |
| EPI_ISL_1906302, EPI_ISL_1906303 | Michigan Department of Health and Human Services, Bureau of Laboratories | Michigan Department of Health and Human Services, Bureau of Laboratories | Blankenship HM, Riner D, Soehnlen MK |
| EPI_ISL_1906304 | Quest Diagnostics Incorporated | Centers for Disease Control and Prevention Division of Viral Diseases, Pathogen Discovery | Dakota Howard, Dhwani Batra, Peter W. Cook, Kara Moser, Adrian Paskey, Jason Caravas, Benjamin Rambo-Martin, Shatavia Morrison, Christopher Gulvick, Scott Sammons, Yvette Unoarumhi, Darlene Wagner, Matthew Schmerer, S. H. Rosenthal, A. Gerasimova, R. M. Kagan, B. Anderson, M. Hua, Y. Liu, L.E. Bernstein, K.E. Livingston, A. Perez, I. A. Shlyakhter, R. V. Rolando, R. Owen, P. Tanpaiboon, F. Lacbawan, Clinton R. Paden, Duncan MacCannell |
| EPI_ISL_1906305, EPI_ISL_1906306, EPI_ISL_1906307 | Michigan Department of Health and Human Services, Bureau of Laboratories | Michigan Department of Health and Human Services, Bureau of Laboratories | Blankenship HM, Riner D, Soehnlen MK |
| EPI_ISL_1906308, EPI_ISL_1906310 | Quest Diagnostics Incorporated | Centers for Disease Control and Prevention Division of Viral Diseases, Pathogen Discovery | Dakota Howard, Dhwani Batra, Peter W. Cook, Kara Moser, Adrian Paskey, Jason Caravas, Benjamin Rambo-Martin, Shatavia Morrison, Christopher Gulvick, Scott Sammons, Yvette Unoarumhi, Darlene Wagner, Matthew Schmerer, S. H. Rosenthal, A. Gerasimova, R. M. Kagan, B. Anderson, M. Hua, Y. Liu, L.E. Bernstein, K.E. Livingston, A. Perez, I. A. Shlyakhter, R. V. Rolando, R. Owen, P. Tanpaiboon, F. Lacbawan, Clinton R. Paden, Duncan MacCannell |
| EPI_ISL_1906311 | Michigan Department of Health and Human Services, Bureau of Laboratories | Michigan Department of Health and Human Services, Bureau of Laboratories | Blankenship HM, Riner D, Soehnlen MK |
| EPI_ISL_1906312 | Quest Diagnostics Incorporated | Centers for Disease Control and Prevention Division of Viral Diseases, Pathogen Discovery | Dakota Howard, Dhwani Batra, Peter W. Cook, Kara Moser, Adrian Paskey, Jason Caravas, Benjamin Rambo-Martin, Shatavia Morrison, Christopher Gulvick, Scott Sammons, Yvette Unoarumhi, Darlene Wagner, Matthew Schmerer, S. H. Rosenthal, A. Gerasimova, R. M. Kagan, B. Anderson, M. Hua, Y. Liu, L.E. Bernstein, K.E. Livingston, A. Perez, I. A. Shlyakhter, R. V. Rolando, R. Owen, P. Tanpaiboon, F. Lacbawan, Clinton R. Paden, Duncan MacCannell |
| EPI_ISL_1906313 | Michigan Department of Health and Human Services, Bureau of Laboratories | Michigan Department of Health and Human Services, Bureau of Laboratories | Blankenship HM, Riner D, Soehnlen MK |
| EPI_ISL_1906314 | Quest Diagnostics Incorporated | Centers for Disease Control and Prevention Division of Viral Diseases, Pathogen Discovery | Dakota Howard, Dhwani Batra, Peter W. Cook, Kara Moser, Adrian Paskey, Jason Caravas, Benjamin Rambo-Martin, Shatavia Morrison, Christopher Gulvick, Scott Sammons, Yvette Unoarumhi, Darlene Wagner, Matthew Schmerer, S. H. Rosenthal, A. Gerasimova, R. M. Kagan, B. Anderson, M. Hua, Y. Liu, L.E. Bernstein, K.E. Livingston, A. Perez, I. A. Shlyakhter, R. V. Rolando, R. Owen, P. Tanpaiboon, F. Lacbawan, Clinton R. Paden, Duncan MacCannell |
| EPI_ISL_1906315, EPI_ISL_1906317 | Michigan Department of Health and Human Services, Bureau of Laboratories | Michigan Department of Health and Human Services, Bureau of Laboratories | Blankenship HM, Riner D, Soehnlen MK |
| EPI_ISL_1906318, EPI_ISL_1906320 | Quest Diagnostics Incorporated | Centers for Disease Control and Prevention Division of Viral Diseases, Pathogen Discovery | Dakota Howard, Dhwani Batra, Peter W. Cook, Kara Moser, Adrian Paskey, Jason Caravas, Benjamin Rambo-Martin, Shatavia Morrison, Christopher Gulvick, Scott Sammons, Yvette Unoarumhi, Darlene Wagner, Matthew Schmerer, S. H. Rosenthal, A. Gerasimova, R. M. Kagan, B. Anderson, M. Hua, Y. Liu, L.E. Bernstein, K.E. Livingston, A. Perez, I. A. Shlyakhter, R. V. Rolando, R. Owen, P. Tanpaiboon, F. Lacbawan, Clinton R. Paden, Duncan MacCannell |
| EPI_ISL_1906321, EPI_ISL_1906322 | Michigan Department of Health and Human Services, Bureau of Laboratories | Michigan Department of Health and Human Services, Bureau of Laboratories | Blankenship HM, Riner D, Soehnlen MK |
| EPI_ISL_1906323 | Quest Diagnostics Incorporated | Centers for Disease Control and Prevention Division of Viral Diseases, Pathogen Discovery | Dakota Howard, Dhwani Batra, Peter W. Cook, Kara Moser, Adrian Paskey, Jason Caravas, Benjamin Rambo-Martin, Shatavia Morrison, Christopher Gulvick, Scott Sammons, Yvette Unoarumhi, Darlene Wagner, Matthew Schmerer, S. H. Rosenthal, A. Gerasimova, R. M. Kagan, B. Anderson, M. Hua, Y. Liu, L.E. Bernstein, K.E. Livingston, A. Perez, I. A. Shlyakhter, R. V. Rolando, R. Owen, P. Tanpaiboon, F. Lacbawan, Clinton R. Paden, Duncan MacCannell |
| EPI_ISL_1906324 | Michigan Department of Health and Human Services, Bureau of Laboratories | Michigan Department of Health and Human Services, Bureau of Laboratories | Blankenship HM, Riner D, Soehnlen MK |
| EPI_ISL_1906325 | Quest Diagnostics Incorporated | Centers for Disease Control and Prevention Division of Viral Diseases, Pathogen Discovery | Dakota Howard, Dhwani Batra, Peter W. Cook, Kara Moser, Adrian Paskey, Jason Caravas, Benjamin Rambo-Martin, Shatavia Morrison, Christopher Gulvick, Scott Sammons, Yvette Unoarumhi, Darlene Wagner, Matthew Schmerer, S. H. Rosenthal, A. Gerasimova, R. M. Kagan, B. Anderson, M. Hua, Y. Liu, L.E. Bernstein, K.E. Livingston, A. Perez, I. A. Shlyakhter, R. V. Rolando, R. Owen, P. Tanpaiboon, F. Lacbawan, Clinton R. Paden, Duncan MacCannell |

| | | | |
|---|---|---|---|
| EPI_ISL_1906326 | Michigan Department of Health and Human Services, Bureau of Laboratories | Michigan Department of Health and Human Services, Bureau of Laboratories | Blankenship HM, Riner D, Soehnlen MK |
| EPI_ISL_1906327 | Quest Diagnostics Incorporated | Centers for Disease Control and Prevention Division of Viral Diseases, Pathogen Discovery | Dakota Howard, Dhwani Batra, Peter W. Cook, Kara Moser, Adrian Paskey, Jason Caravas, Benjamin Rambo-Martin, Shatavia Morrison, Christopher Gulvick, Scott Sammons, Yvette Unoarumhi, Darlene Wagner, Matthew Schmerer, S. H. Rosenthal, A. Gerasimova, R. M. Kagan, B. Anderson, M. Hua, Y. Liu, L.E. Bernstein, K.E. Livingston, A. Perez, I. A. Shlyakhter, R. V. Rolando, R. Owen, P. Tanpaiboon, F. Lacbawan, Clinton R. Paden, Duncan MacCannell |
| EPI_ISL_1906328 | Michigan Department of Health and Human Services, Bureau of Laboratories | Michigan Department of Health and Human Services, Bureau of Laboratories | Blankenship HM, Riner D, Soehnlen MK |
| EPI_ISL_1906329 | Quest Diagnostics Incorporated | Centers for Disease Control and Prevention Division of Viral Diseases, Pathogen Discovery | Dakota Howard, Dhwani Batra, Peter W. Cook, Kara Moser, Adrian Paskey, Jason Caravas, Benjamin Rambo-Martin, Shatavia Morrison, Christopher Gulvick, Scott Sammons, Yvette Unoarumhi, Darlene Wagner, Matthew Schmerer, S. H. Rosenthal, A. Gerasimova, R. M. Kagan, B. Anderson, M. Hua, Y. Liu, L.E. Bernstein, K.E. Livingston, A. Perez, I. A. Shlyakhter, R. V. Rolando, R. Owen, P. Tanpaiboon, F. Lacbawan, Clinton R. Paden, Duncan MacCannell |
| EPI_ISL_1906330, EPI_ISL_1906331 | Michigan Department of Health and Human Services, Bureau of Laboratories | Michigan Department of Health and Human Services, Bureau of Laboratories | Blankenship HM, Riner D, Soehnlen MK |
| EPI_ISL_1906332 | Quest Diagnostics Incorporated | Centers for Disease Control and Prevention Division of Viral Diseases, Pathogen Discovery | Dakota Howard, Dhwani Batra, Peter W. Cook, Kara Moser, Adrian Paskey, Jason Caravas, Benjamin Rambo-Martin, Shatavia Morrison, Christopher Gulvick, Scott Sammons, Yvette Unoarumhi, Darlene Wagner, Matthew Schmerer, S. H. Rosenthal, A. Gerasimova, R. M. Kagan, B. Anderson, M. Hua, Y. Liu, L.E. Bernstein, K.E. Livingston, A. Perez, I. A. Shlyakhter, R. V. Rolando, R. Owen, P. Tanpaiboon, F. Lacbawan, Clinton R. Paden, Duncan MacCannell |
| EPI_ISL_1906333 | Michigan Department of Health and Human Services, Bureau of Laboratories | Michigan Department of Health and Human Services, Bureau of Laboratories | Blankenship HM, Riner D, Soehnlen MK |
| EPI_ISL_1906334 | Quest Diagnostics Incorporated | Centers for Disease Control and Prevention Division of Viral Diseases, Pathogen Discovery | Dakota Howard, Dhwani Batra, Peter W. Cook, Kara Moser, Adrian Paskey, Jason Caravas, Benjamin Rambo-Martin, Shatavia Morrison, Christopher Gulvick, Scott Sammons, Yvette Unoarumhi, Darlene Wagner, Matthew Schmerer, S. H. Rosenthal, A. Gerasimova, R. M. Kagan, B. Anderson, M. Hua, Y. Liu, L.E. Bernstein, K.E. Livingston, A. Perez, I. A. Shlyakhter, R. V. Rolando, R. Owen, P. Tanpaiboon, F. Lacbawan, Clinton R. Paden, Duncan MacCannell |
| EPI_ISL_1906335 | Michigan Department of Health and Human Services, Bureau of Laboratories | Michigan Department of Health and Human Services, Bureau of Laboratories | Blankenship HM, Riner D, Soehnlen MK |
| EPI_ISL_1906336, EPI_ISL_1906338 | Quest Diagnostics Incorporated | Centers for Disease Control and Prevention Division of Viral Diseases, Pathogen Discovery | Dakota Howard, Dhwani Batra, Peter W. Cook, Kara Moser, Adrian Paskey, Jason Caravas, Benjamin Rambo-Martin, Shatavia Morrison, Christopher Gulvick, Scott Sammons, Yvette Unoarumhi, Darlene Wagner, Matthew Schmerer, S. H. Rosenthal, A. Gerasimova, R. M. Kagan, B. Anderson, M. Hua, Y. Liu, L.E. Bernstein, K.E. Livingston, A. Perez, I. A. Shlyakhter, R. V. Rolando, R. Owen, P. Tanpaiboon, F. Lacbawan, Clinton R. Paden, Duncan MacCannell |
| EPI_ISL_1906339 | Michigan Department of Health and Human Services, Bureau of Laboratories | Michigan Department of Health and Human Services, Bureau of Laboratories | Blankenship HM, Riner D, Soehnlen MK |
| EPI_ISL_1906340 | Quest Diagnostics Incorporated | Centers for Disease Control and Prevention Division of Viral Diseases, Pathogen Discovery | Dakota Howard, Dhwani Batra, Peter W. Cook, Kara Moser, Adrian Paskey, Jason Caravas, Benjamin Rambo-Martin, Shatavia Morrison, Christopher Gulvick, Scott Sammons, Yvette Unoarumhi, Darlene Wagner, Matthew Schmerer, S. H. Rosenthal, A. Gerasimova, R. M. Kagan, B. Anderson, M. Hua, Y. Liu, L.E. Bernstein, K.E. Livingston, A. Perez, I. A. Shlyakhter, R. V. Rolando, R. Owen, P. Tanpaiboon, F. Lacbawan, Clinton R. Paden, Duncan MacCannell |
| EPI_ISL_1906341, EPI_ISL_1906342 | Michigan Department of Health and Human Services, Bureau of Laboratories | Michigan Department of Health and Human Services, Bureau of Laboratories | Blankenship HM, Riner D, Soehnlen MK |
| EPI_ISL_1906343 | Quest Diagnostics Incorporated | Centers for Disease Control and Prevention Division of Viral Diseases, Pathogen Discovery | Dakota Howard, Dhwani Batra, Peter W. Cook, Kara Moser, Adrian Paskey, Jason Caravas, Benjamin Rambo-Martin, Shatavia Morrison, Christopher Gulvick, Scott Sammons, Yvette Unoarumhi, Darlene Wagner, Matthew Schmerer, S. H. Rosenthal, A. Gerasimova, R. M. Kagan, B. Anderson, M. Hua, Y. Liu, L.E. Bernstein, K.E. Livingston, A. Perez, I. A. Shlyakhter, R. V. Rolando, R. Owen, P. Tanpaiboon, F. Lacbawan, Clinton R. Paden, Duncan MacCannell |
| EPI_ISL_1906344 | Michigan Department of Health and Human Services, Bureau of Laboratories | Michigan Department of Health and Human Services, Bureau of Laboratories | Blankenship HM, Riner D, Soehnlen MK |
| EPI_ISL_1906345 | Quest Diagnostics Incorporated | Centers for Disease Control and Prevention Division of Viral Diseases, Pathogen Discovery | Dakota Howard, Dhwani Batra, Peter W. Cook, Kara Moser, Adrian Paskey, Jason Caravas, Benjamin Rambo-Martin, Shatavia Morrison, Christopher Gulvick, Scott Sammons, Yvette Unoarumhi, Darlene Wagner, Matthew Schmerer, S. H. Rosenthal, A. Gerasimova, R. M. Kagan, B. Anderson, M. Hua, Y. Liu, L.E. Bernstein, K.E. Livingston, A. Perez, I. A. Shlyakhter, R. V. Rolando, R. Owen, P. Tanpaiboon, F. Lacbawan, Clinton R. Paden, Duncan MacCannell |
| EPI_ISL_1906346 | Michigan Department of Health and Human Services, Bureau of Laboratories | Michigan Department of Health and Human Services, Bureau of Laboratories | Blankenship HM, Riner D, Soehnlen MK |
| EPI_ISL_1906347 | Quest Diagnostics Incorporated | Centers for Disease Control and Prevention Division of Viral Diseases, Pathogen Discovery | Dakota Howard, Dhwani Batra, Peter W. Cook, Kara Moser, Adrian Paskey, Jason Caravas, Benjamin Rambo-Martin, Shatavia Morrison, Christopher Gulvick, Scott Sammons, Yvette Unoarumhi, Darlene Wagner, Matthew Schmerer, S. H. Rosenthal, A. Gerasimova, R. M. Kagan, B. Anderson, M. Hua, Y. Liu, L.E. Bernstein, K.E. Livingston, A. Perez, I. A. Shlyakhter, R. V. Rolando, R. Owen, P. Tanpaiboon, F. Lacbawan, Clinton R. Paden, Duncan MacCannell |
| EPI_ISL_1906348 | Michigan Department of Health and Human Services, Bureau of Laboratories | Michigan Department of Health and Human Services, Bureau of Laboratories | Blankenship HM, Riner D, Soehnlen MK |
| EPI_ISL_1906349 | Quest Diagnostics Incorporated | Centers for Disease Control and Prevention Division of Viral Diseases, Pathogen Discovery | Dakota Howard, Dhwani Batra, Peter W. Cook, Kara Moser, Adrian Paskey, Jason Caravas, Benjamin Rambo-Martin, Shatavia Morrison, Christopher Gulvick, Scott Sammons, Yvette Unoarumhi, Darlene Wagner, Matthew Schmerer, S. H. Rosenthal, A. Gerasimova, R. M. Kagan, B. Anderson, M. Hua, Y. Liu, L.E. Bernstein, K.E. Livingston, A. Perez, I. A. Shlyakhter, R. V. Rolando, R. Owen, P. Tanpaiboon, F. Lacbawan, Clinton R. Paden, Duncan MacCannell |
| EPI_ISL_1906350, EPI_ISL_1906351, EPI_ISL_1906352 | Michigan Department of Health and Human Services, Bureau of Laboratories | Michigan Department of Health and Human Services, Bureau of Laboratories | Blankenship HM, Riner D, Soehnlen MK |
| EPI_ISL_1906353 | Quest Diagnostics Incorporated | Centers for Disease Control and Prevention Division of Viral Diseases, Pathogen Discovery | Dakota Howard, Dhwani Batra, Peter W. Cook, Kara Moser, Adrian Paskey, Jason Caravas, Benjamin Rambo-Martin, Shatavia Morrison, Christopher Gulvick, Scott Sammons, Yvette Unoarumhi, Darlene Wagner, Matthew Schmerer, S. H. Rosenthal, A. Gerasimova, R. M. Kagan, B. Anderson, M. Hua, Y. Liu, L.E. Bernstein, K.E. Livingston, A. Perez, I. A. Shlyakhter, R. V. Rolando, R. Owen, P. Tanpaiboon, F. Lacbawan, Clinton R. Paden, Duncan MacCannell |
| EPI_ISL_1906354, EPI_ISL_1906355 | Michigan Department of Health and Human Services, Bureau of Laboratories | Michigan Department of Health and Human Services, Bureau of Laboratories | Blankenship HM, Riner D, Soehnlen MK |
| EPI_ISL_1906356 | Quest Diagnostics Incorporated | Centers for Disease Control and Prevention Division of Viral Diseases, Pathogen Discovery | Dakota Howard, Dhwani Batra, Peter W. Cook, Kara Moser, Adrian Paskey, Jason Caravas, Benjamin Rambo-Martin, Shatavia Morrison, Christopher Gulvick, Scott Sammons, Yvette Unoarumhi, Darlene Wagner, Matthew Schmerer, S. H. Rosenthal, A. Gerasimova, R. M. Kagan, B. Anderson, M. Hua, Y. Liu, L.E. Bernstein, K.E. Livingston, A. Perez, I. A. Shlyakhter, R. V. Rolando, R. Owen, P. Tanpaiboon, F. Lacbawan, Clinton R. Paden, Duncan MacCannell |
| EPI_ISL_1906357, EPI_ISL_1906358 | Michigan Department of Health and Human Services, Bureau of Laboratories | Michigan Department of Health and Human Services, Bureau of Laboratories | Blankenship HM, Riner D, Soehnlen MK |
| EPI_ISL_1906359 | Quest Diagnostics Incorporated | Centers for Disease Control and Prevention Division of Viral Diseases, Pathogen Discovery | Dakota Howard, Dhwani Batra, Peter W. Cook, Kara Moser, Adrian Paskey, Jason Caravas, Benjamin Rambo-Martin, Shatavia Morrison, Christopher Gulvick, Scott Sammons, Yvette Unoarumhi, Darlene Wagner, Matthew Schmerer, S. H. Rosenthal, A. Gerasimova, R. M. Kagan, B. Anderson, M. Hua, Y. Liu, L.E. Bernstein, K.E. Livingston, A. Perez, I. A. Shlyakhter, R. V. Rolando, R. Owen, P. Tanpaiboon, F. Lacbawan, Clinton R. Paden, Duncan MacCannell |
| EPI_ISL_1906360 | Michigan Department of Health and Human Services, Bureau of Laboratories | Michigan Department of Health and Human Services, Bureau of Laboratories | Blankenship HM, Riner D, Soehnlen MK |
| EPI_ISL_1906361 | Quest Diagnostics Incorporated | Centers for Disease Control and Prevention Division of Viral Diseases, Pathogen Discovery | Dakota Howard, Dhwani Batra, Peter W. Cook, Kara Moser, Adrian Paskey, Jason Caravas, Benjamin Rambo-Martin, Shatavia Morrison, Christopher Gulvick, Scott Sammons, Yvette Unoarumhi, Darlene Wagner, Matthew Schmerer, S. H. Rosenthal, A. Gerasimova, R. M. Kagan, B. Anderson, M. Hua, Y. Liu, L.E. Bernstein, K.E. Livingston, A. Perez, I. A. Shlyakhter, R. V. Rolando, R. Owen, P. Tanpaiboon, F. Lacbawan, Clinton R. Paden, Duncan MacCannell |
| EPI_ISL_1906362 | Michigan Department of Health and Human Services, Bureau of Laboratories | Michigan Department of Health and Human Services, Bureau of Laboratories | Blankenship HM, Riner D, Soehnlen MK |
| EPI_ISL_1906363 | Quest Diagnostics Incorporated | Centers for Disease Control and Prevention Division of Viral Diseases, Pathogen Discovery | Dakota Howard, Dhwani Batra, Peter W. Cook, Kara Moser, Adrian Paskey, Jason Caravas, Benjamin Rambo-Martin, Shatavia Morrison, Christopher Gulvick, Scott Sammons, Yvette Unoarumhi, Darlene Wagner, Matthew Schmerer, S. H. Rosenthal, A. Gerasimova, R. M. Kagan, B. Anderson, M. Hua, Y. Liu, L.E. Bernstein, K.E. Livingston, A. Perez, I. A. Shlyakhter, R. V. Rolando, R. Owen, P. Tanpaiboon, F. Lacbawan, Clinton R. Paden, Duncan MacCannell |
| EPI_ISL_1906364, EPI_ISL_1906365 | Michigan Department of Health and Human Services, Bureau of Laboratories | Michigan Department of Health and Human Services, Bureau of Laboratories | Blankenship HM, Riner D, Soehnlen MK |
| EPI_ISL_1906366 | Quest Diagnostics Incorporated | Centers for Disease Control and Prevention Division of Viral Diseases, Pathogen Discovery | Dakota Howard, Dhwani Batra, Peter W. Cook, Kara Moser, Adrian Paskey, Jason Caravas, Benjamin Rambo-Martin, Shatavia Morrison, Christopher Gulvick, Scott Sammons, Yvette Unoarumhi, Darlene Wagner, Matthew Schmerer, S. H. Rosenthal, A. Gerasimova, R. M. Kagan, B. Anderson, M. Hua, Y. Liu, L.E. Bernstein, K.E. Livingston, A. Perez, I. A. Shlyakhter, R. V. Rolando, R. Owen, P. Tanpaiboon, F. Lacbawan, Clinton R. Paden, Duncan MacCannell |
| EPI_ISL_1906367 | Michigan Department of Health and Human Services, Bureau of Laboratories | Michigan Department of Health and Human Services, Bureau of Laboratories | Blankenship HM, Riner D, Soehnlen MK |
| EPI_ISL_1906368 | Quest Diagnostics Incorporated | Centers for Disease Control and Prevention Division of Viral Diseases, Pathogen Discovery | Dakota Howard, Dhwani Batra, Peter W. Cook, Kara Moser, Adrian Paskey, Jason Caravas, Benjamin Rambo-Martin, Shatavia Morrison, Christopher Gulvick, Scott Sammons, Yvette Unoarumhi, Darlene Wagner, Matthew Schmerer, S. H. Rosenthal, A. Gerasimova, R. M. Kagan, B. Anderson, M. Hua, Y. Liu, L.E. Bernstein, K.E. Livingston, A. Perez, I. A. Shlyakhter, R. V. Rolando, R. Owen, P. Tanpaiboon, F. Lacbawan, Clinton R. Paden, Duncan MacCannell |
| EPI_ISL_1906369 | Michigan Department of Health and Human Services, Bureau of Laboratories | Michigan Department of Health and Human Services, Bureau of Laboratories | Blankenship HM, Riner D, Soehnlen MK |
| EPI_ISL_1906370 | Quest Diagnostics Incorporated | Centers for Disease Control and Prevention Division of Viral Diseases, Pathogen Discovery | Dakota Howard, Dhwani Batra, Peter W. Cook, Kara Moser, Adrian Paskey, Jason Caravas, Benjamin Rambo-Martin, Shatavia Morrison, Christopher Gulvick, Scott Sammons, Yvette Unoarumhi, Darlene Wagner, Matthew Schmerer, S. H. Rosenthal, A. Gerasimova, R. M. Kagan, B. Anderson, M. Hua, Y. Liu, L.E. Bernstein, K.E. Livingston, A. Perez, I. A. Shlyakhter, R. V. Rolando, R. Owen, P. Tanpaiboon, F. Lacbawan, Clinton R. Paden, Duncan MacCannell |
| EPI_ISL_1906371 | Michigan Department of Health and Human Services, Bureau of Laboratories | Michigan Department of Health and Human Services, Bureau of Laboratories | Blankenship HM, Riner D, Soehnlen MK |
| EPI_ISL_1906372 | Quest Diagnostics Incorporated | Centers for Disease Control and Prevention | Dakota Howard, Dhwani Batra, Peter W. Cook, Kara Moser, Adrian Paskey, Jason Caravas, Benjamin Rambo-Martin, Shatavia Morrison, Christopher Gulvick, Scott Sammons, Yvette Unoarumhi, Darlene Wagner, Matthew Schmerer, S. H. Rosenthal, A. Gerasimova, R. M. Kagan, B. |

| | | | |
|---|---|---|---|
| | | Division of Viral Diseases, Pathogen Discovery | Anderson, M. Hua, Y. Liu, L.E. Bernstein, K.E. Livingston, A. Perez, I. A. Shlyakhter, R. V. Rolando, R. Owen, P. Tanpaiboon, F. Lacbawan, Clinton R. Paden, Duncan MacCannell |
| EPI_ISL_1906373 | Michigan Department of Health and Human Services, Bureau of Laboratories | Michigan Department of Health and Human Services, Bureau of Laboratories | Blankenship HM, Riner D, Soehnlen MK |
| EPI_ISL_1906374 | Quest Diagnostics Incorporated | Centers for Disease Control and Prevention Division of Viral Diseases, Pathogen Discovery | Dakota Howard, Dhwani Batra, Peter W. Cook, Kara Moser, Adrian Paskey, Jason Caravas, Benjamin Rambo-Martin, Shatavia Morrison, Christopher Gulvick, Scott Sammons, Yvette Unoarumhi, Darlene Wagner, Matthew Schmerer, S. H. Rosenthal, A. Gerasimova, R. M. Kagan, B. Anderson, M. Hua, Y. Liu, L.E. Bernstein, K.E. Livingston, A. Perez, I. A. Shlyakhter, R. V. Rolando, R. Owen, P. Tanpaiboon, F. Lacbawan, Clinton R. Paden, Duncan MacCannell |
| EPI_ISL_1906375 | Michigan Department of Health and Human Services, Bureau of Laboratories | Michigan Department of Health and Human Services, Bureau of Laboratories | Blankenship HM, Riner D, Soehnlen MK |
| EPI_ISL_1906376 | Quest Diagnostics Incorporated | Centers for Disease Control and Prevention Division of Viral Diseases, Pathogen Discovery | Dakota Howard, Dhwani Batra, Peter W. Cook, Kara Moser, Adrian Paskey, Jason Caravas, Benjamin Rambo-Martin, Shatavia Morrison, Christopher Gulvick, Scott Sammons, Yvette Unoarumhi, Darlene Wagner, Matthew Schmerer, S. H. Rosenthal, A. Gerasimova, R. M. Kagan, B. Anderson, M. Hua, Y. Liu, L.E. Bernstein, K.E. Livingston, A. Perez, I. A. Shlyakhter, R. V. Rolando, R. Owen, P. Tanpaiboon, F. Lacbawan, Clinton R. Paden, Duncan MacCannell |
| EPI_ISL_1906377, EPI_ISL_1906378 | Michigan Department of Health and Human Services, Bureau of Laboratories | Michigan Department of Health and Human Services, Bureau of Laboratories | Blankenship HM, Riner D, Soehnlen MK |
| EPI_ISL_1906379 | Quest Diagnostics Incorporated | Centers for Disease Control and Prevention Division of Viral Diseases, Pathogen Discovery | Dakota Howard, Dhwani Batra, Peter W. Cook, Kara Moser, Adrian Paskey, Jason Caravas, Benjamin Rambo-Martin, Shatavia Morrison, Christopher Gulvick, Scott Sammons, Yvette Unoarumhi, Darlene Wagner, Matthew Schmerer, S. H. Rosenthal, A. Gerasimova, R. M. Kagan, B. Anderson, M. Hua, Y. Liu, L.E. Bernstein, K.E. Livingston, A. Perez, I. A. Shlyakhter, R. V. Rolando, R. Owen, P. Tanpaiboon, F. Lacbawan, Clinton R. Paden, Duncan MacCannell |
| EPI_ISL_1906380, EPI_ISL_1906381 | Michigan Department of Health and Human Services, Bureau of Laboratories | Michigan Department of Health and Human Services, Bureau of Laboratories | Blankenship HM, Riner D, Soehnlen MK |
| EPI_ISL_1906382 | Quest Diagnostics Incorporated | Centers for Disease Control and Prevention Division of Viral Diseases, Pathogen Discovery | Dakota Howard, Dhwani Batra, Peter W. Cook, Kara Moser, Adrian Paskey, Jason Caravas, Benjamin Rambo-Martin, Shatavia Morrison, Christopher Gulvick, Scott Sammons, Yvette Unoarumhi, Darlene Wagner, Matthew Schmerer, S. H. Rosenthal, A. Gerasimova, R. M. Kagan, B. Anderson, M. Hua, Y. Liu, L.E. Bernstein, K.E. Livingston, A. Perez, I. A. Shlyakhter, R. V. Rolando, R. Owen, P. Tanpaiboon, F. Lacbawan, Clinton R. Paden, Duncan MacCannell |
| EPI_ISL_1906383, EPI_ISL_1906384 | Michigan Department of Health and Human Services, Bureau of Laboratories | Michigan Department of Health and Human Services, Bureau of Laboratories | Blankenship HM, Riner D, Soehnlen MK |
| EPI_ISL_1906385 | Quest Diagnostics Incorporated | Centers for Disease Control and Prevention Division of Viral Diseases, Pathogen Discovery | Dakota Howard, Dhwani Batra, Peter W. Cook, Kara Moser, Adrian Paskey, Jason Caravas, Benjamin Rambo-Martin, Shatavia Morrison, Christopher Gulvick, Scott Sammons, Yvette Unoarumhi, Darlene Wagner, Matthew Schmerer, S. H. Rosenthal, A. Gerasimova, R. M. Kagan, B. Anderson, M. Hua, Y. Liu, L.E. Bernstein, K.E. Livingston, A. Perez, I. A. Shlyakhter, R. V. Rolando, R. Owen, P. Tanpaiboon, F. Lacbawan, Clinton R. Paden, Duncan MacCannell |
| EPI_ISL_1906386 | Michigan Department of Health and Human Services, Bureau of Laboratories | Michigan Department of Health and Human Services, Bureau of Laboratories | Blankenship HM, Riner D, Soehnlen MK |
| EPI_ISL_1906387 | Quest Diagnostics Incorporated | Centers for Disease Control and Prevention Division of Viral Diseases, Pathogen Discovery | Dakota Howard, Dhwani Batra, Peter W. Cook, Kara Moser, Adrian Paskey, Jason Caravas, Benjamin Rambo-Martin, Shatavia Morrison, Christopher Gulvick, Scott Sammons, Yvette Unoarumhi, Darlene Wagner, Matthew Schmerer, S. H. Rosenthal, A. Gerasimova, R. M. Kagan, B. Anderson, M. Hua, Y. Liu, L.E. Bernstein, K.E. Livingston, A. Perez, I. A. Shlyakhter, R. V. Rolando, R. Owen, P. Tanpaiboon, F. Lacbawan, Clinton R. Paden, Duncan MacCannell |
| EPI_ISL_1906388 | Michigan Department of Health and Human Services, Bureau of Laboratories | Michigan Department of Health and Human Services, Bureau of Laboratories | Blankenship HM, Riner D, Soehnlen MK |
| EPI_ISL_1906389 | Quest Diagnostics Incorporated | Centers for Disease Control and Prevention Division of Viral Diseases, Pathogen Discovery | Dakota Howard, Dhwani Batra, Peter W. Cook, Kara Moser, Adrian Paskey, Jason Caravas, Benjamin Rambo-Martin, Shatavia Morrison, Christopher Gulvick, Scott Sammons, Yvette Unoarumhi, Darlene Wagner, Matthew Schmerer, S. H. Rosenthal, A. Gerasimova, R. M. Kagan, B. Anderson, M. Hua, Y. Liu, L.E. Bernstein, K.E. Livingston, A. Perez, I. A. Shlyakhter, R. V. Rolando, R. Owen, P. Tanpaiboon, F. Lacbawan, Clinton R. Paden, Duncan MacCannell |
| EPI_ISL_1906390, EPI_ISL_1906391 | Michigan Department of Health and Human Services, Bureau of Laboratories | Michigan Department of Health and Human Services, Bureau of Laboratories | Blankenship HM, Riner D, Soehnlen MK |
| EPI_ISL_1906392 | Quest Diagnostics Incorporated | Centers for Disease Control and Prevention Division of Viral Diseases, Pathogen Discovery | Dakota Howard, Dhwani Batra, Peter W. Cook, Kara Moser, Adrian Paskey, Jason Caravas, Benjamin Rambo-Martin, Shatavia Morrison, Christopher Gulvick, Scott Sammons, Yvette Unoarumhi, Darlene Wagner, Matthew Schmerer, S. H. Rosenthal, A. Gerasimova, R. M. Kagan, B. Anderson, M. Hua, Y. Liu, L.E. Bernstein, K.E. Livingston, A. Perez, I. A. Shlyakhter, R. V. Rolando, R. Owen, P. Tanpaiboon, F. Lacbawan, Clinton R. Paden, Duncan MacCannell |
| EPI_ISL_1906394 | Michigan Department of Health and Human Services, Bureau of Laboratories | Michigan Department of Health and Human Services, Bureau of Laboratories | Blankenship HM, Riner D, Soehnlen MK |
| EPI_ISL_1906395, EPI_ISL_1906397 | Quest Diagnostics Incorporated | Centers for Disease Control and Prevention Division of Viral Diseases, Pathogen Discovery | Dakota Howard, Dhwani Batra, Peter W. Cook, Kara Moser, Adrian Paskey, Jason Caravas, Benjamin Rambo-Martin, Shatavia Morrison, Christopher Gulvick, Scott Sammons, Yvette Unoarumhi, Darlene Wagner, Matthew Schmerer, S. H. Rosenthal, A. Gerasimova, R. M. Kagan, B. Anderson, M. Hua, Y. Liu, L.E. Bernstein, K.E. Livingston, A. Perez, I. A. Shlyakhter, R. V. Rolando, R. Owen, P. Tanpaiboon, F. Lacbawan, Clinton R. Paden, Duncan MacCannell |
| EPI_ISL_1906398 | Michigan Department of Health and Human Services, Bureau of Laboratories | Michigan Department of Health and Human Services, Bureau of Laboratories | Blankenship HM, Riner D, Soehnlen MK |
| EPI_ISL_1906400 | Quest Diagnostics Incorporated | Centers for Disease Control and Prevention Division of Viral Diseases, Pathogen Discovery | Dakota Howard, Dhwani Batra, Peter W. Cook, Kara Moser, Adrian Paskey, Jason Caravas, Benjamin Rambo-Martin, Shatavia Morrison, Christopher Gulvick, Scott Sammons, Yvette Unoarumhi, Darlene Wagner, Matthew Schmerer, S. H. Rosenthal, A. Gerasimova, R. M. Kagan, B. Anderson, M. Hua, Y. Liu, L.E. Bernstein, K.E. Livingston, A. Perez, I. A. Shlyakhter, R. V. Rolando, R. Owen, P. Tanpaiboon, F. Lacbawan, Clinton R. Paden, Duncan MacCannell |
| EPI_ISL_1906401 | Michigan Department of Health and Human Services, Bureau of Laboratories | Michigan Department of Health and Human Services, Bureau of Laboratories | Blankenship HM, Riner D, Soehnlen MK |
| EPI_ISL_1906402, EPI_ISL_1906404 | Quest Diagnostics Incorporated | Centers for Disease Control and Prevention Division of Viral Diseases, Pathogen Discovery | Dakota Howard, Dhwani Batra, Peter W. Cook, Kara Moser, Adrian Paskey, Jason Caravas, Benjamin Rambo-Martin, Shatavia Morrison, Christopher Gulvick, Scott Sammons, Yvette Unoarumhi, Darlene Wagner, Matthew Schmerer, S. H. Rosenthal, A. Gerasimova, R. M. Kagan, B. Anderson, M. Hua, Y. Liu, L.E. Bernstein, K.E. Livingston, A. Perez, I. A. Shlyakhter, R. V. Rolando, R. Owen, P. Tanpaiboon, F. Lacbawan, Clinton R. Paden, Duncan MacCannell |
| EPI_ISL_1906406 | Michigan Department of Health and Human Services, Bureau of Laboratories | Michigan Department of Health and Human Services, Bureau of Laboratories | Blankenship HM, Riner D, Soehnlen MK |
| EPI_ISL_1906407, EPI_ISL_1906409, EPI_ISL_1906411, EPI_ISL_1906413 | Quest Diagnostics Incorporated | Centers for Disease Control and Prevention Division of Viral Diseases, Pathogen Discovery | Dakota Howard, Dhwani Batra, Peter W. Cook, Kara Moser, Adrian Paskey, Jason Caravas, Benjamin Rambo-Martin, Shatavia Morrison, Christopher Gulvick, Scott Sammons, Yvette Unoarumhi, Darlene Wagner, Matthew Schmerer, S. H. Rosenthal, A. Gerasimova, R. M. Kagan, B. Anderson, M. Hua, Y. Liu, L.E. Bernstein, K.E. Livingston, A. Perez, I. A. Shlyakhter, R. V. Rolando, R. Owen, P. Tanpaiboon, F. Lacbawan, Clinton R. Paden, Duncan MacCannell |
| EPI_ISL_1906416 | Michigan Department of Health and Human Services, Bureau of Laboratories | Michigan Department of Health and Human Services, Bureau of Laboratories | Blankenship HM, Riner D, Soehnlen MK |
| EPI_ISL_1906417 | Quest Diagnostics Incorporated | Centers for Disease Control and Prevention Division of Viral Diseases, Pathogen Discovery | Dakota Howard, Dhwani Batra, Peter W. Cook, Kara Moser, Adrian Paskey, Jason Caravas, Benjamin Rambo-Martin, Shatavia Morrison, Christopher Gulvick, Scott Sammons, Yvette Unoarumhi, Darlene Wagner, Matthew Schmerer, S. H. Rosenthal, A. Gerasimova, R. M. Kagan, B. Anderson, M. Hua, Y. Liu, L.E. Bernstein, K.E. Livingston, A. Perez, I. A. Shlyakhter, R. V. Rolando, R. Owen, P. Tanpaiboon, F. Lacbawan, Clinton R. Paden, Duncan MacCannell |
| EPI_ISL_1906419 | Michigan Department of Health and Human Services, Bureau of Laboratories | Michigan Department of Health and Human Services, Bureau of Laboratories | Blankenship HM, Riner D, Soehnlen MK |
| EPI_ISL_1906420, EPI_ISL_1906422, EPI_ISL_1906424 | Quest Diagnostics Incorporated | Centers for Disease Control and Prevention Division of Viral Diseases, Pathogen Discovery | Dakota Howard, Dhwani Batra, Peter W. Cook, Kara Moser, Adrian Paskey, Jason Caravas, Benjamin Rambo-Martin, Shatavia Morrison, Christopher Gulvick, Scott Sammons, Yvette Unoarumhi, Darlene Wagner, Matthew Schmerer, S. H. Rosenthal, A. Gerasimova, R. M. Kagan, B. Anderson, M. Hua, Y. Liu, L.E. Bernstein, K.E. Livingston, A. Perez, I. A. Shlyakhter, R. V. Rolando, R. Owen, P. Tanpaiboon, F. Lacbawan, Clinton R. Paden, Duncan MacCannell |
| EPI_ISL_1906425 | Michigan Department of Health and Human Services, Bureau of Laboratories | Michigan Department of Health and Human Services, Bureau of Laboratories | Blankenship HM, Riner D, Soehnlen MK |
| EPI_ISL_1906426 | Quest Diagnostics Incorporated | Centers for Disease Control and Prevention Division of Viral Diseases, Pathogen Discovery | Dakota Howard, Dhwani Batra, Peter W. Cook, Kara Moser, Adrian Paskey, Jason Caravas, Benjamin Rambo-Martin, Shatavia Morrison, Christopher Gulvick, Scott Sammons, Yvette Unoarumhi, Darlene Wagner, Matthew Schmerer, S. H. Rosenthal, A. Gerasimova, R. M. Kagan, B. Anderson, M. Hua, Y. Liu, L.E. Bernstein, K.E. Livingston, A. Perez, I. A. Shlyakhter, R. V. Rolando, R. Owen, P. Tanpaiboon, F. Lacbawan, Clinton R. Paden, Duncan MacCannell |
| EPI_ISL_1906427 | Michigan Department of Health and Human Services, Bureau of Laboratories | Michigan Department of Health and Human Services, Bureau of Laboratories | Blankenship HM, Riner D, Soehnlen MK |
| EPI_ISL_1906428 | Quest Diagnostics Incorporated | Centers for Disease Control and Prevention Division of Viral Diseases, Pathogen Discovery | Dakota Howard, Dhwani Batra, Peter W. Cook, Kara Moser, Adrian Paskey, Jason Caravas, Benjamin Rambo-Martin, Shatavia Morrison, Christopher Gulvick, Scott Sammons, Yvette Unoarumhi, Darlene Wagner, Matthew Schmerer, S. H. Rosenthal, A. Gerasimova, R. M. Kagan, B. Anderson, M. Hua, Y. Liu, L.E. Bernstein, K.E. Livingston, A. Perez, I. A. Shlyakhter, R. V. Rolando, R. Owen, P. Tanpaiboon, F. Lacbawan, Clinton R. Paden, Duncan MacCannell |
| EPI_ISL_1906430 | Michigan Department of Health and Human Services, Bureau of Laboratories | Michigan Department of Health and Human Services, Bureau of Laboratories | Blankenship HM, Riner D, Soehnlen MK |
| EPI_ISL_1906431 | Quest Diagnostics Incorporated | Centers for Disease Control and Prevention Division of Viral Diseases, Pathogen Discovery | Dakota Howard, Dhwani Batra, Peter W. Cook, Kara Moser, Adrian Paskey, Jason Caravas, Benjamin Rambo-Martin, Shatavia Morrison, Christopher Gulvick, Scott Sammons, Yvette Unoarumhi, Darlene Wagner, Matthew Schmerer, S. H. Rosenthal, A. Gerasimova, R. M. Kagan, B. Anderson, M. Hua, Y. Liu, L.E. Bernstein, K.E. Livingston, A. Perez, I. A. Shlyakhter, R. V. Rolando, R. Owen, P. Tanpaiboon, F. Lacbawan, Clinton R. Paden, Duncan MacCannell |
| EPI_ISL_1906432 | Michigan Department of Health and Human Services, Bureau of Laboratories | Michigan Department of Health and Human Services, Bureau of Laboratories | Blankenship HM, Riner D, Soehnlen MK |
| EPI_ISL_1906433 | Quest Diagnostics Incorporated | Centers for Disease Control and Prevention Division of Viral Diseases, Pathogen Discovery | Dakota Howard, Dhwani Batra, Peter W. Cook, Kara Moser, Adrian Paskey, Jason Caravas, Benjamin Rambo-Martin, Shatavia Morrison, Christopher Gulvick, Scott Sammons, Yvette Unoarumhi, Darlene Wagner, Matthew Schmerer, S. H. Rosenthal, A. Gerasimova, R. M. Kagan, B. Anderson, M. Hua, Y. Liu, L.E. Bernstein, K.E. Livingston, A. Perez, I. A. Shlyakhter, R. V. Rolando, R. Owen, P. Tanpaiboon, F. Lacbawan, Clinton R. Paden, Duncan MacCannell |
| EPI_ISL_1906434 | Michigan Department of Health and | Michigan Department of Health and Human | Blankenship HM, Riner D, Soehnlen MK |

| | | | |
|---|---|---|---|
| | Human Services, Bureau of Laboratories | Services, Bureau of Laboratories | |
| EPI_ISL_1906435 | Quest Diagnostics Incorporated | Centers for Disease Control and Prevention Division of Viral Diseases, Pathogen Discovery | Dakota Howard, Dhwani Batra, Peter W. Cook, Kara Moser, Adrian Paskey, Jason Caravas, Benjamin Rambo-Martin, Shatavia Morrison, Christopher Gulvick, Scott Sammons, Yvette Unoarumhi, Darlene Wagner, Matthew Schmerer, S. H. Rosenthal, A. Gerasimova, R. M. Kagan, B. Anderson, M. Hua, Y. Liu, L.E. Bernstein, K.E. Livingston, A. Perez, I. A. Shlyakhter, R. V. Rolando, R. Owen, P. Tanpaiboon, F. Lacbawan, Clinton R. Paden, Duncan MacCannell |
| EPI_ISL_1906436 | Michigan Department of Health and Human Services, Bureau of Laboratories | Michigan Department of Health and Human Services, Bureau of Laboratories | Blankenship HM, Riner D, Soehnlen MK |
| EPI_ISL_1906437 | Quest Diagnostics Incorporated | Centers for Disease Control and Prevention Division of Viral Diseases, Pathogen Discovery | Dakota Howard, Dhwani Batra, Peter W. Cook, Kara Moser, Adrian Paskey, Jason Caravas, Benjamin Rambo-Martin, Shatavia Morrison, Christopher Gulvick, Scott Sammons, Yvette Unoarumhi, Darlene Wagner, Matthew Schmerer, S. H. Rosenthal, A. Gerasimova, R. M. Kagan, B. Anderson, M. Hua, Y. Liu, L.E. Bernstein, K.E. Livingston, A. Perez, I. A. Shlyakhter, R. V. Rolando, R. Owen, P. Tanpaiboon, F. Lacbawan, Clinton R. Paden, Duncan MacCannell |
| EPI_ISL_1906438, EPI_ISL_1906439 | Michigan Department of Health and Human Services, Bureau of Laboratories | Michigan Department of Health and Human Services, Bureau of Laboratories | Blankenship HM, Riner D, Soehnlen MK |
| EPI_ISL_1906440 | Quest Diagnostics Incorporated | Centers for Disease Control and Prevention Division of Viral Diseases, Pathogen Discovery | Dakota Howard, Dhwani Batra, Peter W. Cook, Kara Moser, Adrian Paskey, Jason Caravas, Benjamin Rambo-Martin, Shatavia Morrison, Christopher Gulvick, Scott Sammons, Yvette Unoarumhi, Darlene Wagner, Matthew Schmerer, S. H. Rosenthal, A. Gerasimova, R. M. Kagan, B. Anderson, M. Hua, Y. Liu, L.E. Bernstein, K.E. Livingston, A. Perez, I. A. Shlyakhter, R. V. Rolando, R. Owen, P. Tanpaiboon, F. Lacbawan, Clinton R. Paden, Duncan MacCannell |
| EPI_ISL_1906441 | Michigan Department of Health and Human Services, Bureau of Laboratories | Michigan Department of Health and Human Services, Bureau of Laboratories | Blankenship HM, Riner D, Soehnlen MK |
| EPI_ISL_1906442 | Quest Diagnostics Incorporated | Centers for Disease Control and Prevention Division of Viral Diseases, Pathogen Discovery | Dakota Howard, Dhwani Batra, Peter W. Cook, Kara Moser, Adrian Paskey, Jason Caravas, Benjamin Rambo-Martin, Shatavia Morrison, Christopher Gulvick, Scott Sammons, Yvette Unoarumhi, Darlene Wagner, Matthew Schmerer, S. H. Rosenthal, A. Gerasimova, R. M. Kagan, B. Anderson, M. Hua, Y. Liu, L.E. Bernstein, K.E. Livingston, A. Perez, I. A. Shlyakhter, R. V. Rolando, R. Owen, P. Tanpaiboon, F. Lacbawan, Clinton R. Paden, Duncan MacCannell |
| EPI_ISL_1906443 | Michigan Department of Health and Human Services, Bureau of Laboratories | Michigan Department of Health and Human Services, Bureau of Laboratories | Blankenship HM, Riner D, Soehnlen MK |
| EPI_ISL_1906444 | Quest Diagnostics Incorporated | Centers for Disease Control and Prevention Division of Viral Diseases, Pathogen Discovery | Dakota Howard, Dhwani Batra, Peter W. Cook, Kara Moser, Adrian Paskey, Jason Caravas, Benjamin Rambo-Martin, Shatavia Morrison, Christopher Gulvick, Scott Sammons, Yvette Unoarumhi, Darlene Wagner, Matthew Schmerer, S. H. Rosenthal, A. Gerasimova, R. M. Kagan, B. Anderson, M. Hua, Y. Liu, L.E. Bernstein, K.E. Livingston, A. Perez, I. A. Shlyakhter, R. V. Rolando, R. Owen, P. Tanpaiboon, F. Lacbawan, Clinton R. Paden, Duncan MacCannell |
| EPI_ISL_1906445, EPI_ISL_1906446, EPI_ISL_1906447 | Michigan Department of Health and Human Services, Bureau of Laboratories | Michigan Department of Health and Human Services, Bureau of Laboratories | Blankenship HM, Riner D, Soehnlen MK |
| EPI_ISL_1906448 | Quest Diagnostics Incorporated | Centers for Disease Control and Prevention Division of Viral Diseases, Pathogen Discovery | Dakota Howard, Dhwani Batra, Peter W. Cook, Kara Moser, Adrian Paskey, Jason Caravas, Benjamin Rambo-Martin, Shatavia Morrison, Christopher Gulvick, Scott Sammons, Yvette Unoarumhi, Darlene Wagner, Matthew Schmerer, S. H. Rosenthal, A. Gerasimova, R. M. Kagan, B. Anderson, M. Hua, Y. Liu, L.E. Bernstein, K.E. Livingston, A. Perez, I. A. Shlyakhter, R. V. Rolando, R. Owen, P. Tanpaiboon, F. Lacbawan, Clinton R. Paden, Duncan MacCannell |
| EPI_ISL_1906449 | Michigan Department of Health and Human Services, Bureau of Laboratories | Michigan Department of Health and Human Services, Bureau of Laboratories | Blankenship HM, Riner D, Soehnlen MK |
| EPI_ISL_1906450 | Quest Diagnostics Incorporated | Centers for Disease Control and Prevention Division of Viral Diseases, Pathogen Discovery | Dakota Howard, Dhwani Batra, Peter W. Cook, Kara Moser, Adrian Paskey, Jason Caravas, Benjamin Rambo-Martin, Shatavia Morrison, Christopher Gulvick, Scott Sammons, Yvette Unoarumhi, Darlene Wagner, Matthew Schmerer, S. H. Rosenthal, A. Gerasimova, R. M. Kagan, B. Anderson, M. Hua, Y. Liu, L.E. Bernstein, K.E. Livingston, A. Perez, I. A. Shlyakhter, R. V. Rolando, R. Owen, P. Tanpaiboon, F. Lacbawan, Clinton R. Paden, Duncan MacCannell |
| EPI_ISL_1906451 | Michigan Department of Health and Human Services, Bureau of Laboratories | Michigan Department of Health and Human Services, Bureau of Laboratories | Blankenship HM, Riner D, Soehnlen MK |
| EPI_ISL_1906452 | Quest Diagnostics Incorporated | Centers for Disease Control and Prevention Division of Viral Diseases, Pathogen Discovery | Dakota Howard, Dhwani Batra, Peter W. Cook, Kara Moser, Adrian Paskey, Jason Caravas, Benjamin Rambo-Martin, Shatavia Morrison, Christopher Gulvick, Scott Sammons, Yvette Unoarumhi, Darlene Wagner, Matthew Schmerer, S. H. Rosenthal, A. Gerasimova, R. M. Kagan, B. Anderson, M. Hua, Y. Liu, L.E. Bernstein, K.E. Livingston, A. Perez, I. A. Shlyakhter, R. V. Rolando, R. Owen, P. Tanpaiboon, F. Lacbawan, Clinton R. Paden, Duncan MacCannell |
| EPI_ISL_1906453, EPI_ISL_1906454 | Michigan Department of Health and Human Services, Bureau of Laboratories | Michigan Department of Health and Human Services, Bureau of Laboratories | Blankenship HM, Riner D, Soehnlen MK |
| EPI_ISL_1906455 | Quest Diagnostics Incorporated | Centers for Disease Control and Prevention Division of Viral Diseases, Pathogen Discovery | Dakota Howard, Dhwani Batra, Peter W. Cook, Kara Moser, Adrian Paskey, Jason Caravas, Benjamin Rambo-Martin, Shatavia Morrison, Christopher Gulvick, Scott Sammons, Yvette Unoarumhi, Darlene Wagner, Matthew Schmerer, S. H. Rosenthal, A. Gerasimova, R. M. Kagan, B. Anderson, M. Hua, Y. Liu, L.E. Bernstein, K.E. Livingston, A. Perez, I. A. Shlyakhter, R. V. Rolando, R. Owen, P. Tanpaiboon, F. Lacbawan, Clinton R. Paden, Duncan MacCannell |
| EPI_ISL_1906456, EPI_ISL_1906457 | Michigan Department of Health and Human Services, Bureau of Laboratories | Michigan Department of Health and Human Services, Bureau of Laboratories | Blankenship HM, Riner D, Soehnlen MK |
| EPI_ISL_1906458 | Quest Diagnostics Incorporated | Centers for Disease Control and Prevention Division of Viral Diseases, Pathogen Discovery | Dakota Howard, Dhwani Batra, Peter W. Cook, Kara Moser, Adrian Paskey, Jason Caravas, Benjamin Rambo-Martin, Shatavia Morrison, Christopher Gulvick, Scott Sammons, Yvette Unoarumhi, Darlene Wagner, Matthew Schmerer, S. H. Rosenthal, A. Gerasimova, R. M. Kagan, B. Anderson, M. Hua, Y. Liu, L.E. Bernstein, K.E. Livingston, A. Perez, I. A. Shlyakhter, R. V. Rolando, R. Owen, P. Tanpaiboon, F. Lacbawan, Clinton R. Paden, Duncan MacCannell |
| EPI_ISL_1906459, EPI_ISL_1906460 | Michigan Department of Health and Human Services, Bureau of Laboratories | Michigan Department of Health and Human Services, Bureau of Laboratories | Blankenship HM, Riner D, Soehnlen MK |
| EPI_ISL_1906461 | Quest Diagnostics Incorporated | Centers for Disease Control and Prevention Division of Viral Diseases, Pathogen Discovery | Dakota Howard, Dhwani Batra, Peter W. Cook, Kara Moser, Adrian Paskey, Jason Caravas, Benjamin Rambo-Martin, Shatavia Morrison, Christopher Gulvick, Scott Sammons, Yvette Unoarumhi, Darlene Wagner, Matthew Schmerer, S. H. Rosenthal, A. Gerasimova, R. M. Kagan, B. Anderson, M. Hua, Y. Liu, L.E. Bernstein, K.E. Livingston, A. Perez, I. A. Shlyakhter, R. V. Rolando, R. Owen, P. Tanpaiboon, F. Lacbawan, Clinton R. Paden, Duncan MacCannell |
| EPI_ISL_1906462 | Michigan Department of Health and Human Services, Bureau of Laboratories | Michigan Department of Health and Human Services, Bureau of Laboratories | Blankenship HM, Riner D, Soehnlen MK |
| EPI_ISL_1906463 | Quest Diagnostics Incorporated | Centers for Disease Control and Prevention Division of Viral Diseases, Pathogen Discovery | Dakota Howard, Dhwani Batra, Peter W. Cook, Kara Moser, Adrian Paskey, Jason Caravas, Benjamin Rambo-Martin, Shatavia Morrison, Christopher Gulvick, Scott Sammons, Yvette Unoarumhi, Darlene Wagner, Matthew Schmerer, S. H. Rosenthal, A. Gerasimova, R. M. Kagan, B. Anderson, M. Hua, Y. Liu, L.E. Bernstein, K.E. Livingston, A. Perez, I. A. Shlyakhter, R. V. Rolando, R. Owen, P. Tanpaiboon, F. Lacbawan, Clinton R. Paden, Duncan MacCannell |
| EPI_ISL_1906465 | Michigan Department of Health and Human Services, Bureau of Laboratories | Michigan Department of Health and Human Services, Bureau of Laboratories | Blankenship HM, Riner D, Soehnlen MK |
| EPI_ISL_1906466 | Quest Diagnostics Incorporated | Centers for Disease Control and Prevention Division of Viral Diseases, Pathogen Discovery | Dakota Howard, Dhwani Batra, Peter W. Cook, Kara Moser, Adrian Paskey, Jason Caravas, Benjamin Rambo-Martin, Shatavia Morrison, Christopher Gulvick, Scott Sammons, Yvette Unoarumhi, Darlene Wagner, Matthew Schmerer, S. H. Rosenthal, A. Gerasimova, R. M. Kagan, B. Anderson, M. Hua, Y. Liu, L.E. Bernstein, K.E. Livingston, A. Perez, I. A. Shlyakhter, R. V. Rolando, R. Owen, P. Tanpaiboon, F. Lacbawan, Clinton R. Paden, Duncan MacCannell |
| EPI_ISL_1906467, EPI_ISL_1906468, EPI_ISL_1906469 | Michigan Department of Health and Human Services, Bureau of Laboratories | Michigan Department of Health and Human Services, Bureau of Laboratories | Blankenship HM, Riner D, Soehnlen MK |
| EPI_ISL_1906470 | Quest Diagnostics Incorporated | Centers for Disease Control and Prevention Division of Viral Diseases, Pathogen Discovery | Dakota Howard, Dhwani Batra, Peter W. Cook, Kara Moser, Adrian Paskey, Jason Caravas, Benjamin Rambo-Martin, Shatavia Morrison, Christopher Gulvick, Scott Sammons, Yvette Unoarumhi, Darlene Wagner, Matthew Schmerer, S. H. Rosenthal, A. Gerasimova, R. M. Kagan, B. Anderson, M. Hua, Y. Liu, L.E. Bernstein, K.E. Livingston, A. Perez, I. A. Shlyakhter, R. V. Rolando, R. Owen, P. Tanpaiboon, F. Lacbawan, Clinton R. Paden, Duncan MacCannell |
| EPI_ISL_1906471 | Michigan Department of Health and Human Services, Bureau of Laboratories | Michigan Department of Health and Human Services, Bureau of Laboratories | Blankenship HM, Riner D, Soehnlen MK |
| EPI_ISL_1906472 | Quest Diagnostics Incorporated | Centers for Disease Control and Prevention Division of Viral Diseases, Pathogen Discovery | Dakota Howard, Dhwani Batra, Peter W. Cook, Kara Moser, Adrian Paskey, Jason Caravas, Benjamin Rambo-Martin, Shatavia Morrison, Christopher Gulvick, Scott Sammons, Yvette Unoarumhi, Darlene Wagner, Matthew Schmerer, S. H. Rosenthal, A. Gerasimova, R. M. Kagan, B. Anderson, M. Hua, Y. Liu, L.E. Bernstein, K.E. Livingston, A. Perez, I. A. Shlyakhter, R. V. Rolando, R. Owen, P. Tanpaiboon, F. Lacbawan, Clinton R. Paden, Duncan MacCannell |
| EPI_ISL_1906473 | Michigan Department of Health and Human Services, Bureau of Laboratories | Michigan Department of Health and Human Services, Bureau of Laboratories | Blankenship HM, Riner D, Soehnlen MK |
| EPI_ISL_1906474 | Quest Diagnostics Incorporated | Centers for Disease Control and Prevention Division of Viral Diseases, Pathogen Discovery | Dakota Howard, Dhwani Batra, Peter W. Cook, Kara Moser, Adrian Paskey, Jason Caravas, Benjamin Rambo-Martin, Shatavia Morrison, Christopher Gulvick, Scott Sammons, Yvette Unoarumhi, Darlene Wagner, Matthew Schmerer, S. H. Rosenthal, A. Gerasimova, R. M. Kagan, B. Anderson, M. Hua, Y. Liu, L.E. Bernstein, K.E. Livingston, A. Perez, I. A. Shlyakhter, R. V. Rolando, R. Owen, P. Tanpaiboon, F. Lacbawan, Clinton R. Paden, Duncan MacCannell |
| EPI_ISL_1906475 | Michigan Department of Health and Human Services, Bureau of Laboratories | Michigan Department of Health and Human Services, Bureau of Laboratories | Blankenship HM, Riner D, Soehnlen MK |
| EPI_ISL_1906476 | Quest Diagnostics Incorporated | Centers for Disease Control and Prevention Division of Viral Diseases, Pathogen Discovery | Dakota Howard, Dhwani Batra, Peter W. Cook, Kara Moser, Adrian Paskey, Jason Caravas, Benjamin Rambo-Martin, Shatavia Morrison, Christopher Gulvick, Scott Sammons, Yvette Unoarumhi, Darlene Wagner, Matthew Schmerer, S. H. Rosenthal, A. Gerasimova, R. M. Kagan, B. Anderson, M. Hua, Y. Liu, L.E. Bernstein, K.E. Livingston, A. Perez, I. A. Shlyakhter, R. V. Rolando, R. Owen, P. Tanpaiboon, F. Lacbawan, Clinton R. Paden, Duncan MacCannell |
| EPI_ISL_1906477 | Michigan Department of Health and Human Services, Bureau of Laboratories | Michigan Department of Health and Human Services, Bureau of Laboratories | Blankenship HM, Riner D, Soehnlen MK |
| EPI_ISL_1906478 | Quest Diagnostics Incorporated | Centers for Disease Control and Prevention Division of Viral Diseases, Pathogen Discovery | Dakota Howard, Dhwani Batra, Peter W. Cook, Kara Moser, Adrian Paskey, Jason Caravas, Benjamin Rambo-Martin, Shatavia Morrison, Christopher Gulvick, Scott Sammons, Yvette Unoarumhi, Darlene Wagner, Matthew Schmerer, S. H. Rosenthal, A. Gerasimova, R. M. Kagan, B. Anderson, M. Hua, Y. Liu, L.E. Bernstein, K.E. Livingston, A. Perez, I. A. Shlyakhter, R. V. Rolando, R. Owen, P. Tanpaiboon, F. Lacbawan, Clinton R. Paden, Duncan MacCannell |
| EPI_ISL_1906479 | Michigan Department of Health and Human Services, Bureau of Laboratories | Michigan Department of Health and Human Services, Bureau of Laboratories | Blankenship HM, Riner D, Soehnlen MK |

| | | | |
|---|---|---|---|
| EPI_ISL_1906480 | Quest Diagnostics Incorporated | Centers for Disease Control and Prevention Division of Viral Diseases, Pathogen Discovery | Dakota Howard, Dhwani Batra, Peter W. Cook, Kara Moser, Adrian Paskey, Jason Caravas, Benjamin Rambo-Martin, Shatavia Morrison, Christopher Gulvick, Scott Sammons, Yvette Unoarumhi, Darlene Wagner, Matthew Schmerer, S. H. Rosenthal, A. Gerasimova, R. M. Kagan, B. Anderson, M. Hua, Y. Liu, L.E. Bernstein, K.E. Livingston, A. Perez, I. A. Shlyakhter, R. V. Rolando, R. Owen, P. Tanpaiboon, F. Lacbawan, Clinton R. Paden, Duncan MacCannell |
| EPI_ISL_1906481 | Michigan Department of Health and Human Services, Bureau of Laboratories | Michigan Department of Health and Human Services, Bureau of Laboratories | Blankenship HM, Riner D, Soehnlen MK |
| EPI_ISL_1906482 | Quest Diagnostics Incorporated | Centers for Disease Control and Prevention Division of Viral Diseases, Pathogen Discovery | Dakota Howard, Dhwani Batra, Peter W. Cook, Kara Moser, Adrian Paskey, Jason Caravas, Benjamin Rambo-Martin, Shatavia Morrison, Christopher Gulvick, Scott Sammons, Yvette Unoarumhi, Darlene Wagner, Matthew Schmerer, S. H. Rosenthal, A. Gerasimova, R. M. Kagan, B. Anderson, M. Hua, Y. Liu, L.E. Bernstein, K.E. Livingston, A. Perez, I. A. Shlyakhter, R. V. Rolando, R. Owen, P. Tanpaiboon, F. Lacbawan, Clinton R. Paden, Duncan MacCannell |
| EPI_ISL_1906483 | Michigan Department of Health and Human Services, Bureau of Laboratories | Michigan Department of Health and Human Services, Bureau of Laboratories | Blankenship HM, Riner D, Soehnlen MK |
| EPI_ISL_1906484 | Quest Diagnostics Incorporated | Centers for Disease Control and Prevention Division of Viral Diseases, Pathogen Discovery | Dakota Howard, Dhwani Batra, Peter W. Cook, Kara Moser, Adrian Paskey, Jason Caravas, Benjamin Rambo-Martin, Shatavia Morrison, Christopher Gulvick, Scott Sammons, Yvette Unoarumhi, Darlene Wagner, Matthew Schmerer, S. H. Rosenthal, A. Gerasimova, R. M. Kagan, B. Anderson, M. Hua, Y. Liu, L.E. Bernstein, K.E. Livingston, A. Perez, I. A. Shlyakhter, R. V. Rolando, R. Owen, P. Tanpaiboon, F. Lacbawan, Clinton R. Paden, Duncan MacCannell |
| EPI_ISL_1906485 | Michigan Department of Health and Human Services, Bureau of Laboratories | Michigan Department of Health and Human Services, Bureau of Laboratories | Blankenship HM, Riner D, Soehnlen MK |
| EPI_ISL_1906486 | Quest Diagnostics Incorporated | Centers for Disease Control and Prevention Division of Viral Diseases, Pathogen Discovery | Dakota Howard, Dhwani Batra, Peter W. Cook, Kara Moser, Adrian Paskey, Jason Caravas, Benjamin Rambo-Martin, Shatavia Morrison, Christopher Gulvick, Scott Sammons, Yvette Unoarumhi, Darlene Wagner, Matthew Schmerer, S. H. Rosenthal, A. Gerasimova, R. M. Kagan, B. Anderson, M. Hua, Y. Liu, L.E. Bernstein, K.E. Livingston, A. Perez, I. A. Shlyakhter, R. V. Rolando, R. Owen, P. Tanpaiboon, F. Lacbawan, Clinton R. Paden, Duncan MacCannell |
| EPI_ISL_1906487, EPI_ISL_1906488 | Michigan Department of Health and Human Services, Bureau of Laboratories | Michigan Department of Health and Human Services, Bureau of Laboratories | Blankenship HM, Riner D, Soehnlen MK |
| EPI_ISL_1906489 | Quest Diagnostics Incorporated | Centers for Disease Control and Prevention Division of Viral Diseases, Pathogen Discovery | Dakota Howard, Dhwani Batra, Peter W. Cook, Kara Moser, Adrian Paskey, Jason Caravas, Benjamin Rambo-Martin, Shatavia Morrison, Christopher Gulvick, Scott Sammons, Yvette Unoarumhi, Darlene Wagner, Matthew Schmerer, S. H. Rosenthal, A. Gerasimova, R. M. Kagan, B. Anderson, M. Hua, Y. Liu, L.E. Bernstein, K.E. Livingston, A. Perez, I. A. Shlyakhter, R. V. Rolando, R. Owen, P. Tanpaiboon, F. Lacbawan, Clinton R. Paden, Duncan MacCannell |
| EPI_ISL_1906490 | Michigan Department of Health and Human Services, Bureau of Laboratories | Michigan Department of Health and Human Services, Bureau of Laboratories | Blankenship HM, Riner D, Soehnlen MK |
| EPI_ISL_1906491 | Quest Diagnostics Incorporated | Centers for Disease Control and Prevention Division of Viral Diseases, Pathogen Discovery | Dakota Howard, Dhwani Batra, Peter W. Cook, Kara Moser, Adrian Paskey, Jason Caravas, Benjamin Rambo-Martin, Shatavia Morrison, Christopher Gulvick, Scott Sammons, Yvette Unoarumhi, Darlene Wagner, Matthew Schmerer, S. H. Rosenthal, A. Gerasimova, R. M. Kagan, B. Anderson, M. Hua, Y. Liu, L.E. Bernstein, K.E. Livingston, A. Perez, I. A. Shlyakhter, R. V. Rolando, R. Owen, P. Tanpaiboon, F. Lacbawan, Clinton R. Paden, Duncan MacCannell |
| EPI_ISL_1906492, EPI_ISL_1906493 | Michigan Department of Health and Human Services, Bureau of Laboratories | Michigan Department of Health and Human Services, Bureau of Laboratories | Blankenship HM, Riner D, Soehnlen MK |
| EPI_ISL_1906494, EPI_ISL_1906495 | Quest Diagnostics Incorporated | Centers for Disease Control and Prevention Division of Viral Diseases, Pathogen Discovery | Dakota Howard, Dhwani Batra, Peter W. Cook, Kara Moser, Adrian Paskey, Jason Caravas, Benjamin Rambo-Martin, Shatavia Morrison, Christopher Gulvick, Scott Sammons, Yvette Unoarumhi, Darlene Wagner, Matthew Schmerer, S. H. Rosenthal, A. Gerasimova, R. M. Kagan, B. Anderson, M. Hua, Y. Liu, L.E. Bernstein, K.E. Livingston, A. Perez, I. A. Shlyakhter, R. V. Rolando, R. Owen, P. Tanpaiboon, F. Lacbawan, Clinton R. Paden, Duncan MacCannell |
| EPI_ISL_1906496, EPI_ISL_1906497, EPI_ISL_1906498 | Michigan Department of Health and Human Services, Bureau of Laboratories | Michigan Department of Health and Human Services, Bureau of Laboratories | Blankenship HM, Riner D, Soehnlen MK |
| EPI_ISL_1906500 | Quest Diagnostics Incorporated | Centers for Disease Control and Prevention Division of Viral Diseases, Pathogen Discovery | Dakota Howard, Dhwani Batra, Peter W. Cook, Kara Moser, Adrian Paskey, Jason Caravas, Benjamin Rambo-Martin, Shatavia Morrison, Christopher Gulvick, Scott Sammons, Yvette Unoarumhi, Darlene Wagner, Matthew Schmerer, S. H. Rosenthal, A. Gerasimova, R. M. Kagan, B. Anderson, M. Hua, Y. Liu, L.E. Bernstein, K.E. Livingston, A. Perez, I. A. Shlyakhter, R. V. Rolando, R. Owen, P. Tanpaiboon, F. Lacbawan, Clinton R. Paden, Duncan MacCannell |
| EPI_ISL_1906501 | Michigan Department of Health and Human Services, Bureau of Laboratories | Michigan Department of Health and Human Services, Bureau of Laboratories | Blankenship HM, Riner D, Soehnlen MK |
| EPI_ISL_1906502 | Quest Diagnostics Incorporated | Centers for Disease Control and Prevention Division of Viral Diseases, Pathogen Discovery | Dakota Howard, Dhwani Batra, Peter W. Cook, Kara Moser, Adrian Paskey, Jason Caravas, Benjamin Rambo-Martin, Shatavia Morrison, Christopher Gulvick, Scott Sammons, Yvette Unoarumhi, Darlene Wagner, Matthew Schmerer, S. H. Rosenthal, A. Gerasimova, R. M. Kagan, B. Anderson, M. Hua, Y. Liu, L.E. Bernstein, K.E. Livingston, A. Perez, I. A. Shlyakhter, R. V. Rolando, R. Owen, P. Tanpaiboon, F. Lacbawan, Clinton R. Paden, Duncan MacCannell |
| EPI_ISL_1906503 | Michigan Department of Health and Human Services, Bureau of Laboratories | Michigan Department of Health and Human Services, Bureau of Laboratories | Blankenship HM, Riner D, Soehnlen MK |
| EPI_ISL_1906504 | Quest Diagnostics Incorporated | Centers for Disease Control and Prevention Division of Viral Diseases, Pathogen Discovery | Dakota Howard, Dhwani Batra, Peter W. Cook, Kara Moser, Adrian Paskey, Jason Caravas, Benjamin Rambo-Martin, Shatavia Morrison, Christopher Gulvick, Scott Sammons, Yvette Unoarumhi, Darlene Wagner, Matthew Schmerer, S. H. Rosenthal, A. Gerasimova, R. M. Kagan, B. Anderson, M. Hua, Y. Liu, L.E. Bernstein, K.E. Livingston, A. Perez, I. A. Shlyakhter, R. V. Rolando, R. Owen, P. Tanpaiboon, F. Lacbawan, Clinton R. Paden, Duncan MacCannell |
| EPI_ISL_1906505, EPI_ISL_1906506 | Michigan Department of Health and Human Services, Bureau of Laboratories | Michigan Department of Health and Human Services, Bureau of Laboratories | Blankenship HM, Riner D, Soehnlen MK |
| EPI_ISL_1906507 | Quest Diagnostics Incorporated | Centers for Disease Control and Prevention Division of Viral Diseases, Pathogen Discovery | Dakota Howard, Dhwani Batra, Peter W. Cook, Kara Moser, Adrian Paskey, Jason Caravas, Benjamin Rambo-Martin, Shatavia Morrison, Christopher Gulvick, Scott Sammons, Yvette Unoarumhi, Darlene Wagner, Matthew Schmerer, S. H. Rosenthal, A. Gerasimova, R. M. Kagan, B. Anderson, M. Hua, Y. Liu, L.E. Bernstein, K.E. Livingston, A. Perez, I. A. Shlyakhter, R. V. Rolando, R. Owen, P. Tanpaiboon, F. Lacbawan, Clinton R. Paden, Duncan MacCannell |
| EPI_ISL_1906508 | Michigan Department of Health and Human Services, Bureau of Laboratories | Michigan Department of Health and Human Services, Bureau of Laboratories | Blankenship HM, Riner D, Soehnlen MK |
| EPI_ISL_1906509 | Quest Diagnostics Incorporated | Centers for Disease Control and Prevention Division of Viral Diseases, Pathogen Discovery | Dakota Howard, Dhwani Batra, Peter W. Cook, Kara Moser, Adrian Paskey, Jason Caravas, Benjamin Rambo-Martin, Shatavia Morrison, Christopher Gulvick, Scott Sammons, Yvette Unoarumhi, Darlene Wagner, Matthew Schmerer, S. H. Rosenthal, A. Gerasimova, R. M. Kagan, B. Anderson, M. Hua, Y. Liu, L.E. Bernstein, K.E. Livingston, A. Perez, I. A. Shlyakhter, R. V. Rolando, R. Owen, P. Tanpaiboon, F. Lacbawan, Clinton R. Paden, Duncan MacCannell |
| EPI_ISL_1906510 | Michigan Department of Health and Human Services, Bureau of Laboratories | Michigan Department of Health and Human Services, Bureau of Laboratories | Blankenship HM, Riner D, Soehnlen MK |
| EPI_ISL_1906511 | Quest Diagnostics Incorporated | Centers for Disease Control and Prevention Division of Viral Diseases, Pathogen Discovery | Dakota Howard, Dhwani Batra, Peter W. Cook, Kara Moser, Adrian Paskey, Jason Caravas, Benjamin Rambo-Martin, Shatavia Morrison, Christopher Gulvick, Scott Sammons, Yvette Unoarumhi, Darlene Wagner, Matthew Schmerer, S. H. Rosenthal, A. Gerasimova, R. M. Kagan, B. Anderson, M. Hua, Y. Liu, L.E. Bernstein, K.E. Livingston, A. Perez, I. A. Shlyakhter, R. V. Rolando, R. Owen, P. Tanpaiboon, F. Lacbawan, Clinton R. Paden, Duncan MacCannell |
| EPI_ISL_1906512, EPI_ISL_1906513 | Michigan Department of Health and Human Services, Bureau of Laboratories | Michigan Department of Health and Human Services, Bureau of Laboratories | Blankenship HM, Riner D, Soehnlen MK |
| EPI_ISL_1906514 | Quest Diagnostics Incorporated | Centers for Disease Control and Prevention Division of Viral Diseases, Pathogen Discovery | Dakota Howard, Dhwani Batra, Peter W. Cook, Kara Moser, Adrian Paskey, Jason Caravas, Benjamin Rambo-Martin, Shatavia Morrison, Christopher Gulvick, Scott Sammons, Yvette Unoarumhi, Darlene Wagner, Matthew Schmerer, S. H. Rosenthal, A. Gerasimova, R. M. Kagan, B. Anderson, M. Hua, Y. Liu, L.E. Bernstein, K.E. Livingston, A. Perez, I. A. Shlyakhter, R. V. Rolando, R. Owen, P. Tanpaiboon, F. Lacbawan, Clinton R. Paden, Duncan MacCannell |
| EPI_ISL_1906515 | Michigan Department of Health and Human Services, Bureau of Laboratories | Michigan Department of Health and Human Services, Bureau of Laboratories | Blankenship HM, Riner D, Soehnlen MK |
| EPI_ISL_1906516 | Quest Diagnostics Incorporated | Centers for Disease Control and Prevention Division of Viral Diseases, Pathogen Discovery | Dakota Howard, Dhwani Batra, Peter W. Cook, Kara Moser, Adrian Paskey, Jason Caravas, Benjamin Rambo-Martin, Shatavia Morrison, Christopher Gulvick, Scott Sammons, Yvette Unoarumhi, Darlene Wagner, Matthew Schmerer, S. H. Rosenthal, A. Gerasimova, R. M. Kagan, B. Anderson, M. Hua, Y. Liu, L.E. Bernstein, K.E. Livingston, A. Perez, I. A. Shlyakhter, R. V. Rolando, R. Owen, P. Tanpaiboon, F. Lacbawan, Clinton R. Paden, Duncan MacCannell |
| EPI_ISL_1906517 | Michigan Department of Health and Human Services, Bureau of Laboratories | Michigan Department of Health and Human Services, Bureau of Laboratories | Blankenship HM, Riner D, Soehnlen MK |
| EPI_ISL_1906518 | Quest Diagnostics Incorporated | Centers for Disease Control and Prevention Division of Viral Diseases, Pathogen Discovery | Dakota Howard, Dhwani Batra, Peter W. Cook, Kara Moser, Adrian Paskey, Jason Caravas, Benjamin Rambo-Martin, Shatavia Morrison, Christopher Gulvick, Scott Sammons, Yvette Unoarumhi, Darlene Wagner, Matthew Schmerer, S. H. Rosenthal, A. Gerasimova, R. M. Kagan, B. Anderson, M. Hua, Y. Liu, L.E. Bernstein, K.E. Livingston, A. Perez, I. A. Shlyakhter, R. V. Rolando, R. Owen, P. Tanpaiboon, F. Lacbawan, Clinton R. Paden, Duncan MacCannell |
| EPI_ISL_1906519 | Michigan Department of Health and Human Services, Bureau of Laboratories | Michigan Department of Health and Human Services, Bureau of Laboratories | Blankenship HM, Riner D, Soehnlen MK |
| EPI_ISL_1906520 | Quest Diagnostics Incorporated | Centers for Disease Control and Prevention Division of Viral Diseases, Pathogen Discovery | Dakota Howard, Dhwani Batra, Peter W. Cook, Kara Moser, Adrian Paskey, Jason Caravas, Benjamin Rambo-Martin, Shatavia Morrison, Christopher Gulvick, Scott Sammons, Yvette Unoarumhi, Darlene Wagner, Matthew Schmerer, S. H. Rosenthal, A. Gerasimova, R. M. Kagan, B. Anderson, M. Hua, Y. Liu, L.E. Bernstein, K.E. Livingston, A. Perez, I. A. Shlyakhter, R. V. Rolando, R. Owen, P. Tanpaiboon, F. Lacbawan, Clinton R. Paden, Duncan MacCannell |
| EPI_ISL_1906521, EPI_ISL_1906522 | Michigan Department of Health and Human Services, Bureau of Laboratories | Michigan Department of Health and Human Services, Bureau of Laboratories | Blankenship HM, Riner D, Soehnlen MK |
| EPI_ISL_1906523 | Quest Diagnostics Incorporated | Centers for Disease Control and Prevention Division of Viral Diseases, Pathogen Discovery | Dakota Howard, Dhwani Batra, Peter W. Cook, Kara Moser, Adrian Paskey, Jason Caravas, Benjamin Rambo-Martin, Shatavia Morrison, Christopher Gulvick, Scott Sammons, Yvette Unoarumhi, Darlene Wagner, Matthew Schmerer, S. H. Rosenthal, A. Gerasimova, R. M. Kagan, B. Anderson, M. Hua, Y. Liu, L.E. Bernstein, K.E. Livingston, A. Perez, I. A. Shlyakhter, R. V. Rolando, R. Owen, P. Tanpaiboon, F. Lacbawan, Clinton R. Paden, Duncan MacCannell |
| EPI_ISL_1906524, EPI_ISL_1906525 | Michigan Department of Health and Human Services, Bureau of Laboratories | Michigan Department of Health and Human Services, Bureau of Laboratories | Blankenship HM, Riner D, Soehnlen MK |
| EPI_ISL_1906526, EPI_ISL_1906528, EPI_ISL_1906530 | Quest Diagnostics Incorporated | Centers for Disease Control and Prevention Division of Viral Diseases, Pathogen Discovery | Dakota Howard, Dhwani Batra, Peter W. Cook, Kara Moser, Adrian Paskey, Jason Caravas, Benjamin Rambo-Martin, Shatavia Morrison, Christopher Gulvick, Scott Sammons, Yvette Unoarumhi, Darlene Wagner, Matthew Schmerer, S. H. Rosenthal, A. Gerasimova, R. M. Kagan, B. Anderson, M. Hua, Y. Liu, L.E. Bernstein, K.E. Livingston, A. Perez, I. A. Shlyakhter, R. V. Rolando, R. Owen, P. Tanpaiboon, F. Lacbawan, Clinton R. Paden, Duncan MacCannell |

| | | | |
|---|---|---|---|
| EPI_ISL_1906531 | Michigan Department of Health and Human Services, Bureau of Laboratories | Michigan Department of Health and Human Services, Bureau of Laboratories | Blankenship HM, Riner D, Soehnlen MK |
| EPI_ISL_1906532 | Quest Diagnostics Incorporated | Centers for Disease Control and Prevention Division of Viral Diseases, Pathogen Discovery | Dakota Howard, Dhwani Batra, Peter W. Cook, Kara Moser, Adrian Paskey, Jason Caravas, Benjamin Rambo-Martin, Shatavia Morrison, Christopher Gulvick, Scott Sammons, Yvette Unoarumhi, Darlene Wagner, Matthew Schmerer, S. H. Rosenthal, A. Gerasimova, R. M. Kagan, B. Anderson, M. Hua, Y. Liu, L.E. Bernstein, K.E. Livingston, A. Perez, I. A. Shlyakhter, R. V. Rolando, R. Owen, P. Tanpaiboon, F. Lacbawan, Clinton R. Paden, Duncan MacCannell |
| EPI_ISL_1906533 | Michigan Department of Health and Human Services, Bureau of Laboratories | Michigan Department of Health and Human Services, Bureau of Laboratories | Blankenship HM, Riner D, Soehnlen MK |
| EPI_ISL_1906534, EPI_ISL_1906536 | Quest Diagnostics Incorporated | Centers for Disease Control and Prevention Division of Viral Diseases, Pathogen Discovery | Dakota Howard, Dhwani Batra, Peter W. Cook, Kara Moser, Adrian Paskey, Jason Caravas, Benjamin Rambo-Martin, Shatavia Morrison, Christopher Gulvick, Scott Sammons, Yvette Unoarumhi, Darlene Wagner, Matthew Schmerer, S. H. Rosenthal, A. Gerasimova, R. M. Kagan, B. Anderson, M. Hua, Y. Liu, L.E. Bernstein, K.E. Livingston, A. Perez, I. A. Shlyakhter, R. V. Rolando, R. Owen, P. Tanpaiboon, F. Lacbawan, Clinton R. Paden, Duncan MacCannell |
| EPI_ISL_1906537 | Michigan Department of Health and Human Services, Bureau of Laboratories | Michigan Department of Health and Human Services, Bureau of Laboratories | Blankenship HM, Riner D, Soehnlen MK |
| EPI_ISL_1906538 | Quest Diagnostics Incorporated | Centers for Disease Control and Prevention Division of Viral Diseases, Pathogen Discovery | Dakota Howard, Dhwani Batra, Peter W. Cook, Kara Moser, Adrian Paskey, Jason Caravas, Benjamin Rambo-Martin, Shatavia Morrison, Christopher Gulvick, Scott Sammons, Yvette Unoarumhi, Darlene Wagner, Matthew Schmerer, S. H. Rosenthal, A. Gerasimova, R. M. Kagan, B. Anderson, M. Hua, Y. Liu, L.E. Bernstein, K.E. Livingston, A. Perez, I. A. Shlyakhter, R. V. Rolando, R. Owen, P. Tanpaiboon, F. Lacbawan, Clinton R. Paden, Duncan MacCannell |
| EPI_ISL_1906539 | Michigan Department of Health and Human Services, Bureau of Laboratories | Michigan Department of Health and Human Services, Bureau of Laboratories | Blankenship HM, Riner D, Soehnlen MK |
| EPI_ISL_1906540 | Quest Diagnostics Incorporated | Centers for Disease Control and Prevention Division of Viral Diseases, Pathogen Discovery | Dakota Howard, Dhwani Batra, Peter W. Cook, Kara Moser, Adrian Paskey, Jason Caravas, Benjamin Rambo-Martin, Shatavia Morrison, Christopher Gulvick, Scott Sammons, Yvette Unoarumhi, Darlene Wagner, Matthew Schmerer, S. H. Rosenthal, A. Gerasimova, R. M. Kagan, B. Anderson, M. Hua, Y. Liu, L.E. Bernstein, K.E. Livingston, A. Perez, I. A. Shlyakhter, R. V. Rolando, R. Owen, P. Tanpaiboon, F. Lacbawan, Clinton R. Paden, Duncan MacCannell |
| EPI_ISL_1906541 | Michigan Department of Health and Human Services, Bureau of Laboratories | Michigan Department of Health and Human Services, Bureau of Laboratories | Blankenship HM, Riner D, Soehnlen MK |
| EPI_ISL_1906542 | Quest Diagnostics Incorporated | Centers for Disease Control and Prevention Division of Viral Diseases, Pathogen Discovery | Dakota Howard, Dhwani Batra, Peter W. Cook, Kara Moser, Adrian Paskey, Jason Caravas, Benjamin Rambo-Martin, Shatavia Morrison, Christopher Gulvick, Scott Sammons, Yvette Unoarumhi, Darlene Wagner, Matthew Schmerer, S. H. Rosenthal, A. Gerasimova, R. M. Kagan, B. Anderson, M. Hua, Y. Liu, L.E. Bernstein, K.E. Livingston, A. Perez, I. A. Shlyakhter, R. V. Rolando, R. Owen, P. Tanpaiboon, F. Lacbawan, Clinton R. Paden, Duncan MacCannell |
| EPI_ISL_1906543, EPI_ISL_1906544 | Michigan Department of Health and Human Services, Bureau of Laboratories | Michigan Department of Health and Human Services, Bureau of Laboratories | Blankenship HM, Riner D, Soehnlen MK |
| EPI_ISL_1906545, EPI_ISL_1906546 | Quest Diagnostics Incorporated | Centers for Disease Control and Prevention Division of Viral Diseases, Pathogen Discovery | Dakota Howard, Dhwani Batra, Peter W. Cook, Kara Moser, Adrian Paskey, Jason Caravas, Benjamin Rambo-Martin, Shatavia Morrison, Christopher Gulvick, Scott Sammons, Yvette Unoarumhi, Darlene Wagner, Matthew Schmerer, S. H. Rosenthal, A. Gerasimova, R. M. Kagan, B. Anderson, M. Hua, Y. Liu, L.E. Bernstein, K.E. Livingston, A. Perez, I. A. Shlyakhter, R. V. Rolando, R. Owen, P. Tanpaiboon, F. Lacbawan, Clinton R. Paden, Duncan MacCannell |
| EPI_ISL_1906547, EPI_ISL_1906548 | Michigan Department of Health and Human Services, Bureau of Laboratories | Michigan Department of Health and Human Services, Bureau of Laboratories | Blankenship HM, Riner D, Soehnlen MK |
| EPI_ISL_1906549 | Quest Diagnostics Incorporated | Centers for Disease Control and Prevention Division of Viral Diseases, Pathogen Discovery | Dakota Howard, Dhwani Batra, Peter W. Cook, Kara Moser, Adrian Paskey, Jason Caravas, Benjamin Rambo-Martin, Shatavia Morrison, Christopher Gulvick, Scott Sammons, Yvette Unoarumhi, Darlene Wagner, Matthew Schmerer, S. H. Rosenthal, A. Gerasimova, R. M. Kagan, B. Anderson, M. Hua, Y. Liu, L.E. Bernstein, K.E. Livingston, A. Perez, I. A. Shlyakhter, R. V. Rolando, R. Owen, P. Tanpaiboon, F. Lacbawan, Clinton R. Paden, Duncan MacCannell |
| EPI_ISL_1906550 | Michigan Department of Health and Human Services, Bureau of Laboratories | Michigan Department of Health and Human Services, Bureau of Laboratories | Blankenship HM, Riner D, Soehnlen MK |
| EPI_ISL_1906551 | Quest Diagnostics Incorporated | Centers for Disease Control and Prevention Division of Viral Diseases, Pathogen Discovery | Dakota Howard, Dhwani Batra, Peter W. Cook, Kara Moser, Adrian Paskey, Jason Caravas, Benjamin Rambo-Martin, Shatavia Morrison, Christopher Gulvick, Scott Sammons, Yvette Unoarumhi, Darlene Wagner, Matthew Schmerer, S. H. Rosenthal, A. Gerasimova, R. M. Kagan, B. Anderson, M. Hua, Y. Liu, L.E. Bernstein, K.E. Livingston, A. Perez, I. A. Shlyakhter, R. V. Rolando, R. Owen, P. Tanpaiboon, F. Lacbawan, Clinton R. Paden, Duncan MacCannell |
| EPI_ISL_1906552 | Michigan Department of Health and Human Services, Bureau of Laboratories | Michigan Department of Health and Human Services, Bureau of Laboratories | Blankenship HM, Riner D, Soehnlen MK |
| EPI_ISL_1906553 | Quest Diagnostics Incorporated | Centers for Disease Control and Prevention Division of Viral Diseases, Pathogen Discovery | Dakota Howard, Dhwani Batra, Peter W. Cook, Kara Moser, Adrian Paskey, Jason Caravas, Benjamin Rambo-Martin, Shatavia Morrison, Christopher Gulvick, Scott Sammons, Yvette Unoarumhi, Darlene Wagner, Matthew Schmerer, S. H. Rosenthal, A. Gerasimova, R. M. Kagan, B. Anderson, M. Hua, Y. Liu, L.E. Bernstein, K.E. Livingston, A. Perez, I. A. Shlyakhter, R. V. Rolando, R. Owen, P. Tanpaiboon, F. Lacbawan, Clinton R. Paden, Duncan MacCannell |
| EPI_ISL_1906554 | Michigan Department of Health and Human Services, Bureau of Laboratories | Michigan Department of Health and Human Services, Bureau of Laboratories | Blankenship HM, Riner D, Soehnlen MK |
| EPI_ISL_1906555 | Quest Diagnostics Incorporated | Centers for Disease Control and Prevention Division of Viral Diseases, Pathogen Discovery | Dakota Howard, Dhwani Batra, Peter W. Cook, Kara Moser, Adrian Paskey, Jason Caravas, Benjamin Rambo-Martin, Shatavia Morrison, Christopher Gulvick, Scott Sammons, Yvette Unoarumhi, Darlene Wagner, Matthew Schmerer, S. H. Rosenthal, A. Gerasimova, R. M. Kagan, B. Anderson, M. Hua, Y. Liu, L.E. Bernstein, K.E. Livingston, A. Perez, I. A. Shlyakhter, R. V. Rolando, R. Owen, P. Tanpaiboon, F. Lacbawan, Clinton R. Paden, Duncan MacCannell |
| EPI_ISL_1906556 | Michigan Department of Health and Human Services, Bureau of Laboratories | Michigan Department of Health and Human Services, Bureau of Laboratories | Blankenship HM, Riner D, Soehnlen MK |
| EPI_ISL_1906557 | Quest Diagnostics Incorporated | Centers for Disease Control and Prevention Division of Viral Diseases, Pathogen Discovery | Dakota Howard, Dhwani Batra, Peter W. Cook, Kara Moser, Adrian Paskey, Jason Caravas, Benjamin Rambo-Martin, Shatavia Morrison, Christopher Gulvick, Scott Sammons, Yvette Unoarumhi, Darlene Wagner, Matthew Schmerer, S. H. Rosenthal, A. Gerasimova, R. M. Kagan, B. Anderson, M. Hua, Y. Liu, L.E. Bernstein, K.E. Livingston, A. Perez, I. A. Shlyakhter, R. V. Rolando, R. Owen, P. Tanpaiboon, F. Lacbawan, Clinton R. Paden, Duncan MacCannell |
| EPI_ISL_1906558, EPI_ISL_1906559 | Michigan Department of Health and Human Services, Bureau of Laboratories | Michigan Department of Health and Human Services, Bureau of Laboratories | Blankenship HM, Riner D, Soehnlen MK |
| EPI_ISL_1906560 | Quest Diagnostics Incorporated | Centers for Disease Control and Prevention Division of Viral Diseases, Pathogen Discovery | Dakota Howard, Dhwani Batra, Peter W. Cook, Kara Moser, Adrian Paskey, Jason Caravas, Benjamin Rambo-Martin, Shatavia Morrison, Christopher Gulvick, Scott Sammons, Yvette Unoarumhi, Darlene Wagner, Matthew Schmerer, S. H. Rosenthal, A. Gerasimova, R. M. Kagan, B. Anderson, M. Hua, Y. Liu, L.E. Bernstein, K.E. Livingston, A. Perez, I. A. Shlyakhter, R. V. Rolando, R. Owen, P. Tanpaiboon, F. Lacbawan, Clinton R. Paden, Duncan MacCannell |
| EPI_ISL_1906561 | Michigan Department of Health and Human Services, Bureau of Laboratories | Michigan Department of Health and Human Services, Bureau of Laboratories | Blankenship HM, Riner D, Soehnlen MK |
| EPI_ISL_1906562 | Quest Diagnostics Incorporated | Centers for Disease Control and Prevention Division of Viral Diseases, Pathogen Discovery | Dakota Howard, Dhwani Batra, Peter W. Cook, Kara Moser, Adrian Paskey, Jason Caravas, Benjamin Rambo-Martin, Shatavia Morrison, Christopher Gulvick, Scott Sammons, Yvette Unoarumhi, Darlene Wagner, Matthew Schmerer, S. H. Rosenthal, A. Gerasimova, R. M. Kagan, B. Anderson, M. Hua, Y. Liu, L.E. Bernstein, K.E. Livingston, A. Perez, I. A. Shlyakhter, R. V. Rolando, R. Owen, P. Tanpaiboon, F. Lacbawan, Clinton R. Paden, Duncan MacCannell |
| EPI_ISL_1906563 | Michigan Department of Health and Human Services, Bureau of Laboratories | Michigan Department of Health and Human Services, Bureau of Laboratories | Blankenship HM, Riner D, Soehnlen MK |
| EPI_ISL_1906564 | Quest Diagnostics Incorporated | Centers for Disease Control and Prevention Division of Viral Diseases, Pathogen Discovery | Dakota Howard, Dhwani Batra, Peter W. Cook, Kara Moser, Adrian Paskey, Jason Caravas, Benjamin Rambo-Martin, Shatavia Morrison, Christopher Gulvick, Scott Sammons, Yvette Unoarumhi, Darlene Wagner, Matthew Schmerer, S. H. Rosenthal, A. Gerasimova, R. M. Kagan, B. Anderson, M. Hua, Y. Liu, L.E. Bernstein, K.E. Livingston, A. Perez, I. A. Shlyakhter, R. V. Rolando, R. Owen, P. Tanpaiboon, F. Lacbawan, Clinton R. Paden, Duncan MacCannell |
| EPI_ISL_1906566 | Michigan Department of Health and Human Services, Bureau of Laboratories | Michigan Department of Health and Human Services, Bureau of Laboratories | Blankenship HM, Riner D, Soehnlen MK |
| EPI_ISL_1906567 | Quest Diagnostics Incorporated | Centers for Disease Control and Prevention Division of Viral Diseases, Pathogen Discovery | Dakota Howard, Dhwani Batra, Peter W. Cook, Kara Moser, Adrian Paskey, Jason Caravas, Benjamin Rambo-Martin, Shatavia Morrison, Christopher Gulvick, Scott Sammons, Yvette Unoarumhi, Darlene Wagner, Matthew Schmerer, S. H. Rosenthal, A. Gerasimova, R. M. Kagan, B. Anderson, M. Hua, Y. Liu, L.E. Bernstein, K.E. Livingston, A. Perez, I. A. Shlyakhter, R. V. Rolando, R. Owen, P. Tanpaiboon, F. Lacbawan, Clinton R. Paden, Duncan MacCannell |
| EPI_ISL_1906568, EPI_ISL_1906569, EPI_ISL_1906570 | Michigan Department of Health and Human Services, Bureau of Laboratories | Michigan Department of Health and Human Services, Bureau of Laboratories | Blankenship HM, Riner D, Soehnlen MK |
| EPI_ISL_1906571 | Quest Diagnostics Incorporated | Centers for Disease Control and Prevention Division of Viral Diseases, Pathogen Discovery | Dakota Howard, Dhwani Batra, Peter W. Cook, Kara Moser, Adrian Paskey, Jason Caravas, Benjamin Rambo-Martin, Shatavia Morrison, Christopher Gulvick, Scott Sammons, Yvette Unoarumhi, Darlene Wagner, Matthew Schmerer, S. H. Rosenthal, A. Gerasimova, R. M. Kagan, B. Anderson, M. Hua, Y. Liu, L.E. Bernstein, K.E. Livingston, A. Perez, I. A. Shlyakhter, R. V. Rolando, R. Owen, P. Tanpaiboon, F. Lacbawan, Clinton R. Paden, Duncan MacCannell |
| EPI_ISL_1906572 | Michigan Department of Health and Human Services, Bureau of Laboratories | Michigan Department of Health and Human Services, Bureau of Laboratories | Blankenship HM, Riner D, Soehnlen MK |
| EPI_ISL_1906573 | Quest Diagnostics Incorporated | Centers for Disease Control and Prevention Division of Viral Diseases, Pathogen Discovery | Dakota Howard, Dhwani Batra, Peter W. Cook, Kara Moser, Adrian Paskey, Jason Caravas, Benjamin Rambo-Martin, Shatavia Morrison, Christopher Gulvick, Scott Sammons, Yvette Unoarumhi, Darlene Wagner, Matthew Schmerer, S. H. Rosenthal, A. Gerasimova, R. M. Kagan, B. Anderson, M. Hua, Y. Liu, L.E. Bernstein, K.E. Livingston, A. Perez, I. A. Shlyakhter, R. V. Rolando, R. Owen, P. Tanpaiboon, F. Lacbawan, Clinton R. Paden, Duncan MacCannell |
| EPI_ISL_1906574 | Michigan Department of Health and Human Services, Bureau of Laboratories | Michigan Department of Health and Human Services, Bureau of Laboratories | Blankenship HM, Riner D, Soehnlen MK |
| EPI_ISL_1906575 | Quest Diagnostics Incorporated | Centers for Disease Control and Prevention Division of Viral Diseases, Pathogen Discovery | Dakota Howard, Dhwani Batra, Peter W. Cook, Kara Moser, Adrian Paskey, Jason Caravas, Benjamin Rambo-Martin, Shatavia Morrison, Christopher Gulvick, Scott Sammons, Yvette Unoarumhi, Darlene Wagner, Matthew Schmerer, S. H. Rosenthal, A. Gerasimova, R. M. Kagan, B. Anderson, M. Hua, Y. Liu, L.E. Bernstein, K.E. Livingston, A. Perez, I. A. Shlyakhter, R. V. Rolando, R. Owen, P. Tanpaiboon, F. Lacbawan, Clinton R. Paden, Duncan MacCannell |
| EPI_ISL_1906576 | Michigan Department of Health and Human Services, Bureau of Laboratories | Michigan Department of Health and Human Services, Bureau of Laboratories | Blankenship HM, Riner D, Soehnlen MK |

| | | | |
|---|---|---|---|
| EPI_ISL_1906577 | Quest Diagnostics Incorporated | Centers for Disease Control and Prevention Division of Viral Diseases, Pathogen Discovery | Dakota Howard, Dhwani Batra, Peter W. Cook, Kara Moser, Adrian Paskey, Jason Caravas, Benjamin Rambo-Martin, Shatavia Morrison, Christopher Gulvick, Scott Sammons, Yvette Unoarumhi, Darlene Wagner, Matthew Schmerer, S. H. Rosenthal, A. Gerasimova, R. M. Kagan, B. Anderson, M. Hua, Y. Liu, L.E. Bernstein, K.E. Livingston, A. Perez, I. A. Shlyakhter, R. V. Rolando, R. Owen, P. Tanpaiboon, F. Lacbawan, Clinton R. Paden, Duncan MacCannell |
| EPI_ISL_1906578, EPI_ISL_1906579 | Michigan Department of Health and Human Services, Bureau of Laboratories | Michigan Department of Health and Human Services, Bureau of Laboratories | Blankenship HM, Riner D, Soehnlen MK |
| EPI_ISL_1906580 | Quest Diagnostics Incorporated | Centers for Disease Control and Prevention Division of Viral Diseases, Pathogen Discovery | Dakota Howard, Dhwani Batra, Peter W. Cook, Kara Moser, Adrian Paskey, Jason Caravas, Benjamin Rambo-Martin, Shatavia Morrison, Christopher Gulvick, Scott Sammons, Yvette Unoarumhi, Darlene Wagner, Matthew Schmerer, S. H. Rosenthal, A. Gerasimova, R. M. Kagan, B. Anderson, M. Hua, Y. Liu, L.E. Bernstein, K.E. Livingston, A. Perez, I. A. Shlyakhter, R. V. Rolando, R. Owen, P. Tanpaiboon, F. Lacbawan, Clinton R. Paden, Duncan MacCannell |
| EPI_ISL_1906581 | Michigan Department of Health and Human Services, Bureau of Laboratories | Michigan Department of Health and Human Services, Bureau of Laboratories | Blankenship HM, Riner D, Soehnlen MK |
| EPI_ISL_1906582 | Quest Diagnostics Incorporated | Centers for Disease Control and Prevention Division of Viral Diseases, Pathogen Discovery | Dakota Howard, Dhwani Batra, Peter W. Cook, Kara Moser, Adrian Paskey, Jason Caravas, Benjamin Rambo-Martin, Shatavia Morrison, Christopher Gulvick, Scott Sammons, Yvette Unoarumhi, Darlene Wagner, Matthew Schmerer, S. H. Rosenthal, A. Gerasimova, R. M. Kagan, B. Anderson, M. Hua, Y. Liu, L.E. Bernstein, K.E. Livingston, A. Perez, I. A. Shlyakhter, R. V. Rolando, R. Owen, P. Tanpaiboon, F. Lacbawan, Clinton R. Paden, Duncan MacCannell |
| EPI_ISL_1906583 | Michigan Department of Health and Human Services, Bureau of Laboratories | Michigan Department of Health and Human Services, Bureau of Laboratories | Blankenship HM, Riner D, Soehnlen MK |
| EPI_ISL_1906584 | Quest Diagnostics Incorporated | Centers for Disease Control and Prevention Division of Viral Diseases, Pathogen Discovery | Dakota Howard, Dhwani Batra, Peter W. Cook, Kara Moser, Adrian Paskey, Jason Caravas, Benjamin Rambo-Martin, Shatavia Morrison, Christopher Gulvick, Scott Sammons, Yvette Unoarumhi, Darlene Wagner, Matthew Schmerer, S. H. Rosenthal, A. Gerasimova, R. M. Kagan, B. Anderson, M. Hua, Y. Liu, L.E. Bernstein, K.E. Livingston, A. Perez, I. A. Shlyakhter, R. V. Rolando, R. Owen, P. Tanpaiboon, F. Lacbawan, Clinton R. Paden, Duncan MacCannell |
| EPI_ISL_1906585 | Michigan Department of Health and Human Services, Bureau of Laboratories | Michigan Department of Health and Human Services, Bureau of Laboratories | Blankenship HM, Riner D, Soehnlen MK |
| EPI_ISL_1906586 | Quest Diagnostics Incorporated | Centers for Disease Control and Prevention Division of Viral Diseases, Pathogen Discovery | Dakota Howard, Dhwani Batra, Peter W. Cook, Kara Moser, Adrian Paskey, Jason Caravas, Benjamin Rambo-Martin, Shatavia Morrison, Christopher Gulvick, Scott Sammons, Yvette Unoarumhi, Darlene Wagner, Matthew Schmerer, S. H. Rosenthal, A. Gerasimova, R. M. Kagan, B. Anderson, M. Hua, Y. Liu, L.E. Bernstein, K.E. Livingston, A. Perez, I. A. Shlyakhter, R. V. Rolando, R. Owen, P. Tanpaiboon, F. Lacbawan, Clinton R. Paden, Duncan MacCannell |
| EPI_ISL_1906587, EPI_ISL_1906588 | Michigan Department of Health and Human Services, Bureau of Laboratories | Michigan Department of Health and Human Services, Bureau of Laboratories | Blankenship HM, Riner D, Soehnlen MK |
| EPI_ISL_1906589 | Quest Diagnostics Incorporated | Centers for Disease Control and Prevention Division of Viral Diseases, Pathogen Discovery | Dakota Howard, Dhwani Batra, Peter W. Cook, Kara Moser, Adrian Paskey, Jason Caravas, Benjamin Rambo-Martin, Shatavia Morrison, Christopher Gulvick, Scott Sammons, Yvette Unoarumhi, Darlene Wagner, Matthew Schmerer, S. H. Rosenthal, A. Gerasimova, R. M. Kagan, B. Anderson, M. Hua, Y. Liu, L.E. Bernstein, K.E. Livingston, A. Perez, I. A. Shlyakhter, R. V. Rolando, R. Owen, P. Tanpaiboon, F. Lacbawan, Clinton R. Paden, Duncan MacCannell |
| EPI_ISL_1906590 | Michigan Department of Health and Human Services, Bureau of Laboratories | Michigan Department of Health and Human Services, Bureau of Laboratories | Blankenship HM, Riner D, Soehnlen MK |
| EPI_ISL_1906591 | Quest Diagnostics Incorporated | Centers for Disease Control and Prevention Division of Viral Diseases, Pathogen Discovery | Dakota Howard, Dhwani Batra, Peter W. Cook, Kara Moser, Adrian Paskey, Jason Caravas, Benjamin Rambo-Martin, Shatavia Morrison, Christopher Gulvick, Scott Sammons, Yvette Unoarumhi, Darlene Wagner, Matthew Schmerer, S. H. Rosenthal, A. Gerasimova, R. M. Kagan, B. Anderson, M. Hua, Y. Liu, L.E. Bernstein, K.E. Livingston, A. Perez, I. A. Shlyakhter, R. V. Rolando, R. Owen, P. Tanpaiboon, F. Lacbawan, Clinton R. Paden, Duncan MacCannell |
| EPI_ISL_1906592 | Michigan Department of Health and Human Services, Bureau of Laboratories | Michigan Department of Health and Human Services, Bureau of Laboratories | Blankenship HM, Riner D, Soehnlen MK |
| EPI_ISL_1906593 | Quest Diagnostics Incorporated | Centers for Disease Control and Prevention Division of Viral Diseases, Pathogen Discovery | Dakota Howard, Dhwani Batra, Peter W. Cook, Kara Moser, Adrian Paskey, Jason Caravas, Benjamin Rambo-Martin, Shatavia Morrison, Christopher Gulvick, Scott Sammons, Yvette Unoarumhi, Darlene Wagner, Matthew Schmerer, S. H. Rosenthal, A. Gerasimova, R. M. Kagan, B. Anderson, M. Hua, Y. Liu, L.E. Bernstein, K.E. Livingston, A. Perez, I. A. Shlyakhter, R. V. Rolando, R. Owen, P. Tanpaiboon, F. Lacbawan, Clinton R. Paden, Duncan MacCannell |
| EPI_ISL_1906594 | Michigan Department of Health and Human Services, Bureau of Laboratories | Michigan Department of Health and Human Services, Bureau of Laboratories | Blankenship HM, Riner D, Soehnlen MK |
| EPI_ISL_1906595 | Quest Diagnostics Incorporated | Centers for Disease Control and Prevention Division of Viral Diseases, Pathogen Discovery | Dakota Howard, Dhwani Batra, Peter W. Cook, Kara Moser, Adrian Paskey, Jason Caravas, Benjamin Rambo-Martin, Shatavia Morrison, Christopher Gulvick, Scott Sammons, Yvette Unoarumhi, Darlene Wagner, Matthew Schmerer, S. H. Rosenthal, A. Gerasimova, R. M. Kagan, B. Anderson, M. Hua, Y. Liu, L.E. Bernstein, K.E. Livingston, A. Perez, I. A. Shlyakhter, R. V. Rolando, R. Owen, P. Tanpaiboon, F. Lacbawan, Clinton R. Paden, Duncan MacCannell |
| EPI_ISL_1906596, EPI_ISL_1906597 | Michigan Department of Health and Human Services, Bureau of Laboratories | Michigan Department of Health and Human Services, Bureau of Laboratories | Blankenship HM, Riner D, Soehnlen MK |
| EPI_ISL_1906598 | Quest Diagnostics Incorporated | Centers for Disease Control and Prevention Division of Viral Diseases, Pathogen Discovery | Dakota Howard, Dhwani Batra, Peter W. Cook, Kara Moser, Adrian Paskey, Jason Caravas, Benjamin Rambo-Martin, Shatavia Morrison, Christopher Gulvick, Scott Sammons, Yvette Unoarumhi, Darlene Wagner, Matthew Schmerer, S. H. Rosenthal, A. Gerasimova, R. M. Kagan, B. Anderson, M. Hua, Y. Liu, L.E. Bernstein, K.E. Livingston, A. Perez, I. A. Shlyakhter, R. V. Rolando, R. Owen, P. Tanpaiboon, F. Lacbawan, Clinton R. Paden, Duncan MacCannell |
| EPI_ISL_1906599 | Michigan Department of Health and Human Services, Bureau of Laboratories | Michigan Department of Health and Human Services, Bureau of Laboratories | Blankenship HM, Riner D, Soehnlen MK |
| EPI_ISL_1906600 | Quest Diagnostics Incorporated | Centers for Disease Control and Prevention Division of Viral Diseases, Pathogen Discovery | Dakota Howard, Dhwani Batra, Peter W. Cook, Kara Moser, Adrian Paskey, Jason Caravas, Benjamin Rambo-Martin, Shatavia Morrison, Christopher Gulvick, Scott Sammons, Yvette Unoarumhi, Darlene Wagner, Matthew Schmerer, S. H. Rosenthal, A. Gerasimova, R. M. Kagan, B. Anderson, M. Hua, Y. Liu, L.E. Bernstein, K.E. Livingston, A. Perez, I. A. Shlyakhter, R. V. Rolando, R. Owen, P. Tanpaiboon, F. Lacbawan, Clinton R. Paden, Duncan MacCannell |
| EPI_ISL_1906601 | Michigan Department of Health and Human Services, Bureau of Laboratories | Michigan Department of Health and Human Services, Bureau of Laboratories | Blankenship HM, Riner D, Soehnlen MK |
| EPI_ISL_1906602 | Quest Diagnostics Incorporated | Centers for Disease Control and Prevention Division of Viral Diseases, Pathogen Discovery | Dakota Howard, Dhwani Batra, Peter W. Cook, Kara Moser, Adrian Paskey, Jason Caravas, Benjamin Rambo-Martin, Shatavia Morrison, Christopher Gulvick, Scott Sammons, Yvette Unoarumhi, Darlene Wagner, Matthew Schmerer, S. H. Rosenthal, A. Gerasimova, R. M. Kagan, B. Anderson, M. Hua, Y. Liu, L.E. Bernstein, K.E. Livingston, A. Perez, I. A. Shlyakhter, R. V. Rolando, R. Owen, P. Tanpaiboon, F. Lacbawan, Clinton R. Paden, Duncan MacCannell |
| EPI_ISL_1906603 | Michigan Department of Health and Human Services, Bureau of Laboratories | Michigan Department of Health and Human Services, Bureau of Laboratories | Blankenship HM, Riner D, Soehnlen MK |
| EPI_ISL_1906604 | Quest Diagnostics Incorporated | Centers for Disease Control and Prevention Division of Viral Diseases, Pathogen Discovery | Dakota Howard, Dhwani Batra, Peter W. Cook, Kara Moser, Adrian Paskey, Jason Caravas, Benjamin Rambo-Martin, Shatavia Morrison, Christopher Gulvick, Scott Sammons, Yvette Unoarumhi, Darlene Wagner, Matthew Schmerer, S. H. Rosenthal, A. Gerasimova, R. M. Kagan, B. Anderson, M. Hua, Y. Liu, L.E. Bernstein, K.E. Livingston, A. Perez, I. A. Shlyakhter, R. V. Rolando, R. Owen, P. Tanpaiboon, F. Lacbawan, Clinton R. Paden, Duncan MacCannell |
| EPI_ISL_1906605 | Michigan Department of Health and Human Services, Bureau of Laboratories | Michigan Department of Health and Human Services, Bureau of Laboratories | Blankenship HM, Riner D, Soehnlen MK |
| EPI_ISL_1906606 | Quest Diagnostics Incorporated | Centers for Disease Control and Prevention Division of Viral Diseases, Pathogen Discovery | Dakota Howard, Dhwani Batra, Peter W. Cook, Kara Moser, Adrian Paskey, Jason Caravas, Benjamin Rambo-Martin, Shatavia Morrison, Christopher Gulvick, Scott Sammons, Yvette Unoarumhi, Darlene Wagner, Matthew Schmerer, S. H. Rosenthal, A. Gerasimova, R. M. Kagan, B. Anderson, M. Hua, Y. Liu, L.E. Bernstein, K.E. Livingston, A. Perez, I. A. Shlyakhter, R. V. Rolando, R. Owen, P. Tanpaiboon, F. Lacbawan, Clinton R. Paden, Duncan MacCannell |
| EPI_ISL_1906607, EPI_ISL_1906608 | Michigan Department of Health and Human Services, Bureau of Laboratories | Michigan Department of Health and Human Services, Bureau of Laboratories | Blankenship HM, Riner D, Soehnlen MK |
| EPI_ISL_1906609 | Quest Diagnostics Incorporated | Centers for Disease Control and Prevention Division of Viral Diseases, Pathogen Discovery | Dakota Howard, Dhwani Batra, Peter W. Cook, Kara Moser, Adrian Paskey, Jason Caravas, Benjamin Rambo-Martin, Shatavia Morrison, Christopher Gulvick, Scott Sammons, Yvette Unoarumhi, Darlene Wagner, Matthew Schmerer, S. H. Rosenthal, A. Gerasimova, R. M. Kagan, B. Anderson, M. Hua, Y. Liu, L.E. Bernstein, K.E. Livingston, A. Perez, I. A. Shlyakhter, R. V. Rolando, R. Owen, P. Tanpaiboon, F. Lacbawan, Clinton R. Paden, Duncan MacCannell |
| EPI_ISL_1906610 | Michigan Department of Health and Human Services, Bureau of Laboratories | Michigan Department of Health and Human Services, Bureau of Laboratories | Blankenship HM, Riner D, Soehnlen MK |
| EPI_ISL_1906611 | Quest Diagnostics Incorporated | Centers for Disease Control and Prevention Division of Viral Diseases, Pathogen Discovery | Dakota Howard, Dhwani Batra, Peter W. Cook, Kara Moser, Adrian Paskey, Jason Caravas, Benjamin Rambo-Martin, Shatavia Morrison, Christopher Gulvick, Scott Sammons, Yvette Unoarumhi, Darlene Wagner, Matthew Schmerer, S. H. Rosenthal, A. Gerasimova, R. M. Kagan, B. Anderson, M. Hua, Y. Liu, L.E. Bernstein, K.E. Livingston, A. Perez, I. A. Shlyakhter, R. V. Rolando, R. Owen, P. Tanpaiboon, F. Lacbawan, Clinton R. Paden, Duncan MacCannell |
| EPI_ISL_1906612 | Michigan Department of Health and Human Services, Bureau of Laboratories | Michigan Department of Health and Human Services, Bureau of Laboratories | Blankenship HM, Riner D, Soehnlen MK |
| EPI_ISL_1906613 | Quest Diagnostics Incorporated | Centers for Disease Control and Prevention Division of Viral Diseases, Pathogen Discovery | Dakota Howard, Dhwani Batra, Peter W. Cook, Kara Moser, Adrian Paskey, Jason Caravas, Benjamin Rambo-Martin, Shatavia Morrison, Christopher Gulvick, Scott Sammons, Yvette Unoarumhi, Darlene Wagner, Matthew Schmerer, S. H. Rosenthal, A. Gerasimova, R. M. Kagan, B. Anderson, M. Hua, Y. Liu, L.E. Bernstein, K.E. Livingston, A. Perez, I. A. Shlyakhter, R. V. Rolando, R. Owen, P. Tanpaiboon, F. Lacbawan, Clinton R. Paden, Duncan MacCannell |
| EPI_ISL_1906614 | Michigan Department of Health and Human Services, Bureau of Laboratories | Michigan Department of Health and Human Services, Bureau of Laboratories | Blankenship HM, Riner D, Soehnlen MK |
| EPI_ISL_1906615 | Quest Diagnostics Incorporated | Centers for Disease Control and Prevention Division of Viral Diseases, Pathogen Discovery | Dakota Howard, Dhwani Batra, Peter W. Cook, Kara Moser, Adrian Paskey, Jason Caravas, Benjamin Rambo-Martin, Shatavia Morrison, Christopher Gulvick, Scott Sammons, Yvette Unoarumhi, Darlene Wagner, Matthew Schmerer, S. H. Rosenthal, A. Gerasimova, R. M. Kagan, B. Anderson, M. Hua, Y. Liu, L.E. Bernstein, K.E. Livingston, A. Perez, I. A. Shlyakhter, R. V. Rolando, R. Owen, P. Tanpaiboon, F. Lacbawan, Clinton R. Paden, Duncan MacCannell |
| EPI_ISL_1906616 | Michigan Department of Health and Human Services, Bureau of Laboratories | Michigan Department of Health and Human Services, Bureau of Laboratories | Blankenship HM, Riner D, Soehnlen MK |
| EPI_ISL_1906617 | Quest Diagnostics Incorporated | Centers for Disease Control and Prevention Division of Viral Diseases, Pathogen Discovery | Dakota Howard, Dhwani Batra, Peter W. Cook, Kara Moser, Adrian Paskey, Jason Caravas, Benjamin Rambo-Martin, Shatavia Morrison, Christopher Gulvick, Scott Sammons, Yvette Unoarumhi, Darlene Wagner, Matthew Schmerer, S. H. Rosenthal, A. Gerasimova, R. M. Kagan, B. Anderson, M. Hua, Y. Liu, L.E. Bernstein, K.E. Livingston, A. Perez, I. A. Shlyakhter, R. V. Rolando, R. Owen, P. Tanpaiboon, F. Lacbawan, Clinton R. Paden, Duncan MacCannell |

| | | | |
|---|---|---|---|
| EPI_ISL_1906618 | Michigan Department of Health and Human Services, Bureau of Laboratories | Michigan Department of Health and Human Services, Bureau of Laboratories | Blankenship HM, Riner D, Soehnlen MK |
| EPI_ISL_1906619 | Quest Diagnostics Incorporated | Centers for Disease Control and Prevention Division of Viral Diseases, Pathogen Discovery | Dakota Howard, Dhwani Batra, Peter W. Cook, Kara Moser, Adrian Paskey, Jason Caravas, Benjamin Rambo-Martin, Shatavia Morrison, Christopher Gulvick, Scott Sammons, Yvette Unoarumhi, Darlene Wagner, Matthew Schmerer, S. H. Rosenthal, A. Gerasimova, R. M. Kagan, B. Anderson, M. Hua, Y. Liu, L.E. Bernstein, K.E. Livingston, A. Perez, I. A. Shlyakhter, R. V. Rolando, R. Owen, P. Tanpaiboon, F. Lacbawan, Clinton R. Paden, Duncan MacCannell |
| EPI_ISL_1906620 | Michigan Department of Health and Human Services, Bureau of Laboratories | Michigan Department of Health and Human Services, Bureau of Laboratories | Blankenship HM, Riner D, Soehnlen MK |
| EPI_ISL_1906621 | Quest Diagnostics Incorporated | Centers for Disease Control and Prevention Division of Viral Diseases, Pathogen Discovery | Dakota Howard, Dhwani Batra, Peter W. Cook, Kara Moser, Adrian Paskey, Jason Caravas, Benjamin Rambo-Martin, Shatavia Morrison, Christopher Gulvick, Scott Sammons, Yvette Unoarumhi, Darlene Wagner, Matthew Schmerer, S. H. Rosenthal, A. Gerasimova, R. M. Kagan, B. Anderson, M. Hua, Y. Liu, L.E. Bernstein, K.E. Livingston, A. Perez, I. A. Shlyakhter, R. V. Rolando, R. Owen, P. Tanpaiboon, F. Lacbawan, Clinton R. Paden, Duncan MacCannell |
| EPI_ISL_1906622 | Michigan Department of Health and Human Services, Bureau of Laboratories | Michigan Department of Health and Human Services, Bureau of Laboratories | Blankenship HM, Riner D, Soehnlen MK |
| EPI_ISL_1906623 | Quest Diagnostics Incorporated | Centers for Disease Control and Prevention Division of Viral Diseases, Pathogen Discovery | Dakota Howard, Dhwani Batra, Peter W. Cook, Kara Moser, Adrian Paskey, Jason Caravas, Benjamin Rambo-Martin, Shatavia Morrison, Christopher Gulvick, Scott Sammons, Yvette Unoarumhi, Darlene Wagner, Matthew Schmerer, S. H. Rosenthal, A. Gerasimova, R. M. Kagan, B. Anderson, M. Hua, Y. Liu, L.E. Bernstein, K.E. Livingston, A. Perez, I. A. Shlyakhter, R. V. Rolando, R. Owen, P. Tanpaiboon, F. Lacbawan, Clinton R. Paden, Duncan MacCannell |
| EPI_ISL_1906624 | Michigan Department of Health and Human Services, Bureau of Laboratories | Michigan Department of Health and Human Services, Bureau of Laboratories | Blankenship HM, Riner D, Soehnlen MK |
| EPI_ISL_1906625 | Quest Diagnostics Incorporated | Centers for Disease Control and Prevention Division of Viral Diseases, Pathogen Discovery | Dakota Howard, Dhwani Batra, Peter W. Cook, Kara Moser, Adrian Paskey, Jason Caravas, Benjamin Rambo-Martin, Shatavia Morrison, Christopher Gulvick, Scott Sammons, Yvette Unoarumhi, Darlene Wagner, Matthew Schmerer, S. H. Rosenthal, A. Gerasimova, R. M. Kagan, B. Anderson, M. Hua, Y. Liu, L.E. Bernstein, K.E. Livingston, A. Perez, I. A. Shlyakhter, R. V. Rolando, R. Owen, P. Tanpaiboon, F. Lacbawan, Clinton R. Paden, Duncan MacCannell |
| EPI_ISL_1906626, EPI_ISL_1906627 | Michigan Department of Health and Human Services, Bureau of Laboratories | Michigan Department of Health and Human Services, Bureau of Laboratories | Blankenship HM, Riner D, Soehnlen MK |
| EPI_ISL_1906628 | Quest Diagnostics Incorporated | Centers for Disease Control and Prevention Division of Viral Diseases, Pathogen Discovery | Dakota Howard, Dhwani Batra, Peter W. Cook, Kara Moser, Adrian Paskey, Jason Caravas, Benjamin Rambo-Martin, Shatavia Morrison, Christopher Gulvick, Scott Sammons, Yvette Unoarumhi, Darlene Wagner, Matthew Schmerer, S. H. Rosenthal, A. Gerasimova, R. M. Kagan, B. Anderson, M. Hua, Y. Liu, L.E. Bernstein, K.E. Livingston, A. Perez, I. A. Shlyakhter, R. V. Rolando, R. Owen, P. Tanpaiboon, F. Lacbawan, Clinton R. Paden, Duncan MacCannell |
| EPI_ISL_1906629, EPI_ISL_1906631 | Michigan Department of Health and Human Services, Bureau of Laboratories | Michigan Department of Health and Human Services, Bureau of Laboratories | Blankenship HM, Riner D, Soehnlen MK |
| EPI_ISL_1906632 | Quest Diagnostics Incorporated | Centers for Disease Control and Prevention Division of Viral Diseases, Pathogen Discovery | Dakota Howard, Dhwani Batra, Peter W. Cook, Kara Moser, Adrian Paskey, Jason Caravas, Benjamin Rambo-Martin, Shatavia Morrison, Christopher Gulvick, Scott Sammons, Yvette Unoarumhi, Darlene Wagner, Matthew Schmerer, S. H. Rosenthal, A. Gerasimova, R. M. Kagan, B. Anderson, M. Hua, Y. Liu, L.E. Bernstein, K.E. Livingston, A. Perez, I. A. Shlyakhter, R. V. Rolando, R. Owen, P. Tanpaiboon, F. Lacbawan, Clinton R. Paden, Duncan MacCannell |
| EPI_ISL_1906633 | Michigan Department of Health and Human Services, Bureau of Laboratories | Michigan Department of Health and Human Services, Bureau of Laboratories | Blankenship HM, Riner D, Soehnlen MK |
| EPI_ISL_1906634 | Quest Diagnostics Incorporated | Centers for Disease Control and Prevention Division of Viral Diseases, Pathogen Discovery | Dakota Howard, Dhwani Batra, Peter W. Cook, Kara Moser, Adrian Paskey, Jason Caravas, Benjamin Rambo-Martin, Shatavia Morrison, Christopher Gulvick, Scott Sammons, Yvette Unoarumhi, Darlene Wagner, Matthew Schmerer, S. H. Rosenthal, A. Gerasimova, R. M. Kagan, B. Anderson, M. Hua, Y. Liu, L.E. Bernstein, K.E. Livingston, A. Perez, I. A. Shlyakhter, R. V. Rolando, R. Owen, P. Tanpaiboon, F. Lacbawan, Clinton R. Paden, Duncan MacCannell |
| EPI_ISL_1906635, EPI_ISL_1906636 | Michigan Department of Health and Human Services, Bureau of Laboratories | Michigan Department of Health and Human Services, Bureau of Laboratories | Blankenship HM, Riner D, Soehnlen MK |
| EPI_ISL_1906637 | Quest Diagnostics Incorporated | Centers for Disease Control and Prevention Division of Viral Diseases, Pathogen Discovery | Dakota Howard, Dhwani Batra, Peter W. Cook, Kara Moser, Adrian Paskey, Jason Caravas, Benjamin Rambo-Martin, Shatavia Morrison, Christopher Gulvick, Scott Sammons, Yvette Unoarumhi, Darlene Wagner, Matthew Schmerer, S. H. Rosenthal, A. Gerasimova, R. M. Kagan, B. Anderson, M. Hua, Y. Liu, L.E. Bernstein, K.E. Livingston, A. Perez, I. A. Shlyakhter, R. V. Rolando, R. Owen, P. Tanpaiboon, F. Lacbawan, Clinton R. Paden, Duncan MacCannell |
| EPI_ISL_1906638 | Michigan Department of Health and Human Services, Bureau of Laboratories | Michigan Department of Health and Human Services, Bureau of Laboratories | Blankenship HM, Riner D, Soehnlen MK |
| EPI_ISL_1906639 | Quest Diagnostics Incorporated | Centers for Disease Control and Prevention Division of Viral Diseases, Pathogen Discovery | Dakota Howard, Dhwani Batra, Peter W. Cook, Kara Moser, Adrian Paskey, Jason Caravas, Benjamin Rambo-Martin, Shatavia Morrison, Christopher Gulvick, Scott Sammons, Yvette Unoarumhi, Darlene Wagner, Matthew Schmerer, S. H. Rosenthal, A. Gerasimova, R. M. Kagan, B. Anderson, M. Hua, Y. Liu, L.E. Bernstein, K.E. Livingston, A. Perez, I. A. Shlyakhter, R. V. Rolando, R. Owen, P. Tanpaiboon, F. Lacbawan, Clinton R. Paden, Duncan MacCannell |
| EPI_ISL_1906640 | Michigan Department of Health and Human Services, Bureau of Laboratories | Michigan Department of Health and Human Services, Bureau of Laboratories | Blankenship HM, Riner D, Soehnlen MK |
| EPI_ISL_1906641 | Quest Diagnostics Incorporated | Centers for Disease Control and Prevention Division of Viral Diseases, Pathogen Discovery | Dakota Howard, Dhwani Batra, Peter W. Cook, Kara Moser, Adrian Paskey, Jason Caravas, Benjamin Rambo-Martin, Shatavia Morrison, Christopher Gulvick, Scott Sammons, Yvette Unoarumhi, Darlene Wagner, Matthew Schmerer, S. H. Rosenthal, A. Gerasimova, R. M. Kagan, B. Anderson, M. Hua, Y. Liu, L.E. Bernstein, K.E. Livingston, A. Perez, I. A. Shlyakhter, R. V. Rolando, R. Owen, P. Tanpaiboon, F. Lacbawan, Clinton R. Paden, Duncan MacCannell |
| EPI_ISL_1906642, EPI_ISL_1906643, EPI_ISL_1906645 | Michigan Department of Health and Human Services, Bureau of Laboratories | Michigan Department of Health and Human Services, Bureau of Laboratories | Blankenship HM, Riner D, Soehnlen MK |
| EPI_ISL_1906646 | Quest Diagnostics Incorporated | Centers for Disease Control and Prevention Division of Viral Diseases, Pathogen Discovery | Dakota Howard, Dhwani Batra, Peter W. Cook, Kara Moser, Adrian Paskey, Jason Caravas, Benjamin Rambo-Martin, Shatavia Morrison, Christopher Gulvick, Scott Sammons, Yvette Unoarumhi, Darlene Wagner, Matthew Schmerer, S. H. Rosenthal, A. Gerasimova, R. M. Kagan, B. Anderson, M. Hua, Y. Liu, L.E. Bernstein, K.E. Livingston, A. Perez, I. A. Shlyakhter, R. V. Rolando, R. Owen, P. Tanpaiboon, F. Lacbawan, Clinton R. Paden, Duncan MacCannell |
| EPI_ISL_1906647, EPI_ISL_1906648, EPI_ISL_1906649 | Michigan Department of Health and Human Services, Bureau of Laboratories | Michigan Department of Health and Human Services, Bureau of Laboratories | Blankenship HM, Riner D, Soehnlen MK |
| EPI_ISL_1906650 | Quest Diagnostics Incorporated | Centers for Disease Control and Prevention Division of Viral Diseases, Pathogen Discovery | Dakota Howard, Dhwani Batra, Peter W. Cook, Kara Moser, Adrian Paskey, Jason Caravas, Benjamin Rambo-Martin, Shatavia Morrison, Christopher Gulvick, Scott Sammons, Yvette Unoarumhi, Darlene Wagner, Matthew Schmerer, S. H. Rosenthal, A. Gerasimova, R. M. Kagan, B. Anderson, M. Hua, Y. Liu, L.E. Bernstein, K.E. Livingston, A. Perez, I. A. Shlyakhter, R. V. Rolando, R. Owen, P. Tanpaiboon, F. Lacbawan, Clinton R. Paden, Duncan MacCannell |
| EPI_ISL_1906651, EPI_ISL_1906652, EPI_ISL_1906653 | Michigan Department of Health and Human Services, Bureau of Laboratories | Michigan Department of Health and Human Services, Bureau of Laboratories | Blankenship HM, Riner D, Soehnlen MK |
| EPI_ISL_1906654 | Quest Diagnostics Incorporated | Centers for Disease Control and Prevention Division of Viral Diseases, Pathogen Discovery | Dakota Howard, Dhwani Batra, Peter W. Cook, Kara Moser, Adrian Paskey, Jason Caravas, Benjamin Rambo-Martin, Shatavia Morrison, Christopher Gulvick, Scott Sammons, Yvette Unoarumhi, Darlene Wagner, Matthew Schmerer, S. H. Rosenthal, A. Gerasimova, R. M. Kagan, B. Anderson, M. Hua, Y. Liu, L.E. Bernstein, K.E. Livingston, A. Perez, I. A. Shlyakhter, R. V. Rolando, R. Owen, P. Tanpaiboon, F. Lacbawan, Clinton R. Paden, Duncan MacCannell |
| EPI_ISL_1906655, EPI_ISL_1906656 | Michigan Department of Health and Human Services, Bureau of Laboratories | Michigan Department of Health and Human Services, Bureau of Laboratories | Blankenship HM, Riner D, Soehnlen MK |
| EPI_ISL_1906657 | Quest Diagnostics Incorporated | Centers for Disease Control and Prevention Division of Viral Diseases, Pathogen Discovery | Dakota Howard, Dhwani Batra, Peter W. Cook, Kara Moser, Adrian Paskey, Jason Caravas, Benjamin Rambo-Martin, Shatavia Morrison, Christopher Gulvick, Scott Sammons, Yvette Unoarumhi, Darlene Wagner, Matthew Schmerer, S. H. Rosenthal, A. Gerasimova, R. M. Kagan, B. Anderson, M. Hua, Y. Liu, L.E. Bernstein, K.E. Livingston, A. Perez, I. A. Shlyakhter, R. V. Rolando, R. Owen, P. Tanpaiboon, F. Lacbawan, Clinton R. Paden, Duncan MacCannell |
| EPI_ISL_1906658 | Michigan Department of Health and Human Services, Bureau of Laboratories | Michigan Department of Health and Human Services, Bureau of Laboratories | Blankenship HM, Riner D, Soehnlen MK |
| EPI_ISL_1906659 | Quest Diagnostics Incorporated | Centers for Disease Control and Prevention Division of Viral Diseases, Pathogen Discovery | Dakota Howard, Dhwani Batra, Peter W. Cook, Kara Moser, Adrian Paskey, Jason Caravas, Benjamin Rambo-Martin, Shatavia Morrison, Christopher Gulvick, Scott Sammons, Yvette Unoarumhi, Darlene Wagner, Matthew Schmerer, S. H. Rosenthal, A. Gerasimova, R. M. Kagan, B. Anderson, M. Hua, Y. Liu, L.E. Bernstein, K.E. Livingston, A. Perez, I. A. Shlyakhter, R. V. Rolando, R. Owen, P. Tanpaiboon, F. Lacbawan, Clinton R. Paden, Duncan MacCannell |
| EPI_ISL_1906660 | Michigan Department of Health and Human Services, Bureau of Laboratories | Michigan Department of Health and Human Services, Bureau of Laboratories | Blankenship HM, Riner D, Soehnlen MK |
| EPI_ISL_1906661 | Quest Diagnostics Incorporated | Centers for Disease Control and Prevention Division of Viral Diseases, Pathogen Discovery | Dakota Howard, Dhwani Batra, Peter W. Cook, Kara Moser, Adrian Paskey, Jason Caravas, Benjamin Rambo-Martin, Shatavia Morrison, Christopher Gulvick, Scott Sammons, Yvette Unoarumhi, Darlene Wagner, Matthew Schmerer, S. H. Rosenthal, A. Gerasimova, R. M. Kagan, B. Anderson, M. Hua, Y. Liu, L.E. Bernstein, K.E. Livingston, A. Perez, I. A. Shlyakhter, R. V. Rolando, R. Owen, P. Tanpaiboon, F. Lacbawan, Clinton R. Paden, Duncan MacCannell |
| EPI_ISL_1906662 | Michigan Department of Health and Human Services, Bureau of Laboratories | Michigan Department of Health and Human Services, Bureau of Laboratories | Blankenship HM, Riner D, Soehnlen MK |
| EPI_ISL_1906663 | Quest Diagnostics Incorporated | Centers for Disease Control and Prevention Division of Viral Diseases, Pathogen Discovery | Dakota Howard, Dhwani Batra, Peter W. Cook, Kara Moser, Adrian Paskey, Jason Caravas, Benjamin Rambo-Martin, Shatavia Morrison, Christopher Gulvick, Scott Sammons, Yvette Unoarumhi, Darlene Wagner, Matthew Schmerer, S. H. Rosenthal, A. Gerasimova, R. M. Kagan, B. Anderson, M. Hua, Y. Liu, L.E. Bernstein, K.E. Livingston, A. Perez, I. A. Shlyakhter, R. V. Rolando, R. Owen, P. Tanpaiboon, F. Lacbawan, Clinton R. Paden, Duncan MacCannell |
| EPI_ISL_1906664 | Michigan Department of Health and Human Services, Bureau of Laboratories | Michigan Department of Health and Human Services, Bureau of Laboratories | Blankenship HM, Riner D, Soehnlen MK |
| EPI_ISL_1906665 | Quest Diagnostics Incorporated | Centers for Disease Control and Prevention Division of Viral Diseases, Pathogen Discovery | Dakota Howard, Dhwani Batra, Peter W. Cook, Kara Moser, Adrian Paskey, Jason Caravas, Benjamin Rambo-Martin, Shatavia Morrison, Christopher Gulvick, Scott Sammons, Yvette Unoarumhi, Darlene Wagner, Matthew Schmerer, S. H. Rosenthal, A. Gerasimova, R. M. Kagan, B. Anderson, M. Hua, Y. Liu, L.E. Bernstein, K.E. Livingston, A. Perez, I. A. Shlyakhter, R. V. Rolando, R. Owen, P. Tanpaiboon, F. Lacbawan, Clinton R. Paden, Duncan MacCannell |
| EPI_ISL_1906666 | Michigan Department of Health and | Michigan Department of Health and Human | Blankenship HM, Riner D, Soehnlen MK |

| | | | |
|---|---|---|---|
| | Human Services, Bureau of Laboratories | Services, Bureau of Laboratories | |
| EPI_ISL_1906668 | Quest Diagnostics Incorporated | Centers for Disease Control and Prevention Division of Viral Diseases, Pathogen Discovery | Dakota Howard, Dhwani Batra, Peter W. Cook, Kara Moser, Adrian Paskey, Jason Caravas, Benjamin Rambo-Martin, Shatavia Morrison, Christopher Gulvick, Scott Sammons, Yvette Unoarumhi, Darlene Wagner, Matthew Schmerer, S. H. Rosenthal, A. Gerasimova, R. M. Kagan, B. Anderson, M. Hua, Y. Liu, L.E. Bernstein, K.E. Livingston, A. Perez, I. A. Shlyakhter, R. V. Rolando, R. Owen, P. Tanpaiboon, F. Lacbawan, Clinton R. Paden, Duncan MacCannell |
| EPI_ISL_1906669 | Michigan Department of Health and Human Services, Bureau of Laboratories | Michigan Department of Health and Human Services, Bureau of Laboratories | Blankenship HM, Riner D, Soehnlen MK |
| EPI_ISL_1906670, EPI_ISL_1906672 | Quest Diagnostics Incorporated | Centers for Disease Control and Prevention Division of Viral Diseases, Pathogen Discovery | Dakota Howard, Dhwani Batra, Peter W. Cook, Kara Moser, Adrian Paskey, Jason Caravas, Benjamin Rambo-Martin, Shatavia Morrison, Christopher Gulvick, Scott Sammons, Yvette Unoarumhi, Darlene Wagner, Matthew Schmerer, S. H. Rosenthal, A. Gerasimova, R. M. Kagan, B. Anderson, M. Hua, Y. Liu, L.E. Bernstein, K.E. Livingston, A. Perez, I. A. Shlyakhter, R. V. Rolando, R. Owen, P. Tanpaiboon, F. Lacbawan, Clinton R. Paden, Duncan MacCannell |
| EPI_ISL_1906673 | Michigan Department of Health and Human Services, Bureau of Laboratories | Michigan Department of Health and Human Services, Bureau of Laboratories | Blankenship HM, Riner D, Soehnlen MK |
| EPI_ISL_1906674 | Quest Diagnostics Incorporated | Centers for Disease Control and Prevention Division of Viral Diseases, Pathogen Discovery | Dakota Howard, Dhwani Batra, Peter W. Cook, Kara Moser, Adrian Paskey, Jason Caravas, Benjamin Rambo-Martin, Shatavia Morrison, Christopher Gulvick, Scott Sammons, Yvette Unoarumhi, Darlene Wagner, Matthew Schmerer, S. H. Rosenthal, A. Gerasimova, R. M. Kagan, B. Anderson, M. Hua, Y. Liu, L.E. Bernstein, K.E. Livingston, A. Perez, I. A. Shlyakhter, R. V. Rolando, R. Owen, P. Tanpaiboon, F. Lacbawan, Clinton R. Paden, Duncan MacCannell |
| EPI_ISL_1906675, EPI_ISL_1906676 | Michigan Department of Health and Human Services, Bureau of Laboratories | Michigan Department of Health and Human Services, Bureau of Laboratories | Blankenship HM, Riner D, Soehnlen MK |
| EPI_ISL_1906677 | Quest Diagnostics Incorporated | Centers for Disease Control and Prevention Division of Viral Diseases, Pathogen Discovery | Dakota Howard, Dhwani Batra, Peter W. Cook, Kara Moser, Adrian Paskey, Jason Caravas, Benjamin Rambo-Martin, Shatavia Morrison, Christopher Gulvick, Scott Sammons, Yvette Unoarumhi, Darlene Wagner, Matthew Schmerer, S. H. Rosenthal, A. Gerasimova, R. M. Kagan, B. Anderson, M. Hua, Y. Liu, L.E. Bernstein, K.E. Livingston, A. Perez, I. A. Shlyakhter, R. V. Rolando, R. Owen, P. Tanpaiboon, F. Lacbawan, Clinton R. Paden, Duncan MacCannell |
| EPI_ISL_1906678 | Michigan Department of Health and Human Services, Bureau of Laboratories | Michigan Department of Health and Human Services, Bureau of Laboratories | Blankenship HM, Riner D, Soehnlen MK |
| EPI_ISL_1906679 | Quest Diagnostics Incorporated | Centers for Disease Control and Prevention Division of Viral Diseases, Pathogen Discovery | Dakota Howard, Dhwani Batra, Peter W. Cook, Kara Moser, Adrian Paskey, Jason Caravas, Benjamin Rambo-Martin, Shatavia Morrison, Christopher Gulvick, Scott Sammons, Yvette Unoarumhi, Darlene Wagner, Matthew Schmerer, S. H. Rosenthal, A. Gerasimova, R. M. Kagan, B. Anderson, M. Hua, Y. Liu, L.E. Bernstein, K.E. Livingston, A. Perez, I. A. Shlyakhter, R. V. Rolando, R. Owen, P. Tanpaiboon, F. Lacbawan, Clinton R. Paden, Duncan MacCannell |
| EPI_ISL_1906680 | Michigan Department of Health and Human Services, Bureau of Laboratories | Michigan Department of Health and Human Services, Bureau of Laboratories | Blankenship HM, Riner D, Soehnlen MK |
| EPI_ISL_1906681 | Quest Diagnostics Incorporated | Centers for Disease Control and Prevention Division of Viral Diseases, Pathogen Discovery | Dakota Howard, Dhwani Batra, Peter W. Cook, Kara Moser, Adrian Paskey, Jason Caravas, Benjamin Rambo-Martin, Shatavia Morrison, Christopher Gulvick, Scott Sammons, Yvette Unoarumhi, Darlene Wagner, Matthew Schmerer, S. H. Rosenthal, A. Gerasimova, R. M. Kagan, B. Anderson, M. Hua, Y. Liu, L.E. Bernstein, K.E. Livingston, A. Perez, I. A. Shlyakhter, R. V. Rolando, R. Owen, P. Tanpaiboon, F. Lacbawan, Clinton R. Paden, Duncan MacCannell |
| EPI_ISL_1906682 | Michigan Department of Health and Human Services, Bureau of Laboratories | Michigan Department of Health and Human Services, Bureau of Laboratories | Blankenship HM, Riner D, Soehnlen MK |
| EPI_ISL_1906684 | Quest Diagnostics Incorporated | Centers for Disease Control and Prevention Division of Viral Diseases, Pathogen Discovery | Dakota Howard, Dhwani Batra, Peter W. Cook, Kara Moser, Adrian Paskey, Jason Caravas, Benjamin Rambo-Martin, Shatavia Morrison, Christopher Gulvick, Scott Sammons, Yvette Unoarumhi, Darlene Wagner, Matthew Schmerer, S. H. Rosenthal, A. Gerasimova, R. M. Kagan, B. Anderson, M. Hua, Y. Liu, L.E. Bernstein, K.E. Livingston, A. Perez, I. A. Shlyakhter, R. V. Rolando, R. Owen, P. Tanpaiboon, F. Lacbawan, Clinton R. Paden, Duncan MacCannell |
| EPI_ISL_1906685 | Michigan Department of Health and Human Services, Bureau of Laboratories | Michigan Department of Health and Human Services, Bureau of Laboratories | Blankenship HM, Riner D, Soehnlen MK |
| EPI_ISL_1906686 | Quest Diagnostics Incorporated | Centers for Disease Control and Prevention Division of Viral Diseases, Pathogen Discovery | Dakota Howard, Dhwani Batra, Peter W. Cook, Kara Moser, Adrian Paskey, Jason Caravas, Benjamin Rambo-Martin, Shatavia Morrison, Christopher Gulvick, Scott Sammons, Yvette Unoarumhi, Darlene Wagner, Matthew Schmerer, S. H. Rosenthal, A. Gerasimova, R. M. Kagan, B. Anderson, M. Hua, Y. Liu, L.E. Bernstein, K.E. Livingston, A. Perez, I. A. Shlyakhter, R. V. Rolando, R. Owen, P. Tanpaiboon, F. Lacbawan, Clinton R. Paden, Duncan MacCannell |
| EPI_ISL_1906687, EPI_ISL_1906688, EPI_ISL_1906689 | Michigan Department of Health and Human Services, Bureau of Laboratories | Michigan Department of Health and Human Services, Bureau of Laboratories | Blankenship HM, Riner D, Soehnlen MK |
| EPI_ISL_1906690 | Quest Diagnostics Incorporated | Centers for Disease Control and Prevention Division of Viral Diseases, Pathogen Discovery | Dakota Howard, Dhwani Batra, Peter W. Cook, Kara Moser, Adrian Paskey, Jason Caravas, Benjamin Rambo-Martin, Shatavia Morrison, Christopher Gulvick, Scott Sammons, Yvette Unoarumhi, Darlene Wagner, Matthew Schmerer, S. H. Rosenthal, A. Gerasimova, R. M. Kagan, B. Anderson, M. Hua, Y. Liu, L.E. Bernstein, K.E. Livingston, A. Perez, I. A. Shlyakhter, R. V. Rolando, R. Owen, P. Tanpaiboon, F. Lacbawan, Clinton R. Paden, Duncan MacCannell |
| EPI_ISL_1906691 | Michigan Department of Health and Human Services, Bureau of Laboratories | Michigan Department of Health and Human Services, Bureau of Laboratories | Blankenship HM, Riner D, Soehnlen MK |
| EPI_ISL_1906692 | Quest Diagnostics Incorporated | Centers for Disease Control and Prevention Division of Viral Diseases, Pathogen Discovery | Dakota Howard, Dhwani Batra, Peter W. Cook, Kara Moser, Adrian Paskey, Jason Caravas, Benjamin Rambo-Martin, Shatavia Morrison, Christopher Gulvick, Scott Sammons, Yvette Unoarumhi, Darlene Wagner, Matthew Schmerer, S. H. Rosenthal, A. Gerasimova, R. M. Kagan, B. Anderson, M. Hua, Y. Liu, L.E. Bernstein, K.E. Livingston, A. Perez, I. A. Shlyakhter, R. V. Rolando, R. Owen, P. Tanpaiboon, F. Lacbawan, Clinton R. Paden, Duncan MacCannell |
| EPI_ISL_1906693 | Michigan Department of Health and Human Services, Bureau of Laboratories | Michigan Department of Health and Human Services, Bureau of Laboratories | Blankenship HM, Riner D, Soehnlen MK |
| EPI_ISL_1906694 | Quest Diagnostics Incorporated | Centers for Disease Control and Prevention Division of Viral Diseases, Pathogen Discovery | Dakota Howard, Dhwani Batra, Peter W. Cook, Kara Moser, Adrian Paskey, Jason Caravas, Benjamin Rambo-Martin, Shatavia Morrison, Christopher Gulvick, Scott Sammons, Yvette Unoarumhi, Darlene Wagner, Matthew Schmerer, S. H. Rosenthal, A. Gerasimova, R. M. Kagan, B. Anderson, M. Hua, Y. Liu, L.E. Bernstein, K.E. Livingston, A. Perez, I. A. Shlyakhter, R. V. Rolando, R. Owen, P. Tanpaiboon, F. Lacbawan, Clinton R. Paden, Duncan MacCannell |
| EPI_ISL_1906695 | Michigan Department of Health and Human Services, Bureau of Laboratories | Michigan Department of Health and Human Services, Bureau of Laboratories | Blankenship HM, Riner D, Soehnlen MK |
| EPI_ISL_1906696 | Quest Diagnostics Incorporated | Centers for Disease Control and Prevention Division of Viral Diseases, Pathogen Discovery | Dakota Howard, Dhwani Batra, Peter W. Cook, Kara Moser, Adrian Paskey, Jason Caravas, Benjamin Rambo-Martin, Shatavia Morrison, Christopher Gulvick, Scott Sammons, Yvette Unoarumhi, Darlene Wagner, Matthew Schmerer, S. H. Rosenthal, A. Gerasimova, R. M. Kagan, B. Anderson, M. Hua, Y. Liu, L.E. Bernstein, K.E. Livingston, A. Perez, I. A. Shlyakhter, R. V. Rolando, R. Owen, P. Tanpaiboon, F. Lacbawan, Clinton R. Paden, Duncan MacCannell |
| EPI_ISL_1906697, EPI_ISL_1906698 | Michigan Department of Health and Human Services, Bureau of Laboratories | Michigan Department of Health and Human Services, Bureau of Laboratories | Blankenship HM, Riner D, Soehnlen MK |
| EPI_ISL_1906699 | Quest Diagnostics Incorporated | Centers for Disease Control and Prevention Division of Viral Diseases, Pathogen Discovery | Dakota Howard, Dhwani Batra, Peter W. Cook, Kara Moser, Adrian Paskey, Jason Caravas, Benjamin Rambo-Martin, Shatavia Morrison, Christopher Gulvick, Scott Sammons, Yvette Unoarumhi, Darlene Wagner, Matthew Schmerer, S. H. Rosenthal, A. Gerasimova, R. M. Kagan, B. Anderson, M. Hua, Y. Liu, L.E. Bernstein, K.E. Livingston, A. Perez, I. A. Shlyakhter, R. V. Rolando, R. Owen, P. Tanpaiboon, F. Lacbawan, Clinton R. Paden, Duncan MacCannell |
| EPI_ISL_1906700, EPI_ISL_1906701, EPI_ISL_1906702 | Michigan Department of Health and Human Services, Bureau of Laboratories | Michigan Department of Health and Human Services, Bureau of Laboratories | Blankenship HM, Riner D, Soehnlen MK |
| EPI_ISL_1906703 | Quest Diagnostics Incorporated | Centers for Disease Control and Prevention Division of Viral Diseases, Pathogen Discovery | Dakota Howard, Dhwani Batra, Peter W. Cook, Kara Moser, Adrian Paskey, Jason Caravas, Benjamin Rambo-Martin, Shatavia Morrison, Christopher Gulvick, Scott Sammons, Yvette Unoarumhi, Darlene Wagner, Matthew Schmerer, S. H. Rosenthal, A. Gerasimova, R. M. Kagan, B. Anderson, M. Hua, Y. Liu, L.E. Bernstein, K.E. Livingston, A. Perez, I. A. Shlyakhter, R. V. Rolando, R. Owen, P. Tanpaiboon, F. Lacbawan, Clinton R. Paden, Duncan MacCannell |
| EPI_ISL_1906704 | Michigan Department of Health and Human Services, Bureau of Laboratories | Michigan Department of Health and Human Services, Bureau of Laboratories | Blankenship HM, Riner D, Soehnlen MK |
| EPI_ISL_1906705 | Quest Diagnostics Incorporated | Centers for Disease Control and Prevention Division of Viral Diseases, Pathogen Discovery | Dakota Howard, Dhwani Batra, Peter W. Cook, Kara Moser, Adrian Paskey, Jason Caravas, Benjamin Rambo-Martin, Shatavia Morrison, Christopher Gulvick, Scott Sammons, Yvette Unoarumhi, Darlene Wagner, Matthew Schmerer, S. H. Rosenthal, A. Gerasimova, R. M. Kagan, B. Anderson, M. Hua, Y. Liu, L.E. Bernstein, K.E. Livingston, A. Perez, I. A. Shlyakhter, R. V. Rolando, R. Owen, P. Tanpaiboon, F. Lacbawan, Clinton R. Paden, Duncan MacCannell |
| EPI_ISL_1906706, EPI_ISL_1906707, EPI_ISL_1906708 | Michigan Department of Health and Human Services, Bureau of Laboratories | Michigan Department of Health and Human Services, Bureau of Laboratories | Blankenship HM, Riner D, Soehnlen MK |
| EPI_ISL_1906709 | Quest Diagnostics Incorporated | Centers for Disease Control and Prevention Division of Viral Diseases, Pathogen Discovery | Dakota Howard, Dhwani Batra, Peter W. Cook, Kara Moser, Adrian Paskey, Jason Caravas, Benjamin Rambo-Martin, Shatavia Morrison, Christopher Gulvick, Scott Sammons, Yvette Unoarumhi, Darlene Wagner, Matthew Schmerer, S. H. Rosenthal, A. Gerasimova, R. M. Kagan, B. Anderson, M. Hua, Y. Liu, L.E. Bernstein, K.E. Livingston, A. Perez, I. A. Shlyakhter, R. V. Rolando, R. Owen, P. Tanpaiboon, F. Lacbawan, Clinton R. Paden, Duncan MacCannell |
| EPI_ISL_1906710, EPI_ISL_1906711 | Michigan Department of Health and Human Services, Bureau of Laboratories | Michigan Department of Health and Human Services, Bureau of Laboratories | Blankenship HM, Riner D, Soehnlen MK |
| EPI_ISL_1906712 | Quest Diagnostics Incorporated | Centers for Disease Control and Prevention Division of Viral Diseases, Pathogen Discovery | Dakota Howard, Dhwani Batra, Peter W. Cook, Kara Moser, Adrian Paskey, Jason Caravas, Benjamin Rambo-Martin, Shatavia Morrison, Christopher Gulvick, Scott Sammons, Yvette Unoarumhi, Darlene Wagner, Matthew Schmerer, S. H. Rosenthal, A. Gerasimova, R. M. Kagan, B. Anderson, M. Hua, Y. Liu, L.E. Bernstein, K.E. Livingston, A. Perez, I. A. Shlyakhter, R. V. Rolando, R. Owen, P. Tanpaiboon, F. Lacbawan, Clinton R. Paden, Duncan MacCannell |
| EPI_ISL_1906713 | Michigan Department of Health and Human Services, Bureau of Laboratories | Michigan Department of Health and Human Services, Bureau of Laboratories | Blankenship HM, Riner D, Soehnlen MK |
| EPI_ISL_1906714, EPI_ISL_1906716, EPI_ISL_1906718 | Quest Diagnostics Incorporated | Centers for Disease Control and Prevention Division of Viral Diseases, Pathogen Discovery | Dakota Howard, Dhwani Batra, Peter W. Cook, Kara Moser, Adrian Paskey, Jason Caravas, Benjamin Rambo-Martin, Shatavia Morrison, Christopher Gulvick, Scott Sammons, Yvette Unoarumhi, Darlene Wagner, Matthew Schmerer, S. H. Rosenthal, A. Gerasimova, R. M. Kagan, B. Anderson, M. Hua, Y. Liu, L.E. Bernstein, K.E. Livingston, A. Perez, I. A. Shlyakhter, R. V. Rolando, R. Owen, P. Tanpaiboon, F. Lacbawan, Clinton R. Paden, Duncan MacCannell |
| EPI_ISL_1906719, EPI_ISL_1906720 | Michigan Department of Health and Human Services, Bureau of Laboratories | Michigan Department of Health and Human Services, Bureau of Laboratories | Blankenship HM, Riner D, Soehnlen MK |

| | | | |
|---|---|---|---|
| EPI_ISL_1906721 | Quest Diagnostics Incorporated | Centers for Disease Control and Prevention Division of Viral Diseases, Pathogen Discovery | Dakota Howard, Dhwani Batra, Peter W. Cook, Kara Moser, Adrian Paskey, Jason Caravas, Benjamin Rambo-Martin, Shatavia Morrison, Christopher Gulvick, Scott Sammons, Yvette Unoarumhi, Darlene Wagner, Matthew Schmerer, S. H. Rosenthal, A. Gerasimova, R. M. Kagan, B. Anderson, M. Hua, Y. Liu, L.E. Bernstein, K.E. Livingston, A. Perez, I. A. Shlyakhter, R. V. Rolando, R. Owen, P. Tanpaiboon, F. Lacbawan, Clinton R. Paden, Duncan MacCannell |
| EPI_ISL_1906722 | Michigan Department of Health and Human Services, Bureau of Laboratories | Michigan Department of Health and Human Services, Bureau of Laboratories | Blankenship HM, Riner D, Soehnlen MK |
| EPI_ISL_1906723 | Quest Diagnostics Incorporated | Centers for Disease Control and Prevention Division of Viral Diseases, Pathogen Discovery | Dakota Howard, Dhwani Batra, Peter W. Cook, Kara Moser, Adrian Paskey, Jason Caravas, Benjamin Rambo-Martin, Shatavia Morrison, Christopher Gulvick, Scott Sammons, Yvette Unoarumhi, Darlene Wagner, Matthew Schmerer, S. H. Rosenthal, A. Gerasimova, R. M. Kagan, B. Anderson, M. Hua, Y. Liu, L.E. Bernstein, K.E. Livingston, A. Perez, I. A. Shlyakhter, R. V. Rolando, R. Owen, P. Tanpaiboon, F. Lacbawan, Clinton R. Paden, Duncan MacCannell |
| EPI_ISL_1906724, EPI_ISL_1906725, EPI_ISL_1906726 | Michigan Department of Health and Human Services, Bureau of Laboratories | Michigan Department of Health and Human Services, Bureau of Laboratories | Blankenship HM, Riner D, Soehnlen MK |
| EPI_ISL_1906727 | Quest Diagnostics Incorporated | Centers for Disease Control and Prevention Division of Viral Diseases, Pathogen Discovery | Dakota Howard, Dhwani Batra, Peter W. Cook, Kara Moser, Adrian Paskey, Jason Caravas, Benjamin Rambo-Martin, Shatavia Morrison, Christopher Gulvick, Scott Sammons, Yvette Unoarumhi, Darlene Wagner, Matthew Schmerer, S. H. Rosenthal, A. Gerasimova, R. M. Kagan, B. Anderson, M. Hua, Y. Liu, L.E. Bernstein, K.E. Livingston, A. Perez, I. A. Shlyakhter, R. V. Rolando, R. Owen, P. Tanpaiboon, F. Lacbawan, Clinton R. Paden, Duncan MacCannell |
| EPI_ISL_1906728 | Michigan Department of Health and Human Services, Bureau of Laboratories | Michigan Department of Health and Human Services, Bureau of Laboratories | Blankenship HM, Riner D, Soehnlen MK |
| EPI_ISL_1906729 | Quest Diagnostics Incorporated | Centers for Disease Control and Prevention Division of Viral Diseases, Pathogen Discovery | Dakota Howard, Dhwani Batra, Peter W. Cook, Kara Moser, Adrian Paskey, Jason Caravas, Benjamin Rambo-Martin, Shatavia Morrison, Christopher Gulvick, Scott Sammons, Yvette Unoarumhi, Darlene Wagner, Matthew Schmerer, S. H. Rosenthal, A. Gerasimova, R. M. Kagan, B. Anderson, M. Hua, Y. Liu, L.E. Bernstein, K.E. Livingston, A. Perez, I. A. Shlyakhter, R. V. Rolando, R. Owen, P. Tanpaiboon, F. Lacbawan, Clinton R. Paden, Duncan MacCannell |
| EPI_ISL_1906730 | Michigan Department of Health and Human Services, Bureau of Laboratories | Michigan Department of Health and Human Services, Bureau of Laboratories | Blankenship HM, Riner D, Soehnlen MK |
| EPI_ISL_1906731 | Quest Diagnostics Incorporated | Centers for Disease Control and Prevention Division of Viral Diseases, Pathogen Discovery | Dakota Howard, Dhwani Batra, Peter W. Cook, Kara Moser, Adrian Paskey, Jason Caravas, Benjamin Rambo-Martin, Shatavia Morrison, Christopher Gulvick, Scott Sammons, Yvette Unoarumhi, Darlene Wagner, Matthew Schmerer, S. H. Rosenthal, A. Gerasimova, R. M. Kagan, B. Anderson, M. Hua, Y. Liu, L.E. Bernstein, K.E. Livingston, A. Perez, I. A. Shlyakhter, R. V. Rolando, R. Owen, P. Tanpaiboon, F. Lacbawan, Clinton R. Paden, Duncan MacCannell |
| EPI_ISL_1906732, EPI_ISL_1906733 | Michigan Department of Health and Human Services, Bureau of Laboratories | Michigan Department of Health and Human Services, Bureau of Laboratories | Blankenship HM, Riner D, Soehnlen MK |
| EPI_ISL_1906734 | Quest Diagnostics Incorporated | Centers for Disease Control and Prevention Division of Viral Diseases, Pathogen Discovery | Dakota Howard, Dhwani Batra, Peter W. Cook, Kara Moser, Adrian Paskey, Jason Caravas, Benjamin Rambo-Martin, Shatavia Morrison, Christopher Gulvick, Scott Sammons, Yvette Unoarumhi, Darlene Wagner, Matthew Schmerer, S. H. Rosenthal, A. Gerasimova, R. M. Kagan, B. Anderson, M. Hua, Y. Liu, L.E. Bernstein, K.E. Livingston, A. Perez, I. A. Shlyakhter, R. V. Rolando, R. Owen, P. Tanpaiboon, F. Lacbawan, Clinton R. Paden, Duncan MacCannell |
| EPI_ISL_1906735 | Michigan Department of Health and Human Services, Bureau of Laboratories | Michigan Department of Health and Human Services, Bureau of Laboratories | Blankenship HM, Riner D, Soehnlen MK |
| EPI_ISL_1906736 | Quest Diagnostics Incorporated | Centers for Disease Control and Prevention Division of Viral Diseases, Pathogen Discovery | Dakota Howard, Dhwani Batra, Peter W. Cook, Kara Moser, Adrian Paskey, Jason Caravas, Benjamin Rambo-Martin, Shatavia Morrison, Christopher Gulvick, Scott Sammons, Yvette Unoarumhi, Darlene Wagner, Matthew Schmerer, S. H. Rosenthal, A. Gerasimova, R. M. Kagan, B. Anderson, M. Hua, Y. Liu, L.E. Bernstein, K.E. Livingston, A. Perez, I. A. Shlyakhter, R. V. Rolando, R. Owen, P. Tanpaiboon, F. Lacbawan, Clinton R. Paden, Duncan MacCannell |
| EPI_ISL_1906737 | Michigan Department of Health and Human Services, Bureau of Laboratories | Michigan Department of Health and Human Services, Bureau of Laboratories | Blankenship HM, Riner D, Soehnlen MK |
| EPI_ISL_1906738 | Quest Diagnostics Incorporated | Centers for Disease Control and Prevention Division of Viral Diseases, Pathogen Discovery | Dakota Howard, Dhwani Batra, Peter W. Cook, Kara Moser, Adrian Paskey, Jason Caravas, Benjamin Rambo-Martin, Shatavia Morrison, Christopher Gulvick, Scott Sammons, Yvette Unoarumhi, Darlene Wagner, Matthew Schmerer, S. H. Rosenthal, A. Gerasimova, R. M. Kagan, B. Anderson, M. Hua, Y. Liu, L.E. Bernstein, K.E. Livingston, A. Perez, I. A. Shlyakhter, R. V. Rolando, R. Owen, P. Tanpaiboon, F. Lacbawan, Clinton R. Paden, Duncan MacCannell |
| EPI_ISL_1906739 | Michigan Department of Health and Human Services, Bureau of Laboratories | Michigan Department of Health and Human Services, Bureau of Laboratories | Blankenship HM, Riner D, Soehnlen MK |
| EPI_ISL_1906740 | Quest Diagnostics Incorporated | Centers for Disease Control and Prevention Division of Viral Diseases, Pathogen Discovery | Dakota Howard, Dhwani Batra, Peter W. Cook, Kara Moser, Adrian Paskey, Jason Caravas, Benjamin Rambo-Martin, Shatavia Morrison, Christopher Gulvick, Scott Sammons, Yvette Unoarumhi, Darlene Wagner, Matthew Schmerer, S. H. Rosenthal, A. Gerasimova, R. M. Kagan, B. Anderson, M. Hua, Y. Liu, L.E. Bernstein, K.E. Livingston, A. Perez, I. A. Shlyakhter, R. V. Rolando, R. Owen, P. Tanpaiboon, F. Lacbawan, Clinton R. Paden, Duncan MacCannell |
| EPI_ISL_1906741 | Michigan Department of Health and Human Services, Bureau of Laboratories | Michigan Department of Health and Human Services, Bureau of Laboratories | Blankenship HM, Riner D, Soehnlen MK |
| EPI_ISL_1906742 | Quest Diagnostics Incorporated | Centers for Disease Control and Prevention Division of Viral Diseases, Pathogen Discovery | Dakota Howard, Dhwani Batra, Peter W. Cook, Kara Moser, Adrian Paskey, Jason Caravas, Benjamin Rambo-Martin, Shatavia Morrison, Christopher Gulvick, Scott Sammons, Yvette Unoarumhi, Darlene Wagner, Matthew Schmerer, S. H. Rosenthal, A. Gerasimova, R. M. Kagan, B. Anderson, M. Hua, Y. Liu, L.E. Bernstein, K.E. Livingston, A. Perez, I. A. Shlyakhter, R. V. Rolando, R. Owen, P. Tanpaiboon, F. Lacbawan, Clinton R. Paden, Duncan MacCannell |
| EPI_ISL_1906743 | Michigan Department of Health and Human Services, Bureau of Laboratories | Michigan Department of Health and Human Services, Bureau of Laboratories | Blankenship HM, Riner D, Soehnlen MK |
| EPI_ISL_1906744 | Quest Diagnostics Incorporated | Centers for Disease Control and Prevention Division of Viral Diseases, Pathogen Discovery | Dakota Howard, Dhwani Batra, Peter W. Cook, Kara Moser, Adrian Paskey, Jason Caravas, Benjamin Rambo-Martin, Shatavia Morrison, Christopher Gulvick, Scott Sammons, Yvette Unoarumhi, Darlene Wagner, Matthew Schmerer, S. H. Rosenthal, A. Gerasimova, R. M. Kagan, B. Anderson, M. Hua, Y. Liu, L.E. Bernstein, K.E. Livingston, A. Perez, I. A. Shlyakhter, R. V. Rolando, R. Owen, P. Tanpaiboon, F. Lacbawan, Clinton R. Paden, Duncan MacCannell |
| EPI_ISL_1906745, EPI_ISL_1906746 | Michigan Department of Health and Human Services, Bureau of Laboratories | Michigan Department of Health and Human Services, Bureau of Laboratories | Blankenship HM, Riner D, Soehnlen MK |
| EPI_ISL_1906747 | Quest Diagnostics Incorporated | Centers for Disease Control and Prevention Division of Viral Diseases, Pathogen Discovery | Dakota Howard, Dhwani Batra, Peter W. Cook, Kara Moser, Adrian Paskey, Jason Caravas, Benjamin Rambo-Martin, Shatavia Morrison, Christopher Gulvick, Scott Sammons, Yvette Unoarumhi, Darlene Wagner, Matthew Schmerer, S. H. Rosenthal, A. Gerasimova, R. M. Kagan, B. Anderson, M. Hua, Y. Liu, L.E. Bernstein, K.E. Livingston, A. Perez, I. A. Shlyakhter, R. V. Rolando, R. Owen, P. Tanpaiboon, F. Lacbawan, Clinton R. Paden, Duncan MacCannell |
| EPI_ISL_1906748 | Michigan Department of Health and Human Services, Bureau of Laboratories | Michigan Department of Health and Human Services, Bureau of Laboratories | Blankenship HM, Riner D, Soehnlen MK |
| EPI_ISL_1906749 | Quest Diagnostics Incorporated | Centers for Disease Control and Prevention Division of Viral Diseases, Pathogen Discovery | Dakota Howard, Dhwani Batra, Peter W. Cook, Kara Moser, Adrian Paskey, Jason Caravas, Benjamin Rambo-Martin, Shatavia Morrison, Christopher Gulvick, Scott Sammons, Yvette Unoarumhi, Darlene Wagner, Matthew Schmerer, S. H. Rosenthal, A. Gerasimova, R. M. Kagan, B. Anderson, M. Hua, Y. Liu, L.E. Bernstein, K.E. Livingston, A. Perez, I. A. Shlyakhter, R. V. Rolando, R. Owen, P. Tanpaiboon, F. Lacbawan, Clinton R. Paden, Duncan MacCannell |
| EPI_ISL_1906750 | Michigan Department of Health and Human Services, Bureau of Laboratories | Michigan Department of Health and Human Services, Bureau of Laboratories | Blankenship HM, Riner D, Soehnlen MK |
| EPI_ISL_1906751 | Quest Diagnostics Incorporated | Centers for Disease Control and Prevention Division of Viral Diseases, Pathogen Discovery | Dakota Howard, Dhwani Batra, Peter W. Cook, Kara Moser, Adrian Paskey, Jason Caravas, Benjamin Rambo-Martin, Shatavia Morrison, Christopher Gulvick, Scott Sammons, Yvette Unoarumhi, Darlene Wagner, Matthew Schmerer, S. H. Rosenthal, A. Gerasimova, R. M. Kagan, B. Anderson, M. Hua, Y. Liu, L.E. Bernstein, K.E. Livingston, A. Perez, I. A. Shlyakhter, R. V. Rolando, R. Owen, P. Tanpaiboon, F. Lacbawan, Clinton R. Paden, Duncan MacCannell |
| EPI_ISL_1906752 | Michigan Department of Health and Human Services, Bureau of Laboratories | Michigan Department of Health and Human Services, Bureau of Laboratories | Blankenship HM, Riner D, Soehnlen MK |
| EPI_ISL_1906753 | Quest Diagnostics Incorporated | Centers for Disease Control and Prevention Division of Viral Diseases, Pathogen Discovery | Dakota Howard, Dhwani Batra, Peter W. Cook, Kara Moser, Adrian Paskey, Jason Caravas, Benjamin Rambo-Martin, Shatavia Morrison, Christopher Gulvick, Scott Sammons, Yvette Unoarumhi, Darlene Wagner, Matthew Schmerer, S. H. Rosenthal, A. Gerasimova, R. M. Kagan, B. Anderson, M. Hua, Y. Liu, L.E. Bernstein, K.E. Livingston, A. Perez, I. A. Shlyakhter, R. V. Rolando, R. Owen, P. Tanpaiboon, F. Lacbawan, Clinton R. Paden, Duncan MacCannell |
| EPI_ISL_1906754 | Michigan Department of Health and Human Services, Bureau of Laboratories | Michigan Department of Health and Human Services, Bureau of Laboratories | Blankenship HM, Riner D, Soehnlen MK |
| EPI_ISL_1906755 | Quest Diagnostics Incorporated | Centers for Disease Control and Prevention Division of Viral Diseases, Pathogen Discovery | Dakota Howard, Dhwani Batra, Peter W. Cook, Kara Moser, Adrian Paskey, Jason Caravas, Benjamin Rambo-Martin, Shatavia Morrison, Christopher Gulvick, Scott Sammons, Yvette Unoarumhi, Darlene Wagner, Matthew Schmerer, S. H. Rosenthal, A. Gerasimova, R. M. Kagan, B. Anderson, M. Hua, Y. Liu, L.E. Bernstein, K.E. Livingston, A. Perez, I. A. Shlyakhter, R. V. Rolando, R. Owen, P. Tanpaiboon, F. Lacbawan, Clinton R. Paden, Duncan MacCannell |
| EPI_ISL_1906756 | Michigan Department of Health and Human Services, Bureau of Laboratories | Michigan Department of Health and Human Services, Bureau of Laboratories | Blankenship HM, Riner D, Soehnlen MK |
| EPI_ISL_1906757 | Quest Diagnostics Incorporated | Centers for Disease Control and Prevention Division of Viral Diseases, Pathogen Discovery | Dakota Howard, Dhwani Batra, Peter W. Cook, Kara Moser, Adrian Paskey, Jason Caravas, Benjamin Rambo-Martin, Shatavia Morrison, Christopher Gulvick, Scott Sammons, Yvette Unoarumhi, Darlene Wagner, Matthew Schmerer, S. H. Rosenthal, A. Gerasimova, R. M. Kagan, B. Anderson, M. Hua, Y. Liu, L.E. Bernstein, K.E. Livingston, A. Perez, I. A. Shlyakhter, R. V. Rolando, R. Owen, P. Tanpaiboon, F. Lacbawan, Clinton R. Paden, Duncan MacCannell |
| EPI_ISL_1906758 | Michigan Department of Health and Human Services, Bureau of Laboratories | Michigan Department of Health and Human Services, Bureau of Laboratories | Blankenship HM, Riner D, Soehnlen MK |
| EPI_ISL_1906759 | Quest Diagnostics Incorporated | Centers for Disease Control and Prevention Division of Viral Diseases, Pathogen Discovery | Dakota Howard, Dhwani Batra, Peter W. Cook, Kara Moser, Adrian Paskey, Jason Caravas, Benjamin Rambo-Martin, Shatavia Morrison, Christopher Gulvick, Scott Sammons, Yvette Unoarumhi, Darlene Wagner, Matthew Schmerer, S. H. Rosenthal, A. Gerasimova, R. M. Kagan, B. Anderson, M. Hua, Y. Liu, L.E. Bernstein, K.E. Livingston, A. Perez, I. A. Shlyakhter, R. V. Rolando, R. Owen, P. Tanpaiboon, F. Lacbawan, Clinton R. Paden, Duncan MacCannell |
| EPI_ISL_1906760 | Michigan Department of Health and Human Services, Bureau of Laboratories | Michigan Department of Health and Human Services, Bureau of Laboratories | Blankenship HM, Riner D, Soehnlen MK |
| EPI_ISL_1906761 | Quest Diagnostics Incorporated | Centers for Disease Control and Prevention Division of Viral Diseases, Pathogen Discovery | Dakota Howard, Dhwani Batra, Peter W. Cook, Kara Moser, Adrian Paskey, Jason Caravas, Benjamin Rambo-Martin, Shatavia Morrison, Christopher Gulvick, Scott Sammons, Yvette Unoarumhi, Darlene Wagner, Matthew Schmerer, S. H. Rosenthal, A. Gerasimova, R. M. Kagan, B. Anderson, M. Hua, Y. Liu, L.E. Bernstein, K.E. Livingston, A. Perez, I. A. Shlyakhter, R. V. Rolando, R. Owen, P. Tanpaiboon, F. Lacbawan, Clinton R. Paden, Duncan MacCannell |

| | | | |
|---|---|---|---|
| EPI_ISL_1906762 | Michigan Department of Health and Human Services, Bureau of Laboratories | Michigan Department of Health and Human Services, Bureau of Laboratories | Blankenship HM, Riner D, Soehnlen MK |
| EPI_ISL_1906763 | Quest Diagnostics Incorporated | Centers for Disease Control and Prevention Division of Viral Diseases, Pathogen Discovery | Dakota Howard, Dhwani Batra, Peter W. Cook, Kara Moser, Adrian Paskey, Jason Caravas, Benjamin Rambo-Martin, Shatavia Morrison, Christopher Gulvick, Scott Sammons, Yvette Unoarumhi, Darlene Wagner, Matthew Schmerer, S. H. Rosenthal, A. Gerasimova, R. M. Kagan, B. Anderson, M. Hua, Y. Liu, L.E. Bernstein, K.E. Livingston, A. Perez, I. A. Shlyakhter, R. V. Rolando, R. Owen, P. Tanpaiboon, F. Lacbawan, Clinton R. Paden, Duncan MacCannell |
| EPI_ISL_1906764, EPI_ISL_1906765 | Michigan Department of Health and Human Services, Bureau of Laboratories | Michigan Department of Health and Human Services, Bureau of Laboratories | Blankenship HM, Riner D, Soehnlen MK |
| EPI_ISL_1906766 | Quest Diagnostics Incorporated | Centers for Disease Control and Prevention Division of Viral Diseases, Pathogen Discovery | Dakota Howard, Dhwani Batra, Peter W. Cook, Kara Moser, Adrian Paskey, Jason Caravas, Benjamin Rambo-Martin, Shatavia Morrison, Christopher Gulvick, Scott Sammons, Yvette Unoarumhi, Darlene Wagner, Matthew Schmerer, S. H. Rosenthal, A. Gerasimova, R. M. Kagan, B. Anderson, M. Hua, Y. Liu, L.E. Bernstein, K.E. Livingston, A. Perez, I. A. Shlyakhter, R. V. Rolando, R. Owen, P. Tanpaiboon, F. Lacbawan, Clinton R. Paden, Duncan MacCannell |
| EPI_ISL_1906767 | Michigan Department of Health and Human Services, Bureau of Laboratories | Michigan Department of Health and Human Services, Bureau of Laboratories | Blankenship HM, Riner D, Soehnlen MK |
| EPI_ISL_1906768 | Quest Diagnostics Incorporated | Centers for Disease Control and Prevention Division of Viral Diseases, Pathogen Discovery | Dakota Howard, Dhwani Batra, Peter W. Cook, Kara Moser, Adrian Paskey, Jason Caravas, Benjamin Rambo-Martin, Shatavia Morrison, Christopher Gulvick, Scott Sammons, Yvette Unoarumhi, Darlene Wagner, Matthew Schmerer, S. H. Rosenthal, A. Gerasimova, R. M. Kagan, B. Anderson, M. Hua, Y. Liu, L.E. Bernstein, K.E. Livingston, A. Perez, I. A. Shlyakhter, R. V. Rolando, R. Owen, P. Tanpaiboon, F. Lacbawan, Clinton R. Paden, Duncan MacCannell |
| EPI_ISL_1906769 | Michigan Department of Health and Human Services, Bureau of Laboratories | Michigan Department of Health and Human Services, Bureau of Laboratories | Blankenship HM, Riner D, Soehnlen MK |
| EPI_ISL_1906770 | Quest Diagnostics Incorporated | Centers for Disease Control and Prevention Division of Viral Diseases, Pathogen Discovery | Dakota Howard, Dhwani Batra, Peter W. Cook, Kara Moser, Adrian Paskey, Jason Caravas, Benjamin Rambo-Martin, Shatavia Morrison, Christopher Gulvick, Scott Sammons, Yvette Unoarumhi, Darlene Wagner, Matthew Schmerer, S. H. Rosenthal, A. Gerasimova, R. M. Kagan, B. Anderson, M. Hua, Y. Liu, L.E. Bernstein, K.E. Livingston, A. Perez, I. A. Shlyakhter, R. V. Rolando, R. Owen, P. Tanpaiboon, F. Lacbawan, Clinton R. Paden, Duncan MacCannell |
| EPI_ISL_1906771 | Michigan Department of Health and Human Services, Bureau of Laboratories | Michigan Department of Health and Human Services, Bureau of Laboratories | Blankenship HM, Riner D, Soehnlen MK |
| EPI_ISL_1906772 | Quest Diagnostics Incorporated | Centers for Disease Control and Prevention Division of Viral Diseases, Pathogen Discovery | Dakota Howard, Dhwani Batra, Peter W. Cook, Kara Moser, Adrian Paskey, Jason Caravas, Benjamin Rambo-Martin, Shatavia Morrison, Christopher Gulvick, Scott Sammons, Yvette Unoarumhi, Darlene Wagner, Matthew Schmerer, S. H. Rosenthal, A. Gerasimova, R. M. Kagan, B. Anderson, M. Hua, Y. Liu, L.E. Bernstein, K.E. Livingston, A. Perez, I. A. Shlyakhter, R. V. Rolando, R. Owen, P. Tanpaiboon, F. Lacbawan, Clinton R. Paden, Duncan MacCannell |
| EPI_ISL_1906773 | Michigan Department of Health and Human Services, Bureau of Laboratories | Michigan Department of Health and Human Services, Bureau of Laboratories | Blankenship HM, Riner D, Soehnlen MK |
| EPI_ISL_1906774 | Quest Diagnostics Incorporated | Centers for Disease Control and Prevention Division of Viral Diseases, Pathogen Discovery | Dakota Howard, Dhwani Batra, Peter W. Cook, Kara Moser, Adrian Paskey, Jason Caravas, Benjamin Rambo-Martin, Shatavia Morrison, Christopher Gulvick, Scott Sammons, Yvette Unoarumhi, Darlene Wagner, Matthew Schmerer, S. H. Rosenthal, A. Gerasimova, R. M. Kagan, B. Anderson, M. Hua, Y. Liu, L.E. Bernstein, K.E. Livingston, A. Perez, I. A. Shlyakhter, R. V. Rolando, R. Owen, P. Tanpaiboon, F. Lacbawan, Clinton R. Paden, Duncan MacCannell |
| EPI_ISL_1906775, EPI_ISL_1906776 | Michigan Department of Health and Human Services, Bureau of Laboratories | Michigan Department of Health and Human Services, Bureau of Laboratories | Blankenship HM, Riner D, Soehnlen MK |
| EPI_ISL_1906777, EPI_ISL_1906778 | Quest Diagnostics Incorporated | Centers for Disease Control and Prevention Division of Viral Diseases, Pathogen Discovery | Dakota Howard, Dhwani Batra, Peter W. Cook, Kara Moser, Adrian Paskey, Jason Caravas, Benjamin Rambo-Martin, Shatavia Morrison, Christopher Gulvick, Scott Sammons, Yvette Unoarumhi, Darlene Wagner, Matthew Schmerer, S. H. Rosenthal, A. Gerasimova, R. M. Kagan, B. Anderson, M. Hua, Y. Liu, L.E. Bernstein, K.E. Livingston, A. Perez, I. A. Shlyakhter, R. V. Rolando, R. Owen, P. Tanpaiboon, F. Lacbawan, Clinton R. Paden, Duncan MacCannell |
| EPI_ISL_1906779, EPI_ISL_1906780 | Michigan Department of Health and Human Services, Bureau of Laboratories | Michigan Department of Health and Human Services, Bureau of Laboratories | Blankenship HM, Riner D, Soehnlen MK |
| EPI_ISL_1906781 | Quest Diagnostics Incorporated | Centers for Disease Control and Prevention Division of Viral Diseases, Pathogen Discovery | Dakota Howard, Dhwani Batra, Peter W. Cook, Kara Moser, Adrian Paskey, Jason Caravas, Benjamin Rambo-Martin, Shatavia Morrison, Christopher Gulvick, Scott Sammons, Yvette Unoarumhi, Darlene Wagner, Matthew Schmerer, S. H. Rosenthal, A. Gerasimova, R. M. Kagan, B. Anderson, M. Hua, Y. Liu, L.E. Bernstein, K.E. Livingston, A. Perez, I. A. Shlyakhter, R. V. Rolando, R. Owen, P. Tanpaiboon, F. Lacbawan, Clinton R. Paden, Duncan MacCannell |
| EPI_ISL_1906782, EPI_ISL_1906783 | Michigan Department of Health and Human Services, Bureau of Laboratories | Michigan Department of Health and Human Services, Bureau of Laboratories | Blankenship HM, Riner D, Soehnlen MK |
| EPI_ISL_1906784 | Quest Diagnostics Incorporated | Centers for Disease Control and Prevention Division of Viral Diseases, Pathogen Discovery | Dakota Howard, Dhwani Batra, Peter W. Cook, Kara Moser, Adrian Paskey, Jason Caravas, Benjamin Rambo-Martin, Shatavia Morrison, Christopher Gulvick, Scott Sammons, Yvette Unoarumhi, Darlene Wagner, Matthew Schmerer, S. H. Rosenthal, A. Gerasimova, R. M. Kagan, B. Anderson, M. Hua, Y. Liu, L.E. Bernstein, K.E. Livingston, A. Perez, I. A. Shlyakhter, R. V. Rolando, R. Owen, P. Tanpaiboon, F. Lacbawan, Clinton R. Paden, Duncan MacCannell |
| EPI_ISL_1906785 | Michigan Department of Health and Human Services, Bureau of Laboratories | Michigan Department of Health and Human Services, Bureau of Laboratories | Blankenship HM, Riner D, Soehnlen MK |
| EPI_ISL_1906786 | Quest Diagnostics Incorporated | Centers for Disease Control and Prevention Division of Viral Diseases, Pathogen Discovery | Dakota Howard, Dhwani Batra, Peter W. Cook, Kara Moser, Adrian Paskey, Jason Caravas, Benjamin Rambo-Martin, Shatavia Morrison, Christopher Gulvick, Scott Sammons, Yvette Unoarumhi, Darlene Wagner, Matthew Schmerer, S. H. Rosenthal, A. Gerasimova, R. M. Kagan, B. Anderson, M. Hua, Y. Liu, L.E. Bernstein, K.E. Livingston, A. Perez, I. A. Shlyakhter, R. V. Rolando, R. Owen, P. Tanpaiboon, F. Lacbawan, Clinton R. Paden, Duncan MacCannell |
| EPI_ISL_1906787 | Michigan Department of Health and Human Services, Bureau of Laboratories | Michigan Department of Health and Human Services, Bureau of Laboratories | Blankenship HM, Riner D, Soehnlen MK |
| EPI_ISL_1906788 | Quest Diagnostics Incorporated | Centers for Disease Control and Prevention Division of Viral Diseases, Pathogen Discovery | Dakota Howard, Dhwani Batra, Peter W. Cook, Kara Moser, Adrian Paskey, Jason Caravas, Benjamin Rambo-Martin, Shatavia Morrison, Christopher Gulvick, Scott Sammons, Yvette Unoarumhi, Darlene Wagner, Matthew Schmerer, S. H. Rosenthal, A. Gerasimova, R. M. Kagan, B. Anderson, M. Hua, Y. Liu, L.E. Bernstein, K.E. Livingston, A. Perez, I. A. Shlyakhter, R. V. Rolando, R. Owen, P. Tanpaiboon, F. Lacbawan, Clinton R. Paden, Duncan MacCannell |
| EPI_ISL_1906789 | Michigan Department of Health and Human Services, Bureau of Laboratories | Michigan Department of Health and Human Services, Bureau of Laboratories | Blankenship HM, Riner D, Soehnlen MK |
| EPI_ISL_1906790 | Quest Diagnostics Incorporated | Centers for Disease Control and Prevention Division of Viral Diseases, Pathogen Discovery | Dakota Howard, Dhwani Batra, Peter W. Cook, Kara Moser, Adrian Paskey, Jason Caravas, Benjamin Rambo-Martin, Shatavia Morrison, Christopher Gulvick, Scott Sammons, Yvette Unoarumhi, Darlene Wagner, Matthew Schmerer, S. H. Rosenthal, A. Gerasimova, R. M. Kagan, B. Anderson, M. Hua, Y. Liu, L.E. Bernstein, K.E. Livingston, A. Perez, I. A. Shlyakhter, R. V. Rolando, R. Owen, P. Tanpaiboon, F. Lacbawan, Clinton R. Paden, Duncan MacCannell |
| EPI_ISL_1906791, EPI_ISL_1906792, EPI_ISL_1906793 | Michigan Department of Health and Human Services, Bureau of Laboratories | Michigan Department of Health and Human Services, Bureau of Laboratories | Blankenship HM, Riner D, Soehnlen MK |
| EPI_ISL_1906794 | Quest Diagnostics Incorporated | Centers for Disease Control and Prevention Division of Viral Diseases, Pathogen Discovery | Dakota Howard, Dhwani Batra, Peter W. Cook, Kara Moser, Adrian Paskey, Jason Caravas, Benjamin Rambo-Martin, Shatavia Morrison, Christopher Gulvick, Scott Sammons, Yvette Unoarumhi, Darlene Wagner, Matthew Schmerer, S. H. Rosenthal, A. Gerasimova, R. M. Kagan, B. Anderson, M. Hua, Y. Liu, L.E. Bernstein, K.E. Livingston, A. Perez, I. A. Shlyakhter, R. V. Rolando, R. Owen, P. Tanpaiboon, F. Lacbawan, Clinton R. Paden, Duncan MacCannell |
| EPI_ISL_1906795, EPI_ISL_1906796 | Michigan Department of Health and Human Services, Bureau of Laboratories | Michigan Department of Health and Human Services, Bureau of Laboratories | Blankenship HM, Riner D, Soehnlen MK |
| EPI_ISL_1906797 | Quest Diagnostics Incorporated | Centers for Disease Control and Prevention Division of Viral Diseases, Pathogen Discovery | Dakota Howard, Dhwani Batra, Peter W. Cook, Kara Moser, Adrian Paskey, Jason Caravas, Benjamin Rambo-Martin, Shatavia Morrison, Christopher Gulvick, Scott Sammons, Yvette Unoarumhi, Darlene Wagner, Matthew Schmerer, S. H. Rosenthal, A. Gerasimova, R. M. Kagan, B. Anderson, M. Hua, Y. Liu, L.E. Bernstein, K.E. Livingston, A. Perez, I. A. Shlyakhter, R. V. Rolando, R. Owen, P. Tanpaiboon, F. Lacbawan, Clinton R. Paden, Duncan MacCannell |
| EPI_ISL_1906798 | Michigan Department of Health and Human Services, Bureau of Laboratories | Michigan Department of Health and Human Services, Bureau of Laboratories | Blankenship HM, Riner D, Soehnlen MK |
| EPI_ISL_1906799 | Quest Diagnostics Incorporated | Centers for Disease Control and Prevention Division of Viral Diseases, Pathogen Discovery | Dakota Howard, Dhwani Batra, Peter W. Cook, Kara Moser, Adrian Paskey, Jason Caravas, Benjamin Rambo-Martin, Shatavia Morrison, Christopher Gulvick, Scott Sammons, Yvette Unoarumhi, Darlene Wagner, Matthew Schmerer, S. H. Rosenthal, A. Gerasimova, R. M. Kagan, B. Anderson, M. Hua, Y. Liu, L.E. Bernstein, K.E. Livingston, A. Perez, I. A. Shlyakhter, R. V. Rolando, R. Owen, P. Tanpaiboon, F. Lacbawan, Clinton R. Paden, Duncan MacCannell |
| EPI_ISL_1906800 | Michigan Department of Health and Human Services, Bureau of Laboratories | Michigan Department of Health and Human Services, Bureau of Laboratories | Blankenship HM, Riner D, Soehnlen MK |
| EPI_ISL_1906801 | Quest Diagnostics Incorporated | Centers for Disease Control and Prevention Division of Viral Diseases, Pathogen Discovery | Dakota Howard, Dhwani Batra, Peter W. Cook, Kara Moser, Adrian Paskey, Jason Caravas, Benjamin Rambo-Martin, Shatavia Morrison, Christopher Gulvick, Scott Sammons, Yvette Unoarumhi, Darlene Wagner, Matthew Schmerer, S. H. Rosenthal, A. Gerasimova, R. M. Kagan, B. Anderson, M. Hua, Y. Liu, L.E. Bernstein, K.E. Livingston, A. Perez, I. A. Shlyakhter, R. V. Rolando, R. Owen, P. Tanpaiboon, F. Lacbawan, Clinton R. Paden, Duncan MacCannell |
| EPI_ISL_1906802, EPI_ISL_1906803 | Michigan Department of Health and Human Services, Bureau of Laboratories | Michigan Department of Health and Human Services, Bureau of Laboratories | Blankenship HM, Riner D, Soehnlen MK |
| EPI_ISL_1906804 | Quest Diagnostics Incorporated | Centers for Disease Control and Prevention Division of Viral Diseases, Pathogen Discovery | Dakota Howard, Dhwani Batra, Peter W. Cook, Kara Moser, Adrian Paskey, Jason Caravas, Benjamin Rambo-Martin, Shatavia Morrison, Christopher Gulvick, Scott Sammons, Yvette Unoarumhi, Darlene Wagner, Matthew Schmerer, S. H. Rosenthal, A. Gerasimova, R. M. Kagan, B. Anderson, M. Hua, Y. Liu, L.E. Bernstein, K.E. Livingston, A. Perez, I. A. Shlyakhter, R. V. Rolando, R. Owen, P. Tanpaiboon, F. Lacbawan, Clinton R. Paden, Duncan MacCannell |
| EPI_ISL_1906805 | Michigan Department of Health and Human Services, Bureau of Laboratories | Michigan Department of Health and Human Services, Bureau of Laboratories | Blankenship HM, Riner D, Soehnlen MK |
| EPI_ISL_1906806 | Quest Diagnostics Incorporated | Centers for Disease Control and Prevention Division of Viral Diseases, Pathogen Discovery | Dakota Howard, Dhwani Batra, Peter W. Cook, Kara Moser, Adrian Paskey, Jason Caravas, Benjamin Rambo-Martin, Shatavia Morrison, Christopher Gulvick, Scott Sammons, Yvette Unoarumhi, Darlene Wagner, Matthew Schmerer, S. H. Rosenthal, A. Gerasimova, R. M. Kagan, B. Anderson, M. Hua, Y. Liu, L.E. Bernstein, K.E. Livingston, A. Perez, I. A. Shlyakhter, R. V. Rolando, R. Owen, P. Tanpaiboon, F. Lacbawan, Clinton R. Paden, Duncan MacCannell |
| EPI_ISL_1906807 | Michigan Department of Health and Human Services, Bureau of Laboratories | Michigan Department of Health and Human Services, Bureau of Laboratories | Blankenship HM, Riner D, Soehnlen MK |
| EPI_ISL_1906808 | Quest Diagnostics Incorporated | Centers for Disease Control and Prevention | Dakota Howard, Dhwani Batra, Peter W. Cook, Kara Moser, Adrian Paskey, Jason Caravas, Benjamin Rambo-Martin, Shatavia Morrison, Christopher Gulvick, Scott Sammons, Yvette Unoarumhi, Darlene Wagner, Matthew Schmerer, S. H. Rosenthal, A. Gerasimova, R. M. Kagan, B. |

| | | | |
|---|---|---|---|
| | | Division of Viral Diseases, Pathogen Discovery | Anderson, M. Hua, Y. Liu, L.E. Bernstein, K.E. Livingston, A. Perez, I. A. Shlyakhter, R. V. Rolando, R. Owen, P. Tanpaiboon, F. Lacbawan, Clinton R. Paden, Duncan MacCannell |
| EPI_ISL_1906809 | Michigan Department of Health and Human Services, Bureau of Laboratories | Michigan Department of Health and Human Services, Bureau of Laboratories | Blankenship HM, Riner D, Soehnlen MK |
| EPI_ISL_1906810 | Quest Diagnostics Incorporated | Centers for Disease Control and Prevention Division of Viral Diseases, Pathogen Discovery | Dakota Howard, Dhwani Batra, Peter W. Cook, Kara Moser, Adrian Paskey, Jason Caravas, Benjamin Rambo-Martin, Shatavia Morrison, Christopher Gulvick, Scott Sammons, Yvette Unoarumhi, Darlene Wagner, Matthew Schmerer, S. H. Rosenthal, A. Gerasimova, R. M. Kagan, B. Anderson, M. Hua, Y. Liu, L.E. Bernstein, K.E. Livingston, A. Perez, I. A. Shlyakhter, R. V. Rolando, R. Owen, P. Tanpaiboon, F. Lacbawan, Clinton R. Paden, Duncan MacCannell |
| EPI_ISL_1906811 | Michigan Department of Health and Human Services, Bureau of Laboratories | Michigan Department of Health and Human Services, Bureau of Laboratories | Blankenship HM, Riner D, Soehnlen MK |
| EPI_ISL_1906812 | Quest Diagnostics Incorporated | Centers for Disease Control and Prevention Division of Viral Diseases, Pathogen Discovery | Dakota Howard, Dhwani Batra, Peter W. Cook, Kara Moser, Adrian Paskey, Jason Caravas, Benjamin Rambo-Martin, Shatavia Morrison, Christopher Gulvick, Scott Sammons, Yvette Unoarumhi, Darlene Wagner, Matthew Schmerer, S. H. Rosenthal, A. Gerasimova, R. M. Kagan, B. Anderson, M. Hua, Y. Liu, L.E. Bernstein, K.E. Livingston, A. Perez, I. A. Shlyakhter, R. V. Rolando, R. Owen, P. Tanpaiboon, F. Lacbawan, Clinton R. Paden, Duncan MacCannell |
| EPI_ISL_1906813 | Michigan Department of Health and Human Services, Bureau of Laboratories | Michigan Department of Health and Human Services, Bureau of Laboratories | Blankenship HM, Riner D, Soehnlen MK |
| EPI_ISL_1906814 | Quest Diagnostics Incorporated | Centers for Disease Control and Prevention Division of Viral Diseases, Pathogen Discovery | Dakota Howard, Dhwani Batra, Peter W. Cook, Kara Moser, Adrian Paskey, Jason Caravas, Benjamin Rambo-Martin, Shatavia Morrison, Christopher Gulvick, Scott Sammons, Yvette Unoarumhi, Darlene Wagner, Matthew Schmerer, S. H. Rosenthal, A. Gerasimova, R. M. Kagan, B. Anderson, M. Hua, Y. Liu, L.E. Bernstein, K.E. Livingston, A. Perez, I. A. Shlyakhter, R. V. Rolando, R. Owen, P. Tanpaiboon, F. Lacbawan, Clinton R. Paden, Duncan MacCannell |
| EPI_ISL_1906815, EPI_ISL_1906816 | Michigan Department of Health and Human Services, Bureau of Laboratories | Michigan Department of Health and Human Services, Bureau of Laboratories | Blankenship HM, Riner D, Soehnlen MK |
| EPI_ISL_1906817 | Quest Diagnostics Incorporated | Centers for Disease Control and Prevention Division of Viral Diseases, Pathogen Discovery | Dakota Howard, Dhwani Batra, Peter W. Cook, Kara Moser, Adrian Paskey, Jason Caravas, Benjamin Rambo-Martin, Shatavia Morrison, Christopher Gulvick, Scott Sammons, Yvette Unoarumhi, Darlene Wagner, Matthew Schmerer, S. H. Rosenthal, A. Gerasimova, R. M. Kagan, B. Anderson, M. Hua, Y. Liu, L.E. Bernstein, K.E. Livingston, A. Perez, I. A. Shlyakhter, R. V. Rolando, R. Owen, P. Tanpaiboon, F. Lacbawan, Clinton R. Paden, Duncan MacCannell |
| EPI_ISL_1906818 | Michigan Department of Health and Human Services, Bureau of Laboratories | Michigan Department of Health and Human Services, Bureau of Laboratories | Blankenship HM, Riner D, Soehnlen MK |
| EPI_ISL_1906819 | Quest Diagnostics Incorporated | Centers for Disease Control and Prevention Division of Viral Diseases, Pathogen Discovery | Dakota Howard, Dhwani Batra, Peter W. Cook, Kara Moser, Adrian Paskey, Jason Caravas, Benjamin Rambo-Martin, Shatavia Morrison, Christopher Gulvick, Scott Sammons, Yvette Unoarumhi, Darlene Wagner, Matthew Schmerer, S. H. Rosenthal, A. Gerasimova, R. M. Kagan, B. Anderson, M. Hua, Y. Liu, L.E. Bernstein, K.E. Livingston, A. Perez, I. A. Shlyakhter, R. V. Rolando, R. Owen, P. Tanpaiboon, F. Lacbawan, Clinton R. Paden, Duncan MacCannell |
| EPI_ISL_1906820 | Michigan Department of Health and Human Services, Bureau of Laboratories | Michigan Department of Health and Human Services, Bureau of Laboratories | Blankenship HM, Riner D, Soehnlen MK |
| EPI_ISL_1906821 | Quest Diagnostics Incorporated | Centers for Disease Control and Prevention Division of Viral Diseases, Pathogen Discovery | Dakota Howard, Dhwani Batra, Peter W. Cook, Kara Moser, Adrian Paskey, Jason Caravas, Benjamin Rambo-Martin, Shatavia Morrison, Christopher Gulvick, Scott Sammons, Yvette Unoarumhi, Darlene Wagner, Matthew Schmerer, S. H. Rosenthal, A. Gerasimova, R. M. Kagan, B. Anderson, M. Hua, Y. Liu, L.E. Bernstein, K.E. Livingston, A. Perez, I. A. Shlyakhter, R. V. Rolando, R. Owen, P. Tanpaiboon, F. Lacbawan, Clinton R. Paden, Duncan MacCannell |
| EPI_ISL_1906822 | Michigan Department of Health and Human Services, Bureau of Laboratories | Michigan Department of Health and Human Services, Bureau of Laboratories | Blankenship HM, Riner D, Soehnlen MK |
| EPI_ISL_1906823 | Quest Diagnostics Incorporated | Centers for Disease Control and Prevention Division of Viral Diseases, Pathogen Discovery | Dakota Howard, Dhwani Batra, Peter W. Cook, Kara Moser, Adrian Paskey, Jason Caravas, Benjamin Rambo-Martin, Shatavia Morrison, Christopher Gulvick, Scott Sammons, Yvette Unoarumhi, Darlene Wagner, Matthew Schmerer, S. H. Rosenthal, A. Gerasimova, R. M. Kagan, B. Anderson, M. Hua, Y. Liu, L.E. Bernstein, K.E. Livingston, A. Perez, I. A. Shlyakhter, R. V. Rolando, R. Owen, P. Tanpaiboon, F. Lacbawan, Clinton R. Paden, Duncan MacCannell |
| EPI_ISL_1906824 | Michigan Department of Health and Human Services, Bureau of Laboratories | Michigan Department of Health and Human Services, Bureau of Laboratories | Blankenship HM, Riner D, Soehnlen MK |
| EPI_ISL_1906825 | Quest Diagnostics Incorporated | Centers for Disease Control and Prevention Division of Viral Diseases, Pathogen Discovery | Dakota Howard, Dhwani Batra, Peter W. Cook, Kara Moser, Adrian Paskey, Jason Caravas, Benjamin Rambo-Martin, Shatavia Morrison, Christopher Gulvick, Scott Sammons, Yvette Unoarumhi, Darlene Wagner, Matthew Schmerer, S. H. Rosenthal, A. Gerasimova, R. M. Kagan, B. Anderson, M. Hua, Y. Liu, L.E. Bernstein, K.E. Livingston, A. Perez, I. A. Shlyakhter, R. V. Rolando, R. Owen, P. Tanpaiboon, F. Lacbawan, Clinton R. Paden, Duncan MacCannell |
| EPI_ISL_1906826, EPI_ISL_1906827 | Michigan Department of Health and Human Services, Bureau of Laboratories | Michigan Department of Health and Human Services, Bureau of Laboratories | Blankenship HM, Riner D, Soehnlen MK |
| EPI_ISL_1906828 | Quest Diagnostics Incorporated | Centers for Disease Control and Prevention Division of Viral Diseases, Pathogen Discovery | Dakota Howard, Dhwani Batra, Peter W. Cook, Kara Moser, Adrian Paskey, Jason Caravas, Benjamin Rambo-Martin, Shatavia Morrison, Christopher Gulvick, Scott Sammons, Yvette Unoarumhi, Darlene Wagner, Matthew Schmerer, S. H. Rosenthal, A. Gerasimova, R. M. Kagan, B. Anderson, M. Hua, Y. Liu, L.E. Bernstein, K.E. Livingston, A. Perez, I. A. Shlyakhter, R. V. Rolando, R. Owen, P. Tanpaiboon, F. Lacbawan, Clinton R. Paden, Duncan MacCannell |
| EPI_ISL_1906829 | Michigan Department of Health and Human Services, Bureau of Laboratories | Michigan Department of Health and Human Services, Bureau of Laboratories | Blankenship HM, Riner D, Soehnlen MK |
| EPI_ISL_1906830 | Quest Diagnostics Incorporated | Centers for Disease Control and Prevention Division of Viral Diseases, Pathogen Discovery | Dakota Howard, Dhwani Batra, Peter W. Cook, Kara Moser, Adrian Paskey, Jason Caravas, Benjamin Rambo-Martin, Shatavia Morrison, Christopher Gulvick, Scott Sammons, Yvette Unoarumhi, Darlene Wagner, Matthew Schmerer, S. H. Rosenthal, A. Gerasimova, R. M. Kagan, B. Anderson, M. Hua, Y. Liu, L.E. Bernstein, K.E. Livingston, A. Perez, I. A. Shlyakhter, R. V. Rolando, R. Owen, P. Tanpaiboon, F. Lacbawan, Clinton R. Paden, Duncan MacCannell |
| EPI_ISL_1906831 | Michigan Department of Health and Human Services, Bureau of Laboratories | Michigan Department of Health and Human Services, Bureau of Laboratories | Blankenship HM, Riner D, Soehnlen MK |
| EPI_ISL_1906832 | Quest Diagnostics Incorporated | Centers for Disease Control and Prevention Division of Viral Diseases, Pathogen Discovery | Dakota Howard, Dhwani Batra, Peter W. Cook, Kara Moser, Adrian Paskey, Jason Caravas, Benjamin Rambo-Martin, Shatavia Morrison, Christopher Gulvick, Scott Sammons, Yvette Unoarumhi, Darlene Wagner, Matthew Schmerer, S. H. Rosenthal, A. Gerasimova, R. M. Kagan, B. Anderson, M. Hua, Y. Liu, L.E. Bernstein, K.E. Livingston, A. Perez, I. A. Shlyakhter, R. V. Rolando, R. Owen, P. Tanpaiboon, F. Lacbawan, Clinton R. Paden, Duncan MacCannell |
| EPI_ISL_1906833 | Michigan Department of Health and Human Services, Bureau of Laboratories | Michigan Department of Health and Human Services, Bureau of Laboratories | Blankenship HM, Riner D, Soehnlen MK |
| EPI_ISL_1906834 | Quest Diagnostics Incorporated | Centers for Disease Control and Prevention Division of Viral Diseases, Pathogen Discovery | Dakota Howard, Dhwani Batra, Peter W. Cook, Kara Moser, Adrian Paskey, Jason Caravas, Benjamin Rambo-Martin, Shatavia Morrison, Christopher Gulvick, Scott Sammons, Yvette Unoarumhi, Darlene Wagner, Matthew Schmerer, S. H. Rosenthal, A. Gerasimova, R. M. Kagan, B. Anderson, M. Hua, Y. Liu, L.E. Bernstein, K.E. Livingston, A. Perez, I. A. Shlyakhter, R. V. Rolando, R. Owen, P. Tanpaiboon, F. Lacbawan, Clinton R. Paden, Duncan MacCannell |
| EPI_ISL_1906835 | Michigan Department of Health and Human Services, Bureau of Laboratories | Michigan Department of Health and Human Services, Bureau of Laboratories | Blankenship HM, Riner D, Soehnlen MK |
| EPI_ISL_1906836 | Quest Diagnostics Incorporated | Centers for Disease Control and Prevention Division of Viral Diseases, Pathogen Discovery | Dakota Howard, Dhwani Batra, Peter W. Cook, Kara Moser, Adrian Paskey, Jason Caravas, Benjamin Rambo-Martin, Shatavia Morrison, Christopher Gulvick, Scott Sammons, Yvette Unoarumhi, Darlene Wagner, Matthew Schmerer, S. H. Rosenthal, A. Gerasimova, R. M. Kagan, B. Anderson, M. Hua, Y. Liu, L.E. Bernstein, K.E. Livingston, A. Perez, I. A. Shlyakhter, R. V. Rolando, R. Owen, P. Tanpaiboon, F. Lacbawan, Clinton R. Paden, Duncan MacCannell |
| EPI_ISL_1906837 | Michigan Department of Health and Human Services, Bureau of Laboratories | Michigan Department of Health and Human Services, Bureau of Laboratories | Blankenship HM, Riner D, Soehnlen MK |
| EPI_ISL_1906838 | Quest Diagnostics Incorporated | Centers for Disease Control and Prevention Division of Viral Diseases, Pathogen Discovery | Dakota Howard, Dhwani Batra, Peter W. Cook, Kara Moser, Adrian Paskey, Jason Caravas, Benjamin Rambo-Martin, Shatavia Morrison, Christopher Gulvick, Scott Sammons, Yvette Unoarumhi, Darlene Wagner, Matthew Schmerer, S. H. Rosenthal, A. Gerasimova, R. M. Kagan, B. Anderson, M. Hua, Y. Liu, L.E. Bernstein, K.E. Livingston, A. Perez, I. A. Shlyakhter, R. V. Rolando, R. Owen, P. Tanpaiboon, F. Lacbawan, Clinton R. Paden, Duncan MacCannell |
| EPI_ISL_1906839, EPI_ISL_1906840, EPI_ISL_1906841 | Michigan Department of Health and Human Services, Bureau of Laboratories | Michigan Department of Health and Human Services, Bureau of Laboratories | Blankenship HM, Riner D, Soehnlen MK |
| EPI_ISL_1906842 | Quest Diagnostics Incorporated | Centers for Disease Control and Prevention Division of Viral Diseases, Pathogen Discovery | Dakota Howard, Dhwani Batra, Peter W. Cook, Kara Moser, Adrian Paskey, Jason Caravas, Benjamin Rambo-Martin, Shatavia Morrison, Christopher Gulvick, Scott Sammons, Yvette Unoarumhi, Darlene Wagner, Matthew Schmerer, S. H. Rosenthal, A. Gerasimova, R. M. Kagan, B. Anderson, M. Hua, Y. Liu, L.E. Bernstein, K.E. Livingston, A. Perez, I. A. Shlyakhter, R. V. Rolando, R. Owen, P. Tanpaiboon, F. Lacbawan, Clinton R. Paden, Duncan MacCannell |
| EPI_ISL_1906843 | Michigan Department of Health and Human Services, Bureau of Laboratories | Michigan Department of Health and Human Services, Bureau of Laboratories | Blankenship HM, Riner D, Soehnlen MK |
| EPI_ISL_1906844, EPI_ISL_1906845, EPI_ISL_1906846, EPI_ISL_1906847, EPI_ISL_1906848, EPI_ISL_1906849, EPI_ISL_1906850, EPI_ISL_1906851, EPI_ISL_1906852, EPI_ISL_1906853, EPI_ISL_1906854, EPI_ISL_1906855, EPI_ISL_1906856, EPI_ISL_1906857, EPI_ISL_1906858, EPI_ISL_1906859, EPI_ISL_1906860, EPI_ISL_1906861, EPI_ISL_1906862, EPI_ISL_1906863, EPI_ISL_1906864, EPI_ISL_1906865, EPI_ISL_1906866, EPI_ISL_1906867, EPI_ISL_1906868, EPI_ISL_1906869, EPI_ISL_1906870, EPI_ISL_1906871, EPI_ISL_1906872, EPI_ISL_1906873, EPI_ISL_1906874, EPI_ISL_1906875, EPI_ISL_1906876, EPI_ISL_1906877, EPI_ISL_1906878, EPI_ISL_1906879, EPI_ISL_1906880, EPI_ISL_1906881, EPI_ISL_1906882, EPI_ISL_1906883 | | | |
| see above | Quest Diagnostics Incorporated | Centers for Disease Control and Prevention Division of Viral Diseases, Pathogen Discovery | Dakota Howard, Dhwani Batra, Peter W. Cook, Kara Moser, Adrian Paskey, Jason Caravas, Benjamin Rambo-Martin, Shatavia Morrison, Christopher Gulvick, Scott Sammons, Yvette Unoarumhi, Darlene Wagner, Matthew Schmerer, S. H. Rosenthal, A. Gerasimova, R. M. Kagan, B. Anderson, M. Hua, Y. Liu, L.E. Bernstein, K.E. Livingston, A. Perez, I. A. Shlyakhter, R. V. Rolando, R. Owen, P. Tanpaiboon, F. Lacbawan, Clinton R. Paden, Duncan MacCannell |
| EPI_ISL_1906884, EPI_ISL_1906885, EPI_ISL_1906886, EPI_ISL_1906887, EPI_ISL_1906888, EPI_ISL_1906889, EPI_ISL_1906890, EPI_ISL_1906891, EPI_ISL_1906892, EPI_ISL_1906893, EPI_ISL_1906894, EPI_ISL_1906895, EPI_ISL_1906896, EPI_ISL_1906897, EPI_ISL_1906898, EPI_ISL_1906899, EPI_ISL_1906900, EPI_ISL_1906901, EPI_ISL_1906902, EPI_ISL_1906903, EPI_ISL_1906904, EPI_ISL_1906905, EPI_ISL_1906906, EPI_ISL_1906907, EPI_ISL_1906908, EPI_ISL_1906909, EPI_ISL_1906910, EPI_ISL_1906911, EPI_ISL_1906912, EPI_ISL_1906913, EPI_ISL_1906914, EPI_ISL_1906915, EPI_ISL_1906916, EPI_ISL_1906917, EPI_ISL_1906918, EPI_ISL_1906919, EPI_ISL_1906920, EPI_ISL_1906921, EPI_ISL_1906922, EPI_ISL_1906923, EPI_ISL_1906924, EPI_ISL_1906925, EPI_ISL_1906926, EPI_ISL_1906927, EPI_ISL_1906928, EPI_ISL_1906929, EPI_ISL_1906930, EPI_ISL_1906931, EPI_ISL_1906932, EPI_ISL_1906933, EPI_ISL_1906934, EPI_ISL_1906935, EPI_ISL_1906936, EPI_ISL_1906937, EPI_ISL_1906938, EPI_ISL_1906939, EPI_ISL_1906940, EPI_ISL_1906941, EPI_ISL_1906942, EPI_ISL_1906943, EPI_ISL_1906944, EPI_ISL_1906945, EPI_ISL_1906946, EPI_ISL_1906947, EPI_ISL_1906948, EPI_ISL_1906949, EPI_ISL_1906950, EPI_ISL_1906951, EPI_ISL_1906952, EPI_ISL_1906953, EPI_ISL_1906954, EPI_ISL_1906955, EPI_ISL_1906956, EPI_ISL_1906957, EPI_ISL_1906958, EPI_ISL_1906959, EPI_ISL_1906960, EPI_ISL_1906961, EPI_ISL_1906962, EPI_ISL_1906963, EPI_ISL_1906964, EPI_ISL_1906965, EPI_ISL_1906966, EPI_ISL_1906967, EPI_ISL_1906968, EPI_ISL_1906969, EPI_ISL_1906970, EPI_ISL_1906971, EPI_ISL_1906972, EPI_ISL_1906973, EPI_ISL_1906974, EPI_ISL_1906975, EPI_ISL_1906976, EPI_ISL_1906977, EPI_ISL_1906978, EPI_ISL_1906979, EPI_ISL_1906980, EPI_ISL_1906981, EPI_ISL_1906982, EPI_ISL_1906983, EPI_ISL_1906984, EPI_ISL_1906985, EPI_ISL_1906986, EPI_ISL_1906987, EPI_ISL_1906988, EPI_ISL_1906989, EPI_ISL_1906990, EPI_ISL_1906991, EPI_ISL_1906992, EPI_ISL_1906993, EPI_ISL_1906994, EPI_ISL_1906995, EPI_ISL_1906996, EPI_ISL_1906997, EPI_ISL_1906998, EPI_ISL_1906999, EPI_ISL_1907000, EPI_ISL_1907001, EPI_ISL_1907002, EPI_ISL_1907003, EPI_ISL_1907004, EPI_ISL_1907005, EPI_ISL_1907006, EPI_ISL_1907007, EPI_ISL_1907008, EPI_ISL_1907009, EPI_ISL_1907010, EPI_ISL_1907011, EPI_ISL_1907012, EPI_ISL_1907013, EPI_ISL_1907014, EPI_ISL_1907015, EPI_ISL_1907016, EPI_ISL_1907017, EPI_ISL_1907018, EPI_ISL_1907019, EPI_ISL_1907020, EPI_ISL_1907021, EPI_ISL_1907022, EPI_ISL_1907023, EPI_ISL_1907024, EPI_ISL_1907025, EPI_ISL_1907026, EPI_ISL_1907027, EPI_ISL_1907028, EPI_ISL_1907029, EPI_ISL_1907030, EPI_ISL_1907031, EPI_ISL_1907032, EPI_ISL_1907033, EPI_ISL_1907034, EPI_ISL_1907036, EPI_ISL_1907037, EPI_ISL_1907038, EPI_ISL_1907039, EPI_ISL_1907040, EPI_ISL_1907041, EPI_ISL_1907042, EPI_ISL_1907043, EPI_ISL_1907044, EPI_ISL_1907045, EPI_ISL_1907046, EPI_ISL_1907047, EPI_ISL_1907048, EPI_ISL_1907049, EPI_ISL_1907050, EPI_ISL_1907051, EPI_ISL_1907052, EPI_ISL_1907053, EPI_ISL_1907054, EPI_ISL_1907055, EPI_ISL_1907056, EPI_ISL_1907057, EPI_ISL_1907058, EPI_ISL_1907059, EPI_ISL_1907060, EPI_ISL_1907061, EPI_ISL_1907062, EPI_ISL_1907063, EPI_ISL_1907064, EPI_ISL_1907065, EPI_ISL_1907066, EPI_ISL_1907067, EPI_ISL_1907068, EPI_ISL_1907069, EPI_ISL_1907070, EPI_ISL_1907071, EPI_ISL_1907072, EPI_ISL_1907073, EPI_ISL_1907074, EPI_ISL_1907075, EPI_ISL_1907076, EPI_ISL_1907077, EPI_ISL_1907078, EPI_ISL_1907079, EPI_ISL_1907080, EPI_ISL_1907081, EPI_ISL_1907082, EPI_ISL_1907083, EPI_ISL_1907084, EPI_ISL_1907085, EPI_ISL_1907086, EPI_ISL_1907087, EPI_ISL_1907088, EPI_ISL_1907089, EPI_ISL_1907090, EPI_ISL_1907091, EPI_ISL_1907092, EPI_ISL_1907093, EPI_ISL_1907094, EPI_ISL_1907095, EPI_ISL_1907096, EPI_ISL_1907097, EPI_ISL_1907098, EPI_ISL_1907099, EPI_ISL_1907100, EPI_ISL_1907101, EPI_ISL_1907102, EPI_ISL_1907103, EPI_ISL_1907104, | | | |

| | | | |
|---|---|---|---|
| EPI_ISL_1907105, EPI_ISL_1907106, EPI_ISL_1907107, EPI_ISL_1907108, EPI_ISL_1907109, EPI_ISL_1907110, EPI_ISL_1907111, EPI_ISL_1907112, EPI_ISL_1907113, EPI_ISL_1907114, EPI_ISL_1907115, EPI_ISL_1907116, EPI_ISL_1907117, EPI_ISL_1907118, EPI_ISL_1907119, EPI_ISL_1907120, EPI_ISL_1907121, EPI_ISL_1907122, EPI_ISL_1907123, EPI_ISL_1907124, EPI_ISL_1907125, EPI_ISL_1907126, EPI_ISL_1907127, EPI_ISL_1907128, EPI_ISL_1907129, EPI_ISL_1907130, EPI_ISL_1907131, EPI_ISL_1907132, EPI_ISL_1907134, EPI_ISL_1907135, EPI_ISL_1907136, EPI_ISL_1907137, EPI_ISL_1907138, EPI_ISL_1907139, EPI_ISL_1907140, EPI_ISL_1907141, EPI_ISL_1907142, EPI_ISL_1907143, EPI_ISL_1907144, EPI_ISL_1907145, EPI_ISL_1907146, EPI_ISL_1907147, EPI_ISL_1907148, EPI_ISL_1907149, EPI_ISL_1907151, EPI_ISL_1907152, EPI_ISL_1907153, EPI_ISL_1907154, EPI_ISL_1907156, EPI_ISL_1907157, EPI_ISL_1907158, EPI_ISL_1907159, EPI_ISL_1907160, EPI_ISL_1907161, EPI_ISL_1907162, EPI_ISL_1907163, EPI_ISL_1907164, EPI_ISL_1907165, EPI_ISL_1907166, EPI_ISL_1907167, EPI_ISL_1907168, EPI_ISL_1907169, EPI_ISL_1907170, EPI_ISL_1907171, EPI_ISL_1907172, EPI_ISL_1907173, EPI_ISL_1907174, EPI_ISL_1907175, EPI_ISL_1907176, EPI_ISL_1907177, EPI_ISL_1907179, EPI_ISL_1907180, EPI_ISL_1907181, EPI_ISL_1907182, EPI_ISL_1907183, EPI_ISL_1907184, EPI_ISL_1907185, EPI_ISL_1907186, EPI_ISL_1907187, EPI_ISL_1907188, EPI_ISL_1907189, EPI_ISL_1907190, EPI_ISL_1907191, EPI_ISL_1907192, EPI_ISL_1907193, EPI_ISL_1907194, EPI_ISL_1907195, EPI_ISL_1907196, EPI_ISL_1907197, EPI_ISL_1907198, EPI_ISL_1907199, EPI_ISL_1907200, EPI_ISL_1907201, EPI_ISL_1907202, EPI_ISL_1907203, EPI_ISL_1907204, EPI_ISL_1907205, EPI_ISL_1907206, EPI_ISL_1907207, EPI_ISL_1907208, EPI_ISL_1907209, EPI_ISL_1907210, EPI_ISL_1907211, EPI_ISL_1907212, EPI_ISL_1907213, EPI_ISL_1907214, EPI_ISL_1907215, EPI_ISL_1907216, EPI_ISL_1907217, EPI_ISL_1907218, EPI_ISL_1907219, EPI_ISL_1907220, EPI_ISL_1907221, EPI_ISL_1907222, EPI_ISL_1907223, EPI_ISL_1907224, EPI_ISL_1907225, EPI_ISL_1907226, EPI_ISL_1907227, EPI_ISL_1907228, EPI_ISL_1907229, EPI_ISL_1907230, EPI_ISL_1907231, EPI_ISL_1907232, EPI_ISL_1907233, EPI_ISL_1907234, EPI_ISL_1907235, EPI_ISL_1907236, EPI_ISL_1907237, EPI_ISL_1907238, EPI_ISL_1907239, EPI_ISL_1907240, EPI_ISL_1907241, EPI_ISL_1907242, EPI_ISL_1907243, EPI_ISL_1907244, EPI_ISL_1907245, EPI_ISL_1907246, EPI_ISL_1907247, EPI_ISL_1907248, EPI_ISL_1907249, EPI_ISL_1907250, EPI_ISL_1907251, EPI_ISL_1907253, EPI_ISL_1907254, EPI_ISL_1907255, EPI_ISL_1907256, EPI_ISL_1907257, EPI_ISL_1907259, EPI_ISL_1907260, EPI_ISL_1907261, EPI_ISL_1907262, EPI_ISL_1907263, EPI_ISL_1907264, EPI_ISL_1907265, EPI_ISL_1907266, EPI_ISL_1907267, EPI_ISL_1907268, EPI_ISL_1907269, EPI_ISL_1907270, EPI_ISL_1907271, EPI_ISL_1907272, EPI_ISL_1907273, EPI_ISL_1907274, EPI_ISL_1907275, EPI_ISL_1907276, EPI_ISL_1907277, EPI_ISL_1907278, EPI_ISL_1907279, EPI_ISL_1907280, EPI_ISL_1907281, EPI_ISL_1907282, EPI_ISL_1907283, EPI_ISL_1907284, EPI_ISL_1907285, EPI_ISL_1907286, EPI_ISL_1907287, EPI_ISL_1907288, EPI_ISL_1907289, EPI_ISL_1907290, EPI_ISL_1907291, EPI_ISL_1907292, EPI_ISL_1907293, EPI_ISL_1907294, EPI_ISL_1907295, EPI_ISL_1907296, EPI_ISL_1907297, EPI_ISL_1907298, EPI_ISL_1907299, EPI_ISL_1907300, EPI_ISL_1907301, EPI_ISL_1907302, EPI_ISL_1907303, EPI_ISL_1907304, EPI_ISL_1907305, EPI_ISL_1907306, EPI_ISL_1907307, EPI_ISL_1907308, EPI_ISL_1907309, EPI_ISL_1907310, EPI_ISL_1907311, EPI_ISL_1907312, EPI_ISL_1907313, EPI_ISL_1907314, EPI_ISL_1907315, EPI_ISL_1907316, EPI_ISL_1907317, EPI_ISL_1907318, EPI_ISL_1907319, EPI_ISL_1907320, EPI_ISL_1907321, EPI_ISL_1907322, EPI_ISL_1907323, EPI_ISL_1907324, EPI_ISL_1907325, EPI_ISL_1907326, EPI_ISL_1907327, EPI_ISL_1907328, EPI_ISL_1907329, EPI_ISL_1907330, EPI_ISL_1907331, EPI_ISL_1907332, EPI_ISL_1907333, EPI_ISL_1907334, EPI_ISL_1907335, EPI_ISL_1907336, EPI_ISL_1907337, EPI_ISL_1907338, EPI_ISL_1907339, EPI_ISL_1907340, EPI_ISL_1907341, EPI_ISL_1907342, EPI_ISL_1907343, EPI_ISL_1907344, EPI_ISL_1907345, EPI_ISL_1907346, EPI_ISL_1907347, EPI_ISL_1907348, EPI_ISL_1907349, EPI_ISL_1907350, EPI_ISL_1907351, EPI_ISL_1907352, EPI_ISL_1907353, EPI_ISL_1907354, EPI_ISL_1907355, EPI_ISL_1907356, EPI_ISL_1907357, EPI_ISL_1907358, EPI_ISL_1907359, EPI_ISL_1907360, EPI_ISL_1907361, EPI_ISL_1907363, EPI_ISL_1907364, EPI_ISL_1907365, EPI_ISL_1907366, EPI_ISL_1907367, EPI_ISL_1907368, EPI_ISL_1907369, EPI_ISL_1907370, EPI_ISL_1907371, EPI_ISL_1907372, EPI_ISL_1907373, EPI_ISL_1907374, EPI_ISL_1907375, EPI_ISL_1907376, EPI_ISL_1907377, EPI_ISL_1907378, EPI_ISL_1907379, EPI_ISL_1907380, EPI_ISL_1907381, EPI_ISL_1907382, EPI_ISL_1907383, EPI_ISL_1907384, EPI_ISL_1907385, EPI_ISL_1907386, EPI_ISL_1907387, EPI_ISL_1907388, EPI_ISL_1907389, EPI_ISL_1907390, EPI_ISL_1907391, EPI_ISL_1907392, EPI_ISL_1907393, EPI_ISL_1907394, EPI_ISL_1907395, EPI_ISL_1907396, EPI_ISL_1907397, EPI_ISL_1907398, EPI_ISL_1907399, EPI_ISL_1907400, EPI_ISL_1907401, EPI_ISL_1907402, EPI_ISL_1907403, EPI_ISL_1907404, EPI_ISL_1907405, EPI_ISL_1907406, EPI_ISL_1907407, EPI_ISL_1907408, EPI_ISL_1907409, EPI_ISL_1907410, EPI_ISL_1907411, EPI_ISL_1907412, EPI_ISL_1907413, EPI_ISL_1907414, EPI_ISL_1907415, EPI_ISL_1907416, EPI_ISL_1907417, EPI_ISL_1907418, EPI_ISL_1907419, EPI_ISL_1907420, EPI_ISL_1907421, EPI_ISL_1907422, EPI_ISL_1907423, EPI_ISL_1907424, EPI_ISL_1907425, EPI_ISL_1907426, EPI_ISL_1907427, EPI_ISL_1907428, EPI_ISL_1907429, EPI_ISL_1907430, EPI_ISL_1907431, EPI_ISL_1907432, EPI_ISL_1907433, EPI_ISL_1907434, EPI_ISL_1907435, EPI_ISL_1907436, EPI_ISL_1907437, EPI_ISL_1907438, EPI_ISL_1907439, EPI_ISL_1907440, EPI_ISL_1907441, EPI_ISL_1907442, EPI_ISL_1907443, EPI_ISL_1907444, EPI_ISL_1907445, EPI_ISL_1907446, EPI_ISL_1907447, EPI_ISL_1907448, EPI_ISL_1907449, EPI_ISL_1907450, EPI_ISL_1907451, EPI_ISL_1907452, EPI_ISL_1907453, EPI_ISL_1907454, EPI_ISL_1907455, EPI_ISL_1907456, EPI_ISL_1907457, EPI_ISL_1907458, EPI_ISL_1907459, EPI_ISL_1907460, EPI_ISL_1907461, EPI_ISL_1907462, EPI_ISL_1907463, EPI_ISL_1907464, EPI_ISL_1907465, EPI_ISL_1907466, EPI_ISL_1907467, EPI_ISL_1907468, EPI_ISL_1907469, EPI_ISL_1907470, EPI_ISL_1907471, EPI_ISL_1907472, EPI_ISL_1907473, EPI_ISL_1907474, EPI_ISL_1907475, EPI_ISL_1907476, EPI_ISL_1907477, EPI_ISL_1907478, EPI_ISL_1907479, EPI_ISL_1907480, EPI_ISL_1907481, EPI_ISL_1907482, EPI_ISL_1907483, EPI_ISL_1907484, EPI_ISL_1907485, EPI_ISL_1907486, EPI_ISL_1907487, EPI_ISL_1907488, EPI_ISL_1907489, EPI_ISL_1907490, EPI_ISL_1907491, EPI_ISL_1907492, EPI_ISL_1907493, EPI_ISL_1907494, EPI_ISL_1907495, EPI_ISL_1907497, EPI_ISL_1907498, EPI_ISL_1907499, EPI_ISL_1907500, EPI_ISL_1907502, EPI_ISL_1907503, EPI_ISL_1907504, EPI_ISL_1907505, EPI_ISL_1907507, EPI_ISL_1907508, EPI_ISL_1907509, EPI_ISL_1907510, EPI_ISL_1907511, EPI_ISL_1907512, EPI_ISL_1907513, EPI_ISL_1907514, EPI_ISL_1907515, EPI_ISL_1907516, EPI_ISL_1907517, EPI_ISL_1907518, EPI_ISL_1907519, EPI_ISL_1907520, EPI_ISL_1907521, EPI_ISL_1907522, EPI_ISL_1907523, EPI_ISL_1907524, EPI_ISL_1907525, EPI_ISL_1907526, EPI_ISL_1907527, EPI_ISL_1907528, EPI_ISL_1907529, EPI_ISL_1907530, EPI_ISL_1907531, EPI_ISL_1907532, EPI_ISL_1907533, EPI_ISL_1907534, EPI_ISL_1907535, EPI_ISL_1907536, EPI_ISL_1907537, EPI_ISL_1907538, EPI_ISL_1907539, EPI_ISL_1907540, EPI_ISL_1907541, EPI_ISL_1907542, EPI_ISL_1907543, EPI_ISL_1907544, EPI_ISL_1907545, EPI_ISL_1907546, EPI_ISL_1907547, EPI_ISL_1907548, EPI_ISL_1907549, EPI_ISL_1907550, EPI_ISL_1907551, EPI_ISL_1907552, EPI_ISL_1907553, EPI_ISL_1907554, EPI_ISL_1907555, EPI_ISL_1907556, EPI_ISL_1907557, EPI_ISL_1907558, EPI_ISL_1907559, EPI_ISL_1907560, EPI_ISL_1907561, EPI_ISL_1907562, EPI_ISL_1907563, EPI_ISL_1907564, EPI_ISL_1907565, EPI_ISL_1907566, EPI_ISL_1907567, EPI_ISL_1907568, EPI_ISL_1907569, EPI_ISL_1907570, EPI_ISL_1907571, EPI_ISL_1907572, EPI_ISL_1907573, EPI_ISL_1907574, EPI_ISL_1907575, EPI_ISL_1907576, EPI_ISL_1907577, EPI_ISL_1907578, EPI_ISL_1907579, EPI_ISL_1907580, EPI_ISL_1907582, EPI_ISL_1907583, EPI_ISL_1907584, EPI_ISL_1907585, EPI_ISL_1907586, EPI_ISL_1907587, EPI_ISL_1907588, EPI_ISL_1907589, EPI_ISL_1907590, EPI_ISL_1907591, EPI_ISL_1907592, EPI_ISL_1907593, EPI_ISL_1907594, EPI_ISL_1907595, EPI_ISL_1907596, EPI_ISL_1907597, EPI_ISL_1907598, EPI_ISL_1907599, EPI_ISL_1907600, EPI_ISL_1907601, EPI_ISL_1907602, EPI_ISL_1907603, EPI_ISL_1907604, EPI_ISL_1907605, EPI_ISL_1907606, EPI_ISL_1907607, EPI_ISL_1907608, EPI_ISL_1907609, EPI_ISL_1907610, EPI_ISL_1907611, EPI_ISL_1907612, EPI_ISL_1907613, EPI_ISL_1907614, EPI_ISL_1907615, EPI_ISL_1907616, EPI_ISL_1907617, EPI_ISL_1907618, EPI_ISL_1907619, EPI_ISL_1907620, EPI_ISL_1907621, EPI_ISL_1907622, EPI_ISL_1907623, EPI_ISL_1907624, EPI_ISL_1907625, EPI_ISL_1907626, EPI_ISL_1907627, EPI_ISL_1907628, EPI_ISL_1907629, EPI_ISL_1907630, EPI_ISL_1907631, EPI_ISL_1907632, EPI_ISL_1907633, EPI_ISL_1907634, EPI_ISL_1907635, EPI_ISL_1907636, EPI_ISL_1907637, EPI_ISL_1907638, EPI_ISL_1907639, EPI_ISL_1907640, EPI_ISL_1907641, EPI_ISL_1907642, EPI_ISL_1907643, EPI_ISL_1907644, EPI_ISL_1907645, EPI_ISL_1907646, EPI_ISL_1907647, EPI_ISL_1907648, EPI_ISL_1907649, EPI_ISL_1907650, EPI_ISL_1907651, EPI_ISL_1907652, EPI_ISL_1907653, EPI_ISL_1907654, EPI_ISL_1907655, EPI_ISL_1907656, EPI_ISL_1907657, EPI_ISL_1907658, EPI_ISL_1907659, EPI_ISL_1907660, EPI_ISL_1907661, EPI_ISL_1907662, EPI_ISL_1907663, EPI_ISL_1907664, EPI_ISL_1907665, EPI_ISL_1907666, EPI_ISL_1907667, EPI_ISL_1907668, EPI_ISL_1907669, EPI_ISL_1907670, EPI_ISL_1907671, EPI_ISL_1907672, EPI_ISL_1907673, EPI_ISL_1907674, EPI_ISL_1907675, EPI_ISL_1907676, EPI_ISL_1907677, EPI_ISL_1907678, EPI_ISL_1907679, EPI_ISL_1907680, EPI_ISL_1907681, EPI_ISL_1907682, EPI_ISL_1907683, EPI_ISL_1907684, EPI_ISL_1907685, EPI_ISL_1907686, EPI_ISL_1907687, EPI_ISL_1907688, EPI_ISL_1907689, EPI_ISL_1907690, EPI_ISL_1907691, EPI_ISL_1907692, EPI_ISL_1907693, EPI_ISL_1907694, EPI_ISL_1907695, EPI_ISL_1907696, EPI_ISL_1907697, EPI_ISL_1907698, EPI_ISL_1907699, EPI_ISL_1907700, EPI_ISL_1907701, EPI_ISL_1907702, EPI_ISL_1907703, EPI_ISL_1907704, EPI_ISL_1907705, EPI_ISL_1907706, EPI_ISL_1907707, EPI_ISL_1907708, EPI_ISL_1907709, EPI_ISL_1907710, EPI_ISL_1907711, EPI_ISL_1907712, EPI_ISL_1907713, EPI_ISL_1907714, EPI_ISL_1907715, EPI_ISL_1907716, EPI_ISL_1907717, EPI_ISL_1907718, EPI_ISL_1907719, EPI_ISL_1907720, EPI_ISL_1907721, EPI_ISL_1907722, EPI_ISL_1907723, EPI_ISL_1907724, EPI_ISL_1907725, EPI_ISL_1907726, EPI_ISL_1907727, EPI_ISL_1907728, EPI_ISL_1907729, EPI_ISL_1907730, EPI_ISL_1907731, EPI_ISL_1907732, EPI_ISL_1907733, EPI_ISL_1907734, EPI_ISL_1907735, EPI_ISL_1907736, EPI_ISL_1907737, EPI_ISL_1907738, EPI_ISL_1907739, EPI_ISL_1907740, EPI_ISL_1907741, EPI_ISL_1907742, EPI_ISL_1907743, EPI_ISL_1907744, EPI_ISL_1907745, EPI_ISL_1907746, EPI_ISL_1907747, EPI_ISL_1907748, EPI_ISL_1907749, EPI_ISL_1907750, EPI_ISL_1907751, EPI_ISL_1907752, EPI_ISL_1907753, EPI_ISL_1907754, EPI_ISL_1907755, EPI_ISL_1907756, EPI_ISL_1907757, EPI_ISL_1907758, EPI_ISL_1907759, EPI_ISL_1907760, EPI_ISL_1907761, EPI_ISL_1907762, EPI_ISL_1907763, EPI_ISL_1907764, EPI_ISL_1907765, EPI_ISL_1907766, EPI_ISL_1907767, EPI_ISL_1907768, EPI_ISL_1907769, EPI_ISL_1907770, EPI_ISL_1907771, EPI_ISL_1907772, EPI_ISL_1907773, EPI_ISL_1907774, EPI_ISL_1907775, EPI_ISL_1907776, EPI_ISL_1907777, EPI_ISL_1907778, EPI_ISL_1907779, EPI_ISL_1907780, EPI_ISL_1907781, EPI_ISL_1907782, EPI_ISL_1907783, EPI_ISL_1907784, EPI_ISL_1907785, EPI_ISL_1907786, EPI_ISL_1907787, EPI_ISL_1907788, EPI_ISL_1907789, EPI_ISL_1907790, EPI_ISL_1907791, EPI_ISL_1907792, EPI_ISL_1907793, EPI_ISL_1907794, EPI_ISL_1907795, EPI_ISL_1907796, EPI_ISL_1907797, EPI_ISL_1907798, EPI_ISL_1907799, EPI_ISL_1907800, EPI_ISL_1907801, EPI_ISL_1907802, EPI_ISL_1907803, EPI_ISL_1907804, EPI_ISL_1907805, EPI_ISL_1907806, EPI_ISL_1907807, EPI_ISL_1907808, EPI_ISL_1907809, EPI_ISL_1907810, EPI_ISL_1907811, EPI_ISL_1907812, EPI_ISL_1907813, EPI_ISL_1907814, EPI_ISL_1907815, EPI_ISL_1907816, EPI_ISL_1907817, EPI_ISL_1907818, EPI_ISL_1907819, EPI_ISL_1907820, EPI_ISL_1907821, EPI_ISL_1907822, EPI_ISL_1907823, EPI_ISL_1907824, EPI_ISL_1907825, EPI_ISL_1907826, EPI_ISL_1907827, EPI_ISL_1907828, EPI_ISL_1907829, EPI_ISL_1907830, EPI_ISL_1907831, EPI_ISL_1907832, EPI_ISL_1907833, EPI_ISL_1907834, EPI_ISL_1907835, EPI_ISL_1907836, EPI_ISL_1907837, EPI_ISL_1907838, EPI_ISL_1907839, EPI_ISL_1907840, EPI_ISL_1907841, EPI_ISL_1907842, EPI_ISL_1907843, EPI_ISL_1907844, EPI_ISL_1907845, EPI_ISL_1907846, EPI_ISL_1907847, EPI_ISL_1907848, EPI_ISL_1907849, EPI_ISL_1907850, EPI_ISL_1907851, EPI_ISL_1907852, EPI_ISL_1907853, EPI_ISL_1907854, EPI_ISL_1907855, EPI_ISL_1907856, EPI_ISL_1907857, EPI_ISL_1907858, EPI_ISL_1907859, EPI_ISL_1907860, EPI_ISL_1907861, EPI_ISL_1907862, EPI_ISL_1907863, EPI_ISL_1907864, EPI_ISL_1907865, EPI_ISL_1907866, EPI_ISL_1907867, EPI_ISL_1907868, EPI_ISL_1907869, EPI_ISL_1907870, EPI_ISL_1907871, EPI_ISL_1907872, EPI_ISL_1907873, EPI_ISL_1907874, EPI_ISL_1907875, EPI_ISL_1907876, EPI_ISL_1907877, EPI_ISL_1907878, EPI_ISL_1907879, EPI_ISL_1907880, EPI_ISL_1907881, EPI_ISL_1907882, EPI_ISL_1907883, EPI_ISL_1907884, EPI_ISL_1907885, EPI_ISL_1907886, EPI_ISL_1907887, EPI_ISL_1907888, EPI_ISL_1907889, EPI_ISL_1907890, EPI_ISL_1907891, EPI_ISL_1907892, EPI_ISL_1907893, EPI_ISL_1907894, EPI_ISL_1907895, EPI_ISL_1907896, EPI_ISL_1907897, EPI_ISL_1907898, EPI_ISL_1907899, EPI_ISL_1907900, EPI_ISL_1907901, EPI_ISL_1907902, EPI_ISL_1907903, EPI_ISL_1907904, EPI_ISL_1907905, EPI_ISL_1907906, EPI_ISL_1907907, EPI_ISL_1907908, EPI_ISL_1907909, EPI_ISL_1907910, EPI_ISL_1907911, EPI_ISL_1907912, EPI_ISL_1907913, EPI_ISL_1907914, EPI_ISL_1907915, EPI_ISL_1907916, EPI_ISL_1907917, EPI_ISL_1907918, EPI_ISL_1907919, EPI_ISL_1907920, EPI_ISL_1907921, EPI_ISL_1907922, EPI_ISL_1907923, EPI_ISL_1907924, EPI_ISL_1907925, EPI_ISL_1907926, EPI_ISL_1907927, EPI_ISL_1907928, EPI_ISL_1907929, EPI_ISL_1907931, EPI_ISL_1907932, EPI_ISL_1907933, EPI_ISL_1907934, EPI_ISL_1907935, EPI_ISL_1907936, EPI_ISL_1907937, EPI_ISL_1907938, EPI_ISL_1907939, EPI_ISL_1907940, EPI_ISL_1907941, EPI_ISL_1907942, EPI_ISL_1907943, EPI_ISL_1907944, EPI_ISL_1907945, EPI_ISL_1907946, EPI_ISL_1907947, EPI_ISL_1907948, EPI_ISL_1907949, EPI_ISL_1907950, EPI_ISL_1907951, EPI_ISL_1907952, EPI_ISL_1907953, EPI_ISL_1907954, EPI_ISL_1907955, EPI_ISL_1907956, EPI_ISL_1907957, EPI_ISL_1907958, EPI_ISL_1907959, EPI_ISL_1907960, EPI_ISL_1907962, EPI_ISL_1907963, EPI_ISL_1907964, EPI_ISL_1907965, EPI_ISL_1907966, EPI_ISL_1907967, EPI_ISL_1907968, EPI_ISL_1907969, EPI_ISL_1907970, EPI_ISL_1907971, EPI_ISL_1907972, EPI_ISL_1907973, EPI_ISL_1907974, EPI_ISL_1907975, EPI_ISL_1907976, EPI_ISL_1907977, EPI_ISL_1907978, EPI_ISL_1907979, EPI_ISL_1907980, EPI_ISL_1907981, EPI_ISL_1907982, EPI_ISL_1907983, EPI_ISL_1907984, EPI_ISL_1907985, EPI_ISL_1907986, EPI_ISL_1907987, EPI_ISL_1907988, EPI_ISL_1907989, EPI_ISL_1907990, EPI_ISL_1907991, EPI_ISL_1907992, EPI_ISL_1907993, EPI_ISL_1907994, EPI_ISL_1907995, EPI_ISL_1907996, EPI_ISL_1907997, EPI_ISL_1907998, EPI_ISL_1907999, EPI_ISL_1908000, EPI_ISL_1908001, EPI_ISL_1908002, EPI_ISL_1908003, EPI_ISL_1908004, EPI_ISL_1908005, EPI_ISL_1908006, EPI_ISL_1908007, EPI_ISL_1908008, EPI_ISL_1908009, EPI_ISL_1908010, EPI_ISL_1908011, EPI_ISL_1908012, EPI_ISL_1908013, EPI_ISL_1908014, EPI_ISL_1908015, EPI_ISL_1908016, EPI_ISL_1908017, EPI_ISL_1908018, EPI_ISL_1908019, EPI_ISL_1908020, EPI_ISL_1908021, EPI_ISL_1908022, EPI_ISL_1908023, EPI_ISL_1908024, EPI_ISL_1908025, EPI_ISL_1908026, EPI_ISL_1908027, EPI_ISL_1908028, EPI_ISL_1908029, EPI_ISL_1908030, EPI_ISL_1908031, EPI_ISL_1908032, EPI_ISL_1908033, EPI_ISL_1908034, EPI_ISL_1908035, EPI_ISL_1908036, EPI_ISL_1908037, EPI_ISL_1908038, EPI_ISL_1908039, EPI_ISL_1908040, EPI_ISL_1908041, EPI_ISL_1908042, EPI_ISL_1908043, EPI_ISL_1908044, EPI_ISL_1908045, EPI_ISL_1908046, EPI_ISL_1908047, EPI_ISL_1908048, EPI_ISL_1908049, EPI_ISL_1908050, EPI_ISL_1908051, EPI_ISL_1908052, EPI_ISL_1908053, EPI_ISL_1908054, EPI_ISL_1908055, EPI_ISL_1908056, EPI_ISL_1908057, EPI_ISL_1908058, EPI_ISL_1908059, EPI_ISL_1908060, EPI_ISL_1908061, EPI_ISL_1908062, EPI_ISL_1908063, EPI_ISL_1908064, EPI_ISL_1908065, EPI_ISL_1908066, EPI_ISL_1908067, EPI_ISL_1908068, EPI_ISL_1908069, EPI_ISL_1908070, EPI_ISL_1908071, EPI_ISL_1908072, EPI_ISL_1908073, EPI_ISL_1908074, EPI_ISL_1908075, EPI_ISL_1908076, EPI_ISL_1908077, EPI_ISL_1908078, EPI_ISL_1908079, EPI_ISL_1908080, EPI_ISL_1908081, EPI_ISL_1908082, EPI_ISL_1908083, EPI_ISL_1908084, EPI_ISL_1908085, EPI_ISL_1908086, EPI_ISL_1908087, EPI_ISL_1908088, EPI_ISL_1908089, EPI_ISL_1908090, EPI_ISL_1908091, EPI_ISL_1908092, EPI_ISL_1908093, EPI_ISL_1908094, EPI_ISL_1908095, EPI_ISL_1908096, EPI_ISL_1908097, EPI_ISL_1908098, EPI_ISL_1908099, EPI_ISL_1908100, EPI_ISL_1908101, EPI_ISL_1908102, EPI_ISL_1908103, EPI_ISL_1908104, EPI_ISL_1908105, EPI_ISL_1908106, EPI_ISL_1908107, EPI_ISL_1908108, EPI_ISL_1908109, EPI_ISL_1908110, EPI_ISL_1908111, EPI_ISL_1908112, EPI_ISL_1908113, EPI_ISL_1908114, EPI_ISL_1908115, EPI_ISL_1908116, EPI_ISL_1908117, EPI_ISL_1908118, EPI_ISL_1908119, EPI_ISL_1908120, EPI_ISL_1908121, EPI_ISL_1908122, EPI_ISL_1908123, EPI_ISL_1908124, EPI_ISL_1908125, EPI_ISL_1908126, EPI_ISL_1908127, EPI_ISL_1908128, EPI_ISL_1908129, EPI_ISL_1908130, EPI_ISL_1908131, EPI_ISL_1908132, EPI_ISL_1908133, EPI_ISL_1908134, EPI_ISL_1908135, EPI_ISL_1908136, EPI_ISL_1908137, EPI_ISL_1908138, EPI_ISL_1908139, EPI_ISL_1908140, EPI_ISL_1908141, EPI_ISL_1908142, EPI_ISL_1908143, EPI_ISL_1908144, EPI_ISL_1908145, EPI_ISL_1908146 | | | |
| see above | Helix/Illumina | Centers for Disease Control and Prevention Division of Viral Diseases, Pathogen Discovery | Dakota Howard, Dhwani Batra, Peter W. Cook, Kara Moser, Adrian Paskey, Jason Caravas, Benjamin Rambo-Martin, Shatavia Morrison, Christopher Gulvick, Scott Sammons, Yvette Unoarumhi, Darlene Wagner, Matthew Schmerer, Eileen de Feo, Jan Antico, Christine Tran, Matthew Tolentino, Shannon Wickline, Kim Gietzen, Brad Sickler, Jingtao Liu, Eric Allen, Phil Febbo, Nicole L. Washington, Simon White, Geraint Levan, Kelly Schiabor Barrett, Elizabeth Cirulli, Alexandre Bolze, Ary Ascencio, Charlotte Rivera-Garcia, Ryan Cho, Jason Nguyen, Sherry Wang, Jimmy Ramirez, Tyler Cassens, Efren Sandoval, Magnus Isaksson, William Lee, David Becker, Marc Laurent, James Lu, Clinton R. Paden, Duncan MacCannell |
| EPI_ISL_1908435, EPI_ISL_1908436, EPI_ISL_1908437, EPI_ISL_1908438, EPI_ISL_1908439, EPI_ISL_1908440, EPI_ISL_1908441, EPI_ISL_1908442, EPI_ISL_1908443, EPI_ISL_1908444, EPI_ISL_1908445, EPI_ISL_1908446, EPI_ISL_1908447, EPI_ISL_1908448, EPI_ISL_1908449, EPI_ISL_1908450, EPI_ISL_1908451, EPI_ISL_1908452, EPI_ISL_1908453, EPI_ISL_1908454, EPI_ISL_1908455, EPI_ISL_1908456, EPI_ISL_1908457, EPI_ISL_1908458, EPI_ISL_1908459, EPI_ISL_1908460, EPI_ISL_1908461, EPI_ISL_1908462, EPI_ISL_1908463, EPI_ISL_1908464, EPI_ISL_1908465, EPI_ISL_1908466, EPI_ISL_1908467, EPI_ISL_1908468, EPI_ISL_1908469, EPI_ISL_1908470, EPI_ISL_1908471, EPI_ISL_1908472, EPI_ISL_1908473, EPI_ISL_1908474, EPI_ISL_1908475, EPI_ISL_1908476, EPI_ISL_1908477, EPI_ISL_1908478, EPI_ISL_1908479, EPI_ISL_1908480, EPI_ISL_1908481, EPI_ISL_1908482, EPI_ISL_1908483, EPI_ISL_1908484, EPI_ISL_1908485, EPI_ISL_1908486, EPI_ISL_1908487, EPI_ISL_1908488, EPI_ISL_1908489, EPI_ISL_1908490, EPI_ISL_1908491, EPI_ISL_1908492, EPI_ISL_1908493, EPI_ISL_1908494, EPI_ISL_1908495, EPI_ISL_1908496, EPI_ISL_1908497, EPI_ISL_1908498, EPI_ISL_1908499, EPI_ISL_1908500, EPI_ISL_1908501, EPI_ISL_1908502, EPI_ISL_1908503, EPI_ISL_1908504, EPI_ISL_1908505, EPI_ISL_1908506, EPI_ISL_1908507, EPI_ISL_1908508, EPI_ISL_1908509, EPI_ISL_1908510, EPI_ISL_1908511, EPI_ISL_1908512, EPI_ISL_1908513, EPI_ISL_1908514, EPI_ISL_1908515, EPI_ISL_1908516, EPI_ISL_1908517, EPI_ISL_1908518, EPI_ISL_1908519, EPI_ISL_1908520, EPI_ISL_1908521, EPI_ISL_1908522, EPI_ISL_1908523, EPI_ISL_1908524, EPI_ISL_1908525, EPI_ISL_1908526 | | | |
| see above | MD PHL | MD PHL | Maryland Department of Health Laboratories Administration |
| EPI_ISL_1908899, EPI_ISL_1908901, EPI_ISL_1908902, EPI_ISL_1908903, EPI_ISL_1908904, EPI_ISL_1908905, EPI_ISL_1908906, EPI_ISL_1908909, EPI_ISL_1908910, EPI_ISL_1908912, EPI_ISL_1908915, EPI_ISL_1908916, EPI_ISL_1908917, EPI_ISL_1908918, EPI_ISL_1908919, EPI_ISL_1908921, EPI_ISL_1908922, EPI_ISL_1908923, EPI_ISL_1908924, EPI_ISL_1908925, EPI_ISL_1908926, EPI_ISL_1908928, EPI_ISL_1908929, EPI_ISL_1908930, EPI_ISL_1908931, EPI_ISL_1908932, EPI_ISL_1908933, EPI_ISL_1908934, EPI_ISL_1908935, EPI_ISL_1908936, EPI_ISL_1908937, EPI_ISL_1908938, EPI_ISL_1908939, EPI_ISL_1908940, EPI_ISL_1908941, EPI_ISL_1908943, EPI_ISL_1908945, EPI_ISL_1908946, EPI_ISL_1908947, EPI_ISL_1908948, EPI_ISL_1908949, EPI_ISL_1908950, EPI_ISL_1908951, EPI_ISL_1908952, EPI_ISL_1908953, EPI_ISL_1908954, EPI_ISL_1908955, EPI_ISL_1908956, EPI_ISL_1908957, EPI_ISL_1908958, EPI_ISL_1908959, EPI_ISL_1908961, EPI_ISL_1908962, EPI_ISL_1908963, EPI_ISL_1908965, EPI_ISL_1908966, EPI_ISL_1908968, EPI_ISL_1908969, EPI_ISL_1908970, EPI_ISL_1908971, EPI_ISL_1908972, EPI_ISL_1908975, EPI_ISL_1908977, EPI_ISL_1908978, EPI_ISL_1908979, EPI_ISL_1908980, EPI_ISL_1908982, EPI_ISL_1908983, EPI_ISL_1908984, EPI_ISL_1908985, EPI_ISL_1908986, EPI_ISL_1908987, EPI_ISL_1908988, EPI_ISL_1908989, EPI_ISL_1908990, EPI_ISL_1908991, EPI_ISL_1908992, EPI_ISL_1908993, EPI_ISL_1908994, EPI_ISL_1908995, EPI_ISL_1908996, EPI_ISL_1908997, EPI_ISL_1908998, EPI_ISL_1908999, EPI_ISL_1909000, EPI_ISL_1909001, EPI_ISL_1909002, EPI_ISL_1909003, EPI_ISL_1909004, EPI_ISL_1909005, EPI_ISL_1909006, EPI_ISL_1909009, EPI_ISL_1909010, EPI_ISL_1909011, EPI_ISL_1909012, EPI_ISL_1909013, EPI_ISL_1909014, EPI_ISL_1909015, EPI_ISL_1909016, EPI_ISL_1909017, EPI_ISL_1909018, EPI_ISL_1909019, EPI_ISL_1909020, EPI_ISL_1909021, EPI_ISL_1909022, EPI_ISL_1909023, EPI_ISL_1909024, EPI_ISL_1909025, EPI_ISL_1909026, EPI_ISL_1909027, EPI_ISL_1909028, EPI_ISL_1909029, EPI_ISL_1909030, EPI_ISL_1909032, EPI_ISL_1909033, EPI_ISL_1909034, EPI_ISL_1909035, EPI_ISL_1909036, EPI_ISL_1909037, EPI_ISL_1909038, EPI_ISL_1909039, EPI_ISL_1909040, EPI_ISL_1909041, EPI_ISL_1909042, EPI_ISL_1909043, EPI_ISL_1909044, EPI_ISL_1909045, EPI_ISL_1909046, EPI_ISL_1909047, EPI_ISL_1909048, EPI_ISL_1909049, EPI_ISL_1909050, EPI_ISL_1909051, EPI_ISL_1909052, EPI_ISL_1909053, EPI_ISL_1909054, EPI_ISL_1909055, EPI_ISL_1909056, EPI_ISL_1909058, EPI_ISL_1909059, EPI_ISL_1909060, EPI_ISL_1909061, EPI_ISL_1909063, EPI_ISL_1909064, EPI_ISL_1909066, EPI_ISL_1909067, EPI_ISL_1909068, EPI_ISL_1909069, EPI_ISL_1909071, EPI_ISL_1909072, EPI_ISL_1909073, EPI_ISL_1909074, EPI_ISL_1909076, EPI_ISL_1909077, EPI_ISL_1909079, EPI_ISL_1909080, EPI_ISL_1909081, EPI_ISL_1909082, EPI_ISL_1909083, EPI_ISL_1909085, EPI_ISL_1909086, EPI_ISL_1909087, EPI_ISL_1909089, EPI_ISL_1909090, EPI_ISL_1909091 | | | |
| see above | DPHL | Delaware Public Health Lab | Rebecca Savage |
| EPI_ISL_1909096, EPI_ISL_1909097, EPI_ISL_1909098, EPI_ISL_1909100, EPI_ISL_1909101, EPI_ISL_1909103, EPI_ISL_1909106, EPI_ISL_1909107, EPI_ISL_1909108, EPI_ISL_1909110, EPI_ISL_1909111, EPI_ISL_1909112, EPI_ISL_1909113, EPI_ISL_1909114, EPI_ISL_1909115, EPI_ISL_1909116, EPI_ISL_1909117, EPI_ISL_1909118, EPI_ISL_1909119, EPI_ISL_1909120 | | | |
| see above | Wyoming Public Health Laboratory | Wyoming Public Health Laboratory | Jim Mildenberger, Taylor Fearing, Channing Weber, Ashley Norberg, Chayse Rowley, Marley Goetz, Brian Dominguez, Elliot Thomasson, Brittany Oher, Sam Britz, Cari Sloma, and Rob Christensen |
| EPI_ISL_1909240 | University of Michigan Clinical Microbiology Laboratory | Lauring Lab, University of Michigan, Department of Microbiology and Immunology | Valesano |
| EPI_ISL_1909245, EPI_ISL_1909248 | Montana Public Health Laboratory | Montana Public Health Laboratory | Joy Ritter, Michelle Mozer, Carrie Biskupiak, Deborah Gibson |
| EPI_ISL_1909259, EPI_ISL_1909263, EPI_ISL_1909265, EPI_ISL_1909268, EPI_ISL_1909270, EPI_ISL_1909271, EPI_ISL_1909276, EPI_ISL_1909279, EPI_ISL_1909282 | Sonora Quest Laboratories | TGen North | "Jolene Bowers, Heather Centner, Chris French, Hayley Yaglom, Ashlyn Pfeiffer, Darrin Lemmer, Dave Engelthaler, The Arizona COVID Genomics Union (ACGU)" |
| EPI_ISL_1909286, EPI_ISL_1909287 | TGen North | TGen North | "Jolene Bowers, Heather Centner, Chris French, Hayley Yaglom, Ashlyn Pfeiffer, Darrin Lemmer, Dave Engelthaler, The Arizona COVID Genomics Union (ACGU)" |
| EPI_ISL_1909288, EPI_ISL_1909289, EPI_ISL_1909292, EPI_ISL_1909293, EPI_ISL_1909294, EPI_ISL_1909295, EPI_ISL_1909296, EPI_ISL_1909300, EPI_ISL_1909303, EPI_ISL_1909304, EPI_ISL_1909305, EPI_ISL_1909306, EPI_ISL_1909307, EPI_ISL_1909308, EPI_ISL_1909309, EPI_ISL_1909310, EPI_ISL_1909313 | | | |
| see above | Sonora Quest Laboratories | TGen North | "Jolene Bowers, Heather Centner, Chris French, Hayley Yaglom, Ashlyn Pfeiffer, Darrin Lemmer, Dave Engelthaler, The Arizona COVID Genomics Union (ACGU)" |
| EPI_ISL_1909316 | TGen North | TGen North | "Jolene Bowers, Heather Centner, Chris French, Hayley Yaglom, Ashlyn Pfeiffer, Darrin Lemmer, Dave Engelthaler, The Arizona COVID Genomics Union (ACGU)" |
| EPI_ISL_1909317, EPI_ISL_1909318, EPI_ISL_1909319, EPI_ISL_1909320, EPI_ISL_1909322, EPI_ISL_1909328, EPI_ISL_1909329, EPI_ISL_1909331, EPI_ISL_1909334 | Sonora Quest Laboratories | TGen North | "Jolene Bowers, Heather Centner, Chris French, Hayley Yaglom, Ashlyn Pfeiffer, Darrin Lemmer, Dave Engelthaler, The Arizona COVID Genomics Union (ACGU)" |
| EPI_ISL_1909335 | TGen North | TGen North | "Jolene Bowers, Heather Centner, Chris French, Hayley Yaglom, Ashlyn Pfeiffer, Darrin Lemmer, Dave Engelthaler, The Arizona COVID Genomics Union (ACGU)" |
| EPI_ISL_1909336, EPI_ISL_1909339, EPI_ISL_1909341, EPI_ISL_1909342, EPI_ISL_1909345, EPI_ISL_1909346, EPI_ISL_1909348, EPI_ISL_1909350, EPI_ISL_1909352, EPI_ISL_1909353, EPI_ISL_1909354, EPI_ISL_1909355, EPI_ISL_1909356, EPI_ISL_1909357, EPI_ISL_1909361, EPI_ISL_1909363, EPI_ISL_1909365, EPI_ISL_1909367, EPI_ISL_1909369, EPI_ISL_1909371, EPI_ISL_1909372, EPI_ISL_1909376, EPI_ISL_1909382, EPI_ISL_1909383, EPI_ISL_1909384, EPI_ISL_1909385, EPI_ISL_1909387 | | | |
| see above | Sonora Quest Laboratories | TGen North | "Jolene Bowers, Heather Centner, Chris French, Hayley Yaglom, Ashlyn Pfeiffer, Darrin Lemmer, Dave Engelthaler, The Arizona COVID Genomics Union (ACGU)" |
| EPI_ISL_1909389 | TGen North | TGen North | "Jolene Bowers, Heather Centner, Chris French, Hayley Yaglom, Ashlyn Pfeiffer, Darrin Lemmer, Dave Engelthaler, The Arizona COVID Genomics Union (ACGU)" |
| EPI_ISL_1909391, EPI_ISL_1909393 | Sonora Quest Laboratories | TGen North | "Jolene Bowers, Heather Centner, Chris French, Hayley Yaglom, Ashlyn Pfeiffer, Darrin Lemmer, Dave Engelthaler, The Arizona COVID Genomics Union (ACGU)" |

| | | | |
|---|---|---|---|
| EPI_ISL_1909394 | TGen North | TGen North | "Jolene Bowers, Heather Centner, Chris French, Hayley Yaglom, Ashlyn Pfeiffer, Darrin Lemmer, Dave Engelthaler, The Arizona COVID Genomics Union (ACGU)" |
| EPI_ISL_1909395, EPI_ISL_1909398, EPI_ISL_1909401, EPI_ISL_1909403, EPI_ISL_1909406, EPI_ISL_1909408, EPI_ISL_1909411, EPI_ISL_1909412, EPI_ISL_1909413, EPI_ISL_1909414, EPI_ISL_1909415, EPI_ISL_1909419 | | | |
| see above | Sonora Quest Laboratories | TGen North | "Jolene Bowers, Heather Centner, Chris French, Hayley Yaglom, Ashlyn Pfeiffer, Darrin Lemmer, Dave Engelthaler, The Arizona COVID Genomics Union (ACGU)" |
| EPI_ISL_1909422 | TGen North | TGen North | "Jolene Bowers, Heather Centner, Chris French, Hayley Yaglom, Ashlyn Pfeiffer, Darrin Lemmer, Dave Engelthaler, The Arizona COVID Genomics Union (ACGU)" |
| EPI_ISL_1909425, EPI_ISL_1909426, EPI_ISL_1909427, EPI_ISL_1909431, EPI_ISL_1909432, EPI_ISL_1909434 | Sonora Quest Laboratories | TGen North | "Jolene Bowers, Heather Centner, Chris French, Hayley Yaglom, Ashlyn Pfeiffer, Darrin Lemmer, Dave Engelthaler, The Arizona COVID Genomics Union (ACGU)" |
| EPI_ISL_1909436 | TGen North | TGen North | "Jolene Bowers, Heather Centner, Chris French, Hayley Yaglom, Ashlyn Pfeiffer, Darrin Lemmer, Dave Engelthaler, The Arizona COVID Genomics Union (ACGU)" |
| EPI_ISL_1909441, EPI_ISL_1909442, EPI_ISL_1909443, EPI_ISL_1909444, EPI_ISL_1909445, EPI_ISL_1909446, EPI_ISL_1909448, EPI_ISL_1909449, EPI_ISL_1909450, EPI_ISL_1909451 | Sonora Quest Laboratories | TGen North | "Jolene Bowers, Heather Centner, Chris French, Hayley Yaglom, Ashlyn Pfeiffer, Darrin Lemmer, Dave Engelthaler, The Arizona COVID Genomics Union (ACGU)" |
| EPI_ISL_1909452 | TGen North | TGen North | "Jolene Bowers, Heather Centner, Chris French, Hayley Yaglom, Ashlyn Pfeiffer, Darrin Lemmer, Dave Engelthaler, The Arizona COVID Genomics Union (ACGU)" |
| EPI_ISL_1909453, EPI_ISL_1909455, EPI_ISL_1909457, EPI_ISL_1909463, EPI_ISL_1909465, EPI_ISL_1909466, EPI_ISL_1909467, EPI_ISL_1909469, EPI_ISL_1909474, EPI_ISL_1909475, EPI_ISL_1909480 | | | |
| see above | Sonora Quest Laboratories | TGen North | "Jolene Bowers, Heather Centner, Chris French, Hayley Yaglom, Ashlyn Pfeiffer, Darrin Lemmer, Dave Engelthaler, The Arizona COVID Genomics Union (ACGU)" |
| EPI_ISL_1909482 | TGen North | TGen North | "Jolene Bowers, Heather Centner, Chris French, Hayley Yaglom, Ashlyn Pfeiffer, Darrin Lemmer, Dave Engelthaler, The Arizona COVID Genomics Union (ACGU)" |
| EPI_ISL_1909484, EPI_ISL_1909485, EPI_ISL_1909488, EPI_ISL_1909490, EPI_ISL_1909492, EPI_ISL_1909494, EPI_ISL_1909495, EPI_ISL_1909496, EPI_ISL_1909497, EPI_ISL_1909498, EPI_ISL_1909499, EPI_ISL_1909500, EPI_ISL_1909502, EPI_ISL_1909503, EPI_ISL_1909504, EPI_ISL_1909505, EPI_ISL_1909507, EPI_ISL_1909508, EPI_ISL_1909509, EPI_ISL_1909512, EPI_ISL_1909514, EPI_ISL_1909515, EPI_ISL_1909516, EPI_ISL_1909517, EPI_ISL_1909518, EPI_ISL_1909519, EPI_ISL_1909521, EPI_ISL_1909523, EPI_ISL_1909527, EPI_ISL_1909528, EPI_ISL_1909535, EPI_ISL_1909536, EPI_ISL_1909537, EPI_ISL_1909538, EPI_ISL_1909542 | | | |
| see above | Sonora Quest Laboratories | TGen North | "Jolene Bowers, Heather Centner, Chris French, Hayley Yaglom, Ashlyn Pfeiffer, Darrin Lemmer, Dave Engelthaler, The Arizona COVID Genomics Union (ACGU)" |
| EPI_ISL_1909545 | TGen North | TGen North | "Jolene Bowers, Heather Centner, Chris French, Hayley Yaglom, Ashlyn Pfeiffer, Darrin Lemmer, Dave Engelthaler, The Arizona COVID Genomics Union (ACGU)" |
| EPI_ISL_1909547, EPI_ISL_1909550 | Sonora Quest Laboratories | TGen North | "Jolene Bowers, Heather Centner, Chris French, Hayley Yaglom, Ashlyn Pfeiffer, Darrin Lemmer, Dave Engelthaler, The Arizona COVID Genomics Union (ACGU)" |
| EPI_ISL_1909556 | TGen North | TGen North | "Jolene Bowers, Heather Centner, Chris French, Hayley Yaglom, Ashlyn Pfeiffer, Darrin Lemmer, Dave Engelthaler, The Arizona COVID Genomics Union (ACGU)" |
| EPI_ISL_1909559, EPI_ISL_1909567, EPI_ISL_1909570, EPI_ISL_1909571, EPI_ISL_1909579, EPI_ISL_1909581, EPI_ISL_1909582, EPI_ISL_1909583, EPI_ISL_1909584, EPI_ISL_1909589, EPI_ISL_1909591, EPI_ISL_1909597 | | | |
| see above | Sonora Quest Laboratories | TGen North | "Jolene Bowers, Heather Centner, Chris French, Hayley Yaglom, Ashlyn Pfeiffer, Darrin Lemmer, Dave Engelthaler, The Arizona COVID Genomics Union (ACGU)" |
| EPI_ISL_1909598 | TGen North | TGen North | "Jolene Bowers, Heather Centner, Chris French, Hayley Yaglom, Ashlyn Pfeiffer, Darrin Lemmer, Dave Engelthaler, The Arizona COVID Genomics Union (ACGU)" |
| EPI_ISL_1909599, EPI_ISL_1909601, EPI_ISL_1909604, EPI_ISL_1909605, EPI_ISL_1909606, EPI_ISL_1909609 | Sonora Quest Laboratories | TGen North | "Jolene Bowers, Heather Centner, Chris French, Hayley Yaglom, Ashlyn Pfeiffer, Darrin Lemmer, Dave Engelthaler, The Arizona COVID Genomics Union (ACGU)" |
| EPI_ISL_1909618, EPI_ISL_1909621 | TGen North | TGen North | "Jolene Bowers, Heather Centner, Chris French, Hayley Yaglom, Ashlyn Pfeiffer, Darrin Lemmer, Dave Engelthaler, The Arizona COVID Genomics Union (ACGU)" |
| EPI_ISL_1909624, EPI_ISL_1909625, EPI_ISL_1909629, EPI_ISL_1909631, EPI_ISL_1909633 | Sonora Quest Laboratories | TGen North | "Jolene Bowers, Heather Centner, Chris French, Hayley Yaglom, Ashlyn Pfeiffer, Darrin Lemmer, Dave Engelthaler, The Arizona COVID Genomics Union (ACGU)" |
| EPI_ISL_1909634 | TGen North | TGen North | "Jolene Bowers, Heather Centner, Chris French, Hayley Yaglom, Ashlyn Pfeiffer, Darrin Lemmer, Dave Engelthaler, The Arizona COVID Genomics Union (ACGU)" |
| EPI_ISL_1909639, EPI_ISL_1909640, EPI_ISL_1909641 | Sonora Quest Laboratories | TGen North | "Jolene Bowers, Heather Centner, Chris French, Hayley Yaglom, Ashlyn Pfeiffer, Darrin Lemmer, Dave Engelthaler, The Arizona COVID Genomics Union (ACGU)" |
| EPI_ISL_1909645 | TGen North | TGen North | "Jolene Bowers, Heather Centner, Chris French, Hayley Yaglom, Ashlyn Pfeiffer, Darrin Lemmer, Dave Engelthaler, The Arizona COVID Genomics Union (ACGU)" |
| EPI_ISL_1909651, EPI_ISL_1909653, EPI_ISL_1909655, EPI_ISL_1909656, EPI_ISL_1909657, EPI_ISL_1909658, EPI_ISL_1909659, EPI_ISL_1909660, EPI_ISL_1909664, EPI_ISL_1909665, EPI_ISL_1909670, EPI_ISL_1909673, EPI_ISL_1909675, EPI_ISL_1909678, EPI_ISL_1909679, EPI_ISL_1909680, EPI_ISL_1909681 | | | |
| see above | Sonora Quest Laboratories | TGen North | "Jolene Bowers, Heather Centner, Chris French, Hayley Yaglom, Ashlyn Pfeiffer, Darrin Lemmer, Dave Engelthaler, The Arizona COVID Genomics Union (ACGU)" |
| EPI_ISL_1909683 | TGen North | TGen North | "Jolene Bowers, Heather Centner, Chris French, Hayley Yaglom, Ashlyn Pfeiffer, Darrin Lemmer, Dave Engelthaler, The Arizona COVID Genomics Union (ACGU)" |
| EPI_ISL_1909686, EPI_ISL_1909688, EPI_ISL_1909690, EPI_ISL_1909692, EPI_ISL_1909693, EPI_ISL_1909697, EPI_ISL_1909702 | Sonora Quest Laboratories | TGen North | "Jolene Bowers, Heather Centner, Chris French, Hayley Yaglom, Ashlyn Pfeiffer, Darrin Lemmer, Dave Engelthaler, The Arizona COVID Genomics Union (ACGU)" |
| EPI_ISL_1909704 | TGen North | TGen North | "Jolene Bowers, Heather Centner, Chris French, Hayley Yaglom, Ashlyn Pfeiffer, Darrin Lemmer, Dave Engelthaler, The Arizona COVID Genomics Union (ACGU)" |
| EPI_ISL_1909707 | Sonora Quest Laboratories | TGen North | "Jolene Bowers, Heather Centner, Chris French, Hayley Yaglom, Ashlyn Pfeiffer, Darrin Lemmer, Dave Engelthaler, The Arizona COVID Genomics Union (ACGU)" |
| EPI_ISL_1909708 | TGen North | TGen North | "Jolene Bowers, Heather Centner, Chris French, Hayley Yaglom, Ashlyn Pfeiffer, Darrin Lemmer, Dave Engelthaler, The Arizona COVID Genomics Union (ACGU)" |
| EPI_ISL_1909709, EPI_ISL_1909710, EPI_ISL_1909711, EPI_ISL_1909713, EPI_ISL_1909717, EPI_ISL_1909718, EPI_ISL_1909719, EPI_ISL_1909723, EPI_ISL_1909726, EPI_ISL_1909729, EPI_ISL_1909732, EPI_ISL_1909737, EPI_ISL_1909738, EPI_ISL_1909742, EPI_ISL_1909747, EPI_ISL_1909749, EPI_ISL_1909751, EPI_ISL_1909752, EPI_ISL_1909755, EPI_ISL_1909757, EPI_ISL_1909760, EPI_ISL_1909761, EPI_ISL_1909762, EPI_ISL_1909765, EPI_ISL_1909767, EPI_ISL_1909771, EPI_ISL_1909773, EPI_ISL_1909774, EPI_ISL_1909775, EPI_ISL_1909776 | | | |
| see above | Sonora Quest Laboratories | TGen North | "Jolene Bowers, Heather Centner, Chris French, Hayley Yaglom, Ashlyn Pfeiffer, Darrin Lemmer, Dave Engelthaler, The Arizona COVID Genomics Union (ACGU)" |
| EPI_ISL_1909778, EPI_ISL_1909779 | TGen North | TGen North | "Jolene Bowers, Heather Centner, Chris French, Hayley Yaglom, Ashlyn Pfeiffer, Darrin Lemmer, Dave Engelthaler, The Arizona COVID Genomics Union (ACGU)" |
| EPI_ISL_1910451, EPI_ISL_1910452, EPI_ISL_1910454, EPI_ISL_1910455, EPI_ISL_1910456, EPI_ISL_1910457, EPI_ISL_1910458, EPI_ISL_1910459, EPI_ISL_1910460, EPI_ISL_1910461, EPI_ISL_1910462, EPI_ISL_1910463, EPI_ISL_1910464, EPI_ISL_1910465, EPI_ISL_1910466 | | | |
| see above | County of San Diego Health and Human Services Agency | Pathogen Discovery, Respiratory Viruses Branch, Division of Viral Diseases, Centers for Disease Control and Prevention | Anna Kelleher, Ying Tao, Yan Li, Jing Zhang, Brian Lynch, Krista Queen, Anna Uehara, Peter Cook, Han Jia Justin Ng, Rachel Marine, Clinton R. Paden, Dhwani Batra, Haibin Wang, Tara Coalter, Jasmine Padilla, Morgan Davis, Mili Sheth, Sarah Nobles, Mark Burroughs, Justin Lee, Adam Retchless, Suxiang Tong |
| EPI_ISL_1910629, EPI_ISL_1910630, EPI_ISL_1910631, EPI_ISL_1910632, EPI_ISL_1910633, EPI_ISL_1910634, EPI_ISL_1910635, EPI_ISL_1910636, EPI_ISL_1910637, EPI_ISL_1910638, EPI_ISL_1910639, EPI_ISL_1910640, EPI_ISL_1910641, EPI_ISL_1910642, EPI_ISL_1910643, EPI_ISL_1910644, EPI_ISL_1910645, EPI_ISL_1910646, EPI_ISL_1910647, EPI_ISL_1910648, EPI_ISL_1910649, EPI_ISL_1910650, EPI_ISL_1910651, EPI_ISL_1910652, EPI_ISL_1910653, EPI_ISL_1910654, EPI_ISL_1910655, EPI_ISL_1910656, EPI_ISL_1910657, EPI_ISL_1910658, EPI_ISL_1910659, EPI_ISL_1910660, EPI_ISL_1910661, EPI_ISL_1910662, EPI_ISL_1910663, EPI_ISL_1910664, EPI_ISL_1910665, EPI_ISL_1910666, EPI_ISL_1910667, EPI_ISL_1910668, EPI_ISL_1910669, EPI_ISL_1910670, EPI_ISL_1910671, EPI_ISL_1910672, EPI_ISL_1910673, EPI_ISL_1910674, EPI_ISL_1910675, EPI_ISL_1910676, EPI_ISL_1910677, EPI_ISL_1910678, EPI_ISL_1910679, EPI_ISL_1910680, EPI_ISL_1910681, EPI_ISL_1910682, EPI_ISL_1910683, EPI_ISL_1910684, EPI_ISL_1910685, EPI_ISL_1910686, EPI_ISL_1910687, EPI_ISL_1910688, EPI_ISL_1910689, EPI_ISL_1910690, EPI_ISL_1910691, EPI_ISL_1910692, EPI_ISL_1910693, EPI_ISL_1910694, EPI_ISL_1910695, EPI_ISL_1910696, EPI_ISL_1910697, EPI_ISL_1910698, EPI_ISL_1910699, EPI_ISL_1910700, EPI_ISL_1910701, EPI_ISL_1910702, EPI_ISL_1910703, EPI_ISL_1910704, EPI_ISL_1910705, EPI_ISL_1910706, EPI_ISL_1910707, EPI_ISL_1910708, EPI_ISL_1910709, EPI_ISL_1910710, EPI_ISL_1910711, EPI_ISL_1910712, EPI_ISL_1910713, EPI_ISL_1910714, EPI_ISL_1910715, EPI_ISL_1910716, EPI_ISL_1910717, EPI_ISL_1910718, EPI_ISL_1910719, EPI_ISL_1910720, EPI_ISL_1910721, EPI_ISL_1910722, EPI_ISL_1910723, EPI_ISL_1910724, EPI_ISL_1910725, EPI_ISL_1910726, EPI_ISL_1910727, EPI_ISL_1910728, EPI_ISL_1910729, EPI_ISL_1910730, EPI_ISL_1910731, EPI_ISL_1910732, EPI_ISL_1910733, EPI_ISL_1910734, EPI_ISL_1910735, EPI_ISL_1910736, EPI_ISL_1910737, EPI_ISL_1910738, EPI_ISL_1910739, EPI_ISL_1910740, EPI_ISL_1910741, EPI_ISL_1910742, EPI_ISL_1910743, EPI_ISL_1910744, EPI_ISL_1910745, EPI_ISL_1910746, EPI_ISL_1910747, EPI_ISL_1910748, EPI_ISL_1910749, EPI_ISL_1910750, EPI_ISL_1910751, EPI_ISL_1910752, EPI_ISL_1910753, EPI_ISL_1910754 | | | |
| see above | Utah Public Health Laboratory | Utah Public Health Laboratory | Erin L. Young, Kelly F. Oakeson, Tara Gallagher |
| EPI_ISL_1910762, EPI_ISL_1910763, EPI_ISL_1910764, EPI_ISL_1910765, EPI_ISL_1910766, EPI_ISL_1910767, EPI_ISL_1910768, EPI_ISL_1910769, EPI_ISL_1910770, EPI_ISL_1910771, EPI_ISL_1910772, EPI_ISL_1910773, EPI_ISL_1910774, EPI_ISL_1910775, EPI_ISL_1910776, EPI_ISL_1910777, EPI_ISL_1910778, EPI_ISL_1910779, EPI_ISL_1910781, EPI_ISL_1910783, EPI_ISL_1910784, EPI_ISL_1910785, EPI_ISL_1910788, EPI_ISL_1910790, EPI_ISL_1910791, EPI_ISL_1910793, EPI_ISL_1910794, EPI_ISL_1910795, EPI_ISL_1910796, EPI_ISL_1910799, EPI_ISL_1910800, EPI_ISL_1910802, EPI_ISL_1910804, EPI_ISL_1910806, EPI_ISL_1910807, EPI_ISL_1910808, EPI_ISL_1910810, EPI_ISL_1910811, EPI_ISL_1910812, EPI_ISL_1910813, EPI_ISL_1910815, EPI_ISL_1910816, EPI_ISL_1910817, EPI_ISL_1910818, EPI_ISL_1910820, EPI_ISL_1910821, EPI_ISL_1910823, EPI_ISL_1910824, EPI_ISL_1910825, EPI_ISL_1910826 | | | |
| see above | Arizona State University | Arizona State University | Peter T. Skidmore, LaRinda A. Holland, Matthew F. Smith, Rabia Maqsood, Nicholas J. Mellor, Joy M. Blain, Valerie Harris, Joshua LaBaer, Vel Murugan, Efrem S. Lim |
| EPI_ISL_1910827, EPI_ISL_1910828, EPI_ISL_1910829, EPI_ISL_1910830, EPI_ISL_1910831, EPI_ISL_1910832, EPI_ISL_1910833, EPI_ISL_1910834, EPI_ISL_1910835, EPI_ISL_1910836, EPI_ISL_1910837, EPI_ISL_1910838, EPI_ISL_1910839, EPI_ISL_1910840, EPI_ISL_1910841, EPI_ISL_1910842, EPI_ISL_1910843, EPI_ISL_1910844, EPI_ISL_1910845, EPI_ISL_1910846, EPI_ISL_1910847, EPI_ISL_1910848, EPI_ISL_1910849, EPI_ISL_1910850, EPI_ISL_1910851, EPI_ISL_1910852, EPI_ISL_1910853, EPI_ISL_1910854, EPI_ISL_1910855 | | | |
| see above | University of Michigan Clinical Microbiology Laboratory | Lauring Lab, University of Michigan, Department of Microbiology and Immunology | Valesano |
| EPI_ISL_1911005, | Notre Dame COVID Response and | Notre Dame Genomics & Bioinformatics Core | Michael Pfrender, Joseph Sarro, Melissa Stephens |

| | | | |
|---|---|---|---|
| EPI_ISL_1911007 | Testing Laboratory | Facility | |
| EPI_ISL_1911013, EPI_ISL_1911014, EPI_ISL_1911015, EPI_ISL_1911016, EPI_ISL_1911017, EPI_ISL_1911018, EPI_ISL_1911019, EPI_ISL_1911020, EPI_ISL_1911021, EPI_ISL_1911022, EPI_ISL_1911023, EPI_ISL_1911024, EPI_ISL_1911025, EPI_ISL_1911026, EPI_ISL_1911027, EPI_ISL_1911028, EPI_ISL_1911029, EPI_ISL_1911030, EPI_ISL_1911031, EPI_ISL_1911032, EPI_ISL_1911033, EPI_ISL_1911034, EPI_ISL_1911035, EPI_ISL_1911036, EPI_ISL_1911037, EPI_ISL_1911038, EPI_ISL_1911039, EPI_ISL_1911040, EPI_ISL_1911041, EPI_ISL_1911042, EPI_ISL_1911043, EPI_ISL_1911044, EPI_ISL_1911045, EPI_ISL_1911046, EPI_ISL_1911047, EPI_ISL_1911048, EPI_ISL_1911049, EPI_ISL_1911050, EPI_ISL_1911051, EPI_ISL_1911052, EPI_ISL_1911053, EPI_ISL_1911054, EPI_ISL_1911055, EPI_ISL_1911056, EPI_ISL_1911057, EPI_ISL_1911058, EPI_ISL_1911059, EPI_ISL_1911060, EPI_ISL_1911061, EPI_ISL_1911062, EPI_ISL_1911063, EPI_ISL_1911064, EPI_ISL_1911065, EPI_ISL_1911066, EPI_ISL_1911067, EPI_ISL_1911068, EPI_ISL_1911069, EPI_ISL_1911070, EPI_ISL_1911071, EPI_ISL_1911072, EPI_ISL_1911073, EPI_ISL_1911074, EPI_ISL_1911075, EPI_ISL_1911076, EPI_ISL_1911077, EPI_ISL_1911078, EPI_ISL_1911079, EPI_ISL_1911080, EPI_ISL_1911081, EPI_ISL_1911082, EPI_ISL_1911084, EPI_ISL_1911085, EPI_ISL_1911086, EPI_ISL_1911087, EPI_ISL_1911088, EPI_ISL_1911089, EPI_ISL_1911090, EPI_ISL_1911091, EPI_ISL_1911092, EPI_ISL_1911093, EPI_ISL_1911094, EPI_ISL_1911095, EPI_ISL_1911096, EPI_ISL_1911097, EPI_ISL_1911098, EPI_ISL_1911099, EPI_ISL_1911100, EPI_ISL_1911101 | | | |
| see above | The Ohio State University Applied Microbiology Services Laboratory | The Ohio State University Applied Microbiology Services Laboratory | Seth A. Faith PhD |
| EPI_ISL_1911102, EPI_ISL_1911103, EPI_ISL_1911104, EPI_ISL_1911106, EPI_ISL_1911107, EPI_ISL_1911108, EPI_ISL_1911109, EPI_ISL_1911110, EPI_ISL_1911111 | Maine Health and Environmental Testing Laboratory | Maine Health and Environmental Testing Laboratory | Matluk,N., Dewey,H., Martha,J., Meak,S., Grieser,H.,Barter,M., Lynch,R., Munger,H. and Tewhey,R. |
| EPI_ISL_1911198, EPI_ISL_1911199, EPI_ISL_1911200, EPI_ISL_1911201 | Naval Infectious Diseases Diagnostic Laboratory | Naval Medical Research Center Biological Defense Research Directorate | Logan Voegtly, Catherine Arnold, Bishwo Adhikari, Francisco Malagon Bautista, Andrea Luquette, Gregory Rice, Andrew Bennett, Kyle Long, Lindsay Glang, Michael Deschenes, Megan Schilling, Victor Sugiharto, Regina Cer, Kimberly Bishop-Lilly |
| EPI_ISL_1911202, EPI_ISL_1911203, EPI_ISL_1911204, EPI_ISL_1911205, EPI_ISL_1911206, EPI_ISL_1911207, EPI_ISL_1911208, EPI_ISL_1911209, EPI_ISL_1911210 | Naval Health Clinic Annapolis | Naval Medical Research Center Biological Defense Research Directorate | Logan Voegtly, Catherine Arnold, Bishwo Adhikari, Francisco Malagon Bautista, Andrea Luquette, Gregory Rice, Andrew Bennett, Kyle Long, Lindsay Glang, Michael Deschenes, Regina Cer, Kimberly Bishop-Lilly |
| EPI_ISL_1911211, EPI_ISL_1911212, EPI_ISL_1911213, EPI_ISL_1911214, EPI_ISL_1911215, EPI_ISL_1911216, EPI_ISL_1911217, EPI_ISL_1911218, EPI_ISL_1911219, EPI_ISL_1911220, EPI_ISL_1911221, EPI_ISL_1911222, EPI_ISL_1911223, EPI_ISL_1911224, EPI_ISL_1911225, EPI_ISL_1911226, EPI_ISL_1911227, EPI_ISL_1911228, EPI_ISL_1911229, EPI_ISL_1911230, EPI_ISL_1911231, EPI_ISL_1911232, EPI_ISL_1911233, EPI_ISL_1911234, EPI_ISL_1911235, EPI_ISL_1911236, EPI_ISL_1911237, EPI_ISL_1911238, EPI_ISL_1911239, EPI_ISL_1911240, EPI_ISL_1911241, EPI_ISL_1911242, EPI_ISL_1911243, EPI_ISL_1911244, EPI_ISL_1911245, EPI_ISL_1911246, EPI_ISL_1911247, EPI_ISL_1911248, EPI_ISL_1911249, EPI_ISL_1911251, EPI_ISL_1911252 | | | |
| see above | Naval Infectious Diseases Diagnostic Laboratory | Naval Medical Research Center Biological Defense Research Directorate | Logan Voegtly, Catherine Arnold, Bishwo Adhikari, Francisco Malagon Bautista, Andrea Luquette, Gregory Rice, Andrew Bennett, Kyle Long, Lindsay Glang, Michael Deschenes, Megan Schilling, Victor Sugiharto, Regina Cer, Kimberly Bishop-Lilly |
| EPI_ISL_1911253, EPI_ISL_1911254, EPI_ISL_1911255, EPI_ISL_1911256, EPI_ISL_1911260, EPI_ISL_1911261, EPI_ISL_1911262, EPI_ISL_1911263, EPI_ISL_1911265, EPI_ISL_1911267, EPI_ISL_1911268, EPI_ISL_1911273, EPI_ISL_1911276, EPI_ISL_1911277, EPI_ISL_1911279, EPI_ISL_1911281, EPI_ISL_1911285, EPI_ISL_1911286, EPI_ISL_1911288, EPI_ISL_1911290, EPI_ISL_1911291, EPI_ISL_1911293, EPI_ISL_1911296, EPI_ISL_1911297, EPI_ISL_1911298, EPI_ISL_1911300, EPI_ISL_1911301, EPI_ISL_1911302, EPI_ISL_1911305, EPI_ISL_1911306, EPI_ISL_1911307, EPI_ISL_1911310 | | | |
| see above | LESPNL | LESPNL | (in alphabetical order) Consuelo Treviño-Garza, Eduardo Isaac de la Rosa-Moreno, Else del Carmen Garcia-Garcia, Gloria Alejandra Jasso-de la Peña, Manuel Enrique de la O-Cavazos, Olín Medina-Chávez, Yulianna Mayre Cordero-Cruz |
| EPI_ISL_1911349, EPI_ISL_1911350, EPI_ISL_1911351, EPI_ISL_1911352, EPI_ISL_1911353, EPI_ISL_1911354, EPI_ISL_1911355, EPI_ISL_1911356, EPI_ISL_1911357, EPI_ISL_1911358, EPI_ISL_1911359, EPI_ISL_1911360, EPI_ISL_1911361, EPI_ISL_1911362, EPI_ISL_1911363, EPI_ISL_1911364, EPI_ISL_1911365, EPI_ISL_1911366, EPI_ISL_1911367, EPI_ISL_1911368 | | | |
| see above | CDPH, Viral and Rickettsial Disease Laboratory | Pathogen Discovery, Respiratory Viruses Branch, Division of Viral Diseases, Centers for Disease Control and Prevention | Anna Kelleher, Ying Tao, Yan Li, Jing Zhang, Brian Lynch, Krista Queen, Anna Uehara, Peter Cook, Han Jia Justin Ng, Rachel Marine, Clinton R. Paden, Dhwani Batra, Haibin Wang, Tara Coalter, Jasmine Padilla, Morgan Davis, Mili Sheth, Sarah Nobles, Mark Burroughs, Justin Lee, Adam Retchless, Suxiang Tong |
| EPI_ISL_1911369, EPI_ISL_1911370, EPI_ISL_1911371, EPI_ISL_1911372, EPI_ISL_1911373, EPI_ISL_1911374, EPI_ISL_1911375, EPI_ISL_1911376, EPI_ISL_1911377, EPI_ISL_1911378, EPI_ISL_1911379, EPI_ISL_1911380 | | | |
| see above | Nebraska Public Health Laboratory | NPHL COVID-19 Response Team | NPHL COVID-19 Response Team |
| EPI_ISL_1911384, EPI_ISL_1911385, EPI_ISL_1911386, EPI_ISL_1911388, EPI_ISL_1911389, EPI_ISL_1911390, EPI_ISL_1911391, EPI_ISL_1911393, EPI_ISL_1911394, EPI_ISL_1911395, EPI_ISL_1911396, EPI_ISL_1911397, EPI_ISL_1911398, EPI_ISL_1911399, EPI_ISL_1911400, EPI_ISL_1911401, EPI_ISL_1911402, EPI_ISL_1911403, EPI_ISL_1911404, EPI_ISL_1911405, EPI_ISL_1911406, EPI_ISL_1911407, EPI_ISL_1911408, EPI_ISL_1911409, EPI_ISL_1911410, EPI_ISL_1911411, EPI_ISL_1911412, EPI_ISL_1911413, EPI_ISL_1911414, EPI_ISL_1911415, EPI_ISL_1911416, EPI_ISL_1911417, EPI_ISL_1911418, EPI_ISL_1911419, EPI_ISL_1911420, EPI_ISL_1911421, EPI_ISL_1911422 | | | |
| see above | Wyoming Public Health Laboratory | Wyoming Public Health Laboratory | Jim Mildenberger, Wanda Manley, Noah Hull, Taylor Fearing, Lynette Gumbleton, Channing Weber, Ashley Norberg, Chayse Rowley, Marley Goetz, Brian Dominguez, Elliot Thomasson, Sam Britz, Cari Sloma, and Rob Christensen |
| EPI_ISL_1911509, EPI_ISL_1911510 | Kansas Health and Environmental Lab | Kansas Health and Environmental Lab | Mike Grose, Jonathan Barnell, Ben Olsen, and Phil Adam |
| EPI_ISL_1911511, EPI_ISL_1911513 | Cytocheck Laboratory | Kansas Health and Environmental Lab | Mike Grose, Jonathan Barnell, Ben Olsen, and Phil Adam |
| EPI_ISL_1911515 | Kansas Health and Environmental Lab | Kansas Health and Environmental Lab | Mike Grose, Jonathan Barnell, Ben Olsen, and Phil Adam |
| EPI_ISL_1911516, EPI_ISL_1911517, EPI_ISL_1911518, EPI_ISL_1911519, EPI_ISL_1911521, EPI_ISL_1911522, EPI_ISL_1911523, EPI_ISL_1911524, EPI_ISL_1911525 | Arizona State University | Arizona State University | Rabia Maqsood, LaRinda A. Holland, Matthew F. Smith, Peter T. Skidmore, Nicholas J. Mellor, Joy M. Blain, Valerie Harris, Joshua LaBaer, Vel Murugan, Efrem S. Lim |
| EPI_ISL_1911902, EPI_ISL_1911903, EPI_ISL_1911904, EPI_ISL_1911905, EPI_ISL_1911906, EPI_ISL_1911907, EPI_ISL_1911908, EPI_ISL_1911909, EPI_ISL_1911910, EPI_ISL_1911911, EPI_ISL_1911912, EPI_ISL_1911913, EPI_ISL_1911914, EPI_ISL_1911915, EPI_ISL_1911916, EPI_ISL_1911917, EPI_ISL_1911918, EPI_ISL_1911919, EPI_ISL_1911920, EPI_ISL_1911921, EPI_ISL_1911922, EPI_ISL_1911923, EPI_ISL_1911924, EPI_ISL_1911925, EPI_ISL_1911926, EPI_ISL_1911927, EPI_ISL_1911928, EPI_ISL_1911929, EPI_ISL_1911930, EPI_ISL_1911931, EPI_ISL_1911932, EPI_ISL_1911933, EPI_ISL_1911934, EPI_ISL_1911935, EPI_ISL_1911936, EPI_ISL_1911937, EPI_ISL_1911938, EPI_ISL_1911939, EPI_ISL_1911940, EPI_ISL_1911941, EPI_ISL_1911942, EPI_ISL_1911943, EPI_ISL_1911944, EPI_ISL_1911945, EPI_ISL_1911946, EPI_ISL_1911947, EPI_ISL_1911948, EPI_ISL_1911949, EPI_ISL_1911950, EPI_ISL_1911951, EPI_ISL_1911952, EPI_ISL_1911953, EPI_ISL_1911954, EPI_ISL_1911955, EPI_ISL_1911956 | | | |
| see above | Baylor Scott & White-Temple | Baylor Scott & White-Temple | Ari Rao, Linden Morales, Kimberly Walker, Marcus Volz, Shelby Hendrickson |
| EPI_ISL_1911967, EPI_ISL_1911969, EPI_ISL_1911974, EPI_ISL_1911975, EPI_ISL_1911976, EPI_ISL_1911979, EPI_ISL_1911980, EPI_ISL_1911982, EPI_ISL_1911983, EPI_ISL_1911985, EPI_ISL_1911986 | | | |
| see above | Wyoming Public Health Laboratory | Wyoming Public Health Laboratory | Jim Mildenberger, Taylor Fearing, Channing Weber, Ashley Norberg, Chayse Rowley, Marley Goetz, Brian Dominguez, Elliot Thomasson, Brittany Oher, Sam Britz, Cari Sloma, and Rob Christensen |
| EPI_ISL_1912791, EPI_ISL_1912792, EPI_ISL_1912793, EPI_ISL_1912794, EPI_ISL_1912795, EPI_ISL_1912796, EPI_ISL_1912797, EPI_ISL_1912798, EPI_ISL_1912799, EPI_ISL_1912800, EPI_ISL_1912801, EPI_ISL_1912802, EPI_ISL_1912803, EPI_ISL_1912804, EPI_ISL_1912805, EPI_ISL_1912806, EPI_ISL_1912807, EPI_ISL_1912808, EPI_ISL_1912809, EPI_ISL_1912810, EPI_ISL_1912811, EPI_ISL_1912812, EPI_ISL_1912813, EPI_ISL_1912814, EPI_ISL_1912815, EPI_ISL_1912816, EPI_ISL_1912817, EPI_ISL_1912818, EPI_ISL_1912819, EPI_ISL_1912820, EPI_ISL_1912821, EPI_ISL_1912822, EPI_ISL_1912823, EPI_ISL_1912824, EPI_ISL_1912825, EPI_ISL_1912826, EPI_ISL_1912827, EPI_ISL_1912828, EPI_ISL_1912829, EPI_ISL_1912830, EPI_ISL_1912831, EPI_ISL_1912832, EPI_ISL_1912833, EPI_ISL_1912834, EPI_ISL_1912835, EPI_ISL_1912836, EPI_ISL_1912837, EPI_ISL_1912838, EPI_ISL_1912839, EPI_ISL_1912840, EPI_ISL_1912841, EPI_ISL_1912842, EPI_ISL_1912843, EPI_ISL_1912844, EPI_ISL_1912845, EPI_ISL_1912846, EPI_ISL_1912847, EPI_ISL_1912848, EPI_ISL_1912849, EPI_ISL_1912850, EPI_ISL_1912851, EPI_ISL_1912852, EPI_ISL_1912853, EPI_ISL_1912854, EPI_ISL_1912855, EPI_ISL_1912856, EPI_ISL_1912857, EPI_ISL_1912858, EPI_ISL_1912859, EPI_ISL_1912860, EPI_ISL_1912861, EPI_ISL_1912862, EPI_ISL_1912863, EPI_ISL_1912864, EPI_ISL_1912865, EPI_ISL_1912866, EPI_ISL_1912867, EPI_ISL_1912868, EPI_ISL_1912869, EPI_ISL_1912870, EPI_ISL_1912871, EPI_ISL_1912872, EPI_ISL_1912873, EPI_ISL_1912874, EPI_ISL_1912875, EPI_ISL_1912876, EPI_ISL_1912877, EPI_ISL_1912878, EPI_ISL_1912879, EPI_ISL_1912880, EPI_ISL_1912881, EPI_ISL_1912882, EPI_ISL_1912883, EPI_ISL_1912884, EPI_ISL_1912885, EPI_ISL_1912886, EPI_ISL_1912887, EPI_ISL_1912888, EPI_ISL_1912889, EPI_ISL_1912890, EPI_ISL_1912891, EPI_ISL_1912892, EPI_ISL_1912893, EPI_ISL_1912894, EPI_ISL_1912895, EPI_ISL_1912896, EPI_ISL_1912897, EPI_ISL_1912898, EPI_ISL_1912899, EPI_ISL_1912900, EPI_ISL_1912901, EPI_ISL_1912902, EPI_ISL_1912903, EPI_ISL_1912904, EPI_ISL_1912905, EPI_ISL_1912906, EPI_ISL_1912907, EPI_ISL_1912908, EPI_ISL_1912909, EPI_ISL_1912910, EPI_ISL_1912911, EPI_ISL_1912912, EPI_ISL_1912913, EPI_ISL_1912914, EPI_ISL_1912915, EPI_ISL_1912916, EPI_ISL_1912918, EPI_ISL_1912919, EPI_ISL_1912920, EPI_ISL_1912921, EPI_ISL_1912922, EPI_ISL_1912923, EPI_ISL_1912924, EPI_ISL_1912925, EPI_ISL_1912926, EPI_ISL_1912927, EPI_ISL_1912928, EPI_ISL_1912929, EPI_ISL_1912930, EPI_ISL_1912931, EPI_ISL_1912932, EPI_ISL_1912933, EPI_ISL_1912934 | | | |
| see above | HMH | New York Genome Center | Michael Zody, Andre Corvelo, Dayna M. Oschwald, Samantha Fennessey, Tom Maniatis, Liang Chen, Jose Mediavilla, Marcus Cunningham, Kaelea Composto, Kar Chow, David Perlin, Barry Kreiswirth |
| EPI_ISL_1912938 | LESP Queretaro | Instituto de Diagnostico y Referencia Epidemiologicos (INDRE) | Claudia Wong-Arambula, Abril Rodriguez-Maldonado, Vanessa Rivero-Arredondo, Ariadna Medina-Benitez, Joaquin Quiroz-Mercado, Sergio Rangel-Guerrero, Natividad Cruz-Ortiz, Tatiana Nunez-Garcia, Gisela Barrera-Badillo, Lucia Hernandez-Rivas, Irma Lopez-Martinez, Ernesto Ramirez-Gonzalez. |
| EPI_ISL_1912939 | LESP Nuevo Leon | Instituto de Diagnostico y Referencia Epidemiologicos (INDRE) | Claudia Wong-Arambula, Abril Rodriguez-Maldonado, Vanessa Rivero-Arredondo, Ariadna Medina-Benitez, Joaquin Quiroz-Mercado, Sergio Rangel-Guerrero, Natividad Cruz-Ortiz, Tatiana Nunez-Garcia, Gisela Barrera-Badillo, Lucia Hernandez-Rivas, Irma Lopez-Martinez, Ernesto Ramirez-Gonzalez. |
| EPI_ISL_1912940, EPI_ISL_1912941 | LESP Queretaro | Instituto de Diagnostico y Referencia Epidemiologicos (INDRE) | Claudia Wong-Arambula, Abril Rodriguez-Maldonado, Vanessa Rivero-Arredondo, Ariadna Medina-Benitez, Joaquin Quiroz-Mercado, Sergio Rangel-Guerrero, Natividad Cruz-Ortiz, Tatiana Nunez-Garcia, Gisela Barrera-Badillo, Lucia Hernandez-Rivas, Irma Lopez-Martinez, Ernesto Ramirez-Gonzalez. |
| EPI_ISL_1912942, EPI_ISL_1912943, EPI_ISL_1912944, EPI_ISL_1912945 | LESP Tamaulipas | Instituto de Diagnostico y Referencia Epidemiologicos (INDRE) | Claudia Wong-Arambula, Abril Rodriguez-Maldonado, Vanessa Rivero-Arredondo, Ariadna Medina-Benitez, Joaquin Quiroz-Mercado, Sergio Rangel-Guerrero, Natividad Cruz-Ortiz, Tatiana Nunez-Garcia, Gisela Barrera-Badillo, Lucia Hernandez-Rivas, Irma Lopez-Martinez, Ernesto Ramirez-Gonzalez. |
| EPI_ISL_1912946 | LESP Veracruz | Instituto de Diagnostico y Referencia Epidemiologicos (INDRE) | Claudia Wong-Arambula, Abril Rodriguez-Maldonado, Vanessa Rivero-Arredondo, Ariadna Medina-Benitez, Joaquin Quiroz-Mercado, Sergio Rangel-Guerrero, Natividad Cruz-Ortiz, Tatiana Nunez-Garcia, Gisela Barrera-Badillo, Lucia Hernandez-Rivas, Irma Lopez-Martinez, Ernesto Ramirez-Gonzalez. |
| EPI_ISL_1912947, EPI_ISL_1912948, EPI_ISL_1912949, EPI_ISL_1912950, EPI_ISL_1912951, EPI_ISL_1912952, EPI_ISL_1912953, EPI_ISL_1912954, EPI_ISL_1912955, EPI_ISL_1912956, EPI_ISL_1912957, EPI_ISL_1912958, EPI_ISL_1912959, EPI_ISL_1912960, EPI_ISL_1912961, EPI_ISL_1912962, EPI_ISL_1912963, EPI_ISL_1912964, EPI_ISL_1912965, EPI_ISL_1912966, EPI_ISL_1912967, EPI_ISL_1912968, EPI_ISL_1912969, EPI_ISL_1912970, EPI_ISL_1912971, EPI_ISL_1912972, EPI_ISL_1912973, EPI_ISL_1912974, EPI_ISL_1912975, EPI_ISL_1912976, EPI_ISL_1912977, EPI_ISL_1912978, EPI_ISL_1912979 | | | |
| see above | Poplar Healthcare | Colleen B. Jonsson | Walter Reichard, Mariah Taylor, Joythi Parvathareddy, Jacob Nichols, Surekha Surendranathan, Colleen Jonsson |
| EPI_ISL_1912980, EPI_ISL_1912981, EPI_ISL_1912982, EPI_ISL_1912983, EPI_ISL_1912984, EPI_ISL_1912985, EPI_ISL_1912986, EPI_ISL_1912987, EPI_ISL_1912988, EPI_ISL_1912989, EPI_ISL_1912990, EPI_ISL_1912991, EPI_ISL_1912992, EPI_ISL_1912993, EPI_ISL_1912994, EPI_ISL_1912995, EPI_ISL_1912996, EPI_ISL_1912997, EPI_ISL_1912998, EPI_ISL_1912999 | | | |
| see above | American Esoteric Laboratories | Colleen B. Jonsson | Walter Reichard, Mariah Taylor, Joythi Parvathareddy, Jacob Nichols, Surekha Surendranathan, Colleen Jonsson |
| EPI_ISL_1913000, EPI_ISL_1913001, EPI_ISL_1913002, EPI_ISL_1913003, EPI_ISL_1913004 | Montana Public Health Laboratory | Montana Public Health Laboratory | Joy Ritter, Michelle Mozer, Carrie Biskupiak, Deborah Gibson |

| | | | |
|---|---|---|---|
| EPI_ISL_1913007, EPI_ISL_1913011, EPI_ISL_1913012, EPI_ISL_1913013 | Wyoming Public Health Laboratory | Wyoming Public Health Laboratory | Jim Mildenberger, Wanda Manley, Noah Hull, Taylor Fearing, Lynette Gumbleton, Channing Weber, Ashley Norberg, Chayse Rowley, Marley Goetz, Brian Dominguez, Elliot Thomasson, Sam Britz, Cari Sloma, and Rob Christensen |
| EPI_ISL_1913215 | Santa Clara Valley Medical Center | Santa Clara County public Health Laboratory | Santa Clara County Public Health Department |
| EPI_ISL_1913216 | Santa Clara County Public Health Laboratory | Santa Clara County public Health Laboratory | Santa Clara County Public Health Department |
| EPI_ISL_1913217 | KAISER SAN JOSE | Santa Clara County public Health Laboratory | Santa Clara County Public Health Department |
| EPI_ISL_1913218 | Santa Clara Valley Medical Center | Santa Clara County public Health Laboratory | Santa Clara County Public Health Department |
| EPI_ISL_1913225 | KAISER SAN JOSE | Santa Clara County public Health Laboratory | Santa Clara County Public Health Department |
| EPI_ISL_1913226, EPI_ISL_1913227, EPI_ISL_1913228, EPI_ISL_1913229, EPI_ISL_1913230, EPI_ISL_1913231, EPI_ISL_1913233, EPI_ISL_1913234, EPI_ISL_1913235, EPI_ISL_1913236, EPI_ISL_1913237, EPI_ISL_1913238, EPI_ISL_1913239, EPI_ISL_1913241, EPI_ISL_1913242, EPI_ISL_1913245, EPI_ISL_1913247, EPI_ISL_1913248, EPI_ISL_1913249 | | | |
| see above | Santa Clara Valley Medical Center | Santa Clara County public Health Laboratory | Santa Clara County Public Health Department |
| EPI_ISL_1913251 | Santa Clara County Public Health Laboratory | Santa Clara County public Health Laboratory | Santa Clara County Public Health Department |
| EPI_ISL_1913256, EPI_ISL_1913257, EPI_ISL_1913258, EPI_ISL_1913259, EPI_ISL_1913260, EPI_ISL_1913261, EPI_ISL_1913262, EPI_ISL_1913263, EPI_ISL_1913264, EPI_ISL_1913265 | LSUHS Emerging Viral Threat Laboratory | Microbial Genome Sequencing Center | Jeremy P. Kamil, Jennifer L. Carroll, Rona S. Scott, Gregory L. Ware, Alexander Mijalis, Maarten Van Diest, Andrew D. Yurochko, Christopher G. Kevil, Daniel J. Snyder, Vaughn S. Cooper, John A. Vanchiere |
| EPI_ISL_1913266 | LSUHS Emerging Viral Threat Laboratory | Microbial Genome Sequencing Center | Jennifer L. Carroll, Jeremy P. Kamil, Rona S. Scott, Gregory L. Ware, Alexander Mijalis, Maarten Van Diest, Andrew D. Yurochko, Christopher G. Kevil, Daniel J. Snyder, Vaughn S. Cooper, John A. Vanchiere |
| EPI_ISL_1913267 | LSUHS Emerging Viral Threat Laboratory | Microbial Genome Sequencing Center | Maarten Van Diest, Jeremy P. Kamil, Jennifer L. Carroll, Rona S. Scott, Gregory L. Ware, Alexander Mijalis, Andrew D. Yurochko, Christopher G. Kevil, Daniel J. Snyder, Vaughn S. Cooper, John A. Vanchiere |
| EPI_ISL_1914663, EPI_ISL_1914664 | University of Michigan Clinical Microbiology Laboratory | Lauring Lab, University of Michigan, Department of Microbiology and Immunology | Valesano |
| EPI_ISL_1915100, EPI_ISL_1915101, EPI_ISL_1915102, EPI_ISL_1915103 | Florida Bureau of Public Health Laboratories | Florida Bureau of Public Health Laboratories | Sarah Schmedes, Jason Blanton |
| EPI_ISL_1915604 | The Ohio State University Applied Microbiology Services Laboratory | The Ohio State University Applied Microbiology Services Laboratory | Seth A. Faith PhD |
| EPI_ISL_1918401, EPI_ISL_1918403 | Arizona State University | Arizona State University | Peter T. Skidmore, LaRinda A. Holland, Matthew F. Smith, Rabia Maqsood, Nicholas J. Mellor, Joy M. Blain, Valerie Harris, Joshua LaBaer, Vel Murugan, Efrem S. Lim |
| EPI_ISL_1918404 | UNC Charlotte COVID-19 Testing Lab | UNC Charlotte Environmental Monitoring Laboratory | Cynthia Gibas, Kevin Lambirth, Angelica Martins, Jannatul Ferdous, Visva Barua, Jessica Schlueter |
| EPI_ISL_1918405, EPI_ISL_1918406, EPI_ISL_1918407, EPI_ISL_1918408, EPI_ISL_1918410, EPI_ISL_1918411, EPI_ISL_1918412, EPI_ISL_1918413, EPI_ISL_1918414, EPI_ISL_1918415, EPI_ISL_1918418, EPI_ISL_1918419, EPI_ISL_1918420, EPI_ISL_1918421, EPI_ISL_1918422, EPI_ISL_1918423, EPI_ISL_1918424, EPI_ISL_1918425, EPI_ISL_1918426, EPI_ISL_1918427, EPI_ISL_1918428, EPI_ISL_1918429, EPI_ISL_1918430, EPI_ISL_1918431, EPI_ISL_1918432, EPI_ISL_1918433, EPI_ISL_1918434, EPI_ISL_1918435, EPI_ISL_1918436, EPI_ISL_1918437, EPI_ISL_1918438, EPI_ISL_1918439, EPI_ISL_1918440, EPI_ISL_1918441, EPI_ISL_1918442, EPI_ISL_1918443, EPI_ISL_1918444, EPI_ISL_1918445, EPI_ISL_1918446, EPI_ISL_1918447, EPI_ISL_1918448, EPI_ISL_1918449, EPI_ISL_1918450, EPI_ISL_1918451, EPI_ISL_1918452, EPI_ISL_1918453, EPI_ISL_1918454, EPI_ISL_1918455, EPI_ISL_1918456, EPI_ISL_1918457, EPI_ISL_1918458, EPI_ISL_1918459, EPI_ISL_1918460, EPI_ISL_1918461, EPI_ISL_1918462, EPI_ISL_1918463, EPI_ISL_1918464, EPI_ISL_1918465, EPI_ISL_1918468, EPI_ISL_1918469, EPI_ISL_1918470, EPI_ISL_1918471, EPI_ISL_1918472, EPI_ISL_1918473, EPI_ISL_1918474, EPI_ISL_1918475, EPI_ISL_1918476, EPI_ISL_1918477, EPI_ISL_1918478, EPI_ISL_1918479, EPI_ISL_1918480, EPI_ISL_1918481, EPI_ISL_1918482, EPI_ISL_1918483, EPI_ISL_1918484, EPI_ISL_1918485, EPI_ISL_1918486, EPI_ISL_1918487, EPI_ISL_1918488, EPI_ISL_1918489, EPI_ISL_1918490, EPI_ISL_1918491, EPI_ISL_1918492, EPI_ISL_1918493, EPI_ISL_1918494, EPI_ISL_1918495, EPI_ISL_1918496, EPI_ISL_1918497, EPI_ISL_1918498, EPI_ISL_1918499, EPI_ISL_1918500, EPI_ISL_1918501, EPI_ISL_1918502, EPI_ISL_1918503, EPI_ISL_1918504, EPI_ISL_1918505, EPI_ISL_1918506, EPI_ISL_1918507, EPI_ISL_1918508, EPI_ISL_1918509, EPI_ISL_1918510, EPI_ISL_1918511, EPI_ISL_1918512, EPI_ISL_1918513, EPI_ISL_1918514, EPI_ISL_1918515, EPI_ISL_1918516, EPI_ISL_1918517, EPI_ISL_1918518, EPI_ISL_1918519, EPI_ISL_1918520, EPI_ISL_1918521, EPI_ISL_1918522, EPI_ISL_1918523, EPI_ISL_1918524, EPI_ISL_1918526, EPI_ISL_1918527, EPI_ISL_1918528, EPI_ISL_1918529, EPI_ISL_1918530, EPI_ISL_1918531, EPI_ISL_1918532, EPI_ISL_1918533, EPI_ISL_1918534, EPI_ISL_1918535, EPI_ISL_1918536, EPI_ISL_1918537, EPI_ISL_1918538, EPI_ISL_1918540, EPI_ISL_1918541, EPI_ISL_1918542, EPI_ISL_1918543, EPI_ISL_1918544, EPI_ISL_1918545, EPI_ISL_1918546, EPI_ISL_1918547, EPI_ISL_1918548, EPI_ISL_1918549, EPI_ISL_1918550, EPI_ISL_1918551, EPI_ISL_1918552, EPI_ISL_1918553, EPI_ISL_1918554, EPI_ISL_1918555, EPI_ISL_1918556, EPI_ISL_1918557, EPI_ISL_1918558, EPI_ISL_1918559, EPI_ISL_1918560, EPI_ISL_1918561, EPI_ISL_1918562, EPI_ISL_1918563, EPI_ISL_1918564, EPI_ISL_1918565, EPI_ISL_1918566, EPI_ISL_1918567, EPI_ISL_1918568, EPI_ISL_1918569, EPI_ISL_1918570, EPI_ISL_1918571, EPI_ISL_1918572, EPI_ISL_1918573, EPI_ISL_1918574, EPI_ISL_1918575, EPI_ISL_1918576, EPI_ISL_1918577, EPI_ISL_1918578, EPI_ISL_1918579, EPI_ISL_1918580, EPI_ISL_1918581, EPI_ISL_1918582, EPI_ISL_1918583, EPI_ISL_1918584, EPI_ISL_1918585, EPI_ISL_1918586, EPI_ISL_1918587, EPI_ISL_1918588, EPI_ISL_1918589, EPI_ISL_1918590, EPI_ISL_1918591, EPI_ISL_1918592, EPI_ISL_1918593, EPI_ISL_1918594, EPI_ISL_1918595, EPI_ISL_1918596, EPI_ISL_1918597, EPI_ISL_1918598, EPI_ISL_1918599, EPI_ISL_1918600, EPI_ISL_1918601, EPI_ISL_1918602, EPI_ISL_1918603, EPI_ISL_1918604, EPI_ISL_1918605, EPI_ISL_1918606, EPI_ISL_1918607, EPI_ISL_1918608, EPI_ISL_1918609, EPI_ISL_1918610, EPI_ISL_1918611, EPI_ISL_1918612, EPI_ISL_1918613, EPI_ISL_1918614, EPI_ISL_1918615, EPI_ISL_1918616, EPI_ISL_1918617, EPI_ISL_1918618, EPI_ISL_1918619, EPI_ISL_1918620, EPI_ISL_1918621, EPI_ISL_1918622, EPI_ISL_1918623, EPI_ISL_1918624, EPI_ISL_1918625, EPI_ISL_1918626, EPI_ISL_1918627, EPI_ISL_1918628, EPI_ISL_1918629, EPI_ISL_1918630, EPI_ISL_1918631, EPI_ISL_1918632, EPI_ISL_1918633, EPI_ISL_1918634, EPI_ISL_1918635, EPI_ISL_1918636, EPI_ISL_1918637, EPI_ISL_1918638, EPI_ISL_1918639, EPI_ISL_1918640, EPI_ISL_1918641, EPI_ISL_1918642, EPI_ISL_1918643, EPI_ISL_1918644, EPI_ISL_1918645, EPI_ISL_1918646, EPI_ISL_1918647, EPI_ISL_1918648, EPI_ISL_1918649, EPI_ISL_1918650, EPI_ISL_1918651, EPI_ISL_1918652, EPI_ISL_1918653, EPI_ISL_1918654, EPI_ISL_1918655, EPI_ISL_1918656, EPI_ISL_1918657, EPI_ISL_1918658, EPI_ISL_1918659, EPI_ISL_1918660, EPI_ISL_1918661, EPI_ISL_1918662, EPI_ISL_1918663, EPI_ISL_1918664, EPI_ISL_1918665, EPI_ISL_1918666, EPI_ISL_1918667, EPI_ISL_1918668, EPI_ISL_1918669, EPI_ISL_1918670, EPI_ISL_1918671, EPI_ISL_1918672, EPI_ISL_1918673, EPI_ISL_1918674, EPI_ISL_1918675, EPI_ISL_1918676, EPI_ISL_1918677, EPI_ISL_1918678, EPI_ISL_1918679, EPI_ISL_1918680, EPI_ISL_1918681, EPI_ISL_1918682, EPI_ISL_1918683, EPI_ISL_1918684, EPI_ISL_1918685, EPI_ISL_1918686, EPI_ISL_1918687, EPI_ISL_1918688, EPI_ISL_1918689, EPI_ISL_1918690, EPI_ISL_1918691, EPI_ISL_1918692, EPI_ISL_1918693, EPI_ISL_1918694, EPI_ISL_1918695, EPI_ISL_1918696, EPI_ISL_1918697, EPI_ISL_1918698, EPI_ISL_1918699, EPI_ISL_1918700, EPI_ISL_1918701, EPI_ISL_1918702, EPI_ISL_1918703, EPI_ISL_1918704, EPI_ISL_1918705, EPI_ISL_1918706, EPI_ISL_1918707, EPI_ISL_1918708, EPI_ISL_1918709, EPI_ISL_1918710, EPI_ISL_1918711, EPI_ISL_1918712, EPI_ISL_1918713, EPI_ISL_1918714, EPI_ISL_1918715, EPI_ISL_1918716, EPI_ISL_1918717, EPI_ISL_1918718, EPI_ISL_1918719, EPI_ISL_1918720, EPI_ISL_1918721, EPI_ISL_1918722, EPI_ISL_1918723, EPI_ISL_1918724, EPI_ISL_1918725, EPI_ISL_1918726, EPI_ISL_1918727, EPI_ISL_1918728, EPI_ISL_1918729, EPI_ISL_1918730, EPI_ISL_1918731, EPI_ISL_1918732, EPI_ISL_1918733, EPI_ISL_1918735, EPI_ISL_1918736, EPI_ISL_1918737, EPI_ISL_1918738, EPI_ISL_1918739, EPI_ISL_1918740, EPI_ISL_1918741, EPI_ISL_1918742, EPI_ISL_1918743, EPI_ISL_1918744, EPI_ISL_1918745, EPI_ISL_1918746, EPI_ISL_1918747, EPI_ISL_1918748, EPI_ISL_1918749, EPI_ISL_1918750, EPI_ISL_1918751, EPI_ISL_1918752, EPI_ISL_1918753, EPI_ISL_1918754, EPI_ISL_1918755, EPI_ISL_1918756, EPI_ISL_1918757, EPI_ISL_1918758, EPI_ISL_1918759, EPI_ISL_1918760, EPI_ISL_1918761, EPI_ISL_1918762, EPI_ISL_1918763, EPI_ISL_1918764, EPI_ISL_1918765, EPI_ISL_1918766, EPI_ISL_1918767, EPI_ISL_1918768, EPI_ISL_1918769, EPI_ISL_1918770, EPI_ISL_1918771, EPI_ISL_1918772, EPI_ISL_1918773, EPI_ISL_1918774, EPI_ISL_1918775, EPI_ISL_1918776, EPI_ISL_1918777, EPI_ISL_1918778, EPI_ISL_1918779, EPI_ISL_1918781, EPI_ISL_1918782, EPI_ISL_1918783, EPI_ISL_1918784, EPI_ISL_1918786, EPI_ISL_1918787, EPI_ISL_1918788, EPI_ISL_1918789, EPI_ISL_1918790, EPI_ISL_1918791, EPI_ISL_1918792, EPI_ISL_1918793, EPI_ISL_1918794, EPI_ISL_1918795, EPI_ISL_1918796, EPI_ISL_1918797, EPI_ISL_1918798, EPI_ISL_1918799, EPI_ISL_1918800, EPI_ISL_1918801, EPI_ISL_1918802, EPI_ISL_1918803, EPI_ISL_1918804, EPI_ISL_1918805, EPI_ISL_1918806, EPI_ISL_1918807, EPI_ISL_1918808, EPI_ISL_1918809, EPI_ISL_1918810, EPI_ISL_1918811, EPI_ISL_1918812, EPI_ISL_1918813, EPI_ISL_1918814, EPI_ISL_1918815, EPI_ISL_1918816, EPI_ISL_1918817, EPI_ISL_1918818, EPI_ISL_1918819, EPI_ISL_1918820, EPI_ISL_1918821, EPI_ISL_1918822, EPI_ISL_1918823, EPI_ISL_1918824, EPI_ISL_1918825, EPI_ISL_1918826, EPI_ISL_1918827, EPI_ISL_1918828, EPI_ISL_1918829, EPI_ISL_1918830, EPI_ISL_1918831, EPI_ISL_1918832, EPI_ISL_1918833, EPI_ISL_1918834, EPI_ISL_1918835, EPI_ISL_1918836, EPI_ISL_1918837, EPI_ISL_1918838, EPI_ISL_1918839, EPI_ISL_1918840, EPI_ISL_1918841, EPI_ISL_1918842, EPI_ISL_1918843, EPI_ISL_1918844, EPI_ISL_1918845, EPI_ISL_1918846, EPI_ISL_1918847, EPI_ISL_1918848, EPI_ISL_1918849, EPI_ISL_1918850, EPI_ISL_1918851, EPI_ISL_1918852, EPI_ISL_1918853, EPI_ISL_1918854, EPI_ISL_1918855, EPI_ISL_1918856, EPI_ISL_1918857, EPI_ISL_1918858, EPI_ISL_1918859, EPI_ISL_1918860, EPI_ISL_1918861, EPI_ISL_1918862, EPI_ISL_1918863, EPI_ISL_1918864, EPI_ISL_1918865, EPI_ISL_1918866, EPI_ISL_1918867, EPI_ISL_1918868, EPI_ISL_1918869, EPI_ISL_1918870, EPI_ISL_1918871, EPI_ISL_1918872, EPI_ISL_1918873, EPI_ISL_1918874, EPI_ISL_1918875, EPI_ISL_1918876, EPI_ISL_1918877, EPI_ISL_1918878, EPI_ISL_1918879, EPI_ISL_1918880, EPI_ISL_1918881, EPI_ISL_1918882, EPI_ISL_1918883, EPI_ISL_1918884, EPI_ISL_1918885, EPI_ISL_1918886, EPI_ISL_1918887, EPI_ISL_1918888, EPI_ISL_1918889, EPI_ISL_1918890, EPI_ISL_1918891, EPI_ISL_1918892, EPI_ISL_1918893, EPI_ISL_1918894, EPI_ISL_1918895, EPI_ISL_1918896, EPI_ISL_1918897, EPI_ISL_1918898, EPI_ISL_1918899, EPI_ISL_1918900, EPI_ISL_1918901, EPI_ISL_1918902, EPI_ISL_1918903, EPI_ISL_1918904, EPI_ISL_1918905, EPI_ISL_1918906, EPI_ISL_1918907, EPI_ISL_1918908, EPI_ISL_1918909, EPI_ISL_1918910, EPI_ISL_1918911, EPI_ISL_1918912, EPI_ISL_1918913, EPI_ISL_1918914, EPI_ISL_1918915, EPI_ISL_1918916, EPI_ISL_1918917, EPI_ISL_1918918, EPI_ISL_1918919, EPI_ISL_1918920, EPI_ISL_1918921, EPI_ISL_1918922, EPI_ISL_1918923, EPI_ISL_1918924, EPI_ISL_1918925, EPI_ISL_1918926, EPI_ISL_1918927, EPI_ISL_1918928, EPI_ISL_1918929, EPI_ISL_1918930, EPI_ISL_1918931, EPI_ISL_1918932, EPI_ISL_1918933, EPI_ISL_1918934, EPI_ISL_1918935, EPI_ISL_1918937, EPI_ISL_1918938, EPI_ISL_1918939, EPI_ISL_1918940, EPI_ISL_1918941, EPI_ISL_1918942, EPI_ISL_1918943, EPI_ISL_1918944, EPI_ISL_1918945, EPI_ISL_1918946, EPI_ISL_1918947, EPI_ISL_1918948, EPI_ISL_1918949, EPI_ISL_1918950, EPI_ISL_1918951, EPI_ISL_1918952, EPI_ISL_1918953, EPI_ISL_1918954, EPI_ISL_1918955, EPI_ISL_1918956, EPI_ISL_1918957, EPI_ISL_1918958, EPI_ISL_1918959, EPI_ISL_1918960, EPI_ISL_1918961, EPI_ISL_1918962, EPI_ISL_1918963, EPI_ISL_1918964, EPI_ISL_1918965, EPI_ISL_1918966, EPI_ISL_1918967, EPI_ISL_1918968, EPI_ISL_1918969, EPI_ISL_1918970, EPI_ISL_1918971, EPI_ISL_1918972, EPI_ISL_1918973, EPI_ISL_1918974, EPI_ISL_1918975, EPI_ISL_1918976, EPI_ISL_1918977, EPI_ISL_1918978, EPI_ISL_1918979, EPI_ISL_1918980, EPI_ISL_1918981, EPI_ISL_1918982, EPI_ISL_1918983, EPI_ISL_1918984, EPI_ISL_1918985, EPI_ISL_1918986, EPI_ISL_1918987, EPI_ISL_1918988, EPI_ISL_1918989, EPI_ISL_1918990 | | | |
| see above | Broad Institute Clinical Research Sequencing Platform | Infectious Disease Program, Broad Institute of Harvard and MIT | Siddle,K.J., Adams,G., Pearlman,L., Gladden-Young,A., Vicente,G., Blumenstiel,B., DeFelice,M., Lee,M., McGovern,S., Lagerborg,K., Rudy,M., DeRuff,K., Carter,A., Normandin,E., Bauer,M., Reilly,S., Tomkins-Tinch,C., Loreth,C., Chaluvadi,S., Meldrim,J., Granger,B., Lemieux,J.E., Birren,B.W., Sabeti,P.C., Larkin,K., Dodge,S., Lennon,N., Madoff,L., Brown,C., Gallagher,G., Smole,S., Park,D.J., Gabriel,S., and MacInnis,B.L. |
| EPI_ISL_1918991, EPI_ISL_1918992 | Flow Health | Infectious Disease Program, Broad Institute of Harvard and MIT | Lemieux,J.E., Siddle,K.J., Adams,G., Gladden-Young,A., Lagerborg,K., Rudy,M., DeRuff,K., Carter,A., Normandin,E., Bauer,M., Reilly,S., Tomkins-Tinch,C., Loreth,C., Chaluvadi,S., Birren,B.W., Gallagher,G., Smole,S., Park,D.J., MacInnis,B.L., and Sabeti,P.C. |
| EPI_ISL_1918993 | Warrior Diagnostics Laboratory | Infectious Disease Program, Broad Institute of Harvard and MIT | Lemieux,J.E., Siddle,K.J., Marshall,J., ONeill,M., Bronson,A., Adams,G., Gladden-Young,A., Lagerborg,K., Rudy,M., DeRuff,K., Carter,A., Normandin,E., Bauer,M., Reilly,S., Tomkins-Tinch,C., Loreth,C., Chaluvadi,S., Birren,B.W., Gallagher,G., Smole,S., Park,D.J., MacInnis,B.L., and Sabeti,P.C. |
| EPI_ISL_1918996, EPI_ISL_1919000, EPI_ISL_1919001, EPI_ISL_1919002, EPI_ISL_1919003, EPI_ISL_1919004, EPI_ISL_1919005, EPI_ISL_1919006, EPI_ISL_1919007, EPI_ISL_1919008, EPI_ISL_1919009, EPI_ISL_1919010, EPI_ISL_1919013, EPI_ISL_1919014, EPI_ISL_1919015, EPI_ISL_1919016, EPI_ISL_1919017, EPI_ISL_1919018, EPI_ISL_1919020, EPI_ISL_1919021, EPI_ISL_1919022, EPI_ISL_1919023, EPI_ISL_1919024, EPI_ISL_1919025, EPI_ISL_1919026, EPI_ISL_1919027, EPI_ISL_1919028, EPI_ISL_1919029, EPI_ISL_1919030, EPI_ISL_1919031, EPI_ISL_1919033, EPI_ISL_1919034, EPI_ISL_1919036, EPI_ISL_1919037, EPI_ISL_1919038, EPI_ISL_1919039, EPI_ISL_1919040, EPI_ISL_1919041, EPI_ISL_1919042, EPI_ISL_1919044, EPI_ISL_1919045, EPI_ISL_1919046, EPI_ISL_1919047, EPI_ISL_1919048, EPI_ISL_1919049, EPI_ISL_1919050, EPI_ISL_1919052, EPI_ISL_1919053, EPI_ISL_1919054, EPI_ISL_1919055, EPI_ISL_1919056, EPI_ISL_1919057, EPI_ISL_1919058, EPI_ISL_1919059, EPI_ISL_1919060, EPI_ISL_1919062, EPI_ISL_1919063, EPI_ISL_1919064, EPI_ISL_1919065, EPI_ISL_1919066, EPI_ISL_1919067, EPI_ISL_1919069, EPI_ISL_1919070, EPI_ISL_1919071, EPI_ISL_1919072, EPI_ISL_1919073, EPI_ISL_1919074, EPI_ISL_1919075, EPI_ISL_1919076, EPI_ISL_1919077, EPI_ISL_1919078, EPI_ISL_1919079, EPI_ISL_1919080, EPI_ISL_1919081, EPI_ISL_1919082, EPI_ISL_1919083, EPI_ISL_1919084, EPI_ISL_1919085, EPI_ISL_1919086, EPI_ISL_1919087, EPI_ISL_1919088, EPI_ISL_1919089, EPI_ISL_1919090, EPI_ISL_1919091, EPI_ISL_1919092, EPI_ISL_1919093, EPI_ISL_1919094, EPI_ISL_1919095, EPI_ISL_1919097, EPI_ISL_1919098, EPI_ISL_1919099, EPI_ISL_1919100, EPI_ISL_1919101, EPI_ISL_1919102, EPI_ISL_1919103, EPI_ISL_1919104, EPI_ISL_1919105, EPI_ISL_1919106, EPI_ISL_1919107, EPI_ISL_1919110, EPI_ISL_1919111, EPI_ISL_1919112, EPI_ISL_1919113, EPI_ISL_1919114, EPI_ISL_1919115, EPI_ISL_1919116, EPI_ISL_1919117, EPI_ISL_1919118, EPI_ISL_1919119, EPI_ISL_1919120, EPI_ISL_1919121, EPI_ISL_1919122, EPI_ISL_1919123, EPI_ISL_1919124, EPI_ISL_1919125, EPI_ISL_1919126, EPI_ISL_1919127, EPI_ISL_1919128, EPI_ISL_1919129 | | | |
| see above | Broad Institute Clinical Research Sequencing Platform | Infectious Disease Program, Broad Institute of Harvard and MIT | Siddle,K.J., Adams,G., Pearlman,L., Gladden-Young,A., Vicente,G., Blumenstiel,B., DeFelice,M., Lee,M., McGovern,S., Lagerborg,K., Rudy,M., DeRuff,K., Carter,A., Normandin,E., Bauer,M., Reilly,S., Tomkins-Tinch,C., Loreth,C., Chaluvadi,S., Meldrim,J., Granger,B., Lemieux,J.E., Birren,B.W., Sabeti,P.C., Larkin,K., Dodge,S., Lennon,N., Madoff,L., Brown,C., Gallagher,G., Smole,S., Park,D.J., Gabriel,S., and MacInnis,B.L. |
| EPI_ISL_1919130, EPI_ISL_1919131, EPI_ISL_1919132, EPI_ISL_1919133, EPI_ISL_1919134, EPI_ISL_1919135, EPI_ISL_1919136, EPI_ISL_1919137, EPI_ISL_1919138, EPI_ISL_1919139, EPI_ISL_1919140, EPI_ISL_1919141, EPI_ISL_1919142, EPI_ISL_1919143, EPI_ISL_1919144, EPI_ISL_1919145, EPI_ISL_1919146, EPI_ISL_1919147, EPI_ISL_1919148, EPI_ISL_1919149, EPI_ISL_1919150, EPI_ISL_1919151, EPI_ISL_1919152, EPI_ISL_1919153, EPI_ISL_1919154, EPI_ISL_1919155, EPI_ISL_1919156, EPI_ISL_1919157, EPI_ISL_1919158, EPI_ISL_1919159, EPI_ISL_1919160, EPI_ISL_1919161, EPI_ISL_1919162, EPI_ISL_1919163, EPI_ISL_1919164, EPI_ISL_1919165, EPI_ISL_1919166, EPI_ISL_1919167, EPI_ISL_1919168, EPI_ISL_1919169, EPI_ISL_1919170, EPI_ISL_1919171, EPI_ISL_1919172, EPI_ISL_1919173, EPI_ISL_1919174, EPI_ISL_1919175, EPI_ISL_1919176, EPI_ISL_1919177, EPI_ISL_1919178, EPI_ISL_1919179, EPI_ISL_1919180, EPI_ISL_1919181, EPI_ISL_1919182, EPI_ISL_1919183, EPI_ISL_1919184, EPI_ISL_1919185, EPI_ISL_1919186, EPI_ISL_1919187, EPI_ISL_1919188, EPI_ISL_1919189, EPI_ISL_1919190, EPI_ISL_1919191, EPI_ISL_1919192, EPI_ISL_1919193, EPI_ISL_1919194, EPI_ISL_1919195, EPI_ISL_1919196, EPI_ISL_1919197, EPI_ISL_1919198, EPI_ISL_1919199, EPI_ISL_1919200, EPI_ISL_1919201, EPI_ISL_1919202, EPI_ISL_1919203, EPI_ISL_1919204, EPI_ISL_1919205, EPI_ISL_1919206, EPI_ISL_1919207, EPI_ISL_1919208, EPI_ISL_1919209, EPI_ISL_1919210, EPI_ISL_1919211, EPI_ISL_1919212, EPI_ISL_1919213, EPI_ISL_1919214, EPI_ISL_1919215, EPI_ISL_1919216, EPI_ISL_1919217, EPI_ISL_1919218, EPI_ISL_1919219, EPI_ISL_1919220, EPI_ISL_1919221, EPI_ISL_1919222, EPI_ISL_1919223, EPI_ISL_1919224, EPI_ISL_1919225, EPI_ISL_1919226, EPI_ISL_1919227, EPI_ISL_1919228, EPI_ISL_1919229, EPI_ISL_1919230, EPI_ISL_1919231, EPI_ISL_1919232, EPI_ISL_1919233, EPI_ISL_1919234, EPI_ISL_1919235, EPI_ISL_1919236, EPI_ISL_1919237, EPI_ISL_1919238, EPI_ISL_1919239, EPI_ISL_1919240, EPI_ISL_1919241, EPI_ISL_1919242, EPI_ISL_1919243, EPI_ISL_1919244, EPI_ISL_1919245, EPI_ISL_1919246, EPI_ISL_1919247, EPI_ISL_1919248, EPI_ISL_1919249 | | | |
| see above | Rhode Island Department of Health | Infectious Disease Program, Broad Institute of Harvard and MIT | Siddle,K.J., Azevedo,K., Miller,A., Adams,G., Pearlman,L., Gladden-Young,A., Lagerborg,K., Rudy,M., DeRuff,K., Carter,A., Normandin,E., Bauer,M., Reilly,S., Tomkins-Tinch,C., Loreth,C., Chaluvadi,S., Lemieux,J.E., Birren,B.W., Sabeti,P.C., Huard,R., King,E., Park,D.J., and MacInnis,B.L. |
| EPI_ISL_1919250, EPI_ISL_1919251, EPI_ISL_1919252, EPI_ISL_1919253, EPI_ISL_1919254, EPI_ISL_1919255, EPI_ISL_1919256, EPI_ISL_1919257, EPI_ISL_1919258, EPI_ISL_1919259, EPI_ISL_1919260, EPI_ISL_1919261, EPI_ISL_1919262, EPI_ISL_1919263, EPI_ISL_1919264, EPI_ISL_1919265, EPI_ISL_1919267, EPI_ISL_1919268, EPI_ISL_1919269, EPI_ISL_1919270, EPI_ISL_1919271 | | | |
| see above | Broad Institute Clinical Research Sequencing Platform | Infectious Disease Program, Broad Institute of Harvard and MIT | Siddle,K.J., Adams,G., Pearlman,L., Gladden-Young,A., Vicente,G., Blumenstiel,B., DeFelice,M., Lee,M., McGovern,S., Lagerborg,K., Rudy,M., DeRuff,K., Carter,A., Normandin,E., Bauer,M., Reilly,S., Tomkins-Tinch,C., Loreth,C., Chaluvadi,S., Meldrim,J., Granger,B., Lemieux,J.E., Birren,B.W., Sabeti,P.C., Larkin,K., Dodge,S., Lennon,N., Madoff,L., Brown,C., Gallagher,G., Smole,S., Park,D.J., Gabriel,S., and MacInnis,B.L. |
| EPI_ISL_1919576, EPI_ISL_1919577, EPI_ISL_1919578, EPI_ISL_1919580, EPI_ISL_1919581, EPI_ISL_1919587, EPI_ISL_1919588, EPI_ISL_1919592, EPI_ISL_1919595, EPI_ISL_1919596, EPI_ISL_1919597, EPI_ISL_1919598, EPI_ISL_1919599 | | | |
| see above | Wyoming Public Health Laboratory | Wyoming Public Health Laboratory | Jim Mildenberger, Wanda Manley, Noah Hull, Taylor Fearing, Lynette Gumbleton, Channing Weber, Ashley Norberg, Chayse Rowley, Marley Goetz, Brian Dominguez, Elliot Thomasson, Sam Britz, Cari Sloma, and Rob Christensen |
| EPI_ISL_1919759, EPI_ISL_1919760, EPI_ISL_1919762, EPI_ISL_1919765, EPI_ISL_1919766, EPI_ISL_1919767, EPI_ISL_1919769, EPI_ISL_1919770, EPI_ISL_1919771, EPI_ISL_1919772, EPI_ISL_1919773, EPI_ISL_1919774, EPI_ISL_1919775, EPI_ISL_1919776, EPI_ISL_1919777, EPI_ISL_1919778, EPI_ISL_1919779, EPI_ISL_1919780, EPI_ISL_1919781, EPI_ISL_1919782, EPI_ISL_1919783, EPI_ISL_1919784, EPI_ISL_1919785, EPI_ISL_1919786, EPI_ISL_1919787, EPI_ISL_1919788, EPI_ISL_1919789, EPI_ISL_1919791, EPI_ISL_1919792, EPI_ISL_1919793, EPI_ISL_1919795, EPI_ISL_1919796, EPI_ISL_1919797, EPI_ISL_1919798, EPI_ISL_1919799, EPI_ISL_1919800, EPI_ISL_1919801, EPI_ISL_1919802, EPI_ISL_1919803, EPI_ISL_1919805, EPI_ISL_1919806, EPI_ISL_1919807, EPI_ISL_1919808, EPI_ISL_1919809, EPI_ISL_1919810, EPI_ISL_1919811 | | | |

| | | | |
|---|---|---|---|
| see above | North Dakota Department of Health, Public Health Laboratory | North Dakota Department of Health, Public Health Laboratory | Lisa Wingerter |
| EPI_ISL_1919864, EPI_ISL_1919865, EPI_ISL_1919866, EPI_ISL_1919867, EPI_ISL_1919868, EPI_ISL_1919869, EPI_ISL_1919870, EPI_ISL_1919871, EPI_ISL_1919872, EPI_ISL_1919873, EPI_ISL_1919874, EPI_ISL_1919875, EPI_ISL_1919876, EPI_ISL_1919877, EPI_ISL_1919878, EPI_ISL_1919879, EPI_ISL_1919880, EPI_ISL_1919881, EPI_ISL_1919882, EPI_ISL_1919883, EPI_ISL_1919884, EPI_ISL_1919885, EPI_ISL_1919886, EPI_ISL_1919887, EPI_ISL_1919888, EPI_ISL_1919889, EPI_ISL_1919890, EPI_ISL_1919891, EPI_ISL_1919892, EPI_ISL_1919893, EPI_ISL_1919894, EPI_ISL_1919895, EPI_ISL_1919897, EPI_ISL_1919898, EPI_ISL_1919899, EPI_ISL_1919900, EPI_ISL_1919901, EPI_ISL_1919902, EPI_ISL_1919903, EPI_ISL_1919904, EPI_ISL_1919905, EPI_ISL_1919906, EPI_ISL_1919907, EPI_ISL_1919908, EPI_ISL_1919909, EPI_ISL_1919910, EPI_ISL_1919911, EPI_ISL_1919912, EPI_ISL_1919913, EPI_ISL_1919914, EPI_ISL_1919915, EPI_ISL_1919916, EPI_ISL_1919917, EPI_ISL_1919918, EPI_ISL_1919919, EPI_ISL_1919920, EPI_ISL_1919922, EPI_ISL_1919923, EPI_ISL_1919924, EPI_ISL_1919925, EPI_ISL_1919926, EPI_ISL_1919927, EPI_ISL_1919928, EPI_ISL_1919929, EPI_ISL_1919930, EPI_ISL_1919931, EPI_ISL_1919932, EPI_ISL_1919934, EPI_ISL_1919935, EPI_ISL_1919936, EPI_ISL_1919937, EPI_ISL_1919938, EPI_ISL_1919939, EPI_ISL_1919940, EPI_ISL_1919941, EPI_ISL_1919942, EPI_ISL_1919943 | | | |
| see above | University of Michigan Clinical Microbiology Laboratory | Lauring Lab, University of Michigan, Department of Microbiology and Immunology | Valesano |
| EPI_ISL_1919988, EPI_ISL_1919989 | Northwell Health Laboratories | Wadsworth Center, New York State Department of Health | Kirsten St. George, Daryl M. Lamson, Alexis Russell, Matthew Shudt, Melissa A Leisner, Jonathan Plitnick, Catharine Prussing, Navjot Singh, John Kelly, Erasmus Schneider, Erica Lasek-Nesselquist |
| EPI_ISL_1919990 | Suny Upstate Medical University | Wadsworth Center, New York State Department of Health | Kirsten St. George, Daryl M. Lamson, Alexis Russell, Matthew Shudt, Melissa A Leisner, Jonathan Plitnick, Catharine Prussing, Navjot Singh, John Kelly, Erasmus Schneider, Erica Lasek-Nesselquist |
| EPI_ISL_1919991, EPI_ISL_1919992 | Stony Brook University Hospital | Wadsworth Center, New York State Department of Health | Kirsten St. George, Daryl M. Lamson, Alexis Russell, Matthew Shudt, Melissa A Leisner, Jonathan Plitnick, Catharine Prussing, Navjot Singh, John Kelly, Erasmus Schneider, Erica Lasek-Nesselquist |
| EPI_ISL_1919993, EPI_ISL_1919994, EPI_ISL_1919995, EPI_ISL_1919996 | Northwell Health Laboratories | Wadsworth Center, New York State Department of Health | Kirsten St. George, Daryl M. Lamson, Alexis Russell, Matthew Shudt, Melissa A Leisner, Jonathan Plitnick, Catharine Prussing, Navjot Singh, John Kelly, Erasmus Schneider, Erica Lasek-Nesselquist |
| EPI_ISL_1919997, EPI_ISL_1919998, EPI_ISL_1919999, EPI_ISL_1920000, EPI_ISL_1920001, EPI_ISL_1920002, EPI_ISL_1920003, EPI_ISL_1920004, EPI_ISL_1920005, EPI_ISL_1920006 | Montefiore Medical Center Laboratories | Wadsworth Center, New York State Department of Health | Kirsten St. George, Daryl M. Lamson, Alexis Russell, Matthew Shudt, Melissa A Leisner, Jonathan Plitnick, Catharine Prussing, Navjot Singh, John Kelly, Erasmus Schneider, Erica Lasek-Nesselquist |
| EPI_ISL_1920007 | Westchester Medical Center | Wadsworth Center, New York State Department of Health | Kirsten St. George, Daryl M. Lamson, Alexis Russell, Matthew Shudt, Melissa A Leisner, Jonathan Plitnick, Catharine Prussing, Navjot Singh, John Kelly, Erasmus Schneider, Erica Lasek-Nesselquist |
| EPI_ISL_1920008 | Northwell Health Laboratories | Wadsworth Center, New York State Department of Health | Kirsten St. George, Daryl M. Lamson, Alexis Russell, Matthew Shudt, Melissa A Leisner, Jonathan Plitnick, Catharine Prussing, Navjot Singh, John Kelly, Erasmus Schneider, Erica Lasek-Nesselquist |
| EPI_ISL_1920009 | Westchester Medical Center | Wadsworth Center, New York State Department of Health | Kirsten St. George, Daryl M. Lamson, Alexis Russell, Matthew Shudt, Melissa A Leisner, Jonathan Plitnick, Catharine Prussing, Navjot Singh, John Kelly, Erasmus Schneider, Erica Lasek-Nesselquist |
| EPI_ISL_1920010, EPI_ISL_1920011, EPI_ISL_1920012, EPI_ISL_1920013, EPI_ISL_1920014, EPI_ISL_1920015, EPI_ISL_1920016, EPI_ISL_1920017, EPI_ISL_1920018, EPI_ISL_1920019, EPI_ISL_1920020, EPI_ISL_1920021, EPI_ISL_1920022, EPI_ISL_1920023, EPI_ISL_1920024, EPI_ISL_1920025, EPI_ISL_1920026, EPI_ISL_1920027, EPI_ISL_1920028 | | | |
| see above | Northwell Health Laboratories | Wadsworth Center, New York State Department of Health | Kirsten St. George, Daryl M. Lamson, Alexis Russell, Matthew Shudt, Melissa A Leisner, Jonathan Plitnick, Catharine Prussing, Navjot Singh, John Kelly, Erasmus Schneider, Erica Lasek-Nesselquist |
| EPI_ISL_1920029 | St. Francis Hospital Laboratory | Wadsworth Center, New York State Department of Health | Kirsten St. George, Daryl M. Lamson, Alexis Russell, Matthew Shudt, Melissa A Leisner, Jonathan Plitnick, Catharine Prussing, Navjot Singh, John Kelly, Erasmus Schneider, Erica Lasek-Nesselquist |
| EPI_ISL_1920030 | Stony Brook University Hospital | Wadsworth Center, New York State Department of Health | Kirsten St. George, Daryl M. Lamson, Alexis Russell, Matthew Shudt, Melissa A Leisner, Jonathan Plitnick, Catharine Prussing, Navjot Singh, John Kelly, Erasmus Schneider, Erica Lasek-Nesselquist |
| EPI_ISL_1920031, EPI_ISL_1920032, EPI_ISL_1920033, EPI_ISL_1920034 | Northwell Health Laboratories | Wadsworth Center, New York State Department of Health | Kirsten St. George, Daryl M. Lamson, Alexis Russell, Matthew Shudt, Melissa A Leisner, Jonathan Plitnick, Catharine Prussing, Navjot Singh, John Kelly, Erasmus Schneider, Erica Lasek-Nesselquist |
| EPI_ISL_1920035 | Stony Brook University Hospital | Wadsworth Center, New York State Department of Health | Kirsten St. George, Daryl M. Lamson, Alexis Russell, Matthew Shudt, Melissa A Leisner, Jonathan Plitnick, Catharine Prussing, Navjot Singh, John Kelly, Erasmus Schneider, Erica Lasek-Nesselquist |
| EPI_ISL_1920036, EPI_ISL_1920037, EPI_ISL_1920038, EPI_ISL_1920039, EPI_ISL_1920040 | Northwell Health Laboratories | Wadsworth Center, New York State Department of Health | Kirsten St. George, Daryl M. Lamson, Alexis Russell, Matthew Shudt, Melissa A Leisner, Jonathan Plitnick, Catharine Prussing, Navjot Singh, John Kelly, Erasmus Schneider, Erica Lasek-Nesselquist |
| EPI_ISL_1920041 | Stony Brook University Hospital | Wadsworth Center, New York State Department of Health | Kirsten St. George, Daryl M. Lamson, Alexis Russell, Matthew Shudt, Melissa A Leisner, Jonathan Plitnick, Catharine Prussing, Navjot Singh, John Kelly, Erasmus Schneider, Erica Lasek-Nesselquist |
| EPI_ISL_1920042, EPI_ISL_1920043 | Northwell Health Laboratories | Wadsworth Center, New York State Department of Health | Kirsten St. George, Daryl M. Lamson, Alexis Russell, Matthew Shudt, Melissa A Leisner, Jonathan Plitnick, Catharine Prussing, Navjot Singh, John Kelly, Erasmus Schneider, Erica Lasek-Nesselquist |
| EPI_ISL_1920044 | Stony Brook University Hospital | Wadsworth Center, New York State Department of Health | Kirsten St. George, Daryl M. Lamson, Alexis Russell, Matthew Shudt, Melissa A Leisner, Jonathan Plitnick, Catharine Prussing, Navjot Singh, John Kelly, Erasmus Schneider, Erica Lasek-Nesselquist |
| EPI_ISL_1920045 | Northwell Health Laboratories | Wadsworth Center, New York State Department of Health | Kirsten St. George, Daryl M. Lamson, Alexis Russell, Matthew Shudt, Melissa A Leisner, Jonathan Plitnick, Catharine Prussing, Navjot Singh, John Kelly, Erasmus Schneider, Erica Lasek-Nesselquist |
| EPI_ISL_1920046, EPI_ISL_1920047, EPI_ISL_1920048, EPI_ISL_1920049 | Stony Brook University Hospital | Wadsworth Center, New York State Department of Health | Kirsten St. George, Daryl M. Lamson, Alexis Russell, Matthew Shudt, Melissa A Leisner, Jonathan Plitnick, Catharine Prussing, Navjot Singh, John Kelly, Erasmus Schneider, Erica Lasek-Nesselquist |
| EPI_ISL_1920050, EPI_ISL_1920051, EPI_ISL_1920052, EPI_ISL_1920053, EPI_ISL_1920054, EPI_ISL_1920055, EPI_ISL_1920056, EPI_ISL_1920057, EPI_ISL_1920058, EPI_ISL_1920059, EPI_ISL_1920060 | | | |
| see above | Albany Medical Center | Wadsworth Center, New York State Department of Health | Kirsten St. George, Daryl M. Lamson, Alexis Russell, Matthew Shudt, Melissa A Leisner, Jonathan Plitnick, Catharine Prussing, Navjot Singh, John Kelly, Erasmus Schneider, Erica Lasek-Nesselquist |
| EPI_ISL_1920061 | Memorial Sloan Kettering Cancer Center | Wadsworth Center, New York State Department of Health | Kirsten St. George, Daryl M. Lamson, Alexis Russell, Matthew Shudt, Melissa A Leisner, Jonathan Plitnick, Catharine Prussing, Navjot Singh, John Kelly, Erasmus Schneider, Erica Lasek-Nesselquist |
| EPI_ISL_1920062, EPI_ISL_1920063, EPI_ISL_1920064, EPI_ISL_1920065 | Westchester Medical Center | Wadsworth Center, New York State Department of Health | Kirsten St. George, Daryl M. Lamson, Alexis Russell, Matthew Shudt, Melissa A Leisner, Jonathan Plitnick, Catharine Prussing, Navjot Singh, John Kelly, Erasmus Schneider, Erica Lasek-Nesselquist |
| EPI_ISL_1920066, EPI_ISL_1920067, EPI_ISL_1920068, EPI_ISL_1920069 | URMC LABS | Wadsworth Center, New York State Department of Health | Kirsten St. George, Daryl M. Lamson, Alexis Russell, Matthew Shudt, Melissa A Leisner, Jonathan Plitnick, Catharine Prussing, Navjot Singh, John Kelly, Erasmus Schneider, Erica Lasek-Nesselquist |
| EPI_ISL_1920070, EPI_ISL_1920071, EPI_ISL_1920072, EPI_ISL_1920073 | NYC Pandemic Response Lab | Wadsworth Center, New York State Department of Health | Kirsten St. George, Daryl M. Lamson, Alexis Russell, Matthew Shudt, Melissa A Leisner, Jonathan Plitnick, Catharine Prussing, Navjot Singh, John Kelly, Erasmus Schneider, Erica Lasek-Nesselquist |
| EPI_ISL_1920074, EPI_ISL_1920075 | Montefiore Medical Center Laboratories | Wadsworth Center, New York State Department of Health | Kirsten St. George, Daryl M. Lamson, Alexis Russell, Matthew Shudt, Melissa A Leisner, Jonathan Plitnick, Catharine Prussing, Navjot Singh, John Kelly, Erasmus Schneider, Erica Lasek-Nesselquist |
| EPI_ISL_1920076 | Memorial Sloan Kettering Cancer Center | Wadsworth Center, New York State Department of Health | Kirsten St. George, Daryl M. Lamson, Alexis Russell, Matthew Shudt, Melissa A Leisner, Jonathan Plitnick, Catharine Prussing, Navjot Singh, John Kelly, Erasmus Schneider, Erica Lasek-Nesselquist |
| EPI_ISL_1920077 | Montefiore Medical Center Laboratories | Wadsworth Center, New York State Department of Health | Kirsten St. George, Daryl M. Lamson, Alexis Russell, Matthew Shudt, Melissa A Leisner, Jonathan Plitnick, Catharine Prussing, Navjot Singh, John Kelly, Erasmus Schneider, Erica Lasek-Nesselquist |
| EPI_ISL_1920078 | Columbia University Irving Medical Center | Wadsworth Center, New York State Department of Health | Kirsten St. George, Daryl M. Lamson, Alexis Russell, Matthew Shudt, Melissa A Leisner, Jonathan Plitnick, Catharine Prussing, Navjot Singh, John Kelly, Erasmus Schneider, Erica Lasek-Nesselquist |
| EPI_ISL_1920079, EPI_ISL_1920080 | NYC Pandemic Response Lab | Wadsworth Center, New York State Department of Health | Kirsten St. George, Daryl M. Lamson, Alexis Russell, Matthew Shudt, Melissa A Leisner, Jonathan Plitnick, Catharine Prussing, Navjot Singh, John Kelly, Erasmus Schneider, Erica Lasek-Nesselquist |
| EPI_ISL_1920081, EPI_ISL_1920082 | Westchester Medical Center | Wadsworth Center, New York State Department of Health | Kirsten St. George, Daryl M. Lamson, Alexis Russell, Matthew Shudt, Melissa A Leisner, Jonathan Plitnick, Catharine Prussing, Navjot Singh, John Kelly, Erasmus Schneider, Erica Lasek-Nesselquist |
| EPI_ISL_1920083 | NYC Pandemic Response Lab | Wadsworth Center, New York State | Kirsten St. George, Daryl M. Lamson, Alexis Russell, Matthew Shudt, Melissa A Leisner, Jonathan Plitnick, Catharine Prussing, Navjot Singh, John Kelly, Erasmus Schneider, Erica Lasek-Nesselquist |

| | | | |
|---|---|---|---|
| | | Department of Health | |
| EPI_ISL_1920084, EPI_ISL_1920085, EPI_ISL_1920086 | Montefiore Medical Center Laboratories | Wadsworth Center, New York State Department of Health | Kirsten St. George, Daryl M. Lamson, Alexis Russell, Matthew Shudt, Melissa A Leisner, Jonathan Plitnick, Catharine Prussing, Navjot Singh, John Kelly, Erasmus Schneider, Erica Lasek-Nesselquist |
| EPI_ISL_1920087, EPI_ISL_1920088 | NYC Pandemic Response Lab | Wadsworth Center, New York State Department of Health | Kirsten St. George, Daryl M. Lamson, Alexis Russell, Matthew Shudt, Melissa A Leisner, Jonathan Plitnick, Catharine Prussing, Navjot Singh, John Kelly, Erasmus Schneider, Erica Lasek-Nesselquist |
| EPI_ISL_1920089, EPI_ISL_1920090, EPI_ISL_1920091, EPI_ISL_1920092 | Montefiore Medical Center Laboratories | Wadsworth Center, New York State Department of Health | Kirsten St. George, Daryl M. Lamson, Alexis Russell, Matthew Shudt, Melissa A Leisner, Jonathan Plitnick, Catharine Prussing, Navjot Singh, John Kelly, Erasmus Schneider, Erica Lasek-Nesselquist |
| EPI_ISL_1920093 | Westchester Medical Center | Wadsworth Center, New York State Department of Health | Kirsten St. George, Daryl M. Lamson, Alexis Russell, Matthew Shudt, Melissa A Leisner, Jonathan Plitnick, Catharine Prussing, Navjot Singh, John Kelly, Erasmus Schneider, Erica Lasek-Nesselquist |
| EPI_ISL_1920094 | Columbia University Irving Medical Center | Wadsworth Center, New York State Department of Health | Kirsten St. George, Daryl M. Lamson, Alexis Russell, Matthew Shudt, Melissa A Leisner, Jonathan Plitnick, Catharine Prussing, Navjot Singh, John Kelly, Erasmus Schneider, Erica Lasek-Nesselquist |
| EPI_ISL_1920095 | NYC Pandemic Response Lab | Wadsworth Center, New York State Department of Health | Kirsten St. George, Daryl M. Lamson, Alexis Russell, Matthew Shudt, Melissa A Leisner, Jonathan Plitnick, Catharine Prussing, Navjot Singh, John Kelly, Erasmus Schneider, Erica Lasek-Nesselquist |
| EPI_ISL_1920096 | Montefiore Medical Center Laboratories | Wadsworth Center, New York State Department of Health | Kirsten St. George, Daryl M. Lamson, Alexis Russell, Matthew Shudt, Melissa A Leisner, Jonathan Plitnick, Catharine Prussing, Navjot Singh, John Kelly, Erasmus Schneider, Erica Lasek-Nesselquist |
| EPI_ISL_1920097 | New York Presbyterian Hospital | Wadsworth Center, New York State Department of Health | Kirsten St. George, Daryl M. Lamson, Alexis Russell, Matthew Shudt, Melissa A Leisner, Jonathan Plitnick, Catharine Prussing, Navjot Singh, John Kelly, Erasmus Schneider, Erica Lasek-Nesselquist |
| EPI_ISL_1920098, EPI_ISL_1920099, EPI_ISL_1920100 | Tempus Labs Inc | Wadsworth Center, New York State Department of Health | Kirsten St. George, Daryl M. Lamson, Alexis Russell, Matthew Shudt, Melissa A Leisner, Jonathan Plitnick, Catharine Prussing, Navjot Singh, John Kelly, Erasmus Schneider, Erica Lasek-Nesselquist |
| EPI_ISL_1920101 | Northwell Health Laboratories | Wadsworth Center, New York State Department of Health | Kirsten St. George, Daryl M. Lamson, Alexis Russell, Matthew Shudt, Melissa A Leisner, Jonathan Plitnick, Catharine Prussing, Navjot Singh, John Kelly, Erasmus Schneider, Erica Lasek-Nesselquist |
| EPI_ISL_1920102, EPI_ISL_1920103 | Tempus Labs Inc | Wadsworth Center, New York State Department of Health | Kirsten St. George, Daryl M. Lamson, Alexis Russell, Matthew Shudt, Melissa A Leisner, Jonathan Plitnick, Catharine Prussing, Navjot Singh, John Kelly, Erasmus Schneider, Erica Lasek-Nesselquist |
| EPI_ISL_1920104, EPI_ISL_1920105 | Albany Medical Center | Wadsworth Center, New York State Department of Health | Kirsten St. George, Daryl M. Lamson, Alexis Russell, Matthew Shudt, Melissa A Leisner, Jonathan Plitnick, Catharine Prussing, Navjot Singh, John Kelly, Erasmus Schneider, Erica Lasek-Nesselquist |
| EPI_ISL_1920106, EPI_ISL_1920107 | Tempus Labs Inc | Wadsworth Center, New York State Department of Health | Kirsten St. George, Daryl M. Lamson, Alexis Russell, Matthew Shudt, Melissa A Leisner, Jonathan Plitnick, Catharine Prussing, Navjot Singh, John Kelly, Erasmus Schneider, Erica Lasek-Nesselquist |
| EPI_ISL_1920108, EPI_ISL_1920109, EPI_ISL_1920110, EPI_ISL_1920111, EPI_ISL_1920112 | Albany Medical Center | Wadsworth Center, New York State Department of Health | Kirsten St. George, Daryl M. Lamson, Alexis Russell, Matthew Shudt, Melissa A Leisner, Jonathan Plitnick, Catharine Prussing, Navjot Singh, John Kelly, Erasmus Schneider, Erica Lasek-Nesselquist |
| EPI_ISL_1920113, EPI_ISL_1920114 | Wadsworth Center, New York State Department of Health | Wadsworth Center, New York State Department of Health | Kirsten St. George, Daryl M. Lamson, Alexis Russell, Matthew Shudt, Melissa A Leisner, Jonathan Plitnick, Catharine Prussing, Navjot Singh, John Kelly, Erasmus Schneider, Erica Lasek-Nesselquist |
| EPI_ISL_1920115, EPI_ISL_1920116, EPI_ISL_1920117, EPI_ISL_1920118 | Tempus Labs Inc | Wadsworth Center, New York State Department of Health | Kirsten St. George, Daryl M. Lamson, Alexis Russell, Matthew Shudt, Melissa A Leisner, Jonathan Plitnick, Catharine Prussing, Navjot Singh, John Kelly, Erasmus Schneider, Erica Lasek-Nesselquist |
| EPI_ISL_1920119, EPI_ISL_1920120 | Glens Falls Hospital Laboratory | Wadsworth Center, New York State Department of Health | Kirsten St. George, Daryl M. Lamson, Alexis Russell, Matthew Shudt, Melissa A Leisner, Jonathan Plitnick, Catharine Prussing, Navjot Singh, John Kelly, Erasmus Schneider, Erica Lasek-Nesselquist |
| EPI_ISL_1920121 | Wadsworth Center, New York State Department of Health | Wadsworth Center, New York State Department of Health | Kirsten St. George, Daryl M. Lamson, Alexis Russell, Matthew Shudt, Melissa A Leisner, Jonathan Plitnick, Catharine Prussing, Navjot Singh, John Kelly, Erasmus Schneider, Erica Lasek-Nesselquist |
| EPI_ISL_1920122 | Pulaski Urgent Care | Wadsworth Center, New York State Department of Health | Kirsten St. George, Daryl M. Lamson, Alexis Russell, Matthew Shudt, Melissa A Leisner, Jonathan Plitnick, Catharine Prussing, Navjot Singh, John Kelly, Erasmus Schneider, Erica Lasek-Nesselquist |
| EPI_ISL_1920123 | Tempus Labs Inc | Wadsworth Center, New York State Department of Health | Kirsten St. George, Daryl M. Lamson, Alexis Russell, Matthew Shudt, Melissa A Leisner, Jonathan Plitnick, Catharine Prussing, Navjot Singh, John Kelly, Erasmus Schneider, Erica Lasek-Nesselquist |
| EPI_ISL_1920124, EPI_ISL_1920125, EPI_ISL_1920126, EPI_ISL_1920127, EPI_ISL_1920128, EPI_ISL_1920129, EPI_ISL_1920130, EPI_ISL_1920131 | URMC LABS | Wadsworth Center, New York State Department of Health | Kirsten St. George, Daryl M. Lamson, Alexis Russell, Matthew Shudt, Melissa A Leisner, Jonathan Plitnick, Catharine Prussing, Navjot Singh, John Kelly, Erasmus Schneider, Erica Lasek-Nesselquist |
| EPI_ISL_1920132 | Northwell Health Laboratories | Wadsworth Center, New York State Department of Health | Kirsten St. George, Daryl M. Lamson, Alexis Russell, Matthew Shudt, Melissa A Leisner, Jonathan Plitnick, Catharine Prussing, Navjot Singh, John Kelly, Erasmus Schneider, Erica Lasek-Nesselquist |
| EPI_ISL_1920133, EPI_ISL_1920134, EPI_ISL_1920135, EPI_ISL_1920136 | New York Presbyterian Hospital | Wadsworth Center, New York State Department of Health | Kirsten St. George, Daryl M. Lamson, Alexis Russell, Matthew Shudt, Melissa A Leisner, Jonathan Plitnick, Catharine Prussing, Navjot Singh, John Kelly, Erasmus Schneider, Erica Lasek-Nesselquist |
| EPI_ISL_1920137 | Columbia University Irving Medical Center | Wadsworth Center, New York State Department of Health | Kirsten St. George, Daryl M. Lamson, Alexis Russell, Matthew Shudt, Melissa A Leisner, Jonathan Plitnick, Catharine Prussing, Navjot Singh, John Kelly, Erasmus Schneider, Erica Lasek-Nesselquist |
| EPI_ISL_1920138 | New York Presbyterian Hospital | Wadsworth Center, New York State Department of Health | Kirsten St. George, Daryl M. Lamson, Alexis Russell, Matthew Shudt, Melissa A Leisner, Jonathan Plitnick, Catharine Prussing, Navjot Singh, John Kelly, Erasmus Schneider, Erica Lasek-Nesselquist |
| EPI_ISL_1920139, EPI_ISL_1920140, EPI_ISL_1920141, EPI_ISL_1920142, EPI_ISL_1920143, EPI_ISL_1920144, EPI_ISL_1920145, EPI_ISL_1920146 | Columbia University Irving Medical Center | Wadsworth Center, New York State Department of Health | Kirsten St. George, Daryl M. Lamson, Alexis Russell, Matthew Shudt, Melissa A Leisner, Jonathan Plitnick, Catharine Prussing, Navjot Singh, John Kelly, Erasmus Schneider, Erica Lasek-Nesselquist |
| EPI_ISL_1920147 | New York Presbyterian Hospital | Wadsworth Center, New York State Department of Health | Kirsten St. George, Daryl M. Lamson, Alexis Russell, Matthew Shudt, Melissa A Leisner, Jonathan Plitnick, Catharine Prussing, Navjot Singh, John Kelly, Erasmus Schneider, Erica Lasek-Nesselquist |
| EPI_ISL_1920148, EPI_ISL_1920149 | Columbia University Irving Medical Center | Wadsworth Center, New York State Department of Health | Kirsten St. George, Daryl M. Lamson, Alexis Russell, Matthew Shudt, Melissa A Leisner, Jonathan Plitnick, Catharine Prussing, Navjot Singh, John Kelly, Erasmus Schneider, Erica Lasek-Nesselquist |
| EPI_ISL_1920150 | New York Presbyterian Hospital | Wadsworth Center, New York State Department of Health | Kirsten St. George, Daryl M. Lamson, Alexis Russell, Matthew Shudt, Melissa A Leisner, Jonathan Plitnick, Catharine Prussing, Navjot Singh, John Kelly, Erasmus Schneider, Erica Lasek-Nesselquist |
| EPI_ISL_1920151 | Northwell Health Laboratories | Wadsworth Center, New York State Department of Health | Kirsten St. George, Daryl M. Lamson, Alexis Russell, Matthew Shudt, Melissa A Leisner, Jonathan Plitnick, Catharine Prussing, Navjot Singh, John Kelly, Erasmus Schneider, Erica Lasek-Nesselquist |
| EPI_ISL_1920152, EPI_ISL_1920153 | Westchester Medical Center | Wadsworth Center, New York State Department of Health | Kirsten St. George, Daryl M. Lamson, Alexis Russell, Matthew Shudt, Melissa A Leisner, Jonathan Plitnick, Catharine Prussing, Navjot Singh, John Kelly, Erasmus Schneider, Erica Lasek-Nesselquist |
| EPI_ISL_1920154 | Columbia University Irving Medical Center | Wadsworth Center, New York State Department of Health | Kirsten St. George, Daryl M. Lamson, Alexis Russell, Matthew Shudt, Melissa A Leisner, Jonathan Plitnick, Catharine Prussing, Navjot Singh, John Kelly, Erasmus Schneider, Erica Lasek-Nesselquist |
| EPI_ISL_1920155 | Westchester Medical Center | Wadsworth Center, New York State Department of Health | Kirsten St. George, Daryl M. Lamson, Alexis Russell, Matthew Shudt, Melissa A Leisner, Jonathan Plitnick, Catharine Prussing, Navjot Singh, John Kelly, Erasmus Schneider, Erica Lasek-Nesselquist |
| EPI_ISL_1920156 | Columbia University Irving Medical Center | Wadsworth Center, New York State Department of Health | Kirsten St. George, Daryl M. Lamson, Alexis Russell, Matthew Shudt, Melissa A Leisner, Jonathan Plitnick, Catharine Prussing, Navjot Singh, John Kelly, Erasmus Schneider, Erica Lasek-Nesselquist |
| EPI_ISL_1920157 | Memorial Sloan Kettering Cancer Center | Wadsworth Center, New York State Department of Health | Kirsten St. George, Daryl M. Lamson, Alexis Russell, Matthew Shudt, Melissa A Leisner, Jonathan Plitnick, Catharine Prussing, Navjot Singh, John Kelly, Erasmus Schneider, Erica Lasek-Nesselquist |

| | | | |
|---|---|---|---|
| EPI_ISL_1920158 | Westchester Medical Center | Wadsworth Center, New York State Department of Health | Kirsten St. George, Daryl M. Lamson, Alexis Russell, Matthew Shudt, Melissa A Leisner, Jonathan Plitnick, Catharine Prussing, Navjot Singh, John Kelly, Erasmus Schneider, Erica Lasek-Nesselquist |
| EPI_ISL_1920159 | Montefiore Medical Center Laboratories | Wadsworth Center, New York State Department of Health | Kirsten St. George, Daryl M. Lamson, Alexis Russell, Matthew Shudt, Melissa A Leisner, Jonathan Plitnick, Catharine Prussing, Navjot Singh, John Kelly, Erasmus Schneider, Erica Lasek-Nesselquist |
| EPI_ISL_1920160 | Columbia University Irving Medical Center | Wadsworth Center, New York State Department of Health | Kirsten St. George, Daryl M. Lamson, Alexis Russell, Matthew Shudt, Melissa A Leisner, Jonathan Plitnick, Catharine Prussing, Navjot Singh, John Kelly, Erasmus Schneider, Erica Lasek-Nesselquist |
| EPI_ISL_1920161 | Montefiore Medical Center Laboratories | Wadsworth Center, New York State Department of Health | Kirsten St. George, Daryl M. Lamson, Alexis Russell, Matthew Shudt, Melissa A Leisner, Jonathan Plitnick, Catharine Prussing, Navjot Singh, John Kelly, Erasmus Schneider, Erica Lasek-Nesselquist |
| EPI_ISL_1920162 | New York Presbyterian Hospital | Wadsworth Center, New York State Department of Health | Kirsten St. George, Daryl M. Lamson, Alexis Russell, Matthew Shudt, Melissa A Leisner, Jonathan Plitnick, Catharine Prussing, Navjot Singh, John Kelly, Erasmus Schneider, Erica Lasek-Nesselquist |
| EPI_ISL_1920163 | NYC Pandemic Response Lab | Wadsworth Center, New York State Department of Health | Kirsten St. George, Daryl M. Lamson, Alexis Russell, Matthew Shudt, Melissa A Leisner, Jonathan Plitnick, Catharine Prussing, Navjot Singh, John Kelly, Erasmus Schneider, Erica Lasek-Nesselquist |
| EPI_ISL_1920164, EPI_ISL_1920165 | New York Presbyterian Hospital | Wadsworth Center, New York State Department of Health | Kirsten St. George, Daryl M. Lamson, Alexis Russell, Matthew Shudt, Melissa A Leisner, Jonathan Plitnick, Catharine Prussing, Navjot Singh, John Kelly, Erasmus Schneider, Erica Lasek-Nesselquist |
| EPI_ISL_1920166 | Northwell Health Laboratories | Wadsworth Center, New York State Department of Health | Kirsten St. George, Daryl M. Lamson, Alexis Russell, Matthew Shudt, Melissa A Leisner, Jonathan Plitnick, Catharine Prussing, Navjot Singh, John Kelly, Erasmus Schneider, Erica Lasek-Nesselquist |
| EPI_ISL_1920167 | Westchester Medical Center | Wadsworth Center, New York State Department of Health | Kirsten St. George, Daryl M. Lamson, Alexis Russell, Matthew Shudt, Melissa A Leisner, Jonathan Plitnick, Catharine Prussing, Navjot Singh, John Kelly, Erasmus Schneider, Erica Lasek-Nesselquist |
| EPI_ISL_1920168, EPI_ISL_1920169, EPI_ISL_1920170 | Northwell Health Laboratories | Wadsworth Center, New York State Department of Health | Kirsten St. George, Daryl M. Lamson, Alexis Russell, Matthew Shudt, Melissa A Leisner, Jonathan Plitnick, Catharine Prussing, Navjot Singh, John Kelly, Erasmus Schneider, Erica Lasek-Nesselquist |
| EPI_ISL_1920171 | New York Presbyterian Hospital | Wadsworth Center, New York State Department of Health | Kirsten St. George, Daryl M. Lamson, Alexis Russell, Matthew Shudt, Melissa A Leisner, Jonathan Plitnick, Catharine Prussing, Navjot Singh, John Kelly, Erasmus Schneider, Erica Lasek-Nesselquist |
| EPI_ISL_1920172, EPI_ISL_1920173, EPI_ISL_1920174 | Westchester Medical Center | Wadsworth Center, New York State Department of Health | Kirsten St. George, Daryl M. Lamson, Alexis Russell, Matthew Shudt, Melissa A Leisner, Jonathan Plitnick, Catharine Prussing, Navjot Singh, John Kelly, Erasmus Schneider, Erica Lasek-Nesselquist |
| EPI_ISL_1920510, EPI_ISL_1920511, EPI_ISL_1920512, EPI_ISL_1920513, EPI_ISL_1920514, EPI_ISL_1920515, EPI_ISL_1920516, EPI_ISL_1920518, EPI_ISL_1920520, EPI_ISL_1920521, EPI_ISL_1920522, EPI_ISL_1920523, EPI_ISL_1920524, EPI_ISL_1920526, EPI_ISL_1920527, EPI_ISL_1920528, EPI_ISL_1920529, EPI_ISL_1920530, EPI_ISL_1920531, EPI_ISL_1920532, EPI_ISL_1920533, EPI_ISL_1920534, EPI_ISL_1920535, EPI_ISL_1920536, EPI_ISL_1920537, EPI_ISL_1920538, EPI_ISL_1920539, EPI_ISL_1920540, EPI_ISL_1920541, EPI_ISL_1920542, EPI_ISL_1920543, EPI_ISL_1920544, EPI_ISL_1920545, EPI_ISL_1920546, EPI_ISL_1920547, EPI_ISL_1920548, EPI_ISL_1920549, EPI_ISL_1920550, EPI_ISL_1920551, EPI_ISL_1920552, EPI_ISL_1920553, EPI_ISL_1920554, EPI_ISL_1920556, EPI_ISL_1920557, EPI_ISL_1920559, EPI_ISL_1920560 | | | |
| see above | North Dakota Department of Health, Public Health Laboratory | North Dakota Department of Health, Public Health Laboratory | Lisa Wingerter |
| EPI_ISL_1920836, EPI_ISL_1920837, EPI_ISL_1920838, EPI_ISL_1920839, EPI_ISL_1920840, EPI_ISL_1920841, EPI_ISL_1920842, EPI_ISL_1920843, EPI_ISL_1920845, EPI_ISL_1920849, EPI_ISL_1920852, EPI_ISL_1920853, EPI_ISL_1920854, EPI_ISL_1920855, EPI_ISL_1920856, EPI_ISL_1920861, EPI_ISL_1920862, EPI_ISL_1920864, EPI_ISL_1920867, EPI_ISL_1920871, EPI_ISL_1920873, EPI_ISL_1920875, EPI_ISL_1920876, EPI_ISL_1920878, EPI_ISL_1920879, EPI_ISL_1920881, EPI_ISL_1920882, EPI_ISL_1920883, EPI_ISL_1920884, EPI_ISL_1920885, EPI_ISL_1920886, EPI_ISL_1920887, EPI_ISL_1920890, EPI_ISL_1920891, EPI_ISL_1920892, EPI_ISL_1920896, EPI_ISL_1920897, EPI_ISL_1920898, EPI_ISL_1920899, EPI_ISL_1920900 | | | |
| see above | Santa Clara County Public Health Laboratory | Santa Clara County Public Health Laboratory | Santa Clara County Public Health Department |
| EPI_ISL_1920902, EPI_ISL_1920903, EPI_ISL_1920904, EPI_ISL_1920905, EPI_ISL_1920906, EPI_ISL_1920907, EPI_ISL_1920908, EPI_ISL_1920909, EPI_ISL_1920911, EPI_ISL_1920912, EPI_ISL_1920913, EPI_ISL_1920914, EPI_ISL_1920915, EPI_ISL_1920916, EPI_ISL_1920918, EPI_ISL_1920919, EPI_ISL_1920920, EPI_ISL_1920921, EPI_ISL_1920922, EPI_ISL_1920923, EPI_ISL_1920924, EPI_ISL_1920925, EPI_ISL_1920926, EPI_ISL_1920927, EPI_ISL_1920928, EPI_ISL_1920929, EPI_ISL_1920930, EPI_ISL_1920931, EPI_ISL_1920932, EPI_ISL_1920933, EPI_ISL_1920934, EPI_ISL_1920935, EPI_ISL_1920936, EPI_ISL_1920937, EPI_ISL_1920938, EPI_ISL_1920939, EPI_ISL_1920940, EPI_ISL_1920941, EPI_ISL_1920943, EPI_ISL_1920944, EPI_ISL_1920945, EPI_ISL_1920946, EPI_ISL_1920947, EPI_ISL_1920948, EPI_ISL_1920949, EPI_ISL_1920950, EPI_ISL_1920951, EPI_ISL_1920952, EPI_ISL_1920953, EPI_ISL_1920954, EPI_ISL_1920955, EPI_ISL_1920956, EPI_ISL_1920957, EPI_ISL_1920958, EPI_ISL_1920959, EPI_ISL_1920960, EPI_ISL_1920961 | | | |
| see above | WVT Laboratory, Corvallis, Oregon | OSU Center for Genome Research and Biocomputing | Oregon State University TRACE Project |
| EPI_ISL_1920963, EPI_ISL_1920964, EPI_ISL_1920965, EPI_ISL_1920966, EPI_ISL_1920967, EPI_ISL_1920968, EPI_ISL_1920969, EPI_ISL_1920970, EPI_ISL_1920971, EPI_ISL_1920972, EPI_ISL_1920973, EPI_ISL_1920974, EPI_ISL_1920975, EPI_ISL_1920976, EPI_ISL_1920977, EPI_ISL_1920978, EPI_ISL_1920979, EPI_ISL_1920980, EPI_ISL_1920981, EPI_ISL_1920982, EPI_ISL_1920983, EPI_ISL_1920984, EPI_ISL_1920985, EPI_ISL_1920986, EPI_ISL_1920987, EPI_ISL_1920988, EPI_ISL_1920989, EPI_ISL_1920990, EPI_ISL_1920991, EPI_ISL_1920992, EPI_ISL_1920993, EPI_ISL_1920994, EPI_ISL_1920995, EPI_ISL_1920996 | | | |
| see above | Alaska State Virology Laboratory | Alaska State Virology Laboratory | Stephanie DeRonde, Elva House, Jacob Zidek, Lisa Smith, Ph.D., Jack Chen, Ph.D. |
| EPI_ISL_1920997, EPI_ISL_1920998, EPI_ISL_1920999, EPI_ISL_1921000, EPI_ISL_1921001, EPI_ISL_1921002, EPI_ISL_1921003, EPI_ISL_1921004, EPI_ISL_1921005, EPI_ISL_1921006, EPI_ISL_1921007, EPI_ISL_1921008, EPI_ISL_1921009, EPI_ISL_1921010 | | | |
| see above | URMC LABS | Wadsworth Center, New York State Department of Health | Kirsten St. George, Daryl M. Lamson, Alexis Russell, Matthew Shudt, Melissa A Leisner, Jonathan Plitnick, Catharine Prussing, Navjot Singh, John Kelly, Erasmus Schneider, Erica Lasek-Nesselquist |
| EPI_ISL_1921011, EPI_ISL_1921012, EPI_ISL_1921013, EPI_ISL_1921014, EPI_ISL_1921015, EPI_ISL_1921016, EPI_ISL_1921017 | SUNY Upstate Medical University | Wadsworth Center, New York State Department of Health | Kirsten St. George, Daryl M. Lamson, Alexis Russell, Matthew Shudt, Melissa A Leisner, Jonathan Plitnick, Catharine Prussing, Navjot Singh, John Kelly, Erasmus Schneider, Erica Lasek-Nesselquist |
| EPI_ISL_1921018 | Albany Medical Center | Wadsworth Center, New York State Department of Health | Kirsten St. George, Daryl M. Lamson, Alexis Russell, Matthew Shudt, Melissa A Leisner, Jonathan Plitnick, Catharine Prussing, Navjot Singh, John Kelly, Erasmus Schneider, Erica Lasek-Nesselquist |
| EPI_ISL_1921019, EPI_ISL_1921020, EPI_ISL_1921021 | SUNY Upstate Medical University | Wadsworth Center, New York State Department of Health | Kirsten St. George, Daryl M. Lamson, Alexis Russell, Matthew Shudt, Melissa A Leisner, Jonathan Plitnick, Catharine Prussing, Navjot Singh, John Kelly, Erasmus Schneider, Erica Lasek-Nesselquist |
| EPI_ISL_1921022 | Albany Medical Center | Wadsworth Center, New York State Department of Health | Kirsten St. George, Daryl M. Lamson, Alexis Russell, Matthew Shudt, Melissa A Leisner, Jonathan Plitnick, Catharine Prussing, Navjot Singh, John Kelly, Erasmus Schneider, Erica Lasek-Nesselquist |
| EPI_ISL_1921023, EPI_ISL_1921024, EPI_ISL_1921025, EPI_ISL_1921026, EPI_ISL_1921027 | SUNY Upstate Medical University | Wadsworth Center, New York State Department of Health | Kirsten St. George, Daryl M. Lamson, Alexis Russell, Matthew Shudt, Melissa A Leisner, Jonathan Plitnick, Catharine Prussing, Navjot Singh, John Kelly, Erasmus Schneider, Erica Lasek-Nesselquist |
| EPI_ISL_1921028, EPI_ISL_1921029, EPI_ISL_1921030, EPI_ISL_1921031, EPI_ISL_1921032, EPI_ISL_1921033, EPI_ISL_1921034, EPI_ISL_1921035, EPI_ISL_1921036 | Albany Medical Center | Wadsworth Center, New York State Department of Health | Kirsten St. George, Daryl M. Lamson, Alexis Russell, Matthew Shudt, Melissa A Leisner, Jonathan Plitnick, Catharine Prussing, Navjot Singh, John Kelly, Erasmus Schneider, Erica Lasek-Nesselquist |
| EPI_ISL_1921037 | Glens Falls Hospital Laboratory | Wadsworth Center, New York State Department of Health | Kirsten St. George, Daryl M. Lamson, Alexis Russell, Matthew Shudt, Melissa A Leisner, Jonathan Plitnick, Catharine Prussing, Navjot Singh, John Kelly, Erasmus Schneider, Erica Lasek-Nesselquist |
| EPI_ISL_1921038, EPI_ISL_1921039 | SUNY Upstate Medical University | Wadsworth Center, New York State Department of Health | Kirsten St. George, Daryl M. Lamson, Alexis Russell, Matthew Shudt, Melissa A Leisner, Jonathan Plitnick, Catharine Prussing, Navjot Singh, John Kelly, Erasmus Schneider, Erica Lasek-Nesselquist |
| EPI_ISL_1921040, EPI_ISL_1921041, EPI_ISL_1921042, EPI_ISL_1921043, EPI_ISL_1921044, EPI_ISL_1921045, EPI_ISL_1921046, EPI_ISL_1921047, EPI_ISL_1921048, EPI_ISL_1921049, EPI_ISL_1921050, EPI_ISL_1921051, EPI_ISL_1921052, EPI_ISL_1921053, EPI_ISL_1921054, EPI_ISL_1921055, EPI_ISL_1921056, EPI_ISL_1921057, EPI_ISL_1921058, EPI_ISL_1921059, EPI_ISL_1921060, EPI_ISL_1921061, EPI_ISL_1921062, EPI_ISL_1921063, EPI_ISL_1921064, EPI_ISL_1921065, EPI_ISL_1921066, EPI_ISL_1921067, EPI_ISL_1921068, EPI_ISL_1921069, EPI_ISL_1921070, EPI_ISL_1921071, EPI_ISL_1921072, EPI_ISL_1921073, EPI_ISL_1921074, EPI_ISL_1921075, EPI_ISL_1921076, EPI_ISL_1921077, EPI_ISL_1921078, EPI_ISL_1921079 | | | |
| see above | URMC LABS | Wadsworth Center, New York State Department of Health | Kirsten St. George, Daryl M. Lamson, Alexis Russell, Matthew Shudt, Melissa A Leisner, Jonathan Plitnick, Catharine Prussing, Navjot Singh, John Kelly, Erasmus Schneider, Erica Lasek-Nesselquist |
| EPI_ISL_1921080, EPI_ISL_1921081, EPI_ISL_1921082, EPI_ISL_1921083, EPI_ISL_1921084, EPI_ISL_1921085, EPI_ISL_1921086, EPI_ISL_1921087, EPI_ISL_1921088, EPI_ISL_1921089, EPI_ISL_1921090, EPI_ISL_1921091, EPI_ISL_1921092, EPI_ISL_1921093, EPI_ISL_1921094, EPI_ISL_1921095, EPI_ISL_1921096, EPI_ISL_1921097, EPI_ISL_1921098, EPI_ISL_1921099, EPI_ISL_1921100, EPI_ISL_1921101, EPI_ISL_1921102, EPI_ISL_1921103, EPI_ISL_1921104, EPI_ISL_1921105, EPI_ISL_1921106, EPI_ISL_1921107, EPI_ISL_1921108, EPI_ISL_1921109, EPI_ISL_1921110, EPI_ISL_1921111, EPI_ISL_1921112, EPI_ISL_1921113, EPI_ISL_1921114, EPI_ISL_1921115, EPI_ISL_1921116, EPI_ISL_1921117, EPI_ISL_1921118, EPI_ISL_1921119, EPI_ISL_1921120, EPI_ISL_1921121, EPI_ISL_1921122, EPI_ISL_1921123, EPI_ISL_1921124, EPI_ISL_1921125, EPI_ISL_1921126, EPI_ISL_1921127, EPI_ISL_1921128, EPI_ISL_1921129, EPI_ISL_1921130, EPI_ISL_1921131, EPI_ISL_1921132, EPI_ISL_1921133, EPI_ISL_1921134, EPI_ISL_1921135, EPI_ISL_1921136, EPI_ISL_1921137, EPI_ISL_1921138, EPI_ISL_1921139, EPI_ISL_1921140, EPI_ISL_1921141, EPI_ISL_1921142, EPI_ISL_1921143, EPI_ISL_1921144, EPI_ISL_1921145, EPI_ISL_1921146, EPI_ISL_1921147, EPI_ISL_1921148, EPI_ISL_1921149, EPI_ISL_1921150, EPI_ISL_1921151, EPI_ISL_1921152, EPI_ISL_1921153, EPI_ISL_1921154, EPI_ISL_1921155, EPI_ISL_1921156, EPI_ISL_1921157, EPI_ISL_1921158, EPI_ISL_1921159, EPI_ISL_1921160, EPI_ISL_1921161, EPI_ISL_1921162, EPI_ISL_1921163, EPI_ISL_1921164 | | | |
| see above | UCSC Genomics Institute | UCSC Genomics Institute | Mark Akeson, Ikenna Anigbogu, Eric Beraut, Isabel Bjork, Molly Cassatt-Johnstone, Terren Chang, Russell Corbett-Detig, Namrita Dhillon, Maximilian Haeussler, David Haussler, Angie Hinrichs, Miten Jain, Joshua Kapp, A. Marm Kilpatrick, Jakob McBroome, Hugh Olsen, Jeremy Sanford, Beth Shapiro, Michael Stone, Bryan Thornlow, Yatish Turakhia, Ciara Wanket |
| EPI_ISL_1921740, EPI_ISL_1921742, EPI_ISL_1921743, EPI_ISL_1921744, EPI_ISL_1921746, EPI_ISL_1921747, EPI_ISL_1921748, EPI_ISL_1921749, EPI_ISL_1921750, EPI_ISL_1921751, EPI_ISL_1921752, EPI_ISL_1921753, EPI_ISL_1921754, EPI_ISL_1921755, EPI_ISL_1921756, EPI_ISL_1921757, EPI_ISL_1921758, EPI_ISL_1921759, EPI_ISL_1921760, EPI_ISL_1921761, EPI_ISL_1921762 | | | |
| see above | Wyoming Public Health Laboratory | Wyoming Public Health Laboratory | Jim Mildenberger, Taylor Fearing, Channing Weber, Ashley Norberg, Chayse Rowley, Marley Goetz, Brian Dominguez, Elliot Thomasson, Brittany Oher, Sam Britz, Cari Sloma, and Rob Christensen |
| EPI_ISL_1921860, EPI_ISL_1921861 | Kansas Health and Environmental Lab | Kansas Health and Environmental Lab | Mike Grose, Jonathan Barnell, Ben Olsen, and Phil Adam |
| EPI_ISL_1921862, EPI_ISL_1921863, EPI_ISL_1921864, EPI_ISL_1921865, EPI_ISL_1921866, EPI_ISL_1921867, EPI_ISL_1921868, EPI_ISL_1921869, EPI_ISL_1921870, EPI_ISL_1921871, EPI_ISL_1921872, EPI_ISL_1921874, EPI_ISL_1921876, EPI_ISL_1921877 | | | |

| | | | |
|---|---|---|---|
| see above | Wichita State University - Molecular Diagnostics Lab | Kansas Health and Environmental Lab | Mike Grose, Jonathan Barnell, Ben Olsen, and Phil Adam |
| EPI_ISL_1921880, EPI_ISL_1921881, EPI_ISL_1921882, EPI_ISL_1921883, EPI_ISL_1921884, EPI_ISL_1921885, EPI_ISL_1921886, EPI_ISL_1921887, EPI_ISL_1921888, EPI_ISL_1921889, EPI_ISL_1921890, EPI_ISL_1921891 | | | |
| see above | Monterey County Public Health Laboratory | Monterey County Public Health Laboratory | Monterey County Public Health Laboratory |
| EPI_ISL_1921897, EPI_ISL_1921899, EPI_ISL_1921900, EPI_ISL_1921901, EPI_ISL_1921902, EPI_ISL_1921903, EPI_ISL_1921904, EPI_ISL_1921905, EPI_ISL_1921906, EPI_ISL_1921907, EPI_ISL_1921908, EPI_ISL_1921909, EPI_ISL_1921910, EPI_ISL_1921911, EPI_ISL_1921912, EPI_ISL_1921913, EPI_ISL_1921914, EPI_ISL_1921915, EPI_ISL_1921916, EPI_ISL_1921919, EPI_ISL_1921920, EPI_ISL_1921921, EPI_ISL_1921922, EPI_ISL_1921923, EPI_ISL_1921924, EPI_ISL_1921925, EPI_ISL_1921926, EPI_ISL_1921928, EPI_ISL_1921930, EPI_ISL_1921933, EPI_ISL_1921935, EPI_ISL_1921936, EPI_ISL_1921937, EPI_ISL_1921938, EPI_ISL_1921939, EPI_ISL_1921940, EPI_ISL_1921941, EPI_ISL_1921942, EPI_ISL_1921943, EPI_ISL_1921944, EPI_ISL_1921945, EPI_ISL_1921946, EPI_ISL_1921947, EPI_ISL_1921948, EPI_ISL_1921949, EPI_ISL_1921950, EPI_ISL_1921951, EPI_ISL_1921952, EPI_ISL_1921953, EPI_ISL_1921955, EPI_ISL_1921956, EPI_ISL_1921957, EPI_ISL_1921958, EPI_ISL_1921959, EPI_ISL_1921960, EPI_ISL_1921961, EPI_ISL_1921963, EPI_ISL_1921964, EPI_ISL_1921966, EPI_ISL_1921967, EPI_ISL_1921968, EPI_ISL_1921969, EPI_ISL_1921970, EPI_ISL_1921971, EPI_ISL_1921972, EPI_ISL_1921973, EPI_ISL_1921974, EPI_ISL_1921975, EPI_ISL_1921976, EPI_ISL_1921977, EPI_ISL_1921978, EPI_ISL_1921979, EPI_ISL_1921980, EPI_ISL_1921981, EPI_ISL_1921982, EPI_ISL_1921983, EPI_ISL_1921984, EPI_ISL_1921985, EPI_ISL_1921986, EPI_ISL_1921987, EPI_ISL_1921989, EPI_ISL_1921990, EPI_ISL_1921991, EPI_ISL_1921993, EPI_ISL_1921994, EPI_ISL_1921995, EPI_ISL_1921996, EPI_ISL_1921997, EPI_ISL_1921998, EPI_ISL_1921999, EPI_ISL_1922001, EPI_ISL_1922002 | | | |
| see above | Illinois Department of Public Health | Gagnon Lab, Southern Illinois University | Keith Gagnon |
| EPI_ISL_1922042, EPI_ISL_1922043, EPI_ISL_1922044, EPI_ISL_1922045, EPI_ISL_1922046, EPI_ISL_1922047, EPI_ISL_1922048, EPI_ISL_1922049, EPI_ISL_1922050, EPI_ISL_1922051, EPI_ISL_1922052, EPI_ISL_1922053, EPI_ISL_1922054, EPI_ISL_1922055, EPI_ISL_1922056, EPI_ISL_1922057, EPI_ISL_1922058, EPI_ISL_1922126, EPI_ISL_1922127, EPI_ISL_1922128, EPI_ISL_1922129, EPI_ISL_1922130, EPI_ISL_1922131, EPI_ISL_1922132, EPI_ISL_1922133, EPI_ISL_1922134, EPI_ISL_1922135 | | | |
| see above | GA Department of Public Health | GA Department of Public Health | Stacy Reeves, Jonathan Edwards, Cynthia Dixey, Tonia Parrott, Aliyah Fields, Taylor Smith |
| EPI_ISL_1922520, EPI_ISL_1922521, EPI_ISL_1922522, EPI_ISL_1922523, EPI_ISL_1922524, EPI_ISL_1922525, EPI_ISL_1922526, EPI_ISL_1922527, EPI_ISL_1922528, EPI_ISL_1922529, EPI_ISL_1922530, EPI_ISL_1922531, EPI_ISL_1922532, EPI_ISL_1922533, EPI_ISL_1922534, EPI_ISL_1922535, EPI_ISL_1922536, EPI_ISL_1922537, EPI_ISL_1922538, EPI_ISL_1922539, EPI_ISL_1922540, EPI_ISL_1922541, EPI_ISL_1922542, EPI_ISL_1922543, EPI_ISL_1922544, EPI_ISL_1922545, EPI_ISL_1922546, EPI_ISL_1922547, EPI_ISL_1922548, EPI_ISL_1922549, EPI_ISL_1922550, EPI_ISL_1922551, EPI_ISL_1922552, EPI_ISL_1922553, EPI_ISL_1922554, EPI_ISL_1922555, EPI_ISL_1922556, EPI_ISL_1922557, EPI_ISL_1922558, EPI_ISL_1922559, EPI_ISL_1922560, EPI_ISL_1922561, EPI_ISL_1922562, EPI_ISL_1922563, EPI_ISL_1922564, EPI_ISL_1922565, EPI_ISL_1922566, EPI_ISL_1922567, EPI_ISL_1922568, EPI_ISL_1922569, EPI_ISL_1922571, EPI_ISL_1922572, EPI_ISL_1922573, EPI_ISL_1922574, EPI_ISL_1922575, EPI_ISL_1922576, EPI_ISL_1922578, EPI_ISL_1922579, EPI_ISL_1922580, EPI_ISL_1922581, EPI_ISL_1922582, EPI_ISL_1922583, EPI_ISL_1922584, EPI_ISL_1922585, EPI_ISL_1922586, EPI_ISL_1922587, EPI_ISL_1922588, EPI_ISL_1922589, EPI_ISL_1922590, EPI_ISL_1922591, EPI_ISL_1922592, EPI_ISL_1922593, EPI_ISL_1922594, EPI_ISL_1922595, EPI_ISL_1922596, EPI_ISL_1922597, EPI_ISL_1922598, EPI_ISL_1922599, EPI_ISL_1922600, EPI_ISL_1922601, EPI_ISL_1922602, EPI_ISL_1922603, EPI_ISL_1922604, EPI_ISL_1922605, EPI_ISL_1922606, EPI_ISL_1922607, EPI_ISL_1922608, EPI_ISL_1922609, EPI_ISL_1922610, EPI_ISL_1922611, EPI_ISL_1922612, EPI_ISL_1922613, EPI_ISL_1922614, EPI_ISL_1922615, EPI_ISL_1922616, EPI_ISL_1922617, EPI_ISL_1922618, EPI_ISL_1922619, EPI_ISL_1922620, EPI_ISL_1922621, EPI_ISL_1922622, EPI_ISL_1922623, EPI_ISL_1922624, EPI_ISL_1922625, EPI_ISL_1922626, EPI_ISL_1922627, EPI_ISL_1922628, EPI_ISL_1922629, EPI_ISL_1922630, EPI_ISL_1922631, EPI_ISL_1922632, EPI_ISL_1922633, EPI_ISL_1922634, EPI_ISL_1922635, EPI_ISL_1922636, EPI_ISL_1922637, EPI_ISL_1922638, EPI_ISL_1922639, EPI_ISL_1922640, EPI_ISL_1922641, EPI_ISL_1922642, EPI_ISL_1922643, EPI_ISL_1922644, EPI_ISL_1922645, EPI_ISL_1922646, EPI_ISL_1922647, EPI_ISL_1922648, EPI_ISL_1922649, EPI_ISL_1922650, EPI_ISL_1922651, EPI_ISL_1922652, EPI_ISL_1922653, EPI_ISL_1922654, EPI_ISL_1922655, EPI_ISL_1922656, EPI_ISL_1922657, EPI_ISL_1922658, EPI_ISL_1922659, EPI_ISL_1922660, EPI_ISL_1922661, EPI_ISL_1922662, EPI_ISL_1922663, EPI_ISL_1922664, EPI_ISL_1922665, EPI_ISL_1922666, EPI_ISL_1922667, EPI_ISL_1922668, EPI_ISL_1922669, EPI_ISL_1922670, EPI_ISL_1922671, EPI_ISL_1922672, EPI_ISL_1922673, EPI_ISL_1922674, EPI_ISL_1922675, EPI_ISL_1922676, EPI_ISL_1922677, EPI_ISL_1922678, EPI_ISL_1922679, EPI_ISL_1922680, EPI_ISL_1922681, EPI_ISL_1922682, EPI_ISL_1922683, EPI_ISL_1922684, EPI_ISL_1922685, EPI_ISL_1922686, EPI_ISL_1922687, EPI_ISL_1922688, EPI_ISL_1922689, EPI_ISL_1922690, EPI_ISL_1922691, EPI_ISL_1922692, EPI_ISL_1922693, EPI_ISL_1922694 | | | |
| see above | Maryland Genomics, Institute for Genome Sciences, University of Maryland School of Medicine | Maryland Genomics, Institute for Genome Sciences, University of Maryland School of Medicine | Tallon, Luke J; Sadzewicz, Lisa D; Humphrys, Mike; Ott, Sandra; Roussey, Holly; Mehta, Aditya; Vavikolanu, Kranthi; Fraser, Claire M; Ravel, Jacques |
| EPI_ISL_1922695, EPI_ISL_1922696 | Kansas Health and Environmental Lab | Kansas Health and Environmental Lab | Mike Grose, Jonathan Barnell, Ben Olsen, and Phil Adam |
| EPI_ISL_1922697, EPI_ISL_1922698, EPI_ISL_1922699, EPI_ISL_1922700, EPI_ISL_1922701, EPI_ISL_1922702, EPI_ISL_1922703 | Wichita State University - Molecular Diagnostics Lab | Kansas Health and Environmental Lab | Mike Grose, Jonathan Barnell, Ben Olsen, and Phil Adam |
| EPI_ISL_1922704 | Clinical Reference Laboratory | Kansas Health and Environmental Lab | Mike Grose, Jonathan Barnell, Ben Olsen, and Phil Adam |
| EPI_ISL_1922706 | Kansas Health and Environmental Lab | Kansas Health and Environmental Lab | Mike Grose, Jonathan Barnell, Ben Olsen, and Phil Adam |
| EPI_ISL_1922707, EPI_ISL_1922708 | Wichita State University - Molecular Diagnostics Lab | Kansas Health and Environmental Lab | Mike Grose, Jonathan Barnell, Ben Olsen, and Phil Adam |
| EPI_ISL_1922710, EPI_ISL_1922711, EPI_ISL_1922712, EPI_ISL_1922713, EPI_ISL_1922714, EPI_ISL_1922717, EPI_ISL_1922718, EPI_ISL_1922719, EPI_ISL_1922720, EPI_ISL_1922721, EPI_ISL_1922722, EPI_ISL_1922723, EPI_ISL_1922724, EPI_ISL_1922725, EPI_ISL_1922726, EPI_ISL_1922727, EPI_ISL_1922728, EPI_ISL_1922729, EPI_ISL_1922730, EPI_ISL_1922731, EPI_ISL_1922732, EPI_ISL_1922733, EPI_ISL_1922735, EPI_ISL_1922736, EPI_ISL_1922737, EPI_ISL_1922738, EPI_ISL_1922740, EPI_ISL_1922742, EPI_ISL_1922743, EPI_ISL_1922744, EPI_ISL_1922745, EPI_ISL_1922747, EPI_ISL_1922749, EPI_ISL_1922750, EPI_ISL_1922751, EPI_ISL_1922752, EPI_ISL_1922753, EPI_ISL_1922754, EPI_ISL_1922755, EPI_ISL_1922756, EPI_ISL_1922757, EPI_ISL_1922758, EPI_ISL_1922759, EPI_ISL_1922761, EPI_ISL_1922762, EPI_ISL_1922763, EPI_ISL_1922764, EPI_ISL_1922765, EPI_ISL_1922766, EPI_ISL_1922767, EPI_ISL_1922768, EPI_ISL_1922770, EPI_ISL_1922773, EPI_ISL_1922774, EPI_ISL_1922775, EPI_ISL_1922776, EPI_ISL_1922777, EPI_ISL_1922778, EPI_ISL_1922779, EPI_ISL_1922780, EPI_ISL_1922781, EPI_ISL_1922782, EPI_ISL_1922783, EPI_ISL_1922784, EPI_ISL_1922785, EPI_ISL_1922786, EPI_ISL_1922787, EPI_ISL_1922788, EPI_ISL_1922789, EPI_ISL_1922790, EPI_ISL_1922791, EPI_ISL_1922792, EPI_ISL_1922793, EPI_ISL_1922794, EPI_ISL_1922795, EPI_ISL_1922796, EPI_ISL_1922797, EPI_ISL_1922799, EPI_ISL_1922800, EPI_ISL_1922801, EPI_ISL_1922803, EPI_ISL_1922804, EPI_ISL_1922808, EPI_ISL_1922810, EPI_ISL_1922812, EPI_ISL_1922814, EPI_ISL_1922816, EPI_ISL_1922817, EPI_ISL_1922818, EPI_ISL_1922824, EPI_ISL_1922825, EPI_ISL_1922826, EPI_ISL_1922827, EPI_ISL_1922829, EPI_ISL_1922831, EPI_ISL_1922832, EPI_ISL_1922833, EPI_ISL_1922836, EPI_ISL_1922841, EPI_ISL_1922842, EPI_ISL_1922843, EPI_ISL_1922844, EPI_ISL_1922847, EPI_ISL_1922851, EPI_ISL_1922852, EPI_ISL_1922854, EPI_ISL_1922855, EPI_ISL_1922857, EPI_ISL_1922861, EPI_ISL_1922865, EPI_ISL_1922868, EPI_ISL_1922869, EPI_ISL_1922870, EPI_ISL_1922871 | | | |
| see above | NYU Langone Health | Departments of Pathology and Medicine, New York University School of Medicine | Adriana Heguy, Dacia Dimartino, Emily Guzman, Christian Marier, Peter Meyn, Sitharam Ramaswami, Gael Westby, Paul Zappile, Yutong Zhang, Paolo Cotzia, Guiqing Wang |
| EPI_ISL_1922873, EPI_ISL_1922874, EPI_ISL_1922875, EPI_ISL_1922876, EPI_ISL_1922877 | University of Michigan Clinical Microbiology Laboratory | Lauring Lab, University of Michigan, Department of Microbiology and Immunology | Valesano |
| EPI_ISL_1922893, EPI_ISL_1922894, EPI_ISL_1922895, EPI_ISL_1922896, EPI_ISL_1922897, EPI_ISL_1922898, EPI_ISL_1922899, EPI_ISL_1922900, EPI_ISL_1922901, EPI_ISL_1922902, EPI_ISL_1922903, EPI_ISL_1922904, EPI_ISL_1922905, EPI_ISL_1922906, EPI_ISL_1922907, EPI_ISL_1922908, EPI_ISL_1922909, EPI_ISL_1922910, EPI_ISL_1922911, EPI_ISL_1922912, EPI_ISL_1922913, EPI_ISL_1922914, EPI_ISL_1922915, EPI_ISL_1922916, EPI_ISL_1922917, EPI_ISL_1922918, EPI_ISL_1922919, EPI_ISL_1922920, EPI_ISL_1922921, EPI_ISL_1922922, EPI_ISL_1922923, EPI_ISL_1922924, EPI_ISL_1922925, EPI_ISL_1922926, EPI_ISL_1922927, EPI_ISL_1922928, EPI_ISL_1922929, EPI_ISL_1922930, EPI_ISL_1922931, EPI_ISL_1922932, EPI_ISL_1922933, EPI_ISL_1922934, EPI_ISL_1922935, EPI_ISL_1922937, EPI_ISL_1922938, EPI_ISL_1922939, EPI_ISL_1922940, EPI_ISL_1922941, EPI_ISL_1922942, EPI_ISL_1922943, EPI_ISL_1922944, EPI_ISL_1922945, EPI_ISL_1922946, EPI_ISL_1922947, EPI_ISL_1922948, EPI_ISL_1922949, EPI_ISL_1922950, EPI_ISL_1922951, EPI_ISL_1922952, EPI_ISL_1922953, EPI_ISL_1922954, EPI_ISL_1922956, EPI_ISL_1922957, EPI_ISL_1922958, EPI_ISL_1922959, EPI_ISL_1922960, EPI_ISL_1922961, EPI_ISL_1922962, EPI_ISL_1922963, EPI_ISL_1922964, EPI_ISL_1922965, EPI_ISL_1922966, EPI_ISL_1922967, EPI_ISL_1922968, EPI_ISL_1922969, EPI_ISL_1922970, EPI_ISL_1922971, EPI_ISL_1922972, EPI_ISL_1922973, EPI_ISL_1922974, EPI_ISL_1922975, EPI_ISL_1922976, EPI_ISL_1922977, EPI_ISL_1922978, EPI_ISL_1922979, EPI_ISL_1922980, EPI_ISL_1922981, EPI_ISL_1922982, EPI_ISL_1922983, EPI_ISL_1922984, EPI_ISL_1922985, EPI_ISL_1922986, EPI_ISL_1922987, EPI_ISL_1922988, EPI_ISL_1922989, EPI_ISL_1922990, EPI_ISL_1922991, EPI_ISL_1922992, EPI_ISL_1922993, EPI_ISL_1922994, EPI_ISL_1922995, EPI_ISL_1922996, EPI_ISL_1922997, EPI_ISL_1922998, EPI_ISL_1922999, EPI_ISL_1923001, EPI_ISL_1923002, EPI_ISL_1923003, EPI_ISL_1923004, EPI_ISL_1923005, EPI_ISL_1923006, EPI_ISL_1923007, EPI_ISL_1923008, EPI_ISL_1923009, EPI_ISL_1923010, EPI_ISL_1923011, EPI_ISL_1923012, EPI_ISL_1923013, EPI_ISL_1923014, EPI_ISL_1923015, EPI_ISL_1923016, EPI_ISL_1923017, EPI_ISL_1923018, EPI_ISL_1923019, EPI_ISL_1923020, EPI_ISL_1923021, EPI_ISL_1923022, EPI_ISL_1923023, EPI_ISL_1923024, EPI_ISL_1923025, EPI_ISL_1923026, EPI_ISL_1923027, EPI_ISL_1923028, EPI_ISL_1923029, EPI_ISL_1923030, EPI_ISL_1923031, EPI_ISL_1923032, EPI_ISL_1923033, EPI_ISL_1923034, EPI_ISL_1923035, EPI_ISL_1923036, EPI_ISL_1923037, EPI_ISL_1923038, EPI_ISL_1923039, EPI_ISL_1923040, EPI_ISL_1923042, EPI_ISL_1923043, EPI_ISL_1923044, EPI_ISL_1923045, EPI_ISL_1923046, EPI_ISL_1923047, EPI_ISL_1923048, EPI_ISL_1923049, EPI_ISL_1923050, EPI_ISL_1923051, EPI_ISL_1923052, EPI_ISL_1923053, EPI_ISL_1923054, EPI_ISL_1923055, EPI_ISL_1923056, EPI_ISL_1923057, EPI_ISL_1923058, EPI_ISL_1923059, EPI_ISL_1923060, EPI_ISL_1923061, EPI_ISL_1923062, EPI_ISL_1923063, EPI_ISL_1923064, EPI_ISL_1923065, EPI_ISL_1923066, EPI_ISL_1923067, EPI_ISL_1923068, EPI_ISL_1923069, EPI_ISL_1923070, EPI_ISL_1923071, EPI_ISL_1923072, EPI_ISL_1923073, EPI_ISL_1923074, EPI_ISL_1923075, EPI_ISL_1923076, EPI_ISL_1923077, EPI_ISL_1923078, EPI_ISL_1923079, EPI_ISL_1923080, EPI_ISL_1923081, EPI_ISL_1923082, EPI_ISL_1923083, EPI_ISL_1923084, EPI_ISL_1923085, EPI_ISL_1923086, EPI_ISL_1923087, EPI_ISL_1923088, EPI_ISL_1923089, EPI_ISL_1923090, EPI_ISL_1923091, EPI_ISL_1923092, EPI_ISL_1923093, EPI_ISL_1923094, EPI_ISL_1923095, EPI_ISL_1923096, EPI_ISL_1923098, EPI_ISL_1923099, EPI_ISL_1923100, EPI_ISL_1923101, EPI_ISL_1923102, EPI_ISL_1923103, EPI_ISL_1923104, EPI_ISL_1923105, EPI_ISL_1923106, EPI_ISL_1923107, EPI_ISL_1923108, EPI_ISL_1923109, EPI_ISL_1923110, EPI_ISL_1923111, EPI_ISL_1923112, EPI_ISL_1923113, EPI_ISL_1923114, EPI_ISL_1923115, EPI_ISL_1923116, EPI_ISL_1923117, EPI_ISL_1923118, EPI_ISL_1923119, EPI_ISL_1923120, EPI_ISL_1923121, EPI_ISL_1923122, EPI_ISL_1923123, EPI_ISL_1923124, EPI_ISL_1923125, EPI_ISL_1923126, EPI_ISL_1923127, EPI_ISL_1923128, EPI_ISL_1923129, EPI_ISL_1923130, EPI_ISL_1923131, EPI_ISL_1923132, EPI_ISL_1923133, EPI_ISL_1923134, EPI_ISL_1923135, EPI_ISL_1923136, EPI_ISL_1923138, EPI_ISL_1923139, EPI_ISL_1923140, EPI_ISL_1923141, EPI_ISL_1923142, EPI_ISL_1923143, EPI_ISL_1923144, EPI_ISL_1923145, EPI_ISL_1923146, EPI_ISL_1923147, EPI_ISL_1923148, EPI_ISL_1923149, EPI_ISL_1923150, EPI_ISL_1923151, EPI_ISL_1923152, EPI_ISL_1923153, EPI_ISL_1923154, EPI_ISL_1923155, EPI_ISL_1923156, EPI_ISL_1923157, EPI_ISL_1923158, EPI_ISL_1923159, EPI_ISL_1923160, EPI_ISL_1923161, EPI_ISL_1923162, EPI_ISL_1923163, EPI_ISL_1923164, EPI_ISL_1923165 | | | |
| see above | Aegis Sciences Corporation | Centers for Disease Control and Prevention Division of Viral Diseases, Pathogen Discovery | Dakota Howard, Dhwani Batra, Peter W. Cook, Kara Moser, Adrian Paskey, Jason Caravas, Benjamin Rambo-Martin, Shatavia Morrison, Christopher Gulvick, Scott Sammons, Yvette Unoarumhi, Darlene Wagner, Matthew Schmerer, Cyndi Clark, Patrick Campbell, Rob Case, Vikramsinha Ghorpade, Holly Houdeshell, Ola Kvalvaag, Dillon Nall, Ethan Sanders, Alec Vest, Shaun Westlund, Matthew Hardison, Clinton R. Paden, Duncan MacCannell |
| EPI_ISL_1923166, EPI_ISL_1923167, EPI_ISL_1923168, EPI_ISL_1923169, EPI_ISL_1923170, EPI_ISL_1923171, EPI_ISL_1923172, EPI_ISL_1923173, EPI_ISL_1923174, EPI_ISL_1923175, EPI_ISL_1923176, EPI_ISL_1923177, EPI_ISL_1923178, EPI_ISL_1923179, EPI_ISL_1923180, EPI_ISL_1923181, EPI_ISL_1923182, EPI_ISL_1923183, EPI_ISL_1923184, EPI_ISL_1923185, EPI_ISL_1923186, EPI_ISL_1923187, EPI_ISL_1923188, EPI_ISL_1923189, EPI_ISL_1923190, EPI_ISL_1923191, EPI_ISL_1923192, EPI_ISL_1923193, EPI_ISL_1923194, EPI_ISL_1923195, EPI_ISL_1923196, EPI_ISL_1923197, EPI_ISL_1923198, EPI_ISL_1923199, EPI_ISL_1923200, EPI_ISL_1923201, EPI_ISL_1923202, EPI_ISL_1923203, EPI_ISL_1923204, EPI_ISL_1923205, EPI_ISL_1923206, EPI_ISL_1923207, EPI_ISL_1923208, EPI_ISL_1923209, EPI_ISL_1923210, EPI_ISL_1923211, EPI_ISL_1923212, EPI_ISL_1923213, EPI_ISL_1923214, EPI_ISL_1923215, EPI_ISL_1923216, EPI_ISL_1923217, EPI_ISL_1923218, EPI_ISL_1923219, EPI_ISL_1923220, EPI_ISL_1923221, EPI_ISL_1923222, EPI_ISL_1923223, EPI_ISL_1923224, EPI_ISL_1923225, EPI_ISL_1923226, EPI_ISL_1923227, EPI_ISL_1923228, EPI_ISL_1923229, EPI_ISL_1923230, EPI_ISL_1923231, EPI_ISL_1923232, EPI_ISL_1923233, EPI_ISL_1923234, EPI_ISL_1923235, EPI_ISL_1923236, EPI_ISL_1923237, EPI_ISL_1923238, EPI_ISL_1923239, EPI_ISL_1923240, EPI_ISL_1923241, EPI_ISL_1923242, EPI_ISL_1923243, EPI_ISL_1923244, EPI_ISL_1923245, EPI_ISL_1923246, EPI_ISL_1923247, EPI_ISL_1923248, EPI_ISL_1923249, EPI_ISL_1923250, EPI_ISL_1923251, EPI_ISL_1923252, EPI_ISL_1923253, EPI_ISL_1923254, EPI_ISL_1923255, EPI_ISL_1923256, EPI_ISL_1923257, EPI_ISL_1923258, EPI_ISL_1923259, EPI_ISL_1923260, EPI_ISL_1923261, EPI_ISL_1923262, EPI_ISL_1923263, EPI_ISL_1923264, EPI_ISL_1923265, EPI_ISL_1923266, EPI_ISL_1923267, EPI_ISL_1923268, EPI_ISL_1923269, EPI_ISL_1923270, EPI_ISL_1923271, EPI_ISL_1923272, EPI_ISL_1923273, EPI_ISL_1923274, EPI_ISL_1923275, EPI_ISL_1923276, EPI_ISL_1923277, EPI_ISL_1923278, EPI_ISL_1923279, EPI_ISL_1923280, EPI_ISL_1923281, EPI_ISL_1923282, EPI_ISL_1923283, EPI_ISL_1923284, EPI_ISL_1923285, EPI_ISL_1923286, EPI_ISL_1923287, EPI_ISL_1923288, EPI_ISL_1923289, EPI_ISL_1923290, EPI_ISL_1923291, EPI_ISL_1923292, EPI_ISL_1923293, EPI_ISL_1923294, EPI_ISL_1923295, EPI_ISL_1923296, EPI_ISL_1923297, EPI_ISL_1923298, EPI_ISL_1923299, EPI_ISL_1923300, EPI_ISL_1923301, EPI_ISL_1923302, EPI_ISL_1923303, EPI_ISL_1923304, EPI_ISL_1923305, EPI_ISL_1923306, EPI_ISL_1923307, EPI_ISL_1923308, EPI_ISL_1923309, EPI_ISL_1923310, EPI_ISL_1923311, EPI_ISL_1923312, EPI_ISL_1923313, EPI_ISL_1923314, EPI_ISL_1923315, EPI_ISL_1923316, EPI_ISL_1923317, EPI_ISL_1923318, EPI_ISL_1923319, EPI_ISL_1923320, EPI_ISL_1923321, EPI_ISL_1923322, EPI_ISL_1923323, EPI_ISL_1923324, EPI_ISL_1923325, EPI_ISL_1923326, EPI_ISL_1923327, EPI_ISL_1923328, EPI_ISL_1923329, EPI_ISL_1923330, EPI_ISL_1923331, EPI_ISL_1923332, EPI_ISL_1923333, EPI_ISL_1923334, EPI_ISL_1923335, EPI_ISL_1923336, EPI_ISL_1923337, EPI_ISL_1923338, EPI_ISL_1923339, EPI_ISL_1923340, EPI_ISL_1923341, EPI_ISL_1923342, EPI_ISL_1923343, EPI_ISL_1923344, EPI_ISL_1923345, EPI_ISL_1923346, EPI_ISL_1923347, EPI_ISL_1923348, EPI_ISL_1923349, EPI_ISL_1923350, EPI_ISL_1923351, EPI_ISL_1923352, EPI_ISL_1923353, EPI_ISL_1923354, EPI_ISL_1923355, EPI_ISL_1923356, EPI_ISL_1923357, EPI_ISL_1923358, EPI_ISL_1923359, EPI_ISL_1923360, EPI_ISL_1923361, EPI_ISL_1923362, EPI_ISL_1923363, EPI_ISL_1923364, EPI_ISL_1923365, EPI_ISL_1923366, EPI_ISL_1923367, EPI_ISL_1923368, EPI_ISL_1923369, EPI_ISL_1923370, EPI_ISL_1923371, EPI_ISL_1923372, EPI_ISL_1923373, EPI_ISL_1923374, EPI_ISL_1923375, EPI_ISL_1923376, EPI_ISL_1923377, EPI_ISL_1923378, EPI_ISL_1923379, EPI_ISL_1923380, EPI_ISL_1923381, EPI_ISL_1923382, EPI_ISL_1923383, EPI_ISL_1923384, EPI_ISL_1923385, EPI_ISL_1923386, EPI_ISL_1923387, EPI_ISL_1923388, EPI_ISL_1923389, EPI_ISL_1923390, EPI_ISL_1923391, EPI_ISL_1923392, EPI_ISL_1923393, EPI_ISL_1923394, EPI_ISL_1923395, EPI_ISL_1923396, EPI_ISL_1923397, EPI_ISL_1923398, EPI_ISL_1923399, EPI_ISL_1923400, EPI_ISL_1923401, EPI_ISL_1923402, EPI_ISL_1923403, EPI_ISL_1923404, EPI_ISL_1923405, EPI_ISL_1923406, EPI_ISL_1923407, EPI_ISL_1923408, EPI_ISL_1923409, EPI_ISL_1923410, EPI_ISL_1923411, EPI_ISL_1923412, EPI_ISL_1923413, EPI_ISL_1923414, EPI_ISL_1923415, EPI_ISL_1923416, EPI_ISL_1923417, EPI_ISL_1923418, EPI_ISL_1923419, EPI_ISL_1923420, EPI_ISL_1923421, EPI_ISL_1923422, EPI_ISL_1923423, EPI_ISL_1923424, EPI_ISL_1923425, EPI_ISL_1923426, EPI_ISL_1923427, EPI_ISL_1923428, EPI_ISL_1923429, EPI_ISL_1923430, EPI_ISL_1923431, EPI_ISL_1923432, EPI_ISL_1923433, EPI_ISL_1923434, EPI_ISL_1923435, EPI_ISL_1923436, EPI_ISL_1923437, EPI_ISL_1923438, EPI_ISL_1923439, EPI_ISL_1923440, EPI_ISL_1923441, EPI_ISL_1923442, EPI_ISL_1923443, EPI_ISL_1923444, EPI_ISL_1923445, EPI_ISL_1923446, EPI_ISL_1923447, EPI_ISL_1923448, EPI_ISL_1923449, EPI_ISL_1923450, EPI_ISL_1923451, EPI_ISL_1923452, EPI_ISL_1923453, EPI_ISL_1923454, EPI_ISL_1923455, EPI_ISL_1923456, EPI_ISL_1923457, EPI_ISL_1923458, EPI_ISL_1923459, EPI_ISL_1923460, EPI_ISL_1923461, EPI_ISL_1923462, EPI_ISL_1923463, EPI_ISL_1923464, EPI_ISL_1923465, EPI_ISL_1923466, EPI_ISL_1923467, EPI_ISL_1923468, EPI_ISL_1923469, EPI_ISL_1923470, EPI_ISL_1923471, EPI_ISL_1923472, EPI_ISL_1923473, EPI_ISL_1923474, EPI_ISL_1923475, EPI_ISL_1923476, EPI_ISL_1923477, EPI_ISL_1923478, EPI_ISL_1923479, EPI_ISL_1923480, EPI_ISL_1923481, EPI_ISL_1923482, EPI_ISL_1923483, EPI_ISL_1923484, EPI_ISL_1923485, EPI_ISL_1923486, EPI_ISL_1923487, EPI_ISL_1923488, EPI_ISL_1923489, EPI_ISL_1923490, EPI_ISL_1923491, EPI_ISL_1923492, EPI_ISL_1923493, EPI_ISL_1923494, EPI_ISL_1923495, EPI_ISL_1923496, EPI_ISL_1923497, EPI_ISL_1923499, EPI_ISL_1923500, EPI_ISL_1923501, EPI_ISL_1923502, EPI_ISL_1923503, EPI_ISL_1923504, EPI_ISL_1923505, EPI_ISL_1923506, EPI_ISL_1923507, EPI_ISL_1923508, EPI_ISL_1923509, EPI_ISL_1923510, EPI_ISL_1923511, EPI_ISL_1923512, EPI_ISL_1923513, EPI_ISL_1923514, EPI_ISL_1923515, EPI_ISL_1923516, EPI_ISL_1923517, EPI_ISL_1923518, EPI_ISL_1923519, EPI_ISL_1923520, EPI_ISL_1923521, EPI_ISL_1923522, EPI_ISL_1923523, EPI_ISL_1923524, EPI_ISL_1923525, EPI_ISL_1923526, EPI_ISL_1923527, EPI_ISL_1923528, EPI_ISL_1923529, EPI_ISL_1923530, EPI_ISL_1923531, EPI_ISL_1923533, EPI_ISL_1923536, EPI_ISL_1923539, EPI_ISL_1923542, EPI_ISL_1923545, EPI_ISL_1923548, EPI_ISL_1923551, EPI_ISL_1923555, EPI_ISL_1923558, EPI_ISL_1923561, EPI_ISL_1923564, EPI_ISL_1923567, EPI_ISL_1923568, EPI_ISL_1923571, EPI_ISL_1923574, EPI_ISL_1923577, EPI_ISL_1923580, EPI_ISL_1923582, EPI_ISL_1923585, EPI_ISL_1923588, EPI_ISL_1923591, EPI_ISL_1923594, EPI_ISL_1923597, EPI_ISL_1923600, EPI_ISL_1923603, EPI_ISL_1923604, EPI_ISL_1923605, EPI_ISL_1923606, EPI_ISL_1923607, EPI_ISL_1923608, EPI_ISL_1923609, EPI_ISL_1923610, EPI_ISL_1923611, EPI_ISL_1923612, EPI_ISL_1923613, EPI_ISL_1923614, EPI_ISL_1923615, EPI_ISL_1923616, EPI_ISL_1923617, EPI_ISL_1923618, EPI_ISL_1923619, EPI_ISL_1923620, EPI_ISL_1923621, EPI_ISL_1923622, EPI_ISL_1923623, EPI_ISL_1923624, EPI_ISL_1923625, EPI_ISL_1923626, EPI_ISL_1923627, EPI_ISL_1923628, EPI_ISL_1923629, EPI_ISL_1923630, EPI_ISL_1923631, EPI_ISL_1923632, EPI_ISL_1923633, EPI_ISL_1923634, EPI_ISL_1923635, EPI_ISL_1923636, EPI_ISL_1923637, EPI_ISL_1923638, EPI_ISL_1923639, EPI_ISL_1923640, EPI_ISL_1923641, EPI_ISL_1923642, EPI_ISL_1923643, EPI_ISL_1923644, EPI_ISL_1923645, EPI_ISL_1923646, EPI_ISL_1923647, EPI_ISL_1923648, EPI_ISL_1923649, EPI_ISL_1923650, EPI_ISL_1923651, EPI_ISL_1923652, EPI_ISL_1923653, EPI_ISL_1923654, EPI_ISL_1923655, EPI_ISL_1923656, EPI_ISL_1923657, EPI_ISL_1923658, EPI_ISL_1923659, EPI_ISL_1923660, EPI_ISL_1923661, EPI_ISL_1923662, EPI_ISL_1923663, EPI_ISL_1923664, EPI_ISL_1923666, EPI_ISL_1923667, EPI_ISL_1923668, EPI_ISL_1923671, EPI_ISL_1923672, EPI_ISL_1923673, EPI_ISL_1923674, EPI_ISL_1923675, EPI_ISL_1923676, EPI_ISL_1923677, EPI_ISL_1923678, EPI_ISL_1923679, EPI_ISL_1923680, EPI_ISL_1923681, EPI_ISL_1923682, EPI_ISL_1923683, EPI_ISL_1923684, EPI_ISL_1923685, EPI_ISL_1923686, EPI_ISL_1923687, EPI_ISL_1923688, EPI_ISL_1923689, EPI_ISL_1923690, EPI_ISL_1923691, EPI_ISL_1923692, EPI_ISL_1923693, EPI_ISL_1923694, EPI_ISL_1923695, EPI_ISL_1923696, EPI_ISL_1923697, EPI_ISL_1923698, EPI_ISL_1923699, EPI_ISL_1923700, EPI_ISL_1923701, EPI_ISL_1923702, EPI_ISL_1923703, EPI_ISL_1923704, EPI_ISL_1923705, EPI_ISL_1923706, EPI_ISL_1923707, EPI_ISL_1923708, EPI_ISL_1923709, EPI_ISL_1923710, EPI_ISL_1923711, EPI_ISL_1923712, EPI_ISL_1923713, EPI_ISL_1923714, EPI_ISL_1923715, EPI_ISL_1923716, EPI_ISL_1923717, EPI_ISL_1923718, EPI_ISL_1923719, EPI_ISL_1923720, EPI_ISL_1923721, EPI_ISL_1923722, EPI_ISL_1923723, EPI_ISL_1923724, EPI_ISL_1923725, EPI_ISL_1923726, EPI_ISL_1923727, EPI_ISL_1923728, EPI_ISL_1923729, EPI_ISL_1923730, EPI_ISL_1923731, EPI_ISL_1923732, EPI_ISL_1923733, EPI_ISL_1923734, EPI_ISL_1923735, EPI_ISL_1923736, EPI_ISL_1923737, EPI_ISL_1923738, EPI_ISL_1923739, EPI_ISL_1923740, EPI_ISL_1923741, EPI_ISL_1923742, EPI_ISL_1923744, EPI_ISL_1923745, EPI_ISL_1923746, EPI_ISL_1923747, EPI_ISL_1923748, EPI_ISL_1923749, EPI_ISL_1923750, EPI_ISL_1923751, EPI_ISL_1923752, EPI_ISL_1923753, EPI_ISL_1923754, EPI_ISL_1923755, EPI_ISL_1923756, EPI_ISL_1923757, EPI_ISL_1923758, EPI_ISL_1923759, EPI_ISL_1923760, EPI_ISL_1923761, EPI_ISL_1923762, EPI_ISL_1923763, EPI_ISL_1923764, EPI_ISL_1923765, EPI_ISL_1923766, EPI_ISL_1923768, EPI_ISL_1923769, EPI_ISL_1923770, EPI_ISL_1923771, EPI_ISL_1923772, EPI_ISL_1923773, EPI_ISL_1923774, EPI_ISL_1923775, EPI_ISL_1923776, EPI_ISL_1923777, EPI_ISL_1923778, EPI_ISL_1923779, EPI_ISL_1923780, EPI_ISL_1923781, EPI_ISL_1923782, EPI_ISL_1923783, EPI_ISL_1923784, EPI_ISL_1923785, EPI_ISL_1923786, EPI_ISL_1923787, EPI_ISL_1923788, EPI_ISL_1923789, EPI_ISL_1923790, EPI_ISL_1923791, EPI_ISL_1923792, EPI_ISL_1923793, EPI_ISL_1923794, EPI_ISL_1923795, EPI_ISL_1923796, EPI_ISL_1923797, EPI_ISL_1923798, EPI_ISL_1923799, EPI_ISL_1923800, EPI_ISL_1923801, EPI_ISL_1923802, EPI_ISL_1923803, EPI_ISL_1923804, EPI_ISL_1923805, EPI_ISL_1923806, EPI_ISL_1923807, EPI_ISL_1923808, EPI_ISL_1923809, EPI_ISL_1923810, EPI_ISL_1923811, EPI_ISL_1923812, EPI_ISL_1923813, EPI_ISL_1923814, EPI_ISL_1923815, EPI_ISL_1923816, EPI_ISL_1923817, EPI_ISL_1923818, EPI_ISL_1923819, EPI_ISL_1923820, EPI_ISL_1923821, EPI_ISL_1923822, EPI_ISL_1923823, EPI_ISL_1923824, EPI_ISL_1923825, EPI_ISL_1923826, EPI_ISL_1923827, EPI_ISL_1923828, EPI_ISL_1923829, EPI_ISL_1923830, EPI_ISL_1923831, EPI_ISL_1923832, EPI_ISL_1923833, EPI_ISL_1923834, EPI_ISL_1923835, EPI_ISL_1923836, EPI_ISL_1923837, EPI_ISL_1923838, EPI_ISL_1923839, EPI_ISL_1923841, EPI_ISL_1923842, EPI_ISL_1923843, EPI_ISL_1923844, EPI_ISL_1923845, EPI_ISL_1923846, EPI_ISL_1923847, EPI_ISL_1923848, EPI_ISL_1923849, EPI_ISL_1923850, EPI_ISL_1923851, EPI_ISL_1923852, EPI_ISL_1923853, EPI_ISL_1923854, EPI_ISL_1923855, EPI_ISL_1923856, EPI_ISL_1923857, EPI_ISL_1923858, EPI_ISL_1923859, EPI_ISL_1923860, EPI_ISL_1923861, EPI_ISL_1923862, EPI_ISL_1923863, EPI_ISL_1923864, EPI_ISL_1923865, EPI_ISL_1923866, EPI_ISL_1923867, EPI_ISL_1923868, EPI_ISL_1923869, EPI_ISL_1923870, EPI_ISL_1923871, EPI_ISL_1923873, EPI_ISL_1923874, EPI_ISL_1923875, EPI_ISL_1923876, EPI_ISL_1923877, EPI_ISL_1923878, EPI_ISL_1923879, EPI_ISL_1923880, EPI_ISL_1923881, EPI_ISL_1923882, EPI_ISL_1923883, EPI_ISL_1923884, EPI_ISL_1923885, EPI_ISL_1923886, EPI_ISL_1923887, EPI_ISL_1923888, EPI_ISL_1923889, EPI_ISL_1923890, EPI_ISL_1923891, EPI_ISL_1923892, EPI_ISL_1923893, EPI_ISL_1923894, EPI_ISL_1923895, EPI_ISL_1923896, EPI_ISL_1923897, EPI_ISL_1923898, EPI_ISL_1923899, EPI_ISL_1923900, EPI_ISL_1923901, EPI_ISL_1923902, EPI_ISL_1923903, EPI_ISL_1923904, EPI_ISL_1923905, EPI_ISL_1923906, EPI_ISL_1923907, EPI_ISL_1923908, EPI_ISL_1923909, EPI_ISL_1923910, EPI_ISL_1923911, EPI_ISL_1923912, EPI_ISL_1923913, EPI_ISL_1923914, EPI_ISL_1923915, EPI_ISL_1923916, EPI_ISL_1923917, EPI_ISL_1923918, EPI_ISL_1923919, EPI_ISL_1923920, EPI_ISL_1923921, EPI_ISL_1923922, EPI_ISL_1923923, EPI_ISL_1923924, EPI_ISL_1923925, EPI_ISL_1923926, EPI_ISL_1923927, EPI_ISL_1923928, EPI_ISL_1923929, EPI_ISL_1923930, EPI_ISL_1923931, EPI_ISL_1923932, EPI_ISL_1923933, EPI_ISL_1923934, EPI_ISL_1923935, EPI_ISL_1923936, EPI_ISL_1923937, EPI_ISL_1923938, EPI_ISL_1923939, EPI_ISL_1923940, EPI_ISL_1923941, EPI_ISL_1923942, EPI_ISL_1923943, EPI_ISL_1923944, EPI_ISL_1923945, EPI_ISL_1923946, EPI_ISL_1923947, EPI_ISL_1923948, EPI_ISL_1923949, EPI_ISL_1923950, EPI_ISL_1923951, EPI_ISL_1923952, EPI_ISL_1923953, EPI_ISL_1923954, EPI_ISL_1923955, EPI_ISL_1923956, EPI_ISL_1923957, EPI_ISL_1923958, EPI_ISL_1923959, EPI_ISL_1923960, EPI_ISL_1923961, EPI_ISL_1923962, EPI_ISL_1923963, EPI_ISL_1923964, EPI_ISL_1923965, EPI_ISL_1923966, EPI_ISL_1923967, EPI_ISL_1923968, EPI_ISL_1923969, EPI_ISL_1923970, EPI_ISL_1923971, EPI_ISL_1923972, EPI_ISL_1923973, EPI_ISL_1923974, EPI_ISL_1923975, EPI_ISL_1923976, EPI_ISL_1923977, EPI_ISL_1923978, EPI_ISL_1923979, EPI_ISL_1923980, EPI_ISL_1923981, EPI_ISL_1923982, EPI_ISL_1923983, EPI_ISL_1923984, EPI_ISL_1923985, EPI_ISL_1923986, EPI_ISL_1923987, EPI_ISL_1923988, EPI_ISL_1923989, EPI_ISL_1923990, | | | |

| | | | |
|---|---|---|---|
| EPI_ISL_1923991, EPI_ISL_1923992, EPI_ISL_1923993, EPI_ISL_1923994, EPI_ISL_1923995, EPI_ISL_1923996, EPI_ISL_1923997, EPI_ISL_1923998, EPI_ISL_1923999, EPI_ISL_1924000, EPI_ISL_1924001, EPI_ISL_1924002, EPI_ISL_1924003, EPI_ISL_1924004, EPI_ISL_1924005, EPI_ISL_1924006, EPI_ISL_1924007, EPI_ISL_1924008, EPI_ISL_1924009, EPI_ISL_1924010, EPI_ISL_1924011, EPI_ISL_1924012, EPI_ISL_1924013, EPI_ISL_1924014, EPI_ISL_1924015, EPI_ISL_1924016, EPI_ISL_1924017, EPI_ISL_1924018, EPI_ISL_1924019, EPI_ISL_1924020, EPI_ISL_1924021, EPI_ISL_1924022, EPI_ISL_1924023, EPI_ISL_1924024, EPI_ISL_1924025, EPI_ISL_1924026, EPI_ISL_1924027, EPI_ISL_1924028, EPI_ISL_1924029, EPI_ISL_1924030, EPI_ISL_1924031, EPI_ISL_1924032, EPI_ISL_1924033, EPI_ISL_1924034, EPI_ISL_1924035, EPI_ISL_1924036, EPI_ISL_1924037, EPI_ISL_1924038, EPI_ISL_1924039, EPI_ISL_1924040, EPI_ISL_1924041, EPI_ISL_1924042, EPI_ISL_1924043, EPI_ISL_1924044, EPI_ISL_1924045, EPI_ISL_1924046, EPI_ISL_1924047, EPI_ISL_1924048, EPI_ISL_1924049, EPI_ISL_1924051, EPI_ISL_1924052, EPI_ISL_1924053, EPI_ISL_1924054, EPI_ISL_1924055, EPI_ISL_1924056, EPI_ISL_1924057, EPI_ISL_1924058, EPI_ISL_1924059, EPI_ISL_1924060, EPI_ISL_1924061, EPI_ISL_1924062, EPI_ISL_1924063, EPI_ISL_1924064, EPI_ISL_1924065, EPI_ISL_1924066, EPI_ISL_1924067, EPI_ISL_1924068, EPI_ISL_1924069, EPI_ISL_1924070, EPI_ISL_1924071, EPI_ISL_1924072, EPI_ISL_1924073, EPI_ISL_1924074, EPI_ISL_1924075, EPI_ISL_1924076, EPI_ISL_1924077, EPI_ISL_1924078, EPI_ISL_1924079, EPI_ISL_1924080, EPI_ISL_1924081, EPI_ISL_1924082, EPI_ISL_1924083, EPI_ISL_1924084, EPI_ISL_1924085, EPI_ISL_1924086, EPI_ISL_1924087, EPI_ISL_1924088, EPI_ISL_1924089, EPI_ISL_1924090, EPI_ISL_1924091, EPI_ISL_1924092, EPI_ISL_1924093, EPI_ISL_1924094, EPI_ISL_1924095, EPI_ISL_1924096, EPI_ISL_1924097, EPI_ISL_1924098, EPI_ISL_1924099, EPI_ISL_1924100, EPI_ISL_1924101, EPI_ISL_1924102, EPI_ISL_1924103, EPI_ISL_1924104, EPI_ISL_1924105, EPI_ISL_1924106, EPI_ISL_1924107, EPI_ISL_1924108, EPI_ISL_1924109, EPI_ISL_1924110, EPI_ISL_1924111, EPI_ISL_1924112, EPI_ISL_1924113, EPI_ISL_1924114, EPI_ISL_1924115, EPI_ISL_1924116, EPI_ISL_1924117, EPI_ISL_1924118, EPI_ISL_1924119, EPI_ISL_1924120, EPI_ISL_1924121, EPI_ISL_1924122, EPI_ISL_1924123, EPI_ISL_1924124, EPI_ISL_1924125, EPI_ISL_1924126, EPI_ISL_1924127, EPI_ISL_1924128, EPI_ISL_1924129, EPI_ISL_1924130, EPI_ISL_1924131, EPI_ISL_1924132, EPI_ISL_1924133, EPI_ISL_1924134, EPI_ISL_1924135, EPI_ISL_1924136, EPI_ISL_1924137, EPI_ISL_1924138, EPI_ISL_1924139, EPI_ISL_1924140, EPI_ISL_1924141, EPI_ISL_1924142, EPI_ISL_1924143, EPI_ISL_1924144, EPI_ISL_1924145, EPI_ISL_1924146, EPI_ISL_1924147, EPI_ISL_1924148, EPI_ISL_1924149, EPI_ISL_1924150, EPI_ISL_1924151, EPI_ISL_1924152, EPI_ISL_1924153, EPI_ISL_1924154, EPI_ISL_1924155, EPI_ISL_1924156, EPI_ISL_1924157, EPI_ISL_1924158, EPI_ISL_1924159, EPI_ISL_1924160, EPI_ISL_1924161, EPI_ISL_1924162, EPI_ISL_1924163, EPI_ISL_1924164, EPI_ISL_1924165, EPI_ISL_1924166, EPI_ISL_1924167, EPI_ISL_1924168, EPI_ISL_1924169, EPI_ISL_1924170, EPI_ISL_1924171, EPI_ISL_1924172, EPI_ISL_1924173, EPI_ISL_1924174, EPI_ISL_1924175, EPI_ISL_1924176, EPI_ISL_1924177, EPI_ISL_1924178, EPI_ISL_1924179, EPI_ISL_1924180, EPI_ISL_1924181, EPI_ISL_1924182, EPI_ISL_1924183, EPI_ISL_1924184, EPI_ISL_1924185, EPI_ISL_1924186, EPI_ISL_1924187, EPI_ISL_1924188, EPI_ISL_1924189, EPI_ISL_1924190, EPI_ISL_1924191, EPI_ISL_1924192, EPI_ISL_1924193, EPI_ISL_1924194, EPI_ISL_1924195, EPI_ISL_1924196, EPI_ISL_1924197, EPI_ISL_1924198, EPI_ISL_1924199, EPI_ISL_1924200, EPI_ISL_1924201, EPI_ISL_1924202, EPI_ISL_1924203, EPI_ISL_1924204, EPI_ISL_1924205, EPI_ISL_1924206, EPI_ISL_1924207, EPI_ISL_1924208, EPI_ISL_1924209, EPI_ISL_1924210, EPI_ISL_1924211, EPI_ISL_1924212, EPI_ISL_1924213, EPI_ISL_1924214, EPI_ISL_1924215, EPI_ISL_1924216, EPI_ISL_1924217, EPI_ISL_1924218, EPI_ISL_1924219, EPI_ISL_1924220, EPI_ISL_1924221, EPI_ISL_1924222, EPI_ISL_1924223, EPI_ISL_1924224, EPI_ISL_1924225, EPI_ISL_1924226, EPI_ISL_1924227, EPI_ISL_1924228, EPI_ISL_1924229, EPI_ISL_1924230, EPI_ISL_1924231, EPI_ISL_1924232, EPI_ISL_1924233, EPI_ISL_1924234, EPI_ISL_1924235, EPI_ISL_1924236, EPI_ISL_1924237, EPI_ISL_1924238, EPI_ISL_1924239, EPI_ISL_1924240, EPI_ISL_1924241, EPI_ISL_1924242, EPI_ISL_1924243, EPI_ISL_1924244, EPI_ISL_1924245, EPI_ISL_1924246, EPI_ISL_1924247, EPI_ISL_1924248, EPI_ISL_1924249, EPI_ISL_1924250, EPI_ISL_1924251, EPI_ISL_1924252, EPI_ISL_1924253, EPI_ISL_1924254, EPI_ISL_1924255, EPI_ISL_1924256, EPI_ISL_1924257, EPI_ISL_1924258, EPI_ISL_1924259, EPI_ISL_1924260, EPI_ISL_1924261, EPI_ISL_1924262, EPI_ISL_1924263, EPI_ISL_1924264, EPI_ISL_1924265, EPI_ISL_1924266, EPI_ISL_1924267, EPI_ISL_1924268, EPI_ISL_1924270, EPI_ISL_1924271, EPI_ISL_1924272, EPI_ISL_1924273, EPI_ISL_1924274, EPI_ISL_1924275, EPI_ISL_1924276, EPI_ISL_1924277 | | | |
| see above | Helix/Illumina | Centers for Disease Control and Prevention Division of Viral Diseases, Pathogen Discovery | Dakota Howard, Dhwani Batra, Peter W. Cook, Kara Moser, Adrian Paskey, Jason Caravas, Benjamin Rambo-Martin, Shatavia Morrison, Christopher Gulvick, Scott Sammons, Yvette Unoarumhi, Darlene Wagner, Matthew Schmerer, Eileen de Feo, Jan Antico, Christine Tran, Matthew Tolentino, Shannon Wickline, Kim Gietzen, Brad Sickler, Jingtao Liu, Eric Allen, Phil Febbo, Nicole L. Washington, Simon White, Geraint Levan, Kelly Schiabor Barrett, Elizabeth Cirulli, Alexandre Bolze, Ary Ascencio, Charlotte Rivera-Garcia, Ryan Cho, Jason Nguyen, Sherry Wang, Jimmy Ramirez, Tyler Cassens, Efren Sandoval, Magnus Isaksson, William Lee, David Becker, Marc Laurent, James Lu, Clinton R. Paden, Duncan MacCannell |
| EPI_ISL_1924278, EPI_ISL_1924279, EPI_ISL_1924280, EPI_ISL_1924281, EPI_ISL_1924282, EPI_ISL_1924283, EPI_ISL_1924284, EPI_ISL_1924285, EPI_ISL_1924286, EPI_ISL_1924287, EPI_ISL_1924288, EPI_ISL_1924289, EPI_ISL_1924290, EPI_ISL_1924291, EPI_ISL_1924292, EPI_ISL_1924293, EPI_ISL_1924294, EPI_ISL_1924295, EPI_ISL_1924296, EPI_ISL_1924297, EPI_ISL_1924298, EPI_ISL_1924299, EPI_ISL_1924300, EPI_ISL_1924301, EPI_ISL_1924302, EPI_ISL_1924303, EPI_ISL_1924304, EPI_ISL_1924305, EPI_ISL_1924306, EPI_ISL_1924307, EPI_ISL_1924308, EPI_ISL_1924309, EPI_ISL_1924310, EPI_ISL_1924311, EPI_ISL_1924312, EPI_ISL_1924313, EPI_ISL_1924314, EPI_ISL_1924315, EPI_ISL_1924316, EPI_ISL_1924317, EPI_ISL_1924318, EPI_ISL_1924319, EPI_ISL_1924320, EPI_ISL_1924321, EPI_ISL_1924322, EPI_ISL_1924323, EPI_ISL_1924324, EPI_ISL_1924325, EPI_ISL_1924326, EPI_ISL_1924327, EPI_ISL_1924328, EPI_ISL_1924329, EPI_ISL_1924330, EPI_ISL_1924331, EPI_ISL_1924332, EPI_ISL_1924333, EPI_ISL_1924334, EPI_ISL_1924335, EPI_ISL_1924336, EPI_ISL_1924337, EPI_ISL_1924338, EPI_ISL_1924339, EPI_ISL_1924340, EPI_ISL_1924341, EPI_ISL_1924342, EPI_ISL_1924343, EPI_ISL_1924344, EPI_ISL_1924345, EPI_ISL_1924346, EPI_ISL_1924347, EPI_ISL_1924348, EPI_ISL_1924349, EPI_ISL_1924350, EPI_ISL_1924351, EPI_ISL_1924352, EPI_ISL_1924353, EPI_ISL_1924354, EPI_ISL_1924355, EPI_ISL_1924356, EPI_ISL_1924357, EPI_ISL_1924358, EPI_ISL_1924359, EPI_ISL_1924360, EPI_ISL_1924361, EPI_ISL_1924362, EPI_ISL_1924363, EPI_ISL_1924364, EPI_ISL_1924365, EPI_ISL_1924366, EPI_ISL_1924367, EPI_ISL_1924368, EPI_ISL_1924369, EPI_ISL_1924370, EPI_ISL_1924371, EPI_ISL_1924372, EPI_ISL_1924373, EPI_ISL_1924374, EPI_ISL_1924375, EPI_ISL_1924376, EPI_ISL_1924377, EPI_ISL_1924378, EPI_ISL_1924379, EPI_ISL_1924380, EPI_ISL_1924381, EPI_ISL_1924382, EPI_ISL_1924383, EPI_ISL_1924384, EPI_ISL_1924385, EPI_ISL_1924386, EPI_ISL_1924387, EPI_ISL_1924388, EPI_ISL_1924389, EPI_ISL_1924390, EPI_ISL_1924391, EPI_ISL_1924392, EPI_ISL_1924393, EPI_ISL_1924394, EPI_ISL_1924395, EPI_ISL_1924396, EPI_ISL_1924397, EPI_ISL_1924398, EPI_ISL_1924399, EPI_ISL_1924400, EPI_ISL_1924401, EPI_ISL_1924402, EPI_ISL_1924403, EPI_ISL_1924404, EPI_ISL_1924405, EPI_ISL_1924406, EPI_ISL_1924407, EPI_ISL_1924408, EPI_ISL_1924409, EPI_ISL_1924410, EPI_ISL_1924411, EPI_ISL_1924412, EPI_ISL_1924413, EPI_ISL_1924414, EPI_ISL_1924415, EPI_ISL_1924416, EPI_ISL_1924417, EPI_ISL_1924418, EPI_ISL_1924419, EPI_ISL_1924420, EPI_ISL_1924421, EPI_ISL_1924422, EPI_ISL_1924423, EPI_ISL_1924424, EPI_ISL_1924425, EPI_ISL_1924426, EPI_ISL_1924427, EPI_ISL_1924428, EPI_ISL_1924429, EPI_ISL_1924430, EPI_ISL_1924431, EPI_ISL_1924432, EPI_ISL_1924433, EPI_ISL_1924434, EPI_ISL_1924435, EPI_ISL_1924436, EPI_ISL_1924437, EPI_ISL_1924438, EPI_ISL_1924439, EPI_ISL_1924440, EPI_ISL_1924441, EPI_ISL_1924442, EPI_ISL_1924443, EPI_ISL_1924444, EPI_ISL_1924445, EPI_ISL_1924446, EPI_ISL_1924447, EPI_ISL_1924448, EPI_ISL_1924449, EPI_ISL_1924450, EPI_ISL_1924451, EPI_ISL_1924452, EPI_ISL_1924453, EPI_ISL_1924454, EPI_ISL_1924455, EPI_ISL_1924456, EPI_ISL_1924457, EPI_ISL_1924458, EPI_ISL_1924459, EPI_ISL_1924460, EPI_ISL_1924461, EPI_ISL_1924462, EPI_ISL_1924463, EPI_ISL_1924464, EPI_ISL_1924465, EPI_ISL_1924466, EPI_ISL_1924467, EPI_ISL_1924468, EPI_ISL_1924469, EPI_ISL_1924470, EPI_ISL_1924471, EPI_ISL_1924472, EPI_ISL_1924473, EPI_ISL_1924474, EPI_ISL_1924475, EPI_ISL_1924476, EPI_ISL_1924477, EPI_ISL_1924478, EPI_ISL_1924479, EPI_ISL_1924480, EPI_ISL_1924481, EPI_ISL_1924482, EPI_ISL_1924483, EPI_ISL_1924484, EPI_ISL_1924485, EPI_ISL_1924486, EPI_ISL_1924487, EPI_ISL_1924488, EPI_ISL_1924489, EPI_ISL_1924490, EPI_ISL_1924491, EPI_ISL_1924492, EPI_ISL_1924493, EPI_ISL_1924494, EPI_ISL_1924495, EPI_ISL_1924496, EPI_ISL_1924497, EPI_ISL_1924498, EPI_ISL_1924499, EPI_ISL_1924500, EPI_ISL_1924501, EPI_ISL_1924502, EPI_ISL_1924503, EPI_ISL_1924504, EPI_ISL_1924505, EPI_ISL_1924506, EPI_ISL_1924507, EPI_ISL_1924508, EPI_ISL_1924509, EPI_ISL_1924510, EPI_ISL_1924511, EPI_ISL_1924512, EPI_ISL_1924513, EPI_ISL_1924514, EPI_ISL_1924515, EPI_ISL_1924516, EPI_ISL_1924517, EPI_ISL_1924518, EPI_ISL_1924519, EPI_ISL_1924520, EPI_ISL_1924521, EPI_ISL_1924522, EPI_ISL_1924523, EPI_ISL_1924524, EPI_ISL_1924525, EPI_ISL_1924526, EPI_ISL_1924527, EPI_ISL_1924528, EPI_ISL_1924529, EPI_ISL_1924530, EPI_ISL_1924531, EPI_ISL_1924532, EPI_ISL_1924533, EPI_ISL_1924534, EPI_ISL_1924535, EPI_ISL_1924536, EPI_ISL_1924537, EPI_ISL_1924538, EPI_ISL_1924539, EPI_ISL_1924540, EPI_ISL_1924541, EPI_ISL_1924542, EPI_ISL_1924543, EPI_ISL_1924544, EPI_ISL_1924545, EPI_ISL_1924546, EPI_ISL_1924547, EPI_ISL_1924548, EPI_ISL_1924549, EPI_ISL_1924550, EPI_ISL_1924551, EPI_ISL_1924552, EPI_ISL_1924553, EPI_ISL_1924554, EPI_ISL_1924555, EPI_ISL_1924556, EPI_ISL_1924557, EPI_ISL_1924558, EPI_ISL_1924559, EPI_ISL_1924560, EPI_ISL_1924561, EPI_ISL_1924562, EPI_ISL_1924563, EPI_ISL_1924564, EPI_ISL_1924565, EPI_ISL_1924566, EPI_ISL_1924567, EPI_ISL_1924568, EPI_ISL_1924569, EPI_ISL_1924570, EPI_ISL_1924571, EPI_ISL_1924572, EPI_ISL_1924573, EPI_ISL_1924574, EPI_ISL_1924575, EPI_ISL_1924576, EPI_ISL_1924577, EPI_ISL_1924578, EPI_ISL_1924579, EPI_ISL_1924580, EPI_ISL_1924581 | | | |
| see above | Quest Diagnostics Incorporated | Centers for Disease Control and Prevention Division of Viral Diseases, Pathogen Discovery | Dakota Howard, Dhwani Batra, Peter W. Cook, Kara Moser, Adrian Paskey, Jason Caravas, Benjamin Rambo-Martin, Shatavia Morrison, Christopher Gulvick, Scott Sammons, Yvette Unoarumhi, Darlene Wagner, Matthew Schmerer, S. H. Rosenthal, A. Gerasimova, R. M. Kagan, B. Anderson, M. Hua, Y. Liu, L.E. Bernstein, K.E. Livingston, A. Perez, I. A. Shlyakhter, R. V. Rolando, R. Owen, P. Tanpaiboon, F. Lacbawan, Clinton R. Paden, Duncan MacCannell |
| EPI_ISL_1924582 | Orange County Public Health Lab | Chan-Zuckerberg Biohub | CZB Cliahub Consortium |
| EPI_ISL_1924583, EPI_ISL_1924584, EPI_ISL_1924585, EPI_ISL_1924586, EPI_ISL_1924587, EPI_ISL_1924588, EPI_ISL_1924589, EPI_ISL_1924591, EPI_ISL_1924594, EPI_ISL_1924595 | CA DPH Viral and Rickettsial Disease Laboratory | Chan-Zuckerberg Biohub | CZB Cliahub Consortium |
| EPI_ISL_1924596 | Orange County Public Health Lab | Chan-Zuckerberg Biohub | CZB Cliahub Consortium |
| EPI_ISL_1924597, EPI_ISL_1924598 | CA DPH Viral and Rickettsial Disease Laboratory | Chan-Zuckerberg Biohub | CZB Cliahub Consortium |
| EPI_ISL_1924600 | Santa Clara County Public Health Laboratory | Chan-Zuckerberg Biohub | CZB Cliahub Consortium |
| EPI_ISL_1924601, EPI_ISL_1924603, EPI_ISL_1924606, EPI_ISL_1924607, EPI_ISL_1924608, EPI_ISL_1924609, EPI_ISL_1924610, EPI_ISL_1924611, EPI_ISL_1924612, EPI_ISL_1924613, EPI_ISL_1924614, EPI_ISL_1924615, EPI_ISL_1924617, EPI_ISL_1924620, EPI_ISL_1924621, EPI_ISL_1924622, EPI_ISL_1924623, EPI_ISL_1924624, EPI_ISL_1924625, EPI_ISL_1924626, EPI_ISL_1924627, EPI_ISL_1924628 | | | |
| see above | CA DPH Viral and Rickettsial Disease Laboratory | Chan-Zuckerberg Biohub | CZB Cliahub Consortium |
| EPI_ISL_1924629 | Orange County Public Health Lab | Chan-Zuckerberg Biohub | CZB Cliahub Consortium |
| EPI_ISL_1924630 | CHILDRENS HOSPITAL OF NORCAL | Santa Clara County public Health Laboratory | Santa Clara County Public Health Department |
| EPI_ISL_1924631, EPI_ISL_1924632 | Santa Clara Valley Medical Center | Santa Clara County public Health Laboratory | Santa Clara County Public Health Department |
| EPI_ISL_1924633, EPI_ISL_1924634, EPI_ISL_1924636, EPI_ISL_1924638, EPI_ISL_1924639, EPI_ISL_1924640, EPI_ISL_1924642, EPI_ISL_1924643, EPI_ISL_1924644, EPI_ISL_1924645, EPI_ISL_1924646, EPI_ISL_1924648 | | | |
| see above | Stanford Clinical Virology Laboratory | Santa Clara County public Health Laboratory | Santa Clara County Public Health Department |
| EPI_ISL_1924649, EPI_ISL_1924650, EPI_ISL_1924651, EPI_ISL_1924652, EPI_ISL_1924653, EPI_ISL_1924654, EPI_ISL_1924655, EPI_ISL_1924656, EPI_ISL_1924657, EPI_ISL_1924658, EPI_ISL_1924659, EPI_ISL_1924660, EPI_ISL_1924661, EPI_ISL_1924662, EPI_ISL_1924663, EPI_ISL_1924664, EPI_ISL_1924665, EPI_ISL_1924666, EPI_ISL_1924667, EPI_ISL_1924668, EPI_ISL_1924669, EPI_ISL_1924670, EPI_ISL_1924671, EPI_ISL_1924672, EPI_ISL_1924673, EPI_ISL_1924674, EPI_ISL_1924675, EPI_ISL_1924676, EPI_ISL_1924677, EPI_ISL_1924678, EPI_ISL_1924679, EPI_ISL_1924680, EPI_ISL_1924681, EPI_ISL_1924682, EPI_ISL_1924683, EPI_ISL_1924684, EPI_ISL_1924685, EPI_ISL_1924686, EPI_ISL_1924687, EPI_ISL_1924688, EPI_ISL_1924689, EPI_ISL_1924690, EPI_ISL_1924691, EPI_ISL_1924692, EPI_ISL_1924693, EPI_ISL_1924694, EPI_ISL_1924695, EPI_ISL_1924696, EPI_ISL_1924697, EPI_ISL_1924698, EPI_ISL_1924699, EPI_ISL_1924700, EPI_ISL_1924701, EPI_ISL_1924702, EPI_ISL_1924703, EPI_ISL_1924704, EPI_ISL_1924705, EPI_ISL_1924706, EPI_ISL_1924707, EPI_ISL_1924708, EPI_ISL_1924709, EPI_ISL_1924710, EPI_ISL_1924711, EPI_ISL_1924712, EPI_ISL_1924713, EPI_ISL_1924714, EPI_ISL_1924715, EPI_ISL_1924716, EPI_ISL_1924717, EPI_ISL_1924718, EPI_ISL_1924719, EPI_ISL_1924720, EPI_ISL_1924721, EPI_ISL_1924722, EPI_ISL_1924723, EPI_ISL_1924724, EPI_ISL_1924725, EPI_ISL_1924726, EPI_ISL_1924727, EPI_ISL_1924728, EPI_ISL_1924729, EPI_ISL_1924730, EPI_ISL_1924731, EPI_ISL_1924732, EPI_ISL_1924733, EPI_ISL_1924734, EPI_ISL_1924735, EPI_ISL_1924736, EPI_ISL_1924737, EPI_ISL_1924738, EPI_ISL_1924739, EPI_ISL_1924740, EPI_ISL_1924741, EPI_ISL_1924742, EPI_ISL_1924743, EPI_ISL_1924744, EPI_ISL_1924745, EPI_ISL_1924746, EPI_ISL_1924747, EPI_ISL_1924748, EPI_ISL_1924749, EPI_ISL_1924750, EPI_ISL_1924751, EPI_ISL_1924752, EPI_ISL_1924753, EPI_ISL_1924754, EPI_ISL_1924755, EPI_ISL_1924756, EPI_ISL_1924757, EPI_ISL_1924758, EPI_ISL_1924759, EPI_ISL_1924760, EPI_ISL_1924761, EPI_ISL_1924762, EPI_ISL_1924763, EPI_ISL_1924764, EPI_ISL_1924765, EPI_ISL_1924766, EPI_ISL_1924767, EPI_ISL_1924768, EPI_ISL_1924769, EPI_ISL_1924770, EPI_ISL_1924771, EPI_ISL_1924772, EPI_ISL_1924773, EPI_ISL_1924774, EPI_ISL_1924775, EPI_ISL_1924776, EPI_ISL_1924777, EPI_ISL_1924778, EPI_ISL_1924779, EPI_ISL_1924780, EPI_ISL_1924781, EPI_ISL_1924782, EPI_ISL_1924783, EPI_ISL_1924784, EPI_ISL_1924785, EPI_ISL_1924786, EPI_ISL_1924787, EPI_ISL_1924788, EPI_ISL_1924789, EPI_ISL_1924790, EPI_ISL_1924791, EPI_ISL_1924792, EPI_ISL_1924793, EPI_ISL_1924794, EPI_ISL_1924795, EPI_ISL_1924796, EPI_ISL_1924797, EPI_ISL_1924798, EPI_ISL_1924799, EPI_ISL_1924800, EPI_ISL_1924801, EPI_ISL_1924802, EPI_ISL_1924803, EPI_ISL_1924804, EPI_ISL_1924805, EPI_ISL_1924806, EPI_ISL_1924807, EPI_ISL_1924808, EPI_ISL_1924809, EPI_ISL_1924810, EPI_ISL_1924811, EPI_ISL_1924812, EPI_ISL_1924813, EPI_ISL_1924814, EPI_ISL_1924815, EPI_ISL_1924816, EPI_ISL_1924817, EPI_ISL_1924818, EPI_ISL_1924819, EPI_ISL_1924820, EPI_ISL_1924821, EPI_ISL_1924822, EPI_ISL_1924823, EPI_ISL_1924824, EPI_ISL_1924825, EPI_ISL_1924826, EPI_ISL_1924827, EPI_ISL_1924828, EPI_ISL_1924829, EPI_ISL_1924830, EPI_ISL_1924831, EPI_ISL_1924832, EPI_ISL_1924833, EPI_ISL_1924834, EPI_ISL_1924835, EPI_ISL_1924836, EPI_ISL_1924837, EPI_ISL_1924838, EPI_ISL_1924839, EPI_ISL_1924840, EPI_ISL_1924841, EPI_ISL_1924842, EPI_ISL_1924843, EPI_ISL_1924844, EPI_ISL_1924845, EPI_ISL_1924846, EPI_ISL_1924847, EPI_ISL_1924848, EPI_ISL_1924849, EPI_ISL_1924850, EPI_ISL_1924851, EPI_ISL_1924852, EPI_ISL_1924853, EPI_ISL_1924854, EPI_ISL_1924855, EPI_ISL_1924856, EPI_ISL_1924857, EPI_ISL_1924858, EPI_ISL_1924859, EPI_ISL_1924860, EPI_ISL_1924861, EPI_ISL_1924862, EPI_ISL_1924863, EPI_ISL_1924864, EPI_ISL_1924865, EPI_ISL_1924866, EPI_ISL_1924867, EPI_ISL_1924868, EPI_ISL_1924869, EPI_ISL_1924870, EPI_ISL_1924871, EPI_ISL_1924872, EPI_ISL_1924873, EPI_ISL_1924874, EPI_ISL_1924875, EPI_ISL_1924876, EPI_ISL_1924877, EPI_ISL_1924878, EPI_ISL_1924879, EPI_ISL_1924881, EPI_ISL_1924882, EPI_ISL_1924883, EPI_ISL_1924884, EPI_ISL_1924885, EPI_ISL_1924886, EPI_ISL_1924887, EPI_ISL_1924888, EPI_ISL_1924889, EPI_ISL_1924890, EPI_ISL_1924891, EPI_ISL_1924892, EPI_ISL_1924893, EPI_ISL_1924894, EPI_ISL_1924895, EPI_ISL_1924896, EPI_ISL_1924897, EPI_ISL_1924898, EPI_ISL_1924899, EPI_ISL_1924900, EPI_ISL_1924901, EPI_ISL_1924902, EPI_ISL_1924903, EPI_ISL_1924904, EPI_ISL_1924905, EPI_ISL_1924906, EPI_ISL_1924907, EPI_ISL_1924908, EPI_ISL_1924909, EPI_ISL_1924910, EPI_ISL_1924911, EPI_ISL_1924912, EPI_ISL_1924913, EPI_ISL_1924914, EPI_ISL_1924915, EPI_ISL_1924916, EPI_ISL_1924917, EPI_ISL_1924918, EPI_ISL_1924919, EPI_ISL_1924920, EPI_ISL_1924921, EPI_ISL_1924922, EPI_ISL_1924923, EPI_ISL_1924924, EPI_ISL_1924925, EPI_ISL_1924926, EPI_ISL_1924927, EPI_ISL_1924928, EPI_ISL_1924929, EPI_ISL_1924930, EPI_ISL_1924931, EPI_ISL_1924932, EPI_ISL_1924933, EPI_ISL_1924934, EPI_ISL_1924935, EPI_ISL_1924936, EPI_ISL_1924937, EPI_ISL_1924938, EPI_ISL_1924939, EPI_ISL_1924940, EPI_ISL_1924941, EPI_ISL_1924942, EPI_ISL_1924943, EPI_ISL_1924944, EPI_ISL_1924945, EPI_ISL_1924946, EPI_ISL_1924947, EPI_ISL_1924948, EPI_ISL_1924949, EPI_ISL_1924950, EPI_ISL_1924951, EPI_ISL_1924952, EPI_ISL_1924953, EPI_ISL_1924954, EPI_ISL_1924955, EPI_ISL_1924956, EPI_ISL_1924957, EPI_ISL_1924958, EPI_ISL_1924959, EPI_ISL_1924960, EPI_ISL_1924961, EPI_ISL_1924962, EPI_ISL_1924963, EPI_ISL_1924964, EPI_ISL_1924965, EPI_ISL_1924966, EPI_ISL_1924967, EPI_ISL_1924968, EPI_ISL_1924969, EPI_ISL_1924970, EPI_ISL_1924971, EPI_ISL_1924972, EPI_ISL_1924973, EPI_ISL_1924974, EPI_ISL_1924975, EPI_ISL_1924976, EPI_ISL_1924977, EPI_ISL_1924978, EPI_ISL_1924979, EPI_ISL_1924980, EPI_ISL_1924981, EPI_ISL_1924982, EPI_ISL_1924983, EPI_ISL_1924984, EPI_ISL_1924985, EPI_ISL_1924986, EPI_ISL_1924987, EPI_ISL_1924988, EPI_ISL_1924989, EPI_ISL_1924990, EPI_ISL_1924991, EPI_ISL_1924992, EPI_ISL_1924993, EPI_ISL_1924994, EPI_ISL_1924995, EPI_ISL_1924996, EPI_ISL_1924997, EPI_ISL_1924998, EPI_ISL_1924999, EPI_ISL_1925000, EPI_ISL_1925001, EPI_ISL_1925002, EPI_ISL_1925003, EPI_ISL_1925004, EPI_ISL_1925005, EPI_ISL_1925006, EPI_ISL_1925007, EPI_ISL_1925008, EPI_ISL_1925009, EPI_ISL_1925010, EPI_ISL_1925011, EPI_ISL_1925012, EPI_ISL_1925013, EPI_ISL_1925014, EPI_ISL_1925015, EPI_ISL_1925016, EPI_ISL_1925017, EPI_ISL_1925018, EPI_ISL_1925019, EPI_ISL_1925020, EPI_ISL_1925021, EPI_ISL_1925022, EPI_ISL_1925023, EPI_ISL_1925024, EPI_ISL_1925025, EPI_ISL_1925026, EPI_ISL_1925027, EPI_ISL_1925028, EPI_ISL_1925029, EPI_ISL_1925030, EPI_ISL_1925031, EPI_ISL_1925032, EPI_ISL_1925033, EPI_ISL_1925034, EPI_ISL_1925035, EPI_ISL_1925036, EPI_ISL_1925037, EPI_ISL_1925038, EPI_ISL_1925039, EPI_ISL_1925040, EPI_ISL_1925041, EPI_ISL_1925042, EPI_ISL_1925043, EPI_ISL_1925044, EPI_ISL_1925045, EPI_ISL_1925046, EPI_ISL_1925047, EPI_ISL_1925048, EPI_ISL_1925049, EPI_ISL_1925050, EPI_ISL_1925051, EPI_ISL_1925052, EPI_ISL_1925053, EPI_ISL_1925054, EPI_ISL_1925055, EPI_ISL_1925056, EPI_ISL_1925057, EPI_ISL_1925058, EPI_ISL_1925059, EPI_ISL_1925060, EPI_ISL_1925061, EPI_ISL_1925062, EPI_ISL_1925063, EPI_ISL_1925064, EPI_ISL_1925065, EPI_ISL_1925066, EPI_ISL_1925067, EPI_ISL_1925068, EPI_ISL_1925069, EPI_ISL_1925070, EPI_ISL_1925071, EPI_ISL_1925072, EPI_ISL_1925073, EPI_ISL_1925074, EPI_ISL_1925075, EPI_ISL_1925076 | | | |
| see above | Quest Diagnostics Incorporated | Centers for Disease Control and Prevention Division of Viral Diseases, Pathogen Discovery | Dakota Howard, Dhwani Batra, Peter W. Cook, Kara Moser, Adrian Paskey, Jason Caravas, Benjamin Rambo-Martin, Shatavia Morrison, Christopher Gulvick, Scott Sammons, Yvette Unoarumhi, Darlene Wagner, Matthew Schmerer, S. H. Rosenthal, A. Gerasimova, R. M. Kagan, B. Anderson, M. Hua, Y. Liu, L.E. Bernstein, K.E. Livingston, A. Perez, I. A. Shlyakhter, R. V. Rolando, R. Owen, P. Tanpaiboon, F. Lacbawan, Clinton R. Paden, Duncan MacCannell |
| EPI_ISL_1925077, EPI_ISL_1925079, EPI_ISL_1925080, EPI_ISL_1925081, EPI_ISL_1925082, EPI_ISL_1925083, EPI_ISL_1925084, EPI_ISL_1925085, EPI_ISL_1925086, EPI_ISL_1925087, EPI_ISL_1925088, EPI_ISL_1925089, EPI_ISL_1925090, EPI_ISL_1925091, EPI_ISL_1925092, EPI_ISL_1925093, EPI_ISL_1925094, EPI_ISL_1925095, EPI_ISL_1925096, EPI_ISL_1925097, EPI_ISL_1925098, EPI_ISL_1925099, EPI_ISL_1925100, EPI_ISL_1925101, EPI_ISL_1925102, EPI_ISL_1925103, EPI_ISL_1925104, EPI_ISL_1925105, EPI_ISL_1925106, EPI_ISL_1925107, EPI_ISL_1925108, EPI_ISL_1925109, EPI_ISL_1925110, EPI_ISL_1925111, EPI_ISL_1925112, EPI_ISL_1925113, EPI_ISL_1925114, EPI_ISL_1925115, EPI_ISL_1925116, EPI_ISL_1925117, EPI_ISL_1925118, EPI_ISL_1925119, EPI_ISL_1925120, EPI_ISL_1925121, EPI_ISL_1925122, EPI_ISL_1925123, EPI_ISL_1925124, EPI_ISL_1925125, EPI_ISL_1925126, EPI_ISL_1925127, EPI_ISL_1925128, EPI_ISL_1925129, EPI_ISL_1925130, EPI_ISL_1925131, EPI_ISL_1925132, EPI_ISL_1925133, EPI_ISL_1925134, EPI_ISL_1925135, EPI_ISL_1925136, EPI_ISL_1925137, EPI_ISL_1925138, EPI_ISL_1925139, EPI_ISL_1925140, EPI_ISL_1925141, EPI_ISL_1925142, EPI_ISL_1925143, EPI_ISL_1925144, EPI_ISL_1925145, EPI_ISL_1925146, EPI_ISL_1925147, EPI_ISL_1925148, EPI_ISL_1925149, EPI_ISL_1925150, EPI_ISL_1925151, EPI_ISL_1925152, EPI_ISL_1925153, EPI_ISL_1925154, EPI_ISL_1925155, EPI_ISL_1925156, EPI_ISL_1925157, EPI_ISL_1925158, EPI_ISL_1925159, EPI_ISL_1925160, EPI_ISL_1925161, EPI_ISL_1925162, EPI_ISL_1925163, EPI_ISL_1925164, EPI_ISL_1925165, EPI_ISL_1925166, EPI_ISL_1925167, EPI_ISL_1925168, EPI_ISL_1925169, EPI_ISL_1925170, EPI_ISL_1925171, EPI_ISL_1925172, EPI_ISL_1925173, EPI_ISL_1925174, EPI_ISL_1925175, EPI_ISL_1925176, EPI_ISL_1925177, EPI_ISL_1925178, EPI_ISL_1925179, EPI_ISL_1925180, EPI_ISL_1925181, EPI_ISL_1925182, EPI_ISL_1925183, EPI_ISL_1925184, EPI_ISL_1925185, EPI_ISL_1925186, EPI_ISL_1925187, EPI_ISL_1925188, EPI_ISL_1925189, EPI_ISL_1925190, EPI_ISL_1925191, EPI_ISL_1925192, EPI_ISL_1925193, EPI_ISL_1925194, EPI_ISL_1925195, EPI_ISL_1925196, EPI_ISL_1925197, EPI_ISL_1925198, EPI_ISL_1925199, EPI_ISL_1925200, EPI_ISL_1925201, EPI_ISL_1925202, EPI_ISL_1925203, EPI_ISL_1925204, EPI_ISL_1925205, EPI_ISL_1925206, EPI_ISL_1925207, EPI_ISL_1925208, EPI_ISL_1925209, EPI_ISL_1925210, EPI_ISL_1925211, EPI_ISL_1925212, EPI_ISL_1925213, EPI_ISL_1925214, EPI_ISL_1925215, EPI_ISL_1925216, EPI_ISL_1925217, EPI_ISL_1925218, EPI_ISL_1925219, EPI_ISL_1925220, EPI_ISL_1925221, EPI_ISL_1925222, EPI_ISL_1925223, EPI_ISL_1925224, EPI_ISL_1925225, EPI_ISL_1925226, EPI_ISL_1925227, EPI_ISL_1925228, EPI_ISL_1925229, EPI_ISL_1925230, EPI_ISL_1925231, EPI_ISL_1925232, EPI_ISL_1925233, EPI_ISL_1925234, EPI_ISL_1925235, EPI_ISL_1925236, EPI_ISL_1925237, EPI_ISL_1925238, EPI_ISL_1925239, EPI_ISL_1925240, EPI_ISL_1925241, EPI_ISL_1925242, EPI_ISL_1925243, EPI_ISL_1925244, EPI_ISL_1925245, EPI_ISL_1925246, EPI_ISL_1925247, EPI_ISL_1925248, EPI_ISL_1925249, EPI_ISL_1925250, EPI_ISL_1925251, EPI_ISL_1925252, EPI_ISL_1925253, EPI_ISL_1925254, EPI_ISL_1925255, EPI_ISL_1925256, EPI_ISL_1925257, EPI_ISL_1925259, EPI_ISL_1925260, EPI_ISL_1925261, EPI_ISL_1925262, EPI_ISL_1925263, EPI_ISL_1925264, EPI_ISL_1925265, EPI_ISL_1925266, EPI_ISL_1925267, EPI_ISL_1925268, EPI_ISL_1925269, EPI_ISL_1925270, EPI_ISL_1925271, EPI_ISL_1925272, EPI_ISL_1925273, EPI_ISL_1925274, EPI_ISL_1925275, EPI_ISL_1925276, EPI_ISL_1925277, EPI_ISL_1925278, EPI_ISL_1925279, EPI_ISL_1925280, EPI_ISL_1925281, EPI_ISL_1925282, EPI_ISL_1925283, EPI_ISL_1925284, EPI_ISL_1925285, EPI_ISL_1925286, EPI_ISL_1925287, EPI_ISL_1925288, EPI_ISL_1925289, EPI_ISL_1925290, EPI_ISL_1925291, EPI_ISL_1925292, EPI_ISL_1925293, EPI_ISL_1925294, EPI_ISL_1925295, EPI_ISL_1925296, EPI_ISL_1925297, EPI_ISL_1925298, EPI_ISL_1925299, EPI_ISL_1925300, EPI_ISL_1925301, EPI_ISL_1925302, EPI_ISL_1925303, EPI_ISL_1925304, EPI_ISL_1925305, EPI_ISL_1925306, EPI_ISL_1925307, EPI_ISL_1925308, EPI_ISL_1925309, EPI_ISL_1925310, EPI_ISL_1925311, EPI_ISL_1925312, EPI_ISL_1925313, EPI_ISL_1925314, EPI_ISL_1925315, EPI_ISL_1925316, EPI_ISL_1925317, EPI_ISL_1925318, EPI_ISL_1925319, EPI_ISL_1925320, EPI_ISL_1925321, EPI_ISL_1925322, EPI_ISL_1925324, EPI_ISL_1925325, EPI_ISL_1925326, EPI_ISL_1925327, EPI_ISL_1925328, EPI_ISL_1925329, EPI_ISL_1925330, EPI_ISL_1925331, EPI_ISL_1925332, EPI_ISL_1925333, EPI_ISL_1925334, EPI_ISL_1925335, EPI_ISL_1925336, EPI_ISL_1925337, EPI_ISL_1925338, EPI_ISL_1925339, EPI_ISL_1925340, EPI_ISL_1925341, EPI_ISL_1925342, EPI_ISL_1925343, EPI_ISL_1925344, EPI_ISL_1925345, EPI_ISL_1925346, EPI_ISL_1925347, EPI_ISL_1925348, EPI_ISL_1925349, EPI_ISL_1925350, EPI_ISL_1925351, EPI_ISL_1925352, EPI_ISL_1925353, EPI_ISL_1925354, EPI_ISL_1925355, EPI_ISL_1925356, EPI_ISL_1925357, EPI_ISL_1925358, EPI_ISL_1925359, EPI_ISL_1925360, EPI_ISL_1925361, EPI_ISL_1925362, EPI_ISL_1925363, EPI_ISL_1925364, EPI_ISL_1925365, EPI_ISL_1925366, EPI_ISL_1925367, EPI_ISL_1925368, EPI_ISL_1925369, EPI_ISL_1925370, EPI_ISL_1925371, EPI_ISL_1925372, EPI_ISL_1925373, EPI_ISL_1925374, EPI_ISL_1925375, EPI_ISL_1925376, EPI_ISL_1925377, EPI_ISL_1925378, EPI_ISL_1925379, EPI_ISL_1925380, EPI_ISL_1925381, EPI_ISL_1925382, EPI_ISL_1925383, EPI_ISL_1925384, EPI_ISL_1925385, EPI_ISL_1925386, EPI_ISL_1925387, EPI_ISL_1925388, EPI_ISL_1925389, EPI_ISL_1925390, EPI_ISL_1925391, EPI_ISL_1925392, EPI_ISL_1925393, EPI_ISL_1925394, EPI_ISL_1925395, EPI_ISL_1925396, EPI_ISL_1925397, EPI_ISL_1925398, EPI_ISL_1925399, EPI_ISL_1925400, EPI_ISL_1925401, EPI_ISL_1925402, EPI_ISL_1925403, EPI_ISL_1925404, EPI_ISL_1925405, EPI_ISL_1925406, EPI_ISL_1925407, EPI_ISL_1925408, EPI_ISL_1925409, EPI_ISL_1925410, EPI_ISL_1925411, EPI_ISL_1925412, EPI_ISL_1925413, EPI_ISL_1925414, EPI_ISL_1925415, EPI_ISL_1925416, EPI_ISL_1925417, EPI_ISL_1925418, EPI_ISL_1925419, EPI_ISL_1925420, EPI_ISL_1925421, EPI_ISL_1925422, EPI_ISL_1925423, EPI_ISL_1925424, EPI_ISL_1925425, EPI_ISL_1925426, EPI_ISL_1925427, EPI_ISL_1925428, EPI_ISL_1925429, EPI_ISL_1925430, EPI_ISL_1925431, EPI_ISL_1925432, EPI_ISL_1925433, EPI_ISL_1925434, EPI_ISL_1925435, EPI_ISL_1925436, EPI_ISL_1925437, EPI_ISL_1925438, EPI_ISL_1925439, EPI_ISL_1925440, EPI_ISL_1925441, EPI_ISL_1925442, EPI_ISL_1925443, EPI_ISL_1925444, EPI_ISL_1925445, EPI_ISL_1925446, EPI_ISL_1925447, EPI_ISL_1925448, EPI_ISL_1925449, EPI_ISL_1925450, EPI_ISL_1925451, EPI_ISL_1925452, EPI_ISL_1925453, EPI_ISL_1925454, EPI_ISL_1925455, EPI_ISL_1925456, EPI_ISL_1925457, EPI_ISL_1925458, EPI_ISL_1925459, EPI_ISL_1925460, EPI_ISL_1925461, EPI_ISL_1925462, EPI_ISL_1925463, EPI_ISL_1925464, EPI_ISL_1925465, EPI_ISL_1925466, EPI_ISL_1925467, EPI_ISL_1925468, EPI_ISL_1925469, EPI_ISL_1925470, EPI_ISL_1925471, EPI_ISL_1925472, EPI_ISL_1925473, EPI_ISL_1925474, EPI_ISL_1925475, EPI_ISL_1925476, EPI_ISL_1925478, EPI_ISL_1925479, EPI_ISL_1925480, EPI_ISL_1925481, EPI_ISL_1925482, EPI_ISL_1925483, EPI_ISL_1925484, EPI_ISL_1925485, EPI_ISL_1925486, EPI_ISL_1925487, EPI_ISL_1925488, EPI_ISL_1925489, EPI_ISL_1925490, EPI_ISL_1925491, EPI_ISL_1925492, EPI_ISL_1925493, EPI_ISL_1925494, EPI_ISL_1925495, EPI_ISL_1925496, EPI_ISL_1925497, EPI_ISL_1925498, EPI_ISL_1925499, EPI_ISL_1925500, EPI_ISL_1925502, EPI_ISL_1925503, EPI_ISL_1925504, EPI_ISL_1925505, EPI_ISL_1925507, EPI_ISL_1925508, EPI_ISL_1925509, EPI_ISL_1925510, EPI_ISL_1925511, EPI_ISL_1925512, EPI_ISL_1925513, EPI_ISL_1925514, EPI_ISL_1925515, EPI_ISL_1925516, EPI_ISL_1925517, EPI_ISL_1925518, EPI_ISL_1925519, EPI_ISL_1925520, EPI_ISL_1925521, EPI_ISL_1925522, EPI_ISL_1925523, EPI_ISL_1925524, EPI_ISL_1925525, EPI_ISL_1925526, EPI_ISL_1925527, EPI_ISL_1925528, EPI_ISL_1925529, EPI_ISL_1925530, EPI_ISL_1925531, EPI_ISL_1925532, EPI_ISL_1925533, EPI_ISL_1925534, EPI_ISL_1925535, EPI_ISL_1925536, EPI_ISL_1925537, EPI_ISL_1925538, EPI_ISL_1925539, EPI_ISL_1925540, EPI_ISL_1925541, EPI_ISL_1925542, EPI_ISL_1925543, EPI_ISL_1925544, | | | |

EPI_ISL_1925545, EPI_ISL_1925546, EPI_ISL_1925547, EPI_ISL_1925548, EPI_ISL_1925549, EPI_ISL_1925550, EPI_ISL_1925551, EPI_ISL_1925552, EPI_ISL_1925553, EPI_ISL_1925554, EPI_ISL_1925555, EPI_ISL_1925556, EPI_ISL_1925557, EPI_ISL_1925558, EPI_ISL_1925559, EPI_ISL_1925560, EPI_ISL_1925561, EPI_ISL_1925562, EPI_ISL_1925563, EPI_ISL_1925564, EPI_ISL_1925565, EPI_ISL_1925566, EPI_ISL_1925567, EPI_ISL_1925568, EPI_ISL_1925569, EPI_ISL_1925570, EPI_ISL_1925571, EPI_ISL_1925572, EPI_ISL_1925573, EPI_ISL_1925574, EPI_ISL_1925575, EPI_ISL_1925576, EPI_ISL_1925577, EPI_ISL_1925578, EPI_ISL_1925579, EPI_ISL_1925580, EPI_ISL_1925581, EPI_ISL_1925582, EPI_ISL_1925583, EPI_ISL_1925584, EPI_ISL_1925586, EPI_ISL_1925587, EPI_ISL_1925588, EPI_ISL_1925589, EPI_ISL_1925590, EPI_ISL_1925591, EPI_ISL_1925592, EPI_ISL_1925593, EPI_ISL_1925594, EPI_ISL_1925595, EPI_ISL_1925596, EPI_ISL_1925597, EPI_ISL_1925598, EPI_ISL_1925600, EPI_ISL_1925601, EPI_ISL_1925602, EPI_ISL_1925603, EPI_ISL_1925604, EPI_ISL_1925605, EPI_ISL_1925606, EPI_ISL_1925607, EPI_ISL_1925608, EPI_ISL_1925609, EPI_ISL_1925610, EPI_ISL_1925611, EPI_ISL_1925612, EPI_ISL_1925613, EPI_ISL_1925614, EPI_ISL_1925615, EPI_ISL_1925616, EPI_ISL_1925617

| | | | |
|---|---|---|---|
| see above | Aegis Sciences Corporation | Centers for Disease Control and Prevention Division of Viral Diseases, Pathogen Discovery | Dakota Howard, Dhwani Batra, Peter W. Cook, Kara Moser, Adrian Paskey, Jason Caravas, Benjamin Rambo-Martin, Shatavia Morrison, Christopher Gulvick, Scott Sammons, Yvette Unoarumhi, Darlene Wagner, Matthew Schmerer, Cyndi Clark, Patrick Campbell, Rob Case, Vikramsinha Ghorpade, Holly Houdeshell, Ola Kvalvaag, Dillon Nall, Ethan Sanders, Alec Vest, Shaun Westlund, Matthew Hardison, Clinton R. Paden, Duncan MacCannell |

EPI_ISL_1925618, EPI_ISL_1925619, EPI_ISL_1925620, EPI_ISL_1925621, EPI_ISL_1925622, EPI_ISL_1925623, EPI_ISL_1925625, EPI_ISL_1925626, EPI_ISL_1925627, EPI_ISL_1925628, EPI_ISL_1925629, EPI_ISL_1925630, EPI_ISL_1925633, EPI_ISL_1925634, EPI_ISL_1925635, EPI_ISL_1925636, EPI_ISL_1925637, EPI_ISL_1925639, EPI_ISL_1925640, EPI_ISL_1925641, EPI_ISL_1925642, EPI_ISL_1925643, EPI_ISL_1925644, EPI_ISL_1925645, EPI_ISL_1925646, EPI_ISL_1925647, EPI_ISL_1925648, EPI_ISL_1925650, EPI_ISL_1925651, EPI_ISL_1925652, EPI_ISL_1925653, EPI_ISL_1925654, EPI_ISL_1925655, EPI_ISL_1925656, EPI_ISL_1925657, EPI_ISL_1925658, EPI_ISL_1925659, EPI_ISL_1925661, EPI_ISL_1925663, EPI_ISL_1925664, EPI_ISL_1925665, EPI_ISL_1925666, EPI_ISL_1925667, EPI_ISL_1925668, EPI_ISL_1925670, EPI_ISL_1925671, EPI_ISL_1925672, EPI_ISL_1925673, EPI_ISL_1925674, EPI_ISL_1925675, EPI_ISL_1925676, EPI_ISL_1925677, EPI_ISL_1925678, EPI_ISL_1925680, EPI_ISL_1925681, EPI_ISL_1925682, EPI_ISL_1925683, EPI_ISL_1925684, EPI_ISL_1925686, EPI_ISL_1925687, EPI_ISL_1925688, EPI_ISL_1925689, EPI_ISL_1925694, EPI_ISL_1925695, EPI_ISL_1925697, EPI_ISL_1925699, EPI_ISL_1925700, EPI_ISL_1925701, EPI_ISL_1925703, EPI_ISL_1925704, EPI_ISL_1925705, EPI_ISL_1925707, EPI_ISL_1925708, EPI_ISL_1925710, EPI_ISL_1925713, EPI_ISL_1925714, EPI_ISL_1925717, EPI_ISL_1925718, EPI_ISL_1925719, EPI_ISL_1925720, EPI_ISL_1925721, EPI_ISL_1925722, EPI_ISL_1925723, EPI_ISL_1925724, EPI_ISL_1925725, EPI_ISL_1925727, EPI_ISL_1925728, EPI_ISL_1925729, EPI_ISL_1925730, EPI_ISL_1925731, EPI_ISL_1925732, EPI_ISL_1925733, EPI_ISL_1925734, EPI_ISL_1925735, EPI_ISL_1925736, EPI_ISL_1925737, EPI_ISL_1925738, EPI_ISL_1925739, EPI_ISL_1925740, EPI_ISL_1925741, EPI_ISL_1925742, EPI_ISL_1925743, EPI_ISL_1925744, EPI_ISL_1925746, EPI_ISL_1925750, EPI_ISL_1925751, EPI_ISL_1925752, EPI_ISL_1925754, EPI_ISL_1925755, EPI_ISL_1925757, EPI_ISL_1925758, EPI_ISL_1925759, EPI_ISL_1925760, EPI_ISL_1925762, EPI_ISL_1925763, EPI_ISL_1925764, EPI_ISL_1925765, EPI_ISL_1925767, EPI_ISL_1925770, EPI_ISL_1925771, EPI_ISL_1925772, EPI_ISL_1925774, EPI_ISL_1925775, EPI_ISL_1925776, EPI_ISL_1925777, EPI_ISL_1925778, EPI_ISL_1925779, EPI_ISL_1925780, EPI_ISL_1925781, EPI_ISL_1925782, EPI_ISL_1925784, EPI_ISL_1925785, EPI_ISL_1925786, EPI_ISL_1925788, EPI_ISL_1925789, EPI_ISL_1925790, EPI_ISL_1925791, EPI_ISL_1925792, EPI_ISL_1925794, EPI_ISL_1925795, EPI_ISL_1925796, EPI_ISL_1925799

| | | | |
|---|---|---|---|
| see above | Laboratory Corporation of America | Centers for Disease Control and Prevention Division of Viral Diseases, Pathogen Discovery | Dakota Howard, Dhwani Batra, Peter W. Cook, Kara Moser, Adrian Paskey, Jason Caravas, Benjamin Rambo-Martin, Shatavia Morrison, Christopher Gulvick, Scott Sammons, Yvette Unoarumhi, Darlene Wagner, Matthew Schmerer, Minoo Agarwal, Eyad Almasri, Debbie Boles, Ayla Burns, Nuthawin Charoensri, Oren Cohen, Susan Countryman, Mary Ann Cristobal, Bobbi Croy, Suzanne Dale, Hrushikesh Deshmukh, Amanda Douglas, Vincent Drouillon, Marcia Eisenberg, Howard Engler, Rama Ghatti, Prashant Gupta, Susan Hicks, Jake Humphrey, Lax Iyer, Manoj Jain, Mohan Kolli, Brian Krueger, Tim Kuphal, Stanley Letovsky, Michael Levandoski, Craig Lukasik, Jonathan Meltzer, Brian Norvell, Mindy Nye, Scott Parker, Christos Petropoulos, John Pruitt, Steven Ragan, Scott Ryan, Mike Sapeta, Jana Schroth, Suresh Babu Selvaraju, Goran Stevovic, Amanda Suchanek, Andrea Throop, Lyndon Tilson, Thomas Urban, Joe Voshell, Kimberly Wagner, Jonathan Williams, Mary Williamson, Qian Zeng, Tricia Zwiefelhofer, Clinton R. Paden, Duncan MacCannell |
| EPI_ISL_1925804 | Interpath Laboratory | Seattle Flu Study | Deborah A. Nickerson, Chris D. Frazar, Jover Lee, Benjamin Pelle, Erica Ryke, Matthew Richardson, Amanda Adler, Elisabeth Brandstetter, Peter D. Han, Kairsten Fay, Misja Ilcisin, Kirsten Lacombe, Thomas R. Sibley, Melissa Truong, Caitlin R. Wolf, Romesh Gautom, Geoff Melly, Brian Hiatt, Philip Dykema, Scott Lindquist, Michael Boeckh, Janet A. Englund, Michael Famulare, Barry R. Lutz, Mark J. Rieder, Lea M. Starita, Matthew Thompson, Helen Y. Chu, Jay Shendure, Trevor Bedford |
| EPI_ISL_1925805, EPI_ISL_1925808 | Laboratory Corporation of America | Centers for Disease Control and Prevention Division of Viral Diseases, Pathogen Discovery | Dakota Howard, Dhwani Batra, Peter W. Cook, Kara Moser, Adrian Paskey, Jason Caravas, Benjamin Rambo-Martin, Shatavia Morrison, Christopher Gulvick, Scott Sammons, Yvette Unoarumhi, Darlene Wagner, Matthew Schmerer, Minoo Agarwal, Eyad Almasri, Debbie Boles, Ayla Burns, Nuthawin Charoensri, Oren Cohen, Susan Countryman, Mary Ann Cristobal, Bobbi Croy, Suzanne Dale, Hrushikesh Deshmukh, Amanda Douglas, Vincent Drouillon, Marcia Eisenberg, Howard Engler, Rama Ghatti, Prashant Gupta, Susan Hicks, Jake Humphrey, Lax Iyer, Manoj Jain, Mohan Kolli, Brian Krueger, Tim Kuphal, Stanley Letovsky, Michael Levandoski, Craig Lukasik, Jonathan Meltzer, Brian Norvell, Mindy Nye, Scott Parker, Christos Petropoulos, John Pruitt, Steven Ragan, Scott Ryan, Mike Sapeta, Jana Schroth, Suresh Babu Selvaraju, Goran Stevovic, Amanda Suchanek, Andrea Throop, Lyndon Tilson, Thomas Urban, Joe Voshell, Kimberly Wagner, Jonathan Williams, Mary Williamson, Qian Zeng, Tricia Zwiefelhofer, Clinton R. Paden, Duncan MacCannell |
| EPI_ISL_1925809, EPI_ISL_1925810, EPI_ISL_1925812 | Interpath Laboratory | Seattle Flu Study | Deborah A. Nickerson, Chris D. Frazar, Jover Lee, Benjamin Pelle, Erica Ryke, Matthew Richardson, Amanda Adler, Elisabeth Brandstetter, Peter D. Han, Kairsten Fay, Misja Ilcisin, Kirsten Lacombe, Thomas R. Sibley, Melissa Truong, Caitlin R. Wolf, Romesh Gautom, Geoff Melly, Brian Hiatt, Philip Dykema, Scott Lindquist, Michael Boeckh, Janet A. Englund, Michael Famulare, Barry R. Lutz, Mark J. Rieder, Lea M. Starita, Matthew Thompson, Helen Y. Chu, Jay Shendure, Trevor Bedford |
| EPI_ISL_1925814 | Laboratory Corporation of America | Centers for Disease Control and Prevention Division of Viral Diseases, Pathogen Discovery | Dakota Howard, Dhwani Batra, Peter W. Cook, Kara Moser, Adrian Paskey, Jason Caravas, Benjamin Rambo-Martin, Shatavia Morrison, Christopher Gulvick, Scott Sammons, Yvette Unoarumhi, Darlene Wagner, Matthew Schmerer, Minoo Agarwal, Eyad Almasri, Debbie Boles, Ayla Burns, Nuthawin Charoensri, Oren Cohen, Susan Countryman, Mary Ann Cristobal, Bobbi Croy, Suzanne Dale, Hrushikesh Deshmukh, Amanda Douglas, Vincent Drouillon, Marcia Eisenberg, Howard Engler, Rama Ghatti, Prashant Gupta, Susan Hicks, Jake Humphrey, Lax Iyer, Manoj Jain, Mohan Kolli, Brian Krueger, Tim Kuphal, Stanley Letovsky, Michael Levandoski, Craig Lukasik, Jonathan Meltzer, Brian Norvell, Mindy Nye, Scott Parker, Christos Petropoulos, John Pruitt, Steven Ragan, Scott Ryan, Mike Sapeta, Jana Schroth, Suresh Babu Selvaraju, Goran Stevovic, Amanda Suchanek, Andrea Throop, Lyndon Tilson, Thomas Urban, Joe Voshell, Kimberly Wagner, Jonathan Williams, Mary Williamson, Qian Zeng, Tricia Zwiefelhofer, Clinton R. Paden, Duncan MacCannell |
| EPI_ISL_1925817 | Interpath Laboratory | Seattle Flu Study | Deborah A. Nickerson, Chris D. Frazar, Jover Lee, Benjamin Pelle, Erica Ryke, Matthew Richardson, Amanda Adler, Elisabeth Brandstetter, Peter D. Han, Kairsten Fay, Misja Ilcisin, Kirsten Lacombe, Thomas R. Sibley, Melissa Truong, Caitlin R. Wolf, Romesh Gautom, Geoff Melly, Brian Hiatt, Philip Dykema, Scott Lindquist, Michael Boeckh, Janet A. Englund, Michael Famulare, Barry R. Lutz, Mark J. Rieder, Lea M. Starita, Matthew Thompson, Helen Y. Chu, Jay Shendure, Trevor Bedford |
| EPI_ISL_1925818 | Laboratory Corporation of America | Centers for Disease Control and Prevention Division of Viral Diseases, Pathogen Discovery | Dakota Howard, Dhwani Batra, Peter W. Cook, Kara Moser, Adrian Paskey, Jason Caravas, Benjamin Rambo-Martin, Shatavia Morrison, Christopher Gulvick, Scott Sammons, Yvette Unoarumhi, Darlene Wagner, Matthew Schmerer, Minoo Agarwal, Eyad Almasri, Debbie Boles, Ayla Burns, Nuthawin Charoensri, Oren Cohen, Susan Countryman, Mary Ann Cristobal, Bobbi Croy, Suzanne Dale, Hrushikesh Deshmukh, Amanda Douglas, Vincent Drouillon, Marcia Eisenberg, Howard Engler, Rama Ghatti, Prashant Gupta, Susan Hicks, Jake Humphrey, Lax Iyer, Manoj Jain, Mohan Kolli, Brian Krueger, Tim Kuphal, Stanley Letovsky, Michael Levandoski, Craig Lukasik, Jonathan Meltzer, Brian Norvell, Mindy Nye, Scott Parker, Christos Petropoulos, John Pruitt, Steven Ragan, Scott Ryan, Mike Sapeta, Jana Schroth, Suresh Babu Selvaraju, Goran Stevovic, Amanda Suchanek, Andrea Throop, Lyndon Tilson, Thomas Urban, Joe Voshell, Kimberly Wagner, Jonathan Williams, Mary Williamson, Qian Zeng, Tricia Zwiefelhofer, Clinton R. Paden, Duncan MacCannell |
| EPI_ISL_1925819, EPI_ISL_1925820 | Interpath Laboratory | Seattle Flu Study | Deborah A. Nickerson, Chris D. Frazar, Jover Lee, Benjamin Pelle, Erica Ryke, Matthew Richardson, Amanda Adler, Elisabeth Brandstetter, Peter D. Han, Kairsten Fay, Misja Ilcisin, Kirsten Lacombe, Thomas R. Sibley, Melissa Truong, Caitlin R. Wolf, Romesh Gautom, Geoff Melly, Brian Hiatt, Philip Dykema, Scott Lindquist, Michael Boeckh, Janet A. Englund, Michael Famulare, Barry R. Lutz, Mark J. Rieder, Lea M. Starita, Matthew Thompson, Helen Y. Chu, Jay Shendure, Trevor Bedford |
| EPI_ISL_1925821 | Laboratory Corporation of America | Centers for Disease Control and Prevention Division of Viral Diseases, Pathogen Discovery | Dakota Howard, Dhwani Batra, Peter W. Cook, Kara Moser, Adrian Paskey, Jason Caravas, Benjamin Rambo-Martin, Shatavia Morrison, Christopher Gulvick, Scott Sammons, Yvette Unoarumhi, Darlene Wagner, Matthew Schmerer, Minoo Agarwal, Eyad Almasri, Debbie Boles, Ayla Burns, Nuthawin Charoensri, Oren Cohen, Susan Countryman, Mary Ann Cristobal, Bobbi Croy, Suzanne Dale, Hrushikesh Deshmukh, Amanda Douglas, Vincent Drouillon, Marcia Eisenberg, Howard Engler, Rama Ghatti, Prashant Gupta, Susan Hicks, Jake Humphrey, Lax Iyer, Manoj Jain, Mohan Kolli, Brian Krueger, Tim Kuphal, Stanley Letovsky, Michael Levandoski, Craig Lukasik, Jonathan Meltzer, Brian Norvell, Mindy Nye, Scott Parker, Christos Petropoulos, John Pruitt, Steven Ragan, Scott Ryan, Mike Sapeta, Jana Schroth, Suresh Babu Selvaraju, Goran Stevovic, Amanda Suchanek, Andrea Throop, Lyndon Tilson, Thomas Urban, Joe Voshell, Kimberly Wagner, Jonathan Williams, Mary Williamson, Qian Zeng, Tricia Zwiefelhofer, Clinton R. Paden, Duncan MacCannell |
| EPI_ISL_1925823 | Interpath Laboratory | Seattle Flu Study | Deborah A. Nickerson, Chris D. Frazar, Jover Lee, Benjamin Pelle, Erica Ryke, Matthew Richardson, Amanda Adler, Elisabeth Brandstetter, Peter D. Han, Kairsten Fay, Misja Ilcisin, Kirsten Lacombe, Thomas R. Sibley, Melissa Truong, Caitlin R. Wolf, Romesh Gautom, Geoff Melly, Brian Hiatt, Philip Dykema, Scott Lindquist, Michael Boeckh, Janet A. Englund, Michael Famulare, Barry R. Lutz, Mark J. Rieder, Lea M. Starita, Matthew Thompson, Helen Y. Chu, Jay Shendure, Trevor Bedford |
| EPI_ISL_1925824 | Laboratory Corporation of America | Centers for Disease Control and Prevention Division of Viral Diseases, Pathogen Discovery | Dakota Howard, Dhwani Batra, Peter W. Cook, Kara Moser, Adrian Paskey, Jason Caravas, Benjamin Rambo-Martin, Shatavia Morrison, Christopher Gulvick, Scott Sammons, Yvette Unoarumhi, Darlene Wagner, Matthew Schmerer, Minoo Agarwal, Eyad Almasri, Debbie Boles, Ayla Burns, Nuthawin Charoensri, Oren Cohen, Susan Countryman, Mary Ann Cristobal, Bobbi Croy, Suzanne Dale, Hrushikesh Deshmukh, Amanda Douglas, Vincent Drouillon, Marcia Eisenberg, Howard Engler, Rama Ghatti, Prashant Gupta, Susan Hicks, Jake Humphrey, Lax Iyer, Manoj Jain, Mohan Kolli, Brian Krueger, Tim Kuphal, Stanley Letovsky, Michael Levandoski, Craig Lukasik, Jonathan Meltzer, Brian Norvell, Mindy Nye, Scott Parker, Christos Petropoulos, John Pruitt, Steven Ragan, Scott Ryan, Mike Sapeta, Jana Schroth, Suresh Babu Selvaraju, Goran Stevovic, Amanda Suchanek, Andrea Throop, Lyndon Tilson, Thomas Urban, Joe Voshell, Kimberly Wagner, Jonathan Williams, Mary Williamson, Qian Zeng, Tricia Zwiefelhofer, Clinton R. Paden, Duncan MacCannell |
| EPI_ISL_1925826 | Interpath Laboratory | Seattle Flu Study | Deborah A. Nickerson, Chris D. Frazar, Jover Lee, Benjamin Pelle, Erica Ryke, Matthew Richardson, Amanda Adler, Elisabeth Brandstetter, Peter D. Han, Kairsten Fay, Misja Ilcisin, Kirsten Lacombe, Thomas R. Sibley, Melissa Truong, Caitlin R. Wolf, Romesh Gautom, Geoff Melly, Brian Hiatt, Philip Dykema, Scott Lindquist, Michael Boeckh, Janet A. Englund, Michael Famulare, Barry R. Lutz, Mark J. Rieder, Lea M. Starita, Matthew Thompson, Helen Y. Chu, Jay Shendure, Trevor Bedford |
| EPI_ISL_1925827 | Laboratory Corporation of America | Centers for Disease Control and Prevention Division of Viral Diseases, Pathogen Discovery | Dakota Howard, Dhwani Batra, Peter W. Cook, Kara Moser, Adrian Paskey, Jason Caravas, Benjamin Rambo-Martin, Shatavia Morrison, Christopher Gulvick, Scott Sammons, Yvette Unoarumhi, Darlene Wagner, Matthew Schmerer, Minoo Agarwal, Eyad Almasri, Debbie Boles, Ayla Burns, Nuthawin Charoensri, Oren Cohen, Susan Countryman, Mary Ann Cristobal, Bobbi Croy, Suzanne Dale, Hrushikesh Deshmukh, Amanda Douglas, Vincent Drouillon, Marcia Eisenberg, Howard Engler, Rama Ghatti, Prashant Gupta, Susan Hicks, Jake Humphrey, Lax Iyer, Manoj Jain, Mohan Kolli, Brian Krueger, Tim Kuphal, Stanley Letovsky, Michael Levandoski, Craig Lukasik, Jonathan Meltzer, Brian Norvell, Mindy Nye, Scott Parker, Christos Petropoulos, John Pruitt, Steven Ragan, Scott Ryan, Mike Sapeta, Jana Schroth, Suresh Babu Selvaraju, Goran Stevovic, Amanda Suchanek, Andrea Throop, Lyndon Tilson, Thomas Urban, Joe Voshell, Kimberly Wagner, Jonathan Williams, Mary Williamson, Qian Zeng, Tricia Zwiefelhofer, Clinton R. Paden, Duncan MacCannell |
| EPI_ISL_1925828 | Interpath Laboratory | Seattle Flu Study | Deborah A. Nickerson, Chris D. Frazar, Jover Lee, Benjamin Pelle, Erica Ryke, Matthew Richardson, Amanda Adler, Elisabeth Brandstetter, Peter D. Han, Kairsten Fay, Misja Ilcisin, Kirsten Lacombe, Thomas R. Sibley, Melissa Truong, Caitlin R. Wolf, Romesh Gautom, Geoff Melly, Brian Hiatt, Philip Dykema, Scott Lindquist, Michael Boeckh, Janet A. Englund, Michael Famulare, Barry R. Lutz, Mark J. Rieder, Lea M. Starita, Matthew Thompson, Helen Y. Chu, Jay Shendure, Trevor Bedford |
| EPI_ISL_1925830 | Laboratory Corporation of America | Centers for Disease Control and Prevention Division of Viral Diseases, Pathogen Discovery | Dakota Howard, Dhwani Batra, Peter W. Cook, Kara Moser, Adrian Paskey, Jason Caravas, Benjamin Rambo-Martin, Shatavia Morrison, Christopher Gulvick, Scott Sammons, Yvette Unoarumhi, Darlene Wagner, Matthew Schmerer, Minoo Agarwal, Eyad Almasri, Debbie Boles, Ayla Burns, Nuthawin Charoensri, Oren Cohen, Susan Countryman, Mary Ann Cristobal, Bobbi Croy, Suzanne Dale, Hrushikesh Deshmukh, Amanda Douglas, Vincent Drouillon, Marcia Eisenberg, Howard Engler, Rama Ghatti, Prashant Gupta, Susan Hicks, Jake Humphrey, Lax Iyer, Manoj Jain, Mohan Kolli, Brian Krueger, Tim Kuphal, Stanley Letovsky, Michael Levandoski, Craig Lukasik, Jonathan Meltzer, Brian Norvell, Mindy Nye, Scott Parker, Christos Petropoulos, John Pruitt, Steven Ragan, Scott Ryan, Mike Sapeta, Jana Schroth, Suresh Babu Selvaraju, Goran Stevovic, Amanda Suchanek, Andrea Throop, Lyndon Tilson, Thomas Urban, Joe Voshell, Kimberly Wagner, Jonathan Williams, Mary Williamson, Qian Zeng, Tricia Zwiefelhofer, Clinton R. Paden, Duncan MacCannell |
| EPI_ISL_1925833 | Interpath Laboratory | Seattle Flu Study | Deborah A. Nickerson, Chris D. Frazar, Jover Lee, Benjamin Pelle, Erica Ryke, Matthew Richardson, Amanda Adler, Elisabeth Brandstetter, Peter D. Han, Kairsten Fay, Misja Ilcisin, Kirsten Lacombe, Thomas R. Sibley, Melissa Truong, Caitlin R. Wolf, Romesh Gautom, Geoff Melly, Brian Hiatt, Philip Dykema, Scott Lindquist, Michael Boeckh, Janet A. Englund, Michael Famulare, Barry R. Lutz, Mark J. Rieder, Lea M. Starita, Matthew Thompson, Helen Y. Chu, Jay Shendure, Trevor Bedford |
| EPI_ISL_1925834 | Laboratory Corporation of America | Centers for Disease Control and Prevention Division of Viral Diseases, Pathogen Discovery | Dakota Howard, Dhwani Batra, Peter W. Cook, Kara Moser, Adrian Paskey, Jason Caravas, Benjamin Rambo-Martin, Shatavia Morrison, Christopher Gulvick, Scott Sammons, Yvette Unoarumhi, Darlene Wagner, Matthew Schmerer, Minoo Agarwal, Eyad Almasri, Debbie Boles, Ayla Burns, Nuthawin Charoensri, Oren Cohen, Susan Countryman, Mary Ann Cristobal, Bobbi Croy, Suzanne Dale, Hrushikesh Deshmukh, Amanda Douglas, Vincent Drouillon, Marcia Eisenberg, Howard Engler, Rama Ghatti, Prashant Gupta, Susan Hicks, Jake Humphrey, Lax Iyer, Manoj Jain, Mohan Kolli, Brian Krueger, Tim Kuphal, Stanley Letovsky, Michael Levandoski, Craig Lukasik, Jonathan Meltzer, Brian Norvell, Mindy Nye, Scott Parker, Christos Petropoulos, John Pruitt, Steven Ragan, Scott Ryan, Mike Sapeta, Jana Schroth, Suresh Babu Selvaraju, Goran Stevovic, Amanda Suchanek, Andrea Throop, Lyndon Tilson, Thomas Urban, Joe Voshell, Kimberly Wagner, Jonathan Williams, Mary Williamson, Qian Zeng, Tricia Zwiefelhofer, Clinton R. Paden, Duncan MacCannell |
| EPI_ISL_1925836 | Interpath Laboratory | Seattle Flu Study | Deborah A. Nickerson, Chris D. Frazar, Jover Lee, Benjamin Pelle, Erica Ryke, Matthew Richardson, Amanda Adler, Elisabeth Brandstetter, Peter D. Han, Kairsten Fay, Misja Ilcisin, Kirsten Lacombe, Thomas R. Sibley, Melissa Truong, Caitlin R. Wolf, Romesh Gautom, Geoff Melly, Brian Hiatt, Philip Dykema, Scott Lindquist, Michael Boeckh, Janet A. Englund, Michael Famulare, Barry R. Lutz, Mark J. Rieder, Lea M. Starita, Matthew Thompson, Helen Y. Chu, Jay Shendure, Trevor Bedford |
| EPI_ISL_1925837, EPI_ISL_1925840, EPI_ISL_1925846 | Laboratory Corporation of America | Centers for Disease Control and Prevention Division of Viral Diseases, Pathogen Discovery | Dakota Howard, Dhwani Batra, Peter W. Cook, Kara Moser, Adrian Paskey, Jason Caravas, Benjamin Rambo-Martin, Shatavia Morrison, Christopher Gulvick, Scott Sammons, Yvette Unoarumhi, Darlene Wagner, Matthew Schmerer, Minoo Agarwal, Eyad Almasri, Debbie Boles, Ayla Burns, Nuthawin Charoensri, Oren Cohen, Susan Countryman, Mary Ann Cristobal, Bobbi Croy, Suzanne Dale, Hrushikesh Deshmukh, Amanda Douglas, Vincent Drouillon, Marcia Eisenberg, Howard Engler, Rama Ghatti, Prashant Gupta, Susan Hicks, Jake Humphrey, Lax Iyer, Manoj Jain, Mohan Kolli, Brian Krueger, Tim Kuphal, Stanley Letovsky, Michael Levandoski, Craig Lukasik, Jonathan Meltzer, Brian Norvell, Mindy Nye, Scott Parker, Christos Petropoulos, John Pruitt, Steven Ragan, Scott Ryan, Mike Sapeta, Jana Schroth, Suresh Babu Selvaraju, Goran Stevovic, Amanda Suchanek, Andrea Throop, Lyndon Tilson, Thomas Urban, Joe Voshell, Kimberly Wagner, Jonathan Williams, Mary Williamson, Qian Zeng, Tricia Zwiefelhofer, Clinton R. Paden, Duncan MacCannell |
| EPI_ISL_1925847 | Interpath Laboratory | Seattle Flu Study | Deborah A. Nickerson, Chris D. Frazar, Jover Lee, Benjamin Pelle, Erica Ryke, Matthew Richardson, Amanda Adler, Elisabeth Brandstetter, Peter D. Han, Kairsten Fay, Misja Ilcisin, Kirsten Lacombe, Thomas R. Sibley, Melissa Truong, Caitlin R. Wolf, Romesh Gautom, Geoff Melly, Brian Hiatt, Philip Dykema, Scott Lindquist, Michael Boeckh, Janet A. Englund, Michael Famulare, Barry R. Lutz, Mark J. Rieder, Lea M. Starita, Matthew Thompson, Helen Y. Chu, Jay Shendure, Trevor Bedford |
| EPI_ISL_1925849 | Laboratory Corporation of America | Centers for Disease Control and Prevention Division of Viral Diseases, Pathogen Discovery | Dakota Howard, Dhwani Batra, Peter W. Cook, Kara Moser, Adrian Paskey, Jason Caravas, Benjamin Rambo-Martin, Shatavia Morrison, Christopher Gulvick, Scott Sammons, Yvette Unoarumhi, Darlene Wagner, Matthew Schmerer, Minoo Agarwal, Eyad Almasri, Debbie Boles, Ayla Burns, Nuthawin Charoensri, Oren Cohen, Susan Countryman, Mary Ann Cristobal, Bobbi Croy, Suzanne Dale, Hrushikesh Deshmukh, Amanda Douglas, Vincent Drouillon, Marcia Eisenberg, Howard Engler, Rama Ghatti, Prashant Gupta, Susan Hicks, Jake Humphrey, Lax Iyer, Manoj Jain, Mohan Kolli, Brian Krueger, Tim Kuphal, Stanley Letovsky, Michael Levandoski, Craig Lukasik, Jonathan Meltzer, Brian Norvell, Mindy Nye, Scott Parker, Christos Petropoulos, John Pruitt, Steven Ragan, Scott Ryan, Mike Sapeta, Jana Schroth, Suresh Babu Selvaraju, Goran Stevovic, Amanda Suchanek, Andrea Throop, Lyndon Tilson, Thomas Urban, Joe Voshell, Kimberly Wagner, Jonathan Williams, Mary Williamson, Qian Zeng, Tricia Zwiefelhofer, Clinton R. Paden, Duncan MacCannell |
| EPI_ISL_1925850, EPI_ISL_1925851 | Interpath Laboratory | Seattle Flu Study | Deborah A. Nickerson, Chris D. Frazar, Jover Lee, Benjamin Pelle, Erica Ryke, Matthew Richardson, Amanda Adler, Elisabeth Brandstetter, Peter D. Han, Kairsten Fay, Misja Ilcisin, Kirsten Lacombe, Thomas R. Sibley, Melissa Truong, Caitlin R. Wolf, Romesh Gautom, Geoff Melly, Brian Hiatt, Philip Dykema, Scott Lindquist, Michael Boeckh, Janet A. Englund, Michael Famulare, Barry R. Lutz, Mark J. Rieder, Lea M. Starita, Matthew Thompson, Helen Y. Chu, Jay Shendure, Trevor Bedford |
| EPI_ISL_1925852 | Laboratory Corporation of America | Centers for Disease Control and Prevention Division of Viral Diseases, Pathogen Discovery | Dakota Howard, Dhwani Batra, Peter W. Cook, Kara Moser, Adrian Paskey, Jason Caravas, Benjamin Rambo-Martin, Shatavia Morrison, Christopher Gulvick, Scott Sammons, Yvette Unoarumhi, Darlene Wagner, Matthew Schmerer, Minoo Agarwal, Eyad Almasri, Debbie Boles, Ayla Burns, Nuthawin Charoensri, Oren Cohen, Susan Countryman, Mary Ann Cristobal, Bobbi Croy, Suzanne Dale, Hrushikesh Deshmukh, Amanda Douglas, Vincent Drouillon, Marcia Eisenberg, Howard Engler, Rama Ghatti, Prashant Gupta, Susan Hicks, Jake Humphrey, Lax Iyer, Manoj Jain, Mohan Kolli, Brian Krueger, Tim Kuphal, Stanley Letovsky, Michael Levandoski, Craig Lukasik, Jonathan Meltzer, Brian Norvell, Mindy Nye, Scott Parker, Christos Petropoulos, John Pruitt, Steven Ragan, Scott Ryan, Mike Sapeta, Jana Schroth, Suresh Babu Selvaraju, Goran Stevovic, Amanda Suchanek, Andrea Throop, Lyndon Tilson, Thomas Urban, Joe Voshell, Kimberly Wagner, Jonathan Williams, Mary Williamson, Qian Zeng, Tricia Zwiefelhofer, Clinton R. Paden, Duncan MacCannell |
| EPI_ISL_1925853, EPI_ISL_1925854 | Interpath Laboratory | Seattle Flu Study | Deborah A. Nickerson, Chris D. Frazar, Jover Lee, Benjamin Pelle, Erica Ryke, Matthew Richardson, Amanda Adler, Elisabeth Brandstetter, Peter D. Han, Kairsten Fay, Misja Ilcisin, Kirsten Lacombe, Thomas R. Sibley, Melissa Truong, Caitlin R. Wolf, Romesh Gautom, Geoff Melly, Brian Hiatt, Philip Dykema, Scott Lindquist, Michael Boeckh, Janet A. Englund, Michael Famulare, Barry R. Lutz, Mark J. Rieder, Lea M. Starita, Matthew Thompson, Helen Y. Chu, Jay Shendure, Trevor Bedford |
| EPI_ISL_1925856 | Laboratory Corporation of America | Centers for Disease Control and Prevention Division of Viral Diseases, Pathogen Discovery | Dakota Howard, Dhwani Batra, Peter W. Cook, Kara Moser, Adrian Paskey, Jason Caravas, Benjamin Rambo-Martin, Shatavia Morrison, Christopher Gulvick, Scott Sammons, Yvette Unoarumhi, Darlene Wagner, Matthew Schmerer, Minoo Agarwal, Eyad Almasri, Debbie Boles, Ayla Burns, Nuthawin Charoensri, Oren Cohen, Susan Countryman, Mary Ann Cristobal, Bobbi Croy, Suzanne Dale, Hrushikesh Deshmukh, Amanda Douglas, Vincent Drouillon, Marcia Eisenberg, Howard Engler, Rama Ghatti, Prashant Gupta, Susan Hicks, Jake Humphrey, Lax Iyer, Manoj Jain, Mohan Kolli, Brian Krueger, Tim Kuphal, Stanley Letovsky, Michael Levandoski, Craig Lukasik, Jonathan Meltzer, Brian Norvell, Mindy Nye, Scott Parker, Christos Petropoulos, John Pruitt, Steven Ragan, Scott Ryan, Mike Sapeta, Jana Schroth, Suresh Babu Selvaraju, Goran Stevovic, Amanda Suchanek, Andrea Throop, Lyndon Tilson, Thomas Urban, Joe Voshell, Kimberly Wagner, Jonathan Williams, Mary Williamson, Qian Zeng, Tricia Zwiefelhofer, Clinton R. Paden, Duncan MacCannell |
| EPI_ISL_1925857, EPI_ISL_1925858 | Interpath Laboratory | Seattle Flu Study | Deborah A. Nickerson, Chris D. Frazar, Jover Lee, Benjamin Pelle, Erica Ryke, Matthew Richardson, Amanda Adler, Elisabeth Brandstetter, Peter D. Han, Kairsten Fay, Misja Ilcisin, Kirsten Lacombe, Thomas R. Sibley, Melissa Truong, Caitlin R. Wolf, Romesh Gautom, Geoff Melly, Brian Hiatt, Philip Dykema, Scott Lindquist, Michael Boeckh, Janet A. Englund, Michael Famulare, Barry R. Lutz, Mark J. Rieder, Lea M. Starita, Matthew Thompson, Helen Y. Chu, Jay Shendure, Trevor Bedford |
| EPI_ISL_1925859 | Laboratory Corporation of America | Centers for Disease Control and Prevention Division of Viral Diseases, Pathogen Discovery | Dakota Howard, Dhwani Batra, Peter W. Cook, Kara Moser, Adrian Paskey, Jason Caravas, Benjamin Rambo-Martin, Shatavia Morrison, Christopher Gulvick, Scott Sammons, Yvette Unoarumhi, Darlene Wagner, Matthew Schmerer, Minoo Agarwal, Eyad Almasri, Debbie Boles, Ayla Burns, Nuthawin Charoensri, Oren Cohen, Susan Countryman, Mary Ann Cristobal, Bobbi Croy, Suzanne Dale, Hrushikesh Deshmukh, Amanda Douglas, Vincent Drouillon, Marcia Eisenberg, Howard Engler, Rama Ghatti, Prashant Gupta, Susan Hicks, Jake Humphrey, Lax Iyer, Manoj Jain, Mohan Kolli, Brian Krueger, Tim Kuphal, Stanley Letovsky, Michael Levandoski, Craig Lukasik, Jonathan Meltzer, Brian Norvell, Mindy Nye, Scott Parker, Christos Petropoulos, John Pruitt, Steven Ragan, Scott Ryan, Mike Sapeta, Jana Schroth, Suresh Babu Selvaraju, Goran Stevovic, Amanda Suchanek, Andrea Throop, Lyndon Tilson, Thomas Urban, Joe Voshell, Kimberly Wagner, Jonathan Williams, Mary Williamson, Qian Zeng, Tricia Zwiefelhofer, Clinton R. Paden, Duncan MacCannell |

| | | | |
|---|---|---|---|
| EPI_ISL_1925860, EPI_ISL_1925861 | Interpath Laboratory | Seattle Flu Study | Deborah A. Nickerson, Chris D. Frazar, Jover Lee, Benjamin Pelle, Erica Ryke, Matthew Richardson, Amanda Adler, Elisabeth Brandstetter, Peter D. Han, Kairsten Fay, Misja Ilcisin, Kirsten Lacombe, Thomas R. Sibley, Melissa Truong, Caitlin R. Wolf, Romesh Gautom, Geoff Melly, Brian Hiatt, Philip Dykema, Scott Lindquist, Michael Boeckh, Janet A. Englund, Michael Famulare, Barry R. Lutz, Mark J. Rieder, Lea M. Starita, Matthew Thompson, Helen Y. Chu, Jay Shendure, Trevor Bedford |
| EPI_ISL_1925862 | Laboratory Corporation of America | Centers for Disease Control and Prevention Division of Viral Diseases, Pathogen Discovery | Dakota Howard, Dhwani Batra, Peter W. Cook, Kara Moser, Adrian Paskey, Jason Caravas, Benjamin Rambo-Martin, Shatavia Morrison, Christopher Gulvick, Scott Sammons, Yvette Unoarumhi, Darlene Wagner, Matthew Schmerer, Minoo Agarwal, Eyad Almasri, Debbie Boles, Ayla Burns, Nuthawin Charoensri, Oren Cohen, Susan Countryman, Mary Ann Cristobal, Bobbi Croy, Suzanne Dale, Hrushikesh Deshmukh, Amanda Douglas, Vincent Drouillon, Marcia Eisenberg, Howard Engler, Rama Ghatti, Prashant Gupta, Susan Hicks, Jake Humphrey, Lax Iyer, Manoj Jain, Mohan Kolli, Brian Krueger, Tim Kuphal, Stanley Letovsky, Michael Levandoski, Craig Lukasik, Jonathan Meltzer, Brian Norvell, Mindy Nye, Scott Parker, Christos Petropoulos, John Pruitt, Steven Ragan, Scott Ryan, Mike Sapeta, Jana Schroth, Suresh Babu Selvaraju, Goran Stevovic, Amanda Suchanek, Andrea Throop, Lyndon Tilson, Thomas Urban, Joe Voshell, Kimberly Wagner, Jonathan Williams, Mary Williamson, Qian Zeng, Tricia Zwiefelhofer, Clinton R. Paden, Duncan MacCannell |
| EPI_ISL_1925866, EPI_ISL_1925867, EPI_ISL_1925869 | Interpath Laboratory | Seattle Flu Study | Deborah A. Nickerson, Chris D. Frazar, Jover Lee, Benjamin Pelle, Erica Ryke, Matthew Richardson, Amanda Adler, Elisabeth Brandstetter, Peter D. Han, Kairsten Fay, Misja Ilcisin, Kirsten Lacombe, Thomas R. Sibley, Melissa Truong, Caitlin R. Wolf, Romesh Gautom, Geoff Melly, Brian Hiatt, Philip Dykema, Scott Lindquist, Michael Boeckh, Janet A. Englund, Michael Famulare, Barry R. Lutz, Mark J. Rieder, Lea M. Starita, Matthew Thompson, Helen Y. Chu, Jay Shendure, Trevor Bedford |
| EPI_ISL_1925871 | Laboratory Corporation of America | Centers for Disease Control and Prevention Division of Viral Diseases, Pathogen Discovery | Dakota Howard, Dhwani Batra, Peter W. Cook, Kara Moser, Adrian Paskey, Jason Caravas, Benjamin Rambo-Martin, Shatavia Morrison, Christopher Gulvick, Scott Sammons, Yvette Unoarumhi, Darlene Wagner, Matthew Schmerer, Minoo Agarwal, Eyad Almasri, Debbie Boles, Ayla Burns, Nuthawin Charoensri, Oren Cohen, Susan Countryman, Mary Ann Cristobal, Bobbi Croy, Suzanne Dale, Hrushikesh Deshmukh, Amanda Douglas, Vincent Drouillon, Marcia Eisenberg, Howard Engler, Rama Ghatti, Prashant Gupta, Susan Hicks, Jake Humphrey, Lax Iyer, Manoj Jain, Mohan Kolli, Brian Krueger, Tim Kuphal, Stanley Letovsky, Michael Levandoski, Craig Lukasik, Jonathan Meltzer, Brian Norvell, Mindy Nye, Scott Parker, Christos Petropoulos, John Pruitt, Steven Ragan, Scott Ryan, Mike Sapeta, Jana Schroth, Suresh Babu Selvaraju, Goran Stevovic, Amanda Suchanek, Andrea Throop, Lyndon Tilson, Thomas Urban, Joe Voshell, Kimberly Wagner, Jonathan Williams, Mary Williamson, Qian Zeng, Tricia Zwiefelhofer, Clinton R. Paden, Duncan MacCannell |
| EPI_ISL_1925875, EPI_ISL_1925876, EPI_ISL_1925879, EPI_ISL_1925880 | Interpath Laboratory | Seattle Flu Study | Deborah A. Nickerson, Chris D. Frazar, Jover Lee, Benjamin Pelle, Erica Ryke, Matthew Richardson, Amanda Adler, Elisabeth Brandstetter, Peter D. Han, Kairsten Fay, Misja Ilcisin, Kirsten Lacombe, Thomas R. Sibley, Melissa Truong, Caitlin R. Wolf, Romesh Gautom, Geoff Melly, Brian Hiatt, Philip Dykema, Scott Lindquist, Michael Boeckh, Janet A. Englund, Michael Famulare, Barry R. Lutz, Mark J. Rieder, Lea M. Starita, Matthew Thompson, Helen Y. Chu, Jay Shendure, Trevor Bedford |
| EPI_ISL_1925881 | Laboratory Corporation of America | Centers for Disease Control and Prevention Division of Viral Diseases, Pathogen Discovery | Dakota Howard, Dhwani Batra, Peter W. Cook, Kara Moser, Adrian Paskey, Jason Caravas, Benjamin Rambo-Martin, Shatavia Morrison, Christopher Gulvick, Scott Sammons, Yvette Unoarumhi, Darlene Wagner, Matthew Schmerer, Minoo Agarwal, Eyad Almasri, Debbie Boles, Ayla Burns, Nuthawin Charoensri, Oren Cohen, Susan Countryman, Mary Ann Cristobal, Bobbi Croy, Suzanne Dale, Hrushikesh Deshmukh, Amanda Douglas, Vincent Drouillon, Marcia Eisenberg, Howard Engler, Rama Ghatti, Prashant Gupta, Susan Hicks, Jake Humphrey, Lax Iyer, Manoj Jain, Mohan Kolli, Brian Krueger, Tim Kuphal, Stanley Letovsky, Michael Levandoski, Craig Lukasik, Jonathan Meltzer, Brian Norvell, Mindy Nye, Scott Parker, Christos Petropoulos, John Pruitt, Steven Ragan, Scott Ryan, Mike Sapeta, Jana Schroth, Suresh Babu Selvaraju, Goran Stevovic, Amanda Suchanek, Andrea Throop, Lyndon Tilson, Thomas Urban, Joe Voshell, Kimberly Wagner, Jonathan Williams, Mary Williamson, Qian Zeng, Tricia Zwiefelhofer, Clinton R. Paden, Duncan MacCannell |
| EPI_ISL_1925882, EPI_ISL_1925885, EPI_ISL_1925886, EPI_ISL_1925889 | Interpath Laboratory | Seattle Flu Study | Deborah A. Nickerson, Chris D. Frazar, Jover Lee, Benjamin Pelle, Erica Ryke, Matthew Richardson, Amanda Adler, Elisabeth Brandstetter, Peter D. Han, Kairsten Fay, Misja Ilcisin, Kirsten Lacombe, Thomas R. Sibley, Melissa Truong, Caitlin R. Wolf, Romesh Gautom, Geoff Melly, Brian Hiatt, Philip Dykema, Scott Lindquist, Michael Boeckh, Janet A. Englund, Michael Famulare, Barry R. Lutz, Mark J. Rieder, Lea M. Starita, Matthew Thompson, Helen Y. Chu, Jay Shendure, Trevor Bedford |
| EPI_ISL_1925890 | Laboratory Corporation of America | Centers for Disease Control and Prevention Division of Viral Diseases, Pathogen Discovery | Dakota Howard, Dhwani Batra, Peter W. Cook, Kara Moser, Adrian Paskey, Jason Caravas, Benjamin Rambo-Martin, Shatavia Morrison, Christopher Gulvick, Scott Sammons, Yvette Unoarumhi, Darlene Wagner, Matthew Schmerer, Minoo Agarwal, Eyad Almasri, Debbie Boles, Ayla Burns, Nuthawin Charoensri, Oren Cohen, Susan Countryman, Mary Ann Cristobal, Bobbi Croy, Suzanne Dale, Hrushikesh Deshmukh, Amanda Douglas, Vincent Drouillon, Marcia Eisenberg, Howard Engler, Rama Ghatti, Prashant Gupta, Susan Hicks, Jake Humphrey, Lax Iyer, Manoj Jain, Mohan Kolli, Brian Krueger, Tim Kuphal, Stanley Letovsky, Michael Levandoski, Craig Lukasik, Jonathan Meltzer, Brian Norvell, Mindy Nye, Scott Parker, Christos Petropoulos, John Pruitt, Steven Ragan, Scott Ryan, Mike Sapeta, Jana Schroth, Suresh Babu Selvaraju, Goran Stevovic, Amanda Suchanek, Andrea Throop, Lyndon Tilson, Thomas Urban, Joe Voshell, Kimberly Wagner, Jonathan Williams, Mary Williamson, Qian Zeng, Tricia Zwiefelhofer, Clinton R. Paden, Duncan MacCannell |
| EPI_ISL_1925892 | Interpath Laboratory | Seattle Flu Study | Deborah A. Nickerson, Chris D. Frazar, Jover Lee, Benjamin Pelle, Erica Ryke, Matthew Richardson, Amanda Adler, Elisabeth Brandstetter, Peter D. Han, Kairsten Fay, Misja Ilcisin, Kirsten Lacombe, Thomas R. Sibley, Melissa Truong, Caitlin R. Wolf, Romesh Gautom, Geoff Melly, Brian Hiatt, Philip Dykema, Scott Lindquist, Michael Boeckh, Janet A. Englund, Michael Famulare, Barry R. Lutz, Mark J. Rieder, Lea M. Starita, Matthew Thompson, Helen Y. Chu, Jay Shendure, Trevor Bedford |
| EPI_ISL_1925893 | Laboratory Corporation of America | Centers for Disease Control and Prevention Division of Viral Diseases, Pathogen Discovery | Dakota Howard, Dhwani Batra, Peter W. Cook, Kara Moser, Adrian Paskey, Jason Caravas, Benjamin Rambo-Martin, Shatavia Morrison, Christopher Gulvick, Scott Sammons, Yvette Unoarumhi, Darlene Wagner, Matthew Schmerer, Minoo Agarwal, Eyad Almasri, Debbie Boles, Ayla Burns, Nuthawin Charoensri, Oren Cohen, Susan Countryman, Mary Ann Cristobal, Bobbi Croy, Suzanne Dale, Hrushikesh Deshmukh, Amanda Douglas, Vincent Drouillon, Marcia Eisenberg, Howard Engler, Rama Ghatti, Prashant Gupta, Susan Hicks, Jake Humphrey, Lax Iyer, Manoj Jain, Mohan Kolli, Brian Krueger, Tim Kuphal, Stanley Letovsky, Michael Levandoski, Craig Lukasik, Jonathan Meltzer, Brian Norvell, Mindy Nye, Scott Parker, Christos Petropoulos, John Pruitt, Steven Ragan, Scott Ryan, Mike Sapeta, Jana Schroth, Suresh Babu Selvaraju, Goran Stevovic, Amanda Suchanek, Andrea Throop, Lyndon Tilson, Thomas Urban, Joe Voshell, Kimberly Wagner, Jonathan Williams, Mary Williamson, Qian Zeng, Tricia Zwiefelhofer, Clinton R. Paden, Duncan MacCannell |
| EPI_ISL_1925895, EPI_ISL_1925897 | Interpath Laboratory | Seattle Flu Study | Deborah A. Nickerson, Chris D. Frazar, Jover Lee, Benjamin Pelle, Erica Ryke, Matthew Richardson, Amanda Adler, Elisabeth Brandstetter, Peter D. Han, Kairsten Fay, Misja Ilcisin, Kirsten Lacombe, Thomas R. Sibley, Melissa Truong, Caitlin R. Wolf, Romesh Gautom, Geoff Melly, Brian Hiatt, Philip Dykema, Scott Lindquist, Michael Boeckh, Janet A. Englund, Michael Famulare, Barry R. Lutz, Mark J. Rieder, Lea M. Starita, Matthew Thompson, Helen Y. Chu, Jay Shendure, Trevor Bedford |
| EPI_ISL_1925899 | Laboratory Corporation of America | Centers for Disease Control and Prevention Division of Viral Diseases, Pathogen Discovery | Dakota Howard, Dhwani Batra, Peter W. Cook, Kara Moser, Adrian Paskey, Jason Caravas, Benjamin Rambo-Martin, Shatavia Morrison, Christopher Gulvick, Scott Sammons, Yvette Unoarumhi, Darlene Wagner, Matthew Schmerer, Minoo Agarwal, Eyad Almasri, Debbie Boles, Ayla Burns, Nuthawin Charoensri, Oren Cohen, Susan Countryman, Mary Ann Cristobal, Bobbi Croy, Suzanne Dale, Hrushikesh Deshmukh, Amanda Douglas, Vincent Drouillon, Marcia Eisenberg, Howard Engler, Rama Ghatti, Prashant Gupta, Susan Hicks, Jake Humphrey, Lax Iyer, Manoj Jain, Mohan Kolli, Brian Krueger, Tim Kuphal, Stanley Letovsky, Michael Levandoski, Craig Lukasik, Jonathan Meltzer, Brian Norvell, Mindy Nye, Scott Parker, Christos Petropoulos, John Pruitt, Steven Ragan, Scott Ryan, Mike Sapeta, Jana Schroth, Suresh Babu Selvaraju, Goran Stevovic, Amanda Suchanek, Andrea Throop, Lyndon Tilson, Thomas Urban, Joe Voshell, Kimberly Wagner, Jonathan Williams, Mary Williamson, Qian Zeng, Tricia Zwiefelhofer, Clinton R. Paden, Duncan MacCannell |
| EPI_ISL_1925900, EPI_ISL_1925903, EPI_ISL_1925904 | Interpath Laboratory | Seattle Flu Study | Deborah A. Nickerson, Chris D. Frazar, Jover Lee, Benjamin Pelle, Erica Ryke, Matthew Richardson, Amanda Adler, Elisabeth Brandstetter, Peter D. Han, Kairsten Fay, Misja Ilcisin, Kirsten Lacombe, Thomas R. Sibley, Melissa Truong, Caitlin R. Wolf, Romesh Gautom, Geoff Melly, Brian Hiatt, Philip Dykema, Scott Lindquist, Michael Boeckh, Janet A. Englund, Michael Famulare, Barry R. Lutz, Mark J. Rieder, Lea M. Starita, Matthew Thompson, Helen Y. Chu, Jay Shendure, Trevor Bedford |
| EPI_ISL_1925905 | Laboratory Corporation of America | Centers for Disease Control and Prevention Division of Viral Diseases, Pathogen Discovery | Dakota Howard, Dhwani Batra, Peter W. Cook, Kara Moser, Adrian Paskey, Jason Caravas, Benjamin Rambo-Martin, Shatavia Morrison, Christopher Gulvick, Scott Sammons, Yvette Unoarumhi, Darlene Wagner, Matthew Schmerer, Minoo Agarwal, Eyad Almasri, Debbie Boles, Ayla Burns, Nuthawin Charoensri, Oren Cohen, Susan Countryman, Mary Ann Cristobal, Bobbi Croy, Suzanne Dale, Hrushikesh Deshmukh, Amanda Douglas, Vincent Drouillon, Marcia Eisenberg, Howard Engler, Rama Ghatti, Prashant Gupta, Susan Hicks, Jake Humphrey, Lax Iyer, Manoj Jain, Mohan Kolli, Brian Krueger, Tim Kuphal, Stanley Letovsky, Michael Levandoski, Craig Lukasik, Jonathan Meltzer, Brian Norvell, Mindy Nye, Scott Parker, Christos Petropoulos, John Pruitt, Steven Ragan, Scott Ryan, Mike Sapeta, Jana Schroth, Suresh Babu Selvaraju, Goran Stevovic, Amanda Suchanek, Andrea Throop, Lyndon Tilson, Thomas Urban, Joe Voshell, Kimberly Wagner, Jonathan Williams, Mary Williamson, Qian Zeng, Tricia Zwiefelhofer, Clinton R. Paden, Duncan MacCannell |
| EPI_ISL_1925906, EPI_ISL_1925907 | Interpath Laboratory | Seattle Flu Study | Deborah A. Nickerson, Chris D. Frazar, Jover Lee, Benjamin Pelle, Erica Ryke, Matthew Richardson, Amanda Adler, Elisabeth Brandstetter, Peter D. Han, Kairsten Fay, Misja Ilcisin, Kirsten Lacombe, Thomas R. Sibley, Melissa Truong, Caitlin R. Wolf, Romesh Gautom, Geoff Melly, Brian Hiatt, Philip Dykema, Scott Lindquist, Michael Boeckh, Janet A. Englund, Michael Famulare, Barry R. Lutz, Mark J. Rieder, Lea M. Starita, Matthew Thompson, Helen Y. Chu, Jay Shendure, Trevor Bedford |
| EPI_ISL_1925908 | Laboratory Corporation of America | Centers for Disease Control and Prevention Division of Viral Diseases, Pathogen Discovery | Dakota Howard, Dhwani Batra, Peter W. Cook, Kara Moser, Adrian Paskey, Jason Caravas, Benjamin Rambo-Martin, Shatavia Morrison, Christopher Gulvick, Scott Sammons, Yvette Unoarumhi, Darlene Wagner, Matthew Schmerer, Minoo Agarwal, Eyad Almasri, Debbie Boles, Ayla Burns, Nuthawin Charoensri, Oren Cohen, Susan Countryman, Mary Ann Cristobal, Bobbi Croy, Suzanne Dale, Hrushikesh Deshmukh, Amanda Douglas, Vincent Drouillon, Marcia Eisenberg, Howard Engler, Rama Ghatti, Prashant Gupta, Susan Hicks, Jake Humphrey, Lax Iyer, Manoj Jain, Mohan Kolli, Brian Krueger, Tim Kuphal, Stanley Letovsky, Michael Levandoski, Craig Lukasik, Jonathan Meltzer, Brian Norvell, Mindy Nye, Scott Parker, Christos Petropoulos, John Pruitt, Steven Ragan, Scott Ryan, Mike Sapeta, Jana Schroth, Suresh Babu Selvaraju, Goran Stevovic, Amanda Suchanek, Andrea Throop, Lyndon Tilson, Thomas Urban, Joe Voshell, Kimberly Wagner, Jonathan Williams, Mary Williamson, Qian Zeng, Tricia Zwiefelhofer, Clinton R. Paden, Duncan MacCannell |
| EPI_ISL_1925909, EPI_ISL_1925911 | Interpath Laboratory | Seattle Flu Study | Deborah A. Nickerson, Chris D. Frazar, Jover Lee, Benjamin Pelle, Erica Ryke, Matthew Richardson, Amanda Adler, Elisabeth Brandstetter, Peter D. Han, Kairsten Fay, Misja Ilcisin, Kirsten Lacombe, Thomas R. Sibley, Melissa Truong, Caitlin R. Wolf, Romesh Gautom, Geoff Melly, Brian Hiatt, Philip Dykema, Scott Lindquist, Michael Boeckh, Janet A. Englund, Michael Famulare, Barry R. Lutz, Mark J. Rieder, Lea M. Starita, Matthew Thompson, Helen Y. Chu, Jay Shendure, Trevor Bedford |
| EPI_ISL_1925912 | Laboratory Corporation of America | Centers for Disease Control and Prevention Division of Viral Diseases, Pathogen Discovery | Dakota Howard, Dhwani Batra, Peter W. Cook, Kara Moser, Adrian Paskey, Jason Caravas, Benjamin Rambo-Martin, Shatavia Morrison, Christopher Gulvick, Scott Sammons, Yvette Unoarumhi, Darlene Wagner, Matthew Schmerer, Minoo Agarwal, Eyad Almasri, Debbie Boles, Ayla Burns, Nuthawin Charoensri, Oren Cohen, Susan Countryman, Mary Ann Cristobal, Bobbi Croy, Suzanne Dale, Hrushikesh Deshmukh, Amanda Douglas, Vincent Drouillon, Marcia Eisenberg, Howard Engler, Rama Ghatti, Prashant Gupta, Susan Hicks, Jake Humphrey, Lax Iyer, Manoj Jain, Mohan Kolli, Brian Krueger, Tim Kuphal, Stanley Letovsky, Michael Levandoski, Craig Lukasik, Jonathan Meltzer, Brian Norvell, Mindy Nye, Scott Parker, Christos Petropoulos, John Pruitt, Steven Ragan, Scott Ryan, Mike Sapeta, Jana Schroth, Suresh Babu Selvaraju, Goran Stevovic, Amanda Suchanek, Andrea Throop, Lyndon Tilson, Thomas Urban, Joe Voshell, Kimberly Wagner, Jonathan Williams, Mary Williamson, Qian Zeng, Tricia Zwiefelhofer, Clinton R. Paden, Duncan MacCannell |
| EPI_ISL_1925913, EPI_ISL_1925914 | Interpath Laboratory | Seattle Flu Study | Deborah A. Nickerson, Chris D. Frazar, Jover Lee, Benjamin Pelle, Erica Ryke, Matthew Richardson, Amanda Adler, Elisabeth Brandstetter, Peter D. Han, Kairsten Fay, Misja Ilcisin, Kirsten Lacombe, Thomas R. Sibley, Melissa Truong, Caitlin R. Wolf, Romesh Gautom, Geoff Melly, Brian Hiatt, Philip Dykema, Scott Lindquist, Michael Boeckh, Janet A. Englund, Michael Famulare, Barry R. Lutz, Mark J. Rieder, Lea M. Starita, Matthew Thompson, Helen Y. Chu, Jay Shendure, Trevor Bedford |
| EPI_ISL_1925915 | Laboratory Corporation of America | Centers for Disease Control and Prevention Division of Viral Diseases, Pathogen Discovery | Dakota Howard, Dhwani Batra, Peter W. Cook, Kara Moser, Adrian Paskey, Jason Caravas, Benjamin Rambo-Martin, Shatavia Morrison, Christopher Gulvick, Scott Sammons, Yvette Unoarumhi, Darlene Wagner, Matthew Schmerer, Minoo Agarwal, Eyad Almasri, Debbie Boles, Ayla Burns, Nuthawin Charoensri, Oren Cohen, Susan Countryman, Mary Ann Cristobal, Bobbi Croy, Suzanne Dale, Hrushikesh Deshmukh, Amanda Douglas, Vincent Drouillon, Marcia Eisenberg, Howard Engler, Rama Ghatti, Prashant Gupta, Susan Hicks, Jake Humphrey, Lax Iyer, Manoj Jain, Mohan Kolli, Brian Krueger, Tim Kuphal, Stanley Letovsky, Michael Levandoski, Craig Lukasik, Jonathan Meltzer, Brian Norvell, Mindy Nye, Scott Parker, Christos Petropoulos, John Pruitt, Steven Ragan, Scott Ryan, Mike Sapeta, Jana Schroth, Suresh Babu Selvaraju, Goran Stevovic, Amanda Suchanek, Andrea Throop, Lyndon Tilson, Thomas Urban, Joe Voshell, Kimberly Wagner, Jonathan Williams, Mary Williamson, Qian Zeng, Tricia Zwiefelhofer, Clinton R. Paden, Duncan MacCannell |
| EPI_ISL_1925916, EPI_ISL_1925917 | Interpath Laboratory | Seattle Flu Study | Deborah A. Nickerson, Chris D. Frazar, Jover Lee, Benjamin Pelle, Erica Ryke, Matthew Richardson, Amanda Adler, Elisabeth Brandstetter, Peter D. Han, Kairsten Fay, Misja Ilcisin, Kirsten Lacombe, Thomas R. Sibley, Melissa Truong, Caitlin R. Wolf, Romesh Gautom, Geoff Melly, Brian Hiatt, Philip Dykema, Scott Lindquist, Michael Boeckh, Janet A. Englund, Michael Famulare, Barry R. Lutz, Mark J. Rieder, Lea M. Starita, Matthew Thompson, Helen Y. Chu, Jay Shendure, Trevor Bedford |
| EPI_ISL_1925918 | Laboratory Corporation of America | Centers for Disease Control and Prevention Division of Viral Diseases, Pathogen Discovery | Dakota Howard, Dhwani Batra, Peter W. Cook, Kara Moser, Adrian Paskey, Jason Caravas, Benjamin Rambo-Martin, Shatavia Morrison, Christopher Gulvick, Scott Sammons, Yvette Unoarumhi, Darlene Wagner, Matthew Schmerer, Minoo Agarwal, Eyad Almasri, Debbie Boles, Ayla Burns, Nuthawin Charoensri, Oren Cohen, Susan Countryman, Mary Ann Cristobal, Bobbi Croy, Suzanne Dale, Hrushikesh Deshmukh, Amanda Douglas, Vincent Drouillon, Marcia Eisenberg, Howard Engler, Rama Ghatti, Prashant Gupta, Susan Hicks, Jake Humphrey, Lax Iyer, Manoj Jain, Mohan Kolli, Brian Krueger, Tim Kuphal, Stanley Letovsky, Michael Levandoski, Craig Lukasik, Jonathan Meltzer, Brian Norvell, Mindy Nye, Scott Parker, Christos Petropoulos, John Pruitt, Steven Ragan, Scott Ryan, Mike Sapeta, Jana Schroth, Suresh Babu Selvaraju, Goran Stevovic, Amanda Suchanek, Andrea Throop, Lyndon Tilson, Thomas Urban, Joe Voshell, Kimberly Wagner, Jonathan Williams, Mary Williamson, Qian Zeng, Tricia Zwiefelhofer, Clinton R. Paden, Duncan MacCannell |
| EPI_ISL_1925919, EPI_ISL_1925920 | Interpath Laboratory | Seattle Flu Study | Deborah A. Nickerson, Chris D. Frazar, Jover Lee, Benjamin Pelle, Erica Ryke, Matthew Richardson, Amanda Adler, Elisabeth Brandstetter, Peter D. Han, Kairsten Fay, Misja Ilcisin, Kirsten Lacombe, Thomas R. Sibley, Melissa Truong, Caitlin R. Wolf, Romesh Gautom, Geoff Melly, Brian Hiatt, Philip Dykema, Scott Lindquist, Michael Boeckh, Janet A. Englund, Michael Famulare, Barry R. Lutz, Mark J. Rieder, Lea M. Starita, Matthew Thompson, Helen Y. Chu, Jay Shendure, Trevor Bedford |
| EPI_ISL_1925921 | Laboratory Corporation of America | Centers for Disease Control and Prevention Division of Viral Diseases, Pathogen Discovery | Dakota Howard, Dhwani Batra, Peter W. Cook, Kara Moser, Adrian Paskey, Jason Caravas, Benjamin Rambo-Martin, Shatavia Morrison, Christopher Gulvick, Scott Sammons, Yvette Unoarumhi, Darlene Wagner, Matthew Schmerer, Minoo Agarwal, Eyad Almasri, Debbie Boles, Ayla Burns, Nuthawin Charoensri, Oren Cohen, Susan Countryman, Mary Ann Cristobal, Bobbi Croy, Suzanne Dale, Hrushikesh Deshmukh, Amanda Douglas, Vincent Drouillon, Marcia Eisenberg, Howard Engler, Rama Ghatti, Prashant Gupta, Susan Hicks, Jake Humphrey, Lax Iyer, Manoj Jain, Mohan Kolli, Brian Krueger, Tim Kuphal, Stanley Letovsky, Michael Levandoski, Craig Lukasik, Jonathan Meltzer, Brian Norvell, Mindy Nye, Scott Parker, Christos Petropoulos, John Pruitt, Steven Ragan, Scott Ryan, Mike Sapeta, Jana Schroth, Suresh Babu Selvaraju, Goran Stevovic, Amanda Suchanek, Andrea Throop, Lyndon Tilson, Thomas Urban, Joe Voshell, Kimberly Wagner, Jonathan Williams, Mary Williamson, Qian Zeng, Tricia Zwiefelhofer, Clinton R. Paden, Duncan MacCannell |
| EPI_ISL_1925922, EPI_ISL_1925923 | Interpath Laboratory | Seattle Flu Study | Deborah A. Nickerson, Chris D. Frazar, Jover Lee, Benjamin Pelle, Erica Ryke, Matthew Richardson, Amanda Adler, Elisabeth Brandstetter, Peter D. Han, Kairsten Fay, Misja Ilcisin, Kirsten Lacombe, Thomas R. Sibley, Melissa Truong, Caitlin R. Wolf, Romesh Gautom, Geoff Melly, Brian Hiatt, Philip Dykema, Scott Lindquist, Michael Boeckh, Janet A. Englund, Michael Famulare, Barry R. Lutz, Mark J. Rieder, Lea M. Starita, Matthew Thompson, Helen Y. Chu, Jay Shendure, Trevor Bedford |
| EPI_ISL_1925924 | Laboratory Corporation of America | Centers for Disease Control and Prevention Division of Viral Diseases, Pathogen Discovery | Dakota Howard, Dhwani Batra, Peter W. Cook, Kara Moser, Adrian Paskey, Jason Caravas, Benjamin Rambo-Martin, Shatavia Morrison, Christopher Gulvick, Scott Sammons, Yvette Unoarumhi, Darlene Wagner, Matthew Schmerer, Minoo Agarwal, Eyad Almasri, Debbie Boles, Ayla Burns, Nuthawin Charoensri, Oren Cohen, Susan Countryman, Mary Ann Cristobal, Bobbi Croy, Suzanne Dale, Hrushikesh Deshmukh, Amanda Douglas, Vincent Drouillon, Marcia Eisenberg, Howard Engler, Rama Ghatti, Prashant Gupta, Susan Hicks, Jake Humphrey, Lax Iyer, Manoj Jain, Mohan Kolli, Brian Krueger, Tim Kuphal, Stanley Letovsky, Michael Levandoski, Craig Lukasik, Jonathan Meltzer, Brian Norvell, Mindy Nye, Scott Parker, Christos Petropoulos, John Pruitt, Steven Ragan, Scott Ryan, Mike Sapeta, Jana Schroth, Suresh Babu Selvaraju, Goran Stevovic, Amanda Suchanek, Andrea Throop, Lyndon Tilson, Thomas Urban, Joe Voshell, Kimberly Wagner, Jonathan Williams, Mary Williamson, Qian Zeng, Tricia Zwiefelhofer, Clinton R. Paden, Duncan MacCannell |
| EPI_ISL_1925926 | Interpath Laboratory | Seattle Flu Study | Deborah A. Nickerson, Chris D. Frazar, Jover Lee, Benjamin Pelle, Erica Ryke, Matthew Richardson, Amanda Adler, Elisabeth Brandstetter, Peter D. Han, Kairsten Fay, Misja Ilcisin, Kirsten Lacombe, Thomas R. Sibley, Melissa Truong, Caitlin R. Wolf, Romesh Gautom, Geoff Melly, Brian Hiatt, Philip Dykema, Scott Lindquist, Michael Boeckh, Janet A. Englund, Michael Famulare, Barry R. Lutz, Mark J. Rieder, Lea M. Starita, Matthew Thompson, Helen Y. Chu, Jay Shendure, Trevor Bedford |
| EPI_ISL_1925927 | Laboratory Corporation of America | Centers for Disease Control and Prevention Division of Viral Diseases, Pathogen Discovery | Dakota Howard, Dhwani Batra, Peter W. Cook, Kara Moser, Adrian Paskey, Jason Caravas, Benjamin Rambo-Martin, Shatavia Morrison, Christopher Gulvick, Scott Sammons, Yvette Unoarumhi, Darlene Wagner, Matthew Schmerer, Minoo Agarwal, Eyad Almasri, Debbie Boles, Ayla Burns, Nuthawin Charoensri, Oren Cohen, Susan Countryman, Mary Ann Cristobal, Bobbi Croy, Suzanne Dale, Hrushikesh Deshmukh, Amanda Douglas, Vincent Drouillon, Marcia Eisenberg, Howard Engler, Rama Ghatti, Prashant Gupta, Susan Hicks, Jake Humphrey, Lax Iyer, Manoj Jain, Mohan Kolli, Brian Krueger, Tim Kuphal, Stanley Letovsky, Michael Levandoski, Craig Lukasik, Jonathan Meltzer, Brian Norvell, Mindy Nye, Scott Parker, Christos Petropoulos, John Pruitt, Steven Ragan, Scott Ryan, Mike Sapeta, Jana Schroth, Suresh Babu Selvaraju, Goran Stevovic, Amanda Suchanek, Andrea Throop, Lyndon Tilson, Thomas Urban, Joe Voshell, Kimberly Wagner, Jonathan Williams, Mary Williamson, Qian Zeng, Tricia Zwiefelhofer, Clinton R. Paden, Duncan MacCannell |
| EPI_ISL_1925928, EPI_ISL_1925929 | Interpath Laboratory | Seattle Flu Study | Deborah A. Nickerson, Chris D. Frazar, Jover Lee, Benjamin Pelle, Erica Ryke, Matthew Richardson, Amanda Adler, Elisabeth Brandstetter, Peter D. Han, Kairsten Fay, Misja Ilcisin, Kirsten Lacombe, Thomas R. Sibley, Melissa Truong, Caitlin R. Wolf, Romesh Gautom, Geoff Melly, Brian Hiatt, Philip Dykema, Scott Lindquist, Michael Boeckh, Janet A. Englund, Michael Famulare, Barry R. Lutz, Mark J. Rieder, Lea M. Starita, Matthew Thompson, Helen Y. Chu, Jay Shendure, Trevor Bedford |
| EPI_ISL_1925930, EPI_ISL_1925933 | Laboratory Corporation of America | Centers for Disease Control and Prevention Division of Viral Diseases, Pathogen Discovery | Dakota Howard, Dhwani Batra, Peter W. Cook, Kara Moser, Adrian Paskey, Jason Caravas, Benjamin Rambo-Martin, Shatavia Morrison, Christopher Gulvick, Scott Sammons, Yvette Unoarumhi, Darlene Wagner, Matthew Schmerer, Minoo Agarwal, Eyad Almasri, Debbie Boles, Ayla Burns, Nuthawin Charoensri, Oren Cohen, Susan Countryman, Mary Ann Cristobal, Bobbi Croy, Suzanne Dale, Hrushikesh Deshmukh, Amanda Douglas, Vincent Drouillon, Marcia Eisenberg, Howard Engler, Rama Ghatti, Prashant Gupta, Susan Hicks, Jake Humphrey, Lax Iyer, Manoj Jain, Mohan Kolli, Brian Krueger, Tim Kuphal, Stanley Letovsky, Michael Levandoski, Craig Lukasik, Jonathan Meltzer, Brian Norvell, Mindy Nye, Scott Parker, Christos Petropoulos, John Pruitt, Steven Ragan, Scott Ryan, Mike Sapeta, Jana Schroth, Suresh Babu Selvaraju, Goran Stevovic, Amanda Suchanek, Andrea Throop, Lyndon Tilson, Thomas Urban, Joe Voshell, Kimberly Wagner, Jonathan Williams, Mary Williamson, Qian Zeng, Tricia Zwiefelhofer, Clinton R. Paden, Duncan MacCannell |

| | | | |
|---|---|---|---|
| EPI_ISL_1925934, EPI_ISL_1925935 | Interpath Laboratory | Seattle Flu Study | Deborah A. Nickerson, Chris D. Frazar, Jover Lee, Benjamin Pelle, Erica Ryke, Matthew Richardson, Amanda Adler, Elisabeth Brandstetter, Peter D. Han, Kairsten Fay, Misja Ilcisin, Kirsten Lacombe, Thomas R. Sibley, Melissa Truong, Caitlin R. Wolf, Romesh Gautom, Geoff Melly, Brian Hiatt, Philip Dykema, Scott Lindquist, Michael Boeckh, Janet A. Englund, Michael Famulare, Barry R. Lutz, Mark J. Rieder, Lea M. Starita, Matthew Thompson, Helen Y. Chu, Jay Shendure, Trevor Bedford |
| EPI_ISL_1925936 | Laboratory Corporation of America | Centers for Disease Control and Prevention Division of Viral Diseases, Pathogen Discovery | Dakota Howard, Dhwani Batra, Peter W. Cook, Kara Moser, Adrian Paskey, Jason Caravas, Benjamin Rambo-Martin, Shatavia Morrison, Christopher Gulvick, Scott Sammons, Yvette Unoarumhi, Darlene Wagner, Matthew Schmerer, Minoo Agarwal, Eyad Almasri, Debbie Boles, Ayla Burns, Nuthawin Charoensri, Oren Cohen, Susan Countryman, Mary Ann Cristobal, Bobbi Croy, Suzanne Dale, Hrushikesh Deshmukh, Amanda Douglas, Vincent Drouillon, Marcia Eisenberg, Howard Engler, Rama Ghatti, Prashant Gupta, Susan Hicks, Jake Humphrey, Lax Iyer, Manoj Jain, Mohan Kolli, Brian Krueger, Tim Kuphal, Stanley Letovsky, Michael Levandoski, Craig Lukasik, Jonathan Meltzer, Brian Norvell, Mindy Nye, Scott Parker, Christos Petropoulos, John Pruitt, Steven Ragan, Scott Ryan, Mike Sapeta, Jana Schroth, Suresh Babu Selvaraju, Goran Stevovic, Amanda Suchanek, Andrea Throop, Lyndon Tilson, Thomas Urban, Joe Voshell, Kimberly Wagner, Jonathan Williams, Mary Williamson, Qian Zeng, Tricia Zwiefelhofer, Clinton R. Paden, Duncan MacCannell |
| EPI_ISL_1925937, EPI_ISL_1925938 | Interpath Laboratory | Seattle Flu Study | Deborah A. Nickerson, Chris D. Frazar, Jover Lee, Benjamin Pelle, Erica Ryke, Matthew Richardson, Amanda Adler, Elisabeth Brandstetter, Peter D. Han, Kairsten Fay, Misja Ilcisin, Kirsten Lacombe, Thomas R. Sibley, Melissa Truong, Caitlin R. Wolf, Romesh Gautom, Geoff Melly, Brian Hiatt, Philip Dykema, Scott Lindquist, Michael Boeckh, Janet A. Englund, Michael Famulare, Barry R. Lutz, Mark J. Rieder, Lea M. Starita, Matthew Thompson, Helen Y. Chu, Jay Shendure, Trevor Bedford |
| EPI_ISL_1925939 | Laboratory Corporation of America | Centers for Disease Control and Prevention Division of Viral Diseases, Pathogen Discovery | Dakota Howard, Dhwani Batra, Peter W. Cook, Kara Moser, Adrian Paskey, Jason Caravas, Benjamin Rambo-Martin, Shatavia Morrison, Christopher Gulvick, Scott Sammons, Yvette Unoarumhi, Darlene Wagner, Matthew Schmerer, Minoo Agarwal, Eyad Almasri, Debbie Boles, Ayla Burns, Nuthawin Charoensri, Oren Cohen, Susan Countryman, Mary Ann Cristobal, Bobbi Croy, Suzanne Dale, Hrushikesh Deshmukh, Amanda Douglas, Vincent Drouillon, Marcia Eisenberg, Howard Engler, Rama Ghatti, Prashant Gupta, Susan Hicks, Jake Humphrey, Lax Iyer, Manoj Jain, Mohan Kolli, Brian Krueger, Tim Kuphal, Stanley Letovsky, Michael Levandoski, Craig Lukasik, Jonathan Meltzer, Brian Norvell, Mindy Nye, Scott Parker, Christos Petropoulos, John Pruitt, Steven Ragan, Scott Ryan, Mike Sapeta, Jana Schroth, Suresh Babu Selvaraju, Goran Stevovic, Amanda Suchanek, Andrea Throop, Lyndon Tilson, Thomas Urban, Joe Voshell, Kimberly Wagner, Jonathan Williams, Mary Williamson, Qian Zeng, Tricia Zwiefelhofer, Clinton R. Paden, Duncan MacCannell |
| EPI_ISL_1925941, EPI_ISL_1925943, EPI_ISL_1925944 | Interpath Laboratory | Seattle Flu Study | Deborah A. Nickerson, Chris D. Frazar, Jover Lee, Benjamin Pelle, Erica Ryke, Matthew Richardson, Amanda Adler, Elisabeth Brandstetter, Peter D. Han, Kairsten Fay, Misja Ilcisin, Kirsten Lacombe, Thomas R. Sibley, Melissa Truong, Caitlin R. Wolf, Romesh Gautom, Geoff Melly, Brian Hiatt, Philip Dykema, Scott Lindquist, Michael Boeckh, Janet A. Englund, Michael Famulare, Barry R. Lutz, Mark J. Rieder, Lea M. Starita, Matthew Thompson, Helen Y. Chu, Jay Shendure, Trevor Bedford |
| EPI_ISL_1925948 | Laboratory Corporation of America | Centers for Disease Control and Prevention Division of Viral Diseases, Pathogen Discovery | Dakota Howard, Dhwani Batra, Peter W. Cook, Kara Moser, Adrian Paskey, Jason Caravas, Benjamin Rambo-Martin, Shatavia Morrison, Christopher Gulvick, Scott Sammons, Yvette Unoarumhi, Darlene Wagner, Matthew Schmerer, Minoo Agarwal, Eyad Almasri, Debbie Boles, Ayla Burns, Nuthawin Charoensri, Oren Cohen, Susan Countryman, Mary Ann Cristobal, Bobbi Croy, Suzanne Dale, Hrushikesh Deshmukh, Amanda Douglas, Vincent Drouillon, Marcia Eisenberg, Howard Engler, Rama Ghatti, Prashant Gupta, Susan Hicks, Jake Humphrey, Lax Iyer, Manoj Jain, Mohan Kolli, Brian Krueger, Tim Kuphal, Stanley Letovsky, Michael Levandoski, Craig Lukasik, Jonathan Meltzer, Brian Norvell, Mindy Nye, Scott Parker, Christos Petropoulos, John Pruitt, Steven Ragan, Scott Ryan, Mike Sapeta, Jana Schroth, Suresh Babu Selvaraju, Goran Stevovic, Amanda Suchanek, Andrea Throop, Lyndon Tilson, Thomas Urban, Joe Voshell, Kimberly Wagner, Jonathan Williams, Mary Williamson, Qian Zeng, Tricia Zwiefelhofer, Clinton R. Paden, Duncan MacCannell |
| EPI_ISL_1925950, EPI_ISL_1925951, EPI_ISL_1925953, EPI_ISL_1925954 | Interpath Laboratory | Seattle Flu Study | Deborah A. Nickerson, Chris D. Frazar, Jover Lee, Benjamin Pelle, Erica Ryke, Matthew Richardson, Amanda Adler, Elisabeth Brandstetter, Peter D. Han, Kairsten Fay, Misja Ilcisin, Kirsten Lacombe, Thomas R. Sibley, Melissa Truong, Caitlin R. Wolf, Romesh Gautom, Geoff Melly, Brian Hiatt, Philip Dykema, Scott Lindquist, Michael Boeckh, Janet A. Englund, Michael Famulare, Barry R. Lutz, Mark J. Rieder, Lea M. Starita, Matthew Thompson, Helen Y. Chu, Jay Shendure, Trevor Bedford |
| EPI_ISL_1925955 | Laboratory Corporation of America | Centers for Disease Control and Prevention Division of Viral Diseases, Pathogen Discovery | Dakota Howard, Dhwani Batra, Peter W. Cook, Kara Moser, Adrian Paskey, Jason Caravas, Benjamin Rambo-Martin, Shatavia Morrison, Christopher Gulvick, Scott Sammons, Yvette Unoarumhi, Darlene Wagner, Matthew Schmerer, Minoo Agarwal, Eyad Almasri, Debbie Boles, Ayla Burns, Nuthawin Charoensri, Oren Cohen, Susan Countryman, Mary Ann Cristobal, Bobbi Croy, Suzanne Dale, Hrushikesh Deshmukh, Amanda Douglas, Vincent Drouillon, Marcia Eisenberg, Howard Engler, Rama Ghatti, Prashant Gupta, Susan Hicks, Jake Humphrey, Lax Iyer, Manoj Jain, Mohan Kolli, Brian Krueger, Tim Kuphal, Stanley Letovsky, Michael Levandoski, Craig Lukasik, Jonathan Meltzer, Brian Norvell, Mindy Nye, Scott Parker, Christos Petropoulos, John Pruitt, Steven Ragan, Scott Ryan, Mike Sapeta, Jana Schroth, Suresh Babu Selvaraju, Goran Stevovic, Amanda Suchanek, Andrea Throop, Lyndon Tilson, Thomas Urban, Joe Voshell, Kimberly Wagner, Jonathan Williams, Mary Williamson, Qian Zeng, Tricia Zwiefelhofer, Clinton R. Paden, Duncan MacCannell |
| EPI_ISL_1925957 | Interpath Laboratory | Seattle Flu Study | Deborah A. Nickerson, Chris D. Frazar, Jover Lee, Benjamin Pelle, Erica Ryke, Matthew Richardson, Amanda Adler, Elisabeth Brandstetter, Peter D. Han, Kairsten Fay, Misja Ilcisin, Kirsten Lacombe, Thomas R. Sibley, Melissa Truong, Caitlin R. Wolf, Romesh Gautom, Geoff Melly, Brian Hiatt, Philip Dykema, Scott Lindquist, Michael Boeckh, Janet A. Englund, Michael Famulare, Barry R. Lutz, Mark J. Rieder, Lea M. Starita, Matthew Thompson, Helen Y. Chu, Jay Shendure, Trevor Bedford |
| EPI_ISL_1925958 | Laboratory Corporation of America | Centers for Disease Control and Prevention Division of Viral Diseases, Pathogen Discovery | Dakota Howard, Dhwani Batra, Peter W. Cook, Kara Moser, Adrian Paskey, Jason Caravas, Benjamin Rambo-Martin, Shatavia Morrison, Christopher Gulvick, Scott Sammons, Yvette Unoarumhi, Darlene Wagner, Matthew Schmerer, Minoo Agarwal, Eyad Almasri, Debbie Boles, Ayla Burns, Nuthawin Charoensri, Oren Cohen, Susan Countryman, Mary Ann Cristobal, Bobbi Croy, Suzanne Dale, Hrushikesh Deshmukh, Amanda Douglas, Vincent Drouillon, Marcia Eisenberg, Howard Engler, Rama Ghatti, Prashant Gupta, Susan Hicks, Jake Humphrey, Lax Iyer, Manoj Jain, Mohan Kolli, Brian Krueger, Tim Kuphal, Stanley Letovsky, Michael Levandoski, Craig Lukasik, Jonathan Meltzer, Brian Norvell, Mindy Nye, Scott Parker, Christos Petropoulos, John Pruitt, Steven Ragan, Scott Ryan, Mike Sapeta, Jana Schroth, Suresh Babu Selvaraju, Goran Stevovic, Amanda Suchanek, Andrea Throop, Lyndon Tilson, Thomas Urban, Joe Voshell, Kimberly Wagner, Jonathan Williams, Mary Williamson, Qian Zeng, Tricia Zwiefelhofer, Clinton R. Paden, Duncan MacCannell |
| EPI_ISL_1925960 | Interpath Laboratory | Seattle Flu Study | Deborah A. Nickerson, Chris D. Frazar, Jover Lee, Benjamin Pelle, Erica Ryke, Matthew Richardson, Amanda Adler, Elisabeth Brandstetter, Peter D. Han, Kairsten Fay, Misja Ilcisin, Kirsten Lacombe, Thomas R. Sibley, Melissa Truong, Caitlin R. Wolf, Romesh Gautom, Geoff Melly, Brian Hiatt, Philip Dykema, Scott Lindquist, Michael Boeckh, Janet A. Englund, Michael Famulare, Barry R. Lutz, Mark J. Rieder, Lea M. Starita, Matthew Thompson, Helen Y. Chu, Jay Shendure, Trevor Bedford |
| EPI_ISL_1925961 | Laboratory Corporation of America | Centers for Disease Control and Prevention Division of Viral Diseases, Pathogen Discovery | Dakota Howard, Dhwani Batra, Peter W. Cook, Kara Moser, Adrian Paskey, Jason Caravas, Benjamin Rambo-Martin, Shatavia Morrison, Christopher Gulvick, Scott Sammons, Yvette Unoarumhi, Darlene Wagner, Matthew Schmerer, Minoo Agarwal, Eyad Almasri, Debbie Boles, Ayla Burns, Nuthawin Charoensri, Oren Cohen, Susan Countryman, Mary Ann Cristobal, Bobbi Croy, Suzanne Dale, Hrushikesh Deshmukh, Amanda Douglas, Vincent Drouillon, Marcia Eisenberg, Howard Engler, Rama Ghatti, Prashant Gupta, Susan Hicks, Jake Humphrey, Lax Iyer, Manoj Jain, Mohan Kolli, Brian Krueger, Tim Kuphal, Stanley Letovsky, Michael Levandoski, Craig Lukasik, Jonathan Meltzer, Brian Norvell, Mindy Nye, Scott Parker, Christos Petropoulos, John Pruitt, Steven Ragan, Scott Ryan, Mike Sapeta, Jana Schroth, Suresh Babu Selvaraju, Goran Stevovic, Amanda Suchanek, Andrea Throop, Lyndon Tilson, Thomas Urban, Joe Voshell, Kimberly Wagner, Jonathan Williams, Mary Williamson, Qian Zeng, Tricia Zwiefelhofer, Clinton R. Paden, Duncan MacCannell |
| EPI_ISL_1925962, EPI_ISL_1925963, EPI_ISL_1925965, EPI_ISL_1925966 | Interpath Laboratory | Seattle Flu Study | Deborah A. Nickerson, Chris D. Frazar, Jover Lee, Benjamin Pelle, Erica Ryke, Matthew Richardson, Amanda Adler, Elisabeth Brandstetter, Peter D. Han, Kairsten Fay, Misja Ilcisin, Kirsten Lacombe, Thomas R. Sibley, Melissa Truong, Caitlin R. Wolf, Romesh Gautom, Geoff Melly, Brian Hiatt, Philip Dykema, Scott Lindquist, Michael Boeckh, Janet A. Englund, Michael Famulare, Barry R. Lutz, Mark J. Rieder, Lea M. Starita, Matthew Thompson, Helen Y. Chu, Jay Shendure, Trevor Bedford |
| EPI_ISL_1925967 | Laboratory Corporation of America | Centers for Disease Control and Prevention Division of Viral Diseases, Pathogen Discovery | Dakota Howard, Dhwani Batra, Peter W. Cook, Kara Moser, Adrian Paskey, Jason Caravas, Benjamin Rambo-Martin, Shatavia Morrison, Christopher Gulvick, Scott Sammons, Yvette Unoarumhi, Darlene Wagner, Matthew Schmerer, Minoo Agarwal, Eyad Almasri, Debbie Boles, Ayla Burns, Nuthawin Charoensri, Oren Cohen, Susan Countryman, Mary Ann Cristobal, Bobbi Croy, Suzanne Dale, Hrushikesh Deshmukh, Amanda Douglas, Vincent Drouillon, Marcia Eisenberg, Howard Engler, Rama Ghatti, Prashant Gupta, Susan Hicks, Jake Humphrey, Lax Iyer, Manoj Jain, Mohan Kolli, Brian Krueger, Tim Kuphal, Stanley Letovsky, Michael Levandoski, Craig Lukasik, Jonathan Meltzer, Brian Norvell, Mindy Nye, Scott Parker, Christos Petropoulos, John Pruitt, Steven Ragan, Scott Ryan, Mike Sapeta, Jana Schroth, Suresh Babu Selvaraju, Goran Stevovic, Amanda Suchanek, Andrea Throop, Lyndon Tilson, Thomas Urban, Joe Voshell, Kimberly Wagner, Jonathan Williams, Mary Williamson, Qian Zeng, Tricia Zwiefelhofer, Clinton R. Paden, Duncan MacCannell |
| EPI_ISL_1925968, EPI_ISL_1925969 | Interpath Laboratory | Seattle Flu Study | Deborah A. Nickerson, Chris D. Frazar, Jover Lee, Benjamin Pelle, Erica Ryke, Matthew Richardson, Amanda Adler, Elisabeth Brandstetter, Peter D. Han, Kairsten Fay, Misja Ilcisin, Kirsten Lacombe, Thomas R. Sibley, Melissa Truong, Caitlin R. Wolf, Romesh Gautom, Geoff Melly, Brian Hiatt, Philip Dykema, Scott Lindquist, Michael Boeckh, Janet A. Englund, Michael Famulare, Barry R. Lutz, Mark J. Rieder, Lea M. Starita, Matthew Thompson, Helen Y. Chu, Jay Shendure, Trevor Bedford |
| EPI_ISL_1925970, EPI_ISL_1925974 | Laboratory Corporation of America | Centers for Disease Control and Prevention Division of Viral Diseases, Pathogen Discovery | Dakota Howard, Dhwani Batra, Peter W. Cook, Kara Moser, Adrian Paskey, Jason Caravas, Benjamin Rambo-Martin, Shatavia Morrison, Christopher Gulvick, Scott Sammons, Yvette Unoarumhi, Darlene Wagner, Matthew Schmerer, Minoo Agarwal, Eyad Almasri, Debbie Boles, Ayla Burns, Nuthawin Charoensri, Oren Cohen, Susan Countryman, Mary Ann Cristobal, Bobbi Croy, Suzanne Dale, Hrushikesh Deshmukh, Amanda Douglas, Vincent Drouillon, Marcia Eisenberg, Howard Engler, Rama Ghatti, Prashant Gupta, Susan Hicks, Jake Humphrey, Lax Iyer, Manoj Jain, Mohan Kolli, Brian Krueger, Tim Kuphal, Stanley Letovsky, Michael Levandoski, Craig Lukasik, Jonathan Meltzer, Brian Norvell, Mindy Nye, Scott Parker, Christos Petropoulos, John Pruitt, Steven Ragan, Scott Ryan, Mike Sapeta, Jana Schroth, Suresh Babu Selvaraju, Goran Stevovic, Amanda Suchanek, Andrea Throop, Lyndon Tilson, Thomas Urban, Joe Voshell, Kimberly Wagner, Jonathan Williams, Mary Williamson, Qian Zeng, Tricia Zwiefelhofer, Clinton R. Paden, Duncan MacCannell |
| EPI_ISL_1925976 | Northwest Laboratories | Seattle Flu Study | Deborah A. Nickerson, Chris D. Frazar, Jover Lee, Benjamin Pelle, Erica Ryke, Matthew Richardson, Amanda Adler, Elisabeth Brandstetter, Peter D. Han, Kairsten Fay, Misja Ilcisin, Kirsten Lacombe, Thomas R. Sibley, Melissa Truong, Caitlin R. Wolf, Romesh Gautom, Geoff Melly, Brian Hiatt, Philip Dykema, Scott Lindquist, Michael Boeckh, Janet A. Englund, Michael Famulare, Barry R. Lutz, Mark J. Rieder, Lea M. Starita, Matthew Thompson, Helen Y. Chu, Jay Shendure, Trevor Bedford |
| EPI_ISL_1925977 | Laboratory Corporation of America | Centers for Disease Control and Prevention Division of Viral Diseases, Pathogen Discovery | Dakota Howard, Dhwani Batra, Peter W. Cook, Kara Moser, Adrian Paskey, Jason Caravas, Benjamin Rambo-Martin, Shatavia Morrison, Christopher Gulvick, Scott Sammons, Yvette Unoarumhi, Darlene Wagner, Matthew Schmerer, Minoo Agarwal, Eyad Almasri, Debbie Boles, Ayla Burns, Nuthawin Charoensri, Oren Cohen, Susan Countryman, Mary Ann Cristobal, Bobbi Croy, Suzanne Dale, Hrushikesh Deshmukh, Amanda Douglas, Vincent Drouillon, Marcia Eisenberg, Howard Engler, Rama Ghatti, Prashant Gupta, Susan Hicks, Jake Humphrey, Lax Iyer, Manoj Jain, Mohan Kolli, Brian Krueger, Tim Kuphal, Stanley Letovsky, Michael Levandoski, Craig Lukasik, Jonathan Meltzer, Brian Norvell, Mindy Nye, Scott Parker, Christos Petropoulos, John Pruitt, Steven Ragan, Scott Ryan, Mike Sapeta, Jana Schroth, Suresh Babu Selvaraju, Goran Stevovic, Amanda Suchanek, Andrea Throop, Lyndon Tilson, Thomas Urban, Joe Voshell, Kimberly Wagner, Jonathan Williams, Mary Williamson, Qian Zeng, Tricia Zwiefelhofer, Clinton R. Paden, Duncan MacCannell |
| EPI_ISL_1925978, EPI_ISL_1925979 | Northwest Laboratories | Seattle Flu Study | Deborah A. Nickerson, Chris D. Frazar, Jover Lee, Benjamin Pelle, Erica Ryke, Matthew Richardson, Amanda Adler, Elisabeth Brandstetter, Peter D. Han, Kairsten Fay, Misja Ilcisin, Kirsten Lacombe, Thomas R. Sibley, Melissa Truong, Caitlin R. Wolf, Romesh Gautom, Geoff Melly, Brian Hiatt, Philip Dykema, Scott Lindquist, Michael Boeckh, Janet A. Englund, Michael Famulare, Barry R. Lutz, Mark J. Rieder, Lea M. Starita, Matthew Thompson, Helen Y. Chu, Jay Shendure, Trevor Bedford |
| EPI_ISL_1925980 | Laboratory Corporation of America | Centers for Disease Control and Prevention Division of Viral Diseases, Pathogen Discovery | Dakota Howard, Dhwani Batra, Peter W. Cook, Kara Moser, Adrian Paskey, Jason Caravas, Benjamin Rambo-Martin, Shatavia Morrison, Christopher Gulvick, Scott Sammons, Yvette Unoarumhi, Darlene Wagner, Matthew Schmerer, Minoo Agarwal, Eyad Almasri, Debbie Boles, Ayla Burns, Nuthawin Charoensri, Oren Cohen, Susan Countryman, Mary Ann Cristobal, Bobbi Croy, Suzanne Dale, Hrushikesh Deshmukh, Amanda Douglas, Vincent Drouillon, Marcia Eisenberg, Howard Engler, Rama Ghatti, Prashant Gupta, Susan Hicks, Jake Humphrey, Lax Iyer, Manoj Jain, Mohan Kolli, Brian Krueger, Tim Kuphal, Stanley Letovsky, Michael Levandoski, Craig Lukasik, Jonathan Meltzer, Brian Norvell, Mindy Nye, Scott Parker, Christos Petropoulos, John Pruitt, Steven Ragan, Scott Ryan, Mike Sapeta, Jana Schroth, Suresh Babu Selvaraju, Goran Stevovic, Amanda Suchanek, Andrea Throop, Lyndon Tilson, Thomas Urban, Joe Voshell, Kimberly Wagner, Jonathan Williams, Mary Williamson, Qian Zeng, Tricia Zwiefelhofer, Clinton R. Paden, Duncan MacCannell |
| EPI_ISL_1925981, EPI_ISL_1925982, EPI_ISL_1925985 | Northwest Laboratories | Seattle Flu Study | Deborah A. Nickerson, Chris D. Frazar, Jover Lee, Benjamin Pelle, Erica Ryke, Matthew Richardson, Amanda Adler, Elisabeth Brandstetter, Peter D. Han, Kairsten Fay, Misja Ilcisin, Kirsten Lacombe, Thomas R. Sibley, Melissa Truong, Caitlin R. Wolf, Romesh Gautom, Geoff Melly, Brian Hiatt, Philip Dykema, Scott Lindquist, Michael Boeckh, Janet A. Englund, Michael Famulare, Barry R. Lutz, Mark J. Rieder, Lea M. Starita, Matthew Thompson, Helen Y. Chu, Jay Shendure, Trevor Bedford |
| EPI_ISL_1925986 | Laboratory Corporation of America | Centers for Disease Control and Prevention Division of Viral Diseases, Pathogen Discovery | Dakota Howard, Dhwani Batra, Peter W. Cook, Kara Moser, Adrian Paskey, Jason Caravas, Benjamin Rambo-Martin, Shatavia Morrison, Christopher Gulvick, Scott Sammons, Yvette Unoarumhi, Darlene Wagner, Matthew Schmerer, Minoo Agarwal, Eyad Almasri, Debbie Boles, Ayla Burns, Nuthawin Charoensri, Oren Cohen, Susan Countryman, Mary Ann Cristobal, Bobbi Croy, Suzanne Dale, Hrushikesh Deshmukh, Amanda Douglas, Vincent Drouillon, Marcia Eisenberg, Howard Engler, Rama Ghatti, Prashant Gupta, Susan Hicks, Jake Humphrey, Lax Iyer, Manoj Jain, Mohan Kolli, Brian Krueger, Tim Kuphal, Stanley Letovsky, Michael Levandoski, Craig Lukasik, Jonathan Meltzer, Brian Norvell, Mindy Nye, Scott Parker, Christos Petropoulos, John Pruitt, Steven Ragan, Scott Ryan, Mike Sapeta, Jana Schroth, Suresh Babu Selvaraju, Goran Stevovic, Amanda Suchanek, Andrea Throop, Lyndon Tilson, Thomas Urban, Joe Voshell, Kimberly Wagner, Jonathan Williams, Mary Williamson, Qian Zeng, Tricia Zwiefelhofer, Clinton R. Paden, Duncan MacCannell |
| EPI_ISL_1925987 | Northwest Laboratories | Seattle Flu Study | Deborah A. Nickerson, Chris D. Frazar, Jover Lee, Benjamin Pelle, Erica Ryke, Matthew Richardson, Amanda Adler, Elisabeth Brandstetter, Peter D. Han, Kairsten Fay, Misja Ilcisin, Kirsten Lacombe, Thomas R. Sibley, Melissa Truong, Caitlin R. Wolf, Romesh Gautom, Geoff Melly, Brian Hiatt, Philip Dykema, Scott Lindquist, Michael Boeckh, Janet A. Englund, Michael Famulare, Barry R. Lutz, Mark J. Rieder, Lea M. Starita, Matthew Thompson, Helen Y. Chu, Jay Shendure, Trevor Bedford |
| EPI_ISL_1925989 | Laboratory Corporation of America | Centers for Disease Control and Prevention Division of Viral Diseases, Pathogen Discovery | Dakota Howard, Dhwani Batra, Peter W. Cook, Kara Moser, Adrian Paskey, Jason Caravas, Benjamin Rambo-Martin, Shatavia Morrison, Christopher Gulvick, Scott Sammons, Yvette Unoarumhi, Darlene Wagner, Matthew Schmerer, Minoo Agarwal, Eyad Almasri, Debbie Boles, Ayla Burns, Nuthawin Charoensri, Oren Cohen, Susan Countryman, Mary Ann Cristobal, Bobbi Croy, Suzanne Dale, Hrushikesh Deshmukh, Amanda Douglas, Vincent Drouillon, Marcia Eisenberg, Howard Engler, Rama Ghatti, Prashant Gupta, Susan Hicks, Jake Humphrey, Lax Iyer, Manoj Jain, Mohan Kolli, Brian Krueger, Tim Kuphal, Stanley Letovsky, Michael Levandoski, Craig Lukasik, Jonathan Meltzer, Brian Norvell, Mindy Nye, Scott Parker, Christos Petropoulos, John Pruitt, Steven Ragan, Scott Ryan, Mike Sapeta, Jana Schroth, Suresh Babu Selvaraju, Goran Stevovic, Amanda Suchanek, Andrea Throop, Lyndon Tilson, Thomas Urban, Joe Voshell, Kimberly Wagner, Jonathan Williams, Mary Williamson, Qian Zeng, Tricia Zwiefelhofer, Clinton R. Paden, Duncan MacCannell |
| EPI_ISL_1925990, EPI_ISL_1925991 | Northwest Laboratories | Seattle Flu Study | Deborah A. Nickerson, Chris D. Frazar, Jover Lee, Benjamin Pelle, Erica Ryke, Matthew Richardson, Amanda Adler, Elisabeth Brandstetter, Peter D. Han, Kairsten Fay, Misja Ilcisin, Kirsten Lacombe, Thomas R. Sibley, Melissa Truong, Caitlin R. Wolf, Romesh Gautom, Geoff Melly, Brian Hiatt, Philip Dykema, Scott Lindquist, Michael Boeckh, Janet A. Englund, Michael Famulare, Barry R. Lutz, Mark J. Rieder, Lea M. Starita, Matthew Thompson, Helen Y. Chu, Jay Shendure, Trevor Bedford |
| EPI_ISL_1925992 | Laboratory Corporation of America | Centers for Disease Control and Prevention Division of Viral Diseases, Pathogen Discovery | Dakota Howard, Dhwani Batra, Peter W. Cook, Kara Moser, Adrian Paskey, Jason Caravas, Benjamin Rambo-Martin, Shatavia Morrison, Christopher Gulvick, Scott Sammons, Yvette Unoarumhi, Darlene Wagner, Matthew Schmerer, Minoo Agarwal, Eyad Almasri, Debbie Boles, Ayla Burns, Nuthawin Charoensri, Oren Cohen, Susan Countryman, Mary Ann Cristobal, Bobbi Croy, Suzanne Dale, Hrushikesh Deshmukh, Amanda Douglas, Vincent Drouillon, Marcia Eisenberg, Howard Engler, Rama Ghatti, Prashant Gupta, Susan Hicks, Jake Humphrey, Lax Iyer, Manoj Jain, Mohan Kolli, Brian Krueger, Tim Kuphal, Stanley Letovsky, Michael Levandoski, Craig Lukasik, Jonathan Meltzer, Brian Norvell, Mindy Nye, Scott Parker, Christos Petropoulos, John Pruitt, Steven Ragan, Scott Ryan, Mike Sapeta, Jana Schroth, Suresh Babu Selvaraju, Goran Stevovic, Amanda Suchanek, Andrea Throop, Lyndon Tilson, Thomas Urban, Joe Voshell, Kimberly Wagner, Jonathan Williams, Mary Williamson, Qian Zeng, Tricia Zwiefelhofer, Clinton R. Paden, Duncan MacCannell |
| EPI_ISL_1925993, EPI_ISL_1925994 | Northwest Laboratories | Seattle Flu Study | Deborah A. Nickerson, Chris D. Frazar, Jover Lee, Benjamin Pelle, Erica Ryke, Matthew Richardson, Amanda Adler, Elisabeth Brandstetter, Peter D. Han, Kairsten Fay, Misja Ilcisin, Kirsten Lacombe, Thomas R. Sibley, Melissa Truong, Caitlin R. Wolf, Romesh Gautom, Geoff Melly, Brian Hiatt, Philip Dykema, Scott Lindquist, Michael Boeckh, Janet A. Englund, Michael Famulare, Barry R. Lutz, Mark J. Rieder, Lea M. Starita, Matthew Thompson, Helen Y. Chu, Jay Shendure, Trevor Bedford |
| EPI_ISL_1925995, EPI_ISL_1925998 | Laboratory Corporation of America | Centers for Disease Control and Prevention Division of Viral Diseases, Pathogen Discovery | Dakota Howard, Dhwani Batra, Peter W. Cook, Kara Moser, Adrian Paskey, Jason Caravas, Benjamin Rambo-Martin, Shatavia Morrison, Christopher Gulvick, Scott Sammons, Yvette Unoarumhi, Darlene Wagner, Matthew Schmerer, Minoo Agarwal, Eyad Almasri, Debbie Boles, Ayla Burns, Nuthawin Charoensri, Oren Cohen, Susan Countryman, Mary Ann Cristobal, Bobbi Croy, Suzanne Dale, Hrushikesh Deshmukh, Amanda Douglas, Vincent Drouillon, Marcia Eisenberg, Howard Engler, Rama Ghatti, Prashant Gupta, Susan Hicks, Jake Humphrey, Lax Iyer, Manoj Jain, Mohan Kolli, Brian Krueger, Tim Kuphal, Stanley Letovsky, Michael Levandoski, Craig Lukasik, Jonathan Meltzer, Brian Norvell, Mindy Nye, Scott Parker, Christos Petropoulos, John Pruitt, Steven Ragan, Scott Ryan, Mike Sapeta, Jana Schroth, Suresh Babu Selvaraju, Goran Stevovic, Amanda Suchanek, Andrea Throop, Lyndon Tilson, Thomas Urban, Joe Voshell, Kimberly Wagner, Jonathan Williams, Mary Williamson, Qian Zeng, Tricia Zwiefelhofer, Clinton R. Paden, Duncan MacCannell |
| EPI_ISL_1925999 | Northwest Laboratories | Seattle Flu Study | Deborah A. Nickerson, Chris D. Frazar, Jover Lee, Benjamin Pelle, Erica Ryke, Matthew Richardson, Amanda Adler, Elisabeth Brandstetter, Peter D. Han, Kairsten Fay, Misja Ilcisin, Kirsten Lacombe, Thomas R. Sibley, Melissa Truong, Caitlin R. Wolf, Romesh Gautom, Geoff Melly, Brian Hiatt, Philip Dykema, Scott Lindquist, Michael Boeckh, Janet A. Englund, Michael Famulare, Barry R. Lutz, Mark J. Rieder, Lea M. Starita, Matthew Thompson, Helen Y. Chu, Jay Shendure, Trevor Bedford |
| EPI_ISL_1926005 | Laboratory Corporation of America | Centers for Disease Control and Prevention Division of Viral Diseases, Pathogen Discovery | Dakota Howard, Dhwani Batra, Peter W. Cook, Kara Moser, Adrian Paskey, Jason Caravas, Benjamin Rambo-Martin, Shatavia Morrison, Christopher Gulvick, Scott Sammons, Yvette Unoarumhi, Darlene Wagner, Matthew Schmerer, Minoo Agarwal, Eyad Almasri, Debbie Boles, Ayla Burns, Nuthawin Charoensri, Oren Cohen, Susan Countryman, Mary Ann Cristobal, Bobbi Croy, Suzanne Dale, Hrushikesh Deshmukh, Amanda Douglas, Vincent Drouillon, Marcia Eisenberg, Howard Engler, Rama Ghatti, Prashant Gupta, Susan Hicks, Jake Humphrey, Lax Iyer, Manoj Jain, Mohan Kolli, Brian Krueger, Tim Kuphal, Stanley Letovsky, Michael Levandoski, Craig Lukasik, Jonathan Meltzer, Brian Norvell, Mindy Nye, Scott Parker, Christos Petropoulos, John Pruitt, Steven Ragan, Scott Ryan, Mike Sapeta, Jana Schroth, Suresh Babu Selvaraju, Goran Stevovic, Amanda Suchanek, Andrea Throop, Lyndon Tilson, Thomas Urban, Joe Voshell, Kimberly Wagner, Jonathan Williams, Mary Williamson, Qian Zeng, Tricia Zwiefelhofer, Clinton R. Paden, Duncan MacCannell |

| | | | |
|---|---|---|---|
| EPI_ISL_1926007 | Northwest Laboratories | Seattle Flu Study | Deborah A. Nickerson, Chris D. Frazar, Jover Lee, Benjamin Pelle, Erica Ryke, Matthew Richardson, Amanda Adler, Elisabeth Brandstetter, Peter D. Han, Kairsten Fay, Misja Ilcisin, Kirsten Lacombe, Thomas R. Sibley, Melissa Truong, Caitlin R. Wolf, Romesh Gautom, Geoff Melly, Brian Hiatt, Philip Dykema, Scott Lindquist, Michael Boeckh, Janet A. Englund, Michael Famulare, Barry R. Lutz, Mark J. Rieder, Lea M. Starita, Matthew Thompson, Helen Y. Chu, Jay Shendure, Trevor Bedford |
| EPI_ISL_1926008 | Laboratory Corporation of America | Centers for Disease Control and Prevention Division of Viral Diseases, Pathogen Discovery | Dakota Howard, Dhwani Batra, Peter W. Cook, Kara Moser, Adrian Paskey, Jason Caravas, Benjamin Rambo-Martin, Shatavia Morrison, Christopher Gulvick, Scott Sammons, Yvette Unoarumhi, Darlene Wagner, Matthew Schmerer, Minoo Agarwal, Eyad Almasri, Debbie Boles, Ayla Burns, Nuthawin Charoensri, Oren Cohen, Susan Countryman, Mary Ann Cristobal, Bobbi Croy, Suzanne Dale, Hrushikesh Deshmukh, Amanda Douglas, Vincent Drouillon, Marcia Eisenberg, Howard Engler, Rama Ghatti, Prashant Gupta, Susan Hicks, Jake Humphrey, Lax Iyer, Manoj Jain, Mohan Kolli, Brian Krueger, Tim Kuphal, Stanley Letovsky, Michael Levandoski, Craig Lukasik, Jonathan Meltzer, Brian Norvell, Mindy Nye, Scott Parker, Christos Petropoulos, John Pruitt, Steven Ragan, Scott Ryan, Mike Sapeta, Jana Schroth, Suresh Babu Selvaraju, Goran Stevovic, Amanda Suchanek, Andrea Throop, Lyndon Tilson, Thomas Urban, Joe Voshell, Kimberly Wagner, Jonathan Williams, Mary Williamson, Qian Zeng, Tricia Zwiefelhofer, Clinton R. Paden, Duncan MacCannell |
| EPI_ISL_1926010, EPI_ISL_1926011 | Northwest Laboratories | Seattle Flu Study | Deborah A. Nickerson, Chris D. Frazar, Jover Lee, Benjamin Pelle, Erica Ryke, Matthew Richardson, Amanda Adler, Elisabeth Brandstetter, Peter D. Han, Kairsten Fay, Misja Ilcisin, Kirsten Lacombe, Thomas R. Sibley, Melissa Truong, Caitlin R. Wolf, Romesh Gautom, Geoff Melly, Brian Hiatt, Philip Dykema, Scott Lindquist, Michael Boeckh, Janet A. Englund, Michael Famulare, Barry R. Lutz, Mark J. Rieder, Lea M. Starita, Matthew Thompson, Helen Y. Chu, Jay Shendure, Trevor Bedford |
| EPI_ISL_1926012 | Laboratory Corporation of America | Centers for Disease Control and Prevention Division of Viral Diseases, Pathogen Discovery | Dakota Howard, Dhwani Batra, Peter W. Cook, Kara Moser, Adrian Paskey, Jason Caravas, Benjamin Rambo-Martin, Shatavia Morrison, Christopher Gulvick, Scott Sammons, Yvette Unoarumhi, Darlene Wagner, Matthew Schmerer, Minoo Agarwal, Eyad Almasri, Debbie Boles, Ayla Burns, Nuthawin Charoensri, Oren Cohen, Susan Countryman, Mary Ann Cristobal, Bobbi Croy, Suzanne Dale, Hrushikesh Deshmukh, Amanda Douglas, Vincent Drouillon, Marcia Eisenberg, Howard Engler, Rama Ghatti, Prashant Gupta, Susan Hicks, Jake Humphrey, Lax Iyer, Manoj Jain, Mohan Kolli, Brian Krueger, Tim Kuphal, Stanley Letovsky, Michael Levandoski, Craig Lukasik, Jonathan Meltzer, Brian Norvell, Mindy Nye, Scott Parker, Christos Petropoulos, John Pruitt, Steven Ragan, Scott Ryan, Mike Sapeta, Jana Schroth, Suresh Babu Selvaraju, Goran Stevovic, Amanda Suchanek, Andrea Throop, Lyndon Tilson, Thomas Urban, Joe Voshell, Kimberly Wagner, Jonathan Williams, Mary Williamson, Qian Zeng, Tricia Zwiefelhofer, Clinton R. Paden, Duncan MacCannell |
| EPI_ISL_1926013 | Northwest Laboratories | Seattle Flu Study | Deborah A. Nickerson, Chris D. Frazar, Jover Lee, Benjamin Pelle, Erica Ryke, Matthew Richardson, Amanda Adler, Elisabeth Brandstetter, Peter D. Han, Kairsten Fay, Misja Ilcisin, Kirsten Lacombe, Thomas R. Sibley, Melissa Truong, Caitlin R. Wolf, Romesh Gautom, Geoff Melly, Brian Hiatt, Philip Dykema, Scott Lindquist, Michael Boeckh, Janet A. Englund, Michael Famulare, Barry R. Lutz, Mark J. Rieder, Lea M. Starita, Matthew Thompson, Helen Y. Chu, Jay Shendure, Trevor Bedford |
| EPI_ISL_1926015, EPI_ISL_1926018 | Laboratory Corporation of America | Centers for Disease Control and Prevention Division of Viral Diseases, Pathogen Discovery | Dakota Howard, Dhwani Batra, Peter W. Cook, Kara Moser, Adrian Paskey, Jason Caravas, Benjamin Rambo-Martin, Shatavia Morrison, Christopher Gulvick, Scott Sammons, Yvette Unoarumhi, Darlene Wagner, Matthew Schmerer, Minoo Agarwal, Eyad Almasri, Debbie Boles, Ayla Burns, Nuthawin Charoensri, Oren Cohen, Susan Countryman, Mary Ann Cristobal, Bobbi Croy, Suzanne Dale, Hrushikesh Deshmukh, Amanda Douglas, Vincent Drouillon, Marcia Eisenberg, Howard Engler, Rama Ghatti, Prashant Gupta, Susan Hicks, Jake Humphrey, Lax Iyer, Manoj Jain, Mohan Kolli, Brian Krueger, Tim Kuphal, Stanley Letovsky, Michael Levandoski, Craig Lukasik, Jonathan Meltzer, Brian Norvell, Mindy Nye, Scott Parker, Christos Petropoulos, John Pruitt, Steven Ragan, Scott Ryan, Mike Sapeta, Jana Schroth, Suresh Babu Selvaraju, Goran Stevovic, Amanda Suchanek, Andrea Throop, Lyndon Tilson, Thomas Urban, Joe Voshell, Kimberly Wagner, Jonathan Williams, Mary Williamson, Qian Zeng, Tricia Zwiefelhofer, Clinton R. Paden, Duncan MacCannell |
| EPI_ISL_1926019 | Northwest Laboratories | Seattle Flu Study | Deborah A. Nickerson, Chris D. Frazar, Jover Lee, Benjamin Pelle, Erica Ryke, Matthew Richardson, Amanda Adler, Elisabeth Brandstetter, Peter D. Han, Kairsten Fay, Misja Ilcisin, Kirsten Lacombe, Thomas R. Sibley, Melissa Truong, Caitlin R. Wolf, Romesh Gautom, Geoff Melly, Brian Hiatt, Philip Dykema, Scott Lindquist, Michael Boeckh, Janet A. Englund, Michael Famulare, Barry R. Lutz, Mark J. Rieder, Lea M. Starita, Matthew Thompson, Helen Y. Chu, Jay Shendure, Trevor Bedford |
| EPI_ISL_1926021 | Laboratory Corporation of America | Centers for Disease Control and Prevention Division of Viral Diseases, Pathogen Discovery | Dakota Howard, Dhwani Batra, Peter W. Cook, Kara Moser, Adrian Paskey, Jason Caravas, Benjamin Rambo-Martin, Shatavia Morrison, Christopher Gulvick, Scott Sammons, Yvette Unoarumhi, Darlene Wagner, Matthew Schmerer, Minoo Agarwal, Eyad Almasri, Debbie Boles, Ayla Burns, Nuthawin Charoensri, Oren Cohen, Susan Countryman, Mary Ann Cristobal, Bobbi Croy, Suzanne Dale, Hrushikesh Deshmukh, Amanda Douglas, Vincent Drouillon, Marcia Eisenberg, Howard Engler, Rama Ghatti, Prashant Gupta, Susan Hicks, Jake Humphrey, Lax Iyer, Manoj Jain, Mohan Kolli, Brian Krueger, Tim Kuphal, Stanley Letovsky, Michael Levandoski, Craig Lukasik, Jonathan Meltzer, Brian Norvell, Mindy Nye, Scott Parker, Christos Petropoulos, John Pruitt, Steven Ragan, Scott Ryan, Mike Sapeta, Jana Schroth, Suresh Babu Selvaraju, Goran Stevovic, Amanda Suchanek, Andrea Throop, Lyndon Tilson, Thomas Urban, Joe Voshell, Kimberly Wagner, Jonathan Williams, Mary Williamson, Qian Zeng, Tricia Zwiefelhofer, Clinton R. Paden, Duncan MacCannell |
| EPI_ISL_1926022, EPI_ISL_1926023 | Northwest Laboratories | Seattle Flu Study | Deborah A. Nickerson, Chris D. Frazar, Jover Lee, Benjamin Pelle, Erica Ryke, Matthew Richardson, Amanda Adler, Elisabeth Brandstetter, Peter D. Han, Kairsten Fay, Misja Ilcisin, Kirsten Lacombe, Thomas R. Sibley, Melissa Truong, Caitlin R. Wolf, Romesh Gautom, Geoff Melly, Brian Hiatt, Philip Dykema, Scott Lindquist, Michael Boeckh, Janet A. Englund, Michael Famulare, Barry R. Lutz, Mark J. Rieder, Lea M. Starita, Matthew Thompson, Helen Y. Chu, Jay Shendure, Trevor Bedford |
| EPI_ISL_1926024 | Laboratory Corporation of America | Centers for Disease Control and Prevention Division of Viral Diseases, Pathogen Discovery | Dakota Howard, Dhwani Batra, Peter W. Cook, Kara Moser, Adrian Paskey, Jason Caravas, Benjamin Rambo-Martin, Shatavia Morrison, Christopher Gulvick, Scott Sammons, Yvette Unoarumhi, Darlene Wagner, Matthew Schmerer, Minoo Agarwal, Eyad Almasri, Debbie Boles, Ayla Burns, Nuthawin Charoensri, Oren Cohen, Susan Countryman, Mary Ann Cristobal, Bobbi Croy, Suzanne Dale, Hrushikesh Deshmukh, Amanda Douglas, Vincent Drouillon, Marcia Eisenberg, Howard Engler, Rama Ghatti, Prashant Gupta, Susan Hicks, Jake Humphrey, Lax Iyer, Manoj Jain, Mohan Kolli, Brian Krueger, Tim Kuphal, Stanley Letovsky, Michael Levandoski, Craig Lukasik, Jonathan Meltzer, Brian Norvell, Mindy Nye, Scott Parker, Christos Petropoulos, John Pruitt, Steven Ragan, Scott Ryan, Mike Sapeta, Jana Schroth, Suresh Babu Selvaraju, Goran Stevovic, Amanda Suchanek, Andrea Throop, Lyndon Tilson, Thomas Urban, Joe Voshell, Kimberly Wagner, Jonathan Williams, Mary Williamson, Qian Zeng, Tricia Zwiefelhofer, Clinton R. Paden, Duncan MacCannell |
| EPI_ISL_1926025, EPI_ISL_1926026 | Northwest Laboratories | Seattle Flu Study | Deborah A. Nickerson, Chris D. Frazar, Jover Lee, Benjamin Pelle, Erica Ryke, Matthew Richardson, Amanda Adler, Elisabeth Brandstetter, Peter D. Han, Kairsten Fay, Misja Ilcisin, Kirsten Lacombe, Thomas R. Sibley, Melissa Truong, Caitlin R. Wolf, Romesh Gautom, Geoff Melly, Brian Hiatt, Philip Dykema, Scott Lindquist, Michael Boeckh, Janet A. Englund, Michael Famulare, Barry R. Lutz, Mark J. Rieder, Lea M. Starita, Matthew Thompson, Helen Y. Chu, Jay Shendure, Trevor Bedford |
| EPI_ISL_1926027 | Laboratory Corporation of America | Centers for Disease Control and Prevention Division of Viral Diseases, Pathogen Discovery | Dakota Howard, Dhwani Batra, Peter W. Cook, Kara Moser, Adrian Paskey, Jason Caravas, Benjamin Rambo-Martin, Shatavia Morrison, Christopher Gulvick, Scott Sammons, Yvette Unoarumhi, Darlene Wagner, Matthew Schmerer, Minoo Agarwal, Eyad Almasri, Debbie Boles, Ayla Burns, Nuthawin Charoensri, Oren Cohen, Susan Countryman, Mary Ann Cristobal, Bobbi Croy, Suzanne Dale, Hrushikesh Deshmukh, Amanda Douglas, Vincent Drouillon, Marcia Eisenberg, Howard Engler, Rama Ghatti, Prashant Gupta, Susan Hicks, Jake Humphrey, Lax Iyer, Manoj Jain, Mohan Kolli, Brian Krueger, Tim Kuphal, Stanley Letovsky, Michael Levandoski, Craig Lukasik, Jonathan Meltzer, Brian Norvell, Mindy Nye, Scott Parker, Christos Petropoulos, John Pruitt, Steven Ragan, Scott Ryan, Mike Sapeta, Jana Schroth, Suresh Babu Selvaraju, Goran Stevovic, Amanda Suchanek, Andrea Throop, Lyndon Tilson, Thomas Urban, Joe Voshell, Kimberly Wagner, Jonathan Williams, Mary Williamson, Qian Zeng, Tricia Zwiefelhofer, Clinton R. Paden, Duncan MacCannell |
| EPI_ISL_1926028, EPI_ISL_1926029 | Northwest Laboratories | Seattle Flu Study | Deborah A. Nickerson, Chris D. Frazar, Jover Lee, Benjamin Pelle, Erica Ryke, Matthew Richardson, Amanda Adler, Elisabeth Brandstetter, Peter D. Han, Kairsten Fay, Misja Ilcisin, Kirsten Lacombe, Thomas R. Sibley, Melissa Truong, Caitlin R. Wolf, Romesh Gautom, Geoff Melly, Brian Hiatt, Philip Dykema, Scott Lindquist, Michael Boeckh, Janet A. Englund, Michael Famulare, Barry R. Lutz, Mark J. Rieder, Lea M. Starita, Matthew Thompson, Helen Y. Chu, Jay Shendure, Trevor Bedford |
| EPI_ISL_1926030 | Laboratory Corporation of America | Centers for Disease Control and Prevention Division of Viral Diseases, Pathogen Discovery | Dakota Howard, Dhwani Batra, Peter W. Cook, Kara Moser, Adrian Paskey, Jason Caravas, Benjamin Rambo-Martin, Shatavia Morrison, Christopher Gulvick, Scott Sammons, Yvette Unoarumhi, Darlene Wagner, Matthew Schmerer, Minoo Agarwal, Eyad Almasri, Debbie Boles, Ayla Burns, Nuthawin Charoensri, Oren Cohen, Susan Countryman, Mary Ann Cristobal, Bobbi Croy, Suzanne Dale, Hrushikesh Deshmukh, Amanda Douglas, Vincent Drouillon, Marcia Eisenberg, Howard Engler, Rama Ghatti, Prashant Gupta, Susan Hicks, Jake Humphrey, Lax Iyer, Manoj Jain, Mohan Kolli, Brian Krueger, Tim Kuphal, Stanley Letovsky, Michael Levandoski, Craig Lukasik, Jonathan Meltzer, Brian Norvell, Mindy Nye, Scott Parker, Christos Petropoulos, John Pruitt, Steven Ragan, Scott Ryan, Mike Sapeta, Jana Schroth, Suresh Babu Selvaraju, Goran Stevovic, Amanda Suchanek, Andrea Throop, Lyndon Tilson, Thomas Urban, Joe Voshell, Kimberly Wagner, Jonathan Williams, Mary Williamson, Qian Zeng, Tricia Zwiefelhofer, Clinton R. Paden, Duncan MacCannell |
| EPI_ISL_1926031, EPI_ISL_1926032 | Northwest Laboratories | Seattle Flu Study | Deborah A. Nickerson, Chris D. Frazar, Jover Lee, Benjamin Pelle, Erica Ryke, Matthew Richardson, Amanda Adler, Elisabeth Brandstetter, Peter D. Han, Kairsten Fay, Misja Ilcisin, Kirsten Lacombe, Thomas R. Sibley, Melissa Truong, Caitlin R. Wolf, Romesh Gautom, Geoff Melly, Brian Hiatt, Philip Dykema, Scott Lindquist, Michael Boeckh, Janet A. Englund, Michael Famulare, Barry R. Lutz, Mark J. Rieder, Lea M. Starita, Matthew Thompson, Helen Y. Chu, Jay Shendure, Trevor Bedford |
| EPI_ISL_1926033 | Laboratory Corporation of America | Centers for Disease Control and Prevention Division of Viral Diseases, Pathogen Discovery | Dakota Howard, Dhwani Batra, Peter W. Cook, Kara Moser, Adrian Paskey, Jason Caravas, Benjamin Rambo-Martin, Shatavia Morrison, Christopher Gulvick, Scott Sammons, Yvette Unoarumhi, Darlene Wagner, Matthew Schmerer, Minoo Agarwal, Eyad Almasri, Debbie Boles, Ayla Burns, Nuthawin Charoensri, Oren Cohen, Susan Countryman, Mary Ann Cristobal, Bobbi Croy, Suzanne Dale, Hrushikesh Deshmukh, Amanda Douglas, Vincent Drouillon, Marcia Eisenberg, Howard Engler, Rama Ghatti, Prashant Gupta, Susan Hicks, Jake Humphrey, Lax Iyer, Manoj Jain, Mohan Kolli, Brian Krueger, Tim Kuphal, Stanley Letovsky, Michael Levandoski, Craig Lukasik, Jonathan Meltzer, Brian Norvell, Mindy Nye, Scott Parker, Christos Petropoulos, John Pruitt, Steven Ragan, Scott Ryan, Mike Sapeta, Jana Schroth, Suresh Babu Selvaraju, Goran Stevovic, Amanda Suchanek, Andrea Throop, Lyndon Tilson, Thomas Urban, Joe Voshell, Kimberly Wagner, Jonathan Williams, Mary Williamson, Qian Zeng, Tricia Zwiefelhofer, Clinton R. Paden, Duncan MacCannell |
| EPI_ISL_1926034 | Northwest Laboratories | Seattle Flu Study | Deborah A. Nickerson, Chris D. Frazar, Jover Lee, Benjamin Pelle, Erica Ryke, Matthew Richardson, Amanda Adler, Elisabeth Brandstetter, Peter D. Han, Kairsten Fay, Misja Ilcisin, Kirsten Lacombe, Thomas R. Sibley, Melissa Truong, Caitlin R. Wolf, Romesh Gautom, Geoff Melly, Brian Hiatt, Philip Dykema, Scott Lindquist, Michael Boeckh, Janet A. Englund, Michael Famulare, Barry R. Lutz, Mark J. Rieder, Lea M. Starita, Matthew Thompson, Helen Y. Chu, Jay Shendure, Trevor Bedford |
| EPI_ISL_1926036 | Laboratory Corporation of America | Centers for Disease Control and Prevention Division of Viral Diseases, Pathogen Discovery | Dakota Howard, Dhwani Batra, Peter W. Cook, Kara Moser, Adrian Paskey, Jason Caravas, Benjamin Rambo-Martin, Shatavia Morrison, Christopher Gulvick, Scott Sammons, Yvette Unoarumhi, Darlene Wagner, Matthew Schmerer, Minoo Agarwal, Eyad Almasri, Debbie Boles, Ayla Burns, Nuthawin Charoensri, Oren Cohen, Susan Countryman, Mary Ann Cristobal, Bobbi Croy, Suzanne Dale, Hrushikesh Deshmukh, Amanda Douglas, Vincent Drouillon, Marcia Eisenberg, Howard Engler, Rama Ghatti, Prashant Gupta, Susan Hicks, Jake Humphrey, Lax Iyer, Manoj Jain, Mohan Kolli, Brian Krueger, Tim Kuphal, Stanley Letovsky, Michael Levandoski, Craig Lukasik, Jonathan Meltzer, Brian Norvell, Mindy Nye, Scott Parker, Christos Petropoulos, John Pruitt, Steven Ragan, Scott Ryan, Mike Sapeta, Jana Schroth, Suresh Babu Selvaraju, Goran Stevovic, Amanda Suchanek, Andrea Throop, Lyndon Tilson, Thomas Urban, Joe Voshell, Kimberly Wagner, Jonathan Williams, Mary Williamson, Qian Zeng, Tricia Zwiefelhofer, Clinton R. Paden, Duncan MacCannell |
| EPI_ISL_1926037, EPI_ISL_1926038 | Northwest Laboratories | Seattle Flu Study | Deborah A. Nickerson, Chris D. Frazar, Jover Lee, Benjamin Pelle, Erica Ryke, Matthew Richardson, Amanda Adler, Elisabeth Brandstetter, Peter D. Han, Kairsten Fay, Misja Ilcisin, Kirsten Lacombe, Thomas R. Sibley, Melissa Truong, Caitlin R. Wolf, Romesh Gautom, Geoff Melly, Brian Hiatt, Philip Dykema, Scott Lindquist, Michael Boeckh, Janet A. Englund, Michael Famulare, Barry R. Lutz, Mark J. Rieder, Lea M. Starita, Matthew Thompson, Helen Y. Chu, Jay Shendure, Trevor Bedford |
| EPI_ISL_1926039, EPI_ISL_1926042 | Laboratory Corporation of America | Centers for Disease Control and Prevention Division of Viral Diseases, Pathogen Discovery | Dakota Howard, Dhwani Batra, Peter W. Cook, Kara Moser, Adrian Paskey, Jason Caravas, Benjamin Rambo-Martin, Shatavia Morrison, Christopher Gulvick, Scott Sammons, Yvette Unoarumhi, Darlene Wagner, Matthew Schmerer, Minoo Agarwal, Eyad Almasri, Debbie Boles, Ayla Burns, Nuthawin Charoensri, Oren Cohen, Susan Countryman, Mary Ann Cristobal, Bobbi Croy, Suzanne Dale, Hrushikesh Deshmukh, Amanda Douglas, Vincent Drouillon, Marcia Eisenberg, Howard Engler, Rama Ghatti, Prashant Gupta, Susan Hicks, Jake Humphrey, Lax Iyer, Manoj Jain, Mohan Kolli, Brian Krueger, Tim Kuphal, Stanley Letovsky, Michael Levandoski, Craig Lukasik, Jonathan Meltzer, Brian Norvell, Mindy Nye, Scott Parker, Christos Petropoulos, John Pruitt, Steven Ragan, Scott Ryan, Mike Sapeta, Jana Schroth, Suresh Babu Selvaraju, Goran Stevovic, Amanda Suchanek, Andrea Throop, Lyndon Tilson, Thomas Urban, Joe Voshell, Kimberly Wagner, Jonathan Williams, Mary Williamson, Qian Zeng, Tricia Zwiefelhofer, Clinton R. Paden, Duncan MacCannell |
| EPI_ISL_1926043, EPI_ISL_1926044 | Northwest Laboratories | Seattle Flu Study | Deborah A. Nickerson, Chris D. Frazar, Jover Lee, Benjamin Pelle, Erica Ryke, Matthew Richardson, Amanda Adler, Elisabeth Brandstetter, Peter D. Han, Kairsten Fay, Misja Ilcisin, Kirsten Lacombe, Thomas R. Sibley, Melissa Truong, Caitlin R. Wolf, Romesh Gautom, Geoff Melly, Brian Hiatt, Philip Dykema, Scott Lindquist, Michael Boeckh, Janet A. Englund, Michael Famulare, Barry R. Lutz, Mark J. Rieder, Lea M. Starita, Matthew Thompson, Helen Y. Chu, Jay Shendure, Trevor Bedford |
| EPI_ISL_1926045, EPI_ISL_1926048 | Laboratory Corporation of America | Centers for Disease Control and Prevention Division of Viral Diseases, Pathogen Discovery | Dakota Howard, Dhwani Batra, Peter W. Cook, Kara Moser, Adrian Paskey, Jason Caravas, Benjamin Rambo-Martin, Shatavia Morrison, Christopher Gulvick, Scott Sammons, Yvette Unoarumhi, Darlene Wagner, Matthew Schmerer, Minoo Agarwal, Eyad Almasri, Debbie Boles, Ayla Burns, Nuthawin Charoensri, Oren Cohen, Susan Countryman, Mary Ann Cristobal, Bobbi Croy, Suzanne Dale, Hrushikesh Deshmukh, Amanda Douglas, Vincent Drouillon, Marcia Eisenberg, Howard Engler, Rama Ghatti, Prashant Gupta, Susan Hicks, Jake Humphrey, Lax Iyer, Manoj Jain, Mohan Kolli, Brian Krueger, Tim Kuphal, Stanley Letovsky, Michael Levandoski, Craig Lukasik, Jonathan Meltzer, Brian Norvell, Mindy Nye, Scott Parker, Christos Petropoulos, John Pruitt, Steven Ragan, Scott Ryan, Mike Sapeta, Jana Schroth, Suresh Babu Selvaraju, Goran Stevovic, Amanda Suchanek, Andrea Throop, Lyndon Tilson, Thomas Urban, Joe Voshell, Kimberly Wagner, Jonathan Williams, Mary Williamson, Qian Zeng, Tricia Zwiefelhofer, Clinton R. Paden, Duncan MacCannell |
| EPI_ISL_1926050 | Northwest Laboratories | Seattle Flu Study | Deborah A. Nickerson, Chris D. Frazar, Jover Lee, Benjamin Pelle, Erica Ryke, Matthew Richardson, Amanda Adler, Elisabeth Brandstetter, Peter D. Han, Kairsten Fay, Misja Ilcisin, Kirsten Lacombe, Thomas R. Sibley, Melissa Truong, Caitlin R. Wolf, Romesh Gautom, Geoff Melly, Brian Hiatt, Philip Dykema, Scott Lindquist, Michael Boeckh, Janet A. Englund, Michael Famulare, Barry R. Lutz, Mark J. Rieder, Lea M. Starita, Matthew Thompson, Helen Y. Chu, Jay Shendure, Trevor Bedford |
| EPI_ISL_1926051 | Laboratory Corporation of America | Centers for Disease Control and Prevention Division of Viral Diseases, Pathogen Discovery | Dakota Howard, Dhwani Batra, Peter W. Cook, Kara Moser, Adrian Paskey, Jason Caravas, Benjamin Rambo-Martin, Shatavia Morrison, Christopher Gulvick, Scott Sammons, Yvette Unoarumhi, Darlene Wagner, Matthew Schmerer, Minoo Agarwal, Eyad Almasri, Debbie Boles, Ayla Burns, Nuthawin Charoensri, Oren Cohen, Susan Countryman, Mary Ann Cristobal, Bobbi Croy, Suzanne Dale, Hrushikesh Deshmukh, Amanda Douglas, Vincent Drouillon, Marcia Eisenberg, Howard Engler, Rama Ghatti, Prashant Gupta, Susan Hicks, Jake Humphrey, Lax Iyer, Manoj Jain, Mohan Kolli, Brian Krueger, Tim Kuphal, Stanley Letovsky, Michael Levandoski, Craig Lukasik, Jonathan Meltzer, Brian Norvell, Mindy Nye, Scott Parker, Christos Petropoulos, John Pruitt, Steven Ragan, Scott Ryan, Mike Sapeta, Jana Schroth, Suresh Babu Selvaraju, Goran Stevovic, Amanda Suchanek, Andrea Throop, Lyndon Tilson, Thomas Urban, Joe Voshell, Kimberly Wagner, Jonathan Williams, Mary Williamson, Qian Zeng, Tricia Zwiefelhofer, Clinton R. Paden, Duncan MacCannell |
| EPI_ISL_1926055, EPI_ISL_1926057 | Northwest Laboratories | Seattle Flu Study | Deborah A. Nickerson, Chris D. Frazar, Jover Lee, Benjamin Pelle, Erica Ryke, Matthew Richardson, Amanda Adler, Elisabeth Brandstetter, Peter D. Han, Kairsten Fay, Misja Ilcisin, Kirsten Lacombe, Thomas R. Sibley, Melissa Truong, Caitlin R. Wolf, Romesh Gautom, Geoff Melly, Brian Hiatt, Philip Dykema, Scott Lindquist, Michael Boeckh, Janet A. Englund, Michael Famulare, Barry R. Lutz, Mark J. Rieder, Lea M. Starita, Matthew Thompson, Helen Y. Chu, Jay Shendure, Trevor Bedford |
| EPI_ISL_1926058 | Laboratory Corporation of America | Centers for Disease Control and Prevention Division of Viral Diseases, Pathogen Discovery | Dakota Howard, Dhwani Batra, Peter W. Cook, Kara Moser, Adrian Paskey, Jason Caravas, Benjamin Rambo-Martin, Shatavia Morrison, Christopher Gulvick, Scott Sammons, Yvette Unoarumhi, Darlene Wagner, Matthew Schmerer, Minoo Agarwal, Eyad Almasri, Debbie Boles, Ayla Burns, Nuthawin Charoensri, Oren Cohen, Susan Countryman, Mary Ann Cristobal, Bobbi Croy, Suzanne Dale, Hrushikesh Deshmukh, Amanda Douglas, Vincent Drouillon, Marcia Eisenberg, Howard Engler, Rama Ghatti, Prashant Gupta, Susan Hicks, Jake Humphrey, Lax Iyer, Manoj Jain, Mohan Kolli, Brian Krueger, Tim Kuphal, Stanley Letovsky, Michael Levandoski, Craig Lukasik, Jonathan Meltzer, Brian Norvell, Mindy Nye, Scott Parker, Christos Petropoulos, John Pruitt, Steven Ragan, Scott Ryan, Mike Sapeta, Jana Schroth, Suresh Babu Selvaraju, Goran Stevovic, Amanda Suchanek, Andrea Throop, Lyndon Tilson, Thomas Urban, Joe Voshell, Kimberly Wagner, Jonathan Williams, Mary Williamson, Qian Zeng, Tricia Zwiefelhofer, Clinton R. Paden, Duncan MacCannell |
| EPI_ISL_1926059, EPI_ISL_1926060 | Northwest Laboratories | Seattle Flu Study | Deborah A. Nickerson, Chris D. Frazar, Jover Lee, Benjamin Pelle, Erica Ryke, Matthew Richardson, Amanda Adler, Elisabeth Brandstetter, Peter D. Han, Kairsten Fay, Misja Ilcisin, Kirsten Lacombe, Thomas R. Sibley, Melissa Truong, Caitlin R. Wolf, Romesh Gautom, Geoff Melly, Brian Hiatt, Philip Dykema, Scott Lindquist, Michael Boeckh, Janet A. Englund, Michael Famulare, Barry R. Lutz, Mark J. Rieder, Lea M. Starita, Matthew Thompson, Helen Y. Chu, Jay Shendure, Trevor Bedford |
| EPI_ISL_1926061 | Laboratory Corporation of America | Centers for Disease Control and Prevention Division of Viral Diseases, Pathogen Discovery | Dakota Howard, Dhwani Batra, Peter W. Cook, Kara Moser, Adrian Paskey, Jason Caravas, Benjamin Rambo-Martin, Shatavia Morrison, Christopher Gulvick, Scott Sammons, Yvette Unoarumhi, Darlene Wagner, Matthew Schmerer, Minoo Agarwal, Eyad Almasri, Debbie Boles, Ayla Burns, Nuthawin Charoensri, Oren Cohen, Susan Countryman, Mary Ann Cristobal, Bobbi Croy, Suzanne Dale, Hrushikesh Deshmukh, Amanda Douglas, Vincent Drouillon, Marcia Eisenberg, Howard Engler, Rama Ghatti, Prashant Gupta, Susan Hicks, Jake Humphrey, Lax Iyer, Manoj Jain, Mohan Kolli, Brian Krueger, Tim Kuphal, Stanley Letovsky, Michael Levandoski, Craig Lukasik, Jonathan Meltzer, Brian Norvell, Mindy Nye, Scott Parker, Christos Petropoulos, John Pruitt, Steven Ragan, Scott Ryan, Mike Sapeta, Jana Schroth, Suresh Babu Selvaraju, Goran Stevovic, Amanda Suchanek, Andrea Throop, Lyndon Tilson, Thomas Urban, Joe Voshell, Kimberly Wagner, Jonathan Williams, Mary Williamson, Qian Zeng, Tricia Zwiefelhofer, Clinton R. Paden, Duncan MacCannell |
| EPI_ISL_1926062, EPI_ISL_1926063 | Northwest Laboratories | Seattle Flu Study | Deborah A. Nickerson, Chris D. Frazar, Jover Lee, Benjamin Pelle, Erica Ryke, Matthew Richardson, Amanda Adler, Elisabeth Brandstetter, Peter D. Han, Kairsten Fay, Misja Ilcisin, Kirsten Lacombe, Thomas R. Sibley, Melissa Truong, Caitlin R. Wolf, Romesh Gautom, Geoff Melly, Brian Hiatt, Philip Dykema, Scott Lindquist, Michael Boeckh, Janet A. Englund, Michael Famulare, Barry R. Lutz, Mark J. Rieder, Lea M. Starita, Matthew Thompson, Helen Y. Chu, Jay Shendure, Trevor Bedford |
| EPI_ISL_1926064 | Laboratory Corporation of America | Centers for Disease Control and Prevention Division of Viral Diseases, Pathogen Discovery | Dakota Howard, Dhwani Batra, Peter W. Cook, Kara Moser, Adrian Paskey, Jason Caravas, Benjamin Rambo-Martin, Shatavia Morrison, Christopher Gulvick, Scott Sammons, Yvette Unoarumhi, Darlene Wagner, Matthew Schmerer, Minoo Agarwal, Eyad Almasri, Debbie Boles, Ayla Burns, Nuthawin Charoensri, Oren Cohen, Susan Countryman, Mary Ann Cristobal, Bobbi Croy, Suzanne Dale, Hrushikesh Deshmukh, Amanda Douglas, Vincent Drouillon, Marcia Eisenberg, Howard Engler, Rama Ghatti, Prashant Gupta, Susan Hicks, Jake Humphrey, Lax Iyer, Manoj Jain, Mohan Kolli, Brian Krueger, Tim Kuphal, Stanley Letovsky, Michael Levandoski, Craig Lukasik, Jonathan Meltzer, Brian Norvell, Mindy Nye, Scott Parker, Christos Petropoulos, John Pruitt, Steven Ragan, Scott Ryan, Mike Sapeta, Jana Schroth, Suresh Babu Selvaraju, Goran Stevovic, Amanda Suchanek, Andrea Throop, Lyndon Tilson, Thomas Urban, Joe Voshell, Kimberly Wagner, Jonathan Williams, Mary Williamson, Qian Zeng, Tricia Zwiefelhofer, Clinton R. Paden, Duncan MacCannell |
| EPI_ISL_1926065, EPI_ISL_1926066, EPI_ISL_1926068, EPI_ISL_1926069 | Northwest Laboratories | Seattle Flu Study | Deborah A. Nickerson, Chris D. Frazar, Jover Lee, Benjamin Pelle, Erica Ryke, Matthew Richardson, Amanda Adler, Elisabeth Brandstetter, Peter D. Han, Kairsten Fay, Misja Ilcisin, Kirsten Lacombe, Thomas R. Sibley, Melissa Truong, Caitlin R. Wolf, Romesh Gautom, Geoff Melly, Brian Hiatt, Philip Dykema, Scott Lindquist, Michael Boeckh, Janet A. Englund, Michael Famulare, Barry R. Lutz, Mark J. Rieder, Lea M. Starita, Matthew Thompson, Helen Y. Chu, Jay Shendure, Trevor Bedford |
| EPI_ISL_1926070 | Laboratory Corporation of America | Centers for Disease Control and Prevention Division of Viral Diseases, Pathogen Discovery | Dakota Howard, Dhwani Batra, Peter W. Cook, Kara Moser, Adrian Paskey, Jason Caravas, Benjamin Rambo-Martin, Shatavia Morrison, Christopher Gulvick, Scott Sammons, Yvette Unoarumhi, Darlene Wagner, Matthew Schmerer, Minoo Agarwal, Eyad Almasri, Debbie Boles, Ayla Burns, Nuthawin Charoensri, Oren Cohen, Susan Countryman, Mary Ann Cristobal, Bobbi Croy, Suzanne Dale, Hrushikesh Deshmukh, Amanda Douglas, Vincent Drouillon, Marcia Eisenberg, Howard Engler, Rama Ghatti, Prashant Gupta, Susan Hicks, Jake Humphrey, Lax Iyer, Manoj Jain, Mohan Kolli, Brian Krueger, Tim Kuphal, Stanley Letovsky, Michael Levandoski, Craig Lukasik, Jonathan Meltzer, Brian Norvell, Mindy Nye, Scott Parker, Christos Petropoulos, John Pruitt, Steven Ragan, Scott Ryan, Mike Sapeta, Jana Schroth, Suresh Babu Selvaraju, Goran Stevovic, Amanda |

| | | | |
|---|---|---|---|
| | | | Suchanek, Andrea Throop, Lyndon Tilson, Thomas Urban, Joe Voshell, Kimberly Wagner, Jonathan Williams, Mary Williamson, Qian Zeng, Tricia Zwiefelhofer, Clinton R. Paden, Duncan MacCannell |
| EPI_ISL_1926076 | Northwest Laboratories | Seattle Flu Study | Deborah A. Nickerson, Chris D. Frazar, Jover Lee, Benjamin Pelle, Erica Ryke, Matthew Richardson, Amanda Adler, Elisabeth Brandstetter, Peter D. Han, Kairsten Fay, Misja Ilcisin, Kirsten Lacombe, Thomas R. Sibley, Melissa Truong, Caitlin R. Wolf, Romesh Gautom, Geoff Melly, Brian Hiatt, Philip Dykema, Scott Lindquist, Michael Boeckh, Janet A. Englund, Michael Famulare, Barry R. Lutz, Mark J. Rieder, Lea M. Starita, Matthew Thompson, Helen Y. Chu, Jay Shendure, Trevor Bedford |
| EPI_ISL_1926077 | Laboratory Corporation of America | Centers for Disease Control and Prevention Division of Viral Diseases, Pathogen Discovery | Dakota Howard, Dhwani Batra, Peter W. Cook, Kara Moser, Adrian Paskey, Jason Caravas, Benjamin Rambo-Martin, Shatavia Morrison, Christopher Gulvick, Scott Sammons, Yvette Unoarumhi, Darlene Wagner, Matthew Schmerer, Minoo Agarwal, Eyad Almasri, Debbie Boles, Ayla Burns, Nuthawin Charoensri, Oren Cohen, Susan Countryman, Mary Ann Cristobal, Bobbi Croy, Suzanne Dale, Hrushikesh Deshmukh, Amanda Douglas, Vincent Drouillon, Marcia Eisenberg, Howard Engler, Rama Ghatti, Prashant Gupta, Susan Hicks, Jake Humphrey, Lax Iyer, Manoj Jain, Mohan Kolli, Brian Krueger, Tim Kuphal, Stanley Letovsky, Michael Levandoski, Craig Lukasik, Jonathan Meltzer, Brian Norvell, Mindy Nye, Scott Parker, Christos Petropoulos, John Pruitt, Steven Ragan, Scott Ryan, Mike Sapeta, Jana Schroth, Suresh Babu Selvaraju, Goran Stevovic, Amanda Suchanek, Andrea Throop, Lyndon Tilson, Thomas Urban, Joe Voshell, Kimberly Wagner, Jonathan Williams, Mary Williamson, Qian Zeng, Tricia Zwiefelhofer, Clinton R. Paden, Duncan MacCannell |
| EPI_ISL_1926078 | Northwest Laboratories | Seattle Flu Study | Deborah A. Nickerson, Chris D. Frazar, Jover Lee, Benjamin Pelle, Erica Ryke, Matthew Richardson, Amanda Adler, Elisabeth Brandstetter, Peter D. Han, Kairsten Fay, Misja Ilcisin, Kirsten Lacombe, Thomas R. Sibley, Melissa Truong, Caitlin R. Wolf, Romesh Gautom, Geoff Melly, Brian Hiatt, Philip Dykema, Scott Lindquist, Michael Boeckh, Janet A. Englund, Michael Famulare, Barry R. Lutz, Mark J. Rieder, Lea M. Starita, Matthew Thompson, Helen Y. Chu, Jay Shendure, Trevor Bedford |
| EPI_ISL_1926080 | Laboratory Corporation of America | Centers for Disease Control and Prevention Division of Viral Diseases, Pathogen Discovery | Dakota Howard, Dhwani Batra, Peter W. Cook, Kara Moser, Adrian Paskey, Jason Caravas, Benjamin Rambo-Martin, Shatavia Morrison, Christopher Gulvick, Scott Sammons, Yvette Unoarumhi, Darlene Wagner, Matthew Schmerer, Minoo Agarwal, Eyad Almasri, Debbie Boles, Ayla Burns, Nuthawin Charoensri, Oren Cohen, Susan Countryman, Mary Ann Cristobal, Bobbi Croy, Suzanne Dale, Hrushikesh Deshmukh, Amanda Douglas, Vincent Drouillon, Marcia Eisenberg, Howard Engler, Rama Ghatti, Prashant Gupta, Susan Hicks, Jake Humphrey, Lax Iyer, Manoj Jain, Mohan Kolli, Brian Krueger, Tim Kuphal, Stanley Letovsky, Michael Levandoski, Craig Lukasik, Jonathan Meltzer, Brian Norvell, Mindy Nye, Scott Parker, Christos Petropoulos, John Pruitt, Steven Ragan, Scott Ryan, Mike Sapeta, Jana Schroth, Suresh Babu Selvaraju, Goran Stevovic, Amanda Suchanek, Andrea Throop, Lyndon Tilson, Thomas Urban, Joe Voshell, Kimberly Wagner, Jonathan Williams, Mary Williamson, Qian Zeng, Tricia Zwiefelhofer, Clinton R. Paden, Duncan MacCannell |
| EPI_ISL_1926082 | Northwest Laboratories | Seattle Flu Study | Deborah A. Nickerson, Chris D. Frazar, Jover Lee, Benjamin Pelle, Erica Ryke, Matthew Richardson, Amanda Adler, Elisabeth Brandstetter, Peter D. Han, Kairsten Fay, Misja Ilcisin, Kirsten Lacombe, Thomas R. Sibley, Melissa Truong, Caitlin R. Wolf, Romesh Gautom, Geoff Melly, Brian Hiatt, Philip Dykema, Scott Lindquist, Michael Boeckh, Janet A. Englund, Michael Famulare, Barry R. Lutz, Mark J. Rieder, Lea M. Starita, Matthew Thompson, Helen Y. Chu, Jay Shendure, Trevor Bedford |
| EPI_ISL_1926083, EPI_ISL_1926086 | Laboratory Corporation of America | Centers for Disease Control and Prevention Division of Viral Diseases, Pathogen Discovery | Dakota Howard, Dhwani Batra, Peter W. Cook, Kara Moser, Adrian Paskey, Jason Caravas, Benjamin Rambo-Martin, Shatavia Morrison, Christopher Gulvick, Scott Sammons, Yvette Unoarumhi, Darlene Wagner, Matthew Schmerer, Minoo Agarwal, Eyad Almasri, Debbie Boles, Ayla Burns, Nuthawin Charoensri, Oren Cohen, Susan Countryman, Mary Ann Cristobal, Bobbi Croy, Suzanne Dale, Hrushikesh Deshmukh, Amanda Douglas, Vincent Drouillon, Marcia Eisenberg, Howard Engler, Rama Ghatti, Prashant Gupta, Susan Hicks, Jake Humphrey, Lax Iyer, Manoj Jain, Mohan Kolli, Brian Krueger, Tim Kuphal, Stanley Letovsky, Michael Levandoski, Craig Lukasik, Jonathan Meltzer, Brian Norvell, Mindy Nye, Scott Parker, Christos Petropoulos, John Pruitt, Steven Ragan, Scott Ryan, Mike Sapeta, Jana Schroth, Suresh Babu Selvaraju, Goran Stevovic, Amanda Suchanek, Andrea Throop, Lyndon Tilson, Thomas Urban, Joe Voshell, Kimberly Wagner, Jonathan Williams, Mary Williamson, Qian Zeng, Tricia Zwiefelhofer, Clinton R. Paden, Duncan MacCannell |
| EPI_ISL_1926087 | Northwest Laboratories | Seattle Flu Study | Deborah A. Nickerson, Chris D. Frazar, Jover Lee, Benjamin Pelle, Erica Ryke, Matthew Richardson, Amanda Adler, Elisabeth Brandstetter, Peter D. Han, Kairsten Fay, Misja Ilcisin, Kirsten Lacombe, Thomas R. Sibley, Melissa Truong, Caitlin R. Wolf, Romesh Gautom, Geoff Melly, Brian Hiatt, Philip Dykema, Scott Lindquist, Michael Boeckh, Janet A. Englund, Michael Famulare, Barry R. Lutz, Mark J. Rieder, Lea M. Starita, Matthew Thompson, Helen Y. Chu, Jay Shendure, Trevor Bedford |
| EPI_ISL_1926089 | Laboratory Corporation of America | Centers for Disease Control and Prevention Division of Viral Diseases, Pathogen Discovery | Dakota Howard, Dhwani Batra, Peter W. Cook, Kara Moser, Adrian Paskey, Jason Caravas, Benjamin Rambo-Martin, Shatavia Morrison, Christopher Gulvick, Scott Sammons, Yvette Unoarumhi, Darlene Wagner, Matthew Schmerer, Minoo Agarwal, Eyad Almasri, Debbie Boles, Ayla Burns, Nuthawin Charoensri, Oren Cohen, Susan Countryman, Mary Ann Cristobal, Bobbi Croy, Suzanne Dale, Hrushikesh Deshmukh, Amanda Douglas, Vincent Drouillon, Marcia Eisenberg, Howard Engler, Rama Ghatti, Prashant Gupta, Susan Hicks, Jake Humphrey, Lax Iyer, Manoj Jain, Mohan Kolli, Brian Krueger, Tim Kuphal, Stanley Letovsky, Michael Levandoski, Craig Lukasik, Jonathan Meltzer, Brian Norvell, Mindy Nye, Scott Parker, Christos Petropoulos, John Pruitt, Steven Ragan, Scott Ryan, Mike Sapeta, Jana Schroth, Suresh Babu Selvaraju, Goran Stevovic, Amanda Suchanek, Andrea Throop, Lyndon Tilson, Thomas Urban, Joe Voshell, Kimberly Wagner, Jonathan Williams, Mary Williamson, Qian Zeng, Tricia Zwiefelhofer, Clinton R. Paden, Duncan MacCannell |
| EPI_ISL_1926090, EPI_ISL_1926091 | Northwest Laboratories | Seattle Flu Study | Deborah A. Nickerson, Chris D. Frazar, Jover Lee, Benjamin Pelle, Erica Ryke, Matthew Richardson, Amanda Adler, Elisabeth Brandstetter, Peter D. Han, Kairsten Fay, Misja Ilcisin, Kirsten Lacombe, Thomas R. Sibley, Melissa Truong, Caitlin R. Wolf, Romesh Gautom, Geoff Melly, Brian Hiatt, Philip Dykema, Scott Lindquist, Michael Boeckh, Janet A. Englund, Michael Famulare, Barry R. Lutz, Mark J. Rieder, Lea M. Starita, Matthew Thompson, Helen Y. Chu, Jay Shendure, Trevor Bedford |
| EPI_ISL_1926093 | Laboratory Corporation of America | Centers for Disease Control and Prevention Division of Viral Diseases, Pathogen Discovery | Dakota Howard, Dhwani Batra, Peter W. Cook, Kara Moser, Adrian Paskey, Jason Caravas, Benjamin Rambo-Martin, Shatavia Morrison, Christopher Gulvick, Scott Sammons, Yvette Unoarumhi, Darlene Wagner, Matthew Schmerer, Minoo Agarwal, Eyad Almasri, Debbie Boles, Ayla Burns, Nuthawin Charoensri, Oren Cohen, Susan Countryman, Mary Ann Cristobal, Bobbi Croy, Suzanne Dale, Hrushikesh Deshmukh, Amanda Douglas, Vincent Drouillon, Marcia Eisenberg, Howard Engler, Rama Ghatti, Prashant Gupta, Susan Hicks, Jake Humphrey, Lax Iyer, Manoj Jain, Mohan Kolli, Brian Krueger, Tim Kuphal, Stanley Letovsky, Michael Levandoski, Craig Lukasik, Jonathan Meltzer, Brian Norvell, Mindy Nye, Scott Parker, Christos Petropoulos, John Pruitt, Steven Ragan, Scott Ryan, Mike Sapeta, Jana Schroth, Suresh Babu Selvaraju, Goran Stevovic, Amanda Suchanek, Andrea Throop, Lyndon Tilson, Thomas Urban, Joe Voshell, Kimberly Wagner, Jonathan Williams, Mary Williamson, Qian Zeng, Tricia Zwiefelhofer, Clinton R. Paden, Duncan MacCannell |
| EPI_ISL_1926095 | Altius Institute | Seattle Flu Study | Deborah A. Nickerson, Chris D. Frazar, Jover Lee, Benjamin Pelle, Erica Ryke, Matthew Richardson, Amanda Adler, Elisabeth Brandstetter, Peter D. Han, Kairsten Fay, Misja Ilcisin, Kirsten Lacombe, Thomas R. Sibley, Melissa Truong, Caitlin R. Wolf, Ryan Alexander, Daniel Bates, Rebecca Bruders, Stephanie DeBaun, Clem Green, Muhammad Halimun, Jessica Halow, Kneshay Harper, Matt Hartman, Andrew Meuser, Alex Nguyen, Truong Nguyen, Sofia Olsson, Sadie Patraw, Hannah Petersen, Tobias Ragoczy, Joshua Richards, Jacob Rodriguez, John Stamatoyannopoulos, Julia Wald, Olivia Waltner, Michael Boeckh, Janet A. Englund, Michael Famulare, Barry R. Lutz, Mark J. Rieder, Lea M. Starita, Matthew Thompson, Helen Y. Chu, Jay Shendure, Trevor Bedford |
| EPI_ISL_1926096 | Laboratory Corporation of America | Centers for Disease Control and Prevention Division of Viral Diseases, Pathogen Discovery | Dakota Howard, Dhwani Batra, Peter W. Cook, Kara Moser, Adrian Paskey, Jason Caravas, Benjamin Rambo-Martin, Shatavia Morrison, Christopher Gulvick, Scott Sammons, Yvette Unoarumhi, Darlene Wagner, Matthew Schmerer, Minoo Agarwal, Eyad Almasri, Debbie Boles, Ayla Burns, Nuthawin Charoensri, Oren Cohen, Susan Countryman, Mary Ann Cristobal, Bobbi Croy, Suzanne Dale, Hrushikesh Deshmukh, Amanda Douglas, Vincent Drouillon, Marcia Eisenberg, Howard Engler, Rama Ghatti, Prashant Gupta, Susan Hicks, Jake Humphrey, Lax Iyer, Manoj Jain, Mohan Kolli, Brian Krueger, Tim Kuphal, Stanley Letovsky, Michael Levandoski, Craig Lukasik, Jonathan Meltzer, Brian Norvell, Mindy Nye, Scott Parker, Christos Petropoulos, John Pruitt, Steven Ragan, Scott Ryan, Mike Sapeta, Jana Schroth, Suresh Babu Selvaraju, Goran Stevovic, Amanda Suchanek, Andrea Throop, Lyndon Tilson, Thomas Urban, Joe Voshell, Kimberly Wagner, Jonathan Williams, Mary Williamson, Qian Zeng, Tricia Zwiefelhofer, Clinton R. Paden, Duncan MacCannell |
| EPI_ISL_1926097, EPI_ISL_1926098 | Altius Institute | Seattle Flu Study | Deborah A. Nickerson, Chris D. Frazar, Jover Lee, Benjamin Pelle, Erica Ryke, Matthew Richardson, Amanda Adler, Elisabeth Brandstetter, Peter D. Han, Kairsten Fay, Misja Ilcisin, Kirsten Lacombe, Thomas R. Sibley, Melissa Truong, Caitlin R. Wolf, Ryan Alexander, Daniel Bates, Rebecca Bruders, Stephanie DeBaun, Clem Green, Muhammad Halimun, Jessica Halow, Kneshay Harper, Matt Hartman, Andrew Meuser, Alex Nguyen, Truong Nguyen, Sofia Olsson, Sadie Patraw, Hannah Petersen, Tobias Ragoczy, Joshua Richards, Jacob Rodriguez, John Stamatoyannopoulos, Julia Wald, Olivia Waltner, Michael Boeckh, Janet A. Englund, Michael Famulare, Barry R. Lutz, Mark J. Rieder, Lea M. Starita, Matthew Thompson, Helen Y. Chu, Jay Shendure, Trevor Bedford |
| EPI_ISL_1926099 | Laboratory Corporation of America | Centers for Disease Control and Prevention Division of Viral Diseases, Pathogen Discovery | Dakota Howard, Dhwani Batra, Peter W. Cook, Kara Moser, Adrian Paskey, Jason Caravas, Benjamin Rambo-Martin, Shatavia Morrison, Christopher Gulvick, Scott Sammons, Yvette Unoarumhi, Darlene Wagner, Matthew Schmerer, Minoo Agarwal, Eyad Almasri, Debbie Boles, Ayla Burns, Nuthawin Charoensri, Oren Cohen, Susan Countryman, Mary Ann Cristobal, Bobbi Croy, Suzanne Dale, Hrushikesh Deshmukh, Amanda Douglas, Vincent Drouillon, Marcia Eisenberg, Howard Engler, Rama Ghatti, Prashant Gupta, Susan Hicks, Jake Humphrey, Lax Iyer, Manoj Jain, Mohan Kolli, Brian Krueger, Tim Kuphal, Stanley Letovsky, Michael Levandoski, Craig Lukasik, Jonathan Meltzer, Brian Norvell, Mindy Nye, Scott Parker, Christos Petropoulos, John Pruitt, Steven Ragan, Scott Ryan, Mike Sapeta, Jana Schroth, Suresh Babu Selvaraju, Goran Stevovic, Amanda Suchanek, Andrea Throop, Lyndon Tilson, Thomas Urban, Joe Voshell, Kimberly Wagner, Jonathan Williams, Mary Williamson, Qian Zeng, Tricia Zwiefelhofer, Clinton R. Paden, Duncan MacCannell |
| EPI_ISL_1926100, EPI_ISL_1926103, EPI_ISL_1926104 | Altius Institute | Seattle Flu Study | Deborah A. Nickerson, Chris D. Frazar, Jover Lee, Benjamin Pelle, Erica Ryke, Matthew Richardson, Amanda Adler, Elisabeth Brandstetter, Peter D. Han, Kairsten Fay, Misja Ilcisin, Kirsten Lacombe, Thomas R. Sibley, Melissa Truong, Caitlin R. Wolf, Ryan Alexander, Daniel Bates, Rebecca Bruders, Stephanie DeBaun, Clem Green, Muhammad Halimun, Jessica Halow, Kneshay Harper, Matt Hartman, Andrew Meuser, Alex Nguyen, Truong Nguyen, Sofia Olsson, Sadie Patraw, Hannah Petersen, Tobias Ragoczy, Joshua Richards, Jacob Rodriguez, John Stamatoyannopoulos, Julia Wald, Olivia Waltner, Michael Boeckh, Janet A. Englund, Michael Famulare, Barry R. Lutz, Mark J. Rieder, Lea M. Starita, Matthew Thompson, Helen Y. Chu, Jay Shendure, Trevor Bedford |
| EPI_ISL_1926105 | Laboratory Corporation of America | Centers for Disease Control and Prevention Division of Viral Diseases, Pathogen Discovery | Dakota Howard, Dhwani Batra, Peter W. Cook, Kara Moser, Adrian Paskey, Jason Caravas, Benjamin Rambo-Martin, Shatavia Morrison, Christopher Gulvick, Scott Sammons, Yvette Unoarumhi, Darlene Wagner, Matthew Schmerer, Minoo Agarwal, Eyad Almasri, Debbie Boles, Ayla Burns, Nuthawin Charoensri, Oren Cohen, Susan Countryman, Mary Ann Cristobal, Bobbi Croy, Suzanne Dale, Hrushikesh Deshmukh, Amanda Douglas, Vincent Drouillon, Marcia Eisenberg, Howard Engler, Rama Ghatti, Prashant Gupta, Susan Hicks, Jake Humphrey, Lax Iyer, Manoj Jain, Mohan Kolli, Brian Krueger, Tim Kuphal, Stanley Letovsky, Michael Levandoski, Craig Lukasik, Jonathan Meltzer, Brian Norvell, Mindy Nye, Scott Parker, Christos Petropoulos, John Pruitt, Steven Ragan, Scott Ryan, Mike Sapeta, Jana Schroth, Suresh Babu Selvaraju, Goran Stevovic, Amanda Suchanek, Andrea Throop, Lyndon Tilson, Thomas Urban, Joe Voshell, Kimberly Wagner, Jonathan Williams, Mary Williamson, Qian Zeng, Tricia Zwiefelhofer, Clinton R. Paden, Duncan MacCannell |
| EPI_ISL_1926106, EPI_ISL_1926107 | Altius Institute | Seattle Flu Study | Deborah A. Nickerson, Chris D. Frazar, Jover Lee, Benjamin Pelle, Erica Ryke, Matthew Richardson, Amanda Adler, Elisabeth Brandstetter, Peter D. Han, Kairsten Fay, Misja Ilcisin, Kirsten Lacombe, Thomas R. Sibley, Melissa Truong, Caitlin R. Wolf, Ryan Alexander, Daniel Bates, Rebecca Bruders, Stephanie DeBaun, Clem Green, Muhammad Halimun, Jessica Halow, Kneshay Harper, Matt Hartman, Andrew Meuser, Alex Nguyen, Truong Nguyen, Sofia Olsson, Sadie Patraw, Hannah Petersen, Tobias Ragoczy, Joshua Richards, Jacob Rodriguez, John Stamatoyannopoulos, Julia Wald, Olivia Waltner, Michael Boeckh, Janet A. Englund, Michael Famulare, Barry R. Lutz, Mark J. Rieder, Lea M. Starita, Matthew Thompson, Helen Y. Chu, Jay Shendure, Trevor Bedford |
| EPI_ISL_1926108 | Laboratory Corporation of America | Centers for Disease Control and Prevention Division of Viral Diseases, Pathogen Discovery | Dakota Howard, Dhwani Batra, Peter W. Cook, Kara Moser, Adrian Paskey, Jason Caravas, Benjamin Rambo-Martin, Shatavia Morrison, Christopher Gulvick, Scott Sammons, Yvette Unoarumhi, Darlene Wagner, Matthew Schmerer, Minoo Agarwal, Eyad Almasri, Debbie Boles, Ayla Burns, Nuthawin Charoensri, Oren Cohen, Susan Countryman, Mary Ann Cristobal, Bobbi Croy, Suzanne Dale, Hrushikesh Deshmukh, Amanda Douglas, Vincent Drouillon, Marcia Eisenberg, Howard Engler, Rama Ghatti, Prashant Gupta, Susan Hicks, Jake Humphrey, Lax Iyer, Manoj Jain, Mohan Kolli, Brian Krueger, Tim Kuphal, Stanley Letovsky, Michael Levandoski, Craig Lukasik, Jonathan Meltzer, Brian Norvell, Mindy Nye, Scott Parker, Christos Petropoulos, John Pruitt, Steven Ragan, Scott Ryan, Mike Sapeta, Jana Schroth, Suresh Babu Selvaraju, Goran Stevovic, Amanda Suchanek, Andrea Throop, Lyndon Tilson, Thomas Urban, Joe Voshell, Kimberly Wagner, Jonathan Williams, Mary Williamson, Qian Zeng, Tricia Zwiefelhofer, Clinton R. Paden, Duncan MacCannell |
| EPI_ISL_1926109, EPI_ISL_1926110, EPI_ISL_1926113 | Altius Institute | Seattle Flu Study | Deborah A. Nickerson, Chris D. Frazar, Jover Lee, Benjamin Pelle, Erica Ryke, Matthew Richardson, Amanda Adler, Elisabeth Brandstetter, Peter D. Han, Kairsten Fay, Misja Ilcisin, Kirsten Lacombe, Thomas R. Sibley, Melissa Truong, Caitlin R. Wolf, Ryan Alexander, Daniel Bates, Rebecca Bruders, Stephanie DeBaun, Clem Green, Muhammad Halimun, Jessica Halow, Kneshay Harper, Matt Hartman, Andrew Meuser, Alex Nguyen, Truong Nguyen, Sofia Olsson, Sadie Patraw, Hannah Petersen, Tobias Ragoczy, Joshua Richards, Jacob Rodriguez, John Stamatoyannopoulos, Julia Wald, Olivia Waltner, Michael Boeckh, Janet A. Englund, Michael Famulare, Barry R. Lutz, Mark J. Rieder, Lea M. Starita, Matthew Thompson, Helen Y. Chu, Jay Shendure, Trevor Bedford |
| EPI_ISL_1926114 | Laboratory Corporation of America | Centers for Disease Control and Prevention Division of Viral Diseases, Pathogen Discovery | Dakota Howard, Dhwani Batra, Peter W. Cook, Kara Moser, Adrian Paskey, Jason Caravas, Benjamin Rambo-Martin, Shatavia Morrison, Christopher Gulvick, Scott Sammons, Yvette Unoarumhi, Darlene Wagner, Matthew Schmerer, Minoo Agarwal, Eyad Almasri, Debbie Boles, Ayla Burns, Nuthawin Charoensri, Oren Cohen, Susan Countryman, Mary Ann Cristobal, Bobbi Croy, Suzanne Dale, Hrushikesh Deshmukh, Amanda Douglas, Vincent Drouillon, Marcia Eisenberg, Howard Engler, Rama Ghatti, Prashant Gupta, Susan Hicks, Jake Humphrey, Lax Iyer, Manoj Jain, Mohan Kolli, Brian Krueger, Tim Kuphal, Stanley Letovsky, Michael Levandoski, Craig Lukasik, Jonathan Meltzer, Brian Norvell, Mindy Nye, Scott Parker, Christos Petropoulos, John Pruitt, Steven Ragan, Scott Ryan, Mike Sapeta, Jana Schroth, Suresh Babu Selvaraju, Goran Stevovic, Amanda Suchanek, Andrea Throop, Lyndon Tilson, Thomas Urban, Joe Voshell, Kimberly Wagner, Jonathan Williams, Mary Williamson, Qian Zeng, Tricia Zwiefelhofer, Clinton R. Paden, Duncan MacCannell |
| EPI_ISL_1926115, EPI_ISL_1926116 | Altius Institute | Seattle Flu Study | Deborah A. Nickerson, Chris D. Frazar, Jover Lee, Benjamin Pelle, Erica Ryke, Matthew Richardson, Amanda Adler, Elisabeth Brandstetter, Peter D. Han, Kairsten Fay, Misja Ilcisin, Kirsten Lacombe, Thomas R. Sibley, Melissa Truong, Caitlin R. Wolf, Ryan Alexander, Daniel Bates, Rebecca Bruders, Stephanie DeBaun, Clem Green, Muhammad Halimun, Jessica Halow, Kneshay Harper, Matt Hartman, Andrew Meuser, Alex Nguyen, Truong Nguyen, Sofia Olsson, Sadie Patraw, Hannah Petersen, Tobias Ragoczy, Joshua Richards, Jacob Rodriguez, John Stamatoyannopoulos, Julia Wald, Olivia Waltner, Michael Boeckh, Janet A. Englund, Michael Famulare, Barry R. Lutz, Mark J. Rieder, Lea M. Starita, Matthew Thompson, Helen Y. Chu, Jay Shendure, Trevor Bedford |
| EPI_ISL_1926117, EPI_ISL_1926120, EPI_ISL_1926122 | Laboratory Corporation of America | Centers for Disease Control and Prevention Division of Viral Diseases, Pathogen Discovery | Dakota Howard, Dhwani Batra, Peter W. Cook, Kara Moser, Adrian Paskey, Jason Caravas, Benjamin Rambo-Martin, Shatavia Morrison, Christopher Gulvick, Scott Sammons, Yvette Unoarumhi, Darlene Wagner, Matthew Schmerer, Minoo Agarwal, Eyad Almasri, Debbie Boles, Ayla Burns, Nuthawin Charoensri, Oren Cohen, Susan Countryman, Mary Ann Cristobal, Bobbi Croy, Suzanne Dale, Hrushikesh Deshmukh, Amanda Douglas, Vincent Drouillon, Marcia Eisenberg, Howard Engler, Rama Ghatti, Prashant Gupta, Susan Hicks, Jake Humphrey, Lax Iyer, Manoj Jain, Mohan Kolli, Brian Krueger, Tim Kuphal, Stanley Letovsky, Michael Levandoski, Craig Lukasik, Jonathan Meltzer, Brian Norvell, Mindy Nye, Scott Parker, Christos Petropoulos, John Pruitt, Steven Ragan, Scott Ryan, Mike Sapeta, Jana Schroth, Suresh Babu Selvaraju, Goran Stevovic, Amanda Suchanek, Andrea Throop, Lyndon Tilson, Thomas Urban, Joe Voshell, Kimberly Wagner, Jonathan Williams, Mary Williamson, Qian Zeng, Tricia Zwiefelhofer, Clinton R. Paden, Duncan MacCannell |
| EPI_ISL_1926123, EPI_ISL_1926125 | Altius Institute | Seattle Flu Study | Deborah A. Nickerson, Chris D. Frazar, Jover Lee, Benjamin Pelle, Erica Ryke, Matthew Richardson, Amanda Adler, Elisabeth Brandstetter, Peter D. Han, Kairsten Fay, Misja Ilcisin, Kirsten Lacombe, Thomas R. Sibley, Melissa Truong, Caitlin R. Wolf, Ryan Alexander, Daniel Bates, Rebecca Bruders, Stephanie DeBaun, Clem Green, Muhammad Halimun, Jessica Halow, Kneshay Harper, Matt Hartman, Andrew Meuser, Alex Nguyen, Truong Nguyen, Sofia Olsson, Sadie Patraw, Hannah Petersen, Tobias Ragoczy, Joshua Richards, Jacob Rodriguez, John Stamatoyannopoulos, Julia Wald, Olivia Waltner, Michael Boeckh, Janet A. Englund, Michael Famulare, Barry R. Lutz, Mark J. Rieder, Lea M. Starita, Matthew Thompson, Helen Y. Chu, Jay Shendure, Trevor Bedford |
| EPI_ISL_1926126 | Laboratory Corporation of America | Centers for Disease Control and Prevention Division of Viral Diseases, Pathogen Discovery | Dakota Howard, Dhwani Batra, Peter W. Cook, Kara Moser, Adrian Paskey, Jason Caravas, Benjamin Rambo-Martin, Shatavia Morrison, Christopher Gulvick, Scott Sammons, Yvette Unoarumhi, Darlene Wagner, Matthew Schmerer, Minoo Agarwal, Eyad Almasri, Debbie Boles, Ayla Burns, Nuthawin Charoensri, Oren Cohen, Susan Countryman, Mary Ann Cristobal, Bobbi Croy, Suzanne Dale, Hrushikesh Deshmukh, Amanda Douglas, Vincent Drouillon, Marcia Eisenberg, Howard Engler, Rama Ghatti, Prashant Gupta, Susan Hicks, Jake Humphrey, Lax Iyer, Manoj Jain, Mohan Kolli, Brian Krueger, Tim Kuphal, Stanley Letovsky, Michael Levandoski, Craig Lukasik, Jonathan Meltzer, Brian Norvell, Mindy Nye, Scott Parker, Christos Petropoulos, John Pruitt, Steven Ragan, Scott Ryan, Mike Sapeta, Jana Schroth, Suresh Babu Selvaraju, Goran Stevovic, Amanda Suchanek, Andrea Throop, Lyndon Tilson, Thomas Urban, Joe Voshell, Kimberly Wagner, Jonathan Williams, Mary Williamson, Qian Zeng, Tricia Zwiefelhofer, Clinton R. Paden, Duncan MacCannell |
| EPI_ISL_1926127 | Altius Institute | Seattle Flu Study | Deborah A. Nickerson, Chris D. Frazar, Jover Lee, Benjamin Pelle, Erica Ryke, Matthew Richardson, Amanda Adler, Elisabeth Brandstetter, Peter D. Han, Kairsten Fay, Misja Ilcisin, Kirsten Lacombe, Thomas R. Sibley, Melissa Truong, Caitlin R. Wolf, Ryan Alexander, Daniel Bates, Rebecca Bruders, Stephanie DeBaun, Clem Green, Muhammad Halimun, Jessica Halow, Kneshay Harper, Matt Hartman, Andrew Meuser, Alex Nguyen, Truong Nguyen, Sofia Olsson, Sadie Patraw, Hannah Petersen, Tobias Ragoczy, Joshua Richards, Jacob Rodriguez, John Stamatoyannopoulos, Julia Wald, Olivia Waltner, Michael Boeckh, Janet A. Englund, Michael Famulare, Barry R. Lutz, Mark J. Rieder, Lea M. Starita, Matthew Thompson, Helen Y. Chu, Jay Shendure, Trevor Bedford |
| EPI_ISL_1926129 | Laboratory Corporation of America | Centers for Disease Control and Prevention Division of Viral Diseases, Pathogen Discovery | Dakota Howard, Dhwani Batra, Peter W. Cook, Kara Moser, Adrian Paskey, Jason Caravas, Benjamin Rambo-Martin, Shatavia Morrison, Christopher Gulvick, Scott Sammons, Yvette Unoarumhi, Darlene Wagner, Matthew Schmerer, Minoo Agarwal, Eyad Almasri, Debbie Boles, Ayla Burns, Nuthawin Charoensri, Oren Cohen, Susan Countryman, Mary Ann Cristobal, Bobbi Croy, Suzanne Dale, Hrushikesh Deshmukh, Amanda Douglas, Vincent Drouillon, Marcia Eisenberg, Howard Engler, Rama Ghatti, Prashant Gupta, Susan Hicks, Jake Humphrey, Lax Iyer, Manoj Jain, Mohan Kolli, Brian Krueger, Tim Kuphal, Stanley Letovsky, Michael Levandoski, Craig Lukasik, Jonathan Meltzer, Brian Norvell, Mindy Nye, Scott Parker, Christos Petropoulos, John Pruitt, Steven Ragan, Scott Ryan, Mike Sapeta, Jana Schroth, Suresh Babu Selvaraju, Goran Stevovic, Amanda Suchanek, Andrea Throop, Lyndon Tilson, Thomas Urban, Joe Voshell, Kimberly Wagner, Jonathan Williams, Mary Williamson, Qian Zeng, Tricia Zwiefelhofer, Clinton R. Paden, Duncan MacCannell |
| EPI_ISL_1926133 | Altius Institute | Seattle Flu Study | Deborah A. Nickerson, Chris D. Frazar, Jover Lee, Benjamin Pelle, Erica Ryke, Matthew Richardson, Amanda Adler, Elisabeth Brandstetter, Peter D. Han, Kairsten Fay, Misja Ilcisin, Kirsten Lacombe, Thomas R. Sibley, Melissa Truong, Caitlin R. Wolf, Ryan Alexander, Daniel Bates, Rebecca Bruders, Stephanie DeBaun, Clem Green, Muhammad Halimun, Jessica Halow, Kneshay Harper, Matt Hartman, Andrew Meuser, Alex Nguyen, Truong Nguyen, Sofia Olsson, Sadie Patraw, Hannah Petersen, Tobias Ragoczy, Joshua Richards, Jacob Rodriguez, John Stamatoyannopoulos, Julia Wald, Olivia Waltner, Michael Boeckh, Janet A. Englund, Michael Famulare, Barry R. Lutz, Mark J. Rieder, Lea M. Starita, Matthew Thompson, Helen Y. Chu, Jay Shendure, Trevor Bedford |
| EPI_ISL_1926135 | Laboratory Corporation of America | Centers for Disease Control and Prevention Division of Viral Diseases, Pathogen Discovery | Dakota Howard, Dhwani Batra, Peter W. Cook, Kara Moser, Adrian Paskey, Jason Caravas, Benjamin Rambo-Martin, Shatavia Morrison, Christopher Gulvick, Scott Sammons, Yvette Unoarumhi, Darlene Wagner, Matthew Schmerer, Minoo Agarwal, Eyad Almasri, Debbie Boles, Ayla Burns, Nuthawin Charoensri, Oren Cohen, Susan Countryman, Mary Ann Cristobal, Bobbi Croy, Suzanne Dale, Hrushikesh Deshmukh, Amanda Douglas, Vincent Drouillon, Marcia Eisenberg, Howard Engler, Rama Ghatti, Prashant Gupta, Susan Hicks, Jake Humphrey, Lax Iyer, Manoj Jain, Mohan Kolli, Brian Krueger, Tim Kuphal, Stanley Letovsky, Michael Levandoski, Craig Lukasik, Jonathan Meltzer, Brian Norvell, Mindy Nye, Scott Parker, Christos Petropoulos, John Pruitt, Steven Ragan, Scott Ryan, Mike Sapeta, Jana Schroth, Suresh Babu Selvaraju, Goran Stevovic, Amanda Suchanek, Andrea Throop, Lyndon Tilson, Thomas Urban, Joe Voshell, Kimberly Wagner, Jonathan Williams, Mary Williamson, Qian Zeng, Tricia Zwiefelhofer, Clinton R. Paden, Duncan MacCannell |
| EPI_ISL_1926136, EPI_ISL_1926137 | Altius Institute | Seattle Flu Study | Deborah A. Nickerson, Chris D. Frazar, Jover Lee, Benjamin Pelle, Erica Ryke, Matthew Richardson, Amanda Adler, Elisabeth Brandstetter, Peter D. Han, Kairsten Fay, Misja Ilcisin, Kirsten Lacombe, Thomas R. Sibley, Melissa Truong, Caitlin R. Wolf, Ryan Alexander, Daniel Bates, Rebecca Bruders, Stephanie DeBaun, Clem Green, Muhammad Halimun, Jessica Halow, Kneshay Harper, Matt Hartman, Andrew Meuser, Alex Nguyen, Truong Nguyen, Sofia Olsson, Sadie Patraw, Hannah Petersen, Tobias Ragoczy, Joshua Richards, Jacob Rodriguez, John Stamatoyannopoulos, Julia Wald, Olivia Waltner, Michael Boeckh, Janet A. Englund, Michael Famulare, Barry R. Lutz, Mark J. Rieder, Lea M. Starita, Matthew Thompson, Helen Y. Chu, Jay Shendure, Trevor Bedford |

| | | | |
|---|---|---|---|
| EPI_ISL_1926138, EPI_ISL_1926141 | Laboratory Corporation of America | Centers for Disease Control and Prevention Division of Viral Diseases, Pathogen Discovery | Dakota Howard, Dhwani Batra, Peter W. Cook, Kara Moser, Adrian Paskey, Jason Caravas, Benjamin Rambo-Martin, Shatavia Morrison, Christopher Gulvick, Scott Sammons, Yvette Unoarumhi, Darlene Wagner, Matthew Schmerer, Minoo Agarwal, Eyad Almasri, Debbie Boles, Ayla Burns, Nuthawin Charoensri, Oren Cohen, Susan Countryman, Mary Ann Cristobal, Bobbi Croy, Suzanne Dale, Hrushikesh Deshmukh, Amanda Douglas, Vincent Drouillon, Marcia Eisenberg, Howard Engler, Rama Ghatti, Prashant Gupta, Susan Hicks, Jake Humphrey, Lax Iyer, Manoj Jain, Mohan Kolli, Brian Krueger, Tim Kuphal, Stanley Letovsky, Michael Levandoski, Craig Lukasik, Jonathan Meltzer, Brian Norvell, Mindy Nye, Scott Parker, Christos Petropoulos, John Pruitt, Steven Ragan, Scott Ryan, Mike Sapeta, Jana Schroth, Suresh Babu Selvaraju, Goran Stevovic, Amanda Suchanek, Andrea Throop, Lyndon Tilson, Thomas Urban, Joe Voshell, Kimberly Wagner, Jonathan Williams, Mary Williamson, Qian Zeng, Tricia Zwiefelhofer, Clinton R. Paden, Duncan MacCannell |
| EPI_ISL_1926142, EPI_ISL_1926143, EPI_ISL_1926144, EPI_ISL_1926146, EPI_ISL_1926147 | Altius Institute | Seattle Flu Study | Deborah A. Nickerson, Chris D. Frazar, Jover Lee, Benjamin Pelle, Erica Ryke, Matthew Richardson, Amanda Adler, Elisabeth Brandstetter, Peter D. Han, Kairsten Fay, Misja Ilcisin, Kirsten Lacombe, Thomas R. Sibley, Melissa Truong, Caitlin R. Wolf, Ryan Alexander, Daniel Bates, Rebecca Bruders, Stephanie DeBaun, Clem Green, Muhammad Halimun, Jessica Halow, Kneshay Harper, Matt Hartman, Andrew Meuser, Alex Nguyen, Truong Nguyen, Sofia Olsson, Sadie Patraw, Hannah Petersen, Tobias Ragoczy, Joshua Richards, Jacob Rodriguez, John Stamatoyannopoulos, Julia Wald, Olivia Waltner, Michael Boeckh, Janet A. Englund, Michael Famulare, Barry R. Lutz, Mark J. Rieder, Lea M. Starita, Matthew Thompson, Helen Y. Chu, Jay Shendure, Trevor Bedford |
| EPI_ISL_1926148, EPI_ISL_1926151 | Laboratory Corporation of America | Centers for Disease Control and Prevention Division of Viral Diseases, Pathogen Discovery | Dakota Howard, Dhwani Batra, Peter W. Cook, Kara Moser, Adrian Paskey, Jason Caravas, Benjamin Rambo-Martin, Shatavia Morrison, Christopher Gulvick, Scott Sammons, Yvette Unoarumhi, Darlene Wagner, Matthew Schmerer, Minoo Agarwal, Eyad Almasri, Debbie Boles, Ayla Burns, Nuthawin Charoensri, Oren Cohen, Susan Countryman, Mary Ann Cristobal, Bobbi Croy, Suzanne Dale, Hrushikesh Deshmukh, Amanda Douglas, Vincent Drouillon, Marcia Eisenberg, Howard Engler, Rama Ghatti, Prashant Gupta, Susan Hicks, Jake Humphrey, Lax Iyer, Manoj Jain, Mohan Kolli, Brian Krueger, Tim Kuphal, Stanley Letovsky, Michael Levandoski, Craig Lukasik, Jonathan Meltzer, Brian Norvell, Mindy Nye, Scott Parker, Christos Petropoulos, John Pruitt, Steven Ragan, Scott Ryan, Mike Sapeta, Jana Schroth, Suresh Babu Selvaraju, Goran Stevovic, Amanda Suchanek, Andrea Throop, Lyndon Tilson, Thomas Urban, Joe Voshell, Kimberly Wagner, Jonathan Williams, Mary Williamson, Qian Zeng, Tricia Zwiefelhofer, Clinton R. Paden, Duncan MacCannell |
| EPI_ISL_1926152 | Altius Institute | Seattle Flu Study | Deborah A. Nickerson, Chris D. Frazar, Jover Lee, Benjamin Pelle, Erica Ryke, Matthew Richardson, Amanda Adler, Elisabeth Brandstetter, Peter D. Han, Kairsten Fay, Misja Ilcisin, Kirsten Lacombe, Thomas R. Sibley, Melissa Truong, Caitlin R. Wolf, Ryan Alexander, Daniel Bates, Rebecca Bruders, Stephanie DeBaun, Clem Green, Muhammad Halimun, Jessica Halow, Kneshay Harper, Matt Hartman, Andrew Meuser, Alex Nguyen, Truong Nguyen, Sofia Olsson, Sadie Patraw, Hannah Petersen, Tobias Ragoczy, Joshua Richards, Jacob Rodriguez, John Stamatoyannopoulos, Julia Wald, Olivia Waltner, Michael Boeckh, Janet A. Englund, Michael Famulare, Barry R. Lutz, Mark J. Rieder, Lea M. Starita, Matthew Thompson, Helen Y. Chu, Jay Shendure, Trevor Bedford |
| EPI_ISL_1926153 | Laboratory Corporation of America | Centers for Disease Control and Prevention Division of Viral Diseases, Pathogen Discovery | Dakota Howard, Dhwani Batra, Peter W. Cook, Kara Moser, Adrian Paskey, Jason Caravas, Benjamin Rambo-Martin, Shatavia Morrison, Christopher Gulvick, Scott Sammons, Yvette Unoarumhi, Darlene Wagner, Matthew Schmerer, Minoo Agarwal, Eyad Almasri, Debbie Boles, Ayla Burns, Nuthawin Charoensri, Oren Cohen, Susan Countryman, Mary Ann Cristobal, Bobbi Croy, Suzanne Dale, Hrushikesh Deshmukh, Amanda Douglas, Vincent Drouillon, Marcia Eisenberg, Howard Engler, Rama Ghatti, Prashant Gupta, Susan Hicks, Jake Humphrey, Lax Iyer, Manoj Jain, Mohan Kolli, Brian Krueger, Tim Kuphal, Stanley Letovsky, Michael Levandoski, Craig Lukasik, Jonathan Meltzer, Brian Norvell, Mindy Nye, Scott Parker, Christos Petropoulos, John Pruitt, Steven Ragan, Scott Ryan, Mike Sapeta, Jana Schroth, Suresh Babu Selvaraju, Goran Stevovic, Amanda Suchanek, Andrea Throop, Lyndon Tilson, Thomas Urban, Joe Voshell, Kimberly Wagner, Jonathan Williams, Mary Williamson, Qian Zeng, Tricia Zwiefelhofer, Clinton R. Paden, Duncan MacCannell |
| EPI_ISL_1926156 | Altius Institute | Seattle Flu Study | Deborah A. Nickerson, Chris D. Frazar, Jover Lee, Benjamin Pelle, Erica Ryke, Matthew Richardson, Amanda Adler, Elisabeth Brandstetter, Peter D. Han, Kairsten Fay, Misja Ilcisin, Kirsten Lacombe, Thomas R. Sibley, Melissa Truong, Caitlin R. Wolf, Ryan Alexander, Daniel Bates, Rebecca Bruders, Stephanie DeBaun, Clem Green, Muhammad Halimun, Jessica Halow, Kneshay Harper, Matt Hartman, Andrew Meuser, Alex Nguyen, Truong Nguyen, Sofia Olsson, Sadie Patraw, Hannah Petersen, Tobias Ragoczy, Joshua Richards, Jacob Rodriguez, John Stamatoyannopoulos, Julia Wald, Olivia Waltner, Michael Boeckh, Janet A. Englund, Michael Famulare, Barry R. Lutz, Mark J. Rieder, Lea M. Starita, Matthew Thompson, Helen Y. Chu, Jay Shendure, Trevor Bedford |
| EPI_ISL_1926157 | Laboratory Corporation of America | Centers for Disease Control and Prevention Division of Viral Diseases, Pathogen Discovery | Dakota Howard, Dhwani Batra, Peter W. Cook, Kara Moser, Adrian Paskey, Jason Caravas, Benjamin Rambo-Martin, Shatavia Morrison, Christopher Gulvick, Scott Sammons, Yvette Unoarumhi, Darlene Wagner, Matthew Schmerer, Minoo Agarwal, Eyad Almasri, Debbie Boles, Ayla Burns, Nuthawin Charoensri, Oren Cohen, Susan Countryman, Mary Ann Cristobal, Bobbi Croy, Suzanne Dale, Hrushikesh Deshmukh, Amanda Douglas, Vincent Drouillon, Marcia Eisenberg, Howard Engler, Rama Ghatti, Prashant Gupta, Susan Hicks, Jake Humphrey, Lax Iyer, Manoj Jain, Mohan Kolli, Brian Krueger, Tim Kuphal, Stanley Letovsky, Michael Levandoski, Craig Lukasik, Jonathan Meltzer, Brian Norvell, Mindy Nye, Scott Parker, Christos Petropoulos, John Pruitt, Steven Ragan, Scott Ryan, Mike Sapeta, Jana Schroth, Suresh Babu Selvaraju, Goran Stevovic, Amanda Suchanek, Andrea Throop, Lyndon Tilson, Thomas Urban, Joe Voshell, Kimberly Wagner, Jonathan Williams, Mary Williamson, Qian Zeng, Tricia Zwiefelhofer, Clinton R. Paden, Duncan MacCannell |
| EPI_ISL_1926159 | Altius Institute | Seattle Flu Study | Deborah A. Nickerson, Chris D. Frazar, Jover Lee, Benjamin Pelle, Erica Ryke, Matthew Richardson, Amanda Adler, Elisabeth Brandstetter, Peter D. Han, Kairsten Fay, Misja Ilcisin, Kirsten Lacombe, Thomas R. Sibley, Melissa Truong, Caitlin R. Wolf, Ryan Alexander, Daniel Bates, Rebecca Bruders, Stephanie DeBaun, Clem Green, Muhammad Halimun, Jessica Halow, Kneshay Harper, Matt Hartman, Andrew Meuser, Alex Nguyen, Truong Nguyen, Sofia Olsson, Sadie Patraw, Hannah Petersen, Tobias Ragoczy, Joshua Richards, Jacob Rodriguez, John Stamatoyannopoulos, Julia Wald, Olivia Waltner, Michael Boeckh, Janet A. Englund, Michael Famulare, Barry R. Lutz, Mark J. Rieder, Lea M. Starita, Matthew Thompson, Helen Y. Chu, Jay Shendure, Trevor Bedford |
| EPI_ISL_1926160 | Laboratory Corporation of America | Centers for Disease Control and Prevention Division of Viral Diseases, Pathogen Discovery | Dakota Howard, Dhwani Batra, Peter W. Cook, Kara Moser, Adrian Paskey, Jason Caravas, Benjamin Rambo-Martin, Shatavia Morrison, Christopher Gulvick, Scott Sammons, Yvette Unoarumhi, Darlene Wagner, Matthew Schmerer, Minoo Agarwal, Eyad Almasri, Debbie Boles, Ayla Burns, Nuthawin Charoensri, Oren Cohen, Susan Countryman, Mary Ann Cristobal, Bobbi Croy, Suzanne Dale, Hrushikesh Deshmukh, Amanda Douglas, Vincent Drouillon, Marcia Eisenberg, Howard Engler, Rama Ghatti, Prashant Gupta, Susan Hicks, Jake Humphrey, Lax Iyer, Manoj Jain, Mohan Kolli, Brian Krueger, Tim Kuphal, Stanley Letovsky, Michael Levandoski, Craig Lukasik, Jonathan Meltzer, Brian Norvell, Mindy Nye, Scott Parker, Christos Petropoulos, John Pruitt, Steven Ragan, Scott Ryan, Mike Sapeta, Jana Schroth, Suresh Babu Selvaraju, Goran Stevovic, Amanda Suchanek, Andrea Throop, Lyndon Tilson, Thomas Urban, Joe Voshell, Kimberly Wagner, Jonathan Williams, Mary Williamson, Qian Zeng, Tricia Zwiefelhofer, Clinton R. Paden, Duncan MacCannell |
| EPI_ISL_1926161, EPI_ISL_1926162 | Altius Institute | Seattle Flu Study | Deborah A. Nickerson, Chris D. Frazar, Jover Lee, Benjamin Pelle, Erica Ryke, Matthew Richardson, Amanda Adler, Elisabeth Brandstetter, Peter D. Han, Kairsten Fay, Misja Ilcisin, Kirsten Lacombe, Thomas R. Sibley, Melissa Truong, Caitlin R. Wolf, Ryan Alexander, Daniel Bates, Rebecca Bruders, Stephanie DeBaun, Clem Green, Muhammad Halimun, Jessica Halow, Kneshay Harper, Matt Hartman, Andrew Meuser, Alex Nguyen, Truong Nguyen, Sofia Olsson, Sadie Patraw, Hannah Petersen, Tobias Ragoczy, Joshua Richards, Jacob Rodriguez, John Stamatoyannopoulos, Julia Wald, Olivia Waltner, Michael Boeckh, Janet A. Englund, Michael Famulare, Barry R. Lutz, Mark J. Rieder, Lea M. Starita, Matthew Thompson, Helen Y. Chu, Jay Shendure, Trevor Bedford |
| EPI_ISL_1926163 | Laboratory Corporation of America | Centers for Disease Control and Prevention Division of Viral Diseases, Pathogen Discovery | Dakota Howard, Dhwani Batra, Peter W. Cook, Kara Moser, Adrian Paskey, Jason Caravas, Benjamin Rambo-Martin, Shatavia Morrison, Christopher Gulvick, Scott Sammons, Yvette Unoarumhi, Darlene Wagner, Matthew Schmerer, Minoo Agarwal, Eyad Almasri, Debbie Boles, Ayla Burns, Nuthawin Charoensri, Oren Cohen, Susan Countryman, Mary Ann Cristobal, Bobbi Croy, Suzanne Dale, Hrushikesh Deshmukh, Amanda Douglas, Vincent Drouillon, Marcia Eisenberg, Howard Engler, Rama Ghatti, Prashant Gupta, Susan Hicks, Jake Humphrey, Lax Iyer, Manoj Jain, Mohan Kolli, Brian Krueger, Tim Kuphal, Stanley Letovsky, Michael Levandoski, Craig Lukasik, Jonathan Meltzer, Brian Norvell, Mindy Nye, Scott Parker, Christos Petropoulos, John Pruitt, Steven Ragan, Scott Ryan, Mike Sapeta, Jana Schroth, Suresh Babu Selvaraju, Goran Stevovic, Amanda Suchanek, Andrea Throop, Lyndon Tilson, Thomas Urban, Joe Voshell, Kimberly Wagner, Jonathan Williams, Mary Williamson, Qian Zeng, Tricia Zwiefelhofer, Clinton R. Paden, Duncan MacCannell |
| EPI_ISL_1926165 | Altius Institute | Seattle Flu Study | Deborah A. Nickerson, Chris D. Frazar, Jover Lee, Benjamin Pelle, Erica Ryke, Matthew Richardson, Amanda Adler, Elisabeth Brandstetter, Peter D. Han, Kairsten Fay, Misja Ilcisin, Kirsten Lacombe, Thomas R. Sibley, Melissa Truong, Caitlin R. Wolf, Ryan Alexander, Daniel Bates, Rebecca Bruders, Stephanie DeBaun, Clem Green, Muhammad Halimun, Jessica Halow, Kneshay Harper, Matt Hartman, Andrew Meuser, Alex Nguyen, Truong Nguyen, Sofia Olsson, Sadie Patraw, Hannah Petersen, Tobias Ragoczy, Joshua Richards, Jacob Rodriguez, John Stamatoyannopoulos, Julia Wald, Olivia Waltner, Michael Boeckh, Janet A. Englund, Michael Famulare, Barry R. Lutz, Mark J. Rieder, Lea M. Starita, Matthew Thompson, Helen Y. Chu, Jay Shendure, Trevor Bedford |
| EPI_ISL_1926166 | Laboratory Corporation of America | Centers for Disease Control and Prevention Division of Viral Diseases, Pathogen Discovery | Dakota Howard, Dhwani Batra, Peter W. Cook, Kara Moser, Adrian Paskey, Jason Caravas, Benjamin Rambo-Martin, Shatavia Morrison, Christopher Gulvick, Scott Sammons, Yvette Unoarumhi, Darlene Wagner, Matthew Schmerer, Minoo Agarwal, Eyad Almasri, Debbie Boles, Ayla Burns, Nuthawin Charoensri, Oren Cohen, Susan Countryman, Mary Ann Cristobal, Bobbi Croy, Suzanne Dale, Hrushikesh Deshmukh, Amanda Douglas, Vincent Drouillon, Marcia Eisenberg, Howard Engler, Rama Ghatti, Prashant Gupta, Susan Hicks, Jake Humphrey, Lax Iyer, Manoj Jain, Mohan Kolli, Brian Krueger, Tim Kuphal, Stanley Letovsky, Michael Levandoski, Craig Lukasik, Jonathan Meltzer, Brian Norvell, Mindy Nye, Scott Parker, Christos Petropoulos, John Pruitt, Steven Ragan, Scott Ryan, Mike Sapeta, Jana Schroth, Suresh Babu Selvaraju, Goran Stevovic, Amanda Suchanek, Andrea Throop, Lyndon Tilson, Thomas Urban, Joe Voshell, Kimberly Wagner, Jonathan Williams, Mary Williamson, Qian Zeng, Tricia Zwiefelhofer, Clinton R. Paden, Duncan MacCannell |
| EPI_ISL_1926167 | Altius Institute | Seattle Flu Study | Deborah A. Nickerson, Chris D. Frazar, Jover Lee, Benjamin Pelle, Erica Ryke, Matthew Richardson, Amanda Adler, Elisabeth Brandstetter, Peter D. Han, Kairsten Fay, Misja Ilcisin, Kirsten Lacombe, Thomas R. Sibley, Melissa Truong, Caitlin R. Wolf, Ryan Alexander, Daniel Bates, Rebecca Bruders, Stephanie DeBaun, Clem Green, Muhammad Halimun, Jessica Halow, Kneshay Harper, Matt Hartman, Andrew Meuser, Alex Nguyen, Truong Nguyen, Sofia Olsson, Sadie Patraw, Hannah Petersen, Tobias Ragoczy, Joshua Richards, Jacob Rodriguez, John Stamatoyannopoulos, Julia Wald, Olivia Waltner, Michael Boeckh, Janet A. Englund, Michael Famulare, Barry R. Lutz, Mark J. Rieder, Lea M. Starita, Matthew Thompson, Helen Y. Chu, Jay Shendure, Trevor Bedford |
| EPI_ISL_1926169 | Laboratory Corporation of America | Centers for Disease Control and Prevention Division of Viral Diseases, Pathogen Discovery | Dakota Howard, Dhwani Batra, Peter W. Cook, Kara Moser, Adrian Paskey, Jason Caravas, Benjamin Rambo-Martin, Shatavia Morrison, Christopher Gulvick, Scott Sammons, Yvette Unoarumhi, Darlene Wagner, Matthew Schmerer, Minoo Agarwal, Eyad Almasri, Debbie Boles, Ayla Burns, Nuthawin Charoensri, Oren Cohen, Susan Countryman, Mary Ann Cristobal, Bobbi Croy, Suzanne Dale, Hrushikesh Deshmukh, Amanda Douglas, Vincent Drouillon, Marcia Eisenberg, Howard Engler, Rama Ghatti, Prashant Gupta, Susan Hicks, Jake Humphrey, Lax Iyer, Manoj Jain, Mohan Kolli, Brian Krueger, Tim Kuphal, Stanley Letovsky, Michael Levandoski, Craig Lukasik, Jonathan Meltzer, Brian Norvell, Mindy Nye, Scott Parker, Christos Petropoulos, John Pruitt, Steven Ragan, Scott Ryan, Mike Sapeta, Jana Schroth, Suresh Babu Selvaraju, Goran Stevovic, Amanda Suchanek, Andrea Throop, Lyndon Tilson, Thomas Urban, Joe Voshell, Kimberly Wagner, Jonathan Williams, Mary Williamson, Qian Zeng, Tricia Zwiefelhofer, Clinton R. Paden, Duncan MacCannell |
| EPI_ISL_1926170 | Altius Institute | Seattle Flu Study | Deborah A. Nickerson, Chris D. Frazar, Jover Lee, Benjamin Pelle, Erica Ryke, Matthew Richardson, Amanda Adler, Elisabeth Brandstetter, Peter D. Han, Kairsten Fay, Misja Ilcisin, Kirsten Lacombe, Thomas R. Sibley, Melissa Truong, Caitlin R. Wolf, Ryan Alexander, Daniel Bates, Rebecca Bruders, Stephanie DeBaun, Clem Green, Muhammad Halimun, Jessica Halow, Kneshay Harper, Matt Hartman, Andrew Meuser, Alex Nguyen, Truong Nguyen, Sofia Olsson, Sadie Patraw, Hannah Petersen, Tobias Ragoczy, Joshua Richards, Jacob Rodriguez, John Stamatoyannopoulos, Julia Wald, Olivia Waltner, Michael Boeckh, Janet A. Englund, Michael Famulare, Barry R. Lutz, Mark J. Rieder, Lea M. Starita, Matthew Thompson, Helen Y. Chu, Jay Shendure, Trevor Bedford |
| EPI_ISL_1926172 | Laboratory Corporation of America | Centers for Disease Control and Prevention Division of Viral Diseases, Pathogen Discovery | Dakota Howard, Dhwani Batra, Peter W. Cook, Kara Moser, Adrian Paskey, Jason Caravas, Benjamin Rambo-Martin, Shatavia Morrison, Christopher Gulvick, Scott Sammons, Yvette Unoarumhi, Darlene Wagner, Matthew Schmerer, Minoo Agarwal, Eyad Almasri, Debbie Boles, Ayla Burns, Nuthawin Charoensri, Oren Cohen, Susan Countryman, Mary Ann Cristobal, Bobbi Croy, Suzanne Dale, Hrushikesh Deshmukh, Amanda Douglas, Vincent Drouillon, Marcia Eisenberg, Howard Engler, Rama Ghatti, Prashant Gupta, Susan Hicks, Jake Humphrey, Lax Iyer, Manoj Jain, Mohan Kolli, Brian Krueger, Tim Kuphal, Stanley Letovsky, Michael Levandoski, Craig Lukasik, Jonathan Meltzer, Brian Norvell, Mindy Nye, Scott Parker, Christos Petropoulos, John Pruitt, Steven Ragan, Scott Ryan, Mike Sapeta, Jana Schroth, Suresh Babu Selvaraju, Goran Stevovic, Amanda Suchanek, Andrea Throop, Lyndon Tilson, Thomas Urban, Joe Voshell, Kimberly Wagner, Jonathan Williams, Mary Williamson, Qian Zeng, Tricia Zwiefelhofer, Clinton R. Paden, Duncan MacCannell |
| EPI_ISL_1926173, EPI_ISL_1926174 | Altius Institute | Seattle Flu Study | Deborah A. Nickerson, Chris D. Frazar, Jover Lee, Benjamin Pelle, Erica Ryke, Matthew Richardson, Amanda Adler, Elisabeth Brandstetter, Peter D. Han, Kairsten Fay, Misja Ilcisin, Kirsten Lacombe, Thomas R. Sibley, Melissa Truong, Caitlin R. Wolf, Ryan Alexander, Daniel Bates, Rebecca Bruders, Stephanie DeBaun, Clem Green, Muhammad Halimun, Jessica Halow, Kneshay Harper, Matt Hartman, Andrew Meuser, Alex Nguyen, Truong Nguyen, Sofia Olsson, Sadie Patraw, Hannah Petersen, Tobias Ragoczy, Joshua Richards, Jacob Rodriguez, John Stamatoyannopoulos, Julia Wald, Olivia Waltner, Michael Boeckh, Janet A. Englund, Michael Famulare, Barry R. Lutz, Mark J. Rieder, Lea M. Starita, Matthew Thompson, Helen Y. Chu, Jay Shendure, Trevor Bedford |
| EPI_ISL_1926175 | Laboratory Corporation of America | Centers for Disease Control and Prevention Division of Viral Diseases, Pathogen Discovery | Dakota Howard, Dhwani Batra, Peter W. Cook, Kara Moser, Adrian Paskey, Jason Caravas, Benjamin Rambo-Martin, Shatavia Morrison, Christopher Gulvick, Scott Sammons, Yvette Unoarumhi, Darlene Wagner, Matthew Schmerer, Minoo Agarwal, Eyad Almasri, Debbie Boles, Ayla Burns, Nuthawin Charoensri, Oren Cohen, Susan Countryman, Mary Ann Cristobal, Bobbi Croy, Suzanne Dale, Hrushikesh Deshmukh, Amanda Douglas, Vincent Drouillon, Marcia Eisenberg, Howard Engler, Rama Ghatti, Prashant Gupta, Susan Hicks, Jake Humphrey, Lax Iyer, Manoj Jain, Mohan Kolli, Brian Krueger, Tim Kuphal, Stanley Letovsky, Michael Levandoski, Craig Lukasik, Jonathan Meltzer, Brian Norvell, Mindy Nye, Scott Parker, Christos Petropoulos, John Pruitt, Steven Ragan, Scott Ryan, Mike Sapeta, Jana Schroth, Suresh Babu Selvaraju, Goran Stevovic, Amanda Suchanek, Andrea Throop, Lyndon Tilson, Thomas Urban, Joe Voshell, Kimberly Wagner, Jonathan Williams, Mary Williamson, Qian Zeng, Tricia Zwiefelhofer, Clinton R. Paden, Duncan MacCannell |
| EPI_ISL_1926177, EPI_ISL_1926179 | Altius Institute | Seattle Flu Study | Deborah A. Nickerson, Chris D. Frazar, Jover Lee, Benjamin Pelle, Erica Ryke, Matthew Richardson, Amanda Adler, Elisabeth Brandstetter, Peter D. Han, Kairsten Fay, Misja Ilcisin, Kirsten Lacombe, Thomas R. Sibley, Melissa Truong, Caitlin R. Wolf, Ryan Alexander, Daniel Bates, Rebecca Bruders, Stephanie DeBaun, Clem Green, Muhammad Halimun, Jessica Halow, Kneshay Harper, Matt Hartman, Andrew Meuser, Alex Nguyen, Truong Nguyen, Sofia Olsson, Sadie Patraw, Hannah Petersen, Tobias Ragoczy, Joshua Richards, Jacob Rodriguez, John Stamatoyannopoulos, Julia Wald, Olivia Waltner, Michael Boeckh, Janet A. Englund, Michael Famulare, Barry R. Lutz, Mark J. Rieder, Lea M. Starita, Matthew Thompson, Helen Y. Chu, Jay Shendure, Trevor Bedford |
| EPI_ISL_1926181 | Laboratory Corporation of America | Centers for Disease Control and Prevention Division of Viral Diseases, Pathogen Discovery | Dakota Howard, Dhwani Batra, Peter W. Cook, Kara Moser, Adrian Paskey, Jason Caravas, Benjamin Rambo-Martin, Shatavia Morrison, Christopher Gulvick, Scott Sammons, Yvette Unoarumhi, Darlene Wagner, Matthew Schmerer, Minoo Agarwal, Eyad Almasri, Debbie Boles, Ayla Burns, Nuthawin Charoensri, Oren Cohen, Susan Countryman, Mary Ann Cristobal, Bobbi Croy, Suzanne Dale, Hrushikesh Deshmukh, Amanda Douglas, Vincent Drouillon, Marcia Eisenberg, Howard Engler, Rama Ghatti, Prashant Gupta, Susan Hicks, Jake Humphrey, Lax Iyer, Manoj Jain, Mohan Kolli, Brian Krueger, Tim Kuphal, Stanley Letovsky, Michael Levandoski, Craig Lukasik, Jonathan Meltzer, Brian Norvell, Mindy Nye, Scott Parker, Christos Petropoulos, John Pruitt, Steven Ragan, Scott Ryan, Mike Sapeta, Jana Schroth, Suresh Babu Selvaraju, Goran Stevovic, Amanda Suchanek, Andrea Throop, Lyndon Tilson, Thomas Urban, Joe Voshell, Kimberly Wagner, Jonathan Williams, Mary Williamson, Qian Zeng, Tricia Zwiefelhofer, Clinton R. Paden, Duncan MacCannell |
| EPI_ISL_1926182, EPI_ISL_1926183, EPI_ISL_1926184 | Altius Institute | Seattle Flu Study | Deborah A. Nickerson, Chris D. Frazar, Jover Lee, Benjamin Pelle, Erica Ryke, Matthew Richardson, Amanda Adler, Elisabeth Brandstetter, Peter D. Han, Kairsten Fay, Misja Ilcisin, Kirsten Lacombe, Thomas R. Sibley, Melissa Truong, Caitlin R. Wolf, Ryan Alexander, Daniel Bates, Rebecca Bruders, Stephanie DeBaun, Clem Green, Muhammad Halimun, Jessica Halow, Kneshay Harper, Matt Hartman, Andrew Meuser, Alex Nguyen, Truong Nguyen, Sofia Olsson, Sadie Patraw, Hannah Petersen, Tobias Ragoczy, Joshua Richards, Jacob Rodriguez, John Stamatoyannopoulos, Julia Wald, Olivia Waltner, Michael Boeckh, Janet A. Englund, Michael Famulare, Barry R. Lutz, Mark J. Rieder, Lea M. Starita, Matthew Thompson, Helen Y. Chu, Jay Shendure, Trevor Bedford |
| EPI_ISL_1926185 | Laboratory Corporation of America | Centers for Disease Control and Prevention Division of Viral Diseases, Pathogen Discovery | Dakota Howard, Dhwani Batra, Peter W. Cook, Kara Moser, Adrian Paskey, Jason Caravas, Benjamin Rambo-Martin, Shatavia Morrison, Christopher Gulvick, Scott Sammons, Yvette Unoarumhi, Darlene Wagner, Matthew Schmerer, Minoo Agarwal, Eyad Almasri, Debbie Boles, Ayla Burns, Nuthawin Charoensri, Oren Cohen, Susan Countryman, Mary Ann Cristobal, Bobbi Croy, Suzanne Dale, Hrushikesh Deshmukh, Amanda Douglas, Vincent Drouillon, Marcia Eisenberg, Howard Engler, Rama Ghatti, Prashant Gupta, Susan Hicks, Jake Humphrey, Lax Iyer, Manoj Jain, Mohan Kolli, Brian Krueger, Tim Kuphal, Stanley Letovsky, Michael Levandoski, Craig Lukasik, Jonathan Meltzer, Brian Norvell, Mindy Nye, Scott Parker, Christos Petropoulos, John Pruitt, Steven Ragan, Scott Ryan, Mike Sapeta, Jana Schroth, Suresh Babu Selvaraju, Goran Stevovic, Amanda Suchanek, Andrea Throop, Lyndon Tilson, Thomas Urban, Joe Voshell, Kimberly Wagner, Jonathan Williams, Mary Williamson, Qian Zeng, Tricia Zwiefelhofer, Clinton R. Paden, Duncan MacCannell |
| EPI_ISL_1926186 | Altius Institute | Seattle Flu Study | Deborah A. Nickerson, Chris D. Frazar, Jover Lee, Benjamin Pelle, Erica Ryke, Matthew Richardson, Amanda Adler, Elisabeth Brandstetter, Peter D. Han, Kairsten Fay, Misja Ilcisin, Kirsten Lacombe, Thomas R. Sibley, Melissa Truong, Caitlin R. Wolf, Ryan Alexander, Daniel Bates, Rebecca Bruders, Stephanie DeBaun, Clem Green, Muhammad Halimun, Jessica Halow, Kneshay Harper, Matt Hartman, Andrew Meuser, Alex Nguyen, Truong Nguyen, Sofia Olsson, Sadie Patraw, Hannah Petersen, Tobias Ragoczy, Joshua Richards, Jacob Rodriguez, John Stamatoyannopoulos, Julia Wald, Olivia Waltner, Michael Boeckh, Janet A. Englund, Michael Famulare, Barry R. Lutz, Mark J. Rieder, Lea M. Starita, Matthew Thompson, Helen Y. Chu, Jay Shendure, Trevor Bedford |
| EPI_ISL_1926188 | Laboratory Corporation of America | Centers for Disease Control and Prevention Division of Viral Diseases, Pathogen Discovery | Dakota Howard, Dhwani Batra, Peter W. Cook, Kara Moser, Adrian Paskey, Jason Caravas, Benjamin Rambo-Martin, Shatavia Morrison, Christopher Gulvick, Scott Sammons, Yvette Unoarumhi, Darlene Wagner, Matthew Schmerer, Minoo Agarwal, Eyad Almasri, Debbie Boles, Ayla Burns, Nuthawin Charoensri, Oren Cohen, Susan Countryman, Mary Ann Cristobal, Bobbi Croy, Suzanne Dale, Hrushikesh Deshmukh, Amanda Douglas, Vincent Drouillon, Marcia Eisenberg, Howard Engler, Rama Ghatti, Prashant Gupta, Susan Hicks, Jake Humphrey, Lax Iyer, Manoj Jain, Mohan Kolli, Brian Krueger, Tim Kuphal, Stanley Letovsky, Michael Levandoski, Craig Lukasik, Jonathan Meltzer, Brian Norvell, Mindy Nye, Scott Parker, Christos Petropoulos, John Pruitt, Steven Ragan, Scott Ryan, Mike Sapeta, Jana Schroth, Suresh Babu Selvaraju, Goran Stevovic, Amanda Suchanek, Andrea Throop, Lyndon Tilson, Thomas Urban, Joe Voshell, Kimberly Wagner, Jonathan Williams, Mary Williamson, Qian Zeng, Tricia Zwiefelhofer, Clinton R. Paden, Duncan MacCannell |
| EPI_ISL_1926189, EPI_ISL_1926190 | Altius Institute | Seattle Flu Study | Deborah A. Nickerson, Chris D. Frazar, Jover Lee, Benjamin Pelle, Erica Ryke, Matthew Richardson, Amanda Adler, Elisabeth Brandstetter, Peter D. Han, Kairsten Fay, Misja Ilcisin, Kirsten Lacombe, Thomas R. Sibley, Melissa Truong, Caitlin R. Wolf, Ryan Alexander, Daniel Bates, Rebecca Bruders, Stephanie DeBaun, Clem Green, Muhammad Halimun, Jessica Halow, Kneshay Harper, Matt Hartman, Andrew Meuser, Alex Nguyen, Truong Nguyen, Sofia Olsson, Sadie Patraw, Hannah Petersen, Tobias Ragoczy, Joshua Richards, Jacob Rodriguez, John Stamatoyannopoulos, Julia Wald, Olivia Waltner, Michael Boeckh, Janet A. Englund, Michael Famulare, Barry R. Lutz, Mark J. Rieder, Lea M. Starita, Matthew Thompson, Helen Y. Chu, Jay Shendure, Trevor Bedford |
| EPI_ISL_1926191 | Laboratory Corporation of America | Centers for Disease Control and Prevention Division of Viral Diseases, Pathogen Discovery | Dakota Howard, Dhwani Batra, Peter W. Cook, Kara Moser, Adrian Paskey, Jason Caravas, Benjamin Rambo-Martin, Shatavia Morrison, Christopher Gulvick, Scott Sammons, Yvette Unoarumhi, Darlene Wagner, Matthew Schmerer, Minoo Agarwal, Eyad Almasri, Debbie Boles, Ayla Burns, Nuthawin Charoensri, Oren Cohen, Susan Countryman, Mary Ann Cristobal, Bobbi Croy, Suzanne Dale, Hrushikesh Deshmukh, Amanda Douglas, Vincent Drouillon, Marcia Eisenberg, Howard Engler, Rama Ghatti, Prashant Gupta, Susan Hicks, Jake Humphrey, Lax Iyer, Manoj Jain, Mohan Kolli, Brian Krueger, Tim Kuphal, Stanley Letovsky, Michael Levandoski, Craig Lukasik, Jonathan Meltzer, Brian Norvell, Mindy Nye, Scott Parker, Christos Petropoulos, John Pruitt, Steven Ragan, Scott Ryan, Mike Sapeta, Jana Schroth, Suresh Babu Selvaraju, Goran Stevovic, Amanda Suchanek, Andrea Throop, Lyndon Tilson, Thomas Urban, Joe Voshell, Kimberly Wagner, Jonathan Williams, Mary Williamson, Qian Zeng, Tricia Zwiefelhofer, Clinton R. Paden, Duncan MacCannell |
| EPI_ISL_1926192 | Altius Institute | Seattle Flu Study | Deborah A. Nickerson, Chris D. Frazar, Jover Lee, Benjamin Pelle, Erica Ryke, Matthew Richardson, Amanda Adler, Elisabeth Brandstetter, Peter D. Han, Kairsten Fay, Misja Ilcisin, Kirsten Lacombe, Thomas R. Sibley, Melissa Truong, Caitlin R. Wolf, Ryan Alexander, Daniel Bates, Rebecca |

| | | | |
|---|---|---|---|
| | | | Bruders, Stephanie DeBaun, Clem Green, Muhammad Halimun, Jessica Halow, Kneshay Harper, Matt Hartman, Andrew Meuser, Alex Nguyen, Truong Nguyen, Sofia Olsson, Sadie Patraw, Hannah Petersen, Tobias Ragoczy, Joshua Richards, Jacob Rodriguez, John Stamatoyannopoulos, Julia Wald, Olivia Waltner, Michael Boeckh, Janet A. Englund, Michael Famulare, Barry R. Lutz, Mark J. Rieder, Lea M. Starita, Matthew Thompson, Helen Y. Chu, Jay Shendure, Trevor Bedford |
| EPI_ISL_1926194 | Laboratory Corporation of America | Centers for Disease Control and Prevention Division of Viral Diseases, Pathogen Discovery | Dakota Howard, Dhwani Batra, Peter W. Cook, Kara Moser, Adrian Paskey, Jason Caravas, Benjamin Rambo-Martin, Shatavia Morrison, Christopher Gulvick, Scott Sammons, Yvette Unoarumhi, Darlene Wagner, Matthew Schmerer, Minoo Agarwal, Eyad Almasri, Debbie Boles, Ayla Burns, Nuthawin Charoensri, Oren Cohen, Susan Countryman, Mary Ann Cristobal, Bobbi Croy, Suzanne Dale, Hrushikesh Deshmukh, Amanda Douglas, Vincent Drouillon, Marcia Eisenberg, Howard Engler, Rama Ghatti, Prashant Gupta, Susan Hicks, Jake Humphrey, Lax Iyer, Manoj Jain, Mohan Kolli, Brian Krueger, Tim Kuphal, Stanley Letovsky, Michael Levandoski, Craig Lukasik, Jonathan Meltzer, Brian Norvell, Mindy Nye, Scott Parker, Christos Petropoulos, John Pruitt, Steven Ragan, Scott Ryan, Mike Sapeta, Jana Schroth, Suresh Babu Selvaraju, Goran Stevovic, Amanda Suchanek, Andrea Throop, Lyndon Tilson, Thomas Urban, Joe Voshell, Kimberly Wagner, Jonathan Williams, Mary Williamson, Qian Zeng, Tricia Zwiefelhofer, Clinton R. Paden, Duncan MacCannell |
| EPI_ISL_1926195, EPI_ISL_1926196 | Altius Institute | Seattle Flu Study | Deborah A. Nickerson, Chris D. Frazar, Jover Lee, Benjamin Pelle, Erica Ryke, Matthew Richardson, Amanda Adler, Elisabeth Brandstetter, Peter D. Han, Kairsten Fay, Misja Ilcisin, Kirsten Lacombe, Thomas R. Sibley, Melissa Truong, Caitlin R. Wolf, Ryan Alexander, Daniel Bates, Rebecca Bruders, Stephanie DeBaun, Clem Green, Muhammad Halimun, Jessica Halow, Kneshay Harper, Matt Hartman, Andrew Meuser, Alex Nguyen, Truong Nguyen, Sofia Olsson, Sadie Patraw, Hannah Petersen, Tobias Ragoczy, Joshua Richards, Jacob Rodriguez, John Stamatoyannopoulos, Julia Wald, Olivia Waltner, Michael Boeckh, Janet A. Englund, Michael Famulare, Barry R. Lutz, Mark J. Rieder, Lea M. Starita, Matthew Thompson, Helen Y. Chu, Jay Shendure, Trevor Bedford |
| EPI_ISL_1926197, EPI_ISL_1926200 | Laboratory Corporation of America | Centers for Disease Control and Prevention Division of Viral Diseases, Pathogen Discovery | Dakota Howard, Dhwani Batra, Peter W. Cook, Kara Moser, Adrian Paskey, Jason Caravas, Benjamin Rambo-Martin, Shatavia Morrison, Christopher Gulvick, Scott Sammons, Yvette Unoarumhi, Darlene Wagner, Matthew Schmerer, Minoo Agarwal, Eyad Almasri, Debbie Boles, Ayla Burns, Nuthawin Charoensri, Oren Cohen, Susan Countryman, Mary Ann Cristobal, Bobbi Croy, Suzanne Dale, Hrushikesh Deshmukh, Amanda Douglas, Vincent Drouillon, Marcia Eisenberg, Howard Engler, Rama Ghatti, Prashant Gupta, Susan Hicks, Jake Humphrey, Lax Iyer, Manoj Jain, Mohan Kolli, Brian Krueger, Tim Kuphal, Stanley Letovsky, Michael Levandoski, Craig Lukasik, Jonathan Meltzer, Brian Norvell, Mindy Nye, Scott Parker, Christos Petropoulos, John Pruitt, Steven Ragan, Scott Ryan, Mike Sapeta, Jana Schroth, Suresh Babu Selvaraju, Goran Stevovic, Amanda Suchanek, Andrea Throop, Lyndon Tilson, Thomas Urban, Joe Voshell, Kimberly Wagner, Jonathan Williams, Mary Williamson, Qian Zeng, Tricia Zwiefelhofer, Clinton R. Paden, Duncan MacCannell |
| EPI_ISL_1926202 | Altius Institute | Seattle Flu Study | Deborah A. Nickerson, Chris D. Frazar, Jover Lee, Benjamin Pelle, Erica Ryke, Matthew Richardson, Amanda Adler, Elisabeth Brandstetter, Peter D. Han, Kairsten Fay, Misja Ilcisin, Kirsten Lacombe, Thomas R. Sibley, Melissa Truong, Caitlin R. Wolf, Ryan Alexander, Daniel Bates, Rebecca Bruders, Stephanie DeBaun, Clem Green, Muhammad Halimun, Jessica Halow, Kneshay Harper, Matt Hartman, Andrew Meuser, Alex Nguyen, Truong Nguyen, Sofia Olsson, Sadie Patraw, Hannah Petersen, Tobias Ragoczy, Joshua Richards, Jacob Rodriguez, John Stamatoyannopoulos, Julia Wald, Olivia Waltner, Michael Boeckh, Janet A. Englund, Michael Famulare, Barry R. Lutz, Mark J. Rieder, Lea M. Starita, Matthew Thompson, Helen Y. Chu, Jay Shendure, Trevor Bedford |
| EPI_ISL_1926203 | Laboratory Corporation of America | Centers for Disease Control and Prevention Division of Viral Diseases, Pathogen Discovery | Dakota Howard, Dhwani Batra, Peter W. Cook, Kara Moser, Adrian Paskey, Jason Caravas, Benjamin Rambo-Martin, Shatavia Morrison, Christopher Gulvick, Scott Sammons, Yvette Unoarumhi, Darlene Wagner, Matthew Schmerer, Minoo Agarwal, Eyad Almasri, Debbie Boles, Ayla Burns, Nuthawin Charoensri, Oren Cohen, Susan Countryman, Mary Ann Cristobal, Bobbi Croy, Suzanne Dale, Hrushikesh Deshmukh, Amanda Douglas, Vincent Drouillon, Marcia Eisenberg, Howard Engler, Rama Ghatti, Prashant Gupta, Susan Hicks, Jake Humphrey, Lax Iyer, Manoj Jain, Mohan Kolli, Brian Krueger, Tim Kuphal, Stanley Letovsky, Michael Levandoski, Craig Lukasik, Jonathan Meltzer, Brian Norvell, Mindy Nye, Scott Parker, Christos Petropoulos, John Pruitt, Steven Ragan, Scott Ryan, Mike Sapeta, Jana Schroth, Suresh Babu Selvaraju, Goran Stevovic, Amanda Suchanek, Andrea Throop, Lyndon Tilson, Thomas Urban, Joe Voshell, Kimberly Wagner, Jonathan Williams, Mary Williamson, Qian Zeng, Tricia Zwiefelhofer, Clinton R. Paden, Duncan MacCannell |
| EPI_ISL_1926205 | Altius Institute | Seattle Flu Study | Deborah A. Nickerson, Chris D. Frazar, Jover Lee, Benjamin Pelle, Erica Ryke, Matthew Richardson, Amanda Adler, Elisabeth Brandstetter, Peter D. Han, Kairsten Fay, Misja Ilcisin, Kirsten Lacombe, Thomas R. Sibley, Melissa Truong, Caitlin R. Wolf, Ryan Alexander, Daniel Bates, Rebecca Bruders, Stephanie DeBaun, Clem Green, Muhammad Halimun, Jessica Halow, Kneshay Harper, Matt Hartman, Andrew Meuser, Alex Nguyen, Truong Nguyen, Sofia Olsson, Sadie Patraw, Hannah Petersen, Tobias Ragoczy, Joshua Richards, Jacob Rodriguez, John Stamatoyannopoulos, Julia Wald, Olivia Waltner, Michael Boeckh, Janet A. Englund, Michael Famulare, Barry R. Lutz, Mark J. Rieder, Lea M. Starita, Matthew Thompson, Helen Y. Chu, Jay Shendure, Trevor Bedford |
| EPI_ISL_1926206 | Laboratory Corporation of America | Centers for Disease Control and Prevention Division of Viral Diseases, Pathogen Discovery | Dakota Howard, Dhwani Batra, Peter W. Cook, Kara Moser, Adrian Paskey, Jason Caravas, Benjamin Rambo-Martin, Shatavia Morrison, Christopher Gulvick, Scott Sammons, Yvette Unoarumhi, Darlene Wagner, Matthew Schmerer, Minoo Agarwal, Eyad Almasri, Debbie Boles, Ayla Burns, Nuthawin Charoensri, Oren Cohen, Susan Countryman, Mary Ann Cristobal, Bobbi Croy, Suzanne Dale, Hrushikesh Deshmukh, Amanda Douglas, Vincent Drouillon, Marcia Eisenberg, Howard Engler, Rama Ghatti, Prashant Gupta, Susan Hicks, Jake Humphrey, Lax Iyer, Manoj Jain, Mohan Kolli, Brian Krueger, Tim Kuphal, Stanley Letovsky, Michael Levandoski, Craig Lukasik, Jonathan Meltzer, Brian Norvell, Mindy Nye, Scott Parker, Christos Petropoulos, John Pruitt, Steven Ragan, Scott Ryan, Mike Sapeta, Jana Schroth, Suresh Babu Selvaraju, Goran Stevovic, Amanda Suchanek, Andrea Throop, Lyndon Tilson, Thomas Urban, Joe Voshell, Kimberly Wagner, Jonathan Williams, Mary Williamson, Qian Zeng, Tricia Zwiefelhofer, Clinton R. Paden, Duncan MacCannell |
| EPI_ISL_1926208 | Altius Institute | Seattle Flu Study | Deborah A. Nickerson, Chris D. Frazar, Jover Lee, Benjamin Pelle, Erica Ryke, Matthew Richardson, Amanda Adler, Elisabeth Brandstetter, Peter D. Han, Kairsten Fay, Misja Ilcisin, Kirsten Lacombe, Thomas R. Sibley, Melissa Truong, Caitlin R. Wolf, Ryan Alexander, Daniel Bates, Rebecca Bruders, Stephanie DeBaun, Clem Green, Muhammad Halimun, Jessica Halow, Kneshay Harper, Matt Hartman, Andrew Meuser, Alex Nguyen, Truong Nguyen, Sofia Olsson, Sadie Patraw, Hannah Petersen, Tobias Ragoczy, Joshua Richards, Jacob Rodriguez, John Stamatoyannopoulos, Julia Wald, Olivia Waltner, Michael Boeckh, Janet A. Englund, Michael Famulare, Barry R. Lutz, Mark J. Rieder, Lea M. Starita, Matthew Thompson, Helen Y. Chu, Jay Shendure, Trevor Bedford |
| EPI_ISL_1926209 | Laboratory Corporation of America | Centers for Disease Control and Prevention Division of Viral Diseases, Pathogen Discovery | Dakota Howard, Dhwani Batra, Peter W. Cook, Kara Moser, Adrian Paskey, Jason Caravas, Benjamin Rambo-Martin, Shatavia Morrison, Christopher Gulvick, Scott Sammons, Yvette Unoarumhi, Darlene Wagner, Matthew Schmerer, Minoo Agarwal, Eyad Almasri, Debbie Boles, Ayla Burns, Nuthawin Charoensri, Oren Cohen, Susan Countryman, Mary Ann Cristobal, Bobbi Croy, Suzanne Dale, Hrushikesh Deshmukh, Amanda Douglas, Vincent Drouillon, Marcia Eisenberg, Howard Engler, Rama Ghatti, Prashant Gupta, Susan Hicks, Jake Humphrey, Lax Iyer, Manoj Jain, Mohan Kolli, Brian Krueger, Tim Kuphal, Stanley Letovsky, Michael Levandoski, Craig Lukasik, Jonathan Meltzer, Brian Norvell, Mindy Nye, Scott Parker, Christos Petropoulos, John Pruitt, Steven Ragan, Scott Ryan, Mike Sapeta, Jana Schroth, Suresh Babu Selvaraju, Goran Stevovic, Amanda Suchanek, Andrea Throop, Lyndon Tilson, Thomas Urban, Joe Voshell, Kimberly Wagner, Jonathan Williams, Mary Williamson, Qian Zeng, Tricia Zwiefelhofer, Clinton R. Paden, Duncan MacCannell |
| EPI_ISL_1926211, EPI_ISL_1926213, EPI_ISL_1926214 | Altius Institute | Seattle Flu Study | Deborah A. Nickerson, Chris D. Frazar, Jover Lee, Benjamin Pelle, Erica Ryke, Matthew Richardson, Amanda Adler, Elisabeth Brandstetter, Peter D. Han, Kairsten Fay, Misja Ilcisin, Kirsten Lacombe, Thomas R. Sibley, Melissa Truong, Caitlin R. Wolf, Ryan Alexander, Daniel Bates, Rebecca Bruders, Stephanie DeBaun, Clem Green, Muhammad Halimun, Jessica Halow, Kneshay Harper, Matt Hartman, Andrew Meuser, Alex Nguyen, Truong Nguyen, Sofia Olsson, Sadie Patraw, Hannah Petersen, Tobias Ragoczy, Joshua Richards, Jacob Rodriguez, John Stamatoyannopoulos, Julia Wald, Olivia Waltner, Michael Boeckh, Janet A. Englund, Michael Famulare, Barry R. Lutz, Mark J. Rieder, Lea M. Starita, Matthew Thompson, Helen Y. Chu, Jay Shendure, Trevor Bedford |
| EPI_ISL_1926215, EPI_ISL_1926218 | Laboratory Corporation of America | Centers for Disease Control and Prevention Division of Viral Diseases, Pathogen Discovery | Dakota Howard, Dhwani Batra, Peter W. Cook, Kara Moser, Adrian Paskey, Jason Caravas, Benjamin Rambo-Martin, Shatavia Morrison, Christopher Gulvick, Scott Sammons, Yvette Unoarumhi, Darlene Wagner, Matthew Schmerer, Minoo Agarwal, Eyad Almasri, Debbie Boles, Ayla Burns, Nuthawin Charoensri, Oren Cohen, Susan Countryman, Mary Ann Cristobal, Bobbi Croy, Suzanne Dale, Hrushikesh Deshmukh, Amanda Douglas, Vincent Drouillon, Marcia Eisenberg, Howard Engler, Rama Ghatti, Prashant Gupta, Susan Hicks, Jake Humphrey, Lax Iyer, Manoj Jain, Mohan Kolli, Brian Krueger, Tim Kuphal, Stanley Letovsky, Michael Levandoski, Craig Lukasik, Jonathan Meltzer, Brian Norvell, Mindy Nye, Scott Parker, Christos Petropoulos, John Pruitt, Steven Ragan, Scott Ryan, Mike Sapeta, Jana Schroth, Suresh Babu Selvaraju, Goran Stevovic, Amanda Suchanek, Andrea Throop, Lyndon Tilson, Thomas Urban, Joe Voshell, Kimberly Wagner, Jonathan Williams, Mary Williamson, Qian Zeng, Tricia Zwiefelhofer, Clinton R. Paden, Duncan MacCannell |
| EPI_ISL_1926220, EPI_ISL_1926222, EPI_ISL_1926224 | Altius Institute | Seattle Flu Study | Deborah A. Nickerson, Chris D. Frazar, Jover Lee, Benjamin Pelle, Erica Ryke, Matthew Richardson, Amanda Adler, Elisabeth Brandstetter, Peter D. Han, Kairsten Fay, Misja Ilcisin, Kirsten Lacombe, Thomas R. Sibley, Melissa Truong, Caitlin R. Wolf, Ryan Alexander, Daniel Bates, Rebecca Bruders, Stephanie DeBaun, Clem Green, Muhammad Halimun, Jessica Halow, Kneshay Harper, Matt Hartman, Andrew Meuser, Alex Nguyen, Truong Nguyen, Sofia Olsson, Sadie Patraw, Hannah Petersen, Tobias Ragoczy, Joshua Richards, Jacob Rodriguez, John Stamatoyannopoulos, Julia Wald, Olivia Waltner, Michael Boeckh, Janet A. Englund, Michael Famulare, Barry R. Lutz, Mark J. Rieder, Lea M. Starita, Matthew Thompson, Helen Y. Chu, Jay Shendure, Trevor Bedford |
| EPI_ISL_1926225 | Laboratory Corporation of America | Centers for Disease Control and Prevention Division of Viral Diseases, Pathogen Discovery | Dakota Howard, Dhwani Batra, Peter W. Cook, Kara Moser, Adrian Paskey, Jason Caravas, Benjamin Rambo-Martin, Shatavia Morrison, Christopher Gulvick, Scott Sammons, Yvette Unoarumhi, Darlene Wagner, Matthew Schmerer, Minoo Agarwal, Eyad Almasri, Debbie Boles, Ayla Burns, Nuthawin Charoensri, Oren Cohen, Susan Countryman, Mary Ann Cristobal, Bobbi Croy, Suzanne Dale, Hrushikesh Deshmukh, Amanda Douglas, Vincent Drouillon, Marcia Eisenberg, Howard Engler, Rama Ghatti, Prashant Gupta, Susan Hicks, Jake Humphrey, Lax Iyer, Manoj Jain, Mohan Kolli, Brian Krueger, Tim Kuphal, Stanley Letovsky, Michael Levandoski, Craig Lukasik, Jonathan Meltzer, Brian Norvell, Mindy Nye, Scott Parker, Christos Petropoulos, John Pruitt, Steven Ragan, Scott Ryan, Mike Sapeta, Jana Schroth, Suresh Babu Selvaraju, Goran Stevovic, Amanda Suchanek, Andrea Throop, Lyndon Tilson, Thomas Urban, Joe Voshell, Kimberly Wagner, Jonathan Williams, Mary Williamson, Qian Zeng, Tricia Zwiefelhofer, Clinton R. Paden, Duncan MacCannell |
| EPI_ISL_1926226 | Altius Institute | Seattle Flu Study | Deborah A. Nickerson, Chris D. Frazar, Jover Lee, Benjamin Pelle, Erica Ryke, Matthew Richardson, Amanda Adler, Elisabeth Brandstetter, Peter D. Han, Kairsten Fay, Misja Ilcisin, Kirsten Lacombe, Thomas R. Sibley, Melissa Truong, Caitlin R. Wolf, Ryan Alexander, Daniel Bates, Rebecca Bruders, Stephanie DeBaun, Clem Green, Muhammad Halimun, Jessica Halow, Kneshay Harper, Matt Hartman, Andrew Meuser, Alex Nguyen, Truong Nguyen, Sofia Olsson, Sadie Patraw, Hannah Petersen, Tobias Ragoczy, Joshua Richards, Jacob Rodriguez, John Stamatoyannopoulos, Julia Wald, Olivia Waltner, Michael Boeckh, Janet A. Englund, Michael Famulare, Barry R. Lutz, Mark J. Rieder, Lea M. Starita, Matthew Thompson, Helen Y. Chu, Jay Shendure, Trevor Bedford |

EPI_ISL_1926227, EPI_ISL_1926228, EPI_ISL_1926229, EPI_ISL_1926230, EPI_ISL_1926231, EPI_ISL_1926232, EPI_ISL_1926233, EPI_ISL_1926234, EPI_ISL_1926235, EPI_ISL_1926237, EPI_ISL_1926238, EPI_ISL_1926239, EPI_ISL_1926240, EPI_ISL_1926241, EPI_ISL_1926242, EPI_ISL_1926245, EPI_ISL_1926246, EPI_ISL_1926247, EPI_ISL_1926248, EPI_ISL_1926249, EPI_ISL_1926250, EPI_ISL_1926251, EPI_ISL_1926252, EPI_ISL_1926253, EPI_ISL_1926254, EPI_ISL_1926255, EPI_ISL_1926256, EPI_ISL_1926258, EPI_ISL_1926259, EPI_ISL_1926260, EPI_ISL_1926261, EPI_ISL_1926262, EPI_ISL_1926263, EPI_ISL_1926264, EPI_ISL_1926265, EPI_ISL_1926266, EPI_ISL_1926267, EPI_ISL_1926268, EPI_ISL_1926269, EPI_ISL_1926271, EPI_ISL_1926272, EPI_ISL_1926273, EPI_ISL_1926274, EPI_ISL_1926276, EPI_ISL_1926278, EPI_ISL_1926279, EPI_ISL_1926280, EPI_ISL_1926281, EPI_ISL_1926282, EPI_ISL_1926283, EPI_ISL_1926284, EPI_ISL_1926285, EPI_ISL_1926288, EPI_ISL_1926289, EPI_ISL_1926290, EPI_ISL_1926291, EPI_ISL_1926292, EPI_ISL_1926293, EPI_ISL_1926294, EPI_ISL_1926295, EPI_ISL_1926297, EPI_ISL_1926298, EPI_ISL_1926299, EPI_ISL_1926300, EPI_ISL_1926301, EPI_ISL_1926302, EPI_ISL_1926303, EPI_ISL_1926304, EPI_ISL_1926305, EPI_ISL_1926307, EPI_ISL_1926308, EPI_ISL_1926309, EPI_ISL_1926310, EPI_ISL_1926311, EPI_ISL_1926312, EPI_ISL_1926313, EPI_ISL_1926315, EPI_ISL_1926316, EPI_ISL_1926317, EPI_ISL_1926318, EPI_ISL_1926320, EPI_ISL_1926322, EPI_ISL_1926323, EPI_ISL_1926324, EPI_ISL_1926325, EPI_ISL_1926327, EPI_ISL_1926328, EPI_ISL_1926329, EPI_ISL_1926330, EPI_ISL_1926331, EPI_ISL_1926332, EPI_ISL_1926333, EPI_ISL_1926334, EPI_ISL_1926335, EPI_ISL_1926336, EPI_ISL_1926338, EPI_ISL_1926339, EPI_ISL_1926341, EPI_ISL_1926342, EPI_ISL_1926343, EPI_ISL_1926344, EPI_ISL_1926345, EPI_ISL_1926346, EPI_ISL_1926347, EPI_ISL_1926348, EPI_ISL_1926349, EPI_ISL_1926350, EPI_ISL_1926352, EPI_ISL_1926353, EPI_ISL_1926354, EPI_ISL_1926355, EPI_ISL_1926356, EPI_ISL_1926357, EPI_ISL_1926358, EPI_ISL_1926359, EPI_ISL_1926360, EPI_ISL_1926361, EPI_ISL_1926362, EPI_ISL_1926363, EPI_ISL_1926364, EPI_ISL_1926365, EPI_ISL_1926366, EPI_ISL_1926367, EPI_ISL_1926368, EPI_ISL_1926370, EPI_ISL_1926371, EPI_ISL_1926373, EPI_ISL_1926374, EPI_ISL_1926375, EPI_ISL_1926376, EPI_ISL_1926377, EPI_ISL_1926378, EPI_ISL_1926380, EPI_ISL_1926382, EPI_ISL_1926383, EPI_ISL_1926384, EPI_ISL_1926385, EPI_ISL_1926386, EPI_ISL_1926387, EPI_ISL_1926388, EPI_ISL_1926389, EPI_ISL_1926390, EPI_ISL_1926391, EPI_ISL_1926392, EPI_ISL_1926393, EPI_ISL_1926394, EPI_ISL_1926395, EPI_ISL_1926396, EPI_ISL_1926397, EPI_ISL_1926399, EPI_ISL_1926400, EPI_ISL_1926401, EPI_ISL_1926402, EPI_ISL_1926403, EPI_ISL_1926404, EPI_ISL_1926405, EPI_ISL_1926407, EPI_ISL_1926408, EPI_ISL_1926409, EPI_ISL_1926410, EPI_ISL_1926411, EPI_ISL_1926412, EPI_ISL_1926413, EPI_ISL_1926414, EPI_ISL_1926416, EPI_ISL_1926417, EPI_ISL_1926419, EPI_ISL_1926420, EPI_ISL_1926421, EPI_ISL_1926422, EPI_ISL_1926423, EPI_ISL_1926424, EPI_ISL_1926426, EPI_ISL_1926428, EPI_ISL_1926429, EPI_ISL_1926430, EPI_ISL_1926431, EPI_ISL_1926432, EPI_ISL_1926433, EPI_ISL_1926434, EPI_ISL_1926435, EPI_ISL_1926436, EPI_ISL_1926437, EPI_ISL_1926438, EPI_ISL_1926439, EPI_ISL_1926440, EPI_ISL_1926441, EPI_ISL_1926442, EPI_ISL_1926443, EPI_ISL_1926444, EPI_ISL_1926445, EPI_ISL_1926446, EPI_ISL_1926447, EPI_ISL_1926448, EPI_ISL_1926449, EPI_ISL_1926451, EPI_ISL_1926452, EPI_ISL_1926453, EPI_ISL_1926454, EPI_ISL_1926455, EPI_ISL_1926457, EPI_ISL_1926458, EPI_ISL_1926460, EPI_ISL_1926462, EPI_ISL_1926463, EPI_ISL_1926464, EPI_ISL_1926465, EPI_ISL_1926466, EPI_ISL_1926468, EPI_ISL_1926469, EPI_ISL_1926471, EPI_ISL_1926472, EPI_ISL_1926473, EPI_ISL_1926474, EPI_ISL_1926475, EPI_ISL_1926476, EPI_ISL_1926477, EPI_ISL_1926478, EPI_ISL_1926479, EPI_ISL_1926480, EPI_ISL_1926481, EPI_ISL_1926483, EPI_ISL_1926485, EPI_ISL_1926486, EPI_ISL_1926487, EPI_ISL_1926488, EPI_ISL_1926490, EPI_ISL_1926492, EPI_ISL_1926493, EPI_ISL_1926496, EPI_ISL_1926497, EPI_ISL_1926498, EPI_ISL_1926499, EPI_ISL_1926500, EPI_ISL_1926501, EPI_ISL_1926502, EPI_ISL_1926503, EPI_ISL_1926504, EPI_ISL_1926505, EPI_ISL_1926506, EPI_ISL_1926507, EPI_ISL_1926509, EPI_ISL_1926510, EPI_ISL_1926511, EPI_ISL_1926512, EPI_ISL_1926514, EPI_ISL_1926515, EPI_ISL_1926516, EPI_ISL_1926517, EPI_ISL_1926518, EPI_ISL_1926519, EPI_ISL_1926520, EPI_ISL_1926521, EPI_ISL_1926522, EPI_ISL_1926523, EPI_ISL_1926524, EPI_ISL_1926525, EPI_ISL_1926527, EPI_ISL_1926528, EPI_ISL_1926529, EPI_ISL_1926531, EPI_ISL_1926532, EPI_ISL_1926533, EPI_ISL_1926534, EPI_ISL_1926536, EPI_ISL_1926538, EPI_ISL_1926539, EPI_ISL_1926540, EPI_ISL_1926541, EPI_ISL_1926542, EPI_ISL_1926543, EPI_ISL_1926544, EPI_ISL_1926545, EPI_ISL_1926546, EPI_ISL_1926548, EPI_ISL_1926549, EPI_ISL_1926550, EPI_ISL_1926551, EPI_ISL_1926552, EPI_ISL_1926553, EPI_ISL_1926554, EPI_ISL_1926555, EPI_ISL_1926556, EPI_ISL_1926557, EPI_ISL_1926558, EPI_ISL_1926559, EPI_ISL_1926560, EPI_ISL_1926561, EPI_ISL_1926562, EPI_ISL_1926564, EPI_ISL_1926565, EPI_ISL_1926566, EPI_ISL_1926567, EPI_ISL_1926568, EPI_ISL_1926570, EPI_ISL_1926572, EPI_ISL_1926573, EPI_ISL_1926574, EPI_ISL_1926577, EPI_ISL_1926578, EPI_ISL_1926579, EPI_ISL_1926580, EPI_ISL_1926581, EPI_ISL_1926582, EPI_ISL_1926583, EPI_ISL_1926584, EPI_ISL_1926585, EPI_ISL_1926586, EPI_ISL_1926587, EPI_ISL_1926588, EPI_ISL_1926589, EPI_ISL_1926590, EPI_ISL_1926591, EPI_ISL_1926592, EPI_ISL_1926593, EPI_ISL_1926595, EPI_ISL_1926596, EPI_ISL_1926597, EPI_ISL_1926598, EPI_ISL_1926599, EPI_ISL_1926600, EPI_ISL_1926601, EPI_ISL_1926603, EPI_ISL_1926605, EPI_ISL_1926606, EPI_ISL_1926607, EPI_ISL_1926609, EPI_ISL_1926610, EPI_ISL_1926611, EPI_ISL_1926612, EPI_ISL_1926613, EPI_ISL_1926614, EPI_ISL_1926615, EPI_ISL_1926616, EPI_ISL_1926617, EPI_ISL_1926618, EPI_ISL_1926619, EPI_ISL_1926620, EPI_ISL_1926621, EPI_ISL_1926622, EPI_ISL_1926623, EPI_ISL_1926624, EPI_ISL_1926625, EPI_ISL_1926626, EPI_ISL_1926627, EPI_ISL_1926628, EPI_ISL_1926629, EPI_ISL_1926630, EPI_ISL_1926631, EPI_ISL_1926632, EPI_ISL_1926633, EPI_ISL_1926634, EPI_ISL_1926635, EPI_ISL_1926637, EPI_ISL_1926638, EPI_ISL_1926639, EPI_ISL_1926642, EPI_ISL_1926643, EPI_ISL_1926644, EPI_ISL_1926645, EPI_ISL_1926646, EPI_ISL_1926647, EPI_ISL_1926648, EPI_ISL_1926649, EPI_ISL_1926650, EPI_ISL_1926651, EPI_ISL_1926652, EPI_ISL_1926653, EPI_ISL_1926654, EPI_ISL_1926655, EPI_ISL_1926656, EPI_ISL_1926657, EPI_ISL_1926658, EPI_ISL_1926659, EPI_ISL_1926660, EPI_ISL_1926661, EPI_ISL_1926663, EPI_ISL_1926664, EPI_ISL_1926665, EPI_ISL_1926666, EPI_ISL_1926667, EPI_ISL_1926668, EPI_ISL_1926670, EPI_ISL_1926671, EPI_ISL_1926672, EPI_ISL_1926673, EPI_ISL_1926674, EPI_ISL_1926675, EPI_ISL_1926677, EPI_ISL_1926678, EPI_ISL_1926679, EPI_ISL_1926680, EPI_ISL_1926681, EPI_ISL_1926682, EPI_ISL_1926683, EPI_ISL_1926684, EPI_ISL_1926685, EPI_ISL_1926686, EPI_ISL_1926687, EPI_ISL_1926688, EPI_ISL_1926689, EPI_ISL_1926690, EPI_ISL_1926691, EPI_ISL_1926692, EPI_ISL_1926693, EPI_ISL_1926694, EPI_ISL_1926695, EPI_ISL_1926698, EPI_ISL_1926699, EPI_ISL_1926700, EPI_ISL_1926701, EPI_ISL_1926702, EPI_ISL_1926703, EPI_ISL_1926704, EPI_ISL_1926705, EPI_ISL_1926707, EPI_ISL_1926708, EPI_ISL_1926710, EPI_ISL_1926712, EPI_ISL_1926713, EPI_ISL_1926714, EPI_ISL_1926715, EPI_ISL_1926716, EPI_ISL_1926717, EPI_ISL_1926718, EPI_ISL_1926719, EPI_ISL_1926720, EPI_ISL_1926721, EPI_ISL_1926723, EPI_ISL_1926725, EPI_ISL_1926726, EPI_ISL_1926727, EPI_ISL_1926728, EPI_ISL_1926729, EPI_ISL_1926731, EPI_ISL_1926732, EPI_ISL_1926733, EPI_ISL_1926734, EPI_ISL_1926735, EPI_ISL_1926736, EPI_ISL_1926737, EPI_ISL_1926738, EPI_ISL_1926739, EPI_ISL_1926740, EPI_ISL_1926741, EPI_ISL_1926743, EPI_ISL_1926744, EPI_ISL_1926745, EPI_ISL_1926746, EPI_ISL_1926748, EPI_ISL_1926749, EPI_ISL_1926750, EPI_ISL_1926751, EPI_ISL_1926752, EPI_ISL_1926753, EPI_ISL_1926754, EPI_ISL_1926756, EPI_ISL_1926757, EPI_ISL_1926758, EPI_ISL_1926760, EPI_ISL_1926761, EPI_ISL_1926762, EPI_ISL_1926763, EPI_ISL_1926764, EPI_ISL_1926765, EPI_ISL_1926766, EPI_ISL_1926769, EPI_ISL_1926770, EPI_ISL_1926771, EPI_ISL_1926772, EPI_ISL_1926773, EPI_ISL_1926774, EPI_ISL_1926775, EPI_ISL_1926776, EPI_ISL_1926777, EPI_ISL_1926778, EPI_ISL_1926779, EPI_ISL_1926783, EPI_ISL_1926784, EPI_ISL_1926785, EPI_ISL_1926787, EPI_ISL_1926788, EPI_ISL_1926789, EPI_ISL_1926790, EPI_ISL_1926791, EPI_ISL_1926792, EPI_ISL_1926793, EPI_ISL_1926794, EPI_ISL_1926795, EPI_ISL_1926796, EPI_ISL_1926797, EPI_ISL_1926799, EPI_ISL_1926800, EPI_ISL_1926801, EPI_ISL_1926802, EPI_ISL_1926803, EPI_ISL_1926804, EPI_ISL_1926805, EPI_ISL_1926806, EPI_ISL_1926807, EPI_ISL_1926808, EPI_ISL_1926811, EPI_ISL_1926812, EPI_ISL_1926815, EPI_ISL_1926816, EPI_ISL_1926817, EPI_ISL_1926818, EPI_ISL_1926819, EPI_ISL_1926821, EPI_ISL_1926823, EPI_ISL_1926824, EPI_ISL_1926825, EPI_ISL_1926826, EPI_ISL_1926827, EPI_ISL_1926828, EPI_ISL_1926829, EPI_ISL_1926831, EPI_ISL_1926834, EPI_ISL_1926835, EPI_ISL_1926836, EPI_ISL_1926838, EPI_ISL_1926839, EPI_ISL_1926840, EPI_ISL_1926841, EPI_ISL_1926844, EPI_ISL_1926845, EPI_ISL_1926846, EPI_ISL_1926849, EPI_ISL_1926850, EPI_ISL_1926851, EPI_ISL_1926853, EPI_ISL_1926854, EPI_ISL_1926855, EPI_ISL_1926856, EPI_ISL_1926859, EPI_ISL_1926863, EPI_ISL_1926864, EPI_ISL_1926866, EPI_ISL_1926867, EPI_ISL_1926868, EPI_ISL_1926869, EPI_ISL_1926870, EPI_ISL_1926871, EPI_ISL_1926872, EPI_ISL_1926873, EPI_ISL_1926876, EPI_ISL_1926879, EPI_ISL_1926886, EPI_ISL_1926889, EPI_ISL_1926892, EPI_ISL_1926895, EPI_ISL_1926897, EPI_ISL_1926900, EPI_ISL_1926907, EPI_ISL_1926910, EPI_ISL_1926916, EPI_ISL_1926920, EPI_ISL_1926923, EPI_ISL_1926935, EPI_ISL_1926940, EPI_ISL_1926943, EPI_ISL_1926950, EPI_ISL_1926953, EPI_ISL_1926956, EPI_ISL_1926959, EPI_ISL_1926968, EPI_ISL_1926974, EPI_ISL_1926977, EPI_ISL_1926986, EPI_ISL_1926989, EPI_ISL_1926995, EPI_ISL_1926998, EPI_ISL_1927001, EPI_ISL_1927003, EPI_ISL_1927006, EPI_ISL_1927009, EPI_ISL_1927012, EPI_ISL_1927015, EPI_ISL_1927018, EPI_ISL_1927021, EPI_ISL_1927025, EPI_ISL_1927028, EPI_ISL_1927031, EPI_ISL_1927033, EPI_ISL_1927043, EPI_ISL_1927052, EPI_ISL_1927055, EPI_ISL_1927057, EPI_ISL_1927063, EPI_ISL_1927072, EPI_ISL_1927075, EPI_ISL_1927081, EPI_ISL_1927085, EPI_ISL_1927088, EPI_ISL_1927094, EPI_ISL_1927097, EPI_ISL_1927103, EPI_ISL_1927106, EPI_ISL_1927115, EPI_ISL_1927118, EPI_ISL_1927121, EPI_ISL_1927124, EPI_ISL_1927128, EPI_ISL_1927134, EPI_ISL_1927137, EPI_ISL_1927142, EPI_ISL_1927149, EPI_ISL_1927152, EPI_ISL_1927158, EPI_ISL_1927161, EPI_ISL_1927164, EPI_ISL_1927170, EPI_ISL_1927173, EPI_ISL_1927176, EPI_ISL_1927185, EPI_ISL_1927188, EPI_ISL_1927191, EPI_ISL_1927194, EPI_ISL_1927198, EPI_ISL_1927201, EPI_ISL_1927206, EPI_ISL_1927210, EPI_ISL_1927216, EPI_ISL_1927219, EPI_ISL_1927222, EPI_ISL_1927225, EPI_ISL_1927228, EPI_ISL_1927231, EPI_ISL_1927234, EPI_ISL_1927237, EPI_ISL_1927240, EPI_ISL_1927243, EPI_ISL_1927253, EPI_ISL_1927256, EPI_ISL_1927259, EPI_ISL_1927265, EPI_ISL_1927274, EPI_ISL_1927277, EPI_ISL_1927281, EPI_ISL_1927284, EPI_ISL_1927287, EPI_ISL_1927290, EPI_ISL_1927292, EPI_ISL_1927295, EPI_ISL_1927298, EPI_ISL_1927301, EPI_ISL_1927304, EPI_ISL_1927307, EPI_ISL_1927313, EPI_ISL_1927323, EPI_ISL_1927326, EPI_ISL_1927338, EPI_ISL_1927350, EPI_ISL_1927353, EPI_ISL_1927359, EPI_ISL_1927363, EPI_ISL_1927366, EPI_ISL_1927369, EPI_ISL_1927372, EPI_ISL_1927375, EPI_ISL_1927385, EPI_ISL_1927387, EPI_ISL_1927393, EPI_ISL_1927396, EPI_ISL_1927399, EPI_ISL_1927402, EPI_ISL_1927406, EPI_ISL_1927409, EPI_ISL_1927415, EPI_ISL_1927418, EPI_ISL_1927421, EPI_ISL_1927425, EPI_ISL_1927434, EPI_ISL_1927437, EPI_ISL_1927440, EPI_ISL_1927443, EPI_ISL_1927446, EPI_ISL_1927452, EPI_ISL_1927455, EPI_ISL_1927458, EPI_ISL_1927461, EPI_ISL_1927464, EPI_ISL_1927467, EPI_ISL_1927470, EPI_ISL_1927474, EPI_ISL_1927480, EPI_ISL_1927483, EPI_ISL_1927486, EPI_ISL_1927493, EPI_ISL_1927502, EPI_ISL_1927504, EPI_ISL_1927510, EPI_ISL_1927513, EPI_ISL_1927516, EPI_ISL_1927519, EPI_ISL_1927526, EPI_ISL_1927529, EPI_ISL_1927535, EPI_ISL_1927541, EPI_ISL_1927544, EPI_ISL_1927548, EPI_ISL_1927551, EPI_ISL_1927557, EPI_ISL_1927560, EPI_ISL_1927562, EPI_ISL_1927565, EPI_ISL_1927568, EPI_ISL_1927572, EPI_ISL_1927574, EPI_ISL_1927577, EPI_ISL_1927580, EPI_ISL_1927587, EPI_ISL_1927590, EPI_ISL_1927593, EPI_ISL_1927597, EPI_ISL_1927603, EPI_ISL_1927607, EPI_ISL_1927610, EPI_ISL_1927613, EPI_ISL_1927616, EPI_ISL_1927622, EPI_ISL_1927625, EPI_ISL_1927628, EPI_ISL_1927631, EPI_ISL_1927634, EPI_ISL_1927636, EPI_ISL_1927639, EPI_ISL_1927642, EPI_ISL_1927646, EPI_ISL_1927648, EPI_ISL_1927651, EPI_ISL_1927654, EPI_ISL_1927657, EPI_ISL_1927661, EPI_ISL_1927664, EPI_ISL_1927666, EPI_ISL_1927669, EPI_ISL_1927672, EPI_ISL_1927679, EPI_ISL_1927682, EPI_ISL_1927685, EPI_ISL_1927688, EPI_ISL_1927691, EPI_ISL_1927694, EPI_ISL_1927697, EPI_ISL_1927700, EPI_ISL_1927709, EPI_ISL_1927712, EPI_ISL_1927715, EPI_ISL_1927718, EPI_ISL_1927721, EPI_ISL_1927724, EPI_ISL_1927728, EPI_ISL_1927731, EPI_ISL_1927734, EPI_ISL_1927737, EPI_ISL_1927740, EPI_ISL_1927743, EPI_ISL_1927747, EPI_ISL_1927750, EPI_ISL_1927754, EPI_ISL_1927757, EPI_ISL_1927760, EPI_ISL_1927763, EPI_ISL_1927766, EPI_ISL_1927769, EPI_ISL_1927772, EPI_ISL_1927778, EPI_ISL_1927781, EPI_ISL_1927784, EPI_ISL_1927787, EPI_ISL_1927790, EPI_ISL_1927793, EPI_ISL_1927796, EPI_ISL_1927799, EPI_ISL_1927802, EPI_ISL_1927804, EPI_ISL_1927807, EPI_ISL_1927810, EPI_ISL_1927817, EPI_ISL_1927820, EPI_ISL_1927823, EPI_ISL_1927826, EPI_ISL_1927829, EPI_ISL_1927832, EPI_ISL_1927835, EPI_ISL_1927838, EPI_ISL_1927841, EPI_ISL_1927844, EPI_ISL_1927847, EPI_ISL_1927850, EPI_ISL_1927853, EPI_ISL_1927859, EPI_ISL_1927862, EPI_ISL_1927865, EPI_ISL_1927868, EPI_ISL_1927871, EPI_ISL_1927874, EPI_ISL_1927877, EPI_ISL_1927880, EPI_ISL_1927883, EPI_ISL_1927892, EPI_ISL_1927899, EPI_ISL_1927905, EPI_ISL_1927911, EPI_ISL_1927914, EPI_ISL_1927917, EPI_ISL_1927920, EPI_ISL_1927923, EPI_ISL_1927926, EPI_ISL_1927929, EPI_ISL_1927932, EPI_ISL_1927934, EPI_ISL_1927938, EPI_ISL_1927941, EPI_ISL_1927943, EPI_ISL_1927947, EPI_ISL_1927950, EPI_ISL_1927956, EPI_ISL_1927959, EPI_ISL_1927962, EPI_ISL_1927965, EPI_ISL_1927968, EPI_ISL_1927971, EPI_ISL_1927974, EPI_ISL_1927977, EPI_ISL_1927980, EPI_ISL_1927983, EPI_ISL_1927986, EPI_ISL_1927989, EPI_ISL_1927992, EPI_ISL_1927995, EPI_ISL_1927998, EPI_ISL_1928001, EPI_ISL_1928004, EPI_ISL_1928013, EPI_ISL_1928016, EPI_ISL_1928019, EPI_ISL_1928022, EPI_ISL_1928025, EPI_ISL_1928028, EPI_ISL_1928034, EPI_ISL_1928037, EPI_ISL_1928040, EPI_ISL_1928043, EPI_ISL_1928046, EPI_ISL_1928049, EPI_ISL_1928052, EPI_ISL_1928055, EPI_ISL_1928058, EPI_ISL_1928061, EPI_ISL_1928067, EPI_ISL_1928070, EPI_ISL_1928073, EPI_ISL_1928076, EPI_ISL_1928079, EPI_ISL_1928082, EPI_ISL_1928085, EPI_ISL_1928088, EPI_ISL_1928091, EPI_ISL_1928097, EPI_ISL_1928103, EPI_ISL_1928106, EPI_ISL_1928109, EPI_ISL_1928111, EPI_ISL_1928115, EPI_ISL_1928118, EPI_ISL_1928121, EPI_ISL_1928124, EPI_ISL_1928127, EPI_ISL_1928130, EPI_ISL_1928133, EPI_ISL_1928135, EPI_ISL_1928139, EPI_ISL_1928142, EPI_ISL_1928145, EPI_ISL_1928148, EPI_ISL_1928151, EPI_ISL_1928154, EPI_ISL_1928157, EPI_ISL_1928160, EPI_ISL_1928163, EPI_ISL_1928170, EPI_ISL_1928172, EPI_ISL_1928176, EPI_ISL_1928179, EPI_ISL_1928182, EPI_ISL_1928185, EPI_ISL_1928188, EPI_ISL_1928191, EPI_ISL_1928194, EPI_ISL_1928197, EPI_ISL_1928200, EPI_ISL_1928204, EPI_ISL_1928210, EPI_ISL_1928213, EPI_ISL_1928216, EPI_ISL_1928219, EPI_ISL_1928222, EPI_ISL_1928224, EPI_ISL_1928227, EPI_ISL_1928231, EPI_ISL_1928234, EPI_ISL_1928237, EPI_ISL_1928240, EPI_ISL_1928243, EPI_ISL_1928246, EPI_ISL_1928249, EPI_ISL_1928252, EPI_ISL_1928255, EPI_ISL_1928262, EPI_ISL_1928265, EPI_ISL_1928268, EPI_ISL_1928271, EPI_ISL_1928274, EPI_ISL_1928277, EPI_ISL_1928280, EPI_ISL_1928282, EPI_ISL_1928289, EPI_ISL_1928292, EPI_ISL_1928295, EPI_ISL_1928298, EPI_ISL_1928301, EPI_ISL_1928307, EPI_ISL_1928310, EPI_ISL_1928314, EPI_ISL_1928317, EPI_ISL_1928319, EPI_ISL_1928322, EPI_ISL_1928326, EPI_ISL_1928329, EPI_ISL_1928332, EPI_ISL_1928335, EPI_ISL_1928338, EPI_ISL_1928341, EPI_ISL_1928345, EPI_ISL_1928347, EPI_ISL_1928357, EPI_ISL_1928366, EPI_ISL_1928369, EPI_ISL_1928371, EPI_ISL_1928372, EPI_ISL_1928373, EPI_ISL_1928374, EPI_ISL_1928376, EPI_ISL_1928377, EPI_ISL_1928379, EPI_ISL_1928380, EPI_ISL_1928381, EPI_ISL_1928382, EPI_ISL_1928384, EPI_ISL_1928386, EPI_ISL_1928390, EPI_ISL_1928393, EPI_ISL_1928397, EPI_ISL_1928400, EPI_ISL_1928403, EPI_ISL_1928406, EPI_ISL_1928409, EPI_ISL_1928412, EPI_ISL_1928415, EPI_ISL_1928418, EPI_ISL_1928421, EPI_ISL_1928424, EPI_ISL_1928428, EPI_ISL_1928431, EPI_ISL_1928434, EPI_ISL_1928437, EPI_ISL_1928440, EPI_ISL_1928443, EPI_ISL_1928446, EPI_ISL_1928452, EPI_ISL_1928455, EPI_ISL_1928458, EPI_ISL_1928461, EPI_ISL_1928470, EPI_ISL_1928473, EPI_ISL_1928476, EPI_ISL_1928479, EPI_ISL_1928482, EPI_ISL_1928486, EPI_ISL_1928489, EPI_ISL_1928492, EPI_ISL_1928495, EPI_ISL_1928501, EPI_ISL_1928504, EPI_ISL_1928508, EPI_ISL_1928511, EPI_ISL_1928514, EPI_ISL_1928516, EPI_ISL_1928519, EPI_ISL_1928523, EPI_ISL_1928526, EPI_ISL_1928529, EPI_ISL_1928532, EPI_ISL_1928535, EPI_ISL_1928542, EPI_ISL_1928545, EPI_ISL_1928548, EPI_ISL_1928551, EPI_ISL_1928554, EPI_ISL_1928557, EPI_ISL_1928560, EPI_ISL_1928564, EPI_ISL_1928567, EPI_ISL_1928570, EPI_ISL_1928574, EPI_ISL_1928577, EPI_ISL_1928580, EPI_ISL_1928582, EPI_ISL_1928585, EPI_ISL_1928589, EPI_ISL_1928591, EPI_ISL_1928594, EPI_ISL_1928597, EPI_ISL_1928600, EPI_ISL_1928604, EPI_ISL_1928606, EPI_ISL_1928609, EPI_ISL_1928612, EPI_ISL_1928616, EPI_ISL_1928619, EPI_ISL_1928621, EPI_ISL_1928625, EPI_ISL_1928628, EPI_ISL_1928630, EPI_ISL_1928634, EPI_ISL_1928637, EPI_ISL_1928640, EPI_ISL_1928643, EPI_ISL_1928649, EPI_ISL_1928652, EPI_ISL_1928653, EPI_ISL_1928654, EPI_ISL_1928655, EPI_ISL_1928656, EPI_ISL_1928659, EPI_ISL_1928660, EPI_ISL_1928661, EPI_ISL_1928662, EPI_ISL_1928663, EPI_ISL_1928664, EPI_ISL_1928665, EPI_ISL_1928666, EPI_ISL_1928667, EPI_ISL_1928668, EPI_ISL_1928669, EPI_ISL_1928670, EPI_ISL_1928671, EPI_ISL_1928672, EPI_ISL_1928673, EPI_ISL_1928674, EPI_ISL_1928675, EPI_ISL_1928676, EPI_ISL_1928677, EPI_ISL_1928678, EPI_ISL_1928680, EPI_ISL_1928681, EPI_ISL_1928682, EPI_ISL_1928683, EPI_ISL_1928684, EPI_ISL_1928685, EPI_ISL_1928686, EPI_ISL_1928687, EPI_ISL_1928688, EPI_ISL_1928689, EPI_ISL_1928690, EPI_ISL_1928691, EPI_ISL_1928692, EPI_ISL_1928693, EPI_ISL_1928694, EPI_ISL_1928695, EPI_ISL_1928696, EPI_ISL_1928697, EPI_ISL_1928698, EPI_ISL_1928699, EPI_ISL_1928700, EPI_ISL_1928701, EPI_ISL_1928702, EPI_ISL_1928703, EPI_ISL_1928704, EPI_ISL_1928705, EPI_ISL_1928707, EPI_ISL_1928708, EPI_ISL_1928710, EPI_ISL_1928711, EPI_ISL_1928712, EPI_ISL_1928713, EPI_ISL_1928714, EPI_ISL_1928715, EPI_ISL_1928716, EPI_ISL_1928718, EPI_ISL_1928720, EPI_ISL_1928721, EPI_ISL_1928722, EPI_ISL_1928724, EPI_ISL_1928726, EPI_ISL_1928727, EPI_ISL_1928729, EPI_ISL_1928730, EPI_ISL_1928731, EPI_ISL_1928733, EPI_ISL_1928734, EPI_ISL_1928735, EPI_ISL_1928736, EPI_ISL_1928737, EPI_ISL_1928738, EPI_ISL_1928739, EPI_ISL_1928741, EPI_ISL_1928742, EPI_ISL_1928743, EPI_ISL_1928745, EPI_ISL_1928746, EPI_ISL_1928748, EPI_ISL_1928750, EPI_ISL_1928751, EPI_ISL_1928752, EPI_ISL_1928753, EPI_ISL_1928756, EPI_ISL_1928757, EPI_ISL_1928759, EPI_ISL_1928761, EPI_ISL_1928762, EPI_ISL_1928766, EPI_ISL_1928770, EPI_ISL_1928771, EPI_ISL_1928772, EPI_ISL_1928773, EPI_ISL_1928775, EPI_ISL_1928776, EPI_ISL_1928778, EPI_ISL_1928781, EPI_ISL_1928782, EPI_ISL_1928783, EPI_ISL_1928784, EPI_ISL_1928785, EPI_ISL_1928787, EPI_ISL_1928788, EPI_ISL_1928789, EPI_ISL_1928792, EPI_ISL_1928793, EPI_ISL_1928794, EPI_ISL_1928795, EPI_ISL_1928796, EPI_ISL_1928798, EPI_ISL_1928802, EPI_ISL_1928803, EPI_ISL_1928806, EPI_ISL_1928809, EPI_ISL_1928811, EPI_ISL_1928814, EPI_ISL_1928815, EPI_ISL_1928816, EPI_ISL_1928818, EPI_ISL_1928819, EPI_ISL_1928820, EPI_ISL_1928821, EPI_ISL_1928822, EPI_ISL_1928823, EPI_ISL_1928824, EPI_ISL_1928828, EPI_ISL_1928832, EPI_ISL_1928833, EPI_ISL_1928835, EPI_ISL_1928839, EPI_ISL_1928842,

| | | | |
|---|---|---|---|
| EPI_ISL_1928843, EPI_ISL_1928844, EPI_ISL_1928845, EPI_ISL_1928846, EPI_ISL_1928847, EPI_ISL_1928849, EPI_ISL_1928850, EPI_ISL_1928852, EPI_ISL_1928853, EPI_ISL_1928855, EPI_ISL_1928856, EPI_ISL_1928858, EPI_ISL_1928859, EPI_ISL_1928860, EPI_ISL_1928861, EPI_ISL_1928864, EPI_ISL_1928865, EPI_ISL_1928867, EPI_ISL_1928868, EPI_ISL_1928869, EPI_ISL_1928870, EPI_ISL_1928871, EPI_ISL_1928872, EPI_ISL_1928874, EPI_ISL_1928876, EPI_ISL_1928877, EPI_ISL_1928879, EPI_ISL_1928881, EPI_ISL_1928883, EPI_ISL_1928885, EPI_ISL_1928887, EPI_ISL_1928888, EPI_ISL_1928889, EPI_ISL_1928890, EPI_ISL_1928891, EPI_ISL_1928894, EPI_ISL_1928897, EPI_ISL_1928898, EPI_ISL_1928900, EPI_ISL_1928902, EPI_ISL_1928903, EPI_ISL_1928904, EPI_ISL_1928905, EPI_ISL_1928907, EPI_ISL_1928908, EPI_ISL_1928909, EPI_ISL_1928910, EPI_ISL_1928914, EPI_ISL_1928915, EPI_ISL_1928916, EPI_ISL_1928917, EPI_ISL_1928919, EPI_ISL_1928920, EPI_ISL_1928921, EPI_ISL_1928922, EPI_ISL_1928923, EPI_ISL_1928928, EPI_ISL_1928929, EPI_ISL_1928931, EPI_ISL_1928932, EPI_ISL_1928934, EPI_ISL_1928936, EPI_ISL_1928937, EPI_ISL_1928938, EPI_ISL_1928940, EPI_ISL_1928941, EPI_ISL_1928943, EPI_ISL_1928945, EPI_ISL_1928946, EPI_ISL_1928948, EPI_ISL_1928949, EPI_ISL_1928950, EPI_ISL_1928951, EPI_ISL_1928952, EPI_ISL_1928953, EPI_ISL_1928954, EPI_ISL_1928955, EPI_ISL_1928957, EPI_ISL_1928958, EPI_ISL_1928963, EPI_ISL_1928964, EPI_ISL_1928965, EPI_ISL_1928966, EPI_ISL_1928969, EPI_ISL_1928970, EPI_ISL_1928971, EPI_ISL_1928972, EPI_ISL_1928973, EPI_ISL_1928976, EPI_ISL_1928977, EPI_ISL_1928978, EPI_ISL_1928979, EPI_ISL_1928980, EPI_ISL_1928983, EPI_ISL_1928986, EPI_ISL_1928987, EPI_ISL_1928988, EPI_ISL_1928989, EPI_ISL_1928990, EPI_ISL_1928991, EPI_ISL_1928992, EPI_ISL_1928993, EPI_ISL_1928994, EPI_ISL_1928996, EPI_ISL_1928997, EPI_ISL_1928999, EPI_ISL_1929004, EPI_ISL_1929006, EPI_ISL_1929007, EPI_ISL_1929008, EPI_ISL_1929009, EPI_ISL_1929011, EPI_ISL_1929012, EPI_ISL_1929013, EPI_ISL_1929014, EPI_ISL_1929015, EPI_ISL_1929016, EPI_ISL_1929017, EPI_ISL_1929018, EPI_ISL_1929019, EPI_ISL_1929021, EPI_ISL_1929022, EPI_ISL_1929023, EPI_ISL_1929024, EPI_ISL_1929025, EPI_ISL_1929027, EPI_ISL_1929031, EPI_ISL_1929032, EPI_ISL_1929033, EPI_ISL_1929034, EPI_ISL_1929035, EPI_ISL_1929036, EPI_ISL_1929037, EPI_ISL_1929038, EPI_ISL_1929039, EPI_ISL_1929040, EPI_ISL_1929041, EPI_ISL_1929042, EPI_ISL_1929044, EPI_ISL_1929045, EPI_ISL_1929047, EPI_ISL_1929048, EPI_ISL_1929049, EPI_ISL_1929050, EPI_ISL_1929051, EPI_ISL_1929052, EPI_ISL_1929053, EPI_ISL_1929055, EPI_ISL_1929057, EPI_ISL_1929059, EPI_ISL_1929060, EPI_ISL_1929062, EPI_ISL_1929063, EPI_ISL_1929064, EPI_ISL_1929065, EPI_ISL_1929067, EPI_ISL_1929068, EPI_ISL_1929069, EPI_ISL_1929070, EPI_ISL_1929071, EPI_ISL_1929072, EPI_ISL_1929075, EPI_ISL_1929078, EPI_ISL_1929081, EPI_ISL_1929084, EPI_ISL_1929087, EPI_ISL_1929090, EPI_ISL_1929093, EPI_ISL_1929097, EPI_ISL_1929103, EPI_ISL_1929105, EPI_ISL_1929108, EPI_ISL_1929111, EPI_ISL_1929114, EPI_ISL_1929118, EPI_ISL_1929120, EPI_ISL_1929123, EPI_ISL_1929126, EPI_ISL_1929129, EPI_ISL_1929137, EPI_ISL_1929140, EPI_ISL_1929146, EPI_ISL_1929149, EPI_ISL_1929152, EPI_ISL_1929154, EPI_ISL_1929157, EPI_ISL_1929161, EPI_ISL_1929163, EPI_ISL_1929166, EPI_ISL_1929169, EPI_ISL_1929172, EPI_ISL_1929178, EPI_ISL_1929181, EPI_ISL_1929184, EPI_ISL_1929187, EPI_ISL_1929190, EPI_ISL_1929192, EPI_ISL_1929195, EPI_ISL_1929198, EPI_ISL_1929201, EPI_ISL_1929204, EPI_ISL_1929207, EPI_ISL_1929210, EPI_ISL_1929213, EPI_ISL_1929215, EPI_ISL_1929218, EPI_ISL_1929222, EPI_ISL_1929224, EPI_ISL_1929230, EPI_ISL_1929232, EPI_ISL_1929235, EPI_ISL_1929238, EPI_ISL_1929242, EPI_ISL_1929245, EPI_ISL_1929248, EPI_ISL_1929251, EPI_ISL_1929255, EPI_ISL_1929257, EPI_ISL_1929260, EPI_ISL_1929263, EPI_ISL_1929267, EPI_ISL_1929270, EPI_ISL_1929273, EPI_ISL_1929276, EPI_ISL_1929279, EPI_ISL_1929282, EPI_ISL_1929289, EPI_ISL_1929292, EPI_ISL_1929295, EPI_ISL_1929298, EPI_ISL_1929301, EPI_ISL_1929304, EPI_ISL_1929308, EPI_ISL_1929311, EPI_ISL_1929314, EPI_ISL_1929317, EPI_ISL_1929320, EPI_ISL_1929323, EPI_ISL_1929325, EPI_ISL_1929332, EPI_ISL_1929335, EPI_ISL_1929338, EPI_ISL_1929341, EPI_ISL_1929350, EPI_ISL_1929356, EPI_ISL_1929359, EPI_ISL_1929362, EPI_ISL_1929365, EPI_ISL_1929367, EPI_ISL_1929370, EPI_ISL_1929374, EPI_ISL_1929376, EPI_ISL_1929379, EPI_ISL_1929382, EPI_ISL_1929385, EPI_ISL_1929388, EPI_ISL_1929394, EPI_ISL_1929397, EPI_ISL_1929400, EPI_ISL_1929403, EPI_ISL_1929406, EPI_ISL_1929409, EPI_ISL_1929412, EPI_ISL_1929415, EPI_ISL_1929418, EPI_ISL_1929421, EPI_ISL_1929425, EPI_ISL_1929431, EPI_ISL_1929434, EPI_ISL_1929437, EPI_ISL_1929442, EPI_ISL_1929445, EPI_ISL_1929448, EPI_ISL_1929451, EPI_ISL_1929454, EPI_ISL_1929457, EPI_ISL_1929460, EPI_ISL_1929466, EPI_ISL_1929469, EPI_ISL_1929472, EPI_ISL_1929478, EPI_ISL_1929481, EPI_ISL_1929484, EPI_ISL_1929487, EPI_ISL_1929490, EPI_ISL_1929493, EPI_ISL_1929496, EPI_ISL_1929499, EPI_ISL_1929502, EPI_ISL_1929505, EPI_ISL_1929508, EPI_ISL_1929514, EPI_ISL_1929517, EPI_ISL_1929520, EPI_ISL_1929523, EPI_ISL_1929526, EPI_ISL_1929529, EPI_ISL_1929536, EPI_ISL_1929538, EPI_ISL_1929542, EPI_ISL_1929544, EPI_ISL_1929547, EPI_ISL_1929553, EPI_ISL_1929556, EPI_ISL_1929559, EPI_ISL_1929562, EPI_ISL_1929565, EPI_ISL_1929568, EPI_ISL_1929571, EPI_ISL_1929574, EPI_ISL_1929577, EPI_ISL_1929580, EPI_ISL_1929583, EPI_ISL_1929586, EPI_ISL_1929589, EPI_ISL_1929592, EPI_ISL_1929595, EPI_ISL_1929598, EPI_ISL_1929601, EPI_ISL_1929604, EPI_ISL_1929607, EPI_ISL_1929610, EPI_ISL_1929613, EPI_ISL_1929619, EPI_ISL_1929622, EPI_ISL_1929625, EPI_ISL_1929628, EPI_ISL_1929631, EPI_ISL_1929634, EPI_ISL_1929640, EPI_ISL_1929644, EPI_ISL_1929647, EPI_ISL_1929650, EPI_ISL_1929653, EPI_ISL_1929660, EPI_ISL_1929663, EPI_ISL_1929666, EPI_ISL_1929669, EPI_ISL_1929672, EPI_ISL_1929676, EPI_ISL_1929679, EPI_ISL_1929682, EPI_ISL_1929685, EPI_ISL_1929688, EPI_ISL_1929691, EPI_ISL_1929694, EPI_ISL_1929697, EPI_ISL_1929700, EPI_ISL_1929703, EPI_ISL_1929706, EPI_ISL_1929712, EPI_ISL_1929715, EPI_ISL_1929718, EPI_ISL_1929721, EPI_ISL_1929724, EPI_ISL_1929726, EPI_ISL_1929729, EPI_ISL_1929733, EPI_ISL_1929736, EPI_ISL_1929738, EPI_ISL_1929741, EPI_ISL_1929744, EPI_ISL_1929747, EPI_ISL_1929750, EPI_ISL_1929756, EPI_ISL_1929759, EPI_ISL_1929762, EPI_ISL_1929765, EPI_ISL_1929767, EPI_ISL_1929773, EPI_ISL_1929776, EPI_ISL_1929779, EPI_ISL_1929782, EPI_ISL_1929786, EPI_ISL_1929788, EPI_ISL_1929791, EPI_ISL_1929794, EPI_ISL_1929798, EPI_ISL_1929801, EPI_ISL_1929804, EPI_ISL_1929807, EPI_ISL_1929810, EPI_ISL_1929813, EPI_ISL_1929816, EPI_ISL_1929819, EPI_ISL_1929825, EPI_ISL_1929828, EPI_ISL_1929830, EPI_ISL_1929833, EPI_ISL_1929836, EPI_ISL_1929839, EPI_ISL_1929842, EPI_ISL_1929846, EPI_ISL_1929850, EPI_ISL_1929853, EPI_ISL_1929856, EPI_ISL_1929859, EPI_ISL_1929864, EPI_ISL_1929870, EPI_ISL_1929873, EPI_ISL_1929876, EPI_ISL_1929879, EPI_ISL_1929882, EPI_ISL_1929885, EPI_ISL_1929896, EPI_ISL_1929902, EPI_ISL_1929905, EPI_ISL_1929908, EPI_ISL_1929911, EPI_ISL_1929914, EPI_ISL_1929917, EPI_ISL_1929920, EPI_ISL_1929926, EPI_ISL_1929929, EPI_ISL_1929932, EPI_ISL_1929935, EPI_ISL_1929941, EPI_ISL_1929944, EPI_ISL_1929947, EPI_ISL_1929950, EPI_ISL_1929953, EPI_ISL_1929956, EPI_ISL_1929961, EPI_ISL_1929965, EPI_ISL_1929971, EPI_ISL_1929974, EPI_ISL_1929977, EPI_ISL_1929980, EPI_ISL_1929984, EPI_ISL_1929987, EPI_ISL_1929990, EPI_ISL_1929994, EPI_ISL_1929996, EPI_ISL_1929999, EPI_ISL_1930003, EPI_ISL_1930006, EPI_ISL_1930009, EPI_ISL_1930012, EPI_ISL_1930014, EPI_ISL_1930017, EPI_ISL_1930020, EPI_ISL_1930028, EPI_ISL_1930031, EPI_ISL_1930034, EPI_ISL_1930037, EPI_ISL_1930040, EPI_ISL_1930043, EPI_ISL_1930045, EPI_ISL_1930048, EPI_ISL_1930051, EPI_ISL_1930054, EPI_ISL_1930057, EPI_ISL_1930060, EPI_ISL_1930062, EPI_ISL_1930066, EPI_ISL_1930068, EPI_ISL_1930071, EPI_ISL_1930074, EPI_ISL_1930077, EPI_ISL_1930081, EPI_ISL_1930084, EPI_ISL_1930087, EPI_ISL_1930089, EPI_ISL_1930092, EPI_ISL_1930095, EPI_ISL_1930097, EPI_ISL_1930100, EPI_ISL_1930103, EPI_ISL_1930106, EPI_ISL_1930109, EPI_ISL_1930112, EPI_ISL_1930115, EPI_ISL_1930117, EPI_ISL_1930120, EPI_ISL_1930126, EPI_ISL_1930129, EPI_ISL_1930131, EPI_ISL_1930134, EPI_ISL_1930137, EPI_ISL_1930140, EPI_ISL_1930143, EPI_ISL_1930145, EPI_ISL_1930153, EPI_ISL_1930156, EPI_ISL_1930158, EPI_ISL_1930162, EPI_ISL_1930165, EPI_ISL_1930168, EPI_ISL_1930170, EPI_ISL_1930173, EPI_ISL_1930179, EPI_ISL_1930184, EPI_ISL_1930187, EPI_ISL_1930190, EPI_ISL_1930193, EPI_ISL_1930198, EPI_ISL_1930201, EPI_ISL_1930207, EPI_ISL_1930210, EPI_ISL_1930216, EPI_ISL_1930219, EPI_ISL_1930222, EPI_ISL_1930225, EPI_ISL_1930228, EPI_ISL_1930231, EPI_ISL_1930234, EPI_ISL_1930239, EPI_ISL_1930242, EPI_ISL_1930244, EPI_ISL_1930247, EPI_ISL_1930250, EPI_ISL_1930255, EPI_ISL_1930261, EPI_ISL_1930264, EPI_ISL_1930267, EPI_ISL_1930270, EPI_ISL_1930273, EPI_ISL_1930276, EPI_ISL_1930278, EPI_ISL_1930281, EPI_ISL_1930284, EPI_ISL_1930287, EPI_ISL_1930290, EPI_ISL_1930292, EPI_ISL_1930295, EPI_ISL_1930298, EPI_ISL_1930301, EPI_ISL_1930304, EPI_ISL_1930307, EPI_ISL_1930313, EPI_ISL_1930315, EPI_ISL_1930318, EPI_ISL_1930321, EPI_ISL_1930324, EPI_ISL_1930327, EPI_ISL_1930330, EPI_ISL_1930333, EPI_ISL_1930335, EPI_ISL_1930339, EPI_ISL_1930343, EPI_ISL_1930346, EPI_ISL_1930350, EPI_ISL_1930353, EPI_ISL_1930356, EPI_ISL_1930358, EPI_ISL_1930361, EPI_ISL_1930365, EPI_ISL_1930368, EPI_ISL_1930371, EPI_ISL_1930374, EPI_ISL_1930377, EPI_ISL_1930386, EPI_ISL_1930388, EPI_ISL_1930391, EPI_ISL_1930394, EPI_ISL_1930400, EPI_ISL_1930407, EPI_ISL_1930409, EPI_ISL_1930412, EPI_ISL_1930415, EPI_ISL_1930418, EPI_ISL_1930421, EPI_ISL_1930424, EPI_ISL_1930427, EPI_ISL_1930430, EPI_ISL_1930436, EPI_ISL_1930439, EPI_ISL_1930442, EPI_ISL_1930446, EPI_ISL_1930449, EPI_ISL_1930452, EPI_ISL_1930455, EPI_ISL_1930458, EPI_ISL_1930461, EPI_ISL_1930467, EPI_ISL_1930470, EPI_ISL_1930473, EPI_ISL_1930475, EPI_ISL_1930478, EPI_ISL_1930481, EPI_ISL_1930484, EPI_ISL_1930487, EPI_ISL_1930491, EPI_ISL_1930494, EPI_ISL_1930497, EPI_ISL_1930500, EPI_ISL_1930502, EPI_ISL_1930505, EPI_ISL_1930508, EPI_ISL_1930511, EPI_ISL_1930513, EPI_ISL_1930517, EPI_ISL_1930519, EPI_ISL_1930523, EPI_ISL_1930525, EPI_ISL_1930528, EPI_ISL_1930531, EPI_ISL_1930533, EPI_ISL_1930536, EPI_ISL_1930539, EPI_ISL_1930542, EPI_ISL_1930545, EPI_ISL_1930551, EPI_ISL_1930554, EPI_ISL_1930557, EPI_ISL_1930560, EPI_ISL_1930563, EPI_ISL_1930566, EPI_ISL_1930569, EPI_ISL_1930572, EPI_ISL_1930575, EPI_ISL_1930576, EPI_ISL_1930577, EPI_ISL_1930578, EPI_ISL_1930579, EPI_ISL_1930580, EPI_ISL_1930582, EPI_ISL_1930583, EPI_ISL_1930584, EPI_ISL_1930585, EPI_ISL_1930586, EPI_ISL_1930588, EPI_ISL_1930589, EPI_ISL_1930590, EPI_ISL_1930591, EPI_ISL_1930593, EPI_ISL_1930594, EPI_ISL_1930595, EPI_ISL_1930596, EPI_ISL_1930597, EPI_ISL_1930598, EPI_ISL_1930599, EPI_ISL_1930600, EPI_ISL_1930601, EPI_ISL_1930602, EPI_ISL_1930604, EPI_ISL_1930606, EPI_ISL_1930609, EPI_ISL_1930612, EPI_ISL_1930615, EPI_ISL_1930618, EPI_ISL_1930621, EPI_ISL_1930625, EPI_ISL_1930628, EPI_ISL_1930631, EPI_ISL_1930634, EPI_ISL_1930637, EPI_ISL_1930640, EPI_ISL_1930643, EPI_ISL_1930650, EPI_ISL_1930653, EPI_ISL_1930656, EPI_ISL_1930659, EPI_ISL_1930662, EPI_ISL_1930665, EPI_ISL_1930668, EPI_ISL_1930669, EPI_ISL_1930670, EPI_ISL_1930671, EPI_ISL_1930672, EPI_ISL_1930673, EPI_ISL_1930674, EPI_ISL_1930675, EPI_ISL_1930677, EPI_ISL_1930678, EPI_ISL_1930679, EPI_ISL_1930681, EPI_ISL_1930682, EPI_ISL_1930683, EPI_ISL_1930684, EPI_ISL_1930687, EPI_ISL_1930688, EPI_ISL_1930689, EPI_ISL_1930690, EPI_ISL_1930692, EPI_ISL_1930693, EPI_ISL_1930695, EPI_ISL_1930697, EPI_ISL_1930698, EPI_ISL_1930699, EPI_ISL_1930700, EPI_ISL_1930701, EPI_ISL_1930702, EPI_ISL_1930705, EPI_ISL_1930706, EPI_ISL_1930711, EPI_ISL_1930713, EPI_ISL_1930715, EPI_ISL_1930716, EPI_ISL_1930717, EPI_ISL_1930718, EPI_ISL_1930719, EPI_ISL_1930720, EPI_ISL_1930721, EPI_ISL_1930723, EPI_ISL_1930724, EPI_ISL_1930725, EPI_ISL_1930727, EPI_ISL_1930728, EPI_ISL_1930729, EPI_ISL_1930730, EPI_ISL_1930731, EPI_ISL_1930733, EPI_ISL_1930734, EPI_ISL_1930738, EPI_ISL_1930739, EPI_ISL_1930742, EPI_ISL_1930743, EPI_ISL_1930745, EPI_ISL_1930746, EPI_ISL_1930747, EPI_ISL_1930748, EPI_ISL_1930749, EPI_ISL_1930750, EPI_ISL_1930751, EPI_ISL_1930753, EPI_ISL_1930754, EPI_ISL_1930755, EPI_ISL_1930756, EPI_ISL_1930758, EPI_ISL_1930759, EPI_ISL_1930761, EPI_ISL_1930762, EPI_ISL_1930764, EPI_ISL_1930765, EPI_ISL_1930766, EPI_ISL_1930768, EPI_ISL_1930770, EPI_ISL_1930771, EPI_ISL_1930772, EPI_ISL_1930773, EPI_ISL_1930774, EPI_ISL_1930776, EPI_ISL_1930777, EPI_ISL_1930778, EPI_ISL_1930779, EPI_ISL_1930780, EPI_ISL_1930781, EPI_ISL_1930782, EPI_ISL_1930783, EPI_ISL_1930784, EPI_ISL_1930785, EPI_ISL_1930787, EPI_ISL_1930788, EPI_ISL_1930789, EPI_ISL_1930790, EPI_ISL_1930791, EPI_ISL_1930793, EPI_ISL_1930794, EPI_ISL_1930795, EPI_ISL_1930797, EPI_ISL_1930798, EPI_ISL_1930799, EPI_ISL_1930800, EPI_ISL_1930801, EPI_ISL_1930802, EPI_ISL_1930803, EPI_ISL_1930804, EPI_ISL_1930806, EPI_ISL_1930809, EPI_ISL_1930810, EPI_ISL_1930811, EPI_ISL_1930812, EPI_ISL_1930813, EPI_ISL_1930815, EPI_ISL_1930816, EPI_ISL_1930817, EPI_ISL_1930818, EPI_ISL_1930819, EPI_ISL_1930820, EPI_ISL_1930822, EPI_ISL_1930823, EPI_ISL_1930825, EPI_ISL_1930826, EPI_ISL_1930827, EPI_ISL_1930828, EPI_ISL_1930829, EPI_ISL_1930830, EPI_ISL_1930832, EPI_ISL_1930833, EPI_ISL_1930834, EPI_ISL_1930835, EPI_ISL_1930836, EPI_ISL_1930837, EPI_ISL_1930839, EPI_ISL_1930840, EPI_ISL_1930841, EPI_ISL_1930842, EPI_ISL_1930843, EPI_ISL_1930844, EPI_ISL_1930845, EPI_ISL_1930846, EPI_ISL_1930848, EPI_ISL_1930850, EPI_ISL_1930851, EPI_ISL_1930852, EPI_ISL_1930853, EPI_ISL_1930854, EPI_ISL_1930857, EPI_ISL_1930858, EPI_ISL_1930859, EPI_ISL_1930860, EPI_ISL_1930861, EPI_ISL_1930862, EPI_ISL_1930865, EPI_ISL_1930868, EPI_ISL_1930871, EPI_ISL_1930874, EPI_ISL_1930877, EPI_ISL_1930880, EPI_ISL_1930886, EPI_ISL_1930888, EPI_ISL_1930892, EPI_ISL_1930895, EPI_ISL_1930898, EPI_ISL_1930904, EPI_ISL_1930907, EPI_ISL_1930916, EPI_ISL_1930919, EPI_ISL_1930922, EPI_ISL_1930928, EPI_ISL_1930931, EPI_ISL_1930937, EPI_ISL_1930940, EPI_ISL_1930943, EPI_ISL_1930946, EPI_ISL_1930949, EPI_ISL_1930955, EPI_ISL_1930958, EPI_ISL_1930964, EPI_ISL_1930967, EPI_ISL_1930970, EPI_ISL_1930977, EPI_ISL_1930980, EPI_ISL_1930986, EPI_ISL_1930989, EPI_ISL_1930992, EPI_ISL_1930995, EPI_ISL_1930998, EPI_ISL_1931001, EPI_ISL_1931003, EPI_ISL_1931010, EPI_ISL_1931013, EPI_ISL_1931016, EPI_ISL_1931019, EPI_ISL_1931022, EPI_ISL_1931024, EPI_ISL_1931028, EPI_ISL_1931031, EPI_ISL_1931034, EPI_ISL_1931040, EPI_ISL_1931042, EPI_ISL_1931046, EPI_ISL_1931049, EPI_ISL_1931052, EPI_ISL_1931055, EPI_ISL_1931058, EPI_ISL_1931061, EPI_ISL_1931064, EPI_ISL_1931067, EPI_ISL_1931070, EPI_ISL_1931073, EPI_ISL_1931076, EPI_ISL_1931079, EPI_ISL_1931082, EPI_ISL_1931089, EPI_ISL_1931092, EPI_ISL_1931098, EPI_ISL_1931112, EPI_ISL_1931115, EPI_ISL_1931117, EPI_ISL_1931119, EPI_ISL_1931122, EPI_ISL_1931123, EPI_ISL_1931124, EPI_ISL_1931126, EPI_ISL_1931127, EPI_ISL_1931129, EPI_ISL_1931130, EPI_ISL_1931131, EPI_ISL_1931133, EPI_ISL_1931136, EPI_ISL_1931139, EPI_ISL_1931140, EPI_ISL_1931142, EPI_ISL_1931143, EPI_ISL_1931145, EPI_ISL_1931146, EPI_ISL_1931151, EPI_ISL_1931158, EPI_ISL_1931160, EPI_ISL_1931163, EPI_ISL_1931167, EPI_ISL_1931168, EPI_ISL_1931171, EPI_ISL_1931173, EPI_ISL_1931177, EPI_ISL_1931183, EPI_ISL_1931185, EPI_ISL_1931186, EPI_ISL_1931189, EPI_ISL_1931190, EPI_ISL_1931192, EPI_ISL_1931195, EPI_ISL_1931196, EPI_ISL_1931198, EPI_ISL_1931201, EPI_ISL_1931202, EPI_ISL_1931205, EPI_ISL_1931208, EPI_ISL_1931209, EPI_ISL_1931211, EPI_ISL_1931213, EPI_ISL_1931215, EPI_ISL_1931217, EPI_ISL_1931218, EPI_ISL_1931220, EPI_ISL_1931221, EPI_ISL_1931223, EPI_ISL_1931224, EPI_ISL_1931227, EPI_ISL_1931229, EPI_ISL_1931230, EPI_ISL_1931232, EPI_ISL_1931235, EPI_ISL_1931238, EPI_ISL_1931241, EPI_ISL_1931244, EPI_ISL_1931246, EPI_ISL_1931247, EPI_ISL_1931250, EPI_ISL_1931251, EPI_ISL_1931256, EPI_ISL_1931257, EPI_ISL_1931260, EPI_ISL_1931261, EPI_ISL_1931264, EPI_ISL_1931266, EPI_ISL_1931267, EPI_ISL_1931268, EPI_ISL_1931270, EPI_ISL_1931272, EPI_ISL_1931273, EPI_ISL_1931275, EPI_ISL_1931276, EPI_ISL_1931278, EPI_ISL_1931279, EPI_ISL_1931281, EPI_ISL_1931282, EPI_ISL_1931285, EPI_ISL_1931287, EPI_ISL_1931288, EPI_ISL_1931290, EPI_ISL_1931291, EPI_ISL_1931294, EPI_ISL_1931296, EPI_ISL_1931300, EPI_ISL_1931302, EPI_ISL_1931304, EPI_ISL_1931306, EPI_ISL_1931307, EPI_ISL_1931311, EPI_ISL_1931312, EPI_ISL_1931315, EPI_ISL_1931318, EPI_ISL_1931319, EPI_ISL_1931321, EPI_ISL_1931322, EPI_ISL_1931324, EPI_ISL_1931328, EPI_ISL_1931331, EPI_ISL_1931334, EPI_ISL_1931337, EPI_ISL_1931340, EPI_ISL_1931346, EPI_ISL_1931351, EPI_ISL_1931354, EPI_ISL_1931357, EPI_ISL_1931361, EPI_ISL_1931367, EPI_ISL_1931373, EPI_ISL_1931379, EPI_ISL_1931384, EPI_ISL_1931387, EPI_ISL_1931390, EPI_ISL_1931391, EPI_ISL_1931394, EPI_ISL_1931397, EPI_ISL_1931400, EPI_ISL_1931406, EPI_ISL_1931409, EPI_ISL_1931412, EPI_ISL_1931418, EPI_ISL_1931423, EPI_ISL_1931425, EPI_ISL_1931428, EPI_ISL_1931440, EPI_ISL_1931443, EPI_ISL_1931445, EPI_ISL_1931448, EPI_ISL_1931451, EPI_ISL_1931454, EPI_ISL_1931457, EPI_ISL_1931460, EPI_ISL_1931463, EPI_ISL_1931471, EPI_ISL_1931476, EPI_ISL_1931479, EPI_ISL_1931481, EPI_ISL_1931484, EPI_ISL_1931487, EPI_ISL_1931490, EPI_ISL_1931492, EPI_ISL_1931495, EPI_ISL_1931498, EPI_ISL_1931500, EPI_ISL_1931501, EPI_ISL_1931504, EPI_ISL_1931507, EPI_ISL_1931510, EPI_ISL_1931513, EPI_ISL_1931518, EPI_ISL_1931521, EPI_ISL_1931522, EPI_ISL_1931523, EPI_ISL_1931532, EPI_ISL_1931537, EPI_ISL_1931540, EPI_ISL_1931543, EPI_ISL_1931545, EPI_ISL_1931554, EPI_ISL_1931557, EPI_ISL_1931559, EPI_ISL_1931560, EPI_ISL_1931562, EPI_ISL_1931563, EPI_ISL_1931565, EPI_ISL_1931566, EPI_ISL_1931569, EPI_ISL_1931570, EPI_ISL_1931572, EPI_ISL_1931573, EPI_ISL_1931574, EPI_ISL_1931576, EPI_ISL_1931577, EPI_ISL_1931580, EPI_ISL_1931583, EPI_ISL_1931591, EPI_ISL_1931594, EPI_ISL_1931598, EPI_ISL_1931601, EPI_ISL_1931604, EPI_ISL_1931606, EPI_ISL_1931609, EPI_ISL_1931612, EPI_ISL_1931615, EPI_ISL_1931616, EPI_ISL_1931617, EPI_ISL_1931620, EPI_ISL_1931622, EPI_ISL_1931626, EPI_ISL_1931629, EPI_ISL_1931630, EPI_ISL_1931632, EPI_ISL_1931633, EPI_ISL_1931634, EPI_ISL_1931637, EPI_ISL_1931639, EPI_ISL_1931642, EPI_ISL_1931650, EPI_ISL_1931654, EPI_ISL_1931656, EPI_ISL_1931659, EPI_ISL_1931661, EPI_ISL_1931662, EPI_ISL_1931663, EPI_ISL_1931665, EPI_ISL_1931666, EPI_ISL_1931668, EPI_ISL_1931669, EPI_ISL_1931672, EPI_ISL_1931673, EPI_ISL_1931675, EPI_ISL_1931676, EPI_ISL_1931678, EPI_ISL_1931680, EPI_ISL_1931685, EPI_ISL_1931688, EPI_ISL_1931691, EPI_ISL_1931694, EPI_ISL_1931697, EPI_ISL_1931700, EPI_ISL_1931706, EPI_ISL_1931710, EPI_ISL_1931711, EPI_ISL_1931713, EPI_ISL_1931716, EPI_ISL_1931718, EPI_ISL_1931720, EPI_ISL_1931723, EPI_ISL_1931726, EPI_ISL_1931728, EPI_ISL_1931731, EPI_ISL_1931734, EPI_ISL_1931737, EPI_ISL_1931740, EPI_ISL_1931745, EPI_ISL_1931748, EPI_ISL_1931751, EPI_ISL_1931754, EPI_ISL_1931755, EPI_ISL_1931758, EPI_ISL_1931759, EPI_ISL_1931762, EPI_ISL_1931763, EPI_ISL_1931766, EPI_ISL_1931769, EPI_ISL_1931771, EPI_ISL_1931774, EPI_ISL_1931777, EPI_ISL_1931780, EPI_ISL_1931783, EPI_ISL_1931785, EPI_ISL_1931788, EPI_ISL_1931791, EPI_ISL_1931793, EPI_ISL_1931796, EPI_ISL_1931799, EPI_ISL_1931802, EPI_ISL_1931805, EPI_ISL_1931807, EPI_ISL_1931808, EPI_ISL_1931809, EPI_ISL_1931811, EPI_ISL_1931812, EPI_ISL_1931814, EPI_ISL_1931816, EPI_ISL_1931818, EPI_ISL_1931819, EPI_ISL_1931821, EPI_ISL_1931822, EPI_ISL_1931824, EPI_ISL_1931825, EPI_ISL_1931826, EPI_ISL_1931828, EPI_ISL_1931829, EPI_ISL_1931831, EPI_ISL_1931832, EPI_ISL_1931834, EPI_ISL_1931837, EPI_ISL_1931839, EPI_ISL_1931842, EPI_ISL_1931848, EPI_ISL_1931851, EPI_ISL_1931857, EPI_ISL_1931860, EPI_ISL_1931863, EPI_ISL_1931866, EPI_ISL_1931869, EPI_ISL_1931870, EPI_ISL_1931871, EPI_ISL_1931873, EPI_ISL_1931877, EPI_ISL_1931883, EPI_ISL_1931885, EPI_ISL_1931888, EPI_ISL_1931889, EPI_ISL_1931893, EPI_ISL_1931896, EPI_ISL_1931899, EPI_ISL_1931900, EPI_ISL_1931906, EPI_ISL_1931908, EPI_ISL_1931911, EPI_ISL_1931917, EPI_ISL_1931920, EPI_ISL_1931923, EPI_ISL_1931925, EPI_ISL_1931927, EPI_ISL_1931928, EPI_ISL_1931930, EPI_ISL_1931932, EPI_ISL_1931935, EPI_ISL_1931938, EPI_ISL_1931943, EPI_ISL_1931946, EPI_ISL_1931950, EPI_ISL_1931952, EPI_ISL_1931953, EPI_ISL_1931954, EPI_ISL_1931956, EPI_ISL_1931957, EPI_ISL_1931958, EPI_ISL_1931960, EPI_ISL_1931961, EPI_ISL_1931962, EPI_ISL_1931964, EPI_ISL_1931973, EPI_ISL_1931975, EPI_ISL_1931978, EPI_ISL_1931979, EPI_ISL_1931981, EPI_ISL_1931982, EPI_ISL_1931983, EPI_ISL_1931985, EPI_ISL_1931986, EPI_ISL_1931987, EPI_ISL_1931989, EPI_ISL_1931990, EPI_ISL_1931991, EPI_ISL_1931994, EPI_ISL_1931999, EPI_ISL_1932000, EPI_ISL_1932003, EPI_ISL_1932006, EPI_ISL_1932009, EPI_ISL_1932012, EPI_ISL_1932015, EPI_ISL_1932018, EPI_ISL_1932026, EPI_ISL_1932040, EPI_ISL_1932045, EPI_ISL_1932050, EPI_ISL_1932053, EPI_ISL_1932054 | | | |
| see above | Laboratory Corporation of America | Centers for Disease Control and Prevention Division of Viral Diseases, Pathogen Discovery | Dakota Howard, Dhwani Batra, Peter W. Cook, Kara Moser, Adrian Paskey, Jason Caravas, Benjamin Rambo-Martin, Shatavia Morrison, Christopher Gulvick, Scott Sammons, Yvette Unoarumhi, Darlene Wagner, Matthew Schmerer, Minoo Agarwal, Eyad Almasri, Debbie Boles, Ayla Burns, Nuthawin Charoensri, Oren Cohen, Susan Countryman, Mary Ann Cristobal, Bobbi Croy, Suzanne Dale, Hrushikesh Deshmukh, Amanda Douglas, Vincent Drouillon, Marcia Eisenberg, Howard Engler, Rama Ghatti, Prashant Gupta, Susan Hicks, Jake Humphrey, Lax Iyer, Manoj Jain, Mohan Kolli, Brian Krueger, Tim Kuphal, Stanley Letovsky, Michael Levandoski, Craig Lukasik, Jonathan Meltzer, Brian Norvell, Mindy Nye, Scott Parker, Christos Petropoulos, John Pruitt, Steven Ragan, Scott Ryan, Mike Sapeta, Jana Schroth, Suresh Babu Selvaraju, Goran Stevovic, Amanda Suchanek, Andrea Throop, Lyndon Tilson, Thomas Urban, Joe Voshell, Kimberly Wagner, Jonathan Williams, Mary Williamson, Qian Zeng, Tricia Zwiefelhofer, Clinton R. Paden, Duncan MacCannell |
| EPI_ISL_1935443, EPI_ISL_1935444, EPI_ISL_1935445, EPI_ISL_1935446, EPI_ISL_1935447, EPI_ISL_1935448, EPI_ISL_1935449, EPI_ISL_1935450, EPI_ISL_1935451, EPI_ISL_1935452, EPI_ISL_1935453, EPI_ISL_1935454, EPI_ISL_1935455, EPI_ISL_1935456, EPI_ISL_1935457, EPI_ISL_1935458, EPI_ISL_1935459, EPI_ISL_1935460, EPI_ISL_1935461, EPI_ISL_1935462, EPI_ISL_1935463, EPI_ISL_1935464, EPI_ISL_1935465, EPI_ISL_1935466, EPI_ISL_1935467, EPI_ISL_1935468, EPI_ISL_1935469, EPI_ISL_1935470, EPI_ISL_1935471, EPI_ISL_1935472, EPI_ISL_1935473, EPI_ISL_1935474, EPI_ISL_1935475, EPI_ISL_1935476, EPI_ISL_1935477, EPI_ISL_1935478, EPI_ISL_1935479, EPI_ISL_1935480, EPI_ISL_1935481, EPI_ISL_1935482, EPI_ISL_1935483, EPI_ISL_1935484, EPI_ISL_1935485, EPI_ISL_1935486, EPI_ISL_1935487, EPI_ISL_1935488, EPI_ISL_1935489, EPI_ISL_1935490, EPI_ISL_1935491, EPI_ISL_1935492, EPI_ISL_1935493, EPI_ISL_1935494, EPI_ISL_1935495, EPI_ISL_1935496, EPI_ISL_1935497, EPI_ISL_1935498, EPI_ISL_1935499, EPI_ISL_1935500, EPI_ISL_1935501 | | | |
| see above | Minnesota Department of Health, Public Health Laboratory | Minnesota Department of Health, Public Health Laboratory | Alexandra Lorentz, Jacob Garfin, Matt Plumb, and Xiong Wang |
| EPI_ISL_1935502, EPI_ISL_1935503 | M Health Fairview | Minnesota Department of Health, Public Health Laboratory | Alexandra Lorentz, Jacob Garfin, Matt Plumb, and Xiong Wang |
| EPI_ISL_1935505, EPI_ISL_1935506, EPI_ISL_1935507, EPI_ISL_1935508, EPI_ISL_1935509, EPI_ISL_1935510, EPI_ISL_1935511, EPI_ISL_1935512, EPI_ISL_1935513, EPI_ISL_1935514, EPI_ISL_1935515, EPI_ISL_1935516, EPI_ISL_1935517, EPI_ISL_1935518 | | | |
| see above | Minnesota Department of Health, Public Health Laboratory | Minnesota Department of Health, Public Health Laboratory | Alexandra Lorentz, Jacob Garfin, Matt Plumb, and Xiong Wang |
| EPI_ISL_1935556, EPI_ISL_1935557, EPI_ISL_1935558, EPI_ISL_1935559, EPI_ISL_1935560, EPI_ISL_1935561, EPI_ISL_1935562, EPI_ISL_1935563, EPI_ISL_1935564, EPI_ISL_1935565, EPI_ISL_1935566, EPI_ISL_1935567, EPI_ISL_1935568, EPI_ISL_1935569, EPI_ISL_1935570, EPI_ISL_1935571, EPI_ISL_1935572, EPI_ISL_1935573, EPI_ISL_1935574 | | | |
| see above | Quest Diagnostics Incorporated | Centers for Disease Control and Prevention Division of Viral Diseases, Pathogen Discovery | Dakota Howard, Dhwani Batra, Peter W. Cook, Kara Moser, Adrian Paskey, Jason Caravas, Benjamin Rambo-Martin, Shatavia Morrison, Christopher Gulvick, Scott Sammons, Yvette Unoarumhi, Darlene Wagner, Matthew Schmerer, S. H. Rosenthal, A. Gerasimova, R. M. Kagan, B. Anderson, M. Hua, Y. Liu, L.E. Bernstein, K.E. Livingston, A. Perez, I. A. Shlyakhter, R. V. Rolando, R. Owen, P. Tanpaiboon, F. Lacbawan, Clinton R. Paden, Duncan MacCannell |
| EPI_ISL_1935575 | UVA Clinical Microbiology Lab | Mathers Lab | Hardik I. Parikh, Katie E. Barry, Amy J. Mathers |
| EPI_ISL_1935576 | Quest Diagnostics Incorporated | Centers for Disease Control and Prevention Division of Viral Diseases, Pathogen Discovery | Dakota Howard, Dhwani Batra, Peter W. Cook, Kara Moser, Adrian Paskey, Jason Caravas, Benjamin Rambo-Martin, Shatavia Morrison, Christopher Gulvick, Scott Sammons, Yvette Unoarumhi, Darlene Wagner, Matthew Schmerer, S. H. Rosenthal, A. Gerasimova, R. M. Kagan, B. Anderson, M. Hua, Y. Liu, L.E. Bernstein, K.E. Livingston, A. Perez, I. A. Shlyakhter, R. V. Rolando, R. Owen, P. Tanpaiboon, F. Lacbawan, Clinton R. Paden, Duncan MacCannell |
| EPI_ISL_1935577 | UVA Clinical Microbiology Lab | Mathers Lab | Hardik I. Parikh, Katie E. Barry, Amy J. Mathers |
| EPI_ISL_1935578 | Quest Diagnostics Incorporated | Centers for Disease Control and Prevention Division of Viral Diseases, Pathogen Discovery | Dakota Howard, Dhwani Batra, Peter W. Cook, Kara Moser, Adrian Paskey, Jason Caravas, Benjamin Rambo-Martin, Shatavia Morrison, Christopher Gulvick, Scott Sammons, Yvette Unoarumhi, Darlene Wagner, Matthew Schmerer, S. H. Rosenthal, A. Gerasimova, R. M. Kagan, B. Anderson, M. Hua, Y. Liu, L.E. Bernstein, K.E. Livingston, A. Perez, I. A. Shlyakhter, R. V. Rolando, R. Owen, P. Tanpaiboon, F. Lacbawan, Clinton R. Paden, Duncan MacCannell |
| EPI_ISL_1935579, EPI_ISL_1935580 | UVA Clinical Microbiology Lab | Mathers Lab | Hardik I. Parikh, Katie E. Barry, Amy J. Mathers |
| EPI_ISL_1935581, EPI_ISL_1935582, EPI_ISL_1935583 | Quest Diagnostics Incorporated | Centers for Disease Control and Prevention Division of Viral Diseases, Pathogen Discovery | Dakota Howard, Dhwani Batra, Peter W. Cook, Kara Moser, Adrian Paskey, Jason Caravas, Benjamin Rambo-Martin, Shatavia Morrison, Christopher Gulvick, Scott Sammons, Yvette Unoarumhi, Darlene Wagner, Matthew Schmerer, S. H. Rosenthal, A. Gerasimova, R. M. Kagan, B. Anderson, M. Hua, Y. Liu, L.E. Bernstein, K.E. Livingston, A. Perez, I. A. Shlyakhter, R. V. Rolando, R. Owen, P. Tanpaiboon, F. Lacbawan, Clinton R. Paden, Duncan MacCannell |
| EPI_ISL_1935584, EPI_ISL_1935585, EPI_ISL_1935586, EPI_ISL_1935587, EPI_ISL_1935588, EPI_ISL_1935589, EPI_ISL_1935590, EPI_ISL_1935591, EPI_ISL_1935592, EPI_ISL_1935593, EPI_ISL_1935594, EPI_ISL_1935595, EPI_ISL_1935596, EPI_ISL_1935597 | | | |
| see above | Helix/Illumina | Centers for Disease Control and Prevention Division of Viral Diseases, Pathogen Discovery | Dakota Howard, Dhwani Batra, Peter W. Cook, Kara Moser, Adrian Paskey, Jason Caravas, Benjamin Rambo-Martin, Shatavia Morrison, Christopher Gulvick, Scott Sammons, Yvette Unoarumhi, Darlene Wagner, Matthew Schmerer, Eileen de Feo, Jan Antico, Christine Tran, Matthew Tolentino, Shannon Wickline, Kim Gietzen, Brad Sickler, Jingtao Liu, Eric Allen, Phil Febbo, Nicole L. Washington, Simon White, Geraint Levan, Kelly Schiabor Barrett, Elizabeth Cirulli, Alexandre Bolze, Ary Ascencio, Charlotte Rivera-Garcia, Ryan Cho, Jason Nguyen, Sherry Wang, Jimmy Ramirez, Tyler Cassens, Efren Sandoval, Magnus Isaksson, William Lee, David Becker, Marc Laurent, James Lu, Clinton R. Paden, Duncan MacCannell |
| EPI_ISL_1935607 | ALBANY MEDICAL CENTER | Wadsworth Center, New York State Department of Health | Kirsten St. George, Daryl M. Lamson, Alexis Russell, Matthew Shudt, Melissa A Leisner, Jonathan Plitnick, Catharine Prussing, Navjot Singh, John Kelly, Erasmus Schneider, Erica Lasek-Nesselquist |
| EPI_ISL_1935608, EPI_ISL_1935609 | NORTHWELL HEALTH LABORATORIES | Wadsworth Center, New York State Department of Health | Kirsten St. George, Daryl M. Lamson, Alexis Russell, Matthew Shudt, Melissa A Leisner, Jonathan Plitnick, Catharine Prussing, Navjot Singh, John Kelly, Erasmus Schneider, Erica Lasek-Nesselquist |
| EPI_ISL_1935610 | MONTEFIORE MEDICAL CENTER LABORATORIES | Wadsworth Center, New York State Department of Health | Kirsten St. George, Daryl M. Lamson, Alexis Russell, Matthew Shudt, Melissa A Leisner, Jonathan Plitnick, Catharine Prussing, Navjot Singh, John Kelly, Erasmus Schneider, Erica Lasek-Nesselquist |
| EPI_ISL_1935611 | URMC LABS | Wadsworth Center, New York State Department of Health | Kirsten St. George, Daryl M. Lamson, Alexis Russell, Matthew Shudt, Melissa A Leisner, Jonathan Plitnick, Catharine Prussing, Navjot Singh, John Kelly, Erasmus Schneider, Erica Lasek-Nesselquist |
| EPI_ISL_1935612 | WESTCHESTER MEDICAL CENTER | Wadsworth Center, New York State | Kirsten St. George, Daryl M. Lamson, Alexis Russell, Matthew Shudt, Melissa A Leisner, Jonathan Plitnick, Catharine Prussing, Navjot Singh, John Kelly, Erasmus Schneider, Erica Lasek-Nesselquist |

| | | | |
|---|---|---|---|
| | | Department of Health | |
| EPI_ISL_1935613 | TEMPUS LABS INC | Wadsworth Center, New York State Department of Health | Kirsten St. George, Daryl M. Lamson, Alexis Russell, Matthew Shudt, Melissa A Leisner, Jonathan Plitnick, Catharine Prussing, Navjot Singh, John Kelly, Erasmus Schneider, Erica Lasek-Nesselquist |
| EPI_ISL_1935614 | WESTCHESTER MEDICAL CENTER | Wadsworth Center, New York State Department of Health | Kirsten St. George, Daryl M. Lamson, Alexis Russell, Matthew Shudt, Melissa A Leisner, Jonathan Plitnick, Catharine Prussing, Navjot Singh, John Kelly, Erasmus Schneider, Erica Lasek-Nesselquist |
| EPI_ISL_1935615 | MONTEFIORE MEDICAL CENTER LABORATORIES | Wadsworth Center, New York State Department of Health | Kirsten St. George, Daryl M. Lamson, Alexis Russell, Matthew Shudt, Melissa A Leisner, Jonathan Plitnick, Catharine Prussing, Navjot Singh, John Kelly, Erasmus Schneider, Erica Lasek-Nesselquist |
| EPI_ISL_1935616 | URMC LABS | Wadsworth Center, New York State Department of Health | Kirsten St. George, Daryl M. Lamson, Alexis Russell, Matthew Shudt, Melissa A Leisner, Jonathan Plitnick, Catharine Prussing, Navjot Singh, John Kelly, Erasmus Schneider, Erica Lasek-Nesselquist |
| EPI_ISL_1935617 | BOSTON HEART DIAGNOSTICS CORP | Wadsworth Center, New York State Department of Health | Kirsten St. George, Daryl M. Lamson, Alexis Russell, Matthew Shudt, Melissa A Leisner, Jonathan Plitnick, Catharine Prussing, Navjot Singh, John Kelly, Erasmus Schneider, Erica Lasek-Nesselquist |
| EPI_ISL_1935618 | URMC LABS | Wadsworth Center, New York State Department of Health | Kirsten St. George, Daryl M. Lamson, Alexis Russell, Matthew Shudt, Melissa A Leisner, Jonathan Plitnick, Catharine Prussing, Navjot Singh, John Kelly, Erasmus Schneider, Erica Lasek-Nesselquist |
| EPI_ISL_1935619 | NYC Pandemic Response Lab | Wadsworth Center, New York State Department of Health | Kirsten St. George, Daryl M. Lamson, Alexis Russell, Matthew Shudt, Melissa A Leisner, Jonathan Plitnick, Catharine Prussing, Navjot Singh, John Kelly, Erasmus Schneider, Erica Lasek-Nesselquist |
| EPI_ISL_1935620 | BOSTON HEART DIAGNOSTICS CORP | Wadsworth Center, New York State Department of Health | Kirsten St. George, Daryl M. Lamson, Alexis Russell, Matthew Shudt, Melissa A Leisner, Jonathan Plitnick, Catharine Prussing, Navjot Singh, John Kelly, Erasmus Schneider, Erica Lasek-Nesselquist |
| EPI_ISL_1935621 | NORTHWELL HEALTH LABORATORIES | Wadsworth Center, New York State Department of Health | Kirsten St. George, Daryl M. Lamson, Alexis Russell, Matthew Shudt, Melissa A Leisner, Jonathan Plitnick, Catharine Prussing, Navjot Singh, John Kelly, Erasmus Schneider, Erica Lasek-Nesselquist |
| EPI_ISL_1935622, EPI_ISL_1935623 | URMC LABS | Wadsworth Center, New York State Department of Health | Kirsten St. George, Daryl M. Lamson, Alexis Russell, Matthew Shudt, Melissa A Leisner, Jonathan Plitnick, Catharine Prussing, Navjot Singh, John Kelly, Erasmus Schneider, Erica Lasek-Nesselquist |
| EPI_ISL_1935624 | NORTHWELL HEALTH LABORATORIES | Wadsworth Center, New York State Department of Health | Kirsten St. George, Daryl M. Lamson, Alexis Russell, Matthew Shudt, Melissa A Leisner, Jonathan Plitnick, Catharine Prussing, Navjot Singh, John Kelly, Erasmus Schneider, Erica Lasek-Nesselquist |
| EPI_ISL_1935625 | Columbia University Irving Medical Center | Wadsworth Center, New York State Department of Health | Kirsten St. George, Daryl M. Lamson, Alexis Russell, Matthew Shudt, Melissa A Leisner, Jonathan Plitnick, Catharine Prussing, Navjot Singh, John Kelly, Erasmus Schneider, Erica Lasek-Nesselquist |
| EPI_ISL_1935626 | NORTHWELL HEALTH LABORATORIES | Wadsworth Center, New York State Department of Health | Kirsten St. George, Daryl M. Lamson, Alexis Russell, Matthew Shudt, Melissa A Leisner, Jonathan Plitnick, Catharine Prussing, Navjot Singh, John Kelly, Erasmus Schneider, Erica Lasek-Nesselquist |
| EPI_ISL_1935627 | TEMPUS LABS INC | Wadsworth Center, New York State Department of Health | Kirsten St. George, Daryl M. Lamson, Alexis Russell, Matthew Shudt, Melissa A Leisner, Jonathan Plitnick, Catharine Prussing, Navjot Singh, John Kelly, Erasmus Schneider, Erica Lasek-Nesselquist |
| EPI_ISL_1935628 | NYC Pandemic Response Lab | Wadsworth Center, New York State Department of Health | Kirsten St. George, Daryl M. Lamson, Alexis Russell, Matthew Shudt, Melissa A Leisner, Jonathan Plitnick, Catharine Prussing, Navjot Singh, John Kelly, Erasmus Schneider, Erica Lasek-Nesselquist |
| EPI_ISL_1935629 | NORTHWELL HEALTH LABORATORIES | Wadsworth Center, New York State Department of Health | Kirsten St. George, Daryl M. Lamson, Alexis Russell, Matthew Shudt, Melissa A Leisner, Jonathan Plitnick, Catharine Prussing, Navjot Singh, John Kelly, Erasmus Schneider, Erica Lasek-Nesselquist |
| EPI_ISL_1935630 | ALBANY MEDICAL CENTER | Wadsworth Center, New York State Department of Health | Kirsten St. George, Daryl M. Lamson, Alexis Russell, Matthew Shudt, Melissa A Leisner, Jonathan Plitnick, Catharine Prussing, Navjot Singh, John Kelly, Erasmus Schneider, Erica Lasek-Nesselquist |
| EPI_ISL_1935631 | NYC Pandemic Response Lab | Wadsworth Center, New York State Department of Health | Kirsten St. George, Daryl M. Lamson, Alexis Russell, Matthew Shudt, Melissa A Leisner, Jonathan Plitnick, Catharine Prussing, Navjot Singh, John Kelly, Erasmus Schneider, Erica Lasek-Nesselquist |
| EPI_ISL_1935632, EPI_ISL_1935633 | TEMPUS LABS INC | Wadsworth Center, New York State Department of Health | Kirsten St. George, Daryl M. Lamson, Alexis Russell, Matthew Shudt, Melissa A Leisner, Jonathan Plitnick, Catharine Prussing, Navjot Singh, John Kelly, Erasmus Schneider, Erica Lasek-Nesselquist |
| EPI_ISL_1935634, EPI_ISL_1935635 | URMC LABS | Wadsworth Center, New York State Department of Health | Kirsten St. George, Daryl M. Lamson, Alexis Russell, Matthew Shudt, Melissa A Leisner, Jonathan Plitnick, Catharine Prussing, Navjot Singh, John Kelly, Erasmus Schneider, Erica Lasek-Nesselquist |
| EPI_ISL_1935636 | NYC Pandemic Response Lab | Wadsworth Center, New York State Department of Health | Kirsten St. George, Daryl M. Lamson, Alexis Russell, Matthew Shudt, Melissa A Leisner, Jonathan Plitnick, Catharine Prussing, Navjot Singh, John Kelly, Erasmus Schneider, Erica Lasek-Nesselquist |
| EPI_ISL_1935637 | NORTHWELL HEALTH LABORATORIES | Wadsworth Center, New York State Department of Health | Kirsten St. George, Daryl M. Lamson, Alexis Russell, Matthew Shudt, Melissa A Leisner, Jonathan Plitnick, Catharine Prussing, Navjot Singh, John Kelly, Erasmus Schneider, Erica Lasek-Nesselquist |
| EPI_ISL_1935638 | ALBANY MEDICAL CENTER | Wadsworth Center, New York State Department of Health | Kirsten St. George, Daryl M. Lamson, Alexis Russell, Matthew Shudt, Melissa A Leisner, Jonathan Plitnick, Catharine Prussing, Navjot Singh, John Kelly, Erasmus Schneider, Erica Lasek-Nesselquist |
| EPI_ISL_1935639, EPI_ISL_1935640, EPI_ISL_1935641, EPI_ISL_1935642 | SUNY UPSTATE MEDICAL UNIVERSITY | Wadsworth Center, New York State Department of Health | Kirsten St. George, Daryl M. Lamson, Alexis Russell, Matthew Shudt, Melissa A Leisner, Jonathan Plitnick, Catharine Prussing, Navjot Singh, John Kelly, Erasmus Schneider, Erica Lasek-Nesselquist |
| EPI_ISL_1935643 | MONTEFIORE MEDICAL CENTER LABORATORIES | Wadsworth Center, New York State Department of Health | Kirsten St. George, Daryl M. Lamson, Alexis Russell, Matthew Shudt, Melissa A Leisner, Jonathan Plitnick, Catharine Prussing, Navjot Singh, John Kelly, Erasmus Schneider, Erica Lasek-Nesselquist |
| EPI_ISL_1935644 | ALBANY MEDICAL CENTER | Wadsworth Center, New York State Department of Health | Kirsten St. George, Daryl M. Lamson, Alexis Russell, Matthew Shudt, Melissa A Leisner, Jonathan Plitnick, Catharine Prussing, Navjot Singh, John Kelly, Erasmus Schneider, Erica Lasek-Nesselquist |
| EPI_ISL_1935645 | URMC LABS | Wadsworth Center, New York State Department of Health | Kirsten St. George, Daryl M. Lamson, Alexis Russell, Matthew Shudt, Melissa A Leisner, Jonathan Plitnick, Catharine Prussing, Navjot Singh, John Kelly, Erasmus Schneider, Erica Lasek-Nesselquist |
| EPI_ISL_1935646, EPI_ISL_1935647 | SUNY UPSTATE MEDICAL UNIVERSITY | Wadsworth Center, New York State Department of Health | Kirsten St. George, Daryl M. Lamson, Alexis Russell, Matthew Shudt, Melissa A Leisner, Jonathan Plitnick, Catharine Prussing, Navjot Singh, John Kelly, Erasmus Schneider, Erica Lasek-Nesselquist |
| EPI_ISL_1935648 | URMC LABS | Wadsworth Center, New York State Department of Health | Kirsten St. George, Daryl M. Lamson, Alexis Russell, Matthew Shudt, Melissa A Leisner, Jonathan Plitnick, Catharine Prussing, Navjot Singh, John Kelly, Erasmus Schneider, Erica Lasek-Nesselquist |
| EPI_ISL_1935649, EPI_ISL_1935650, EPI_ISL_1935651 | SUNY UPSTATE MEDICAL UNIVERSITY | Wadsworth Center, New York State Department of Health | Kirsten St. George, Daryl M. Lamson, Alexis Russell, Matthew Shudt, Melissa A Leisner, Jonathan Plitnick, Catharine Prussing, Navjot Singh, John Kelly, Erasmus Schneider, Erica Lasek-Nesselquist |
| EPI_ISL_1935652 | URMC LABS | Wadsworth Center, New York State Department of Health | Kirsten St. George, Daryl M. Lamson, Alexis Russell, Matthew Shudt, Melissa A Leisner, Jonathan Plitnick, Catharine Prussing, Navjot Singh, John Kelly, Erasmus Schneider, Erica Lasek-Nesselquist |
| EPI_ISL_1936146, EPI_ISL_1936147, EPI_ISL_1936148, EPI_ISL_1936149, EPI_ISL_1936150, EPI_ISL_1936151, EPI_ISL_1936152, EPI_ISL_1936153, EPI_ISL_1936154, EPI_ISL_1936155, EPI_ISL_1936156, EPI_ISL_1936157, EPI_ISL_1936158, EPI_ISL_1936159, EPI_ISL_1936160, EPI_ISL_1936161, EPI_ISL_1936162, EPI_ISL_1936163, EPI_ISL_1936164, EPI_ISL_1936165, EPI_ISL_1936166, EPI_ISL_1936167, EPI_ISL_1936168, EPI_ISL_1936169, EPI_ISL_1936170, EPI_ISL_1936171, EPI_ISL_1936172, EPI_ISL_1936173, EPI_ISL_1936174, EPI_ISL_1936175, EPI_ISL_1936176, EPI_ISL_1936177, EPI_ISL_1936178, EPI_ISL_1936179, EPI_ISL_1936180, EPI_ISL_1936181, EPI_ISL_1936182, EPI_ISL_1936183, EPI_ISL_1936184, EPI_ISL_1936185, EPI_ISL_1936186, EPI_ISL_1936187, EPI_ISL_1936188, EPI_ISL_1936189, EPI_ISL_1936190, EPI_ISL_1936191, EPI_ISL_1936192, EPI_ISL_1936193 | | | |
| see above | MD PHL | MD PHL | Maryland Department of Health Laboratories Administration |
| EPI_ISL_1936238, EPI_ISL_1936239, EPI_ISL_1936295 | Ohio Department of Health Laboratory | Ohio Department of Health Laboratory | Holmes, Jennifer; Eric Brandt, Keoni Omura, Glen McGillivary, Caitlin McDonnell, Jade Mowery, Stephanie Mccracken, Tyler Payne, Kirtana Ramadugu, Erica Leasure, Brent Lee, Kelsey Florek, Heather Blankenship, Allison Black, Quanta Brown, and Tammy Bannerman |
| EPI_ISL_1937024, EPI_ISL_1937025, EPI_ISL_1937026, EPI_ISL_1937027, EPI_ISL_1937028, EPI_ISL_1937029, EPI_ISL_1937030, EPI_ISL_1937031, EPI_ISL_1937032, EPI_ISL_1937033, EPI_ISL_1937035, EPI_ISL_1937038, EPI_ISL_1937039, EPI_ISL_1937040, EPI_ISL_1937041, EPI_ISL_1937042, EPI_ISL_1937043, EPI_ISL_1937044, EPI_ISL_1937045, EPI_ISL_1937047, EPI_ISL_1937048, EPI_ISL_1937049, EPI_ISL_1937050, EPI_ISL_1937052, EPI_ISL_1937054, EPI_ISL_1937055, EPI_ISL_1937059, EPI_ISL_1937061, EPI_ISL_1937062, EPI_ISL_1937063, EPI_ISL_1937064, EPI_ISL_1937065, EPI_ISL_1937066, EPI_ISL_1937067, EPI_ISL_1937069, EPI_ISL_1937070, EPI_ISL_1937071, EPI_ISL_1937072, EPI_ISL_1937075, EPI_ISL_1937076, EPI_ISL_1937077, EPI_ISL_1937078, EPI_ISL_1937079, EPI_ISL_1937080, EPI_ISL_1937082, EPI_ISL_1937084, EPI_ISL_1937085, EPI_ISL_1937086, EPI_ISL_1937087, EPI_ISL_1937088, EPI_ISL_1937089, EPI_ISL_1937090, EPI_ISL_1937093, EPI_ISL_1937096, EPI_ISL_1937097, EPI_ISL_1937098, EPI_ISL_1937099, EPI_ISL_1937100, EPI_ISL_1937101, EPI_ISL_1937102, EPI_ISL_1937103, EPI_ISL_1937104, EPI_ISL_1937105, EPI_ISL_1937106, EPI_ISL_1937107, EPI_ISL_1937108, EPI_ISL_1937110, EPI_ISL_1937111, EPI_ISL_1937112, EPI_ISL_1937113, EPI_ISL_1937114, EPI_ISL_1937115, EPI_ISL_1937117, EPI_ISL_1937118, EPI_ISL_1937119, EPI_ISL_1937123, EPI_ISL_1937124, EPI_ISL_1937125, EPI_ISL_1937126, EPI_ISL_1937127, EPI_ISL_1937129, EPI_ISL_1937130, EPI_ISL_1937131, EPI_ISL_1937132, EPI_ISL_1937133, EPI_ISL_1937134, EPI_ISL_1937135, EPI_ISL_1937136, EPI_ISL_1937139, EPI_ISL_1937140, EPI_ISL_1937141, EPI_ISL_1937142, EPI_ISL_1937143, EPI_ISL_1937144, EPI_ISL_1937145, EPI_ISL_1937146, EPI_ISL_1937147, EPI_ISL_1937148, EPI_ISL_1937149, EPI_ISL_1937150, EPI_ISL_1937151, EPI_ISL_1937152, EPI_ISL_1937154, EPI_ISL_1937156, EPI_ISL_1937157, EPI_ISL_1937158, EPI_ISL_1937159, EPI_ISL_1937160, EPI_ISL_1937161, EPI_ISL_1937162, EPI_ISL_1937163, EPI_ISL_1937164, EPI_ISL_1937166, EPI_ISL_1937168, EPI_ISL_1937169, EPI_ISL_1937170, EPI_ISL_1937173, EPI_ISL_1937175, EPI_ISL_1937176, EPI_ISL_1937177, EPI_ISL_1937179, EPI_ISL_1937181, EPI_ISL_1937182, EPI_ISL_1937183, EPI_ISL_1937184, EPI_ISL_1937185, EPI_ISL_1937186, EPI_ISL_1937187, EPI_ISL_1937188, EPI_ISL_1937191, EPI_ISL_1937193, EPI_ISL_1937194, EPI_ISL_1937195, EPI_ISL_1937201, EPI_ISL_1937204, EPI_ISL_1937205, EPI_ISL_1937207, EPI_ISL_1937208, EPI_ISL_1937209, EPI_ISL_1937210, EPI_ISL_1937211, EPI_ISL_1937213, EPI_ISL_1937214, EPI_ISL_1937216, EPI_ISL_1937217, EPI_ISL_1937218, EPI_ISL_1937219, EPI_ISL_1937221, EPI_ISL_1937222, EPI_ISL_1937224, EPI_ISL_1937225, EPI_ISL_1937226, EPI_ISL_1937227, EPI_ISL_1937228, EPI_ISL_1937229, EPI_ISL_1937230, EPI_ISL_1937231, EPI_ISL_1937232, EPI_ISL_1937234, EPI_ISL_1937236, EPI_ISL_1937237, EPI_ISL_1937238, EPI_ISL_1937239, EPI_ISL_1937241, EPI_ISL_1937242, EPI_ISL_1937243, EPI_ISL_1937244, EPI_ISL_1937245, EPI_ISL_1937246, EPI_ISL_1937249, EPI_ISL_1937250, EPI_ISL_1937251, EPI_ISL_1937253, EPI_ISL_1937254, EPI_ISL_1937255, EPI_ISL_1937256, EPI_ISL_1937257, EPI_ISL_1937258, EPI_ISL_1937259, EPI_ISL_1937260, EPI_ISL_1937261, EPI_ISL_1937262, EPI_ISL_1937263, EPI_ISL_1937265, EPI_ISL_1937266, EPI_ISL_1937267, EPI_ISL_1937268, EPI_ISL_1937269, EPI_ISL_1937270, EPI_ISL_1937271, EPI_ISL_1937272, EPI_ISL_1937273, EPI_ISL_1937274, EPI_ISL_1937275, EPI_ISL_1937276, EPI_ISL_1937277, EPI_ISL_1937279, EPI_ISL_1937280, EPI_ISL_1937281, EPI_ISL_1937283, EPI_ISL_1937284, EPI_ISL_1937285, EPI_ISL_1937286, EPI_ISL_1937287, EPI_ISL_1937288, EPI_ISL_1937289, EPI_ISL_1937290, EPI_ISL_1937291, EPI_ISL_1937293, EPI_ISL_1937294, EPI_ISL_1937295, EPI_ISL_1937296, EPI_ISL_1937297, EPI_ISL_1937298, EPI_ISL_1937299, EPI_ISL_1937300, EPI_ISL_1937301, EPI_ISL_1937302, EPI_ISL_1937303, EPI_ISL_1937304, EPI_ISL_1937305, EPI_ISL_1937306, EPI_ISL_1937307, EPI_ISL_1937308, EPI_ISL_1937309, EPI_ISL_1937310, EPI_ISL_1937311, EPI_ISL_1937313, EPI_ISL_1937314, EPI_ISL_1937315, EPI_ISL_1937316, EPI_ISL_1937317, EPI_ISL_1937318, EPI_ISL_1937320, EPI_ISL_1937321, EPI_ISL_1937322, EPI_ISL_1937323, EPI_ISL_1937324, EPI_ISL_1937326, EPI_ISL_1937327, EPI_ISL_1937328, EPI_ISL_1937329, EPI_ISL_1937332, EPI_ISL_1937334, EPI_ISL_1937335, EPI_ISL_1937336, EPI_ISL_1937337, EPI_ISL_1937338, EPI_ISL_1937339, EPI_ISL_1937342, EPI_ISL_1937343, EPI_ISL_1937344, EPI_ISL_1937346, EPI_ISL_1937347, EPI_ISL_1937348, EPI_ISL_1937349, EPI_ISL_1937352, EPI_ISL_1937353, EPI_ISL_1937356, EPI_ISL_1937357, EPI_ISL_1937359, EPI_ISL_1937360, EPI_ISL_1937361, EPI_ISL_1937362, EPI_ISL_1937363, EPI_ISL_1937365, EPI_ISL_1937366, EPI_ISL_1937368, EPI_ISL_1937369, EPI_ISL_1937370, EPI_ISL_1937371, EPI_ISL_1937372, EPI_ISL_1937374, EPI_ISL_1937375, EPI_ISL_1937376, EPI_ISL_1937377, EPI_ISL_1937378, EPI_ISL_1937379, EPI_ISL_1937381, EPI_ISL_1937382, EPI_ISL_1937383, EPI_ISL_1937386, EPI_ISL_1937387, EPI_ISL_1937388, EPI_ISL_1937389, EPI_ISL_1937390, EPI_ISL_1937391, EPI_ISL_1937392, EPI_ISL_1937393, EPI_ISL_1937394, EPI_ISL_1937396, EPI_ISL_1937397, EPI_ISL_1937399, EPI_ISL_1937400, EPI_ISL_1937401, EPI_ISL_1937403, EPI_ISL_1937404, EPI_ISL_1937405, EPI_ISL_1937408, EPI_ISL_1937409, EPI_ISL_1937410, EPI_ISL_1937411, EPI_ISL_1937412, EPI_ISL_1937414, EPI_ISL_1937415, EPI_ISL_1937417, EPI_ISL_1937418, EPI_ISL_1937420, EPI_ISL_1937422, EPI_ISL_1937423, EPI_ISL_1937424, EPI_ISL_1937425, EPI_ISL_1937426, EPI_ISL_1937427, EPI_ISL_1937428, EPI_ISL_1937429, EPI_ISL_1937430, EPI_ISL_1937431, EPI_ISL_1937433, EPI_ISL_1937434, EPI_ISL_1937437, EPI_ISL_1937438, EPI_ISL_1937440, EPI_ISL_1937441, EPI_ISL_1937442, EPI_ISL_1937443, EPI_ISL_1937444, EPI_ISL_1937446, EPI_ISL_1937448, EPI_ISL_1937449 | | | |
| see above | Fulgent Genetics | Fulgent Genetics | Harry Gao, Mickey Li, John Gao, Joseph Fierro, Benafsh Sapra, Becky Tsai, Yan Meng, Doreen Ng, James Xie |
| EPI_ISL_1937450 | PR Public Health Lab | Centers for Disease Control and Prevention Division of Viral Diseases, Pathogen Discovery | Mili Sheth, Sarah Nobles, Jasmine Padilla, Mark Burroughs, Shoshona Le, Katie Dillon, Peter Cook, Clinton R. Paden, Dhwani Batra, Krista Queen, Kristen Knipe, Dakota Howard, Yvette Unoarumhi, Darlene Wagner, Matthew Schmerer, Ben L. Rambo-Martin, Kristine Lacek, Sam Shepard, Alison Laufer Halpin, Dave Wentworth, Vivien Dugan, Suxiang Tong, Justin Lee |
| EPI_ISL_1937451 | Fulgent Genetics | Fulgent Genetics | Harry Gao, Mickey Li, John Gao, Joseph Fierro, Benafsh Sapra, Becky Tsai, Yan Meng, Doreen Ng, James Xie |

| | | | |
|---|---|---|---|
| EPI_ISL_1937452 | PR Public Health Lab | Centers for Disease Control and Prevention Division of Viral Diseases, Pathogen Discovery | Mili Sheth, Sarah Nobles, Jasmine Padilla, Mark Burroughs, Shoshona Le, Katie Dillon, Peter Cook, Clinton R. Paden, Dhwani Batra, Krista Queen, Kristen Knipe, Dakota Howard, Yvette Unoarumhi, Darlene Wagner, Matthew Schmerer, Ben L. Rambo-Martin, Kristine Lacek, Sam Shepard, Alison Laufer Halpin, Dave Wentworth, Vivien Dugan, Suxiang Tong, Justin Lee |
| EPI_ISL_1937453 | Fulgent Genetics | Fulgent Genetics | Harry Gao, Mickey Li, John Gao, Joseph Fierro, Benafsh Sapra, Becky Tsai, Yan Meng, Doreen Ng, James Xie |
| EPI_ISL_1937454, EPI_ISL_1937455 | PR Public Health Lab | Centers for Disease Control and Prevention Division of Viral Diseases, Pathogen Discovery | Mili Sheth, Sarah Nobles, Jasmine Padilla, Mark Burroughs, Shoshona Le, Katie Dillon, Peter Cook, Clinton R. Paden, Dhwani Batra, Krista Queen, Kristen Knipe, Dakota Howard, Yvette Unoarumhi, Darlene Wagner, Matthew Schmerer, Ben L. Rambo-Martin, Kristine Lacek, Sam Shepard, Alison Laufer Halpin, Dave Wentworth, Vivien Dugan, Suxiang Tong, Justin Lee |
| EPI_ISL_1937456 | Fulgent Genetics | Fulgent Genetics | Harry Gao, Mickey Li, John Gao, Joseph Fierro, Benafsh Sapra, Becky Tsai, Yan Meng, Doreen Ng, James Xie |
| EPI_ISL_1937457 | PR Public Health Lab | Centers for Disease Control and Prevention Division of Viral Diseases, Pathogen Discovery | Mili Sheth, Sarah Nobles, Jasmine Padilla, Mark Burroughs, Shoshona Le, Katie Dillon, Peter Cook, Clinton R. Paden, Dhwani Batra, Krista Queen, Kristen Knipe, Dakota Howard, Yvette Unoarumhi, Darlene Wagner, Matthew Schmerer, Ben L. Rambo-Martin, Kristine Lacek, Sam Shepard, Alison Laufer Halpin, Dave Wentworth, Vivien Dugan, Suxiang Tong, Justin Lee |
| EPI_ISL_1937458 | Fulgent Genetics | Fulgent Genetics | Harry Gao, Mickey Li, John Gao, Joseph Fierro, Benafsh Sapra, Becky Tsai, Yan Meng, Doreen Ng, James Xie |
| EPI_ISL_1937459, EPI_ISL_1937460 | PR Public Health Lab | Centers for Disease Control and Prevention Division of Viral Diseases, Pathogen Discovery | Mili Sheth, Sarah Nobles, Jasmine Padilla, Mark Burroughs, Shoshona Le, Katie Dillon, Peter Cook, Clinton R. Paden, Dhwani Batra, Krista Queen, Kristen Knipe, Dakota Howard, Yvette Unoarumhi, Darlene Wagner, Matthew Schmerer, Ben L. Rambo-Martin, Kristine Lacek, Sam Shepard, Alison Laufer Halpin, Dave Wentworth, Vivien Dugan, Suxiang Tong, Justin Lee |
| EPI_ISL_1937461 | Fulgent Genetics | Fulgent Genetics | Harry Gao, Mickey Li, John Gao, Joseph Fierro, Benafsh Sapra, Becky Tsai, Yan Meng, Doreen Ng, James Xie |
| EPI_ISL_1937462 | Philadelphia Department of Public Health | Centers for Disease Control and Prevention Division of Viral Diseases, Pathogen Discovery | Mili Sheth, Sarah Nobles, Jasmine Padilla, Mark Burroughs, Shoshona Le, Katie Dillon, Peter Cook, Clinton R. Paden, Dhwani Batra, Krista Queen, Kristen Knipe, Dakota Howard, Yvette Unoarumhi, Darlene Wagner, Matthew Schmerer, Ben L. Rambo-Martin, Kristine Lacek, Sam Shepard, Alison Laufer Halpin, Dave Wentworth, Vivien Dugan, Suxiang Tong, Justin Lee |
| EPI_ISL_1937463 | Fulgent Genetics | Fulgent Genetics | Harry Gao, Mickey Li, John Gao, Joseph Fierro, Benafsh Sapra, Becky Tsai, Yan Meng, Doreen Ng, James Xie |
| EPI_ISL_1937464 | Philadelphia Department of Public Health | Centers for Disease Control and Prevention Division of Viral Diseases, Pathogen Discovery | Mili Sheth, Sarah Nobles, Jasmine Padilla, Mark Burroughs, Shoshona Le, Katie Dillon, Peter Cook, Clinton R. Paden, Dhwani Batra, Krista Queen, Kristen Knipe, Dakota Howard, Yvette Unoarumhi, Darlene Wagner, Matthew Schmerer, Ben L. Rambo-Martin, Kristine Lacek, Sam Shepard, Alison Laufer Halpin, Dave Wentworth, Vivien Dugan, Suxiang Tong, Justin Lee |
| EPI_ISL_1937465 | Fulgent Genetics | Fulgent Genetics | Harry Gao, Mickey Li, John Gao, Joseph Fierro, Benafsh Sapra, Becky Tsai, Yan Meng, Doreen Ng, James Xie |
| EPI_ISL_1937466, EPI_ISL_1937467, EPI_ISL_1937469 | Philadelphia Department of Public Health | Centers for Disease Control and Prevention Division of Viral Diseases, Pathogen Discovery | Mili Sheth, Sarah Nobles, Jasmine Padilla, Mark Burroughs, Shoshona Le, Katie Dillon, Peter Cook, Clinton R. Paden, Dhwani Batra, Krista Queen, Kristen Knipe, Dakota Howard, Yvette Unoarumhi, Darlene Wagner, Matthew Schmerer, Ben L. Rambo-Martin, Kristine Lacek, Sam Shepard, Alison Laufer Halpin, Dave Wentworth, Vivien Dugan, Suxiang Tong, Justin Lee |
| EPI_ISL_1937470 | Fulgent Genetics | Fulgent Genetics | Harry Gao, Mickey Li, John Gao, Joseph Fierro, Benafsh Sapra, Becky Tsai, Yan Meng, Doreen Ng, James Xie |
| EPI_ISL_1937471, EPI_ISL_1937472, EPI_ISL_1937474, EPI_ISL_1937476, EPI_ISL_1937477, EPI_ISL_1937479 | TX-Houston Health Department Laboratory | Centers for Disease Control and Prevention Division of Viral Diseases, Pathogen Discovery | Mili Sheth, Sarah Nobles, Jasmine Padilla, Mark Burroughs, Shoshona Le, Katie Dillon, Peter Cook, Clinton R. Paden, Dhwani Batra, Krista Queen, Kristen Knipe, Dakota Howard, Yvette Unoarumhi, Darlene Wagner, Matthew Schmerer, Ben L. Rambo-Martin, Kristine Lacek, Sam Shepard, Alison Laufer Halpin, Dave Wentworth, Vivien Dugan, Suxiang Tong, Justin Lee |
| EPI_ISL_1937480 | Fulgent Genetics | Fulgent Genetics | Harry Gao, Mickey Li, John Gao, Joseph Fierro, Benafsh Sapra, Becky Tsai, Yan Meng, Doreen Ng, James Xie |
| EPI_ISL_1937481, EPI_ISL_1937483 | TX-Houston Health Department Laboratory | Centers for Disease Control and Prevention Division of Viral Diseases, Pathogen Discovery | Mili Sheth, Sarah Nobles, Jasmine Padilla, Mark Burroughs, Shoshona Le, Katie Dillon, Peter Cook, Clinton R. Paden, Dhwani Batra, Krista Queen, Kristen Knipe, Dakota Howard, Yvette Unoarumhi, Darlene Wagner, Matthew Schmerer, Ben L. Rambo-Martin, Kristine Lacek, Sam Shepard, Alison Laufer Halpin, Dave Wentworth, Vivien Dugan, Suxiang Tong, Justin Lee |
| EPI_ISL_1937484 | Fulgent Genetics | Fulgent Genetics | Harry Gao, Mickey Li, John Gao, Joseph Fierro, Benafsh Sapra, Becky Tsai, Yan Meng, Doreen Ng, James Xie |
| EPI_ISL_1937485, EPI_ISL_1937486 | OH Department of Health Laboratory | Centers for Disease Control and Prevention Division of Viral Diseases, Pathogen Discovery | Mili Sheth, Sarah Nobles, Jasmine Padilla, Mark Burroughs, Shoshona Le, Katie Dillon, Peter Cook, Clinton R. Paden, Dhwani Batra, Krista Queen, Kristen Knipe, Dakota Howard, Yvette Unoarumhi, Darlene Wagner, Matthew Schmerer, Ben L. Rambo-Martin, Kristine Lacek, Sam Shepard, Alison Laufer Halpin, Dave Wentworth, Vivien Dugan, Suxiang Tong, Justin Lee |
| EPI_ISL_1937487 | Fulgent Genetics | Fulgent Genetics | Harry Gao, Mickey Li, John Gao, Joseph Fierro, Benafsh Sapra, Becky Tsai, Yan Meng, Doreen Ng, James Xie |
| EPI_ISL_1937488, EPI_ISL_1937490, EPI_ISL_1937491, EPI_ISL_1937493 | OH Department of Health Laboratory | Centers for Disease Control and Prevention Division of Viral Diseases, Pathogen Discovery | Mili Sheth, Sarah Nobles, Jasmine Padilla, Mark Burroughs, Shoshona Le, Katie Dillon, Peter Cook, Clinton R. Paden, Dhwani Batra, Krista Queen, Kristen Knipe, Dakota Howard, Yvette Unoarumhi, Darlene Wagner, Matthew Schmerer, Ben L. Rambo-Martin, Kristine Lacek, Sam Shepard, Alison Laufer Halpin, Dave Wentworth, Vivien Dugan, Suxiang Tong, Justin Lee |
| EPI_ISL_1937494 | Fulgent Genetics | Fulgent Genetics | Harry Gao, Mickey Li, John Gao, Joseph Fierro, Benafsh Sapra, Becky Tsai, Yan Meng, Doreen Ng, James Xie |
| EPI_ISL_1937495 | OH Department of Health Laboratory | Centers for Disease Control and Prevention Division of Viral Diseases, Pathogen Discovery | Mili Sheth, Sarah Nobles, Jasmine Padilla, Mark Burroughs, Shoshona Le, Katie Dillon, Peter Cook, Clinton R. Paden, Dhwani Batra, Krista Queen, Kristen Knipe, Dakota Howard, Yvette Unoarumhi, Darlene Wagner, Matthew Schmerer, Ben L. Rambo-Martin, Kristine Lacek, Sam Shepard, Alison Laufer Halpin, Dave Wentworth, Vivien Dugan, Suxiang Tong, Justin Lee |
| EPI_ISL_1937496 | Fulgent Genetics | Fulgent Genetics | Harry Gao, Mickey Li, John Gao, Joseph Fierro, Benafsh Sapra, Becky Tsai, Yan Meng, Doreen Ng, James Xie |
| EPI_ISL_1937497, EPI_ISL_1937498 | OH Department of Health Laboratory | Centers for Disease Control and Prevention Division of Viral Diseases, Pathogen Discovery | Mili Sheth, Sarah Nobles, Jasmine Padilla, Mark Burroughs, Shoshona Le, Katie Dillon, Peter Cook, Clinton R. Paden, Dhwani Batra, Krista Queen, Kristen Knipe, Dakota Howard, Yvette Unoarumhi, Darlene Wagner, Matthew Schmerer, Ben L. Rambo-Martin, Kristine Lacek, Sam Shepard, Alison Laufer Halpin, Dave Wentworth, Vivien Dugan, Suxiang Tong, Justin Lee |
| EPI_ISL_1937499 | Fulgent Genetics | Fulgent Genetics | Harry Gao, Mickey Li, John Gao, Joseph Fierro, Benafsh Sapra, Becky Tsai, Yan Meng, Doreen Ng, James Xie |
| EPI_ISL_1937500, EPI_ISL_1937502, EPI_ISL_1937503 | OH Department of Health Laboratory | Centers for Disease Control and Prevention Division of Viral Diseases, Pathogen Discovery | Mili Sheth, Sarah Nobles, Jasmine Padilla, Mark Burroughs, Shoshona Le, Katie Dillon, Peter Cook, Clinton R. Paden, Dhwani Batra, Krista Queen, Kristen Knipe, Dakota Howard, Yvette Unoarumhi, Darlene Wagner, Matthew Schmerer, Ben L. Rambo-Martin, Kristine Lacek, Sam Shepard, Alison Laufer Halpin, Dave Wentworth, Vivien Dugan, Suxiang Tong, Justin Lee |
| EPI_ISL_1937504 | Fulgent Genetics | Fulgent Genetics | Harry Gao, Mickey Li, John Gao, Joseph Fierro, Benafsh Sapra, Becky Tsai, Yan Meng, Doreen Ng, James Xie |
| EPI_ISL_1937505 | NE Public Health Laboratory | Centers for Disease Control and Prevention Division of Viral Diseases, Pathogen Discovery | Mili Sheth, Sarah Nobles, Jasmine Padilla, Mark Burroughs, Shoshona Le, Katie Dillon, Peter Cook, Clinton R. Paden, Dhwani Batra, Krista Queen, Kristen Knipe, Dakota Howard, Yvette Unoarumhi, Darlene Wagner, Matthew Schmerer, Ben L. Rambo-Martin, Kristine Lacek, Sam Shepard, Alison Laufer Halpin, Dave Wentworth, Vivien Dugan, Suxiang Tong, Justin Lee |
| EPI_ISL_1937506 | Fulgent Genetics | Fulgent Genetics | Harry Gao, Mickey Li, John Gao, Joseph Fierro, Benafsh Sapra, Becky Tsai, Yan Meng, Doreen Ng, James Xie |
| EPI_ISL_1937507, EPI_ISL_1937508 | NE Public Health Laboratory | Centers for Disease Control and Prevention Division of Viral Diseases, Pathogen Discovery | Mili Sheth, Sarah Nobles, Jasmine Padilla, Mark Burroughs, Shoshona Le, Katie Dillon, Peter Cook, Clinton R. Paden, Dhwani Batra, Krista Queen, Kristen Knipe, Dakota Howard, Yvette Unoarumhi, Darlene Wagner, Matthew Schmerer, Ben L. Rambo-Martin, Kristine Lacek, Sam Shepard, Alison Laufer Halpin, Dave Wentworth, Vivien Dugan, Suxiang Tong, Justin Lee |
| EPI_ISL_1937509 | Fulgent Genetics | Fulgent Genetics | Harry Gao, Mickey Li, John Gao, Joseph Fierro, Benafsh Sapra, Becky Tsai, Yan Meng, Doreen Ng, James Xie |
| EPI_ISL_1937510, EPI_ISL_1937512, EPI_ISL_1937513, EPI_ISL_1937515 | NE Public Health Laboratory | Centers for Disease Control and Prevention Division of Viral Diseases, Pathogen Discovery | Mili Sheth, Sarah Nobles, Jasmine Padilla, Mark Burroughs, Shoshona Le, Katie Dillon, Peter Cook, Clinton R. Paden, Dhwani Batra, Krista Queen, Kristen Knipe, Dakota Howard, Yvette Unoarumhi, Darlene Wagner, Matthew Schmerer, Ben L. Rambo-Martin, Kristine Lacek, Sam Shepard, Alison Laufer Halpin, Dave Wentworth, Vivien Dugan, Suxiang Tong, Justin Lee |
| EPI_ISL_1937516 | Fulgent Genetics | Fulgent Genetics | Harry Gao, Mickey Li, John Gao, Joseph Fierro, Benafsh Sapra, Becky Tsai, Yan Meng, Doreen Ng, James Xie |
| EPI_ISL_1937517 | NE Public Health Laboratory | Centers for Disease Control and Prevention Division of Viral Diseases, Pathogen Discovery | Mili Sheth, Sarah Nobles, Jasmine Padilla, Mark Burroughs, Shoshona Le, Katie Dillon, Peter Cook, Clinton R. Paden, Dhwani Batra, Krista Queen, Kristen Knipe, Dakota Howard, Yvette Unoarumhi, Darlene Wagner, Matthew Schmerer, Ben L. Rambo-Martin, Kristine Lacek, Sam Shepard, Alison Laufer Halpin, Dave Wentworth, Vivien Dugan, Suxiang Tong, Justin Lee |
| EPI_ISL_1937518 | Fulgent Genetics | Fulgent Genetics | Harry Gao, Mickey Li, John Gao, Joseph Fierro, Benafsh Sapra, Becky Tsai, Yan Meng, Doreen Ng, James Xie |
| EPI_ISL_1937519, EPI_ISL_1937520 | NE Public Health Laboratory | Centers for Disease Control and Prevention Division of Viral Diseases, Pathogen Discovery | Mili Sheth, Sarah Nobles, Jasmine Padilla, Mark Burroughs, Shoshona Le, Katie Dillon, Peter Cook, Clinton R. Paden, Dhwani Batra, Krista Queen, Kristen Knipe, Dakota Howard, Yvette Unoarumhi, Darlene Wagner, Matthew Schmerer, Ben L. Rambo-Martin, Kristine Lacek, Sam Shepard, Alison Laufer Halpin, Dave Wentworth, Vivien Dugan, Suxiang Tong, Justin Lee |
| EPI_ISL_1937521 | Fulgent Genetics | Fulgent Genetics | Harry Gao, Mickey Li, John Gao, Joseph Fierro, Benafsh Sapra, Becky Tsai, Yan Meng, Doreen Ng, James Xie |
| EPI_ISL_1937522, EPI_ISL_1937523, EPI_ISL_1937525 | NE Public Health Laboratory | Centers for Disease Control and Prevention Division of Viral Diseases, Pathogen Discovery | Mili Sheth, Sarah Nobles, Jasmine Padilla, Mark Burroughs, Shoshona Le, Katie Dillon, Peter Cook, Clinton R. Paden, Dhwani Batra, Krista Queen, Kristen Knipe, Dakota Howard, Yvette Unoarumhi, Darlene Wagner, Matthew Schmerer, Ben L. Rambo-Martin, Kristine Lacek, Sam Shepard, Alison Laufer Halpin, Dave Wentworth, Vivien Dugan, Suxiang Tong, Justin Lee |
| EPI_ISL_1937526 | Fulgent Genetics | Fulgent Genetics | Harry Gao, Mickey Li, John Gao, Joseph Fierro, Benafsh Sapra, Becky Tsai, Yan Meng, Doreen Ng, James Xie |
| EPI_ISL_1937527 | ND Dept. of Health Laboratory Services-Microbiology | Centers for Disease Control and Prevention Division of Viral Diseases, Pathogen Discovery | Mili Sheth, Sarah Nobles, Jasmine Padilla, Mark Burroughs, Shoshona Le, Katie Dillon, Peter Cook, Clinton R. Paden, Dhwani Batra, Krista Queen, Kristen Knipe, Dakota Howard, Yvette Unoarumhi, Darlene Wagner, Matthew Schmerer, Ben L. Rambo-Martin, Kristine Lacek, Sam Shepard, Alison Laufer Halpin, Dave Wentworth, Vivien Dugan, Suxiang Tong, Justin Lee |
| EPI_ISL_1937528 | Fulgent Genetics | Fulgent Genetics | Harry Gao, Mickey Li, John Gao, Joseph Fierro, Benafsh Sapra, Becky Tsai, Yan Meng, Doreen Ng, James Xie |
| EPI_ISL_1937529, EPI_ISL_1937530 | ND Dept. of Health Laboratory Services-Microbiology | Centers for Disease Control and Prevention Division of Viral Diseases, Pathogen | Mili Sheth, Sarah Nobles, Jasmine Padilla, Mark Burroughs, Shoshona Le, Katie Dillon, Peter Cook, Clinton R. Paden, Dhwani Batra, Krista Queen, Kristen Knipe, Dakota Howard, Yvette Unoarumhi, Darlene Wagner, Matthew Schmerer, Ben L. Rambo-Martin, Kristine Lacek, Sam Shepard, Alison Laufer Halpin, Dave Wentworth, Vivien Dugan, Suxiang Tong, Justin Lee |

| | | | |
|---|---|---|---|
| | | Discovery | |
| EPI_ISL_1937531 | Fulgent Genetics | Fulgent Genetics | Harry Gao, Mickey Li, John Gao, Joseph Fierro, Benafsh Sapra, Becky Tsai, Yan Meng, Doreen Ng, James Xie |
| EPI_ISL_1937532 | ND Dept. of Health Laboratory Services-Microbiology | Centers for Disease Control and Prevention Division of Viral Diseases, Pathogen Discovery | Mili Sheth, Sarah Nobles, Jasmine Padilla, Mark Burroughs, Shoshona Le, Katie Dillon, Peter Cook, Clinton R. Paden, Dhwani Batra, Krista Queen, Kristen Knipe, Dakota Howard, Yvette Unoarumhi, Darlene Wagner, Matthew Schmerer, Ben L. Rambo-Martin, Kristine Lacek, Sam Shepard, Alison Laufer Halpin, Dave Wentworth, Vivien Dugan, Suxiang Tong, Justin Lee |
| EPI_ISL_1937533 | Fulgent Genetics | Fulgent Genetics | Harry Gao, Mickey Li, John Gao, Joseph Fierro, Benafsh Sapra, Becky Tsai, Yan Meng, Doreen Ng, James Xie |
| EPI_ISL_1937534 | ND Dept. of Health Laboratory Services-Microbiology | Centers for Disease Control and Prevention Division of Viral Diseases, Pathogen Discovery | Mili Sheth, Sarah Nobles, Jasmine Padilla, Mark Burroughs, Shoshona Le, Katie Dillon, Peter Cook, Clinton R. Paden, Dhwani Batra, Krista Queen, Kristen Knipe, Dakota Howard, Yvette Unoarumhi, Darlene Wagner, Matthew Schmerer, Ben L. Rambo-Martin, Kristine Lacek, Sam Shepard, Alison Laufer Halpin, Dave Wentworth, Vivien Dugan, Suxiang Tong, Justin Lee |
| EPI_ISL_1937535 | NC State Laboratory of Public Health | Centers for Disease Control and Prevention Division of Viral Diseases, Pathogen Discovery | Mili Sheth, Sarah Nobles, Jasmine Padilla, Mark Burroughs, Shoshona Le, Katie Dillon, Peter Cook, Clinton R. Paden, Dhwani Batra, Krista Queen, Kristen Knipe, Dakota Howard, Yvette Unoarumhi, Darlene Wagner, Matthew Schmerer, Ben L. Rambo-Martin, Kristine Lacek, Sam Shepard, Alison Laufer Halpin, Dave Wentworth, Vivien Dugan, Suxiang Tong, Justin Lee |
| EPI_ISL_1937536 | Fulgent Genetics | Fulgent Genetics | Harry Gao, Mickey Li, John Gao, Joseph Fierro, Benafsh Sapra, Becky Tsai, Yan Meng, Doreen Ng, James Xie |
| EPI_ISL_1937537 | NC State Laboratory of Public Health | Centers for Disease Control and Prevention Division of Viral Diseases, Pathogen Discovery | Mili Sheth, Sarah Nobles, Jasmine Padilla, Mark Burroughs, Shoshona Le, Katie Dillon, Peter Cook, Clinton R. Paden, Dhwani Batra, Krista Queen, Kristen Knipe, Dakota Howard, Yvette Unoarumhi, Darlene Wagner, Matthew Schmerer, Ben L. Rambo-Martin, Kristine Lacek, Sam Shepard, Alison Laufer Halpin, Dave Wentworth, Vivien Dugan, Suxiang Tong, Justin Lee |
| EPI_ISL_1937538 | Fulgent Genetics | Fulgent Genetics | Harry Gao, Mickey Li, John Gao, Joseph Fierro, Benafsh Sapra, Becky Tsai, Yan Meng, Doreen Ng, James Xie |
| EPI_ISL_1937539, EPI_ISL_1937540 | NC State Laboratory of Public Health | Centers for Disease Control and Prevention Division of Viral Diseases, Pathogen Discovery | Mili Sheth, Sarah Nobles, Jasmine Padilla, Mark Burroughs, Shoshona Le, Katie Dillon, Peter Cook, Clinton R. Paden, Dhwani Batra, Krista Queen, Kristen Knipe, Dakota Howard, Yvette Unoarumhi, Darlene Wagner, Matthew Schmerer, Ben L. Rambo-Martin, Kristine Lacek, Sam Shepard, Alison Laufer Halpin, Dave Wentworth, Vivien Dugan, Suxiang Tong, Justin Lee |
| EPI_ISL_1937541 | Fulgent Genetics | Fulgent Genetics | Harry Gao, Mickey Li, John Gao, Joseph Fierro, Benafsh Sapra, Becky Tsai, Yan Meng, Doreen Ng, James Xie |
| EPI_ISL_1937542 | NC State Laboratory of Public Health | Centers for Disease Control and Prevention Division of Viral Diseases, Pathogen Discovery | Mili Sheth, Sarah Nobles, Jasmine Padilla, Mark Burroughs, Shoshona Le, Katie Dillon, Peter Cook, Clinton R. Paden, Dhwani Batra, Krista Queen, Kristen Knipe, Dakota Howard, Yvette Unoarumhi, Darlene Wagner, Matthew Schmerer, Ben L. Rambo-Martin, Kristine Lacek, Sam Shepard, Alison Laufer Halpin, Dave Wentworth, Vivien Dugan, Suxiang Tong, Justin Lee |
| EPI_ISL_1937543 | Fulgent Genetics | Fulgent Genetics | Harry Gao, Mickey Li, John Gao, Joseph Fierro, Benafsh Sapra, Becky Tsai, Yan Meng, Doreen Ng, James Xie |
| EPI_ISL_1937544 | NC State Laboratory of Public Health | Centers for Disease Control and Prevention Division of Viral Diseases, Pathogen Discovery | Mili Sheth, Sarah Nobles, Jasmine Padilla, Mark Burroughs, Shoshona Le, Katie Dillon, Peter Cook, Clinton R. Paden, Dhwani Batra, Krista Queen, Kristen Knipe, Dakota Howard, Yvette Unoarumhi, Darlene Wagner, Matthew Schmerer, Ben L. Rambo-Martin, Kristine Lacek, Sam Shepard, Alison Laufer Halpin, Dave Wentworth, Vivien Dugan, Suxiang Tong, Justin Lee |
| EPI_ISL_1937545 | Fulgent Genetics | Fulgent Genetics | Harry Gao, Mickey Li, John Gao, Joseph Fierro, Benafsh Sapra, Becky Tsai, Yan Meng, Doreen Ng, James Xie |
| EPI_ISL_1937546 | NC State Laboratory of Public Health | Centers for Disease Control and Prevention Division of Viral Diseases, Pathogen Discovery | Mili Sheth, Sarah Nobles, Jasmine Padilla, Mark Burroughs, Shoshona Le, Katie Dillon, Peter Cook, Clinton R. Paden, Dhwani Batra, Krista Queen, Kristen Knipe, Dakota Howard, Yvette Unoarumhi, Darlene Wagner, Matthew Schmerer, Ben L. Rambo-Martin, Kristine Lacek, Sam Shepard, Alison Laufer Halpin, Dave Wentworth, Vivien Dugan, Suxiang Tong, Justin Lee |
| EPI_ISL_1937547 | MO State Public Health Laboratory | Centers for Disease Control and Prevention Division of Viral Diseases, Pathogen Discovery | Mili Sheth, Sarah Nobles, Jasmine Padilla, Mark Burroughs, Shoshona Le, Katie Dillon, Peter Cook, Clinton R. Paden, Dhwani Batra, Krista Queen, Kristen Knipe, Dakota Howard, Yvette Unoarumhi, Darlene Wagner, Matthew Schmerer, Ben L. Rambo-Martin, Kristine Lacek, Sam Shepard, Alison Laufer Halpin, Dave Wentworth, Vivien Dugan, Suxiang Tong, Justin Lee |
| EPI_ISL_1937548 | Fulgent Genetics | Fulgent Genetics | Harry Gao, Mickey Li, John Gao, Joseph Fierro, Benafsh Sapra, Becky Tsai, Yan Meng, Doreen Ng, James Xie |
| EPI_ISL_1937549 | "MN PHL Division, Minnesota Department of Health" | Centers for Disease Control and Prevention Division of Viral Diseases, Pathogen Discovery | Mili Sheth, Sarah Nobles, Jasmine Padilla, Mark Burroughs, Shoshona Le, Katie Dillon, Peter Cook, Clinton R. Paden, Dhwani Batra, Krista Queen, Kristen Knipe, Dakota Howard, Yvette Unoarumhi, Darlene Wagner, Matthew Schmerer, Ben L. Rambo-Martin, Kristine Lacek, Sam Shepard, Alison Laufer Halpin, Dave Wentworth, Vivien Dugan, Suxiang Tong, Justin Lee |
| EPI_ISL_1937550 | Fulgent Genetics | Fulgent Genetics | Harry Gao, Mickey Li, John Gao, Joseph Fierro, Benafsh Sapra, Becky Tsai, Yan Meng, Doreen Ng, James Xie |
| EPI_ISL_1937551, EPI_ISL_1937552 | "MN PHL Division, Minnesota Department of Health" | Centers for Disease Control and Prevention Division of Viral Diseases, Pathogen Discovery | Mili Sheth, Sarah Nobles, Jasmine Padilla, Mark Burroughs, Shoshona Le, Katie Dillon, Peter Cook, Clinton R. Paden, Dhwani Batra, Krista Queen, Kristen Knipe, Dakota Howard, Yvette Unoarumhi, Darlene Wagner, Matthew Schmerer, Ben L. Rambo-Martin, Kristine Lacek, Sam Shepard, Alison Laufer Halpin, Dave Wentworth, Vivien Dugan, Suxiang Tong, Justin Lee |
| EPI_ISL_1937553 | Fulgent Genetics | Fulgent Genetics | Harry Gao, Mickey Li, John Gao, Joseph Fierro, Benafsh Sapra, Becky Tsai, Yan Meng, Doreen Ng, James Xie |
| EPI_ISL_1937554 | "MN PHL Division, Minnesota Department of Health" | Centers for Disease Control and Prevention Division of Viral Diseases, Pathogen Discovery | Mili Sheth, Sarah Nobles, Jasmine Padilla, Mark Burroughs, Shoshona Le, Katie Dillon, Peter Cook, Clinton R. Paden, Dhwani Batra, Krista Queen, Kristen Knipe, Dakota Howard, Yvette Unoarumhi, Darlene Wagner, Matthew Schmerer, Ben L. Rambo-Martin, Kristine Lacek, Sam Shepard, Alison Laufer Halpin, Dave Wentworth, Vivien Dugan, Suxiang Tong, Justin Lee |
| EPI_ISL_1937555 | Fulgent Genetics | Fulgent Genetics | Harry Gao, Mickey Li, John Gao, Joseph Fierro, Benafsh Sapra, Becky Tsai, Yan Meng, Doreen Ng, James Xie |
| EPI_ISL_1937556, EPI_ISL_1937558, EPI_ISL_1937559 | "MN PHL Division, Minnesota Department of Health" | Centers for Disease Control and Prevention Division of Viral Diseases, Pathogen Discovery | Mili Sheth, Sarah Nobles, Jasmine Padilla, Mark Burroughs, Shoshona Le, Katie Dillon, Peter Cook, Clinton R. Paden, Dhwani Batra, Krista Queen, Kristen Knipe, Dakota Howard, Yvette Unoarumhi, Darlene Wagner, Matthew Schmerer, Ben L. Rambo-Martin, Kristine Lacek, Sam Shepard, Alison Laufer Halpin, Dave Wentworth, Vivien Dugan, Suxiang Tong, Justin Lee |
| EPI_ISL_1937560 | Fulgent Genetics | Fulgent Genetics | Harry Gao, Mickey Li, John Gao, Joseph Fierro, Benafsh Sapra, Becky Tsai, Yan Meng, Doreen Ng, James Xie |
| EPI_ISL_1937561, EPI_ISL_1937562 | MT Public Health Laboratory | Centers for Disease Control and Prevention Division of Viral Diseases, Pathogen Discovery | Mili Sheth, Sarah Nobles, Jasmine Padilla, Mark Burroughs, Shoshona Le, Katie Dillon, Peter Cook, Clinton R. Paden, Dhwani Batra, Krista Queen, Kristen Knipe, Dakota Howard, Yvette Unoarumhi, Darlene Wagner, Matthew Schmerer, Ben L. Rambo-Martin, Kristine Lacek, Sam Shepard, Alison Laufer Halpin, Dave Wentworth, Vivien Dugan, Suxiang Tong, Justin Lee |
| EPI_ISL_1937563 | Fulgent Genetics | Fulgent Genetics | Harry Gao, Mickey Li, John Gao, Joseph Fierro, Benafsh Sapra, Becky Tsai, Yan Meng, Doreen Ng, James Xie |
| EPI_ISL_1937564 | MT Public Health Laboratory | Centers for Disease Control and Prevention Division of Viral Diseases, Pathogen Discovery | Mili Sheth, Sarah Nobles, Jasmine Padilla, Mark Burroughs, Shoshona Le, Katie Dillon, Peter Cook, Clinton R. Paden, Dhwani Batra, Krista Queen, Kristen Knipe, Dakota Howard, Yvette Unoarumhi, Darlene Wagner, Matthew Schmerer, Ben L. Rambo-Martin, Kristine Lacek, Sam Shepard, Alison Laufer Halpin, Dave Wentworth, Vivien Dugan, Suxiang Tong, Justin Lee |
| EPI_ISL_1937566, EPI_ISL_1937567, EPI_ISL_1937569 | VA-Division of Consolidated Laboratory Services | Centers for Disease Control and Prevention Division of Viral Diseases, Pathogen Discovery | Mili Sheth, Sarah Nobles, Jasmine Padilla, Mark Burroughs, Shoshona Le, Katie Dillon, Peter Cook, Clinton R. Paden, Dhwani Batra, Krista Queen, Kristen Knipe, Dakota Howard, Yvette Unoarumhi, Darlene Wagner, Matthew Schmerer, Ben L. Rambo-Martin, Kristine Lacek, Sam Shepard, Alison Laufer Halpin, Dave Wentworth, Vivien Dugan, Suxiang Tong, Justin Lee |
| EPI_ISL_1937570 | Fulgent Genetics | Fulgent Genetics | Harry Gao, Mickey Li, John Gao, Joseph Fierro, Benafsh Sapra, Becky Tsai, Yan Meng, Doreen Ng, James Xie |
| EPI_ISL_1937571, EPI_ISL_1937572, EPI_ISL_1937574 | VA-Division of Consolidated Laboratory Services | Centers for Disease Control and Prevention Division of Viral Diseases, Pathogen Discovery | Mili Sheth, Sarah Nobles, Jasmine Padilla, Mark Burroughs, Shoshona Le, Katie Dillon, Peter Cook, Clinton R. Paden, Dhwani Batra, Krista Queen, Kristen Knipe, Dakota Howard, Yvette Unoarumhi, Darlene Wagner, Matthew Schmerer, Ben L. Rambo-Martin, Kristine Lacek, Sam Shepard, Alison Laufer Halpin, Dave Wentworth, Vivien Dugan, Suxiang Tong, Justin Lee |
| EPI_ISL_1937575 | Fulgent Genetics | Fulgent Genetics | Harry Gao, Mickey Li, John Gao, Joseph Fierro, Benafsh Sapra, Becky Tsai, Yan Meng, Doreen Ng, James Xie |
| EPI_ISL_1937576, EPI_ISL_1937578, EPI_ISL_1937579 | VA-Division of Consolidated Laboratory Services | Centers for Disease Control and Prevention Division of Viral Diseases, Pathogen Discovery | Mili Sheth, Sarah Nobles, Jasmine Padilla, Mark Burroughs, Shoshona Le, Katie Dillon, Peter Cook, Clinton R. Paden, Dhwani Batra, Krista Queen, Kristen Knipe, Dakota Howard, Yvette Unoarumhi, Darlene Wagner, Matthew Schmerer, Ben L. Rambo-Martin, Kristine Lacek, Sam Shepard, Alison Laufer Halpin, Dave Wentworth, Vivien Dugan, Suxiang Tong, Justin Lee |
| EPI_ISL_1937580 | Fulgent Genetics | Fulgent Genetics | Harry Gao, Mickey Li, John Gao, Joseph Fierro, Benafsh Sapra, Becky Tsai, Yan Meng, Doreen Ng, James Xie |
| EPI_ISL_1937581 | VA-Division of Consolidated Laboratory Services | Centers for Disease Control and Prevention Division of Viral Diseases, Pathogen Discovery | Mili Sheth, Sarah Nobles, Jasmine Padilla, Mark Burroughs, Shoshona Le, Katie Dillon, Peter Cook, Clinton R. Paden, Dhwani Batra, Krista Queen, Kristen Knipe, Dakota Howard, Yvette Unoarumhi, Darlene Wagner, Matthew Schmerer, Ben L. Rambo-Martin, Kristine Lacek, Sam Shepard, Alison Laufer Halpin, Dave Wentworth, Vivien Dugan, Suxiang Tong, Justin Lee |
| EPI_ISL_1937582 | Fulgent Genetics | Fulgent Genetics | Harry Gao, Mickey Li, John Gao, Joseph Fierro, Benafsh Sapra, Becky Tsai, Yan Meng, Doreen Ng, James Xie |
| EPI_ISL_1937583 | VA-Division of Consolidated Laboratory Services | Centers for Disease Control and Prevention Division of Viral Diseases, Pathogen Discovery | Mili Sheth, Sarah Nobles, Jasmine Padilla, Mark Burroughs, Shoshona Le, Katie Dillon, Peter Cook, Clinton R. Paden, Dhwani Batra, Krista Queen, Kristen Knipe, Dakota Howard, Yvette Unoarumhi, Darlene Wagner, Matthew Schmerer, Ben L. Rambo-Martin, Kristine Lacek, Sam Shepard, Alison Laufer Halpin, Dave Wentworth, Vivien Dugan, Suxiang Tong, Justin Lee |
| EPI_ISL_1937584 | Fulgent Genetics | Fulgent Genetics | Harry Gao, Mickey Li, John Gao, Joseph Fierro, Benafsh Sapra, Becky Tsai, Yan Meng, Doreen Ng, James Xie |
| EPI_ISL_1937585, EPI_ISL_1937586 | VA-Division of Consolidated Laboratory Services | Centers for Disease Control and Prevention Division of Viral Diseases, Pathogen Discovery | Mili Sheth, Sarah Nobles, Jasmine Padilla, Mark Burroughs, Shoshona Le, Katie Dillon, Peter Cook, Clinton R. Paden, Dhwani Batra, Krista Queen, Kristen Knipe, Dakota Howard, Yvette Unoarumhi, Darlene Wagner, Matthew Schmerer, Ben L. Rambo-Martin, Kristine Lacek, Sam Shepard, Alison Laufer Halpin, Dave Wentworth, Vivien Dugan, Suxiang Tong, Justin Lee |
| EPI_ISL_1937587 | Fulgent Genetics | Fulgent Genetics | Harry Gao, Mickey Li, John Gao, Joseph Fierro, Benafsh Sapra, Becky Tsai, Yan Meng, Doreen Ng, James Xie |
| EPI_ISL_1937588, EPI_ISL_1937589, EPI_ISL_1937591 | VA-Division of Consolidated Laboratory Services | Centers for Disease Control and Prevention Division of Viral Diseases, Pathogen Discovery | Mili Sheth, Sarah Nobles, Jasmine Padilla, Mark Burroughs, Shoshona Le, Katie Dillon, Peter Cook, Clinton R. Paden, Dhwani Batra, Krista Queen, Kristen Knipe, Dakota Howard, Yvette Unoarumhi, Darlene Wagner, Matthew Schmerer, Ben L. Rambo-Martin, Kristine Lacek, Sam Shepard, Alison Laufer Halpin, Dave Wentworth, Vivien Dugan, Suxiang Tong, Justin Lee |
| EPI_ISL_1937592 | Fulgent Genetics | Fulgent Genetics | Harry Gao, Mickey Li, John Gao, Joseph Fierro, Benafsh Sapra, Becky Tsai, Yan Meng, Doreen Ng, James Xie |
| EPI_ISL_1937593 | VA-Division of Consolidated Laboratory Services | Centers for Disease Control and Prevention Division of Viral Diseases, Pathogen Discovery | Mili Sheth, Sarah Nobles, Jasmine Padilla, Mark Burroughs, Shoshona Le, Katie Dillon, Peter Cook, Clinton R. Paden, Dhwani Batra, Krista Queen, Kristen Knipe, Dakota Howard, Yvette Unoarumhi, Darlene Wagner, Matthew Schmerer, Ben L. Rambo-Martin, Kristine Lacek, Sam Shepard, Alison Laufer Halpin, Dave Wentworth, Vivien Dugan, Suxiang Tong, Justin Lee |
| EPI_ISL_1937594 | Fulgent Genetics | Fulgent Genetics | Harry Gao, Mickey Li, John Gao, Joseph Fierro, Benafsh Sapra, Becky Tsai, Yan Meng, Doreen Ng, James Xie |

| | | | |
|---|---|---|---|
| EPI_ISL_1937595 | VA-Division of Consolidated Laboratory Services | Centers for Disease Control and Prevention Division of Viral Diseases, Pathogen Discovery | Mili Sheth, Sarah Nobles, Jasmine Padilla, Mark Burroughs, Shoshona Le, Katie Dillon, Peter Cook, Clinton R. Paden, Dhwani Batra, Krista Queen, Kristen Knipe, Dakota Howard, Yvette Unoarumhi, Darlene Wagner, Matthew Schmerer, Ben L. Rambo-Martin, Kristine Lacek, Sam Shepard, Alison Laufer Halpin, Dave Wentworth, Vivien Dugan, Suxiang Tong, Justin Lee |
| EPI_ISL_1937596, EPI_ISL_1937598 | NYC Department of Health and Mental Hygiene | Centers for Disease Control and Prevention Division of Viral Diseases, Pathogen Discovery | Mili Sheth, Sarah Nobles, Jasmine Padilla, Mark Burroughs, Shoshona Le, Katie Dillon, Peter Cook, Clinton R. Paden, Dhwani Batra, Krista Queen, Kristen Knipe, Dakota Howard, Yvette Unoarumhi, Darlene Wagner, Matthew Schmerer, Ben L. Rambo-Martin, Kristine Lacek, Sam Shepard, Alison Laufer Halpin, Dave Wentworth, Vivien Dugan, Suxiang Tong, Justin Lee |
| EPI_ISL_1937599 | Fulgent Genetics | Fulgent Genetics | Harry Gao, Mickey Li, John Gao, Joseph Fierro, Benafsh Sapra, Becky Tsai, Yan Meng, Doreen Ng, James Xie |
| EPI_ISL_1937600, EPI_ISL_1937601, EPI_ISL_1937603 | NYC Department of Health and Mental Hygiene | Centers for Disease Control and Prevention Division of Viral Diseases, Pathogen Discovery | Mili Sheth, Sarah Nobles, Jasmine Padilla, Mark Burroughs, Shoshona Le, Katie Dillon, Peter Cook, Clinton R. Paden, Dhwani Batra, Krista Queen, Kristen Knipe, Dakota Howard, Yvette Unoarumhi, Darlene Wagner, Matthew Schmerer, Ben L. Rambo-Martin, Kristine Lacek, Sam Shepard, Alison Laufer Halpin, Dave Wentworth, Vivien Dugan, Suxiang Tong, Justin Lee |
| EPI_ISL_1937604 | Fulgent Genetics | Fulgent Genetics | Harry Gao, Mickey Li, John Gao, Joseph Fierro, Benafsh Sapra, Becky Tsai, Yan Meng, Doreen Ng, James Xie |
| EPI_ISL_1937605, EPI_ISL_1937606 | NYC Department of Health and Mental Hygiene | Centers for Disease Control and Prevention Division of Viral Diseases, Pathogen Discovery | Mili Sheth, Sarah Nobles, Jasmine Padilla, Mark Burroughs, Shoshona Le, Katie Dillon, Peter Cook, Clinton R. Paden, Dhwani Batra, Krista Queen, Kristen Knipe, Dakota Howard, Yvette Unoarumhi, Darlene Wagner, Matthew Schmerer, Ben L. Rambo-Martin, Kristine Lacek, Sam Shepard, Alison Laufer Halpin, Dave Wentworth, Vivien Dugan, Suxiang Tong, Justin Lee |
| EPI_ISL_1937607 | Fulgent Genetics | Fulgent Genetics | Harry Gao, Mickey Li, John Gao, Joseph Fierro, Benafsh Sapra, Becky Tsai, Yan Meng, Doreen Ng, James Xie |
| EPI_ISL_1937608 | NYC Department of Health and Mental Hygiene | Centers for Disease Control and Prevention Division of Viral Diseases, Pathogen Discovery | Mili Sheth, Sarah Nobles, Jasmine Padilla, Mark Burroughs, Shoshona Le, Katie Dillon, Peter Cook, Clinton R. Paden, Dhwani Batra, Krista Queen, Kristen Knipe, Dakota Howard, Yvette Unoarumhi, Darlene Wagner, Matthew Schmerer, Ben L. Rambo-Martin, Kristine Lacek, Sam Shepard, Alison Laufer Halpin, Dave Wentworth, Vivien Dugan, Suxiang Tong, Justin Lee |
| EPI_ISL_1937609 | Fulgent Genetics | Fulgent Genetics | Harry Gao, Mickey Li, John Gao, Joseph Fierro, Benafsh Sapra, Becky Tsai, Yan Meng, Doreen Ng, James Xie |
| EPI_ISL_1937610, EPI_ISL_1937611 | NYC Department of Health and Mental Hygiene | Centers for Disease Control and Prevention Division of Viral Diseases, Pathogen Discovery | Mili Sheth, Sarah Nobles, Jasmine Padilla, Mark Burroughs, Shoshona Le, Katie Dillon, Peter Cook, Clinton R. Paden, Dhwani Batra, Krista Queen, Kristen Knipe, Dakota Howard, Yvette Unoarumhi, Darlene Wagner, Matthew Schmerer, Ben L. Rambo-Martin, Kristine Lacek, Sam Shepard, Alison Laufer Halpin, Dave Wentworth, Vivien Dugan, Suxiang Tong, Justin Lee |
| EPI_ISL_1937612 | Fulgent Genetics | Fulgent Genetics | Harry Gao, Mickey Li, John Gao, Joseph Fierro, Benafsh Sapra, Becky Tsai, Yan Meng, Doreen Ng, James Xie |
| EPI_ISL_1937613 | NYC Department of Health and Mental Hygiene | Centers for Disease Control and Prevention Division of Viral Diseases, Pathogen Discovery | Mili Sheth, Sarah Nobles, Jasmine Padilla, Mark Burroughs, Shoshona Le, Katie Dillon, Peter Cook, Clinton R. Paden, Dhwani Batra, Krista Queen, Kristen Knipe, Dakota Howard, Yvette Unoarumhi, Darlene Wagner, Matthew Schmerer, Ben L. Rambo-Martin, Kristine Lacek, Sam Shepard, Alison Laufer Halpin, Dave Wentworth, Vivien Dugan, Suxiang Tong, Justin Lee |
| EPI_ISL_1937614 | Fulgent Genetics | Fulgent Genetics | Harry Gao, Mickey Li, John Gao, Joseph Fierro, Benafsh Sapra, Becky Tsai, Yan Meng, Doreen Ng, James Xie |
| EPI_ISL_1937615, EPI_ISL_1937616 | NYC Department of Health and Mental Hygiene | Centers for Disease Control and Prevention Division of Viral Diseases, Pathogen Discovery | Mili Sheth, Sarah Nobles, Jasmine Padilla, Mark Burroughs, Shoshona Le, Katie Dillon, Peter Cook, Clinton R. Paden, Dhwani Batra, Krista Queen, Kristen Knipe, Dakota Howard, Yvette Unoarumhi, Darlene Wagner, Matthew Schmerer, Ben L. Rambo-Martin, Kristine Lacek, Sam Shepard, Alison Laufer Halpin, Dave Wentworth, Vivien Dugan, Suxiang Tong, Justin Lee |
| EPI_ISL_1937617 | Fulgent Genetics | Fulgent Genetics | Harry Gao, Mickey Li, John Gao, Joseph Fierro, Benafsh Sapra, Becky Tsai, Yan Meng, Doreen Ng, James Xie |
| EPI_ISL_1937618 | NYC Department of Health and Mental Hygiene | Centers for Disease Control and Prevention Division of Viral Diseases, Pathogen Discovery | Mili Sheth, Sarah Nobles, Jasmine Padilla, Mark Burroughs, Shoshona Le, Katie Dillon, Peter Cook, Clinton R. Paden, Dhwani Batra, Krista Queen, Kristen Knipe, Dakota Howard, Yvette Unoarumhi, Darlene Wagner, Matthew Schmerer, Ben L. Rambo-Martin, Kristine Lacek, Sam Shepard, Alison Laufer Halpin, Dave Wentworth, Vivien Dugan, Suxiang Tong, Justin Lee |
| EPI_ISL_1937619 | Fulgent Genetics | Fulgent Genetics | Harry Gao, Mickey Li, John Gao, Joseph Fierro, Benafsh Sapra, Becky Tsai, Yan Meng, Doreen Ng, James Xie |
| EPI_ISL_1937620, EPI_ISL_1937622, EPI_ISL_1937623 | NYC Department of Health and Mental Hygiene | Centers for Disease Control and Prevention Division of Viral Diseases, Pathogen Discovery | Mili Sheth, Sarah Nobles, Jasmine Padilla, Mark Burroughs, Shoshona Le, Katie Dillon, Peter Cook, Clinton R. Paden, Dhwani Batra, Krista Queen, Kristen Knipe, Dakota Howard, Yvette Unoarumhi, Darlene Wagner, Matthew Schmerer, Ben L. Rambo-Martin, Kristine Lacek, Sam Shepard, Alison Laufer Halpin, Dave Wentworth, Vivien Dugan, Suxiang Tong, Justin Lee |
| EPI_ISL_1937624 | Fulgent Genetics | Fulgent Genetics | Harry Gao, Mickey Li, John Gao, Joseph Fierro, Benafsh Sapra, Becky Tsai, Yan Meng, Doreen Ng, James Xie |
| EPI_ISL_1937625, EPI_ISL_1937626, EPI_ISL_1937627, EPI_ISL_1937628 | NJ Public Health and Environmental Laboratories | Centers for Disease Control and Prevention Division of Viral Diseases, Pathogen Discovery | Mili Sheth, Sarah Nobles, Jasmine Padilla, Mark Burroughs, Shoshona Le, Katie Dillon, Peter Cook, Clinton R. Paden, Dhwani Batra, Krista Queen, Kristen Knipe, Dakota Howard, Yvette Unoarumhi, Darlene Wagner, Matthew Schmerer, Ben L. Rambo-Martin, Kristine Lacek, Sam Shepard, Alison Laufer Halpin, Dave Wentworth, Vivien Dugan, Suxiang Tong, Justin Lee |
| EPI_ISL_1937629 | Fulgent Genetics | Fulgent Genetics | Harry Gao, Mickey Li, John Gao, Joseph Fierro, Benafsh Sapra, Becky Tsai, Yan Meng, Doreen Ng, James Xie |
| EPI_ISL_1937630, EPI_ISL_1937631 | NJ Public Health and Environmental Laboratories | Centers for Disease Control and Prevention Division of Viral Diseases, Pathogen Discovery | Mili Sheth, Sarah Nobles, Jasmine Padilla, Mark Burroughs, Shoshona Le, Katie Dillon, Peter Cook, Clinton R. Paden, Dhwani Batra, Krista Queen, Kristen Knipe, Dakota Howard, Yvette Unoarumhi, Darlene Wagner, Matthew Schmerer, Ben L. Rambo-Martin, Kristine Lacek, Sam Shepard, Alison Laufer Halpin, Dave Wentworth, Vivien Dugan, Suxiang Tong, Justin Lee |
| EPI_ISL_1937632 | Fulgent Genetics | Fulgent Genetics | Harry Gao, Mickey Li, John Gao, Joseph Fierro, Benafsh Sapra, Becky Tsai, Yan Meng, Doreen Ng, James Xie |
| EPI_ISL_1937633 | NJ Public Health and Environmental Laboratories | Centers for Disease Control and Prevention Division of Viral Diseases, Pathogen Discovery | Mili Sheth, Sarah Nobles, Jasmine Padilla, Mark Burroughs, Shoshona Le, Katie Dillon, Peter Cook, Clinton R. Paden, Dhwani Batra, Krista Queen, Kristen Knipe, Dakota Howard, Yvette Unoarumhi, Darlene Wagner, Matthew Schmerer, Ben L. Rambo-Martin, Kristine Lacek, Sam Shepard, Alison Laufer Halpin, Dave Wentworth, Vivien Dugan, Suxiang Tong, Justin Lee |
| EPI_ISL_1937634 | Fulgent Genetics | Fulgent Genetics | Harry Gao, Mickey Li, John Gao, Joseph Fierro, Benafsh Sapra, Becky Tsai, Yan Meng, Doreen Ng, James Xie |
| EPI_ISL_1937635, EPI_ISL_1937636 | NJ Public Health and Environmental Laboratories | Centers for Disease Control and Prevention Division of Viral Diseases, Pathogen Discovery | Mili Sheth, Sarah Nobles, Jasmine Padilla, Mark Burroughs, Shoshona Le, Katie Dillon, Peter Cook, Clinton R. Paden, Dhwani Batra, Krista Queen, Kristen Knipe, Dakota Howard, Yvette Unoarumhi, Darlene Wagner, Matthew Schmerer, Ben L. Rambo-Martin, Kristine Lacek, Sam Shepard, Alison Laufer Halpin, Dave Wentworth, Vivien Dugan, Suxiang Tong, Justin Lee |
| EPI_ISL_1937637 | Fulgent Genetics | Fulgent Genetics | Harry Gao, Mickey Li, John Gao, Joseph Fierro, Benafsh Sapra, Becky Tsai, Yan Meng, Doreen Ng, James Xie |
| EPI_ISL_1937638 | NJ Public Health and Environmental Laboratories | Centers for Disease Control and Prevention Division of Viral Diseases, Pathogen Discovery | Mili Sheth, Sarah Nobles, Jasmine Padilla, Mark Burroughs, Shoshona Le, Katie Dillon, Peter Cook, Clinton R. Paden, Dhwani Batra, Krista Queen, Kristen Knipe, Dakota Howard, Yvette Unoarumhi, Darlene Wagner, Matthew Schmerer, Ben L. Rambo-Martin, Kristine Lacek, Sam Shepard, Alison Laufer Halpin, Dave Wentworth, Vivien Dugan, Suxiang Tong, Justin Lee |
| EPI_ISL_1937639 | Fulgent Genetics | Fulgent Genetics | Harry Gao, Mickey Li, John Gao, Joseph Fierro, Benafsh Sapra, Becky Tsai, Yan Meng, Doreen Ng, James Xie |
| EPI_ISL_1937640, EPI_ISL_1937641 | NJ Public Health and Environmental Laboratories | Centers for Disease Control and Prevention Division of Viral Diseases, Pathogen Discovery | Mili Sheth, Sarah Nobles, Jasmine Padilla, Mark Burroughs, Shoshona Le, Katie Dillon, Peter Cook, Clinton R. Paden, Dhwani Batra, Krista Queen, Kristen Knipe, Dakota Howard, Yvette Unoarumhi, Darlene Wagner, Matthew Schmerer, Ben L. Rambo-Martin, Kristine Lacek, Sam Shepard, Alison Laufer Halpin, Dave Wentworth, Vivien Dugan, Suxiang Tong, Justin Lee |
| EPI_ISL_1937642 | Fulgent Genetics | Fulgent Genetics | Harry Gao, Mickey Li, John Gao, Joseph Fierro, Benafsh Sapra, Becky Tsai, Yan Meng, Doreen Ng, James Xie |
| EPI_ISL_1937643 | NJ Public Health and Environmental Laboratories | Centers for Disease Control and Prevention Division of Viral Diseases, Pathogen Discovery | Mili Sheth, Sarah Nobles, Jasmine Padilla, Mark Burroughs, Shoshona Le, Katie Dillon, Peter Cook, Clinton R. Paden, Dhwani Batra, Krista Queen, Kristen Knipe, Dakota Howard, Yvette Unoarumhi, Darlene Wagner, Matthew Schmerer, Ben L. Rambo-Martin, Kristine Lacek, Sam Shepard, Alison Laufer Halpin, Dave Wentworth, Vivien Dugan, Suxiang Tong, Justin Lee |
| EPI_ISL_1937644 | Fulgent Genetics | Fulgent Genetics | Harry Gao, Mickey Li, John Gao, Joseph Fierro, Benafsh Sapra, Becky Tsai, Yan Meng, Doreen Ng, James Xie |
| EPI_ISL_1937645 | GA Department of Public Health Laboratory | Centers for Disease Control and Prevention Division of Viral Diseases, Pathogen Discovery | Mili Sheth, Sarah Nobles, Jasmine Padilla, Mark Burroughs, Shoshona Le, Katie Dillon, Peter Cook, Clinton R. Paden, Dhwani Batra, Krista Queen, Kristen Knipe, Dakota Howard, Yvette Unoarumhi, Darlene Wagner, Matthew Schmerer, Ben L. Rambo-Martin, Kristine Lacek, Sam Shepard, Alison Laufer Halpin, Dave Wentworth, Vivien Dugan, Suxiang Tong, Justin Lee |
| EPI_ISL_1937646 | Fulgent Genetics | Fulgent Genetics | Harry Gao, Mickey Li, John Gao, Joseph Fierro, Benafsh Sapra, Becky Tsai, Yan Meng, Doreen Ng, James Xie |
| EPI_ISL_1937647, EPI_ISL_1937648, EPI_ISL_1937650 | GA Department of Public Health Laboratory | Centers for Disease Control and Prevention Division of Viral Diseases, Pathogen Discovery | Mili Sheth, Sarah Nobles, Jasmine Padilla, Mark Burroughs, Shoshona Le, Katie Dillon, Peter Cook, Clinton R. Paden, Dhwani Batra, Krista Queen, Kristen Knipe, Dakota Howard, Yvette Unoarumhi, Darlene Wagner, Matthew Schmerer, Ben L. Rambo-Martin, Kristine Lacek, Sam Shepard, Alison Laufer Halpin, Dave Wentworth, Vivien Dugan, Suxiang Tong, Justin Lee |
| EPI_ISL_1937651 | Fulgent Genetics | Fulgent Genetics | Harry Gao, Mickey Li, John Gao, Joseph Fierro, Benafsh Sapra, Becky Tsai, Yan Meng, Doreen Ng, James Xie |
| EPI_ISL_1937652 | GA Department of Public Health Laboratory | Centers for Disease Control and Prevention Division of Viral Diseases, Pathogen Discovery | Mili Sheth, Sarah Nobles, Jasmine Padilla, Mark Burroughs, Shoshona Le, Katie Dillon, Peter Cook, Clinton R. Paden, Dhwani Batra, Krista Queen, Kristen Knipe, Dakota Howard, Yvette Unoarumhi, Darlene Wagner, Matthew Schmerer, Ben L. Rambo-Martin, Kristine Lacek, Sam Shepard, Alison Laufer Halpin, Dave Wentworth, Vivien Dugan, Suxiang Tong, Justin Lee |
| EPI_ISL_1937653 | Fulgent Genetics | Fulgent Genetics | Harry Gao, Mickey Li, John Gao, Joseph Fierro, Benafsh Sapra, Becky Tsai, Yan Meng, Doreen Ng, James Xie |
| EPI_ISL_1937654, EPI_ISL_1937655 | GA Department of Public Health Laboratory | Centers for Disease Control and Prevention Division of Viral Diseases, Pathogen Discovery | Mili Sheth, Sarah Nobles, Jasmine Padilla, Mark Burroughs, Shoshona Le, Katie Dillon, Peter Cook, Clinton R. Paden, Dhwani Batra, Krista Queen, Kristen Knipe, Dakota Howard, Yvette Unoarumhi, Darlene Wagner, Matthew Schmerer, Ben L. Rambo-Martin, Kristine Lacek, Sam Shepard, Alison Laufer Halpin, Dave Wentworth, Vivien Dugan, Suxiang Tong, Justin Lee |
| EPI_ISL_1937656 | Fulgent Genetics | Fulgent Genetics | Harry Gao, Mickey Li, John Gao, Joseph Fierro, Benafsh Sapra, Becky Tsai, Yan Meng, Doreen Ng, James Xie |
| EPI_ISL_1937657 | GA Department of Public Health Laboratory | Centers for Disease Control and Prevention Division of Viral Diseases, Pathogen Discovery | Mili Sheth, Sarah Nobles, Jasmine Padilla, Mark Burroughs, Shoshona Le, Katie Dillon, Peter Cook, Clinton R. Paden, Dhwani Batra, Krista Queen, Kristen Knipe, Dakota Howard, Yvette Unoarumhi, Darlene Wagner, Matthew Schmerer, Ben L. Rambo-Martin, Kristine Lacek, Sam Shepard, Alison Laufer Halpin, Dave Wentworth, Vivien Dugan, Suxiang Tong, Justin Lee |
| EPI_ISL_1937658 | Fulgent Genetics | Fulgent Genetics | Harry Gao, Mickey Li, John Gao, Joseph Fierro, Benafsh Sapra, Becky Tsai, Yan Meng, Doreen Ng, James Xie |
| EPI_ISL_1937659 | GA Department of Public Health Laboratory | Centers for Disease Control and Prevention Division of Viral Diseases, Pathogen | Mili Sheth, Sarah Nobles, Jasmine Padilla, Mark Burroughs, Shoshona Le, Katie Dillon, Peter Cook, Clinton R. Paden, Dhwani Batra, Krista Queen, Kristen Knipe, Dakota Howard, Yvette Unoarumhi, Darlene Wagner, Matthew Schmerer, Ben L. Rambo-Martin, Kristine Lacek, Sam Shepard, Alison Laufer Halpin, Dave Wentworth, Vivien Dugan, Suxiang Tong, Justin Lee |

| | | | |
|---|---|---|---|
| | | Discovery | |
| EPI_ISL_1937660 | Fulgent Genetics | Fulgent Genetics | Harry Gao, Mickey Li, John Gao, Joseph Fierro, Benafsh Sapra, Becky Tsai, Yan Meng, Doreen Ng, James Xie |
| EPI_ISL_1937661, EPI_ISL_1937662 | GA Department of Public Health Laboratory | Centers for Disease Control and Prevention Division of Viral Diseases, Pathogen Discovery | Mili Sheth, Sarah Nobles, Jasmine Padilla, Mark Burroughs, Shoshona Le, Katie Dillon, Peter Cook, Clinton R. Paden, Dhwani Batra, Krista Queen, Kristen Knipe, Dakota Howard, Yvette Unoarumhi, Darlene Wagner, Matthew Schmerer, Ben L. Rambo-Martin, Kristine Lacek, Sam Shepard, Alison Laufer Halpin, Dave Wentworth, Vivien Dugan, Suxiang Tong, Justin Lee |
| EPI_ISL_1937663 | Fulgent Genetics | Fulgent Genetics | Harry Gao, Mickey Li, John Gao, Joseph Fierro, Benafsh Sapra, Becky Tsai, Yan Meng, Doreen Ng, James Xie |
| EPI_ISL_1937664 | GA Department of Public Health Laboratory | Centers for Disease Control and Prevention Division of Viral Diseases, Pathogen Discovery | Mili Sheth, Sarah Nobles, Jasmine Padilla, Mark Burroughs, Shoshona Le, Katie Dillon, Peter Cook, Clinton R. Paden, Dhwani Batra, Krista Queen, Kristen Knipe, Dakota Howard, Yvette Unoarumhi, Darlene Wagner, Matthew Schmerer, Ben L. Rambo-Martin, Kristine Lacek, Sam Shepard, Alison Laufer Halpin, Dave Wentworth, Vivien Dugan, Suxiang Tong, Justin Lee |
| EPI_ISL_1937665 | Fulgent Genetics | Fulgent Genetics | Harry Gao, Mickey Li, John Gao, Joseph Fierro, Benafsh Sapra, Becky Tsai, Yan Meng, Doreen Ng, James Xie |
| EPI_ISL_1937666, EPI_ISL_1937667 | GA Department of Public Health Laboratory | Centers for Disease Control and Prevention Division of Viral Diseases, Pathogen Discovery | Mili Sheth, Sarah Nobles, Jasmine Padilla, Mark Burroughs, Shoshona Le, Katie Dillon, Peter Cook, Clinton R. Paden, Dhwani Batra, Krista Queen, Kristen Knipe, Dakota Howard, Yvette Unoarumhi, Darlene Wagner, Matthew Schmerer, Ben L. Rambo-Martin, Kristine Lacek, Sam Shepard, Alison Laufer Halpin, Dave Wentworth, Vivien Dugan, Suxiang Tong, Justin Lee |
| EPI_ISL_1937668 | Fulgent Genetics | Fulgent Genetics | Harry Gao, Mickey Li, John Gao, Joseph Fierro, Benafsh Sapra, Becky Tsai, Yan Meng, Doreen Ng, James Xie |
| EPI_ISL_1937669 | GA Department of Public Health Laboratory | Centers for Disease Control and Prevention Division of Viral Diseases, Pathogen Discovery | Mili Sheth, Sarah Nobles, Jasmine Padilla, Mark Burroughs, Shoshona Le, Katie Dillon, Peter Cook, Clinton R. Paden, Dhwani Batra, Krista Queen, Kristen Knipe, Dakota Howard, Yvette Unoarumhi, Darlene Wagner, Matthew Schmerer, Ben L. Rambo-Martin, Kristine Lacek, Sam Shepard, Alison Laufer Halpin, Dave Wentworth, Vivien Dugan, Suxiang Tong, Justin Lee |
| EPI_ISL_1937670 | Fulgent Genetics | Fulgent Genetics | Harry Gao, Mickey Li, John Gao, Joseph Fierro, Benafsh Sapra, Becky Tsai, Yan Meng, Doreen Ng, James Xie |
| EPI_ISL_1937671 | GA Department of Public Health Laboratory | Centers for Disease Control and Prevention Division of Viral Diseases, Pathogen Discovery | Mili Sheth, Sarah Nobles, Jasmine Padilla, Mark Burroughs, Shoshona Le, Katie Dillon, Peter Cook, Clinton R. Paden, Dhwani Batra, Krista Queen, Kristen Knipe, Dakota Howard, Yvette Unoarumhi, Darlene Wagner, Matthew Schmerer, Ben L. Rambo-Martin, Kristine Lacek, Sam Shepard, Alison Laufer Halpin, Dave Wentworth, Vivien Dugan, Suxiang Tong, Justin Lee |
| EPI_ISL_1937672 | Fulgent Genetics | Fulgent Genetics | Harry Gao, Mickey Li, John Gao, Joseph Fierro, Benafsh Sapra, Becky Tsai, Yan Meng, Doreen Ng, James Xie |
| EPI_ISL_1937673, EPI_ISL_1937674 | GA Department of Public Health Laboratory | Centers for Disease Control and Prevention Division of Viral Diseases, Pathogen Discovery | Mili Sheth, Sarah Nobles, Jasmine Padilla, Mark Burroughs, Shoshona Le, Katie Dillon, Peter Cook, Clinton R. Paden, Dhwani Batra, Krista Queen, Kristen Knipe, Dakota Howard, Yvette Unoarumhi, Darlene Wagner, Matthew Schmerer, Ben L. Rambo-Martin, Kristine Lacek, Sam Shepard, Alison Laufer Halpin, Dave Wentworth, Vivien Dugan, Suxiang Tong, Justin Lee |
| EPI_ISL_1937675 | Fulgent Genetics | Fulgent Genetics | Harry Gao, Mickey Li, John Gao, Joseph Fierro, Benafsh Sapra, Becky Tsai, Yan Meng, Doreen Ng, James Xie |
| EPI_ISL_1937676 | GA Department of Public Health Laboratory | Centers for Disease Control and Prevention Division of Viral Diseases, Pathogen Discovery | Mili Sheth, Sarah Nobles, Jasmine Padilla, Mark Burroughs, Shoshona Le, Katie Dillon, Peter Cook, Clinton R. Paden, Dhwani Batra, Krista Queen, Kristen Knipe, Dakota Howard, Yvette Unoarumhi, Darlene Wagner, Matthew Schmerer, Ben L. Rambo-Martin, Kristine Lacek, Sam Shepard, Alison Laufer Halpin, Dave Wentworth, Vivien Dugan, Suxiang Tong, Justin Lee |
| EPI_ISL_1937677 | Fulgent Genetics | Fulgent Genetics | Harry Gao, Mickey Li, John Gao, Joseph Fierro, Benafsh Sapra, Becky Tsai, Yan Meng, Doreen Ng, James Xie |
| EPI_ISL_1937678, EPI_ISL_1937679 | MI - Michigan Department of Health and Human Services - Bureau of Laboratories | Centers for Disease Control and Prevention Division of Viral Diseases, Pathogen Discovery | Mili Sheth, Sarah Nobles, Jasmine Padilla, Mark Burroughs, Shoshona Le, Katie Dillon, Peter Cook, Clinton R. Paden, Dhwani Batra, Krista Queen, Kristen Knipe, Dakota Howard, Yvette Unoarumhi, Darlene Wagner, Matthew Schmerer, Ben L. Rambo-Martin, Kristine Lacek, Sam Shepard, Alison Laufer Halpin, Dave Wentworth, Vivien Dugan, Suxiang Tong, Justin Lee |
| EPI_ISL_1937680 | Fulgent Genetics | Fulgent Genetics | Harry Gao, Mickey Li, John Gao, Joseph Fierro, Benafsh Sapra, Becky Tsai, Yan Meng, Doreen Ng, James Xie |
| EPI_ISL_1937681, EPI_ISL_1937682 | MI - Michigan Department of Health and Human Services - Bureau of Laboratories | Centers for Disease Control and Prevention Division of Viral Diseases, Pathogen Discovery | Mili Sheth, Sarah Nobles, Jasmine Padilla, Mark Burroughs, Shoshona Le, Katie Dillon, Peter Cook, Clinton R. Paden, Dhwani Batra, Krista Queen, Kristen Knipe, Dakota Howard, Yvette Unoarumhi, Darlene Wagner, Matthew Schmerer, Ben L. Rambo-Martin, Kristine Lacek, Sam Shepard, Alison Laufer Halpin, Dave Wentworth, Vivien Dugan, Suxiang Tong, Justin Lee |
| EPI_ISL_1937683 | Fulgent Genetics | Fulgent Genetics | Harry Gao, Mickey Li, John Gao, Joseph Fierro, Benafsh Sapra, Becky Tsai, Yan Meng, Doreen Ng, James Xie |
| EPI_ISL_1937684, EPI_ISL_1937685 | MI - Michigan Department of Health and Human Services - Bureau of Laboratories | Centers for Disease Control and Prevention Division of Viral Diseases, Pathogen Discovery | Mili Sheth, Sarah Nobles, Jasmine Padilla, Mark Burroughs, Shoshona Le, Katie Dillon, Peter Cook, Clinton R. Paden, Dhwani Batra, Krista Queen, Kristen Knipe, Dakota Howard, Yvette Unoarumhi, Darlene Wagner, Matthew Schmerer, Ben L. Rambo-Martin, Kristine Lacek, Sam Shepard, Alison Laufer Halpin, Dave Wentworth, Vivien Dugan, Suxiang Tong, Justin Lee |
| EPI_ISL_1937686 | Fulgent Genetics | Fulgent Genetics | Harry Gao, Mickey Li, John Gao, Joseph Fierro, Benafsh Sapra, Becky Tsai, Yan Meng, Doreen Ng, James Xie |
| EPI_ISL_1937687 | MI - Michigan Department of Health and Human Services - Bureau of Laboratories | Centers for Disease Control and Prevention Division of Viral Diseases, Pathogen Discovery | Mili Sheth, Sarah Nobles, Jasmine Padilla, Mark Burroughs, Shoshona Le, Katie Dillon, Peter Cook, Clinton R. Paden, Dhwani Batra, Krista Queen, Kristen Knipe, Dakota Howard, Yvette Unoarumhi, Darlene Wagner, Matthew Schmerer, Ben L. Rambo-Martin, Kristine Lacek, Sam Shepard, Alison Laufer Halpin, Dave Wentworth, Vivien Dugan, Suxiang Tong, Justin Lee |
| EPI_ISL_1937688 | Fulgent Genetics | Fulgent Genetics | Harry Gao, Mickey Li, John Gao, Joseph Fierro, Benafsh Sapra, Becky Tsai, Yan Meng, Doreen Ng, James Xie |
| EPI_ISL_1937689, EPI_ISL_1937690 | MI - Michigan Department of Health and Human Services - Bureau of Laboratories | Centers for Disease Control and Prevention Division of Viral Diseases, Pathogen Discovery | Mili Sheth, Sarah Nobles, Jasmine Padilla, Mark Burroughs, Shoshona Le, Katie Dillon, Peter Cook, Clinton R. Paden, Dhwani Batra, Krista Queen, Kristen Knipe, Dakota Howard, Yvette Unoarumhi, Darlene Wagner, Matthew Schmerer, Ben L. Rambo-Martin, Kristine Lacek, Sam Shepard, Alison Laufer Halpin, Dave Wentworth, Vivien Dugan, Suxiang Tong, Justin Lee |
| EPI_ISL_1937691 | Fulgent Genetics | Fulgent Genetics | Harry Gao, Mickey Li, John Gao, Joseph Fierro, Benafsh Sapra, Becky Tsai, Yan Meng, Doreen Ng, James Xie |
| EPI_ISL_1937692 | MI - Michigan Department of Health and Human Services - Bureau of Laboratories | Centers for Disease Control and Prevention Division of Viral Diseases, Pathogen Discovery | Mili Sheth, Sarah Nobles, Jasmine Padilla, Mark Burroughs, Shoshona Le, Katie Dillon, Peter Cook, Clinton R. Paden, Dhwani Batra, Krista Queen, Kristen Knipe, Dakota Howard, Yvette Unoarumhi, Darlene Wagner, Matthew Schmerer, Ben L. Rambo-Martin, Kristine Lacek, Sam Shepard, Alison Laufer Halpin, Dave Wentworth, Vivien Dugan, Suxiang Tong, Justin Lee |
| EPI_ISL_1937693 | Fulgent Genetics | Fulgent Genetics | Harry Gao, Mickey Li, John Gao, Joseph Fierro, Benafsh Sapra, Becky Tsai, Yan Meng, Doreen Ng, James Xie |
| EPI_ISL_1937694 | WY Public Health Laboratory | Centers for Disease Control and Prevention Division of Viral Diseases, Pathogen Discovery | Mili Sheth, Sarah Nobles, Jasmine Padilla, Mark Burroughs, Shoshona Le, Katie Dillon, Peter Cook, Clinton R. Paden, Dhwani Batra, Krista Queen, Kristen Knipe, Dakota Howard, Yvette Unoarumhi, Darlene Wagner, Matthew Schmerer, Ben L. Rambo-Martin, Kristine Lacek, Sam Shepard, Alison Laufer Halpin, Dave Wentworth, Vivien Dugan, Suxiang Tong, Justin Lee |
| EPI_ISL_1937695 | Fulgent Genetics | Fulgent Genetics | Harry Gao, Mickey Li, John Gao, Joseph Fierro, Benafsh Sapra, Becky Tsai, Yan Meng, Doreen Ng, James Xie |
| EPI_ISL_1937696, EPI_ISL_1937697 | WY Public Health Laboratory | Centers for Disease Control and Prevention Division of Viral Diseases, Pathogen Discovery | Mili Sheth, Sarah Nobles, Jasmine Padilla, Mark Burroughs, Shoshona Le, Katie Dillon, Peter Cook, Clinton R. Paden, Dhwani Batra, Krista Queen, Kristen Knipe, Dakota Howard, Yvette Unoarumhi, Darlene Wagner, Matthew Schmerer, Ben L. Rambo-Martin, Kristine Lacek, Sam Shepard, Alison Laufer Halpin, Dave Wentworth, Vivien Dugan, Suxiang Tong, Justin Lee |
| EPI_ISL_1937698 | Fulgent Genetics | Fulgent Genetics | Harry Gao, Mickey Li, John Gao, Joseph Fierro, Benafsh Sapra, Becky Tsai, Yan Meng, Doreen Ng, James Xie |
| EPI_ISL_1937699 | WY Public Health Laboratory | Centers for Disease Control and Prevention Division of Viral Diseases, Pathogen Discovery | Mili Sheth, Sarah Nobles, Jasmine Padilla, Mark Burroughs, Shoshona Le, Katie Dillon, Peter Cook, Clinton R. Paden, Dhwani Batra, Krista Queen, Kristen Knipe, Dakota Howard, Yvette Unoarumhi, Darlene Wagner, Matthew Schmerer, Ben L. Rambo-Martin, Kristine Lacek, Sam Shepard, Alison Laufer Halpin, Dave Wentworth, Vivien Dugan, Suxiang Tong, Justin Lee |
| EPI_ISL_1937700 | Fulgent Genetics | Fulgent Genetics | Harry Gao, Mickey Li, John Gao, Joseph Fierro, Benafsh Sapra, Becky Tsai, Yan Meng, Doreen Ng, James Xie |
| EPI_ISL_1937701 | WY Public Health Laboratory | Centers for Disease Control and Prevention Division of Viral Diseases, Pathogen Discovery | Mili Sheth, Sarah Nobles, Jasmine Padilla, Mark Burroughs, Shoshona Le, Katie Dillon, Peter Cook, Clinton R. Paden, Dhwani Batra, Krista Queen, Kristen Knipe, Dakota Howard, Yvette Unoarumhi, Darlene Wagner, Matthew Schmerer, Ben L. Rambo-Martin, Kristine Lacek, Sam Shepard, Alison Laufer Halpin, Dave Wentworth, Vivien Dugan, Suxiang Tong, Justin Lee |
| EPI_ISL_1937702 | Fulgent Genetics | Fulgent Genetics | Harry Gao, Mickey Li, John Gao, Joseph Fierro, Benafsh Sapra, Becky Tsai, Yan Meng, Doreen Ng, James Xie |
| EPI_ISL_1937703 | WY Public Health Laboratory | Centers for Disease Control and Prevention Division of Viral Diseases, Pathogen Discovery | Mili Sheth, Sarah Nobles, Jasmine Padilla, Mark Burroughs, Shoshona Le, Katie Dillon, Peter Cook, Clinton R. Paden, Dhwani Batra, Krista Queen, Kristen Knipe, Dakota Howard, Yvette Unoarumhi, Darlene Wagner, Matthew Schmerer, Ben L. Rambo-Martin, Kristine Lacek, Sam Shepard, Alison Laufer Halpin, Dave Wentworth, Vivien Dugan, Suxiang Tong, Justin Lee |
| EPI_ISL_1937704 | MD DOH Laboratories Administration | Centers for Disease Control and Prevention Division of Viral Diseases, Pathogen Discovery | Mili Sheth, Sarah Nobles, Jasmine Padilla, Mark Burroughs, Shoshona Le, Katie Dillon, Peter Cook, Clinton R. Paden, Dhwani Batra, Krista Queen, Kristen Knipe, Dakota Howard, Yvette Unoarumhi, Darlene Wagner, Matthew Schmerer, Ben L. Rambo-Martin, Kristine Lacek, Sam Shepard, Alison Laufer Halpin, Dave Wentworth, Vivien Dugan, Suxiang Tong, Justin Lee |
| EPI_ISL_1937705 | Fulgent Genetics | Fulgent Genetics | Harry Gao, Mickey Li, John Gao, Joseph Fierro, Benafsh Sapra, Becky Tsai, Yan Meng, Doreen Ng, James Xie |
| EPI_ISL_1937706, EPI_ISL_1937708, EPI_ISL_1937709 | MD DOH Laboratories Administration | Centers for Disease Control and Prevention Division of Viral Diseases, Pathogen Discovery | Mili Sheth, Sarah Nobles, Jasmine Padilla, Mark Burroughs, Shoshona Le, Katie Dillon, Peter Cook, Clinton R. Paden, Dhwani Batra, Krista Queen, Kristen Knipe, Dakota Howard, Yvette Unoarumhi, Darlene Wagner, Matthew Schmerer, Ben L. Rambo-Martin, Kristine Lacek, Sam Shepard, Alison Laufer Halpin, Dave Wentworth, Vivien Dugan, Suxiang Tong, Justin Lee |
| EPI_ISL_1937710 | Fulgent Genetics | Fulgent Genetics | Harry Gao, Mickey Li, John Gao, Joseph Fierro, Benafsh Sapra, Becky Tsai, Yan Meng, Doreen Ng, James Xie |
| EPI_ISL_1937711 | MD DOH Laboratories Administration | Centers for Disease Control and Prevention Division of Viral Diseases, Pathogen Discovery | Mili Sheth, Sarah Nobles, Jasmine Padilla, Mark Burroughs, Shoshona Le, Katie Dillon, Peter Cook, Clinton R. Paden, Dhwani Batra, Krista Queen, Kristen Knipe, Dakota Howard, Yvette Unoarumhi, Darlene Wagner, Matthew Schmerer, Ben L. Rambo-Martin, Kristine Lacek, Sam Shepard, Alison Laufer Halpin, Dave Wentworth, Vivien Dugan, Suxiang Tong, Justin Lee |
| EPI_ISL_1937712 | Fulgent Genetics | Fulgent Genetics | Harry Gao, Mickey Li, John Gao, Joseph Fierro, Benafsh Sapra, Becky Tsai, Yan Meng, Doreen Ng, James Xie |
| EPI_ISL_1937713, EPI_ISL_1937714 | MD DOH Laboratories Administration | Centers for Disease Control and Prevention Division of Viral Diseases, Pathogen Discovery | Mili Sheth, Sarah Nobles, Jasmine Padilla, Mark Burroughs, Shoshona Le, Katie Dillon, Peter Cook, Clinton R. Paden, Dhwani Batra, Krista Queen, Kristen Knipe, Dakota Howard, Yvette Unoarumhi, Darlene Wagner, Matthew Schmerer, Ben L. Rambo-Martin, Kristine Lacek, Sam Shepard, Alison Laufer Halpin, Dave Wentworth, Vivien Dugan, Suxiang Tong, Justin Lee |
| EPI_ISL_1937715 | Fulgent Genetics | Fulgent Genetics | Harry Gao, Mickey Li, John Gao, Joseph Fierro, Benafsh Sapra, Becky Tsai, Yan Meng, Doreen Ng, James Xie |
| EPI_ISL_1937716 | MD DOH Laboratories Administration | Centers for Disease Control and Prevention | Mili Sheth, Sarah Nobles, Jasmine Padilla, Mark Burroughs, Shoshona Le, Katie Dillon, Peter Cook, Clinton R. Paden, Dhwani Batra, Krista Queen, Kristen Knipe, Dakota Howard, Yvette Unoarumhi, Darlene Wagner, Matthew Schmerer, Ben L. Rambo-Martin, Kristine Lacek, Sam Shepard, |

| | | | |
|---|---|---|---|
| | | Division of Viral Diseases, Pathogen Discovery | Alison Laufer Halpin, Dave Wentworth, Vivien Dugan, Suxiang Tong, Justin Lee |
| EPI_ISL_1937717 | Fulgent Genetics | Fulgent Genetics | Harry Gao, Mickey Li, John Gao, Joseph Fierro, Benafsh Sapra, Becky Tsai, Yan Meng, Doreen Ng, James Xie |
| EPI_ISL_1937718, EPI_ISL_1937719 | MD DOH Laboratories Administration | Centers for Disease Control and Prevention Division of Viral Diseases, Pathogen Discovery | Mili Sheth, Sarah Nobles, Jasmine Padilla, Mark Burroughs, Shoshona Le, Katie Dillon, Peter Cook, Clinton R. Paden, Dhwani Batra, Krista Queen, Kristen Knipe, Dakota Howard, Yvette Unoarumhi, Darlene Wagner, Matthew Schmerer, Ben L. Rambo-Martin, Kristine Lacek, Sam Shepard, Alison Laufer Halpin, Dave Wentworth, Vivien Dugan, Suxiang Tong, Justin Lee |
| EPI_ISL_1937720 | Fulgent Genetics | Fulgent Genetics | Harry Gao, Mickey Li, John Gao, Joseph Fierro, Benafsh Sapra, Becky Tsai, Yan Meng, Doreen Ng, James Xie |
| EPI_ISL_1937721 | MD DOH Laboratories Administration | Centers for Disease Control and Prevention Division of Viral Diseases, Pathogen Discovery | Mili Sheth, Sarah Nobles, Jasmine Padilla, Mark Burroughs, Shoshona Le, Katie Dillon, Peter Cook, Clinton R. Paden, Dhwani Batra, Krista Queen, Kristen Knipe, Dakota Howard, Yvette Unoarumhi, Darlene Wagner, Matthew Schmerer, Ben L. Rambo-Martin, Kristine Lacek, Sam Shepard, Alison Laufer Halpin, Dave Wentworth, Vivien Dugan, Suxiang Tong, Justin Lee |
| EPI_ISL_1937722 | Fulgent Genetics | Fulgent Genetics | Harry Gao, Mickey Li, John Gao, Joseph Fierro, Benafsh Sapra, Becky Tsai, Yan Meng, Doreen Ng, James Xie |
| EPI_ISL_1937723, EPI_ISL_1937724 | MD DOH Laboratories Administration | Centers for Disease Control and Prevention Division of Viral Diseases, Pathogen Discovery | Mili Sheth, Sarah Nobles, Jasmine Padilla, Mark Burroughs, Shoshona Le, Katie Dillon, Peter Cook, Clinton R. Paden, Dhwani Batra, Krista Queen, Kristen Knipe, Dakota Howard, Yvette Unoarumhi, Darlene Wagner, Matthew Schmerer, Ben L. Rambo-Martin, Kristine Lacek, Sam Shepard, Alison Laufer Halpin, Dave Wentworth, Vivien Dugan, Suxiang Tong, Justin Lee |
| EPI_ISL_1937725 | Fulgent Genetics | Fulgent Genetics | Harry Gao, Mickey Li, John Gao, Joseph Fierro, Benafsh Sapra, Becky Tsai, Yan Meng, Doreen Ng, James Xie |
| EPI_ISL_1937726 | MD DOH Laboratories Administration | Centers for Disease Control and Prevention Division of Viral Diseases, Pathogen Discovery | Mili Sheth, Sarah Nobles, Jasmine Padilla, Mark Burroughs, Shoshona Le, Katie Dillon, Peter Cook, Clinton R. Paden, Dhwani Batra, Krista Queen, Kristen Knipe, Dakota Howard, Yvette Unoarumhi, Darlene Wagner, Matthew Schmerer, Ben L. Rambo-Martin, Kristine Lacek, Sam Shepard, Alison Laufer Halpin, Dave Wentworth, Vivien Dugan, Suxiang Tong, Justin Lee |
| EPI_ISL_1937727 | Fulgent Genetics | Fulgent Genetics | Harry Gao, Mickey Li, John Gao, Joseph Fierro, Benafsh Sapra, Becky Tsai, Yan Meng, Doreen Ng, James Xie |
| EPI_ISL_1937728 | MD DOH Laboratories Administration | Centers for Disease Control and Prevention Division of Viral Diseases, Pathogen Discovery | Mili Sheth, Sarah Nobles, Jasmine Padilla, Mark Burroughs, Shoshona Le, Katie Dillon, Peter Cook, Clinton R. Paden, Dhwani Batra, Krista Queen, Kristen Knipe, Dakota Howard, Yvette Unoarumhi, Darlene Wagner, Matthew Schmerer, Ben L. Rambo-Martin, Kristine Lacek, Sam Shepard, Alison Laufer Halpin, Dave Wentworth, Vivien Dugan, Suxiang Tong, Justin Lee |
| EPI_ISL_1937729 | Fulgent Genetics | Fulgent Genetics | Harry Gao, Mickey Li, John Gao, Joseph Fierro, Benafsh Sapra, Becky Tsai, Yan Meng, Doreen Ng, James Xie |
| EPI_ISL_1937730, EPI_ISL_1937731 | MD DOH Laboratories Administration | Centers for Disease Control and Prevention Division of Viral Diseases, Pathogen Discovery | Mili Sheth, Sarah Nobles, Jasmine Padilla, Mark Burroughs, Shoshona Le, Katie Dillon, Peter Cook, Clinton R. Paden, Dhwani Batra, Krista Queen, Kristen Knipe, Dakota Howard, Yvette Unoarumhi, Darlene Wagner, Matthew Schmerer, Ben L. Rambo-Martin, Kristine Lacek, Sam Shepard, Alison Laufer Halpin, Dave Wentworth, Vivien Dugan, Suxiang Tong, Justin Lee |
| EPI_ISL_1937732 | Fulgent Genetics | Fulgent Genetics | Harry Gao, Mickey Li, John Gao, Joseph Fierro, Benafsh Sapra, Becky Tsai, Yan Meng, Doreen Ng, James Xie |
| EPI_ISL_1937733 | MD DOH Laboratories Administration | Centers for Disease Control and Prevention Division of Viral Diseases, Pathogen Discovery | Mili Sheth, Sarah Nobles, Jasmine Padilla, Mark Burroughs, Shoshona Le, Katie Dillon, Peter Cook, Clinton R. Paden, Dhwani Batra, Krista Queen, Kristen Knipe, Dakota Howard, Yvette Unoarumhi, Darlene Wagner, Matthew Schmerer, Ben L. Rambo-Martin, Kristine Lacek, Sam Shepard, Alison Laufer Halpin, Dave Wentworth, Vivien Dugan, Suxiang Tong, Justin Lee |
| EPI_ISL_1937734 | Fulgent Genetics | Fulgent Genetics | Harry Gao, Mickey Li, John Gao, Joseph Fierro, Benafsh Sapra, Becky Tsai, Yan Meng, Doreen Ng, James Xie |
| EPI_ISL_1937735, EPI_ISL_1937736 | IA State Hygienic Laboratory | Centers for Disease Control and Prevention Division of Viral Diseases, Pathogen Discovery | Mili Sheth, Sarah Nobles, Jasmine Padilla, Mark Burroughs, Shoshona Le, Katie Dillon, Peter Cook, Clinton R. Paden, Dhwani Batra, Krista Queen, Kristen Knipe, Dakota Howard, Yvette Unoarumhi, Darlene Wagner, Matthew Schmerer, Ben L. Rambo-Martin, Kristine Lacek, Sam Shepard, Alison Laufer Halpin, Dave Wentworth, Vivien Dugan, Suxiang Tong, Justin Lee |
| EPI_ISL_1937737 | Fulgent Genetics | Fulgent Genetics | Harry Gao, Mickey Li, John Gao, Joseph Fierro, Benafsh Sapra, Becky Tsai, Yan Meng, Doreen Ng, James Xie |
| EPI_ISL_1937738, EPI_ISL_1937739, EPI_ISL_1937740 | IA State Hygienic Laboratory | Centers for Disease Control and Prevention Division of Viral Diseases, Pathogen Discovery | Mili Sheth, Sarah Nobles, Jasmine Padilla, Mark Burroughs, Shoshona Le, Katie Dillon, Peter Cook, Clinton R. Paden, Dhwani Batra, Krista Queen, Kristen Knipe, Dakota Howard, Yvette Unoarumhi, Darlene Wagner, Matthew Schmerer, Ben L. Rambo-Martin, Kristine Lacek, Sam Shepard, Alison Laufer Halpin, Dave Wentworth, Vivien Dugan, Suxiang Tong, Justin Lee |
| EPI_ISL_1937741 | Fulgent Genetics | Fulgent Genetics | Harry Gao, Mickey Li, John Gao, Joseph Fierro, Benafsh Sapra, Becky Tsai, Yan Meng, Doreen Ng, James Xie |
| EPI_ISL_1937742 | IA State Hygienic Laboratory | Centers for Disease Control and Prevention Division of Viral Diseases, Pathogen Discovery | Mili Sheth, Sarah Nobles, Jasmine Padilla, Mark Burroughs, Shoshona Le, Katie Dillon, Peter Cook, Clinton R. Paden, Dhwani Batra, Krista Queen, Kristen Knipe, Dakota Howard, Yvette Unoarumhi, Darlene Wagner, Matthew Schmerer, Ben L. Rambo-Martin, Kristine Lacek, Sam Shepard, Alison Laufer Halpin, Dave Wentworth, Vivien Dugan, Suxiang Tong, Justin Lee |
| EPI_ISL_1937743 | Fulgent Genetics | Fulgent Genetics | Harry Gao, Mickey Li, John Gao, Joseph Fierro, Benafsh Sapra, Becky Tsai, Yan Meng, Doreen Ng, James Xie |
| EPI_ISL_1937744 | IA State Hygienic Laboratory | Centers for Disease Control and Prevention Division of Viral Diseases, Pathogen Discovery | Mili Sheth, Sarah Nobles, Jasmine Padilla, Mark Burroughs, Shoshona Le, Katie Dillon, Peter Cook, Clinton R. Paden, Dhwani Batra, Krista Queen, Kristen Knipe, Dakota Howard, Yvette Unoarumhi, Darlene Wagner, Matthew Schmerer, Ben L. Rambo-Martin, Kristine Lacek, Sam Shepard, Alison Laufer Halpin, Dave Wentworth, Vivien Dugan, Suxiang Tong, Justin Lee |
| EPI_ISL_1937745 | Fulgent Genetics | Fulgent Genetics | Harry Gao, Mickey Li, John Gao, Joseph Fierro, Benafsh Sapra, Becky Tsai, Yan Meng, Doreen Ng, James Xie |
| EPI_ISL_1937746, EPI_ISL_1937747, EPI_ISL_1937749 | IA State Hygienic Laboratory | Centers for Disease Control and Prevention Division of Viral Diseases, Pathogen Discovery | Mili Sheth, Sarah Nobles, Jasmine Padilla, Mark Burroughs, Shoshona Le, Katie Dillon, Peter Cook, Clinton R. Paden, Dhwani Batra, Krista Queen, Kristen Knipe, Dakota Howard, Yvette Unoarumhi, Darlene Wagner, Matthew Schmerer, Ben L. Rambo-Martin, Kristine Lacek, Sam Shepard, Alison Laufer Halpin, Dave Wentworth, Vivien Dugan, Suxiang Tong, Justin Lee |
| EPI_ISL_1937750 | Fulgent Genetics | Fulgent Genetics | Harry Gao, Mickey Li, John Gao, Joseph Fierro, Benafsh Sapra, Becky Tsai, Yan Meng, Doreen Ng, James Xie |
| EPI_ISL_1937751, EPI_ISL_1937752 | SC Dept of Health and Env. Control-Bureau of Laboratories | Centers for Disease Control and Prevention Division of Viral Diseases, Pathogen Discovery | Mili Sheth, Sarah Nobles, Jasmine Padilla, Mark Burroughs, Shoshona Le, Katie Dillon, Peter Cook, Clinton R. Paden, Dhwani Batra, Krista Queen, Kristen Knipe, Dakota Howard, Yvette Unoarumhi, Darlene Wagner, Matthew Schmerer, Ben L. Rambo-Martin, Kristine Lacek, Sam Shepard, Alison Laufer Halpin, Dave Wentworth, Vivien Dugan, Suxiang Tong, Justin Lee |
| EPI_ISL_1937753 | Fulgent Genetics | Fulgent Genetics | Harry Gao, Mickey Li, John Gao, Joseph Fierro, Benafsh Sapra, Becky Tsai, Yan Meng, Doreen Ng, James Xie |
| EPI_ISL_1937754 | SC Dept of Health and Env. Control-Bureau of Laboratories | Centers for Disease Control and Prevention Division of Viral Diseases, Pathogen Discovery | Mili Sheth, Sarah Nobles, Jasmine Padilla, Mark Burroughs, Shoshona Le, Katie Dillon, Peter Cook, Clinton R. Paden, Dhwani Batra, Krista Queen, Kristen Knipe, Dakota Howard, Yvette Unoarumhi, Darlene Wagner, Matthew Schmerer, Ben L. Rambo-Martin, Kristine Lacek, Sam Shepard, Alison Laufer Halpin, Dave Wentworth, Vivien Dugan, Suxiang Tong, Justin Lee |
| EPI_ISL_1937755 | Fulgent Genetics | Fulgent Genetics | Harry Gao, Mickey Li, John Gao, Joseph Fierro, Benafsh Sapra, Becky Tsai, Yan Meng, Doreen Ng, James Xie |
| EPI_ISL_1937756 | SC Dept of Health and Env. Control-Bureau of Laboratories | Centers for Disease Control and Prevention Division of Viral Diseases, Pathogen Discovery | Mili Sheth, Sarah Nobles, Jasmine Padilla, Mark Burroughs, Shoshona Le, Katie Dillon, Peter Cook, Clinton R. Paden, Dhwani Batra, Krista Queen, Kristen Knipe, Dakota Howard, Yvette Unoarumhi, Darlene Wagner, Matthew Schmerer, Ben L. Rambo-Martin, Kristine Lacek, Sam Shepard, Alison Laufer Halpin, Dave Wentworth, Vivien Dugan, Suxiang Tong, Justin Lee |
| EPI_ISL_1937757 | Fulgent Genetics | Fulgent Genetics | Harry Gao, Mickey Li, John Gao, Joseph Fierro, Benafsh Sapra, Becky Tsai, Yan Meng, Doreen Ng, James Xie |
| EPI_ISL_1937758, EPI_ISL_1937759 | SC Dept of Health and Env. Control-Bureau of Laboratories | Centers for Disease Control and Prevention Division of Viral Diseases, Pathogen Discovery | Mili Sheth, Sarah Nobles, Jasmine Padilla, Mark Burroughs, Shoshona Le, Katie Dillon, Peter Cook, Clinton R. Paden, Dhwani Batra, Krista Queen, Kristen Knipe, Dakota Howard, Yvette Unoarumhi, Darlene Wagner, Matthew Schmerer, Ben L. Rambo-Martin, Kristine Lacek, Sam Shepard, Alison Laufer Halpin, Dave Wentworth, Vivien Dugan, Suxiang Tong, Justin Lee |
| EPI_ISL_1937760 | Fulgent Genetics | Fulgent Genetics | Harry Gao, Mickey Li, John Gao, Joseph Fierro, Benafsh Sapra, Becky Tsai, Yan Meng, Doreen Ng, James Xie |
| EPI_ISL_1937761, EPI_ISL_1937763, EPI_ISL_1937764 | SC Dept of Health and Env. Control-Bureau of Laboratories | Centers for Disease Control and Prevention Division of Viral Diseases, Pathogen Discovery | Mili Sheth, Sarah Nobles, Jasmine Padilla, Mark Burroughs, Shoshona Le, Katie Dillon, Peter Cook, Clinton R. Paden, Dhwani Batra, Krista Queen, Kristen Knipe, Dakota Howard, Yvette Unoarumhi, Darlene Wagner, Matthew Schmerer, Ben L. Rambo-Martin, Kristine Lacek, Sam Shepard, Alison Laufer Halpin, Dave Wentworth, Vivien Dugan, Suxiang Tong, Justin Lee |
| EPI_ISL_1937765 | Fulgent Genetics | Fulgent Genetics | Harry Gao, Mickey Li, John Gao, Joseph Fierro, Benafsh Sapra, Becky Tsai, Yan Meng, Doreen Ng, James Xie |
| EPI_ISL_1937766 | IL Dept. of Public Health Springfield Laboratory | Centers for Disease Control and Prevention Division of Viral Diseases, Pathogen Discovery | Mili Sheth, Sarah Nobles, Jasmine Padilla, Mark Burroughs, Shoshona Le, Katie Dillon, Peter Cook, Clinton R. Paden, Dhwani Batra, Krista Queen, Kristen Knipe, Dakota Howard, Yvette Unoarumhi, Darlene Wagner, Matthew Schmerer, Ben L. Rambo-Martin, Kristine Lacek, Sam Shepard, Alison Laufer Halpin, Dave Wentworth, Vivien Dugan, Suxiang Tong, Justin Lee |
| EPI_ISL_1937767 | Fulgent Genetics | Fulgent Genetics | Harry Gao, Mickey Li, John Gao, Joseph Fierro, Benafsh Sapra, Becky Tsai, Yan Meng, Doreen Ng, James Xie |
| EPI_ISL_1937768 | IL Dept. of Public Health Springfield Laboratory | Centers for Disease Control and Prevention Division of Viral Diseases, Pathogen Discovery | Mili Sheth, Sarah Nobles, Jasmine Padilla, Mark Burroughs, Shoshona Le, Katie Dillon, Peter Cook, Clinton R. Paden, Dhwani Batra, Krista Queen, Kristen Knipe, Dakota Howard, Yvette Unoarumhi, Darlene Wagner, Matthew Schmerer, Ben L. Rambo-Martin, Kristine Lacek, Sam Shepard, Alison Laufer Halpin, Dave Wentworth, Vivien Dugan, Suxiang Tong, Justin Lee |
| EPI_ISL_1937769 | Fulgent Genetics | Fulgent Genetics | Harry Gao, Mickey Li, John Gao, Joseph Fierro, Benafsh Sapra, Becky Tsai, Yan Meng, Doreen Ng, James Xie |
| EPI_ISL_1937770, EPI_ISL_1937771 | IL Dept. of Public Health Springfield Laboratory | Centers for Disease Control and Prevention Division of Viral Diseases, Pathogen Discovery | Mili Sheth, Sarah Nobles, Jasmine Padilla, Mark Burroughs, Shoshona Le, Katie Dillon, Peter Cook, Clinton R. Paden, Dhwani Batra, Krista Queen, Kristen Knipe, Dakota Howard, Yvette Unoarumhi, Darlene Wagner, Matthew Schmerer, Ben L. Rambo-Martin, Kristine Lacek, Sam Shepard, Alison Laufer Halpin, Dave Wentworth, Vivien Dugan, Suxiang Tong, Justin Lee |
| EPI_ISL_1937772 | Fulgent Genetics | Fulgent Genetics | Harry Gao, Mickey Li, John Gao, Joseph Fierro, Benafsh Sapra, Becky Tsai, Yan Meng, Doreen Ng, James Xie |
| EPI_ISL_1937773 | IL Dept. of Public Health Springfield Laboratory | Centers for Disease Control and Prevention Division of Viral Diseases, Pathogen Discovery | Mili Sheth, Sarah Nobles, Jasmine Padilla, Mark Burroughs, Shoshona Le, Katie Dillon, Peter Cook, Clinton R. Paden, Dhwani Batra, Krista Queen, Kristen Knipe, Dakota Howard, Yvette Unoarumhi, Darlene Wagner, Matthew Schmerer, Ben L. Rambo-Martin, Kristine Lacek, Sam Shepard, Alison Laufer Halpin, Dave Wentworth, Vivien Dugan, Suxiang Tong, Justin Lee |
| EPI_ISL_1937774 | Fulgent Genetics | Fulgent Genetics | Harry Gao, Mickey Li, John Gao, Joseph Fierro, Benafsh Sapra, Becky Tsai, Yan Meng, Doreen Ng, James Xie |
| EPI_ISL_1937775 | IL Dept. of Public Health Springfield Laboratory | Centers for Disease Control and Prevention Division of Viral Diseases, Pathogen Discovery | Mili Sheth, Sarah Nobles, Jasmine Padilla, Mark Burroughs, Shoshona Le, Katie Dillon, Peter Cook, Clinton R. Paden, Dhwani Batra, Krista Queen, Kristen Knipe, Dakota Howard, Yvette Unoarumhi, Darlene Wagner, Matthew Schmerer, Ben L. Rambo-Martin, Kristine Lacek, Sam Shepard, Alison Laufer Halpin, Dave Wentworth, Vivien Dugan, Suxiang Tong, Justin Lee |

| | | | |
|---|---|---|---|
| EPI_ISL_1937776 | Fulgent Genetics | Fulgent Genetics | Harry Gao, Mickey Li, John Gao, Joseph Fierro, Benafsh Sapra, Becky Tsai, Yan Meng, Doreen Ng, James Xie |
| EPI_ISL_1937777, EPI_ISL_1937778 | IL Dept. of Public Health Springfield Laboratory | Centers for Disease Control and Prevention Division of Viral Diseases, Pathogen Discovery | Mili Sheth, Sarah Nobles, Jasmine Padilla, Mark Burroughs, Shoshona Le, Katie Dillon, Peter Cook, Clinton R. Paden, Dhwani Batra, Krista Queen, Kristen Knipe, Dakota Howard, Yvette Unoarumhi, Darlene Wagner, Matthew Schmerer, Ben L. Rambo-Martin, Kristine Lacek, Sam Shepard, Alison Laufer Halpin, Dave Wentworth, Vivien Dugan, Suxiang Tong, Justin Lee |
| EPI_ISL_1937779 | Fulgent Genetics | Fulgent Genetics | Harry Gao, Mickey Li, John Gao, Joseph Fierro, Benafsh Sapra, Becky Tsai, Yan Meng, Doreen Ng, James Xie |
| EPI_ISL_1937780, EPI_ISL_1937781, EPI_ISL_1937782, EPI_ISL_1937783, EPI_ISL_1937784, EPI_ISL_1937785 | IL Dept. of Public Health Springfield Laboratory | Centers for Disease Control and Prevention Division of Viral Diseases, Pathogen Discovery | Mili Sheth, Sarah Nobles, Jasmine Padilla, Mark Burroughs, Shoshona Le, Katie Dillon, Peter Cook, Clinton R. Paden, Dhwani Batra, Krista Queen, Kristen Knipe, Dakota Howard, Yvette Unoarumhi, Darlene Wagner, Matthew Schmerer, Ben L. Rambo-Martin, Kristine Lacek, Sam Shepard, Alison Laufer Halpin, Dave Wentworth, Vivien Dugan, Suxiang Tong, Justin Lee |
| EPI_ISL_1937786, EPI_ISL_1937787, EPI_ISL_1937788, EPI_ISL_1937789, EPI_ISL_1937790, EPI_ISL_1937791, EPI_ISL_1937792, EPI_ISL_1937793, EPI_ISL_1937794 | "HI Dept. of Health, State Laboratories Division" | Centers for Disease Control and Prevention Division of Viral Diseases, Pathogen Discovery | Mili Sheth, Sarah Nobles, Jasmine Padilla, Mark Burroughs, Shoshona Le, Katie Dillon, Peter Cook, Clinton R. Paden, Dhwani Batra, Krista Queen, Kristen Knipe, Dakota Howard, Yvette Unoarumhi, Darlene Wagner, Matthew Schmerer, Ben L. Rambo-Martin, Kristine Lacek, Sam Shepard, Alison Laufer Halpin, Dave Wentworth, Vivien Dugan, Suxiang Tong, Justin Lee |
| EPI_ISL_1937795, EPI_ISL_1937796, EPI_ISL_1937797, EPI_ISL_1937798 | IN State Department of Health Laboratory Services | Centers for Disease Control and Prevention Division of Viral Diseases, Pathogen Discovery | Mili Sheth, Sarah Nobles, Jasmine Padilla, Mark Burroughs, Shoshona Le, Katie Dillon, Peter Cook, Clinton R. Paden, Dhwani Batra, Krista Queen, Kristen Knipe, Dakota Howard, Yvette Unoarumhi, Darlene Wagner, Matthew Schmerer, Ben L. Rambo-Martin, Kristine Lacek, Sam Shepard, Alison Laufer Halpin, Dave Wentworth, Vivien Dugan, Suxiang Tong, Justin Lee |
| EPI_ISL_1937799, EPI_ISL_1937800, EPI_ISL_1937801, EPI_ISL_1937802, EPI_ISL_1937803, EPI_ISL_1937804, EPI_ISL_1937805 | WI State Laboratory of Hygiene | Centers for Disease Control and Prevention Division of Viral Diseases, Pathogen Discovery | Mili Sheth, Sarah Nobles, Jasmine Padilla, Mark Burroughs, Shoshona Le, Katie Dillon, Peter Cook, Clinton R. Paden, Dhwani Batra, Krista Queen, Kristen Knipe, Dakota Howard, Yvette Unoarumhi, Darlene Wagner, Matthew Schmerer, Ben L. Rambo-Martin, Kristine Lacek, Sam Shepard, Alison Laufer Halpin, Dave Wentworth, Vivien Dugan, Suxiang Tong, Justin Lee |
| EPI_ISL_1937806, EPI_ISL_1937807, EPI_ISL_1937808, EPI_ISL_1937809, EPI_ISL_1937810, EPI_ISL_1937811 | VT Dept. of Health Laboratory | Centers for Disease Control and Prevention Division of Viral Diseases, Pathogen Discovery | Mili Sheth, Sarah Nobles, Jasmine Padilla, Mark Burroughs, Shoshona Le, Katie Dillon, Peter Cook, Clinton R. Paden, Dhwani Batra, Krista Queen, Kristen Knipe, Dakota Howard, Yvette Unoarumhi, Darlene Wagner, Matthew Schmerer, Ben L. Rambo-Martin, Kristine Lacek, Sam Shepard, Alison Laufer Halpin, Dave Wentworth, Vivien Dugan, Suxiang Tong, Justin Lee |
| EPI_ISL_1937812, EPI_ISL_1937813, EPI_ISL_1937814, EPI_ISL_1937815, EPI_ISL_1937816, EPI_ISL_1937817, EPI_ISL_1937818, EPI_ISL_1937819, EPI_ISL_1937820, EPI_ISL_1937822, EPI_ISL_1937823, EPI_ISL_1937824, EPI_ISL_1937825, EPI_ISL_1937826, EPI_ISL_1937827, EPI_ISL_1937828, EPI_ISL_1937829 | | | |
| see above | CA-Los Angeles County Public Health Laboratory | Centers for Disease Control and Prevention Division of Viral Diseases, Pathogen Discovery | Mili Sheth, Sarah Nobles, Jasmine Padilla, Mark Burroughs, Shoshona Le, Katie Dillon, Peter Cook, Clinton R. Paden, Dhwani Batra, Krista Queen, Kristen Knipe, Dakota Howard, Yvette Unoarumhi, Darlene Wagner, Matthew Schmerer, Ben L. Rambo-Martin, Kristine Lacek, Sam Shepard, Alison Laufer Halpin, Dave Wentworth, Vivien Dugan, Suxiang Tong, Justin Lee |
| EPI_ISL_1937830, EPI_ISL_1937831, EPI_ISL_1937832, EPI_ISL_1937833 | UT-Unified State Labs: Public Health Utah DOH | Centers for Disease Control and Prevention Division of Viral Diseases, Pathogen Discovery | Mili Sheth, Sarah Nobles, Jasmine Padilla, Mark Burroughs, Shoshona Le, Katie Dillon, Peter Cook, Clinton R. Paden, Dhwani Batra, Krista Queen, Kristen Knipe, Dakota Howard, Yvette Unoarumhi, Darlene Wagner, Matthew Schmerer, Ben L. Rambo-Martin, Kristine Lacek, Sam Shepard, Alison Laufer Halpin, Dave Wentworth, Vivien Dugan, Suxiang Tong, Justin Lee |
| EPI_ISL_1937834, EPI_ISL_1937835, EPI_ISL_1937836, EPI_ISL_1937837, EPI_ISL_1937838, EPI_ISL_1937839, EPI_ISL_1937840 | DC Public Health Lab/ Dept. of Forensic Sciences | Centers for Disease Control and Prevention Division of Viral Diseases, Pathogen Discovery | Mili Sheth, Sarah Nobles, Jasmine Padilla, Mark Burroughs, Shoshona Le, Katie Dillon, Peter Cook, Clinton R. Paden, Dhwani Batra, Krista Queen, Kristen Knipe, Dakota Howard, Yvette Unoarumhi, Darlene Wagner, Matthew Schmerer, Ben L. Rambo-Martin, Kristine Lacek, Sam Shepard, Alison Laufer Halpin, Dave Wentworth, Vivien Dugan, Suxiang Tong, Justin Lee |
| EPI_ISL_1937841, EPI_ISL_1937842, EPI_ISL_1937843, EPI_ISL_1937844, EPI_ISL_1937845, EPI_ISL_1937846, EPI_ISL_1937847, EPI_ISL_1937848, EPI_ISL_1937849 | NH Dept. of Health and Human Services Public Health Labs | Centers for Disease Control and Prevention Division of Viral Diseases, Pathogen Discovery | Mili Sheth, Sarah Nobles, Jasmine Padilla, Mark Burroughs, Shoshona Le, Katie Dillon, Peter Cook, Clinton R. Paden, Dhwani Batra, Krista Queen, Kristen Knipe, Dakota Howard, Yvette Unoarumhi, Darlene Wagner, Matthew Schmerer, Ben L. Rambo-Martin, Kristine Lacek, Sam Shepard, Alison Laufer Halpin, Dave Wentworth, Vivien Dugan, Suxiang Tong, Justin Lee |
| EPI_ISL_1937850, EPI_ISL_1937851, EPI_ISL_1937852, EPI_ISL_1937853, EPI_ISL_1937854, EPI_ISL_1937856, EPI_ISL_1937858, EPI_ISL_1937859, EPI_ISL_1937860, EPI_ISL_1937861, EPI_ISL_1937862, EPI_ISL_1937863, EPI_ISL_1937864, EPI_ISL_1937865, EPI_ISL_1937866, EPI_ISL_1937867, EPI_ISL_1937868, EPI_ISL_1937869, EPI_ISL_1937870, EPI_ISL_1937871, EPI_ISL_1937872, EPI_ISL_1937873, EPI_ISL_1937874, EPI_ISL_1937875, EPI_ISL_1937876, EPI_ISL_1937877, EPI_ISL_1937879, EPI_ISL_1937880 | | | |
| see above | Fulgent Genetics | Fulgent Genetics | Harry Gao, Mickey Li, John Gao, Joseph Fierro, Benafsh Sapra, Becky Tsai, Yan Meng, Doreen Ng, James Xie |
| EPI_ISL_1937893, EPI_ISL_1937894, EPI_ISL_1937896, EPI_ISL_1937897, EPI_ISL_1937898, EPI_ISL_1937899, EPI_ISL_1937900, EPI_ISL_1937901, EPI_ISL_1937902, EPI_ISL_1937903, EPI_ISL_1937904, EPI_ISL_1937905, EPI_ISL_1937906, EPI_ISL_1937907, EPI_ISL_1937908, EPI_ISL_1937909, EPI_ISL_1937910, EPI_ISL_1937911, EPI_ISL_1937912, EPI_ISL_1937913, EPI_ISL_1937914, EPI_ISL_1937915, EPI_ISL_1937916, EPI_ISL_1937917, EPI_ISL_1937918, EPI_ISL_1937919, EPI_ISL_1937920, EPI_ISL_1937921, EPI_ISL_1937922, EPI_ISL_1937923, EPI_ISL_1937924, EPI_ISL_1937925, EPI_ISL_1937926, EPI_ISL_1937927, EPI_ISL_1937928, EPI_ISL_1937929, EPI_ISL_1937930, EPI_ISL_1937931, EPI_ISL_1937932, EPI_ISL_1937933, EPI_ISL_1937934, EPI_ISL_1937935, EPI_ISL_1937936, EPI_ISL_1937937, EPI_ISL_1937938, EPI_ISL_1937939, EPI_ISL_1937940, EPI_ISL_1937941, EPI_ISL_1937942, EPI_ISL_1937943, EPI_ISL_1937944, EPI_ISL_1937945, EPI_ISL_1937946, EPI_ISL_1937947, EPI_ISL_1937948, EPI_ISL_1937949, EPI_ISL_1937950, EPI_ISL_1937951, EPI_ISL_1937952, EPI_ISL_1937953, EPI_ISL_1937954, EPI_ISL_1937955, EPI_ISL_1937956, EPI_ISL_1937957, EPI_ISL_1937958, EPI_ISL_1937959, EPI_ISL_1937960, EPI_ISL_1937961, EPI_ISL_1937962, EPI_ISL_1937963, EPI_ISL_1937964, EPI_ISL_1937965, EPI_ISL_1937966, EPI_ISL_1937967, EPI_ISL_1937968, EPI_ISL_1937969, EPI_ISL_1937970, EPI_ISL_1937971, EPI_ISL_1937972, EPI_ISL_1937973, EPI_ISL_1937975, EPI_ISL_1937976, EPI_ISL_1937977, EPI_ISL_1937978, EPI_ISL_1937979, EPI_ISL_1937980, EPI_ISL_1937981, EPI_ISL_1937982, EPI_ISL_1937983, EPI_ISL_1937984, EPI_ISL_1937985, EPI_ISL_1937986, EPI_ISL_1937987, EPI_ISL_1937988, EPI_ISL_1937989, EPI_ISL_1937990, EPI_ISL_1937991, EPI_ISL_1937992, EPI_ISL_1937994, EPI_ISL_1937995, EPI_ISL_1937997, EPI_ISL_1937998, EPI_ISL_1937999, EPI_ISL_1938000, EPI_ISL_1938001, EPI_ISL_1938002, EPI_ISL_1938003, EPI_ISL_1938004, EPI_ISL_1938005, EPI_ISL_1938006, EPI_ISL_1938007, EPI_ISL_1938008, EPI_ISL_1938009, EPI_ISL_1938010, EPI_ISL_1938011, EPI_ISL_1938012, EPI_ISL_1938013, EPI_ISL_1938014, EPI_ISL_1938015, EPI_ISL_1938016, EPI_ISL_1938017, EPI_ISL_1938018, EPI_ISL_1938020, EPI_ISL_1938021, EPI_ISL_1938022, EPI_ISL_1938023, EPI_ISL_1938024, EPI_ISL_1938025, EPI_ISL_1938026, EPI_ISL_1938027, EPI_ISL_1938028, EPI_ISL_1938029, EPI_ISL_1938030, EPI_ISL_1938031, EPI_ISL_1938032, EPI_ISL_1938033, EPI_ISL_1938034, EPI_ISL_1938035, EPI_ISL_1938036, EPI_ISL_1938037, EPI_ISL_1938038, EPI_ISL_1938039, EPI_ISL_1938040, EPI_ISL_1938041, EPI_ISL_1938042, EPI_ISL_1938043, EPI_ISL_1938044, EPI_ISL_1938045, EPI_ISL_1938046, EPI_ISL_1938047, EPI_ISL_1938048, EPI_ISL_1938049, EPI_ISL_1938050, EPI_ISL_1938051, EPI_ISL_1938052, EPI_ISL_1938053, EPI_ISL_1938054, EPI_ISL_1938055, EPI_ISL_1938056, EPI_ISL_1938057, EPI_ISL_1938058, EPI_ISL_1938059, EPI_ISL_1938060, EPI_ISL_1938061, EPI_ISL_1938062, EPI_ISL_1938063, EPI_ISL_1938064, EPI_ISL_1938065, EPI_ISL_1938066, EPI_ISL_1938067, EPI_ISL_1938068, EPI_ISL_1938069, EPI_ISL_1938070, EPI_ISL_1938071, EPI_ISL_1938072, EPI_ISL_1938073, EPI_ISL_1938074, EPI_ISL_1938075, EPI_ISL_1938076, EPI_ISL_1938077, EPI_ISL_1938078, EPI_ISL_1938079, EPI_ISL_1938080, EPI_ISL_1938081, EPI_ISL_1938082, EPI_ISL_1938083, EPI_ISL_1938084, EPI_ISL_1938085, EPI_ISL_1938086, EPI_ISL_1938087, EPI_ISL_1938088, EPI_ISL_1938089, EPI_ISL_1938090, EPI_ISL_1938091, EPI_ISL_1938092, EPI_ISL_1938093, EPI_ISL_1938094, EPI_ISL_1938095, EPI_ISL_1938096, EPI_ISL_1938097, EPI_ISL_1938098, EPI_ISL_1938099, EPI_ISL_1938100, EPI_ISL_1938101, EPI_ISL_1938102, EPI_ISL_1938103, EPI_ISL_1938104, EPI_ISL_1938105, EPI_ISL_1938106, EPI_ISL_1938107, EPI_ISL_1938108, EPI_ISL_1938109, EPI_ISL_1938110, EPI_ISL_1938111, EPI_ISL_1938112, EPI_ISL_1938113, EPI_ISL_1938114, EPI_ISL_1938115, EPI_ISL_1938116, EPI_ISL_1938118, EPI_ISL_1938119, EPI_ISL_1938120, EPI_ISL_1938121, EPI_ISL_1938122, EPI_ISL_1938123, EPI_ISL_1938124, EPI_ISL_1938125, EPI_ISL_1938126, EPI_ISL_1938128, EPI_ISL_1938129, EPI_ISL_1938130, EPI_ISL_1938131, EPI_ISL_1938132, EPI_ISL_1938134, EPI_ISL_1938135, EPI_ISL_1938136, EPI_ISL_1938137, EPI_ISL_1938138, EPI_ISL_1938139, EPI_ISL_1938140, EPI_ISL_1938141, EPI_ISL_1938142, EPI_ISL_1938143, EPI_ISL_1938144, EPI_ISL_1938145, EPI_ISL_1938146, EPI_ISL_1938147, EPI_ISL_1938148, EPI_ISL_1938149, EPI_ISL_1938150, EPI_ISL_1938151, EPI_ISL_1938152, EPI_ISL_1938153, EPI_ISL_1938154, EPI_ISL_1938155, EPI_ISL_1938156, EPI_ISL_1938157, EPI_ISL_1938158, EPI_ISL_1938159, EPI_ISL_1938160, EPI_ISL_1938161, EPI_ISL_1938162, EPI_ISL_1938163, EPI_ISL_1938164, EPI_ISL_1938165, EPI_ISL_1938166, EPI_ISL_1938167, EPI_ISL_1938168, EPI_ISL_1938169, EPI_ISL_1938170, EPI_ISL_1938171, EPI_ISL_1938172, EPI_ISL_1938173, EPI_ISL_1938175, EPI_ISL_1938176, EPI_ISL_1938177, EPI_ISL_1938178, EPI_ISL_1938179, EPI_ISL_1938180, EPI_ISL_1938181, EPI_ISL_1938182, EPI_ISL_1938183, EPI_ISL_1938184, EPI_ISL_1938185, EPI_ISL_1938186, EPI_ISL_1938187, EPI_ISL_1938188, EPI_ISL_1938189, EPI_ISL_1938190, EPI_ISL_1938191, EPI_ISL_1938192, EPI_ISL_1938193, EPI_ISL_1938194, EPI_ISL_1938195, EPI_ISL_1938196, EPI_ISL_1938197, EPI_ISL_1938198, EPI_ISL_1938199, EPI_ISL_1938200, EPI_ISL_1938201, EPI_ISL_1938202, EPI_ISL_1938203, EPI_ISL_1938204, EPI_ISL_1938205, EPI_ISL_1938206, EPI_ISL_1938207, EPI_ISL_1938208, EPI_ISL_1938210, EPI_ISL_1938211, EPI_ISL_1938212, EPI_ISL_1938213, EPI_ISL_1938214, EPI_ISL_1938215, EPI_ISL_1938216, EPI_ISL_1938217, EPI_ISL_1938218, EPI_ISL_1938219, EPI_ISL_1938220, EPI_ISL_1938221, EPI_ISL_1938222, EPI_ISL_1938223, EPI_ISL_1938224, EPI_ISL_1938225, EPI_ISL_1938226, EPI_ISL_1938227, EPI_ISL_1938228, EPI_ISL_1938229, EPI_ISL_1938230, EPI_ISL_1938232, EPI_ISL_1938233, EPI_ISL_1938234, EPI_ISL_1938235, EPI_ISL_1938236, EPI_ISL_1938237, EPI_ISL_1938238, EPI_ISL_1938239, EPI_ISL_1938240, EPI_ISL_1938241, EPI_ISL_1938242, EPI_ISL_1938243, EPI_ISL_1938244, EPI_ISL_1938245, EPI_ISL_1938246, EPI_ISL_1938247, EPI_ISL_1938248, EPI_ISL_1938249, EPI_ISL_1938250, EPI_ISL_1938251, EPI_ISL_1938252, EPI_ISL_1938253, EPI_ISL_1938254, EPI_ISL_1938255, EPI_ISL_1938256, EPI_ISL_1938257, EPI_ISL_1938258, EPI_ISL_1938259, EPI_ISL_1938260, EPI_ISL_1938261, EPI_ISL_1938262, EPI_ISL_1938263, EPI_ISL_1938264, EPI_ISL_1938265, EPI_ISL_1938266, EPI_ISL_1938267, EPI_ISL_1938268, EPI_ISL_1938269, EPI_ISL_1938270, EPI_ISL_1938271, EPI_ISL_1938272, EPI_ISL_1938273, EPI_ISL_1938274, EPI_ISL_1938275, EPI_ISL_1938276, EPI_ISL_1938277, EPI_ISL_1938278, EPI_ISL_1938279, EPI_ISL_1938280, EPI_ISL_1938281, EPI_ISL_1938282, EPI_ISL_1938283, EPI_ISL_1938284, EPI_ISL_1938285, EPI_ISL_1938286, EPI_ISL_1938287, EPI_ISL_1938288, EPI_ISL_1938289, EPI_ISL_1938290, EPI_ISL_1938291, EPI_ISL_1938292, EPI_ISL_1938293, EPI_ISL_1938294, EPI_ISL_1938295, EPI_ISL_1938296, EPI_ISL_1938297, EPI_ISL_1938298, EPI_ISL_1938299, EPI_ISL_1938300, EPI_ISL_1938301, EPI_ISL_1938302, EPI_ISL_1938303, EPI_ISL_1938304, EPI_ISL_1938305, EPI_ISL_1938306 | | | |
| see above | UNC Charlotte COVID-19 Testing Lab | UNC Charlotte Environmental Monitoring Laboratory | Cynthia Gibas, Kevin Lambirth, Angelica Martins, Jannatul Ferdous, Visva Barua, Jessica Schlueter |
| EPI_ISL_1938482, EPI_ISL_1938483, EPI_ISL_1938484, EPI_ISL_1938485, EPI_ISL_1938486, EPI_ISL_1938487, EPI_ISL_1938488, EPI_ISL_1938489, EPI_ISL_1938490, EPI_ISL_1938492, EPI_ISL_1938493, EPI_ISL_1938494, EPI_ISL_1938495, EPI_ISL_1938496, EPI_ISL_1938498, EPI_ISL_1938500, EPI_ISL_1938501, EPI_ISL_1938502, EPI_ISL_1938503, EPI_ISL_1938504, EPI_ISL_1938505, EPI_ISL_1938506, EPI_ISL_1938507, EPI_ISL_1938508, EPI_ISL_1938509, EPI_ISL_1938510, EPI_ISL_1938511, EPI_ISL_1938512, EPI_ISL_1938513, EPI_ISL_1938514, EPI_ISL_1938515, EPI_ISL_1938516, EPI_ISL_1938517, EPI_ISL_1938518, EPI_ISL_1938520, EPI_ISL_1938521, EPI_ISL_1938524, EPI_ISL_1938525, EPI_ISL_1938526, EPI_ISL_1938527, EPI_ISL_1938528, EPI_ISL_1938529, EPI_ISL_1938530, EPI_ISL_1938531, EPI_ISL_1938532, EPI_ISL_1938533, EPI_ISL_1938534, EPI_ISL_1938535, EPI_ISL_1938537, EPI_ISL_1938538, EPI_ISL_1938539, EPI_ISL_1938540, EPI_ISL_1938541, EPI_ISL_1938542, EPI_ISL_1938543, EPI_ISL_1938544, EPI_ISL_1938545, EPI_ISL_1938546, EPI_ISL_1938547, EPI_ISL_1938548, EPI_ISL_1938550, EPI_ISL_1938551, EPI_ISL_1938552, EPI_ISL_1938553, EPI_ISL_1938554, EPI_ISL_1938555, EPI_ISL_1938556, EPI_ISL_1938557, EPI_ISL_1938558, EPI_ISL_1938559, EPI_ISL_1938560, EPI_ISL_1938561, EPI_ISL_1938562, EPI_ISL_1938563, EPI_ISL_1938564, EPI_ISL_1938565, EPI_ISL_1938566, EPI_ISL_1938567, EPI_ISL_1938568, EPI_ISL_1938569, EPI_ISL_1938570, EPI_ISL_1938571, EPI_ISL_1938572, EPI_ISL_1938574, EPI_ISL_1938575, EPI_ISL_1938576, EPI_ISL_1938577, EPI_ISL_1938578, EPI_ISL_1938579, EPI_ISL_1938580, EPI_ISL_1938581, EPI_ISL_1938582, EPI_ISL_1938583, EPI_ISL_1938584, EPI_ISL_1938585, EPI_ISL_1938586, EPI_ISL_1938587, EPI_ISL_1938588, EPI_ISL_1938589, EPI_ISL_1938590, EPI_ISL_1938591, EPI_ISL_1938592, EPI_ISL_1938593, EPI_ISL_1938594, EPI_ISL_1938595, EPI_ISL_1938596, EPI_ISL_1938597, EPI_ISL_1938598, EPI_ISL_1938599, EPI_ISL_1938600, EPI_ISL_1938601, EPI_ISL_1938602, EPI_ISL_1938603, EPI_ISL_1938604, EPI_ISL_1938605, EPI_ISL_1938606, EPI_ISL_1938607, EPI_ISL_1938608, EPI_ISL_1938609, EPI_ISL_1938610, EPI_ISL_1938611, EPI_ISL_1938612, EPI_ISL_1938613, EPI_ISL_1938614, EPI_ISL_1938615, EPI_ISL_1938616, EPI_ISL_1938617, EPI_ISL_1938618, EPI_ISL_1938619, EPI_ISL_1938620, EPI_ISL_1938621, EPI_ISL_1938622, EPI_ISL_1938623, EPI_ISL_1938624, EPI_ISL_1938626, EPI_ISL_1938627, EPI_ISL_1938628, EPI_ISL_1938629, EPI_ISL_1938630, EPI_ISL_1938631, EPI_ISL_1938632, EPI_ISL_1938633, EPI_ISL_1938634, EPI_ISL_1938635, EPI_ISL_1938636, EPI_ISL_1938637, EPI_ISL_1938638, EPI_ISL_1938639, EPI_ISL_1938640, EPI_ISL_1938641, EPI_ISL_1938642, EPI_ISL_1938643, EPI_ISL_1938644, EPI_ISL_1938645, EPI_ISL_1938646, EPI_ISL_1938647, EPI_ISL_1938648, EPI_ISL_1938649, EPI_ISL_1938650, EPI_ISL_1938651, EPI_ISL_1938652, EPI_ISL_1938653, EPI_ISL_1938654, EPI_ISL_1938655, EPI_ISL_1938656, EPI_ISL_1938657, EPI_ISL_1938658, EPI_ISL_1938659, EPI_ISL_1938661, EPI_ISL_1938662, EPI_ISL_1938663, EPI_ISL_1938664, EPI_ISL_1938665, EPI_ISL_1938666, EPI_ISL_1938667, EPI_ISL_1938669, EPI_ISL_1938670, EPI_ISL_1938671 | | | |
| see above | Illinois Department of Public Health - Springfield Lab | Illinois Department of Public Health - Springfield Lab | Bryan Sim, Gordon McCall |
| EPI_ISL_1938672, EPI_ISL_1938673, EPI_ISL_1938674, EPI_ISL_1938675, EPI_ISL_1938676, EPI_ISL_1938678, EPI_ISL_1938679, EPI_ISL_1938680, EPI_ISL_1938681, EPI_ISL_1938682, EPI_ISL_1938683, EPI_ISL_1938685, EPI_ISL_1938686, EPI_ISL_1938687, EPI_ISL_1938689, EPI_ISL_1938690, EPI_ISL_1938691, EPI_ISL_1938692, EPI_ISL_1938693, EPI_ISL_1938694, EPI_ISL_1938695, EPI_ISL_1938696, EPI_ISL_1938697, EPI_ISL_1938698, EPI_ISL_1938699, EPI_ISL_1938700, EPI_ISL_1938701, EPI_ISL_1938702, EPI_ISL_1938703 | | | |
| see above | Wyoming Public Health Laboratory | Wyoming Public Health Laboratory | Jim Mildenberger, Wanda Manley, Noah Hull, Taylor Fearing, Lynette Gumbleton, Channing Weber, Ashley Norberg, Chayse Rowley, Marley Goetz, Brian Dominguez, Elliot Thomasson, Sam Britz, Cari Sloma, and Rob Christensen |
| EPI_ISL_1938795, EPI_ISL_1938799, EPI_ISL_1938800, EPI_ISL_1938801, EPI_ISL_1938804, EPI_ISL_1938805, EPI_ISL_1938806, EPI_ISL_1938807, EPI_ISL_1938808, EPI_ISL_1938809, EPI_ISL_1938810, EPI_ISL_1938811, EPI_ISL_1938812, EPI_ISL_1938815, EPI_ISL_1938816, EPI_ISL_1938817, EPI_ISL_1938887, EPI_ISL_1938888, EPI_ISL_1938889, EPI_ISL_1938890, EPI_ISL_1938891, EPI_ISL_1938892, | | | |

| | | | |
|---|---|---|---|
| EPI_ISL_1938894, EPI_ISL_1938895, EPI_ISL_1938896, EPI_ISL_1938897, EPI_ISL_1938898, EPI_ISL_1938899, EPI_ISL_1938900, EPI_ISL_1938902, EPI_ISL_1938903, EPI_ISL_1938904, EPI_ISL_1938905, EPI_ISL_1938907 | | | |
| see above | Wyoming Public Health Laboratory | Wyoming Public Health Laboratory | Jim Mildenberger, Taylor Fearing, Channing Weber, Ashley Norberg, Chayse Rowley, Marley Goetz, Brian Dominguez, Elliot Thomasson, Brittany Oher, Sam Britz, Cari Sloma, and Rob Christensen |
| EPI_ISL_1938908, EPI_ISL_1938909 | Wyoming Public Health Laboratory | Wyoming Public Health Laboratory | Jim Mildenberger, Wanda Manley, Noah Hull, Taylor Fearing, Lynette Gumbleton, Channing Weber, Ashley Norberg, Chayse Rowley, Marley Goetz, Brian Dominguez, Elliot Thomasson, Sam Britz, Cari Sloma, and Rob Christensen |
| EPI_ISL_1939201, EPI_ISL_1939202, EPI_ISL_1939203, EPI_ISL_1939204, EPI_ISL_1939205, EPI_ISL_1939206, EPI_ISL_1939207, EPI_ISL_1939291, EPI_ISL_1939292, EPI_ISL_1939293, EPI_ISL_1939294, EPI_ISL_1939296, EPI_ISL_1939297, EPI_ISL_1939298, EPI_ISL_1939299, EPI_ISL_1939300, EPI_ISL_1939301, EPI_ISL_1939302, EPI_ISL_1939303, EPI_ISL_1939305, EPI_ISL_1939306, EPI_ISL_1939308, EPI_ISL_1939309, EPI_ISL_1939310, EPI_ISL_1939385, EPI_ISL_1939386, EPI_ISL_1939387, EPI_ISL_1939388, EPI_ISL_1939389, EPI_ISL_1939390, EPI_ISL_1939391, EPI_ISL_1939392, EPI_ISL_1939393, EPI_ISL_1939395, EPI_ISL_1939396, EPI_ISL_1939397, EPI_ISL_1939398, EPI_ISL_1939399, EPI_ISL_1939400, EPI_ISL_1939401, EPI_ISL_1939402, EPI_ISL_1939403, EPI_ISL_1939404, EPI_ISL_1939405 | | | |
| see above | Wyoming Public Health Laboratory | Wyoming Public Health Laboratory | Jim Mildenberger, Taylor Fearing, Channing Weber, Ashley Norberg, Chayse Rowley, Marley Goetz, Brian Dominguez, Elliot Thomasson, Brittany Oher, Sam Britz, Cari Sloma, and Rob Christensen |
| EPI_ISL_1939407, EPI_ISL_1939408, EPI_ISL_1939409, EPI_ISL_1939410, EPI_ISL_1939412, EPI_ISL_1939413, EPI_ISL_1939414, EPI_ISL_1939415, EPI_ISL_1939416, EPI_ISL_1939417, EPI_ISL_1939418, EPI_ISL_1939420, EPI_ISL_1939421, EPI_ISL_1939422, EPI_ISL_1939423, EPI_ISL_1939424, EPI_ISL_1939425, EPI_ISL_1939426, EPI_ISL_1939427, EPI_ISL_1939428, EPI_ISL_1939429, EPI_ISL_1939430, EPI_ISL_1939431, EPI_ISL_1939432, EPI_ISL_1939433, EPI_ISL_1939434, EPI_ISL_1939437, EPI_ISL_1939438, EPI_ISL_1939439, EPI_ISL_1939440, EPI_ISL_1939441, EPI_ISL_1939442, EPI_ISL_1939443, EPI_ISL_1939444, EPI_ISL_1939445, EPI_ISL_1939446, EPI_ISL_1939447, EPI_ISL_1939449, EPI_ISL_1939450, EPI_ISL_1939451, EPI_ISL_1939452, EPI_ISL_1939453, EPI_ISL_1939454, EPI_ISL_1939455, EPI_ISL_1939456, EPI_ISL_1939457, EPI_ISL_1939458, EPI_ISL_1939459, EPI_ISL_1939460, EPI_ISL_1939461, EPI_ISL_1939462, EPI_ISL_1939463, EPI_ISL_1939464, EPI_ISL_1939466, EPI_ISL_1939467, EPI_ISL_1939469, EPI_ISL_1939470, EPI_ISL_1939472, EPI_ISL_1939473 | | | |
| see above | North Dakota Department of Health, Public Health Laboratory | North Dakota Department of Health, Public Health Laboratory | Lisa Wingerter |
| EPI_ISL_1940143, EPI_ISL_1940144, EPI_ISL_1940145, EPI_ISL_1940146, EPI_ISL_1940147, EPI_ISL_1940148, EPI_ISL_1940149, EPI_ISL_1940150, EPI_ISL_1940151, EPI_ISL_1940152, EPI_ISL_1940153, EPI_ISL_1940154, EPI_ISL_1940155, EPI_ISL_1940156, EPI_ISL_1940158, EPI_ISL_1940159, EPI_ISL_1940160 | | | |
| see above | Wichita State University - Molecular Diagnostics Lab | Kansas Health and Environmental Lab | Mike Grose, Jonathan Barnell, Ben Olsen, and Phil Adam |
| EPI_ISL_1941584, EPI_ISL_1941585, EPI_ISL_1941586, EPI_ISL_1941587, EPI_ISL_1941588, EPI_ISL_1941589, EPI_ISL_1941594, EPI_ISL_1941595, EPI_ISL_1941597, EPI_ISL_1941599, EPI_ISL_1941602, EPI_ISL_1941604, EPI_ISL_1941613, EPI_ISL_1941619, EPI_ISL_1941620, EPI_ISL_1941622, EPI_ISL_1941624, EPI_ISL_1941625, EPI_ISL_1941626, EPI_ISL_1941628, EPI_ISL_1941629, EPI_ISL_1941631, EPI_ISL_1941633, EPI_ISL_1941639, EPI_ISL_1941646 | | | |
| see above | IN State Department of Health Laboratory Services | IN State Department of Health Laboratory Services | Cassandra Campion, Jamie Yeadon, Brian Pope, Lixia Liu, Kyle Brownlee, Melissa Hindenlang, Mark Glazier |
| EPI_ISL_1941714, EPI_ISL_1941717, EPI_ISL_1941720, EPI_ISL_1941722, EPI_ISL_1941724, EPI_ISL_1941726, EPI_ISL_1941729, EPI_ISL_1941732 | Sonic Reference Laboratory | Sonic Reference Laboratory | "Robert Moller, Bijaya Dhakal, Krishna Maddula, John Woody, Cora Lahey, Ellen Pizarek" |
| EPI_ISL_1942137, EPI_ISL_1942138, EPI_ISL_1942139, EPI_ISL_1942140, EPI_ISL_1942141, EPI_ISL_1942142, EPI_ISL_1942143, EPI_ISL_1942144, EPI_ISL_1942145, EPI_ISL_1942146, EPI_ISL_1942147, EPI_ISL_1942148, EPI_ISL_1942149, EPI_ISL_1942150 | | | |
| see above | Aegis Sciences Corporation | Centers for Disease Control and Prevention Division of Viral Diseases, Pathogen Discovery | Dakota Howard, Dhwani Batra, Peter W. Cook, Kara Moser, Adrian Paskey, Jason Caravas, Benjamin Rambo-Martin, Shatavia Morrison, Christopher Gulvick, Scott Sammons, Yvette Unoarumhi, Darlene Wagner, Matthew Schmerer, Cyndi Clark, Patrick Campbell, Rob Case, Vikramsinha Ghorpade, Holly Houdeshell, Ola Kvalvaag, Dillon Nall, Ethan Sanders, Alec Vest, Shaun Westlund, Matthew Hardison, Clinton R. Paden, Duncan MacCannell |
| EPI_ISL_1942151, EPI_ISL_1942152, EPI_ISL_1942153, EPI_ISL_1942154, EPI_ISL_1942155, EPI_ISL_1942156, EPI_ISL_1942157, EPI_ISL_1942158, EPI_ISL_1942159, EPI_ISL_1942160, EPI_ISL_1942161, EPI_ISL_1942162, EPI_ISL_1942163, EPI_ISL_1942164, EPI_ISL_1942165, EPI_ISL_1942166, EPI_ISL_1942167, EPI_ISL_1942168 | | | |
| see above | Helix/Illumina | Centers for Disease Control and Prevention Division of Viral Diseases, Pathogen Discovery | Dakota Howard, Dhwani Batra, Peter W. Cook, Kara Moser, Adrian Paskey, Jason Caravas, Benjamin Rambo-Martin, Shatavia Morrison, Christopher Gulvick, Scott Sammons, Yvette Unoarumhi, Darlene Wagner, Matthew Schmerer, Eileen de Feo, Jan Antico, Christine Tran, Matthew Tolentino, Shannon Wickline, Kim Gietzen, Brad Sickler, Jingtao Liu, Eric Allen, Phil Febbo, Nicole L. Washington, Simon White, Geraint Levan, Kelly Schiabor Barrett, Elizabeth Cirulli, Alexandre Bolze, Ary Ascencio, Charlotte Rivera-Garcia, Ryan Cho, Jason Nguyen, Sherry Wang, Jimmy Ramirez, Tyler Cassens, Efren Sandoval, Magnus Isaksson, William Lee, David Becker, Marc Laurent, James Lu, Clinton R. Paden, Duncan MacCannell |
| EPI_ISL_1942169, EPI_ISL_1942170, EPI_ISL_1942171, EPI_ISL_1942172, EPI_ISL_1942173, EPI_ISL_1942174, EPI_ISL_1942175, EPI_ISL_1942176, EPI_ISL_1942177, EPI_ISL_1942178, EPI_ISL_1942179, EPI_ISL_1942180, EPI_ISL_1942181, EPI_ISL_1942182, EPI_ISL_1942183, EPI_ISL_1942184, EPI_ISL_1942185, EPI_ISL_1942186, EPI_ISL_1942187, EPI_ISL_1942188, EPI_ISL_1942189, EPI_ISL_1942190, EPI_ISL_1942191, EPI_ISL_1942192, EPI_ISL_1942193, EPI_ISL_1942194, EPI_ISL_1942195, EPI_ISL_1942196, EPI_ISL_1942197, EPI_ISL_1942198, EPI_ISL_1942199, EPI_ISL_1942200, EPI_ISL_1942201, EPI_ISL_1942202, EPI_ISL_1942203, EPI_ISL_1942204, EPI_ISL_1942205, EPI_ISL_1942206, EPI_ISL_1942207, EPI_ISL_1942208, EPI_ISL_1942209, EPI_ISL_1942210, EPI_ISL_1942211, EPI_ISL_1942212, EPI_ISL_1942213, EPI_ISL_1942214, EPI_ISL_1942215, EPI_ISL_1942216 | | | |
| see above | Quest Diagnostics Incorporated | Centers for Disease Control and Prevention Division of Viral Diseases, Pathogen Discovery | Dakota Howard, Dhwani Batra, Peter W. Cook, Kara Moser, Adrian Paskey, Jason Caravas, Benjamin Rambo-Martin, Shatavia Morrison, Christopher Gulvick, Scott Sammons, Yvette Unoarumhi, Darlene Wagner, Matthew Schmerer, S. H. Rosenthal, A. Gerasimova, R. M. Kagan, B. Anderson, M. Hua, Y. Liu, L.E. Bernstein, K.E. Livingston, A. Perez, I. A. Shlyakhter, R. V. Rolando, R. Owen, P. Tanpaiboon, F. Lacbawan, Clinton R. Paden, Duncan MacCannell |
| EPI_ISL_1942217, EPI_ISL_1942218, EPI_ISL_1942219, EPI_ISL_1942220, EPI_ISL_1942221, EPI_ISL_1942222, EPI_ISL_1942223, EPI_ISL_1942224, EPI_ISL_1942225, EPI_ISL_1942226, EPI_ISL_1942227, EPI_ISL_1942228, EPI_ISL_1942229, EPI_ISL_1942230, EPI_ISL_1942231, EPI_ISL_1942232, EPI_ISL_1942233, EPI_ISL_1942234, EPI_ISL_1942235 | | | |
| see above | Aegis Sciences Corporation | Centers for Disease Control and Prevention Division of Viral Diseases, Pathogen Discovery | Dakota Howard, Dhwani Batra, Peter W. Cook, Kara Moser, Adrian Paskey, Jason Caravas, Benjamin Rambo-Martin, Shatavia Morrison, Christopher Gulvick, Scott Sammons, Yvette Unoarumhi, Darlene Wagner, Matthew Schmerer, Cyndi Clark, Patrick Campbell, Rob Case, Vikramsinha Ghorpade, Holly Houdeshell, Ola Kvalvaag, Dillon Nall, Ethan Sanders, Alec Vest, Shaun Westlund, Matthew Hardison, Clinton R. Paden, Duncan MacCannell |
| EPI_ISL_1942236, EPI_ISL_1942238, EPI_ISL_1942239, EPI_ISL_1942240 | Laboratory Corporation of America | Centers for Disease Control and Prevention Division of Viral Diseases, Pathogen Discovery | Dakota Howard, Dhwani Batra, Peter W. Cook, Kara Moser, Adrian Paskey, Jason Caravas, Benjamin Rambo-Martin, Shatavia Morrison, Christopher Gulvick, Scott Sammons, Yvette Unoarumhi, Darlene Wagner, Matthew Schmerer, Minoo Agarwal, Eyad Almasri, Debbie Boles, Ayla Burns, Nuthawin Charoensri, Oren Cohen, Susan Countryman, Mary Ann Cristobal, Bobbi Croy, Suzanne Dale, Hrushikesh Deshmukh, Amanda Douglas, Vincent Drouillon, Marcia Eisenberg, Howard Engler, Rama Ghatti, Prashant Gupta, Susan Hicks, Jake Humphrey, Lax Iyer, Manoj Jain, Mohan Kolli, Brian Krueger, Tim Kuphal, Stanley Letovsky, Michael Levandoski, Craig Lukasik, Jonathan Meltzer, Brian Norvell, Mindy Nye, Scott Parker, Christos Petropoulos, John Pruitt, Steven Ragan, Scott Ryan, Mike Sapeta, Jana Schroth, Suresh Babu Selvaraju, Goran Stevovic, Amanda Suchanek, Andrea Throop, Lyndon Tilson, Thomas Urban, Joe Voshell, Kimberly Wagner, Jonathan Williams, Mary Williamson, Qian Zeng, Tricia Zwiefelhofer, Clinton R. Paden, Duncan MacCannell |
| EPI_ISL_1960000, EPI_ISL_1960001, EPI_ISL_1960002, EPI_ISL_1960003, EPI_ISL_1960004, EPI_ISL_1960005, EPI_ISL_1960006, EPI_ISL_1960007, EPI_ISL_1960008, EPI_ISL_1960009, EPI_ISL_1960010, EPI_ISL_1960011, EPI_ISL_1960012, EPI_ISL_1960014, EPI_ISL_1960015, EPI_ISL_1960016, EPI_ISL_1960017, EPI_ISL_1960018, EPI_ISL_1960019, EPI_ISL_1960020 | | | |
| see above | NJDOH, Public Health and Environmental Laboratories | NJ_PHEL | Lindsey Bodnar, Shiv K. Verma, Jacquelyn Deverell, Matthew Scarnati, Dana Woell, Byeong Jeong |
| EPI_ISL_1960025, EPI_ISL_1960031, EPI_ISL_1960032, EPI_ISL_1960035 | Interpath Laboratory | Seattle Flu Study | Deborah A. Nickerson, Chris D. Frazar, Jover Lee, Benjamin Pelle, Erica Ryke, Matthew Richardson, Amanda Adler, Elisabeth Brandstetter, Peter D. Han, Kairsten Fay, Misja Ilcisin, Kirsten Lacombe, Thomas R. Sibley, Melissa Truong, Caitlin R. Wolf, Romesh Gautom, Geoff Melly, Brian Hiatt, Philip Dykema, Scott Lindquist, Michael Boeckh, Janet A. Englund, Michael Famulare, Barry R. Lutz, Mark J. Rieder, Lea M. Starita, Matthew Thompson, Helen Y. Chu, Jay Shendure, Trevor Bedford |
| EPI_ISL_1960036 | Northwest Laboratories | Seattle Flu Study | Deborah A. Nickerson, Chris D. Frazar, Jover Lee, Benjamin Pelle, Erica Ryke, Matthew Richardson, Amanda Adler, Elisabeth Brandstetter, Peter D. Han, Kairsten Fay, Misja Ilcisin, Kirsten Lacombe, Thomas R. Sibley, Melissa Truong, Caitlin R. Wolf, Romesh Gautom, Geoff Melly, Brian Hiatt, Philip Dykema, Scott Lindquist, Michael Boeckh, Janet A. Englund, Michael Famulare, Barry R. Lutz, Mark J. Rieder, Lea M. Starita, Matthew Thompson, Helen Y. Chu, Jay Shendure, Trevor Bedford |
| EPI_ISL_1960037 | Altius Institute | Seattle Flu Study | Deborah A. Nickerson, Chris D. Frazar, Jover Lee, Benjamin Pelle, Erica Ryke, Matthew Richardson, Amanda Adler, Elisabeth Brandstetter, Peter D. Han, Kairsten Fay, Misja Ilcisin, Kirsten Lacombe, Thomas R. Sibley, Melissa Truong, Caitlin R. Wolf, Ryan Alexander, Daniel Bates, Rebecca Bruders, Stephanie DeBaun, Clem Green, Muhammad Halimun, Jessica Halow, Kneshay Harper, Matt Hartman, Andrew Meuser, Alex Nguyen, Truong Nguyen, Sofia Olsson, Sadie Patraw, Hannah Petersen, Tobias Ragoczy, Joshua Richards, Jacob Rodriguez, John Stamatoyannopoulos, Julia Wald, Olivia Waltner, Michael Boeckh, Janet A. Englund, Michael Famulare, Barry R. Lutz, Mark J. Rieder, Lea M. Starita, Matthew Thompson, Helen Y. Chu, Jay Shendure, Trevor Bedford |
| EPI_ISL_1960083, EPI_ISL_1960084, EPI_ISL_1960085, EPI_ISL_1960086, EPI_ISL_1960087, EPI_ISL_1960088, EPI_ISL_1960089, EPI_ISL_1960090, EPI_ISL_1960091, EPI_ISL_1960092, EPI_ISL_1960093, EPI_ISL_1960094, EPI_ISL_1960095 | | | |
| see above | Maryland Genomics, Institute for Genome Sciences, University of Maryland School of Medicine | Maryland Genomics, Institute for Genome Sciences, University of Maryland School of Medicine | Tallon, Luke J; Sadzewicz, Lisa D; Humphrys, Mike; Ott, Sandra; Roussey, Holly; Mehta, Aditya; Vavikolanu, Kranthi; Fraser, Claire M; Ravel, Jacques |
| EPI_ISL_1960096, EPI_ISL_1960097, EPI_ISL_1960098, EPI_ISL_1960099, EPI_ISL_1960100, EPI_ISL_1960101, EPI_ISL_1960102, EPI_ISL_1960103, EPI_ISL_1960104, EPI_ISL_1960105, EPI_ISL_1960106, EPI_ISL_1960107, EPI_ISL_1960108, EPI_ISL_1960109, EPI_ISL_1960110, EPI_ISL_1960111, EPI_ISL_1960112, EPI_ISL_1960113, EPI_ISL_1960114, EPI_ISL_1960115, EPI_ISL_1960116, EPI_ISL_1960117, EPI_ISL_1960118, EPI_ISL_1960119, EPI_ISL_1960120, EPI_ISL_1960121, EPI_ISL_1960122, EPI_ISL_1960123, EPI_ISL_1960124, EPI_ISL_1960125, EPI_ISL_1960126, EPI_ISL_1960127 | | | |
| see above | Gundersen Clinical Microbiology Laboratory | Kabara Cancer Research Institute | Craig S. Richmond, Paraic A. Kenny |
| EPI_ISL_1960128, EPI_ISL_1960129, EPI_ISL_1960130 | Maryland Genomics, Institute for Genome Sciences, University of Maryland School of Medicine | Maryland Genomics, Institute for Genome Sciences, University of Maryland School of Medicine | Tallon, Luke J; Sadzewicz, Lisa D; Humphrys, Mike; Ott, Sandra; Roussey, Holly; Mehta, Aditya; Vavikolanu, Kranthi; Fraser, Claire M; Ravel, Jacques |
| EPI_ISL_1960907, EPI_ISL_1960910, EPI_ISL_1960911, EPI_ISL_1960912, EPI_ISL_1960913, EPI_ISL_1960914, EPI_ISL_1960916, EPI_ISL_1960920, EPI_ISL_1960921, EPI_ISL_1960922, EPI_ISL_1960929, EPI_ISL_1960930, EPI_ISL_1960931, EPI_ISL_1960933, EPI_ISL_1960934, EPI_ISL_1960936, EPI_ISL_1960937, EPI_ISL_1960939, EPI_ISL_1960941, EPI_ISL_1960942, EPI_ISL_1960943 | | | |
| see above | Willis-Knighton Medical Center Hospital Laboratory | LSUHS Emerging Viral Threat Laboratory | Gregory L. Ware, Alexander Mijalis, Jeremy P. Kamil, Jennifer L. Carroll, Maarten Van Diest, Rona S. Scott, Andrew D. Yurochko, Christopher G. Kevil, John A. Vanchiere |
| EPI_ISL_1960964, EPI_ISL_1960965, EPI_ISL_1960966, EPI_ISL_1960967, EPI_ISL_1960968, EPI_ISL_1960969, EPI_ISL_1960970, EPI_ISL_1960973 | Willis-Knighton Medical Center Hospital Laboratory | LSUHS Emerging Viral Threat Laboratory | Alexander Mijalis, Gregory L. Ware,Jeremy P. Kamil, Jennifer L. Carroll, Maarten Van Diest, Rona S. Scott, Andrew D. Yurochko, Christopher G. Kevil, John A. Vanchiere |
| EPI_ISL_1960974, EPI_ISL_1960975, EPI_ISL_1960976, EPI_ISL_1960977, EPI_ISL_1960978, EPI_ISL_1960979, EPI_ISL_1960980, EPI_ISL_1960981, EPI_ISL_1960982, EPI_ISL_1960983, EPI_ISL_1960984 | | | |
| see above | Willis-Knighton Medical Center Hospital Laboratory | LSUHS Emerging Viral Threat Laboratory | Gregory L. Ware, Alexander Mijalis, Jeremy P. Kamil, Jennifer L. Carroll, Maarten Van Diest, Rona S. Scott, Andrew D. Yurochko, Christopher G. Kevil, John A. Vanchiere |
| EPI_ISL_1960985, EPI_ISL_1960986, EPI_ISL_1960987, EPI_ISL_1960988, EPI_ISL_1960989, EPI_ISL_1960990, EPI_ISL_1960991, EPI_ISL_1960993, | Willis-Knighton Medical Center Hospital Laboratory | LSUHS Emerging Viral Threat Laboratory | Jennifer L. Carroll, Gregory L. Ware, Alexander Mijalis, Jeremy P. Kamil, Maarten Van Diest, Rona S. Scott, Andrew D. Yurochko, Christopher G. Kevil, John A. Vanchiere |

| | | | |
|---|---|---|---|
| EPI_ISL_1960994, EPI_ISL_1960995 | | | |
| EPI_ISL_1960997, EPI_ISL_1960998, EPI_ISL_1960999 | Willis-Knighton Medical Center Hospital Laboratory | LSUHS Emerging Viral Threat Laboratory | Jeremy P. Kamil, Gregory L. Ware, Alexander Mijalis, Jennifer L. Carroll, Maarten Van Diest, Rona S. Scott, Andrew D. Yurochko, Christopher G. Kevil, John A. Vanchiere |
| EPI_ISL_1961001, EPI_ISL_1961002, EPI_ISL_1961003, EPI_ISL_1961004, EPI_ISL_1961005, EPI_ISL_1961006, EPI_ISL_1961007, EPI_ISL_1961008 | Willis-Knighton Medical Center Hospital Laboratory | LSUHS Emerging Viral Threat Laboratory | Rona S. Scott, Gregory L. Ware, Alexander Mijalis, Jeremy P. Kamil, Jennifer L. Carroll, Maarten Van Diest, Andrew D. Yurochko, Christopher G. Kevil, John A. Vanchiere |
| EPI_ISL_1961009, EPI_ISL_1961010, EPI_ISL_1961011, EPI_ISL_1961012 | Wichita State University - Molecular Diagnostics Lab | Kansas Health and Environmental Lab | Mike Grose, Jonathan Barnell, Ben Olsen, and Phil Adam |
| EPI_ISL_1961013, EPI_ISL_1961014 | Kansas Health and Environmental Lab | Kansas Health and Environmental Lab | Mike Grose, Jonathan Barnell, Ben Olsen, and Phil Adam |
| EPI_ISL_1961015, EPI_ISL_1961016, EPI_ISL_1961017, EPI_ISL_1961018, EPI_ISL_1961019 | Wichita State University - Molecular Diagnostics Lab | Kansas Health and Environmental Lab | Mike Grose, Jonathan Barnell, Ben Olsen, and Phil Adam |
| EPI_ISL_1961020, EPI_ISL_1961021, EPI_ISL_1961022, EPI_ISL_1961023, EPI_ISL_1961024, EPI_ISL_1961025, EPI_ISL_1961026, EPI_ISL_1961027, EPI_ISL_1961028, EPI_ISL_1961029, EPI_ISL_1961030, EPI_ISL_1961031, EPI_ISL_1961032, EPI_ISL_1961033, EPI_ISL_1961034 | | | |
| see above | Houston Health Dept. | Houston Health Dept. | Ryker Penn, Pamela Brown, Adolpho Lara, Yanlai Lai |
| EPI_ISL_1961036, EPI_ISL_1961037, EPI_ISL_1961040, EPI_ISL_1961041, EPI_ISL_1961042, EPI_ISL_1961043, EPI_ISL_1961044, EPI_ISL_1961045 | Nevada State Public Health Laboratory | Nevada State Public Health Laboratory | Andrew Gorzalski, Mark Pandori |
| EPI_ISL_1961046, EPI_ISL_1961047 | Arizona State Public Health Laboratory | Arizona State Public Health Laboratory | Trung Huynh, Jessica Escobar, Katherine Fullerton, Nobuko Fukushima, Stacy White, Linda Getsinger, Victor Waddell |
| EPI_ISL_1961053, EPI_ISL_1961054, EPI_ISL_1961055, EPI_ISL_1961056, EPI_ISL_1961057, EPI_ISL_1961058, EPI_ISL_1961059, EPI_ISL_1961060, EPI_ISL_1961061, EPI_ISL_1961062, EPI_ISL_1961063, EPI_ISL_1961064, EPI_ISL_1961065, EPI_ISL_1961066, EPI_ISL_1961067, EPI_ISL_1961068, EPI_ISL_1961069 | | | |
| see above | Oregon State University TRACE Project | OSU Center for Genome Research and Biocomputing | Oregon State University TRACE Project |
| EPI_ISL_1961070, EPI_ISL_1961071, EPI_ISL_1961072, EPI_ISL_1961076, EPI_ISL_1961078, EPI_ISL_1961079, EPI_ISL_1961080, EPI_ISL_1961081, EPI_ISL_1961082, EPI_ISL_1961083, EPI_ISL_1961084, EPI_ISL_1961085, EPI_ISL_1961086, EPI_ISL_1961087, EPI_ISL_1961088, EPI_ISL_1961089, EPI_ISL_1961090, EPI_ISL_1961091, EPI_ISL_1961092, EPI_ISL_1961093, EPI_ISL_1961094, EPI_ISL_1961095, EPI_ISL_1961096, EPI_ISL_1961097, EPI_ISL_1961098, EPI_ISL_1961099, EPI_ISL_1961100, EPI_ISL_1961101, EPI_ISL_1961102, EPI_ISL_1961103, EPI_ISL_1961104, EPI_ISL_1961105, EPI_ISL_1961106, EPI_ISL_1961107, EPI_ISL_1961108, EPI_ISL_1961109, EPI_ISL_1961110, EPI_ISL_1961111, EPI_ISL_1961112, EPI_ISL_1961113, EPI_ISL_1961114, EPI_ISL_1961115, EPI_ISL_1961116, EPI_ISL_1961117, EPI_ISL_1961118, EPI_ISL_1961120, EPI_ISL_1961121, EPI_ISL_1961122, EPI_ISL_1961123, EPI_ISL_1961124, EPI_ISL_1961125, EPI_ISL_1961126, EPI_ISL_1961127, EPI_ISL_1961128, EPI_ISL_1961129, EPI_ISL_1961130, EPI_ISL_1961131, EPI_ISL_1961132, EPI_ISL_1961133, EPI_ISL_1961134, EPI_ISL_1961135, EPI_ISL_1961136 | | | |
| see above | TXDSHS | TXDSHS | Rashmi Tuladhar, Bonnie Oh, Jenny Zhang, Maliha Rahman, Mayela Pedrueza, Anita Pokharel, Lorraine Rodriguez, Myong Koag, Chun Wang, Rachel Lee, Grace Kubin |
| EPI_ISL_1963645, EPI_ISL_1963646, EPI_ISL_1963647, EPI_ISL_1963648, EPI_ISL_1963649, EPI_ISL_1963650, EPI_ISL_1963651, EPI_ISL_1963652, EPI_ISL_1963653, EPI_ISL_1963654, EPI_ISL_1963655, EPI_ISL_1963656, EPI_ISL_1963657, EPI_ISL_1963658, EPI_ISL_1963659, EPI_ISL_1963660, EPI_ISL_1963661, EPI_ISL_1963662, EPI_ISL_1963663, EPI_ISL_1963664, EPI_ISL_1963665, EPI_ISL_1963666, EPI_ISL_1963667, EPI_ISL_1963668, EPI_ISL_1963669, EPI_ISL_1963670, EPI_ISL_1963671, EPI_ISL_1963672, EPI_ISL_1963673, EPI_ISL_1963674, EPI_ISL_1963675, EPI_ISL_1963676, EPI_ISL_1963677, EPI_ISL_1963678, EPI_ISL_1963679, EPI_ISL_1963680, EPI_ISL_1963681, EPI_ISL_1963682, EPI_ISL_1963683, EPI_ISL_1963684, EPI_ISL_1963685, EPI_ISL_1963686, EPI_ISL_1963687, EPI_ISL_1963688, EPI_ISL_1963689, EPI_ISL_1963690, EPI_ISL_1963691, EPI_ISL_1963692, EPI_ISL_1963693, EPI_ISL_1963694, EPI_ISL_1963695, EPI_ISL_1963696, EPI_ISL_1963697, EPI_ISL_1963698, EPI_ISL_1963699, EPI_ISL_1963700, EPI_ISL_1963701, EPI_ISL_1963703, EPI_ISL_1963704, EPI_ISL_1963705, EPI_ISL_1963706, EPI_ISL_1963707, EPI_ISL_1963708, EPI_ISL_1963709, EPI_ISL_1963710, EPI_ISL_1963711, EPI_ISL_1963712, EPI_ISL_1963713, EPI_ISL_1963714, EPI_ISL_1963715, EPI_ISL_1963716, EPI_ISL_1963717, EPI_ISL_1963718, EPI_ISL_1963719, EPI_ISL_1963720, EPI_ISL_1963721, EPI_ISL_1963722, EPI_ISL_1963723, EPI_ISL_1963724, EPI_ISL_1963725, EPI_ISL_1963726, EPI_ISL_1963727, EPI_ISL_1963728, EPI_ISL_1963729, EPI_ISL_1963730, EPI_ISL_1963731, EPI_ISL_1963732, EPI_ISL_1963733, EPI_ISL_1963734, EPI_ISL_1963735, EPI_ISL_1963736, EPI_ISL_1963738, EPI_ISL_1963739, EPI_ISL_1963740, EPI_ISL_1963741, EPI_ISL_1963742, EPI_ISL_1963743, EPI_ISL_1963744, EPI_ISL_1963745, EPI_ISL_1963746, EPI_ISL_1963747, EPI_ISL_1963748, EPI_ISL_1963749, EPI_ISL_1963750, EPI_ISL_1963751, EPI_ISL_1963752, EPI_ISL_1963753, EPI_ISL_1963754, EPI_ISL_1963755, EPI_ISL_1963756, EPI_ISL_1963757, EPI_ISL_1963758, EPI_ISL_1963759, EPI_ISL_1963760, EPI_ISL_1963761, EPI_ISL_1963762, EPI_ISL_1963763, EPI_ISL_1963764, EPI_ISL_1963765, EPI_ISL_1963766, EPI_ISL_1963767, EPI_ISL_1963768, EPI_ISL_1963769, EPI_ISL_1963770, EPI_ISL_1963771, EPI_ISL_1963772, EPI_ISL_1963773, EPI_ISL_1963774, EPI_ISL_1963775, EPI_ISL_1963776, EPI_ISL_1963777, EPI_ISL_1963778, EPI_ISL_1963779, EPI_ISL_1963780, EPI_ISL_1963781, EPI_ISL_1963782, EPI_ISL_1963783, EPI_ISL_1963784, EPI_ISL_1963785, EPI_ISL_1963786, EPI_ISL_1963787, EPI_ISL_1963788, EPI_ISL_1963789, EPI_ISL_1963790, EPI_ISL_1963791, EPI_ISL_1963792, EPI_ISL_1963793, EPI_ISL_1963794, EPI_ISL_1963795, EPI_ISL_1963796, EPI_ISL_1963797, EPI_ISL_1963798, EPI_ISL_1963799, EPI_ISL_1963800, EPI_ISL_1963801, EPI_ISL_1963802, EPI_ISL_1963803, EPI_ISL_1963804, EPI_ISL_1963805, EPI_ISL_1963806, EPI_ISL_1963807, EPI_ISL_1963808, EPI_ISL_1963809, EPI_ISL_1963810, EPI_ISL_1963811, EPI_ISL_1963812, EPI_ISL_1963813, EPI_ISL_1963814, EPI_ISL_1963815, EPI_ISL_1963816, EPI_ISL_1963817, EPI_ISL_1963818, EPI_ISL_1963819, EPI_ISL_1963820, EPI_ISL_1963821, EPI_ISL_1963822, EPI_ISL_1963823, EPI_ISL_1963824, EPI_ISL_1963825, EPI_ISL_1963826 | | | |
| see above | Yale Clinical Virology Lab | Grubaugh Lab - Yale School of Public Health | Joseph Fauver, Mallery Breban, Isabel Ott, Tara Alpert, Mary Petrone, Anderson Brito, Chantal Vogels, Annie Watkins, Chaney Kalinich, Jessica Rothman, Marie L. Landry, Nathan Grubaugh |
| EPI_ISL_1964153, EPI_ISL_1964154, EPI_ISL_1964155, EPI_ISL_1964156, EPI_ISL_1964157, EPI_ISL_1964158, EPI_ISL_1964159, EPI_ISL_1964160, EPI_ISL_1964161, EPI_ISL_1964162, EPI_ISL_1964163, EPI_ISL_1964164, EPI_ISL_1964165, EPI_ISL_1964166, EPI_ISL_1964167, EPI_ISL_1964168, EPI_ISL_1964169, EPI_ISL_1964170, EPI_ISL_1964171, EPI_ISL_1964172, EPI_ISL_1964173, EPI_ISL_1964174, EPI_ISL_1964175, EPI_ISL_1964176, EPI_ISL_1964177, EPI_ISL_1964178 | | | |
| see above | Oregon State Public Health Laboratory | Oregon State Public Health Laboratory | Rafia Razzaque, Eugene Yeboah, Vanda Makris, Laura Tsaknaridis, John Fontana and Shane Sevey |
| EPI_ISL_1964224, EPI_ISL_1964225, EPI_ISL_1964226, EPI_ISL_1964227 | Kansas Health and Environmental Lab | Kansas Health and Environmental Lab | Mike Grose, Jonathan Barnell, Ben Olsen, and Phil Adam |
| EPI_ISL_1964228, EPI_ISL_1964229, EPI_ISL_1964230, EPI_ISL_1964231, EPI_ISL_1964232, EPI_ISL_1964233, EPI_ISL_1964234, EPI_ISL_1964235, EPI_ISL_1964236, EPI_ISL_1964237 | Kansas Health and Environmental Lab | Wichita State University - Molecular Diagnostics Lab | Mike Grose, Jonathan Barnell, Ben Olsen, and Phil Adam |
| EPI_ISL_1964969, EPI_ISL_1964970, EPI_ISL_1964971, EPI_ISL_1964972, EPI_ISL_1964974, EPI_ISL_1964975, EPI_ISL_1964976, EPI_ISL_1964977, EPI_ISL_1964978, EPI_ISL_1964979, EPI_ISL_1964981, EPI_ISL_1964982, EPI_ISL_1964983, EPI_ISL_1964985, EPI_ISL_1964986, EPI_ISL_1964988, EPI_ISL_1964990, EPI_ISL_1964991, EPI_ISL_1964992, EPI_ISL_1964993, EPI_ISL_1964994, EPI_ISL_1964995, EPI_ISL_1964997, EPI_ISL_1964998, EPI_ISL_1964999, EPI_ISL_1965000, EPI_ISL_1965001, EPI_ISL_1965002, EPI_ISL_1965003, EPI_ISL_1965004, EPI_ISL_1965005, EPI_ISL_1965006, EPI_ISL_1965007, EPI_ISL_1965008, EPI_ISL_1965009, EPI_ISL_1965010, EPI_ISL_1965011, EPI_ISL_1965012, EPI_ISL_1965013, EPI_ISL_1965015, EPI_ISL_1965016, EPI_ISL_1965017, EPI_ISL_1965019, EPI_ISL_1965020, EPI_ISL_1965021, EPI_ISL_1965022, EPI_ISL_1965023, EPI_ISL_1965024, EPI_ISL_1965025, EPI_ISL_1965026, EPI_ISL_1965027, EPI_ISL_1965028, EPI_ISL_1965029, EPI_ISL_1965031, EPI_ISL_1965032, EPI_ISL_1965033, EPI_ISL_1965039, EPI_ISL_1965041, EPI_ISL_1965042, EPI_ISL_1965045, EPI_ISL_1965046, EPI_ISL_1965047, EPI_ISL_1965048, EPI_ISL_1965049, EPI_ISL_1965050, EPI_ISL_1965051, EPI_ISL_1965052, EPI_ISL_1965053, EPI_ISL_1965054, EPI_ISL_1965055, EPI_ISL_1965056, EPI_ISL_1965057, EPI_ISL_1965058, EPI_ISL_1965059, EPI_ISL_1965060, EPI_ISL_1965061, EPI_ISL_1965062, EPI_ISL_1965064, EPI_ISL_1965065, EPI_ISL_1965066, EPI_ISL_1965067, EPI_ISL_1965071, EPI_ISL_1965072, EPI_ISL_1965073, EPI_ISL_1965074, EPI_ISL_1965076, EPI_ISL_1965077, EPI_ISL_1965078, EPI_ISL_1965079, EPI_ISL_1965080, EPI_ISL_1965081, EPI_ISL_1965082, EPI_ISL_1965083, EPI_ISL_1965084, EPI_ISL_1965085, EPI_ISL_1965086, EPI_ISL_1965087, EPI_ISL_1965088, EPI_ISL_1965089, EPI_ISL_1965090, EPI_ISL_1965091, EPI_ISL_1965092, EPI_ISL_1965093, EPI_ISL_1965094, EPI_ISL_1965095, EPI_ISL_1965096, EPI_ISL_1965097, EPI_ISL_1965098, EPI_ISL_1965099, EPI_ISL_1965100, EPI_ISL_1965101, EPI_ISL_1965102, EPI_ISL_1965103, EPI_ISL_1965104, EPI_ISL_1965105, EPI_ISL_1965106, EPI_ISL_1965107, EPI_ISL_1965108, EPI_ISL_1965109, EPI_ISL_1965110, EPI_ISL_1965111, EPI_ISL_1965112, EPI_ISL_1965113, EPI_ISL_1965114, EPI_ISL_1965115, EPI_ISL_1965116, EPI_ISL_1965117, EPI_ISL_1965118 | | | |
| see above | Illinois Department of Public Health | Illinois Department of Public Health - Chicago Lab | Vineet K. Dhiman, Ira Heimler, Joel Price |
| EPI_ISL_1965723, EPI_ISL_1965724, EPI_ISL_1965725, EPI_ISL_1965726, EPI_ISL_1965727, EPI_ISL_1965729, EPI_ISL_1965730, EPI_ISL_1965731, EPI_ISL_1965732, EPI_ISL_1965733, EPI_ISL_1965734, EPI_ISL_1965735, EPI_ISL_1965736, EPI_ISL_1965737, EPI_ISL_1965738, EPI_ISL_1965739, EPI_ISL_1965740, EPI_ISL_1965742, EPI_ISL_1965743, EPI_ISL_1965744, EPI_ISL_1965745, EPI_ISL_1965746, EPI_ISL_1965747, EPI_ISL_1965748, EPI_ISL_1965749, EPI_ISL_1965750, EPI_ISL_1965751, EPI_ISL_1965752, EPI_ISL_1965753, EPI_ISL_1965754, EPI_ISL_1965755, EPI_ISL_1965756, EPI_ISL_1965757, EPI_ISL_1965758, EPI_ISL_1965759, EPI_ISL_1965760, EPI_ISL_1965761, EPI_ISL_1965762, EPI_ISL_1965763, EPI_ISL_1965764, EPI_ISL_1965765, EPI_ISL_1965766, EPI_ISL_1965767, EPI_ISL_1965768, EPI_ISL_1965769, EPI_ISL_1965770, EPI_ISL_1965773, EPI_ISL_1965774, EPI_ISL_1965776, EPI_ISL_1965777, EPI_ISL_1965778, EPI_ISL_1965779, EPI_ISL_1965780, EPI_ISL_1965781, EPI_ISL_1965782, EPI_ISL_1965783, EPI_ISL_1965784, EPI_ISL_1965785, EPI_ISL_1965786, EPI_ISL_1965787, EPI_ISL_1965788, EPI_ISL_1965790, EPI_ISL_1965791, EPI_ISL_1965793, EPI_ISL_1965794, EPI_ISL_1965798, EPI_ISL_1965800, EPI_ISL_1965802, EPI_ISL_1965803, EPI_ISL_1965804, EPI_ISL_1965805, EPI_ISL_1965807, EPI_ISL_1965811, EPI_ISL_1965812, EPI_ISL_1965814, EPI_ISL_1965815, EPI_ISL_1965816, EPI_ISL_1965817, EPI_ISL_1965818, EPI_ISL_1965819, EPI_ISL_1965820, EPI_ISL_1965821, EPI_ISL_1965822, EPI_ISL_1965823, EPI_ISL_1965824, EPI_ISL_1965825, EPI_ISL_1965826, EPI_ISL_1965827, EPI_ISL_1965828, EPI_ISL_1965829, EPI_ISL_1965830, EPI_ISL_1965831, EPI_ISL_1965832, EPI_ISL_1965833, EPI_ISL_1965834, EPI_ISL_1965835, EPI_ISL_1965836, EPI_ISL_1965837, EPI_ISL_1965838, EPI_ISL_1965839, EPI_ISL_1965841, EPI_ISL_1965842, EPI_ISL_1965843, EPI_ISL_1965844, EPI_ISL_1965845, EPI_ISL_1965846, EPI_ISL_1965847, EPI_ISL_1965849, EPI_ISL_1965850, EPI_ISL_1965851, EPI_ISL_1965854, EPI_ISL_1965855, EPI_ISL_1965856, EPI_ISL_1965857, EPI_ISL_1965858, EPI_ISL_1965859, EPI_ISL_1965860, EPI_ISL_1965861, EPI_ISL_1965862, EPI_ISL_1965863, EPI_ISL_1965864, EPI_ISL_1965865, EPI_ISL_1965866, EPI_ISL_1965867, EPI_ISL_1965868, EPI_ISL_1965869, EPI_ISL_1965871, EPI_ISL_1965872, EPI_ISL_1965873, EPI_ISL_1965875, EPI_ISL_1965876, EPI_ISL_1965877, EPI_ISL_1965879, EPI_ISL_1965881, EPI_ISL_1965882, EPI_ISL_1965883, EPI_ISL_1965885, EPI_ISL_1965886, EPI_ISL_1965887, EPI_ISL_1965888, EPI_ISL_1965891, EPI_ISL_1965893, EPI_ISL_1965894, EPI_ISL_1965896, EPI_ISL_1965897, EPI_ISL_1965898, EPI_ISL_1965899, EPI_ISL_1965900, EPI_ISL_1965901, EPI_ISL_1965902, EPI_ISL_1965903, EPI_ISL_1965904, EPI_ISL_1965906, EPI_ISL_1965907, EPI_ISL_1965908, EPI_ISL_1965910, EPI_ISL_1965911, EPI_ISL_1965913, EPI_ISL_1965914, EPI_ISL_1965915, EPI_ISL_1965916, EPI_ISL_1965918, EPI_ISL_1965921, EPI_ISL_1965922, EPI_ISL_1965923, EPI_ISL_1965924, EPI_ISL_1965926, EPI_ISL_1965927, EPI_ISL_1965928, EPI_ISL_1965929, EPI_ISL_1965930, EPI_ISL_1965932, EPI_ISL_1965933, EPI_ISL_1965934, EPI_ISL_1965935, EPI_ISL_1965936, EPI_ISL_1965937, EPI_ISL_1965938, EPI_ISL_1965939, EPI_ISL_1965940, EPI_ISL_1965942, EPI_ISL_1965943, EPI_ISL_1965944, EPI_ISL_1965945, EPI_ISL_1965946, EPI_ISL_1965947, EPI_ISL_1965948, EPI_ISL_1965949, EPI_ISL_1965950, EPI_ISL_1965951, EPI_ISL_1965952, EPI_ISL_1965953, EPI_ISL_1965954, EPI_ISL_1965956, EPI_ISL_1965957, EPI_ISL_1965958, EPI_ISL_1965960, EPI_ISL_1965961, EPI_ISL_1965963, EPI_ISL_1965965, EPI_ISL_1965966, EPI_ISL_1965967, EPI_ISL_1965968, EPI_ISL_1965969, EPI_ISL_1965970, EPI_ISL_1965971, EPI_ISL_1965972, EPI_ISL_1965973, EPI_ISL_1965974, EPI_ISL_1965975, EPI_ISL_1965976, EPI_ISL_1965977, EPI_ISL_1965978, EPI_ISL_1965980, EPI_ISL_1965981, EPI_ISL_1965982, EPI_ISL_1965983, EPI_ISL_1965984, EPI_ISL_1965985, EPI_ISL_1965986, EPI_ISL_1965987, EPI_ISL_1965988, EPI_ISL_1965989, EPI_ISL_1965990, EPI_ISL_1965991, EPI_ISL_1965992, EPI_ISL_1965994, EPI_ISL_1965995, EPI_ISL_1965996, EPI_ISL_1965997, EPI_ISL_1965998, EPI_ISL_1965999, EPI_ISL_1966001, EPI_ISL_1966003, EPI_ISL_1966005, EPI_ISL_1966006, EPI_ISL_1966007, EPI_ISL_1966008, EPI_ISL_1966011, EPI_ISL_1966012, EPI_ISL_1966013, EPI_ISL_1966014, EPI_ISL_1966015, EPI_ISL_1966016, EPI_ISL_1966017, EPI_ISL_1966018, EPI_ISL_1966019, EPI_ISL_1966020, EPI_ISL_1966021, EPI_ISL_1966022, EPI_ISL_1966024, EPI_ISL_1966025, EPI_ISL_1966026, EPI_ISL_1966027, EPI_ISL_1966028, EPI_ISL_1966029, EPI_ISL_1966030, EPI_ISL_1966031, EPI_ISL_1966032, EPI_ISL_1966034, EPI_ISL_1966035, EPI_ISL_1966036, EPI_ISL_1966037, EPI_ISL_1966038, EPI_ISL_1966039, EPI_ISL_1966040, EPI_ISL_1966042, EPI_ISL_1966044, EPI_ISL_1966046, EPI_ISL_1966047, EPI_ISL_1966049, EPI_ISL_1966050, EPI_ISL_1966052, EPI_ISL_1966053, EPI_ISL_1966056, EPI_ISL_1966058 | | | |
| see above | Clinical Microbiology Laboratory, NewYork Presbyterian Hospital/Columbia University Irving Medical Center | Uhlemann Laboratory, Columbia University Irving Medical Center | Medini K. Annavajhala, Anne Kelley, Maya Tagliavia, Anne-Catrin Uhlemann |
| EPI_ISL_1967476, EPI_ISL_1967478, EPI_ISL_1967479, EPI_ISL_1967481, | Wyoming Public Health Laboratory | Wyoming Public Health Laboratory | Jim Mildenberger, Taylor Fearing, Channing Weber, Ashley Norberg, Chayse Rowley, Marley Goetz, Brian Dominguez, Elliot Thomasson, Brittany Oher, Sam Britz, Cari Sloma, and Rob Christensen |

| | | | |
|---|---|---|---|
| EPI_ISL_1967488 | | | |
| EPI_ISL_1967491, EPI_ISL_1967492, EPI_ISL_1967493, EPI_ISL_1967494, EPI_ISL_1967495, EPI_ISL_1967496, EPI_ISL_1967497, EPI_ISL_1967498, EPI_ISL_1967499, EPI_ISL_1967500, EPI_ISL_1967501, EPI_ISL_1967502, EPI_ISL_1967503, EPI_ISL_1967504, EPI_ISL_1967505, EPI_ISL_1967506, EPI_ISL_1967507, EPI_ISL_1967508, EPI_ISL_1967509, EPI_ISL_1967510, EPI_ISL_1967511, EPI_ISL_1967512, EPI_ISL_1967513, EPI_ISL_1967514, EPI_ISL_1967515, EPI_ISL_1967516, EPI_ISL_1967517, EPI_ISL_1967518, EPI_ISL_1967519, EPI_ISL_1967520, EPI_ISL_1967521, EPI_ISL_1967522, EPI_ISL_1967523, EPI_ISL_1967524, EPI_ISL_1967525, EPI_ISL_1967526, EPI_ISL_1967527, EPI_ISL_1967528, EPI_ISL_1967529, EPI_ISL_1967530, EPI_ISL_1967531, EPI_ISL_1967532, EPI_ISL_1967533, EPI_ISL_1967534, EPI_ISL_1967535, EPI_ISL_1967536, EPI_ISL_1967537, EPI_ISL_1967538, EPI_ISL_1967539, EPI_ISL_1967540, EPI_ISL_1967541, EPI_ISL_1967542, EPI_ISL_1967543, EPI_ISL_1967544, EPI_ISL_1967545, EPI_ISL_1967546, EPI_ISL_1967547, EPI_ISL_1967548, EPI_ISL_1967549, EPI_ISL_1967550, EPI_ISL_1967551, EPI_ISL_1967552, EPI_ISL_1967553, EPI_ISL_1967554, EPI_ISL_1967555, EPI_ISL_1967556, EPI_ISL_1967557, EPI_ISL_1967558, EPI_ISL_1967559, EPI_ISL_1967560, EPI_ISL_1967561, EPI_ISL_1967562, EPI_ISL_1967563, EPI_ISL_1967564, EPI_ISL_1967565, EPI_ISL_1967566, EPI_ISL_1967567, EPI_ISL_1967568, EPI_ISL_1967569, EPI_ISL_1967570, EPI_ISL_1967571, EPI_ISL_1967572, EPI_ISL_1967573, EPI_ISL_1967574, EPI_ISL_1967575, EPI_ISL_1967576, EPI_ISL_1967577, EPI_ISL_1967578, EPI_ISL_1967579, EPI_ISL_1967580, EPI_ISL_1967581, EPI_ISL_1967582, EPI_ISL_1967583, EPI_ISL_1967584, EPI_ISL_1967585, EPI_ISL_1967586, EPI_ISL_1967587, EPI_ISL_1967588, EPI_ISL_1967589, EPI_ISL_1967590, EPI_ISL_1967591, EPI_ISL_1967592, EPI_ISL_1967593, EPI_ISL_1967594, EPI_ISL_1967595, EPI_ISL_1967596, EPI_ISL_1967597, EPI_ISL_1967598, EPI_ISL_1967599, EPI_ISL_1967600, EPI_ISL_1967601, EPI_ISL_1967602, EPI_ISL_1967603, EPI_ISL_1967604, EPI_ISL_1967605, EPI_ISL_1967606, EPI_ISL_1967607, EPI_ISL_1967608, EPI_ISL_1967609, EPI_ISL_1967610, EPI_ISL_1967611, EPI_ISL_1967612, EPI_ISL_1967613, EPI_ISL_1967614, EPI_ISL_1967615, EPI_ISL_1967616, EPI_ISL_1967617, EPI_ISL_1967618, EPI_ISL_1967619, EPI_ISL_1967620, EPI_ISL_1967621, EPI_ISL_1967622, EPI_ISL_1967623, EPI_ISL_1967624, EPI_ISL_1967625, EPI_ISL_1967626, EPI_ISL_1967627, EPI_ISL_1967628, EPI_ISL_1967629, EPI_ISL_1967630, EPI_ISL_1967631, EPI_ISL_1967632, EPI_ISL_1967633, EPI_ISL_1967634, EPI_ISL_1967635, EPI_ISL_1967636, EPI_ISL_1967637, EPI_ISL_1967638, EPI_ISL_1967639, EPI_ISL_1967640, EPI_ISL_1967641, EPI_ISL_1967642, EPI_ISL_1967643, EPI_ISL_1967644, EPI_ISL_1967645, EPI_ISL_1967646, EPI_ISL_1967647, EPI_ISL_1967648, EPI_ISL_1967649, EPI_ISL_1967650, EPI_ISL_1967651, EPI_ISL_1967652, EPI_ISL_1967653, EPI_ISL_1967654, EPI_ISL_1967655, EPI_ISL_1967656, EPI_ISL_1967657, EPI_ISL_1967658, EPI_ISL_1967659, EPI_ISL_1967660, EPI_ISL_1967661 | | | |
| see above | Maryland Genomics, Institute for Genome Sciences, University of Maryland School of Medicine | Maryland Genomics, Institute for Genome Sciences, University of Maryland School of Medicine | Tallon, Luke J; Sadzewicz, Lisa D; Humphrys, Mike; Ott, Sandra; Roussey, Holly; Mehta, Aditya; Vavikolanu, Kranthi; Fraser, Claire M; Ravel, Jacques |
| EPI_ISL_1967662, EPI_ISL_1967663, EPI_ISL_1967664, EPI_ISL_1967665, EPI_ISL_1967666, EPI_ISL_1967667, EPI_ISL_1967668, EPI_ISL_1967669, EPI_ISL_1967670, EPI_ISL_1967671, EPI_ISL_1967672, EPI_ISL_1967673, EPI_ISL_1967674, EPI_ISL_1967675, EPI_ISL_1967676, EPI_ISL_1967677, EPI_ISL_1967678, EPI_ISL_1967679, EPI_ISL_1967680, EPI_ISL_1967681, EPI_ISL_1967682, EPI_ISL_1967683, EPI_ISL_1967684 | | | |
| see above | Weill Cornell Medicine | New York Genome Center | Michael Zody, Andre Corvelo, Dayna M. Oschwald, Samantha Fennessey, Tom Maniatis, Melissa Cushing, Olivier Elemento, Margaret Elizabeth Ross, Chris Mason, Priya Velu, Hanna Rennert, Arryn Craney, Lars F Westblade |
| EPI_ISL_1967685, EPI_ISL_1967686, EPI_ISL_1967687, EPI_ISL_1967688 | Stony Brook Medicine | New York Genome Center | Michael Zody, Andre Corvelo, Dayna M. Oschwald, Samantha Fennessey, Tom Maniatis , Sarah A. Halbohn, Alaa B. Salaheldin, Sarah A. Malik, Josue M. Flores Acuna, Dimitri Gnatenko, Silvia G. Spitzer, Margaret M. McGovern, Eric D. Spitzer, Richard R. Kew, Kenneth R. Shroyer, Jiang Chen |
| EPI_ISL_1967689, EPI_ISL_1967690, EPI_ISL_1967691, EPI_ISL_1967692, EPI_ISL_1967693, EPI_ISL_1967694, EPI_ISL_1967695 | Stony Brook Medicine | New York Genome Center | Michael Zody, Andre Corvelo, Dayna M. Oschwald, Samantha Fennessey, Tom Maniatis , Sarah A. Halbohn, Alaa B. Salaheldin, Sarah A. Malik, Josue M. Flores Acuna, Dimitri Gnatenko, Silvia G. Spitzer, Margaret M. McGovern, Eric D. Spitzer, Richard R. Kew, Kenneth R. Shroyer, Jiang Chen, Susan V. Donelan |
| EPI_ISL_1967708, EPI_ISL_1967709, EPI_ISL_1967710, EPI_ISL_1967711, EPI_ISL_1967712, EPI_ISL_1967713, EPI_ISL_1967714, EPI_ISL_1967715, EPI_ISL_1967716, EPI_ISL_1967717, EPI_ISL_1967718, EPI_ISL_1967719, EPI_ISL_1967720, EPI_ISL_1967721, EPI_ISL_1967722, EPI_ISL_1967723, EPI_ISL_1967724 | | | |
| see above | Oregon State Public Health Laboratory | Oregon State Public Health Laboratory | Rafia Razzaque, Eugene Yeboah, Vanda Makris, Laura Tsaknaridis, John Fontana and Shane Sevey |
| EPI_ISL_1967903, EPI_ISL_1967904, EPI_ISL_1967905, EPI_ISL_1967906, EPI_ISL_1967907, EPI_ISL_1967908, EPI_ISL_1967909, EPI_ISL_1967910, EPI_ISL_1967911, EPI_ISL_1967912, EPI_ISL_1967913, EPI_ISL_1967914, EPI_ISL_1967915, EPI_ISL_1967916, EPI_ISL_1967917, EPI_ISL_1967918, EPI_ISL_1967919, EPI_ISL_1967920, EPI_ISL_1967921, EPI_ISL_1967922, EPI_ISL_1967923, EPI_ISL_1967924, EPI_ISL_1967925, EPI_ISL_1967926, EPI_ISL_1967927, EPI_ISL_1967928, EPI_ISL_1967929, EPI_ISL_1967930, EPI_ISL_1967931, EPI_ISL_1967932, EPI_ISL_1967933, EPI_ISL_1967934, EPI_ISL_1967935, EPI_ISL_1967936, EPI_ISL_1967937, EPI_ISL_1967938, EPI_ISL_1967939, EPI_ISL_1967940, EPI_ISL_1967941 | | | |
| see above | Alaska State Virology Laboratory | Alaska State Virology Laboratory | Stephanie DeRonde, Elva House, Jacob Zidek, Lisa Smith, Ph.D., Jack Chen, Ph.D. |
| EPI_ISL_1967942, EPI_ISL_1967943, EPI_ISL_1967944, EPI_ISL_1967945, EPI_ISL_1967946, EPI_ISL_1967947, EPI_ISL_1967948, EPI_ISL_1967949, EPI_ISL_1967950, EPI_ISL_1967951, EPI_ISL_1967952, EPI_ISL_1967953, EPI_ISL_1967954, EPI_ISL_1967955, EPI_ISL_1967956, EPI_ISL_1967957, EPI_ISL_1967958, EPI_ISL_1967959, EPI_ISL_1967960, EPI_ISL_1967961, EPI_ISL_1967962, EPI_ISL_1967963, EPI_ISL_1967964, EPI_ISL_1967965, EPI_ISL_1967966, EPI_ISL_1967967, EPI_ISL_1967968, EPI_ISL_1967969, EPI_ISL_1967970, EPI_ISL_1967971, EPI_ISL_1967972, EPI_ISL_1967973, EPI_ISL_1967974, EPI_ISL_1967975, EPI_ISL_1967976, EPI_ISL_1967977, EPI_ISL_1967978, EPI_ISL_1967979, EPI_ISL_1967980, EPI_ISL_1967981, EPI_ISL_1967982, EPI_ISL_1967983, EPI_ISL_1967984, EPI_ISL_1967985, EPI_ISL_1967986, EPI_ISL_1967987, EPI_ISL_1967988, EPI_ISL_1967989, EPI_ISL_1967990, EPI_ISL_1967991, EPI_ISL_1967992, EPI_ISL_1967993, EPI_ISL_1967994, EPI_ISL_1967995, EPI_ISL_1967996, EPI_ISL_1967997, EPI_ISL_1967998, EPI_ISL_1967999, EPI_ISL_1968000, EPI_ISL_1968001, EPI_ISL_1968002, EPI_ISL_1968003, EPI_ISL_1968004, EPI_ISL_1968005, EPI_ISL_1968006, EPI_ISL_1968007, EPI_ISL_1968008, EPI_ISL_1968009, EPI_ISL_1968010, EPI_ISL_1968011, EPI_ISL_1968012, EPI_ISL_1968013, EPI_ISL_1968014, EPI_ISL_1968015, EPI_ISL_1968016, EPI_ISL_1968017, EPI_ISL_1968018, EPI_ISL_1968019, EPI_ISL_1968020, EPI_ISL_1968021, EPI_ISL_1968022, EPI_ISL_1968023, EPI_ISL_1968024, EPI_ISL_1968025, EPI_ISL_1968027, EPI_ISL_1968028, EPI_ISL_1968029, EPI_ISL_1968030, EPI_ISL_1968031, EPI_ISL_1968032, EPI_ISL_1968033, EPI_ISL_1968034, EPI_ISL_1968035, EPI_ISL_1968036, EPI_ISL_1968037, EPI_ISL_1968038, EPI_ISL_1968039, EPI_ISL_1968040, EPI_ISL_1968041, EPI_ISL_1968042, EPI_ISL_1968043, EPI_ISL_1968044, EPI_ISL_1968045, EPI_ISL_1968046, EPI_ISL_1968047, EPI_ISL_1968048, EPI_ISL_1968049, EPI_ISL_1968050, EPI_ISL_1968051, EPI_ISL_1968052, EPI_ISL_1968053, EPI_ISL_1968054, EPI_ISL_1968055, EPI_ISL_1968056, EPI_ISL_1968057, EPI_ISL_1968058, EPI_ISL_1968059, EPI_ISL_1968060, EPI_ISL_1968061, EPI_ISL_1968062, EPI_ISL_1968063, EPI_ISL_1968064, EPI_ISL_1968065, EPI_ISL_1968066, EPI_ISL_1968067, EPI_ISL_1968068, EPI_ISL_1968069, EPI_ISL_1968070, EPI_ISL_1968071, EPI_ISL_1968072, EPI_ISL_1968073, EPI_ISL_1968074, EPI_ISL_1968075, EPI_ISL_1968076, EPI_ISL_1968077, EPI_ISL_1968078, EPI_ISL_1968079, EPI_ISL_1968080, EPI_ISL_1968081, EPI_ISL_1968082, EPI_ISL_1968083, EPI_ISL_1968084, EPI_ISL_1968085, EPI_ISL_1968086, EPI_ISL_1968087, EPI_ISL_1968088, EPI_ISL_1968089, EPI_ISL_1968090, EPI_ISL_1968091, EPI_ISL_1968092, EPI_ISL_1968093, EPI_ISL_1968094, EPI_ISL_1968095, EPI_ISL_1968096, EPI_ISL_1968097, EPI_ISL_1968098, EPI_ISL_1968099, EPI_ISL_1968100, EPI_ISL_1968101, EPI_ISL_1968102, EPI_ISL_1968103, EPI_ISL_1968104, EPI_ISL_1968105, EPI_ISL_1968106, EPI_ISL_1968107, EPI_ISL_1968108, EPI_ISL_1968109, EPI_ISL_1968110, EPI_ISL_1968111, EPI_ISL_1968112, EPI_ISL_1968113, EPI_ISL_1968114, EPI_ISL_1968115, EPI_ISL_1968116, EPI_ISL_1968117, EPI_ISL_1968118, EPI_ISL_1968119, EPI_ISL_1968120, EPI_ISL_1968121, EPI_ISL_1968122, EPI_ISL_1968123, EPI_ISL_1968124, EPI_ISL_1968125, EPI_ISL_1968126, EPI_ISL_1968127, EPI_ISL_1968128, EPI_ISL_1968129, EPI_ISL_1968130, EPI_ISL_1968131, EPI_ISL_1968132, EPI_ISL_1968133, EPI_ISL_1968134, EPI_ISL_1968135, EPI_ISL_1968136, EPI_ISL_1968137, EPI_ISL_1968138, EPI_ISL_1968139, EPI_ISL_1968140, EPI_ISL_1968141, EPI_ISL_1968142, EPI_ISL_1968143, EPI_ISL_1968144, EPI_ISL_1968145, EPI_ISL_1968146, EPI_ISL_1968147, EPI_ISL_1968148, EPI_ISL_1968149, EPI_ISL_1968150, EPI_ISL_1968151, EPI_ISL_1968152, EPI_ISL_1968153, EPI_ISL_1968154, EPI_ISL_1968155, EPI_ISL_1968156, EPI_ISL_1968157, EPI_ISL_1968158, EPI_ISL_1968159, EPI_ISL_1968160, EPI_ISL_1968161, EPI_ISL_1968162, EPI_ISL_1968163, EPI_ISL_1968164, EPI_ISL_1968165, EPI_ISL_1968166, EPI_ISL_1968167, EPI_ISL_1968168, EPI_ISL_1968169, EPI_ISL_1968170, EPI_ISL_1968171, EPI_ISL_1968172, EPI_ISL_1968173, EPI_ISL_1968174, EPI_ISL_1968175, EPI_ISL_1968176, EPI_ISL_1968177, EPI_ISL_1968178, EPI_ISL_1968179, EPI_ISL_1968180, EPI_ISL_1968181, EPI_ISL_1968182, EPI_ISL_1968183, EPI_ISL_1968184, EPI_ISL_1968185, EPI_ISL_1968186, EPI_ISL_1968187, EPI_ISL_1968188, EPI_ISL_1968189, EPI_ISL_1968190, EPI_ISL_1968191, EPI_ISL_1968192, EPI_ISL_1968193, EPI_ISL_1968194, EPI_ISL_1968195, EPI_ISL_1968196, EPI_ISL_1968197, EPI_ISL_1968198, EPI_ISL_1968199, EPI_ISL_1968200, EPI_ISL_1968201, EPI_ISL_1968202, EPI_ISL_1968203, EPI_ISL_1968204, EPI_ISL_1968205, EPI_ISL_1968206, EPI_ISL_1968207, EPI_ISL_1968208, EPI_ISL_1968209, EPI_ISL_1968210, EPI_ISL_1968211, EPI_ISL_1968212, EPI_ISL_1968213, EPI_ISL_1968214, EPI_ISL_1968215, EPI_ISL_1968216, EPI_ISL_1968217, EPI_ISL_1968218, EPI_ISL_1968219, EPI_ISL_1968220, EPI_ISL_1968221, EPI_ISL_1968222, EPI_ISL_1968223, EPI_ISL_1968224, EPI_ISL_1968225, EPI_ISL_1968226, EPI_ISL_1968227, EPI_ISL_1968228, EPI_ISL_1968229, EPI_ISL_1968230, EPI_ISL_1968231, EPI_ISL_1968232, EPI_ISL_1968233, EPI_ISL_1968234, EPI_ISL_1968235, EPI_ISL_1968236, EPI_ISL_1968237, EPI_ISL_1968238, EPI_ISL_1968239, EPI_ISL_1968240, EPI_ISL_1968241, EPI_ISL_1968242, EPI_ISL_1968243, EPI_ISL_1968244, EPI_ISL_1968245, EPI_ISL_1968246, EPI_ISL_1968247, EPI_ISL_1968248, EPI_ISL_1968249, EPI_ISL_1968250, EPI_ISL_1968251, EPI_ISL_1968252, EPI_ISL_1968253, EPI_ISL_1968254, EPI_ISL_1968255, EPI_ISL_1968256, EPI_ISL_1968257, EPI_ISL_1968258, EPI_ISL_1968259, EPI_ISL_1968260, EPI_ISL_1968262, EPI_ISL_1968263, EPI_ISL_1968264, EPI_ISL_1968265, EPI_ISL_1968266, EPI_ISL_1968267, EPI_ISL_1968268, EPI_ISL_1968269, EPI_ISL_1968270, EPI_ISL_1968271, EPI_ISL_1968272, EPI_ISL_1968273, EPI_ISL_1968274, EPI_ISL_1968275, EPI_ISL_1968276, EPI_ISL_1968277, EPI_ISL_1968278, EPI_ISL_1968279, EPI_ISL_1968280, EPI_ISL_1968281, EPI_ISL_1968282, EPI_ISL_1968283, EPI_ISL_1968284, EPI_ISL_1968285, EPI_ISL_1968286, EPI_ISL_1968287, EPI_ISL_1968288, EPI_ISL_1968289, EPI_ISL_1968290, EPI_ISL_1968291, EPI_ISL_1968292, EPI_ISL_1968293, EPI_ISL_1968294, EPI_ISL_1968295, EPI_ISL_1968296, EPI_ISL_1968297, EPI_ISL_1968298, EPI_ISL_1968299, EPI_ISL_1968300, EPI_ISL_1968301, EPI_ISL_1968302, EPI_ISL_1968303, EPI_ISL_1968304, EPI_ISL_1968305, EPI_ISL_1968306, EPI_ISL_1968307, EPI_ISL_1968308, EPI_ISL_1968309, EPI_ISL_1968310, EPI_ISL_1968311, EPI_ISL_1968312, EPI_ISL_1968313, EPI_ISL_1968314, EPI_ISL_1968315, EPI_ISL_1968316, EPI_ISL_1968317, EPI_ISL_1968318, EPI_ISL_1968319, EPI_ISL_1968320, EPI_ISL_1968321, EPI_ISL_1968322, EPI_ISL_1968323, EPI_ISL_1968324, EPI_ISL_1968325, EPI_ISL_1968326, EPI_ISL_1968327, EPI_ISL_1968328, EPI_ISL_1968329, EPI_ISL_1968330, EPI_ISL_1968331, EPI_ISL_1968332, EPI_ISL_1968333, EPI_ISL_1968334, EPI_ISL_1968335, EPI_ISL_1968336, EPI_ISL_1968337, EPI_ISL_1968338, EPI_ISL_1968339, EPI_ISL_1968340, EPI_ISL_1968341, EPI_ISL_1968342, EPI_ISL_1968343, EPI_ISL_1968344, EPI_ISL_1968345, EPI_ISL_1968346, EPI_ISL_1968347, EPI_ISL_1968348, EPI_ISL_1968349, EPI_ISL_1968350, EPI_ISL_1968351, EPI_ISL_1968352, EPI_ISL_1968353, EPI_ISL_1968354, EPI_ISL_1968355, EPI_ISL_1968356, EPI_ISL_1968357, EPI_ISL_1968358, EPI_ISL_1968359, EPI_ISL_1968360, EPI_ISL_1968361, EPI_ISL_1968363, EPI_ISL_1968364, EPI_ISL_1968365, EPI_ISL_1968366, EPI_ISL_1968367, EPI_ISL_1968368, EPI_ISL_1968369, EPI_ISL_1968370, EPI_ISL_1968371, EPI_ISL_1968372, EPI_ISL_1968373, EPI_ISL_1968374, EPI_ISL_1968375, EPI_ISL_1968376, EPI_ISL_1968377, EPI_ISL_1968378, EPI_ISL_1968379, EPI_ISL_1968380, EPI_ISL_1968381, EPI_ISL_1968382, EPI_ISL_1968383, EPI_ISL_1968384, EPI_ISL_1968385, EPI_ISL_1968386, EPI_ISL_1968387, EPI_ISL_1968388, EPI_ISL_1968389, EPI_ISL_1968390, EPI_ISL_1968391, EPI_ISL_1968392, EPI_ISL_1968393, EPI_ISL_1968394, EPI_ISL_1968395, EPI_ISL_1968396, EPI_ISL_1968397, EPI_ISL_1968398, EPI_ISL_1968399, EPI_ISL_1968400, EPI_ISL_1968401, EPI_ISL_1968402, EPI_ISL_1968403, EPI_ISL_1968404, EPI_ISL_1968405, EPI_ISL_1968406, EPI_ISL_1968407, EPI_ISL_1968408, EPI_ISL_1968409, EPI_ISL_1968410, EPI_ISL_1968411, EPI_ISL_1968412, EPI_ISL_1968413, EPI_ISL_1968414, EPI_ISL_1968415, EPI_ISL_1968416, EPI_ISL_1968417, EPI_ISL_1968418, EPI_ISL_1968419, EPI_ISL_1968420, EPI_ISL_1968421, EPI_ISL_1968422, EPI_ISL_1968423, EPI_ISL_1968424, EPI_ISL_1968425, EPI_ISL_1968426, EPI_ISL_1968427, EPI_ISL_1968428, EPI_ISL_1968429, EPI_ISL_1968430, EPI_ISL_1968431, EPI_ISL_1968432, EPI_ISL_1968433, EPI_ISL_1968434, EPI_ISL_1968435, EPI_ISL_1968436, EPI_ISL_1968437, EPI_ISL_1968438, EPI_ISL_1968439, EPI_ISL_1968440, EPI_ISL_1968441, EPI_ISL_1968442, EPI_ISL_1968443, EPI_ISL_1968444, EPI_ISL_1968445, EPI_ISL_1968446, EPI_ISL_1968447, EPI_ISL_1968448, EPI_ISL_1968449, EPI_ISL_1968450, EPI_ISL_1968451, EPI_ISL_1968452, EPI_ISL_1968453, EPI_ISL_1968454, EPI_ISL_1968455, EPI_ISL_1968456, EPI_ISL_1968457, EPI_ISL_1968458, EPI_ISL_1968459, EPI_ISL_1968460, EPI_ISL_1968461, EPI_ISL_1968462, EPI_ISL_1968463, EPI_ISL_1968464, EPI_ISL_1968465, EPI_ISL_1968466, EPI_ISL_1968467, EPI_ISL_1968468, EPI_ISL_1968469, EPI_ISL_1968470, EPI_ISL_1968471, EPI_ISL_1968472, EPI_ISL_1968473, EPI_ISL_1968474, EPI_ISL_1968475, EPI_ISL_1968476, EPI_ISL_1968477, EPI_ISL_1968478, EPI_ISL_1968479, EPI_ISL_1968480, EPI_ISL_1968481, EPI_ISL_1968482, EPI_ISL_1968483, EPI_ISL_1968484, EPI_ISL_1968485, EPI_ISL_1968486, EPI_ISL_1968487, EPI_ISL_1968488, EPI_ISL_1968489, EPI_ISL_1968490, EPI_ISL_1968491, EPI_ISL_1968492, EPI_ISL_1968493, EPI_ISL_1968494, EPI_ISL_1968495, EPI_ISL_1968496, EPI_ISL_1968497, EPI_ISL_1968498, EPI_ISL_1968499, EPI_ISL_1968500, EPI_ISL_1968501, EPI_ISL_1968502, EPI_ISL_1968503, EPI_ISL_1968504, EPI_ISL_1968505, EPI_ISL_1968506, EPI_ISL_1968507, EPI_ISL_1968508, EPI_ISL_1968509, EPI_ISL_1968510, EPI_ISL_1968511, EPI_ISL_1968512, EPI_ISL_1968513, EPI_ISL_1968514, EPI_ISL_1968515, EPI_ISL_1968516, EPI_ISL_1968517, EPI_ISL_1968518, EPI_ISL_1968519, EPI_ISL_1968520, EPI_ISL_1968521, EPI_ISL_1968522, EPI_ISL_1968523, EPI_ISL_1968524, EPI_ISL_1968525, EPI_ISL_1968526, EPI_ISL_1968527, EPI_ISL_1968528, EPI_ISL_1968529, EPI_ISL_1968530, EPI_ISL_1968531, EPI_ISL_1968532, EPI_ISL_1968533, EPI_ISL_1968534, EPI_ISL_1968535, EPI_ISL_1968536, EPI_ISL_1968537, EPI_ISL_1968538, EPI_ISL_1968539, EPI_ISL_1968540, EPI_ISL_1968541, EPI_ISL_1968542, EPI_ISL_1968543, EPI_ISL_1968544, EPI_ISL_1968545, EPI_ISL_1968546, EPI_ISL_1968547, EPI_ISL_1968548, EPI_ISL_1968549, EPI_ISL_1968550, EPI_ISL_1968551, EPI_ISL_1968552, EPI_ISL_1968553, EPI_ISL_1968554, EPI_ISL_1968555, EPI_ISL_1968556, EPI_ISL_1968557, EPI_ISL_1968558, EPI_ISL_1968559, EPI_ISL_1968560 | | | |
| see above | Broad Institute Clinical Research Sequencing Platform | Infectious Disease Program, Broad Institute of Harvard and MIT | Siddle,K.J., Adams,G., Pearlman,L., Gladden-Young,A., Vicente,G., Blumenstiel,B., DeFelice,M., Lee,M., McGovern,S., Lagerborg,K., Rudy,M., DeRuff,K., Carter,A., Normandin,E., Bauer,M., Reilly,S., Tomkins-Tinch,C., Loreth,C., Chaluvadi,S., Meldrim,J., Granger,B., Lemieux,J.E., Birren,B.W., Sabeti,P.C., Larkin,K., Dodge,S., Lennon,N., Madoff,L., Brown,C., Gallagher,G., Smole,S., Park,D.J., Gabriel,S., and MacInnis,B.L. |
| EPI_ISL_1968561, EPI_ISL_1968562, EPI_ISL_1968563, EPI_ISL_1968564, EPI_ISL_1968565, EPI_ISL_1968566, EPI_ISL_1968567, EPI_ISL_1968568, EPI_ISL_1968569, EPI_ISL_1968570, EPI_ISL_1968571, EPI_ISL_1968572, EPI_ISL_1968573, EPI_ISL_1968574, EPI_ISL_1968575, EPI_ISL_1968576, EPI_ISL_1968577, EPI_ISL_1968578, EPI_ISL_1968579, EPI_ISL_1968580, EPI_ISL_1968581, EPI_ISL_1968582, EPI_ISL_1968583, EPI_ISL_1968584, EPI_ISL_1968585, EPI_ISL_1968586, EPI_ISL_1968587, EPI_ISL_1968588, EPI_ISL_1968589, EPI_ISL_1968590, EPI_ISL_1968591, EPI_ISL_1968592, EPI_ISL_1968593, EPI_ISL_1968594, EPI_ISL_1968595, EPI_ISL_1968596, EPI_ISL_1968597, EPI_ISL_1968598, EPI_ISL_1968599, EPI_ISL_1968600, EPI_ISL_1968601, EPI_ISL_1968602, EPI_ISL_1968603, EPI_ISL_1968604, EPI_ISL_1968605, EPI_ISL_1968606, EPI_ISL_1968607, EPI_ISL_1968608, EPI_ISL_1968609, EPI_ISL_1968610, EPI_ISL_1968611, EPI_ISL_1968612, EPI_ISL_1968613, EPI_ISL_1968614, EPI_ISL_1968615, EPI_ISL_1968616, EPI_ISL_1968617, EPI_ISL_1968618, EPI_ISL_1968619, EPI_ISL_1968620, EPI_ISL_1968621, EPI_ISL_1968622, EPI_ISL_1968623, EPI_ISL_1968624, EPI_ISL_1968625, EPI_ISL_1968626, EPI_ISL_1968627 | | | |
| see above | Weill Cornell Medicine | New York Genome Center | Michael Zody, Andre Corvelo, Dayna M. Oschwald, Samantha Fennessey, Tom Maniatis, Melissa Cushing, Olivier Elemento, Margaret Elizabeth Ross, Chris Mason, Priya Velu, Hanna Rennert, Arryn Craney, Lars F Westblade |
| EPI_ISL_1968661 | County of San Diego Health and Human Services Agency | Pathogen Discovery, Respiratory Viruses Branch, Division of Viral Diseases, Centers for Disease Control and Prevention | Anna Kelleher, Ying Tao, Yan Li, Jing Zhang, Brian Lynch, Krista Queen, Anna Uehara, Peter Cook, Han Jia Justin Ng, Rachel Marine, Clinton R. Paden, Dhwani Batra, Haibin Wang, Tara Coalter, Jasmine Padilla, Morgan Davis, Mili Sheth, Sarah Nobles, Mark Burroughs, Justin Lee, Adam Retchless, Suxiang Tong |
| EPI_ISL_1968662, EPI_ISL_1968663 | GA Department of Public Health Laboratory | Pathogen Discovery, Respiratory Viruses Branch, Division of Viral Diseases, Centers for Disease Control and Prevention | Anna Kelleher, Ying Tao, Yan Li, Jing Zhang, Brian Lynch, Krista Queen, Anna Uehara, Peter Cook, Han Jia Justin Ng, Rachel Marine, Clinton R. Paden, Dhwani Batra, Haibin Wang, Tara Coalter, Jasmine Padilla, Morgan Davis, Mili Sheth, Sarah Nobles, Mark Burroughs, Justin Lee, Adam Retchless, Suxiang Tong |
| EPI_ISL_1968664, EPI_ISL_1968665 | TN Division of Laboratory Services | Pathogen Discovery, Respiratory Viruses Branch, Division of Viral Diseases, Centers for Disease Control and Prevention | Anna Kelleher, Ying Tao, Yan Li, Jing Zhang, Brian Lynch, Krista Queen, Anna Uehara, Peter Cook, Han Jia Justin Ng, Rachel Marine, Clinton R. Paden, Dhwani Batra, Haibin Wang, Tara Coalter, Jasmine Padilla, Morgan Davis, Mili Sheth, Sarah Nobles, Mark Burroughs, Justin Lee, Adam Retchless, Suxiang Tong |
| EPI_ISL_1968666 | NC State Laboratory of Public Health | Pathogen Discovery, Respiratory Viruses Branch, Division of Viral Diseases, Centers for Disease Control and Prevention | Anna Kelleher, Ying Tao, Yan Li, Jing Zhang, Brian Lynch, Krista Queen, Anna Uehara, Peter Cook, Han Jia Justin Ng, Rachel Marine, Clinton R. Paden, Dhwani Batra, Haibin Wang, Tara Coalter, Jasmine Padilla, Morgan Davis, Mili Sheth, Sarah Nobles, Mark Burroughs, Justin Lee, Adam Retchless, Suxiang Tong |
| EPI_ISL_1968717, EPI_ISL_1968718, EPI_ISL_1968719 | NYP-WCM | New York Genome Center | Michael Zody, Andre Corvelo, Dayna M. Oschwald, Samantha Fennessey, Tom Maniatis, Melissa Cushing, Olivier Elemento, Margaret Elizabeth Ross, Chris Mason, Priya Velu, Hanna Rennert, Arryn Craney, Lars F Westblade |
| EPI_ISL_1968720, EPI_ISL_1968721 | Hackensack Meridian Health | New York Genome Center | Michael Zody, Andre Corvelo, Dayna M. Oschwald, Samantha Fennessey, Tom Maniatis, Liang Chen, Jose Mediavilla, Marcus Cunningham, Kaelea Composto, Kar Chow, David Perlin, Barry Kreiswirth |
| EPI_ISL_1968722, EPI_ISL_1968723, EPI_ISL_1968724, EPI_ISL_1968725 | NYP-WCM | New York Genome Center | Michael Zody, Andre Corvelo, Dayna M. Oschwald, Samantha Fennessey, Tom Maniatis, Melissa Cushing, Olivier Elemento, Margaret Elizabeth Ross, Chris Mason, Priya Velu, Hanna Rennert, Arryn Craney, Lars F Westblade |
| EPI_ISL_1969000 | Idaho Bureau of Laboratories | Center for Global Health, University of New Mexico Health Sciences Center | Daryl Domman, Kurt Schwalm, Valerie Morley, Matthew Burns, Robert Voermans, Christopher Ball, Darrell Dinwiddie |
| EPI_ISL_1969001, EPI_ISL_1969002, EPI_ISL_1969003, EPI_ISL_1969004, EPI_ISL_1969005, EPI_ISL_1969006, EPI_ISL_1969007, EPI_ISL_1969008, EPI_ISL_1969010, EPI_ISL_1969011, EPI_ISL_1969012, EPI_ISL_1969013, EPI_ISL_1969014, EPI_ISL_1969015, EPI_ISL_1969016, EPI_ISL_1969017, EPI_ISL_1969018, EPI_ISL_1969019, EPI_ISL_1969020, EPI_ISL_1969021, EPI_ISL_1969022, EPI_ISL_1969023, EPI_ISL_1969024, EPI_ISL_1969025, EPI_ISL_1969026, EPI_ISL_1969027, EPI_ISL_1969028, EPI_ISL_1969029, EPI_ISL_1969030, EPI_ISL_1969031, EPI_ISL_1969032, EPI_ISL_1969033, EPI_ISL_1969034, EPI_ISL_1969035, EPI_ISL_1969036, EPI_ISL_1969037, EPI_ISL_1969038, EPI_ISL_1969039, EPI_ISL_1969041, EPI_ISL_1969042, EPI_ISL_1969043, EPI_ISL_1969044, EPI_ISL_1969045, EPI_ISL_1969046, EPI_ISL_1969047, EPI_ISL_1969048, EPI_ISL_1969050, EPI_ISL_1969051, EPI_ISL_1969052, EPI_ISL_1969053, EPI_ISL_1969054, EPI_ISL_1969055, EPI_ISL_1969057, EPI_ISL_1969058, EPI_ISL_1969059, EPI_ISL_1969061, EPI_ISL_1969062, EPI_ISL_1969063, EPI_ISL_1969064, EPI_ISL_1969065, EPI_ISL_1969066, EPI_ISL_1969067, EPI_ISL_1969068, EPI_ISL_1969069, EPI_ISL_1969070, EPI_ISL_1969071, EPI_ISL_1969072, EPI_ISL_1969073, EPI_ISL_1969074, EPI_ISL_1969075, EPI_ISL_1969076, EPI_ISL_1969077 | | | |
| see above | NJDOH, Public Health and Environmental Laboratories | NJ_PHEL | Lindsey Bodnar, Shiv K. Verma, Jacquelyn Deverell, Matthew Scarnati, Dana Woell, Byeong Jeong |
| EPI_ISL_1969205, EPI_ISL_1969206, EPI_ISL_1969207, EPI_ISL_1969208, EPI_ISL_1969209, EPI_ISL_1969210, EPI_ISL_1969211, EPI_ISL_1969212, EPI_ISL_1969213, EPI_ISL_1969214, EPI_ISL_1969215, EPI_ISL_1969216, EPI_ISL_1969217, EPI_ISL_1969218, EPI_ISL_1969219, EPI_ISL_1969220, EPI_ISL_1969221, EPI_ISL_1969222, EPI_ISL_1969223, EPI_ISL_1969224, EPI_ISL_1969225, EPI_ISL_1969226 | | | |
| see above | Oregon State Public Health Laboratory | Oregon State Public Health Laboratory | Rafia Razzaque, Eugene Yeboah, Vanda Makris, Laura Tsaknaridis, John Fontana and Shane Sevey |
| EPI_ISL_1969227, EPI_ISL_1969228, EPI_ISL_1969229, EPI_ISL_1969230, EPI_ISL_1969231, EPI_ISL_1969232, EPI_ISL_1969233, EPI_ISL_1969234, EPI_ISL_1969235, EPI_ISL_1969236, EPI_ISL_1969238 | | | |

| | | | |
|---|---|---|---|
| see above | Wyoming Public Health Laboratory | Wyoming Public Health Laboratory | Jim Mildenberger, Taylor Fearing, Channing Weber, Ashley Norberg, Chayse Rowley, Marley Goetz, Brian Dominguez, Elliot Thomasson, Brittany Oher, Sam Britz, Cari Sloma, and Rob Christensen |
| EPI_ISL_1969251, EPI_ISL_1969252, EPI_ISL_1969253, EPI_ISL_1969254, EPI_ISL_1969255, EPI_ISL_1969256, EPI_ISL_1969257, EPI_ISL_1969258, EPI_ISL_1969259, EPI_ISL_1969260, EPI_ISL_1969261, EPI_ISL_1969262, EPI_ISL_1969263, EPI_ISL_1969264, EPI_ISL_1969265, EPI_ISL_1969266, EPI_ISL_1969267, EPI_ISL_1969268, EPI_ISL_1969269, EPI_ISL_1969270 | | | |
| see above | San Diego County Public Health Laboratory | Andersen lab at Scripps Research | SEARCH Alliance San Diego with Tracy Basler, Jovan Shephard, Brett Austin |
| EPI_ISL_1969271, EPI_ISL_1969272, EPI_ISL_1969273, EPI_ISL_1969274, EPI_ISL_1969275, EPI_ISL_1969276, EPI_ISL_1969277, EPI_ISL_1969278, EPI_ISL_1969279, EPI_ISL_1969280, EPI_ISL_1969281 | | | |
| see above | Scripps Medical Laboratory | Andersen lab at Scripps Research | SEARCH Alliance San Diego with Michael Quigley, Ellen Stefanski, Ian Mchardy |
| EPI_ISL_1969282, EPI_ISL_1969283, EPI_ISL_1969284, EPI_ISL_1969285, EPI_ISL_1969286, EPI_ISL_1969287, EPI_ISL_1969288, EPI_ISL_1969289, EPI_ISL_1969290, EPI_ISL_1969291, EPI_ISL_1969292, EPI_ISL_1969293, EPI_ISL_1969294, EPI_ISL_1969295, EPI_ISL_1969296, EPI_ISL_1969297, EPI_ISL_1969298, EPI_ISL_1969299, EPI_ISL_1969300, EPI_ISL_1969301, EPI_ISL_1969302, EPI_ISL_1969303, EPI_ISL_1969304, EPI_ISL_1969305, EPI_ISL_1969306, EPI_ISL_1969307, EPI_ISL_1969308, EPI_ISL_1969309, EPI_ISL_1969310, EPI_ISL_1969311 | | | |
| see above | San Diego County Public Health Laboratory | Andersen lab at Scripps Research | SEARCH Alliance San Diego with Tracy Basler, Jovan Shephard, Brett Austin |
| EPI_ISL_1969312, EPI_ISL_1969313 | Sharp HealthCare Laboratory | Andersen lab at Scripps Research | SEARCH Alliance San Diego with Aaron Harding, Jacquelyn Berumen, Cathy Woerle, Liam McGinnis, Art Mendoza, Omid Bakhtar |
| EPI_ISL_1969314 | Scripps Medical Laboratory | Andersen lab at Scripps Research | SEARCH Alliance San Diego with Michael Quigley, Ellen Stefanski, Ian Mchardy |
| EPI_ISL_1969315, EPI_ISL_1969316, EPI_ISL_1969317, EPI_ISL_1969318 | Sharp HealthCare Laboratory | Andersen lab at Scripps Research | SEARCH Alliance San Diego with Aaron Harding, Jacquelyn Berumen, Cathy Woerle, Liam McGinnis, Art Mendoza, Omid Bakhtar |
| EPI_ISL_1969319, EPI_ISL_1969320, EPI_ISL_1969321, EPI_ISL_1969322, EPI_ISL_1969323, EPI_ISL_1969324, EPI_ISL_1969325, EPI_ISL_1969326 | San Diego County Public Health Laboratory | Andersen lab at Scripps Research | SEARCH Alliance San Diego with Tracy Basler, Jovan Shephard, Brett Austin |
| EPI_ISL_1969327, EPI_ISL_1969328, EPI_ISL_1969329, EPI_ISL_1969330, EPI_ISL_1969331, EPI_ISL_1969332, EPI_ISL_1969333, EPI_ISL_1969334, EPI_ISL_1969335, EPI_ISL_1969336, EPI_ISL_1969337, EPI_ISL_1969338, EPI_ISL_1969339, EPI_ISL_1969340, EPI_ISL_1969341, EPI_ISL_1969342, EPI_ISL_1969343, EPI_ISL_1969344, EPI_ISL_1969345, EPI_ISL_1969346, EPI_ISL_1969347, EPI_ISL_1969348, EPI_ISL_1969349, EPI_ISL_1969350, EPI_ISL_1969351, EPI_ISL_1969352, EPI_ISL_1969353, EPI_ISL_1969354, EPI_ISL_1969355, EPI_ISL_1969356, EPI_ISL_1969357, EPI_ISL_1969358, EPI_ISL_1969359, EPI_ISL_1969360, EPI_ISL_1969361, EPI_ISL_1969362, EPI_ISL_1969363, EPI_ISL_1969364, EPI_ISL_1969365, EPI_ISL_1969366, EPI_ISL_1969367, EPI_ISL_1969368, EPI_ISL_1969369, EPI_ISL_1969370 | | | |
| see above | Scripps Medical Laboratory | Andersen lab at Scripps Research | SEARCH Alliance San Diego with Michael Quigley, Ellen Stefanski, Ian Mchardy |
| EPI_ISL_1969371, EPI_ISL_1969372, EPI_ISL_1969373, EPI_ISL_1969374, EPI_ISL_1969375, EPI_ISL_1969376, EPI_ISL_1969377, EPI_ISL_1969378, EPI_ISL_1969379, EPI_ISL_1969380, EPI_ISL_1969381, EPI_ISL_1969382, EPI_ISL_1969383, EPI_ISL_1969384, EPI_ISL_1969385, EPI_ISL_1969386, EPI_ISL_1969387, EPI_ISL_1969388, EPI_ISL_1969389, EPI_ISL_1969390, EPI_ISL_1969391, EPI_ISL_1969392, EPI_ISL_1969393, EPI_ISL_1969394, EPI_ISL_1969395, EPI_ISL_1969396, EPI_ISL_1969397, EPI_ISL_1969398, EPI_ISL_1969399, EPI_ISL_1969400, EPI_ISL_1969401, EPI_ISL_1969402, EPI_ISL_1969403, EPI_ISL_1969404, EPI_ISL_1969405, EPI_ISL_1969406, EPI_ISL_1969407, EPI_ISL_1969408, EPI_ISL_1969409, EPI_ISL_1969410, EPI_ISL_1969411, EPI_ISL_1969412, EPI_ISL_1969413, EPI_ISL_1969414, EPI_ISL_1969415, EPI_ISL_1969416, EPI_ISL_1969417, EPI_ISL_1969418, EPI_ISL_1969419, EPI_ISL_1969420, EPI_ISL_1969421, EPI_ISL_1969422, EPI_ISL_1969423, EPI_ISL_1969424, EPI_ISL_1969425, EPI_ISL_1969426, EPI_ISL_1969427, EPI_ISL_1969428, EPI_ISL_1969429, EPI_ISL_1969430, EPI_ISL_1969431, EPI_ISL_1969432, EPI_ISL_1969433, EPI_ISL_1969434, EPI_ISL_1969435, EPI_ISL_1969436, EPI_ISL_1969437, EPI_ISL_1969438, EPI_ISL_1969439, EPI_ISL_1969440, EPI_ISL_1969441, EPI_ISL_1969442, EPI_ISL_1969443, EPI_ISL_1969444, EPI_ISL_1969445, EPI_ISL_1969446, EPI_ISL_1969447, EPI_ISL_1969448, EPI_ISL_1969449, EPI_ISL_1969450, EPI_ISL_1969451, EPI_ISL_1969452, EPI_ISL_1969453, EPI_ISL_1969454, EPI_ISL_1969455, EPI_ISL_1969456, EPI_ISL_1969457, EPI_ISL_1969458, EPI_ISL_1969459, EPI_ISL_1969460, EPI_ISL_1969461, EPI_ISL_1969462, EPI_ISL_1969463, EPI_ISL_1969464, EPI_ISL_1969465, EPI_ISL_1969466, EPI_ISL_1969467, EPI_ISL_1969468, EPI_ISL_1969469, EPI_ISL_1969470, EPI_ISL_1969471, EPI_ISL_1969472, EPI_ISL_1969473, EPI_ISL_1969474, EPI_ISL_1969475, EPI_ISL_1969476, EPI_ISL_1969477, EPI_ISL_1969478, EPI_ISL_1969479, EPI_ISL_1969480, EPI_ISL_1969481, EPI_ISL_1969482, EPI_ISL_1969483, EPI_ISL_1969484, EPI_ISL_1969485, EPI_ISL_1969486, EPI_ISL_1969487, EPI_ISL_1969488, EPI_ISL_1969489, EPI_ISL_1969490, EPI_ISL_1969491, EPI_ISL_1969492, EPI_ISL_1969493, EPI_ISL_1969494, EPI_ISL_1969495, EPI_ISL_1969496, EPI_ISL_1969497, EPI_ISL_1969498, EPI_ISL_1969499, EPI_ISL_1969500, EPI_ISL_1969501, EPI_ISL_1969502, EPI_ISL_1969503, EPI_ISL_1969504, EPI_ISL_1969505, EPI_ISL_1969506 | | | |
| see above | San Diego County Public Health Laboratory | Andersen lab at Scripps Research | SEARCH Alliance San Diego with Ashleigh Murphy, Jovan Shephard, Brett Austin |
| EPI_ISL_1969507, EPI_ISL_1969508, EPI_ISL_1969509, EPI_ISL_1969510, EPI_ISL_1969511, EPI_ISL_1969512, EPI_ISL_1969513, EPI_ISL_1969514, EPI_ISL_1969515, EPI_ISL_1969516, EPI_ISL_1969517, EPI_ISL_1969518, EPI_ISL_1969519, EPI_ISL_1969520, EPI_ISL_1969521, EPI_ISL_1969522, EPI_ISL_1969523, EPI_ISL_1969524, EPI_ISL_1969525, EPI_ISL_1969526, EPI_ISL_1969527, EPI_ISL_1969528, EPI_ISL_1969529, EPI_ISL_1969530, EPI_ISL_1969531, EPI_ISL_1969532, EPI_ISL_1969533, EPI_ISL_1969534, EPI_ISL_1969535, EPI_ISL_1969536, EPI_ISL_1969537, EPI_ISL_1969538, EPI_ISL_1969539, EPI_ISL_1969540, EPI_ISL_1969541, EPI_ISL_1969542, EPI_ISL_1969543, EPI_ISL_1969544, EPI_ISL_1969545, EPI_ISL_1969546, EPI_ISL_1969547, EPI_ISL_1969548, EPI_ISL_1969549, EPI_ISL_1969550, EPI_ISL_1969551, EPI_ISL_1969552, EPI_ISL_1969553 | | | |
| see above | Scripps Medical Laboratory | Andersen lab at Scripps Research | SEARCH Alliance San Diego with Michael Quigley, Ellen Stefanski, Ian Mchardy |
| EPI_ISL_1969554, EPI_ISL_1969555, EPI_ISL_1969556, EPI_ISL_1969557, EPI_ISL_1969558, EPI_ISL_1969559, EPI_ISL_1969560, EPI_ISL_1969561, EPI_ISL_1969562, EPI_ISL_1969563, EPI_ISL_1969564, EPI_ISL_1969565, EPI_ISL_1969566, EPI_ISL_1969567, EPI_ISL_1969568, EPI_ISL_1969569, EPI_ISL_1969570, EPI_ISL_1969571, EPI_ISL_1969572, EPI_ISL_1969573, EPI_ISL_1969574, EPI_ISL_1969575, EPI_ISL_1969576, EPI_ISL_1969577, EPI_ISL_1969578, EPI_ISL_1969579, EPI_ISL_1969580, EPI_ISL_1969581, EPI_ISL_1969582, EPI_ISL_1969583, EPI_ISL_1969584, EPI_ISL_1969585, EPI_ISL_1969586, EPI_ISL_1969587, EPI_ISL_1969588, EPI_ISL_1969589, EPI_ISL_1969590, EPI_ISL_1969591, EPI_ISL_1969592, EPI_ISL_1969593, EPI_ISL_1969594, EPI_ISL_1969595, EPI_ISL_1969596, EPI_ISL_1969597, EPI_ISL_1969598, EPI_ISL_1969599, EPI_ISL_1969600, EPI_ISL_1969601, EPI_ISL_1969602, EPI_ISL_1969603, EPI_ISL_1969604, EPI_ISL_1969605, EPI_ISL_1969606, EPI_ISL_1969607, EPI_ISL_1969608, EPI_ISL_1969609, EPI_ISL_1969610, EPI_ISL_1969611, EPI_ISL_1969612, EPI_ISL_1969613, EPI_ISL_1969614, EPI_ISL_1969615, EPI_ISL_1969616, EPI_ISL_1969617, EPI_ISL_1969618, EPI_ISL_1969619, EPI_ISL_1969620, EPI_ISL_1969621, EPI_ISL_1969622, EPI_ISL_1969623, EPI_ISL_1969624, EPI_ISL_1969625, EPI_ISL_1969626, EPI_ISL_1969627, EPI_ISL_1969628, EPI_ISL_1969629, EPI_ISL_1969630, EPI_ISL_1969631, EPI_ISL_1969632, EPI_ISL_1969633, EPI_ISL_1969634, EPI_ISL_1969635, EPI_ISL_1969636, EPI_ISL_1969637, EPI_ISL_1969638, EPI_ISL_1969639, EPI_ISL_1969640, EPI_ISL_1969641, EPI_ISL_1969642, EPI_ISL_1969643, EPI_ISL_1969644, EPI_ISL_1969645, EPI_ISL_1969646, EPI_ISL_1969647, EPI_ISL_1969648, EPI_ISL_1969649, EPI_ISL_1969650, EPI_ISL_1969651, EPI_ISL_1969652, EPI_ISL_1969653, EPI_ISL_1969654, EPI_ISL_1969655, EPI_ISL_1969656, EPI_ISL_1969657, EPI_ISL_1969658, EPI_ISL_1969659, EPI_ISL_1969660, EPI_ISL_1969661, EPI_ISL_1969662, EPI_ISL_1969663, EPI_ISL_1969664, EPI_ISL_1969665, EPI_ISL_1969666 | | | |
| see above | Sharp HealthCare Laboratory | Andersen lab at Scripps Research | SEARCH Alliance San Diego with Aaron Harding, Jacquelyn Berumen, Cathy Woerle, Liam McGinnis, Art Mendoza, Omid Bakhtar |
| EPI_ISL_1969693, EPI_ISL_1969694, EPI_ISL_1969695, EPI_ISL_1969696, EPI_ISL_1969697, EPI_ISL_1969698, EPI_ISL_1969699, EPI_ISL_1969700, EPI_ISL_1969701, EPI_ISL_1969702, EPI_ISL_1969703, EPI_ISL_1969704, EPI_ISL_1969705, EPI_ISL_1969706, EPI_ISL_1969707, EPI_ISL_1969708, EPI_ISL_1969709, EPI_ISL_1969710, EPI_ISL_1969711, EPI_ISL_1969712, EPI_ISL_1969713, EPI_ISL_1969714, EPI_ISL_1969715, EPI_ISL_1969716, EPI_ISL_1969717, EPI_ISL_1969718, EPI_ISL_1969719, EPI_ISL_1969720, EPI_ISL_1969721, EPI_ISL_1969722 | | | |
| see above | Wyoming Public Health Laboratory | Wyoming Public Health Laboratory | Jim Mildenberger, Taylor Fearing, Channing Weber, Ashley Norberg, Chayse Rowley, Marley Goetz, Brian Dominguez, Elliot Thomasson, Brittany Oher, Sam Britz, Cari Sloma, and Rob Christensen |
| EPI_ISL_1969723, EPI_ISL_1969724, EPI_ISL_1969725, EPI_ISL_1969726, EPI_ISL_1969727, EPI_ISL_1969728, EPI_ISL_1969729, EPI_ISL_1969730, EPI_ISL_1969731 | Wyoming Public Health Laboratory | Wyoming Public Health Laboratory | Jim Mildenberger, Wanda Manley, Noah Hull, Taylor Fearing, Lynette Gumbleton, Channing Weber, Ashley Norberg, Chayse Rowley, Marley Goetz, Brian Dominguez, Elliot Thomasson, Sam Britz, Cari Sloma, and Rob Christensen |
| EPI_ISL_1969732, EPI_ISL_1969733, EPI_ISL_1969734, EPI_ISL_1969735, EPI_ISL_1969736, EPI_ISL_1969737, EPI_ISL_1969738, EPI_ISL_1969739, EPI_ISL_1969740, EPI_ISL_1969741, EPI_ISL_1969742, EPI_ISL_1969744, EPI_ISL_1969745, EPI_ISL_1969746, EPI_ISL_1969747, EPI_ISL_1969748, EPI_ISL_1969749, EPI_ISL_1969750, EPI_ISL_1969751 | | | |
| see above | Wyoming Public Health Laboratory | Wyoming Public Health Laboratory | Jim Mildenberger, Taylor Fearing, Channing Weber, Ashley Norberg, Chayse Rowley, Marley Goetz, Brian Dominguez, Elliot Thomasson, Brittany Oher, Sam Britz, Cari Sloma, and Rob Christensen |
| EPI_ISL_1970634, EPI_ISL_1970635, EPI_ISL_1970636, EPI_ISL_1970637, EPI_ISL_1970638, EPI_ISL_1970639, EPI_ISL_1970641, EPI_ISL_1970642, EPI_ISL_1970643, EPI_ISL_1970644, EPI_ISL_1970645, EPI_ISL_1970646, EPI_ISL_1970647, EPI_ISL_1970648, EPI_ISL_1970649, EPI_ISL_1970650, EPI_ISL_1970651, EPI_ISL_1970652, EPI_ISL_1970653, EPI_ISL_1970654, EPI_ISL_1970655, EPI_ISL_1970656, EPI_ISL_1970657, EPI_ISL_1970658, EPI_ISL_1970659, EPI_ISL_1970660, EPI_ISL_1970661, EPI_ISL_1970662, EPI_ISL_1970663, EPI_ISL_1970664, EPI_ISL_1970665, EPI_ISL_1970666, EPI_ISL_1970667, EPI_ISL_1970668, EPI_ISL_1970669, EPI_ISL_1970670, EPI_ISL_1970671, EPI_ISL_1970672, EPI_ISL_1970673, EPI_ISL_1970674, EPI_ISL_1970675, EPI_ISL_1970676, EPI_ISL_1970677, EPI_ISL_1970678, EPI_ISL_1970679, EPI_ISL_1970680, EPI_ISL_1970681, EPI_ISL_1970682, EPI_ISL_1970683, EPI_ISL_1970684, EPI_ISL_1970685, EPI_ISL_1970686 | | | |
| see above | The Jackson Laboratory | The Jackson Laboratory | Bergeron D, Renzette N, Adams M, Omerza G, Kelly K, Li L |
| EPI_ISL_1970836, EPI_ISL_1970837 | Centro de Investigación en Enfermedades Infecciosas (CIENI), Instituto Nacional de Enfermedades Respiratorias (INER) | Centro de Investigación en Enfermedades Infecciosas (CIENI), Instituto Nacional de Enfermedades Respiratorias (INER) | Celia Boukadida, Carlos F. Arias, Santiago Ávila-Ríos, Eduardo Becerril-Vargas, Ricardo Grande, Alejandra Hernández-Terán, Pavel Isa, José Arturo Martínez-Orozco, Margarita Matías-Florentino, Fidencio Mejía-Nepomuceno, Mario Mújica-Sánchez, Hector Esteban Paz-Juárez, Francisco Pulido, Jorge Salas-Hernández, Alejandro Sanchez-Flores, Blanca Taboada, Joel Armando Vázquez-Pérez and Consorcio Mexicano de Vigilancia Genómica (CoViGen-Mex) |
| EPI_ISL_1970838, EPI_ISL_1970839, EPI_ISL_1970840, EPI_ISL_1970841, EPI_ISL_1970842, EPI_ISL_1970843, EPI_ISL_1970844, EPI_ISL_1970845 | Instituto Nacional de Medicina Genomica | Centro de Investigación en Enfermedades Infecciosas (CIENI), Instituto Nacional de Enfermedades Respiratorias (INER) | Matias-Florentino M, Perez-Garcia M, Hidalgo-Miranda A, Mendoza-Vargas A, Reyes-Grajeda JP, Cedro-Tanda A, Arriaga-Canon C, Herrera-Montalvo LA, Reyes-Teran G, Avila-Rios S, Boukadida C |
| EPI_ISL_1970846, EPI_ISL_1970847, EPI_ISL_1970848, EPI_ISL_1970849, EPI_ISL_1970850, EPI_ISL_1970851, EPI_ISL_1970852, EPI_ISL_1970853, EPI_ISL_1970854, EPI_ISL_1970855, EPI_ISL_1970856, EPI_ISL_1970857, EPI_ISL_1970858, EPI_ISL_1970859, EPI_ISL_1970860, EPI_ISL_1970862, EPI_ISL_1970864, EPI_ISL_1970865, EPI_ISL_1970866, EPI_ISL_1970868, EPI_ISL_1970869, EPI_ISL_1970870, EPI_ISL_1970871, EPI_ISL_1970872, EPI_ISL_1970873, EPI_ISL_1970874, EPI_ISL_1970875, EPI_ISL_1970876, EPI_ISL_1970877, EPI_ISL_1970878, EPI_ISL_1970879, EPI_ISL_1970880, EPI_ISL_1970881, EPI_ISL_1970882, EPI_ISL_1970883, EPI_ISL_1970884, EPI_ISL_1970886, EPI_ISL_1970887, EPI_ISL_1970888, EPI_ISL_1970889, EPI_ISL_1970890, EPI_ISL_1970891, EPI_ISL_1970892, EPI_ISL_1970893 | | | |
| see above | Curative | New Mexico Department of Health Scientific Laboratory | Ellie Johnson, Anastacia Griego-Fisher, D'eldra Malone, Jennifer Benoit, Linda Salazar, Ratheesh Rajan, Mark Willmon |
| EPI_ISL_1970894, EPI_ISL_1970895, EPI_ISL_1970896, EPI_ISL_1970897, EPI_ISL_1970898, EPI_ISL_1970899, EPI_ISL_1970900, EPI_ISL_1970901, EPI_ISL_1970902, EPI_ISL_1970903, EPI_ISL_1970904, EPI_ISL_1970905, EPI_ISL_1970906, EPI_ISL_1970907, EPI_ISL_1970908, EPI_ISL_1970909, EPI_ISL_1970910, EPI_ISL_1970911, EPI_ISL_1970912, EPI_ISL_1970913, EPI_ISL_1970914, EPI_ISL_1970915, EPI_ISL_1970917, EPI_ISL_1970918, EPI_ISL_1970919 | | | |
| see above | New Mexico Department of Health Scientific Laboratory | New Mexico Department of Health Scientific Laboratory | Ellie Johnson, Anastacia Griego-Fisher, D'eldra Malone, Jennifer Benoit, Linda Salazar, Ratheesh Rajan, Mark Willmon |
| EPI_ISL_1970920, EPI_ISL_1970921, EPI_ISL_1970922, EPI_ISL_1970923, EPI_ISL_1970924, EPI_ISL_1970925, EPI_ISL_1970926, EPI_ISL_1970927 | San Juan Regional Medical Center | New Mexico Department of Health Scientific Laboratory | Ellie Johnson, Anastacia Griego-Fisher, D'eldra Malone, Jennifer Benoit, Linda Salazar, Ratheesh Rajan, Mark Willmon |

| | | | |
|---|---|---|---|
| EPI_ISL_1970928, EPI_ISL_1970929, EPI_ISL_1970930, EPI_ISL_1970931, EPI_ISL_1970932, EPI_ISL_1970933 | TriCore Reference Laboratories | New Mexico Department of Health Scientific Laboratory | Ellie Johnson, Anastacia Griego-Fisher, D'eldra Malone, Jennifer Benoit, Linda Salazar, Ratheesh Rajan, Mark Willmon |
| EPI_ISL_1970934, EPI_ISL_1970935, EPI_ISL_1970936 | New Mexico Department of Health Scientific Laboratory | New Mexico Department of Health Scientific Laboratory | Ellie Johnson, Anastacia Griego-Fisher, D'eldra Malone, Jennifer Benoit, Linda Salazar, Ratheesh Rajan, Mark Willmon |
| EPI_ISL_1970937 | TriCore Reference Laboratories | New Mexico Department of Health Scientific Laboratory | Ellie Johnson, Anastacia Griego-Fisher, D'eldra Malone, Jennifer Benoit, Linda Salazar, Ratheesh Rajan, Mark Willmon |
| EPI_ISL_1970938 | New Mexico Department of Health Scientific Laboratory | New Mexico Department of Health Scientific Laboratory | Ellie Johnson, Anastacia Griego-Fisher, D'eldra Malone, Jennifer Benoit, Linda Salazar, Ratheesh Rajan, Mark Willmon |
| EPI_ISL_1970940 | PHS/IHS Hospital-Jicarilla Service Unit | New Mexico Department of Health Scientific Laboratory | Ellie Johnson, Anastacia Griego-Fisher, D'eldra Malone, Jennifer Benoit, Linda Salazar, Ratheesh Rajan, Mark Willmon |
| EPI_ISL_1970942, EPI_ISL_1970944, EPI_ISL_1970945, EPI_ISL_1970946, EPI_ISL_1970947 | New Mexico Department of Health Scientific Laboratory | New Mexico Department of Health Scientific Laboratory | Ellie Johnson, Anastacia Griego-Fisher, D'eldra Malone, Jennifer Benoit, Linda Salazar, Ratheesh Rajan, Mark Willmon |
| EPI_ISL_1970948, EPI_ISL_1970949 | TriCore Reference Laboratory | New Mexico Department of Health Scientific Laboratory | Ellie Johnson, Anastacia Griego-Fisher, D'eldra Malone, Jennifer Benoit, Linda Salazar, Ratheesh Rajan, Mark Willmon |
| EPI_ISL_1970951, EPI_ISL_1970952, EPI_ISL_1970953, EPI_ISL_1970954, EPI_ISL_1970955 | San Juan Regional Medical Center | New Mexico Department of Health Scientific Laboratory | Ellie Johnson, Anastacia Griego-Fisher, D'eldra Malone, Jennifer Benoit, Linda Salazar, Ratheesh Rajan, Mark Willmon |
| EPI_ISL_1970958, EPI_ISL_1970959, EPI_ISL_1970960, EPI_ISL_1970961, EPI_ISL_1970962 | New Mexico Department of Health Scientific Laboratory | New Mexico Department of Health Scientific Laboratory | Ellie Johnson, Anastacia Griego-Fisher, D'eldra Malone, Jennifer Benoit, Linda Salazar, Ratheesh Rajan, Mark Willmon |
| EPI_ISL_1970963 | San Juan Regional Medical Center | New Mexico Department of Health Scientific Laboratory | Ellie Johnson, Anastacia Griego-Fisher, D'eldra Malone, Jennifer Benoit, Linda Salazar, Ratheesh Rajan, Mark Willmon |
| EPI_ISL_1970964, EPI_ISL_1970965, EPI_ISL_1970966, EPI_ISL_1970967, EPI_ISL_1970969, EPI_ISL_1970970, EPI_ISL_1970971, EPI_ISL_1970972, EPI_ISL_1970974 | New Mexico Department of Health Scientific Laboratory | New Mexico Department of Health Scientific Laboratory | Ellie Johnson, Anastacia Griego-Fisher, D'eldra Malone, Jennifer Benoit, Linda Salazar, Ratheesh Rajan, Mark Willmon |
| EPI_ISL_1970975, EPI_ISL_1970976, EPI_ISL_1970977, EPI_ISL_1970978, EPI_ISL_1970979, EPI_ISL_1970980, EPI_ISL_1970981, EPI_ISL_1970982, EPI_ISL_1970983, EPI_ISL_1970984 | San Juan Regional Medical Center | New Mexico Department of Health Scientific Laboratory | Ellie Johnson, Anastacia Griego-Fisher, D'eldra Malone, Jennifer Benoit, Linda Salazar, Ratheesh Rajan, Mark Willmon |
| EPI_ISL_1971088, EPI_ISL_1971089, EPI_ISL_1971090, EPI_ISL_1971091, EPI_ISL_1971092, EPI_ISL_1971093, EPI_ISL_1971094, EPI_ISL_1971095, EPI_ISL_1971096, EPI_ISL_1971097, EPI_ISL_1971098, EPI_ISL_1971099, EPI_ISL_1971100, EPI_ISL_1971101, EPI_ISL_1971102, EPI_ISL_1971103, EPI_ISL_1971104, EPI_ISL_1971105, EPI_ISL_1971106, EPI_ISL_1971107, EPI_ISL_1971108, EPI_ISL_1971109, EPI_ISL_1971110, EPI_ISL_1971111, EPI_ISL_1971112, EPI_ISL_1971113, EPI_ISL_1971114, EPI_ISL_1971115, EPI_ISL_1971116, EPI_ISL_1971117, EPI_ISL_1971118, EPI_ISL_1971119, EPI_ISL_1971120, EPI_ISL_1971121, EPI_ISL_1971122, EPI_ISL_1971123, EPI_ISL_1971124, EPI_ISL_1971125, EPI_ISL_1971126, EPI_ISL_1971127, EPI_ISL_1971128, EPI_ISL_1971129, EPI_ISL_1971130, EPI_ISL_1971131, EPI_ISL_1971132, EPI_ISL_1971133, EPI_ISL_1971134, EPI_ISL_1971135, EPI_ISL_1971136, EPI_ISL_1971137, EPI_ISL_1971138, EPI_ISL_1971139, EPI_ISL_1971140, EPI_ISL_1971141, EPI_ISL_1971142, EPI_ISL_1971143, EPI_ISL_1971144, EPI_ISL_1971145, EPI_ISL_1971146, EPI_ISL_1971147, EPI_ISL_1971148, EPI_ISL_1971149, EPI_ISL_1971150, EPI_ISL_1971151, EPI_ISL_1971152, EPI_ISL_1971153, EPI_ISL_1971154, EPI_ISL_1971155, EPI_ISL_1971156, EPI_ISL_1971157, EPI_ISL_1971158, EPI_ISL_1971159, EPI_ISL_1971160, EPI_ISL_1971161, EPI_ISL_1971162, EPI_ISL_1971163, EPI_ISL_1971164, EPI_ISL_1971165, EPI_ISL_1971166, EPI_ISL_1971167, EPI_ISL_1971168, EPI_ISL_1971169, EPI_ISL_1971170, EPI_ISL_1971171, EPI_ISL_1971172, EPI_ISL_1971173, EPI_ISL_1971174, EPI_ISL_1971175, EPI_ISL_1971176, EPI_ISL_1971177, EPI_ISL_1971178, EPI_ISL_1971179, EPI_ISL_1971180, EPI_ISL_1971181, EPI_ISL_1971182, EPI_ISL_1971183, EPI_ISL_1971184, EPI_ISL_1971185, EPI_ISL_1971186, EPI_ISL_1971187, EPI_ISL_1971188, EPI_ISL_1971189, EPI_ISL_1971190, EPI_ISL_1971191, EPI_ISL_1971192, EPI_ISL_1971193, EPI_ISL_1971194, EPI_ISL_1971195, EPI_ISL_1971196, EPI_ISL_1971197, EPI_ISL_1971198, EPI_ISL_1971199, EPI_ISL_1971200, EPI_ISL_1971201, EPI_ISL_1971202, EPI_ISL_1971203, EPI_ISL_1971204, EPI_ISL_1971205, EPI_ISL_1971206, EPI_ISL_1971207, EPI_ISL_1971208, EPI_ISL_1971209, EPI_ISL_1971210, EPI_ISL_1971211, EPI_ISL_1971212, EPI_ISL_1971213, EPI_ISL_1971214, EPI_ISL_1971215, EPI_ISL_1971216, EPI_ISL_1971217, EPI_ISL_1971218, EPI_ISL_1971219, EPI_ISL_1971220, EPI_ISL_1971221, EPI_ISL_1971222, EPI_ISL_1971223, EPI_ISL_1971224, EPI_ISL_1971225, EPI_ISL_1971226, EPI_ISL_1971227, EPI_ISL_1971228, EPI_ISL_1971229, EPI_ISL_1971230, EPI_ISL_1971231 | | | |
| see above | Weill Cornell Medicine | New York Genome Center | Michael Zody, Andre Corvelo, Dayna M. Oschwald, Samantha Fennessey, Tom Maniatis, Melissa Cushing, Olivier Elemento, Margaret Elizabeth Ross, Chris Mason, Priya Velu, Hanna Rennert, Arryn Craney, Lars F Westblade |
| EPI_ISL_1971232, EPI_ISL_1971233, EPI_ISL_1971234, EPI_ISL_1971235, EPI_ISL_1971236, EPI_ISL_1971237, EPI_ISL_1971238, EPI_ISL_1971239, EPI_ISL_1971240, EPI_ISL_1971241, EPI_ISL_1971242, EPI_ISL_1971243, EPI_ISL_1971244, EPI_ISL_1971245, EPI_ISL_1971246, EPI_ISL_1971247, EPI_ISL_1971248, EPI_ISL_1971249, EPI_ISL_1971250, EPI_ISL_1971251, EPI_ISL_1971252, EPI_ISL_1971253, EPI_ISL_1971254, EPI_ISL_1971255, EPI_ISL_1971256, EPI_ISL_1971257, EPI_ISL_1971258, EPI_ISL_1971259, EPI_ISL_1971260, EPI_ISL_1971261, EPI_ISL_1971262, EPI_ISL_1971263, EPI_ISL_1971264, EPI_ISL_1971265, EPI_ISL_1971266, EPI_ISL_1971267, EPI_ISL_1971268, EPI_ISL_1971269, EPI_ISL_1971270, EPI_ISL_1971271, EPI_ISL_1971272, EPI_ISL_1971273, EPI_ISL_1971274, EPI_ISL_1971275, EPI_ISL_1971276, EPI_ISL_1971277, EPI_ISL_1971278, EPI_ISL_1971279, EPI_ISL_1971280, EPI_ISL_1971281, EPI_ISL_1971282, EPI_ISL_1971283, EPI_ISL_1971284, EPI_ISL_1971285, EPI_ISL_1971286, EPI_ISL_1971287, EPI_ISL_1971288, EPI_ISL_1971289, EPI_ISL_1971290, EPI_ISL_1971291, EPI_ISL_1971292, EPI_ISL_1971293, EPI_ISL_1971294, EPI_ISL_1971295, EPI_ISL_1971296, EPI_ISL_1971297, EPI_ISL_1971298, EPI_ISL_1971299, EPI_ISL_1971300, EPI_ISL_1971301, EPI_ISL_1971302, EPI_ISL_1971303, EPI_ISL_1971304, EPI_ISL_1971305, EPI_ISL_1971306, EPI_ISL_1971307, EPI_ISL_1971308, EPI_ISL_1971309, EPI_ISL_1971310, EPI_ISL_1971311, EPI_ISL_1971312, EPI_ISL_1971313, EPI_ISL_1971314, EPI_ISL_1971315, EPI_ISL_1971316, EPI_ISL_1971317, EPI_ISL_1971318, EPI_ISL_1971319, EPI_ISL_1971320, EPI_ISL_1971321, EPI_ISL_1971322, EPI_ISL_1971323, EPI_ISL_1971324, EPI_ISL_1971325, EPI_ISL_1971326, EPI_ISL_1971327, EPI_ISL_1971328, EPI_ISL_1971329, EPI_ISL_1971330, EPI_ISL_1971331, EPI_ISL_1971332, EPI_ISL_1971333, EPI_ISL_1971334, EPI_ISL_1971335, EPI_ISL_1971336, EPI_ISL_1971337, EPI_ISL_1971338, EPI_ISL_1971339, EPI_ISL_1971340, EPI_ISL_1971341, EPI_ISL_1971342, EPI_ISL_1971343, EPI_ISL_1971344, EPI_ISL_1971345, EPI_ISL_1971346, EPI_ISL_1971347, EPI_ISL_1971348, EPI_ISL_1971349, EPI_ISL_1971350, EPI_ISL_1971351, EPI_ISL_1971352, EPI_ISL_1971353, EPI_ISL_1971354, EPI_ISL_1971355, EPI_ISL_1971356, EPI_ISL_1971357, EPI_ISL_1971358, EPI_ISL_1971359, EPI_ISL_1971360, EPI_ISL_1971361, EPI_ISL_1971362, EPI_ISL_1971363, EPI_ISL_1971364, EPI_ISL_1971365, EPI_ISL_1971366, EPI_ISL_1971367 | | | |
| see above | Hackensack Medical Center | New York Genome Center | Michael Zody, Andre Corvelo, Dayna M. Oschwald, Samantha Fennessey, Tom Maniatis, Liang Chen, Jose Mediavilla, Marcus Cunningham, Kaelea Composto, Kar Chow, David Perlin, Barry Kreiswirth |
| EPI_ISL_1971369, EPI_ISL_1971370, EPI_ISL_1971371, EPI_ISL_1971372, EPI_ISL_1971373, EPI_ISL_1971374, EPI_ISL_1971375, EPI_ISL_1971376, EPI_ISL_1971377, EPI_ISL_1971378, EPI_ISL_1971379, EPI_ISL_1971380, EPI_ISL_1971381, EPI_ISL_1971382, EPI_ISL_1971383, EPI_ISL_1971384, EPI_ISL_1971385, EPI_ISL_1971386, EPI_ISL_1971387, EPI_ISL_1971388, EPI_ISL_1971389, EPI_ISL_1971390, EPI_ISL_1971391, EPI_ISL_1971392, EPI_ISL_1971393, EPI_ISL_1971394, EPI_ISL_1971395, EPI_ISL_1971396, EPI_ISL_1971397, EPI_ISL_1971398, EPI_ISL_1971399, EPI_ISL_1971400, EPI_ISL_1971401, EPI_ISL_1971402, EPI_ISL_1971403, EPI_ISL_1971404, EPI_ISL_1971405, EPI_ISL_1971406 | | | |
| see above | Weill Cornell Medicine | New York Genome Center | Michael Zody, Andre Corvelo, Dayna M. Oschwald, Samantha Fennessey, Tom Maniatis, Melissa Cushing, Olivier Elemento, Margaret Elizabeth Ross, Chris Mason, Priya Velu, Hanna Rennert, Arryn Craney, Lars F Westblade |
| EPI_ISL_1971407, EPI_ISL_1971408, EPI_ISL_1971409, EPI_ISL_1971410, EPI_ISL_1971411, EPI_ISL_1971412, EPI_ISL_1971413, EPI_ISL_1971414, EPI_ISL_1971415, EPI_ISL_1971416, EPI_ISL_1971417, EPI_ISL_1971418, EPI_ISL_1971419, EPI_ISL_1971420, EPI_ISL_1971421, EPI_ISL_1971422, EPI_ISL_1971423, EPI_ISL_1971424, EPI_ISL_1971425, EPI_ISL_1971426, EPI_ISL_1971427, EPI_ISL_1971428, EPI_ISL_1971429, EPI_ISL_1971430, EPI_ISL_1971431, EPI_ISL_1971432, EPI_ISL_1971433, EPI_ISL_1971434, EPI_ISL_1971435, EPI_ISL_1971436, EPI_ISL_1971437, EPI_ISL_1971438, EPI_ISL_1971439, EPI_ISL_1971440, EPI_ISL_1971441, EPI_ISL_1971442, EPI_ISL_1971443, EPI_ISL_1971444, EPI_ISL_1971445, EPI_ISL_1971446, EPI_ISL_1971447, EPI_ISL_1971448, EPI_ISL_1971449, EPI_ISL_1971450, EPI_ISL_1971460, EPI_ISL_1971461, EPI_ISL_1971462, EPI_ISL_1971463, EPI_ISL_1971464, EPI_ISL_1971465, EPI_ISL_1971466, EPI_ISL_1971467, EPI_ISL_1971468, EPI_ISL_1971469, EPI_ISL_1971470 | | | |
| see above | TXDSHS | TXDSHS | Rashmi Tuladhar, Bonnie Oh, Jenny Zhang, Maliha Rahman, Mayela Pedrueza, Anita Pokharel, Karen Bobier, Lorraine Rodriguez, Myong Koag, Chun Wang, Rachel Lee, Grace Kubin |
| EPI_ISL_1971507, EPI_ISL_1971508, EPI_ISL_1971509, EPI_ISL_1971510, EPI_ISL_1971511, EPI_ISL_1971512, EPI_ISL_1971513, EPI_ISL_1971514, EPI_ISL_1971515, EPI_ISL_1971516, EPI_ISL_1971518, EPI_ISL_1971519, EPI_ISL_1971520, EPI_ISL_1971521, EPI_ISL_1971522, EPI_ISL_1971523, EPI_ISL_1971524, EPI_ISL_1971525, EPI_ISL_1971526, EPI_ISL_1971527, EPI_ISL_1971528, EPI_ISL_1971529, EPI_ISL_1971530, EPI_ISL_1971531, EPI_ISL_1971532, EPI_ISL_1971533, EPI_ISL_1971534, EPI_ISL_1971535, EPI_ISL_1971536, EPI_ISL_1971537, EPI_ISL_1971538, EPI_ISL_1971539, EPI_ISL_1971540, EPI_ISL_1971541, EPI_ISL_1971542, EPI_ISL_1971543, EPI_ISL_1971544, EPI_ISL_1971545, EPI_ISL_1971546, EPI_ISL_1971547, EPI_ISL_1971548, EPI_ISL_1971549, EPI_ISL_1971550, EPI_ISL_1971551, EPI_ISL_1971552, EPI_ISL_1971553, EPI_ISL_1971554, EPI_ISL_1971555, EPI_ISL_1971556, EPI_ISL_1971557, EPI_ISL_1971558, EPI_ISL_1971559, EPI_ISL_1971560, EPI_ISL_1971561, EPI_ISL_1971562, EPI_ISL_1971563, EPI_ISL_1971564, EPI_ISL_1971565, EPI_ISL_1971566, EPI_ISL_1971567, EPI_ISL_1971568, EPI_ISL_1971569, EPI_ISL_1971570, EPI_ISL_1971571, EPI_ISL_1971572, EPI_ISL_1971573, EPI_ISL_1971574, EPI_ISL_1971575, EPI_ISL_1971576, EPI_ISL_1971577, EPI_ISL_1971578, EPI_ISL_1971579, EPI_ISL_1971580, EPI_ISL_1971581, EPI_ISL_1971582, EPI_ISL_1971583, EPI_ISL_1971584, EPI_ISL_1971585, EPI_ISL_1971586, EPI_ISL_1971587, EPI_ISL_1971588, EPI_ISL_1971589, EPI_ISL_1971590, EPI_ISL_1971591, EPI_ISL_1971592, EPI_ISL_1971593, EPI_ISL_1971594, EPI_ISL_1971595, EPI_ISL_1971596, EPI_ISL_1971597, EPI_ISL_1971598, EPI_ISL_1971599, EPI_ISL_1971600, EPI_ISL_1971601, EPI_ISL_1971602, EPI_ISL_1971603, EPI_ISL_1971604, EPI_ISL_1971605, EPI_ISL_1971606, EPI_ISL_1971607, EPI_ISL_1971608, EPI_ISL_1971609, EPI_ISL_1971610, EPI_ISL_1971611, EPI_ISL_1971612, EPI_ISL_1971613, EPI_ISL_1971614, EPI_ISL_1971615, EPI_ISL_1971616, EPI_ISL_1971617, EPI_ISL_1971618, EPI_ISL_1971619, EPI_ISL_1971620, EPI_ISL_1971621, EPI_ISL_1971622, EPI_ISL_1971623, EPI_ISL_1971624, EPI_ISL_1971625, EPI_ISL_1971626, EPI_ISL_1971627, EPI_ISL_1971628, EPI_ISL_1971629, EPI_ISL_1971630, EPI_ISL_1971631, EPI_ISL_1971632, EPI_ISL_1971633, EPI_ISL_1971635, EPI_ISL_1971636, EPI_ISL_1971637, EPI_ISL_1971638, EPI_ISL_1971639, EPI_ISL_1971640, EPI_ISL_1971641, EPI_ISL_1971642, EPI_ISL_1971643, EPI_ISL_1971644, EPI_ISL_1971645, EPI_ISL_1971646, EPI_ISL_1971647, EPI_ISL_1971648, EPI_ISL_1971649, EPI_ISL_1971651, EPI_ISL_1971652, EPI_ISL_1971653, EPI_ISL_1971654, EPI_ISL_1971655, EPI_ISL_1971656, EPI_ISL_1971657, EPI_ISL_1971658, EPI_ISL_1971659, EPI_ISL_1971660, EPI_ISL_1971661, EPI_ISL_1971662, EPI_ISL_1971663, EPI_ISL_1971664, EPI_ISL_1971665, EPI_ISL_1971666, EPI_ISL_1971667, EPI_ISL_1971668, EPI_ISL_1971669, EPI_ISL_1971670, EPI_ISL_1971671, EPI_ISL_1971673, EPI_ISL_1971674, EPI_ISL_1971675, EPI_ISL_1971676, EPI_ISL_1971677, EPI_ISL_1971678, EPI_ISL_1971679, EPI_ISL_1971680, EPI_ISL_1971681, EPI_ISL_1971682, EPI_ISL_1971683, EPI_ISL_1971684, EPI_ISL_1971685, EPI_ISL_1971686, EPI_ISL_1971687, EPI_ISL_1971688, EPI_ISL_1971689, EPI_ISL_1971690, EPI_ISL_1971691, EPI_ISL_1971692, EPI_ISL_1971693, EPI_ISL_1971694, EPI_ISL_1971695, EPI_ISL_1971696, EPI_ISL_1971697, EPI_ISL_1971698, EPI_ISL_1971699, EPI_ISL_1971700, EPI_ISL_1971701, EPI_ISL_1971702, EPI_ISL_1971703, EPI_ISL_1971704, EPI_ISL_1971705, EPI_ISL_1971706, EPI_ISL_1971707, EPI_ISL_1971708, EPI_ISL_1971709, EPI_ISL_1971710, EPI_ISL_1971712, EPI_ISL_1971713, EPI_ISL_1971714, EPI_ISL_1971715, EPI_ISL_1971716, EPI_ISL_1971717, EPI_ISL_1971718, EPI_ISL_1971719, EPI_ISL_1971720, EPI_ISL_1971721, EPI_ISL_1971722, EPI_ISL_1971723, EPI_ISL_1971724, EPI_ISL_1971725, EPI_ISL_1971726, EPI_ISL_1971727, EPI_ISL_1971728, EPI_ISL_1971729, EPI_ISL_1971730, EPI_ISL_1971731, EPI_ISL_1971732, EPI_ISL_1971733, EPI_ISL_1971734, EPI_ISL_1971735, EPI_ISL_1971736, EPI_ISL_1971737, EPI_ISL_1971738, EPI_ISL_1971739, EPI_ISL_1971740, EPI_ISL_1971741, EPI_ISL_1971742, EPI_ISL_1971744, EPI_ISL_1971745, EPI_ISL_1971746, EPI_ISL_1971747, EPI_ISL_1971748, EPI_ISL_1971749, EPI_ISL_1971750, EPI_ISL_1971751, EPI_ISL_1971752, EPI_ISL_1971753, EPI_ISL_1971754, EPI_ISL_1971755, EPI_ISL_1971756, EPI_ISL_1971757, EPI_ISL_1971758, EPI_ISL_1971759, EPI_ISL_1971760, EPI_ISL_1971761, EPI_ISL_1971762, EPI_ISL_1971763, EPI_ISL_1971764, EPI_ISL_1971765, EPI_ISL_1971766, EPI_ISL_1971768, EPI_ISL_1971769, EPI_ISL_1971770, EPI_ISL_1971771, EPI_ISL_1971772, EPI_ISL_1971773, EPI_ISL_1971774, EPI_ISL_1971775, EPI_ISL_1971776, EPI_ISL_1971777, EPI_ISL_1971778, EPI_ISL_1971779, EPI_ISL_1971780, EPI_ISL_1971781, EPI_ISL_1971782, EPI_ISL_1971783, EPI_ISL_1971784, EPI_ISL_1971785, EPI_ISL_1971786, EPI_ISL_1971788, EPI_ISL_1971789, EPI_ISL_1971790, EPI_ISL_1971791, EPI_ISL_1971792, EPI_ISL_1971793, EPI_ISL_1971794, EPI_ISL_1971795, EPI_ISL_1971796, EPI_ISL_1971797, EPI_ISL_1971798, EPI_ISL_1971799, EPI_ISL_1971800, EPI_ISL_1971801, EPI_ISL_1971802, EPI_ISL_1971803, EPI_ISL_1971804, EPI_ISL_1971805, EPI_ISL_1971806, EPI_ISL_1971807, EPI_ISL_1971808, EPI_ISL_1971809, EPI_ISL_1971810, EPI_ISL_1971811, EPI_ISL_1971812, EPI_ISL_1971813, EPI_ISL_1971814, EPI_ISL_1971815, EPI_ISL_1971816, EPI_ISL_1971817, EPI_ISL_1971818, EPI_ISL_1971819, EPI_ISL_1971820, EPI_ISL_1971821, EPI_ISL_1971822, EPI_ISL_1971823, EPI_ISL_1971824, EPI_ISL_1971825, EPI_ISL_1971826, EPI_ISL_1971827, EPI_ISL_1971828, EPI_ISL_1971829, EPI_ISL_1971830, EPI_ISL_1971831, EPI_ISL_1971832, EPI_ISL_1971833, EPI_ISL_1971834, EPI_ISL_1971835, EPI_ISL_1971836, EPI_ISL_1971837, EPI_ISL_1971838, EPI_ISL_1971839, EPI_ISL_1971840, EPI_ISL_1971841, EPI_ISL_1971842, EPI_ISL_1971843, EPI_ISL_1971844, EPI_ISL_1971845, EPI_ISL_1971846, EPI_ISL_1971847, EPI_ISL_1971848, EPI_ISL_1971849, EPI_ISL_1971850, EPI_ISL_1971851, EPI_ISL_1971852, EPI_ISL_1971853, EPI_ISL_1971854, EPI_ISL_1971855, EPI_ISL_1971856, EPI_ISL_1971857, EPI_ISL_1971858, EPI_ISL_1971859, EPI_ISL_1971860, EPI_ISL_1971862, EPI_ISL_1971863, EPI_ISL_1971864, EPI_ISL_1971865, EPI_ISL_1971866, EPI_ISL_1971867, EPI_ISL_1971868, EPI_ISL_1971869, EPI_ISL_1971870, EPI_ISL_1971871, EPI_ISL_1971872, EPI_ISL_1971873, EPI_ISL_1971874, EPI_ISL_1971875, EPI_ISL_1971876, EPI_ISL_1971877, EPI_ISL_1971878, EPI_ISL_1971879, EPI_ISL_1971880, EPI_ISL_1971881, EPI_ISL_1971882, EPI_ISL_1971883, EPI_ISL_1971884, EPI_ISL_1971885, EPI_ISL_1971886, EPI_ISL_1971887, EPI_ISL_1971888, EPI_ISL_1971889, EPI_ISL_1971890, EPI_ISL_1971891, EPI_ISL_1971892, EPI_ISL_1971893, EPI_ISL_1971894, EPI_ISL_1971895, EPI_ISL_1971896, EPI_ISL_1971897, EPI_ISL_1971898, EPI_ISL_1971899, EPI_ISL_1971900, EPI_ISL_1971901, EPI_ISL_1971902, EPI_ISL_1971903, EPI_ISL_1971904, EPI_ISL_1971905, EPI_ISL_1971906, EPI_ISL_1971907, EPI_ISL_1971908, EPI_ISL_1971909, EPI_ISL_1971910, EPI_ISL_1971911, EPI_ISL_1971912, EPI_ISL_1971913, EPI_ISL_1971914, EPI_ISL_1971915, EPI_ISL_1971916, EPI_ISL_1971917, EPI_ISL_1971918, EPI_ISL_1971919, EPI_ISL_1971920, EPI_ISL_1971921, EPI_ISL_1971922, EPI_ISL_1971923, EPI_ISL_1971924, EPI_ISL_1971925, EPI_ISL_1971926, EPI_ISL_1971927, EPI_ISL_1971928, EPI_ISL_1971929, EPI_ISL_1971930, EPI_ISL_1971931, EPI_ISL_1971932, EPI_ISL_1971933, EPI_ISL_1971934, EPI_ISL_1971935, EPI_ISL_1971936, EPI_ISL_1971937, EPI_ISL_1971938, EPI_ISL_1971939, EPI_ISL_1971940, EPI_ISL_1971941, EPI_ISL_1971942, EPI_ISL_1971943, EPI_ISL_1971944, EPI_ISL_1971945, EPI_ISL_1971946, EPI_ISL_1971947, EPI_ISL_1971949, EPI_ISL_1971950, EPI_ISL_1971951, EPI_ISL_1971952, EPI_ISL_1971953, EPI_ISL_1971954, EPI_ISL_1971955, EPI_ISL_1971956, EPI_ISL_1971957, EPI_ISL_1971958, EPI_ISL_1971959, EPI_ISL_1971960, EPI_ISL_1971961, EPI_ISL_1971962, EPI_ISL_1971963, EPI_ISL_1971964, EPI_ISL_1971965, EPI_ISL_1971966, EPI_ISL_1971967, EPI_ISL_1971968, EPI_ISL_1971969, EPI_ISL_1971970, EPI_ISL_1971971, EPI_ISL_1971972, EPI_ISL_1971973, EPI_ISL_1971974, EPI_ISL_1971975, EPI_ISL_1971976, EPI_ISL_1971977, EPI_ISL_1971978, EPI_ISL_1971979, EPI_ISL_1971980, EPI_ISL_1971981, EPI_ISL_1971982, EPI_ISL_1971983, EPI_ISL_1971984, EPI_ISL_1971985, EPI_ISL_1971986, EPI_ISL_1971987, EPI_ISL_1971988, EPI_ISL_1971989, EPI_ISL_1971990, EPI_ISL_1971991, EPI_ISL_1971992, EPI_ISL_1971993, EPI_ISL_1971994, EPI_ISL_1971995, EPI_ISL_1971996, EPI_ISL_1971997, EPI_ISL_1971998, EPI_ISL_1971999, EPI_ISL_1972000, EPI_ISL_1972001, EPI_ISL_1972002, EPI_ISL_1972003, EPI_ISL_1972004, EPI_ISL_1972005, EPI_ISL_1972006, EPI_ISL_1972007, EPI_ISL_1972008, EPI_ISL_1972009, EPI_ISL_1972010, EPI_ISL_1972011, EPI_ISL_1972012, EPI_ISL_1972013, EPI_ISL_1972014, EPI_ISL_1972015, EPI_ISL_1972016, EPI_ISL_1972017, EPI_ISL_1972019, EPI_ISL_1972020, EPI_ISL_1972021, EPI_ISL_1972022, EPI_ISL_1972023, EPI_ISL_1972024, EPI_ISL_1972025, EPI_ISL_1972026, EPI_ISL_1972027, EPI_ISL_1972028, EPI_ISL_1972029, EPI_ISL_1972030, EPI_ISL_1972031, EPI_ISL_1972032, EPI_ISL_1972033, EPI_ISL_1972034, EPI_ISL_1972035, EPI_ISL_1972036, EPI_ISL_1972037, EPI_ISL_1972039, EPI_ISL_1972040, EPI_ISL_1972041, EPI_ISL_1972042, EPI_ISL_1972043, EPI_ISL_1972044, EPI_ISL_1972045, EPI_ISL_1972046, EPI_ISL_1972047, EPI_ISL_1972048, EPI_ISL_1972049, EPI_ISL_1972050, EPI_ISL_1972051, EPI_ISL_1972052, EPI_ISL_1972053, EPI_ISL_1972054, EPI_ISL_1972055, EPI_ISL_1972056, EPI_ISL_1972057, EPI_ISL_1972058, EPI_ISL_1972059, EPI_ISL_1972060, EPI_ISL_1972061, EPI_ISL_1972062, EPI_ISL_1972063, EPI_ISL_1972064, EPI_ISL_1972065, EPI_ISL_1972066, EPI_ISL_1972067, EPI_ISL_1972068, | | | |

| | | | |
|---|---|---|---|
| EPI_ISL_1972069, EPI_ISL_1972070, EPI_ISL_1972071, EPI_ISL_1972072, EPI_ISL_1972073, EPI_ISL_1972074, EPI_ISL_1972075, EPI_ISL_1972076, EPI_ISL_1972077, EPI_ISL_1972079, EPI_ISL_1972080, EPI_ISL_1972081, EPI_ISL_1972082, EPI_ISL_1972083, EPI_ISL_1972084, EPI_ISL_1972085, EPI_ISL_1972086, EPI_ISL_1972087, EPI_ISL_1972088, EPI_ISL_1972089, EPI_ISL_1972090, EPI_ISL_1972091, EPI_ISL_1972092, EPI_ISL_1972093, EPI_ISL_1972094, EPI_ISL_1972095, EPI_ISL_1972096, EPI_ISL_1972097, EPI_ISL_1972098, EPI_ISL_1972099, EPI_ISL_1972100, EPI_ISL_1972101 | | | |
| see above | Broad Institute Clinical Research Sequencing Platform | Infectious Disease Program, Broad Institute of Harvard and MIT | Siddle,K.J., Adams,G., Pearlman,L., Gladden-Young,A., Vicente,G., Blumenstiel,B., DeFelice,M., Lee,M., McGovern,S., Lagerborg,K., Rudy,M., DeRuff,K., Carter,A., Normandin,E., Bauer,M., Reilly,S., Tomkins-Tinch,C., Loreth,C., Chaluvadi,S., Meldrim,J., Granger,B., Lemieux,J.E., Birren,B.W., Sabeti,P.C., Larkin,K., Dodge,S., Lennon,N., Madoff,L., Brown,C., Gallagher,G., Smole,S., Park,D.J., Gabriel,S., and MacInnis,B.L. |
| EPI_ISL_1972238, EPI_ISL_1972239, EPI_ISL_1972240, EPI_ISL_1972241, EPI_ISL_1972242, EPI_ISL_1972243, EPI_ISL_1972244, EPI_ISL_1972245, EPI_ISL_1972246, EPI_ISL_1972247, EPI_ISL_1972248, EPI_ISL_1972249, EPI_ISL_1972250, EPI_ISL_1972251, EPI_ISL_1972252, EPI_ISL_1972253, EPI_ISL_1972254, EPI_ISL_1972255, EPI_ISL_1972256, EPI_ISL_1972257, EPI_ISL_1972258, EPI_ISL_1972259, EPI_ISL_1972260, EPI_ISL_1972261, EPI_ISL_1972262, EPI_ISL_1972263, EPI_ISL_1972264, EPI_ISL_1972265, EPI_ISL_1972266, EPI_ISL_1972267, EPI_ISL_1972268, EPI_ISL_1972269, EPI_ISL_1972270, EPI_ISL_1972271, EPI_ISL_1972273, EPI_ISL_1972274, EPI_ISL_1972275, EPI_ISL_1972276, EPI_ISL_1972277, EPI_ISL_1972278, EPI_ISL_1972279, EPI_ISL_1972280, EPI_ISL_1972281, EPI_ISL_1972282, EPI_ISL_1972283, EPI_ISL_1972284, EPI_ISL_1972285, EPI_ISL_1972286, EPI_ISL_1972287, EPI_ISL_1972288, EPI_ISL_1972289, EPI_ISL_1972290, EPI_ISL_1972291, EPI_ISL_1972292, EPI_ISL_1972293, EPI_ISL_1972294, EPI_ISL_1972295, EPI_ISL_1972296, EPI_ISL_1972297, EPI_ISL_1972298, EPI_ISL_1972299, EPI_ISL_1972300, EPI_ISL_1972301, EPI_ISL_1972302, EPI_ISL_1972303, EPI_ISL_1972304 | | | |
| see above | Baylor Scott & White-Temple | Baylor Scott & White-Temple | Ari Rao, Linden Morales, Kimberly Walker, Marcus Volz, Shelby Hendrickson |
| EPI_ISL_1972328, EPI_ISL_1972329, EPI_ISL_1972330, EPI_ISL_1972331, EPI_ISL_1972332, EPI_ISL_1972333, EPI_ISL_1972334 | Hospital Margarita Maza de Juárez | Microbial Genomics Laboratory | Bruno Gomez-Gil, Julissa Enciso-Ibarra, Alejandra Garcia-Gasca, Daniel Fregoso-Rueda |
| EPI_ISL_1972335, EPI_ISL_1972336, EPI_ISL_1972337 | Imágenes Diagnósticas de Mazatlán | Microbial Genomics Laboratory | Bruno Gomez-Gil, Julissa Enciso-Ibarra, Alejandra Garcia-Gasca, Daniel Fregoso-Rueda |
| EPI_ISL_1972338, EPI_ISL_1972339, EPI_ISL_1972340, EPI_ISL_1972341, EPI_ISL_1972342, EPI_ISL_1972570 | Hospital Sharp | Microbial Genomics Laboratory | Bruno Gomez-Gil, Julissa Enciso-Ibarra, Alejandra Garcia-Gasca, Daniel Fregoso-Rueda |
| EPI_ISL_1972949, EPI_ISL_1972950, EPI_ISL_1972952, EPI_ISL_1972953, EPI_ISL_1972954, EPI_ISL_1972955, EPI_ISL_1972956, EPI_ISL_1972957, EPI_ISL_1972958, EPI_ISL_1972959, EPI_ISL_1972960, EPI_ISL_1972961, EPI_ISL_1972962, EPI_ISL_1972963, EPI_ISL_1972964, EPI_ISL_1972965, EPI_ISL_1972966, EPI_ISL_1972967, EPI_ISL_1972968, EPI_ISL_1972969, EPI_ISL_1972970, EPI_ISL_1972971, EPI_ISL_1972973, EPI_ISL_1972974, EPI_ISL_1972975, EPI_ISL_1972976, EPI_ISL_1972977, EPI_ISL_1972978, EPI_ISL_1972979, EPI_ISL_1972980, EPI_ISL_1972981, EPI_ISL_1972982, EPI_ISL_1972985, EPI_ISL_1972986, EPI_ISL_1972987, EPI_ISL_1972988, EPI_ISL_1972989, EPI_ISL_1972990, EPI_ISL_1972991, EPI_ISL_1972992, EPI_ISL_1972993, EPI_ISL_1972994, EPI_ISL_1972995, EPI_ISL_1972996, EPI_ISL_1972997, EPI_ISL_1972998, EPI_ISL_1972999, EPI_ISL_1973000, EPI_ISL_1973001, EPI_ISL_1973002, EPI_ISL_1973003, EPI_ISL_1973004, EPI_ISL_1973006, EPI_ISL_1973007, EPI_ISL_1973008, EPI_ISL_1973009, EPI_ISL_1973010, EPI_ISL_1973011, EPI_ISL_1973012, EPI_ISL_1973013, EPI_ISL_1973014, EPI_ISL_1973015, EPI_ISL_1973016, EPI_ISL_1973017, EPI_ISL_1973018, EPI_ISL_1973019, EPI_ISL_1973020, EPI_ISL_1973021, EPI_ISL_1973022, EPI_ISL_1973023, EPI_ISL_1973024, EPI_ISL_1973025, EPI_ISL_1973026, EPI_ISL_1973027, EPI_ISL_1973028, EPI_ISL_1973029, EPI_ISL_1973030, EPI_ISL_1973031, EPI_ISL_1973032, EPI_ISL_1973033, EPI_ISL_1973034, EPI_ISL_1973035, EPI_ISL_1973036, EPI_ISL_1973037, EPI_ISL_1973038, EPI_ISL_1973039, EPI_ISL_1973040, EPI_ISL_1973041, EPI_ISL_1973042, EPI_ISL_1973043, EPI_ISL_1973044, EPI_ISL_1973045, EPI_ISL_1973046, EPI_ISL_1973047, EPI_ISL_1973048, EPI_ISL_1973049, EPI_ISL_1973050, EPI_ISL_1973051, EPI_ISL_1973052, EPI_ISL_1973053, EPI_ISL_1973054, EPI_ISL_1973055, EPI_ISL_1973056, EPI_ISL_1973057, EPI_ISL_1973058, EPI_ISL_1973059, EPI_ISL_1973060, EPI_ISL_1973061, EPI_ISL_1973062, EPI_ISL_1973063, EPI_ISL_1973064, EPI_ISL_1973065, EPI_ISL_1973066, EPI_ISL_1973067, EPI_ISL_1973068, EPI_ISL_1973069, EPI_ISL_1973070, EPI_ISL_1973071, EPI_ISL_1973072, EPI_ISL_1973073, EPI_ISL_1973074, EPI_ISL_1973075, EPI_ISL_1973076, EPI_ISL_1973077, EPI_ISL_1973078, EPI_ISL_1973079, EPI_ISL_1973080, EPI_ISL_1973081, EPI_ISL_1973082, EPI_ISL_1973083, EPI_ISL_1973084, EPI_ISL_1973085, EPI_ISL_1973086, EPI_ISL_1973087, EPI_ISL_1973088, EPI_ISL_1973089, EPI_ISL_1973090 | | | |
| see above | Maine Health and Environmental Testing Laboratory | Tewhey Lab, The Jackson Laboratory | Matluk,N., Dewey,H., Iosue,F., Barter,M., Lynch,R., Munger,H. and Tewhey,R. |
| EPI_ISL_1973330, EPI_ISL_1973331, EPI_ISL_1973332, EPI_ISL_1973333, EPI_ISL_1973334, EPI_ISL_1973335, EPI_ISL_1973336, EPI_ISL_1973337, EPI_ISL_1973338, EPI_ISL_1973339, EPI_ISL_1973340, EPI_ISL_1973341, EPI_ISL_1973342, EPI_ISL_1973343, EPI_ISL_1973344, EPI_ISL_1973345, EPI_ISL_1973346, EPI_ISL_1973347, EPI_ISL_1973348, EPI_ISL_1973349, EPI_ISL_1973350, EPI_ISL_1973351, EPI_ISL_1973352, EPI_ISL_1973353, EPI_ISL_1973354, EPI_ISL_1973355, EPI_ISL_1973356, EPI_ISL_1973357, EPI_ISL_1973358, EPI_ISL_1973359, EPI_ISL_1973360, EPI_ISL_1973361, EPI_ISL_1973362, EPI_ISL_1973363, EPI_ISL_1973364, EPI_ISL_1973365, EPI_ISL_1973366, EPI_ISL_1973367, EPI_ISL_1973368, EPI_ISL_1973369, EPI_ISL_1973370, EPI_ISL_1973371, EPI_ISL_1973372, EPI_ISL_1973373, EPI_ISL_1973374, EPI_ISL_1973375, EPI_ISL_1973376, EPI_ISL_1973377, EPI_ISL_1973378, EPI_ISL_1973379, EPI_ISL_1973380, EPI_ISL_1973381, EPI_ISL_1973382, EPI_ISL_1973383, EPI_ISL_1973384, EPI_ISL_1973385, EPI_ISL_1973386, EPI_ISL_1973387, EPI_ISL_1973388, EPI_ISL_1973389, EPI_ISL_1973390, EPI_ISL_1973391, EPI_ISL_1973392, EPI_ISL_1973393, EPI_ISL_1973394, EPI_ISL_1973395, EPI_ISL_1973396, EPI_ISL_1973398, EPI_ISL_1973399, EPI_ISL_1973401, EPI_ISL_1973402, EPI_ISL_1973403, EPI_ISL_1973404, EPI_ISL_1973405, EPI_ISL_1973406, EPI_ISL_1973407, EPI_ISL_1973408, EPI_ISL_1973409, EPI_ISL_1973410, EPI_ISL_1973411, EPI_ISL_1973412, EPI_ISL_1973413, EPI_ISL_1973414, EPI_ISL_1973415, EPI_ISL_1973416, EPI_ISL_1973417, EPI_ISL_1973418, EPI_ISL_1973419, EPI_ISL_1973421, EPI_ISL_1973422, EPI_ISL_1973423, EPI_ISL_1973424, EPI_ISL_1973425, EPI_ISL_1973426, EPI_ISL_1973427, EPI_ISL_1973428, EPI_ISL_1973429, EPI_ISL_1973431, EPI_ISL_1973432, EPI_ISL_1973433, EPI_ISL_1973434, EPI_ISL_1973435, EPI_ISL_1973436, EPI_ISL_1973437, EPI_ISL_1973438, EPI_ISL_1973439, EPI_ISL_1973440, EPI_ISL_1973441, EPI_ISL_1973442, EPI_ISL_1973443, EPI_ISL_1973444, EPI_ISL_1973445, EPI_ISL_1973446, EPI_ISL_1973447, EPI_ISL_1973448, EPI_ISL_1973449, EPI_ISL_1973450, EPI_ISL_1973451, EPI_ISL_1973452, EPI_ISL_1973453, EPI_ISL_1973454, EPI_ISL_1973455, EPI_ISL_1973456, EPI_ISL_1973457, EPI_ISL_1973458, EPI_ISL_1973459, EPI_ISL_1973460, EPI_ISL_1973461, EPI_ISL_1973462, EPI_ISL_1973463, EPI_ISL_1973464, EPI_ISL_1973465, EPI_ISL_1973466, EPI_ISL_1973467, EPI_ISL_1973468, EPI_ISL_1973469, EPI_ISL_1973470, EPI_ISL_1973471, EPI_ISL_1973472, EPI_ISL_1973473, EPI_ISL_1973474, EPI_ISL_1973476, EPI_ISL_1973477, EPI_ISL_1973478, EPI_ISL_1973479, EPI_ISL_1973480, EPI_ISL_1973481, EPI_ISL_1973482, EPI_ISL_1973483, EPI_ISL_1973484, EPI_ISL_1973485, EPI_ISL_1973486, EPI_ISL_1973487, EPI_ISL_1973488, EPI_ISL_1973489, EPI_ISL_1973490, EPI_ISL_1973491, EPI_ISL_1973492, EPI_ISL_1973493, EPI_ISL_1973494, EPI_ISL_1973495, EPI_ISL_1973496, EPI_ISL_1973497, EPI_ISL_1973498, EPI_ISL_1973499, EPI_ISL_1973500, EPI_ISL_1973501, EPI_ISL_1973502, EPI_ISL_1973503, EPI_ISL_1973504, EPI_ISL_1973505, EPI_ISL_1973506, EPI_ISL_1973508, EPI_ISL_1973509, EPI_ISL_1973510, EPI_ISL_1973511, EPI_ISL_1973512, EPI_ISL_1973513, EPI_ISL_1973514, EPI_ISL_1973515, EPI_ISL_1973516, EPI_ISL_1973517, EPI_ISL_1973518, EPI_ISL_1973519, EPI_ISL_1973520, EPI_ISL_1973521, EPI_ISL_1973522, EPI_ISL_1973523, EPI_ISL_1973524, EPI_ISL_1973525, EPI_ISL_1973526, EPI_ISL_1973527, EPI_ISL_1973528, EPI_ISL_1973529, EPI_ISL_1973530, EPI_ISL_1973531, EPI_ISL_1973532, EPI_ISL_1973533, EPI_ISL_1973534, EPI_ISL_1973535, EPI_ISL_1973537, EPI_ISL_1973538, EPI_ISL_1973539, EPI_ISL_1973540, EPI_ISL_1973541, EPI_ISL_1973542, EPI_ISL_1973543, EPI_ISL_1973544, EPI_ISL_1973545, EPI_ISL_1973546, EPI_ISL_1973547, EPI_ISL_1973548, EPI_ISL_1973549, EPI_ISL_1973551, EPI_ISL_1973552, EPI_ISL_1973553, EPI_ISL_1973554, EPI_ISL_1973555 | | | |
| see above | Johns Hopkins Hospital Department of Pathology | Johns Hopkins Hospital Department of Pathology | C. Paul Morris, Chun Huai Luo, Adannaya Amadi, Nicholas Gallagher, Matthew Schwartz, Heba H. Mostafa |
| EPI_ISL_1989678, EPI_ISL_1989679, EPI_ISL_1989680, EPI_ISL_1989681, EPI_ISL_1989682, EPI_ISL_1989683, EPI_ISL_1989684, EPI_ISL_1989685, EPI_ISL_1989686, EPI_ISL_1989687, EPI_ISL_1989688, EPI_ISL_1989689, EPI_ISL_1989690, EPI_ISL_1989691, EPI_ISL_1989692, EPI_ISL_1989693, EPI_ISL_1989694, EPI_ISL_1989695, EPI_ISL_1989696, EPI_ISL_1989697, EPI_ISL_1989698, EPI_ISL_1989699, EPI_ISL_1989700, EPI_ISL_1989701, EPI_ISL_1989702, EPI_ISL_1989703, EPI_ISL_1989704, EPI_ISL_1989705, EPI_ISL_1989706, EPI_ISL_1989707, EPI_ISL_1989708, EPI_ISL_1989709, EPI_ISL_1989710, EPI_ISL_1989711, EPI_ISL_1989712, EPI_ISL_1989713, EPI_ISL_1989714, EPI_ISL_1989715, EPI_ISL_1989716, EPI_ISL_1989717, EPI_ISL_1989718, EPI_ISL_1989719, EPI_ISL_1989720, EPI_ISL_1989721, EPI_ISL_1989722, EPI_ISL_1989723, EPI_ISL_1989724, EPI_ISL_1989725, EPI_ISL_1989726, EPI_ISL_1989727, EPI_ISL_1989728, EPI_ISL_1989729, EPI_ISL_1989730, EPI_ISL_1989731, EPI_ISL_1989732, EPI_ISL_1989733, EPI_ISL_1989734, EPI_ISL_1989735, EPI_ISL_1989736, EPI_ISL_1989737, EPI_ISL_1989738, EPI_ISL_1989739, EPI_ISL_1989740, EPI_ISL_1989741, EPI_ISL_1989742, EPI_ISL_1989743, EPI_ISL_1989744, EPI_ISL_1989745, EPI_ISL_1989746, EPI_ISL_1989747, EPI_ISL_1989748, EPI_ISL_1989749, EPI_ISL_1989750, EPI_ISL_1989751, EPI_ISL_1989752, EPI_ISL_1989753, EPI_ISL_1989754, EPI_ISL_1989755, EPI_ISL_1989756, EPI_ISL_1989757, EPI_ISL_1989758, EPI_ISL_1989759, EPI_ISL_1989760, EPI_ISL_1989761, EPI_ISL_1989762, EPI_ISL_1989763, EPI_ISL_1989764, EPI_ISL_1989765, EPI_ISL_1989766, EPI_ISL_1989767, EPI_ISL_1989768, EPI_ISL_1989769, EPI_ISL_1989770, EPI_ISL_1989771, EPI_ISL_1989772, EPI_ISL_1989773, EPI_ISL_1989774, EPI_ISL_1989775, EPI_ISL_1989776, EPI_ISL_1989777, EPI_ISL_1989778, EPI_ISL_1989779, EPI_ISL_1989780, EPI_ISL_1989781, EPI_ISL_1989782, EPI_ISL_1989783, EPI_ISL_1989784, EPI_ISL_1989785, EPI_ISL_1989786, EPI_ISL_1989787, EPI_ISL_1989788, EPI_ISL_1989789, EPI_ISL_1989790, EPI_ISL_1989791, EPI_ISL_1989792, EPI_ISL_1989793, EPI_ISL_1989794, EPI_ISL_1989795, EPI_ISL_1989796, EPI_ISL_1989797, EPI_ISL_1989798, EPI_ISL_1989799, EPI_ISL_1989800, EPI_ISL_1989801, EPI_ISL_1989802, EPI_ISL_1989803, EPI_ISL_1989804, EPI_ISL_1989805, EPI_ISL_1989806, EPI_ISL_1989807, EPI_ISL_1989808, EPI_ISL_1989809, EPI_ISL_1989810, EPI_ISL_1989811, EPI_ISL_1989812, EPI_ISL_1989813, EPI_ISL_1989814, EPI_ISL_1989815, EPI_ISL_1989816, EPI_ISL_1989817, EPI_ISL_1989818, EPI_ISL_1989819, EPI_ISL_1989820, EPI_ISL_1989821, EPI_ISL_1989822, EPI_ISL_1989823, EPI_ISL_1989824, EPI_ISL_1989825, EPI_ISL_1989826, EPI_ISL_1989827, EPI_ISL_1989828, EPI_ISL_1989829, EPI_ISL_1989830, EPI_ISL_1989831, EPI_ISL_1989832, EPI_ISL_1989833, EPI_ISL_1989834, EPI_ISL_1989835, EPI_ISL_1989836, EPI_ISL_1989837, EPI_ISL_1989838, EPI_ISL_1989839, EPI_ISL_1989840, EPI_ISL_1989841, EPI_ISL_1989842, EPI_ISL_1989843, EPI_ISL_1989844, EPI_ISL_1989845, EPI_ISL_1989846, EPI_ISL_1989847, EPI_ISL_1989848, EPI_ISL_1989849, EPI_ISL_1989850, EPI_ISL_1989851, EPI_ISL_1989852, EPI_ISL_1989853, EPI_ISL_1989854, EPI_ISL_1989855, EPI_ISL_1989856, EPI_ISL_1989857, EPI_ISL_1989858, EPI_ISL_1989859, EPI_ISL_1989860, EPI_ISL_1989861, EPI_ISL_1989862, EPI_ISL_1989863, EPI_ISL_1989864, EPI_ISL_1989865, EPI_ISL_1989866, EPI_ISL_1989867, EPI_ISL_1989868, EPI_ISL_1989869, EPI_ISL_1989870, EPI_ISL_1989871, EPI_ISL_1989872, EPI_ISL_1989873, EPI_ISL_1989874, EPI_ISL_1989875, EPI_ISL_1989876, EPI_ISL_1989877, EPI_ISL_1989878, EPI_ISL_1989879, EPI_ISL_1989880, EPI_ISL_1989881, EPI_ISL_1989882, EPI_ISL_1989883, EPI_ISL_1989884, EPI_ISL_1989885, EPI_ISL_1989886, EPI_ISL_1989887, EPI_ISL_1989888, EPI_ISL_1989889, EPI_ISL_1989890, EPI_ISL_1989891, EPI_ISL_1989892, EPI_ISL_1989893, EPI_ISL_1989894, EPI_ISL_1989895, EPI_ISL_1989896, EPI_ISL_1989897, EPI_ISL_1989898, EPI_ISL_1989899, EPI_ISL_1989900, EPI_ISL_1989901, EPI_ISL_1989902, EPI_ISL_1989903, EPI_ISL_1989904, EPI_ISL_1989905, EPI_ISL_1989906, EPI_ISL_1989907, EPI_ISL_1989908, EPI_ISL_1989909, EPI_ISL_1989910, EPI_ISL_1989911, EPI_ISL_1989912, EPI_ISL_1989913, EPI_ISL_1989914, EPI_ISL_1989915, EPI_ISL_1989916, EPI_ISL_1989917, EPI_ISL_1989918, EPI_ISL_1989919, EPI_ISL_1989920, EPI_ISL_1989921, EPI_ISL_1989922, EPI_ISL_1989923, EPI_ISL_1989924, EPI_ISL_1989925, EPI_ISL_1989926, EPI_ISL_1989927, EPI_ISL_1989928, EPI_ISL_1989929, EPI_ISL_1989930, EPI_ISL_1989931, EPI_ISL_1989932, EPI_ISL_1989933, EPI_ISL_1989934, EPI_ISL_1989935, EPI_ISL_1989936, EPI_ISL_1989937, EPI_ISL_1989938, EPI_ISL_1989939, EPI_ISL_1989940, EPI_ISL_1989941, EPI_ISL_1989942, EPI_ISL_1989943, EPI_ISL_1989944, EPI_ISL_1989945, EPI_ISL_1989946, EPI_ISL_1989947, EPI_ISL_1989948, EPI_ISL_1989949, EPI_ISL_1989950, EPI_ISL_1989951, EPI_ISL_1989952, EPI_ISL_1989953, EPI_ISL_1989954, EPI_ISL_1989955, EPI_ISL_1989956, EPI_ISL_1989957, EPI_ISL_1989958, EPI_ISL_1989959, EPI_ISL_1989960, EPI_ISL_1989961, EPI_ISL_1989962, EPI_ISL_1989963, EPI_ISL_1989964, EPI_ISL_1989965, EPI_ISL_1989966, EPI_ISL_1989967, EPI_ISL_1989968, EPI_ISL_1989969, EPI_ISL_1989970, EPI_ISL_1989971, EPI_ISL_1989972, EPI_ISL_1989973, EPI_ISL_1989974, EPI_ISL_1989975 | | | |
| see above | Quest Diagnostics Incorporated | Centers for Disease Control and Prevention Division of Viral Diseases, Pathogen Discovery | Dakota Howard, Dhwani Batra, Peter W. Cook, Kara Moser, Adrian Paskey, Jason Caravas, Benjamin Rambo-Martin, Shatavia Morrison, Christopher Gulvick, Scott Sammons, Yvette Unoarumhi, Darlene Wagner, Matthew Schmerer, S. H. Rosenthal, A. Gerasimova, R. M. Kagan, B. Anderson, M. Hua, Y. Liu, L.E. Bernstein, K.E. Livingston, A. Perez, I. A. Shlyakhter, R. V. Rolando, R. Owen, P. Tanpaiboon, F. Lacbawan, Clinton R. Paden, Duncan MacCannell |
| EPI_ISL_1989976 | PR Public Health Lab | Centers for Disease Control and Prevention Division of Viral Diseases, Pathogen Discovery | Mili Sheth, Sarah Nobles, Jasmine Padilla, Mark Burroughs, Shoshona Le, Katie Dillon, Peter Cook, Clinton R. Paden, Dhwani Batra, Krista Queen, Kristen Knipe, Dakota Howard, Yvette Unoarumhi, Darlene Wagner, Matthew Schmerer, Ben L. Rambo-Martin, Kristine Lacek, Sam Shepard, Alison Laufer Halpin, Dave Wentworth, Vivien Dugan, Suxiang Tong, Justin Lee |
| EPI_ISL_1989977 | IL Dept. of Public Health Springfield Laboratory | Centers for Disease Control and Prevention Division of Viral Diseases, Pathogen Discovery | Mili Sheth, Sarah Nobles, Jasmine Padilla, Mark Burroughs, Shoshona Le, Katie Dillon, Peter Cook, Clinton R. Paden, Dhwani Batra, Krista Queen, Kristen Knipe, Dakota Howard, Yvette Unoarumhi, Darlene Wagner, Matthew Schmerer, Ben L. Rambo-Martin, Kristine Lacek, Sam Shepard, Alison Laufer Halpin, Dave Wentworth, Vivien Dugan, Suxiang Tong, Justin Lee |
| EPI_ISL_1989978 | WI State Laboratory of Hygiene | Centers for Disease Control and Prevention Division of Viral Diseases, Pathogen Discovery | Mili Sheth, Sarah Nobles, Jasmine Padilla, Mark Burroughs, Shoshona Le, Katie Dillon, Peter Cook, Clinton R. Paden, Dhwani Batra, Krista Queen, Kristen Knipe, Dakota Howard, Yvette Unoarumhi, Darlene Wagner, Matthew Schmerer, Ben L. Rambo-Martin, Kristine Lacek, Sam Shepard, Alison Laufer Halpin, Dave Wentworth, Vivien Dugan, Suxiang Tong, Justin Lee |
| EPI_ISL_1989979 | DC Public Health Lab/ Dept. of Forensic Sciences | Centers for Disease Control and Prevention Division of Viral Diseases, Pathogen Discovery | Mili Sheth, Sarah Nobles, Jasmine Padilla, Mark Burroughs, Shoshona Le, Katie Dillon, Peter Cook, Clinton R. Paden, Dhwani Batra, Krista Queen, Kristen Knipe, Dakota Howard, Yvette Unoarumhi, Darlene Wagner, Matthew Schmerer, Ben L. Rambo-Martin, Kristine Lacek, Sam Shepard, Alison Laufer Halpin, Dave Wentworth, Vivien Dugan, Suxiang Tong, Justin Lee |
| EPI_ISL_1989980, EPI_ISL_1989981, EPI_ISL_1989982, EPI_ISL_1989983, EPI_ISL_1989984, EPI_ISL_1989985, EPI_ISL_1989986, EPI_ISL_1989987, EPI_ISL_1989988, EPI_ISL_1989989, EPI_ISL_1989990, EPI_ISL_1989991, EPI_ISL_1989992, EPI_ISL_1989993, EPI_ISL_1989994, EPI_ISL_1989995, EPI_ISL_1989996, EPI_ISL_1989997, EPI_ISL_1989998, EPI_ISL_1989999, EPI_ISL_1990000, EPI_ISL_1990001, EPI_ISL_1990002, EPI_ISL_1990003 | | | |
| see above | Quest Diagnostics Incorporated | Centers for Disease Control and Prevention Division of Viral Diseases, Pathogen Discovery | Dakota Howard, Dhwani Batra, Peter W. Cook, Kara Moser, Adrian Paskey, Jason Caravas, Benjamin Rambo-Martin, Shatavia Morrison, Christopher Gulvick, Scott Sammons, Yvette Unoarumhi, Darlene Wagner, Matthew Schmerer, S. H. Rosenthal, A. Gerasimova, R. M. Kagan, B. Anderson, M. Hua, Y. Liu, L.E. Bernstein, K.E. Livingston, A. Perez, I. A. Shlyakhter, R. V. Rolando, R. Owen, P. Tanpaiboon, F. Lacbawan, Clinton R. Paden, Duncan MacCannell |
| EPI_ISL_1990051 | Laboratory Corporation of America | Centers for Disease Control and Prevention Division of Viral Diseases, Pathogen Discovery | Dakota Howard, Dhwani Batra, Peter W. Cook, Kara Moser, Adrian Paskey, Jason Caravas, Benjamin Rambo-Martin, Shatavia Morrison, Christopher Gulvick, Scott Sammons, Yvette Unoarumhi, Darlene Wagner, Matthew Schmerer, Minoo Agarwal, Eyad Almasri, Debbie Boles, Ayla Burns, Nuthawin Charoensri, Oren Cohen, Susan Countryman, Mary Ann Cristobal, Bobbi Croy, Suzanne Dale, Hrushikesh Deshmukh, Amanda Douglas, Vincent Drouillon, Marcia Eisenberg, Howard Engler, Rama Ghatti, Prashant Gupta, Susan Hicks, Jake Humphrey, Lax Iyer, Manoj Jain, Mohan Kolli, Brian Krueger, Tim Kuphal, Stanley Letovsky, Michael Levandoski, Craig Lukasik, Jonathan Meltzer, Brian Norvell, Mindy Nye, Scott Parker, Christos Petropoulos, John Pruitt, Steven Ragan, Scott Ryan, Mike Sapeta, Jana Schroth, Suresh Babu Selvaraju, Goran Stevovic, Amanda Suchanek, Andrea Throop, Lyndon Tilson, Thomas Urban, Joe Voshell, Kimberly Wagner, Jonathan Williams, Mary Williamson, Qian Zeng, Tricia Zwiefelhofer, Clinton R. Paden, Duncan MacCannell |
| EPI_ISL_1990052, EPI_ISL_1990139, EPI_ISL_1990140, EPI_ISL_1990141, EPI_ISL_1990142, EPI_ISL_1990143, EPI_ISL_1990144, EPI_ISL_1990145, EPI_ISL_1990146, EPI_ISL_1990147, EPI_ISL_1990148, EPI_ISL_1990149, EPI_ISL_1990150, EPI_ISL_1990151, EPI_ISL_1990152, EPI_ISL_1990153, EPI_ISL_1990154, EPI_ISL_1990155, EPI_ISL_1990156, EPI_ISL_1990157, EPI_ISL_1990158, EPI_ISL_1990159, EPI_ISL_1990160, EPI_ISL_1990161, EPI_ISL_1990162, EPI_ISL_1990163, EPI_ISL_1990164, EPI_ISL_1990165, EPI_ISL_1990166, EPI_ISL_1990167, EPI_ISL_1990168, EPI_ISL_1990169, EPI_ISL_1990170, EPI_ISL_1990171, EPI_ISL_1990172, EPI_ISL_1990173, EPI_ISL_1990174, EPI_ISL_1990175, EPI_ISL_1990176, EPI_ISL_1990177, EPI_ISL_1990178, EPI_ISL_1990179, EPI_ISL_1990180, EPI_ISL_1990181, EPI_ISL_1990182, EPI_ISL_1990183, EPI_ISL_1990184, EPI_ISL_1990185, EPI_ISL_1990186, EPI_ISL_1990187, EPI_ISL_1990188, EPI_ISL_1990189, EPI_ISL_1990190, EPI_ISL_1990191, EPI_ISL_1990192, EPI_ISL_1990193, EPI_ISL_1990194, EPI_ISL_1990195, EPI_ISL_1990196, EPI_ISL_1990197, EPI_ISL_1990198, EPI_ISL_1990199, EPI_ISL_1990200, EPI_ISL_1990201, EPI_ISL_1990202, EPI_ISL_1990203, EPI_ISL_1990204, EPI_ISL_1990205, EPI_ISL_1990206, EPI_ISL_1990207, EPI_ISL_1990208, EPI_ISL_1990209, EPI_ISL_1990210, EPI_ISL_1990211, EPI_ISL_1990212, EPI_ISL_1990213, EPI_ISL_1990214, EPI_ISL_1990215, EPI_ISL_1990216, EPI_ISL_1990217, EPI_ISL_1990218, EPI_ISL_1990219, EPI_ISL_1990220, EPI_ISL_1990221, EPI_ISL_1990222, EPI_ISL_1990223, EPI_ISL_1990224, EPI_ISL_1990225, EPI_ISL_1990226, EPI_ISL_1990227, EPI_ISL_1990228, EPI_ISL_1990229, EPI_ISL_1990230, EPI_ISL_1990231, EPI_ISL_1990232, EPI_ISL_1990233, EPI_ISL_1990234, EPI_ISL_1990235, EPI_ISL_1990236, EPI_ISL_1990237, EPI_ISL_1990238, EPI_ISL_1990239, EPI_ISL_1990240, EPI_ISL_1990241, EPI_ISL_1990242, EPI_ISL_1990243, EPI_ISL_1990244, EPI_ISL_1990245, EPI_ISL_1990246, EPI_ISL_1990247, EPI_ISL_1990248, EPI_ISL_1990249, EPI_ISL_1990250, EPI_ISL_1990251, EPI_ISL_1990252, EPI_ISL_1990253, EPI_ISL_1990254, EPI_ISL_1990255, EPI_ISL_1990256, EPI_ISL_1990257, EPI_ISL_1990258, EPI_ISL_1990259, EPI_ISL_1990260, EPI_ISL_1990261, EPI_ISL_1990262, EPI_ISL_1990263, EPI_ISL_1990264, EPI_ISL_1990265, EPI_ISL_1990266, EPI_ISL_1990267, EPI_ISL_1990268, EPI_ISL_1990269, EPI_ISL_1990270, EPI_ISL_1990271, EPI_ISL_1990272, EPI_ISL_1990273, EPI_ISL_1990274, EPI_ISL_1990275, EPI_ISL_1990276, EPI_ISL_1990277, EPI_ISL_1990278, EPI_ISL_1990279, EPI_ISL_1990280, EPI_ISL_1990281, EPI_ISL_1990282, EPI_ISL_1990283, EPI_ISL_1990284, EPI_ISL_1990285, EPI_ISL_1990286, EPI_ISL_1990287, EPI_ISL_1990288, EPI_ISL_1990289, EPI_ISL_1990290, EPI_ISL_1990291, EPI_ISL_1990292, EPI_ISL_1990293, EPI_ISL_1990294, EPI_ISL_1990295, EPI_ISL_1990296, EPI_ISL_1990297, EPI_ISL_1990298, EPI_ISL_1990299, EPI_ISL_1990300, EPI_ISL_1990301, EPI_ISL_1990302, EPI_ISL_1990303, EPI_ISL_1990304, EPI_ISL_1990305, EPI_ISL_1990306, EPI_ISL_1990307, EPI_ISL_1990308, EPI_ISL_1990309, EPI_ISL_1990310, EPI_ISL_1990311, EPI_ISL_1990312, EPI_ISL_1990313, EPI_ISL_1990314, EPI_ISL_1990315, EPI_ISL_1990316, EPI_ISL_1990317, EPI_ISL_1990318, EPI_ISL_1990319, EPI_ISL_1990320, EPI_ISL_1990321, EPI_ISL_1990322, EPI_ISL_1990323, EPI_ISL_1990324, EPI_ISL_1990325, EPI_ISL_1990326, EPI_ISL_1990327, EPI_ISL_1990328, EPI_ISL_1990329, EPI_ISL_1990330, EPI_ISL_1990331, EPI_ISL_1990332, EPI_ISL_1990333, EPI_ISL_1990334, EPI_ISL_1990335, EPI_ISL_1990336, EPI_ISL_1990337, EPI_ISL_1990338, EPI_ISL_1990339, EPI_ISL_1990340, EPI_ISL_1990341, EPI_ISL_1990342, EPI_ISL_1990343, EPI_ISL_1990344, EPI_ISL_1990345, EPI_ISL_1990346, EPI_ISL_1990347, EPI_ISL_1990348, EPI_ISL_1990349, EPI_ISL_1990350, EPI_ISL_1990351, EPI_ISL_1990352, EPI_ISL_1990353, EPI_ISL_1990354, EPI_ISL_1990355, EPI_ISL_1990356, EPI_ISL_1990357, EPI_ISL_1990358, EPI_ISL_1990359, EPI_ISL_1990360, EPI_ISL_1990361, EPI_ISL_1990362, EPI_ISL_1990363, EPI_ISL_1990364, EPI_ISL_1990365, EPI_ISL_1990366, EPI_ISL_1990367, EPI_ISL_1990368, EPI_ISL_1990369, EPI_ISL_1990370, EPI_ISL_1990371, EPI_ISL_1990372, EPI_ISL_1990373, EPI_ISL_1990374, EPI_ISL_1990375, EPI_ISL_1990376, EPI_ISL_1990377, EPI_ISL_1990378, EPI_ISL_1990379, EPI_ISL_1990380, EPI_ISL_1990381, EPI_ISL_1990382, EPI_ISL_1990383, EPI_ISL_1990384, EPI_ISL_1990385, EPI_ISL_1990386, EPI_ISL_1990387, EPI_ISL_1990388, EPI_ISL_1990389, EPI_ISL_1990390, EPI_ISL_1990391, EPI_ISL_1990392, EPI_ISL_1990393, EPI_ISL_1990394, EPI_ISL_1990395, EPI_ISL_1990396, EPI_ISL_1990397, EPI_ISL_1990398, EPI_ISL_1990399, EPI_ISL_1990400, EPI_ISL_1990401, EPI_ISL_1990402, EPI_ISL_1990403, EPI_ISL_1990404, EPI_ISL_1990405, EPI_ISL_1990406, EPI_ISL_1990407, EPI_ISL_1990408, EPI_ISL_1990409, EPI_ISL_1990410, EPI_ISL_1990411, EPI_ISL_1990412, EPI_ISL_1990413, EPI_ISL_1990414, EPI_ISL_1990415, EPI_ISL_1990416, EPI_ISL_1990417, EPI_ISL_1990418, EPI_ISL_1990419, EPI_ISL_1990420, EPI_ISL_1990421, EPI_ISL_1990422, EPI_ISL_1990423, EPI_ISL_1990424, EPI_ISL_1990425, EPI_ISL_1990426, EPI_ISL_1990427, EPI_ISL_1990428, EPI_ISL_1990429, EPI_ISL_1990430, EPI_ISL_1990431, EPI_ISL_1990432, EPI_ISL_1990433, EPI_ISL_1990434, EPI_ISL_1990435, EPI_ISL_1990436, EPI_ISL_1990437, EPI_ISL_1990438, EPI_ISL_1990439, EPI_ISL_1990440, EPI_ISL_1990441, EPI_ISL_1990442, EPI_ISL_1990443, EPI_ISL_1990444, EPI_ISL_1990445, EPI_ISL_1990446, EPI_ISL_1990447, EPI_ISL_1990448, EPI_ISL_1990449, EPI_ISL_1990450, EPI_ISL_1990451, EPI_ISL_1990452, EPI_ISL_1990453, EPI_ISL_1990454, EPI_ISL_1990455, EPI_ISL_1990456, EPI_ISL_1990457, EPI_ISL_1990458, EPI_ISL_1990459, EPI_ISL_1990460, EPI_ISL_1990461, EPI_ISL_1990462, EPI_ISL_1990463, EPI_ISL_1990464, EPI_ISL_1990465, EPI_ISL_1990466, EPI_ISL_1990467, EPI_ISL_1990468, EPI_ISL_1990469, EPI_ISL_1990470, EPI_ISL_1990471, EPI_ISL_1990472, EPI_ISL_1990473, EPI_ISL_1990474, EPI_ISL_1990475, EPI_ISL_1990476, EPI_ISL_1990477, EPI_ISL_1990478, EPI_ISL_1990479, EPI_ISL_1990480, EPI_ISL_1990481, EPI_ISL_1990482, EPI_ISL_1990483, EPI_ISL_1990484, EPI_ISL_1990485, EPI_ISL_1990486, EPI_ISL_1990487, EPI_ISL_1990488, EPI_ISL_1990489, EPI_ISL_1990490, EPI_ISL_1990491, EPI_ISL_1990492, EPI_ISL_1990493, EPI_ISL_1990494, EPI_ISL_1990495, EPI_ISL_1990496, EPI_ISL_1990497, EPI_ISL_1990498, EPI_ISL_1990499, EPI_ISL_1990500, EPI_ISL_1990501, EPI_ISL_1990502, EPI_ISL_1990503, EPI_ISL_1990504, EPI_ISL_1990505, EPI_ISL_1990506, EPI_ISL_1990507, EPI_ISL_1990508, EPI_ISL_1990509, EPI_ISL_1990510, EPI_ISL_1990511, EPI_ISL_1990512, EPI_ISL_1990513, EPI_ISL_1990514, EPI_ISL_1990515, EPI_ISL_1990516, EPI_ISL_1990517, EPI_ISL_1990518, EPI_ISL_1990519, EPI_ISL_1990520, EPI_ISL_1990521, EPI_ISL_1990522, EPI_ISL_1990523, EPI_ISL_1990524, EPI_ISL_1990525, EPI_ISL_1990526, EPI_ISL_1990527, EPI_ISL_1990528, EPI_ISL_1990529, EPI_ISL_1990530, EPI_ISL_1990531, EPI_ISL_1990532, EPI_ISL_1990533, | | | |

EPI_ISL_1990534, EPI_ISL_1990535, EPI_ISL_1990536, EPI_ISL_1990537, EPI_ISL_1990538, EPI_ISL_1990539, EPI_ISL_1990540, EPI_ISL_1990541, EPI_ISL_1990542, EPI_ISL_1990543

| | | | |
|---|---|---|---|
| see above | Quest Diagnostics Incorporated | Centers for Disease Control and Prevention Division of Viral Diseases, Pathogen Discovery | Dakota Howard, Dhwani Batra, Peter W. Cook, Kara Moser, Adrian Paskey, Jason Caravas, Benjamin Rambo-Martin, Shatavia Morrison, Christopher Gulvick, Scott Sammons, Yvette Unoarumhi, Darlene Wagner, Matthew Schmerer, S. H. Rosenthal, A. Gerasimova, R. M. Kagan, B. Anderson, M. Hua, Y. Liu, L.E. Bernstein, K.E. Livingston, A. Perez, I. A. Shlyakhter, R. V. Rolando, R. Owen, P. Tanpaiboon, F. Lacbawan, Clinton R. Paden, Duncan MacCannell |

EPI_ISL_1990544, EPI_ISL_1990545, EPI_ISL_1990546, EPI_ISL_1990547, EPI_ISL_1990548, EPI_ISL_1990549, EPI_ISL_1990550, EPI_ISL_1990551, EPI_ISL_1990552, EPI_ISL_1990553, EPI_ISL_1990554, EPI_ISL_1990555, EPI_ISL_1990556, EPI_ISL_1990557, EPI_ISL_1990558, EPI_ISL_1990559, EPI_ISL_1990560, EPI_ISL_1990561, EPI_ISL_1990562, EPI_ISL_1990563, EPI_ISL_1990564, EPI_ISL_1990565, EPI_ISL_1990566, EPI_ISL_1990567, EPI_ISL_1990568, EPI_ISL_1990569, EPI_ISL_1990570, EPI_ISL_1990571, EPI_ISL_1990572, EPI_ISL_1990573, EPI_ISL_1990574, EPI_ISL_1990575, EPI_ISL_1990576, EPI_ISL_1990577, EPI_ISL_1990578, EPI_ISL_1990579, EPI_ISL_1990580, EPI_ISL_1990581, EPI_ISL_1990582, EPI_ISL_1990583, EPI_ISL_1990584, EPI_ISL_1990585, EPI_ISL_1990586, EPI_ISL_1990587, EPI_ISL_1990588, EPI_ISL_1990589, EPI_ISL_1990590, EPI_ISL_1990591, EPI_ISL_1990592, EPI_ISL_1990593, EPI_ISL_1990594, EPI_ISL_1990595, EPI_ISL_1990596, EPI_ISL_1990597, EPI_ISL_1990598, EPI_ISL_1990599, EPI_ISL_1990600, EPI_ISL_1990601, EPI_ISL_1990602, EPI_ISL_1990603, EPI_ISL_1990604, EPI_ISL_1990605, EPI_ISL_1990606, EPI_ISL_1990607, EPI_ISL_1990608, EPI_ISL_1990609, EPI_ISL_1990610, EPI_ISL_1990611, EPI_ISL_1990612, EPI_ISL_1990613, EPI_ISL_1990614, EPI_ISL_1990615, EPI_ISL_1990616, EPI_ISL_1990617, EPI_ISL_1990618, EPI_ISL_1990619, EPI_ISL_1990620, EPI_ISL_1990621, EPI_ISL_1990622, EPI_ISL_1990623, EPI_ISL_1990624, EPI_ISL_1990625, EPI_ISL_1990626, EPI_ISL_1990627, EPI_ISL_1990628, EPI_ISL_1990629, EPI_ISL_1990630, EPI_ISL_1990631, EPI_ISL_1990632, EPI_ISL_1990633, EPI_ISL_1990634, EPI_ISL_1990635, EPI_ISL_1990636, EPI_ISL_1990637, EPI_ISL_1990638, EPI_ISL_1990639, EPI_ISL_1990640, EPI_ISL_1990641, EPI_ISL_1990642, EPI_ISL_1990643, EPI_ISL_1990644, EPI_ISL_1990645, EPI_ISL_1990646, EPI_ISL_1990647, EPI_ISL_1990648, EPI_ISL_1990649, EPI_ISL_1990650, EPI_ISL_1990654, EPI_ISL_1990657, EPI_ISL_1990661, EPI_ISL_1990664, EPI_ISL_1990668, EPI_ISL_1990671, EPI_ISL_1990675, EPI_ISL_1990678, EPI_ISL_1990682, EPI_ISL_1990686, EPI_ISL_1990689, EPI_ISL_1990693, EPI_ISL_1990697, EPI_ISL_1990700, EPI_ISL_1990704, EPI_ISL_1990707, EPI_ISL_1990710, EPI_ISL_1990711, EPI_ISL_1990712, EPI_ISL_1990713, EPI_ISL_1990714, EPI_ISL_1990715, EPI_ISL_1990716, EPI_ISL_1990717, EPI_ISL_1990718, EPI_ISL_1990719, EPI_ISL_1990720, EPI_ISL_1990721, EPI_ISL_1990722, EPI_ISL_1990723, EPI_ISL_1990724, EPI_ISL_1990725, EPI_ISL_1990726, EPI_ISL_1990727, EPI_ISL_1990728, EPI_ISL_1990729, EPI_ISL_1990730, EPI_ISL_1990731, EPI_ISL_1990732, EPI_ISL_1990733, EPI_ISL_1990734, EPI_ISL_1990735, EPI_ISL_1990736, EPI_ISL_1990737, EPI_ISL_1990738, EPI_ISL_1990739, EPI_ISL_1990740, EPI_ISL_1990741, EPI_ISL_1990742, EPI_ISL_1990743, EPI_ISL_1990744, EPI_ISL_1990745, EPI_ISL_1990746, EPI_ISL_1990747, EPI_ISL_1990748, EPI_ISL_1990749, EPI_ISL_1990750, EPI_ISL_1990751, EPI_ISL_1990752, EPI_ISL_1990753, EPI_ISL_1990754, EPI_ISL_1990755, EPI_ISL_1990756, EPI_ISL_1990757, EPI_ISL_1990758, EPI_ISL_1990759, EPI_ISL_1990760, EPI_ISL_1990761, EPI_ISL_1990762, EPI_ISL_1990763, EPI_ISL_1990764, EPI_ISL_1990765, EPI_ISL_1990766, EPI_ISL_1990767, EPI_ISL_1990768, EPI_ISL_1990769, EPI_ISL_1990770, EPI_ISL_1990771, EPI_ISL_1990772, EPI_ISL_1990773, EPI_ISL_1990774, EPI_ISL_1990775, EPI_ISL_1990776, EPI_ISL_1990777, EPI_ISL_1990778, EPI_ISL_1990779, EPI_ISL_1990780, EPI_ISL_1990781, EPI_ISL_1990782, EPI_ISL_1990783, EPI_ISL_1990784, EPI_ISL_1990785, EPI_ISL_1990786, EPI_ISL_1990787, EPI_ISL_1990788, EPI_ISL_1990789, EPI_ISL_1990790, EPI_ISL_1990791, EPI_ISL_1990792, EPI_ISL_1990793, EPI_ISL_1990794, EPI_ISL_1990795, EPI_ISL_1990796, EPI_ISL_1990797, EPI_ISL_1990798, EPI_ISL_1990799, EPI_ISL_1990800, EPI_ISL_1990801, EPI_ISL_1990802, EPI_ISL_1990803, EPI_ISL_1990804, EPI_ISL_1990805, EPI_ISL_1990806, EPI_ISL_1990807, EPI_ISL_1990808, EPI_ISL_1990809, EPI_ISL_1990810, EPI_ISL_1990811, EPI_ISL_1990812, EPI_ISL_1990813, EPI_ISL_1990814, EPI_ISL_1990815, EPI_ISL_1990816, EPI_ISL_1990817, EPI_ISL_1990818, EPI_ISL_1990819, EPI_ISL_1990820, EPI_ISL_1990821, EPI_ISL_1990822, EPI_ISL_1990823, EPI_ISL_1990824, EPI_ISL_1990825, EPI_ISL_1990826, EPI_ISL_1990827, EPI_ISL_1990828, EPI_ISL_1990829, EPI_ISL_1990830, EPI_ISL_1990831, EPI_ISL_1990832, EPI_ISL_1990833, EPI_ISL_1990834, EPI_ISL_1990835, EPI_ISL_1990836, EPI_ISL_1990837, EPI_ISL_1990838, EPI_ISL_1990839, EPI_ISL_1990840, EPI_ISL_1990841, EPI_ISL_1990842, EPI_ISL_1990843, EPI_ISL_1990844, EPI_ISL_1990845, EPI_ISL_1990846, EPI_ISL_1990847, EPI_ISL_1990848, EPI_ISL_1990849, EPI_ISL_1990850, EPI_ISL_1990851, EPI_ISL_1990852, EPI_ISL_1990853, EPI_ISL_1990854, EPI_ISL_1990855, EPI_ISL_1990856, EPI_ISL_1990857, EPI_ISL_1990858, EPI_ISL_1990859, EPI_ISL_1990860, EPI_ISL_1990861, EPI_ISL_1990862, EPI_ISL_1990863, EPI_ISL_1990864, EPI_ISL_1990865, EPI_ISL_1990866, EPI_ISL_1990867, EPI_ISL_1990868, EPI_ISL_1990869, EPI_ISL_1990870, EPI_ISL_1990871, EPI_ISL_1990872, EPI_ISL_1990873, EPI_ISL_1990874, EPI_ISL_1990875, EPI_ISL_1990876, EPI_ISL_1990877, EPI_ISL_1990878, EPI_ISL_1990879, EPI_ISL_1990880, EPI_ISL_1990881, EPI_ISL_1990882, EPI_ISL_1990883, EPI_ISL_1990884, EPI_ISL_1990885, EPI_ISL_1990886, EPI_ISL_1990887, EPI_ISL_1990888, EPI_ISL_1990889, EPI_ISL_1990890, EPI_ISL_1990891, EPI_ISL_1990892, EPI_ISL_1990893, EPI_ISL_1990894, EPI_ISL_1990895, EPI_ISL_1990896, EPI_ISL_1990897, EPI_ISL_1990898, EPI_ISL_1990899, EPI_ISL_1990900, EPI_ISL_1990901, EPI_ISL_1990902, EPI_ISL_1990903, EPI_ISL_1990904, EPI_ISL_1990905, EPI_ISL_1990906, EPI_ISL_1990907, EPI_ISL_1990908, EPI_ISL_1990909, EPI_ISL_1990910, EPI_ISL_1990911, EPI_ISL_1990912, EPI_ISL_1990913, EPI_ISL_1990914, EPI_ISL_1990915, EPI_ISL_1990916, EPI_ISL_1990917, EPI_ISL_1990918, EPI_ISL_1990919, EPI_ISL_1990920, EPI_ISL_1990921, EPI_ISL_1990922, EPI_ISL_1990923, EPI_ISL_1990924, EPI_ISL_1990925, EPI_ISL_1990926, EPI_ISL_1990927, EPI_ISL_1990928, EPI_ISL_1990929, EPI_ISL_1990930, EPI_ISL_1990931, EPI_ISL_1990932, EPI_ISL_1990933, EPI_ISL_1990934, EPI_ISL_1990935, EPI_ISL_1990936, EPI_ISL_1990937, EPI_ISL_1990938, EPI_ISL_1990939, EPI_ISL_1990940, EPI_ISL_1990941, EPI_ISL_1990942, EPI_ISL_1990943, EPI_ISL_1990944, EPI_ISL_1990945, EPI_ISL_1990946, EPI_ISL_1990947, EPI_ISL_1990948, EPI_ISL_1990949, EPI_ISL_1990950, EPI_ISL_1990951, EPI_ISL_1990952, EPI_ISL_1990954, EPI_ISL_1990955, EPI_ISL_1990956, EPI_ISL_1990957, EPI_ISL_1990958, EPI_ISL_1990959, EPI_ISL_1990960, EPI_ISL_1990961, EPI_ISL_1990962, EPI_ISL_1990963, EPI_ISL_1990964, EPI_ISL_1990965, EPI_ISL_1990966, EPI_ISL_1990967, EPI_ISL_1990968, EPI_ISL_1990969, EPI_ISL_1990970, EPI_ISL_1990971, EPI_ISL_1990972, EPI_ISL_1990973, EPI_ISL_1990974, EPI_ISL_1990975, EPI_ISL_1990976, EPI_ISL_1990977, EPI_ISL_1990978, EPI_ISL_1990979, EPI_ISL_1990980, EPI_ISL_1990981, EPI_ISL_1990982, EPI_ISL_1990983, EPI_ISL_1990984, EPI_ISL_1990985, EPI_ISL_1990986, EPI_ISL_1990987, EPI_ISL_1990988, EPI_ISL_1990989, EPI_ISL_1990990, EPI_ISL_1990991, EPI_ISL_1990992, EPI_ISL_1990993, EPI_ISL_1990994, EPI_ISL_1990995, EPI_ISL_1990996, EPI_ISL_1990997, EPI_ISL_1990998, EPI_ISL_1990999, EPI_ISL_1991000, EPI_ISL_1991001, EPI_ISL_1991002, EPI_ISL_1991003, EPI_ISL_1991004, EPI_ISL_1991005, EPI_ISL_1991006, EPI_ISL_1991007, EPI_ISL_1991008, EPI_ISL_1991009, EPI_ISL_1991010, EPI_ISL_1991011, EPI_ISL_1991012, EPI_ISL_1991013, EPI_ISL_1991014, EPI_ISL_1991015, EPI_ISL_1991016, EPI_ISL_1991017, EPI_ISL_1991018, EPI_ISL_1991019, EPI_ISL_1991020, EPI_ISL_1991021, EPI_ISL_1991022, EPI_ISL_1991023, EPI_ISL_1991024, EPI_ISL_1991025, EPI_ISL_1991026, EPI_ISL_1991027, EPI_ISL_1991028, EPI_ISL_1991029, EPI_ISL_1991030, EPI_ISL_1991031, EPI_ISL_1991032, EPI_ISL_1991033, EPI_ISL_1991034, EPI_ISL_1991035, EPI_ISL_1991036, EPI_ISL_1991037, EPI_ISL_1991038, EPI_ISL_1991039, EPI_ISL_1991040, EPI_ISL_1991041, EPI_ISL_1991042, EPI_ISL_1991043, EPI_ISL_1991044, EPI_ISL_1991045, EPI_ISL_1991046, EPI_ISL_1991047, EPI_ISL_1991048, EPI_ISL_1991049, EPI_ISL_1991050, EPI_ISL_1991051, EPI_ISL_1991052, EPI_ISL_1991053, EPI_ISL_1991054, EPI_ISL_1991055, EPI_ISL_1991056, EPI_ISL_1991057, EPI_ISL_1991058, EPI_ISL_1991059, EPI_ISL_1991060, EPI_ISL_1991061, EPI_ISL_1991062, EPI_ISL_1991063, EPI_ISL_1991064, EPI_ISL_1991065, EPI_ISL_1991066, EPI_ISL_1991067, EPI_ISL_1991068, EPI_ISL_1991069, EPI_ISL_1991071, EPI_ISL_1991072, EPI_ISL_1991073, EPI_ISL_1991074, EPI_ISL_1991075, EPI_ISL_1991076, EPI_ISL_1991077, EPI_ISL_1991078, EPI_ISL_1991079, EPI_ISL_1991080, EPI_ISL_1991081, EPI_ISL_1991082, EPI_ISL_1991083, EPI_ISL_1991084, EPI_ISL_1991085, EPI_ISL_1991086, EPI_ISL_1991087, EPI_ISL_1991088, EPI_ISL_1991089, EPI_ISL_1991090, EPI_ISL_1991091, EPI_ISL_1991092, EPI_ISL_1991093, EPI_ISL_1991094, EPI_ISL_1991095, EPI_ISL_1991096, EPI_ISL_1991097, EPI_ISL_1991098, EPI_ISL_1991099, EPI_ISL_1991100, EPI_ISL_1991101, EPI_ISL_1991102, EPI_ISL_1991103, EPI_ISL_1991104, EPI_ISL_1991105, EPI_ISL_1991106, EPI_ISL_1991107, EPI_ISL_1991108, EPI_ISL_1991109, EPI_ISL_1991110, EPI_ISL_1991111, EPI_ISL_1991112, EPI_ISL_1991113, EPI_ISL_1991114, EPI_ISL_1991115, EPI_ISL_1991116, EPI_ISL_1991117, EPI_ISL_1991118, EPI_ISL_1991119, EPI_ISL_1991120, EPI_ISL_1991121, EPI_ISL_1991122, EPI_ISL_1991123, EPI_ISL_1991124, EPI_ISL_1991125, EPI_ISL_1991126, EPI_ISL_1991127, EPI_ISL_1991128, EPI_ISL_1991129, EPI_ISL_1991130, EPI_ISL_1991131, EPI_ISL_1991132, EPI_ISL_1991133, EPI_ISL_1991134, EPI_ISL_1991135, EPI_ISL_1991136, EPI_ISL_1991137, EPI_ISL_1991138, EPI_ISL_1991139, EPI_ISL_1991140, EPI_ISL_1991141, EPI_ISL_1991142, EPI_ISL_1991143, EPI_ISL_1991144, EPI_ISL_1991145, EPI_ISL_1991146, EPI_ISL_1991147, EPI_ISL_1991148, EPI_ISL_1991149, EPI_ISL_1991150, EPI_ISL_1991151, EPI_ISL_1991152, EPI_ISL_1991153, EPI_ISL_1991154, EPI_ISL_1991155, EPI_ISL_1991156, EPI_ISL_1991157, EPI_ISL_1991158, EPI_ISL_1991159, EPI_ISL_1991160, EPI_ISL_1991161, EPI_ISL_1991162, EPI_ISL_1991163, EPI_ISL_1991164, EPI_ISL_1991165, EPI_ISL_1991166, EPI_ISL_1991167, EPI_ISL_1991168, EPI_ISL_1991169, EPI_ISL_1991170, EPI_ISL_1991171, EPI_ISL_1991172, EPI_ISL_1991173, EPI_ISL_1991174, EPI_ISL_1991175, EPI_ISL_1991176, EPI_ISL_1991177, EPI_ISL_1991178, EPI_ISL_1991179, EPI_ISL_1991180, EPI_ISL_1991181, EPI_ISL_1991182, EPI_ISL_1991183, EPI_ISL_1991184, EPI_ISL_1991185, EPI_ISL_1991186, EPI_ISL_1991187, EPI_ISL_1991188, EPI_ISL_1991189, EPI_ISL_1991190, EPI_ISL_1991191, EPI_ISL_1991192, EPI_ISL_1991193, EPI_ISL_1991194, EPI_ISL_1991195, EPI_ISL_1991196, EPI_ISL_1991197, EPI_ISL_1991198, EPI_ISL_1991199, EPI_ISL_1991200, EPI_ISL_1991201, EPI_ISL_1991202, EPI_ISL_1991203, EPI_ISL_1991204, EPI_ISL_1991205, EPI_ISL_1991206, EPI_ISL_1991207, EPI_ISL_1991208, EPI_ISL_1991209, EPI_ISL_1991210, EPI_ISL_1991211, EPI_ISL_1991212, EPI_ISL_1991213, EPI_ISL_1991214, EPI_ISL_1991215, EPI_ISL_1991216, EPI_ISL_1991217, EPI_ISL_1991218, EPI_ISL_1991219, EPI_ISL_1991220, EPI_ISL_1991221, EPI_ISL_1991222, EPI_ISL_1991223, EPI_ISL_1991224, EPI_ISL_1991225, EPI_ISL_1991226, EPI_ISL_1991227, EPI_ISL_1991228, EPI_ISL_1991229, EPI_ISL_1991230, EPI_ISL_1991231, EPI_ISL_1991232, EPI_ISL_1991233, EPI_ISL_1991234, EPI_ISL_1991235, EPI_ISL_1991236, EPI_ISL_1991237, EPI_ISL_1991238, EPI_ISL_1991239, EPI_ISL_1991240, EPI_ISL_1991241, EPI_ISL_1991242, EPI_ISL_1991243, EPI_ISL_1991244, EPI_ISL_1991245, EPI_ISL_1991246, EPI_ISL_1991247, EPI_ISL_1991248, EPI_ISL_1991249, EPI_ISL_1991250, EPI_ISL_1991251, EPI_ISL_1991252, EPI_ISL_1991253, EPI_ISL_1991254, EPI_ISL_1991255, EPI_ISL_1991256, EPI_ISL_1991257, EPI_ISL_1991258, EPI_ISL_1991259, EPI_ISL_1991260, EPI_ISL_1991261, EPI_ISL_1991262, EPI_ISL_1991263, EPI_ISL_1991264, EPI_ISL_1991265, EPI_ISL_1991266, EPI_ISL_1991267, EPI_ISL_1991268, EPI_ISL_1991269, EPI_ISL_1991270, EPI_ISL_1991271, EPI_ISL_1991272, EPI_ISL_1991273, EPI_ISL_1991274, EPI_ISL_1991275, EPI_ISL_1991276, EPI_ISL_1991277, EPI_ISL_1991278, EPI_ISL_1991279, EPI_ISL_1991280, EPI_ISL_1991281, EPI_ISL_1991282, EPI_ISL_1991283, EPI_ISL_1991284, EPI_ISL_1991285, EPI_ISL_1991286, EPI_ISL_1991287, EPI_ISL_1991288, EPI_ISL_1991289, EPI_ISL_1991290, EPI_ISL_1991291, EPI_ISL_1991292, EPI_ISL_1991293, EPI_ISL_1991294, EPI_ISL_1991295, EPI_ISL_1991296, EPI_ISL_1991297, EPI_ISL_1991298, EPI_ISL_1991299, EPI_ISL_1991300, EPI_ISL_1991301, EPI_ISL_1991302, EPI_ISL_1991303, EPI_ISL_1991304, EPI_ISL_1991305, EPI_ISL_1991306, EPI_ISL_1991307, EPI_ISL_1991308, EPI_ISL_1991309, EPI_ISL_1991310, EPI_ISL_1991311, EPI_ISL_1991312, EPI_ISL_1991313, EPI_ISL_1991314, EPI_ISL_1991315, EPI_ISL_1991316, EPI_ISL_1991317, EPI_ISL_1991318, EPI_ISL_1991319, EPI_ISL_1991320, EPI_ISL_1991321, EPI_ISL_1991322, EPI_ISL_1991323, EPI_ISL_1991324, EPI_ISL_1991325, EPI_ISL_1991326, EPI_ISL_1991327, EPI_ISL_1991328, EPI_ISL_1991329, EPI_ISL_1991330, EPI_ISL_1991331, EPI_ISL_1991332, EPI_ISL_1991333, EPI_ISL_1991334, EPI_ISL_1991335, EPI_ISL_1991336, EPI_ISL_1991337, EPI_ISL_1991338, EPI_ISL_1991339, EPI_ISL_1991340, EPI_ISL_1991341, EPI_ISL_1991342, EPI_ISL_1991343, EPI_ISL_1991344, EPI_ISL_1991345, EPI_ISL_1991346, EPI_ISL_1991347, EPI_ISL_1991348, EPI_ISL_1991349, EPI_ISL_1991350, EPI_ISL_1991351, EPI_ISL_1991352, EPI_ISL_1991353, EPI_ISL_1991354, EPI_ISL_1991355, EPI_ISL_1991356, EPI_ISL_1991357, EPI_ISL_1991358, EPI_ISL_1991359, EPI_ISL_1991360, EPI_ISL_1991361, EPI_ISL_1991362, EPI_ISL_1991363, EPI_ISL_1991364, EPI_ISL_1991365, EPI_ISL_1991366, EPI_ISL_1991367, EPI_ISL_1991368, EPI_ISL_1991369, EPI_ISL_1991370, EPI_ISL_1991371, EPI_ISL_1991372, EPI_ISL_1991373, EPI_ISL_1991374, EPI_ISL_1991375, EPI_ISL_1991376, EPI_ISL_1991377, EPI_ISL_1991378, EPI_ISL_1991379, EPI_ISL_1991380, EPI_ISL_1991381, EPI_ISL_1991382, EPI_ISL_1991383, EPI_ISL_1991384, EPI_ISL_1991385, EPI_ISL_1991386, EPI_ISL_1991387, EPI_ISL_1991388, EPI_ISL_1991389, EPI_ISL_1991390, EPI_ISL_1991391, EPI_ISL_1991392, EPI_ISL_1991393, EPI_ISL_1991394, EPI_ISL_1991395, EPI_ISL_1991396, EPI_ISL_1991397, EPI_ISL_1991398, EPI_ISL_1991399, EPI_ISL_1991400, EPI_ISL_1991401, EPI_ISL_1991402, EPI_ISL_1991403, EPI_ISL_1991404, EPI_ISL_1991405, EPI_ISL_1991406, EPI_ISL_1991407, EPI_ISL_1991408, EPI_ISL_1991409, EPI_ISL_1991410, EPI_ISL_1991411, EPI_ISL_1991412, EPI_ISL_1991413, EPI_ISL_1991414, EPI_ISL_1991415, EPI_ISL_1991416, EPI_ISL_1991417, EPI_ISL_1991418, EPI_ISL_1991419, EPI_ISL_1991421, EPI_ISL_1991422, EPI_ISL_1991423, EPI_ISL_1991424, EPI_ISL_1991425, EPI_ISL_1991426, EPI_ISL_1991427, EPI_ISL_1991428, EPI_ISL_1991429, EPI_ISL_1991430, EPI_ISL_1991431, EPI_ISL_1991432, EPI_ISL_1991433, EPI_ISL_1991434, EPI_ISL_1991435, EPI_ISL_1991436, EPI_ISL_1991437, EPI_ISL_1991438, EPI_ISL_1991439, EPI_ISL_1991440, EPI_ISL_1991441, EPI_ISL_1991442, EPI_ISL_1991443, EPI_ISL_1991444, EPI_ISL_1991445, EPI_ISL_1991446, EPI_ISL_1991447, EPI_ISL_1991448, EPI_ISL_1991449, EPI_ISL_1991450, EPI_ISL_1991451, EPI_ISL_1991452, EPI_ISL_1991453, EPI_ISL_1991454, EPI_ISL_1991455, EPI_ISL_1991456, EPI_ISL_1991457, EPI_ISL_1991458, EPI_ISL_1991459, EPI_ISL_1991460, EPI_ISL_1991461, EPI_ISL_1991462, EPI_ISL_1991463, EPI_ISL_1991464, EPI_ISL_1991465, EPI_ISL_1991466, EPI_ISL_1991467, EPI_ISL_1991468, EPI_ISL_1991469, EPI_ISL_1991470, EPI_ISL_1991471, EPI_ISL_1991472, EPI_ISL_1991473, EPI_ISL_1991474, EPI_ISL_1991475, EPI_ISL_1991476, EPI_ISL_1991477, EPI_ISL_1991478, EPI_ISL_1991479, EPI_ISL_1991480, EPI_ISL_1991481, EPI_ISL_1991482, EPI_ISL_1991483, EPI_ISL_1991484, EPI_ISL_1991485, EPI_ISL_1991486, EPI_ISL_1991487, EPI_ISL_1991488, EPI_ISL_1991489, EPI_ISL_1991491, EPI_ISL_1991492, EPI_ISL_1991493, EPI_ISL_1991494, EPI_ISL_1991495, EPI_ISL_1991496, EPI_ISL_1991497, EPI_ISL_1991498, EPI_ISL_1991499, EPI_ISL_1991500, EPI_ISL_1991501, EPI_ISL_1991502, EPI_ISL_1991503, EPI_ISL_1991504, EPI_ISL_1991505, EPI_ISL_1991506, EPI_ISL_1991507, EPI_ISL_1991508, EPI_ISL_1991509, EPI_ISL_1991510, EPI_ISL_1991511, EPI_ISL_1991512, EPI_ISL_1991514, EPI_ISL_1991515, EPI_ISL_1991516, EPI_ISL_1991518, EPI_ISL_1991519, EPI_ISL_1991520, EPI_ISL_1991521, EPI_ISL_1991522, EPI_ISL_1991523, EPI_ISL_1991524, EPI_ISL_1991525, EPI_ISL_1991526, EPI_ISL_1991527, EPI_ISL_1991529, EPI_ISL_1991530, EPI_ISL_1991531, EPI_ISL_1991532, EPI_ISL_1991533, EPI_ISL_1991534, EPI_ISL_1991535, EPI_ISL_1991536, EPI_ISL_1991537, EPI_ISL_1991538, EPI_ISL_1991539, EPI_ISL_1991540, EPI_ISL_1991541, EPI_ISL_1991543, EPI_ISL_1991544, EPI_ISL_1991545, EPI_ISL_1991546, EPI_ISL_1991547, EPI_ISL_1991548, EPI_ISL_1991549, EPI_ISL_1991550, EPI_ISL_1991551, EPI_ISL_1991552, EPI_ISL_1991553, EPI_ISL_1991554, EPI_ISL_1991555, EPI_ISL_1991556, EPI_ISL_1991557, EPI_ISL_1991558, EPI_ISL_1991559, EPI_ISL_1991560, EPI_ISL_1991561, EPI_ISL_1991562, EPI_ISL_1991563, EPI_ISL_1991564, EPI_ISL_1991565, EPI_ISL_1991566, EPI_ISL_1991567, EPI_ISL_1991568, EPI_ISL_1991569, EPI_ISL_1991570, EPI_ISL_1991571, EPI_ISL_1991572, EPI_ISL_1991573, EPI_ISL_1991574, EPI_ISL_1991575, EPI_ISL_1991576, EPI_ISL_1991577, EPI_ISL_1991578, EPI_ISL_1991579, EPI_ISL_1991580, EPI_ISL_1991581, EPI_ISL_1991582, EPI_ISL_1991583, EPI_ISL_1991584, EPI_ISL_1991585, EPI_ISL_1991586, EPI_ISL_1991588, EPI_ISL_1991589, EPI_ISL_1991590, EPI_ISL_1991591, EPI_ISL_1991592, EPI_ISL_1991593, EPI_ISL_1991594, EPI_ISL_1991595, EPI_ISL_1991596, EPI_ISL_1991598, EPI_ISL_1991601, EPI_ISL_1991605, EPI_ISL_1991608, EPI_ISL_1991612, EPI_ISL_1991615, EPI_ISL_1991619, EPI_ISL_1991622, EPI_ISL_1991625, EPI_ISL_1991629, EPI_ISL_1991633, EPI_ISL_1991636, EPI_ISL_1991639, EPI_ISL_1991642, EPI_ISL_1991649, EPI_ISL_1991653, EPI_ISL_1991657, EPI_ISL_1991658, EPI_ISL_1991659, EPI_ISL_1991660, EPI_ISL_1991661, EPI_ISL_1991662, EPI_ISL_1991663, EPI_ISL_1991664, EPI_ISL_1991665, EPI_ISL_1991666, EPI_ISL_1991667, EPI_ISL_1991668, EPI_ISL_1991669, EPI_ISL_1991670, EPI_ISL_1991671, EPI_ISL_1991672, EPI_ISL_1991673, EPI_ISL_1991674, EPI_ISL_1991675, EPI_ISL_1991676, EPI_ISL_1991677, EPI_ISL_1991678, EPI_ISL_1991679, EPI_ISL_1991680, EPI_ISL_1991681, EPI_ISL_1991682, EPI_ISL_1991683, EPI_ISL_1991684, EPI_ISL_1991685, EPI_ISL_1991686, EPI_ISL_1991687, EPI_ISL_1991688, EPI_ISL_1991689, EPI_ISL_1991690, EPI_ISL_1991691, EPI_ISL_1991692, EPI_ISL_1991693, EPI_ISL_1991694, EPI_ISL_1991695, EPI_ISL_1991696, EPI_ISL_1991697, EPI_ISL_1991698, EPI_ISL_1991699, EPI_ISL_1991700, EPI_ISL_1991701, EPI_ISL_1991702, EPI_ISL_1991703, EPI_ISL_1991704, EPI_ISL_1991705, EPI_ISL_1991706, EPI_ISL_1991707, EPI_ISL_1991708, EPI_ISL_1991709, EPI_ISL_1991710, EPI_ISL_1991711, EPI_ISL_1991712, EPI_ISL_1991713, EPI_ISL_1991714, EPI_ISL_1991715, EPI_ISL_1991716, EPI_ISL_1991717, EPI_ISL_1991718, EPI_ISL_1991719, EPI_ISL_1991720, EPI_ISL_1991721, EPI_ISL_1991722, EPI_ISL_1991723, EPI_ISL_1991724, EPI_ISL_1991725, EPI_ISL_1991726, EPI_ISL_1991727, EPI_ISL_1991728, EPI_ISL_1991729, EPI_ISL_1991730, EPI_ISL_1991731, EPI_ISL_1991732, EPI_ISL_1991733, EPI_ISL_1991734, EPI_ISL_1991735, EPI_ISL_1991736, EPI_ISL_1991737, EPI_ISL_1991738, EPI_ISL_1991739, EPI_ISL_1991740, EPI_ISL_1991741, EPI_ISL_1991742, EPI_ISL_1991743, EPI_ISL_1991744, EPI_ISL_1991745, EPI_ISL_1991746, EPI_ISL_1991747, EPI_ISL_1991748, EPI_ISL_1991749, EPI_ISL_1991750, EPI_ISL_1991751, EPI_ISL_1991752, EPI_ISL_1991754, EPI_ISL_1991755, EPI_ISL_1991756, EPI_ISL_1991757, EPI_ISL_1991758, EPI_ISL_1991759, EPI_ISL_1991760, EPI_ISL_1991761, EPI_ISL_1991762, EPI_ISL_1991763, EPI_ISL_1991764, EPI_ISL_1991765, EPI_ISL_1991766, EPI_ISL_1991767, EPI_ISL_1991768, EPI_ISL_1991769, EPI_ISL_1991770, EPI_ISL_1991771, EPI_ISL_1991772, EPI_ISL_1991773, EPI_ISL_1991774, EPI_ISL_1991775, EPI_ISL_1991776, EPI_ISL_1991777, EPI_ISL_1991778, EPI_ISL_1991779, EPI_ISL_1991780, EPI_ISL_1991781, EPI_ISL_1991782, EPI_ISL_1991783, EPI_ISL_1991784, EPI_ISL_1991785, EPI_ISL_1991786, EPI_ISL_1991787, EPI_ISL_1991788, EPI_ISL_1991789, EPI_ISL_1991790, EPI_ISL_1991791, EPI_ISL_1991792, EPI_ISL_1991793, EPI_ISL_1991794, EPI_ISL_1991795, EPI_ISL_1991796, EPI_ISL_1991797, EPI_ISL_1991798, EPI_ISL_1991799, EPI_ISL_1991800, EPI_ISL_1991801, EPI_ISL_1991802, EPI_ISL_1991803, EPI_ISL_1991804, EPI_ISL_1991805, EPI_ISL_1991806, EPI_ISL_1991807, EPI_ISL_1991808, EPI_ISL_1991809, EPI_ISL_1991810, EPI_ISL_1991811, EPI_ISL_1991812, EPI_ISL_1991813, EPI_ISL_1991814, EPI_ISL_1991815, EPI_ISL_1991816, EPI_ISL_1991817, EPI_ISL_1991818, EPI_ISL_1991819, EPI_ISL_1991820, EPI_ISL_1991821, EPI_ISL_1991822, EPI_ISL_1991823, EPI_ISL_1991824, EPI_ISL_1991825, EPI_ISL_1991827, EPI_ISL_1991828, EPI_ISL_1991829, EPI_ISL_1991830, EPI_ISL_1991831, EPI_ISL_1991832, EPI_ISL_1991833, EPI_ISL_1991834, EPI_ISL_1991835, EPI_ISL_1991836, EPI_ISL_1991837, EPI_ISL_1991838, EPI_ISL_1991839, EPI_ISL_1991840, EPI_ISL_1991841, EPI_ISL_1991842, EPI_ISL_1991843

| | | | |
|---|---|---|---|
| see above | Helix/Illumina | Centers for Disease Control and Prevention Division of Viral Diseases, Pathogen Discovery | Dakota Howard, Dhwani Batra, Peter W. Cook, Kara Moser, Adrian Paskey, Jason Caravas, Benjamin Rambo-Martin, Shatavia Morrison, Christopher Gulvick, Scott Sammons, Yvette Unoarumhi, Darlene Wagner, Matthew Schmerer, Eileen de Feo, Jan Antico, Christine Tran, Matthew Tolentino, Shannon Wickline, Kim Gietzen, Brad Sickler, Jingtao Liu, Eric Allen, Phil Febbo, Nicole L. Washington, Simon White, Geraint Levan, Kelly Schiabor Barrett, Elizabeth Cirulli, Alexandre Bolze, Ary Ascencio, Charlotte Rivera-Garcia, Ryan Cho, Jason Nguyen, Sherry Wang, Jimmy Ramirez, Tyler Cassens, Efren Sandoval, Magnus Isaksson, William Lee, David Becker, Marc Laurent, James Lu, Clinton R. Paden, Duncan MacCannell |

EPI_ISL_1992018, EPI_ISL_1992019, EPI_ISL_1992020, EPI_ISL_1992021, EPI_ISL_1992023, EPI_ISL_1992024, EPI_ISL_1992025, EPI_ISL_1992026, EPI_ISL_1992027, EPI_ISL_1992028, EPI_ISL_1992029, EPI_ISL_1992030, EPI_ISL_1992031, EPI_ISL_1992032, EPI_ISL_1992033, EPI_ISL_1992034, EPI_ISL_1992035, EPI_ISL_1992036, EPI_ISL_1992037, EPI_ISL_1992038, EPI_ISL_1992039, EPI_ISL_1992040, EPI_ISL_1992041, EPI_ISL_1992042, EPI_ISL_1992043, EPI_ISL_1992045, EPI_ISL_1992046, EPI_ISL_1992047, EPI_ISL_1992048, EPI_ISL_1992049, EPI_ISL_1992050, EPI_ISL_1992051, EPI_ISL_1992052, EPI_ISL_1992053, EPI_ISL_1992054, EPI_ISL_1992055, EPI_ISL_1992056, EPI_ISL_1992057, EPI_ISL_1992058, EPI_ISL_1992059, EPI_ISL_1992060, EPI_ISL_1992061, EPI_ISL_1992062, EPI_ISL_1992063, EPI_ISL_1992064, EPI_ISL_1992065, EPI_ISL_1992066, EPI_ISL_1992067, EPI_ISL_1992068, EPI_ISL_1992069, EPI_ISL_1992070, EPI_ISL_1992071, EPI_ISL_1992072, EPI_ISL_1992074, EPI_ISL_1992075, EPI_ISL_1992077, EPI_ISL_1992079, EPI_ISL_1992080, EPI_ISL_1992081, EPI_ISL_1992082, EPI_ISL_1992083, EPI_ISL_1992084, EPI_ISL_1992085, EPI_ISL_1992086, EPI_ISL_1992087, EPI_ISL_1992088, EPI_ISL_1992089, EPI_ISL_1992090, EPI_ISL_1992091, EPI_ISL_1992092, EPI_ISL_1992093, EPI_ISL_1992094, EPI_ISL_1992095, EPI_ISL_1992096, EPI_ISL_1992097, EPI_ISL_1992098, EPI_ISL_1992099, EPI_ISL_1992101, EPI_ISL_1992102, EPI_ISL_1992103, EPI_ISL_1992104, EPI_ISL_1992105, EPI_ISL_1992106, EPI_ISL_1992107, EPI_ISL_1992108, EPI_ISL_1992109, EPI_ISL_1992110, EPI_ISL_1992111, EPI_ISL_1992112, EPI_ISL_1992113, EPI_ISL_1992115, EPI_ISL_1992116, EPI_ISL_1992117, EPI_ISL_1992118, EPI_ISL_1992119, EPI_ISL_1992120, EPI_ISL_1992121, EPI_ISL_1992122, EPI_ISL_1992123, EPI_ISL_1992124, EPI_ISL_1992125, EPI_ISL_1992126, EPI_ISL_1992127, EPI_ISL_1992128, EPI_ISL_1992129, EPI_ISL_1992130, EPI_ISL_1992131, EPI_ISL_1992132, EPI_ISL_1992133, EPI_ISL_1992134, EPI_ISL_1992136, EPI_ISL_1992137, EPI_ISL_1992138, EPI_ISL_1992139, EPI_ISL_1992140, EPI_ISL_1992141, EPI_ISL_1992142, EPI_ISL_1992143, EPI_ISL_1992144, EPI_ISL_1992145, EPI_ISL_1992146, EPI_ISL_1992147, EPI_ISL_1992148, EPI_ISL_1992149, EPI_ISL_1992150, EPI_ISL_1992151, EPI_ISL_1992152, EPI_ISL_1992153, EPI_ISL_1992154, EPI_ISL_1992155, EPI_ISL_1992156, EPI_ISL_1992157, EPI_ISL_1992158, EPI_ISL_1992159, EPI_ISL_1992160, EPI_ISL_1992161, EPI_ISL_1992163, EPI_ISL_1992165, EPI_ISL_1992166, EPI_ISL_1992167, EPI_ISL_1992168, EPI_ISL_1992169, EPI_ISL_1992170, EPI_ISL_1992171, EPI_ISL_1992172, EPI_ISL_1992173, EPI_ISL_1992174, EPI_ISL_1992175, EPI_ISL_1992176, EPI_ISL_1992177, EPI_ISL_1992178, EPI_ISL_1992179, EPI_ISL_1992180, EPI_ISL_1992181, EPI_ISL_1992182, EPI_ISL_1992183, EPI_ISL_1992184, EPI_ISL_1992185, EPI_ISL_1992186, EPI_ISL_1992187, EPI_ISL_1992188, EPI_ISL_1992189, EPI_ISL_1992190, EPI_ISL_1992191, EPI_ISL_1992192, EPI_ISL_1992193, EPI_ISL_1992194, EPI_ISL_1992195, EPI_ISL_1992196, EPI_ISL_1992198, EPI_ISL_1992199, EPI_ISL_1992200, EPI_ISL_1992201, EPI_ISL_1992202, EPI_ISL_1992203, EPI_ISL_1992204, EPI_ISL_1992205, EPI_ISL_1992206, EPI_ISL_1992207, EPI_ISL_1992208, EPI_ISL_1992209, EPI_ISL_1992211, EPI_ISL_1992212, EPI_ISL_1992213, EPI_ISL_1992214, EPI_ISL_1992215, EPI_ISL_1992216, EPI_ISL_1992217, EPI_ISL_1992218, EPI_ISL_1992219, EPI_ISL_1992220, EPI_ISL_1992221, EPI_ISL_1992222, EPI_ISL_1992223, EPI_ISL_1992224, EPI_ISL_1992225, EPI_ISL_1992227, EPI_ISL_1992228, EPI_ISL_1992229, EPI_ISL_1992230, EPI_ISL_1992231, EPI_ISL_1992232, EPI_ISL_1992233, EPI_ISL_1992234, EPI_ISL_1992235, EPI_ISL_1992236, EPI_ISL_1992237, EPI_ISL_1992238, EPI_ISL_1992239, EPI_ISL_1992240, EPI_ISL_1992243, EPI_ISL_1992244, EPI_ISL_1992245, EPI_ISL_1992247, EPI_ISL_1992248, EPI_ISL_1992249, EPI_ISL_1992250, EPI_ISL_1992252, EPI_ISL_1992254, EPI_ISL_1992255, EPI_ISL_1992256, EPI_ISL_1992257, EPI_ISL_1992258, EPI_ISL_1992259, EPI_ISL_1992260, EPI_ISL_1992261, EPI_ISL_1992262, EPI_ISL_1992263, EPI_ISL_1992264, EPI_ISL_1992266, EPI_ISL_1992267, EPI_ISL_1992268, EPI_ISL_1992269, EPI_ISL_1992270, EPI_ISL_1992271, EPI_ISL_1992272, EPI_ISL_1992273, EPI_ISL_1992274, EPI_ISL_1992275, EPI_ISL_1992276, EPI_ISL_1992278, EPI_ISL_1992279, EPI_ISL_1992280, EPI_ISL_1992281, EPI_ISL_1992282, EPI_ISL_1992284, EPI_ISL_1992285, EPI_ISL_1992286, EPI_ISL_1992287, EPI_ISL_1992288, EPI_ISL_1992289, EPI_ISL_1992290, EPI_ISL_1992291, EPI_ISL_1992292, EPI_ISL_1992293, EPI_ISL_1992294, EPI_ISL_1992295, EPI_ISL_1992296, EPI_ISL_1992297, EPI_ISL_1992298, EPI_ISL_1992299, EPI_ISL_1992300, EPI_ISL_1992301, EPI_ISL_1992302, EPI_ISL_1992303, EPI_ISL_1992304, EPI_ISL_1992305, EPI_ISL_1992306, EPI_ISL_1992307, EPI_ISL_1992308, EPI_ISL_1992309, EPI_ISL_1992310, EPI_ISL_1992311, EPI_ISL_1992312, EPI_ISL_1992313, EPI_ISL_1992314, EPI_ISL_1992315, EPI_ISL_1992316, EPI_ISL_1992317, EPI_ISL_1992318, EPI_ISL_1992320, EPI_ISL_1992321, EPI_ISL_1992322, EPI_ISL_1992323, EPI_ISL_1992324, EPI_ISL_1992325, EPI_ISL_1992326, EPI_ISL_1992327, EPI_ISL_1992328, EPI_ISL_1992329, EPI_ISL_1992330, EPI_ISL_1992331, EPI_ISL_1992332, EPI_ISL_1992333, EPI_ISL_1992334, EPI_ISL_1992335, EPI_ISL_1992337, EPI_ISL_1992338, EPI_ISL_1992339, EPI_ISL_1992340, EPI_ISL_1992341, EPI_ISL_1992342, EPI_ISL_1992343, EPI_ISL_1992345, EPI_ISL_1992347, EPI_ISL_1992348, EPI_ISL_1992349, EPI_ISL_1992350, EPI_ISL_1992351, EPI_ISL_1992352, EPI_ISL_1992353, EPI_ISL_1992354, EPI_ISL_1992355, EPI_ISL_1992356, EPI_ISL_1992357, EPI_ISL_1992358, EPI_ISL_1992359, EPI_ISL_1992360, EPI_ISL_1992361, EPI_ISL_1992362, EPI_ISL_1992363, EPI_ISL_1992364, EPI_ISL_1992365, EPI_ISL_1992366, EPI_ISL_1992367, EPI_ISL_1992368, EPI_ISL_1992369, EPI_ISL_1992370, EPI_ISL_1992371, EPI_ISL_1992372, EPI_ISL_1992373, EPI_ISL_1992374, EPI_ISL_1992375, EPI_ISL_1992376, EPI_ISL_1992377, EPI_ISL_1992378, EPI_ISL_1992379, EPI_ISL_1992380, EPI_ISL_1992381, EPI_ISL_1992382, EPI_ISL_1992383, EPI_ISL_1992384, EPI_ISL_1992385, EPI_ISL_1992386, EPI_ISL_1992387, EPI_ISL_1992388, EPI_ISL_1992389, EPI_ISL_1992390, EPI_ISL_1992391, EPI_ISL_1992392, EPI_ISL_1992393, EPI_ISL_1992394, EPI_ISL_1992395, EPI_ISL_1992396, EPI_ISL_1992398, EPI_ISL_1992400, EPI_ISL_1992401, EPI_ISL_1992402, EPI_ISL_1992404, EPI_ISL_1992405, EPI_ISL_1992406, EPI_ISL_1992408, EPI_ISL_1992409, EPI_ISL_1992410, EPI_ISL_1992411, EPI_ISL_1992412, EPI_ISL_1992413, EPI_ISL_1992414, EPI_ISL_1992416, EPI_ISL_1992417, EPI_ISL_1992418, EPI_ISL_1992419, EPI_ISL_1992420, EPI_ISL_1992421, EPI_ISL_1992422, EPI_ISL_1992423, EPI_ISL_1992424, EPI_ISL_1992425, EPI_ISL_1992426, EPI_ISL_1992427, EPI_ISL_1992428, EPI_ISL_1992429, EPI_ISL_1992430, EPI_ISL_1992431, EPI_ISL_1992432, EPI_ISL_1992433, EPI_ISL_1992434, EPI_ISL_1992435, EPI_ISL_1992436, EPI_ISL_1992437, EPI_ISL_1992439, EPI_ISL_1992440, EPI_ISL_1992442, EPI_ISL_1992444, EPI_ISL_1992445, EPI_ISL_1992446, EPI_ISL_1992447, EPI_ISL_1992448, EPI_ISL_1992449, EPI_ISL_1992450, EPI_ISL_1992451, EPI_ISL_1992452, EPI_ISL_1992453, EPI_ISL_1992454, EPI_ISL_1992455, EPI_ISL_1992456, EPI_ISL_1992457, EPI_ISL_1992458, EPI_ISL_1992459, EPI_ISL_1992460, EPI_ISL_1992462, EPI_ISL_1992463, EPI_ISL_1992464, EPI_ISL_1992465, EPI_ISL_1992466, EPI_ISL_1992467, EPI_ISL_1992468, EPI_ISL_1992469, EPI_ISL_1992470, EPI_ISL_1992471, EPI_ISL_1992472, EPI_ISL_1992473, EPI_ISL_1992474, EPI_ISL_1992475, EPI_ISL_1992476, EPI_ISL_1992477, EPI_ISL_1992478, EPI_ISL_1992480, EPI_ISL_1992481, EPI_ISL_1992482, EPI_ISL_1992484, EPI_ISL_1992485, EPI_ISL_1992486, EPI_ISL_1992487, EPI_ISL_1992488, EPI_ISL_1992489, EPI_ISL_1992490, EPI_ISL_1992491, EPI_ISL_1992492, EPI_ISL_1992493, EPI_ISL_1992494, EPI_ISL_1992495, EPI_ISL_1992496, EPI_ISL_1992497, EPI_ISL_1992498, EPI_ISL_1992499, EPI_ISL_1992500, EPI_ISL_1992501, EPI_ISL_1992502, EPI_ISL_1992503, EPI_ISL_1992504, EPI_ISL_1992505, EPI_ISL_1992507, EPI_ISL_1992508, EPI_ISL_1992509, EPI_ISL_1992510, EPI_ISL_1992511, EPI_ISL_1992512, EPI_ISL_1992513, EPI_ISL_1992515, EPI_ISL_1992516, EPI_ISL_1992517, EPI_ISL_1992518, EPI_ISL_1992519, EPI_ISL_1992520, EPI_ISL_1992521, EPI_ISL_1992522, EPI_ISL_1992523, EPI_ISL_1992524, EPI_ISL_1992525, EPI_ISL_1992526, EPI_ISL_1992527, EPI_ISL_1992528, EPI_ISL_1992529, EPI_ISL_1992530, EPI_ISL_1992531, EPI_ISL_1992532, EPI_ISL_1992533, EPI_ISL_1992534, EPI_ISL_1992535, EPI_ISL_1992536, EPI_ISL_1992537, EPI_ISL_1992538, EPI_ISL_1992539, EPI_ISL_1992541, EPI_ISL_1992542, EPI_ISL_1992543, EPI_ISL_1992544, EPI_ISL_1992545, EPI_ISL_1992547, EPI_ISL_1992549, EPI_ISL_1992550, EPI_ISL_1992551, EPI_ISL_1992552, EPI_ISL_1992553, EPI_ISL_1992554, EPI_ISL_1992555, EPI_ISL_1992556, EPI_ISL_1992557, EPI_ISL_1992558, EPI_ISL_1992559, EPI_ISL_1992560, EPI_ISL_1992561, EPI_ISL_1992562, EPI_ISL_1992563, EPI_ISL_1992565, EPI_ISL_1992566, EPI_ISL_1992568, EPI_ISL_1992569, EPI_ISL_1992571, EPI_ISL_1992572, EPI_ISL_1992573, EPI_ISL_1992574, EPI_ISL_1992575, EPI_ISL_1992577, EPI_ISL_1992578, EPI_ISL_1992579, EPI_ISL_1992580, EPI_ISL_1992581, EPI_ISL_1992582, EPI_ISL_1992584, EPI_ISL_1992585, EPI_ISL_1992586, EPI_ISL_1992587, EPI_ISL_1992588, EPI_ISL_1992589, EPI_ISL_1992590, EPI_ISL_1992591, EPI_ISL_1992592, EPI_ISL_1992593, EPI_ISL_1992594, EPI_ISL_1992595, EPI_ISL_1992596, EPI_ISL_1992597, EPI_ISL_1992598, EPI_ISL_1992600, EPI_ISL_1992601, EPI_ISL_1992602, EPI_ISL_1992603, EPI_ISL_1992604, EPI_ISL_1992606, EPI_ISL_1992607, EPI_ISL_1992608, EPI_ISL_1992609, EPI_ISL_1992611, EPI_ISL_1992612, EPI_ISL_1992613, EPI_ISL_1992614, EPI_ISL_1992615, EPI_ISL_1992616, EPI_ISL_1992617, EPI_ISL_1992618, EPI_ISL_1992619, EPI_ISL_1992620, EPI_ISL_1992622, EPI_ISL_1992624, EPI_ISL_1992625, EPI_ISL_1992626, EPI_ISL_1992627, EPI_ISL_1992628, EPI_ISL_1992630, EPI_ISL_1992632, EPI_ISL_1992633, EPI_ISL_1992634, EPI_ISL_1992635, EPI_ISL_1992636, EPI_ISL_1992637, EPI_ISL_1992638, EPI_ISL_1992639, EPI_ISL_1992640, EPI_ISL_1992642, EPI_ISL_1992643, EPI_ISL_1992646, EPI_ISL_1992648, EPI_ISL_1992650, EPI_ISL_1992651, EPI_ISL_1992652, EPI_ISL_1992653, EPI_ISL_1992654, EPI_ISL_1992657, EPI_ISL_1992658, EPI_ISL_1992660, EPI_ISL_1992661, EPI_ISL_1992662, EPI_ISL_1992663, EPI_ISL_1992665, EPI_ISL_1992666, EPI_ISL_1992667, EPI_ISL_1992668, EPI_ISL_1992669, EPI_ISL_1992670, EPI_ISL_1992672, EPI_ISL_1992673, EPI_ISL_1992674, EPI_ISL_1992675, EPI_ISL_1992676, EPI_ISL_1992678, EPI_ISL_1992679, EPI_ISL_1992680, EPI_ISL_1992681, EPI_ISL_1992682, EPI_ISL_1992683, EPI_ISL_1992684, EPI_ISL_1992685, EPI_ISL_1992686, EPI_ISL_1992687, EPI_ISL_1992688, EPI_ISL_1992689, EPI_ISL_1992690, EPI_ISL_1992691, EPI_ISL_1992692, EPI_ISL_1992693, EPI_ISL_1992694, EPI_ISL_1992696, EPI_ISL_1992697, EPI_ISL_1992698, EPI_ISL_1992699, EPI_ISL_1992701, EPI_ISL_1992703, EPI_ISL_1992704, EPI_ISL_1992705, EPI_ISL_1992706, EPI_ISL_1992707, EPI_ISL_1992708, EPI_ISL_1992709, EPI_ISL_1992710, EPI_ISL_1992711, EPI_ISL_1992712, EPI_ISL_1992713, EPI_ISL_1992714, EPI_ISL_1992715, EPI_ISL_1992716, EPI_ISL_1992717, EPI_ISL_1992718, EPI_ISL_1992719, EPI_ISL_1992720, EPI_ISL_1992721, EPI_ISL_1992723, EPI_ISL_1992725, EPI_ISL_1992726, EPI_ISL_1992727, EPI_ISL_1992729, EPI_ISL_1992730, EPI_ISL_1992731, EPI_ISL_1992732, EPI_ISL_1992733, EPI_ISL_1992737, EPI_ISL_1992738, EPI_ISL_1992739, EPI_ISL_1992740, EPI_ISL_1992741, EPI_ISL_1992742, EPI_ISL_1992743, EPI_ISL_1992744, EPI_ISL_1992746, EPI_ISL_1992747, EPI_ISL_1992749, EPI_ISL_1992750, EPI_ISL_1992751, EPI_ISL_1992752, EPI_ISL_1992753, EPI_ISL_1992754, EPI_ISL_1992755, EPI_ISL_1992756, EPI_ISL_1992757, EPI_ISL_1992758, EPI_ISL_1992759, EPI_ISL_1992760, EPI_ISL_1992761, EPI_ISL_1992762, EPI_ISL_1992763, EPI_ISL_1992764, EPI_ISL_1992765, EPI_ISL_1992766, EPI_ISL_1992767, EPI_ISL_1992768, EPI_ISL_1992769, EPI_ISL_1992770, EPI_ISL_1992771, EPI_ISL_1992775, EPI_ISL_1992776, EPI_ISL_1992777, EPI_ISL_1992778, EPI_ISL_1992779, EPI_ISL_1992780, EPI_ISL_1992781, EPI_ISL_1992782, EPI_ISL_1992783, EPI_ISL_1992784, EPI_ISL_1992785, EPI_ISL_1992786, EPI_ISL_1992787, EPI_ISL_1992788, EPI_ISL_1992790, EPI_ISL_1992791, EPI_ISL_1992792, EPI_ISL_1992793, EPI_ISL_1992794, EPI_ISL_1992795, EPI_ISL_1992796, EPI_ISL_1992797, EPI_ISL_1992798, EPI_ISL_1992799, EPI_ISL_1992800, EPI_ISL_1992801, EPI_ISL_1992802, EPI_ISL_1992803, EPI_ISL_1992804, EPI_ISL_1992805, EPI_ISL_1992807, EPI_ISL_1992808, EPI_ISL_1992809, EPI_ISL_1992810, EPI_ISL_1992811, EPI_ISL_1992812, EPI_ISL_1992813, EPI_ISL_1992814, EPI_ISL_1992815, EPI_ISL_1992816, EPI_ISL_1992817, EPI_ISL_1992818, EPI_ISL_1992819, EPI_ISL_1992820, EPI_ISL_1992821, EPI_ISL_1992822, EPI_ISL_1992823, EPI_ISL_1992824, EPI_ISL_1992826, EPI_ISL_1992827, EPI_ISL_1992829, EPI_ISL_1992830, EPI_ISL_1992831, EPI_ISL_1992832, EPI_ISL_1992833, EPI_ISL_1992834, EPI_ISL_1992835, EPI_ISL_1992836, EPI_ISL_1992837, EPI_ISL_1992838, EPI_ISL_1992839, EPI_ISL_1992841, EPI_ISL_1992842, EPI_ISL_1992843, EPI_ISL_1992844, EPI_ISL_1992845, EPI_ISL_1992846, EPI_ISL_1992847, EPI_ISL_1992848, EPI_ISL_1992849, EPI_ISL_1992850, EPI_ISL_1992851, EPI_ISL_1992855, EPI_ISL_1992862, EPI_ISL_1992865, EPI_ISL_1992869, EPI_ISL_1992873, EPI_ISL_1992876, EPI_ISL_1992881, EPI_ISL_1992884, EPI_ISL_1992888, EPI_ISL_1992892, EPI_ISL_1992896, EPI_ISL_1992900, EPI_ISL_1992907, EPI_ISL_1992910, EPI_ISL_1992912, EPI_ISL_1992914, EPI_ISL_1992915, EPI_ISL_1992916, EPI_ISL_1992917, EPI_ISL_1992919, EPI_ISL_1992920, EPI_ISL_1992921, EPI_ISL_1992922, EPI_ISL_1992923, EPI_ISL_1992924, EPI_ISL_1992925, EPI_ISL_1992926, EPI_ISL_1992927, EPI_ISL_1992928, EPI_ISL_1992929, EPI_ISL_1992930, EPI_ISL_1992931, EPI_ISL_1992932, EPI_ISL_1992933, EPI_ISL_1992934, EPI_ISL_1992935, EPI_ISL_1992936, EPI_ISL_1992937, EPI_ISL_1992938, EPI_ISL_1992939, EPI_ISL_1992941, EPI_ISL_1992942, EPI_ISL_1992943, EPI_ISL_1992944, EPI_ISL_1992945, EPI_ISL_1992947, EPI_ISL_1992949, EPI_ISL_1992950, EPI_ISL_1992951, EPI_ISL_1992952, EPI_ISL_1992953, EPI_ISL_1992954, EPI_ISL_1992955, EPI_ISL_1992956, EPI_ISL_1992957, EPI_ISL_1992958, EPI_ISL_1992959, EPI_ISL_1992961, EPI_ISL_1992962, EPI_ISL_1992963, EPI_ISL_1992964, EPI_ISL_1992965, EPI_ISL_1992966, EPI_ISL_1992968, EPI_ISL_1992969, EPI_ISL_1992970, EPI_ISL_1992971, EPI_ISL_1992972, EPI_ISL_1992973, EPI_ISL_1992974, EPI_ISL_1992975, EPI_ISL_1992976, EPI_ISL_1992977, EPI_ISL_1992978, EPI_ISL_1992979, EPI_ISL_1992980, EPI_ISL_1992981, EPI_ISL_1992983, EPI_ISL_1992984, EPI_ISL_1992985, EPI_ISL_1992986, EPI_ISL_1992987, EPI_ISL_1992988, EPI_ISL_1992989, EPI_ISL_1992990, EPI_ISL_1992991, EPI_ISL_1992992, EPI_ISL_1992993, EPI_ISL_1992994, EPI_ISL_1992995, EPI_ISL_1992996, EPI_ISL_1992997, EPI_ISL_1992998, EPI_ISL_1992999, EPI_ISL_1993001, EPI_ISL_1993002, EPI_ISL_1993003, EPI_ISL_1993004, EPI_ISL_1993005, EPI_ISL_1993006, EPI_ISL_1993008, EPI_ISL_1993009, EPI_ISL_1993010, EPI_ISL_1993011, EPI_ISL_1993012, EPI_ISL_1993013, EPI_ISL_1993014, EPI_ISL_1993015, EPI_ISL_1993016, EPI_ISL_1993017, EPI_ISL_1993018, EPI_ISL_1993019, EPI_ISL_1993020, EPI_ISL_1993021, EPI_ISL_1993022, EPI_ISL_1993023, EPI_ISL_1993024, EPI_ISL_1993025, EPI_ISL_1993026, EPI_ISL_1993028, EPI_ISL_1993029, EPI_ISL_1993030, EPI_ISL_1993031, EPI_ISL_1993032, EPI_ISL_1993033, EPI_ISL_1993034, EPI_ISL_1993035, EPI_ISL_1993037, EPI_ISL_1993039, EPI_ISL_1993040, EPI_ISL_1993041, EPI_ISL_1993042, EPI_ISL_1993043, EPI_ISL_1993044, EPI_ISL_1993046, EPI_ISL_1993047, EPI_ISL_1993048, EPI_ISL_1993049, EPI_ISL_1993050, EPI_ISL_1993051, EPI_ISL_1993052, EPI_ISL_1993053, EPI_ISL_1993056, EPI_ISL_1993057, EPI_ISL_1993058, EPI_ISL_1993059, EPI_ISL_1993060, EPI_ISL_1993061, EPI_ISL_1993062, EPI_ISL_1993063, EPI_ISL_1993064, EPI_ISL_1993065, EPI_ISL_1993066, EPI_ISL_1993067, EPI_ISL_1993068, EPI_ISL_1993069, EPI_ISL_1993070, EPI_ISL_1993071, EPI_ISL_1993072, EPI_ISL_1993073, EPI_ISL_1993074, EPI_ISL_1993075, EPI_ISL_1993076, EPI_ISL_1993077, EPI_ISL_1993078, EPI_ISL_1993079, EPI_ISL_1993080, EPI_ISL_1993081, EPI_ISL_1993082, EPI_ISL_1993083, EPI_ISL_1993084, EPI_ISL_1993085, EPI_ISL_1993086, EPI_ISL_1993087, EPI_ISL_1993088, EPI_ISL_1993089, EPI_ISL_1993092, EPI_ISL_1993093, EPI_ISL_1993094, EPI_ISL_1993095, EPI_ISL_1993096, EPI_ISL_1993097, EPI_ISL_1993098, EPI_ISL_1993099, EPI_ISL_1993100, EPI_ISL_1993101, EPI_ISL_1993102, EPI_ISL_1993103, EPI_ISL_1993104, EPI_ISL_1993105, EPI_ISL_1993107, EPI_ISL_1993108, EPI_ISL_1993110, EPI_ISL_1993111, EPI_ISL_1993112, EPI_ISL_1993113, EPI_ISL_1993114, EPI_ISL_1993115, EPI_ISL_1993116, EPI_ISL_1993117, EPI_ISL_1993118, EPI_ISL_1993119, EPI_ISL_1993120, EPI_ISL_1993121, EPI_ISL_1993122, EPI_ISL_1993124, EPI_ISL_1993125, EPI_ISL_1993126, EPI_ISL_1993128, EPI_ISL_1993129, EPI_ISL_1993130, EPI_ISL_1993131, EPI_ISL_1993132, EPI_ISL_1993133, EPI_ISL_1993134, EPI_ISL_1993136, EPI_ISL_1993137, EPI_ISL_1993139, EPI_ISL_1993140, EPI_ISL_1993141, EPI_ISL_1993142, EPI_ISL_1993143, EPI_ISL_1993144, EPI_ISL_1993145, EPI_ISL_1993147, EPI_ISL_1993148, EPI_ISL_1993149, EPI_ISL_1993150, EPI_ISL_1993151, EPI_ISL_1993152, EPI_ISL_1993153, EPI_ISL_1993154, EPI_ISL_1993155, EPI_ISL_1993156, EPI_ISL_1993157, EPI_ISL_1993158, EPI_ISL_1993160, EPI_ISL_1993161, EPI_ISL_1993162, EPI_ISL_1993163, EPI_ISL_1993164, EPI_ISL_1993165, EPI_ISL_1993166, EPI_ISL_1993167, EPI_ISL_1993168, EPI_ISL_1993169, EPI_ISL_1993170, EPI_ISL_1993171, EPI_ISL_1993172, EPI_ISL_1993173, EPI_ISL_1993174, EPI_ISL_1993175, EPI_ISL_1993176, EPI_ISL_1993177, EPI_ISL_1993178, EPI_ISL_1993179, EPI_ISL_1993180, EPI_ISL_1993181, EPI_ISL_1993182, EPI_ISL_1993183, EPI_ISL_1993184, EPI_ISL_1993185, EPI_ISL_1993186, EPI_ISL_1993187, EPI_ISL_1993188, EPI_ISL_1993189, EPI_ISL_1993190, EPI_ISL_1993191, EPI_ISL_1993192, EPI_ISL_1993193, EPI_ISL_1993194, EPI_ISL_1993195, EPI_ISL_1993196, EPI_ISL_1993197, EPI_ISL_1993198, EPI_ISL_1993199, EPI_ISL_1993200, EPI_ISL_1993202, EPI_ISL_1993203, EPI_ISL_1993204, EPI_ISL_1993205, EPI_ISL_1993206, EPI_ISL_1993208,

EPI_ISL_1993209, EPI_ISL_1993210, EPI_ISL_1993211, EPI_ISL_1993212, EPI_ISL_1993213, EPI_ISL_1993214, EPI_ISL_1993215, EPI_ISL_1993216, EPI_ISL_1993217, EPI_ISL_1993218, EPI_ISL_1993219, EPI_ISL_1993220, EPI_ISL_1993221, EPI_ISL_1993222, EPI_ISL_1993223, EPI_ISL_1993224, EPI_ISL_1993225, EPI_ISL_1993227, EPI_ISL_1993228, EPI_ISL_1993229, EPI_ISL_1993230, EPI_ISL_1993231, EPI_ISL_1993232, EPI_ISL_1993233, EPI_ISL_1993234, EPI_ISL_1993235, EPI_ISL_1993236, EPI_ISL_1993237, EPI_ISL_1993238, EPI_ISL_1993239, EPI_ISL_1993240, EPI_ISL_1993241, EPI_ISL_1993242, EPI_ISL_1993243, EPI_ISL_1993244, EPI_ISL_1993245, EPI_ISL_1993246, EPI_ISL_1993247, EPI_ISL_1993248, EPI_ISL_1993249, EPI_ISL_1993250, EPI_ISL_1993251, EPI_ISL_1993252, EPI_ISL_1993253, EPI_ISL_1993254, EPI_ISL_1993255, EPI_ISL_1993256, EPI_ISL_1993257, EPI_ISL_1993258, EPI_ISL_1993259, EPI_ISL_1993260, EPI_ISL_1993261, EPI_ISL_1993262, EPI_ISL_1993263, EPI_ISL_1993264, EPI_ISL_1993265, EPI_ISL_1993266, EPI_ISL_1993267, EPI_ISL_1993268, EPI_ISL_1993269, EPI_ISL_1993270, EPI_ISL_1993271, EPI_ISL_1993272, EPI_ISL_1993273, EPI_ISL_1993274, EPI_ISL_1993275, EPI_ISL_1993276, EPI_ISL_1993277, EPI_ISL_1993278, EPI_ISL_1993279, EPI_ISL_1993280, EPI_ISL_1993281, EPI_ISL_1993282, EPI_ISL_1993283, EPI_ISL_1993284, EPI_ISL_1993288, EPI_ISL_1993289, EPI_ISL_1993290, EPI_ISL_1993291, EPI_ISL_1993293, EPI_ISL_1993294, EPI_ISL_1993295, EPI_ISL_1993296, EPI_ISL_1993297, EPI_ISL_1993298, EPI_ISL_1993299, EPI_ISL_1993300, EPI_ISL_1993301, EPI_ISL_1993302, EPI_ISL_1993303, EPI_ISL_1993304, EPI_ISL_1993305, EPI_ISL_1993306, EPI_ISL_1993307, EPI_ISL_1993309, EPI_ISL_1993311, EPI_ISL_1993312, EPI_ISL_1993314, EPI_ISL_1993315, EPI_ISL_1993316, EPI_ISL_1993318, EPI_ISL_1993319, EPI_ISL_1993320, EPI_ISL_1993321, EPI_ISL_1993323, EPI_ISL_1993324, EPI_ISL_1993325, EPI_ISL_1993326, EPI_ISL_1993327, EPI_ISL_1993328, EPI_ISL_1993329, EPI_ISL_1993330, EPI_ISL_1993331, EPI_ISL_1993333, EPI_ISL_1993335, EPI_ISL_1993336, EPI_ISL_1993337, EPI_ISL_1993338, EPI_ISL_1993340, EPI_ISL_1993341, EPI_ISL_1993342, EPI_ISL_1993343, EPI_ISL_1993344, EPI_ISL_1993345, EPI_ISL_1993346, EPI_ISL_1993347, EPI_ISL_1993348, EPI_ISL_1993350, EPI_ISL_1993351, EPI_ISL_1993352, EPI_ISL_1993353, EPI_ISL_1993354, EPI_ISL_1993355, EPI_ISL_1993356, EPI_ISL_1993357, EPI_ISL_1993360, EPI_ISL_1993361, EPI_ISL_1993362, EPI_ISL_1993363, EPI_ISL_1993364, EPI_ISL_1993365, EPI_ISL_1993366, EPI_ISL_1993367, EPI_ISL_1993368, EPI_ISL_1993369, EPI_ISL_1993370, EPI_ISL_1993371, EPI_ISL_1993372, EPI_ISL_1993373, EPI_ISL_1993374

see above | Fulgent Genetics | Centers for Disease Control and Prevention Division of Viral Diseases, Pathogen Discovery | Dakota Howard, Dhwani Batra, Peter W. Cook, Kara Moser, Adrian Paskey, Jason Caravas, Benjamin Rambo-Martin, Shatavia Morrison, Christopher Gulvick, Scott Sammons, Yvette Unoarumhi, Darlene Wagner, Matthew Schmerer, Harry Gao, Mickey Li, John Gao, Joseph Fierro, Benafsh Sapra, Becky Tsai, Yan Meng, Doreen Ng, James Xie, Clinton R. Paden, Duncan MacCannell

EPI_ISL_1994064, EPI_ISL_1994065, EPI_ISL_1994066, EPI_ISL_1994067, EPI_ISL_1994068, EPI_ISL_1994069, EPI_ISL_1994070, EPI_ISL_1994072, EPI_ISL_1994073, EPI_ISL_1994074, EPI_ISL_1994075, EPI_ISL_1994076, EPI_ISL_1994079, EPI_ISL_1994080, EPI_ISL_1994081, EPI_ISL_1994082, EPI_ISL_1994084, EPI_ISL_1994085, EPI_ISL_1994086, EPI_ISL_1994087, EPI_ISL_1994088, EPI_ISL_1994089, EPI_ISL_1994090, EPI_ISL_1994091, EPI_ISL_1994092, EPI_ISL_1994093, EPI_ISL_1994094, EPI_ISL_1994095, EPI_ISL_1994096, EPI_ISL_1994097, EPI_ISL_1994098, EPI_ISL_1994099, EPI_ISL_1994100, EPI_ISL_1994101, EPI_ISL_1994102, EPI_ISL_1994103, EPI_ISL_1994105, EPI_ISL_1994106, EPI_ISL_1994107, EPI_ISL_1994108, EPI_ISL_1994109, EPI_ISL_1994112, EPI_ISL_1994113, EPI_ISL_1994114, EPI_ISL_1994115, EPI_ISL_1994116, EPI_ISL_1994117, EPI_ISL_1994119, EPI_ISL_1994120, EPI_ISL_1994121, EPI_ISL_1994122, EPI_ISL_1994123, EPI_ISL_1994125, EPI_ISL_1994127, EPI_ISL_1994128, EPI_ISL_1994129, EPI_ISL_1994130, EPI_ISL_1994131, EPI_ISL_1994132, EPI_ISL_1994133, EPI_ISL_1994134, EPI_ISL_1994135, EPI_ISL_1994136, EPI_ISL_1994137, EPI_ISL_1994139, EPI_ISL_1994140, EPI_ISL_1994141, EPI_ISL_1994142, EPI_ISL_1994143, EPI_ISL_1994144, EPI_ISL_1994146, EPI_ISL_1994147, EPI_ISL_1994148, EPI_ISL_1994149, EPI_ISL_1994151, EPI_ISL_1994152, EPI_ISL_1994153, EPI_ISL_1994154, EPI_ISL_1994155, EPI_ISL_1994156, EPI_ISL_1994157, EPI_ISL_1994158, EPI_ISL_1994159, EPI_ISL_1994160, EPI_ISL_1994161, EPI_ISL_1994162, EPI_ISL_1994163, EPI_ISL_1994164, EPI_ISL_1994165, EPI_ISL_1994166, EPI_ISL_1994167, EPI_ISL_1994168, EPI_ISL_1994169, EPI_ISL_1994171, EPI_ISL_1994172, EPI_ISL_1994175, EPI_ISL_1994176, EPI_ISL_1994177, EPI_ISL_1994178, EPI_ISL_1994179, EPI_ISL_1994180, EPI_ISL_1994181, EPI_ISL_1994182, EPI_ISL_1994183, EPI_ISL_1994184, EPI_ISL_1994185, EPI_ISL_1994186, EPI_ISL_1994187, EPI_ISL_1994188, EPI_ISL_1994189, EPI_ISL_1994191, EPI_ISL_1994193, EPI_ISL_1994196, EPI_ISL_1994197, EPI_ISL_1994198, EPI_ISL_1994199, EPI_ISL_1994200, EPI_ISL_1994201, EPI_ISL_1994202, EPI_ISL_1994203, EPI_ISL_1994204, EPI_ISL_1994205, EPI_ISL_1994206, EPI_ISL_1994207, EPI_ISL_1994208, EPI_ISL_1994209, EPI_ISL_1994210, EPI_ISL_1994211, EPI_ISL_1994212, EPI_ISL_1994213, EPI_ISL_1994214, EPI_ISL_1994215, EPI_ISL_1994216, EPI_ISL_1994217, EPI_ISL_1994218, EPI_ISL_1994219, EPI_ISL_1994220, EPI_ISL_1994221, EPI_ISL_1994222, EPI_ISL_1994223, EPI_ISL_1994224, EPI_ISL_1994225, EPI_ISL_1994226, EPI_ISL_1994227, EPI_ISL_1994228, EPI_ISL_1994229, EPI_ISL_1994230, EPI_ISL_1994231, EPI_ISL_1994232, EPI_ISL_1994233, EPI_ISL_1994234, EPI_ISL_1994235, EPI_ISL_1994236, EPI_ISL_1994237, EPI_ISL_1994238, EPI_ISL_1994239, EPI_ISL_1994240, EPI_ISL_1994241, EPI_ISL_1994242, EPI_ISL_1994243, EPI_ISL_1994244, EPI_ISL_1994245, EPI_ISL_1994246, EPI_ISL_1994247, EPI_ISL_1994248, EPI_ISL_1994249, EPI_ISL_1994250, EPI_ISL_1994251, EPI_ISL_1994252, EPI_ISL_1994253, EPI_ISL_1994254, EPI_ISL_1994255, EPI_ISL_1994256, EPI_ISL_1994257, EPI_ISL_1994258, EPI_ISL_1994259, EPI_ISL_1994260, EPI_ISL_1994261, EPI_ISL_1994262, EPI_ISL_1994263, EPI_ISL_1994265, EPI_ISL_1994266, EPI_ISL_1994267, EPI_ISL_1994268, EPI_ISL_1994269, EPI_ISL_1994270, EPI_ISL_1994271, EPI_ISL_1994272, EPI_ISL_1994274, EPI_ISL_1994275, EPI_ISL_1994276, EPI_ISL_1994277, EPI_ISL_1994279, EPI_ISL_1994280, EPI_ISL_1994281, EPI_ISL_1994282, EPI_ISL_1994283, EPI_ISL_1994284, EPI_ISL_1994285, EPI_ISL_1994286, EPI_ISL_1994287, EPI_ISL_1994288, EPI_ISL_1994289, EPI_ISL_1994290, EPI_ISL_1994291, EPI_ISL_1994292, EPI_ISL_1994293, EPI_ISL_1994294, EPI_ISL_1994295, EPI_ISL_1994296, EPI_ISL_1994297, EPI_ISL_1994298, EPI_ISL_1994299, EPI_ISL_1994300, EPI_ISL_1994301, EPI_ISL_1994302, EPI_ISL_1994303, EPI_ISL_1994304, EPI_ISL_1994305, EPI_ISL_1994307, EPI_ISL_1994308, EPI_ISL_1994309, EPI_ISL_1994310, EPI_ISL_1994311, EPI_ISL_1994312, EPI_ISL_1994313, EPI_ISL_1994314, EPI_ISL_1994315, EPI_ISL_1994316, EPI_ISL_1994317, EPI_ISL_1994318, EPI_ISL_1994319, EPI_ISL_1994321, EPI_ISL_1994322, EPI_ISL_1994323, EPI_ISL_1994324, EPI_ISL_1994325, EPI_ISL_1994326, EPI_ISL_1994327, EPI_ISL_1994328, EPI_ISL_1994329, EPI_ISL_1994330, EPI_ISL_1994331, EPI_ISL_1994332, EPI_ISL_1994333, EPI_ISL_1994334, EPI_ISL_1994335, EPI_ISL_1994336, EPI_ISL_1994337, EPI_ISL_1994338, EPI_ISL_1994339, EPI_ISL_1994340, EPI_ISL_1994341, EPI_ISL_1994342, EPI_ISL_1994344, EPI_ISL_1994345, EPI_ISL_1994346, EPI_ISL_1994347, EPI_ISL_1994348, EPI_ISL_1994349, EPI_ISL_1994350, EPI_ISL_1994351, EPI_ISL_1994352, EPI_ISL_1994353, EPI_ISL_1994354, EPI_ISL_1994355, EPI_ISL_1994356, EPI_ISL_1994357, EPI_ISL_1994358, EPI_ISL_1994359, EPI_ISL_1994360, EPI_ISL_1994361, EPI_ISL_1994362, EPI_ISL_1994363, EPI_ISL_1994364, EPI_ISL_1994366, EPI_ISL_1994368, EPI_ISL_1994369, EPI_ISL_1994371, EPI_ISL_1994372, EPI_ISL_1994373, EPI_ISL_1994374, EPI_ISL_1994375, EPI_ISL_1994376, EPI_ISL_1994377, EPI_ISL_1994378, EPI_ISL_1994379, EPI_ISL_1994380, EPI_ISL_1994381, EPI_ISL_1994382, EPI_ISL_1994383, EPI_ISL_1994384, EPI_ISL_1994385, EPI_ISL_1994386, EPI_ISL_1994387, EPI_ISL_1994388, EPI_ISL_1994389, EPI_ISL_1994390, EPI_ISL_1994391, EPI_ISL_1994392, EPI_ISL_1994393, EPI_ISL_1994394, EPI_ISL_1994395, EPI_ISL_1994396, EPI_ISL_1994397, EPI_ISL_1994398, EPI_ISL_1994399, EPI_ISL_1994400, EPI_ISL_1994401, EPI_ISL_1994402, EPI_ISL_1994403, EPI_ISL_1994404, EPI_ISL_1994405, EPI_ISL_1994406, EPI_ISL_1994407, EPI_ISL_1994408, EPI_ISL_1994409, EPI_ISL_1994410, EPI_ISL_1994411, EPI_ISL_1994412, EPI_ISL_1994413, EPI_ISL_1994414, EPI_ISL_1994416, EPI_ISL_1994417, EPI_ISL_1994418, EPI_ISL_1994419, EPI_ISL_1994421, EPI_ISL_1994422, EPI_ISL_1994424, EPI_ISL_1994425, EPI_ISL_1994426, EPI_ISL_1994427, EPI_ISL_1994428, EPI_ISL_1994429, EPI_ISL_1994430, EPI_ISL_1994431, EPI_ISL_1994432, EPI_ISL_1994433, EPI_ISL_1994434, EPI_ISL_1994435, EPI_ISL_1994436, EPI_ISL_1994437, EPI_ISL_1994438, EPI_ISL_1994439, EPI_ISL_1994440, EPI_ISL_1994441, EPI_ISL_1994442, EPI_ISL_1994443, EPI_ISL_1994444, EPI_ISL_1994445, EPI_ISL_1994446, EPI_ISL_1994447, EPI_ISL_1994448, EPI_ISL_1994449, EPI_ISL_1994450, EPI_ISL_1994452, EPI_ISL_1994453, EPI_ISL_1994454, EPI_ISL_1994455, EPI_ISL_1994456, EPI_ISL_1994457, EPI_ISL_1994458, EPI_ISL_1994459, EPI_ISL_1994460, EPI_ISL_1994461, EPI_ISL_1994462, EPI_ISL_1994463, EPI_ISL_1994464, EPI_ISL_1994465, EPI_ISL_1994466, EPI_ISL_1994468, EPI_ISL_1994469, EPI_ISL_1994470, EPI_ISL_1994471, EPI_ISL_1994472, EPI_ISL_1994473, EPI_ISL_1994474, EPI_ISL_1994475, EPI_ISL_1994476, EPI_ISL_1994477, EPI_ISL_1994478, EPI_ISL_1994479, EPI_ISL_1994480, EPI_ISL_1994482, EPI_ISL_1994483, EPI_ISL_1994484, EPI_ISL_1994489, EPI_ISL_1994490, EPI_ISL_1994491, EPI_ISL_1994492, EPI_ISL_1994494, EPI_ISL_1994496, EPI_ISL_1994498, EPI_ISL_1994499, EPI_ISL_1994500, EPI_ISL_1994501, EPI_ISL_1994502, EPI_ISL_1994503, EPI_ISL_1994504, EPI_ISL_1994505, EPI_ISL_1994506, EPI_ISL_1994508, EPI_ISL_1994509, EPI_ISL_1994510, EPI_ISL_1994511, EPI_ISL_1994512, EPI_ISL_1994513, EPI_ISL_1994514, EPI_ISL_1994516, EPI_ISL_1994517, EPI_ISL_1994518, EPI_ISL_1994519, EPI_ISL_1994520, EPI_ISL_1994521, EPI_ISL_1994522, EPI_ISL_1994523, EPI_ISL_1994524, EPI_ISL_1994525, EPI_ISL_1994526, EPI_ISL_1994527, EPI_ISL_1994528, EPI_ISL_1994529, EPI_ISL_1994530, EPI_ISL_1994531, EPI_ISL_1994532, EPI_ISL_1994533, EPI_ISL_1994534, EPI_ISL_1994535, EPI_ISL_1994536, EPI_ISL_1994537, EPI_ISL_1994538, EPI_ISL_1994539, EPI_ISL_1994540, EPI_ISL_1994541, EPI_ISL_1994542, EPI_ISL_1994609, EPI_ISL_1994610, EPI_ISL_1994611, EPI_ISL_1994612, EPI_ISL_1994613, EPI_ISL_1994614, EPI_ISL_1994615, EPI_ISL_1994616, EPI_ISL_1994617, EPI_ISL_1994618, EPI_ISL_1994619, EPI_ISL_1994620, EPI_ISL_1994621, EPI_ISL_1994622, EPI_ISL_1994623, EPI_ISL_1994624, EPI_ISL_1994625, EPI_ISL_1994626, EPI_ISL_1994627, EPI_ISL_1994628, EPI_ISL_1994629, EPI_ISL_1994630, EPI_ISL_1994631, EPI_ISL_1994632, EPI_ISL_1994633, EPI_ISL_1994634, EPI_ISL_1994635, EPI_ISL_1994636, EPI_ISL_1994637, EPI_ISL_1994638, EPI_ISL_1994639, EPI_ISL_1994640, EPI_ISL_1994641, EPI_ISL_1994642, EPI_ISL_1994643, EPI_ISL_1994644, EPI_ISL_1994645, EPI_ISL_1994646, EPI_ISL_1994647, EPI_ISL_1994648, EPI_ISL_1994649, EPI_ISL_1994650, EPI_ISL_1994651, EPI_ISL_1994652, EPI_ISL_1994653, EPI_ISL_1994655, EPI_ISL_1994656, EPI_ISL_1994657, EPI_ISL_1994658, EPI_ISL_1994659, EPI_ISL_1994660, EPI_ISL_1994661, EPI_ISL_1994662, EPI_ISL_1994663, EPI_ISL_1994664, EPI_ISL_1994665, EPI_ISL_1994666, EPI_ISL_1994667, EPI_ISL_1994668, EPI_ISL_1994669, EPI_ISL_1994671, EPI_ISL_1994672, EPI_ISL_1994673, EPI_ISL_1994674, EPI_ISL_1994675, EPI_ISL_1994677, EPI_ISL_1994679, EPI_ISL_1994680, EPI_ISL_1994681, EPI_ISL_1994682, EPI_ISL_1994683, EPI_ISL_1994684, EPI_ISL_1994685, EPI_ISL_1994686, EPI_ISL_1994687, EPI_ISL_1994688, EPI_ISL_1994689, EPI_ISL_1994690, EPI_ISL_1994691, EPI_ISL_1994692, EPI_ISL_1994694, EPI_ISL_1994695, EPI_ISL_1994696, EPI_ISL_1994697, EPI_ISL_1994698, EPI_ISL_1994699, EPI_ISL_1994700, EPI_ISL_1994701, EPI_ISL_1994702, EPI_ISL_1994703, EPI_ISL_1994704, EPI_ISL_1994705, EPI_ISL_1994706, EPI_ISL_1994707, EPI_ISL_1994708, EPI_ISL_1994709, EPI_ISL_1994710, EPI_ISL_1994711, EPI_ISL_1994712, EPI_ISL_1994713, EPI_ISL_1994714, EPI_ISL_1994715, EPI_ISL_1994716, EPI_ISL_1994717, EPI_ISL_1994718, EPI_ISL_1994719, EPI_ISL_1994720, EPI_ISL_1994721, EPI_ISL_1994722, EPI_ISL_1994723, EPI_ISL_1994724, EPI_ISL_1994725, EPI_ISL_1994726, EPI_ISL_1994727, EPI_ISL_1994729, EPI_ISL_1994730, EPI_ISL_1994731, EPI_ISL_1994732, EPI_ISL_1994733, EPI_ISL_1994734, EPI_ISL_1994735, EPI_ISL_1994736, EPI_ISL_1994737, EPI_ISL_1994738, EPI_ISL_1994739, EPI_ISL_1994740, EPI_ISL_1994741, EPI_ISL_1994742, EPI_ISL_1994743, EPI_ISL_1994744, EPI_ISL_1994745, EPI_ISL_1994746, EPI_ISL_1994747, EPI_ISL_1994748, EPI_ISL_1994749, EPI_ISL_1994752, EPI_ISL_1994753, EPI_ISL_1994754, EPI_ISL_1994755, EPI_ISL_1994757, EPI_ISL_1994758, EPI_ISL_1994759, EPI_ISL_1994760, EPI_ISL_1994761, EPI_ISL_1994762, EPI_ISL_1994763, EPI_ISL_1994764, EPI_ISL_1994765, EPI_ISL_1994766, EPI_ISL_1994767, EPI_ISL_1994768, EPI_ISL_1994769, EPI_ISL_1994770, EPI_ISL_1994772, EPI_ISL_1994773, EPI_ISL_1994774, EPI_ISL_1994775, EPI_ISL_1994776, EPI_ISL_1994777, EPI_ISL_1994778, EPI_ISL_1994779, EPI_ISL_1994780, EPI_ISL_1994781, EPI_ISL_1994782, EPI_ISL_1994783, EPI_ISL_1994785, EPI_ISL_1994787, EPI_ISL_1994788, EPI_ISL_1994789, EPI_ISL_1994790, EPI_ISL_1994791, EPI_ISL_1994792, EPI_ISL_1994793, EPI_ISL_1994794, EPI_ISL_1994795, EPI_ISL_1994796, EPI_ISL_1994797, EPI_ISL_1994798, EPI_ISL_1994799, EPI_ISL_1994800, EPI_ISL_1994801, EPI_ISL_1994802, EPI_ISL_1994803, EPI_ISL_1994804, EPI_ISL_1994805, EPI_ISL_1994809, EPI_ISL_1994810, EPI_ISL_1994811, EPI_ISL_1994812, EPI_ISL_1994813, EPI_ISL_1994814, EPI_ISL_1994815, EPI_ISL_1994816, EPI_ISL_1994817, EPI_ISL_1994818, EPI_ISL_1994819, EPI_ISL_1994820, EPI_ISL_1994821, EPI_ISL_1994822, EPI_ISL_1994824, EPI_ISL_1994825, EPI_ISL_1994826, EPI_ISL_1994827, EPI_ISL_1994828, EPI_ISL_1994829, EPI_ISL_1994830, EPI_ISL_1994831, EPI_ISL_1994832, EPI_ISL_1994833, EPI_ISL_1994834, EPI_ISL_1994835, EPI_ISL_1994836, EPI_ISL_1994837, EPI_ISL_1994838, EPI_ISL_1994839, EPI_ISL_1994840, EPI_ISL_1994841, EPI_ISL_1994842, EPI_ISL_1994843, EPI_ISL_1994844, EPI_ISL_1994846, EPI_ISL_1994847, EPI_ISL_1994848, EPI_ISL_1994849, EPI_ISL_1994850, EPI_ISL_1994851, EPI_ISL_1994852, EPI_ISL_1994853, EPI_ISL_1994854, EPI_ISL_1994855, EPI_ISL_1994857, EPI_ISL_1994858, EPI_ISL_1994859, EPI_ISL_1994860, EPI_ISL_1994861, EPI_ISL_1994862, EPI_ISL_1994864, EPI_ISL_1994865, EPI_ISL_1994867, EPI_ISL_1994868, EPI_ISL_1994869, EPI_ISL_1994870, EPI_ISL_1994871, EPI_ISL_1994872

see above | Michigan Department of Health and Human Services, Bureau of Laboratories | Michigan Department of Health and Human Services, Bureau of Laboratories | Blankenship HM, Riner D, Soehnlen MK

EPI_ISL_1994873, EPI_ISL_1994876, EPI_ISL_1994878, EPI_ISL_1994879, EPI_ISL_1994880, EPI_ISL_1994881, EPI_ISL_1994882, EPI_ISL_1994883, EPI_ISL_1994885, EPI_ISL_1994886, EPI_ISL_1994887, EPI_ISL_1994888, EPI_ISL_1994890, EPI_ISL_1994891, EPI_ISL_1994892, EPI_ISL_1994894, EPI_ISL_1994895, EPI_ISL_1994896, EPI_ISL_1994898, EPI_ISL_1994900, EPI_ISL_1994901, EPI_ISL_1994902, EPI_ISL_1994903, EPI_ISL_1994904, EPI_ISL_1994905, EPI_ISL_1994906, EPI_ISL_1994907, EPI_ISL_1994908, EPI_ISL_1994909, EPI_ISL_1994910, EPI_ISL_1994911, EPI_ISL_1994912, EPI_ISL_1994913, EPI_ISL_1994914, EPI_ISL_1994915, EPI_ISL_1994916, EPI_ISL_1994917, EPI_ISL_1994918, EPI_ISL_1994920

see above | IN State Department of Health Laboratory Services | IN State Department of Health Laboratory Services | Cassandra Campion, Jamie Yeadon, Brian Pope, Lixia Liu, Kyle Brownlee, Melissa Hindenlang, Mark Glazier

EPI_ISL_1994963, EPI_ISL_1994964, EPI_ISL_1994965, EPI_ISL_1994966, EPI_ISL_1994967, EPI_ISL_1994968, EPI_ISL_1994969, EPI_ISL_1994970, EPI_ISL_1994971, EPI_ISL_1994972, EPI_ISL_1994973, EPI_ISL_1994974, EPI_ISL_1994975, EPI_ISL_1994976, EPI_ISL_1994977, EPI_ISL_1994978, EPI_ISL_1994979, EPI_ISL_1994980, EPI_ISL_1994981, EPI_ISL_1994982, EPI_ISL_1994983, EPI_ISL_1994984, EPI_ISL_1994985, EPI_ISL_1994986, EPI_ISL_1994987, EPI_ISL_1994988, EPI_ISL_1994989, EPI_ISL_1994990, EPI_ISL_1994991, EPI_ISL_1994992, EPI_ISL_1994993, EPI_ISL_1994994, EPI_ISL_1994995, EPI_ISL_1994996, EPI_ISL_1994997, EPI_ISL_1994998, EPI_ISL_1994999, EPI_ISL_1995000, EPI_ISL_1995001, EPI_ISL_1995002, EPI_ISL_1995003, EPI_ISL_1995004, EPI_ISL_1995005, EPI_ISL_1995006, EPI_ISL_1995007, EPI_ISL_1995008, EPI_ISL_1995009, EPI_ISL_1995010, EPI_ISL_1995011, EPI_ISL_1995012, EPI_ISL_1995013, EPI_ISL_1995014, EPI_ISL_1995015, EPI_ISL_1995016, EPI_ISL_1995017, EPI_ISL_1995018, EPI_ISL_1995019, EPI_ISL_1995020, EPI_ISL_1995021, EPI_ISL_1995022, EPI_ISL_1995023, EPI_ISL_1995024, EPI_ISL_1995025, EPI_ISL_1995026, EPI_ISL_1995027, EPI_ISL_1995028, EPI_ISL_1995029, EPI_ISL_1995030, EPI_ISL_1995031, EPI_ISL_1995032, EPI_ISL_1995033, EPI_ISL_1995034, EPI_ISL_1995035, EPI_ISL_1995036, EPI_ISL_1995037, EPI_ISL_1995038, EPI_ISL_1995039, EPI_ISL_1995040, EPI_ISL_1995041, EPI_ISL_1995042, EPI_ISL_1995043, EPI_ISL_1995044, EPI_ISL_1995045, EPI_ISL_1995046, EPI_ISL_1995047, EPI_ISL_1995048, EPI_ISL_1995049, EPI_ISL_1995050, EPI_ISL_1995051, EPI_ISL_1995052, EPI_ISL_1995053, EPI_ISL_1995054, EPI_ISL_1995055, EPI_ISL_1995056, EPI_ISL_1995057, EPI_ISL_1995058, EPI_ISL_1995059, EPI_ISL_1995060, EPI_ISL_1995061, EPI_ISL_1995062, EPI_ISL_1995063, EPI_ISL_1995064, EPI_ISL_1995065, EPI_ISL_1995066, EPI_ISL_1995067, EPI_ISL_1995068, EPI_ISL_1995069, EPI_ISL_1995070, EPI_ISL_1995071, EPI_ISL_1995072, EPI_ISL_1995073, EPI_ISL_1995074, EPI_ISL_1995075, EPI_ISL_1995076, EPI_ISL_1995077, EPI_ISL_1995078, EPI_ISL_1995079, EPI_ISL_1995080, EPI_ISL_1995081, EPI_ISL_1995082, EPI_ISL_1995083, EPI_ISL_1995084, EPI_ISL_1995085, EPI_ISL_1995086, EPI_ISL_1995087, EPI_ISL_1995088, EPI_ISL_1995089, EPI_ISL_1995090, EPI_ISL_1995092, EPI_ISL_1995093, EPI_ISL_1995094, EPI_ISL_1995095, EPI_ISL_1995096, EPI_ISL_1995097, EPI_ISL_1995098, EPI_ISL_1995100, EPI_ISL_1995101, EPI_ISL_1995102, EPI_ISL_1995103, EPI_ISL_1995104, EPI_ISL_1995105, EPI_ISL_1995106, EPI_ISL_1995107, EPI_ISL_1995108, EPI_ISL_1995109, EPI_ISL_1995110, EPI_ISL_1995111, EPI_ISL_1995112, EPI_ISL_1995113, EPI_ISL_1995114, EPI_ISL_1995115, EPI_ISL_1995116, EPI_ISL_1995117, EPI_ISL_1995118, EPI_ISL_1995119, EPI_ISL_1995120, EPI_ISL_1995121, EPI_ISL_1995122, EPI_ISL_1995123, EPI_ISL_1995124, EPI_ISL_1995125, EPI_ISL_1995126, EPI_ISL_1995127, EPI_ISL_1995128, EPI_ISL_1995129, EPI_ISL_1995130, EPI_ISL_1995131, EPI_ISL_1995132, EPI_ISL_1995133, EPI_ISL_1995134, EPI_ISL_1995135, EPI_ISL_1995136, EPI_ISL_1995137, EPI_ISL_1995138, EPI_ISL_1995139, EPI_ISL_1995140, EPI_ISL_1995141, EPI_ISL_1995142, EPI_ISL_1995143, EPI_ISL_1995144, EPI_ISL_1995145, EPI_ISL_1995146, EPI_ISL_1995147, EPI_ISL_1995148, EPI_ISL_1995149, EPI_ISL_1995150, EPI_ISL_1995151, EPI_ISL_1995152, EPI_ISL_1995155, EPI_ISL_1995156, EPI_ISL_1995157, EPI_ISL_1995158, EPI_ISL_1995159, EPI_ISL_1995160, EPI_ISL_1995161, EPI_ISL_1995162, EPI_ISL_1995163, EPI_ISL_1995164, EPI_ISL_1995165, EPI_ISL_1995166, EPI_ISL_1995167, EPI_ISL_1995168, EPI_ISL_1995169, EPI_ISL_1995170, EPI_ISL_1995171, EPI_ISL_1995172, EPI_ISL_1995173, EPI_ISL_1995174, EPI_ISL_1995175, EPI_ISL_1995176, EPI_ISL_1995177, EPI_ISL_1995178, EPI_ISL_1995179, EPI_ISL_1995180, EPI_ISL_1995181, EPI_ISL_1995182, EPI_ISL_1995183, EPI_ISL_1995184, EPI_ISL_1995185, EPI_ISL_1995186, EPI_ISL_1995187, EPI_ISL_1995188, EPI_ISL_1995189, EPI_ISL_1995190, EPI_ISL_1995191, EPI_ISL_1995192, EPI_ISL_1995193, EPI_ISL_1995194, EPI_ISL_1995195, EPI_ISL_1995196, EPI_ISL_1995197, EPI_ISL_1995198, EPI_ISL_1995199, EPI_ISL_1995200, EPI_ISL_1995201, EPI_ISL_1995202, EPI_ISL_1995203, EPI_ISL_1995204, EPI_ISL_1995205, EPI_ISL_1995206, EPI_ISL_1995207, EPI_ISL_1995208, EPI_ISL_1995209, EPI_ISL_1995210, EPI_ISL_1995211, EPI_ISL_1995212, EPI_ISL_1995213, EPI_ISL_1995214, EPI_ISL_1995215, EPI_ISL_1995216, EPI_ISL_1995217, EPI_ISL_1995218, EPI_ISL_1995219, EPI_ISL_1995220, EPI_ISL_1995221, EPI_ISL_1995222, EPI_ISL_1995224, EPI_ISL_1995225, EPI_ISL_1995226, EPI_ISL_1995228, EPI_ISL_1995229, EPI_ISL_1995230, EPI_ISL_1995231, EPI_ISL_1995232, EPI_ISL_1995233, EPI_ISL_1995234, EPI_ISL_1995235, EPI_ISL_1995236, EPI_ISL_1995237, EPI_ISL_1995238, EPI_ISL_1995239, EPI_ISL_1995240, EPI_ISL_1995241, EPI_ISL_1995242, EPI_ISL_1995243, EPI_ISL_1995244, EPI_ISL_1995245, EPI_ISL_1995246, EPI_ISL_1995247, EPI_ISL_1995248, EPI_ISL_1995249, EPI_ISL_1995250, EPI_ISL_1995251, EPI_ISL_1995252, EPI_ISL_1995253, EPI_ISL_1995254, EPI_ISL_1995255, EPI_ISL_1995257, EPI_ISL_1995258, EPI_ISL_1995259, EPI_ISL_1995260, EPI_ISL_1995261, EPI_ISL_1995262, EPI_ISL_1995263, EPI_ISL_1995264, EPI_ISL_1995266, EPI_ISL_1995267, EPI_ISL_1995268, EPI_ISL_1995269, EPI_ISL_1995270, EPI_ISL_1995271, EPI_ISL_1995272, EPI_ISL_1995273, EPI_ISL_1995274, EPI_ISL_1995275, EPI_ISL_1995276, EPI_ISL_1995277, EPI_ISL_1995278, EPI_ISL_1995279, EPI_ISL_1995280, EPI_ISL_1995281, EPI_ISL_1995282, EPI_ISL_1995283, EPI_ISL_1995284, EPI_ISL_1995285, EPI_ISL_1995286, EPI_ISL_1995287, EPI_ISL_1995288, EPI_ISL_1995289, EPI_ISL_1995290, EPI_ISL_1995291, EPI_ISL_1995292, EPI_ISL_1995293, EPI_ISL_1995294, EPI_ISL_1995295, EPI_ISL_1995296, EPI_ISL_1995297, EPI_ISL_1995298, EPI_ISL_1995299, EPI_ISL_1995300, EPI_ISL_1995301, EPI_ISL_1995302, EPI_ISL_1995303, EPI_ISL_1995304, EPI_ISL_1995305, EPI_ISL_1995306, EPI_ISL_1995307, EPI_ISL_1995308, EPI_ISL_1995309, EPI_ISL_1995310, EPI_ISL_1995311, EPI_ISL_1995312, EPI_ISL_1995313, EPI_ISL_1995314, EPI_ISL_1995315, EPI_ISL_1995316, EPI_ISL_1995317, EPI_ISL_1995318, EPI_ISL_1995319, EPI_ISL_1995320, EPI_ISL_1995321, EPI_ISL_1995322, EPI_ISL_1995323, EPI_ISL_1995324, EPI_ISL_1995325, EPI_ISL_1995326, EPI_ISL_1995327, EPI_ISL_1995328, EPI_ISL_1995329, EPI_ISL_1995330, EPI_ISL_1995331, EPI_ISL_1995332, EPI_ISL_1995333, EPI_ISL_1995334, EPI_ISL_1995335, EPI_ISL_1995336, EPI_ISL_1995337, EPI_ISL_1995338, EPI_ISL_1995339, EPI_ISL_1995340, EPI_ISL_1995341, EPI_ISL_1995342, EPI_ISL_1995343, EPI_ISL_1995345, EPI_ISL_1995346, EPI_ISL_1995347, EPI_ISL_1995348, EPI_ISL_1995349, EPI_ISL_1995351, EPI_ISL_1995352, EPI_ISL_1995353, EPI_ISL_1995354, EPI_ISL_1995355, EPI_ISL_1995356, EPI_ISL_1995357, EPI_ISL_1995358, EPI_ISL_1995359, EPI_ISL_1995360, EPI_ISL_1995361, EPI_ISL_1995362, EPI_ISL_1995363, EPI_ISL_1995365, EPI_ISL_1995366, EPI_ISL_1995367, EPI_ISL_1995368, EPI_ISL_1995369, EPI_ISL_1995370, EPI_ISL_1995371, EPI_ISL_1995372, EPI_ISL_1995373, EPI_ISL_1995375, EPI_ISL_1995376, EPI_ISL_1995377, EPI_ISL_1995378, EPI_ISL_1995379, EPI_ISL_1995380, EPI_ISL_1995381, EPI_ISL_1995382, EPI_ISL_1995383, EPI_ISL_1995384, EPI_ISL_1995385, EPI_ISL_1995386, EPI_ISL_1995387, EPI_ISL_1995388, EPI_ISL_1995389, EPI_ISL_1995390, EPI_ISL_1995391, EPI_ISL_1995392, EPI_ISL_1995393, EPI_ISL_1995394, EPI_ISL_1995395, EPI_ISL_1995396, EPI_ISL_1995397, EPI_ISL_1995398, EPI_ISL_1995399, EPI_ISL_1995400, EPI_ISL_1995401, EPI_ISL_1995402, EPI_ISL_1995403, EPI_ISL_1995404, EPI_ISL_1995405, EPI_ISL_1995406, EPI_ISL_1995407, EPI_ISL_1995408, EPI_ISL_1995409, EPI_ISL_1995410, EPI_ISL_1995411, EPI_ISL_1995412, EPI_ISL_1995413, EPI_ISL_1995414, EPI_ISL_1995415, EPI_ISL_1995416, EPI_ISL_1995417, EPI_ISL_1995418, EPI_ISL_1995419, EPI_ISL_1995420, EPI_ISL_1995421, EPI_ISL_1995422, EPI_ISL_1995423, EPI_ISL_1995424, EPI_ISL_1995425, EPI_ISL_1995426, EPI_ISL_1995427, EPI_ISL_1995428, EPI_ISL_1995429, EPI_ISL_1995430, EPI_ISL_1995431, EPI_ISL_1995432, EPI_ISL_1995433, EPI_ISL_1995434, EPI_ISL_1995435, EPI_ISL_1995436, EPI_ISL_1995437, EPI_ISL_1995438, EPI_ISL_1995439, EPI_ISL_1995440, EPI_ISL_1995441, EPI_ISL_1995442, EPI_ISL_1995443, EPI_ISL_1995444, EPI_ISL_1995445, EPI_ISL_1995446, EPI_ISL_1995447, EPI_ISL_1995448, EPI_ISL_1995449, EPI_ISL_1995450, EPI_ISL_1995451, EPI_ISL_1995452, EPI_ISL_1995453, EPI_ISL_1995454, EPI_ISL_1995456, EPI_ISL_1995457, EPI_ISL_1995458, EPI_ISL_1995459, EPI_ISL_1995460, EPI_ISL_1995461, EPI_ISL_1995462, EPI_ISL_1995463, EPI_ISL_1995464, EPI_ISL_1995465, EPI_ISL_1995466, EPI_ISL_1995467, EPI_ISL_1995468, EPI_ISL_1995469, EPI_ISL_1995470, EPI_ISL_1995471, EPI_ISL_1995472, EPI_ISL_1995473, EPI_ISL_1995474, EPI_ISL_1995475, EPI_ISL_1995476, EPI_ISL_1995477, EPI_ISL_1995478, EPI_ISL_1995479, EPI_ISL_1995480, EPI_ISL_1995481, EPI_ISL_1995482, EPI_ISL_1995483, EPI_ISL_1995484, EPI_ISL_1995485, EPI_ISL_1995486, EPI_ISL_1995487, EPI_ISL_1995488, EPI_ISL_1995489, EPI_ISL_1995490, EPI_ISL_1995491, EPI_ISL_1995492, EPI_ISL_1995493, EPI_ISL_1995494, EPI_ISL_1995495, EPI_ISL_1995496, EPI_ISL_1995497, EPI_ISL_1995498, EPI_ISL_1995499, EPI_ISL_1995500, EPI_ISL_1995501, EPI_ISL_1995502, EPI_ISL_1995503, EPI_ISL_1995504, EPI_ISL_1995505, EPI_ISL_1995506, EPI_ISL_1995507, EPI_ISL_1995508, EPI_ISL_1995509, EPI_ISL_1995510, EPI_ISL_1995511, EPI_ISL_1995512, EPI_ISL_1995513, EPI_ISL_1995514, EPI_ISL_1995515, EPI_ISL_1995516, EPI_ISL_1995517, EPI_ISL_1995518, EPI_ISL_1995519, EPI_ISL_1995520, EPI_ISL_1995521, EPI_ISL_1995522, EPI_ISL_1995523, EPI_ISL_1995524, EPI_ISL_1995525, EPI_ISL_1995526, EPI_ISL_1995527, EPI_ISL_1995528, EPI_ISL_1995529, EPI_ISL_1995530, EPI_ISL_1995531, EPI_ISL_1995532, EPI_ISL_1995533, EPI_ISL_1995534, EPI_ISL_1995535, EPI_ISL_1995536, EPI_ISL_1995537, EPI_ISL_1995538, EPI_ISL_1995539, EPI_ISL_1995540, EPI_ISL_1995541, EPI_ISL_1995542, EPI_ISL_1995543, EPI_ISL_1995544, EPI_ISL_1995545, EPI_ISL_1995546, EPI_ISL_1995548, EPI_ISL_1995549, EPI_ISL_1995550, EPI_ISL_1995551, EPI_ISL_1995552, EPI_ISL_1995554, EPI_ISL_1995555, EPI_ISL_1995556, EPI_ISL_1995557, EPI_ISL_1995558, EPI_ISL_1995559, EPI_ISL_1995560, EPI_ISL_1995561, EPI_ISL_1995562, EPI_ISL_1995563, EPI_ISL_1995564, EPI_ISL_1995565, EPI_ISL_1995566, EPI_ISL_1995567, EPI_ISL_1995568, EPI_ISL_1995569, EPI_ISL_1995570, EPI_ISL_1995571, EPI_ISL_1995572, EPI_ISL_1995573, EPI_ISL_1995574, EPI_ISL_1995575, EPI_ISL_1995577, EPI_ISL_1995578, EPI_ISL_1995579, EPI_ISL_1995580, EPI_ISL_1995581, EPI_ISL_1995582, EPI_ISL_1995583, EPI_ISL_1995584, EPI_ISL_1995585, EPI_ISL_1995586, EPI_ISL_1995587, EPI_ISL_1995588, EPI_ISL_1995589, EPI_ISL_1995590, EPI_ISL_1995591, EPI_ISL_1995592, EPI_ISL_1995593, EPI_ISL_1995594, EPI_ISL_1995595, EPI_ISL_1995596, EPI_ISL_1995597, EPI_ISL_1995598, EPI_ISL_1995599, EPI_ISL_1995600, EPI_ISL_1995601, EPI_ISL_1995602, EPI_ISL_1995603, EPI_ISL_1995604, EPI_ISL_1995605, EPI_ISL_1995606, EPI_ISL_1995607, EPI_ISL_1995608, EPI_ISL_1995609, EPI_ISL_1995610, EPI_ISL_1995611, EPI_ISL_1995612, EPI_ISL_1995613, EPI_ISL_1995614, EPI_ISL_1995615, EPI_ISL_1995616, EPI_ISL_1995617, EPI_ISL_1995618, EPI_ISL_1995619, EPI_ISL_1995620, EPI_ISL_1995621, EPI_ISL_1995622, EPI_ISL_1995623, EPI_ISL_1995624, EPI_ISL_1995625, EPI_ISL_1995626, EPI_ISL_1995627, EPI_ISL_1995628, EPI_ISL_1995629, EPI_ISL_1995630, EPI_ISL_1995631, EPI_ISL_1995632, EPI_ISL_1995633, EPI_ISL_1995634, EPI_ISL_1995635, EPI_ISL_1995636, EPI_ISL_1995637, EPI_ISL_1995638, EPI_ISL_1995639, EPI_ISL_1995640, EPI_ISL_1995641, EPI_ISL_1995643, EPI_ISL_1995644, EPI_ISL_1995645, EPI_ISL_1995646, EPI_ISL_1995647, EPI_ISL_1995648, EPI_ISL_1995649, EPI_ISL_1995650, EPI_ISL_1995651, EPI_ISL_1995652, EPI_ISL_1995653, EPI_ISL_1995654, EPI_ISL_1995655, EPI_ISL_1995656, EPI_ISL_1995657, EPI_ISL_1995658, EPI_ISL_1995659, EPI_ISL_1995660, EPI_ISL_1995661, EPI_ISL_1995662, EPI_ISL_1995663, EPI_ISL_1995664, EPI_ISL_1995665, EPI_ISL_1995666, EPI_ISL_1995667, EPI_ISL_1995668, EPI_ISL_1995669, EPI_ISL_1995670, EPI_ISL_1995671, EPI_ISL_1995672, EPI_ISL_1995673, EPI_ISL_1995674, EPI_ISL_1995675, EPI_ISL_1995676, EPI_ISL_1995677, EPI_ISL_1995678, EPI_ISL_1995679, EPI_ISL_1995680, EPI_ISL_1995681, EPI_ISL_1995682, EPI_ISL_1995683, EPI_ISL_1995684, EPI_ISL_1995685, EPI_ISL_1995686, EPI_ISL_1995688, EPI_ISL_1995689, EPI_ISL_1995690, EPI_ISL_1995691, EPI_ISL_1995692, EPI_ISL_1995693, EPI_ISL_1995694, EPI_ISL_1995695, EPI_ISL_1995696, EPI_ISL_1995697, EPI_ISL_1995698, EPI_ISL_1995699, EPI_ISL_1995700, EPI_ISL_1995701, EPI_ISL_1995702, EPI_ISL_1995703, EPI_ISL_1995704, EPI_ISL_1995705, EPI_ISL_1995706, EPI_ISL_1995707, EPI_ISL_1995708, EPI_ISL_1995709, EPI_ISL_1995710, EPI_ISL_1995711, EPI_ISL_1995712, EPI_ISL_1995713, EPI_ISL_1995714, EPI_ISL_1995715, EPI_ISL_1995716, EPI_ISL_1995718, EPI_ISL_1995719, EPI_ISL_1995720, EPI_ISL_1995721, EPI_ISL_1995722, EPI_ISL_1995723, EPI_ISL_1995724, EPI_ISL_1995725, EPI_ISL_1995726, EPI_ISL_1995727, EPI_ISL_1995728, EPI_ISL_1995729, EPI_ISL_1995730, EPI_ISL_1995731, EPI_ISL_1995732, EPI_ISL_1995733, EPI_ISL_1995734, EPI_ISL_1995736, EPI_ISL_1995737, EPI_ISL_1995738, EPI_ISL_1995739, EPI_ISL_1995740, EPI_ISL_1995741, EPI_ISL_1995742, EPI_ISL_1995743, EPI_ISL_1995744, EPI_ISL_1995745, EPI_ISL_1995746, EPI_ISL_1995747, EPI_ISL_1995748, EPI_ISL_1995749, EPI_ISL_1995750, EPI_ISL_1995751, EPI_ISL_1995752, EPI_ISL_1995753, EPI_ISL_1995754, EPI_ISL_1995755, EPI_ISL_1995756, EPI_ISL_1995757, EPI_ISL_1995758, EPI_ISL_1995759, EPI_ISL_1995760, EPI_ISL_1995761, EPI_ISL_1995762, EPI_ISL_1995763, EPI_ISL_1995764, EPI_ISL_1995765, EPI_ISL_1995766, EPI_ISL_1995767, EPI_ISL_1995768, EPI_ISL_1995769, EPI_ISL_1995770, EPI_ISL_1995771, EPI_ISL_1995772, EPI_ISL_1995773, EPI_ISL_1995774, EPI_ISL_1995775, EPI_ISL_1995776, EPI_ISL_1995777, EPI_ISL_1995778, EPI_ISL_1995779, EPI_ISL_1995780, EPI_ISL_1995781, EPI_ISL_1995782, EPI_ISL_1995783, EPI_ISL_1995784, EPI_ISL_1995785, EPI_ISL_1995786, EPI_ISL_1995787, EPI_ISL_1995788, EPI_ISL_1995789, EPI_ISL_1995790, EPI_ISL_1995791, EPI_ISL_1995792, EPI_ISL_1995793, EPI_ISL_1995794, EPI_ISL_1995795, EPI_ISL_1995796, EPI_ISL_1995797, EPI_ISL_1995798, EPI_ISL_1995799, EPI_ISL_1995800, EPI_ISL_1995801, EPI_ISL_1995802, EPI_ISL_1995803, EPI_ISL_1995804, EPI_ISL_1995805, EPI_ISL_1995806, EPI_ISL_1995807, EPI_ISL_1995808, EPI_ISL_1995809, EPI_ISL_1995810, EPI_ISL_1995811, EPI_ISL_1995812, EPI_ISL_1995813, EPI_ISL_1995814, EPI_ISL_1995815, EPI_ISL_1995816, EPI_ISL_1995817, EPI_ISL_1995818, EPI_ISL_1995819, EPI_ISL_1995820, EPI_ISL_1995821, EPI_ISL_1995822, EPI_ISL_1995823, EPI_ISL_1995824, EPI_ISL_1995825, EPI_ISL_1995826, EPI_ISL_1995827, EPI_ISL_1995828, EPI_ISL_1995829, EPI_ISL_1995830, EPI_ISL_1995831, EPI_ISL_1995832, EPI_ISL_1995833, EPI_ISL_1995834, EPI_ISL_1995835, EPI_ISL_1995836, EPI_ISL_1995837, EPI_ISL_1995838, EPI_ISL_1995839, EPI_ISL_1995840, EPI_ISL_1995841, EPI_ISL_1995842, EPI_ISL_1995843, EPI_ISL_1995844, EPI_ISL_1995845, EPI_ISL_1995846, EPI_ISL_1995847, EPI_ISL_1995848, EPI_ISL_1995849, EPI_ISL_1995850, EPI_ISL_1995851, EPI_ISL_1995852, EPI_ISL_1995853, EPI_ISL_1995854, EPI_ISL_1995855, EPI_ISL_1995856, EPI_ISL_1995857, EPI_ISL_1995858, EPI_ISL_1995859, EPI_ISL_1995860, EPI_ISL_1995861, EPI_ISL_1995862, EPI_ISL_1995863, EPI_ISL_1995864, EPI_ISL_1995865, EPI_ISL_1995866, EPI_ISL_1995868, EPI_ISL_1995869, EPI_ISL_1995870, EPI_ISL_1995871, EPI_ISL_1995872, EPI_ISL_1995873, EPI_ISL_1995874, EPI_ISL_1995875, EPI_ISL_1995876, EPI_ISL_1995877, EPI_ISL_1995878, EPI_ISL_1995879, EPI_ISL_1995880, EPI_ISL_1995881, EPI_ISL_1995882, EPI_ISL_1995883, EPI_ISL_1995884, EPI_ISL_1995885, EPI_ISL_1995887, EPI_ISL_1995888, EPI_ISL_1995889, EPI_ISL_1995891, EPI_ISL_1995892, EPI_ISL_1995893, EPI_ISL_1995894, EPI_ISL_1995895, EPI_ISL_1995896, EPI_ISL_1995897, EPI_ISL_1995898, EPI_ISL_1995899, EPI_ISL_1995900, EPI_ISL_1995901, EPI_ISL_1995902, EPI_ISL_1995903, EPI_ISL_1995904, EPI_ISL_1995905, EPI_ISL_1995906, EPI_ISL_1995907, EPI_ISL_1995908, EPI_ISL_1995909, EPI_ISL_1995910, EPI_ISL_1995911, EPI_ISL_1995912, EPI_ISL_1995913, EPI_ISL_1995914, EPI_ISL_1995915, EPI_ISL_1995916, EPI_ISL_1995917, EPI_ISL_1995918, EPI_ISL_1995919, EPI_ISL_1995920, EPI_ISL_1995921, EPI_ISL_1995922, EPI_ISL_1995923, EPI_ISL_1995924, EPI_ISL_1995925, EPI_ISL_1995926, EPI_ISL_1995927, EPI_ISL_1995928, EPI_ISL_1995929, EPI_ISL_1995930, EPI_ISL_1995931, EPI_ISL_1995932, EPI_ISL_1995933, EPI_ISL_1995934, EPI_ISL_1995935, EPI_ISL_1995936, EPI_ISL_1995937, EPI_ISL_1995938, EPI_ISL_1995939, EPI_ISL_1995940, EPI_ISL_1995941, EPI_ISL_1995942, EPI_ISL_1995943, EPI_ISL_1995944, EPI_ISL_1995945, EPI_ISL_1995946, EPI_ISL_1995947, EPI_ISL_1995948, EPI_ISL_1995949, EPI_ISL_1995950, EPI_ISL_1995951, EPI_ISL_1995952, EPI_ISL_1995953, EPI_ISL_1995954, EPI_ISL_1995955, EPI_ISL_1995956, EPI_ISL_1995957, EPI_ISL_1995958, EPI_ISL_1995962, EPI_ISL_1995963, EPI_ISL_1995964, EPI_ISL_1995965, EPI_ISL_1995966, EPI_ISL_1995967, EPI_ISL_1995968, EPI_ISL_1995970, EPI_ISL_1995971, EPI_ISL_1995972, EPI_ISL_1995973, EPI_ISL_1995974, EPI_ISL_1995975, EPI_ISL_1995976, EPI_ISL_1995977, EPI_ISL_1995978, EPI_ISL_1995979, EPI_ISL_1995980, EPI_ISL_1995981, EPI_ISL_1995982, EPI_ISL_1995983, EPI_ISL_1995984, EPI_ISL_1995985, EPI_ISL_1995986, EPI_ISL_1995987, EPI_ISL_1995988, EPI_ISL_1995989, EPI_ISL_1995990, EPI_ISL_1995991, EPI_ISL_1995992, EPI_ISL_1995993, EPI_ISL_1995994, EPI_ISL_1995995, EPI_ISL_1995996, EPI_ISL_1995997, EPI_ISL_1995998, EPI_ISL_1995999, EPI_ISL_1996000, EPI_ISL_1996001, EPI_ISL_1996002, EPI_ISL_1996003, EPI_ISL_1996004, EPI_ISL_1996005, EPI_ISL_1996006, EPI_ISL_1996008, EPI_ISL_1996009, EPI_ISL_1996010, EPI_ISL_1996011, EPI_ISL_1996012, EPI_ISL_1996013, EPI_ISL_1996014, EPI_ISL_1996015, EPI_ISL_1996016, EPI_ISL_1996017, EPI_ISL_1996018, EPI_ISL_1996019, EPI_ISL_1996020, EPI_ISL_1996021, EPI_ISL_1996022, EPI_ISL_1996023, EPI_ISL_1996024, EPI_ISL_1996025, EPI_ISL_1996026, EPI_ISL_1996027, EPI_ISL_1996028, EPI_ISL_1996029, EPI_ISL_1996030, EPI_ISL_1996031, EPI_ISL_1996032, EPI_ISL_1996033, EPI_ISL_1996034, EPI_ISL_1996035, EPI_ISL_1996036, EPI_ISL_1996037, EPI_ISL_1996038, EPI_ISL_1996039, EPI_ISL_1996040, EPI_ISL_1996041, EPI_ISL_1996042, EPI_ISL_1996043, EPI_ISL_1996044, EPI_ISL_1996045, EPI_ISL_1996046, EPI_ISL_1996047, EPI_ISL_1996048, EPI_ISL_1996049, EPI_ISL_1996050, EPI_ISL_1996051, EPI_ISL_1996052, EPI_ISL_1996053, EPI_ISL_1996054, EPI_ISL_1996055, EPI_ISL_1996056, EPI_ISL_1996057, EPI_ISL_1996058, EPI_ISL_1996059, EPI_ISL_1996060, EPI_ISL_1996061, EPI_ISL_1996062, EPI_ISL_1996063, EPI_ISL_1996064, EPI_ISL_1996065, EPI_ISL_1996066, EPI_ISL_1996067, EPI_ISL_1996068, EPI_ISL_1996069, EPI_ISL_1996070, EPI_ISL_1996071, EPI_ISL_1996072, EPI_ISL_1996073, EPI_ISL_1996074, EPI_ISL_1996075, EPI_ISL_1996076, EPI_ISL_1996077, EPI_ISL_1996078, EPI_ISL_1996079, EPI_ISL_1996081, EPI_ISL_1996082, EPI_ISL_1996083, EPI_ISL_1996084, EPI_ISL_1996085, EPI_ISL_1996086, EPI_ISL_1996087, EPI_ISL_1996088, EPI_ISL_1996089, EPI_ISL_1996090, EPI_ISL_1996091, EPI_ISL_1996092, EPI_ISL_1996093, EPI_ISL_1996094, EPI_ISL_1996095, EPI_ISL_1996096, EPI_ISL_1996097, EPI_ISL_1996098, EPI_ISL_1996099, EPI_ISL_1996100, EPI_ISL_1996101, EPI_ISL_1996102, EPI_ISL_1996103, EPI_ISL_1996104, EPI_ISL_1996105, EPI_ISL_1996106, EPI_ISL_1996107, EPI_ISL_1996108, EPI_ISL_1996109, EPI_ISL_1996110, EPI_ISL_1996111, EPI_ISL_1996112, EPI_ISL_1996113, EPI_ISL_1996114, EPI_ISL_1996115, EPI_ISL_1996116, EPI_ISL_1996117, EPI_ISL_1996118, EPI_ISL_1996119, EPI_ISL_1996120, EPI_ISL_1996121, EPI_ISL_1996122, EPI_ISL_1996123, EPI_ISL_1996124, EPI_ISL_1996125, EPI_ISL_1996126, EPI_ISL_1996127, EPI_ISL_1996128, EPI_ISL_1996129, EPI_ISL_1996130, EPI_ISL_1996131, EPI_ISL_1996132, EPI_ISL_1996133, EPI_ISL_1996134, EPI_ISL_1996135, EPI_ISL_1996136, EPI_ISL_1996137, EPI_ISL_1996138, EPI_ISL_1996139, EPI_ISL_1996140, EPI_ISL_1996141, EPI_ISL_1996142, EPI_ISL_1996143, EPI_ISL_1996144, EPI_ISL_1996145, EPI_ISL_1996146, EPI_ISL_1996147, EPI_ISL_1996148, EPI_ISL_1996149, EPI_ISL_1996150, EPI_ISL_1996152, EPI_ISL_1996153, EPI_ISL_1996154, EPI_ISL_1996155, EPI_ISL_1996156, EPI_ISL_1996157, EPI_ISL_1996159, EPI_ISL_1996160, EPI_ISL_1996161, EPI_ISL_1996162, EPI_ISL_1996164, EPI_ISL_1996165, EPI_ISL_1996166, EPI_ISL_1996167, EPI_ISL_1996168, EPI_ISL_1996169, EPI_ISL_1996170, EPI_ISL_1996171, EPI_ISL_1996172, EPI_ISL_1996173, EPI_ISL_1996175, EPI_ISL_1996176, EPI_ISL_1996177, EPI_ISL_1996178, EPI_ISL_1996179, EPI_ISL_1996180, EPI_ISL_1996181, EPI_ISL_1996182, EPI_ISL_1996183, EPI_ISL_1996184, EPI_ISL_1996185, EPI_ISL_1996186, EPI_ISL_1996187, EPI_ISL_1996188, EPI_ISL_1996189, EPI_ISL_1996190, EPI_ISL_1996191, EPI_ISL_1996192, EPI_ISL_1996193, EPI_ISL_1996194, EPI_ISL_1996195, EPI_ISL_1996196, EPI_ISL_1996197, EPI_ISL_1996198, EPI_ISL_1996199, EPI_ISL_1996200, EPI_ISL_1996201, EPI_ISL_1996202, EPI_ISL_1996203, EPI_ISL_1996204, EPI_ISL_1996205, EPI_ISL_1996206, EPI_ISL_1996207, EPI_ISL_1996208, EPI_ISL_1996209, EPI_ISL_1996210, EPI_ISL_1996211, EPI_ISL_1996212, EPI_ISL_1996213, EPI_ISL_1996215, EPI_ISL_1996216, EPI_ISL_1996217, EPI_ISL_1996218, EPI_ISL_1996219, EPI_ISL_1996220, EPI_ISL_1996221, EPI_ISL_1996222, EPI_ISL_1996223, EPI_ISL_1996224, EPI_ISL_1996225, EPI_ISL_1996226, EPI_ISL_1996227, EPI_ISL_1996228, EPI_ISL_1996229, EPI_ISL_1996230, EPI_ISL_1996231, EPI_ISL_1996232, EPI_ISL_1996233, EPI_ISL_1996234, EPI_ISL_1996235, EPI_ISL_1996236, EPI_ISL_1996237, EPI_ISL_1996238, EPI_ISL_1996239, EPI_ISL_1996240, EPI_ISL_1996241, EPI_ISL_1996242, EPI_ISL_1996243, EPI_ISL_1996244, EPI_ISL_1996245, EPI_ISL_1996246, EPI_ISL_1996247, EPI_ISL_1996248, EPI_ISL_1996249, EPI_ISL_1996250, EPI_ISL_1996251, EPI_ISL_1996252, EPI_ISL_1996253, EPI_ISL_1996254, EPI_ISL_1996255, EPI_ISL_1996256, EPI_ISL_1996257, EPI_ISL_1996258, EPI_ISL_1996259, EPI_ISL_1996260, EPI_ISL_1996261, EPI_ISL_1996262, EPI_ISL_1996263, EPI_ISL_1996265, EPI_ISL_1996266, EPI_ISL_1996267, EPI_ISL_1996268, EPI_ISL_1996269, EPI_ISL_1996270, EPI_ISL_1996271, EPI_ISL_1996272, EPI_ISL_1996273, EPI_ISL_1996274, EPI_ISL_1996275, EPI_ISL_1996276, EPI_ISL_1996277, EPI_ISL_1996278, EPI_ISL_1996279, EPI_ISL_1996280, EPI_ISL_1996282, EPI_ISL_1996283, EPI_ISL_1996285, EPI_ISL_1996286, EPI_ISL_1996287, EPI_ISL_1996291, EPI_ISL_1996293, EPI_ISL_1996294, EPI_ISL_1996295, EPI_ISL_1996297, EPI_ISL_1996298, EPI_ISL_1996300, EPI_ISL_1996301, EPI_ISL_1996303, EPI_ISL_1996304, EPI_ISL_1996305, EPI_ISL_1996306, EPI_ISL_1996307, EPI_ISL_1996308, EPI_ISL_1996309, EPI_ISL_1996310, EPI_ISL_1996311, EPI_ISL_1996312, EPI_ISL_1996313, EPI_ISL_1996314, EPI_ISL_1996315, EPI_ISL_1996316, EPI_ISL_1996317, EPI_ISL_1996318, EPI_ISL_1996319,

EPI_ISL_1996320, EPI_ISL_1996321, EPI_ISL_1996322, EPI_ISL_1996323, EPI_ISL_1996324, EPI_ISL_1996325, EPI_ISL_1996326, EPI_ISL_1996327, EPI_ISL_1996328, EPI_ISL_1996329, EPI_ISL_1996330, EPI_ISL_1996331, EPI_ISL_1996332, EPI_ISL_1996333, EPI_ISL_1996334, EPI_ISL_1996335, EPI_ISL_1996337, EPI_ISL_1996338, EPI_ISL_1996339, EPI_ISL_1996340, EPI_ISL_1996341, EPI_ISL_1996342,
EPI_ISL_1996343, EPI_ISL_1996344, EPI_ISL_1996345, EPI_ISL_1996346, EPI_ISL_1996347, EPI_ISL_1996348, EPI_ISL_1996349, EPI_ISL_1996350, EPI_ISL_1996351, EPI_ISL_1996352, EPI_ISL_1996353, EPI_ISL_1996354, EPI_ISL_1996355, EPI_ISL_1996356, EPI_ISL_1996357, EPI_ISL_1996358, EPI_ISL_1996359, EPI_ISL_1996360, EPI_ISL_1996361, EPI_ISL_1996362, EPI_ISL_1996363, EPI_ISL_1996364,
EPI_ISL_1996365, EPI_ISL_1996366, EPI_ISL_1996368, EPI_ISL_1996369, EPI_ISL_1996370, EPI_ISL_1996371, EPI_ISL_1996372, EPI_ISL_1996373, EPI_ISL_1996374, EPI_ISL_1996375, EPI_ISL_1996376, EPI_ISL_1996377, EPI_ISL_1996378, EPI_ISL_1996379, EPI_ISL_1996380, EPI_ISL_1996381, EPI_ISL_1996382, EPI_ISL_1996383, EPI_ISL_1996384, EPI_ISL_1996385, EPI_ISL_1996386, EPI_ISL_1996387,
EPI_ISL_1996388, EPI_ISL_1996389, EPI_ISL_1996390, EPI_ISL_1996391, EPI_ISL_1996392, EPI_ISL_1996393, EPI_ISL_1996394, EPI_ISL_1996395, EPI_ISL_1996396, EPI_ISL_1996397, EPI_ISL_1996398, EPI_ISL_1996399, EPI_ISL_1996400, EPI_ISL_1996401, EPI_ISL_1996402, EPI_ISL_1996405, EPI_ISL_1996406, EPI_ISL_1996407, EPI_ISL_1996408, EPI_ISL_1996409, EPI_ISL_1996410, EPI_ISL_1996411,
EPI_ISL_1996412, EPI_ISL_1996413, EPI_ISL_1996414, EPI_ISL_1996415, EPI_ISL_1996416, EPI_ISL_1996417, EPI_ISL_1996418, EPI_ISL_1996419, EPI_ISL_1996420, EPI_ISL_1996421, EPI_ISL_1996422, EPI_ISL_1996423, EPI_ISL_1996424, EPI_ISL_1996425, EPI_ISL_1996427, EPI_ISL_1996428, EPI_ISL_1996429, EPI_ISL_1996430, EPI_ISL_1996431, EPI_ISL_1996432, EPI_ISL_1996433, EPI_ISL_1996434,
EPI_ISL_1996435, EPI_ISL_1996436, EPI_ISL_1996437, EPI_ISL_1996438, EPI_ISL_1996439, EPI_ISL_1996440, EPI_ISL_1996441, EPI_ISL_1996442, EPI_ISL_1996443, EPI_ISL_1996444, EPI_ISL_1996445, EPI_ISL_1996446, EPI_ISL_1996447, EPI_ISL_1996448, EPI_ISL_1996449, EPI_ISL_1996450, EPI_ISL_1996451, EPI_ISL_1996452, EPI_ISL_1996453, EPI_ISL_1996454, EPI_ISL_1996455, EPI_ISL_1996456,
EPI_ISL_1996457, EPI_ISL_1996458, EPI_ISL_1996459, EPI_ISL_1996460, EPI_ISL_1996461, EPI_ISL_1996462, EPI_ISL_1996463, EPI_ISL_1996464, EPI_ISL_1996465, EPI_ISL_1996466, EPI_ISL_1996467, EPI_ISL_1996468, EPI_ISL_1996469, EPI_ISL_1996470, EPI_ISL_1996471, EPI_ISL_1996472, EPI_ISL_1996473, EPI_ISL_1996474, EPI_ISL_1996475, EPI_ISL_1996476, EPI_ISL_1996477, EPI_ISL_1996478,
EPI_ISL_1996479, EPI_ISL_1996480, EPI_ISL_1996481, EPI_ISL_1996482, EPI_ISL_1996483, EPI_ISL_1996484, EPI_ISL_1996485, EPI_ISL_1996486, EPI_ISL_1996487, EPI_ISL_1996488, EPI_ISL_1996489, EPI_ISL_1996490, EPI_ISL_1996491, EPI_ISL_1996492, EPI_ISL_1996493, EPI_ISL_1996494, EPI_ISL_1996495, EPI_ISL_1996496, EPI_ISL_1996497, EPI_ISL_1996498, EPI_ISL_1996499, EPI_ISL_1996500,
EPI_ISL_1996501, EPI_ISL_1996502, EPI_ISL_1996503, EPI_ISL_1996504, EPI_ISL_1996505, EPI_ISL_1996506, EPI_ISL_1996507, EPI_ISL_1996508, EPI_ISL_1996509, EPI_ISL_1996510, EPI_ISL_1996511, EPI_ISL_1996512, EPI_ISL_1996513, EPI_ISL_1996514, EPI_ISL_1996515, EPI_ISL_1996516, EPI_ISL_1996517, EPI_ISL_1996518, EPI_ISL_1996519, EPI_ISL_1996520, EPI_ISL_1996521, EPI_ISL_1996522,
EPI_ISL_1996523, EPI_ISL_1996524, EPI_ISL_1996525, EPI_ISL_1996526, EPI_ISL_1996527, EPI_ISL_1996528, EPI_ISL_1996529, EPI_ISL_1996530, EPI_ISL_1996531, EPI_ISL_1996532, EPI_ISL_1996533, EPI_ISL_1996534, EPI_ISL_1996535, EPI_ISL_1996536, EPI_ISL_1996537, EPI_ISL_1996538, EPI_ISL_1996539, EPI_ISL_1996540, EPI_ISL_1996541, EPI_ISL_1996542, EPI_ISL_1996543, EPI_ISL_1996544,
EPI_ISL_1996545, EPI_ISL_1996546, EPI_ISL_1996547, EPI_ISL_1996548, EPI_ISL_1996549, EPI_ISL_1996550, EPI_ISL_1996551, EPI_ISL_1996552, EPI_ISL_1996553, EPI_ISL_1996554, EPI_ISL_1996555, EPI_ISL_1996556, EPI_ISL_1996557, EPI_ISL_1996558, EPI_ISL_1996559, EPI_ISL_1996560, EPI_ISL_1996561, EPI_ISL_1996562, EPI_ISL_1996563, EPI_ISL_1996564, EPI_ISL_1996565, EPI_ISL_1996566,
EPI_ISL_1996567, EPI_ISL_1996568, EPI_ISL_1996569, EPI_ISL_1996570, EPI_ISL_1996572, EPI_ISL_1996573, EPI_ISL_1996574, EPI_ISL_1996575, EPI_ISL_1996576, EPI_ISL_1996577, EPI_ISL_1996578, EPI_ISL_1996579, EPI_ISL_1996580, EPI_ISL_1996581, EPI_ISL_1996582, EPI_ISL_1996583, EPI_ISL_1996584, EPI_ISL_1996585, EPI_ISL_1996586, EPI_ISL_1996587, EPI_ISL_1996588, EPI_ISL_1996589,
EPI_ISL_1996590, EPI_ISL_1996591, EPI_ISL_1996592, EPI_ISL_1996593, EPI_ISL_1996594, EPI_ISL_1996595, EPI_ISL_1996596, EPI_ISL_1996597, EPI_ISL_1996598, EPI_ISL_1996599, EPI_ISL_1996600, EPI_ISL_1996601, EPI_ISL_1996602, EPI_ISL_1996603, EPI_ISL_1996604, EPI_ISL_1996605, EPI_ISL_1996606, EPI_ISL_1996607, EPI_ISL_1996608, EPI_ISL_1996609, EPI_ISL_1996610, EPI_ISL_1996611,
EPI_ISL_1996612, EPI_ISL_1996613, EPI_ISL_1996614, EPI_ISL_1996615, EPI_ISL_1996616, EPI_ISL_1996617, EPI_ISL_1996618, EPI_ISL_1996619, EPI_ISL_1996620, EPI_ISL_1996621, EPI_ISL_1996622, EPI_ISL_1996623, EPI_ISL_1996624, EPI_ISL_1996625, EPI_ISL_1996626, EPI_ISL_1996627, EPI_ISL_1996628, EPI_ISL_1996629, EPI_ISL_1996630, EPI_ISL_1996631, EPI_ISL_1996632, EPI_ISL_1996633,
EPI_ISL_1996634, EPI_ISL_1996635, EPI_ISL_1996636, EPI_ISL_1996637, EPI_ISL_1996638, EPI_ISL_1996639, EPI_ISL_1996640, EPI_ISL_1996641, EPI_ISL_1996642, EPI_ISL_1996643, EPI_ISL_1996644, EPI_ISL_1996645, EPI_ISL_1996646, EPI_ISL_1996647, EPI_ISL_1996648, EPI_ISL_1996649, EPI_ISL_1996650, EPI_ISL_1996651, EPI_ISL_1996652, EPI_ISL_1996653, EPI_ISL_1996654, EPI_ISL_1996655,
EPI_ISL_1996656, EPI_ISL_1996657, EPI_ISL_1996658, EPI_ISL_1996659, EPI_ISL_1996660, EPI_ISL_1996661, EPI_ISL_1996662, EPI_ISL_1996663, EPI_ISL_1996664, EPI_ISL_1996665, EPI_ISL_1996666, EPI_ISL_1996667, EPI_ISL_1996668, EPI_ISL_1996669, EPI_ISL_1996670, EPI_ISL_1996671, EPI_ISL_1996672, EPI_ISL_1996673, EPI_ISL_1996674, EPI_ISL_1996675, EPI_ISL_1996676, EPI_ISL_1996677,
EPI_ISL_1996678, EPI_ISL_1996679, EPI_ISL_1996680, EPI_ISL_1996681, EPI_ISL_1996682, EPI_ISL_1996683, EPI_ISL_1996684, EPI_ISL_1996685, EPI_ISL_1996686, EPI_ISL_1996687, EPI_ISL_1996688, EPI_ISL_1996689, EPI_ISL_1996690, EPI_ISL_1996691, EPI_ISL_1996692, EPI_ISL_1996693, EPI_ISL_1996694, EPI_ISL_1996695, EPI_ISL_1996696, EPI_ISL_1996697, EPI_ISL_1996698, EPI_ISL_1996699,
EPI_ISL_1996700, EPI_ISL_1996701, EPI_ISL_1996702, EPI_ISL_1996703, EPI_ISL_1996704, EPI_ISL_1996705, EPI_ISL_1996706, EPI_ISL_1996707, EPI_ISL_1996708, EPI_ISL_1996709, EPI_ISL_1996710, EPI_ISL_1996711, EPI_ISL_1996712, EPI_ISL_1996713, EPI_ISL_1996714, EPI_ISL_1996715, EPI_ISL_1996716, EPI_ISL_1996717, EPI_ISL_1996718, EPI_ISL_1996719, EPI_ISL_1996720, EPI_ISL_1996721,
EPI_ISL_1996722, EPI_ISL_1996723, EPI_ISL_1996724, EPI_ISL_1996725, EPI_ISL_1996726, EPI_ISL_1996727, EPI_ISL_1996728, EPI_ISL_1996729, EPI_ISL_1996730, EPI_ISL_1996731, EPI_ISL_1996732, EPI_ISL_1996733, EPI_ISL_1996734, EPI_ISL_1996735, EPI_ISL_1996736, EPI_ISL_1996737, EPI_ISL_1996738, EPI_ISL_1996739, EPI_ISL_1996740, EPI_ISL_1996741, EPI_ISL_1996742, EPI_ISL_1996743,
EPI_ISL_1996744, EPI_ISL_1996745, EPI_ISL_1996746, EPI_ISL_1996747, EPI_ISL_1996748, EPI_ISL_1996749, EPI_ISL_1996750, EPI_ISL_1996752, EPI_ISL_1996753, EPI_ISL_1996754, EPI_ISL_1996755, EPI_ISL_1996756, EPI_ISL_1996757, EPI_ISL_1996759, EPI_ISL_1996760, EPI_ISL_1996761, EPI_ISL_1996762, EPI_ISL_1996763, EPI_ISL_1996764, EPI_ISL_1996765, EPI_ISL_1996766, EPI_ISL_1996767,
EPI_ISL_1996768, EPI_ISL_1996769, EPI_ISL_1996770, EPI_ISL_1996771, EPI_ISL_1996772, EPI_ISL_1996773, EPI_ISL_1996774, EPI_ISL_1996775, EPI_ISL_1996776, EPI_ISL_1996777, EPI_ISL_1996778, EPI_ISL_1996779, EPI_ISL_1996780, EPI_ISL_1996781, EPI_ISL_1996782, EPI_ISL_1996783, EPI_ISL_1996784, EPI_ISL_1996785, EPI_ISL_1996786, EPI_ISL_1996787, EPI_ISL_1996788, EPI_ISL_1996789,
EPI_ISL_1996790, EPI_ISL_1996791, EPI_ISL_1996792, EPI_ISL_1996793, EPI_ISL_1996794, EPI_ISL_1996795, EPI_ISL_1996796, EPI_ISL_1996797, EPI_ISL_1996798, EPI_ISL_1996799, EPI_ISL_1996800, EPI_ISL_1996801, EPI_ISL_1996802, EPI_ISL_1996803, EPI_ISL_1996804, EPI_ISL_1996805, EPI_ISL_1996806, EPI_ISL_1996807, EPI_ISL_1996808, EPI_ISL_1996809, EPI_ISL_1996810, EPI_ISL_1996811,
EPI_ISL_1996812, EPI_ISL_1996813, EPI_ISL_1996814, EPI_ISL_1996815, EPI_ISL_1996816, EPI_ISL_1996817, EPI_ISL_1996818, EPI_ISL_1996819, EPI_ISL_1996820, EPI_ISL_1996821, EPI_ISL_1996822, EPI_ISL_1996823, EPI_ISL_1996824, EPI_ISL_1996825, EPI_ISL_1996826, EPI_ISL_1996827, EPI_ISL_1996828, EPI_ISL_1996829, EPI_ISL_1996830, EPI_ISL_1996831, EPI_ISL_1996832, EPI_ISL_1996833,
EPI_ISL_1996834, EPI_ISL_1996835, EPI_ISL_1996836, EPI_ISL_1996837, EPI_ISL_1996838, EPI_ISL_1996839, EPI_ISL_1996840, EPI_ISL_1996841, EPI_ISL_1996842, EPI_ISL_1996843, EPI_ISL_1996844, EPI_ISL_1996845, EPI_ISL_1996846, EPI_ISL_1996847, EPI_ISL_1996848, EPI_ISL_1996849, EPI_ISL_1996850, EPI_ISL_1996851, EPI_ISL_1996852, EPI_ISL_1996853, EPI_ISL_1996854, EPI_ISL_1996855,
EPI_ISL_1996856, EPI_ISL_1996858, EPI_ISL_1996859, EPI_ISL_1996860, EPI_ISL_1996861, EPI_ISL_1996862, EPI_ISL_1996863, EPI_ISL_1996864, EPI_ISL_1996865, EPI_ISL_1996866, EPI_ISL_1996867, EPI_ISL_1996868, EPI_ISL_1996869, EPI_ISL_1996870, EPI_ISL_1996871, EPI_ISL_1996872, EPI_ISL_1996873, EPI_ISL_1996874, EPI_ISL_1996875, EPI_ISL_1996876, EPI_ISL_1996877, EPI_ISL_1996878,
EPI_ISL_1996879, EPI_ISL_1996880, EPI_ISL_1996881, EPI_ISL_1996882, EPI_ISL_1996883, EPI_ISL_1996884, EPI_ISL_1996885, EPI_ISL_1996886, EPI_ISL_1996887, EPI_ISL_1996888, EPI_ISL_1996889, EPI_ISL_1996890, EPI_ISL_1996891, EPI_ISL_1996892, EPI_ISL_1996893, EPI_ISL_1996894, EPI_ISL_1996895, EPI_ISL_1996896, EPI_ISL_1996897, EPI_ISL_1996898, EPI_ISL_1996899, EPI_ISL_1996900,
EPI_ISL_1996901, EPI_ISL_1996902, EPI_ISL_1996903, EPI_ISL_1996904, EPI_ISL_1996905, EPI_ISL_1996906, EPI_ISL_1996907, EPI_ISL_1996908, EPI_ISL_1996909, EPI_ISL_1996910, EPI_ISL_1996911, EPI_ISL_1996912, EPI_ISL_1996913, EPI_ISL_1996914, EPI_ISL_1996915, EPI_ISL_1996916, EPI_ISL_1996917, EPI_ISL_1996918, EPI_ISL_1996919, EPI_ISL_1996920, EPI_ISL_1996921, EPI_ISL_1996922,
EPI_ISL_1996923, EPI_ISL_1996924, EPI_ISL_1996925, EPI_ISL_1996926, EPI_ISL_1996927, EPI_ISL_1996928, EPI_ISL_1996929, EPI_ISL_1996930, EPI_ISL_1996931, EPI_ISL_1996932, EPI_ISL_1996933, EPI_ISL_1996934, EPI_ISL_1996935, EPI_ISL_1996936, EPI_ISL_1996937, EPI_ISL_1996938, EPI_ISL_1996939, EPI_ISL_1996940, EPI_ISL_1996941, EPI_ISL_1996942, EPI_ISL_1996943, EPI_ISL_1996944,
EPI_ISL_1996945, EPI_ISL_1996946, EPI_ISL_1996947, EPI_ISL_1996948, EPI_ISL_1996949, EPI_ISL_1996950, EPI_ISL_1996951, EPI_ISL_1996952, EPI_ISL_1996953, EPI_ISL_1996954, EPI_ISL_1996955, EPI_ISL_1996956, EPI_ISL_1996957, EPI_ISL_1996958, EPI_ISL_1996959, EPI_ISL_1996960, EPI_ISL_1996961, EPI_ISL_1996962, EPI_ISL_1996963, EPI_ISL_1996964, EPI_ISL_1996965, EPI_ISL_1996966,
EPI_ISL_1996967, EPI_ISL_1996968, EPI_ISL_1996969, EPI_ISL_1996970, EPI_ISL_1996971, EPI_ISL_1996972, EPI_ISL_1996973, EPI_ISL_1996974, EPI_ISL_1996975, EPI_ISL_1996977, EPI_ISL_1996978, EPI_ISL_1996979, EPI_ISL_1996980, EPI_ISL_1996981, EPI_ISL_1996982, EPI_ISL_1996983, EPI_ISL_1996984, EPI_ISL_1996985, EPI_ISL_1996986, EPI_ISL_1996987, EPI_ISL_1996988, EPI_ISL_1996989,
EPI_ISL_1996990, EPI_ISL_1996991, EPI_ISL_1996992, EPI_ISL_1996993, EPI_ISL_1996994, EPI_ISL_1996995, EPI_ISL_1996996, EPI_ISL_1996998, EPI_ISL_1996999, EPI_ISL_1997000, EPI_ISL_1997001, EPI_ISL_1997002, EPI_ISL_1997003, EPI_ISL_1997004, EPI_ISL_1997005, EPI_ISL_1997006, EPI_ISL_1997007, EPI_ISL_1997008, EPI_ISL_1997009, EPI_ISL_1997010, EPI_ISL_1997011, EPI_ISL_1997012,
EPI_ISL_1997013, EPI_ISL_1997014, EPI_ISL_1997015, EPI_ISL_1997016, EPI_ISL_1997017, EPI_ISL_1997018, EPI_ISL_1997019, EPI_ISL_1997020, EPI_ISL_1997021, EPI_ISL_1997022, EPI_ISL_1997023, EPI_ISL_1997024, EPI_ISL_1997025, EPI_ISL_1997026, EPI_ISL_1997027, EPI_ISL_1997028, EPI_ISL_1997029, EPI_ISL_1997030, EPI_ISL_1997031, EPI_ISL_1997032, EPI_ISL_1997033, EPI_ISL_1997034,
EPI_ISL_1997035, EPI_ISL_1997036, EPI_ISL_1997037, EPI_ISL_1997038, EPI_ISL_1997039, EPI_ISL_1997040, EPI_ISL_1997041, EPI_ISL_1997042, EPI_ISL_1997043, EPI_ISL_1997044, EPI_ISL_1997045, EPI_ISL_1997046, EPI_ISL_1997047, EPI_ISL_1997049, EPI_ISL_1997050, EPI_ISL_1997051, EPI_ISL_1997052, EPI_ISL_1997053, EPI_ISL_1997054, EPI_ISL_1997055, EPI_ISL_1997056, EPI_ISL_1997057,
EPI_ISL_1997058, EPI_ISL_1997059, EPI_ISL_1997060, EPI_ISL_1997061, EPI_ISL_1997062, EPI_ISL_1997063, EPI_ISL_1997064, EPI_ISL_1997065, EPI_ISL_1997066, EPI_ISL_1997067, EPI_ISL_1997068, EPI_ISL_1997069, EPI_ISL_1997070, EPI_ISL_1997071, EPI_ISL_1997072, EPI_ISL_1997073, EPI_ISL_1997074, EPI_ISL_1997075, EPI_ISL_1997076, EPI_ISL_1997077, EPI_ISL_1997078, EPI_ISL_1997079,
EPI_ISL_1997080, EPI_ISL_1997081, EPI_ISL_1997082, EPI_ISL_1997083, EPI_ISL_1997084, EPI_ISL_1997085, EPI_ISL_1997086, EPI_ISL_1997088, EPI_ISL_1997089, EPI_ISL_1997090, EPI_ISL_1997091, EPI_ISL_1997092, EPI_ISL_1997093, EPI_ISL_1997094, EPI_ISL_1997095, EPI_ISL_1997096, EPI_ISL_1997097, EPI_ISL_1997098, EPI_ISL_1997099, EPI_ISL_1997100, EPI_ISL_1997101, EPI_ISL_1997102,
EPI_ISL_1997103, EPI_ISL_1997104, EPI_ISL_1997105, EPI_ISL_1997106, EPI_ISL_1997107, EPI_ISL_1997108, EPI_ISL_1997109, EPI_ISL_1997110, EPI_ISL_1997111, EPI_ISL_1997112, EPI_ISL_1997113, EPI_ISL_1997114, EPI_ISL_1997115, EPI_ISL_1997116, EPI_ISL_1997117, EPI_ISL_1997118, EPI_ISL_1997119, EPI_ISL_1997120, EPI_ISL_1997121, EPI_ISL_1997122, EPI_ISL_1997123, EPI_ISL_1997124,
EPI_ISL_1997125, EPI_ISL_1997126, EPI_ISL_1997127, EPI_ISL_1997128, EPI_ISL_1997129, EPI_ISL_1997130, EPI_ISL_1997131, EPI_ISL_1997132, EPI_ISL_1997133, EPI_ISL_1997134, EPI_ISL_1997135, EPI_ISL_1997136, EPI_ISL_1997137, EPI_ISL_1997138, EPI_ISL_1997139, EPI_ISL_1997140, EPI_ISL_1997141, EPI_ISL_1997142, EPI_ISL_1997143, EPI_ISL_1997144, EPI_ISL_1997145, EPI_ISL_1997146,
EPI_ISL_1997147, EPI_ISL_1997148, EPI_ISL_1997149, EPI_ISL_1997150, EPI_ISL_1997151, EPI_ISL_1997152, EPI_ISL_1997153, EPI_ISL_1997154, EPI_ISL_1997155, EPI_ISL_1997156, EPI_ISL_1997157, EPI_ISL_1997158, EPI_ISL_1997159, EPI_ISL_1997160, EPI_ISL_1997161, EPI_ISL_1997162, EPI_ISL_1997163, EPI_ISL_1997164, EPI_ISL_1997165, EPI_ISL_1997166, EPI_ISL_1997167, EPI_ISL_1997168,
EPI_ISL_1997169, EPI_ISL_1997170, EPI_ISL_1997171, EPI_ISL_1997172, EPI_ISL_1997173, EPI_ISL_1997174, EPI_ISL_1997175, EPI_ISL_1997176, EPI_ISL_1997177, EPI_ISL_1997179, EPI_ISL_1997180, EPI_ISL_1997181, EPI_ISL_1997182, EPI_ISL_1997183, EPI_ISL_1997184, EPI_ISL_1997185, EPI_ISL_1997186, EPI_ISL_1997187, EPI_ISL_1997188, EPI_ISL_1997189, EPI_ISL_1997190, EPI_ISL_1997191,
EPI_ISL_1997192, EPI_ISL_1997193, EPI_ISL_1997194, EPI_ISL_1997195, EPI_ISL_1997196, EPI_ISL_1997197, EPI_ISL_1997198, EPI_ISL_1997199, EPI_ISL_1997200, EPI_ISL_1997202, EPI_ISL_1997203, EPI_ISL_1997204, EPI_ISL_1997205, EPI_ISL_1997206, EPI_ISL_1997207, EPI_ISL_1997208, EPI_ISL_1997209, EPI_ISL_1997210, EPI_ISL_1997211, EPI_ISL_1997212, EPI_ISL_1997213, EPI_ISL_1997214,
EPI_ISL_1997215, EPI_ISL_1997216, EPI_ISL_1997217, EPI_ISL_1997218, EPI_ISL_1997219, EPI_ISL_1997220, EPI_ISL_1997221, EPI_ISL_1997222, EPI_ISL_1997223, EPI_ISL_1997224, EPI_ISL_1997225, EPI_ISL_1997226, EPI_ISL_1997227, EPI_ISL_1997228, EPI_ISL_1997229, EPI_ISL_1997230, EPI_ISL_1997231, EPI_ISL_1997232, EPI_ISL_1997233, EPI_ISL_1997234, EPI_ISL_1997235, EPI_ISL_1997236,
EPI_ISL_1997237, EPI_ISL_1997238, EPI_ISL_1997239, EPI_ISL_1997240, EPI_ISL_1997241, EPI_ISL_1997242, EPI_ISL_1997243, EPI_ISL_1997244, EPI_ISL_1997245, EPI_ISL_1997246, EPI_ISL_1997247, EPI_ISL_1997248, EPI_ISL_1997249, EPI_ISL_1997250, EPI_ISL_1997251, EPI_ISL_1997252, EPI_ISL_1997253, EPI_ISL_1997254, EPI_ISL_1997255, EPI_ISL_1997256, EPI_ISL_1997257, EPI_ISL_1997258,
EPI_ISL_1997259, EPI_ISL_1997260, EPI_ISL_1997261, EPI_ISL_1997262, EPI_ISL_1997263, EPI_ISL_1997264, EPI_ISL_1997265, EPI_ISL_1997266, EPI_ISL_1997267, EPI_ISL_1997268, EPI_ISL_1997269, EPI_ISL_1997270, EPI_ISL_1997271, EPI_ISL_1997272, EPI_ISL_1997273, EPI_ISL_1997274, EPI_ISL_1997275, EPI_ISL_1997276, EPI_ISL_1997277, EPI_ISL_1997278, EPI_ISL_1997279, EPI_ISL_1997280,
EPI_ISL_1997281, EPI_ISL_1997282, EPI_ISL_1997283, EPI_ISL_1997284, EPI_ISL_1997285, EPI_ISL_1997286, EPI_ISL_1997287, EPI_ISL_1997288, EPI_ISL_1997289, EPI_ISL_1997290, EPI_ISL_1997291, EPI_ISL_1997292, EPI_ISL_1997293, EPI_ISL_1997294, EPI_ISL_1997295, EPI_ISL_1997296, EPI_ISL_1997298, EPI_ISL_1997299, EPI_ISL_1997300, EPI_ISL_1997301, EPI_ISL_1997302, EPI_ISL_1997303,
EPI_ISL_1997304, EPI_ISL_1997305, EPI_ISL_1997306, EPI_ISL_1997307, EPI_ISL_1997308, EPI_ISL_1997309, EPI_ISL_1997310, EPI_ISL_1997311, EPI_ISL_1997312, EPI_ISL_1997313, EPI_ISL_1997314, EPI_ISL_1997315, EPI_ISL_1997316, EPI_ISL_1997317, EPI_ISL_1997318, EPI_ISL_1997319, EPI_ISL_1997320, EPI_ISL_1997321, EPI_ISL_1997322, EPI_ISL_1997323, EPI_ISL_1997324, EPI_ISL_1997325,
EPI_ISL_1997326, EPI_ISL_1997327, EPI_ISL_1997328, EPI_ISL_1997329, EPI_ISL_1997330, EPI_ISL_1997331, EPI_ISL_1997332, EPI_ISL_1997333, EPI_ISL_1997334, EPI_ISL_1997335, EPI_ISL_1997336, EPI_ISL_1997337, EPI_ISL_1997338, EPI_ISL_1997339, EPI_ISL_1997340, EPI_ISL_1997341, EPI_ISL_1997342, EPI_ISL_1997343, EPI_ISL_1997344, EPI_ISL_1997345, EPI_ISL_1997346, EPI_ISL_1997347,
EPI_ISL_1997348, EPI_ISL_1997349, EPI_ISL_1997350, EPI_ISL_1997351, EPI_ISL_1997352, EPI_ISL_1997353, EPI_ISL_1997354, EPI_ISL_1997355, EPI_ISL_1997356, EPI_ISL_1997357, EPI_ISL_1997358, EPI_ISL_1997359, EPI_ISL_1997360, EPI_ISL_1997361, EPI_ISL_1997362, EPI_ISL_1997363, EPI_ISL_1997364, EPI_ISL_1997365, EPI_ISL_1997366, EPI_ISL_1997367, EPI_ISL_1997368, EPI_ISL_1997369,
EPI_ISL_1997370, EPI_ISL_1997371, EPI_ISL_1997372, EPI_ISL_1997373, EPI_ISL_1997374, EPI_ISL_1997375, EPI_ISL_1997376, EPI_ISL_1997377, EPI_ISL_1997378, EPI_ISL_1997379, EPI_ISL_1997381, EPI_ISL_1997382, EPI_ISL_1997383, EPI_ISL_1997384, EPI_ISL_1997385, EPI_ISL_1997386, EPI_ISL_1997387, EPI_ISL_1997388, EPI_ISL_1997389, EPI_ISL_1997390, EPI_ISL_1997391, EPI_ISL_1997392,
EPI_ISL_1997393, EPI_ISL_1997394, EPI_ISL_1997395, EPI_ISL_1997396, EPI_ISL_1997397, EPI_ISL_1997398, EPI_ISL_1997400, EPI_ISL_1997401, EPI_ISL_1997402, EPI_ISL_1997403, EPI_ISL_1997404, EPI_ISL_1997405, EPI_ISL_1997406, EPI_ISL_1997407, EPI_ISL_1997408, EPI_ISL_1997409, EPI_ISL_1997410, EPI_ISL_1997411, EPI_ISL_1997412, EPI_ISL_1997413, EPI_ISL_1997415, EPI_ISL_1997416,
EPI_ISL_1997417, EPI_ISL_1997418, EPI_ISL_1997419, EPI_ISL_1997420, EPI_ISL_1997421, EPI_ISL_1997422, EPI_ISL_1997423, EPI_ISL_1997424, EPI_ISL_1997425, EPI_ISL_1997427, EPI_ISL_1997428, EPI_ISL_1997429, EPI_ISL_1997430, EPI_ISL_1997431, EPI_ISL_1997432, EPI_ISL_1997433, EPI_ISL_1997434, EPI_ISL_1997435, EPI_ISL_1997436, EPI_ISL_1997437, EPI_ISL_1997438, EPI_ISL_1997439,
EPI_ISL_1997440, EPI_ISL_1997441, EPI_ISL_1997442, EPI_ISL_1997443, EPI_ISL_1997444, EPI_ISL_1997445, EPI_ISL_1997446, EPI_ISL_1997447, EPI_ISL_1997448, EPI_ISL_1997449, EPI_ISL_1997450, EPI_ISL_1997451, EPI_ISL_1997452, EPI_ISL_1997453, EPI_ISL_1997454, EPI_ISL_1997455, EPI_ISL_1997456, EPI_ISL_1997457, EPI_ISL_1997458, EPI_ISL_1997459, EPI_ISL_1997460, EPI_ISL_1997461,
EPI_ISL_1997462, EPI_ISL_1997463, EPI_ISL_1997464, EPI_ISL_1997465, EPI_ISL_1997466, EPI_ISL_1997467, EPI_ISL_1997468, EPI_ISL_1997469, EPI_ISL_1997470, EPI_ISL_1997471, EPI_ISL_1997472, EPI_ISL_1997473, EPI_ISL_1997474, EPI_ISL_1997475, EPI_ISL_1997476, EPI_ISL_1997477, EPI_ISL_1997478, EPI_ISL_1997479, EPI_ISL_1997480, EPI_ISL_1997481, EPI_ISL_1997482, EPI_ISL_1997483,
EPI_ISL_1997484, EPI_ISL_1997485, EPI_ISL_1997486, EPI_ISL_1997487, EPI_ISL_1997488, EPI_ISL_1997489, EPI_ISL_1997490, EPI_ISL_1997491, EPI_ISL_1997492, EPI_ISL_1997493, EPI_ISL_1997494, EPI_ISL_1997495, EPI_ISL_1997496, EPI_ISL_1997497, EPI_ISL_1997499, EPI_ISL_1997500, EPI_ISL_1997501, EPI_ISL_1997502, EPI_ISL_1997504, EPI_ISL_1997505, EPI_ISL_1997506, EPI_ISL_1997507,
EPI_ISL_1997508, EPI_ISL_1997509, EPI_ISL_1997510, EPI_ISL_1997511, EPI_ISL_1997512, EPI_ISL_1997513, EPI_ISL_1997514, EPI_ISL_1997515, EPI_ISL_1997516, EPI_ISL_1997517, EPI_ISL_1997518, EPI_ISL_1997519, EPI_ISL_1997520, EPI_ISL_1997521, EPI_ISL_1997523, EPI_ISL_1997524, EPI_ISL_1997525, EPI_ISL_1997526, EPI_ISL_1997527, EPI_ISL_1997528, EPI_ISL_1997529, EPI_ISL_1997530,
EPI_ISL_1997531, EPI_ISL_1997532, EPI_ISL_1997533, EPI_ISL_1997534, EPI_ISL_1997535, EPI_ISL_1997536, EPI_ISL_1997537, EPI_ISL_1997538, EPI_ISL_1997540, EPI_ISL_1997541, EPI_ISL_1997542, EPI_ISL_1997543, EPI_ISL_1997544, EPI_ISL_1997545, EPI_ISL_1997546, EPI_ISL_1997547, EPI_ISL_1997548, EPI_ISL_1997549, EPI_ISL_1997550, EPI_ISL_1997551, EPI_ISL_1997552, EPI_ISL_1997553,
EPI_ISL_1997554, EPI_ISL_1997555, EPI_ISL_1997556, EPI_ISL_1997557, EPI_ISL_1997558, EPI_ISL_1997559, EPI_ISL_1997560, EPI_ISL_1997561, EPI_ISL_1997562, EPI_ISL_1997563, EPI_ISL_1997564, EPI_ISL_1997565, EPI_ISL_1997566, EPI_ISL_1997567, EPI_ISL_1997568, EPI_ISL_1997569, EPI_ISL_1997570, EPI_ISL_1997572, EPI_ISL_1997573, EPI_ISL_1997574, EPI_ISL_1997575, EPI_ISL_1997576,
EPI_ISL_1997577, EPI_ISL_1997578, EPI_ISL_1997579, EPI_ISL_1997580, EPI_ISL_1997581, EPI_ISL_1997582, EPI_ISL_1997583, EPI_ISL_1997584, EPI_ISL_1997585, EPI_ISL_1997586, EPI_ISL_1997587, EPI_ISL_1997588, EPI_ISL_1997589, EPI_ISL_1997590, EPI_ISL_1997591, EPI_ISL_1997592, EPI_ISL_1997593, EPI_ISL_1997594, EPI_ISL_1997595, EPI_ISL_1997596, EPI_ISL_1997597, EPI_ISL_1997598,
EPI_ISL_1997599, EPI_ISL_1997600, EPI_ISL_1997601, EPI_ISL_1997602, EPI_ISL_1997603, EPI_ISL_1997604, EPI_ISL_1997605, EPI_ISL_1997606, EPI_ISL_1997607, EPI_ISL_1997608, EPI_ISL_1997609, EPI_ISL_1997610, EPI_ISL_1997611, EPI_ISL_1997612, EPI_ISL_1997613, EPI_ISL_1997614, EPI_ISL_1997615, EPI_ISL_1997616, EPI_ISL_1997617, EPI_ISL_1997618, EPI_ISL_1997619, EPI_ISL_1997620,
EPI_ISL_1997621, EPI_ISL_1997622, EPI_ISL_1997623, EPI_ISL_1997624, EPI_ISL_1997625, EPI_ISL_1997626, EPI_ISL_1997627, EPI_ISL_1997628, EPI_ISL_1997629, EPI_ISL_1997630, EPI_ISL_1997631, EPI_ISL_1997632, EPI_ISL_1997633, EPI_ISL_1997634, EPI_ISL_1997635, EPI_ISL_1997636, EPI_ISL_1997637, EPI_ISL_1997638, EPI_ISL_1997639, EPI_ISL_1997640, EPI_ISL_1997641, EPI_ISL_1997642,
EPI_ISL_1997643, EPI_ISL_1997644, EPI_ISL_1997645, EPI_ISL_1997646, EPI_ISL_1997647, EPI_ISL_1997648, EPI_ISL_1997649, EPI_ISL_1997650, EPI_ISL_1997651, EPI_ISL_1997652, EPI_ISL_1997653, EPI_ISL_1997654, EPI_ISL_1997655, EPI_ISL_1997656, EPI_ISL_1997657, EPI_ISL_1997658, EPI_ISL_1997659, EPI_ISL_1997660, EPI_ISL_1997661, EPI_ISL_1997662, EPI_ISL_1997663, EPI_ISL_1997664,
EPI_ISL_1997665, EPI_ISL_1997666, EPI_ISL_1997667, EPI_ISL_1997668, EPI_ISL_1997669, EPI_ISL_1997670, EPI_ISL_1997671, EPI_ISL_1997672, EPI_ISL_1997673, EPI_ISL_1997674, EPI_ISL_1997675, EPI_ISL_1997676, EPI_ISL_1997677, EPI_ISL_1997678, EPI_ISL_1997679, EPI_ISL_1997680, EPI_ISL_1997681, EPI_ISL_1997682, EPI_ISL_1997683, EPI_ISL_1997684, EPI_ISL_1997685, EPI_ISL_1997686,
EPI_ISL_1997687, EPI_ISL_1997688, EPI_ISL_1997689, EPI_ISL_1997690, EPI_ISL_1997691, EPI_ISL_1997692, EPI_ISL_1997693, EPI_ISL_1997694, EPI_ISL_1997695, EPI_ISL_1997696, EPI_ISL_1997698, EPI_ISL_1997699, EPI_ISL_1997700, EPI_ISL_1997701, EPI_ISL_1997702, EPI_ISL_1997703, EPI_ISL_1997704, EPI_ISL_1997705, EPI_ISL_1997706, EPI_ISL_1997707, EPI_ISL_1997708, EPI_ISL_1997709,
EPI_ISL_1997710, EPI_ISL_1997711, EPI_ISL_1997712, EPI_ISL_1997713, EPI_ISL_1997714, EPI_ISL_1997715, EPI_ISL_1997716, EPI_ISL_1997717, EPI_ISL_1997718, EPI_ISL_1997719, EPI_ISL_1997720, EPI_ISL_1997721, EPI_ISL_1997722, EPI_ISL_1997723, EPI_ISL_1997724, EPI_ISL_1997725, EPI_ISL_1997726, EPI_ISL_1997727, EPI_ISL_1997728, EPI_ISL_1997730, EPI_ISL_1997731, EPI_ISL_1997732,
EPI_ISL_1997733, EPI_ISL_1997734, EPI_ISL_1997735, EPI_ISL_1997736, EPI_ISL_1997737, EPI_ISL_1997738, EPI_ISL_1997739, EPI_ISL_1997740, EPI_ISL_1997741, EPI_ISL_1997742, EPI_ISL_1997743, EPI_ISL_1997744, EPI_ISL_1997745, EPI_ISL_1997747, EPI_ISL_1997748, EPI_ISL_1997749, EPI_ISL_1997750, EPI_ISL_1997751, EPI_ISL_1997752, EPI_ISL_1997753, EPI_ISL_1997754, EPI_ISL_1997755,
EPI_ISL_1997756, EPI_ISL_1997757, EPI_ISL_1997758, EPI_ISL_1997759, EPI_ISL_1997760, EPI_ISL_1997761, EPI_ISL_1997762, EPI_ISL_1997763, EPI_ISL_1997764, EPI_ISL_1997765, EPI_ISL_1997766, EPI_ISL_1997767, EPI_ISL_1997768, EPI_ISL_1997769, EPI_ISL_1997770, EPI_ISL_1997771, EPI_ISL_1997772, EPI_ISL_1997773, EPI_ISL_1997774, EPI_ISL_1997775, EPI_ISL_1997776, EPI_ISL_1997777,
EPI_ISL_1997778, EPI_ISL_1997779, EPI_ISL_1997780, EPI_ISL_1997781, EPI_ISL_1997782, EPI_ISL_1997783, EPI_ISL_1997784, EPI_ISL_1997785, EPI_ISL_1997786, EPI_ISL_1997787, EPI_ISL_1997788, EPI_ISL_1997789, EPI_ISL_1997790, EPI_ISL_1997791, EPI_ISL_1997792, EPI_ISL_1997793, EPI_ISL_1997794, EPI_ISL_1997795, EPI_ISL_1997796, EPI_ISL_1997797, EPI_ISL_1997798, EPI_ISL_1997799,
EPI_ISL_1997800, EPI_ISL_1997801, EPI_ISL_1997802, EPI_ISL_1997803, EPI_ISL_1997804, EPI_ISL_1997805, EPI_ISL_1997806, EPI_ISL_1997807, EPI_ISL_1997808, EPI_ISL_1997809, EPI_ISL_1997810, EPI_ISL_1997811, EPI_ISL_1997812, EPI_ISL_1997813, EPI_ISL_1997814, EPI_ISL_1997815, EPI_ISL_1997816, EPI_ISL_1997817, EPI_ISL_1997818, EPI_ISL_1997819, EPI_ISL_1997820, EPI_ISL_1997821,
EPI_ISL_1997822, EPI_ISL_1997823, EPI_ISL_1997824, EPI_ISL_1997825, EPI_ISL_1997826, EPI_ISL_1997827, EPI_ISL_1997828, EPI_ISL_1997829, EPI_ISL_1997830, EPI_ISL_1997831, EPI_ISL_1997832, EPI_ISL_1997833, EPI_ISL_1997834, EPI_ISL_1997835, EPI_ISL_1997836, EPI_ISL_1997837, EPI_ISL_1997838, EPI_ISL_1997839, EPI_ISL_1997840, EPI_ISL_1997841, EPI_ISL_1997842, EPI_ISL_1997843,
EPI_ISL_1997844, EPI_ISL_1997845, EPI_ISL_1997846, EPI_ISL_1997847, EPI_ISL_1997848, EPI_ISL_1997849, EPI_ISL_1997850, EPI_ISL_1997851, EPI_ISL_1997852, EPI_ISL_1997853, EPI_ISL_1997854, EPI_ISL_1997855, EPI_ISL_1997856, EPI_ISL_1997857, EPI_ISL_1997858, EPI_ISL_1997859, EPI_ISL_1997860, EPI_ISL_1997861, EPI_ISL_1997862, EPI_ISL_1997863, EPI_ISL_1997864, EPI_ISL_1997865,
EPI_ISL_1997866, EPI_ISL_1997868, EPI_ISL_1997869, EPI_ISL_1997870, EPI_ISL_1997871, EPI_ISL_1997872, EPI_ISL_1997873, EPI_ISL_1997874, EPI_ISL_1997875, EPI_ISL_1997876, EPI_ISL_1997877, EPI_ISL_1997878, EPI_ISL_1997879, EPI_ISL_1997880, EPI_ISL_1997881, EPI_ISL_1997882, EPI_ISL_1997883, EPI_ISL_1997884, EPI_ISL_1997885, EPI_ISL_1997886, EPI_ISL_1997887, EPI_ISL_1997888,
EPI_ISL_1997889, EPI_ISL_1997890, EPI_ISL_1997891, EPI_ISL_1997892, EPI_ISL_1997893, EPI_ISL_1997894, EPI_ISL_1997895, EPI_ISL_1997896, EPI_ISL_1997897, EPI_ISL_1997898, EPI_ISL_1997900, EPI_ISL_1997901, EPI_ISL_1997902, EPI_ISL_1997903, EPI_ISL_1997904, EPI_ISL_1997905, EPI_ISL_1997906, EPI_ISL_1997907, EPI_ISL_1997908, EPI_ISL_1997909, EPI_ISL_1997910, EPI_ISL_1997911,
EPI_ISL_1997912, EPI_ISL_1997913, EPI_ISL_1997914, EPI_ISL_1997915, EPI_ISL_1997916, EPI_ISL_1997917, EPI_ISL_1997918, EPI_ISL_1997919, EPI_ISL_1997920, EPI_ISL_1997921, EPI_ISL_1997922, EPI_ISL_1997923, EPI_ISL_1997924, EPI_ISL_1997925, EPI_ISL_1997926, EPI_ISL_1997927, EPI_ISL_1997928, EPI_ISL_1997929, EPI_ISL_1997930, EPI_ISL_1997931, EPI_ISL_1997932, EPI_ISL_1997933,
EPI_ISL_1997934, EPI_ISL_1997935, EPI_ISL_1997936, EPI_ISL_1997937, EPI_ISL_1997938, EPI_ISL_1997939, EPI_ISL_1997940, EPI_ISL_1997941, EPI_ISL_1997942, EPI_ISL_1997943, EPI_ISL_1997944, EPI_ISL_1997945, EPI_ISL_1997946, EPI_ISL_1997947, EPI_ISL_1997948, EPI_ISL_1997950, EPI_ISL_1997951, EPI_ISL_1997952, EPI_ISL_1997953, EPI_ISL_1997954, EPI_ISL_1997955, EPI_ISL_1997956,
EPI_ISL_1997957, EPI_ISL_1997958, EPI_ISL_1997959, EPI_ISL_1997960, EPI_ISL_1997961, EPI_ISL_1997962, EPI_ISL_1997963, EPI_ISL_1997964, EPI_ISL_1997965, EPI_ISL_1997966, EPI_ISL_1997967, EPI_ISL_1997968, EPI_ISL_1997969, EPI_ISL_1997970, EPI_ISL_1997971, EPI_ISL_1997972, EPI_ISL_1997973, EPI_ISL_1997974, EPI_ISL_1997975, EPI_ISL_1997976, EPI_ISL_1997977, EPI_ISL_1997978,
EPI_ISL_1997979, EPI_ISL_1997980, EPI_ISL_1997981, EPI_ISL_1997982, EPI_ISL_1997983, EPI_ISL_1997984, EPI_ISL_1997985, EPI_ISL_1997986, EPI_ISL_1997987, EPI_ISL_1997989, EPI_ISL_1997990, EPI_ISL_1997991, EPI_ISL_1997992, EPI_ISL_1997993, EPI_ISL_1997994, EPI_ISL_1997995, EPI_ISL_1997996, EPI_ISL_1997998, EPI_ISL_1997999, EPI_ISL_1998000, EPI_ISL_1998001, EPI_ISL_1998002,
EPI_ISL_1998003, EPI_ISL_1998004, EPI_ISL_1998005, EPI_ISL_1998006, EPI_ISL_1998007, EPI_ISL_1998008, EPI_ISL_1998009, EPI_ISL_1998010, EPI_ISL_1998011, EPI_ISL_1998012, EPI_ISL_1998013, EPI_ISL_1998014, EPI_ISL_1998015, EPI_ISL_1998016, EPI_ISL_1998017, EPI_ISL_1998018, EPI_ISL_1998019, EPI_ISL_1998020, EPI_ISL_1998021, EPI_ISL_1998022, EPI_ISL_1998023, EPI_ISL_1998024,
EPI_ISL_1998025, EPI_ISL_1998026, EPI_ISL_1998027, EPI_ISL_1998028, EPI_ISL_1998029, EPI_ISL_1998031, EPI_ISL_1998032, EPI_ISL_1998033, EPI_ISL_1998034, EPI_ISL_1998035, EPI_ISL_1998037, EPI_ISL_1998038, EPI_ISL_1998039, EPI_ISL_1998040, EPI_ISL_1998041, EPI_ISL_1998043, EPI_ISL_1998044, EPI_ISL_1998045, EPI_ISL_1998046, EPI_ISL_1998047, EPI_ISL_1998048, EPI_ISL_1998049,
EPI_ISL_1998050, EPI_ISL_1998051, EPI_ISL_1998052, EPI_ISL_1998053, EPI_ISL_1998054, EPI_ISL_1998055, EPI_ISL_1998056, EPI_ISL_1998057, EPI_ISL_1998058, EPI_ISL_1998059, EPI_ISL_1998060, EPI_ISL_1998061, EPI_ISL_1998063, EPI_ISL_1998064, EPI_ISL_1998065, EPI_ISL_1998066, EPI_ISL_1998067, EPI_ISL_1998068, EPI_ISL_1998069, EPI_ISL_1998070, EPI_ISL_1998071, EPI_ISL_1998072,
EPI_ISL_1998073, EPI_ISL_1998074, EPI_ISL_1998075, EPI_ISL_1998076, EPI_ISL_1998077, EPI_ISL_1998078, EPI_ISL_1998079, EPI_ISL_1998080, EPI_ISL_1998081, EPI_ISL_1998082, EPI_ISL_1998084, EPI_ISL_1998085, EPI_ISL_1998086, EPI_ISL_1998087, EPI_ISL_1998088, EPI_ISL_1998089, EPI_ISL_1998090, EPI_ISL_1998091, EPI_ISL_1998092, EPI_ISL_1998093, EPI_ISL_1998094, EPI_ISL_1998095,
EPI_ISL_1998096, EPI_ISL_1998097, EPI_ISL_1998098, EPI_ISL_1998099, EPI_ISL_1998100, EPI_ISL_1998101, EPI_ISL_1998102, EPI_ISL_1998103, EPI_ISL_1998104, EPI_ISL_1998105, EPI_ISL_1998106, EPI_ISL_1998107, EPI_ISL_1998108, EPI_ISL_1998109, EPI_ISL_1998110, EPI_ISL_1998111, EPI_ISL_1998112, EPI_ISL_1998113, EPI_ISL_1998114, EPI_ISL_1998115, EPI_ISL_1998116, EPI_ISL_1998117,
EPI_ISL_1998118, EPI_ISL_1998119, EPI_ISL_1998120, EPI_ISL_1998121, EPI_ISL_1998122, EPI_ISL_1998123, EPI_ISL_1998124, EPI_ISL_1998125, EPI_ISL_1998126, EPI_ISL_1998127, EPI_ISL_1998128, EPI_ISL_1998129, EPI_ISL_1998130, EPI_ISL_1998131, EPI_ISL_1998132, EPI_ISL_1998133, EPI_ISL_1998134, EPI_ISL_1998135, EPI_ISL_1998136, EPI_ISL_1998137, EPI_ISL_1998138, EPI_ISL_1998139,
EPI_ISL_1998140, EPI_ISL_1998141, EPI_ISL_1998142, EPI_ISL_1998143, EPI_ISL_1998145, EPI_ISL_1998146, EPI_ISL_1998147, EPI_ISL_1998148, EPI_ISL_1998149, EPI_ISL_1998150, EPI_ISL_1998151, EPI_ISL_1998152, EPI_ISL_1998153, EPI_ISL_1998154, EPI_ISL_1998155, EPI_ISL_1998156, EPI_ISL_1998158, EPI_ISL_1998159, EPI_ISL_1998160, EPI_ISL_1998161, EPI_ISL_1998162, EPI_ISL_1998163,
EPI_ISL_1998164, EPI_ISL_1998165, EPI_ISL_1998166, EPI_ISL_1998167, EPI_ISL_1998168, EPI_ISL_1998169, EPI_ISL_1998170, EPI_ISL_1998171, EPI_ISL_1998172, EPI_ISL_1998173, EPI_ISL_1998175, EPI_ISL_1998176, EPI_ISL_1998177, EPI_ISL_1998178, EPI_ISL_1998179, EPI_ISL_1998180, EPI_ISL_1998181, EPI_ISL_1998182, EPI_ISL_1998183, EPI_ISL_1998184, EPI_ISL_1998185, EPI_ISL_1998186,
EPI_ISL_1998187, EPI_ISL_1998188, EPI_ISL_1998189, EPI_ISL_1998190, EPI_ISL_1998191, EPI_ISL_1998193, EPI_ISL_1998194, EPI_ISL_1998195, EPI_ISL_1998196, EPI_ISL_1998197, EPI_ISL_1998198, EPI_ISL_1998199, EPI_ISL_1998200, EPI_ISL_1998201, EPI_ISL_1998202, EPI_ISL_1998203, EPI_ISL_1998204, EPI_ISL_1998205, EPI_ISL_1998206, EPI_ISL_1998207, EPI_ISL_1998208, EPI_ISL_1998209,
EPI_ISL_1998210, EPI_ISL_1998211, EPI_ISL_1998212, EPI_ISL_1998213, EPI_ISL_1998214, EPI_ISL_1998215, EPI_ISL_1998216, EPI_ISL_1998217, EPI_ISL_1998218, EPI_ISL_1998219, EPI_ISL_1998220, EPI_ISL_1998221, EPI_ISL_1998222, EPI_ISL_1998223, EPI_ISL_1998224, EPI_ISL_1998225, EPI_ISL_1998226, EPI_ISL_1998227, EPI_ISL_1998228, EPI_ISL_1998229, EPI_ISL_1998230, EPI_ISL_1998231,
EPI_ISL_1998232, EPI_ISL_1998233, EPI_ISL_1998234, EPI_ISL_1998235, EPI_ISL_1998236, EPI_ISL_1998237, EPI_ISL_1998238, EPI_ISL_1998239, EPI_ISL_1998240, EPI_ISL_1998241, EPI_ISL_1998242, EPI_ISL_1998243, EPI_ISL_1998244, EPI_ISL_1998245, EPI_ISL_1998246, EPI_ISL_1998247, EPI_ISL_1998248, EPI_ISL_1998250, EPI_ISL_1998251, EPI_ISL_1998252, EPI_ISL_1998253, EPI_ISL_1998254,
EPI_ISL_1998255, EPI_ISL_1998256, EPI_ISL_1998257, EPI_ISL_1998258, EPI_ISL_1998259, EPI_ISL_1998260, EPI_ISL_1998261, EPI_ISL_1998262, EPI_ISL_1998263, EPI_ISL_1998264, EPI_ISL_1998265, EPI_ISL_1998266, EPI_ISL_1998267, EPI_ISL_1998268, EPI_ISL_1998269, EPI_ISL_1998270, EPI_ISL_1998271, EPI_ISL_1998272, EPI_ISL_1998273, EPI_ISL_1998274, EPI_ISL_1998275, EPI_ISL_1998276,
EPI_ISL_1998277, EPI_ISL_1998278, EPI_ISL_1998279, EPI_ISL_1998280, EPI_ISL_1998281, EPI_ISL_1998282, EPI_ISL_1998283, EPI_ISL_1998284, EPI_ISL_1998285, EPI_ISL_1998286, EPI_ISL_1998287, EPI_ISL_1998288, EPI_ISL_1998289, EPI_ISL_1998290, EPI_ISL_1998291, EPI_ISL_1998292, EPI_ISL_1998293, EPI_ISL_1998294, EPI_ISL_1998295, EPI_ISL_1998296, EPI_ISL_1998297, EPI_ISL_1998298,
EPI_ISL_1998299, EPI_ISL_1998300, EPI_ISL_1998301, EPI_ISL_1998302, EPI_ISL_1998303, EPI_ISL_1998304, EPI_ISL_1998305, EPI_ISL_1998306, EPI_ISL_1998307, EPI_ISL_1998308, EPI_ISL_1998309, EPI_ISL_1998310, EPI_ISL_1998311, EPI_ISL_1998312, EPI_ISL_1998313, EPI_ISL_1998314, EPI_ISL_1998315, EPI_ISL_1998316, EPI_ISL_1998317, EPI_ISL_1998318, EPI_ISL_1998319, EPI_ISL_1998320,
EPI_ISL_1998321, EPI_ISL_1998322, EPI_ISL_1998323, EPI_ISL_1998324, EPI_ISL_1998325, EPI_ISL_1998326, EPI_ISL_1998327, EPI_ISL_1998328, EPI_ISL_1998329, EPI_ISL_1998330, EPI_ISL_1998331, EPI_ISL_1998332, EPI_ISL_1998333, EPI_ISL_1998334, EPI_ISL_1998335, EPI_ISL_1998336, EPI_ISL_1998337, EPI_ISL_1998338, EPI_ISL_1998339, EPI_ISL_1998340, EPI_ISL_1998341, EPI_ISL_1998342,
EPI_ISL_1998343, EPI_ISL_1998344, EPI_ISL_1998345, EPI_ISL_1998346, EPI_ISL_1998348, EPI_ISL_1998349, EPI_ISL_1998350, EPI_ISL_1998351, EPI_ISL_1998352, EPI_ISL_1998353, EPI_ISL_1998354, EPI_ISL_1998355, EPI_ISL_1998356, EPI_ISL_1998357, EPI_ISL_1998358, EPI_ISL_1998359, EPI_ISL_1998360, EPI_ISL_1998361, EPI_ISL_1998362, EPI_ISL_1998363, EPI_ISL_1998364, EPI_ISL_1998365,
EPI_ISL_1998366, EPI_ISL_1998367, EPI_ISL_1998368, EPI_ISL_1998369, EPI_ISL_1998370, EPI_ISL_1998371, EPI_ISL_1998372, EPI_ISL_1998373, EPI_ISL_1998374, EPI_ISL_1998375, EPI_ISL_1998378, EPI_ISL_1998379, EPI_ISL_1998380, EPI_ISL_1998381, EPI_ISL_1998382, EPI_ISL_1998383, EPI_ISL_1998384, EPI_ISL_1998385, EPI_ISL_1998386, EPI_ISL_1998387, EPI_ISL_1998388, EPI_ISL_1998389,
EPI_ISL_1998390, EPI_ISL_1998391, EPI_ISL_1998392, EPI_ISL_1998393, EPI_ISL_1998394, EPI_ISL_1998395, EPI_ISL_1998396, EPI_ISL_1998397, EPI_ISL_1998398, EPI_ISL_1998399, EPI_ISL_1998400, EPI_ISL_1998401, EPI_ISL_1998402, EPI_ISL_1998403, EPI_ISL_1998404, EPI_ISL_1998405, EPI_ISL_1998406, EPI_ISL_1998407, EPI_ISL_1998408, EPI_ISL_1998409, EPI_ISL_1998410, EPI_ISL_1998411,
EPI_ISL_1998412, EPI_ISL_1998413, EPI_ISL_1998414, EPI_ISL_1998415, EPI_ISL_1998416, EPI_ISL_1998417, EPI_ISL_1998419, EPI_ISL_1998420, EPI_ISL_1998421, EPI_ISL_1998422, EPI_ISL_1998423, EPI_ISL_1998424, EPI_ISL_1998425, EPI_ISL_1998426, EPI_ISL_1998427, EPI_ISL_1998428, EPI_ISL_1998429, EPI_ISL_1998430, EPI_ISL_1998431, EPI_ISL_1998432, EPI_ISL_1998433, EPI_ISL_1998434,
EPI_ISL_1998435, EPI_ISL_1998436, EPI_ISL_1998437, EPI_ISL_1998438, EPI_ISL_1998439, EPI_ISL_1998440, EPI_ISL_1998441, EPI_ISL_1998442, EPI_ISL_1998443, EPI_ISL_1998444, EPI_ISL_1998445, EPI_ISL_1998446, EPI_ISL_1998447, EPI_ISL_1998448, EPI_ISL_1998449, EPI_ISL_1998450, EPI_ISL_1998451, EPI_ISL_1998452, EPI_ISL_1998453, EPI_ISL_1998454, EPI_ISL_1998455, EPI_ISL_1998456,
EPI_ISL_1998457, EPI_ISL_1998458, EPI_ISL_1998459, EPI_ISL_1998460, EPI_ISL_1998461, EPI_ISL_1998462, EPI_ISL_1998463, EPI_ISL_1998464, EPI_ISL_1998465, EPI_ISL_1998466, EPI_ISL_1998467, EPI_ISL_1998468, EPI_ISL_1998469, EPI_ISL_1998470, EPI_ISL_1998471, EPI_ISL_1998472, EPI_ISL_1998473, EPI_ISL_1998474, EPI_ISL_1998475, EPI_ISL_1998476, EPI_ISL_1998477, EPI_ISL_1998479,
EPI_ISL_1998480, EPI_ISL_1998481, EPI_ISL_1998482, EPI_ISL_1998483, EPI_ISL_1998484, EPI_ISL_1998485, EPI_ISL_1998486, EPI_ISL_1998487, EPI_ISL_1998488, EPI_ISL_1998489, EPI_ISL_1998490, EPI_ISL_1998491, EPI_ISL_1998492, EPI_ISL_1998493, EPI_ISL_1998494, EPI_ISL_1998495, EPI_ISL_1998496, EPI_ISL_1998497, EPI_ISL_1998498, EPI_ISL_1998499, EPI_ISL_1998500, EPI_ISL_1998501,
EPI_ISL_1998502, EPI_ISL_1998503, EPI_ISL_1998504, EPI_ISL_1998505, EPI_ISL_1998506, EPI_ISL_1998507, EPI_ISL_1998508, EPI_ISL_1998509, EPI_ISL_1998510, EPI_ISL_1998511, EPI_ISL_1998512, EPI_ISL_1998513, EPI_ISL_1998514, EPI_ISL_1998515, EPI_ISL_1998516, EPI_ISL_1998517, EPI_ISL_1998518, EPI_ISL_1998519, EPI_ISL_1998520, EPI_ISL_1998521, EPI_ISL_1998522, EPI_ISL_1998523,
EPI_ISL_1998524, EPI_ISL_1998525, EPI_ISL_1998526, EPI_ISL_1998527, EPI_ISL_1998528, EPI_ISL_1998530, EPI_ISL_1998531, EPI_ISL_1998532, EPI_ISL_1998533, EPI_ISL_1998534, EPI_ISL_1998535, EPI_ISL_1998536, EPI_ISL_1998537, EPI_ISL_1998538, EPI_ISL_1998539, EPI_ISL_1998540, EPI_ISL_1998541, EPI_ISL_1998543, EPI_ISL_1998544, EPI_ISL_1998545, EPI_ISL_1998546, EPI_ISL_1998547,
EPI_ISL_1998548, EPI_ISL_1998549, EPI_ISL_1998550, EPI_ISL_1998551, EPI_ISL_1998552, EPI_ISL_1998553, EPI_ISL_1998554, EPI_ISL_1998555, EPI_ISL_1998556, EPI_ISL_1998557, EPI_ISL_1998558, EPI_ISL_1998559, EPI_ISL_1998560, EPI_ISL_1998561, EPI_ISL_1998562, EPI_ISL_1998563, EPI_ISL_1998564, EPI_ISL_1998565, EPI_ISL_1998566, EPI_ISL_1998567, EPI_ISL_1998568, EPI_ISL_1998569,
EPI_ISL_1998570, EPI_ISL_1998571, EPI_ISL_1998572, EPI_ISL_1998573, EPI_ISL_1998574, EPI_ISL_1998575, EPI_ISL_1998576, EPI_ISL_1998578, EPI_ISL_1998579, EPI_ISL_1998580, EPI_ISL_1998581, EPI_ISL_1998582, EPI_ISL_1998583, EPI_ISL_1998584, EPI_ISL_1998585, EPI_ISL_1998586, EPI_ISL_1998587, EPI_ISL_1998588, EPI_ISL_1998589, EPI_ISL_1998590, EPI_ISL_1998591, EPI_ISL_1998592,
EPI_ISL_1998593, EPI_ISL_1998594, EPI_ISL_1998595, EPI_ISL_1998596, EPI_ISL_1998597, EPI_ISL_1998598, EPI_ISL_1998599, EPI_ISL_1998600, EPI_ISL_1998601, EPI_ISL_1998602, EPI_ISL_1998603, EPI_ISL_1998604, EPI_ISL_1998605, EPI_ISL_1998606, EPI_ISL_1998607, EPI_ISL_1998608, EPI_ISL_1998609, EPI_ISL_1998610, EPI_ISL_1998611, EPI_ISL_1998612, EPI_ISL_1998613, EPI_ISL_1998614,
EPI_ISL_1998616, EPI_ISL_1998617, EPI_ISL_1998618, EPI_ISL_1998619, EPI_ISL_1998620, EPI_ISL_1998621, EPI_ISL_1998622, EPI_ISL_1998623, EPI_ISL_1998624, EPI_ISL_1998625, EPI_ISL_1998626, EPI_ISL_1998627, EPI_ISL_1998628, EPI_ISL_1998629, EPI_ISL_1998630, EPI_ISL_1998631, EPI_ISL_1998632, EPI_ISL_1998633, EPI_ISL_1998634, EPI_ISL_1998635, EPI_ISL_1998636, EPI_ISL_1998637,
EPI_ISL_1998638, EPI_ISL_1998639, EPI_ISL_1998640, EPI_ISL_1998641, EPI_ISL_1998642, EPI_ISL_1998643, EPI_ISL_1998644, EPI_ISL_1998645, EPI_ISL_1998646, EPI_ISL_1998647, EPI_ISL_1998648, EPI_ISL_1998649, EPI_ISL_1998650, EPI_ISL_1998652, EPI_ISL_1998653, EPI_ISL_1998654, EPI_ISL_1998655, EPI_ISL_1998656, EPI_ISL_1998657, EPI_ISL_1998658, EPI_ISL_1998659, EPI_ISL_1998660,
EPI_ISL_1998661, EPI_ISL_1998662, EPI_ISL_1998663, EPI_ISL_1998664, EPI_ISL_1998665, EPI_ISL_1998666, EPI_ISL_1998667, EPI_ISL_1998668, EPI_ISL_1998669, EPI_ISL_1998670, EPI_ISL_1998671, EPI_ISL_1998672, EPI_ISL_1998673, EPI_ISL_1998674, EPI_ISL_1998675, EPI_ISL_1998676, EPI_ISL_1998677, EPI_ISL_1998678, EPI_ISL_1998679, EPI_ISL_1998680, EPI_ISL_1998681, EPI_ISL_1998682,
EPI_ISL_1998683, EPI_ISL_1998684, EPI_ISL_1998685, EPI_ISL_1998686, EPI_ISL_1998687, EPI_ISL_1998688, EPI_ISL_1998689, EPI_ISL_1998690, EPI_ISL_1998691, EPI_ISL_1998692, EPI_ISL_1998693, EPI_ISL_1998694, EPI_ISL_1998695, EPI_ISL_1998696, EPI_ISL_1998697, EPI_ISL_1998698, EPI_ISL_1998699, EPI_ISL_1998700, EPI_ISL_1998701, EPI_ISL_1998702, EPI_ISL_1998703, EPI_ISL_1998704,
EPI_ISL_1998705, EPI_ISL_1998706, EPI_ISL_1998707, EPI_ISL_1998708, EPI_ISL_1998709, EPI_ISL_1998710, EPI_ISL_1998711, EPI_ISL_1998712, EPI_ISL_1998713, EPI_ISL_1998714, EPI_ISL_1998715, EPI_ISL_1998716, EPI_ISL_1998717, EPI_ISL_1998718, EPI_ISL_1998719, EPI_ISL_1998720, EPI_ISL_1998721, EPI_ISL_1998722, EPI_ISL_1998723, EPI_ISL_1998724, EPI_ISL_1998725, EPI_ISL_1998726,
EPI_ISL_1998727, EPI_ISL_1998728, EPI_ISL_1998729, EPI_ISL_1998730, EPI_ISL_1998731, EPI_ISL_1998732, EPI_ISL_1998733, EPI_ISL_1998734, EPI_ISL_1998735, EPI_ISL_1998736, EPI_ISL_1998737, EPI_ISL_1998739, EPI_ISL_1998740, EPI_ISL_1998741, EPI_ISL_1998742, EPI_ISL_1998743, EPI_ISL_1998744, EPI_ISL_1998745, EPI_ISL_1998746, EPI_ISL_1998747, EPI_ISL_1998748, EPI_ISL_1998749,
EPI_ISL_1998750, EPI_ISL_1998751, EPI_ISL_1998752, EPI_ISL_1998753, EPI_ISL_1998754, EPI_ISL_1998755, EPI_ISL_1998756, EPI_ISL_1998757, EPI_ISL_1998758, EPI_ISL_1998759, EPI_ISL_1998760, EPI_ISL_1998761, EPI_ISL_1998762, EPI_ISL_1998763, EPI_ISL_1998764, EPI_ISL_1998765, EPI_ISL_1998766, EPI_ISL_1998767, EPI_ISL_1998768, EPI_ISL_1998769, EPI_ISL_1998770, EPI_ISL_1998771,
EPI_ISL_1998772, EPI_ISL_1998773, EPI_ISL_1998774, EPI_ISL_1998775, EPI_ISL_1998776, EPI_ISL_1998777, EPI_ISL_1998778, EPI_ISL_1998779, EPI_ISL_1998780, EPI_ISL_1998781, EPI_ISL_1998782, EPI_ISL_1998783, EPI_ISL_1998784, EPI_ISL_1998785, EPI_ISL_1998786, EPI_ISL_1998787, EPI_ISL_1998788, EPI_ISL_1998789, EPI_ISL_1998790, EPI_ISL_1998791, EPI_ISL_1998792, EPI_ISL_1998793,

EPI_ISL_1998794, EPI_ISL_1998795, EPI_ISL_1998796, EPI_ISL_1998797, EPI_ISL_1998798, EPI_ISL_1998799, EPI_ISL_1998800, EPI_ISL_1998801, EPI_ISL_1998802, EPI_ISL_1998804, EPI_ISL_1998805, EPI_ISL_1998806, EPI_ISL_1998807, EPI_ISL_1998808, EPI_ISL_1998809, EPI_ISL_1998810, EPI_ISL_1998811, EPI_ISL_1998812, EPI_ISL_1998813, EPI_ISL_1998814, EPI_ISL_1998815, EPI_ISL_1998816,
EPI_ISL_1998817, EPI_ISL_1998818, EPI_ISL_1998819, EPI_ISL_1998820, EPI_ISL_1998821, EPI_ISL_1998822, EPI_ISL_1998823, EPI_ISL_1998824, EPI_ISL_1998825, EPI_ISL_1998826, EPI_ISL_1998827, EPI_ISL_1998828, EPI_ISL_1998829, EPI_ISL_1998830, EPI_ISL_1998831, EPI_ISL_1998832, EPI_ISL_1998833, EPI_ISL_1998834, EPI_ISL_1998835, EPI_ISL_1998836, EPI_ISL_1998837, EPI_ISL_1998838,
EPI_ISL_1998839, EPI_ISL_1998840, EPI_ISL_1998841, EPI_ISL_1998842, EPI_ISL_1998843, EPI_ISL_1998844, EPI_ISL_1998845, EPI_ISL_1998846, EPI_ISL_1998847, EPI_ISL_1998848, EPI_ISL_1998849, EPI_ISL_1998850, EPI_ISL_1998851, EPI_ISL_1998852, EPI_ISL_1998853, EPI_ISL_1998854, EPI_ISL_1998855, EPI_ISL_1998856, EPI_ISL_1998857, EPI_ISL_1998858, EPI_ISL_1998859, EPI_ISL_1998860,
EPI_ISL_1998861, EPI_ISL_1998862, EPI_ISL_1998863, EPI_ISL_1998864, EPI_ISL_1998865, EPI_ISL_1998866, EPI_ISL_1998867, EPI_ISL_1998868, EPI_ISL_1998869, EPI_ISL_1998870, EPI_ISL_1998871, EPI_ISL_1998872, EPI_ISL_1998873, EPI_ISL_1998874, EPI_ISL_1998875, EPI_ISL_1998876, EPI_ISL_1998877, EPI_ISL_1998878, EPI_ISL_1998879, EPI_ISL_1998880, EPI_ISL_1998881, EPI_ISL_1998882,
EPI_ISL_1998883, EPI_ISL_1998884, EPI_ISL_1998885, EPI_ISL_1998886, EPI_ISL_1998887, EPI_ISL_1998888, EPI_ISL_1998889, EPI_ISL_1998890, EPI_ISL_1998891, EPI_ISL_1998892, EPI_ISL_1998893, EPI_ISL_1998894, EPI_ISL_1998895, EPI_ISL_1998896, EPI_ISL_1998897, EPI_ISL_1998898, EPI_ISL_1998900, EPI_ISL_1998901, EPI_ISL_1998902, EPI_ISL_1998903, EPI_ISL_1998904, EPI_ISL_1998905,
EPI_ISL_1998906, EPI_ISL_1998907, EPI_ISL_1998908, EPI_ISL_1998909, EPI_ISL_1998910, EPI_ISL_1998911, EPI_ISL_1998912, EPI_ISL_1998913, EPI_ISL_1998914, EPI_ISL_1998915, EPI_ISL_1998916, EPI_ISL_1998917, EPI_ISL_1998918, EPI_ISL_1998919, EPI_ISL_1998920, EPI_ISL_1998921, EPI_ISL_1998922, EPI_ISL_1998923, EPI_ISL_1998924, EPI_ISL_1998925, EPI_ISL_1998926, EPI_ISL_1998927,
EPI_ISL_1998928, EPI_ISL_1998929, EPI_ISL_1998930, EPI_ISL_1998931, EPI_ISL_1998932, EPI_ISL_1998933, EPI_ISL_1998934, EPI_ISL_1998936, EPI_ISL_1998937, EPI_ISL_1998938, EPI_ISL_1998939, EPI_ISL_1998940, EPI_ISL_1998941, EPI_ISL_1998942, EPI_ISL_1998943, EPI_ISL_1998944, EPI_ISL_1998945, EPI_ISL_1998946, EPI_ISL_1998947, EPI_ISL_1998948, EPI_ISL_1998949, EPI_ISL_1998950,
EPI_ISL_1998951, EPI_ISL_1998952, EPI_ISL_1998953, EPI_ISL_1998954, EPI_ISL_1998955, EPI_ISL_1998956, EPI_ISL_1998957, EPI_ISL_1998958, EPI_ISL_1998959, EPI_ISL_1998960, EPI_ISL_1998961, EPI_ISL_1998962, EPI_ISL_1998963, EPI_ISL_1998964, EPI_ISL_1998965, EPI_ISL_1998966, EPI_ISL_1998967, EPI_ISL_1998968, EPI_ISL_1998969, EPI_ISL_1998970, EPI_ISL_1998971, EPI_ISL_1998972,
EPI_ISL_1998973, EPI_ISL_1998974, EPI_ISL_1998975, EPI_ISL_1998976, EPI_ISL_1998977, EPI_ISL_1998978, EPI_ISL_1998979, EPI_ISL_1998980, EPI_ISL_1998981, EPI_ISL_1998982, EPI_ISL_1998983, EPI_ISL_1998984, EPI_ISL_1998985, EPI_ISL_1998986, EPI_ISL_1998987, EPI_ISL_1998988, EPI_ISL_1998989, EPI_ISL_1998990, EPI_ISL_1998991, EPI_ISL_1998992, EPI_ISL_1998993, EPI_ISL_1998994,
EPI_ISL_1998995, EPI_ISL_1998996, EPI_ISL_1998997, EPI_ISL_1998998, EPI_ISL_1998999, EPI_ISL_1999000, EPI_ISL_1999001, EPI_ISL_1999002, EPI_ISL_1999003, EPI_ISL_1999004, EPI_ISL_1999005, EPI_ISL_1999006, EPI_ISL_1999007, EPI_ISL_1999008, EPI_ISL_1999009, EPI_ISL_1999010, EPI_ISL_1999011, EPI_ISL_1999012, EPI_ISL_1999013, EPI_ISL_1999014, EPI_ISL_1999015, EPI_ISL_1999016,
EPI_ISL_1999017, EPI_ISL_1999018, EPI_ISL_1999019, EPI_ISL_1999020, EPI_ISL_1999021, EPI_ISL_1999022, EPI_ISL_1999023, EPI_ISL_1999024, EPI_ISL_1999025, EPI_ISL_1999026, EPI_ISL_1999027, EPI_ISL_1999028, EPI_ISL_1999029, EPI_ISL_1999030, EPI_ISL_1999031, EPI_ISL_1999032, EPI_ISL_1999033, EPI_ISL_1999034, EPI_ISL_1999035, EPI_ISL_1999036, EPI_ISL_1999037, EPI_ISL_1999038,
EPI_ISL_1999039, EPI_ISL_1999040, EPI_ISL_1999041, EPI_ISL_1999042, EPI_ISL_1999043, EPI_ISL_1999044, EPI_ISL_1999045, EPI_ISL_1999046, EPI_ISL_1999047, EPI_ISL_1999048, EPI_ISL_1999049, EPI_ISL_1999050, EPI_ISL_1999051, EPI_ISL_1999052, EPI_ISL_1999053, EPI_ISL_1999054, EPI_ISL_1999055, EPI_ISL_1999056, EPI_ISL_1999057, EPI_ISL_1999058, EPI_ISL_1999059, EPI_ISL_1999060,
EPI_ISL_1999061, EPI_ISL_1999062, EPI_ISL_1999063, EPI_ISL_1999064, EPI_ISL_1999065, EPI_ISL_1999066, EPI_ISL_1999067, EPI_ISL_1999068, EPI_ISL_1999069, EPI_ISL_1999070, EPI_ISL_1999071, EPI_ISL_1999072, EPI_ISL_1999073, EPI_ISL_1999074, EPI_ISL_1999075, EPI_ISL_1999076, EPI_ISL_1999077, EPI_ISL_1999079, EPI_ISL_1999080, EPI_ISL_1999081, EPI_ISL_1999082, EPI_ISL_1999083,
EPI_ISL_1999084, EPI_ISL_1999085, EPI_ISL_1999086, EPI_ISL_1999087, EPI_ISL_1999088, EPI_ISL_1999089, EPI_ISL_1999090, EPI_ISL_1999091, EPI_ISL_1999092, EPI_ISL_1999093, EPI_ISL_1999094, EPI_ISL_1999095, EPI_ISL_1999096, EPI_ISL_1999097, EPI_ISL_1999098, EPI_ISL_1999099, EPI_ISL_1999100, EPI_ISL_1999101, EPI_ISL_1999102, EPI_ISL_1999104, EPI_ISL_1999105, EPI_ISL_1999106,
EPI_ISL_1999107, EPI_ISL_1999108, EPI_ISL_1999109, EPI_ISL_1999110, EPI_ISL_1999111, EPI_ISL_1999112, EPI_ISL_1999113, EPI_ISL_1999114, EPI_ISL_1999115, EPI_ISL_1999116, EPI_ISL_1999117, EPI_ISL_1999118, EPI_ISL_1999119, EPI_ISL_1999120, EPI_ISL_1999121, EPI_ISL_1999122, EPI_ISL_1999123, EPI_ISL_1999124, EPI_ISL_1999125, EPI_ISL_1999126, EPI_ISL_1999127, EPI_ISL_1999128,
EPI_ISL_1999129, EPI_ISL_1999130, EPI_ISL_1999131, EPI_ISL_1999132, EPI_ISL_1999133, EPI_ISL_1999134, EPI_ISL_1999135, EPI_ISL_1999136, EPI_ISL_1999137, EPI_ISL_1999138, EPI_ISL_1999139, EPI_ISL_1999140, EPI_ISL_1999141, EPI_ISL_1999142, EPI_ISL_1999143, EPI_ISL_1999144, EPI_ISL_1999145, EPI_ISL_1999146, EPI_ISL_1999147, EPI_ISL_1999148, EPI_ISL_1999149, EPI_ISL_1999150,
EPI_ISL_1999151, EPI_ISL_1999152, EPI_ISL_1999153, EPI_ISL_1999154, EPI_ISL_1999155, EPI_ISL_1999156, EPI_ISL_1999157, EPI_ISL_1999158, EPI_ISL_1999159, EPI_ISL_1999160, EPI_ISL_1999161, EPI_ISL_1999162, EPI_ISL_1999163, EPI_ISL_1999164, EPI_ISL_1999165, EPI_ISL_1999166, EPI_ISL_1999167, EPI_ISL_1999169, EPI_ISL_1999170, EPI_ISL_1999171, EPI_ISL_1999172, EPI_ISL_1999173,
EPI_ISL_1999174, EPI_ISL_1999175, EPI_ISL_1999176, EPI_ISL_1999177, EPI_ISL_1999178, EPI_ISL_1999179, EPI_ISL_1999180, EPI_ISL_1999181, EPI_ISL_1999182, EPI_ISL_1999183, EPI_ISL_1999184, EPI_ISL_1999185, EPI_ISL_1999186, EPI_ISL_1999187, EPI_ISL_1999189, EPI_ISL_1999190, EPI_ISL_1999191, EPI_ISL_1999192, EPI_ISL_1999193, EPI_ISL_1999194, EPI_ISL_1999195, EPI_ISL_1999196,
EPI_ISL_1999197, EPI_ISL_1999198, EPI_ISL_1999199, EPI_ISL_1999201, EPI_ISL_1999202, EPI_ISL_1999203, EPI_ISL_1999204, EPI_ISL_1999205, EPI_ISL_1999206, EPI_ISL_1999207, EPI_ISL_1999208, EPI_ISL_1999209, EPI_ISL_1999210, EPI_ISL_1999211, EPI_ISL_1999212, EPI_ISL_1999213, EPI_ISL_1999214, EPI_ISL_1999215, EPI_ISL_1999216, EPI_ISL_1999217, EPI_ISL_1999218, EPI_ISL_1999219,
EPI_ISL_1999220, EPI_ISL_1999221, EPI_ISL_1999222, EPI_ISL_1999223, EPI_ISL_1999224, EPI_ISL_1999225, EPI_ISL_1999226, EPI_ISL_1999227, EPI_ISL_1999228, EPI_ISL_1999229, EPI_ISL_1999230, EPI_ISL_1999231, EPI_ISL_1999232, EPI_ISL_1999233, EPI_ISL_1999234, EPI_ISL_1999235, EPI_ISL_1999236, EPI_ISL_1999237, EPI_ISL_1999238, EPI_ISL_1999239, EPI_ISL_1999240, EPI_ISL_1999241,
EPI_ISL_1999242, EPI_ISL_1999243, EPI_ISL_1999244, EPI_ISL_1999245, EPI_ISL_1999246, EPI_ISL_1999247, EPI_ISL_1999248, EPI_ISL_1999249, EPI_ISL_1999250, EPI_ISL_1999251, EPI_ISL_1999252, EPI_ISL_1999253, EPI_ISL_1999254, EPI_ISL_1999255, EPI_ISL_1999256, EPI_ISL_1999257, EPI_ISL_1999258, EPI_ISL_1999259, EPI_ISL_1999260, EPI_ISL_1999261, EPI_ISL_1999262, EPI_ISL_1999263,
EPI_ISL_1999264, EPI_ISL_1999265, EPI_ISL_1999266, EPI_ISL_1999267, EPI_ISL_1999268, EPI_ISL_1999269, EPI_ISL_1999270, EPI_ISL_1999271, EPI_ISL_1999272, EPI_ISL_1999273, EPI_ISL_1999274, EPI_ISL_1999275, EPI_ISL_1999276, EPI_ISL_1999277, EPI_ISL_1999278, EPI_ISL_1999279, EPI_ISL_1999280, EPI_ISL_1999281, EPI_ISL_1999282, EPI_ISL_1999283, EPI_ISL_1999284, EPI_ISL_1999285,
EPI_ISL_1999286, EPI_ISL_1999287, EPI_ISL_1999288, EPI_ISL_1999289, EPI_ISL_1999290, EPI_ISL_1999291, EPI_ISL_1999292, EPI_ISL_1999293, EPI_ISL_1999294, EPI_ISL_1999295, EPI_ISL_1999296, EPI_ISL_1999297, EPI_ISL_1999298, EPI_ISL_1999299, EPI_ISL_1999300, EPI_ISL_1999301, EPI_ISL_1999302, EPI_ISL_1999303, EPI_ISL_1999304, EPI_ISL_1999305, EPI_ISL_1999306, EPI_ISL_1999307,
EPI_ISL_1999308, EPI_ISL_1999309, EPI_ISL_1999310, EPI_ISL_1999311, EPI_ISL_1999312, EPI_ISL_1999313, EPI_ISL_1999314, EPI_ISL_1999315, EPI_ISL_1999316, EPI_ISL_1999317, EPI_ISL_1999318, EPI_ISL_1999319, EPI_ISL_1999320, EPI_ISL_1999321, EPI_ISL_1999322, EPI_ISL_1999323, EPI_ISL_1999324, EPI_ISL_1999325, EPI_ISL_1999326, EPI_ISL_1999327, EPI_ISL_1999328, EPI_ISL_1999329,
EPI_ISL_1999330, EPI_ISL_1999331, EPI_ISL_1999332, EPI_ISL_1999333, EPI_ISL_1999334, EPI_ISL_1999335, EPI_ISL_1999336, EPI_ISL_1999337, EPI_ISL_1999338, EPI_ISL_1999339, EPI_ISL_1999340, EPI_ISL_1999341, EPI_ISL_1999342, EPI_ISL_1999343, EPI_ISL_1999344, EPI_ISL_1999345, EPI_ISL_1999346, EPI_ISL_1999347, EPI_ISL_1999348, EPI_ISL_1999349, EPI_ISL_1999350, EPI_ISL_1999351,
EPI_ISL_1999352, EPI_ISL_1999353, EPI_ISL_1999354, EPI_ISL_1999355, EPI_ISL_1999356, EPI_ISL_1999357, EPI_ISL_1999358, EPI_ISL_1999359, EPI_ISL_1999360, EPI_ISL_1999361, EPI_ISL_1999362, EPI_ISL_1999363, EPI_ISL_1999364, EPI_ISL_1999365, EPI_ISL_1999366, EPI_ISL_1999367, EPI_ISL_1999368, EPI_ISL_1999369, EPI_ISL_1999370, EPI_ISL_1999371, EPI_ISL_1999372, EPI_ISL_1999373,
EPI_ISL_1999374, EPI_ISL_1999375, EPI_ISL_1999376, EPI_ISL_1999377, EPI_ISL_1999378, EPI_ISL_1999379, EPI_ISL_1999380, EPI_ISL_1999381, EPI_ISL_1999382, EPI_ISL_1999383, EPI_ISL_1999384, EPI_ISL_1999385, EPI_ISL_1999386, EPI_ISL_1999387, EPI_ISL_1999388, EPI_ISL_1999389, EPI_ISL_1999390, EPI_ISL_1999391, EPI_ISL_1999392, EPI_ISL_1999393, EPI_ISL_1999394, EPI_ISL_1999395,
EPI_ISL_1999397, EPI_ISL_1999398, EPI_ISL_1999399, EPI_ISL_1999400, EPI_ISL_1999401, EPI_ISL_1999402, EPI_ISL_1999403, EPI_ISL_1999404, EPI_ISL_1999405, EPI_ISL_1999406, EPI_ISL_1999407, EPI_ISL_1999408, EPI_ISL_1999409, EPI_ISL_1999410, EPI_ISL_1999411, EPI_ISL_1999412, EPI_ISL_1999413, EPI_ISL_1999414, EPI_ISL_1999415, EPI_ISL_1999416, EPI_ISL_1999417, EPI_ISL_1999418,
EPI_ISL_1999419, EPI_ISL_1999420, EPI_ISL_1999421, EPI_ISL_1999422, EPI_ISL_1999424, EPI_ISL_1999425, EPI_ISL_1999426, EPI_ISL_1999427, EPI_ISL_1999428, EPI_ISL_1999429, EPI_ISL_1999430, EPI_ISL_1999431, EPI_ISL_1999432, EPI_ISL_1999433, EPI_ISL_1999434, EPI_ISL_1999435, EPI_ISL_1999436, EPI_ISL_1999437, EPI_ISL_1999438, EPI_ISL_1999439, EPI_ISL_1999441, EPI_ISL_1999442,
EPI_ISL_1999443, EPI_ISL_1999444, EPI_ISL_1999445, EPI_ISL_1999446, EPI_ISL_1999447, EPI_ISL_1999448, EPI_ISL_1999449, EPI_ISL_1999450, EPI_ISL_1999451, EPI_ISL_1999452, EPI_ISL_1999453, EPI_ISL_1999454, EPI_ISL_1999455, EPI_ISL_1999456, EPI_ISL_1999457, EPI_ISL_1999458, EPI_ISL_1999459, EPI_ISL_1999460, EPI_ISL_1999461, EPI_ISL_1999462, EPI_ISL_1999463, EPI_ISL_1999464,
EPI_ISL_1999465, EPI_ISL_1999466, EPI_ISL_1999467, EPI_ISL_1999468, EPI_ISL_1999469, EPI_ISL_1999470, EPI_ISL_1999471, EPI_ISL_1999472, EPI_ISL_1999473, EPI_ISL_1999474, EPI_ISL_1999475, EPI_ISL_1999476, EPI_ISL_1999478, EPI_ISL_1999479, EPI_ISL_1999480, EPI_ISL_1999481, EPI_ISL_1999482, EPI_ISL_1999483, EPI_ISL_1999484, EPI_ISL_1999485, EPI_ISL_1999486, EPI_ISL_1999487,
EPI_ISL_1999488, EPI_ISL_1999489, EPI_ISL_1999490, EPI_ISL_1999491, EPI_ISL_1999492, EPI_ISL_1999493, EPI_ISL_1999494, EPI_ISL_1999495, EPI_ISL_1999496, EPI_ISL_1999497, EPI_ISL_1999498, EPI_ISL_1999499, EPI_ISL_1999500, EPI_ISL_1999501, EPI_ISL_1999502, EPI_ISL_1999503, EPI_ISL_1999504, EPI_ISL_1999505, EPI_ISL_1999506, EPI_ISL_1999507, EPI_ISL_1999508, EPI_ISL_1999509,
EPI_ISL_1999510, EPI_ISL_1999511, EPI_ISL_1999512, EPI_ISL_1999513, EPI_ISL_1999515, EPI_ISL_1999516, EPI_ISL_1999518, EPI_ISL_1999519, EPI_ISL_1999520, EPI_ISL_1999521, EPI_ISL_1999522, EPI_ISL_1999523, EPI_ISL_1999524, EPI_ISL_1999525, EPI_ISL_1999526, EPI_ISL_1999527, EPI_ISL_1999528, EPI_ISL_1999529, EPI_ISL_1999530, EPI_ISL_1999531, EPI_ISL_1999532, EPI_ISL_1999533,
EPI_ISL_1999534, EPI_ISL_1999535, EPI_ISL_1999536, EPI_ISL_1999537, EPI_ISL_1999538, EPI_ISL_1999539, EPI_ISL_1999540, EPI_ISL_1999541, EPI_ISL_1999542, EPI_ISL_1999543, EPI_ISL_1999544, EPI_ISL_1999545, EPI_ISL_1999546, EPI_ISL_1999547, EPI_ISL_1999548, EPI_ISL_1999549, EPI_ISL_1999550, EPI_ISL_1999551, EPI_ISL_1999553, EPI_ISL_1999554, EPI_ISL_1999555, EPI_ISL_1999556,
EPI_ISL_1999557, EPI_ISL_1999558, EPI_ISL_1999559, EPI_ISL_1999560, EPI_ISL_1999561, EPI_ISL_1999562, EPI_ISL_1999563, EPI_ISL_1999564, EPI_ISL_1999565, EPI_ISL_1999566, EPI_ISL_1999567, EPI_ISL_1999568, EPI_ISL_1999569, EPI_ISL_1999570, EPI_ISL_1999571, EPI_ISL_1999572, EPI_ISL_1999573, EPI_ISL_1999574, EPI_ISL_1999575, EPI_ISL_1999576, EPI_ISL_1999577, EPI_ISL_1999578,
EPI_ISL_1999579, EPI_ISL_1999580, EPI_ISL_1999581, EPI_ISL_1999582, EPI_ISL_1999583, EPI_ISL_1999584, EPI_ISL_1999585, EPI_ISL_1999586, EPI_ISL_1999587, EPI_ISL_1999588, EPI_ISL_1999589, EPI_ISL_1999590, EPI_ISL_1999591, EPI_ISL_1999592, EPI_ISL_1999593, EPI_ISL_1999594, EPI_ISL_1999595, EPI_ISL_1999596, EPI_ISL_1999597, EPI_ISL_1999598, EPI_ISL_1999599, EPI_ISL_1999600,
EPI_ISL_1999601, EPI_ISL_1999602, EPI_ISL_1999603, EPI_ISL_1999604, EPI_ISL_1999605, EPI_ISL_1999606, EPI_ISL_1999607, EPI_ISL_1999608, EPI_ISL_1999609, EPI_ISL_1999610, EPI_ISL_1999611, EPI_ISL_1999612, EPI_ISL_1999613, EPI_ISL_1999614, EPI_ISL_1999615, EPI_ISL_1999616, EPI_ISL_1999617, EPI_ISL_1999618, EPI_ISL_1999619, EPI_ISL_1999620, EPI_ISL_1999621, EPI_ISL_1999622,
EPI_ISL_1999623, EPI_ISL_1999624, EPI_ISL_1999625, EPI_ISL_1999626, EPI_ISL_1999627, EPI_ISL_1999628, EPI_ISL_1999629, EPI_ISL_1999630, EPI_ISL_1999631, EPI_ISL_1999632, EPI_ISL_1999633, EPI_ISL_1999634, EPI_ISL_1999635, EPI_ISL_1999636, EPI_ISL_1999637, EPI_ISL_1999639, EPI_ISL_1999640, EPI_ISL_1999641, EPI_ISL_1999643, EPI_ISL_1999644, EPI_ISL_1999645, EPI_ISL_1999647,
EPI_ISL_1999648, EPI_ISL_1999650, EPI_ISL_1999651, EPI_ISL_1999653, EPI_ISL_1999654, EPI_ISL_1999656, EPI_ISL_1999657, EPI_ISL_1999658, EPI_ISL_1999660, EPI_ISL_1999661, EPI_ISL_1999663, EPI_ISL_1999664, EPI_ISL_1999666, EPI_ISL_1999667, EPI_ISL_1999668, EPI_ISL_1999670, EPI_ISL_1999671, EPI_ISL_1999673, EPI_ISL_1999674, EPI_ISL_1999676, EPI_ISL_1999677, EPI_ISL_1999679,
EPI_ISL_1999680, EPI_ISL_1999682, EPI_ISL_1999683, EPI_ISL_1999686, EPI_ISL_1999687, EPI_ISL_1999689, EPI_ISL_1999690, EPI_ISL_1999691, EPI_ISL_1999693, EPI_ISL_1999695, EPI_ISL_1999696, EPI_ISL_1999697, EPI_ISL_1999699, EPI_ISL_1999700, EPI_ISL_1999702, EPI_ISL_1999703, EPI_ISL_1999705, EPI_ISL_1999706, EPI_ISL_1999707, EPI_ISL_1999709, EPI_ISL_1999710, EPI_ISL_1999712,
EPI_ISL_1999713, EPI_ISL_1999715, EPI_ISL_1999716, EPI_ISL_1999717, EPI_ISL_1999719, EPI_ISL_1999720, EPI_ISL_1999722, EPI_ISL_1999723, EPI_ISL_1999725, EPI_ISL_1999726, EPI_ISL_1999728, EPI_ISL_1999729, EPI_ISL_1999730, EPI_ISL_1999732, EPI_ISL_1999733, EPI_ISL_1999735, EPI_ISL_1999736, EPI_ISL_1999738, EPI_ISL_1999739, EPI_ISL_1999741, EPI_ISL_1999742, EPI_ISL_1999743,
EPI_ISL_1999745, EPI_ISL_1999746, EPI_ISL_1999748, EPI_ISL_1999749, EPI_ISL_1999751, EPI_ISL_1999752, EPI_ISL_1999753, EPI_ISL_1999755, EPI_ISL_1999756, EPI_ISL_1999758, EPI_ISL_1999759, EPI_ISL_1999761, EPI_ISL_1999762, EPI_ISL_1999764, EPI_ISL_1999765, EPI_ISL_1999767, EPI_ISL_1999768, EPI_ISL_1999769, EPI_ISL_1999771, EPI_ISL_1999772, EPI_ISL_1999773, EPI_ISL_1999775,
EPI_ISL_1999778, EPI_ISL_1999779, EPI_ISL_1999780, EPI_ISL_1999784, EPI_ISL_1999787, EPI_ISL_1999789, EPI_ISL_1999790, EPI_ISL_1999792, EPI_ISL_1999793, EPI_ISL_1999795, EPI_ISL_1999796, EPI_ISL_1999798, EPI_ISL_1999799, EPI_ISL_1999801, EPI_ISL_1999802, EPI_ISL_1999804, EPI_ISL_1999805, EPI_ISL_1999807, EPI_ISL_1999808, EPI_ISL_1999810, EPI_ISL_1999813, EPI_ISL_1999814,
EPI_ISL_1999816, EPI_ISL_1999817, EPI_ISL_1999819, EPI_ISL_1999820, EPI_ISL_1999822, EPI_ISL_1999823, EPI_ISL_1999825, EPI_ISL_1999826, EPI_ISL_1999827, EPI_ISL_1999829, EPI_ISL_1999830, EPI_ISL_1999832, EPI_ISL_1999833, EPI_ISL_1999836, EPI_ISL_1999838, EPI_ISL_1999839, EPI_ISL_1999841, EPI_ISL_1999842, EPI_ISL_1999845, EPI_ISL_1999849, EPI_ISL_1999852, EPI_ISL_1999856,
EPI_ISL_1999859, EPI_ISL_1999863, EPI_ISL_1999865, EPI_ISL_1999866, EPI_ISL_1999867, EPI_ISL_1999868, EPI_ISL_1999869, EPI_ISL_1999870, EPI_ISL_1999871, EPI_ISL_1999872, EPI_ISL_1999873, EPI_ISL_1999874, EPI_ISL_1999875, EPI_ISL_1999876, EPI_ISL_1999877, EPI_ISL_1999878, EPI_ISL_1999879, EPI_ISL_1999880, EPI_ISL_1999881, EPI_ISL_1999882, EPI_ISL_1999883, EPI_ISL_1999884,
EPI_ISL_1999885, EPI_ISL_1999886, EPI_ISL_1999887, EPI_ISL_1999888, EPI_ISL_1999889, EPI_ISL_1999890, EPI_ISL_1999891, EPI_ISL_1999892, EPI_ISL_1999893, EPI_ISL_1999894, EPI_ISL_1999895, EPI_ISL_1999896, EPI_ISL_1999897, EPI_ISL_1999898, EPI_ISL_1999899, EPI_ISL_1999900, EPI_ISL_1999901, EPI_ISL_1999902, EPI_ISL_1999903, EPI_ISL_1999904, EPI_ISL_1999905, EPI_ISL_1999906,
EPI_ISL_1999907, EPI_ISL_1999908, EPI_ISL_1999909, EPI_ISL_1999910, EPI_ISL_1999911, EPI_ISL_1999912, EPI_ISL_1999913, EPI_ISL_1999914, EPI_ISL_1999915, EPI_ISL_1999916, EPI_ISL_1999917, EPI_ISL_1999918, EPI_ISL_1999919, EPI_ISL_1999920, EPI_ISL_1999921, EPI_ISL_1999922, EPI_ISL_1999923, EPI_ISL_1999925, EPI_ISL_1999926, EPI_ISL_1999927, EPI_ISL_1999928, EPI_ISL_1999929,
EPI_ISL_1999930, EPI_ISL_1999931, EPI_ISL_1999932, EPI_ISL_1999933, EPI_ISL_1999934, EPI_ISL_1999935, EPI_ISL_1999936, EPI_ISL_1999937, EPI_ISL_1999938, EPI_ISL_1999939, EPI_ISL_1999940, EPI_ISL_1999941, EPI_ISL_1999942, EPI_ISL_1999943, EPI_ISL_1999944, EPI_ISL_1999945, EPI_ISL_1999947, EPI_ISL_1999948, EPI_ISL_1999949, EPI_ISL_1999950, EPI_ISL_1999951, EPI_ISL_1999952,
EPI_ISL_1999953, EPI_ISL_1999954, EPI_ISL_1999955, EPI_ISL_1999956, EPI_ISL_1999957, EPI_ISL_1999958, EPI_ISL_1999959, EPI_ISL_1999960, EPI_ISL_1999961, EPI_ISL_1999962, EPI_ISL_1999963, EPI_ISL_1999964, EPI_ISL_1999965, EPI_ISL_1999966, EPI_ISL_1999968, EPI_ISL_1999969, EPI_ISL_1999970, EPI_ISL_1999971, EPI_ISL_1999972, EPI_ISL_1999973, EPI_ISL_1999974, EPI_ISL_1999975,
EPI_ISL_1999976, EPI_ISL_1999977, EPI_ISL_1999978, EPI_ISL_1999979, EPI_ISL_1999980, EPI_ISL_1999981, EPI_ISL_1999982, EPI_ISL_1999983, EPI_ISL_1999984, EPI_ISL_1999985, EPI_ISL_1999986, EPI_ISL_1999987, EPI_ISL_1999988, EPI_ISL_1999989, EPI_ISL_1999990, EPI_ISL_1999991, EPI_ISL_1999992, EPI_ISL_1999993, EPI_ISL_1999994, EPI_ISL_1999995, EPI_ISL_1999996, EPI_ISL_1999997,
EPI_ISL_2000000, EPI_ISL_2000004, EPI_ISL_2000007, EPI_ISL_2000014, EPI_ISL_2000018, EPI_ISL_2000021, EPI_ISL_2000025, EPI_ISL_2000028, EPI_ISL_2000032, EPI_ISL_2000035, EPI_ISL_2000038, EPI_ISL_2000042, EPI_ISL_2000045, EPI_ISL_2000049, EPI_ISL_2000052, EPI_ISL_2000056, EPI_ISL_2000059, EPI_ISL_2000064, EPI_ISL_2000067, EPI_ISL_2000071, EPI_ISL_2000074, EPI_ISL_2000078,
EPI_ISL_2000082, EPI_ISL_2000086, EPI_ISL_2000090, EPI_ISL_2000093, EPI_ISL_2000097, EPI_ISL_2000100, EPI_ISL_2000104, EPI_ISL_2000107, EPI_ISL_2000111, EPI_ISL_2000116, EPI_ISL_2000119, EPI_ISL_2000123, EPI_ISL_2000126, EPI_ISL_2000129, EPI_ISL_2000130, EPI_ISL_2000131, EPI_ISL_2000132, EPI_ISL_2000133, EPI_ISL_2000134, EPI_ISL_2000135, EPI_ISL_2000136, EPI_ISL_2000137,
EPI_ISL_2000138, EPI_ISL_2000139, EPI_ISL_2000140, EPI_ISL_2000141, EPI_ISL_2000142, EPI_ISL_2000143, EPI_ISL_2000144, EPI_ISL_2000145, EPI_ISL_2000146, EPI_ISL_2000147, EPI_ISL_2000148, EPI_ISL_2000149, EPI_ISL_2000150, EPI_ISL_2000151, EPI_ISL_2000152, EPI_ISL_2000153, EPI_ISL_2000154, EPI_ISL_2000155, EPI_ISL_2000156, EPI_ISL_2000157, EPI_ISL_2000158, EPI_ISL_2000159,
EPI_ISL_2000160, EPI_ISL_2000161, EPI_ISL_2000162, EPI_ISL_2000163, EPI_ISL_2000164, EPI_ISL_2000165, EPI_ISL_2000166, EPI_ISL_2000167, EPI_ISL_2000168, EPI_ISL_2000169, EPI_ISL_2000170, EPI_ISL_2000171, EPI_ISL_2000172, EPI_ISL_2000173, EPI_ISL_2000174, EPI_ISL_2000175, EPI_ISL_2000176, EPI_ISL_2000177, EPI_ISL_2000178, EPI_ISL_2000179, EPI_ISL_2000180, EPI_ISL_2000181,
EPI_ISL_2000182, EPI_ISL_2000183, EPI_ISL_2000184, EPI_ISL_2000185, EPI_ISL_2000186, EPI_ISL_2000187, EPI_ISL_2000188, EPI_ISL_2000189, EPI_ISL_2000190, EPI_ISL_2000191, EPI_ISL_2000192, EPI_ISL_2000193, EPI_ISL_2000194, EPI_ISL_2000195, EPI_ISL_2000196, EPI_ISL_2000197, EPI_ISL_2000198, EPI_ISL_2000199, EPI_ISL_2000200, EPI_ISL_2000201, EPI_ISL_2000202, EPI_ISL_2000203,
EPI_ISL_2000205, EPI_ISL_2000206, EPI_ISL_2000207, EPI_ISL_2000208, EPI_ISL_2000209, EPI_ISL_2000210, EPI_ISL_2000211, EPI_ISL_2000212, EPI_ISL_2000213, EPI_ISL_2000214, EPI_ISL_2000215, EPI_ISL_2000216, EPI_ISL_2000217, EPI_ISL_2000218, EPI_ISL_2000219, EPI_ISL_2000220, EPI_ISL_2000221, EPI_ISL_2000222, EPI_ISL_2000223, EPI_ISL_2000224, EPI_ISL_2000225, EPI_ISL_2000226,
EPI_ISL_2000227, EPI_ISL_2000228, EPI_ISL_2000229, EPI_ISL_2000230, EPI_ISL_2000231, EPI_ISL_2000232, EPI_ISL_2000233, EPI_ISL_2000234, EPI_ISL_2000235, EPI_ISL_2000236, EPI_ISL_2000237, EPI_ISL_2000238, EPI_ISL_2000239, EPI_ISL_2000240, EPI_ISL_2000242, EPI_ISL_2000243, EPI_ISL_2000244, EPI_ISL_2000245, EPI_ISL_2000246, EPI_ISL_2000247, EPI_ISL_2000248, EPI_ISL_2000249,
EPI_ISL_2000250, EPI_ISL_2000251, EPI_ISL_2000252, EPI_ISL_2000253, EPI_ISL_2000255, EPI_ISL_2000256, EPI_ISL_2000257, EPI_ISL_2000259, EPI_ISL_2000260, EPI_ISL_2000262, EPI_ISL_2000263, EPI_ISL_2000264, EPI_ISL_2000265, EPI_ISL_2000266, EPI_ISL_2000267, EPI_ISL_2000268, EPI_ISL_2000269, EPI_ISL_2000270, EPI_ISL_2000271, EPI_ISL_2000272, EPI_ISL_2000273, EPI_ISL_2000274,
EPI_ISL_2000275, EPI_ISL_2000276, EPI_ISL_2000277, EPI_ISL_2000278, EPI_ISL_2000279, EPI_ISL_2000280, EPI_ISL_2000281, EPI_ISL_2000282, EPI_ISL_2000283, EPI_ISL_2000284, EPI_ISL_2000285, EPI_ISL_2000286, EPI_ISL_2000287, EPI_ISL_2000288, EPI_ISL_2000289, EPI_ISL_2000290, EPI_ISL_2000291, EPI_ISL_2000292, EPI_ISL_2000293, EPI_ISL_2000294, EPI_ISL_2000295, EPI_ISL_2000296,
EPI_ISL_2000297, EPI_ISL_2000298, EPI_ISL_2000299, EPI_ISL_2000300, EPI_ISL_2000301, EPI_ISL_2000302, EPI_ISL_2000303, EPI_ISL_2000304, EPI_ISL_2000305, EPI_ISL_2000306, EPI_ISL_2000307, EPI_ISL_2000308, EPI_ISL_2000309, EPI_ISL_2000310, EPI_ISL_2000311, EPI_ISL_2000312, EPI_ISL_2000313, EPI_ISL_2000314, EPI_ISL_2000315, EPI_ISL_2000316, EPI_ISL_2000317, EPI_ISL_2000318,
EPI_ISL_2000319, EPI_ISL_2000320, EPI_ISL_2000321, EPI_ISL_2000322, EPI_ISL_2000323, EPI_ISL_2000324, EPI_ISL_2000325, EPI_ISL_2000326, EPI_ISL_2000327, EPI_ISL_2000328, EPI_ISL_2000329, EPI_ISL_2000330, EPI_ISL_2000331, EPI_ISL_2000332, EPI_ISL_2000333, EPI_ISL_2000334, EPI_ISL_2000336, EPI_ISL_2000337, EPI_ISL_2000338, EPI_ISL_2000339, EPI_ISL_2000340, EPI_ISL_2000341,
EPI_ISL_2000342, EPI_ISL_2000343, EPI_ISL_2000344, EPI_ISL_2000345, EPI_ISL_2000346, EPI_ISL_2000347, EPI_ISL_2000348, EPI_ISL_2000349, EPI_ISL_2000350, EPI_ISL_2000351, EPI_ISL_2000352, EPI_ISL_2000353, EPI_ISL_2000354, EPI_ISL_2000355, EPI_ISL_2000356, EPI_ISL_2000357, EPI_ISL_2000358, EPI_ISL_2000359, EPI_ISL_2000360, EPI_ISL_2000361, EPI_ISL_2000362, EPI_ISL_2000363,
EPI_ISL_2000364, EPI_ISL_2000365, EPI_ISL_2000366, EPI_ISL_2000367, EPI_ISL_2000368, EPI_ISL_2000369, EPI_ISL_2000370, EPI_ISL_2000371, EPI_ISL_2000372, EPI_ISL_2000373, EPI_ISL_2000374, EPI_ISL_2000375, EPI_ISL_2000376, EPI_ISL_2000377, EPI_ISL_2000378, EPI_ISL_2000379, EPI_ISL_2000380, EPI_ISL_2000381, EPI_ISL_2000382, EPI_ISL_2000383, EPI_ISL_2000384, EPI_ISL_2000385,
EPI_ISL_2000386, EPI_ISL_2000387, EPI_ISL_2000388, EPI_ISL_2000389, EPI_ISL_2000390, EPI_ISL_2000391, EPI_ISL_2000392, EPI_ISL_2000393, EPI_ISL_2000394, EPI_ISL_2000395, EPI_ISL_2000396, EPI_ISL_2000397, EPI_ISL_2000398, EPI_ISL_2000399, EPI_ISL_2000400, EPI_ISL_2000401, EPI_ISL_2000402, EPI_ISL_2000403, EPI_ISL_2000404, EPI_ISL_2000405, EPI_ISL_2000406, EPI_ISL_2000407,
EPI_ISL_2000408, EPI_ISL_2000409, EPI_ISL_2000410, EPI_ISL_2000411, EPI_ISL_2000412, EPI_ISL_2000413, EPI_ISL_2000414, EPI_ISL_2000415, EPI_ISL_2000416, EPI_ISL_2000417, EPI_ISL_2000418, EPI_ISL_2000419, EPI_ISL_2000420, EPI_ISL_2000421, EPI_ISL_2000422, EPI_ISL_2000423, EPI_ISL_2000424, EPI_ISL_2000425, EPI_ISL_2000426, EPI_ISL_2000427, EPI_ISL_2000428, EPI_ISL_2000429,
EPI_ISL_2000430, EPI_ISL_2000431, EPI_ISL_2000432, EPI_ISL_2000433, EPI_ISL_2000434, EPI_ISL_2000435, EPI_ISL_2000436, EPI_ISL_2000437, EPI_ISL_2000438, EPI_ISL_2000439, EPI_ISL_2000440, EPI_ISL_2000441, EPI_ISL_2000442, EPI_ISL_2000443, EPI_ISL_2000444, EPI_ISL_2000445, EPI_ISL_2000446, EPI_ISL_2000447, EPI_ISL_2000448, EPI_ISL_2000449, EPI_ISL_2000450, EPI_ISL_2000451,
EPI_ISL_2000452, EPI_ISL_2000453, EPI_ISL_2000454, EPI_ISL_2000455, EPI_ISL_2000456, EPI_ISL_2000457, EPI_ISL_2000458, EPI_ISL_2000459, EPI_ISL_2000460, EPI_ISL_2000461, EPI_ISL_2000462, EPI_ISL_2000463, EPI_ISL_2000464, EPI_ISL_2000465, EPI_ISL_2000466, EPI_ISL_2000467, EPI_ISL_2000468, EPI_ISL_2000469, EPI_ISL_2000470, EPI_ISL_2000471, EPI_ISL_2000472, EPI_ISL_2000473,
EPI_ISL_2000474, EPI_ISL_2000475, EPI_ISL_2000476, EPI_ISL_2000477, EPI_ISL_2000478, EPI_ISL_2000479, EPI_ISL_2000480, EPI_ISL_2000481, EPI_ISL_2000482, EPI_ISL_2000483, EPI_ISL_2000484, EPI_ISL_2000485, EPI_ISL_2000486, EPI_ISL_2000487, EPI_ISL_2000488, EPI_ISL_2000489, EPI_ISL_2000490, EPI_ISL_2000491, EPI_ISL_2000492, EPI_ISL_2000493, EPI_ISL_2000494, EPI_ISL_2000495,
EPI_ISL_2000496, EPI_ISL_2000497, EPI_ISL_2000498, EPI_ISL_2000499, EPI_ISL_2000500, EPI_ISL_2000501, EPI_ISL_2000502, EPI_ISL_2000504, EPI_ISL_2000505, EPI_ISL_2000506, EPI_ISL_2000507, EPI_ISL_2000508, EPI_ISL_2000509, EPI_ISL_2000510, EPI_ISL_2000511, EPI_ISL_2000512, EPI_ISL_2000513, EPI_ISL_2000514, EPI_ISL_2000515, EPI_ISL_2000516, EPI_ISL_2000517, EPI_ISL_2000518,
EPI_ISL_2000519, EPI_ISL_2000520, EPI_ISL_2000521, EPI_ISL_2000522, EPI_ISL_2000523, EPI_ISL_2000524, EPI_ISL_2000525, EPI_ISL_2000526, EPI_ISL_2000527, EPI_ISL_2000528, EPI_ISL_2000529, EPI_ISL_2000530, EPI_ISL_2000531, EPI_ISL_2000532, EPI_ISL_2000533, EPI_ISL_2000534, EPI_ISL_2000535, EPI_ISL_2000536, EPI_ISL_2000537, EPI_ISL_2000538, EPI_ISL_2000539, EPI_ISL_2000540,
EPI_ISL_2000541, EPI_ISL_2000542, EPI_ISL_2000543, EPI_ISL_2000544, EPI_ISL_2000545, EPI_ISL_2000546, EPI_ISL_2000547, EPI_ISL_2000548, EPI_ISL_2000549, EPI_ISL_2000550

| | | | |
|---|---|---|---|
| see above | Aegis Sciences Corporation | Centers for Disease Control and Prevention Division of Viral Diseases, Pathogen Discovery | Dakota Howard, Dhwani Batra, Peter W. Cook, Kara Moser, Adrian Paskey, Jason Caravas, Benjamin Rambo-Martin, Shatavia Morrison, Christopher Gulvick, Scott Sammons, Yvette Unoarumhi, Darlene Wagner, Matthew Schmerer, Cyndi Clark, Patrick Campbell, Rob Case, Vikramsinha Ghorpade, Holly Houdeshell, Ola Kvalvaag, Dillon Nall, Ethan Sanders, Alec Vest, Shaun Westlund, Matthew Hardison, Clinton R. Paden, Duncan MacCannell |

EPI_ISL_2000573, EPI_ISL_2000574, EPI_ISL_2000575, EPI_ISL_2000576, EPI_ISL_2000577, EPI_ISL_2000578, EPI_ISL_2000579, EPI_ISL_2000580, EPI_ISL_2000581, EPI_ISL_2000582, EPI_ISL_2000583, EPI_ISL_2000584, EPI_ISL_2000585, EPI_ISL_2000586, EPI_ISL_2000587, EPI_ISL_2000588, EPI_ISL_2000589, EPI_ISL_2000590, EPI_ISL_2000591, EPI_ISL_2000592, EPI_ISL_2000593, EPI_ISL_2000594,
EPI_ISL_2000595, EPI_ISL_2000596, EPI_ISL_2000597, EPI_ISL_2000598

| | | | |
|---|---|---|---|
| see above | Rami Kantor lab | Rami Kantor lab | Josephine Darpolor, Vlad Novitsky, Mark Howison, Richard Huard, Ewa King, Rami Kantor |

EPI_ISL_2000599, EPI_ISL_2000600, EPI_ISL_2000601, EPI_ISL_2000602, EPI_ISL_2000603, EPI_ISL_2000604, EPI_ISL_2000605, EPI_ISL_2000606, EPI_ISL_2000607, EPI_ISL_2000608, EPI_ISL_2000609, EPI_ISL_2000610, EPI_ISL_2000611, EPI_ISL_2000612, EPI_ISL_2000613, EPI_ISL_2000614, EPI_ISL_2000615, EPI_ISL_2000616

| | | | |
|---|---|---|---|
| see above | ISDH | Quantigen Biosciences | Paul J. Childress |

EPI_ISL_2000766, EPI_ISL_2000767, EPI_ISL_2000768, EPI_ISL_2000769, EPI_ISL_2000770, EPI_ISL_2000771, EPI_ISL_2000772, EPI_ISL_2000773, EPI_ISL_2000774, EPI_ISL_2000775, EPI_ISL_2000776, EPI_ISL_2000777, EPI_ISL_2000778, EPI_ISL_2000779, EPI_ISL_2000780, EPI_ISL_2000781, EPI_ISL_2000782, EPI_ISL_2000783, EPI_ISL_2000784, EPI_ISL_2000785, EPI_ISL_2000786, EPI_ISL_2000787,
EPI_ISL_2000788, EPI_ISL_2000789, EPI_ISL_2000790, EPI_ISL_2000791, EPI_ISL_2000792, EPI_ISL_2000793, EPI_ISL_2000794, EPI_ISL_2000795, EPI_ISL_2000796, EPI_ISL_2000797, EPI_ISL_2000798, EPI_ISL_2000799, EPI_ISL_2000800, EPI_ISL_2000801, EPI_ISL_2000802, EPI_ISL_2000803, EPI_ISL_2000804, EPI_ISL_2000805, EPI_ISL_2000806, EPI_ISL_2000807, EPI_ISL_2000808, EPI_ISL_2000809,
EPI_ISL_2000810, EPI_ISL_2000811, EPI_ISL_2000812, EPI_ISL_2000813, EPI_ISL_2000814, EPI_ISL_2000815, EPI_ISL_2000816, EPI_ISL_2000817, EPI_ISL_2000818, EPI_ISL_2000819, EPI_ISL_2000820, EPI_ISL_2000821, EPI_ISL_2000822, EPI_ISL_2000823, EPI_ISL_2000824, EPI_ISL_2000825, EPI_ISL_2000826, EPI_ISL_2000827, EPI_ISL_2000828, EPI_ISL_2000829, EPI_ISL_2000830, EPI_ISL_2000831,
EPI_ISL_2000832, EPI_ISL_2000833, EPI_ISL_2000834, EPI_ISL_2000835, EPI_ISL_2000836, EPI_ISL_2000837, EPI_ISL_2000838, EPI_ISL_2000839, EPI_ISL_2000841, EPI_ISL_2000843, EPI_ISL_2000844, EPI_ISL_2000845, EPI_ISL_2000846, EPI_ISL_2000847, EPI_ISL_2000848, EPI_ISL_2000849, EPI_ISL_2000850, EPI_ISL_2000851, EPI_ISL_2000853, EPI_ISL_2000854, EPI_ISL_2000855, EPI_ISL_2000856,
EPI_ISL_2000857, EPI_ISL_2000858, EPI_ISL_2000859

| | | | |
|---|---|---|---|
| see above | Illinois Department of Public Health - Springfield Lab | Illinois Department of Public Health - Springfield Lab | Bryan Sim, Gordon McCall |

EPI_ISL_2000866, EPI_ISL_2000868, EPI_ISL_2000869, EPI_ISL_2000873, EPI_ISL_2000874, EPI_ISL_2000877, EPI_ISL_2000878, EPI_ISL_2000880, EPI_ISL_2000885, EPI_ISL_2000886, EPI_ISL_2000887, EPI_ISL_2000889, EPI_ISL_2000890, EPI_ISL_2000892, EPI_ISL_2000895, EPI_ISL_2000896, EPI_ISL_2000899, EPI_ISL_2000900, EPI_ISL_2000903, EPI_ISL_2000904, EPI_ISL_2000907, EPI_ISL_2000908,
EPI_ISL_2000909, EPI_ISL_2000910, EPI_ISL_2000913

| | | | |
|---|---|---|---|
| see above | TGen North | TGen North | "Jolene Bowers, Heather Centner, Chris French, Hayley Yaglom, Ashlyn Pfeiffer, Darrin Lemmer, Dave Engelthaler, The Arizona COVID Genomics Union (ACGU)" |
| EPI_ISL_2000914 | Sonora Quest Laboratories | TGen North | "Jolene Bowers, Heather Centner, Chris French, Hayley Yaglom, Ashlyn Pfeiffer, Darrin Lemmer, Dave Engelthaler, The Arizona COVID Genomics Union (ACGU)" |

EPI_ISL_2000916, EPI_ISL_2000920, EPI_ISL_2000926, EPI_ISL_2000928, EPI_ISL_2000931, EPI_ISL_2000932, EPI_ISL_2000933, EPI_ISL_2000936, EPI_ISL_2000938, EPI_ISL_2000939, EPI_ISL_2000941, EPI_ISL_2000944, EPI_ISL_2000948, EPI_ISL_2000949, EPI_ISL_2000951, EPI_ISL_2000952, EPI_ISL_2000956, EPI_ISL_2000957, EPI_ISL_2000964, EPI_ISL_2000965, EPI_ISL_2000967, EPI_ISL_2000968,
EPI_ISL_2000970, EPI_ISL_2000971, EPI_ISL_2000972, EPI_ISL_2000974, EPI_ISL_2000975, EPI_ISL_2000979, EPI_ISL_2000980, EPI_ISL_2000981, EPI_ISL_2000986, EPI_ISL_2000990, EPI_ISL_2000992, EPI_ISL_2000995, EPI_ISL_2000997, EPI_ISL_2000999, EPI_ISL_2001000, EPI_ISL_2001001, EPI_ISL_2001002, EPI_ISL_2001006, EPI_ISL_2001007, EPI_ISL_2001010, EPI_ISL_2001011, EPI_ISL_2001017,
EPI_ISL_2001021, EPI_ISL_2001023, EPI_ISL_2001024

| | | | |
|---|---|---|---|
| see above | TGen North | TGen North | "Jolene Bowers, Heather Centner, Chris French, Hayley Yaglom, Ashlyn Pfeiffer, Darrin Lemmer, Dave Engelthaler, The Arizona COVID Genomics Union (ACGU)" |
| EPI_ISL_2001029, EPI_ISL_2001032, EPI_ISL_2001034, EPI_ISL_2001036, EPI_ISL_2001039 | Monterey County Public Health Laboratory | Monterey County Public Health Laboratory | Monterey County Public Health Laboratory |
| EPI_ISL_2001050 | Hospital Sharp | Microbial Genomics Laboratory | Bruno Gomez-Gil, Julissa Enciso-Ibarra, Alejandra Garcia-Gasca, Daniel Fregoso-Rueda |

EPI_ISL_2001214, EPI_ISL_2001215, EPI_ISL_2001216, EPI_ISL_2001217, EPI_ISL_2001218, EPI_ISL_2001219, EPI_ISL_2001220, EPI_ISL_2001221, EPI_ISL_2001222, EPI_ISL_2001223, EPI_ISL_2001226, EPI_ISL_2001227, EPI_ISL_2001229, EPI_ISL_2001230, EPI_ISL_2001231, EPI_ISL_2001232, EPI_ISL_2001233, EPI_ISL_2001234, EPI_ISL_2001235, EPI_ISL_2001236, EPI_ISL_2001237, EPI_ISL_2001238,
EPI_ISL_2001239, EPI_ISL_2001240, EPI_ISL_2001241, EPI_ISL_2001242, EPI_ISL_2001244, EPI_ISL_2001245, EPI_ISL_2001246, EPI_ISL_2001247, EPI_ISL_2001248, EPI_ISL_2001249, EPI_ISL_2001250, EPI_ISL_2001251, EPI_ISL_2001252, EPI_ISL_2001253, EPI_ISL_2001254, EPI_ISL_2001255, EPI_ISL_2001256, EPI_ISL_2001257, EPI_ISL_2001258, EPI_ISL_2001259, EPI_ISL_2001260, EPI_ISL_2001261,
EPI_ISL_2001262, EPI_ISL_2001263, EPI_ISL_2001264, EPI_ISL_2001265, EPI_ISL_2001267, EPI_ISL_2001268, EPI_ISL_2001269, EPI_ISL_2001270, EPI_ISL_2001271, EPI_ISL_2001272, EPI_ISL_2001273, EPI_ISL_2001274, EPI_ISL_2001275, EPI_ISL_2001276, EPI_ISL_2001277, EPI_ISL_2001278, EPI_ISL_2001279, EPI_ISL_2001280, EPI_ISL_2001282, EPI_ISL_2001283, EPI_ISL_2001284, EPI_ISL_2001285,
EPI_ISL_2001286, EPI_ISL_2001287, EPI_ISL_2001289, EPI_ISL_2001290, EPI_ISL_2001292, EPI_ISL_2001293, EPI_ISL_2001294, EPI_ISL_2001295, EPI_ISL_2001296

| | | | |
|---|---|---|---|
| see above | University of Michigan Clinical Microbiology Laboratory | Lauring Lab, University of Michigan, Department of Microbiology and Immunology | Valesano |
| EPI_ISL_2001657, EPI_ISL_2001658 | Wyoming Public Health Laboratory | Wyoming Public Health Laboratory | Jim Mildenberger, Wanda Manley, Noah Hull, Taylor Fearing, Lynette Gumbleton, Channing Weber, Ashley Norberg, Chayse Rowley, Marley Goetz, Brian Dominguez, Elliot Thomasson, Sam Britz, Cari Sloma, and Rob Christensen |
| EPI_ISL_2002139, EPI_ISL_2002140, EPI_ISL_2002141, EPI_ISL_2002142, EPI_ISL_2002143, EPI_ISL_2002144, EPI_ISL_2002145, EPI_ISL_2002146, EPI_ISL_2002147, EPI_ISL_2002148, EPI_ISL_2002149, EPI_ISL_2002150, EPI_ISL_2002151, EPI_ISL_2002152, EPI_ISL_2002153, EPI_ISL_2002154, EPI_ISL_2002155, EPI_ISL_2002156, EPI_ISL_2002157, EPI_ISL_2002158, EPI_ISL_2002159, EPI_ISL_2002160, EPI_ISL_2002161, EPI_ISL_2002163, EPI_ISL_2002164, EPI_ISL_2002165, EPI_ISL_2002166, EPI_ISL_2002167, EPI_ISL_2002168, EPI_ISL_2002169, EPI_ISL_2002170, EPI_ISL_2002171, EPI_ISL_2002172, EPI_ISL_2002173, EPI_ISL_2002174, EPI_ISL_2002175, EPI_ISL_2002176, EPI_ISL_2002177, EPI_ISL_2002178, EPI_ISL_2002179, EPI_ISL_2002180, EPI_ISL_2002181, EPI_ISL_2002182, EPI_ISL_2002183, EPI_ISL_2002184, EPI_ISL_2002185, EPI_ISL_2002186, EPI_ISL_2002187, EPI_ISL_2002188, EPI_ISL_2002189, EPI_ISL_2002190, EPI_ISL_2002191, EPI_ISL_2002192, EPI_ISL_2002193, EPI_ISL_2002194, EPI_ISL_2002195, EPI_ISL_2002196, EPI_ISL_2002197, EPI_ISL_2002198, EPI_ISL_2002199, EPI_ISL_2002200, EPI_ISL_2002201, EPI_ISL_2002202, EPI_ISL_2002203, EPI_ISL_2002204, EPI_ISL_2002205, EPI_ISL_2002206, EPI_ISL_2002207, EPI_ISL_2002208, EPI_ISL_2002209, EPI_ISL_2002210, EPI_ISL_2002211, EPI_ISL_2002212, EPI_ISL_2002213, EPI_ISL_2002214, EPI_ISL_2002215, EPI_ISL_2002216, EPI_ISL_2002217, EPI_ISL_2002218, EPI_ISL_2002219, EPI_ISL_2002220, EPI_ISL_2002221, EPI_ISL_2002222, EPI_ISL_2002223, EPI_ISL_2002224, EPI_ISL_2002225, EPI_ISL_2002226, EPI_ISL_2002227, EPI_ISL_2002228, EPI_ISL_2002229, EPI_ISL_2002230, EPI_ISL_2002231, EPI_ISL_2002232, EPI_ISL_2002233, EPI_ISL_2002234, EPI_ISL_2002235, EPI_ISL_2002236, EPI_ISL_2002237, EPI_ISL_2002238, EPI_ISL_2002239, EPI_ISL_2002240, EPI_ISL_2002241, EPI_ISL_2002242, EPI_ISL_2002243, EPI_ISL_2002244, EPI_ISL_2002245, EPI_ISL_2002246, EPI_ISL_2002247, EPI_ISL_2002248, EPI_ISL_2002249, EPI_ISL_2002250, EPI_ISL_2002251, EPI_ISL_2002252, EPI_ISL_2002253, EPI_ISL_2002254, EPI_ISL_2002255, EPI_ISL_2002256, EPI_ISL_2002257, EPI_ISL_2002258, EPI_ISL_2002259, EPI_ISL_2002261, EPI_ISL_2002262, EPI_ISL_2002263, EPI_ISL_2002264, EPI_ISL_2002265, EPI_ISL_2002266, EPI_ISL_2002267, EPI_ISL_2002268, EPI_ISL_2002269, EPI_ISL_2002270, EPI_ISL_2002271, EPI_ISL_2002272, EPI_ISL_2002274, EPI_ISL_2002275, EPI_ISL_2002276, EPI_ISL_2002277, EPI_ISL_2002278, EPI_ISL_2002279, EPI_ISL_2002280, EPI_ISL_2002281, EPI_ISL_2002282, EPI_ISL_2002283, EPI_ISL_2002284, EPI_ISL_2002285, EPI_ISL_2002286, EPI_ISL_2002287, EPI_ISL_2002288, EPI_ISL_2002289, EPI_ISL_2002290, EPI_ISL_2002291, EPI_ISL_2002292, EPI_ISL_2002293, EPI_ISL_2002294, EPI_ISL_2002295, EPI_ISL_2002296, EPI_ISL_2002297, EPI_ISL_2002298, EPI_ISL_2002299, EPI_ISL_2002300, EPI_ISL_2002301, EPI_ISL_2002302, EPI_ISL_2002304, EPI_ISL_2002305, EPI_ISL_2002306, EPI_ISL_2002307, EPI_ISL_2002309, EPI_ISL_2002310, EPI_ISL_2002311, EPI_ISL_2002312, EPI_ISL_2002313, EPI_ISL_2002314, EPI_ISL_2002315, EPI_ISL_2002316, EPI_ISL_2002317, EPI_ISL_2002318, EPI_ISL_2002319, EPI_ISL_2002320, EPI_ISL_2002321, EPI_ISL_2002322, EPI_ISL_2002323, EPI_ISL_2002324, EPI_ISL_2002325, EPI_ISL_2002326, EPI_ISL_2002327, EPI_ISL_2002328, EPI_ISL_2002329, EPI_ISL_2002330, EPI_ISL_2002331, EPI_ISL_2002332, EPI_ISL_2002333, EPI_ISL_2002334, EPI_ISL_2002335, EPI_ISL_2002336, EPI_ISL_2002337, EPI_ISL_2002338, EPI_ISL_2002339, EPI_ISL_2002340, EPI_ISL_2002341, EPI_ISL_2002342, EPI_ISL_2002343, EPI_ISL_2002344, EPI_ISL_2002345, EPI_ISL_2002346, EPI_ISL_2002347, EPI_ISL_2002348, EPI_ISL_2002349, EPI_ISL_2002350, EPI_ISL_2002351, EPI_ISL_2002352, EPI_ISL_2002353, EPI_ISL_2002354, EPI_ISL_2002355, EPI_ISL_2002356, EPI_ISL_2002357, EPI_ISL_2002358, EPI_ISL_2002359, EPI_ISL_2002360, EPI_ISL_2002361, EPI_ISL_2002362, EPI_ISL_2002363, EPI_ISL_2002364, EPI_ISL_2002365, EPI_ISL_2002366, EPI_ISL_2002367, EPI_ISL_2002368, EPI_ISL_2002369, EPI_ISL_2002370, EPI_ISL_2002371, EPI_ISL_2002372, EPI_ISL_2002373, EPI_ISL_2002374, EPI_ISL_2002375, EPI_ISL_2002376, EPI_ISL_2002377, EPI_ISL_2002378, EPI_ISL_2002379, EPI_ISL_2002380, EPI_ISL_2002381, EPI_ISL_2002382, EPI_ISL_2002383, EPI_ISL_2002384, EPI_ISL_2002385, EPI_ISL_2002386, EPI_ISL_2002387, EPI_ISL_2002388, EPI_ISL_2002389, EPI_ISL_2002390, EPI_ISL_2002391, EPI_ISL_2002392, EPI_ISL_2002393, EPI_ISL_2002394, EPI_ISL_2002395, EPI_ISL_2002396, EPI_ISL_2002397, EPI_ISL_2002398, EPI_ISL_2002399, EPI_ISL_2002400, EPI_ISL_2002401, EPI_ISL_2002402, EPI_ISL_2002403, EPI_ISL_2002404, EPI_ISL_2002405, EPI_ISL_2002406, EPI_ISL_2002407, EPI_ISL_2002408, EPI_ISL_2002409, EPI_ISL_2002410, EPI_ISL_2002411, EPI_ISL_2002412, EPI_ISL_2002413, EPI_ISL_2002414, EPI_ISL_2002415, EPI_ISL_2002416, EPI_ISL_2002417, EPI_ISL_2002418, EPI_ISL_2002419, EPI_ISL_2002420, EPI_ISL_2002421, EPI_ISL_2002422, EPI_ISL_2002423, EPI_ISL_2002424, EPI_ISL_2002425, EPI_ISL_2002426, EPI_ISL_2002427, EPI_ISL_2002428, EPI_ISL_2002429, EPI_ISL_2002430, EPI_ISL_2002431, EPI_ISL_2002432, EPI_ISL_2002433, EPI_ISL_2002434, EPI_ISL_2002435, EPI_ISL_2002436, EPI_ISL_2002437, EPI_ISL_2002438, EPI_ISL_2002439, EPI_ISL_2002440, EPI_ISL_2002441, EPI_ISL_2002442, EPI_ISL_2002443, EPI_ISL_2002444, EPI_ISL_2002445, EPI_ISL_2002446, EPI_ISL_2002447, EPI_ISL_2002448, EPI_ISL_2002449, EPI_ISL_2002450, EPI_ISL_2002451, EPI_ISL_2002452, EPI_ISL_2002453, EPI_ISL_2002454, EPI_ISL_2002455, EPI_ISL_2002456, EPI_ISL_2002457, EPI_ISL_2002458, EPI_ISL_2002459, EPI_ISL_2002460, EPI_ISL_2002461, EPI_ISL_2002462, EPI_ISL_2002463, EPI_ISL_2002464, EPI_ISL_2002465, EPI_ISL_2002466, EPI_ISL_2002467, EPI_ISL_2002468, EPI_ISL_2002469, EPI_ISL_2002470, EPI_ISL_2002471, EPI_ISL_2002472, EPI_ISL_2002473, EPI_ISL_2002474, EPI_ISL_2002475, EPI_ISL_2002476, EPI_ISL_2002477, EPI_ISL_2002478, EPI_ISL_2002479, EPI_ISL_2002480, EPI_ISL_2002481 | | | |
| see above | UW Virology Lab | UW Virology Lab | Pavitra Roychoudhury, Hong Xie, Lasata Shrestha, Tien V. Nguyen, Shah Mohamed Bakhash, Michelle Lin, Noah R. Baker, Sean Ellis, Meei-Li Huang, Keith R Jerome, Alexander Greninger |
| EPI_ISL_2002484, EPI_ISL_2002485, EPI_ISL_2002486, EPI_ISL_2002487, EPI_ISL_2002488, EPI_ISL_2002489, EPI_ISL_2002490, EPI_ISL_2002491, EPI_ISL_2002492 | Kansas Health and Environmental Lab | Kansas Health and Environmental Lab | Mike Grose, Jonathan Barnell, Ben Olsen, and Phil Adam |
| EPI_ISL_2002493, EPI_ISL_2002494, EPI_ISL_2002495, EPI_ISL_2002496, EPI_ISL_2002497, EPI_ISL_2002498, EPI_ISL_2002499, EPI_ISL_2002500, EPI_ISL_2002501, EPI_ISL_2002502, EPI_ISL_2002503, EPI_ISL_2002504, EPI_ISL_2002505, EPI_ISL_2002506, EPI_ISL_2002507, EPI_ISL_2002508, EPI_ISL_2002509, EPI_ISL_2002510, EPI_ISL_2002511, EPI_ISL_2002512, EPI_ISL_2002513, EPI_ISL_2002514, EPI_ISL_2002515, EPI_ISL_2002516, EPI_ISL_2002517, EPI_ISL_2002518, EPI_ISL_2002519, EPI_ISL_2002520, EPI_ISL_2002521, EPI_ISL_2002522, EPI_ISL_2002523, EPI_ISL_2002524, EPI_ISL_2002525, EPI_ISL_2002526, EPI_ISL_2002527, EPI_ISL_2002528, EPI_ISL_2002529, EPI_ISL_2002530, EPI_ISL_2002531, EPI_ISL_2002532, EPI_ISL_2002533, EPI_ISL_2002534, EPI_ISL_2002535, EPI_ISL_2002536, EPI_ISL_2002537, EPI_ISL_2002538, EPI_ISL_2002539, EPI_ISL_2002540, EPI_ISL_2002541, EPI_ISL_2002542, EPI_ISL_2002543, EPI_ISL_2002544, EPI_ISL_2002545, EPI_ISL_2002546, EPI_ISL_2002547, EPI_ISL_2002548, EPI_ISL_2002549, EPI_ISL_2002550, EPI_ISL_2002551, EPI_ISL_2002552, EPI_ISL_2002553, EPI_ISL_2002554, EPI_ISL_2002555, EPI_ISL_2002556, EPI_ISL_2002557, EPI_ISL_2002558, EPI_ISL_2002559, EPI_ISL_2002560, EPI_ISL_2002561, EPI_ISL_2002562, EPI_ISL_2002563, EPI_ISL_2002564, EPI_ISL_2002565, EPI_ISL_2002566, EPI_ISL_2002567, EPI_ISL_2002568, EPI_ISL_2002569, EPI_ISL_2002570, EPI_ISL_2002571, EPI_ISL_2002572, EPI_ISL_2002573, EPI_ISL_2002574, EPI_ISL_2002575, EPI_ISL_2002576, EPI_ISL_2002577 | | | |
| see above | UW Virology Lab | UW Virology Lab | Pavitra Roychoudhury, Hong Xie, Lasata Shrestha, Tien V. Nguyen, Shah Mohamed Bakhash, Michelle Lin, Noah R. Baker, Sean Ellis, Meei-Li Huang, Keith R Jerome, Alexander Greninger |
| EPI_ISL_2002578, EPI_ISL_2002579, EPI_ISL_2002580, EPI_ISL_2002582, EPI_ISL_2002583, EPI_ISL_2002584, EPI_ISL_2002585, EPI_ISL_2002586, EPI_ISL_2002587, EPI_ISL_2002588, EPI_ISL_2002589, EPI_ISL_2002590, EPI_ISL_2002591, EPI_ISL_2002592, EPI_ISL_2002593, EPI_ISL_2002594, EPI_ISL_2002595, EPI_ISL_2002596, EPI_ISL_2002597 | | | |
| see above | Kansas Health and Environmental Lab | Kansas Health and Environmental Lab | Mike Grose, Jonathan Barnell, Ben Olsen, and Phil Adam |
| EPI_ISL_2002689, EPI_ISL_2002690, EPI_ISL_2002691, EPI_ISL_2002692, EPI_ISL_2002693, EPI_ISL_2002694, EPI_ISL_2002695, EPI_ISL_2002696, EPI_ISL_2002697, EPI_ISL_2002698, EPI_ISL_2002699, EPI_ISL_2002700, EPI_ISL_2002701, EPI_ISL_2002702, EPI_ISL_2002703, EPI_ISL_2002704, EPI_ISL_2002705, EPI_ISL_2002706, EPI_ISL_2002707, EPI_ISL_2002708, EPI_ISL_2002709, EPI_ISL_2002710, EPI_ISL_2002711, EPI_ISL_2002712, EPI_ISL_2002713, EPI_ISL_2002714, EPI_ISL_2002715, EPI_ISL_2002716, EPI_ISL_2002717, EPI_ISL_2002718, EPI_ISL_2002719, EPI_ISL_2002720, EPI_ISL_2002721, EPI_ISL_2002722, EPI_ISL_2002723, EPI_ISL_2002724, EPI_ISL_2002725, EPI_ISL_2002726, EPI_ISL_2002727, EPI_ISL_2002728, EPI_ISL_2002729, EPI_ISL_2002730, EPI_ISL_2002731, EPI_ISL_2002732, EPI_ISL_2002733, EPI_ISL_2002734, EPI_ISL_2002735, EPI_ISL_2002736, EPI_ISL_2002737, EPI_ISL_2002738, EPI_ISL_2002739, EPI_ISL_2002740, EPI_ISL_2002741, EPI_ISL_2002742, EPI_ISL_2002743, EPI_ISL_2002744, EPI_ISL_2002745, EPI_ISL_2002746, EPI_ISL_2002747, EPI_ISL_2002749, EPI_ISL_2002750, EPI_ISL_2002751, EPI_ISL_2002752, EPI_ISL_2002753, EPI_ISL_2002754, EPI_ISL_2002755, EPI_ISL_2002756, EPI_ISL_2002757, EPI_ISL_2002759, EPI_ISL_2002760, EPI_ISL_2002761, EPI_ISL_2002762, EPI_ISL_2002763, EPI_ISL_2002764, EPI_ISL_2002765, EPI_ISL_2002766, EPI_ISL_2002767, EPI_ISL_2002768, EPI_ISL_2002769, EPI_ISL_2002770, EPI_ISL_2002771, EPI_ISL_2002772, EPI_ISL_2002773, EPI_ISL_2002775, EPI_ISL_2002776, EPI_ISL_2002777, EPI_ISL_2002778, EPI_ISL_2002779, EPI_ISL_2002780, EPI_ISL_2002781, EPI_ISL_2002782, EPI_ISL_2002783, EPI_ISL_2002784, EPI_ISL_2002785, EPI_ISL_2002786, EPI_ISL_2002787, EPI_ISL_2002788, EPI_ISL_2002789, EPI_ISL_2002790, EPI_ISL_2002791, EPI_ISL_2002792, EPI_ISL_2002793, EPI_ISL_2002794, EPI_ISL_2002795, EPI_ISL_2002796, EPI_ISL_2002797, EPI_ISL_2002798, EPI_ISL_2002800, EPI_ISL_2002801, EPI_ISL_2002802, EPI_ISL_2002803, EPI_ISL_2002804, EPI_ISL_2002805, EPI_ISL_2002806, EPI_ISL_2002807, EPI_ISL_2002808, EPI_ISL_2002809, EPI_ISL_2002810, EPI_ISL_2002811, EPI_ISL_2002812, EPI_ISL_2002813, EPI_ISL_2002814, EPI_ISL_2002815, EPI_ISL_2002816, EPI_ISL_2002817, EPI_ISL_2002818, EPI_ISL_2002819, EPI_ISL_2002820, EPI_ISL_2002821, EPI_ISL_2002822, EPI_ISL_2002823, EPI_ISL_2002824, EPI_ISL_2002825, EPI_ISL_2002826, EPI_ISL_2002827, EPI_ISL_2002828, EPI_ISL_2002829, EPI_ISL_2002830, EPI_ISL_2002831, EPI_ISL_2002832, EPI_ISL_2002833, EPI_ISL_2002834, EPI_ISL_2002835, EPI_ISL_2002836, EPI_ISL_2002837, EPI_ISL_2002838, EPI_ISL_2002839, EPI_ISL_2002840, EPI_ISL_2002841, EPI_ISL_2002842, EPI_ISL_2002843, EPI_ISL_2002844, EPI_ISL_2002845, EPI_ISL_2002846, EPI_ISL_2002847, EPI_ISL_2002848, EPI_ISL_2002849, EPI_ISL_2002850, EPI_ISL_2002851, EPI_ISL_2002852, EPI_ISL_2002853, EPI_ISL_2002854, EPI_ISL_2002855, EPI_ISL_2002856, EPI_ISL_2002857, EPI_ISL_2002858, EPI_ISL_2002859, EPI_ISL_2002860, EPI_ISL_2002861, EPI_ISL_2002862, EPI_ISL_2002863, EPI_ISL_2002864, EPI_ISL_2002865, EPI_ISL_2002866, EPI_ISL_2002867, EPI_ISL_2002868, EPI_ISL_2002869, EPI_ISL_2002870, EPI_ISL_2002871, EPI_ISL_2002872, EPI_ISL_2002873, EPI_ISL_2002874, EPI_ISL_2002875, EPI_ISL_2002876, EPI_ISL_2002877, EPI_ISL_2002878, EPI_ISL_2002879, EPI_ISL_2002880, EPI_ISL_2002881, EPI_ISL_2002882, EPI_ISL_2002884, EPI_ISL_2002885, EPI_ISL_2002886, EPI_ISL_2002887, EPI_ISL_2002888, EPI_ISL_2002889, EPI_ISL_2002890, EPI_ISL_2002891, EPI_ISL_2002892, EPI_ISL_2002893, EPI_ISL_2002894, EPI_ISL_2002895, EPI_ISL_2002897, EPI_ISL_2002898, EPI_ISL_2002899, EPI_ISL_2002900, EPI_ISL_2002901, EPI_ISL_2002902, EPI_ISL_2002903, EPI_ISL_2002904, EPI_ISL_2002905, EPI_ISL_2002906, EPI_ISL_2002907, EPI_ISL_2002908, EPI_ISL_2002909, EPI_ISL_2002910, EPI_ISL_2002911, EPI_ISL_2002912, EPI_ISL_2002913, EPI_ISL_2002914, EPI_ISL_2002915, EPI_ISL_2002916, EPI_ISL_2002917, EPI_ISL_2002918, EPI_ISL_2002919, EPI_ISL_2002920, EPI_ISL_2002921, EPI_ISL_2002922, EPI_ISL_2002923, EPI_ISL_2002924, EPI_ISL_2002925, EPI_ISL_2002927, EPI_ISL_2002928, EPI_ISL_2002929, EPI_ISL_2002930, EPI_ISL_2002931, EPI_ISL_2002932, EPI_ISL_2002933, EPI_ISL_2002934, EPI_ISL_2002935, EPI_ISL_2002936, EPI_ISL_2002937, EPI_ISL_2002938, EPI_ISL_2002939, EPI_ISL_2002940 | | | |
| see above | UW Virology Lab | UW Virology Lab | Pavitra Roychoudhury, Hong Xie, Lasata Shrestha, Tien V. Nguyen, Shah Mohamed Bakhash, Michelle Lin, Noah R. Baker, Sean Ellis, Meei-Li Huang, Keith R Jerome, Alexander Greninger |
| EPI_ISL_2003171, EPI_ISL_2003172, EPI_ISL_2003173, EPI_ISL_2003174, EPI_ISL_2003175, EPI_ISL_2003176, EPI_ISL_2003177, EPI_ISL_2003178, EPI_ISL_2003179, EPI_ISL_2003180, EPI_ISL_2003181, EPI_ISL_2003182, EPI_ISL_2003183, EPI_ISL_2003185, EPI_ISL_2003186, EPI_ISL_2003187, EPI_ISL_2003188, EPI_ISL_2003189, EPI_ISL_2003190, EPI_ISL_2003191, EPI_ISL_2003192, EPI_ISL_2003193, EPI_ISL_2003194, EPI_ISL_2003195 | | | |
| see above | Kansas Health and Environmental Lab | Kansas Health and Environmental Lab | Mike Grose, Jonathan Barnell, Ben Olsen, and Phil Adam |
| EPI_ISL_2003439, EPI_ISL_2003440, EPI_ISL_2003441, EPI_ISL_2003442, EPI_ISL_2003443, EPI_ISL_2003444, EPI_ISL_2003445, EPI_ISL_2003446, EPI_ISL_2003447, EPI_ISL_2003448, EPI_ISL_2003449, EPI_ISL_2003450, EPI_ISL_2003451, EPI_ISL_2003452, EPI_ISL_2003453, EPI_ISL_2003454, EPI_ISL_2003455, EPI_ISL_2003456, EPI_ISL_2003457, EPI_ISL_2003458, EPI_ISL_2003460, EPI_ISL_2003461, EPI_ISL_2003462, EPI_ISL_2003463, EPI_ISL_2003464, EPI_ISL_2003465, EPI_ISL_2003466, EPI_ISL_2003467, EPI_ISL_2003468, EPI_ISL_2003469, EPI_ISL_2003470, EPI_ISL_2003471, EPI_ISL_2003472, EPI_ISL_2003473, EPI_ISL_2003474, EPI_ISL_2003475, EPI_ISL_2003476, EPI_ISL_2003477, EPI_ISL_2003478, EPI_ISL_2003479, EPI_ISL_2003480, EPI_ISL_2003481, EPI_ISL_2003482, EPI_ISL_2003483, EPI_ISL_2003484, EPI_ISL_2003485, EPI_ISL_2003486, EPI_ISL_2003487, EPI_ISL_2003488, EPI_ISL_2003489, EPI_ISL_2003490, EPI_ISL_2003491, EPI_ISL_2003492, EPI_ISL_2003493, EPI_ISL_2003494, EPI_ISL_2003495, EPI_ISL_2003496, EPI_ISL_2003497 | | | |
| see above | Minnesota Department of Health, Public Health Laboratory | Minnesota Department of Health, Public Health Laboratory | Alexandra Lorentz, Jacob Garfin, Matt Plumb, and Xiong Wang |
| EPI_ISL_2003498, EPI_ISL_2003499, EPI_ISL_2003500, EPI_ISL_2003501, EPI_ISL_2003502, EPI_ISL_2003503, EPI_ISL_2003504, EPI_ISL_2003505, EPI_ISL_2003506, EPI_ISL_2003507, EPI_ISL_2003509, EPI_ISL_2003510, EPI_ISL_2003511, EPI_ISL_2003512, EPI_ISL_2003513, EPI_ISL_2003515, EPI_ISL_2003517, EPI_ISL_2003518, EPI_ISL_2003519, EPI_ISL_2003520, EPI_ISL_2003522, EPI_ISL_2003523, EPI_ISL_2003524, EPI_ISL_2003525, EPI_ISL_2003527, EPI_ISL_2003528, EPI_ISL_2003529, EPI_ISL_2003531, EPI_ISL_2003532, EPI_ISL_2003535, EPI_ISL_2003536, EPI_ISL_2003537, EPI_ISL_2003538, EPI_ISL_2003539, EPI_ISL_2003540, EPI_ISL_2003542, EPI_ISL_2003543, EPI_ISL_2003544, EPI_ISL_2003545, EPI_ISL_2003546, EPI_ISL_2003547, EPI_ISL_2003548, EPI_ISL_2003549, EPI_ISL_2003550, EPI_ISL_2003556, EPI_ISL_2003558, EPI_ISL_2003559, EPI_ISL_2003560, EPI_ISL_2003561, EPI_ISL_2003562, EPI_ISL_2003563, EPI_ISL_2003564, EPI_ISL_2003565, EPI_ISL_2003566, EPI_ISL_2003567, EPI_ISL_2003568, EPI_ISL_2003569, EPI_ISL_2003570, EPI_ISL_2003571, EPI_ISL_2003572, EPI_ISL_2003573, EPI_ISL_2003574, EPI_ISL_2003575, EPI_ISL_2003577, EPI_ISL_2003578, EPI_ISL_2003579, EPI_ISL_2003580, EPI_ISL_2003581, EPI_ISL_2003582, EPI_ISL_2003583, EPI_ISL_2003584, EPI_ISL_2003585, EPI_ISL_2003586, EPI_ISL_2003587, EPI_ISL_2003588, EPI_ISL_2003589, EPI_ISL_2003590, EPI_ISL_2003591, EPI_ISL_2003592, EPI_ISL_2003593, EPI_ISL_2003594, EPI_ISL_2003595, EPI_ISL_2003596, EPI_ISL_2003597, EPI_ISL_2003598, EPI_ISL_2003599, EPI_ISL_2003600, EPI_ISL_2003601, EPI_ISL_2003602, EPI_ISL_2003603, EPI_ISL_2003604, EPI_ISL_2003605, EPI_ISL_2003606, EPI_ISL_2003607, EPI_ISL_2003608, EPI_ISL_2003609, EPI_ISL_2003610, EPI_ISL_2003611, EPI_ISL_2003612, EPI_ISL_2003613, EPI_ISL_2003614, EPI_ISL_2003615, EPI_ISL_2003616, EPI_ISL_2003617, EPI_ISL_2003618, EPI_ISL_2003619, EPI_ISL_2003620 | | | |
| see above | Virginia Division of Consolidated Laboratory Services | Virginia Division of Consolidated Laboratory Services | Virginia DCLS |
| EPI_ISL_2003621, EPI_ISL_2003622, EPI_ISL_2003623, EPI_ISL_2003624, EPI_ISL_2003625, EPI_ISL_2003626, EPI_ISL_2003627, EPI_ISL_2003628, EPI_ISL_2003629, EPI_ISL_2003630, EPI_ISL_2003631, EPI_ISL_2003632, EPI_ISL_2003633, EPI_ISL_2003634, EPI_ISL_2003635, EPI_ISL_2003636, EPI_ISL_2003637, EPI_ISL_2003638, EPI_ISL_2003639, EPI_ISL_2003640, EPI_ISL_2003641, EPI_ISL_2003642, EPI_ISL_2003643, EPI_ISL_2003644, EPI_ISL_2003645, EPI_ISL_2003646, EPI_ISL_2003648, EPI_ISL_2003649, EPI_ISL_2003650, EPI_ISL_2003651, EPI_ISL_2003652, EPI_ISL_2003653, EPI_ISL_2003654 | | | |
| see above | University of Michigan Clinical Microbiology Laboratory | Lauring Lab, University of Michigan, Department of Microbiology and Immunology | Valesano |
| EPI_ISL_2003655, EPI_ISL_2003656, EPI_ISL_2003657, EPI_ISL_2003658, EPI_ISL_2003659, EPI_ISL_2003660, EPI_ISL_2003661, EPI_ISL_2003662, EPI_ISL_2003663, EPI_ISL_2003664, EPI_ISL_2003665, EPI_ISL_2003666, EPI_ISL_2003667 | | | |
| see above | Kansas Health and Environmental Lab | Kansas Health and Environmental Lab | Mike Grose, Jonathan Barnell, Ben Olsen, and Phil Adam |
| EPI_ISL_2003668, EPI_ISL_2003669, EPI_ISL_2003670, EPI_ISL_2003671, EPI_ISL_2003672, EPI_ISL_2003673, EPI_ISL_2003674, EPI_ISL_2003675, EPI_ISL_2003676, EPI_ISL_2003677, EPI_ISL_2003678, EPI_ISL_2003679, EPI_ISL_2003680, EPI_ISL_2003681, EPI_ISL_2003682, EPI_ISL_2003683, EPI_ISL_2003684, EPI_ISL_2003685, EPI_ISL_2003686, EPI_ISL_2003687, EPI_ISL_2003689, EPI_ISL_2003690, EPI_ISL_2003691, EPI_ISL_2003692, EPI_ISL_2003693, EPI_ISL_2003694, EPI_ISL_2003695, EPI_ISL_2003697, EPI_ISL_2003698, EPI_ISL_2003699, EPI_ISL_2003700, EPI_ISL_2003701, EPI_ISL_2003702, EPI_ISL_2003703, EPI_ISL_2003704, EPI_ISL_2003705 | | | |
| see above | Virginia Division of Consolidated Laboratory Services | Virginia Division of Consolidated Laboratory Services | Virginia DCLS |
| EPI_ISL_2003779, EPI_ISL_2003780, EPI_ISL_2003781, EPI_ISL_2003782, EPI_ISL_2003783, EPI_ISL_2003784, EPI_ISL_2003785, EPI_ISL_2003786, EPI_ISL_2003787, EPI_ISL_2003788, EPI_ISL_2003789, EPI_ISL_2003790, EPI_ISL_2003791, EPI_ISL_2003792, EPI_ISL_2003793, EPI_ISL_2003794, EPI_ISL_2003795, EPI_ISL_2003796, EPI_ISL_2003797, EPI_ISL_2003798, EPI_ISL_2003799 | | | |
| see above | Kansas Health and Environmental Lab | Kansas Health and Environmental Lab | Mike Grose, Jonathan Barnell, Ben Olsen, and Phil Adam |
| EPI_ISL_2003800, EPI_ISL_2003801, EPI_ISL_2003802, EPI_ISL_2003803, EPI_ISL_2003804, EPI_ISL_2003805, EPI_ISL_2003806, EPI_ISL_2003807, EPI_ISL_2003808, EPI_ISL_2003809, EPI_ISL_2003810, EPI_ISL_2003811, EPI_ISL_2003812, EPI_ISL_2003813, EPI_ISL_2003814, EPI_ISL_2003815, EPI_ISL_2003816, EPI_ISL_2003817, EPI_ISL_2003818, EPI_ISL_2003819, EPI_ISL_2003820, EPI_ISL_2003821, EPI_ISL_2003822, EPI_ISL_2003823, EPI_ISL_2003824, EPI_ISL_2003825, EPI_ISL_2003826, EPI_ISL_2003827, EPI_ISL_2003828, EPI_ISL_2003829, EPI_ISL_2003830, EPI_ISL_2003831, EPI_ISL_2003832, EPI_ISL_2003833, EPI_ISL_2003834, EPI_ISL_2003835, EPI_ISL_2003836, EPI_ISL_2003837, EPI_ISL_2003838, EPI_ISL_2003839, EPI_ISL_2003840, EPI_ISL_2003841, EPI_ISL_2003842, EPI_ISL_2003843, EPI_ISL_2003844, EPI_ISL_2003845, EPI_ISL_2003846, EPI_ISL_2003847, EPI_ISL_2003848, EPI_ISL_2003849, EPI_ISL_2003850, EPI_ISL_2003851, EPI_ISL_2003852, EPI_ISL_2003853, EPI_ISL_2003854, EPI_ISL_2003855, EPI_ISL_2003856, EPI_ISL_2003857, EPI_ISL_2003858, EPI_ISL_2003859, EPI_ISL_2003860, EPI_ISL_2003861, EPI_ISL_2003862, EPI_ISL_2003863, EPI_ISL_2003864, EPI_ISL_2003865, EPI_ISL_2003866, EPI_ISL_2003867, EPI_ISL_2003868, EPI_ISL_2003869, EPI_ISL_2003870, EPI_ISL_2003871, EPI_ISL_2003872, EPI_ISL_2003873, EPI_ISL_2003874, EPI_ISL_2003875, EPI_ISL_2003876, EPI_ISL_2003877, EPI_ISL_2003878, EPI_ISL_2003879, EPI_ISL_2003880, EPI_ISL_2003881, EPI_ISL_2003882, EPI_ISL_2003883, EPI_ISL_2003884, EPI_ISL_2003885, EPI_ISL_2003886, EPI_ISL_2003887, EPI_ISL_2003888, EPI_ISL_2003889, EPI_ISL_2003890, EPI_ISL_2003891, EPI_ISL_2003892, EPI_ISL_2003893, EPI_ISL_2003894, EPI_ISL_2003895, EPI_ISL_2003896, EPI_ISL_2003897, EPI_ISL_2003898, EPI_ISL_2003899, EPI_ISL_2003900, EPI_ISL_2003901, EPI_ISL_2003902, EPI_ISL_2003903, EPI_ISL_2003904, EPI_ISL_2003905, EPI_ISL_2003906, EPI_ISL_2003907, EPI_ISL_2003908, EPI_ISL_2003909, EPI_ISL_2003910, EPI_ISL_2003911, EPI_ISL_2003912, EPI_ISL_2003913, EPI_ISL_2003914, EPI_ISL_2003915, EPI_ISL_2003916, EPI_ISL_2003917, EPI_ISL_2003918, EPI_ISL_2003919, EPI_ISL_2003920, EPI_ISL_2003921, EPI_ISL_2003922, EPI_ISL_2003923, EPI_ISL_2003924, EPI_ISL_2003925, EPI_ISL_2003926, EPI_ISL_2003927, EPI_ISL_2003928, EPI_ISL_2003929, EPI_ISL_2003930, EPI_ISL_2003931, EPI_ISL_2003932, EPI_ISL_2003933, EPI_ISL_2003934, EPI_ISL_2003935, EPI_ISL_2003936, EPI_ISL_2003937, EPI_ISL_2003938, EPI_ISL_2003939, EPI_ISL_2003940, EPI_ISL_2003941, EPI_ISL_2003942, EPI_ISL_2003943, EPI_ISL_2003944, EPI_ISL_2003945, EPI_ISL_2003946, EPI_ISL_2003947, EPI_ISL_2003948, EPI_ISL_2003949, EPI_ISL_2003950, EPI_ISL_2003951, EPI_ISL_2003952, EPI_ISL_2003953, EPI_ISL_2003954, EPI_ISL_2003955, EPI_ISL_2003956, EPI_ISL_2003957, EPI_ISL_2003958, EPI_ISL_2003959, EPI_ISL_2003960, EPI_ISL_2003961, EPI_ISL_2003962, EPI_ISL_2003963, EPI_ISL_2003964, EPI_ISL_2003965, EPI_ISL_2003966, EPI_ISL_2003967, EPI_ISL_2003968, EPI_ISL_2003969, EPI_ISL_2003970, EPI_ISL_2003971, EPI_ISL_2003972, EPI_ISL_2003973, EPI_ISL_2003974, EPI_ISL_2003975, EPI_ISL_2003976, EPI_ISL_2003977, EPI_ISL_2003978, EPI_ISL_2003979 | | | |
| see above | Hospital of the University of Pennsylvania Molecular Pathology Lab | Bushman Lab - University of Pennsylvania | John Everett, Kyle Rodino, Shantan Reddy, Pascha Hokama, Aoife M. Roche, Young Hwang, Abigail Glascock, Scott Sherrill-Mix, Samantha A. Whiteside, Jevon Graham-Wooten, Layla A. Khatib, Ayannah S. Fitzgerald, Arupa Ganguly, Mike Feldman, Brendan Kelly, Ronald G. Collman and Frederic Bushman |
| EPI_ISL_2004079, EPI_ISL_2004080, EPI_ISL_2004081, EPI_ISL_2004082, EPI_ISL_2004083, EPI_ISL_2004084, EPI_ISL_2004085, EPI_ISL_2004086, EPI_ISL_2004087, | GA Department of Public Health | GA Department of Public Health | Stacy Reeves, Jonathan Edwards, Cynthia Dixey, Tonia Parrott, Aliyah Fields, Taylor Smith |

| | | | |
|---|---|---|---|
| EPI_ISL_2004088 | | | |
| EPI_ISL_2006723, EPI_ISL_2006724, EPI_ISL_2006725, EPI_ISL_2006726, EPI_ISL_2006727, EPI_ISL_2006728, EPI_ISL_2006729, EPI_ISL_2006730, EPI_ISL_2006731, EPI_ISL_2006732, EPI_ISL_2006733, EPI_ISL_2006734, EPI_ISL_2006735, EPI_ISL_2006736, EPI_ISL_2006737, EPI_ISL_2006738, EPI_ISL_2006739, EPI_ISL_2006740, EPI_ISL_2006741, EPI_ISL_2006742, EPI_ISL_2006743, EPI_ISL_2006744, EPI_ISL_2006745, EPI_ISL_2006746, EPI_ISL_2006747, EPI_ISL_2006748, EPI_ISL_2006749, EPI_ISL_2006750, EPI_ISL_2006751, EPI_ISL_2006752, EPI_ISL_2006753, EPI_ISL_2006754, EPI_ISL_2006755, EPI_ISL_2006756, EPI_ISL_2006757, EPI_ISL_2006758, EPI_ISL_2006759, EPI_ISL_2006760, EPI_ISL_2006761, EPI_ISL_2006762, EPI_ISL_2006763, EPI_ISL_2006764, EPI_ISL_2006765, EPI_ISL_2006766, EPI_ISL_2006767, EPI_ISL_2006768, EPI_ISL_2006769, EPI_ISL_2006770, EPI_ISL_2006771, EPI_ISL_2006772, EPI_ISL_2006773, EPI_ISL_2006774, EPI_ISL_2006775, EPI_ISL_2006776, EPI_ISL_2006777, EPI_ISL_2006778, EPI_ISL_2006779, EPI_ISL_2006780, EPI_ISL_2006781, EPI_ISL_2006782, EPI_ISL_2006783, EPI_ISL_2006784, EPI_ISL_2006785, EPI_ISL_2006786, EPI_ISL_2006787, EPI_ISL_2006788, EPI_ISL_2006789, EPI_ISL_2006790, EPI_ISL_2006791, EPI_ISL_2006792, EPI_ISL_2006793, EPI_ISL_2006794, EPI_ISL_2006796, EPI_ISL_2006797 | | | |
| see above | Moderna Inc. | Moderna Inc. | Yamuna Paila; Groves Dixon; Rolando Pajon |
| EPI_ISL_2006872, EPI_ISL_2006873, EPI_ISL_2006874, EPI_ISL_2006875, EPI_ISL_2006876, EPI_ISL_2006879, EPI_ISL_2006880, EPI_ISL_2006881, EPI_ISL_2006882, EPI_ISL_2006883, EPI_ISL_2006884, EPI_ISL_2006885, EPI_ISL_2006886, EPI_ISL_2006887, EPI_ISL_2006888, EPI_ISL_2006889, EPI_ISL_2006890, EPI_ISL_2006891, EPI_ISL_2006892, EPI_ISL_2006893, EPI_ISL_2006894, EPI_ISL_2006895, EPI_ISL_2006898, EPI_ISL_2006899 | | | |
| see above | South Dakota Public Health Laboratory | South Dakota Public Health Laboratory | Jacob Garfin and Chris Carlson |
| EPI_ISL_2006900, EPI_ISL_2006901, EPI_ISL_2006902, EPI_ISL_2006903, EPI_ISL_2006904, EPI_ISL_2006905, EPI_ISL_2006906, EPI_ISL_2006907, EPI_ISL_2006908, EPI_ISL_2006909, EPI_ISL_2006910, EPI_ISL_2006911, EPI_ISL_2006912, EPI_ISL_2006913, EPI_ISL_2006914, EPI_ISL_2006915, EPI_ISL_2006916, EPI_ISL_2006917, EPI_ISL_2006918, EPI_ISL_2006919, EPI_ISL_2006920, EPI_ISL_2006921, EPI_ISL_2006922, EPI_ISL_2006923, EPI_ISL_2006924 | | | |
| see above | Montana Public Health Laboratory | Montana Public Health Laboratory | Joy Ritter, Michelle Mozer, Carrie Biskupiak, Deborah Gibson |
| EPI_ISL_2006928, EPI_ISL_2006929, EPI_ISL_2006930, EPI_ISL_2006931, EPI_ISL_2006932, EPI_ISL_2006933, EPI_ISL_2006934, EPI_ISL_2006935, EPI_ISL_2006936, EPI_ISL_2006937, EPI_ISL_2006938, EPI_ISL_2006939, EPI_ISL_2006940, EPI_ISL_2006941, EPI_ISL_2006942, EPI_ISL_2006943, EPI_ISL_2006944, EPI_ISL_2006945, EPI_ISL_2006946, EPI_ISL_2006947, EPI_ISL_2006948, EPI_ISL_2006949, EPI_ISL_2006950, EPI_ISL_2006951, EPI_ISL_2006952, EPI_ISL_2006953, EPI_ISL_2006954, EPI_ISL_2006955, EPI_ISL_2006956, EPI_ISL_2006957, EPI_ISL_2006958, EPI_ISL_2006959, EPI_ISL_2006960, EPI_ISL_2006961, EPI_ISL_2006963, EPI_ISL_2006964, EPI_ISL_2006965, EPI_ISL_2006966, EPI_ISL_2006968, EPI_ISL_2006969, EPI_ISL_2006970, EPI_ISL_2006973, EPI_ISL_2006975, EPI_ISL_2006976, EPI_ISL_2006978, EPI_ISL_2006979, EPI_ISL_2006980, EPI_ISL_2006981, EPI_ISL_2006982, EPI_ISL_2006983, EPI_ISL_2006984, EPI_ISL_2006985, EPI_ISL_2006986, EPI_ISL_2006987, EPI_ISL_2006989, EPI_ISL_2006990, EPI_ISL_2006991, EPI_ISL_2006992, EPI_ISL_2006993, EPI_ISL_2006994, EPI_ISL_2006995, EPI_ISL_2006996, EPI_ISL_2006997, EPI_ISL_2006998, EPI_ISL_2006999, EPI_ISL_2007000, EPI_ISL_2007001, EPI_ISL_2007002, EPI_ISL_2007003, EPI_ISL_2007004, EPI_ISL_2007005, EPI_ISL_2007006, EPI_ISL_2007008, EPI_ISL_2007009 | | | |
| see above | The Ohio State University Applied Microbiology Services Laboratory | The Ohio State University Applied Microbiology Services Laboratory | Seth A. Faith PhD |
| EPI_ISL_2007088, EPI_ISL_2007089, EPI_ISL_2007090, EPI_ISL_2007091, EPI_ISL_2007092, EPI_ISL_2007093, EPI_ISL_2007094, EPI_ISL_2007095, EPI_ISL_2007096, EPI_ISL_2007097, EPI_ISL_2007098, EPI_ISL_2007099, EPI_ISL_2007100, EPI_ISL_2007101, EPI_ISL_2007102, EPI_ISL_2007103, EPI_ISL_2007105, EPI_ISL_2007119, EPI_ISL_2007120, EPI_ISL_2007121, EPI_ISL_2007122, EPI_ISL_2007123, EPI_ISL_2007124, EPI_ISL_2007125, EPI_ISL_2007126, EPI_ISL_2007127, EPI_ISL_2007128, EPI_ISL_2007129, EPI_ISL_2007130, EPI_ISL_2007131, EPI_ISL_2007132, EPI_ISL_2007133, EPI_ISL_2007134, EPI_ISL_2007135, EPI_ISL_2007136, EPI_ISL_2007137, EPI_ISL_2007138, EPI_ISL_2007139, EPI_ISL_2007140, EPI_ISL_2007141, EPI_ISL_2007142, EPI_ISL_2007143, EPI_ISL_2007144, EPI_ISL_2007145, EPI_ISL_2007146, EPI_ISL_2007147 | | | |
| see above | Kansas Health and Environmental Lab | Kansas Health and Environmental Lab | Mike Grose, Jonathan Barnell, Ben Olsen, and Phil Adam |
| EPI_ISL_2007148, EPI_ISL_2007149 | Rami Kantor lab | Rami Kantor lab | Josephine Darpolor, Vlad Novitsky, Mark Howison, Richard Huard, Ewa King, Rami Kantor |
| EPI_ISL_2007150, EPI_ISL_2007151, EPI_ISL_2007152, EPI_ISL_2007153, EPI_ISL_2007154, EPI_ISL_2007155, EPI_ISL_2007156, EPI_ISL_2007157, EPI_ISL_2007158, EPI_ISL_2007159, EPI_ISL_2007160, EPI_ISL_2007173, EPI_ISL_2007174, EPI_ISL_2007175, EPI_ISL_2007176, EPI_ISL_2007177, EPI_ISL_2007178, EPI_ISL_2007179, EPI_ISL_2007180, EPI_ISL_2007181, EPI_ISL_2007182, EPI_ISL_2007183, EPI_ISL_2007184, EPI_ISL_2007185, EPI_ISL_2007186, EPI_ISL_2007187, EPI_ISL_2007188, EPI_ISL_2007189, EPI_ISL_2007190, EPI_ISL_2007191, EPI_ISL_2007192, EPI_ISL_2007193 | | | |
| see above | Arizona State University | Arizona State University | Peter T. Skidmore, LaRinda A. Holland, Matthew F. Smith, Rabia Maqsood, Nicholas J. Mellor, Joy M. Blain, Valerie Harris, Joshua LaBaer, Vel Murugan, Efrem S. Lim |
| EPI_ISL_2007459, EPI_ISL_2007460 | Minnesota Department of Health, Public Health Laboratory | Minnesota Department of Health, Public Health Laboratory | Alexandra Lorentz, Jacob Garfin, Matt Plumb, and Xiong Wang |
| EPI_ISL_2007461, EPI_ISL_2007462, EPI_ISL_2007463, EPI_ISL_2007464, EPI_ISL_2007465, EPI_ISL_2007466, EPI_ISL_2007467, EPI_ISL_2007468, EPI_ISL_2007469, EPI_ISL_2007470 | University of Michigan Clinical Microbiology Laboratory | Lauring Lab, University of Michigan, Department of Microbiology and Immunology | Valesano |
| EPI_ISL_2007785, EPI_ISL_2007788, EPI_ISL_2007794, EPI_ISL_2007795, EPI_ISL_2007796, EPI_ISL_2007799 | Wyoming Public Health Laboratory | Wyoming Public Health Laboratory | Jim Mildenberger, Taylor Fearing, Channing Weber, Ashley Norberg, Chayse Rowley, Marley Goetz, Brian Dominguez, Elliot Thomasson, Brittany Oher, Sam Britz, Cari Sloma, and Rob Christensen |
| EPI_ISL_2007801, EPI_ISL_2007802, EPI_ISL_2007803, EPI_ISL_2007804, EPI_ISL_2007805, EPI_ISL_2007806, EPI_ISL_2007807, EPI_ISL_2007808, EPI_ISL_2007809, EPI_ISL_2007810, EPI_ISL_2007811, EPI_ISL_2007812, EPI_ISL_2007813, EPI_ISL_2007814, EPI_ISL_2007815, EPI_ISL_2007816, EPI_ISL_2007817, EPI_ISL_2007818, EPI_ISL_2007819, EPI_ISL_2007820, EPI_ISL_2007821, EPI_ISL_2007822, EPI_ISL_2007823, EPI_ISL_2007824, EPI_ISL_2007829, EPI_ISL_2007830, EPI_ISL_2007831, EPI_ISL_2007832, EPI_ISL_2007833, EPI_ISL_2007834 | | | |
| see above | University of Oregon COVID-19 MAP Laboratory | University of Oregon Genomics and Cell Characterization Core Facility (GC3F) | Douglas Turnbull, Ariana White, Jeff Bishop, Jason Carriere, Jason Sydes, Peter Batzel |
| EPI_ISL_2007835, EPI_ISL_2007836, EPI_ISL_2007837, EPI_ISL_2007838, EPI_ISL_2007839, EPI_ISL_2007840, EPI_ISL_2007841, EPI_ISL_2007842, EPI_ISL_2007843, EPI_ISL_2007844, EPI_ISL_2007845, EPI_ISL_2007846, EPI_ISL_2007847, EPI_ISL_2007848, EPI_ISL_2007849, EPI_ISL_2007850, EPI_ISL_2007851, EPI_ISL_2007852, EPI_ISL_2007853, EPI_ISL_2007854, EPI_ISL_2007855, EPI_ISL_2007856, EPI_ISL_2007857, EPI_ISL_2007859, EPI_ISL_2007860, EPI_ISL_2007861, EPI_ISL_2007862, EPI_ISL_2007864, EPI_ISL_2007865, EPI_ISL_2007866, EPI_ISL_2007867, EPI_ISL_2007868, EPI_ISL_2007869, EPI_ISL_2007870, EPI_ISL_2007871, EPI_ISL_2007872, EPI_ISL_2007873, EPI_ISL_2007874, EPI_ISL_2007875, EPI_ISL_2007876, EPI_ISL_2007877, EPI_ISL_2007878, EPI_ISL_2007880, EPI_ISL_2007881, EPI_ISL_2007882, EPI_ISL_2007883, EPI_ISL_2007884, EPI_ISL_2007885, EPI_ISL_2007886, EPI_ISL_2007887, EPI_ISL_2007888, EPI_ISL_2007889, EPI_ISL_2007890, EPI_ISL_2007891, EPI_ISL_2007892, EPI_ISL_2007893, EPI_ISL_2007895, EPI_ISL_2007896, EPI_ISL_2007897, EPI_ISL_2007899, EPI_ISL_2007900, EPI_ISL_2007901, EPI_ISL_2007902, EPI_ISL_2007903, EPI_ISL_2007904, EPI_ISL_2007905, EPI_ISL_2007906, EPI_ISL_2007907, EPI_ISL_2007908, EPI_ISL_2007909, EPI_ISL_2007910, EPI_ISL_2007911, EPI_ISL_2007912, EPI_ISL_2007913, EPI_ISL_2007914, EPI_ISL_2007915, EPI_ISL_2007916, EPI_ISL_2007917, EPI_ISL_2007918, EPI_ISL_2007919, EPI_ISL_2007920, EPI_ISL_2007921, EPI_ISL_2007922, EPI_ISL_2007923, EPI_ISL_2007925, EPI_ISL_2007926, EPI_ISL_2007927, EPI_ISL_2007928, EPI_ISL_2007929, EPI_ISL_2007930, EPI_ISL_2007931, EPI_ISL_2007932, EPI_ISL_2007933, EPI_ISL_2007934, EPI_ISL_2007935, EPI_ISL_2007936, EPI_ISL_2007937, EPI_ISL_2007940, EPI_ISL_2007941, EPI_ISL_2007942, EPI_ISL_2007943, EPI_ISL_2007944, EPI_ISL_2007945, EPI_ISL_2007946, EPI_ISL_2007947, EPI_ISL_2007948, EPI_ISL_2007949, EPI_ISL_2007951, EPI_ISL_2007953, EPI_ISL_2007954, EPI_ISL_2007955, EPI_ISL_2007956, EPI_ISL_2007957, EPI_ISL_2007958, EPI_ISL_2007959, EPI_ISL_2007960, EPI_ISL_2007961, EPI_ISL_2007962, EPI_ISL_2007963, EPI_ISL_2007965, EPI_ISL_2007966, EPI_ISL_2007967, EPI_ISL_2007968, EPI_ISL_2007969, EPI_ISL_2007970, EPI_ISL_2007971, EPI_ISL_2007972, EPI_ISL_2007973, EPI_ISL_2007974, EPI_ISL_2007975, EPI_ISL_2007976, EPI_ISL_2007977, EPI_ISL_2007978, EPI_ISL_2007979, EPI_ISL_2007980, EPI_ISL_2007981, EPI_ISL_2007982, EPI_ISL_2007983, EPI_ISL_2007984, EPI_ISL_2007985, EPI_ISL_2007986, EPI_ISL_2007987, EPI_ISL_2007988, EPI_ISL_2007989, EPI_ISL_2007990, EPI_ISL_2007991, EPI_ISL_2007992, EPI_ISL_2007993, EPI_ISL_2007994, EPI_ISL_2007995, EPI_ISL_2007996, EPI_ISL_2007997, EPI_ISL_2007998, EPI_ISL_2007999, EPI_ISL_2008000, EPI_ISL_2008001, EPI_ISL_2008002, EPI_ISL_2008003, EPI_ISL_2008004, EPI_ISL_2008005, EPI_ISL_2008006, EPI_ISL_2008007, EPI_ISL_2008008, EPI_ISL_2008009, EPI_ISL_2008010, EPI_ISL_2008011, EPI_ISL_2008012, EPI_ISL_2008013, EPI_ISL_2008014, EPI_ISL_2008015, EPI_ISL_2008016, EPI_ISL_2008017, EPI_ISL_2008018, EPI_ISL_2008019, EPI_ISL_2008020, EPI_ISL_2008021, EPI_ISL_2008022, EPI_ISL_2008023, EPI_ISL_2008024, EPI_ISL_2008025, EPI_ISL_2008027, EPI_ISL_2008028, EPI_ISL_2008029, EPI_ISL_2008030, EPI_ISL_2008031, EPI_ISL_2008032, EPI_ISL_2008033, EPI_ISL_2008034, EPI_ISL_2008035, EPI_ISL_2008036 | | | |
| see above | State Laboratories Division, Hawaii State Department of Health | State Laboratories Division, Hawaii State Department of Health | Pamela O'Brien, Drew Kuwazaki, Ayana Garnet, Razvan Sultana, Edward Desmond |
| EPI_ISL_2008038, EPI_ISL_2008039, EPI_ISL_2008040, EPI_ISL_2008041, EPI_ISL_2008042, EPI_ISL_2008043, EPI_ISL_2008044, EPI_ISL_2008045, EPI_ISL_2008046, EPI_ISL_2008047, EPI_ISL_2008048, EPI_ISL_2008049, EPI_ISL_2008050, EPI_ISL_2008051, EPI_ISL_2008052, EPI_ISL_2008053, EPI_ISL_2008054, EPI_ISL_2008055, EPI_ISL_2008056, EPI_ISL_2008057, EPI_ISL_2008058, EPI_ISL_2008059, EPI_ISL_2008060, EPI_ISL_2008061, EPI_ISL_2008062, EPI_ISL_2008063, EPI_ISL_2008064, EPI_ISL_2008065, EPI_ISL_2008066, EPI_ISL_2008067, EPI_ISL_2008068, EPI_ISL_2008069, EPI_ISL_2008070, EPI_ISL_2008071, EPI_ISL_2008072, EPI_ISL_2008073, EPI_ISL_2008074, EPI_ISL_2008075, EPI_ISL_2008076, EPI_ISL_2008077, EPI_ISL_2008078, EPI_ISL_2008079, EPI_ISL_2008080, EPI_ISL_2008081, EPI_ISL_2008082, EPI_ISL_2008083, EPI_ISL_2008084, EPI_ISL_2008085, EPI_ISL_2008086, EPI_ISL_2008087, EPI_ISL_2008088, EPI_ISL_2008089, EPI_ISL_2008090, EPI_ISL_2008091, EPI_ISL_2008092, EPI_ISL_2008093, EPI_ISL_2008094, EPI_ISL_2008095, EPI_ISL_2008096, EPI_ISL_2008097, EPI_ISL_2008098, EPI_ISL_2008099, EPI_ISL_2008100, EPI_ISL_2008101, EPI_ISL_2008102, EPI_ISL_2008103, EPI_ISL_2008104, EPI_ISL_2008105, EPI_ISL_2008106, EPI_ISL_2008107, EPI_ISL_2008108, EPI_ISL_2008109, EPI_ISL_2008110, EPI_ISL_2008111, EPI_ISL_2008112, EPI_ISL_2008113, EPI_ISL_2008114, EPI_ISL_2008115, EPI_ISL_2008116 | | | |
| see above | Hackensack Medical Center | New York Genome Center | Michael Zody, Andre Corvelo, Dayna M. Oschwald, Samantha Fennessey, Tom Maniatis, Liang Chen, Jose Mediavilla, Marcus Cunningham, Kaelea Composto, Kar Chow, David Perlin, Barry Kreiswirth |
| EPI_ISL_2008117, EPI_ISL_2008118, EPI_ISL_2008119, EPI_ISL_2008120, EPI_ISL_2008121, EPI_ISL_2008122, EPI_ISL_2008123, EPI_ISL_2008124, EPI_ISL_2008125, EPI_ISL_2008126, EPI_ISL_2008127, EPI_ISL_2008128, EPI_ISL_2008129, EPI_ISL_2008130, EPI_ISL_2008131, EPI_ISL_2008132, EPI_ISL_2008133, EPI_ISL_2008134, EPI_ISL_2008135, EPI_ISL_2008136, EPI_ISL_2008137, EPI_ISL_2008138, EPI_ISL_2008139, EPI_ISL_2008140, EPI_ISL_2008141, EPI_ISL_2008142, EPI_ISL_2008143, EPI_ISL_2008144, EPI_ISL_2008145, EPI_ISL_2008146, EPI_ISL_2008147, EPI_ISL_2008148, EPI_ISL_2008149, EPI_ISL_2008150, EPI_ISL_2008151, EPI_ISL_2008152, EPI_ISL_2008153, EPI_ISL_2008154, EPI_ISL_2008155, EPI_ISL_2008156, EPI_ISL_2008157, EPI_ISL_2008158, EPI_ISL_2008159, EPI_ISL_2008160, EPI_ISL_2008161, EPI_ISL_2008162, EPI_ISL_2008163, EPI_ISL_2008164, EPI_ISL_2008165, EPI_ISL_2008166, EPI_ISL_2008167, EPI_ISL_2008168, EPI_ISL_2008169, EPI_ISL_2008170, EPI_ISL_2008171, EPI_ISL_2008172, EPI_ISL_2008173, EPI_ISL_2008174, EPI_ISL_2008175, EPI_ISL_2008176, EPI_ISL_2008177, EPI_ISL_2008178, EPI_ISL_2008179, EPI_ISL_2008180, EPI_ISL_2008181, EPI_ISL_2008182, EPI_ISL_2008183, EPI_ISL_2008184, EPI_ISL_2008185, EPI_ISL_2008186, EPI_ISL_2008187, EPI_ISL_2008188, EPI_ISL_2008189, EPI_ISL_2008190, EPI_ISL_2008191, EPI_ISL_2008192, EPI_ISL_2008193, EPI_ISL_2008194, EPI_ISL_2008195, EPI_ISL_2008196, EPI_ISL_2008197, EPI_ISL_2008198, EPI_ISL_2008199, EPI_ISL_2008200, EPI_ISL_2008201, EPI_ISL_2008202, EPI_ISL_2008203, EPI_ISL_2008204, EPI_ISL_2008205, EPI_ISL_2008206, EPI_ISL_2008207, EPI_ISL_2008208, EPI_ISL_2008209, EPI_ISL_2008210, EPI_ISL_2008211, EPI_ISL_2008212, EPI_ISL_2008213, EPI_ISL_2008214, EPI_ISL_2008215, EPI_ISL_2008216, EPI_ISL_2008217, EPI_ISL_2008218, EPI_ISL_2008219, EPI_ISL_2008220, EPI_ISL_2008221, EPI_ISL_2008222, EPI_ISL_2008223, EPI_ISL_2008224, EPI_ISL_2008225, EPI_ISL_2008226, EPI_ISL_2008227, EPI_ISL_2008228, EPI_ISL_2008229, EPI_ISL_2008230, EPI_ISL_2008231, EPI_ISL_2008232, EPI_ISL_2008233, EPI_ISL_2008234, EPI_ISL_2008235, EPI_ISL_2008236, EPI_ISL_2008237, EPI_ISL_2008238, EPI_ISL_2008239, EPI_ISL_2008240, EPI_ISL_2008241, EPI_ISL_2008242, EPI_ISL_2008243, EPI_ISL_2008244, EPI_ISL_2008245, EPI_ISL_2008246, EPI_ISL_2008247, EPI_ISL_2008248, EPI_ISL_2008249, EPI_ISL_2008250, EPI_ISL_2008251, EPI_ISL_2008252, EPI_ISL_2008253, EPI_ISL_2008254, EPI_ISL_2008255, EPI_ISL_2008256, EPI_ISL_2008257, EPI_ISL_2008258, EPI_ISL_2008259, EPI_ISL_2008260, EPI_ISL_2008261, EPI_ISL_2008262, EPI_ISL_2008263, EPI_ISL_2008264, EPI_ISL_2008265, EPI_ISL_2008266, EPI_ISL_2008267, EPI_ISL_2008268, EPI_ISL_2008269, EPI_ISL_2008270, EPI_ISL_2008271, EPI_ISL_2008272, EPI_ISL_2008273, EPI_ISL_2008274, EPI_ISL_2008275, EPI_ISL_2008276, EPI_ISL_2008277, EPI_ISL_2008278, EPI_ISL_2008279, EPI_ISL_2008280, EPI_ISL_2008281, EPI_ISL_2008282, EPI_ISL_2008283, EPI_ISL_2008284, EPI_ISL_2008285, EPI_ISL_2008286, EPI_ISL_2008287, EPI_ISL_2008288, EPI_ISL_2008289, EPI_ISL_2008290, EPI_ISL_2008291, EPI_ISL_2008292, EPI_ISL_2008293, EPI_ISL_2008294, EPI_ISL_2008295, EPI_ISL_2008296, EPI_ISL_2008297, EPI_ISL_2008298, EPI_ISL_2008299, EPI_ISL_2008300, EPI_ISL_2008301, EPI_ISL_2008302, EPI_ISL_2008303, EPI_ISL_2008304, EPI_ISL_2008305, EPI_ISL_2008306, EPI_ISL_2008307, EPI_ISL_2008308, EPI_ISL_2008309, EPI_ISL_2008310, EPI_ISL_2008311, EPI_ISL_2008312, EPI_ISL_2008313, EPI_ISL_2008314, EPI_ISL_2008335, EPI_ISL_2008336, EPI_ISL_2008337, EPI_ISL_2008338, EPI_ISL_2008339, EPI_ISL_2008340, EPI_ISL_2008341, EPI_ISL_2008342, EPI_ISL_2008343, EPI_ISL_2008344, EPI_ISL_2008345, EPI_ISL_2008346, EPI_ISL_2008347, EPI_ISL_2008348, EPI_ISL_2008349, EPI_ISL_2008350, EPI_ISL_2008351, EPI_ISL_2008352, EPI_ISL_2008353, EPI_ISL_2008354, EPI_ISL_2008355, EPI_ISL_2008356, EPI_ISL_2008357, EPI_ISL_2008358, EPI_ISL_2008359, EPI_ISL_2008360, EPI_ISL_2008361, EPI_ISL_2008362, EPI_ISL_2008363, EPI_ISL_2008364, EPI_ISL_2008365, EPI_ISL_2008366, EPI_ISL_2008367, EPI_ISL_2008368, EPI_ISL_2008369, EPI_ISL_2008370, EPI_ISL_2008371, EPI_ISL_2008372, EPI_ISL_2008373, EPI_ISL_2008374, EPI_ISL_2008375, EPI_ISL_2008376, EPI_ISL_2008377, EPI_ISL_2008378, EPI_ISL_2008379, EPI_ISL_2008380, EPI_ISL_2008381, EPI_ISL_2008382, EPI_ISL_2008383, EPI_ISL_2008384, EPI_ISL_2008385, EPI_ISL_2008386, EPI_ISL_2008387, EPI_ISL_2008388, EPI_ISL_2008389, EPI_ISL_2008390, EPI_ISL_2008391, EPI_ISL_2008392, EPI_ISL_2008393, EPI_ISL_2008394, EPI_ISL_2008395, EPI_ISL_2008396, EPI_ISL_2008397, EPI_ISL_2008398, EPI_ISL_2008399, EPI_ISL_2008400 | | | |
| see above | Massachusetts State Public Health Laboratory | Massachusetts State Public Health Laboratory | Andrew Lang, Timelia Fink, Glen Gallagher, Sandra Smole |
| EPI_ISL_2008401, EPI_ISL_2008402, EPI_ISL_2008403, EPI_ISL_2008404, EPI_ISL_2008405, EPI_ISL_2008406, EPI_ISL_2008407, EPI_ISL_2008408, EPI_ISL_2008409, EPI_ISL_2008410, EPI_ISL_2008411, EPI_ISL_2008412, EPI_ISL_2008413, EPI_ISL_2008414, EPI_ISL_2008415, EPI_ISL_2008416, EPI_ISL_2008417, EPI_ISL_2008418, EPI_ISL_2008419, EPI_ISL_2008420, EPI_ISL_2008421, EPI_ISL_2008422, EPI_ISL_2008423, EPI_ISL_2008424, EPI_ISL_2008425, EPI_ISL_2008426, EPI_ISL_2008427, EPI_ISL_2008428, EPI_ISL_2008429, EPI_ISL_2008430, EPI_ISL_2008431, EPI_ISL_2008432, EPI_ISL_2008433, EPI_ISL_2008434, EPI_ISL_2008435, EPI_ISL_2008436, EPI_ISL_2008437, EPI_ISL_2008438, EPI_ISL_2008439, EPI_ISL_2008440, EPI_ISL_2008441, EPI_ISL_2008443, EPI_ISL_2008444, EPI_ISL_2008445, EPI_ISL_2008446, EPI_ISL_2008447, EPI_ISL_2008449, EPI_ISL_2008450, EPI_ISL_2008451, EPI_ISL_2008452, EPI_ISL_2008454, EPI_ISL_2008455, EPI_ISL_2008456, EPI_ISL_2008457, EPI_ISL_2008458, EPI_ISL_2008459, EPI_ISL_2008460, EPI_ISL_2008462, EPI_ISL_2008463, EPI_ISL_2008465, EPI_ISL_2008466, EPI_ISL_2008467, EPI_ISL_2008468, EPI_ISL_2008469, EPI_ISL_2008470, EPI_ISL_2008471, EPI_ISL_2008472, EPI_ISL_2008474, EPI_ISL_2008475, EPI_ISL_2008476, EPI_ISL_2008477, EPI_ISL_2008478, EPI_ISL_2008479, EPI_ISL_2008480, EPI_ISL_2008481, EPI_ISL_2008482, EPI_ISL_2008483, EPI_ISL_2008484, EPI_ISL_2008485, EPI_ISL_2008486, EPI_ISL_2008487, EPI_ISL_2008488, EPI_ISL_2008489, EPI_ISL_2008490, EPI_ISL_2008491, EPI_ISL_2008492, EPI_ISL_2008493, EPI_ISL_2008495, EPI_ISL_2008496, EPI_ISL_2008497, EPI_ISL_2008498, EPI_ISL_2008499, EPI_ISL_2008500, EPI_ISL_2008501, EPI_ISL_2008502, EPI_ISL_2008504, EPI_ISL_2008505, EPI_ISL_2008506, EPI_ISL_2008507, EPI_ISL_2008508, EPI_ISL_2008509, EPI_ISL_2008510, EPI_ISL_2008511, EPI_ISL_2008512, EPI_ISL_2008513, EPI_ISL_2008514, EPI_ISL_2008515, EPI_ISL_2008516, EPI_ISL_2008517, EPI_ISL_2008518, EPI_ISL_2008519, EPI_ISL_2008520, EPI_ISL_2008521, EPI_ISL_2008522, EPI_ISL_2008523, EPI_ISL_2008524, EPI_ISL_2008525, EPI_ISL_2008526, EPI_ISL_2008527, EPI_ISL_2008528, EPI_ISL_2008529, EPI_ISL_2008530, EPI_ISL_2008531, EPI_ISL_2008532, EPI_ISL_2008533, EPI_ISL_2008534, EPI_ISL_2008535, EPI_ISL_2008536, EPI_ISL_2008537, EPI_ISL_2008538, EPI_ISL_2008539, EPI_ISL_2008540, EPI_ISL_2008541, EPI_ISL_2008542, EPI_ISL_2008543, EPI_ISL_2008544, EPI_ISL_2008545, EPI_ISL_2008546, EPI_ISL_2008548, EPI_ISL_2008549, EPI_ISL_2008550, EPI_ISL_2008551, EPI_ISL_2008553, EPI_ISL_2008554, EPI_ISL_2008558, EPI_ISL_2008559, EPI_ISL_2008560, EPI_ISL_2008561, EPI_ISL_2008562, EPI_ISL_2008563, EPI_ISL_2008564, EPI_ISL_2008565, EPI_ISL_2008566, EPI_ISL_2008568, EPI_ISL_2008569, EPI_ISL_2008570, EPI_ISL_2008571, EPI_ISL_2008572, EPI_ISL_2008574, EPI_ISL_2008575, EPI_ISL_2008576, EPI_ISL_2008577, EPI_ISL_2008578, EPI_ISL_2008579, EPI_ISL_2008580, EPI_ISL_2008581, EPI_ISL_2008582, EPI_ISL_2008583, EPI_ISL_2008584, EPI_ISL_2008585, EPI_ISL_2008587, EPI_ISL_2008588, EPI_ISL_2008589, EPI_ISL_2008590, EPI_ISL_2008591, EPI_ISL_2008592, EPI_ISL_2008593, EPI_ISL_2008594, EPI_ISL_2008595, EPI_ISL_2008596, EPI_ISL_2008597, EPI_ISL_2008598, EPI_ISL_2008599, EPI_ISL_2008600, EPI_ISL_2008601, EPI_ISL_2008602, EPI_ISL_2008603, EPI_ISL_2008604, EPI_ISL_2008605, EPI_ISL_2008606, EPI_ISL_2008607, EPI_ISL_2008608, EPI_ISL_2008609, EPI_ISL_2008610, EPI_ISL_2008611, EPI_ISL_2008612, EPI_ISL_2008613, EPI_ISL_2008614, EPI_ISL_2008615, EPI_ISL_2008616, EPI_ISL_2008617, EPI_ISL_2008618, EPI_ISL_2008619, EPI_ISL_2008620, EPI_ISL_2008621, EPI_ISL_2008622, EPI_ISL_2008624, EPI_ISL_2008625, EPI_ISL_2008626, EPI_ISL_2008627, EPI_ISL_2008628, EPI_ISL_2008629, EPI_ISL_2008630, EPI_ISL_2008631, EPI_ISL_2008632, EPI_ISL_2008633, EPI_ISL_2008634, EPI_ISL_2008635, EPI_ISL_2008636, EPI_ISL_2008637, EPI_ISL_2008639, EPI_ISL_2008641, EPI_ISL_2008642, EPI_ISL_2008643, EPI_ISL_2008644, EPI_ISL_2008645, EPI_ISL_2008646, EPI_ISL_2008647, EPI_ISL_2008648, EPI_ISL_2008649, EPI_ISL_2008651, EPI_ISL_2008652, EPI_ISL_2008653, EPI_ISL_2008654, EPI_ISL_2008655, EPI_ISL_2008656, EPI_ISL_2008657, EPI_ISL_2008658, EPI_ISL_2008659, EPI_ISL_2008660, EPI_ISL_2008661, EPI_ISL_2008662, EPI_ISL_2008663, EPI_ISL_2008664, EPI_ISL_2008665, EPI_ISL_2008666, EPI_ISL_2008667, EPI_ISL_2008668, EPI_ISL_2008669, EPI_ISL_2008670, EPI_ISL_2008671, EPI_ISL_2008672, EPI_ISL_2008673, EPI_ISL_2008674, EPI_ISL_2008675, EPI_ISL_2008676, EPI_ISL_2008677, EPI_ISL_2008678, EPI_ISL_2008680, EPI_ISL_2008681, EPI_ISL_2008682, EPI_ISL_2008684, EPI_ISL_2008685, EPI_ISL_2008686, EPI_ISL_2008687, EPI_ISL_2008688, EPI_ISL_2008689, EPI_ISL_2008691, EPI_ISL_2008692, EPI_ISL_2008693, EPI_ISL_2008694, EPI_ISL_2008695, EPI_ISL_2008696, EPI_ISL_2008697, EPI_ISL_2008698, EPI_ISL_2008699, EPI_ISL_2008700, EPI_ISL_2008701, EPI_ISL_2008702, EPI_ISL_2008703, EPI_ISL_2008705, EPI_ISL_2008706, EPI_ISL_2008707, EPI_ISL_2008708, EPI_ISL_2008709, EPI_ISL_2008711, EPI_ISL_2008712, EPI_ISL_2008713, EPI_ISL_2008714, EPI_ISL_2008715, EPI_ISL_2008716, EPI_ISL_2008717, EPI_ISL_2008718, EPI_ISL_2008719, EPI_ISL_2008720, EPI_ISL_2008722, EPI_ISL_2008723, EPI_ISL_2008724, EPI_ISL_2008725, EPI_ISL_2008726, EPI_ISL_2008727, EPI_ISL_2008728, EPI_ISL_2008729, EPI_ISL_2008730, EPI_ISL_2008732, EPI_ISL_2008733, EPI_ISL_2008734, EPI_ISL_2008735, EPI_ISL_2008736, EPI_ISL_2008738, EPI_ISL_2008739, EPI_ISL_2008740, EPI_ISL_2008741, EPI_ISL_2008742, EPI_ISL_2008743, EPI_ISL_2008744, EPI_ISL_2008745, EPI_ISL_2008746, EPI_ISL_2008747, EPI_ISL_2008749, EPI_ISL_2008750, EPI_ISL_2008752, EPI_ISL_2008754, EPI_ISL_2008755, EPI_ISL_2008756, EPI_ISL_2008757, EPI_ISL_2008759, EPI_ISL_2008761, EPI_ISL_2008762, EPI_ISL_2008763, EPI_ISL_2008764, EPI_ISL_2008765, EPI_ISL_2008767, EPI_ISL_2008768, EPI_ISL_2008770, EPI_ISL_2008771, EPI_ISL_2008772, EPI_ISL_2008773, EPI_ISL_2008776, EPI_ISL_2008778, EPI_ISL_2008779, EPI_ISL_2008780, EPI_ISL_2008781, EPI_ISL_2008783, EPI_ISL_2008784, EPI_ISL_2008785, EPI_ISL_2008787, EPI_ISL_2008788, EPI_ISL_2008789, EPI_ISL_2008791, EPI_ISL_2008792, EPI_ISL_2008793, EPI_ISL_2008795, EPI_ISL_2008796, EPI_ISL_2008797, EPI_ISL_2008798, EPI_ISL_2008799, EPI_ISL_2008800, EPI_ISL_2008801, EPI_ISL_2008802, EPI_ISL_2008803, EPI_ISL_2008804, EPI_ISL_2008805, EPI_ISL_2008806, EPI_ISL_2008808, EPI_ISL_2008812, EPI_ISL_2008813, EPI_ISL_2008815, EPI_ISL_2008817, EPI_ISL_2008818, EPI_ISL_2008819, EPI_ISL_2008820, EPI_ISL_2008821, EPI_ISL_2008822, EPI_ISL_2008823, EPI_ISL_2008824, EPI_ISL_2008825, EPI_ISL_2008826, EPI_ISL_2008827, EPI_ISL_2008828, EPI_ISL_2008829, EPI_ISL_2008830, EPI_ISL_2008831, EPI_ISL_2008832, EPI_ISL_2008834, EPI_ISL_2008835, EPI_ISL_2008836, EPI_ISL_2008837, EPI_ISL_2008838, EPI_ISL_2008839, EPI_ISL_2008840, EPI_ISL_2008841, EPI_ISL_2008842, EPI_ISL_2008843, EPI_ISL_2008844, EPI_ISL_2008845, EPI_ISL_2008847, EPI_ISL_2008848, EPI_ISL_2008849, EPI_ISL_2008850, EPI_ISL_2008851, EPI_ISL_2008852, EPI_ISL_2008853, EPI_ISL_2008854, EPI_ISL_2008855, EPI_ISL_2008856, EPI_ISL_2008857, EPI_ISL_2008858, EPI_ISL_2008859, EPI_ISL_2008860, EPI_ISL_2008861, EPI_ISL_2008862, EPI_ISL_2008863, EPI_ISL_2008864, EPI_ISL_2008865, EPI_ISL_2008866, EPI_ISL_2008867, EPI_ISL_2008868, EPI_ISL_2008869, EPI_ISL_2008870, EPI_ISL_2008871, EPI_ISL_2008872, EPI_ISL_2008873, EPI_ISL_2008875, EPI_ISL_2008876, EPI_ISL_2008877, EPI_ISL_2008878, EPI_ISL_2008879, EPI_ISL_2008880, EPI_ISL_2008881, EPI_ISL_2008882, EPI_ISL_2008883, EPI_ISL_2008884, EPI_ISL_2008885, EPI_ISL_2008886, EPI_ISL_2008887, EPI_ISL_2008889, EPI_ISL_2008890, EPI_ISL_2008891, EPI_ISL_2008892, | | | |

| | | | |
|---|---|---|---|
| EPI_ISL_2008893, EPI_ISL_2008895, EPI_ISL_2008896, EPI_ISL_2008897, EPI_ISL_2008898, EPI_ISL_2008899, EPI_ISL_2008900, EPI_ISL_2008901, EPI_ISL_2008902, EPI_ISL_2008903, EPI_ISL_2008904, EPI_ISL_2008905 | | | |
| see above | Florida Bureau of Public Health Laboratories | Florida Bureau of Public Health Laboratories | Sarah Schmedes, Jason Blanton |
| EPI_ISL_2008970, EPI_ISL_2008971, EPI_ISL_2008972, EPI_ISL_2008973, EPI_ISL_2008974, EPI_ISL_2008975, EPI_ISL_2008976, EPI_ISL_2008977 | Wyoming Public Health Laboratory | Wyoming Public Health Laboratory | Jim Mildenberger, Taylor Fearing, Channing Weber, Ashley Norberg, Chayse Rowley, Marley Goetz, Brian Dominguez, Elliot Thomasson, Brittany Oher, Sam Britz, Cari Sloma, and Rob Christensen |
| EPI_ISL_2008978, EPI_ISL_2008979, EPI_ISL_2008980, EPI_ISL_2008981, EPI_ISL_2008982, EPI_ISL_2008983, EPI_ISL_2008984, EPI_ISL_2008985, EPI_ISL_2008986, EPI_ISL_2008987, EPI_ISL_2008988, EPI_ISL_2008989, EPI_ISL_2008990, EPI_ISL_2008991, EPI_ISL_2008992, EPI_ISL_2008993, EPI_ISL_2008994, EPI_ISL_2008995, EPI_ISL_2008996, EPI_ISL_2008997, EPI_ISL_2008998, EPI_ISL_2008999, EPI_ISL_2009000, EPI_ISL_2009001, EPI_ISL_2009002, EPI_ISL_2009003, EPI_ISL_2009004, EPI_ISL_2009005, EPI_ISL_2009006, EPI_ISL_2009007, EPI_ISL_2009008, EPI_ISL_2009009, EPI_ISL_2009010, EPI_ISL_2009011, EPI_ISL_2009012, EPI_ISL_2009013, EPI_ISL_2009014 | | | |
| see above | Alaska State Virology Laboratory | Alaska State Virology Laboratory | Stephanie DeRonde, Elva House, Jacob Zidek, Lisa Smith, Ph.D., Jack Chen, Ph.D. |
| EPI_ISL_2009015, EPI_ISL_2009016, EPI_ISL_2009017, EPI_ISL_2009018, EPI_ISL_2009019, EPI_ISL_2009020, EPI_ISL_2009021, EPI_ISL_2009022, EPI_ISL_2009023, EPI_ISL_2009024, EPI_ISL_2009025, EPI_ISL_2009026, EPI_ISL_2009027, EPI_ISL_2009028, EPI_ISL_2009029 | | | |
| see above | Maryland Genomics, Institute for Genome Sciences, University of Maryland School of Medicine | Maryland Genomics, Institute for Genome Sciences, University of Maryland School of Medicine | Tallon, Luke J; Sadzewicz, Lisa D; Humphrys, Mike; Ott, Sandra; Roussey, Holly; Mehta, Aditya; Vavikolanu, Kranthi; Fraser, Claire M; Ravel, Jacques |
| EPI_ISL_2009037 | Wyoming Public Health Laboratory | Wyoming Public Health Laboratory | Jim Mildenberger, Taylor Fearing, Channing Weber, Ashley Norberg, Chayse Rowley, Marley Goetz, Brian Dominguez, Elliot Thomasson, Brittany Oher, Sam Britz, Cari Sloma, and Rob Christensen |
| EPI_ISL_2009049 | Stanford Health Care | Stanford University School of Medicine, Clinical Virology Laboratory | Fumiko Yamamoto, Malaya K. Sahoo, Mamdouh Sibai, Daniel Solis, Michelle Verghese, ChunHong Huang, James Zehnder, and Benjamin A. Pinsky |
| EPI_ISL_2009050, EPI_ISL_2009051, EPI_ISL_2009052 | Stanford Health Care | Stanford University School of Medicine, Clinical Virology Laboratory | Michelle Verghese, ChunHong Huang, Mamdouh Sibai, Daniel Solis, Fumiko Yamamoto, Malaya K. Sahoo, James Zehnder, and Benjamin A. Pinsky |
| EPI_ISL_2009053, EPI_ISL_2009054, EPI_ISL_2009055, EPI_ISL_2009056, EPI_ISL_2009057 | Stanford Health Care | Stanford University School of Medicine, Clinical Virology Laboratory | Mamdouh Sibai, Daniel Solis, Fumiko Yamamoto, Malaya K. Sahoo, Michelle Verghese, ChunHong Huang, James Zehnder, and Benjamin A. Pinsky |
| EPI_ISL_2009070, EPI_ISL_2009071 | Stanford Health Care | Stanford University School of Medicine, Clinical Virology Laboratory | Michelle Verghese, ChunHong Huang, Mamdouh Sibai, Daniel Solis, Fumiko Yamamoto, Malaya K. Sahoo, James Zehnder, and Benjamin A. Pinsky |
| EPI_ISL_2009073, EPI_ISL_2009074, EPI_ISL_2009075, EPI_ISL_2009076, EPI_ISL_2009077, EPI_ISL_2009078, EPI_ISL_2009079, EPI_ISL_2009080, EPI_ISL_2009081, EPI_ISL_2009082, EPI_ISL_2009083, EPI_ISL_2009088, EPI_ISL_2009089, EPI_ISL_2009090, EPI_ISL_2009091, EPI_ISL_2009092, EPI_ISL_2009093, EPI_ISL_2009094, EPI_ISL_2009095, EPI_ISL_2009099, EPI_ISL_2009100, EPI_ISL_2009101, EPI_ISL_2009102, EPI_ISL_2009103, EPI_ISL_2009104, EPI_ISL_2009105, EPI_ISL_2009106, EPI_ISL_2009107, EPI_ISL_2009108, EPI_ISL_2009109, EPI_ISL_2009110, EPI_ISL_2009111, EPI_ISL_2009112, EPI_ISL_2009113, EPI_ISL_2009114, EPI_ISL_2009115 | | | |
| see above | Wyoming Public Health Laboratory | Wyoming Public Health Laboratory | Jim Mildenberger, Taylor Fearing, Channing Weber, Ashley Norberg, Chayse Rowley, Marley Goetz, Brian Dominguez, Elliot Thomasson, Brittany Oher, Sam Britz, Cari Sloma, and Rob Christensen |
| EPI_ISL_2009117, EPI_ISL_2009118, EPI_ISL_2009119, EPI_ISL_2009120, EPI_ISL_2009121, EPI_ISL_2009122, EPI_ISL_2009123, EPI_ISL_2009124, EPI_ISL_2009125, EPI_ISL_2009126, EPI_ISL_2009127, EPI_ISL_2009128, EPI_ISL_2009129, EPI_ISL_2009130, EPI_ISL_2009131, EPI_ISL_2009132, EPI_ISL_2009133, EPI_ISL_2009134, EPI_ISL_2009135, EPI_ISL_2009136, EPI_ISL_2009137, EPI_ISL_2009138, EPI_ISL_2009139, EPI_ISL_2009140, EPI_ISL_2009141, EPI_ISL_2009142, EPI_ISL_2009143, EPI_ISL_2009165, EPI_ISL_2009166, EPI_ISL_2009167, EPI_ISL_2009168, EPI_ISL_2009169, EPI_ISL_2009170, EPI_ISL_2009171, EPI_ISL_2009172, EPI_ISL_2009173, EPI_ISL_2009174, EPI_ISL_2009175, EPI_ISL_2009176, EPI_ISL_2009177, EPI_ISL_2009178, EPI_ISL_2009179, EPI_ISL_2009180, EPI_ISL_2009181, EPI_ISL_2009182, EPI_ISL_2009183, EPI_ISL_2009184, EPI_ISL_2009185 | | | |
| see above | MSK Microbiology Lab | MSK Microbiology Lab | Esther Babady, Tracy McMillen, Krupa Jani |
| EPI_ISL_2009186, EPI_ISL_2009187, EPI_ISL_2009188, EPI_ISL_2009189, EPI_ISL_2009190, EPI_ISL_2009191, EPI_ISL_2009192 | LSUHS Emerging Viral Threat Laboratory | LSUHS Emerging Viral Threat Laboratory | Gregory L. Ware, Alexander Mijalis, Jeremy P. Kamil, Jennifer L. Carroll, Maarten Van Diest, Rona S. Scott, Andrew D. Yurochko, Christopher G. Kevil, John A. Vanchiere |
| EPI_ISL_2009306 | Northwestern Memorial Hospital | Northwestern University - Ozer Lab | Ramon Lorenzo-Redondo, Lacy M. Simons, Taylor J. Dean, Chad J. Achenbach, Lawrence J. Jennings, Chao Qi, Michael G. Ison, Judd F. Hultquist, Egon A. Ozer |
| EPI_ISL_2009307 | Lurie Children's Hospital of Chicago | Northwestern University - Ozer Lab | Ramon Lorenzo-Redondo, Lacy M. Simons, Taylor J. Dean, Michael G. Ison, Xiaotian, Zheng, William J. Muller, Larry K. Kociolek, Judd F. Hultquist, Egon A. Ozer |
| EPI_ISL_2009308, EPI_ISL_2009309, EPI_ISL_2009310, EPI_ISL_2009311, EPI_ISL_2009312, EPI_ISL_2009313, EPI_ISL_2009314, EPI_ISL_2009315, EPI_ISL_2009316, EPI_ISL_2009317, EPI_ISL_2009318, EPI_ISL_2009319, EPI_ISL_2009320, EPI_ISL_2009321, EPI_ISL_2009322, EPI_ISL_2009323, EPI_ISL_2009324, EPI_ISL_2009325, EPI_ISL_2009326, EPI_ISL_2009327, EPI_ISL_2009328, EPI_ISL_2009329, EPI_ISL_2009330, EPI_ISL_2009331, EPI_ISL_2009332, EPI_ISL_2009333, EPI_ISL_2009334, EPI_ISL_2009335, EPI_ISL_2009336, EPI_ISL_2009337, EPI_ISL_2009338, EPI_ISL_2009339, EPI_ISL_2009340, EPI_ISL_2009341, EPI_ISL_2009342, EPI_ISL_2009343, EPI_ISL_2009344, EPI_ISL_2009345, EPI_ISL_2009346, EPI_ISL_2009347, EPI_ISL_2009348, EPI_ISL_2009349, EPI_ISL_2009350, EPI_ISL_2009351, EPI_ISL_2009352, EPI_ISL_2009353, EPI_ISL_2009354, EPI_ISL_2009355, EPI_ISL_2009356, EPI_ISL_2009357, EPI_ISL_2009358, EPI_ISL_2009359, EPI_ISL_2009360, EPI_ISL_2009361, EPI_ISL_2009362, EPI_ISL_2009363, EPI_ISL_2009364, EPI_ISL_2009365, EPI_ISL_2009366, EPI_ISL_2009367 | | | |
| see above | Northwestern Memorial Hospital | Northwestern University - Ozer Lab | Ramon Lorenzo-Redondo, Lacy M. Simons, Taylor J. Dean, Chad J. Achenbach, Lawrence J. Jennings, Chao Qi, Michael G. Ison, Judd F. Hultquist, Egon A. Ozer |
| EPI_ISL_2009368, EPI_ISL_2009369, EPI_ISL_2009370, EPI_ISL_2009371, EPI_ISL_2009372, EPI_ISL_2009373, EPI_ISL_2009374, EPI_ISL_2009375, EPI_ISL_2009376, EPI_ISL_2009377, EPI_ISL_2009378, EPI_ISL_2009379, EPI_ISL_2009380, EPI_ISL_2009382, EPI_ISL_2009383, EPI_ISL_2009384, EPI_ISL_2009385, EPI_ISL_2009386, EPI_ISL_2009387, EPI_ISL_2009388, EPI_ISL_2009389, EPI_ISL_2009390, EPI_ISL_2009391, EPI_ISL_2009392, EPI_ISL_2009393, EPI_ISL_2009394, EPI_ISL_2009395, EPI_ISL_2009396, EPI_ISL_2009397, EPI_ISL_2009398, EPI_ISL_2009399, EPI_ISL_2009400, EPI_ISL_2009401 | | | |
| see above | Lurie Children's Hospital of Chicago | Northwestern University - Ozer Lab | Ramon Lorenzo-Redondo, Lacy M. Simons, Taylor J. Dean, Michael G. Ison, Xiaotian, Zheng, William J. Muller, Larry K. Kociolek, Judd F. Hultquist, Egon A. Ozer |
| EPI_ISL_2009402, EPI_ISL_2009403, EPI_ISL_2009404, EPI_ISL_2009405 | Moderna Inc. | Moderna Inc. | Yamuna Paila; Groves Dixon; Rolando Pajon |
| EPI_ISL_2009406 | Lurie Children's Hospital of Chicago | Northwestern University - Ozer Lab | Ramon Lorenzo-Redondo, Lacy M. Simons, Taylor J. Dean, Michael G. Ison, Xiaotian, Zheng, William J. Muller, Larry K. Kociolek, Judd F. Hultquist, Egon A. Ozer |
| EPI_ISL_2009407, EPI_ISL_2009408 | Moderna Inc. | Moderna Inc. | Yamuna Paila; Groves Dixon; Rolando Pajon |
| EPI_ISL_2009409 | Lurie Children's Hospital of Chicago | Northwestern University - Ozer Lab | Ramon Lorenzo-Redondo, Lacy M. Simons, Taylor J. Dean, Michael G. Ison, Xiaotian, Zheng, William J. Muller, Larry K. Kociolek, Judd F. Hultquist, Egon A. Ozer |
| EPI_ISL_2009410, EPI_ISL_2009411, EPI_ISL_2009412, EPI_ISL_2009413, EPI_ISL_2009414, EPI_ISL_2009415, EPI_ISL_2009416, EPI_ISL_2009417, EPI_ISL_2009418 | Moderna Inc. | Moderna Inc. | Yamuna Paila; Groves Dixon; Rolando Pajon |
| EPI_ISL_2009419 | Lurie Children's Hospital of Chicago | Northwestern University - Ozer Lab | Ramon Lorenzo-Redondo, Lacy M. Simons, Taylor J. Dean, Michael G. Ison, Xiaotian, Zheng, William J. Muller, Larry K. Kociolek, Judd F. Hultquist, Egon A. Ozer |
| EPI_ISL_2009420, EPI_ISL_2009421, EPI_ISL_2009422 | Moderna Inc. | Moderna Inc. | Yamuna Paila; Groves Dixon; Rolando Pajon |
| EPI_ISL_2009423 | Lurie Children's Hospital of Chicago | Northwestern University - Ozer Lab | Ramon Lorenzo-Redondo, Lacy M. Simons, Taylor J. Dean, Michael G. Ison, Xiaotian, Zheng, William J. Muller, Larry K. Kociolek, Judd F. Hultquist, Egon A. Ozer |
| EPI_ISL_2009424, EPI_ISL_2009425 | Moderna Inc. | Moderna Inc. | Yamuna Paila; Groves Dixon; Rolando Pajon |
| EPI_ISL_2009426 | Lurie Children's Hospital of Chicago | Northwestern University - Ozer Lab | Ramon Lorenzo-Redondo, Lacy M. Simons, Taylor J. Dean, Michael G. Ison, Xiaotian, Zheng, William J. Muller, Larry K. Kociolek, Judd F. Hultquist, Egon A. Ozer |
| EPI_ISL_2009427, EPI_ISL_2009428, EPI_ISL_2009429, EPI_ISL_2009430, EPI_ISL_2009431, EPI_ISL_2009432, EPI_ISL_2009433, EPI_ISL_2009434 | Moderna Inc. | Moderna Inc. | Yamuna Paila; Groves Dixon; Rolando Pajon |
| EPI_ISL_2009435 | Lurie Children's Hospital of Chicago | Northwestern University - Ozer Lab | Ramon Lorenzo-Redondo, Lacy M. Simons, Taylor J. Dean, Michael G. Ison, Xiaotian, Zheng, William J. Muller, Larry K. Kociolek, Judd F. Hultquist, Egon A. Ozer |
| EPI_ISL_2009436, EPI_ISL_2009437, EPI_ISL_2009438, EPI_ISL_2009439, EPI_ISL_2009440, EPI_ISL_2009441, EPI_ISL_2009442, | Moderna Inc. | Moderna Inc. | Yamuna Paila; Groves Dixon; Rolando Pajon |

| | | | |
|---|---|---|---|
| EPI_ISL_2009443,<br>EPI_ISL_2009444 | | | |
| EPI_ISL_2009445 | Lurie Children's Hospital of Chicago | Northwestern University - Ozer Lab | Ramon Lorenzo-Redondo, Lacy M. Simons, Taylor J. Dean, Michael G. Ison, Xiaotian, Zheng, William J. Muller, Larry K. Kociolek, Judd F. Hultquist, Egon A. Ozer |
| EPI_ISL_2009446,<br>EPI_ISL_2009447,<br>EPI_ISL_2009448 | Moderna Inc. | Moderna Inc. | Yamuna Paila; Groves Dixon; Rolando Pajon |
| EPI_ISL_2009449 | Lurie Children's Hospital of Chicago | Northwestern University - Ozer Lab | Ramon Lorenzo-Redondo, Lacy M. Simons, Taylor J. Dean, Michael G. Ison, Xiaotian, Zheng, William J. Muller, Larry K. Kociolek, Judd F. Hultquist, Egon A. Ozer |
| EPI_ISL_2009450,<br>EPI_ISL_2009451,<br>EPI_ISL_2009452,<br>EPI_ISL_2009453,<br>EPI_ISL_2009454,<br>EPI_ISL_2009455,<br>EPI_ISL_2009456,<br>EPI_ISL_2009457 | Moderna Inc. | Moderna Inc. | Yamuna Paila; Groves Dixon; Rolando Pajon |
| EPI_ISL_2009458 | Lurie Children's Hospital of Chicago | Northwestern University - Ozer Lab | Ramon Lorenzo-Redondo, Lacy M. Simons, Taylor J. Dean, Michael G. Ison, Xiaotian, Zheng, William J. Muller, Larry K. Kociolek, Judd F. Hultquist, Egon A. Ozer |
| EPI_ISL_2009459,<br>EPI_ISL_2009460,<br>EPI_ISL_2009461,<br>EPI_ISL_2009462 | Moderna Inc. | Moderna Inc. | Yamuna Paila; Groves Dixon; Rolando Pajon |
| EPI_ISL_2009463 | Lurie Children's Hospital of Chicago | Northwestern University - Ozer Lab | Ramon Lorenzo-Redondo, Lacy M. Simons, Taylor J. Dean, Michael G. Ison, Xiaotian, Zheng, William J. Muller, Larry K. Kociolek, Judd F. Hultquist, Egon A. Ozer |
| EPI_ISL_2009464,<br>EPI_ISL_2009465,<br>EPI_ISL_2009466,<br>EPI_ISL_2009467,<br>EPI_ISL_2009468,<br>EPI_ISL_2009469,<br>EPI_ISL_2009470 | Moderna Inc. | Moderna Inc. | Yamuna Paila; Groves Dixon; Rolando Pajon |
| EPI_ISL_2009471 | Lurie Children's Hospital of Chicago | Northwestern University - Ozer Lab | Ramon Lorenzo-Redondo, Lacy M. Simons, Taylor J. Dean, Michael G. Ison, Xiaotian, Zheng, William J. Muller, Larry K. Kociolek, Judd F. Hultquist, Egon A. Ozer |
| EPI_ISL_2009472,<br>EPI_ISL_2009473,<br>EPI_ISL_2009474,<br>EPI_ISL_2009475,<br>EPI_ISL_2009476,<br>EPI_ISL_2009477,<br>EPI_ISL_2009478,<br>EPI_ISL_2009479 | Moderna Inc. | Moderna Inc. | Yamuna Paila; Groves Dixon; Rolando Pajon |
| EPI_ISL_2009480 | Lurie Children's Hospital of Chicago | Northwestern University - Ozer Lab | Ramon Lorenzo-Redondo, Lacy M. Simons, Taylor J. Dean, Michael G. Ison, Xiaotian, Zheng, William J. Muller, Larry K. Kociolek, Judd F. Hultquist, Egon A. Ozer |
| EPI_ISL_2009481,<br>EPI_ISL_2009482,<br>EPI_ISL_2009483 | Moderna Inc. | Moderna Inc. | Yamuna Paila; Groves Dixon; Rolando Pajon |
| EPI_ISL_2009484 | Lurie Children's Hospital of Chicago | Northwestern University - Ozer Lab | Ramon Lorenzo-Redondo, Lacy M. Simons, Taylor J. Dean, Michael G. Ison, Xiaotian, Zheng, William J. Muller, Larry K. Kociolek, Judd F. Hultquist, Egon A. Ozer |
| EPI_ISL_2009485,<br>EPI_ISL_2009486,<br>EPI_ISL_2009487 | Moderna Inc. | Moderna Inc. | Yamuna Paila; Groves Dixon; Rolando Pajon |
| EPI_ISL_2009488 | Lurie Children's Hospital of Chicago | Northwestern University - Ozer Lab | Ramon Lorenzo-Redondo, Lacy M. Simons, Taylor J. Dean, Michael G. Ison, Xiaotian, Zheng, William J. Muller, Larry K. Kociolek, Judd F. Hultquist, Egon A. Ozer |
| EPI_ISL_2009489,<br>EPI_ISL_2009490 | Moderna Inc. | Moderna Inc. | Yamuna Paila; Groves Dixon; Rolando Pajon |
| EPI_ISL_2009491 | Lurie Children's Hospital of Chicago | Northwestern University - Ozer Lab | Ramon Lorenzo-Redondo, Lacy M. Simons, Taylor J. Dean, Michael G. Ison, Xiaotian, Zheng, William J. Muller, Larry K. Kociolek, Judd F. Hultquist, Egon A. Ozer |
| EPI_ISL_2009492,<br>EPI_ISL_2009493 | Moderna Inc. | Moderna Inc. | Yamuna Paila; Groves Dixon; Rolando Pajon |
| EPI_ISL_2009494 | Lurie Children's Hospital of Chicago | Northwestern University - Ozer Lab | Ramon Lorenzo-Redondo, Lacy M. Simons, Taylor J. Dean, Michael G. Ison, Xiaotian, Zheng, William J. Muller, Larry K. Kociolek, Judd F. Hultquist, Egon A. Ozer |
| EPI_ISL_2009495,<br>EPI_ISL_2009496,<br>EPI_ISL_2009497,<br>EPI_ISL_2009498,<br>EPI_ISL_2009499,<br>EPI_ISL_2009500,<br>EPI_ISL_2009501,<br>EPI_ISL_2009502,<br>EPI_ISL_2009503 | Moderna Inc. | Moderna Inc. | Yamuna Paila; Groves Dixon; Rolando Pajon |
| EPI_ISL_2009504 | Lurie Children's Hospital of Chicago | Northwestern University - Ozer Lab | Ramon Lorenzo-Redondo, Lacy M. Simons, Taylor J. Dean, Michael G. Ison, Xiaotian, Zheng, William J. Muller, Larry K. Kociolek, Judd F. Hultquist, Egon A. Ozer |
| EPI_ISL_2009505,<br>EPI_ISL_2009506 | Moderna Inc. | Moderna Inc. | Yamuna Paila; Groves Dixon; Rolando Pajon |
| EPI_ISL_2009507, EPI_ISL_2009508, EPI_ISL_2009509, EPI_ISL_2009510, EPI_ISL_2009511, EPI_ISL_2009512, EPI_ISL_2009513, EPI_ISL_2009514, EPI_ISL_2009515, EPI_ISL_2009516, EPI_ISL_2009517, EPI_ISL_2009518, EPI_ISL_2009519, EPI_ISL_2009520, EPI_ISL_2009521, EPI_ISL_2009522, EPI_ISL_2009523, EPI_ISL_2009524, EPI_ISL_2009525, EPI_ISL_2009526, EPI_ISL_2009527, EPI_ISL_2009528 | | | |
| see above | Lurie Children's Hospital of Chicago | Northwestern University - Ozer Lab | Ramon Lorenzo-Redondo, Lacy M. Simons, Taylor J. Dean, Michael G. Ison, Xiaotian, Zheng, William J. Muller, Larry K. Kociolek, Judd F. Hultquist, Egon A. Ozer |
| EPI_ISL_2009665, EPI_ISL_2009666, EPI_ISL_2009667, EPI_ISL_2009668, EPI_ISL_2009669, EPI_ISL_2009670, EPI_ISL_2009671, EPI_ISL_2009672, EPI_ISL_2009673, EPI_ISL_2009674, EPI_ISL_2009675, EPI_ISL_2009676, EPI_ISL_2009677, EPI_ISL_2009678, EPI_ISL_2009680, EPI_ISL_2009681, EPI_ISL_2009682, EPI_ISL_2009683, EPI_ISL_2009684, EPI_ISL_2009685, EPI_ISL_2009686, EPI_ISL_2009687,<br>EPI_ISL_2009688, EPI_ISL_2009689, EPI_ISL_2009690, EPI_ISL_2009691, EPI_ISL_2009692, EPI_ISL_2009693, EPI_ISL_2009694, EPI_ISL_2009695, EPI_ISL_2009696, EPI_ISL_2009697, EPI_ISL_2009698, EPI_ISL_2009699, EPI_ISL_2009700, EPI_ISL_2009701, EPI_ISL_2009702, EPI_ISL_2009703, EPI_ISL_2009704, EPI_ISL_2009705, EPI_ISL_2009706, EPI_ISL_2009707, EPI_ISL_2009708, EPI_ISL_2009709,<br>EPI_ISL_2009710, EPI_ISL_2009711, EPI_ISL_2009712, EPI_ISL_2009713, EPI_ISL_2009714, EPI_ISL_2009715, EPI_ISL_2009716, EPI_ISL_2009717, EPI_ISL_2009718, EPI_ISL_2009719, EPI_ISL_2009720, EPI_ISL_2009721, EPI_ISL_2009722, EPI_ISL_2009723, EPI_ISL_2009724, EPI_ISL_2009725, EPI_ISL_2009726, EPI_ISL_2009727, EPI_ISL_2009728, EPI_ISL_2009729, EPI_ISL_2009730, EPI_ISL_2009731,<br>EPI_ISL_2009732, EPI_ISL_2009733, EPI_ISL_2009734, EPI_ISL_2009735, EPI_ISL_2009736, EPI_ISL_2009737, EPI_ISL_2009738, EPI_ISL_2009739, EPI_ISL_2009740, EPI_ISL_2009741, EPI_ISL_2009742, EPI_ISL_2009743, EPI_ISL_2009744, EPI_ISL_2009745, EPI_ISL_2009746, EPI_ISL_2009747, EPI_ISL_2009748, EPI_ISL_2009749, EPI_ISL_2009750, EPI_ISL_2009751, EPI_ISL_2009752, EPI_ISL_2009753,<br>EPI_ISL_2009754, EPI_ISL_2009755, EPI_ISL_2009756, EPI_ISL_2009757, EPI_ISL_2009758, EPI_ISL_2009759, EPI_ISL_2009760, EPI_ISL_2009761, EPI_ISL_2009762, EPI_ISL_2009763, EPI_ISL_2009764, EPI_ISL_2009765, EPI_ISL_2009766, EPI_ISL_2009767, EPI_ISL_2009768, EPI_ISL_2009769, EPI_ISL_2009770, EPI_ISL_2009771, EPI_ISL_2009772, EPI_ISL_2009773, EPI_ISL_2009774, EPI_ISL_2009775,<br>EPI_ISL_2009776, EPI_ISL_2009777, EPI_ISL_2009778, EPI_ISL_2009779, EPI_ISL_2009780, EPI_ISL_2009781, EPI_ISL_2009782, EPI_ISL_2009783, EPI_ISL_2009784, EPI_ISL_2009785, EPI_ISL_2009786, EPI_ISL_2009787, EPI_ISL_2009788, EPI_ISL_2009789, EPI_ISL_2009790, EPI_ISL_2009791, EPI_ISL_2009792, EPI_ISL_2009793, EPI_ISL_2009794, EPI_ISL_2009795, EPI_ISL_2009796, EPI_ISL_2009797,<br>EPI_ISL_2009798, EPI_ISL_2009799, EPI_ISL_2009800, EPI_ISL_2009801, EPI_ISL_2009802, EPI_ISL_2009803, EPI_ISL_2009804, EPI_ISL_2009805, EPI_ISL_2009806, EPI_ISL_2009807, EPI_ISL_2009808, EPI_ISL_2009809, EPI_ISL_2009810, EPI_ISL_2009811, EPI_ISL_2009812, EPI_ISL_2009813, EPI_ISL_2009814, EPI_ISL_2009815, EPI_ISL_2009816, EPI_ISL_2009817, EPI_ISL_2009818, EPI_ISL_2009819,<br>EPI_ISL_2009820, EPI_ISL_2009821, EPI_ISL_2009822, EPI_ISL_2009823, EPI_ISL_2009824, EPI_ISL_2009825, EPI_ISL_2009826, EPI_ISL_2009827, EPI_ISL_2009828, EPI_ISL_2009829, EPI_ISL_2009830, EPI_ISL_2009831, EPI_ISL_2009832, EPI_ISL_2009833, EPI_ISL_2009834, EPI_ISL_2009835, EPI_ISL_2009836, EPI_ISL_2009837, EPI_ISL_2009838, EPI_ISL_2009839, EPI_ISL_2009840, EPI_ISL_2009841,<br>EPI_ISL_2009842, EPI_ISL_2009843, EPI_ISL_2009844, EPI_ISL_2009845, EPI_ISL_2009846, EPI_ISL_2009847, EPI_ISL_2009848, EPI_ISL_2009849, EPI_ISL_2009850, EPI_ISL_2009851, EPI_ISL_2009852, EPI_ISL_2009853, EPI_ISL_2009854, EPI_ISL_2009855, EPI_ISL_2009856, EPI_ISL_2009857, EPI_ISL_2009858, EPI_ISL_2009859, EPI_ISL_2009860, EPI_ISL_2009861, EPI_ISL_2009862, EPI_ISL_2009863,<br>EPI_ISL_2009864, EPI_ISL_2009865, EPI_ISL_2009866, EPI_ISL_2009867, EPI_ISL_2009868, EPI_ISL_2009869, EPI_ISL_2009870, EPI_ISL_2009871, EPI_ISL_2009872, EPI_ISL_2009873, EPI_ISL_2009874, EPI_ISL_2009875, EPI_ISL_2009876, EPI_ISL_2009877, EPI_ISL_2009878, EPI_ISL_2009879, EPI_ISL_2009880, EPI_ISL_2009881, EPI_ISL_2009882, EPI_ISL_2009883, EPI_ISL_2009884, EPI_ISL_2009885,<br>EPI_ISL_2009886, EPI_ISL_2009887, EPI_ISL_2009888, EPI_ISL_2009889, EPI_ISL_2009890, EPI_ISL_2009891, EPI_ISL_2009892, EPI_ISL_2009893, EPI_ISL_2009894, EPI_ISL_2009895, EPI_ISL_2009896, EPI_ISL_2009897, EPI_ISL_2009898, EPI_ISL_2009899, EPI_ISL_2009900, EPI_ISL_2009901, EPI_ISL_2009902, EPI_ISL_2009903, EPI_ISL_2009904, EPI_ISL_2009905, EPI_ISL_2009906, EPI_ISL_2009907,<br>EPI_ISL_2009908, EPI_ISL_2009909, EPI_ISL_2009910, EPI_ISL_2009911, EPI_ISL_2009912, EPI_ISL_2009913, EPI_ISL_2009914, EPI_ISL_2009915, EPI_ISL_2009916, EPI_ISL_2009917, EPI_ISL_2009918, EPI_ISL_2009919, EPI_ISL_2009920, EPI_ISL_2009921, EPI_ISL_2009922, EPI_ISL_2009923, EPI_ISL_2009924, EPI_ISL_2009925, EPI_ISL_2009926, EPI_ISL_2009927, EPI_ISL_2009928, EPI_ISL_2009929,<br>EPI_ISL_2009930, EPI_ISL_2009931, EPI_ISL_2009932, EPI_ISL_2009933, EPI_ISL_2009934, EPI_ISL_2009935, EPI_ISL_2009936, EPI_ISL_2009937, EPI_ISL_2009938, EPI_ISL_2009939, EPI_ISL_2009940, EPI_ISL_2009941, EPI_ISL_2009942, EPI_ISL_2009943, EPI_ISL_2009944, EPI_ISL_2009945, EPI_ISL_2009946, EPI_ISL_2009947, EPI_ISL_2009948, EPI_ISL_2009949, EPI_ISL_2009950, EPI_ISL_2009951,<br>EPI_ISL_2009952, EPI_ISL_2009953, EPI_ISL_2009954, EPI_ISL_2009955, EPI_ISL_2009956, EPI_ISL_2009957, EPI_ISL_2009958, EPI_ISL_2009959, EPI_ISL_2009960, EPI_ISL_2009961, EPI_ISL_2009962, EPI_ISL_2009963, EPI_ISL_2009964, EPI_ISL_2009965, EPI_ISL_2009966, EPI_ISL_2009967, EPI_ISL_2009968, EPI_ISL_2009969, EPI_ISL_2009970, EPI_ISL_2009971, EPI_ISL_2009972, EPI_ISL_2009973,<br>EPI_ISL_2009974, EPI_ISL_2009975, EPI_ISL_2009976, EPI_ISL_2009977, EPI_ISL_2009978, EPI_ISL_2009979, EPI_ISL_2009980, EPI_ISL_2009981, EPI_ISL_2009982, EPI_ISL_2009983, EPI_ISL_2009984, EPI_ISL_2009985, EPI_ISL_2009986, EPI_ISL_2009987, EPI_ISL_2009988, EPI_ISL_2009989, EPI_ISL_2009990, EPI_ISL_2009991, EPI_ISL_2009992, EPI_ISL_2009993, EPI_ISL_2009994, EPI_ISL_2009995,<br>EPI_ISL_2009996, EPI_ISL_2009997, EPI_ISL_2009998, EPI_ISL_2009999, EPI_ISL_2010000, EPI_ISL_2010001, EPI_ISL_2010002, EPI_ISL_2010003, EPI_ISL_2010004, EPI_ISL_2010005, EPI_ISL_2010006, EPI_ISL_2010007, EPI_ISL_2010008, EPI_ISL_2010009, EPI_ISL_2010010, EPI_ISL_2010011, EPI_ISL_2010012, EPI_ISL_2010013, EPI_ISL_2010014, EPI_ISL_2010015, EPI_ISL_2010016, EPI_ISL_2010017,<br>EPI_ISL_2010018, EPI_ISL_2010019, EPI_ISL_2010020, EPI_ISL_2010021, EPI_ISL_2010022, EPI_ISL_2010023, EPI_ISL_2010024, EPI_ISL_2010025, EPI_ISL_2010026, EPI_ISL_2010027, EPI_ISL_2010028, EPI_ISL_2010029, EPI_ISL_2010030, EPI_ISL_2010031, EPI_ISL_2010032, EPI_ISL_2010033, EPI_ISL_2010034, EPI_ISL_2010035, EPI_ISL_2010036, EPI_ISL_2010037, EPI_ISL_2010038, EPI_ISL_2010039,<br>EPI_ISL_2010040, EPI_ISL_2010041, EPI_ISL_2010042, EPI_ISL_2010043, EPI_ISL_2010044, EPI_ISL_2010045, EPI_ISL_2010046, EPI_ISL_2010047, EPI_ISL_2010048, EPI_ISL_2010049, EPI_ISL_2010050, EPI_ISL_2010051, EPI_ISL_2010052, EPI_ISL_2010053, EPI_ISL_2010054, EPI_ISL_2010055, EPI_ISL_2010056, EPI_ISL_2010057, EPI_ISL_2010058, EPI_ISL_2010059, EPI_ISL_2010060, EPI_ISL_2010061,<br>EPI_ISL_2010062, EPI_ISL_2010063, EPI_ISL_2010064, EPI_ISL_2010065, EPI_ISL_2010066, EPI_ISL_2010067, EPI_ISL_2010068, EPI_ISL_2010069, EPI_ISL_2010070, EPI_ISL_2010071, EPI_ISL_2010072, EPI_ISL_2010073, EPI_ISL_2010074, EPI_ISL_2010075, EPI_ISL_2010076, EPI_ISL_2010077, EPI_ISL_2010078, EPI_ISL_2010079, EPI_ISL_2010080, EPI_ISL_2010081, EPI_ISL_2010082, EPI_ISL_2010083,<br>EPI_ISL_2010084, EPI_ISL_2010085, EPI_ISL_2010086, EPI_ISL_2010087, EPI_ISL_2010088, EPI_ISL_2010089, EPI_ISL_2010090, EPI_ISL_2010091, EPI_ISL_2010092, EPI_ISL_2010093, EPI_ISL_2010094, EPI_ISL_2010095, EPI_ISL_2010096, EPI_ISL_2010097, EPI_ISL_2010098, EPI_ISL_2010099, EPI_ISL_2010100, EPI_ISL_2010101, EPI_ISL_2010102, EPI_ISL_2010103, EPI_ISL_2010104, EPI_ISL_2010105,<br>EPI_ISL_2010106, EPI_ISL_2010107, EPI_ISL_2010108, EPI_ISL_2010109, EPI_ISL_2010110, EPI_ISL_2010111, EPI_ISL_2010112, EPI_ISL_2010113, EPI_ISL_2010114, EPI_ISL_2010115, EPI_ISL_2010116, EPI_ISL_2010117, EPI_ISL_2010118, EPI_ISL_2010119, EPI_ISL_2010120, EPI_ISL_2010121, EPI_ISL_2010122, EPI_ISL_2010123, EPI_ISL_2010124, EPI_ISL_2010125, EPI_ISL_2010126, EPI_ISL_2010127,<br>EPI_ISL_2010128, EPI_ISL_2010129, EPI_ISL_2010130, EPI_ISL_2010131, EPI_ISL_2010132, EPI_ISL_2010133, EPI_ISL_2010134, EPI_ISL_2010135, EPI_ISL_2010136, EPI_ISL_2010137, EPI_ISL_2010138, EPI_ISL_2010139, EPI_ISL_2010140, EPI_ISL_2010141, EPI_ISL_2010142, EPI_ISL_2010143, EPI_ISL_2010144, EPI_ISL_2010145, EPI_ISL_2010146, EPI_ISL_2010147, EPI_ISL_2010148, EPI_ISL_2010149,<br>EPI_ISL_2010150, EPI_ISL_2010151, EPI_ISL_2010152, EPI_ISL_2010153, EPI_ISL_2010154, EPI_ISL_2010155, EPI_ISL_2010156, EPI_ISL_2010157, EPI_ISL_2010158, EPI_ISL_2010159, EPI_ISL_2010160, EPI_ISL_2010161, EPI_ISL_2010162, EPI_ISL_2010163, EPI_ISL_2010164, EPI_ISL_2010165, EPI_ISL_2010166, EPI_ISL_2010167, EPI_ISL_2010168, EPI_ISL_2010169, EPI_ISL_2010170, EPI_ISL_2010171,<br>EPI_ISL_2010172, EPI_ISL_2010173, EPI_ISL_2010174, EPI_ISL_2010175, EPI_ISL_2010176, EPI_ISL_2010177, EPI_ISL_2010178, EPI_ISL_2010179, EPI_ISL_2010180, EPI_ISL_2010181, EPI_ISL_2010182, EPI_ISL_2010183, EPI_ISL_2010184, EPI_ISL_2010185, EPI_ISL_2010186, EPI_ISL_2010187, EPI_ISL_2010188, EPI_ISL_2010189, EPI_ISL_2010190, EPI_ISL_2010191, EPI_ISL_2010192, EPI_ISL_2010193,<br>EPI_ISL_2010194, EPI_ISL_2010195, EPI_ISL_2010196, EPI_ISL_2010197, EPI_ISL_2010198, EPI_ISL_2010199, EPI_ISL_2010200, EPI_ISL_2010201, EPI_ISL_2010202, EPI_ISL_2010203, EPI_ISL_2010204, EPI_ISL_2010205, EPI_ISL_2010206, EPI_ISL_2010207, EPI_ISL_2010208, EPI_ISL_2010209, EPI_ISL_2010210, EPI_ISL_2010211, EPI_ISL_2010212, EPI_ISL_2010213, EPI_ISL_2010214, EPI_ISL_2010215,<br>EPI_ISL_2010216, EPI_ISL_2010217, EPI_ISL_2010218, EPI_ISL_2010219, EPI_ISL_2010220, EPI_ISL_2010221, EPI_ISL_2010222, EPI_ISL_2010223, EPI_ISL_2010224, EPI_ISL_2010225, EPI_ISL_2010226, EPI_ISL_2010227, EPI_ISL_2010228, EPI_ISL_2010229, EPI_ISL_2010230, EPI_ISL_2010231, EPI_ISL_2010232, EPI_ISL_2010233, EPI_ISL_2010234, EPI_ISL_2010235, EPI_ISL_2010236, EPI_ISL_2010237,<br>EPI_ISL_2010238, EPI_ISL_2010239, EPI_ISL_2010240, EPI_ISL_2010241, EPI_ISL_2010242, EPI_ISL_2010243, EPI_ISL_2010244, EPI_ISL_2010245, EPI_ISL_2010246, EPI_ISL_2010247, EPI_ISL_2010248, EPI_ISL_2010249, EPI_ISL_2010250, EPI_ISL_2010251, EPI_ISL_2010252, EPI_ISL_2010253, EPI_ISL_2010254, EPI_ISL_2010255, EPI_ISL_2010256, EPI_ISL_2010257, EPI_ISL_2010258, EPI_ISL_2010259,<br>EPI_ISL_2010260, EPI_ISL_2010261, EPI_ISL_2010262, EPI_ISL_2010263, EPI_ISL_2010264, EPI_ISL_2010265, EPI_ISL_2010266, EPI_ISL_2010267, EPI_ISL_2010268, EPI_ISL_2010269, EPI_ISL_2010270, EPI_ISL_2010271, EPI_ISL_2010272, EPI_ISL_2010273, EPI_ISL_2010274, EPI_ISL_2010275, EPI_ISL_2010276, EPI_ISL_2010277, EPI_ISL_2010278, EPI_ISL_2010279, EPI_ISL_2010280, EPI_ISL_2010281,<br>EPI_ISL_2010282, EPI_ISL_2010283, EPI_ISL_2010284, EPI_ISL_2010285, EPI_ISL_2010286, EPI_ISL_2010287, EPI_ISL_2010288, EPI_ISL_2010289, EPI_ISL_2010290, EPI_ISL_2010291, EPI_ISL_2010292, EPI_ISL_2010293, EPI_ISL_2010294, EPI_ISL_2010295, EPI_ISL_2010296, EPI_ISL_2010297, EPI_ISL_2010298, EPI_ISL_2010299, EPI_ISL_2010300, EPI_ISL_2010301, EPI_ISL_2010302, EPI_ISL_2010303,<br>EPI_ISL_2010304, EPI_ISL_2010305, EPI_ISL_2010306, EPI_ISL_2010307, EPI_ISL_2010308, EPI_ISL_2010309, EPI_ISL_2010310, EPI_ISL_2010311, EPI_ISL_2010312, EPI_ISL_2010313, EPI_ISL_2010314, EPI_ISL_2010315, EPI_ISL_2010316, EPI_ISL_2010317, EPI_ISL_2010318, EPI_ISL_2010319, EPI_ISL_2010320, EPI_ISL_2010321, EPI_ISL_2010322, EPI_ISL_2010323, EPI_ISL_2010324, EPI_ISL_2010325,<br>EPI_ISL_2010326, EPI_ISL_2010327, EPI_ISL_2010328, EPI_ISL_2010329, EPI_ISL_2010330, EPI_ISL_2010331, EPI_ISL_2010332, EPI_ISL_2010333, EPI_ISL_2010334, EPI_ISL_2010335, EPI_ISL_2010336, EPI_ISL_2010337, EPI_ISL_2010338, EPI_ISL_2010339, EPI_ISL_2010340, EPI_ISL_2010341, EPI_ISL_2010342, EPI_ISL_2010343, EPI_ISL_2010344, EPI_ISL_2010345, EPI_ISL_2010346, EPI_ISL_2010347,<br>EPI_ISL_2010348, EPI_ISL_2010349, EPI_ISL_2010350, EPI_ISL_2010351, EPI_ISL_2010352, EPI_ISL_2010353, EPI_ISL_2010354, EPI_ISL_2010355, EPI_ISL_2010356, EPI_ISL_2010357, EPI_ISL_2010358, EPI_ISL_2010359, EPI_ISL_2010360, EPI_ISL_2010361, EPI_ISL_2010362, EPI_ISL_2010363, EPI_ISL_2010364, EPI_ISL_2010365, EPI_ISL_2010366, EPI_ISL_2010367, EPI_ISL_2010368, EPI_ISL_2010369,<br>EPI_ISL_2010370, EPI_ISL_2010371, EPI_ISL_2010372, EPI_ISL_2010373, EPI_ISL_2010374, EPI_ISL_2010375, EPI_ISL_2010376, EPI_ISL_2010377, EPI_ISL_2010378, EPI_ISL_2010379, EPI_ISL_2010380, EPI_ISL_2010381, EPI_ISL_2010382, EPI_ISL_2010383, EPI_ISL_2010384, EPI_ISL_2010385, EPI_ISL_2010386, EPI_ISL_2010387, EPI_ISL_2010388, EPI_ISL_2010389, EPI_ISL_2010390, EPI_ISL_2010391, | | | |

| | | | |
|---|---|---|---|
| EPI_ISL_2010392, EPI_ISL_2010393, EPI_ISL_2010394, EPI_ISL_2010395, EPI_ISL_2010396, EPI_ISL_2010397, EPI_ISL_2010398, EPI_ISL_2010399, EPI_ISL_2010400, EPI_ISL_2010401, EPI_ISL_2010402, EPI_ISL_2010403, EPI_ISL_2010404, EPI_ISL_2010405, EPI_ISL_2010406, EPI_ISL_2010407, EPI_ISL_2010408, EPI_ISL_2010409, EPI_ISL_2010410, EPI_ISL_2010411, EPI_ISL_2010412, EPI_ISL_2010413, EPI_ISL_2010414, EPI_ISL_2010415, EPI_ISL_2010416, EPI_ISL_2010417, EPI_ISL_2010418, EPI_ISL_2010419, EPI_ISL_2010420, EPI_ISL_2010421, EPI_ISL_2010422, EPI_ISL_2010423, EPI_ISL_2010424, EPI_ISL_2010425, EPI_ISL_2010426, EPI_ISL_2010427, EPI_ISL_2010428, EPI_ISL_2010429, EPI_ISL_2010430, EPI_ISL_2010431, EPI_ISL_2010432, EPI_ISL_2010433, EPI_ISL_2010434, EPI_ISL_2010435, EPI_ISL_2010436, EPI_ISL_2010437, EPI_ISL_2010438, EPI_ISL_2010439, EPI_ISL_2010440, EPI_ISL_2010441, EPI_ISL_2010442, EPI_ISL_2010443, EPI_ISL_2010444, EPI_ISL_2010445, EPI_ISL_2010446, EPI_ISL_2010447, EPI_ISL_2010448, EPI_ISL_2010449, EPI_ISL_2010450, EPI_ISL_2010451, EPI_ISL_2010452, EPI_ISL_2010453, EPI_ISL_2010454, EPI_ISL_2010455, EPI_ISL_2010456, EPI_ISL_2010457, EPI_ISL_2010458, EPI_ISL_2010459, EPI_ISL_2010460, EPI_ISL_2010461, EPI_ISL_2010462, EPI_ISL_2010463, EPI_ISL_2010464, EPI_ISL_2010465, EPI_ISL_2010466, EPI_ISL_2010467, EPI_ISL_2010468, EPI_ISL_2010469, EPI_ISL_2010470, EPI_ISL_2010471, EPI_ISL_2010472, EPI_ISL_2010473, EPI_ISL_2010474, EPI_ISL_2010475, EPI_ISL_2010476, EPI_ISL_2010477, EPI_ISL_2010478, EPI_ISL_2010479, EPI_ISL_2010480, EPI_ISL_2010481, EPI_ISL_2010482, EPI_ISL_2010483, EPI_ISL_2010484, EPI_ISL_2010485, EPI_ISL_2010486, EPI_ISL_2010487, EPI_ISL_2010488, EPI_ISL_2010489, EPI_ISL_2010490, EPI_ISL_2010491, EPI_ISL_2010492, EPI_ISL_2010493, EPI_ISL_2010494, EPI_ISL_2010495, EPI_ISL_2010496, EPI_ISL_2010497, EPI_ISL_2010498, EPI_ISL_2010499, EPI_ISL_2010500, EPI_ISL_2010501, EPI_ISL_2010502, EPI_ISL_2010503, EPI_ISL_2010504, EPI_ISL_2010505, EPI_ISL_2010506, EPI_ISL_2010507, EPI_ISL_2010508, EPI_ISL_2010509, EPI_ISL_2010510, EPI_ISL_2010511, EPI_ISL_2010512, EPI_ISL_2010513, EPI_ISL_2010514, EPI_ISL_2010515, EPI_ISL_2010516, EPI_ISL_2010517, EPI_ISL_2010518, EPI_ISL_2010519, EPI_ISL_2010520, EPI_ISL_2010521, EPI_ISL_2010522, EPI_ISL_2010523, EPI_ISL_2010524, EPI_ISL_2010525, EPI_ISL_2010526, EPI_ISL_2010527, EPI_ISL_2010528, EPI_ISL_2010529, EPI_ISL_2010530, EPI_ISL_2010531, EPI_ISL_2010532, EPI_ISL_2010533, EPI_ISL_2010534, EPI_ISL_2010535, EPI_ISL_2010536, EPI_ISL_2010537, EPI_ISL_2010538, EPI_ISL_2010539, EPI_ISL_2010540, EPI_ISL_2010541, EPI_ISL_2010542, EPI_ISL_2010543, EPI_ISL_2010544, EPI_ISL_2010545, EPI_ISL_2010546, EPI_ISL_2010547, EPI_ISL_2010548, EPI_ISL_2010549, EPI_ISL_2010550, EPI_ISL_2010551, EPI_ISL_2010552, EPI_ISL_2010553, EPI_ISL_2010554, EPI_ISL_2010555, EPI_ISL_2010556, EPI_ISL_2010557, EPI_ISL_2010558, EPI_ISL_2010559, EPI_ISL_2010560, EPI_ISL_2010561, EPI_ISL_2010562, EPI_ISL_2010563, EPI_ISL_2010564, EPI_ISL_2010565, EPI_ISL_2010566, EPI_ISL_2010567, EPI_ISL_2010568, EPI_ISL_2010569, EPI_ISL_2010570, EPI_ISL_2010571, EPI_ISL_2010572, EPI_ISL_2010573, EPI_ISL_2010574, EPI_ISL_2010575, EPI_ISL_2010576, EPI_ISL_2010577, EPI_ISL_2010578, EPI_ISL_2010579, EPI_ISL_2010580, EPI_ISL_2010581, EPI_ISL_2010582, EPI_ISL_2010583, EPI_ISL_2010584, EPI_ISL_2010585, EPI_ISL_2010586, EPI_ISL_2010587, EPI_ISL_2010588, EPI_ISL_2010589, EPI_ISL_2010590, EPI_ISL_2010591, EPI_ISL_2010592, EPI_ISL_2010593, EPI_ISL_2010594, EPI_ISL_2010595, EPI_ISL_2010596, EPI_ISL_2010597, EPI_ISL_2010598, EPI_ISL_2010599, EPI_ISL_2010600, EPI_ISL_2010601, EPI_ISL_2010602, EPI_ISL_2010603, EPI_ISL_2010604, EPI_ISL_2010605, EPI_ISL_2010606, EPI_ISL_2010607, EPI_ISL_2010608, EPI_ISL_2010609, EPI_ISL_2010610, EPI_ISL_2010611, EPI_ISL_2010612, EPI_ISL_2010613, EPI_ISL_2010614, EPI_ISL_2010615, EPI_ISL_2010616, EPI_ISL_2010617, EPI_ISL_2010618, EPI_ISL_2010619, EPI_ISL_2010620, EPI_ISL_2010621, EPI_ISL_2010622, EPI_ISL_2010623, EPI_ISL_2010624, EPI_ISL_2010625, EPI_ISL_2010626, EPI_ISL_2010627, EPI_ISL_2010628, EPI_ISL_2010629, EPI_ISL_2010630, EPI_ISL_2010631, EPI_ISL_2010632, EPI_ISL_2010633, EPI_ISL_2010634, EPI_ISL_2010635, EPI_ISL_2010636, EPI_ISL_2010637, EPI_ISL_2010638, EPI_ISL_2010639, EPI_ISL_2010640, EPI_ISL_2010641, EPI_ISL_2010642, EPI_ISL_2010643, EPI_ISL_2010644, EPI_ISL_2010645, EPI_ISL_2010646, EPI_ISL_2010647, EPI_ISL_2010648, EPI_ISL_2010649, EPI_ISL_2010650, EPI_ISL_2010651, EPI_ISL_2010652, EPI_ISL_2010653, EPI_ISL_2010654, EPI_ISL_2010655, EPI_ISL_2010656, EPI_ISL_2010657, EPI_ISL_2010658, EPI_ISL_2010659, EPI_ISL_2010660, EPI_ISL_2010661, EPI_ISL_2010662, EPI_ISL_2010663, EPI_ISL_2010664, EPI_ISL_2010665, EPI_ISL_2010666, EPI_ISL_2010667, EPI_ISL_2010668, EPI_ISL_2010669, EPI_ISL_2010670, EPI_ISL_2010671, EPI_ISL_2010672, EPI_ISL_2010673, EPI_ISL_2010674, EPI_ISL_2010675, EPI_ISL_2010676, EPI_ISL_2010677, EPI_ISL_2010678, EPI_ISL_2010679, EPI_ISL_2010680, EPI_ISL_2010681, EPI_ISL_2010682, EPI_ISL_2010683, EPI_ISL_2010684, EPI_ISL_2010685, EPI_ISL_2010686, EPI_ISL_2010687, EPI_ISL_2010688, EPI_ISL_2010689, EPI_ISL_2010690, EPI_ISL_2010691, EPI_ISL_2010692, EPI_ISL_2010693, EPI_ISL_2010694, EPI_ISL_2010695, EPI_ISL_2010696, EPI_ISL_2010697, EPI_ISL_2010698, EPI_ISL_2010699, EPI_ISL_2010700, EPI_ISL_2010701, EPI_ISL_2010702, EPI_ISL_2010703, EPI_ISL_2010704, EPI_ISL_2010705, EPI_ISL_2010706, EPI_ISL_2010707, EPI_ISL_2010708, EPI_ISL_2010709, EPI_ISL_2010710, EPI_ISL_2010711, EPI_ISL_2010712, EPI_ISL_2010713, EPI_ISL_2010714, EPI_ISL_2010715, EPI_ISL_2010716, EPI_ISL_2010717, EPI_ISL_2010718, EPI_ISL_2010719, EPI_ISL_2010720, EPI_ISL_2010721, EPI_ISL_2010722, EPI_ISL_2010723, EPI_ISL_2010724, EPI_ISL_2010725, EPI_ISL_2010726, EPI_ISL_2010727, EPI_ISL_2010728, EPI_ISL_2010729, EPI_ISL_2010730, EPI_ISL_2010731, EPI_ISL_2010732, EPI_ISL_2010733, EPI_ISL_2010734, EPI_ISL_2010735, EPI_ISL_2010736, EPI_ISL_2010737, EPI_ISL_2010738, EPI_ISL_2010739, EPI_ISL_2010740, EPI_ISL_2010741, EPI_ISL_2010742, EPI_ISL_2010743, EPI_ISL_2010744, EPI_ISL_2010745, EPI_ISL_2010746, EPI_ISL_2010747, EPI_ISL_2010748, EPI_ISL_2010749, EPI_ISL_2010750, EPI_ISL_2010751, EPI_ISL_2010752, EPI_ISL_2010753, EPI_ISL_2010754, EPI_ISL_2010755, EPI_ISL_2010756, EPI_ISL_2010757, EPI_ISL_2010758, EPI_ISL_2010759, EPI_ISL_2010760, EPI_ISL_2010761, EPI_ISL_2010762, EPI_ISL_2010763, EPI_ISL_2010764, EPI_ISL_2010765, EPI_ISL_2010766, EPI_ISL_2010767, EPI_ISL_2010768, EPI_ISL_2010769, EPI_ISL_2010770, EPI_ISL_2010771, EPI_ISL_2010772, EPI_ISL_2010773, EPI_ISL_2010774, EPI_ISL_2010775, EPI_ISL_2010776, EPI_ISL_2010777, EPI_ISL_2010778, EPI_ISL_2010779, EPI_ISL_2010780, EPI_ISL_2010781, EPI_ISL_2010782, EPI_ISL_2010783, EPI_ISL_2010784, EPI_ISL_2010785, EPI_ISL_2010786, EPI_ISL_2010787, EPI_ISL_2010788, EPI_ISL_2010789, EPI_ISL_2010790, EPI_ISL_2010791, EPI_ISL_2010792, EPI_ISL_2010793, EPI_ISL_2010794, EPI_ISL_2010795, EPI_ISL_2010796, EPI_ISL_2010797, EPI_ISL_2010798, EPI_ISL_2010799, EPI_ISL_2010800, EPI_ISL_2010801, EPI_ISL_2010802, EPI_ISL_2010803, EPI_ISL_2010804, EPI_ISL_2010805, EPI_ISL_2010806, EPI_ISL_2010807, EPI_ISL_2010808, EPI_ISL_2010809, EPI_ISL_2010810, EPI_ISL_2010811, EPI_ISL_2010812, EPI_ISL_2010813, EPI_ISL_2010814, EPI_ISL_2010815, EPI_ISL_2010816, EPI_ISL_2010817, EPI_ISL_2010818, EPI_ISL_2010819, EPI_ISL_2010820, EPI_ISL_2010821, EPI_ISL_2010822, EPI_ISL_2010823, EPI_ISL_2010824, EPI_ISL_2010825, EPI_ISL_2010826, EPI_ISL_2010827, EPI_ISL_2010828, EPI_ISL_2010829, EPI_ISL_2010830, EPI_ISL_2010831, EPI_ISL_2010832, EPI_ISL_2010833, EPI_ISL_2010834, EPI_ISL_2010835, EPI_ISL_2010836, EPI_ISL_2010837, EPI_ISL_2010838, EPI_ISL_2010839, EPI_ISL_2010840, EPI_ISL_2010841, EPI_ISL_2010842, EPI_ISL_2010843, EPI_ISL_2010844, EPI_ISL_2010845, EPI_ISL_2010846, EPI_ISL_2010847, EPI_ISL_2010848, EPI_ISL_2010849, EPI_ISL_2010850, EPI_ISL_2010851, EPI_ISL_2010852, EPI_ISL_2010853, EPI_ISL_2010854, EPI_ISL_2010855, EPI_ISL_2010856, EPI_ISL_2010857, EPI_ISL_2010858, EPI_ISL_2010859, EPI_ISL_2010860, EPI_ISL_2010861, EPI_ISL_2010862, EPI_ISL_2010863, EPI_ISL_2010864, EPI_ISL_2010865, EPI_ISL_2010866, EPI_ISL_2010867, EPI_ISL_2010868, EPI_ISL_2010869, EPI_ISL_2010870, EPI_ISL_2010871, EPI_ISL_2010872, EPI_ISL_2010873, EPI_ISL_2010874, EPI_ISL_2010875, EPI_ISL_2010876, EPI_ISL_2010877, EPI_ISL_2010878, EPI_ISL_2010879, EPI_ISL_2010880, EPI_ISL_2010881, EPI_ISL_2010882, EPI_ISL_2010883, EPI_ISL_2010884, EPI_ISL_2010885, EPI_ISL_2010886, EPI_ISL_2010887, EPI_ISL_2010888 | | | |
| see above | Helix/Illumina | Centers for Disease Control and Prevention Division of Viral Diseases, Pathogen Discovery | Dakota Howard, Dhwani Batra, Peter W. Cook, Kara Moser, Adrian Paskey, Jason Caravas, Benjamin Rambo-Martin, Shatavia Morrison, Christopher Gulvick, Scott Sammons, Yvette Unoarumhi, Darlene Wagner, Matthew Schmerer, Eileen de Feo, Jan Antico, Christine Tran, Matthew Tolentino, Shannon Wickline, Kim Gietzen, Brad Sickler, Jingtao Liu, Eric Allen, Phil Febbo, Nicole L. Washington, Simon White, Geraint Levan, Kelly Schiabor Barrett, Elizabeth Cirulli, Alexandre Bolze, Ary Ascencio, Charlotte Rivera-Garcia, Ryan Cho, Jason Nguyen, Sherry Wang, Jimmy Ramirez, Tyler Cassens, Efren Sandoval, Magnus Isaksson, William Lee, David Becker, Marc Laurent, James Lu, Clinton R. Paden, Duncan MacCannell |
| EPI_ISL_2010889, EPI_ISL_2010890, EPI_ISL_2010891, EPI_ISL_2010892, EPI_ISL_2010893, EPI_ISL_2010894, EPI_ISL_2010895, EPI_ISL_2010896, EPI_ISL_2010897, EPI_ISL_2010898, EPI_ISL_2010899, EPI_ISL_2010900, EPI_ISL_2010901, EPI_ISL_2010902, EPI_ISL_2010903, EPI_ISL_2010904, EPI_ISL_2010905, EPI_ISL_2010906, EPI_ISL_2010907, EPI_ISL_2010908, EPI_ISL_2010909, EPI_ISL_2010910, EPI_ISL_2010911, EPI_ISL_2010912, EPI_ISL_2010913, EPI_ISL_2010914, EPI_ISL_2010915, EPI_ISL_2010916, EPI_ISL_2010917, EPI_ISL_2010918, EPI_ISL_2010919, EPI_ISL_2010920, EPI_ISL_2010921, EPI_ISL_2010922, EPI_ISL_2010923, EPI_ISL_2010924, EPI_ISL_2010925, EPI_ISL_2010926, EPI_ISL_2010927, EPI_ISL_2010928, EPI_ISL_2010929, EPI_ISL_2010930, EPI_ISL_2010931, EPI_ISL_2010932, EPI_ISL_2010933, EPI_ISL_2010934, EPI_ISL_2010935, EPI_ISL_2010936, EPI_ISL_2010937, EPI_ISL_2010938, EPI_ISL_2010939, EPI_ISL_2010940, EPI_ISL_2010941, EPI_ISL_2010942, EPI_ISL_2010943, EPI_ISL_2010944, EPI_ISL_2010945, EPI_ISL_2010946, EPI_ISL_2010947, EPI_ISL_2010948, EPI_ISL_2010949, EPI_ISL_2010950, EPI_ISL_2010951, EPI_ISL_2010952, EPI_ISL_2010953, EPI_ISL_2010954, EPI_ISL_2010955, EPI_ISL_2010956, EPI_ISL_2010957, EPI_ISL_2010958, EPI_ISL_2010959, EPI_ISL_2010960, EPI_ISL_2010961, EPI_ISL_2010962, EPI_ISL_2010963, EPI_ISL_2010964, EPI_ISL_2010965, EPI_ISL_2010966, EPI_ISL_2010967, EPI_ISL_2010968, EPI_ISL_2010969, EPI_ISL_2010970, EPI_ISL_2010971, EPI_ISL_2010972, EPI_ISL_2010973, EPI_ISL_2010974, EPI_ISL_2010975, EPI_ISL_2010976, EPI_ISL_2010977, EPI_ISL_2010978, EPI_ISL_2010979, EPI_ISL_2010980, EPI_ISL_2010981, EPI_ISL_2010982, EPI_ISL_2010983, EPI_ISL_2010984, EPI_ISL_2010985, EPI_ISL_2010986, EPI_ISL_2010987, EPI_ISL_2010988, EPI_ISL_2010989, EPI_ISL_2010990, EPI_ISL_2010991, EPI_ISL_2010992, EPI_ISL_2010993, EPI_ISL_2010994, EPI_ISL_2010995, EPI_ISL_2010996, EPI_ISL_2010997, EPI_ISL_2010998, EPI_ISL_2010999, EPI_ISL_2011000, EPI_ISL_2011001, EPI_ISL_2011002, EPI_ISL_2011003, EPI_ISL_2011004, EPI_ISL_2011005, EPI_ISL_2011006, EPI_ISL_2011007, EPI_ISL_2011008, EPI_ISL_2011009, EPI_ISL_2011010, EPI_ISL_2011011, EPI_ISL_2011012, EPI_ISL_2011013, EPI_ISL_2011014, EPI_ISL_2011015, EPI_ISL_2011016, EPI_ISL_2011017, EPI_ISL_2011018, EPI_ISL_2011019, EPI_ISL_2011020, EPI_ISL_2011021, EPI_ISL_2011022, EPI_ISL_2011023, EPI_ISL_2011024, EPI_ISL_2011025, EPI_ISL_2011026, EPI_ISL_2011027, EPI_ISL_2011028, EPI_ISL_2011029, EPI_ISL_2011030, EPI_ISL_2011031, EPI_ISL_2011032, EPI_ISL_2011033, EPI_ISL_2011034, EPI_ISL_2011035, EPI_ISL_2011036, EPI_ISL_2011037, EPI_ISL_2011038, EPI_ISL_2011039, EPI_ISL_2011040, EPI_ISL_2011041, EPI_ISL_2011042, EPI_ISL_2011043, EPI_ISL_2011044, EPI_ISL_2011045, EPI_ISL_2011046, EPI_ISL_2011047, EPI_ISL_2011048, EPI_ISL_2011049, EPI_ISL_2011050, EPI_ISL_2011051, EPI_ISL_2011052, EPI_ISL_2011053, EPI_ISL_2011054, EPI_ISL_2011055, EPI_ISL_2011056, EPI_ISL_2011057, EPI_ISL_2011058, EPI_ISL_2011059, EPI_ISL_2011060, EPI_ISL_2011061, EPI_ISL_2011062, EPI_ISL_2011063, EPI_ISL_2011064, EPI_ISL_2011065, EPI_ISL_2011066, EPI_ISL_2011067, EPI_ISL_2011068, EPI_ISL_2011069, EPI_ISL_2011070, EPI_ISL_2011071, EPI_ISL_2011072, EPI_ISL_2011073, EPI_ISL_2011074, EPI_ISL_2011075, EPI_ISL_2011076, EPI_ISL_2011077, EPI_ISL_2011078, EPI_ISL_2011079, EPI_ISL_2011080, EPI_ISL_2011081, EPI_ISL_2011082, EPI_ISL_2011083, EPI_ISL_2011084, EPI_ISL_2011085, EPI_ISL_2011086, EPI_ISL_2011087, EPI_ISL_2011088, EPI_ISL_2011089, EPI_ISL_2011090, EPI_ISL_2011091, EPI_ISL_2011092, EPI_ISL_2011093, EPI_ISL_2011094, EPI_ISL_2011095, EPI_ISL_2011096, EPI_ISL_2011097, EPI_ISL_2011098, EPI_ISL_2011099, EPI_ISL_2011100, EPI_ISL_2011101, EPI_ISL_2011102, EPI_ISL_2011103, EPI_ISL_2011104, EPI_ISL_2011105, EPI_ISL_2011106, EPI_ISL_2011107, EPI_ISL_2011108, EPI_ISL_2011109, EPI_ISL_2011110, EPI_ISL_2011111, EPI_ISL_2011112, EPI_ISL_2011113, EPI_ISL_2011114, EPI_ISL_2011115, EPI_ISL_2011116, EPI_ISL_2011117, EPI_ISL_2011118, EPI_ISL_2011119, EPI_ISL_2011120, EPI_ISL_2011121, EPI_ISL_2011122, EPI_ISL_2011123, EPI_ISL_2011124, EPI_ISL_2011125, EPI_ISL_2011126, EPI_ISL_2011127, EPI_ISL_2011128, EPI_ISL_2011129, EPI_ISL_2011130, EPI_ISL_2011131, EPI_ISL_2011132, EPI_ISL_2011133, EPI_ISL_2011134, EPI_ISL_2011135, EPI_ISL_2011136, EPI_ISL_2011137, EPI_ISL_2011138, EPI_ISL_2011139, EPI_ISL_2011140, EPI_ISL_2011141, EPI_ISL_2011142, EPI_ISL_2011143, EPI_ISL_2011144, EPI_ISL_2011145, EPI_ISL_2011146, EPI_ISL_2011147, EPI_ISL_2011148, EPI_ISL_2011149, EPI_ISL_2011150, EPI_ISL_2011151, EPI_ISL_2011152, EPI_ISL_2011153, EPI_ISL_2011154, EPI_ISL_2011155, EPI_ISL_2011156, EPI_ISL_2011157, EPI_ISL_2011158, EPI_ISL_2011159, EPI_ISL_2011160, EPI_ISL_2011161, EPI_ISL_2011162, EPI_ISL_2011163, EPI_ISL_2011164, EPI_ISL_2011165, EPI_ISL_2011166, EPI_ISL_2011167, EPI_ISL_2011168, EPI_ISL_2011169, EPI_ISL_2011170, EPI_ISL_2011171, EPI_ISL_2011172, EPI_ISL_2011173, EPI_ISL_2011174, EPI_ISL_2011175, EPI_ISL_2011176, EPI_ISL_2011177, EPI_ISL_2011178, EPI_ISL_2011179, EPI_ISL_2011180, EPI_ISL_2011181, EPI_ISL_2011182, EPI_ISL_2011183, EPI_ISL_2011184, EPI_ISL_2011185, EPI_ISL_2011186, EPI_ISL_2011187, EPI_ISL_2011188, EPI_ISL_2011189, EPI_ISL_2011190, EPI_ISL_2011191, EPI_ISL_2011192, EPI_ISL_2011193, EPI_ISL_2011194, EPI_ISL_2011195, EPI_ISL_2011196, EPI_ISL_2011197, EPI_ISL_2011198, EPI_ISL_2011199, EPI_ISL_2011200, EPI_ISL_2011201, EPI_ISL_2011202, EPI_ISL_2011203, EPI_ISL_2011204, EPI_ISL_2011205, EPI_ISL_2011206, EPI_ISL_2011207, EPI_ISL_2011208, EPI_ISL_2011209, EPI_ISL_2011210, EPI_ISL_2011211, EPI_ISL_2011212, EPI_ISL_2011213, EPI_ISL_2011214, EPI_ISL_2011215, EPI_ISL_2011216, EPI_ISL_2011217, EPI_ISL_2011218, EPI_ISL_2011219, EPI_ISL_2011220, EPI_ISL_2011221, EPI_ISL_2011222, EPI_ISL_2011223, EPI_ISL_2011224, EPI_ISL_2011225, EPI_ISL_2011226, EPI_ISL_2011227, EPI_ISL_2011228, EPI_ISL_2011229, EPI_ISL_2011230, EPI_ISL_2011231, EPI_ISL_2011232, EPI_ISL_2011233, EPI_ISL_2011234, EPI_ISL_2011235, EPI_ISL_2011236, EPI_ISL_2011237, EPI_ISL_2011238, EPI_ISL_2011239, EPI_ISL_2011240, EPI_ISL_2011241, EPI_ISL_2011242, EPI_ISL_2011243, EPI_ISL_2011244, EPI_ISL_2011245, EPI_ISL_2011246, EPI_ISL_2011247, EPI_ISL_2011248 | | | |
| see above | Quest Diagnostics Incorporated | Centers for Disease Control and Prevention Division of Viral Diseases, Pathogen Discovery | Dakota Howard, Dhwani Batra, Peter W. Cook, Kara Moser, Adrian Paskey, Jason Caravas, Benjamin Rambo-Martin, Shatavia Morrison, Christopher Gulvick, Scott Sammons, Yvette Unoarumhi, Darlene Wagner, Matthew Schmerer, S. H. Rosenthal, A. Gerasimova, R. M. Kagan, B. Anderson, M. Hua, Y. Liu, L.E. Bernstein, K.E. Livingston, A. Perez, I. A. Shlyakhter, R. V. Rolando, R. Owen, P. Tanpaiboon, F. Lacbawan, Clinton R. Paden, Duncan MacCannell |
| EPI_ISL_2011249, EPI_ISL_2011250 | Seattle Flu Study | Seattle Flu Study | Deborah A. Nickerson, Chris D. Frazar, Jover Lee, Benjamin Pelle, Erica Ryke, Matthew Richardson, Amanda Adler, Elisabeth Brandstetter, Peter D. Han, Kairsten Fay, Misja Ilcisin, Kirsten Lacombe, Thomas R. Sibley, Melissa Truong, Caitlin R. Wolf, Karen Cowgill, Stephanie Schrag, Jeff Duchin, Michael Boeckh, Janet A. Englund, Michael Famulare, Barry R. Lutz, Mark J. Rieder, Lea M. Starita, Matthew Thompson, Helen Y. Chu, Trevor Bedford, Jay Shendure |
| EPI_ISL_2011251 | Quest Diagnostics Incorporated | Centers for Disease Control and Prevention Division of Viral Diseases, Pathogen Discovery | Dakota Howard, Dhwani Batra, Peter W. Cook, Kara Moser, Adrian Paskey, Jason Caravas, Benjamin Rambo-Martin, Shatavia Morrison, Christopher Gulvick, Scott Sammons, Yvette Unoarumhi, Darlene Wagner, Matthew Schmerer, S. H. Rosenthal, A. Gerasimova, R. M. Kagan, B. Anderson, M. Hua, Y. Liu, L.E. Bernstein, K.E. Livingston, A. Perez, I. A. Shlyakhter, R. V. Rolando, R. Owen, P. Tanpaiboon, F. Lacbawan, Clinton R. Paden, Duncan MacCannell |
| EPI_ISL_2011252, EPI_ISL_2011254 | Seattle Flu Study | Seattle Flu Study | Deborah A. Nickerson, Chris D. Frazar, Jover Lee, Benjamin Pelle, Erica Ryke, Matthew Richardson, Amanda Adler, Elisabeth Brandstetter, Peter D. Han, Kairsten Fay, Misja Ilcisin, Kirsten Lacombe, Thomas R. Sibley, Melissa Truong, Caitlin R. Wolf, Michael Boeckh, Janet A. Englund, Michael Famulare, Barry R. Lutz, Mark J. Rieder, Lea M. Starita, Matthew Thompson, Jay Shendure, Trevor Bedford, Helen Y. Chu |
| EPI_ISL_2011255 | Quest Diagnostics Incorporated | Centers for Disease Control and Prevention Division of Viral Diseases, Pathogen Discovery | Dakota Howard, Dhwani Batra, Peter W. Cook, Kara Moser, Adrian Paskey, Jason Caravas, Benjamin Rambo-Martin, Shatavia Morrison, Christopher Gulvick, Scott Sammons, Yvette Unoarumhi, Darlene Wagner, Matthew Schmerer, S. H. Rosenthal, A. Gerasimova, R. M. Kagan, B. Anderson, M. Hua, Y. Liu, L.E. Bernstein, K.E. Livingston, A. Perez, I. A. Shlyakhter, R. V. Rolando, R. Owen, P. Tanpaiboon, F. Lacbawan, Clinton R. Paden, Duncan MacCannell |
| EPI_ISL_2011256 | Seattle Flu Study | Seattle Flu Study | Deborah A. Nickerson, Chris D. Frazar, Jover Lee, Benjamin Pelle, Erica Ryke, Matthew Richardson, Amanda Adler, Elisabeth Brandstetter, Peter D. Han, Kairsten Fay, Misja Ilcisin, Kirsten Lacombe, Thomas R. Sibley, Melissa Truong, Caitlin R. Wolf, Michael Boeckh, Janet A. Englund, Michael Famulare, Barry R. Lutz, Mark J. Rieder, Lea M. Starita, Matthew Thompson, Jay Shendure, Trevor Bedford, Helen Y. Chu |
| EPI_ISL_2011258, EPI_ISL_2011262 | Quest Diagnostics Incorporated | Centers for Disease Control and Prevention Division of Viral Diseases, Pathogen Discovery | Dakota Howard, Dhwani Batra, Peter W. Cook, Kara Moser, Adrian Paskey, Jason Caravas, Benjamin Rambo-Martin, Shatavia Morrison, Christopher Gulvick, Scott Sammons, Yvette Unoarumhi, Darlene Wagner, Matthew Schmerer, S. H. Rosenthal, A. Gerasimova, R. M. Kagan, B. Anderson, M. Hua, Y. Liu, L.E. Bernstein, K.E. Livingston, A. Perez, I. A. Shlyakhter, R. V. Rolando, R. Owen, P. Tanpaiboon, F. Lacbawan, Clinton R. Paden, Duncan MacCannell |
| EPI_ISL_2011263, EPI_ISL_2011265 | Seattle Flu Study | Seattle Flu Study | Deborah A. Nickerson, Chris D. Frazar, Jover Lee, Benjamin Pelle, Erica Ryke, Matthew Richardson, Amanda Adler, Elisabeth Brandstetter, Peter D. Han, Kairsten Fay, Misja Ilcisin, Kirsten Lacombe, Thomas R. Sibley, Melissa Truong, Caitlin R. Wolf, Michael Boeckh, Janet A. Englund, Michael Famulare, Barry R. Lutz, Mark J. Rieder, Lea M. Starita, Matthew Thompson, Jay Shendure, Trevor Bedford, Helen Y. Chu |
| EPI_ISL_2011266 | Quest Diagnostics Incorporated | Centers for Disease Control and Prevention Division of Viral Diseases, Pathogen Discovery | Dakota Howard, Dhwani Batra, Peter W. Cook, Kara Moser, Adrian Paskey, Jason Caravas, Benjamin Rambo-Martin, Shatavia Morrison, Christopher Gulvick, Scott Sammons, Yvette Unoarumhi, Darlene Wagner, Matthew Schmerer, S. H. Rosenthal, A. Gerasimova, R. M. Kagan, B. Anderson, M. Hua, Y. Liu, L.E. Bernstein, K.E. Livingston, A. Perez, I. A. Shlyakhter, R. V. Rolando, R. Owen, P. Tanpaiboon, F. Lacbawan, Clinton R. Paden, Duncan MacCannell |
| EPI_ISL_2011267, EPI_ISL_2011268, EPI_ISL_2011269 | Seattle Flu Study | Seattle Flu Study | Deborah A. Nickerson, Chris D. Frazar, Jover Lee, Benjamin Pelle, Erica Ryke, Matthew Richardson, Amanda Adler, Elisabeth Brandstetter, Peter D. Han, Kairsten Fay, Misja Ilcisin, Kirsten Lacombe, Thomas R. Sibley, Melissa Truong, Caitlin R. Wolf, Karen Cowgill, Stephanie Schrag, Jeff Duchin, Michael Boeckh, Janet A. Englund, Michael Famulare, Barry R. Lutz, Mark J. Rieder, Lea M. Starita, Matthew Thompson, Helen Y. Chu, Trevor Bedford, Jay Shendure |
| EPI_ISL_2011270 | Quest Diagnostics Incorporated | Centers for Disease Control and Prevention Division of Viral Diseases, Pathogen Discovery | Dakota Howard, Dhwani Batra, Peter W. Cook, Kara Moser, Adrian Paskey, Jason Caravas, Benjamin Rambo-Martin, Shatavia Morrison, Christopher Gulvick, Scott Sammons, Yvette Unoarumhi, Darlene Wagner, Matthew Schmerer, S. H. Rosenthal, A. Gerasimova, R. M. Kagan, B. Anderson, M. Hua, Y. Liu, L.E. Bernstein, K.E. Livingston, A. Perez, I. A. Shlyakhter, R. V. Rolando, R. Owen, P. Tanpaiboon, F. Lacbawan, Clinton R. Paden, Duncan MacCannell |
| EPI_ISL_2011273 | Seattle Flu Study | Seattle Flu Study | Deborah A. Nickerson, Chris D. Frazar, Jover Lee, Benjamin Pelle, Erica Ryke, Matthew Richardson, Amanda Adler, Elisabeth Brandstetter, Peter D. Han, Kairsten Fay, Misja Ilcisin, Kirsten Lacombe, Thomas R. Sibley, Melissa Truong, Caitlin R. Wolf, Michael Boeckh, Janet A. Englund, Michael Famulare, Barry R. Lutz, Mark J. Rieder, Lea M. Starita, Matthew Thompson, Jay Shendure, Trevor Bedford, Helen Y. Chu |
| EPI_ISL_2011274 | Quest Diagnostics Incorporated | Centers for Disease Control and Prevention Division of Viral Diseases, Pathogen Discovery | Dakota Howard, Dhwani Batra, Peter W. Cook, Kara Moser, Adrian Paskey, Jason Caravas, Benjamin Rambo-Martin, Shatavia Morrison, Christopher Gulvick, Scott Sammons, Yvette Unoarumhi, Darlene Wagner, Matthew Schmerer, S. H. Rosenthal, A. Gerasimova, R. M. Kagan, B. Anderson, M. Hua, Y. Liu, L.E. Bernstein, K.E. Livingston, A. Perez, I. A. Shlyakhter, R. V. Rolando, R. Owen, P. Tanpaiboon, F. Lacbawan, Clinton R. Paden, Duncan MacCannell |
| EPI_ISL_2011275 | Seattle Flu Study | Seattle Flu Study | Deborah A. Nickerson, Chris D. Frazar, Jover Lee, Benjamin Pelle, Erica Ryke, Matthew Richardson, Amanda Adler, Elisabeth Brandstetter, Peter D. Han, Kairsten Fay, Misja Ilcisin, Kirsten Lacombe, Thomas R. Sibley, Melissa Truong, Caitlin R. Wolf, Michael Boeckh, Janet A. Englund, Michael Famulare, Barry R. Lutz, Mark J. Rieder, Lea M. Starita, Matthew Thompson, Jay Shendure, Trevor Bedford, Helen Y. Chu |
| EPI_ISL_2011276 | Seattle Flu Study | Seattle Flu Study | Deborah A. Nickerson, Chris D. Frazar, Jover Lee, Benjamin Pelle, Erica Ryke, Matthew Richardson, Amanda Adler, Elisabeth Brandstetter, Peter D. Han, Kairsten Fay, Misja Ilcisin, Kirsten Lacombe, Thomas R. Sibley, Melissa Truong, Caitlin R. Wolf, Karen Cowgill, Stephanie Schrag, Jeff Duchin, Michael Boeckh, Janet A. Englund, Michael Famulare, Barry R. Lutz, Mark J. Rieder, Lea M. Starita, Matthew Thompson, Helen Y. Chu, Trevor Bedford, Jay Shendure |
| EPI_ISL_2011277, EPI_ISL_2011278 | Seattle Flu Study | Seattle Flu Study | Deborah A. Nickerson, Chris D. Frazar, Jover Lee, Benjamin Pelle, Erica Ryke, Matthew Richardson, Amanda Adler, Elisabeth Brandstetter, Peter D. Han, Kairsten Fay, Misja Ilcisin, Kirsten Lacombe, Thomas R. Sibley, Melissa Truong, Caitlin R. Wolf, Michael Boeckh, Janet A. Englund, Michael Famulare, Barry R. Lutz, Mark J. Rieder, Lea M. Starita, Matthew Thompson, Jay Shendure, Trevor Bedford, Helen Y. Chu |
| EPI_ISL_2011279 | Interpath Laboratory | Seattle Flu Study | Deborah A. Nickerson, Chris D. Frazar, Jover Lee, Benjamin Pelle, Erica Ryke, Matthew Richardson, Amanda Adler, Elisabeth Brandstetter, Peter D. Han, Kairsten Fay, Misja Ilcisin, Kirsten Lacombe, Thomas R. Sibley, Melissa Truong, Caitlin R. Wolf, Romesh Gautom, Geoff Melly, Brian Hiatt, Philip Dykema, Scott Lindquist, Michael Boeckh, Janet A. Englund, Michael Famulare, Barry R. Lutz, Mark J. Rieder, Lea M. Starita, Matthew Thompson, Helen Y. Chu, Jay Shendure, Trevor Bedford |
| EPI_ISL_2011280 | Quest Diagnostics Incorporated | Centers for Disease Control and Prevention Division of Viral Diseases, Pathogen Discovery | Dakota Howard, Dhwani Batra, Peter W. Cook, Kara Moser, Adrian Paskey, Jason Caravas, Benjamin Rambo-Martin, Shatavia Morrison, Christopher Gulvick, Scott Sammons, Yvette Unoarumhi, Darlene Wagner, Matthew Schmerer, S. H. Rosenthal, A. Gerasimova, R. M. Kagan, B. Anderson, M. Hua, Y. Liu, L.E. Bernstein, K.E. Livingston, A. Perez, I. A. Shlyakhter, R. V. Rolando, R. Owen, P. Tanpaiboon, F. Lacbawan, Clinton R. Paden, Duncan MacCannell |
| EPI_ISL_2011281, EPI_ISL_2011282, EPI_ISL_2011283, EPI_ISL_2011284 | Interpath Laboratory | Seattle Flu Study | Deborah A. Nickerson, Chris D. Frazar, Jover Lee, Benjamin Pelle, Erica Ryke, Matthew Richardson, Amanda Adler, Elisabeth Brandstetter, Peter D. Han, Kairsten Fay, Misja Ilcisin, Kirsten Lacombe, Thomas R. Sibley, Melissa Truong, Caitlin R. Wolf, Romesh Gautom, Geoff Melly, Brian Hiatt, Philip Dykema, Scott Lindquist, Michael Boeckh, Janet A. Englund, Michael Famulare, Barry R. Lutz, Mark J. Rieder, Lea M. Starita, Matthew Thompson, Helen Y. Chu, Jay Shendure, Trevor Bedford |
| EPI_ISL_2011285, EPI_ISL_2011289 | Quest Diagnostics Incorporated | Centers for Disease Control and Prevention Division of Viral Diseases, Pathogen Discovery | Dakota Howard, Dhwani Batra, Peter W. Cook, Kara Moser, Adrian Paskey, Jason Caravas, Benjamin Rambo-Martin, Shatavia Morrison, Christopher Gulvick, Scott Sammons, Yvette Unoarumhi, Darlene Wagner, Matthew Schmerer, S. H. Rosenthal, A. Gerasimova, R. M. Kagan, B. Anderson, M. Hua, Y. Liu, L.E. Bernstein, K.E. Livingston, A. Perez, I. A. Shlyakhter, R. V. Rolando, R. Owen, P. Tanpaiboon, F. Lacbawan, Clinton R. Paden, Duncan MacCannell |
| EPI_ISL_2011290, EPI_ISL_2011291 | Seattle Flu Study | Seattle Flu Study | Deborah A. Nickerson, Chris D. Frazar, Jover Lee, Benjamin Pelle, Erica Ryke, Matthew Richardson, Amanda Adler, Elisabeth Brandstetter, Peter D. Han, Kairsten Fay, Misja Ilcisin, Kirsten Lacombe, Thomas R. Sibley, Melissa Truong, Caitlin R. Wolf, Karen Cowgill, Stephanie Schrag, Jeff Duchin, Michael Boeckh, Janet A. Englund, Michael Famulare, Barry R. Lutz, Mark J. Rieder, Lea M. Starita, Matthew Thompson, Helen Y. Chu, Trevor Bedford, Jay Shendure |
| EPI_ISL_2011292 | Quest Diagnostics Incorporated | Centers for Disease Control and Prevention Division of Viral Diseases, Pathogen | Dakota Howard, Dhwani Batra, Peter W. Cook, Kara Moser, Adrian Paskey, Jason Caravas, Benjamin Rambo-Martin, Shatavia Morrison, Christopher Gulvick, Scott Sammons, Yvette Unoarumhi, Darlene Wagner, Matthew Schmerer, S. H. Rosenthal, A. Gerasimova, R. M. Kagan, B. Anderson, M. Hua, Y. Liu, L.E. Bernstein, K.E. Livingston, A. Perez, I. A. Shlyakhter, R. V. Rolando, R. Owen, P. Tanpaiboon, F. Lacbawan, Clinton R. Paden, Duncan MacCannell |

| | | | |
|---|---|---|---|
| | | Discovery | |
| EPI_ISL_2011293, EPI_ISL_2011294 | Seattle Flu Study | Seattle Flu Study | Deborah A. Nickerson, Chris D. Frazar, Jover Lee, Benjamin Pelle, Erica Ryke, Matthew Richardson, Amanda Adler, Elisabeth Brandstetter, Peter D. Han, Kairsten Fay, Misja Ilcisin, Kirsten Lacombe, Thomas R. Sibley, Melissa Truong, Caitlin R. Wolf, Michael Boeckh, Janet A. Englund, Michael Famulare, Barry R. Lutz, Mark J. Rieder, Lea M. Starita, Matthew Thompson, Jay Shendure, Trevor Bedford, Helen Y. Chu |
| EPI_ISL_2011295 | Quest Diagnostics Incorporated | Centers for Disease Control and Prevention Division of Viral Diseases, Pathogen Discovery | Dakota Howard, Dhwani Batra, Peter W. Cook, Kara Moser, Adrian Paskey, Jason Caravas, Benjamin Rambo-Martin, Shatavia Morrison, Christopher Gulvick, Scott Sammons, Yvette Unoarumhi, Darlene Wagner, Matthew Schmerer, S. H. Rosenthal, A. Gerasimova, R. M. Kagan, B. Anderson, M. Hua, Y. Liu, L.E. Bernstein, K.E. Livingston, A. Perez, I. A. Shlyakhter, R. V. Rolando, R. Owen, P. Tanpaiboon, F. Lacbawan, Clinton R. Paden, Duncan MacCannell |
| EPI_ISL_2011296 | Seattle Flu Study | Seattle Flu Study | Deborah A. Nickerson, Chris D. Frazar, Jover Lee, Benjamin Pelle, Erica Ryke, Matthew Richardson, Amanda Adler, Elisabeth Brandstetter, Peter D. Han, Kairsten Fay, Misja Ilcisin, Kirsten Lacombe, Thomas R. Sibley, Melissa Truong, Caitlin R. Wolf, Michael Boeckh, Janet A. Englund, Michael Famulare, Barry R. Lutz, Mark J. Rieder, Lea M. Starita, Matthew Thompson, Jay Shendure, Trevor Bedford, Helen Y. Chu |
| EPI_ISL_2011297 | Seattle Flu Study | Seattle Flu Study | Deborah A. Nickerson, Chris D. Frazar, Jover Lee, Benjamin Pelle, Erica Ryke, Matthew Richardson, Amanda Adler, Elisabeth Brandstetter, Peter D. Han, Kairsten Fay, Misja Ilcisin, Kirsten Lacombe, Thomas R. Sibley, Melissa Truong, Caitlin R. Wolf, Karen Cowgill, Stephanie Schrag, Jeff Duchin, Michael Boeckh, Janet A. Englund, Michael Famulare, Barry R. Lutz, Mark J. Rieder, Lea M. Starita, Matthew Thompson, Helen Y. Chu, Trevor Bedford, Jay Shendure |
| EPI_ISL_2011299 | Quest Diagnostics Incorporated | Centers for Disease Control and Prevention Division of Viral Diseases, Pathogen Discovery | Dakota Howard, Dhwani Batra, Peter W. Cook, Kara Moser, Adrian Paskey, Jason Caravas, Benjamin Rambo-Martin, Shatavia Morrison, Christopher Gulvick, Scott Sammons, Yvette Unoarumhi, Darlene Wagner, Matthew Schmerer, S. H. Rosenthal, A. Gerasimova, R. M. Kagan, B. Anderson, M. Hua, Y. Liu, L.E. Bernstein, K.E. Livingston, A. Perez, I. A. Shlyakhter, R. V. Rolando, R. Owen, P. Tanpaiboon, F. Lacbawan, Clinton R. Paden, Duncan MacCannell |
| EPI_ISL_2011300 | Seattle Flu Study | Seattle Flu Study | Deborah A. Nickerson, Chris D. Frazar, Jover Lee, Benjamin Pelle, Erica Ryke, Matthew Richardson, Amanda Adler, Elisabeth Brandstetter, Peter D. Han, Kairsten Fay, Misja Ilcisin, Kirsten Lacombe, Thomas R. Sibley, Melissa Truong, Caitlin R. Wolf, Karen Cowgill, Stephanie Schrag, Jeff Duchin, Michael Boeckh, Janet A. Englund, Michael Famulare, Barry R. Lutz, Mark J. Rieder, Lea M. Starita, Matthew Thompson, Helen Y. Chu, Trevor Bedford, Jay Shendure |
| EPI_ISL_2011301 | Seattle Flu Study | Seattle Flu Study | Deborah A. Nickerson, Chris D. Frazar, Jover Lee, Benjamin Pelle, Erica Ryke, Matthew Richardson, Amanda Adler, Elisabeth Brandstetter, Peter D. Han, Kairsten Fay, Misja Ilcisin, Kirsten Lacombe, Thomas R. Sibley, Melissa Truong, Caitlin R. Wolf, Michael Boeckh, Janet A. Englund, Michael Famulare, Barry R. Lutz, Mark J. Rieder, Lea M. Starita, Matthew Thompson, Jay Shendure, Trevor Bedford, Helen Y. Chu |
| EPI_ISL_2011302 | Seattle Flu Study | Seattle Flu Study | Deborah A. Nickerson, Chris D. Frazar, Jover Lee, Benjamin Pelle, Erica Ryke, Matthew Richardson, Amanda Adler, Elisabeth Brandstetter, Peter D. Han, Kairsten Fay, Misja Ilcisin, Kirsten Lacombe, Thomas R. Sibley, Melissa Truong, Caitlin R. Wolf, Karen Cowgill, Stephanie Schrag, Jeff Duchin, Michael Boeckh, Janet A. Englund, Michael Famulare, Barry R. Lutz, Mark J. Rieder, Lea M. Starita, Matthew Thompson, Helen Y. Chu, Trevor Bedford, Jay Shendure |
| EPI_ISL_2011303 | Quest Diagnostics Incorporated | Centers for Disease Control and Prevention Division of Viral Diseases, Pathogen Discovery | Dakota Howard, Dhwani Batra, Peter W. Cook, Kara Moser, Adrian Paskey, Jason Caravas, Benjamin Rambo-Martin, Shatavia Morrison, Christopher Gulvick, Scott Sammons, Yvette Unoarumhi, Darlene Wagner, Matthew Schmerer, S. H. Rosenthal, A. Gerasimova, R. M. Kagan, B. Anderson, M. Hua, Y. Liu, L.E. Bernstein, K.E. Livingston, A. Perez, I. A. Shlyakhter, R. V. Rolando, R. Owen, P. Tanpaiboon, F. Lacbawan, Clinton R. Paden, Duncan MacCannell |
| EPI_ISL_2011304, EPI_ISL_2011305 | Seattle Flu Study | Seattle Flu Study | Deborah A. Nickerson, Chris D. Frazar, Jover Lee, Benjamin Pelle, Erica Ryke, Matthew Richardson, Amanda Adler, Elisabeth Brandstetter, Peter D. Han, Kairsten Fay, Misja Ilcisin, Kirsten Lacombe, Thomas R. Sibley, Melissa Truong, Caitlin R. Wolf, Karen Cowgill, Stephanie Schrag, Jeff Duchin, Michael Boeckh, Janet A. Englund, Michael Famulare, Barry R. Lutz, Mark J. Rieder, Lea M. Starita, Matthew Thompson, Helen Y. Chu, Trevor Bedford, Jay Shendure |
| EPI_ISL_2011306, EPI_ISL_2011307 | Seattle Flu Study | Seattle Flu Study | Deborah A. Nickerson, Chris D. Frazar, Jover Lee, Benjamin Pelle, Erica Ryke, Matthew Richardson, Amanda Adler, Elisabeth Brandstetter, Peter D. Han, Kairsten Fay, Misja Ilcisin, Kirsten Lacombe, Thomas R. Sibley, Melissa Truong, Caitlin R. Wolf, Michael Boeckh, Janet A. Englund, Michael Famulare, Barry R. Lutz, Mark J. Rieder, Lea M. Starita, Matthew Thompson, Jay Shendure, Trevor Bedford, Helen Y. Chu |
| EPI_ISL_2011308 | Quest Diagnostics Incorporated | Centers for Disease Control and Prevention Division of Viral Diseases, Pathogen Discovery | Dakota Howard, Dhwani Batra, Peter W. Cook, Kara Moser, Adrian Paskey, Jason Caravas, Benjamin Rambo-Martin, Shatavia Morrison, Christopher Gulvick, Scott Sammons, Yvette Unoarumhi, Darlene Wagner, Matthew Schmerer, S. H. Rosenthal, A. Gerasimova, R. M. Kagan, B. Anderson, M. Hua, Y. Liu, L.E. Bernstein, K.E. Livingston, A. Perez, I. A. Shlyakhter, R. V. Rolando, R. Owen, P. Tanpaiboon, F. Lacbawan, Clinton R. Paden, Duncan MacCannell |
| EPI_ISL_2011309, EPI_ISL_2011311 | Seattle Flu Study | Seattle Flu Study | Deborah A. Nickerson, Chris D. Frazar, Jover Lee, Benjamin Pelle, Erica Ryke, Matthew Richardson, Amanda Adler, Elisabeth Brandstetter, Peter D. Han, Kairsten Fay, Misja Ilcisin, Kirsten Lacombe, Thomas R. Sibley, Melissa Truong, Caitlin R. Wolf, Michael Boeckh, Janet A. Englund, Michael Famulare, Barry R. Lutz, Mark J. Rieder, Lea M. Starita, Matthew Thompson, Jay Shendure, Trevor Bedford, Helen Y. Chu |
| EPI_ISL_2011312 | Quest Diagnostics Incorporated | Centers for Disease Control and Prevention Division of Viral Diseases, Pathogen Discovery | Dakota Howard, Dhwani Batra, Peter W. Cook, Kara Moser, Adrian Paskey, Jason Caravas, Benjamin Rambo-Martin, Shatavia Morrison, Christopher Gulvick, Scott Sammons, Yvette Unoarumhi, Darlene Wagner, Matthew Schmerer, S. H. Rosenthal, A. Gerasimova, R. M. Kagan, B. Anderson, M. Hua, Y. Liu, L.E. Bernstein, K.E. Livingston, A. Perez, I. A. Shlyakhter, R. V. Rolando, R. Owen, P. Tanpaiboon, F. Lacbawan, Clinton R. Paden, Duncan MacCannell |
| EPI_ISL_2011313 | Seattle Flu Study | Seattle Flu Study | Deborah A. Nickerson, Chris D. Frazar, Jover Lee, Benjamin Pelle, Erica Ryke, Matthew Richardson, Amanda Adler, Elisabeth Brandstetter, Peter D. Han, Kairsten Fay, Misja Ilcisin, Kirsten Lacombe, Thomas R. Sibley, Melissa Truong, Caitlin R. Wolf, Karen Cowgill, Stephanie Schrag, Jeff Duchin, Michael Boeckh, Janet A. Englund, Michael Famulare, Barry R. Lutz, Mark J. Rieder, Lea M. Starita, Matthew Thompson, Helen Y. Chu, Trevor Bedford, Jay Shendure |
| EPI_ISL_2011316 | Quest Diagnostics Incorporated | Centers for Disease Control and Prevention Division of Viral Diseases, Pathogen Discovery | Dakota Howard, Dhwani Batra, Peter W. Cook, Kara Moser, Adrian Paskey, Jason Caravas, Benjamin Rambo-Martin, Shatavia Morrison, Christopher Gulvick, Scott Sammons, Yvette Unoarumhi, Darlene Wagner, Matthew Schmerer, S. H. Rosenthal, A. Gerasimova, R. M. Kagan, B. Anderson, M. Hua, Y. Liu, L.E. Bernstein, K.E. Livingston, A. Perez, I. A. Shlyakhter, R. V. Rolando, R. Owen, P. Tanpaiboon, F. Lacbawan, Clinton R. Paden, Duncan MacCannell |
| EPI_ISL_2011318, EPI_ISL_2011319, EPI_ISL_2011320 | Seattle Flu Study | Seattle Flu Study | Deborah A. Nickerson, Chris D. Frazar, Jover Lee, Benjamin Pelle, Erica Ryke, Matthew Richardson, Amanda Adler, Elisabeth Brandstetter, Peter D. Han, Kairsten Fay, Misja Ilcisin, Kirsten Lacombe, Thomas R. Sibley, Melissa Truong, Caitlin R. Wolf, Karen Cowgill, Stephanie Schrag, Jeff Duchin, Michael Boeckh, Janet A. Englund, Michael Famulare, Barry R. Lutz, Mark J. Rieder, Lea M. Starita, Matthew Thompson, Helen Y. Chu, Trevor Bedford, Jay Shendure |
| EPI_ISL_2011321, EPI_ISL_2011325 | Quest Diagnostics Incorporated | Centers for Disease Control and Prevention Division of Viral Diseases, Pathogen Discovery | Dakota Howard, Dhwani Batra, Peter W. Cook, Kara Moser, Adrian Paskey, Jason Caravas, Benjamin Rambo-Martin, Shatavia Morrison, Christopher Gulvick, Scott Sammons, Yvette Unoarumhi, Darlene Wagner, Matthew Schmerer, S. H. Rosenthal, A. Gerasimova, R. M. Kagan, B. Anderson, M. Hua, Y. Liu, L.E. Bernstein, K.E. Livingston, A. Perez, I. A. Shlyakhter, R. V. Rolando, R. Owen, P. Tanpaiboon, F. Lacbawan, Clinton R. Paden, Duncan MacCannell |
| EPI_ISL_2011326, EPI_ISL_2011327, EPI_ISL_2011328 | Seattle Flu Study | Seattle Flu Study | Deborah A. Nickerson, Chris D. Frazar, Jover Lee, Benjamin Pelle, Erica Ryke, Matthew Richardson, Amanda Adler, Elisabeth Brandstetter, Peter D. Han, Kairsten Fay, Misja Ilcisin, Kirsten Lacombe, Thomas R. Sibley, Melissa Truong, Caitlin R. Wolf, Karen Cowgill, Stephanie Schrag, Jeff Duchin, Michael Boeckh, Janet A. Englund, Michael Famulare, Barry R. Lutz, Mark J. Rieder, Lea M. Starita, Matthew Thompson, Helen Y. Chu, Trevor Bedford, Jay Shendure |
| EPI_ISL_2011329 | Quest Diagnostics Incorporated | Centers for Disease Control and Prevention Division of Viral Diseases, Pathogen Discovery | Dakota Howard, Dhwani Batra, Peter W. Cook, Kara Moser, Adrian Paskey, Jason Caravas, Benjamin Rambo-Martin, Shatavia Morrison, Christopher Gulvick, Scott Sammons, Yvette Unoarumhi, Darlene Wagner, Matthew Schmerer, S. H. Rosenthal, A. Gerasimova, R. M. Kagan, B. Anderson, M. Hua, Y. Liu, L.E. Bernstein, K.E. Livingston, A. Perez, I. A. Shlyakhter, R. V. Rolando, R. Owen, P. Tanpaiboon, F. Lacbawan, Clinton R. Paden, Duncan MacCannell |
| EPI_ISL_2011330 | Seattle Flu Study | Seattle Flu Study | Deborah A. Nickerson, Chris D. Frazar, Jover Lee, Benjamin Pelle, Erica Ryke, Matthew Richardson, Amanda Adler, Elisabeth Brandstetter, Peter D. Han, Kairsten Fay, Misja Ilcisin, Kirsten Lacombe, Thomas R. Sibley, Melissa Truong, Caitlin R. Wolf, Michael Boeckh, Janet A. Englund, Michael Famulare, Barry R. Lutz, Mark J. Rieder, Lea M. Starita, Matthew Thompson, Jay Shendure, Trevor Bedford, Helen Y. Chu |
| EPI_ISL_2011331 | Atlas Genomics | Seattle Flu Study | Deborah A. Nickerson, Chris D. Frazar, Jover Lee, Benjamin Pelle, Erica Ryke, Matthew Richardson, Amanda Adler, Elisabeth Brandstetter, Peter D. Han, Kairsten Fay, Misja Ilcisin, Kirsten Lacombe, Thomas R. Sibley, Melissa Truong, Caitlin R. Wolf, Romesh Gautom, Geoff Melly, Brian Hiatt, Philip Dykema, Scott Lindquist, Michael Boeckh, Janet A. Englund, Michael Famulare, Barry R. Lutz, Mark J. Rieder, Lea M. Starita, Matthew Thompson, Helen Y. Chu, Jay Shendure, Trevor Bedford |
| EPI_ISL_2011332 | Quest Diagnostics Incorporated | Centers for Disease Control and Prevention Division of Viral Diseases, Pathogen Discovery | Dakota Howard, Dhwani Batra, Peter W. Cook, Kara Moser, Adrian Paskey, Jason Caravas, Benjamin Rambo-Martin, Shatavia Morrison, Christopher Gulvick, Scott Sammons, Yvette Unoarumhi, Darlene Wagner, Matthew Schmerer, S. H. Rosenthal, A. Gerasimova, R. M. Kagan, B. Anderson, M. Hua, Y. Liu, L.E. Bernstein, K.E. Livingston, A. Perez, I. A. Shlyakhter, R. V. Rolando, R. Owen, P. Tanpaiboon, F. Lacbawan, Clinton R. Paden, Duncan MacCannell |
| EPI_ISL_2011334, EPI_ISL_2011335 | Atlas Genomics | Seattle Flu Study | Deborah A. Nickerson, Chris D. Frazar, Jover Lee, Benjamin Pelle, Erica Ryke, Matthew Richardson, Amanda Adler, Elisabeth Brandstetter, Peter D. Han, Kairsten Fay, Misja Ilcisin, Kirsten Lacombe, Thomas R. Sibley, Melissa Truong, Caitlin R. Wolf, Romesh Gautom, Geoff Melly, Brian Hiatt, Philip Dykema, Scott Lindquist, Michael Boeckh, Janet A. Englund, Michael Famulare, Barry R. Lutz, Mark J. Rieder, Lea M. Starita, Matthew Thompson, Helen Y. Chu, Jay Shendure, Trevor Bedford |
| EPI_ISL_2011336 | Quest Diagnostics Incorporated | Centers for Disease Control and Prevention Division of Viral Diseases, Pathogen Discovery | Dakota Howard, Dhwani Batra, Peter W. Cook, Kara Moser, Adrian Paskey, Jason Caravas, Benjamin Rambo-Martin, Shatavia Morrison, Christopher Gulvick, Scott Sammons, Yvette Unoarumhi, Darlene Wagner, Matthew Schmerer, S. H. Rosenthal, A. Gerasimova, R. M. Kagan, B. Anderson, M. Hua, Y. Liu, L.E. Bernstein, K.E. Livingston, A. Perez, I. A. Shlyakhter, R. V. Rolando, R. Owen, P. Tanpaiboon, F. Lacbawan, Clinton R. Paden, Duncan MacCannell |
| EPI_ISL_2011337, EPI_ISL_2011338 | Atlas Genomics | Seattle Flu Study | Deborah A. Nickerson, Chris D. Frazar, Jover Lee, Benjamin Pelle, Erica Ryke, Matthew Richardson, Amanda Adler, Elisabeth Brandstetter, Peter D. Han, Kairsten Fay, Misja Ilcisin, Kirsten Lacombe, Thomas R. Sibley, Melissa Truong, Caitlin R. Wolf, Romesh Gautom, Geoff Melly, Brian Hiatt, Philip Dykema, Scott Lindquist, Michael Boeckh, Janet A. Englund, Michael Famulare, Barry R. Lutz, Mark J. Rieder, Lea M. Starita, Matthew Thompson, Helen Y. Chu, Jay Shendure, Trevor Bedford |
| EPI_ISL_2011340 | Quest Diagnostics Incorporated | Centers for Disease Control and Prevention Division of Viral Diseases, Pathogen Discovery | Dakota Howard, Dhwani Batra, Peter W. Cook, Kara Moser, Adrian Paskey, Jason Caravas, Benjamin Rambo-Martin, Shatavia Morrison, Christopher Gulvick, Scott Sammons, Yvette Unoarumhi, Darlene Wagner, Matthew Schmerer, S. H. Rosenthal, A. Gerasimova, R. M. Kagan, B. Anderson, M. Hua, Y. Liu, L.E. Bernstein, K.E. Livingston, A. Perez, I. A. Shlyakhter, R. V. Rolando, R. Owen, P. Tanpaiboon, F. Lacbawan, Clinton R. Paden, Duncan MacCannell |
| EPI_ISL_2011341, EPI_ISL_2011342 | Atlas Genomics | Seattle Flu Study | Deborah A. Nickerson, Chris D. Frazar, Jover Lee, Benjamin Pelle, Erica Ryke, Matthew Richardson, Amanda Adler, Elisabeth Brandstetter, Peter D. Han, Kairsten Fay, Misja Ilcisin, Kirsten Lacombe, Thomas R. Sibley, Melissa Truong, Caitlin R. Wolf, Romesh Gautom, Geoff Melly, Brian Hiatt, Philip Dykema, Scott Lindquist, Michael Boeckh, Janet A. Englund, Michael Famulare, Barry R. Lutz, Mark J. Rieder, Lea M. Starita, Matthew Thompson, Helen Y. Chu, Jay Shendure, Trevor Bedford |
| EPI_ISL_2011343 | Quest Diagnostics Incorporated | Centers for Disease Control and Prevention Division of Viral Diseases, Pathogen Discovery | Dakota Howard, Dhwani Batra, Peter W. Cook, Kara Moser, Adrian Paskey, Jason Caravas, Benjamin Rambo-Martin, Shatavia Morrison, Christopher Gulvick, Scott Sammons, Yvette Unoarumhi, Darlene Wagner, Matthew Schmerer, S. H. Rosenthal, A. Gerasimova, R. M. Kagan, B. Anderson, M. Hua, Y. Liu, L.E. Bernstein, K.E. Livingston, A. Perez, I. A. Shlyakhter, R. V. Rolando, R. Owen, P. Tanpaiboon, F. Lacbawan, Clinton R. Paden, Duncan MacCannell |
| EPI_ISL_2011345, EPI_ISL_2011346 | Atlas Genomics | Seattle Flu Study | Deborah A. Nickerson, Chris D. Frazar, Jover Lee, Benjamin Pelle, Erica Ryke, Matthew Richardson, Amanda Adler, Elisabeth Brandstetter, Peter D. Han, Kairsten Fay, Misja Ilcisin, Kirsten Lacombe, Thomas R. Sibley, Melissa Truong, Caitlin R. Wolf, Romesh Gautom, Geoff Melly, Brian Hiatt, Philip Dykema, Scott Lindquist, Michael Boeckh, Janet A. Englund, Michael Famulare, Barry R. Lutz, Mark J. Rieder, Lea M. Starita, Matthew Thompson, Helen Y. Chu, Jay Shendure, Trevor Bedford |
| EPI_ISL_2011347 | Quest Diagnostics Incorporated | Centers for Disease Control and Prevention Division of Viral Diseases, Pathogen Discovery | Dakota Howard, Dhwani Batra, Peter W. Cook, Kara Moser, Adrian Paskey, Jason Caravas, Benjamin Rambo-Martin, Shatavia Morrison, Christopher Gulvick, Scott Sammons, Yvette Unoarumhi, Darlene Wagner, Matthew Schmerer, S. H. Rosenthal, A. Gerasimova, R. M. Kagan, B. Anderson, M. Hua, Y. Liu, L.E. Bernstein, K.E. Livingston, A. Perez, I. A. Shlyakhter, R. V. Rolando, R. Owen, P. Tanpaiboon, F. Lacbawan, Clinton R. Paden, Duncan MacCannell |
| EPI_ISL_2011348, EPI_ISL_2011349, EPI_ISL_2011350 | Atlas Genomics | Seattle Flu Study | Deborah A. Nickerson, Chris D. Frazar, Jover Lee, Benjamin Pelle, Erica Ryke, Matthew Richardson, Amanda Adler, Elisabeth Brandstetter, Peter D. Han, Kairsten Fay, Misja Ilcisin, Kirsten Lacombe, Thomas R. Sibley, Melissa Truong, Caitlin R. Wolf, Romesh Gautom, Geoff Melly, Brian Hiatt, Philip Dykema, Scott Lindquist, Michael Boeckh, Janet A. Englund, Michael Famulare, Barry R. Lutz, Mark J. Rieder, Lea M. Starita, Matthew Thompson, Helen Y. Chu, Jay Shendure, Trevor Bedford |
| EPI_ISL_2011351 | Quest Diagnostics Incorporated | Centers for Disease Control and Prevention Division of Viral Diseases, Pathogen Discovery | Dakota Howard, Dhwani Batra, Peter W. Cook, Kara Moser, Adrian Paskey, Jason Caravas, Benjamin Rambo-Martin, Shatavia Morrison, Christopher Gulvick, Scott Sammons, Yvette Unoarumhi, Darlene Wagner, Matthew Schmerer, S. H. Rosenthal, A. Gerasimova, R. M. Kagan, B. Anderson, M. Hua, Y. Liu, L.E. Bernstein, K.E. Livingston, A. Perez, I. A. Shlyakhter, R. V. Rolando, R. Owen, P. Tanpaiboon, F. Lacbawan, Clinton R. Paden, Duncan MacCannell |
| EPI_ISL_2011352 | Atlas Genomics | Seattle Flu Study | Deborah A. Nickerson, Chris D. Frazar, Jover Lee, Benjamin Pelle, Erica Ryke, Matthew Richardson, Amanda Adler, Elisabeth Brandstetter, Peter D. Han, Kairsten Fay, Misja Ilcisin, Kirsten Lacombe, Thomas R. Sibley, Melissa Truong, Caitlin R. Wolf, Romesh Gautom, Geoff Melly, Brian Hiatt, Philip Dykema, Scott Lindquist, Michael Boeckh, Janet A. Englund, Michael Famulare, Barry R. Lutz, Mark J. Rieder, Lea M. Starita, Matthew Thompson, Helen Y. Chu, Jay Shendure, Trevor Bedford |
| EPI_ISL_2011354 | Quest Diagnostics Incorporated | Centers for Disease Control and Prevention Division of Viral Diseases, Pathogen Discovery | Dakota Howard, Dhwani Batra, Peter W. Cook, Kara Moser, Adrian Paskey, Jason Caravas, Benjamin Rambo-Martin, Shatavia Morrison, Christopher Gulvick, Scott Sammons, Yvette Unoarumhi, Darlene Wagner, Matthew Schmerer, S. H. Rosenthal, A. Gerasimova, R. M. Kagan, B. Anderson, M. Hua, Y. Liu, L.E. Bernstein, K.E. Livingston, A. Perez, I. A. Shlyakhter, R. V. Rolando, R. Owen, P. Tanpaiboon, F. Lacbawan, Clinton R. Paden, Duncan MacCannell |
| EPI_ISL_2011355, EPI_ISL_2011356, EPI_ISL_2011357 | Atlas Genomics | Seattle Flu Study | Deborah A. Nickerson, Chris D. Frazar, Jover Lee, Benjamin Pelle, Erica Ryke, Matthew Richardson, Amanda Adler, Elisabeth Brandstetter, Peter D. Han, Kairsten Fay, Misja Ilcisin, Kirsten Lacombe, Thomas R. Sibley, Melissa Truong, Caitlin R. Wolf, Romesh Gautom, Geoff Melly, Brian Hiatt, Philip Dykema, Scott Lindquist, Michael Boeckh, Janet A. Englund, Michael Famulare, Barry R. Lutz, Mark J. Rieder, Lea M. Starita, Matthew Thompson, Helen Y. Chu, Jay Shendure, Trevor Bedford |
| EPI_ISL_2011358 | Quest Diagnostics Incorporated | Centers for Disease Control and Prevention | Dakota Howard, Dhwani Batra, Peter W. Cook, Kara Moser, Adrian Paskey, Jason Caravas, Benjamin Rambo-Martin, Shatavia Morrison, Christopher Gulvick, Scott Sammons, Yvette Unoarumhi, Darlene Wagner, Matthew Schmerer, S. H. Rosenthal, A. Gerasimova, R. M. Kagan, B. |

| | | | |
|---|---|---|---|
| | | Division of Viral Diseases, Pathogen Discovery | Anderson, M. Hua, Y. Liu, L.E. Bernstein, K.E. Livingston, A. Perez, I. A. Shlyakhter, R. V. Rolando, R. Owen, P. Tanpaiboon, F. Lacbawan, Clinton R. Paden, Duncan MacCannell |
| EPI_ISL_2011361 | Atlas Genomics | Seattle Flu Study | Deborah A. Nickerson, Chris D. Frazar, Jover Lee, Benjamin Pelle, Erica Ryke, Matthew Richardson, Amanda Adler, Elisabeth Brandstetter, Peter D. Han, Kairsten Fay, Misja Ilcisin, Kirsten Lacombe, Thomas R. Sibley, Melissa Truong, Caitlin R. Wolf, Romesh Gautom, Geoff Melly, Brian Hiatt, Philip Dykema, Scott Lindquist, Michael Boeckh, Janet A. Englund, Michael Famulare, Barry R. Lutz, Mark J. Rieder, Lea M. Starita, Matthew Thompson, Helen Y. Chu, Jay Shendure, Trevor Bedford |
| EPI_ISL_2011362 | Quest Diagnostics Incorporated | Centers for Disease Control and Prevention Division of Viral Diseases, Pathogen Discovery | Dakota Howard, Dhwani Batra, Peter W. Cook, Kara Moser, Adrian Paskey, Jason Caravas, Benjamin Rambo-Martin, Shatavia Morrison, Christopher Gulvick, Scott Sammons, Yvette Unoarumhi, Darlene Wagner, Matthew Schmerer, S. H. Rosenthal, A. Gerasimova, R. M. Kagan, B. Anderson, M. Hua, Y. Liu, L.E. Bernstein, K.E. Livingston, A. Perez, I. A. Shlyakhter, R. V. Rolando, R. Owen, P. Tanpaiboon, F. Lacbawan, Clinton R. Paden, Duncan MacCannell |
| EPI_ISL_2011363 | Atlas Genomics | Seattle Flu Study | Deborah A. Nickerson, Chris D. Frazar, Jover Lee, Benjamin Pelle, Erica Ryke, Matthew Richardson, Amanda Adler, Elisabeth Brandstetter, Peter D. Han, Kairsten Fay, Misja Ilcisin, Kirsten Lacombe, Thomas R. Sibley, Melissa Truong, Caitlin R. Wolf, Romesh Gautom, Geoff Melly, Brian Hiatt, Philip Dykema, Scott Lindquist, Michael Boeckh, Janet A. Englund, Michael Famulare, Barry R. Lutz, Mark J. Rieder, Lea M. Starita, Matthew Thompson, Helen Y. Chu, Jay Shendure, Trevor Bedford |
| EPI_ISL_2011365 | Quest Diagnostics Incorporated | Centers for Disease Control and Prevention Division of Viral Diseases, Pathogen Discovery | Dakota Howard, Dhwani Batra, Peter W. Cook, Kara Moser, Adrian Paskey, Jason Caravas, Benjamin Rambo-Martin, Shatavia Morrison, Christopher Gulvick, Scott Sammons, Yvette Unoarumhi, Darlene Wagner, Matthew Schmerer, S. H. Rosenthal, A. Gerasimova, R. M. Kagan, B. Anderson, M. Hua, Y. Liu, L.E. Bernstein, K.E. Livingston, A. Perez, I. A. Shlyakhter, R. V. Rolando, R. Owen, P. Tanpaiboon, F. Lacbawan, Clinton R. Paden, Duncan MacCannell |
| EPI_ISL_2011366, EPI_ISL_2011367, EPI_ISL_2011368 | Atlas Genomics | Seattle Flu Study | Deborah A. Nickerson, Chris D. Frazar, Jover Lee, Benjamin Pelle, Erica Ryke, Matthew Richardson, Amanda Adler, Elisabeth Brandstetter, Peter D. Han, Kairsten Fay, Misja Ilcisin, Kirsten Lacombe, Thomas R. Sibley, Melissa Truong, Caitlin R. Wolf, Romesh Gautom, Geoff Melly, Brian Hiatt, Philip Dykema, Scott Lindquist, Michael Boeckh, Janet A. Englund, Michael Famulare, Barry R. Lutz, Mark J. Rieder, Lea M. Starita, Matthew Thompson, Helen Y. Chu, Jay Shendure, Trevor Bedford |
| EPI_ISL_2011375, EPI_ISL_2011379 | Quest Diagnostics Incorporated | Centers for Disease Control and Prevention Division of Viral Diseases, Pathogen Discovery | Dakota Howard, Dhwani Batra, Peter W. Cook, Kara Moser, Adrian Paskey, Jason Caravas, Benjamin Rambo-Martin, Shatavia Morrison, Christopher Gulvick, Scott Sammons, Yvette Unoarumhi, Darlene Wagner, Matthew Schmerer, S. H. Rosenthal, A. Gerasimova, R. M. Kagan, B. Anderson, M. Hua, Y. Liu, L.E. Bernstein, K.E. Livingston, A. Perez, I. A. Shlyakhter, R. V. Rolando, R. Owen, P. Tanpaiboon, F. Lacbawan, Clinton R. Paden, Duncan MacCannell |
| EPI_ISL_2011380, EPI_ISL_2011381 | Atlas Genomics | Seattle Flu Study | Deborah A. Nickerson, Chris D. Frazar, Jover Lee, Benjamin Pelle, Erica Ryke, Matthew Richardson, Amanda Adler, Elisabeth Brandstetter, Peter D. Han, Kairsten Fay, Misja Ilcisin, Kirsten Lacombe, Thomas R. Sibley, Melissa Truong, Caitlin R. Wolf, Romesh Gautom, Geoff Melly, Brian Hiatt, Philip Dykema, Scott Lindquist, Michael Boeckh, Janet A. Englund, Michael Famulare, Barry R. Lutz, Mark J. Rieder, Lea M. Starita, Matthew Thompson, Helen Y. Chu, Jay Shendure, Trevor Bedford |
| EPI_ISL_2011383 | Quest Diagnostics Incorporated | Centers for Disease Control and Prevention Division of Viral Diseases, Pathogen Discovery | Dakota Howard, Dhwani Batra, Peter W. Cook, Kara Moser, Adrian Paskey, Jason Caravas, Benjamin Rambo-Martin, Shatavia Morrison, Christopher Gulvick, Scott Sammons, Yvette Unoarumhi, Darlene Wagner, Matthew Schmerer, S. H. Rosenthal, A. Gerasimova, R. M. Kagan, B. Anderson, M. Hua, Y. Liu, L.E. Bernstein, K.E. Livingston, A. Perez, I. A. Shlyakhter, R. V. Rolando, R. Owen, P. Tanpaiboon, F. Lacbawan, Clinton R. Paden, Duncan MacCannell |
| EPI_ISL_2011385, EPI_ISL_2011387 | Atlas Genomics | Seattle Flu Study | Deborah A. Nickerson, Chris D. Frazar, Jover Lee, Benjamin Pelle, Erica Ryke, Matthew Richardson, Amanda Adler, Elisabeth Brandstetter, Peter D. Han, Kairsten Fay, Misja Ilcisin, Kirsten Lacombe, Thomas R. Sibley, Melissa Truong, Caitlin R. Wolf, Romesh Gautom, Geoff Melly, Brian Hiatt, Philip Dykema, Scott Lindquist, Michael Boeckh, Janet A. Englund, Michael Famulare, Barry R. Lutz, Mark J. Rieder, Lea M. Starita, Matthew Thompson, Helen Y. Chu, Jay Shendure, Trevor Bedford |
| EPI_ISL_2011388, EPI_ISL_2011392, EPI_ISL_2011395 | Quest Diagnostics Incorporated | Centers for Disease Control and Prevention Division of Viral Diseases, Pathogen Discovery | Dakota Howard, Dhwani Batra, Peter W. Cook, Kara Moser, Adrian Paskey, Jason Caravas, Benjamin Rambo-Martin, Shatavia Morrison, Christopher Gulvick, Scott Sammons, Yvette Unoarumhi, Darlene Wagner, Matthew Schmerer, S. H. Rosenthal, A. Gerasimova, R. M. Kagan, B. Anderson, M. Hua, Y. Liu, L.E. Bernstein, K.E. Livingston, A. Perez, I. A. Shlyakhter, R. V. Rolando, R. Owen, P. Tanpaiboon, F. Lacbawan, Clinton R. Paden, Duncan MacCannell |
| EPI_ISL_2011397 | Atlas Genomics | Seattle Flu Study | Deborah A. Nickerson, Chris D. Frazar, Jover Lee, Benjamin Pelle, Erica Ryke, Matthew Richardson, Amanda Adler, Elisabeth Brandstetter, Peter D. Han, Kairsten Fay, Misja Ilcisin, Kirsten Lacombe, Thomas R. Sibley, Melissa Truong, Caitlin R. Wolf, Romesh Gautom, Geoff Melly, Brian Hiatt, Philip Dykema, Scott Lindquist, Michael Boeckh, Janet A. Englund, Michael Famulare, Barry R. Lutz, Mark J. Rieder, Lea M. Starita, Matthew Thompson, Helen Y. Chu, Jay Shendure, Trevor Bedford |
| EPI_ISL_2011399 | Quest Diagnostics Incorporated | Centers for Disease Control and Prevention Division of Viral Diseases, Pathogen Discovery | Dakota Howard, Dhwani Batra, Peter W. Cook, Kara Moser, Adrian Paskey, Jason Caravas, Benjamin Rambo-Martin, Shatavia Morrison, Christopher Gulvick, Scott Sammons, Yvette Unoarumhi, Darlene Wagner, Matthew Schmerer, S. H. Rosenthal, A. Gerasimova, R. M. Kagan, B. Anderson, M. Hua, Y. Liu, L.E. Bernstein, K.E. Livingston, A. Perez, I. A. Shlyakhter, R. V. Rolando, R. Owen, P. Tanpaiboon, F. Lacbawan, Clinton R. Paden, Duncan MacCannell |
| EPI_ISL_2011400, EPI_ISL_2011401 | Atlas Genomics | Seattle Flu Study | Deborah A. Nickerson, Chris D. Frazar, Jover Lee, Benjamin Pelle, Erica Ryke, Matthew Richardson, Amanda Adler, Elisabeth Brandstetter, Peter D. Han, Kairsten Fay, Misja Ilcisin, Kirsten Lacombe, Thomas R. Sibley, Melissa Truong, Caitlin R. Wolf, Romesh Gautom, Geoff Melly, Brian Hiatt, Philip Dykema, Scott Lindquist, Michael Boeckh, Janet A. Englund, Michael Famulare, Barry R. Lutz, Mark J. Rieder, Lea M. Starita, Matthew Thompson, Helen Y. Chu, Jay Shendure, Trevor Bedford |
| EPI_ISL_2011402 | Quest Diagnostics Incorporated | Centers for Disease Control and Prevention Division of Viral Diseases, Pathogen Discovery | Dakota Howard, Dhwani Batra, Peter W. Cook, Kara Moser, Adrian Paskey, Jason Caravas, Benjamin Rambo-Martin, Shatavia Morrison, Christopher Gulvick, Scott Sammons, Yvette Unoarumhi, Darlene Wagner, Matthew Schmerer, S. H. Rosenthal, A. Gerasimova, R. M. Kagan, B. Anderson, M. Hua, Y. Liu, L.E. Bernstein, K.E. Livingston, A. Perez, I. A. Shlyakhter, R. V. Rolando, R. Owen, P. Tanpaiboon, F. Lacbawan, Clinton R. Paden, Duncan MacCannell |
| EPI_ISL_2011403, EPI_ISL_2011404, EPI_ISL_2011405 | Atlas Genomics | Seattle Flu Study | Deborah A. Nickerson, Chris D. Frazar, Jover Lee, Benjamin Pelle, Erica Ryke, Matthew Richardson, Amanda Adler, Elisabeth Brandstetter, Peter D. Han, Kairsten Fay, Misja Ilcisin, Kirsten Lacombe, Thomas R. Sibley, Melissa Truong, Caitlin R. Wolf, Romesh Gautom, Geoff Melly, Brian Hiatt, Philip Dykema, Scott Lindquist, Michael Boeckh, Janet A. Englund, Michael Famulare, Barry R. Lutz, Mark J. Rieder, Lea M. Starita, Matthew Thompson, Helen Y. Chu, Jay Shendure, Trevor Bedford |
| EPI_ISL_2011406 | Quest Diagnostics Incorporated | Centers for Disease Control and Prevention Division of Viral Diseases, Pathogen Discovery | Dakota Howard, Dhwani Batra, Peter W. Cook, Kara Moser, Adrian Paskey, Jason Caravas, Benjamin Rambo-Martin, Shatavia Morrison, Christopher Gulvick, Scott Sammons, Yvette Unoarumhi, Darlene Wagner, Matthew Schmerer, S. H. Rosenthal, A. Gerasimova, R. M. Kagan, B. Anderson, M. Hua, Y. Liu, L.E. Bernstein, K.E. Livingston, A. Perez, I. A. Shlyakhter, R. V. Rolando, R. Owen, P. Tanpaiboon, F. Lacbawan, Clinton R. Paden, Duncan MacCannell |
| EPI_ISL_2011408 | Atlas Genomics | Seattle Flu Study | Deborah A. Nickerson, Chris D. Frazar, Jover Lee, Benjamin Pelle, Erica Ryke, Matthew Richardson, Amanda Adler, Elisabeth Brandstetter, Peter D. Han, Kairsten Fay, Misja Ilcisin, Kirsten Lacombe, Thomas R. Sibley, Melissa Truong, Caitlin R. Wolf, Romesh Gautom, Geoff Melly, Brian Hiatt, Philip Dykema, Scott Lindquist, Michael Boeckh, Janet A. Englund, Michael Famulare, Barry R. Lutz, Mark J. Rieder, Lea M. Starita, Matthew Thompson, Helen Y. Chu, Jay Shendure, Trevor Bedford |
| EPI_ISL_2011409 | Quest Diagnostics Incorporated | Centers for Disease Control and Prevention Division of Viral Diseases, Pathogen Discovery | Dakota Howard, Dhwani Batra, Peter W. Cook, Kara Moser, Adrian Paskey, Jason Caravas, Benjamin Rambo-Martin, Shatavia Morrison, Christopher Gulvick, Scott Sammons, Yvette Unoarumhi, Darlene Wagner, Matthew Schmerer, S. H. Rosenthal, A. Gerasimova, R. M. Kagan, B. Anderson, M. Hua, Y. Liu, L.E. Bernstein, K.E. Livingston, A. Perez, I. A. Shlyakhter, R. V. Rolando, R. Owen, P. Tanpaiboon, F. Lacbawan, Clinton R. Paden, Duncan MacCannell |
| EPI_ISL_2011410, EPI_ISL_2011411, EPI_ISL_2011412 | Atlas Genomics | Seattle Flu Study | Deborah A. Nickerson, Chris D. Frazar, Jover Lee, Benjamin Pelle, Erica Ryke, Matthew Richardson, Amanda Adler, Elisabeth Brandstetter, Peter D. Han, Kairsten Fay, Misja Ilcisin, Kirsten Lacombe, Thomas R. Sibley, Melissa Truong, Caitlin R. Wolf, Romesh Gautom, Geoff Melly, Brian Hiatt, Philip Dykema, Scott Lindquist, Michael Boeckh, Janet A. Englund, Michael Famulare, Barry R. Lutz, Mark J. Rieder, Lea M. Starita, Matthew Thompson, Helen Y. Chu, Jay Shendure, Trevor Bedford |
| EPI_ISL_2011413 | Quest Diagnostics Incorporated | Centers for Disease Control and Prevention Division of Viral Diseases, Pathogen Discovery | Dakota Howard, Dhwani Batra, Peter W. Cook, Kara Moser, Adrian Paskey, Jason Caravas, Benjamin Rambo-Martin, Shatavia Morrison, Christopher Gulvick, Scott Sammons, Yvette Unoarumhi, Darlene Wagner, Matthew Schmerer, S. H. Rosenthal, A. Gerasimova, R. M. Kagan, B. Anderson, M. Hua, Y. Liu, L.E. Bernstein, K.E. Livingston, A. Perez, I. A. Shlyakhter, R. V. Rolando, R. Owen, P. Tanpaiboon, F. Lacbawan, Clinton R. Paden, Duncan MacCannell |
| EPI_ISL_2011415, EPI_ISL_2011416 | Atlas Genomics | Seattle Flu Study | Deborah A. Nickerson, Chris D. Frazar, Jover Lee, Benjamin Pelle, Erica Ryke, Matthew Richardson, Amanda Adler, Elisabeth Brandstetter, Peter D. Han, Kairsten Fay, Misja Ilcisin, Kirsten Lacombe, Thomas R. Sibley, Melissa Truong, Caitlin R. Wolf, Romesh Gautom, Geoff Melly, Brian Hiatt, Philip Dykema, Scott Lindquist, Michael Boeckh, Janet A. Englund, Michael Famulare, Barry R. Lutz, Mark J. Rieder, Lea M. Starita, Matthew Thompson, Helen Y. Chu, Jay Shendure, Trevor Bedford |
| EPI_ISL_2011418, EPI_ISL_2011420, EPI_ISL_2011423 | Quest Diagnostics Incorporated | Centers for Disease Control and Prevention Division of Viral Diseases, Pathogen Discovery | Dakota Howard, Dhwani Batra, Peter W. Cook, Kara Moser, Adrian Paskey, Jason Caravas, Benjamin Rambo-Martin, Shatavia Morrison, Christopher Gulvick, Scott Sammons, Yvette Unoarumhi, Darlene Wagner, Matthew Schmerer, S. H. Rosenthal, A. Gerasimova, R. M. Kagan, B. Anderson, M. Hua, Y. Liu, L.E. Bernstein, K.E. Livingston, A. Perez, I. A. Shlyakhter, R. V. Rolando, R. Owen, P. Tanpaiboon, F. Lacbawan, Clinton R. Paden, Duncan MacCannell |
| EPI_ISL_2011425 | Seattle Flu Study | Seattle Flu Study | Deborah A. Nickerson, Chris D. Frazar, Jover Lee, Benjamin Pelle, Erica Ryke, Matthew Richardson, Amanda Adler, Elisabeth Brandstetter, Peter D. Han, Kairsten Fay, Misja Ilcisin, Kirsten Lacombe, Thomas R. Sibley, Melissa Truong, Caitlin R. Wolf, Karen Cowgill, Stephanie Schrag, Jeff Duchin, Michael Boeckh, Janet A. Englund, Michael Famulare, Barry R. Lutz, Mark J. Rieder, Lea M. Starita, Matthew Thompson, Helen Y. Chu, Trevor Bedford, Jay Shendure |
| EPI_ISL_2011426 | Quest Diagnostics Incorporated | Centers for Disease Control and Prevention Division of Viral Diseases, Pathogen Discovery | Dakota Howard, Dhwani Batra, Peter W. Cook, Kara Moser, Adrian Paskey, Jason Caravas, Benjamin Rambo-Martin, Shatavia Morrison, Christopher Gulvick, Scott Sammons, Yvette Unoarumhi, Darlene Wagner, Matthew Schmerer, S. H. Rosenthal, A. Gerasimova, R. M. Kagan, B. Anderson, M. Hua, Y. Liu, L.E. Bernstein, K.E. Livingston, A. Perez, I. A. Shlyakhter, R. V. Rolando, R. Owen, P. Tanpaiboon, F. Lacbawan, Clinton R. Paden, Duncan MacCannell |
| EPI_ISL_2011427, EPI_ISL_2011429 | Seattle Flu Study | Seattle Flu Study | Deborah A. Nickerson, Chris D. Frazar, Jover Lee, Benjamin Pelle, Erica Ryke, Matthew Richardson, Amanda Adler, Elisabeth Brandstetter, Peter D. Han, Kairsten Fay, Misja Ilcisin, Kirsten Lacombe, Thomas R. Sibley, Melissa Truong, Caitlin R. Wolf, Karen Cowgill, Stephanie Schrag, Jeff Duchin, Michael Boeckh, Janet A. Englund, Michael Famulare, Barry R. Lutz, Mark J. Rieder, Lea M. Starita, Matthew Thompson, Helen Y. Chu, Trevor Bedford, Jay Shendure |
| EPI_ISL_2011430 | Quest Diagnostics Incorporated | Centers for Disease Control and Prevention Division of Viral Diseases, Pathogen Discovery | Dakota Howard, Dhwani Batra, Peter W. Cook, Kara Moser, Adrian Paskey, Jason Caravas, Benjamin Rambo-Martin, Shatavia Morrison, Christopher Gulvick, Scott Sammons, Yvette Unoarumhi, Darlene Wagner, Matthew Schmerer, S. H. Rosenthal, A. Gerasimova, R. M. Kagan, B. Anderson, M. Hua, Y. Liu, L.E. Bernstein, K.E. Livingston, A. Perez, I. A. Shlyakhter, R. V. Rolando, R. Owen, P. Tanpaiboon, F. Lacbawan, Clinton R. Paden, Duncan MacCannell |
| EPI_ISL_2011432 | Seattle Flu Study | Seattle Flu Study | Deborah A. Nickerson, Chris D. Frazar, Jover Lee, Benjamin Pelle, Erica Ryke, Matthew Richardson, Amanda Adler, Elisabeth Brandstetter, Peter D. Han, Kairsten Fay, Misja Ilcisin, Kirsten Lacombe, Thomas R. Sibley, Melissa Truong, Caitlin R. Wolf, Karen Cowgill, Stephanie Schrag, Jeff Duchin, Michael Boeckh, Janet A. Englund, Michael Famulare, Barry R. Lutz, Mark J. Rieder, Lea M. Starita, Matthew Thompson, Helen Y. Chu, Trevor Bedford, Jay Shendure |
| EPI_ISL_2011433 | Quest Diagnostics Incorporated | Centers for Disease Control and Prevention Division of Viral Diseases, Pathogen Discovery | Dakota Howard, Dhwani Batra, Peter W. Cook, Kara Moser, Adrian Paskey, Jason Caravas, Benjamin Rambo-Martin, Shatavia Morrison, Christopher Gulvick, Scott Sammons, Yvette Unoarumhi, Darlene Wagner, Matthew Schmerer, S. H. Rosenthal, A. Gerasimova, R. M. Kagan, B. Anderson, M. Hua, Y. Liu, L.E. Bernstein, K.E. Livingston, A. Perez, I. A. Shlyakhter, R. V. Rolando, R. Owen, P. Tanpaiboon, F. Lacbawan, Clinton R. Paden, Duncan MacCannell |
| EPI_ISL_2011434, EPI_ISL_2011435 | Seattle Flu Study | Seattle Flu Study | Deborah A. Nickerson, Chris D. Frazar, Jover Lee, Benjamin Pelle, Erica Ryke, Matthew Richardson, Amanda Adler, Elisabeth Brandstetter, Peter D. Han, Kairsten Fay, Misja Ilcisin, Kirsten Lacombe, Thomas R. Sibley, Melissa Truong, Caitlin R. Wolf, Karen Cowgill, Stephanie Schrag, Jeff Duchin, Michael Boeckh, Janet A. Englund, Michael Famulare, Barry R. Lutz, Mark J. Rieder, Lea M. Starita, Matthew Thompson, Helen Y. Chu, Trevor Bedford, Jay Shendure |
| EPI_ISL_2011436 | Quest Diagnostics Incorporated | Centers for Disease Control and Prevention Division of Viral Diseases, Pathogen Discovery | Dakota Howard, Dhwani Batra, Peter W. Cook, Kara Moser, Adrian Paskey, Jason Caravas, Benjamin Rambo-Martin, Shatavia Morrison, Christopher Gulvick, Scott Sammons, Yvette Unoarumhi, Darlene Wagner, Matthew Schmerer, S. H. Rosenthal, A. Gerasimova, R. M. Kagan, B. Anderson, M. Hua, Y. Liu, L.E. Bernstein, K.E. Livingston, A. Perez, I. A. Shlyakhter, R. V. Rolando, R. Owen, P. Tanpaiboon, F. Lacbawan, Clinton R. Paden, Duncan MacCannell |
| EPI_ISL_2011437, EPI_ISL_2011438 | Seattle Flu Study | Seattle Flu Study | Deborah A. Nickerson, Chris D. Frazar, Jover Lee, Benjamin Pelle, Erica Ryke, Matthew Richardson, Amanda Adler, Elisabeth Brandstetter, Peter D. Han, Kairsten Fay, Misja Ilcisin, Kirsten Lacombe, Thomas R. Sibley, Melissa Truong, Caitlin R. Wolf, Karen Cowgill, Stephanie Schrag, Jeff Duchin, Michael Boeckh, Janet A. Englund, Michael Famulare, Barry R. Lutz, Mark J. Rieder, Lea M. Starita, Matthew Thompson, Helen Y. Chu, Trevor Bedford, Jay Shendure |
| EPI_ISL_2011439 | Quest Diagnostics Incorporated | Centers for Disease Control and Prevention Division of Viral Diseases, Pathogen Discovery | Dakota Howard, Dhwani Batra, Peter W. Cook, Kara Moser, Adrian Paskey, Jason Caravas, Benjamin Rambo-Martin, Shatavia Morrison, Christopher Gulvick, Scott Sammons, Yvette Unoarumhi, Darlene Wagner, Matthew Schmerer, S. H. Rosenthal, A. Gerasimova, R. M. Kagan, B. Anderson, M. Hua, Y. Liu, L.E. Bernstein, K.E. Livingston, A. Perez, I. A. Shlyakhter, R. V. Rolando, R. Owen, P. Tanpaiboon, F. Lacbawan, Clinton R. Paden, Duncan MacCannell |
| EPI_ISL_2011443 | Seattle Flu Study | Seattle Flu Study | Deborah A. Nickerson, Chris D. Frazar, Jover Lee, Benjamin Pelle, Erica Ryke, Matthew Richardson, Amanda Adler, Elisabeth Brandstetter, Peter D. Han, Kairsten Fay, Misja Ilcisin, Kirsten Lacombe, Thomas R. Sibley, Melissa Truong, Caitlin R. Wolf, Karen Cowgill, Stephanie Schrag, Jeff Duchin, Michael Boeckh, Janet A. Englund, Michael Famulare, Barry R. Lutz, Mark J. Rieder, Lea M. Starita, Matthew Thompson, Helen Y. Chu, Trevor Bedford, Jay Shendure |
| EPI_ISL_2011444 | Quest Diagnostics Incorporated | Centers for Disease Control and Prevention Division of Viral Diseases, Pathogen Discovery | Dakota Howard, Dhwani Batra, Peter W. Cook, Kara Moser, Adrian Paskey, Jason Caravas, Benjamin Rambo-Martin, Shatavia Morrison, Christopher Gulvick, Scott Sammons, Yvette Unoarumhi, Darlene Wagner, Matthew Schmerer, S. H. Rosenthal, A. Gerasimova, R. M. Kagan, B. Anderson, M. Hua, Y. Liu, L.E. Bernstein, K.E. Livingston, A. Perez, I. A. Shlyakhter, R. V. Rolando, R. Owen, P. Tanpaiboon, F. Lacbawan, Clinton R. Paden, Duncan MacCannell |
| EPI_ISL_2011445 | Seattle Flu Study | Seattle Flu Study | Deborah A. Nickerson, Chris D. Frazar, Jover Lee, Benjamin Pelle, Erica Ryke, Matthew Richardson, Amanda Adler, Elisabeth Brandstetter, Peter D. Han, Kairsten Fay, Misja Ilcisin, Kirsten Lacombe, Thomas R. Sibley, Melissa Truong, Caitlin R. Wolf, Karen Cowgill, Stephanie Schrag, Jeff Duchin, Michael Boeckh, Janet A. Englund, Michael Famulare, Barry R. Lutz, Mark J. Rieder, Lea M. Starita, Matthew Thompson, Helen Y. Chu, Trevor Bedford, Jay Shendure |
| EPI_ISL_2011447 | Quest Diagnostics Incorporated | Centers for Disease Control and Prevention | Dakota Howard, Dhwani Batra, Peter W. Cook, Kara Moser, Adrian Paskey, Jason Caravas, Benjamin Rambo-Martin, Shatavia Morrison, Christopher Gulvick, Scott Sammons, Yvette Unoarumhi, Darlene Wagner, Matthew Schmerer, S. H. Rosenthal, A. Gerasimova, R. M. Kagan, B. |

| | | | |
|---|---|---|---|
| | | Division of Viral Diseases, Pathogen Discovery | Anderson, M. Hua, Y. Liu, L.E. Bernstein, K.E. Livingston, A. Perez, I. A. Shlyakhter, R. V. Rolando, R. Owen, P. Tanpaiboon, F. Lacbawan, Clinton R. Paden, Duncan MacCannell |
| EPI_ISL_2011448, EPI_ISL_2011450 | Seattle Flu Study | Seattle Flu Study | Deborah A. Nickerson, Chris D. Frazar, Jover Lee, Benjamin Pelle, Matthew Richardson, Kairsten Fay, Misja Ilcisin, Kirsten Lacombe, Thomas R. Sibley, Melissa Truong, Caitlin R. Wolf, Karen Cowgill, Stephanie Schrag, Jeff Duchin, Michael Boeckh, Janet A. Englund, Michael Famulare, Barry R. Lutz, Mark J. Rieder, Lea M. Starita, Matthew Thompson, Helen Y. Chu, Trevor Bedford, Jay Shendure |
| EPI_ISL_2011451 | Quest Diagnostics Incorporated | Centers for Disease Control and Prevention Division of Viral Diseases, Pathogen Discovery | Dakota Howard, Dhwani Batra, Peter W. Cook, Kara Moser, Adrian Paskey, Jason Caravas, Benjamin Rambo-Martin, Shatavia Morrison, Christopher Gulvick, Scott Sammons, Yvette Unoarumhi, Darlene Wagner, Matthew Schmerer, S. H. Rosenthal, A. Gerasimova, R. M. Kagan, B. Anderson, M. Hua, Y. Liu, L.E. Bernstein, K.E. Livingston, A. Perez, I. A. Shlyakhter, R. V. Rolando, R. Owen, P. Tanpaiboon, F. Lacbawan, Clinton R. Paden, Duncan MacCannell |
| EPI_ISL_2011452 | Incyte Diagnostics Spokane | Seattle Flu Study | Deborah A. Nickerson, Chris D. Frazar, Jover Lee, Benjamin Pelle, Erica Ryke, Matthew Richardson, Amanda Adler, Elisabeth Brandstetter, Peter D. Han, Kairsten Fay, Misja Ilcisin, Kirsten Lacombe, Thomas R. Sibley, Melissa Truong, Caitlin R. Wolf, Romesh Gautom, Geoff Melly, Brian Hiatt, Philip Dykema, Scott Lindquist, Michael Boeckh, Janet A. Englund, Michael Famulare, Barry R. Lutz, Mark J. Rieder, Lea M. Starita, Matthew Thompson, Helen Y. Chu, Jay Shendure, Trevor Bedford |
| EPI_ISL_2011454 | Quest Diagnostics Incorporated | Centers for Disease Control and Prevention Division of Viral Diseases, Pathogen Discovery | Dakota Howard, Dhwani Batra, Peter W. Cook, Kara Moser, Adrian Paskey, Jason Caravas, Benjamin Rambo-Martin, Shatavia Morrison, Christopher Gulvick, Scott Sammons, Yvette Unoarumhi, Darlene Wagner, Matthew Schmerer, S. H. Rosenthal, A. Gerasimova, R. M. Kagan, B. Anderson, M. Hua, Y. Liu, L.E. Bernstein, K.E. Livingston, A. Perez, I. A. Shlyakhter, R. V. Rolando, R. Owen, P. Tanpaiboon, F. Lacbawan, Clinton R. Paden, Duncan MacCannell |
| EPI_ISL_2011455, EPI_ISL_2011456, EPI_ISL_2011457 | Incyte Diagnostics Spokane | Seattle Flu Study | Deborah A. Nickerson, Chris D. Frazar, Jover Lee, Benjamin Pelle, Erica Ryke, Matthew Richardson, Amanda Adler, Elisabeth Brandstetter, Peter D. Han, Kairsten Fay, Misja Ilcisin, Kirsten Lacombe, Thomas R. Sibley, Melissa Truong, Caitlin R. Wolf, Romesh Gautom, Geoff Melly, Brian Hiatt, Philip Dykema, Scott Lindquist, Michael Boeckh, Janet A. Englund, Michael Famulare, Barry R. Lutz, Mark J. Rieder, Lea M. Starita, Matthew Thompson, Helen Y. Chu, Jay Shendure, Trevor Bedford |
| EPI_ISL_2011458 | Quest Diagnostics Incorporated | Centers for Disease Control and Prevention Division of Viral Diseases, Pathogen Discovery | Dakota Howard, Dhwani Batra, Peter W. Cook, Kara Moser, Adrian Paskey, Jason Caravas, Benjamin Rambo-Martin, Shatavia Morrison, Christopher Gulvick, Scott Sammons, Yvette Unoarumhi, Darlene Wagner, Matthew Schmerer, S. H. Rosenthal, A. Gerasimova, R. M. Kagan, B. Anderson, M. Hua, Y. Liu, L.E. Bernstein, K.E. Livingston, A. Perez, I. A. Shlyakhter, R. V. Rolando, R. Owen, P. Tanpaiboon, F. Lacbawan, Clinton R. Paden, Duncan MacCannell |
| EPI_ISL_2011459, EPI_ISL_2011461 | Incyte Diagnostics Spokane | Seattle Flu Study | Deborah A. Nickerson, Chris D. Frazar, Jover Lee, Benjamin Pelle, Erica Ryke, Matthew Richardson, Amanda Adler, Elisabeth Brandstetter, Peter D. Han, Kairsten Fay, Misja Ilcisin, Kirsten Lacombe, Thomas R. Sibley, Melissa Truong, Caitlin R. Wolf, Romesh Gautom, Geoff Melly, Brian Hiatt, Philip Dykema, Scott Lindquist, Michael Boeckh, Janet A. Englund, Michael Famulare, Barry R. Lutz, Mark J. Rieder, Lea M. Starita, Matthew Thompson, Helen Y. Chu, Jay Shendure, Trevor Bedford |
| EPI_ISL_2011462 | Quest Diagnostics Incorporated | Centers for Disease Control and Prevention Division of Viral Diseases, Pathogen Discovery | Dakota Howard, Dhwani Batra, Peter W. Cook, Kara Moser, Adrian Paskey, Jason Caravas, Benjamin Rambo-Martin, Shatavia Morrison, Christopher Gulvick, Scott Sammons, Yvette Unoarumhi, Darlene Wagner, Matthew Schmerer, S. H. Rosenthal, A. Gerasimova, R. M. Kagan, B. Anderson, M. Hua, Y. Liu, L.E. Bernstein, K.E. Livingston, A. Perez, I. A. Shlyakhter, R. V. Rolando, R. Owen, P. Tanpaiboon, F. Lacbawan, Clinton R. Paden, Duncan MacCannell |
| EPI_ISL_2011463, EPI_ISL_2011464 | Incyte Diagnostics Spokane | Seattle Flu Study | Deborah A. Nickerson, Chris D. Frazar, Jover Lee, Benjamin Pelle, Erica Ryke, Matthew Richardson, Amanda Adler, Elisabeth Brandstetter, Peter D. Han, Kairsten Fay, Misja Ilcisin, Kirsten Lacombe, Thomas R. Sibley, Melissa Truong, Caitlin R. Wolf, Romesh Gautom, Geoff Melly, Brian Hiatt, Philip Dykema, Scott Lindquist, Michael Boeckh, Janet A. Englund, Michael Famulare, Barry R. Lutz, Mark J. Rieder, Lea M. Starita, Matthew Thompson, Helen Y. Chu, Jay Shendure, Trevor Bedford |
| EPI_ISL_2011465 | Quest Diagnostics Incorporated | Centers for Disease Control and Prevention Division of Viral Diseases, Pathogen Discovery | Dakota Howard, Dhwani Batra, Peter W. Cook, Kara Moser, Adrian Paskey, Jason Caravas, Benjamin Rambo-Martin, Shatavia Morrison, Christopher Gulvick, Scott Sammons, Yvette Unoarumhi, Darlene Wagner, Matthew Schmerer, S. H. Rosenthal, A. Gerasimova, R. M. Kagan, B. Anderson, M. Hua, Y. Liu, L.E. Bernstein, K.E. Livingston, A. Perez, I. A. Shlyakhter, R. V. Rolando, R. Owen, P. Tanpaiboon, F. Lacbawan, Clinton R. Paden, Duncan MacCannell |
| EPI_ISL_2011467 | Incyte Diagnostics Spokane | Seattle Flu Study | Deborah A. Nickerson, Chris D. Frazar, Jover Lee, Benjamin Pelle, Erica Ryke, Matthew Richardson, Amanda Adler, Elisabeth Brandstetter, Peter D. Han, Kairsten Fay, Misja Ilcisin, Kirsten Lacombe, Thomas R. Sibley, Melissa Truong, Caitlin R. Wolf, Romesh Gautom, Geoff Melly, Brian Hiatt, Philip Dykema, Scott Lindquist, Michael Boeckh, Janet A. Englund, Michael Famulare, Barry R. Lutz, Mark J. Rieder, Lea M. Starita, Matthew Thompson, Helen Y. Chu, Jay Shendure, Trevor Bedford |
| EPI_ISL_2011468 | Quest Diagnostics Incorporated | Centers for Disease Control and Prevention Division of Viral Diseases, Pathogen Discovery | Dakota Howard, Dhwani Batra, Peter W. Cook, Kara Moser, Adrian Paskey, Jason Caravas, Benjamin Rambo-Martin, Shatavia Morrison, Christopher Gulvick, Scott Sammons, Yvette Unoarumhi, Darlene Wagner, Matthew Schmerer, S. H. Rosenthal, A. Gerasimova, R. M. Kagan, B. Anderson, M. Hua, Y. Liu, L.E. Bernstein, K.E. Livingston, A. Perez, I. A. Shlyakhter, R. V. Rolando, R. Owen, P. Tanpaiboon, F. Lacbawan, Clinton R. Paden, Duncan MacCannell |
| EPI_ISL_2011470 | Incyte Diagnostics Spokane | Seattle Flu Study | Deborah A. Nickerson, Chris D. Frazar, Jover Lee, Benjamin Pelle, Erica Ryke, Matthew Richardson, Amanda Adler, Elisabeth Brandstetter, Peter D. Han, Kairsten Fay, Misja Ilcisin, Kirsten Lacombe, Thomas R. Sibley, Melissa Truong, Caitlin R. Wolf, Romesh Gautom, Geoff Melly, Brian Hiatt, Philip Dykema, Scott Lindquist, Michael Boeckh, Janet A. Englund, Michael Famulare, Barry R. Lutz, Mark J. Rieder, Lea M. Starita, Matthew Thompson, Helen Y. Chu, Jay Shendure, Trevor Bedford |
| EPI_ISL_2011472 | Quest Diagnostics Incorporated | Centers for Disease Control and Prevention Division of Viral Diseases, Pathogen Discovery | Dakota Howard, Dhwani Batra, Peter W. Cook, Kara Moser, Adrian Paskey, Jason Caravas, Benjamin Rambo-Martin, Shatavia Morrison, Christopher Gulvick, Scott Sammons, Yvette Unoarumhi, Darlene Wagner, Matthew Schmerer, S. H. Rosenthal, A. Gerasimova, R. M. Kagan, B. Anderson, M. Hua, Y. Liu, L.E. Bernstein, K.E. Livingston, A. Perez, I. A. Shlyakhter, R. V. Rolando, R. Owen, P. Tanpaiboon, F. Lacbawan, Clinton R. Paden, Duncan MacCannell |
| EPI_ISL_2011473, EPI_ISL_2011474, EPI_ISL_2011475 | Incyte Diagnostics Spokane | Seattle Flu Study | Deborah A. Nickerson, Chris D. Frazar, Jover Lee, Benjamin Pelle, Erica Ryke, Matthew Richardson, Amanda Adler, Elisabeth Brandstetter, Peter D. Han, Kairsten Fay, Misja Ilcisin, Kirsten Lacombe, Thomas R. Sibley, Melissa Truong, Caitlin R. Wolf, Romesh Gautom, Geoff Melly, Brian Hiatt, Philip Dykema, Scott Lindquist, Michael Boeckh, Janet A. Englund, Michael Famulare, Barry R. Lutz, Mark J. Rieder, Lea M. Starita, Matthew Thompson, Helen Y. Chu, Jay Shendure, Trevor Bedford |
| EPI_ISL_2011476 | Quest Diagnostics Incorporated | Centers for Disease Control and Prevention Division of Viral Diseases, Pathogen Discovery | Dakota Howard, Dhwani Batra, Peter W. Cook, Kara Moser, Adrian Paskey, Jason Caravas, Benjamin Rambo-Martin, Shatavia Morrison, Christopher Gulvick, Scott Sammons, Yvette Unoarumhi, Darlene Wagner, Matthew Schmerer, S. H. Rosenthal, A. Gerasimova, R. M. Kagan, B. Anderson, M. Hua, Y. Liu, L.E. Bernstein, K.E. Livingston, A. Perez, I. A. Shlyakhter, R. V. Rolando, R. Owen, P. Tanpaiboon, F. Lacbawan, Clinton R. Paden, Duncan MacCannell |
| EPI_ISL_2011478, EPI_ISL_2011479 | Incyte Diagnostics Spokane | Seattle Flu Study | Deborah A. Nickerson, Chris D. Frazar, Jover Lee, Benjamin Pelle, Erica Ryke, Matthew Richardson, Amanda Adler, Elisabeth Brandstetter, Peter D. Han, Kairsten Fay, Misja Ilcisin, Kirsten Lacombe, Thomas R. Sibley, Melissa Truong, Caitlin R. Wolf, Romesh Gautom, Geoff Melly, Brian Hiatt, Philip Dykema, Scott Lindquist, Michael Boeckh, Janet A. Englund, Michael Famulare, Barry R. Lutz, Mark J. Rieder, Lea M. Starita, Matthew Thompson, Helen Y. Chu, Jay Shendure, Trevor Bedford |
| EPI_ISL_2011480 | Quest Diagnostics Incorporated | Centers for Disease Control and Prevention Division of Viral Diseases, Pathogen Discovery | Dakota Howard, Dhwani Batra, Peter W. Cook, Kara Moser, Adrian Paskey, Jason Caravas, Benjamin Rambo-Martin, Shatavia Morrison, Christopher Gulvick, Scott Sammons, Yvette Unoarumhi, Darlene Wagner, Matthew Schmerer, S. H. Rosenthal, A. Gerasimova, R. M. Kagan, B. Anderson, M. Hua, Y. Liu, L.E. Bernstein, K.E. Livingston, A. Perez, I. A. Shlyakhter, R. V. Rolando, R. Owen, P. Tanpaiboon, F. Lacbawan, Clinton R. Paden, Duncan MacCannell |
| EPI_ISL_2011482 | Incyte Diagnostics Spokane | Seattle Flu Study | Deborah A. Nickerson, Chris D. Frazar, Jover Lee, Benjamin Pelle, Erica Ryke, Matthew Richardson, Amanda Adler, Elisabeth Brandstetter, Peter D. Han, Kairsten Fay, Misja Ilcisin, Kirsten Lacombe, Thomas R. Sibley, Melissa Truong, Caitlin R. Wolf, Romesh Gautom, Geoff Melly, Brian Hiatt, Philip Dykema, Scott Lindquist, Michael Boeckh, Janet A. Englund, Michael Famulare, Barry R. Lutz, Mark J. Rieder, Lea M. Starita, Matthew Thompson, Helen Y. Chu, Jay Shendure, Trevor Bedford |
| EPI_ISL_2011483 | Quest Diagnostics Incorporated | Centers for Disease Control and Prevention Division of Viral Diseases, Pathogen Discovery | Dakota Howard, Dhwani Batra, Peter W. Cook, Kara Moser, Adrian Paskey, Jason Caravas, Benjamin Rambo-Martin, Shatavia Morrison, Christopher Gulvick, Scott Sammons, Yvette Unoarumhi, Darlene Wagner, Matthew Schmerer, S. H. Rosenthal, A. Gerasimova, R. M. Kagan, B. Anderson, M. Hua, Y. Liu, L.E. Bernstein, K.E. Livingston, A. Perez, I. A. Shlyakhter, R. V. Rolando, R. Owen, P. Tanpaiboon, F. Lacbawan, Clinton R. Paden, Duncan MacCannell |
| EPI_ISL_2011484, EPI_ISL_2011486 | Incyte Diagnostics Spokane | Seattle Flu Study | Deborah A. Nickerson, Chris D. Frazar, Jover Lee, Benjamin Pelle, Erica Ryke, Matthew Richardson, Amanda Adler, Elisabeth Brandstetter, Peter D. Han, Kairsten Fay, Misja Ilcisin, Kirsten Lacombe, Thomas R. Sibley, Melissa Truong, Caitlin R. Wolf, Romesh Gautom, Geoff Melly, Brian Hiatt, Philip Dykema, Scott Lindquist, Michael Boeckh, Janet A. Englund, Michael Famulare, Barry R. Lutz, Mark J. Rieder, Lea M. Starita, Matthew Thompson, Helen Y. Chu, Jay Shendure, Trevor Bedford |
| EPI_ISL_2011487 | Quest Diagnostics Incorporated | Centers for Disease Control and Prevention Division of Viral Diseases, Pathogen Discovery | Dakota Howard, Dhwani Batra, Peter W. Cook, Kara Moser, Adrian Paskey, Jason Caravas, Benjamin Rambo-Martin, Shatavia Morrison, Christopher Gulvick, Scott Sammons, Yvette Unoarumhi, Darlene Wagner, Matthew Schmerer, S. H. Rosenthal, A. Gerasimova, R. M. Kagan, B. Anderson, M. Hua, Y. Liu, L.E. Bernstein, K.E. Livingston, A. Perez, I. A. Shlyakhter, R. V. Rolando, R. Owen, P. Tanpaiboon, F. Lacbawan, Clinton R. Paden, Duncan MacCannell |
| EPI_ISL_2011488, EPI_ISL_2011489, EPI_ISL_2011490 | Incyte Diagnostics Spokane | Seattle Flu Study | Deborah A. Nickerson, Chris D. Frazar, Jover Lee, Benjamin Pelle, Erica Ryke, Matthew Richardson, Amanda Adler, Elisabeth Brandstetter, Peter D. Han, Kairsten Fay, Misja Ilcisin, Kirsten Lacombe, Thomas R. Sibley, Melissa Truong, Caitlin R. Wolf, Romesh Gautom, Geoff Melly, Brian Hiatt, Philip Dykema, Scott Lindquist, Michael Boeckh, Janet A. Englund, Michael Famulare, Barry R. Lutz, Mark J. Rieder, Lea M. Starita, Matthew Thompson, Helen Y. Chu, Jay Shendure, Trevor Bedford |
| EPI_ISL_2011491 | Quest Diagnostics Incorporated | Centers for Disease Control and Prevention Division of Viral Diseases, Pathogen Discovery | Dakota Howard, Dhwani Batra, Peter W. Cook, Kara Moser, Adrian Paskey, Jason Caravas, Benjamin Rambo-Martin, Shatavia Morrison, Christopher Gulvick, Scott Sammons, Yvette Unoarumhi, Darlene Wagner, Matthew Schmerer, S. H. Rosenthal, A. Gerasimova, R. M. Kagan, B. Anderson, M. Hua, Y. Liu, L.E. Bernstein, K.E. Livingston, A. Perez, I. A. Shlyakhter, R. V. Rolando, R. Owen, P. Tanpaiboon, F. Lacbawan, Clinton R. Paden, Duncan MacCannell |
| EPI_ISL_2011492, EPI_ISL_2011493, EPI_ISL_2011494 | Incyte Diagnostics Spokane | Seattle Flu Study | Deborah A. Nickerson, Chris D. Frazar, Jover Lee, Benjamin Pelle, Erica Ryke, Matthew Richardson, Amanda Adler, Elisabeth Brandstetter, Peter D. Han, Kairsten Fay, Misja Ilcisin, Kirsten Lacombe, Thomas R. Sibley, Melissa Truong, Caitlin R. Wolf, Romesh Gautom, Geoff Melly, Brian Hiatt, Philip Dykema, Scott Lindquist, Michael Boeckh, Janet A. Englund, Michael Famulare, Barry R. Lutz, Mark J. Rieder, Lea M. Starita, Matthew Thompson, Helen Y. Chu, Jay Shendure, Trevor Bedford |
| EPI_ISL_2011495 | Quest Diagnostics Incorporated | Centers for Disease Control and Prevention Division of Viral Diseases, Pathogen Discovery | Dakota Howard, Dhwani Batra, Peter W. Cook, Kara Moser, Adrian Paskey, Jason Caravas, Benjamin Rambo-Martin, Shatavia Morrison, Christopher Gulvick, Scott Sammons, Yvette Unoarumhi, Darlene Wagner, Matthew Schmerer, S. H. Rosenthal, A. Gerasimova, R. M. Kagan, B. Anderson, M. Hua, Y. Liu, L.E. Bernstein, K.E. Livingston, A. Perez, I. A. Shlyakhter, R. V. Rolando, R. Owen, P. Tanpaiboon, F. Lacbawan, Clinton R. Paden, Duncan MacCannell |
| EPI_ISL_2011497 | Incyte Diagnostics Spokane | Seattle Flu Study | Deborah A. Nickerson, Chris D. Frazar, Jover Lee, Benjamin Pelle, Erica Ryke, Matthew Richardson, Amanda Adler, Elisabeth Brandstetter, Peter D. Han, Kairsten Fay, Misja Ilcisin, Kirsten Lacombe, Thomas R. Sibley, Melissa Truong, Caitlin R. Wolf, Romesh Gautom, Geoff Melly, Brian Hiatt, Philip Dykema, Scott Lindquist, Michael Boeckh, Janet A. Englund, Michael Famulare, Barry R. Lutz, Mark J. Rieder, Lea M. Starita, Matthew Thompson, Helen Y. Chu, Jay Shendure, Trevor Bedford |
| EPI_ISL_2011498 | Quest Diagnostics Incorporated | Centers for Disease Control and Prevention Division of Viral Diseases, Pathogen Discovery | Dakota Howard, Dhwani Batra, Peter W. Cook, Kara Moser, Adrian Paskey, Jason Caravas, Benjamin Rambo-Martin, Shatavia Morrison, Christopher Gulvick, Scott Sammons, Yvette Unoarumhi, Darlene Wagner, Matthew Schmerer, S. H. Rosenthal, A. Gerasimova, R. M. Kagan, B. Anderson, M. Hua, Y. Liu, L.E. Bernstein, K.E. Livingston, A. Perez, I. A. Shlyakhter, R. V. Rolando, R. Owen, P. Tanpaiboon, F. Lacbawan, Clinton R. Paden, Duncan MacCannell |
| EPI_ISL_2011499, EPI_ISL_2011500 | Incyte Diagnostics Spokane | Seattle Flu Study | Deborah A. Nickerson, Chris D. Frazar, Jover Lee, Benjamin Pelle, Erica Ryke, Matthew Richardson, Amanda Adler, Elisabeth Brandstetter, Peter D. Han, Kairsten Fay, Misja Ilcisin, Kirsten Lacombe, Thomas R. Sibley, Melissa Truong, Caitlin R. Wolf, Romesh Gautom, Geoff Melly, Brian Hiatt, Philip Dykema, Scott Lindquist, Michael Boeckh, Janet A. Englund, Michael Famulare, Barry R. Lutz, Mark J. Rieder, Lea M. Starita, Matthew Thompson, Helen Y. Chu, Jay Shendure, Trevor Bedford |
| EPI_ISL_2011502 | Quest Diagnostics Incorporated | Centers for Disease Control and Prevention Division of Viral Diseases, Pathogen Discovery | Dakota Howard, Dhwani Batra, Peter W. Cook, Kara Moser, Adrian Paskey, Jason Caravas, Benjamin Rambo-Martin, Shatavia Morrison, Christopher Gulvick, Scott Sammons, Yvette Unoarumhi, Darlene Wagner, Matthew Schmerer, S. H. Rosenthal, A. Gerasimova, R. M. Kagan, B. Anderson, M. Hua, Y. Liu, L.E. Bernstein, K.E. Livingston, A. Perez, I. A. Shlyakhter, R. V. Rolando, R. Owen, P. Tanpaiboon, F. Lacbawan, Clinton R. Paden, Duncan MacCannell |
| EPI_ISL_2011504 | Incyte Diagnostics Spokane | Seattle Flu Study | Deborah A. Nickerson, Chris D. Frazar, Jover Lee, Benjamin Pelle, Erica Ryke, Matthew Richardson, Amanda Adler, Elisabeth Brandstetter, Peter D. Han, Kairsten Fay, Misja Ilcisin, Kirsten Lacombe, Thomas R. Sibley, Melissa Truong, Caitlin R. Wolf, Romesh Gautom, Geoff Melly, Brian Hiatt, Philip Dykema, Scott Lindquist, Michael Boeckh, Janet A. Englund, Michael Famulare, Barry R. Lutz, Mark J. Rieder, Lea M. Starita, Matthew Thompson, Helen Y. Chu, Jay Shendure, Trevor Bedford |
| EPI_ISL_2011505 | Quest Diagnostics Incorporated | Centers for Disease Control and Prevention Division of Viral Diseases, Pathogen Discovery | Dakota Howard, Dhwani Batra, Peter W. Cook, Kara Moser, Adrian Paskey, Jason Caravas, Benjamin Rambo-Martin, Shatavia Morrison, Christopher Gulvick, Scott Sammons, Yvette Unoarumhi, Darlene Wagner, Matthew Schmerer, S. H. Rosenthal, A. Gerasimova, R. M. Kagan, B. Anderson, M. Hua, Y. Liu, L.E. Bernstein, K.E. Livingston, A. Perez, I. A. Shlyakhter, R. V. Rolando, R. Owen, P. Tanpaiboon, F. Lacbawan, Clinton R. Paden, Duncan MacCannell |
| EPI_ISL_2011506, EPI_ISL_2011507 | Incyte Diagnostics Spokane | Seattle Flu Study | Deborah A. Nickerson, Chris D. Frazar, Jover Lee, Benjamin Pelle, Erica Ryke, Matthew Richardson, Amanda Adler, Elisabeth Brandstetter, Peter D. Han, Kairsten Fay, Misja Ilcisin, Kirsten Lacombe, Thomas R. Sibley, Melissa Truong, Caitlin R. Wolf, Romesh Gautom, Geoff Melly, Brian Hiatt, Philip Dykema, Scott Lindquist, Michael Boeckh, Janet A. Englund, Michael Famulare, Barry R. Lutz, Mark J. Rieder, Lea M. Starita, Matthew Thompson, Helen Y. Chu, Jay Shendure, Trevor Bedford |
| EPI_ISL_2011508 | Quest Diagnostics Incorporated | Centers for Disease Control and Prevention Division of Viral Diseases, Pathogen Discovery | Dakota Howard, Dhwani Batra, Peter W. Cook, Kara Moser, Adrian Paskey, Jason Caravas, Benjamin Rambo-Martin, Shatavia Morrison, Christopher Gulvick, Scott Sammons, Yvette Unoarumhi, Darlene Wagner, Matthew Schmerer, S. H. Rosenthal, A. Gerasimova, R. M. Kagan, B. Anderson, M. Hua, Y. Liu, L.E. Bernstein, K.E. Livingston, A. Perez, I. A. Shlyakhter, R. V. Rolando, R. Owen, P. Tanpaiboon, F. Lacbawan, Clinton R. Paden, Duncan MacCannell |
| EPI_ISL_2011509, EPI_ISL_2011510, EPI_ISL_2011511 | Incyte Diagnostics Spokane | Seattle Flu Study | Deborah A. Nickerson, Chris D. Frazar, Jover Lee, Benjamin Pelle, Erica Ryke, Matthew Richardson, Amanda Adler, Elisabeth Brandstetter, Peter D. Han, Kairsten Fay, Misja Ilcisin, Kirsten Lacombe, Thomas R. Sibley, Melissa Truong, Caitlin R. Wolf, Romesh Gautom, Geoff Melly, Brian Hiatt, Philip Dykema, Scott Lindquist, Michael Boeckh, Janet A. Englund, Michael Famulare, Barry R. Lutz, Mark J. Rieder, Lea M. Starita, Matthew Thompson, Helen Y. Chu, Jay Shendure, Trevor Bedford |

| | | | |
|---|---|---|---|
| EPI_ISL_2011512 | Quest Diagnostics Incorporated | Centers for Disease Control and Prevention Division of Viral Diseases, Pathogen Discovery | Dakota Howard, Dhwani Batra, Peter W. Cook, Kara Moser, Adrian Paskey, Jason Caravas, Benjamin Rambo-Martin, Shatavia Morrison, Christopher Gulvick, Scott Sammons, Yvette Unoarumhi, Darlene Wagner, Matthew Schmerer, S. H. Rosenthal, A. Gerasimova, R. M. Kagan, B. Anderson, M. Hua, Y. Liu, L.E. Bernstein, K.E. Livingston, A. Perez, I. A. Shlyakhter, R. V. Rolando, R. Owen, P. Tanpaiboon, F. Lacbawan, Clinton R. Paden, Duncan MacCannell |
| EPI_ISL_2011514 | Incyte Diagnostics Spokane | Seattle Flu Study | Deborah A. Nickerson, Chris D. Frazar, Jover Lee, Benjamin Pelle, Erica Ryke, Matthew Richardson, Amanda Adler, Elisabeth Brandstetter, Peter D. Han, Kairsten Fay, Misja Ilcisin, Kirsten Lacombe, Thomas R. Sibley, Melissa Truong, Caitlin R. Wolf, Romesh Gautom, Geoff Melly, Brian Hiatt, Philip Dykema, Scott Lindquist, Michael Boeckh, Janet A. Englund, Michael Famulare, Barry R. Lutz, Mark J. Rieder, Lea M. Starita, Matthew Thompson, Helen Y. Chu, Jay Shendure, Trevor Bedford |
| EPI_ISL_2011515 | Quest Diagnostics Incorporated | Centers for Disease Control and Prevention Division of Viral Diseases, Pathogen Discovery | Dakota Howard, Dhwani Batra, Peter W. Cook, Kara Moser, Adrian Paskey, Jason Caravas, Benjamin Rambo-Martin, Shatavia Morrison, Christopher Gulvick, Scott Sammons, Yvette Unoarumhi, Darlene Wagner, Matthew Schmerer, S. H. Rosenthal, A. Gerasimova, R. M. Kagan, B. Anderson, M. Hua, Y. Liu, L.E. Bernstein, K.E. Livingston, A. Perez, I. A. Shlyakhter, R. V. Rolando, R. Owen, P. Tanpaiboon, F. Lacbawan, Clinton R. Paden, Duncan MacCannell |
| EPI_ISL_2011516, EPI_ISL_2011517, EPI_ISL_2011518 | Incyte Diagnostics Spokane | Seattle Flu Study | Deborah A. Nickerson, Chris D. Frazar, Jover Lee, Benjamin Pelle, Erica Ryke, Matthew Richardson, Amanda Adler, Elisabeth Brandstetter, Peter D. Han, Kairsten Fay, Misja Ilcisin, Kirsten Lacombe, Thomas R. Sibley, Melissa Truong, Caitlin R. Wolf, Romesh Gautom, Geoff Melly, Brian Hiatt, Philip Dykema, Scott Lindquist, Michael Boeckh, Janet A. Englund, Michael Famulare, Barry R. Lutz, Mark J. Rieder, Lea M. Starita, Matthew Thompson, Helen Y. Chu, Jay Shendure, Trevor Bedford |
| EPI_ISL_2011519 | Quest Diagnostics Incorporated | Centers for Disease Control and Prevention Division of Viral Diseases, Pathogen Discovery | Dakota Howard, Dhwani Batra, Peter W. Cook, Kara Moser, Adrian Paskey, Jason Caravas, Benjamin Rambo-Martin, Shatavia Morrison, Christopher Gulvick, Scott Sammons, Yvette Unoarumhi, Darlene Wagner, Matthew Schmerer, S. H. Rosenthal, A. Gerasimova, R. M. Kagan, B. Anderson, M. Hua, Y. Liu, L.E. Bernstein, K.E. Livingston, A. Perez, I. A. Shlyakhter, R. V. Rolando, R. Owen, P. Tanpaiboon, F. Lacbawan, Clinton R. Paden, Duncan MacCannell |
| EPI_ISL_2011520 | Incyte Diagnostics Spokane | Seattle Flu Study | Deborah A. Nickerson, Chris D. Frazar, Jover Lee, Benjamin Pelle, Erica Ryke, Matthew Richardson, Amanda Adler, Elisabeth Brandstetter, Peter D. Han, Kairsten Fay, Misja Ilcisin, Kirsten Lacombe, Thomas R. Sibley, Melissa Truong, Caitlin R. Wolf, Romesh Gautom, Geoff Melly, Brian Hiatt, Philip Dykema, Scott Lindquist, Michael Boeckh, Janet A. Englund, Michael Famulare, Barry R. Lutz, Mark J. Rieder, Lea M. Starita, Matthew Thompson, Helen Y. Chu, Jay Shendure, Trevor Bedford |
| EPI_ISL_2011522 | Quest Diagnostics Incorporated | Centers for Disease Control and Prevention Division of Viral Diseases, Pathogen Discovery | Dakota Howard, Dhwani Batra, Peter W. Cook, Kara Moser, Adrian Paskey, Jason Caravas, Benjamin Rambo-Martin, Shatavia Morrison, Christopher Gulvick, Scott Sammons, Yvette Unoarumhi, Darlene Wagner, Matthew Schmerer, S. H. Rosenthal, A. Gerasimova, R. M. Kagan, B. Anderson, M. Hua, Y. Liu, L.E. Bernstein, K.E. Livingston, A. Perez, I. A. Shlyakhter, R. V. Rolando, R. Owen, P. Tanpaiboon, F. Lacbawan, Clinton R. Paden, Duncan MacCannell |
| EPI_ISL_2011523 | Incyte Diagnostics Spokane | Seattle Flu Study | Deborah A. Nickerson, Chris D. Frazar, Jover Lee, Benjamin Pelle, Erica Ryke, Matthew Richardson, Amanda Adler, Elisabeth Brandstetter, Peter D. Han, Kairsten Fay, Misja Ilcisin, Kirsten Lacombe, Thomas R. Sibley, Melissa Truong, Caitlin R. Wolf, Romesh Gautom, Geoff Melly, Brian Hiatt, Philip Dykema, Scott Lindquist, Michael Boeckh, Janet A. Englund, Michael Famulare, Barry R. Lutz, Mark J. Rieder, Lea M. Starita, Matthew Thompson, Helen Y. Chu, Jay Shendure, Trevor Bedford |
| EPI_ISL_2011526 | Quest Diagnostics Incorporated | Centers for Disease Control and Prevention Division of Viral Diseases, Pathogen Discovery | Dakota Howard, Dhwani Batra, Peter W. Cook, Kara Moser, Adrian Paskey, Jason Caravas, Benjamin Rambo-Martin, Shatavia Morrison, Christopher Gulvick, Scott Sammons, Yvette Unoarumhi, Darlene Wagner, Matthew Schmerer, S. H. Rosenthal, A. Gerasimova, R. M. Kagan, B. Anderson, M. Hua, Y. Liu, L.E. Bernstein, K.E. Livingston, A. Perez, I. A. Shlyakhter, R. V. Rolando, R. Owen, P. Tanpaiboon, F. Lacbawan, Clinton R. Paden, Duncan MacCannell |
| EPI_ISL_2011527 | Incyte Diagnostics Spokane | Seattle Flu Study | Deborah A. Nickerson, Chris D. Frazar, Jover Lee, Benjamin Pelle, Erica Ryke, Matthew Richardson, Amanda Adler, Elisabeth Brandstetter, Peter D. Han, Kairsten Fay, Misja Ilcisin, Kirsten Lacombe, Thomas R. Sibley, Melissa Truong, Caitlin R. Wolf, Romesh Gautom, Geoff Melly, Brian Hiatt, Philip Dykema, Scott Lindquist, Michael Boeckh, Janet A. Englund, Michael Famulare, Barry R. Lutz, Mark J. Rieder, Lea M. Starita, Matthew Thompson, Helen Y. Chu, Jay Shendure, Trevor Bedford |
| EPI_ISL_2011529 | Quest Diagnostics Incorporated | Centers for Disease Control and Prevention Division of Viral Diseases, Pathogen Discovery | Dakota Howard, Dhwani Batra, Peter W. Cook, Kara Moser, Adrian Paskey, Jason Caravas, Benjamin Rambo-Martin, Shatavia Morrison, Christopher Gulvick, Scott Sammons, Yvette Unoarumhi, Darlene Wagner, Matthew Schmerer, S. H. Rosenthal, A. Gerasimova, R. M. Kagan, B. Anderson, M. Hua, Y. Liu, L.E. Bernstein, K.E. Livingston, A. Perez, I. A. Shlyakhter, R. V. Rolando, R. Owen, P. Tanpaiboon, F. Lacbawan, Clinton R. Paden, Duncan MacCannell |
| EPI_ISL_2011530 | Incyte Diagnostics Spokane | Seattle Flu Study | Deborah A. Nickerson, Chris D. Frazar, Jover Lee, Benjamin Pelle, Erica Ryke, Matthew Richardson, Amanda Adler, Elisabeth Brandstetter, Peter D. Han, Kairsten Fay, Misja Ilcisin, Kirsten Lacombe, Thomas R. Sibley, Melissa Truong, Caitlin R. Wolf, Romesh Gautom, Geoff Melly, Brian Hiatt, Philip Dykema, Scott Lindquist, Michael Boeckh, Janet A. Englund, Michael Famulare, Barry R. Lutz, Mark J. Rieder, Lea M. Starita, Matthew Thompson, Helen Y. Chu, Jay Shendure, Trevor Bedford |
| EPI_ISL_2011533 | Quest Diagnostics Incorporated | Centers for Disease Control and Prevention Division of Viral Diseases, Pathogen Discovery | Dakota Howard, Dhwani Batra, Peter W. Cook, Kara Moser, Adrian Paskey, Jason Caravas, Benjamin Rambo-Martin, Shatavia Morrison, Christopher Gulvick, Scott Sammons, Yvette Unoarumhi, Darlene Wagner, Matthew Schmerer, S. H. Rosenthal, A. Gerasimova, R. M. Kagan, B. Anderson, M. Hua, Y. Liu, L.E. Bernstein, K.E. Livingston, A. Perez, I. A. Shlyakhter, R. V. Rolando, R. Owen, P. Tanpaiboon, F. Lacbawan, Clinton R. Paden, Duncan MacCannell |
| EPI_ISL_2011534 | Incyte Diagnostics Spokane | Seattle Flu Study | Deborah A. Nickerson, Chris D. Frazar, Jover Lee, Benjamin Pelle, Erica Ryke, Matthew Richardson, Amanda Adler, Elisabeth Brandstetter, Peter D. Han, Kairsten Fay, Misja Ilcisin, Kirsten Lacombe, Thomas R. Sibley, Melissa Truong, Caitlin R. Wolf, Romesh Gautom, Geoff Melly, Brian Hiatt, Philip Dykema, Scott Lindquist, Michael Boeckh, Janet A. Englund, Michael Famulare, Barry R. Lutz, Mark J. Rieder, Lea M. Starita, Matthew Thompson, Helen Y. Chu, Jay Shendure, Trevor Bedford |
| EPI_ISL_2011536 | Quest Diagnostics Incorporated | Centers for Disease Control and Prevention Division of Viral Diseases, Pathogen Discovery | Dakota Howard, Dhwani Batra, Peter W. Cook, Kara Moser, Adrian Paskey, Jason Caravas, Benjamin Rambo-Martin, Shatavia Morrison, Christopher Gulvick, Scott Sammons, Yvette Unoarumhi, Darlene Wagner, Matthew Schmerer, S. H. Rosenthal, A. Gerasimova, R. M. Kagan, B. Anderson, M. Hua, Y. Liu, L.E. Bernstein, K.E. Livingston, A. Perez, I. A. Shlyakhter, R. V. Rolando, R. Owen, P. Tanpaiboon, F. Lacbawan, Clinton R. Paden, Duncan MacCannell |
| EPI_ISL_2011537, EPI_ISL_2011538 | Incyte Diagnostics Spokane | Seattle Flu Study | Deborah A. Nickerson, Chris D. Frazar, Jover Lee, Benjamin Pelle, Erica Ryke, Matthew Richardson, Amanda Adler, Elisabeth Brandstetter, Peter D. Han, Kairsten Fay, Misja Ilcisin, Kirsten Lacombe, Thomas R. Sibley, Melissa Truong, Caitlin R. Wolf, Romesh Gautom, Geoff Melly, Brian Hiatt, Philip Dykema, Scott Lindquist, Michael Boeckh, Janet A. Englund, Michael Famulare, Barry R. Lutz, Mark J. Rieder, Lea M. Starita, Matthew Thompson, Helen Y. Chu, Jay Shendure, Trevor Bedford |
| EPI_ISL_2011540 | Quest Diagnostics Incorporated | Centers for Disease Control and Prevention Division of Viral Diseases, Pathogen Discovery | Dakota Howard, Dhwani Batra, Peter W. Cook, Kara Moser, Adrian Paskey, Jason Caravas, Benjamin Rambo-Martin, Shatavia Morrison, Christopher Gulvick, Scott Sammons, Yvette Unoarumhi, Darlene Wagner, Matthew Schmerer, S. H. Rosenthal, A. Gerasimova, R. M. Kagan, B. Anderson, M. Hua, Y. Liu, L.E. Bernstein, K.E. Livingston, A. Perez, I. A. Shlyakhter, R. V. Rolando, R. Owen, P. Tanpaiboon, F. Lacbawan, Clinton R. Paden, Duncan MacCannell |
| EPI_ISL_2011542, EPI_ISL_2011543 | Incyte Diagnostics Spokane | Seattle Flu Study | Deborah A. Nickerson, Chris D. Frazar, Jover Lee, Benjamin Pelle, Erica Ryke, Matthew Richardson, Amanda Adler, Elisabeth Brandstetter, Peter D. Han, Kairsten Fay, Misja Ilcisin, Kirsten Lacombe, Thomas R. Sibley, Melissa Truong, Caitlin R. Wolf, Romesh Gautom, Geoff Melly, Brian Hiatt, Philip Dykema, Scott Lindquist, Michael Boeckh, Janet A. Englund, Michael Famulare, Barry R. Lutz, Mark J. Rieder, Lea M. Starita, Matthew Thompson, Helen Y. Chu, Jay Shendure, Trevor Bedford |
| EPI_ISL_2011544 | Quest Diagnostics Incorporated | Centers for Disease Control and Prevention Division of Viral Diseases, Pathogen Discovery | Dakota Howard, Dhwani Batra, Peter W. Cook, Kara Moser, Adrian Paskey, Jason Caravas, Benjamin Rambo-Martin, Shatavia Morrison, Christopher Gulvick, Scott Sammons, Yvette Unoarumhi, Darlene Wagner, Matthew Schmerer, S. H. Rosenthal, A. Gerasimova, R. M. Kagan, B. Anderson, M. Hua, Y. Liu, L.E. Bernstein, K.E. Livingston, A. Perez, I. A. Shlyakhter, R. V. Rolando, R. Owen, P. Tanpaiboon, F. Lacbawan, Clinton R. Paden, Duncan MacCannell |
| EPI_ISL_2011545, EPI_ISL_2011547 | Incyte Diagnostics Spokane | Seattle Flu Study | Deborah A. Nickerson, Chris D. Frazar, Jover Lee, Benjamin Pelle, Erica Ryke, Matthew Richardson, Amanda Adler, Elisabeth Brandstetter, Peter D. Han, Kairsten Fay, Misja Ilcisin, Kirsten Lacombe, Thomas R. Sibley, Melissa Truong, Caitlin R. Wolf, Romesh Gautom, Geoff Melly, Brian Hiatt, Philip Dykema, Scott Lindquist, Michael Boeckh, Janet A. Englund, Michael Famulare, Barry R. Lutz, Mark J. Rieder, Lea M. Starita, Matthew Thompson, Helen Y. Chu, Jay Shendure, Trevor Bedford |
| EPI_ISL_2011548 | Quest Diagnostics Incorporated | Centers for Disease Control and Prevention Division of Viral Diseases, Pathogen Discovery | Dakota Howard, Dhwani Batra, Peter W. Cook, Kara Moser, Adrian Paskey, Jason Caravas, Benjamin Rambo-Martin, Shatavia Morrison, Christopher Gulvick, Scott Sammons, Yvette Unoarumhi, Darlene Wagner, Matthew Schmerer, S. H. Rosenthal, A. Gerasimova, R. M. Kagan, B. Anderson, M. Hua, Y. Liu, L.E. Bernstein, K.E. Livingston, A. Perez, I. A. Shlyakhter, R. V. Rolando, R. Owen, P. Tanpaiboon, F. Lacbawan, Clinton R. Paden, Duncan MacCannell |
| EPI_ISL_2011549, EPI_ISL_2011550 | Incyte Diagnostics Spokane | Seattle Flu Study | Deborah A. Nickerson, Chris D. Frazar, Jover Lee, Benjamin Pelle, Erica Ryke, Matthew Richardson, Amanda Adler, Elisabeth Brandstetter, Peter D. Han, Kairsten Fay, Misja Ilcisin, Kirsten Lacombe, Thomas R. Sibley, Melissa Truong, Caitlin R. Wolf, Romesh Gautom, Geoff Melly, Brian Hiatt, Philip Dykema, Scott Lindquist, Michael Boeckh, Janet A. Englund, Michael Famulare, Barry R. Lutz, Mark J. Rieder, Lea M. Starita, Matthew Thompson, Helen Y. Chu, Jay Shendure, Trevor Bedford |
| EPI_ISL_2011551 | Quest Diagnostics Incorporated | Centers for Disease Control and Prevention Division of Viral Diseases, Pathogen Discovery | Dakota Howard, Dhwani Batra, Peter W. Cook, Kara Moser, Adrian Paskey, Jason Caravas, Benjamin Rambo-Martin, Shatavia Morrison, Christopher Gulvick, Scott Sammons, Yvette Unoarumhi, Darlene Wagner, Matthew Schmerer, S. H. Rosenthal, A. Gerasimova, R. M. Kagan, B. Anderson, M. Hua, Y. Liu, L.E. Bernstein, K.E. Livingston, A. Perez, I. A. Shlyakhter, R. V. Rolando, R. Owen, P. Tanpaiboon, F. Lacbawan, Clinton R. Paden, Duncan MacCannell |
| EPI_ISL_2011553 | Incyte Diagnostics Spokane | Seattle Flu Study | Deborah A. Nickerson, Chris D. Frazar, Jover Lee, Benjamin Pelle, Erica Ryke, Matthew Richardson, Amanda Adler, Elisabeth Brandstetter, Peter D. Han, Kairsten Fay, Misja Ilcisin, Kirsten Lacombe, Thomas R. Sibley, Melissa Truong, Caitlin R. Wolf, Romesh Gautom, Geoff Melly, Brian Hiatt, Philip Dykema, Scott Lindquist, Michael Boeckh, Janet A. Englund, Michael Famulare, Barry R. Lutz, Mark J. Rieder, Lea M. Starita, Matthew Thompson, Helen Y. Chu, Jay Shendure, Trevor Bedford |
| EPI_ISL_2011554 | Quest Diagnostics Incorporated | Centers for Disease Control and Prevention Division of Viral Diseases, Pathogen Discovery | Dakota Howard, Dhwani Batra, Peter W. Cook, Kara Moser, Adrian Paskey, Jason Caravas, Benjamin Rambo-Martin, Shatavia Morrison, Christopher Gulvick, Scott Sammons, Yvette Unoarumhi, Darlene Wagner, Matthew Schmerer, S. H. Rosenthal, A. Gerasimova, R. M. Kagan, B. Anderson, M. Hua, Y. Liu, L.E. Bernstein, K.E. Livingston, A. Perez, I. A. Shlyakhter, R. V. Rolando, R. Owen, P. Tanpaiboon, F. Lacbawan, Clinton R. Paden, Duncan MacCannell |
| EPI_ISL_2011555, EPI_ISL_2011556 | Incyte Diagnostics Spokane | Seattle Flu Study | Deborah A. Nickerson, Chris D. Frazar, Jover Lee, Benjamin Pelle, Erica Ryke, Matthew Richardson, Amanda Adler, Elisabeth Brandstetter, Peter D. Han, Kairsten Fay, Misja Ilcisin, Kirsten Lacombe, Thomas R. Sibley, Melissa Truong, Caitlin R. Wolf, Romesh Gautom, Geoff Melly, Brian Hiatt, Philip Dykema, Scott Lindquist, Michael Boeckh, Janet A. Englund, Michael Famulare, Barry R. Lutz, Mark J. Rieder, Lea M. Starita, Matthew Thompson, Helen Y. Chu, Jay Shendure, Trevor Bedford |
| EPI_ISL_2011558 | Quest Diagnostics Incorporated | Centers for Disease Control and Prevention Division of Viral Diseases, Pathogen Discovery | Dakota Howard, Dhwani Batra, Peter W. Cook, Kara Moser, Adrian Paskey, Jason Caravas, Benjamin Rambo-Martin, Shatavia Morrison, Christopher Gulvick, Scott Sammons, Yvette Unoarumhi, Darlene Wagner, Matthew Schmerer, S. H. Rosenthal, A. Gerasimova, R. M. Kagan, B. Anderson, M. Hua, Y. Liu, L.E. Bernstein, K.E. Livingston, A. Perez, I. A. Shlyakhter, R. V. Rolando, R. Owen, P. Tanpaiboon, F. Lacbawan, Clinton R. Paden, Duncan MacCannell |
| EPI_ISL_2011559, EPI_ISL_2011560, EPI_ISL_2011561 | Incyte Diagnostics Spokane | Seattle Flu Study | Deborah A. Nickerson, Chris D. Frazar, Jover Lee, Benjamin Pelle, Erica Ryke, Matthew Richardson, Amanda Adler, Elisabeth Brandstetter, Peter D. Han, Kairsten Fay, Misja Ilcisin, Kirsten Lacombe, Thomas R. Sibley, Melissa Truong, Caitlin R. Wolf, Romesh Gautom, Geoff Melly, Brian Hiatt, Philip Dykema, Scott Lindquist, Michael Boeckh, Janet A. Englund, Michael Famulare, Barry R. Lutz, Mark J. Rieder, Lea M. Starita, Matthew Thompson, Helen Y. Chu, Jay Shendure, Trevor Bedford |
| EPI_ISL_2011562 | Quest Diagnostics Incorporated | Centers for Disease Control and Prevention Division of Viral Diseases, Pathogen Discovery | Dakota Howard, Dhwani Batra, Peter W. Cook, Kara Moser, Adrian Paskey, Jason Caravas, Benjamin Rambo-Martin, Shatavia Morrison, Christopher Gulvick, Scott Sammons, Yvette Unoarumhi, Darlene Wagner, Matthew Schmerer, S. H. Rosenthal, A. Gerasimova, R. M. Kagan, B. Anderson, M. Hua, Y. Liu, L.E. Bernstein, K.E. Livingston, A. Perez, I. A. Shlyakhter, R. V. Rolando, R. Owen, P. Tanpaiboon, F. Lacbawan, Clinton R. Paden, Duncan MacCannell |
| EPI_ISL_2011563, EPI_ISL_2011564 | Incyte Diagnostics Spokane | Seattle Flu Study | Deborah A. Nickerson, Chris D. Frazar, Jover Lee, Benjamin Pelle, Erica Ryke, Matthew Richardson, Amanda Adler, Elisabeth Brandstetter, Peter D. Han, Kairsten Fay, Misja Ilcisin, Kirsten Lacombe, Thomas R. Sibley, Melissa Truong, Caitlin R. Wolf, Romesh Gautom, Geoff Melly, Brian Hiatt, Philip Dykema, Scott Lindquist, Michael Boeckh, Janet A. Englund, Michael Famulare, Barry R. Lutz, Mark J. Rieder, Lea M. Starita, Matthew Thompson, Helen Y. Chu, Jay Shendure, Trevor Bedford |
| EPI_ISL_2011565 | Quest Diagnostics Incorporated | Centers for Disease Control and Prevention Division of Viral Diseases, Pathogen Discovery | Dakota Howard, Dhwani Batra, Peter W. Cook, Kara Moser, Adrian Paskey, Jason Caravas, Benjamin Rambo-Martin, Shatavia Morrison, Christopher Gulvick, Scott Sammons, Yvette Unoarumhi, Darlene Wagner, Matthew Schmerer, S. H. Rosenthal, A. Gerasimova, R. M. Kagan, B. Anderson, M. Hua, Y. Liu, L.E. Bernstein, K.E. Livingston, A. Perez, I. A. Shlyakhter, R. V. Rolando, R. Owen, P. Tanpaiboon, F. Lacbawan, Clinton R. Paden, Duncan MacCannell |
| EPI_ISL_2011566, EPI_ISL_2011567, EPI_ISL_2011568 | Incyte Diagnostics Spokane | Seattle Flu Study | Deborah A. Nickerson, Chris D. Frazar, Jover Lee, Benjamin Pelle, Erica Ryke, Matthew Richardson, Amanda Adler, Elisabeth Brandstetter, Peter D. Han, Kairsten Fay, Misja Ilcisin, Kirsten Lacombe, Thomas R. Sibley, Melissa Truong, Caitlin R. Wolf, Romesh Gautom, Geoff Melly, Brian Hiatt, Philip Dykema, Scott Lindquist, Michael Boeckh, Janet A. Englund, Michael Famulare, Barry R. Lutz, Mark J. Rieder, Lea M. Starita, Matthew Thompson, Helen Y. Chu, Jay Shendure, Trevor Bedford |
| EPI_ISL_2011569 | Quest Diagnostics Incorporated | Centers for Disease Control and Prevention Division of Viral Diseases, Pathogen Discovery | Dakota Howard, Dhwani Batra, Peter W. Cook, Kara Moser, Adrian Paskey, Jason Caravas, Benjamin Rambo-Martin, Shatavia Morrison, Christopher Gulvick, Scott Sammons, Yvette Unoarumhi, Darlene Wagner, Matthew Schmerer, S. H. Rosenthal, A. Gerasimova, R. M. Kagan, B. Anderson, M. Hua, Y. Liu, L.E. Bernstein, K.E. Livingston, A. Perez, I. A. Shlyakhter, R. V. Rolando, R. Owen, P. Tanpaiboon, F. Lacbawan, Clinton R. Paden, Duncan MacCannell |
| EPI_ISL_2011570 | Incyte Diagnostics Spokane | Seattle Flu Study | Deborah A. Nickerson, Chris D. Frazar, Jover Lee, Benjamin Pelle, Erica Ryke, Matthew Richardson, Amanda Adler, Elisabeth Brandstetter, Peter D. Han, Kairsten Fay, Misja Ilcisin, Kirsten Lacombe, Thomas R. Sibley, Melissa Truong, Caitlin R. Wolf, Romesh Gautom, Geoff Melly, Brian Hiatt, Philip Dykema, Scott Lindquist, Michael Boeckh, Janet A. Englund, Michael Famulare, Barry R. Lutz, Mark J. Rieder, Lea M. Starita, Matthew Thompson, Helen Y. Chu, Jay Shendure, Trevor Bedford |
| EPI_ISL_2011571 | Seattle Flu Study | Seattle Flu Study | Deborah A. Nickerson, Chris D. Frazar, Jover Lee, Benjamin Pelle, Erica Ryke, Matthew Richardson, Amanda Adler, Elisabeth Brandstetter, Peter D. Han, Kairsten Fay, Misja Ilcisin, Kirsten Lacombe, Thomas R. Sibley, Melissa Truong, Caitlin R. Wolf, Michael Boeckh, Janet A. Englund, Michael Famulare, Barry R. Lutz, Mark J. Rieder, Lea M. Starita, Matthew Thompson, Jay Shendure, Trevor Bedford, Helen Y. Chu |
| EPI_ISL_2011572 | Quest Diagnostics Incorporated | Centers for Disease Control and Prevention Division of Viral Diseases, Pathogen Discovery | Dakota Howard, Dhwani Batra, Peter W. Cook, Kara Moser, Adrian Paskey, Jason Caravas, Benjamin Rambo-Martin, Shatavia Morrison, Christopher Gulvick, Scott Sammons, Yvette Unoarumhi, Darlene Wagner, Matthew Schmerer, S. H. Rosenthal, A. Gerasimova, R. M. Kagan, B. Anderson, M. Hua, Y. Liu, L.E. Bernstein, K.E. Livingston, A. Perez, I. A. Shlyakhter, R. V. Rolando, R. Owen, P. Tanpaiboon, F. Lacbawan, Clinton R. Paden, Duncan MacCannell |
| EPI_ISL_2011574 | Seattle Flu Study | Seattle Flu Study | Deborah A. Nickerson, Chris D. Frazar, Jover Lee, Benjamin Pelle, Erica Ryke, Matthew Richardson, Amanda Adler, Elisabeth Brandstetter, Peter D. Han, Kairsten Fay, Misja Ilcisin, Kirsten Lacombe, Thomas R. Sibley, Melissa Truong, Caitlin R. Wolf, Michael Boeckh, Janet A. Englund, |

| | | | |
|---|---|---|---|
| | | | Michael Famulare, Barry R. Lutz, Mark J. Rieder, Lea M. Starita, Matthew Thompson, Jay Shendure, Trevor Bedford, Helen Y. Chu |
| EPI_ISL_2011575 | Quest Diagnostics Incorporated | Centers for Disease Control and Prevention Division of Viral Diseases, Pathogen Discovery | Dakota Howard, Dhwani Batra, Peter W. Cook, Kara Moser, Adrian Paskey, Jason Caravas, Benjamin Rambo-Martin, Shatavia Morrison, Christopher Gulvick, Scott Sammons, Yvette Unoarumhi, Darlene Wagner, Matthew Schmerer, S. H. Rosenthal, A. Gerasimova, R. M. Kagan, B. Anderson, M. Hua, Y. Liu, L.E. Bernstein, K.E. Livingston, A. Perez, I. A. Shlyakhter, R. V. Rolando, R. Owen, P. Tanpaiboon, F. Lacbawan, Clinton R. Paden, Duncan MacCannell |
| EPI_ISL_2011576, EPI_ISL_2011577 | Seattle Flu Study | Seattle Flu Study | Deborah A. Nickerson, Chris D. Frazar, Jover Lee, Benjamin Pelle, Erica Ryke, Matthew Richardson, Amanda Adler, Elisabeth Brandstetter, Peter D. Han, Kairsten Fay, Misja Ilcisin, Kirsten Lacombe, Thomas R. Sibley, Melissa Truong, Caitlin R. Wolf, Michael Boeckh, Janet A. Englund, Michael Famulare, Barry R. Lutz, Mark J. Rieder, Lea M. Starita, Matthew Thompson, Jay Shendure, Trevor Bedford, Helen Y. Chu |
| EPI_ISL_2011578 | Quest Diagnostics Incorporated | Centers for Disease Control and Prevention Division of Viral Diseases, Pathogen Discovery | Dakota Howard, Dhwani Batra, Peter W. Cook, Kara Moser, Adrian Paskey, Jason Caravas, Benjamin Rambo-Martin, Shatavia Morrison, Christopher Gulvick, Scott Sammons, Yvette Unoarumhi, Darlene Wagner, Matthew Schmerer, S. H. Rosenthal, A. Gerasimova, R. M. Kagan, B. Anderson, M. Hua, Y. Liu, L.E. Bernstein, K.E. Livingston, A. Perez, I. A. Shlyakhter, R. V. Rolando, R. Owen, P. Tanpaiboon, F. Lacbawan, Clinton R. Paden, Duncan MacCannell |
| EPI_ISL_2011580, EPI_ISL_2011582 | Seattle Flu Study | Seattle Flu Study | Deborah A. Nickerson, Chris D. Frazar, Jover Lee, Benjamin Pelle, Erica Ryke, Matthew Richardson, Amanda Adler, Elisabeth Brandstetter, Peter D. Han, Kairsten Fay, Misja Ilcisin, Kirsten Lacombe, Thomas R. Sibley, Melissa Truong, Caitlin R. Wolf, Michael Boeckh, Janet A. Englund, Michael Famulare, Barry R. Lutz, Mark J. Rieder, Lea M. Starita, Matthew Thompson, Jay Shendure, Trevor Bedford, Helen Y. Chu |
| EPI_ISL_2011583 | Quest Diagnostics Incorporated | Centers for Disease Control and Prevention Division of Viral Diseases, Pathogen Discovery | Dakota Howard, Dhwani Batra, Peter W. Cook, Kara Moser, Adrian Paskey, Jason Caravas, Benjamin Rambo-Martin, Shatavia Morrison, Christopher Gulvick, Scott Sammons, Yvette Unoarumhi, Darlene Wagner, Matthew Schmerer, S. H. Rosenthal, A. Gerasimova, R. M. Kagan, B. Anderson, M. Hua, Y. Liu, L.E. Bernstein, K.E. Livingston, A. Perez, I. A. Shlyakhter, R. V. Rolando, R. Owen, P. Tanpaiboon, F. Lacbawan, Clinton R. Paden, Duncan MacCannell |
| EPI_ISL_2011586, EPI_ISL_2011588 | Seattle Flu Study | Seattle Flu Study | Deborah A. Nickerson, Chris D. Frazar, Jover Lee, Benjamin Pelle, Erica Ryke, Matthew Richardson, Amanda Adler, Elisabeth Brandstetter, Peter D. Han, Kairsten Fay, Misja Ilcisin, Kirsten Lacombe, Thomas R. Sibley, Melissa Truong, Caitlin R. Wolf, Michael Boeckh, Janet A. Englund, Michael Famulare, Barry R. Lutz, Mark J. Rieder, Lea M. Starita, Matthew Thompson, Jay Shendure, Trevor Bedford, Helen Y. Chu |
| EPI_ISL_2011589, EPI_ISL_2011592 | Quest Diagnostics Incorporated | Centers for Disease Control and Prevention Division of Viral Diseases, Pathogen Discovery | Dakota Howard, Dhwani Batra, Peter W. Cook, Kara Moser, Adrian Paskey, Jason Caravas, Benjamin Rambo-Martin, Shatavia Morrison, Christopher Gulvick, Scott Sammons, Yvette Unoarumhi, Darlene Wagner, Matthew Schmerer, S. H. Rosenthal, A. Gerasimova, R. M. Kagan, B. Anderson, M. Hua, Y. Liu, L.E. Bernstein, K.E. Livingston, A. Perez, I. A. Shlyakhter, R. V. Rolando, R. Owen, P. Tanpaiboon, F. Lacbawan, Clinton R. Paden, Duncan MacCannell |
| EPI_ISL_2011594 | Seattle Flu Study | Seattle Flu Study | Deborah A. Nickerson, Chris D. Frazar, Jover Lee, Benjamin Pelle, Erica Ryke, Matthew Richardson, Amanda Adler, Elisabeth Brandstetter, Peter D. Han, Kairsten Fay, Misja Ilcisin, Kirsten Lacombe, Thomas R. Sibley, Melissa Truong, Caitlin R. Wolf, Michael Boeckh, Janet A. Englund, Michael Famulare, Barry R. Lutz, Mark J. Rieder, Lea M. Starita, Matthew Thompson, Jay Shendure, Trevor Bedford, Helen Y. Chu |
| EPI_ISL_2011596 | Quest Diagnostics Incorporated | Centers for Disease Control and Prevention Division of Viral Diseases, Pathogen Discovery | Dakota Howard, Dhwani Batra, Peter W. Cook, Kara Moser, Adrian Paskey, Jason Caravas, Benjamin Rambo-Martin, Shatavia Morrison, Christopher Gulvick, Scott Sammons, Yvette Unoarumhi, Darlene Wagner, Matthew Schmerer, S. H. Rosenthal, A. Gerasimova, R. M. Kagan, B. Anderson, M. Hua, Y. Liu, L.E. Bernstein, K.E. Livingston, A. Perez, I. A. Shlyakhter, R. V. Rolando, R. Owen, P. Tanpaiboon, F. Lacbawan, Clinton R. Paden, Duncan MacCannell |
| EPI_ISL_2011597, EPI_ISL_2011598 | Seattle Flu Study | Seattle Flu Study | Deborah A. Nickerson, Chris D. Frazar, Jover Lee, Benjamin Pelle, Erica Ryke, Matthew Richardson, Amanda Adler, Elisabeth Brandstetter, Peter D. Han, Kairsten Fay, Misja Ilcisin, Kirsten Lacombe, Thomas R. Sibley, Melissa Truong, Caitlin R. Wolf, Michael Boeckh, Janet A. Englund, Michael Famulare, Barry R. Lutz, Mark J. Rieder, Lea M. Starita, Matthew Thompson, Jay Shendure, Trevor Bedford, Helen Y. Chu |
| EPI_ISL_2011600 | Quest Diagnostics Incorporated | Centers for Disease Control and Prevention Division of Viral Diseases, Pathogen Discovery | Dakota Howard, Dhwani Batra, Peter W. Cook, Kara Moser, Adrian Paskey, Jason Caravas, Benjamin Rambo-Martin, Shatavia Morrison, Christopher Gulvick, Scott Sammons, Yvette Unoarumhi, Darlene Wagner, Matthew Schmerer, S. H. Rosenthal, A. Gerasimova, R. M. Kagan, B. Anderson, M. Hua, Y. Liu, L.E. Bernstein, K.E. Livingston, A. Perez, I. A. Shlyakhter, R. V. Rolando, R. Owen, P. Tanpaiboon, F. Lacbawan, Clinton R. Paden, Duncan MacCannell |
| EPI_ISL_2011601 | Seattle Flu Study | Seattle Flu Study | Deborah A. Nickerson, Chris D. Frazar, Jover Lee, Benjamin Pelle, Erica Ryke, Matthew Richardson, Amanda Adler, Elisabeth Brandstetter, Peter D. Han, Kairsten Fay, Misja Ilcisin, Kirsten Lacombe, Thomas R. Sibley, Melissa Truong, Caitlin R. Wolf, Michael Boeckh, Janet A. Englund, Michael Famulare, Barry R. Lutz, Mark J. Rieder, Lea M. Starita, Matthew Thompson, Jay Shendure, Trevor Bedford, Helen Y. Chu |
| EPI_ISL_2011603 | Quest Diagnostics Incorporated | Centers for Disease Control and Prevention Division of Viral Diseases, Pathogen Discovery | Dakota Howard, Dhwani Batra, Peter W. Cook, Kara Moser, Adrian Paskey, Jason Caravas, Benjamin Rambo-Martin, Shatavia Morrison, Christopher Gulvick, Scott Sammons, Yvette Unoarumhi, Darlene Wagner, Matthew Schmerer, S. H. Rosenthal, A. Gerasimova, R. M. Kagan, B. Anderson, M. Hua, Y. Liu, L.E. Bernstein, K.E. Livingston, A. Perez, I. A. Shlyakhter, R. V. Rolando, R. Owen, P. Tanpaiboon, F. Lacbawan, Clinton R. Paden, Duncan MacCannell |
| EPI_ISL_2011604, EPI_ISL_2011605 | Seattle Flu Study | Seattle Flu Study | Deborah A. Nickerson, Chris D. Frazar, Jover Lee, Benjamin Pelle, Erica Ryke, Matthew Richardson, Amanda Adler, Elisabeth Brandstetter, Peter D. Han, Kairsten Fay, Misja Ilcisin, Kirsten Lacombe, Thomas R. Sibley, Melissa Truong, Caitlin R. Wolf, Michael Boeckh, Janet A. Englund, Michael Famulare, Barry R. Lutz, Mark J. Rieder, Lea M. Starita, Matthew Thompson, Jay Shendure, Trevor Bedford, Helen Y. Chu |
| EPI_ISL_2011606 | Quest Diagnostics Incorporated | Centers for Disease Control and Prevention Division of Viral Diseases, Pathogen Discovery | Dakota Howard, Dhwani Batra, Peter W. Cook, Kara Moser, Adrian Paskey, Jason Caravas, Benjamin Rambo-Martin, Shatavia Morrison, Christopher Gulvick, Scott Sammons, Yvette Unoarumhi, Darlene Wagner, Matthew Schmerer, S. H. Rosenthal, A. Gerasimova, R. M. Kagan, B. Anderson, M. Hua, Y. Liu, L.E. Bernstein, K.E. Livingston, A. Perez, I. A. Shlyakhter, R. V. Rolando, R. Owen, P. Tanpaiboon, F. Lacbawan, Clinton R. Paden, Duncan MacCannell |
| EPI_ISL_2011608, EPI_ISL_2011609 | Incyte Diagnostics Spokane | Seattle Flu Study | Deborah A. Nickerson, Chris D. Frazar, Jover Lee, Benjamin Pelle, Erica Ryke, Matthew Richardson, Amanda Adler, Elisabeth Brandstetter, Peter D. Han, Kairsten Fay, Misja Ilcisin, Kirsten Lacombe, Thomas R. Sibley, Melissa Truong, Caitlin R. Wolf, Romesh Gautom, Geoff Melly, Brian Hiatt, Philip Dykema, Scott Lindquist, Michael Boeckh, Janet A. Englund, Michael Famulare, Barry R. Lutz, Mark J. Rieder, Lea M. Starita, Matthew Thompson, Helen Y. Chu, Jay Shendure, Trevor Bedford |
| EPI_ISL_2011610 | Quest Diagnostics Incorporated | Centers for Disease Control and Prevention Division of Viral Diseases, Pathogen Discovery | Dakota Howard, Dhwani Batra, Peter W. Cook, Kara Moser, Adrian Paskey, Jason Caravas, Benjamin Rambo-Martin, Shatavia Morrison, Christopher Gulvick, Scott Sammons, Yvette Unoarumhi, Darlene Wagner, Matthew Schmerer, S. H. Rosenthal, A. Gerasimova, R. M. Kagan, B. Anderson, M. Hua, Y. Liu, L.E. Bernstein, K.E. Livingston, A. Perez, I. A. Shlyakhter, R. V. Rolando, R. Owen, P. Tanpaiboon, F. Lacbawan, Clinton R. Paden, Duncan MacCannell |
| EPI_ISL_2011612, EPI_ISL_2011614, EPI_ISL_2011615, EPI_ISL_2011616, EPI_ISL_2011617, EPI_ISL_2011618, EPI_ISL_2011619, EPI_ISL_2011621, EPI_ISL_2011623, EPI_ISL_2011625, EPI_ISL_2011626, EPI_ISL_2011629, EPI_ISL_2011630, EPI_ISL_2011631, EPI_ISL_2011633 | | | |
| see above | Incyte Diagnostics Spokane | Seattle Flu Study | Deborah A. Nickerson, Chris D. Frazar, Jover Lee, Benjamin Pelle, Erica Ryke, Matthew Richardson, Amanda Adler, Elisabeth Brandstetter, Peter D. Han, Kairsten Fay, Misja Ilcisin, Kirsten Lacombe, Thomas R. Sibley, Melissa Truong, Caitlin R. Wolf, Romesh Gautom, Geoff Melly, Brian Hiatt, Philip Dykema, Scott Lindquist, Michael Boeckh, Janet A. Englund, Michael Famulare, Barry R. Lutz, Mark J. Rieder, Lea M. Starita, Matthew Thompson, Helen Y. Chu, Jay Shendure, Trevor Bedford |
| EPI_ISL_2011634 | Quest Diagnostics Incorporated | Centers for Disease Control and Prevention Division of Viral Diseases, Pathogen Discovery | Dakota Howard, Dhwani Batra, Peter W. Cook, Kara Moser, Adrian Paskey, Jason Caravas, Benjamin Rambo-Martin, Shatavia Morrison, Christopher Gulvick, Scott Sammons, Yvette Unoarumhi, Darlene Wagner, Matthew Schmerer, S. H. Rosenthal, A. Gerasimova, R. M. Kagan, B. Anderson, M. Hua, Y. Liu, L.E. Bernstein, K.E. Livingston, A. Perez, I. A. Shlyakhter, R. V. Rolando, R. Owen, P. Tanpaiboon, F. Lacbawan, Clinton R. Paden, Duncan MacCannell |
| EPI_ISL_2011635, EPI_ISL_2011636, EPI_ISL_2011637 | Incyte Diagnostics Spokane | Seattle Flu Study | Deborah A. Nickerson, Chris D. Frazar, Jover Lee, Benjamin Pelle, Erica Ryke, Matthew Richardson, Amanda Adler, Elisabeth Brandstetter, Peter D. Han, Kairsten Fay, Misja Ilcisin, Kirsten Lacombe, Thomas R. Sibley, Melissa Truong, Caitlin R. Wolf, Romesh Gautom, Geoff Melly, Brian Hiatt, Philip Dykema, Scott Lindquist, Michael Boeckh, Janet A. Englund, Michael Famulare, Barry R. Lutz, Mark J. Rieder, Lea M. Starita, Matthew Thompson, Helen Y. Chu, Jay Shendure, Trevor Bedford |
| EPI_ISL_2011638 | Quest Diagnostics Incorporated | Centers for Disease Control and Prevention Division of Viral Diseases, Pathogen Discovery | Dakota Howard, Dhwani Batra, Peter W. Cook, Kara Moser, Adrian Paskey, Jason Caravas, Benjamin Rambo-Martin, Shatavia Morrison, Christopher Gulvick, Scott Sammons, Yvette Unoarumhi, Darlene Wagner, Matthew Schmerer, S. H. Rosenthal, A. Gerasimova, R. M. Kagan, B. Anderson, M. Hua, Y. Liu, L.E. Bernstein, K.E. Livingston, A. Perez, I. A. Shlyakhter, R. V. Rolando, R. Owen, P. Tanpaiboon, F. Lacbawan, Clinton R. Paden, Duncan MacCannell |
| EPI_ISL_2011639 | Incyte Diagnostics Spokane | Seattle Flu Study | Deborah A. Nickerson, Chris D. Frazar, Jover Lee, Benjamin Pelle, Erica Ryke, Matthew Richardson, Amanda Adler, Elisabeth Brandstetter, Peter D. Han, Kairsten Fay, Misja Ilcisin, Kirsten Lacombe, Thomas R. Sibley, Melissa Truong, Caitlin R. Wolf, Romesh Gautom, Geoff Melly, Brian Hiatt, Philip Dykema, Scott Lindquist, Michael Boeckh, Janet A. Englund, Michael Famulare, Barry R. Lutz, Mark J. Rieder, Lea M. Starita, Matthew Thompson, Helen Y. Chu, Jay Shendure, Trevor Bedford |
| EPI_ISL_2011642 | Quest Diagnostics Incorporated | Centers for Disease Control and Prevention Division of Viral Diseases, Pathogen Discovery | Dakota Howard, Dhwani Batra, Peter W. Cook, Kara Moser, Adrian Paskey, Jason Caravas, Benjamin Rambo-Martin, Shatavia Morrison, Christopher Gulvick, Scott Sammons, Yvette Unoarumhi, Darlene Wagner, Matthew Schmerer, S. H. Rosenthal, A. Gerasimova, R. M. Kagan, B. Anderson, M. Hua, Y. Liu, L.E. Bernstein, K.E. Livingston, A. Perez, I. A. Shlyakhter, R. V. Rolando, R. Owen, P. Tanpaiboon, F. Lacbawan, Clinton R. Paden, Duncan MacCannell |
| EPI_ISL_2011643 | Incyte Diagnostics Spokane | Seattle Flu Study | Deborah A. Nickerson, Chris D. Frazar, Jover Lee, Benjamin Pelle, Erica Ryke, Matthew Richardson, Amanda Adler, Elisabeth Brandstetter, Peter D. Han, Kairsten Fay, Misja Ilcisin, Kirsten Lacombe, Thomas R. Sibley, Melissa Truong, Caitlin R. Wolf, Romesh Gautom, Geoff Melly, Brian Hiatt, Philip Dykema, Scott Lindquist, Michael Boeckh, Janet A. Englund, Michael Famulare, Barry R. Lutz, Mark J. Rieder, Lea M. Starita, Matthew Thompson, Helen Y. Chu, Jay Shendure, Trevor Bedford |
| EPI_ISL_2011644, EPI_ISL_2011645, EPI_ISL_2011646, EPI_ISL_2011647, EPI_ISL_2011648, EPI_ISL_2011649, EPI_ISL_2011650, EPI_ISL_2011651, EPI_ISL_2011652, EPI_ISL_2011653, EPI_ISL_2011654, EPI_ISL_2011655, EPI_ISL_2011656, EPI_ISL_2011657, EPI_ISL_2011658, EPI_ISL_2011659, EPI_ISL_2011660, EPI_ISL_2011661, EPI_ISL_2011662, EPI_ISL_2011663, EPI_ISL_2011664, EPI_ISL_2011665, EPI_ISL_2011666, EPI_ISL_2011667, EPI_ISL_2011668, EPI_ISL_2011669, EPI_ISL_2011670, EPI_ISL_2011671, EPI_ISL_2011672, EPI_ISL_2011673, EPI_ISL_2011674, EPI_ISL_2011675, EPI_ISL_2011676, EPI_ISL_2011677, EPI_ISL_2011678, EPI_ISL_2011679, EPI_ISL_2011680, EPI_ISL_2011681, EPI_ISL_2011682 | | | |
| see above | Quest Diagnostics Incorporated | Centers for Disease Control and Prevention Division of Viral Diseases, Pathogen Discovery | Dakota Howard, Dhwani Batra, Peter W. Cook, Kara Moser, Adrian Paskey, Jason Caravas, Benjamin Rambo-Martin, Shatavia Morrison, Christopher Gulvick, Scott Sammons, Yvette Unoarumhi, Darlene Wagner, Matthew Schmerer, S. H. Rosenthal, A. Gerasimova, R. M. Kagan, B. Anderson, M. Hua, Y. Liu, L.E. Bernstein, K.E. Livingston, A. Perez, I. A. Shlyakhter, R. V. Rolando, R. Owen, P. Tanpaiboon, F. Lacbawan, Clinton R. Paden, Duncan MacCannell |
| EPI_ISL_2011688 | DC Public Health Lab/ Dept. of Forensic Sciences | DC Public Health Lab/ Dept. of Forensic Sciences | Janis Doss, Scott Nguyen, Elizabeth Zelaya, Sarah Scott, Connie Maza, Monica Mann, Brittany Hamilton, David Payne, Jocelyn Hauser |
| EPI_ISL_2011689, EPI_ISL_2011690, EPI_ISL_2011691, EPI_ISL_2011692, EPI_ISL_2011693, EPI_ISL_2011694, EPI_ISL_2011695, EPI_ISL_2011696, EPI_ISL_2011697, EPI_ISL_2011698, EPI_ISL_2011700, EPI_ISL_2011701, EPI_ISL_2011702, EPI_ISL_2011703, EPI_ISL_2011704, EPI_ISL_2011705, EPI_ISL_2011706, EPI_ISL_2011707, EPI_ISL_2011708, EPI_ISL_2011709, EPI_ISL_2011710, EPI_ISL_2011711, EPI_ISL_2011712, EPI_ISL_2011713, EPI_ISL_2011714, EPI_ISL_2011715, EPI_ISL_2011716, EPI_ISL_2011717, EPI_ISL_2011718 | | | |
| see above | Moderna Inc. | Moderna Inc. | Yamuna Paila; Groves Dixon; Rolando Pajon |
| EPI_ISL_2013054, EPI_ISL_2013055, EPI_ISL_2013056, EPI_ISL_2013057, EPI_ISL_2013058, EPI_ISL_2013059, EPI_ISL_2013060, EPI_ISL_2013061, EPI_ISL_2013062, EPI_ISL_2013063, EPI_ISL_2013064, EPI_ISL_2013065, EPI_ISL_2013066, EPI_ISL_2013067, EPI_ISL_2013068, EPI_ISL_2013069, EPI_ISL_2013070, EPI_ISL_2013071, EPI_ISL_2013072, EPI_ISL_2013073, EPI_ISL_2013074, EPI_ISL_2013075, EPI_ISL_2013076, EPI_ISL_2013077, EPI_ISL_2013078, EPI_ISL_2013079, EPI_ISL_2013080, EPI_ISL_2013081, EPI_ISL_2013082, EPI_ISL_2013083, EPI_ISL_2013084, EPI_ISL_2013085, EPI_ISL_2013086, EPI_ISL_2013087, EPI_ISL_2013089, EPI_ISL_2013090, EPI_ISL_2013091, EPI_ISL_2013092, EPI_ISL_2013093, EPI_ISL_2013094, EPI_ISL_2013095, EPI_ISL_2013096, EPI_ISL_2013097, EPI_ISL_2013098, EPI_ISL_2013099, EPI_ISL_2013100, EPI_ISL_2013101, EPI_ISL_2013102, EPI_ISL_2013103, EPI_ISL_2013104, EPI_ISL_2013105, EPI_ISL_2013106, EPI_ISL_2013107, EPI_ISL_2013108, EPI_ISL_2013109, EPI_ISL_2013110, EPI_ISL_2013111, EPI_ISL_2013112, EPI_ISL_2013113, EPI_ISL_2013114, EPI_ISL_2013115, EPI_ISL_2013116, EPI_ISL_2013117, EPI_ISL_2013118, EPI_ISL_2013119, EPI_ISL_2013120, EPI_ISL_2013121, EPI_ISL_2013122, EPI_ISL_2013123, EPI_ISL_2013124, EPI_ISL_2013125, EPI_ISL_2013126, EPI_ISL_2013127, EPI_ISL_2013128, EPI_ISL_2013129, EPI_ISL_2013130, EPI_ISL_2013131, EPI_ISL_2013132, EPI_ISL_2013133, EPI_ISL_2013134, EPI_ISL_2013135, EPI_ISL_2013136, EPI_ISL_2013137, EPI_ISL_2013138, EPI_ISL_2013139, EPI_ISL_2013140, EPI_ISL_2013141, EPI_ISL_2013142, EPI_ISL_2013143, EPI_ISL_2013144, EPI_ISL_2013145, EPI_ISL_2013146, EPI_ISL_2013147, EPI_ISL_2013148, EPI_ISL_2013150, EPI_ISL_2013151, EPI_ISL_2013152, EPI_ISL_2013153, EPI_ISL_2013154, EPI_ISL_2013155, EPI_ISL_2013156, EPI_ISL_2013157, EPI_ISL_2013158, EPI_ISL_2013159, EPI_ISL_2013160, EPI_ISL_2013161, EPI_ISL_2013162, EPI_ISL_2013163, EPI_ISL_2013164, EPI_ISL_2013165, EPI_ISL_2013166, EPI_ISL_2013167, EPI_ISL_2013168, EPI_ISL_2013169, EPI_ISL_2013170, EPI_ISL_2013171, EPI_ISL_2013172, EPI_ISL_2013173, EPI_ISL_2013174, EPI_ISL_2013175, EPI_ISL_2013176, EPI_ISL_2013177, EPI_ISL_2013178, EPI_ISL_2013179, EPI_ISL_2013180, EPI_ISL_2013181, EPI_ISL_2013182, EPI_ISL_2013183, EPI_ISL_2013184, EPI_ISL_2013185, EPI_ISL_2013186, EPI_ISL_2013187, EPI_ISL_2013188, EPI_ISL_2013189, EPI_ISL_2013190, EPI_ISL_2013191, EPI_ISL_2013192, EPI_ISL_2013193, EPI_ISL_2013194, EPI_ISL_2013195, EPI_ISL_2013196, EPI_ISL_2013197, EPI_ISL_2013198, EPI_ISL_2013199, EPI_ISL_2013200, EPI_ISL_2013201, EPI_ISL_2013202, EPI_ISL_2013203, EPI_ISL_2013204, EPI_ISL_2013205, EPI_ISL_2013206, EPI_ISL_2013207, EPI_ISL_2013208, EPI_ISL_2013209, EPI_ISL_2013210, EPI_ISL_2013211, EPI_ISL_2013212, EPI_ISL_2013213, EPI_ISL_2013214, EPI_ISL_2013215, EPI_ISL_2013216, EPI_ISL_2013217, EPI_ISL_2013218, EPI_ISL_2013219, EPI_ISL_2013221, EPI_ISL_2013222, EPI_ISL_2013223, EPI_ISL_2013224, EPI_ISL_2013225, EPI_ISL_2013226, EPI_ISL_2013227, EPI_ISL_2013228, EPI_ISL_2013229, EPI_ISL_2013230, EPI_ISL_2013231, EPI_ISL_2013232, EPI_ISL_2013233, EPI_ISL_2013234, EPI_ISL_2013235, EPI_ISL_2013236, EPI_ISL_2013237, EPI_ISL_2013238, EPI_ISL_2013239, EPI_ISL_2013240, EPI_ISL_2013241, EPI_ISL_2013242, EPI_ISL_2013243, EPI_ISL_2013244, EPI_ISL_2013245, EPI_ISL_2013246, EPI_ISL_2013247, EPI_ISL_2013248, EPI_ISL_2013249, EPI_ISL_2013250, EPI_ISL_2013251, EPI_ISL_2013252, EPI_ISL_2013253, EPI_ISL_2013254, EPI_ISL_2013255, EPI_ISL_2013256, EPI_ISL_2013257, EPI_ISL_2013258, EPI_ISL_2013259, EPI_ISL_2013260, EPI_ISL_2013261, EPI_ISL_2013262, EPI_ISL_2013263, EPI_ISL_2013264, EPI_ISL_2013265, EPI_ISL_2013266, EPI_ISL_2013267, EPI_ISL_2013268, EPI_ISL_2013269, EPI_ISL_2013271, EPI_ISL_2013272, EPI_ISL_2013273, EPI_ISL_2013274, EPI_ISL_2013275, EPI_ISL_2013276, EPI_ISL_2013277, EPI_ISL_2013278, EPI_ISL_2013279, EPI_ISL_2013280, EPI_ISL_2013281, EPI_ISL_2013282, EPI_ISL_2013283, EPI_ISL_2013284, EPI_ISL_2013285, EPI_ISL_2013286, EPI_ISL_2013287, EPI_ISL_2013288, EPI_ISL_2013289, EPI_ISL_2013290, EPI_ISL_2013291, EPI_ISL_2013292, EPI_ISL_2013293, EPI_ISL_2013294, EPI_ISL_2013295, EPI_ISL_2013296, EPI_ISL_2013297, EPI_ISL_2013298, EPI_ISL_2013299, EPI_ISL_2013300, EPI_ISL_2013301, EPI_ISL_2013302, EPI_ISL_2013303, EPI_ISL_2013304, EPI_ISL_2013305, EPI_ISL_2013306, EPI_ISL_2013307, EPI_ISL_2013308, EPI_ISL_2013309, EPI_ISL_2013311, EPI_ISL_2013312, EPI_ISL_2013313, EPI_ISL_2013314, EPI_ISL_2013315, EPI_ISL_2013316, EPI_ISL_2013317, EPI_ISL_2013318, EPI_ISL_2013319, EPI_ISL_2013320, EPI_ISL_2013321, EPI_ISL_2013322, EPI_ISL_2013324, EPI_ISL_2013327, EPI_ISL_2013328, EPI_ISL_2013329, EPI_ISL_2013330, EPI_ISL_2013331, EPI_ISL_2013333, EPI_ISL_2013334, EPI_ISL_2013336, EPI_ISL_2013337, EPI_ISL_2013338, EPI_ISL_2013339, EPI_ISL_2013340, EPI_ISL_2013341, EPI_ISL_2013342, EPI_ISL_2013343, EPI_ISL_2013345, EPI_ISL_2013346, EPI_ISL_2013347, EPI_ISL_2013348, EPI_ISL_2013349, EPI_ISL_2013350, EPI_ISL_2013351, EPI_ISL_2013352, EPI_ISL_2013353, EPI_ISL_2013354, EPI_ISL_2013355, EPI_ISL_2013356, EPI_ISL_2013357, EPI_ISL_2013358, EPI_ISL_2013359, EPI_ISL_2013360, EPI_ISL_2013361, EPI_ISL_2013362, EPI_ISL_2013363, EPI_ISL_2013365, EPI_ISL_2013366, EPI_ISL_2013367, EPI_ISL_2013368, EPI_ISL_2013369, EPI_ISL_2013370, EPI_ISL_2013371, EPI_ISL_2013372, EPI_ISL_2013373, EPI_ISL_2013374, EPI_ISL_2013375, EPI_ISL_2013376, EPI_ISL_2013377, EPI_ISL_2013378, EPI_ISL_2013379, EPI_ISL_2013380, EPI_ISL_2013381, EPI_ISL_2013382, EPI_ISL_2013383, EPI_ISL_2013384, EPI_ISL_2013385, EPI_ISL_2013386, EPI_ISL_2013387, EPI_ISL_2013388, EPI_ISL_2013389, EPI_ISL_2013390, EPI_ISL_2013391, EPI_ISL_2013392, EPI_ISL_2013393, EPI_ISL_2013394, EPI_ISL_2013395, EPI_ISL_2013396, EPI_ISL_2013397, EPI_ISL_2013398, EPI_ISL_2013399, EPI_ISL_2013400, EPI_ISL_2013401, EPI_ISL_2013402, EPI_ISL_2013404, EPI_ISL_2013405, EPI_ISL_2013406, EPI_ISL_2013407, EPI_ISL_2013408, EPI_ISL_2013409, EPI_ISL_2013410, EPI_ISL_2013411, EPI_ISL_2013412, EPI_ISL_2013413, EPI_ISL_2013414, EPI_ISL_2013415, EPI_ISL_2013416, EPI_ISL_2013417, EPI_ISL_2013418, EPI_ISL_2013419, EPI_ISL_2013420, EPI_ISL_2013421, EPI_ISL_2013422, EPI_ISL_2013423, EPI_ISL_2013424, EPI_ISL_2013425, EPI_ISL_2013426, EPI_ISL_2013427, EPI_ISL_2013428, EPI_ISL_2013429, EPI_ISL_2013430, EPI_ISL_2013431, EPI_ISL_2013432, EPI_ISL_2013433, EPI_ISL_2013434, EPI_ISL_2013435, EPI_ISL_2013436, EPI_ISL_2013437, EPI_ISL_2013438, EPI_ISL_2013439, EPI_ISL_2013440, EPI_ISL_2013441, EPI_ISL_2013442, EPI_ISL_2013443, EPI_ISL_2013444, EPI_ISL_2013445, EPI_ISL_2013446, EPI_ISL_2013447, EPI_ISL_2013448, EPI_ISL_2013449, EPI_ISL_2013450, EPI_ISL_2013451, EPI_ISL_2013452, EPI_ISL_2013453, EPI_ISL_2013454, EPI_ISL_2013455, EPI_ISL_2013456, EPI_ISL_2013457, EPI_ISL_2013458, EPI_ISL_2013459, EPI_ISL_2013460, EPI_ISL_2013461, EPI_ISL_2013462, EPI_ISL_2013463, EPI_ISL_2013464, EPI_ISL_2013465, EPI_ISL_2013466, EPI_ISL_2013467, EPI_ISL_2013468, EPI_ISL_2013469, EPI_ISL_2013470, EPI_ISL_2013471, EPI_ISL_2013472, EPI_ISL_2013473, EPI_ISL_2013474, EPI_ISL_2013475, EPI_ISL_2013476, EPI_ISL_2013477, EPI_ISL_2013478, EPI_ISL_2013479, EPI_ISL_2013480, EPI_ISL_2013482, EPI_ISL_2013483, EPI_ISL_2013484, EPI_ISL_2013485, EPI_ISL_2013486, EPI_ISL_2013487, EPI_ISL_2013488, EPI_ISL_2013489, EPI_ISL_2013490, EPI_ISL_2013491, EPI_ISL_2013492, EPI_ISL_2013493, EPI_ISL_2013494, EPI_ISL_2013496, EPI_ISL_2013497, EPI_ISL_2013498, EPI_ISL_2013501, EPI_ISL_2013502, EPI_ISL_2013503, EPI_ISL_2013504, EPI_ISL_2013505, EPI_ISL_2013506, EPI_ISL_2013507, EPI_ISL_2013508, EPI_ISL_2013510, EPI_ISL_2013511, EPI_ISL_2013512, EPI_ISL_2013513, EPI_ISL_2013514, EPI_ISL_2013515, EPI_ISL_2013516, EPI_ISL_2013517, EPI_ISL_2013518, EPI_ISL_2013519, EPI_ISL_2013520, EPI_ISL_2013521, EPI_ISL_2013522, EPI_ISL_2013523, EPI_ISL_2013524, EPI_ISL_2013525, EPI_ISL_2013526, EPI_ISL_2013527, EPI_ISL_2013528, EPI_ISL_2013529, EPI_ISL_2013530, EPI_ISL_2013531, EPI_ISL_2013532, EPI_ISL_2013533, EPI_ISL_2013534, EPI_ISL_2013535, EPI_ISL_2013536, EPI_ISL_2013537, EPI_ISL_2013538, EPI_ISL_2013539, EPI_ISL_2013540, EPI_ISL_2013541, EPI_ISL_2013542, EPI_ISL_2013543, EPI_ISL_2013544, EPI_ISL_2013545, EPI_ISL_2013546, EPI_ISL_2013547, EPI_ISL_2013548, EPI_ISL_2013549, EPI_ISL_2013550, EPI_ISL_2013551, EPI_ISL_2013552, EPI_ISL_2013553, EPI_ISL_2013554, EPI_ISL_2013555, EPI_ISL_2013556, EPI_ISL_2013557, EPI_ISL_2013558, EPI_ISL_2013559, EPI_ISL_2013560, EPI_ISL_2013561, EPI_ISL_2013562, EPI_ISL_2013563, EPI_ISL_2013564, EPI_ISL_2013565, EPI_ISL_2013566, EPI_ISL_2013567, EPI_ISL_2013568, EPI_ISL_2013569, EPI_ISL_2013570, EPI_ISL_2013571, EPI_ISL_2013572, EPI_ISL_2013573, EPI_ISL_2013574, EPI_ISL_2013575, EPI_ISL_2013576, EPI_ISL_2013577, EPI_ISL_2013578, EPI_ISL_2013579, EPI_ISL_2013580, EPI_ISL_2013581, EPI_ISL_2013582, EPI_ISL_2013583, EPI_ISL_2013584, EPI_ISL_2013585, EPI_ISL_2013586, EPI_ISL_2013587, EPI_ISL_2013588, EPI_ISL_2013589, EPI_ISL_2013590, EPI_ISL_2013591, EPI_ISL_2013592, EPI_ISL_2013593, EPI_ISL_2013594, EPI_ISL_2013595, EPI_ISL_2013596, EPI_ISL_2013597, EPI_ISL_2013599, EPI_ISL_2013600, | | | |

| | | | |
|---|---|---|---|
| EPI_ISL_2013601, EPI_ISL_2013602, EPI_ISL_2013603, EPI_ISL_2013604, EPI_ISL_2013605, EPI_ISL_2013607, EPI_ISL_2013608, EPI_ISL_2013609, EPI_ISL_2013610, EPI_ISL_2013611, EPI_ISL_2013612, EPI_ISL_2013613, EPI_ISL_2013614, EPI_ISL_2013615, EPI_ISL_2013616, EPI_ISL_2013617, EPI_ISL_2013618, EPI_ISL_2013619, EPI_ISL_2013620, EPI_ISL_2013621, EPI_ISL_2013622, EPI_ISL_2013623, EPI_ISL_2013624, EPI_ISL_2013625, EPI_ISL_2013627, EPI_ISL_2013628, EPI_ISL_2013629, EPI_ISL_2013630, EPI_ISL_2013631, EPI_ISL_2013632, EPI_ISL_2013633, EPI_ISL_2013634, EPI_ISL_2013635, EPI_ISL_2013636, EPI_ISL_2013637, EPI_ISL_2013638, EPI_ISL_2013639, EPI_ISL_2013640, EPI_ISL_2013641, EPI_ISL_2013642, EPI_ISL_2013643, EPI_ISL_2013644, EPI_ISL_2013645, EPI_ISL_2013646, EPI_ISL_2013647, EPI_ISL_2013648, EPI_ISL_2013649, EPI_ISL_2013650, EPI_ISL_2013651, EPI_ISL_2013652, EPI_ISL_2013653, EPI_ISL_2013654, EPI_ISL_2013655, EPI_ISL_2013656, EPI_ISL_2013657, EPI_ISL_2013658, EPI_ISL_2013659, EPI_ISL_2013660, EPI_ISL_2013661, EPI_ISL_2013662, EPI_ISL_2013663, EPI_ISL_2013664, EPI_ISL_2013665, EPI_ISL_2013666, EPI_ISL_2013667, EPI_ISL_2013668, EPI_ISL_2013669 | | | |
| see above | Infinity Biologix | Centers for Disease Control and Prevention Division of Viral Diseases, Pathogen Discovery | Dakota Howard, Dhwani Batra, Peter W. Cook, Kara Moser, Adrian Paskey, Jason Caravas, Benjamin Rambo-Martin, Shatavia Morrison, Christopher Gulvick, Scott Sammons, Yvette Unoarumhi, Darlene Wagner, Matthew Schmerer, Christian Bixby, Yihe Wang, Jonathan Schultz, Chirayu Goswami, Russ Hager, Robin Grimwood, Clinton R. Paden, Duncan MacCannell |
| EPI_ISL_2013670 | UW Virology Lab | UW Virology Lab | Pavitra Roychoudhury, Hong Xie, Lasata Shrestha, Tien V. Nguyen, Shah Mohamed Bakhash, Michelle Lin, Noah R. Baker, Ricardo Perez, Sean Ellis, Nathan Breit, Robert J. Livingston, Meei-Li Huang, Keith R Jerome, Patrick Mathias, Alexander Greninger |
| EPI_ISL_2013671 | Infinity Biologix | Centers for Disease Control and Prevention Division of Viral Diseases, Pathogen Discovery | Dakota Howard, Dhwani Batra, Peter W. Cook, Kara Moser, Adrian Paskey, Jason Caravas, Benjamin Rambo-Martin, Shatavia Morrison, Christopher Gulvick, Scott Sammons, Yvette Unoarumhi, Darlene Wagner, Matthew Schmerer, Christian Bixby, Yihe Wang, Jonathan Schultz, Chirayu Goswami, Russ Hager, Robin Grimwood, Clinton R. Paden, Duncan MacCannell |
| EPI_ISL_2013672 | UW Virology Lab | UW Virology Lab | Pavitra Roychoudhury, Hong Xie, Lasata Shrestha, Tien V. Nguyen, Shah Mohamed Bakhash, Michelle Lin, Noah R. Baker, Ricardo Perez, Sean Ellis, Nathan Breit, Robert J. Livingston, Meei-Li Huang, Keith R Jerome, Patrick Mathias, Alexander Greninger |
| EPI_ISL_2013673, EPI_ISL_2013674, EPI_ISL_2013675, EPI_ISL_2013676, EPI_ISL_2013677, EPI_ISL_2013678, EPI_ISL_2013679, EPI_ISL_2013680, EPI_ISL_2013681, EPI_ISL_2013682, EPI_ISL_2013683, EPI_ISL_2013684, EPI_ISL_2013686, EPI_ISL_2013687, EPI_ISL_2013688, EPI_ISL_2013689, EPI_ISL_2013691, EPI_ISL_2013692, EPI_ISL_2013693, EPI_ISL_2013694, EPI_ISL_2013695, EPI_ISL_2013696, EPI_ISL_2013697, EPI_ISL_2013698, EPI_ISL_2013699, EPI_ISL_2013700, EPI_ISL_2013701, EPI_ISL_2013702, EPI_ISL_2013703, EPI_ISL_2013704, EPI_ISL_2013705, EPI_ISL_2013706, EPI_ISL_2013707, EPI_ISL_2013708, EPI_ISL_2013709, EPI_ISL_2013710, EPI_ISL_2013711, EPI_ISL_2013712, EPI_ISL_2013713, EPI_ISL_2013714, EPI_ISL_2013715, EPI_ISL_2013716, EPI_ISL_2013717, EPI_ISL_2013718, EPI_ISL_2013719, EPI_ISL_2013720, EPI_ISL_2013721, EPI_ISL_2013722, EPI_ISL_2013724, EPI_ISL_2013725, EPI_ISL_2013726, EPI_ISL_2013727, EPI_ISL_2013728, EPI_ISL_2013729, EPI_ISL_2013730, EPI_ISL_2013731, EPI_ISL_2013732, EPI_ISL_2013733, EPI_ISL_2013734, EPI_ISL_2013735, EPI_ISL_2013736, EPI_ISL_2013737, EPI_ISL_2013738, EPI_ISL_2013739, EPI_ISL_2013740, EPI_ISL_2013741, EPI_ISL_2013742, EPI_ISL_2013743, EPI_ISL_2013744, EPI_ISL_2013745, EPI_ISL_2013746, EPI_ISL_2013748, EPI_ISL_2013749, EPI_ISL_2013750, EPI_ISL_2013751, EPI_ISL_2013752, EPI_ISL_2013753, EPI_ISL_2013754, EPI_ISL_2013755, EPI_ISL_2013756, EPI_ISL_2013757, EPI_ISL_2013758, EPI_ISL_2013759, EPI_ISL_2013760, EPI_ISL_2013761, EPI_ISL_2013762, EPI_ISL_2013763, EPI_ISL_2013764, EPI_ISL_2013765, EPI_ISL_2013766, EPI_ISL_2013767, EPI_ISL_2013768, EPI_ISL_2013769, EPI_ISL_2013770, EPI_ISL_2013771, EPI_ISL_2013772, EPI_ISL_2013773, EPI_ISL_2013774, EPI_ISL_2013775, EPI_ISL_2013776, EPI_ISL_2013777, EPI_ISL_2013778, EPI_ISL_2013779, EPI_ISL_2013780, EPI_ISL_2013781, EPI_ISL_2013782, EPI_ISL_2013783, EPI_ISL_2013784, EPI_ISL_2013785, EPI_ISL_2013786, EPI_ISL_2013787, EPI_ISL_2013788, EPI_ISL_2013789, EPI_ISL_2013790, EPI_ISL_2013791, EPI_ISL_2013792, EPI_ISL_2013793, EPI_ISL_2013794, EPI_ISL_2013795, EPI_ISL_2013796, EPI_ISL_2013797, EPI_ISL_2013798, EPI_ISL_2013799, EPI_ISL_2013800, EPI_ISL_2013801, EPI_ISL_2013802, EPI_ISL_2013803, EPI_ISL_2013804, EPI_ISL_2013805, EPI_ISL_2013806, EPI_ISL_2013807, EPI_ISL_2013808, EPI_ISL_2013809, EPI_ISL_2013810, EPI_ISL_2013811, EPI_ISL_2013812, EPI_ISL_2013813, EPI_ISL_2013814, EPI_ISL_2013815, EPI_ISL_2013816, EPI_ISL_2013817, EPI_ISL_2013818, EPI_ISL_2013819, EPI_ISL_2013820, EPI_ISL_2013821, EPI_ISL_2013822, EPI_ISL_2013823, EPI_ISL_2013824, EPI_ISL_2013825, EPI_ISL_2013826, EPI_ISL_2013827, EPI_ISL_2013828, EPI_ISL_2013829, EPI_ISL_2013830, EPI_ISL_2013831, EPI_ISL_2013832, EPI_ISL_2013833, EPI_ISL_2013834, EPI_ISL_2013835, EPI_ISL_2013836, EPI_ISL_2013837, EPI_ISL_2013838, EPI_ISL_2013839, EPI_ISL_2013840, EPI_ISL_2013841, EPI_ISL_2013842, EPI_ISL_2013843, EPI_ISL_2013844, EPI_ISL_2013845, EPI_ISL_2013846, EPI_ISL_2013847, EPI_ISL_2013848, EPI_ISL_2013849, EPI_ISL_2013850, EPI_ISL_2013851, EPI_ISL_2013852, EPI_ISL_2013853, EPI_ISL_2013856, EPI_ISL_2013857, EPI_ISL_2013858, EPI_ISL_2013859, EPI_ISL_2013860, EPI_ISL_2013861, EPI_ISL_2013862, EPI_ISL_2013863, EPI_ISL_2013864, EPI_ISL_2013865, EPI_ISL_2013867, EPI_ISL_2013868, EPI_ISL_2013869, EPI_ISL_2013870, EPI_ISL_2013871, EPI_ISL_2013872, EPI_ISL_2013873, EPI_ISL_2013874, EPI_ISL_2013875, EPI_ISL_2013876, EPI_ISL_2013877, EPI_ISL_2013878, EPI_ISL_2013879, EPI_ISL_2013880, EPI_ISL_2013881, EPI_ISL_2013882, EPI_ISL_2013883, EPI_ISL_2013884, EPI_ISL_2013885, EPI_ISL_2013886, EPI_ISL_2013887, EPI_ISL_2013888, EPI_ISL_2013889, EPI_ISL_2013890, EPI_ISL_2013891, EPI_ISL_2013893, EPI_ISL_2013894, EPI_ISL_2013895, EPI_ISL_2013896, EPI_ISL_2013897, EPI_ISL_2013898, EPI_ISL_2013899, EPI_ISL_2013900, EPI_ISL_2013901, EPI_ISL_2013902, EPI_ISL_2013903, EPI_ISL_2013904, EPI_ISL_2013905, EPI_ISL_2013907, EPI_ISL_2013908, EPI_ISL_2013909, EPI_ISL_2013910, EPI_ISL_2013911, EPI_ISL_2013912, EPI_ISL_2013913, EPI_ISL_2013914, EPI_ISL_2013915, EPI_ISL_2013916, EPI_ISL_2013917, EPI_ISL_2013918, EPI_ISL_2013919, EPI_ISL_2013920, EPI_ISL_2013921, EPI_ISL_2013922, EPI_ISL_2013923, EPI_ISL_2013924, EPI_ISL_2013925, EPI_ISL_2013926, EPI_ISL_2013927, EPI_ISL_2013928, EPI_ISL_2013929, EPI_ISL_2013930, EPI_ISL_2013931, EPI_ISL_2013932, EPI_ISL_2013933, EPI_ISL_2013934, EPI_ISL_2013935, EPI_ISL_2013936, EPI_ISL_2013937, EPI_ISL_2013938, EPI_ISL_2013939, EPI_ISL_2013940, EPI_ISL_2013941, EPI_ISL_2013942, EPI_ISL_2013943, EPI_ISL_2013944, EPI_ISL_2013945, EPI_ISL_2013946, EPI_ISL_2013947, EPI_ISL_2013948, EPI_ISL_2013949, EPI_ISL_2013950, EPI_ISL_2013951, EPI_ISL_2013952, EPI_ISL_2013953, EPI_ISL_2013954, EPI_ISL_2013955, EPI_ISL_2013956, EPI_ISL_2013957, EPI_ISL_2013958, EPI_ISL_2013959, EPI_ISL_2013960, EPI_ISL_2013961, EPI_ISL_2013962, EPI_ISL_2013963, EPI_ISL_2013964, EPI_ISL_2013965, EPI_ISL_2013966, EPI_ISL_2013967, EPI_ISL_2013968, EPI_ISL_2013969, EPI_ISL_2013970, EPI_ISL_2013971, EPI_ISL_2013973, EPI_ISL_2013974, EPI_ISL_2013975, EPI_ISL_2013976, EPI_ISL_2013977, EPI_ISL_2013978, EPI_ISL_2013979, EPI_ISL_2013980, EPI_ISL_2013981, EPI_ISL_2013982, EPI_ISL_2013983, EPI_ISL_2013984, EPI_ISL_2013985, EPI_ISL_2013986, EPI_ISL_2013987, EPI_ISL_2013988, EPI_ISL_2013989, EPI_ISL_2013990, EPI_ISL_2013991, EPI_ISL_2013992, EPI_ISL_2013993, EPI_ISL_2013994, EPI_ISL_2013995, EPI_ISL_2013996, EPI_ISL_2013997, EPI_ISL_2013998, EPI_ISL_2013999, EPI_ISL_2014000, EPI_ISL_2014001, EPI_ISL_2014002, EPI_ISL_2014003, EPI_ISL_2014004, EPI_ISL_2014005, EPI_ISL_2014006, EPI_ISL_2014007, EPI_ISL_2014008, EPI_ISL_2014009, EPI_ISL_2014010, EPI_ISL_2014011, EPI_ISL_2014012, EPI_ISL_2014013, EPI_ISL_2014014, EPI_ISL_2014015, EPI_ISL_2014016, EPI_ISL_2014017, EPI_ISL_2014018, EPI_ISL_2014019, EPI_ISL_2014020, EPI_ISL_2014021, EPI_ISL_2014022, EPI_ISL_2014023, EPI_ISL_2014024, EPI_ISL_2014025, EPI_ISL_2014026, EPI_ISL_2014027, EPI_ISL_2014028, EPI_ISL_2014029, EPI_ISL_2014030, EPI_ISL_2014031, EPI_ISL_2014032, EPI_ISL_2014033, EPI_ISL_2014034, EPI_ISL_2014035, EPI_ISL_2014036, EPI_ISL_2014037, EPI_ISL_2014038, EPI_ISL_2014039, EPI_ISL_2014040, EPI_ISL_2014041, EPI_ISL_2014042, EPI_ISL_2014043, EPI_ISL_2014045, EPI_ISL_2014046, EPI_ISL_2014047, EPI_ISL_2014048, EPI_ISL_2014049, EPI_ISL_2014050, EPI_ISL_2014051, EPI_ISL_2014052, EPI_ISL_2014053, EPI_ISL_2014054, EPI_ISL_2014055, EPI_ISL_2014056, EPI_ISL_2014057, EPI_ISL_2014058, EPI_ISL_2014059, EPI_ISL_2014060, EPI_ISL_2014061, EPI_ISL_2014062, EPI_ISL_2014063, EPI_ISL_2014064, EPI_ISL_2014065, EPI_ISL_2014066, EPI_ISL_2014067, EPI_ISL_2014068, EPI_ISL_2014069, EPI_ISL_2014070, EPI_ISL_2014071, EPI_ISL_2014072, EPI_ISL_2014073, EPI_ISL_2014074, EPI_ISL_2014075, EPI_ISL_2014076, EPI_ISL_2014077, EPI_ISL_2014078, EPI_ISL_2014079, EPI_ISL_2014080, EPI_ISL_2014081, EPI_ISL_2014082, EPI_ISL_2014083, EPI_ISL_2014084, EPI_ISL_2014086, EPI_ISL_2014087, EPI_ISL_2014088, EPI_ISL_2014089, EPI_ISL_2014090, EPI_ISL_2014091, EPI_ISL_2014092, EPI_ISL_2014093, EPI_ISL_2014094, EPI_ISL_2014095, EPI_ISL_2014096, EPI_ISL_2014097, EPI_ISL_2014098, EPI_ISL_2014099, EPI_ISL_2014100, EPI_ISL_2014101, EPI_ISL_2014102, EPI_ISL_2014103, EPI_ISL_2014104, EPI_ISL_2014105, EPI_ISL_2014106, EPI_ISL_2014107, EPI_ISL_2014108, EPI_ISL_2014109, EPI_ISL_2014110, EPI_ISL_2014111, EPI_ISL_2014112, EPI_ISL_2014113, EPI_ISL_2014114, EPI_ISL_2014115, EPI_ISL_2014116, EPI_ISL_2014117, EPI_ISL_2014118, EPI_ISL_2014119, EPI_ISL_2014120, EPI_ISL_2014121, EPI_ISL_2014122, EPI_ISL_2014123, EPI_ISL_2014124, EPI_ISL_2014125, EPI_ISL_2014126, EPI_ISL_2014127, EPI_ISL_2014128, EPI_ISL_2014129, EPI_ISL_2014130, EPI_ISL_2014134, EPI_ISL_2014136, EPI_ISL_2014137, EPI_ISL_2014138, EPI_ISL_2014139, EPI_ISL_2014140, EPI_ISL_2014141, EPI_ISL_2014142, EPI_ISL_2014143, EPI_ISL_2014144, EPI_ISL_2014145, EPI_ISL_2014146, EPI_ISL_2014147, EPI_ISL_2014148, EPI_ISL_2014149, EPI_ISL_2014150, EPI_ISL_2014151, EPI_ISL_2014152, EPI_ISL_2014153, EPI_ISL_2014154, EPI_ISL_2014155, EPI_ISL_2014156, EPI_ISL_2014157, EPI_ISL_2014158, EPI_ISL_2014159, EPI_ISL_2014160, EPI_ISL_2014161, EPI_ISL_2014162, EPI_ISL_2014163, EPI_ISL_2014164, EPI_ISL_2014165, EPI_ISL_2014166, EPI_ISL_2014167, EPI_ISL_2014168, EPI_ISL_2014169, EPI_ISL_2014170, EPI_ISL_2014171, EPI_ISL_2014172, EPI_ISL_2014173, EPI_ISL_2014174, EPI_ISL_2014175, EPI_ISL_2014176, EPI_ISL_2014177, EPI_ISL_2014178, EPI_ISL_2014179, EPI_ISL_2014180, EPI_ISL_2014181, EPI_ISL_2014182, EPI_ISL_2014183, EPI_ISL_2014184, EPI_ISL_2014185, EPI_ISL_2014186, EPI_ISL_2014187, EPI_ISL_2014188, EPI_ISL_2014189, EPI_ISL_2014190, EPI_ISL_2014191, EPI_ISL_2014192, EPI_ISL_2014193, EPI_ISL_2014194, EPI_ISL_2014195, EPI_ISL_2014196, EPI_ISL_2014197, EPI_ISL_2014198, EPI_ISL_2014200, EPI_ISL_2014201, EPI_ISL_2014202, EPI_ISL_2014203, EPI_ISL_2014204, EPI_ISL_2014205, EPI_ISL_2014206, EPI_ISL_2014207, EPI_ISL_2014208, EPI_ISL_2014209, EPI_ISL_2014210, EPI_ISL_2014211, EPI_ISL_2014212, EPI_ISL_2014213, EPI_ISL_2014214, EPI_ISL_2014215, EPI_ISL_2014217, EPI_ISL_2014218, EPI_ISL_2014219, EPI_ISL_2014220, EPI_ISL_2014221, EPI_ISL_2014222, EPI_ISL_2014223, EPI_ISL_2014224, EPI_ISL_2014225, EPI_ISL_2014226, EPI_ISL_2014227, EPI_ISL_2014228, EPI_ISL_2014229, EPI_ISL_2014230, EPI_ISL_2014231, EPI_ISL_2014232, EPI_ISL_2014233, EPI_ISL_2014234, EPI_ISL_2014235, EPI_ISL_2014236, EPI_ISL_2014237, EPI_ISL_2014238, EPI_ISL_2014239, EPI_ISL_2014240, EPI_ISL_2014241, EPI_ISL_2014242, EPI_ISL_2014243, EPI_ISL_2014244, EPI_ISL_2014245, EPI_ISL_2014246, EPI_ISL_2014247, EPI_ISL_2014248, EPI_ISL_2014249, EPI_ISL_2014250, EPI_ISL_2014251, EPI_ISL_2014252, EPI_ISL_2014253, EPI_ISL_2014254, EPI_ISL_2014255, EPI_ISL_2014256, EPI_ISL_2014257, EPI_ISL_2014259, EPI_ISL_2014260, EPI_ISL_2014261, EPI_ISL_2014262, EPI_ISL_2014263, EPI_ISL_2014264, EPI_ISL_2014265, EPI_ISL_2014266, EPI_ISL_2014267, EPI_ISL_2014268, EPI_ISL_2014269, EPI_ISL_2014270, EPI_ISL_2014271, EPI_ISL_2014272, EPI_ISL_2014273, EPI_ISL_2014275, EPI_ISL_2014276, EPI_ISL_2014277, EPI_ISL_2014278, EPI_ISL_2014279, EPI_ISL_2014280, EPI_ISL_2014281, EPI_ISL_2014282, EPI_ISL_2014283, EPI_ISL_2014284, EPI_ISL_2014285, EPI_ISL_2014286, EPI_ISL_2014287, EPI_ISL_2014288, EPI_ISL_2014289, EPI_ISL_2014290, EPI_ISL_2014291, EPI_ISL_2014292, EPI_ISL_2014293, EPI_ISL_2014294, EPI_ISL_2014295, EPI_ISL_2014296, EPI_ISL_2014298, EPI_ISL_2014299, EPI_ISL_2014300, EPI_ISL_2014301, EPI_ISL_2014302, EPI_ISL_2014303, EPI_ISL_2014304, EPI_ISL_2014305, EPI_ISL_2014306, EPI_ISL_2014307, EPI_ISL_2014308, EPI_ISL_2014309, EPI_ISL_2014310, EPI_ISL_2014311, EPI_ISL_2014312, EPI_ISL_2014313, EPI_ISL_2014314, EPI_ISL_2014315, EPI_ISL_2014316, EPI_ISL_2014317, EPI_ISL_2014318, EPI_ISL_2014319, EPI_ISL_2014320, EPI_ISL_2014321, EPI_ISL_2014322, EPI_ISL_2014323, EPI_ISL_2014324, EPI_ISL_2014325, EPI_ISL_2014326, EPI_ISL_2014327, EPI_ISL_2014328, EPI_ISL_2014329, EPI_ISL_2014330, EPI_ISL_2014331, EPI_ISL_2014332, EPI_ISL_2014333, EPI_ISL_2014334, EPI_ISL_2014335, EPI_ISL_2014336, EPI_ISL_2014337, EPI_ISL_2014338, EPI_ISL_2014339, EPI_ISL_2014340, EPI_ISL_2014341, EPI_ISL_2014342, EPI_ISL_2014343, EPI_ISL_2014344, EPI_ISL_2014345, EPI_ISL_2014346, EPI_ISL_2014347, EPI_ISL_2014348, EPI_ISL_2014349, EPI_ISL_2014350, EPI_ISL_2014351, EPI_ISL_2014352, EPI_ISL_2014353, EPI_ISL_2014354, EPI_ISL_2014356, EPI_ISL_2014357, EPI_ISL_2014358, EPI_ISL_2014359, EPI_ISL_2014360, EPI_ISL_2014361, EPI_ISL_2014362, EPI_ISL_2014363, EPI_ISL_2014364, EPI_ISL_2014365, EPI_ISL_2014366, EPI_ISL_2014367, EPI_ISL_2014368, EPI_ISL_2014369, EPI_ISL_2014370, EPI_ISL_2014371, EPI_ISL_2014372, EPI_ISL_2014373, EPI_ISL_2014374, EPI_ISL_2014375, EPI_ISL_2014376, EPI_ISL_2014377, EPI_ISL_2014378, EPI_ISL_2014379, EPI_ISL_2014380, EPI_ISL_2014381, EPI_ISL_2014382, EPI_ISL_2014383, EPI_ISL_2014384, EPI_ISL_2014385, EPI_ISL_2014386, EPI_ISL_2014387, EPI_ISL_2014388, EPI_ISL_2014389, EPI_ISL_2014391, EPI_ISL_2014392, EPI_ISL_2014393, EPI_ISL_2014394, EPI_ISL_2014395, EPI_ISL_2014396, EPI_ISL_2014397, EPI_ISL_2014398, EPI_ISL_2014400, EPI_ISL_2014401, EPI_ISL_2014402, EPI_ISL_2014403, EPI_ISL_2014404, EPI_ISL_2014405, EPI_ISL_2014406, EPI_ISL_2014407, EPI_ISL_2014408, EPI_ISL_2014409, EPI_ISL_2014410, EPI_ISL_2014411, EPI_ISL_2014412, EPI_ISL_2014413, EPI_ISL_2014414, EPI_ISL_2014415, EPI_ISL_2014416, EPI_ISL_2014417, EPI_ISL_2014418, EPI_ISL_2014419, EPI_ISL_2014420, EPI_ISL_2014422, EPI_ISL_2014423, EPI_ISL_2014424, EPI_ISL_2014425, EPI_ISL_2014426, EPI_ISL_2014427, EPI_ISL_2014428, EPI_ISL_2014429, EPI_ISL_2014430, EPI_ISL_2014431, EPI_ISL_2014432, EPI_ISL_2014433, EPI_ISL_2014434, EPI_ISL_2014435, EPI_ISL_2014436, EPI_ISL_2014437, EPI_ISL_2014439, EPI_ISL_2014440, EPI_ISL_2014441, EPI_ISL_2014442, EPI_ISL_2014443, EPI_ISL_2014444, EPI_ISL_2014445, EPI_ISL_2014446, EPI_ISL_2014447, EPI_ISL_2014448, EPI_ISL_2014449, EPI_ISL_2014450, EPI_ISL_2014451, EPI_ISL_2014452, EPI_ISL_2014453, EPI_ISL_2014454, EPI_ISL_2014455, EPI_ISL_2014456, EPI_ISL_2014457, EPI_ISL_2014458, EPI_ISL_2014459, EPI_ISL_2014460, EPI_ISL_2014461, EPI_ISL_2014462, EPI_ISL_2014463, EPI_ISL_2014464, EPI_ISL_2014465, EPI_ISL_2014466, EPI_ISL_2014467, EPI_ISL_2014468, EPI_ISL_2014470, EPI_ISL_2014471, EPI_ISL_2014472, EPI_ISL_2014473, EPI_ISL_2014474, EPI_ISL_2014475, EPI_ISL_2014477, EPI_ISL_2014478, EPI_ISL_2014479, EPI_ISL_2014480, EPI_ISL_2014481, EPI_ISL_2014482, EPI_ISL_2014483, EPI_ISL_2014484, EPI_ISL_2014486, EPI_ISL_2014487, EPI_ISL_2014488, EPI_ISL_2014489, EPI_ISL_2014490, EPI_ISL_2014491, EPI_ISL_2014492, EPI_ISL_2014493, EPI_ISL_2014494, EPI_ISL_2014495, EPI_ISL_2014496, EPI_ISL_2014497, EPI_ISL_2014498, EPI_ISL_2014499, EPI_ISL_2014500, EPI_ISL_2014501, EPI_ISL_2014502, EPI_ISL_2014503, EPI_ISL_2014504, EPI_ISL_2014505, EPI_ISL_2014506, EPI_ISL_2014507, EPI_ISL_2014508, EPI_ISL_2014509, EPI_ISL_2014510, EPI_ISL_2014511, EPI_ISL_2014512, EPI_ISL_2014513, EPI_ISL_2014514, EPI_ISL_2014515, EPI_ISL_2014516, EPI_ISL_2014517, EPI_ISL_2014518, EPI_ISL_2014519, EPI_ISL_2014520, EPI_ISL_2014521, EPI_ISL_2014522, EPI_ISL_2014523, EPI_ISL_2014524, EPI_ISL_2014525, EPI_ISL_2014526, EPI_ISL_2014527, EPI_ISL_2014528, EPI_ISL_2014530, EPI_ISL_2014531, EPI_ISL_2014532, EPI_ISL_2014533, EPI_ISL_2014534, EPI_ISL_2014535, EPI_ISL_2014536, EPI_ISL_2014537, EPI_ISL_2014538, EPI_ISL_2014539, EPI_ISL_2014540, EPI_ISL_2014541, EPI_ISL_2014543, EPI_ISL_2014544, EPI_ISL_2014545, EPI_ISL_2014546, EPI_ISL_2014547, EPI_ISL_2014548, EPI_ISL_2014549, EPI_ISL_2014550, EPI_ISL_2014551, EPI_ISL_2014552, EPI_ISL_2014553, EPI_ISL_2014554, EPI_ISL_2014555, EPI_ISL_2014556, EPI_ISL_2014557, EPI_ISL_2014559, EPI_ISL_2014560, EPI_ISL_2014561, EPI_ISL_2014562, EPI_ISL_2014563, EPI_ISL_2014564, EPI_ISL_2014566, EPI_ISL_2014567, EPI_ISL_2014568, EPI_ISL_2014569, EPI_ISL_2014570, EPI_ISL_2014571, EPI_ISL_2014572, EPI_ISL_2014573, EPI_ISL_2014574, EPI_ISL_2014575, EPI_ISL_2014576, EPI_ISL_2014578, EPI_ISL_2014579, EPI_ISL_2014580, EPI_ISL_2014581, EPI_ISL_2014582, EPI_ISL_2014583, EPI_ISL_2014584, EPI_ISL_2014585, EPI_ISL_2014586, EPI_ISL_2014587, EPI_ISL_2014588, EPI_ISL_2014590, EPI_ISL_2014591, EPI_ISL_2014592, EPI_ISL_2014593, EPI_ISL_2014594, EPI_ISL_2014595, EPI_ISL_2014596, EPI_ISL_2014597, EPI_ISL_2014598, EPI_ISL_2014599, EPI_ISL_2014600, EPI_ISL_2014601, EPI_ISL_2014602, EPI_ISL_2014603, EPI_ISL_2014604, EPI_ISL_2014606, EPI_ISL_2014607, EPI_ISL_2014608, EPI_ISL_2014609, EPI_ISL_2014610, EPI_ISL_2014611, EPI_ISL_2014612, EPI_ISL_2014613, EPI_ISL_2014614, EPI_ISL_2014615, EPI_ISL_2014616, EPI_ISL_2014617, EPI_ISL_2014618, EPI_ISL_2014619, EPI_ISL_2014620, EPI_ISL_2014621, EPI_ISL_2014622, EPI_ISL_2014623, EPI_ISL_2014624, EPI_ISL_2014625, EPI_ISL_2014626, EPI_ISL_2014627, EPI_ISL_2014628, EPI_ISL_2014629, EPI_ISL_2014630, EPI_ISL_2014631, EPI_ISL_2014632, EPI_ISL_2014633, EPI_ISL_2014634, EPI_ISL_2014635, EPI_ISL_2014636, EPI_ISL_2014637, EPI_ISL_2014639, EPI_ISL_2014640, EPI_ISL_2014641, EPI_ISL_2014642, EPI_ISL_2014643, EPI_ISL_2014644, EPI_ISL_2014645, EPI_ISL_2014646, EPI_ISL_2014647, EPI_ISL_2014648, EPI_ISL_2014649, EPI_ISL_2014650, EPI_ISL_2014651, EPI_ISL_2014652, EPI_ISL_2014653, EPI_ISL_2014654, EPI_ISL_2014655, EPI_ISL_2014656, EPI_ISL_2014657, EPI_ISL_2014658, EPI_ISL_2014659, EPI_ISL_2014660, EPI_ISL_2014661, EPI_ISL_2014662, EPI_ISL_2014663, EPI_ISL_2014664, EPI_ISL_2014665, EPI_ISL_2014666, EPI_ISL_2014667, EPI_ISL_2014668, EPI_ISL_2014669, EPI_ISL_2014670, EPI_ISL_2014671, EPI_ISL_2014673, EPI_ISL_2014674, EPI_ISL_2014675, EPI_ISL_2014676, EPI_ISL_2014677, EPI_ISL_2014678, EPI_ISL_2014679, EPI_ISL_2014681, EPI_ISL_2014682, EPI_ISL_2014683, EPI_ISL_2014684, EPI_ISL_2014685, EPI_ISL_2014686, EPI_ISL_2014687, EPI_ISL_2014688, EPI_ISL_2014690, EPI_ISL_2014691, EPI_ISL_2014692, EPI_ISL_2014693, EPI_ISL_2014694, EPI_ISL_2014695, EPI_ISL_2014697, EPI_ISL_2014698, EPI_ISL_2014699, EPI_ISL_2014700, EPI_ISL_2014701, EPI_ISL_2014702, EPI_ISL_2014703, EPI_ISL_2014704, EPI_ISL_2014705, EPI_ISL_2014706, EPI_ISL_2014707, EPI_ISL_2014708, EPI_ISL_2014709, EPI_ISL_2014710, EPI_ISL_2014711, EPI_ISL_2014712, EPI_ISL_2014713, EPI_ISL_2014714, EPI_ISL_2014715, EPI_ISL_2014716, EPI_ISL_2014717, EPI_ISL_2014718, EPI_ISL_2014719, EPI_ISL_2014720, EPI_ISL_2014721, EPI_ISL_2014722, EPI_ISL_2014723, EPI_ISL_2014724, EPI_ISL_2014725, EPI_ISL_2014727, EPI_ISL_2014728, EPI_ISL_2014729, EPI_ISL_2014730, EPI_ISL_2014732, EPI_ISL_2014734, EPI_ISL_2014735, EPI_ISL_2014736, EPI_ISL_2014737, EPI_ISL_2014738, EPI_ISL_2014739, EPI_ISL_2014740, EPI_ISL_2014741, EPI_ISL_2014742, EPI_ISL_2014743, EPI_ISL_2014744, EPI_ISL_2014745, EPI_ISL_2014746, EPI_ISL_2014747, EPI_ISL_2014748, EPI_ISL_2014749, EPI_ISL_2014750, EPI_ISL_2014751, EPI_ISL_2014752, EPI_ISL_2014753, EPI_ISL_2014754, EPI_ISL_2014755, EPI_ISL_2014756, EPI_ISL_2014757, EPI_ISL_2014758, EPI_ISL_2014759, EPI_ISL_2014760, EPI_ISL_2014761, EPI_ISL_2014762, EPI_ISL_2014764, EPI_ISL_2014765, EPI_ISL_2014766, EPI_ISL_2014767, EPI_ISL_2014768, EPI_ISL_2014769, EPI_ISL_2014770, EPI_ISL_2014771, EPI_ISL_2014773, EPI_ISL_2014774, EPI_ISL_2014775, EPI_ISL_2014776, EPI_ISL_2014777, EPI_ISL_2014778, EPI_ISL_2014780, EPI_ISL_2014781, EPI_ISL_2014782, EPI_ISL_2014783, EPI_ISL_2014784, EPI_ISL_2014785, EPI_ISL_2014786, EPI_ISL_2014787, EPI_ISL_2014788, EPI_ISL_2014789, EPI_ISL_2014790, EPI_ISL_2014791, EPI_ISL_2014792, EPI_ISL_2014793, EPI_ISL_2014794, EPI_ISL_2014795, EPI_ISL_2014796, EPI_ISL_2014797, EPI_ISL_2014798, EPI_ISL_2014799, EPI_ISL_2014800, EPI_ISL_2014801, EPI_ISL_2014802, EPI_ISL_2014803, EPI_ISL_2014804, EPI_ISL_2014805, EPI_ISL_2014806, EPI_ISL_2014807, EPI_ISL_2014808, EPI_ISL_2014809, EPI_ISL_2014811, EPI_ISL_2014812, EPI_ISL_2014813, EPI_ISL_2014814, EPI_ISL_2014815, EPI_ISL_2014816, EPI_ISL_2014817, EPI_ISL_2014818, EPI_ISL_2014820, EPI_ISL_2014821, EPI_ISL_2014822, EPI_ISL_2014823, EPI_ISL_2014824, EPI_ISL_2014825, EPI_ISL_2014826, EPI_ISL_2014827, EPI_ISL_2014828, EPI_ISL_2014830, EPI_ISL_2014831, EPI_ISL_2014832, EPI_ISL_2014833, EPI_ISL_2014834, EPI_ISL_2014835, EPI_ISL_2014836, EPI_ISL_2014837, EPI_ISL_2014838, EPI_ISL_2014839, EPI_ISL_2014841, EPI_ISL_2014842, EPI_ISL_2014843, EPI_ISL_2014844, EPI_ISL_2014846, EPI_ISL_2014847, EPI_ISL_2014848, EPI_ISL_2014849, EPI_ISL_2014851, EPI_ISL_2014852 | | | |
| see above | Infinity Biologix | Centers for Disease Control and Prevention Division of Viral Diseases, Pathogen Discovery | Dakota Howard, Dhwani Batra, Peter W. Cook, Kara Moser, Adrian Paskey, Jason Caravas, Benjamin Rambo-Martin, Shatavia Morrison, Christopher Gulvick, Scott Sammons, Yvette Unoarumhi, Darlene Wagner, Matthew Schmerer, Christian Bixby, Yihe Wang, Jonathan Schultz, Chirayu Goswami, Russ Hager, Robin Grimwood, Clinton R. Paden, Duncan MacCannell |
| EPI_ISL_2015144, EPI_ISL_2015145, EPI_ISL_2015146 | Minnesota Department of Health, Public Health Laboratory | Minnesota Department of Health, Public Health Laboratory | Alexandra Lorentz, Jacob Garfin, Matt Plumb, and Xiong Wang |
| EPI_ISL_2015147 | Centracare Laboratory Services | Minnesota Department of Health, Public Health Laboratory | Alexandra Lorentz, Jacob Garfin, Matt Plumb, and Xiong Wang |
| EPI_ISL_2015148, EPI_ISL_2015149, EPI_ISL_2015150, EPI_ISL_2015151, EPI_ISL_2015152, EPI_ISL_2015153, EPI_ISL_2015154, EPI_ISL_2015155, EPI_ISL_2015156, EPI_ISL_2015157, EPI_ISL_2015158, EPI_ISL_2015159, EPI_ISL_2015160, EPI_ISL_2015161, EPI_ISL_2015162, EPI_ISL_2015163, EPI_ISL_2015164, EPI_ISL_2015165, EPI_ISL_2015166, EPI_ISL_2015167, EPI_ISL_2015168, EPI_ISL_2015169, EPI_ISL_2015170, EPI_ISL_2015171, EPI_ISL_2015172, EPI_ISL_2015173, EPI_ISL_2015174, EPI_ISL_2015175, EPI_ISL_2015176, EPI_ISL_2015177, EPI_ISL_2015178, EPI_ISL_2015179, EPI_ISL_2015180, EPI_ISL_2015181, EPI_ISL_2015182, EPI_ISL_2015183, EPI_ISL_2015184, EPI_ISL_2015185, EPI_ISL_2015186, EPI_ISL_2015187, EPI_ISL_2015188, EPI_ISL_2015189 | | | |
| see above | Minnesota Department of Health, Public Health Laboratory | Minnesota Department of Health, Public Health Laboratory | Alexandra Lorentz, Jacob Garfin, Matt Plumb, and Xiong Wang |
| EPI_ISL_2015190 | Centracare Laboratory Services | Minnesota Department of Health, Public Health Laboratory | Alexandra Lorentz, Jacob Garfin, Matt Plumb, and Xiong Wang |
| EPI_ISL_2015191, EPI_ISL_2015192 | M Health Fairview | Minnesota Department of Health, Public Health Laboratory | Alexandra Lorentz, Jacob Garfin, Matt Plumb, and Xiong Wang |
| EPI_ISL_2015193, EPI_ISL_2015194, EPI_ISL_2015195, EPI_ISL_2015196, EPI_ISL_2015197, EPI_ISL_2015198, EPI_ISL_2015199, EPI_ISL_2015200, EPI_ISL_2015201, EPI_ISL_2015202, EPI_ISL_2015203, EPI_ISL_2015204, EPI_ISL_2015205 | | | |
| see above | Minnesota Department of Health, Public Health Laboratory | Minnesota Department of Health, Public Health Laboratory | Alexandra Lorentz, Jacob Garfin, Matt Plumb, and Xiong Wang |
| EPI_ISL_2015206 | Centracare Laboratory Services | Minnesota Department of Health, Public Health Laboratory | Alexandra Lorentz, Jacob Garfin, Matt Plumb, and Xiong Wang |
| EPI_ISL_2015211, EPI_ISL_2015212, EPI_ISL_2015213, EPI_ISL_2015214, EPI_ISL_2015215, EPI_ISL_2015216, EPI_ISL_2015217, EPI_ISL_2015218, EPI_ISL_2015219, EPI_ISL_2015220, EPI_ISL_2015221, EPI_ISL_2015222 | | | |
| see above | Minnesota Department of Health, Public Health Laboratory | Minnesota Department of Health, Public Health Laboratory | Alexandra Lorentz, Jacob Garfin, Matt Plumb, and Xiong Wang |
| EPI_ISL_2015223, EPI_ISL_2015224, EPI_ISL_2015226 | Wyoming Public Health Laboratory | Wyoming Public Health Laboratory | Jim Mildenberger, Wanda Manley, Noah Hull, Taylor Fearing, Lynette Gumbleton, Channing Weber, Ashley Norberg, Chayse Rowley, Marley Goetz, Brian Dominguez, Elliot Thomasson, Sam Britz, Cari Sloma, and Rob Christensen |
| EPI_ISL_2015270 | Wyoming Public Health Laboratory | Wyoming Public Health Laboratory | Jim Mildenberger, Taylor Fearing, Channing Weber, Ashley Norberg, Chayse Rowley, Marley Goetz, Brian Dominguez, Elliot Thomasson, Brittany Oher, Sam Britz, Cari Sloma, and Rob Christensen |
| EPI_ISL_2015300 | Wyoming Public Health Laboratory | Wyoming Public Health Laboratory | Jim Mildenberger, Wanda Manley, Noah Hull, Taylor Fearing, Lynette Gumbleton, Channing Weber, Ashley Norberg, Chayse Rowley, Marley Goetz, Brian Dominguez, Elliot Thomasson, Sam Britz, Cari Sloma, and Rob Christensen |
| EPI_ISL_2015332, EPI_ISL_2015335, EPI_ISL_2015336, EPI_ISL_2015337, EPI_ISL_2015338, EPI_ISL_2015339, EPI_ISL_2015340, EPI_ISL_2015341, EPI_ISL_2015342, EPI_ISL_2015343, EPI_ISL_2015344, EPI_ISL_2015345, EPI_ISL_2015346, EPI_ISL_2015347, EPI_ISL_2015349, EPI_ISL_2015351, EPI_ISL_2015353, EPI_ISL_2015354, EPI_ISL_2015355, EPI_ISL_2015359, EPI_ISL_2015360, EPI_ISL_2015361, EPI_ISL_2015362, EPI_ISL_2015363, EPI_ISL_2015364, EPI_ISL_2015365, EPI_ISL_2015366, EPI_ISL_2015367, EPI_ISL_2015368, EPI_ISL_2015370, EPI_ISL_2015371, EPI_ISL_2015372, EPI_ISL_2015373, EPI_ISL_2015374, EPI_ISL_2015375, EPI_ISL_2015379, EPI_ISL_2015380, EPI_ISL_2015381, EPI_ISL_2015382, EPI_ISL_2015385, EPI_ISL_2015386, EPI_ISL_2015387, EPI_ISL_2015388, EPI_ISL_2015389, EPI_ISL_2015390, EPI_ISL_2015392, EPI_ISL_2015393, EPI_ISL_2015394, EPI_ISL_2015396, EPI_ISL_2015398, EPI_ISL_2015399, EPI_ISL_2015401, EPI_ISL_2015402, EPI_ISL_2015403, EPI_ISL_2015404, EPI_ISL_2015405, EPI_ISL_2015406, EPI_ISL_2015407, EPI_ISL_2015408, EPI_ISL_2015410, EPI_ISL_2015411, EPI_ISL_2015412, EPI_ISL_2015413, EPI_ISL_2015414, EPI_ISL_2015415, EPI_ISL_2015416, EPI_ISL_2015418, EPI_ISL_2015419, EPI_ISL_2015420, EPI_ISL_2015421, EPI_ISL_2015422, EPI_ISL_2015423, EPI_ISL_2015425, EPI_ISL_2015426, EPI_ISL_2015427, EPI_ISL_2015428, EPI_ISL_2015429, EPI_ISL_2015430, EPI_ISL_2015432, EPI_ISL_2015433, EPI_ISL_2015434, EPI_ISL_2015436, EPI_ISL_2015437, EPI_ISL_2015438, EPI_ISL_2015439, EPI_ISL_2015440, EPI_ISL_2015441, EPI_ISL_2015442, EPI_ISL_2015443, EPI_ISL_2015444, EPI_ISL_2015445, EPI_ISL_2015446, EPI_ISL_2015447, EPI_ISL_2015448, EPI_ISL_2015449, EPI_ISL_2015451, EPI_ISL_2015452, EPI_ISL_2015453, EPI_ISL_2015454, EPI_ISL_2015455, EPI_ISL_2015456, EPI_ISL_2015457, EPI_ISL_2015458, EPI_ISL_2015459, EPI_ISL_2015460, EPI_ISL_2015461, EPI_ISL_2015462, EPI_ISL_2015463, EPI_ISL_2015464, EPI_ISL_2015465 | | | |
| see above | Arizona State University | Arizona State University | Peter T. Skidmore, LaRinda A. Holland, Matthew F. Smith, Rabia Maqsood, Nicholas J. Mellor, Joy M. Blain, Valerie Harris, Joshua LaBaer, Vel Murugan, Efrem S. Lim |
| EPI_ISL_2016188, EPI_ISL_2016189, EPI_ISL_2016190, EPI_ISL_2016191, EPI_ISL_2016192, EPI_ISL_2016193, EPI_ISL_2016194, EPI_ISL_2016195, EPI_ISL_2016196, EPI_ISL_2016197, EPI_ISL_2016198, EPI_ISL_2016199, EPI_ISL_2016200, EPI_ISL_2016202, EPI_ISL_2016203, EPI_ISL_2016204, EPI_ISL_2016205, EPI_ISL_2016207, EPI_ISL_2016208, EPI_ISL_2016209, EPI_ISL_2016210, EPI_ISL_2016211, | | | |

| | | | |
|---|---|---|---|
| EPI_ISL_2016212, EPI_ISL_2016213, EPI_ISL_2016214, EPI_ISL_2016215, EPI_ISL_2016216, EPI_ISL_2016217, EPI_ISL_2016218, EPI_ISL_2016219, EPI_ISL_2016220, EPI_ISL_2016221, EPI_ISL_2016222, EPI_ISL_2016224, EPI_ISL_2016225, EPI_ISL_2016226, EPI_ISL_2016227, EPI_ISL_2016228, EPI_ISL_2016229, EPI_ISL_2016230, EPI_ISL_2016231, EPI_ISL_2016232, EPI_ISL_2016233, EPI_ISL_2016234, EPI_ISL_2016235, EPI_ISL_2016236, EPI_ISL_2016237, EPI_ISL_2016238, EPI_ISL_2016239, EPI_ISL_2016240, EPI_ISL_2016241, EPI_ISL_2016242, EPI_ISL_2016243, EPI_ISL_2016244, EPI_ISL_2016245, EPI_ISL_2016246, EPI_ISL_2016247, EPI_ISL_2016248, EPI_ISL_2016249, EPI_ISL_2016250, EPI_ISL_2016251, EPI_ISL_2016252, EPI_ISL_2016253, EPI_ISL_2016254, EPI_ISL_2016255, EPI_ISL_2016256, EPI_ISL_2016258, EPI_ISL_2016259, EPI_ISL_2016260, EPI_ISL_2016261, EPI_ISL_2016262, EPI_ISL_2016264, EPI_ISL_2016265, EPI_ISL_2016266, EPI_ISL_2016267, EPI_ISL_2016268, EPI_ISL_2016269, EPI_ISL_2016270, EPI_ISL_2016271, EPI_ISL_2016272, EPI_ISL_2016274, EPI_ISL_2016275, EPI_ISL_2016276, EPI_ISL_2016277, EPI_ISL_2016278, EPI_ISL_2016279, EPI_ISL_2016280, EPI_ISL_2016281, EPI_ISL_2016282, EPI_ISL_2016283, EPI_ISL_2016284, EPI_ISL_2016285, EPI_ISL_2016286, EPI_ISL_2016287, EPI_ISL_2016288, EPI_ISL_2016289, EPI_ISL_2016290, EPI_ISL_2016291, EPI_ISL_2016292, EPI_ISL_2016293, EPI_ISL_2016294, EPI_ISL_2016295, EPI_ISL_2016296, EPI_ISL_2016297, EPI_ISL_2016298, EPI_ISL_2016299, EPI_ISL_2016300 | | | |
| see above | Pathology and Laboratory Medicine Institute, Cleveland Clinic, Ohio, USA | Pathology and Laboratory Medicine Institute, Cleveland Clinic, Ohio, USA | Jennifer Starbuck, Joy Nakitandwe, Kristen McDonnell, David Plunkett, Zheng Jin Tu, Jay Brock, Yu-Wei Cheng, Gary Procop, Daniel Rhoads, Daniel H. Farkas, David Bosler |
| EPI_ISL_2016384, EPI_ISL_2016385, EPI_ISL_2016386, EPI_ISL_2016389, EPI_ISL_2016390, EPI_ISL_2016392, EPI_ISL_2016393, EPI_ISL_2016394, EPI_ISL_2016400, EPI_ISL_2016401, EPI_ISL_2016402, EPI_ISL_2016404, EPI_ISL_2016405, EPI_ISL_2016406, EPI_ISL_2016408, EPI_ISL_2016409, EPI_ISL_2016410, EPI_ISL_2016411, EPI_ISL_2016413, EPI_ISL_2016415, EPI_ISL_2016417, EPI_ISL_2016418, EPI_ISL_2016419, EPI_ISL_2016420, EPI_ISL_2016421, EPI_ISL_2016422, EPI_ISL_2016423, EPI_ISL_2016424, EPI_ISL_2016425, EPI_ISL_2016426, EPI_ISL_2016427, EPI_ISL_2016429, EPI_ISL_2016430, EPI_ISL_2016431, EPI_ISL_2016432, EPI_ISL_2016434, EPI_ISL_2016435, EPI_ISL_2016437, EPI_ISL_2016439 | | | |
| see above | Broad Institute Clinical Research Sequencing Platform | Infectious Disease Program, Broad Institute of Harvard and MIT | Siddle,K.J., Adams,G., Pearlman,L., Gladden-Young,A., Vicente,G., Blumenstiel,B., DeFelice,M., Lee,M., McGovern,S., Lagerborg,K., Rudy,M., DeRuff,K., Carter,A., Normandin,E., Bauer,M., Reilly,S., Tomkins-Tinch,C., Loreth,C., Chaluvadi,S., Meldrim,J., Granger,B., Lemieux,J.E., Birren,B.W., Sabeti,P.C., Larkin,K., Dodge,S., Lennon,N., Madoff,L., Brown,C., Gallagher,G., Smole,S., Park,D.J., Gabriel,S., and MacInnis,B.L. |
| EPI_ISL_2016440, EPI_ISL_2016441, EPI_ISL_2016442, EPI_ISL_2016443, EPI_ISL_2016444, EPI_ISL_2016445, EPI_ISL_2016446, EPI_ISL_2016447, EPI_ISL_2016448, EPI_ISL_2016449, EPI_ISL_2016450, EPI_ISL_2016451, EPI_ISL_2016452, EPI_ISL_2016453, EPI_ISL_2016454, EPI_ISL_2016455, EPI_ISL_2016456, EPI_ISL_2016457, EPI_ISL_2016458, EPI_ISL_2016459, EPI_ISL_2016461, EPI_ISL_2016462, EPI_ISL_2016463, EPI_ISL_2016464, EPI_ISL_2016465, EPI_ISL_2016466, EPI_ISL_2016467, EPI_ISL_2016468, EPI_ISL_2016469, EPI_ISL_2016470, EPI_ISL_2016471, EPI_ISL_2016472, EPI_ISL_2016473, EPI_ISL_2016474, EPI_ISL_2016475, EPI_ISL_2016476, EPI_ISL_2016477, EPI_ISL_2016478, EPI_ISL_2016479, EPI_ISL_2016480, EPI_ISL_2016481, EPI_ISL_2016482, EPI_ISL_2016483, EPI_ISL_2016484, EPI_ISL_2016485, EPI_ISL_2016486 | | | |
| see above | Massachusetts General Hospital | Infectious Disease Program, Broad Institute of Harvard and MIT | Lemieux,J.E., Siddle,K.J., Shaw,B., Adams,G., Pierce,V., Turbett,S., Anahtar,M., Branda,J., Slater,D., Harris,J., Lin,A.E., Gladden-Young,A., Lagerborg,K., Rudy,M., DeRuff,K., Carter,A., Normandin,E., Bauer,M., Reilly,S., Tomkins-Tinch,C., Loreth,C., Chaluvadi,S., Neumann,A., Cusick,C., Chapman,S.B., Gnirke,A., Flowers,K., Cerrato,F., Birren,B.W., Gallagher,G., Smole,S., Park,D.J., MacInnis,B.L., Ryan,E., LaRocque,R., Rosenberg,E. and Sabeti,P.C. |
| EPI_ISL_2016487, EPI_ISL_2016488, EPI_ISL_2016489, EPI_ISL_2016490, EPI_ISL_2016491, EPI_ISL_2016492, EPI_ISL_2016493, EPI_ISL_2016494, EPI_ISL_2016495, EPI_ISL_2016497, EPI_ISL_2016498, EPI_ISL_2016500, EPI_ISL_2016501, EPI_ISL_2016502, EPI_ISL_2016503, EPI_ISL_2016504, EPI_ISL_2016505, EPI_ISL_2016506, EPI_ISL_2016507, EPI_ISL_2016508, EPI_ISL_2016509, EPI_ISL_2016510, EPI_ISL_2016511, EPI_ISL_2016512, EPI_ISL_2016513, EPI_ISL_2016514, EPI_ISL_2016515, EPI_ISL_2016516, EPI_ISL_2016517, EPI_ISL_2016518, EPI_ISL_2016519, EPI_ISL_2016520, EPI_ISL_2016521, EPI_ISL_2016522, EPI_ISL_2016523, EPI_ISL_2016524, EPI_ISL_2016525, EPI_ISL_2016526, EPI_ISL_2016527, EPI_ISL_2016528, EPI_ISL_2016529, EPI_ISL_2016530, EPI_ISL_2016531, EPI_ISL_2016532, EPI_ISL_2016534, EPI_ISL_2016535, EPI_ISL_2016536, EPI_ISL_2016537, EPI_ISL_2016538, EPI_ISL_2016539, EPI_ISL_2016540, EPI_ISL_2016541, EPI_ISL_2016542, EPI_ISL_2016543, EPI_ISL_2016544, EPI_ISL_2016545, EPI_ISL_2016546, EPI_ISL_2016547, EPI_ISL_2016548, EPI_ISL_2016549, EPI_ISL_2016550, EPI_ISL_2016551, EPI_ISL_2016552, EPI_ISL_2016553, EPI_ISL_2016554, EPI_ISL_2016555, EPI_ISL_2016556, EPI_ISL_2016557, EPI_ISL_2016558, EPI_ISL_2016559, EPI_ISL_2016560, EPI_ISL_2016561, EPI_ISL_2016562, EPI_ISL_2016563, EPI_ISL_2016564, EPI_ISL_2016565, EPI_ISL_2016566, EPI_ISL_2016567, EPI_ISL_2016568, EPI_ISL_2016569, EPI_ISL_2016570, EPI_ISL_2016571, EPI_ISL_2016572, EPI_ISL_2016573, EPI_ISL_2016574, EPI_ISL_2016575, EPI_ISL_2016576, EPI_ISL_2016577, EPI_ISL_2016578, EPI_ISL_2016579, EPI_ISL_2016580, EPI_ISL_2016581, EPI_ISL_2016582, EPI_ISL_2016583, EPI_ISL_2016584, EPI_ISL_2016585, EPI_ISL_2016586, EPI_ISL_2016587, EPI_ISL_2016589, EPI_ISL_2016590, EPI_ISL_2016591, EPI_ISL_2016592, EPI_ISL_2016594, EPI_ISL_2016595, EPI_ISL_2016597, EPI_ISL_2016598, EPI_ISL_2016599, EPI_ISL_2016600, EPI_ISL_2016601, EPI_ISL_2016602, EPI_ISL_2016603, EPI_ISL_2016604, EPI_ISL_2016606, EPI_ISL_2016607, EPI_ISL_2016608, EPI_ISL_2016609, EPI_ISL_2016610, EPI_ISL_2016611, EPI_ISL_2016612, EPI_ISL_2016613, EPI_ISL_2016614, EPI_ISL_2016615, EPI_ISL_2016616 | | | |
| see above | Rhode Island Department of Health | Infectious Disease Program, Broad Institute of Harvard and MIT | Siddle,K.J., Azevedo,K., Miller,A., Adams,G., Pearlman,L., Gladden-Young,A., Lagerborg,K., Rudy,M., DeRuff,K., Carter,A., Normandin,E., Bauer,M., Reilly,S., Tomkins-Tinch,C., Loreth,C., Chaluvadi,S., Lemieux,J.E., Birren,B.W., Sabeti,P.C., Huard,R., King,E., Park,D.J., and MacInnis,B.L. |
| EPI_ISL_2017872, EPI_ISL_2017873, EPI_ISL_2017874, EPI_ISL_2017875, EPI_ISL_2017876, EPI_ISL_2017877, EPI_ISL_2017878, EPI_ISL_2017879, EPI_ISL_2017880, EPI_ISL_2017881, EPI_ISL_2017882, EPI_ISL_2017883, EPI_ISL_2017884, EPI_ISL_2017885, EPI_ISL_2017886, EPI_ISL_2017887, EPI_ISL_2017888, EPI_ISL_2017890, EPI_ISL_2017891, EPI_ISL_2017892, EPI_ISL_2017893, EPI_ISL_2017894, EPI_ISL_2017895, EPI_ISL_2017896, EPI_ISL_2017897, EPI_ISL_2017898, EPI_ISL_2017899, EPI_ISL_2017900, EPI_ISL_2017901, EPI_ISL_2017902, EPI_ISL_2017904, EPI_ISL_2017905, EPI_ISL_2017906, EPI_ISL_2017907, EPI_ISL_2017908, EPI_ISL_2017909, EPI_ISL_2017911, EPI_ISL_2017912, EPI_ISL_2017913, EPI_ISL_2017915, EPI_ISL_2017916, EPI_ISL_2017917, EPI_ISL_2017918, EPI_ISL_2017920, EPI_ISL_2017921, EPI_ISL_2017922, EPI_ISL_2017923, EPI_ISL_2017924, EPI_ISL_2017925, EPI_ISL_2017926, EPI_ISL_2017927, EPI_ISL_2017928, EPI_ISL_2017929, EPI_ISL_2017930, EPI_ISL_2017931, EPI_ISL_2017932, EPI_ISL_2017933, EPI_ISL_2017934, EPI_ISL_2017935, EPI_ISL_2017936, EPI_ISL_2017937, EPI_ISL_2017938, EPI_ISL_2017939, EPI_ISL_2017940, EPI_ISL_2017941 | | | |
| see above | TXDSHS | TXDSHS | Rashmi Tuladhar, Bonnie Oh, Jenny Zhang, Maliha Rahman, Mayela Pedrueza, Anita Pokharel, Karen Bobier, Lorraine Rodriguez, Myong Koag, Chun Wang, Rachel Lee, Grace Kubin |
| EPI_ISL_2017942, EPI_ISL_2017943, EPI_ISL_2017945, EPI_ISL_2017948, EPI_ISL_2017950, EPI_ISL_2017951, EPI_ISL_2017955, EPI_ISL_2017956, EPI_ISL_2017957, EPI_ISL_2017958, EPI_ISL_2017960, EPI_ISL_2017962, EPI_ISL_2017964, EPI_ISL_2017965, EPI_ISL_2017966, EPI_ISL_2017969, EPI_ISL_2017975, EPI_ISL_2017977, EPI_ISL_2017978, EPI_ISL_2017979, EPI_ISL_2017980, EPI_ISL_2017981, EPI_ISL_2017982, EPI_ISL_2017983, EPI_ISL_2017985, EPI_ISL_2017986, EPI_ISL_2017987, EPI_ISL_2017988, EPI_ISL_2017992, EPI_ISL_2017993, EPI_ISL_2017995, EPI_ISL_2017996, EPI_ISL_2017997, EPI_ISL_2017999, EPI_ISL_2018000, EPI_ISL_2018001, EPI_ISL_2018002, EPI_ISL_2018003, EPI_ISL_2018004, EPI_ISL_2018005, EPI_ISL_2018006, EPI_ISL_2018007, EPI_ISL_2018009, EPI_ISL_2018010, EPI_ISL_2018012, EPI_ISL_2018013, EPI_ISL_2018015, EPI_ISL_2018016, EPI_ISL_2018018, EPI_ISL_2018019, EPI_ISL_2018020, EPI_ISL_2018021, EPI_ISL_2018022, EPI_ISL_2018025, EPI_ISL_2018028, EPI_ISL_2018029, EPI_ISL_2018030, EPI_ISL_2018031, EPI_ISL_2018032, EPI_ISL_2018033, EPI_ISL_2018036, EPI_ISL_2018037, EPI_ISL_2018039, EPI_ISL_2018040, EPI_ISL_2018041, EPI_ISL_2018044, EPI_ISL_2018045, EPI_ISL_2018046, EPI_ISL_2018047, EPI_ISL_2018048, EPI_ISL_2018049, EPI_ISL_2018050, EPI_ISL_2018052, EPI_ISL_2018053, EPI_ISL_2018054, EPI_ISL_2018055, EPI_ISL_2018056, EPI_ISL_2018060, EPI_ISL_2018061, EPI_ISL_2018062, EPI_ISL_2018063, EPI_ISL_2018064, EPI_ISL_2018065, EPI_ISL_2018066, EPI_ISL_2018067, EPI_ISL_2018068, EPI_ISL_2018069, EPI_ISL_2018070, EPI_ISL_2018071, EPI_ISL_2018072, EPI_ISL_2018073, EPI_ISL_2018074, EPI_ISL_2018075, EPI_ISL_2018076, EPI_ISL_2018077, EPI_ISL_2018078, EPI_ISL_2018079, EPI_ISL_2018080, EPI_ISL_2018082, EPI_ISL_2018083, EPI_ISL_2018084, EPI_ISL_2018085, EPI_ISL_2018086, EPI_ISL_2018089, EPI_ISL_2018090, EPI_ISL_2018091, EPI_ISL_2018092, EPI_ISL_2018093, EPI_ISL_2018094, EPI_ISL_2018098, EPI_ISL_2018099, EPI_ISL_2018100, EPI_ISL_2018101, EPI_ISL_2018102, EPI_ISL_2018103, EPI_ISL_2018104, EPI_ISL_2018105 | | | |
| see above | Wyoming Public Health Laboratory | Wyoming Public Health Laboratory | Jim Mildenberger, Taylor Fearing, Channing Weber, Ashley Norberg, Chayse Rowley, Marley Goetz, Brian Dominguez, Elliot Thomasson, Brittany Oher, Sam Britz, Cari Sloma, and Rob Christensen |
| EPI_ISL_2018106, EPI_ISL_2018107, EPI_ISL_2018108, EPI_ISL_2018109, EPI_ISL_2018110, EPI_ISL_2018111, EPI_ISL_2018112, EPI_ISL_2018113, EPI_ISL_2018114, EPI_ISL_2018115, EPI_ISL_2018116, EPI_ISL_2018117, EPI_ISL_2018118, EPI_ISL_2018119, EPI_ISL_2018120, EPI_ISL_2018121, EPI_ISL_2018122, EPI_ISL_2018123, EPI_ISL_2018124, EPI_ISL_2018125, EPI_ISL_2018126, EPI_ISL_2018127, EPI_ISL_2018128, EPI_ISL_2018129, EPI_ISL_2018130, EPI_ISL_2018131, EPI_ISL_2018132, EPI_ISL_2018133, EPI_ISL_2018134, EPI_ISL_2018135, EPI_ISL_2018136, EPI_ISL_2018137, EPI_ISL_2018138, EPI_ISL_2018139, EPI_ISL_2018140, EPI_ISL_2018141, EPI_ISL_2018142, EPI_ISL_2018143, EPI_ISL_2018144, EPI_ISL_2018145, EPI_ISL_2018146, EPI_ISL_2018147, EPI_ISL_2018148, EPI_ISL_2018149, EPI_ISL_2018150, EPI_ISL_2018151, EPI_ISL_2018152, EPI_ISL_2018154, EPI_ISL_2018155, EPI_ISL_2018156, EPI_ISL_2018157, EPI_ISL_2018158, EPI_ISL_2018159, EPI_ISL_2018160, EPI_ISL_2018161, EPI_ISL_2018162, EPI_ISL_2018163, EPI_ISL_2018164, EPI_ISL_2018165 | | | |
| see above | Minnesota Department of Health, Public Health Laboratory | Minnesota Department of Health, Public Health Laboratory | Alexandra Lorentz, Jacob Garfin, Matt Plumb, and Xiong Wang |
| EPI_ISL_2018166, EPI_ISL_2018167, EPI_ISL_2018168, EPI_ISL_2018169, EPI_ISL_2018170, EPI_ISL_2018171, EPI_ISL_2018172 | FL Bureau of Public Health Laboratories-Miami | Centers for Disease Control and Prevention Division of Viral Diseases, Pathogen Discovery | Mili Sheth, Sarah Nobles, Jasmine Padilla, Mark Burroughs, Shoshona Le, Katie Dillon, Peter Cook, Clinton R. Paden, Dhwani Batra, Krista Queen, Kristen Knipe, Dakota Howard, Yvette Unoarumhi, Darlene Wagner, Matthew Schmerer, Ben L. Rambo-Martin, Kristine Lacek, Sam Shepard, Alison Laufer Halpin, Dave Wentworth, Vivien Dugan, Suxiang Tong, Justin Lee |
| EPI_ISL_2018173, EPI_ISL_2018174, EPI_ISL_2018175, EPI_ISL_2018176, EPI_ISL_2018177, EPI_ISL_2018178, EPI_ISL_2018179, EPI_ISL_2018180, EPI_ISL_2018181 | NM Dept. Health, Scientific Laboratory Division | Centers for Disease Control and Prevention Division of Viral Diseases, Pathogen Discovery | Mili Sheth, Sarah Nobles, Jasmine Padilla, Mark Burroughs, Shoshona Le, Katie Dillon, Peter Cook, Clinton R. Paden, Dhwani Batra, Krista Queen, Kristen Knipe, Dakota Howard, Yvette Unoarumhi, Darlene Wagner, Matthew Schmerer, Ben L. Rambo-Martin, Kristine Lacek, Sam Shepard, Alison Laufer Halpin, Dave Wentworth, Vivien Dugan, Suxiang Tong, Justin Lee |
| EPI_ISL_2018182, EPI_ISL_2018183, EPI_ISL_2018184, EPI_ISL_2018185, EPI_ISL_2018186, EPI_ISL_2018187, EPI_ISL_2018188, EPI_ISL_2018189, EPI_ISL_2018190, EPI_ISL_2018191, EPI_ISL_2018192, EPI_ISL_2018193, EPI_ISL_2018194, EPI_ISL_2018195, EPI_ISL_2018196 | | | |
| see above | ID Bureau of Laboratories | Centers for Disease Control and Prevention Division of Viral Diseases, Pathogen Discovery | Mili Sheth, Sarah Nobles, Jasmine Padilla, Mark Burroughs, Shoshona Le, Katie Dillon, Peter Cook, Clinton R. Paden, Dhwani Batra, Krista Queen, Kristen Knipe, Dakota Howard, Yvette Unoarumhi, Darlene Wagner, Matthew Schmerer, Ben L. Rambo-Martin, Kristine Lacek, Sam Shepard, Alison Laufer Halpin, Dave Wentworth, Vivien Dugan, Suxiang Tong, Justin Lee |
| EPI_ISL_2018197, EPI_ISL_2018198, EPI_ISL_2018199, EPI_ISL_2018200, EPI_ISL_2018201, EPI_ISL_2018202, EPI_ISL_2018203, EPI_ISL_2018204, EPI_ISL_2018205 | AR Dept. of Health-PHL, Molecular Diagnostics | Centers for Disease Control and Prevention Division of Viral Diseases, Pathogen Discovery | Mili Sheth, Sarah Nobles, Jasmine Padilla, Mark Burroughs, Shoshona Le, Katie Dillon, Peter Cook, Clinton R. Paden, Dhwani Batra, Krista Queen, Kristen Knipe, Dakota Howard, Yvette Unoarumhi, Darlene Wagner, Matthew Schmerer, Ben L. Rambo-Martin, Kristine Lacek, Sam Shepard, Alison Laufer Halpin, Dave Wentworth, Vivien Dugan, Suxiang Tong, Justin Lee |
| EPI_ISL_2018206, EPI_ISL_2018207, EPI_ISL_2018208, EPI_ISL_2018209, EPI_ISL_2018210, EPI_ISL_2018211, EPI_ISL_2018212, EPI_ISL_2018213, EPI_ISL_2018214, EPI_ISL_2018215, EPI_ISL_2018216, EPI_ISL_2018217, EPI_ISL_2018218 | | | |
| see above | SD Public Health Laboratory | Centers for Disease Control and Prevention Division of Viral Diseases, Pathogen Discovery | Mili Sheth, Sarah Nobles, Jasmine Padilla, Mark Burroughs, Shoshona Le, Katie Dillon, Peter Cook, Clinton R. Paden, Dhwani Batra, Krista Queen, Kristen Knipe, Dakota Howard, Yvette Unoarumhi, Darlene Wagner, Matthew Schmerer, Ben L. Rambo-Martin, Kristine Lacek, Sam Shepard, Alison Laufer Halpin, Dave Wentworth, Vivien Dugan, Suxiang Tong, Justin Lee |
| EPI_ISL_2018219, EPI_ISL_2018220, EPI_ISL_2018221, EPI_ISL_2018222, EPI_ISL_2018223, EPI_ISL_2018224, EPI_ISL_2018225, EPI_ISL_2018226, EPI_ISL_2018227, EPI_ISL_2018228 | MA State Public Health Laboratory | Centers for Disease Control and Prevention Division of Viral Diseases, Pathogen Discovery | Mili Sheth, Sarah Nobles, Jasmine Padilla, Mark Burroughs, Shoshona Le, Katie Dillon, Peter Cook, Clinton R. Paden, Dhwani Batra, Krista Queen, Kristen Knipe, Dakota Howard, Yvette Unoarumhi, Darlene Wagner, Matthew Schmerer, Ben L. Rambo-Martin, Kristine Lacek, Sam Shepard, Alison Laufer Halpin, Dave Wentworth, Vivien Dugan, Suxiang Tong, Justin Lee |
| EPI_ISL_2018229 | MN PHL Division, Minnesota Department of Health | Centers for Disease Control and Prevention Division of Viral Diseases, Pathogen Discovery | Mili Sheth, Sarah Nobles, Jasmine Padilla, Mark Burroughs, Shoshona Le, Katie Dillon, Peter Cook, Clinton R. Paden, Dhwani Batra, Krista Queen, Kristen Knipe, Dakota Howard, Yvette Unoarumhi, Darlene Wagner, Matthew Schmerer, Ben L. Rambo-Martin, Kristine Lacek, Sam Shepard, Alison Laufer Halpin, Dave Wentworth, Vivien Dugan, Suxiang Tong, Justin Lee |
| EPI_ISL_2018230, EPI_ISL_2018231, EPI_ISL_2018232, EPI_ISL_2018233 | WA State Department of Health | Centers for Disease Control and Prevention Division of Viral Diseases, Pathogen Discovery | Mili Sheth, Sarah Nobles, Jasmine Padilla, Mark Burroughs, Shoshona Le, Katie Dillon, Peter Cook, Clinton R. Paden, Dhwani Batra, Krista Queen, Kristen Knipe, Dakota Howard, Yvette Unoarumhi, Darlene Wagner, Matthew Schmerer, Ben L. Rambo-Martin, Kristine Lacek, Sam Shepard, Alison Laufer Halpin, Dave Wentworth, Vivien Dugan, Suxiang Tong, Justin Lee |
| EPI_ISL_2018234, EPI_ISL_2018235, EPI_ISL_2018236, EPI_ISL_2018237, EPI_ISL_2018238, EPI_ISL_2018239, EPI_ISL_2018240, EPI_ISL_2018241, EPI_ISL_2018242 | FL Bur. of Public Health Laboratories-Jacksonville | Centers for Disease Control and Prevention Division of Viral Diseases, Pathogen Discovery | Mili Sheth, Sarah Nobles, Jasmine Padilla, Mark Burroughs, Shoshona Le, Katie Dillon, Peter Cook, Clinton R. Paden, Dhwani Batra, Krista Queen, Kristen Knipe, Dakota Howard, Yvette Unoarumhi, Darlene Wagner, Matthew Schmerer, Ben L. Rambo-Martin, Kristine Lacek, Sam Shepard, Alison Laufer Halpin, Dave Wentworth, Vivien Dugan, Suxiang Tong, Justin Lee |

| | | | |
|---|---|---|---|
| EPI_ISL_2018243, EPI_ISL_2018244, EPI_ISL_2018245, EPI_ISL_2018246 | NYSDOH Wadsworth Center, Virology Lab | Centers for Disease Control and Prevention Division of Viral Diseases, Pathogen Discovery | Mili Sheth, Sarah Nobles, Jasmine Padilla, Mark Burroughs, Shoshona Le, Katie Dillon, Peter Cook, Clinton R. Paden, Dhwani Batra, Krista Queen, Kristen Knipe, Dakota Howard, Yvette Unoarumhi, Darlene Wagner, Matthew Schmerer, Ben L. Rambo-Martin, Kristine Lacek, Sam Shepard, Alison Laufer Halpin, Dave Wentworth, Vivien Dugan, Suxiang Tong, Justin Lee |
| EPI_ISL_2018247, EPI_ISL_2018248, EPI_ISL_2018249, EPI_ISL_2018250, EPI_ISL_2018251, EPI_ISL_2018252, EPI_ISL_2018253, EPI_ISL_2018254, EPI_ISL_2018255, EPI_ISL_2018256, EPI_ISL_2018257, EPI_ISL_2018258, EPI_ISL_2018259, EPI_ISL_2018260, EPI_ISL_2018261, EPI_ISL_2018262, EPI_ISL_2018263 | | | |
| see above | AK State Public Health Lab, State Health Department | Centers for Disease Control and Prevention Division of Viral Diseases, Pathogen Discovery | Mili Sheth, Sarah Nobles, Jasmine Padilla, Mark Burroughs, Shoshona Le, Katie Dillon, Peter Cook, Clinton R. Paden, Dhwani Batra, Krista Queen, Kristen Knipe, Dakota Howard, Yvette Unoarumhi, Darlene Wagner, Matthew Schmerer, Ben L. Rambo-Martin, Kristine Lacek, Sam Shepard, Alison Laufer Halpin, Dave Wentworth, Vivien Dugan, Suxiang Tong, Justin Lee |
| EPI_ISL_2018264 | TX DSHS, Lab Services Section MC 1947 | Centers for Disease Control and Prevention Division of Viral Diseases, Pathogen Discovery | Mili Sheth, Sarah Nobles, Jasmine Padilla, Mark Burroughs, Shoshona Le, Katie Dillon, Peter Cook, Clinton R. Paden, Dhwani Batra, Krista Queen, Kristen Knipe, Dakota Howard, Yvette Unoarumhi, Darlene Wagner, Matthew Schmerer, Ben L. Rambo-Martin, Kristine Lacek, Sam Shepard, Alison Laufer Halpin, Dave Wentworth, Vivien Dugan, Suxiang Tong, Justin Lee |
| EPI_ISL_2018265, EPI_ISL_2018266, EPI_ISL_2018267, EPI_ISL_2018268, EPI_ISL_2018269, EPI_ISL_2018270, EPI_ISL_2018271, EPI_ISL_2018272, EPI_ISL_2018273, EPI_ISL_2018274, EPI_ISL_2018275, EPI_ISL_2018276 | | | |
| see above | AL Dept. of Public Health Bureau of Clinical Laboratories | Centers for Disease Control and Prevention Division of Viral Diseases, Pathogen Discovery | Mili Sheth, Sarah Nobles, Jasmine Padilla, Mark Burroughs, Shoshona Le, Katie Dillon, Peter Cook, Clinton R. Paden, Dhwani Batra, Krista Queen, Kristen Knipe, Dakota Howard, Yvette Unoarumhi, Darlene Wagner, Matthew Schmerer, Ben L. Rambo-Martin, Kristine Lacek, Sam Shepard, Alison Laufer Halpin, Dave Wentworth, Vivien Dugan, Suxiang Tong, Justin Lee |
| EPI_ISL_2018277, EPI_ISL_2018278, EPI_ISL_2018279, EPI_ISL_2018280, EPI_ISL_2018281, EPI_ISL_2018282, EPI_ISL_2018283 | DE Public Health Laboratory | Centers for Disease Control and Prevention Division of Viral Diseases, Pathogen Discovery | Mili Sheth, Sarah Nobles, Jasmine Padilla, Mark Burroughs, Shoshona Le, Katie Dillon, Peter Cook, Clinton R. Paden, Dhwani Batra, Krista Queen, Kristen Knipe, Dakota Howard, Yvette Unoarumhi, Darlene Wagner, Matthew Schmerer, Ben L. Rambo-Martin, Kristine Lacek, Sam Shepard, Alison Laufer Halpin, Dave Wentworth, Vivien Dugan, Suxiang Tong, Justin Lee |
| EPI_ISL_2018284, EPI_ISL_2018285, EPI_ISL_2018286, EPI_ISL_2018287, EPI_ISL_2018288, EPI_ISL_2018289, EPI_ISL_2018290, EPI_ISL_2018291, EPI_ISL_2018292, EPI_ISL_2018293, EPI_ISL_2018294, EPI_ISL_2018295, EPI_ISL_2018296, EPI_ISL_2018297, EPI_ISL_2018298, EPI_ISL_2018299, EPI_ISL_2018300, EPI_ISL_2018301 | | | |
| see above | FL Bureau of Public Health Laboratories-Tampa | Centers for Disease Control and Prevention Division of Viral Diseases, Pathogen Discovery | Mili Sheth, Sarah Nobles, Jasmine Padilla, Mark Burroughs, Shoshona Le, Katie Dillon, Peter Cook, Clinton R. Paden, Dhwani Batra, Krista Queen, Kristen Knipe, Dakota Howard, Yvette Unoarumhi, Darlene Wagner, Matthew Schmerer, Ben L. Rambo-Martin, Kristine Lacek, Sam Shepard, Alison Laufer Halpin, Dave Wentworth, Vivien Dugan, Suxiang Tong, Justin Lee |
| EPI_ISL_2018302, EPI_ISL_2018303, EPI_ISL_2018304, EPI_ISL_2018305, EPI_ISL_2018306, EPI_ISL_2018307, EPI_ISL_2018308, EPI_ISL_2018309 | SC Dept of Health and Env. Control-Bureau of Laboratories | Centers for Disease Control and Prevention Division of Viral Diseases, Pathogen Discovery | Mili Sheth, Sarah Nobles, Jasmine Padilla, Mark Burroughs, Shoshona Le, Katie Dillon, Peter Cook, Clinton R. Paden, Dhwani Batra, Krista Queen, Kristen Knipe, Dakota Howard, Yvette Unoarumhi, Darlene Wagner, Matthew Schmerer, Ben L. Rambo-Martin, Kristine Lacek, Sam Shepard, Alison Laufer Halpin, Dave Wentworth, Vivien Dugan, Suxiang Tong, Justin Lee |
| EPI_ISL_2018310, EPI_ISL_2018311, EPI_ISL_2018312, EPI_ISL_2018313, EPI_ISL_2018314, EPI_ISL_2018315, EPI_ISL_2018316, EPI_ISL_2018317, EPI_ISL_2018318, EPI_ISL_2018319, EPI_ISL_2018320, EPI_ISL_2018321, EPI_ISL_2018322, EPI_ISL_2018323, EPI_ISL_2018324, EPI_ISL_2018325 | | | |
| see above | CDPH, Viral and Rickettsial Disease Laboratory | Centers for Disease Control and Prevention Division of Viral Diseases, Pathogen Discovery | Mili Sheth, Sarah Nobles, Jasmine Padilla, Mark Burroughs, Shoshona Le, Katie Dillon, Peter Cook, Clinton R. Paden, Dhwani Batra, Krista Queen, Kristen Knipe, Dakota Howard, Yvette Unoarumhi, Darlene Wagner, Matthew Schmerer, Ben L. Rambo-Martin, Kristine Lacek, Sam Shepard, Alison Laufer Halpin, Dave Wentworth, Vivien Dugan, Suxiang Tong, Justin Lee |
| EPI_ISL_2018326, EPI_ISL_2018327, EPI_ISL_2018328, EPI_ISL_2018329, EPI_ISL_2018330 | WY Public Health Laboratory | Centers for Disease Control and Prevention Division of Viral Diseases, Pathogen Discovery | Mili Sheth, Sarah Nobles, Jasmine Padilla, Mark Burroughs, Shoshona Le, Katie Dillon, Peter Cook, Clinton R. Paden, Dhwani Batra, Krista Queen, Kristen Knipe, Dakota Howard, Yvette Unoarumhi, Darlene Wagner, Matthew Schmerer, Ben L. Rambo-Martin, Kristine Lacek, Sam Shepard, Alison Laufer Halpin, Dave Wentworth, Vivien Dugan, Suxiang Tong, Justin Lee |
| EPI_ISL_2018331, EPI_ISL_2018332, EPI_ISL_2018333, EPI_ISL_2018334, EPI_ISL_2018335, EPI_ISL_2018336, EPI_ISL_2018337, EPI_ISL_2018338, EPI_ISL_2018339, EPI_ISL_2018340, EPI_ISL_2018341, EPI_ISL_2018342, EPI_ISL_2018343, EPI_ISL_2018344, EPI_ISL_2018345, EPI_ISL_2018346, EPI_ISL_2018347, EPI_ISL_2018348, EPI_ISL_2018349, EPI_ISL_2018350, EPI_ISL_2018351, EPI_ISL_2018352, EPI_ISL_2018353, EPI_ISL_2018354, EPI_ISL_2018355 | | | |
| see above | RI State Health Laboratories | Centers for Disease Control and Prevention Division of Viral Diseases, Pathogen Discovery | Mili Sheth, Sarah Nobles, Jasmine Padilla, Mark Burroughs, Shoshona Le, Katie Dillon, Peter Cook, Clinton R. Paden, Dhwani Batra, Krista Queen, Kristen Knipe, Dakota Howard, Yvette Unoarumhi, Darlene Wagner, Matthew Schmerer, Ben L. Rambo-Martin, Kristine Lacek, Sam Shepard, Alison Laufer Halpin, Dave Wentworth, Vivien Dugan, Suxiang Tong, Justin Lee |
| EPI_ISL_2018356 | AZ SPHL, Arizona Department of Health Services | Centers for Disease Control and Prevention Division of Viral Diseases, Pathogen Discovery | Mili Sheth, Sarah Nobles, Jasmine Padilla, Mark Burroughs, Shoshona Le, Katie Dillon, Peter Cook, Clinton R. Paden, Dhwani Batra, Krista Queen, Kristen Knipe, Dakota Howard, Yvette Unoarumhi, Darlene Wagner, Matthew Schmerer, Ben L. Rambo-Martin, Kristine Lacek, Sam Shepard, Alison Laufer Halpin, Dave Wentworth, Vivien Dugan, Suxiang Tong, Justin Lee |
| EPI_ISL_2018357, EPI_ISL_2018358, EPI_ISL_2018359, EPI_ISL_2018360, EPI_ISL_2018361, EPI_ISL_2018362, EPI_ISL_2018363 | MT Public Health Laboratory | Centers for Disease Control and Prevention Division of Viral Diseases, Pathogen Discovery | Mili Sheth, Sarah Nobles, Jasmine Padilla, Mark Burroughs, Shoshona Le, Katie Dillon, Peter Cook, Clinton R. Paden, Dhwani Batra, Krista Queen, Kristen Knipe, Dakota Howard, Yvette Unoarumhi, Darlene Wagner, Matthew Schmerer, Ben L. Rambo-Martin, Kristine Lacek, Sam Shepard, Alison Laufer Halpin, Dave Wentworth, Vivien Dugan, Suxiang Tong, Justin Lee |
| EPI_ISL_2018364, EPI_ISL_2018365, EPI_ISL_2018366, EPI_ISL_2018367, EPI_ISL_2018368, EPI_ISL_2018369, EPI_ISL_2018370, EPI_ISL_2018371, EPI_ISL_2018372, EPI_ISL_2018373, EPI_ISL_2018374, EPI_ISL_2018375, EPI_ISL_2018376, EPI_ISL_2018377, EPI_ISL_2018378 | | | |
| see above | CO Dept. of Public Health and Environment, Lab Services Division | Centers for Disease Control and Prevention Division of Viral Diseases, Pathogen Discovery | Mili Sheth, Sarah Nobles, Jasmine Padilla, Mark Burroughs, Shoshona Le, Katie Dillon, Peter Cook, Clinton R. Paden, Dhwani Batra, Krista Queen, Kristen Knipe, Dakota Howard, Yvette Unoarumhi, Darlene Wagner, Matthew Schmerer, Ben L. Rambo-Martin, Kristine Lacek, Sam Shepard, Alison Laufer Halpin, Dave Wentworth, Vivien Dugan, Suxiang Tong, Justin Lee |
| EPI_ISL_2018379, EPI_ISL_2018380, EPI_ISL_2018381, EPI_ISL_2018382, EPI_ISL_2018383, EPI_ISL_2018384, EPI_ISL_2018385 | LA Office of Public Health Laboratories | Centers for Disease Control and Prevention Division of Viral Diseases, Pathogen Discovery | Mili Sheth, Sarah Nobles, Jasmine Padilla, Mark Burroughs, Shoshona Le, Katie Dillon, Peter Cook, Clinton R. Paden, Dhwani Batra, Krista Queen, Kristen Knipe, Dakota Howard, Yvette Unoarumhi, Darlene Wagner, Matthew Schmerer, Ben L. Rambo-Martin, Kristine Lacek, Sam Shepard, Alison Laufer Halpin, Dave Wentworth, Vivien Dugan, Suxiang Tong, Justin Lee |
| EPI_ISL_2018386, EPI_ISL_2018387, EPI_ISL_2018388, EPI_ISL_2018389, EPI_ISL_2018390, EPI_ISL_2018391 | UT-Unified State Labs: Public Health Utah DOH | Centers for Disease Control and Prevention Division of Viral Diseases, Pathogen Discovery | Mili Sheth, Sarah Nobles, Jasmine Padilla, Mark Burroughs, Shoshona Le, Katie Dillon, Peter Cook, Clinton R. Paden, Dhwani Batra, Krista Queen, Kristen Knipe, Dakota Howard, Yvette Unoarumhi, Darlene Wagner, Matthew Schmerer, Ben L. Rambo-Martin, Kristine Lacek, Sam Shepard, Alison Laufer Halpin, Dave Wentworth, Vivien Dugan, Suxiang Tong, Justin Lee |
| EPI_ISL_2018392, EPI_ISL_2018393, EPI_ISL_2018394, EPI_ISL_2018395, EPI_ISL_2018396, EPI_ISL_2018397, EPI_ISL_2018398, EPI_ISL_2018399, EPI_ISL_2018400, EPI_ISL_2018401 | DC Public Health Lab/ Dept. of Forensic Sciences | Centers for Disease Control and Prevention Division of Viral Diseases, Pathogen Discovery | Mili Sheth, Sarah Nobles, Jasmine Padilla, Mark Burroughs, Shoshona Le, Katie Dillon, Peter Cook, Clinton R. Paden, Dhwani Batra, Krista Queen, Kristen Knipe, Dakota Howard, Yvette Unoarumhi, Darlene Wagner, Matthew Schmerer, Ben L. Rambo-Martin, Kristine Lacek, Sam Shepard, Alison Laufer Halpin, Dave Wentworth, Vivien Dugan, Suxiang Tong, Justin Lee |
| EPI_ISL_2018402, EPI_ISL_2018403, EPI_ISL_2018404, EPI_ISL_2018405, EPI_ISL_2018406, EPI_ISL_2018407, EPI_ISL_2018408, EPI_ISL_2018409, EPI_ISL_2018410, EPI_ISL_2018411, EPI_ISL_2018412, EPI_ISL_2018413, EPI_ISL_2018414, EPI_ISL_2018415, EPI_ISL_2018416, EPI_ISL_2018417 | | | |
| see above | KS Health and Environmental Laboratories | Centers for Disease Control and Prevention Division of Viral Diseases, Pathogen Discovery | Mili Sheth, Sarah Nobles, Jasmine Padilla, Mark Burroughs, Shoshona Le, Katie Dillon, Peter Cook, Clinton R. Paden, Dhwani Batra, Krista Queen, Kristen Knipe, Dakota Howard, Yvette Unoarumhi, Darlene Wagner, Matthew Schmerer, Ben L. Rambo-Martin, Kristine Lacek, Sam Shepard, Alison Laufer Halpin, Dave Wentworth, Vivien Dugan, Suxiang Tong, Justin Lee |
| EPI_ISL_2018418, EPI_ISL_2018419, EPI_ISL_2018420, EPI_ISL_2018421, EPI_ISL_2018422, EPI_ISL_2018423, EPI_ISL_2018424, EPI_ISL_2018425, EPI_ISL_2018426, EPI_ISL_2018427, EPI_ISL_2018428, EPI_ISL_2018429, EPI_ISL_2018430, EPI_ISL_2018431, EPI_ISL_2018432, EPI_ISL_2018433 | | | |
| see above | NYC Department of Health and Mental Hygiene | Centers for Disease Control and Prevention Division of Viral Diseases, Pathogen Discovery | Mili Sheth, Sarah Nobles, Jasmine Padilla, Mark Burroughs, Shoshona Le, Katie Dillon, Peter Cook, Clinton R. Paden, Dhwani Batra, Krista Queen, Kristen Knipe, Dakota Howard, Yvette Unoarumhi, Darlene Wagner, Matthew Schmerer, Ben L. Rambo-Martin, Kristine Lacek, Sam Shepard, Alison Laufer Halpin, Dave Wentworth, Vivien Dugan, Suxiang Tong, Justin Lee |
| EPI_ISL_2018434, EPI_ISL_2018435, EPI_ISL_2018436, EPI_ISL_2018437, EPI_ISL_2018438, | GA Department of Public Health Laboratory | Centers for Disease Control and Prevention Division of Viral Diseases, Pathogen Discovery | Mili Sheth, Sarah Nobles, Jasmine Padilla, Mark Burroughs, Shoshona Le, Katie Dillon, Peter Cook, Clinton R. Paden, Dhwani Batra, Krista Queen, Kristen Knipe, Dakota Howard, Yvette Unoarumhi, Darlene Wagner, Matthew Schmerer, Ben L. Rambo-Martin, Kristine Lacek, Sam Shepard, Alison Laufer Halpin, Dave Wentworth, Vivien Dugan, Suxiang Tong, Justin Lee |

| | | | |
|---|---|---|---|
| EPI_ISL_2018439, EPI_ISL_2018440, EPI_ISL_2018441, EPI_ISL_2018442 | | | |
| EPI_ISL_2018444 | M Health Fairview | Minnesota Department of Health, Public Health Laboratory | Alexandra Lorentz, Jacob Garfin, Matt Plumb, and Xiong Wang |
| EPI_ISL_2018445, EPI_ISL_2018446, EPI_ISL_2018447, EPI_ISL_2018448, EPI_ISL_2018449, EPI_ISL_2018450, EPI_ISL_2018451, EPI_ISL_2018452, EPI_ISL_2018453, EPI_ISL_2018454, EPI_ISL_2018455, EPI_ISL_2018456, EPI_ISL_2018457, EPI_ISL_2018458, EPI_ISL_2018459, EPI_ISL_2018460, EPI_ISL_2018461, EPI_ISL_2018462, EPI_ISL_2018463, EPI_ISL_2018464, EPI_ISL_2018465, EPI_ISL_2018466, EPI_ISL_2018467, EPI_ISL_2018468, EPI_ISL_2018469, EPI_ISL_2018470, EPI_ISL_2018471, EPI_ISL_2018472, EPI_ISL_2018473, EPI_ISL_2018474, EPI_ISL_2018475 | | | |
| see above | Minnesota Department of Health, Public Health Laboratory | Minnesota Department of Health, Public Health Laboratory | Alexandra Lorentz, Jacob Garfin, Matt Plumb, and Xiong Wang |
| EPI_ISL_2018476 | M Health Fairview | Minnesota Department of Health, Public Health Laboratory | Alexandra Lorentz, Jacob Garfin, Matt Plumb, and Xiong Wang |
| EPI_ISL_2018477, EPI_ISL_2018478, EPI_ISL_2018480, EPI_ISL_2018481, EPI_ISL_2018482, EPI_ISL_2018483, EPI_ISL_2018484, EPI_ISL_2018485, EPI_ISL_2018486, EPI_ISL_2018487, EPI_ISL_2018488, EPI_ISL_2018489 | | | |
| see above | Minnesota Department of Health, Public Health Laboratory | Minnesota Department of Health, Public Health Laboratory | Alexandra Lorentz, Jacob Garfin, Matt Plumb, and Xiong Wang |
| EPI_ISL_2018490, EPI_ISL_2018492, EPI_ISL_2018493, EPI_ISL_2018494, EPI_ISL_2018495, EPI_ISL_2018496, EPI_ISL_2018497, EPI_ISL_2018498, EPI_ISL_2018499, EPI_ISL_2018500, EPI_ISL_2018501, EPI_ISL_2018502, EPI_ISL_2018503, EPI_ISL_2018504, EPI_ISL_2018505, EPI_ISL_2018506 | | | |
| see above | Mayo Clinic & Mayo Clinic Laboratories | Minnesota Department of Health, Public Health Laboratory | Alexandra Lorentz, Jacob Garfin, Matt Plumb, and Xiong Wang |
| EPI_ISL_2018508, EPI_ISL_2018509, EPI_ISL_2018510, EPI_ISL_2018511, EPI_ISL_2018512, EPI_ISL_2018513, EPI_ISL_2018514, EPI_ISL_2018515, EPI_ISL_2018516, EPI_ISL_2018517 | M Health Fairview | Minnesota Department of Health, Public Health Laboratory | Alexandra Lorentz, Jacob Garfin, Matt Plumb, and Xiong Wang |
| EPI_ISL_2018545, EPI_ISL_2018546, EPI_ISL_2018547, EPI_ISL_2018548, EPI_ISL_2018549, EPI_ISL_2018550, EPI_ISL_2018551, EPI_ISL_2018552, EPI_ISL_2018553, EPI_ISL_2018554, EPI_ISL_2018555, EPI_ISL_2018556, EPI_ISL_2018557, EPI_ISL_2018558, EPI_ISL_2018559, EPI_ISL_2018560, EPI_ISL_2018561, EPI_ISL_2018562, EPI_ISL_2018563, EPI_ISL_2018564, EPI_ISL_2018565, EPI_ISL_2018566, EPI_ISL_2018567, EPI_ISL_2018568, EPI_ISL_2018569, EPI_ISL_2018570, EPI_ISL_2018571, EPI_ISL_2018572, EPI_ISL_2018573, EPI_ISL_2018574, EPI_ISL_2018575, EPI_ISL_2018576, EPI_ISL_2018577, EPI_ISL_2018578, EPI_ISL_2018579, EPI_ISL_2018580, EPI_ISL_2018581, EPI_ISL_2018582, EPI_ISL_2018583, EPI_ISL_2018584, EPI_ISL_2018585, EPI_ISL_2018586, EPI_ISL_2018587, EPI_ISL_2018588, EPI_ISL_2018589, EPI_ISL_2018590, EPI_ISL_2018591, EPI_ISL_2018592, EPI_ISL_2018593, EPI_ISL_2018594, EPI_ISL_2018595, EPI_ISL_2018596, EPI_ISL_2018597, EPI_ISL_2018598, EPI_ISL_2018599, EPI_ISL_2018600, EPI_ISL_2018601, EPI_ISL_2018602, EPI_ISL_2018603, EPI_ISL_2018604, EPI_ISL_2018605, EPI_ISL_2018606, EPI_ISL_2018607, EPI_ISL_2018608, EPI_ISL_2018609, EPI_ISL_2018610, EPI_ISL_2018611, EPI_ISL_2018612, EPI_ISL_2018613, EPI_ISL_2018614, EPI_ISL_2018615, EPI_ISL_2018616, EPI_ISL_2018617, EPI_ISL_2018618, EPI_ISL_2018619, EPI_ISL_2018620, EPI_ISL_2018621, EPI_ISL_2018622, EPI_ISL_2018623, EPI_ISL_2018624, EPI_ISL_2018626, EPI_ISL_2018627, EPI_ISL_2018628, EPI_ISL_2018629, EPI_ISL_2018630, EPI_ISL_2018631, EPI_ISL_2018632, EPI_ISL_2018633, EPI_ISL_2018634, EPI_ISL_2018635, EPI_ISL_2018636, EPI_ISL_2018637, EPI_ISL_2018638, EPI_ISL_2018639, EPI_ISL_2018640, EPI_ISL_2018641, EPI_ISL_2018642, EPI_ISL_2018643, EPI_ISL_2018644, EPI_ISL_2018645, EPI_ISL_2018646, EPI_ISL_2018647, EPI_ISL_2018648, EPI_ISL_2018649, EPI_ISL_2018650, EPI_ISL_2018651, EPI_ISL_2018652, EPI_ISL_2018653, EPI_ISL_2018654, EPI_ISL_2018655, EPI_ISL_2018656, EPI_ISL_2018657, EPI_ISL_2018658, EPI_ISL_2018659, EPI_ISL_2018660, EPI_ISL_2018661, EPI_ISL_2018662, EPI_ISL_2018663, EPI_ISL_2018664, EPI_ISL_2018665, EPI_ISL_2018666, EPI_ISL_2018667, EPI_ISL_2018668, EPI_ISL_2018669, EPI_ISL_2018670, EPI_ISL_2018671, EPI_ISL_2018672, EPI_ISL_2018673, EPI_ISL_2018674, EPI_ISL_2018675, EPI_ISL_2018676, EPI_ISL_2018677, EPI_ISL_2018678, EPI_ISL_2018680, EPI_ISL_2018681, EPI_ISL_2018682, EPI_ISL_2018683, EPI_ISL_2018684, EPI_ISL_2018685, EPI_ISL_2018686, EPI_ISL_2018688, EPI_ISL_2018689, EPI_ISL_2018690, EPI_ISL_2018691, EPI_ISL_2018692, EPI_ISL_2018693, EPI_ISL_2018694, EPI_ISL_2018695, EPI_ISL_2018696, EPI_ISL_2018697, EPI_ISL_2018698, EPI_ISL_2018699, EPI_ISL_2018700, EPI_ISL_2018701, EPI_ISL_2018702, EPI_ISL_2018703, EPI_ISL_2018704, EPI_ISL_2018705, EPI_ISL_2018706, EPI_ISL_2018707, EPI_ISL_2018708, EPI_ISL_2018709, EPI_ISL_2018710, EPI_ISL_2018711, EPI_ISL_2018712, EPI_ISL_2018713, EPI_ISL_2018714, EPI_ISL_2018715, EPI_ISL_2018716, EPI_ISL_2018717, EPI_ISL_2018718, EPI_ISL_2018719, EPI_ISL_2018720, EPI_ISL_2018721, EPI_ISL_2018722, EPI_ISL_2018723, EPI_ISL_2018724, EPI_ISL_2018725, EPI_ISL_2018726, EPI_ISL_2018727, EPI_ISL_2018728, EPI_ISL_2018729, EPI_ISL_2018730, EPI_ISL_2018731, EPI_ISL_2018732, EPI_ISL_2018733, EPI_ISL_2018734, EPI_ISL_2018735, EPI_ISL_2018736, EPI_ISL_2018737, EPI_ISL_2018738, EPI_ISL_2018739, EPI_ISL_2018740, EPI_ISL_2018741, EPI_ISL_2018742, EPI_ISL_2018743, EPI_ISL_2018744, EPI_ISL_2018745, EPI_ISL_2018746, EPI_ISL_2018747, EPI_ISL_2018748, EPI_ISL_2018749, EPI_ISL_2018750, EPI_ISL_2018751, EPI_ISL_2018752, EPI_ISL_2018753, EPI_ISL_2018754, EPI_ISL_2018755, EPI_ISL_2018756, EPI_ISL_2018757, EPI_ISL_2018758, EPI_ISL_2018759, EPI_ISL_2018760, EPI_ISL_2018761, EPI_ISL_2018762, EPI_ISL_2018763, EPI_ISL_2018764, EPI_ISL_2018765, EPI_ISL_2018766, EPI_ISL_2018767, EPI_ISL_2018768, EPI_ISL_2018769, EPI_ISL_2018770, EPI_ISL_2018771, EPI_ISL_2018772, EPI_ISL_2018773, EPI_ISL_2018774, EPI_ISL_2018775, EPI_ISL_2018776, EPI_ISL_2018777, EPI_ISL_2018778, EPI_ISL_2018779, EPI_ISL_2018780, EPI_ISL_2018781, EPI_ISL_2018782, EPI_ISL_2018783, EPI_ISL_2018784, EPI_ISL_2018785, EPI_ISL_2018787, EPI_ISL_2018788, EPI_ISL_2018789, EPI_ISL_2018790, EPI_ISL_2018791, EPI_ISL_2018792, EPI_ISL_2018793, EPI_ISL_2018794, EPI_ISL_2018795, EPI_ISL_2018796, EPI_ISL_2018797, EPI_ISL_2018798, EPI_ISL_2018800, EPI_ISL_2018801, EPI_ISL_2018802, EPI_ISL_2018803, EPI_ISL_2018804, EPI_ISL_2018805, EPI_ISL_2018806, EPI_ISL_2018807, EPI_ISL_2018808, EPI_ISL_2018809, EPI_ISL_2018810, EPI_ISL_2018811, EPI_ISL_2018812, EPI_ISL_2018813, EPI_ISL_2018814, EPI_ISL_2018815, EPI_ISL_2018816, EPI_ISL_2018817, EPI_ISL_2018818, EPI_ISL_2018819, EPI_ISL_2018820, EPI_ISL_2018821, EPI_ISL_2018822, EPI_ISL_2018823, EPI_ISL_2018824, EPI_ISL_2018825, EPI_ISL_2018826, EPI_ISL_2018827, EPI_ISL_2018828, EPI_ISL_2018829, EPI_ISL_2018830, EPI_ISL_2018831, EPI_ISL_2018832, EPI_ISL_2018833, EPI_ISL_2018834, EPI_ISL_2018835, EPI_ISL_2018836, EPI_ISL_2018837, EPI_ISL_2018838, EPI_ISL_2018839, EPI_ISL_2018840, EPI_ISL_2018841, EPI_ISL_2018842, EPI_ISL_2018843, EPI_ISL_2018844, EPI_ISL_2018845, EPI_ISL_2018846, EPI_ISL_2018847, EPI_ISL_2018848, EPI_ISL_2018849, EPI_ISL_2018850, EPI_ISL_2018851, EPI_ISL_2018853, EPI_ISL_2018854, EPI_ISL_2018855, EPI_ISL_2018857, EPI_ISL_2018858, EPI_ISL_2018859, EPI_ISL_2018860, EPI_ISL_2018861, EPI_ISL_2018862, EPI_ISL_2018863, EPI_ISL_2018864, EPI_ISL_2018865, EPI_ISL_2018866, EPI_ISL_2018867, EPI_ISL_2018868, EPI_ISL_2018869, EPI_ISL_2018870, EPI_ISL_2018871, EPI_ISL_2018872, EPI_ISL_2018873, EPI_ISL_2018874, EPI_ISL_2018875, EPI_ISL_2018876, EPI_ISL_2018877, EPI_ISL_2018878, EPI_ISL_2018879, EPI_ISL_2018880, EPI_ISL_2018881, EPI_ISL_2018882, EPI_ISL_2018883, EPI_ISL_2018884, EPI_ISL_2018885, EPI_ISL_2018886, EPI_ISL_2018887, EPI_ISL_2018889, EPI_ISL_2018890, EPI_ISL_2018892, EPI_ISL_2018893, EPI_ISL_2018895, EPI_ISL_2018899, EPI_ISL_2018900, EPI_ISL_2018901, EPI_ISL_2018902, EPI_ISL_2018903, EPI_ISL_2018904, EPI_ISL_2018905, EPI_ISL_2018909, EPI_ISL_2018910, EPI_ISL_2018911, EPI_ISL_2018912, EPI_ISL_2018913, EPI_ISL_2018914, EPI_ISL_2018915, EPI_ISL_2018916, EPI_ISL_2018917, EPI_ISL_2018918, EPI_ISL_2018919, EPI_ISL_2018920, EPI_ISL_2018921 | | | |
| see above | Wisconsin State Laboratory of Hygiene Communicable Disease Division | Wisconsin State Laboratory of Hygiene Communicable Disease Division | Abigail C. Shockey, Alicia J. Mooney, Erika M. Hanson, Tonya Danz, Richard Griesser, Sara Wagner, Kelsey R. Florek |
| EPI_ISL_2020688, EPI_ISL_2020689, EPI_ISL_2020690, EPI_ISL_2020691, EPI_ISL_2020692, EPI_ISL_2020693, EPI_ISL_2020694, EPI_ISL_2020695, EPI_ISL_2020696, EPI_ISL_2020697, EPI_ISL_2020698, EPI_ISL_2020699, EPI_ISL_2020700, EPI_ISL_2020701, EPI_ISL_2020702, EPI_ISL_2020703, EPI_ISL_2020704, EPI_ISL_2020705, EPI_ISL_2020706, EPI_ISL_2020707, EPI_ISL_2020708, EPI_ISL_2020709, EPI_ISL_2020710, EPI_ISL_2020711, EPI_ISL_2020712, EPI_ISL_2020713, EPI_ISL_2020714, EPI_ISL_2020716, EPI_ISL_2020717, EPI_ISL_2020718, EPI_ISL_2020719, EPI_ISL_2020720, EPI_ISL_2020721, EPI_ISL_2020722, EPI_ISL_2020723, EPI_ISL_2020724, EPI_ISL_2020725, EPI_ISL_2020726, EPI_ISL_2020727, EPI_ISL_2020728, EPI_ISL_2020729, EPI_ISL_2020730, EPI_ISL_2020731, EPI_ISL_2020732, EPI_ISL_2020733, EPI_ISL_2020734, EPI_ISL_2020735, EPI_ISL_2020736, EPI_ISL_2020737, EPI_ISL_2020738, EPI_ISL_2020739, EPI_ISL_2020740, EPI_ISL_2020741, EPI_ISL_2020742, EPI_ISL_2020743, EPI_ISL_2020744, EPI_ISL_2020745, EPI_ISL_2020746, EPI_ISL_2020747, EPI_ISL_2020748, EPI_ISL_2020749, EPI_ISL_2020750, EPI_ISL_2020751, EPI_ISL_2020752, EPI_ISL_2020753, EPI_ISL_2020754, EPI_ISL_2020755, EPI_ISL_2020756, EPI_ISL_2020757, EPI_ISL_2020758, EPI_ISL_2020759, EPI_ISL_2020760, EPI_ISL_2020761, EPI_ISL_2020762, EPI_ISL_2020763, EPI_ISL_2020764, EPI_ISL_2020765, EPI_ISL_2020766, EPI_ISL_2020767, EPI_ISL_2020768, EPI_ISL_2020769, EPI_ISL_2020770, EPI_ISL_2020771, EPI_ISL_2020772, EPI_ISL_2020773, EPI_ISL_2020774, EPI_ISL_2020775, EPI_ISL_2020776, EPI_ISL_2020777, EPI_ISL_2020778, EPI_ISL_2020779, EPI_ISL_2020780, EPI_ISL_2020781, EPI_ISL_2020782, EPI_ISL_2020783, EPI_ISL_2020784, EPI_ISL_2020785, EPI_ISL_2020786, EPI_ISL_2020787, EPI_ISL_2020788, EPI_ISL_2020789, EPI_ISL_2020790, EPI_ISL_2020791, EPI_ISL_2020792, EPI_ISL_2020793, EPI_ISL_2020794, EPI_ISL_2020795, EPI_ISL_2020796, EPI_ISL_2020797, EPI_ISL_2020798, EPI_ISL_2020799, EPI_ISL_2020800, EPI_ISL_2020801, EPI_ISL_2020802, EPI_ISL_2020803, EPI_ISL_2020804, EPI_ISL_2020805, EPI_ISL_2020806, EPI_ISL_2020807, EPI_ISL_2020808, EPI_ISL_2020809, EPI_ISL_2020810, EPI_ISL_2020811, EPI_ISL_2020812, EPI_ISL_2020813, EPI_ISL_2020814, EPI_ISL_2020815, EPI_ISL_2020816, EPI_ISL_2020817, EPI_ISL_2020818, EPI_ISL_2020819, EPI_ISL_2020820, EPI_ISL_2020821, EPI_ISL_2020822, EPI_ISL_2020823, EPI_ISL_2020824, EPI_ISL_2020825, EPI_ISL_2020826, EPI_ISL_2020827, EPI_ISL_2020828, EPI_ISL_2020829, EPI_ISL_2020830, EPI_ISL_2020831, EPI_ISL_2020832, EPI_ISL_2020833, EPI_ISL_2020834, EPI_ISL_2020835, EPI_ISL_2020836, EPI_ISL_2020838, EPI_ISL_2020839, EPI_ISL_2020840, EPI_ISL_2020841, EPI_ISL_2020842, EPI_ISL_2020843, EPI_ISL_2020844, EPI_ISL_2020845, EPI_ISL_2020846, EPI_ISL_2020847, EPI_ISL_2020848, EPI_ISL_2020849, EPI_ISL_2020850, EPI_ISL_2020851, EPI_ISL_2020852, EPI_ISL_2020853, EPI_ISL_2020854, EPI_ISL_2020855, EPI_ISL_2020856, EPI_ISL_2020857, EPI_ISL_2020858, EPI_ISL_2020859, EPI_ISL_2020860, EPI_ISL_2020861 | | | |
| see above | Yale Clinical Virology Lab | Yale Center for Genomic Analysis | Shrikant Mane, Kaya Bilguvar, Curt Scharfe, Irina Tikhonova, Brooke Sullivan |
| EPI_ISL_2020866, EPI_ISL_2020867, EPI_ISL_2020868, EPI_ISL_2020869, EPI_ISL_2020870, EPI_ISL_2020871, EPI_ISL_2020872, EPI_ISL_2020873, EPI_ISL_2020874, EPI_ISL_2020875, EPI_ISL_2020876, EPI_ISL_2020877 | | | |
| see above | Wyoming Public Health Laboratory | Wyoming Public Health Laboratory | Jim Mildenberger, Taylor Fearing, Channing Weber, Ashley Norberg, Chayse Rowley, Marley Goetz, Brian Dominguez, Elliot Thomasson, Brittany Oher, Sam Britz, Cari Sloma, and Rob Christensen |
| EPI_ISL_2020965, EPI_ISL_2020966, EPI_ISL_2020967, EPI_ISL_2020968, EPI_ISL_2020969, EPI_ISL_2020970, EPI_ISL_2020971, EPI_ISL_2020972, EPI_ISL_2020973, EPI_ISL_2020974, EPI_ISL_2020975, EPI_ISL_2020976, EPI_ISL_2020977, EPI_ISL_2020978, EPI_ISL_2020979, EPI_ISL_2020980, EPI_ISL_2020981, EPI_ISL_2020982, EPI_ISL_2020983, EPI_ISL_2020984, EPI_ISL_2020985, EPI_ISL_2020986, EPI_ISL_2020987, EPI_ISL_2020988, EPI_ISL_2020989, EPI_ISL_2020990, EPI_ISL_2020992 | | | |
| see above | UW Virology Lab | UW Virology Lab | Pavitra Roychoudhury, Hong Xie, Lasata Shrestha, Tien V. Nguyen, Shah Mohamed Bakhash, Michelle Lin, Noah R. Baker, Ricardo Perez, Sean Ellis, Nathan Breit, Robert J. Livingston, Meei-Li Huang, Keith R Jerome, Patrick Mathias, Alexander Greninger |
| EPI_ISL_2020993, EPI_ISL_2020994, EPI_ISL_2020995, EPI_ISL_2020998, EPI_ISL_2020999, EPI_ISL_2021000, EPI_ISL_2021002, EPI_ISL_2021003, EPI_ISL_2021004, EPI_ISL_2021005, EPI_ISL_2021006, EPI_ISL_2021007, EPI_ISL_2021008, EPI_ISL_2021009, EPI_ISL_2021010, EPI_ISL_2021011, EPI_ISL_2021012, EPI_ISL_2021017, EPI_ISL_2021018, EPI_ISL_2021019, EPI_ISL_2021020, EPI_ISL_2021021, EPI_ISL_2021022, EPI_ISL_2021025, EPI_ISL_2021026, EPI_ISL_2021027 | | | |
| see above | Wyoming Public Health Laboratory | Wyoming Public Health Laboratory | Jim Mildenberger, Taylor Fearing, Channing Weber, Ashley Norberg, Chayse Rowley, Marley Goetz, Brian Dominguez, Elliot Thomasson, Brittany Oher, Sam Britz, Cari Sloma, and Rob Christensen |
| EPI_ISL_2021032, EPI_ISL_2021034, EPI_ISL_2021037, EPI_ISL_2021038 | Wyoming Public Health Laboratory | Wyoming Public Health Laboratory | Jim Mildenberger, Wanda Manley, Noah Hull, Taylor Fearing, Lynette Gumbleton, Channing Weber, Ashley Norberg, Chayse Rowley, Marley Goetz, Brian Dominguez, Elliot Thomasson, Sam Britz, Cari Sloma, and Rob Christensen |
| EPI_ISL_2023050, EPI_ISL_2023051, EPI_ISL_2023052, EPI_ISL_2023053, EPI_ISL_2023054, EPI_ISL_2023055, EPI_ISL_2023056, EPI_ISL_2023057, EPI_ISL_2023058, EPI_ISL_2023059, EPI_ISL_2023060, EPI_ISL_2023061, EPI_ISL_2023062, EPI_ISL_2023063, EPI_ISL_2023064, EPI_ISL_2023065, EPI_ISL_2023066, EPI_ISL_2023067, EPI_ISL_2023068, EPI_ISL_2023069, EPI_ISL_2023070, EPI_ISL_2023071, EPI_ISL_2023072, EPI_ISL_2023073, EPI_ISL_2023074, EPI_ISL_2023075, EPI_ISL_2023076, EPI_ISL_2023077, EPI_ISL_2023078, EPI_ISL_2023079, EPI_ISL_2023080, EPI_ISL_2023081, EPI_ISL_2023082, EPI_ISL_2023083, EPI_ISL_2023084, EPI_ISL_2023085, EPI_ISL_2023086, EPI_ISL_2023087, EPI_ISL_2023088, EPI_ISL_2023089, EPI_ISL_2023090, EPI_ISL_2023091, EPI_ISL_2023092, EPI_ISL_2023093, EPI_ISL_2023094, EPI_ISL_2023095, EPI_ISL_2023096, EPI_ISL_2023097, EPI_ISL_2023098, EPI_ISL_2023099, EPI_ISL_2023100, EPI_ISL_2023101, EPI_ISL_2023102, EPI_ISL_2023103, EPI_ISL_2023104, EPI_ISL_2023105, EPI_ISL_2023106, EPI_ISL_2023107, EPI_ISL_2023108, EPI_ISL_2023109, EPI_ISL_2023110, EPI_ISL_2023111, EPI_ISL_2023112, EPI_ISL_2023113, EPI_ISL_2023114, EPI_ISL_2023115, EPI_ISL_2023116, EPI_ISL_2023117, EPI_ISL_2023118, EPI_ISL_2023119, EPI_ISL_2023120, EPI_ISL_2023121, EPI_ISL_2023122, EPI_ISL_2023123, EPI_ISL_2023124, EPI_ISL_2023125, EPI_ISL_2023126, EPI_ISL_2023127, EPI_ISL_2023128, EPI_ISL_2023129, EPI_ISL_2023130, EPI_ISL_2023131, EPI_ISL_2023132, EPI_ISL_2023133, EPI_ISL_2023135, EPI_ISL_2023136, EPI_ISL_2023138, EPI_ISL_2023139, EPI_ISL_2023140, EPI_ISL_2023141, EPI_ISL_2023142, EPI_ISL_2023143, EPI_ISL_2023144, EPI_ISL_2023145, EPI_ISL_2023146, EPI_ISL_2023148, EPI_ISL_2023149, EPI_ISL_2023150, EPI_ISL_2023151, EPI_ISL_2023152, EPI_ISL_2023153, EPI_ISL_2023154, EPI_ISL_2023155, EPI_ISL_2023156, EPI_ISL_2023157, EPI_ISL_2023158, EPI_ISL_2023159, EPI_ISL_2023160, EPI_ISL_2023161, EPI_ISL_2023162, EPI_ISL_2023163, EPI_ISL_2023164, EPI_ISL_2023165, EPI_ISL_2023166, EPI_ISL_2023167, EPI_ISL_2023168, EPI_ISL_2023169, EPI_ISL_2023170, EPI_ISL_2023171, EPI_ISL_2023172, EPI_ISL_2023173, EPI_ISL_2023174, EPI_ISL_2023175, EPI_ISL_2023176, EPI_ISL_2023177, EPI_ISL_2023178, EPI_ISL_2023179, EPI_ISL_2023180, EPI_ISL_2023181, EPI_ISL_2023182, EPI_ISL_2023183, EPI_ISL_2023184, EPI_ISL_2023185, EPI_ISL_2023186, EPI_ISL_2023187, EPI_ISL_2023188, EPI_ISL_2023189, EPI_ISL_2023190, EPI_ISL_2023191, EPI_ISL_2023192, EPI_ISL_2023193, EPI_ISL_2023194, EPI_ISL_2023195, EPI_ISL_2023196, EPI_ISL_2023197, EPI_ISL_2023198, EPI_ISL_2023199, EPI_ISL_2023200, EPI_ISL_2023201, EPI_ISL_2023203, EPI_ISL_2023204, EPI_ISL_2023205, EPI_ISL_2023206, EPI_ISL_2023207, EPI_ISL_2023208, EPI_ISL_2023209, EPI_ISL_2023210, EPI_ISL_2023211, EPI_ISL_2023212, EPI_ISL_2023213 | | | |
| see above | Yale Clinical Virology Lab | Grubaugh Lab - Yale School of Public Health | Joseph Fauver, Mallery Breban, Isabel Ott, Tara Alpert, Mary Petrone, Anderson Brito, Chantal Vogels, Annie Watkins, Chaney Kalinich, Jessica Rothman, Marie L. Landry, Nathan Grubaugh |
| EPI_ISL_2023214, EPI_ISL_2023215, EPI_ISL_2023216, EPI_ISL_2023217 | Yale Pathology Lab | Grubaugh Lab - Yale School of Public Health | Joseph Fauver, Mallery Breban, Isabel Ott, Tara Alpert, Mary Petrone, Anderson Brito, Chantal Vogels, Annie Watkins, Chaney Kalinich, Jessica Rothman, Jianhui Wang, Chen Liu, Pei Hui, Nathan Grubaugh |
| EPI_ISL_2023218, EPI_ISL_2023219, EPI_ISL_2023220, EPI_ISL_2023222, EPI_ISL_2023223, EPI_ISL_2023226, EPI_ISL_2023227, EPI_ISL_2023228, EPI_ISL_2023229, EPI_ISL_2023230, EPI_ISL_2023231, EPI_ISL_2023232, EPI_ISL_2023233, EPI_ISL_2023234, EPI_ISL_2023235, EPI_ISL_2023236, EPI_ISL_2023237, EPI_ISL_2023238, EPI_ISL_2023239, EPI_ISL_2023240, EPI_ISL_2023241, EPI_ISL_2023243, EPI_ISL_2023244, EPI_ISL_2023246, EPI_ISL_2023247, EPI_ISL_2023248, EPI_ISL_2023249, EPI_ISL_2023250, EPI_ISL_2023252, EPI_ISL_2023253, EPI_ISL_2023254, EPI_ISL_2023255, EPI_ISL_2023256, EPI_ISL_2023257, EPI_ISL_2023258, EPI_ISL_2023259, EPI_ISL_2023260, EPI_ISL_2023261, EPI_ISL_2023262, EPI_ISL_2023263, EPI_ISL_2023264, EPI_ISL_2023265, EPI_ISL_2023266, EPI_ISL_2023267, EPI_ISL_2023268, EPI_ISL_2023270, EPI_ISL_2023271, EPI_ISL_2023272, EPI_ISL_2023274, EPI_ISL_2023275, EPI_ISL_2023276, EPI_ISL_2023277 | | | |
| see above | Utah Public Health Laboratory | Utah Public Health Laboratory | Erin L. Young, Kelly F. Oakeson, Tara Gallagher |
| EPI_ISL_2023282 | Wyoming Public Health Laboratory | Wyoming Public Health Laboratory | Jim Mildenberger, Wanda Manley, Noah Hull, Taylor Fearing, Lynette Gumbleton, Channing Weber, Ashley Norberg, Chayse Rowley, Marley Goetz, Brian Dominguez, Elliot Thomasson, Sam Britz, Cari Sloma, and Rob Christensen |
| EPI_ISL_2023283, EPI_ISL_2023284, EPI_ISL_2023285, EPI_ISL_2023286, EPI_ISL_2023287, EPI_ISL_2023288, EPI_ISL_2023289, EPI_ISL_2023290, EPI_ISL_2023291, EPI_ISL_2023292, EPI_ISL_2023293, EPI_ISL_2023294, EPI_ISL_2023295, EPI_ISL_2023296, EPI_ISL_2023297, EPI_ISL_2023298 | | | |
| see above | North Carolina State Laboratory of Public Health (NCSLPH) | North Carolina State Laboratory of Public Health (NCSLPH) | Chase K, Miller MC, Greene S, Glover W |
| EPI_ISL_2023300, EPI_ISL_2023301, EPI_ISL_2023302, EPI_ISL_2023303, EPI_ISL_2023304, EPI_ISL_2023305, EPI_ISL_2023306, EPI_ISL_2023307, EPI_ISL_2023308, EPI_ISL_2023310, EPI_ISL_2023311, EPI_ISL_2023312, EPI_ISL_2023313, EPI_ISL_2023314, EPI_ISL_2023315, EPI_ISL_2023316, EPI_ISL_2023317, EPI_ISL_2023318 | | | |
| see above | Kansas Health and Environmental Lab | Kansas Health and Environmental Lab | Mike Grose, Jonathan Barnell, Ben Olsen, and Phil Adam |
| EPI_ISL_2023319, EPI_ISL_2023320, EPI_ISL_2023321, EPI_ISL_2023322 | Wichita State University - Molecular Diagnostics Lab | Kansas Health and Environmental Lab | Mike Grose, Jonathan Barnell, Ben Olsen, and Phil Adam |
| EPI_ISL_2023323, | Clinical Reference Laboratory | Kansas Health and Environmental Lab | Mike Grose, Jonathan Barnell, Ben Olsen, and Phil Adam |

| | | | |
|---|---|---|---|
| EPI_ISL_2023324, EPI_ISL_2023325, EPI_ISL_2023326 | | | |
| EPI_ISL_2023327, EPI_ISL_2023329, EPI_ISL_2023330 | Kansas Health and Environmental Lab | Kansas Health and Environmental Lab | Mike Grose, Jonathan Barnell, Ben Olsen, and Phil Adam |
| EPI_ISL_2023331, EPI_ISL_2023332, EPI_ISL_2023333, EPI_ISL_2023334, EPI_ISL_2023335, EPI_ISL_2023336, EPI_ISL_2023337, EPI_ISL_2023338, EPI_ISL_2023340, EPI_ISL_2023341, EPI_ISL_2023342, EPI_ISL_2023343, EPI_ISL_2023344, EPI_ISL_2023345, EPI_ISL_2023346, EPI_ISL_2023347, EPI_ISL_2023350, EPI_ISL_2023351, EPI_ISL_2023352, EPI_ISL_2023354 | | | |
| see above | Wichita State University - Molecular Diagnostics Lab | Kansas Health and Environmental Lab | Mike Grose, Jonathan Barnell, Ben Olsen, and Phil Adam |
| EPI_ISL_2023355, EPI_ISL_2023356, EPI_ISL_2023357, EPI_ISL_2023358, EPI_ISL_2023359, EPI_ISL_2023360, EPI_ISL_2023361, EPI_ISL_2023362, EPI_ISL_2023363, EPI_ISL_2023364, EPI_ISL_2023365, EPI_ISL_2023366, EPI_ISL_2023367, EPI_ISL_2023368, EPI_ISL_2023369, EPI_ISL_2023370, EPI_ISL_2023371, EPI_ISL_2023372, EPI_ISL_2023373, EPI_ISL_2023374, EPI_ISL_2023375, EPI_ISL_2023376, EPI_ISL_2023377, EPI_ISL_2023378, EPI_ISL_2023379, EPI_ISL_2023380, EPI_ISL_2023381, EPI_ISL_2023382 | | | |
| see above | Montana Public Health Laboratory | Montana Public Health Laboratory | Joy Ritter, Michelle Mozer, Carrie Biskupiak, Deborah Gibson |
| EPI_ISL_2023408, EPI_ISL_2023409, EPI_ISL_2023410, EPI_ISL_2023411, EPI_ISL_2023412, EPI_ISL_2023413, EPI_ISL_2023414, EPI_ISL_2023415, EPI_ISL_2023416, EPI_ISL_2023417, EPI_ISL_2023418, EPI_ISL_2023419, EPI_ISL_2023420, EPI_ISL_2023421, EPI_ISL_2023422, EPI_ISL_2023423, EPI_ISL_2023424, EPI_ISL_2023425, EPI_ISL_2023426, EPI_ISL_2023427, EPI_ISL_2023428 | | | |
| see above | Nebraska Public Health Laboratory | NPHL COVID-19 Response Team | NPHL COVID-19 Response Team |
| EPI_ISL_2023450, EPI_ISL_2023451 | Tampa General Hospital Esoteric Lab | Tampa General Hospital Esoteric Research & Development Lab | Grant Vestal, Deanna Becker, Dominic Uy, Vicki Healer, Amorce Lima, Suzane Silbert |
| EPI_ISL_2023452, EPI_ISL_2023453, EPI_ISL_2023454, EPI_ISL_2023455, EPI_ISL_2023456, EPI_ISL_2023457, EPI_ISL_2023458, EPI_ISL_2023459, EPI_ISL_2023460, EPI_ISL_2023461, EPI_ISL_2023462, EPI_ISL_2023463, EPI_ISL_2023464, EPI_ISL_2023465, EPI_ISL_2023466, EPI_ISL_2023467, EPI_ISL_2023468, EPI_ISL_2023469, EPI_ISL_2023470, EPI_ISL_2023471, EPI_ISL_2023472, EPI_ISL_2023473, EPI_ISL_2023474, EPI_ISL_2023475, EPI_ISL_2023476, EPI_ISL_2023477, EPI_ISL_2023478, EPI_ISL_2023479, EPI_ISL_2023480, EPI_ISL_2023481, EPI_ISL_2023482, EPI_ISL_2023483, EPI_ISL_2023484, EPI_ISL_2023485, EPI_ISL_2023486, EPI_ISL_2023487, EPI_ISL_2023488, EPI_ISL_2023489, EPI_ISL_2023490, EPI_ISL_2023491, EPI_ISL_2023492, EPI_ISL_2023493 | | | |
| see above | Oregon State Public Health Laboratory | Oregon State Public Health Laboratory | Rafia Razzaque, Eugene Yeboah, Vanda Makris, Laura Tsaknaridis, John Fontana and Shane Sevey |
| EPI_ISL_2024490, EPI_ISL_2024492, EPI_ISL_2024494, EPI_ISL_2024496, EPI_ISL_2024498, EPI_ISL_2024499, EPI_ISL_2024500, EPI_ISL_2024501, EPI_ISL_2024503, EPI_ISL_2024505, EPI_ISL_2024507, EPI_ISL_2024509, EPI_ISL_2024511, EPI_ISL_2024513, EPI_ISL_2024515, EPI_ISL_2024545 | | | |
| see above | Arizona State University | Arizona State University | Peter T. Skidmore, LaRinda A. Holland, Matthew F. Smith, Rabia Maqsood, Nicholas J. Mellor, Joy M. Blain, Valerie Harris, Joshua LaBaer, Vel Murugan, Efrem S. Lim |
| EPI_ISL_2025779 | Seattle Flu Study | Seattle Flu Study | Deborah A. Nickerson, Chris D. Frazar, Jover Lee, Benjamin Pelle, Erica Ryke, Matthew Richardson, Amanda Adler, Elisabeth Brandstetter, Peter D. Han, Kairsten Fay, Misja Ilcisin, Kirsten Lacombe, Thomas R. Sibley, Melissa Truong, Caitlin R. Wolf, Michael Boeckh, Janet A. Englund, Michael Famulare, Barry R. Lutz, Mark J. Rieder, Lea M. Starita, Matthew Thompson, Jay Shendure, Trevor Bedford, Helen Y. Chu |
| EPI_ISL_2025781, EPI_ISL_2025783, EPI_ISL_2025786 | Incyte Diagnostics Spokane | Seattle Flu Study | Deborah A. Nickerson, Chris D. Frazar, Jover Lee, Benjamin Pelle, Erica Ryke, Matthew Richardson, Amanda Adler, Elisabeth Brandstetter, Peter D. Han, Kairsten Fay, Misja Ilcisin, Kirsten Lacombe, Thomas R. Sibley, Melissa Truong, Caitlin R. Wolf, Romesh Gautom, Geoff Melly, Brian Hiatt, Philip Dykema, Scott Lindquist, Michael Boeckh, Janet A. Englund, Michael Famulare, Barry R. Lutz, Mark J. Rieder, Lea M. Starita, Matthew Thompson, Helen Y. Chu, Jay Shendure, Trevor Bedford |
| EPI_ISL_2026394 | Hospital Margarita Maza de Juárez | Microbial Genomics Laboratory | Bruno Gomez-Gil, Julissa Enciso-Ibarra, Alejandra Garcia-Gasca, Daniel Fregoso-Rueda |
| EPI_ISL_2026396, EPI_ISL_2026398 | Hospital Sharp | Microbial Genomics Laboratory | Bruno Gomez-Gil, Julissa Enciso-Ibarra, Alejandra Garcia-Gasca, Daniel Fregoso-Rueda |
| EPI_ISL_2026400, EPI_ISL_2026402, EPI_ISL_2026404 | Hospital Margarita Maza de Juárez | Microbial Genomics Laboratory | Bruno Gomez-Gil, Julissa Enciso-Ibarra, Alejandra Garcia-Gasca, Daniel Fregoso-Rueda |
| EPI_ISL_2026406 | Imágenes Diagnósticas de Mazatlán | Microbial Genomics Laboratory | Bruno Gomez-Gil, Julissa Enciso-Ibarra, Alejandra Garcia-Gasca, Daniel Fregoso-Rueda |
| EPI_ISL_2026407 | Hospital Sharp | Microbial Genomics Laboratory | Bruno Gomez-Gil, Julissa Enciso-Ibarra, Alejandra Garcia-Gasca, Daniel Fregoso-Rueda |
| EPI_ISL_2026409, EPI_ISL_2026411, EPI_ISL_2026413, EPI_ISL_2026415, EPI_ISL_2026417 | Hospital Margarita Maza de Juárez | Microbial Genomics Laboratory | Bruno Gomez-Gil, Julissa Enciso-Ibarra, Alejandra Garcia-Gasca, Daniel Fregoso-Rueda |
| EPI_ISL_2026418, EPI_ISL_2026420, EPI_ISL_2026432, EPI_ISL_2026434 | Hospital Sharp | Microbial Genomics Laboratory | Bruno Gomez-Gil, Julissa Enciso-Ibarra, Alejandra Garcia-Gasca, Daniel Fregoso-Rueda |
| EPI_ISL_2026436, EPI_ISL_2026437, EPI_ISL_2026440 | Hospital Margarita Maza de Juárez | Microbial Genomics Laboratory | Bruno Gomez-Gil, Julissa Enciso-Ibarra, Alejandra Garcia-Gasca, Daniel Fregoso-Rueda |
| EPI_ISL_2026442, EPI_ISL_2026444 | Hospital Sharp | Microbial Genomics Laboratory | Bruno Gomez-Gil, Julissa Enciso-Ibarra, Alejandra Garcia-Gasca, Daniel Fregoso-Rueda |
| EPI_ISL_2026446, EPI_ISL_2026448, EPI_ISL_2026450, EPI_ISL_2026452, EPI_ISL_2026454 | Hospital Margarita Maza de Juárez | Microbial Genomics Laboratory | Bruno Gomez-Gil, Julissa Enciso-Ibarra, Alejandra Garcia-Gasca, Daniel Fregoso-Rueda |
| EPI_ISL_2026456 | Hospital Sharp | Microbial Genomics Laboratory | Bruno Gomez-Gil, Julissa Enciso-Ibarra, Alejandra Garcia-Gasca, Daniel Fregoso-Rueda |
| EPI_ISL_2026458 | Hospital Margarita Maza de Juárez | Microbial Genomics Laboratory | Bruno Gomez-Gil, Julissa Enciso-Ibarra, Alejandra Garcia-Gasca, Daniel Fregoso-Rueda |
| EPI_ISL_2026460, EPI_ISL_2026462 | Hospital Sharp | Microbial Genomics Laboratory | Bruno Gomez-Gil, Julissa Enciso-Ibarra, Alejandra Garcia-Gasca, Daniel Fregoso-Rueda |
| EPI_ISL_2027133, EPI_ISL_2027136, EPI_ISL_2027138 | Stanford Health Care | Stanford University School of Medicine, Clinical Virology Laboratory | Malaya K. Sahoo, Fumiko Yamamoto, Daniel Solis, Mamdouh Sibai, Bernadette Troung, Becky Jiang, James Zehnder, and Benjamin A. Pinsky |
| EPI_ISL_2027144 | Stanford Health Care | Stanford University School of Medicine, Clinical Virology Laboratory | Mamdouh Sibai, Daniel Solis, Becky Jiang, Fumiko Yamamoto, Michelle Verghese, ChunHong Huang, James Zehnder, Malaya K. Sahoo, and Benjamin A. Pinsky |
| EPI_ISL_2027147 | Stanford Health Care | Stanford University School of Medicine, Clinical Virology Laboratory | Malaya K. Sahoo, Fumiko Yamamoto, Daniel Solis, Mamdouh Sibai, Bernadette Troung, Becky Jiang, James Zehnder, and Benjamin A. Pinsky |
| EPI_ISL_2027150 | Stanford Health Care | Stanford University School of Medicine, Clinical Virology Laboratory | Mamdouh Sibai, Daniel Solis, Becky Jiang, Fumiko Yamamoto, Michelle Verghese, ChunHong Huang, James Zehnder, Malaya K. Sahoo, and Benjamin A. Pinsky |
| EPI_ISL_2027153, EPI_ISL_2027157 | Stanford Health Care | Stanford University School of Medicine, Clinical Virology Laboratory | Daniel Solis, Mamdouh Sibai, Fumiko Yamamoto, Becky Jiang, Michelle Verghese, ChunHong Huang, James Zehnder, Malaya K. Sahoo, and Benjamin A. Pinsky |
| EPI_ISL_2027160, EPI_ISL_2027164 | Stanford Health Care | Stanford University School of Medicine, Clinical Virology Laboratory | Fumiko Yamamoto, Malaya K. Sahoo, Mamdouh Sibai, Daniel Solis, Michelle Verghese, ChunHong Huang, James Zehnder, and Benjamin A. Pinsky |
| EPI_ISL_2027167 | Stanford Health Care | Stanford University School of Medicine, Clinical Virology Laboratory | Malaya K. Sahoo, Fumiko Yamamoto, Daniel Solis, Mamdouh Sibai, Bernadette Troung, Becky Jiang, James Zehnder, and Benjamin A. Pinsky |
| EPI_ISL_2027171, EPI_ISL_2027174, EPI_ISL_2027177, EPI_ISL_2027178 | Stanford Health Care | Stanford University School of Medicine, Clinical Virology Laboratory | Mamdouh Sibai, Daniel Solis, Becky Jiang, Fumiko Yamamoto, Michelle Verghese, ChunHong Huang, James Zehnder, Malaya K. Sahoo, and Benjamin A. Pinsky |
| EPI_ISL_2027183, EPI_ISL_2027185, EPI_ISL_2027187, EPI_ISL_2027191, EPI_ISL_2027195, EPI_ISL_2027198, EPI_ISL_2027202 | Stanford Health Care | Stanford University School of Medicine, Clinical Virology Laboratory | Daniel Solis, Mamdouh Sibai, Fumiko Yamamoto, Becky Jiang, Michelle Verghese, ChunHong Huang, James Zehnder, Malaya K. Sahoo, and Benjamin A. Pinsky |
| EPI_ISL_2027205, EPI_ISL_2027208, EPI_ISL_2027212, EPI_ISL_2027215, EPI_ISL_2027218 | Stanford Health Care | Stanford University School of Medicine, Clinical Virology Laboratory | Fumiko Yamamoto, Malaya K. Sahoo, Mamdouh Sibai, Daniel Solis, Michelle Verghese, ChunHong Huang, James Zehnder, and Benjamin A. Pinsky |
| EPI_ISL_2027221, EPI_ISL_2027225, EPI_ISL_2027228 | Stanford Health Care | Stanford University School of Medicine, Clinical Virology Laboratory | Malaya K. Sahoo, Fumiko Yamamoto, Daniel Solis, Mamdouh Sibai, Bernadette Troung, Becky Jiang, James Zehnder, and Benjamin A. Pinsky |
| EPI_ISL_2028101, EPI_ISL_2028103, | Pathology and Laboratory Medicine Institute, Cleveland Clinic, Ohio, USA | Pathology and Laboratory Medicine Institute, Cleveland Clinic, Ohio, USA | Jessica Spildener, Joy Nakitandwe, Kristen McDonnell, David Plunkett, Zheng Jin Tu, Jay Brock, Yu-Wei Cheng, Gary Procop, Daniel Rhoads, Daniel H. Farkas, David Bosler |

| | | | |
|---|---|---|---|
| EPI_ISL_2028105 | | | |
| EPI_ISL_2030444, EPI_ISL_2030445, EPI_ISL_2030446, EPI_ISL_2030447, EPI_ISL_2030448, EPI_ISL_2030449, EPI_ISL_2030450, EPI_ISL_2030451, EPI_ISL_2030452, EPI_ISL_2030453, EPI_ISL_2030454, EPI_ISL_2030455, EPI_ISL_2030456, EPI_ISL_2030457 | | | |
| see above | Nebraska Public Health Laboratory | NPHL COVID-19 Response Team | NPHL COVID-19 Response Team |
| EPI_ISL_2031271, EPI_ISL_2031272, EPI_ISL_2031273, EPI_ISL_2031274, EPI_ISL_2031275, EPI_ISL_2031276, EPI_ISL_2031277, EPI_ISL_2031278, EPI_ISL_2031279, EPI_ISL_2031280, EPI_ISL_2031281, EPI_ISL_2031282, EPI_ISL_2031283, EPI_ISL_2031284, EPI_ISL_2031285, EPI_ISL_2031286, EPI_ISL_2031287, EPI_ISL_2031288, EPI_ISL_2031289, EPI_ISL_2031290, EPI_ISL_2031291, EPI_ISL_2031292, EPI_ISL_2031293, EPI_ISL_2031294, EPI_ISL_2031295, EPI_ISL_2031296, EPI_ISL_2031297, EPI_ISL_2031298, EPI_ISL_2031299, EPI_ISL_2031300, EPI_ISL_2031301, EPI_ISL_2031302, EPI_ISL_2031303, EPI_ISL_2031304, EPI_ISL_2031305, EPI_ISL_2031306, EPI_ISL_2031307, EPI_ISL_2031308, EPI_ISL_2031309, EPI_ISL_2031310, EPI_ISL_2031311, EPI_ISL_2031312, EPI_ISL_2031313, EPI_ISL_2031314, EPI_ISL_2031315, EPI_ISL_2031316, EPI_ISL_2031317, EPI_ISL_2031318, EPI_ISL_2031319, EPI_ISL_2031320, EPI_ISL_2031321, EPI_ISL_2031322, EPI_ISL_2031323, EPI_ISL_2031324, EPI_ISL_2031325, EPI_ISL_2031326, EPI_ISL_2031327, EPI_ISL_2031328, EPI_ISL_2031329, EPI_ISL_2031330, EPI_ISL_2031331, EPI_ISL_2031332, EPI_ISL_2031333, EPI_ISL_2031334, EPI_ISL_2031335, EPI_ISL_2031336, EPI_ISL_2031337, EPI_ISL_2031338, EPI_ISL_2031339, EPI_ISL_2031340, EPI_ISL_2031341, EPI_ISL_2031342, EPI_ISL_2031343, EPI_ISL_2031344, EPI_ISL_2031345, EPI_ISL_2031346, EPI_ISL_2031347, EPI_ISL_2031348, EPI_ISL_2031349, EPI_ISL_2031350, EPI_ISL_2031351, EPI_ISL_2031352, EPI_ISL_2031353, EPI_ISL_2031354, EPI_ISL_2031355, EPI_ISL_2031356, EPI_ISL_2031357, EPI_ISL_2031358, EPI_ISL_2031359, EPI_ISL_2031360, EPI_ISL_2031361, EPI_ISL_2031362, EPI_ISL_2031363, EPI_ISL_2031364, EPI_ISL_2031365, EPI_ISL_2031366, EPI_ISL_2031367, EPI_ISL_2031368, EPI_ISL_2031369, EPI_ISL_2031370, EPI_ISL_2031371, EPI_ISL_2031372, EPI_ISL_2031373, EPI_ISL_2031374, EPI_ISL_2031375, EPI_ISL_2031376, EPI_ISL_2031377, EPI_ISL_2031378, EPI_ISL_2031379, EPI_ISL_2031380, EPI_ISL_2031381, EPI_ISL_2031382, EPI_ISL_2031383, EPI_ISL_2031384, EPI_ISL_2031385, EPI_ISL_2031386, EPI_ISL_2031387, EPI_ISL_2031388, EPI_ISL_2031389, EPI_ISL_2031390, EPI_ISL_2031391, EPI_ISL_2031392, EPI_ISL_2031393, EPI_ISL_2031394, EPI_ISL_2031395, EPI_ISL_2031396, EPI_ISL_2031397, EPI_ISL_2031398, EPI_ISL_2031399, EPI_ISL_2031400, EPI_ISL_2031401, EPI_ISL_2031402, EPI_ISL_2031403, EPI_ISL_2031404, EPI_ISL_2031405, EPI_ISL_2031406, EPI_ISL_2031407, EPI_ISL_2031408, EPI_ISL_2031409, EPI_ISL_2031410, EPI_ISL_2031411, EPI_ISL_2031412, EPI_ISL_2031413, EPI_ISL_2031414, EPI_ISL_2031415, EPI_ISL_2031416, EPI_ISL_2031417, EPI_ISL_2031418, EPI_ISL_2031419, EPI_ISL_2031420, EPI_ISL_2031421, EPI_ISL_2031422, EPI_ISL_2031423, EPI_ISL_2031424, EPI_ISL_2031425, EPI_ISL_2031426, EPI_ISL_2031427, EPI_ISL_2031428, EPI_ISL_2031429, EPI_ISL_2031430, EPI_ISL_2031431, EPI_ISL_2031432, EPI_ISL_2031433, EPI_ISL_2031434, EPI_ISL_2031435, EPI_ISL_2031436, EPI_ISL_2031437, EPI_ISL_2031438, EPI_ISL_2031439, EPI_ISL_2031440, EPI_ISL_2031441, EPI_ISL_2031442, EPI_ISL_2031443, EPI_ISL_2031444, EPI_ISL_2031445, EPI_ISL_2031446, EPI_ISL_2031447, EPI_ISL_2031448, EPI_ISL_2031449, EPI_ISL_2031450, EPI_ISL_2031451, EPI_ISL_2031452, EPI_ISL_2031453, EPI_ISL_2031454, EPI_ISL_2031455, EPI_ISL_2031456, EPI_ISL_2031457, EPI_ISL_2031458, EPI_ISL_2031459, EPI_ISL_2031460, EPI_ISL_2031461, EPI_ISL_2031462, EPI_ISL_2031480, EPI_ISL_2031481, EPI_ISL_2031482, EPI_ISL_2031483, EPI_ISL_2031484, EPI_ISL_2031485, EPI_ISL_2031486, EPI_ISL_2031487, EPI_ISL_2031488, EPI_ISL_2031489, EPI_ISL_2031490, EPI_ISL_2031491, EPI_ISL_2031492, EPI_ISL_2031493, EPI_ISL_2031494, EPI_ISL_2031495, EPI_ISL_2031496, EPI_ISL_2031497, EPI_ISL_2031498, EPI_ISL_2031499, EPI_ISL_2031500, EPI_ISL_2031501, EPI_ISL_2031502, EPI_ISL_2031503, EPI_ISL_2031504, EPI_ISL_2031505, EPI_ISL_2031506, EPI_ISL_2031507, EPI_ISL_2031508, EPI_ISL_2031509, EPI_ISL_2031510, EPI_ISL_2031511, EPI_ISL_2031512, EPI_ISL_2031513, EPI_ISL_2031514, EPI_ISL_2031515, EPI_ISL_2031516, EPI_ISL_2031517, EPI_ISL_2031518, EPI_ISL_2031519, EPI_ISL_2031520, EPI_ISL_2031521, EPI_ISL_2031522, EPI_ISL_2031523, EPI_ISL_2031524, EPI_ISL_2031525, EPI_ISL_2031526, EPI_ISL_2031527, EPI_ISL_2031528, EPI_ISL_2031529, EPI_ISL_2031530, EPI_ISL_2031531, EPI_ISL_2031532, EPI_ISL_2031533, EPI_ISL_2031534, EPI_ISL_2031535, EPI_ISL_2031536, EPI_ISL_2031537, EPI_ISL_2031538, EPI_ISL_2031539, EPI_ISL_2031540, EPI_ISL_2031541, EPI_ISL_2031542, EPI_ISL_2031543, EPI_ISL_2031544, EPI_ISL_2031545, EPI_ISL_2031546, EPI_ISL_2031547, EPI_ISL_2031548, EPI_ISL_2031549, EPI_ISL_2031550, EPI_ISL_2031551, EPI_ISL_2031552, EPI_ISL_2031553, EPI_ISL_2031554, EPI_ISL_2031555, EPI_ISL_2031556, EPI_ISL_2031557, EPI_ISL_2031558, EPI_ISL_2031559, EPI_ISL_2031560, EPI_ISL_2031561, EPI_ISL_2031562, EPI_ISL_2031563, EPI_ISL_2031564, EPI_ISL_2031565, EPI_ISL_2031566, EPI_ISL_2031567, EPI_ISL_2031568, EPI_ISL_2031569, EPI_ISL_2031570, EPI_ISL_2031571, EPI_ISL_2031572, EPI_ISL_2031573, EPI_ISL_2031574, EPI_ISL_2031575, EPI_ISL_2031576, EPI_ISL_2031577, EPI_ISL_2031578, EPI_ISL_2031579, EPI_ISL_2031580, EPI_ISL_2031581, EPI_ISL_2031582, EPI_ISL_2031583, EPI_ISL_2031584, EPI_ISL_2031585, EPI_ISL_2031586, EPI_ISL_2031587, EPI_ISL_2031588, EPI_ISL_2031589, EPI_ISL_2031590, EPI_ISL_2031591, EPI_ISL_2031592 | | | |
| see above | UW Virology Lab | UW Virology Lab | Pavitra Roychoudhury, Hong Xie, Lasata Shrestha, Tien V. Nguyen, Shah Mohamed Bakhash, Michelle Lin, Noah R. Baker, Sean Ellis, Meei-Li Huang, Keith R Jerome, Alexander Greninger |
| EPI_ISL_2031593, EPI_ISL_2031594, EPI_ISL_2031595, EPI_ISL_2031596, EPI_ISL_2031598, EPI_ISL_2031599, EPI_ISL_2031600, EPI_ISL_2031601, EPI_ISL_2031602, EPI_ISL_2031603, EPI_ISL_2031604, EPI_ISL_2031605, EPI_ISL_2031606, EPI_ISL_2031607, EPI_ISL_2031608, EPI_ISL_2031609, EPI_ISL_2031610, EPI_ISL_2031612, EPI_ISL_2031613, EPI_ISL_2031614, EPI_ISL_2031615, EPI_ISL_2031616, EPI_ISL_2031617, EPI_ISL_2031618, EPI_ISL_2031619, EPI_ISL_2031620, EPI_ISL_2031621, EPI_ISL_2031622, EPI_ISL_2031623, EPI_ISL_2031624, EPI_ISL_2031625, EPI_ISL_2031626, EPI_ISL_2031627, EPI_ISL_2031628, EPI_ISL_2031629, EPI_ISL_2031630, EPI_ISL_2031631, EPI_ISL_2031632, EPI_ISL_2031633, EPI_ISL_2031634, EPI_ISL_2031635 | | | |
| see above | Idaho Bureau of Laboratories | Idaho Bureau of Laboratories | R. Beukelman, Matthew Charles Burns, Aimee Ceniseros, Robert L. Voermans, Christopher Ball |
| EPI_ISL_2031636, EPI_ISL_2031637, EPI_ISL_2031638, EPI_ISL_2031639, EPI_ISL_2031640 | University of Utah, School of Medicine | Pathogen Discovery, Respiratory Viruses Branch, Division of Viral Diseases, Centers for Disease Control and Prevention | Ying Tao, Yan Li, Jing Zhang, Anna Kelleher, Brian Lynch, Krista Queen, Anna Uehara, Peter Cook, Han Jia Justin Ng, Rachel Marine, Clinton R. Paden, Dhwani Batra, Haibin Wang, Tara Coalter, Jasmine Padilla, Morgan Davis, Mili Sheth, Sarah Nobles, Mark Burroughs, Justin Lee, Adam Retchless, Suxiang Tong |
| EPI_ISL_2031641 | Johns Hopkins University | Pathogen Discovery, Respiratory Viruses Branch, Division of Viral Diseases, Centers for Disease Control and Prevention | Ying Tao, Yan Li, Jing Zhang, Anna Kelleher, Brian Lynch, Krista Queen, Anna Uehara, Peter Cook, Han Jia Justin Ng, Rachel Marine, Clinton R. Paden, Dhwani Batra, Haibin Wang, Tara Coalter, Jasmine Padilla, Morgan Davis, Mili Sheth, Sarah Nobles, Mark Burroughs, Justin Lee, Adam Retchless, Suxiang Tong |
| EPI_ISL_2031642, EPI_ISL_2031643 | Columbia University | Pathogen Discovery, Respiratory Viruses Branch, Division of Viral Diseases, Centers for Disease Control and Prevention | Ying Tao, Yan Li, Jing Zhang, Anna Kelleher, Brian Lynch, Krista Queen, Anna Uehara, Peter Cook, Han Jia Justin Ng, Rachel Marine, Clinton R. Paden, Dhwani Batra, Haibin Wang, Tara Coalter, Jasmine Padilla, Morgan Davis, Mili Sheth, Sarah Nobles, Mark Burroughs, Justin Lee, Adam Retchless, Suxiang Tong |
| EPI_ISL_2031644 | University of Utah, School of Medicine | Pathogen Discovery, Respiratory Viruses Branch, Division of Viral Diseases, Centers for Disease Control and Prevention | Ying Tao, Yan Li, Jing Zhang, Anna Kelleher, Brian Lynch, Krista Queen, Anna Uehara, Peter Cook, Han Jia Justin Ng, Rachel Marine, Clinton R. Paden, Dhwani Batra, Haibin Wang, Tara Coalter, Jasmine Padilla, Morgan Davis, Mili Sheth, Sarah Nobles, Mark Burroughs, Justin Lee, Adam Retchless, Suxiang Tong |
| EPI_ISL_2031645, EPI_ISL_2031646 | Johns Hopkins University | Pathogen Discovery, Respiratory Viruses Branch, Division of Viral Diseases, Centers for Disease Control and Prevention | Ying Tao, Yan Li, Jing Zhang, Anna Kelleher, Brian Lynch, Krista Queen, Anna Uehara, Peter Cook, Han Jia Justin Ng, Rachel Marine, Clinton R. Paden, Dhwani Batra, Haibin Wang, Tara Coalter, Jasmine Padilla, Morgan Davis, Mili Sheth, Sarah Nobles, Mark Burroughs, Justin Lee, Adam Retchless, Suxiang Tong |
| EPI_ISL_2031647 | Columbia University | Pathogen Discovery, Respiratory Viruses Branch, Division of Viral Diseases, Centers for Disease Control and Prevention | Ying Tao, Yan Li, Jing Zhang, Anna Kelleher, Brian Lynch, Krista Queen, Anna Uehara, Peter Cook, Han Jia Justin Ng, Rachel Marine, Clinton R. Paden, Dhwani Batra, Haibin Wang, Tara Coalter, Jasmine Padilla, Morgan Davis, Mili Sheth, Sarah Nobles, Mark Burroughs, Justin Lee, Adam Retchless, Suxiang Tong |
| EPI_ISL_2031648, EPI_ISL_2031649, EPI_ISL_2031650, EPI_ISL_2031651 | AR Dept. of Health-PHL, Molecular Diagnostics | Pathogen Discovery, Respiratory Viruses Branch, Division of Viral Diseases, Centers for Disease Control and Prevention | Ying Tao, Yan Li, Jing Zhang, Anna Kelleher, Brian Lynch, Krista Queen, Anna Uehara, Peter Cook, Han Jia Justin Ng, Rachel Marine, Clinton R. Paden, Dhwani Batra, Haibin Wang, Tara Coalter, Jasmine Padilla, Morgan Davis, Mili Sheth, Sarah Nobles, Mark Burroughs, Justin Lee, Adam Retchless, Suxiang Tong |
| EPI_ISL_2031652, EPI_ISL_2031653, EPI_ISL_2031654, EPI_ISL_2031655 | NC State Laboratory of Public Health | Pathogen Discovery, Respiratory Viruses Branch, Division of Viral Diseases, Centers for Disease Control and Prevention | Ying Tao, Yan Li, Jing Zhang, Anna Kelleher, Brian Lynch, Krista Queen, Anna Uehara, Peter Cook, Han Jia Justin Ng, Rachel Marine, Clinton R. Paden, Dhwani Batra, Haibin Wang, Tara Coalter, Jasmine Padilla, Morgan Davis, Mili Sheth, Sarah Nobles, Mark Burroughs, Justin Lee, Adam Retchless, Suxiang Tong |
| EPI_ISL_2031656, EPI_ISL_2031657, EPI_ISL_2031658, EPI_ISL_2031659, EPI_ISL_2031660, EPI_ISL_2031661 | OK Public Health Laboratory, Oklahoma State DOH | Pathogen Discovery, Respiratory Viruses Branch, Division of Viral Diseases, Centers for Disease Control and Prevention | Ying Tao, Yan Li, Jing Zhang, Anna Kelleher, Brian Lynch, Krista Queen, Anna Uehara, Peter Cook, Han Jia Justin Ng, Rachel Marine, Clinton R. Paden, Dhwani Batra, Haibin Wang, Tara Coalter, Jasmine Padilla, Morgan Davis, Mili Sheth, Sarah Nobles, Mark Burroughs, Justin Lee, Adam Retchless, Suxiang Tong |
| EPI_ISL_2031663, EPI_ISL_2031664, EPI_ISL_2031665 | GA Department of Public Health Laboratory | Pathogen Discovery, Respiratory Viruses Branch, Division of Viral Diseases, Centers for Disease Control and Prevention | Ying Tao, Yan Li, Jing Zhang, Anna Kelleher, Brian Lynch, Krista Queen, Anna Uehara, Peter Cook, Han Jia Justin Ng, Rachel Marine, Clinton R. Paden, Dhwani Batra, Haibin Wang, Tara Coalter, Jasmine Padilla, Morgan Davis, Mili Sheth, Sarah Nobles, Mark Burroughs, Justin Lee, Adam Retchless, Suxiang Tong |
| EPI_ISL_2031667 | AL Dept. of Public Health, Bureau of Clinical Laboratories | Pathogen Discovery, Respiratory Viruses Branch, Division of Viral Diseases, Centers for Disease Control and Prevention | Ying Tao, Yan Li, Jing Zhang, Anna Kelleher, Brian Lynch, Krista Queen, Anna Uehara, Peter Cook, Han Jia Justin Ng, Rachel Marine, Clinton R. Paden, Dhwani Batra, Haibin Wang, Tara Coalter, Jasmine Padilla, Morgan Davis, Mili Sheth, Sarah Nobles, Mark Burroughs, Justin Lee, Adam Retchless, Suxiang Tong |
| EPI_ISL_2031668, EPI_ISL_2031669, EPI_ISL_2031670, EPI_ISL_2031671, EPI_ISL_2031672 | TN Division of Laboratory Services | Pathogen Discovery, Respiratory Viruses Branch, Division of Viral Diseases, Centers for Disease Control and Prevention | Ying Tao, Yan Li, Jing Zhang, Anna Kelleher, Brian Lynch, Krista Queen, Anna Uehara, Peter Cook, Han Jia Justin Ng, Rachel Marine, Clinton R. Paden, Dhwani Batra, Haibin Wang, Tara Coalter, Jasmine Padilla, Morgan Davis, Mili Sheth, Sarah Nobles, Mark Burroughs, Justin Lee, Adam Retchless, Suxiang Tong |
| EPI_ISL_2031763 | Johns Hopkins University | Pathogen Discovery, Respiratory Viruses Branch, Division of Viral Diseases, Centers for Disease Control and Prevention | Ying Tao, Yan Li, Jing Zhang, Anna Kelleher, Brian Lynch, Krista Queen, Anna Uehara, Peter Cook, Han Jia Justin Ng, Rachel Marine, Clinton R. Paden, Dhwani Batra, Haibin Wang, Tara Coalter, Jasmine Padilla, Morgan Davis, Mili Sheth, Sarah Nobles, Mark Burroughs, Justin Lee, Adam Retchless, Suxiang Tong |
| EPI_ISL_2031764, EPI_ISL_2031765, EPI_ISL_2031766, EPI_ISL_2031767, EPI_ISL_2031768, EPI_ISL_2031769, EPI_ISL_2031770, EPI_ISL_2031771 | Weill Cornell Medicine | New York Genome Center | Michael Zody, Andre Corvelo, Dayna M. Oschwald, Samantha Fennessey, Tom Maniatis, Melissa Cushing, Olivier Elemento, Margaret Elizabeth Ross, Chris Mason, Priya Velu, Hanna Rennert, Arryn Craney, Lars F Westblade |
| EPI_ISL_2031772, EPI_ISL_2031774, EPI_ISL_2031775, EPI_ISL_2031776, EPI_ISL_2031777, EPI_ISL_2031778, EPI_ISL_2031780, EPI_ISL_2031781, EPI_ISL_2031782, EPI_ISL_2031783, EPI_ISL_2031784, EPI_ISL_2031785, EPI_ISL_2031786, EPI_ISL_2031787, EPI_ISL_2031788, EPI_ISL_2031789, EPI_ISL_2031790, EPI_ISL_2031791, EPI_ISL_2031792, EPI_ISL_2031793, EPI_ISL_2031794, EPI_ISL_2031795, EPI_ISL_2031796, EPI_ISL_2031797, EPI_ISL_2031798, EPI_ISL_2031799, EPI_ISL_2031800, EPI_ISL_2031801, EPI_ISL_2031802, EPI_ISL_2031803, EPI_ISL_2031804, EPI_ISL_2031805, EPI_ISL_2031806, EPI_ISL_2031807, EPI_ISL_2031808, EPI_ISL_2031809, EPI_ISL_2031810, EPI_ISL_2031811, EPI_ISL_2031812, EPI_ISL_2031813, EPI_ISL_2031814, EPI_ISL_2031815, EPI_ISL_2031816, EPI_ISL_2031817, EPI_ISL_2031818, EPI_ISL_2031819, EPI_ISL_2031820, EPI_ISL_2031821, EPI_ISL_2031822, EPI_ISL_2031823, EPI_ISL_2031824, EPI_ISL_2031825, EPI_ISL_2031826, EPI_ISL_2031827, EPI_ISL_2031828, EPI_ISL_2031829, EPI_ISL_2031830, EPI_ISL_2031831, EPI_ISL_2031832, EPI_ISL_2031833, EPI_ISL_2031834, EPI_ISL_2031835, EPI_ISL_2031836, EPI_ISL_2031837, EPI_ISL_2031838, EPI_ISL_2031839, EPI_ISL_2031840, EPI_ISL_2031841, EPI_ISL_2031843, EPI_ISL_2031844, EPI_ISL_2031845, EPI_ISL_2031846, EPI_ISL_2031847, EPI_ISL_2031848, EPI_ISL_2031850, EPI_ISL_2031851, EPI_ISL_2031852, EPI_ISL_2031853, EPI_ISL_2031854, EPI_ISL_2031855, EPI_ISL_2031856 | | | |
| see above | UCSC Genomics Institute | UCSC Genomics Institute | Mark Akeson, Ikenna Anigbogu, Eric Beraut, Isabel Bjork, Molly Cassatt-Johnstone, Terren Chang, Russell Corbett-Detig, Namrita Dhillon, Maximilian Haeussler, David Haussler, Angie Hinrichs, Miten Jain, Joshua Kapp, A. Marm Kilpatrick, Jakob McBroome, Hugh Olsen, Jeremy Sanford, Beth Shapiro, Michael Stone, Bryan Thornlow, Yatish Turakhia, Ciara Wanket |
| EPI_ISL_2031858 | VA Medical Center | Los Angeles County Public Health Laboratories | P. Hemarajata et al. |
| EPI_ISL_2031860 | Flow Diagnostics Lab | Los Angeles County Public Health Laboratories | P. Hemarajata et al. |
| EPI_ISL_2031979, EPI_ISL_2031980, EPI_ISL_2031981, EPI_ISL_2031982, EPI_ISL_2031983, EPI_ISL_2031984, EPI_ISL_2031985, EPI_ISL_2031986, EPI_ISL_2031987, EPI_ISL_2031988, EPI_ISL_2031989, EPI_ISL_2031990, EPI_ISL_2031991, EPI_ISL_2031992, EPI_ISL_2031993, EPI_ISL_2031994, EPI_ISL_2031995, EPI_ISL_2031996, EPI_ISL_2031997 | | | |
| see above | Los Angeles County Public Health Laboratories | Los Angeles County Public Health Laboratories | P. Hemarajata et al. |
| EPI_ISL_2032397, EPI_ISL_2032399, EPI_ISL_2032401, EPI_ISL_2032403, EPI_ISL_2032407, EPI_ISL_2032410, EPI_ISL_2032412, EPI_ISL_2032414, EPI_ISL_2032418, EPI_ISL_2032422, EPI_ISL_2032427, EPI_ISL_2032431, EPI_ISL_2032433, EPI_ISL_2032435, EPI_ISL_2032437 | | | |
| see above | Nevada State Public Health Laboratory | Nevada State Public Health Laboratory | Andrew Gorzalski, Mark Pandori |
| EPI_ISL_2034694, EPI_ISL_2034695, EPI_ISL_2034696, EPI_ISL_2034697, EPI_ISL_2034698, EPI_ISL_2034699, EPI_ISL_2034700, EPI_ISL_2034701, EPI_ISL_2034702, EPI_ISL_2034703, EPI_ISL_2034704, EPI_ISL_2034705, EPI_ISL_2034706, EPI_ISL_2034707, EPI_ISL_2034708, EPI_ISL_2034710 | | | |
| see above | Connecticut DPH | Yale Center for Genomic Analysis | Joseph Fauver, Mallery Breban, Isabel Ott, Tara Alpert, Mary Petrone, Anderson Brito, Chantal Vogels, Annie Watkins, Chaney Kalinich, Jessica Rothman, Anthony Muyombwe, Randy Downing, Jafar Razeq, Stephen M. Bart, Nathan Grubaugh, Shrikant Mane, Kaya Bilguvar, Curt Scharfe, Irina Tikhonova, Brooke Sullivan |

| | | | |
|---|---|---|---|
| EPI_ISL_2034712 | Quest Diagnostics Incorporated | Centers for Disease Control and Prevention Division of Viral Diseases, Pathogen Discovery | Dakota Howard, Dhwani Batra, Peter W. Cook, Kara Moser, Adrian Paskey, Jason Caravas, Benjamin Rambo-Martin, Shatavia Morrison, Christopher Gulvick, Scott Sammons, Yvette Unoarumhi, Darlene Wagner, Matthew Schmerer, S. H. Rosenthal, A. Gerasimova, R. M. Kagan, B. Anderson, M. Hua, Y. Liu, L.E. Bernstein, K.E. Livingston, A. Perez, I. A. Shlyakhter, R. V. Rolando, R. Owen, P. Tanpaiboon, F. Lacbawan, Clinton R. Paden, Duncan MacCannell |
| EPI_ISL_2034713 | Connecticut DPH | Yale Center for Genomic Analysis | Joseph Fauver, Mallery Breban, Isabel Ott, Tara Alpert, Mary Petrone, Anderson Brito, Chantal Vogels, Annie Watkins, Chaney Kalinich, Jessica Rothman, Anthony Muyombwe, Randy Downing, Jafar Razeq, Stephen M. Bart, Nathan Grubaugh, Shrikant Mane, Kaya Bilguvar, Curt Scharfe, Irina Tikhonova, Brooke Sullivan |
| EPI_ISL_2034714 | Quest Diagnostics Incorporated | Centers for Disease Control and Prevention Division of Viral Diseases, Pathogen Discovery | Dakota Howard, Dhwani Batra, Peter W. Cook, Kara Moser, Adrian Paskey, Jason Caravas, Benjamin Rambo-Martin, Shatavia Morrison, Christopher Gulvick, Scott Sammons, Yvette Unoarumhi, Darlene Wagner, Matthew Schmerer, S. H. Rosenthal, A. Gerasimova, R. M. Kagan, B. Anderson, M. Hua, Y. Liu, L.E. Bernstein, K.E. Livingston, A. Perez, I. A. Shlyakhter, R. V. Rolando, R. Owen, P. Tanpaiboon, F. Lacbawan, Clinton R. Paden, Duncan MacCannell |
| EPI_ISL_2034715 | Connecticut DPH | Yale Center for Genomic Analysis | Joseph Fauver, Mallery Breban, Isabel Ott, Tara Alpert, Mary Petrone, Anderson Brito, Chantal Vogels, Annie Watkins, Chaney Kalinich, Jessica Rothman, Anthony Muyombwe, Randy Downing, Jafar Razeq, Stephen M. Bart, Nathan Grubaugh, Shrikant Mane, Kaya Bilguvar, Curt Scharfe, Irina Tikhonova, Brooke Sullivan |
| EPI_ISL_2034716, EPI_ISL_2034718, EPI_ISL_2034720 | Quest Diagnostics Incorporated | Centers for Disease Control and Prevention Division of Viral Diseases, Pathogen Discovery | Dakota Howard, Dhwani Batra, Peter W. Cook, Kara Moser, Adrian Paskey, Jason Caravas, Benjamin Rambo-Martin, Shatavia Morrison, Christopher Gulvick, Scott Sammons, Yvette Unoarumhi, Darlene Wagner, Matthew Schmerer, S. H. Rosenthal, A. Gerasimova, R. M. Kagan, B. Anderson, M. Hua, Y. Liu, L.E. Bernstein, K.E. Livingston, A. Perez, I. A. Shlyakhter, R. V. Rolando, R. Owen, P. Tanpaiboon, F. Lacbawan, Clinton R. Paden, Duncan MacCannell |
| EPI_ISL_2034721 | Connecticut DPH | Yale Center for Genomic Analysis | Joseph Fauver, Mallery Breban, Isabel Ott, Tara Alpert, Mary Petrone, Anderson Brito, Chantal Vogels, Annie Watkins, Chaney Kalinich, Jessica Rothman, Anthony Muyombwe, Randy Downing, Jafar Razeq, Stephen M. Bart, Nathan Grubaugh, Shrikant Mane, Kaya Bilguvar, Curt Scharfe, Irina Tikhonova, Brooke Sullivan |
| EPI_ISL_2034722 | Quest Diagnostics Incorporated | Centers for Disease Control and Prevention Division of Viral Diseases, Pathogen Discovery | Dakota Howard, Dhwani Batra, Peter W. Cook, Kara Moser, Adrian Paskey, Jason Caravas, Benjamin Rambo-Martin, Shatavia Morrison, Christopher Gulvick, Scott Sammons, Yvette Unoarumhi, Darlene Wagner, Matthew Schmerer, S. H. Rosenthal, A. Gerasimova, R. M. Kagan, B. Anderson, M. Hua, Y. Liu, L.E. Bernstein, K.E. Livingston, A. Perez, I. A. Shlyakhter, R. V. Rolando, R. Owen, P. Tanpaiboon, F. Lacbawan, Clinton R. Paden, Duncan MacCannell |
| EPI_ISL_2034723 | Connecticut DPH | Yale Center for Genomic Analysis | Joseph Fauver, Mallery Breban, Isabel Ott, Tara Alpert, Mary Petrone, Anderson Brito, Chantal Vogels, Annie Watkins, Chaney Kalinich, Jessica Rothman, Anthony Muyombwe, Randy Downing, Jafar Razeq, Stephen M. Bart, Nathan Grubaugh, Shrikant Mane, Kaya Bilguvar, Curt Scharfe, Irina Tikhonova, Brooke Sullivan |
| EPI_ISL_2034724 | Quest Diagnostics Incorporated | Centers for Disease Control and Prevention Division of Viral Diseases, Pathogen Discovery | Dakota Howard, Dhwani Batra, Peter W. Cook, Kara Moser, Adrian Paskey, Jason Caravas, Benjamin Rambo-Martin, Shatavia Morrison, Christopher Gulvick, Scott Sammons, Yvette Unoarumhi, Darlene Wagner, Matthew Schmerer, S. H. Rosenthal, A. Gerasimova, R. M. Kagan, B. Anderson, M. Hua, Y. Liu, L.E. Bernstein, K.E. Livingston, A. Perez, I. A. Shlyakhter, R. V. Rolando, R. Owen, P. Tanpaiboon, F. Lacbawan, Clinton R. Paden, Duncan MacCannell |
| EPI_ISL_2034725, EPI_ISL_2034726 | Connecticut DPH | Yale Center for Genomic Analysis | Joseph Fauver, Mallery Breban, Isabel Ott, Tara Alpert, Mary Petrone, Anderson Brito, Chantal Vogels, Annie Watkins, Chaney Kalinich, Jessica Rothman, Anthony Muyombwe, Randy Downing, Jafar Razeq, Stephen M. Bart, Nathan Grubaugh, Shrikant Mane, Kaya Bilguvar, Curt Scharfe, Irina Tikhonova, Brooke Sullivan |
| EPI_ISL_2034727 | Quest Diagnostics Incorporated | Centers for Disease Control and Prevention Division of Viral Diseases, Pathogen Discovery | Dakota Howard, Dhwani Batra, Peter W. Cook, Kara Moser, Adrian Paskey, Jason Caravas, Benjamin Rambo-Martin, Shatavia Morrison, Christopher Gulvick, Scott Sammons, Yvette Unoarumhi, Darlene Wagner, Matthew Schmerer, S. H. Rosenthal, A. Gerasimova, R. M. Kagan, B. Anderson, M. Hua, Y. Liu, L.E. Bernstein, K.E. Livingston, A. Perez, I. A. Shlyakhter, R. V. Rolando, R. Owen, P. Tanpaiboon, F. Lacbawan, Clinton R. Paden, Duncan MacCannell |
| EPI_ISL_2034728 | Connecticut DPH | Yale Center for Genomic Analysis | Joseph Fauver, Mallery Breban, Isabel Ott, Tara Alpert, Mary Petrone, Anderson Brito, Chantal Vogels, Annie Watkins, Chaney Kalinich, Jessica Rothman, Anthony Muyombwe, Randy Downing, Jafar Razeq, Stephen M. Bart, Nathan Grubaugh, Shrikant Mane, Kaya Bilguvar, Curt Scharfe, Irina Tikhonova, Brooke Sullivan |
| EPI_ISL_2034729 | Quest Diagnostics Incorporated | Centers for Disease Control and Prevention Division of Viral Diseases, Pathogen Discovery | Dakota Howard, Dhwani Batra, Peter W. Cook, Kara Moser, Adrian Paskey, Jason Caravas, Benjamin Rambo-Martin, Shatavia Morrison, Christopher Gulvick, Scott Sammons, Yvette Unoarumhi, Darlene Wagner, Matthew Schmerer, S. H. Rosenthal, A. Gerasimova, R. M. Kagan, B. Anderson, M. Hua, Y. Liu, L.E. Bernstein, K.E. Livingston, A. Perez, I. A. Shlyakhter, R. V. Rolando, R. Owen, P. Tanpaiboon, F. Lacbawan, Clinton R. Paden, Duncan MacCannell |
| EPI_ISL_2034730, EPI_ISL_2034731 | Connecticut DPH | Yale Center for Genomic Analysis | Joseph Fauver, Mallery Breban, Isabel Ott, Tara Alpert, Mary Petrone, Anderson Brito, Chantal Vogels, Annie Watkins, Chaney Kalinich, Jessica Rothman, Anthony Muyombwe, Randy Downing, Jafar Razeq, Stephen M. Bart, Nathan Grubaugh, Shrikant Mane, Kaya Bilguvar, Curt Scharfe, Irina Tikhonova, Brooke Sullivan |
| EPI_ISL_2034732 | Quest Diagnostics Incorporated | Centers for Disease Control and Prevention Division of Viral Diseases, Pathogen Discovery | Dakota Howard, Dhwani Batra, Peter W. Cook, Kara Moser, Adrian Paskey, Jason Caravas, Benjamin Rambo-Martin, Shatavia Morrison, Christopher Gulvick, Scott Sammons, Yvette Unoarumhi, Darlene Wagner, Matthew Schmerer, S. H. Rosenthal, A. Gerasimova, R. M. Kagan, B. Anderson, M. Hua, Y. Liu, L.E. Bernstein, K.E. Livingston, A. Perez, I. A. Shlyakhter, R. V. Rolando, R. Owen, P. Tanpaiboon, F. Lacbawan, Clinton R. Paden, Duncan MacCannell |
| EPI_ISL_2034733, EPI_ISL_2034735 | Connecticut DPH | Yale Center for Genomic Analysis | Joseph Fauver, Mallery Breban, Isabel Ott, Tara Alpert, Mary Petrone, Anderson Brito, Chantal Vogels, Annie Watkins, Chaney Kalinich, Jessica Rothman, Anthony Muyombwe, Randy Downing, Jafar Razeq, Stephen M. Bart, Nathan Grubaugh, Shrikant Mane, Kaya Bilguvar, Curt Scharfe, Irina Tikhonova, Brooke Sullivan |
| EPI_ISL_2034736 | Quest Diagnostics Incorporated | Centers for Disease Control and Prevention Division of Viral Diseases, Pathogen Discovery | Dakota Howard, Dhwani Batra, Peter W. Cook, Kara Moser, Adrian Paskey, Jason Caravas, Benjamin Rambo-Martin, Shatavia Morrison, Christopher Gulvick, Scott Sammons, Yvette Unoarumhi, Darlene Wagner, Matthew Schmerer, S. H. Rosenthal, A. Gerasimova, R. M. Kagan, B. Anderson, M. Hua, Y. Liu, L.E. Bernstein, K.E. Livingston, A. Perez, I. A. Shlyakhter, R. V. Rolando, R. Owen, P. Tanpaiboon, F. Lacbawan, Clinton R. Paden, Duncan MacCannell |
| EPI_ISL_2034737, EPI_ISL_2034738 | Connecticut DPH | Yale Center for Genomic Analysis | Joseph Fauver, Mallery Breban, Isabel Ott, Tara Alpert, Mary Petrone, Anderson Brito, Chantal Vogels, Annie Watkins, Chaney Kalinich, Jessica Rothman, Anthony Muyombwe, Randy Downing, Jafar Razeq, Stephen M. Bart, Nathan Grubaugh, Shrikant Mane, Kaya Bilguvar, Curt Scharfe, Irina Tikhonova, Brooke Sullivan |
| EPI_ISL_2034739, EPI_ISL_2034740 | Quest Diagnostics Incorporated | Centers for Disease Control and Prevention Division of Viral Diseases, Pathogen Discovery | Dakota Howard, Dhwani Batra, Peter W. Cook, Kara Moser, Adrian Paskey, Jason Caravas, Benjamin Rambo-Martin, Shatavia Morrison, Christopher Gulvick, Scott Sammons, Yvette Unoarumhi, Darlene Wagner, Matthew Schmerer, S. H. Rosenthal, A. Gerasimova, R. M. Kagan, B. Anderson, M. Hua, Y. Liu, L.E. Bernstein, K.E. Livingston, A. Perez, I. A. Shlyakhter, R. V. Rolando, R. Owen, P. Tanpaiboon, F. Lacbawan, Clinton R. Paden, Duncan MacCannell |
| EPI_ISL_2034741 | Connecticut DPH | Yale Center for Genomic Analysis | Joseph Fauver, Mallery Breban, Isabel Ott, Tara Alpert, Mary Petrone, Anderson Brito, Chantal Vogels, Annie Watkins, Chaney Kalinich, Jessica Rothman, Anthony Muyombwe, Randy Downing, Jafar Razeq, Stephen M. Bart, Nathan Grubaugh, Shrikant Mane, Kaya Bilguvar, Curt Scharfe, Irina Tikhonova, Brooke Sullivan |
| EPI_ISL_2034742 | Quest Diagnostics Incorporated | Centers for Disease Control and Prevention Division of Viral Diseases, Pathogen Discovery | Dakota Howard, Dhwani Batra, Peter W. Cook, Kara Moser, Adrian Paskey, Jason Caravas, Benjamin Rambo-Martin, Shatavia Morrison, Christopher Gulvick, Scott Sammons, Yvette Unoarumhi, Darlene Wagner, Matthew Schmerer, S. H. Rosenthal, A. Gerasimova, R. M. Kagan, B. Anderson, M. Hua, Y. Liu, L.E. Bernstein, K.E. Livingston, A. Perez, I. A. Shlyakhter, R. V. Rolando, R. Owen, P. Tanpaiboon, F. Lacbawan, Clinton R. Paden, Duncan MacCannell |
| EPI_ISL_2034743 | Connecticut DPH | Yale Center for Genomic Analysis | Joseph Fauver, Mallery Breban, Isabel Ott, Tara Alpert, Mary Petrone, Anderson Brito, Chantal Vogels, Annie Watkins, Chaney Kalinich, Jessica Rothman, Anthony Muyombwe, Randy Downing, Jafar Razeq, Stephen M. Bart, Nathan Grubaugh, Shrikant Mane, Kaya Bilguvar, Curt Scharfe, Irina Tikhonova, Brooke Sullivan |
| EPI_ISL_2034744 | Quest Diagnostics Incorporated | Centers for Disease Control and Prevention Division of Viral Diseases, Pathogen Discovery | Dakota Howard, Dhwani Batra, Peter W. Cook, Kara Moser, Adrian Paskey, Jason Caravas, Benjamin Rambo-Martin, Shatavia Morrison, Christopher Gulvick, Scott Sammons, Yvette Unoarumhi, Darlene Wagner, Matthew Schmerer, S. H. Rosenthal, A. Gerasimova, R. M. Kagan, B. Anderson, M. Hua, Y. Liu, L.E. Bernstein, K.E. Livingston, A. Perez, I. A. Shlyakhter, R. V. Rolando, R. Owen, P. Tanpaiboon, F. Lacbawan, Clinton R. Paden, Duncan MacCannell |
| EPI_ISL_2034745 | Connecticut DPH | Yale Center for Genomic Analysis | Joseph Fauver, Mallery Breban, Isabel Ott, Tara Alpert, Mary Petrone, Anderson Brito, Chantal Vogels, Annie Watkins, Chaney Kalinich, Jessica Rothman, Anthony Muyombwe, Randy Downing, Jafar Razeq, Stephen M. Bart, Nathan Grubaugh, Shrikant Mane, Kaya Bilguvar, Curt Scharfe, Irina Tikhonova, Brooke Sullivan |
| EPI_ISL_2034746 | Quest Diagnostics Incorporated | Centers for Disease Control and Prevention Division of Viral Diseases, Pathogen Discovery | Dakota Howard, Dhwani Batra, Peter W. Cook, Kara Moser, Adrian Paskey, Jason Caravas, Benjamin Rambo-Martin, Shatavia Morrison, Christopher Gulvick, Scott Sammons, Yvette Unoarumhi, Darlene Wagner, Matthew Schmerer, S. H. Rosenthal, A. Gerasimova, R. M. Kagan, B. Anderson, M. Hua, Y. Liu, L.E. Bernstein, K.E. Livingston, A. Perez, I. A. Shlyakhter, R. V. Rolando, R. Owen, P. Tanpaiboon, F. Lacbawan, Clinton R. Paden, Duncan MacCannell |
| EPI_ISL_2034747, EPI_ISL_2034748 | Connecticut DPH | Yale Center for Genomic Analysis | Joseph Fauver, Mallery Breban, Isabel Ott, Tara Alpert, Mary Petrone, Anderson Brito, Chantal Vogels, Annie Watkins, Chaney Kalinich, Jessica Rothman, Anthony Muyombwe, Randy Downing, Jafar Razeq, Stephen M. Bart, Nathan Grubaugh, Shrikant Mane, Kaya Bilguvar, Curt Scharfe, Irina Tikhonova, Brooke Sullivan |
| EPI_ISL_2034749 | Quest Diagnostics Incorporated | Centers for Disease Control and Prevention Division of Viral Diseases, Pathogen Discovery | Dakota Howard, Dhwani Batra, Peter W. Cook, Kara Moser, Adrian Paskey, Jason Caravas, Benjamin Rambo-Martin, Shatavia Morrison, Christopher Gulvick, Scott Sammons, Yvette Unoarumhi, Darlene Wagner, Matthew Schmerer, S. H. Rosenthal, A. Gerasimova, R. M. Kagan, B. Anderson, M. Hua, Y. Liu, L.E. Bernstein, K.E. Livingston, A. Perez, I. A. Shlyakhter, R. V. Rolando, R. Owen, P. Tanpaiboon, F. Lacbawan, Clinton R. Paden, Duncan MacCannell |
| EPI_ISL_2034750 | Connecticut DPH | Yale Center for Genomic Analysis | Joseph Fauver, Mallery Breban, Isabel Ott, Tara Alpert, Mary Petrone, Anderson Brito, Chantal Vogels, Annie Watkins, Chaney Kalinich, Jessica Rothman, Anthony Muyombwe, Randy Downing, Jafar Razeq, Stephen M. Bart, Nathan Grubaugh, Shrikant Mane, Kaya Bilguvar, Curt Scharfe, Irina Tikhonova, Brooke Sullivan |
| EPI_ISL_2034751 | Quest Diagnostics Incorporated | Centers for Disease Control and Prevention Division of Viral Diseases, Pathogen Discovery | Dakota Howard, Dhwani Batra, Peter W. Cook, Kara Moser, Adrian Paskey, Jason Caravas, Benjamin Rambo-Martin, Shatavia Morrison, Christopher Gulvick, Scott Sammons, Yvette Unoarumhi, Darlene Wagner, Matthew Schmerer, S. H. Rosenthal, A. Gerasimova, R. M. Kagan, B. Anderson, M. Hua, Y. Liu, L.E. Bernstein, K.E. Livingston, A. Perez, I. A. Shlyakhter, R. V. Rolando, R. Owen, P. Tanpaiboon, F. Lacbawan, Clinton R. Paden, Duncan MacCannell |
| EPI_ISL_2034752, EPI_ISL_2034753 | Connecticut DPH | Yale Center for Genomic Analysis | Joseph Fauver, Mallery Breban, Isabel Ott, Tara Alpert, Mary Petrone, Anderson Brito, Chantal Vogels, Annie Watkins, Chaney Kalinich, Jessica Rothman, Anthony Muyombwe, Randy Downing, Jafar Razeq, Stephen M. Bart, Nathan Grubaugh, Shrikant Mane, Kaya Bilguvar, Curt Scharfe, Irina Tikhonova, Brooke Sullivan |
| EPI_ISL_2034754 | Yale Clinical Virology Lab | Yale Center for Genomic Analysis | Shrikant Mane, Kaya Bilguvar, Curt Scharfe, Irina Tikhonova, Brooke Sullivan |
| EPI_ISL_2034755 | Quest Diagnostics Incorporated | Centers for Disease Control and Prevention Division of Viral Diseases, Pathogen Discovery | Dakota Howard, Dhwani Batra, Peter W. Cook, Kara Moser, Adrian Paskey, Jason Caravas, Benjamin Rambo-Martin, Shatavia Morrison, Christopher Gulvick, Scott Sammons, Yvette Unoarumhi, Darlene Wagner, Matthew Schmerer, S. H. Rosenthal, A. Gerasimova, R. M. Kagan, B. Anderson, M. Hua, Y. Liu, L.E. Bernstein, K.E. Livingston, A. Perez, I. A. Shlyakhter, R. V. Rolando, R. Owen, P. Tanpaiboon, F. Lacbawan, Clinton R. Paden, Duncan MacCannell |
| EPI_ISL_2034756 | Yale Clinical Virology Lab | Yale Center for Genomic Analysis | Shrikant Mane, Kaya Bilguvar, Curt Scharfe, Irina Tikhonova, Brooke Sullivan |
| EPI_ISL_2034758 | Quest Diagnostics Incorporated | Centers for Disease Control and Prevention Division of Viral Diseases, Pathogen Discovery | Dakota Howard, Dhwani Batra, Peter W. Cook, Kara Moser, Adrian Paskey, Jason Caravas, Benjamin Rambo-Martin, Shatavia Morrison, Christopher Gulvick, Scott Sammons, Yvette Unoarumhi, Darlene Wagner, Matthew Schmerer, S. H. Rosenthal, A. Gerasimova, R. M. Kagan, B. Anderson, M. Hua, Y. Liu, L.E. Bernstein, K.E. Livingston, A. Perez, I. A. Shlyakhter, R. V. Rolando, R. Owen, P. Tanpaiboon, F. Lacbawan, Clinton R. Paden, Duncan MacCannell |
| EPI_ISL_2034759, EPI_ISL_2034761 | Yale Clinical Virology Lab | Yale Center for Genomic Analysis | Shrikant Mane, Kaya Bilguvar, Curt Scharfe, Irina Tikhonova, Brooke Sullivan |
| EPI_ISL_2034762 | Quest Diagnostics Incorporated | Centers for Disease Control and Prevention Division of Viral Diseases, Pathogen Discovery | Dakota Howard, Dhwani Batra, Peter W. Cook, Kara Moser, Adrian Paskey, Jason Caravas, Benjamin Rambo-Martin, Shatavia Morrison, Christopher Gulvick, Scott Sammons, Yvette Unoarumhi, Darlene Wagner, Matthew Schmerer, S. H. Rosenthal, A. Gerasimova, R. M. Kagan, B. Anderson, M. Hua, Y. Liu, L.E. Bernstein, K.E. Livingston, A. Perez, I. A. Shlyakhter, R. V. Rolando, R. Owen, P. Tanpaiboon, F. Lacbawan, Clinton R. Paden, Duncan MacCannell |
| EPI_ISL_2034763, EPI_ISL_2034764, EPI_ISL_2034765 | Yale Clinical Virology Lab | Yale Center for Genomic Analysis | Shrikant Mane, Kaya Bilguvar, Curt Scharfe, Irina Tikhonova, Brooke Sullivan |
| EPI_ISL_2034766, EPI_ISL_2034769 | Quest Diagnostics Incorporated | Centers for Disease Control and Prevention Division of Viral Diseases, Pathogen Discovery | Dakota Howard, Dhwani Batra, Peter W. Cook, Kara Moser, Adrian Paskey, Jason Caravas, Benjamin Rambo-Martin, Shatavia Morrison, Christopher Gulvick, Scott Sammons, Yvette Unoarumhi, Darlene Wagner, Matthew Schmerer, S. H. Rosenthal, A. Gerasimova, R. M. Kagan, B. Anderson, M. Hua, Y. Liu, L.E. Bernstein, K.E. Livingston, A. Perez, I. A. Shlyakhter, R. V. Rolando, R. Owen, P. Tanpaiboon, F. Lacbawan, Clinton R. Paden, Duncan MacCannell |

| | | | |
|---|---|---|---|
| EPI_ISL_2034770, EPI_ISL_2034771, EPI_ISL_2034772 | Yale Clinical Virology Lab | Yale Center for Genomic Analysis | Shrikant Mane, Kaya Bilguvar, Curt Scharfe, Irina Tikhonova, Brooke Sullivan |
| EPI_ISL_2034773 | Quest Diagnostics Incorporated | Centers for Disease Control and Prevention Division of Viral Diseases, Pathogen Discovery | Dakota Howard, Dhwani Batra, Peter W. Cook, Kara Moser, Adrian Paskey, Jason Caravas, Benjamin Rambo-Martin, Shatavia Morrison, Christopher Gulvick, Scott Sammons, Yvette Unoarumhi, Darlene Wagner, Matthew Schmerer, S. H. Rosenthal, A. Gerasimova, R. M. Kagan, B. Anderson, M. Hua, Y. Liu, L.E. Bernstein, K.E. Livingston, A. Perez, I. A. Shlyakhter, R. V. Rolando, R. Owen, P. Tanpaiboon, F. Lacbawan, Clinton R. Paden, Duncan MacCannell |
| EPI_ISL_2034774, EPI_ISL_2034775 | Yale Clinical Virology Lab | Yale Center for Genomic Analysis | Shrikant Mane, Kaya Bilguvar, Curt Scharfe, Irina Tikhonova, Brooke Sullivan |
| EPI_ISL_2034776 | Quest Diagnostics Incorporated | Centers for Disease Control and Prevention Division of Viral Diseases, Pathogen Discovery | Dakota Howard, Dhwani Batra, Peter W. Cook, Kara Moser, Adrian Paskey, Jason Caravas, Benjamin Rambo-Martin, Shatavia Morrison, Christopher Gulvick, Scott Sammons, Yvette Unoarumhi, Darlene Wagner, Matthew Schmerer, S. H. Rosenthal, A. Gerasimova, R. M. Kagan, B. Anderson, M. Hua, Y. Liu, L.E. Bernstein, K.E. Livingston, A. Perez, I. A. Shlyakhter, R. V. Rolando, R. Owen, P. Tanpaiboon, F. Lacbawan, Clinton R. Paden, Duncan MacCannell |
| EPI_ISL_2034777, EPI_ISL_2034778 | Yale Clinical Virology Lab | Yale Center for Genomic Analysis | Shrikant Mane, Kaya Bilguvar, Curt Scharfe, Irina Tikhonova, Brooke Sullivan |
| EPI_ISL_2034779 | Quest Diagnostics Incorporated | Centers for Disease Control and Prevention Division of Viral Diseases, Pathogen Discovery | Dakota Howard, Dhwani Batra, Peter W. Cook, Kara Moser, Adrian Paskey, Jason Caravas, Benjamin Rambo-Martin, Shatavia Morrison, Christopher Gulvick, Scott Sammons, Yvette Unoarumhi, Darlene Wagner, Matthew Schmerer, S. H. Rosenthal, A. Gerasimova, R. M. Kagan, B. Anderson, M. Hua, Y. Liu, L.E. Bernstein, K.E. Livingston, A. Perez, I. A. Shlyakhter, R. V. Rolando, R. Owen, P. Tanpaiboon, F. Lacbawan, Clinton R. Paden, Duncan MacCannell |
| EPI_ISL_2034780, EPI_ISL_2034781 | Yale Clinical Virology Lab | Yale Center for Genomic Analysis | Shrikant Mane, Kaya Bilguvar, Curt Scharfe, Irina Tikhonova, Brooke Sullivan |
| EPI_ISL_2034782 | Quest Diagnostics Incorporated | Centers for Disease Control and Prevention Division of Viral Diseases, Pathogen Discovery | Dakota Howard, Dhwani Batra, Peter W. Cook, Kara Moser, Adrian Paskey, Jason Caravas, Benjamin Rambo-Martin, Shatavia Morrison, Christopher Gulvick, Scott Sammons, Yvette Unoarumhi, Darlene Wagner, Matthew Schmerer, S. H. Rosenthal, A. Gerasimova, R. M. Kagan, B. Anderson, M. Hua, Y. Liu, L.E. Bernstein, K.E. Livingston, A. Perez, I. A. Shlyakhter, R. V. Rolando, R. Owen, P. Tanpaiboon, F. Lacbawan, Clinton R. Paden, Duncan MacCannell |
| EPI_ISL_2034783, EPI_ISL_2034784 | Yale Clinical Virology Lab | Yale Center for Genomic Analysis | Shrikant Mane, Kaya Bilguvar, Curt Scharfe, Irina Tikhonova, Brooke Sullivan |
| EPI_ISL_2034785 | Quest Diagnostics Incorporated | Centers for Disease Control and Prevention Division of Viral Diseases, Pathogen Discovery | Dakota Howard, Dhwani Batra, Peter W. Cook, Kara Moser, Adrian Paskey, Jason Caravas, Benjamin Rambo-Martin, Shatavia Morrison, Christopher Gulvick, Scott Sammons, Yvette Unoarumhi, Darlene Wagner, Matthew Schmerer, S. H. Rosenthal, A. Gerasimova, R. M. Kagan, B. Anderson, M. Hua, Y. Liu, L.E. Bernstein, K.E. Livingston, A. Perez, I. A. Shlyakhter, R. V. Rolando, R. Owen, P. Tanpaiboon, F. Lacbawan, Clinton R. Paden, Duncan MacCannell |
| EPI_ISL_2034786, EPI_ISL_2034787, EPI_ISL_2034788 | Yale Clinical Virology Lab | Yale Center for Genomic Analysis | Shrikant Mane, Kaya Bilguvar, Curt Scharfe, Irina Tikhonova, Brooke Sullivan |
| EPI_ISL_2034789 | Quest Diagnostics Incorporated | Centers for Disease Control and Prevention Division of Viral Diseases, Pathogen Discovery | Dakota Howard, Dhwani Batra, Peter W. Cook, Kara Moser, Adrian Paskey, Jason Caravas, Benjamin Rambo-Martin, Shatavia Morrison, Christopher Gulvick, Scott Sammons, Yvette Unoarumhi, Darlene Wagner, Matthew Schmerer, S. H. Rosenthal, A. Gerasimova, R. M. Kagan, B. Anderson, M. Hua, Y. Liu, L.E. Bernstein, K.E. Livingston, A. Perez, I. A. Shlyakhter, R. V. Rolando, R. Owen, P. Tanpaiboon, F. Lacbawan, Clinton R. Paden, Duncan MacCannell |
| EPI_ISL_2034790, EPI_ISL_2034791, EPI_ISL_2034792 | Yale Clinical Virology Lab | Yale Center for Genomic Analysis | Shrikant Mane, Kaya Bilguvar, Curt Scharfe, Irina Tikhonova, Brooke Sullivan |
| EPI_ISL_2034793 | Quest Diagnostics Incorporated | Centers for Disease Control and Prevention Division of Viral Diseases, Pathogen Discovery | Dakota Howard, Dhwani Batra, Peter W. Cook, Kara Moser, Adrian Paskey, Jason Caravas, Benjamin Rambo-Martin, Shatavia Morrison, Christopher Gulvick, Scott Sammons, Yvette Unoarumhi, Darlene Wagner, Matthew Schmerer, S. H. Rosenthal, A. Gerasimova, R. M. Kagan, B. Anderson, M. Hua, Y. Liu, L.E. Bernstein, K.E. Livingston, A. Perez, I. A. Shlyakhter, R. V. Rolando, R. Owen, P. Tanpaiboon, F. Lacbawan, Clinton R. Paden, Duncan MacCannell |
| EPI_ISL_2034794, EPI_ISL_2034795 | Yale Clinical Virology Lab | Yale Center for Genomic Analysis | Shrikant Mane, Kaya Bilguvar, Curt Scharfe, Irina Tikhonova, Brooke Sullivan |
| EPI_ISL_2034796 | Quest Diagnostics Incorporated | Centers for Disease Control and Prevention Division of Viral Diseases, Pathogen Discovery | Dakota Howard, Dhwani Batra, Peter W. Cook, Kara Moser, Adrian Paskey, Jason Caravas, Benjamin Rambo-Martin, Shatavia Morrison, Christopher Gulvick, Scott Sammons, Yvette Unoarumhi, Darlene Wagner, Matthew Schmerer, S. H. Rosenthal, A. Gerasimova, R. M. Kagan, B. Anderson, M. Hua, Y. Liu, L.E. Bernstein, K.E. Livingston, A. Perez, I. A. Shlyakhter, R. V. Rolando, R. Owen, P. Tanpaiboon, F. Lacbawan, Clinton R. Paden, Duncan MacCannell |
| EPI_ISL_2034797, EPI_ISL_2034798 | Yale Clinical Virology Lab | Yale Center for Genomic Analysis | Shrikant Mane, Kaya Bilguvar, Curt Scharfe, Irina Tikhonova, Brooke Sullivan |
| EPI_ISL_2034799 | Quest Diagnostics Incorporated | Centers for Disease Control and Prevention Division of Viral Diseases, Pathogen Discovery | Dakota Howard, Dhwani Batra, Peter W. Cook, Kara Moser, Adrian Paskey, Jason Caravas, Benjamin Rambo-Martin, Shatavia Morrison, Christopher Gulvick, Scott Sammons, Yvette Unoarumhi, Darlene Wagner, Matthew Schmerer, S. H. Rosenthal, A. Gerasimova, R. M. Kagan, B. Anderson, M. Hua, Y. Liu, L.E. Bernstein, K.E. Livingston, A. Perez, I. A. Shlyakhter, R. V. Rolando, R. Owen, P. Tanpaiboon, F. Lacbawan, Clinton R. Paden, Duncan MacCannell |
| EPI_ISL_2034800, EPI_ISL_2034801, EPI_ISL_2034802, EPI_ISL_2034803 | Yale Clinical Virology Lab | Yale Center for Genomic Analysis | Shrikant Mane, Kaya Bilguvar, Curt Scharfe, Irina Tikhonova, Brooke Sullivan |
| EPI_ISL_2034804 | Quest Diagnostics Incorporated | Centers for Disease Control and Prevention Division of Viral Diseases, Pathogen Discovery | Dakota Howard, Dhwani Batra, Peter W. Cook, Kara Moser, Adrian Paskey, Jason Caravas, Benjamin Rambo-Martin, Shatavia Morrison, Christopher Gulvick, Scott Sammons, Yvette Unoarumhi, Darlene Wagner, Matthew Schmerer, S. H. Rosenthal, A. Gerasimova, R. M. Kagan, B. Anderson, M. Hua, Y. Liu, L.E. Bernstein, K.E. Livingston, A. Perez, I. A. Shlyakhter, R. V. Rolando, R. Owen, P. Tanpaiboon, F. Lacbawan, Clinton R. Paden, Duncan MacCannell |
| EPI_ISL_2034805, EPI_ISL_2034806, EPI_ISL_2034807 | Yale Clinical Virology Lab | Yale Center for Genomic Analysis | Shrikant Mane, Kaya Bilguvar, Curt Scharfe, Irina Tikhonova, Brooke Sullivan |
| EPI_ISL_2034808, EPI_ISL_2034811 | Quest Diagnostics Incorporated | Centers for Disease Control and Prevention Division of Viral Diseases, Pathogen Discovery | Dakota Howard, Dhwani Batra, Peter W. Cook, Kara Moser, Adrian Paskey, Jason Caravas, Benjamin Rambo-Martin, Shatavia Morrison, Christopher Gulvick, Scott Sammons, Yvette Unoarumhi, Darlene Wagner, Matthew Schmerer, S. H. Rosenthal, A. Gerasimova, R. M. Kagan, B. Anderson, M. Hua, Y. Liu, L.E. Bernstein, K.E. Livingston, A. Perez, I. A. Shlyakhter, R. V. Rolando, R. Owen, P. Tanpaiboon, F. Lacbawan, Clinton R. Paden, Duncan MacCannell |
| EPI_ISL_2034812, EPI_ISL_2034813, EPI_ISL_2034814 | Yale Clinical Virology Lab | Yale Center for Genomic Analysis | Shrikant Mane, Kaya Bilguvar, Curt Scharfe, Irina Tikhonova, Brooke Sullivan |
| EPI_ISL_2034815 | Quest Diagnostics Incorporated | Centers for Disease Control and Prevention Division of Viral Diseases, Pathogen Discovery | Dakota Howard, Dhwani Batra, Peter W. Cook, Kara Moser, Adrian Paskey, Jason Caravas, Benjamin Rambo-Martin, Shatavia Morrison, Christopher Gulvick, Scott Sammons, Yvette Unoarumhi, Darlene Wagner, Matthew Schmerer, S. H. Rosenthal, A. Gerasimova, R. M. Kagan, B. Anderson, M. Hua, Y. Liu, L.E. Bernstein, K.E. Livingston, A. Perez, I. A. Shlyakhter, R. V. Rolando, R. Owen, P. Tanpaiboon, F. Lacbawan, Clinton R. Paden, Duncan MacCannell |
| EPI_ISL_2034816 | Yale Clinical Virology Lab | Yale Center for Genomic Analysis | Shrikant Mane, Kaya Bilguvar, Curt Scharfe, Irina Tikhonova, Brooke Sullivan |
| EPI_ISL_2034817 | Quest Diagnostics Incorporated | Centers for Disease Control and Prevention Division of Viral Diseases, Pathogen Discovery | Dakota Howard, Dhwani Batra, Peter W. Cook, Kara Moser, Adrian Paskey, Jason Caravas, Benjamin Rambo-Martin, Shatavia Morrison, Christopher Gulvick, Scott Sammons, Yvette Unoarumhi, Darlene Wagner, Matthew Schmerer, S. H. Rosenthal, A. Gerasimova, R. M. Kagan, B. Anderson, M. Hua, Y. Liu, L.E. Bernstein, K.E. Livingston, A. Perez, I. A. Shlyakhter, R. V. Rolando, R. Owen, P. Tanpaiboon, F. Lacbawan, Clinton R. Paden, Duncan MacCannell |
| EPI_ISL_2034818, EPI_ISL_2034819, EPI_ISL_2034820, EPI_ISL_2034821 | Yale Clinical Virology Lab | Yale Center for Genomic Analysis | Shrikant Mane, Kaya Bilguvar, Curt Scharfe, Irina Tikhonova, Brooke Sullivan |
| EPI_ISL_2034822 | Quest Diagnostics Incorporated | Centers for Disease Control and Prevention Division of Viral Diseases, Pathogen Discovery | Dakota Howard, Dhwani Batra, Peter W. Cook, Kara Moser, Adrian Paskey, Jason Caravas, Benjamin Rambo-Martin, Shatavia Morrison, Christopher Gulvick, Scott Sammons, Yvette Unoarumhi, Darlene Wagner, Matthew Schmerer, S. H. Rosenthal, A. Gerasimova, R. M. Kagan, B. Anderson, M. Hua, Y. Liu, L.E. Bernstein, K.E. Livingston, A. Perez, I. A. Shlyakhter, R. V. Rolando, R. Owen, P. Tanpaiboon, F. Lacbawan, Clinton R. Paden, Duncan MacCannell |
| EPI_ISL_2034823, EPI_ISL_2034824 | Yale Clinical Virology Lab | Yale Center for Genomic Analysis | Shrikant Mane, Kaya Bilguvar, Curt Scharfe, Irina Tikhonova, Brooke Sullivan |
| EPI_ISL_2034825 | Quest Diagnostics Incorporated | Centers for Disease Control and Prevention Division of Viral Diseases, Pathogen Discovery | Dakota Howard, Dhwani Batra, Peter W. Cook, Kara Moser, Adrian Paskey, Jason Caravas, Benjamin Rambo-Martin, Shatavia Morrison, Christopher Gulvick, Scott Sammons, Yvette Unoarumhi, Darlene Wagner, Matthew Schmerer, S. H. Rosenthal, A. Gerasimova, R. M. Kagan, B. Anderson, M. Hua, Y. Liu, L.E. Bernstein, K.E. Livingston, A. Perez, I. A. Shlyakhter, R. V. Rolando, R. Owen, P. Tanpaiboon, F. Lacbawan, Clinton R. Paden, Duncan MacCannell |
| EPI_ISL_2034826, EPI_ISL_2034827, EPI_ISL_2034828 | Yale Clinical Virology Lab | Yale Center for Genomic Analysis | Shrikant Mane, Kaya Bilguvar, Curt Scharfe, Irina Tikhonova, Brooke Sullivan |
| EPI_ISL_2034829 | Quest Diagnostics Incorporated | Centers for Disease Control and Prevention Division of Viral Diseases, Pathogen Discovery | Dakota Howard, Dhwani Batra, Peter W. Cook, Kara Moser, Adrian Paskey, Jason Caravas, Benjamin Rambo-Martin, Shatavia Morrison, Christopher Gulvick, Scott Sammons, Yvette Unoarumhi, Darlene Wagner, Matthew Schmerer, S. H. Rosenthal, A. Gerasimova, R. M. Kagan, B. Anderson, M. Hua, Y. Liu, L.E. Bernstein, K.E. Livingston, A. Perez, I. A. Shlyakhter, R. V. Rolando, R. Owen, P. Tanpaiboon, F. Lacbawan, Clinton R. Paden, Duncan MacCannell |
| EPI_ISL_2034830, EPI_ISL_2034831, EPI_ISL_2034832 | Yale Clinical Virology Lab | Yale Center for Genomic Analysis | Shrikant Mane, Kaya Bilguvar, Curt Scharfe, Irina Tikhonova, Brooke Sullivan |
| EPI_ISL_2034833 | Quest Diagnostics Incorporated | Centers for Disease Control and Prevention Division of Viral Diseases, Pathogen Discovery | Dakota Howard, Dhwani Batra, Peter W. Cook, Kara Moser, Adrian Paskey, Jason Caravas, Benjamin Rambo-Martin, Shatavia Morrison, Christopher Gulvick, Scott Sammons, Yvette Unoarumhi, Darlene Wagner, Matthew Schmerer, S. H. Rosenthal, A. Gerasimova, R. M. Kagan, B. Anderson, M. Hua, Y. Liu, L.E. Bernstein, K.E. Livingston, A. Perez, I. A. Shlyakhter, R. V. Rolando, R. Owen, P. Tanpaiboon, F. Lacbawan, Clinton R. Paden, Duncan MacCannell |

| | | | |
|---|---|---|---|
| EPI_ISL_2034834, EPI_ISL_2034835 | Yale Clinical Virology Lab | Yale Center for Genomic Analysis | Shrikant Mane, Kaya Bilguvar, Curt Scharfe, Irina Tikhonova, Brooke Sullivan |
| EPI_ISL_2034836, EPI_ISL_2034840 | Quest Diagnostics Incorporated | Centers for Disease Control and Prevention Division of Viral Diseases, Pathogen Discovery | Dakota Howard, Dhwani Batra, Peter W. Cook, Kara Moser, Adrian Paskey, Jason Caravas, Benjamin Rambo-Martin, Shatavia Morrison, Christopher Gulvick, Scott Sammons, Yvette Unoarumhi, Darlene Wagner, Matthew Schmerer, S. H. Rosenthal, A. Gerasimova, R. M. Kagan, B. Anderson, M. Hua, Y. Liu, L.E. Bernstein, K.E. Livingston, A. Perez, I. A. Shlyakhter, R. V. Rolando, R. Owen, P. Tanpaiboon, F. Lacbawan, Clinton R. Paden, Duncan MacCannell |
| EPI_ISL_2034841, EPI_ISL_2034842, EPI_ISL_2034843 | Yale Clinical Virology Lab | Yale Center for Genomic Analysis | Shrikant Mane, Kaya Bilguvar, Curt Scharfe, Irina Tikhonova, Brooke Sullivan |
| EPI_ISL_2034844 | Quest Diagnostics Incorporated | Centers for Disease Control and Prevention Division of Viral Diseases, Pathogen Discovery | Dakota Howard, Dhwani Batra, Peter W. Cook, Kara Moser, Adrian Paskey, Jason Caravas, Benjamin Rambo-Martin, Shatavia Morrison, Christopher Gulvick, Scott Sammons, Yvette Unoarumhi, Darlene Wagner, Matthew Schmerer, S. H. Rosenthal, A. Gerasimova, R. M. Kagan, B. Anderson, M. Hua, Y. Liu, L.E. Bernstein, K.E. Livingston, A. Perez, I. A. Shlyakhter, R. V. Rolando, R. Owen, P. Tanpaiboon, F. Lacbawan, Clinton R. Paden, Duncan MacCannell |
| EPI_ISL_2034845, EPI_ISL_2034847, EPI_ISL_2034848, EPI_ISL_2034850, EPI_ISL_2034851, EPI_ISL_2034852, EPI_ISL_2034853, EPI_ISL_2034854, EPI_ISL_2034855, EPI_ISL_2034856, EPI_ISL_2034857, EPI_ISL_2034858, EPI_ISL_2034859, EPI_ISL_2034860, EPI_ISL_2034861, EPI_ISL_2034862, EPI_ISL_2034863, EPI_ISL_2034864, EPI_ISL_2034865, EPI_ISL_2034866, EPI_ISL_2034867, EPI_ISL_2034868, EPI_ISL_2034870 | | | |
| see above | Yale Clinical Virology Lab | Yale Center for Genomic Analysis | Shrikant Mane, Kaya Bilguvar, Curt Scharfe, Irina Tikhonova, Brooke Sullivan |
| EPI_ISL_2034871 | Helix/Illumina | Centers for Disease Control and Prevention Division of Viral Diseases, Pathogen Discovery | Dakota Howard, Dhwani Batra, Peter W. Cook, Kara Moser, Adrian Paskey, Jason Caravas, Benjamin Rambo-Martin, Shatavia Morrison, Christopher Gulvick, Scott Sammons, Yvette Unoarumhi, Darlene Wagner, Matthew Schmerer, Eileen de Feo, Jan Antico, Christine Tran, Matthew Tolentino, Shannon Wickline, Kim Gietzen, Brad Sickler, Jingtao Liu, Eric Allen, Phil Febbo, Nicole L. Washington, Simon White, Geraint Levan, Kelly Schiabor Barrett, Elizabeth Cirulli, Alexandre Bolze, Ary Ascencio, Charlotte Rivera-Garcia, Ryan Cho, Jason Nguyen, Sherry Wang, Jimmy Ramirez, Tyler Cassens, Efren Sandoval, Magnus Isaksson, William Lee, David Becker, Marc Laurent, James Lu, Clinton R. Paden, Duncan MacCannell |
| EPI_ISL_2034872, EPI_ISL_2034873 | Yale Clinical Virology Lab | Yale Center for Genomic Analysis | Shrikant Mane, Kaya Bilguvar, Curt Scharfe, Irina Tikhonova, Brooke Sullivan |
| EPI_ISL_2034874 | Helix/Illumina | Centers for Disease Control and Prevention Division of Viral Diseases, Pathogen Discovery | Dakota Howard, Dhwani Batra, Peter W. Cook, Kara Moser, Adrian Paskey, Jason Caravas, Benjamin Rambo-Martin, Shatavia Morrison, Christopher Gulvick, Scott Sammons, Yvette Unoarumhi, Darlene Wagner, Matthew Schmerer, Eileen de Feo, Jan Antico, Christine Tran, Matthew Tolentino, Shannon Wickline, Kim Gietzen, Brad Sickler, Jingtao Liu, Eric Allen, Phil Febbo, Nicole L. Washington, Simon White, Geraint Levan, Kelly Schiabor Barrett, Elizabeth Cirulli, Alexandre Bolze, Ary Ascencio, Charlotte Rivera-Garcia, Ryan Cho, Jason Nguyen, Sherry Wang, Jimmy Ramirez, Tyler Cassens, Efren Sandoval, Magnus Isaksson, William Lee, David Becker, Marc Laurent, James Lu, Clinton R. Paden, Duncan MacCannell |
| EPI_ISL_2034875, EPI_ISL_2034877 | Yale Clinical Virology Lab | Yale Center for Genomic Analysis | Shrikant Mane, Kaya Bilguvar, Curt Scharfe, Irina Tikhonova, Brooke Sullivan |
| EPI_ISL_2034878 | Helix/Illumina | Centers for Disease Control and Prevention Division of Viral Diseases, Pathogen Discovery | Dakota Howard, Dhwani Batra, Peter W. Cook, Kara Moser, Adrian Paskey, Jason Caravas, Benjamin Rambo-Martin, Shatavia Morrison, Christopher Gulvick, Scott Sammons, Yvette Unoarumhi, Darlene Wagner, Matthew Schmerer, Eileen de Feo, Jan Antico, Christine Tran, Matthew Tolentino, Shannon Wickline, Kim Gietzen, Brad Sickler, Jingtao Liu, Eric Allen, Phil Febbo, Nicole L. Washington, Simon White, Geraint Levan, Kelly Schiabor Barrett, Elizabeth Cirulli, Alexandre Bolze, Ary Ascencio, Charlotte Rivera-Garcia, Ryan Cho, Jason Nguyen, Sherry Wang, Jimmy Ramirez, Tyler Cassens, Efren Sandoval, Magnus Isaksson, William Lee, David Becker, Marc Laurent, James Lu, Clinton R. Paden, Duncan MacCannell |
| EPI_ISL_2034879, EPI_ISL_2034880 | Yale Clinical Virology Lab | Yale Center for Genomic Analysis | Shrikant Mane, Kaya Bilguvar, Curt Scharfe, Irina Tikhonova, Brooke Sullivan |
| EPI_ISL_2034881 | Helix/Illumina | Centers for Disease Control and Prevention Division of Viral Diseases, Pathogen Discovery | Dakota Howard, Dhwani Batra, Peter W. Cook, Kara Moser, Adrian Paskey, Jason Caravas, Benjamin Rambo-Martin, Shatavia Morrison, Christopher Gulvick, Scott Sammons, Yvette Unoarumhi, Darlene Wagner, Matthew Schmerer, Eileen de Feo, Jan Antico, Christine Tran, Matthew Tolentino, Shannon Wickline, Kim Gietzen, Brad Sickler, Jingtao Liu, Eric Allen, Phil Febbo, Nicole L. Washington, Simon White, Geraint Levan, Kelly Schiabor Barrett, Elizabeth Cirulli, Alexandre Bolze, Ary Ascencio, Charlotte Rivera-Garcia, Ryan Cho, Jason Nguyen, Sherry Wang, Jimmy Ramirez, Tyler Cassens, Efren Sandoval, Magnus Isaksson, William Lee, David Becker, Marc Laurent, James Lu, Clinton R. Paden, Duncan MacCannell |
| EPI_ISL_2034882, EPI_ISL_2034883 | Yale Clinical Virology Lab | Yale Center for Genomic Analysis | Shrikant Mane, Kaya Bilguvar, Curt Scharfe, Irina Tikhonova, Brooke Sullivan |
| EPI_ISL_2034884 | Helix/Illumina | Centers for Disease Control and Prevention Division of Viral Diseases, Pathogen Discovery | Dakota Howard, Dhwani Batra, Peter W. Cook, Kara Moser, Adrian Paskey, Jason Caravas, Benjamin Rambo-Martin, Shatavia Morrison, Christopher Gulvick, Scott Sammons, Yvette Unoarumhi, Darlene Wagner, Matthew Schmerer, Eileen de Feo, Jan Antico, Christine Tran, Matthew Tolentino, Shannon Wickline, Kim Gietzen, Brad Sickler, Jingtao Liu, Eric Allen, Phil Febbo, Nicole L. Washington, Simon White, Geraint Levan, Kelly Schiabor Barrett, Elizabeth Cirulli, Alexandre Bolze, Ary Ascencio, Charlotte Rivera-Garcia, Ryan Cho, Jason Nguyen, Sherry Wang, Jimmy Ramirez, Tyler Cassens, Efren Sandoval, Magnus Isaksson, William Lee, David Becker, Marc Laurent, James Lu, Clinton R. Paden, Duncan MacCannell |
| EPI_ISL_2034886, EPI_ISL_2034887 | Yale Clinical Virology Lab | Yale Center for Genomic Analysis | Shrikant Mane, Kaya Bilguvar, Curt Scharfe, Irina Tikhonova, Brooke Sullivan |
| EPI_ISL_2034888 | Helix/Illumina | Centers for Disease Control and Prevention Division of Viral Diseases, Pathogen Discovery | Dakota Howard, Dhwani Batra, Peter W. Cook, Kara Moser, Adrian Paskey, Jason Caravas, Benjamin Rambo-Martin, Shatavia Morrison, Christopher Gulvick, Scott Sammons, Yvette Unoarumhi, Darlene Wagner, Matthew Schmerer, Eileen de Feo, Jan Antico, Christine Tran, Matthew Tolentino, Shannon Wickline, Kim Gietzen, Brad Sickler, Jingtao Liu, Eric Allen, Phil Febbo, Nicole L. Washington, Simon White, Geraint Levan, Kelly Schiabor Barrett, Elizabeth Cirulli, Alexandre Bolze, Ary Ascencio, Charlotte Rivera-Garcia, Ryan Cho, Jason Nguyen, Sherry Wang, Jimmy Ramirez, Tyler Cassens, Efren Sandoval, Magnus Isaksson, William Lee, David Becker, Marc Laurent, James Lu, Clinton R. Paden, Duncan MacCannell |
| EPI_ISL_2034889 | Yale Clinical Virology Lab | Yale Center for Genomic Analysis | Shrikant Mane, Kaya Bilguvar, Curt Scharfe, Irina Tikhonova, Brooke Sullivan |
| EPI_ISL_2034890 | Helix/Illumina | Centers for Disease Control and Prevention Division of Viral Diseases, Pathogen Discovery | Dakota Howard, Dhwani Batra, Peter W. Cook, Kara Moser, Adrian Paskey, Jason Caravas, Benjamin Rambo-Martin, Shatavia Morrison, Christopher Gulvick, Scott Sammons, Yvette Unoarumhi, Darlene Wagner, Matthew Schmerer, Eileen de Feo, Jan Antico, Christine Tran, Matthew Tolentino, Shannon Wickline, Kim Gietzen, Brad Sickler, Jingtao Liu, Eric Allen, Phil Febbo, Nicole L. Washington, Simon White, Geraint Levan, Kelly Schiabor Barrett, Elizabeth Cirulli, Alexandre Bolze, Ary Ascencio, Charlotte Rivera-Garcia, Ryan Cho, Jason Nguyen, Sherry Wang, Jimmy Ramirez, Tyler Cassens, Efren Sandoval, Magnus Isaksson, William Lee, David Becker, Marc Laurent, James Lu, Clinton R. Paden, Duncan MacCannell |
| EPI_ISL_2034891, EPI_ISL_2034892, EPI_ISL_2034893 | Yale Clinical Virology Lab | Yale Center for Genomic Analysis | Shrikant Mane, Kaya Bilguvar, Curt Scharfe, Irina Tikhonova, Brooke Sullivan |
| EPI_ISL_2034894 | Helix/Illumina | Centers for Disease Control and Prevention Division of Viral Diseases, Pathogen Discovery | Dakota Howard, Dhwani Batra, Peter W. Cook, Kara Moser, Adrian Paskey, Jason Caravas, Benjamin Rambo-Martin, Shatavia Morrison, Christopher Gulvick, Scott Sammons, Yvette Unoarumhi, Darlene Wagner, Matthew Schmerer, Eileen de Feo, Jan Antico, Christine Tran, Matthew Tolentino, Shannon Wickline, Kim Gietzen, Brad Sickler, Jingtao Liu, Eric Allen, Phil Febbo, Nicole L. Washington, Simon White, Geraint Levan, Kelly Schiabor Barrett, Elizabeth Cirulli, Alexandre Bolze, Ary Ascencio, Charlotte Rivera-Garcia, Ryan Cho, Jason Nguyen, Sherry Wang, Jimmy Ramirez, Tyler Cassens, Efren Sandoval, Magnus Isaksson, William Lee, David Becker, Marc Laurent, James Lu, Clinton R. Paden, Duncan MacCannell |
| EPI_ISL_2034895, EPI_ISL_2034896, EPI_ISL_2034897 | Yale Clinical Virology Lab | Yale Center for Genomic Analysis | Shrikant Mane, Kaya Bilguvar, Curt Scharfe, Irina Tikhonova, Brooke Sullivan |
| EPI_ISL_2034898 | Helix/Illumina | Centers for Disease Control and Prevention Division of Viral Diseases, Pathogen Discovery | Dakota Howard, Dhwani Batra, Peter W. Cook, Kara Moser, Adrian Paskey, Jason Caravas, Benjamin Rambo-Martin, Shatavia Morrison, Christopher Gulvick, Scott Sammons, Yvette Unoarumhi, Darlene Wagner, Matthew Schmerer, Eileen de Feo, Jan Antico, Christine Tran, Matthew Tolentino, Shannon Wickline, Kim Gietzen, Brad Sickler, Jingtao Liu, Eric Allen, Phil Febbo, Nicole L. Washington, Simon White, Geraint Levan, Kelly Schiabor Barrett, Elizabeth Cirulli, Alexandre Bolze, Ary Ascencio, Charlotte Rivera-Garcia, Ryan Cho, Jason Nguyen, Sherry Wang, Jimmy Ramirez, Tyler Cassens, Efren Sandoval, Magnus Isaksson, William Lee, David Becker, Marc Laurent, James Lu, Clinton R. Paden, Duncan MacCannell |
| EPI_ISL_2034899, EPI_ISL_2034900 | Yale Clinical Virology Lab | Yale Center for Genomic Analysis | Shrikant Mane, Kaya Bilguvar, Curt Scharfe, Irina Tikhonova, Brooke Sullivan |
| EPI_ISL_2034901 | Helix/Illumina | Centers for Disease Control and Prevention Division of Viral Diseases, Pathogen Discovery | Dakota Howard, Dhwani Batra, Peter W. Cook, Kara Moser, Adrian Paskey, Jason Caravas, Benjamin Rambo-Martin, Shatavia Morrison, Christopher Gulvick, Scott Sammons, Yvette Unoarumhi, Darlene Wagner, Matthew Schmerer, Eileen de Feo, Jan Antico, Christine Tran, Matthew Tolentino, Shannon Wickline, Kim Gietzen, Brad Sickler, Jingtao Liu, Eric Allen, Phil Febbo, Nicole L. Washington, Simon White, Geraint Levan, Kelly Schiabor Barrett, Elizabeth Cirulli, Alexandre Bolze, Ary Ascencio, Charlotte Rivera-Garcia, Ryan Cho, Jason Nguyen, Sherry Wang, Jimmy Ramirez, Tyler Cassens, Efren Sandoval, Magnus Isaksson, William Lee, David Becker, Marc Laurent, James Lu, Clinton R. Paden, Duncan MacCannell |
| EPI_ISL_2034902, EPI_ISL_2034903, EPI_ISL_2034904 | Yale Clinical Virology Lab | Yale Center for Genomic Analysis | Shrikant Mane, Kaya Bilguvar, Curt Scharfe, Irina Tikhonova, Brooke Sullivan |
| EPI_ISL_2034905 | Helix/Illumina | Centers for Disease Control and Prevention Division of Viral Diseases, Pathogen Discovery | Dakota Howard, Dhwani Batra, Peter W. Cook, Kara Moser, Adrian Paskey, Jason Caravas, Benjamin Rambo-Martin, Shatavia Morrison, Christopher Gulvick, Scott Sammons, Yvette Unoarumhi, Darlene Wagner, Matthew Schmerer, Eileen de Feo, Jan Antico, Christine Tran, Matthew Tolentino, Shannon Wickline, Kim Gietzen, Brad Sickler, Jingtao Liu, Eric Allen, Phil Febbo, Nicole L. Washington, Simon White, Geraint Levan, Kelly Schiabor Barrett, Elizabeth Cirulli, Alexandre Bolze, Ary Ascencio, Charlotte Rivera-Garcia, Ryan Cho, Jason Nguyen, Sherry Wang, Jimmy Ramirez, Tyler Cassens, Efren Sandoval, Magnus Isaksson, William Lee, David Becker, Marc Laurent, James Lu, Clinton R. Paden, Duncan MacCannell |
| EPI_ISL_2034906, EPI_ISL_2034907, EPI_ISL_2034908 | Yale Clinical Virology Lab | Yale Center for Genomic Analysis | Shrikant Mane, Kaya Bilguvar, Curt Scharfe, Irina Tikhonova, Brooke Sullivan |
| EPI_ISL_2034909 | Helix/Illumina | Centers for Disease Control and Prevention Division of Viral Diseases, Pathogen Discovery | Dakota Howard, Dhwani Batra, Peter W. Cook, Kara Moser, Adrian Paskey, Jason Caravas, Benjamin Rambo-Martin, Shatavia Morrison, Christopher Gulvick, Scott Sammons, Yvette Unoarumhi, Darlene Wagner, Matthew Schmerer, Eileen de Feo, Jan Antico, Christine Tran, Matthew Tolentino, Shannon Wickline, Kim Gietzen, Brad Sickler, Jingtao Liu, Eric Allen, Phil Febbo, Nicole L. Washington, Simon White, Geraint Levan, Kelly Schiabor Barrett, Elizabeth Cirulli, Alexandre Bolze, Ary Ascencio, Charlotte Rivera-Garcia, Ryan Cho, Jason Nguyen, Sherry Wang, Jimmy Ramirez, Tyler Cassens, Efren Sandoval, Magnus Isaksson, William Lee, David Becker, Marc Laurent, James Lu, Clinton R. Paden, Duncan MacCannell |
| EPI_ISL_2034910, EPI_ISL_2034911 | Yale Clinical Virology Lab | Yale Center for Genomic Analysis | Shrikant Mane, Kaya Bilguvar, Curt Scharfe, Irina Tikhonova, Brooke Sullivan |
| EPI_ISL_2034912 | Helix/Illumina | Centers for Disease Control and Prevention Division of Viral Diseases, Pathogen Discovery | Dakota Howard, Dhwani Batra, Peter W. Cook, Kara Moser, Adrian Paskey, Jason Caravas, Benjamin Rambo-Martin, Shatavia Morrison, Christopher Gulvick, Scott Sammons, Yvette Unoarumhi, Darlene Wagner, Matthew Schmerer, Eileen de Feo, Jan Antico, Christine Tran, Matthew Tolentino, Shannon Wickline, Kim Gietzen, Brad Sickler, Jingtao Liu, Eric Allen, Phil Febbo, Nicole L. Washington, Simon White, Geraint Levan, Kelly Schiabor Barrett, Elizabeth Cirulli, Alexandre Bolze, Ary Ascencio, Charlotte Rivera-Garcia, Ryan Cho, Jason Nguyen, Sherry Wang, Jimmy Ramirez, Tyler Cassens, Efren Sandoval, Magnus Isaksson, William Lee, David Becker, Marc Laurent, James Lu, Clinton R. Paden, Duncan MacCannell |
| EPI_ISL_2034913, EPI_ISL_2034914 | Yale Clinical Virology Lab | Yale Center for Genomic Analysis | Shrikant Mane, Kaya Bilguvar, Curt Scharfe, Irina Tikhonova, Brooke Sullivan |
| EPI_ISL_2034915 | Helix/Illumina | Centers for Disease Control and Prevention Division of Viral Diseases, Pathogen Discovery | Dakota Howard, Dhwani Batra, Peter W. Cook, Kara Moser, Adrian Paskey, Jason Caravas, Benjamin Rambo-Martin, Shatavia Morrison, Christopher Gulvick, Scott Sammons, Yvette Unoarumhi, Darlene Wagner, Matthew Schmerer, Eileen de Feo, Jan Antico, Christine Tran, Matthew Tolentino, Shannon Wickline, Kim Gietzen, Brad Sickler, Jingtao Liu, Eric Allen, Phil Febbo, Nicole L. Washington, Simon White, Geraint Levan, Kelly Schiabor Barrett, Elizabeth Cirulli, Alexandre Bolze, Ary Ascencio, Charlotte Rivera-Garcia, Ryan Cho, Jason Nguyen, Sherry Wang, Jimmy Ramirez, Tyler Cassens, Efren Sandoval, Magnus Isaksson, William Lee, David Becker, Marc Laurent, James Lu, Clinton R. Paden, Duncan MacCannell |
| EPI_ISL_2034916, EPI_ISL_2034917, EPI_ISL_2034918 | Yale Clinical Virology Lab | Yale Center for Genomic Analysis | Shrikant Mane, Kaya Bilguvar, Curt Scharfe, Irina Tikhonova, Brooke Sullivan |
| EPI_ISL_2034919 | Helix/Illumina | Centers for Disease Control and Prevention Division of Viral Diseases, Pathogen Discovery | Dakota Howard, Dhwani Batra, Peter W. Cook, Kara Moser, Adrian Paskey, Jason Caravas, Benjamin Rambo-Martin, Shatavia Morrison, Christopher Gulvick, Scott Sammons, Yvette Unoarumhi, Darlene Wagner, Matthew Schmerer, Eileen de Feo, Jan Antico, Christine Tran, Matthew Tolentino, Shannon Wickline, Kim Gietzen, Brad Sickler, Jingtao Liu, Eric Allen, Phil Febbo, Nicole L. Washington, Simon White, Geraint Levan, Kelly Schiabor Barrett, Elizabeth Cirulli, Alexandre Bolze, Ary Ascencio, Charlotte Rivera-Garcia, Ryan Cho, Jason Nguyen, Sherry Wang, Jimmy Ramirez, Tyler Cassens, Efren Sandoval, Magnus Isaksson, William Lee, David Becker, Marc Laurent, James Lu, Clinton R. Paden, Duncan MacCannell |
| EPI_ISL_2034920, EPI_ISL_2034921 | Yale Clinical Virology Lab | Yale Center for Genomic Analysis | Shrikant Mane, Kaya Bilguvar, Curt Scharfe, Irina Tikhonova, Brooke Sullivan |

| | | | |
|---|---|---|---|
| EPI_ISL_2034922 | Helix/Illumina | Centers for Disease Control and Prevention Division of Viral Diseases, Pathogen Discovery | Dakota Howard, Dhwani Batra, Peter W. Cook, Kara Moser, Adrian Paskey, Jason Caravas, Benjamin Rambo-Martin, Shatavia Morrison, Christopher Gulvick, Scott Sammons, Yvette Unoarumhi, Darlene Wagner, Matthew Schmerer, Eileen de Feo, Jan Antico, Christine Tran, Matthew Tolentino, Shannon Wickline, Kim Gietzen, Brad Sickler, Jingtao Liu, Eric Allen, Phil Febbo, Nicole L. Washington, Simon White, Geraint Levan, Kelly Schiabor Barrett, Elizabeth Cirulli, Alexandre Bolze, Ary Ascencio, Charlotte Rivera-Garcia, Ryan Cho, Jason Nguyen, Sherry Wang, Jimmy Ramirez, Tyler Cassens, Efren Sandoval, Magnus Isaksson, William Lee, David Becker, Marc Laurent, James Lu, Clinton R. Paden, Duncan MacCannell |
| EPI_ISL_2034923, EPI_ISL_2034924, EPI_ISL_2034925 | Yale Clinical Virology Lab | Yale Center for Genomic Analysis | Shrikant Mane, Kaya Bilguvar, Curt Scharfe, Irina Tikhonova, Brooke Sullivan |
| EPI_ISL_2034926 | Helix/Illumina | Centers for Disease Control and Prevention Division of Viral Diseases, Pathogen Discovery | Dakota Howard, Dhwani Batra, Peter W. Cook, Kara Moser, Adrian Paskey, Jason Caravas, Benjamin Rambo-Martin, Shatavia Morrison, Christopher Gulvick, Scott Sammons, Yvette Unoarumhi, Darlene Wagner, Matthew Schmerer, Eileen de Feo, Jan Antico, Christine Tran, Matthew Tolentino, Shannon Wickline, Kim Gietzen, Brad Sickler, Jingtao Liu, Eric Allen, Phil Febbo, Nicole L. Washington, Simon White, Geraint Levan, Kelly Schiabor Barrett, Elizabeth Cirulli, Alexandre Bolze, Ary Ascencio, Charlotte Rivera-Garcia, Ryan Cho, Jason Nguyen, Sherry Wang, Jimmy Ramirez, Tyler Cassens, Efren Sandoval, Magnus Isaksson, William Lee, David Becker, Marc Laurent, James Lu, Clinton R. Paden, Duncan MacCannell |
| EPI_ISL_2034927, EPI_ISL_2034928, EPI_ISL_2034929 | Yale Clinical Virology Lab | Yale Center for Genomic Analysis | Shrikant Mane, Kaya Bilguvar, Curt Scharfe, Irina Tikhonova, Brooke Sullivan |
| EPI_ISL_2034930 | Helix/Illumina | Centers for Disease Control and Prevention Division of Viral Diseases, Pathogen Discovery | Dakota Howard, Dhwani Batra, Peter W. Cook, Kara Moser, Adrian Paskey, Jason Caravas, Benjamin Rambo-Martin, Shatavia Morrison, Christopher Gulvick, Scott Sammons, Yvette Unoarumhi, Darlene Wagner, Matthew Schmerer, Eileen de Feo, Jan Antico, Christine Tran, Matthew Tolentino, Shannon Wickline, Kim Gietzen, Brad Sickler, Jingtao Liu, Eric Allen, Phil Febbo, Nicole L. Washington, Simon White, Geraint Levan, Kelly Schiabor Barrett, Elizabeth Cirulli, Alexandre Bolze, Ary Ascencio, Charlotte Rivera-Garcia, Ryan Cho, Jason Nguyen, Sherry Wang, Jimmy Ramirez, Tyler Cassens, Efren Sandoval, Magnus Isaksson, William Lee, David Becker, Marc Laurent, James Lu, Clinton R. Paden, Duncan MacCannell |
| EPI_ISL_2034931, EPI_ISL_2034932 | Yale Clinical Virology Lab | Yale Center for Genomic Analysis | Shrikant Mane, Kaya Bilguvar, Curt Scharfe, Irina Tikhonova, Brooke Sullivan |
| EPI_ISL_2034933 | Helix/Illumina | Centers for Disease Control and Prevention Division of Viral Diseases, Pathogen Discovery | Dakota Howard, Dhwani Batra, Peter W. Cook, Kara Moser, Adrian Paskey, Jason Caravas, Benjamin Rambo-Martin, Shatavia Morrison, Christopher Gulvick, Scott Sammons, Yvette Unoarumhi, Darlene Wagner, Matthew Schmerer, Eileen de Feo, Jan Antico, Christine Tran, Matthew Tolentino, Shannon Wickline, Kim Gietzen, Brad Sickler, Jingtao Liu, Eric Allen, Phil Febbo, Nicole L. Washington, Simon White, Geraint Levan, Kelly Schiabor Barrett, Elizabeth Cirulli, Alexandre Bolze, Ary Ascencio, Charlotte Rivera-Garcia, Ryan Cho, Jason Nguyen, Sherry Wang, Jimmy Ramirez, Tyler Cassens, Efren Sandoval, Magnus Isaksson, William Lee, David Becker, Marc Laurent, James Lu, Clinton R. Paden, Duncan MacCannell |
| EPI_ISL_2034934, EPI_ISL_2034935, EPI_ISL_2034936 | Yale Clinical Virology Lab | Yale Center for Genomic Analysis | Shrikant Mane, Kaya Bilguvar, Curt Scharfe, Irina Tikhonova, Brooke Sullivan |
| EPI_ISL_2034937 | Helix/Illumina | Centers for Disease Control and Prevention Division of Viral Diseases, Pathogen Discovery | Dakota Howard, Dhwani Batra, Peter W. Cook, Kara Moser, Adrian Paskey, Jason Caravas, Benjamin Rambo-Martin, Shatavia Morrison, Christopher Gulvick, Scott Sammons, Yvette Unoarumhi, Darlene Wagner, Matthew Schmerer, Eileen de Feo, Jan Antico, Christine Tran, Matthew Tolentino, Shannon Wickline, Kim Gietzen, Brad Sickler, Jingtao Liu, Eric Allen, Phil Febbo, Nicole L. Washington, Simon White, Geraint Levan, Kelly Schiabor Barrett, Elizabeth Cirulli, Alexandre Bolze, Ary Ascencio, Charlotte Rivera-Garcia, Ryan Cho, Jason Nguyen, Sherry Wang, Jimmy Ramirez, Tyler Cassens, Efren Sandoval, Magnus Isaksson, William Lee, David Becker, Marc Laurent, James Lu, Clinton R. Paden, Duncan MacCannell |
| EPI_ISL_2034938, EPI_ISL_2034939 | Yale Clinical Virology Lab | Yale Center for Genomic Analysis | Shrikant Mane, Kaya Bilguvar, Curt Scharfe, Irina Tikhonova, Brooke Sullivan |
| EPI_ISL_2034940 | Helix/Illumina | Centers for Disease Control and Prevention Division of Viral Diseases, Pathogen Discovery | Dakota Howard, Dhwani Batra, Peter W. Cook, Kara Moser, Adrian Paskey, Jason Caravas, Benjamin Rambo-Martin, Shatavia Morrison, Christopher Gulvick, Scott Sammons, Yvette Unoarumhi, Darlene Wagner, Matthew Schmerer, Eileen de Feo, Jan Antico, Christine Tran, Matthew Tolentino, Shannon Wickline, Kim Gietzen, Brad Sickler, Jingtao Liu, Eric Allen, Phil Febbo, Nicole L. Washington, Simon White, Geraint Levan, Kelly Schiabor Barrett, Elizabeth Cirulli, Alexandre Bolze, Ary Ascencio, Charlotte Rivera-Garcia, Ryan Cho, Jason Nguyen, Sherry Wang, Jimmy Ramirez, Tyler Cassens, Efren Sandoval, Magnus Isaksson, William Lee, David Becker, Marc Laurent, James Lu, Clinton R. Paden, Duncan MacCannell |
| EPI_ISL_2034941, EPI_ISL_2034942 | Yale Clinical Virology Lab | Yale Center for Genomic Analysis | Shrikant Mane, Kaya Bilguvar, Curt Scharfe, Irina Tikhonova, Brooke Sullivan |
| EPI_ISL_2034943 | Helix/Illumina | Centers for Disease Control and Prevention Division of Viral Diseases, Pathogen Discovery | Dakota Howard, Dhwani Batra, Peter W. Cook, Kara Moser, Adrian Paskey, Jason Caravas, Benjamin Rambo-Martin, Shatavia Morrison, Christopher Gulvick, Scott Sammons, Yvette Unoarumhi, Darlene Wagner, Matthew Schmerer, Eileen de Feo, Jan Antico, Christine Tran, Matthew Tolentino, Shannon Wickline, Kim Gietzen, Brad Sickler, Jingtao Liu, Eric Allen, Phil Febbo, Nicole L. Washington, Simon White, Geraint Levan, Kelly Schiabor Barrett, Elizabeth Cirulli, Alexandre Bolze, Ary Ascencio, Charlotte Rivera-Garcia, Ryan Cho, Jason Nguyen, Sherry Wang, Jimmy Ramirez, Tyler Cassens, Efren Sandoval, Magnus Isaksson, William Lee, David Becker, Marc Laurent, James Lu, Clinton R. Paden, Duncan MacCannell |
| EPI_ISL_2034944, EPI_ISL_2034945, EPI_ISL_2034946 | Yale Clinical Virology Lab | Yale Center for Genomic Analysis | Shrikant Mane, Kaya Bilguvar, Curt Scharfe, Irina Tikhonova, Brooke Sullivan |
| EPI_ISL_2034947 | Helix/Illumina | Centers for Disease Control and Prevention Division of Viral Diseases, Pathogen Discovery | Dakota Howard, Dhwani Batra, Peter W. Cook, Kara Moser, Adrian Paskey, Jason Caravas, Benjamin Rambo-Martin, Shatavia Morrison, Christopher Gulvick, Scott Sammons, Yvette Unoarumhi, Darlene Wagner, Matthew Schmerer, Eileen de Feo, Jan Antico, Christine Tran, Matthew Tolentino, Shannon Wickline, Kim Gietzen, Brad Sickler, Jingtao Liu, Eric Allen, Phil Febbo, Nicole L. Washington, Simon White, Geraint Levan, Kelly Schiabor Barrett, Elizabeth Cirulli, Alexandre Bolze, Ary Ascencio, Charlotte Rivera-Garcia, Ryan Cho, Jason Nguyen, Sherry Wang, Jimmy Ramirez, Tyler Cassens, Efren Sandoval, Magnus Isaksson, William Lee, David Becker, Marc Laurent, James Lu, Clinton R. Paden, Duncan MacCannell |
| EPI_ISL_2034948, EPI_ISL_2034949 | Yale Clinical Virology Lab | Yale Center for Genomic Analysis | Shrikant Mane, Kaya Bilguvar, Curt Scharfe, Irina Tikhonova, Brooke Sullivan |
| EPI_ISL_2034950 | Helix/Illumina | Centers for Disease Control and Prevention Division of Viral Diseases, Pathogen Discovery | Dakota Howard, Dhwani Batra, Peter W. Cook, Kara Moser, Adrian Paskey, Jason Caravas, Benjamin Rambo-Martin, Shatavia Morrison, Christopher Gulvick, Scott Sammons, Yvette Unoarumhi, Darlene Wagner, Matthew Schmerer, Eileen de Feo, Jan Antico, Christine Tran, Matthew Tolentino, Shannon Wickline, Kim Gietzen, Brad Sickler, Jingtao Liu, Eric Allen, Phil Febbo, Nicole L. Washington, Simon White, Geraint Levan, Kelly Schiabor Barrett, Elizabeth Cirulli, Alexandre Bolze, Ary Ascencio, Charlotte Rivera-Garcia, Ryan Cho, Jason Nguyen, Sherry Wang, Jimmy Ramirez, Tyler Cassens, Efren Sandoval, Magnus Isaksson, William Lee, David Becker, Marc Laurent, James Lu, Clinton R. Paden, Duncan MacCannell |
| EPI_ISL_2034951, EPI_ISL_2034952 | Yale Clinical Virology Lab | Yale Center for Genomic Analysis | Shrikant Mane, Kaya Bilguvar, Curt Scharfe, Irina Tikhonova, Brooke Sullivan |
| EPI_ISL_2034953 | Helix/Illumina | Centers for Disease Control and Prevention Division of Viral Diseases, Pathogen Discovery | Dakota Howard, Dhwani Batra, Peter W. Cook, Kara Moser, Adrian Paskey, Jason Caravas, Benjamin Rambo-Martin, Shatavia Morrison, Christopher Gulvick, Scott Sammons, Yvette Unoarumhi, Darlene Wagner, Matthew Schmerer, Eileen de Feo, Jan Antico, Christine Tran, Matthew Tolentino, Shannon Wickline, Kim Gietzen, Brad Sickler, Jingtao Liu, Eric Allen, Phil Febbo, Nicole L. Washington, Simon White, Geraint Levan, Kelly Schiabor Barrett, Elizabeth Cirulli, Alexandre Bolze, Ary Ascencio, Charlotte Rivera-Garcia, Ryan Cho, Jason Nguyen, Sherry Wang, Jimmy Ramirez, Tyler Cassens, Efren Sandoval, Magnus Isaksson, William Lee, David Becker, Marc Laurent, James Lu, Clinton R. Paden, Duncan MacCannell |
| EPI_ISL_2034954, EPI_ISL_2034955 | Yale Clinical Virology Lab | Yale Center for Genomic Analysis | Shrikant Mane, Kaya Bilguvar, Curt Scharfe, Irina Tikhonova, Brooke Sullivan |
| EPI_ISL_2034956 | Helix/Illumina | Centers for Disease Control and Prevention Division of Viral Diseases, Pathogen Discovery | Dakota Howard, Dhwani Batra, Peter W. Cook, Kara Moser, Adrian Paskey, Jason Caravas, Benjamin Rambo-Martin, Shatavia Morrison, Christopher Gulvick, Scott Sammons, Yvette Unoarumhi, Darlene Wagner, Matthew Schmerer, Eileen de Feo, Jan Antico, Christine Tran, Matthew Tolentino, Shannon Wickline, Kim Gietzen, Brad Sickler, Jingtao Liu, Eric Allen, Phil Febbo, Nicole L. Washington, Simon White, Geraint Levan, Kelly Schiabor Barrett, Elizabeth Cirulli, Alexandre Bolze, Ary Ascencio, Charlotte Rivera-Garcia, Ryan Cho, Jason Nguyen, Sherry Wang, Jimmy Ramirez, Tyler Cassens, Efren Sandoval, Magnus Isaksson, William Lee, David Becker, Marc Laurent, James Lu, Clinton R. Paden, Duncan MacCannell |
| EPI_ISL_2034957, EPI_ISL_2034958, EPI_ISL_2034959 | Yale Clinical Virology Lab | Yale Center for Genomic Analysis | Shrikant Mane, Kaya Bilguvar, Curt Scharfe, Irina Tikhonova, Brooke Sullivan |
| EPI_ISL_2034960 | Helix/Illumina | Centers for Disease Control and Prevention Division of Viral Diseases, Pathogen Discovery | Dakota Howard, Dhwani Batra, Peter W. Cook, Kara Moser, Adrian Paskey, Jason Caravas, Benjamin Rambo-Martin, Shatavia Morrison, Christopher Gulvick, Scott Sammons, Yvette Unoarumhi, Darlene Wagner, Matthew Schmerer, Eileen de Feo, Jan Antico, Christine Tran, Matthew Tolentino, Shannon Wickline, Kim Gietzen, Brad Sickler, Jingtao Liu, Eric Allen, Phil Febbo, Nicole L. Washington, Simon White, Geraint Levan, Kelly Schiabor Barrett, Elizabeth Cirulli, Alexandre Bolze, Ary Ascencio, Charlotte Rivera-Garcia, Ryan Cho, Jason Nguyen, Sherry Wang, Jimmy Ramirez, Tyler Cassens, Efren Sandoval, Magnus Isaksson, William Lee, David Becker, Marc Laurent, James Lu, Clinton R. Paden, Duncan MacCannell |
| EPI_ISL_2034961, EPI_ISL_2034962, EPI_ISL_2034963 | Yale Clinical Virology Lab | Yale Center for Genomic Analysis | Shrikant Mane, Kaya Bilguvar, Curt Scharfe, Irina Tikhonova, Brooke Sullivan |
| EPI_ISL_2034964 | Helix/Illumina | Centers for Disease Control and Prevention Division of Viral Diseases, Pathogen Discovery | Dakota Howard, Dhwani Batra, Peter W. Cook, Kara Moser, Adrian Paskey, Jason Caravas, Benjamin Rambo-Martin, Shatavia Morrison, Christopher Gulvick, Scott Sammons, Yvette Unoarumhi, Darlene Wagner, Matthew Schmerer, Eileen de Feo, Jan Antico, Christine Tran, Matthew Tolentino, Shannon Wickline, Kim Gietzen, Brad Sickler, Jingtao Liu, Eric Allen, Phil Febbo, Nicole L. Washington, Simon White, Geraint Levan, Kelly Schiabor Barrett, Elizabeth Cirulli, Alexandre Bolze, Ary Ascencio, Charlotte Rivera-Garcia, Ryan Cho, Jason Nguyen, Sherry Wang, Jimmy Ramirez, Tyler Cassens, Efren Sandoval, Magnus Isaksson, William Lee, David Becker, Marc Laurent, James Lu, Clinton R. Paden, Duncan MacCannell |
| EPI_ISL_2034965, EPI_ISL_2034966 | Yale Clinical Virology Lab | Yale Center for Genomic Analysis | Shrikant Mane, Kaya Bilguvar, Curt Scharfe, Irina Tikhonova, Brooke Sullivan |
| EPI_ISL_2034967 | Helix/Illumina | Centers for Disease Control and Prevention Division of Viral Diseases, Pathogen Discovery | Dakota Howard, Dhwani Batra, Peter W. Cook, Kara Moser, Adrian Paskey, Jason Caravas, Benjamin Rambo-Martin, Shatavia Morrison, Christopher Gulvick, Scott Sammons, Yvette Unoarumhi, Darlene Wagner, Matthew Schmerer, Eileen de Feo, Jan Antico, Christine Tran, Matthew Tolentino, Shannon Wickline, Kim Gietzen, Brad Sickler, Jingtao Liu, Eric Allen, Phil Febbo, Nicole L. Washington, Simon White, Geraint Levan, Kelly Schiabor Barrett, Elizabeth Cirulli, Alexandre Bolze, Ary Ascencio, Charlotte Rivera-Garcia, Ryan Cho, Jason Nguyen, Sherry Wang, Jimmy Ramirez, Tyler Cassens, Efren Sandoval, Magnus Isaksson, William Lee, David Becker, Marc Laurent, James Lu, Clinton R. Paden, Duncan MacCannell |
| EPI_ISL_2034968, EPI_ISL_2034969, EPI_ISL_2034970 | Yale Clinical Virology Lab | Yale Center for Genomic Analysis | Shrikant Mane, Kaya Bilguvar, Curt Scharfe, Irina Tikhonova, Brooke Sullivan |
| EPI_ISL_2034971 | Helix/Illumina | Centers for Disease Control and Prevention Division of Viral Diseases, Pathogen Discovery | Dakota Howard, Dhwani Batra, Peter W. Cook, Kara Moser, Adrian Paskey, Jason Caravas, Benjamin Rambo-Martin, Shatavia Morrison, Christopher Gulvick, Scott Sammons, Yvette Unoarumhi, Darlene Wagner, Matthew Schmerer, Eileen de Feo, Jan Antico, Christine Tran, Matthew Tolentino, Shannon Wickline, Kim Gietzen, Brad Sickler, Jingtao Liu, Eric Allen, Phil Febbo, Nicole L. Washington, Simon White, Geraint Levan, Kelly Schiabor Barrett, Elizabeth Cirulli, Alexandre Bolze, Ary Ascencio, Charlotte Rivera-Garcia, Ryan Cho, Jason Nguyen, Sherry Wang, Jimmy Ramirez, Tyler Cassens, Efren Sandoval, Magnus Isaksson, William Lee, David Becker, Marc Laurent, James Lu, Clinton R. Paden, Duncan MacCannell |
| EPI_ISL_2034972, EPI_ISL_2034973, EPI_ISL_2034974 | Yale Clinical Virology Lab | Yale Center for Genomic Analysis | Shrikant Mane, Kaya Bilguvar, Curt Scharfe, Irina Tikhonova, Brooke Sullivan |
| EPI_ISL_2034975 | Helix/Illumina | Centers for Disease Control and Prevention Division of Viral Diseases, Pathogen Discovery | Dakota Howard, Dhwani Batra, Peter W. Cook, Kara Moser, Adrian Paskey, Jason Caravas, Benjamin Rambo-Martin, Shatavia Morrison, Christopher Gulvick, Scott Sammons, Yvette Unoarumhi, Darlene Wagner, Matthew Schmerer, Eileen de Feo, Jan Antico, Christine Tran, Matthew Tolentino, Shannon Wickline, Kim Gietzen, Brad Sickler, Jingtao Liu, Eric Allen, Phil Febbo, Nicole L. Washington, Simon White, Geraint Levan, Kelly Schiabor Barrett, Elizabeth Cirulli, Alexandre Bolze, Ary Ascencio, Charlotte Rivera-Garcia, Ryan Cho, Jason Nguyen, Sherry Wang, Jimmy Ramirez, Tyler Cassens, Efren Sandoval, Magnus Isaksson, William Lee, David Becker, Marc Laurent, James Lu, Clinton R. Paden, Duncan MacCannell |
| EPI_ISL_2034976, EPI_ISL_2034977, EPI_ISL_2034978, EPI_ISL_2034979, EPI_ISL_2034980, EPI_ISL_2034981, EPI_ISL_2034982, EPI_ISL_2034983, EPI_ISL_2034984, EPI_ISL_2034985, EPI_ISL_2034986, EPI_ISL_2034987, EPI_ISL_2034988, EPI_ISL_2034989, EPI_ISL_2034990, EPI_ISL_2034991, EPI_ISL_2034992, EPI_ISL_2034993, EPI_ISL_2034994, EPI_ISL_2034995 | | | |
| see above | Yale Clinical Virology Lab | Yale Center for Genomic Analysis | Shrikant Mane, Kaya Bilguvar, Curt Scharfe, Irina Tikhonova, Brooke Sullivan |
| EPI_ISL_2034996 | Fulgent Genetics | Centers for Disease Control and Prevention Division of Viral Diseases, Pathogen Discovery | Dakota Howard, Dhwani Batra, Peter W. Cook, Kara Moser, Adrian Paskey, Jason Caravas, Benjamin Rambo-Martin, Shatavia Morrison, Christopher Gulvick, Scott Sammons, Yvette Unoarumhi, Darlene Wagner, Matthew Schmerer, Harry Gao, Mickey Li, John Gao, Joseph Fierro, Benafsh Sapra, Becky Tsai, Yan Meng, Doreen Ng, James Xie, Clinton R. Paden, Duncan MacCannell |
| EPI_ISL_2034997, EPI_ISL_2034998, EPI_ISL_2034999 | Yale Clinical Virology Lab | Yale Center for Genomic Analysis | Shrikant Mane, Kaya Bilguvar, Curt Scharfe, Irina Tikhonova, Brooke Sullivan |
| EPI_ISL_2035000 | Fulgent Genetics | Centers for Disease Control and Prevention | Dakota Howard, Dhwani Batra, Peter W. Cook, Kara Moser, Adrian Paskey, Jason Caravas, Benjamin Rambo-Martin, Shatavia Morrison, Christopher Gulvick, Scott Sammons, Yvette Unoarumhi, Darlene Wagner, Matthew Schmerer, Harry Gao, Mickey Li, John Gao, Joseph Fierro, Benafsh |

| | | | |
|---|---|---|---|
| | | Division of Viral Diseases, Pathogen Discovery | Sapra, Becky Tsai, Yan Meng, Doreen Ng, James Xie, Clinton R. Paden, Duncan MacCannell |
| EPI_ISL_2035001, EPI_ISL_2035002, EPI_ISL_2035003 | Yale Clinical Virology Lab | Yale Center for Genomic Analysis | Shrikant Mane, Kaya Bilguvar, Curt Scharfe, Irina Tikhonova, Brooke Sullivan |
| EPI_ISL_2035004 | Fulgent Genetics | Centers for Disease Control and Prevention Division of Viral Diseases, Pathogen Discovery | Dakota Howard, Dhwani Batra, Peter W. Cook, Kara Moser, Adrian Paskey, Jason Caravas, Benjamin Rambo-Martin, Shatavia Morrison, Christopher Gulvick, Scott Sammons, Yvette Unoarumhi, Darlene Wagner, Matthew Schmerer, Harry Gao, Mickey Li, John Gao, Joseph Fierro, Benafsh Sapra, Becky Tsai, Yan Meng, Doreen Ng, James Xie, Clinton R. Paden, Duncan MacCannell |
| EPI_ISL_2035005, EPI_ISL_2035006 | Yale Clinical Virology Lab | Yale Center for Genomic Analysis | Shrikant Mane, Kaya Bilguvar, Curt Scharfe, Irina Tikhonova, Brooke Sullivan |
| EPI_ISL_2035007 | Fulgent Genetics | Centers for Disease Control and Prevention Division of Viral Diseases, Pathogen Discovery | Dakota Howard, Dhwani Batra, Peter W. Cook, Kara Moser, Adrian Paskey, Jason Caravas, Benjamin Rambo-Martin, Shatavia Morrison, Christopher Gulvick, Scott Sammons, Yvette Unoarumhi, Darlene Wagner, Matthew Schmerer, Harry Gao, Mickey Li, John Gao, Joseph Fierro, Benafsh Sapra, Becky Tsai, Yan Meng, Doreen Ng, James Xie, Clinton R. Paden, Duncan MacCannell |
| EPI_ISL_2035008, EPI_ISL_2035009, EPI_ISL_2035010, EPI_ISL_2035011, EPI_ISL_2035012, EPI_ISL_2035013, EPI_ISL_2035014, EPI_ISL_2035015, EPI_ISL_2035016, EPI_ISL_2035017, EPI_ISL_2035018, EPI_ISL_2035019, EPI_ISL_2035020, EPI_ISL_2035022, EPI_ISL_2035023, EPI_ISL_2035026, EPI_ISL_2035027, EPI_ISL_2035028, EPI_ISL_2035029, EPI_ISL_2035030 | | | |
| see above | Yale Clinical Virology Lab | Yale Center for Genomic Analysis | Shrikant Mane, Kaya Bilguvar, Curt Scharfe, Irina Tikhonova, Brooke Sullivan |
| EPI_ISL_2035031 | Aegis Sciences Corporation | Centers for Disease Control and Prevention Division of Viral Diseases, Pathogen Discovery | Dakota Howard, Dhwani Batra, Peter W. Cook, Kara Moser, Adrian Paskey, Jason Caravas, Benjamin Rambo-Martin, Shatavia Morrison, Christopher Gulvick, Scott Sammons, Yvette Unoarumhi, Darlene Wagner, Matthew Schmerer, Cyndi Clark, Patrick Campbell, Rob Case, Vikramsinha Ghorpade, Holly Houdeshell, Ola Kvalvaag, Dillon Nall, Ethan Sanders, Alec Vest, Shaun Westlund, Matthew Hardison, Clinton R. Paden, Duncan MacCannell |
| EPI_ISL_2035032, EPI_ISL_2035033 | Yale Clinical Virology Lab | Yale Center for Genomic Analysis | Shrikant Mane, Kaya Bilguvar, Curt Scharfe, Irina Tikhonova, Brooke Sullivan |
| EPI_ISL_2035034 | Aegis Sciences Corporation | Centers for Disease Control and Prevention Division of Viral Diseases, Pathogen Discovery | Dakota Howard, Dhwani Batra, Peter W. Cook, Kara Moser, Adrian Paskey, Jason Caravas, Benjamin Rambo-Martin, Shatavia Morrison, Christopher Gulvick, Scott Sammons, Yvette Unoarumhi, Darlene Wagner, Matthew Schmerer, Cyndi Clark, Patrick Campbell, Rob Case, Vikramsinha Ghorpade, Holly Houdeshell, Ola Kvalvaag, Dillon Nall, Ethan Sanders, Alec Vest, Shaun Westlund, Matthew Hardison, Clinton R. Paden, Duncan MacCannell |
| EPI_ISL_2035035 | Yale Clinical Virology Lab | Yale Center for Genomic Analysis | Shrikant Mane, Kaya Bilguvar, Curt Scharfe, Irina Tikhonova, Brooke Sullivan |
| EPI_ISL_2035036 | Aegis Sciences Corporation | Centers for Disease Control and Prevention Division of Viral Diseases, Pathogen Discovery | Dakota Howard, Dhwani Batra, Peter W. Cook, Kara Moser, Adrian Paskey, Jason Caravas, Benjamin Rambo-Martin, Shatavia Morrison, Christopher Gulvick, Scott Sammons, Yvette Unoarumhi, Darlene Wagner, Matthew Schmerer, Cyndi Clark, Patrick Campbell, Rob Case, Vikramsinha Ghorpade, Holly Houdeshell, Ola Kvalvaag, Dillon Nall, Ethan Sanders, Alec Vest, Shaun Westlund, Matthew Hardison, Clinton R. Paden, Duncan MacCannell |
| EPI_ISL_2035037, EPI_ISL_2035038 | Yale Clinical Virology Lab | Yale Center for Genomic Analysis | Shrikant Mane, Kaya Bilguvar, Curt Scharfe, Irina Tikhonova, Brooke Sullivan |
| EPI_ISL_2035039 | Aegis Sciences Corporation | Centers for Disease Control and Prevention Division of Viral Diseases, Pathogen Discovery | Dakota Howard, Dhwani Batra, Peter W. Cook, Kara Moser, Adrian Paskey, Jason Caravas, Benjamin Rambo-Martin, Shatavia Morrison, Christopher Gulvick, Scott Sammons, Yvette Unoarumhi, Darlene Wagner, Matthew Schmerer, Cyndi Clark, Patrick Campbell, Rob Case, Vikramsinha Ghorpade, Holly Houdeshell, Ola Kvalvaag, Dillon Nall, Ethan Sanders, Alec Vest, Shaun Westlund, Matthew Hardison, Clinton R. Paden, Duncan MacCannell |
| EPI_ISL_2035040 | Yale Clinical Virology Lab | Yale Center for Genomic Analysis | Shrikant Mane, Kaya Bilguvar, Curt Scharfe, Irina Tikhonova, Brooke Sullivan |
| EPI_ISL_2035041 | Aegis Sciences Corporation | Centers for Disease Control and Prevention Division of Viral Diseases, Pathogen Discovery | Dakota Howard, Dhwani Batra, Peter W. Cook, Kara Moser, Adrian Paskey, Jason Caravas, Benjamin Rambo-Martin, Shatavia Morrison, Christopher Gulvick, Scott Sammons, Yvette Unoarumhi, Darlene Wagner, Matthew Schmerer, Cyndi Clark, Patrick Campbell, Rob Case, Vikramsinha Ghorpade, Holly Houdeshell, Ola Kvalvaag, Dillon Nall, Ethan Sanders, Alec Vest, Shaun Westlund, Matthew Hardison, Clinton R. Paden, Duncan MacCannell |
| EPI_ISL_2035042 | Yale Clinical Virology Lab | Yale Center for Genomic Analysis | Shrikant Mane, Kaya Bilguvar, Curt Scharfe, Irina Tikhonova, Brooke Sullivan |
| EPI_ISL_2035043 | Aegis Sciences Corporation | Centers for Disease Control and Prevention Division of Viral Diseases, Pathogen Discovery | Dakota Howard, Dhwani Batra, Peter W. Cook, Kara Moser, Adrian Paskey, Jason Caravas, Benjamin Rambo-Martin, Shatavia Morrison, Christopher Gulvick, Scott Sammons, Yvette Unoarumhi, Darlene Wagner, Matthew Schmerer, Cyndi Clark, Patrick Campbell, Rob Case, Vikramsinha Ghorpade, Holly Houdeshell, Ola Kvalvaag, Dillon Nall, Ethan Sanders, Alec Vest, Shaun Westlund, Matthew Hardison, Clinton R. Paden, Duncan MacCannell |
| EPI_ISL_2035044, EPI_ISL_2035045 | Yale Clinical Virology Lab | Yale Center for Genomic Analysis | Shrikant Mane, Kaya Bilguvar, Curt Scharfe, Irina Tikhonova, Brooke Sullivan |
| EPI_ISL_2035046 | Aegis Sciences Corporation | Centers for Disease Control and Prevention Division of Viral Diseases, Pathogen Discovery | Dakota Howard, Dhwani Batra, Peter W. Cook, Kara Moser, Adrian Paskey, Jason Caravas, Benjamin Rambo-Martin, Shatavia Morrison, Christopher Gulvick, Scott Sammons, Yvette Unoarumhi, Darlene Wagner, Matthew Schmerer, Cyndi Clark, Patrick Campbell, Rob Case, Vikramsinha Ghorpade, Holly Houdeshell, Ola Kvalvaag, Dillon Nall, Ethan Sanders, Alec Vest, Shaun Westlund, Matthew Hardison, Clinton R. Paden, Duncan MacCannell |
| EPI_ISL_2035047, EPI_ISL_2035048 | Yale Clinical Virology Lab | Yale Center for Genomic Analysis | Shrikant Mane, Kaya Bilguvar, Curt Scharfe, Irina Tikhonova, Brooke Sullivan |
| EPI_ISL_2035049 | Aegis Sciences Corporation | Centers for Disease Control and Prevention Division of Viral Diseases, Pathogen Discovery | Dakota Howard, Dhwani Batra, Peter W. Cook, Kara Moser, Adrian Paskey, Jason Caravas, Benjamin Rambo-Martin, Shatavia Morrison, Christopher Gulvick, Scott Sammons, Yvette Unoarumhi, Darlene Wagner, Matthew Schmerer, Cyndi Clark, Patrick Campbell, Rob Case, Vikramsinha Ghorpade, Holly Houdeshell, Ola Kvalvaag, Dillon Nall, Ethan Sanders, Alec Vest, Shaun Westlund, Matthew Hardison, Clinton R. Paden, Duncan MacCannell |
| EPI_ISL_2035050, EPI_ISL_2035051 | Yale Clinical Virology Lab | Yale Center for Genomic Analysis | Shrikant Mane, Kaya Bilguvar, Curt Scharfe, Irina Tikhonova, Brooke Sullivan |
| EPI_ISL_2035052, EPI_ISL_2035053 | Aegis Sciences Corporation | Centers for Disease Control and Prevention Division of Viral Diseases, Pathogen Discovery | Dakota Howard, Dhwani Batra, Peter W. Cook, Kara Moser, Adrian Paskey, Jason Caravas, Benjamin Rambo-Martin, Shatavia Morrison, Christopher Gulvick, Scott Sammons, Yvette Unoarumhi, Darlene Wagner, Matthew Schmerer, Cyndi Clark, Patrick Campbell, Rob Case, Vikramsinha Ghorpade, Holly Houdeshell, Ola Kvalvaag, Dillon Nall, Ethan Sanders, Alec Vest, Shaun Westlund, Matthew Hardison, Clinton R. Paden, Duncan MacCannell |
| EPI_ISL_2035054, EPI_ISL_2035055 | Yale Clinical Virology Lab | Yale Center for Genomic Analysis | Shrikant Mane, Kaya Bilguvar, Curt Scharfe, Irina Tikhonova, Brooke Sullivan |
| EPI_ISL_2035056 | Aegis Sciences Corporation | Centers for Disease Control and Prevention Division of Viral Diseases, Pathogen Discovery | Dakota Howard, Dhwani Batra, Peter W. Cook, Kara Moser, Adrian Paskey, Jason Caravas, Benjamin Rambo-Martin, Shatavia Morrison, Christopher Gulvick, Scott Sammons, Yvette Unoarumhi, Darlene Wagner, Matthew Schmerer, Cyndi Clark, Patrick Campbell, Rob Case, Vikramsinha Ghorpade, Holly Houdeshell, Ola Kvalvaag, Dillon Nall, Ethan Sanders, Alec Vest, Shaun Westlund, Matthew Hardison, Clinton R. Paden, Duncan MacCannell |
| EPI_ISL_2035057, EPI_ISL_2035058 | Yale Clinical Virology Lab | Yale Center for Genomic Analysis | Shrikant Mane, Kaya Bilguvar, Curt Scharfe, Irina Tikhonova, Brooke Sullivan |
| EPI_ISL_2035059 | Aegis Sciences Corporation | Centers for Disease Control and Prevention Division of Viral Diseases, Pathogen Discovery | Dakota Howard, Dhwani Batra, Peter W. Cook, Kara Moser, Adrian Paskey, Jason Caravas, Benjamin Rambo-Martin, Shatavia Morrison, Christopher Gulvick, Scott Sammons, Yvette Unoarumhi, Darlene Wagner, Matthew Schmerer, Cyndi Clark, Patrick Campbell, Rob Case, Vikramsinha Ghorpade, Holly Houdeshell, Ola Kvalvaag, Dillon Nall, Ethan Sanders, Alec Vest, Shaun Westlund, Matthew Hardison, Clinton R. Paden, Duncan MacCannell |
| EPI_ISL_2035060, EPI_ISL_2035061, EPI_ISL_2035062 | Yale Clinical Virology Lab | Yale Center for Genomic Analysis | Shrikant Mane, Kaya Bilguvar, Curt Scharfe, Irina Tikhonova, Brooke Sullivan |
| EPI_ISL_2035063 | Aegis Sciences Corporation | Centers for Disease Control and Prevention Division of Viral Diseases, Pathogen Discovery | Dakota Howard, Dhwani Batra, Peter W. Cook, Kara Moser, Adrian Paskey, Jason Caravas, Benjamin Rambo-Martin, Shatavia Morrison, Christopher Gulvick, Scott Sammons, Yvette Unoarumhi, Darlene Wagner, Matthew Schmerer, Cyndi Clark, Patrick Campbell, Rob Case, Vikramsinha Ghorpade, Holly Houdeshell, Ola Kvalvaag, Dillon Nall, Ethan Sanders, Alec Vest, Shaun Westlund, Matthew Hardison, Clinton R. Paden, Duncan MacCannell |
| EPI_ISL_2035064 | Yale Clinical Virology Lab | Yale Center for Genomic Analysis | Shrikant Mane, Kaya Bilguvar, Curt Scharfe, Irina Tikhonova, Brooke Sullivan |
| EPI_ISL_2035065 | Aegis Sciences Corporation | Centers for Disease Control and Prevention Division of Viral Diseases, Pathogen Discovery | Dakota Howard, Dhwani Batra, Peter W. Cook, Kara Moser, Adrian Paskey, Jason Caravas, Benjamin Rambo-Martin, Shatavia Morrison, Christopher Gulvick, Scott Sammons, Yvette Unoarumhi, Darlene Wagner, Matthew Schmerer, Cyndi Clark, Patrick Campbell, Rob Case, Vikramsinha Ghorpade, Holly Houdeshell, Ola Kvalvaag, Dillon Nall, Ethan Sanders, Alec Vest, Shaun Westlund, Matthew Hardison, Clinton R. Paden, Duncan MacCannell |
| EPI_ISL_2035066 | Yale Clinical Virology Lab | Yale Center for Genomic Analysis | Shrikant Mane, Kaya Bilguvar, Curt Scharfe, Irina Tikhonova, Brooke Sullivan |
| EPI_ISL_2035067 | Aegis Sciences Corporation | Centers for Disease Control and Prevention Division of Viral Diseases, Pathogen Discovery | Dakota Howard, Dhwani Batra, Peter W. Cook, Kara Moser, Adrian Paskey, Jason Caravas, Benjamin Rambo-Martin, Shatavia Morrison, Christopher Gulvick, Scott Sammons, Yvette Unoarumhi, Darlene Wagner, Matthew Schmerer, Cyndi Clark, Patrick Campbell, Rob Case, Vikramsinha Ghorpade, Holly Houdeshell, Ola Kvalvaag, Dillon Nall, Ethan Sanders, Alec Vest, Shaun Westlund, Matthew Hardison, Clinton R. Paden, Duncan MacCannell |
| EPI_ISL_2035068 | Yale Clinical Virology Lab | Yale Center for Genomic Analysis | Shrikant Mane, Kaya Bilguvar, Curt Scharfe, Irina Tikhonova, Brooke Sullivan |
| EPI_ISL_2035069 | Aegis Sciences Corporation | Centers for Disease Control and Prevention Division of Viral Diseases, Pathogen Discovery | Dakota Howard, Dhwani Batra, Peter W. Cook, Kara Moser, Adrian Paskey, Jason Caravas, Benjamin Rambo-Martin, Shatavia Morrison, Christopher Gulvick, Scott Sammons, Yvette Unoarumhi, Darlene Wagner, Matthew Schmerer, Cyndi Clark, Patrick Campbell, Rob Case, Vikramsinha Ghorpade, Holly Houdeshell, Ola Kvalvaag, Dillon Nall, Ethan Sanders, Alec Vest, Shaun Westlund, Matthew Hardison, Clinton R. Paden, Duncan MacCannell |
| EPI_ISL_2035070 | Yale Clinical Virology Lab | Yale Center for Genomic Analysis | Shrikant Mane, Kaya Bilguvar, Curt Scharfe, Irina Tikhonova, Brooke Sullivan |
| EPI_ISL_2035071 | Aegis Sciences Corporation | Centers for Disease Control and Prevention Division of Viral Diseases, Pathogen Discovery | Dakota Howard, Dhwani Batra, Peter W. Cook, Kara Moser, Adrian Paskey, Jason Caravas, Benjamin Rambo-Martin, Shatavia Morrison, Christopher Gulvick, Scott Sammons, Yvette Unoarumhi, Darlene Wagner, Matthew Schmerer, Cyndi Clark, Patrick Campbell, Rob Case, Vikramsinha Ghorpade, Holly Houdeshell, Ola Kvalvaag, Dillon Nall, Ethan Sanders, Alec Vest, Shaun Westlund, Matthew Hardison, Clinton R. Paden, Duncan MacCannell |
| EPI_ISL_2035072 | Yale Clinical Virology Lab | Yale Center for Genomic Analysis | Shrikant Mane, Kaya Bilguvar, Curt Scharfe, Irina Tikhonova, Brooke Sullivan |
| EPI_ISL_2035073 | Aegis Sciences Corporation | Centers for Disease Control and Prevention Division of Viral Diseases, Pathogen Discovery | Dakota Howard, Dhwani Batra, Peter W. Cook, Kara Moser, Adrian Paskey, Jason Caravas, Benjamin Rambo-Martin, Shatavia Morrison, Christopher Gulvick, Scott Sammons, Yvette Unoarumhi, Darlene Wagner, Matthew Schmerer, Cyndi Clark, Patrick Campbell, Rob Case, Vikramsinha Ghorpade, Holly Houdeshell, Ola Kvalvaag, Dillon Nall, Ethan Sanders, Alec Vest, Shaun Westlund, Matthew Hardison, Clinton R. Paden, Duncan MacCannell |
| EPI_ISL_2035074 | Yale Clinical Virology Lab | Yale Center for Genomic Analysis | Shrikant Mane, Kaya Bilguvar, Curt Scharfe, Irina Tikhonova, Brooke Sullivan |

| | | | |
|---|---|---|---|
| EPI_ISL_2035075 | Aegis Sciences Corporation | Centers for Disease Control and Prevention Division of Viral Diseases, Pathogen Discovery | Dakota Howard, Dhwani Batra, Peter W. Cook, Kara Moser, Adrian Paskey, Jason Caravas, Benjamin Rambo-Martin, Shatavia Morrison, Christopher Gulvick, Scott Sammons, Yvette Unoarumhi, Darlene Wagner, Matthew Schmerer, Cyndi Clark, Patrick Campbell, Rob Case, Vikramsinha Ghorpade, Holly Houdeshell, Ola Kvalvaag, Dillon Nall, Ethan Sanders, Alec Vest, Shaun Westlund, Matthew Hardison, Clinton R. Paden, Duncan MacCannell |
| EPI_ISL_2035076, EPI_ISL_2035077 | Yale Clinical Virology Lab | Yale Center for Genomic Analysis | Shrikant Mane, Kaya Bilguvar, Curt Scharfe, Irina Tikhonova, Brooke Sullivan |
| EPI_ISL_2035078 | Aegis Sciences Corporation | Centers for Disease Control and Prevention Division of Viral Diseases, Pathogen Discovery | Dakota Howard, Dhwani Batra, Peter W. Cook, Kara Moser, Adrian Paskey, Jason Caravas, Benjamin Rambo-Martin, Shatavia Morrison, Christopher Gulvick, Scott Sammons, Yvette Unoarumhi, Darlene Wagner, Matthew Schmerer, Cyndi Clark, Patrick Campbell, Rob Case, Vikramsinha Ghorpade, Holly Houdeshell, Ola Kvalvaag, Dillon Nall, Ethan Sanders, Alec Vest, Shaun Westlund, Matthew Hardison, Clinton R. Paden, Duncan MacCannell |
| EPI_ISL_2035079, EPI_ISL_2035080 | Yale Clinical Virology Lab | Yale Center for Genomic Analysis | Shrikant Mane, Kaya Bilguvar, Curt Scharfe, Irina Tikhonova, Brooke Sullivan |
| EPI_ISL_2035081 | Aegis Sciences Corporation | Centers for Disease Control and Prevention Division of Viral Diseases, Pathogen Discovery | Dakota Howard, Dhwani Batra, Peter W. Cook, Kara Moser, Adrian Paskey, Jason Caravas, Benjamin Rambo-Martin, Shatavia Morrison, Christopher Gulvick, Scott Sammons, Yvette Unoarumhi, Darlene Wagner, Matthew Schmerer, Cyndi Clark, Patrick Campbell, Rob Case, Vikramsinha Ghorpade, Holly Houdeshell, Ola Kvalvaag, Dillon Nall, Ethan Sanders, Alec Vest, Shaun Westlund, Matthew Hardison, Clinton R. Paden, Duncan MacCannell |
| EPI_ISL_2035082, EPI_ISL_2035083 | Yale Clinical Virology Lab | Yale Center for Genomic Analysis | Shrikant Mane, Kaya Bilguvar, Curt Scharfe, Irina Tikhonova, Brooke Sullivan |
| EPI_ISL_2035084 | Aegis Sciences Corporation | Centers for Disease Control and Prevention Division of Viral Diseases, Pathogen Discovery | Dakota Howard, Dhwani Batra, Peter W. Cook, Kara Moser, Adrian Paskey, Jason Caravas, Benjamin Rambo-Martin, Shatavia Morrison, Christopher Gulvick, Scott Sammons, Yvette Unoarumhi, Darlene Wagner, Matthew Schmerer, Cyndi Clark, Patrick Campbell, Rob Case, Vikramsinha Ghorpade, Holly Houdeshell, Ola Kvalvaag, Dillon Nall, Ethan Sanders, Alec Vest, Shaun Westlund, Matthew Hardison, Clinton R. Paden, Duncan MacCannell |
| EPI_ISL_2035085 | Yale Clinical Virology Lab | Yale Center for Genomic Analysis | Shrikant Mane, Kaya Bilguvar, Curt Scharfe, Irina Tikhonova, Brooke Sullivan |
| EPI_ISL_2035086 | Aegis Sciences Corporation | Centers for Disease Control and Prevention Division of Viral Diseases, Pathogen Discovery | Dakota Howard, Dhwani Batra, Peter W. Cook, Kara Moser, Adrian Paskey, Jason Caravas, Benjamin Rambo-Martin, Shatavia Morrison, Christopher Gulvick, Scott Sammons, Yvette Unoarumhi, Darlene Wagner, Matthew Schmerer, Cyndi Clark, Patrick Campbell, Rob Case, Vikramsinha Ghorpade, Holly Houdeshell, Ola Kvalvaag, Dillon Nall, Ethan Sanders, Alec Vest, Shaun Westlund, Matthew Hardison, Clinton R. Paden, Duncan MacCannell |
| EPI_ISL_2035087, EPI_ISL_2035088 | Yale Clinical Virology Lab | Yale Center for Genomic Analysis | Shrikant Mane, Kaya Bilguvar, Curt Scharfe, Irina Tikhonova, Brooke Sullivan |
| EPI_ISL_2035089 | Aegis Sciences Corporation | Centers for Disease Control and Prevention Division of Viral Diseases, Pathogen Discovery | Dakota Howard, Dhwani Batra, Peter W. Cook, Kara Moser, Adrian Paskey, Jason Caravas, Benjamin Rambo-Martin, Shatavia Morrison, Christopher Gulvick, Scott Sammons, Yvette Unoarumhi, Darlene Wagner, Matthew Schmerer, Cyndi Clark, Patrick Campbell, Rob Case, Vikramsinha Ghorpade, Holly Houdeshell, Ola Kvalvaag, Dillon Nall, Ethan Sanders, Alec Vest, Shaun Westlund, Matthew Hardison, Clinton R. Paden, Duncan MacCannell |
| EPI_ISL_2035090, EPI_ISL_2035091 | Yale Clinical Virology Lab | Yale Center for Genomic Analysis | Shrikant Mane, Kaya Bilguvar, Curt Scharfe, Irina Tikhonova, Brooke Sullivan |
| EPI_ISL_2035092 | Aegis Sciences Corporation | Centers for Disease Control and Prevention Division of Viral Diseases, Pathogen Discovery | Dakota Howard, Dhwani Batra, Peter W. Cook, Kara Moser, Adrian Paskey, Jason Caravas, Benjamin Rambo-Martin, Shatavia Morrison, Christopher Gulvick, Scott Sammons, Yvette Unoarumhi, Darlene Wagner, Matthew Schmerer, Cyndi Clark, Patrick Campbell, Rob Case, Vikramsinha Ghorpade, Holly Houdeshell, Ola Kvalvaag, Dillon Nall, Ethan Sanders, Alec Vest, Shaun Westlund, Matthew Hardison, Clinton R. Paden, Duncan MacCannell |
| EPI_ISL_2035093 | Yale Clinical Virology Lab | Yale Center for Genomic Analysis | Shrikant Mane, Kaya Bilguvar, Curt Scharfe, Irina Tikhonova, Brooke Sullivan |
| EPI_ISL_2035094 | Aegis Sciences Corporation | Centers for Disease Control and Prevention Division of Viral Diseases, Pathogen Discovery | Dakota Howard, Dhwani Batra, Peter W. Cook, Kara Moser, Adrian Paskey, Jason Caravas, Benjamin Rambo-Martin, Shatavia Morrison, Christopher Gulvick, Scott Sammons, Yvette Unoarumhi, Darlene Wagner, Matthew Schmerer, Cyndi Clark, Patrick Campbell, Rob Case, Vikramsinha Ghorpade, Holly Houdeshell, Ola Kvalvaag, Dillon Nall, Ethan Sanders, Alec Vest, Shaun Westlund, Matthew Hardison, Clinton R. Paden, Duncan MacCannell |
| EPI_ISL_2035095 | Yale Clinical Virology Lab | Yale Center for Genomic Analysis | Shrikant Mane, Kaya Bilguvar, Curt Scharfe, Irina Tikhonova, Brooke Sullivan |
| EPI_ISL_2035096 | Aegis Sciences Corporation | Centers for Disease Control and Prevention Division of Viral Diseases, Pathogen Discovery | Dakota Howard, Dhwani Batra, Peter W. Cook, Kara Moser, Adrian Paskey, Jason Caravas, Benjamin Rambo-Martin, Shatavia Morrison, Christopher Gulvick, Scott Sammons, Yvette Unoarumhi, Darlene Wagner, Matthew Schmerer, Cyndi Clark, Patrick Campbell, Rob Case, Vikramsinha Ghorpade, Holly Houdeshell, Ola Kvalvaag, Dillon Nall, Ethan Sanders, Alec Vest, Shaun Westlund, Matthew Hardison, Clinton R. Paden, Duncan MacCannell |
| EPI_ISL_2035098 | Yale Clinical Virology Lab | Yale Center for Genomic Analysis | Shrikant Mane, Kaya Bilguvar, Curt Scharfe, Irina Tikhonova, Brooke Sullivan |
| EPI_ISL_2035099 | Aegis Sciences Corporation | Centers for Disease Control and Prevention Division of Viral Diseases, Pathogen Discovery | Dakota Howard, Dhwani Batra, Peter W. Cook, Kara Moser, Adrian Paskey, Jason Caravas, Benjamin Rambo-Martin, Shatavia Morrison, Christopher Gulvick, Scott Sammons, Yvette Unoarumhi, Darlene Wagner, Matthew Schmerer, Cyndi Clark, Patrick Campbell, Rob Case, Vikramsinha Ghorpade, Holly Houdeshell, Ola Kvalvaag, Dillon Nall, Ethan Sanders, Alec Vest, Shaun Westlund, Matthew Hardison, Clinton R. Paden, Duncan MacCannell |
| EPI_ISL_2035100, EPI_ISL_2035101 | Yale Clinical Virology Lab | Yale Center for Genomic Analysis | Shrikant Mane, Kaya Bilguvar, Curt Scharfe, Irina Tikhonova, Brooke Sullivan |
| EPI_ISL_2035102, EPI_ISL_2035103, EPI_ISL_2035104, EPI_ISL_2035105, EPI_ISL_2035106, EPI_ISL_2035107, EPI_ISL_2035108, EPI_ISL_2035109, EPI_ISL_2035110, EPI_ISL_2035111, EPI_ISL_2035112, EPI_ISL_2035113, EPI_ISL_2035114, EPI_ISL_2035115, EPI_ISL_2035116, EPI_ISL_2035117, EPI_ISL_2035118, EPI_ISL_2035119, EPI_ISL_2035120, EPI_ISL_2035121, EPI_ISL_2035122, EPI_ISL_2035123, EPI_ISL_2035124, EPI_ISL_2035125, EPI_ISL_2035126, EPI_ISL_2035127, EPI_ISL_2035128, EPI_ISL_2035129, EPI_ISL_2035130, EPI_ISL_2035131, EPI_ISL_2035132, EPI_ISL_2035133, EPI_ISL_2035135, EPI_ISL_2035136, EPI_ISL_2035137, EPI_ISL_2035138, EPI_ISL_2035139, EPI_ISL_2035140, EPI_ISL_2035141, EPI_ISL_2035142, EPI_ISL_2035143, EPI_ISL_2035144, EPI_ISL_2035145, EPI_ISL_2035146, EPI_ISL_2035148, EPI_ISL_2035149, EPI_ISL_2035150, EPI_ISL_2035151, EPI_ISL_2035152, EPI_ISL_2035153, EPI_ISL_2035154, EPI_ISL_2035155, EPI_ISL_2035156, EPI_ISL_2035157, EPI_ISL_2035158, EPI_ISL_2035159, EPI_ISL_2035160, EPI_ISL_2035161, EPI_ISL_2035162, EPI_ISL_2035163, EPI_ISL_2035164, EPI_ISL_2035165, EPI_ISL_2035166, EPI_ISL_2035167, EPI_ISL_2035168, EPI_ISL_2035169, EPI_ISL_2035170, EPI_ISL_2035171, EPI_ISL_2035172, EPI_ISL_2035173, EPI_ISL_2035174, EPI_ISL_2035175, EPI_ISL_2035176, EPI_ISL_2035177, EPI_ISL_2035178, EPI_ISL_2035179, EPI_ISL_2035180, EPI_ISL_2035181, EPI_ISL_2035182, EPI_ISL_2035183, EPI_ISL_2035184, EPI_ISL_2035185, EPI_ISL_2035186, EPI_ISL_2035187, EPI_ISL_2035188, EPI_ISL_2035189, EPI_ISL_2035190, EPI_ISL_2035191, EPI_ISL_2035192, EPI_ISL_2035193, EPI_ISL_2035194, EPI_ISL_2035195, EPI_ISL_2035196, EPI_ISL_2035197, EPI_ISL_2035198, EPI_ISL_2035199, EPI_ISL_2035200, EPI_ISL_2035201, EPI_ISL_2035202, EPI_ISL_2035203, EPI_ISL_2035204, EPI_ISL_2035205, EPI_ISL_2035206, EPI_ISL_2035207, EPI_ISL_2035208, EPI_ISL_2035209, EPI_ISL_2035210, EPI_ISL_2035211, EPI_ISL_2035212, EPI_ISL_2035213, EPI_ISL_2035214, EPI_ISL_2035215, EPI_ISL_2035216, EPI_ISL_2035217, EPI_ISL_2035218, EPI_ISL_2035219, EPI_ISL_2035220, EPI_ISL_2035221, EPI_ISL_2035222, EPI_ISL_2035223, EPI_ISL_2035224, EPI_ISL_2035225, EPI_ISL_2035226, EPI_ISL_2035227, EPI_ISL_2035228, EPI_ISL_2035229, EPI_ISL_2035230, EPI_ISL_2035231, EPI_ISL_2035232, EPI_ISL_2035233, EPI_ISL_2035234, EPI_ISL_2035235, EPI_ISL_2035236, EPI_ISL_2035237, EPI_ISL_2035238, EPI_ISL_2035239, EPI_ISL_2035240, EPI_ISL_2035241, EPI_ISL_2035242, EPI_ISL_2035243, EPI_ISL_2035244, EPI_ISL_2035245, EPI_ISL_2035246, EPI_ISL_2035247, EPI_ISL_2035248, EPI_ISL_2035249, EPI_ISL_2035250, EPI_ISL_2035251, EPI_ISL_2035253, EPI_ISL_2035254, EPI_ISL_2035255, EPI_ISL_2035256, EPI_ISL_2035257, EPI_ISL_2035258, EPI_ISL_2035259, EPI_ISL_2035260, EPI_ISL_2035261, EPI_ISL_2035262, EPI_ISL_2035263, EPI_ISL_2035264 | | | |
| see above | Aegis Sciences Corporation | Centers for Disease Control and Prevention Division of Viral Diseases, Pathogen Discovery | Dakota Howard, Dhwani Batra, Peter W. Cook, Kara Moser, Adrian Paskey, Jason Caravas, Benjamin Rambo-Martin, Shatavia Morrison, Christopher Gulvick, Scott Sammons, Yvette Unoarumhi, Darlene Wagner, Matthew Schmerer, Cyndi Clark, Patrick Campbell, Rob Case, Vikramsinha Ghorpade, Holly Houdeshell, Ola Kvalvaag, Dillon Nall, Ethan Sanders, Alec Vest, Shaun Westlund, Matthew Hardison, Clinton R. Paden, Duncan MacCannell |
| EPI_ISL_2035266, EPI_ISL_2035267, EPI_ISL_2035268, EPI_ISL_2035269, EPI_ISL_2035270, EPI_ISL_2035271, EPI_ISL_2035272, EPI_ISL_2035273, EPI_ISL_2035274, EPI_ISL_2035275, EPI_ISL_2035276, EPI_ISL_2035277, EPI_ISL_2035278, EPI_ISL_2035279, EPI_ISL_2035280, EPI_ISL_2035281, EPI_ISL_2035282, EPI_ISL_2035284, EPI_ISL_2035285, EPI_ISL_2035286, EPI_ISL_2035287, EPI_ISL_2035288, EPI_ISL_2035289, EPI_ISL_2035290, EPI_ISL_2035291, EPI_ISL_2035292, EPI_ISL_2035293, EPI_ISL_2035294, EPI_ISL_2035295, EPI_ISL_2035296, EPI_ISL_2035297, EPI_ISL_2035298, EPI_ISL_2035299, EPI_ISL_2035300, EPI_ISL_2035301 | | | |
| see above | Helix/Illumina | Centers for Disease Control and Prevention Division of Viral Diseases, Pathogen Discovery | Dakota Howard, Dhwani Batra, Peter W. Cook, Kara Moser, Adrian Paskey, Jason Caravas, Benjamin Rambo-Martin, Shatavia Morrison, Christopher Gulvick, Scott Sammons, Yvette Unoarumhi, Darlene Wagner, Matthew Schmerer, Eileen de Feo, Jan Antico, Christine Tran, Matthew Tolentino, Shannon Wickline, Kim Gietzen, Brad Sickler, Jingtao Liu, Eric Allen, Phil Febbo, Nicole L. Washington, Simon White, Geraint Levan, Kelly Schiabor Barrett, Elizabeth Cirulli, Alexandre Bolze, Ary Ascencio, Charlotte Rivera-Garcia, Ryan Cho, Jason Nguyen, Sherry Wang, Jimmy Ramirez, Tyler Cassens, Efren Sandoval, Magnus Isaksson, William Lee, David Becker, Marc Laurent, James Lu, Clinton R. Paden, Duncan MacCannell |
| EPI_ISL_2035302, EPI_ISL_2035303, EPI_ISL_2035304, EPI_ISL_2035305, EPI_ISL_2035306, EPI_ISL_2035307, EPI_ISL_2035308, EPI_ISL_2035309, EPI_ISL_2035310, EPI_ISL_2035311, EPI_ISL_2035312, EPI_ISL_2035313, EPI_ISL_2035314, EPI_ISL_2035315, EPI_ISL_2035316, EPI_ISL_2035317, EPI_ISL_2035318, EPI_ISL_2035319, EPI_ISL_2035320, EPI_ISL_2035321, EPI_ISL_2035322, EPI_ISL_2035323, EPI_ISL_2035324, EPI_ISL_2035325, EPI_ISL_2035326, EPI_ISL_2035327, EPI_ISL_2035328, EPI_ISL_2035329, EPI_ISL_2035330, EPI_ISL_2035331, EPI_ISL_2035332, EPI_ISL_2035333, EPI_ISL_2035334, EPI_ISL_2035335, EPI_ISL_2035336, EPI_ISL_2035337, EPI_ISL_2035338, EPI_ISL_2035339, EPI_ISL_2035340, EPI_ISL_2035341, EPI_ISL_2035342, EPI_ISL_2035343, EPI_ISL_2035344, EPI_ISL_2035345, EPI_ISL_2035346, EPI_ISL_2035347, EPI_ISL_2035348, EPI_ISL_2035349, EPI_ISL_2035350, EPI_ISL_2035351, EPI_ISL_2035352, EPI_ISL_2035353, EPI_ISL_2035354, EPI_ISL_2035355, EPI_ISL_2035356, EPI_ISL_2035357, EPI_ISL_2035358, EPI_ISL_2035359, EPI_ISL_2035360, EPI_ISL_2035361, EPI_ISL_2035362, EPI_ISL_2035363, EPI_ISL_2035364 | | | |
| see above | Quest Diagnostics Incorporated | Centers for Disease Control and Prevention Division of Viral Diseases, Pathogen Discovery | Dakota Howard, Dhwani Batra, Peter W. Cook, Kara Moser, Adrian Paskey, Jason Caravas, Benjamin Rambo-Martin, Shatavia Morrison, Christopher Gulvick, Scott Sammons, Yvette Unoarumhi, Darlene Wagner, Matthew Schmerer, S. H. Rosenthal, A. Gerasimova, R. M. Kagan, B. Anderson, M. Hua, Y. Liu, L.E. Bernstein, K.E. Livingston, A. Perez, I. A. Shlyakhter, R. V. Rolando, R. Owen, P. Tanpaiboon, F. Lacbawan, Clinton R. Paden, Duncan MacCannell |
| EPI_ISL_2035475, EPI_ISL_2035476, EPI_ISL_2035477, EPI_ISL_2035478, EPI_ISL_2035480, EPI_ISL_2035481, EPI_ISL_2035482, EPI_ISL_2035483, EPI_ISL_2035484, EPI_ISL_2035485, EPI_ISL_2035486, EPI_ISL_2035487, EPI_ISL_2035488, EPI_ISL_2035489, EPI_ISL_2035490, EPI_ISL_2035491, EPI_ISL_2035492, EPI_ISL_2035493, EPI_ISL_2035494, EPI_ISL_2035495, EPI_ISL_2035496, EPI_ISL_2035497, EPI_ISL_2035498, EPI_ISL_2035499, EPI_ISL_2035500, EPI_ISL_2035501, EPI_ISL_2035502, EPI_ISL_2035503, EPI_ISL_2035504, EPI_ISL_2035505, EPI_ISL_2035506, EPI_ISL_2035507, EPI_ISL_2035508, EPI_ISL_2035509, EPI_ISL_2035510, EPI_ISL_2035511, EPI_ISL_2035512, EPI_ISL_2035513, EPI_ISL_2035514, EPI_ISL_2035515, EPI_ISL_2035516, EPI_ISL_2035517, EPI_ISL_2035518, EPI_ISL_2035519, EPI_ISL_2035520, EPI_ISL_2035521, EPI_ISL_2035522, EPI_ISL_2035523, EPI_ISL_2035524, EPI_ISL_2035525, EPI_ISL_2035526, EPI_ISL_2035527, EPI_ISL_2035528, EPI_ISL_2035529, EPI_ISL_2035530, EPI_ISL_2035531, EPI_ISL_2035532, EPI_ISL_2035533, EPI_ISL_2035534, EPI_ISL_2035535, EPI_ISL_2035536, EPI_ISL_2035537, EPI_ISL_2035538, EPI_ISL_2035539, EPI_ISL_2035540, EPI_ISL_2035541, EPI_ISL_2035542, EPI_ISL_2035543, EPI_ISL_2035544, EPI_ISL_2035545, EPI_ISL_2035546, EPI_ISL_2035547, EPI_ISL_2035548, EPI_ISL_2035549, EPI_ISL_2035550, EPI_ISL_2035551, EPI_ISL_2035552, EPI_ISL_2035553, EPI_ISL_2035554, EPI_ISL_2035555, EPI_ISL_2035556, EPI_ISL_2035557, EPI_ISL_2035558, EPI_ISL_2035564, EPI_ISL_2035565, EPI_ISL_2035566, EPI_ISL_2035567, EPI_ISL_2035568, EPI_ISL_2035570, EPI_ISL_2035571, EPI_ISL_2035573, EPI_ISL_2035574, EPI_ISL_2035575, EPI_ISL_2035576, EPI_ISL_2035577, EPI_ISL_2035578, EPI_ISL_2035579, EPI_ISL_2035580, EPI_ISL_2035581, EPI_ISL_2035582, EPI_ISL_2035583, EPI_ISL_2035584, EPI_ISL_2035585, EPI_ISL_2035586, EPI_ISL_2035587, EPI_ISL_2035588, EPI_ISL_2035589, EPI_ISL_2035590, EPI_ISL_2035591, EPI_ISL_2035592, EPI_ISL_2035593, EPI_ISL_2035594, EPI_ISL_2035595, EPI_ISL_2035596, EPI_ISL_2035597, EPI_ISL_2035598, EPI_ISL_2035599, EPI_ISL_2035601, EPI_ISL_2035602, EPI_ISL_2035603, EPI_ISL_2035604, EPI_ISL_2035605, EPI_ISL_2035606, EPI_ISL_2035607, EPI_ISL_2035608, EPI_ISL_2035610, EPI_ISL_2035611, EPI_ISL_2035612, EPI_ISL_2035613, EPI_ISL_2035614, EPI_ISL_2035615, EPI_ISL_2035616, EPI_ISL_2035617, EPI_ISL_2035618, EPI_ISL_2035619, EPI_ISL_2035620, EPI_ISL_2035621, EPI_ISL_2035622, EPI_ISL_2035624, EPI_ISL_2035625, EPI_ISL_2035626, EPI_ISL_2035627, EPI_ISL_2035628, EPI_ISL_2035629, EPI_ISL_2035630, EPI_ISL_2035631, EPI_ISL_2035632, EPI_ISL_2035633, EPI_ISL_2035634, EPI_ISL_2035635, EPI_ISL_2035637 | | | |
| see above | Missouri State Public Health Laboratory | Missouri State Public Health Laboratory | Matthew Sinn, Joshua Barry, Ashley New |
| EPI_ISL_2035639, EPI_ISL_2035640, EPI_ISL_2035641, EPI_ISL_2035642, EPI_ISL_2035643, EPI_ISL_2035644, EPI_ISL_2035645, EPI_ISL_2035646, EPI_ISL_2035647, EPI_ISL_2035648, EPI_ISL_2035649, EPI_ISL_2035650, EPI_ISL_2035651, EPI_ISL_2035652, EPI_ISL_2035653, EPI_ISL_2035654, EPI_ISL_2035655, EPI_ISL_2035657, EPI_ISL_2035658, EPI_ISL_2035661, EPI_ISL_2035663, EPI_ISL_2035664, EPI_ISL_2035665, EPI_ISL_2035667, EPI_ISL_2035668, EPI_ISL_2035669, EPI_ISL_2035670, EPI_ISL_2035671, EPI_ISL_2035672, EPI_ISL_2035674, EPI_ISL_2035675, EPI_ISL_2035676, EPI_ISL_2035677, EPI_ISL_2035678, EPI_ISL_2035679, EPI_ISL_2035680, EPI_ISL_2035681, EPI_ISL_2035682, EPI_ISL_2035683, EPI_ISL_2035684, EPI_ISL_2035685, EPI_ISL_2035686, EPI_ISL_2035687, EPI_ISL_2035688, EPI_ISL_2035690, EPI_ISL_2035691, EPI_ISL_2035692, EPI_ISL_2035693, EPI_ISL_2035694, EPI_ISL_2035699, EPI_ISL_2035700, EPI_ISL_2035701, EPI_ISL_2035702, EPI_ISL_2035703, EPI_ISL_2035704, EPI_ISL_2035705, EPI_ISL_2035709, EPI_ISL_2035711, EPI_ISL_2035712, EPI_ISL_2035717 | | | |
| see above | OHSU Lab Services Molecular Microbiology Lab | Oregon SARS-CoV-2 Genome Sequencing Center | Brendan L. O'Connell, Sally Grindstaff, Sonia Acharya, Cierra LaBlanc, Kayla Carter, Ruth V. Nichols, Alec J. Hirsch, Donna Hansel, Guang Fan, Xuan Qin, Daniel N. Streblow, William B. Messer, Andrew C. Adey, Benjamin N. Bimber, Brian J. O'Roak |
| EPI_ISL_2035989 | Centracare Laboratory Services | Minnesota Department of Health, Public Health Laboratory | Alexandra Lorentz, Jacob Garfin, Matt Plumb, and Xiong Wang |
| EPI_ISL_2035990 | M Health Fairview | Minnesota Department of Health, Public Health Laboratory | Alexandra Lorentz, Jacob Garfin, Matt Plumb, and Xiong Wang |
| EPI_ISL_2035991 | Centracare Laboratory Services | Minnesota Department of Health, Public Health Laboratory | Alexandra Lorentz, Jacob Garfin, Matt Plumb, and Xiong Wang |
| EPI_ISL_2035992, EPI_ISL_2035993, EPI_ISL_2035994 | M Health Fairview | Minnesota Department of Health, Public Health Laboratory | Alexandra Lorentz, Jacob Garfin, Matt Plumb, and Xiong Wang |
| EPI_ISL_2035995 | Centracare Laboratory Services | Minnesota Department of Health, Public Health Laboratory | Alexandra Lorentz, Jacob Garfin, Matt Plumb, and Xiong Wang |
| EPI_ISL_2035996 | M Health Fairview | Minnesota Department of Health, Public Health Laboratory | Alexandra Lorentz, Jacob Garfin, Matt Plumb, and Xiong Wang |

| | | | |
|---|---|---|---|
| EPI_ISL_2035997 | Centracare Laboratory Services | Minnesota Department of Health, Public Health Laboratory | Alexandra Lorentz, Jacob Garfin, Matt Plumb, and Xiong Wang |
| EPI_ISL_2035998, EPI_ISL_2035999 | M Health Fairview | Minnesota Department of Health, Public Health Laboratory | Alexandra Lorentz, Jacob Garfin, Matt Plumb, and Xiong Wang |
| EPI_ISL_2036000 | Centracare Laboratory Services | Minnesota Department of Health, Public Health Laboratory | Alexandra Lorentz, Jacob Garfin, Matt Plumb, and Xiong Wang |
| EPI_ISL_2036001, EPI_ISL_2036002, EPI_ISL_2036003, EPI_ISL_2036004 | M Health Fairview | Minnesota Department of Health, Public Health Laboratory | Alexandra Lorentz, Jacob Garfin, Matt Plumb, and Xiong Wang |
| EPI_ISL_2036005 | Centracare Laboratory Services | Minnesota Department of Health, Public Health Laboratory | Alexandra Lorentz, Jacob Garfin, Matt Plumb, and Xiong Wang |
| EPI_ISL_2036006, EPI_ISL_2036007, EPI_ISL_2036008, EPI_ISL_2036009, EPI_ISL_2036010, EPI_ISL_2036011, EPI_ISL_2036012, EPI_ISL_2036013, EPI_ISL_2036014, EPI_ISL_2036015, EPI_ISL_2036016, EPI_ISL_2036017, EPI_ISL_2036018, EPI_ISL_2036019, EPI_ISL_2036020, EPI_ISL_2036021, EPI_ISL_2036022 | | | |
| see above | M Health Fairview | Minnesota Department of Health, Public Health Laboratory | Alexandra Lorentz, Jacob Garfin, Matt Plumb, and Xiong Wang |
| EPI_ISL_2036023, EPI_ISL_2036024, EPI_ISL_2036025 | Centracare Laboratory Services | Minnesota Department of Health, Public Health Laboratory | Alexandra Lorentz, Jacob Garfin, Matt Plumb, and Xiong Wang |
| EPI_ISL_2036026, EPI_ISL_2036027, EPI_ISL_2036028, EPI_ISL_2036029, EPI_ISL_2036030, EPI_ISL_2036031, EPI_ISL_2036032, EPI_ISL_2036033, EPI_ISL_2036034, EPI_ISL_2036035, EPI_ISL_2036036, EPI_ISL_2036037, EPI_ISL_2036038, EPI_ISL_2036039, EPI_ISL_2036040, EPI_ISL_2036041, EPI_ISL_2036042, EPI_ISL_2036043, EPI_ISL_2036044, EPI_ISL_2036045, EPI_ISL_2036046, EPI_ISL_2036047, EPI_ISL_2036048, EPI_ISL_2036049, EPI_ISL_2036050, EPI_ISL_2036051, EPI_ISL_2036052, EPI_ISL_2036053, EPI_ISL_2036054, EPI_ISL_2036055, EPI_ISL_2036056, EPI_ISL_2036057, EPI_ISL_2036058, EPI_ISL_2036059, EPI_ISL_2036060, EPI_ISL_2036061, EPI_ISL_2036062, EPI_ISL_2036063, EPI_ISL_2036064, EPI_ISL_2036065, EPI_ISL_2036066, EPI_ISL_2036067, EPI_ISL_2036068, EPI_ISL_2036069, EPI_ISL_2036070, EPI_ISL_2036071, EPI_ISL_2036072 | | | |
| see above | M Health Fairview | Minnesota Department of Health, Public Health Laboratory | Alexandra Lorentz, Jacob Garfin, Matt Plumb, and Xiong Wang |
| EPI_ISL_2036073 | Centracare Laboratory Services | Minnesota Department of Health, Public Health Laboratory | Alexandra Lorentz, Jacob Garfin, Matt Plumb, and Xiong Wang |
| EPI_ISL_2036074 | M Health Fairview | Minnesota Department of Health, Public Health Laboratory | Alexandra Lorentz, Jacob Garfin, Matt Plumb, and Xiong Wang |
| EPI_ISL_2036075 | Centracare Laboratory Services | Minnesota Department of Health, Public Health Laboratory | Alexandra Lorentz, Jacob Garfin, Matt Plumb, and Xiong Wang |
| EPI_ISL_2036076 | M Health Fairview | Minnesota Department of Health, Public Health Laboratory | Alexandra Lorentz, Jacob Garfin, Matt Plumb, and Xiong Wang |
| EPI_ISL_2036079 | WVDHHR - Office of Laboratory Services | Pathogen Discovery, Respiratory Viruses Branch, Division of Viral Diseases, Centers for Disease Control and Prevention | Ying Tao, Yan Li, Jing Zhang, Anna Kelleher, Brian Lynch, Krista Queen, Anna Uehara, Peter Cook, Han Jia Justin Ng, Rachel Marine, Clinton R. Paden, Dhwani Batra, Haibin Wang, Tara Coalter, Jasmine Padilla, Morgan Davis, Mili Sheth, Sarah Nobles, Mark Burroughs, Justin Lee, Adam Retchless, Suxiang Tong |
| EPI_ISL_2036275 | University of Oregon COVID-19 MAP Laboratory | University of Oregon Genomics and Cell Characterization Core Facility (GC3F) | Douglas Turnbull, Ariana White, Jeff Bishop, Jason Carriere, Jason Sydes, Peter Batzel |
| EPI_ISL_2036747, EPI_ISL_2036748, EPI_ISL_2036749, EPI_ISL_2036750, EPI_ISL_2036751, EPI_ISL_2036752, EPI_ISL_2036753, EPI_ISL_2036754, EPI_ISL_2036757, EPI_ISL_2036758, EPI_ISL_2036759, EPI_ISL_2036760, EPI_ISL_2036761 | | | |
| see above | CT Department of Public Health | CT Department of Public Health | Claire_Pearson,Tu_N_Nguyen |
| EPI_ISL_2036945, EPI_ISL_2036946, EPI_ISL_2036947, EPI_ISL_2036948, EPI_ISL_2036949, EPI_ISL_2036950, EPI_ISL_2036951, EPI_ISL_2036952, EPI_ISL_2036953, EPI_ISL_2036954, EPI_ISL_2036955, EPI_ISL_2036956, EPI_ISL_2036957, EPI_ISL_2036958, EPI_ISL_2036959, EPI_ISL_2036960, EPI_ISL_2036961, EPI_ISL_2036962, EPI_ISL_2036963 | | | |
| see above | DC Public Health Lab/ Dept. of Forensic Sciences | DC Public Health Lab/ Dept. of Forensic Sciences | Janis Doss, Scott Nguyen, Elizabeth Zelaya, Sarah Scott, Connie Maza, Monica Mann, Brittany Hamilton, David Payne, Jocelyn Hauser |
| EPI_ISL_2036964, EPI_ISL_2036965, EPI_ISL_2036966, EPI_ISL_2036967, EPI_ISL_2036968, EPI_ISL_2036969, EPI_ISL_2036971, EPI_ISL_2036972, EPI_ISL_2036973, EPI_ISL_2036974, EPI_ISL_2036976, EPI_ISL_2036977, EPI_ISL_2036978, EPI_ISL_2036979, EPI_ISL_2036980, EPI_ISL_2036981, EPI_ISL_2036982, EPI_ISL_2036983, EPI_ISL_2036984, EPI_ISL_2036985, EPI_ISL_2036986, EPI_ISL_2036987, EPI_ISL_2036988, EPI_ISL_2036989, EPI_ISL_2036990, EPI_ISL_2036992, EPI_ISL_2036993, EPI_ISL_2036994, EPI_ISL_2036995, EPI_ISL_2036996, EPI_ISL_2036997, EPI_ISL_2036998, EPI_ISL_2036999, EPI_ISL_2037000, EPI_ISL_2037001, EPI_ISL_2037002, EPI_ISL_2037003, EPI_ISL_2037004, EPI_ISL_2037005, EPI_ISL_2037006, EPI_ISL_2037007, EPI_ISL_2037008, EPI_ISL_2037009, EPI_ISL_2037010, EPI_ISL_2037011, EPI_ISL_2037012, EPI_ISL_2037013, EPI_ISL_2037014, EPI_ISL_2037015, EPI_ISL_2037016, EPI_ISL_2037017, EPI_ISL_2037018, EPI_ISL_2037019, EPI_ISL_2037020, EPI_ISL_2037021, EPI_ISL_2037022, EPI_ISL_2037024, EPI_ISL_2037025, EPI_ISL_2037026, EPI_ISL_2037027, EPI_ISL_2037029, EPI_ISL_2037030, EPI_ISL_2037032, EPI_ISL_2037033, EPI_ISL_2037034, EPI_ISL_2037035, EPI_ISL_2037036, EPI_ISL_2037037, EPI_ISL_2037038, EPI_ISL_2037039, EPI_ISL_2037040, EPI_ISL_2037041, EPI_ISL_2037042, EPI_ISL_2037043, EPI_ISL_2037044, EPI_ISL_2037045, EPI_ISL_2037046, EPI_ISL_2037047, EPI_ISL_2037048, EPI_ISL_2037049, EPI_ISL_2037050, EPI_ISL_2037051, EPI_ISL_2037052, EPI_ISL_2037053, EPI_ISL_2037054, EPI_ISL_2037055, EPI_ISL_2037056, EPI_ISL_2037057, EPI_ISL_2037058, EPI_ISL_2037059, EPI_ISL_2037060, EPI_ISL_2037061, EPI_ISL_2037062, EPI_ISL_2037064, EPI_ISL_2037065, EPI_ISL_2037067, EPI_ISL_2037068, EPI_ISL_2037069, EPI_ISL_2037070, EPI_ISL_2037071, EPI_ISL_2037072, EPI_ISL_2037075, EPI_ISL_2037076, EPI_ISL_2037077, EPI_ISL_2037078, EPI_ISL_2037079, EPI_ISL_2037080, EPI_ISL_2037081, EPI_ISL_2037082, EPI_ISL_2037083, EPI_ISL_2037084, EPI_ISL_2037085, EPI_ISL_2037086, EPI_ISL_2037087, EPI_ISL_2037088, EPI_ISL_2037089, EPI_ISL_2037090, EPI_ISL_2037091, EPI_ISL_2037092, EPI_ISL_2037093, EPI_ISL_2037095, EPI_ISL_2037097, EPI_ISL_2037099, EPI_ISL_2037100, EPI_ISL_2037101, EPI_ISL_2037102, EPI_ISL_2037103, EPI_ISL_2037104, EPI_ISL_2037105, EPI_ISL_2037106, EPI_ISL_2037107, EPI_ISL_2037108, EPI_ISL_2037109, EPI_ISL_2037110, EPI_ISL_2037111, EPI_ISL_2037112, EPI_ISL_2037114, EPI_ISL_2037115, EPI_ISL_2037116, EPI_ISL_2037117, EPI_ISL_2037119, EPI_ISL_2037120, EPI_ISL_2037121, EPI_ISL_2037122, EPI_ISL_2037123, EPI_ISL_2037124, EPI_ISL_2037126, EPI_ISL_2037127, EPI_ISL_2037129, EPI_ISL_2037130, EPI_ISL_2037132, EPI_ISL_2037133, EPI_ISL_2037134, EPI_ISL_2037135, EPI_ISL_2037137, EPI_ISL_2037138, EPI_ISL_2037139, EPI_ISL_2037140, EPI_ISL_2037141, EPI_ISL_2037142, EPI_ISL_2037143, EPI_ISL_2037145, EPI_ISL_2037146, EPI_ISL_2037148, EPI_ISL_2037149, EPI_ISL_2037150, EPI_ISL_2037151, EPI_ISL_2037152, EPI_ISL_2037153, EPI_ISL_2037154, EPI_ISL_2037155, EPI_ISL_2037156, EPI_ISL_2037157, EPI_ISL_2037158, EPI_ISL_2037159, EPI_ISL_2037160, EPI_ISL_2037161, EPI_ISL_2037162, EPI_ISL_2037163, EPI_ISL_2037165, EPI_ISL_2037166, EPI_ISL_2037167, EPI_ISL_2037168, EPI_ISL_2037169, EPI_ISL_2037170, EPI_ISL_2037171, EPI_ISL_2037172, EPI_ISL_2037173, EPI_ISL_2037174, EPI_ISL_2037175, EPI_ISL_2037176, EPI_ISL_2037177, EPI_ISL_2037178, EPI_ISL_2037180, EPI_ISL_2037181, EPI_ISL_2037182, EPI_ISL_2037183, EPI_ISL_2037184, EPI_ISL_2037185, EPI_ISL_2037186, EPI_ISL_2037187, EPI_ISL_2037189, EPI_ISL_2037190, EPI_ISL_2037191, EPI_ISL_2037192, EPI_ISL_2037193, EPI_ISL_2037194, EPI_ISL_2037195, EPI_ISL_2037196, EPI_ISL_2037197, EPI_ISL_2037198, EPI_ISL_2037199, EPI_ISL_2037200, EPI_ISL_2037201, EPI_ISL_2037202, EPI_ISL_2037204, EPI_ISL_2037206, EPI_ISL_2037207, EPI_ISL_2037208, EPI_ISL_2037211, EPI_ISL_2037212, EPI_ISL_2037213, EPI_ISL_2037214, EPI_ISL_2037215, EPI_ISL_2037216, EPI_ISL_2037217, EPI_ISL_2037218, EPI_ISL_2037219, EPI_ISL_2037220, EPI_ISL_2037221, EPI_ISL_2037222, EPI_ISL_2037224, EPI_ISL_2037225, EPI_ISL_2037226, EPI_ISL_2037227, EPI_ISL_2037228, EPI_ISL_2037231, EPI_ISL_2037232, EPI_ISL_2037233, EPI_ISL_2037234, EPI_ISL_2037235, EPI_ISL_2037236, EPI_ISL_2037237, EPI_ISL_2037238, EPI_ISL_2037239, EPI_ISL_2037240, EPI_ISL_2037242, EPI_ISL_2037243, EPI_ISL_2037244, EPI_ISL_2037245, EPI_ISL_2037246, EPI_ISL_2037247, EPI_ISL_2037248, EPI_ISL_2037249, EPI_ISL_2037250, EPI_ISL_2037251, EPI_ISL_2037252, EPI_ISL_2037253, EPI_ISL_2037254, EPI_ISL_2037256, EPI_ISL_2037257, EPI_ISL_2037258, EPI_ISL_2037260, EPI_ISL_2037261, EPI_ISL_2037262, EPI_ISL_2037263, EPI_ISL_2037265, EPI_ISL_2037266, EPI_ISL_2037267, EPI_ISL_2037268, EPI_ISL_2037269, EPI_ISL_2037270, EPI_ISL_2037271, EPI_ISL_2037272, EPI_ISL_2037274, EPI_ISL_2037275, EPI_ISL_2037276, EPI_ISL_2037277, EPI_ISL_2037278, EPI_ISL_2037279, EPI_ISL_2037280, EPI_ISL_2037281, EPI_ISL_2037282, EPI_ISL_2037283, EPI_ISL_2037284, EPI_ISL_2037285, EPI_ISL_2037286, EPI_ISL_2037287, EPI_ISL_2037288, EPI_ISL_2037289, EPI_ISL_2037291, EPI_ISL_2037292, EPI_ISL_2037293, EPI_ISL_2037294, EPI_ISL_2037295, EPI_ISL_2037296, EPI_ISL_2037297, EPI_ISL_2037298, EPI_ISL_2037299, EPI_ISL_2037300, EPI_ISL_2037301, EPI_ISL_2037303, EPI_ISL_2037305, EPI_ISL_2037306, EPI_ISL_2037307, EPI_ISL_2037308, EPI_ISL_2037309, EPI_ISL_2037310, EPI_ISL_2037311, EPI_ISL_2037312, EPI_ISL_2037314, EPI_ISL_2037315, EPI_ISL_2037316, EPI_ISL_2037317, EPI_ISL_2037318, EPI_ISL_2037319, EPI_ISL_2037321, EPI_ISL_2037322, EPI_ISL_2037324, EPI_ISL_2037325, EPI_ISL_2037326, EPI_ISL_2037327, EPI_ISL_2037328, EPI_ISL_2037329, EPI_ISL_2037330, EPI_ISL_2037331, EPI_ISL_2037332, EPI_ISL_2037333, EPI_ISL_2037334, EPI_ISL_2037335, EPI_ISL_2037336, EPI_ISL_2037337, EPI_ISL_2037338, EPI_ISL_2037339, EPI_ISL_2037340, EPI_ISL_2037341, EPI_ISL_2037342, EPI_ISL_2037343, EPI_ISL_2037344, EPI_ISL_2037345, EPI_ISL_2037346, EPI_ISL_2037347, EPI_ISL_2037349, EPI_ISL_2037350, EPI_ISL_2037351, EPI_ISL_2037352, EPI_ISL_2037353, EPI_ISL_2037354, EPI_ISL_2037355, EPI_ISL_2037357, EPI_ISL_2037358, EPI_ISL_2037359, EPI_ISL_2037362, EPI_ISL_2037363, EPI_ISL_2037364, EPI_ISL_2037365, EPI_ISL_2037366, EPI_ISL_2037367, EPI_ISL_2037368, EPI_ISL_2037369, EPI_ISL_2037370, EPI_ISL_2037371, EPI_ISL_2037372, EPI_ISL_2037373, EPI_ISL_2037374, EPI_ISL_2037375, EPI_ISL_2037376, EPI_ISL_2037377, EPI_ISL_2037378, EPI_ISL_2037379, EPI_ISL_2037380, EPI_ISL_2037381, EPI_ISL_2037382, EPI_ISL_2037383, EPI_ISL_2037384, EPI_ISL_2037385, EPI_ISL_2037386, EPI_ISL_2037387, EPI_ISL_2037388 | | | |
| see above | Pandemic Response Lab - NYC | Pandemic Response Lab, R&D | Henry Lee, Michael Hammerling, Melissa Hopkins, Cybill del Castillo, Shinyoung Clair Kang, William Ward, Pradeep Bugga, Sol Rey, Dylan Law, Katharine Nelson, Haiping Hao, Jon Laurent |
| EPI_ISL_2037389, EPI_ISL_2037390, EPI_ISL_2037391, EPI_ISL_2037392, EPI_ISL_2037393, EPI_ISL_2037394, EPI_ISL_2037395, EPI_ISL_2037398 | Kansas Health and Environmental Lab | Kansas Health and Environmental Lab | Mike Grose, Jonathan Barnell, Ben Olsen, and Phil Adam |
| EPI_ISL_2037400, EPI_ISL_2037401, EPI_ISL_2037402, EPI_ISL_2037403, EPI_ISL_2037404 | Cytocheck Laboratory | Kansas Health and Environmental Lab | Mike Grose, Jonathan Barnell, Ben Olsen, and Phil Adam |
| EPI_ISL_2037405, EPI_ISL_2037406, EPI_ISL_2037407, EPI_ISL_2037408, EPI_ISL_2037409, EPI_ISL_2037410, EPI_ISL_2037411, EPI_ISL_2037413 | Wichita State University - Molecular Diagnostics Lab | Kansas Health and Environmental Lab | Mike Grose, Jonathan Barnell, Ben Olsen, and Phil Adam |
| EPI_ISL_2037444, EPI_ISL_2037445, EPI_ISL_2037446, EPI_ISL_2037447, EPI_ISL_2037448, EPI_ISL_2037449, EPI_ISL_2037451, EPI_ISL_2037452, EPI_ISL_2037453, EPI_ISL_2037454, EPI_ISL_2037455, EPI_ISL_2037456, EPI_ISL_2037457, EPI_ISL_2037458, EPI_ISL_2037459, EPI_ISL_2037460, EPI_ISL_2037461, EPI_ISL_2037463, EPI_ISL_2037464, EPI_ISL_2037465, EPI_ISL_2037466, EPI_ISL_2037467, EPI_ISL_2037468, EPI_ISL_2037469, EPI_ISL_2037470, EPI_ISL_2037471, EPI_ISL_2037472, EPI_ISL_2037473, EPI_ISL_2037474, EPI_ISL_2037475, EPI_ISL_2037476, EPI_ISL_2037477, EPI_ISL_2037479, EPI_ISL_2037480, EPI_ISL_2037481, EPI_ISL_2037482, EPI_ISL_2037483, EPI_ISL_2037484, EPI_ISL_2037485, EPI_ISL_2037486, EPI_ISL_2037487, EPI_ISL_2037488, EPI_ISL_2037489, EPI_ISL_2037490, EPI_ISL_2037491, EPI_ISL_2037492, EPI_ISL_2037493, EPI_ISL_2037494, EPI_ISL_2037495, EPI_ISL_2037496, EPI_ISL_2037497, EPI_ISL_2037498, EPI_ISL_2037499, EPI_ISL_2037500, EPI_ISL_2037501, EPI_ISL_2037502, EPI_ISL_2037503, EPI_ISL_2037504, EPI_ISL_2037505, EPI_ISL_2037506, EPI_ISL_2037507, EPI_ISL_2037508, EPI_ISL_2037509, EPI_ISL_2037510, EPI_ISL_2037511, EPI_ISL_2037512, EPI_ISL_2037513, EPI_ISL_2037514, EPI_ISL_2037515, EPI_ISL_2037516, EPI_ISL_2037517, EPI_ISL_2037518, EPI_ISL_2037519, EPI_ISL_2037520, EPI_ISL_2037521, EPI_ISL_2037522, EPI_ISL_2037523, EPI_ISL_2037524, EPI_ISL_2037525, EPI_ISL_2037526, EPI_ISL_2037527, EPI_ISL_2037528, EPI_ISL_2037529, EPI_ISL_2037530, EPI_ISL_2037531, EPI_ISL_2037532, EPI_ISL_2037533, EPI_ISL_2037534, EPI_ISL_2037535, EPI_ISL_2037536, EPI_ISL_2037537, EPI_ISL_2037538, EPI_ISL_2037539, EPI_ISL_2037540, EPI_ISL_2037541, EPI_ISL_2037542, EPI_ISL_2037543, EPI_ISL_2037544, EPI_ISL_2037545, EPI_ISL_2037547, EPI_ISL_2037548, EPI_ISL_2037549, EPI_ISL_2037550, EPI_ISL_2037552, EPI_ISL_2037553, EPI_ISL_2037554, EPI_ISL_2037555, EPI_ISL_2037556, EPI_ISL_2037557, EPI_ISL_2037558, EPI_ISL_2037559, EPI_ISL_2037560, EPI_ISL_2037561, EPI_ISL_2037562, EPI_ISL_2037563, EPI_ISL_2037564, EPI_ISL_2037565, EPI_ISL_2037566, EPI_ISL_2037567, EPI_ISL_2037568, EPI_ISL_2037569, EPI_ISL_2037570, EPI_ISL_2037571, EPI_ISL_2037572, EPI_ISL_2037573, EPI_ISL_2037574, EPI_ISL_2037575, EPI_ISL_2037576, EPI_ISL_2037577, EPI_ISL_2037578, EPI_ISL_2037579, EPI_ISL_2037580, EPI_ISL_2037581, EPI_ISL_2037582, EPI_ISL_2037583, EPI_ISL_2037584, EPI_ISL_2037585, EPI_ISL_2037586, EPI_ISL_2037587, EPI_ISL_2037588, EPI_ISL_2037589, EPI_ISL_2037590, EPI_ISL_2037591, EPI_ISL_2037592, EPI_ISL_2037593, EPI_ISL_2037594, EPI_ISL_2037595, EPI_ISL_2037596, EPI_ISL_2037597, EPI_ISL_2037598, EPI_ISL_2037599, EPI_ISL_2037600, EPI_ISL_2037601, EPI_ISL_2037602, EPI_ISL_2037603, EPI_ISL_2037604, EPI_ISL_2037605, EPI_ISL_2037606, EPI_ISL_2037607, EPI_ISL_2037608, EPI_ISL_2037609, EPI_ISL_2037610, EPI_ISL_2037611, EPI_ISL_2037612, EPI_ISL_2037613, EPI_ISL_2037614, EPI_ISL_2037615, EPI_ISL_2037617, EPI_ISL_2037618, EPI_ISL_2037619, EPI_ISL_2037620, EPI_ISL_2037621, EPI_ISL_2037622, EPI_ISL_2037624, EPI_ISL_2037625, EPI_ISL_2037626, EPI_ISL_2037627, EPI_ISL_2037628, EPI_ISL_2037629, EPI_ISL_2037630, EPI_ISL_2037632, EPI_ISL_2037633, EPI_ISL_2037634, EPI_ISL_2037635, EPI_ISL_2037636, EPI_ISL_2037637, EPI_ISL_2037638, EPI_ISL_2037639, EPI_ISL_2037640, EPI_ISL_2037641, EPI_ISL_2037642, EPI_ISL_2037643, EPI_ISL_2037644, EPI_ISL_2037645, EPI_ISL_2037646, EPI_ISL_2037647, EPI_ISL_2037648, EPI_ISL_2037650, EPI_ISL_2037651, EPI_ISL_2037652, EPI_ISL_2037653, EPI_ISL_2037654, EPI_ISL_2037655, EPI_ISL_2037656, EPI_ISL_2037657, EPI_ISL_2037658, EPI_ISL_2037659, EPI_ISL_2037660, EPI_ISL_2037661, EPI_ISL_2037662, EPI_ISL_2037663, EPI_ISL_2037664, EPI_ISL_2037665, EPI_ISL_2037666, EPI_ISL_2037667, EPI_ISL_2037668, EPI_ISL_2037669, EPI_ISL_2037670, EPI_ISL_2037671, EPI_ISL_2037672, EPI_ISL_2037673, EPI_ISL_2037674, EPI_ISL_2037675, EPI_ISL_2037676, EPI_ISL_2037678, EPI_ISL_2037679, EPI_ISL_2037680, EPI_ISL_2037682, EPI_ISL_2037683, EPI_ISL_2037684, EPI_ISL_2037685, EPI_ISL_2037686, EPI_ISL_2037687, EPI_ISL_2037688, EPI_ISL_2037689, EPI_ISL_2037690, EPI_ISL_2037691, EPI_ISL_2037692, EPI_ISL_2037693, EPI_ISL_2037694, EPI_ISL_2037695, EPI_ISL_2037696, EPI_ISL_2037697, EPI_ISL_2037698, EPI_ISL_2037699, EPI_ISL_2037700, EPI_ISL_2037701, EPI_ISL_2037702, EPI_ISL_2037703, EPI_ISL_2037704, EPI_ISL_2037705, EPI_ISL_2037707, EPI_ISL_2037708, EPI_ISL_2037709, EPI_ISL_2037710 | | | |
| see above | UW Virology Lab | UW Virology Lab | Pavitra Roychoudhury, Hong Xie, Lasata Shrestha, Tien V. Nguyen, Shah Mohamed Bakhash, Michelle Lin, Noah R. Baker, Sean Ellis, Meei-Li Huang, Keith R Jerome, Alexander Greninger |
| EPI_ISL_2037711, EPI_ISL_2037712, EPI_ISL_2037713, EPI_ISL_2037714, EPI_ISL_2037717, EPI_ISL_2037718, EPI_ISL_2037719, EPI_ISL_2037720, EPI_ISL_2037721, EPI_ISL_2037722, EPI_ISL_2037723, EPI_ISL_2037724, EPI_ISL_2037725, EPI_ISL_2037727, EPI_ISL_2037728, EPI_ISL_2037729, EPI_ISL_2037730, EPI_ISL_2037731, EPI_ISL_2037732, EPI_ISL_2037733, EPI_ISL_2037734, EPI_ISL_2037736, EPI_ISL_2037738, EPI_ISL_2037739, EPI_ISL_2037740, EPI_ISL_2037741, EPI_ISL_2037742, EPI_ISL_2037744, EPI_ISL_2037745, EPI_ISL_2037746, EPI_ISL_2037747, EPI_ISL_2037748, EPI_ISL_2037749, EPI_ISL_2037750, EPI_ISL_2037751, EPI_ISL_2037752, EPI_ISL_2037753, EPI_ISL_2037754, EPI_ISL_2037756, EPI_ISL_2037757, EPI_ISL_2037758, EPI_ISL_2037759, EPI_ISL_2037760, EPI_ISL_2037761, EPI_ISL_2037762, EPI_ISL_2037763, EPI_ISL_2037765, EPI_ISL_2037766, EPI_ISL_2037767, EPI_ISL_2037768, EPI_ISL_2037770, EPI_ISL_2037771, EPI_ISL_2037773, EPI_ISL_2037774, EPI_ISL_2037776, EPI_ISL_2037777, EPI_ISL_2037778, EPI_ISL_2037779, EPI_ISL_2037780, EPI_ISL_2037782, EPI_ISL_2037784, EPI_ISL_2037785, EPI_ISL_2037786, EPI_ISL_2037787, EPI_ISL_2037788, EPI_ISL_2037789, EPI_ISL_2037790, EPI_ISL_2037791, EPI_ISL_2037792, EPI_ISL_2037793, EPI_ISL_2037794, EPI_ISL_2037795, EPI_ISL_2037796, EPI_ISL_2037798, EPI_ISL_2037800, EPI_ISL_2037801, EPI_ISL_2037802, EPI_ISL_2037803, EPI_ISL_2037804, EPI_ISL_2037805, EPI_ISL_2037806, EPI_ISL_2037807, EPI_ISL_2037808, EPI_ISL_2037809, EPI_ISL_2037810, EPI_ISL_2037811, EPI_ISL_2037812, EPI_ISL_2037813, | | | |

| | | | |
|---|---|---|---|
| EPI_ISL_2037814, EPI_ISL_2037815, EPI_ISL_2037817, EPI_ISL_2037818, EPI_ISL_2037819, EPI_ISL_2037820, EPI_ISL_2037821, EPI_ISL_2037822, EPI_ISL_2037823, EPI_ISL_2037824, EPI_ISL_2037825, EPI_ISL_2037826, EPI_ISL_2037827, EPI_ISL_2037828, EPI_ISL_2037829, EPI_ISL_2037830, EPI_ISL_2037831, EPI_ISL_2037832, EPI_ISL_2037833, EPI_ISL_2037834, EPI_ISL_2037835, EPI_ISL_2037836, EPI_ISL_2037837, EPI_ISL_2037838, EPI_ISL_2037839, EPI_ISL_2037840, EPI_ISL_2037841, EPI_ISL_2037842, EPI_ISL_2037844, EPI_ISL_2037845, EPI_ISL_2037846, EPI_ISL_2037847, EPI_ISL_2037848, EPI_ISL_2037849, EPI_ISL_2037850, EPI_ISL_2037851, EPI_ISL_2037852, EPI_ISL_2037853, EPI_ISL_2037855, EPI_ISL_2037856, EPI_ISL_2037857, EPI_ISL_2037858, EPI_ISL_2037859, EPI_ISL_2037860, EPI_ISL_2037861, EPI_ISL_2037862, EPI_ISL_2037863, EPI_ISL_2037864, EPI_ISL_2037866, EPI_ISL_2037867, EPI_ISL_2037868, EPI_ISL_2037869, EPI_ISL_2037870, EPI_ISL_2037871, EPI_ISL_2037872, EPI_ISL_2037873, EPI_ISL_2037874, EPI_ISL_2037875, EPI_ISL_2037877, EPI_ISL_2037878, EPI_ISL_2037879, EPI_ISL_2037880, EPI_ISL_2037881, EPI_ISL_2037882, EPI_ISL_2037885, EPI_ISL_2037886, EPI_ISL_2037887, EPI_ISL_2037888, EPI_ISL_2037889, EPI_ISL_2037890, EPI_ISL_2037891, EPI_ISL_2037892, EPI_ISL_2037893, EPI_ISL_2037894, EPI_ISL_2037895, EPI_ISL_2037896, EPI_ISL_2037897, EPI_ISL_2037899, EPI_ISL_2037900, EPI_ISL_2037901, EPI_ISL_2037902, EPI_ISL_2037903, EPI_ISL_2037904, EPI_ISL_2037905, EPI_ISL_2037906, EPI_ISL_2037908, EPI_ISL_2037909, EPI_ISL_2037910, EPI_ISL_2037912, EPI_ISL_2037913, EPI_ISL_2037914, EPI_ISL_2037915, EPI_ISL_2037917, EPI_ISL_2037918, EPI_ISL_2037919, EPI_ISL_2037920, EPI_ISL_2037921, EPI_ISL_2037922, EPI_ISL_2037923, EPI_ISL_2037924, EPI_ISL_2037925, EPI_ISL_2037926, EPI_ISL_2037927, EPI_ISL_2037928, EPI_ISL_2037929, EPI_ISL_2037930, EPI_ISL_2037931, EPI_ISL_2037932, EPI_ISL_2037933, EPI_ISL_2037934, EPI_ISL_2037935, EPI_ISL_2037936, EPI_ISL_2037937, EPI_ISL_2037938, EPI_ISL_2037939, EPI_ISL_2037940, EPI_ISL_2037941, EPI_ISL_2037942, EPI_ISL_2037943, EPI_ISL_2037944, EPI_ISL_2037945, EPI_ISL_2037947, EPI_ISL_2037948, EPI_ISL_2037950, EPI_ISL_2037951, EPI_ISL_2037952, EPI_ISL_2037953, EPI_ISL_2037954, EPI_ISL_2037955, EPI_ISL_2037956, EPI_ISL_2037957, EPI_ISL_2037958, EPI_ISL_2037959, EPI_ISL_2037960, EPI_ISL_2037961, EPI_ISL_2037962, EPI_ISL_2037963, EPI_ISL_2037964, EPI_ISL_2037965, EPI_ISL_2037966, EPI_ISL_2037967, EPI_ISL_2037968, EPI_ISL_2037969, EPI_ISL_2037970, EPI_ISL_2037971, EPI_ISL_2037972, EPI_ISL_2037973, EPI_ISL_2037974, EPI_ISL_2037975, EPI_ISL_2037976, EPI_ISL_2037978, EPI_ISL_2037980, EPI_ISL_2037981, EPI_ISL_2037982, EPI_ISL_2037983, EPI_ISL_2037984, EPI_ISL_2037985, EPI_ISL_2037986, EPI_ISL_2037988, EPI_ISL_2037989, EPI_ISL_2037990, EPI_ISL_2037991, EPI_ISL_2037992, EPI_ISL_2037993, EPI_ISL_2037994, EPI_ISL_2037995, EPI_ISL_2037996, EPI_ISL_2037997, EPI_ISL_2037998, EPI_ISL_2037999, EPI_ISL_2038002, EPI_ISL_2038003, EPI_ISL_2038004, EPI_ISL_2038005, EPI_ISL_2038007, EPI_ISL_2038008, EPI_ISL_2038009, EPI_ISL_2038010, EPI_ISL_2038011, EPI_ISL_2038012, EPI_ISL_2038013, EPI_ISL_2038014, EPI_ISL_2038015, EPI_ISL_2038016, EPI_ISL_2038017, EPI_ISL_2038018, EPI_ISL_2038019, EPI_ISL_2038020, EPI_ISL_2038021, EPI_ISL_2038022, EPI_ISL_2038023, EPI_ISL_2038024, EPI_ISL_2038026, EPI_ISL_2038028, EPI_ISL_2038030, EPI_ISL_2038032, EPI_ISL_2038033, EPI_ISL_2038035, EPI_ISL_2038036, EPI_ISL_2038037, EPI_ISL_2038038, EPI_ISL_2038039, EPI_ISL_2038040, EPI_ISL_2038041, EPI_ISL_2038042, EPI_ISL_2038044, EPI_ISL_2038046, EPI_ISL_2038047, EPI_ISL_2038048, EPI_ISL_2038049, EPI_ISL_2038050, EPI_ISL_2038051, EPI_ISL_2038052, EPI_ISL_2038053, EPI_ISL_2038054, EPI_ISL_2038055, EPI_ISL_2038056, EPI_ISL_2038057, EPI_ISL_2038058, EPI_ISL_2038059, EPI_ISL_2038061, EPI_ISL_2038062, EPI_ISL_2038063, EPI_ISL_2038065, EPI_ISL_2038066, EPI_ISL_2038067, EPI_ISL_2038068, EPI_ISL_2038069, EPI_ISL_2038070, EPI_ISL_2038071, EPI_ISL_2038072, EPI_ISL_2038073, EPI_ISL_2038075, EPI_ISL_2038076, EPI_ISL_2038078, EPI_ISL_2038079, EPI_ISL_2038080, EPI_ISL_2038081, EPI_ISL_2038082, EPI_ISL_2038083, EPI_ISL_2038084, EPI_ISL_2038085, EPI_ISL_2038087, EPI_ISL_2038088, EPI_ISL_2038089, EPI_ISL_2038090, EPI_ISL_2038091, EPI_ISL_2038092, EPI_ISL_2038094, EPI_ISL_2038095, EPI_ISL_2038096, EPI_ISL_2038097, EPI_ISL_2038098, EPI_ISL_2038099, EPI_ISL_2038100, EPI_ISL_2038101, EPI_ISL_2038102, EPI_ISL_2038103, EPI_ISL_2038104, EPI_ISL_2038105, EPI_ISL_2038106, EPI_ISL_2038107, EPI_ISL_2038108, EPI_ISL_2038109, EPI_ISL_2038110, EPI_ISL_2038111, EPI_ISL_2038112, EPI_ISL_2038113, EPI_ISL_2038114, EPI_ISL_2038115, EPI_ISL_2038116, EPI_ISL_2038117, EPI_ISL_2038119, EPI_ISL_2038121, EPI_ISL_2038122, EPI_ISL_2038123, EPI_ISL_2038124, EPI_ISL_2038125, EPI_ISL_2038126, EPI_ISL_2038127, EPI_ISL_2038129, EPI_ISL_2038131, EPI_ISL_2038132, EPI_ISL_2038133, EPI_ISL_2038134, EPI_ISL_2038135, EPI_ISL_2038136, EPI_ISL_2038137, EPI_ISL_2038138, EPI_ISL_2038139, EPI_ISL_2038140, EPI_ISL_2038141, EPI_ISL_2038142, EPI_ISL_2038143, EPI_ISL_2038144, EPI_ISL_2038145, EPI_ISL_2038146, EPI_ISL_2038148, EPI_ISL_2038149, EPI_ISL_2038150, EPI_ISL_2038151, EPI_ISL_2038153, EPI_ISL_2038154, EPI_ISL_2038155, EPI_ISL_2038156, EPI_ISL_2038157, EPI_ISL_2038158, EPI_ISL_2038159, EPI_ISL_2038160, EPI_ISL_2038161, EPI_ISL_2038162, EPI_ISL_2038163, EPI_ISL_2038164, EPI_ISL_2038165, EPI_ISL_2038166, EPI_ISL_2038167, EPI_ISL_2038168, EPI_ISL_2038169, EPI_ISL_2038170, EPI_ISL_2038171, EPI_ISL_2038173, EPI_ISL_2038174, EPI_ISL_2038175, EPI_ISL_2038176, EPI_ISL_2038177, EPI_ISL_2038178, EPI_ISL_2038179, EPI_ISL_2038180, EPI_ISL_2038181, EPI_ISL_2038182, EPI_ISL_2038183, EPI_ISL_2038184, EPI_ISL_2038185, EPI_ISL_2038186, EPI_ISL_2038187, EPI_ISL_2038188, EPI_ISL_2038189, EPI_ISL_2038190, EPI_ISL_2038191, EPI_ISL_2038192, EPI_ISL_2038193, EPI_ISL_2038194, EPI_ISL_2038195, EPI_ISL_2038196, EPI_ISL_2038197, EPI_ISL_2038198, EPI_ISL_2038199, EPI_ISL_2038200, EPI_ISL_2038201, EPI_ISL_2038202, EPI_ISL_2038203, EPI_ISL_2038206, EPI_ISL_2038207, EPI_ISL_2038208, EPI_ISL_2038210, EPI_ISL_2038211, EPI_ISL_2038212, EPI_ISL_2038213, EPI_ISL_2038214, EPI_ISL_2038215, EPI_ISL_2038216, EPI_ISL_2038217, EPI_ISL_2038218, EPI_ISL_2038219, EPI_ISL_2038220, EPI_ISL_2038221, EPI_ISL_2038222, EPI_ISL_2038223, EPI_ISL_2038224, EPI_ISL_2038225, EPI_ISL_2038226, EPI_ISL_2038227, EPI_ISL_2038228, EPI_ISL_2038229, EPI_ISL_2038230, EPI_ISL_2038231, EPI_ISL_2038232, EPI_ISL_2038233, EPI_ISL_2038234, EPI_ISL_2038235, EPI_ISL_2038236, EPI_ISL_2038237, EPI_ISL_2038238, EPI_ISL_2038239, EPI_ISL_2038240, EPI_ISL_2038241, EPI_ISL_2038242, EPI_ISL_2038243, EPI_ISL_2038244, EPI_ISL_2038245, EPI_ISL_2038246, EPI_ISL_2038247, EPI_ISL_2038248, EPI_ISL_2038249, EPI_ISL_2038250, EPI_ISL_2038251, EPI_ISL_2038254, EPI_ISL_2038255, EPI_ISL_2038257, EPI_ISL_2038258, EPI_ISL_2038259, EPI_ISL_2038260, EPI_ISL_2038261, EPI_ISL_2038262, EPI_ISL_2038263, EPI_ISL_2038264, EPI_ISL_2038265, EPI_ISL_2038266, EPI_ISL_2038267, EPI_ISL_2038268, EPI_ISL_2038269, EPI_ISL_2038270, EPI_ISL_2038271, EPI_ISL_2038272, EPI_ISL_2038273, EPI_ISL_2038274, EPI_ISL_2038275, EPI_ISL_2038276, EPI_ISL_2038277, EPI_ISL_2038279, EPI_ISL_2038280, EPI_ISL_2038281, EPI_ISL_2038282, EPI_ISL_2038283, EPI_ISL_2038284, EPI_ISL_2038285, EPI_ISL_2038286, EPI_ISL_2038287, EPI_ISL_2038288, EPI_ISL_2038289, EPI_ISL_2038290, EPI_ISL_2038291, EPI_ISL_2038292, EPI_ISL_2038293, EPI_ISL_2038294, EPI_ISL_2038295, EPI_ISL_2038296, EPI_ISL_2038297, EPI_ISL_2038299, EPI_ISL_2038301, EPI_ISL_2038302, EPI_ISL_2038303, EPI_ISL_2038304, EPI_ISL_2038305, EPI_ISL_2038306, EPI_ISL_2038307, EPI_ISL_2038308, EPI_ISL_2038309, EPI_ISL_2038310, EPI_ISL_2038311, EPI_ISL_2038312, EPI_ISL_2038313, EPI_ISL_2038314, EPI_ISL_2038315, EPI_ISL_2038316, EPI_ISL_2038317, EPI_ISL_2038318, EPI_ISL_2038319, EPI_ISL_2038320, EPI_ISL_2038321, EPI_ISL_2038322, EPI_ISL_2038323, EPI_ISL_2038324, EPI_ISL_2038325, EPI_ISL_2038326, EPI_ISL_2038327, EPI_ISL_2038328, EPI_ISL_2038329, EPI_ISL_2038330, EPI_ISL_2038331, EPI_ISL_2038332, EPI_ISL_2038333, EPI_ISL_2038334, EPI_ISL_2038335, EPI_ISL_2038336, EPI_ISL_2038337, EPI_ISL_2038338, EPI_ISL_2038339, EPI_ISL_2038340, EPI_ISL_2038342, EPI_ISL_2038343, EPI_ISL_2038344, EPI_ISL_2038346, EPI_ISL_2038347, EPI_ISL_2038348, EPI_ISL_2038349, EPI_ISL_2038350, EPI_ISL_2038351, EPI_ISL_2038353, EPI_ISL_2038354, EPI_ISL_2038355, EPI_ISL_2038356, EPI_ISL_2038357, EPI_ISL_2038360, EPI_ISL_2038361, EPI_ISL_2038362, EPI_ISL_2038363, EPI_ISL_2038364, EPI_ISL_2038365, EPI_ISL_2038366, EPI_ISL_2038367, EPI_ISL_2038368, EPI_ISL_2038370, EPI_ISL_2038372, EPI_ISL_2038373, EPI_ISL_2038375, EPI_ISL_2038376, EPI_ISL_2038378, EPI_ISL_2038379, EPI_ISL_2038380, EPI_ISL_2038382, EPI_ISL_2038383, EPI_ISL_2038384, EPI_ISL_2038386, EPI_ISL_2038387, EPI_ISL_2038388, EPI_ISL_2038389, EPI_ISL_2038390, EPI_ISL_2038393, EPI_ISL_2038394, EPI_ISL_2038395, EPI_ISL_2038396, EPI_ISL_2038397, EPI_ISL_2038398, EPI_ISL_2038399, EPI_ISL_2038400, EPI_ISL_2038401, EPI_ISL_2038402, EPI_ISL_2038403, EPI_ISL_2038404, EPI_ISL_2038405, EPI_ISL_2038406, EPI_ISL_2038408, EPI_ISL_2038409, EPI_ISL_2038410, EPI_ISL_2038411, EPI_ISL_2038413, EPI_ISL_2038415, EPI_ISL_2038416, EPI_ISL_2038417, EPI_ISL_2038418, EPI_ISL_2038419, EPI_ISL_2038420, EPI_ISL_2038421, EPI_ISL_2038422, EPI_ISL_2038423, EPI_ISL_2038424, EPI_ISL_2038425, EPI_ISL_2038426, EPI_ISL_2038427, EPI_ISL_2038428, EPI_ISL_2038429, EPI_ISL_2038430, EPI_ISL_2038431, EPI_ISL_2038432, EPI_ISL_2038433, EPI_ISL_2038434, EPI_ISL_2038435, EPI_ISL_2038436, EPI_ISL_2038437, EPI_ISL_2038438, EPI_ISL_2038439, EPI_ISL_2038440, EPI_ISL_2038441, EPI_ISL_2038442, EPI_ISL_2038443, EPI_ISL_2038444, EPI_ISL_2038445, EPI_ISL_2038446, EPI_ISL_2038447, EPI_ISL_2038448, EPI_ISL_2038449, EPI_ISL_2038450, EPI_ISL_2038451, EPI_ISL_2038452, EPI_ISL_2038453, EPI_ISL_2038454, EPI_ISL_2038455, EPI_ISL_2038456, EPI_ISL_2038457, EPI_ISL_2038458, EPI_ISL_2038459, EPI_ISL_2038460, EPI_ISL_2038461, EPI_ISL_2038462, EPI_ISL_2038463, EPI_ISL_2038464, EPI_ISL_2038465, EPI_ISL_2038466, EPI_ISL_2038467, EPI_ISL_2038468, EPI_ISL_2038469, EPI_ISL_2038470, EPI_ISL_2038471, EPI_ISL_2038473, EPI_ISL_2038474, EPI_ISL_2038475, EPI_ISL_2038476, EPI_ISL_2038477, EPI_ISL_2038478, EPI_ISL_2038479, EPI_ISL_2038480, EPI_ISL_2038481, EPI_ISL_2038483, EPI_ISL_2038484, EPI_ISL_2038485, EPI_ISL_2038486, EPI_ISL_2038487, EPI_ISL_2038488, EPI_ISL_2038489, EPI_ISL_2038490, EPI_ISL_2038491, EPI_ISL_2038492, EPI_ISL_2038493, EPI_ISL_2038495, EPI_ISL_2038496, EPI_ISL_2038497, EPI_ISL_2038498, EPI_ISL_2038499, EPI_ISL_2038500, EPI_ISL_2038501, EPI_ISL_2038503, EPI_ISL_2038504, EPI_ISL_2038505, EPI_ISL_2038506, EPI_ISL_2038507, EPI_ISL_2038508, EPI_ISL_2038509, EPI_ISL_2038510, EPI_ISL_2038511, EPI_ISL_2038512, EPI_ISL_2038513, EPI_ISL_2038514, EPI_ISL_2038515, EPI_ISL_2038516, EPI_ISL_2038517, EPI_ISL_2038518, EPI_ISL_2038519, EPI_ISL_2038520, EPI_ISL_2038521, EPI_ISL_2038522, EPI_ISL_2038523, EPI_ISL_2038524, EPI_ISL_2038525, EPI_ISL_2038526, EPI_ISL_2038527, EPI_ISL_2038528, EPI_ISL_2038529, EPI_ISL_2038530, EPI_ISL_2038531, EPI_ISL_2038533, EPI_ISL_2038534, EPI_ISL_2038535, EPI_ISL_2038536, EPI_ISL_2038537, EPI_ISL_2038538, EPI_ISL_2038539, EPI_ISL_2038540, EPI_ISL_2038541, EPI_ISL_2038542, EPI_ISL_2038544, EPI_ISL_2038545, EPI_ISL_2038546, EPI_ISL_2038547, EPI_ISL_2038548, EPI_ISL_2038549, EPI_ISL_2038552, EPI_ISL_2038553, EPI_ISL_2038554, EPI_ISL_2038555, EPI_ISL_2038556, EPI_ISL_2038557, EPI_ISL_2038558, EPI_ISL_2038559, EPI_ISL_2038560, EPI_ISL_2038561, EPI_ISL_2038562, EPI_ISL_2038563, EPI_ISL_2038564, EPI_ISL_2038565, EPI_ISL_2038566, EPI_ISL_2038567, EPI_ISL_2038568, EPI_ISL_2038569, EPI_ISL_2038570, EPI_ISL_2038571, EPI_ISL_2038572, EPI_ISL_2038573, EPI_ISL_2038574, EPI_ISL_2038575, EPI_ISL_2038576, EPI_ISL_2038577, EPI_ISL_2038578, EPI_ISL_2038579, EPI_ISL_2038580, EPI_ISL_2038581, EPI_ISL_2038582, EPI_ISL_2038583, EPI_ISL_2038584, EPI_ISL_2038587, EPI_ISL_2038588, EPI_ISL_2038589, EPI_ISL_2038590, EPI_ISL_2038591, EPI_ISL_2038592, EPI_ISL_2038593, EPI_ISL_2038595, EPI_ISL_2038596, EPI_ISL_2038597, EPI_ISL_2038598, EPI_ISL_2038599, EPI_ISL_2038600, EPI_ISL_2038601, EPI_ISL_2038603, EPI_ISL_2038604, EPI_ISL_2038605, EPI_ISL_2038607, EPI_ISL_2038608, EPI_ISL_2038609, EPI_ISL_2038610, EPI_ISL_2038611, EPI_ISL_2038612, EPI_ISL_2038613, EPI_ISL_2038614, EPI_ISL_2038615, EPI_ISL_2038616, EPI_ISL_2038617, EPI_ISL_2038618, EPI_ISL_2038619, EPI_ISL_2038620, EPI_ISL_2038621, EPI_ISL_2038622, EPI_ISL_2038624, EPI_ISL_2038625, EPI_ISL_2038626, EPI_ISL_2038627, EPI_ISL_2038628, EPI_ISL_2038629, EPI_ISL_2038630, EPI_ISL_2038631, EPI_ISL_2038632, EPI_ISL_2038633, EPI_ISL_2038634, EPI_ISL_2038637, EPI_ISL_2038638, EPI_ISL_2038639, EPI_ISL_2038641, EPI_ISL_2038642, EPI_ISL_2038643, EPI_ISL_2038644, EPI_ISL_2038645, EPI_ISL_2038647, EPI_ISL_2038648, EPI_ISL_2038649, EPI_ISL_2038650, EPI_ISL_2038651, EPI_ISL_2038652, EPI_ISL_2038653, EPI_ISL_2038654, EPI_ISL_2038655, EPI_ISL_2038656, EPI_ISL_2038657, EPI_ISL_2038658, EPI_ISL_2038659, EPI_ISL_2038660, EPI_ISL_2038662, EPI_ISL_2038663, EPI_ISL_2038665, EPI_ISL_2038666, EPI_ISL_2038668, EPI_ISL_2038669, EPI_ISL_2038670, EPI_ISL_2038671, EPI_ISL_2038673, EPI_ISL_2038674, EPI_ISL_2038675, EPI_ISL_2038676, EPI_ISL_2038677, EPI_ISL_2038678, EPI_ISL_2038679, EPI_ISL_2038680, EPI_ISL_2038681, EPI_ISL_2038683, EPI_ISL_2038684, EPI_ISL_2038685, EPI_ISL_2038686, EPI_ISL_2038687, EPI_ISL_2038688, EPI_ISL_2038689, EPI_ISL_2038690, EPI_ISL_2038691, EPI_ISL_2038692, EPI_ISL_2038693, EPI_ISL_2038694, EPI_ISL_2038695, EPI_ISL_2038697, EPI_ISL_2038698, EPI_ISL_2038699, EPI_ISL_2038700, EPI_ISL_2038701, EPI_ISL_2038702, EPI_ISL_2038703, EPI_ISL_2038704, EPI_ISL_2038705, EPI_ISL_2038706, EPI_ISL_2038708, EPI_ISL_2038709, EPI_ISL_2038710, EPI_ISL_2038711, EPI_ISL_2038712, EPI_ISL_2038713, EPI_ISL_2038715, EPI_ISL_2038716, EPI_ISL_2038717, EPI_ISL_2038718, EPI_ISL_2038719, EPI_ISL_2038720, EPI_ISL_2038722, EPI_ISL_2038723, EPI_ISL_2038724, EPI_ISL_2038725, EPI_ISL_2038726, EPI_ISL_2038727, EPI_ISL_2038728, EPI_ISL_2038729, EPI_ISL_2038730, EPI_ISL_2038731, EPI_ISL_2038732, EPI_ISL_2038733, EPI_ISL_2038734, EPI_ISL_2038735, EPI_ISL_2038736, EPI_ISL_2038737, EPI_ISL_2038738, EPI_ISL_2038739, EPI_ISL_2038740, EPI_ISL_2038741, EPI_ISL_2038742, EPI_ISL_2038743, EPI_ISL_2038744, EPI_ISL_2038745, EPI_ISL_2038746, EPI_ISL_2038747, EPI_ISL_2038748, EPI_ISL_2038749, EPI_ISL_2038750, EPI_ISL_2038751, EPI_ISL_2038752, EPI_ISL_2038753, EPI_ISL_2038754, EPI_ISL_2038755, EPI_ISL_2038756, EPI_ISL_2038757, EPI_ISL_2038758, EPI_ISL_2038759, EPI_ISL_2038760, EPI_ISL_2038761, EPI_ISL_2038762, EPI_ISL_2038763, EPI_ISL_2038764, EPI_ISL_2038765, EPI_ISL_2038766, EPI_ISL_2038767, EPI_ISL_2038768, EPI_ISL_2038769, EPI_ISL_2038770, EPI_ISL_2038771, EPI_ISL_2038772, EPI_ISL_2038773, EPI_ISL_2038775, EPI_ISL_2038778, EPI_ISL_2038779, EPI_ISL_2038780, EPI_ISL_2038782, EPI_ISL_2038783, EPI_ISL_2038784, EPI_ISL_2038785, EPI_ISL_2038786, EPI_ISL_2038788, EPI_ISL_2038789, EPI_ISL_2038790, EPI_ISL_2038791, EPI_ISL_2038792, EPI_ISL_2038793, EPI_ISL_2038794, EPI_ISL_2038795, EPI_ISL_2038796, EPI_ISL_2038797, EPI_ISL_2038798 | | | |
| see above | Fulgent Genetics | Centers for Disease Control and Prevention Division of Viral Diseases, Pathogen Discovery | Dakota Howard, Dhwani Batra, Peter W. Cook, Kara Moser, Adrian Paskey, Jason Caravas, Benjamin Rambo-Martin, Shatavia Morrison, Christopher Gulvick, Scott Sammons, Yvette Unoarumhi, Darlene Wagner, Matthew Schmerer, Harry Gao, Mickey Li, John Gao, Joseph Fierro, Benafsh Sapra, Becky Tsai, Yan Meng, Doreen Ng, James Xie, Clinton R. Paden, Duncan MacCannell |
| EPI_ISL_2038799, EPI_ISL_2038800, EPI_ISL_2038801, EPI_ISL_2038802 | Minnesota Department of Health, Public Health Laboratory | Minnesota Department of Health, Public Health Laboratory | Alexandra Lorentz, Jacob Garfin, Matt Plumb, and Xiong Wang |
| EPI_ISL_2038803 | Premier Medical Lab | Minnesota Department of Health, Public Health Laboratory | Alexandra Lorentz, Jacob Garfin, Matt Plumb, and Xiong Wang |
| EPI_ISL_2038804, EPI_ISL_2038805, EPI_ISL_2038806, EPI_ISL_2038807, EPI_ISL_2038808, EPI_ISL_2038809, EPI_ISL_2038810, EPI_ISL_2038811, EPI_ISL_2038812, EPI_ISL_2038813, EPI_ISL_2038814, EPI_ISL_2038815, EPI_ISL_2038816, EPI_ISL_2038817, EPI_ISL_2038818 | | | |
| see above | Minnesota Department of Health, Public Health Laboratory | Minnesota Department of Health, Public Health Laboratory | Alexandra Lorentz, Jacob Garfin, Matt Plumb, and Xiong Wang |
| EPI_ISL_2038819 | M Health Fairview | Minnesota Department of Health, Public Health Laboratory | Alexandra Lorentz, Jacob Garfin, Matt Plumb, and Xiong Wang |
| EPI_ISL_2038820, EPI_ISL_2038821, EPI_ISL_2038822, EPI_ISL_2038823, EPI_ISL_2038824, EPI_ISL_2038825, EPI_ISL_2038826, EPI_ISL_2038827, EPI_ISL_2038828, EPI_ISL_2038829, EPI_ISL_2038830, EPI_ISL_2038831, EPI_ISL_2038832, EPI_ISL_2038833, EPI_ISL_2038834, EPI_ISL_2038835, EPI_ISL_2038836 | | | |
| see above | Minnesota Department of Health, Public Health Laboratory | Minnesota Department of Health, Public Health Laboratory | Alexandra Lorentz, Jacob Garfin, Matt Plumb, and Xiong Wang |
| EPI_ISL_2038837, EPI_ISL_2038838, EPI_ISL_2038839, EPI_ISL_2038840 | M Health Fairview | Minnesota Department of Health, Public Health Laboratory | Alexandra Lorentz, Jacob Garfin, Matt Plumb, and Xiong Wang |
| EPI_ISL_2038841, EPI_ISL_2038842, EPI_ISL_2038843, EPI_ISL_2038844, EPI_ISL_2038845, EPI_ISL_2038846, EPI_ISL_2038847, EPI_ISL_2038848, EPI_ISL_2038849, EPI_ISL_2038850 | Minnesota Department of Health, Public Health Laboratory | Minnesota Department of Health, Public Health Laboratory | Alexandra Lorentz, Jacob Garfin, Matt Plumb, and Xiong Wang |
| EPI_ISL_2038851, EPI_ISL_2038852, EPI_ISL_2038853 | M Health Fairview | Minnesota Department of Health, Public Health Laboratory | Alexandra Lorentz, Jacob Garfin, Matt Plumb, and Xiong Wang |
| EPI_ISL_2038854, EPI_ISL_2038860, EPI_ISL_2038861, EPI_ISL_2038862, EPI_ISL_2038863, EPI_ISL_2038864, EPI_ISL_2038865 | Minnesota Department of Health, Public Health Laboratory | Minnesota Department of Health, Public Health Laboratory | Alexandra Lorentz, Jacob Garfin, Matt Plumb, and Xiong Wang |
| EPI_ISL_2038969, EPI_ISL_2038970, EPI_ISL_2038971, EPI_ISL_2038972, EPI_ISL_2038973, EPI_ISL_2038974, EPI_ISL_2038975, EPI_ISL_2038976, EPI_ISL_2038977, EPI_ISL_2038978, EPI_ISL_2038979, EPI_ISL_2038980, EPI_ISL_2038981, EPI_ISL_2038982, EPI_ISL_2038983, EPI_ISL_2038984, EPI_ISL_2038985, EPI_ISL_2038986, EPI_ISL_2038987, EPI_ISL_2038988, EPI_ISL_2038989, EPI_ISL_2038990, EPI_ISL_2038991, EPI_ISL_2038992, EPI_ISL_2038993, EPI_ISL_2038994, EPI_ISL_2038996, EPI_ISL_2038997, EPI_ISL_2038998, EPI_ISL_2038999, EPI_ISL_2039000, EPI_ISL_2039001, EPI_ISL_2039002, EPI_ISL_2039003, EPI_ISL_2039004, EPI_ISL_2039005, EPI_ISL_2039006, EPI_ISL_2039007, EPI_ISL_2039008, EPI_ISL_2039009, EPI_ISL_2039010, EPI_ISL_2039011, EPI_ISL_2039012, EPI_ISL_2039013, EPI_ISL_2039014, EPI_ISL_2039015, EPI_ISL_2039016, EPI_ISL_2039017, EPI_ISL_2039018, EPI_ISL_2039019, EPI_ISL_2039020, EPI_ISL_2039021, EPI_ISL_2039022, EPI_ISL_2039023, EPI_ISL_2039024, EPI_ISL_2039025, EPI_ISL_2039026, EPI_ISL_2039027, EPI_ISL_2039028, EPI_ISL_2039029, EPI_ISL_2039030, EPI_ISL_2039031, EPI_ISL_2039032, EPI_ISL_2039033, EPI_ISL_2039034, EPI_ISL_2039035, EPI_ISL_2039036, EPI_ISL_2039037, EPI_ISL_2039038, EPI_ISL_2039040, EPI_ISL_2039041, EPI_ISL_2039042, EPI_ISL_2039043, EPI_ISL_2039044, EPI_ISL_2039045, EPI_ISL_2039046, EPI_ISL_2039047, EPI_ISL_2039048, EPI_ISL_2039049, EPI_ISL_2039050, EPI_ISL_2039051, EPI_ISL_2039052, EPI_ISL_2039053, EPI_ISL_2039054, EPI_ISL_2039055, EPI_ISL_2039056, EPI_ISL_2039057, EPI_ISL_2039058, EPI_ISL_2039059, EPI_ISL_2039060, EPI_ISL_2039061, EPI_ISL_2039062, EPI_ISL_2039063, EPI_ISL_2039064, EPI_ISL_2039065, EPI_ISL_2039066, EPI_ISL_2039067, EPI_ISL_2039068, EPI_ISL_2039069, EPI_ISL_2039070, EPI_ISL_2039072, EPI_ISL_2039074, EPI_ISL_2039075, EPI_ISL_2039076, EPI_ISL_2039077, EPI_ISL_2039078, EPI_ISL_2039079, EPI_ISL_2039080, EPI_ISL_2039081, EPI_ISL_2039082, EPI_ISL_2039083, EPI_ISL_2039084, EPI_ISL_2039085, EPI_ISL_2039086, EPI_ISL_2039087, EPI_ISL_2039088, EPI_ISL_2039089, EPI_ISL_2039090, EPI_ISL_2039091, EPI_ISL_2039092, EPI_ISL_2039093, EPI_ISL_2039094, EPI_ISL_2039095, EPI_ISL_2039096, EPI_ISL_2039097, EPI_ISL_2039098, EPI_ISL_2039099, EPI_ISL_2039101, EPI_ISL_2039102, EPI_ISL_2039103, EPI_ISL_2039104, EPI_ISL_2039105, EPI_ISL_2039106, EPI_ISL_2039107, EPI_ISL_2039108, EPI_ISL_2039109, EPI_ISL_2039110, EPI_ISL_2039111, EPI_ISL_2039112, EPI_ISL_2039113, EPI_ISL_2039114, EPI_ISL_2039115, EPI_ISL_2039116, EPI_ISL_2039117, EPI_ISL_2039118, EPI_ISL_2039119, EPI_ISL_2039120, EPI_ISL_2039121, EPI_ISL_2039122, EPI_ISL_2039123, EPI_ISL_2039124, EPI_ISL_2039125, EPI_ISL_2039126, EPI_ISL_2039127, EPI_ISL_2039128, EPI_ISL_2039129, EPI_ISL_2039130, EPI_ISL_2039131, EPI_ISL_2039132, EPI_ISL_2039133, EPI_ISL_2039134, EPI_ISL_2039135, EPI_ISL_2039136, EPI_ISL_2039137, EPI_ISL_2039138, EPI_ISL_2039139, EPI_ISL_2039140, EPI_ISL_2039141, EPI_ISL_2039142, EPI_ISL_2039143, EPI_ISL_2039144, EPI_ISL_2039145, EPI_ISL_2039146, EPI_ISL_2039147, EPI_ISL_2039148, EPI_ISL_2039149, EPI_ISL_2039150, EPI_ISL_2039151, EPI_ISL_2039152, EPI_ISL_2039153, EPI_ISL_2039154, EPI_ISL_2039155, EPI_ISL_2039157, EPI_ISL_2039158, EPI_ISL_2039159, EPI_ISL_2039160, EPI_ISL_2039161, EPI_ISL_2039162, EPI_ISL_2039163, EPI_ISL_2039164, EPI_ISL_2039165, EPI_ISL_2039166, EPI_ISL_2039168, EPI_ISL_2039169, EPI_ISL_2039170, EPI_ISL_2039171, EPI_ISL_2039172, EPI_ISL_2039173, EPI_ISL_2039174, EPI_ISL_2039175, EPI_ISL_2039176, EPI_ISL_2039177, EPI_ISL_2039179, EPI_ISL_2039180, EPI_ISL_2039181, EPI_ISL_2039182, EPI_ISL_2039183, EPI_ISL_2039184, EPI_ISL_2039185, EPI_ISL_2039186, EPI_ISL_2039187, EPI_ISL_2039188, EPI_ISL_2039189, EPI_ISL_2039190, EPI_ISL_2039191, EPI_ISL_2039192, EPI_ISL_2039193, EPI_ISL_2039194, EPI_ISL_2039195, EPI_ISL_2039196, EPI_ISL_2039197, EPI_ISL_2039199, EPI_ISL_2039200, EPI_ISL_2039201 | | | |
| see above | Aegis Sciences Corporation | Centers for Disease Control and Prevention Division of Viral Diseases, Pathogen Discovery | Dakota Howard, Dhwani Batra, Peter W. Cook, Kara Moser, Adrian Paskey, Jason Caravas, Benjamin Rambo-Martin, Shatavia Morrison, Christopher Gulvick, Scott Sammons, Yvette Unoarumhi, Darlene Wagner, Matthew Schmerer, Cyndi Clark, Patrick Campbell, Rob Case, Vikramsinha Ghorpade, Holly Houdeshell, Ola Kvalvaag, Dillon Nall, Ethan Sanders, Alec Vest, Shaun Westlund, Matthew Hardison, Clinton R. Paden, Duncan MacCannell |
| EPI_ISL_2039202 | Kentucky State Public Health Lab | Kentucky State Public Health Lab | Anjayya Tummala, Karim George, Joshua Tobias, William Grooms, Vaneet Arora, Matthew Johnson, Rachel Zinner, Rhonda Lucas |
| EPI_ISL_2039203 | Aegis Sciences Corporation | Centers for Disease Control and Prevention Division of Viral Diseases, Pathogen Discovery | Dakota Howard, Dhwani Batra, Peter W. Cook, Kara Moser, Adrian Paskey, Jason Caravas, Benjamin Rambo-Martin, Shatavia Morrison, Christopher Gulvick, Scott Sammons, Yvette Unoarumhi, Darlene Wagner, Matthew Schmerer, Cyndi Clark, Patrick Campbell, Rob Case, Vikramsinha Ghorpade, Holly Houdeshell, Ola Kvalvaag, Dillon Nall, Ethan Sanders, Alec Vest, Shaun Westlund, Matthew Hardison, Clinton R. Paden, Duncan MacCannell |
| EPI_ISL_2039204, EPI_ISL_2039205 | Kentucky State Public Health Lab | Kentucky State Public Health Lab | Anjayya Tummala, Karim George, Joshua Tobias, William Grooms, Vaneet Arora, Matthew Johnson, Rachel Zinner, Rhonda Lucas |

| | | | |
|---|---|---|---|
| EPI_ISL_2039206 | Aegis Sciences Corporation | Centers for Disease Control and Prevention Division of Viral Diseases, Pathogen Discovery | Dakota Howard, Dhwani Batra, Peter W. Cook, Kara Moser, Adrian Paskey, Jason Caravas, Benjamin Rambo-Martin, Shatavia Morrison, Christopher Gulvick, Scott Sammons, Yvette Unoarumhi, Darlene Wagner, Matthew Schmerer, Cyndi Clark, Patrick Campbell, Rob Case, Vikramsinha Ghorpade, Holly Houdeshell, Ola Kvalvaag, Dillon Nall, Ethan Sanders, Alec Vest, Shaun Westlund, Matthew Hardison, Clinton R. Paden, Duncan MacCannell |
| EPI_ISL_2039207, EPI_ISL_2039208 | Kentucky State Public Health Lab | Kentucky State Public Health Lab | Anjayya Tummala, Karim George, Joshua Tobias, William Grooms, Vaneet Arora, Matthew Johnson, Rachel Zinner, Rhonda Lucas |
| EPI_ISL_2039209 | Aegis Sciences Corporation | Centers for Disease Control and Prevention Division of Viral Diseases, Pathogen Discovery | Dakota Howard, Dhwani Batra, Peter W. Cook, Kara Moser, Adrian Paskey, Jason Caravas, Benjamin Rambo-Martin, Shatavia Morrison, Christopher Gulvick, Scott Sammons, Yvette Unoarumhi, Darlene Wagner, Matthew Schmerer, Cyndi Clark, Patrick Campbell, Rob Case, Vikramsinha Ghorpade, Holly Houdeshell, Ola Kvalvaag, Dillon Nall, Ethan Sanders, Alec Vest, Shaun Westlund, Matthew Hardison, Clinton R. Paden, Duncan MacCannell |
| EPI_ISL_2039210, EPI_ISL_2039211, EPI_ISL_2039212 | Kentucky State Public Health Lab | Kentucky State Public Health Lab | Anjayya Tummala, Karim George, Joshua Tobias, William Grooms, Vaneet Arora, Matthew Johnson, Rachel Zinner, Rhonda Lucas |
| EPI_ISL_2039213 | Aegis Sciences Corporation | Centers for Disease Control and Prevention Division of Viral Diseases, Pathogen Discovery | Dakota Howard, Dhwani Batra, Peter W. Cook, Kara Moser, Adrian Paskey, Jason Caravas, Benjamin Rambo-Martin, Shatavia Morrison, Christopher Gulvick, Scott Sammons, Yvette Unoarumhi, Darlene Wagner, Matthew Schmerer, Cyndi Clark, Patrick Campbell, Rob Case, Vikramsinha Ghorpade, Holly Houdeshell, Ola Kvalvaag, Dillon Nall, Ethan Sanders, Alec Vest, Shaun Westlund, Matthew Hardison, Clinton R. Paden, Duncan MacCannell |
| EPI_ISL_2039214, EPI_ISL_2039215 | Kentucky State Public Health Lab | Kentucky State Public Health Lab | Anjayya Tummala, Karim George, Joshua Tobias, William Grooms, Vaneet Arora, Matthew Johnson, Rachel Zinner, Rhonda Lucas |
| EPI_ISL_2039216 | Aegis Sciences Corporation | Centers for Disease Control and Prevention Division of Viral Diseases, Pathogen Discovery | Dakota Howard, Dhwani Batra, Peter W. Cook, Kara Moser, Adrian Paskey, Jason Caravas, Benjamin Rambo-Martin, Shatavia Morrison, Christopher Gulvick, Scott Sammons, Yvette Unoarumhi, Darlene Wagner, Matthew Schmerer, Cyndi Clark, Patrick Campbell, Rob Case, Vikramsinha Ghorpade, Holly Houdeshell, Ola Kvalvaag, Dillon Nall, Ethan Sanders, Alec Vest, Shaun Westlund, Matthew Hardison, Clinton R. Paden, Duncan MacCannell |
| EPI_ISL_2039217, EPI_ISL_2039218, EPI_ISL_2039219 | Kentucky State Public Health Lab | Kentucky State Public Health Lab | Anjayya Tummala, Karim George, Joshua Tobias, William Grooms, Vaneet Arora, Matthew Johnson, Rachel Zinner, Rhonda Lucas |
| EPI_ISL_2039220 | Aegis Sciences Corporation | Centers for Disease Control and Prevention Division of Viral Diseases, Pathogen Discovery | Dakota Howard, Dhwani Batra, Peter W. Cook, Kara Moser, Adrian Paskey, Jason Caravas, Benjamin Rambo-Martin, Shatavia Morrison, Christopher Gulvick, Scott Sammons, Yvette Unoarumhi, Darlene Wagner, Matthew Schmerer, Cyndi Clark, Patrick Campbell, Rob Case, Vikramsinha Ghorpade, Holly Houdeshell, Ola Kvalvaag, Dillon Nall, Ethan Sanders, Alec Vest, Shaun Westlund, Matthew Hardison, Clinton R. Paden, Duncan MacCannell |
| EPI_ISL_2039221 | Kentucky State Public Health Lab | Kentucky State Public Health Lab | Anjayya Tummala, Karim George, Joshua Tobias, William Grooms, Vaneet Arora, Matthew Johnson, Rachel Zinner, Rhonda Lucas |
| EPI_ISL_2039222, EPI_ISL_2039223, EPI_ISL_2039225, EPI_ISL_2039227, EPI_ISL_2039228, EPI_ISL_2039229, EPI_ISL_2039230, EPI_ISL_2039233, EPI_ISL_2039234, EPI_ISL_2039235, EPI_ISL_2039236, EPI_ISL_2039237, EPI_ISL_2039239, EPI_ISL_2039240, EPI_ISL_2039241, EPI_ISL_2039242, EPI_ISL_2039243, EPI_ISL_2039244, EPI_ISL_2039245, EPI_ISL_2039246, EPI_ISL_2039247, EPI_ISL_2039248, EPI_ISL_2039249, EPI_ISL_2039250, EPI_ISL_2039251, EPI_ISL_2039252, EPI_ISL_2039253, EPI_ISL_2039254, EPI_ISL_2039255, EPI_ISL_2039256, EPI_ISL_2039257, EPI_ISL_2039258, EPI_ISL_2039259, EPI_ISL_2039260, EPI_ISL_2039261, EPI_ISL_2039262, EPI_ISL_2039263, EPI_ISL_2039264, EPI_ISL_2039265, EPI_ISL_2039266, EPI_ISL_2039267, EPI_ISL_2039268, EPI_ISL_2039269, EPI_ISL_2039270, EPI_ISL_2039271, EPI_ISL_2039272, EPI_ISL_2039273, EPI_ISL_2039274, EPI_ISL_2039275, EPI_ISL_2039276, EPI_ISL_2039277, EPI_ISL_2039278, EPI_ISL_2039279, EPI_ISL_2039280, EPI_ISL_2039281, EPI_ISL_2039282, EPI_ISL_2039283, EPI_ISL_2039284, EPI_ISL_2039285, EPI_ISL_2039286, EPI_ISL_2039287, EPI_ISL_2039288, EPI_ISL_2039289, EPI_ISL_2039290, EPI_ISL_2039291, EPI_ISL_2039292, EPI_ISL_2039293, EPI_ISL_2039294, EPI_ISL_2039295, EPI_ISL_2039296, EPI_ISL_2039297, EPI_ISL_2039298, EPI_ISL_2039299, EPI_ISL_2039300, EPI_ISL_2039301, EPI_ISL_2039302, EPI_ISL_2039303, EPI_ISL_2039304, EPI_ISL_2039305, EPI_ISL_2039306, EPI_ISL_2039307, EPI_ISL_2039308, EPI_ISL_2039309, EPI_ISL_2039310, EPI_ISL_2039311, EPI_ISL_2039312, EPI_ISL_2039313, EPI_ISL_2039314, EPI_ISL_2039315, EPI_ISL_2039316, EPI_ISL_2039317, EPI_ISL_2039318, EPI_ISL_2039319, EPI_ISL_2039322, EPI_ISL_2039325, EPI_ISL_2039328, EPI_ISL_2039331, EPI_ISL_2039332, EPI_ISL_2039333, EPI_ISL_2039334, EPI_ISL_2039335, EPI_ISL_2039336, EPI_ISL_2039337, EPI_ISL_2039338, EPI_ISL_2039339, EPI_ISL_2039340, EPI_ISL_2039341, EPI_ISL_2039342, EPI_ISL_2039343, EPI_ISL_2039344, EPI_ISL_2039345, EPI_ISL_2039346, EPI_ISL_2039347, EPI_ISL_2039348, EPI_ISL_2039349, EPI_ISL_2039350, EPI_ISL_2039351, EPI_ISL_2039352, EPI_ISL_2039353, EPI_ISL_2039354, EPI_ISL_2039355, EPI_ISL_2039356, EPI_ISL_2039357, EPI_ISL_2039358, EPI_ISL_2039359, EPI_ISL_2039360, EPI_ISL_2039361, EPI_ISL_2039362, EPI_ISL_2039363, EPI_ISL_2039364, EPI_ISL_2039365, EPI_ISL_2039366, EPI_ISL_2039367, EPI_ISL_2039368, EPI_ISL_2039369, EPI_ISL_2039370, EPI_ISL_2039371, EPI_ISL_2039372, EPI_ISL_2039373, EPI_ISL_2039374, EPI_ISL_2039375, EPI_ISL_2039376, EPI_ISL_2039377, EPI_ISL_2039378, EPI_ISL_2039379, EPI_ISL_2039380, EPI_ISL_2039381, EPI_ISL_2039382, EPI_ISL_2039383, EPI_ISL_2039384, EPI_ISL_2039385, EPI_ISL_2039386, EPI_ISL_2039387, EPI_ISL_2039388, EPI_ISL_2039389, EPI_ISL_2039390, EPI_ISL_2039391, EPI_ISL_2039392, EPI_ISL_2039393, EPI_ISL_2039395, EPI_ISL_2039396, EPI_ISL_2039397, EPI_ISL_2039398, EPI_ISL_2039399, EPI_ISL_2039400, EPI_ISL_2039401, EPI_ISL_2039402, EPI_ISL_2039403, EPI_ISL_2039404, EPI_ISL_2039405, EPI_ISL_2039406, EPI_ISL_2039407, EPI_ISL_2039408, EPI_ISL_2039409, EPI_ISL_2039410, EPI_ISL_2039411, EPI_ISL_2039412, EPI_ISL_2039413, EPI_ISL_2039414, EPI_ISL_2039415, EPI_ISL_2039416, EPI_ISL_2039417, EPI_ISL_2039418, EPI_ISL_2039419, EPI_ISL_2039420, EPI_ISL_2039421, EPI_ISL_2039422, EPI_ISL_2039423, EPI_ISL_2039424, EPI_ISL_2039425, EPI_ISL_2039426, EPI_ISL_2039427, EPI_ISL_2039428, EPI_ISL_2039429, EPI_ISL_2039430, EPI_ISL_2039431, EPI_ISL_2039432, EPI_ISL_2039434, EPI_ISL_2039435, EPI_ISL_2039436, EPI_ISL_2039437, EPI_ISL_2039438, EPI_ISL_2039439, EPI_ISL_2039440, EPI_ISL_2039441, EPI_ISL_2039442, EPI_ISL_2039443, EPI_ISL_2039444, EPI_ISL_2039445, EPI_ISL_2039446, EPI_ISL_2039447, EPI_ISL_2039448, EPI_ISL_2039449, EPI_ISL_2039450, EPI_ISL_2039451, EPI_ISL_2039452, EPI_ISL_2039453, EPI_ISL_2039454, EPI_ISL_2039455, EPI_ISL_2039456, EPI_ISL_2039457, EPI_ISL_2039458, EPI_ISL_2039459, EPI_ISL_2039460, EPI_ISL_2039461, EPI_ISL_2039462, EPI_ISL_2039463, EPI_ISL_2039464, EPI_ISL_2039465, EPI_ISL_2039466, EPI_ISL_2039467, EPI_ISL_2039468, EPI_ISL_2039469, EPI_ISL_2039470, EPI_ISL_2039471, EPI_ISL_2039472, EPI_ISL_2039473, EPI_ISL_2039474, EPI_ISL_2039475, EPI_ISL_2039476, EPI_ISL_2039477, EPI_ISL_2039478, EPI_ISL_2039479, EPI_ISL_2039480, EPI_ISL_2039481, EPI_ISL_2039482, EPI_ISL_2039483, EPI_ISL_2039484, EPI_ISL_2039485, EPI_ISL_2039486, EPI_ISL_2039487, EPI_ISL_2039488, EPI_ISL_2039489, EPI_ISL_2039491 | | | |
| see above | Aegis Sciences Corporation | Centers for Disease Control and Prevention Division of Viral Diseases, Pathogen Discovery | Dakota Howard, Dhwani Batra, Peter W. Cook, Kara Moser, Adrian Paskey, Jason Caravas, Benjamin Rambo-Martin, Shatavia Morrison, Christopher Gulvick, Scott Sammons, Yvette Unoarumhi, Darlene Wagner, Matthew Schmerer, Cyndi Clark, Patrick Campbell, Rob Case, Vikramsinha Ghorpade, Holly Houdeshell, Ola Kvalvaag, Dillon Nall, Ethan Sanders, Alec Vest, Shaun Westlund, Matthew Hardison, Clinton R. Paden, Duncan MacCannell |
| EPI_ISL_2039493 | Wyoming Public Health Laboratory | Wyoming Public Health Laboratory | Jim Mildenberger, Taylor Fearing, Channing Weber, Ashley Norberg, Chayse Rowley, Marley Goetz, Brian Dominguez, Elliot Thomasson, Brittany Oher, Sam Britz, Cari Sloma, and Rob Christensen |
| EPI_ISL_2039494 | Aegis Sciences Corporation | Centers for Disease Control and Prevention Division of Viral Diseases, Pathogen Discovery | Dakota Howard, Dhwani Batra, Peter W. Cook, Kara Moser, Adrian Paskey, Jason Caravas, Benjamin Rambo-Martin, Shatavia Morrison, Christopher Gulvick, Scott Sammons, Yvette Unoarumhi, Darlene Wagner, Matthew Schmerer, Cyndi Clark, Patrick Campbell, Rob Case, Vikramsinha Ghorpade, Holly Houdeshell, Ola Kvalvaag, Dillon Nall, Ethan Sanders, Alec Vest, Shaun Westlund, Matthew Hardison, Clinton R. Paden, Duncan MacCannell |
| EPI_ISL_2039496, EPI_ISL_2039497, EPI_ISL_2039498, EPI_ISL_2039500, EPI_ISL_2039502 | Wyoming Public Health Laboratory | Wyoming Public Health Laboratory | Jim Mildenberger, Taylor Fearing, Channing Weber, Ashley Norberg, Chayse Rowley, Marley Goetz, Brian Dominguez, Elliot Thomasson, Brittany Oher, Sam Britz, Cari Sloma, and Rob Christensen |
| EPI_ISL_2039503 | Aegis Sciences Corporation | Centers for Disease Control and Prevention Division of Viral Diseases, Pathogen Discovery | Dakota Howard, Dhwani Batra, Peter W. Cook, Kara Moser, Adrian Paskey, Jason Caravas, Benjamin Rambo-Martin, Shatavia Morrison, Christopher Gulvick, Scott Sammons, Yvette Unoarumhi, Darlene Wagner, Matthew Schmerer, Cyndi Clark, Patrick Campbell, Rob Case, Vikramsinha Ghorpade, Holly Houdeshell, Ola Kvalvaag, Dillon Nall, Ethan Sanders, Alec Vest, Shaun Westlund, Matthew Hardison, Clinton R. Paden, Duncan MacCannell |
| EPI_ISL_2039505, EPI_ISL_2039506 | Wyoming Public Health Laboratory | Wyoming Public Health Laboratory | Jim Mildenberger, Taylor Fearing, Channing Weber, Ashley Norberg, Chayse Rowley, Marley Goetz, Brian Dominguez, Elliot Thomasson, Brittany Oher, Sam Britz, Cari Sloma, and Rob Christensen |
| EPI_ISL_2039507 | Aegis Sciences Corporation | Centers for Disease Control and Prevention Division of Viral Diseases, Pathogen Discovery | Dakota Howard, Dhwani Batra, Peter W. Cook, Kara Moser, Adrian Paskey, Jason Caravas, Benjamin Rambo-Martin, Shatavia Morrison, Christopher Gulvick, Scott Sammons, Yvette Unoarumhi, Darlene Wagner, Matthew Schmerer, Cyndi Clark, Patrick Campbell, Rob Case, Vikramsinha Ghorpade, Holly Houdeshell, Ola Kvalvaag, Dillon Nall, Ethan Sanders, Alec Vest, Shaun Westlund, Matthew Hardison, Clinton R. Paden, Duncan MacCannell |
| EPI_ISL_2039508, EPI_ISL_2039510 | Wyoming Public Health Laboratory | Wyoming Public Health Laboratory | Jim Mildenberger, Taylor Fearing, Channing Weber, Ashley Norberg, Chayse Rowley, Marley Goetz, Brian Dominguez, Elliot Thomasson, Brittany Oher, Sam Britz, Cari Sloma, and Rob Christensen |
| EPI_ISL_2039511, EPI_ISL_2039514, EPI_ISL_2039516, EPI_ISL_2039517, EPI_ISL_2039518, EPI_ISL_2039519, EPI_ISL_2039520, EPI_ISL_2039521, EPI_ISL_2039522, EPI_ISL_2039523, EPI_ISL_2039524, EPI_ISL_2039525, EPI_ISL_2039526, EPI_ISL_2039527, EPI_ISL_2039528, EPI_ISL_2039529, EPI_ISL_2039530, EPI_ISL_2039531, EPI_ISL_2039532, EPI_ISL_2039533, EPI_ISL_2039534, EPI_ISL_2039535, EPI_ISL_2039536, EPI_ISL_2039537, EPI_ISL_2039538, EPI_ISL_2039539, EPI_ISL_2039540, EPI_ISL_2039541, EPI_ISL_2039542, EPI_ISL_2039543, EPI_ISL_2039544, EPI_ISL_2039545, EPI_ISL_2039546, EPI_ISL_2039547, EPI_ISL_2039548, EPI_ISL_2039549, EPI_ISL_2039550, EPI_ISL_2039551, EPI_ISL_2039552, EPI_ISL_2039553, EPI_ISL_2039554, EPI_ISL_2039555, EPI_ISL_2039556, EPI_ISL_2039557, EPI_ISL_2039558, EPI_ISL_2039559, EPI_ISL_2039560, EPI_ISL_2039561, EPI_ISL_2039562, EPI_ISL_2039563, EPI_ISL_2039564, EPI_ISL_2039565, EPI_ISL_2039566, EPI_ISL_2039567, EPI_ISL_2039568, EPI_ISL_2039569, EPI_ISL_2039570, EPI_ISL_2039571, EPI_ISL_2039572, EPI_ISL_2039574, EPI_ISL_2039575, EPI_ISL_2039576, EPI_ISL_2039577, EPI_ISL_2039578, EPI_ISL_2039579, EPI_ISL_2039581, EPI_ISL_2039582, EPI_ISL_2039583, EPI_ISL_2039584, EPI_ISL_2039585, EPI_ISL_2039586, EPI_ISL_2039587, EPI_ISL_2039588, EPI_ISL_2039589, EPI_ISL_2039590, EPI_ISL_2039591, EPI_ISL_2039592, EPI_ISL_2039594, EPI_ISL_2039595, EPI_ISL_2039596, EPI_ISL_2039597, EPI_ISL_2039598, EPI_ISL_2039599, EPI_ISL_2039600, EPI_ISL_2039601, EPI_ISL_2039602, EPI_ISL_2039603, EPI_ISL_2039604, EPI_ISL_2039605, EPI_ISL_2039606, EPI_ISL_2039607, EPI_ISL_2039608, EPI_ISL_2039609, EPI_ISL_2039610, EPI_ISL_2039611, EPI_ISL_2039612, EPI_ISL_2039613, EPI_ISL_2039614, EPI_ISL_2039615, EPI_ISL_2039616, EPI_ISL_2039617, EPI_ISL_2039618, EPI_ISL_2039619, EPI_ISL_2039620, EPI_ISL_2039621, EPI_ISL_2039622, EPI_ISL_2039623, EPI_ISL_2039624, EPI_ISL_2039625, EPI_ISL_2039626, EPI_ISL_2039627, EPI_ISL_2039628, EPI_ISL_2039629, EPI_ISL_2039630, EPI_ISL_2039631, EPI_ISL_2039632, EPI_ISL_2039633, EPI_ISL_2039634, EPI_ISL_2039635, EPI_ISL_2039636, EPI_ISL_2039637, EPI_ISL_2039638, EPI_ISL_2039639, EPI_ISL_2039640, EPI_ISL_2039641, EPI_ISL_2039642, EPI_ISL_2039643, EPI_ISL_2039644, EPI_ISL_2039645, EPI_ISL_2039646, EPI_ISL_2039647, EPI_ISL_2039648, EPI_ISL_2039649, EPI_ISL_2039650, EPI_ISL_2039651, EPI_ISL_2039652, EPI_ISL_2039653, EPI_ISL_2039654, EPI_ISL_2039655, EPI_ISL_2039656, EPI_ISL_2039657, EPI_ISL_2039658, EPI_ISL_2039659, EPI_ISL_2039660, EPI_ISL_2039661, EPI_ISL_2039662, EPI_ISL_2039663, EPI_ISL_2039664, EPI_ISL_2039665, EPI_ISL_2039666, EPI_ISL_2039667, EPI_ISL_2039668, EPI_ISL_2039669, EPI_ISL_2039670, EPI_ISL_2039671, EPI_ISL_2039672, EPI_ISL_2039673, EPI_ISL_2039674, EPI_ISL_2039675, EPI_ISL_2039676, EPI_ISL_2039677, EPI_ISL_2039678, EPI_ISL_2039679, EPI_ISL_2039680, EPI_ISL_2039681, EPI_ISL_2039682, EPI_ISL_2039683, EPI_ISL_2039684, EPI_ISL_2039685, EPI_ISL_2039686, EPI_ISL_2039687, EPI_ISL_2039688, EPI_ISL_2039689, EPI_ISL_2039690, EPI_ISL_2039691, EPI_ISL_2039692, EPI_ISL_2039693, EPI_ISL_2039694, EPI_ISL_2039695, EPI_ISL_2039696, EPI_ISL_2039697, EPI_ISL_2039698, EPI_ISL_2039699, EPI_ISL_2039700, EPI_ISL_2039701, EPI_ISL_2039702, EPI_ISL_2039703, EPI_ISL_2039704, EPI_ISL_2039705, EPI_ISL_2039706, EPI_ISL_2039707, EPI_ISL_2039708, EPI_ISL_2039710, EPI_ISL_2039711, EPI_ISL_2039712, EPI_ISL_2039713, EPI_ISL_2039714, EPI_ISL_2039715, EPI_ISL_2039716, EPI_ISL_2039717, EPI_ISL_2039718, EPI_ISL_2039719, EPI_ISL_2039720, EPI_ISL_2039721, EPI_ISL_2039722, EPI_ISL_2039723, EPI_ISL_2039724, EPI_ISL_2039725, EPI_ISL_2039726, EPI_ISL_2039727, EPI_ISL_2039728, EPI_ISL_2039729, EPI_ISL_2039730, EPI_ISL_2039731, EPI_ISL_2039732, EPI_ISL_2039733, EPI_ISL_2039734, EPI_ISL_2039735, EPI_ISL_2039736, EPI_ISL_2039737, EPI_ISL_2039738, EPI_ISL_2039739, EPI_ISL_2039740, EPI_ISL_2039741, EPI_ISL_2039742, EPI_ISL_2039743, EPI_ISL_2039745, EPI_ISL_2039747, EPI_ISL_2039748, EPI_ISL_2039749, EPI_ISL_2039750, EPI_ISL_2039751, EPI_ISL_2039752, EPI_ISL_2039753, EPI_ISL_2039754, EPI_ISL_2039755, EPI_ISL_2039756, EPI_ISL_2039758, EPI_ISL_2039759, EPI_ISL_2039760, EPI_ISL_2039761, EPI_ISL_2039762, EPI_ISL_2039763, EPI_ISL_2039764, EPI_ISL_2039765, EPI_ISL_2039766, EPI_ISL_2039767, EPI_ISL_2039768, EPI_ISL_2039769, EPI_ISL_2039770, EPI_ISL_2039771, EPI_ISL_2039772, EPI_ISL_2039773, EPI_ISL_2039774, EPI_ISL_2039775, EPI_ISL_2039776, EPI_ISL_2039777, EPI_ISL_2039778, EPI_ISL_2039779, EPI_ISL_2039780, EPI_ISL_2039781, EPI_ISL_2039782, EPI_ISL_2039783, EPI_ISL_2039785, EPI_ISL_2039786, EPI_ISL_2039788, EPI_ISL_2039789, EPI_ISL_2039790, EPI_ISL_2039791, EPI_ISL_2039792, EPI_ISL_2039793, EPI_ISL_2039794, EPI_ISL_2039795, EPI_ISL_2039796, EPI_ISL_2039797, EPI_ISL_2039798, EPI_ISL_2039800, EPI_ISL_2039801, EPI_ISL_2039802, EPI_ISL_2039804, EPI_ISL_2039805, EPI_ISL_2039806, EPI_ISL_2039807, EPI_ISL_2039808, EPI_ISL_2039809, EPI_ISL_2039810, EPI_ISL_2039811, EPI_ISL_2039812, EPI_ISL_2039813, EPI_ISL_2039814, EPI_ISL_2039815, EPI_ISL_2039816, EPI_ISL_2039817, EPI_ISL_2039818, EPI_ISL_2039819, EPI_ISL_2039820, EPI_ISL_2039821, EPI_ISL_2039822, EPI_ISL_2039823, EPI_ISL_2039824, EPI_ISL_2039825, EPI_ISL_2039826, EPI_ISL_2039827, EPI_ISL_2039828, EPI_ISL_2039829, EPI_ISL_2039831, EPI_ISL_2039832, EPI_ISL_2039833, EPI_ISL_2039834, EPI_ISL_2039835, EPI_ISL_2039836, EPI_ISL_2039838, EPI_ISL_2039839, EPI_ISL_2039840, EPI_ISL_2039841, EPI_ISL_2039842, EPI_ISL_2039843, EPI_ISL_2039844, EPI_ISL_2039845, EPI_ISL_2039846, EPI_ISL_2039847, EPI_ISL_2039848, EPI_ISL_2039849, EPI_ISL_2039850, EPI_ISL_2039851, EPI_ISL_2039853, EPI_ISL_2039854, EPI_ISL_2039855, EPI_ISL_2039856, EPI_ISL_2039857, EPI_ISL_2039858, EPI_ISL_2039859, EPI_ISL_2039860, EPI_ISL_2039861, EPI_ISL_2039862, EPI_ISL_2039863, EPI_ISL_2039864, EPI_ISL_2039865, EPI_ISL_2039866, EPI_ISL_2039867, EPI_ISL_2039869, EPI_ISL_2039870, EPI_ISL_2039871, EPI_ISL_2039872, EPI_ISL_2039873, EPI_ISL_2039874, EPI_ISL_2039875, EPI_ISL_2039876, EPI_ISL_2039877, EPI_ISL_2039878, EPI_ISL_2039879, EPI_ISL_2039880, EPI_ISL_2039881, EPI_ISL_2039882, EPI_ISL_2039883, EPI_ISL_2039884, EPI_ISL_2039885, EPI_ISL_2039886, EPI_ISL_2039887, EPI_ISL_2039888, EPI_ISL_2039889, EPI_ISL_2039890, EPI_ISL_2039891, EPI_ISL_2039892, EPI_ISL_2039893, EPI_ISL_2039894, EPI_ISL_2039895, EPI_ISL_2039896, EPI_ISL_2039897, EPI_ISL_2039898, EPI_ISL_2039899, EPI_ISL_2039900, EPI_ISL_2039901, EPI_ISL_2039902, EPI_ISL_2039903, EPI_ISL_2039904, EPI_ISL_2039905, EPI_ISL_2039906, EPI_ISL_2039907, EPI_ISL_2039908, EPI_ISL_2039909, EPI_ISL_2039910, EPI_ISL_2039911, EPI_ISL_2039912, EPI_ISL_2039913, EPI_ISL_2039914, EPI_ISL_2039915, EPI_ISL_2039916, EPI_ISL_2039917, EPI_ISL_2039918, EPI_ISL_2039920, EPI_ISL_2039921, EPI_ISL_2039922, EPI_ISL_2039923, EPI_ISL_2039924, EPI_ISL_2039925, EPI_ISL_2039926, EPI_ISL_2039927, EPI_ISL_2039928, EPI_ISL_2039929, EPI_ISL_2039930, EPI_ISL_2039931, EPI_ISL_2039932, EPI_ISL_2039933, EPI_ISL_2039934, EPI_ISL_2039935, EPI_ISL_2039936, EPI_ISL_2039937, EPI_ISL_2039938, EPI_ISL_2039939, EPI_ISL_2039940, EPI_ISL_2039941, EPI_ISL_2039942, EPI_ISL_2039943, EPI_ISL_2039944, EPI_ISL_2039945, EPI_ISL_2039946, EPI_ISL_2039947, EPI_ISL_2039948, EPI_ISL_2039949, EPI_ISL_2039950, EPI_ISL_2039951, EPI_ISL_2039952, EPI_ISL_2039953, EPI_ISL_2039954, EPI_ISL_2039955, EPI_ISL_2039956, EPI_ISL_2039957, EPI_ISL_2039958, EPI_ISL_2039959, EPI_ISL_2039960, EPI_ISL_2039961, EPI_ISL_2039962, EPI_ISL_2039963, EPI_ISL_2039964, EPI_ISL_2039965, EPI_ISL_2039966, EPI_ISL_2039967, EPI_ISL_2039968, EPI_ISL_2039969, EPI_ISL_2039970, EPI_ISL_2039971, EPI_ISL_2039972, EPI_ISL_2039973, EPI_ISL_2039974, EPI_ISL_2039975, EPI_ISL_2039976, EPI_ISL_2039977, EPI_ISL_2039978, EPI_ISL_2039979, EPI_ISL_2039980, EPI_ISL_2039981, EPI_ISL_2039982, EPI_ISL_2039983, EPI_ISL_2039984, EPI_ISL_2039985, EPI_ISL_2039986, EPI_ISL_2039987, EPI_ISL_2039988, EPI_ISL_2039989, EPI_ISL_2039990, EPI_ISL_2039991, EPI_ISL_2039992, EPI_ISL_2039993, EPI_ISL_2039994, EPI_ISL_2039995, EPI_ISL_2039996, EPI_ISL_2039997, EPI_ISL_2039998, EPI_ISL_2039999, EPI_ISL_2040000, EPI_ISL_2040001, EPI_ISL_2040002, EPI_ISL_2040003, EPI_ISL_2040004, EPI_ISL_2040005, EPI_ISL_2040006, EPI_ISL_2040007, EPI_ISL_2040008, EPI_ISL_2040009, EPI_ISL_2040010, EPI_ISL_2040011, EPI_ISL_2040012, EPI_ISL_2040013, EPI_ISL_2040014, EPI_ISL_2040015, EPI_ISL_2040016, EPI_ISL_2040017, EPI_ISL_2040018, EPI_ISL_2040019, EPI_ISL_2040020, EPI_ISL_2040021, EPI_ISL_2040022, EPI_ISL_2040023, EPI_ISL_2040025, EPI_ISL_2040026, EPI_ISL_2040027, EPI_ISL_2040028, EPI_ISL_2040029, EPI_ISL_2040030, EPI_ISL_2040031, EPI_ISL_2040032, EPI_ISL_2040033, EPI_ISL_2040034, EPI_ISL_2040035, EPI_ISL_2040036, EPI_ISL_2040037, EPI_ISL_2040038, EPI_ISL_2040039, EPI_ISL_2040040, EPI_ISL_2040041, EPI_ISL_2040042, EPI_ISL_2040043, EPI_ISL_2040044, EPI_ISL_2040045, EPI_ISL_2040047, EPI_ISL_2040048, EPI_ISL_2040049, EPI_ISL_2040050, EPI_ISL_2040051, EPI_ISL_2040053, EPI_ISL_2040054, EPI_ISL_2040055, EPI_ISL_2040056, EPI_ISL_2040057, EPI_ISL_2040058, EPI_ISL_2040059, EPI_ISL_2040060, EPI_ISL_2040061, EPI_ISL_2040062, EPI_ISL_2040063, EPI_ISL_2040064, EPI_ISL_2040065, EPI_ISL_2040066, EPI_ISL_2040067, EPI_ISL_2040068, EPI_ISL_2040069, EPI_ISL_2040070, EPI_ISL_2040071, EPI_ISL_2040072, EPI_ISL_2040073, EPI_ISL_2040074, EPI_ISL_2040075, EPI_ISL_2040076, EPI_ISL_2040077, EPI_ISL_2040078, EPI_ISL_2040079, EPI_ISL_2040080, EPI_ISL_2040081, EPI_ISL_2040082, EPI_ISL_2040083, EPI_ISL_2040084, EPI_ISL_2040085, EPI_ISL_2040086, EPI_ISL_2040087, EPI_ISL_2040088, EPI_ISL_2040089, EPI_ISL_2040090, EPI_ISL_2040091, EPI_ISL_2040092, EPI_ISL_2040093, EPI_ISL_2040094, EPI_ISL_2040095, EPI_ISL_2040096, EPI_ISL_2040097, EPI_ISL_2040098, EPI_ISL_2040099, EPI_ISL_2040100, EPI_ISL_2040101, EPI_ISL_2040102, EPI_ISL_2040103, EPI_ISL_2040104, EPI_ISL_2040105, EPI_ISL_2040106, EPI_ISL_2040107, EPI_ISL_2040108, EPI_ISL_2040109, EPI_ISL_2040110, EPI_ISL_2040111, EPI_ISL_2040112, EPI_ISL_2040113, EPI_ISL_2040114, EPI_ISL_2040115, EPI_ISL_2040116, EPI_ISL_2040117, EPI_ISL_2040118, EPI_ISL_2040119, EPI_ISL_2040120, EPI_ISL_2040121, EPI_ISL_2040122, EPI_ISL_2040123, EPI_ISL_2040124, EPI_ISL_2040125, EPI_ISL_2040126, EPI_ISL_2040127, EPI_ISL_2040128, EPI_ISL_2040129, EPI_ISL_2040130, EPI_ISL_2040131, EPI_ISL_2040132, EPI_ISL_2040133, EPI_ISL_2040134, EPI_ISL_2040135, EPI_ISL_2040136, EPI_ISL_2040137, EPI_ISL_2040138, EPI_ISL_2040139, EPI_ISL_2040140, EPI_ISL_2040141, EPI_ISL_2040142, EPI_ISL_2040143, EPI_ISL_2040144, EPI_ISL_2040145, EPI_ISL_2040146, EPI_ISL_2040147, EPI_ISL_2040148, EPI_ISL_2040149, EPI_ISL_2040150, EPI_ISL_2040151, EPI_ISL_2040152, EPI_ISL_2040153, EPI_ISL_2040154, EPI_ISL_2040155, EPI_ISL_2040156, EPI_ISL_2040157, EPI_ISL_2040158, EPI_ISL_2040159, EPI_ISL_2040160, EPI_ISL_2040161, EPI_ISL_2040162, EPI_ISL_2040163, EPI_ISL_2040164, EPI_ISL_2040165, EPI_ISL_2040166, EPI_ISL_2040167, EPI_ISL_2040168, EPI_ISL_2040169, EPI_ISL_2040170, EPI_ISL_2040171, EPI_ISL_2040172, EPI_ISL_2040173, EPI_ISL_2040174, EPI_ISL_2040175, EPI_ISL_2040176, EPI_ISL_2040177, EPI_ISL_2040178, EPI_ISL_2040179, EPI_ISL_2040180, EPI_ISL_2040181, EPI_ISL_2040182, EPI_ISL_2040183, EPI_ISL_2040184, EPI_ISL_2040185, EPI_ISL_2040186, EPI_ISL_2040187, EPI_ISL_2040188, EPI_ISL_2040189, EPI_ISL_2040190, EPI_ISL_2040191, EPI_ISL_2040192, EPI_ISL_2040193, EPI_ISL_2040194, EPI_ISL_2040195, EPI_ISL_2040196, EPI_ISL_2040197, EPI_ISL_2040198, EPI_ISL_2040199, EPI_ISL_2040200, EPI_ISL_2040201, EPI_ISL_2040203, EPI_ISL_2040204, EPI_ISL_2040205, EPI_ISL_2040206, EPI_ISL_2040207, EPI_ISL_2040208, EPI_ISL_2040209, EPI_ISL_2040210, EPI_ISL_2040211, EPI_ISL_2040212, EPI_ISL_2040213, EPI_ISL_2040214, EPI_ISL_2040215, EPI_ISL_2040216, EPI_ISL_2040217, EPI_ISL_2040218, EPI_ISL_2040219, EPI_ISL_2040220, EPI_ISL_2040221, EPI_ISL_2040222, EPI_ISL_2040223, EPI_ISL_2040224, EPI_ISL_2040225, EPI_ISL_2040228, EPI_ISL_2040231, EPI_ISL_2040232, EPI_ISL_2040233, EPI_ISL_2040234, EPI_ISL_2040235, EPI_ISL_2040236, EPI_ISL_2040237, EPI_ISL_2040238, EPI_ISL_2040239, EPI_ISL_2040240, EPI_ISL_2040241, EPI_ISL_2040243, EPI_ISL_2040244, EPI_ISL_2040245, EPI_ISL_2040246, EPI_ISL_2040247, EPI_ISL_2040248, EPI_ISL_2040249, EPI_ISL_2040251, EPI_ISL_2040252, EPI_ISL_2040253, EPI_ISL_2040254, EPI_ISL_2040255, EPI_ISL_2040256, EPI_ISL_2040257, EPI_ISL_2040258, EPI_ISL_2040259, EPI_ISL_2040260, EPI_ISL_2040262, EPI_ISL_2040263, EPI_ISL_2040264, EPI_ISL_2040265, EPI_ISL_2040266, EPI_ISL_2040267, EPI_ISL_2040268, EPI_ISL_2040269, EPI_ISL_2040270, EPI_ISL_2040271, EPI_ISL_2040272, EPI_ISL_2040274, EPI_ISL_2040275, EPI_ISL_2040276, EPI_ISL_2040277, EPI_ISL_2040278, EPI_ISL_2040279, EPI_ISL_2040280, EPI_ISL_2040281, EPI_ISL_2040282, EPI_ISL_2040283, EPI_ISL_2040284, EPI_ISL_2040285, EPI_ISL_2040286, EPI_ISL_2040287, EPI_ISL_2040288, EPI_ISL_2040289, EPI_ISL_2040290, EPI_ISL_2040291, EPI_ISL_2040292, EPI_ISL_2040293, EPI_ISL_2040294, EPI_ISL_2040295, EPI_ISL_2040296, EPI_ISL_2040297, EPI_ISL_2040298, EPI_ISL_2040299, EPI_ISL_2040300, EPI_ISL_2040301, EPI_ISL_2040302, EPI_ISL_2040303, EPI_ISL_2040304, EPI_ISL_2040305, EPI_ISL_2040306, EPI_ISL_2040307, EPI_ISL_2040309, EPI_ISL_2040310, EPI_ISL_2040311, EPI_ISL_2040312, EPI_ISL_2040313, EPI_ISL_2040314, EPI_ISL_2040315, EPI_ISL_2040316, EPI_ISL_2040317, EPI_ISL_2040318, EPI_ISL_2040319, EPI_ISL_2040320, EPI_ISL_2040321, EPI_ISL_2040322, EPI_ISL_2040323, EPI_ISL_2040324, EPI_ISL_2040325, EPI_ISL_2040326, EPI_ISL_2040327, EPI_ISL_2040328, EPI_ISL_2040329, EPI_ISL_2040330, EPI_ISL_2040331, EPI_ISL_2040332, EPI_ISL_2040333, EPI_ISL_2040334, EPI_ISL_2040335, EPI_ISL_2040336, EPI_ISL_2040337, EPI_ISL_2040338, EPI_ISL_2040339, EPI_ISL_2040340, EPI_ISL_2040341, EPI_ISL_2040342, EPI_ISL_2040343, EPI_ISL_2040344, EPI_ISL_2040345, EPI_ISL_2040346, EPI_ISL_2040347, EPI_ISL_2040348, EPI_ISL_2040349, EPI_ISL_2040350, EPI_ISL_2040351, EPI_ISL_2040352, EPI_ISL_2040353, EPI_ISL_2040354, EPI_ISL_2040355, EPI_ISL_2040356, EPI_ISL_2040357, EPI_ISL_2040358, EPI_ISL_2040359, EPI_ISL_2040360, EPI_ISL_2040361, EPI_ISL_2040362, EPI_ISL_2040363, EPI_ISL_2040364, EPI_ISL_2040365, EPI_ISL_2040366, EPI_ISL_2040367, EPI_ISL_2040368, EPI_ISL_2040369, EPI_ISL_2040370, EPI_ISL_2040372, EPI_ISL_2040373, EPI_ISL_2040374, EPI_ISL_2040375, EPI_ISL_2040376, EPI_ISL_2040377, EPI_ISL_2040378, EPI_ISL_2040379, EPI_ISL_2040380, EPI_ISL_2040381, EPI_ISL_2040382, EPI_ISL_2040383, EPI_ISL_2040384, EPI_ISL_2040385, EPI_ISL_2040386, EPI_ISL_2040387, EPI_ISL_2040388, EPI_ISL_2040389, EPI_ISL_2040390, EPI_ISL_2040391, EPI_ISL_2040392, EPI_ISL_2040393, EPI_ISL_2040394, EPI_ISL_2040395, EPI_ISL_2040396, EPI_ISL_2040397, EPI_ISL_2040398, EPI_ISL_2040399, EPI_ISL_2040400, EPI_ISL_2040402, EPI_ISL_2040403, EPI_ISL_2040404, EPI_ISL_2040405, EPI_ISL_2040406, EPI_ISL_2040407, EPI_ISL_2040408, EPI_ISL_2040409, EPI_ISL_2040410, EPI_ISL_2040411, EPI_ISL_2040412, EPI_ISL_2040413, EPI_ISL_2040414, EPI_ISL_2040415, EPI_ISL_2040416, EPI_ISL_2040417, EPI_ISL_2040418, EPI_ISL_2040419, EPI_ISL_2040420, EPI_ISL_2040422, EPI_ISL_2040423, EPI_ISL_2040424, EPI_ISL_2040425, EPI_ISL_2040426, EPI_ISL_2040427, EPI_ISL_2040429, EPI_ISL_2040430, EPI_ISL_2040431, EPI_ISL_2040432, EPI_ISL_2040433, EPI_ISL_2040434, EPI_ISL_2040435, EPI_ISL_2040436, EPI_ISL_2040437, EPI_ISL_2040438, EPI_ISL_2040439, EPI_ISL_2040440, EPI_ISL_2040441, EPI_ISL_2040442, EPI_ISL_2040443, EPI_ISL_2040444, EPI_ISL_2040445, EPI_ISL_2040446, EPI_ISL_2040447, EPI_ISL_2040448, | | | |

EPI_ISL_2040449, EPI_ISL_2040450, EPI_ISL_2040451, EPI_ISL_2040452, EPI_ISL_2040453, EPI_ISL_2040454, EPI_ISL_2040455, EPI_ISL_2040456, EPI_ISL_2040457, EPI_ISL_2040458, EPI_ISL_2040459, EPI_ISL_2040460, EPI_ISL_2040461, EPI_ISL_2040462, EPI_ISL_2040463, EPI_ISL_2040464, EPI_ISL_2040465, EPI_ISL_2040466, EPI_ISL_2040467, EPI_ISL_2040468, EPI_ISL_2040469, EPI_ISL_2040470, EPI_ISL_2040471, EPI_ISL_2040472, EPI_ISL_2040473, EPI_ISL_2040474, EPI_ISL_2040475, EPI_ISL_2040476, EPI_ISL_2040477, EPI_ISL_2040479, EPI_ISL_2040480, EPI_ISL_2040481, EPI_ISL_2040482, EPI_ISL_2040483, EPI_ISL_2040484, EPI_ISL_2040485, EPI_ISL_2040486, EPI_ISL_2040487, EPI_ISL_2040488, EPI_ISL_2040489, EPI_ISL_2040490, EPI_ISL_2040491, EPI_ISL_2040493, EPI_ISL_2040494, EPI_ISL_2040495, EPI_ISL_2040496, EPI_ISL_2040497, EPI_ISL_2040498, EPI_ISL_2040499, EPI_ISL_2040500, EPI_ISL_2040501, EPI_ISL_2040502, EPI_ISL_2040503, EPI_ISL_2040504, EPI_ISL_2040505, EPI_ISL_2040506, EPI_ISL_2040507, EPI_ISL_2040508, EPI_ISL_2040509, EPI_ISL_2040510, EPI_ISL_2040511, EPI_ISL_2040512, EPI_ISL_2040513, EPI_ISL_2040514, EPI_ISL_2040515, EPI_ISL_2040516, EPI_ISL_2040517, EPI_ISL_2040518, EPI_ISL_2040519, EPI_ISL_2040520, EPI_ISL_2040521, EPI_ISL_2040522, EPI_ISL_2040523, EPI_ISL_2040524, EPI_ISL_2040525, EPI_ISL_2040526, EPI_ISL_2040527, EPI_ISL_2040528, EPI_ISL_2040529, EPI_ISL_2040530, EPI_ISL_2040531, EPI_ISL_2040532, EPI_ISL_2040533, EPI_ISL_2040534, EPI_ISL_2040535, EPI_ISL_2040536, EPI_ISL_2040537, EPI_ISL_2040538, EPI_ISL_2040539, EPI_ISL_2040540, EPI_ISL_2040541, EPI_ISL_2040542, EPI_ISL_2040543, EPI_ISL_2040544, EPI_ISL_2040545, EPI_ISL_2040546, EPI_ISL_2040547, EPI_ISL_2040548, EPI_ISL_2040549, EPI_ISL_2040550, EPI_ISL_2040551, EPI_ISL_2040552, EPI_ISL_2040553, EPI_ISL_2040554, EPI_ISL_2040555, EPI_ISL_2040556, EPI_ISL_2040557, EPI_ISL_2040558, EPI_ISL_2040559, EPI_ISL_2040560, EPI_ISL_2040561, EPI_ISL_2040562, EPI_ISL_2040563, EPI_ISL_2040564, EPI_ISL_2040565, EPI_ISL_2040566, EPI_ISL_2040567, EPI_ISL_2040568, EPI_ISL_2040569, EPI_ISL_2040570, EPI_ISL_2040571, EPI_ISL_2040572, EPI_ISL_2040573, EPI_ISL_2040574, EPI_ISL_2040577, EPI_ISL_2040581, EPI_ISL_2040585, EPI_ISL_2040588, EPI_ISL_2040590, EPI_ISL_2040591, EPI_ISL_2040592, EPI_ISL_2040593, EPI_ISL_2040594, EPI_ISL_2040595, EPI_ISL_2040596, EPI_ISL_2040597, EPI_ISL_2040598, EPI_ISL_2040599, EPI_ISL_2040600, EPI_ISL_2040601, EPI_ISL_2040602, EPI_ISL_2040603, EPI_ISL_2040604, EPI_ISL_2040605, EPI_ISL_2040606, EPI_ISL_2040607, EPI_ISL_2040608, EPI_ISL_2040609, EPI_ISL_2040610, EPI_ISL_2040611, EPI_ISL_2040612, EPI_ISL_2040613, EPI_ISL_2040614, EPI_ISL_2040615, EPI_ISL_2040616, EPI_ISL_2040617, EPI_ISL_2040618, EPI_ISL_2040619, EPI_ISL_2040620, EPI_ISL_2040621, EPI_ISL_2040622, EPI_ISL_2040623, EPI_ISL_2040624, EPI_ISL_2040625, EPI_ISL_2040626, EPI_ISL_2040627, EPI_ISL_2040628, EPI_ISL_2040629, EPI_ISL_2040630, EPI_ISL_2040631, EPI_ISL_2040632, EPI_ISL_2040633, EPI_ISL_2040634, EPI_ISL_2040635, EPI_ISL_2040636, EPI_ISL_2040637, EPI_ISL_2040639, EPI_ISL_2040640, EPI_ISL_2040641, EPI_ISL_2040642, EPI_ISL_2040643, EPI_ISL_2040644, EPI_ISL_2040645, EPI_ISL_2040646, EPI_ISL_2040647, EPI_ISL_2040648, EPI_ISL_2040649, EPI_ISL_2040650, EPI_ISL_2040651, EPI_ISL_2040652, EPI_ISL_2040653, EPI_ISL_2040654, EPI_ISL_2040655, EPI_ISL_2040656, EPI_ISL_2040657, EPI_ISL_2040658, EPI_ISL_2040659, EPI_ISL_2040660, EPI_ISL_2040661, EPI_ISL_2040662, EPI_ISL_2040663, EPI_ISL_2040664, EPI_ISL_2040665, EPI_ISL_2040666, EPI_ISL_2040667, EPI_ISL_2040668, EPI_ISL_2040669, EPI_ISL_2040670, EPI_ISL_2040671, EPI_ISL_2040672, EPI_ISL_2040673, EPI_ISL_2040674, EPI_ISL_2040675, EPI_ISL_2040676, EPI_ISL_2040677, EPI_ISL_2040678, EPI_ISL_2040679, EPI_ISL_2040680, EPI_ISL_2040681, EPI_ISL_2040682, EPI_ISL_2040683, EPI_ISL_2040684, EPI_ISL_2040685, EPI_ISL_2040686, EPI_ISL_2040687, EPI_ISL_2040688, EPI_ISL_2040689, EPI_ISL_2040690, EPI_ISL_2040691, EPI_ISL_2040693, EPI_ISL_2040694, EPI_ISL_2040695, EPI_ISL_2040696, EPI_ISL_2040697, EPI_ISL_2040698, EPI_ISL_2040699, EPI_ISL_2040700, EPI_ISL_2040701, EPI_ISL_2040702, EPI_ISL_2040703, EPI_ISL_2040704, EPI_ISL_2040705, EPI_ISL_2040706, EPI_ISL_2040707, EPI_ISL_2040708, EPI_ISL_2040709, EPI_ISL_2040711, EPI_ISL_2040712, EPI_ISL_2040713, EPI_ISL_2040714, EPI_ISL_2040715, EPI_ISL_2040716, EPI_ISL_2040717, EPI_ISL_2040719, EPI_ISL_2040720, EPI_ISL_2040721, EPI_ISL_2040722, EPI_ISL_2040723, EPI_ISL_2040724, EPI_ISL_2040725, EPI_ISL_2040726, EPI_ISL_2040727, EPI_ISL_2040728, EPI_ISL_2040730, EPI_ISL_2040731, EPI_ISL_2040732, EPI_ISL_2040733, EPI_ISL_2040734, EPI_ISL_2040735, EPI_ISL_2040736, EPI_ISL_2040737, EPI_ISL_2040738, EPI_ISL_2040739, EPI_ISL_2040740, EPI_ISL_2040741, EPI_ISL_2040742, EPI_ISL_2040743, EPI_ISL_2040744, EPI_ISL_2040745, EPI_ISL_2040746, EPI_ISL_2040747, EPI_ISL_2040748, EPI_ISL_2040749, EPI_ISL_2040750, EPI_ISL_2040751, EPI_ISL_2040752, EPI_ISL_2040753, EPI_ISL_2040754, EPI_ISL_2040755, EPI_ISL_2040756, EPI_ISL_2040757, EPI_ISL_2040758, EPI_ISL_2040759, EPI_ISL_2040760, EPI_ISL_2040761, EPI_ISL_2040762, EPI_ISL_2040763, EPI_ISL_2040764, EPI_ISL_2040765, EPI_ISL_2040766, EPI_ISL_2040767, EPI_ISL_2040768, EPI_ISL_2040769, EPI_ISL_2040770, EPI_ISL_2040771, EPI_ISL_2040772, EPI_ISL_2040773, EPI_ISL_2040774, EPI_ISL_2040775, EPI_ISL_2040776, EPI_ISL_2040777, EPI_ISL_2040778, EPI_ISL_2040779, EPI_ISL_2040780, EPI_ISL_2040781, EPI_ISL_2040782, EPI_ISL_2040783, EPI_ISL_2040784, EPI_ISL_2040785, EPI_ISL_2040786, EPI_ISL_2040787, EPI_ISL_2040788, EPI_ISL_2040789, EPI_ISL_2040790, EPI_ISL_2040791, EPI_ISL_2040792, EPI_ISL_2040793, EPI_ISL_2040794, EPI_ISL_2040795, EPI_ISL_2040796, EPI_ISL_2040797, EPI_ISL_2040798, EPI_ISL_2040799, EPI_ISL_2040800, EPI_ISL_2040801, EPI_ISL_2040802, EPI_ISL_2040803, EPI_ISL_2040804, EPI_ISL_2040805, EPI_ISL_2040806, EPI_ISL_2040807, EPI_ISL_2040808, EPI_ISL_2040809, EPI_ISL_2040811, EPI_ISL_2040812, EPI_ISL_2040813, EPI_ISL_2040814, EPI_ISL_2040815, EPI_ISL_2040816, EPI_ISL_2040817, EPI_ISL_2040818, EPI_ISL_2040819, EPI_ISL_2040820, EPI_ISL_2040822, EPI_ISL_2040823, EPI_ISL_2040824, EPI_ISL_2040825, EPI_ISL_2040826, EPI_ISL_2040827, EPI_ISL_2040828, EPI_ISL_2040830, EPI_ISL_2040831, EPI_ISL_2040832, EPI_ISL_2040833, EPI_ISL_2040834, EPI_ISL_2040835, EPI_ISL_2040836, EPI_ISL_2040837, EPI_ISL_2040838, EPI_ISL_2040839, EPI_ISL_2040840, EPI_ISL_2040841, EPI_ISL_2040842, EPI_ISL_2040843, EPI_ISL_2040844, EPI_ISL_2040845, EPI_ISL_2040846, EPI_ISL_2040847, EPI_ISL_2040848, EPI_ISL_2040849, EPI_ISL_2040850, EPI_ISL_2040851, EPI_ISL_2040852, EPI_ISL_2040853, EPI_ISL_2040854, EPI_ISL_2040855, EPI_ISL_2040856, EPI_ISL_2040857, EPI_ISL_2040858, EPI_ISL_2040859, EPI_ISL_2040860, EPI_ISL_2040861, EPI_ISL_2040862, EPI_ISL_2040863, EPI_ISL_2040864, EPI_ISL_2040865, EPI_ISL_2040866, EPI_ISL_2040867, EPI_ISL_2040868, EPI_ISL_2040869, EPI_ISL_2040870, EPI_ISL_2040871, EPI_ISL_2040872, EPI_ISL_2040873, EPI_ISL_2040874, EPI_ISL_2040875, EPI_ISL_2040876, EPI_ISL_2040877, EPI_ISL_2040878, EPI_ISL_2040879, EPI_ISL_2040880, EPI_ISL_2040881, EPI_ISL_2040882, EPI_ISL_2040883, EPI_ISL_2040884, EPI_ISL_2040885, EPI_ISL_2040886, EPI_ISL_2040887, EPI_ISL_2040888, EPI_ISL_2040889, EPI_ISL_2040890, EPI_ISL_2040891, EPI_ISL_2040892, EPI_ISL_2040893, EPI_ISL_2040894, EPI_ISL_2040895, EPI_ISL_2040896, EPI_ISL_2040897, EPI_ISL_2040898, EPI_ISL_2040899, EPI_ISL_2040900, EPI_ISL_2040901, EPI_ISL_2040902, EPI_ISL_2040903, EPI_ISL_2040904, EPI_ISL_2040905, EPI_ISL_2040906, EPI_ISL_2040907, EPI_ISL_2040908, EPI_ISL_2040909, EPI_ISL_2040910, EPI_ISL_2040911, EPI_ISL_2040912, EPI_ISL_2040913, EPI_ISL_2040914, EPI_ISL_2040915, EPI_ISL_2040916, EPI_ISL_2040917, EPI_ISL_2040918, EPI_ISL_2040919, EPI_ISL_2040920, EPI_ISL_2040921, EPI_ISL_2040922, EPI_ISL_2040923, EPI_ISL_2040924, EPI_ISL_2040925, EPI_ISL_2040926, EPI_ISL_2040927, EPI_ISL_2040928, EPI_ISL_2040929, EPI_ISL_2040930, EPI_ISL_2040931, EPI_ISL_2040932, EPI_ISL_2040933, EPI_ISL_2040934, EPI_ISL_2040935, EPI_ISL_2040936, EPI_ISL_2040937, EPI_ISL_2040938, EPI_ISL_2040939, EPI_ISL_2040940, EPI_ISL_2040941, EPI_ISL_2040942, EPI_ISL_2040943, EPI_ISL_2040944, EPI_ISL_2040945, EPI_ISL_2040946, EPI_ISL_2040947, EPI_ISL_2040948, EPI_ISL_2040949, EPI_ISL_2040950, EPI_ISL_2040951, EPI_ISL_2040952, EPI_ISL_2040953, EPI_ISL_2040954, EPI_ISL_2040955, EPI_ISL_2040956, EPI_ISL_2040957, EPI_ISL_2040958, EPI_ISL_2040959, EPI_ISL_2040960, EPI_ISL_2040961, EPI_ISL_2040962, EPI_ISL_2040963, EPI_ISL_2040964, EPI_ISL_2040965, EPI_ISL_2040966, EPI_ISL_2040967, EPI_ISL_2040968, EPI_ISL_2040969, EPI_ISL_2040970, EPI_ISL_2040971, EPI_ISL_2040972, EPI_ISL_2040973, EPI_ISL_2040974, EPI_ISL_2040975, EPI_ISL_2040976, EPI_ISL_2040977, EPI_ISL_2040978, EPI_ISL_2040979, EPI_ISL_2040980, EPI_ISL_2040981, EPI_ISL_2040982, EPI_ISL_2040983, EPI_ISL_2040984, EPI_ISL_2040985, EPI_ISL_2040986, EPI_ISL_2040987, EPI_ISL_2040988, EPI_ISL_2040989, EPI_ISL_2040990, EPI_ISL_2040991, EPI_ISL_2040992, EPI_ISL_2040993, EPI_ISL_2040994, EPI_ISL_2040995, EPI_ISL_2040996, EPI_ISL_2040997, EPI_ISL_2040998, EPI_ISL_2040999, EPI_ISL_2041000, EPI_ISL_2041001, EPI_ISL_2041002, EPI_ISL_2041003, EPI_ISL_2041004, EPI_ISL_2041005, EPI_ISL_2041006, EPI_ISL_2041007, EPI_ISL_2041008, EPI_ISL_2041009, EPI_ISL_2041010, EPI_ISL_2041011, EPI_ISL_2041012, EPI_ISL_2041013, EPI_ISL_2041014, EPI_ISL_2041015, EPI_ISL_2041016, EPI_ISL_2041017, EPI_ISL_2041018, EPI_ISL_2041019, EPI_ISL_2041020, EPI_ISL_2041021, EPI_ISL_2041022, EPI_ISL_2041023, EPI_ISL_2041024, EPI_ISL_2041025, EPI_ISL_2041026, EPI_ISL_2041027, EPI_ISL_2041028, EPI_ISL_2041029, EPI_ISL_2041030, EPI_ISL_2041031, EPI_ISL_2041032, EPI_ISL_2041033, EPI_ISL_2041034, EPI_ISL_2041035, EPI_ISL_2041036, EPI_ISL_2041037, EPI_ISL_2041038, EPI_ISL_2041039, EPI_ISL_2041040, EPI_ISL_2041041, EPI_ISL_2041042, EPI_ISL_2041043, EPI_ISL_2041044, EPI_ISL_2041045, EPI_ISL_2041046, EPI_ISL_2041047, EPI_ISL_2041048, EPI_ISL_2041049, EPI_ISL_2041050, EPI_ISL_2041051, EPI_ISL_2041052, EPI_ISL_2041053, EPI_ISL_2041054, EPI_ISL_2041055, EPI_ISL_2041056, EPI_ISL_2041057, EPI_ISL_2041058, EPI_ISL_2041059, EPI_ISL_2041060, EPI_ISL_2041061, EPI_ISL_2041062, EPI_ISL_2041063, EPI_ISL_2041064, EPI_ISL_2041065, EPI_ISL_2041066, EPI_ISL_2041067, EPI_ISL_2041068, EPI_ISL_2041069, EPI_ISL_2041070, EPI_ISL_2041071, EPI_ISL_2041072, EPI_ISL_2041073, EPI_ISL_2041074, EPI_ISL_2041075, EPI_ISL_2041076, EPI_ISL_2041077, EPI_ISL_2041078, EPI_ISL_2041079, EPI_ISL_2041080, EPI_ISL_2041081, EPI_ISL_2041083, EPI_ISL_2041084, EPI_ISL_2041085, EPI_ISL_2041086, EPI_ISL_2041087, EPI_ISL_2041088, EPI_ISL_2041089, EPI_ISL_2041090, EPI_ISL_2041091, EPI_ISL_2041092, EPI_ISL_2041093, EPI_ISL_2041094, EPI_ISL_2041095, EPI_ISL_2041096, EPI_ISL_2041097, EPI_ISL_2041098, EPI_ISL_2041099, EPI_ISL_2041100, EPI_ISL_2041101, EPI_ISL_2041102, EPI_ISL_2041103, EPI_ISL_2041104, EPI_ISL_2041105, EPI_ISL_2041106, EPI_ISL_2041107, EPI_ISL_2041108, EPI_ISL_2041109, EPI_ISL_2041110, EPI_ISL_2041111, EPI_ISL_2041112, EPI_ISL_2041113, EPI_ISL_2041114, EPI_ISL_2041115, EPI_ISL_2041116, EPI_ISL_2041117, EPI_ISL_2041118, EPI_ISL_2041119, EPI_ISL_2041120, EPI_ISL_2041121, EPI_ISL_2041122, EPI_ISL_2041123, EPI_ISL_2041124, EPI_ISL_2041125, EPI_ISL_2041126, EPI_ISL_2041127, EPI_ISL_2041128, EPI_ISL_2041129, EPI_ISL_2041130, EPI_ISL_2041131, EPI_ISL_2041132, EPI_ISL_2041133, EPI_ISL_2041134, EPI_ISL_2041135, EPI_ISL_2041136, EPI_ISL_2041137, EPI_ISL_2041138, EPI_ISL_2041139, EPI_ISL_2041140, EPI_ISL_2041141, EPI_ISL_2041142, EPI_ISL_2041143, EPI_ISL_2041144, EPI_ISL_2041145, EPI_ISL_2041146, EPI_ISL_2041147, EPI_ISL_2041148, EPI_ISL_2041149, EPI_ISL_2041150, EPI_ISL_2041151, EPI_ISL_2041152, EPI_ISL_2041153, EPI_ISL_2041154, EPI_ISL_2041155, EPI_ISL_2041156, EPI_ISL_2041157, EPI_ISL_2041158, EPI_ISL_2041159, EPI_ISL_2041160, EPI_ISL_2041161, EPI_ISL_2041162, EPI_ISL_2041163, EPI_ISL_2041164, EPI_ISL_2041165, EPI_ISL_2041166, EPI_ISL_2041167, EPI_ISL_2041168, EPI_ISL_2041169, EPI_ISL_2041170, EPI_ISL_2041171, EPI_ISL_2041172, EPI_ISL_2041173, EPI_ISL_2041174, EPI_ISL_2041175, EPI_ISL_2041176, EPI_ISL_2041177, EPI_ISL_2041178, EPI_ISL_2041179, EPI_ISL_2041181, EPI_ISL_2041182, EPI_ISL_2041183, EPI_ISL_2041184, EPI_ISL_2041185, EPI_ISL_2041186, EPI_ISL_2041187, EPI_ISL_2041188, EPI_ISL_2041189, EPI_ISL_2041190, EPI_ISL_2041191, EPI_ISL_2041192, EPI_ISL_2041193, EPI_ISL_2041194, EPI_ISL_2041195, EPI_ISL_2041196, EPI_ISL_2041197, EPI_ISL_2041198, EPI_ISL_2041199, EPI_ISL_2041200, EPI_ISL_2041201, EPI_ISL_2041202, EPI_ISL_2041203, EPI_ISL_2041204, EPI_ISL_2041205, EPI_ISL_2041206, EPI_ISL_2041207, EPI_ISL_2041208, EPI_ISL_2041209, EPI_ISL_2041210, EPI_ISL_2041211, EPI_ISL_2041212, EPI_ISL_2041213, EPI_ISL_2041215, EPI_ISL_2041216, EPI_ISL_2041217, EPI_ISL_2041218, EPI_ISL_2041219, EPI_ISL_2041220, EPI_ISL_2041221, EPI_ISL_2041222, EPI_ISL_2041223, EPI_ISL_2041224, EPI_ISL_2041225, EPI_ISL_2041226, EPI_ISL_2041227, EPI_ISL_2041228, EPI_ISL_2041229, EPI_ISL_2041230, EPI_ISL_2041231, EPI_ISL_2041232, EPI_ISL_2041233, EPI_ISL_2041234, EPI_ISL_2041235, EPI_ISL_2041236, EPI_ISL_2041237, EPI_ISL_2041238, EPI_ISL_2041239, EPI_ISL_2041240, EPI_ISL_2041241, EPI_ISL_2041242, EPI_ISL_2041243, EPI_ISL_2041244, EPI_ISL_2041245, EPI_ISL_2041246, EPI_ISL_2041247, EPI_ISL_2041250, EPI_ISL_2041251, EPI_ISL_2041252, EPI_ISL_2041253, EPI_ISL_2041254, EPI_ISL_2041255, EPI_ISL_2041256, EPI_ISL_2041257, EPI_ISL_2041258, EPI_ISL_2041259, EPI_ISL_2041260, EPI_ISL_2041261, EPI_ISL_2041262, EPI_ISL_2041263, EPI_ISL_2041264, EPI_ISL_2041265, EPI_ISL_2041266, EPI_ISL_2041267, EPI_ISL_2041268, EPI_ISL_2041269, EPI_ISL_2041270, EPI_ISL_2041271, EPI_ISL_2041272, EPI_ISL_2041273, EPI_ISL_2041274, EPI_ISL_2041275, EPI_ISL_2041276, EPI_ISL_2041277, EPI_ISL_2041278, EPI_ISL_2041279, EPI_ISL_2041280, EPI_ISL_2041281, EPI_ISL_2041282, EPI_ISL_2041283, EPI_ISL_2041284, EPI_ISL_2041285, EPI_ISL_2041286, EPI_ISL_2041287, EPI_ISL_2041288, EPI_ISL_2041289, EPI_ISL_2041290, EPI_ISL_2041291, EPI_ISL_2041292, EPI_ISL_2041293, EPI_ISL_2041294, EPI_ISL_2041295, EPI_ISL_2041296, EPI_ISL_2041297, EPI_ISL_2041298, EPI_ISL_2041299, EPI_ISL_2041300, EPI_ISL_2041301, EPI_ISL_2041302, EPI_ISL_2041303, EPI_ISL_2041304, EPI_ISL_2041305, EPI_ISL_2041306, EPI_ISL_2041307, EPI_ISL_2041308, EPI_ISL_2041309, EPI_ISL_2041310, EPI_ISL_2041311, EPI_ISL_2041312, EPI_ISL_2041313, EPI_ISL_2041314, EPI_ISL_2041315, EPI_ISL_2041316, EPI_ISL_2041317, EPI_ISL_2041318, EPI_ISL_2041319, EPI_ISL_2041320, EPI_ISL_2041321, EPI_ISL_2041322, EPI_ISL_2041323, EPI_ISL_2041324, EPI_ISL_2041325, EPI_ISL_2041326, EPI_ISL_2041327, EPI_ISL_2041328, EPI_ISL_2041329, EPI_ISL_2041330, EPI_ISL_2041331, EPI_ISL_2041332, EPI_ISL_2041333, EPI_ISL_2041334, EPI_ISL_2041335, EPI_ISL_2041336, EPI_ISL_2041337, EPI_ISL_2041338, EPI_ISL_2041339, EPI_ISL_2041340, EPI_ISL_2041341, EPI_ISL_2041342, EPI_ISL_2041344, EPI_ISL_2041345, EPI_ISL_2041346, EPI_ISL_2041347, EPI_ISL_2041348, EPI_ISL_2041349, EPI_ISL_2041350, EPI_ISL_2041351, EPI_ISL_2041354, EPI_ISL_2041355, EPI_ISL_2041356, EPI_ISL_2041357, EPI_ISL_2041358, EPI_ISL_2041359, EPI_ISL_2041360, EPI_ISL_2041361, EPI_ISL_2041362, EPI_ISL_2041363, EPI_ISL_2041364, EPI_ISL_2041366, EPI_ISL_2041367, EPI_ISL_2041368, EPI_ISL_2041369, EPI_ISL_2041370, EPI_ISL_2041371, EPI_ISL_2041372, EPI_ISL_2041373, EPI_ISL_2041375, EPI_ISL_2041376, EPI_ISL_2041377, EPI_ISL_2041378, EPI_ISL_2041379, EPI_ISL_2041380, EPI_ISL_2041381, EPI_ISL_2041382, EPI_ISL_2041383, EPI_ISL_2041384, EPI_ISL_2041385, EPI_ISL_2041386, EPI_ISL_2041387, EPI_ISL_2041388, EPI_ISL_2041389, EPI_ISL_2041390, EPI_ISL_2041391, EPI_ISL_2041392, EPI_ISL_2041393, EPI_ISL_2041394, EPI_ISL_2041395, EPI_ISL_2041396, EPI_ISL_2041397, EPI_ISL_2041398, EPI_ISL_2041399, EPI_ISL_2041400, EPI_ISL_2041401, EPI_ISL_2041402, EPI_ISL_2041403, EPI_ISL_2041404, EPI_ISL_2041405, EPI_ISL_2041406, EPI_ISL_2041407, EPI_ISL_2041408, EPI_ISL_2041409, EPI_ISL_2041410, EPI_ISL_2041411, EPI_ISL_2041412, EPI_ISL_2041413, EPI_ISL_2041415, EPI_ISL_2041416, EPI_ISL_2041417, EPI_ISL_2041418, EPI_ISL_2041419, EPI_ISL_2041420, EPI_ISL_2041421, EPI_ISL_2041422, EPI_ISL_2041424, EPI_ISL_2041425, EPI_ISL_2041426, EPI_ISL_2041427, EPI_ISL_2041428, EPI_ISL_2041429, EPI_ISL_2041430, EPI_ISL_2041431, EPI_ISL_2041432, EPI_ISL_2041433, EPI_ISL_2041434, EPI_ISL_2041435, EPI_ISL_2041436, EPI_ISL_2041437, EPI_ISL_2041438, EPI_ISL_2041439, EPI_ISL_2041440, EPI_ISL_2041441, EPI_ISL_2041442, EPI_ISL_2041443, EPI_ISL_2041444, EPI_ISL_2041445, EPI_ISL_2041446, EPI_ISL_2041447, EPI_ISL_2041448, EPI_ISL_2041449, EPI_ISL_2041450, EPI_ISL_2041451, EPI_ISL_2041452, EPI_ISL_2041453, EPI_ISL_2041454, EPI_ISL_2041455, EPI_ISL_2041456, EPI_ISL_2041457, EPI_ISL_2041458, EPI_ISL_2041459, EPI_ISL_2041460, EPI_ISL_2041461, EPI_ISL_2041462, EPI_ISL_2041463, EPI_ISL_2041464, EPI_ISL_2041465, EPI_ISL_2041466, EPI_ISL_2041467, EPI_ISL_2041468, EPI_ISL_2041469, EPI_ISL_2041470, EPI_ISL_2041472, EPI_ISL_2041473, EPI_ISL_2041474, EPI_ISL_2041475, EPI_ISL_2041476, EPI_ISL_2041477, EPI_ISL_2041478, EPI_ISL_2041479, EPI_ISL_2041480, EPI_ISL_2041481, EPI_ISL_2041482, EPI_ISL_2041483, EPI_ISL_2041484, EPI_ISL_2041485, EPI_ISL_2041486, EPI_ISL_2041487, EPI_ISL_2041488, EPI_ISL_2041489, EPI_ISL_2041490, EPI_ISL_2041491, EPI_ISL_2041492, EPI_ISL_2041493, EPI_ISL_2041494, EPI_ISL_2041495, EPI_ISL_2041496, EPI_ISL_2041497, EPI_ISL_2041498, EPI_ISL_2041499, EPI_ISL_2041500, EPI_ISL_2041501, EPI_ISL_2041502, EPI_ISL_2041503, EPI_ISL_2041504, EPI_ISL_2041505, EPI_ISL_2041506, EPI_ISL_2041507, EPI_ISL_2041508, EPI_ISL_2041509, EPI_ISL_2041510, EPI_ISL_2041511, EPI_ISL_2041512, EPI_ISL_2041513, EPI_ISL_2041514, EPI_ISL_2041515, EPI_ISL_2041516, EPI_ISL_2041517, EPI_ISL_2041518, EPI_ISL_2041519, EPI_ISL_2041520, EPI_ISL_2041521, EPI_ISL_2041522, EPI_ISL_2041523, EPI_ISL_2041524, EPI_ISL_2041525, EPI_ISL_2041526, EPI_ISL_2041527, EPI_ISL_2041528, EPI_ISL_2041529, EPI_ISL_2041530, EPI_ISL_2041531, EPI_ISL_2041532, EPI_ISL_2041533, EPI_ISL_2041534, EPI_ISL_2041535, EPI_ISL_2041536, EPI_ISL_2041537, EPI_ISL_2041538, EPI_ISL_2041539, EPI_ISL_2041540, EPI_ISL_2041541, EPI_ISL_2041542, EPI_ISL_2041543, EPI_ISL_2041544, EPI_ISL_2041545, EPI_ISL_2041546, EPI_ISL_2041547, EPI_ISL_2041548, EPI_ISL_2041549, EPI_ISL_2041550, EPI_ISL_2041551, EPI_ISL_2041553, EPI_ISL_2041554, EPI_ISL_2041555, EPI_ISL_2041556, EPI_ISL_2041557, EPI_ISL_2041558, EPI_ISL_2041559, EPI_ISL_2041560, EPI_ISL_2041561, EPI_ISL_2041562, EPI_ISL_2041563, EPI_ISL_2041564, EPI_ISL_2041566, EPI_ISL_2041567, EPI_ISL_2041568, EPI_ISL_2041569, EPI_ISL_2041570, EPI_ISL_2041571, EPI_ISL_2041572, EPI_ISL_2041573, EPI_ISL_2041574, EPI_ISL_2041575, EPI_ISL_2041576, EPI_ISL_2041577, EPI_ISL_2041579, EPI_ISL_2041580, EPI_ISL_2041581, EPI_ISL_2041582, EPI_ISL_2041583, EPI_ISL_2041584, EPI_ISL_2041585, EPI_ISL_2041586, EPI_ISL_2041587, EPI_ISL_2041588, EPI_ISL_2041589, EPI_ISL_2041590, EPI_ISL_2041592, EPI_ISL_2041593, EPI_ISL_2041594, EPI_ISL_2041596, EPI_ISL_2041597, EPI_ISL_2041598, EPI_ISL_2041599, EPI_ISL_2041600, EPI_ISL_2041601, EPI_ISL_2041602, EPI_ISL_2041603, EPI_ISL_2041604, EPI_ISL_2041605, EPI_ISL_2041606, EPI_ISL_2041607, EPI_ISL_2041608, EPI_ISL_2041609, EPI_ISL_2041610, EPI_ISL_2041611, EPI_ISL_2041612, EPI_ISL_2041613, EPI_ISL_2041614, EPI_ISL_2041615, EPI_ISL_2041616, EPI_ISL_2041617, EPI_ISL_2041618, EPI_ISL_2041619, EPI_ISL_2041620, EPI_ISL_2041621, EPI_ISL_2041622, EPI_ISL_2041623, EPI_ISL_2041624, EPI_ISL_2041625, EPI_ISL_2041626, EPI_ISL_2041627, EPI_ISL_2041628, EPI_ISL_2041629, EPI_ISL_2041630, EPI_ISL_2041631, EPI_ISL_2041632, EPI_ISL_2041633, EPI_ISL_2041634, EPI_ISL_2041635, EPI_ISL_2041636, EPI_ISL_2041637, EPI_ISL_2041638, EPI_ISL_2041639, EPI_ISL_2041640, EPI_ISL_2041641, EPI_ISL_2041642, EPI_ISL_2041643, EPI_ISL_2041644, EPI_ISL_2041645, EPI_ISL_2041646, EPI_ISL_2041648, EPI_ISL_2041649, EPI_ISL_2041650, EPI_ISL_2041651, EPI_ISL_2041652, EPI_ISL_2041653, EPI_ISL_2041654, EPI_ISL_2041655, EPI_ISL_2041656, EPI_ISL_2041657, EPI_ISL_2041658, EPI_ISL_2041659, EPI_ISL_2041660, EPI_ISL_2041661, EPI_ISL_2041662, EPI_ISL_2041663, EPI_ISL_2041664, EPI_ISL_2041665, EPI_ISL_2041666, EPI_ISL_2041667, EPI_ISL_2041668, EPI_ISL_2041669, EPI_ISL_2041670, EPI_ISL_2041671, EPI_ISL_2041672, EPI_ISL_2041673, EPI_ISL_2041674, EPI_ISL_2041675, EPI_ISL_2041676, EPI_ISL_2041677, EPI_ISL_2041678, EPI_ISL_2041679, EPI_ISL_2041680, EPI_ISL_2041681, EPI_ISL_2041682, EPI_ISL_2041683, EPI_ISL_2041684, EPI_ISL_2041685, EPI_ISL_2041686, EPI_ISL_2041687, EPI_ISL_2041688, EPI_ISL_2041689, EPI_ISL_2041690, EPI_ISL_2041691, EPI_ISL_2041692, EPI_ISL_2041693, EPI_ISL_2041694, EPI_ISL_2041695, EPI_ISL_2041696, EPI_ISL_2041697, EPI_ISL_2041698, EPI_ISL_2041699, EPI_ISL_2041700, EPI_ISL_2041701, EPI_ISL_2041702, EPI_ISL_2041703, EPI_ISL_2041704, EPI_ISL_2041705, EPI_ISL_2041706, EPI_ISL_2041707, EPI_ISL_2041708, EPI_ISL_2041709, EPI_ISL_2041710, EPI_ISL_2041711, EPI_ISL_2041712, EPI_ISL_2041713, EPI_ISL_2041714, EPI_ISL_2041715, EPI_ISL_2041716, EPI_ISL_2041717, EPI_ISL_2041718, EPI_ISL_2041719, EPI_ISL_2041720, EPI_ISL_2041721, EPI_ISL_2041722, EPI_ISL_2041723, EPI_ISL_2041724, EPI_ISL_2041725, EPI_ISL_2041726, EPI_ISL_2041727, EPI_ISL_2041728, EPI_ISL_2041729, EPI_ISL_2041730, EPI_ISL_2041731, EPI_ISL_2041733, EPI_ISL_2041734, EPI_ISL_2041735, EPI_ISL_2041736, EPI_ISL_2041737, EPI_ISL_2041738, EPI_ISL_2041739, EPI_ISL_2041740, EPI_ISL_2041741, EPI_ISL_2041742, EPI_ISL_2041743, EPI_ISL_2041744, EPI_ISL_2041745, EPI_ISL_2041746, EPI_ISL_2041748, EPI_ISL_2041749, EPI_ISL_2041750, EPI_ISL_2041751, EPI_ISL_2041752, EPI_ISL_2041753, EPI_ISL_2041754, EPI_ISL_2041755, EPI_ISL_2041756, EPI_ISL_2041757, EPI_ISL_2041758, EPI_ISL_2041759, EPI_ISL_2041760, EPI_ISL_2041761, EPI_ISL_2041762, EPI_ISL_2041763, EPI_ISL_2041764, EPI_ISL_2041765, EPI_ISL_2041766, EPI_ISL_2041767, EPI_ISL_2041768, EPI_ISL_2041769, EPI_ISL_2041770, EPI_ISL_2041771, EPI_ISL_2041772, EPI_ISL_2041773, EPI_ISL_2041774, EPI_ISL_2041775, EPI_ISL_2041776, EPI_ISL_2041777, EPI_ISL_2041778, EPI_ISL_2041779, EPI_ISL_2041780, EPI_ISL_2041781, EPI_ISL_2041782, EPI_ISL_2041783, EPI_ISL_2041785, EPI_ISL_2041786, EPI_ISL_2041787, EPI_ISL_2041788, EPI_ISL_2041789, EPI_ISL_2041790, EPI_ISL_2041791, EPI_ISL_2041792, EPI_ISL_2041793, EPI_ISL_2041794, EPI_ISL_2041795, EPI_ISL_2041796, EPI_ISL_2041797, EPI_ISL_2041798, EPI_ISL_2041799, EPI_ISL_2041800, EPI_ISL_2041801, EPI_ISL_2041802, EPI_ISL_2041803, EPI_ISL_2041804, EPI_ISL_2041805, EPI_ISL_2041806, EPI_ISL_2041807, EPI_ISL_2041808, EPI_ISL_2041809, EPI_ISL_2041810, EPI_ISL_2041811, EPI_ISL_2041812, EPI_ISL_2041813, EPI_ISL_2041814, EPI_ISL_2041815, EPI_ISL_2041816, EPI_ISL_2041817, EPI_ISL_2041818, EPI_ISL_2041819, EPI_ISL_2041820, EPI_ISL_2041821, EPI_ISL_2041822, EPI_ISL_2041823, EPI_ISL_2041824, EPI_ISL_2041825, EPI_ISL_2041826, EPI_ISL_2041827, EPI_ISL_2041828, EPI_ISL_2041829, EPI_ISL_2041831, EPI_ISL_2041832, EPI_ISL_2041833, EPI_ISL_2041834, EPI_ISL_2041835, EPI_ISL_2041836, EPI_ISL_2041837, EPI_ISL_2041838, EPI_ISL_2041839, EPI_ISL_2041840, EPI_ISL_2041841, EPI_ISL_2041842, EPI_ISL_2041843, EPI_ISL_2041845, EPI_ISL_2041846, EPI_ISL_2041847, EPI_ISL_2041848, EPI_ISL_2041849, EPI_ISL_2041850, EPI_ISL_2041851, EPI_ISL_2041852, EPI_ISL_2041853, EPI_ISL_2041854, EPI_ISL_2041855, EPI_ISL_2041856, EPI_ISL_2041857, EPI_ISL_2041858, EPI_ISL_2041859, EPI_ISL_2041860, EPI_ISL_2041861, EPI_ISL_2041862, EPI_ISL_2041863, EPI_ISL_2041864, EPI_ISL_2041865, EPI_ISL_2041866, EPI_ISL_2041867, EPI_ISL_2041868, EPI_ISL_2041869, EPI_ISL_2041870, EPI_ISL_2041871, EPI_ISL_2041872, EPI_ISL_2041873, EPI_ISL_2041874, EPI_ISL_2041875, EPI_ISL_2041876, EPI_ISL_2041877, EPI_ISL_2041878, EPI_ISL_2041879, EPI_ISL_2041880, EPI_ISL_2041881, EPI_ISL_2041883, EPI_ISL_2041884, EPI_ISL_2041885, EPI_ISL_2041886, EPI_ISL_2041887, EPI_ISL_2041888, EPI_ISL_2041890, EPI_ISL_2041891, EPI_ISL_2041892, EPI_ISL_2041894, EPI_ISL_2041895, EPI_ISL_2041896, EPI_ISL_2041897, EPI_ISL_2041898, EPI_ISL_2041899, EPI_ISL_2041900, EPI_ISL_2041901, EPI_ISL_2041902, EPI_ISL_2041903, EPI_ISL_2041904, EPI_ISL_2041905, EPI_ISL_2041906, EPI_ISL_2041907, EPI_ISL_2041908, EPI_ISL_2041909, EPI_ISL_2041910, EPI_ISL_2041911, EPI_ISL_2041912, EPI_ISL_2041913, EPI_ISL_2041914, EPI_ISL_2041915, EPI_ISL_2041916, EPI_ISL_2041917, EPI_ISL_2041918, EPI_ISL_2041919, EPI_ISL_2041920, EPI_ISL_2041921, EPI_ISL_2041922, EPI_ISL_2041923, EPI_ISL_2041924, EPI_ISL_2041925, EPI_ISL_2041926, EPI_ISL_2041927, EPI_ISL_2041928, EPI_ISL_2041929, EPI_ISL_2041930, EPI_ISL_2041931, EPI_ISL_2041932, EPI_ISL_2041933, EPI_ISL_2041935, EPI_ISL_2041936, EPI_ISL_2041937, EPI_ISL_2041938, EPI_ISL_2041939, EPI_ISL_2041940, EPI_ISL_2041941, EPI_ISL_2041942, EPI_ISL_2041943, EPI_ISL_2041944, EPI_ISL_2041945, EPI_ISL_2041946, EPI_ISL_2041947, EPI_ISL_2041948, EPI_ISL_2041949, EPI_ISL_2041950, EPI_ISL_2041951, EPI_ISL_2041952, EPI_ISL_2041953, EPI_ISL_2041954, EPI_ISL_2041955, EPI_ISL_2041956, EPI_ISL_2041957, EPI_ISL_2041958, EPI_ISL_2041959, EPI_ISL_2041960, EPI_ISL_2041961, EPI_ISL_2041962, EPI_ISL_2041963, EPI_ISL_2041964, EPI_ISL_2041965, EPI_ISL_2041966, EPI_ISL_2041967, EPI_ISL_2041968, EPI_ISL_2041969, EPI_ISL_2041970, EPI_ISL_2041971, EPI_ISL_2041972, EPI_ISL_2041973, EPI_ISL_2041974, EPI_ISL_2041976, EPI_ISL_2041977, EPI_ISL_2041978, EPI_ISL_2041979, EPI_ISL_2041980, EPI_ISL_2041981, EPI_ISL_2041982, EPI_ISL_2041983, EPI_ISL_2041984, EPI_ISL_2041985, EPI_ISL_2041986, EPI_ISL_2041987, EPI_ISL_2041988, EPI_ISL_2041989, EPI_ISL_2041990, EPI_ISL_2041991, EPI_ISL_2041992, EPI_ISL_2041993, EPI_ISL_2041994, EPI_ISL_2041995, EPI_ISL_2041996, EPI_ISL_2041997, EPI_ISL_2041998, EPI_ISL_2041999, EPI_ISL_2042000, EPI_ISL_2042001, EPI_ISL_2042002, EPI_ISL_2042003, EPI_ISL_2042004, EPI_ISL_2042005, EPI_ISL_2042006, EPI_ISL_2042007, EPI_ISL_2042008, EPI_ISL_2042009, EPI_ISL_2042010, EPI_ISL_2042011, EPI_ISL_2042012, EPI_ISL_2042013, EPI_ISL_2042014, EPI_ISL_2042015, EPI_ISL_2042016, EPI_ISL_2042017, EPI_ISL_2042018, EPI_ISL_2042019, EPI_ISL_2042020, EPI_ISL_2042021, EPI_ISL_2042022, EPI_ISL_2042023, EPI_ISL_2042024, EPI_ISL_2042025, EPI_ISL_2042026, EPI_ISL_2042027, EPI_ISL_2042028, EPI_ISL_2042029, EPI_ISL_2042030, EPI_ISL_2042031, EPI_ISL_2042032, EPI_ISL_2042033, EPI_ISL_2042034, EPI_ISL_2042035, EPI_ISL_2042036, EPI_ISL_2042037, EPI_ISL_2042038, EPI_ISL_2042039, EPI_ISL_2042040, EPI_ISL_2042041, EPI_ISL_2042042, EPI_ISL_2042043, EPI_ISL_2042044, EPI_ISL_2042045, EPI_ISL_2042046, EPI_ISL_2042047, EPI_ISL_2042048, EPI_ISL_2042049, EPI_ISL_2042050, EPI_ISL_2042051, EPI_ISL_2042052, EPI_ISL_2042053, EPI_ISL_2042054, EPI_ISL_2042055, EPI_ISL_2042056, EPI_ISL_2042057, EPI_ISL_2042058, EPI_ISL_2042059, EPI_ISL_2042060, EPI_ISL_2042061, EPI_ISL_2042063, EPI_ISL_2042064, EPI_ISL_2042065, EPI_ISL_2042066, EPI_ISL_2042067, EPI_ISL_2042068, EPI_ISL_2042069, EPI_ISL_2042070, EPI_ISL_2042071, EPI_ISL_2042072, EPI_ISL_2042073, EPI_ISL_2042074, EPI_ISL_2042075, EPI_ISL_2042076, EPI_ISL_2042077, EPI_ISL_2042078, EPI_ISL_2042079, EPI_ISL_2042080, EPI_ISL_2042081, EPI_ISL_2042082, EPI_ISL_2042083, EPI_ISL_2042084, EPI_ISL_2042085, EPI_ISL_2042086, EPI_ISL_2042088, EPI_ISL_2042089, EPI_ISL_2042091, EPI_ISL_2042092, EPI_ISL_2042093, EPI_ISL_2042094, EPI_ISL_2042095, EPI_ISL_2042096, EPI_ISL_2042097, EPI_ISL_2042098, EPI_ISL_2042099, EPI_ISL_2042100, EPI_ISL_2042101, EPI_ISL_2042102, EPI_ISL_2042103, EPI_ISL_2042104, EPI_ISL_2042105, EPI_ISL_2042106, EPI_ISL_2042107, EPI_ISL_2042108, EPI_ISL_2042109, EPI_ISL_2042110, EPI_ISL_2042111, EPI_ISL_2042112, EPI_ISL_2042113, EPI_ISL_2042114, EPI_ISL_2042115, EPI_ISL_2042116, EPI_ISL_2042117, EPI_ISL_2042118, EPI_ISL_2042119, EPI_ISL_2042121, EPI_ISL_2042122, EPI_ISL_2042123, EPI_ISL_2042125, EPI_ISL_2042126, EPI_ISL_2042127, EPI_ISL_2042128, EPI_ISL_2042129, EPI_ISL_2042130, EPI_ISL_2042131, EPI_ISL_2042132, EPI_ISL_2042133, EPI_ISL_2042134, EPI_ISL_2042135, EPI_ISL_2042136, EPI_ISL_2042137, EPI_ISL_2042138, EPI_ISL_2042139, EPI_ISL_2042140, EPI_ISL_2042141, EPI_ISL_2042142, EPI_ISL_2042143, EPI_ISL_2042144, EPI_ISL_2042145, EPI_ISL_2042146, EPI_ISL_2042147, EPI_ISL_2042148, EPI_ISL_2042149, EPI_ISL_2042150, EPI_ISL_2042151, EPI_ISL_2042152, EPI_ISL_2042153, EPI_ISL_2042154, EPI_ISL_2042155, EPI_ISL_2042156, EPI_ISL_2042157, EPI_ISL_2042158, EPI_ISL_2042159, EPI_ISL_2042160, EPI_ISL_2042161, EPI_ISL_2042162, EPI_ISL_2042163, EPI_ISL_2042164, EPI_ISL_2042165, EPI_ISL_2042166, EPI_ISL_2042167, EPI_ISL_2042168, EPI_ISL_2042169, EPI_ISL_2042170, EPI_ISL_2042171, EPI_ISL_2042172, EPI_ISL_2042173, EPI_ISL_2042174, EPI_ISL_2042175, EPI_ISL_2042176, EPI_ISL_2042177, EPI_ISL_2042178, EPI_ISL_2042179, EPI_ISL_2042180, EPI_ISL_2042181, EPI_ISL_2042182, EPI_ISL_2042183, EPI_ISL_2042184, EPI_ISL_2042185, EPI_ISL_2042186, EPI_ISL_2042187, EPI_ISL_2042188, EPI_ISL_2042189, EPI_ISL_2042190, EPI_ISL_2042191, EPI_ISL_2042192, EPI_ISL_2042193, EPI_ISL_2042194, EPI_ISL_2042195, EPI_ISL_2042196, EPI_ISL_2042197, EPI_ISL_2042198, EPI_ISL_2042199, EPI_ISL_2042200, EPI_ISL_2042201, EPI_ISL_2042202, EPI_ISL_2042203, EPI_ISL_2042204, EPI_ISL_2042205, EPI_ISL_2042206, EPI_ISL_2042207, EPI_ISL_2042208, EPI_ISL_2042209, EPI_ISL_2042210, EPI_ISL_2042211, EPI_ISL_2042212, EPI_ISL_2042213, EPI_ISL_2042214, EPI_ISL_2042215, EPI_ISL_2042216, EPI_ISL_2042217, EPI_ISL_2042218, EPI_ISL_2042219, EPI_ISL_2042220, EPI_ISL_2042221, EPI_ISL_2042222, EPI_ISL_2042223, EPI_ISL_2042225, EPI_ISL_2042226, EPI_ISL_2042227, EPI_ISL_2042228, EPI_ISL_2042229, EPI_ISL_2042230, EPI_ISL_2042231, EPI_ISL_2042233, EPI_ISL_2042234, EPI_ISL_2042235, EPI_ISL_2042236, EPI_ISL_2042237, EPI_ISL_2042238, EPI_ISL_2042239, EPI_ISL_2042240, EPI_ISL_2042241, EPI_ISL_2042242, EPI_ISL_2042243, EPI_ISL_2042244, EPI_ISL_2042245, EPI_ISL_2042246, EPI_ISL_2042247, EPI_ISL_2042248, EPI_ISL_2042249, EPI_ISL_2042250, EPI_ISL_2042251, EPI_ISL_2042252, EPI_ISL_2042253, EPI_ISL_2042254, EPI_ISL_2042255, EPI_ISL_2042256, EPI_ISL_2042257, EPI_ISL_2042258, EPI_ISL_2042259, EPI_ISL_2042260, EPI_ISL_2042261, EPI_ISL_2042262, EPI_ISL_2042263, EPI_ISL_2042264, EPI_ISL_2042265, EPI_ISL_2042266, EPI_ISL_2042267, EPI_ISL_2042268, EPI_ISL_2042269, EPI_ISL_2042270, EPI_ISL_2042271, EPI_ISL_2042272, EPI_ISL_2042274, EPI_ISL_2042275, EPI_ISL_2042276, EPI_ISL_2042277, EPI_ISL_2042278, EPI_ISL_2042279, EPI_ISL_2042280, EPI_ISL_2042281, EPI_ISL_2042282, EPI_ISL_2042283, EPI_ISL_2042284, EPI_ISL_2042285, EPI_ISL_2042286, EPI_ISL_2042287, EPI_ISL_2042288, EPI_ISL_2042289, EPI_ISL_2042290, EPI_ISL_2042291, EPI_ISL_2042292, EPI_ISL_2042293, EPI_ISL_2042294, EPI_ISL_2042295, EPI_ISL_2042296, EPI_ISL_2042297, EPI_ISL_2042298, EPI_ISL_2042299, EPI_ISL_2042300, EPI_ISL_2042301, EPI_ISL_2042303, EPI_ISL_2042304, EPI_ISL_2042305, EPI_ISL_2042306, EPI_ISL_2042307, EPI_ISL_2042308, EPI_ISL_2042309, EPI_ISL_2042310, EPI_ISL_2042311, EPI_ISL_2042312, EPI_ISL_2042313, EPI_ISL_2042314, EPI_ISL_2042315, EPI_ISL_2042317, EPI_ISL_2042318, EPI_ISL_2042319, EPI_ISL_2042320, EPI_ISL_2042321, EPI_ISL_2042322, EPI_ISL_2042323, EPI_ISL_2042324, EPI_ISL_2042325, EPI_ISL_2042326, EPI_ISL_2042327, EPI_ISL_2042328, EPI_ISL_2042329, EPI_ISL_2042330, EPI_ISL_2042331, EPI_ISL_2042332, EPI_ISL_2042333, EPI_ISL_2042334, EPI_ISL_2042335, EPI_ISL_2042336, EPI_ISL_2042337, EPI_ISL_2042338, EPI_ISL_2042339, EPI_ISL_2042340, EPI_ISL_2042341, EPI_ISL_2042342, EPI_ISL_2042343, EPI_ISL_2042344, EPI_ISL_2042345, EPI_ISL_2042346, EPI_ISL_2042347, EPI_ISL_2042348, EPI_ISL_2042349, EPI_ISL_2042350, EPI_ISL_2042351, EPI_ISL_2042353, EPI_ISL_2042354, EPI_ISL_2042355, EPI_ISL_2042356, EPI_ISL_2042357, EPI_ISL_2042358, EPI_ISL_2042359, EPI_ISL_2042360, EPI_ISL_2042361, EPI_ISL_2042362, EPI_ISL_2042363, EPI_ISL_2042364, EPI_ISL_2042365, EPI_ISL_2042366, EPI_ISL_2042367, EPI_ISL_2042368, EPI_ISL_2042369, EPI_ISL_2042371, EPI_ISL_2042372, EPI_ISL_2042373, EPI_ISL_2042374, EPI_ISL_2042375, EPI_ISL_2042376, EPI_ISL_2042377, EPI_ISL_2042378, EPI_ISL_2042379, EPI_ISL_2042380, EPI_ISL_2042381, EPI_ISL_2042382, EPI_ISL_2042383, EPI_ISL_2042384, EPI_ISL_2042385, EPI_ISL_2042387, EPI_ISL_2042388, EPI_ISL_2042389, EPI_ISL_2042390, EPI_ISL_2042391, EPI_ISL_2042392, EPI_ISL_2042393, EPI_ISL_2042394, EPI_ISL_2042395, EPI_ISL_2042396, EPI_ISL_2042397, EPI_ISL_2042399, EPI_ISL_2042400, EPI_ISL_2042401, EPI_ISL_2042402, EPI_ISL_2042403, EPI_ISL_2042404, EPI_ISL_2042405, EPI_ISL_2042406, EPI_ISL_2042407

| | | | |
|---|---|---|---|
| see above | Aegis Sciences Corporation | Centers for Disease Control and Prevention Division of Viral Diseases, Pathogen Discovery | Dakota Howard, Dhwani Batra, Peter W. Cook, Kara Moser, Adrian Paskey, Jason Caravas, Benjamin Rambo-Martin, Shatavia Morrison, Christopher Gulvick, Scott Sammons, Yvette Unoarumhi, Darlene Wagner, Matthew Schmerer, Cyndi Clark, Patrick Campbell, Rob Case, Vikramsinha Ghorpade, Holly Houdeshell, Ola Kvalvaag, Dillon Nall, Ethan Sanders, Alec Vest, Shaun Westlund, Matthew Hardison, Clinton R. Paden, Duncan MacCannell |
| EPI_ISL_2042408, EPI_ISL_2042409, EPI_ISL_2042410, EPI_ISL_2042411 | TN Division of Laboratory Services | Pathogen Discovery, Respiratory Viruses Branch, Division of Viral Diseases, Centers for Disease Control and Prevention | Ying Tao, Yan Li, Jing Zhang, Anna Kelleher, Brian Lynch, Krista Queen, Anna Uehara, Peter Cook, Han Jia Justin Ng, Rachel Marine, Clinton R. Paden, Dhwani Batra, Haibin Wang, Tara Coalter, Jasmine Padilla, Morgan Davis, Mili Sheth, Sarah Nobles, Mark Burroughs, Justin Lee, Adam Retchless, Suxiang Tong |
| EPI_ISL_2042412, EPI_ISL_2042413, EPI_ISL_2042414 | OK Public Health Laboratory, Oklahoma State DOH | Pathogen Discovery, Respiratory Viruses Branch, Division of Viral Diseases, Centers for Disease Control and Prevention | Ying Tao, Yan Li, Jing Zhang, Anna Kelleher, Brian Lynch, Krista Queen, Anna Uehara, Peter Cook, Han Jia Justin Ng, Rachel Marine, Clinton R. Paden, Dhwani Batra, Haibin Wang, Tara Coalter, Jasmine Padilla, Morgan Davis, Mili Sheth, Sarah Nobles, Mark Burroughs, Justin Lee, Adam Retchless, Suxiang Tong |
| EPI_ISL_2042415 | NC State Laboratory of Public Health | Pathogen Discovery, Respiratory Viruses Branch, Division of Viral Diseases, Centers for Disease Control and Prevention | Ying Tao, Yan Li, Jing Zhang, Anna Kelleher, Brian Lynch, Krista Queen, Anna Uehara, Peter Cook, Han Jia Justin Ng, Rachel Marine, Clinton R. Paden, Dhwani Batra, Haibin Wang, Tara Coalter, Jasmine Padilla, Morgan Davis, Mili Sheth, Sarah Nobles, Mark Burroughs, Justin Lee, Adam Retchless, Suxiang Tong |
| EPI_ISL_2042416, EPI_ISL_2042417 | WVDHHR - Office of Laboratory Services | Pathogen Discovery, Respiratory Viruses Branch, Division of Viral Diseases, Centers for Disease Control and Prevention | Ying Tao, Yan Li, Jing Zhang, Anna Kelleher, Brian Lynch, Krista Queen, Anna Uehara, Peter Cook, Han Jia Justin Ng, Rachel Marine, Clinton R. Paden, Dhwani Batra, Haibin Wang, Tara Coalter, Jasmine Padilla, Morgan Davis, Mili Sheth, Sarah Nobles, Mark Burroughs, Justin Lee, Adam Retchless, Suxiang Tong |
| EPI_ISL_2042418 | NH Dept. of Health and Human Services Public Health Labs | Pathogen Discovery, Respiratory Viruses Branch, Division of Viral Diseases, Centers for Disease Control and Prevention | Ying Tao, Yan Li, Jing Zhang, Anna Kelleher, Brian Lynch, Krista Queen, Anna Uehara, Peter Cook, Han Jia Justin Ng, Rachel Marine, Clinton R. Paden, Dhwani Batra, Haibin Wang, Tara Coalter, Jasmine Padilla, Morgan Davis, Mili Sheth, Sarah Nobles, Mark Burroughs, Justin Lee, Adam Retchless, Suxiang Tong |

EPI_ISL_2042419, EPI_ISL_2042420, EPI_ISL_2042421, EPI_ISL_2042422, EPI_ISL_2042423, EPI_ISL_2042424, EPI_ISL_2042425, EPI_ISL_2042426, EPI_ISL_2042427, EPI_ISL_2042428, EPI_ISL_2042429, EPI_ISL_2042430, EPI_ISL_2042431, EPI_ISL_2042432, EPI_ISL_2042434, EPI_ISL_2042435, EPI_ISL_2042436, EPI_ISL_2042437, EPI_ISL_2042438, EPI_ISL_2042439, EPI_ISL_2042440, EPI_ISL_2042441,

EPI_ISL_2042442, EPI_ISL_2042443, EPI_ISL_2042444, EPI_ISL_2042445, EPI_ISL_2042446, EPI_ISL_2042447, EPI_ISL_2042448, EPI_ISL_2042449, EPI_ISL_2042450, EPI_ISL_2042452, EPI_ISL_2042453, EPI_ISL_2042454, EPI_ISL_2042455, EPI_ISL_2042456, EPI_ISL_2042457, EPI_ISL_2042459, EPI_ISL_2042460, EPI_ISL_2042461, EPI_ISL_2042462, EPI_ISL_2042463, EPI_ISL_2042464, EPI_ISL_2042465, EPI_ISL_2042466, EPI_ISL_2042467, EPI_ISL_2042468, EPI_ISL_2042469, EPI_ISL_2042470, EPI_ISL_2042471, EPI_ISL_2042472, EPI_ISL_2042473, EPI_ISL_2042474, EPI_ISL_2042475, EPI_ISL_2042476, EPI_ISL_2042477, EPI_ISL_2042478, EPI_ISL_2042479, EPI_ISL_2042480, EPI_ISL_2042481, EPI_ISL_2042482, EPI_ISL_2042483, EPI_ISL_2042484, EPI_ISL_2042485, EPI_ISL_2042486, EPI_ISL_2042487, EPI_ISL_2042488, EPI_ISL_2042489, EPI_ISL_2042490, EPI_ISL_2042491, EPI_ISL_2042492, EPI_ISL_2042493, EPI_ISL_2042494, EPI_ISL_2042495, EPI_ISL_2042496, EPI_ISL_2042497, EPI_ISL_2042498, EPI_ISL_2042499, EPI_ISL_2042500, EPI_ISL_2042501, EPI_ISL_2042502, EPI_ISL_2042503, EPI_ISL_2042504, EPI_ISL_2042505, EPI_ISL_2042506, EPI_ISL_2042507, EPI_ISL_2042508, EPI_ISL_2042509, EPI_ISL_2042510, EPI_ISL_2042511, EPI_ISL_2042512, EPI_ISL_2042513, EPI_ISL_2042514, EPI_ISL_2042515, EPI_ISL_2042516, EPI_ISL_2042517, EPI_ISL_2042518, EPI_ISL_2042519, EPI_ISL_2042520, EPI_ISL_2042521, EPI_ISL_2042522, EPI_ISL_2042523, EPI_ISL_2042524, EPI_ISL_2042525, EPI_ISL_2042526, EPI_ISL_2042527, EPI_ISL_2042528, EPI_ISL_2042529, EPI_ISL_2042530, EPI_ISL_2042531, EPI_ISL_2042532, EPI_ISL_2042533, EPI_ISL_2042534, EPI_ISL_2042535, EPI_ISL_2042536, EPI_ISL_2042537, EPI_ISL_2042538, EPI_ISL_2042539, EPI_ISL_2042540, EPI_ISL_2042541, EPI_ISL_2042542, EPI_ISL_2042543, EPI_ISL_2042544, EPI_ISL_2042545, EPI_ISL_2042546, EPI_ISL_2042547, EPI_ISL_2042548, EPI_ISL_2042549, EPI_ISL_2042550, EPI_ISL_2042551, EPI_ISL_2042552, EPI_ISL_2042553, EPI_ISL_2042554, EPI_ISL_2042555, EPI_ISL_2042556, EPI_ISL_2042557, EPI_ISL_2042558, EPI_ISL_2042559, EPI_ISL_2042560, EPI_ISL_2042561, EPI_ISL_2042562, EPI_ISL_2042563, EPI_ISL_2042564, EPI_ISL_2042565, EPI_ISL_2042566, EPI_ISL_2042567, EPI_ISL_2042568, EPI_ISL_2042569, EPI_ISL_2042570, EPI_ISL_2042571, EPI_ISL_2042572, EPI_ISL_2042573, EPI_ISL_2042574, EPI_ISL_2042575, EPI_ISL_2042576, EPI_ISL_2042577, EPI_ISL_2042578, EPI_ISL_2042579, EPI_ISL_2042580, EPI_ISL_2042581, EPI_ISL_2042582, EPI_ISL_2042583, EPI_ISL_2042584, EPI_ISL_2042585, EPI_ISL_2042586, EPI_ISL_2042587, EPI_ISL_2042588, EPI_ISL_2042589, EPI_ISL_2042590, EPI_ISL_2042591, EPI_ISL_2042592, EPI_ISL_2042593, EPI_ISL_2042594, EPI_ISL_2042595, EPI_ISL_2042596, EPI_ISL_2042597, EPI_ISL_2042598, EPI_ISL_2042599, EPI_ISL_2042600, EPI_ISL_2042601, EPI_ISL_2042602, EPI_ISL_2042603, EPI_ISL_2042604, EPI_ISL_2042605, EPI_ISL_2042606, EPI_ISL_2042607, EPI_ISL_2042608, EPI_ISL_2042609, EPI_ISL_2042610, EPI_ISL_2042611, EPI_ISL_2042612, EPI_ISL_2042613, EPI_ISL_2042614, EPI_ISL_2042615, EPI_ISL_2042616, EPI_ISL_2042617, EPI_ISL_2042618, EPI_ISL_2042619, EPI_ISL_2042620, EPI_ISL_2042621, EPI_ISL_2042622, EPI_ISL_2042623, EPI_ISL_2042624, EPI_ISL_2042625, EPI_ISL_2042626, EPI_ISL_2042627, EPI_ISL_2042629, EPI_ISL_2042630, EPI_ISL_2042631, EPI_ISL_2042632, EPI_ISL_2042633, EPI_ISL_2042634, EPI_ISL_2042635, EPI_ISL_2042637, EPI_ISL_2042638, EPI_ISL_2042639, EPI_ISL_2042640, EPI_ISL_2042641, EPI_ISL_2042642, EPI_ISL_2042643, EPI_ISL_2042644, EPI_ISL_2042645, EPI_ISL_2042646, EPI_ISL_2042647, EPI_ISL_2042648, EPI_ISL_2042649, EPI_ISL_2042650, EPI_ISL_2042651, EPI_ISL_2042652, EPI_ISL_2042653, EPI_ISL_2042654, EPI_ISL_2042655, EPI_ISL_2042656, EPI_ISL_2042657, EPI_ISL_2042658, EPI_ISL_2042659, EPI_ISL_2042660, EPI_ISL_2042661, EPI_ISL_2042662, EPI_ISL_2042663, EPI_ISL_2042664, EPI_ISL_2042665, EPI_ISL_2042666, EPI_ISL_2042667, EPI_ISL_2042668, EPI_ISL_2042669, EPI_ISL_2042670, EPI_ISL_2042671, EPI_ISL_2042672, EPI_ISL_2042673, EPI_ISL_2042674, EPI_ISL_2042675, EPI_ISL_2042676, EPI_ISL_2042677, EPI_ISL_2042678, EPI_ISL_2042679, EPI_ISL_2042680, EPI_ISL_2042681, EPI_ISL_2042682, EPI_ISL_2042683, EPI_ISL_2042684, EPI_ISL_2042685, EPI_ISL_2042686, EPI_ISL_2042687, EPI_ISL_2042688, EPI_ISL_2042689, EPI_ISL_2042690, EPI_ISL_2042691, EPI_ISL_2042692, EPI_ISL_2042693, EPI_ISL_2042694, EPI_ISL_2042695, EPI_ISL_2042696, EPI_ISL_2042697, EPI_ISL_2042698, EPI_ISL_2042699, EPI_ISL_2042700, EPI_ISL_2042701, EPI_ISL_2042702, EPI_ISL_2042703, EPI_ISL_2042704, EPI_ISL_2042705, EPI_ISL_2042706, EPI_ISL_2042707, EPI_ISL_2042708, EPI_ISL_2042709, EPI_ISL_2042710, EPI_ISL_2042711, EPI_ISL_2042712, EPI_ISL_2042713, EPI_ISL_2042714, EPI_ISL_2042715, EPI_ISL_2042716, EPI_ISL_2042717, EPI_ISL_2042718, EPI_ISL_2042719, EPI_ISL_2042720, EPI_ISL_2042721, EPI_ISL_2042722, EPI_ISL_2042723, EPI_ISL_2042724, EPI_ISL_2042725, EPI_ISL_2042726, EPI_ISL_2042727, EPI_ISL_2042728, EPI_ISL_2042729, EPI_ISL_2042730, EPI_ISL_2042731, EPI_ISL_2042732, EPI_ISL_2042733, EPI_ISL_2042734, EPI_ISL_2042735, EPI_ISL_2042736, EPI_ISL_2042737, EPI_ISL_2042738, EPI_ISL_2042739, EPI_ISL_2042740, EPI_ISL_2042741, EPI_ISL_2042742, EPI_ISL_2042743, EPI_ISL_2042744, EPI_ISL_2042745, EPI_ISL_2042746, EPI_ISL_2042747, EPI_ISL_2042748, EPI_ISL_2042749, EPI_ISL_2042750, EPI_ISL_2042751, EPI_ISL_2042752, EPI_ISL_2042753, EPI_ISL_2042755, EPI_ISL_2042756, EPI_ISL_2042757, EPI_ISL_2042758, EPI_ISL_2042759, EPI_ISL_2042760, EPI_ISL_2042761, EPI_ISL_2042762, EPI_ISL_2042763, EPI_ISL_2042764, EPI_ISL_2042765, EPI_ISL_2042766, EPI_ISL_2042767, EPI_ISL_2042768, EPI_ISL_2042770, EPI_ISL_2042771, EPI_ISL_2042772, EPI_ISL_2042773, EPI_ISL_2042774, EPI_ISL_2042775, EPI_ISL_2042776, EPI_ISL_2042777, EPI_ISL_2042778, EPI_ISL_2042779, EPI_ISL_2042781, EPI_ISL_2042782, EPI_ISL_2042783, EPI_ISL_2042784, EPI_ISL_2042785, EPI_ISL_2042786, EPI_ISL_2042787, EPI_ISL_2042788, EPI_ISL_2042789, EPI_ISL_2042790, EPI_ISL_2042791, EPI_ISL_2042792, EPI_ISL_2042793, EPI_ISL_2042794, EPI_ISL_2042795, EPI_ISL_2042796, EPI_ISL_2042797, EPI_ISL_2042798, EPI_ISL_2042799, EPI_ISL_2042800, EPI_ISL_2042801, EPI_ISL_2042802, EPI_ISL_2042803, EPI_ISL_2042804, EPI_ISL_2042805, EPI_ISL_2042806, EPI_ISL_2042807, EPI_ISL_2042808, EPI_ISL_2042809, EPI_ISL_2042810, EPI_ISL_2042811, EPI_ISL_2042812, EPI_ISL_2042813, EPI_ISL_2042814, EPI_ISL_2042815, EPI_ISL_2042816, EPI_ISL_2042817, EPI_ISL_2042818, EPI_ISL_2042819, EPI_ISL_2042820, EPI_ISL_2042823, EPI_ISL_2042824, EPI_ISL_2042825, EPI_ISL_2042826, EPI_ISL_2042827, EPI_ISL_2042829, EPI_ISL_2042831, EPI_ISL_2042832, EPI_ISL_2042833, EPI_ISL_2042834, EPI_ISL_2042835, EPI_ISL_2042836, EPI_ISL_2042837, EPI_ISL_2042838, EPI_ISL_2042840, EPI_ISL_2042841, EPI_ISL_2042842, EPI_ISL_2042843, EPI_ISL_2042844, EPI_ISL_2042845, EPI_ISL_2042846, EPI_ISL_2042847, EPI_ISL_2042848, EPI_ISL_2042849, EPI_ISL_2042850, EPI_ISL_2042851, EPI_ISL_2042852, EPI_ISL_2042853, EPI_ISL_2042854, EPI_ISL_2042855, EPI_ISL_2042856, EPI_ISL_2042857, EPI_ISL_2042858, EPI_ISL_2042859, EPI_ISL_2042860, EPI_ISL_2042861, EPI_ISL_2042863, EPI_ISL_2042864, EPI_ISL_2042865, EPI_ISL_2042866, EPI_ISL_2042867, EPI_ISL_2042868, EPI_ISL_2042869, EPI_ISL_2042870, EPI_ISL_2042871, EPI_ISL_2042872, EPI_ISL_2042873, EPI_ISL_2042874, EPI_ISL_2042875, EPI_ISL_2042876, EPI_ISL_2042877, EPI_ISL_2042878, EPI_ISL_2042879, EPI_ISL_2042880, EPI_ISL_2042881, EPI_ISL_2042882, EPI_ISL_2042883, EPI_ISL_2042884, EPI_ISL_2042885, EPI_ISL_2042886, EPI_ISL_2042887, EPI_ISL_2042888, EPI_ISL_2042889, EPI_ISL_2042890, EPI_ISL_2042891, EPI_ISL_2042892, EPI_ISL_2042893, EPI_ISL_2042894, EPI_ISL_2042895, EPI_ISL_2042896, EPI_ISL_2042897, EPI_ISL_2042898, EPI_ISL_2042899, EPI_ISL_2042900, EPI_ISL_2042901, EPI_ISL_2042902, EPI_ISL_2042903, EPI_ISL_2042904, EPI_ISL_2042905, EPI_ISL_2042906, EPI_ISL_2042907, EPI_ISL_2042908, EPI_ISL_2042909, EPI_ISL_2042910, EPI_ISL_2042911, EPI_ISL_2042912, EPI_ISL_2042913, EPI_ISL_2042914, EPI_ISL_2042915, EPI_ISL_2042916, EPI_ISL_2042917, EPI_ISL_2042918, EPI_ISL_2042919, EPI_ISL_2042920, EPI_ISL_2042921, EPI_ISL_2042922, EPI_ISL_2042923, EPI_ISL_2042924, EPI_ISL_2042925, EPI_ISL_2042926, EPI_ISL_2042927, EPI_ISL_2042928, EPI_ISL_2042929, EPI_ISL_2042930, EPI_ISL_2042931, EPI_ISL_2042932, EPI_ISL_2042933, EPI_ISL_2042934, EPI_ISL_2042935, EPI_ISL_2042936, EPI_ISL_2042937, EPI_ISL_2042938, EPI_ISL_2042939, EPI_ISL_2042940, EPI_ISL_2042941, EPI_ISL_2042942, EPI_ISL_2042943, EPI_ISL_2042944, EPI_ISL_2042945, EPI_ISL_2042946, EPI_ISL_2042948, EPI_ISL_2042949, EPI_ISL_2042950, EPI_ISL_2042951, EPI_ISL_2042952, EPI_ISL_2042953, EPI_ISL_2042954, EPI_ISL_2042955, EPI_ISL_2042956, EPI_ISL_2042957, EPI_ISL_2042958, EPI_ISL_2042959, EPI_ISL_2042960, EPI_ISL_2042961, EPI_ISL_2042962, EPI_ISL_2042963, EPI_ISL_2042964, EPI_ISL_2042965, EPI_ISL_2042966, EPI_ISL_2042967, EPI_ISL_2042968, EPI_ISL_2042969, EPI_ISL_2042970, EPI_ISL_2042971, EPI_ISL_2042972, EPI_ISL_2042973, EPI_ISL_2042974, EPI_ISL_2042975, EPI_ISL_2042976, EPI_ISL_2042977, EPI_ISL_2042978, EPI_ISL_2042979, EPI_ISL_2042980, EPI_ISL_2042981, EPI_ISL_2042982, EPI_ISL_2042983, EPI_ISL_2042984, EPI_ISL_2042985, EPI_ISL_2042986, EPI_ISL_2042987, EPI_ISL_2042988, EPI_ISL_2042989, EPI_ISL_2042990, EPI_ISL_2042991, EPI_ISL_2042992, EPI_ISL_2042993, EPI_ISL_2042994, EPI_ISL_2042995, EPI_ISL_2042996, EPI_ISL_2042997, EPI_ISL_2042998, EPI_ISL_2042999, EPI_ISL_2043000, EPI_ISL_2043001, EPI_ISL_2043002, EPI_ISL_2043003, EPI_ISL_2043004, EPI_ISL_2043005, EPI_ISL_2043006, EPI_ISL_2043007, EPI_ISL_2043008, EPI_ISL_2043009, EPI_ISL_2043010, EPI_ISL_2043011, EPI_ISL_2043012, EPI_ISL_2043013, EPI_ISL_2043014, EPI_ISL_2043015, EPI_ISL_2043016, EPI_ISL_2043017, EPI_ISL_2043018, EPI_ISL_2043019, EPI_ISL_2043020, EPI_ISL_2043021, EPI_ISL_2043022, EPI_ISL_2043023, EPI_ISL_2043024, EPI_ISL_2043025, EPI_ISL_2043026, EPI_ISL_2043027, EPI_ISL_2043028, EPI_ISL_2043029, EPI_ISL_2043030, EPI_ISL_2043031, EPI_ISL_2043032, EPI_ISL_2043034, EPI_ISL_2043035, EPI_ISL_2043036, EPI_ISL_2043037, EPI_ISL_2043038, EPI_ISL_2043039, EPI_ISL_2043040, EPI_ISL_2043041, EPI_ISL_2043042, EPI_ISL_2043043, EPI_ISL_2043044, EPI_ISL_2043045, EPI_ISL_2043046, EPI_ISL_2043047, EPI_ISL_2043048, EPI_ISL_2043049, EPI_ISL_2043051, EPI_ISL_2043052, EPI_ISL_2043053, EPI_ISL_2043054, EPI_ISL_2043055, EPI_ISL_2043056, EPI_ISL_2043057, EPI_ISL_2043058, EPI_ISL_2043059, EPI_ISL_2043060, EPI_ISL_2043061, EPI_ISL_2043062, EPI_ISL_2043063, EPI_ISL_2043064, EPI_ISL_2043065, EPI_ISL_2043066, EPI_ISL_2043067, EPI_ISL_2043068, EPI_ISL_2043069, EPI_ISL_2043070, EPI_ISL_2043071, EPI_ISL_2043072, EPI_ISL_2043073, EPI_ISL_2043074, EPI_ISL_2043075, EPI_ISL_2043076, EPI_ISL_2043077, EPI_ISL_2043078, EPI_ISL_2043079, EPI_ISL_2043080, EPI_ISL_2043081, EPI_ISL_2043082, EPI_ISL_2043083, EPI_ISL_2043084, EPI_ISL_2043085, EPI_ISL_2043086, EPI_ISL_2043087, EPI_ISL_2043089, EPI_ISL_2043091, EPI_ISL_2043092, EPI_ISL_2043093, EPI_ISL_2043094, EPI_ISL_2043095, EPI_ISL_2043096, EPI_ISL_2043097, EPI_ISL_2043098, EPI_ISL_2043099, EPI_ISL_2043100, EPI_ISL_2043101, EPI_ISL_2043102, EPI_ISL_2043103, EPI_ISL_2043104, EPI_ISL_2043105, EPI_ISL_2043106, EPI_ISL_2043107, EPI_ISL_2043108, EPI_ISL_2043109, EPI_ISL_2043110, EPI_ISL_2043111, EPI_ISL_2043112, EPI_ISL_2043113, EPI_ISL_2043114, EPI_ISL_2043115, EPI_ISL_2043116, EPI_ISL_2043117, EPI_ISL_2043118, EPI_ISL_2043119, EPI_ISL_2043120, EPI_ISL_2043121, EPI_ISL_2043122, EPI_ISL_2043123, EPI_ISL_2043124, EPI_ISL_2043125, EPI_ISL_2043126, EPI_ISL_2043127, EPI_ISL_2043128, EPI_ISL_2043129, EPI_ISL_2043130, EPI_ISL_2043131, EPI_ISL_2043132, EPI_ISL_2043133, EPI_ISL_2043134, EPI_ISL_2043135, EPI_ISL_2043136, EPI_ISL_2043137, EPI_ISL_2043138, EPI_ISL_2043139, EPI_ISL_2043140, EPI_ISL_2043141, EPI_ISL_2043142, EPI_ISL_2043143, EPI_ISL_2043144, EPI_ISL_2043145, EPI_ISL_2043146, EPI_ISL_2043147, EPI_ISL_2043148, EPI_ISL_2043149, EPI_ISL_2043150, EPI_ISL_2043151, EPI_ISL_2043152, EPI_ISL_2043153, EPI_ISL_2043154, EPI_ISL_2043156, EPI_ISL_2043157, EPI_ISL_2043158, EPI_ISL_2043159, EPI_ISL_2043160, EPI_ISL_2043161, EPI_ISL_2043162, EPI_ISL_2043163, EPI_ISL_2043164, EPI_ISL_2043165, EPI_ISL_2043166, EPI_ISL_2043167, EPI_ISL_2043168, EPI_ISL_2043169, EPI_ISL_2043170, EPI_ISL_2043171, EPI_ISL_2043172, EPI_ISL_2043173, EPI_ISL_2043174, EPI_ISL_2043175, EPI_ISL_2043176, EPI_ISL_2043177, EPI_ISL_2043178, EPI_ISL_2043179, EPI_ISL_2043180, EPI_ISL_2043181, EPI_ISL_2043182, EPI_ISL_2043183, EPI_ISL_2043184, EPI_ISL_2043185, EPI_ISL_2043186, EPI_ISL_2043187, EPI_ISL_2043188, EPI_ISL_2043189, EPI_ISL_2043190, EPI_ISL_2043191, EPI_ISL_2043192, EPI_ISL_2043193, EPI_ISL_2043194, EPI_ISL_2043195, EPI_ISL_2043196, EPI_ISL_2043197, EPI_ISL_2043198, EPI_ISL_2043199, EPI_ISL_2043200, EPI_ISL_2043201, EPI_ISL_2043202, EPI_ISL_2043204, EPI_ISL_2043205, EPI_ISL_2043206, EPI_ISL_2043207, EPI_ISL_2043208, EPI_ISL_2043209, EPI_ISL_2043210, EPI_ISL_2043211, EPI_ISL_2043213, EPI_ISL_2043214, EPI_ISL_2043215, EPI_ISL_2043216, EPI_ISL_2043217, EPI_ISL_2043218, EPI_ISL_2043219, EPI_ISL_2043220, EPI_ISL_2043221, EPI_ISL_2043222, EPI_ISL_2043223, EPI_ISL_2043224, EPI_ISL_2043225, EPI_ISL_2043226, EPI_ISL_2043227, EPI_ISL_2043228, EPI_ISL_2043229, EPI_ISL_2043230, EPI_ISL_2043231, EPI_ISL_2043232, EPI_ISL_2043233, EPI_ISL_2043234, EPI_ISL_2043235, EPI_ISL_2043236, EPI_ISL_2043237, EPI_ISL_2043238, EPI_ISL_2043239, EPI_ISL_2043240, EPI_ISL_2043241, EPI_ISL_2043242, EPI_ISL_2043243, EPI_ISL_2043244, EPI_ISL_2043245, EPI_ISL_2043246, EPI_ISL_2043247, EPI_ISL_2043248, EPI_ISL_2043249, EPI_ISL_2043250, EPI_ISL_2043251, EPI_ISL_2043252, EPI_ISL_2043253, EPI_ISL_2043254, EPI_ISL_2043255, EPI_ISL_2043256, EPI_ISL_2043257, EPI_ISL_2043258, EPI_ISL_2043259, EPI_ISL_2043260, EPI_ISL_2043261, EPI_ISL_2043262, EPI_ISL_2043263, EPI_ISL_2043264, EPI_ISL_2043265, EPI_ISL_2043266, EPI_ISL_2043267, EPI_ISL_2043268, EPI_ISL_2043269, EPI_ISL_2043270, EPI_ISL_2043271, EPI_ISL_2043272, EPI_ISL_2043273, EPI_ISL_2043274, EPI_ISL_2043275, EPI_ISL_2043276, EPI_ISL_2043277, EPI_ISL_2043278, EPI_ISL_2043279, EPI_ISL_2043280, EPI_ISL_2043281, EPI_ISL_2043282, EPI_ISL_2043283, EPI_ISL_2043284, EPI_ISL_2043285, EPI_ISL_2043286, EPI_ISL_2043287, EPI_ISL_2043288, EPI_ISL_2043289, EPI_ISL_2043291, EPI_ISL_2043292, EPI_ISL_2043293, EPI_ISL_2043294, EPI_ISL_2043295, EPI_ISL_2043296, EPI_ISL_2043297, EPI_ISL_2043298, EPI_ISL_2043299, EPI_ISL_2043300, EPI_ISL_2043301, EPI_ISL_2043302, EPI_ISL_2043303, EPI_ISL_2043304, EPI_ISL_2043305, EPI_ISL_2043306, EPI_ISL_2043307, EPI_ISL_2043308, EPI_ISL_2043309, EPI_ISL_2043310, EPI_ISL_2043311, EPI_ISL_2043312, EPI_ISL_2043313, EPI_ISL_2043314, EPI_ISL_2043315, EPI_ISL_2043316, EPI_ISL_2043317, EPI_ISL_2043318, EPI_ISL_2043319, EPI_ISL_2043320, EPI_ISL_2043321, EPI_ISL_2043322, EPI_ISL_2043323, EPI_ISL_2043324, EPI_ISL_2043325, EPI_ISL_2043326, EPI_ISL_2043327, EPI_ISL_2043328, EPI_ISL_2043329, EPI_ISL_2043330, EPI_ISL_2043331, EPI_ISL_2043332, EPI_ISL_2043333, EPI_ISL_2043334, EPI_ISL_2043335, EPI_ISL_2043336, EPI_ISL_2043337, EPI_ISL_2043338, EPI_ISL_2043339, EPI_ISL_2043340, EPI_ISL_2043341, EPI_ISL_2043342, EPI_ISL_2043343, EPI_ISL_2043344, EPI_ISL_2043345, EPI_ISL_2043346, EPI_ISL_2043347, EPI_ISL_2043348, EPI_ISL_2043349, EPI_ISL_2043350, EPI_ISL_2043351, EPI_ISL_2043352, EPI_ISL_2043353, EPI_ISL_2043354, EPI_ISL_2043355, EPI_ISL_2043356, EPI_ISL_2043357, EPI_ISL_2043358, EPI_ISL_2043359, EPI_ISL_2043360, EPI_ISL_2043361, EPI_ISL_2043362, EPI_ISL_2043363, EPI_ISL_2043364, EPI_ISL_2043365, EPI_ISL_2043366, EPI_ISL_2043367, EPI_ISL_2043368, EPI_ISL_2043369, EPI_ISL_2043370, EPI_ISL_2043371, EPI_ISL_2043372, EPI_ISL_2043373, EPI_ISL_2043374, EPI_ISL_2043375, EPI_ISL_2043376, EPI_ISL_2043377, EPI_ISL_2043378, EPI_ISL_2043379, EPI_ISL_2043380, EPI_ISL_2043381, EPI_ISL_2043382, EPI_ISL_2043383, EPI_ISL_2043384, EPI_ISL_2043385, EPI_ISL_2043386, EPI_ISL_2043387, EPI_ISL_2043388, EPI_ISL_2043389, EPI_ISL_2043390, EPI_ISL_2043391, EPI_ISL_2043392, EPI_ISL_2043393, EPI_ISL_2043394, EPI_ISL_2043395, EPI_ISL_2043396, EPI_ISL_2043397, EPI_ISL_2043398, EPI_ISL_2043399, EPI_ISL_2043400, EPI_ISL_2043401, EPI_ISL_2043402, EPI_ISL_2043403, EPI_ISL_2043404, EPI_ISL_2043405, EPI_ISL_2043406, EPI_ISL_2043407, EPI_ISL_2043408, EPI_ISL_2043409, EPI_ISL_2043410, EPI_ISL_2043411, EPI_ISL_2043412, EPI_ISL_2043413, EPI_ISL_2043414, EPI_ISL_2043415, EPI_ISL_2043416, EPI_ISL_2043417, EPI_ISL_2043418, EPI_ISL_2043419, EPI_ISL_2043420, EPI_ISL_2043421, EPI_ISL_2043422, EPI_ISL_2043423, EPI_ISL_2043424, EPI_ISL_2043425, EPI_ISL_2043426, EPI_ISL_2043427, EPI_ISL_2043428, EPI_ISL_2043429, EPI_ISL_2043430, EPI_ISL_2043431, EPI_ISL_2043432, EPI_ISL_2043433, EPI_ISL_2043434, EPI_ISL_2043435, EPI_ISL_2043436, EPI_ISL_2043437, EPI_ISL_2043438, EPI_ISL_2043439, EPI_ISL_2043440, EPI_ISL_2043441, EPI_ISL_2043442, EPI_ISL_2043443, EPI_ISL_2043444, EPI_ISL_2043445, EPI_ISL_2043446, EPI_ISL_2043447, EPI_ISL_2043448, EPI_ISL_2043449, EPI_ISL_2043450, EPI_ISL_2043451, EPI_ISL_2043452, EPI_ISL_2043453, EPI_ISL_2043454, EPI_ISL_2043455, EPI_ISL_2043456, EPI_ISL_2043457, EPI_ISL_2043458, EPI_ISL_2043459, EPI_ISL_2043460, EPI_ISL_2043461, EPI_ISL_2043462, EPI_ISL_2043463, EPI_ISL_2043464, EPI_ISL_2043465, EPI_ISL_2043466, EPI_ISL_2043467, EPI_ISL_2043468, EPI_ISL_2043469, EPI_ISL_2043470, EPI_ISL_2043471, EPI_ISL_2043473, EPI_ISL_2043474, EPI_ISL_2043475, EPI_ISL_2043476, EPI_ISL_2043477, EPI_ISL_2043478, EPI_ISL_2043479, EPI_ISL_2043480, EPI_ISL_2043481, EPI_ISL_2043483, EPI_ISL_2043484, EPI_ISL_2043485, EPI_ISL_2043486, EPI_ISL_2043487, EPI_ISL_2043488, EPI_ISL_2043489, EPI_ISL_2043490, EPI_ISL_2043491, EPI_ISL_2043492, EPI_ISL_2043493, EPI_ISL_2043494, EPI_ISL_2043496, EPI_ISL_2043497, EPI_ISL_2043498, EPI_ISL_2043499, EPI_ISL_2043500, EPI_ISL_2043501, EPI_ISL_2043502, EPI_ISL_2043503, EPI_ISL_2043504, EPI_ISL_2043505, EPI_ISL_2043506, EPI_ISL_2043507, EPI_ISL_2043508, EPI_ISL_2043509, EPI_ISL_2043510, EPI_ISL_2043511, EPI_ISL_2043512, EPI_ISL_2043513, EPI_ISL_2043514, EPI_ISL_2043515, EPI_ISL_2043516, EPI_ISL_2043517, EPI_ISL_2043518, EPI_ISL_2043519, EPI_ISL_2043521, EPI_ISL_2043522, EPI_ISL_2043523, EPI_ISL_2043524, EPI_ISL_2043525, EPI_ISL_2043526, EPI_ISL_2043527, EPI_ISL_2043529, EPI_ISL_2043530, EPI_ISL_2043531, EPI_ISL_2043532, EPI_ISL_2043533, EPI_ISL_2043534, EPI_ISL_2043535, EPI_ISL_2043536, EPI_ISL_2043537, EPI_ISL_2043538, EPI_ISL_2043539, EPI_ISL_2043540, EPI_ISL_2043541, EPI_ISL_2043542, EPI_ISL_2043543, EPI_ISL_2043544, EPI_ISL_2043545, EPI_ISL_2043546, EPI_ISL_2043547, EPI_ISL_2043548, EPI_ISL_2043549

see above | Aegis Sciences Corporation | Centers for Disease Control and Prevention Division of Viral Diseases, Pathogen Discovery | Dakota Howard, Dhwani Batra, Peter W. Cook, Kara Moser, Adrian Paskey, Jason Caravas, Benjamin Rambo-Martin, Shatavia Morrison, Christopher Gulvick, Scott Sammons, Yvette Unoarumhi, Darlene Wagner, Matthew Schmerer, Cyndi Clark, Patrick Campbell, Rob Case, Vikramsinha Ghorpade, Holly Houdeshell, Ola Kvalvaag, Dillon Nall, Ethan Sanders, Alec Vest, Shaun Westlund, Matthew Hardison, Clinton R. Paden, Duncan MacCannell

EPI_ISL_2043550, EPI_ISL_2043551, EPI_ISL_2043552, EPI_ISL_2043553, EPI_ISL_2043554, EPI_ISL_2043555, EPI_ISL_2043556, EPI_ISL_2043557, EPI_ISL_2043558, EPI_ISL_2043559, EPI_ISL_2043560, EPI_ISL_2043561, EPI_ISL_2043562, EPI_ISL_2043563, EPI_ISL_2043564, EPI_ISL_2043565, EPI_ISL_2043566, EPI_ISL_2043567, EPI_ISL_2043568, EPI_ISL_2043569, EPI_ISL_2043570, EPI_ISL_2043571, EPI_ISL_2043572, EPI_ISL_2043573, EPI_ISL_2043574, EPI_ISL_2043575, EPI_ISL_2043576, EPI_ISL_2043577, EPI_ISL_2043578, EPI_ISL_2043579, EPI_ISL_2043580, EPI_ISL_2043581, EPI_ISL_2043582, EPI_ISL_2043583, EPI_ISL_2043584, EPI_ISL_2043585, EPI_ISL_2043586, EPI_ISL_2043587, EPI_ISL_2043588, EPI_ISL_2043589, EPI_ISL_2043590, EPI_ISL_2043591, EPI_ISL_2043592, EPI_ISL_2043593, EPI_ISL_2043594, EPI_ISL_2043595, EPI_ISL_2043596, EPI_ISL_2043597, EPI_ISL_2043599, EPI_ISL_2043600, EPI_ISL_2043601, EPI_ISL_2043602, EPI_ISL_2043603, EPI_ISL_2043604, EPI_ISL_2043605, EPI_ISL_2043606, EPI_ISL_2043607, EPI_ISL_2043608, EPI_ISL_2043609, EPI_ISL_2043610, EPI_ISL_2043611, EPI_ISL_2043612, EPI_ISL_2043613, EPI_ISL_2043614, EPI_ISL_2043615, EPI_ISL_2043616, EPI_ISL_2043617, EPI_ISL_2043618, EPI_ISL_2043619, EPI_ISL_2043623, EPI_ISL_2043624, EPI_ISL_2043625, EPI_ISL_2043626, EPI_ISL_2043627, EPI_ISL_2043628, EPI_ISL_2043630, EPI_ISL_2043631, EPI_ISL_2043632, EPI_ISL_2043633, EPI_ISL_2043635, EPI_ISL_2043636, EPI_ISL_2043638, EPI_ISL_2043641, EPI_ISL_2043642, EPI_ISL_2043644, EPI_ISL_2043645, EPI_ISL_2043646, EPI_ISL_2043647, EPI_ISL_2043648, EPI_ISL_2043649, EPI_ISL_2043650, EPI_ISL_2043652, EPI_ISL_2043654, EPI_ISL_2043656, EPI_ISL_2043657, EPI_ISL_2043659, EPI_ISL_2043660, EPI_ISL_2043662, EPI_ISL_2043664, EPI_ISL_2043665, EPI_ISL_2043666, EPI_ISL_2043667, EPI_ISL_2043668, EPI_ISL_2043669, EPI_ISL_2043670, EPI_ISL_2043673, EPI_ISL_2043674, EPI_ISL_2043675, EPI_ISL_2043676, EPI_ISL_2043677, EPI_ISL_2043678, EPI_ISL_2043679, EPI_ISL_2043681, EPI_ISL_2043682, EPI_ISL_2043684, EPI_ISL_2043685, EPI_ISL_2043686, EPI_ISL_2043687, EPI_ISL_2043688, EPI_ISL_2043689, EPI_ISL_2043690, EPI_ISL_2043691, EPI_ISL_2043692, EPI_ISL_2043693, EPI_ISL_2043694, EPI_ISL_2043695, EPI_ISL_2043697, EPI_ISL_2043698, EPI_ISL_2043699, EPI_ISL_2043700, EPI_ISL_2043701, EPI_ISL_2043702, EPI_ISL_2043703, EPI_ISL_2043704, EPI_ISL_2043705, EPI_ISL_2043706, EPI_ISL_2043707, EPI_ISL_2043709, EPI_ISL_2043710, EPI_ISL_2043711, EPI_ISL_2043712, EPI_ISL_2043713, EPI_ISL_2043714, EPI_ISL_2043716, EPI_ISL_2043717, EPI_ISL_2043718, EPI_ISL_2043719, EPI_ISL_2043720, EPI_ISL_2043721, EPI_ISL_2043722, EPI_ISL_2043723, EPI_ISL_2043724, EPI_ISL_2043725, EPI_ISL_2043727, EPI_ISL_2043728, EPI_ISL_2043729, EPI_ISL_2043730, EPI_ISL_2043731, EPI_ISL_2043732, EPI_ISL_2043733, EPI_ISL_2043734, EPI_ISL_2043735, EPI_ISL_2043736, EPI_ISL_2043737, EPI_ISL_2043738, EPI_ISL_2043739, EPI_ISL_2043740, EPI_ISL_2043741, EPI_ISL_2043742, EPI_ISL_2043743, EPI_ISL_2043744, EPI_ISL_2043745, EPI_ISL_2043746, EPI_ISL_2043747, EPI_ISL_2043748, EPI_ISL_2043749, EPI_ISL_2043750, EPI_ISL_2043751, EPI_ISL_2043752, EPI_ISL_2043753, EPI_ISL_2043754, EPI_ISL_2043755, EPI_ISL_2043756, EPI_ISL_2043757, EPI_ISL_2043758, EPI_ISL_2043759, EPI_ISL_2043760, EPI_ISL_2043761, EPI_ISL_2043762, EPI_ISL_2043763, EPI_ISL_2043764, EPI_ISL_2043765, EPI_ISL_2043766, EPI_ISL_2043767, EPI_ISL_2043768, EPI_ISL_2043769, EPI_ISL_2043770, EPI_ISL_2043771, EPI_ISL_2043772, EPI_ISL_2043773, EPI_ISL_2043774, EPI_ISL_2043775, EPI_ISL_2043777, EPI_ISL_2043778, EPI_ISL_2043779, EPI_ISL_2043780, EPI_ISL_2043781, EPI_ISL_2043782, EPI_ISL_2043783, EPI_ISL_2043784, EPI_ISL_2043785, EPI_ISL_2043786, EPI_ISL_2043787, EPI_ISL_2043788, EPI_ISL_2043789, EPI_ISL_2043790, EPI_ISL_2043791, EPI_ISL_2043792, EPI_ISL_2043795, EPI_ISL_2043796, EPI_ISL_2043797, EPI_ISL_2043798, EPI_ISL_2043799, EPI_ISL_2043800, EPI_ISL_2043801, EPI_ISL_2043802, EPI_ISL_2043803, EPI_ISL_2043804, EPI_ISL_2043805, EPI_ISL_2043806, EPI_ISL_2043808, EPI_ISL_2043809, EPI_ISL_2043810, EPI_ISL_2043811, EPI_ISL_2043812, EPI_ISL_2043813, EPI_ISL_2043814, EPI_ISL_2043816, EPI_ISL_2043817, EPI_ISL_2043818, EPI_ISL_2043819, EPI_ISL_2043820, EPI_ISL_2043822, EPI_ISL_2043823, EPI_ISL_2043824, EPI_ISL_2043825, EPI_ISL_2043826, EPI_ISL_2043827, EPI_ISL_2043828, EPI_ISL_2043829, EPI_ISL_2043830, EPI_ISL_2043831, EPI_ISL_2043832, EPI_ISL_2043833, EPI_ISL_2043834, EPI_ISL_2043835, EPI_ISL_2043836, EPI_ISL_2043837, EPI_ISL_2043838, EPI_ISL_2043839, EPI_ISL_2043840, EPI_ISL_2043841, EPI_ISL_2043842, EPI_ISL_2043843, EPI_ISL_2043844, EPI_ISL_2043845, EPI_ISL_2043846, EPI_ISL_2043847, EPI_ISL_2043849, EPI_ISL_2043851, EPI_ISL_2043852, EPI_ISL_2043853, EPI_ISL_2043854, EPI_ISL_2043855, EPI_ISL_2043856, EPI_ISL_2043857, EPI_ISL_2043858, EPI_ISL_2043859, EPI_ISL_2043860, EPI_ISL_2043861, EPI_ISL_2043863, EPI_ISL_2043864, EPI_ISL_2043865, EPI_ISL_2043867, EPI_ISL_2043868, EPI_ISL_2043869, EPI_ISL_2043870, EPI_ISL_2043871, EPI_ISL_2043872, EPI_ISL_2043873, EPI_ISL_2043874, EPI_ISL_2043875, EPI_ISL_2043876, EPI_ISL_2043877, EPI_ISL_2043878, EPI_ISL_2043879, EPI_ISL_2043880, EPI_ISL_2043881, EPI_ISL_2043882, EPI_ISL_2043885, EPI_ISL_2043886, EPI_ISL_2043888, EPI_ISL_2043889, EPI_ISL_2043890, EPI_ISL_2043891, EPI_ISL_2043892, EPI_ISL_2043893, EPI_ISL_2043894, EPI_ISL_2043895, EPI_ISL_2043896, EPI_ISL_2043897, EPI_ISL_2043898, EPI_ISL_2043901, EPI_ISL_2043903, EPI_ISL_2043904, EPI_ISL_2043905, EPI_ISL_2043907, EPI_ISL_2043908, EPI_ISL_2043909, EPI_ISL_2043910, EPI_ISL_2043911, EPI_ISL_2043912, EPI_ISL_2043913, EPI_ISL_2043914, EPI_ISL_2043916, EPI_ISL_2043917, EPI_ISL_2043918, EPI_ISL_2043919, EPI_ISL_2043920, EPI_ISL_2043921, EPI_ISL_2043922, EPI_ISL_2043923, EPI_ISL_2043924, EPI_ISL_2043926, EPI_ISL_2043927, EPI_ISL_2043928, EPI_ISL_2043929, EPI_ISL_2043930, EPI_ISL_2043931, EPI_ISL_2043932, EPI_ISL_2043933, EPI_ISL_2043934, EPI_ISL_2043935, EPI_ISL_2043936, EPI_ISL_2043937, EPI_ISL_2043938, EPI_ISL_2043939, EPI_ISL_2043940, EPI_ISL_2043941, EPI_ISL_2043942, EPI_ISL_2043943, EPI_ISL_2043944, EPI_ISL_2043945, EPI_ISL_2043946, EPI_ISL_2043947, EPI_ISL_2043948, EPI_ISL_2043949, EPI_ISL_2043950, EPI_ISL_2043951, EPI_ISL_2043952, EPI_ISL_2043953, EPI_ISL_2043954, EPI_ISL_2043955, EPI_ISL_2043956, EPI_ISL_2043957, EPI_ISL_2043958, EPI_ISL_2043959, EPI_ISL_2043960, EPI_ISL_2043961, EPI_ISL_2043962, EPI_ISL_2043963, EPI_ISL_2043964, EPI_ISL_2043965, EPI_ISL_2043966, EPI_ISL_2043967, EPI_ISL_2043968, EPI_ISL_2043969, EPI_ISL_2043970, EPI_ISL_2043971, EPI_ISL_2043972, EPI_ISL_2043973, EPI_ISL_2043974, EPI_ISL_2043976, EPI_ISL_2043977, EPI_ISL_2043978, EPI_ISL_2043979, EPI_ISL_2043980, EPI_ISL_2043981, EPI_ISL_2043982, EPI_ISL_2043983, EPI_ISL_2043984, EPI_ISL_2043985, EPI_ISL_2043986, EPI_ISL_2043987, EPI_ISL_2043988, EPI_ISL_2043989, EPI_ISL_2043991, EPI_ISL_2043992, EPI_ISL_2043993, EPI_ISL_2043994, EPI_ISL_2043995, EPI_ISL_2043996, EPI_ISL_2043997, EPI_ISL_2043998, EPI_ISL_2043999, EPI_ISL_2044000

see above | Fulgent Genetics | Centers for Disease Control and Prevention Division of Viral Diseases, Pathogen Discovery | Dakota Howard, Dhwani Batra, Peter W. Cook, Kara Moser, Adrian Paskey, Jason Caravas, Benjamin Rambo-Martin, Shatavia Morrison, Christopher Gulvick, Scott Sammons, Yvette Unoarumhi, Darlene Wagner, Matthew Schmerer, Harry Gao, Mickey Li, John Gao, Joseph Fierro, Benafsh Sapra, Becky Tsai, Yan Meng, Doreen Ng, James Xie, Clinton R. Paden, Duncan MacCannell

EPI_ISL_2044006, EPI_ISL_2044007, EPI_ISL_2044008, EPI_ISL_2044009, EPI_ISL_2044010, EPI_ISL_2044011, EPI_ISL_2044013, EPI_ISL_2044014, EPI_ISL_2044015, EPI_ISL_2044016, EPI_ISL_2044018, EPI_ISL_2044019, EPI_ISL_2044020, EPI_ISL_2044022, EPI_ISL_2044024, EPI_ISL_2044025, EPI_ISL_2044026, EPI_ISL_2044030, EPI_ISL_2044032, EPI_ISL_2044033, EPI_ISL_2044034, EPI_ISL_2044035, EPI_ISL_2044036, EPI_ISL_2044037, EPI_ISL_2044038, EPI_ISL_2044039, EPI_ISL_2044040, EPI_ISL_2044042, EPI_ISL_2044043, EPI_ISL_2044044, EPI_ISL_2044045, EPI_ISL_2044046, EPI_ISL_2044048, EPI_ISL_2044051, EPI_ISL_2044052, EPI_ISL_2044053, EPI_ISL_2044055, EPI_ISL_2044056, EPI_ISL_2044057, EPI_ISL_2044058, EPI_ISL_2044059, EPI_ISL_2044060, EPI_ISL_2044061, EPI_ISL_2044062, EPI_ISL_2044064, EPI_ISL_2044065, EPI_ISL_2044067, EPI_ISL_2044069, EPI_ISL_2044070, EPI_ISL_2044074, EPI_ISL_2044077, EPI_ISL_2044078, EPI_ISL_2044079, EPI_ISL_2044080, EPI_ISL_2044081, EPI_ISL_2044082, EPI_ISL_2044088, EPI_ISL_2044089, EPI_ISL_2044090, EPI_ISL_2044092, EPI_ISL_2044093, EPI_ISL_2044094, EPI_ISL_2044095, EPI_ISL_2044096, EPI_ISL_2044098, EPI_ISL_2044099, EPI_ISL_2044100, EPI_ISL_2044101, EPI_ISL_2044102, EPI_ISL_2044103, EPI_ISL_2044105, EPI_ISL_2044106, EPI_ISL_2044107, EPI_ISL_2044108, EPI_ISL_2044110, EPI_ISL_2044111, EPI_ISL_2044112, EPI_ISL_2044114, EPI_ISL_2044115, EPI_ISL_2044117, EPI_ISL_2044118, EPI_ISL_2044119, EPI_ISL_2044120, EPI_ISL_2044121, EPI_ISL_2044122, EPI_ISL_2044126, EPI_ISL_2044128, EPI_ISL_2044129, EPI_ISL_2044130, EPI_ISL_2044131, EPI_ISL_2044132, EPI_ISL_2044133, EPI_ISL_2044136, EPI_ISL_2044137, EPI_ISL_2044138, EPI_ISL_2044140, EPI_ISL_2044141, EPI_ISL_2044143, EPI_ISL_2044144, EPI_ISL_2044145, EPI_ISL_2044149, EPI_ISL_2044150, EPI_ISL_2044151, EPI_ISL_2044152, EPI_ISL_2044153, EPI_ISL_2044155, EPI_ISL_2044156, EPI_ISL_2044157, EPI_ISL_2044158, EPI_ISL_2044159, EPI_ISL_2044160, EPI_ISL_2044161, EPI_ISL_2044163, EPI_ISL_2044164, EPI_ISL_2044165, EPI_ISL_2044166, EPI_ISL_2044168, EPI_ISL_2044169, EPI_ISL_2044170, EPI_ISL_2044171, EPI_ISL_2044173, EPI_ISL_2044174, EPI_ISL_2044175, EPI_ISL_2044176, EPI_ISL_2044177, EPI_ISL_2044179, EPI_ISL_2044181, EPI_ISL_2044182, EPI_ISL_2044184, EPI_ISL_2044185, EPI_ISL_2044188, EPI_ISL_2044192, EPI_ISL_2044193, EPI_ISL_2044194, EPI_ISL_2044195, EPI_ISL_2044196, EPI_ISL_2044198, EPI_ISL_2044199, EPI_ISL_2044200, EPI_ISL_2044202, EPI_ISL_2044203, EPI_ISL_2044204, EPI_ISL_2044205, EPI_ISL_2044207, EPI_ISL_2044208, EPI_ISL_2044209, EPI_ISL_2044211, EPI_ISL_2044212, EPI_ISL_2044213, EPI_ISL_2044214, EPI_ISL_2044216, EPI_ISL_2044219, EPI_ISL_2044220, EPI_ISL_2044223, EPI_ISL_2044224, EPI_ISL_2044226, EPI_ISL_2044227, EPI_ISL_2044228, EPI_ISL_2044229, EPI_ISL_2044230, EPI_ISL_2044232, EPI_ISL_2044233, EPI_ISL_2044238, EPI_ISL_2044239, EPI_ISL_2044241, EPI_ISL_2044242, EPI_ISL_2044243, EPI_ISL_2044244, EPI_ISL_2044245, EPI_ISL_2044247, EPI_ISL_2044249, EPI_ISL_2044250, EPI_ISL_2044253, EPI_ISL_2044254, EPI_ISL_2044256, EPI_ISL_2044260, EPI_ISL_2044261, EPI_ISL_2044262, EPI_ISL_2044263, EPI_ISL_2044265, EPI_ISL_2044266, EPI_ISL_2044267, EPI_ISL_2044269, EPI_ISL_2044270, EPI_ISL_2044271, EPI_ISL_2044272, EPI_ISL_2044273, EPI_ISL_2044274, EPI_ISL_2044275, EPI_ISL_2044276, EPI_ISL_2044277, EPI_ISL_2044278, EPI_ISL_2044279, EPI_ISL_2044281, EPI_ISL_2044282, EPI_ISL_2044283, EPI_ISL_2044284, EPI_ISL_2044285, EPI_ISL_2044286, EPI_ISL_2044287, EPI_ISL_2044288, EPI_ISL_2044289, EPI_ISL_2044290, EPI_ISL_2044291, EPI_ISL_2044292, EPI_ISL_2044293, EPI_ISL_2044294, EPI_ISL_2044295, EPI_ISL_2044297, EPI_ISL_2044299, EPI_ISL_2044301, EPI_ISL_2044302, EPI_ISL_2044303, EPI_ISL_2044304, EPI_ISL_2044305, EPI_ISL_2044306, EPI_ISL_2044307, EPI_ISL_2044308, EPI_ISL_2044310, EPI_ISL_2044311, EPI_ISL_2044312, EPI_ISL_2044313, EPI_ISL_2044314, EPI_ISL_2044315, EPI_ISL_2044316, EPI_ISL_2044317, EPI_ISL_2044318, EPI_ISL_2044319, EPI_ISL_2044320, EPI_ISL_2044321, EPI_ISL_2044322, EPI_ISL_2044323, EPI_ISL_2044325, EPI_ISL_2044326, EPI_ISL_2044328, EPI_ISL_2044329, EPI_ISL_2044330, EPI_ISL_2044331, EPI_ISL_2044332, EPI_ISL_2044333, EPI_ISL_2044334, EPI_ISL_2044335, EPI_ISL_2044338, EPI_ISL_2044339, EPI_ISL_2044341, EPI_ISL_2044342, EPI_ISL_2044343, EPI_ISL_2044344, EPI_ISL_2044345, EPI_ISL_2044346, EPI_ISL_2044347, EPI_ISL_2044350, EPI_ISL_2044351, EPI_ISL_2044352, EPI_ISL_2044353, EPI_ISL_2044354, EPI_ISL_2044355, EPI_ISL_2044357, EPI_ISL_2044359, EPI_ISL_2044360, EPI_ISL_2044361, EPI_ISL_2044362, EPI_ISL_2044363, EPI_ISL_2044365, EPI_ISL_2044366, EPI_ISL_2044367, EPI_ISL_2044368, EPI_ISL_2044371, EPI_ISL_2044372, EPI_ISL_2044373, EPI_ISL_2044374, EPI_ISL_2044376, EPI_ISL_2044377, EPI_ISL_2044378, EPI_ISL_2044379, EPI_ISL_2044380, EPI_ISL_2044381, EPI_ISL_2044382, EPI_ISL_2044383, EPI_ISL_2044384, EPI_ISL_2044386, EPI_ISL_2044391, EPI_ISL_2044393, EPI_ISL_2044395, EPI_ISL_2044396, EPI_ISL_2044398, EPI_ISL_2044399, EPI_ISL_2044400, EPI_ISL_2044401, EPI_ISL_2044402, EPI_ISL_2044403, EPI_ISL_2044404, EPI_ISL_2044405, EPI_ISL_2044406, EPI_ISL_2044407, EPI_ISL_2044408, EPI_ISL_2044413, EPI_ISL_2044414, EPI_ISL_2044416, EPI_ISL_2044417, EPI_ISL_2044418, EPI_ISL_2044419, EPI_ISL_2044420, EPI_ISL_2044421, EPI_ISL_2044423, EPI_ISL_2044425, EPI_ISL_2044428, EPI_ISL_2044429, EPI_ISL_2044430, EPI_ISL_2044431, EPI_ISL_2044432, EPI_ISL_2044433, EPI_ISL_2044434, EPI_ISL_2044435, EPI_ISL_2044436, EPI_ISL_2044437, EPI_ISL_2044438, EPI_ISL_2044439, EPI_ISL_2044440, EPI_ISL_2044442, EPI_ISL_2044443, EPI_ISL_2044444, EPI_ISL_2044445, EPI_ISL_2044446, EPI_ISL_2044448, EPI_ISL_2044449, EPI_ISL_2044450, EPI_ISL_2044451, EPI_ISL_2044452, EPI_ISL_2044453, EPI_ISL_2044454, EPI_ISL_2044455, EPI_ISL_2044456, EPI_ISL_2044458, EPI_ISL_2044459, EPI_ISL_2044460, EPI_ISL_2044461, EPI_ISL_2044462, EPI_ISL_2044463, EPI_ISL_2044464, EPI_ISL_2044465, EPI_ISL_2044469, EPI_ISL_2044470, EPI_ISL_2044471, EPI_ISL_2044472, EPI_ISL_2044473, EPI_ISL_2044474, EPI_ISL_2044475, EPI_ISL_2044476, EPI_ISL_2044477, EPI_ISL_2044478, EPI_ISL_2044479, EPI_ISL_2044482, EPI_ISL_2044483, EPI_ISL_2044484, EPI_ISL_2044485, EPI_ISL_2044486, EPI_ISL_2044487, EPI_ISL_2044489, EPI_ISL_2044491, EPI_ISL_2044492, EPI_ISL_2044494, EPI_ISL_2044495, EPI_ISL_2044496, EPI_ISL_2044497, EPI_ISL_2044499, EPI_ISL_2044500, EPI_ISL_2044501, EPI_ISL_2044502, EPI_ISL_2044503, EPI_ISL_2044504, EPI_ISL_2044505, EPI_ISL_2044506, EPI_ISL_2044508, EPI_ISL_2044509, EPI_ISL_2044510, EPI_ISL_2044511, EPI_ISL_2044512, EPI_ISL_2044513, EPI_ISL_2044515, EPI_ISL_2044516, EPI_ISL_2044517, EPI_ISL_2044518, EPI_ISL_2044519, EPI_ISL_2044521, EPI_ISL_2044522, EPI_ISL_2044523, EPI_ISL_2044524, EPI_ISL_2044526, EPI_ISL_2044527, EPI_ISL_2044528, EPI_ISL_2044529, EPI_ISL_2044530, EPI_ISL_2044531, EPI_ISL_2044532, EPI_ISL_2044533, EPI_ISL_2044534, EPI_ISL_2044535, EPI_ISL_2044536, EPI_ISL_2044537, EPI_ISL_2044538, EPI_ISL_2044540, EPI_ISL_2044541, EPI_ISL_2044542, EPI_ISL_2044543, EPI_ISL_2044545, EPI_ISL_2044546, EPI_ISL_2044547, EPI_ISL_2044549, EPI_ISL_2044550, EPI_ISL_2044552, EPI_ISL_2044553, EPI_ISL_2044554, EPI_ISL_2044555, EPI_ISL_2044558, EPI_ISL_2044559, EPI_ISL_2044560, EPI_ISL_2044561, EPI_ISL_2044562, EPI_ISL_2044563, EPI_ISL_2044564, EPI_ISL_2044565, EPI_ISL_2044566, EPI_ISL_2044568, EPI_ISL_2044570, EPI_ISL_2044573, EPI_ISL_2044574, EPI_ISL_2044575, EPI_ISL_2044576, EPI_ISL_2044578, EPI_ISL_2044580, EPI_ISL_2044583, EPI_ISL_2044585, EPI_ISL_2044586, EPI_ISL_2044588, EPI_ISL_2044589, EPI_ISL_2044590, EPI_ISL_2044591, EPI_ISL_2044592, EPI_ISL_2044593, EPI_ISL_2044594, EPI_ISL_2044595, EPI_ISL_2044596, EPI_ISL_2044599, EPI_ISL_2044600, EPI_ISL_2044601, EPI_ISL_2044602, EPI_ISL_2044604, EPI_ISL_2044605, EPI_ISL_2044606, EPI_ISL_2044607, EPI_ISL_2044609, EPI_ISL_2044610, EPI_ISL_2044611, EPI_ISL_2044612, EPI_ISL_2044613, EPI_ISL_2044615, EPI_ISL_2044616, EPI_ISL_2044617, EPI_ISL_2044618, EPI_ISL_2044619, EPI_ISL_2044620, EPI_ISL_2044621, EPI_ISL_2044622, EPI_ISL_2044623, EPI_ISL_2044624, EPI_ISL_2044626, EPI_ISL_2044627, EPI_ISL_2044629, EPI_ISL_2044631, EPI_ISL_2044632, EPI_ISL_2044635, EPI_ISL_2044636, EPI_ISL_2044637, EPI_ISL_2044638, EPI_ISL_2044639, EPI_ISL_2044640, EPI_ISL_2044641, EPI_ISL_2044642, EPI_ISL_2044643, EPI_ISL_2044645, EPI_ISL_2044646, EPI_ISL_2044647, EPI_ISL_2044648, EPI_ISL_2044650, EPI_ISL_2044652, EPI_ISL_2044653, EPI_ISL_2044654, EPI_ISL_2044655, EPI_ISL_2044656, EPI_ISL_2044657, EPI_ISL_2044659, EPI_ISL_2044660, EPI_ISL_2044661, EPI_ISL_2044662, EPI_ISL_2044663, EPI_ISL_2044664, EPI_ISL_2044666, EPI_ISL_2044667, EPI_ISL_2044668, EPI_ISL_2044669, EPI_ISL_2044671, EPI_ISL_2044673, EPI_ISL_2044675, EPI_ISL_2044676, EPI_ISL_2044678, EPI_ISL_2044679, EPI_ISL_2044680, EPI_ISL_2044681, EPI_ISL_2044682, EPI_ISL_2044683, EPI_ISL_2044684, EPI_ISL_2044685, EPI_ISL_2044686, EPI_ISL_2044687, EPI_ISL_2044689, EPI_ISL_2044690, EPI_ISL_2044691, EPI_ISL_2044692, EPI_ISL_2044693, EPI_ISL_2044695, EPI_ISL_2044696, EPI_ISL_2044697, EPI_ISL_2044698, EPI_ISL_2044699, EPI_ISL_2044700, EPI_ISL_2044701, EPI_ISL_2044702, EPI_ISL_2044703, EPI_ISL_2044705, EPI_ISL_2044706, EPI_ISL_2044707, EPI_ISL_2044709, EPI_ISL_2044710, EPI_ISL_2044712, EPI_ISL_2044713, EPI_ISL_2044714, EPI_ISL_2044716, EPI_ISL_2044717, EPI_ISL_2044718, EPI_ISL_2044720, EPI_ISL_2044721, EPI_ISL_2044724, EPI_ISL_2044725, EPI_ISL_2044726, EPI_ISL_2044727, EPI_ISL_2044728, EPI_ISL_2044730, EPI_ISL_2044731, EPI_ISL_2044733, EPI_ISL_2044736, EPI_ISL_2044737, EPI_ISL_2044738, EPI_ISL_2044741, EPI_ISL_2044743, EPI_ISL_2044744, EPI_ISL_2044745, EPI_ISL_2044746, EPI_ISL_2044747, EPI_ISL_2044748, EPI_ISL_2044750, EPI_ISL_2044751, EPI_ISL_2044752, EPI_ISL_2044754, EPI_ISL_2044755, EPI_ISL_2044756, EPI_ISL_2044758, EPI_ISL_2044759, EPI_ISL_2044760, EPI_ISL_2044761, EPI_ISL_2044763, EPI_ISL_2044764, EPI_ISL_2044765, EPI_ISL_2044766, EPI_ISL_2044767, EPI_ISL_2044768, EPI_ISL_2044769, EPI_ISL_2044770, EPI_ISL_2044771, EPI_ISL_2044774, EPI_ISL_2044775, EPI_ISL_2044776, EPI_ISL_2044777, EPI_ISL_2044778, EPI_ISL_2044779, EPI_ISL_2044780, EPI_ISL_2044781, EPI_ISL_2044782, EPI_ISL_2044783, EPI_ISL_2044784, EPI_ISL_2044786, EPI_ISL_2044787, EPI_ISL_2044788, EPI_ISL_2044789, EPI_ISL_2044790, EPI_ISL_2044791, EPI_ISL_2044792, EPI_ISL_2044793, EPI_ISL_2044794, EPI_ISL_2044795, EPI_ISL_2044796, EPI_ISL_2044797, EPI_ISL_2044798, EPI_ISL_2044800, EPI_ISL_2044801, EPI_ISL_2044802, EPI_ISL_2044803, EPI_ISL_2044804, EPI_ISL_2044805, EPI_ISL_2044806, EPI_ISL_2044807, EPI_ISL_2044809, EPI_ISL_2044810, EPI_ISL_2044811, EPI_ISL_2044812, EPI_ISL_2044813, EPI_ISL_2044814, EPI_ISL_2044815, EPI_ISL_2044816, EPI_ISL_2044817, EPI_ISL_2044818, EPI_ISL_2044819, EPI_ISL_2044820, EPI_ISL_2044821, EPI_ISL_2044822, EPI_ISL_2044823, EPI_ISL_2044824, EPI_ISL_2044825, EPI_ISL_2044826, EPI_ISL_2044827, EPI_ISL_2044828, EPI_ISL_2044831, EPI_ISL_2044832, EPI_ISL_2044833, EPI_ISL_2044834, EPI_ISL_2044835, EPI_ISL_2044837, EPI_ISL_2044838, EPI_ISL_2044839, EPI_ISL_2044840, EPI_ISL_2044841, EPI_ISL_2044842, EPI_ISL_2044843, EPI_ISL_2044845, EPI_ISL_2044847, EPI_ISL_2044848, EPI_ISL_2044849, EPI_ISL_2044850, EPI_ISL_2044852, EPI_ISL_2044853, EPI_ISL_2044854, EPI_ISL_2044855, EPI_ISL_2044856, EPI_ISL_2044859, EPI_ISL_2044860, EPI_ISL_2044864, EPI_ISL_2044865, EPI_ISL_2044866, EPI_ISL_2044868, EPI_ISL_2044869, EPI_ISL_2044870, EPI_ISL_2044872, EPI_ISL_2044873, EPI_ISL_2044875, EPI_ISL_2044877, EPI_ISL_2044878, EPI_ISL_2044881, EPI_ISL_2044884, EPI_ISL_2044885, EPI_ISL_2044886, EPI_ISL_2044887, EPI_ISL_2044891, EPI_ISL_2044892, EPI_ISL_2044893, EPI_ISL_2044896, EPI_ISL_2044897, EPI_ISL_2044898, EPI_ISL_2044899, EPI_ISL_2044901, EPI_ISL_2044902, EPI_ISL_2044903, EPI_ISL_2044904, EPI_ISL_2044905, EPI_ISL_2044906, EPI_ISL_2044908, EPI_ISL_2044909, EPI_ISL_2044911, EPI_ISL_2044913, EPI_ISL_2044915, EPI_ISL_2044916, EPI_ISL_2044917, EPI_ISL_2044918, EPI_ISL_2044919, EPI_ISL_2044921, EPI_ISL_2044923, EPI_ISL_2044924, EPI_ISL_2044925, EPI_ISL_2044928, EPI_ISL_2044931, EPI_ISL_2044933, EPI_ISL_2044934, EPI_ISL_2044938, EPI_ISL_2044939, EPI_ISL_2044943, EPI_ISL_2044945, EPI_ISL_2044946, EPI_ISL_2044947, EPI_ISL_2044948, EPI_ISL_2044949, EPI_ISL_2044952, EPI_ISL_2044953, EPI_ISL_2044954, EPI_ISL_2044955, EPI_ISL_2044956, EPI_ISL_2044957, EPI_ISL_2044958, EPI_ISL_2044959, EPI_ISL_2044960, EPI_ISL_2044961, EPI_ISL_2044963, EPI_ISL_2044964, EPI_ISL_2044965, EPI_ISL_2044966, EPI_ISL_2044967, EPI_ISL_2044969, EPI_ISL_2044970, EPI_ISL_2044971, EPI_ISL_2044972, EPI_ISL_2044976, EPI_ISL_2044977, EPI_ISL_2044978, EPI_ISL_2044979, EPI_ISL_2044981, EPI_ISL_2044982, EPI_ISL_2044983, EPI_ISL_2044984, EPI_ISL_2044986, EPI_ISL_2044988, EPI_ISL_2044990, EPI_ISL_2044991, EPI_ISL_2044992, EPI_ISL_2044996, EPI_ISL_2044997, EPI_ISL_2044998, EPI_ISL_2044999, EPI_ISL_2045000, EPI_ISL_2045001, EPI_ISL_2045003,

EPI_ISL_2045006, EPI_ISL_2045007, EPI_ISL_2045008, EPI_ISL_2045009, EPI_ISL_2045010, EPI_ISL_2045011, EPI_ISL_2045012, EPI_ISL_2045015, EPI_ISL_2045018, EPI_ISL_2045019, EPI_ISL_2045020, EPI_ISL_2045021, EPI_ISL_2045022, EPI_ISL_2045023, EPI_ISL_2045024, EPI_ISL_2045025, EPI_ISL_2045026, EPI_ISL_2045027, EPI_ISL_2045028, EPI_ISL_2045029, EPI_ISL_2045030, EPI_ISL_2045031, EPI_ISL_2045032, EPI_ISL_2045035, EPI_ISL_2045036, EPI_ISL_2045037, EPI_ISL_2045038, EPI_ISL_2045041, EPI_ISL_2045042, EPI_ISL_2045043, EPI_ISL_2045045, EPI_ISL_2045046, EPI_ISL_2045047, EPI_ISL_2045048, EPI_ISL_2045049, EPI_ISL_2045050, EPI_ISL_2045051, EPI_ISL_2045052, EPI_ISL_2045053, EPI_ISL_2045054, EPI_ISL_2045056, EPI_ISL_2045057, EPI_ISL_2045058, EPI_ISL_2045059, EPI_ISL_2045061, EPI_ISL_2045062, EPI_ISL_2045065, EPI_ISL_2045066, EPI_ISL_2045067, EPI_ISL_2045069, EPI_ISL_2045071, EPI_ISL_2045072, EPI_ISL_2045073, EPI_ISL_2045074, EPI_ISL_2045076, EPI_ISL_2045077, EPI_ISL_2045078, EPI_ISL_2045079, EPI_ISL_2045080, EPI_ISL_2045085, EPI_ISL_2045087, EPI_ISL_2045088, EPI_ISL_2045089, EPI_ISL_2045091, EPI_ISL_2045092, EPI_ISL_2045093, EPI_ISL_2045094, EPI_ISL_2045095, EPI_ISL_2045097, EPI_ISL_2045098, EPI_ISL_2045100, EPI_ISL_2045102, EPI_ISL_2045103, EPI_ISL_2045104, EPI_ISL_2045105, EPI_ISL_2045106, EPI_ISL_2045107, EPI_ISL_2045108, EPI_ISL_2045111, EPI_ISL_2045112, EPI_ISL_2045113, EPI_ISL_2045114, EPI_ISL_2045115, EPI_ISL_2045116, EPI_ISL_2045118, EPI_ISL_2045120, EPI_ISL_2045123, EPI_ISL_2045124, EPI_ISL_2045125, EPI_ISL_2045127, EPI_ISL_2045128, EPI_ISL_2045129, EPI_ISL_2045130, EPI_ISL_2045131, EPI_ISL_2045132, EPI_ISL_2045133, EPI_ISL_2045135, EPI_ISL_2045138, EPI_ISL_2045139, EPI_ISL_2045140, EPI_ISL_2045141, EPI_ISL_2045142, EPI_ISL_2045143, EPI_ISL_2045145, EPI_ISL_2045146, EPI_ISL_2045148, EPI_ISL_2045149, EPI_ISL_2045150, EPI_ISL_2045151, EPI_ISL_2045152, EPI_ISL_2045153, EPI_ISL_2045154, EPI_ISL_2045155, EPI_ISL_2045156, EPI_ISL_2045157, EPI_ISL_2045158, EPI_ISL_2045159, EPI_ISL_2045160, EPI_ISL_2045161, EPI_ISL_2045162, EPI_ISL_2045163, EPI_ISL_2045164, EPI_ISL_2045167, EPI_ISL_2045168, EPI_ISL_2045169, EPI_ISL_2045171, EPI_ISL_2045172, EPI_ISL_2045173, EPI_ISL_2045174, EPI_ISL_2045175, EPI_ISL_2045176, EPI_ISL_2045177, EPI_ISL_2045179, EPI_ISL_2045180, EPI_ISL_2045181, EPI_ISL_2045182, EPI_ISL_2045183, EPI_ISL_2045184, EPI_ISL_2045186, EPI_ISL_2045187, EPI_ISL_2045188, EPI_ISL_2045190, EPI_ISL_2045191, EPI_ISL_2045192, EPI_ISL_2045195, EPI_ISL_2045196, EPI_ISL_2045197, EPI_ISL_2045198, EPI_ISL_2045199, EPI_ISL_2045200, EPI_ISL_2045201, EPI_ISL_2045202, EPI_ISL_2045203, EPI_ISL_2045204, EPI_ISL_2045206, EPI_ISL_2045207, EPI_ISL_2045208, EPI_ISL_2045209, EPI_ISL_2045210, EPI_ISL_2045211, EPI_ISL_2045213, EPI_ISL_2045215, EPI_ISL_2045216, EPI_ISL_2045217, EPI_ISL_2045218, EPI_ISL_2045219, EPI_ISL_2045220, EPI_ISL_2045221, EPI_ISL_2045222, EPI_ISL_2045224, EPI_ISL_2045225, EPI_ISL_2045226, EPI_ISL_2045228, EPI_ISL_2045229, EPI_ISL_2045230, EPI_ISL_2045231, EPI_ISL_2045232, EPI_ISL_2045233, EPI_ISL_2045235, EPI_ISL_2045236, EPI_ISL_2045237, EPI_ISL_2045238, EPI_ISL_2045239, EPI_ISL_2045240, EPI_ISL_2045241, EPI_ISL_2045242, EPI_ISL_2045243, EPI_ISL_2045244, EPI_ISL_2045246, EPI_ISL_2045247, EPI_ISL_2045248, EPI_ISL_2045250, EPI_ISL_2045251, EPI_ISL_2045252, EPI_ISL_2045253, EPI_ISL_2045255, EPI_ISL_2045257, EPI_ISL_2045258, EPI_ISL_2045259, EPI_ISL_2045260, EPI_ISL_2045261, EPI_ISL_2045262, EPI_ISL_2045264, EPI_ISL_2045266, EPI_ISL_2045267, EPI_ISL_2045268, EPI_ISL_2045269, EPI_ISL_2045270, EPI_ISL_2045271, EPI_ISL_2045272, EPI_ISL_2045273, EPI_ISL_2045274, EPI_ISL_2045275, EPI_ISL_2045276, EPI_ISL_2045277, EPI_ISL_2045278, EPI_ISL_2045279, EPI_ISL_2045280, EPI_ISL_2045281, EPI_ISL_2045282, EPI_ISL_2045283, EPI_ISL_2045284, EPI_ISL_2045287, EPI_ISL_2045288, EPI_ISL_2045290, EPI_ISL_2045291, EPI_ISL_2045292, EPI_ISL_2045293, EPI_ISL_2045294, EPI_ISL_2045295, EPI_ISL_2045296, EPI_ISL_2045297, EPI_ISL_2045298, EPI_ISL_2045299, EPI_ISL_2045300, EPI_ISL_2045301, EPI_ISL_2045302, EPI_ISL_2045303, EPI_ISL_2045306, EPI_ISL_2045307, EPI_ISL_2045308, EPI_ISL_2045309, EPI_ISL_2045310, EPI_ISL_2045311, EPI_ISL_2045312, EPI_ISL_2045315, EPI_ISL_2045316, EPI_ISL_2045317, EPI_ISL_2045318, EPI_ISL_2045319, EPI_ISL_2045320, EPI_ISL_2045321, EPI_ISL_2045322, EPI_ISL_2045323, EPI_ISL_2045324, EPI_ISL_2045325, EPI_ISL_2045326, EPI_ISL_2045327, EPI_ISL_2045328, EPI_ISL_2045329, EPI_ISL_2045330, EPI_ISL_2045331, EPI_ISL_2045332, EPI_ISL_2045334, EPI_ISL_2045335, EPI_ISL_2045336, EPI_ISL_2045337, EPI_ISL_2045338, EPI_ISL_2045340, EPI_ISL_2045341, EPI_ISL_2045342, EPI_ISL_2045345, EPI_ISL_2045346, EPI_ISL_2045348, EPI_ISL_2045350, EPI_ISL_2045351, EPI_ISL_2045352, EPI_ISL_2045353, EPI_ISL_2045354, EPI_ISL_2045355, EPI_ISL_2045356, EPI_ISL_2045357, EPI_ISL_2045358, EPI_ISL_2045359, EPI_ISL_2045360, EPI_ISL_2045361, EPI_ISL_2045364, EPI_ISL_2045366, EPI_ISL_2045367, EPI_ISL_2045368, EPI_ISL_2045369, EPI_ISL_2045370, EPI_ISL_2045371, EPI_ISL_2045372, EPI_ISL_2045373, EPI_ISL_2045374, EPI_ISL_2045375, EPI_ISL_2045376, EPI_ISL_2045377, EPI_ISL_2045378, EPI_ISL_2045379, EPI_ISL_2045380, EPI_ISL_2045381, EPI_ISL_2045382, EPI_ISL_2045383, EPI_ISL_2045384, EPI_ISL_2045385, EPI_ISL_2045386, EPI_ISL_2045387, EPI_ISL_2045388, EPI_ISL_2045389, EPI_ISL_2045391, EPI_ISL_2045392, EPI_ISL_2045393, EPI_ISL_2045394, EPI_ISL_2045395, EPI_ISL_2045396, EPI_ISL_2045397, EPI_ISL_2045398, EPI_ISL_2045399, EPI_ISL_2045400, EPI_ISL_2045401, EPI_ISL_2045403, EPI_ISL_2045404, EPI_ISL_2045405, EPI_ISL_2045406, EPI_ISL_2045407, EPI_ISL_2045408, EPI_ISL_2045409, EPI_ISL_2045410, EPI_ISL_2045411, EPI_ISL_2045412, EPI_ISL_2045413, EPI_ISL_2045414, EPI_ISL_2045415, EPI_ISL_2045416, EPI_ISL_2045417, EPI_ISL_2045418, EPI_ISL_2045419, EPI_ISL_2045420, EPI_ISL_2045421, EPI_ISL_2045422, EPI_ISL_2045423, EPI_ISL_2045424, EPI_ISL_2045426, EPI_ISL_2045427, EPI_ISL_2045428, EPI_ISL_2045429, EPI_ISL_2045430, EPI_ISL_2045431, EPI_ISL_2045432, EPI_ISL_2045433, EPI_ISL_2045434, EPI_ISL_2045436, EPI_ISL_2045437, EPI_ISL_2045439, EPI_ISL_2045440, EPI_ISL_2045441, EPI_ISL_2045442, EPI_ISL_2045443, EPI_ISL_2045444, EPI_ISL_2045446, EPI_ISL_2045447, EPI_ISL_2045448, EPI_ISL_2045449, EPI_ISL_2045450, EPI_ISL_2045451, EPI_ISL_2045452, EPI_ISL_2045453, EPI_ISL_2045454, EPI_ISL_2045455, EPI_ISL_2045457, EPI_ISL_2045458, EPI_ISL_2045459, EPI_ISL_2045460, EPI_ISL_2045462, EPI_ISL_2045463, EPI_ISL_2045464, EPI_ISL_2045465, EPI_ISL_2045466, EPI_ISL_2045467, EPI_ISL_2045468, EPI_ISL_2045469, EPI_ISL_2045470, EPI_ISL_2045471, EPI_ISL_2045472, EPI_ISL_2045473, EPI_ISL_2045474, EPI_ISL_2045475, EPI_ISL_2045477, EPI_ISL_2045478, EPI_ISL_2045479, EPI_ISL_2045480, EPI_ISL_2045481, EPI_ISL_2045482, EPI_ISL_2045483, EPI_ISL_2045484, EPI_ISL_2045485, EPI_ISL_2045486, EPI_ISL_2045487, EPI_ISL_2045488, EPI_ISL_2045489, EPI_ISL_2045490, EPI_ISL_2045491, EPI_ISL_2045492, EPI_ISL_2045493, EPI_ISL_2045495, EPI_ISL_2045496, EPI_ISL_2045497, EPI_ISL_2045498, EPI_ISL_2045499, EPI_ISL_2045500, EPI_ISL_2045501, EPI_ISL_2045502, EPI_ISL_2045506, EPI_ISL_2045507, EPI_ISL_2045508, EPI_ISL_2045509, EPI_ISL_2045510, EPI_ISL_2045511, EPI_ISL_2045512, EPI_ISL_2045513, EPI_ISL_2045514, EPI_ISL_2045516, EPI_ISL_2045517, EPI_ISL_2045519, EPI_ISL_2045520, EPI_ISL_2045521, EPI_ISL_2045522, EPI_ISL_2045523, EPI_ISL_2045524, EPI_ISL_2045525, EPI_ISL_2045526, EPI_ISL_2045527, EPI_ISL_2045528, EPI_ISL_2045529, EPI_ISL_2045530, EPI_ISL_2045531, EPI_ISL_2045532, EPI_ISL_2045533, EPI_ISL_2045535, EPI_ISL_2045536, EPI_ISL_2045537, EPI_ISL_2045538, EPI_ISL_2045539, EPI_ISL_2045540, EPI_ISL_2045542, EPI_ISL_2045543, EPI_ISL_2045544, EPI_ISL_2045545, EPI_ISL_2045546, EPI_ISL_2045547, EPI_ISL_2045548, EPI_ISL_2045549, EPI_ISL_2045550, EPI_ISL_2045551, EPI_ISL_2045552, EPI_ISL_2045553, EPI_ISL_2045554, EPI_ISL_2045555, EPI_ISL_2045557, EPI_ISL_2045558, EPI_ISL_2045559, EPI_ISL_2045560, EPI_ISL_2045561, EPI_ISL_2045562, EPI_ISL_2045563, EPI_ISL_2045564, EPI_ISL_2045565, EPI_ISL_2045566, EPI_ISL_2045567, EPI_ISL_2045568, EPI_ISL_2045569, EPI_ISL_2045570, EPI_ISL_2045571, EPI_ISL_2045572, EPI_ISL_2045573, EPI_ISL_2045574, EPI_ISL_2045575, EPI_ISL_2045576, EPI_ISL_2045577, EPI_ISL_2045578, EPI_ISL_2045580, EPI_ISL_2045581, EPI_ISL_2045582, EPI_ISL_2045583, EPI_ISL_2045584, EPI_ISL_2045585, EPI_ISL_2045586, EPI_ISL_2045587, EPI_ISL_2045588, EPI_ISL_2045589, EPI_ISL_2045590, EPI_ISL_2045592, EPI_ISL_2045593, EPI_ISL_2045594, EPI_ISL_2045595, EPI_ISL_2045596, EPI_ISL_2045598, EPI_ISL_2045599, EPI_ISL_2045600, EPI_ISL_2045601, EPI_ISL_2045602, EPI_ISL_2045603, EPI_ISL_2045604, EPI_ISL_2045605, EPI_ISL_2045606, EPI_ISL_2045607, EPI_ISL_2045608, EPI_ISL_2045609, EPI_ISL_2045610, EPI_ISL_2045611, EPI_ISL_2045612, EPI_ISL_2045613, EPI_ISL_2045614, EPI_ISL_2045615, EPI_ISL_2045616, EPI_ISL_2045617, EPI_ISL_2045618, EPI_ISL_2045619, EPI_ISL_2045620, EPI_ISL_2045621, EPI_ISL_2045622, EPI_ISL_2045623, EPI_ISL_2045624, EPI_ISL_2045625, EPI_ISL_2045626, EPI_ISL_2045627, EPI_ISL_2045628, EPI_ISL_2045629, EPI_ISL_2045630, EPI_ISL_2045631, EPI_ISL_2045632, EPI_ISL_2045635, EPI_ISL_2045636, EPI_ISL_2045638, EPI_ISL_2045640, EPI_ISL_2045641, EPI_ISL_2045642, EPI_ISL_2045643, EPI_ISL_2045645, EPI_ISL_2045646, EPI_ISL_2045647, EPI_ISL_2045648, EPI_ISL_2045649, EPI_ISL_2045650, EPI_ISL_2045651, EPI_ISL_2045652, EPI_ISL_2045653, EPI_ISL_2045654, EPI_ISL_2045656, EPI_ISL_2045657, EPI_ISL_2045658, EPI_ISL_2045659, EPI_ISL_2045660, EPI_ISL_2045662, EPI_ISL_2045663, EPI_ISL_2045664, EPI_ISL_2045665, EPI_ISL_2045666, EPI_ISL_2045667, EPI_ISL_2045669, EPI_ISL_2045670, EPI_ISL_2045672, EPI_ISL_2045673, EPI_ISL_2045674, EPI_ISL_2045675, EPI_ISL_2045676, EPI_ISL_2045677, EPI_ISL_2045678, EPI_ISL_2045679, EPI_ISL_2045680, EPI_ISL_2045681, EPI_ISL_2045682, EPI_ISL_2045683, EPI_ISL_2045684, EPI_ISL_2045685, EPI_ISL_2045686, EPI_ISL_2045687, EPI_ISL_2045688, EPI_ISL_2045689, EPI_ISL_2045690, EPI_ISL_2045691, EPI_ISL_2045692, EPI_ISL_2045694, EPI_ISL_2045695, EPI_ISL_2045697, EPI_ISL_2045698, EPI_ISL_2045699, EPI_ISL_2045700, EPI_ISL_2045701, EPI_ISL_2045702, EPI_ISL_2045703, EPI_ISL_2045704, EPI_ISL_2045705, EPI_ISL_2045706, EPI_ISL_2045707, EPI_ISL_2045708, EPI_ISL_2045709, EPI_ISL_2045710, EPI_ISL_2045711, EPI_ISL_2045712, EPI_ISL_2045713, EPI_ISL_2045714, EPI_ISL_2045715, EPI_ISL_2045717, EPI_ISL_2045718, EPI_ISL_2045720, EPI_ISL_2045721, EPI_ISL_2045722, EPI_ISL_2045723, EPI_ISL_2045725, EPI_ISL_2045726, EPI_ISL_2045727, EPI_ISL_2045728, EPI_ISL_2045729, EPI_ISL_2045732, EPI_ISL_2045733, EPI_ISL_2045735, EPI_ISL_2045737, EPI_ISL_2045738, EPI_ISL_2045739, EPI_ISL_2045741, EPI_ISL_2045742, EPI_ISL_2045743, EPI_ISL_2045744, EPI_ISL_2045745, EPI_ISL_2045746, EPI_ISL_2045747, EPI_ISL_2045748, EPI_ISL_2045749, EPI_ISL_2045750, EPI_ISL_2045751, EPI_ISL_2045754, EPI_ISL_2045755, EPI_ISL_2045756, EPI_ISL_2045758, EPI_ISL_2045759, EPI_ISL_2045761, EPI_ISL_2045762, EPI_ISL_2045763, EPI_ISL_2045764, EPI_ISL_2045765, EPI_ISL_2045766, EPI_ISL_2045767, EPI_ISL_2045770, EPI_ISL_2045771, EPI_ISL_2045772, EPI_ISL_2045773, EPI_ISL_2045774, EPI_ISL_2045775, EPI_ISL_2045776, EPI_ISL_2045777, EPI_ISL_2045778, EPI_ISL_2045779, EPI_ISL_2045780, EPI_ISL_2045781, EPI_ISL_2045782, EPI_ISL_2045783, EPI_ISL_2045784, EPI_ISL_2045785, EPI_ISL_2045786, EPI_ISL_2045788, EPI_ISL_2045789, EPI_ISL_2045791, EPI_ISL_2045792, EPI_ISL_2045794, EPI_ISL_2045795, EPI_ISL_2045796, EPI_ISL_2045797, EPI_ISL_2045798, EPI_ISL_2045799, EPI_ISL_2045800, EPI_ISL_2045801, EPI_ISL_2045802, EPI_ISL_2045803, EPI_ISL_2045804, EPI_ISL_2045805, EPI_ISL_2045806, EPI_ISL_2045807, EPI_ISL_2045808, EPI_ISL_2045809, EPI_ISL_2045810, EPI_ISL_2045813, EPI_ISL_2045814, EPI_ISL_2045815, EPI_ISL_2045816, EPI_ISL_2045817, EPI_ISL_2045818, EPI_ISL_2045819, EPI_ISL_2045820, EPI_ISL_2045822, EPI_ISL_2045823, EPI_ISL_2045824, EPI_ISL_2045825, EPI_ISL_2045826, EPI_ISL_2045827, EPI_ISL_2045828, EPI_ISL_2045830, EPI_ISL_2045831, EPI_ISL_2045832, EPI_ISL_2045833, EPI_ISL_2045834, EPI_ISL_2045835, EPI_ISL_2045836, EPI_ISL_2045837, EPI_ISL_2045838, EPI_ISL_2045839, EPI_ISL_2045840, EPI_ISL_2045841, EPI_ISL_2045842, EPI_ISL_2045843, EPI_ISL_2045844, EPI_ISL_2045845, EPI_ISL_2045846, EPI_ISL_2045847, EPI_ISL_2045848, EPI_ISL_2045849, EPI_ISL_2045850, EPI_ISL_2045851, EPI_ISL_2045852, EPI_ISL_2045853, EPI_ISL_2045855, EPI_ISL_2045857, EPI_ISL_2045858, EPI_ISL_2045859, EPI_ISL_2045860, EPI_ISL_2045861, EPI_ISL_2045862, EPI_ISL_2045863, EPI_ISL_2045865, EPI_ISL_2045866, EPI_ISL_2045868, EPI_ISL_2045870, EPI_ISL_2045871, EPI_ISL_2045872, EPI_ISL_2045873, EPI_ISL_2045874, EPI_ISL_2045875, EPI_ISL_2045876, EPI_ISL_2045877, EPI_ISL_2045879, EPI_ISL_2045884, EPI_ISL_2045885, EPI_ISL_2045886, EPI_ISL_2045887, EPI_ISL_2045888, EPI_ISL_2045889, EPI_ISL_2045890, EPI_ISL_2045891, EPI_ISL_2045892, EPI_ISL_2045893, EPI_ISL_2045894, EPI_ISL_2045895, EPI_ISL_2045896, EPI_ISL_2045897, EPI_ISL_2045898, EPI_ISL_2045899, EPI_ISL_2045900, EPI_ISL_2045901, EPI_ISL_2045902, EPI_ISL_2045903, EPI_ISL_2045904, EPI_ISL_2045905, EPI_ISL_2045906, EPI_ISL_2045907, EPI_ISL_2045909, EPI_ISL_2045910, EPI_ISL_2045911, EPI_ISL_2045912, EPI_ISL_2045913, EPI_ISL_2045914, EPI_ISL_2045915, EPI_ISL_2045916, EPI_ISL_2045917, EPI_ISL_2045918, EPI_ISL_2045919, EPI_ISL_2045920, EPI_ISL_2045921, EPI_ISL_2045922, EPI_ISL_2045923, EPI_ISL_2045924, EPI_ISL_2045925, EPI_ISL_2045927, EPI_ISL_2045928, EPI_ISL_2045929, EPI_ISL_2045930, EPI_ISL_2045932, EPI_ISL_2045933, EPI_ISL_2045934, EPI_ISL_2045935, EPI_ISL_2045936, EPI_ISL_2045937, EPI_ISL_2045938, EPI_ISL_2045939, EPI_ISL_2045940, EPI_ISL_2045941, EPI_ISL_2045942, EPI_ISL_2045943, EPI_ISL_2045944, EPI_ISL_2045946, EPI_ISL_2045947, EPI_ISL_2045948, EPI_ISL_2045949, EPI_ISL_2045951, EPI_ISL_2045952, EPI_ISL_2045953, EPI_ISL_2045954, EPI_ISL_2045955, EPI_ISL_2045956, EPI_ISL_2045957, EPI_ISL_2045958, EPI_ISL_2045959, EPI_ISL_2045960, EPI_ISL_2045961, EPI_ISL_2045963, EPI_ISL_2045965, EPI_ISL_2045966, EPI_ISL_2045969, EPI_ISL_2045972, EPI_ISL_2045973, EPI_ISL_2045974, EPI_ISL_2045976, EPI_ISL_2045978, EPI_ISL_2045979, EPI_ISL_2045980, EPI_ISL_2045981, EPI_ISL_2045983, EPI_ISL_2045984, EPI_ISL_2045985, EPI_ISL_2045986, EPI_ISL_2045987, EPI_ISL_2045989, EPI_ISL_2045990, EPI_ISL_2045991, EPI_ISL_2045992, EPI_ISL_2045993, EPI_ISL_2045994, EPI_ISL_2045995, EPI_ISL_2045996, EPI_ISL_2045997, EPI_ISL_2045999, EPI_ISL_2046001, EPI_ISL_2046002, EPI_ISL_2046003, EPI_ISL_2046004, EPI_ISL_2046006, EPI_ISL_2046007, EPI_ISL_2046008, EPI_ISL_2046009, EPI_ISL_2046011, EPI_ISL_2046012, EPI_ISL_2046013, EPI_ISL_2046015, EPI_ISL_2046016, EPI_ISL_2046017, EPI_ISL_2046018, EPI_ISL_2046020, EPI_ISL_2046021, EPI_ISL_2046023, EPI_ISL_2046025, EPI_ISL_2046026, EPI_ISL_2046027, EPI_ISL_2046029, EPI_ISL_2046031, EPI_ISL_2046032, EPI_ISL_2046034, EPI_ISL_2046036, EPI_ISL_2046037, EPI_ISL_2046038, EPI_ISL_2046039, EPI_ISL_2046040, EPI_ISL_2046041, EPI_ISL_2046042, EPI_ISL_2046043, EPI_ISL_2046044, EPI_ISL_2046045, EPI_ISL_2046047, EPI_ISL_2046048, EPI_ISL_2046050, EPI_ISL_2046054, EPI_ISL_2046055, EPI_ISL_2046057, EPI_ISL_2046058, EPI_ISL_2046059, EPI_ISL_2046060, EPI_ISL_2046063, EPI_ISL_2046064, EPI_ISL_2046065, EPI_ISL_2046066, EPI_ISL_2046067, EPI_ISL_2046068, EPI_ISL_2046069, EPI_ISL_2046070, EPI_ISL_2046071, EPI_ISL_2046072, EPI_ISL_2046074, EPI_ISL_2046076, EPI_ISL_2046077, EPI_ISL_2046078, EPI_ISL_2046079, EPI_ISL_2046081, EPI_ISL_2046082, EPI_ISL_2046083, EPI_ISL_2046084, EPI_ISL_2046088, EPI_ISL_2046089, EPI_ISL_2046091, EPI_ISL_2046092, EPI_ISL_2046094, EPI_ISL_2046095, EPI_ISL_2046096, EPI_ISL_2046097, EPI_ISL_2046098, EPI_ISL_2046100, EPI_ISL_2046101, EPI_ISL_2046102, EPI_ISL_2046104, EPI_ISL_2046105, EPI_ISL_2046106, EPI_ISL_2046107, EPI_ISL_2046108, EPI_ISL_2046109, EPI_ISL_2046110, EPI_ISL_2046112, EPI_ISL_2046114, EPI_ISL_2046115, EPI_ISL_2046116, EPI_ISL_2046117, EPI_ISL_2046118, EPI_ISL_2046119, EPI_ISL_2046120, EPI_ISL_2046121, EPI_ISL_2046122, EPI_ISL_2046123, EPI_ISL_2046124, EPI_ISL_2046125, EPI_ISL_2046126, EPI_ISL_2046127, EPI_ISL_2046128, EPI_ISL_2046129, EPI_ISL_2046130, EPI_ISL_2046133, EPI_ISL_2046134, EPI_ISL_2046135, EPI_ISL_2046136, EPI_ISL_2046137, EPI_ISL_2046138, EPI_ISL_2046139, EPI_ISL_2046140, EPI_ISL_2046141, EPI_ISL_2046143, EPI_ISL_2046144, EPI_ISL_2046145, EPI_ISL_2046146, EPI_ISL_2046147, EPI_ISL_2046148, EPI_ISL_2046149, EPI_ISL_2046150, EPI_ISL_2046151, EPI_ISL_2046153, EPI_ISL_2046154, EPI_ISL_2046155, EPI_ISL_2046156, EPI_ISL_2046157, EPI_ISL_2046158, EPI_ISL_2046159, EPI_ISL_2046160, EPI_ISL_2046161, EPI_ISL_2046163, EPI_ISL_2046164, EPI_ISL_2046165, EPI_ISL_2046167, EPI_ISL_2046168, EPI_ISL_2046170, EPI_ISL_2046171, EPI_ISL_2046172, EPI_ISL_2046174, EPI_ISL_2046176, EPI_ISL_2046178, EPI_ISL_2046179, EPI_ISL_2046180, EPI_ISL_2046181, EPI_ISL_2046182, EPI_ISL_2046183, EPI_ISL_2046184, EPI_ISL_2046185, EPI_ISL_2046186, EPI_ISL_2046187, EPI_ISL_2046189, EPI_ISL_2046191, EPI_ISL_2046192, EPI_ISL_2046193, EPI_ISL_2046194, EPI_ISL_2046195, EPI_ISL_2046196, EPI_ISL_2046198, EPI_ISL_2046199, EPI_ISL_2046200, EPI_ISL_2046201, EPI_ISL_2046204, EPI_ISL_2046205, EPI_ISL_2046209, EPI_ISL_2046210, EPI_ISL_2046211, EPI_ISL_2046212, EPI_ISL_2046213, EPI_ISL_2046217, EPI_ISL_2046218, EPI_ISL_2046221, EPI_ISL_2046222, EPI_ISL_2046225, EPI_ISL_2046227, EPI_ISL_2046229, EPI_ISL_2046230, EPI_ISL_2046231, EPI_ISL_2046233, EPI_ISL_2046234, EPI_ISL_2046236, EPI_ISL_2046237, EPI_ISL_2046238, EPI_ISL_2046239, EPI_ISL_2046242, EPI_ISL_2046243, EPI_ISL_2046244, EPI_ISL_2046246, EPI_ISL_2046247, EPI_ISL_2046248, EPI_ISL_2046249, EPI_ISL_2046252, EPI_ISL_2046253, EPI_ISL_2046254, EPI_ISL_2046257, EPI_ISL_2046260, EPI_ISL_2046262, EPI_ISL_2046263, EPI_ISL_2046264, EPI_ISL_2046266, EPI_ISL_2046267, EPI_ISL_2046268, EPI_ISL_2046270, EPI_ISL_2046271, EPI_ISL_2046275, EPI_ISL_2046278, EPI_ISL_2046279, EPI_ISL_2046280, EPI_ISL_2046281, EPI_ISL_2046282, EPI_ISL_2046283, EPI_ISL_2046284, EPI_ISL_2046285, EPI_ISL_2046286, EPI_ISL_2046287, EPI_ISL_2046288, EPI_ISL_2046290, EPI_ISL_2046291, EPI_ISL_2046292, EPI_ISL_2046293, EPI_ISL_2046294, EPI_ISL_2046295, EPI_ISL_2046296, EPI_ISL_2046297, EPI_ISL_2046298, EPI_ISL_2046299, EPI_ISL_2046300, EPI_ISL_2046302, EPI_ISL_2046304, EPI_ISL_2046306, EPI_ISL_2046307, EPI_ISL_2046308, EPI_ISL_2046309, EPI_ISL_2046310, EPI_ISL_2046311, EPI_ISL_2046312, EPI_ISL_2046313, EPI_ISL_2046314, EPI_ISL_2046315, EPI_ISL_2046316, EPI_ISL_2046317, EPI_ISL_2046320, EPI_ISL_2046321, EPI_ISL_2046322, EPI_ISL_2046323, EPI_ISL_2046324, EPI_ISL_2046325, EPI_ISL_2046327, EPI_ISL_2046328, EPI_ISL_2046329, EPI_ISL_2046330, EPI_ISL_2046331, EPI_ISL_2046332, EPI_ISL_2046333, EPI_ISL_2046334, EPI_ISL_2046335, EPI_ISL_2046337, EPI_ISL_2046339, EPI_ISL_2046340, EPI_ISL_2046341, EPI_ISL_2046342, EPI_ISL_2046343, EPI_ISL_2046344, EPI_ISL_2046345, EPI_ISL_2046346, EPI_ISL_2046347, EPI_ISL_2046349, EPI_ISL_2046350, EPI_ISL_2046351, EPI_ISL_2046352, EPI_ISL_2046353, EPI_ISL_2046354, EPI_ISL_2046358, EPI_ISL_2046360, EPI_ISL_2046364, EPI_ISL_2046365, EPI_ISL_2046366, EPI_ISL_2046367, EPI_ISL_2046368, EPI_ISL_2046369, EPI_ISL_2046370, EPI_ISL_2046371, EPI_ISL_2046373, EPI_ISL_2046374, EPI_ISL_2046376, EPI_ISL_2046377, EPI_ISL_2046378, EPI_ISL_2046379, EPI_ISL_2046380, EPI_ISL_2046381, EPI_ISL_2046382, EPI_ISL_2046383, EPI_ISL_2046384, EPI_ISL_2046385, EPI_ISL_2046386, EPI_ISL_2046387, EPI_ISL_2046388, EPI_ISL_2046389, EPI_ISL_2046390, EPI_ISL_2046391, EPI_ISL_2046392, EPI_ISL_2046393, EPI_ISL_2046394, EPI_ISL_2046395, EPI_ISL_2046396, EPI_ISL_2046397, EPI_ISL_2046398, EPI_ISL_2046399, EPI_ISL_2046400, EPI_ISL_2046403, EPI_ISL_2046404, EPI_ISL_2046405, EPI_ISL_2046406, EPI_ISL_2046407, EPI_ISL_2046408, EPI_ISL_2046409, EPI_ISL_2046410, EPI_ISL_2046411, EPI_ISL_2046412, EPI_ISL_2046413, EPI_ISL_2046414, EPI_ISL_2046415, EPI_ISL_2046416, EPI_ISL_2046417, EPI_ISL_2046418, EPI_ISL_2046419, EPI_ISL_2046420, EPI_ISL_2046421, EPI_ISL_2046422, EPI_ISL_2046423, EPI_ISL_2046424, EPI_ISL_2046427, EPI_ISL_2046428, EPI_ISL_2046429, EPI_ISL_2046430, EPI_ISL_2046431, EPI_ISL_2046432, EPI_ISL_2046433, EPI_ISL_2046434, EPI_ISL_2046435, EPI_ISL_2046436, EPI_ISL_2046439, EPI_ISL_2046440, EPI_ISL_2046441, EPI_ISL_2046442, EPI_ISL_2046443, EPI_ISL_2046444, EPI_ISL_2046445, EPI_ISL_2046446, EPI_ISL_2046447, EPI_ISL_2046449, EPI_ISL_2046450, EPI_ISL_2046453, EPI_ISL_2046454, EPI_ISL_2046455, EPI_ISL_2046456, EPI_ISL_2046457, EPI_ISL_2046458, EPI_ISL_2046459, EPI_ISL_2046460, EPI_ISL_2046461, EPI_ISL_2046462, EPI_ISL_2046463, EPI_ISL_2046465, EPI_ISL_2046466, EPI_ISL_2046467, EPI_ISL_2046468, EPI_ISL_2046469, EPI_ISL_2046470, EPI_ISL_2046471, EPI_ISL_2046472, EPI_ISL_2046473, EPI_ISL_2046474, EPI_ISL_2046475, EPI_ISL_2046476, EPI_ISL_2046477, EPI_ISL_2046478, EPI_ISL_2046479, EPI_ISL_2046480, EPI_ISL_2046481, EPI_ISL_2046482, EPI_ISL_2046483, EPI_ISL_2046485, EPI_ISL_2046486, EPI_ISL_2046488, EPI_ISL_2046489, EPI_ISL_2046490, EPI_ISL_2046491, EPI_ISL_2046492, EPI_ISL_2046493, EPI_ISL_2046494, EPI_ISL_2046495, EPI_ISL_2046497, EPI_ISL_2046498, EPI_ISL_2046500, EPI_ISL_2046502, EPI_ISL_2046503, EPI_ISL_2046504, EPI_ISL_2046505, EPI_ISL_2046506, EPI_ISL_2046507, EPI_ISL_2046508, EPI_ISL_2046509, EPI_ISL_2046510, EPI_ISL_2046511, EPI_ISL_2046512, EPI_ISL_2046513, EPI_ISL_2046514, EPI_ISL_2046516, EPI_ISL_2046518, EPI_ISL_2046520, EPI_ISL_2046522, EPI_ISL_2046523, EPI_ISL_2046524, EPI_ISL_2046526, EPI_ISL_2046528, EPI_ISL_2046530, EPI_ISL_2046531, EPI_ISL_2046532, EPI_ISL_2046533, EPI_ISL_2046534, EPI_ISL_2046535, EPI_ISL_2046536, EPI_ISL_2046538, EPI_ISL_2046539, EPI_ISL_2046540, EPI_ISL_2046541, EPI_ISL_2046542, EPI_ISL_2046543, EPI_ISL_2046544, EPI_ISL_2046545, EPI_ISL_2046546, EPI_ISL_2046547, EPI_ISL_2046548, EPI_ISL_2046549, EPI_ISL_2046550, EPI_ISL_2046551, EPI_ISL_2046552, EPI_ISL_2046553, EPI_ISL_2046554, EPI_ISL_2046555, EPI_ISL_2046556, EPI_ISL_2046558, EPI_ISL_2046559, EPI_ISL_2046560, EPI_ISL_2046561, EPI_ISL_2046562, EPI_ISL_2046564, EPI_ISL_2046565, EPI_ISL_2046567, EPI_ISL_2046568, EPI_ISL_2046569, EPI_ISL_2046570, EPI_ISL_2046571, EPI_ISL_2046572, EPI_ISL_2046574, EPI_ISL_2046576, EPI_ISL_2046577, EPI_ISL_2046578, EPI_ISL_2046580, EPI_ISL_2046581, EPI_ISL_2046582, EPI_ISL_2046583, EPI_ISL_2046584, EPI_ISL_2046585, EPI_ISL_2046586, EPI_ISL_2046587, EPI_ISL_2046588, EPI_ISL_2046589, EPI_ISL_2046590, EPI_ISL_2046592, EPI_ISL_2046593, EPI_ISL_2046594, EPI_ISL_2046595, EPI_ISL_2046596, EPI_ISL_2046597, EPI_ISL_2046598, EPI_ISL_2046599, EPI_ISL_2046600, EPI_ISL_2046601, EPI_ISL_2046602, EPI_ISL_2046604, EPI_ISL_2046605, EPI_ISL_2046606, EPI_ISL_2046607, EPI_ISL_2046608, EPI_ISL_2046609, EPI_ISL_2046610, EPI_ISL_2046611, EPI_ISL_2046612, EPI_ISL_2046613, EPI_ISL_2046614, EPI_ISL_2046615, EPI_ISL_2046616, EPI_ISL_2046618, EPI_ISL_2046619, EPI_ISL_2046620, EPI_ISL_2046623, EPI_ISL_2046624, EPI_ISL_2046625, EPI_ISL_2046626, EPI_ISL_2046627, EPI_ISL_2046628, EPI_ISL_2046629, EPI_ISL_2046631, EPI_ISL_2046632, EPI_ISL_2046633, EPI_ISL_2046634, EPI_ISL_2046635, EPI_ISL_2046636, EPI_ISL_2046638, EPI_ISL_2046639, EPI_ISL_2046640, EPI_ISL_2046642, EPI_ISL_2046643, EPI_ISL_2046645, EPI_ISL_2046648, EPI_ISL_2046649, EPI_ISL_2046652, EPI_ISL_2046653, EPI_ISL_2046654, EPI_ISL_2046656, EPI_ISL_2046657, EPI_ISL_2046661, EPI_ISL_2046662, EPI_ISL_2046663, EPI_ISL_2046664, EPI_ISL_2046665, EPI_ISL_2046666, EPI_ISL_2046667, EPI_ISL_2046668, EPI_ISL_2046669, EPI_ISL_2046670, EPI_ISL_2046672, EPI_ISL_2046673, EPI_ISL_2046674, EPI_ISL_2046675, EPI_ISL_2046676, EPI_ISL_2046677, EPI_ISL_2046678, EPI_ISL_2046679, EPI_ISL_2046680, EPI_ISL_2046681, EPI_ISL_2046682, EPI_ISL_2046683, EPI_ISL_2046684, EPI_ISL_2046685, EPI_ISL_2046686, EPI_ISL_2046687, EPI_ISL_2046688, EPI_ISL_2046689, EPI_ISL_2046690, EPI_ISL_2046691, EPI_ISL_2046692, EPI_ISL_2046693, EPI_ISL_2046694, EPI_ISL_2046695, EPI_ISL_2046696, EPI_ISL_2046697, EPI_ISL_2046698, EPI_ISL_2046699, EPI_ISL_2046700, EPI_ISL_2046701, EPI_ISL_2046702, EPI_ISL_2046705, EPI_ISL_2046706, EPI_ISL_2046708, EPI_ISL_2046710, EPI_ISL_2046711, EPI_ISL_2046713, EPI_ISL_2046715, EPI_ISL_2046716, EPI_ISL_2046717, EPI_ISL_2046718, EPI_ISL_2046721, EPI_ISL_2046722, EPI_ISL_2046723, EPI_ISL_2046724, EPI_ISL_2046725, EPI_ISL_2046726, EPI_ISL_2046727, EPI_ISL_2046729, EPI_ISL_2046730, EPI_ISL_2046731, EPI_ISL_2046732, EPI_ISL_2046733, EPI_ISL_2046734, EPI_ISL_2046735, EPI_ISL_2046736, EPI_ISL_2046737, EPI_ISL_2046738, EPI_ISL_2046739, EPI_ISL_2046740, EPI_ISL_2046741, EPI_ISL_2046742, EPI_ISL_2046744, EPI_ISL_2046747, EPI_ISL_2046748, EPI_ISL_2046749, EPI_ISL_2046751, EPI_ISL_2046753, EPI_ISL_2046754, EPI_ISL_2046755, EPI_ISL_2046756, EPI_ISL_2046757, EPI_ISL_2046758, EPI_ISL_2046759, EPI_ISL_2046760, EPI_ISL_2046761, EPI_ISL_2046762, EPI_ISL_2046764, EPI_ISL_2046765, EPI_ISL_2046768, EPI_ISL_2046769, EPI_ISL_2046770, EPI_ISL_2046771, EPI_ISL_2046773, EPI_ISL_2046776, EPI_ISL_2046777, EPI_ISL_2046778, EPI_ISL_2046779, EPI_ISL_2046781, EPI_ISL_2046783, EPI_ISL_2046784, EPI_ISL_2046785, EPI_ISL_2046786, EPI_ISL_2046787, EPI_ISL_2046788, EPI_ISL_2046789, EPI_ISL_2046791, EPI_ISL_2046793, EPI_ISL_2046794, EPI_ISL_2046795, EPI_ISL_2046796, EPI_ISL_2046797, EPI_ISL_2046798, EPI_ISL_2046800, EPI_ISL_2046801, EPI_ISL_2046802, EPI_ISL_2046803, EPI_ISL_2046804, EPI_ISL_2046808, EPI_ISL_2046809, EPI_ISL_2046810, EPI_ISL_2046811, EPI_ISL_2046812, EPI_ISL_2046814, EPI_ISL_2046815, EPI_ISL_2046816, EPI_ISL_2046818, EPI_ISL_2046819, EPI_ISL_2046820, EPI_ISL_2046821, EPI_ISL_2046822, EPI_ISL_2046823, EPI_ISL_2046824, EPI_ISL_2046825, EPI_ISL_2046826, EPI_ISL_2046827

| | | | |
|---|---|---|---|
| see above | Laboratory Corporation of America | Centers for Disease Control and Prevention Division of Viral Diseases, Pathogen Discovery | Dakota Howard, Dhwani Batra, Peter W. Cook, Kara Moser, Adrian Paskey, Jason Caravas, Benjamin Rambo-Martin, Shatavia Morrison, Christopher Gulvick, Scott Sammons, Yvette Unoarumhi, Darlene Wagner, Matthew Schmerer, Minoo Agarwal, Eyad Almasri, Debbie Boles, Ayla Burns, Nuthawin Charoensri, Oren Cohen, Susan Countryman, Mary Ann Cristobal, Bobbi Croy, Suzanne Dale, Hrushikesh Deshmukh, Amanda Douglas, Vincent Drouillon, Marcia Eisenberg, Howard Engler, Rama Ghatti, Prashant Gupta, Susan Hicks, Jake Humphrey, Lax Iyer, Manoj Jain, Mohan Kolli, Brian Krueger, Tim Kuphal, Stanley Letovsky, Michael Levandoski, Craig Lukasik, Jonathan Meltzer, Brian Norvell, Mindy Nye, Scott Parker, Christos Petropoulos, John Pruitt, Steven Ragan, Scott Ryan, Mike Sapeta, Jana Schroth, Suresh Babu Selvaraju, Goran Stevovic, Amanda Suchanek, Andrea Throop, Lyndon Tilson, Thomas Urban, Joe Voshell, Kimberly Wagner, Jonathan Williams, Mary Williamson, Qian Zeng, Tricia Zwiefelhofer, Clinton R. Paden, Duncan MacCannell |

EPI_ISL_2046828, EPI_ISL_2046829, EPI_ISL_2046830, EPI_ISL_2046831, EPI_ISL_2046832, EPI_ISL_2046833, EPI_ISL_2046834, EPI_ISL_2046835, EPI_ISL_2046836, EPI_ISL_2046837, EPI_ISL_2046838, EPI_ISL_2046839, EPI_ISL_2046840, EPI_ISL_2046841, EPI_ISL_2046842, EPI_ISL_2046843, EPI_ISL_2046844, EPI_ISL_2046845, EPI_ISL_2046846, EPI_ISL_2046847, EPI_ISL_2046848, EPI_ISL_2046849, EPI_ISL_2046850, EPI_ISL_2046851, EPI_ISL_2046852, EPI_ISL_2046853, EPI_ISL_2046854, EPI_ISL_2046855, EPI_ISL_2046856, EPI_ISL_2046857, EPI_ISL_2046858, EPI_ISL_2046859, EPI_ISL_2046860, EPI_ISL_2046861, EPI_ISL_2046862, EPI_ISL_2046863, EPI_ISL_2046864, EPI_ISL_2046865, EPI_ISL_2046866, EPI_ISL_2046867, EPI_ISL_2046868, EPI_ISL_2046869, EPI_ISL_2046870, EPI_ISL_2046871, EPI_ISL_2046872, EPI_ISL_2046873, EPI_ISL_2046874, EPI_ISL_2046875, EPI_ISL_2046876, EPI_ISL_2046877, EPI_ISL_2046878, EPI_ISL_2046879, EPI_ISL_2046880, EPI_ISL_2046881, EPI_ISL_2046882, EPI_ISL_2046883, EPI_ISL_2046884, EPI_ISL_2046885, EPI_ISL_2046886, EPI_ISL_2046887, EPI_ISL_2046888, EPI_ISL_2046889, EPI_ISL_2046890, EPI_ISL_2046891, EPI_ISL_2046892, EPI_ISL_2046893, EPI_ISL_2046894, EPI_ISL_2046895, EPI_ISL_2046896, EPI_ISL_2046897, EPI_ISL_2046898, EPI_ISL_2046899, EPI_ISL_2046900, EPI_ISL_2046901, EPI_ISL_2046902, EPI_ISL_2046903, EPI_ISL_2046904, EPI_ISL_2046905, EPI_ISL_2046906, EPI_ISL_2046907, EPI_ISL_2046908, EPI_ISL_2046909, EPI_ISL_2046910, EPI_ISL_2046911, EPI_ISL_2046912, EPI_ISL_2046913, EPI_ISL_2046914, EPI_ISL_2046915, EPI_ISL_2046916, EPI_ISL_2046917, EPI_ISL_2046918, EPI_ISL_2046919, EPI_ISL_2046920, EPI_ISL_2046921, EPI_ISL_2046922, EPI_ISL_2046923, EPI_ISL_2046924, EPI_ISL_2046925, EPI_ISL_2046926, EPI_ISL_2046927, EPI_ISL_2046928, EPI_ISL_2046929, EPI_ISL_2046930, EPI_ISL_2046931, EPI_ISL_2046932, EPI_ISL_2046933, EPI_ISL_2046934, EPI_ISL_2046935, EPI_ISL_2046936, EPI_ISL_2046937, EPI_ISL_2046938, EPI_ISL_2046939, EPI_ISL_2046940, EPI_ISL_2046941, EPI_ISL_2046942, EPI_ISL_2046943, EPI_ISL_2046944, EPI_ISL_2046945, EPI_ISL_2046946, EPI_ISL_2046947, EPI_ISL_2046948, EPI_ISL_2046949, EPI_ISL_2046950, EPI_ISL_2046951, EPI_ISL_2046952, EPI_ISL_2046953, EPI_ISL_2046954, EPI_ISL_2046955, EPI_ISL_2046956, EPI_ISL_2046957, EPI_ISL_2046958, EPI_ISL_2046959, EPI_ISL_2046960, EPI_ISL_2046961, EPI_ISL_2046962, EPI_ISL_2046963, EPI_ISL_2046964, EPI_ISL_2046965, EPI_ISL_2046966, EPI_ISL_2046967, EPI_ISL_2046968, EPI_ISL_2046969, EPI_ISL_2046970, EPI_ISL_2046971, EPI_ISL_2046972, EPI_ISL_2046973, EPI_ISL_2046974, EPI_ISL_2046975, EPI_ISL_2046976, EPI_ISL_2046977, EPI_ISL_2046978, EPI_ISL_2046979, EPI_ISL_2046980, EPI_ISL_2046981, EPI_ISL_2046982, EPI_ISL_2046983, EPI_ISL_2046984, EPI_ISL_2046985, EPI_ISL_2046986, EPI_ISL_2046987, EPI_ISL_2046988, EPI_ISL_2046989, EPI_ISL_2046990, EPI_ISL_2046991, EPI_ISL_2046992, EPI_ISL_2046993, EPI_ISL_2046994, EPI_ISL_2046995, EPI_ISL_2046996, EPI_ISL_2046997, EPI_ISL_2046998, EPI_ISL_2046999, EPI_ISL_2047000, EPI_ISL_2047001, EPI_ISL_2047002, EPI_ISL_2047003, EPI_ISL_2047004, EPI_ISL_2047005, EPI_ISL_2047006, EPI_ISL_2047007, EPI_ISL_2047008, EPI_ISL_2047009, EPI_ISL_2047010, EPI_ISL_2047011, EPI_ISL_2047012, EPI_ISL_2047013, EPI_ISL_2047014, EPI_ISL_2047015, EPI_ISL_2047016, EPI_ISL_2047017, EPI_ISL_2047018, EPI_ISL_2047019, EPI_ISL_2047020, EPI_ISL_2047021, EPI_ISL_2047022, EPI_ISL_2047023, EPI_ISL_2047024, EPI_ISL_2047025, EPI_ISL_2047026, EPI_ISL_2047027, EPI_ISL_2047028, EPI_ISL_2047029, EPI_ISL_2047030, EPI_ISL_2047031, EPI_ISL_2047032, EPI_ISL_2047033, EPI_ISL_2047034, EPI_ISL_2047035, EPI_ISL_2047036, EPI_ISL_2047037, EPI_ISL_2047038, EPI_ISL_2047039, EPI_ISL_2047040, EPI_ISL_2047041, EPI_ISL_2047042, EPI_ISL_2047043, EPI_ISL_2047044, EPI_ISL_2047045, EPI_ISL_2047046, EPI_ISL_2047047, EPI_ISL_2047048, EPI_ISL_2047049, EPI_ISL_2047050, EPI_ISL_2047051, EPI_ISL_2047052, EPI_ISL_2047054, EPI_ISL_2047055, EPI_ISL_2047056, EPI_ISL_2047057, EPI_ISL_2047058, EPI_ISL_2047059, EPI_ISL_2047060, EPI_ISL_2047061, EPI_ISL_2047062, EPI_ISL_2047063, EPI_ISL_2047064, EPI_ISL_2047065, EPI_ISL_2047066, EPI_ISL_2047067, EPI_ISL_2047068, EPI_ISL_2047069, EPI_ISL_2047070, EPI_ISL_2047071, EPI_ISL_2047072, EPI_ISL_2047073, EPI_ISL_2047074, EPI_ISL_2047075, EPI_ISL_2047076, EPI_ISL_2047077, EPI_ISL_2047078, EPI_ISL_2047079, EPI_ISL_2047080, EPI_ISL_2047081, EPI_ISL_2047082, EPI_ISL_2047083, EPI_ISL_2047084, EPI_ISL_2047085, EPI_ISL_2047086, EPI_ISL_2047087, EPI_ISL_2047088, EPI_ISL_2047089, EPI_ISL_2047090, EPI_ISL_2047091, EPI_ISL_2047092, EPI_ISL_2047093, EPI_ISL_2047094, EPI_ISL_2047095, EPI_ISL_2047096, EPI_ISL_2047097, EPI_ISL_2047098, EPI_ISL_2047099, EPI_ISL_2047100, EPI_ISL_2047101, EPI_ISL_2047102, EPI_ISL_2047103, EPI_ISL_2047104, EPI_ISL_2047105, EPI_ISL_2047106, EPI_ISL_2047107, EPI_ISL_2047108, EPI_ISL_2047109, EPI_ISL_2047110, EPI_ISL_2047111, EPI_ISL_2047112, EPI_ISL_2047113, EPI_ISL_2047114, EPI_ISL_2047115, EPI_ISL_2047116, EPI_ISL_2047117, EPI_ISL_2047118, EPI_ISL_2047119, EPI_ISL_2047120, EPI_ISL_2047121, EPI_ISL_2047122, EPI_ISL_2047123, EPI_ISL_2047124, EPI_ISL_2047125, EPI_ISL_2047126, EPI_ISL_2047127, EPI_ISL_2047128, EPI_ISL_2047129, EPI_ISL_2047130, EPI_ISL_2047131, EPI_ISL_2047132, EPI_ISL_2047133, EPI_ISL_2047134, EPI_ISL_2047135, EPI_ISL_2047136, EPI_ISL_2047137, EPI_ISL_2047138, EPI_ISL_2047139, EPI_ISL_2047140, EPI_ISL_2047141, EPI_ISL_2047142, EPI_ISL_2047143, EPI_ISL_2047144, EPI_ISL_2047145, EPI_ISL_2047146, EPI_ISL_2047147, EPI_ISL_2047148, EPI_ISL_2047149, EPI_ISL_2047150, EPI_ISL_2047151, EPI_ISL_2047152, EPI_ISL_2047153, EPI_ISL_2047154, EPI_ISL_2047155, EPI_ISL_2047156, EPI_ISL_2047157, EPI_ISL_2047158, EPI_ISL_2047159, EPI_ISL_2047160, EPI_ISL_2047161, EPI_ISL_2047162, EPI_ISL_2047163, EPI_ISL_2047164, EPI_ISL_2047165, EPI_ISL_2047166, EPI_ISL_2047167, EPI_ISL_2047168, EPI_ISL_2047169, EPI_ISL_2047170, EPI_ISL_2047171, EPI_ISL_2047172, EPI_ISL_2047173, EPI_ISL_2047174, EPI_ISL_2047175, EPI_ISL_2047176, EPI_ISL_2047177, EPI_ISL_2047178, EPI_ISL_2047179, EPI_ISL_2047180, EPI_ISL_2047181, EPI_ISL_2047182, EPI_ISL_2047183, EPI_ISL_2047184, EPI_ISL_2047185, EPI_ISL_2047186, EPI_ISL_2047187, EPI_ISL_2047188, EPI_ISL_2047189, EPI_ISL_2047190, EPI_ISL_2047191, EPI_ISL_2047192, EPI_ISL_2047193, EPI_ISL_2047194, EPI_ISL_2047195, EPI_ISL_2047196, EPI_ISL_2047197, EPI_ISL_2047198, EPI_ISL_2047199, EPI_ISL_2047200, EPI_ISL_2047201, EPI_ISL_2047202, EPI_ISL_2047203, EPI_ISL_2047204, EPI_ISL_2047205, EPI_ISL_2047206, EPI_ISL_2047207, EPI_ISL_2047208, EPI_ISL_2047209, EPI_ISL_2047210, EPI_ISL_2047211, EPI_ISL_2047212, EPI_ISL_2047213, EPI_ISL_2047214, EPI_ISL_2047215, EPI_ISL_2047216, EPI_ISL_2047217, EPI_ISL_2047218, EPI_ISL_2047219, EPI_ISL_2047220, EPI_ISL_2047221, EPI_ISL_2047222, EPI_ISL_2047223, EPI_ISL_2047224, EPI_ISL_2047225, EPI_ISL_2047226, EPI_ISL_2047227, EPI_ISL_2047228, EPI_ISL_2047229, EPI_ISL_2047230, EPI_ISL_2047231, EPI_ISL_2047232, EPI_ISL_2047233, EPI_ISL_2047234, EPI_ISL_2047235, EPI_ISL_2047236, EPI_ISL_2047237, EPI_ISL_2047238, EPI_ISL_2047239, EPI_ISL_2047240, EPI_ISL_2047241, EPI_ISL_2047242, EPI_ISL_2047243, EPI_ISL_2047244, EPI_ISL_2047245, EPI_ISL_2047246, EPI_ISL_2047247, EPI_ISL_2047248, EPI_ISL_2047249, EPI_ISL_2047250, EPI_ISL_2047251, EPI_ISL_2047252, EPI_ISL_2047253, EPI_ISL_2047254, EPI_ISL_2047255, EPI_ISL_2047256, EPI_ISL_2047257, EPI_ISL_2047258, EPI_ISL_2047259, EPI_ISL_2047260, EPI_ISL_2047261, EPI_ISL_2047262, EPI_ISL_2047263, EPI_ISL_2047264, EPI_ISL_2047265, EPI_ISL_2047266, EPI_ISL_2047267, EPI_ISL_2047268, EPI_ISL_2047269, EPI_ISL_2047270, EPI_ISL_2047271, EPI_ISL_2047272, EPI_ISL_2047273, EPI_ISL_2047274, EPI_ISL_2047275, EPI_ISL_2047276, EPI_ISL_2047277, EPI_ISL_2047278, EPI_ISL_2047279, EPI_ISL_2047280

| | | | |
|---|---|---|---|
| see above | Quest Diagnostics Incorporated | Centers for Disease Control and Prevention Division of Viral Diseases, Pathogen Discovery | Dakota Howard, Dhwani Batra, Peter W. Cook, Kara Moser, Adrian Paskey, Jason Caravas, Benjamin Rambo-Martin, Shatavia Morrison, Christopher Gulvick, Scott Sammons, Yvette Unoarumhi, Darlene Wagner, Matthew Schmerer, S. H. Rosenthal, A. Gerasimova, R. M. Kagan, B. Anderson, M. Hua, Y. Liu, L.E. Bernstein, K.E. Livingston, A. Perez, I. A. Shlyakhter, R. V. Rolando, R. Owen, P. Tanpaiboon, F. Lacbawan, Clinton R. Paden, Duncan MacCannell |

EPI_ISL_2047281, EPI_ISL_2047282, EPI_ISL_2047283, EPI_ISL_2047284, EPI_ISL_2047285, EPI_ISL_2047286, EPI_ISL_2047287, EPI_ISL_2047288, EPI_ISL_2047289, EPI_ISL_2047290, EPI_ISL_2047291, EPI_ISL_2047292, EPI_ISL_2047293, EPI_ISL_2047294, EPI_ISL_2047295, EPI_ISL_2047296, EPI_ISL_2047297, EPI_ISL_2047298, EPI_ISL_2047299, EPI_ISL_2047300, EPI_ISL_2047301, EPI_ISL_2047302, EPI_ISL_2047303, EPI_ISL_2047304, EPI_ISL_2047305, EPI_ISL_2047306, EPI_ISL_2047307, EPI_ISL_2047308, EPI_ISL_2047309, EPI_ISL_2047310, EPI_ISL_2047311, EPI_ISL_2047312, EPI_ISL_2047313, EPI_ISL_2047315, EPI_ISL_2047316, EPI_ISL_2047317, EPI_ISL_2047318, EPI_ISL_2047319, EPI_ISL_2047320, EPI_ISL_2047321, EPI_ISL_2047322, EPI_ISL_2047323, EPI_ISL_2047324, EPI_ISL_2047325, EPI_ISL_2047326, EPI_ISL_2047327, EPI_ISL_2047328, EPI_ISL_2047329, EPI_ISL_2047330, EPI_ISL_2047331, EPI_ISL_2047332, EPI_ISL_2047333, EPI_ISL_2047334, EPI_ISL_2047335, EPI_ISL_2047336, EPI_ISL_2047337, EPI_ISL_2047338, EPI_ISL_2047339, EPI_ISL_2047340, EPI_ISL_2047341, EPI_ISL_2047342, EPI_ISL_2047343, EPI_ISL_2047344, EPI_ISL_2047345, EPI_ISL_2047346, EPI_ISL_2047347, EPI_ISL_2047348, EPI_ISL_2047349, EPI_ISL_2047350, EPI_ISL_2047351, EPI_ISL_2047352, EPI_ISL_2047353, EPI_ISL_2047354, EPI_ISL_2047355, EPI_ISL_2047356, EPI_ISL_2047358, EPI_ISL_2047359, EPI_ISL_2047360, EPI_ISL_2047361, EPI_ISL_2047362, EPI_ISL_2047363, EPI_ISL_2047364, EPI_ISL_2047365, EPI_ISL_2047366, EPI_ISL_2047367, EPI_ISL_2047368, EPI_ISL_2047369, EPI_ISL_2047370, EPI_ISL_2047371, EPI_ISL_2047372, EPI_ISL_2047373, EPI_ISL_2047374, EPI_ISL_2047375, EPI_ISL_2047376, EPI_ISL_2047377, EPI_ISL_2047378, EPI_ISL_2047379, EPI_ISL_2047380, EPI_ISL_2047381, EPI_ISL_2047382, EPI_ISL_2047383, EPI_ISL_2047384, EPI_ISL_2047385, EPI_ISL_2047386, EPI_ISL_2047387, EPI_ISL_2047388, EPI_ISL_2047389, EPI_ISL_2047390, EPI_ISL_2047391, EPI_ISL_2047392, EPI_ISL_2047393, EPI_ISL_2047394, EPI_ISL_2047395, EPI_ISL_2047396, EPI_ISL_2047397, EPI_ISL_2047398, EPI_ISL_2047399, EPI_ISL_2047400, EPI_ISL_2047401, EPI_ISL_2047402, EPI_ISL_2047403, EPI_ISL_2047404, EPI_ISL_2047405, EPI_ISL_2047406, EPI_ISL_2047407, EPI_ISL_2047408, EPI_ISL_2047409, EPI_ISL_2047410, EPI_ISL_2047411, EPI_ISL_2047412, EPI_ISL_2047413, EPI_ISL_2047414, EPI_ISL_2047415, EPI_ISL_2047416, EPI_ISL_2047417, EPI_ISL_2047418, EPI_ISL_2047419, EPI_ISL_2047420, EPI_ISL_2047421, EPI_ISL_2047422, EPI_ISL_2047423, EPI_ISL_2047424, EPI_ISL_2047426, EPI_ISL_2047427, EPI_ISL_2047428, EPI_ISL_2047429, EPI_ISL_2047430, EPI_ISL_2047431, EPI_ISL_2047432, EPI_ISL_2047433, EPI_ISL_2047434, EPI_ISL_2047435, EPI_ISL_2047436, EPI_ISL_2047437, EPI_ISL_2047438, EPI_ISL_2047439, EPI_ISL_2047440, EPI_ISL_2047441, EPI_ISL_2047442, EPI_ISL_2047443, EPI_ISL_2047444, EPI_ISL_2047445, EPI_ISL_2047446, EPI_ISL_2047447, EPI_ISL_2047448, EPI_ISL_2047449, EPI_ISL_2047450, EPI_ISL_2047451, EPI_ISL_2047452, EPI_ISL_2047453, EPI_ISL_2047454, EPI_ISL_2047455, EPI_ISL_2047456, EPI_ISL_2047457, EPI_ISL_2047458, EPI_ISL_2047459, EPI_ISL_2047460, EPI_ISL_2047461, EPI_ISL_2047462, EPI_ISL_2047463, EPI_ISL_2047464, EPI_ISL_2047465, EPI_ISL_2047466, EPI_ISL_2047467, EPI_ISL_2047468, EPI_ISL_2047469, EPI_ISL_2047470, EPI_ISL_2047471, EPI_ISL_2047472, EPI_ISL_2047473, EPI_ISL_2047474, EPI_ISL_2047475, EPI_ISL_2047476, EPI_ISL_2047477, EPI_ISL_2047478, EPI_ISL_2047479, EPI_ISL_2047480, EPI_ISL_2047481, EPI_ISL_2047482, EPI_ISL_2047483, EPI_ISL_2047484, EPI_ISL_2047485, EPI_ISL_2047486, EPI_ISL_2047487, EPI_ISL_2047488, EPI_ISL_2047489, EPI_ISL_2047490, EPI_ISL_2047491, EPI_ISL_2047492, EPI_ISL_2047493, EPI_ISL_2047494, EPI_ISL_2047495, EPI_ISL_2047496, EPI_ISL_2047497, EPI_ISL_2047498, EPI_ISL_2047499, EPI_ISL_2047500, EPI_ISL_2047501, EPI_ISL_2047502, EPI_ISL_2047503, EPI_ISL_2047504, EPI_ISL_2047505

| | | | |
|---|---|---|---|
| see above | Infinity Biologix | Centers for Disease Control and Prevention Division of Viral Diseases, Pathogen Discovery | Dakota Howard, Dhwani Batra, Peter W. Cook, Kara Moser, Adrian Paskey, Jason Caravas, Benjamin Rambo-Martin, Shatavia Morrison, Christopher Gulvick, Scott Sammons, Yvette Unoarumhi, Darlene Wagner, Matthew Schmerer, Christian Bixby, Yihe Wang, Jonathan Schultz, Chirayu Goswami, Russ Hager, Robin Grimwood, Clinton R. Paden, Duncan MacCannell |
| EPI_ISL_2047506 | NYC Department of Health and Mental Hygiene | Centers for Disease Control and Prevention Division of Viral Diseases, Pathogen Discovery | Mili Sheth, Sarah Nobles, Jasmine Padilla, Mark Burroughs, Shoshona Le, Katie Dillon, Peter Cook, Clinton R. Paden, Dhwani Batra, Krista Queen, Kristen Knipe, Dakota Howard, Yvette Unoarumhi, Darlene Wagner, Matthew Schmerer, Ben L. Rambo-Martin, Kristine Lacek, Sam Shepard, Alison Laufer Halpin, Dave Wentworth, Vivien Dugan, Suxiang Tong, Justin Lee |
| EPI_ISL_2047617, EPI_ISL_2047618, EPI_ISL_2047619, EPI_ISL_2047620, EPI_ISL_2047621, EPI_ISL_2047622, EPI_ISL_2047623, EPI_ISL_2047624, EPI_ISL_2047625, EPI_ISL_2047626, EPI_ISL_2047627, EPI_ISL_2047628, EPI_ISL_2047629, EPI_ISL_2047630, EPI_ISL_2047631, EPI_ISL_2047632 | | | |
| see above | Wyoming Public Health Laboratory | Wyoming Public Health Laboratory | Jim Mildenberger, Taylor Fearing, Channing Weber, Ashley Norberg, Chayse Rowley, Marley Goetz, Brian Dominguez, Elliot Thomasson, Brittany Oher, Sam Britz, Cari Sloma, and Rob Christensen |
| EPI_ISL_2047874, EPI_ISL_2047875, EPI_ISL_2047876, EPI_ISL_2047878, EPI_ISL_2047879, EPI_ISL_2047880 | Ochsner Health | Bioinfoexperts, LLC | Amy Feehan, David J. Nolan, Rebecca Rose, Sissy Cross, David Moraga Amador, Tong Yang, Luke Caruso, Wayra Navia, Lydia Von Borstel, Xiao Hui Zhou, Julia-Garcia-Diaz, Susanna L. Lamers |
| EPI_ISL_2080001, EPI_ISL_2080002, EPI_ISL_2080003, EPI_ISL_2080004, EPI_ISL_2080005, EPI_ISL_2080007, EPI_ISL_2080008, EPI_ISL_2080010, EPI_ISL_2080011, EPI_ISL_2080012, EPI_ISL_2080015, EPI_ISL_2080017, EPI_ISL_2080018, EPI_ISL_2080019, EPI_ISL_2080020, EPI_ISL_2080021, EPI_ISL_2080022, EPI_ISL_2080023, EPI_ISL_2080025, EPI_ISL_2080026, EPI_ISL_2080027, EPI_ISL_2080028, EPI_ISL_2080029, EPI_ISL_2080030, EPI_ISL_2080031, EPI_ISL_2080032, EPI_ISL_2080034, EPI_ISL_2080036, EPI_ISL_2080037, EPI_ISL_2080038, EPI_ISL_2080039, EPI_ISL_2080040, EPI_ISL_2080041, EPI_ISL_2080042, EPI_ISL_2080043, EPI_ISL_2080044, EPI_ISL_2080045, EPI_ISL_2080047, EPI_ISL_2080048, EPI_ISL_2080049, EPI_ISL_2080050, EPI_ISL_2080051, EPI_ISL_2080052, EPI_ISL_2080053, EPI_ISL_2080054, EPI_ISL_2080055, EPI_ISL_2080056, EPI_ISL_2080057, EPI_ISL_2080058, EPI_ISL_2080059, EPI_ISL_2080060, EPI_ISL_2080061, EPI_ISL_2080062, EPI_ISL_2080063, EPI_ISL_2080064, EPI_ISL_2080065, EPI_ISL_2080066, EPI_ISL_2080067, EPI_ISL_2080068, EPI_ISL_2080069, EPI_ISL_2080070, EPI_ISL_2080071, EPI_ISL_2080073, EPI_ISL_2080074, EPI_ISL_2080075, EPI_ISL_2080076, EPI_ISL_2080077, EPI_ISL_2080078, EPI_ISL_2080079, EPI_ISL_2080080, EPI_ISL_2080081, EPI_ISL_2080082, EPI_ISL_2080083, EPI_ISL_2080084, EPI_ISL_2080085, EPI_ISL_2080086, EPI_ISL_2080087, EPI_ISL_2080088, EPI_ISL_2080089, EPI_ISL_2080091, EPI_ISL_2080092, EPI_ISL_2080093, EPI_ISL_2080094, EPI_ISL_2080095, EPI_ISL_2080096, EPI_ISL_2080097, EPI_ISL_2080098, EPI_ISL_2080099, EPI_ISL_2080100, EPI_ISL_2080101, EPI_ISL_2080102, EPI_ISL_2080103, EPI_ISL_2080105, EPI_ISL_2080106, EPI_ISL_2080107, EPI_ISL_2080109, EPI_ISL_2080110, EPI_ISL_2080111, EPI_ISL_2080112, EPI_ISL_2080113, EPI_ISL_2080114, EPI_ISL_2080115, EPI_ISL_2080116, EPI_ISL_2080117, EPI_ISL_2080118, EPI_ISL_2080121, EPI_ISL_2080122, EPI_ISL_2080123, EPI_ISL_2080124, EPI_ISL_2080125, EPI_ISL_2080127, EPI_ISL_2080129, EPI_ISL_2080131, EPI_ISL_2080132, EPI_ISL_2080133, EPI_ISL_2080134, EPI_ISL_2080135, EPI_ISL_2080136, EPI_ISL_2080137, EPI_ISL_2080138, EPI_ISL_2080139, EPI_ISL_2080140, EPI_ISL_2080141, EPI_ISL_2080142, EPI_ISL_2080143, EPI_ISL_2080146, EPI_ISL_2080147, EPI_ISL_2080148, EPI_ISL_2080149, EPI_ISL_2080150, EPI_ISL_2080151, EPI_ISL_2080152, EPI_ISL_2080155, EPI_ISL_2080156, EPI_ISL_2080157, EPI_ISL_2080158 | | | |
| see above | Illinois Department of Public Health | Gagnon Lab, Southern Illinois University | Keith Gagnon |
| EPI_ISL_2080159, EPI_ISL_2080160, EPI_ISL_2080161, EPI_ISL_2080162, EPI_ISL_2080163, EPI_ISL_2080164, EPI_ISL_2080165, EPI_ISL_2080166, EPI_ISL_2080167, EPI_ISL_2080168, EPI_ISL_2080169, EPI_ISL_2080170, EPI_ISL_2080171, EPI_ISL_2080172, EPI_ISL_2080173, EPI_ISL_2080174, EPI_ISL_2080175, EPI_ISL_2080176, EPI_ISL_2080177, EPI_ISL_2080178, EPI_ISL_2080179, EPI_ISL_2080180, EPI_ISL_2080181, EPI_ISL_2080182, EPI_ISL_2080183, EPI_ISL_2080184, EPI_ISL_2080185, EPI_ISL_2080187, EPI_ISL_2080188, EPI_ISL_2080189, EPI_ISL_2080190, EPI_ISL_2080191, EPI_ISL_2080192, EPI_ISL_2080193, EPI_ISL_2080194, EPI_ISL_2080195, EPI_ISL_2080196, EPI_ISL_2080197, EPI_ISL_2080198, EPI_ISL_2080199, EPI_ISL_2080200, EPI_ISL_2080201, EPI_ISL_2080202, EPI_ISL_2080203, EPI_ISL_2080204, EPI_ISL_2080205, EPI_ISL_2080207, EPI_ISL_2080209, EPI_ISL_2080210, EPI_ISL_2080211, EPI_ISL_2080212, EPI_ISL_2080213, EPI_ISL_2080216, EPI_ISL_2080217, EPI_ISL_2080218, EPI_ISL_2080219, EPI_ISL_2080220, EPI_ISL_2080221, EPI_ISL_2080222, EPI_ISL_2080223, EPI_ISL_2080225, EPI_ISL_2080226, EPI_ISL_2080227, EPI_ISL_2080228, EPI_ISL_2080229, EPI_ISL_2080230, EPI_ISL_2080231, EPI_ISL_2080233, EPI_ISL_2080234, EPI_ISL_2080235, EPI_ISL_2080236, EPI_ISL_2080237, EPI_ISL_2080238, EPI_ISL_2080239, EPI_ISL_2080240, EPI_ISL_2080241, EPI_ISL_2080242, EPI_ISL_2080243, EPI_ISL_2080244, EPI_ISL_2080245, EPI_ISL_2080246, EPI_ISL_2080247, EPI_ISL_2080248, EPI_ISL_2080249 | | | |
| see above | Simple Laboratories | Gagnon Lab, Southern Illinois University | Keith Gagnon |
| EPI_ISL_2080494, EPI_ISL_2080495, EPI_ISL_2080496, EPI_ISL_2080497, EPI_ISL_2080498, EPI_ISL_2080499, EPI_ISL_2080500, EPI_ISL_2080501, EPI_ISL_2080502, EPI_ISL_2080503, EPI_ISL_2080504, EPI_ISL_2080505, EPI_ISL_2080506, EPI_ISL_2080507, EPI_ISL_2080508, EPI_ISL_2080509, EPI_ISL_2080510, EPI_ISL_2080511, EPI_ISL_2080512, EPI_ISL_2080513, EPI_ISL_2080514, EPI_ISL_2080515, EPI_ISL_2080516, EPI_ISL_2080517, EPI_ISL_2080518, EPI_ISL_2080519, EPI_ISL_2080520, EPI_ISL_2080521, EPI_ISL_2080522, EPI_ISL_2080523, EPI_ISL_2080524, EPI_ISL_2080525, EPI_ISL_2080526, EPI_ISL_2080527, EPI_ISL_2080528, EPI_ISL_2080529, EPI_ISL_2080530, EPI_ISL_2080531, EPI_ISL_2080532, EPI_ISL_2080533, EPI_ISL_2080534, EPI_ISL_2080535, EPI_ISL_2080536, EPI_ISL_2080537, EPI_ISL_2080538, EPI_ISL_2080539, EPI_ISL_2080540, EPI_ISL_2080541, EPI_ISL_2080542, EPI_ISL_2080543, EPI_ISL_2080544, EPI_ISL_2080545, EPI_ISL_2080546, EPI_ISL_2080547, EPI_ISL_2080548, EPI_ISL_2080549, EPI_ISL_2080550, EPI_ISL_2080551, EPI_ISL_2080552, EPI_ISL_2080553, EPI_ISL_2080554, EPI_ISL_2080555, EPI_ISL_2080556, EPI_ISL_2080557, EPI_ISL_2080558, EPI_ISL_2080559, EPI_ISL_2080560, EPI_ISL_2080561, EPI_ISL_2080562, EPI_ISL_2080563, EPI_ISL_2080566, EPI_ISL_2080567, EPI_ISL_2080568, EPI_ISL_2080569, EPI_ISL_2080570, EPI_ISL_2080571, EPI_ISL_2080572 | | | |
| see above | Minnesota Department of Health, Public Health Laboratory | Minnesota Department of Health, Public Health Laboratory | Alexandra Lorentz, Jacob Garfin, Matt Plumb, and Xiong Wang |
| EPI_ISL_2080616, EPI_ISL_2080617 | Instituto Nacional de Medicina Genomica | Instituto Nacional de Medicina Genomica | Hidalgo-Miranda A, Mendoza-Vargas A, Reyes-Grajeda JP, Cedro-Tanda A, Cisneros-Villanueva M, Orjuela-Rodríguez M, Alcaraz N, Gonzalez-Barrera D, Rangel-DeLeon D, Miranda-Ortiz H, Rosas-Escobar P, Canseco Mendez JC, Ramirez-Vega O, Munguia-Garza P, Garcia-Cardenas FJ, Gonzalez-Woge MA, Herrera-Montalvo LA |
| EPI_ISL_2080624, EPI_ISL_2080628, EPI_ISL_2080629, EPI_ISL_2080630, EPI_ISL_2080631, EPI_ISL_2080633, EPI_ISL_2080634, EPI_ISL_2080635, EPI_ISL_2080636, EPI_ISL_2080638, EPI_ISL_2080639, EPI_ISL_2080640, EPI_ISL_2080642, EPI_ISL_2080643, EPI_ISL_2080645, EPI_ISL_2080646, EPI_ISL_2080647, EPI_ISL_2080648, EPI_ISL_2080650, EPI_ISL_2080651, EPI_ISL_2080652, EPI_ISL_2080654, EPI_ISL_2080657, EPI_ISL_2080660, EPI_ISL_2080662, EPI_ISL_2080663, EPI_ISL_2080664, EPI_ISL_2080665, EPI_ISL_2080670, EPI_ISL_2080674, EPI_ISL_2080675, EPI_ISL_2080676, EPI_ISL_2080677, EPI_ISL_2080678, EPI_ISL_2080679, EPI_ISL_2080680, EPI_ISL_2080681, EPI_ISL_2080683, EPI_ISL_2080684, EPI_ISL_2080685, EPI_ISL_2080690, EPI_ISL_2080691, EPI_ISL_2080693, EPI_ISL_2080694 | | | |
| see above | Instituto Nacional de Medicina Genomica | Instituto Nacional de Medicina Genomica | Hidalgo-Miranda A, Mendoza-Vargas A, Reyes-Grajeda JP, Cedro-Tanda A, Alcaraz N, Gonzalez-Barrera D, Rangel-DeLeon D, Munguia-Garza P, Ramirez-Vega O, Escobar-Arrazola, M, Herrera-Montalvo LA. |
| EPI_ISL_2080696, EPI_ISL_2080697, EPI_ISL_2080698, EPI_ISL_2080699, EPI_ISL_2080700, EPI_ISL_2080702, EPI_ISL_2080703, EPI_ISL_2080704, EPI_ISL_2080705, EPI_ISL_2080706, EPI_ISL_2080707, EPI_ISL_2080708, EPI_ISL_2080709, EPI_ISL_2080710, EPI_ISL_2080711, EPI_ISL_2080712, EPI_ISL_2080713, EPI_ISL_2080714, EPI_ISL_2080715, EPI_ISL_2080716, EPI_ISL_2080717, EPI_ISL_2080718, EPI_ISL_2080719, EPI_ISL_2080720, EPI_ISL_2080721, EPI_ISL_2080722, EPI_ISL_2080723, EPI_ISL_2080724, EPI_ISL_2080725, EPI_ISL_2080726, EPI_ISL_2080727, EPI_ISL_2080728, EPI_ISL_2080729, EPI_ISL_2080730, EPI_ISL_2080731, EPI_ISL_2080732, EPI_ISL_2080733, EPI_ISL_2080734, EPI_ISL_2080735, EPI_ISL_2080736, EPI_ISL_2080737, EPI_ISL_2080738, EPI_ISL_2080748, EPI_ISL_2080749, EPI_ISL_2080750, EPI_ISL_2080751, EPI_ISL_2080752, EPI_ISL_2080753, EPI_ISL_2080754, EPI_ISL_2080755, EPI_ISL_2080756, EPI_ISL_2080757, EPI_ISL_2080758, EPI_ISL_2080759, EPI_ISL_2080760, EPI_ISL_2080761, EPI_ISL_2080762, EPI_ISL_2080763, EPI_ISL_2080764, EPI_ISL_2080765, EPI_ISL_2080766, EPI_ISL_2080767, EPI_ISL_2080770, EPI_ISL_2080771, EPI_ISL_2080772, EPI_ISL_2080773, EPI_ISL_2080774, EPI_ISL_2080775, EPI_ISL_2080776, EPI_ISL_2080777, EPI_ISL_2080778, EPI_ISL_2080779, EPI_ISL_2080780, EPI_ISL_2080781, EPI_ISL_2080782, EPI_ISL_2080783, EPI_ISL_2080784, EPI_ISL_2080785, EPI_ISL_2080786, EPI_ISL_2080787, EPI_ISL_2080788, EPI_ISL_2080789, EPI_ISL_2080790, EPI_ISL_2080791, EPI_ISL_2080792, EPI_ISL_2080793, EPI_ISL_2080794, EPI_ISL_2080795, EPI_ISL_2080796, EPI_ISL_2080797, EPI_ISL_2080798, EPI_ISL_2080799, EPI_ISL_2080800, EPI_ISL_2080801, EPI_ISL_2080802, EPI_ISL_2080803, EPI_ISL_2080804, EPI_ISL_2080805, EPI_ISL_2080806, EPI_ISL_2080807, EPI_ISL_2080808, EPI_ISL_2080809, EPI_ISL_2080810, EPI_ISL_2080811, EPI_ISL_2080812, EPI_ISL_2080813, EPI_ISL_2080814, EPI_ISL_2080815, EPI_ISL_2080816, EPI_ISL_2080817, EPI_ISL_2080818, EPI_ISL_2080819, EPI_ISL_2080820, EPI_ISL_2080821, EPI_ISL_2080822, EPI_ISL_2080823, EPI_ISL_2080824, EPI_ISL_2080825, EPI_ISL_2080826, EPI_ISL_2080827, EPI_ISL_2080828, EPI_ISL_2080829, EPI_ISL_2080830, EPI_ISL_2080831, EPI_ISL_2080832, EPI_ISL_2080833, EPI_ISL_2080834, EPI_ISL_2080835, EPI_ISL_2080836, EPI_ISL_2080837, EPI_ISL_2080838, EPI_ISL_2080839, EPI_ISL_2080840, EPI_ISL_2080841, EPI_ISL_2080842, EPI_ISL_2080843, EPI_ISL_2080844, EPI_ISL_2080845, EPI_ISL_2080846, EPI_ISL_2080847, EPI_ISL_2080848, EPI_ISL_2080849, EPI_ISL_2080850, EPI_ISL_2080851, EPI_ISL_2080852, EPI_ISL_2080853, EPI_ISL_2080854, EPI_ISL_2080855, EPI_ISL_2080856, EPI_ISL_2080857, EPI_ISL_2080858, EPI_ISL_2080859, EPI_ISL_2080860, EPI_ISL_2080861, EPI_ISL_2080862 | | | |
| see above | OHSU Lab Services Molecular Microbiology Lab | Oregon SARS-CoV-2 Genome Sequencing Center | Brendan L. O'Connell, Sally Grindstaff, Sonia Acharya, Cierra LaBlanc, Kayla Carter, Ruth V. Nichols, Alec J. Hirsch, Donna Hansel, Guang Fan, Xuan Qin, Daniel N. Streblow, William B. Messer, Andrew C. Adey, Benjamin N. Bimber, Brian J. O'Roak |
| EPI_ISL_2080974, EPI_ISL_2080975 | Wyoming Public Health Laboratory | Wyoming Public Health Laboratory | Jim Mildenberger, Wanda Manley, Noah Hull, Taylor Fearing, Lynette Gumbleton, Channing Weber, Ashley Norberg, Chayse Rowley, Marley Goetz, Brian Dominguez, Elliot Thomasson, Sam Britz, Cari Sloma, and Rob Christensen |
| EPI_ISL_2080983, EPI_ISL_2080988, EPI_ISL_2080990, EPI_ISL_2080995 | Wyoming Public Health Laboratory | Wyoming Public Health Laboratory | Jim Mildenberger, Taylor Fearing, Channing Weber, Ashley Norberg, Chayse Rowley, Marley Goetz, Brian Dominguez, Elliot Thomasson, Brittany Oher, Sam Britz, Cari Sloma, and Rob Christensen |
| EPI_ISL_2080997, EPI_ISL_2080998, EPI_ISL_2080999, EPI_ISL_2081000, EPI_ISL_2081001, EPI_ISL_2081002, EPI_ISL_2081003, EPI_ISL_2081004, EPI_ISL_2081005, EPI_ISL_2081006, EPI_ISL_2081007, EPI_ISL_2081008, EPI_ISL_2081009, EPI_ISL_2081010, EPI_ISL_2081011, EPI_ISL_2081012, EPI_ISL_2081013, EPI_ISL_2081014, EPI_ISL_2081015, EPI_ISL_2081016, EPI_ISL_2081017, EPI_ISL_2081018, EPI_ISL_2081019, EPI_ISL_2081020, EPI_ISL_2081021, EPI_ISL_2081022, EPI_ISL_2081023, EPI_ISL_2081024, EPI_ISL_2081025, EPI_ISL_2081026, EPI_ISL_2081027, EPI_ISL_2081028, EPI_ISL_2081029, EPI_ISL_2081030, EPI_ISL_2081031, EPI_ISL_2081032, EPI_ISL_2081033, EPI_ISL_2081034, EPI_ISL_2081035, EPI_ISL_2081036, EPI_ISL_2081037, EPI_ISL_2081038, EPI_ISL_2081039, EPI_ISL_2081040, EPI_ISL_2081041, EPI_ISL_2081042, EPI_ISL_2081043, EPI_ISL_2081044, EPI_ISL_2081045, EPI_ISL_2081046, EPI_ISL_2081047, EPI_ISL_2081048, EPI_ISL_2081049, EPI_ISL_2081050, EPI_ISL_2081051, EPI_ISL_2081052, EPI_ISL_2081053, EPI_ISL_2081054, EPI_ISL_2081055, EPI_ISL_2081056, EPI_ISL_2081057, EPI_ISL_2081058, EPI_ISL_2081059, EPI_ISL_2081060, EPI_ISL_2081061, EPI_ISL_2081062, EPI_ISL_2081063, EPI_ISL_2081064, EPI_ISL_2081065, EPI_ISL_2081066, EPI_ISL_2081067, EPI_ISL_2081068, EPI_ISL_2081069, EPI_ISL_2081070, EPI_ISL_2081071, EPI_ISL_2081072, EPI_ISL_2081073, EPI_ISL_2081074, EPI_ISL_2081075, EPI_ISL_2081076, EPI_ISL_2081077, EPI_ISL_2081078, EPI_ISL_2081079, EPI_ISL_2081080, EPI_ISL_2081081, EPI_ISL_2081082, EPI_ISL_2081083, EPI_ISL_2081084, EPI_ISL_2081085, EPI_ISL_2081086, EPI_ISL_2081087, EPI_ISL_2081088, EPI_ISL_2081089, EPI_ISL_2081090, EPI_ISL_2081091, EPI_ISL_2081092, EPI_ISL_2081093, EPI_ISL_2081094, EPI_ISL_2081095, EPI_ISL_2081096, EPI_ISL_2081097, EPI_ISL_2081098, EPI_ISL_2081099, EPI_ISL_2081100, EPI_ISL_2081101, EPI_ISL_2081102, EPI_ISL_2081103, EPI_ISL_2081104, EPI_ISL_2081105, EPI_ISL_2081106, EPI_ISL_2081107, EPI_ISL_2081108, EPI_ISL_2081109, EPI_ISL_2081110, EPI_ISL_2081111, EPI_ISL_2081112, EPI_ISL_2081113, EPI_ISL_2081114, EPI_ISL_2081115, EPI_ISL_2081116, EPI_ISL_2081117, EPI_ISL_2081118, EPI_ISL_2081119, EPI_ISL_2081120, EPI_ISL_2081121, EPI_ISL_2081122, EPI_ISL_2081123, EPI_ISL_2081124, EPI_ISL_2081125, EPI_ISL_2081126, EPI_ISL_2081127 | | | |
| see above | Hackensack Medical Center | New York Genome Center | Michael Zody, Andre Corvelo, Dayna M. Oschwald, Samantha Fennessey, Tom Maniatis, Liang Chen, Jose Mediavilla, Marcus Cunningham, Kaelea Composto, Kar Chow, David Perlin, Barry Kreiswirth |
| EPI_ISL_2081128, EPI_ISL_2081129, EPI_ISL_2081130, EPI_ISL_2081131, EPI_ISL_2081132, EPI_ISL_2081133, EPI_ISL_2081134, EPI_ISL_2081135, EPI_ISL_2081136, EPI_ISL_2081137, EPI_ISL_2081138, EPI_ISL_2081139, EPI_ISL_2081140, EPI_ISL_2081141, EPI_ISL_2081142, EPI_ISL_2081143, EPI_ISL_2081144, EPI_ISL_2081145, EPI_ISL_2081146, EPI_ISL_2081147, EPI_ISL_2081148, EPI_ISL_2081149, EPI_ISL_2081150, EPI_ISL_2081151, EPI_ISL_2081152, EPI_ISL_2081153, EPI_ISL_2081154, EPI_ISL_2081155, EPI_ISL_2081156, EPI_ISL_2081157, EPI_ISL_2081158, EPI_ISL_2081159, EPI_ISL_2081160, EPI_ISL_2081161, EPI_ISL_2081162, EPI_ISL_2081163, EPI_ISL_2081164, EPI_ISL_2081165, EPI_ISL_2081166, EPI_ISL_2081167, EPI_ISL_2081168 | | | |
| see above | Weill Cornell Medicine | New York Genome Center | Michael Zody, Andre Corvelo, Dayna M. Oschwald, Samantha Fennessey, Tom Maniatis, Melissa Cushing, Olivier Elemento, Margaret Elizabeth Ross, Chris Mason, Priya Velu, Hanna Rennert, Arryn Craney, Lars F Westblade |
| EPI_ISL_2081169, EPI_ISL_2081170, EPI_ISL_2081172, EPI_ISL_2081173, EPI_ISL_2081174, EPI_ISL_2081176, EPI_ISL_2081177, EPI_ISL_2081178, EPI_ISL_2081180, EPI_ISL_2081181, EPI_ISL_2081182, EPI_ISL_2081183, EPI_ISL_2081184, EPI_ISL_2081186, EPI_ISL_2081188, EPI_ISL_2081190 | | | |
| see above | Wyoming Public Health Laboratory | Wyoming Public Health Laboratory | Jim Mildenberger, Taylor Fearing, Channing Weber, Ashley Norberg, Chayse Rowley, Marley Goetz, Brian Dominguez, Elliot Thomasson, Brittany Oher, Sam Britz, Cari Sloma, and Rob Christensen |
| EPI_ISL_2081192, EPI_ISL_2081193, EPI_ISL_2081194, EPI_ISL_2081195, EPI_ISL_2081196, EPI_ISL_2081197, EPI_ISL_2081198, EPI_ISL_2081199, EPI_ISL_2081200, EPI_ISL_2081201, EPI_ISL_2081202, EPI_ISL_2081203, EPI_ISL_2081204, EPI_ISL_2081205, EPI_ISL_2081206, EPI_ISL_2081207, EPI_ISL_2081208, EPI_ISL_2081209, EPI_ISL_2081210, EPI_ISL_2081211, EPI_ISL_2081212, EPI_ISL_2081213, EPI_ISL_2081214, EPI_ISL_2081215, EPI_ISL_2081216, EPI_ISL_2081217, EPI_ISL_2081218, EPI_ISL_2081219, EPI_ISL_2081220, EPI_ISL_2081221, EPI_ISL_2081222, EPI_ISL_2081223, EPI_ISL_2081224 | | | |
| see above | Premier Laboratory Services | BioInfoExperts LLC | Cheryl Metzler, Valerie Murray, Rebecca Rose, David Nolan, Susanna Lamers |
| EPI_ISL_2081230, EPI_ISL_2081231, EPI_ISL_2081232, EPI_ISL_2081233, EPI_ISL_2081235, EPI_ISL_2081236, EPI_ISL_2081237, EPI_ISL_2081241, EPI_ISL_2081242, EPI_ISL_2081243, EPI_ISL_2081244, EPI_ISL_2081245, EPI_ISL_2081247, EPI_ISL_2081248, EPI_ISL_2081249, EPI_ISL_2081250, EPI_ISL_2081251, EPI_ISL_2081252, EPI_ISL_2081253, EPI_ISL_2081254, EPI_ISL_2081255, EPI_ISL_2081256, EPI_ISL_2081257, EPI_ISL_2081258, EPI_ISL_2081259, EPI_ISL_2081260, EPI_ISL_2081261, EPI_ISL_2081264, EPI_ISL_2081265, EPI_ISL_2081266, EPI_ISL_2081267, EPI_ISL_2081268, EPI_ISL_2081269, EPI_ISL_2081270, EPI_ISL_2081272, EPI_ISL_2081273, EPI_ISL_2081274, EPI_ISL_2081275, EPI_ISL_2081276, EPI_ISL_2081277, EPI_ISL_2081278, EPI_ISL_2081280, EPI_ISL_2081281, EPI_ISL_2081282, EPI_ISL_2081283, EPI_ISL_2081284, EPI_ISL_2081285, EPI_ISL_2081289, EPI_ISL_2081290, EPI_ISL_2081291, EPI_ISL_2081292, EPI_ISL_2081293, EPI_ISL_2081296, EPI_ISL_2081297, EPI_ISL_2081298, EPI_ISL_2081299, EPI_ISL_2081300, EPI_ISL_2081301, EPI_ISL_2081303, EPI_ISL_2081304, EPI_ISL_2081305, EPI_ISL_2081306, EPI_ISL_2081307, EPI_ISL_2081308 | | | |
| see above | UCSC Genomics Institute | UCSC Genomics Institute | Mark Akeson, Ikenna Anigbogu, Eric Beraut, Isabel Bjork, Molly Cassatt-Johnstone, Terren Chang, Russell Corbett-Detig, Namrita Dhillon, Maximilian Haeussler, David Haussler, Angie Hinrichs, Miten Jain, Joshua Kapp, A. Marm Kilpatrick, Jakob McBroome, Hugh Olsen, Jeremy Sanford, Beth Shapiro, Michael Stone, Bryan Thornlow, Yatish Turakhia, Ciara Wanket |
| EPI_ISL_2081322, EPI_ISL_2081326, EPI_ISL_2081328, EPI_ISL_2081329, EPI_ISL_2081335, EPI_ISL_2081339, EPI_ISL_2081340, EPI_ISL_2081343, EPI_ISL_2081346, EPI_ISL_2081350, EPI_ISL_2081353 | | | |
| see above | Notre Dame COVID Response and Testing Laboratory | Notre Dame Genomics & Bioinformatics Core Facility | Michael Pfrender, Joseph Sarro, Melissa Stephens |
| EPI_ISL_2081356, EPI_ISL_2081357, EPI_ISL_2081358, EPI_ISL_2081359, EPI_ISL_2081360, EPI_ISL_2081361, EPI_ISL_2081362, EPI_ISL_2081363, EPI_ISL_2081364, EPI_ISL_2081365, EPI_ISL_2081366, EPI_ISL_2081367, EPI_ISL_2081368, EPI_ISL_2081369, EPI_ISL_2081370, EPI_ISL_2081371, EPI_ISL_2081372, EPI_ISL_2081373, EPI_ISL_2081374, EPI_ISL_2081375, EPI_ISL_2081376, EPI_ISL_2081377, EPI_ISL_2081378, EPI_ISL_2081379, EPI_ISL_2081380, EPI_ISL_2081381, EPI_ISL_2081382, EPI_ISL_2081383, EPI_ISL_2081384, EPI_ISL_2081385, EPI_ISL_2081386, EPI_ISL_2081387, EPI_ISL_2081388, EPI_ISL_2081389 | | | |
| see above | Tampa General Hospital Esoteric Lab | Tampa General Hospital Esoteric Research & Development Lab | Grant Vestal, Deanna Becker, Dominic Uy, Vicki Healer, Amorce Lima, Suzane Silbert |
| EPI_ISL_2081390, EPI_ISL_2081391, EPI_ISL_2081392, EPI_ISL_2081393, EPI_ISL_2081394, EPI_ISL_2081395, EPI_ISL_2081396, EPI_ISL_2081397, EPI_ISL_2081398, EPI_ISL_2081399, EPI_ISL_2081400, EPI_ISL_2081401, EPI_ISL_2081402, EPI_ISL_2081403, EPI_ISL_2081404, EPI_ISL_2081405, EPI_ISL_2081406, EPI_ISL_2081407, EPI_ISL_2081408, EPI_ISL_2081409, EPI_ISL_2081410, EPI_ISL_2081411, EPI_ISL_2081412, EPI_ISL_2081413, EPI_ISL_2081414, EPI_ISL_2081415, EPI_ISL_2081416, EPI_ISL_2081417, EPI_ISL_2081418, EPI_ISL_2081419, EPI_ISL_2081420, EPI_ISL_2081421, EPI_ISL_2081422, EPI_ISL_2081423, EPI_ISL_2081424, EPI_ISL_2081425, EPI_ISL_2081426, EPI_ISL_2081427, EPI_ISL_2081428, EPI_ISL_2081429, EPI_ISL_2081430, EPI_ISL_2081431, EPI_ISL_2081432, EPI_ISL_2081433, EPI_ISL_2081434 | | | |
| see above | MD PHL | MD PHL | Maryland Department of Health Laboratories Administration |
| EPI_ISL_2081435, EPI_ISL_2081436, EPI_ISL_2081437, EPI_ISL_2081438, EPI_ISL_2081439, EPI_ISL_2081440, EPI_ISL_2081441, EPI_ISL_2081442, EPI_ISL_2081443, EPI_ISL_2081444, EPI_ISL_2081448, EPI_ISL_2081449, EPI_ISL_2081450, EPI_ISL_2081453, EPI_ISL_2081454, EPI_ISL_2081456, EPI_ISL_2081457, EPI_ISL_2081458, EPI_ISL_2081459, EPI_ISL_2081462, EPI_ISL_2081463, EPI_ISL_2081464, EPI_ISL_2081465, EPI_ISL_2081466, EPI_ISL_2081468, EPI_ISL_2081470, EPI_ISL_2081473, EPI_ISL_2081474, EPI_ISL_2081475, EPI_ISL_2081476, EPI_ISL_2081477, EPI_ISL_2081478, EPI_ISL_2081479, EPI_ISL_2081480, EPI_ISL_2081481, EPI_ISL_2081482, EPI_ISL_2081483, EPI_ISL_2081486, EPI_ISL_2081487, EPI_ISL_2081488, EPI_ISL_2081490, EPI_ISL_2081491, EPI_ISL_2081492, EPI_ISL_2081493, EPI_ISL_2081494, EPI_ISL_2081495, EPI_ISL_2081498, EPI_ISL_2081499, EPI_ISL_2081500, EPI_ISL_2081501, EPI_ISL_2081502, EPI_ISL_2081503, EPI_ISL_2081505, EPI_ISL_2081506, EPI_ISL_2081507, EPI_ISL_2081508, EPI_ISL_2081509, EPI_ISL_2081510, EPI_ISL_2081511 | | | |
| see above | Virginia Division of Consolidated Laboratory Services | Virginia Division of Consolidated Laboratory Services | Virginia DCLS |
| EPI_ISL_2081512, EPI_ISL_2081513, EPI_ISL_2081514, EPI_ISL_2081515, EPI_ISL_2081516, EPI_ISL_2081517, EPI_ISL_2081518, EPI_ISL_2081519, EPI_ISL_2081520, EPI_ISL_2081521, EPI_ISL_2081522, EPI_ISL_2081523, EPI_ISL_2081524, EPI_ISL_2081525, EPI_ISL_2081526 | | | |
| see above | Oregon State Public Health Laboratory | Oregon State Public Health Laboratory | Rafia Razzaque, Eugene Yeboah, Vanda Makris, Laura Tsaknaridis, John Fontana and Shane Sevey |

| | | | |
|---|---|---|---|
| EPI_ISL_2081530, EPI_ISL_2081531, EPI_ISL_2081532, EPI_ISL_2081533, EPI_ISL_2081534, EPI_ISL_2081535, EPI_ISL_2081536, EPI_ISL_2081537, EPI_ISL_2081538, EPI_ISL_2081539, EPI_ISL_2081540, EPI_ISL_2081541, EPI_ISL_2081542, EPI_ISL_2081543, EPI_ISL_2081544, EPI_ISL_2081546, EPI_ISL_2081547, EPI_ISL_2081548, EPI_ISL_2081549, EPI_ISL_2081550, EPI_ISL_2081551, EPI_ISL_2081552, EPI_ISL_2081553, EPI_ISL_2081554, EPI_ISL_2081555, EPI_ISL_2081556, EPI_ISL_2081557, EPI_ISL_2081558, EPI_ISL_2081559, EPI_ISL_2081560, EPI_ISL_2081561, EPI_ISL_2081562, EPI_ISL_2081563, EPI_ISL_2081564, EPI_ISL_2081565, EPI_ISL_2081566, EPI_ISL_2081567, EPI_ISL_2081568, EPI_ISL_2081569, EPI_ISL_2081570, EPI_ISL_2081571, EPI_ISL_2081572, EPI_ISL_2081573, EPI_ISL_2081574, EPI_ISL_2081575, EPI_ISL_2081576, EPI_ISL_2081577, EPI_ISL_2081578, EPI_ISL_2081579, EPI_ISL_2081580, EPI_ISL_2081581, EPI_ISL_2081582, EPI_ISL_2081583, EPI_ISL_2081584, EPI_ISL_2081585, EPI_ISL_2081586, EPI_ISL_2081587, EPI_ISL_2081588, EPI_ISL_2081589, EPI_ISL_2081590, EPI_ISL_2081591, EPI_ISL_2081592, EPI_ISL_2081593, EPI_ISL_2081594, EPI_ISL_2081595, EPI_ISL_2081596, EPI_ISL_2081597, EPI_ISL_2081598, EPI_ISL_2081599, EPI_ISL_2081600, EPI_ISL_2081601, EPI_ISL_2081602, EPI_ISL_2081603, EPI_ISL_2081604, EPI_ISL_2081605, EPI_ISL_2081606, EPI_ISL_2081607, EPI_ISL_2081608, EPI_ISL_2081609, EPI_ISL_2081610, EPI_ISL_2081611, EPI_ISL_2081612, EPI_ISL_2081613 | | | |
| see above | UCSC Genomics Institute | UCSC Genomics Institute | Mark Akeson, Ikenna Anigbogu, Eric Beraut, Isabel Bjork, Molly Cassatt-Johnstone, Terren Chang, Russell Corbett-Detig, Namrita Dhillon, Maximilian Haeussler, David Haussler, Angie Hinrichs, Miten Jain, Joshua Kapp, A. Marm Kilpatrick, Jakob McBroome, Hugh Olsen, Jeremy Sanford, Beth Shapiro, Michael Stone, Bryan Thornlow, Yatish Turakhia, Ciara Wanket |
| EPI_ISL_2081685, EPI_ISL_2081686, EPI_ISL_2081687, EPI_ISL_2081688, EPI_ISL_2081691, EPI_ISL_2081692, EPI_ISL_2081693, EPI_ISL_2081694, EPI_ISL_2081695, EPI_ISL_2081696, EPI_ISL_2081697, EPI_ISL_2081698, EPI_ISL_2081699, EPI_ISL_2081700, EPI_ISL_2081701, EPI_ISL_2081702, EPI_ISL_2081703, EPI_ISL_2081704, EPI_ISL_2081705, EPI_ISL_2081706, EPI_ISL_2081707, EPI_ISL_2081708, EPI_ISL_2081709, EPI_ISL_2081710, EPI_ISL_2081711, EPI_ISL_2081712, EPI_ISL_2081713, EPI_ISL_2081714, EPI_ISL_2081715, EPI_ISL_2081717, EPI_ISL_2081718, EPI_ISL_2081719, EPI_ISL_2081720, EPI_ISL_2081721, EPI_ISL_2081722, EPI_ISL_2081723, EPI_ISL_2081724, EPI_ISL_2081726, EPI_ISL_2081727, EPI_ISL_2081728, EPI_ISL_2081729, EPI_ISL_2081730, EPI_ISL_2081731, EPI_ISL_2081732, EPI_ISL_2081733, EPI_ISL_2081734, EPI_ISL_2081735, EPI_ISL_2081738, EPI_ISL_2081739, EPI_ISL_2081740, EPI_ISL_2081741, EPI_ISL_2081742, EPI_ISL_2081743, EPI_ISL_2081744, EPI_ISL_2081745, EPI_ISL_2081746, EPI_ISL_2081747, EPI_ISL_2081748, EPI_ISL_2081749, EPI_ISL_2081750, EPI_ISL_2081751, EPI_ISL_2081752, EPI_ISL_2081753, EPI_ISL_2081754, EPI_ISL_2081755, EPI_ISL_2081756, EPI_ISL_2081757, EPI_ISL_2081758, EPI_ISL_2081759, EPI_ISL_2081760, EPI_ISL_2081761, EPI_ISL_2081763, EPI_ISL_2081764, EPI_ISL_2081765, EPI_ISL_2081766, EPI_ISL_2081767, EPI_ISL_2081768, EPI_ISL_2081769, EPI_ISL_2081770, EPI_ISL_2081771, EPI_ISL_2081772, EPI_ISL_2081773, EPI_ISL_2081774, EPI_ISL_2081775, EPI_ISL_2081776, EPI_ISL_2081777, EPI_ISL_2081778, EPI_ISL_2081779, EPI_ISL_2081780, EPI_ISL_2081781, EPI_ISL_2081782, EPI_ISL_2081783, EPI_ISL_2081784, EPI_ISL_2081786, EPI_ISL_2081787, EPI_ISL_2081788, EPI_ISL_2081789, EPI_ISL_2081790, EPI_ISL_2081791, EPI_ISL_2081792, EPI_ISL_2081793, EPI_ISL_2081794, EPI_ISL_2081795, EPI_ISL_2081796, EPI_ISL_2081797, EPI_ISL_2081798, EPI_ISL_2081799, EPI_ISL_2081800, EPI_ISL_2081801, EPI_ISL_2081802, EPI_ISL_2081803, EPI_ISL_2081804, EPI_ISL_2081805, EPI_ISL_2081806, EPI_ISL_2081807, EPI_ISL_2081808, EPI_ISL_2081809, EPI_ISL_2081810, EPI_ISL_2081811, EPI_ISL_2081812, EPI_ISL_2081813, EPI_ISL_2081814, EPI_ISL_2081815, EPI_ISL_2081816, EPI_ISL_2081817, EPI_ISL_2081818, EPI_ISL_2081819, EPI_ISL_2081820, EPI_ISL_2081821, EPI_ISL_2081822, EPI_ISL_2081823, EPI_ISL_2081824, EPI_ISL_2081825, EPI_ISL_2081827, EPI_ISL_2081828, EPI_ISL_2081829, EPI_ISL_2081830, EPI_ISL_2081831, EPI_ISL_2081832, EPI_ISL_2081833 | | | |
| see above | Utah Public Health Laboratory | Utah Public Health Laboratory | Erin L. Young, Kelly F. Oakeson, Tara Gallagher |
| EPI_ISL_2082475, EPI_ISL_2082476, EPI_ISL_2082477 | Arizona State Public Health Laboratory | Arizona State Public Health Laboratory | Trung Huynh, Jessica Escobar, Katherine Fullerton, Nobuko Fukushima, Stacy White, Linda Getsinger, Victor Waddell |
| EPI_ISL_2082481, EPI_ISL_2082482, EPI_ISL_2082483, EPI_ISL_2082484, EPI_ISL_2082485, EPI_ISL_2082486, EPI_ISL_2082487, EPI_ISL_2082488, EPI_ISL_2082489, EPI_ISL_2082490, EPI_ISL_2082491, EPI_ISL_2082492, EPI_ISL_2082493, EPI_ISL_2082494, EPI_ISL_2082495, EPI_ISL_2082496, EPI_ISL_2082497, EPI_ISL_2082498, EPI_ISL_2082499, EPI_ISL_2082500, EPI_ISL_2082501, EPI_ISL_2082502, EPI_ISL_2082503, EPI_ISL_2082504, EPI_ISL_2082505, EPI_ISL_2082506, EPI_ISL_2082507, EPI_ISL_2082508, EPI_ISL_2082509, EPI_ISL_2082510, EPI_ISL_2082511, EPI_ISL_2082512, EPI_ISL_2082513, EPI_ISL_2082514, EPI_ISL_2082515, EPI_ISL_2082516, EPI_ISL_2082517, EPI_ISL_2082518, EPI_ISL_2082519, EPI_ISL_2082520, EPI_ISL_2082521, EPI_ISL_2082522, EPI_ISL_2082523, EPI_ISL_2082524, EPI_ISL_2082525, EPI_ISL_2082526, EPI_ISL_2082527, EPI_ISL_2082528, EPI_ISL_2082529, EPI_ISL_2082530, EPI_ISL_2082531, EPI_ISL_2082532, EPI_ISL_2082533, EPI_ISL_2082534, EPI_ISL_2082535, EPI_ISL_2082536, EPI_ISL_2082537, EPI_ISL_2082538, EPI_ISL_2082539, EPI_ISL_2082540, EPI_ISL_2082541, EPI_ISL_2082542, EPI_ISL_2082543, EPI_ISL_2082544, EPI_ISL_2082545, EPI_ISL_2082546, EPI_ISL_2082547, EPI_ISL_2082548, EPI_ISL_2082549, EPI_ISL_2082550, EPI_ISL_2082551, EPI_ISL_2082552, EPI_ISL_2082553, EPI_ISL_2082554, EPI_ISL_2082555, EPI_ISL_2082556, EPI_ISL_2082557, EPI_ISL_2082558, EPI_ISL_2082559, EPI_ISL_2082560, EPI_ISL_2082561, EPI_ISL_2082562, EPI_ISL_2082563, EPI_ISL_2082564, EPI_ISL_2082565, EPI_ISL_2082566, EPI_ISL_2082567, EPI_ISL_2082568, EPI_ISL_2082569, EPI_ISL_2082570, EPI_ISL_2082571, EPI_ISL_2082572, EPI_ISL_2082573, EPI_ISL_2082575, EPI_ISL_2082576, EPI_ISL_2082577, EPI_ISL_2082578, EPI_ISL_2082579, EPI_ISL_2082580, EPI_ISL_2082581, EPI_ISL_2082582, EPI_ISL_2082583, EPI_ISL_2082584, EPI_ISL_2082585, EPI_ISL_2082586, EPI_ISL_2082587, EPI_ISL_2082588, EPI_ISL_2082589, EPI_ISL_2082590, EPI_ISL_2082591, EPI_ISL_2082592, EPI_ISL_2082593, EPI_ISL_2082594, EPI_ISL_2082595, EPI_ISL_2082596, EPI_ISL_2082597, EPI_ISL_2082598, EPI_ISL_2082599, EPI_ISL_2082600, EPI_ISL_2082601, EPI_ISL_2082602, EPI_ISL_2082603, EPI_ISL_2082604, EPI_ISL_2082605, EPI_ISL_2082606, EPI_ISL_2082607, EPI_ISL_2082608, EPI_ISL_2082609, EPI_ISL_2082610, EPI_ISL_2082611, EPI_ISL_2082612, EPI_ISL_2082613, EPI_ISL_2082614, EPI_ISL_2082615, EPI_ISL_2082616, EPI_ISL_2082617, EPI_ISL_2082618, EPI_ISL_2082619, EPI_ISL_2082620, EPI_ISL_2082621, EPI_ISL_2082622, EPI_ISL_2082623, EPI_ISL_2082624, EPI_ISL_2082625, EPI_ISL_2082626, EPI_ISL_2082627, EPI_ISL_2082628, EPI_ISL_2082629, EPI_ISL_2082630, EPI_ISL_2082631, EPI_ISL_2082632, EPI_ISL_2082633, EPI_ISL_2082634, EPI_ISL_2082635, EPI_ISL_2082636, EPI_ISL_2082637, EPI_ISL_2082638, EPI_ISL_2082639, EPI_ISL_2082640, EPI_ISL_2082641, EPI_ISL_2082642, EPI_ISL_2082643, EPI_ISL_2082644, EPI_ISL_2082645, EPI_ISL_2082646, EPI_ISL_2082647, EPI_ISL_2082648, EPI_ISL_2082649, EPI_ISL_2082650, EPI_ISL_2082651, EPI_ISL_2082652, EPI_ISL_2082653, EPI_ISL_2082654, EPI_ISL_2082655, EPI_ISL_2082656, EPI_ISL_2082657, EPI_ISL_2082659, EPI_ISL_2082660, EPI_ISL_2082661, EPI_ISL_2082662, EPI_ISL_2082663, EPI_ISL_2082664, EPI_ISL_2082665, EPI_ISL_2082666, EPI_ISL_2082667, EPI_ISL_2082668, EPI_ISL_2082669, EPI_ISL_2082671, EPI_ISL_2082672, EPI_ISL_2082673, EPI_ISL_2082674, EPI_ISL_2082675, EPI_ISL_2082676, EPI_ISL_2082677, EPI_ISL_2082678, EPI_ISL_2082679, EPI_ISL_2082680, EPI_ISL_2082681, EPI_ISL_2082682, EPI_ISL_2082683, EPI_ISL_2082685, EPI_ISL_2082686, EPI_ISL_2082687, EPI_ISL_2082688, EPI_ISL_2082689, EPI_ISL_2082690, EPI_ISL_2082691, EPI_ISL_2082692, EPI_ISL_2082694, EPI_ISL_2082695, EPI_ISL_2082696, EPI_ISL_2082697, EPI_ISL_2082698, EPI_ISL_2082699, EPI_ISL_2082700, EPI_ISL_2082701, EPI_ISL_2082702, EPI_ISL_2082703, EPI_ISL_2082704, EPI_ISL_2082705 | | | |
| see above | UW Virology Lab | UW Virology Lab | Pavitra Roychoudhury, Hong Xie, Lasata Shrestha, Tien V. Nguyen, Shah Mohamed Bakhash, Michelle Lin, Noah R. Baker, Sean Ellis, Meei-Li Huang, Keith R Jerome, Alexander Greninger |
| EPI_ISL_2082707, EPI_ISL_2082708, EPI_ISL_2082709, EPI_ISL_2082710, EPI_ISL_2082711, EPI_ISL_2082713, EPI_ISL_2082714, EPI_ISL_2082715, EPI_ISL_2082716, EPI_ISL_2082717, EPI_ISL_2082718, EPI_ISL_2082719, EPI_ISL_2082720, EPI_ISL_2082721, EPI_ISL_2082722, EPI_ISL_2082723, EPI_ISL_2082724, EPI_ISL_2082725, EPI_ISL_2082726, EPI_ISL_2082727, EPI_ISL_2082728 | | | |
| see above | Arizona State Public Health Laboratory | Arizona State Public Health Laboratory | Trung Huynh, Jessica Escobar, Katherine Fullerton, Nobuko Fukushima, Stacy White, Linda Getsinger, Victor Waddell |
| EPI_ISL_2082729 | Weill Cornell Medicine | New York Genome Center | Michael Zody, Andre Corvelo, Dayna M. Oschwald, Samantha Fennessey, Tom Maniatis, Melissa Cushing, Olivier Elemento, Margaret Elizabeth Ross, Chris Mason, Priya Velu, Hanna Rennert, Arryn Craney, Lars F Westblade |
| EPI_ISL_2082730, EPI_ISL_2082731, EPI_ISL_2082732, EPI_ISL_2082733, EPI_ISL_2082734, EPI_ISL_2082735, EPI_ISL_2082736, EPI_ISL_2082737, EPI_ISL_2082738, EPI_ISL_2082739, EPI_ISL_2082740, EPI_ISL_2082741, EPI_ISL_2082742, EPI_ISL_2082743, EPI_ISL_2082744, EPI_ISL_2082745, EPI_ISL_2082746, EPI_ISL_2082747, EPI_ISL_2082748, EPI_ISL_2082749, EPI_ISL_2082750, EPI_ISL_2082751, EPI_ISL_2082752, EPI_ISL_2082753, EPI_ISL_2082754, EPI_ISL_2082755, EPI_ISL_2082756, EPI_ISL_2082757, EPI_ISL_2082758, EPI_ISL_2082759, EPI_ISL_2082760, EPI_ISL_2082761, EPI_ISL_2082762, EPI_ISL_2082763, EPI_ISL_2082764, EPI_ISL_2082765, EPI_ISL_2082766, EPI_ISL_2082767, EPI_ISL_2082768, EPI_ISL_2082769, EPI_ISL_2082770, EPI_ISL_2082771, EPI_ISL_2082772, EPI_ISL_2082773, EPI_ISL_2082774, EPI_ISL_2082775, EPI_ISL_2082776, EPI_ISL_2082777, EPI_ISL_2082778, EPI_ISL_2082779, EPI_ISL_2082780, EPI_ISL_2082781, EPI_ISL_2082782, EPI_ISL_2082783, EPI_ISL_2082784, EPI_ISL_2082785, EPI_ISL_2082786, EPI_ISL_2082787, EPI_ISL_2082788, EPI_ISL_2082789, EPI_ISL_2082790, EPI_ISL_2082791, EPI_ISL_2082792, EPI_ISL_2082793, EPI_ISL_2082794, EPI_ISL_2082795, EPI_ISL_2082796, EPI_ISL_2082797, EPI_ISL_2082798, EPI_ISL_2082799, EPI_ISL_2082800, EPI_ISL_2082801, EPI_ISL_2082802, EPI_ISL_2082803, EPI_ISL_2082804, EPI_ISL_2082805, EPI_ISL_2082806, EPI_ISL_2082807, EPI_ISL_2082808, EPI_ISL_2082809, EPI_ISL_2082810, EPI_ISL_2082811, EPI_ISL_2082812, EPI_ISL_2082813, EPI_ISL_2082814, EPI_ISL_2082815, EPI_ISL_2082816, EPI_ISL_2082817, EPI_ISL_2082818, EPI_ISL_2082819, EPI_ISL_2082820, EPI_ISL_2082821, EPI_ISL_2082822, EPI_ISL_2082823, EPI_ISL_2082824, EPI_ISL_2082825, EPI_ISL_2082826, EPI_ISL_2082827, EPI_ISL_2082828, EPI_ISL_2082829, EPI_ISL_2082830, EPI_ISL_2082831, EPI_ISL_2082832, EPI_ISL_2082833, EPI_ISL_2082834, EPI_ISL_2082835, EPI_ISL_2082836, EPI_ISL_2082837, EPI_ISL_2082838, EPI_ISL_2082839, EPI_ISL_2082840, EPI_ISL_2082841, EPI_ISL_2082842, EPI_ISL_2082843, EPI_ISL_2082844, EPI_ISL_2082845, EPI_ISL_2082846, EPI_ISL_2082847, EPI_ISL_2082848, EPI_ISL_2082849, EPI_ISL_2082850, EPI_ISL_2082851, EPI_ISL_2082852, EPI_ISL_2082853, EPI_ISL_2082854, EPI_ISL_2082855, EPI_ISL_2082856, EPI_ISL_2082857, EPI_ISL_2082858, EPI_ISL_2082859, EPI_ISL_2082860, EPI_ISL_2082861, EPI_ISL_2082862, EPI_ISL_2082863, EPI_ISL_2082864, EPI_ISL_2082865, EPI_ISL_2082866, EPI_ISL_2082867, EPI_ISL_2082868, EPI_ISL_2082869, EPI_ISL_2082870, EPI_ISL_2082871, EPI_ISL_2082872, EPI_ISL_2082873, EPI_ISL_2082874, EPI_ISL_2082875, EPI_ISL_2082876, EPI_ISL_2082877, EPI_ISL_2082878, EPI_ISL_2082879, EPI_ISL_2082880, EPI_ISL_2082881, EPI_ISL_2082882, EPI_ISL_2082883, EPI_ISL_2082884, EPI_ISL_2082885, EPI_ISL_2082886, EPI_ISL_2082887, EPI_ISL_2082888, EPI_ISL_2082889, EPI_ISL_2082890, EPI_ISL_2082891, EPI_ISL_2082892, EPI_ISL_2082893, EPI_ISL_2082894, EPI_ISL_2082895, EPI_ISL_2082896, EPI_ISL_2082897, EPI_ISL_2082898, EPI_ISL_2082899, EPI_ISL_2082900, EPI_ISL_2082901, EPI_ISL_2082902 | | | |
| see above | Maryland Genomics, Institute for Genome Sciences, University of Maryland School of Medicine | Maryland Genomics, Institute for Genome Sciences, University of Maryland School of Medicine | Tallon, Luke J; Sadzewicz, Lisa D; Humphrys, Mike; Ott, Sandra; Roussey, Holly; Mehta, Aditya; Vavikolanu, Kranthi; Fraser, Claire M; Ravel, Jacques |
| EPI_ISL_2082903, EPI_ISL_2082905, EPI_ISL_2082906, EPI_ISL_2082907, EPI_ISL_2082908, EPI_ISL_2082909, EPI_ISL_2082910, EPI_ISL_2082911, EPI_ISL_2082912, EPI_ISL_2082913, EPI_ISL_2082914, EPI_ISL_2082915, EPI_ISL_2082916, EPI_ISL_2082917, EPI_ISL_2082918, EPI_ISL_2082919, EPI_ISL_2082920, EPI_ISL_2082921, EPI_ISL_2082922, EPI_ISL_2082923, EPI_ISL_2082924, EPI_ISL_2082925, EPI_ISL_2082926, EPI_ISL_2082927, EPI_ISL_2082928, EPI_ISL_2082929, EPI_ISL_2082930 | | | |
| see above | Arizona State Public Health Laboratory | Arizona State Public Health Laboratory | Trung Huynh, Jessica Escobar, Katherine Fullerton, Nobuko Fukushima, Stacy White, Linda Getsinger, Victor Waddell |
| EPI_ISL_2082937, EPI_ISL_2082938, EPI_ISL_2082939, EPI_ISL_2082940, EPI_ISL_2082941, EPI_ISL_2082942, EPI_ISL_2082943, EPI_ISL_2082945, EPI_ISL_2082946, EPI_ISL_2082947, EPI_ISL_2082948, EPI_ISL_2082949, EPI_ISL_2082950, EPI_ISL_2082951, EPI_ISL_2082952, EPI_ISL_2082953, EPI_ISL_2082954, EPI_ISL_2082955, EPI_ISL_2082956, EPI_ISL_2082957, EPI_ISL_2082958, EPI_ISL_2082959, EPI_ISL_2082960, EPI_ISL_2082961, EPI_ISL_2082962, EPI_ISL_2082963, EPI_ISL_2082965, EPI_ISL_2082966, EPI_ISL_2082967, EPI_ISL_2082968, EPI_ISL_2082969, EPI_ISL_2082970, EPI_ISL_2082971, EPI_ISL_2082972, EPI_ISL_2082973, EPI_ISL_2082974, EPI_ISL_2082975, EPI_ISL_2082976, EPI_ISL_2082977, EPI_ISL_2082978, EPI_ISL_2082979, EPI_ISL_2082980, EPI_ISL_2082982, EPI_ISL_2082983, EPI_ISL_2082984, EPI_ISL_2082985, EPI_ISL_2082986, EPI_ISL_2082987, EPI_ISL_2082988, EPI_ISL_2082989, EPI_ISL_2082990, EPI_ISL_2082991, EPI_ISL_2082992, EPI_ISL_2082993, EPI_ISL_2082994, EPI_ISL_2082996, EPI_ISL_2082997, EPI_ISL_2082998, EPI_ISL_2082999, EPI_ISL_2083000, EPI_ISL_2083001, EPI_ISL_2083002, EPI_ISL_2083003, EPI_ISL_2083004, EPI_ISL_2083005, EPI_ISL_2083006, EPI_ISL_2083007, EPI_ISL_2083008, EPI_ISL_2083011, EPI_ISL_2083012, EPI_ISL_2083013, EPI_ISL_2083014, EPI_ISL_2083015 | | | |
| see above | UCSC Genomics Institute | UCSC Genomics Institute | Mark Akeson, Ikenna Anigbogu, Eric Beraut, Isabel Bjork, Molly Cassatt-Johnstone, Terren Chang, Russell Corbett-Detig, Namrita Dhillon, Maximilian Haeussler, David Haussler, Angie Hinrichs, Miten Jain, Joshua Kapp, A. Marm Kilpatrick, Jakob McBroome, Hugh Olsen, Jeremy Sanford, Beth Shapiro, Michael Stone, Bryan Thornlow, Yatish Turakhia, Ciara Wanket |
| EPI_ISL_2083016, EPI_ISL_2083017, EPI_ISL_2083018, EPI_ISL_2083019, EPI_ISL_2083020, EPI_ISL_2083021, EPI_ISL_2083022, EPI_ISL_2083025, EPI_ISL_2083026, EPI_ISL_2083027, EPI_ISL_2083028, EPI_ISL_2083029, EPI_ISL_2083030, EPI_ISL_2083031, EPI_ISL_2083032 | | | |
| see above | Baylor Scott & White-Temple | Baylor Scott & White-Temple | Ari Rao, Linden Morales, Kimberly Walker, Marcus Volz, Shelby Hendrickson |
| EPI_ISL_2083033 | Baylor Scott & White-Grapevine | Baylor Scott & White-Temple | Ari Rao, Linden Morales, Kimberly Walker, Marcus Volz, Shelby Hendrickson |
| EPI_ISL_2083034, EPI_ISL_2083035, EPI_ISL_2083036, EPI_ISL_2083037, EPI_ISL_2083038, EPI_ISL_2083039, EPI_ISL_2083040, EPI_ISL_2083041, EPI_ISL_2083042, EPI_ISL_2083043, EPI_ISL_2083044, EPI_ISL_2083045, EPI_ISL_2083046, EPI_ISL_2083047, EPI_ISL_2083048, EPI_ISL_2083049, EPI_ISL_2083050, EPI_ISL_2083051, EPI_ISL_2083052, EPI_ISL_2083053, EPI_ISL_2083055, EPI_ISL_2083056, EPI_ISL_2083057, EPI_ISL_2083058, EPI_ISL_2083059, EPI_ISL_2083060, EPI_ISL_2083061, EPI_ISL_2083062, EPI_ISL_2083063, EPI_ISL_2083064, EPI_ISL_2083065, EPI_ISL_2083066, EPI_ISL_2083067, EPI_ISL_2083068, EPI_ISL_2083069, EPI_ISL_2083070, EPI_ISL_2083071, EPI_ISL_2083072, EPI_ISL_2083073, EPI_ISL_2083074, EPI_ISL_2083076, EPI_ISL_2083077, EPI_ISL_2083078, EPI_ISL_2083079, EPI_ISL_2083080, EPI_ISL_2083081, EPI_ISL_2083082, EPI_ISL_2083083, EPI_ISL_2083084, EPI_ISL_2083085, EPI_ISL_2083086 | | | |
| see above | Baylor Scott & White-Temple | Baylor Scott & White-Temple | Ari Rao, Linden Morales, Kimberly Walker, Marcus Volz, Shelby Hendrickson |
| EPI_ISL_2086119, EPI_ISL_2086120, EPI_ISL_2086161 | The Ohio State University Applied Microbiology Services Laboratory | The Ohio State University Applied Microbiology Services Laboratory | Seth A. Faith PhD |
| EPI_ISL_2086248, EPI_ISL_2086249, EPI_ISL_2086250, EPI_ISL_2086251, EPI_ISL_2086252, EPI_ISL_2086253, EPI_ISL_2086254, EPI_ISL_2086255, EPI_ISL_2086256, EPI_ISL_2086257, EPI_ISL_2086258, EPI_ISL_2086259, EPI_ISL_2086260, EPI_ISL_2086261, EPI_ISL_2086263, EPI_ISL_2086264, EPI_ISL_2086265 | | | |
| see above | Kansas Health and Environmental Lab | Kansas Health and Environmental Lab | Mike Grose, Jonathan Barnell, Ben Olsen, and Phil Adam |
| EPI_ISL_2086266 | Weill Cornell Medicine | New York Genome Center | Michael Zody, Andre Corvelo, Dayna M. Oschwald, Samantha Fennessey, Tom Maniatis, Melissa Cushing, Olivier Elemento, Margaret Elizabeth Ross, Chris Mason, Priya Velu, Hanna Rennert, Arryn Craney, Lars F Westblade |
| EPI_ISL_2086269, EPI_ISL_2086270, EPI_ISL_2086271, EPI_ISL_2086272, EPI_ISL_2086273, EPI_ISL_2086274, EPI_ISL_2086275, EPI_ISL_2086276, EPI_ISL_2086277, EPI_ISL_2086278, EPI_ISL_2086279, EPI_ISL_2086280, EPI_ISL_2086281, EPI_ISL_2086282, EPI_ISL_2086283, EPI_ISL_2086284, EPI_ISL_2086285 | | | |
| see above | Hackensack Medical Center | New York Genome Center | Michael Zody, Andre Corvelo, Dayna M. Oschwald, Samantha Fennessey, Tom Maniatis, Liang Chen, Jose Mediavilla, Marcus Cunningham, Kaelea Composto, Kar Chow, David Perlin, Barry Kreiswirth |
| EPI_ISL_2086298, EPI_ISL_2086299 | Arizona State Public Health Laboratory | Arizona State Public Health Laboratory | Trung Huynh, Jessica Escobar, Katherine Fullerton, Nobuko Fukushima, Stacy White, Linda Getsinger, Victor Waddell |
| EPI_ISL_2086300, EPI_ISL_2086301, EPI_ISL_2086302, EPI_ISL_2086303 | Weill Cornell Medicine | New York Genome Center | Michael Zody, Andre Corvelo, Dayna M. Oschwald, Samantha Fennessey, Tom Maniatis, Melissa Cushing, Olivier Elemento, Margaret Elizabeth Ross, Chris Mason, Priya Velu, Hanna Rennert, Arryn Craney, Lars F Westblade |
| EPI_ISL_2086307, EPI_ISL_2086311, EPI_ISL_2086312 | Arizona State Public Health Laboratory | Arizona State Public Health Laboratory | Trung Huynh, Jessica Escobar, Katherine Fullerton, Nobuko Fukushima, Stacy White, Linda Getsinger, Victor Waddell |
| EPI_ISL_2086316, EPI_ISL_2086317, EPI_ISL_2086318 | MD PHL | MD PHL | Maryland Department of Health Laboratories Administration |
| EPI_ISL_2086320, EPI_ISL_2086321, EPI_ISL_2086322 | Arizona State Public Health Laboratory | Arizona State Public Health Laboratory | Trung Huynh, Jessica Escobar, Katherine Fullerton, Nobuko Fukushima, Stacy White, Linda Getsinger, Victor Waddell |
| EPI_ISL_2086323, EPI_ISL_2086324, EPI_ISL_2086325 | IN State Department of Health Laboratory Services | IN State Department of Health Laboratory Services | Cassandra Campion, Jamie Yeadon, Brian Pope, Lixia Liu, Kyle Brownlee, Melissa Hindenlang, Mark Glazier |
| EPI_ISL_2086326, EPI_ISL_2086327, EPI_ISL_2086328, EPI_ISL_2086329, EPI_ISL_2086330, EPI_ISL_2086331, EPI_ISL_2086332, EPI_ISL_2086333, EPI_ISL_2086334, EPI_ISL_2086335, EPI_ISL_2086336, EPI_ISL_2086337, EPI_ISL_2086338, EPI_ISL_2086339, EPI_ISL_2086340, EPI_ISL_2086341, EPI_ISL_2086342, EPI_ISL_2086343, EPI_ISL_2086344, EPI_ISL_2086345, EPI_ISL_2086346, EPI_ISL_2086347, EPI_ISL_2086348, EPI_ISL_2086349, EPI_ISL_2086350, EPI_ISL_2086351, EPI_ISL_2086352, EPI_ISL_2086353, EPI_ISL_2086354, EPI_ISL_2086355, EPI_ISL_2086356, EPI_ISL_2086357, EPI_ISL_2086358, EPI_ISL_2086359, EPI_ISL_2086360, EPI_ISL_2086361, EPI_ISL_2086362, EPI_ISL_2086363, EPI_ISL_2086364, EPI_ISL_2086365, EPI_ISL_2086366, EPI_ISL_2086367, EPI_ISL_2086368, EPI_ISL_2086369, EPI_ISL_2086370, EPI_ISL_2086371, EPI_ISL_2086372, EPI_ISL_2086373, EPI_ISL_2086374, EPI_ISL_2086375, EPI_ISL_2086376, EPI_ISL_2086377, EPI_ISL_2086378, EPI_ISL_2086379, EPI_ISL_2086380, EPI_ISL_2086381, EPI_ISL_2086382, EPI_ISL_2086383, EPI_ISL_2086384, EPI_ISL_2086385, EPI_ISL_2086386, EPI_ISL_2086387, EPI_ISL_2086388, EPI_ISL_2086389, EPI_ISL_2086390, EPI_ISL_2086391, EPI_ISL_2086392, EPI_ISL_2086393, EPI_ISL_2086394, EPI_ISL_2086395, EPI_ISL_2086396, EPI_ISL_2086397, EPI_ISL_2086398, EPI_ISL_2086399, EPI_ISL_2086400, EPI_ISL_2086401, EPI_ISL_2086402, EPI_ISL_2086403, EPI_ISL_2086404, EPI_ISL_2086405, EPI_ISL_2086406, EPI_ISL_2086407, EPI_ISL_2086408, EPI_ISL_2086409, EPI_ISL_2086411, EPI_ISL_2086412, EPI_ISL_2086413, EPI_ISL_2086414, EPI_ISL_2086415, EPI_ISL_2086416, EPI_ISL_2086417, EPI_ISL_2086418, EPI_ISL_2086419, EPI_ISL_2086420, EPI_ISL_2086421, EPI_ISL_2086422 | | | |
| see above | UW Virology Lab | UW Virology Lab | Pavitra Roychoudhury, Hong Xie, Lasata Shrestha, Tien V. Nguyen, Shah Mohamed Bakhash, Michelle Lin, Noah R. Baker, Ricardo Perez, Sean Ellis, Nathan Breit, Robert J. Livingston, Meei-Li Huang, Keith R Jerome, Patrick Mathias, Alexander Greninger |
| EPI_ISL_2086423, EPI_ISL_2086424, EPI_ISL_2086425, EPI_ISL_2086426, EPI_ISL_2086427, EPI_ISL_2086428, EPI_ISL_2086429, EPI_ISL_2086430, EPI_ISL_2086431, EPI_ISL_2086432, EPI_ISL_2086433, EPI_ISL_2086435, EPI_ISL_2086436, EPI_ISL_2086437, EPI_ISL_2086438, EPI_ISL_2086439, EPI_ISL_2086440, EPI_ISL_2086442, EPI_ISL_2086443, EPI_ISL_2086445, EPI_ISL_2086447, EPI_ISL_2086448, EPI_ISL_2086451 | | | |
| see above | Wichita State University - Molecular Diagnostics Lab | Kansas Health and Environmental Lab | Mike Grose, Jonathan Barnell, Ben Olsen, and Phil Adam |

| | | | |
|---|---|---|---|
| EPI_ISL_2086454 | IN State Department of Health Laboratory Services | IN State Department of Health Laboratory Services | Cassandra Campion, Jamie Yeadon, Brian Pope, Lixia Liu, Kyle Brownlee, Melissa Hindenlang, Mark Glazier |
| EPI_ISL_2086457, EPI_ISL_2086458, EPI_ISL_2086459, EPI_ISL_2086461, EPI_ISL_2086462, EPI_ISL_2086464, EPI_ISL_2086466, EPI_ISL_2086467, EPI_ISL_2086468, EPI_ISL_2086469, EPI_ISL_2086471, EPI_ISL_2086472, EPI_ISL_2086473, EPI_ISL_2086474, EPI_ISL_2086475, EPI_ISL_2086476, EPI_ISL_2086477, EPI_ISL_2086478, EPI_ISL_2086479, EPI_ISL_2086480, EPI_ISL_2086481, EPI_ISL_2086482, EPI_ISL_2086483, EPI_ISL_2086484, EPI_ISL_2086485, EPI_ISL_2086486, EPI_ISL_2086487, EPI_ISL_2086489, EPI_ISL_2086490 | | | |
| see above | North Dakota Department of Health, Public Health Laboratory | North Dakota Department of Health, Public Health Laboratory | Lisa Wingerter |
| EPI_ISL_2086491, EPI_ISL_2086492, EPI_ISL_2086493, EPI_ISL_2086494, EPI_ISL_2086495, EPI_ISL_2086496, EPI_ISL_2086497, EPI_ISL_2086498, EPI_ISL_2086499, EPI_ISL_2086500, EPI_ISL_2086501, EPI_ISL_2086502, EPI_ISL_2086503, EPI_ISL_2086504, EPI_ISL_2086505, EPI_ISL_2086506, EPI_ISL_2086507, EPI_ISL_2086508, EPI_ISL_2086509, EPI_ISL_2086510, EPI_ISL_2086511, EPI_ISL_2086512, EPI_ISL_2086513, EPI_ISL_2086514, EPI_ISL_2086515, EPI_ISL_2086516, EPI_ISL_2086517, EPI_ISL_2086518, EPI_ISL_2086520, EPI_ISL_2086521, EPI_ISL_2086522, EPI_ISL_2086523, EPI_ISL_2086524, EPI_ISL_2086525, EPI_ISL_2086526, EPI_ISL_2086527, EPI_ISL_2086528, EPI_ISL_2086529, EPI_ISL_2086530, EPI_ISL_2086531, EPI_ISL_2086532, EPI_ISL_2086533, EPI_ISL_2086534, EPI_ISL_2086535, EPI_ISL_2086536, EPI_ISL_2086537, EPI_ISL_2086538, EPI_ISL_2086539, EPI_ISL_2086540, EPI_ISL_2086541, EPI_ISL_2086542, EPI_ISL_2086543, EPI_ISL_2086544, EPI_ISL_2086545, EPI_ISL_2086546, EPI_ISL_2086547, EPI_ISL_2086548, EPI_ISL_2086550, EPI_ISL_2086551, EPI_ISL_2086552, EPI_ISL_2086553, EPI_ISL_2086554, EPI_ISL_2086555, EPI_ISL_2086556, EPI_ISL_2086557, EPI_ISL_2086558, EPI_ISL_2086559, EPI_ISL_2086560, EPI_ISL_2086561, EPI_ISL_2086562, EPI_ISL_2086563, EPI_ISL_2086564, EPI_ISL_2086566, EPI_ISL_2086567, EPI_ISL_2086568, EPI_ISL_2086569, EPI_ISL_2086570, EPI_ISL_2086571, EPI_ISL_2086572, EPI_ISL_2086573, EPI_ISL_2086574, EPI_ISL_2086575, EPI_ISL_2086576, EPI_ISL_2086577, EPI_ISL_2086578, EPI_ISL_2086579, EPI_ISL_2086580, EPI_ISL_2086581, EPI_ISL_2086582, EPI_ISL_2086583, EPI_ISL_2086584 | | | |
| see above | Illinois Department of Public Health - Springfield Lab | Illinois Department of Public Health - Springfield Lab | Bryan Sim, Gordon McCall |
| EPI_ISL_2086677, EPI_ISL_2086678, EPI_ISL_2086679, EPI_ISL_2086680, EPI_ISL_2086681, EPI_ISL_2086682, EPI_ISL_2086683, EPI_ISL_2086684, EPI_ISL_2086685, EPI_ISL_2086686, EPI_ISL_2086687, EPI_ISL_2086688, EPI_ISL_2086689, EPI_ISL_2086690, EPI_ISL_2086691, EPI_ISL_2086692, EPI_ISL_2086693, EPI_ISL_2086694, EPI_ISL_2086695, EPI_ISL_2086696, EPI_ISL_2086697 | | | |
| see above | Oregon State Public Health Laboratory | Oregon State Public Health Laboratory | Rafia Razzaque, Eugene Yeboah, Vanda Makris, Laura Tsaknaridis, John Fontana and Shane Sevey |
| EPI_ISL_2086768, EPI_ISL_2086769, EPI_ISL_2086770, EPI_ISL_2086771, EPI_ISL_2086774, EPI_ISL_2086775, EPI_ISL_2086776, EPI_ISL_2086777, EPI_ISL_2086778, EPI_ISL_2086779, EPI_ISL_2086780, EPI_ISL_2086781, EPI_ISL_2086782, EPI_ISL_2086783 | | | |
| see above | Atlanta VA Medical Center | Pathogen Discovery, Respiratory Viruses Branch, Division of Viral Diseases, Centers for Disease Control and Prevention | Ying Tao, Yan Li, Jing Zhang, Anna Kelleher, Brian Lynch, Krista Queen, Anna Uehara, Peter Cook, Han Jia Justin Ng, Rachel Marine, Clinton R. Paden, Dhwani Batra, Haibin Wang, Tara Coalter, Jasmine Padilla, Morgan Davis, Mili Sheth, Sarah Nobles, Mark Burroughs, Justin Lee, Adam Retchless, Suxiang Tong |
| EPI_ISL_2087048, EPI_ISL_2087049, EPI_ISL_2087050 | Kansas Health and Environmental Lab | Kansas Health and Environmental Lab | Mike Grose, Jonathan Barnell, Ben Olsen, and Phil Adam |
| EPI_ISL_2087053, EPI_ISL_2087054, EPI_ISL_2087055, EPI_ISL_2087056, EPI_ISL_2087057, EPI_ISL_2087058, EPI_ISL_2087059, EPI_ISL_2087060, EPI_ISL_2087061, EPI_ISL_2087062, EPI_ISL_2087063, EPI_ISL_2087064, EPI_ISL_2087065, EPI_ISL_2087066, EPI_ISL_2087067, EPI_ISL_2087068, EPI_ISL_2087069, EPI_ISL_2087070, EPI_ISL_2087071, EPI_ISL_2087072, EPI_ISL_2087073, EPI_ISL_2087074, EPI_ISL_2087075, EPI_ISL_2087076, EPI_ISL_2087077, EPI_ISL_2087078, EPI_ISL_2087079, EPI_ISL_2087080, EPI_ISL_2087081, EPI_ISL_2087082, EPI_ISL_2087083, EPI_ISL_2087084, EPI_ISL_2087085, EPI_ISL_2087086, EPI_ISL_2087087, EPI_ISL_2087088, EPI_ISL_2087089, EPI_ISL_2087090, EPI_ISL_2087091, EPI_ISL_2087092, EPI_ISL_2087093, EPI_ISL_2087094, EPI_ISL_2087095, EPI_ISL_2087096, EPI_ISL_2087097, EPI_ISL_2087098, EPI_ISL_2087099, EPI_ISL_2087100, EPI_ISL_2087101, EPI_ISL_2087102, EPI_ISL_2087103, EPI_ISL_2087104, EPI_ISL_2087105, EPI_ISL_2087106, EPI_ISL_2087107, EPI_ISL_2087108, EPI_ISL_2087109, EPI_ISL_2087110, EPI_ISL_2087111, EPI_ISL_2087112, EPI_ISL_2087113, EPI_ISL_2087114, EPI_ISL_2087115, EPI_ISL_2087116, EPI_ISL_2087117, EPI_ISL_2087118, EPI_ISL_2087119, EPI_ISL_2087120, EPI_ISL_2087121, EPI_ISL_2087122, EPI_ISL_2087123, EPI_ISL_2087124, EPI_ISL_2087125, EPI_ISL_2087126, EPI_ISL_2087127, EPI_ISL_2087128, EPI_ISL_2087129, EPI_ISL_2087130, EPI_ISL_2087131, EPI_ISL_2087132, EPI_ISL_2087133, EPI_ISL_2087134, EPI_ISL_2087135, EPI_ISL_2087136, EPI_ISL_2087137, EPI_ISL_2087138, EPI_ISL_2087139, EPI_ISL_2087140, EPI_ISL_2087141, EPI_ISL_2087142, EPI_ISL_2087143, EPI_ISL_2087144, EPI_ISL_2087145, EPI_ISL_2087146, EPI_ISL_2087147, EPI_ISL_2087148, EPI_ISL_2087149, EPI_ISL_2087150, EPI_ISL_2087151, EPI_ISL_2087152, EPI_ISL_2087153, EPI_ISL_2087154, EPI_ISL_2087155, EPI_ISL_2087156, EPI_ISL_2087157, EPI_ISL_2087158, EPI_ISL_2087159, EPI_ISL_2087160, EPI_ISL_2087161, EPI_ISL_2087162, EPI_ISL_2087163, EPI_ISL_2087164, EPI_ISL_2087165, EPI_ISL_2087166, EPI_ISL_2087167, EPI_ISL_2087168, EPI_ISL_2087169, EPI_ISL_2087170, EPI_ISL_2087171, EPI_ISL_2087172, EPI_ISL_2087174, EPI_ISL_2087175, EPI_ISL_2087176, EPI_ISL_2087177, EPI_ISL_2087178, EPI_ISL_2087179, EPI_ISL_2087180, EPI_ISL_2087181, EPI_ISL_2087182, EPI_ISL_2087183, EPI_ISL_2087184, EPI_ISL_2087185, EPI_ISL_2087186, EPI_ISL_2087187, EPI_ISL_2087188, EPI_ISL_2087189, EPI_ISL_2087190, EPI_ISL_2087191, EPI_ISL_2087192, EPI_ISL_2087193, EPI_ISL_2087194, EPI_ISL_2087195, EPI_ISL_2087196, EPI_ISL_2087197, EPI_ISL_2087198, EPI_ISL_2087199, EPI_ISL_2087200, EPI_ISL_2087201, EPI_ISL_2087202, EPI_ISL_2087203, EPI_ISL_2087204, EPI_ISL_2087205, EPI_ISL_2087206, EPI_ISL_2087207, EPI_ISL_2087208, EPI_ISL_2087209, EPI_ISL_2087210, EPI_ISL_2087211, EPI_ISL_2087212, EPI_ISL_2087213, EPI_ISL_2087214, EPI_ISL_2087215, EPI_ISL_2087216, EPI_ISL_2087217, EPI_ISL_2087218, EPI_ISL_2087219, EPI_ISL_2087220, EPI_ISL_2087221, EPI_ISL_2087222, EPI_ISL_2087223, EPI_ISL_2087224, EPI_ISL_2087225, EPI_ISL_2087226, EPI_ISL_2087227, EPI_ISL_2087228, EPI_ISL_2087229, EPI_ISL_2087230, EPI_ISL_2087231, EPI_ISL_2087232, EPI_ISL_2087233, EPI_ISL_2087234, EPI_ISL_2087235, EPI_ISL_2087236, EPI_ISL_2087237, EPI_ISL_2087238, EPI_ISL_2087239, EPI_ISL_2087240, EPI_ISL_2087241, EPI_ISL_2087242, EPI_ISL_2087243, EPI_ISL_2087244, EPI_ISL_2087245, EPI_ISL_2087246, EPI_ISL_2087247, EPI_ISL_2087248, EPI_ISL_2087249, EPI_ISL_2087250, EPI_ISL_2087251, EPI_ISL_2087252, EPI_ISL_2087253, EPI_ISL_2087254, EPI_ISL_2087255, EPI_ISL_2087256, EPI_ISL_2087257, EPI_ISL_2087258, EPI_ISL_2087259, EPI_ISL_2087260, EPI_ISL_2087261, EPI_ISL_2087262, EPI_ISL_2087263, EPI_ISL_2087264, EPI_ISL_2087265, EPI_ISL_2087266, EPI_ISL_2087267, EPI_ISL_2087268, EPI_ISL_2087269, EPI_ISL_2087270, EPI_ISL_2087271, EPI_ISL_2087272, EPI_ISL_2087273, EPI_ISL_2087274, EPI_ISL_2087275, EPI_ISL_2087276, EPI_ISL_2087277, EPI_ISL_2087278, EPI_ISL_2087279, EPI_ISL_2087280, EPI_ISL_2087281, EPI_ISL_2087282, EPI_ISL_2087283, EPI_ISL_2087284, EPI_ISL_2087285, EPI_ISL_2087286, EPI_ISL_2087287, EPI_ISL_2087288, EPI_ISL_2087289, EPI_ISL_2087290, EPI_ISL_2087291, EPI_ISL_2087292, EPI_ISL_2087293, EPI_ISL_2087294, EPI_ISL_2087295, EPI_ISL_2087296, EPI_ISL_2087297, EPI_ISL_2087298, EPI_ISL_2087299, EPI_ISL_2087300, EPI_ISL_2087301, EPI_ISL_2087302, EPI_ISL_2087303, EPI_ISL_2087304, EPI_ISL_2087305, EPI_ISL_2087306, EPI_ISL_2087307, EPI_ISL_2087308, EPI_ISL_2087309, EPI_ISL_2087310, EPI_ISL_2087311, EPI_ISL_2087312, EPI_ISL_2087313, EPI_ISL_2087314, EPI_ISL_2087315, EPI_ISL_2087316, EPI_ISL_2087317, EPI_ISL_2087318, EPI_ISL_2087319, EPI_ISL_2087320, EPI_ISL_2087321, EPI_ISL_2087322, EPI_ISL_2087323, EPI_ISL_2087324, EPI_ISL_2087325, EPI_ISL_2087326, EPI_ISL_2087327, EPI_ISL_2087328, EPI_ISL_2087329, EPI_ISL_2087330, EPI_ISL_2087331, EPI_ISL_2087332, EPI_ISL_2087333, EPI_ISL_2087334, EPI_ISL_2087335, EPI_ISL_2087336, EPI_ISL_2087337, EPI_ISL_2087338, EPI_ISL_2087339, EPI_ISL_2087340, EPI_ISL_2087341, EPI_ISL_2087342, EPI_ISL_2087343, EPI_ISL_2087344, EPI_ISL_2087345, EPI_ISL_2087346, EPI_ISL_2087347, EPI_ISL_2087348, EPI_ISL_2087349, EPI_ISL_2087350, EPI_ISL_2087351, EPI_ISL_2087352, EPI_ISL_2087353, EPI_ISL_2087354, EPI_ISL_2087355, EPI_ISL_2087356, EPI_ISL_2087357, EPI_ISL_2087358, EPI_ISL_2087359, EPI_ISL_2087360, EPI_ISL_2087361, EPI_ISL_2087362, EPI_ISL_2087363, EPI_ISL_2087364, EPI_ISL_2087365, EPI_ISL_2087366, EPI_ISL_2087367, EPI_ISL_2087368, EPI_ISL_2087369, EPI_ISL_2087370, EPI_ISL_2087371, EPI_ISL_2087372, EPI_ISL_2087373, EPI_ISL_2087374, EPI_ISL_2087375, EPI_ISL_2087376, EPI_ISL_2087377, EPI_ISL_2087378, EPI_ISL_2087379, EPI_ISL_2087380, EPI_ISL_2087381, EPI_ISL_2087382, EPI_ISL_2087383, EPI_ISL_2087384, EPI_ISL_2087385, EPI_ISL_2087386, EPI_ISL_2087387, EPI_ISL_2087388, EPI_ISL_2087389, EPI_ISL_2087390, EPI_ISL_2087391, EPI_ISL_2087392, EPI_ISL_2087393, EPI_ISL_2087394, EPI_ISL_2087395, EPI_ISL_2087396, EPI_ISL_2087397, EPI_ISL_2087398, EPI_ISL_2087399, EPI_ISL_2087400, EPI_ISL_2087401, EPI_ISL_2087402, EPI_ISL_2087403, EPI_ISL_2087404, EPI_ISL_2087405, EPI_ISL_2087406, EPI_ISL_2087407, EPI_ISL_2087408, EPI_ISL_2087409, EPI_ISL_2087410, EPI_ISL_2087411, EPI_ISL_2087412, EPI_ISL_2087413, EPI_ISL_2087414, EPI_ISL_2087415, EPI_ISL_2087416, EPI_ISL_2087417, EPI_ISL_2087418, EPI_ISL_2087419, EPI_ISL_2087420, EPI_ISL_2087421, EPI_ISL_2087422, EPI_ISL_2087423, EPI_ISL_2087424, EPI_ISL_2087425, EPI_ISL_2087426, EPI_ISL_2087427, EPI_ISL_2087428, EPI_ISL_2087429, EPI_ISL_2087430, EPI_ISL_2087431, EPI_ISL_2087432, EPI_ISL_2087433, EPI_ISL_2087434, EPI_ISL_2087435, EPI_ISL_2087436, EPI_ISL_2087437, EPI_ISL_2087438, EPI_ISL_2087439, EPI_ISL_2087440, EPI_ISL_2087441, EPI_ISL_2087442, EPI_ISL_2087443, EPI_ISL_2087444, EPI_ISL_2087445, EPI_ISL_2087446, EPI_ISL_2087447, EPI_ISL_2087448, EPI_ISL_2087449, EPI_ISL_2087450, EPI_ISL_2087451, EPI_ISL_2087452, EPI_ISL_2087453, EPI_ISL_2087454, EPI_ISL_2087455, EPI_ISL_2087456, EPI_ISL_2087457, EPI_ISL_2087458, EPI_ISL_2087459, EPI_ISL_2087460, EPI_ISL_2087461, EPI_ISL_2087462, EPI_ISL_2087463, EPI_ISL_2087464, EPI_ISL_2087465, EPI_ISL_2087466, EPI_ISL_2087467, EPI_ISL_2087468, EPI_ISL_2087469, EPI_ISL_2087470, EPI_ISL_2087471, EPI_ISL_2087472, EPI_ISL_2087473, EPI_ISL_2087474, EPI_ISL_2087475, EPI_ISL_2087476, EPI_ISL_2087477, EPI_ISL_2087478, EPI_ISL_2087479, EPI_ISL_2087480, EPI_ISL_2087481, EPI_ISL_2087482, EPI_ISL_2087483, EPI_ISL_2087484, EPI_ISL_2087485, EPI_ISL_2087486, EPI_ISL_2087487, EPI_ISL_2087488, EPI_ISL_2087489, EPI_ISL_2087490, EPI_ISL_2087491, EPI_ISL_2087492, EPI_ISL_2087493, EPI_ISL_2087494, EPI_ISL_2087495, EPI_ISL_2087496, EPI_ISL_2087497, EPI_ISL_2087498, EPI_ISL_2087499, EPI_ISL_2087500, EPI_ISL_2087501, EPI_ISL_2087502, EPI_ISL_2087503, EPI_ISL_2087504, EPI_ISL_2087505, EPI_ISL_2087506, EPI_ISL_2087507, EPI_ISL_2087508, EPI_ISL_2087509, EPI_ISL_2087510, EPI_ISL_2087511, EPI_ISL_2087512, EPI_ISL_2087513, EPI_ISL_2087514, EPI_ISL_2087515, EPI_ISL_2087516, EPI_ISL_2087517, EPI_ISL_2087518, EPI_ISL_2087519, EPI_ISL_2087520, EPI_ISL_2087521, EPI_ISL_2087522, EPI_ISL_2087523, EPI_ISL_2087524, EPI_ISL_2087525, EPI_ISL_2087526, EPI_ISL_2087527, EPI_ISL_2087528, EPI_ISL_2087529, EPI_ISL_2087530, EPI_ISL_2087531, EPI_ISL_2087532, EPI_ISL_2087533, EPI_ISL_2087534, EPI_ISL_2087535, EPI_ISL_2087536, EPI_ISL_2087537, EPI_ISL_2087538, EPI_ISL_2087539, EPI_ISL_2087541, EPI_ISL_2087542, EPI_ISL_2087543, EPI_ISL_2087544, EPI_ISL_2087545, EPI_ISL_2087546, EPI_ISL_2087547, EPI_ISL_2087548, EPI_ISL_2087549, EPI_ISL_2087550, EPI_ISL_2087551, EPI_ISL_2087552, EPI_ISL_2087553, EPI_ISL_2087554, EPI_ISL_2087555, EPI_ISL_2087556, EPI_ISL_2087557, EPI_ISL_2087558, EPI_ISL_2087559, EPI_ISL_2087560, EPI_ISL_2087561, EPI_ISL_2087562, EPI_ISL_2087563, EPI_ISL_2087564, EPI_ISL_2087565, EPI_ISL_2087566, EPI_ISL_2087567, EPI_ISL_2087568, EPI_ISL_2087569, EPI_ISL_2087570, EPI_ISL_2087571, EPI_ISL_2087572, EPI_ISL_2087573, EPI_ISL_2087574, EPI_ISL_2087575, EPI_ISL_2087576, EPI_ISL_2087577, EPI_ISL_2087578, EPI_ISL_2087579, EPI_ISL_2087580, EPI_ISL_2087581, EPI_ISL_2087582, EPI_ISL_2087583, EPI_ISL_2087584, EPI_ISL_2087585, EPI_ISL_2087586, EPI_ISL_2087587, EPI_ISL_2087588, EPI_ISL_2087589, EPI_ISL_2087590, EPI_ISL_2087591, EPI_ISL_2087592, EPI_ISL_2087593, EPI_ISL_2087594, EPI_ISL_2087595, EPI_ISL_2087596, EPI_ISL_2087597, EPI_ISL_2087598, EPI_ISL_2087599, EPI_ISL_2087600, EPI_ISL_2087601, EPI_ISL_2087602, EPI_ISL_2087603, EPI_ISL_2087604, EPI_ISL_2087605, EPI_ISL_2087606, EPI_ISL_2087607, EPI_ISL_2087608, EPI_ISL_2087609, EPI_ISL_2087610, EPI_ISL_2087611, EPI_ISL_2087612, EPI_ISL_2087613, EPI_ISL_2087614, EPI_ISL_2087615, EPI_ISL_2087616, EPI_ISL_2087617, EPI_ISL_2087618, EPI_ISL_2087619, EPI_ISL_2087620, EPI_ISL_2087621, EPI_ISL_2087622, EPI_ISL_2087623, EPI_ISL_2087624, EPI_ISL_2087625, EPI_ISL_2087626, EPI_ISL_2087627, EPI_ISL_2087628, EPI_ISL_2087629, EPI_ISL_2087630, EPI_ISL_2087631, EPI_ISL_2087632, EPI_ISL_2087633, EPI_ISL_2087634, EPI_ISL_2087635, EPI_ISL_2087636, EPI_ISL_2087637, EPI_ISL_2087638, EPI_ISL_2087639, EPI_ISL_2087640, EPI_ISL_2087641, EPI_ISL_2087642, EPI_ISL_2087643, EPI_ISL_2087644, EPI_ISL_2087645, EPI_ISL_2087646, EPI_ISL_2087647, EPI_ISL_2087648, EPI_ISL_2087649, EPI_ISL_2087650, EPI_ISL_2087651, EPI_ISL_2087652, EPI_ISL_2087653, EPI_ISL_2087654, EPI_ISL_2087655, EPI_ISL_2087656, EPI_ISL_2087657, EPI_ISL_2087658, EPI_ISL_2087659, EPI_ISL_2087660, EPI_ISL_2087661, EPI_ISL_2087662, EPI_ISL_2087663, EPI_ISL_2087664, EPI_ISL_2087665, EPI_ISL_2087666, EPI_ISL_2087667, EPI_ISL_2087668, EPI_ISL_2087669, EPI_ISL_2087670, EPI_ISL_2087671, EPI_ISL_2087672, EPI_ISL_2087673, EPI_ISL_2087674, EPI_ISL_2087675, EPI_ISL_2087676, EPI_ISL_2087677, EPI_ISL_2087678, EPI_ISL_2087679, EPI_ISL_2087680, EPI_ISL_2087681, EPI_ISL_2087682, EPI_ISL_2087683, EPI_ISL_2087684, EPI_ISL_2087685, EPI_ISL_2087686, EPI_ISL_2087687, EPI_ISL_2087688, EPI_ISL_2087689, EPI_ISL_2087690, EPI_ISL_2087691, EPI_ISL_2087692, EPI_ISL_2087693, EPI_ISL_2087694, EPI_ISL_2087695, EPI_ISL_2087696, EPI_ISL_2087697, EPI_ISL_2087698, EPI_ISL_2087699, EPI_ISL_2087700, EPI_ISL_2087701, EPI_ISL_2087702, EPI_ISL_2087703, EPI_ISL_2087704, EPI_ISL_2087705, EPI_ISL_2087706, EPI_ISL_2087707, EPI_ISL_2087708, EPI_ISL_2087709, EPI_ISL_2087710, EPI_ISL_2087711, EPI_ISL_2087712, EPI_ISL_2087713, EPI_ISL_2087714, EPI_ISL_2087715, EPI_ISL_2087716, EPI_ISL_2087717, EPI_ISL_2087718, EPI_ISL_2087719, EPI_ISL_2087720, EPI_ISL_2087721, EPI_ISL_2087722, EPI_ISL_2087723, EPI_ISL_2087724, EPI_ISL_2087725, EPI_ISL_2087726, EPI_ISL_2087727, EPI_ISL_2087728, EPI_ISL_2087729, EPI_ISL_2087730, EPI_ISL_2087731, EPI_ISL_2087732, EPI_ISL_2087733, EPI_ISL_2087734, EPI_ISL_2087735, EPI_ISL_2087736, EPI_ISL_2087737, EPI_ISL_2087738, EPI_ISL_2087739, EPI_ISL_2087740, EPI_ISL_2087741, EPI_ISL_2087742, EPI_ISL_2087743, EPI_ISL_2087744, EPI_ISL_2087745, EPI_ISL_2087746, EPI_ISL_2087747, EPI_ISL_2087748, EPI_ISL_2087749, EPI_ISL_2087750, EPI_ISL_2087751, EPI_ISL_2087752, EPI_ISL_2087753, EPI_ISL_2087754, EPI_ISL_2087755, EPI_ISL_2087756, EPI_ISL_2087757, EPI_ISL_2087758, EPI_ISL_2087759, EPI_ISL_2087760, EPI_ISL_2087761, EPI_ISL_2087762, EPI_ISL_2087763, EPI_ISL_2087764, EPI_ISL_2087765, EPI_ISL_2087766, EPI_ISL_2087767, EPI_ISL_2087768, EPI_ISL_2087769, EPI_ISL_2087770, EPI_ISL_2087771, EPI_ISL_2087772, EPI_ISL_2087773, EPI_ISL_2087774, EPI_ISL_2087775, EPI_ISL_2087776, EPI_ISL_2087777, EPI_ISL_2087778, EPI_ISL_2087779, EPI_ISL_2087780, EPI_ISL_2087781, EPI_ISL_2087782, EPI_ISL_2087783, EPI_ISL_2087784, EPI_ISL_2087785, EPI_ISL_2087786, EPI_ISL_2087787, EPI_ISL_2087788, EPI_ISL_2087789, EPI_ISL_2087790, EPI_ISL_2087791, EPI_ISL_2087792, EPI_ISL_2087793, EPI_ISL_2087794, EPI_ISL_2087795, EPI_ISL_2087796, EPI_ISL_2087797, EPI_ISL_2087798, EPI_ISL_2087799, EPI_ISL_2087800, EPI_ISL_2087801, EPI_ISL_2087802, EPI_ISL_2087803, EPI_ISL_2087804, EPI_ISL_2087805, EPI_ISL_2087806, EPI_ISL_2087807, EPI_ISL_2087808, EPI_ISL_2087809, EPI_ISL_2087810, EPI_ISL_2087811, EPI_ISL_2087812, EPI_ISL_2087813, EPI_ISL_2087814, EPI_ISL_2087815, EPI_ISL_2087816, EPI_ISL_2087817, EPI_ISL_2087818, EPI_ISL_2087819, EPI_ISL_2087820, EPI_ISL_2087821, EPI_ISL_2087822, EPI_ISL_2087823, EPI_ISL_2087824, EPI_ISL_2087825, EPI_ISL_2087826, EPI_ISL_2087827, EPI_ISL_2087828, EPI_ISL_2087829, EPI_ISL_2087830, EPI_ISL_2087831, EPI_ISL_2087832, EPI_ISL_2087833, EPI_ISL_2087834, EPI_ISL_2087835, EPI_ISL_2087836, EPI_ISL_2087837, EPI_ISL_2087838, EPI_ISL_2087839, EPI_ISL_2087840, EPI_ISL_2087841, EPI_ISL_2087842, EPI_ISL_2087843, EPI_ISL_2087844, EPI_ISL_2087845, EPI_ISL_2087846, EPI_ISL_2087847, EPI_ISL_2087848, EPI_ISL_2087849, EPI_ISL_2087850, EPI_ISL_2087851, EPI_ISL_2087852, EPI_ISL_2087853, EPI_ISL_2087854, EPI_ISL_2087855, EPI_ISL_2087856, EPI_ISL_2087857, EPI_ISL_2087858, EPI_ISL_2087859, EPI_ISL_2087860, EPI_ISL_2087861, EPI_ISL_2087862, EPI_ISL_2087863, EPI_ISL_2087864, EPI_ISL_2087865, EPI_ISL_2087866, EPI_ISL_2087867, EPI_ISL_2087868, EPI_ISL_2087869, EPI_ISL_2087870, EPI_ISL_2087871, EPI_ISL_2087872, EPI_ISL_2087873, EPI_ISL_2087874, EPI_ISL_2087875, EPI_ISL_2087876, EPI_ISL_2087877, EPI_ISL_2087878, EPI_ISL_2087879, EPI_ISL_2087880, EPI_ISL_2087881, EPI_ISL_2087882, EPI_ISL_2087883, EPI_ISL_2087884, EPI_ISL_2087885, EPI_ISL_2087886, EPI_ISL_2087887, EPI_ISL_2087888, EPI_ISL_2087889, EPI_ISL_2087890, EPI_ISL_2087891, EPI_ISL_2087892, EPI_ISL_2087893, EPI_ISL_2087894, EPI_ISL_2087895, EPI_ISL_2087896, EPI_ISL_2087897, EPI_ISL_2087898, EPI_ISL_2087899, EPI_ISL_2087900, EPI_ISL_2087901, EPI_ISL_2087902, EPI_ISL_2087903, EPI_ISL_2087904, EPI_ISL_2087905, EPI_ISL_2087906, EPI_ISL_2087907, EPI_ISL_2087908, EPI_ISL_2087909, EPI_ISL_2087910, EPI_ISL_2087911, EPI_ISL_2087912, EPI_ISL_2087913, EPI_ISL_2087914, EPI_ISL_2087915, EPI_ISL_2087916, EPI_ISL_2087917, EPI_ISL_2087918, EPI_ISL_2087919, EPI_ISL_2087920, EPI_ISL_2087921, EPI_ISL_2087922, EPI_ISL_2087923, EPI_ISL_2087924, EPI_ISL_2087925, EPI_ISL_2087926, EPI_ISL_2087927, EPI_ISL_2087928, EPI_ISL_2087929, EPI_ISL_2087930, EPI_ISL_2087931, EPI_ISL_2087932, EPI_ISL_2087933, EPI_ISL_2087934, EPI_ISL_2087935, EPI_ISL_2087936, EPI_ISL_2087937, EPI_ISL_2087938, EPI_ISL_2087939, EPI_ISL_2087940, EPI_ISL_2087941, EPI_ISL_2087942, EPI_ISL_2087946, EPI_ISL_2087948, EPI_ISL_2087952, EPI_ISL_2087954, EPI_ISL_2087957, EPI_ISL_2087960, EPI_ISL_2087963, EPI_ISL_2087966, EPI_ISL_2087969, EPI_ISL_2087971, EPI_ISL_2087975, EPI_ISL_2087978, EPI_ISL_2087981, EPI_ISL_2087984, EPI_ISL_2087987, EPI_ISL_2087990, EPI_ISL_2087992, EPI_ISL_2087995, EPI_ISL_2087998, EPI_ISL_2088001, EPI_ISL_2088004, EPI_ISL_2088007, EPI_ISL_2088010, EPI_ISL_2088013, EPI_ISL_2088015, EPI_ISL_2088018, EPI_ISL_2088021, EPI_ISL_2088026, EPI_ISL_2088029, EPI_ISL_2088032, EPI_ISL_2088035, EPI_ISL_2088037, EPI_ISL_2088040, EPI_ISL_2088043, EPI_ISL_2088045, EPI_ISL_2088046, EPI_ISL_2088050, EPI_ISL_2088052, EPI_ISL_2088055, EPI_ISL_2088057, EPI_ISL_2088060, EPI_ISL_2088063, EPI_ISL_2088066, EPI_ISL_2088068, EPI_ISL_2088071, EPI_ISL_2088074, EPI_ISL_2088077, EPI_ISL_2088080, EPI_ISL_2088082, EPI_ISL_2088085, EPI_ISL_2088088, EPI_ISL_2088090, EPI_ISL_2088093, EPI_ISL_2088096, EPI_ISL_2088101, EPI_ISL_2088103, EPI_ISL_2088106, EPI_ISL_2088109, EPI_ISL_2088111, EPI_ISL_2088114, EPI_ISL_2088117, EPI_ISL_2088120, EPI_ISL_2088123, EPI_ISL_2088130, EPI_ISL_2088133, EPI_ISL_2088135, EPI_ISL_2088138, EPI_ISL_2088141, EPI_ISL_2088144, EPI_ISL_2088147, EPI_ISL_2088150, EPI_ISL_2088153, EPI_ISL_2088156, EPI_ISL_2088159, EPI_ISL_2088162, EPI_ISL_2088164, EPI_ISL_2088168, EPI_ISL_2088170, EPI_ISL_2088173, EPI_ISL_2088176, EPI_ISL_2088179, EPI_ISL_2088181, EPI_ISL_2088184, EPI_ISL_2088186, EPI_ISL_2088189, EPI_ISL_2088192, EPI_ISL_2088195, EPI_ISL_2088197, EPI_ISL_2088200, EPI_ISL_2088203, EPI_ISL_2088205, EPI_ISL_2088209, EPI_ISL_2088212, EPI_ISL_2088216, EPI_ISL_2088218, EPI_ISL_2088221, EPI_ISL_2088224, EPI_ISL_2088226, EPI_ISL_2088229, EPI_ISL_2088232, EPI_ISL_2088235, EPI_ISL_2088238, EPI_ISL_2088241, EPI_ISL_2088244, EPI_ISL_2088245, EPI_ISL_2088247, EPI_ISL_2088250, EPI_ISL_2088254, EPI_ISL_2088257, EPI_ISL_2088266, EPI_ISL_2088268, EPI_ISL_2088271, EPI_ISL_2088274, EPI_ISL_2088277, EPI_ISL_2088280, EPI_ISL_2088282, EPI_ISL_2088285, EPI_ISL_2088288, EPI_ISL_2088291, EPI_ISL_2088294, EPI_ISL_2088297, EPI_ISL_2088300, EPI_ISL_2088303, EPI_ISL_2088305, EPI_ISL_2088308, EPI_ISL_2088311, EPI_ISL_2088314, EPI_ISL_2088316, EPI_ISL_2088317, EPI_ISL_2088320, EPI_ISL_2088322, EPI_ISL_2088325, EPI_ISL_2088328, EPI_ISL_2088331, EPI_ISL_2088333, EPI_ISL_2088335, EPI_ISL_2088338, EPI_ISL_2088340, EPI_ISL_2088343, EPI_ISL_2088346, EPI_ISL_2088348, EPI_ISL_2088350, EPI_ISL_2088353, EPI_ISL_2088356, EPI_ISL_2088359, EPI_ISL_2088362, EPI_ISL_2088365, EPI_ISL_2088368, EPI_ISL_2088371, EPI_ISL_2088373, EPI_ISL_2088376, EPI_ISL_2088377, EPI_ISL_2088378, EPI_ISL_2088379, EPI_ISL_2088380, EPI_ISL_2088381, EPI_ISL_2088382, EPI_ISL_2088383, EPI_ISL_2088384, EPI_ISL_2088385, EPI_ISL_2088386, EPI_ISL_2088387, EPI_ISL_2088388, EPI_ISL_2088389, EPI_ISL_2088390, EPI_ISL_2088391, EPI_ISL_2088392, EPI_ISL_2088393, EPI_ISL_2088394, EPI_ISL_2088395, EPI_ISL_2088396, EPI_ISL_2088397, EPI_ISL_2088398, EPI_ISL_2088399, EPI_ISL_2088400, EPI_ISL_2088401, EPI_ISL_2088402, EPI_ISL_2088403, EPI_ISL_2088404, EPI_ISL_2088405, EPI_ISL_2088406, EPI_ISL_2088407, EPI_ISL_2088408, EPI_ISL_2088409, EPI_ISL_2088410, EPI_ISL_2088411, EPI_ISL_2088412, EPI_ISL_2088413, EPI_ISL_2088414, EPI_ISL_2088415, EPI_ISL_2088416, EPI_ISL_2088417, EPI_ISL_2088418, EPI_ISL_2088419, EPI_ISL_2088420, EPI_ISL_2088421, EPI_ISL_2088422, EPI_ISL_2088423, EPI_ISL_2088424, EPI_ISL_2088425, EPI_ISL_2088426, EPI_ISL_2088427, EPI_ISL_2088428, EPI_ISL_2088429, EPI_ISL_2088430, EPI_ISL_2088431, EPI_ISL_2088432, EPI_ISL_2088433, EPI_ISL_2088434, EPI_ISL_2088435, EPI_ISL_2088436, EPI_ISL_2088437, EPI_ISL_2088438, EPI_ISL_2088439, EPI_ISL_2088440, EPI_ISL_2088441, EPI_ISL_2088442, EPI_ISL_2088443, EPI_ISL_2088444, EPI_ISL_2088445, EPI_ISL_2088446, EPI_ISL_2088447, EPI_ISL_2088448, EPI_ISL_2088449, EPI_ISL_2088450, EPI_ISL_2088451, EPI_ISL_2088452, EPI_ISL_2088453, EPI_ISL_2088454, EPI_ISL_2088455, EPI_ISL_2088456, EPI_ISL_2088457, EPI_ISL_2088458, EPI_ISL_2088459, EPI_ISL_2088460, EPI_ISL_2088461, EPI_ISL_2088462, EPI_ISL_2088463, EPI_ISL_2088464, EPI_ISL_2088465, EPI_ISL_2088466, EPI_ISL_2088467, EPI_ISL_2088468, EPI_ISL_2088469, EPI_ISL_2088470, EPI_ISL_2088471, EPI_ISL_2088472, EPI_ISL_2088473, EPI_ISL_2088474, EPI_ISL_2088475, EPI_ISL_2088476, EPI_ISL_2088477, EPI_ISL_2088478, EPI_ISL_2088479, EPI_ISL_2088480, EPI_ISL_2088481, EPI_ISL_2088482, EPI_ISL_2088483, EPI_ISL_2088484, EPI_ISL_2088485, EPI_ISL_2088486, EPI_ISL_2088487, EPI_ISL_2088488, EPI_ISL_2088489, EPI_ISL_2088490, EPI_ISL_2088491, EPI_ISL_2088492, EPI_ISL_2088493, EPI_ISL_2088494, EPI_ISL_2088495, EPI_ISL_2088496, EPI_ISL_2088497, EPI_ISL_2088498, EPI_ISL_2088499, EPI_ISL_2088500, EPI_ISL_2088501, EPI_ISL_2088502, EPI_ISL_2088503, EPI_ISL_2088504, EPI_ISL_2088505, EPI_ISL_2088506, EPI_ISL_2088507, EPI_ISL_2088508, EPI_ISL_2088509, EPI_ISL_2088510, EPI_ISL_2088511, EPI_ISL_2088512, EPI_ISL_2088513, EPI_ISL_2088514, EPI_ISL_2088515, EPI_ISL_2088516, EPI_ISL_2088517, EPI_ISL_2088518, EPI_ISL_2088519, EPI_ISL_2088520, EPI_ISL_2088521, EPI_ISL_2088522, EPI_ISL_2088523, EPI_ISL_2088524, EPI_ISL_2088525, EPI_ISL_2088526, EPI_ISL_2088527, EPI_ISL_2088528, EPI_ISL_2088529, EPI_ISL_2088530, EPI_ISL_2088531, EPI_ISL_2088532, EPI_ISL_2088533, EPI_ISL_2088534, EPI_ISL_2088535, EPI_ISL_2088536, EPI_ISL_2088537, EPI_ISL_2088538, EPI_ISL_2088539, EPI_ISL_2088540, EPI_ISL_2088541, EPI_ISL_2088542, EPI_ISL_2088543, EPI_ISL_2088544, EPI_ISL_2088545, EPI_ISL_2088546, EPI_ISL_2088547, EPI_ISL_2088548, EPI_ISL_2088549, EPI_ISL_2088550, EPI_ISL_2088551, EPI_ISL_2088552, EPI_ISL_2088553, EPI_ISL_2088554, EPI_ISL_2088555, EPI_ISL_2088556, EPI_ISL_2088557, EPI_ISL_2088558, EPI_ISL_2088559, EPI_ISL_2088560, EPI_ISL_2088561, EPI_ISL_2088562, EPI_ISL_2088563, EPI_ISL_2088564, EPI_ISL_2088565, EPI_ISL_2088566, EPI_ISL_2088567, EPI_ISL_2088568, EPI_ISL_2088569, EPI_ISL_2088570, EPI_ISL_2088571, EPI_ISL_2088572, EPI_ISL_2088573, EPI_ISL_2088574, EPI_ISL_2088575, EPI_ISL_2088576, EPI_ISL_2088577, EPI_ISL_2088578, EPI_ISL_2088579, EPI_ISL_2088580, EPI_ISL_2088581, EPI_ISL_2088582, EPI_ISL_2088583, EPI_ISL_2088584, EPI_ISL_2088585, EPI_ISL_2088586, EPI_ISL_2088587, EPI_ISL_2088588, EPI_ISL_2088589, EPI_ISL_2088590, EPI_ISL_2088591, EPI_ISL_2088592, EPI_ISL_2088593, EPI_ISL_2088594, EPI_ISL_2088595, EPI_ISL_2088596, EPI_ISL_2088597, EPI_ISL_2088598, EPI_ISL_2088599, EPI_ISL_2088600, EPI_ISL_2088601, EPI_ISL_2088602, EPI_ISL_2088603, EPI_ISL_2088604, EPI_ISL_2088605, EPI_ISL_2088606, EPI_ISL_2088607, EPI_ISL_2088608, EPI_ISL_2088609, EPI_ISL_2088610, EPI_ISL_2088611, EPI_ISL_2088612, EPI_ISL_2088613, EPI_ISL_2088614, EPI_ISL_2088615, EPI_ISL_2088616, EPI_ISL_2088617, EPI_ISL_2088618, EPI_ISL_2088619, EPI_ISL_2088620, EPI_ISL_2088621, EPI_ISL_2088622, EPI_ISL_2088623, EPI_ISL_2088624, EPI_ISL_2088625, EPI_ISL_2088626, EPI_ISL_2088627, EPI_ISL_2088628, EPI_ISL_2088629, EPI_ISL_2088630, EPI_ISL_2088631, EPI_ISL_2088632, EPI_ISL_2088633, EPI_ISL_2088634, EPI_ISL_2088635, EPI_ISL_2088636, EPI_ISL_2088637, EPI_ISL_2088638, EPI_ISL_2088639, EPI_ISL_2088640, EPI_ISL_2088641, EPI_ISL_2088642, EPI_ISL_2088643, EPI_ISL_2088644, EPI_ISL_2088645, EPI_ISL_2088646, EPI_ISL_2088647, EPI_ISL_2088648, EPI_ISL_2088649, EPI_ISL_2088650, EPI_ISL_2088651, EPI_ISL_2088652, EPI_ISL_2088653, EPI_ISL_2088654, EPI_ISL_2088655, EPI_ISL_2088656, EPI_ISL_2088657, EPI_ISL_2088658, EPI_ISL_2088659, EPI_ISL_2088660, EPI_ISL_2088662, EPI_ISL_2088663, EPI_ISL_2088664, EPI_ISL_2088665, EPI_ISL_2088666, EPI_ISL_2088667, EPI_ISL_2088668, EPI_ISL_2088669, EPI_ISL_2088670, EPI_ISL_2088671, EPI_ISL_2088672, EPI_ISL_2088673, EPI_ISL_2088674, EPI_ISL_2088675, EPI_ISL_2088676, EPI_ISL_2088677, EPI_ISL_2088678, EPI_ISL_2088679, EPI_ISL_2088680, EPI_ISL_2088681, EPI_ISL_2088682, EPI_ISL_2088683, EPI_ISL_2088684, EPI_ISL_2088685, EPI_ISL_2088686, EPI_ISL_2088687, EPI_ISL_2088688, EPI_ISL_2088689, EPI_ISL_2088690, EPI_ISL_2088691, EPI_ISL_2088692, EPI_ISL_2088693, EPI_ISL_2088694, EPI_ISL_2088695, EPI_ISL_2088696, EPI_ISL_2088697, EPI_ISL_2088698, EPI_ISL_2088699, EPI_ISL_2088700, EPI_ISL_2088701, EPI_ISL_2088702, EPI_ISL_2088703, EPI_ISL_2088704, EPI_ISL_2088705, EPI_ISL_2088706, EPI_ISL_2088707, EPI_ISL_2088708, EPI_ISL_2088709, EPI_ISL_2088710, EPI_ISL_2088711, EPI_ISL_2088712, EPI_ISL_2088713, EPI_ISL_2088714, EPI_ISL_2088715, EPI_ISL_2088716, EPI_ISL_2088717, EPI_ISL_2088718, EPI_ISL_2088719, EPI_ISL_2088720, EPI_ISL_2088721, EPI_ISL_2088722, EPI_ISL_2088723, EPI_ISL_2088724, EPI_ISL_2088725, EPI_ISL_2088726, EPI_ISL_2088727, EPI_ISL_2088728, EPI_ISL_2088729, EPI_ISL_2088730, EPI_ISL_2088731, EPI_ISL_2088732, EPI_ISL_2088733, EPI_ISL_2088734, EPI_ISL_2088735, EPI_ISL_2088736, EPI_ISL_2088738, EPI_ISL_2088739, EPI_ISL_2088740, EPI_ISL_2088741, EPI_ISL_2088742, EPI_ISL_2088743, EPI_ISL_2088744, EPI_ISL_2088745, EPI_ISL_2088746, EPI_ISL_2088747, EPI_ISL_2088748, EPI_ISL_2088749, EPI_ISL_2088750, EPI_ISL_2088751, EPI_ISL_2088752, EPI_ISL_2088753, EPI_ISL_2088754, EPI_ISL_2088755, EPI_ISL_2088756, EPI_ISL_2088757, EPI_ISL_2088758, EPI_ISL_2088759, EPI_ISL_2088760, EPI_ISL_2088761, EPI_ISL_2088762, EPI_ISL_2088763, EPI_ISL_2088764, EPI_ISL_2088765, EPI_ISL_2088766, EPI_ISL_2088767, EPI_ISL_2088768, EPI_ISL_2088769, EPI_ISL_2088770, EPI_ISL_2088771, EPI_ISL_2088772, EPI_ISL_2088773, EPI_ISL_2088774, EPI_ISL_2088775, EPI_ISL_2088777, EPI_ISL_2088778, EPI_ISL_2088779, EPI_ISL_2088780, EPI_ISL_2088781, EPI_ISL_2088782, EPI_ISL_2088783, EPI_ISL_2088784, EPI_ISL_2088785, EPI_ISL_2088786, EPI_ISL_2088787, EPI_ISL_2088788, EPI_ISL_2088789, EPI_ISL_2088790, EPI_ISL_2088791, EPI_ISL_2088792, EPI_ISL_2088793, EPI_ISL_2088795, EPI_ISL_2088796, EPI_ISL_2088797, EPI_ISL_2088798, EPI_ISL_2088799, EPI_ISL_2088800, EPI_ISL_2088801, EPI_ISL_2088802, EPI_ISL_2088803, EPI_ISL_2088804, EPI_ISL_2088805, EPI_ISL_2088806, EPI_ISL_2088807, EPI_ISL_2088808, EPI_ISL_2088809, EPI_ISL_2088810, EPI_ISL_2088811, EPI_ISL_2088812, EPI_ISL_2088813, EPI_ISL_2088814, EPI_ISL_2088815, EPI_ISL_2088816, EPI_ISL_2088817, EPI_ISL_2088818, EPI_ISL_2088819, EPI_ISL_2088820, EPI_ISL_2088821, EPI_ISL_2088822, EPI_ISL_2088823, EPI_ISL_2088824, EPI_ISL_2088825, EPI_ISL_2088826, EPI_ISL_2088827, EPI_ISL_2088828, EPI_ISL_2088829, EPI_ISL_2088830, EPI_ISL_2088831, EPI_ISL_2088832, EPI_ISL_2088833, EPI_ISL_2088834, EPI_ISL_2088835, EPI_ISL_2088836, EPI_ISL_2088837, EPI_ISL_2088838, EPI_ISL_2088839, EPI_ISL_2088840, EPI_ISL_2088841, EPI_ISL_2088842, EPI_ISL_2088843, EPI_ISL_2088844, EPI_ISL_2088845, EPI_ISL_2088846, EPI_ISL_2088847, EPI_ISL_2088848, EPI_ISL_2088849, EPI_ISL_2088850, EPI_ISL_2088851, EPI_ISL_2088852, EPI_ISL_2088853, EPI_ISL_2088854, EPI_ISL_2088855, EPI_ISL_2088856, EPI_ISL_2088857, EPI_ISL_2088858, EPI_ISL_2088859, EPI_ISL_2088860, EPI_ISL_2088861, EPI_ISL_2088862, EPI_ISL_2088863, EPI_ISL_2088864, EPI_ISL_2088865, EPI_ISL_2088866, EPI_ISL_2088867, EPI_ISL_2088868, EPI_ISL_2088869, EPI_ISL_2088870, EPI_ISL_2088871, EPI_ISL_2088872, EPI_ISL_2088873, EPI_ISL_2088874, EPI_ISL_2088875, EPI_ISL_2088876, EPI_ISL_2088877, EPI_ISL_2088879, EPI_ISL_2088880, EPI_ISL_2088881, EPI_ISL_2088882, EPI_ISL_2088883, EPI_ISL_2088884, EPI_ISL_2088885, EPI_ISL_2088886, EPI_ISL_2088887, EPI_ISL_2088888, EPI_ISL_2088889, EPI_ISL_2088890, EPI_ISL_2088891, EPI_ISL_2088892, EPI_ISL_2088893, EPI_ISL_2088894, EPI_ISL_2088895, EPI_ISL_2088896, EPI_ISL_2088897, EPI_ISL_2088898, EPI_ISL_2088900, EPI_ISL_2088901, EPI_ISL_2088902, EPI_ISL_2088903, EPI_ISL_2088905, EPI_ISL_2088906, EPI_ISL_2088907, EPI_ISL_2088908, EPI_ISL_2088909, EPI_ISL_2088910, EPI_ISL_2088911, EPI_ISL_2088912, EPI_ISL_2088913, EPI_ISL_2088914, EPI_ISL_2088915, EPI_ISL_2088916, EPI_ISL_2088917, EPI_ISL_2088918, EPI_ISL_2088919, EPI_ISL_2088920, EPI_ISL_2088921, EPI_ISL_2088922, EPI_ISL_2088923, EPI_ISL_2088924, EPI_ISL_2088925, EPI_ISL_2088926, EPI_ISL_2088927, EPI_ISL_2088928, EPI_ISL_2088929, EPI_ISL_2088930, EPI_ISL_2088931, EPI_ISL_2088932, EPI_ISL_2088933, EPI_ISL_2088934, EPI_ISL_2088935, EPI_ISL_2088936, EPI_ISL_2088937, EPI_ISL_2088938, EPI_ISL_2088939, EPI_ISL_2088940, EPI_ISL_2088941, EPI_ISL_2088942, EPI_ISL_2088943, EPI_ISL_2088944, EPI_ISL_2088945, EPI_ISL_2088946, EPI_ISL_2088947, EPI_ISL_2088949, EPI_ISL_2088951, EPI_ISL_2088952, EPI_ISL_2088953, EPI_ISL_2088954, EPI_ISL_2088955, EPI_ISL_2088956, EPI_ISL_2088957, EPI_ISL_2088958, EPI_ISL_2088959, EPI_ISL_2088960, EPI_ISL_2088961, EPI_ISL_2088962, EPI_ISL_2088963, EPI_ISL_2088964, EPI_ISL_2088965, EPI_ISL_2088966, EPI_ISL_2088967, EPI_ISL_2088968, EPI_ISL_2088969, EPI_ISL_2088970, EPI_ISL_2088971, EPI_ISL_2088972, EPI_ISL_2088973, EPI_ISL_2088975, EPI_ISL_2088976, EPI_ISL_2088977, EPI_ISL_2088978, EPI_ISL_2088979, EPI_ISL_2088981, EPI_ISL_2088982, EPI_ISL_2088983, EPI_ISL_2088984, EPI_ISL_2088985, EPI_ISL_2088986, EPI_ISL_2088987, EPI_ISL_2088988, EPI_ISL_2088989, EPI_ISL_2088990, EPI_ISL_2088991, EPI_ISL_2088992, EPI_ISL_2088993, EPI_ISL_2088994, EPI_ISL_2088995, EPI_ISL_2088996, EPI_ISL_2088997, EPI_ISL_2088998, EPI_ISL_2088999, EPI_ISL_2089000, EPI_ISL_2089001, EPI_ISL_2089002, EPI_ISL_2089003, EPI_ISL_2089004, EPI_ISL_2089005, EPI_ISL_2089006, EPI_ISL_2089007, EPI_ISL_2089008, EPI_ISL_2089009, EPI_ISL_2089010, EPI_ISL_2089011, EPI_ISL_2089012, EPI_ISL_2089013, EPI_ISL_2089014, EPI_ISL_2089015, EPI_ISL_2089016, EPI_ISL_2089017, EPI_ISL_2089018, EPI_ISL_2089019, EPI_ISL_2089020, EPI_ISL_2089021, EPI_ISL_2089022, EPI_ISL_2089023, EPI_ISL_2089024, EPI_ISL_2089025, EPI_ISL_2089026, EPI_ISL_2089027, EPI_ISL_2089028, EPI_ISL_2089029, EPI_ISL_2089031, EPI_ISL_2089032, EPI_ISL_2089033, EPI_ISL_2089034, EPI_ISL_2089035, EPI_ISL_2089036, EPI_ISL_2089037, EPI_ISL_2089038, EPI_ISL_2089039, EPI_ISL_2089041, EPI_ISL_2089042, EPI_ISL_2089043, EPI_ISL_2089044, EPI_ISL_2089045, EPI_ISL_2089046, EPI_ISL_2089047, EPI_ISL_2089048, EPI_ISL_2089049, EPI_ISL_2089050, EPI_ISL_2089051, EPI_ISL_2089052, EPI_ISL_2089053, EPI_ISL_2089054, EPI_ISL_2089055, EPI_ISL_2089056, EPI_ISL_2089057, EPI_ISL_2089058, EPI_ISL_2089059, EPI_ISL_2089060, EPI_ISL_2089061, EPI_ISL_2089062, EPI_ISL_2089063, EPI_ISL_2089064, EPI_ISL_2089065, EPI_ISL_2089066, EPI_ISL_2089067, EPI_ISL_2089068, EPI_ISL_2089069, EPI_ISL_2089070, EPI_ISL_2089071, EPI_ISL_2089072, EPI_ISL_2089073, EPI_ISL_2089074, EPI_ISL_2089075, EPI_ISL_2089076, EPI_ISL_2089077, EPI_ISL_2089078, EPI_ISL_2089080, EPI_ISL_2089081, EPI_ISL_2089082, EPI_ISL_2089084, EPI_ISL_2089085, EPI_ISL_2089086, EPI_ISL_2089087, EPI_ISL_2089088, EPI_ISL_2089089, EPI_ISL_2089090, EPI_ISL_2089091, EPI_ISL_2089092, EPI_ISL_2089093, EPI_ISL_2089094, EPI_ISL_2089095, EPI_ISL_2089096, EPI_ISL_2089097, EPI_ISL_2089098, EPI_ISL_2089099, EPI_ISL_2089100, EPI_ISL_2089101, EPI_ISL_2089102, EPI_ISL_2089103, EPI_ISL_2089104, EPI_ISL_2089105, EPI_ISL_2089106, EPI_ISL_2089107, EPI_ISL_2089108, EPI_ISL_2089109, EPI_ISL_2089110, EPI_ISL_2089111, EPI_ISL_2089112, EPI_ISL_2089113, EPI_ISL_2089114, EPI_ISL_2089115, EPI_ISL_2089116, EPI_ISL_2089117, EPI_ISL_2089118, EPI_ISL_2089119, EPI_ISL_2089120, EPI_ISL_2089121, EPI_ISL_2089122, EPI_ISL_2089123, EPI_ISL_2089124, EPI_ISL_2089125, EPI_ISL_2089126, EPI_ISL_2089127, EPI_ISL_2089128, EPI_ISL_2089129, EPI_ISL_2089130, EPI_ISL_2089131, EPI_ISL_2089132, EPI_ISL_2089133, EPI_ISL_2089134, EPI_ISL_2089135, EPI_ISL_2089136, EPI_ISL_2089137, EPI_ISL_2089138, EPI_ISL_2089139, EPI_ISL_2089141, EPI_ISL_2089142, EPI_ISL_2089143, EPI_ISL_2089144, EPI_ISL_2089145, EPI_ISL_2089146, EPI_ISL_2089147, EPI_ISL_2089148, EPI_ISL_2089149, EPI_ISL_2089150, EPI_ISL_2089151, EPI_ISL_2089152, EPI_ISL_2089153, EPI_ISL_2089154, EPI_ISL_2089155, EPI_ISL_2089157, EPI_ISL_2089158, EPI_ISL_2089159, EPI_ISL_2089160, EPI_ISL_2089161, EPI_ISL_2089162, EPI_ISL_2089163, EPI_ISL_2089164, EPI_ISL_2089165, EPI_ISL_2089166, EPI_ISL_2089167, EPI_ISL_2089168, EPI_ISL_2089169, EPI_ISL_2089170, EPI_ISL_2089171, EPI_ISL_2089172, EPI_ISL_2089173, EPI_ISL_2089174, EPI_ISL_2089175, EPI_ISL_2089177, EPI_ISL_2089178, EPI_ISL_2089179, EPI_ISL_2089180, EPI_ISL_2089181, | | | |

| | | | |
|---|---|---|---|
| EPI_ISL_2089184, EPI_ISL_2089185, EPI_ISL_2089186, EPI_ISL_2089187, EPI_ISL_2089188, EPI_ISL_2089189, EPI_ISL_2089190, EPI_ISL_2089191, EPI_ISL_2089192, EPI_ISL_2089193, EPI_ISL_2089194, EPI_ISL_2089195, EPI_ISL_2089196, EPI_ISL_2089197, EPI_ISL_2089198, EPI_ISL_2089199, EPI_ISL_2089200, EPI_ISL_2089201, EPI_ISL_2089202, EPI_ISL_2089203, EPI_ISL_2089204, EPI_ISL_2089205, EPI_ISL_2089207, EPI_ISL_2089208, EPI_ISL_2089209, EPI_ISL_2089210, EPI_ISL_2089211, EPI_ISL_2089212, EPI_ISL_2089213, EPI_ISL_2089214, EPI_ISL_2089215, EPI_ISL_2089216, EPI_ISL_2089217, EPI_ISL_2089218, EPI_ISL_2089219, EPI_ISL_2089220, EPI_ISL_2089221, EPI_ISL_2089222, EPI_ISL_2089223, EPI_ISL_2089224, EPI_ISL_2089225, EPI_ISL_2089226, EPI_ISL_2089227, EPI_ISL_2089228, EPI_ISL_2089229, EPI_ISL_2089230, EPI_ISL_2089232, EPI_ISL_2089233, EPI_ISL_2089234, EPI_ISL_2089235, EPI_ISL_2089236, EPI_ISL_2089238, EPI_ISL_2089239, EPI_ISL_2089241, EPI_ISL_2089242, EPI_ISL_2089243, EPI_ISL_2089244, EPI_ISL_2089245, EPI_ISL_2089246, EPI_ISL_2089247, EPI_ISL_2089248, EPI_ISL_2089249, EPI_ISL_2089250, EPI_ISL_2089251, EPI_ISL_2089252, EPI_ISL_2089253, EPI_ISL_2089254, EPI_ISL_2089255, EPI_ISL_2089256, EPI_ISL_2089257, EPI_ISL_2089258, EPI_ISL_2089259, EPI_ISL_2089260, EPI_ISL_2089262, EPI_ISL_2089263, EPI_ISL_2089264, EPI_ISL_2089265, EPI_ISL_2089266, EPI_ISL_2089268, EPI_ISL_2089269, EPI_ISL_2089270, EPI_ISL_2089271, EPI_ISL_2089272, EPI_ISL_2089273, EPI_ISL_2089274, EPI_ISL_2089275, EPI_ISL_2089276, EPI_ISL_2089277, EPI_ISL_2089278, EPI_ISL_2089279, EPI_ISL_2089280, EPI_ISL_2089281, EPI_ISL_2089282, EPI_ISL_2089283, EPI_ISL_2089284, EPI_ISL_2089285, EPI_ISL_2089286, EPI_ISL_2089287, EPI_ISL_2089288, EPI_ISL_2089289, EPI_ISL_2089290, EPI_ISL_2089291, EPI_ISL_2089292, EPI_ISL_2089293, EPI_ISL_2089294, EPI_ISL_2089295, EPI_ISL_2089296, EPI_ISL_2089297, EPI_ISL_2089298, EPI_ISL_2089300, EPI_ISL_2089301, EPI_ISL_2089302, EPI_ISL_2089303, EPI_ISL_2089304, EPI_ISL_2089305, EPI_ISL_2089306, EPI_ISL_2089307, EPI_ISL_2089308, EPI_ISL_2089309, EPI_ISL_2089310, EPI_ISL_2089311, EPI_ISL_2089312, EPI_ISL_2089313, EPI_ISL_2089314, EPI_ISL_2089315, EPI_ISL_2089316, EPI_ISL_2089317, EPI_ISL_2089318, EPI_ISL_2089319, EPI_ISL_2089320, EPI_ISL_2089321, EPI_ISL_2089323, EPI_ISL_2089324, EPI_ISL_2089325, EPI_ISL_2089326, EPI_ISL_2089327, EPI_ISL_2089328, EPI_ISL_2089329, EPI_ISL_2089330, EPI_ISL_2089331, EPI_ISL_2089332, EPI_ISL_2089334, EPI_ISL_2089335, EPI_ISL_2089336, EPI_ISL_2089337, EPI_ISL_2089338, EPI_ISL_2089339, EPI_ISL_2089340, EPI_ISL_2089341, EPI_ISL_2089342, EPI_ISL_2089343, EPI_ISL_2089344, EPI_ISL_2089345, EPI_ISL_2089346, EPI_ISL_2089347, EPI_ISL_2089348, EPI_ISL_2089349, EPI_ISL_2089350, EPI_ISL_2089351, EPI_ISL_2089352, EPI_ISL_2089353, EPI_ISL_2089354, EPI_ISL_2089355, EPI_ISL_2089356, EPI_ISL_2089357, EPI_ISL_2089358, EPI_ISL_2089360, EPI_ISL_2089361, EPI_ISL_2089362, EPI_ISL_2089363, EPI_ISL_2089364, EPI_ISL_2089365, EPI_ISL_2089366, EPI_ISL_2089369, EPI_ISL_2089370, EPI_ISL_2089371, EPI_ISL_2089372, EPI_ISL_2089373, EPI_ISL_2089374, EPI_ISL_2089375, EPI_ISL_2089376, EPI_ISL_2089377, EPI_ISL_2089378, EPI_ISL_2089380, EPI_ISL_2089382, EPI_ISL_2089383, EPI_ISL_2089384, EPI_ISL_2089385, EPI_ISL_2089386, EPI_ISL_2089387, EPI_ISL_2089388, EPI_ISL_2089389, EPI_ISL_2089390, EPI_ISL_2089391, EPI_ISL_2089392, EPI_ISL_2089393, EPI_ISL_2089394, EPI_ISL_2089395, EPI_ISL_2089396, EPI_ISL_2089397, EPI_ISL_2089398, EPI_ISL_2089399, EPI_ISL_2089400, EPI_ISL_2089401, EPI_ISL_2089402, EPI_ISL_2089403, EPI_ISL_2089404, EPI_ISL_2089406, EPI_ISL_2089407, EPI_ISL_2089408, EPI_ISL_2089409, EPI_ISL_2089410, EPI_ISL_2089411, EPI_ISL_2089412, EPI_ISL_2089413, EPI_ISL_2089414, EPI_ISL_2089416, EPI_ISL_2089417, EPI_ISL_2089418, EPI_ISL_2089419, EPI_ISL_2089420, EPI_ISL_2089421, EPI_ISL_2089422, EPI_ISL_2089423, EPI_ISL_2089424, EPI_ISL_2089425, EPI_ISL_2089426, EPI_ISL_2089427, EPI_ISL_2089428, EPI_ISL_2089429, EPI_ISL_2089431, EPI_ISL_2089432, EPI_ISL_2089433, EPI_ISL_2089434, EPI_ISL_2089435, EPI_ISL_2089436, EPI_ISL_2089437, EPI_ISL_2089438, EPI_ISL_2089439, EPI_ISL_2089440, EPI_ISL_2089441, EPI_ISL_2089442, EPI_ISL_2089443, EPI_ISL_2089445, EPI_ISL_2089446, EPI_ISL_2089447, EPI_ISL_2089448, EPI_ISL_2089449, EPI_ISL_2089450, EPI_ISL_2089452 | | | |
| see above | Infinity Biologix | Centers for Disease Control and Prevention Division of Viral Diseases, Pathogen Discovery | Dakota Howard, Dhwani Batra, Peter W. Cook, Kara Moser, Adrian Paskey, Jason Caravas, Benjamin Rambo-Martin, Shatavia Morrison, Christopher Gulvick, Scott Sammons, Yvette Unoarumhi, Darlene Wagner, Matthew Schmerer, Christian Bixby, Yihe Wang, Jonathan Schultz, Chirayu Goswami, Russ Hager, Robin Grimwood, Clinton R. Paden, Duncan MacCannell |
| EPI_ISL_2089453, EPI_ISL_2089454, EPI_ISL_2089455, EPI_ISL_2089456, EPI_ISL_2089457, EPI_ISL_2089459, EPI_ISL_2089460, EPI_ISL_2089461, EPI_ISL_2089462, EPI_ISL_2089463, EPI_ISL_2089464, EPI_ISL_2089465, EPI_ISL_2089466, EPI_ISL_2089467, EPI_ISL_2089468, EPI_ISL_2089469, EPI_ISL_2089470, EPI_ISL_2089471, EPI_ISL_2089472, EPI_ISL_2089473, EPI_ISL_2089474, EPI_ISL_2089475, EPI_ISL_2089476, EPI_ISL_2089477, EPI_ISL_2089478, EPI_ISL_2089479, EPI_ISL_2089480, EPI_ISL_2089481, EPI_ISL_2089482, EPI_ISL_2089483, EPI_ISL_2089484, EPI_ISL_2089486, EPI_ISL_2089487, EPI_ISL_2089488, EPI_ISL_2089489, EPI_ISL_2089490, EPI_ISL_2089491, EPI_ISL_2089492, EPI_ISL_2089493, EPI_ISL_2089494, EPI_ISL_2089495, EPI_ISL_2089496, EPI_ISL_2089497, EPI_ISL_2089498, EPI_ISL_2089499, EPI_ISL_2089500, EPI_ISL_2089501, EPI_ISL_2089502, EPI_ISL_2089503, EPI_ISL_2089504, EPI_ISL_2089505, EPI_ISL_2089506, EPI_ISL_2089507, EPI_ISL_2089508, EPI_ISL_2089509, EPI_ISL_2089510, EPI_ISL_2089511, EPI_ISL_2089512, EPI_ISL_2089513, EPI_ISL_2089514, EPI_ISL_2089515, EPI_ISL_2089517, EPI_ISL_2089518, EPI_ISL_2089519, EPI_ISL_2089520, EPI_ISL_2089521, EPI_ISL_2089522, EPI_ISL_2089523, EPI_ISL_2089524, EPI_ISL_2089525, EPI_ISL_2089526, EPI_ISL_2089527, EPI_ISL_2089528, EPI_ISL_2089529, EPI_ISL_2089531, EPI_ISL_2089532, EPI_ISL_2089533, EPI_ISL_2089534, EPI_ISL_2089535, EPI_ISL_2089536, EPI_ISL_2089537, EPI_ISL_2089538, EPI_ISL_2089539, EPI_ISL_2089540, EPI_ISL_2089541, EPI_ISL_2089542, EPI_ISL_2089543, EPI_ISL_2089544, EPI_ISL_2089545, EPI_ISL_2089546, EPI_ISL_2089547, EPI_ISL_2089548, EPI_ISL_2089549, EPI_ISL_2089550, EPI_ISL_2089551, EPI_ISL_2089552, EPI_ISL_2089554, EPI_ISL_2089555, EPI_ISL_2089556, EPI_ISL_2089557, EPI_ISL_2089558, EPI_ISL_2089559, EPI_ISL_2089560, EPI_ISL_2089561, EPI_ISL_2089562, EPI_ISL_2089563, EPI_ISL_2089564, EPI_ISL_2089565, EPI_ISL_2089566, EPI_ISL_2089567, EPI_ISL_2089568, EPI_ISL_2089569, EPI_ISL_2089570, EPI_ISL_2089571, EPI_ISL_2089572, EPI_ISL_2089573, EPI_ISL_2089575, EPI_ISL_2089576, EPI_ISL_2089577, EPI_ISL_2089578, EPI_ISL_2089579, EPI_ISL_2089580, EPI_ISL_2089581, EPI_ISL_2089582, EPI_ISL_2089583, EPI_ISL_2089584, EPI_ISL_2089585, EPI_ISL_2089586, EPI_ISL_2089587, EPI_ISL_2089588, EPI_ISL_2089590, EPI_ISL_2089591, EPI_ISL_2089592, EPI_ISL_2089593, EPI_ISL_2089594, EPI_ISL_2089595, EPI_ISL_2089596, EPI_ISL_2089597, EPI_ISL_2089598, EPI_ISL_2089599, EPI_ISL_2089600, EPI_ISL_2089601, EPI_ISL_2089602, EPI_ISL_2089603, EPI_ISL_2089604, EPI_ISL_2089605, EPI_ISL_2089606, EPI_ISL_2089607, EPI_ISL_2089608, EPI_ISL_2089609, EPI_ISL_2089610, EPI_ISL_2089611, EPI_ISL_2089612, EPI_ISL_2089613, EPI_ISL_2089614, EPI_ISL_2089615, EPI_ISL_2089616, EPI_ISL_2089617, EPI_ISL_2089618, EPI_ISL_2089619, EPI_ISL_2089620, EPI_ISL_2089621, EPI_ISL_2089622, EPI_ISL_2089623, EPI_ISL_2089624, EPI_ISL_2089625, EPI_ISL_2089626, EPI_ISL_2089627, EPI_ISL_2089628, EPI_ISL_2089629, EPI_ISL_2089630, EPI_ISL_2089631, EPI_ISL_2089632, EPI_ISL_2089633, EPI_ISL_2089634, EPI_ISL_2089635, EPI_ISL_2089636, EPI_ISL_2089637, EPI_ISL_2089638, EPI_ISL_2089639, EPI_ISL_2089640, EPI_ISL_2089641, EPI_ISL_2089642, EPI_ISL_2089643, EPI_ISL_2089644, EPI_ISL_2089645, EPI_ISL_2089646, EPI_ISL_2089648, EPI_ISL_2089649, EPI_ISL_2089650, EPI_ISL_2089651, EPI_ISL_2089652, EPI_ISL_2089653, EPI_ISL_2089654, EPI_ISL_2089655, EPI_ISL_2089656, EPI_ISL_2089657, EPI_ISL_2089658, EPI_ISL_2089659, EPI_ISL_2089660, EPI_ISL_2089661, EPI_ISL_2089662, EPI_ISL_2089663, EPI_ISL_2089664, EPI_ISL_2089665, EPI_ISL_2089666, EPI_ISL_2089667, EPI_ISL_2089668, EPI_ISL_2089670, EPI_ISL_2089671, EPI_ISL_2089672, EPI_ISL_2089673, EPI_ISL_2089674, EPI_ISL_2089675, EPI_ISL_2089676, EPI_ISL_2089677, EPI_ISL_2089678, EPI_ISL_2089679, EPI_ISL_2089680, EPI_ISL_2089681, EPI_ISL_2089682, EPI_ISL_2089683, EPI_ISL_2089684, EPI_ISL_2089685, EPI_ISL_2089686, EPI_ISL_2089687, EPI_ISL_2089688, EPI_ISL_2089689, EPI_ISL_2089690, EPI_ISL_2089691, EPI_ISL_2089692, EPI_ISL_2089693, EPI_ISL_2089694, EPI_ISL_2089695, EPI_ISL_2089697, EPI_ISL_2089698, EPI_ISL_2089699, EPI_ISL_2089700, EPI_ISL_2089701, EPI_ISL_2089702, EPI_ISL_2089703, EPI_ISL_2089704, EPI_ISL_2089705, EPI_ISL_2089706, EPI_ISL_2089707, EPI_ISL_2089708, EPI_ISL_2089709, EPI_ISL_2089710, EPI_ISL_2089711, EPI_ISL_2089712, EPI_ISL_2089713, EPI_ISL_2089714, EPI_ISL_2089715, EPI_ISL_2089716, EPI_ISL_2089717, EPI_ISL_2089718, EPI_ISL_2089719, EPI_ISL_2089720, EPI_ISL_2089721, EPI_ISL_2089722, EPI_ISL_2089723, EPI_ISL_2089724, EPI_ISL_2089725, EPI_ISL_2089726, EPI_ISL_2089727, EPI_ISL_2089728, EPI_ISL_2089729, EPI_ISL_2089730, EPI_ISL_2089731, EPI_ISL_2089732, EPI_ISL_2089733, EPI_ISL_2089734, EPI_ISL_2089735, EPI_ISL_2089736, EPI_ISL_2089737, EPI_ISL_2089738, EPI_ISL_2089739, EPI_ISL_2089740, EPI_ISL_2089741, EPI_ISL_2089742, EPI_ISL_2089743, EPI_ISL_2089744, EPI_ISL_2089745, EPI_ISL_2089746, EPI_ISL_2089747, EPI_ISL_2089748, EPI_ISL_2089749, EPI_ISL_2089751, EPI_ISL_2089752, EPI_ISL_2089753, EPI_ISL_2089754, EPI_ISL_2089755, EPI_ISL_2089756, EPI_ISL_2089757, EPI_ISL_2089758, EPI_ISL_2089759, EPI_ISL_2089760, EPI_ISL_2089761, EPI_ISL_2089763, EPI_ISL_2089764, EPI_ISL_2089765, EPI_ISL_2089766, EPI_ISL_2089767, EPI_ISL_2089768, EPI_ISL_2089769, EPI_ISL_2089770, EPI_ISL_2089771, EPI_ISL_2089772, EPI_ISL_2089773, EPI_ISL_2089774, EPI_ISL_2089775, EPI_ISL_2089776, EPI_ISL_2089777, EPI_ISL_2089778, EPI_ISL_2089779, EPI_ISL_2089780, EPI_ISL_2089781, EPI_ISL_2089782, EPI_ISL_2089783, EPI_ISL_2089784, EPI_ISL_2089785, EPI_ISL_2089786, EPI_ISL_2089788, EPI_ISL_2089789, EPI_ISL_2089791, EPI_ISL_2089792, EPI_ISL_2089793, EPI_ISL_2089795, EPI_ISL_2089796, EPI_ISL_2089797, EPI_ISL_2089798, EPI_ISL_2089799, EPI_ISL_2089800, EPI_ISL_2089801, EPI_ISL_2089802, EPI_ISL_2089803, EPI_ISL_2089804, EPI_ISL_2089805, EPI_ISL_2089806, EPI_ISL_2089807, EPI_ISL_2089808, EPI_ISL_2089809, EPI_ISL_2089810, EPI_ISL_2089811, EPI_ISL_2089812, EPI_ISL_2089813, EPI_ISL_2089814, EPI_ISL_2089815, EPI_ISL_2089816, EPI_ISL_2089817, EPI_ISL_2089818, EPI_ISL_2089819, EPI_ISL_2089820, EPI_ISL_2089821, EPI_ISL_2089822, EPI_ISL_2089823, EPI_ISL_2089824, EPI_ISL_2089825, EPI_ISL_2089826, EPI_ISL_2089827, EPI_ISL_2089828, EPI_ISL_2089829, EPI_ISL_2089830, EPI_ISL_2089832, EPI_ISL_2089833, EPI_ISL_2089834, EPI_ISL_2089835, EPI_ISL_2089836, EPI_ISL_2089837, EPI_ISL_2089838, EPI_ISL_2089839, EPI_ISL_2089840, EPI_ISL_2089841, EPI_ISL_2089842, EPI_ISL_2089843, EPI_ISL_2089844, EPI_ISL_2089845, EPI_ISL_2089846, EPI_ISL_2089847, EPI_ISL_2089848, EPI_ISL_2089849, EPI_ISL_2089850, EPI_ISL_2089851, EPI_ISL_2089852, EPI_ISL_2089853, EPI_ISL_2089854, EPI_ISL_2089855, EPI_ISL_2089856, EPI_ISL_2089857, EPI_ISL_2089858, EPI_ISL_2089859, EPI_ISL_2089860, EPI_ISL_2089861, EPI_ISL_2089863, EPI_ISL_2089864, EPI_ISL_2089865, EPI_ISL_2089866, EPI_ISL_2089867, EPI_ISL_2089868, EPI_ISL_2089869, EPI_ISL_2089870, EPI_ISL_2089871, EPI_ISL_2089872, EPI_ISL_2089874, EPI_ISL_2089875, EPI_ISL_2089876, EPI_ISL_2089877, EPI_ISL_2089878, EPI_ISL_2089879, EPI_ISL_2089880, EPI_ISL_2089881, EPI_ISL_2089882, EPI_ISL_2089883, EPI_ISL_2089884, EPI_ISL_2089886, EPI_ISL_2089887, EPI_ISL_2089888, EPI_ISL_2089889, EPI_ISL_2089890, EPI_ISL_2089891, EPI_ISL_2089892, EPI_ISL_2089893, EPI_ISL_2089894, EPI_ISL_2089895, EPI_ISL_2089896, EPI_ISL_2089897, EPI_ISL_2089898, EPI_ISL_2089899, EPI_ISL_2089900, EPI_ISL_2089901, EPI_ISL_2089902, EPI_ISL_2089903, EPI_ISL_2089904, EPI_ISL_2089905, EPI_ISL_2089906, EPI_ISL_2089907, EPI_ISL_2089908, EPI_ISL_2089909, EPI_ISL_2089910, EPI_ISL_2089912, EPI_ISL_2089913, EPI_ISL_2089914, EPI_ISL_2089915, EPI_ISL_2089916, EPI_ISL_2089917, EPI_ISL_2089918, EPI_ISL_2089919, EPI_ISL_2089920, EPI_ISL_2089921, EPI_ISL_2089922, EPI_ISL_2089923, EPI_ISL_2089924, EPI_ISL_2089925, EPI_ISL_2089926, EPI_ISL_2089927, EPI_ISL_2089928, EPI_ISL_2089929, EPI_ISL_2089930, EPI_ISL_2089931, EPI_ISL_2089932, EPI_ISL_2089933, EPI_ISL_2089934, EPI_ISL_2089935, EPI_ISL_2089936, EPI_ISL_2089937, EPI_ISL_2089938, EPI_ISL_2089939, EPI_ISL_2089940, EPI_ISL_2089941, EPI_ISL_2089942, EPI_ISL_2089943, EPI_ISL_2089944, EPI_ISL_2089946, EPI_ISL_2089947, EPI_ISL_2089948, EPI_ISL_2089950, EPI_ISL_2089951, EPI_ISL_2089952, EPI_ISL_2089953, EPI_ISL_2089954, EPI_ISL_2089955, EPI_ISL_2089956, EPI_ISL_2089957, EPI_ISL_2089958, EPI_ISL_2089959, EPI_ISL_2089960, EPI_ISL_2089961, EPI_ISL_2089962, EPI_ISL_2089963, EPI_ISL_2089964, EPI_ISL_2089965, EPI_ISL_2089966, EPI_ISL_2089967, EPI_ISL_2089968, EPI_ISL_2089969, EPI_ISL_2089970, EPI_ISL_2089971, EPI_ISL_2089972, EPI_ISL_2089973, EPI_ISL_2089974, EPI_ISL_2089975, EPI_ISL_2089976, EPI_ISL_2089977, EPI_ISL_2089978, EPI_ISL_2089979, EPI_ISL_2089980, EPI_ISL_2089981, EPI_ISL_2089982, EPI_ISL_2089983, EPI_ISL_2089984, EPI_ISL_2089985, EPI_ISL_2089986, EPI_ISL_2089987, EPI_ISL_2089988, EPI_ISL_2089989, EPI_ISL_2089990, EPI_ISL_2089991, EPI_ISL_2089992, EPI_ISL_2089993, EPI_ISL_2089994, EPI_ISL_2089995, EPI_ISL_2089996, EPI_ISL_2089997, EPI_ISL_2089998, EPI_ISL_2089999, EPI_ISL_2090000, EPI_ISL_2090001, EPI_ISL_2090002, EPI_ISL_2090003, EPI_ISL_2090004, EPI_ISL_2090006, EPI_ISL_2090007, EPI_ISL_2090008, EPI_ISL_2090009, EPI_ISL_2090010, EPI_ISL_2090011, EPI_ISL_2090012, EPI_ISL_2090013, EPI_ISL_2090014, EPI_ISL_2090015, EPI_ISL_2090016, EPI_ISL_2090017, EPI_ISL_2090018, EPI_ISL_2090019, EPI_ISL_2090020, EPI_ISL_2090021, EPI_ISL_2090022, EPI_ISL_2090023, EPI_ISL_2090024, EPI_ISL_2090025, EPI_ISL_2090026, EPI_ISL_2090027, EPI_ISL_2090028, EPI_ISL_2090029, EPI_ISL_2090030, EPI_ISL_2090031, EPI_ISL_2090033, EPI_ISL_2090034, EPI_ISL_2090035, EPI_ISL_2090036, EPI_ISL_2090037, EPI_ISL_2090038, EPI_ISL_2090039, EPI_ISL_2090040, EPI_ISL_2090041, EPI_ISL_2090042, EPI_ISL_2090043, EPI_ISL_2090044, EPI_ISL_2090045, EPI_ISL_2090046, EPI_ISL_2090047, EPI_ISL_2090048, EPI_ISL_2090049, EPI_ISL_2090050, EPI_ISL_2090051, EPI_ISL_2090052, EPI_ISL_2090053, EPI_ISL_2090054, EPI_ISL_2090055, EPI_ISL_2090056, EPI_ISL_2090057, EPI_ISL_2090058, EPI_ISL_2090059, EPI_ISL_2090060, EPI_ISL_2090061, EPI_ISL_2090062, EPI_ISL_2090063, EPI_ISL_2090064, EPI_ISL_2090065, EPI_ISL_2090066, EPI_ISL_2090067, EPI_ISL_2090068, EPI_ISL_2090069, EPI_ISL_2090070, EPI_ISL_2090071, EPI_ISL_2090072, EPI_ISL_2090073, EPI_ISL_2090074, EPI_ISL_2090075, EPI_ISL_2090076, EPI_ISL_2090077, EPI_ISL_2090078, EPI_ISL_2090079, EPI_ISL_2090080, EPI_ISL_2090081, EPI_ISL_2090082, EPI_ISL_2090083, EPI_ISL_2090084, EPI_ISL_2090085, EPI_ISL_2090086, EPI_ISL_2090087, EPI_ISL_2090088, EPI_ISL_2090089, EPI_ISL_2090090, EPI_ISL_2090091, EPI_ISL_2090092, EPI_ISL_2090093, EPI_ISL_2090094, EPI_ISL_2090095, EPI_ISL_2090096, EPI_ISL_2090097, EPI_ISL_2090098, EPI_ISL_2090099, EPI_ISL_2090100, EPI_ISL_2090101, EPI_ISL_2090102, EPI_ISL_2090103, EPI_ISL_2090104, EPI_ISL_2090105, EPI_ISL_2090106, EPI_ISL_2090107, EPI_ISL_2090108, EPI_ISL_2090109, EPI_ISL_2090110, EPI_ISL_2090111, EPI_ISL_2090112, EPI_ISL_2090113, EPI_ISL_2090114, EPI_ISL_2090115, EPI_ISL_2090116, EPI_ISL_2090117, EPI_ISL_2090118, EPI_ISL_2090119, EPI_ISL_2090120, EPI_ISL_2090121, EPI_ISL_2090122, EPI_ISL_2090123, EPI_ISL_2090124, EPI_ISL_2090125, EPI_ISL_2090126, EPI_ISL_2090127, EPI_ISL_2090128, EPI_ISL_2090129, EPI_ISL_2090131, EPI_ISL_2090132, EPI_ISL_2090134, EPI_ISL_2090135, EPI_ISL_2090136, EPI_ISL_2090137, EPI_ISL_2090138, EPI_ISL_2090139, EPI_ISL_2090140, EPI_ISL_2090141, EPI_ISL_2090142, EPI_ISL_2090143, EPI_ISL_2090144, EPI_ISL_2090145, EPI_ISL_2090146, EPI_ISL_2090147, EPI_ISL_2090148, EPI_ISL_2090149, EPI_ISL_2090150, EPI_ISL_2090151, EPI_ISL_2090152, EPI_ISL_2090153, EPI_ISL_2090154, EPI_ISL_2090155, EPI_ISL_2090156, EPI_ISL_2090157, EPI_ISL_2090158, EPI_ISL_2090159, EPI_ISL_2090160, EPI_ISL_2090161, EPI_ISL_2090162, EPI_ISL_2090163, EPI_ISL_2090164, EPI_ISL_2090165, EPI_ISL_2090166, EPI_ISL_2090167, EPI_ISL_2090168, EPI_ISL_2090169, EPI_ISL_2090170, EPI_ISL_2090171, EPI_ISL_2090172, EPI_ISL_2090173, EPI_ISL_2090174, EPI_ISL_2090175, EPI_ISL_2090176, EPI_ISL_2090177, EPI_ISL_2090178, EPI_ISL_2090179, EPI_ISL_2090180, EPI_ISL_2090181, EPI_ISL_2090182, EPI_ISL_2090183, EPI_ISL_2090184, EPI_ISL_2090185, EPI_ISL_2090186, EPI_ISL_2090187, EPI_ISL_2090188, EPI_ISL_2090189, EPI_ISL_2090190, EPI_ISL_2090191, EPI_ISL_2090192, EPI_ISL_2090193, EPI_ISL_2090194, EPI_ISL_2090195, EPI_ISL_2090196, EPI_ISL_2090197, EPI_ISL_2090198, EPI_ISL_2090199, EPI_ISL_2090200, EPI_ISL_2090202, EPI_ISL_2090203, EPI_ISL_2090204, EPI_ISL_2090205, EPI_ISL_2090206, EPI_ISL_2090207, EPI_ISL_2090208, EPI_ISL_2090209, EPI_ISL_2090210, EPI_ISL_2090211, EPI_ISL_2090212, EPI_ISL_2090213, EPI_ISL_2090214, EPI_ISL_2090215, EPI_ISL_2090216, EPI_ISL_2090218, EPI_ISL_2090219, EPI_ISL_2090220, EPI_ISL_2090221, EPI_ISL_2090222, EPI_ISL_2090223, EPI_ISL_2090224, EPI_ISL_2090225, EPI_ISL_2090226, EPI_ISL_2090227, EPI_ISL_2090228, EPI_ISL_2090229, EPI_ISL_2090230, EPI_ISL_2090231, EPI_ISL_2090232, EPI_ISL_2090233, EPI_ISL_2090234, EPI_ISL_2090236, EPI_ISL_2090237, EPI_ISL_2090238, EPI_ISL_2090239, EPI_ISL_2090240, EPI_ISL_2090241, EPI_ISL_2090242, EPI_ISL_2090243, EPI_ISL_2090244, EPI_ISL_2090245, EPI_ISL_2090246, EPI_ISL_2090247, EPI_ISL_2090248, EPI_ISL_2090249, EPI_ISL_2090250, EPI_ISL_2090251, EPI_ISL_2090252, EPI_ISL_2090253, EPI_ISL_2090254, EPI_ISL_2090255, EPI_ISL_2090256, EPI_ISL_2090257, EPI_ISL_2090258, EPI_ISL_2090259, EPI_ISL_2090260, EPI_ISL_2090261, EPI_ISL_2090262, EPI_ISL_2090263, EPI_ISL_2090264, EPI_ISL_2090265, EPI_ISL_2090266, EPI_ISL_2090267, EPI_ISL_2090268, EPI_ISL_2090269, EPI_ISL_2090270, EPI_ISL_2090271, EPI_ISL_2090272, EPI_ISL_2090273, EPI_ISL_2090274, EPI_ISL_2090275, EPI_ISL_2090276, EPI_ISL_2090277, EPI_ISL_2090279, EPI_ISL_2090280, EPI_ISL_2090281, EPI_ISL_2090282, EPI_ISL_2090283, EPI_ISL_2090284, EPI_ISL_2090285, EPI_ISL_2090286, EPI_ISL_2090287, EPI_ISL_2090288, EPI_ISL_2090289, EPI_ISL_2090291, EPI_ISL_2090292, EPI_ISL_2090293, EPI_ISL_2090294, EPI_ISL_2090295, EPI_ISL_2090296, EPI_ISL_2090297, EPI_ISL_2090298, EPI_ISL_2090299, EPI_ISL_2090300, EPI_ISL_2090301, EPI_ISL_2090302, EPI_ISL_2090303, EPI_ISL_2090304, EPI_ISL_2090305, EPI_ISL_2090306, EPI_ISL_2090307, EPI_ISL_2090308, EPI_ISL_2090309, EPI_ISL_2090310, EPI_ISL_2090311, EPI_ISL_2090312, EPI_ISL_2090313, EPI_ISL_2090314, EPI_ISL_2090315, EPI_ISL_2090317, EPI_ISL_2090318, EPI_ISL_2090319, EPI_ISL_2090320, EPI_ISL_2090321, EPI_ISL_2090322, EPI_ISL_2090323, EPI_ISL_2090324, EPI_ISL_2090325, EPI_ISL_2090326, EPI_ISL_2090327, EPI_ISL_2090328, EPI_ISL_2090329, EPI_ISL_2090330, EPI_ISL_2090332, EPI_ISL_2090333, EPI_ISL_2090334, EPI_ISL_2090335, EPI_ISL_2090336, EPI_ISL_2090337, EPI_ISL_2090338, EPI_ISL_2090339, EPI_ISL_2090340, EPI_ISL_2090341, EPI_ISL_2090342, EPI_ISL_2090343, EPI_ISL_2090344, EPI_ISL_2090345, EPI_ISL_2090346, EPI_ISL_2090347, EPI_ISL_2090348, EPI_ISL_2090349, EPI_ISL_2090350, EPI_ISL_2090351, EPI_ISL_2090352, EPI_ISL_2090353, EPI_ISL_2090354, EPI_ISL_2090355, EPI_ISL_2090356, EPI_ISL_2090357, EPI_ISL_2090358, EPI_ISL_2090359, EPI_ISL_2090360, EPI_ISL_2090361, EPI_ISL_2090362, EPI_ISL_2090363, EPI_ISL_2090364, EPI_ISL_2090365, EPI_ISL_2090366, EPI_ISL_2090367, EPI_ISL_2090368, EPI_ISL_2090369, EPI_ISL_2090370, EPI_ISL_2090371, EPI_ISL_2090372, EPI_ISL_2090373, EPI_ISL_2090374, EPI_ISL_2090375, EPI_ISL_2090376, EPI_ISL_2090377, EPI_ISL_2090378, EPI_ISL_2090379, EPI_ISL_2090380, EPI_ISL_2090381, EPI_ISL_2090382, EPI_ISL_2090383, EPI_ISL_2090384, EPI_ISL_2090385, EPI_ISL_2090386, EPI_ISL_2090387, EPI_ISL_2090388, EPI_ISL_2090389, EPI_ISL_2090391, EPI_ISL_2090392, EPI_ISL_2090393, EPI_ISL_2090394, EPI_ISL_2090395, EPI_ISL_2090396, EPI_ISL_2090397, EPI_ISL_2090398, EPI_ISL_2090399, EPI_ISL_2090400, EPI_ISL_2090401, EPI_ISL_2090402, EPI_ISL_2090403, EPI_ISL_2090404, EPI_ISL_2090405, EPI_ISL_2090406, EPI_ISL_2090407, EPI_ISL_2090408, EPI_ISL_2090409, EPI_ISL_2090410, EPI_ISL_2090411, EPI_ISL_2090412, EPI_ISL_2090413, EPI_ISL_2090414, EPI_ISL_2090415, EPI_ISL_2090416, EPI_ISL_2090417, EPI_ISL_2090418, EPI_ISL_2090419, EPI_ISL_2090420, EPI_ISL_2090421, EPI_ISL_2090422, EPI_ISL_2090423, EPI_ISL_2090424, EPI_ISL_2090425, EPI_ISL_2090426, EPI_ISL_2090427, EPI_ISL_2090428, EPI_ISL_2090429, EPI_ISL_2090430, EPI_ISL_2090431, EPI_ISL_2090432, EPI_ISL_2090433, EPI_ISL_2090434, EPI_ISL_2090435, EPI_ISL_2090436, EPI_ISL_2090437, EPI_ISL_2090438, EPI_ISL_2090439, EPI_ISL_2090440, EPI_ISL_2090441, EPI_ISL_2090442, EPI_ISL_2090443, EPI_ISL_2090444, EPI_ISL_2090445, EPI_ISL_2090446, EPI_ISL_2090447, EPI_ISL_2090448, EPI_ISL_2090449, EPI_ISL_2090450, EPI_ISL_2090451, EPI_ISL_2090452, EPI_ISL_2090453, EPI_ISL_2090454, EPI_ISL_2090455, EPI_ISL_2090456, EPI_ISL_2090457, EPI_ISL_2090458, EPI_ISL_2090459, EPI_ISL_2090460, EPI_ISL_2090464, EPI_ISL_2090465, EPI_ISL_2090466, EPI_ISL_2090467, EPI_ISL_2090468, EPI_ISL_2090469, EPI_ISL_2090470, EPI_ISL_2090471, EPI_ISL_2090472 | | | |
| see above | Aegis Sciences Corporation | Centers for Disease Control and Prevention Division of Viral Diseases, Pathogen Discovery | Dakota Howard, Dhwani Batra, Peter W. Cook, Kara Moser, Adrian Paskey, Jason Caravas, Benjamin Rambo-Martin, Shatavia Morrison, Christopher Gulvick, Scott Sammons, Yvette Unoarumhi, Darlene Wagner, Matthew Schmerer, Cyndi Clark, Patrick Campbell, Rob Case, Vikramsinha Ghorpade, Holly Houdeshell, Ola Kvalvaag, Dillon Nall, Ethan Sanders, Alec Vest, Shaun Westlund, Matthew Hardison, Clinton R. Paden, Duncan MacCannell |
| EPI_ISL_2090473, EPI_ISL_2090474, EPI_ISL_2090475, EPI_ISL_2090476, EPI_ISL_2090477, EPI_ISL_2090478, EPI_ISL_2090479, EPI_ISL_2090480, EPI_ISL_2090481, EPI_ISL_2090482, EPI_ISL_2090483, EPI_ISL_2090484, EPI_ISL_2090485, EPI_ISL_2090486, EPI_ISL_2090487, EPI_ISL_2090488, EPI_ISL_2090489, EPI_ISL_2090490, EPI_ISL_2090491, EPI_ISL_2090492, EPI_ISL_2090493, EPI_ISL_2090494, EPI_ISL_2090495, EPI_ISL_2090496, EPI_ISL_2090497, EPI_ISL_2090498, EPI_ISL_2090499, EPI_ISL_2090500, EPI_ISL_2090501, EPI_ISL_2090502, EPI_ISL_2090503, EPI_ISL_2090505, EPI_ISL_2090506, EPI_ISL_2090507, EPI_ISL_2090508, EPI_ISL_2090509, EPI_ISL_2090510, EPI_ISL_2090511, EPI_ISL_2090512, EPI_ISL_2090513, EPI_ISL_2090514, EPI_ISL_2090515, EPI_ISL_2090516, EPI_ISL_2090517, EPI_ISL_2090518, EPI_ISL_2090519, EPI_ISL_2090520, EPI_ISL_2090521, EPI_ISL_2090522, EPI_ISL_2090523, EPI_ISL_2090524, EPI_ISL_2090525, EPI_ISL_2090526, EPI_ISL_2090527, EPI_ISL_2090528, EPI_ISL_2090529, EPI_ISL_2090530, EPI_ISL_2090531, EPI_ISL_2090532, EPI_ISL_2090533, EPI_ISL_2090534, EPI_ISL_2090535, EPI_ISL_2090536, EPI_ISL_2090537, EPI_ISL_2090538, EPI_ISL_2090539, EPI_ISL_2090540, EPI_ISL_2090541, EPI_ISL_2090542, EPI_ISL_2090543, EPI_ISL_2090544, EPI_ISL_2090545, EPI_ISL_2090546, EPI_ISL_2090547, EPI_ISL_2090548, EPI_ISL_2090549, EPI_ISL_2090550, EPI_ISL_2090551, EPI_ISL_2090552, EPI_ISL_2090553, EPI_ISL_2090554, EPI_ISL_2090555, EPI_ISL_2090556, EPI_ISL_2090557, EPI_ISL_2090558, EPI_ISL_2090559, EPI_ISL_2090560, EPI_ISL_2090561, EPI_ISL_2090562, EPI_ISL_2090563, EPI_ISL_2090564, EPI_ISL_2090565, EPI_ISL_2090566, EPI_ISL_2090567, EPI_ISL_2090568, EPI_ISL_2090569, EPI_ISL_2090570, EPI_ISL_2090571, EPI_ISL_2090572, EPI_ISL_2090573, EPI_ISL_2090574, EPI_ISL_2090575, EPI_ISL_2090576, EPI_ISL_2090577, EPI_ISL_2090578, EPI_ISL_2090579, EPI_ISL_2090580, EPI_ISL_2090581, EPI_ISL_2090582, EPI_ISL_2090583, EPI_ISL_2090584, EPI_ISL_2090585, EPI_ISL_2090586, EPI_ISL_2090587, EPI_ISL_2090588, EPI_ISL_2090589, EPI_ISL_2090590, EPI_ISL_2090591, EPI_ISL_2090592, EPI_ISL_2090593, EPI_ISL_2090594, EPI_ISL_2090595, EPI_ISL_2090596, EPI_ISL_2090597, EPI_ISL_2090598, EPI_ISL_2090599, EPI_ISL_2090600, EPI_ISL_2090601, EPI_ISL_2090602, EPI_ISL_2090603, EPI_ISL_2090604 | | | |
| see above | Quest Diagnostics Incorporated | Centers for Disease Control and Prevention Division of Viral Diseases, Pathogen Discovery | Dakota Howard, Dhwani Batra, Peter W. Cook, Kara Moser, Adrian Paskey, Jason Caravas, Benjamin Rambo-Martin, Shatavia Morrison, Christopher Gulvick, Scott Sammons, Yvette Unoarumhi, Darlene Wagner, Matthew Schmerer, S. H. Rosenthal, A. Gerasimova, R. M. Kagan, B. Anderson, M. Hua, Y. Liu, L.E. Bernstein, K.E. Livingston, A. Perez, I. A. Shlyakhter, R. V. Rolando, R. Owen, P. Tanpaiboon, F. Lacbawan, Clinton R. Paden, Duncan MacCannell |
| EPI_ISL_2090617, EPI_ISL_2090618 | Laboratory Medicine, University of Washington | Laboratory Medicine, University of Washington | Roychoudhury,P., Xie,H., Shrestha,L., Bakhash,S.M., Rachleff,V.M.,Lin,M.J., Baker,N.R., Ellis,S., Sathees,S., Huang,M.-L.,Jerome,K.R. and Greninger,A. |
| EPI_ISL_2090619 | Microbiology, University of Pennsylvania | Microbiology, University of Pennsylvania | Everett,J., Rodino,K., Reddy,S., Hokama,P., Roche,A.M., Hwang,Y.,Glascock,A., Sherrill-Mix,S., Whiteside,S.A., Graham-Wooten,J.,Khatib,L.A., Fitzgerald,A.S., Ganguly,A., Feldman,M., Kelly,B.,Collman,R.G. and Bushman,F. |
| EPI_ISL_2090629, EPI_ISL_2090632, EPI_ISL_2090633, EPI_ISL_2090634, EPI_ISL_2090635, EPI_ISL_2090636, EPI_ISL_2090637, EPI_ISL_2090638, EPI_ISL_2090639, EPI_ISL_2090640, EPI_ISL_2090641, EPI_ISL_2090642, EPI_ISL_2090643, EPI_ISL_2090644, EPI_ISL_2090645, EPI_ISL_2090646, EPI_ISL_2090647 | | | |
| see above | Idaho Bureau of Laboratories | Idaho Bureau of Laboratories | R. Beukelman, Matthew Charles Burns, Aimee Ceniseros, Robert L. Voermans, Christopher Ball |
| EPI_ISL_2090648 | InDRE | Instituto Nacional de Medicina Genomica | Hidalgo-Miranda A, Mendoza-Vargas A, Reyes-Grajeda JP, Cedro-Tanda A,Cisneros-Villanueva M, Orjuela-Rodríguez M, Alcaraz N, Gisela Barrera-Badillo, Irma Lopez-Martinez, Jose Ernesto Ramirez González, Gonzalez-Barrera D, Rangel-DeLeon D, Miranda-Ortiz H, Rosas-Escobar P, Canseco Mendez JC, Munguia-Garza P, Ramirez-Vega O, Garcia-Cardenas FJ, Gonzalez-Woge MA, Herrera-Montalvo LA. |
| EPI_ISL_2090649 | InDRE | Instituto Nacional de Medicina Genomica | Hidalgo-Miranda A, Mendoza-Vargas A, Reyes-Grajeda JP, Cedro-Tanda A, Cisneros-Villanueva M, Orjuela-Rodríguez M, Alcaraz N, Gisela Barrera-Badillo, Irma Lopez-Martinez, Jose Ernesto Ramirez González, Gonzalez-Barrera D, Rangel-DeLeon D, Miranda-Ortiz H, Rosas-Escobar P, Canseco Mendez JC, Munguia-Garza P, Ramirez-Vega O, Garcia-Cardenas FJ, Gonzalez-Woge MA, Herrera-Montalvo LA. |
| EPI_ISL_2090650, EPI_ISL_2090651, EPI_ISL_2090652, EPI_ISL_2090653 | Rush University Medical Center | RIPHL at Rush University Medical Center | Stefan Green, Kevin Kunstman, Max Kolton, Marieta Hyde, Laura Furtado, Felix Araujo Perez |
| EPI_ISL_2090654, EPI_ISL_2090655, EPI_ISL_2090656, EPI_ISL_2090657, EPI_ISL_2090658, EPI_ISL_2090659, EPI_ISL_2090661 | University of Chicago Medicine | RIPHL at Rush University Medical Center | Stefan Green, Kevin Kunstman, Max Kolton, Marieta Hyde, Laura Furtado, Felix Araujo Perez |
| EPI_ISL_2090662, EPI_ISL_2090663, EPI_ISL_2090664, EPI_ISL_2090665, EPI_ISL_2090666, EPI_ISL_2090667, EPI_ISL_2090668, EPI_ISL_2090669, EPI_ISL_2090670, EPI_ISL_2090671, EPI_ISL_2090672, EPI_ISL_2090673, EPI_ISL_2090674, EPI_ISL_2090675, EPI_ISL_2090676, EPI_ISL_2090677, EPI_ISL_2090678, EPI_ISL_2090679, EPI_ISL_2090680, EPI_ISL_2090681, EPI_ISL_2090682, EPI_ISL_2090683, EPI_ISL_2090684, EPI_ISL_2090685, EPI_ISL_2090686 | | | |
| see above | Northwestern Memorial Hospital | RIPHL at Rush University Medical Center | Stefan Green, Kevin Kunstman, Max Kolton, Marieta Hyde, Laura Furtado, Felix Araujo Perez |
| EPI_ISL_2090687 | John H Stroger Jr Hospital of Cook County | RIPHL at Rush University Medical Center | Stefan Green, Kevin Kunstman, Max Kolton, Marieta Hyde, Laura Furtado, Felix Araujo Perez |
| EPI_ISL_2090688, EPI_ISL_2090689, EPI_ISL_2090691, EPI_ISL_2090692, EPI_ISL_2090695, EPI_ISL_2090696, | Wyoming Public Health Laboratory | Wyoming Public Health Laboratory | Jim Mildenberger, Taylor Fearing, Channing Weber, Ashley Norberg, Chayse Rowley, Marley Goetz, Brian Dominguez, Elliot Thomasson, Brittany Oher, Sam Britz, Cari Sloma, and Rob Christensen |

| | | | |
|---|---|---|---|
| EPI_ISL_2090697, EPI_ISL_2090698, EPI_ISL_2090701 | | | |
| EPI_ISL_2090702, EPI_ISL_2090703, EPI_ISL_2090704, EPI_ISL_2090705, EPI_ISL_2090706, EPI_ISL_2090707, EPI_ISL_2090708, EPI_ISL_2090709, EPI_ISL_2090710, EPI_ISL_2090711, EPI_ISL_2090712, EPI_ISL_2090713, EPI_ISL_2090714, EPI_ISL_2090715, EPI_ISL_2090716, EPI_ISL_2090717, EPI_ISL_2090718, EPI_ISL_2090719, EPI_ISL_2090720, EPI_ISL_2090721, EPI_ISL_2090722, EPI_ISL_2090723, EPI_ISL_2090724, EPI_ISL_2090725, EPI_ISL_2090726, EPI_ISL_2090727, EPI_ISL_2090892, EPI_ISL_2090893, EPI_ISL_2090894, EPI_ISL_2090895, EPI_ISL_2090896, EPI_ISL_2090897, EPI_ISL_2090898, EPI_ISL_2090899, EPI_ISL_2090900, EPI_ISL_2090901, EPI_ISL_2090902, EPI_ISL_2090903, EPI_ISL_2090904, EPI_ISL_2090905 | | | |
| see above | Nebraska Public Health Laboratory | NPHL COVID-19 Response Team | NPHL COVID-19 Response Team |
| EPI_ISL_2090907, EPI_ISL_2090908, EPI_ISL_2090909, EPI_ISL_2090911, EPI_ISL_2090912, EPI_ISL_2090915, EPI_ISL_2090916, EPI_ISL_2090918, EPI_ISL_2090919, EPI_ISL_2090920, EPI_ISL_2090921, EPI_ISL_2090923, EPI_ISL_2090924, EPI_ISL_2090925, EPI_ISL_2090926, EPI_ISL_2090927, EPI_ISL_2090929, EPI_ISL_2090930, EPI_ISL_2090931, EPI_ISL_2090932, EPI_ISL_2090934, EPI_ISL_2090935, EPI_ISL_2090936, EPI_ISL_2090937, EPI_ISL_2090938, EPI_ISL_2090939, EPI_ISL_2090941, EPI_ISL_2090942, EPI_ISL_2090948, EPI_ISL_2090950, EPI_ISL_2090951, EPI_ISL_2090954, EPI_ISL_2090956, EPI_ISL_2090957, EPI_ISL_2090959, EPI_ISL_2090960, EPI_ISL_2090961, EPI_ISL_2090962, EPI_ISL_2090964, EPI_ISL_2090969, EPI_ISL_2090971, EPI_ISL_2090972, EPI_ISL_2090973, EPI_ISL_2090975, EPI_ISL_2090976, EPI_ISL_2090978 | | | |
| see above | North Dakota Department of Health, Public Health Laboratory | North Dakota Department of Health, Public Health Laboratory | Lisa Wingerter |
| EPI_ISL_2090980, EPI_ISL_2090981, EPI_ISL_2090982, EPI_ISL_2090983, EPI_ISL_2090984, EPI_ISL_2090985, EPI_ISL_2090986, EPI_ISL_2090987, EPI_ISL_2090989, EPI_ISL_2090990, EPI_ISL_2090991, EPI_ISL_2090993, EPI_ISL_2090994, EPI_ISL_2090995, EPI_ISL_2090996, EPI_ISL_2090997, EPI_ISL_2090998, EPI_ISL_2090999 | | | |
| see above | Wyoming Public Health Laboratory | Wyoming Public Health Laboratory | Jim Mildenberger, Taylor Fearing, Channing Weber, Ashley Norberg, Chayse Rowley, Marley Goetz, Brian Dominguez, Elliot Thomasson, Brittany Oher, Sam Britz, Cari Sloma, and Rob Christensen |
| EPI_ISL_2091002, EPI_ISL_2091003, EPI_ISL_2091004, EPI_ISL_2091005, EPI_ISL_2091006, EPI_ISL_2091007, EPI_ISL_2091008, EPI_ISL_2091009, EPI_ISL_2091010, EPI_ISL_2091011, EPI_ISL_2091012 | | | |
| see above | Nebraska Public Health Laboratory | NPHL COVID-19 Response Team | NPHL COVID-19 Response Team |
| EPI_ISL_2091026, EPI_ISL_2091027, EPI_ISL_2091029, EPI_ISL_2091030, EPI_ISL_2091031, EPI_ISL_2091032, EPI_ISL_2091033, EPI_ISL_2091034, EPI_ISL_2091035, EPI_ISL_2091036, EPI_ISL_2091037, EPI_ISL_2091040, EPI_ISL_2091041, EPI_ISL_2091042, EPI_ISL_2091043, EPI_ISL_2091044, EPI_ISL_2091045, EPI_ISL_2091046, EPI_ISL_2091047, EPI_ISL_2091050, EPI_ISL_2091052 | | | |
| see above | Kansas Health and Environmental Lab | Wichita State University - Molecular Diagnostics Lab | Mike Grose, Jonathan Barnell, Ben Olsen, and Phil Adam |
| EPI_ISL_2091053, EPI_ISL_2091054, EPI_ISL_2091055, EPI_ISL_2091056, EPI_ISL_2091057, EPI_ISL_2091058, EPI_ISL_2091059, EPI_ISL_2091060, EPI_ISL_2091061, EPI_ISL_2091062, EPI_ISL_2091063, EPI_ISL_2091064, EPI_ISL_2091065, EPI_ISL_2091066, EPI_ISL_2091067, EPI_ISL_2091068, EPI_ISL_2091069, EPI_ISL_2091070, EPI_ISL_2091071, EPI_ISL_2091072, EPI_ISL_2091073, EPI_ISL_2091074, EPI_ISL_2091075, EPI_ISL_2091076, EPI_ISL_2091077, EPI_ISL_2091078, EPI_ISL_2091079, EPI_ISL_2091080, EPI_ISL_2091081, EPI_ISL_2091082, EPI_ISL_2091083, EPI_ISL_2091084, EPI_ISL_2091085, EPI_ISL_2091086, EPI_ISL_2091087, EPI_ISL_2091088, EPI_ISL_2091089, EPI_ISL_2091090, EPI_ISL_2091091, EPI_ISL_2091092, EPI_ISL_2091093, EPI_ISL_2091094, EPI_ISL_2091095, EPI_ISL_2091096, EPI_ISL_2091097, EPI_ISL_2091098, EPI_ISL_2091099, EPI_ISL_2091100, EPI_ISL_2091101, EPI_ISL_2091102, EPI_ISL_2091103, EPI_ISL_2091104, EPI_ISL_2091105, EPI_ISL_2091106, EPI_ISL_2091107, EPI_ISL_2091108, EPI_ISL_2091109, EPI_ISL_2091110, EPI_ISL_2091111, EPI_ISL_2091112, EPI_ISL_2091113, EPI_ISL_2091114, EPI_ISL_2091115, EPI_ISL_2091116, EPI_ISL_2091117, EPI_ISL_2091118 | | | |
| see above | University Hospitals Translational Laboratory (UHTL), University Hospitals | University Hospitals Translational Laboratory (UHTL), University Hospitals | Sadri,N., Alouani,D., Song,X. |
| EPI_ISL_2091132 | Unidad de Investigación Biomédica de Zacatecas (UIBZ) | Centro de Investigación en Enfermedades Infecciosas (CIENI), Instituto Nacional de Enfermedades Respiratorias (INER) | Consorcio Mexicano de Vigilancia Genómica (CoViGen-Mex). Authors (in alphabetical order): Julio Elias Alvarado-Yaah, Carlos F. Arias, Santiago Ávila-Ríos, Víctor Hugo Borja-Aburto, Celia Boukadida, Juan Bautista Chale-Dzul, Célida Duque Molina, José Antonio Enciso-Moreno, Gloria Elena Espinosa-Ayala, Fernando Fontove-Herrera, Victor Eduardo Garcia-Arias, Concepción Grajales-Muñiz, Ricardo Grande, Alfredo Herrera-Estrella, Carla Ivón Herrera-Najera, Pavel Isa, Brenda Irasema Maldonado-Meza, Bernardo Martínez-Miguel, Margarita Matías-Florentino, María Guadalupe de Jesús Mireles-Rivera, Gloria María Molina-Salinas, Hector Montoya-Fuentes, José Esteban Muñoz-Medina, José de Jesús Nuñez-Contreras, Alicia Ocaña-Mondragón, Luis Alberto Ochoa-Carrera, Hector Esteban Paz-Juárez, Francisco Pulido, Helen Haydee Fernanda Ramírez-Plascencia, Angel Gustavo Salas-Lais, Alejandro Sanchez-Flores, Clara Esperanza Santacruz-Tinoco, María Guadalupe Santiago-Mauricio, Nelly Sélem-Mojica, Blanca Taboada, Gloria Vazquez |
| EPI_ISL_2091133, EPI_ISL_2091134 | Laboratorio Central de Epidemiología (LCE) | Centro de Investigación en Enfermedades Infecciosas (CIENI), Instituto Nacional de Enfermedades Respiratorias (INER) | Consorcio Mexicano de Vigilancia Genómica (CoViGen-Mex). Authors (in alphabetical order): Julio Elias Alvarado-Yaah, Carlos F. Arias, Santiago Ávila-Ríos, Víctor Hugo Borja-Aburto, Celia Boukadida, Juan Bautista Chale-Dzul, Célida Duque Molina, José Antonio Enciso-Moreno, Gloria Elena Espinosa-Ayala, Fernando Fontove-Herrera, Victor Eduardo Garcia-Arias, Concepción Grajales-Muñiz, Ricardo Grande, Alfredo Herrera-Estrella, Carla Ivón Herrera-Najera, Pavel Isa, Brenda Irasema Maldonado-Meza, Bernardo Martínez-Miguel, Margarita Matías-Florentino, María Guadalupe de Jesús Mireles-Rivera, Gloria María Molina-Salinas, Hector Montoya-Fuentes, José Esteban Muñoz-Medina, José de Jesús Nuñez-Contreras, Alicia Ocaña-Mondragón, Luis Alberto Ochoa-Carrera, Hector Esteban Paz-Juárez, Francisco Pulido, Helen Haydee Fernanda Ramírez-Plascencia, Angel Gustavo Salas-Lais, Alejandro Sanchez-Flores, Clara Esperanza Santacruz-Tinoco, María Guadalupe Santiago-Mauricio, Nelly Sélem-Mojica, Blanca Taboada, Gloria Vazquez |
| EPI_ISL_2091135, EPI_ISL_2091136 | Centro de Investigación Biomédica del Noreste (CIBIN) | Centro de Investigación en Enfermedades Infecciosas (CIENI), Instituto Nacional de Enfermedades Respiratorias (INER) | Consorcio Mexicano de Vigilancia Genómica (CoViGen-Mex). Authors (in alphabetical order): Julio Elias Alvarado-Yaah, Carlos F. Arias, Santiago Ávila-Ríos, Víctor Hugo Borja-Aburto, Celia Boukadida, Juan Bautista Chale-Dzul, Célida Duque Molina, José Antonio Enciso-Moreno, Gloria Elena Espinosa-Ayala, Fernando Fontove-Herrera, Victor Eduardo Garcia-Arias, Concepción Grajales-Muñiz, Ricardo Grande, Alfredo Herrera-Estrella, Carla Ivón Herrera-Najera, Pavel Isa, Brenda Irasema Maldonado-Meza, Bernardo Martínez-Miguel, Margarita Matías-Florentino, María Guadalupe de Jesús Mireles-Rivera, Gloria María Molina-Salinas, Hector Montoya-Fuentes, José Esteban Muñoz-Medina, José de Jesús Nuñez-Contreras, Alicia Ocaña-Mondragón, Luis Alberto Ochoa-Carrera, Hector Esteban Paz-Juárez, Francisco Pulido, Helen Haydee Fernanda Ramírez-Plascencia, Angel Gustavo Salas-Lais, Alejandro Sanchez-Flores, Clara Esperanza Santacruz-Tinoco, María Guadalupe Santiago-Mauricio, Nelly Sélem-Mojica, Blanca Taboada, Gloria Vazquez |
| EPI_ISL_2091137 | Laboratorio Central de Epidemiología (LCE) | Centro de Investigación en Enfermedades Infecciosas (CIENI), Instituto Nacional de Enfermedades Respiratorias (INER) | Consorcio Mexicano de Vigilancia Genómica (CoViGen-Mex). Authors (in alphabetical order): Julio Elias Alvarado-Yaah, Carlos F. Arias, Santiago Ávila-Ríos, Víctor Hugo Borja-Aburto, Celia Boukadida, Juan Bautista Chale-Dzul, Célida Duque Molina, José Antonio Enciso-Moreno, Gloria Elena Espinosa-Ayala, Fernando Fontove-Herrera, Victor Eduardo Garcia-Arias, Concepción Grajales-Muñiz, Ricardo Grande, Alfredo Herrera-Estrella, Carla Ivón Herrera-Najera, Pavel Isa, Brenda Irasema Maldonado-Meza, Bernardo Martínez-Miguel, Margarita Matías-Florentino, María Guadalupe de Jesús Mireles-Rivera, Gloria María Molina-Salinas, Hector Montoya-Fuentes, José Esteban Muñoz-Medina, José de Jesús Nuñez-Contreras, Alicia Ocaña-Mondragón, Luis Alberto Ochoa-Carrera, Hector Esteban Paz-Juárez, Francisco Pulido, Helen Haydee Fernanda Ramírez-Plascencia, Angel Gustavo Salas-Lais, Alejandro Sanchez-Flores, Clara Esperanza Santacruz-Tinoco, María Guadalupe Santiago-Mauricio, Nelly Sélem-Mojica, Blanca Taboada, Gloria Vazquez |
| EPI_ISL_2091138, EPI_ISL_2091139 | Centro de Investigación Biomédica del Noreste (CIBIN) | Centro de Investigación en Enfermedades Infecciosas (CIENI), Instituto Nacional de Enfermedades Respiratorias (INER) | Consorcio Mexicano de Vigilancia Genómica (CoViGen-Mex). Authors (in alphabetical order): Julio Elias Alvarado-Yaah, Carlos F. Arias, Santiago Ávila-Ríos, Víctor Hugo Borja-Aburto, Celia Boukadida, Juan Bautista Chale-Dzul, Célida Duque Molina, José Antonio Enciso-Moreno, Gloria Elena Espinosa-Ayala, Fernando Fontove-Herrera, Victor Eduardo Garcia-Arias, Concepción Grajales-Muñiz, Ricardo Grande, Alfredo Herrera-Estrella, Carla Ivón Herrera-Najera, Pavel Isa, Brenda Irasema Maldonado-Meza, Bernardo Martínez-Miguel, Margarita Matías-Florentino, María Guadalupe de Jesús Mireles-Rivera, Gloria María Molina-Salinas, Hector Montoya-Fuentes, José Esteban Muñoz-Medina, José de Jesús Nuñez-Contreras, Alicia Ocaña-Mondragón, Luis Alberto Ochoa-Carrera, Hector Esteban Paz-Juárez, Francisco Pulido, Helen Haydee Fernanda Ramírez-Plascencia, Angel Gustavo Salas-Lais, Alejandro Sanchez-Flores, Clara Esperanza Santacruz-Tinoco, María Guadalupe Santiago-Mauricio, Nelly Sélem-Mojica, Blanca Taboada, Gloria Vazquez |
| EPI_ISL_2091140, EPI_ISL_2091141, EPI_ISL_2091142 | Centro de Investigación Biomédica de Occidente (CIBO) | Centro de Investigación en Enfermedades Infecciosas (CIENI), Instituto Nacional de Enfermedades Respiratorias (INER) | Consorcio Mexicano de Vigilancia Genómica (CoViGen-Mex). Authors (in alphabetical order): Julio Elias Alvarado-Yaah, Carlos F. Arias, Santiago Ávila-Ríos, Víctor Hugo Borja-Aburto, Celia Boukadida, Juan Bautista Chale-Dzul, Célida Duque Molina, José Antonio Enciso-Moreno, Gloria Elena Espinosa-Ayala, Fernando Fontove-Herrera, Victor Eduardo Garcia-Arias, Concepción Grajales-Muñiz, Ricardo Grande, Alfredo Herrera-Estrella, Carla Ivón Herrera-Najera, Pavel Isa, Brenda Irasema Maldonado-Meza, Bernardo Martínez-Miguel, Margarita Matías-Florentino, María Guadalupe de Jesús Mireles-Rivera, Gloria María Molina-Salinas, Hector Montoya-Fuentes, José Esteban Muñoz-Medina, José de Jesús Nuñez-Contreras, Alicia Ocaña-Mondragón, Luis Alberto Ochoa-Carrera, Hector Esteban Paz-Juárez, Francisco Pulido, Helen Haydee Fernanda Ramírez-Plascencia, Angel Gustavo Salas-Lais, Alejandro Sanchez-Flores, Clara Esperanza Santacruz-Tinoco, María Guadalupe Santiago-Mauricio, Nelly Sélem-Mojica, Blanca Taboada, Gloria Vazquez |
| EPI_ISL_2091143, EPI_ISL_2091144 | Laboratorio Central de Epidemiología (LCE) | Centro de Investigación en Enfermedades Infecciosas (CIENI), Instituto Nacional de Enfermedades Respiratorias (INER) | Consorcio Mexicano de Vigilancia Genómica (CoViGen-Mex). Authors (in alphabetical order): Julio Elias Alvarado-Yaah, Carlos F. Arias, Santiago Ávila-Ríos, Víctor Hugo Borja-Aburto, Celia Boukadida, Juan Bautista Chale-Dzul, Célida Duque Molina, José Antonio Enciso-Moreno, Gloria Elena Espinosa-Ayala, Fernando Fontove-Herrera, Victor Eduardo Garcia-Arias, Concepción Grajales-Muñiz, Ricardo Grande, Alfredo Herrera-Estrella, Carla Ivón Herrera-Najera, Pavel Isa, Brenda Irasema Maldonado-Meza, Bernardo Martínez-Miguel, Margarita Matías-Florentino, María Guadalupe de Jesús Mireles-Rivera, Gloria María Molina-Salinas, Hector Montoya-Fuentes, José Esteban Muñoz-Medina, José de Jesús Nuñez-Contreras, Alicia Ocaña-Mondragón, Luis Alberto Ochoa-Carrera, Hector Esteban Paz-Juárez, Francisco Pulido, Helen Haydee Fernanda Ramírez-Plascencia, Angel Gustavo Salas-Lais, Alejandro Sanchez-Flores, Clara Esperanza Santacruz-Tinoco, María Guadalupe Santiago-Mauricio, Nelly Sélem-Mojica, Blanca Taboada, Gloria Vazquez |
| EPI_ISL_2091145 | Centro de Investigación Biomédica de Occidente (CIBO) | Centro de Investigación en Enfermedades Infecciosas (CIENI), Instituto Nacional de Enfermedades Respiratorias (INER) | Consorcio Mexicano de Vigilancia Genómica (CoViGen-Mex). Authors (in alphabetical order): Julio Elias Alvarado-Yaah, Carlos F. Arias, Santiago Ávila-Ríos, Víctor Hugo Borja-Aburto, Celia Boukadida, Juan Bautista Chale-Dzul, Célida Duque Molina, José Antonio Enciso-Moreno, Gloria Elena Espinosa-Ayala, Fernando Fontove-Herrera, Victor Eduardo Garcia-Arias, Concepción Grajales-Muñiz, Ricardo Grande, Alfredo Herrera-Estrella, Carla Ivón Herrera-Najera, Pavel Isa, Brenda Irasema Maldonado-Meza, Bernardo Martínez-Miguel, Margarita Matías-Florentino, María Guadalupe de Jesús Mireles-Rivera, Gloria María Molina-Salinas, Hector Montoya-Fuentes, José Esteban Muñoz-Medina, José de Jesús Nuñez-Contreras, Alicia Ocaña-Mondragón, Luis Alberto Ochoa-Carrera, Hector Esteban Paz-Juárez, Francisco Pulido, Helen Haydee Fernanda Ramírez-Plascencia, Angel Gustavo Salas-Lais, Alejandro Sanchez-Flores, Clara Esperanza Santacruz-Tinoco, María Guadalupe Santiago-Mauricio, Nelly Sélem-Mojica, Blanca Taboada, Gloria Vazquez |
| EPI_ISL_2091146, EPI_ISL_2091147, EPI_ISL_2091148, EPI_ISL_2091149, EPI_ISL_2091150, EPI_ISL_2091151, EPI_ISL_2091152, EPI_ISL_2091153, EPI_ISL_2091154, EPI_ISL_2091155, EPI_ISL_2091156, EPI_ISL_2091157, EPI_ISL_2091158, EPI_ISL_2091159 | | | |
| see above | Laboratorio Central de Epidemiología (LCE) | Centro de Investigación en Enfermedades Infecciosas (CIENI), Instituto Nacional de Enfermedades Respiratorias (INER) | Consorcio Mexicano de Vigilancia Genómica (CoViGen-Mex). Authors (in alphabetical order): Julio Elias Alvarado-Yaah, Carlos F. Arias, Santiago Ávila-Ríos, Víctor Hugo Borja-Aburto, Celia Boukadida, Juan Bautista Chale-Dzul, Célida Duque Molina, José Antonio Enciso-Moreno, Gloria Elena Espinosa-Ayala, Fernando Fontove-Herrera, Victor Eduardo Garcia-Arias, Concepción Grajales-Muñiz, Ricardo Grande, Alfredo Herrera-Estrella, Carla Ivón Herrera-Najera, Pavel Isa, Brenda Irasema Maldonado-Meza, Bernardo Martínez-Miguel, Margarita Matías-Florentino, María Guadalupe de Jesús Mireles-Rivera, Gloria María Molina-Salinas, Hector Montoya-Fuentes, José Esteban Muñoz-Medina, José de Jesús Nuñez-Contreras, Alicia Ocaña-Mondragón, Luis Alberto Ochoa-Carrera, Hector Esteban Paz-Juárez, Francisco Pulido, Helen Haydee Fernanda Ramírez-Plascencia, Angel Gustavo Salas-Lais, Alejandro Sanchez-Flores, Clara Esperanza Santacruz-Tinoco, María Guadalupe Santiago-Mauricio, Nelly Sélem-Mojica, Blanca Taboada, Gloria Vazquez |
| EPI_ISL_2091160, EPI_ISL_2091161, EPI_ISL_2091162, EPI_ISL_2091163, EPI_ISL_2091164, EPI_ISL_2091165, EPI_ISL_2091166, EPI_ISL_2091167, EPI_ISL_2091168, EPI_ISL_2091169, EPI_ISL_2091170, EPI_ISL_2091171, EPI_ISL_2091172, EPI_ISL_2091173, EPI_ISL_2091174, EPI_ISL_2091175, EPI_ISL_2091176, EPI_ISL_2091177, EPI_ISL_2091178, EPI_ISL_2091179, EPI_ISL_2091180, EPI_ISL_2091182, EPI_ISL_2091183, EPI_ISL_2091184, EPI_ISL_2091185, EPI_ISL_2091186, EPI_ISL_2091187, EPI_ISL_2091188, EPI_ISL_2091189, EPI_ISL_2091190, EPI_ISL_2091191, EPI_ISL_2091192, EPI_ISL_2091193, EPI_ISL_2091194, EPI_ISL_2091195, EPI_ISL_2091196, EPI_ISL_2091197, EPI_ISL_2091198, EPI_ISL_2091199, EPI_ISL_2091200, EPI_ISL_2091201, EPI_ISL_2091202, EPI_ISL_2091203, EPI_ISL_2091204, EPI_ISL_2091205, EPI_ISL_2091206, EPI_ISL_2091207, EPI_ISL_2091208, EPI_ISL_2091209, EPI_ISL_2091210, EPI_ISL_2091211, EPI_ISL_2091212, EPI_ISL_2091213, EPI_ISL_2091214, EPI_ISL_2091215, EPI_ISL_2091216, EPI_ISL_2091217, EPI_ISL_2091218, EPI_ISL_2091219, EPI_ISL_2091220, EPI_ISL_2091221, EPI_ISL_2091222, EPI_ISL_2091223, EPI_ISL_2091224 | | | |
| see above | Centro de Investigación Biomédica del Noreste (CIBIN) | Centro de Investigación en Enfermedades Infecciosas (CIENI), Instituto Nacional de Enfermedades Respiratorias (INER) | Consorcio Mexicano de Vigilancia Genómica (CoViGen-Mex). Authors (in alphabetical order): Julio Elias Alvarado-Yaah, Carlos F. Arias, Santiago Ávila-Ríos, Víctor Hugo Borja-Aburto, Celia Boukadida, Juan Bautista Chale-Dzul, Célida Duque Molina, José Antonio Enciso-Moreno, Gloria Elena Espinosa-Ayala, Fernando Fontove-Herrera, Victor Eduardo Garcia-Arias, Concepción Grajales-Muñiz, Ricardo Grande, Alfredo Herrera-Estrella, Carla Ivón Herrera-Najera, Pavel Isa, Brenda Irasema Maldonado-Meza, Bernardo Martínez-Miguel, Margarita Matías-Florentino, María Guadalupe de Jesús Mireles-Rivera, Gloria María Molina-Salinas, Hector Montoya-Fuentes, José Esteban Muñoz-Medina, José de Jesús Nuñez-Contreras, Alicia Ocaña-Mondragón, Luis Alberto Ochoa-Carrera, Hector Esteban Paz-Juárez, Francisco Pulido, Helen Haydee Fernanda Ramírez-Plascencia, Angel Gustavo Salas-Lais, Alejandro Sanchez-Flores, Clara Esperanza Santacruz-Tinoco, María Guadalupe Santiago-Mauricio, Nelly Sélem-Mojica, Blanca Taboada, Gloria Vazquez |
| EPI_ISL_2091225, EPI_ISL_2091226, EPI_ISL_2091227, EPI_ISL_2091228, EPI_ISL_2091229, EPI_ISL_2091230, EPI_ISL_2091231, EPI_ISL_2091232, EPI_ISL_2091233, EPI_ISL_2091234, EPI_ISL_2091235, EPI_ISL_2091236, EPI_ISL_2091237, EPI_ISL_2091238, EPI_ISL_2091239, EPI_ISL_2091240, EPI_ISL_2091241, EPI_ISL_2091242, EPI_ISL_2091243, EPI_ISL_2091244, EPI_ISL_2091245, EPI_ISL_2091246, EPI_ISL_2091247, EPI_ISL_2091248, EPI_ISL_2091249, EPI_ISL_2091250, EPI_ISL_2091251, EPI_ISL_2091252, EPI_ISL_2091253, EPI_ISL_2091254, EPI_ISL_2091255, EPI_ISL_2091256, EPI_ISL_2091257 | | | |
| see above | Unidad de Investigación Médica de Yucatán (UIMY) | Centro de Investigación en Enfermedades Infecciosas (CIENI), Instituto Nacional de Enfermedades Respiratorias (INER) | Consorcio Mexicano de Vigilancia Genómica (CoViGen-Mex). Authors (in alphabetical order): Julio Elias Alvarado-Yaah, Carlos F. Arias, Santiago Ávila-Ríos, Víctor Hugo Borja-Aburto, Celia Boukadida, Juan Bautista Chale-Dzul, Célida Duque Molina, José Antonio Enciso-Moreno, Gloria Elena Espinosa-Ayala, Fernando Fontove-Herrera, Victor Eduardo Garcia-Arias, Concepción Grajales-Muñiz, Ricardo Grande, Alfredo Herrera-Estrella, Carla Ivón Herrera-Najera, Pavel Isa, Brenda Irasema Maldonado-Meza, Bernardo Martínez-Miguel, Margarita Matías-Florentino, María Guadalupe de Jesús Mireles-Rivera, Gloria María Molina-Salinas, Hector Montoya-Fuentes, José Esteban Muñoz-Medina, José de Jesús Nuñez-Contreras, Alicia Ocaña-Mondragón, Luis Alberto Ochoa-Carrera, Hector Esteban Paz-Juárez, Francisco Pulido, Helen Haydee Fernanda Ramírez-Plascencia, Angel Gustavo Salas-Lais, Alejandro Sanchez-Flores, Clara Esperanza Santacruz-Tinoco, María Guadalupe Santiago-Mauricio, Nelly Sélem-Mojica, Blanca Taboada, Gloria Vazquez |
| EPI_ISL_2091258, EPI_ISL_2091259, EPI_ISL_2091260, EPI_ISL_2091261, EPI_ISL_2091262, EPI_ISL_2091263, EPI_ISL_2091264, EPI_ISL_2091265, EPI_ISL_2091266, EPI_ISL_2091267, EPI_ISL_2091268, EPI_ISL_2091269, EPI_ISL_2091270, EPI_ISL_2091271, EPI_ISL_2091272, EPI_ISL_2091273, EPI_ISL_2091274, EPI_ISL_2091275, EPI_ISL_2091276, EPI_ISL_2091277, EPI_ISL_2091278, EPI_ISL_2091279, EPI_ISL_2091280, EPI_ISL_2091281, EPI_ISL_2091282, EPI_ISL_2091283, EPI_ISL_2091284, EPI_ISL_2091285, EPI_ISL_2091286, EPI_ISL_2091287, EPI_ISL_2091288, EPI_ISL_2091289, EPI_ISL_2091290, EPI_ISL_2091291, EPI_ISL_2091292, EPI_ISL_2091293, EPI_ISL_2091294, EPI_ISL_2091295, EPI_ISL_2091296, EPI_ISL_2091297, EPI_ISL_2091298, EPI_ISL_2091299, EPI_ISL_2091300, EPI_ISL_2091301, EPI_ISL_2091302, EPI_ISL_2091303, EPI_ISL_2091304, EPI_ISL_2091305, EPI_ISL_2091306, EPI_ISL_2091307, EPI_ISL_2091308, EPI_ISL_2091309, EPI_ISL_2091310, EPI_ISL_2091311, EPI_ISL_2091312, EPI_ISL_2091313, EPI_ISL_2091314, EPI_ISL_2091315, EPI_ISL_2091316, EPI_ISL_2091317, EPI_ISL_2091318, EPI_ISL_2091319 | | | |
| see above | Centro de Investigación Biomédica de Occidente (CIBO) | Centro de Investigación en Enfermedades Infecciosas (CIENI), Instituto Nacional de Enfermedades Respiratorias (INER) | Consorcio Mexicano de Vigilancia Genómica (CoViGen-Mex). Authors (in alphabetical order): Julio Elias Alvarado-Yaah, Carlos F. Arias, Santiago Ávila-Ríos, Víctor Hugo Borja-Aburto, Celia Boukadida, Juan Bautista Chale-Dzul, Célida Duque Molina, José Antonio Enciso-Moreno, Gloria Elena Espinosa-Ayala, Fernando Fontove-Herrera, Victor Eduardo Garcia-Arias, Concepción Grajales-Muñiz, Ricardo Grande, Alfredo Herrera-Estrella, Carla Ivón Herrera-Najera, Pavel Isa, Brenda Irasema Maldonado-Meza, Bernardo Martínez-Miguel, Margarita Matías-Florentino, María Guadalupe de Jesús Mireles-Rivera, Gloria María Molina-Salinas, Hector Montoya-Fuentes, José Esteban Muñoz-Medina, José de Jesús Nuñez-Contreras, Alicia Ocaña-Mondragón, Luis Alberto Ochoa-Carrera, Hector Esteban Paz-Juárez, Francisco Pulido, Helen Haydee Fernanda Ramírez-Plascencia, Angel Gustavo Salas-Lais, Alejandro Sanchez-Flores, Clara Esperanza Santacruz-Tinoco, María Guadalupe Santiago-Mauricio, Nelly Sélem-Mojica, Blanca Taboada, Gloria Vazquez |
| EPI_ISL_2091320, EPI_ISL_2091321, EPI_ISL_2091322, EPI_ISL_2091323, EPI_ISL_2091324, EPI_ISL_2091325, EPI_ISL_2091326, EPI_ISL_2091327, EPI_ISL_2091328, EPI_ISL_2091329, EPI_ISL_2091330, EPI_ISL_2091331, EPI_ISL_2091332, EPI_ISL_2091333, EPI_ISL_2091334, EPI_ISL_2091335, EPI_ISL_2091336, EPI_ISL_2091337, EPI_ISL_2091338, EPI_ISL_2091339, EPI_ISL_2091340, EPI_ISL_2091341, EPI_ISL_2091342, EPI_ISL_2091343, EPI_ISL_2091344, EPI_ISL_2091345, EPI_ISL_2091346, EPI_ISL_2091347, EPI_ISL_2091348, EPI_ISL_2091349, EPI_ISL_2091350, EPI_ISL_2091351, EPI_ISL_2091352, EPI_ISL_2091353, EPI_ISL_2091354, EPI_ISL_2091355, EPI_ISL_2091356, EPI_ISL_2091357, EPI_ISL_2091358, EPI_ISL_2091359, EPI_ISL_2091360, EPI_ISL_2091361, EPI_ISL_2091362, EPI_ISL_2091363, EPI_ISL_2091364, EPI_ISL_2091365, EPI_ISL_2091366, EPI_ISL_2091367, EPI_ISL_2091368, EPI_ISL_2091369, EPI_ISL_2091370, EPI_ISL_2091371, EPI_ISL_2091372, EPI_ISL_2091373, EPI_ISL_2091374, EPI_ISL_2091375, EPI_ISL_2091376, EPI_ISL_2091377, EPI_ISL_2091378, EPI_ISL_2091379, EPI_ISL_2091380, EPI_ISL_2091381, EPI_ISL_2091382, EPI_ISL_2091383, EPI_ISL_2091384, EPI_ISL_2091385, EPI_ISL_2091386, EPI_ISL_2091387, EPI_ISL_2091388, EPI_ISL_2091389, EPI_ISL_2091390, EPI_ISL_2091391, EPI_ISL_2091392, EPI_ISL_2091393, EPI_ISL_2091394, EPI_ISL_2091395, EPI_ISL_2091396, EPI_ISL_2091397, EPI_ISL_2091398, EPI_ISL_2091399, EPI_ISL_2091400, EPI_ISL_2091401, EPI_ISL_2091402, EPI_ISL_2091403, EPI_ISL_2091404, EPI_ISL_2091405, EPI_ISL_2091406, EPI_ISL_2091407, EPI_ISL_2091408, EPI_ISL_2091409, EPI_ISL_2091410, EPI_ISL_2091411, EPI_ISL_2091412, EPI_ISL_2091413, EPI_ISL_2091414, EPI_ISL_2091415, EPI_ISL_2091416, EPI_ISL_2091417, EPI_ISL_2091418, EPI_ISL_2091419, EPI_ISL_2091420, EPI_ISL_2091421, EPI_ISL_2091422, EPI_ISL_2091423, EPI_ISL_2091424, EPI_ISL_2091425, EPI_ISL_2091426, EPI_ISL_2091427, EPI_ISL_2091428, EPI_ISL_2091429, EPI_ISL_2091430, EPI_ISL_2091431, EPI_ISL_2091432, EPI_ISL_2091433, EPI_ISL_2091434, EPI_ISL_2091435, EPI_ISL_2091436, EPI_ISL_2091437, EPI_ISL_2091438, EPI_ISL_2091439, EPI_ISL_2091440, EPI_ISL_2091441, EPI_ISL_2091442, EPI_ISL_2091443, EPI_ISL_2091444, EPI_ISL_2091445, EPI_ISL_2091446, EPI_ISL_2091447, EPI_ISL_2091448, EPI_ISL_2091449, EPI_ISL_2091450, EPI_ISL_2091451, EPI_ISL_2091452, EPI_ISL_2091453, EPI_ISL_2091454, EPI_ISL_2091455, EPI_ISL_2091456 | | | |
| see above | Laboratorio Central de Epidemiología (LCE) | Centro de Investigación en Enfermedades Infecciosas (CIENI), Instituto Nacional de Enfermedades Respiratorias (INER) | Consorcio Mexicano de Vigilancia Genómica (CoViGen-Mex). Authors (in alphabetical order): Julio Elias Alvarado-Yaah, Carlos F. Arias, Santiago Ávila-Ríos, Víctor Hugo Borja-Aburto, Celia Boukadida, Juan Bautista Chale-Dzul, Célida Duque Molina, José Antonio Enciso-Moreno, Gloria Elena Espinosa-Ayala, Fernando Fontove-Herrera, Victor Eduardo Garcia-Arias, Concepción Grajales-Muñiz, Ricardo Grande, Alfredo Herrera-Estrella, Carla Ivón Herrera-Najera, Pavel Isa, Brenda Irasema Maldonado-Meza, Bernardo Martínez-Miguel, Margarita Matías-Florentino, María Guadalupe de Jesús Mireles-Rivera, Gloria María Molina-Salinas, Hector Montoya-Fuentes, José Esteban Muñoz-Medina, José de Jesús Nuñez-Contreras, Alicia Ocaña-Mondragón, Luis Alberto Ochoa-Carrera, Hector Esteban Paz-Juárez, Francisco Pulido, Helen Haydee Fernanda Ramírez-Plascencia, Angel Gustavo Salas-Lais, Alejandro Sanchez-Flores, Clara Esperanza Santacruz-Tinoco, María Guadalupe Santiago-Mauricio, Nelly Sélem-Mojica, Blanca Taboada, Gloria Vazquez |
| EPI_ISL_2092757, EPI_ISL_2092758 | Minnesota Department of Health, Public Health Laboratory | Minnesota Department of Health, Public Health Laboratory | Alexandra Lorentz, Jacob Garfin, Matt Plumb, and Xiong Wang |
| EPI_ISL_2092759 | M Health Fairview | Minnesota Department of Health, Public Health Laboratory | Alexandra Lorentz, Jacob Garfin, Matt Plumb, and Xiong Wang |

| | | | |
|---|---|---|---|
| EPI_ISL_2092760 | Minnesota Department of Health, Public Health Laboratory | Minnesota Department of Health, Public Health Laboratory | Alexandra Lorentz, Jacob Garfin, Matt Plumb, and Xiong Wang |
| EPI_ISL_2092761 | Centracare Laboratory Services | Minnesota Department of Health, Public Health Laboratory | Alexandra Lorentz, Jacob Garfin, Matt Plumb, and Xiong Wang |
| EPI_ISL_2092762, EPI_ISL_2092763 | M Health Fairview | Minnesota Department of Health, Public Health Laboratory | Alexandra Lorentz, Jacob Garfin, Matt Plumb, and Xiong Wang |
| EPI_ISL_2092764, EPI_ISL_2092765 | Minnesota Department of Health, Public Health Laboratory | Minnesota Department of Health, Public Health Laboratory | Alexandra Lorentz, Jacob Garfin, Matt Plumb, and Xiong Wang |
| EPI_ISL_2092766 | M Health Fairview | Minnesota Department of Health, Public Health Laboratory | Alexandra Lorentz, Jacob Garfin, Matt Plumb, and Xiong Wang |
| EPI_ISL_2092767, EPI_ISL_2092768, EPI_ISL_2092769, EPI_ISL_2092770, EPI_ISL_2092771, EPI_ISL_2092772, EPI_ISL_2092773, EPI_ISL_2092774, EPI_ISL_2092775, EPI_ISL_2092776 | Minnesota Department of Health, Public Health Laboratory | Minnesota Department of Health, Public Health Laboratory | Alexandra Lorentz, Jacob Garfin, Matt Plumb, and Xiong Wang |
| EPI_ISL_2092777, EPI_ISL_2092778, EPI_ISL_2092779, EPI_ISL_2092780, EPI_ISL_2092781, EPI_ISL_2092782, EPI_ISL_2092783, EPI_ISL_2092784 | Centracare Laboratory Services | Minnesota Department of Health, Public Health Laboratory | Alexandra Lorentz, Jacob Garfin, Matt Plumb, and Xiong Wang |
| EPI_ISL_2092785, EPI_ISL_2092786, EPI_ISL_2092787, EPI_ISL_2092788, EPI_ISL_2092789 | M Health Fairview | Minnesota Department of Health, Public Health Laboratory | Alexandra Lorentz, Jacob Garfin, Matt Plumb, and Xiong Wang |
| EPI_ISL_2092790, EPI_ISL_2092791, EPI_ISL_2092792 | Minnesota Department of Health, Public Health Laboratory | Minnesota Department of Health, Public Health Laboratory | Alexandra Lorentz, Jacob Garfin, Matt Plumb, and Xiong Wang |
| EPI_ISL_2092793, EPI_ISL_2092794, EPI_ISL_2092795, EPI_ISL_2092796, EPI_ISL_2092797 | Centracare Laboratory Services | Minnesota Department of Health, Public Health Laboratory | Alexandra Lorentz, Jacob Garfin, Matt Plumb, and Xiong Wang |
| EPI_ISL_2092798, EPI_ISL_2092799, EPI_ISL_2092802 | M Health Fairview | Minnesota Department of Health, Public Health Laboratory | Alexandra Lorentz, Jacob Garfin, Matt Plumb, and Xiong Wang |
| EPI_ISL_2092803 | Minnesota Department of Health, Public Health Laboratory | Minnesota Department of Health, Public Health Laboratory | Alexandra Lorentz, Jacob Garfin, Matt Plumb, and Xiong Wang |
| EPI_ISL_2092804, EPI_ISL_2092805, EPI_ISL_2092806, EPI_ISL_2092807, EPI_ISL_2092808, EPI_ISL_2092809, EPI_ISL_2092810 | Centracare Laboratory Services | Minnesota Department of Health, Public Health Laboratory | Alexandra Lorentz, Jacob Garfin, Matt Plumb, and Xiong Wang |
| EPI_ISL_2092811 | Minnesota Department of Health, Public Health Laboratory | Minnesota Department of Health, Public Health Laboratory | Alexandra Lorentz, Jacob Garfin, Matt Plumb, and Xiong Wang |
| EPI_ISL_2092812, EPI_ISL_2092813, EPI_ISL_2092814, EPI_ISL_2092815 | Centracare Laboratory Services | Minnesota Department of Health, Public Health Laboratory | Alexandra Lorentz, Jacob Garfin, Matt Plumb, and Xiong Wang |
| EPI_ISL_2092816, EPI_ISL_2092817, EPI_ISL_2092818 | Wichita State University - Molecular Diagnostics Lab | Kansas Health and Environmental Lab | Mike Grose, Jonathan Barnell, Ben Olsen, and Phil Adam |
| EPI_ISL_2092819 | Kansas Health and Environmental Lab | Kansas Health and Environmental Lab | Mike Grose, Jonathan Barnell, Ben Olsen, and Phil Adam |
| EPI_ISL_2092820 | Wichita State University - Molecular Diagnostics Lab | Kansas Health and Environmental Lab | Mike Grose, Jonathan Barnell, Ben Olsen, and Phil Adam |
| EPI_ISL_2092821, EPI_ISL_2092822 | Kansas Health and Environmental Lab | Kansas Health and Environmental Lab | Mike Grose, Jonathan Barnell, Ben Olsen, and Phil Adam |
| EPI_ISL_2092823 | Wichita State University - Molecular Diagnostics Lab | Kansas Health and Environmental Lab | Mike Grose, Jonathan Barnell, Ben Olsen, and Phil Adam |
| EPI_ISL_2092832, EPI_ISL_2092833, EPI_ISL_2092834 | Kansas Health and Environmental Lab | Kansas Health and Environmental Lab | Mike Grose, Jonathan Barnell, Ben Olsen, and Phil Adam |
| EPI_ISL_2092835, EPI_ISL_2092836, EPI_ISL_2092837, EPI_ISL_2092838, EPI_ISL_2092839 | Wichita State University - Molecular Diagnostics Lab | Kansas Health and Environmental Lab | Mike Grose, Jonathan Barnell, Ben Olsen, and Phil Adam |
| EPI_ISL_2092840 | Kansas Health and Environmental Lab | Kansas Health and Environmental Lab | Mike Grose, Jonathan Barnell, Ben Olsen, and Phil Adam |
| EPI_ISL_2092842, EPI_ISL_2092843, EPI_ISL_2092844, EPI_ISL_2092845, EPI_ISL_2092846, EPI_ISL_2092849, EPI_ISL_2092850, EPI_ISL_2092851, EPI_ISL_2092852, EPI_ISL_2092853, EPI_ISL_2092854, EPI_ISL_2092855, EPI_ISL_2092856, EPI_ISL_2092857, EPI_ISL_2092858, EPI_ISL_2092859, EPI_ISL_2092860, EPI_ISL_2092861, EPI_ISL_2092862, EPI_ISL_2092863, EPI_ISL_2092864 | | | |
| see above | UNC Charlotte COVID-19 Testing Lab | UNC Charlotte Environmental Monitoring Laboratory | Cynthia Gibas, Kevin Lambirth, Angelica Martins, Jannatul Ferdous, Visva Barua, Jessica Schlueter |
| EPI_ISL_2094443, EPI_ISL_2094444, EPI_ISL_2094445, EPI_ISL_2094447, EPI_ISL_2094448, EPI_ISL_2094449, EPI_ISL_2094450, EPI_ISL_2094452, EPI_ISL_2094453, EPI_ISL_2094454, EPI_ISL_2094455, EPI_ISL_2094457, EPI_ISL_2094458, EPI_ISL_2094459, EPI_ISL_2094462, EPI_ISL_2094464, EPI_ISL_2094465, EPI_ISL_2094466, EPI_ISL_2094467, EPI_ISL_2094469, EPI_ISL_2094472, EPI_ISL_2094474, EPI_ISL_2094475, EPI_ISL_2094476, EPI_ISL_2094477, EPI_ISL_2094478, EPI_ISL_2094479, EPI_ISL_2094480, EPI_ISL_2094482, EPI_ISL_2094483, EPI_ISL_2094484, EPI_ISL_2094485, EPI_ISL_2094487, EPI_ISL_2094488, EPI_ISL_2094489, EPI_ISL_2094490, EPI_ISL_2094491, EPI_ISL_2094492, EPI_ISL_2094493, EPI_ISL_2094494, EPI_ISL_2094495, EPI_ISL_2094496, EPI_ISL_2094497, EPI_ISL_2094500, EPI_ISL_2094503 | | | |
| see above | North Dakota Department of Health, Public Health Laboratory | North Dakota Department of Health, Public Health Laboratory | Lisa Wingerter |
| EPI_ISL_2094505 | LESP Jalisco | Instituto de Diagnostico y Referencia Epidemiologicos (INDRE) | Claudia Wong-Arambula, Abril Rodriguez-Maldonado, Vanessa Rivero-Arredondo, Ariadna Medina-Benitez, Joaquin Quiroz-Mercado, Sergio Rangel-Guerrero, Natividad Cruz-Ortiz, Tatiana Nunez-Garcia, Gisela Barrera-Badillo, Lucia Hernandez-Rivas, Irma Lopez-Martinez, Ernesto Ramirez-Gonzalez. |
| EPI_ISL_2094506, EPI_ISL_2094507, EPI_ISL_2094508 | LESP San Luis Potosi | Instituto de Diagnostico y Referencia Epidemiologicos (INDRE) | Claudia Wong-Arambula, Abril Rodriguez-Maldonado, Vanessa Rivero-Arredondo, Ariadna Medina-Benitez, Joaquin Quiroz-Mercado, Sergio Rangel-Guerrero, Natividad Cruz-Ortiz, Tatiana Nunez-Garcia, Gisela Barrera-Badillo, Lucia Hernandez-Rivas, Irma Lopez-Martinez, Ernesto Ramirez-Gonzalez. |
| EPI_ISL_2094509, EPI_ISL_2094510, EPI_ISL_2094511, EPI_ISL_2094512 | LESP Campeche | Instituto de Diagnostico y Referencia Epidemiologicos (INDRE) | Claudia Wong-Arambula, Abril Rodriguez-Maldonado, Vanessa Rivero-Arredondo, Ariadna Medina-Benitez, Joaquin Quiroz-Mercado, Sergio Rangel-Guerrero, Natividad Cruz-Ortiz, Tatiana Nunez-Garcia, Gisela Barrera-Badillo, Lucia Hernandez-Rivas, Irma Lopez-Martinez, Ernesto Ramirez-Gonzalez. |
| EPI_ISL_2095693, EPI_ISL_2095694, | Idaho Bureau of Laboratories | Center for Global Health, University of New Mexico Health Sciences Center | Daryl Domman, Kurt Schwalm, Valerie Morley, Matthew Burns, Robert Voermans, Christopher Ball, Darrell Dinwiddie |

| | | | |
|---|---|---|---|
| EPI_ISL_2095695, EPI_ISL_2095697, EPI_ISL_2095699, EPI_ISL_2095700 | | | |
| EPI_ISL_2095753, EPI_ISL_2095754, EPI_ISL_2095755, EPI_ISL_2095756, EPI_ISL_2095757, EPI_ISL_2095758, EPI_ISL_2095759, EPI_ISL_2095761, EPI_ISL_2095762, EPI_ISL_2095763, EPI_ISL_2095765, EPI_ISL_2095766, EPI_ISL_2095767, EPI_ISL_2095768, EPI_ISL_2095770, EPI_ISL_2095773, EPI_ISL_2095775, EPI_ISL_2095776, EPI_ISL_2095778, EPI_ISL_2095781, EPI_ISL_2095782, EPI_ISL_2095783, EPI_ISL_2095784, EPI_ISL_2095786, EPI_ISL_2095787, EPI_ISL_2095788, EPI_ISL_2095789, EPI_ISL_2095790, EPI_ISL_2095791, EPI_ISL_2095793, EPI_ISL_2095794, EPI_ISL_2095795, EPI_ISL_2095796, EPI_ISL_2095797, EPI_ISL_2095798, EPI_ISL_2095800, EPI_ISL_2095801, EPI_ISL_2095802, EPI_ISL_2095804, EPI_ISL_2095805, EPI_ISL_2095806, EPI_ISL_2095807, EPI_ISL_2095808, EPI_ISL_2095810, EPI_ISL_2095812 | | | |
| see above | North Dakota Department of Health, Public Health Laboratory | North Dakota Department of Health, Public Health Laboratory | Lisa Wingerter |
| EPI_ISL_2096054, EPI_ISL_2096055, EPI_ISL_2096056 | Instituto Nacional de Medicina Genomica | Instituto Nacional de Medicina Genomica | Hidalgo-Miranda A, Mendoza-Vargas A, Reyes-Grajeda JP, Cedro-Tanda A, Cisneros-Villanueva M, Orjuela-Rodríguez M, Alcaraz N, Gonzalez-Barrera D, Rangel-DeLeon D, Munguia-Garza P, Ramirez-Vega O, Garcia-Cardenas FJ, Gonzalez-Woge MA, Herrera-Montalvo LA |
| EPI_ISL_2096067, EPI_ISL_2096068, EPI_ISL_2096069, EPI_ISL_2096071, EPI_ISL_2096072, EPI_ISL_2096073, EPI_ISL_2096074, EPI_ISL_2096075, EPI_ISL_2096076, EPI_ISL_2096077, EPI_ISL_2096078, EPI_ISL_2096079, EPI_ISL_2096080, EPI_ISL_2096081, EPI_ISL_2096082, EPI_ISL_2096083, EPI_ISL_2096084, EPI_ISL_2096085, EPI_ISL_2096086, EPI_ISL_2096087, EPI_ISL_2096088, EPI_ISL_2096089, EPI_ISL_2096090, EPI_ISL_2096091, EPI_ISL_2096092, EPI_ISL_2096093, EPI_ISL_2096094, EPI_ISL_2096095, EPI_ISL_2096096, EPI_ISL_2096097, EPI_ISL_2096098, EPI_ISL_2096103, EPI_ISL_2096104, EPI_ISL_2096105, EPI_ISL_2096106, EPI_ISL_2096107, EPI_ISL_2096108, EPI_ISL_2096109, EPI_ISL_2096110, EPI_ISL_2096111, EPI_ISL_2096112, EPI_ISL_2096113, EPI_ISL_2096114, EPI_ISL_2096116, EPI_ISL_2096117, EPI_ISL_2096118, EPI_ISL_2096119, EPI_ISL_2096127, EPI_ISL_2096128, EPI_ISL_2096129, EPI_ISL_2096130, EPI_ISL_2096131 | | | |
| see above | Washington State Department of Health Public Health Laboratories | Washington State Department of Health Public Health Laboratories | Drew MacKellar, Philip Dykema, Denny Russell, Joenice Gonzalez, Hannah Gray, Geoff Melly, Vanessa De Los Santos, Darren Lucas, JohnAric Peterson, Avi Singh, Rebecca Cao |
| EPI_ISL_2096132, EPI_ISL_2096133, EPI_ISL_2096134, EPI_ISL_2096136, EPI_ISL_2096137, EPI_ISL_2096138, EPI_ISL_2096139, EPI_ISL_2096140, EPI_ISL_2096141, EPI_ISL_2096142, EPI_ISL_2096144, EPI_ISL_2096146, EPI_ISL_2096147, EPI_ISL_2096148, EPI_ISL_2096149, EPI_ISL_2096151, EPI_ISL_2096152, EPI_ISL_2096153, EPI_ISL_2096155, EPI_ISL_2096156, EPI_ISL_2096157, EPI_ISL_2096158, EPI_ISL_2096159, EPI_ISL_2096160, EPI_ISL_2096161, EPI_ISL_2096164, EPI_ISL_2096165, EPI_ISL_2096166, EPI_ISL_2096167, EPI_ISL_2096168, EPI_ISL_2096169, EPI_ISL_2096170, EPI_ISL_2096171, EPI_ISL_2096172, EPI_ISL_2096174, EPI_ISL_2096175, EPI_ISL_2096176, EPI_ISL_2096177, EPI_ISL_2096178, EPI_ISL_2096179, EPI_ISL_2096181, EPI_ISL_2096182, EPI_ISL_2096183, EPI_ISL_2096184, EPI_ISL_2096185, EPI_ISL_2096186, EPI_ISL_2096187, EPI_ISL_2096188, EPI_ISL_2096189, EPI_ISL_2096190, EPI_ISL_2096191, EPI_ISL_2096192, EPI_ISL_2096194, EPI_ISL_2096195, EPI_ISL_2096196, EPI_ISL_2096197, EPI_ISL_2096198 | | | |
| see above | North Dakota Department of Health, Public Health Laboratory | North Dakota Department of Health, Public Health Laboratory | Lisa Wingerter |
| EPI_ISL_2096204, EPI_ISL_2096205, EPI_ISL_2096206, EPI_ISL_2096207, EPI_ISL_2096208, EPI_ISL_2096209, EPI_ISL_2096210, EPI_ISL_2096211, EPI_ISL_2096212, EPI_ISL_2096213 | Maryland Genomics, Institute for Genome Sciences, University of Maryland School of Medicine | Maryland Genomics, Institute for Genome Sciences, University of Maryland School of Medicine | Tallon, Luke J; Sadzewicz, Lisa D; Humphrys, Mike; Ott, Sandra; Roussey, Holly; Mehta, Aditya; Vavikolanu, Kranthi; Fraser, Claire M; Ravel, Jacques |
| EPI_ISL_2096217, EPI_ISL_2096218, EPI_ISL_2096219, EPI_ISL_2096220, EPI_ISL_2096221, EPI_ISL_2096222, EPI_ISL_2096223, EPI_ISL_2096224, EPI_ISL_2096225, EPI_ISL_2096226, EPI_ISL_2096227, EPI_ISL_2096228, EPI_ISL_2096229, EPI_ISL_2096230, EPI_ISL_2096231, EPI_ISL_2096232, EPI_ISL_2096233, EPI_ISL_2096234, EPI_ISL_2096235, EPI_ISL_2096236, EPI_ISL_2096237, EPI_ISL_2096238, EPI_ISL_2096239, EPI_ISL_2096240, EPI_ISL_2096241, EPI_ISL_2096242, EPI_ISL_2096243, EPI_ISL_2096244, EPI_ISL_2096245, EPI_ISL_2096246, EPI_ISL_2096247, EPI_ISL_2096248, EPI_ISL_2096249, EPI_ISL_2096250, EPI_ISL_2096251, EPI_ISL_2096252, EPI_ISL_2096253, EPI_ISL_2096254, EPI_ISL_2096255, EPI_ISL_2096257, EPI_ISL_2096258, EPI_ISL_2096259, EPI_ISL_2096260, EPI_ISL_2096261, EPI_ISL_2096262, EPI_ISL_2096263, EPI_ISL_2096264, EPI_ISL_2096265, EPI_ISL_2096266, EPI_ISL_2096267, EPI_ISL_2096268, EPI_ISL_2096269, EPI_ISL_2096270, EPI_ISL_2096271, EPI_ISL_2096272, EPI_ISL_2096273, EPI_ISL_2096274, EPI_ISL_2096275, EPI_ISL_2096277, EPI_ISL_2096278, EPI_ISL_2096279, EPI_ISL_2096280, EPI_ISL_2096281, EPI_ISL_2096282, EPI_ISL_2096283, EPI_ISL_2096284, EPI_ISL_2096285, EPI_ISL_2096286, EPI_ISL_2096287, EPI_ISL_2096288, EPI_ISL_2096289, EPI_ISL_2096290, EPI_ISL_2096291, EPI_ISL_2096292, EPI_ISL_2096293, EPI_ISL_2096294, EPI_ISL_2096295, EPI_ISL_2096296, EPI_ISL_2096297, EPI_ISL_2096298, EPI_ISL_2096300, EPI_ISL_2096301, EPI_ISL_2096302, EPI_ISL_2096303, EPI_ISL_2096304, EPI_ISL_2096305, EPI_ISL_2096306, EPI_ISL_2096307, EPI_ISL_2096308, EPI_ISL_2096309, EPI_ISL_2096310, EPI_ISL_2096311, EPI_ISL_2096312, EPI_ISL_2096313, EPI_ISL_2096314, EPI_ISL_2096315, EPI_ISL_2096316, EPI_ISL_2096317, EPI_ISL_2096318, EPI_ISL_2096319, EPI_ISL_2096320, EPI_ISL_2096321, EPI_ISL_2096322, EPI_ISL_2096323, EPI_ISL_2096324, EPI_ISL_2096325, EPI_ISL_2096326, EPI_ISL_2096327, EPI_ISL_2096328, EPI_ISL_2096329, EPI_ISL_2096330, EPI_ISL_2096331, EPI_ISL_2096332, EPI_ISL_2096333, EPI_ISL_2096334, EPI_ISL_2096335, EPI_ISL_2096336, EPI_ISL_2096337, EPI_ISL_2096338, EPI_ISL_2096339, EPI_ISL_2096340, EPI_ISL_2096341, EPI_ISL_2096342, EPI_ISL_2096343, EPI_ISL_2096344, EPI_ISL_2096345, EPI_ISL_2096346, EPI_ISL_2096347, EPI_ISL_2096348, EPI_ISL_2096350, EPI_ISL_2096351, EPI_ISL_2096352, EPI_ISL_2096353, EPI_ISL_2096354, EPI_ISL_2096355, EPI_ISL_2096356, EPI_ISL_2096357, EPI_ISL_2096358, EPI_ISL_2096359, EPI_ISL_2096360, EPI_ISL_2096361, EPI_ISL_2096362, EPI_ISL_2096363, EPI_ISL_2096364, EPI_ISL_2096365, EPI_ISL_2096366, EPI_ISL_2096367, EPI_ISL_2096368, EPI_ISL_2096369, EPI_ISL_2096370, EPI_ISL_2096371, EPI_ISL_2096372, EPI_ISL_2096374, EPI_ISL_2096375, EPI_ISL_2096376, EPI_ISL_2096377, EPI_ISL_2096378, EPI_ISL_2096379, EPI_ISL_2096380, EPI_ISL_2096381, EPI_ISL_2096382, EPI_ISL_2096383, EPI_ISL_2096384, EPI_ISL_2096385, EPI_ISL_2096386, EPI_ISL_2096387, EPI_ISL_2096388, EPI_ISL_2096389, EPI_ISL_2096390, EPI_ISL_2096391, EPI_ISL_2096392, EPI_ISL_2096393, EPI_ISL_2096394, EPI_ISL_2096395, EPI_ISL_2096396, EPI_ISL_2096397, EPI_ISL_2096398, EPI_ISL_2096399, EPI_ISL_2096400, EPI_ISL_2096401, EPI_ISL_2096402, EPI_ISL_2096403, EPI_ISL_2096404, EPI_ISL_2096405, EPI_ISL_2096406, EPI_ISL_2096407, EPI_ISL_2096408, EPI_ISL_2096409, EPI_ISL_2096410, EPI_ISL_2096411, EPI_ISL_2096412, EPI_ISL_2096413, EPI_ISL_2096414, EPI_ISL_2096415, EPI_ISL_2096416, EPI_ISL_2096417, EPI_ISL_2096418, EPI_ISL_2096419, EPI_ISL_2096420, EPI_ISL_2096421, EPI_ISL_2096422, EPI_ISL_2096423, EPI_ISL_2096424, EPI_ISL_2096425, EPI_ISL_2096426, EPI_ISL_2096427, EPI_ISL_2096428, EPI_ISL_2096429, EPI_ISL_2096430, EPI_ISL_2096431, EPI_ISL_2096432, EPI_ISL_2096433, EPI_ISL_2096434, EPI_ISL_2096435, EPI_ISL_2096436, EPI_ISL_2096437, EPI_ISL_2096438, EPI_ISL_2096439, EPI_ISL_2096440, EPI_ISL_2096441, EPI_ISL_2096442, EPI_ISL_2096443, EPI_ISL_2096444, EPI_ISL_2096445, EPI_ISL_2096446, EPI_ISL_2096447, EPI_ISL_2096448, EPI_ISL_2096449, EPI_ISL_2096450, EPI_ISL_2096451, EPI_ISL_2096452, EPI_ISL_2096453, EPI_ISL_2096454, EPI_ISL_2096455, EPI_ISL_2096456, EPI_ISL_2096457, EPI_ISL_2096459, EPI_ISL_2096460, EPI_ISL_2096461, EPI_ISL_2096462, EPI_ISL_2096463, EPI_ISL_2096464, EPI_ISL_2096465, EPI_ISL_2096466, EPI_ISL_2096467, EPI_ISL_2096468, EPI_ISL_2096469, EPI_ISL_2096470, EPI_ISL_2096471, EPI_ISL_2096472, EPI_ISL_2096473, EPI_ISL_2096474, EPI_ISL_2096475, EPI_ISL_2096476, EPI_ISL_2096477, EPI_ISL_2096478, EPI_ISL_2096479, EPI_ISL_2096480, EPI_ISL_2096481, EPI_ISL_2096482, EPI_ISL_2096483, EPI_ISL_2096484, EPI_ISL_2096485, EPI_ISL_2096486, EPI_ISL_2096487, EPI_ISL_2096488, EPI_ISL_2096489, EPI_ISL_2096490, EPI_ISL_2096491, EPI_ISL_2096492, EPI_ISL_2096493, EPI_ISL_2096494, EPI_ISL_2096495, EPI_ISL_2096496, EPI_ISL_2096497, EPI_ISL_2096498, EPI_ISL_2096499, EPI_ISL_2096500, EPI_ISL_2096501, EPI_ISL_2096502, EPI_ISL_2096503, EPI_ISL_2096504, EPI_ISL_2096505, EPI_ISL_2096506, EPI_ISL_2096507, EPI_ISL_2096508, EPI_ISL_2096509, EPI_ISL_2096510, EPI_ISL_2096511, EPI_ISL_2096512, EPI_ISL_2096513, EPI_ISL_2096514, EPI_ISL_2096515, EPI_ISL_2096516, EPI_ISL_2096517, EPI_ISL_2096518, EPI_ISL_2096519, EPI_ISL_2096520, EPI_ISL_2096521, EPI_ISL_2096522, EPI_ISL_2096523, EPI_ISL_2096524, EPI_ISL_2096525, EPI_ISL_2096526, EPI_ISL_2096527, EPI_ISL_2096528, EPI_ISL_2096529, EPI_ISL_2096530, EPI_ISL_2096531, EPI_ISL_2096532, EPI_ISL_2096533, EPI_ISL_2096534, EPI_ISL_2096535, EPI_ISL_2096536, EPI_ISL_2096537, EPI_ISL_2096538, EPI_ISL_2096539, EPI_ISL_2096540, EPI_ISL_2096541, EPI_ISL_2096542, EPI_ISL_2096543, EPI_ISL_2096544, EPI_ISL_2096545, EPI_ISL_2096546, EPI_ISL_2096547, EPI_ISL_2096548, EPI_ISL_2096549, EPI_ISL_2096550, EPI_ISL_2096551, EPI_ISL_2096552, EPI_ISL_2096553, EPI_ISL_2096554, EPI_ISL_2096555, EPI_ISL_2096556, EPI_ISL_2096557, EPI_ISL_2096558, EPI_ISL_2096559, EPI_ISL_2096560, EPI_ISL_2096561, EPI_ISL_2096562, EPI_ISL_2096563, EPI_ISL_2096564, EPI_ISL_2096565, EPI_ISL_2096566, EPI_ISL_2096567, EPI_ISL_2096568, EPI_ISL_2096569, EPI_ISL_2096570, EPI_ISL_2096571, EPI_ISL_2096572, EPI_ISL_2096573, EPI_ISL_2096574, EPI_ISL_2096575, EPI_ISL_2096576, EPI_ISL_2096577, EPI_ISL_2096578, EPI_ISL_2096579, EPI_ISL_2096580, EPI_ISL_2096581, EPI_ISL_2096582, EPI_ISL_2096583, EPI_ISL_2096584, EPI_ISL_2096585, EPI_ISL_2096586, EPI_ISL_2096587, EPI_ISL_2096588, EPI_ISL_2096589, EPI_ISL_2096590, EPI_ISL_2096591, EPI_ISL_2096592, EPI_ISL_2096593, EPI_ISL_2096594, EPI_ISL_2096595, EPI_ISL_2096596, EPI_ISL_2096597, EPI_ISL_2096598, EPI_ISL_2096599, EPI_ISL_2096600, EPI_ISL_2096601, EPI_ISL_2096602, EPI_ISL_2096603, EPI_ISL_2096604, EPI_ISL_2096605, EPI_ISL_2096606, EPI_ISL_2096607, EPI_ISL_2096608, EPI_ISL_2096609, EPI_ISL_2096610, EPI_ISL_2096611, EPI_ISL_2096612, EPI_ISL_2096613, EPI_ISL_2096614, EPI_ISL_2096615, EPI_ISL_2096616, EPI_ISL_2096617, EPI_ISL_2096618, EPI_ISL_2096619, EPI_ISL_2096620, EPI_ISL_2096621, EPI_ISL_2096622, EPI_ISL_2096623, EPI_ISL_2096624, EPI_ISL_2096625, EPI_ISL_2096626, EPI_ISL_2096627, EPI_ISL_2096628, EPI_ISL_2096629, EPI_ISL_2096630, EPI_ISL_2096631, EPI_ISL_2096632, EPI_ISL_2096633, EPI_ISL_2096635, EPI_ISL_2096636, EPI_ISL_2096637, EPI_ISL_2096638, EPI_ISL_2096639, EPI_ISL_2096640, EPI_ISL_2096641, EPI_ISL_2096642, EPI_ISL_2096643, EPI_ISL_2096644, EPI_ISL_2096645, EPI_ISL_2096646, EPI_ISL_2096647, EPI_ISL_2096648, EPI_ISL_2096649, EPI_ISL_2096650, EPI_ISL_2096651, EPI_ISL_2096652, EPI_ISL_2096653, EPI_ISL_2096654, EPI_ISL_2096655, EPI_ISL_2096656, EPI_ISL_2096657, EPI_ISL_2096658, EPI_ISL_2096659, EPI_ISL_2096660, EPI_ISL_2096661, EPI_ISL_2096662, EPI_ISL_2096663, EPI_ISL_2096664, EPI_ISL_2096665, EPI_ISL_2096666, EPI_ISL_2096667, EPI_ISL_2096668, EPI_ISL_2096669, EPI_ISL_2096670, EPI_ISL_2096671, EPI_ISL_2096672, EPI_ISL_2096673, EPI_ISL_2096674, EPI_ISL_2096675, EPI_ISL_2096676, EPI_ISL_2096677, EPI_ISL_2096678, EPI_ISL_2096679, EPI_ISL_2096680, EPI_ISL_2096681, EPI_ISL_2096682, EPI_ISL_2096683, EPI_ISL_2096684, EPI_ISL_2096685, EPI_ISL_2096686, EPI_ISL_2096687, EPI_ISL_2096688, EPI_ISL_2096689, EPI_ISL_2096690, EPI_ISL_2096691, EPI_ISL_2096692, EPI_ISL_2096693, EPI_ISL_2096694, EPI_ISL_2096695, EPI_ISL_2096696, EPI_ISL_2096697, EPI_ISL_2096698, EPI_ISL_2096699, EPI_ISL_2096700, EPI_ISL_2096701, EPI_ISL_2096702, EPI_ISL_2096703, EPI_ISL_2096704, EPI_ISL_2096705, EPI_ISL_2096706, EPI_ISL_2096707, EPI_ISL_2096708, EPI_ISL_2096709, EPI_ISL_2096710, EPI_ISL_2096711, EPI_ISL_2096712, EPI_ISL_2096713, EPI_ISL_2096714, EPI_ISL_2096715, EPI_ISL_2096716, EPI_ISL_2096717, EPI_ISL_2096718, EPI_ISL_2096719, EPI_ISL_2096720, EPI_ISL_2096721, EPI_ISL_2096722, EPI_ISL_2096723, EPI_ISL_2096724, EPI_ISL_2096725, EPI_ISL_2096727, EPI_ISL_2096728, EPI_ISL_2096729, EPI_ISL_2096730, EPI_ISL_2096731, EPI_ISL_2096732, EPI_ISL_2096733, EPI_ISL_2096734, EPI_ISL_2096735, EPI_ISL_2096736, EPI_ISL_2096737, EPI_ISL_2096738, EPI_ISL_2096739, EPI_ISL_2096740, EPI_ISL_2096741, EPI_ISL_2096742, EPI_ISL_2096743, EPI_ISL_2096744, EPI_ISL_2096745, EPI_ISL_2096746, EPI_ISL_2096747, EPI_ISL_2096748, EPI_ISL_2096749, EPI_ISL_2096750, EPI_ISL_2096751, EPI_ISL_2096752, EPI_ISL_2096753, EPI_ISL_2096754, EPI_ISL_2096755, EPI_ISL_2096756, EPI_ISL_2096757, EPI_ISL_2096758, EPI_ISL_2096759, EPI_ISL_2096760, EPI_ISL_2096761, EPI_ISL_2096762, EPI_ISL_2096763, EPI_ISL_2096764, EPI_ISL_2096765, EPI_ISL_2096766, EPI_ISL_2096767, EPI_ISL_2096768, EPI_ISL_2096769, EPI_ISL_2096770, EPI_ISL_2096771, EPI_ISL_2096772, EPI_ISL_2096773, EPI_ISL_2096774 | | | |
| see above | Broad Institute Clinical Research Sequencing Platform | Infectious Disease Program, Broad Institute of Harvard and MIT | Siddle,K.J., Adams,G., Pearlman,L., Gladden-Young,A., Vicente,G., Blumenstiel,B., DeFelice,M., Lee,M., McGovern,S., Lagerborg,K., Rudy,M., DeRuff,K., Carter,A., Normandin,E., Bauer,M., Reilly,S., Tomkins-Tinch,C., Loreth,C., Chaluvadi,S., Meldrim,J., Granger,B., Lemieux,J.E., Birren,B.W., Sabeti,P.C., Larkin,K., Dodge,S., Lennon,N., Madoff,L., Brown,C., Gallagher,G., Smole,S., Park,D.J., Gabriel,S., and MacInnis,B.L. |
| EPI_ISL_2097172, EPI_ISL_2097173, EPI_ISL_2097174, EPI_ISL_2097175, EPI_ISL_2097176, EPI_ISL_2097177, EPI_ISL_2097178, EPI_ISL_2097179, EPI_ISL_2097180, EPI_ISL_2097181, EPI_ISL_2097182, EPI_ISL_2097183, EPI_ISL_2097184, EPI_ISL_2097185, EPI_ISL_2097186, EPI_ISL_2097187, EPI_ISL_2097188, EPI_ISL_2097189, EPI_ISL_2097190, EPI_ISL_2097192, EPI_ISL_2097193, EPI_ISL_2097195, EPI_ISL_2097197, EPI_ISL_2097198, EPI_ISL_2097199, EPI_ISL_2097200, EPI_ISL_2097201, EPI_ISL_2097202, EPI_ISL_2097203, EPI_ISL_2097204, EPI_ISL_2097205, EPI_ISL_2097206, EPI_ISL_2097207, EPI_ISL_2097209, EPI_ISL_2097210, EPI_ISL_2097211, EPI_ISL_2097212, EPI_ISL_2097213 | | | |
| see above | IN State Department of Health Laboratory Services | IN State Department of Health Laboratory Services | Cassandra Campion, Jamie Yeadon, Brian Pope, Lixia Liu, Kyle Brownlee, Melissa Hindenlang, Mark Glazier |
| EPI_ISL_2097232, EPI_ISL_2097233, EPI_ISL_2097234, EPI_ISL_2097235, EPI_ISL_2097236, EPI_ISL_2097237, EPI_ISL_2097238 | ALBANY MEDICAL CENTER | Wadsworth Center, New York State Department of Health | Kirsten St. George, Daryl M. Lamson, Alexis Russell, Matthew Shudt, Melissa A Leisner, Jonathan Plitnick, Catharine Prussing, Navjot Singh, John Kelly, Erasmus Schneider, Erica Lasek-Nesselquist |
| EPI_ISL_2097239, EPI_ISL_2097240, EPI_ISL_2097241, EPI_ISL_2097242, EPI_ISL_2097243, EPI_ISL_2097244, EPI_ISL_2097245, EPI_ISL_2097246, EPI_ISL_2097247, EPI_ISL_2097248, EPI_ISL_2097249, EPI_ISL_2097250, EPI_ISL_2097251 | | | |
| see above | BOSTON HEART DIAGNOSTICS CORP | Wadsworth Center, New York State Department of Health | Kirsten St. George, Daryl M. Lamson, Alexis Russell, Matthew Shudt, Melissa A Leisner, Jonathan Plitnick, Catharine Prussing, Navjot Singh, John Kelly, Erasmus Schneider, Erica Lasek-Nesselquist |
| EPI_ISL_2097252 | CENTERS LAB NJ, LLC (DBA MEDLABS DIAGNOSTICS) | Wadsworth Center, New York State Department of Health | Kirsten St. George, Daryl M. Lamson, Alexis Russell, Matthew Shudt, Melissa A Leisner, Jonathan Plitnick, Catharine Prussing, Navjot Singh, John Kelly, Erasmus Schneider, Erica Lasek-Nesselquist |
| EPI_ISL_2097253, EPI_ISL_2097254, EPI_ISL_2097255, EPI_ISL_2097256, EPI_ISL_2097257, EPI_ISL_2097258, EPI_ISL_2097259, EPI_ISL_2097260, EPI_ISL_2097261, EPI_ISL_2097262, EPI_ISL_2097263, EPI_ISL_2097264, EPI_ISL_2097265 | | | |
| see above | MONTEFIORE MEDICAL CENTER LABORATORIES | Wadsworth Center, New York State Department of Health | Kirsten St. George, Daryl M. Lamson, Alexis Russell, Matthew Shudt, Melissa A Leisner, Jonathan Plitnick, Catharine Prussing, Navjot Singh, John Kelly, Erasmus Schneider, Erica Lasek-Nesselquist |
| EPI_ISL_2097266, EPI_ISL_2097267, EPI_ISL_2097268, EPI_ISL_2097269, EPI_ISL_2097270, EPI_ISL_2097271, EPI_ISL_2097272, EPI_ISL_2097273, EPI_ISL_2097274, EPI_ISL_2097275, EPI_ISL_2097276, EPI_ISL_2097277, EPI_ISL_2097278, EPI_ISL_2097279, EPI_ISL_2097280, EPI_ISL_2097281, EPI_ISL_2097282, EPI_ISL_2097283, EPI_ISL_2097284, EPI_ISL_2097285, EPI_ISL_2097286, EPI_ISL_2097287, EPI_ISL_2097288, EPI_ISL_2097289, EPI_ISL_2097290, EPI_ISL_2097291, EPI_ISL_2097292, EPI_ISL_2097293, EPI_ISL_2097294 | | | |
| see above | NORTHWELL HEALTH LABORATORIES | Wadsworth Center, New York State Department of Health | Kirsten St. George, Daryl M. Lamson, Alexis Russell, Matthew Shudt, Melissa A Leisner, Jonathan Plitnick, Catharine Prussing, Navjot Singh, John Kelly, Erasmus Schneider, Erica Lasek-Nesselquist |
| EPI_ISL_2097295 | ST. CATHERINE OF SIENA HOSPITAL LABORATORY | Wadsworth Center, New York State Department of Health | Kirsten St. George, Daryl M. Lamson, Alexis Russell, Matthew Shudt, Melissa A Leisner, Jonathan Plitnick, Catharine Prussing, Navjot Singh, John Kelly, Erasmus Schneider, Erica Lasek-Nesselquist |
| EPI_ISL_2097296, EPI_ISL_2097297, EPI_ISL_2097298, EPI_ISL_2097299, EPI_ISL_2097300, EPI_ISL_2097301, EPI_ISL_2097302, EPI_ISL_2097303 | STONY BROOK UNIVERSITY HOSPITAL | Wadsworth Center, New York State Department of Health | Kirsten St. George, Daryl M. Lamson, Alexis Russell, Matthew Shudt, Melissa A Leisner, Jonathan Plitnick, Catharine Prussing, Navjot Singh, John Kelly, Erasmus Schneider, Erica Lasek-Nesselquist |
| EPI_ISL_2097304, EPI_ISL_2097305 | SUNY UPSTATE MEDICAL UNIVERSITY | Wadsworth Center, New York State Department of Health | Kirsten St. George, Daryl M. Lamson, Alexis Russell, Matthew Shudt, Melissa A Leisner, Jonathan Plitnick, Catharine Prussing, Navjot Singh, John Kelly, Erasmus Schneider, Erica Lasek-Nesselquist |
| EPI_ISL_2097306, EPI_ISL_2097307, EPI_ISL_2097308, EPI_ISL_2097309, EPI_ISL_2097310, | WESTCHESTER MEDICAL CENTER | Wadsworth Center, New York State Department of Health | Kirsten St. George, Daryl M. Lamson, Alexis Russell, Matthew Shudt, Melissa A Leisner, Jonathan Plitnick, Catharine Prussing, Navjot Singh, John Kelly, Erasmus Schneider, Erica Lasek-Nesselquist |

| | | | |
|---|---|---|---|
| EPI_ISL_2097311, EPI_ISL_2097312, EPI_ISL_2097313, EPI_ISL_2097314 | | | |
| EPI_ISL_2097317, EPI_ISL_2097318, EPI_ISL_2097319, EPI_ISL_2097320, EPI_ISL_2097321, EPI_ISL_2097322, EPI_ISL_2097323, EPI_ISL_2097324 | Minnesota Department of Health, Public Health Laboratory | Minnesota Department of Health, Public Health Laboratory | Alexandra Lorentz, Jacob Garfin, Matt Plumb, and Xiong Wang |
| EPI_ISL_2097325 | M Health Fairview | Minnesota Department of Health, Public Health Laboratory | Alexandra Lorentz, Jacob Garfin, Matt Plumb, and Xiong Wang |
| EPI_ISL_2097326, EPI_ISL_2097327, EPI_ISL_2097328, EPI_ISL_2097329, EPI_ISL_2097330, EPI_ISL_2097331, EPI_ISL_2097332, EPI_ISL_2097333, EPI_ISL_2097334, EPI_ISL_2097335, EPI_ISL_2097336, EPI_ISL_2097337, EPI_ISL_2097338 | | | |
| see above | Minnesota Department of Health, Public Health Laboratory | Minnesota Department of Health, Public Health Laboratory | Alexandra Lorentz, Jacob Garfin, Matt Plumb, and Xiong Wang |
| EPI_ISL_2097339, EPI_ISL_2097340, EPI_ISL_2097341 | M Health Fairview | Minnesota Department of Health, Public Health Laboratory | Alexandra Lorentz, Jacob Garfin, Matt Plumb, and Xiong Wang |
| EPI_ISL_2097342 | Minnesota Department of Health, Public Health Laboratory | Minnesota Department of Health, Public Health Laboratory | Alexandra Lorentz, Jacob Garfin, Matt Plumb, and Xiong Wang |
| EPI_ISL_2097343 | Helix/Illumina | Centers for Disease Control and Prevention Division of Viral Diseases, Pathogen Discovery | Dakota Howard, Dhwani Batra, Peter W. Cook, Kara Moser, Adrian Paskey, Jason Caravas, Benjamin Rambo-Martin, Shatavia Morrison, Christopher Gulvick, Scott Sammons, Yvette Unoarumhi, Darlene Wagner, Matthew Schmerer, Eileen de Feo, Jan Antico, Christine Tran, Matthew Tolentino, Shannon Wickline, Kim Gietzen, Brad Sickler, Jingtao Liu, Eric Allen, Phil Febbo, Nicole L. Washington, Simon White, Geraint Levan, Kelly Schiabor Barrett, Elizabeth Cirulli, Alexandre Bolze, Ary Ascencio, Charlotte Rivera-Garcia, Ryan Cho, Jason Nguyen, Sherry Wang, Jimmy Ramirez, Tyler Cassens, Efren Sandoval, Magnus Isaksson, William Lee, David Becker, Marc Laurent, James Lu, Clinton R. Paden, Duncan MacCannell |
| EPI_ISL_2097344, EPI_ISL_2097345, EPI_ISL_2097346 | Minnesota Department of Health, Public Health Laboratory | Minnesota Department of Health, Public Health Laboratory | Alexandra Lorentz, Jacob Garfin, Matt Plumb, and Xiong Wang |
| EPI_ISL_2097347 | Helix/Illumina | Centers for Disease Control and Prevention Division of Viral Diseases, Pathogen Discovery | Dakota Howard, Dhwani Batra, Peter W. Cook, Kara Moser, Adrian Paskey, Jason Caravas, Benjamin Rambo-Martin, Shatavia Morrison, Christopher Gulvick, Scott Sammons, Yvette Unoarumhi, Darlene Wagner, Matthew Schmerer, Eileen de Feo, Jan Antico, Christine Tran, Matthew Tolentino, Shannon Wickline, Kim Gietzen, Brad Sickler, Jingtao Liu, Eric Allen, Phil Febbo, Nicole L. Washington, Simon White, Geraint Levan, Kelly Schiabor Barrett, Elizabeth Cirulli, Alexandre Bolze, Ary Ascencio, Charlotte Rivera-Garcia, Ryan Cho, Jason Nguyen, Sherry Wang, Jimmy Ramirez, Tyler Cassens, Efren Sandoval, Magnus Isaksson, William Lee, David Becker, Marc Laurent, James Lu, Clinton R. Paden, Duncan MacCannell |
| EPI_ISL_2097348 | Minnesota Department of Health, Public Health Laboratory | Minnesota Department of Health, Public Health Laboratory | Alexandra Lorentz, Jacob Garfin, Matt Plumb, and Xiong Wang |
| EPI_ISL_2097350, EPI_ISL_2097352, EPI_ISL_2097355, EPI_ISL_2097358 | Helix/Illumina | Centers for Disease Control and Prevention Division of Viral Diseases, Pathogen Discovery | Dakota Howard, Dhwani Batra, Peter W. Cook, Kara Moser, Adrian Paskey, Jason Caravas, Benjamin Rambo-Martin, Shatavia Morrison, Christopher Gulvick, Scott Sammons, Yvette Unoarumhi, Darlene Wagner, Matthew Schmerer, Eileen de Feo, Jan Antico, Christine Tran, Matthew Tolentino, Shannon Wickline, Kim Gietzen, Brad Sickler, Jingtao Liu, Eric Allen, Phil Febbo, Nicole L. Washington, Simon White, Geraint Levan, Kelly Schiabor Barrett, Elizabeth Cirulli, Alexandre Bolze, Ary Ascencio, Charlotte Rivera-Garcia, Ryan Cho, Jason Nguyen, Sherry Wang, Jimmy Ramirez, Tyler Cassens, Efren Sandoval, Magnus Isaksson, William Lee, David Becker, Marc Laurent, James Lu, Clinton R. Paden, Duncan MacCannell |
| EPI_ISL_2097359, EPI_ISL_2097360 | Minnesota Department of Health, Public Health Laboratory | Minnesota Department of Health, Public Health Laboratory | Alexandra Lorentz, Jacob Garfin, Matt Plumb, and Xiong Wang |
| EPI_ISL_2097361 | Helix/Illumina | Centers for Disease Control and Prevention Division of Viral Diseases, Pathogen Discovery | Dakota Howard, Dhwani Batra, Peter W. Cook, Kara Moser, Adrian Paskey, Jason Caravas, Benjamin Rambo-Martin, Shatavia Morrison, Christopher Gulvick, Scott Sammons, Yvette Unoarumhi, Darlene Wagner, Matthew Schmerer, Eileen de Feo, Jan Antico, Christine Tran, Matthew Tolentino, Shannon Wickline, Kim Gietzen, Brad Sickler, Jingtao Liu, Eric Allen, Phil Febbo, Nicole L. Washington, Simon White, Geraint Levan, Kelly Schiabor Barrett, Elizabeth Cirulli, Alexandre Bolze, Ary Ascencio, Charlotte Rivera-Garcia, Ryan Cho, Jason Nguyen, Sherry Wang, Jimmy Ramirez, Tyler Cassens, Efren Sandoval, Magnus Isaksson, William Lee, David Becker, Marc Laurent, James Lu, Clinton R. Paden, Duncan MacCannell |
| EPI_ISL_2097362 | Minnesota Department of Health, Public Health Laboratory | Minnesota Department of Health, Public Health Laboratory | Alexandra Lorentz, Jacob Garfin, Matt Plumb, and Xiong Wang |
| EPI_ISL_2097363 | Helix/Illumina | Centers for Disease Control and Prevention Division of Viral Diseases, Pathogen Discovery | Dakota Howard, Dhwani Batra, Peter W. Cook, Kara Moser, Adrian Paskey, Jason Caravas, Benjamin Rambo-Martin, Shatavia Morrison, Christopher Gulvick, Scott Sammons, Yvette Unoarumhi, Darlene Wagner, Matthew Schmerer, Eileen de Feo, Jan Antico, Christine Tran, Matthew Tolentino, Shannon Wickline, Kim Gietzen, Brad Sickler, Jingtao Liu, Eric Allen, Phil Febbo, Nicole L. Washington, Simon White, Geraint Levan, Kelly Schiabor Barrett, Elizabeth Cirulli, Alexandre Bolze, Ary Ascencio, Charlotte Rivera-Garcia, Ryan Cho, Jason Nguyen, Sherry Wang, Jimmy Ramirez, Tyler Cassens, Efren Sandoval, Magnus Isaksson, William Lee, David Becker, Marc Laurent, James Lu, Clinton R. Paden, Duncan MacCannell |
| EPI_ISL_2097364, EPI_ISL_2097365 | Minnesota Department of Health, Public Health Laboratory | Minnesota Department of Health, Public Health Laboratory | Alexandra Lorentz, Jacob Garfin, Matt Plumb, and Xiong Wang |
| EPI_ISL_2097366 | Helix/Illumina | Centers for Disease Control and Prevention Division of Viral Diseases, Pathogen Discovery | Dakota Howard, Dhwani Batra, Peter W. Cook, Kara Moser, Adrian Paskey, Jason Caravas, Benjamin Rambo-Martin, Shatavia Morrison, Christopher Gulvick, Scott Sammons, Yvette Unoarumhi, Darlene Wagner, Matthew Schmerer, Eileen de Feo, Jan Antico, Christine Tran, Matthew Tolentino, Shannon Wickline, Kim Gietzen, Brad Sickler, Jingtao Liu, Eric Allen, Phil Febbo, Nicole L. Washington, Simon White, Geraint Levan, Kelly Schiabor Barrett, Elizabeth Cirulli, Alexandre Bolze, Ary Ascencio, Charlotte Rivera-Garcia, Ryan Cho, Jason Nguyen, Sherry Wang, Jimmy Ramirez, Tyler Cassens, Efren Sandoval, Magnus Isaksson, William Lee, David Becker, Marc Laurent, James Lu, Clinton R. Paden, Duncan MacCannell |
| EPI_ISL_2097367 | Minnesota Department of Health, Public Health Laboratory | Minnesota Department of Health, Public Health Laboratory | Alexandra Lorentz, Jacob Garfin, Matt Plumb, and Xiong Wang |
| EPI_ISL_2097368 | Helix/Illumina | Centers for Disease Control and Prevention Division of Viral Diseases, Pathogen Discovery | Dakota Howard, Dhwani Batra, Peter W. Cook, Kara Moser, Adrian Paskey, Jason Caravas, Benjamin Rambo-Martin, Shatavia Morrison, Christopher Gulvick, Scott Sammons, Yvette Unoarumhi, Darlene Wagner, Matthew Schmerer, Eileen de Feo, Jan Antico, Christine Tran, Matthew Tolentino, Shannon Wickline, Kim Gietzen, Brad Sickler, Jingtao Liu, Eric Allen, Phil Febbo, Nicole L. Washington, Simon White, Geraint Levan, Kelly Schiabor Barrett, Elizabeth Cirulli, Alexandre Bolze, Ary Ascencio, Charlotte Rivera-Garcia, Ryan Cho, Jason Nguyen, Sherry Wang, Jimmy Ramirez, Tyler Cassens, Efren Sandoval, Magnus Isaksson, William Lee, David Becker, Marc Laurent, James Lu, Clinton R. Paden, Duncan MacCannell |
| EPI_ISL_2097369, EPI_ISL_2097370 | Minnesota Department of Health, Public Health Laboratory | Minnesota Department of Health, Public Health Laboratory | Alexandra Lorentz, Jacob Garfin, Matt Plumb, and Xiong Wang |
| EPI_ISL_2097371 | Helix/Illumina | Centers for Disease Control and Prevention Division of Viral Diseases, Pathogen Discovery | Dakota Howard, Dhwani Batra, Peter W. Cook, Kara Moser, Adrian Paskey, Jason Caravas, Benjamin Rambo-Martin, Shatavia Morrison, Christopher Gulvick, Scott Sammons, Yvette Unoarumhi, Darlene Wagner, Matthew Schmerer, Eileen de Feo, Jan Antico, Christine Tran, Matthew Tolentino, Shannon Wickline, Kim Gietzen, Brad Sickler, Jingtao Liu, Eric Allen, Phil Febbo, Nicole L. Washington, Simon White, Geraint Levan, Kelly Schiabor Barrett, Elizabeth Cirulli, Alexandre Bolze, Ary Ascencio, Charlotte Rivera-Garcia, Ryan Cho, Jason Nguyen, Sherry Wang, Jimmy Ramirez, Tyler Cassens, Efren Sandoval, Magnus Isaksson, William Lee, David Becker, Marc Laurent, James Lu, Clinton R. Paden, Duncan MacCannell |
| EPI_ISL_2097372, EPI_ISL_2097373 | Minnesota Department of Health, Public Health Laboratory | Minnesota Department of Health, Public Health Laboratory | Alexandra Lorentz, Jacob Garfin, Matt Plumb, and Xiong Wang |
| EPI_ISL_2097374 | Helix/Illumina | Centers for Disease Control and Prevention Division of Viral Diseases, Pathogen Discovery | Dakota Howard, Dhwani Batra, Peter W. Cook, Kara Moser, Adrian Paskey, Jason Caravas, Benjamin Rambo-Martin, Shatavia Morrison, Christopher Gulvick, Scott Sammons, Yvette Unoarumhi, Darlene Wagner, Matthew Schmerer, Eileen de Feo, Jan Antico, Christine Tran, Matthew Tolentino, Shannon Wickline, Kim Gietzen, Brad Sickler, Jingtao Liu, Eric Allen, Phil Febbo, Nicole L. Washington, Simon White, Geraint Levan, Kelly Schiabor Barrett, Elizabeth Cirulli, Alexandre Bolze, Ary Ascencio, Charlotte Rivera-Garcia, Ryan Cho, Jason Nguyen, Sherry Wang, Jimmy Ramirez, Tyler Cassens, Efren Sandoval, Magnus Isaksson, William Lee, David Becker, Marc Laurent, James Lu, Clinton R. Paden, Duncan MacCannell |
| EPI_ISL_2097375 | Minnesota Department of Health, Public Health Laboratory | Minnesota Department of Health, Public Health Laboratory | Alexandra Lorentz, Jacob Garfin, Matt Plumb, and Xiong Wang |

EPI_ISL_2097376, EPI_ISL_2097378, EPI_ISL_2097379, EPI_ISL_2097380, EPI_ISL_2097381, EPI_ISL_2097382, EPI_ISL_2097383, EPI_ISL_2097384, EPI_ISL_2097385, EPI_ISL_2097386, EPI_ISL_2097387, EPI_ISL_2097388, EPI_ISL_2097389, EPI_ISL_2097390, EPI_ISL_2097391, EPI_ISL_2097392, EPI_ISL_2097393, EPI_ISL_2097394, EPI_ISL_2097395, EPI_ISL_2097396, EPI_ISL_2097397, EPI_ISL_2097398,
EPI_ISL_2097399, EPI_ISL_2097400, EPI_ISL_2097401, EPI_ISL_2097402, EPI_ISL_2097403, EPI_ISL_2097404, EPI_ISL_2097405, EPI_ISL_2097406, EPI_ISL_2097407, EPI_ISL_2097408, EPI_ISL_2097409, EPI_ISL_2097410, EPI_ISL_2097411, EPI_ISL_2097412, EPI_ISL_2097413, EPI_ISL_2097414, EPI_ISL_2097415, EPI_ISL_2097416, EPI_ISL_2097417, EPI_ISL_2097418, EPI_ISL_2097419, EPI_ISL_2097420,
EPI_ISL_2097421, EPI_ISL_2097422, EPI_ISL_2097423, EPI_ISL_2097424, EPI_ISL_2097425, EPI_ISL_2097426, EPI_ISL_2097427, EPI_ISL_2097428, EPI_ISL_2097429, EPI_ISL_2097430, EPI_ISL_2097431, EPI_ISL_2097432, EPI_ISL_2097433, EPI_ISL_2097434, EPI_ISL_2097435, EPI_ISL_2097436, EPI_ISL_2097437, EPI_ISL_2097438, EPI_ISL_2097439, EPI_ISL_2097440, EPI_ISL_2097441, EPI_ISL_2097442,
EPI_ISL_2097443, EPI_ISL_2097444, EPI_ISL_2097445, EPI_ISL_2097446, EPI_ISL_2097447, EPI_ISL_2097448, EPI_ISL_2097449, EPI_ISL_2097450, EPI_ISL_2097451, EPI_ISL_2097452, EPI_ISL_2097453, EPI_ISL_2097454, EPI_ISL_2097455, EPI_ISL_2097456, EPI_ISL_2097457, EPI_ISL_2097458, EPI_ISL_2097459, EPI_ISL_2097460, EPI_ISL_2097461, EPI_ISL_2097462, EPI_ISL_2097463, EPI_ISL_2097464,
EPI_ISL_2097465, EPI_ISL_2097466, EPI_ISL_2097467, EPI_ISL_2097468, EPI_ISL_2097469, EPI_ISL_2097470, EPI_ISL_2097471, EPI_ISL_2097472, EPI_ISL_2097473, EPI_ISL_2097474, EPI_ISL_2097475, EPI_ISL_2097476, EPI_ISL_2097477, EPI_ISL_2097478, EPI_ISL_2097479, EPI_ISL_2097480, EPI_ISL_2097481, EPI_ISL_2097482, EPI_ISL_2097483, EPI_ISL_2097484, EPI_ISL_2097485, EPI_ISL_2097486,
EPI_ISL_2097487, EPI_ISL_2097488, EPI_ISL_2097489, EPI_ISL_2097490, EPI_ISL_2097491, EPI_ISL_2097492, EPI_ISL_2097493, EPI_ISL_2097494, EPI_ISL_2097495, EPI_ISL_2097496, EPI_ISL_2097497, EPI_ISL_2097498, EPI_ISL_2097499, EPI_ISL_2097500, EPI_ISL_2097501, EPI_ISL_2097502, EPI_ISL_2097503, EPI_ISL_2097504, EPI_ISL_2097505, EPI_ISL_2097506, EPI_ISL_2097507, EPI_ISL_2097508,
EPI_ISL_2097509, EPI_ISL_2097510, EPI_ISL_2097511, EPI_ISL_2097512, EPI_ISL_2097513, EPI_ISL_2097514, EPI_ISL_2097515, EPI_ISL_2097516, EPI_ISL_2097517, EPI_ISL_2097518, EPI_ISL_2097519, EPI_ISL_2097520, EPI_ISL_2097521, EPI_ISL_2097522, EPI_ISL_2097523, EPI_ISL_2097524, EPI_ISL_2097525, EPI_ISL_2097526, EPI_ISL_2097527, EPI_ISL_2097528, EPI_ISL_2097529, EPI_ISL_2097530,
EPI_ISL_2097531, EPI_ISL_2097532, EPI_ISL_2097533, EPI_ISL_2097534, EPI_ISL_2097535, EPI_ISL_2097536, EPI_ISL_2097537, EPI_ISL_2097538, EPI_ISL_2097539, EPI_ISL_2097540, EPI_ISL_2097541, EPI_ISL_2097542, EPI_ISL_2097543, EPI_ISL_2097544, EPI_ISL_2097545, EPI_ISL_2097546, EPI_ISL_2097547, EPI_ISL_2097548, EPI_ISL_2097549, EPI_ISL_2097550, EPI_ISL_2097551, EPI_ISL_2097553,
EPI_ISL_2097554, EPI_ISL_2097555, EPI_ISL_2097556, EPI_ISL_2097557, EPI_ISL_2097558, EPI_ISL_2097559, EPI_ISL_2097560, EPI_ISL_2097561, EPI_ISL_2097562, EPI_ISL_2097563, EPI_ISL_2097564, EPI_ISL_2097566, EPI_ISL_2097567, EPI_ISL_2097568, EPI_ISL_2097569, EPI_ISL_2097570, EPI_ISL_2097571, EPI_ISL_2097572, EPI_ISL_2097573, EPI_ISL_2097574, EPI_ISL_2097575, EPI_ISL_2097576,
EPI_ISL_2097577, EPI_ISL_2097578, EPI_ISL_2097580, EPI_ISL_2097581, EPI_ISL_2097582, EPI_ISL_2097583, EPI_ISL_2097584, EPI_ISL_2097585, EPI_ISL_2097586, EPI_ISL_2097587, EPI_ISL_2097588, EPI_ISL_2097589, EPI_ISL_2097590, EPI_ISL_2097591, EPI_ISL_2097592, EPI_ISL_2097593, EPI_ISL_2097594, EPI_ISL_2097595, EPI_ISL_2097596, EPI_ISL_2097597, EPI_ISL_2097598, EPI_ISL_2097599,
EPI_ISL_2097600, EPI_ISL_2097601, EPI_ISL_2097602, EPI_ISL_2097603, EPI_ISL_2097604, EPI_ISL_2097605, EPI_ISL_2097606, EPI_ISL_2097607, EPI_ISL_2097608, EPI_ISL_2097609, EPI_ISL_2097610, EPI_ISL_2097611, EPI_ISL_2097612, EPI_ISL_2097613, EPI_ISL_2097614, EPI_ISL_2097615, EPI_ISL_2097616, EPI_ISL_2097617, EPI_ISL_2097618, EPI_ISL_2097619, EPI_ISL_2097620, EPI_ISL_2097621,
EPI_ISL_2097622, EPI_ISL_2097623, EPI_ISL_2097624, EPI_ISL_2097625, EPI_ISL_2097626, EPI_ISL_2097627, EPI_ISL_2097628, EPI_ISL_2097629, EPI_ISL_2097630, EPI_ISL_2097631, EPI_ISL_2097632, EPI_ISL_2097633, EPI_ISL_2097634, EPI_ISL_2097635, EPI_ISL_2097636, EPI_ISL_2097637, EPI_ISL_2097638, EPI_ISL_2097639, EPI_ISL_2097640, EPI_ISL_2097642, EPI_ISL_2097643, EPI_ISL_2097644,
EPI_ISL_2097645, EPI_ISL_2097646, EPI_ISL_2097647, EPI_ISL_2097648, EPI_ISL_2097649, EPI_ISL_2097650, EPI_ISL_2097651, EPI_ISL_2097652, EPI_ISL_2097653, EPI_ISL_2097654, EPI_ISL_2097655, EPI_ISL_2097656, EPI_ISL_2097657, EPI_ISL_2097658, EPI_ISL_2097659, EPI_ISL_2097660, EPI_ISL_2097661, EPI_ISL_2097662, EPI_ISL_2097663, EPI_ISL_2097664, EPI_ISL_2097665, EPI_ISL_2097666,
EPI_ISL_2097667, EPI_ISL_2097668, EPI_ISL_2097669, EPI_ISL_2097670, EPI_ISL_2097671, EPI_ISL_2097672, EPI_ISL_2097673, EPI_ISL_2097674, EPI_ISL_2097675, EPI_ISL_2097676, EPI_ISL_2097677, EPI_ISL_2097678, EPI_ISL_2097679, EPI_ISL_2097680, EPI_ISL_2097681, EPI_ISL_2097682, EPI_ISL_2097683, EPI_ISL_2097684, EPI_ISL_2097685, EPI_ISL_2097686, EPI_ISL_2097687, EPI_ISL_2097688,
EPI_ISL_2097689, EPI_ISL_2097690, EPI_ISL_2097691, EPI_ISL_2097692, EPI_ISL_2097693, EPI_ISL_2097695, EPI_ISL_2097696, EPI_ISL_2097697, EPI_ISL_2097698, EPI_ISL_2097699, EPI_ISL_2097700, EPI_ISL_2097701, EPI_ISL_2097702, EPI_ISL_2097703, EPI_ISL_2097704, EPI_ISL_2097705, EPI_ISL_2097706, EPI_ISL_2097707, EPI_ISL_2097709, EPI_ISL_2097710, EPI_ISL_2097711, EPI_ISL_2097712,
EPI_ISL_2097713, EPI_ISL_2097714, EPI_ISL_2097715, EPI_ISL_2097716, EPI_ISL_2097717, EPI_ISL_2097718, EPI_ISL_2097719, EPI_ISL_2097720, EPI_ISL_2097721, EPI_ISL_2097722, EPI_ISL_2097723, EPI_ISL_2097725, EPI_ISL_2097726, EPI_ISL_2097728, EPI_ISL_2097729, EPI_ISL_2097730, EPI_ISL_2097731, EPI_ISL_2097732, EPI_ISL_2097733, EPI_ISL_2097734, EPI_ISL_2097735, EPI_ISL_2097736,
EPI_ISL_2097737, EPI_ISL_2097738, EPI_ISL_2097739, EPI_ISL_2097740, EPI_ISL_2097741, EPI_ISL_2097742, EPI_ISL_2097743, EPI_ISL_2097744, EPI_ISL_2097745, EPI_ISL_2097746, EPI_ISL_2097747, EPI_ISL_2097748, EPI_ISL_2097749, EPI_ISL_2097750, EPI_ISL_2097751, EPI_ISL_2097752, EPI_ISL_2097753, EPI_ISL_2097754, EPI_ISL_2097755, EPI_ISL_2097756, EPI_ISL_2097757, EPI_ISL_2097758,
EPI_ISL_2097759, EPI_ISL_2097760, EPI_ISL_2097761, EPI_ISL_2097762, EPI_ISL_2097763, EPI_ISL_2097764, EPI_ISL_2097765, EPI_ISL_2097767, EPI_ISL_2097768, EPI_ISL_2097769, EPI_ISL_2097770, EPI_ISL_2097771, EPI_ISL_2097772, EPI_ISL_2097773, EPI_ISL_2097774, EPI_ISL_2097775, EPI_ISL_2097776, EPI_ISL_2097777, EPI_ISL_2097778, EPI_ISL_2097779, EPI_ISL_2097780, EPI_ISL_2097781,
EPI_ISL_2097782, EPI_ISL_2097783, EPI_ISL_2097784, EPI_ISL_2097785, EPI_ISL_2097786, EPI_ISL_2097787, EPI_ISL_2097788, EPI_ISL_2097789, EPI_ISL_2097790, EPI_ISL_2097791, EPI_ISL_2097792, EPI_ISL_2097793, EPI_ISL_2097794, EPI_ISL_2097795, EPI_ISL_2097796, EPI_ISL_2097797, EPI_ISL_2097798, EPI_ISL_2097799, EPI_ISL_2097800, EPI_ISL_2097801, EPI_ISL_2097802, EPI_ISL_2097803,
EPI_ISL_2097804, EPI_ISL_2097805, EPI_ISL_2097806, EPI_ISL_2097807, EPI_ISL_2097809, EPI_ISL_2097810, EPI_ISL_2097811, EPI_ISL_2097812, EPI_ISL_2097813, EPI_ISL_2097814, EPI_ISL_2097815, EPI_ISL_2097816, EPI_ISL_2097817, EPI_ISL_2097818, EPI_ISL_2097819, EPI_ISL_2097820, EPI_ISL_2097821, EPI_ISL_2097822, EPI_ISL_2097823, EPI_ISL_2097824, EPI_ISL_2097825, EPI_ISL_2097826,
EPI_ISL_2097827, EPI_ISL_2097828, EPI_ISL_2097829, EPI_ISL_2097830, EPI_ISL_2097831, EPI_ISL_2097832, EPI_ISL_2097833, EPI_ISL_2097834, EPI_ISL_2097835, EPI_ISL_2097836, EPI_ISL_2097837, EPI_ISL_2097838, EPI_ISL_2097839, EPI_ISL_2097840, EPI_ISL_2097841, EPI_ISL_2097842, EPI_ISL_2097843, EPI_ISL_2097844, EPI_ISL_2097845, EPI_ISL_2097846, EPI_ISL_2097847, EPI_ISL_2097848,
EPI_ISL_2097849, EPI_ISL_2097850, EPI_ISL_2097851, EPI_ISL_2097852, EPI_ISL_2097853, EPI_ISL_2097854, EPI_ISL_2097855, EPI_ISL_2097856, EPI_ISL_2097857, EPI_ISL_2097858, EPI_ISL_2097859, EPI_ISL_2097860, EPI_ISL_2097861, EPI_ISL_2097862, EPI_ISL_2097863, EPI_ISL_2097864, EPI_ISL_2097865, EPI_ISL_2097866, EPI_ISL_2097867, EPI_ISL_2097868, EPI_ISL_2097869, EPI_ISL_2097870,
EPI_ISL_2097872, EPI_ISL_2097873, EPI_ISL_2097874, EPI_ISL_2097875, EPI_ISL_2097876, EPI_ISL_2097877, EPI_ISL_2097878, EPI_ISL_2097879, EPI_ISL_2097880, EPI_ISL_2097881, EPI_ISL_2097882, EPI_ISL_2097883, EPI_ISL_2097884, EPI_ISL_2097885, EPI_ISL_2097886, EPI_ISL_2097887, EPI_ISL_2097888, EPI_ISL_2097889, EPI_ISL_2097890, EPI_ISL_2097891, EPI_ISL_2097892, EPI_ISL_2097893,
EPI_ISL_2097894, EPI_ISL_2097895, EPI_ISL_2097896, EPI_ISL_2097897, EPI_ISL_2097898, EPI_ISL_2097899, EPI_ISL_2097900, EPI_ISL_2097901, EPI_ISL_2097902, EPI_ISL_2097903, EPI_ISL_2097904, EPI_ISL_2097905, EPI_ISL_2097906, EPI_ISL_2097907, EPI_ISL_2097908, EPI_ISL_2097909, EPI_ISL_2097910, EPI_ISL_2097911, EPI_ISL_2097912, EPI_ISL_2097913, EPI_ISL_2097914, EPI_ISL_2097915,
EPI_ISL_2097916, EPI_ISL_2097917, EPI_ISL_2097918, EPI_ISL_2097919, EPI_ISL_2097920, EPI_ISL_2097921, EPI_ISL_2097922, EPI_ISL_2097923, EPI_ISL_2097924, EPI_ISL_2097925, EPI_ISL_2097926, EPI_ISL_2097927, EPI_ISL_2097928, EPI_ISL_2097929, EPI_ISL_2097930, EPI_ISL_2097931, EPI_ISL_2097932, EPI_ISL_2097933, EPI_ISL_2097934, EPI_ISL_2097935, EPI_ISL_2097936, EPI_ISL_2097937,
EPI_ISL_2097938, EPI_ISL_2097939, EPI_ISL_2097940, EPI_ISL_2097941, EPI_ISL_2097942, EPI_ISL_2097943, EPI_ISL_2097944, EPI_ISL_2097945, EPI_ISL_2097946, EPI_ISL_2097947, EPI_ISL_2097948, EPI_ISL_2097949, EPI_ISL_2097950, EPI_ISL_2097951, EPI_ISL_2097952, EPI_ISL_2097953, EPI_ISL_2097954, EPI_ISL_2097955, EPI_ISL_2097956, EPI_ISL_2097957, EPI_ISL_2097958, EPI_ISL_2097959,
EPI_ISL_2097960, EPI_ISL_2097961, EPI_ISL_2097962, EPI_ISL_2097963, EPI_ISL_2097964, EPI_ISL_2097965, EPI_ISL_2097966, EPI_ISL_2097967, EPI_ISL_2097968, EPI_ISL_2097969, EPI_ISL_2097970, EPI_ISL_2097971, EPI_ISL_2097972, EPI_ISL_2097973, EPI_ISL_2097974, EPI_ISL_2097975, EPI_ISL_2097976, EPI_ISL_2097977, EPI_ISL_2097978, EPI_ISL_2097979, EPI_ISL_2097980, EPI_ISL_2097981,
EPI_ISL_2097982, EPI_ISL_2097983, EPI_ISL_2097984, EPI_ISL_2097985, EPI_ISL_2097986, EPI_ISL_2097987, EPI_ISL_2097988, EPI_ISL_2097989, EPI_ISL_2097990, EPI_ISL_2097991, EPI_ISL_2097992, EPI_ISL_2097993, EPI_ISL_2097994, EPI_ISL_2097995, EPI_ISL_2097996, EPI_ISL_2097997, EPI_ISL_2097998, EPI_ISL_2097999, EPI_ISL_2098000, EPI_ISL_2098001, EPI_ISL_2098002, EPI_ISL_2098003,
EPI_ISL_2098004, EPI_ISL_2098005, EPI_ISL_2098006, EPI_ISL_2098007, EPI_ISL_2098008, EPI_ISL_2098009, EPI_ISL_2098010, EPI_ISL_2098011, EPI_ISL_2098012, EPI_ISL_2098013, EPI_ISL_2098014, EPI_ISL_2098015, EPI_ISL_2098016, EPI_ISL_2098017, EPI_ISL_2098018, EPI_ISL_2098019, EPI_ISL_2098020, EPI_ISL_2098021, EPI_ISL_2098022, EPI_ISL_2098023, EPI_ISL_2098024, EPI_ISL_2098025,
EPI_ISL_2098026, EPI_ISL_2098027, EPI_ISL_2098028, EPI_ISL_2098029, EPI_ISL_2098030, EPI_ISL_2098031, EPI_ISL_2098032, EPI_ISL_2098033, EPI_ISL_2098034, EPI_ISL_2098035, EPI_ISL_2098036, EPI_ISL_2098037, EPI_ISL_2098038, EPI_ISL_2098039, EPI_ISL_2098040, EPI_ISL_2098041, EPI_ISL_2098042, EPI_ISL_2098043, EPI_ISL_2098044, EPI_ISL_2098045, EPI_ISL_2098046, EPI_ISL_2098047,

EPI_ISL_2098048, EPI_ISL_2098049, EPI_ISL_2098050, EPI_ISL_2098051, EPI_ISL_2098052, EPI_ISL_2098053, EPI_ISL_2098054, EPI_ISL_2098055, EPI_ISL_2098056, EPI_ISL_2098057, EPI_ISL_2098058, EPI_ISL_2098059, EPI_ISL_2098060, EPI_ISL_2098061, EPI_ISL_2098062, EPI_ISL_2098063, EPI_ISL_2098064, EPI_ISL_2098065, EPI_ISL_2098066, EPI_ISL_2098067, EPI_ISL_2098068, EPI_ISL_2098069, EPI_ISL_2098070, EPI_ISL_2098071, EPI_ISL_2098072, EPI_ISL_2098073, EPI_ISL_2098074, EPI_ISL_2098075, EPI_ISL_2098076, EPI_ISL_2098077, EPI_ISL_2098078, EPI_ISL_2098079, EPI_ISL_2098080, EPI_ISL_2098081, EPI_ISL_2098082, EPI_ISL_2098083, EPI_ISL_2098084, EPI_ISL_2098085, EPI_ISL_2098086, EPI_ISL_2098087, EPI_ISL_2098088, EPI_ISL_2098089, EPI_ISL_2098090, EPI_ISL_2098091, EPI_ISL_2098092, EPI_ISL_2098093, EPI_ISL_2098094, EPI_ISL_2098095, EPI_ISL_2098096, EPI_ISL_2098097, EPI_ISL_2098098, EPI_ISL_2098099, EPI_ISL_2098100, EPI_ISL_2098101, EPI_ISL_2098102, EPI_ISL_2098103, EPI_ISL_2098104, EPI_ISL_2098105, EPI_ISL_2098106, EPI_ISL_2098107, EPI_ISL_2098108, EPI_ISL_2098109, EPI_ISL_2098110, EPI_ISL_2098111, EPI_ISL_2098112, EPI_ISL_2098113, EPI_ISL_2098114

| | | | |
|---|---|---|---|
| see above | Helix/Illumina | Centers for Disease Control and Prevention Division of Viral Diseases, Pathogen Discovery | Dakota Howard, Dhwani Batra, Peter W. Cook, Kara Moser, Adrian Paskey, Jason Caravas, Benjamin Rambo-Martin, Shatavia Morrison, Christopher Gulvick, Scott Sammons, Yvette Unoarumhi, Darlene Wagner, Matthew Schmerer, Eileen de Feo, Jan Antico, Christine Tran, Matthew Tolentino, Shannon Wickline, Kim Gietzen, Brad Sickler, Jingtao Liu, Eric Allen, Phil Febbo, Nicole L. Washington, Simon White, Geraint Levan, Kelly Schiabor Barrett, Elizabeth Cirulli, Alexandre Bolze, Ary Ascencio, Charlotte Rivera-Garcia, Ryan Cho, Jason Nguyen, Sherry Wang, Jimmy Ramirez, Tyler Cassens, Efren Sandoval, Magnus Isaksson, William Lee, David Becker, Marc Laurent, James Lu, Clinton R. Paden, Duncan MacCannell |
| EPI_ISL_2098115 | ALBANY MEDICAL CENTER | Wadsworth Center, New York State Department of Health | Kirsten St. George, Daryl M. Lamson, Alexis Russell, Matthew Shudt, Melissa A Leisner, Jonathan Plitnick, Catharine Prussing, Navjot Singh, John Kelly, Erasmus Schneider, Erica Lasek-Nesselquist |
| EPI_ISL_2098116 | Helix/Illumina | Centers for Disease Control and Prevention Division of Viral Diseases, Pathogen Discovery | Dakota Howard, Dhwani Batra, Peter W. Cook, Kara Moser, Adrian Paskey, Jason Caravas, Benjamin Rambo-Martin, Shatavia Morrison, Christopher Gulvick, Scott Sammons, Yvette Unoarumhi, Darlene Wagner, Matthew Schmerer, Eileen de Feo, Jan Antico, Christine Tran, Matthew Tolentino, Shannon Wickline, Kim Gietzen, Brad Sickler, Jingtao Liu, Eric Allen, Phil Febbo, Nicole L. Washington, Simon White, Geraint Levan, Kelly Schiabor Barrett, Elizabeth Cirulli, Alexandre Bolze, Ary Ascencio, Charlotte Rivera-Garcia, Ryan Cho, Jason Nguyen, Sherry Wang, Jimmy Ramirez, Tyler Cassens, Efren Sandoval, Magnus Isaksson, William Lee, David Becker, Marc Laurent, James Lu, Clinton R. Paden, Duncan MacCannell |
| EPI_ISL_2098117 | ALBANY MEDICAL CENTER | Wadsworth Center, New York State Department of Health | Kirsten St. George, Daryl M. Lamson, Alexis Russell, Matthew Shudt, Melissa A Leisner, Jonathan Plitnick, Catharine Prussing, Navjot Singh, John Kelly, Erasmus Schneider, Erica Lasek-Nesselquist |
| EPI_ISL_2098118 | Helix/Illumina | Centers for Disease Control and Prevention Division of Viral Diseases, Pathogen Discovery | Dakota Howard, Dhwani Batra, Peter W. Cook, Kara Moser, Adrian Paskey, Jason Caravas, Benjamin Rambo-Martin, Shatavia Morrison, Christopher Gulvick, Scott Sammons, Yvette Unoarumhi, Darlene Wagner, Matthew Schmerer, Eileen de Feo, Jan Antico, Christine Tran, Matthew Tolentino, Shannon Wickline, Kim Gietzen, Brad Sickler, Jingtao Liu, Eric Allen, Phil Febbo, Nicole L. Washington, Simon White, Geraint Levan, Kelly Schiabor Barrett, Elizabeth Cirulli, Alexandre Bolze, Ary Ascencio, Charlotte Rivera-Garcia, Ryan Cho, Jason Nguyen, Sherry Wang, Jimmy Ramirez, Tyler Cassens, Efren Sandoval, Magnus Isaksson, William Lee, David Becker, Marc Laurent, James Lu, Clinton R. Paden, Duncan MacCannell |
| EPI_ISL_2098119, EPI_ISL_2098120 | ALBANY MEDICAL CENTER | Wadsworth Center, New York State Department of Health | Kirsten St. George, Daryl M. Lamson, Alexis Russell, Matthew Shudt, Melissa A Leisner, Jonathan Plitnick, Catharine Prussing, Navjot Singh, John Kelly, Erasmus Schneider, Erica Lasek-Nesselquist |
| EPI_ISL_2098121 | Helix/Illumina | Centers for Disease Control and Prevention Division of Viral Diseases, Pathogen Discovery | Dakota Howard, Dhwani Batra, Peter W. Cook, Kara Moser, Adrian Paskey, Jason Caravas, Benjamin Rambo-Martin, Shatavia Morrison, Christopher Gulvick, Scott Sammons, Yvette Unoarumhi, Darlene Wagner, Matthew Schmerer, Eileen de Feo, Jan Antico, Christine Tran, Matthew Tolentino, Shannon Wickline, Kim Gietzen, Brad Sickler, Jingtao Liu, Eric Allen, Phil Febbo, Nicole L. Washington, Simon White, Geraint Levan, Kelly Schiabor Barrett, Elizabeth Cirulli, Alexandre Bolze, Ary Ascencio, Charlotte Rivera-Garcia, Ryan Cho, Jason Nguyen, Sherry Wang, Jimmy Ramirez, Tyler Cassens, Efren Sandoval, Magnus Isaksson, William Lee, David Becker, Marc Laurent, James Lu, Clinton R. Paden, Duncan MacCannell |
| EPI_ISL_2098122, EPI_ISL_2098123 | ALBANY MEDICAL CENTER | Wadsworth Center, New York State Department of Health | Kirsten St. George, Daryl M. Lamson, Alexis Russell, Matthew Shudt, Melissa A Leisner, Jonathan Plitnick, Catharine Prussing, Navjot Singh, John Kelly, Erasmus Schneider, Erica Lasek-Nesselquist |
| EPI_ISL_2098124 | Helix/Illumina | Centers for Disease Control and Prevention Division of Viral Diseases, Pathogen Discovery | Dakota Howard, Dhwani Batra, Peter W. Cook, Kara Moser, Adrian Paskey, Jason Caravas, Benjamin Rambo-Martin, Shatavia Morrison, Christopher Gulvick, Scott Sammons, Yvette Unoarumhi, Darlene Wagner, Matthew Schmerer, Eileen de Feo, Jan Antico, Christine Tran, Matthew Tolentino, Shannon Wickline, Kim Gietzen, Brad Sickler, Jingtao Liu, Eric Allen, Phil Febbo, Nicole L. Washington, Simon White, Geraint Levan, Kelly Schiabor Barrett, Elizabeth Cirulli, Alexandre Bolze, Ary Ascencio, Charlotte Rivera-Garcia, Ryan Cho, Jason Nguyen, Sherry Wang, Jimmy Ramirez, Tyler Cassens, Efren Sandoval, Magnus Isaksson, William Lee, David Becker, Marc Laurent, James Lu, Clinton R. Paden, Duncan MacCannell |
| EPI_ISL_2098125 | ALBANY MEDICAL CENTER | Wadsworth Center, New York State Department of Health | Kirsten St. George, Daryl M. Lamson, Alexis Russell, Matthew Shudt, Melissa A Leisner, Jonathan Plitnick, Catharine Prussing, Navjot Singh, John Kelly, Erasmus Schneider, Erica Lasek-Nesselquist |
| EPI_ISL_2098126 | BOSTON HEART DIAGNOSTICS CORP | Wadsworth Center, New York State Department of Health | Kirsten St. George, Daryl M. Lamson, Alexis Russell, Matthew Shudt, Melissa A Leisner, Jonathan Plitnick, Catharine Prussing, Navjot Singh, John Kelly, Erasmus Schneider, Erica Lasek-Nesselquist |
| EPI_ISL_2098127 | Helix/Illumina | Centers for Disease Control and Prevention Division of Viral Diseases, Pathogen Discovery | Dakota Howard, Dhwani Batra, Peter W. Cook, Kara Moser, Adrian Paskey, Jason Caravas, Benjamin Rambo-Martin, Shatavia Morrison, Christopher Gulvick, Scott Sammons, Yvette Unoarumhi, Darlene Wagner, Matthew Schmerer, Eileen de Feo, Jan Antico, Christine Tran, Matthew Tolentino, Shannon Wickline, Kim Gietzen, Brad Sickler, Jingtao Liu, Eric Allen, Phil Febbo, Nicole L. Washington, Simon White, Geraint Levan, Kelly Schiabor Barrett, Elizabeth Cirulli, Alexandre Bolze, Ary Ascencio, Charlotte Rivera-Garcia, Ryan Cho, Jason Nguyen, Sherry Wang, Jimmy Ramirez, Tyler Cassens, Efren Sandoval, Magnus Isaksson, William Lee, David Becker, Marc Laurent, James Lu, Clinton R. Paden, Duncan MacCannell |
| EPI_ISL_2098128 | BOSTON HEART DIAGNOSTICS CORP | Wadsworth Center, New York State Department of Health | Kirsten St. George, Daryl M. Lamson, Alexis Russell, Matthew Shudt, Melissa A Leisner, Jonathan Plitnick, Catharine Prussing, Navjot Singh, John Kelly, Erasmus Schneider, Erica Lasek-Nesselquist |
| EPI_ISL_2098129 | Helix/Illumina | Centers for Disease Control and Prevention Division of Viral Diseases, Pathogen Discovery | Dakota Howard, Dhwani Batra, Peter W. Cook, Kara Moser, Adrian Paskey, Jason Caravas, Benjamin Rambo-Martin, Shatavia Morrison, Christopher Gulvick, Scott Sammons, Yvette Unoarumhi, Darlene Wagner, Matthew Schmerer, Eileen de Feo, Jan Antico, Christine Tran, Matthew Tolentino, Shannon Wickline, Kim Gietzen, Brad Sickler, Jingtao Liu, Eric Allen, Phil Febbo, Nicole L. Washington, Simon White, Geraint Levan, Kelly Schiabor Barrett, Elizabeth Cirulli, Alexandre Bolze, Ary Ascencio, Charlotte Rivera-Garcia, Ryan Cho, Jason Nguyen, Sherry Wang, Jimmy Ramirez, Tyler Cassens, Efren Sandoval, Magnus Isaksson, William Lee, David Becker, Marc Laurent, James Lu, Clinton R. Paden, Duncan MacCannell |
| EPI_ISL_2098130 | MONTEFIORE MEDICAL CENTER LABORATORIES | Wadsworth Center, New York State Department of Health | Kirsten St. George, Daryl M. Lamson, Alexis Russell, Matthew Shudt, Melissa A Leisner, Jonathan Plitnick, Catharine Prussing, Navjot Singh, John Kelly, Erasmus Schneider, Erica Lasek-Nesselquist |
| EPI_ISL_2098131 | Helix/Illumina | Centers for Disease Control and Prevention Division of Viral Diseases, Pathogen Discovery | Dakota Howard, Dhwani Batra, Peter W. Cook, Kara Moser, Adrian Paskey, Jason Caravas, Benjamin Rambo-Martin, Shatavia Morrison, Christopher Gulvick, Scott Sammons, Yvette Unoarumhi, Darlene Wagner, Matthew Schmerer, Eileen de Feo, Jan Antico, Christine Tran, Matthew Tolentino, Shannon Wickline, Kim Gietzen, Brad Sickler, Jingtao Liu, Eric Allen, Phil Febbo, Nicole L. Washington, Simon White, Geraint Levan, Kelly Schiabor Barrett, Elizabeth Cirulli, Alexandre Bolze, Ary Ascencio, Charlotte Rivera-Garcia, Ryan Cho, Jason Nguyen, Sherry Wang, Jimmy Ramirez, Tyler Cassens, Efren Sandoval, Magnus Isaksson, William Lee, David Becker, Marc Laurent, James Lu, Clinton R. Paden, Duncan MacCannell |
| EPI_ISL_2098132 | NORTHWELL HEALTH LABORATORIES | Wadsworth Center, New York State Department of Health | Kirsten St. George, Daryl M. Lamson, Alexis Russell, Matthew Shudt, Melissa A Leisner, Jonathan Plitnick, Catharine Prussing, Navjot Singh, John Kelly, Erasmus Schneider, Erica Lasek-Nesselquist |
| EPI_ISL_2098133 | SARATOGA HOSPITAL LABORATORY | Wadsworth Center, New York State Department of Health | Kirsten St. George, Daryl M. Lamson, Alexis Russell, Matthew Shudt, Melissa A Leisner, Jonathan Plitnick, Catharine Prussing, Navjot Singh, John Kelly, Erasmus Schneider, Erica Lasek-Nesselquist |
| EPI_ISL_2098134 | Helix/Illumina | Centers for Disease Control and Prevention Division of Viral Diseases, Pathogen Discovery | Dakota Howard, Dhwani Batra, Peter W. Cook, Kara Moser, Adrian Paskey, Jason Caravas, Benjamin Rambo-Martin, Shatavia Morrison, Christopher Gulvick, Scott Sammons, Yvette Unoarumhi, Darlene Wagner, Matthew Schmerer, Eileen de Feo, Jan Antico, Christine Tran, Matthew Tolentino, Shannon Wickline, Kim Gietzen, Brad Sickler, Jingtao Liu, Eric Allen, Phil Febbo, Nicole L. Washington, Simon White, Geraint Levan, Kelly Schiabor Barrett, Elizabeth Cirulli, Alexandre Bolze, Ary Ascencio, Charlotte Rivera-Garcia, Ryan Cho, Jason Nguyen, Sherry Wang, Jimmy Ramirez, Tyler Cassens, Efren Sandoval, Magnus Isaksson, William Lee, David Becker, Marc Laurent, James Lu, Clinton R. Paden, Duncan MacCannell |
| EPI_ISL_2098135, EPI_ISL_2098136 | SUNY UPSTATE MEDICAL UNIVERSITY | Wadsworth Center, New York State Department of Health | Kirsten St. George, Daryl M. Lamson, Alexis Russell, Matthew Shudt, Melissa A Leisner, Jonathan Plitnick, Catharine Prussing, Navjot Singh, John Kelly, Erasmus Schneider, Erica Lasek-Nesselquist |
| EPI_ISL_2098137 | Helix/Illumina | Centers for Disease Control and Prevention Division of Viral Diseases, Pathogen Discovery | Dakota Howard, Dhwani Batra, Peter W. Cook, Kara Moser, Adrian Paskey, Jason Caravas, Benjamin Rambo-Martin, Shatavia Morrison, Christopher Gulvick, Scott Sammons, Yvette Unoarumhi, Darlene Wagner, Matthew Schmerer, Eileen de Feo, Jan Antico, Christine Tran, Matthew Tolentino, Shannon Wickline, Kim Gietzen, Brad Sickler, Jingtao Liu, Eric Allen, Phil Febbo, Nicole L. Washington, Simon White, Geraint Levan, Kelly Schiabor Barrett, Elizabeth Cirulli, Alexandre Bolze, Ary Ascencio, Charlotte Rivera-Garcia, Ryan Cho, Jason Nguyen, Sherry Wang, Jimmy Ramirez, Tyler Cassens, Efren Sandoval, Magnus Isaksson, William Lee, David Becker, Marc Laurent, James Lu, Clinton R. Paden, Duncan MacCannell |
| EPI_ISL_2098138 | SUNY UPSTATE MEDICAL UNIVERSITY | Wadsworth Center, New York State Department of Health | Kirsten St. George, Daryl M. Lamson, Alexis Russell, Matthew Shudt, Melissa A Leisner, Jonathan Plitnick, Catharine Prussing, Navjot Singh, John Kelly, Erasmus Schneider, Erica Lasek-Nesselquist |
| EPI_ISL_2098139 | Helix/Illumina | Centers for Disease Control and Prevention Division of Viral Diseases, Pathogen Discovery | Dakota Howard, Dhwani Batra, Peter W. Cook, Kara Moser, Adrian Paskey, Jason Caravas, Benjamin Rambo-Martin, Shatavia Morrison, Christopher Gulvick, Scott Sammons, Yvette Unoarumhi, Darlene Wagner, Matthew Schmerer, Eileen de Feo, Jan Antico, Christine Tran, Matthew Tolentino, Shannon Wickline, Kim Gietzen, Brad Sickler, Jingtao Liu, Eric Allen, Phil Febbo, Nicole L. Washington, Simon White, Geraint Levan, Kelly Schiabor Barrett, Elizabeth Cirulli, Alexandre Bolze, Ary Ascencio, Charlotte Rivera-Garcia, Ryan Cho, Jason Nguyen, Sherry Wang, Jimmy Ramirez, Tyler Cassens, Efren Sandoval, Magnus Isaksson, William Lee, David Becker, Marc Laurent, James Lu, Clinton R. Paden, Duncan MacCannell |
| EPI_ISL_2098140, EPI_ISL_2098141 | SUNY UPSTATE MEDICAL UNIVERSITY | Wadsworth Center, New York State Department of Health | Kirsten St. George, Daryl M. Lamson, Alexis Russell, Matthew Shudt, Melissa A Leisner, Jonathan Plitnick, Catharine Prussing, Navjot Singh, John Kelly, Erasmus Schneider, Erica Lasek-Nesselquist |
| EPI_ISL_2098142 | Helix/Illumina | Centers for Disease Control and Prevention Division of Viral Diseases, Pathogen Discovery | Dakota Howard, Dhwani Batra, Peter W. Cook, Kara Moser, Adrian Paskey, Jason Caravas, Benjamin Rambo-Martin, Shatavia Morrison, Christopher Gulvick, Scott Sammons, Yvette Unoarumhi, Darlene Wagner, Matthew Schmerer, Eileen de Feo, Jan Antico, Christine Tran, Matthew Tolentino, Shannon Wickline, Kim Gietzen, Brad Sickler, Jingtao Liu, Eric Allen, Phil Febbo, Nicole L. Washington, Simon White, Geraint Levan, Kelly Schiabor Barrett, Elizabeth Cirulli, Alexandre Bolze, Ary Ascencio, Charlotte Rivera-Garcia, Ryan Cho, Jason Nguyen, Sherry Wang, Jimmy Ramirez, Tyler Cassens, Efren Sandoval, Magnus Isaksson, William Lee, David Becker, Marc Laurent, James Lu, Clinton R. Paden, Duncan MacCannell |
| EPI_ISL_2098143, EPI_ISL_2098144 | SUNY UPSTATE MEDICAL UNIVERSITY | Wadsworth Center, New York State Department of Health | Kirsten St. George, Daryl M. Lamson, Alexis Russell, Matthew Shudt, Melissa A Leisner, Jonathan Plitnick, Catharine Prussing, Navjot Singh, John Kelly, Erasmus Schneider, Erica Lasek-Nesselquist |
| EPI_ISL_2098145 | Helix/Illumina | Centers for Disease Control and Prevention Division of Viral Diseases, Pathogen Discovery | Dakota Howard, Dhwani Batra, Peter W. Cook, Kara Moser, Adrian Paskey, Jason Caravas, Benjamin Rambo-Martin, Shatavia Morrison, Christopher Gulvick, Scott Sammons, Yvette Unoarumhi, Darlene Wagner, Matthew Schmerer, Eileen de Feo, Jan Antico, Christine Tran, Matthew Tolentino, Shannon Wickline, Kim Gietzen, Brad Sickler, Jingtao Liu, Eric Allen, Phil Febbo, Nicole L. Washington, Simon White, Geraint Levan, Kelly Schiabor Barrett, Elizabeth Cirulli, Alexandre Bolze, Ary Ascencio, Charlotte Rivera-Garcia, Ryan Cho, Jason Nguyen, Sherry Wang, Jimmy Ramirez, Tyler Cassens, Efren Sandoval, Magnus Isaksson, William Lee, David Becker, Marc Laurent, James Lu, Clinton R. Paden, Duncan MacCannell |
| EPI_ISL_2098146, EPI_ISL_2098147 | SUNY UPSTATE MEDICAL UNIVERSITY | Wadsworth Center, New York State Department of Health | Kirsten St. George, Daryl M. Lamson, Alexis Russell, Matthew Shudt, Melissa A Leisner, Jonathan Plitnick, Catharine Prussing, Navjot Singh, John Kelly, Erasmus Schneider, Erica Lasek-Nesselquist |
| EPI_ISL_2098148 | Helix/Illumina | Centers for Disease Control and Prevention Division of Viral Diseases, Pathogen Discovery | Dakota Howard, Dhwani Batra, Peter W. Cook, Kara Moser, Adrian Paskey, Jason Caravas, Benjamin Rambo-Martin, Shatavia Morrison, Christopher Gulvick, Scott Sammons, Yvette Unoarumhi, Darlene Wagner, Matthew Schmerer, Eileen de Feo, Jan Antico, Christine Tran, Matthew Tolentino, Shannon Wickline, Kim Gietzen, Brad Sickler, Jingtao Liu, Eric Allen, Phil Febbo, Nicole L. Washington, Simon White, Geraint Levan, Kelly Schiabor Barrett, Elizabeth Cirulli, Alexandre Bolze, Ary Ascencio, Charlotte Rivera-Garcia, Ryan Cho, Jason Nguyen, Sherry Wang, Jimmy Ramirez, Tyler Cassens, Efren Sandoval, Magnus Isaksson, William Lee, David Becker, Marc Laurent, James Lu, Clinton R. Paden, Duncan MacCannell |
| EPI_ISL_2098149, EPI_ISL_2098150, EPI_ISL_2098151 | SUNY UPSTATE MEDICAL UNIVERSITY | Wadsworth Center, New York State Department of Health | Kirsten St. George, Daryl M. Lamson, Alexis Russell, Matthew Shudt, Melissa A Leisner, Jonathan Plitnick, Catharine Prussing, Navjot Singh, John Kelly, Erasmus Schneider, Erica Lasek-Nesselquist |
| EPI_ISL_2098152 | Helix/Illumina | Centers for Disease Control and Prevention Division of Viral Diseases, Pathogen Discovery | Dakota Howard, Dhwani Batra, Peter W. Cook, Kara Moser, Adrian Paskey, Jason Caravas, Benjamin Rambo-Martin, Shatavia Morrison, Christopher Gulvick, Scott Sammons, Yvette Unoarumhi, Darlene Wagner, Matthew Schmerer, Eileen de Feo, Jan Antico, Christine Tran, Matthew Tolentino, Shannon Wickline, Kim Gietzen, Brad Sickler, Jingtao Liu, Eric Allen, Phil Febbo, Nicole L. Washington, Simon White, Geraint Levan, Kelly Schiabor Barrett, Elizabeth Cirulli, Alexandre Bolze, Ary Ascencio, Charlotte Rivera-Garcia, Ryan Cho, Jason Nguyen, Sherry Wang, Jimmy Ramirez, Tyler Cassens, Efren Sandoval, Magnus Isaksson, William Lee, David Becker, Marc Laurent, James Lu, Clinton R. Paden, Duncan MacCannell |
| EPI_ISL_2098153 | SUNY UPSTATE MEDICAL UNIVERSITY | Wadsworth Center, New York State Department of Health | Kirsten St. George, Daryl M. Lamson, Alexis Russell, Matthew Shudt, Melissa A Leisner, Jonathan Plitnick, Catharine Prussing, Navjot Singh, John Kelly, Erasmus Schneider, Erica Lasek-Nesselquist |
| EPI_ISL_2098154 | Helix/Illumina | Centers for Disease Control and Prevention Division of Viral Diseases, Pathogen Discovery | Dakota Howard, Dhwani Batra, Peter W. Cook, Kara Moser, Adrian Paskey, Jason Caravas, Benjamin Rambo-Martin, Shatavia Morrison, Christopher Gulvick, Scott Sammons, Yvette Unoarumhi, Darlene Wagner, Matthew Schmerer, Eileen de Feo, Jan Antico, Christine Tran, Matthew Tolentino, Shannon Wickline, Kim Gietzen, Brad Sickler, Jingtao Liu, Eric Allen, Phil Febbo, Nicole L. Washington, Simon White, Geraint Levan, Kelly Schiabor Barrett, Elizabeth Cirulli, Alexandre Bolze, Ary Ascencio, Charlotte Rivera-Garcia, Ryan Cho, Jason Nguyen, Sherry Wang, Jimmy Ramirez, Tyler Cassens, Efren Sandoval, Magnus Isaksson, William Lee, David Becker, Marc Laurent, James Lu, Clinton R. Paden, Duncan MacCannell |
| EPI_ISL_2098155, EPI_ISL_2098156 | SUNY UPSTATE MEDICAL UNIVERSITY | Wadsworth Center, New York State Department of Health | Kirsten St. George, Daryl M. Lamson, Alexis Russell, Matthew Shudt, Melissa A Leisner, Jonathan Plitnick, Catharine Prussing, Navjot Singh, John Kelly, Erasmus Schneider, Erica Lasek-Nesselquist |
| EPI_ISL_2098157 | Helix/Illumina | Centers for Disease Control and Prevention Division of Viral Diseases, Pathogen Discovery | Dakota Howard, Dhwani Batra, Peter W. Cook, Kara Moser, Adrian Paskey, Jason Caravas, Benjamin Rambo-Martin, Shatavia Morrison, Christopher Gulvick, Scott Sammons, Yvette Unoarumhi, Darlene Wagner, Matthew Schmerer, Eileen de Feo, Jan Antico, Christine Tran, Matthew Tolentino, Shannon Wickline, Kim Gietzen, Brad Sickler, Jingtao Liu, Eric Allen, Phil Febbo, Nicole L. Washington, Simon White, Geraint Levan, Kelly Schiabor Barrett, Elizabeth Cirulli, Alexandre Bolze, Ary Ascencio, Charlotte Rivera-Garcia, Ryan Cho, Jason Nguyen, Sherry Wang, Jimmy Ramirez, Tyler Cassens, Efren Sandoval, Magnus Isaksson, William Lee, David Becker, Marc Laurent, James Lu, Clinton R. Paden, Duncan MacCannell |
| EPI_ISL_2098158, EPI_ISL_2098159 | SUNY UPSTATE MEDICAL UNIVERSITY | Wadsworth Center, New York State Department of Health | Kirsten St. George, Daryl M. Lamson, Alexis Russell, Matthew Shudt, Melissa A Leisner, Jonathan Plitnick, Catharine Prussing, Navjot Singh, John Kelly, Erasmus Schneider, Erica Lasek-Nesselquist |

| | | | |
|---|---|---|---|
| EPI_ISL_2098160 | Helix/Illumina | Centers for Disease Control and Prevention Division of Viral Diseases, Pathogen Discovery | Dakota Howard, Dhwani Batra, Peter W. Cook, Kara Moser, Adrian Paskey, Jason Caravas, Benjamin Rambo-Martin, Shatavia Morrison, Christopher Gulvick, Scott Sammons, Yvette Unoarumhi, Darlene Wagner, Matthew Schmerer, Eileen de Feo, Jan Antico, Christine Tran, Matthew Tolentino, Shannon Wickline, Kim Gietzen, Brad Sickler, Jingtao Liu, Eric Allen, Phil Febbo, Nicole L. Washington, Simon White, Geraint Levan, Kelly Schiabor Barrett, Elizabeth Cirulli, Alexandre Bolze, Ary Ascencio, Charlotte Rivera-Garcia, Ryan Cho, Jason Nguyen, Sherry Wang, Jimmy Ramirez, Tyler Cassens, Efren Sandoval, Magnus Isaksson, William Lee, David Becker, Marc Laurent, James Lu, Clinton R. Paden, Duncan MacCannell |
| EPI_ISL_2098161, EPI_ISL_2098162 | URMC LABS | Wadsworth Center, New York State Department of Health | Kirsten St. George, Daryl M. Lamson, Alexis Russell, Matthew Shudt, Melissa A Leisner, Jonathan Plitnick, Catharine Prussing, Navjot Singh, John Kelly, Erasmus Schneider, Erica Lasek-Nesselquist |
| EPI_ISL_2098163 | Helix/Illumina | Centers for Disease Control and Prevention Division of Viral Diseases, Pathogen Discovery | Dakota Howard, Dhwani Batra, Peter W. Cook, Kara Moser, Adrian Paskey, Jason Caravas, Benjamin Rambo-Martin, Shatavia Morrison, Christopher Gulvick, Scott Sammons, Yvette Unoarumhi, Darlene Wagner, Matthew Schmerer, Eileen de Feo, Jan Antico, Christine Tran, Matthew Tolentino, Shannon Wickline, Kim Gietzen, Brad Sickler, Jingtao Liu, Eric Allen, Phil Febbo, Nicole L. Washington, Simon White, Geraint Levan, Kelly Schiabor Barrett, Elizabeth Cirulli, Alexandre Bolze, Ary Ascencio, Charlotte Rivera-Garcia, Ryan Cho, Jason Nguyen, Sherry Wang, Jimmy Ramirez, Tyler Cassens, Efren Sandoval, Magnus Isaksson, William Lee, David Becker, Marc Laurent, James Lu, Clinton R. Paden, Duncan MacCannell |
| EPI_ISL_2098164 | URMC LABS | Wadsworth Center, New York State Department of Health | Kirsten St. George, Daryl M. Lamson, Alexis Russell, Matthew Shudt, Melissa A Leisner, Jonathan Plitnick, Catharine Prussing, Navjot Singh, John Kelly, Erasmus Schneider, Erica Lasek-Nesselquist |
| EPI_ISL_2098165 | Helix/Illumina | Centers for Disease Control and Prevention Division of Viral Diseases, Pathogen Discovery | Dakota Howard, Dhwani Batra, Peter W. Cook, Kara Moser, Adrian Paskey, Jason Caravas, Benjamin Rambo-Martin, Shatavia Morrison, Christopher Gulvick, Scott Sammons, Yvette Unoarumhi, Darlene Wagner, Matthew Schmerer, Eileen de Feo, Jan Antico, Christine Tran, Matthew Tolentino, Shannon Wickline, Kim Gietzen, Brad Sickler, Jingtao Liu, Eric Allen, Phil Febbo, Nicole L. Washington, Simon White, Geraint Levan, Kelly Schiabor Barrett, Elizabeth Cirulli, Alexandre Bolze, Ary Ascencio, Charlotte Rivera-Garcia, Ryan Cho, Jason Nguyen, Sherry Wang, Jimmy Ramirez, Tyler Cassens, Efren Sandoval, Magnus Isaksson, William Lee, David Becker, Marc Laurent, James Lu, Clinton R. Paden, Duncan MacCannell |
| EPI_ISL_2098166, EPI_ISL_2098167 | URMC LABS | Wadsworth Center, New York State Department of Health | Kirsten St. George, Daryl M. Lamson, Alexis Russell, Matthew Shudt, Melissa A Leisner, Jonathan Plitnick, Catharine Prussing, Navjot Singh, John Kelly, Erasmus Schneider, Erica Lasek-Nesselquist |
| EPI_ISL_2098168 | Helix/Illumina | Centers for Disease Control and Prevention Division of Viral Diseases, Pathogen Discovery | Dakota Howard, Dhwani Batra, Peter W. Cook, Kara Moser, Adrian Paskey, Jason Caravas, Benjamin Rambo-Martin, Shatavia Morrison, Christopher Gulvick, Scott Sammons, Yvette Unoarumhi, Darlene Wagner, Matthew Schmerer, Eileen de Feo, Jan Antico, Christine Tran, Matthew Tolentino, Shannon Wickline, Kim Gietzen, Brad Sickler, Jingtao Liu, Eric Allen, Phil Febbo, Nicole L. Washington, Simon White, Geraint Levan, Kelly Schiabor Barrett, Elizabeth Cirulli, Alexandre Bolze, Ary Ascencio, Charlotte Rivera-Garcia, Ryan Cho, Jason Nguyen, Sherry Wang, Jimmy Ramirez, Tyler Cassens, Efren Sandoval, Magnus Isaksson, William Lee, David Becker, Marc Laurent, James Lu, Clinton R. Paden, Duncan MacCannell |
| EPI_ISL_2098169, EPI_ISL_2098170 | URMC LABS | Wadsworth Center, New York State Department of Health | Kirsten St. George, Daryl M. Lamson, Alexis Russell, Matthew Shudt, Melissa A Leisner, Jonathan Plitnick, Catharine Prussing, Navjot Singh, John Kelly, Erasmus Schneider, Erica Lasek-Nesselquist |
| EPI_ISL_2098171 | Helix/Illumina | Centers for Disease Control and Prevention Division of Viral Diseases, Pathogen Discovery | Dakota Howard, Dhwani Batra, Peter W. Cook, Kara Moser, Adrian Paskey, Jason Caravas, Benjamin Rambo-Martin, Shatavia Morrison, Christopher Gulvick, Scott Sammons, Yvette Unoarumhi, Darlene Wagner, Matthew Schmerer, Eileen de Feo, Jan Antico, Christine Tran, Matthew Tolentino, Shannon Wickline, Kim Gietzen, Brad Sickler, Jingtao Liu, Eric Allen, Phil Febbo, Nicole L. Washington, Simon White, Geraint Levan, Kelly Schiabor Barrett, Elizabeth Cirulli, Alexandre Bolze, Ary Ascencio, Charlotte Rivera-Garcia, Ryan Cho, Jason Nguyen, Sherry Wang, Jimmy Ramirez, Tyler Cassens, Efren Sandoval, Magnus Isaksson, William Lee, David Becker, Marc Laurent, James Lu, Clinton R. Paden, Duncan MacCannell |
| EPI_ISL_2098172, EPI_ISL_2098173 | URMC LABS | Wadsworth Center, New York State Department of Health | Kirsten St. George, Daryl M. Lamson, Alexis Russell, Matthew Shudt, Melissa A Leisner, Jonathan Plitnick, Catharine Prussing, Navjot Singh, John Kelly, Erasmus Schneider, Erica Lasek-Nesselquist |
| EPI_ISL_2098174 | Helix/Illumina | Centers for Disease Control and Prevention Division of Viral Diseases, Pathogen Discovery | Dakota Howard, Dhwani Batra, Peter W. Cook, Kara Moser, Adrian Paskey, Jason Caravas, Benjamin Rambo-Martin, Shatavia Morrison, Christopher Gulvick, Scott Sammons, Yvette Unoarumhi, Darlene Wagner, Matthew Schmerer, Eileen de Feo, Jan Antico, Christine Tran, Matthew Tolentino, Shannon Wickline, Kim Gietzen, Brad Sickler, Jingtao Liu, Eric Allen, Phil Febbo, Nicole L. Washington, Simon White, Geraint Levan, Kelly Schiabor Barrett, Elizabeth Cirulli, Alexandre Bolze, Ary Ascencio, Charlotte Rivera-Garcia, Ryan Cho, Jason Nguyen, Sherry Wang, Jimmy Ramirez, Tyler Cassens, Efren Sandoval, Magnus Isaksson, William Lee, David Becker, Marc Laurent, James Lu, Clinton R. Paden, Duncan MacCannell |
| EPI_ISL_2098175, EPI_ISL_2098176 | URMC LABS | Wadsworth Center, New York State Department of Health | Kirsten St. George, Daryl M. Lamson, Alexis Russell, Matthew Shudt, Melissa A Leisner, Jonathan Plitnick, Catharine Prussing, Navjot Singh, John Kelly, Erasmus Schneider, Erica Lasek-Nesselquist |
| EPI_ISL_2098177 | Helix/Illumina | Centers for Disease Control and Prevention Division of Viral Diseases, Pathogen Discovery | Dakota Howard, Dhwani Batra, Peter W. Cook, Kara Moser, Adrian Paskey, Jason Caravas, Benjamin Rambo-Martin, Shatavia Morrison, Christopher Gulvick, Scott Sammons, Yvette Unoarumhi, Darlene Wagner, Matthew Schmerer, Eileen de Feo, Jan Antico, Christine Tran, Matthew Tolentino, Shannon Wickline, Kim Gietzen, Brad Sickler, Jingtao Liu, Eric Allen, Phil Febbo, Nicole L. Washington, Simon White, Geraint Levan, Kelly Schiabor Barrett, Elizabeth Cirulli, Alexandre Bolze, Ary Ascencio, Charlotte Rivera-Garcia, Ryan Cho, Jason Nguyen, Sherry Wang, Jimmy Ramirez, Tyler Cassens, Efren Sandoval, Magnus Isaksson, William Lee, David Becker, Marc Laurent, James Lu, Clinton R. Paden, Duncan MacCannell |
| EPI_ISL_2098178 | URMC LABS | Wadsworth Center, New York State Department of Health | Kirsten St. George, Daryl M. Lamson, Alexis Russell, Matthew Shudt, Melissa A Leisner, Jonathan Plitnick, Catharine Prussing, Navjot Singh, John Kelly, Erasmus Schneider, Erica Lasek-Nesselquist |
| EPI_ISL_2098179 | Helix/Illumina | Centers for Disease Control and Prevention Division of Viral Diseases, Pathogen Discovery | Dakota Howard, Dhwani Batra, Peter W. Cook, Kara Moser, Adrian Paskey, Jason Caravas, Benjamin Rambo-Martin, Shatavia Morrison, Christopher Gulvick, Scott Sammons, Yvette Unoarumhi, Darlene Wagner, Matthew Schmerer, Eileen de Feo, Jan Antico, Christine Tran, Matthew Tolentino, Shannon Wickline, Kim Gietzen, Brad Sickler, Jingtao Liu, Eric Allen, Phil Febbo, Nicole L. Washington, Simon White, Geraint Levan, Kelly Schiabor Barrett, Elizabeth Cirulli, Alexandre Bolze, Ary Ascencio, Charlotte Rivera-Garcia, Ryan Cho, Jason Nguyen, Sherry Wang, Jimmy Ramirez, Tyler Cassens, Efren Sandoval, Magnus Isaksson, William Lee, David Becker, Marc Laurent, James Lu, Clinton R. Paden, Duncan MacCannell |
| EPI_ISL_2098180, EPI_ISL_2098181 | URMC LABS | Wadsworth Center, New York State Department of Health | Kirsten St. George, Daryl M. Lamson, Alexis Russell, Matthew Shudt, Melissa A Leisner, Jonathan Plitnick, Catharine Prussing, Navjot Singh, John Kelly, Erasmus Schneider, Erica Lasek-Nesselquist |
| EPI_ISL_2098182 | Helix/Illumina | Centers for Disease Control and Prevention Division of Viral Diseases, Pathogen Discovery | Dakota Howard, Dhwani Batra, Peter W. Cook, Kara Moser, Adrian Paskey, Jason Caravas, Benjamin Rambo-Martin, Shatavia Morrison, Christopher Gulvick, Scott Sammons, Yvette Unoarumhi, Darlene Wagner, Matthew Schmerer, Eileen de Feo, Jan Antico, Christine Tran, Matthew Tolentino, Shannon Wickline, Kim Gietzen, Brad Sickler, Jingtao Liu, Eric Allen, Phil Febbo, Nicole L. Washington, Simon White, Geraint Levan, Kelly Schiabor Barrett, Elizabeth Cirulli, Alexandre Bolze, Ary Ascencio, Charlotte Rivera-Garcia, Ryan Cho, Jason Nguyen, Sherry Wang, Jimmy Ramirez, Tyler Cassens, Efren Sandoval, Magnus Isaksson, William Lee, David Becker, Marc Laurent, James Lu, Clinton R. Paden, Duncan MacCannell |
| EPI_ISL_2098183, EPI_ISL_2098184 | URMC LABS | Wadsworth Center, New York State Department of Health | Kirsten St. George, Daryl M. Lamson, Alexis Russell, Matthew Shudt, Melissa A Leisner, Jonathan Plitnick, Catharine Prussing, Navjot Singh, John Kelly, Erasmus Schneider, Erica Lasek-Nesselquist |
| EPI_ISL_2098185 | Helix/Illumina | Centers for Disease Control and Prevention Division of Viral Diseases, Pathogen Discovery | Dakota Howard, Dhwani Batra, Peter W. Cook, Kara Moser, Adrian Paskey, Jason Caravas, Benjamin Rambo-Martin, Shatavia Morrison, Christopher Gulvick, Scott Sammons, Yvette Unoarumhi, Darlene Wagner, Matthew Schmerer, Eileen de Feo, Jan Antico, Christine Tran, Matthew Tolentino, Shannon Wickline, Kim Gietzen, Brad Sickler, Jingtao Liu, Eric Allen, Phil Febbo, Nicole L. Washington, Simon White, Geraint Levan, Kelly Schiabor Barrett, Elizabeth Cirulli, Alexandre Bolze, Ary Ascencio, Charlotte Rivera-Garcia, Ryan Cho, Jason Nguyen, Sherry Wang, Jimmy Ramirez, Tyler Cassens, Efren Sandoval, Magnus Isaksson, William Lee, David Becker, Marc Laurent, James Lu, Clinton R. Paden, Duncan MacCannell |
| EPI_ISL_2098186, EPI_ISL_2098187 | URMC LABS | Wadsworth Center, New York State Department of Health | Kirsten St. George, Daryl M. Lamson, Alexis Russell, Matthew Shudt, Melissa A Leisner, Jonathan Plitnick, Catharine Prussing, Navjot Singh, John Kelly, Erasmus Schneider, Erica Lasek-Nesselquist |
| EPI_ISL_2098188 | Helix/Illumina | Centers for Disease Control and Prevention Division of Viral Diseases, Pathogen Discovery | Dakota Howard, Dhwani Batra, Peter W. Cook, Kara Moser, Adrian Paskey, Jason Caravas, Benjamin Rambo-Martin, Shatavia Morrison, Christopher Gulvick, Scott Sammons, Yvette Unoarumhi, Darlene Wagner, Matthew Schmerer, Eileen de Feo, Jan Antico, Christine Tran, Matthew Tolentino, Shannon Wickline, Kim Gietzen, Brad Sickler, Jingtao Liu, Eric Allen, Phil Febbo, Nicole L. Washington, Simon White, Geraint Levan, Kelly Schiabor Barrett, Elizabeth Cirulli, Alexandre Bolze, Ary Ascencio, Charlotte Rivera-Garcia, Ryan Cho, Jason Nguyen, Sherry Wang, Jimmy Ramirez, Tyler Cassens, Efren Sandoval, Magnus Isaksson, William Lee, David Becker, Marc Laurent, James Lu, Clinton R. Paden, Duncan MacCannell |
| EPI_ISL_2098189 | URMC LABS | Wadsworth Center, New York State Department of Health | Kirsten St. George, Daryl M. Lamson, Alexis Russell, Matthew Shudt, Melissa A Leisner, Jonathan Plitnick, Catharine Prussing, Navjot Singh, John Kelly, Erasmus Schneider, Erica Lasek-Nesselquist |
| EPI_ISL_2098190 | Helix/Illumina | Centers for Disease Control and Prevention Division of Viral Diseases, Pathogen Discovery | Dakota Howard, Dhwani Batra, Peter W. Cook, Kara Moser, Adrian Paskey, Jason Caravas, Benjamin Rambo-Martin, Shatavia Morrison, Christopher Gulvick, Scott Sammons, Yvette Unoarumhi, Darlene Wagner, Matthew Schmerer, Eileen de Feo, Jan Antico, Christine Tran, Matthew Tolentino, Shannon Wickline, Kim Gietzen, Brad Sickler, Jingtao Liu, Eric Allen, Phil Febbo, Nicole L. Washington, Simon White, Geraint Levan, Kelly Schiabor Barrett, Elizabeth Cirulli, Alexandre Bolze, Ary Ascencio, Charlotte Rivera-Garcia, Ryan Cho, Jason Nguyen, Sherry Wang, Jimmy Ramirez, Tyler Cassens, Efren Sandoval, Magnus Isaksson, William Lee, David Becker, Marc Laurent, James Lu, Clinton R. Paden, Duncan MacCannell |
| EPI_ISL_2098191, EPI_ISL_2098192, EPI_ISL_2098193 | URMC LABS | Wadsworth Center, New York State Department of Health | Kirsten St. George, Daryl M. Lamson, Alexis Russell, Matthew Shudt, Melissa A Leisner, Jonathan Plitnick, Catharine Prussing, Navjot Singh, John Kelly, Erasmus Schneider, Erica Lasek-Nesselquist |
| EPI_ISL_2098194 | Helix/Illumina | Centers for Disease Control and Prevention Division of Viral Diseases, Pathogen Discovery | Dakota Howard, Dhwani Batra, Peter W. Cook, Kara Moser, Adrian Paskey, Jason Caravas, Benjamin Rambo-Martin, Shatavia Morrison, Christopher Gulvick, Scott Sammons, Yvette Unoarumhi, Darlene Wagner, Matthew Schmerer, Eileen de Feo, Jan Antico, Christine Tran, Matthew Tolentino, Shannon Wickline, Kim Gietzen, Brad Sickler, Jingtao Liu, Eric Allen, Phil Febbo, Nicole L. Washington, Simon White, Geraint Levan, Kelly Schiabor Barrett, Elizabeth Cirulli, Alexandre Bolze, Ary Ascencio, Charlotte Rivera-Garcia, Ryan Cho, Jason Nguyen, Sherry Wang, Jimmy Ramirez, Tyler Cassens, Efren Sandoval, Magnus Isaksson, William Lee, David Becker, Marc Laurent, James Lu, Clinton R. Paden, Duncan MacCannell |
| EPI_ISL_2098195, EPI_ISL_2098196 | URMC LABS | Wadsworth Center, New York State Department of Health | Kirsten St. George, Daryl M. Lamson, Alexis Russell, Matthew Shudt, Melissa A Leisner, Jonathan Plitnick, Catharine Prussing, Navjot Singh, John Kelly, Erasmus Schneider, Erica Lasek-Nesselquist |
| EPI_ISL_2098197 | Helix/Illumina | Centers for Disease Control and Prevention Division of Viral Diseases, Pathogen Discovery | Dakota Howard, Dhwani Batra, Peter W. Cook, Kara Moser, Adrian Paskey, Jason Caravas, Benjamin Rambo-Martin, Shatavia Morrison, Christopher Gulvick, Scott Sammons, Yvette Unoarumhi, Darlene Wagner, Matthew Schmerer, Eileen de Feo, Jan Antico, Christine Tran, Matthew Tolentino, Shannon Wickline, Kim Gietzen, Brad Sickler, Jingtao Liu, Eric Allen, Phil Febbo, Nicole L. Washington, Simon White, Geraint Levan, Kelly Schiabor Barrett, Elizabeth Cirulli, Alexandre Bolze, Ary Ascencio, Charlotte Rivera-Garcia, Ryan Cho, Jason Nguyen, Sherry Wang, Jimmy Ramirez, Tyler Cassens, Efren Sandoval, Magnus Isaksson, William Lee, David Becker, Marc Laurent, James Lu, Clinton R. Paden, Duncan MacCannell |
| EPI_ISL_2098198, EPI_ISL_2098199 | URMC LABS | Wadsworth Center, New York State Department of Health | Kirsten St. George, Daryl M. Lamson, Alexis Russell, Matthew Shudt, Melissa A Leisner, Jonathan Plitnick, Catharine Prussing, Navjot Singh, John Kelly, Erasmus Schneider, Erica Lasek-Nesselquist |
| EPI_ISL_2098200 | Helix/Illumina | Centers for Disease Control and Prevention Division of Viral Diseases, Pathogen Discovery | Dakota Howard, Dhwani Batra, Peter W. Cook, Kara Moser, Adrian Paskey, Jason Caravas, Benjamin Rambo-Martin, Shatavia Morrison, Christopher Gulvick, Scott Sammons, Yvette Unoarumhi, Darlene Wagner, Matthew Schmerer, Eileen de Feo, Jan Antico, Christine Tran, Matthew Tolentino, Shannon Wickline, Kim Gietzen, Brad Sickler, Jingtao Liu, Eric Allen, Phil Febbo, Nicole L. Washington, Simon White, Geraint Levan, Kelly Schiabor Barrett, Elizabeth Cirulli, Alexandre Bolze, Ary Ascencio, Charlotte Rivera-Garcia, Ryan Cho, Jason Nguyen, Sherry Wang, Jimmy Ramirez, Tyler Cassens, Efren Sandoval, Magnus Isaksson, William Lee, David Becker, Marc Laurent, James Lu, Clinton R. Paden, Duncan MacCannell |
| EPI_ISL_2098201, EPI_ISL_2098202 | URMC LABS | Wadsworth Center, New York State Department of Health | Kirsten St. George, Daryl M. Lamson, Alexis Russell, Matthew Shudt, Melissa A Leisner, Jonathan Plitnick, Catharine Prussing, Navjot Singh, John Kelly, Erasmus Schneider, Erica Lasek-Nesselquist |
| EPI_ISL_2098203 | Helix/Illumina | Centers for Disease Control and Prevention Division of Viral Diseases, Pathogen Discovery | Dakota Howard, Dhwani Batra, Peter W. Cook, Kara Moser, Adrian Paskey, Jason Caravas, Benjamin Rambo-Martin, Shatavia Morrison, Christopher Gulvick, Scott Sammons, Yvette Unoarumhi, Darlene Wagner, Matthew Schmerer, Eileen de Feo, Jan Antico, Christine Tran, Matthew Tolentino, Shannon Wickline, Kim Gietzen, Brad Sickler, Jingtao Liu, Eric Allen, Phil Febbo, Nicole L. Washington, Simon White, Geraint Levan, Kelly Schiabor Barrett, Elizabeth Cirulli, Alexandre Bolze, Ary Ascencio, Charlotte Rivera-Garcia, Ryan Cho, Jason Nguyen, Sherry Wang, Jimmy Ramirez, Tyler Cassens, Efren Sandoval, Magnus Isaksson, William Lee, David Becker, Marc Laurent, James Lu, Clinton R. Paden, Duncan MacCannell |
| EPI_ISL_2098204 | URMC LABS | Wadsworth Center, New York State Department of Health | Kirsten St. George, Daryl M. Lamson, Alexis Russell, Matthew Shudt, Melissa A Leisner, Jonathan Plitnick, Catharine Prussing, Navjot Singh, John Kelly, Erasmus Schneider, Erica Lasek-Nesselquist |
| EPI_ISL_2098205 | Helix/Illumina | Centers for Disease Control and Prevention Division of Viral Diseases, Pathogen Discovery | Dakota Howard, Dhwani Batra, Peter W. Cook, Kara Moser, Adrian Paskey, Jason Caravas, Benjamin Rambo-Martin, Shatavia Morrison, Christopher Gulvick, Scott Sammons, Yvette Unoarumhi, Darlene Wagner, Matthew Schmerer, Eileen de Feo, Jan Antico, Christine Tran, Matthew Tolentino, Shannon Wickline, Kim Gietzen, Brad Sickler, Jingtao Liu, Eric Allen, Phil Febbo, Nicole L. Washington, Simon White, Geraint Levan, Kelly Schiabor Barrett, Elizabeth Cirulli, Alexandre Bolze, Ary Ascencio, Charlotte Rivera-Garcia, Ryan Cho, Jason Nguyen, Sherry Wang, Jimmy Ramirez, Tyler Cassens, Efren Sandoval, Magnus Isaksson, William Lee, David Becker, Marc Laurent, James Lu, Clinton R. Paden, Duncan MacCannell |
| EPI_ISL_2098206, EPI_ISL_2098208, EPI_ISL_2098209 | URMC LABS | Wadsworth Center, New York State Department of Health | Kirsten St. George, Daryl M. Lamson, Alexis Russell, Matthew Shudt, Melissa A Leisner, Jonathan Plitnick, Catharine Prussing, Navjot Singh, John Kelly, Erasmus Schneider, Erica Lasek-Nesselquist |
| EPI_ISL_2098210 | Helix/Illumina | Centers for Disease Control and Prevention Division of Viral Diseases, Pathogen Discovery | Dakota Howard, Dhwani Batra, Peter W. Cook, Kara Moser, Adrian Paskey, Jason Caravas, Benjamin Rambo-Martin, Shatavia Morrison, Christopher Gulvick, Scott Sammons, Yvette Unoarumhi, Darlene Wagner, Matthew Schmerer, Eileen de Feo, Jan Antico, Christine Tran, Matthew Tolentino, Shannon Wickline, Kim Gietzen, Brad Sickler, Jingtao Liu, Eric Allen, Phil Febbo, Nicole L. Washington, Simon White, Geraint Levan, Kelly Schiabor Barrett, Elizabeth Cirulli, Alexandre Bolze, Ary Ascencio, Charlotte Rivera-Garcia, Ryan Cho, Jason Nguyen, Sherry Wang, Jimmy Ramirez, Tyler Cassens, Efren Sandoval, Magnus Isaksson, William Lee, David Becker, Marc Laurent, James Lu, Clinton R. Paden, Duncan MacCannell |
| EPI_ISL_2098211, EPI_ISL_2098212 | URMC LABS | Wadsworth Center, New York State Department of Health | Kirsten St. George, Daryl M. Lamson, Alexis Russell, Matthew Shudt, Melissa A Leisner, Jonathan Plitnick, Catharine Prussing, Navjot Singh, John Kelly, Erasmus Schneider, Erica Lasek-Nesselquist |
| EPI_ISL_2098213 | Helix/Illumina | Centers for Disease Control and Prevention Division of Viral Diseases, Pathogen | Dakota Howard, Dhwani Batra, Peter W. Cook, Kara Moser, Adrian Paskey, Jason Caravas, Benjamin Rambo-Martin, Shatavia Morrison, Christopher Gulvick, Scott Sammons, Yvette Unoarumhi, Darlene Wagner, Matthew Schmerer, Eileen de Feo, Jan Antico, Christine Tran, Matthew Tolentino, Shannon Wickline, Kim Gietzen, Brad Sickler, Jingtao Liu, Eric Allen, Phil Febbo, Nicole L. Washington, Simon White, Geraint Levan, Kelly Schiabor Barrett, Elizabeth Cirulli, Alexandre Bolze, Ary Ascencio, Charlotte Rivera-Garcia, Ryan Cho, Jason Nguyen, Sherry Wang, Jimmy |

| | | | |
|---|---|---|---|
| | | Discovery | Ramirez, Tyler Cassens, Efren Sandoval, Magnus Isaksson, William Lee, David Becker, Marc Laurent, James Lu, Clinton R. Paden, Duncan MacCannell |
| EPI_ISL_2098214, EPI_ISL_2098215 | URMC LABS | Wadsworth Center, New York State Department of Health | Kirsten St. George, Daryl M. Lamson, Alexis Russell, Matthew Shudt, Melissa A Leisner, Jonathan Plitnick, Catharine Prussing, Navjot Singh, John Kelly, Erasmus Schneider, Erica Lasek-Nesselquist |
| EPI_ISL_2098216 | Helix/Illumina | Centers for Disease Control and Prevention Division of Viral Diseases, Pathogen Discovery | Dakota Howard, Dhwani Batra, Peter W. Cook, Kara Moser, Adrian Paskey, Jason Caravas, Benjamin Rambo-Martin, Shatavia Morrison, Christopher Gulvick, Scott Sammons, Yvette Unoarumhi, Darlene Wagner, Matthew Schmerer, Eileen de Feo, Jan Antico, Christine Tran, Matthew Tolentino, Shannon Wickline, Kim Gietzen, Brad Sickler, Jingtao Liu, Eric Allen, Phil Febbo, Nicole L. Washington, Simon White, Geraint Levan, Kelly Schiabor Barrett, Elizabeth Cirulli, Alexandre Bolze, Ary Ascencio, Charlotte Rivera-Garcia, Ryan Cho, Jason Nguyen, Sherry Wang, Jimmy Ramirez, Tyler Cassens, Efren Sandoval, Magnus Isaksson, William Lee, David Becker, Marc Laurent, James Lu, Clinton R. Paden, Duncan MacCannell |
| EPI_ISL_2098217, EPI_ISL_2098218 | URMC LABS | Wadsworth Center, New York State Department of Health | Kirsten St. George, Daryl M. Lamson, Alexis Russell, Matthew Shudt, Melissa A Leisner, Jonathan Plitnick, Catharine Prussing, Navjot Singh, John Kelly, Erasmus Schneider, Erica Lasek-Nesselquist |
| EPI_ISL_2098219 | Helix/Illumina | Centers for Disease Control and Prevention Division of Viral Diseases, Pathogen Discovery | Dakota Howard, Dhwani Batra, Peter W. Cook, Kara Moser, Adrian Paskey, Jason Caravas, Benjamin Rambo-Martin, Shatavia Morrison, Christopher Gulvick, Scott Sammons, Yvette Unoarumhi, Darlene Wagner, Matthew Schmerer, Eileen de Feo, Jan Antico, Christine Tran, Matthew Tolentino, Shannon Wickline, Kim Gietzen, Brad Sickler, Jingtao Liu, Eric Allen, Phil Febbo, Nicole L. Washington, Simon White, Geraint Levan, Kelly Schiabor Barrett, Elizabeth Cirulli, Alexandre Bolze, Ary Ascencio, Charlotte Rivera-Garcia, Ryan Cho, Jason Nguyen, Sherry Wang, Jimmy Ramirez, Tyler Cassens, Efren Sandoval, Magnus Isaksson, William Lee, David Becker, Marc Laurent, James Lu, Clinton R. Paden, Duncan MacCannell |
| EPI_ISL_2098220 | URMC LABS | Wadsworth Center, New York State Department of Health | Kirsten St. George, Daryl M. Lamson, Alexis Russell, Matthew Shudt, Melissa A Leisner, Jonathan Plitnick, Catharine Prussing, Navjot Singh, John Kelly, Erasmus Schneider, Erica Lasek-Nesselquist |
| EPI_ISL_2098221 | Helix/Illumina | Centers for Disease Control and Prevention Division of Viral Diseases, Pathogen Discovery | Dakota Howard, Dhwani Batra, Peter W. Cook, Kara Moser, Adrian Paskey, Jason Caravas, Benjamin Rambo-Martin, Shatavia Morrison, Christopher Gulvick, Scott Sammons, Yvette Unoarumhi, Darlene Wagner, Matthew Schmerer, Eileen de Feo, Jan Antico, Christine Tran, Matthew Tolentino, Shannon Wickline, Kim Gietzen, Brad Sickler, Jingtao Liu, Eric Allen, Phil Febbo, Nicole L. Washington, Simon White, Geraint Levan, Kelly Schiabor Barrett, Elizabeth Cirulli, Alexandre Bolze, Ary Ascencio, Charlotte Rivera-Garcia, Ryan Cho, Jason Nguyen, Sherry Wang, Jimmy Ramirez, Tyler Cassens, Efren Sandoval, Magnus Isaksson, William Lee, David Becker, Marc Laurent, James Lu, Clinton R. Paden, Duncan MacCannell |
| EPI_ISL_2098222, EPI_ISL_2098223, EPI_ISL_2098224, EPI_ISL_2098225 | URMC LABS | Wadsworth Center, New York State Department of Health | Kirsten St. George, Daryl M. Lamson, Alexis Russell, Matthew Shudt, Melissa A Leisner, Jonathan Plitnick, Catharine Prussing, Navjot Singh, John Kelly, Erasmus Schneider, Erica Lasek-Nesselquist |
| EPI_ISL_2098226 | Helix/Illumina | Centers for Disease Control and Prevention Division of Viral Diseases, Pathogen Discovery | Dakota Howard, Dhwani Batra, Peter W. Cook, Kara Moser, Adrian Paskey, Jason Caravas, Benjamin Rambo-Martin, Shatavia Morrison, Christopher Gulvick, Scott Sammons, Yvette Unoarumhi, Darlene Wagner, Matthew Schmerer, Eileen de Feo, Jan Antico, Christine Tran, Matthew Tolentino, Shannon Wickline, Kim Gietzen, Brad Sickler, Jingtao Liu, Eric Allen, Phil Febbo, Nicole L. Washington, Simon White, Geraint Levan, Kelly Schiabor Barrett, Elizabeth Cirulli, Alexandre Bolze, Ary Ascencio, Charlotte Rivera-Garcia, Ryan Cho, Jason Nguyen, Sherry Wang, Jimmy Ramirez, Tyler Cassens, Efren Sandoval, Magnus Isaksson, William Lee, David Becker, Marc Laurent, James Lu, Clinton R. Paden, Duncan MacCannell |
| EPI_ISL_2098227, EPI_ISL_2098228 | URMC LABS | Wadsworth Center, New York State Department of Health | Kirsten St. George, Daryl M. Lamson, Alexis Russell, Matthew Shudt, Melissa A Leisner, Jonathan Plitnick, Catharine Prussing, Navjot Singh, John Kelly, Erasmus Schneider, Erica Lasek-Nesselquist |
| EPI_ISL_2098229 | Helix/Illumina | Centers for Disease Control and Prevention Division of Viral Diseases, Pathogen Discovery | Dakota Howard, Dhwani Batra, Peter W. Cook, Kara Moser, Adrian Paskey, Jason Caravas, Benjamin Rambo-Martin, Shatavia Morrison, Christopher Gulvick, Scott Sammons, Yvette Unoarumhi, Darlene Wagner, Matthew Schmerer, Eileen de Feo, Jan Antico, Christine Tran, Matthew Tolentino, Shannon Wickline, Kim Gietzen, Brad Sickler, Jingtao Liu, Eric Allen, Phil Febbo, Nicole L. Washington, Simon White, Geraint Levan, Kelly Schiabor Barrett, Elizabeth Cirulli, Alexandre Bolze, Ary Ascencio, Charlotte Rivera-Garcia, Ryan Cho, Jason Nguyen, Sherry Wang, Jimmy Ramirez, Tyler Cassens, Efren Sandoval, Magnus Isaksson, William Lee, David Becker, Marc Laurent, James Lu, Clinton R. Paden, Duncan MacCannell |
| EPI_ISL_2098230, EPI_ISL_2098231 | URMC LABS | Wadsworth Center, New York State Department of Health | Kirsten St. George, Daryl M. Lamson, Alexis Russell, Matthew Shudt, Melissa A Leisner, Jonathan Plitnick, Catharine Prussing, Navjot Singh, John Kelly, Erasmus Schneider, Erica Lasek-Nesselquist |
| EPI_ISL_2098232 | Helix/Illumina | Centers for Disease Control and Prevention Division of Viral Diseases, Pathogen Discovery | Dakota Howard, Dhwani Batra, Peter W. Cook, Kara Moser, Adrian Paskey, Jason Caravas, Benjamin Rambo-Martin, Shatavia Morrison, Christopher Gulvick, Scott Sammons, Yvette Unoarumhi, Darlene Wagner, Matthew Schmerer, Eileen de Feo, Jan Antico, Christine Tran, Matthew Tolentino, Shannon Wickline, Kim Gietzen, Brad Sickler, Jingtao Liu, Eric Allen, Phil Febbo, Nicole L. Washington, Simon White, Geraint Levan, Kelly Schiabor Barrett, Elizabeth Cirulli, Alexandre Bolze, Ary Ascencio, Charlotte Rivera-Garcia, Ryan Cho, Jason Nguyen, Sherry Wang, Jimmy Ramirez, Tyler Cassens, Efren Sandoval, Magnus Isaksson, William Lee, David Becker, Marc Laurent, James Lu, Clinton R. Paden, Duncan MacCannell |
| EPI_ISL_2098233 | WESTCHESTER MEDICAL CENTER | Wadsworth Center, New York State Department of Health | Kirsten St. George, Daryl M. Lamson, Alexis Russell, Matthew Shudt, Melissa A Leisner, Jonathan Plitnick, Catharine Prussing, Navjot Singh, John Kelly, Erasmus Schneider, Erica Lasek-Nesselquist |
| EPI_ISL_2098234, EPI_ISL_2098235, EPI_ISL_2098236, EPI_ISL_2098237, EPI_ISL_2098238, EPI_ISL_2098239, EPI_ISL_2098240, EPI_ISL_2098241, EPI_ISL_2098242, EPI_ISL_2098243, EPI_ISL_2098244, EPI_ISL_2098245, EPI_ISL_2098246, EPI_ISL_2098247, EPI_ISL_2098248, EPI_ISL_2098249, EPI_ISL_2098250, EPI_ISL_2098251, EPI_ISL_2098252, EPI_ISL_2098253, EPI_ISL_2098254, EPI_ISL_2098255, EPI_ISL_2098256, EPI_ISL_2098257, EPI_ISL_2098258, EPI_ISL_2098259, EPI_ISL_2098260, EPI_ISL_2098261, EPI_ISL_2098262, EPI_ISL_2098263, EPI_ISL_2098264, EPI_ISL_2098265, EPI_ISL_2098266, EPI_ISL_2098267 | | | |
| see above | Helix/Illumina | Centers for Disease Control and Prevention Division of Viral Diseases, Pathogen Discovery | Dakota Howard, Dhwani Batra, Peter W. Cook, Kara Moser, Adrian Paskey, Jason Caravas, Benjamin Rambo-Martin, Shatavia Morrison, Christopher Gulvick, Scott Sammons, Yvette Unoarumhi, Darlene Wagner, Matthew Schmerer, Eileen de Feo, Jan Antico, Christine Tran, Matthew Tolentino, Shannon Wickline, Kim Gietzen, Brad Sickler, Jingtao Liu, Eric Allen, Phil Febbo, Nicole L. Washington, Simon White, Geraint Levan, Kelly Schiabor Barrett, Elizabeth Cirulli, Alexandre Bolze, Ary Ascencio, Charlotte Rivera-Garcia, Ryan Cho, Jason Nguyen, Sherry Wang, Jimmy Ramirez, Tyler Cassens, Efren Sandoval, Magnus Isaksson, William Lee, David Becker, Marc Laurent, James Lu, Clinton R. Paden, Duncan MacCannell |
| EPI_ISL_2098268, EPI_ISL_2098269, EPI_ISL_2098270, EPI_ISL_2098271, EPI_ISL_2098272, EPI_ISL_2098273, EPI_ISL_2098274, EPI_ISL_2098275, EPI_ISL_2098276, EPI_ISL_2098277, EPI_ISL_2098278, EPI_ISL_2098279, EPI_ISL_2098280, EPI_ISL_2098281, EPI_ISL_2098282, EPI_ISL_2098283, EPI_ISL_2098284, EPI_ISL_2098285, EPI_ISL_2098286, EPI_ISL_2098287, EPI_ISL_2098288, EPI_ISL_2098289, EPI_ISL_2098290, EPI_ISL_2098291, EPI_ISL_2098292, EPI_ISL_2098293, EPI_ISL_2098294, EPI_ISL_2098295, EPI_ISL_2098296, EPI_ISL_2098297, EPI_ISL_2098298, EPI_ISL_2098299, EPI_ISL_2098300, EPI_ISL_2098301, EPI_ISL_2098302, EPI_ISL_2098303, EPI_ISL_2098304, EPI_ISL_2098305, EPI_ISL_2098306, EPI_ISL_2098307, EPI_ISL_2098308, EPI_ISL_2098309, EPI_ISL_2098310, EPI_ISL_2098311, EPI_ISL_2098312, EPI_ISL_2098313, EPI_ISL_2098314, EPI_ISL_2098315, EPI_ISL_2098316, EPI_ISL_2098317, EPI_ISL_2098318, EPI_ISL_2098319, EPI_ISL_2098320, EPI_ISL_2098321, EPI_ISL_2098322, EPI_ISL_2098323, EPI_ISL_2098324, EPI_ISL_2098325, EPI_ISL_2098326, EPI_ISL_2098327, EPI_ISL_2098328, EPI_ISL_2098329, EPI_ISL_2098330, EPI_ISL_2098331, EPI_ISL_2098332, EPI_ISL_2098333, EPI_ISL_2098334, EPI_ISL_2098335, EPI_ISL_2098336, EPI_ISL_2098337, EPI_ISL_2098338, EPI_ISL_2098339, EPI_ISL_2098340, EPI_ISL_2098341, EPI_ISL_2098342, EPI_ISL_2098343, EPI_ISL_2098344, EPI_ISL_2098345, EPI_ISL_2098346, EPI_ISL_2098347, EPI_ISL_2098348, EPI_ISL_2098349, EPI_ISL_2098350, EPI_ISL_2098351, EPI_ISL_2098352, EPI_ISL_2098353, EPI_ISL_2098354, EPI_ISL_2098355, EPI_ISL_2098356, EPI_ISL_2098357, EPI_ISL_2098358, EPI_ISL_2098359, EPI_ISL_2098360, EPI_ISL_2098361, EPI_ISL_2098362, EPI_ISL_2098363, EPI_ISL_2098364, EPI_ISL_2098365, EPI_ISL_2098366, EPI_ISL_2098367, EPI_ISL_2098368, EPI_ISL_2098369, EPI_ISL_2098370, EPI_ISL_2098371, EPI_ISL_2098372, EPI_ISL_2098373, EPI_ISL_2098374, EPI_ISL_2098375, EPI_ISL_2098376, EPI_ISL_2098377, EPI_ISL_2098378, EPI_ISL_2098379, EPI_ISL_2098380, EPI_ISL_2098381, EPI_ISL_2098382, EPI_ISL_2098383, EPI_ISL_2098384, EPI_ISL_2098385, EPI_ISL_2098386, EPI_ISL_2098387, EPI_ISL_2098388, EPI_ISL_2098389, EPI_ISL_2098390, EPI_ISL_2098391, EPI_ISL_2098392, EPI_ISL_2098393, EPI_ISL_2098394, EPI_ISL_2098395, EPI_ISL_2098396, EPI_ISL_2098397, EPI_ISL_2098398, EPI_ISL_2098399, EPI_ISL_2098400, EPI_ISL_2098401, EPI_ISL_2098402, EPI_ISL_2098403, EPI_ISL_2098404, EPI_ISL_2098405, EPI_ISL_2098406, EPI_ISL_2098407, EPI_ISL_2098408, EPI_ISL_2098409, EPI_ISL_2098410, EPI_ISL_2098411, EPI_ISL_2098412, EPI_ISL_2098413, EPI_ISL_2098414, EPI_ISL_2098415 | | | |
| see above | Quest Diagnostics Incorporated | Centers for Disease Control and Prevention Division of Viral Diseases, Pathogen Discovery | Dakota Howard, Dhwani Batra, Peter W. Cook, Kara Moser, Adrian Paskey, Jason Caravas, Benjamin Rambo-Martin, Shatavia Morrison, Christopher Gulvick, Scott Sammons, Yvette Unoarumhi, Darlene Wagner, Matthew Schmerer, S. H. Rosenthal, A. Gerasimova, R. M. Kagan, B. Anderson, M. Hua, Y. Liu, L.E. Bernstein, K.E. Livingston, A. Perez, I. A. Shlyakhter, R. V. Rolando, R. Owen, P. Tanpaiboon, F. Lacbawan, Clinton R. Paden, Duncan MacCannell |
| EPI_ISL_2098499 | ADIRONDACK MEDICAL CENTER | Wadsworth Center, New York State Department of Health | Kirsten St. George, Daryl M. Lamson, Alexis Russell, Matthew Shudt, Melissa A Leisner, Jonathan Plitnick, Catharine Prussing, Navjot Singh, John Kelly, Erasmus Schneider, Erica Lasek-Nesselquist |
| EPI_ISL_2098500 | ALBANY MEDICAL CENTER | Wadsworth Center, New York State Department of Health | Kirsten St. George, Daryl M. Lamson, Alexis Russell, Matthew Shudt, Melissa A Leisner, Jonathan Plitnick, Catharine Prussing, Navjot Singh, John Kelly, Erasmus Schneider, Erica Lasek-Nesselquist |
| EPI_ISL_2098501, EPI_ISL_2098502, EPI_ISL_2098503, EPI_ISL_2098504, EPI_ISL_2098505, EPI_ISL_2098506, EPI_ISL_2098507, EPI_ISL_2098508, EPI_ISL_2098509, EPI_ISL_2098510, EPI_ISL_2098511, EPI_ISL_2098512, EPI_ISL_2098513, EPI_ISL_2098514, EPI_ISL_2098515, EPI_ISL_2098516, EPI_ISL_2098517, EPI_ISL_2098518, EPI_ISL_2098519, EPI_ISL_2098520, EPI_ISL_2098521, EPI_ISL_2098522, EPI_ISL_2098523, EPI_ISL_2098524, EPI_ISL_2098525, EPI_ISL_2098526, EPI_ISL_2098527, EPI_ISL_2098528, EPI_ISL_2098529, EPI_ISL_2098530, EPI_ISL_2098531, EPI_ISL_2098532, EPI_ISL_2098533, EPI_ISL_2098534, EPI_ISL_2098535, EPI_ISL_2098536, EPI_ISL_2098537, EPI_ISL_2098538, EPI_ISL_2098539, EPI_ISL_2098540, EPI_ISL_2098541, EPI_ISL_2098542, EPI_ISL_2098543, EPI_ISL_2098544, EPI_ISL_2098545, EPI_ISL_2098546, EPI_ISL_2098547, EPI_ISL_2098548, EPI_ISL_2098549, EPI_ISL_2098550, EPI_ISL_2098551, EPI_ISL_2098552, EPI_ISL_2098553, EPI_ISL_2098554, EPI_ISL_2098555, EPI_ISL_2098556, EPI_ISL_2098557, EPI_ISL_2098558, EPI_ISL_2098559, EPI_ISL_2098560, EPI_ISL_2098561, EPI_ISL_2098562, EPI_ISL_2098563, EPI_ISL_2098564, EPI_ISL_2098565, EPI_ISL_2098566, EPI_ISL_2098567, EPI_ISL_2098568, EPI_ISL_2098569, EPI_ISL_2098570, EPI_ISL_2098571, EPI_ISL_2098572, EPI_ISL_2098573, EPI_ISL_2098574, EPI_ISL_2098575, EPI_ISL_2098576, EPI_ISL_2098577, EPI_ISL_2098578, EPI_ISL_2098579, EPI_ISL_2098580, EPI_ISL_2098581, EPI_ISL_2098582, EPI_ISL_2098583, EPI_ISL_2098584, EPI_ISL_2098585, EPI_ISL_2098586, EPI_ISL_2098587 | | | |
| see above | URMC LABS | Wadsworth Center, New York State Department of Health | Kirsten St. George, Daryl M. Lamson, Alexis Russell, Matthew Shudt, Melissa A Leisner, Jonathan Plitnick, Catharine Prussing, Navjot Singh, John Kelly, Erasmus Schneider, Erica Lasek-Nesselquist |
| EPI_ISL_2098588 | IN State Department of Health Laboratory Services | IN State Department of Health Laboratory Services | Cassandra Campion, Jamie Yeadon, Brian Pope, Lixia Liu, Kyle Brownlee, Melissa Hindenlang, Mark Glazier |
| EPI_ISL_2098589, EPI_ISL_2098590, EPI_ISL_2098591, EPI_ISL_2098592, EPI_ISL_2098593, EPI_ISL_2098594, EPI_ISL_2098595, EPI_ISL_2098596, EPI_ISL_2098597, EPI_ISL_2098598, EPI_ISL_2098599 | | | |
| see above | BOSTON HEART DIAGNOSTICS CORP | Wadsworth Center, New York State Department of Health | Kirsten St. George, Daryl M. Lamson, Alexis Russell, Matthew Shudt, Melissa A Leisner, Jonathan Plitnick, Catharine Prussing, Navjot Singh, John Kelly, Erasmus Schneider, Erica Lasek-Nesselquist |
| EPI_ISL_2098600 | DELHI REHABILITATION AND NURSING CENTER | Wadsworth Center, New York State Department of Health | Kirsten St. George, Daryl M. Lamson, Alexis Russell, Matthew Shudt, Melissa A Leisner, Jonathan Plitnick, Catharine Prussing, Navjot Singh, John Kelly, Erasmus Schneider, Erica Lasek-Nesselquist |
| EPI_ISL_2098601, EPI_ISL_2098602, EPI_ISL_2098603, EPI_ISL_2098604, EPI_ISL_2098605, EPI_ISL_2098606, EPI_ISL_2098607, EPI_ISL_2098608, EPI_ISL_2098609, EPI_ISL_2098610, EPI_ISL_2098611, EPI_ISL_2098612, EPI_ISL_2098613, EPI_ISL_2098614, EPI_ISL_2098615, EPI_ISL_2098616, EPI_ISL_2098617, EPI_ISL_2098618, EPI_ISL_2098619, EPI_ISL_2098620, EPI_ISL_2098621 | | | |
| see above | NYC Pandemic Response Lab | Wadsworth Center, New York State Department of Health | Kirsten St. George, Daryl M. Lamson, Alexis Russell, Matthew Shudt, Melissa A Leisner, Jonathan Plitnick, Catharine Prussing, Navjot Singh, John Kelly, Erasmus Schneider, Erica Lasek-Nesselquist |
| EPI_ISL_2098622, EPI_ISL_2098623, EPI_ISL_2098624, EPI_ISL_2098625, EPI_ISL_2098626, EPI_ISL_2098627, EPI_ISL_2098628 | SARATOGA HOSPITAL LABORATORY | Wadsworth Center, New York State Department of Health | Kirsten St. George, Daryl M. Lamson, Alexis Russell, Matthew Shudt, Melissa A Leisner, Jonathan Plitnick, Catharine Prussing, Navjot Singh, John Kelly, Erasmus Schneider, Erica Lasek-Nesselquist |
| EPI_ISL_2098629, EPI_ISL_2098630 | ST. FRANCIS HOSPITAL LABORATORY | Wadsworth Center, New York State Department of Health | Kirsten St. George, Daryl M. Lamson, Alexis Russell, Matthew Shudt, Melissa A Leisner, Jonathan Plitnick, Catharine Prussing, Navjot Singh, John Kelly, Erasmus Schneider, Erica Lasek-Nesselquist |
| EPI_ISL_2098631, EPI_ISL_2098632, EPI_ISL_2098633, EPI_ISL_2098634, EPI_ISL_2098635, EPI_ISL_2098636, EPI_ISL_2098637, EPI_ISL_2098638, EPI_ISL_2098639, EPI_ISL_2098640, EPI_ISL_2098641, EPI_ISL_2098642, EPI_ISL_2098643, EPI_ISL_2098644, EPI_ISL_2098645, EPI_ISL_2098646, EPI_ISL_2098647, EPI_ISL_2098648, EPI_ISL_2098649, EPI_ISL_2098650, EPI_ISL_2098651, EPI_ISL_2098652, EPI_ISL_2098653, EPI_ISL_2098654, EPI_ISL_2098655, EPI_ISL_2098656, EPI_ISL_2098657, EPI_ISL_2098658, EPI_ISL_2098659 | | | |
| see above | SUNY UPSTATE MEDICAL UNIVERSITY | Wadsworth Center, New York State Department of Health | Kirsten St. George, Daryl M. Lamson, Alexis Russell, Matthew Shudt, Melissa A Leisner, Jonathan Plitnick, Catharine Prussing, Navjot Singh, John Kelly, Erasmus Schneider, Erica Lasek-Nesselquist |
| EPI_ISL_2098660, EPI_ISL_2098661, EPI_ISL_2098662, EPI_ISL_2098663, EPI_ISL_2098664, EPI_ISL_2098665 | URMC LABS | Wadsworth Center, New York State Department of Health | Kirsten St. George, Daryl M. Lamson, Alexis Russell, Matthew Shudt, Melissa A Leisner, Jonathan Plitnick, Catharine Prussing, Navjot Singh, John Kelly, Erasmus Schneider, Erica Lasek-Nesselquist |
| EPI_ISL_2098667, EPI_ISL_2098668, EPI_ISL_2098669, EPI_ISL_2098670, EPI_ISL_2098671, EPI_ISL_2098672, EPI_ISL_2098673, EPI_ISL_2098674, EPI_ISL_2098675, EPI_ISL_2098676, EPI_ISL_2098677, EPI_ISL_2098678, EPI_ISL_2098679, EPI_ISL_2098680, EPI_ISL_2098681, EPI_ISL_2098682, EPI_ISL_2098683, EPI_ISL_2098684, EPI_ISL_2098685, EPI_ISL_2098686, EPI_ISL_2098687, EPI_ISL_2098688, EPI_ISL_2098689, EPI_ISL_2098691, EPI_ISL_2098692, EPI_ISL_2098693, EPI_ISL_2098694, EPI_ISL_2098695, EPI_ISL_2098696, EPI_ISL_2098697, EPI_ISL_2098698, EPI_ISL_2098699, EPI_ISL_2098700, EPI_ISL_2098701, EPI_ISL_2098702, EPI_ISL_2098703, EPI_ISL_2098704, EPI_ISL_2098705, EPI_ISL_2098706, EPI_ISL_2098707, EPI_ISL_2098708, EPI_ISL_2098709 | | | |
| see above | Virginia Division of Consolidated Laboratory Services | Virginia Division of Consolidated Laboratory Services | Virginia DCLS |
| EPI_ISL_2098724, EPI_ISL_2098726, EPI_ISL_2098728, EPI_ISL_2098729, EPI_ISL_2098730, EPI_ISL_2098731, EPI_ISL_2098732, EPI_ISL_2098734, EPI_ISL_2098735, EPI_ISL_2098737, EPI_ISL_2098738, EPI_ISL_2098739, EPI_ISL_2098741, EPI_ISL_2098742, EPI_ISL_2098744, EPI_ISL_2098745, EPI_ISL_2098746, EPI_ISL_2098747, EPI_ISL_2098749, EPI_ISL_2098750, EPI_ISL_2098752, EPI_ISL_2098753, EPI_ISL_2098754, EPI_ISL_2098755, EPI_ISL_2098756, EPI_ISL_2098757, EPI_ISL_2098758, EPI_ISL_2098759, EPI_ISL_2098761, EPI_ISL_2098762, EPI_ISL_2098763, EPI_ISL_2098764, EPI_ISL_2098765, EPI_ISL_2098766, EPI_ISL_2098767, EPI_ISL_2098769, EPI_ISL_2098770, EPI_ISL_2098771, EPI_ISL_2098772, EPI_ISL_2098773, EPI_ISL_2098774, EPI_ISL_2098776, EPI_ISL_2098778, EPI_ISL_2098779, | | | |

| | | | |
|---|---|---|---|
| EPI_ISL_2098780, EPI_ISL_2098783, EPI_ISL_2098784, EPI_ISL_2098785, EPI_ISL_2098786, EPI_ISL_2098787, EPI_ISL_2098789, EPI_ISL_2098790, EPI_ISL_2098791, EPI_ISL_2098792, EPI_ISL_2098793, EPI_ISL_2098794, EPI_ISL_2098800, EPI_ISL_2098801, EPI_ISL_2098803, EPI_ISL_2098804, EPI_ISL_2098805, EPI_ISL_2098806, EPI_ISL_2098807, EPI_ISL_2098810, EPI_ISL_2098811, EPI_ISL_2098812, EPI_ISL_2098813, EPI_ISL_2098814, EPI_ISL_2098815, EPI_ISL_2098816, EPI_ISL_2098818, EPI_ISL_2098820, EPI_ISL_2098822, EPI_ISL_2098823, EPI_ISL_2098824, EPI_ISL_2098825, EPI_ISL_2098826, EPI_ISL_2098827, EPI_ISL_2098828, EPI_ISL_2098830, EPI_ISL_2098831, EPI_ISL_2098832, EPI_ISL_2098833, EPI_ISL_2098835, EPI_ISL_2098836, EPI_ISL_2098837, EPI_ISL_2098839 | | | |
| see above | Clinical Molecular Microbiology Laboratory, UNC Hospitals | Jeremy Wang | Jeremy Wang, Alexander Rubinsteyn, Colleen Rice, Jose Luis Torres Castillo, Brianna de la Houssaye, Jason Smedberg, Shawn Hawken, Melissa Miller, Corbin Jones, Robert Hagan |
| EPI_ISL_2099184, EPI_ISL_2099185, EPI_ISL_2099186, EPI_ISL_2099187, EPI_ISL_2099188, EPI_ISL_2099189, EPI_ISL_2099190, EPI_ISL_2099191, EPI_ISL_2099192, EPI_ISL_2099193, EPI_ISL_2099194, EPI_ISL_2099195, EPI_ISL_2099196, EPI_ISL_2099197, EPI_ISL_2099198, EPI_ISL_2099199, EPI_ISL_2099200, EPI_ISL_2099201, EPI_ISL_2099202, EPI_ISL_2099203, EPI_ISL_2099204, EPI_ISL_2099205, EPI_ISL_2099206, EPI_ISL_2099207, EPI_ISL_2099208, EPI_ISL_2099209, EPI_ISL_2099210, EPI_ISL_2099218, EPI_ISL_2099219, EPI_ISL_2099220, EPI_ISL_2099221, EPI_ISL_2099222, EPI_ISL_2099223, EPI_ISL_2099224, EPI_ISL_2099225, EPI_ISL_2099226, EPI_ISL_2099227, EPI_ISL_2099228, EPI_ISL_2099229, EPI_ISL_2099230, EPI_ISL_2099231, EPI_ISL_2099232, EPI_ISL_2099233, EPI_ISL_2099234, EPI_ISL_2099235, EPI_ISL_2099236, EPI_ISL_2099237, EPI_ISL_2099238, EPI_ISL_2099239, EPI_ISL_2099240, EPI_ISL_2099241, EPI_ISL_2099242, EPI_ISL_2099243, EPI_ISL_2099244, EPI_ISL_2099245, EPI_ISL_2099246, EPI_ISL_2099247, EPI_ISL_2099248, EPI_ISL_2099249, EPI_ISL_2099250, EPI_ISL_2099251, EPI_ISL_2099252, EPI_ISL_2099253, EPI_ISL_2099254, EPI_ISL_2099255, EPI_ISL_2099256, EPI_ISL_2099257, EPI_ISL_2099258, EPI_ISL_2099259, EPI_ISL_2099260, EPI_ISL_2099261, EPI_ISL_2099262, EPI_ISL_2099263, EPI_ISL_2099264, EPI_ISL_2099265, EPI_ISL_2099266, EPI_ISL_2099267, EPI_ISL_2099268, EPI_ISL_2099269, EPI_ISL_2099270, EPI_ISL_2099271, EPI_ISL_2099272, EPI_ISL_2099273, EPI_ISL_2099274, EPI_ISL_2099275, EPI_ISL_2099276, EPI_ISL_2099277, EPI_ISL_2099278, EPI_ISL_2099279, EPI_ISL_2099280, EPI_ISL_2099281, EPI_ISL_2099282, EPI_ISL_2099283, EPI_ISL_2099284, EPI_ISL_2099285, EPI_ISL_2099286, EPI_ISL_2099287, EPI_ISL_2099288, EPI_ISL_2099771 | | | |
| see above | Michael E. DeBakey VA Medical Center | Pathogen Discovery, Respiratory Viruses Branch, Division of Viral Diseases, Centers for Disease Control and Prevention | Ying Tao, Yan Li, Jing Zhang, Anna Kelleher, Brian Lynch, Krista Queen, Anna Uehara, Peter Cook, Han Jia Justin Ng, Rachel Marine, Clinton R. Paden, Dhwani Batra, Haibin Wang, Tara Coalter, Jasmine Padilla, Morgan Davis, Mili Sheth, Sarah Nobles, Mark Burroughs, Justin Lee, Adam Retchless, Suxiang Tong |
| EPI_ISL_2100134 | Tampa General Hospital Esoteric Lab | Tampa General Hospital Esoteric Research & Development Lab | Grant Vestal, Deanna Becker, Dominic Uy, Vicki Healer, Amorce Lima, Suzane Silbert |
| EPI_ISL_2100150, EPI_ISL_2100151, EPI_ISL_2100152, EPI_ISL_2100153, EPI_ISL_2100154, EPI_ISL_2100155, EPI_ISL_2100156, EPI_ISL_2100157, EPI_ISL_2100158, EPI_ISL_2100160, EPI_ISL_2100161, EPI_ISL_2100162, EPI_ISL_2100163, EPI_ISL_2100164, EPI_ISL_2100165, EPI_ISL_2100166, EPI_ISL_2100168, EPI_ISL_2100169, EPI_ISL_2100170, EPI_ISL_2100171, EPI_ISL_2100172, EPI_ISL_2100173, EPI_ISL_2100174, EPI_ISL_2100175, EPI_ISL_2100176, EPI_ISL_2100177, EPI_ISL_2100178, EPI_ISL_2100179, EPI_ISL_2100180, EPI_ISL_2100181, EPI_ISL_2100182, EPI_ISL_2100183, EPI_ISL_2100184 | | | |
| see above | DC Public Health Lab/ Dept. of Forensic Sciences | DC Public Health Lab/ Dept. of Forensic Sciences | Janis Doss, Scott Nguyen, Elizabeth Zelaya, Sarah Scott, Connie Maza, Monica Mann, Brittany Hamilton, David Payne, Jocelyn Hauser |
| EPI_ISL_2100224, EPI_ISL_2100225 | NCSLPH | NCSLPH | Chase K, Miller MC, Greene S, Glover W |
| EPI_ISL_2100262, EPI_ISL_2100264, EPI_ISL_2100265, EPI_ISL_2100266, EPI_ISL_2100267, EPI_ISL_2100268 | LSUHS Emerging Viral Threat Laboratory | LSUHS Emerging Viral Threat Laboratory | Gregory L. Ware, Alexander Mijalis, Jeremy P. Kamil, Jennifer L. Carroll, Maarten Van Diest, Rona S. Scott, Andrew D. Yurochko, Christopher G. Kevil, John A. Vanchiere |
| EPI_ISL_2100269, EPI_ISL_2100272, EPI_ISL_2100273 | Atlanta VA Medical Center | Pathogen Discovery, Respiratory Viruses Branch, Division of Viral Diseases, Centers for Disease Control and Prevention | Ying Tao, Yan Li, Jing Zhang, Anna Kelleher, Brian Lynch, Krista Queen, Anna Uehara, Peter Cook, Han Jia Justin Ng, Rachel Marine, Clinton R. Paden, Dhwani Batra, Haibin Wang, Tara Coalter, Jasmine Padilla, Morgan Davis, Mili Sheth, Sarah Nobles, Mark Burroughs, Justin Lee, Adam Retchless, Suxiang Tong |
| EPI_ISL_2100351, EPI_ISL_2100352, EPI_ISL_2100353, EPI_ISL_2100354, EPI_ISL_2100355, EPI_ISL_2100356, EPI_ISL_2100357, EPI_ISL_2100358, EPI_ISL_2100359, EPI_ISL_2100360, EPI_ISL_2100361, EPI_ISL_2100362, EPI_ISL_2100363, EPI_ISL_2100365, EPI_ISL_2100366 | | | |
| see above | Wyoming Public Health Laboratory | Wyoming Public Health Laboratory | Jim Mildenberger, Taylor Fearing, Channing Weber, Ashley Norberg, Chayse Rowley, Marley Goetz, Brian Dominguez, Elliot Thomasson, Brittany Oher, Sam Britz, Cari Sloma, and Rob Christensen |
| EPI_ISL_2100367, EPI_ISL_2100368, EPI_ISL_2100369, EPI_ISL_2100370, EPI_ISL_2100371, EPI_ISL_2100372, EPI_ISL_2100373, EPI_ISL_2100374, EPI_ISL_2100375, EPI_ISL_2100376, EPI_ISL_2100377, EPI_ISL_2100378, EPI_ISL_2100379, EPI_ISL_2100380, EPI_ISL_2100381, EPI_ISL_2100382, EPI_ISL_2100384, EPI_ISL_2100385, EPI_ISL_2100386, EPI_ISL_2100387, EPI_ISL_2100389, EPI_ISL_2100390, EPI_ISL_2100391, EPI_ISL_2100392, EPI_ISL_2100393, EPI_ISL_2100395, EPI_ISL_2100396, EPI_ISL_2100397, EPI_ISL_2100398, EPI_ISL_2100399, EPI_ISL_2100400, EPI_ISL_2100401, EPI_ISL_2100403, EPI_ISL_2100404, EPI_ISL_2100405 | | | |
| see above | University of Michigan Clinical Microbiology Laboratory | Lauring Lab, University of Michigan, Department of Microbiology and Immunology | Valesano |
| EPI_ISL_2100406, EPI_ISL_2100407, EPI_ISL_2100408, EPI_ISL_2100409, EPI_ISL_2100410, EPI_ISL_2100411, EPI_ISL_2100412, EPI_ISL_2100413, EPI_ISL_2100415, EPI_ISL_2100416, EPI_ISL_2100417, EPI_ISL_2100418, EPI_ISL_2100419, EPI_ISL_2100420, EPI_ISL_2100421, EPI_ISL_2100422 | | | |
| see above | Wyoming Public Health Laboratory | Wyoming Public Health Laboratory | Jim Mildenberger, Taylor Fearing, Channing Weber, Ashley Norberg, Chayse Rowley, Marley Goetz, Brian Dominguez, Elliot Thomasson, Brittany Oher, Sam Britz, Cari Sloma, and Rob Christensen |
| EPI_ISL_2100511, EPI_ISL_2100512, EPI_ISL_2100514, EPI_ISL_2100515, EPI_ISL_2100516, EPI_ISL_2100517, EPI_ISL_2100518, EPI_ISL_2100519, EPI_ISL_2100520, EPI_ISL_2100521, EPI_ISL_2100522, EPI_ISL_2100524, EPI_ISL_2100525, EPI_ISL_2100526, EPI_ISL_2100527, EPI_ISL_2100528, EPI_ISL_2100529, EPI_ISL_2100530, EPI_ISL_2100531, EPI_ISL_2100532, EPI_ISL_2100533, EPI_ISL_2100534, EPI_ISL_2100535, EPI_ISL_2100536, EPI_ISL_2100537, EPI_ISL_2100538, EPI_ISL_2100539, EPI_ISL_2100540, EPI_ISL_2100541, EPI_ISL_2100542, EPI_ISL_2100543, EPI_ISL_2100544, EPI_ISL_2100545, EPI_ISL_2100546, EPI_ISL_2100547, EPI_ISL_2100548, EPI_ISL_2100549, EPI_ISL_2100550, EPI_ISL_2100551, EPI_ISL_2100552, EPI_ISL_2100553, EPI_ISL_2100554, EPI_ISL_2100555, EPI_ISL_2100556, EPI_ISL_2100557, EPI_ISL_2100558, EPI_ISL_2100559, EPI_ISL_2100560, EPI_ISL_2100561, EPI_ISL_2100562, EPI_ISL_2100563, EPI_ISL_2100564, EPI_ISL_2100565, EPI_ISL_2100566, EPI_ISL_2100567, EPI_ISL_2100568, EPI_ISL_2100569, EPI_ISL_2100570, EPI_ISL_2100571, EPI_ISL_2100572, EPI_ISL_2100573, EPI_ISL_2100574, EPI_ISL_2100575, EPI_ISL_2100576, EPI_ISL_2100577, EPI_ISL_2100578, EPI_ISL_2100579, EPI_ISL_2100580, EPI_ISL_2100581, EPI_ISL_2100582, EPI_ISL_2100583, EPI_ISL_2100584, EPI_ISL_2100585, EPI_ISL_2100586, EPI_ISL_2100587, EPI_ISL_2100588, EPI_ISL_2100589, EPI_ISL_2100590, EPI_ISL_2100591, EPI_ISL_2100592, EPI_ISL_2100593, EPI_ISL_2100594, EPI_ISL_2100595, EPI_ISL_2100596, EPI_ISL_2100597, EPI_ISL_2100598, EPI_ISL_2100599, EPI_ISL_2100600, EPI_ISL_2100601, EPI_ISL_2100602, EPI_ISL_2100603, EPI_ISL_2100604, EPI_ISL_2100605, EPI_ISL_2100606, EPI_ISL_2100607, EPI_ISL_2100608, EPI_ISL_2100609, EPI_ISL_2100610, EPI_ISL_2100611, EPI_ISL_2100612, EPI_ISL_2100613, EPI_ISL_2100614, EPI_ISL_2100615, EPI_ISL_2100616, EPI_ISL_2100617, EPI_ISL_2100618, EPI_ISL_2100619, EPI_ISL_2100620, EPI_ISL_2100621, EPI_ISL_2100622, EPI_ISL_2100623, EPI_ISL_2100624, EPI_ISL_2100625, EPI_ISL_2100626, EPI_ISL_2100628, EPI_ISL_2100629, EPI_ISL_2100630, EPI_ISL_2100631, EPI_ISL_2100632, EPI_ISL_2100633 | | | |
| see above | Pathology and Laboratory Medicine Institute, Cleveland Clinic, Ohio, USA | Pathology and Laboratory Medicine Institute, Cleveland Clinic, Ohio, USA | Jennifer Starbuck, Joy Nakitandwe, Kristen McDonnell, David Plunkett, Zheng Jin Tu, Jay Brock, Yu-Wei Cheng, Gary Procop, Daniel Rhoads, Daniel H. Farkas, David Bosler |
| EPI_ISL_2100701, EPI_ISL_2100702, EPI_ISL_2100703, EPI_ISL_2100704, EPI_ISL_2100705, EPI_ISL_2100706, EPI_ISL_2100707, EPI_ISL_2100708, EPI_ISL_2100709, EPI_ISL_2100710, EPI_ISL_2100711, EPI_ISL_2100712, EPI_ISL_2100713, EPI_ISL_2100714, EPI_ISL_2100715, EPI_ISL_2100716, EPI_ISL_2100717, EPI_ISL_2100718, EPI_ISL_2100719, EPI_ISL_2100720, EPI_ISL_2100721, EPI_ISL_2100722, EPI_ISL_2100723, EPI_ISL_2100724, EPI_ISL_2100725, EPI_ISL_2100726, EPI_ISL_2100727, EPI_ISL_2100728, EPI_ISL_2100729, EPI_ISL_2100730, EPI_ISL_2100731, EPI_ISL_2100732, EPI_ISL_2100733, EPI_ISL_2100734, EPI_ISL_2100735, EPI_ISL_2100736, EPI_ISL_2100737, EPI_ISL_2100738, EPI_ISL_2100739, EPI_ISL_2100740, EPI_ISL_2100741, EPI_ISL_2100742, EPI_ISL_2100743, EPI_ISL_2100744, EPI_ISL_2100745, EPI_ISL_2100746, EPI_ISL_2100747, EPI_ISL_2100748, EPI_ISL_2100749, EPI_ISL_2100750, EPI_ISL_2100751, EPI_ISL_2100752, EPI_ISL_2100753, EPI_ISL_2100754, EPI_ISL_2100755, EPI_ISL_2100756, EPI_ISL_2100757, EPI_ISL_2100758, EPI_ISL_2100759, EPI_ISL_2100760, EPI_ISL_2100761, EPI_ISL_2100762, EPI_ISL_2100763, EPI_ISL_2100764, EPI_ISL_2100765, EPI_ISL_2100766, EPI_ISL_2100767, EPI_ISL_2100768, EPI_ISL_2100769, EPI_ISL_2100770, EPI_ISL_2100771, EPI_ISL_2100772, EPI_ISL_2100773, EPI_ISL_2100774, EPI_ISL_2100775, EPI_ISL_2100776, EPI_ISL_2100777, EPI_ISL_2100778, EPI_ISL_2100779, EPI_ISL_2100780, EPI_ISL_2100781, EPI_ISL_2100782, EPI_ISL_2100783, EPI_ISL_2100784, EPI_ISL_2100785, EPI_ISL_2100786, EPI_ISL_2100788, EPI_ISL_2100789, EPI_ISL_2100790, EPI_ISL_2100791, EPI_ISL_2100792, EPI_ISL_2100793, EPI_ISL_2100794, EPI_ISL_2100795, EPI_ISL_2100796, EPI_ISL_2100797, EPI_ISL_2100798, EPI_ISL_2100799, EPI_ISL_2100800, EPI_ISL_2100801, EPI_ISL_2100802, EPI_ISL_2100803, EPI_ISL_2100804, EPI_ISL_2100805, EPI_ISL_2100806, EPI_ISL_2100807, EPI_ISL_2100808, EPI_ISL_2100809, EPI_ISL_2100810, EPI_ISL_2100811, EPI_ISL_2100812, EPI_ISL_2100813, EPI_ISL_2100814, EPI_ISL_2100816, EPI_ISL_2100818, EPI_ISL_2100819, EPI_ISL_2100820, EPI_ISL_2100821, EPI_ISL_2100822, EPI_ISL_2100823, EPI_ISL_2100824, EPI_ISL_2100825, EPI_ISL_2100826, EPI_ISL_2100827, EPI_ISL_2100828, EPI_ISL_2100829, EPI_ISL_2100830, EPI_ISL_2100831, EPI_ISL_2100832, EPI_ISL_2100833, EPI_ISL_2100834, EPI_ISL_2100835, EPI_ISL_2100836, EPI_ISL_2100837, EPI_ISL_2100838, EPI_ISL_2100839, EPI_ISL_2100840, EPI_ISL_2100841, EPI_ISL_2100842, EPI_ISL_2100843, EPI_ISL_2100844, EPI_ISL_2100845, EPI_ISL_2100846, EPI_ISL_2100847, EPI_ISL_2100848, EPI_ISL_2100849, EPI_ISL_2100850, EPI_ISL_2100851, EPI_ISL_2100852, EPI_ISL_2100853, EPI_ISL_2100854, EPI_ISL_2100855, EPI_ISL_2100856, EPI_ISL_2100857, EPI_ISL_2100858, EPI_ISL_2100859, EPI_ISL_2100860, EPI_ISL_2100861, EPI_ISL_2100862, EPI_ISL_2100863, EPI_ISL_2100864, EPI_ISL_2100865, EPI_ISL_2100866, EPI_ISL_2100867, EPI_ISL_2100868, EPI_ISL_2100869, EPI_ISL_2100871, EPI_ISL_2100872, EPI_ISL_2100873, EPI_ISL_2100874, EPI_ISL_2100875, EPI_ISL_2100876, EPI_ISL_2100877, EPI_ISL_2100878, EPI_ISL_2100879, EPI_ISL_2100882, EPI_ISL_2100883, EPI_ISL_2100884, EPI_ISL_2100885, EPI_ISL_2100886, EPI_ISL_2100887, EPI_ISL_2100888, EPI_ISL_2100889, EPI_ISL_2100890, EPI_ISL_2100891, EPI_ISL_2100892, EPI_ISL_2100893, EPI_ISL_2100894, EPI_ISL_2100895, EPI_ISL_2100896, EPI_ISL_2100897, EPI_ISL_2100898, EPI_ISL_2100899, EPI_ISL_2100900, EPI_ISL_2100901, EPI_ISL_2100902, EPI_ISL_2100903, EPI_ISL_2100905, EPI_ISL_2100906, EPI_ISL_2100907, EPI_ISL_2100908, EPI_ISL_2100909, EPI_ISL_2100910, EPI_ISL_2100911, EPI_ISL_2100912, EPI_ISL_2100913, EPI_ISL_2100914, EPI_ISL_2100915, EPI_ISL_2100916, EPI_ISL_2100917, EPI_ISL_2100918, EPI_ISL_2100919, EPI_ISL_2100920, EPI_ISL_2100921, EPI_ISL_2100922, EPI_ISL_2100923, EPI_ISL_2100924, EPI_ISL_2100925, EPI_ISL_2100926, EPI_ISL_2100927, EPI_ISL_2100928, EPI_ISL_2100929, EPI_ISL_2100930, EPI_ISL_2100931, EPI_ISL_2100932, EPI_ISL_2100933, EPI_ISL_2100934, EPI_ISL_2100935, EPI_ISL_2100936, EPI_ISL_2100937, EPI_ISL_2100938, EPI_ISL_2100939, EPI_ISL_2100940, EPI_ISL_2100941, EPI_ISL_2100942, EPI_ISL_2100943, EPI_ISL_2100944, EPI_ISL_2100945, EPI_ISL_2100946, EPI_ISL_2100947, EPI_ISL_2100948, EPI_ISL_2100949, EPI_ISL_2100950, EPI_ISL_2100951, EPI_ISL_2100952, EPI_ISL_2100953, EPI_ISL_2100954, EPI_ISL_2100955, EPI_ISL_2100956, EPI_ISL_2100957, EPI_ISL_2100958, EPI_ISL_2100960, EPI_ISL_2100961, EPI_ISL_2100962, EPI_ISL_2100963, EPI_ISL_2100964, EPI_ISL_2100965, EPI_ISL_2100966, EPI_ISL_2100968 | | | |
| see above | The Ohio State University Applied Microbiology Services Laboratory | The Ohio State University Applied Microbiology Services Laboratory | Seth A. Faith PhD |
| EPI_ISL_2100972, EPI_ISL_2100973, EPI_ISL_2100974, EPI_ISL_2100975, EPI_ISL_2100976, EPI_ISL_2100977, EPI_ISL_2100978, EPI_ISL_2100979, EPI_ISL_2100980, EPI_ISL_2100981, EPI_ISL_2100982, EPI_ISL_2100983, EPI_ISL_2100984, EPI_ISL_2100985, EPI_ISL_2100986, EPI_ISL_2100987, EPI_ISL_2100988, EPI_ISL_2100989, EPI_ISL_2100990, EPI_ISL_2100991, EPI_ISL_2100992, EPI_ISL_2100993, EPI_ISL_2100994, EPI_ISL_2100995, EPI_ISL_2100996, EPI_ISL_2100997, EPI_ISL_2100998, EPI_ISL_2100999, EPI_ISL_2101000, EPI_ISL_2101001, EPI_ISL_2101002, EPI_ISL_2101003, EPI_ISL_2101004, EPI_ISL_2101005, EPI_ISL_2101006, EPI_ISL_2101007, EPI_ISL_2101008, EPI_ISL_2101009, EPI_ISL_2101010, EPI_ISL_2101011, EPI_ISL_2101012, EPI_ISL_2101013, EPI_ISL_2101014, EPI_ISL_2101015 | | | |
| see above | Maine HETL | Health and Environmental Testing Laboratory | Matluk,N., Dewey,H., Martha,J., Meak,S., Grieser,H.,Barter,M., Lynch,R., Munger,H. and Tewhey,R. |
| EPI_ISL_2101359, EPI_ISL_2101360, EPI_ISL_2101361, EPI_ISL_2101362, EPI_ISL_2101364 | Wichita State University - Molecular Diagnostics Lab | Kansas Health and Environmental Lab | Mike Grose, Jonathan Barnell, Ben Olsen, and Phil Adam |
| EPI_ISL_2101365, EPI_ISL_2101366, EPI_ISL_2101367, EPI_ISL_2101368, EPI_ISL_2101370 | Kansas Health and Environmental Lab | Kansas Health and Environmental Lab | Mike Grose, Jonathan Barnell, Ben Olsen, and Phil Adam |
| EPI_ISL_2101371 | Wichita State University - Molecular Diagnostics Lab | Kansas Health and Environmental Lab | Mike Grose, Jonathan Barnell, Ben Olsen, and Phil Adam |
| EPI_ISL_2101373, EPI_ISL_2101374, EPI_ISL_2101375, EPI_ISL_2101376, EPI_ISL_2101377, EPI_ISL_2101378, EPI_ISL_2101379 | Kansas Health and Environmental Lab | Kansas Health and Environmental Lab | Mike Grose, Jonathan Barnell, Ben Olsen, and Phil Adam |
| EPI_ISL_2101380 | Clinical Reference Laboratory | Kansas Health and Environmental Lab | Mike Grose, Jonathan Barnell, Ben Olsen, and Phil Adam |
| EPI_ISL_2101381, EPI_ISL_2101382 | Hackensack Medical Center | New York Genome Center | Michael Zody, Andre Corvelo, Dayna M. Oschwald, Samantha Fennessey, Tom Maniatis, Liang Chen, Jose Mediavilla, Marcus Cunningham, Kaelea Composto, Kar Chow, David Perlin, Barry Kreiswirth |
| EPI_ISL_2101749, EPI_ISL_2101750, EPI_ISL_2101751, EPI_ISL_2101752, EPI_ISL_2101753, EPI_ISL_2101754, EPI_ISL_2101755, EPI_ISL_2101756, EPI_ISL_2101757, EPI_ISL_2101758, EPI_ISL_2101759, EPI_ISL_2101760, EPI_ISL_2101761, EPI_ISL_2101762, EPI_ISL_2101763, EPI_ISL_2101764, EPI_ISL_2101765, EPI_ISL_2101766, EPI_ISL_2101767, EPI_ISL_2101768, EPI_ISL_2101769, EPI_ISL_2101770, EPI_ISL_2101771, EPI_ISL_2101772, EPI_ISL_2101773, EPI_ISL_2101774, EPI_ISL_2101775, EPI_ISL_2101776, EPI_ISL_2101777, EPI_ISL_2101778 | | | |
| see above | Alaska State Virology Laboratory | Alaska State Virology Laboratory | Stephanie DeRonde, Elva House, Jacob Zidek, Lisa Smith, Ph.D., Jack Chen, Ph.D. |
| EPI_ISL_2101779, EPI_ISL_2101780, EPI_ISL_2101781, EPI_ISL_2101782 | WLA VAMC | Pathogen Discovery, Respiratory Viruses Branch, Division of Viral Diseases, Centers for Disease Control and Prevention | Ying Tao, Yan Li, Jing Zhang, Anna Kelleher, Brian Lynch, Krista Queen, Anna Uehara, Peter Cook, Han Jia Justin Ng, Rachel Marine, Clinton R. Paden, Dhwani Batra, Haibin Wang, Tara Coalter, Jasmine Padilla, Morgan Davis, Mili Sheth, Sarah Nobles, Mark Burroughs, Justin Lee, Adam Retchless, Suxiang Tong |
| EPI_ISL_2101783, EPI_ISL_2101784, EPI_ISL_2101785 | Michael E. DeBakey VA Medical Center | Pathogen Discovery, Respiratory Viruses Branch, Division of Viral Diseases, Centers for Disease Control and Prevention | Ying Tao, Yan Li, Jing Zhang, Anna Kelleher, Brian Lynch, Krista Queen, Anna Uehara, Peter Cook, Han Jia Justin Ng, Rachel Marine, Clinton R. Paden, Dhwani Batra, Haibin Wang, Tara Coalter, Jasmine Padilla, Morgan Davis, Mili Sheth, Sarah Nobles, Mark Burroughs, Justin Lee, Adam Retchless, Suxiang Tong |
| EPI_ISL_2101786 | WLA VAMC | Pathogen Discovery, Respiratory Viruses | Ying Tao, Yan Li, Jing Zhang, Anna Kelleher, Brian Lynch, Krista Queen, Anna Uehara, Peter Cook, Han Jia Justin Ng, Rachel Marine, Clinton R. Paden, Dhwani Batra, Haibin Wang, Tara Coalter, Jasmine Padilla, Morgan Davis, Mili Sheth, Sarah Nobles, Mark Burroughs, Justin Lee, Adam |

| | | | |
|---|---|---|---|
| | | Branch, Division of Viral Diseases, Centers for Disease Control and Prevention | Retchless, Suxiang Tong |
| EPI_ISL_2101787 | Michael E. DeBakey VA Medical Center | Pathogen Discovery, Respiratory Viruses Branch, Division of Viral Diseases, Centers for Disease Control and Prevention | Ying Tao, Yan Li, Jing Zhang, Anna Kelleher, Brian Lynch, Krista Queen, Anna Uehara, Peter Cook, Han Jia Justin Ng, Rachel Marine, Clinton R. Paden, Dhwani Batra, Haibin Wang, Tara Coalter, Jasmine Padilla, Morgan Davis, Mili Sheth, Sarah Nobles, Mark Burroughs, Justin Lee, Adam Retchless, Suxiang Tong |
| EPI_ISL_2101788, EPI_ISL_2101789, EPI_ISL_2101790 | Bronx Veterans Medical Research Foundation | Pathogen Discovery, Respiratory Viruses Branch, Division of Viral Diseases, Centers for Disease Control and Prevention | Ying Tao, Yan Li, Jing Zhang, Anna Kelleher, Brian Lynch, Krista Queen, Anna Uehara, Peter Cook, Han Jia Justin Ng, Rachel Marine, Clinton R. Paden, Dhwani Batra, Haibin Wang, Tara Coalter, Jasmine Padilla, Morgan Davis, Mili Sheth, Sarah Nobles, Mark Burroughs, Justin Lee, Adam Retchless, Suxiang Tong |
| EPI_ISL_2101791, EPI_ISL_2101792, EPI_ISL_2101793, EPI_ISL_2101794, EPI_ISL_2101795, EPI_ISL_2101796, EPI_ISL_2101797, EPI_ISL_2101798 | Michael E. DeBakey VA Medical Center | Pathogen Discovery, Respiratory Viruses Branch, Division of Viral Diseases, Centers for Disease Control and Prevention | Ying Tao, Yan Li, Jing Zhang, Anna Kelleher, Brian Lynch, Krista Queen, Anna Uehara, Peter Cook, Han Jia Justin Ng, Rachel Marine, Clinton R. Paden, Dhwani Batra, Haibin Wang, Tara Coalter, Jasmine Padilla, Morgan Davis, Mili Sheth, Sarah Nobles, Mark Burroughs, Justin Lee, Adam Retchless, Suxiang Tong |
| EPI_ISL_2101800 | Bronx Veterans Medical Research Foundation | Pathogen Discovery, Respiratory Viruses Branch, Division of Viral Diseases, Centers for Disease Control and Prevention | Ying Tao, Yan Li, Jing Zhang, Anna Kelleher, Brian Lynch, Krista Queen, Anna Uehara, Peter Cook, Han Jia Justin Ng, Rachel Marine, Clinton R. Paden, Dhwani Batra, Haibin Wang, Tara Coalter, Jasmine Padilla, Morgan Davis, Mili Sheth, Sarah Nobles, Mark Burroughs, Justin Lee, Adam Retchless, Suxiang Tong |
| EPI_ISL_2101801 | Michael E. DeBakey VA Medical Center | Pathogen Discovery, Respiratory Viruses Branch, Division of Viral Diseases, Centers for Disease Control and Prevention | Ying Tao, Yan Li, Jing Zhang, Anna Kelleher, Brian Lynch, Krista Queen, Anna Uehara, Peter Cook, Han Jia Justin Ng, Rachel Marine, Clinton R. Paden, Dhwani Batra, Haibin Wang, Tara Coalter, Jasmine Padilla, Morgan Davis, Mili Sheth, Sarah Nobles, Mark Burroughs, Justin Lee, Adam Retchless, Suxiang Tong |
| EPI_ISL_2101802, EPI_ISL_2101803, EPI_ISL_2101804, EPI_ISL_2101805, EPI_ISL_2101806, EPI_ISL_2101807, EPI_ISL_2101808, EPI_ISL_2101809, EPI_ISL_2101810, EPI_ISL_2101811, EPI_ISL_2101812, EPI_ISL_2101813, EPI_ISL_2101814, EPI_ISL_2101815, EPI_ISL_2101816, EPI_ISL_2101817, EPI_ISL_2101818, EPI_ISL_2101819, EPI_ISL_2101820, EPI_ISL_2101821, EPI_ISL_2101822, EPI_ISL_2101823, EPI_ISL_2101824, EPI_ISL_2101825, EPI_ISL_2101826, EPI_ISL_2101827, EPI_ISL_2101828, EPI_ISL_2101829, EPI_ISL_2101830, EPI_ISL_2101831, EPI_ISL_2101832, EPI_ISL_2101833, EPI_ISL_2101834, EPI_ISL_2101835, EPI_ISL_2101836, EPI_ISL_2101837, EPI_ISL_2101838, EPI_ISL_2101839, EPI_ISL_2101840, EPI_ISL_2101841, EPI_ISL_2101842, EPI_ISL_2101843, EPI_ISL_2101844, EPI_ISL_2101845, EPI_ISL_2101846, EPI_ISL_2101847, EPI_ISL_2101848, EPI_ISL_2101849, EPI_ISL_2101850, EPI_ISL_2101851, EPI_ISL_2101852, EPI_ISL_2101853, EPI_ISL_2101854, EPI_ISL_2101855, EPI_ISL_2101856, EPI_ISL_2101857, EPI_ISL_2101858, EPI_ISL_2101859, EPI_ISL_2101860, EPI_ISL_2101861, EPI_ISL_2101862, EPI_ISL_2101863, EPI_ISL_2101864, EPI_ISL_2101865, EPI_ISL_2101866, EPI_ISL_2101867, EPI_ISL_2101868 | | | |
| see above | WLA VAMC | Pathogen Discovery, Respiratory Viruses Branch, Division of Viral Diseases, Centers for Disease Control and Prevention | Ying Tao, Yan Li, Jing Zhang, Anna Kelleher, Brian Lynch, Krista Queen, Anna Uehara, Peter Cook, Han Jia Justin Ng, Rachel Marine, Clinton R. Paden, Dhwani Batra, Haibin Wang, Tara Coalter, Jasmine Padilla, Morgan Davis, Mili Sheth, Sarah Nobles, Mark Burroughs, Justin Lee, Adam Retchless, Suxiang Tong |
| EPI_ISL_2101869 | LESP Jalisco | Instituto de Diagnostico y Referencia Epidemiologicos (INDRE) | Claudia Wong-Arambula, Abril Rodriguez-Maldonado, Vanessa Rivero-Arredondo, Ariadna Medina-Benitez, Joaquin Quiroz-Mercado, Sergio Rangel-Guerrero, Natividad Cruz-Ortiz, Tatiana Nunez-Garcia, Gisela Barrera-Badillo, Lucia Hernandez-Rivas, Irma Lopez-Martinez, Ernesto Ramirez-Gonzalez. |
| EPI_ISL_2101870 | LESP Morelos | Instituto de Diagnostico y Referencia Epidemiologicos (INDRE) | Claudia Wong-Arambula, Abril Rodriguez-Maldonado, Vanessa Rivero-Arredondo, Ariadna Medina-Benitez, Joaquin Quiroz-Mercado, Sergio Rangel-Guerrero, Natividad Cruz-Ortiz, Tatiana Nunez-Garcia, Gisela Barrera-Badillo, Lucia Hernandez-Rivas, Irma Lopez-Martinez, Ernesto Ramirez-Gonzalez. |
| EPI_ISL_2101871, EPI_ISL_2101872, EPI_ISL_2101873, EPI_ISL_2101874, EPI_ISL_2101875, EPI_ISL_2101876, EPI_ISL_2101877, EPI_ISL_2101878 | LESP Chihuahua | Instituto de Diagnostico y Referencia Epidemiologicos (INDRE) | Claudia Wong-Arambula, Abril Rodriguez-Maldonado, Vanessa Rivero-Arredondo, Ariadna Medina-Benitez, Joaquin Quiroz-Mercado, Sergio Rangel-Guerrero, Natividad Cruz-Ortiz, Tatiana Nunez-Garcia, Gisela Barrera-Badillo, Lucia Hernandez-Rivas, Irma Lopez-Martinez, Ernesto Ramirez-Gonzalez. |
| EPI_ISL_2101879 | LESP Quintana Roo | Instituto de Diagnostico y Referencia Epidemiologicos (INDRE) | Claudia Wong-Arambula, Abril Rodriguez-Maldonado, Vanessa Rivero-Arredondo, Ariadna Medina-Benitez, Joaquin Quiroz-Mercado, Sergio Rangel-Guerrero, Natividad Cruz-Ortiz, Tatiana Nunez-Garcia, Gisela Barrera-Badillo, Lucia Hernandez-Rivas, Irma Lopez-Martinez, Ernesto Ramirez-Gonzalez. |
| EPI_ISL_2101880 | LESP Baja California | Instituto de Diagnostico y Referencia Epidemiologicos (INDRE) | Claudia Wong-Arambula, Abril Rodriguez-Maldonado, Vanessa Rivero-Arredondo, Ariadna Medina-Benitez, Joaquin Quiroz-Mercado, Sergio Rangel-Guerrero, Natividad Cruz-Ortiz, Tatiana Nunez-Garcia, Gisela Barrera-Badillo, Lucia Hernandez-Rivas, Irma Lopez-Martinez, Ernesto Ramirez-Gonzalez. |
| EPI_ISL_2101881, EPI_ISL_2101882, EPI_ISL_2101883, EPI_ISL_2101884, EPI_ISL_2101885, EPI_ISL_2101886, EPI_ISL_2101887, EPI_ISL_2101888, EPI_ISL_2101889, EPI_ISL_2101890 | LESP Chihuahua | Instituto de Diagnostico y Referencia Epidemiologicos (INDRE) | Claudia Wong-Arambula, Abril Rodriguez-Maldonado, Vanessa Rivero-Arredondo, Ariadna Medina-Benitez, Joaquin Quiroz-Mercado, Sergio Rangel-Guerrero, Natividad Cruz-Ortiz, Tatiana Nunez-Garcia, Gisela Barrera-Badillo, Lucia Hernandez-Rivas, Irma Lopez-Martinez, Ernesto Ramirez-Gonzalez. |
| EPI_ISL_2101891 | LESP Jalisco | Instituto de Diagnostico y Referencia Epidemiologicos (INDRE) | Claudia Wong-Arambula, Abril Rodriguez-Maldonado, Vanessa Rivero-Arredondo, Ariadna Medina-Benitez, Joaquin Quiroz-Mercado, Sergio Rangel-Guerrero, Natividad Cruz-Ortiz, Tatiana Nunez-Garcia, Gisela Barrera-Badillo, Lucia Hernandez-Rivas, Irma Lopez-Martinez, Ernesto Ramirez-Gonzalez. |
| EPI_ISL_2101892 | LESP Veracruz | Instituto de Diagnostico y Referencia Epidemiologicos (INDRE) | Claudia Wong-Arambula, Abril Rodriguez-Maldonado, Vanessa Rivero-Arredondo, Ariadna Medina-Benitez, Joaquin Quiroz-Mercado, Sergio Rangel-Guerrero, Natividad Cruz-Ortiz, Tatiana Nunez-Garcia, Gisela Barrera-Badillo, Lucia Hernandez-Rivas, Irma Lopez-Martinez, Ernesto Ramirez-Gonzalez. |
| EPI_ISL_2101893, EPI_ISL_2101894 | LESP Guanajuato | Instituto de Diagnostico y Referencia Epidemiologicos (INDRE) | Claudia Wong-Arambula, Abril Rodriguez-Maldonado, Vanessa Rivero-Arredondo, Ariadna Medina-Benitez, Joaquin Quiroz-Mercado, Sergio Rangel-Guerrero, Natividad Cruz-Ortiz, Tatiana Nunez-Garcia, Gisela Barrera-Badillo, Lucia Hernandez-Rivas, Irma Lopez-Martinez, Ernesto Ramirez-Gonzalez. |
| EPI_ISL_2101895 | LESP Jalisco | Instituto de Diagnostico y Referencia Epidemiologicos (INDRE) | Claudia Wong-Arambula, Abril Rodriguez-Maldonado, Vanessa Rivero-Arredondo, Ariadna Medina-Benitez, Joaquin Quiroz-Mercado, Sergio Rangel-Guerrero, Natividad Cruz-Ortiz, Tatiana Nunez-Garcia, Gisela Barrera-Badillo, Lucia Hernandez-Rivas, Irma Lopez-Martinez, Ernesto Ramirez-Gonzalez. |
| EPI_ISL_2101896 | LESP Sinaloa | Instituto de Diagnostico y Referencia Epidemiologicos (INDRE) | Claudia Wong-Arambula, Abril Rodriguez-Maldonado, Vanessa Rivero-Arredondo, Ariadna Medina-Benitez, Joaquin Quiroz-Mercado, Sergio Rangel-Guerrero, Natividad Cruz-Ortiz, Tatiana Nunez-Garcia, Gisela Barrera-Badillo, Lucia Hernandez-Rivas, Irma Lopez-Martinez, Ernesto Ramirez-Gonzalez. |
| EPI_ISL_2101897 | LESP Sonora | Instituto de Diagnostico y Referencia Epidemiologicos (INDRE) | Claudia Wong-Arambula, Abril Rodriguez-Maldonado, Vanessa Rivero-Arredondo, Ariadna Medina-Benitez, Joaquin Quiroz-Mercado, Sergio Rangel-Guerrero, Natividad Cruz-Ortiz, Tatiana Nunez-Garcia, Gisela Barrera-Badillo, Lucia Hernandez-Rivas, Irma Lopez-Martinez, Ernesto Ramirez-Gonzalez. |
| EPI_ISL_2101898, EPI_ISL_2101899 | LESP Quintana Roo | Instituto de Diagnostico y Referencia Epidemiologicos (INDRE) | Claudia Wong-Arambula, Abril Rodriguez-Maldonado, Vanessa Rivero-Arredondo, Ariadna Medina-Benitez, Joaquin Quiroz-Mercado, Sergio Rangel-Guerrero, Natividad Cruz-Ortiz, Tatiana Nunez-Garcia, Gisela Barrera-Badillo, Lucia Hernandez-Rivas, Irma Lopez-Martinez, Ernesto Ramirez-Gonzalez. |
| EPI_ISL_2101900 | LESP Jalisco | Instituto de Diagnostico y Referencia Epidemiologicos (INDRE) | Claudia Wong-Arambula, Abril Rodriguez-Maldonado, Vanessa Rivero-Arredondo, Ariadna Medina-Benitez, Joaquin Quiroz-Mercado, Sergio Rangel-Guerrero, Natividad Cruz-Ortiz, Tatiana Nunez-Garcia, Gisela Barrera-Badillo, Lucia Hernandez-Rivas, Irma Lopez-Martinez, Ernesto Ramirez-Gonzalez. |
| EPI_ISL_2101901 | LESP Coahuila | Instituto de Diagnostico y Referencia Epidemiologicos (INDRE) | Claudia Wong-Arambula, Abril Rodriguez-Maldonado, Vanessa Rivero-Arredondo, Ariadna Medina-Benitez, Joaquin Quiroz-Mercado, Sergio Rangel-Guerrero, Natividad Cruz-Ortiz, Tatiana Nunez-Garcia, Gisela Barrera-Badillo, Lucia Hernandez-Rivas, Irma Lopez-Martinez, Ernesto Ramirez-Gonzalez. |
| EPI_ISL_2101902 | LESP Jalisco /Unidad de Patologia | Instituto de Diagnostico y Referencia Epidemiologicos (INDRE) | Claudia Wong-Arambula, Abril Rodriguez-Maldonado, Vanessa Rivero-Arredondo, Ariadna Medina-Benitez, Joaquin Quiroz-Mercado, Sergio Rangel-Guerrero, Natividad Cruz-Ortiz, Tatiana Nunez-Garcia, Gisela Barrera-Badillo, Lucia Hernandez-Rivas, Irma Lopez-Martinez, Ernesto Ramirez-Gonzalez. |
| EPI_ISL_2101903, EPI_ISL_2101904 | LESP Chihuahua | Instituto de Diagnostico y Referencia Epidemiologicos (INDRE) | Claudia Wong-Arambula, Abril Rodriguez-Maldonado, Vanessa Rivero-Arredondo, Ariadna Medina-Benitez, Joaquin Quiroz-Mercado, Sergio Rangel-Guerrero, Natividad Cruz-Ortiz, Tatiana Nunez-Garcia, Gisela Barrera-Badillo, Lucia Hernandez-Rivas, Irma Lopez-Martinez, Ernesto Ramirez-Gonzalez. |
| EPI_ISL_2101905 | LESP Jalisco | Instituto de Diagnostico y Referencia Epidemiologicos (INDRE) | Claudia Wong-Arambula, Abril Rodriguez-Maldonado, Vanessa Rivero-Arredondo, Ariadna Medina-Benitez, Joaquin Quiroz-Mercado, Sergio Rangel-Guerrero, Natividad Cruz-Ortiz, Tatiana Nunez-Garcia, Gisela Barrera-Badillo, Lucia Hernandez-Rivas, Irma Lopez-Martinez, Ernesto Ramirez-Gonzalez. |
| EPI_ISL_2101906, EPI_ISL_2101907, EPI_ISL_2101908, EPI_ISL_2101909, EPI_ISL_2101910, EPI_ISL_2101911, EPI_ISL_2101912 | LESP Ciudad de Mexico | Instituto de Diagnostico y Referencia Epidemiologicos (INDRE) | Claudia Wong-Arambula, Abril Rodriguez-Maldonado, Vanessa Rivero-Arredondo, Ariadna Medina-Benitez, Joaquin Quiroz-Mercado, Sergio Rangel-Guerrero, Natividad Cruz-Ortiz, Tatiana Nunez-Garcia, Gisela Barrera-Badillo, Lucia Hernandez-Rivas, Irma Lopez-Martinez, Ernesto Ramirez-Gonzalez. |
| EPI_ISL_2101913, EPI_ISL_2101914, EPI_ISL_2101916, EPI_ISL_2101917, EPI_ISL_2101918, EPI_ISL_2101919, EPI_ISL_2101920, EPI_ISL_2101921, EPI_ISL_2101922, EPI_ISL_2101923, EPI_ISL_2101924, EPI_ISL_2101925, EPI_ISL_2101926, EPI_ISL_2101927, EPI_ISL_2101928, EPI_ISL_2101929, EPI_ISL_2101930, EPI_ISL_2101931, EPI_ISL_2101932, EPI_ISL_2101933, EPI_ISL_2101934, EPI_ISL_2101935, EPI_ISL_2101936, EPI_ISL_2101937, EPI_ISL_2101938, EPI_ISL_2101939, EPI_ISL_2101940, EPI_ISL_2101941, EPI_ISL_2101942, EPI_ISL_2101943, EPI_ISL_2101944, EPI_ISL_2101945, EPI_ISL_2101946, EPI_ISL_2101947, EPI_ISL_2101948, EPI_ISL_2101949, EPI_ISL_2101950, EPI_ISL_2101951, EPI_ISL_2101952, EPI_ISL_2101953, EPI_ISL_2101954, EPI_ISL_2101955, EPI_ISL_2101956, EPI_ISL_2101957, EPI_ISL_2101958, EPI_ISL_2101959, EPI_ISL_2101960, EPI_ISL_2101961, EPI_ISL_2101962, EPI_ISL_2101963, EPI_ISL_2101964, EPI_ISL_2101965, EPI_ISL_2101966, EPI_ISL_2101967, EPI_ISL_2101968, EPI_ISL_2101969, EPI_ISL_2101970, EPI_ISL_2101971, EPI_ISL_2101972, EPI_ISL_2101973, EPI_ISL_2101974, EPI_ISL_2101975, EPI_ISL_2101977, EPI_ISL_2101978, EPI_ISL_2101979, EPI_ISL_2101980, EPI_ISL_2101981, EPI_ISL_2101982, EPI_ISL_2101983, EPI_ISL_2101984, EPI_ISL_2101985, EPI_ISL_2101986, EPI_ISL_2101987, EPI_ISL_2101988, EPI_ISL_2101989, EPI_ISL_2101990, EPI_ISL_2101991 | | | |
| see above | Virginia Division of Consolidated Laboratory Services | Virginia Division of Consolidated Laboratory Services | Virginia DCLS |
| EPI_ISL_2101993, EPI_ISL_2101994, EPI_ISL_2101995, EPI_ISL_2101996, EPI_ISL_2101997, | Univeristy of New Mexico Hospital | Center for Global Health, University of New Mexico Health Sciences Center | Daryl Domman, Kurt Schwalm, Valerie Morley, Justin Bacca, Jon Femling, Darrell Dinwiddie |

| | | | |
|---|---|---|---|
| EPI_ISL_2101998 | | | |
| EPI_ISL_2101999, EPI_ISL_2102000, EPI_ISL_2102001, EPI_ISL_2102002, EPI_ISL_2102003, EPI_ISL_2102004, EPI_ISL_2102005, EPI_ISL_2102006, EPI_ISL_2102007, EPI_ISL_2102008, EPI_ISL_2102009, EPI_ISL_2102010, EPI_ISL_2102011, EPI_ISL_2102012, EPI_ISL_2102013, EPI_ISL_2102015, EPI_ISL_2102016, EPI_ISL_2102017, EPI_ISL_2102019, EPI_ISL_2102020, EPI_ISL_2102021, EPI_ISL_2102022, EPI_ISL_2102023, EPI_ISL_2102024, EPI_ISL_2102025, EPI_ISL_2102026, EPI_ISL_2102027, EPI_ISL_2102028 | | | |
| see above | South Dakota Public Health Laboratory | South Dakota Public Health Laboratory | Jacob Garfin and Chris Carlson |
| EPI_ISL_2102291 | LSUHS Emerging Viral Threat Laboratory | LSUHS Emerging Viral Threat Laboratory | Jeremy P. Kamil, Jennifer L. Carroll, Maarten Van Diest, Rona S. Scott, Gregory L. Ware, Alexander Mijalis, Andrew D. Yurochko, Christopher G. Kevil, John A. Vanchiere |
| EPI_ISL_2102292 | LSUHS Emerging Viral Threat Laboratory | LSUHS Emerging Viral Threat Laboratory | Jennifer L. Carroll, Maarten Van Diest, Rona S. Scott, Gregory L. Ware, Alexander Mijalis, Jeremy P. Kamil, Andrew D. Yurochko, Christopher G. Kevil, John A. Vanchiere |
| EPI_ISL_2102293 | LSUHS Emerging Viral Threat Laboratory | LSUHS Emerging Viral Threat Laboratory | Maarten Van Diest, Rona S. Scott, Gregory L. Ware, Alexander Mijalis, Jeremy P. Kamil, Jennifer L. Carroll, Andrew D. Yurochko, Christopher G. Kevil, John A. Vanchiere |
| EPI_ISL_2102294 | LSUHS Emerging Viral Threat Laboratory | LSUHS Emerging Viral Threat Laboratory | Jennifer L. Carroll, Maarten Van Diest, Rona S. Scott, Gregory L. Ware, Alexander Mijalis, Jeremy P. Kamil, Andrew D. Yurochko, Christopher G. Kevil, John A. Vanchiere |
| EPI_ISL_2102295 | LSUHS Emerging Viral Threat Laboratory | LSUHS Emerging Viral Threat Laboratory | Jeremy P. Kamil, Jennifer L. Carroll, Maarten Van Diest, Rona S. Scott, Gregory L. Ware, Alexander Mijalis, Andrew D. Yurochko, Christopher G. Kevil, John A. Vanchiere |
| EPI_ISL_2102296 | LSUHS Emerging Viral Threat Laboratory | LSUHS Emerging Viral Threat Laboratory | Rona S. Scott, Gregory L. Ware, Alexander Mijalis, Jeremy P. Kamil, Jennifer L. Carroll, Maarten Van Diest, Andrew D. Yurochko, Christopher G. Kevil, John A. Vanchiere |
| EPI_ISL_2102297 | LSUHS Emerging Viral Threat Laboratory | LSUHS Emerging Viral Threat Laboratory | Alexander Mijalis, Jeremy P. Kamil, Jennifer L. Carroll, Gregory L. Ware, Maarten Van Diest, Rona S. Scott, Andrew D. Yurochko, Christopher G. Kevil, John A. Vanchiere |
| EPI_ISL_2102298, EPI_ISL_2102299 | LSUHS Emerging Viral Threat Laboratory | LSUHS Emerging Viral Threat Laboratory | Jeremy P. Kamil, Jennifer L. Carroll, Maarten Van Diest, Rona S. Scott, Gregory L. Ware, Alexander Mijalis, Andrew D. Yurochko, Christopher G. Kevil, John A. Vanchiere |
| EPI_ISL_2102300 | LSUHS Emerging Viral Threat Laboratory | LSUHS Emerging Viral Threat Laboratory | Jennifer L. Carroll, Maarten Van Diest, Rona S. Scott, Gregory L. Ware, Alexander Mijalis, Jeremy P. Kamil, Andrew D. Yurochko, Christopher G. Kevil, John A. Vanchiere |
| EPI_ISL_2102301 | LSUHS Emerging Viral Threat Laboratory | LSUHS Emerging Viral Threat Laboratory | Rona S. Scott, Gregory L. Ware, Alexander Mijalis, Jeremy P. Kamil, Jennifer L. Carroll, Maarten Van Diest, Andrew D. Yurochko, Christopher G. Kevil, John A. Vanchiere |
| EPI_ISL_2102302 | LSUHS Emerging Viral Threat Laboratory | LSUHS Emerging Viral Threat Laboratory | Jennifer L. Carroll, Maarten Van Diest, Rona S. Scott, Gregory L. Ware, Alexander Mijalis, Jeremy P. Kamil, Andrew D. Yurochko, Christopher G. Kevil, John A. Vanchiere |
| EPI_ISL_2102303 | LSUHS Emerging Viral Threat Laboratory | LSUHS Emerging Viral Threat Laboratory | Rona S. Scott, Gregory L. Ware, Alexander Mijalis, Jeremy P. Kamil, Jennifer L. Carroll, Maarten Van Diest, Andrew D. Yurochko, Christopher G. Kevil, John A. Vanchiere |
| EPI_ISL_2102304 | LSUHS Emerging Viral Threat Laboratory | LSUHS Emerging Viral Threat Laboratory | Jeremy P. Kamil, Jennifer L. Carroll, Maarten Van Diest, Rona S. Scott, Gregory L. Ware, Alexander Mijalis, Andrew D. Yurochko, Christopher G. Kevil, John A. Vanchiere |
| EPI_ISL_2102305 | LSUHS Emerging Viral Threat Laboratory | LSUHS Emerging Viral Threat Laboratory | Alexander Mijalis, Jeremy P. Kamil, Jennifer L. Carroll, Gregory L. Ware, Maarten Van Diest, Rona S. Scott, Andrew D. Yurochko, Christopher G. Kevil, John A. Vanchiere |
| EPI_ISL_2102306 | LSUHS Emerging Viral Threat Laboratory | LSUHS Emerging Viral Threat Laboratory | Maarten Van Diest, Rona S. Scott, Gregory L. Ware, Alexander Mijalis, Jeremy P. Kamil, Jennifer L. Carroll, Andrew D. Yurochko, Christopher G. Kevil, John A. Vanchiere |
| EPI_ISL_2102307, EPI_ISL_2102308 | LSUHS Emerging Viral Threat Laboratory | LSUHS Emerging Viral Threat Laboratory | Rona S. Scott, Gregory L. Ware, Alexander Mijalis, Jeremy P. Kamil, Jennifer L. Carroll, Maarten Van Diest, Andrew D. Yurochko, Christopher G. Kevil, John A. Vanchiere |
| EPI_ISL_2102309 | LSUHS Emerging Viral Threat Laboratory | LSUHS Emerging Viral Threat Laboratory | Jeremy P. Kamil, Jennifer L. Carroll, Maarten Van Diest, Rona S. Scott, Gregory L. Ware, Alexander Mijalis, Andrew D. Yurochko, Christopher G. Kevil, John A. Vanchiere |
| EPI_ISL_2102310 | LSUHS Emerging Viral Threat Laboratory | LSUHS Emerging Viral Threat Laboratory | Alexander Mijalis, Jeremy P. Kamil, Jennifer L. Carroll, Gregory L. Ware, Maarten Van Diest, Rona S. Scott, Andrew D. Yurochko, Christopher G. Kevil, John A. Vanchiere |
| EPI_ISL_2102311 | LSUHS Emerging Viral Threat Laboratory | LSUHS Emerging Viral Threat Laboratory | Rona S. Scott, Gregory L. Ware, Alexander Mijalis, Jeremy P. Kamil, Jennifer L. Carroll, Maarten Van Diest, Andrew D. Yurochko, Christopher G. Kevil, John A. Vanchiere |
| EPI_ISL_2102312 | LSUHS Emerging Viral Threat Laboratory | LSUHS Emerging Viral Threat Laboratory | Maarten Van Diest, Rona S. Scott, Gregory L. Ware, Alexander Mijalis, Jeremy P. Kamil, Jennifer L. Carroll, Andrew D. Yurochko, Christopher G. Kevil, John A. Vanchiere |
| EPI_ISL_2102313 | LSUHS Emerging Viral Threat Laboratory | LSUHS Emerging Viral Threat Laboratory | Rona S. Scott, Gregory L. Ware, Alexander Mijalis, Jeremy P. Kamil, Jennifer L. Carroll, Maarten Van Diest, Andrew D. Yurochko, Christopher G. Kevil, John A. Vanchiere |
| EPI_ISL_2102314 | LSUHS Emerging Viral Threat Laboratory | LSUHS Emerging Viral Threat Laboratory | Alexander Mijalis, Jeremy P. Kamil, Jennifer L. Carroll, Gregory L. Ware, Maarten Van Diest, Rona S. Scott, Andrew D. Yurochko, Christopher G. Kevil, John A. Vanchiere |
| EPI_ISL_2102332, EPI_ISL_2102333, EPI_ISL_2102334, EPI_ISL_2102335, EPI_ISL_2102336, EPI_ISL_2102337, EPI_ISL_2102338, EPI_ISL_2102339, EPI_ISL_2102340, EPI_ISL_2102341, EPI_ISL_2102342, EPI_ISL_2102343, EPI_ISL_2102344, EPI_ISL_2102345, EPI_ISL_2102346, EPI_ISL_2102347, EPI_ISL_2102348, EPI_ISL_2102349, EPI_ISL_2102350, EPI_ISL_2102351, EPI_ISL_2102352, EPI_ISL_2102353, EPI_ISL_2102354, EPI_ISL_2102355, EPI_ISL_2102356, EPI_ISL_2102357, EPI_ISL_2102358, EPI_ISL_2102359, EPI_ISL_2102360, EPI_ISL_2102361, EPI_ISL_2102362, EPI_ISL_2102363, EPI_ISL_2102364, EPI_ISL_2102365, EPI_ISL_2102366, EPI_ISL_2102367, EPI_ISL_2102368, EPI_ISL_2102369, EPI_ISL_2102370, EPI_ISL_2102371, EPI_ISL_2102372, EPI_ISL_2102373, EPI_ISL_2102374, EPI_ISL_2102375, EPI_ISL_2102376, EPI_ISL_2102377, EPI_ISL_2102378, EPI_ISL_2102379, EPI_ISL_2102380, EPI_ISL_2102381, EPI_ISL_2102382, EPI_ISL_2102383, EPI_ISL_2102384, EPI_ISL_2102385, EPI_ISL_2102386, EPI_ISL_2102387, EPI_ISL_2102388, EPI_ISL_2102389, EPI_ISL_2102390, EPI_ISL_2102391, EPI_ISL_2102392, EPI_ISL_2102393, EPI_ISL_2102394, EPI_ISL_2102395, EPI_ISL_2102396, EPI_ISL_2102397, EPI_ISL_2102398, EPI_ISL_2102399, EPI_ISL_2102400, EPI_ISL_2102401, EPI_ISL_2102402, EPI_ISL_2102403, EPI_ISL_2102404, EPI_ISL_2102405, EPI_ISL_2102406, EPI_ISL_2102407, EPI_ISL_2102408, EPI_ISL_2102409, EPI_ISL_2102410, EPI_ISL_2102411, EPI_ISL_2102412, EPI_ISL_2102413, EPI_ISL_2102414, EPI_ISL_2102415, EPI_ISL_2102416, EPI_ISL_2102417, EPI_ISL_2102418, EPI_ISL_2102419, EPI_ISL_2102420, EPI_ISL_2102421, EPI_ISL_2102422, EPI_ISL_2102423, EPI_ISL_2102424, EPI_ISL_2102425, EPI_ISL_2102426, EPI_ISL_2102427, EPI_ISL_2102428, EPI_ISL_2102429, EPI_ISL_2102430, EPI_ISL_2102431, EPI_ISL_2102432, EPI_ISL_2102433, EPI_ISL_2102434, EPI_ISL_2102435, EPI_ISL_2102436, EPI_ISL_2102437, EPI_ISL_2102438, EPI_ISL_2102439, EPI_ISL_2102440, EPI_ISL_2102441, EPI_ISL_2102442, EPI_ISL_2102443, EPI_ISL_2102444, EPI_ISL_2102445, EPI_ISL_2102446, EPI_ISL_2102447, EPI_ISL_2102448, EPI_ISL_2102449, EPI_ISL_2102450, EPI_ISL_2102451, EPI_ISL_2102452, EPI_ISL_2102453, EPI_ISL_2102454, EPI_ISL_2102455, EPI_ISL_2102456, EPI_ISL_2102457, EPI_ISL_2102458, EPI_ISL_2102459, EPI_ISL_2102460, EPI_ISL_2102461, EPI_ISL_2102462, EPI_ISL_2102463, EPI_ISL_2102464, EPI_ISL_2102465, EPI_ISL_2102466, EPI_ISL_2102467, EPI_ISL_2102468, EPI_ISL_2102479, EPI_ISL_2102480, EPI_ISL_2102481, EPI_ISL_2102482, EPI_ISL_2102483, EPI_ISL_2102484, EPI_ISL_2102485, EPI_ISL_2102486, EPI_ISL_2102487, EPI_ISL_2102488, EPI_ISL_2102489, EPI_ISL_2102490, EPI_ISL_2102491, EPI_ISL_2102492, EPI_ISL_2102493, EPI_ISL_2102494, EPI_ISL_2102495, EPI_ISL_2102496, EPI_ISL_2102497, EPI_ISL_2102498, EPI_ISL_2102499, EPI_ISL_2102500, EPI_ISL_2102501, EPI_ISL_2102502, EPI_ISL_2102503, EPI_ISL_2102504, EPI_ISL_2102505, EPI_ISL_2102506, EPI_ISL_2102507, EPI_ISL_2102508, EPI_ISL_2102509, EPI_ISL_2102510, EPI_ISL_2102511, EPI_ISL_2102512, EPI_ISL_2102513, EPI_ISL_2102514, EPI_ISL_2102515, EPI_ISL_2102516 | | | |
| see above | Maryland Genomics, Institute for Genome Sciences, University of Maryland School of Medicine | Maryland Genomics, Institute for Genome Sciences, University of Maryland School of Medicine | Tallon, Luke J; Sadzewicz, Lisa D; Humphrys, Mike; Ott, Sandra; Roussey, Holly; Mehta, Aditya; Vavikolanu, Kranthi; Fraser, Claire M; Ravel, Jacques |
| EPI_ISL_2102522 | Contra Costa County Public Health Lab | Chan-Zuckerberg Biohub | CZB Cliahub Consortium |
| EPI_ISL_2102523 | CA DPH Viral and Rickettsial Disease Laboratory | Chan-Zuckerberg Biohub | CZB Cliahub Consortium |
| EPI_ISL_2102524, EPI_ISL_2102526 | Contra Costa County Public Health Lab | Chan-Zuckerberg Biohub | CZB Cliahub Consortium |
| EPI_ISL_2102527 | CA DPH Viral and Rickettsial Disease Laboratory | Chan-Zuckerberg Biohub | CZB Cliahub Consortium |
| EPI_ISL_2102528 | Humboldt County Public Health Laboratory | Chan-Zuckerberg Biohub | CZB Cliahub Consortium |
| EPI_ISL_2102529, EPI_ISL_2102530, EPI_ISL_2102531, EPI_ISL_2102532 | CA DPH Viral and Rickettsial Disease Laboratory | Chan-Zuckerberg Biohub | CZB Cliahub Consortium |
| EPI_ISL_2102533 | Humboldt County Public Health Laboratory | Chan-Zuckerberg Biohub | CZB Cliahub Consortium |
| EPI_ISL_2102534 | Contra Costa County Public Health Lab | Chan-Zuckerberg Biohub | CZB Cliahub Consortium |
| EPI_ISL_2102536 | CA DPH Viral and Rickettsial Disease Laboratory | Chan-Zuckerberg Biohub | CZB Cliahub Consortium |
| EPI_ISL_2102537 | Humboldt County Public Health Laboratory | Chan-Zuckerberg Biohub | CZB Cliahub Consortium |
| EPI_ISL_2102538 | Contra Costa County Public Health Lab | Chan-Zuckerberg Biohub | CZB Cliahub Consortium |
| EPI_ISL_2102539, EPI_ISL_2102540 | CA DPH Viral and Rickettsial Disease Laboratory | Chan-Zuckerberg Biohub | CZB Cliahub Consortium |
| EPI_ISL_2102541 | Contra Costa County Public Health Lab | Chan-Zuckerberg Biohub | CZB Cliahub Consortium |
| EPI_ISL_2102542 | CA DPH Viral and Rickettsial Disease Laboratory | Chan-Zuckerberg Biohub | CZB Cliahub Consortium |
| EPI_ISL_2102543 | Contra Costa County Public Health Lab | Chan-Zuckerberg Biohub | CZB Cliahub Consortium |
| EPI_ISL_2102544, EPI_ISL_2102545, EPI_ISL_2102546, EPI_ISL_2102547, EPI_ISL_2102548, | CA DPH Viral and Rickettsial Disease Laboratory | Chan-Zuckerberg Biohub | CZB Cliahub Consortium |

| | | | |
|---|---|---|---|
| EPI_ISL_2102549 | | | |
| EPI_ISL_2102550 | Humboldt County Public Health Laboratory | Chan-Zuckerberg Biohub | CZB Cliahub Consortium |
| EPI_ISL_2102551, EPI_ISL_2102552, EPI_ISL_2102553, EPI_ISL_2102554 | CA DPH Viral and Rickettsial Disease Laboratory | Chan-Zuckerberg Biohub | CZB Cliahub Consortium |
| EPI_ISL_2102555 | Contra Costa County Public Health Lab | Chan-Zuckerberg Biohub | CZB Cliahub Consortium |
| EPI_ISL_2102556, EPI_ISL_2102557, EPI_ISL_2102558, EPI_ISL_2102559, EPI_ISL_2102560 | CA DPH Viral and Rickettsial Disease Laboratory | Chan-Zuckerberg Biohub | CZB Cliahub Consortium |
| EPI_ISL_2102561 | Humboldt County Public Health Laboratory | Chan-Zuckerberg Biohub | CZB Cliahub Consortium |
| EPI_ISL_2102562, EPI_ISL_2102563, EPI_ISL_2102564, EPI_ISL_2102565, EPI_ISL_2102566, EPI_ISL_2102567, EPI_ISL_2102568 | CA DPH Viral and Rickettsial Disease Laboratory | Chan-Zuckerberg Biohub | CZB Cliahub Consortium |
| EPI_ISL_2102569 | Contra Costa County Public Health Lab | Chan-Zuckerberg Biohub | CZB Cliahub Consortium |
| EPI_ISL_2102570, EPI_ISL_2102571, EPI_ISL_2102572, EPI_ISL_2102573, EPI_ISL_2102574, EPI_ISL_2102575, EPI_ISL_2102576, EPI_ISL_2102577, EPI_ISL_2102578 | CA DPH Viral and Rickettsial Disease Laboratory | Chan-Zuckerberg Biohub | CZB Cliahub Consortium |
| EPI_ISL_2102579 | Contra Costa County Public Health Lab | Chan-Zuckerberg Biohub | CZB Cliahub Consortium |
| EPI_ISL_2102581, EPI_ISL_2102582, EPI_ISL_2102583 | CA DPH Viral and Rickettsial Disease Laboratory | Chan-Zuckerberg Biohub | CZB Cliahub Consortium |
| EPI_ISL_2102585 | Humboldt County Public Health Laboratory | Chan-Zuckerberg Biohub | CZB Cliahub Consortium |
| EPI_ISL_2102586, EPI_ISL_2102587 | CA DPH Viral and Rickettsial Disease Laboratory | Chan-Zuckerberg Biohub | CZB Cliahub Consortium |
| EPI_ISL_2102588 | Contra Costa County Public Health Lab | Chan-Zuckerberg Biohub | CZB Cliahub Consortium |
| EPI_ISL_2102589 | CA DPH Viral and Rickettsial Disease Laboratory | Chan-Zuckerberg Biohub | CZB Cliahub Consortium |
| EPI_ISL_2102590, EPI_ISL_2102591 | Contra Costa County Public Health Lab | Chan-Zuckerberg Biohub | CZB Cliahub Consortium |
| EPI_ISL_2102592, EPI_ISL_2102593, EPI_ISL_2102594, EPI_ISL_2102595, EPI_ISL_2102596, EPI_ISL_2102597, EPI_ISL_2102598 | CA DPH Viral and Rickettsial Disease Laboratory | Chan-Zuckerberg Biohub | CZB Cliahub Consortium |
| EPI_ISL_2102604, EPI_ISL_2102626, EPI_ISL_2102658, EPI_ISL_2102675 | Laboratorio Estatal de Salud Publica de Nuevo Leon | Laboratorio Estatal de Salud Publica de Nuevo Leon | (in alphabetical order) Consuelo Treviño-Garza, Eduardo Isaac de la Rosa-Moreno, Else del Carmen Garcia-Garcia, Gloria Alejandra Jasso-de la Peña, Manuel Enrique de la O-Cavazos, Olín Medina-Chávez, Yulianna Mayre Cordero-Cruz |
| EPI_ISL_2102681 | Willis-Knighton Medical Center Hospital Laboratory | LSUHS Emerging Viral Threat Laboratory | Maarten Van Diest, Rona S. Scott, Gregory L. Ware, Alexander Mijalis, Jeremy P. Kamil, Jennifer L. Carroll, Andrew D. Yurochko, Christopher G. Kevil, John A. Vanchiere |
| EPI_ISL_2102683 | Willis-Knighton Medical Center Hospital Laboratory | LSUHS Emerging Viral Threat Laboratory | Jeremy P. Kamil, Jennifer L. Carroll, Maarten Van Diest, Rona S. Scott, Gregory L. Ware, Alexander Mijalis, Andrew D. Yurochko, Christopher G. Kevil, John A. Vanchiere |
| EPI_ISL_2102685 | Willis-Knighton Medical Center Hospital Laboratory | LSUHS Emerging Viral Threat Laboratory | Gregory L. Ware, Alexander Mijalis, Jeremy P. Kamil, Jennifer L. Carroll, Maarten Van Diest, Rona S. Scott, Andrew D. Yurochko, Christopher G. Kevil, John A. Vanchiere |
| EPI_ISL_2102687, EPI_ISL_2102688 | Willis-Knighton Medical Center Hospital Laboratory | LSUHS Emerging Viral Threat Laboratory | Rona S. Scott, Gregory L. Ware, Alexander Mijalis, Jeremy P. Kamil, Jennifer L. Carroll, Maarten Van Diest, Andrew D. Yurochko, Christopher G. Kevil, John A. Vanchiere |
| EPI_ISL_2102693 | Willis-Knighton Medical Center Hospital Laboratory | LSUHS Emerging Viral Threat Laboratory | Jennifer L. Carroll, Maarten Van Diest, Rona S. Scott, Gregory L. Ware, Alexander Mijalis, Jeremy P. Kamil, Andrew D. Yurochko, Christopher G. Kevil, John A. Vanchiere |
| EPI_ISL_2102694 | Willis-Knighton Medical Center Hospital Laboratory | LSUHS Emerging Viral Threat Laboratory | Alexander Mijalis, Jeremy P. Kamil, Jennifer L. Carroll, Maarten Van Diest, Rona S. Scott,Gregory L. Ware, Andrew D. Yurochko, Christopher G. Kevil, John A. Vanchiere |
| EPI_ISL_2102695 | Willis-Knighton Medical Center Hospital Laboratory | LSUHS Emerging Viral Threat Laboratory | Maarten Van Diest, Rona S. Scott, Gregory L. Ware, Alexander Mijalis, Jeremy P. Kamil, Jennifer L. Carroll, Andrew D. Yurochko, Christopher G. Kevil, John A. Vanchiere |
| EPI_ISL_2102698, EPI_ISL_2102700 | Willis-Knighton Medical Center Hospital Laboratory | LSUHS Emerging Viral Threat Laboratory | Rona S. Scott, Gregory L. Ware, Alexander Mijalis, Jeremy P. Kamil, Jennifer L. Carroll, Maarten Van Diest, Andrew D. Yurochko, Christopher G. Kevil, John A. Vanchiere |
| EPI_ISL_2102704 | Willis-Knighton Medical Center Hospital Laboratory | LSUHS Emerging Viral Threat Laboratory | Alexander Mijalis, Jeremy P. Kamil, Jennifer L. Carroll, Maarten Van Diest, Rona S. Scott,Gregory L. Ware, Andrew D. Yurochko, Christopher G. Kevil, John A. Vanchiere |
| EPI_ISL_2102706 | Willis-Knighton Medical Center Hospital Laboratory | LSUHS Emerging Viral Threat Laboratory | Jennifer L. Carroll, Maarten Van Diest, Rona S. Scott, Gregory L. Ware, Alexander Mijalis, Jeremy P. Kamil, Andrew D. Yurochko, Christopher G. Kevil, John A. Vanchiere |
| EPI_ISL_2102708 | Willis-Knighton Medical Center Hospital Laboratory | LSUHS Emerging Viral Threat Laboratory | Alexander Mijalis, Jeremy P. Kamil, Jennifer L. Carroll, Maarten Van Diest, Rona S. Scott,Gregory L. Ware, Andrew D. Yurochko, Christopher G. Kevil, John A. Vanchiere |
| EPI_ISL_2102711 | Willis-Knighton Medical Center Hospital Laboratory | LSUHS Emerging Viral Threat Laboratory | Jennifer L. Carroll, Maarten Van Diest, Rona S. Scott, Gregory L. Ware, Alexander Mijalis, Jeremy P. Kamil, Andrew D. Yurochko, Christopher G. Kevil, John A. Vanchiere |
| EPI_ISL_2102712 | Willis-Knighton Medical Center Hospital Laboratory | LSUHS Emerging Viral Threat Laboratory | Rona S. Scott, Gregory L. Ware, Alexander Mijalis, Jeremy P. Kamil, Jennifer L. Carroll, Maarten Van Diest, Andrew D. Yurochko, Christopher G. Kevil, John A. Vanchiere |
| EPI_ISL_2102713 | Willis-Knighton Medical Center Hospital Laboratory | LSUHS Emerging Viral Threat Laboratory | Alexander Mijalis, Jeremy P. Kamil, Jennifer L. Carroll, Maarten Van Diest, Rona S. Scott,Gregory L. Ware, Andrew D. Yurochko, Christopher G. Kevil, John A. Vanchiere |
| EPI_ISL_2102715, EPI_ISL_2102717 | Willis-Knighton Medical Center Hospital Laboratory | LSUHS Emerging Viral Threat Laboratory | Gregory L. Ware, Alexander Mijalis, Jeremy P. Kamil, Jennifer L. Carroll, Maarten Van Diest, Rona S. Scott, Andrew D. Yurochko, Christopher G. Kevil, John A. Vanchiere |
| EPI_ISL_2102718 | Willis-Knighton Medical Center Hospital Laboratory | LSUHS Emerging Viral Threat Laboratory | Jeremy P. Kamil, Jennifer L. Carroll, Maarten Van Diest, Rona S. Scott, Gregory L. Ware, Alexander Mijalis, Andrew D. Yurochko, Christopher G. Kevil, John A. Vanchiere |
| EPI_ISL_2102719 | Willis-Knighton Medical Center Hospital Laboratory | LSUHS Emerging Viral Threat Laboratory | Maarten Van Diest, Rona S. Scott, Gregory L. Ware, Alexander Mijalis, Jeremy P. Kamil, Jennifer L. Carroll, Andrew D. Yurochko, Christopher G. Kevil, John A. Vanchiere |
| EPI_ISL_2102720 | Willis-Knighton Medical Center Hospital Laboratory | LSUHS Emerging Viral Threat Laboratory | Gregory L. Ware, Alexander Mijalis, Jeremy P. Kamil, Jennifer L. Carroll, Maarten Van Diest, Rona S. Scott, Andrew D. Yurochko, Christopher G. Kevil, John A. Vanchiere |
| EPI_ISL_2102721 | Willis-Knighton Medical Center Hospital Laboratory | LSUHS Emerging Viral Threat Laboratory | Rona S. Scott, Gregory L. Ware, Alexander Mijalis, Jeremy P. Kamil, Jennifer L. Carroll, Maarten Van Diest, Andrew D. Yurochko, Christopher G. Kevil, John A. Vanchiere |

| | | | |
|---|---|---|---|
| EPI_ISL_2102722 | Willis-Knighton Medical Center Hospital Laboratory | LSUHS Emerging Viral Threat Laboratory | Jennifer L. Carroll, Maarten Van Diest, Rona S. Scott, Gregory L. Ware, Alexander Mijalis, Jeremy P. Kamil, Andrew D. Yurochko, Christopher G. Kevil, John A. Vanchiere |
| EPI_ISL_2102724 | Willis-Knighton Medical Center Hospital Laboratory | LSUHS Emerging Viral Threat Laboratory | Jeremy P. Kamil, Jennifer L. Carroll, Maarten Van Diest, Rona S. Scott, Gregory L. Ware, Alexander Mijalis, Andrew D. Yurochko, Christopher G. Kevil, John A. Vanchiere |
| EPI_ISL_2102725 | Willis-Knighton Medical Center Hospital Laboratory | LSUHS Emerging Viral Threat Laboratory | Jennifer L. Carroll, Maarten Van Diest, Rona S. Scott, Gregory L. Ware, Alexander Mijalis, Jeremy P. Kamil, Andrew D. Yurochko, Christopher G. Kevil, John A. Vanchiere |
| EPI_ISL_2102726, EPI_ISL_2102727 | Willis-Knighton Medical Center Hospital Laboratory | LSUHS Emerging Viral Threat Laboratory | Rona S. Scott, Gregory L. Ware, Alexander Mijalis, Jeremy P. Kamil, Jennifer L. Carroll, Maarten Van Diest, Andrew D. Yurochko, Christopher G. Kevil, John A. Vanchiere |
| EPI_ISL_2102728 | Willis-Knighton Medical Center Hospital Laboratory | LSUHS Emerging Viral Threat Laboratory | Gregory L. Ware, Alexander Mijalis, Jeremy P. Kamil, Jennifer L. Carroll, Maarten Van Diest, Rona S. Scott, Andrew D. Yurochko, Christopher G. Kevil, John A. Vanchiere |
| EPI_ISL_2102729, EPI_ISL_2102734 | Willis-Knighton Medical Center Hospital Laboratory | LSUHS Emerging Viral Threat Laboratory | Rona S. Scott, Gregory L. Ware, Alexander Mijalis, Jeremy P. Kamil, Jennifer L. Carroll, Maarten Van Diest, Andrew D. Yurochko, Christopher G. Kevil, John A. Vanchiere |
| EPI_ISL_2102736 | Willis-Knighton Medical Center Hospital Laboratory | LSUHS Emerging Viral Threat Laboratory | Alexander Mijalis, Jeremy P. Kamil, Jennifer L. Carroll, Maarten Van Diest, Rona S. Scott,Gregory L. Ware, Andrew D. Yurochko, Christopher G. Kevil, John A. Vanchiere |
| EPI_ISL_2102737 | Willis-Knighton Medical Center Hospital Laboratory | LSUHS Emerging Viral Threat Laboratory | Jennifer L. Carroll, Maarten Van Diest, Rona S. Scott, Gregory L. Ware, Alexander Mijalis, Jeremy P. Kamil, Andrew D. Yurochko, Christopher G. Kevil, John A. Vanchiere |
| EPI_ISL_2102739 | Willis-Knighton Medical Center Hospital Laboratory | LSUHS Emerging Viral Threat Laboratory | Maarten Van Diest, Rona S. Scott, Gregory L. Ware, Alexander Mijalis, Jeremy P. Kamil, Jennifer L. Carroll, Maarten Van Diest, Andrew D. Yurochko, Christopher G. Kevil, John A. Vanchiere |
| EPI_ISL_2102741 | CA DPH Viral and Rickettsial Disease Laboratory | Chan-Zuckerberg Biohub | CZB Cliahub Consortium |
| EPI_ISL_2102742 | Santa Clara County Public Health Laboratory | Chan-Zuckerberg Biohub | CZB Cliahub Consortium |
| EPI_ISL_2102743 | CA DPH Viral and Rickettsial Disease Laboratory | Chan-Zuckerberg Biohub | CZB Cliahub Consortium |
| EPI_ISL_2102744 | Santa Clara County Public Health Laboratory | Chan-Zuckerberg Biohub | CZB Cliahub Consortium |
| EPI_ISL_2102745, EPI_ISL_2102746 | Contra Costa County Public Health Lab | Chan-Zuckerberg Biohub | CZB Cliahub Consortium |
| EPI_ISL_2102747, EPI_ISL_2102748 | County of San Luis Obispo Public Health Laboratory | Chan-Zuckerberg Biohub | CZB Cliahub Consortium |
| EPI_ISL_2102749 | Contra Costa County Public Health Lab | Chan-Zuckerberg Biohub | CZB Cliahub Consortium |
| EPI_ISL_2102750 | CA DPH Viral and Rickettsial Disease Laboratory | Chan-Zuckerberg Biohub | CZB Cliahub Consortium |
| EPI_ISL_2102751 | Contra Costa County Public Health Lab | Chan-Zuckerberg Biohub | CZB Cliahub Consortium |
| EPI_ISL_2102753 | Humboldt County Public Health Laboratory | Chan-Zuckerberg Biohub | CZB Cliahub Consortium |
| EPI_ISL_2102754 | CA DPH Viral and Rickettsial Disease Laboratory | Chan-Zuckerberg Biohub | CZB Cliahub Consortium |
| EPI_ISL_2102755 | County of San Luis Obispo Public Health Laboratory | Chan-Zuckerberg Biohub | CZB Cliahub Consortium |
| EPI_ISL_2102756 | CA DPH Viral and Rickettsial Disease Laboratory | Chan-Zuckerberg Biohub | CZB Cliahub Consortium |
| EPI_ISL_2102757, EPI_ISL_2102758, EPI_ISL_2102759, EPI_ISL_2102760, EPI_ISL_2102761, EPI_ISL_2102762, EPI_ISL_2102763, EPI_ISL_2102764 | Contra Costa County Public Health Lab | Chan-Zuckerberg Biohub | CZB Cliahub Consortium |
| EPI_ISL_2102765 | CA DPH Viral and Rickettsial Disease Laboratory | Chan-Zuckerberg Biohub | CZB Cliahub Consortium |
| EPI_ISL_2102766 | Santa Clara County Public Health Laboratory | Chan-Zuckerberg Biohub | CZB Cliahub Consortium |
| EPI_ISL_2102767 | Contra Costa County Public Health Lab | Chan-Zuckerberg Biohub | CZB Cliahub Consortium |
| EPI_ISL_2102768 | Santa Clara County Public Health Laboratory | Chan-Zuckerberg Biohub | CZB Cliahub Consortium |
| EPI_ISL_2102769, EPI_ISL_2102770 | Contra Costa County Public Health Lab | Chan-Zuckerberg Biohub | CZB Cliahub Consortium |
| EPI_ISL_2102771 | CA DPH Viral and Rickettsial Disease Laboratory | Chan-Zuckerberg Biohub | CZB Cliahub Consortium |
| EPI_ISL_2102772 | Contra Costa County Public Health Lab | Chan-Zuckerberg Biohub | CZB Cliahub Consortium |
| EPI_ISL_2102773 | CA DPH Viral and Rickettsial Disease Laboratory | Chan-Zuckerberg Biohub | CZB Cliahub Consortium |
| EPI_ISL_2102774 | Santa Clara County Public Health Laboratory | Chan-Zuckerberg Biohub | CZB Cliahub Consortium |
| EPI_ISL_2102775 | CA DPH Viral and Rickettsial Disease Laboratory | Chan-Zuckerberg Biohub | CZB Cliahub Consortium |
| EPI_ISL_2102776 | County of San Luis Obispo Public Health Laboratory | Chan-Zuckerberg Biohub | CZB Cliahub Consortium |
| EPI_ISL_2102777, EPI_ISL_2102778 | CA DPH Viral and Rickettsial Disease Laboratory | Chan-Zuckerberg Biohub | CZB Cliahub Consortium |
| EPI_ISL_2102779 | Santa Clara County Public Health Laboratory | Chan-Zuckerberg Biohub | CZB Cliahub Consortium |
| EPI_ISL_2102780 | CA DPH Viral and Rickettsial Disease Laboratory | Chan-Zuckerberg Biohub | CZB Cliahub Consortium |
| EPI_ISL_2102781 | Santa Clara County Public Health Laboratory | Chan-Zuckerberg Biohub | CZB Cliahub Consortium |
| EPI_ISL_2102782 | Contra Costa County Public Health Lab | Chan-Zuckerberg Biohub | CZB Cliahub Consortium |
| EPI_ISL_2102783 | CA DPH Viral and Rickettsial Disease Laboratory | Chan-Zuckerberg Biohub | CZB Cliahub Consortium |
| EPI_ISL_2102784, EPI_ISL_2102785 | Santa Clara County Public Health Laboratory | Chan-Zuckerberg Biohub | CZB Cliahub Consortium |
| EPI_ISL_2102786, EPI_ISL_2102787 | CA DPH Viral and Rickettsial Disease Laboratory | Chan-Zuckerberg Biohub | CZB Cliahub Consortium |
| EPI_ISL_2102788 | Contra Costa County Public Health Lab | Chan-Zuckerberg Biohub | CZB Cliahub Consortium |
| EPI_ISL_2102789 | CA DPH Viral and Rickettsial Disease Laboratory | Chan-Zuckerberg Biohub | CZB Cliahub Consortium |
| EPI_ISL_2102790 | County of San Luis Obispo Public Health Laboratory | Chan-Zuckerberg Biohub | CZB Cliahub Consortium |

| | | | |
|---|---|---|---|
| EPI_ISL_2102791 | Humboldt County Public Health Laboratory | Chan-Zuckerberg Biohub | CZB Cliahub Consortium |
| EPI_ISL_2102792 | Contra Costa County Public Health Lab | Chan-Zuckerberg Biohub | CZB Cliahub Consortium |
| EPI_ISL_2102793, EPI_ISL_2102794 | CA DPH Viral and Rickettsial Disease Laboratory | Chan-Zuckerberg Biohub | CZB Cliahub Consortium |
| EPI_ISL_2102795, EPI_ISL_2102796 | Contra Costa County Public Health Lab | Chan-Zuckerberg Biohub | CZB Cliahub Consortium |
| EPI_ISL_2102797 | Humboldt County Public Health Laboratory | Chan-Zuckerberg Biohub | CZB Cliahub Consortium |
| EPI_ISL_2102798 | Contra Costa County Public Health Lab | Chan-Zuckerberg Biohub | CZB Cliahub Consortium |
| EPI_ISL_2102799 | CA DPH Viral and Rickettsial Disease Laboratory | Chan-Zuckerberg Biohub | CZB Cliahub Consortium |
| EPI_ISL_2102800, EPI_ISL_2102801 | Contra Costa County Public Health Lab | Chan-Zuckerberg Biohub | CZB Cliahub Consortium |
| EPI_ISL_2102802 | County of San Luis Obispo Public Health Laboratory | Chan-Zuckerberg Biohub | CZB Cliahub Consortium |
| EPI_ISL_2102803 | Santa Clara County Public Health Laboratory | Chan-Zuckerberg Biohub | CZB Cliahub Consortium |
| EPI_ISL_2102804 | Contra Costa County Public Health Lab | Chan-Zuckerberg Biohub | CZB Cliahub Consortium |
| EPI_ISL_2102805 | Santa Clara County Public Health Laboratory | Chan-Zuckerberg Biohub | CZB Cliahub Consortium |
| EPI_ISL_2102806 | Humboldt County Public Health Laboratory | Chan-Zuckerberg Biohub | CZB Cliahub Consortium |
| EPI_ISL_2102807 | CA DPH Viral and Rickettsial Disease Laboratory | Chan-Zuckerberg Biohub | CZB Cliahub Consortium |
| EPI_ISL_2102808 | Humboldt County Public Health Laboratory | Chan-Zuckerberg Biohub | CZB Cliahub Consortium |
| EPI_ISL_2102809 | County of San Luis Obispo Public Health Laboratory | Chan-Zuckerberg Biohub | CZB Cliahub Consortium |
| EPI_ISL_2102810 | Santa Clara County Public Health Laboratory | Chan-Zuckerberg Biohub | CZB Cliahub Consortium |
| EPI_ISL_2102811 | County of San Luis Obispo Public Health Laboratory | Chan-Zuckerberg Biohub | CZB Cliahub Consortium |
| EPI_ISL_2102812, EPI_ISL_2102813 | Santa Clara County Public Health Laboratory | Chan-Zuckerberg Biohub | CZB Cliahub Consortium |
| EPI_ISL_2102814, EPI_ISL_2102815 | Contra Costa County Public Health Lab | Chan-Zuckerberg Biohub | CZB Cliahub Consortium |
| EPI_ISL_2102816 | CA DPH Viral and Rickettsial Disease Laboratory | Chan-Zuckerberg Biohub | CZB Cliahub Consortium |
| EPI_ISL_2102817 | Contra Costa County Public Health Lab | Chan-Zuckerberg Biohub | CZB Cliahub Consortium |
| EPI_ISL_2102818, EPI_ISL_2102819 | CA DPH Viral and Rickettsial Disease Laboratory | Chan-Zuckerberg Biohub | CZB Cliahub Consortium |
| EPI_ISL_2102820 | County of San Luis Obispo Public Health Laboratory | Chan-Zuckerberg Biohub | CZB Cliahub Consortium |
| EPI_ISL_2102821 | Contra Costa County Public Health Lab | Chan-Zuckerberg Biohub | CZB Cliahub Consortium |
| EPI_ISL_2102822 | Humboldt County Public Health Laboratory | Chan-Zuckerberg Biohub | CZB Cliahub Consortium |
| EPI_ISL_2102823 | Santa Clara County Public Health Laboratory | Chan-Zuckerberg Biohub | CZB Cliahub Consortium |
| EPI_ISL_2102824 | Contra Costa County Public Health Lab | Chan-Zuckerberg Biohub | CZB Cliahub Consortium |
| EPI_ISL_2102825 | Santa Clara County Public Health Laboratory | Chan-Zuckerberg Biohub | CZB Cliahub Consortium |
| EPI_ISL_2102826 | Contra Costa County Public Health Lab | Chan-Zuckerberg Biohub | CZB Cliahub Consortium |
| EPI_ISL_2102827 | Santa Clara County Public Health Laboratory | Chan-Zuckerberg Biohub | CZB Cliahub Consortium |
| EPI_ISL_2102828 | Contra Costa County Public Health Lab | Chan-Zuckerberg Biohub | CZB Cliahub Consortium |
| EPI_ISL_2102829 | CA DPH Viral and Rickettsial Disease Laboratory | Chan-Zuckerberg Biohub | CZB Cliahub Consortium |
| EPI_ISL_2102830 | County of San Luis Obispo Public Health Laboratory | Chan-Zuckerberg Biohub | CZB Cliahub Consortium |
| EPI_ISL_2102831 | Santa Clara County Public Health Laboratory | Chan-Zuckerberg Biohub | CZB Cliahub Consortium |
| EPI_ISL_2102832 | Humboldt County Public Health Laboratory | Chan-Zuckerberg Biohub | CZB Cliahub Consortium |
| EPI_ISL_2102833 | Contra Costa County Public Health Lab | Chan-Zuckerberg Biohub | CZB Cliahub Consortium |
| EPI_ISL_2102834, EPI_ISL_2102835 | CA DPH Viral and Rickettsial Disease Laboratory | Chan-Zuckerberg Biohub | CZB Cliahub Consortium |
| EPI_ISL_2102836 | Contra Costa County Public Health Lab | Chan-Zuckerberg Biohub | CZB Cliahub Consortium |
| EPI_ISL_2102837 | County of San Luis Obispo Public Health Laboratory | Chan-Zuckerberg Biohub | CZB Cliahub Consortium |
| EPI_ISL_2102838 | Humboldt County Public Health Laboratory | Chan-Zuckerberg Biohub | CZB Cliahub Consortium |
| EPI_ISL_2102839 | CA DPH Viral and Rickettsial Disease Laboratory | Chan-Zuckerberg Biohub | CZB Cliahub Consortium |
| EPI_ISL_2102840, EPI_ISL_2102841 | Contra Costa County Public Health Lab | Chan-Zuckerberg Biohub | CZB Cliahub Consortium |
| EPI_ISL_2102842 | County of San Luis Obispo Public Health Laboratory | Chan-Zuckerberg Biohub | CZB Cliahub Consortium |
| EPI_ISL_2102843 | Contra Costa County Public Health Lab | Chan-Zuckerberg Biohub | CZB Cliahub Consortium |
| EPI_ISL_2102844, EPI_ISL_2102845 | County of San Luis Obispo Public Health Laboratory | Chan-Zuckerberg Biohub | CZB Cliahub Consortium |
| EPI_ISL_2102846 | CA DPH Viral and Rickettsial Disease Laboratory | Chan-Zuckerberg Biohub | CZB Cliahub Consortium |
| EPI_ISL_2102847 | Humboldt County Public Health Laboratory | Chan-Zuckerberg Biohub | CZB Cliahub Consortium |
| EPI_ISL_2102848, EPI_ISL_2102849, EPI_ISL_2102850 | County of San Luis Obispo Public Health Laboratory | Chan-Zuckerberg Biohub | CZB Cliahub Consortium |

| | | | |
|---|---|---|---|
| EPI_ISL_2102851 | Santa Clara County Public Health Laboratory | Chan-Zuckerberg Biohub | CZB Cliahub Consortium |
| EPI_ISL_2102852, EPI_ISL_2102853 | Humboldt County Public Health Laboratory | Chan-Zuckerberg Biohub | CZB Cliahub Consortium |
| EPI_ISL_2102854 | CA DPH Viral and Rickettsial Disease Laboratory | Chan-Zuckerberg Biohub | CZB Cliahub Consortium |
| EPI_ISL_2102855 | County of San Luis Obispo Public Health Laboratory | Chan-Zuckerberg Biohub | CZB Cliahub Consortium |
| EPI_ISL_2102856 | Contra Costa County Public Health Lab | Chan-Zuckerberg Biohub | CZB Cliahub Consortium |
| EPI_ISL_2102857 | CA DPH Viral and Rickettsial Disease Laboratory | Chan-Zuckerberg Biohub | CZB Cliahub Consortium |
| EPI_ISL_2102858 | Contra Costa County Public Health Lab | Chan-Zuckerberg Biohub | CZB Cliahub Consortium |
| EPI_ISL_2102859 | Santa Clara County Public Health Laboratory | Chan-Zuckerberg Biohub | CZB Cliahub Consortium |
| EPI_ISL_2102860 | CA DPH Viral and Rickettsial Disease Laboratory | Chan-Zuckerberg Biohub | CZB Cliahub Consortium |
| EPI_ISL_2102861, EPI_ISL_2102863 | Contra Costa County Public Health Lab | Chan-Zuckerberg Biohub | CZB Cliahub Consortium |
| EPI_ISL_2102864 | CA DPH Viral and Rickettsial Disease Laboratory | Chan-Zuckerberg Biohub | CZB Cliahub Consortium |
| EPI_ISL_2102865 | Santa Clara County Public Health Laboratory | Chan-Zuckerberg Biohub | CZB Cliahub Consortium |
| EPI_ISL_2102866 | Contra Costa County Public Health Lab | Chan-Zuckerberg Biohub | CZB Cliahub Consortium |
| EPI_ISL_2102867 | CA DPH Viral and Rickettsial Disease Laboratory | Chan-Zuckerberg Biohub | CZB Cliahub Consortium |
| EPI_ISL_2102868 | Contra Costa County Public Health Lab | Chan-Zuckerberg Biohub | CZB Cliahub Consortium |
| EPI_ISL_2102869, EPI_ISL_2102870 | County of San Luis Obispo Public Health Laboratory | Chan-Zuckerberg Biohub | CZB Cliahub Consortium |
| EPI_ISL_2102871 | Santa Clara County Public Health Laboratory | Chan-Zuckerberg Biohub | CZB Cliahub Consortium |
| EPI_ISL_2102872 | Humboldt County Public Health Laboratory | Chan-Zuckerberg Biohub | CZB Cliahub Consortium |
| EPI_ISL_2102873, EPI_ISL_2102874 | CA DPH Viral and Rickettsial Disease Laboratory | Chan-Zuckerberg Biohub | CZB Cliahub Consortium |
| EPI_ISL_2102875, EPI_ISL_2102876 | Santa Clara County Public Health Laboratory | Chan-Zuckerberg Biohub | CZB Cliahub Consortium |
| EPI_ISL_2102877 | CA DPH Viral and Rickettsial Disease Laboratory | Chan-Zuckerberg Biohub | CZB Cliahub Consortium |
| EPI_ISL_2102878 | Santa Clara County Public Health Laboratory | Chan-Zuckerberg Biohub | CZB Cliahub Consortium |
| EPI_ISL_2102879 | CA DPH Viral and Rickettsial Disease Laboratory | Chan-Zuckerberg Biohub | CZB Cliahub Consortium |
| EPI_ISL_2102880 | Contra Costa County Public Health Lab | Chan-Zuckerberg Biohub | CZB Cliahub Consortium |
| EPI_ISL_2102881 | CA DPH Viral and Rickettsial Disease Laboratory | Chan-Zuckerberg Biohub | CZB Cliahub Consortium |
| EPI_ISL_2102882, EPI_ISL_2102883 | Santa Clara County Public Health Laboratory | Chan-Zuckerberg Biohub | CZB Cliahub Consortium |
| EPI_ISL_2102884 | CA DPH Viral and Rickettsial Disease Laboratory | Chan-Zuckerberg Biohub | CZB Cliahub Consortium |
| EPI_ISL_2102885, EPI_ISL_2102886, EPI_ISL_2102887 | Contra Costa County Public Health Lab | Chan-Zuckerberg Biohub | CZB Cliahub Consortium |
| EPI_ISL_2102888 | Humboldt County Public Health Laboratory | Chan-Zuckerberg Biohub | CZB Cliahub Consortium |
| EPI_ISL_2102889, EPI_ISL_2102890 | County of San Luis Obispo Public Health Laboratory | Chan-Zuckerberg Biohub | CZB Cliahub Consortium |
| EPI_ISL_2102891 | Contra Costa County Public Health Lab | Chan-Zuckerberg Biohub | CZB Cliahub Consortium |
| EPI_ISL_2102892 | Santa Clara County Public Health Laboratory | Chan-Zuckerberg Biohub | CZB Cliahub Consortium |
| EPI_ISL_2102893 | County of San Luis Obispo Public Health Laboratory | Chan-Zuckerberg Biohub | CZB Cliahub Consortium |
| EPI_ISL_2102894 | Contra Costa County Public Health Lab | Chan-Zuckerberg Biohub | CZB Cliahub Consortium |
| EPI_ISL_2102895 | Santa Clara County Public Health Laboratory | Chan-Zuckerberg Biohub | CZB Cliahub Consortium |
| EPI_ISL_2102896 | Contra Costa County Public Health Lab | Chan-Zuckerberg Biohub | CZB Cliahub Consortium |
| EPI_ISL_2102897 | CA DPH Viral and Rickettsial Disease Laboratory | Chan-Zuckerberg Biohub | CZB Cliahub Consortium |
| EPI_ISL_2102898, EPI_ISL_2102899 | Santa Clara County Public Health Laboratory | Chan-Zuckerberg Biohub | CZB Cliahub Consortium |
| EPI_ISL_2102900 | CA DPH Viral and Rickettsial Disease Laboratory | Chan-Zuckerberg Biohub | CZB Cliahub Consortium |
| EPI_ISL_2102901 | Contra Costa County Public Health Lab | Chan-Zuckerberg Biohub | CZB Cliahub Consortium |
| EPI_ISL_2102902 | County of San Luis Obispo Public Health Laboratory | Chan-Zuckerberg Biohub | CZB Cliahub Consortium |
| EPI_ISL_2102903 | Humboldt County Public Health Laboratory | Chan-Zuckerberg Biohub | CZB Cliahub Consortium |
| EPI_ISL_2102904 | Santa Clara County Public Health Laboratory | Chan-Zuckerberg Biohub | CZB Cliahub Consortium |
| EPI_ISL_2102905 | County of San Luis Obispo Public Health Laboratory | Chan-Zuckerberg Biohub | CZB Cliahub Consortium |
| EPI_ISL_2102906 | Humboldt County Public Health Laboratory | Chan-Zuckerberg Biohub | CZB Cliahub Consortium |
| EPI_ISL_2102907, EPI_ISL_2102908, EPI_ISL_2102909 | Contra Costa County Public Health Lab | Chan-Zuckerberg Biohub | CZB Cliahub Consortium |
| EPI_ISL_2102910 | CA DPH Viral and Rickettsial Disease Laboratory | Chan-Zuckerberg Biohub | CZB Cliahub Consortium |
| EPI_ISL_2102911 | Santa Clara County Public Health Laboratory | Chan-Zuckerberg Biohub | CZB Cliahub Consortium |

| | | | |
|---|---|---|---|
| EPI_ISL_2102912 | County of San Luis Obispo Public Health Laboratory | Chan-Zuckerberg Biohub | CZB Cliahub Consortium |
| EPI_ISL_2102913, EPI_ISL_2102914 | Santa Clara County Public Health Laboratory | Chan-Zuckerberg Biohub | CZB Cliahub Consortium |
| EPI_ISL_2102915 | CA DPH Viral and Rickettsial Disease Laboratory | Chan-Zuckerberg Biohub | CZB Cliahub Consortium |
| EPI_ISL_2102916, EPI_ISL_2102917, EPI_ISL_2102918, EPI_ISL_2102919 | Contra Costa County Public Health Lab | Chan-Zuckerberg Biohub | CZB Cliahub Consortium |
| EPI_ISL_2102920 | Santa Clara County Public Health Laboratory | Chan-Zuckerberg Biohub | CZB Cliahub Consortium |
| EPI_ISL_2102921 | CA DPH Viral and Rickettsial Disease Laboratory | Chan-Zuckerberg Biohub | CZB Cliahub Consortium |
| EPI_ISL_2102922 | Contra Costa County Public Health Lab | Chan-Zuckerberg Biohub | CZB Cliahub Consortium |
| EPI_ISL_2102923 | Santa Clara County Public Health Laboratory | Chan-Zuckerberg Biohub | CZB Cliahub Consortium |
| EPI_ISL_2102924 | CA DPH Viral and Rickettsial Disease Laboratory | Chan-Zuckerberg Biohub | CZB Cliahub Consortium |
| EPI_ISL_2102925, EPI_ISL_2102926 | County of San Luis Obispo Public Health Laboratory | Chan-Zuckerberg Biohub | CZB Cliahub Consortium |
| EPI_ISL_2102927 | Contra Costa County Public Health Lab | Chan-Zuckerberg Biohub | CZB Cliahub Consortium |
| EPI_ISL_2102928, EPI_ISL_2102929 | CA DPH Viral and Rickettsial Disease Laboratory | Chan-Zuckerberg Biohub | CZB Cliahub Consortium |
| EPI_ISL_2102930, EPI_ISL_2102931, EPI_ISL_2102932 | Contra Costa County Public Health Lab | Chan-Zuckerberg Biohub | CZB Cliahub Consortium |
| EPI_ISL_2102933 | County of San Luis Obispo Public Health Laboratory | Chan-Zuckerberg Biohub | CZB Cliahub Consortium |
| EPI_ISL_2102934 | CA DPH Viral and Rickettsial Disease Laboratory | Chan-Zuckerberg Biohub | CZB Cliahub Consortium |
| EPI_ISL_2102935 | Humboldt County Public Health Laboratory | Chan-Zuckerberg Biohub | CZB Cliahub Consortium |
| EPI_ISL_2102936, EPI_ISL_2102937 | Contra Costa County Public Health Lab | Chan-Zuckerberg Biohub | CZB Cliahub Consortium |
| EPI_ISL_2102938 | County of San Luis Obispo Public Health Laboratory | Chan-Zuckerberg Biohub | CZB Cliahub Consortium |
| EPI_ISL_2102939 | Contra Costa County Public Health Lab | Chan-Zuckerberg Biohub | CZB Cliahub Consortium |
| EPI_ISL_2102940 | CA DPH Viral and Rickettsial Disease Laboratory | Chan-Zuckerberg Biohub | CZB Cliahub Consortium |
| EPI_ISL_2102941 | Santa Clara County Public Health Laboratory | Chan-Zuckerberg Biohub | CZB Cliahub Consortium |
| EPI_ISL_2102942 | CA DPH Viral and Rickettsial Disease Laboratory | Chan-Zuckerberg Biohub | CZB Cliahub Consortium |
| EPI_ISL_2102943 | County of San Luis Obispo Public Health Laboratory | Chan-Zuckerberg Biohub | CZB Cliahub Consortium |
| EPI_ISL_2102944 | CA DPH Viral and Rickettsial Disease Laboratory | Chan-Zuckerberg Biohub | CZB Cliahub Consortium |
| EPI_ISL_2102945, EPI_ISL_2102946 | Contra Costa County Public Health Lab | Chan-Zuckerberg Biohub | CZB Cliahub Consortium |
| EPI_ISL_2102947 | County of San Luis Obispo Public Health Laboratory | Chan-Zuckerberg Biohub | CZB Cliahub Consortium |
| EPI_ISL_2102948 | Humboldt County Public Health Laboratory | Chan-Zuckerberg Biohub | CZB Cliahub Consortium |
| EPI_ISL_2102949 | Contra Costa County Public Health Lab | Chan-Zuckerberg Biohub | CZB Cliahub Consortium |
| EPI_ISL_2102950 | Santa Clara County Public Health Laboratory | Chan-Zuckerberg Biohub | CZB Cliahub Consortium |
| EPI_ISL_2102951, EPI_ISL_2102952 | Contra Costa County Public Health Lab | Chan-Zuckerberg Biohub | CZB Cliahub Consortium |
| EPI_ISL_2102953 | County of San Luis Obispo Public Health Laboratory | Chan-Zuckerberg Biohub | CZB Cliahub Consortium |
| EPI_ISL_2102954 | Contra Costa County Public Health Lab | Chan-Zuckerberg Biohub | CZB Cliahub Consortium |
| EPI_ISL_2102955 | County of San Luis Obispo Public Health Laboratory | Chan-Zuckerberg Biohub | CZB Cliahub Consortium |
| EPI_ISL_2102956 | Santa Clara County Public Health Laboratory | Chan-Zuckerberg Biohub | CZB Cliahub Consortium |
| EPI_ISL_2102957 | CA DPH Viral and Rickettsial Disease Laboratory | Chan-Zuckerberg Biohub | CZB Cliahub Consortium |
| EPI_ISL_2102958 | County of San Luis Obispo Public Health Laboratory | Chan-Zuckerberg Biohub | CZB Cliahub Consortium |
| EPI_ISL_2102960, EPI_ISL_2102961 | Contra Costa County Public Health Lab | Chan-Zuckerberg Biohub | CZB Cliahub Consortium |
| EPI_ISL_2102962 | CA DPH Viral and Rickettsial Disease Laboratory | Chan-Zuckerberg Biohub | CZB Cliahub Consortium |
| EPI_ISL_2102963 | Santa Clara County Public Health Laboratory | Chan-Zuckerberg Biohub | CZB Cliahub Consortium |
| EPI_ISL_2102964 | Humboldt County Public Health Laboratory | Chan-Zuckerberg Biohub | CZB Cliahub Consortium |
| EPI_ISL_2102965 | CA DPH Viral and Rickettsial Disease Laboratory | Chan-Zuckerberg Biohub | CZB Cliahub Consortium |
| EPI_ISL_2102966, EPI_ISL_2102967 | Santa Clara County Public Health Laboratory | Chan-Zuckerberg Biohub | CZB Cliahub Consortium |
| EPI_ISL_2102968 | CA DPH Viral and Rickettsial Disease Laboratory | Chan-Zuckerberg Biohub | CZB Cliahub Consortium |
| EPI_ISL_2102969 | Santa Clara County Public Health Laboratory | Chan-Zuckerberg Biohub | CZB Cliahub Consortium |
| EPI_ISL_2102970 | Contra Costa County Public Health Lab | Chan-Zuckerberg Biohub | CZB Cliahub Consortium |
| EPI_ISL_2102971 | County of San Luis Obispo Public Health Laboratory | Chan-Zuckerberg Biohub | CZB Cliahub Consortium |
| EPI_ISL_2102972, | Contra Costa County Public Health Lab | Chan-Zuckerberg Biohub | CZB Cliahub Consortium |

| | | | |
|---|---|---|---|
| EPI_ISL_2102973 | | | |
| EPI_ISL_2102974 | CA DPH Viral and Rickettsial Disease Laboratory | Chan-Zuckerberg Biohub | CZB Cliahub Consortium |
| EPI_ISL_2102975 | Contra Costa County Public Health Lab | Chan-Zuckerberg Biohub | CZB Cliahub Consortium |
| EPI_ISL_2102976 | County of San Luis Obispo Public Health Laboratory | Chan-Zuckerberg Biohub | CZB Cliahub Consortium |
| EPI_ISL_2102977 | CA DPH Viral and Rickettsial Disease Laboratory | Chan-Zuckerberg Biohub | CZB Cliahub Consortium |
| EPI_ISL_2102978 | County of San Luis Obispo Public Health Laboratory | Chan-Zuckerberg Biohub | CZB Cliahub Consortium |
| EPI_ISL_2102979 | Humboldt County Public Health Laboratory | Chan-Zuckerberg Biohub | CZB Cliahub Consortium |
| EPI_ISL_2102980 | CA DPH Viral and Rickettsial Disease Laboratory | Chan-Zuckerberg Biohub | CZB Cliahub Consortium |
| EPI_ISL_2102981, EPI_ISL_2102982 | Contra Costa County Public Health Lab | Chan-Zuckerberg Biohub | CZB Cliahub Consortium |
| EPI_ISL_2102983, EPI_ISL_2102984 | CA DPH Viral and Rickettsial Disease Laboratory | Chan-Zuckerberg Biohub | CZB Cliahub Consortium |
| EPI_ISL_2102985 | Contra Costa County Public Health Lab | Chan-Zuckerberg Biohub | CZB Cliahub Consortium |
| EPI_ISL_2102986 | Humboldt County Public Health Laboratory | Chan-Zuckerberg Biohub | CZB Cliahub Consortium |
| EPI_ISL_2102987 | CA DPH Viral and Rickettsial Disease Laboratory | Chan-Zuckerberg Biohub | CZB Cliahub Consortium |
| EPI_ISL_2102988 | Contra Costa County Public Health Lab | Chan-Zuckerberg Biohub | CZB Cliahub Consortium |
| EPI_ISL_2102989 | CA DPH Viral and Rickettsial Disease Laboratory | Chan-Zuckerberg Biohub | CZB Cliahub Consortium |
| EPI_ISL_2102990 | Contra Costa County Public Health Lab | Chan-Zuckerberg Biohub | CZB Cliahub Consortium |
| EPI_ISL_2102991 | CA DPH Viral and Rickettsial Disease Laboratory | Chan-Zuckerberg Biohub | CZB Cliahub Consortium |
| EPI_ISL_2102992 | Humboldt County Public Health Laboratory | Chan-Zuckerberg Biohub | CZB Cliahub Consortium |
| EPI_ISL_2102993 | CA DPH Viral and Rickettsial Disease Laboratory | Chan-Zuckerberg Biohub | CZB Cliahub Consortium |
| EPI_ISL_2102994 | County of San Luis Obispo Public Health Laboratory | Chan-Zuckerberg Biohub | CZB Cliahub Consortium |
| EPI_ISL_2102995 | Santa Clara County Public Health Laboratory | Chan-Zuckerberg Biohub | CZB Cliahub Consortium |
| EPI_ISL_2102996, EPI_ISL_2102997 | Contra Costa County Public Health Lab | Chan-Zuckerberg Biohub | CZB Cliahub Consortium |
| EPI_ISL_2102998 | County of San Luis Obispo Public Health Laboratory | Chan-Zuckerberg Biohub | CZB Cliahub Consortium |
| EPI_ISL_2102999, EPI_ISL_2103000, EPI_ISL_2103001, EPI_ISL_2103002 | CA DPH Viral and Rickettsial Disease Laboratory | Chan-Zuckerberg Biohub | CZB Cliahub Consortium |
| EPI_ISL_2103003 | Contra Costa County Public Health Lab | Chan-Zuckerberg Biohub | CZB Cliahub Consortium |
| EPI_ISL_2103004 | CA DPH Viral and Rickettsial Disease Laboratory | Chan-Zuckerberg Biohub | CZB Cliahub Consortium |
| EPI_ISL_2103005 | Contra Costa County Public Health Lab | Chan-Zuckerberg Biohub | CZB Cliahub Consortium |
| EPI_ISL_2103006 | Orange County Public Health Lab | Chan-Zuckerberg Biohub | CZB Cliahub Consortium |
| EPI_ISL_2103007 | Contra Costa County Public Health Lab | Chan-Zuckerberg Biohub | CZB Cliahub Consortium |
| EPI_ISL_2103008 | County of San Luis Obispo Public Health Laboratory | Chan-Zuckerberg Biohub | CZB Cliahub Consortium |
| EPI_ISL_2103009, EPI_ISL_2103010, EPI_ISL_2103011, EPI_ISL_2103012, EPI_ISL_2103013 | Contra Costa County Public Health Lab | Chan-Zuckerberg Biohub | CZB Cliahub Consortium |
| EPI_ISL_2103015 | CA DPH Viral and Rickettsial Disease Laboratory | Chan-Zuckerberg Biohub | CZB Cliahub Consortium |
| EPI_ISL_2103016, EPI_ISL_2103017 | Contra Costa County Public Health Lab | Chan-Zuckerberg Biohub | CZB Cliahub Consortium |
| EPI_ISL_2103018, EPI_ISL_2103019, EPI_ISL_2103020 | CA DPH Viral and Rickettsial Disease Laboratory | Chan-Zuckerberg Biohub | CZB Cliahub Consortium |
| EPI_ISL_2103021 | Santa Clara County Public Health Laboratory | Chan-Zuckerberg Biohub | CZB Cliahub Consortium |
| EPI_ISL_2103022 | Contra Costa County Public Health Lab | Chan-Zuckerberg Biohub | CZB Cliahub Consortium |
| EPI_ISL_2103023 | Santa Clara County Public Health Laboratory | Chan-Zuckerberg Biohub | CZB Cliahub Consortium |
| EPI_ISL_2103024 | Contra Costa County Public Health Lab | Chan-Zuckerberg Biohub | CZB Cliahub Consortium |
| EPI_ISL_2103025 | Orange County Public Health Lab | Chan-Zuckerberg Biohub | CZB Cliahub Consortium |
| EPI_ISL_2103026 | County of San Luis Obispo Public Health Laboratory | Chan-Zuckerberg Biohub | CZB Cliahub Consortium |
| EPI_ISL_2103027 | CA DPH Viral and Rickettsial Disease Laboratory | Chan-Zuckerberg Biohub | CZB Cliahub Consortium |
| EPI_ISL_2103028, EPI_ISL_2103029 | Orange County Public Health Lab | Chan-Zuckerberg Biohub | CZB Cliahub Consortium |
| EPI_ISL_2103030, EPI_ISL_2103031 | Contra Costa County Public Health Lab | Chan-Zuckerberg Biohub | CZB Cliahub Consortium |
| EPI_ISL_2103032 | CA DPH Viral and Rickettsial Disease Laboratory | Chan-Zuckerberg Biohub | CZB Cliahub Consortium |
| EPI_ISL_2103033 | Santa Clara County Public Health Laboratory | Chan-Zuckerberg Biohub | CZB Cliahub Consortium |
| EPI_ISL_2103034 | CA DPH Viral and Rickettsial Disease Laboratory | Chan-Zuckerberg Biohub | CZB Cliahub Consortium |
| EPI_ISL_2103035, EPI_ISL_2103036 | Orange County Public Health Lab | Chan-Zuckerberg Biohub | CZB Cliahub Consortium |
| EPI_ISL_2103037 | Contra Costa County Public Health Lab | Chan-Zuckerberg Biohub | CZB Cliahub Consortium |

| | | | |
|---|---|---|---|
| EPI_ISL_2103038 | CA DPH Viral and Rickettsial Disease Laboratory | Chan-Zuckerberg Biohub | CZB Cliahub Consortium |
| EPI_ISL_2103039, EPI_ISL_2103040, EPI_ISL_2103041 | Contra Costa County Public Health Lab | Chan-Zuckerberg Biohub | CZB Cliahub Consortium |
| EPI_ISL_2103042 | Santa Clara County Public Health Laboratory | Chan-Zuckerberg Biohub | CZB Cliahub Consortium |
| EPI_ISL_2103043 | Contra Costa County Public Health Lab | Chan-Zuckerberg Biohub | CZB Cliahub Consortium |
| EPI_ISL_2103044, EPI_ISL_2103045 | County of San Luis Obispo Public Health Laboratory | Chan-Zuckerberg Biohub | CZB Cliahub Consortium |
| EPI_ISL_2103046, EPI_ISL_2103047 | CA DPH Viral and Rickettsial Disease Laboratory | Chan-Zuckerberg Biohub | CZB Cliahub Consortium |
| EPI_ISL_2103048, EPI_ISL_2103049 | Contra Costa County Public Health Lab | Chan-Zuckerberg Biohub | CZB Cliahub Consortium |
| EPI_ISL_2103050, EPI_ISL_2103051 | CA DPH Viral and Rickettsial Disease Laboratory | Chan-Zuckerberg Biohub | CZB Cliahub Consortium |
| EPI_ISL_2103052 | Contra Costa County Public Health Lab | Chan-Zuckerberg Biohub | CZB Cliahub Consortium |
| EPI_ISL_2103053 | CA DPH Viral and Rickettsial Disease Laboratory | Chan-Zuckerberg Biohub | CZB Cliahub Consortium |
| EPI_ISL_2103054, EPI_ISL_2103055, EPI_ISL_2103056, EPI_ISL_2103057 | Contra Costa County Public Health Lab | Chan-Zuckerberg Biohub | CZB Cliahub Consortium |
| EPI_ISL_2103058, EPI_ISL_2103059 | Santa Clara County Public Health Laboratory | Chan-Zuckerberg Biohub | CZB Cliahub Consortium |
| EPI_ISL_2103060 | Contra Costa County Public Health Lab | Chan-Zuckerberg Biohub | CZB Cliahub Consortium |
| EPI_ISL_2103061 | Orange County Public Health Lab | Chan-Zuckerberg Biohub | CZB Cliahub Consortium |
| EPI_ISL_2103062, EPI_ISL_2103063 | Contra Costa County Public Health Lab | Chan-Zuckerberg Biohub | CZB Cliahub Consortium |
| EPI_ISL_2103064 | CA DPH Viral and Rickettsial Disease Laboratory | Chan-Zuckerberg Biohub | CZB Cliahub Consortium |
| EPI_ISL_2103065 | Contra Costa County Public Health Lab | Chan-Zuckerberg Biohub | CZB Cliahub Consortium |
| EPI_ISL_2103066 | CA DPH Viral and Rickettsial Disease Laboratory | Chan-Zuckerberg Biohub | CZB Cliahub Consortium |
| EPI_ISL_2103067 | Orange County Public Health Lab | Chan-Zuckerberg Biohub | CZB Cliahub Consortium |
| EPI_ISL_2103068, EPI_ISL_2103069, EPI_ISL_2103070 | Contra Costa County Public Health Lab | Chan-Zuckerberg Biohub | CZB Cliahub Consortium |
| EPI_ISL_2103071 | CA DPH Viral and Rickettsial Disease Laboratory | Chan-Zuckerberg Biohub | CZB Cliahub Consortium |
| EPI_ISL_2103072 | Contra Costa County Public Health Lab | Chan-Zuckerberg Biohub | CZB Cliahub Consortium |
| EPI_ISL_2103073 | County of San Luis Obispo Public Health Laboratory | Chan-Zuckerberg Biohub | CZB Cliahub Consortium |
| EPI_ISL_2103074, EPI_ISL_2103075 | CA DPH Viral and Rickettsial Disease Laboratory | Chan-Zuckerberg Biohub | CZB Cliahub Consortium |
| EPI_ISL_2103077, EPI_ISL_2103078, EPI_ISL_2103079, EPI_ISL_2103080 | Contra Costa County Public Health Lab | Chan-Zuckerberg Biohub | CZB Cliahub Consortium |
| EPI_ISL_2103081 | Orange County Public Health Lab | Chan-Zuckerberg Biohub | CZB Cliahub Consortium |
| EPI_ISL_2103082 | Contra Costa County Public Health Lab | Chan-Zuckerberg Biohub | CZB Cliahub Consortium |
| EPI_ISL_2103083 | Santa Clara County Public Health Laboratory | Chan-Zuckerberg Biohub | CZB Cliahub Consortium |
| EPI_ISL_2103084 | Contra Costa County Public Health Lab | Chan-Zuckerberg Biohub | CZB Cliahub Consortium |
| EPI_ISL_2103085, EPI_ISL_2103086 | Orange County Public Health Lab | Chan-Zuckerberg Biohub | CZB Cliahub Consortium |
| EPI_ISL_2103087 | Contra Costa County Public Health Lab | Chan-Zuckerberg Biohub | CZB Cliahub Consortium |
| EPI_ISL_2103088 | CA DPH Viral and Rickettsial Disease Laboratory | Chan-Zuckerberg Biohub | CZB Cliahub Consortium |
| EPI_ISL_2103089 | Santa Clara County Public Health Laboratory | Chan-Zuckerberg Biohub | CZB Cliahub Consortium |
| EPI_ISL_2103090, EPI_ISL_2103091, EPI_ISL_2103092 | County of San Luis Obispo Public Health Laboratory | Chan-Zuckerberg Biohub | CZB Cliahub Consortium |
| EPI_ISL_2103093 | CA DPH Viral and Rickettsial Disease Laboratory | Chan-Zuckerberg Biohub | CZB Cliahub Consortium |
| EPI_ISL_2103094 | Orange County Public Health Lab | Chan-Zuckerberg Biohub | CZB Cliahub Consortium |
| EPI_ISL_2103095 | CA DPH Viral and Rickettsial Disease Laboratory | Chan-Zuckerberg Biohub | CZB Cliahub Consortium |
| EPI_ISL_2103096 | County of San Luis Obispo Public Health Laboratory | Chan-Zuckerberg Biohub | CZB Cliahub Consortium |
| EPI_ISL_2103097 | Contra Costa County Public Health Lab | Chan-Zuckerberg Biohub | CZB Cliahub Consortium |
| EPI_ISL_2103098 | CA DPH Viral and Rickettsial Disease Laboratory | Chan-Zuckerberg Biohub | CZB Cliahub Consortium |
| EPI_ISL_2103099 | Contra Costa County Public Health Lab | Chan-Zuckerberg Biohub | CZB Cliahub Consortium |
| EPI_ISL_2103100 | Orange County Public Health Lab | Chan-Zuckerberg Biohub | CZB Cliahub Consortium |
| EPI_ISL_2103101, EPI_ISL_2103102 | Contra Costa County Public Health Lab | Chan-Zuckerberg Biohub | CZB Cliahub Consortium |
| EPI_ISL_2103103 | CA DPH Viral and Rickettsial Disease Laboratory | Chan-Zuckerberg Biohub | CZB Cliahub Consortium |
| EPI_ISL_2103104, EPI_ISL_2103105, EPI_ISL_2103106 | Contra Costa County Public Health Lab | Chan-Zuckerberg Biohub | CZB Cliahub Consortium |
| EPI_ISL_2103107 | CA DPH Viral and Rickettsial Disease Laboratory | Chan-Zuckerberg Biohub | CZB Cliahub Consortium |
| EPI_ISL_2103108 | Contra Costa County Public Health Lab | Chan-Zuckerberg Biohub | CZB Cliahub Consortium |
| EPI_ISL_2103109 | Santa Clara County Public Health Laboratory | Chan-Zuckerberg Biohub | CZB Cliahub Consortium |

| | | | |
|---|---|---|---|
| EPI_ISL_2103110 | Orange County Public Health Lab | Chan-Zuckerberg Biohub | CZB Cliahub Consortium |
| EPI_ISL_2103111 | Contra Costa County Public Health Lab | Chan-Zuckerberg Biohub | CZB Cliahub Consortium |
| EPI_ISL_2103112 | County of San Luis Obispo Public Health Laboratory | Chan-Zuckerberg Biohub | CZB Cliahub Consortium |
| EPI_ISL_2103113 | Contra Costa County Public Health Lab | Chan-Zuckerberg Biohub | CZB Cliahub Consortium |
| EPI_ISL_2103114, EPI_ISL_2103115 | Orange County Public Health Lab | Chan-Zuckerberg Biohub | CZB Cliahub Consortium |
| EPI_ISL_2103116 | CA DPH Viral and Rickettsial Disease Laboratory | Chan-Zuckerberg Biohub | CZB Cliahub Consortium |
| EPI_ISL_2103117 | Contra Costa County Public Health Lab | Chan-Zuckerberg Biohub | CZB Cliahub Consortium |
| EPI_ISL_2103118, EPI_ISL_2103119, EPI_ISL_2103120 | CA DPH Viral and Rickettsial Disease Laboratory | Chan-Zuckerberg Biohub | CZB Cliahub Consortium |
| EPI_ISL_2103121 | Santa Clara County Public Health Laboratory | Chan-Zuckerberg Biohub | CZB Cliahub Consortium |
| EPI_ISL_2103122 | Orange County Public Health Lab | Chan-Zuckerberg Biohub | CZB Cliahub Consortium |
| EPI_ISL_2103124 | CA DPH Viral and Rickettsial Disease Laboratory | Chan-Zuckerberg Biohub | CZB Cliahub Consortium |
| EPI_ISL_2103125 | Contra Costa County Public Health Lab | Chan-Zuckerberg Biohub | CZB Cliahub Consortium |
| EPI_ISL_2103126 | County of San Luis Obispo Public Health Laboratory | Chan-Zuckerberg Biohub | CZB Cliahub Consortium |
| EPI_ISL_2103127 | Contra Costa County Public Health Lab | Chan-Zuckerberg Biohub | CZB Cliahub Consortium |
| EPI_ISL_2103128, EPI_ISL_2103129 | CA DPH Viral and Rickettsial Disease Laboratory | Chan-Zuckerberg Biohub | CZB Cliahub Consortium |
| EPI_ISL_2103130 | County of San Luis Obispo Public Health Laboratory | Chan-Zuckerberg Biohub | CZB Cliahub Consortium |
| EPI_ISL_2103131 | Contra Costa County Public Health Lab | Chan-Zuckerberg Biohub | CZB Cliahub Consortium |
| EPI_ISL_2103132 | CA DPH Viral and Rickettsial Disease Laboratory | Chan-Zuckerberg Biohub | CZB Cliahub Consortium |
| EPI_ISL_2103133 | County of San Luis Obispo Public Health Laboratory | Chan-Zuckerberg Biohub | CZB Cliahub Consortium |
| EPI_ISL_2103134 | Orange County Public Health Lab | Chan-Zuckerberg Biohub | CZB Cliahub Consortium |
| EPI_ISL_2103135 | CA DPH Viral and Rickettsial Disease Laboratory | Chan-Zuckerberg Biohub | CZB Cliahub Consortium |
| EPI_ISL_2103136 | Contra Costa County Public Health Lab | Chan-Zuckerberg Biohub | CZB Cliahub Consortium |
| EPI_ISL_2103137 | CA DPH Viral and Rickettsial Disease Laboratory | Chan-Zuckerberg Biohub | CZB Cliahub Consortium |
| EPI_ISL_2103138 | Contra Costa County Public Health Lab | Chan-Zuckerberg Biohub | CZB Cliahub Consortium |
| EPI_ISL_2103139, EPI_ISL_2103140 | CA DPH Viral and Rickettsial Disease Laboratory | Chan-Zuckerberg Biohub | CZB Cliahub Consortium |
| EPI_ISL_2103141, EPI_ISL_2103143 | Contra Costa County Public Health Lab | Chan-Zuckerberg Biohub | CZB Cliahub Consortium |
| EPI_ISL_2103144 | Santa Clara County Public Health Laboratory | Chan-Zuckerberg Biohub | CZB Cliahub Consortium |
| EPI_ISL_2103145 | Contra Costa County Public Health Lab | Chan-Zuckerberg Biohub | CZB Cliahub Consortium |
| EPI_ISL_2103146 | Santa Clara County Public Health Laboratory | Chan-Zuckerberg Biohub | CZB Cliahub Consortium |
| EPI_ISL_2103147 | CA DPH Viral and Rickettsial Disease Laboratory | Chan-Zuckerberg Biohub | CZB Cliahub Consortium |
| EPI_ISL_2103150 | Contra Costa County Public Health Lab | Chan-Zuckerberg Biohub | CZB Cliahub Consortium |
| EPI_ISL_2103151 | CA DPH Viral and Rickettsial Disease Laboratory | Chan-Zuckerberg Biohub | CZB Cliahub Consortium |
| EPI_ISL_2103152 | Orange County Public Health Lab | Chan-Zuckerberg Biohub | CZB Cliahub Consortium |
| EPI_ISL_2103153 | County of San Luis Obispo Public Health Laboratory | Chan-Zuckerberg Biohub | CZB Cliahub Consortium |
| EPI_ISL_2103154 | Contra Costa County Public Health Lab | Chan-Zuckerberg Biohub | CZB Cliahub Consortium |
| EPI_ISL_2103155 | CA DPH Viral and Rickettsial Disease Laboratory | Chan-Zuckerberg Biohub | CZB Cliahub Consortium |
| EPI_ISL_2103156, EPI_ISL_2103157 | Contra Costa County Public Health Lab | Chan-Zuckerberg Biohub | CZB Cliahub Consortium |
| EPI_ISL_2103158, EPI_ISL_2103159 | Orange County Public Health Lab | Chan-Zuckerberg Biohub | CZB Cliahub Consortium |
| EPI_ISL_2103160 | CA DPH Viral and Rickettsial Disease Laboratory | Chan-Zuckerberg Biohub | CZB Cliahub Consortium |
| EPI_ISL_2103161 | County of San Luis Obispo Public Health Laboratory | Chan-Zuckerberg Biohub | CZB Cliahub Consortium |
| EPI_ISL_2103162, EPI_ISL_2103163 | CA DPH Viral and Rickettsial Disease Laboratory | Chan-Zuckerberg Biohub | CZB Cliahub Consortium |
| EPI_ISL_2103164 | Contra Costa County Public Health Lab | Chan-Zuckerberg Biohub | CZB Cliahub Consortium |
| EPI_ISL_2103165, EPI_ISL_2103166, EPI_ISL_2103167 | CA DPH Viral and Rickettsial Disease Laboratory | Chan-Zuckerberg Biohub | CZB Cliahub Consortium |
| EPI_ISL_2103168, EPI_ISL_2103169, EPI_ISL_2103170, EPI_ISL_2103171 | Contra Costa County Public Health Lab | Chan-Zuckerberg Biohub | CZB Cliahub Consortium |
| EPI_ISL_2103172 | County of San Luis Obispo Public Health Laboratory | Chan-Zuckerberg Biohub | CZB Cliahub Consortium |
| EPI_ISL_2103173, EPI_ISL_2103174, EPI_ISL_2103175 | Contra Costa County Public Health Lab | Chan-Zuckerberg Biohub | CZB Cliahub Consortium |
| EPI_ISL_2103176 | County of San Luis Obispo Public Health Laboratory | Chan-Zuckerberg Biohub | CZB Cliahub Consortium |
| EPI_ISL_2103177 | Santa Clara County Public Health Laboratory | Chan-Zuckerberg Biohub | CZB Cliahub Consortium |
| EPI_ISL_2103178 | Orange County Public Health Lab | Chan-Zuckerberg Biohub | CZB Cliahub Consortium |
| EPI_ISL_2103179 | Contra Costa County Public Health Lab | Chan-Zuckerberg Biohub | CZB Cliahub Consortium |
| EPI_ISL_2103180, | CA DPH Viral and Rickettsial Disease | Chan-Zuckerberg Biohub | CZB Cliahub Consortium |

| | | | |
|---|---|---|---|
| EPI_ISL_2103181 | Laboratory | | |
| EPI_ISL_2103182, EPI_ISL_2103183, EPI_ISL_2103184, EPI_ISL_2103185, EPI_ISL_2103186 | Contra Costa County Public Health Lab | Chan-Zuckerberg Biohub | CZB Cliahub Consortium |
| EPI_ISL_2103187 | Orange County Public Health Lab | Chan-Zuckerberg Biohub | CZB Cliahub Consortium |
| EPI_ISL_2103188, EPI_ISL_2103189 | County of San Luis Obispo Public Health Laboratory | Chan-Zuckerberg Biohub | CZB Cliahub Consortium |
| EPI_ISL_2103190 | Santa Clara County Public Health Laboratory | Chan-Zuckerberg Biohub | CZB Cliahub Consortium |
| EPI_ISL_2103191 | Contra Costa County Public Health Lab | Chan-Zuckerberg Biohub | CZB Cliahub Consortium |
| EPI_ISL_2103192, EPI_ISL_2103193, EPI_ISL_2103194 | CA DPH Viral and Rickettsial Disease Laboratory | Chan-Zuckerberg Biohub | CZB Cliahub Consortium |
| EPI_ISL_2103195, EPI_ISL_2103196 | Contra Costa County Public Health Lab | Chan-Zuckerberg Biohub | CZB Cliahub Consortium |
| EPI_ISL_2103197 | Orange County Public Health Lab | Chan-Zuckerberg Biohub | CZB Cliahub Consortium |
| EPI_ISL_2103198 | Contra Costa County Public Health Lab | Chan-Zuckerberg Biohub | CZB Cliahub Consortium |
| EPI_ISL_2103199 | Instituto Nacional de Medicina Genomica | Instituto Nacional de Medicina Genomica | Hidalgo-Miranda A, Mendoza-Vargas A, Reyes-Grajeda JP, Cedro-Tanda A, Cisneros-Villanueva M, Orjuela-Rodríguez M, Alcaraz N, Gonzalez-Barrera D, Rangel-DeLeon D, Munguia-Garza P, Ramirez-Vega O, Escobar-Arrazola, M, Herrera-Montalvo LA. |
| EPI_ISL_2103203, EPI_ISL_2103204, EPI_ISL_2103205, EPI_ISL_2103206, EPI_ISL_2103207, EPI_ISL_2103208, EPI_ISL_2103209, EPI_ISL_2103210, EPI_ISL_2103211, EPI_ISL_2103212, EPI_ISL_2103213, EPI_ISL_2103214, EPI_ISL_2103215, EPI_ISL_2103216, EPI_ISL_2103217, EPI_ISL_2103218, EPI_ISL_2103219, EPI_ISL_2103220, EPI_ISL_2103221, EPI_ISL_2103222, EPI_ISL_2103223, EPI_ISL_2103224, EPI_ISL_2103225, EPI_ISL_2103226, EPI_ISL_2103227, EPI_ISL_2103228 | | | |
| see above | Humboldt County Public Health Laboratory | Chan-Zuckerberg Biohub | CZB Cliahub Consortium |
| EPI_ISL_2103229, EPI_ISL_2103230 | UW Virology Lab | UW Virology Lab | Pavitra Roychoudhury, Hong Xie, Lasata Shrestha, Shah Mohamed Bakhash, Michelle Lin, Noah R. Baker, Sean Ellis, Saraswathi Sathees, Meei-Li Huang, Keith R Jerome, Alexander Greninger |
| EPI_ISL_2103252, EPI_ISL_2103254, EPI_ISL_2103255, EPI_ISL_2103259, EPI_ISL_2103261, EPI_ISL_2103262, EPI_ISL_2103263, EPI_ISL_2103265, EPI_ISL_2103266, EPI_ISL_2103268, EPI_ISL_2103269, EPI_ISL_2103272, EPI_ISL_2103274, EPI_ISL_2103275, EPI_ISL_2103276, EPI_ISL_2103277, EPI_ISL_2103279, EPI_ISL_2103280, EPI_ISL_2103281, EPI_ISL_2103282, EPI_ISL_2103284, EPI_ISL_2103285, EPI_ISL_2103286, EPI_ISL_2103287, EPI_ISL_2103288, EPI_ISL_2103289, EPI_ISL_2103290, EPI_ISL_2103291, EPI_ISL_2103292, EPI_ISL_2103293, EPI_ISL_2103294, EPI_ISL_2103295, EPI_ISL_2103296, EPI_ISL_2103297, EPI_ISL_2103298, EPI_ISL_2103299, EPI_ISL_2103300, EPI_ISL_2103301, EPI_ISL_2103303, EPI_ISL_2103304, EPI_ISL_2103305, EPI_ISL_2103306, EPI_ISL_2103307, EPI_ISL_2103308, EPI_ISL_2103309, EPI_ISL_2103310, EPI_ISL_2103311, EPI_ISL_2103312, EPI_ISL_2103314, EPI_ISL_2103315, EPI_ISL_2103316, EPI_ISL_2103317, EPI_ISL_2103318, EPI_ISL_2103319, EPI_ISL_2103320, EPI_ISL_2103321, EPI_ISL_2103322, EPI_ISL_2103323, EPI_ISL_2103325, EPI_ISL_2103326, EPI_ISL_2103328, EPI_ISL_2103329, EPI_ISL_2103330, EPI_ISL_2103331, EPI_ISL_2103332, EPI_ISL_2103333, EPI_ISL_2103335, EPI_ISL_2103336, EPI_ISL_2103337, EPI_ISL_2103338, EPI_ISL_2103339, EPI_ISL_2103340, EPI_ISL_2103341, EPI_ISL_2103342, EPI_ISL_2103343, EPI_ISL_2103345, EPI_ISL_2103346, EPI_ISL_2103347, EPI_ISL_2103348, EPI_ISL_2103349, EPI_ISL_2103350, EPI_ISL_2103351, EPI_ISL_2103352, EPI_ISL_2103353, EPI_ISL_2103354, EPI_ISL_2103355 | | | |
| see above | Johns Hopkins Hospital Department of Pathology | Johns Hopkins Hospital Department of Pathology | C. Paul Morris, Chun Huai Luo, Adannaya Amadi, Matthew Schwartz, Nicholas Gallagher, Heba H. Mostafa |
| EPI_ISL_2103356, EPI_ISL_2103357, EPI_ISL_2103358, EPI_ISL_2103359, EPI_ISL_2103360, EPI_ISL_2103361, EPI_ISL_2103362, EPI_ISL_2103363, EPI_ISL_2103364, EPI_ISL_2103365, EPI_ISL_2103366, EPI_ISL_2103367 | | | |
| see above | Johns Hopkins Hospital Department of Pathology | Johns Hopkins Hospital Department of Pathology | C. Paul Morris, Chun Huai Luo, Adannaya Amadi, Nicholas Gallagher, Matthew Schwartz, Heba H. Mostafa |
| EPI_ISL_2103825, EPI_ISL_2103826, EPI_ISL_2103827, EPI_ISL_2103828, EPI_ISL_2103829, EPI_ISL_2103830, EPI_ISL_2103831, EPI_ISL_2103832, EPI_ISL_2103833, EPI_ISL_2103834, EPI_ISL_2103835, EPI_ISL_2103836, EPI_ISL_2103837, EPI_ISL_2103838, EPI_ISL_2103839, EPI_ISL_2103840, EPI_ISL_2103841, EPI_ISL_2103842, EPI_ISL_2103843, EPI_ISL_2103845, EPI_ISL_2103846, EPI_ISL_2103847, EPI_ISL_2103848, EPI_ISL_2103849, EPI_ISL_2103850, EPI_ISL_2103851, EPI_ISL_2103852, EPI_ISL_2103853, EPI_ISL_2103854, EPI_ISL_2103855, EPI_ISL_2103856, EPI_ISL_2103858, EPI_ISL_2103859, EPI_ISL_2103860, EPI_ISL_2103861, EPI_ISL_2103862, EPI_ISL_2103863, EPI_ISL_2103864, EPI_ISL_2103865, EPI_ISL_2103866, EPI_ISL_2103867, EPI_ISL_2103868, EPI_ISL_2103869, EPI_ISL_2103870, EPI_ISL_2103871, EPI_ISL_2103872, EPI_ISL_2103873, EPI_ISL_2103874, EPI_ISL_2103875, EPI_ISL_2103876, EPI_ISL_2103877, EPI_ISL_2103878, EPI_ISL_2103879, EPI_ISL_2103880, EPI_ISL_2103881, EPI_ISL_2103882, EPI_ISL_2103883, EPI_ISL_2103884, EPI_ISL_2103885, EPI_ISL_2103886, EPI_ISL_2103887, EPI_ISL_2103888, EPI_ISL_2103889, EPI_ISL_2103890, EPI_ISL_2103891, EPI_ISL_2103893, EPI_ISL_2103894, EPI_ISL_2103895, EPI_ISL_2103896, EPI_ISL_2103897, EPI_ISL_2103898, EPI_ISL_2103899, EPI_ISL_2103900, EPI_ISL_2103901, EPI_ISL_2103902, EPI_ISL_2103903, EPI_ISL_2103904, EPI_ISL_2103905, EPI_ISL_2103906, EPI_ISL_2103907, EPI_ISL_2103908, EPI_ISL_2103909, EPI_ISL_2103910, EPI_ISL_2103911, EPI_ISL_2103912, EPI_ISL_2103913, EPI_ISL_2103914, EPI_ISL_2103915, EPI_ISL_2103916, EPI_ISL_2103917, EPI_ISL_2103918, EPI_ISL_2103919, EPI_ISL_2103920, EPI_ISL_2103921, EPI_ISL_2103922, EPI_ISL_2103923, EPI_ISL_2103924, EPI_ISL_2103925, EPI_ISL_2103926, EPI_ISL_2103927, EPI_ISL_2103928, EPI_ISL_2103929, EPI_ISL_2103930, EPI_ISL_2103931, EPI_ISL_2103932, EPI_ISL_2103933, EPI_ISL_2103934, EPI_ISL_2103935, EPI_ISL_2103936, EPI_ISL_2103937, EPI_ISL_2103939, EPI_ISL_2103940, EPI_ISL_2103942, EPI_ISL_2103943, EPI_ISL_2103945, EPI_ISL_2103946, EPI_ISL_2103947, EPI_ISL_2103948, EPI_ISL_2103949, EPI_ISL_2103950, EPI_ISL_2103951, EPI_ISL_2103952, EPI_ISL_2103953, EPI_ISL_2103954, EPI_ISL_2103955, EPI_ISL_2103956, EPI_ISL_2103957, EPI_ISL_2103958, EPI_ISL_2103959, EPI_ISL_2103960, EPI_ISL_2103961, EPI_ISL_2103962, EPI_ISL_2103963, EPI_ISL_2103964, EPI_ISL_2103965, EPI_ISL_2103966, EPI_ISL_2103967, EPI_ISL_2103968, EPI_ISL_2103969, EPI_ISL_2103970, EPI_ISL_2103971, EPI_ISL_2103972, EPI_ISL_2103973, EPI_ISL_2103974, EPI_ISL_2103975, EPI_ISL_2103976, EPI_ISL_2103977, EPI_ISL_2103979, EPI_ISL_2103980, EPI_ISL_2103981, EPI_ISL_2103982, EPI_ISL_2103983, EPI_ISL_2103984, EPI_ISL_2103985, EPI_ISL_2103986, EPI_ISL_2103987, EPI_ISL_2103988, EPI_ISL_2103989, EPI_ISL_2103990, EPI_ISL_2103991, EPI_ISL_2103992, EPI_ISL_2103993, EPI_ISL_2103994, EPI_ISL_2103995, EPI_ISL_2103996, EPI_ISL_2103997, EPI_ISL_2103998, EPI_ISL_2103999, EPI_ISL_2104000, EPI_ISL_2104001, EPI_ISL_2104002, EPI_ISL_2104003, EPI_ISL_2104004, EPI_ISL_2104005, EPI_ISL_2104006, EPI_ISL_2104007, EPI_ISL_2104008, EPI_ISL_2104009, EPI_ISL_2104010, EPI_ISL_2104011, EPI_ISL_2104012, EPI_ISL_2104013, EPI_ISL_2104014, EPI_ISL_2104015, EPI_ISL_2104016, EPI_ISL_2104017, EPI_ISL_2104018, EPI_ISL_2104019, EPI_ISL_2104020, EPI_ISL_2104021, EPI_ISL_2104022, EPI_ISL_2104023, EPI_ISL_2104024, EPI_ISL_2104025, EPI_ISL_2104026, EPI_ISL_2104027, EPI_ISL_2104028, EPI_ISL_2104029, EPI_ISL_2104030, EPI_ISL_2104031, EPI_ISL_2104032, EPI_ISL_2104033, EPI_ISL_2104034, EPI_ISL_2104035, EPI_ISL_2104036, EPI_ISL_2104037, EPI_ISL_2104038, EPI_ISL_2104039, EPI_ISL_2104040, EPI_ISL_2104041, EPI_ISL_2104042, EPI_ISL_2104043, EPI_ISL_2104044, EPI_ISL_2104045, EPI_ISL_2104046, EPI_ISL_2104047, EPI_ISL_2104048, EPI_ISL_2104049, EPI_ISL_2104050, EPI_ISL_2104051, EPI_ISL_2104052, EPI_ISL_2104053, EPI_ISL_2104054, EPI_ISL_2104055, EPI_ISL_2104056, EPI_ISL_2104057, EPI_ISL_2104058, EPI_ISL_2104059, EPI_ISL_2104060, EPI_ISL_2104061, EPI_ISL_2104062, EPI_ISL_2104063, EPI_ISL_2104064, EPI_ISL_2104065, EPI_ISL_2104066, EPI_ISL_2104067, EPI_ISL_2104068, EPI_ISL_2104069, EPI_ISL_2104070, EPI_ISL_2104071, EPI_ISL_2104072, EPI_ISL_2104073, EPI_ISL_2104074, EPI_ISL_2104075, EPI_ISL_2104076, EPI_ISL_2104077, EPI_ISL_2104078, EPI_ISL_2104080, EPI_ISL_2104081, EPI_ISL_2104082, EPI_ISL_2104083, EPI_ISL_2104084, EPI_ISL_2104085, EPI_ISL_2104086, EPI_ISL_2104087, EPI_ISL_2104088, EPI_ISL_2104089, EPI_ISL_2104090, EPI_ISL_2104091, EPI_ISL_2104092, EPI_ISL_2104093, EPI_ISL_2104094, EPI_ISL_2104095, EPI_ISL_2104096, EPI_ISL_2104097, EPI_ISL_2104098, EPI_ISL_2104099, EPI_ISL_2104100, EPI_ISL_2104101, EPI_ISL_2104102, EPI_ISL_2104103, EPI_ISL_2104104, EPI_ISL_2104105, EPI_ISL_2104106, EPI_ISL_2104107, EPI_ISL_2104108, EPI_ISL_2104109, EPI_ISL_2104110, EPI_ISL_2104111, EPI_ISL_2104112, EPI_ISL_2104113, EPI_ISL_2104114, EPI_ISL_2104115, EPI_ISL_2104116, EPI_ISL_2104117, EPI_ISL_2104118, EPI_ISL_2104119, EPI_ISL_2104120, EPI_ISL_2104121, EPI_ISL_2104122, EPI_ISL_2104123, EPI_ISL_2104124, EPI_ISL_2104125, EPI_ISL_2104126, EPI_ISL_2104127, EPI_ISL_2104128, EPI_ISL_2104129, EPI_ISL_2104130, EPI_ISL_2104131, EPI_ISL_2104132, EPI_ISL_2104133, EPI_ISL_2104134, EPI_ISL_2104135, EPI_ISL_2104136, EPI_ISL_2104137, EPI_ISL_2104138, EPI_ISL_2104139, EPI_ISL_2104140, EPI_ISL_2104141, EPI_ISL_2104142, EPI_ISL_2104143, EPI_ISL_2104144, EPI_ISL_2104145, EPI_ISL_2104146, EPI_ISL_2104147, EPI_ISL_2104148, EPI_ISL_2104149, EPI_ISL_2104150, EPI_ISL_2104151, EPI_ISL_2104152, EPI_ISL_2104153, EPI_ISL_2104154, EPI_ISL_2104155, EPI_ISL_2104156, EPI_ISL_2104157, EPI_ISL_2104158, EPI_ISL_2104159, EPI_ISL_2104160, EPI_ISL_2104161, EPI_ISL_2104162, EPI_ISL_2104163, EPI_ISL_2104164, EPI_ISL_2104165, EPI_ISL_2104166, EPI_ISL_2104167, EPI_ISL_2104168, EPI_ISL_2104169, EPI_ISL_2104170, EPI_ISL_2104172, EPI_ISL_2104173, EPI_ISL_2104174, EPI_ISL_2104175, EPI_ISL_2104176, EPI_ISL_2104177, EPI_ISL_2104178, EPI_ISL_2104179, EPI_ISL_2104180, EPI_ISL_2104181, EPI_ISL_2104182, EPI_ISL_2104183, EPI_ISL_2104184, EPI_ISL_2104185, EPI_ISL_2104186, EPI_ISL_2104187, EPI_ISL_2104188, EPI_ISL_2104189, EPI_ISL_2104190, EPI_ISL_2104191, EPI_ISL_2104192, EPI_ISL_2104193, EPI_ISL_2104194, EPI_ISL_2104195, EPI_ISL_2104196, EPI_ISL_2104197, EPI_ISL_2104198, EPI_ISL_2104199, EPI_ISL_2104200, EPI_ISL_2104201, EPI_ISL_2104202, EPI_ISL_2104203, EPI_ISL_2104204, EPI_ISL_2104205, EPI_ISL_2104207, EPI_ISL_2104208, EPI_ISL_2104209, EPI_ISL_2104210, EPI_ISL_2104211, EPI_ISL_2104212, EPI_ISL_2104213, EPI_ISL_2104214, EPI_ISL_2104215, EPI_ISL_2104216, EPI_ISL_2104217, EPI_ISL_2104218, EPI_ISL_2104219, EPI_ISL_2104220, EPI_ISL_2104222, EPI_ISL_2104223, EPI_ISL_2104224, EPI_ISL_2104225, EPI_ISL_2104226, EPI_ISL_2104227, EPI_ISL_2104228, EPI_ISL_2104229, EPI_ISL_2104230, EPI_ISL_2104231, EPI_ISL_2104232, EPI_ISL_2104233, EPI_ISL_2104234, EPI_ISL_2104235, EPI_ISL_2104236, EPI_ISL_2104237, EPI_ISL_2104238, EPI_ISL_2104239, EPI_ISL_2104240, EPI_ISL_2104241, EPI_ISL_2104242, EPI_ISL_2104243, EPI_ISL_2104244, EPI_ISL_2104245, EPI_ISL_2104246, EPI_ISL_2104247, EPI_ISL_2104248, EPI_ISL_2104249, EPI_ISL_2104250, EPI_ISL_2104251, EPI_ISL_2104252, EPI_ISL_2104253, EPI_ISL_2104254, EPI_ISL_2104255, EPI_ISL_2104256, EPI_ISL_2104257, EPI_ISL_2104258, EPI_ISL_2104259, EPI_ISL_2104260, EPI_ISL_2104261, EPI_ISL_2104262, EPI_ISL_2104263, EPI_ISL_2104264, EPI_ISL_2104265, EPI_ISL_2104266, EPI_ISL_2104267, EPI_ISL_2104268, EPI_ISL_2104269, EPI_ISL_2104270, EPI_ISL_2104272, EPI_ISL_2104273, EPI_ISL_2104274, EPI_ISL_2104275, EPI_ISL_2104276, EPI_ISL_2104277, EPI_ISL_2104278, EPI_ISL_2104279, EPI_ISL_2104280, EPI_ISL_2104281, EPI_ISL_2104282, EPI_ISL_2104283, EPI_ISL_2104284, EPI_ISL_2104285, EPI_ISL_2104286, EPI_ISL_2104287, EPI_ISL_2104288, EPI_ISL_2104289, EPI_ISL_2104290, EPI_ISL_2104291, EPI_ISL_2104292, EPI_ISL_2104294, EPI_ISL_2104295, EPI_ISL_2104296, EPI_ISL_2104297, EPI_ISL_2104298, EPI_ISL_2104299, EPI_ISL_2104300, EPI_ISL_2104301, EPI_ISL_2104302, EPI_ISL_2104303, EPI_ISL_2104304, EPI_ISL_2104305, EPI_ISL_2104306, EPI_ISL_2104307, EPI_ISL_2104308, EPI_ISL_2104309, EPI_ISL_2104310, EPI_ISL_2104311, EPI_ISL_2104312, EPI_ISL_2104313, EPI_ISL_2104314, EPI_ISL_2104315, EPI_ISL_2104316, EPI_ISL_2104317, EPI_ISL_2104318, EPI_ISL_2104319, EPI_ISL_2104320, EPI_ISL_2104321, EPI_ISL_2104322, EPI_ISL_2104323, EPI_ISL_2104324, EPI_ISL_2104326, EPI_ISL_2104327, EPI_ISL_2104328, EPI_ISL_2104329, EPI_ISL_2104330, EPI_ISL_2104331, EPI_ISL_2104332, EPI_ISL_2104333, EPI_ISL_2104334, EPI_ISL_2104335, EPI_ISL_2104336, EPI_ISL_2104337, EPI_ISL_2104338, EPI_ISL_2104339, EPI_ISL_2104340, EPI_ISL_2104341, EPI_ISL_2104342, EPI_ISL_2104343, EPI_ISL_2104344, EPI_ISL_2104345, EPI_ISL_2104346, EPI_ISL_2104347, EPI_ISL_2104348, EPI_ISL_2104349, EPI_ISL_2104350, EPI_ISL_2104351, EPI_ISL_2104352, EPI_ISL_2104353, EPI_ISL_2104354, EPI_ISL_2104355, EPI_ISL_2104356, EPI_ISL_2104357, EPI_ISL_2104358, EPI_ISL_2104359, EPI_ISL_2104360, EPI_ISL_2104361, EPI_ISL_2104362, EPI_ISL_2104363, EPI_ISL_2104364, EPI_ISL_2104366, EPI_ISL_2104367, EPI_ISL_2104368, EPI_ISL_2104369, EPI_ISL_2104370, EPI_ISL_2104371, EPI_ISL_2104372, EPI_ISL_2104373, EPI_ISL_2104374, EPI_ISL_2104375, EPI_ISL_2104376, EPI_ISL_2104377, EPI_ISL_2104378, EPI_ISL_2104379, EPI_ISL_2104380, EPI_ISL_2104381, EPI_ISL_2104382, EPI_ISL_2104383, EPI_ISL_2104384, EPI_ISL_2104385, EPI_ISL_2104386, EPI_ISL_2104387, EPI_ISL_2104388, EPI_ISL_2104389, EPI_ISL_2104390, EPI_ISL_2104391, EPI_ISL_2104392, EPI_ISL_2104393, EPI_ISL_2104394, EPI_ISL_2104395, EPI_ISL_2104396, EPI_ISL_2104397, EPI_ISL_2104398, EPI_ISL_2104399, EPI_ISL_2104400, EPI_ISL_2104401, EPI_ISL_2104402, EPI_ISL_2104403, EPI_ISL_2104404, EPI_ISL_2104405, EPI_ISL_2104406, EPI_ISL_2104407, EPI_ISL_2104408, EPI_ISL_2104409, EPI_ISL_2104410 | | | |
| see above | Broad Institute Clinical Research Sequencing Platform | Infectious Disease Program, Broad Institute of Harvard and MIT | Siddle,K.J., Adams,G., Pearlman,L., Gladden-Young,A., Vicente,G., Blumenstiel,B., DeFelice,M., Lee,M., McGovern,S., Lagerborg,K., Rudy,M., DeRuff,K., Carter,A., Normandin,E., Bauer,M., Reilly,S., Tomkins-Tinch,C., Loreth,C., Chaluvadi,S., Meldrim,J., Granger,B., Lemieux,J.E., Birren,B.W., Sabeti,P.C., Larkin,K., Dodge,S., Lennon,N., Madoff,L., Brown,C., Gallagher,G., Smole,S., Park,D.J., Gabriel,S., and MacInnis,B.L. |
| EPI_ISL_2104827, EPI_ISL_2104828, EPI_ISL_2104829, EPI_ISL_2104830, EPI_ISL_2104831, EPI_ISL_2104832, EPI_ISL_2104833, EPI_ISL_2104834, EPI_ISL_2104835, EPI_ISL_2104836, EPI_ISL_2104837, EPI_ISL_2104838, EPI_ISL_2104839, EPI_ISL_2104840, EPI_ISL_2104841, EPI_ISL_2104842, EPI_ISL_2104843, EPI_ISL_2104844, EPI_ISL_2104845, EPI_ISL_2104846, EPI_ISL_2104847, EPI_ISL_2104848, EPI_ISL_2104849, EPI_ISL_2104850, EPI_ISL_2104851, EPI_ISL_2104852, EPI_ISL_2104853, EPI_ISL_2104854, EPI_ISL_2104855, EPI_ISL_2104856, EPI_ISL_2104857, EPI_ISL_2104859, EPI_ISL_2104860, EPI_ISL_2104861, EPI_ISL_2104862, EPI_ISL_2104863, EPI_ISL_2104864, EPI_ISL_2104865, EPI_ISL_2104866, EPI_ISL_2104867, EPI_ISL_2104868, EPI_ISL_2104869, EPI_ISL_2104870, EPI_ISL_2104871, EPI_ISL_2104872, EPI_ISL_2104874, EPI_ISL_2104875, EPI_ISL_2104876, EPI_ISL_2104877, EPI_ISL_2104878, EPI_ISL_2104879, EPI_ISL_2104880, EPI_ISL_2104881, EPI_ISL_2104882, EPI_ISL_2104883, EPI_ISL_2104884, EPI_ISL_2104885, EPI_ISL_2104886, EPI_ISL_2104887, EPI_ISL_2104888, EPI_ISL_2104889, EPI_ISL_2104890, EPI_ISL_2104891, EPI_ISL_2104892, EPI_ISL_2104893, EPI_ISL_2104894, EPI_ISL_2104895, EPI_ISL_2104896, EPI_ISL_2104897, EPI_ISL_2104898, EPI_ISL_2104899, EPI_ISL_2104900, EPI_ISL_2104901, EPI_ISL_2104902, EPI_ISL_2104903, EPI_ISL_2104904, EPI_ISL_2104905, EPI_ISL_2104906, EPI_ISL_2104907, EPI_ISL_2104908, EPI_ISL_2104909, EPI_ISL_2104910, EPI_ISL_2104911, EPI_ISL_2104912, EPI_ISL_2104913, EPI_ISL_2104914, EPI_ISL_2104915, EPI_ISL_2104916, EPI_ISL_2104917, EPI_ISL_2104918, EPI_ISL_2104919, EPI_ISL_2104920, EPI_ISL_2104921, EPI_ISL_2104922, EPI_ISL_2104923, EPI_ISL_2104924, EPI_ISL_2104925, EPI_ISL_2104926, EPI_ISL_2104927, EPI_ISL_2104928, EPI_ISL_2104929, EPI_ISL_2104930, EPI_ISL_2104931, EPI_ISL_2104932, EPI_ISL_2104933, EPI_ISL_2104934, EPI_ISL_2104935, EPI_ISL_2104937, EPI_ISL_2104938, EPI_ISL_2104940, EPI_ISL_2104941, EPI_ISL_2104942, EPI_ISL_2104943, EPI_ISL_2104944, EPI_ISL_2104945, EPI_ISL_2104947, EPI_ISL_2104948, EPI_ISL_2104949, EPI_ISL_2104950, EPI_ISL_2104951, EPI_ISL_2104952, EPI_ISL_2104953, EPI_ISL_2104954, EPI_ISL_2104956, EPI_ISL_2104957, EPI_ISL_2104958, EPI_ISL_2104959, EPI_ISL_2104960, EPI_ISL_2104961, EPI_ISL_2104962, EPI_ISL_2104963, EPI_ISL_2104964, EPI_ISL_2104965, EPI_ISL_2104966, EPI_ISL_2104967, EPI_ISL_2104968, EPI_ISL_2104969, EPI_ISL_2104970, EPI_ISL_2104971, EPI_ISL_2104972, EPI_ISL_2104973, EPI_ISL_2104974, EPI_ISL_2104975, EPI_ISL_2104976, EPI_ISL_2104977, EPI_ISL_2104978, EPI_ISL_2104979, EPI_ISL_2104980, EPI_ISL_2104981, EPI_ISL_2104982, EPI_ISL_2104983, EPI_ISL_2104984, EPI_ISL_2104985, EPI_ISL_2104986, EPI_ISL_2104987, EPI_ISL_2104988, EPI_ISL_2104989, EPI_ISL_2104990, EPI_ISL_2104991, EPI_ISL_2104992, EPI_ISL_2104993, EPI_ISL_2104994, EPI_ISL_2104995, EPI_ISL_2104996, EPI_ISL_2104997, EPI_ISL_2104998, EPI_ISL_2104999, EPI_ISL_2105000, EPI_ISL_2105001, EPI_ISL_2105002, EPI_ISL_2105003, EPI_ISL_2105004, EPI_ISL_2105005, EPI_ISL_2105006, EPI_ISL_2105007, EPI_ISL_2105008, EPI_ISL_2105009, EPI_ISL_2105010, EPI_ISL_2105011, EPI_ISL_2105012, EPI_ISL_2105013, EPI_ISL_2105014, EPI_ISL_2105015, EPI_ISL_2105016, EPI_ISL_2105017, EPI_ISL_2105018, EPI_ISL_2105019, EPI_ISL_2105020, EPI_ISL_2105021, EPI_ISL_2105022, EPI_ISL_2105023, EPI_ISL_2105024, EPI_ISL_2105025, EPI_ISL_2105026, EPI_ISL_2105027, EPI_ISL_2105028, EPI_ISL_2105029, EPI_ISL_2105030, EPI_ISL_2105031, EPI_ISL_2105032, EPI_ISL_2105033, EPI_ISL_2105034, EPI_ISL_2105035, EPI_ISL_2105037, EPI_ISL_2105038, EPI_ISL_2105039, EPI_ISL_2105040, EPI_ISL_2105041, EPI_ISL_2105042, EPI_ISL_2105043, EPI_ISL_2105044, EPI_ISL_2105045, EPI_ISL_2105046, EPI_ISL_2105047, EPI_ISL_2105049, EPI_ISL_2105050, EPI_ISL_2105051, EPI_ISL_2105052, EPI_ISL_2105053, EPI_ISL_2105054, EPI_ISL_2105055, EPI_ISL_2105056, EPI_ISL_2105057, EPI_ISL_2105058, EPI_ISL_2105060, EPI_ISL_2105061, EPI_ISL_2105062, EPI_ISL_2105063, EPI_ISL_2105064, EPI_ISL_2105065, EPI_ISL_2105066, EPI_ISL_2105067, EPI_ISL_2105068, EPI_ISL_2105069, EPI_ISL_2105070, EPI_ISL_2105071, EPI_ISL_2105072, EPI_ISL_2105073, EPI_ISL_2105074, EPI_ISL_2105075, EPI_ISL_2105076, EPI_ISL_2105077, EPI_ISL_2105078, EPI_ISL_2105079, EPI_ISL_2105080, EPI_ISL_2105081, EPI_ISL_2105082, EPI_ISL_2105083, EPI_ISL_2105084, EPI_ISL_2105085, EPI_ISL_2105086, EPI_ISL_2105087, EPI_ISL_2105090, EPI_ISL_2105091, EPI_ISL_2105092, EPI_ISL_2105093, EPI_ISL_2105094, EPI_ISL_2105095, EPI_ISL_2105096, EPI_ISL_2105097 | | | |
| see above | UW Virology Lab | UW Virology Lab | Pavitra Roychoudhury, Hong Xie, Lasata Shrestha, Tien V. Nguyen, Shah Mohamed Bakhash, Michelle Lin, Noah R. Baker, Ricardo Perez, Sean Ellis, Nathan Breit, Robert J. Livingston, Meei-Li Huang, Keith R Jerome, Patrick Mathias, Alexander Greninger |
| EPI_ISL_2105098, EPI_ISL_2105099 | SD Public Health Laboratory | Centers for Disease Control and Prevention Division of Viral Diseases, Pathogen Discovery | Mili Sheth, Sarah Nobles, Jasmine Padilla, Mark Burroughs, Shoshona Le, Katie Dillon, Peter Cook, Clinton R. Paden, Dhwani Batra, Krista Queen, Kristen Knipe, Dakota Howard, Yvette Unoarumhi, Darlene Wagner, Matthew Schmerer, Ben L. Rambo-Martin, Kristine Lacek, Sam Shepard, Alison Laufer Halpin, Dave Wentworth, Vivien Dugan, Suxiang Tong, Justin Lee |
| EPI_ISL_2105100, EPI_ISL_2105101, EPI_ISL_2105102 | RI State Health Laboratories | Centers for Disease Control and Prevention Division of Viral Diseases, Pathogen Discovery | Mili Sheth, Sarah Nobles, Jasmine Padilla, Mark Burroughs, Shoshona Le, Katie Dillon, Peter Cook, Clinton R. Paden, Dhwani Batra, Krista Queen, Kristen Knipe, Dakota Howard, Yvette Unoarumhi, Darlene Wagner, Matthew Schmerer, Ben L. Rambo-Martin, Kristine Lacek, Sam Shepard, Alison Laufer Halpin, Dave Wentworth, Vivien Dugan, Suxiang Tong, Justin Lee |
| EPI_ISL_2105103 | SD Public Health Laboratory | Centers for Disease Control and Prevention Division of Viral Diseases, Pathogen Discovery | Mili Sheth, Sarah Nobles, Jasmine Padilla, Mark Burroughs, Shoshona Le, Katie Dillon, Peter Cook, Clinton R. Paden, Dhwani Batra, Krista Queen, Kristen Knipe, Dakota Howard, Yvette Unoarumhi, Darlene Wagner, Matthew Schmerer, Ben L. Rambo-Martin, Kristine Lacek, Sam Shepard, Alison Laufer Halpin, Dave Wentworth, Vivien Dugan, Suxiang Tong, Justin Lee |
| EPI_ISL_2105104 | MT Public Health Laboratory | Centers for Disease Control and Prevention Division of Viral Diseases, Pathogen Discovery | Mili Sheth, Sarah Nobles, Jasmine Padilla, Mark Burroughs, Shoshona Le, Katie Dillon, Peter Cook, Clinton R. Paden, Dhwani Batra, Krista Queen, Kristen Knipe, Dakota Howard, Yvette Unoarumhi, Darlene Wagner, Matthew Schmerer, Ben L. Rambo-Martin, Kristine Lacek, Sam Shepard, Alison Laufer Halpin, Dave Wentworth, Vivien Dugan, Suxiang Tong, Justin Lee |
| EPI_ISL_2105105 | KY State Public Health Lab | Centers for Disease Control and Prevention Division of Viral Diseases, Pathogen Discovery | Mili Sheth, Sarah Nobles, Jasmine Padilla, Mark Burroughs, Shoshona Le, Katie Dillon, Peter Cook, Clinton R. Paden, Dhwani Batra, Krista Queen, Kristen Knipe, Dakota Howard, Yvette Unoarumhi, Darlene Wagner, Matthew Schmerer, Ben L. Rambo-Martin, Kristine Lacek, Sam Shepard, Alison Laufer Halpin, Dave Wentworth, Vivien Dugan, Suxiang Tong, Justin Lee |
| EPI_ISL_2105106 | RI State Health Laboratories | Centers for Disease Control and Prevention Division of Viral Diseases, Pathogen Discovery | Mili Sheth, Sarah Nobles, Jasmine Padilla, Mark Burroughs, Shoshona Le, Katie Dillon, Peter Cook, Clinton R. Paden, Dhwani Batra, Krista Queen, Kristen Knipe, Dakota Howard, Yvette Unoarumhi, Darlene Wagner, Matthew Schmerer, Ben L. Rambo-Martin, Kristine Lacek, Sam Shepard, Alison Laufer Halpin, Dave Wentworth, Vivien Dugan, Suxiang Tong, Justin Lee |
| EPI_ISL_2105107, EPI_ISL_2105108, | CA-Los Angeles County Public Health Laboratory | Centers for Disease Control and Prevention Division of Viral Diseases, Pathogen | Mili Sheth, Sarah Nobles, Jasmine Padilla, Mark Burroughs, Shoshona Le, Katie Dillon, Peter Cook, Clinton R. Paden, Dhwani Batra, Krista Queen, Kristen Knipe, Dakota Howard, Yvette Unoarumhi, Darlene Wagner, Matthew Schmerer, Ben L. Rambo-Martin, Kristine Lacek, Sam Shepard, Alison Laufer Halpin, Dave Wentworth, Vivien Dugan, Suxiang Tong, Justin Lee |

| | | | |
|---|---|---|---|
| EPI_ISL_2105109 | | Discovery | |
| EPI_ISL_2105110 | IN State Department of Health Laboratory Services | Centers for Disease Control and Prevention Division of Viral Diseases, Pathogen Discovery | Mili Sheth, Sarah Nobles, Jasmine Padilla, Mark Burroughs, Shoshona Le, Katie Dillon, Peter Cook, Clinton R. Paden, Dhwani Batra, Krista Queen, Kristen Knipe, Dakota Howard, Yvette Unoarumhi, Darlene Wagner, Matthew Schmerer, Ben L. Rambo-Martin, Kristine Lacek, Sam Shepard, Alison Laufer Halpin, Dave Wentworth, Vivien Dugan, Suxiang Tong, Justin Lee |
| EPI_ISL_2105111, EPI_ISL_2105112, EPI_ISL_2105113 | RI State Health Laboratories | Centers for Disease Control and Prevention Division of Viral Diseases, Pathogen Discovery | Mili Sheth, Sarah Nobles, Jasmine Padilla, Mark Burroughs, Shoshona Le, Katie Dillon, Peter Cook, Clinton R. Paden, Dhwani Batra, Krista Queen, Kristen Knipe, Dakota Howard, Yvette Unoarumhi, Darlene Wagner, Matthew Schmerer, Ben L. Rambo-Martin, Kristine Lacek, Sam Shepard, Alison Laufer Halpin, Dave Wentworth, Vivien Dugan, Suxiang Tong, Justin Lee |
| EPI_ISL_2105114, EPI_ISL_2105115, EPI_ISL_2105116 | SD Public Health Laboratory | Centers for Disease Control and Prevention Division of Viral Diseases, Pathogen Discovery | Mili Sheth, Sarah Nobles, Jasmine Padilla, Mark Burroughs, Shoshona Le, Katie Dillon, Peter Cook, Clinton R. Paden, Dhwani Batra, Krista Queen, Kristen Knipe, Dakota Howard, Yvette Unoarumhi, Darlene Wagner, Matthew Schmerer, Ben L. Rambo-Martin, Kristine Lacek, Sam Shepard, Alison Laufer Halpin, Dave Wentworth, Vivien Dugan, Suxiang Tong, Justin Lee |
| EPI_ISL_2105117, EPI_ISL_2105118, EPI_ISL_2105119 | RI State Health Laboratories | Centers for Disease Control and Prevention Division of Viral Diseases, Pathogen Discovery | Mili Sheth, Sarah Nobles, Jasmine Padilla, Mark Burroughs, Shoshona Le, Katie Dillon, Peter Cook, Clinton R. Paden, Dhwani Batra, Krista Queen, Kristen Knipe, Dakota Howard, Yvette Unoarumhi, Darlene Wagner, Matthew Schmerer, Ben L. Rambo-Martin, Kristine Lacek, Sam Shepard, Alison Laufer Halpin, Dave Wentworth, Vivien Dugan, Suxiang Tong, Justin Lee |
| EPI_ISL_2105120 | MT Public Health Laboratory | Centers for Disease Control and Prevention Division of Viral Diseases, Pathogen Discovery | Mili Sheth, Sarah Nobles, Jasmine Padilla, Mark Burroughs, Shoshona Le, Katie Dillon, Peter Cook, Clinton R. Paden, Dhwani Batra, Krista Queen, Kristen Knipe, Dakota Howard, Yvette Unoarumhi, Darlene Wagner, Matthew Schmerer, Ben L. Rambo-Martin, Kristine Lacek, Sam Shepard, Alison Laufer Halpin, Dave Wentworth, Vivien Dugan, Suxiang Tong, Justin Lee |
| EPI_ISL_2105121 | RI State Health Laboratories | Centers for Disease Control and Prevention Division of Viral Diseases, Pathogen Discovery | Mili Sheth, Sarah Nobles, Jasmine Padilla, Mark Burroughs, Shoshona Le, Katie Dillon, Peter Cook, Clinton R. Paden, Dhwani Batra, Krista Queen, Kristen Knipe, Dakota Howard, Yvette Unoarumhi, Darlene Wagner, Matthew Schmerer, Ben L. Rambo-Martin, Kristine Lacek, Sam Shepard, Alison Laufer Halpin, Dave Wentworth, Vivien Dugan, Suxiang Tong, Justin Lee |
| EPI_ISL_2105122 | KY State Public Health Lab | Centers for Disease Control and Prevention Division of Viral Diseases, Pathogen Discovery | Mili Sheth, Sarah Nobles, Jasmine Padilla, Mark Burroughs, Shoshona Le, Katie Dillon, Peter Cook, Clinton R. Paden, Dhwani Batra, Krista Queen, Kristen Knipe, Dakota Howard, Yvette Unoarumhi, Darlene Wagner, Matthew Schmerer, Ben L. Rambo-Martin, Kristine Lacek, Sam Shepard, Alison Laufer Halpin, Dave Wentworth, Vivien Dugan, Suxiang Tong, Justin Lee |
| EPI_ISL_2105123 | SD Public Health Laboratory | Centers for Disease Control and Prevention Division of Viral Diseases, Pathogen Discovery | Mili Sheth, Sarah Nobles, Jasmine Padilla, Mark Burroughs, Shoshona Le, Katie Dillon, Peter Cook, Clinton R. Paden, Dhwani Batra, Krista Queen, Kristen Knipe, Dakota Howard, Yvette Unoarumhi, Darlene Wagner, Matthew Schmerer, Ben L. Rambo-Martin, Kristine Lacek, Sam Shepard, Alison Laufer Halpin, Dave Wentworth, Vivien Dugan, Suxiang Tong, Justin Lee |
| EPI_ISL_2105124 | MT Public Health Laboratory | Centers for Disease Control and Prevention Division of Viral Diseases, Pathogen Discovery | Mili Sheth, Sarah Nobles, Jasmine Padilla, Mark Burroughs, Shoshona Le, Katie Dillon, Peter Cook, Clinton R. Paden, Dhwani Batra, Krista Queen, Kristen Knipe, Dakota Howard, Yvette Unoarumhi, Darlene Wagner, Matthew Schmerer, Ben L. Rambo-Martin, Kristine Lacek, Sam Shepard, Alison Laufer Halpin, Dave Wentworth, Vivien Dugan, Suxiang Tong, Justin Lee |
| EPI_ISL_2105125 | SD Public Health Laboratory | Centers for Disease Control and Prevention Division of Viral Diseases, Pathogen Discovery | Mili Sheth, Sarah Nobles, Jasmine Padilla, Mark Burroughs, Shoshona Le, Katie Dillon, Peter Cook, Clinton R. Paden, Dhwani Batra, Krista Queen, Kristen Knipe, Dakota Howard, Yvette Unoarumhi, Darlene Wagner, Matthew Schmerer, Ben L. Rambo-Martin, Kristine Lacek, Sam Shepard, Alison Laufer Halpin, Dave Wentworth, Vivien Dugan, Suxiang Tong, Justin Lee |
| EPI_ISL_2105126 | RI State Health Laboratories | Centers for Disease Control and Prevention Division of Viral Diseases, Pathogen Discovery | Mili Sheth, Sarah Nobles, Jasmine Padilla, Mark Burroughs, Shoshona Le, Katie Dillon, Peter Cook, Clinton R. Paden, Dhwani Batra, Krista Queen, Kristen Knipe, Dakota Howard, Yvette Unoarumhi, Darlene Wagner, Matthew Schmerer, Ben L. Rambo-Martin, Kristine Lacek, Sam Shepard, Alison Laufer Halpin, Dave Wentworth, Vivien Dugan, Suxiang Tong, Justin Lee |
| EPI_ISL_2105127 | IN State Department of Health Laboratory Services | Centers for Disease Control and Prevention Division of Viral Diseases, Pathogen Discovery | Mili Sheth, Sarah Nobles, Jasmine Padilla, Mark Burroughs, Shoshona Le, Katie Dillon, Peter Cook, Clinton R. Paden, Dhwani Batra, Krista Queen, Kristen Knipe, Dakota Howard, Yvette Unoarumhi, Darlene Wagner, Matthew Schmerer, Ben L. Rambo-Martin, Kristine Lacek, Sam Shepard, Alison Laufer Halpin, Dave Wentworth, Vivien Dugan, Suxiang Tong, Justin Lee |
| EPI_ISL_2105128 | CA-Los Angeles County Public Health Laboratory | Centers for Disease Control and Prevention Division of Viral Diseases, Pathogen Discovery | Mili Sheth, Sarah Nobles, Jasmine Padilla, Mark Burroughs, Shoshona Le, Katie Dillon, Peter Cook, Clinton R. Paden, Dhwani Batra, Krista Queen, Kristen Knipe, Dakota Howard, Yvette Unoarumhi, Darlene Wagner, Matthew Schmerer, Ben L. Rambo-Martin, Kristine Lacek, Sam Shepard, Alison Laufer Halpin, Dave Wentworth, Vivien Dugan, Suxiang Tong, Justin Lee |
| EPI_ISL_2105129 | RI State Health Laboratories | Centers for Disease Control and Prevention Division of Viral Diseases, Pathogen Discovery | Mili Sheth, Sarah Nobles, Jasmine Padilla, Mark Burroughs, Shoshona Le, Katie Dillon, Peter Cook, Clinton R. Paden, Dhwani Batra, Krista Queen, Kristen Knipe, Dakota Howard, Yvette Unoarumhi, Darlene Wagner, Matthew Schmerer, Ben L. Rambo-Martin, Kristine Lacek, Sam Shepard, Alison Laufer Halpin, Dave Wentworth, Vivien Dugan, Suxiang Tong, Justin Lee |
| EPI_ISL_2105130 | SD Public Health Laboratory | Centers for Disease Control and Prevention Division of Viral Diseases, Pathogen Discovery | Mili Sheth, Sarah Nobles, Jasmine Padilla, Mark Burroughs, Shoshona Le, Katie Dillon, Peter Cook, Clinton R. Paden, Dhwani Batra, Krista Queen, Kristen Knipe, Dakota Howard, Yvette Unoarumhi, Darlene Wagner, Matthew Schmerer, Ben L. Rambo-Martin, Kristine Lacek, Sam Shepard, Alison Laufer Halpin, Dave Wentworth, Vivien Dugan, Suxiang Tong, Justin Lee |
| EPI_ISL_2105131 | MT Public Health Laboratory | Centers for Disease Control and Prevention Division of Viral Diseases, Pathogen Discovery | Mili Sheth, Sarah Nobles, Jasmine Padilla, Mark Burroughs, Shoshona Le, Katie Dillon, Peter Cook, Clinton R. Paden, Dhwani Batra, Krista Queen, Kristen Knipe, Dakota Howard, Yvette Unoarumhi, Darlene Wagner, Matthew Schmerer, Ben L. Rambo-Martin, Kristine Lacek, Sam Shepard, Alison Laufer Halpin, Dave Wentworth, Vivien Dugan, Suxiang Tong, Justin Lee |
| EPI_ISL_2105132, EPI_ISL_2105133 | RI State Health Laboratories | Centers for Disease Control and Prevention Division of Viral Diseases, Pathogen Discovery | Mili Sheth, Sarah Nobles, Jasmine Padilla, Mark Burroughs, Shoshona Le, Katie Dillon, Peter Cook, Clinton R. Paden, Dhwani Batra, Krista Queen, Kristen Knipe, Dakota Howard, Yvette Unoarumhi, Darlene Wagner, Matthew Schmerer, Ben L. Rambo-Martin, Kristine Lacek, Sam Shepard, Alison Laufer Halpin, Dave Wentworth, Vivien Dugan, Suxiang Tong, Justin Lee |
| EPI_ISL_2105134, EPI_ISL_2105135 | CA-Los Angeles County Public Health Laboratory | Centers for Disease Control and Prevention Division of Viral Diseases, Pathogen Discovery | Mili Sheth, Sarah Nobles, Jasmine Padilla, Mark Burroughs, Shoshona Le, Katie Dillon, Peter Cook, Clinton R. Paden, Dhwani Batra, Krista Queen, Kristen Knipe, Dakota Howard, Yvette Unoarumhi, Darlene Wagner, Matthew Schmerer, Ben L. Rambo-Martin, Kristine Lacek, Sam Shepard, Alison Laufer Halpin, Dave Wentworth, Vivien Dugan, Suxiang Tong, Justin Lee |
| EPI_ISL_2105136, EPI_ISL_2105137, EPI_ISL_2105138 | RI State Health Laboratories | Centers for Disease Control and Prevention Division of Viral Diseases, Pathogen Discovery | Mili Sheth, Sarah Nobles, Jasmine Padilla, Mark Burroughs, Shoshona Le, Katie Dillon, Peter Cook, Clinton R. Paden, Dhwani Batra, Krista Queen, Kristen Knipe, Dakota Howard, Yvette Unoarumhi, Darlene Wagner, Matthew Schmerer, Ben L. Rambo-Martin, Kristine Lacek, Sam Shepard, Alison Laufer Halpin, Dave Wentworth, Vivien Dugan, Suxiang Tong, Justin Lee |
| EPI_ISL_2105139, EPI_ISL_2105140 | MT Public Health Laboratory | Centers for Disease Control and Prevention Division of Viral Diseases, Pathogen Discovery | Mili Sheth, Sarah Nobles, Jasmine Padilla, Mark Burroughs, Shoshona Le, Katie Dillon, Peter Cook, Clinton R. Paden, Dhwani Batra, Krista Queen, Kristen Knipe, Dakota Howard, Yvette Unoarumhi, Darlene Wagner, Matthew Schmerer, Ben L. Rambo-Martin, Kristine Lacek, Sam Shepard, Alison Laufer Halpin, Dave Wentworth, Vivien Dugan, Suxiang Tong, Justin Lee |
| EPI_ISL_2105141, EPI_ISL_2105142 | SD Public Health Laboratory | Centers for Disease Control and Prevention Division of Viral Diseases, Pathogen Discovery | Mili Sheth, Sarah Nobles, Jasmine Padilla, Mark Burroughs, Shoshona Le, Katie Dillon, Peter Cook, Clinton R. Paden, Dhwani Batra, Krista Queen, Kristen Knipe, Dakota Howard, Yvette Unoarumhi, Darlene Wagner, Matthew Schmerer, Ben L. Rambo-Martin, Kristine Lacek, Sam Shepard, Alison Laufer Halpin, Dave Wentworth, Vivien Dugan, Suxiang Tong, Justin Lee |
| EPI_ISL_2105143, EPI_ISL_2105144 | RI State Health Laboratories | Centers for Disease Control and Prevention Division of Viral Diseases, Pathogen Discovery | Mili Sheth, Sarah Nobles, Jasmine Padilla, Mark Burroughs, Shoshona Le, Katie Dillon, Peter Cook, Clinton R. Paden, Dhwani Batra, Krista Queen, Kristen Knipe, Dakota Howard, Yvette Unoarumhi, Darlene Wagner, Matthew Schmerer, Ben L. Rambo-Martin, Kristine Lacek, Sam Shepard, Alison Laufer Halpin, Dave Wentworth, Vivien Dugan, Suxiang Tong, Justin Lee |
| EPI_ISL_2105145, EPI_ISL_2105146 | KY State Public Health Lab | Centers for Disease Control and Prevention Division of Viral Diseases, Pathogen Discovery | Mili Sheth, Sarah Nobles, Jasmine Padilla, Mark Burroughs, Shoshona Le, Katie Dillon, Peter Cook, Clinton R. Paden, Dhwani Batra, Krista Queen, Kristen Knipe, Dakota Howard, Yvette Unoarumhi, Darlene Wagner, Matthew Schmerer, Ben L. Rambo-Martin, Kristine Lacek, Sam Shepard, Alison Laufer Halpin, Dave Wentworth, Vivien Dugan, Suxiang Tong, Justin Lee |
| EPI_ISL_2105147 | MT Public Health Laboratory | Centers for Disease Control and Prevention Division of Viral Diseases, Pathogen Discovery | Mili Sheth, Sarah Nobles, Jasmine Padilla, Mark Burroughs, Shoshona Le, Katie Dillon, Peter Cook, Clinton R. Paden, Dhwani Batra, Krista Queen, Kristen Knipe, Dakota Howard, Yvette Unoarumhi, Darlene Wagner, Matthew Schmerer, Ben L. Rambo-Martin, Kristine Lacek, Sam Shepard, Alison Laufer Halpin, Dave Wentworth, Vivien Dugan, Suxiang Tong, Justin Lee |
| EPI_ISL_2105148 | SD Public Health Laboratory | Centers for Disease Control and Prevention Division of Viral Diseases, Pathogen Discovery | Mili Sheth, Sarah Nobles, Jasmine Padilla, Mark Burroughs, Shoshona Le, Katie Dillon, Peter Cook, Clinton R. Paden, Dhwani Batra, Krista Queen, Kristen Knipe, Dakota Howard, Yvette Unoarumhi, Darlene Wagner, Matthew Schmerer, Ben L. Rambo-Martin, Kristine Lacek, Sam Shepard, Alison Laufer Halpin, Dave Wentworth, Vivien Dugan, Suxiang Tong, Justin Lee |
| EPI_ISL_2105149 | RI State Health Laboratories | Centers for Disease Control and Prevention Division of Viral Diseases, Pathogen Discovery | Mili Sheth, Sarah Nobles, Jasmine Padilla, Mark Burroughs, Shoshona Le, Katie Dillon, Peter Cook, Clinton R. Paden, Dhwani Batra, Krista Queen, Kristen Knipe, Dakota Howard, Yvette Unoarumhi, Darlene Wagner, Matthew Schmerer, Ben L. Rambo-Martin, Kristine Lacek, Sam Shepard, Alison Laufer Halpin, Dave Wentworth, Vivien Dugan, Suxiang Tong, Justin Lee |
| EPI_ISL_2105150 | KY State Public Health Lab | Centers for Disease Control and Prevention Division of Viral Diseases, Pathogen Discovery | Mili Sheth, Sarah Nobles, Jasmine Padilla, Mark Burroughs, Shoshona Le, Katie Dillon, Peter Cook, Clinton R. Paden, Dhwani Batra, Krista Queen, Kristen Knipe, Dakota Howard, Yvette Unoarumhi, Darlene Wagner, Matthew Schmerer, Ben L. Rambo-Martin, Kristine Lacek, Sam Shepard, Alison Laufer Halpin, Dave Wentworth, Vivien Dugan, Suxiang Tong, Justin Lee |
| EPI_ISL_2105151, EPI_ISL_2105152 | MT Public Health Laboratory | Centers for Disease Control and Prevention Division of Viral Diseases, Pathogen Discovery | Mili Sheth, Sarah Nobles, Jasmine Padilla, Mark Burroughs, Shoshona Le, Katie Dillon, Peter Cook, Clinton R. Paden, Dhwani Batra, Krista Queen, Kristen Knipe, Dakota Howard, Yvette Unoarumhi, Darlene Wagner, Matthew Schmerer, Ben L. Rambo-Martin, Kristine Lacek, Sam Shepard, Alison Laufer Halpin, Dave Wentworth, Vivien Dugan, Suxiang Tong, Justin Lee |
| EPI_ISL_2105153 | RI State Health Laboratories | Centers for Disease Control and Prevention Division of Viral Diseases, Pathogen Discovery | Mili Sheth, Sarah Nobles, Jasmine Padilla, Mark Burroughs, Shoshona Le, Katie Dillon, Peter Cook, Clinton R. Paden, Dhwani Batra, Krista Queen, Kristen Knipe, Dakota Howard, Yvette Unoarumhi, Darlene Wagner, Matthew Schmerer, Ben L. Rambo-Martin, Kristine Lacek, Sam Shepard, Alison Laufer Halpin, Dave Wentworth, Vivien Dugan, Suxiang Tong, Justin Lee |
| EPI_ISL_2105154 | MT Public Health Laboratory | Centers for Disease Control and Prevention Division of Viral Diseases, Pathogen Discovery | Mili Sheth, Sarah Nobles, Jasmine Padilla, Mark Burroughs, Shoshona Le, Katie Dillon, Peter Cook, Clinton R. Paden, Dhwani Batra, Krista Queen, Kristen Knipe, Dakota Howard, Yvette Unoarumhi, Darlene Wagner, Matthew Schmerer, Ben L. Rambo-Martin, Kristine Lacek, Sam Shepard, Alison Laufer Halpin, Dave Wentworth, Vivien Dugan, Suxiang Tong, Justin Lee |
| EPI_ISL_2105155 | KY State Public Health Lab | Centers for Disease Control and Prevention Division of Viral Diseases, Pathogen Discovery | Mili Sheth, Sarah Nobles, Jasmine Padilla, Mark Burroughs, Shoshona Le, Katie Dillon, Peter Cook, Clinton R. Paden, Dhwani Batra, Krista Queen, Kristen Knipe, Dakota Howard, Yvette Unoarumhi, Darlene Wagner, Matthew Schmerer, Ben L. Rambo-Martin, Kristine Lacek, Sam Shepard, Alison Laufer Halpin, Dave Wentworth, Vivien Dugan, Suxiang Tong, Justin Lee |
| EPI_ISL_2105156 | SD Public Health Laboratory | Centers for Disease Control and Prevention | Mili Sheth, Sarah Nobles, Jasmine Padilla, Mark Burroughs, Shoshona Le, Katie Dillon, Peter Cook, Clinton R. Paden, Dhwani Batra, Krista Queen, Kristen Knipe, Dakota Howard, Yvette Unoarumhi, Darlene Wagner, Matthew Schmerer, Ben L. Rambo-Martin, Kristine Lacek, Sam Shepard, |

| | | | |
|---|---|---|---|
| | | Division of Viral Diseases, Pathogen Discovery | Alison Laufer Halpin, Dave Wentworth, Vivien Dugan, Suxiang Tong, Justin Lee |
| EPI_ISL_2105157 | IN State Department of Health Laboratory Services | Centers for Disease Control and Prevention Division of Viral Diseases, Pathogen Discovery | Mili Sheth, Sarah Nobles, Jasmine Padilla, Mark Burroughs, Shoshona Le, Katie Dillon, Peter Cook, Clinton R. Paden, Dhwani Batra, Krista Queen, Kristen Knipe, Dakota Howard, Yvette Unoarumhi, Darlene Wagner, Matthew Schmerer, Ben L. Rambo-Martin, Kristine Lacek, Sam Shepard, Alison Laufer Halpin, Dave Wentworth, Vivien Dugan, Suxiang Tong, Justin Lee |
| EPI_ISL_2105158 | CA-Los Angeles County Public Health Laboratory | Centers for Disease Control and Prevention Division of Viral Diseases, Pathogen Discovery | Mili Sheth, Sarah Nobles, Jasmine Padilla, Mark Burroughs, Shoshona Le, Katie Dillon, Peter Cook, Clinton R. Paden, Dhwani Batra, Krista Queen, Kristen Knipe, Dakota Howard, Yvette Unoarumhi, Darlene Wagner, Matthew Schmerer, Ben L. Rambo-Martin, Kristine Lacek, Sam Shepard, Alison Laufer Halpin, Dave Wentworth, Vivien Dugan, Suxiang Tong, Justin Lee |
| EPI_ISL_2105159 | IA State Hygienic Laboratory | Centers for Disease Control and Prevention Division of Viral Diseases, Pathogen Discovery | Mili Sheth, Sarah Nobles, Jasmine Padilla, Mark Burroughs, Shoshona Le, Katie Dillon, Peter Cook, Clinton R. Paden, Dhwani Batra, Krista Queen, Kristen Knipe, Dakota Howard, Yvette Unoarumhi, Darlene Wagner, Matthew Schmerer, Ben L. Rambo-Martin, Kristine Lacek, Sam Shepard, Alison Laufer Halpin, Dave Wentworth, Vivien Dugan, Suxiang Tong, Justin Lee |
| EPI_ISL_2105160 | CT-Dr. Katherine A. Kelley State Public Health Lab | Centers for Disease Control and Prevention Division of Viral Diseases, Pathogen Discovery | Mili Sheth, Sarah Nobles, Jasmine Padilla, Mark Burroughs, Shoshona Le, Katie Dillon, Peter Cook, Clinton R. Paden, Dhwani Batra, Krista Queen, Kristen Knipe, Dakota Howard, Yvette Unoarumhi, Darlene Wagner, Matthew Schmerer, Ben L. Rambo-Martin, Kristine Lacek, Sam Shepard, Alison Laufer Halpin, Dave Wentworth, Vivien Dugan, Suxiang Tong, Justin Lee |
| EPI_ISL_2105161 | NJ Public Health and Environmental Laboratories | Centers for Disease Control and Prevention Division of Viral Diseases, Pathogen Discovery | Mili Sheth, Sarah Nobles, Jasmine Padilla, Mark Burroughs, Shoshona Le, Katie Dillon, Peter Cook, Clinton R. Paden, Dhwani Batra, Krista Queen, Kristen Knipe, Dakota Howard, Yvette Unoarumhi, Darlene Wagner, Matthew Schmerer, Ben L. Rambo-Martin, Kristine Lacek, Sam Shepard, Alison Laufer Halpin, Dave Wentworth, Vivien Dugan, Suxiang Tong, Justin Lee |
| EPI_ISL_2105162 | NH Dept. of Health and Human Services Public Health Labs | Centers for Disease Control and Prevention Division of Viral Diseases, Pathogen Discovery | Mili Sheth, Sarah Nobles, Jasmine Padilla, Mark Burroughs, Shoshona Le, Katie Dillon, Peter Cook, Clinton R. Paden, Dhwani Batra, Krista Queen, Kristen Knipe, Dakota Howard, Yvette Unoarumhi, Darlene Wagner, Matthew Schmerer, Ben L. Rambo-Martin, Kristine Lacek, Sam Shepard, Alison Laufer Halpin, Dave Wentworth, Vivien Dugan, Suxiang Tong, Justin Lee |
| EPI_ISL_2105163 | WA State Department of Health | Centers for Disease Control and Prevention Division of Viral Diseases, Pathogen Discovery | Mili Sheth, Sarah Nobles, Jasmine Padilla, Mark Burroughs, Shoshona Le, Katie Dillon, Peter Cook, Clinton R. Paden, Dhwani Batra, Krista Queen, Kristen Knipe, Dakota Howard, Yvette Unoarumhi, Darlene Wagner, Matthew Schmerer, Ben L. Rambo-Martin, Kristine Lacek, Sam Shepard, Alison Laufer Halpin, Dave Wentworth, Vivien Dugan, Suxiang Tong, Justin Lee |
| EPI_ISL_2105164 | AZ SPHL, Arizona Department of Health Services | Centers for Disease Control and Prevention Division of Viral Diseases, Pathogen Discovery | Mili Sheth, Sarah Nobles, Jasmine Padilla, Mark Burroughs, Shoshona Le, Katie Dillon, Peter Cook, Clinton R. Paden, Dhwani Batra, Krista Queen, Kristen Knipe, Dakota Howard, Yvette Unoarumhi, Darlene Wagner, Matthew Schmerer, Ben L. Rambo-Martin, Kristine Lacek, Sam Shepard, Alison Laufer Halpin, Dave Wentworth, Vivien Dugan, Suxiang Tong, Justin Lee |
| EPI_ISL_2105165 | KS Health and Environmental Laboratories | Centers for Disease Control and Prevention Division of Viral Diseases, Pathogen Discovery | Mili Sheth, Sarah Nobles, Jasmine Padilla, Mark Burroughs, Shoshona Le, Katie Dillon, Peter Cook, Clinton R. Paden, Dhwani Batra, Krista Queen, Kristen Knipe, Dakota Howard, Yvette Unoarumhi, Darlene Wagner, Matthew Schmerer, Ben L. Rambo-Martin, Kristine Lacek, Sam Shepard, Alison Laufer Halpin, Dave Wentworth, Vivien Dugan, Suxiang Tong, Justin Lee |
| EPI_ISL_2105166 | IA State Hygienic Laboratory | Centers for Disease Control and Prevention Division of Viral Diseases, Pathogen Discovery | Mili Sheth, Sarah Nobles, Jasmine Padilla, Mark Burroughs, Shoshona Le, Katie Dillon, Peter Cook, Clinton R. Paden, Dhwani Batra, Krista Queen, Kristen Knipe, Dakota Howard, Yvette Unoarumhi, Darlene Wagner, Matthew Schmerer, Ben L. Rambo-Martin, Kristine Lacek, Sam Shepard, Alison Laufer Halpin, Dave Wentworth, Vivien Dugan, Suxiang Tong, Justin Lee |
| EPI_ISL_2105167 | KS Health and Environmental Laboratories | Centers for Disease Control and Prevention Division of Viral Diseases, Pathogen Discovery | Mili Sheth, Sarah Nobles, Jasmine Padilla, Mark Burroughs, Shoshona Le, Katie Dillon, Peter Cook, Clinton R. Paden, Dhwani Batra, Krista Queen, Kristen Knipe, Dakota Howard, Yvette Unoarumhi, Darlene Wagner, Matthew Schmerer, Ben L. Rambo-Martin, Kristine Lacek, Sam Shepard, Alison Laufer Halpin, Dave Wentworth, Vivien Dugan, Suxiang Tong, Justin Lee |
| EPI_ISL_2105168 | NC State Laboratory of Public Health | Centers for Disease Control and Prevention Division of Viral Diseases, Pathogen Discovery | Mili Sheth, Sarah Nobles, Jasmine Padilla, Mark Burroughs, Shoshona Le, Katie Dillon, Peter Cook, Clinton R. Paden, Dhwani Batra, Krista Queen, Kristen Knipe, Dakota Howard, Yvette Unoarumhi, Darlene Wagner, Matthew Schmerer, Ben L. Rambo-Martin, Kristine Lacek, Sam Shepard, Alison Laufer Halpin, Dave Wentworth, Vivien Dugan, Suxiang Tong, Justin Lee |
| EPI_ISL_2105169 | OH Department of Health Laboratory | Centers for Disease Control and Prevention Division of Viral Diseases, Pathogen Discovery | Mili Sheth, Sarah Nobles, Jasmine Padilla, Mark Burroughs, Shoshona Le, Katie Dillon, Peter Cook, Clinton R. Paden, Dhwani Batra, Krista Queen, Kristen Knipe, Dakota Howard, Yvette Unoarumhi, Darlene Wagner, Matthew Schmerer, Ben L. Rambo-Martin, Kristine Lacek, Sam Shepard, Alison Laufer Halpin, Dave Wentworth, Vivien Dugan, Suxiang Tong, Justin Lee |
| EPI_ISL_2105170 | NJ Public Health and Environmental Laboratories | Centers for Disease Control and Prevention Division of Viral Diseases, Pathogen Discovery | Mili Sheth, Sarah Nobles, Jasmine Padilla, Mark Burroughs, Shoshona Le, Katie Dillon, Peter Cook, Clinton R. Paden, Dhwani Batra, Krista Queen, Kristen Knipe, Dakota Howard, Yvette Unoarumhi, Darlene Wagner, Matthew Schmerer, Ben L. Rambo-Martin, Kristine Lacek, Sam Shepard, Alison Laufer Halpin, Dave Wentworth, Vivien Dugan, Suxiang Tong, Justin Lee |
| EPI_ISL_2105171 | CT-Dr. Katherine A. Kelley State Public Health Lab | Centers for Disease Control and Prevention Division of Viral Diseases, Pathogen Discovery | Mili Sheth, Sarah Nobles, Jasmine Padilla, Mark Burroughs, Shoshona Le, Katie Dillon, Peter Cook, Clinton R. Paden, Dhwani Batra, Krista Queen, Kristen Knipe, Dakota Howard, Yvette Unoarumhi, Darlene Wagner, Matthew Schmerer, Ben L. Rambo-Martin, Kristine Lacek, Sam Shepard, Alison Laufer Halpin, Dave Wentworth, Vivien Dugan, Suxiang Tong, Justin Lee |
| EPI_ISL_2105172 | OH Department of Health Laboratory | Centers for Disease Control and Prevention Division of Viral Diseases, Pathogen Discovery | Mili Sheth, Sarah Nobles, Jasmine Padilla, Mark Burroughs, Shoshona Le, Katie Dillon, Peter Cook, Clinton R. Paden, Dhwani Batra, Krista Queen, Kristen Knipe, Dakota Howard, Yvette Unoarumhi, Darlene Wagner, Matthew Schmerer, Ben L. Rambo-Martin, Kristine Lacek, Sam Shepard, Alison Laufer Halpin, Dave Wentworth, Vivien Dugan, Suxiang Tong, Justin Lee |
| EPI_ISL_2105173 | WVDHHR - Office of Laboratory Services | Centers for Disease Control and Prevention Division of Viral Diseases, Pathogen Discovery | Mili Sheth, Sarah Nobles, Jasmine Padilla, Mark Burroughs, Shoshona Le, Katie Dillon, Peter Cook, Clinton R. Paden, Dhwani Batra, Krista Queen, Kristen Knipe, Dakota Howard, Yvette Unoarumhi, Darlene Wagner, Matthew Schmerer, Ben L. Rambo-Martin, Kristine Lacek, Sam Shepard, Alison Laufer Halpin, Dave Wentworth, Vivien Dugan, Suxiang Tong, Justin Lee |
| EPI_ISL_2105174 | WA State Department of Health | Centers for Disease Control and Prevention Division of Viral Diseases, Pathogen Discovery | Mili Sheth, Sarah Nobles, Jasmine Padilla, Mark Burroughs, Shoshona Le, Katie Dillon, Peter Cook, Clinton R. Paden, Dhwani Batra, Krista Queen, Kristen Knipe, Dakota Howard, Yvette Unoarumhi, Darlene Wagner, Matthew Schmerer, Ben L. Rambo-Martin, Kristine Lacek, Sam Shepard, Alison Laufer Halpin, Dave Wentworth, Vivien Dugan, Suxiang Tong, Justin Lee |
| EPI_ISL_2105175 | NC State Laboratory of Public Health | Centers for Disease Control and Prevention Division of Viral Diseases, Pathogen Discovery | Mili Sheth, Sarah Nobles, Jasmine Padilla, Mark Burroughs, Shoshona Le, Katie Dillon, Peter Cook, Clinton R. Paden, Dhwani Batra, Krista Queen, Kristen Knipe, Dakota Howard, Yvette Unoarumhi, Darlene Wagner, Matthew Schmerer, Ben L. Rambo-Martin, Kristine Lacek, Sam Shepard, Alison Laufer Halpin, Dave Wentworth, Vivien Dugan, Suxiang Tong, Justin Lee |
| EPI_ISL_2105176 | CT-Dr. Katherine A. Kelley State Public Health Lab | Centers for Disease Control and Prevention Division of Viral Diseases, Pathogen Discovery | Mili Sheth, Sarah Nobles, Jasmine Padilla, Mark Burroughs, Shoshona Le, Katie Dillon, Peter Cook, Clinton R. Paden, Dhwani Batra, Krista Queen, Kristen Knipe, Dakota Howard, Yvette Unoarumhi, Darlene Wagner, Matthew Schmerer, Ben L. Rambo-Martin, Kristine Lacek, Sam Shepard, Alison Laufer Halpin, Dave Wentworth, Vivien Dugan, Suxiang Tong, Justin Lee |
| EPI_ISL_2105177 | IA State Hygienic Laboratory | Centers for Disease Control and Prevention Division of Viral Diseases, Pathogen Discovery | Mili Sheth, Sarah Nobles, Jasmine Padilla, Mark Burroughs, Shoshona Le, Katie Dillon, Peter Cook, Clinton R. Paden, Dhwani Batra, Krista Queen, Kristen Knipe, Dakota Howard, Yvette Unoarumhi, Darlene Wagner, Matthew Schmerer, Ben L. Rambo-Martin, Kristine Lacek, Sam Shepard, Alison Laufer Halpin, Dave Wentworth, Vivien Dugan, Suxiang Tong, Justin Lee |
| EPI_ISL_2105178 | NH Dept. of Health and Human Services Public Health Labs | Centers for Disease Control and Prevention Division of Viral Diseases, Pathogen Discovery | Mili Sheth, Sarah Nobles, Jasmine Padilla, Mark Burroughs, Shoshona Le, Katie Dillon, Peter Cook, Clinton R. Paden, Dhwani Batra, Krista Queen, Kristen Knipe, Dakota Howard, Yvette Unoarumhi, Darlene Wagner, Matthew Schmerer, Ben L. Rambo-Martin, Kristine Lacek, Sam Shepard, Alison Laufer Halpin, Dave Wentworth, Vivien Dugan, Suxiang Tong, Justin Lee |
| EPI_ISL_2105179, EPI_ISL_2105180 | NC State Laboratory of Public Health | Centers for Disease Control and Prevention Division of Viral Diseases, Pathogen Discovery | Mili Sheth, Sarah Nobles, Jasmine Padilla, Mark Burroughs, Shoshona Le, Katie Dillon, Peter Cook, Clinton R. Paden, Dhwani Batra, Krista Queen, Kristen Knipe, Dakota Howard, Yvette Unoarumhi, Darlene Wagner, Matthew Schmerer, Ben L. Rambo-Martin, Kristine Lacek, Sam Shepard, Alison Laufer Halpin, Dave Wentworth, Vivien Dugan, Suxiang Tong, Justin Lee |
| EPI_ISL_2105181 | OH Department of Health Laboratory | Centers for Disease Control and Prevention Division of Viral Diseases, Pathogen Discovery | Mili Sheth, Sarah Nobles, Jasmine Padilla, Mark Burroughs, Shoshona Le, Katie Dillon, Peter Cook, Clinton R. Paden, Dhwani Batra, Krista Queen, Kristen Knipe, Dakota Howard, Yvette Unoarumhi, Darlene Wagner, Matthew Schmerer, Ben L. Rambo-Martin, Kristine Lacek, Sam Shepard, Alison Laufer Halpin, Dave Wentworth, Vivien Dugan, Suxiang Tong, Justin Lee |
| EPI_ISL_2105182 | KS Health and Environmental Laboratories | Centers for Disease Control and Prevention Division of Viral Diseases, Pathogen Discovery | Mili Sheth, Sarah Nobles, Jasmine Padilla, Mark Burroughs, Shoshona Le, Katie Dillon, Peter Cook, Clinton R. Paden, Dhwani Batra, Krista Queen, Kristen Knipe, Dakota Howard, Yvette Unoarumhi, Darlene Wagner, Matthew Schmerer, Ben L. Rambo-Martin, Kristine Lacek, Sam Shepard, Alison Laufer Halpin, Dave Wentworth, Vivien Dugan, Suxiang Tong, Justin Lee |
| EPI_ISL_2105183 | IA State Hygienic Laboratory | Centers for Disease Control and Prevention Division of Viral Diseases, Pathogen Discovery | Mili Sheth, Sarah Nobles, Jasmine Padilla, Mark Burroughs, Shoshona Le, Katie Dillon, Peter Cook, Clinton R. Paden, Dhwani Batra, Krista Queen, Kristen Knipe, Dakota Howard, Yvette Unoarumhi, Darlene Wagner, Matthew Schmerer, Ben L. Rambo-Martin, Kristine Lacek, Sam Shepard, Alison Laufer Halpin, Dave Wentworth, Vivien Dugan, Suxiang Tong, Justin Lee |
| EPI_ISL_2105184 | MO State Public Health Laboratory | Centers for Disease Control and Prevention Division of Viral Diseases, Pathogen Discovery | Mili Sheth, Sarah Nobles, Jasmine Padilla, Mark Burroughs, Shoshona Le, Katie Dillon, Peter Cook, Clinton R. Paden, Dhwani Batra, Krista Queen, Kristen Knipe, Dakota Howard, Yvette Unoarumhi, Darlene Wagner, Matthew Schmerer, Ben L. Rambo-Martin, Kristine Lacek, Sam Shepard, Alison Laufer Halpin, Dave Wentworth, Vivien Dugan, Suxiang Tong, Justin Lee |
| EPI_ISL_2105185 | KS Health and Environmental Laboratories | Centers for Disease Control and Prevention Division of Viral Diseases, Pathogen Discovery | Mili Sheth, Sarah Nobles, Jasmine Padilla, Mark Burroughs, Shoshona Le, Katie Dillon, Peter Cook, Clinton R. Paden, Dhwani Batra, Krista Queen, Kristen Knipe, Dakota Howard, Yvette Unoarumhi, Darlene Wagner, Matthew Schmerer, Ben L. Rambo-Martin, Kristine Lacek, Sam Shepard, Alison Laufer Halpin, Dave Wentworth, Vivien Dugan, Suxiang Tong, Justin Lee |
| EPI_ISL_2105186 | PR Public Health Lab | Centers for Disease Control and Prevention Division of Viral Diseases, Pathogen Discovery | Mili Sheth, Sarah Nobles, Jasmine Padilla, Mark Burroughs, Shoshona Le, Katie Dillon, Peter Cook, Clinton R. Paden, Dhwani Batra, Krista Queen, Kristen Knipe, Dakota Howard, Yvette Unoarumhi, Darlene Wagner, Matthew Schmerer, Ben L. Rambo-Martin, Kristine Lacek, Sam Shepard, Alison Laufer Halpin, Dave Wentworth, Vivien Dugan, Suxiang Tong, Justin Lee |
| EPI_ISL_2105187 | IA State Hygienic Laboratory | Centers for Disease Control and Prevention Division of Viral Diseases, Pathogen Discovery | Mili Sheth, Sarah Nobles, Jasmine Padilla, Mark Burroughs, Shoshona Le, Katie Dillon, Peter Cook, Clinton R. Paden, Dhwani Batra, Krista Queen, Kristen Knipe, Dakota Howard, Yvette Unoarumhi, Darlene Wagner, Matthew Schmerer, Ben L. Rambo-Martin, Kristine Lacek, Sam Shepard, Alison Laufer Halpin, Dave Wentworth, Vivien Dugan, Suxiang Tong, Justin Lee |
| EPI_ISL_2105188 | KS Health and Environmental Laboratories | Centers for Disease Control and Prevention Division of Viral Diseases, Pathogen Discovery | Mili Sheth, Sarah Nobles, Jasmine Padilla, Mark Burroughs, Shoshona Le, Katie Dillon, Peter Cook, Clinton R. Paden, Dhwani Batra, Krista Queen, Kristen Knipe, Dakota Howard, Yvette Unoarumhi, Darlene Wagner, Matthew Schmerer, Ben L. Rambo-Martin, Kristine Lacek, Sam Shepard, Alison Laufer Halpin, Dave Wentworth, Vivien Dugan, Suxiang Tong, Justin Lee |
| EPI_ISL_2105189 | IA State Hygienic Laboratory | Centers for Disease Control and Prevention | Mili Sheth, Sarah Nobles, Jasmine Padilla, Mark Burroughs, Shoshona Le, Katie Dillon, Peter Cook, Clinton R. Paden, Dhwani Batra, Krista Queen, Kristen Knipe, Dakota Howard, Yvette Unoarumhi, Darlene Wagner, Matthew Schmerer, Ben L. Rambo-Martin, Kristine Lacek, Sam Shepard, |

| | | | |
|---|---|---|---|
| | | Division of Viral Diseases, Pathogen Discovery | Alison Laufer Halpin, Dave Wentworth, Vivien Dugan, Suxiang Tong, Justin Lee |
| EPI_ISL_2105190 | KS Health and Environmental Laboratories | Centers for Disease Control and Prevention Division of Viral Diseases, Pathogen Discovery | Mili Sheth, Sarah Nobles, Jasmine Padilla, Mark Burroughs, Shoshona Le, Katie Dillon, Peter Cook, Clinton R. Paden, Dhwani Batra, Krista Queen, Kristen Knipe, Dakota Howard, Yvette Unoarumhi, Darlene Wagner, Matthew Schmerer, Ben L. Rambo-Martin, Kristine Lacek, Sam Shepard, Alison Laufer Halpin, Dave Wentworth, Vivien Dugan, Suxiang Tong, Justin Lee |
| EPI_ISL_2105191 | OK Public Health Laboratory, Oklahoma State DOH | Centers for Disease Control and Prevention Division of Viral Diseases, Pathogen Discovery | Mili Sheth, Sarah Nobles, Jasmine Padilla, Mark Burroughs, Shoshona Le, Katie Dillon, Peter Cook, Clinton R. Paden, Dhwani Batra, Krista Queen, Kristen Knipe, Dakota Howard, Yvette Unoarumhi, Darlene Wagner, Matthew Schmerer, Ben L. Rambo-Martin, Kristine Lacek, Sam Shepard, Alison Laufer Halpin, Dave Wentworth, Vivien Dugan, Suxiang Tong, Justin Lee |
| EPI_ISL_2105192 | NC State Laboratory of Public Health | Centers for Disease Control and Prevention Division of Viral Diseases, Pathogen Discovery | Mili Sheth, Sarah Nobles, Jasmine Padilla, Mark Burroughs, Shoshona Le, Katie Dillon, Peter Cook, Clinton R. Paden, Dhwani Batra, Krista Queen, Kristen Knipe, Dakota Howard, Yvette Unoarumhi, Darlene Wagner, Matthew Schmerer, Ben L. Rambo-Martin, Kristine Lacek, Sam Shepard, Alison Laufer Halpin, Dave Wentworth, Vivien Dugan, Suxiang Tong, Justin Lee |
| EPI_ISL_2105193, EPI_ISL_2105194 | NH Dept. of Health and Human Services Public Health Labs | Centers for Disease Control and Prevention Division of Viral Diseases, Pathogen Discovery | Mili Sheth, Sarah Nobles, Jasmine Padilla, Mark Burroughs, Shoshona Le, Katie Dillon, Peter Cook, Clinton R. Paden, Dhwani Batra, Krista Queen, Kristen Knipe, Dakota Howard, Yvette Unoarumhi, Darlene Wagner, Matthew Schmerer, Ben L. Rambo-Martin, Kristine Lacek, Sam Shepard, Alison Laufer Halpin, Dave Wentworth, Vivien Dugan, Suxiang Tong, Justin Lee |
| EPI_ISL_2105196 | OH Department of Health Laboratory | Centers for Disease Control and Prevention Division of Viral Diseases, Pathogen Discovery | Mili Sheth, Sarah Nobles, Jasmine Padilla, Mark Burroughs, Shoshona Le, Katie Dillon, Peter Cook, Clinton R. Paden, Dhwani Batra, Krista Queen, Kristen Knipe, Dakota Howard, Yvette Unoarumhi, Darlene Wagner, Matthew Schmerer, Ben L. Rambo-Martin, Kristine Lacek, Sam Shepard, Alison Laufer Halpin, Dave Wentworth, Vivien Dugan, Suxiang Tong, Justin Lee |
| EPI_ISL_2105197 | IA State Hygienic Laboratory | Centers for Disease Control and Prevention Division of Viral Diseases, Pathogen Discovery | Mili Sheth, Sarah Nobles, Jasmine Padilla, Mark Burroughs, Shoshona Le, Katie Dillon, Peter Cook, Clinton R. Paden, Dhwani Batra, Krista Queen, Kristen Knipe, Dakota Howard, Yvette Unoarumhi, Darlene Wagner, Matthew Schmerer, Ben L. Rambo-Martin, Kristine Lacek, Sam Shepard, Alison Laufer Halpin, Dave Wentworth, Vivien Dugan, Suxiang Tong, Justin Lee |
| EPI_ISL_2105198 | KS Health and Environmental Laboratories | Centers for Disease Control and Prevention Division of Viral Diseases, Pathogen Discovery | Mili Sheth, Sarah Nobles, Jasmine Padilla, Mark Burroughs, Shoshona Le, Katie Dillon, Peter Cook, Clinton R. Paden, Dhwani Batra, Krista Queen, Kristen Knipe, Dakota Howard, Yvette Unoarumhi, Darlene Wagner, Matthew Schmerer, Ben L. Rambo-Martin, Kristine Lacek, Sam Shepard, Alison Laufer Halpin, Dave Wentworth, Vivien Dugan, Suxiang Tong, Justin Lee |
| EPI_ISL_2105199 | PR Public Health Lab | Centers for Disease Control and Prevention Division of Viral Diseases, Pathogen Discovery | Mili Sheth, Sarah Nobles, Jasmine Padilla, Mark Burroughs, Shoshona Le, Katie Dillon, Peter Cook, Clinton R. Paden, Dhwani Batra, Krista Queen, Kristen Knipe, Dakota Howard, Yvette Unoarumhi, Darlene Wagner, Matthew Schmerer, Ben L. Rambo-Martin, Kristine Lacek, Sam Shepard, Alison Laufer Halpin, Dave Wentworth, Vivien Dugan, Suxiang Tong, Justin Lee |
| EPI_ISL_2105200 | NJ Public Health and Environmental Laboratories | Centers for Disease Control and Prevention Division of Viral Diseases, Pathogen Discovery | Mili Sheth, Sarah Nobles, Jasmine Padilla, Mark Burroughs, Shoshona Le, Katie Dillon, Peter Cook, Clinton R. Paden, Dhwani Batra, Krista Queen, Kristen Knipe, Dakota Howard, Yvette Unoarumhi, Darlene Wagner, Matthew Schmerer, Ben L. Rambo-Martin, Kristine Lacek, Sam Shepard, Alison Laufer Halpin, Dave Wentworth, Vivien Dugan, Suxiang Tong, Justin Lee |
| EPI_ISL_2105201 | NC State Laboratory of Public Health | Centers for Disease Control and Prevention Division of Viral Diseases, Pathogen Discovery | Mili Sheth, Sarah Nobles, Jasmine Padilla, Mark Burroughs, Shoshona Le, Katie Dillon, Peter Cook, Clinton R. Paden, Dhwani Batra, Krista Queen, Kristen Knipe, Dakota Howard, Yvette Unoarumhi, Darlene Wagner, Matthew Schmerer, Ben L. Rambo-Martin, Kristine Lacek, Sam Shepard, Alison Laufer Halpin, Dave Wentworth, Vivien Dugan, Suxiang Tong, Justin Lee |
| EPI_ISL_2105202 | OH Department of Health Laboratory | Centers for Disease Control and Prevention Division of Viral Diseases, Pathogen Discovery | Mili Sheth, Sarah Nobles, Jasmine Padilla, Mark Burroughs, Shoshona Le, Katie Dillon, Peter Cook, Clinton R. Paden, Dhwani Batra, Krista Queen, Kristen Knipe, Dakota Howard, Yvette Unoarumhi, Darlene Wagner, Matthew Schmerer, Ben L. Rambo-Martin, Kristine Lacek, Sam Shepard, Alison Laufer Halpin, Dave Wentworth, Vivien Dugan, Suxiang Tong, Justin Lee |
| EPI_ISL_2105203 | NH Dept. of Health and Human Services Public Health Labs | Centers for Disease Control and Prevention Division of Viral Diseases, Pathogen Discovery | Mili Sheth, Sarah Nobles, Jasmine Padilla, Mark Burroughs, Shoshona Le, Katie Dillon, Peter Cook, Clinton R. Paden, Dhwani Batra, Krista Queen, Kristen Knipe, Dakota Howard, Yvette Unoarumhi, Darlene Wagner, Matthew Schmerer, Ben L. Rambo-Martin, Kristine Lacek, Sam Shepard, Alison Laufer Halpin, Dave Wentworth, Vivien Dugan, Suxiang Tong, Justin Lee |
| EPI_ISL_2105204, EPI_ISL_2105205 | CT-Dr. Katherine A. Kelley State Public Health Lab | Centers for Disease Control and Prevention Division of Viral Diseases, Pathogen Discovery | Mili Sheth, Sarah Nobles, Jasmine Padilla, Mark Burroughs, Shoshona Le, Katie Dillon, Peter Cook, Clinton R. Paden, Dhwani Batra, Krista Queen, Kristen Knipe, Dakota Howard, Yvette Unoarumhi, Darlene Wagner, Matthew Schmerer, Ben L. Rambo-Martin, Kristine Lacek, Sam Shepard, Alison Laufer Halpin, Dave Wentworth, Vivien Dugan, Suxiang Tong, Justin Lee |
| EPI_ISL_2105206 | NJ Public Health and Environmental Laboratories | Centers for Disease Control and Prevention Division of Viral Diseases, Pathogen Discovery | Mili Sheth, Sarah Nobles, Jasmine Padilla, Mark Burroughs, Shoshona Le, Katie Dillon, Peter Cook, Clinton R. Paden, Dhwani Batra, Krista Queen, Kristen Knipe, Dakota Howard, Yvette Unoarumhi, Darlene Wagner, Matthew Schmerer, Ben L. Rambo-Martin, Kristine Lacek, Sam Shepard, Alison Laufer Halpin, Dave Wentworth, Vivien Dugan, Suxiang Tong, Justin Lee |
| EPI_ISL_2105207, EPI_ISL_2105208 | OH Department of Health Laboratory | Centers for Disease Control and Prevention Division of Viral Diseases, Pathogen Discovery | Mili Sheth, Sarah Nobles, Jasmine Padilla, Mark Burroughs, Shoshona Le, Katie Dillon, Peter Cook, Clinton R. Paden, Dhwani Batra, Krista Queen, Kristen Knipe, Dakota Howard, Yvette Unoarumhi, Darlene Wagner, Matthew Schmerer, Ben L. Rambo-Martin, Kristine Lacek, Sam Shepard, Alison Laufer Halpin, Dave Wentworth, Vivien Dugan, Suxiang Tong, Justin Lee |
| EPI_ISL_2105209, EPI_ISL_2105210 | NC State Laboratory of Public Health | Centers for Disease Control and Prevention Division of Viral Diseases, Pathogen Discovery | Mili Sheth, Sarah Nobles, Jasmine Padilla, Mark Burroughs, Shoshona Le, Katie Dillon, Peter Cook, Clinton R. Paden, Dhwani Batra, Krista Queen, Kristen Knipe, Dakota Howard, Yvette Unoarumhi, Darlene Wagner, Matthew Schmerer, Ben L. Rambo-Martin, Kristine Lacek, Sam Shepard, Alison Laufer Halpin, Dave Wentworth, Vivien Dugan, Suxiang Tong, Justin Lee |
| EPI_ISL_2105211 | NH Dept. of Health and Human Services Public Health Labs | Centers for Disease Control and Prevention Division of Viral Diseases, Pathogen Discovery | Mili Sheth, Sarah Nobles, Jasmine Padilla, Mark Burroughs, Shoshona Le, Katie Dillon, Peter Cook, Clinton R. Paden, Dhwani Batra, Krista Queen, Kristen Knipe, Dakota Howard, Yvette Unoarumhi, Darlene Wagner, Matthew Schmerer, Ben L. Rambo-Martin, Kristine Lacek, Sam Shepard, Alison Laufer Halpin, Dave Wentworth, Vivien Dugan, Suxiang Tong, Justin Lee |
| EPI_ISL_2105212, EPI_ISL_2105213 | CT-Dr. Katherine A. Kelley State Public Health Lab | Centers for Disease Control and Prevention Division of Viral Diseases, Pathogen Discovery | Mili Sheth, Sarah Nobles, Jasmine Padilla, Mark Burroughs, Shoshona Le, Katie Dillon, Peter Cook, Clinton R. Paden, Dhwani Batra, Krista Queen, Kristen Knipe, Dakota Howard, Yvette Unoarumhi, Darlene Wagner, Matthew Schmerer, Ben L. Rambo-Martin, Kristine Lacek, Sam Shepard, Alison Laufer Halpin, Dave Wentworth, Vivien Dugan, Suxiang Tong, Justin Lee |
| EPI_ISL_2105214 | OH Department of Health Laboratory | Centers for Disease Control and Prevention Division of Viral Diseases, Pathogen Discovery | Mili Sheth, Sarah Nobles, Jasmine Padilla, Mark Burroughs, Shoshona Le, Katie Dillon, Peter Cook, Clinton R. Paden, Dhwani Batra, Krista Queen, Kristen Knipe, Dakota Howard, Yvette Unoarumhi, Darlene Wagner, Matthew Schmerer, Ben L. Rambo-Martin, Kristine Lacek, Sam Shepard, Alison Laufer Halpin, Dave Wentworth, Vivien Dugan, Suxiang Tong, Justin Lee |
| EPI_ISL_2105215 | NC State Laboratory of Public Health | Centers for Disease Control and Prevention Division of Viral Diseases, Pathogen Discovery | Mili Sheth, Sarah Nobles, Jasmine Padilla, Mark Burroughs, Shoshona Le, Katie Dillon, Peter Cook, Clinton R. Paden, Dhwani Batra, Krista Queen, Kristen Knipe, Dakota Howard, Yvette Unoarumhi, Darlene Wagner, Matthew Schmerer, Ben L. Rambo-Martin, Kristine Lacek, Sam Shepard, Alison Laufer Halpin, Dave Wentworth, Vivien Dugan, Suxiang Tong, Justin Lee |
| EPI_ISL_2105216 | NH Dept. of Health and Human Services Public Health Labs | Centers for Disease Control and Prevention Division of Viral Diseases, Pathogen Discovery | Mili Sheth, Sarah Nobles, Jasmine Padilla, Mark Burroughs, Shoshona Le, Katie Dillon, Peter Cook, Clinton R. Paden, Dhwani Batra, Krista Queen, Kristen Knipe, Dakota Howard, Yvette Unoarumhi, Darlene Wagner, Matthew Schmerer, Ben L. Rambo-Martin, Kristine Lacek, Sam Shepard, Alison Laufer Halpin, Dave Wentworth, Vivien Dugan, Suxiang Tong, Justin Lee |
| EPI_ISL_2105679 | Nebraska Public Health Laboratory | NPHL COVID-19 Response Team | NPHL COVID-19 Response Team |
| EPI_ISL_2105680 | LESP Nuevo Leon | Instituto de Diagnostico y Referencia Epidemiologicos (INDRE) | Claudia Wong-Arambula, Abril Rodriguez-Maldonado, Vanessa Rivero-Arredondo, Ariadna Medina-Benitez, Joaquin Quiroz-Mercado, Sergio Rangel-Guerrero, Natividad Cruz-Ortiz, Tatiana Nunez-Garcia, Gisela Barrera-Badillo, Lucia Hernandez-Rivas, Irma Lopez-Martinez, Ernesto Ramirez-Gonzalez. |
| EPI_ISL_2105681, EPI_ISL_2105682, EPI_ISL_2105683, EPI_ISL_2105684, EPI_ISL_2105685, EPI_ISL_2105686, EPI_ISL_2105687 | LESP Guanajuato | Instituto de Diagnostico y Referencia Epidemiologicos (INDRE) | Claudia Wong-Arambula, Abril Rodriguez-Maldonado, Vanessa Rivero-Arredondo, Ariadna Medina-Benitez, Joaquin Quiroz-Mercado, Sergio Rangel-Guerrero, Natividad Cruz-Ortiz, Tatiana Nunez-Garcia, Gisela Barrera-Badillo, Lucia Hernandez-Rivas, Irma Lopez-Martinez, Ernesto Ramirez-Gonzalez. |
| EPI_ISL_2105688 | LESP Baja California | Instituto de Diagnostico y Referencia Epidemiologicos (INDRE) | Claudia Wong-Arambula, Abril Rodriguez-Maldonado, Vanessa Rivero-Arredondo, Ariadna Medina-Benitez, Joaquin Quiroz-Mercado, Sergio Rangel-Guerrero, Natividad Cruz-Ortiz, Tatiana Nunez-Garcia, Gisela Barrera-Badillo, Lucia Hernandez-Rivas, Irma Lopez-Martinez, Ernesto Ramirez-Gonzalez. |
| EPI_ISL_2105689, EPI_ISL_2105690, EPI_ISL_2105691, EPI_ISL_2105692, EPI_ISL_2105693 | LESP Quintana Roo | Instituto de Diagnostico y Referencia Epidemiologicos (INDRE) | Claudia Wong-Arambula, Abril Rodriguez-Maldonado, Vanessa Rivero-Arredondo, Ariadna Medina-Benitez, Joaquin Quiroz-Mercado, Sergio Rangel-Guerrero, Natividad Cruz-Ortiz, Tatiana Nunez-Garcia, Gisela Barrera-Badillo, Lucia Hernandez-Rivas, Irma Lopez-Martinez, Ernesto Ramirez-Gonzalez. |
| EPI_ISL_2105694 | LESP Ciudad de Mexico | Instituto de Diagnostico y Referencia Epidemiologicos (INDRE) | Claudia Wong-Arambula, Abril Rodriguez-Maldonado, Vanessa Rivero-Arredondo, Ariadna Medina-Benitez, Joaquin Quiroz-Mercado, Sergio Rangel-Guerrero, Natividad Cruz-Ortiz, Tatiana Nunez-Garcia, Gisela Barrera-Badillo, Lucia Hernandez-Rivas, Irma Lopez-Martinez, Ernesto Ramirez-Gonzalez. |
| EPI_ISL_2105695, EPI_ISL_2105696, EPI_ISL_2105697, EPI_ISL_2105698, EPI_ISL_2105699, EPI_ISL_2105700, EPI_ISL_2105701, EPI_ISL_2105702, EPI_ISL_2105703, EPI_ISL_2105704, EPI_ISL_2105705, EPI_ISL_2105706, EPI_ISL_2105707, EPI_ISL_2105708, EPI_ISL_2105709, EPI_ISL_2105710, EPI_ISL_2105711, EPI_ISL_2105712, EPI_ISL_2105713, EPI_ISL_2105714, EPI_ISL_2105715 | | | |
| see above | LESP Jalisco | Instituto de Diagnostico y Referencia Epidemiologicos (INDRE) | Claudia Wong-Arambula, Abril Rodriguez-Maldonado, Vanessa Rivero-Arredondo, Ariadna Medina-Benitez, Joaquin Quiroz-Mercado, Sergio Rangel-Guerrero, Natividad Cruz-Ortiz, Tatiana Nunez-Garcia, Gisela Barrera-Badillo, Lucia Hernandez-Rivas, Irma Lopez-Martinez, Ernesto Ramirez-Gonzalez. |
| EPI_ISL_2105716, EPI_ISL_2105717, EPI_ISL_2105718 | LESP Ciudad de Mexico | Instituto de Diagnostico y Referencia Epidemiologicos (INDRE) | Claudia Wong-Arambula, Abril Rodriguez-Maldonado, Vanessa Rivero-Arredondo, Ariadna Medina-Benitez, Joaquin Quiroz-Mercado, Sergio Rangel-Guerrero, Natividad Cruz-Ortiz, Tatiana Nunez-Garcia, Gisela Barrera-Badillo, Lucia Hernandez-Rivas, Irma Lopez-Martinez, Ernesto Ramirez-Gonzalez. |
| EPI_ISL_2105721, EPI_ISL_2105722, EPI_ISL_2105723 | Instituto Nacional de Medicina Genomica | Instituto Nacional de Medicina Genomica | Hidalgo-Miranda A, Cedro-Tanda A, Mendoza-Vargas A, Reyes-Grajeda JP, Gonzalez-Barrera D, Rangel-DeLeon D, Munguia-Garza P, Ramirez-Vega O, Escobar-Arrazola, M, Herrera-Montalvo LA. |
| EPI_ISL_2105724, EPI_ISL_2105725, EPI_ISL_2105726, | Arión Genética | Instituto Nacional de Medicina Genomica | Hidalgo-Miranda A, Cedro-Tanda A, Mendoza-Vargas A, Reyes-Grajeda JP, Gonzalez-Barrera D, Rangel-DeLeon D, Munguia-Garza P, Ramirez-Vega O, Escobar-Arrazola M, Uribe-Figueroa Laura, Roldan-Castillo Magaly, Verea Jazmin, Herrera-Montalvo LA. |

| | | | |
|---|---|---|---|
| EPI_ISL_2105727, EPI_ISL_2105728, EPI_ISL_2105729, EPI_ISL_2105730 | | | |
| EPI_ISL_2105731 | Salud Digna | Instituto Nacional de Medicina Genomica | Hidalgo-Miranda A, Cedro-Tanda A, Mendoza-Vargas A, Reyes-Grajeda JP, Abraham Campos-Romero, Moreno-Camacho José Luis, Rodriguez-Gallegos Jorge, Luna-Ruiz Marco, Gonzalez-Barrera D, Rangel-DeLeon D, Munguia-Garza P, Ramirez-Vega O, Escobar-Arrazola, M, Herrera-Montalvo LA. |
| EPI_ISL_2105732, EPI_ISL_2105733, EPI_ISL_2105734, EPI_ISL_2105735, EPI_ISL_2105736, EPI_ISL_2105737 | Arión Genética | Instituto Nacional de Medicina Genomica | Hidalgo-Miranda A, Cedro-Tanda A, Mendoza-Vargas A, Reyes-Grajeda JP, Gonzalez-Barrera D, Rangel-DeLeon D, Munguia-Garza P, Ramirez-Vega O, Escobar-Arrazola M, Uribe-Figueroa Laura, Roldan-Castillo Magaly, Verea Jazmin, Herrera-Montalvo LA. |
| EPI_ISL_2105738 | Salud Digna | Instituto Nacional de Medicina Genomica | Hidalgo-Miranda A, Cedro-Tanda A, Mendoza-Vargas A, Reyes-Grajeda JP, Abraham Campos-Romero, Moreno-Camacho José Luis, Rodriguez-Gallegos Jorge, Luna-Ruiz Marco, Gonzalez-Barrera D, Rangel-DeLeon D, Munguia-Garza P, Ramirez-Vega O, Escobar-Arrazola, M, Herrera-Montalvo LA. |
| EPI_ISL_2105739, EPI_ISL_2105740, EPI_ISL_2105741 | Arión Genética | Instituto Nacional de Medicina Genomica | Hidalgo-Miranda A, Cedro-Tanda A, Mendoza-Vargas A, Reyes-Grajeda JP, Gonzalez-Barrera D, Rangel-DeLeon D, Munguia-Garza P, Ramirez-Vega O, Escobar-Arrazola M, Uribe-Figueroa Laura, Roldan-Castillo Magaly, Verea Jazmin, Herrera-Montalvo LA. |
| EPI_ISL_2105742, EPI_ISL_2105743, EPI_ISL_2105744, EPI_ISL_2105745, EPI_ISL_2105746, EPI_ISL_2105747, EPI_ISL_2105748, EPI_ISL_2105749, EPI_ISL_2105750, EPI_ISL_2105751, EPI_ISL_2105752, EPI_ISL_2105753, EPI_ISL_2105754, EPI_ISL_2105755, EPI_ISL_2105756, EPI_ISL_2105757, EPI_ISL_2105758, EPI_ISL_2105759, EPI_ISL_2105760, EPI_ISL_2105761, EPI_ISL_2105762, EPI_ISL_2105763, EPI_ISL_2105765, EPI_ISL_2105766, EPI_ISL_2105767, EPI_ISL_2105769, EPI_ISL_2105770, EPI_ISL_2105771, EPI_ISL_2105772, EPI_ISL_2105773, EPI_ISL_2105774, EPI_ISL_2105775, EPI_ISL_2105776, EPI_ISL_2105778, EPI_ISL_2105779, EPI_ISL_2105780, EPI_ISL_2105781, EPI_ISL_2105782, EPI_ISL_2105783, EPI_ISL_2105785, EPI_ISL_2105786, EPI_ISL_2105787, EPI_ISL_2105788, EPI_ISL_2105789, EPI_ISL_2105790, EPI_ISL_2105791, EPI_ISL_2105792, EPI_ISL_2105793, EPI_ISL_2105794, EPI_ISL_2105795, EPI_ISL_2105796, EPI_ISL_2105797, EPI_ISL_2105798, EPI_ISL_2105799, EPI_ISL_2105800, EPI_ISL_2105801, EPI_ISL_2105802, EPI_ISL_2105803, EPI_ISL_2105804, EPI_ISL_2105805, EPI_ISL_2105806, EPI_ISL_2105807, EPI_ISL_2105808, EPI_ISL_2105809, EPI_ISL_2105810, EPI_ISL_2105811, EPI_ISL_2105812, EPI_ISL_2105813, EPI_ISL_2105814, EPI_ISL_2105815, EPI_ISL_2105816, EPI_ISL_2105817, EPI_ISL_2105818, EPI_ISL_2105819, EPI_ISL_2105820, EPI_ISL_2105821, EPI_ISL_2105822, EPI_ISL_2105823, EPI_ISL_2105826, EPI_ISL_2105827, EPI_ISL_2105828, EPI_ISL_2105829, EPI_ISL_2105830, EPI_ISL_2105831, EPI_ISL_2105832, EPI_ISL_2105833, EPI_ISL_2105834, EPI_ISL_2105835, EPI_ISL_2105836, EPI_ISL_2105837, EPI_ISL_2105838, EPI_ISL_2105839, EPI_ISL_2105840, EPI_ISL_2105841, EPI_ISL_2105842, EPI_ISL_2105843, EPI_ISL_2105844, EPI_ISL_2105845, EPI_ISL_2105846, EPI_ISL_2105847, EPI_ISL_2105848, EPI_ISL_2105849, EPI_ISL_2105850, EPI_ISL_2105851, EPI_ISL_2105852, EPI_ISL_2105853, EPI_ISL_2105854, EPI_ISL_2105855, EPI_ISL_2105856, EPI_ISL_2105857, EPI_ISL_2105858, EPI_ISL_2105859, EPI_ISL_2105860, EPI_ISL_2105861, EPI_ISL_2105862, EPI_ISL_2105863, EPI_ISL_2105864, EPI_ISL_2105865, EPI_ISL_2105866, EPI_ISL_2105867, EPI_ISL_2105869, EPI_ISL_2105870, EPI_ISL_2105872, EPI_ISL_2105873, EPI_ISL_2105874 | | | |
| see above | Salud Digna | Instituto Nacional de Medicina Genomica | Hidalgo-Miranda A, Cedro-Tanda A, Mendoza-Vargas A, Reyes-Grajeda JP, Abraham Campos-Romero, Moreno-Camacho José Luis, Rodriguez-Gallegos Jorge, Luna-Ruiz Marco, Gonzalez-Barrera D, Rangel-DeLeon D, Munguia-Garza P, Ramirez-Vega O, Escobar-Arrazola, M, Herrera-Montalvo LA. |
| EPI_ISL_2105884, EPI_ISL_2105885, EPI_ISL_2105886, EPI_ISL_2105887, EPI_ISL_2105888, EPI_ISL_2105889, EPI_ISL_2105890, EPI_ISL_2105891, EPI_ISL_2105892, EPI_ISL_2105893, EPI_ISL_2105894, EPI_ISL_2105895, EPI_ISL_2105896, EPI_ISL_2105897, EPI_ISL_2105898, EPI_ISL_2105899, EPI_ISL_2105900, EPI_ISL_2105901, EPI_ISL_2105902, EPI_ISL_2105903, EPI_ISL_2105904, EPI_ISL_2105905, EPI_ISL_2105906, EPI_ISL_2105907, EPI_ISL_2105908, EPI_ISL_2105909, EPI_ISL_2105910, EPI_ISL_2105911, EPI_ISL_2105912, EPI_ISL_2105913, EPI_ISL_2105914, EPI_ISL_2105915, EPI_ISL_2105916, EPI_ISL_2105917, EPI_ISL_2105918, EPI_ISL_2105919, EPI_ISL_2105920, EPI_ISL_2105921, EPI_ISL_2105922, EPI_ISL_2105923, EPI_ISL_2105924, EPI_ISL_2105925, EPI_ISL_2105926, EPI_ISL_2105927, EPI_ISL_2105928, EPI_ISL_2105929, EPI_ISL_2105930, EPI_ISL_2105931 | | | |
| see above | CSL, Maryland Department of Health | CSL, Maryland Department of Health | Keller,E., Loomis,J., Low,K. |
| EPI_ISL_2105960, EPI_ISL_2105961, EPI_ISL_2105962, EPI_ISL_2105963, EPI_ISL_2105964, EPI_ISL_2105965, EPI_ISL_2105966, EPI_ISL_2105967, EPI_ISL_2105968, EPI_ISL_2105969, EPI_ISL_2105970, EPI_ISL_2105971, EPI_ISL_2105972, EPI_ISL_2105973, EPI_ISL_2105974, EPI_ISL_2105975, EPI_ISL_2105976, EPI_ISL_2105977, EPI_ISL_2105978, EPI_ISL_2105979, EPI_ISL_2105980, EPI_ISL_2105981, EPI_ISL_2105982, EPI_ISL_2105983, EPI_ISL_2105984, EPI_ISL_2105985, EPI_ISL_2105986, EPI_ISL_2105987, EPI_ISL_2105988, EPI_ISL_2105989, EPI_ISL_2105990, EPI_ISL_2105991, EPI_ISL_2105993, EPI_ISL_2105994, EPI_ISL_2105995, EPI_ISL_2105996, EPI_ISL_2105997, EPI_ISL_2105998, EPI_ISL_2105999, EPI_ISL_2106000, EPI_ISL_2106001, EPI_ISL_2106002, EPI_ISL_2106003, EPI_ISL_2106004, EPI_ISL_2106005, EPI_ISL_2106006, EPI_ISL_2106007, EPI_ISL_2106008, EPI_ISL_2106009, EPI_ISL_2106010, EPI_ISL_2106011, EPI_ISL_2106012, EPI_ISL_2106013, EPI_ISL_2106014, EPI_ISL_2106015, EPI_ISL_2106016, EPI_ISL_2106017, EPI_ISL_2106018, EPI_ISL_2106019, EPI_ISL_2106020, EPI_ISL_2106021, EPI_ISL_2106022, EPI_ISL_2106023, EPI_ISL_2106024, EPI_ISL_2106025, EPI_ISL_2106026, EPI_ISL_2106027, EPI_ISL_2106028, EPI_ISL_2106029, EPI_ISL_2106030, EPI_ISL_2106031, EPI_ISL_2106032, EPI_ISL_2106033, EPI_ISL_2106034, EPI_ISL_2106035, EPI_ISL_2106036, EPI_ISL_2106037, EPI_ISL_2106038, EPI_ISL_2106039, EPI_ISL_2106040, EPI_ISL_2106041, EPI_ISL_2106042, EPI_ISL_2106043, EPI_ISL_2106044, EPI_ISL_2106045, EPI_ISL_2106046, EPI_ISL_2106047, EPI_ISL_2106048, EPI_ISL_2106049, EPI_ISL_2106050, EPI_ISL_2106051, EPI_ISL_2106052, EPI_ISL_2106053, EPI_ISL_2106054, EPI_ISL_2106055, EPI_ISL_2106056, EPI_ISL_2106057, EPI_ISL_2106058, EPI_ISL_2106059, EPI_ISL_2106060, EPI_ISL_2106061, EPI_ISL_2106062, EPI_ISL_2106063, EPI_ISL_2106064, EPI_ISL_2106065, EPI_ISL_2106066, EPI_ISL_2106067, EPI_ISL_2106068, EPI_ISL_2106069, EPI_ISL_2106070, EPI_ISL_2106071, EPI_ISL_2106072, EPI_ISL_2106073, EPI_ISL_2106074, EPI_ISL_2106075, EPI_ISL_2106076, EPI_ISL_2106077, EPI_ISL_2106078, EPI_ISL_2106079, EPI_ISL_2106080, EPI_ISL_2106081, EPI_ISL_2106082, EPI_ISL_2106083, EPI_ISL_2106084, EPI_ISL_2106085, EPI_ISL_2106086, EPI_ISL_2106087, EPI_ISL_2106088, EPI_ISL_2106089, EPI_ISL_2106090, EPI_ISL_2106091, EPI_ISL_2106092, EPI_ISL_2106093, EPI_ISL_2106094, EPI_ISL_2106095, EPI_ISL_2106096, EPI_ISL_2106097, EPI_ISL_2106098, EPI_ISL_2106099, EPI_ISL_2106100, EPI_ISL_2106101, EPI_ISL_2106102, EPI_ISL_2106103, EPI_ISL_2106104, EPI_ISL_2106105, EPI_ISL_2106106, EPI_ISL_2106107, EPI_ISL_2106108, EPI_ISL_2106109, EPI_ISL_2106110, EPI_ISL_2106111, EPI_ISL_2106112, EPI_ISL_2106113, EPI_ISL_2106114, EPI_ISL_2106115, EPI_ISL_2106116, EPI_ISL_2106117, EPI_ISL_2106118, EPI_ISL_2106119, EPI_ISL_2106120, EPI_ISL_2106121, EPI_ISL_2106122 | | | |
| see above | Respiratory Viruses Branch, Centers for Disease Control and Prevention | Respiratory Viruses Branch, Centers for Disease Control and Prevention | Howard,D., Batra,D., Cook,P.W., Moser,K., Paskey,A., Caravas,J., Rambo-Martin,B., Morrison,S., Gulvick,C., Sammons,S., Unoarumhi,Y., Wagner,D., Roychoudhury,M.S., Xie,H., Shrestha,L., Nguyen,T.V., Bakhash,S.M., Lin,M., Baker,N.R., Perez,R., Ellis,S., Breit,N., Livingston,R.J., Huang,M.-L., Jerome,K.R., Mathias,P., Greninger,A., Paden,C.R., MacCannell,D. |
| EPI_ISL_2106123, EPI_ISL_2106124, EPI_ISL_2106125, EPI_ISL_2106126, EPI_ISL_2106128, EPI_ISL_2106129, EPI_ISL_2106130, EPI_ISL_2106132, EPI_ISL_2106133, EPI_ISL_2106134, EPI_ISL_2106135, EPI_ISL_2106136, EPI_ISL_2106137, EPI_ISL_2106138, EPI_ISL_2106139, EPI_ISL_2106140, EPI_ISL_2106142, EPI_ISL_2106143, EPI_ISL_2106144 | | | |
| see above | Microbiology Division, SC DHEC | Microbiology Division, SC DHEC | Flores,H., Freeman,J. |
| EPI_ISL_2106145, EPI_ISL_2106146, EPI_ISL_2106147, EPI_ISL_2106148, EPI_ISL_2106149, EPI_ISL_2106150, EPI_ISL_2106151, EPI_ISL_2106152 | Microbiology Division, SC DHEC | Microbiology Division, SC DHEC | Freeman,J. |
| EPI_ISL_2106154, EPI_ISL_2106155, EPI_ISL_2106156, EPI_ISL_2106157, EPI_ISL_2106159, EPI_ISL_2106160, EPI_ISL_2106161, EPI_ISL_2106163, EPI_ISL_2106164, EPI_ISL_2106165, EPI_ISL_2106166, EPI_ISL_2106167, EPI_ISL_2106168, EPI_ISL_2106169, EPI_ISL_2106170 | | | |
| see above | Microbiology Division, SC DHEC | Microbiology Division, SC DHEC | Flores,H., Freeman,J. |
| EPI_ISL_2106171, EPI_ISL_2106172, EPI_ISL_2106173, EPI_ISL_2106174, EPI_ISL_2106175, EPI_ISL_2106176 | Infectious Disease Program, Broad Institute of Harvard and MIT | Infectious Disease Program, Broad Institute of Harvard and MIT | Lemieux,J.E., Siddle,K.J., Shaw,B., Adams,G., Pierce,V., Turbett,S., Anahtar,M., Branda,J., Slater,D., Harris,J., Lin,A.E., Gladden-Young,A., Lagerborg,K., Rudy,M., DeRuff,K., Carter,A., Normandin,E., Bauer,M., Reilly,S., Tomkins-Tinch,C., Loreth,C., Chaluvadi,S., Neumann,A., Cusick,C., Chapman,S.B., Gnirke,A., Flowers,K., Cerrato,F., Birren,B.W., Gallagher,G., Smole,S., Park,D.J., MacInnis,B.L., Ryan,E., LaRocque,R., Rosenberg,E., Sabeti,P.C. |
| EPI_ISL_2106177, EPI_ISL_2106178, EPI_ISL_2106179 | Infectious Diseases, NC SLPH COVID-19 Response Team | Infectious Diseases, NC SLPH COVID-19 Response Team | Chase,K. |
| EPI_ISL_2106180 | Department of General Services Division of Consolidated Laboratory Services, Virginia Division of Consolidated Laboratory Services Sequencing Submission Group | Department of General Services Division of Consolidated Laboratory Services, Virginia Division of Consolidated Laboratory Services Sequencing Submission Group | Fink,L. |
| EPI_ISL_2106181, EPI_ISL_2106182 | Los Angeles County Public Health Laboratories | Los Angeles County Public Health Laboratories | Hemarajata,P. and Green,N. |
| EPI_ISL_2106202, EPI_ISL_2106203, EPI_ISL_2106204, EPI_ISL_2106206, EPI_ISL_2106208 | DPH, Massachusetts State Public Health Lab | DPH, Massachusetts State Public Health Lab | Lang,A.S., Fink,T., Gallagher,G.R., Smole,S.C. |
| EPI_ISL_2106209, EPI_ISL_2106210, EPI_ISL_2106211, EPI_ISL_2106212, EPI_ISL_2106213, EPI_ISL_2106214, EPI_ISL_2106215, EPI_ISL_2106216, EPI_ISL_2106217, EPI_ISL_2106218, EPI_ISL_2106219, EPI_ISL_2106220, EPI_ISL_2106221, EPI_ISL_2106222, EPI_ISL_2106223, EPI_ISL_2106225, EPI_ISL_2106227, EPI_ISL_2106228 | | | |
| see above | Microbiology Division, SC DHEC | Microbiology Division, SC DHEC | Flores,H., Freeman,J. |
| EPI_ISL_2106283, EPI_ISL_2106284, EPI_ISL_2106285, EPI_ISL_2106286, EPI_ISL_2106287, EPI_ISL_2106289, EPI_ISL_2106290, EPI_ISL_2106291, EPI_ISL_2106292, EPI_ISL_2106293, EPI_ISL_2106294, EPI_ISL_2106295, EPI_ISL_2106297, EPI_ISL_2106298, EPI_ISL_2106299, EPI_ISL_2106300, EPI_ISL_2106301, EPI_ISL_2106302, EPI_ISL_2106304, EPI_ISL_2106305, EPI_ISL_2106306, EPI_ISL_2106307, EPI_ISL_2106308, EPI_ISL_2106309, EPI_ISL_2106310, EPI_ISL_2106311, EPI_ISL_2106312, EPI_ISL_2106313, EPI_ISL_2106314, EPI_ISL_2106315, EPI_ISL_2106316, EPI_ISL_2106317, EPI_ISL_2106318, EPI_ISL_2106319, EPI_ISL_2106320, EPI_ISL_2106321, EPI_ISL_2106322, EPI_ISL_2106323, EPI_ISL_2106326, EPI_ISL_2106328, EPI_ISL_2106329, EPI_ISL_2106330, EPI_ISL_2106331, EPI_ISL_2106332, EPI_ISL_2106334, EPI_ISL_2106335, EPI_ISL_2106336, EPI_ISL_2106337, EPI_ISL_2106338, EPI_ISL_2106339, EPI_ISL_2106340, EPI_ISL_2106341, EPI_ISL_2106342, EPI_ISL_2106343, EPI_ISL_2106345, EPI_ISL_2106346, EPI_ISL_2106348, EPI_ISL_2106349, EPI_ISL_2106350, EPI_ISL_2106351, EPI_ISL_2106352, EPI_ISL_2106353, EPI_ISL_2106354, EPI_ISL_2106355, EPI_ISL_2106356, EPI_ISL_2106357, EPI_ISL_2106358, EPI_ISL_2106359, EPI_ISL_2106360, EPI_ISL_2106361, EPI_ISL_2106363, EPI_ISL_2106364, EPI_ISL_2106365, EPI_ISL_2106366, EPI_ISL_2106367, EPI_ISL_2106368, EPI_ISL_2106369, EPI_ISL_2106370, EPI_ISL_2106371, EPI_ISL_2106372, EPI_ISL_2106373, EPI_ISL_2106374, EPI_ISL_2106375, EPI_ISL_2106376, EPI_ISL_2106377, EPI_ISL_2106378, EPI_ISL_2106379, EPI_ISL_2106380, EPI_ISL_2106381, EPI_ISL_2106382, EPI_ISL_2106383, EPI_ISL_2106384, EPI_ISL_2106385, EPI_ISL_2106386, EPI_ISL_2106387, EPI_ISL_2106388, EPI_ISL_2106389, EPI_ISL_2106391, EPI_ISL_2106392, EPI_ISL_2106393, EPI_ISL_2106394, EPI_ISL_2106395, EPI_ISL_2106396, EPI_ISL_2106397, EPI_ISL_2106398, EPI_ISL_2106399, EPI_ISL_2106400, EPI_ISL_2106402, EPI_ISL_2106403, EPI_ISL_2106404, EPI_ISL_2106405, EPI_ISL_2106406, EPI_ISL_2106407, EPI_ISL_2106408, EPI_ISL_2106410, EPI_ISL_2106411, EPI_ISL_2106412, EPI_ISL_2106413, EPI_ISL_2106414, EPI_ISL_2106415, EPI_ISL_2106418, EPI_ISL_2106420, EPI_ISL_2106421, EPI_ISL_2106422, EPI_ISL_2106423, EPI_ISL_2106424, EPI_ISL_2106425, EPI_ISL_2106427, EPI_ISL_2106428, EPI_ISL_2106429, EPI_ISL_2106430, EPI_ISL_2106431, EPI_ISL_2106432, EPI_ISL_2106433, EPI_ISL_2106434, EPI_ISL_2106435, EPI_ISL_2106436, EPI_ISL_2106437, EPI_ISL_2106438, EPI_ISL_2106439, EPI_ISL_2106440, EPI_ISL_2106441, EPI_ISL_2106442, EPI_ISL_2106443, EPI_ISL_2106444, EPI_ISL_2106445, EPI_ISL_2106446, EPI_ISL_2106447, EPI_ISL_2106448, EPI_ISL_2106449, EPI_ISL_2106450, EPI_ISL_2106451, EPI_ISL_2106452, EPI_ISL_2106454, EPI_ISL_2106455, EPI_ISL_2106456, EPI_ISL_2106457, EPI_ISL_2106458, EPI_ISL_2106459, EPI_ISL_2106460, EPI_ISL_2106461, EPI_ISL_2106462, EPI_ISL_2106463, EPI_ISL_2106464, EPI_ISL_2106465, EPI_ISL_2106466, EPI_ISL_2106467, EPI_ISL_2106468, EPI_ISL_2106469, EPI_ISL_2106470, EPI_ISL_2106471, EPI_ISL_2106472, EPI_ISL_2106474, EPI_ISL_2106475, EPI_ISL_2106476, EPI_ISL_2106477, EPI_ISL_2106478, EPI_ISL_2106479, EPI_ISL_2106480, EPI_ISL_2106481, EPI_ISL_2106483, EPI_ISL_2106484, EPI_ISL_2106485, EPI_ISL_2106486, EPI_ISL_2106487, EPI_ISL_2106488, EPI_ISL_2106489, EPI_ISL_2106490, EPI_ISL_2106491, EPI_ISL_2106492, EPI_ISL_2106493, EPI_ISL_2106494, EPI_ISL_2106495, EPI_ISL_2106496, EPI_ISL_2106497, EPI_ISL_2106498, EPI_ISL_2106500, EPI_ISL_2106501, EPI_ISL_2106502, EPI_ISL_2106503, EPI_ISL_2106504, EPI_ISL_2106505, EPI_ISL_2106506, EPI_ISL_2106507, EPI_ISL_2106508, EPI_ISL_2106509, EPI_ISL_2106510, EPI_ISL_2106512, EPI_ISL_2106513, EPI_ISL_2106515, EPI_ISL_2106516, EPI_ISL_2106517, EPI_ISL_2106518, EPI_ISL_2106521, EPI_ISL_2106522, EPI_ISL_2106523, EPI_ISL_2106524, EPI_ISL_2106525, EPI_ISL_2106526, EPI_ISL_2106527, EPI_ISL_2106529, EPI_ISL_2106530, EPI_ISL_2106531, EPI_ISL_2106532, EPI_ISL_2106533, EPI_ISL_2106536, EPI_ISL_2106537, EPI_ISL_2106538, EPI_ISL_2106539, EPI_ISL_2106540, EPI_ISL_2106541, EPI_ISL_2106542, EPI_ISL_2106543, EPI_ISL_2106544, EPI_ISL_2106545, EPI_ISL_2106546, EPI_ISL_2106547, EPI_ISL_2106548, EPI_ISL_2106549, EPI_ISL_2106550, EPI_ISL_2106551, EPI_ISL_2106552, EPI_ISL_2106553, EPI_ISL_2106554, EPI_ISL_2106555, EPI_ISL_2106556, EPI_ISL_2106557, EPI_ISL_2106558, EPI_ISL_2106559, EPI_ISL_2106560, EPI_ISL_2106561, EPI_ISL_2106563, EPI_ISL_2106564, EPI_ISL_2106565, EPI_ISL_2106566, EPI_ISL_2106567, EPI_ISL_2106568, EPI_ISL_2106569, EPI_ISL_2106571, EPI_ISL_2106572, EPI_ISL_2106573, EPI_ISL_2106574, EPI_ISL_2106575, EPI_ISL_2106578, EPI_ISL_2106579, EPI_ISL_2106580, EPI_ISL_2106581, EPI_ISL_2106582, EPI_ISL_2106583, EPI_ISL_2106584, EPI_ISL_2106585, EPI_ISL_2106586, EPI_ISL_2106587, EPI_ISL_2106588, EPI_ISL_2106589, EPI_ISL_2106591, EPI_ISL_2106593 | | | |
| see above | Pandemic Response Lab - NYC | Pandemic Response Lab, R&D | Henry Lee, Michael Hammerling, Melissa Hopkins, Cybill del Castillo, Shinyoung Clair Kang, William Ward, Pradeep Bugga, Sol Rey, Dylan Law, Katharine Nelson, Haiping Hao, Jon Laurent |
| EPI_ISL_2107109, EPI_ISL_2107110 | University of Michigan Clinical Microbiology Laboratory | Lauring Lab, University of Michigan, Department of Microbiology and Immunology | Valesano |
| EPI_ISL_2107350, EPI_ISL_2107351, EPI_ISL_2107352, EPI_ISL_2107353, EPI_ISL_2107354 | Emory Molecular Diagnostics Laboratory, Emory Healthcare | Piantadosi Lab, Emory Department of Pathology | Ahmed Babiker, Anne Piantadosi |
| EPI_ISL_2107355, EPI_ISL_2107356, EPI_ISL_2107357, EPI_ISL_2107358, EPI_ISL_2107359, EPI_ISL_2107360, EPI_ISL_2107361, EPI_ISL_2107362, EPI_ISL_2107363, EPI_ISL_2107364, EPI_ISL_2107365, EPI_ISL_2107366, EPI_ISL_2107367, EPI_ISL_2107368, EPI_ISL_2107369, EPI_ISL_2107370, EPI_ISL_2107371, EPI_ISL_2107373, EPI_ISL_2107374, EPI_ISL_2107375, EPI_ISL_2107376, EPI_ISL_2107377, EPI_ISL_2107378, EPI_ISL_2107379, EPI_ISL_2107380, EPI_ISL_2107381, EPI_ISL_2107382, EPI_ISL_2107383, EPI_ISL_2107384, EPI_ISL_2107385, EPI_ISL_2107386, EPI_ISL_2107387, EPI_ISL_2107388, EPI_ISL_2107389 | | | |
| see above | Grady Memorial Hospital, Clinical | Piantadosi Lab, Emory Department of | Yun F (Wayne) Wang, Anne Piantadosi |

| | | | |
|---|---|---|---|
| | Microbiology Laboratory | Pathology | |
| EPI_ISL_2107390, EPI_ISL_2107391, EPI_ISL_2107392, EPI_ISL_2107393, EPI_ISL_2107394, EPI_ISL_2107395, EPI_ISL_2107396, EPI_ISL_2107397, EPI_ISL_2107398, EPI_ISL_2107399, EPI_ISL_2107400 | | | |
| see above | Emory Molecular Diagnostics Laboratory, Emory Healthcare | Piantadosi Lab, Emory Department of Pathology | Ahmed Babiker, Anne Piantadosi |
| EPI_ISL_2107401, EPI_ISL_2107402, EPI_ISL_2107403, EPI_ISL_2107404 | Grady Memorial Hospital, Clinical Microbiology Laboratory | Piantadosi Lab, Emory Department of Pathology | Yun F (Wayne) Wang, Anne Piantadosi |
| EPI_ISL_2107405, EPI_ISL_2107406 | Emory Molecular Diagnostics Laboratory, Emory Healthcare | Piantadosi Lab, Emory Department of Pathology | Ahmed Babiker, Anne Piantadosi |
| EPI_ISL_2107408 | Grady Memorial Hospital, Clinical Microbiology Laboratory | Piantadosi Lab, Emory Department of Pathology | Yun F (Wayne) Wang, Anne Piantadosi |
| EPI_ISL_2107409, EPI_ISL_2107410, EPI_ISL_2107411, EPI_ISL_2107412, EPI_ISL_2107413, EPI_ISL_2107414, EPI_ISL_2107415, EPI_ISL_2107416, EPI_ISL_2107417, EPI_ISL_2107418, EPI_ISL_2107419, EPI_ISL_2107420, EPI_ISL_2107421, EPI_ISL_2107422, EPI_ISL_2107423, EPI_ISL_2107424, EPI_ISL_2107425, EPI_ISL_2107426, EPI_ISL_2107427, EPI_ISL_2107428, EPI_ISL_2107429, EPI_ISL_2107431 | | | |
| see above | Emory Molecular Diagnostics Laboratory, Emory Healthcare | Piantadosi Lab, Emory Department of Pathology | Ahmed Babiker, Anne Piantadosi |
| EPI_ISL_2107536, EPI_ISL_2107537, EPI_ISL_2107538, EPI_ISL_2107539, EPI_ISL_2107540, EPI_ISL_2107541, EPI_ISL_2107542, EPI_ISL_2107543, EPI_ISL_2107545, EPI_ISL_2107547, EPI_ISL_2107548, EPI_ISL_2107549, EPI_ISL_2107550, EPI_ISL_2107551, EPI_ISL_2107552, EPI_ISL_2107553, EPI_ISL_2107554, EPI_ISL_2107555, EPI_ISL_2107556, EPI_ISL_2107557, EPI_ISL_2107558, EPI_ISL_2107559, EPI_ISL_2107563, EPI_ISL_2107564, EPI_ISL_2107565, EPI_ISL_2107566, EPI_ISL_2107567, EPI_ISL_2107568, EPI_ISL_2107570, EPI_ISL_2107571, EPI_ISL_2107572, EPI_ISL_2107573, EPI_ISL_2107574, EPI_ISL_2107575, EPI_ISL_2107576, EPI_ISL_2107577, EPI_ISL_2107578, EPI_ISL_2107579, EPI_ISL_2107580, EPI_ISL_2107581, EPI_ISL_2107582, EPI_ISL_2107583, EPI_ISL_2107584, EPI_ISL_2107585, EPI_ISL_2107586, EPI_ISL_2107587, EPI_ISL_2107588, EPI_ISL_2107589, EPI_ISL_2107590, EPI_ISL_2107591, EPI_ISL_2107592, EPI_ISL_2107594, EPI_ISL_2107595, EPI_ISL_2107596, EPI_ISL_2107597, EPI_ISL_2107598, EPI_ISL_2107599, EPI_ISL_2107601, EPI_ISL_2107602, EPI_ISL_2107603, EPI_ISL_2107604, EPI_ISL_2107605, EPI_ISL_2107606, EPI_ISL_2107608, EPI_ISL_2107609, EPI_ISL_2107610, EPI_ISL_2107611, EPI_ISL_2107612, EPI_ISL_2107613, EPI_ISL_2107614, EPI_ISL_2107615, EPI_ISL_2107616, EPI_ISL_2107617, EPI_ISL_2107618, EPI_ISL_2107620, EPI_ISL_2107621, EPI_ISL_2107622, EPI_ISL_2107623, EPI_ISL_2107624, EPI_ISL_2107625, EPI_ISL_2107628, EPI_ISL_2107629, EPI_ISL_2107630, EPI_ISL_2107631, EPI_ISL_2107632, EPI_ISL_2107633, EPI_ISL_2107634, EPI_ISL_2107635, EPI_ISL_2107637, EPI_ISL_2107638, EPI_ISL_2107639, EPI_ISL_2107640, EPI_ISL_2107641, EPI_ISL_2107642, EPI_ISL_2107643, EPI_ISL_2107645, EPI_ISL_2107646, EPI_ISL_2107647, EPI_ISL_2107648, EPI_ISL_2107649, EPI_ISL_2107650, EPI_ISL_2107651, EPI_ISL_2107652, EPI_ISL_2107653, EPI_ISL_2107654, EPI_ISL_2107655, EPI_ISL_2107656, EPI_ISL_2107657, EPI_ISL_2107659, EPI_ISL_2107660, EPI_ISL_2107661, EPI_ISL_2107662, EPI_ISL_2107663, EPI_ISL_2107664, EPI_ISL_2107665, EPI_ISL_2107666, EPI_ISL_2107668, EPI_ISL_2107669, EPI_ISL_2107672, EPI_ISL_2107673, EPI_ISL_2107674, EPI_ISL_2107675, EPI_ISL_2107678, EPI_ISL_2107682, EPI_ISL_2107685, EPI_ISL_2107686, EPI_ISL_2107687, EPI_ISL_2107688, EPI_ISL_2107689, EPI_ISL_2107690, EPI_ISL_2107692, EPI_ISL_2107693, EPI_ISL_2107694, EPI_ISL_2107695, EPI_ISL_2107696, EPI_ISL_2107698, EPI_ISL_2107699, EPI_ISL_2107700, EPI_ISL_2107702, EPI_ISL_2107703, EPI_ISL_2107705, EPI_ISL_2107706, EPI_ISL_2107708, EPI_ISL_2107709, EPI_ISL_2107711, EPI_ISL_2107712, EPI_ISL_2107713, EPI_ISL_2107714, EPI_ISL_2107715, EPI_ISL_2107716, EPI_ISL_2107718, EPI_ISL_2107719, EPI_ISL_2107720, EPI_ISL_2107721, EPI_ISL_2107723, EPI_ISL_2107725, EPI_ISL_2107727, EPI_ISL_2107728, EPI_ISL_2107729, EPI_ISL_2107731, EPI_ISL_2107732, EPI_ISL_2107733, EPI_ISL_2107734, EPI_ISL_2107735, EPI_ISL_2107736, EPI_ISL_2107739, EPI_ISL_2107740, EPI_ISL_2107742, EPI_ISL_2107743, EPI_ISL_2107744, EPI_ISL_2107745, EPI_ISL_2107746, EPI_ISL_2107747, EPI_ISL_2107748, EPI_ISL_2107749, EPI_ISL_2107750, EPI_ISL_2107751, EPI_ISL_2107753, EPI_ISL_2107754, EPI_ISL_2107755, EPI_ISL_2107756, EPI_ISL_2107757, EPI_ISL_2107758, EPI_ISL_2107759, EPI_ISL_2107760, EPI_ISL_2107761, EPI_ISL_2107762, EPI_ISL_2107763, EPI_ISL_2107764, EPI_ISL_2107765, EPI_ISL_2107766, EPI_ISL_2107767, EPI_ISL_2107768, EPI_ISL_2107770, EPI_ISL_2107771, EPI_ISL_2107772, EPI_ISL_2107773, EPI_ISL_2107775 | | | |
| see above | Delaware Public Health Lab | Delaware Public Health Lab | Rebecca Savage |
| EPI_ISL_2107776, EPI_ISL_2107777, EPI_ISL_2107778, EPI_ISL_2107779, EPI_ISL_2107780, EPI_ISL_2107781, EPI_ISL_2107782, EPI_ISL_2107783, EPI_ISL_2107784, EPI_ISL_2107785, EPI_ISL_2107786, EPI_ISL_2107787, EPI_ISL_2107788, EPI_ISL_2107789, EPI_ISL_2107790, EPI_ISL_2107791 | | | |
| see above | Department of Laboratory Medicine, Clinical Center, National Institutes of Health | Laboratory of Parasitic Diseases, Systems Genomics Section, National Institute of Allergy and Infectious Diseases, National Institutes of Health | Allison Roder, Stephanie Banakis, Matthew Chung, Jung-ho Youn, Rachel Mercado, Wei Wang, Tara Palmore, Michael Bell, Heike Bailin, Jessica McCormick-Ell, Sanchita Das, Jennifer Kwan, Elodie Ghedin |
| EPI_ISL_2112524, EPI_ISL_2112525, EPI_ISL_2112527 | Epidemiology of Microbial Diseases, Yale School of Public Health | Epidemiology of Microbial Diseases, Yale School of Public Health | Vogels,C.B., Lucas,C., Fauver,J.R., Breban,M.I., Ott,I.M., Alpert,T., Petrone,M.E., Brito,A.F., Watkins,A.E., Kalinich,C.C., Rothman,J.E., Iwasaki,A., Landry,M.L., Grubaugh,N.D. |
| EPI_ISL_2132229, EPI_ISL_2132231, EPI_ISL_2132232, EPI_ISL_2132235, EPI_ISL_2132240, EPI_ISL_2132242, EPI_ISL_2132247 | Clinical Molecular Microbiology Laboratory, UNC Hospitals | Jeremy Wang | Jeremy Wang, Alexander Rubinsteyn, Colleen Rice, Jose Luis Torres Castillo, Brianna de la Houssaye, Jason Smedberg, Shawn Hawken, Melissa Miller, Corbin Jones, Robert Hagan |
| EPI_ISL_2132252, EPI_ISL_2132253, EPI_ISL_2132254, EPI_ISL_2132255, EPI_ISL_2132256, EPI_ISL_2132257, EPI_ISL_2132258, EPI_ISL_2132259, EPI_ISL_2132260, EPI_ISL_2132261, EPI_ISL_2132262, EPI_ISL_2132263, EPI_ISL_2132264, EPI_ISL_2132265, EPI_ISL_2132266, EPI_ISL_2132267, EPI_ISL_2132268, EPI_ISL_2132269, EPI_ISL_2132270, EPI_ISL_2132271, EPI_ISL_2132272, EPI_ISL_2132273, EPI_ISL_2132274, EPI_ISL_2132275, EPI_ISL_2132276, EPI_ISL_2132277, EPI_ISL_2132278, EPI_ISL_2132279, EPI_ISL_2132280, EPI_ISL_2132281, EPI_ISL_2132282, EPI_ISL_2132283, EPI_ISL_2132284, EPI_ISL_2132285, EPI_ISL_2132286, EPI_ISL_2132287, EPI_ISL_2132288, EPI_ISL_2132289, EPI_ISL_2132290, EPI_ISL_2132291, EPI_ISL_2132292, EPI_ISL_2132293, EPI_ISL_2132294, EPI_ISL_2132295, EPI_ISL_2132296, EPI_ISL_2132297, EPI_ISL_2132298, EPI_ISL_2132299, EPI_ISL_2132300, EPI_ISL_2132301, EPI_ISL_2132302, EPI_ISL_2132303, EPI_ISL_2132304, EPI_ISL_2132305, EPI_ISL_2132306, EPI_ISL_2132307, EPI_ISL_2132308, EPI_ISL_2132310, EPI_ISL_2132311, EPI_ISL_2132312, EPI_ISL_2132313, EPI_ISL_2132314, EPI_ISL_2132315, EPI_ISL_2132316, EPI_ISL_2132317, EPI_ISL_2132318, EPI_ISL_2132319, EPI_ISL_2132320, EPI_ISL_2132321, EPI_ISL_2132322, EPI_ISL_2132323, EPI_ISL_2132324, EPI_ISL_2132325, EPI_ISL_2132326, EPI_ISL_2132327, EPI_ISL_2132328, EPI_ISL_2132329, EPI_ISL_2132330, EPI_ISL_2132331, EPI_ISL_2132332, EPI_ISL_2132333, EPI_ISL_2132334, EPI_ISL_2132335, EPI_ISL_2132336, EPI_ISL_2132337, EPI_ISL_2132338, EPI_ISL_2132339, EPI_ISL_2132340, EPI_ISL_2132341, EPI_ISL_2132342, EPI_ISL_2132343, EPI_ISL_2132344, EPI_ISL_2132345, EPI_ISL_2132346, EPI_ISL_2132347, EPI_ISL_2132348, EPI_ISL_2132349, EPI_ISL_2132350, EPI_ISL_2132351, EPI_ISL_2132352, EPI_ISL_2132353, EPI_ISL_2132354, EPI_ISL_2132355, EPI_ISL_2132356, EPI_ISL_2132357, EPI_ISL_2132358, EPI_ISL_2132359, EPI_ISL_2132360, EPI_ISL_2132361, EPI_ISL_2132362, EPI_ISL_2132363, EPI_ISL_2132364, EPI_ISL_2132365, EPI_ISL_2132366, EPI_ISL_2132367, EPI_ISL_2132368, EPI_ISL_2132369, EPI_ISL_2132370, EPI_ISL_2132371, EPI_ISL_2132372, EPI_ISL_2132373, EPI_ISL_2132374, EPI_ISL_2132375, EPI_ISL_2132376, EPI_ISL_2132377, EPI_ISL_2132378, EPI_ISL_2132379, EPI_ISL_2132380, EPI_ISL_2132381, EPI_ISL_2132382, EPI_ISL_2132383, EPI_ISL_2132384, EPI_ISL_2132385, EPI_ISL_2132386, EPI_ISL_2132387, EPI_ISL_2132388, EPI_ISL_2132389, EPI_ISL_2132390, EPI_ISL_2132391, EPI_ISL_2132392, EPI_ISL_2132393, EPI_ISL_2132394, EPI_ISL_2132395, EPI_ISL_2132396, EPI_ISL_2132397, EPI_ISL_2132398, EPI_ISL_2132399, EPI_ISL_2132400, EPI_ISL_2132401, EPI_ISL_2132402, EPI_ISL_2132403, EPI_ISL_2132404, EPI_ISL_2132405, EPI_ISL_2132406, EPI_ISL_2132407, EPI_ISL_2132408, EPI_ISL_2132409, EPI_ISL_2132410, EPI_ISL_2132411, EPI_ISL_2132412, EPI_ISL_2132413, EPI_ISL_2132414, EPI_ISL_2132415, EPI_ISL_2132416, EPI_ISL_2132417, EPI_ISL_2132418, EPI_ISL_2132419, EPI_ISL_2132420, EPI_ISL_2132421, EPI_ISL_2132422, EPI_ISL_2132423, EPI_ISL_2132424, EPI_ISL_2132425, EPI_ISL_2132426, EPI_ISL_2132427, EPI_ISL_2132428, EPI_ISL_2132429, EPI_ISL_2132430, EPI_ISL_2132431, EPI_ISL_2132432, EPI_ISL_2132433, EPI_ISL_2132434, EPI_ISL_2132435, EPI_ISL_2132436, EPI_ISL_2132437, EPI_ISL_2132438, EPI_ISL_2132439, EPI_ISL_2132440, EPI_ISL_2132441, EPI_ISL_2132442, EPI_ISL_2132443, EPI_ISL_2132444, EPI_ISL_2132445, EPI_ISL_2132446, EPI_ISL_2132447, EPI_ISL_2132448, EPI_ISL_2132449, EPI_ISL_2132450, EPI_ISL_2132451, EPI_ISL_2132452, EPI_ISL_2132453, EPI_ISL_2132454, EPI_ISL_2132455, EPI_ISL_2132456, EPI_ISL_2132457, EPI_ISL_2132458, EPI_ISL_2132459, EPI_ISL_2132460, EPI_ISL_2132461, EPI_ISL_2132462, EPI_ISL_2132463, EPI_ISL_2132464, EPI_ISL_2132465, EPI_ISL_2132466, EPI_ISL_2132467, EPI_ISL_2132468, EPI_ISL_2132469, EPI_ISL_2132470, EPI_ISL_2132471, EPI_ISL_2132472, EPI_ISL_2132473, EPI_ISL_2132474, EPI_ISL_2132475, EPI_ISL_2132476, EPI_ISL_2132477, EPI_ISL_2132478, EPI_ISL_2132479, EPI_ISL_2132480, EPI_ISL_2132481, EPI_ISL_2132482, EPI_ISL_2132483, EPI_ISL_2132484, EPI_ISL_2132485, EPI_ISL_2132486, EPI_ISL_2132487, EPI_ISL_2132488, EPI_ISL_2132489, EPI_ISL_2132490, EPI_ISL_2132491, EPI_ISL_2132492, EPI_ISL_2132493, EPI_ISL_2132494, EPI_ISL_2132495, EPI_ISL_2132496, EPI_ISL_2132497, EPI_ISL_2132498, EPI_ISL_2132499, EPI_ISL_2132500, EPI_ISL_2132501, EPI_ISL_2132502, EPI_ISL_2132503, EPI_ISL_2132504, EPI_ISL_2132505, EPI_ISL_2132506, EPI_ISL_2132507, EPI_ISL_2132508, EPI_ISL_2132509, EPI_ISL_2132510, EPI_ISL_2132511, EPI_ISL_2132512, EPI_ISL_2132513, EPI_ISL_2132514, EPI_ISL_2132515, EPI_ISL_2132516, EPI_ISL_2132517, EPI_ISL_2132518, EPI_ISL_2132519, EPI_ISL_2132520, EPI_ISL_2132521, EPI_ISL_2132523, EPI_ISL_2132524, EPI_ISL_2132525, EPI_ISL_2132526, EPI_ISL_2132527, EPI_ISL_2132528, EPI_ISL_2132529, EPI_ISL_2132530, EPI_ISL_2132531, EPI_ISL_2132532 | | | |
| see above | Infinity Biologix | Centers for Disease Control and Prevention Division of Viral Diseases, Pathogen Discovery | Dakota Howard, Dhwani Batra, Peter W. Cook, Kara Moser, Adrian Paskey, Jason Caravas, Benjamin Rambo-Martin, Shatavia Morrison, Christopher Gulvick, Scott Sammons, Yvette Unoarumhi, Darlene Wagner, Matthew Schmerer, Christian Bixby, Yihe Wang, Jonathan Schultz, Chirayu Goswami, Russ Hager, Robin Grimwood, Clinton R. Paden, Duncan MacCannell |
| EPI_ISL_2132533, EPI_ISL_2132534, EPI_ISL_2132535, EPI_ISL_2132537, EPI_ISL_2132538, EPI_ISL_2132539, EPI_ISL_2132540, EPI_ISL_2132541, EPI_ISL_2132542, EPI_ISL_2132543, EPI_ISL_2132544, EPI_ISL_2132545, EPI_ISL_2132546, EPI_ISL_2132547, EPI_ISL_2132549, EPI_ISL_2132550, EPI_ISL_2132551, EPI_ISL_2132552, EPI_ISL_2132553, EPI_ISL_2132555, EPI_ISL_2132556, EPI_ISL_2132557, EPI_ISL_2132558, EPI_ISL_2132559, EPI_ISL_2132560, EPI_ISL_2132561, EPI_ISL_2132562, EPI_ISL_2132563, EPI_ISL_2132565, EPI_ISL_2132567, EPI_ISL_2132568, EPI_ISL_2132569, EPI_ISL_2132570, EPI_ISL_2132571, EPI_ISL_2132572, EPI_ISL_2132573 | | | |
| see above | Aegis Sciences Corporation | Centers for Disease Control and Prevention Division of Viral Diseases, Pathogen Discovery | Dakota Howard, Dhwani Batra, Peter W. Cook, Kara Moser, Adrian Paskey, Jason Caravas, Benjamin Rambo-Martin, Shatavia Morrison, Christopher Gulvick, Scott Sammons, Yvette Unoarumhi, Darlene Wagner, Matthew Schmerer, Cyndi Clark, Patrick Campbell, Rob Case, Vikramsinha Ghorpade, Holly Houdeshell, Ola Kvalvaag, Dillon Nall, Ethan Sanders, Alec Vest, Shaun Westlund, Matthew Hardison, Clinton R. Paden, Duncan MacCannell |
| EPI_ISL_2132574, EPI_ISL_2132575, EPI_ISL_2132576, EPI_ISL_2132577, EPI_ISL_2132578, EPI_ISL_2132579, EPI_ISL_2132580, EPI_ISL_2132581, EPI_ISL_2132582, EPI_ISL_2132583, EPI_ISL_2132584, EPI_ISL_2132585, EPI_ISL_2132586 | | | |
| see above | Quest Diagnostics Incorporated | Centers for Disease Control and Prevention Division of Viral Diseases, Pathogen Discovery | Dakota Howard, Dhwani Batra, Peter W. Cook, Kara Moser, Adrian Paskey, Jason Caravas, Benjamin Rambo-Martin, Shatavia Morrison, Christopher Gulvick, Scott Sammons, Yvette Unoarumhi, Darlene Wagner, Matthew Schmerer, S. H. Rosenthal, A. Gerasimova, R. M. Kagan, B. Anderson, M. Hua, Y. Liu, L.E. Bernstein, K.E. Livingston, A. Perez, I. A. Shlyakhter, R. V. Rolando, R. Owen, P. Tanpaiboon, F. Lacbawan, Clinton R. Paden, Duncan MacCannell |
| EPI_ISL_2132587, EPI_ISL_2132588, EPI_ISL_2132589, EPI_ISL_2132590, EPI_ISL_2132591, EPI_ISL_2132592, EPI_ISL_2132593, EPI_ISL_2132594, EPI_ISL_2132595, EPI_ISL_2132596, EPI_ISL_2132597, EPI_ISL_2132598, EPI_ISL_2132599, EPI_ISL_2132600, EPI_ISL_2132601, EPI_ISL_2132602, EPI_ISL_2132603, EPI_ISL_2132604, EPI_ISL_2132605, EPI_ISL_2132606, EPI_ISL_2132607, EPI_ISL_2132608, EPI_ISL_2132609, EPI_ISL_2132610, EPI_ISL_2132611, EPI_ISL_2132612, EPI_ISL_2132613, EPI_ISL_2132614, EPI_ISL_2132615, EPI_ISL_2132616, EPI_ISL_2132617, EPI_ISL_2132618 | | | |
| see above | Helix/Illumina | Centers for Disease Control and Prevention Division of Viral Diseases, Pathogen Discovery | Dakota Howard, Dhwani Batra, Peter W. Cook, Kara Moser, Adrian Paskey, Jason Caravas, Benjamin Rambo-Martin, Shatavia Morrison, Christopher Gulvick, Scott Sammons, Yvette Unoarumhi, Darlene Wagner, Matthew Schmerer, Eileen de Feo, Jan Antico, Christine Tran, Matthew Tolentino, Shannon Wickline, Kim Gietzen, Brad Sickler, Jingtao Liu, Eric Allen, Phil Febbo, Nicole L. Washington, Simon White, Geraint Levan, Kelly Schiabor Barrett, Elizabeth Cirulli, Alexandre Bolze, Ary Ascencio, Charlotte Rivera-Garcia, Ryan Cho, Jason Nguyen, Sherry Wang, Jimmy Ramirez, Tyler Cassens, Efren Sandoval, Magnus Isaksson, William Lee, David Becker, Marc Laurent, James Lu, Clinton R. Paden, Duncan MacCannell |
| EPI_ISL_2132619, EPI_ISL_2132620, EPI_ISL_2132621, EPI_ISL_2132622, EPI_ISL_2132623, EPI_ISL_2132624, EPI_ISL_2132625, EPI_ISL_2132626, EPI_ISL_2132627, EPI_ISL_2132628, EPI_ISL_2132629, EPI_ISL_2132630, EPI_ISL_2132631, EPI_ISL_2133094, EPI_ISL_2133095, EPI_ISL_2133096, EPI_ISL_2133097, EPI_ISL_2133098, EPI_ISL_2133099, EPI_ISL_2133100, EPI_ISL_2133101, EPI_ISL_2133102, EPI_ISL_2133103, EPI_ISL_2133104, EPI_ISL_2133105, EPI_ISL_2133106, EPI_ISL_2133107, EPI_ISL_2133108, EPI_ISL_2133109, EPI_ISL_2133110, EPI_ISL_2133111, EPI_ISL_2133112, EPI_ISL_2133113, EPI_ISL_2133114, EPI_ISL_2133115, EPI_ISL_2133116, EPI_ISL_2133117, EPI_ISL_2133118, EPI_ISL_2133119, EPI_ISL_2133120, EPI_ISL_2133121, EPI_ISL_2133122, EPI_ISL_2133123, EPI_ISL_2133124, EPI_ISL_2133125, EPI_ISL_2133126, EPI_ISL_2133127, EPI_ISL_2133128, EPI_ISL_2133129, EPI_ISL_2133130, EPI_ISL_2133131, EPI_ISL_2133132, EPI_ISL_2133133, EPI_ISL_2133134, EPI_ISL_2133135, EPI_ISL_2133136, EPI_ISL_2133137, EPI_ISL_2133138, EPI_ISL_2133139, EPI_ISL_2133140, EPI_ISL_2133141, EPI_ISL_2133142, EPI_ISL_2133143, EPI_ISL_2133144, EPI_ISL_2133145, EPI_ISL_2133146, EPI_ISL_2133147, EPI_ISL_2133148, EPI_ISL_2133149, EPI_ISL_2133150, EPI_ISL_2133151, EPI_ISL_2133152, EPI_ISL_2133153, EPI_ISL_2133154, EPI_ISL_2133155, EPI_ISL_2133156, EPI_ISL_2133157, EPI_ISL_2133158, EPI_ISL_2133159, EPI_ISL_2133160, EPI_ISL_2133161, EPI_ISL_2133162, EPI_ISL_2133163, EPI_ISL_2133164, EPI_ISL_2133165, EPI_ISL_2133166, EPI_ISL_2133167, EPI_ISL_2133168, EPI_ISL_2133169, EPI_ISL_2133170, EPI_ISL_2133171, EPI_ISL_2133172, EPI_ISL_2133173, EPI_ISL_2133174, EPI_ISL_2133175, EPI_ISL_2133176, EPI_ISL_2133177, EPI_ISL_2133178, EPI_ISL_2133179, EPI_ISL_2133180, EPI_ISL_2133181, EPI_ISL_2133182, EPI_ISL_2133183, EPI_ISL_2133184, EPI_ISL_2133185, EPI_ISL_2133186, EPI_ISL_2133187, EPI_ISL_2133188, EPI_ISL_2133189, EPI_ISL_2133190, EPI_ISL_2133191, EPI_ISL_2133192, EPI_ISL_2133193, EPI_ISL_2133194, EPI_ISL_2133195, EPI_ISL_2133196, EPI_ISL_2133197, EPI_ISL_2133198, EPI_ISL_2133199, EPI_ISL_2133200, EPI_ISL_2133201, EPI_ISL_2133202, EPI_ISL_2133203, EPI_ISL_2133204, EPI_ISL_2133205, EPI_ISL_2133206, EPI_ISL_2133207, EPI_ISL_2133208, EPI_ISL_2133209, EPI_ISL_2133210, EPI_ISL_2133211, EPI_ISL_2133212, EPI_ISL_2133213, EPI_ISL_2133214, EPI_ISL_2133215, EPI_ISL_2133216, EPI_ISL_2133217, EPI_ISL_2133218, EPI_ISL_2133219, EPI_ISL_2133220, EPI_ISL_2133221, EPI_ISL_2133222, EPI_ISL_2133223, EPI_ISL_2133224, EPI_ISL_2133225, EPI_ISL_2133226, EPI_ISL_2133227, EPI_ISL_2133228, EPI_ISL_2133229, EPI_ISL_2133230, EPI_ISL_2133231, EPI_ISL_2133232, EPI_ISL_2133233, EPI_ISL_2133234, EPI_ISL_2133235, EPI_ISL_2133236, EPI_ISL_2133237, EPI_ISL_2133238, EPI_ISL_2133239, EPI_ISL_2133240, EPI_ISL_2133241, EPI_ISL_2133242, EPI_ISL_2133243, EPI_ISL_2133244, EPI_ISL_2133245, EPI_ISL_2133246, EPI_ISL_2133247, EPI_ISL_2133248, EPI_ISL_2133249, EPI_ISL_2133250, EPI_ISL_2133251, EPI_ISL_2133252, EPI_ISL_2133253, EPI_ISL_2133254, EPI_ISL_2133255, EPI_ISL_2133256, EPI_ISL_2133257, EPI_ISL_2133258, EPI_ISL_2133259, EPI_ISL_2133260, EPI_ISL_2133261, EPI_ISL_2133262, EPI_ISL_2133263, EPI_ISL_2133264, EPI_ISL_2133265, EPI_ISL_2133266, EPI_ISL_2133267, EPI_ISL_2133268, EPI_ISL_2133269, EPI_ISL_2133270, EPI_ISL_2133271, EPI_ISL_2133272, EPI_ISL_2133273, EPI_ISL_2133274, EPI_ISL_2133275, EPI_ISL_2133276, EPI_ISL_2133277, EPI_ISL_2133278, EPI_ISL_2133279, EPI_ISL_2133280, EPI_ISL_2133281, EPI_ISL_2133282, EPI_ISL_2133283, EPI_ISL_2133284, EPI_ISL_2133285, EPI_ISL_2133286, EPI_ISL_2133287, EPI_ISL_2133288, EPI_ISL_2133289, EPI_ISL_2133290, EPI_ISL_2133291, EPI_ISL_2133292, EPI_ISL_2133293, EPI_ISL_2133294, EPI_ISL_2133295, EPI_ISL_2133296, EPI_ISL_2133297, EPI_ISL_2133298, EPI_ISL_2133299, EPI_ISL_2133300, EPI_ISL_2133301, EPI_ISL_2133302, EPI_ISL_2133303, EPI_ISL_2133304, EPI_ISL_2133305, EPI_ISL_2133306, EPI_ISL_2133307, EPI_ISL_2133308, EPI_ISL_2133309, EPI_ISL_2133310, EPI_ISL_2133311, EPI_ISL_2133312, EPI_ISL_2133313, EPI_ISL_2133314, EPI_ISL_2133315, EPI_ISL_2133316, EPI_ISL_2133317, EPI_ISL_2133318, EPI_ISL_2133319, EPI_ISL_2133320, EPI_ISL_2133321, EPI_ISL_2133322, EPI_ISL_2133323, EPI_ISL_2133324, EPI_ISL_2133325, EPI_ISL_2133326, EPI_ISL_2133327, EPI_ISL_2133328, EPI_ISL_2133329, EPI_ISL_2133330, EPI_ISL_2133331, EPI_ISL_2133332, EPI_ISL_2133333, EPI_ISL_2133334, EPI_ISL_2133335, EPI_ISL_2133336, EPI_ISL_2133337, EPI_ISL_2133338, EPI_ISL_2133339, EPI_ISL_2133340, EPI_ISL_2133341, EPI_ISL_2133342, EPI_ISL_2133343, EPI_ISL_2133344 | | | |
| see above | Quest Diagnostics Incorporated | Centers for Disease Control and Prevention Division of Viral Diseases, Pathogen Discovery | Dakota Howard, Dhwani Batra, Peter W. Cook, Kara Moser, Adrian Paskey, Jason Caravas, Benjamin Rambo-Martin, Shatavia Morrison, Christopher Gulvick, Scott Sammons, Yvette Unoarumhi, Darlene Wagner, Matthew Schmerer, S. H. Rosenthal, A. Gerasimova, R. M. Kagan, B. Anderson, M. Hua, Y. Liu, L.E. Bernstein, K.E. Livingston, A. Perez, I. A. Shlyakhter, R. V. Rolando, R. Owen, P. Tanpaiboon, F. Lacbawan, Clinton R. Paden, Duncan MacCannell |
| EPI_ISL_2133345, EPI_ISL_2133346, EPI_ISL_2133347, EPI_ISL_2133348, EPI_ISL_2133349, EPI_ISL_2133351, EPI_ISL_2133352, EPI_ISL_2133353, EPI_ISL_2133354, EPI_ISL_2133355, EPI_ISL_2133356, EPI_ISL_2133357, EPI_ISL_2133358, EPI_ISL_2133359, EPI_ISL_2133360, EPI_ISL_2133361, EPI_ISL_2133362, EPI_ISL_2133363, EPI_ISL_2133364, EPI_ISL_2133365, EPI_ISL_2133366, EPI_ISL_2133367, EPI_ISL_2133368, EPI_ISL_2133369, EPI_ISL_2133370, EPI_ISL_2133371, EPI_ISL_2133373, EPI_ISL_2133375, EPI_ISL_2133376, EPI_ISL_2133377, EPI_ISL_2133378, EPI_ISL_2133379, EPI_ISL_2133380 | | | |
| see above | The Jackson Laboratory | The Jackson Laboratory | Long J, Bergeron D, Renzette N, Adams M, Omerza G, Kelly K, Li L |
| EPI_ISL_2133389, EPI_ISL_2133390, EPI_ISL_2133391, EPI_ISL_2133392, EPI_ISL_2133393, EPI_ISL_2133394, EPI_ISL_2133395, EPI_ISL_2133396, EPI_ISL_2133397, EPI_ISL_2133398, EPI_ISL_2133400, EPI_ISL_2133402, EPI_ISL_2133403, EPI_ISL_2133404, EPI_ISL_2133405, EPI_ISL_2133406, EPI_ISL_2133407, EPI_ISL_2133408, EPI_ISL_2133409, EPI_ISL_2133410, EPI_ISL_2133411, EPI_ISL_2133412, EPI_ISL_2133413, EPI_ISL_2133414, EPI_ISL_2133415, EPI_ISL_2133416, EPI_ISL_2133417, EPI_ISL_2133418, EPI_ISL_2133419, EPI_ISL_2133420, EPI_ISL_2133421, EPI_ISL_2133422, EPI_ISL_2133423, EPI_ISL_2133424, EPI_ISL_2133425, EPI_ISL_2133426, EPI_ISL_2133427, EPI_ISL_2133428, EPI_ISL_2133429, EPI_ISL_2133430, EPI_ISL_2133431, EPI_ISL_2133432, EPI_ISL_2133433, EPI_ISL_2133434, EPI_ISL_2133435, EPI_ISL_2133436, EPI_ISL_2133437, EPI_ISL_2133438, EPI_ISL_2133439, EPI_ISL_2133440, EPI_ISL_2133441, EPI_ISL_2133442, EPI_ISL_2133443, EPI_ISL_2133444, EPI_ISL_2133445, EPI_ISL_2133446, EPI_ISL_2133447, EPI_ISL_2133448, EPI_ISL_2133449, EPI_ISL_2133450, EPI_ISL_2133451, EPI_ISL_2133452, EPI_ISL_2133453, EPI_ISL_2133454, EPI_ISL_2133455, EPI_ISL_2133456, EPI_ISL_2133457, EPI_ISL_2133458, EPI_ISL_2133459, EPI_ISL_2133460, EPI_ISL_2133461, EPI_ISL_2133462, EPI_ISL_2133463, EPI_ISL_2133464, EPI_ISL_2133465, EPI_ISL_2133466, EPI_ISL_2133467, EPI_ISL_2133468, EPI_ISL_2133469, EPI_ISL_2133470, EPI_ISL_2133471, EPI_ISL_2133472, EPI_ISL_2133473, EPI_ISL_2133474, EPI_ISL_2133475, EPI_ISL_2133476, EPI_ISL_2133477, EPI_ISL_2133478, EPI_ISL_2133479, EPI_ISL_2133480, EPI_ISL_2133481, EPI_ISL_2133482, EPI_ISL_2133483, EPI_ISL_2133484, EPI_ISL_2133485, EPI_ISL_2133486, EPI_ISL_2133487, EPI_ISL_2133488, EPI_ISL_2133489, EPI_ISL_2133490, EPI_ISL_2133491, EPI_ISL_2133492, EPI_ISL_2133493, EPI_ISL_2133494, EPI_ISL_2133495, EPI_ISL_2133496, EPI_ISL_2133497, EPI_ISL_2133498, EPI_ISL_2133499, EPI_ISL_2133500, EPI_ISL_2133501, EPI_ISL_2133502, EPI_ISL_2133503, EPI_ISL_2133504, EPI_ISL_2133505, EPI_ISL_2133506, EPI_ISL_2133507, EPI_ISL_2133508, EPI_ISL_2133509, EPI_ISL_2133510, EPI_ISL_2133511, EPI_ISL_2133512, EPI_ISL_2133513, EPI_ISL_2133514, EPI_ISL_2133515, EPI_ISL_2133516, EPI_ISL_2133517, EPI_ISL_2133518, EPI_ISL_2133519, EPI_ISL_2133520, EPI_ISL_2133521, EPI_ISL_2133522, EPI_ISL_2133523, EPI_ISL_2133524, EPI_ISL_2133525, EPI_ISL_2133526, EPI_ISL_2133527, EPI_ISL_2133528, EPI_ISL_2133529, EPI_ISL_2133530, EPI_ISL_2133531, EPI_ISL_2133532, EPI_ISL_2133533, EPI_ISL_2133534, EPI_ISL_2133535, EPI_ISL_2133536, EPI_ISL_2133537, EPI_ISL_2133538, EPI_ISL_2133539, EPI_ISL_2133540, EPI_ISL_2133541, EPI_ISL_2133542, EPI_ISL_2133543, EPI_ISL_2133544, EPI_ISL_2133545, EPI_ISL_2133546, EPI_ISL_2133547, EPI_ISL_2133548, EPI_ISL_2133549, EPI_ISL_2133550, EPI_ISL_2133551, EPI_ISL_2133552, EPI_ISL_2133553, EPI_ISL_2133554, EPI_ISL_2133555, EPI_ISL_2133556, EPI_ISL_2133557, EPI_ISL_2133558, EPI_ISL_2133559, EPI_ISL_2133560, EPI_ISL_2133561, EPI_ISL_2133562, EPI_ISL_2133563, EPI_ISL_2133564, EPI_ISL_2133565, EPI_ISL_2133566, EPI_ISL_2133567, EPI_ISL_2133568, EPI_ISL_2133569, EPI_ISL_2133570, EPI_ISL_2133571, EPI_ISL_2133572, EPI_ISL_2133573, EPI_ISL_2133574, EPI_ISL_2133575, EPI_ISL_2133576, EPI_ISL_2133577, EPI_ISL_2133578, EPI_ISL_2133579, EPI_ISL_2133580, EPI_ISL_2133581, EPI_ISL_2133582, EPI_ISL_2133583, EPI_ISL_2133584, EPI_ISL_2133585, EPI_ISL_2133586, EPI_ISL_2133587, EPI_ISL_2133588, EPI_ISL_2133589, EPI_ISL_2133590, EPI_ISL_2133591, EPI_ISL_2133592, EPI_ISL_2133593, EPI_ISL_2133594, EPI_ISL_2133595, EPI_ISL_2133596, EPI_ISL_2133597, EPI_ISL_2133598, EPI_ISL_2133599, EPI_ISL_2133600, EPI_ISL_2133601, EPI_ISL_2133602, EPI_ISL_2133603, EPI_ISL_2133604, EPI_ISL_2133605, EPI_ISL_2133606, EPI_ISL_2133607, EPI_ISL_2133608, EPI_ISL_2133609, EPI_ISL_2133610, EPI_ISL_2133611, EPI_ISL_2133612, EPI_ISL_2133613, EPI_ISL_2133614, EPI_ISL_2133615, EPI_ISL_2133616, EPI_ISL_2133617, EPI_ISL_2133618, EPI_ISL_2133619, EPI_ISL_2133620 | | | |

| | | | |
|---|---|---|---|
| see above | Quest Diagnostics Incorporated | Centers for Disease Control and Prevention Division of Viral Diseases, Pathogen Discovery | Dakota Howard, Dhwani Batra, Peter W. Cook, Kara Moser, Adrian Paskey, Jason Caravas, Benjamin Rambo-Martin, Shatavia Morrison, Christopher Gulvick, Scott Sammons, Yvette Unoarumhi, Darlene Wagner, Matthew Schmerer, S. H. Rosenthal, A. Gerasimova, R. M. Kagan, B. Anderson, M. Hua, Y. Liu, L.E. Bernstein, K.E. Livingston, A. Perez, I. A. Shlyakhter, R. V. Rolando, R. Owen, P. Tanpaiboon, F. Lacbawan, Clinton R. Paden, Duncan MacCannell |
| EPI_ISL_2133621 | Warrior Diagnostics Laboratory | Infectious Disease Program, Broad Institute of Harvard and MIT | Lemieux,J.E., Siddle,K.J., Marshall,J., ONeill,M., Bronson,A., Adams,G., Gladden-Young,A., Lagerborg,K., Rudy,M., DeRuff,K., Carter,A., Normandin,E., Bauer,M., Reilly,S., Tomkins-Tinch,C., Loreth,C., Chaluvadi,S., Birren,B.W., Gallagher,G., Smole,S., Park,D.J., MacInnis,B.L., and Sabeti,P.C. |
| EPI_ISL_2133622 | Quest Diagnostics Incorporated | Centers for Disease Control and Prevention Division of Viral Diseases, Pathogen Discovery | Dakota Howard, Dhwani Batra, Peter W. Cook, Kara Moser, Adrian Paskey, Jason Caravas, Benjamin Rambo-Martin, Shatavia Morrison, Christopher Gulvick, Scott Sammons, Yvette Unoarumhi, Darlene Wagner, Matthew Schmerer, S. H. Rosenthal, A. Gerasimova, R. M. Kagan, B. Anderson, M. Hua, Y. Liu, L.E. Bernstein, K.E. Livingston, A. Perez, I. A. Shlyakhter, R. V. Rolando, R. Owen, P. Tanpaiboon, F. Lacbawan, Clinton R. Paden, Duncan MacCannell |
| EPI_ISL_2133623 | Warrior Diagnostics Laboratory | Infectious Disease Program, Broad Institute of Harvard and MIT | Lemieux,J.E., Siddle,K.J., Marshall,J., ONeill,M., Bronson,A., Adams,G., Gladden-Young,A., Lagerborg,K., Rudy,M., DeRuff,K., Carter,A., Normandin,E., Bauer,M., Reilly,S., Tomkins-Tinch,C., Loreth,C., Chaluvadi,S., Birren,B.W., Gallagher,G., Smole,S., Park,D.J., MacInnis,B.L., and Sabeti,P.C. |
| EPI_ISL_2133624 | Quest Diagnostics Incorporated | Centers for Disease Control and Prevention Division of Viral Diseases, Pathogen Discovery | Dakota Howard, Dhwani Batra, Peter W. Cook, Kara Moser, Adrian Paskey, Jason Caravas, Benjamin Rambo-Martin, Shatavia Morrison, Christopher Gulvick, Scott Sammons, Yvette Unoarumhi, Darlene Wagner, Matthew Schmerer, S. H. Rosenthal, A. Gerasimova, R. M. Kagan, B. Anderson, M. Hua, Y. Liu, L.E. Bernstein, K.E. Livingston, A. Perez, I. A. Shlyakhter, R. V. Rolando, R. Owen, P. Tanpaiboon, F. Lacbawan, Clinton R. Paden, Duncan MacCannell |
| EPI_ISL_2133625, EPI_ISL_2133626 | Warrior Diagnostics Laboratory | Infectious Disease Program, Broad Institute of Harvard and MIT | Lemieux,J.E., Siddle,K.J., Marshall,J., ONeill,M., Bronson,A., Adams,G., Gladden-Young,A., Lagerborg,K., Rudy,M., DeRuff,K., Carter,A., Normandin,E., Bauer,M., Reilly,S., Tomkins-Tinch,C., Loreth,C., Chaluvadi,S., Birren,B.W., Gallagher,G., Smole,S., Park,D.J., MacInnis,B.L., and Sabeti,P.C. |
| EPI_ISL_2133627 | Quest Diagnostics Incorporated | Centers for Disease Control and Prevention Division of Viral Diseases, Pathogen Discovery | Dakota Howard, Dhwani Batra, Peter W. Cook, Kara Moser, Adrian Paskey, Jason Caravas, Benjamin Rambo-Martin, Shatavia Morrison, Christopher Gulvick, Scott Sammons, Yvette Unoarumhi, Darlene Wagner, Matthew Schmerer, S. H. Rosenthal, A. Gerasimova, R. M. Kagan, B. Anderson, M. Hua, Y. Liu, L.E. Bernstein, K.E. Livingston, A. Perez, I. A. Shlyakhter, R. V. Rolando, R. Owen, P. Tanpaiboon, F. Lacbawan, Clinton R. Paden, Duncan MacCannell |
| EPI_ISL_2133628 | Warrior Diagnostics Laboratory | Infectious Disease Program, Broad Institute of Harvard and MIT | Lemieux,J.E., Siddle,K.J., Marshall,J., ONeill,M., Bronson,A., Adams,G., Gladden-Young,A., Lagerborg,K., Rudy,M., DeRuff,K., Carter,A., Normandin,E., Bauer,M., Reilly,S., Tomkins-Tinch,C., Loreth,C., Chaluvadi,S., Birren,B.W., Gallagher,G., Smole,S., Park,D.J., MacInnis,B.L., and Sabeti,P.C. |
| EPI_ISL_2133629 | Quest Diagnostics Incorporated | Centers for Disease Control and Prevention Division of Viral Diseases, Pathogen Discovery | Dakota Howard, Dhwani Batra, Peter W. Cook, Kara Moser, Adrian Paskey, Jason Caravas, Benjamin Rambo-Martin, Shatavia Morrison, Christopher Gulvick, Scott Sammons, Yvette Unoarumhi, Darlene Wagner, Matthew Schmerer, S. H. Rosenthal, A. Gerasimova, R. M. Kagan, B. Anderson, M. Hua, Y. Liu, L.E. Bernstein, K.E. Livingston, A. Perez, I. A. Shlyakhter, R. V. Rolando, R. Owen, P. Tanpaiboon, F. Lacbawan, Clinton R. Paden, Duncan MacCannell |
| EPI_ISL_2133630 | Rhode Island Department of Health | Infectious Disease Program, Broad Institute of Harvard and MIT | Siddle,K.J., Azevedo,K., Miller,A., Adams,G., Pearlman,L., Gladden-Young,A., Lagerborg,K., Rudy,M., DeRuff,K., Carter,A., Normandin,E., Bauer,M., Reilly,S., Tomkins-Tinch,C., Loreth,C., Chaluvadi,S., Lemieux,J.E., Birren,B.W., Sabeti,P.C., Huard,R., King,E., Park,D.J., and MacInnis,B.L. |
| EPI_ISL_2133631 | Quest Diagnostics Incorporated | Centers for Disease Control and Prevention Division of Viral Diseases, Pathogen Discovery | Dakota Howard, Dhwani Batra, Peter W. Cook, Kara Moser, Adrian Paskey, Jason Caravas, Benjamin Rambo-Martin, Shatavia Morrison, Christopher Gulvick, Scott Sammons, Yvette Unoarumhi, Darlene Wagner, Matthew Schmerer, S. H. Rosenthal, A. Gerasimova, R. M. Kagan, B. Anderson, M. Hua, Y. Liu, L.E. Bernstein, K.E. Livingston, A. Perez, I. A. Shlyakhter, R. V. Rolando, R. Owen, P. Tanpaiboon, F. Lacbawan, Clinton R. Paden, Duncan MacCannell |
| EPI_ISL_2133632, EPI_ISL_2133633 | Rhode Island Department of Health | Infectious Disease Program, Broad Institute of Harvard and MIT | Siddle,K.J., Azevedo,K., Miller,A., Adams,G., Pearlman,L., Gladden-Young,A., Lagerborg,K., Rudy,M., DeRuff,K., Carter,A., Normandin,E., Bauer,M., Reilly,S., Tomkins-Tinch,C., Loreth,C., Chaluvadi,S., Lemieux,J.E., Birren,B.W., Sabeti,P.C., Huard,R., King,E., Park,D.J., and MacInnis,B.L. |
| EPI_ISL_2133634 | Quest Diagnostics Incorporated | Centers for Disease Control and Prevention Division of Viral Diseases, Pathogen Discovery | Dakota Howard, Dhwani Batra, Peter W. Cook, Kara Moser, Adrian Paskey, Jason Caravas, Benjamin Rambo-Martin, Shatavia Morrison, Christopher Gulvick, Scott Sammons, Yvette Unoarumhi, Darlene Wagner, Matthew Schmerer, S. H. Rosenthal, A. Gerasimova, R. M. Kagan, B. Anderson, M. Hua, Y. Liu, L.E. Bernstein, K.E. Livingston, A. Perez, I. A. Shlyakhter, R. V. Rolando, R. Owen, P. Tanpaiboon, F. Lacbawan, Clinton R. Paden, Duncan MacCannell |
| EPI_ISL_2133635 | Rhode Island Department of Health | Infectious Disease Program, Broad Institute of Harvard and MIT | Siddle,K.J., Azevedo,K., Miller,A., Adams,G., Pearlman,L., Gladden-Young,A., Lagerborg,K., Rudy,M., DeRuff,K., Carter,A., Normandin,E., Bauer,M., Reilly,S., Tomkins-Tinch,C., Loreth,C., Chaluvadi,S., Lemieux,J.E., Birren,B.W., Sabeti,P.C., Huard,R., King,E., Park,D.J., and MacInnis,B.L. |
| EPI_ISL_2133636 | Quest Diagnostics Incorporated | Centers for Disease Control and Prevention Division of Viral Diseases, Pathogen Discovery | Dakota Howard, Dhwani Batra, Peter W. Cook, Kara Moser, Adrian Paskey, Jason Caravas, Benjamin Rambo-Martin, Shatavia Morrison, Christopher Gulvick, Scott Sammons, Yvette Unoarumhi, Darlene Wagner, Matthew Schmerer, S. H. Rosenthal, A. Gerasimova, R. M. Kagan, B. Anderson, M. Hua, Y. Liu, L.E. Bernstein, K.E. Livingston, A. Perez, I. A. Shlyakhter, R. V. Rolando, R. Owen, P. Tanpaiboon, F. Lacbawan, Clinton R. Paden, Duncan MacCannell |
| EPI_ISL_2133637 | Rhode Island Department of Health | Infectious Disease Program, Broad Institute of Harvard and MIT | Siddle,K.J., Azevedo,K., Miller,A., Adams,G., Pearlman,L., Gladden-Young,A., Lagerborg,K., Rudy,M., DeRuff,K., Carter,A., Normandin,E., Bauer,M., Reilly,S., Tomkins-Tinch,C., Loreth,C., Chaluvadi,S., Lemieux,J.E., Birren,B.W., Sabeti,P.C., Huard,R., King,E., Park,D.J., and MacInnis,B.L. |
| EPI_ISL_2133638, EPI_ISL_2133639 | Quest Diagnostics Incorporated | Centers for Disease Control and Prevention Division of Viral Diseases, Pathogen Discovery | Dakota Howard, Dhwani Batra, Peter W. Cook, Kara Moser, Adrian Paskey, Jason Caravas, Benjamin Rambo-Martin, Shatavia Morrison, Christopher Gulvick, Scott Sammons, Yvette Unoarumhi, Darlene Wagner, Matthew Schmerer, S. H. Rosenthal, A. Gerasimova, R. M. Kagan, B. Anderson, M. Hua, Y. Liu, L.E. Bernstein, K.E. Livingston, A. Perez, I. A. Shlyakhter, R. V. Rolando, R. Owen, P. Tanpaiboon, F. Lacbawan, Clinton R. Paden, Duncan MacCannell |
| EPI_ISL_2133640 | Rhode Island Department of Health | Infectious Disease Program, Broad Institute of Harvard and MIT | Siddle,K.J., Azevedo,K., Miller,A., Adams,G., Pearlman,L., Gladden-Young,A., Lagerborg,K., Rudy,M., DeRuff,K., Carter,A., Normandin,E., Bauer,M., Reilly,S., Tomkins-Tinch,C., Loreth,C., Chaluvadi,S., Lemieux,J.E., Birren,B.W., Sabeti,P.C., Huard,R., King,E., Park,D.J., and MacInnis,B.L. |
| EPI_ISL_2133641 | Quest Diagnostics Incorporated | Centers for Disease Control and Prevention Division of Viral Diseases, Pathogen Discovery | Dakota Howard, Dhwani Batra, Peter W. Cook, Kara Moser, Adrian Paskey, Jason Caravas, Benjamin Rambo-Martin, Shatavia Morrison, Christopher Gulvick, Scott Sammons, Yvette Unoarumhi, Darlene Wagner, Matthew Schmerer, S. H. Rosenthal, A. Gerasimova, R. M. Kagan, B. Anderson, M. Hua, Y. Liu, L.E. Bernstein, K.E. Livingston, A. Perez, I. A. Shlyakhter, R. V. Rolando, R. Owen, P. Tanpaiboon, F. Lacbawan, Clinton R. Paden, Duncan MacCannell |
| EPI_ISL_2133642 | Rhode Island Department of Health | Infectious Disease Program, Broad Institute of Harvard and MIT | Siddle,K.J., Azevedo,K., Miller,A., Adams,G., Pearlman,L., Gladden-Young,A., Lagerborg,K., Rudy,M., DeRuff,K., Carter,A., Normandin,E., Bauer,M., Reilly,S., Tomkins-Tinch,C., Loreth,C., Chaluvadi,S., Lemieux,J.E., Birren,B.W., Sabeti,P.C., Huard,R., King,E., Park,D.J., and MacInnis,B.L. |
| EPI_ISL_2133643 | Quest Diagnostics Incorporated | Centers for Disease Control and Prevention Division of Viral Diseases, Pathogen Discovery | Dakota Howard, Dhwani Batra, Peter W. Cook, Kara Moser, Adrian Paskey, Jason Caravas, Benjamin Rambo-Martin, Shatavia Morrison, Christopher Gulvick, Scott Sammons, Yvette Unoarumhi, Darlene Wagner, Matthew Schmerer, S. H. Rosenthal, A. Gerasimova, R. M. Kagan, B. Anderson, M. Hua, Y. Liu, L.E. Bernstein, K.E. Livingston, A. Perez, I. A. Shlyakhter, R. V. Rolando, R. Owen, P. Tanpaiboon, F. Lacbawan, Clinton R. Paden, Duncan MacCannell |
| EPI_ISL_2133644 | Rhode Island Department of Health | Infectious Disease Program, Broad Institute of Harvard and MIT | Siddle,K.J., Azevedo,K., Miller,A., Adams,G., Pearlman,L., Gladden-Young,A., Lagerborg,K., Rudy,M., DeRuff,K., Carter,A., Normandin,E., Bauer,M., Reilly,S., Tomkins-Tinch,C., Loreth,C., Chaluvadi,S., Lemieux,J.E., Birren,B.W., Sabeti,P.C., Huard,R., King,E., Park,D.J., and MacInnis,B.L. |
| EPI_ISL_2133645, EPI_ISL_2133646 | Quest Diagnostics Incorporated | Centers for Disease Control and Prevention Division of Viral Diseases, Pathogen Discovery | Dakota Howard, Dhwani Batra, Peter W. Cook, Kara Moser, Adrian Paskey, Jason Caravas, Benjamin Rambo-Martin, Shatavia Morrison, Christopher Gulvick, Scott Sammons, Yvette Unoarumhi, Darlene Wagner, Matthew Schmerer, S. H. Rosenthal, A. Gerasimova, R. M. Kagan, B. Anderson, M. Hua, Y. Liu, L.E. Bernstein, K.E. Livingston, A. Perez, I. A. Shlyakhter, R. V. Rolando, R. Owen, P. Tanpaiboon, F. Lacbawan, Clinton R. Paden, Duncan MacCannell |
| EPI_ISL_2133647 | Rhode Island Department of Health | Infectious Disease Program, Broad Institute of Harvard and MIT | Siddle,K.J., Azevedo,K., Miller,A., Adams,G., Pearlman,L., Gladden-Young,A., Lagerborg,K., Rudy,M., DeRuff,K., Carter,A., Normandin,E., Bauer,M., Reilly,S., Tomkins-Tinch,C., Loreth,C., Chaluvadi,S., Lemieux,J.E., Birren,B.W., Sabeti,P.C., Huard,R., King,E., Park,D.J., and MacInnis,B.L. |
| EPI_ISL_2133648 | Quest Diagnostics Incorporated | Centers for Disease Control and Prevention Division of Viral Diseases, Pathogen Discovery | Dakota Howard, Dhwani Batra, Peter W. Cook, Kara Moser, Adrian Paskey, Jason Caravas, Benjamin Rambo-Martin, Shatavia Morrison, Christopher Gulvick, Scott Sammons, Yvette Unoarumhi, Darlene Wagner, Matthew Schmerer, S. H. Rosenthal, A. Gerasimova, R. M. Kagan, B. Anderson, M. Hua, Y. Liu, L.E. Bernstein, K.E. Livingston, A. Perez, I. A. Shlyakhter, R. V. Rolando, R. Owen, P. Tanpaiboon, F. Lacbawan, Clinton R. Paden, Duncan MacCannell |
| EPI_ISL_2133649 | Rhode Island Department of Health | Infectious Disease Program, Broad Institute of Harvard and MIT | Siddle,K.J., Azevedo,K., Miller,A., Adams,G., Pearlman,L., Gladden-Young,A., Lagerborg,K., Rudy,M., DeRuff,K., Carter,A., Normandin,E., Bauer,M., Reilly,S., Tomkins-Tinch,C., Loreth,C., Chaluvadi,S., Lemieux,J.E., Birren,B.W., Sabeti,P.C., Huard,R., King,E., Park,D.J., and MacInnis,B.L. |
| EPI_ISL_2133650 | Quest Diagnostics Incorporated | Centers for Disease Control and Prevention Division of Viral Diseases, Pathogen Discovery | Dakota Howard, Dhwani Batra, Peter W. Cook, Kara Moser, Adrian Paskey, Jason Caravas, Benjamin Rambo-Martin, Shatavia Morrison, Christopher Gulvick, Scott Sammons, Yvette Unoarumhi, Darlene Wagner, Matthew Schmerer, S. H. Rosenthal, A. Gerasimova, R. M. Kagan, B. Anderson, M. Hua, Y. Liu, L.E. Bernstein, K.E. Livingston, A. Perez, I. A. Shlyakhter, R. V. Rolando, R. Owen, P. Tanpaiboon, F. Lacbawan, Clinton R. Paden, Duncan MacCannell |
| EPI_ISL_2133651, EPI_ISL_2133652 | Rhode Island Department of Health | Infectious Disease Program, Broad Institute of Harvard and MIT | Siddle,K.J., Azevedo,K., Miller,A., Adams,G., Pearlman,L., Gladden-Young,A., Lagerborg,K., Rudy,M., DeRuff,K., Carter,A., Normandin,E., Bauer,M., Reilly,S., Tomkins-Tinch,C., Loreth,C., Chaluvadi,S., Lemieux,J.E., Birren,B.W., Sabeti,P.C., Huard,R., King,E., Park,D.J., and MacInnis,B.L. |
| EPI_ISL_2133653 | Quest Diagnostics Incorporated | Centers for Disease Control and Prevention Division of Viral Diseases, Pathogen Discovery | Dakota Howard, Dhwani Batra, Peter W. Cook, Kara Moser, Adrian Paskey, Jason Caravas, Benjamin Rambo-Martin, Shatavia Morrison, Christopher Gulvick, Scott Sammons, Yvette Unoarumhi, Darlene Wagner, Matthew Schmerer, S. H. Rosenthal, A. Gerasimova, R. M. Kagan, B. Anderson, M. Hua, Y. Liu, L.E. Bernstein, K.E. Livingston, A. Perez, I. A. Shlyakhter, R. V. Rolando, R. Owen, P. Tanpaiboon, F. Lacbawan, Clinton R. Paden, Duncan MacCannell |
| EPI_ISL_2133654 | Rhode Island Department of Health | Infectious Disease Program, Broad Institute of Harvard and MIT | Siddle,K.J., Azevedo,K., Miller,A., Adams,G., Pearlman,L., Gladden-Young,A., Lagerborg,K., Rudy,M., DeRuff,K., Carter,A., Normandin,E., Bauer,M., Reilly,S., Tomkins-Tinch,C., Loreth,C., Chaluvadi,S., Lemieux,J.E., Birren,B.W., Sabeti,P.C., Huard,R., King,E., Park,D.J., and MacInnis,B.L. |
| EPI_ISL_2133655 | Quest Diagnostics Incorporated | Centers for Disease Control and Prevention Division of Viral Diseases, Pathogen Discovery | Dakota Howard, Dhwani Batra, Peter W. Cook, Kara Moser, Adrian Paskey, Jason Caravas, Benjamin Rambo-Martin, Shatavia Morrison, Christopher Gulvick, Scott Sammons, Yvette Unoarumhi, Darlene Wagner, Matthew Schmerer, S. H. Rosenthal, A. Gerasimova, R. M. Kagan, B. Anderson, M. Hua, Y. Liu, L.E. Bernstein, K.E. Livingston, A. Perez, I. A. Shlyakhter, R. V. Rolando, R. Owen, P. Tanpaiboon, F. Lacbawan, Clinton R. Paden, Duncan MacCannell |
| EPI_ISL_2133656 | Rhode Island Department of Health | Infectious Disease Program, Broad Institute of Harvard and MIT | Siddle,K.J., Azevedo,K., Miller,A., Adams,G., Pearlman,L., Gladden-Young,A., Lagerborg,K., Rudy,M., DeRuff,K., Carter,A., Normandin,E., Bauer,M., Reilly,S., Tomkins-Tinch,C., Loreth,C., Chaluvadi,S., Lemieux,J.E., Birren,B.W., Sabeti,P.C., Huard,R., King,E., Park,D.J., and MacInnis,B.L. |
| EPI_ISL_2133657 | Quest Diagnostics Incorporated | Centers for Disease Control and Prevention Division of Viral Diseases, Pathogen Discovery | Dakota Howard, Dhwani Batra, Peter W. Cook, Kara Moser, Adrian Paskey, Jason Caravas, Benjamin Rambo-Martin, Shatavia Morrison, Christopher Gulvick, Scott Sammons, Yvette Unoarumhi, Darlene Wagner, Matthew Schmerer, S. H. Rosenthal, A. Gerasimova, R. M. Kagan, B. Anderson, M. Hua, Y. Liu, L.E. Bernstein, K.E. Livingston, A. Perez, I. A. Shlyakhter, R. V. Rolando, R. Owen, P. Tanpaiboon, F. Lacbawan, Clinton R. Paden, Duncan MacCannell |
| EPI_ISL_2133658 | Rhode Island Department of Health | Infectious Disease Program, Broad Institute of Harvard and MIT | Siddle,K.J., Azevedo,K., Miller,A., Adams,G., Pearlman,L., Gladden-Young,A., Lagerborg,K., Rudy,M., DeRuff,K., Carter,A., Normandin,E., Bauer,M., Reilly,S., Tomkins-Tinch,C., Loreth,C., Chaluvadi,S., Lemieux,J.E., Birren,B.W., Sabeti,P.C., Huard,R., King,E., Park,D.J., and MacInnis,B.L. |
| EPI_ISL_2133659 | Quest Diagnostics Incorporated | Centers for Disease Control and Prevention Division of Viral Diseases, Pathogen Discovery | Dakota Howard, Dhwani Batra, Peter W. Cook, Kara Moser, Adrian Paskey, Jason Caravas, Benjamin Rambo-Martin, Shatavia Morrison, Christopher Gulvick, Scott Sammons, Yvette Unoarumhi, Darlene Wagner, Matthew Schmerer, S. H. Rosenthal, A. Gerasimova, R. M. Kagan, B. Anderson, M. Hua, Y. Liu, L.E. Bernstein, K.E. Livingston, A. Perez, I. A. Shlyakhter, R. V. Rolando, R. Owen, P. Tanpaiboon, F. Lacbawan, Clinton R. Paden, Duncan MacCannell |
| EPI_ISL_2133660 | Rhode Island Department of Health | Infectious Disease Program, Broad Institute of Harvard and MIT | Siddle,K.J., Azevedo,K., Miller,A., Adams,G., Pearlman,L., Gladden-Young,A., Lagerborg,K., Rudy,M., DeRuff,K., Carter,A., Normandin,E., Bauer,M., Reilly,S., Tomkins-Tinch,C., Loreth,C., Chaluvadi,S., Lemieux,J.E., Birren,B.W., Sabeti,P.C., Huard,R., King,E., Park,D.J., and MacInnis,B.L. |
| EPI_ISL_2133661 | Quest Diagnostics Incorporated | Centers for Disease Control and Prevention Division of Viral Diseases, Pathogen Discovery | Dakota Howard, Dhwani Batra, Peter W. Cook, Kara Moser, Adrian Paskey, Jason Caravas, Benjamin Rambo-Martin, Shatavia Morrison, Christopher Gulvick, Scott Sammons, Yvette Unoarumhi, Darlene Wagner, Matthew Schmerer, S. H. Rosenthal, A. Gerasimova, R. M. Kagan, B. Anderson, M. Hua, Y. Liu, L.E. Bernstein, K.E. Livingston, A. Perez, I. A. Shlyakhter, R. V. Rolando, R. Owen, P. Tanpaiboon, F. Lacbawan, Clinton R. Paden, Duncan MacCannell |

| | | | |
|---|---|---|---|
| EPI_ISL_2133662 | Rhode Island Department of Health | Infectious Disease Program, Broad Institute of Harvard and MIT | Siddle,K.J., Azevedo,K., Miller,A., Adams,G., Pearlman,L., Gladden-Young,A., Lagerborg,K., Rudy,M., DeRuff,K., Carter,A., Normandin,E., Bauer,M., Reilly,S., Tomkins-Tinch,C., Loreth,C., Chaluvadi,S., Lemieux,J.E., Birren,B.W., Sabeti,P.C., Huard,R., King,E., Park,D.J., and MacInnis,B.L. |
| EPI_ISL_2133663 | Quest Diagnostics Incorporated | Centers for Disease Control and Prevention Division of Viral Diseases, Pathogen Discovery | Dakota Howard, Dhwani Batra, Peter W. Cook, Kara Moser, Adrian Paskey, Jason Caravas, Benjamin Rambo-Martin, Shatavia Morrison, Christopher Gulvick, Scott Sammons, Yvette Unoarumhi, Darlene Wagner, Matthew Schmerer, S. H. Rosenthal, A. Gerasimova, R. M. Kagan, B. Anderson, M. Hua, Y. Liu, L.E. Bernstein, K.E. Livingston, A. Perez, I. A. Shlyakhter, R. V. Rolando, R. Owen, P. Tanpaiboon, F. Lacbawan, Clinton R. Paden, Duncan MacCannell |
| EPI_ISL_2133664 | Rhode Island Department of Health | Infectious Disease Program, Broad Institute of Harvard and MIT | Siddle,K.J., Azevedo,K., Miller,A., Adams,G., Pearlman,L., Gladden-Young,A., Lagerborg,K., Rudy,M., DeRuff,K., Carter,A., Normandin,E., Bauer,M., Reilly,S., Tomkins-Tinch,C., Loreth,C., Chaluvadi,S., Lemieux,J.E., Birren,B.W., Sabeti,P.C., Huard,R., King,E., Park,D.J., and MacInnis,B.L. |
| EPI_ISL_2133665 | Quest Diagnostics Incorporated | Centers for Disease Control and Prevention Division of Viral Diseases, Pathogen Discovery | Dakota Howard, Dhwani Batra, Peter W. Cook, Kara Moser, Adrian Paskey, Jason Caravas, Benjamin Rambo-Martin, Shatavia Morrison, Christopher Gulvick, Scott Sammons, Yvette Unoarumhi, Darlene Wagner, Matthew Schmerer, S. H. Rosenthal, A. Gerasimova, R. M. Kagan, B. Anderson, M. Hua, Y. Liu, L.E. Bernstein, K.E. Livingston, A. Perez, I. A. Shlyakhter, R. V. Rolando, R. Owen, P. Tanpaiboon, F. Lacbawan, Clinton R. Paden, Duncan MacCannell |
| EPI_ISL_2133666, EPI_ISL_2133667, EPI_ISL_2133668 | Rhode Island Department of Health | Infectious Disease Program, Broad Institute of Harvard and MIT | Siddle,K.J., Azevedo,K., Miller,A., Adams,G., Pearlman,L., Gladden-Young,A., Lagerborg,K., Rudy,M., DeRuff,K., Carter,A., Normandin,E., Bauer,M., Reilly,S., Tomkins-Tinch,C., Loreth,C., Chaluvadi,S., Lemieux,J.E., Birren,B.W., Sabeti,P.C., Huard,R., King,E., Park,D.J., and MacInnis,B.L. |
| EPI_ISL_2133669, EPI_ISL_2133670 | Quest Diagnostics Incorporated | Centers for Disease Control and Prevention Division of Viral Diseases, Pathogen Discovery | Dakota Howard, Dhwani Batra, Peter W. Cook, Kara Moser, Adrian Paskey, Jason Caravas, Benjamin Rambo-Martin, Shatavia Morrison, Christopher Gulvick, Scott Sammons, Yvette Unoarumhi, Darlene Wagner, Matthew Schmerer, S. H. Rosenthal, A. Gerasimova, R. M. Kagan, B. Anderson, M. Hua, Y. Liu, L.E. Bernstein, K.E. Livingston, A. Perez, I. A. Shlyakhter, R. V. Rolando, R. Owen, P. Tanpaiboon, F. Lacbawan, Clinton R. Paden, Duncan MacCannell |
| EPI_ISL_2133671 | Rhode Island Department of Health | Infectious Disease Program, Broad Institute of Harvard and MIT | Siddle,K.J., Azevedo,K., Miller,A., Adams,G., Pearlman,L., Gladden-Young,A., Lagerborg,K., Rudy,M., DeRuff,K., Carter,A., Normandin,E., Bauer,M., Reilly,S., Tomkins-Tinch,C., Loreth,C., Chaluvadi,S., Lemieux,J.E., Birren,B.W., Sabeti,P.C., Huard,R., King,E., Park,D.J., and MacInnis,B.L. |
| EPI_ISL_2133672 | Quest Diagnostics Incorporated | Centers for Disease Control and Prevention Division of Viral Diseases, Pathogen Discovery | Dakota Howard, Dhwani Batra, Peter W. Cook, Kara Moser, Adrian Paskey, Jason Caravas, Benjamin Rambo-Martin, Shatavia Morrison, Christopher Gulvick, Scott Sammons, Yvette Unoarumhi, Darlene Wagner, Matthew Schmerer, S. H. Rosenthal, A. Gerasimova, R. M. Kagan, B. Anderson, M. Hua, Y. Liu, L.E. Bernstein, K.E. Livingston, A. Perez, I. A. Shlyakhter, R. V. Rolando, R. Owen, P. Tanpaiboon, F. Lacbawan, Clinton R. Paden, Duncan MacCannell |
| EPI_ISL_2133673 | Rhode Island Department of Health | Infectious Disease Program, Broad Institute of Harvard and MIT | Siddle,K.J., Azevedo,K., Miller,A., Adams,G., Pearlman,L., Gladden-Young,A., Lagerborg,K., Rudy,M., DeRuff,K., Carter,A., Normandin,E., Bauer,M., Reilly,S., Tomkins-Tinch,C., Loreth,C., Chaluvadi,S., Lemieux,J.E., Birren,B.W., Sabeti,P.C., Huard,R., King,E., Park,D.J., and MacInnis,B.L. |
| EPI_ISL_2133674 | Quest Diagnostics Incorporated | Centers for Disease Control and Prevention Division of Viral Diseases, Pathogen Discovery | Dakota Howard, Dhwani Batra, Peter W. Cook, Kara Moser, Adrian Paskey, Jason Caravas, Benjamin Rambo-Martin, Shatavia Morrison, Christopher Gulvick, Scott Sammons, Yvette Unoarumhi, Darlene Wagner, Matthew Schmerer, S. H. Rosenthal, A. Gerasimova, R. M. Kagan, B. Anderson, M. Hua, Y. Liu, L.E. Bernstein, K.E. Livingston, A. Perez, I. A. Shlyakhter, R. V. Rolando, R. Owen, P. Tanpaiboon, F. Lacbawan, Clinton R. Paden, Duncan MacCannell |
| EPI_ISL_2133675 | Rhode Island Department of Health | Infectious Disease Program, Broad Institute of Harvard and MIT | Siddle,K.J., Azevedo,K., Miller,A., Adams,G., Pearlman,L., Gladden-Young,A., Lagerborg,K., Rudy,M., DeRuff,K., Carter,A., Normandin,E., Bauer,M., Reilly,S., Tomkins-Tinch,C., Loreth,C., Chaluvadi,S., Lemieux,J.E., Birren,B.W., Sabeti,P.C., Huard,R., King,E., Park,D.J., and MacInnis,B.L. |
| EPI_ISL_2133676 | Quest Diagnostics Incorporated | Centers for Disease Control and Prevention Division of Viral Diseases, Pathogen Discovery | Dakota Howard, Dhwani Batra, Peter W. Cook, Kara Moser, Adrian Paskey, Jason Caravas, Benjamin Rambo-Martin, Shatavia Morrison, Christopher Gulvick, Scott Sammons, Yvette Unoarumhi, Darlene Wagner, Matthew Schmerer, S. H. Rosenthal, A. Gerasimova, R. M. Kagan, B. Anderson, M. Hua, Y. Liu, L.E. Bernstein, K.E. Livingston, A. Perez, I. A. Shlyakhter, R. V. Rolando, R. Owen, P. Tanpaiboon, F. Lacbawan, Clinton R. Paden, Duncan MacCannell |
| EPI_ISL_2133677, EPI_ISL_2133678 | Rhode Island Department of Health | Infectious Disease Program, Broad Institute of Harvard and MIT | Siddle,K.J., Azevedo,K., Miller,A., Adams,G., Pearlman,L., Gladden-Young,A., Lagerborg,K., Rudy,M., DeRuff,K., Carter,A., Normandin,E., Bauer,M., Reilly,S., Tomkins-Tinch,C., Loreth,C., Chaluvadi,S., Lemieux,J.E., Birren,B.W., Sabeti,P.C., Huard,R., King,E., Park,D.J., and MacInnis,B.L. |
| EPI_ISL_2133679 | Quest Diagnostics Incorporated | Centers for Disease Control and Prevention Division of Viral Diseases, Pathogen Discovery | Dakota Howard, Dhwani Batra, Peter W. Cook, Kara Moser, Adrian Paskey, Jason Caravas, Benjamin Rambo-Martin, Shatavia Morrison, Christopher Gulvick, Scott Sammons, Yvette Unoarumhi, Darlene Wagner, Matthew Schmerer, S. H. Rosenthal, A. Gerasimova, R. M. Kagan, B. Anderson, M. Hua, Y. Liu, L.E. Bernstein, K.E. Livingston, A. Perez, I. A. Shlyakhter, R. V. Rolando, R. Owen, P. Tanpaiboon, F. Lacbawan, Clinton R. Paden, Duncan MacCannell |
| EPI_ISL_2133680 | Rhode Island Department of Health | Infectious Disease Program, Broad Institute of Harvard and MIT | Siddle,K.J., Azevedo,K., Miller,A., Adams,G., Pearlman,L., Gladden-Young,A., Lagerborg,K., Rudy,M., DeRuff,K., Carter,A., Normandin,E., Bauer,M., Reilly,S., Tomkins-Tinch,C., Loreth,C., Chaluvadi,S., Lemieux,J.E., Birren,B.W., Sabeti,P.C., Huard,R., King,E., Park,D.J., and MacInnis,B.L. |
| EPI_ISL_2133681 | Quest Diagnostics Incorporated | Centers for Disease Control and Prevention Division of Viral Diseases, Pathogen Discovery | Dakota Howard, Dhwani Batra, Peter W. Cook, Kara Moser, Adrian Paskey, Jason Caravas, Benjamin Rambo-Martin, Shatavia Morrison, Christopher Gulvick, Scott Sammons, Yvette Unoarumhi, Darlene Wagner, Matthew Schmerer, S. H. Rosenthal, A. Gerasimova, R. M. Kagan, B. Anderson, M. Hua, Y. Liu, L.E. Bernstein, K.E. Livingston, A. Perez, I. A. Shlyakhter, R. V. Rolando, R. Owen, P. Tanpaiboon, F. Lacbawan, Clinton R. Paden, Duncan MacCannell |
| EPI_ISL_2133682 | Rhode Island Department of Health | Infectious Disease Program, Broad Institute of Harvard and MIT | Siddle,K.J., Azevedo,K., Miller,A., Adams,G., Pearlman,L., Gladden-Young,A., Lagerborg,K., Rudy,M., DeRuff,K., Carter,A., Normandin,E., Bauer,M., Reilly,S., Tomkins-Tinch,C., Loreth,C., Chaluvadi,S., Lemieux,J.E., Birren,B.W., Sabeti,P.C., Huard,R., King,E., Park,D.J., and MacInnis,B.L. |
| EPI_ISL_2133683 | Quest Diagnostics Incorporated | Centers for Disease Control and Prevention Division of Viral Diseases, Pathogen Discovery | Dakota Howard, Dhwani Batra, Peter W. Cook, Kara Moser, Adrian Paskey, Jason Caravas, Benjamin Rambo-Martin, Shatavia Morrison, Christopher Gulvick, Scott Sammons, Yvette Unoarumhi, Darlene Wagner, Matthew Schmerer, S. H. Rosenthal, A. Gerasimova, R. M. Kagan, B. Anderson, M. Hua, Y. Liu, L.E. Bernstein, K.E. Livingston, A. Perez, I. A. Shlyakhter, R. V. Rolando, R. Owen, P. Tanpaiboon, F. Lacbawan, Clinton R. Paden, Duncan MacCannell |
| EPI_ISL_2133684 | Rhode Island Department of Health | Infectious Disease Program, Broad Institute of Harvard and MIT | Siddle,K.J., Azevedo,K., Miller,A., Adams,G., Pearlman,L., Gladden-Young,A., Lagerborg,K., Rudy,M., DeRuff,K., Carter,A., Normandin,E., Bauer,M., Reilly,S., Tomkins-Tinch,C., Loreth,C., Chaluvadi,S., Lemieux,J.E., Birren,B.W., Sabeti,P.C., Huard,R., King,E., Park,D.J., and MacInnis,B.L. |
| EPI_ISL_2133685 | Quest Diagnostics Incorporated | Centers for Disease Control and Prevention Division of Viral Diseases, Pathogen Discovery | Dakota Howard, Dhwani Batra, Peter W. Cook, Kara Moser, Adrian Paskey, Jason Caravas, Benjamin Rambo-Martin, Shatavia Morrison, Christopher Gulvick, Scott Sammons, Yvette Unoarumhi, Darlene Wagner, Matthew Schmerer, S. H. Rosenthal, A. Gerasimova, R. M. Kagan, B. Anderson, M. Hua, Y. Liu, L.E. Bernstein, K.E. Livingston, A. Perez, I. A. Shlyakhter, R. V. Rolando, R. Owen, P. Tanpaiboon, F. Lacbawan, Clinton R. Paden, Duncan MacCannell |
| EPI_ISL_2133686 | Rhode Island Department of Health | Infectious Disease Program, Broad Institute of Harvard and MIT | Siddle,K.J., Azevedo,K., Miller,A., Adams,G., Pearlman,L., Gladden-Young,A., Lagerborg,K., Rudy,M., DeRuff,K., Carter,A., Normandin,E., Bauer,M., Reilly,S., Tomkins-Tinch,C., Loreth,C., Chaluvadi,S., Lemieux,J.E., Birren,B.W., Sabeti,P.C., Huard,R., King,E., Park,D.J., and MacInnis,B.L. |
| EPI_ISL_2133687 | Quest Diagnostics Incorporated | Centers for Disease Control and Prevention Division of Viral Diseases, Pathogen Discovery | Dakota Howard, Dhwani Batra, Peter W. Cook, Kara Moser, Adrian Paskey, Jason Caravas, Benjamin Rambo-Martin, Shatavia Morrison, Christopher Gulvick, Scott Sammons, Yvette Unoarumhi, Darlene Wagner, Matthew Schmerer, S. H. Rosenthal, A. Gerasimova, R. M. Kagan, B. Anderson, M. Hua, Y. Liu, L.E. Bernstein, K.E. Livingston, A. Perez, I. A. Shlyakhter, R. V. Rolando, R. Owen, P. Tanpaiboon, F. Lacbawan, Clinton R. Paden, Duncan MacCannell |
| EPI_ISL_2133688 | Rhode Island Department of Health | Infectious Disease Program, Broad Institute of Harvard and MIT | Siddle,K.J., Azevedo,K., Miller,A., Adams,G., Pearlman,L., Gladden-Young,A., Lagerborg,K., Rudy,M., DeRuff,K., Carter,A., Normandin,E., Bauer,M., Reilly,S., Tomkins-Tinch,C., Loreth,C., Chaluvadi,S., Lemieux,J.E., Birren,B.W., Sabeti,P.C., Huard,R., King,E., Park,D.J., and MacInnis,B.L. |
| EPI_ISL_2133689 | Quest Diagnostics Incorporated | Centers for Disease Control and Prevention Division of Viral Diseases, Pathogen Discovery | Dakota Howard, Dhwani Batra, Peter W. Cook, Kara Moser, Adrian Paskey, Jason Caravas, Benjamin Rambo-Martin, Shatavia Morrison, Christopher Gulvick, Scott Sammons, Yvette Unoarumhi, Darlene Wagner, Matthew Schmerer, S. H. Rosenthal, A. Gerasimova, R. M. Kagan, B. Anderson, M. Hua, Y. Liu, L.E. Bernstein, K.E. Livingston, A. Perez, I. A. Shlyakhter, R. V. Rolando, R. Owen, P. Tanpaiboon, F. Lacbawan, Clinton R. Paden, Duncan MacCannell |
| EPI_ISL_2133690 | Rhode Island Department of Health | Infectious Disease Program, Broad Institute of Harvard and MIT | Siddle,K.J., Azevedo,K., Miller,A., Adams,G., Pearlman,L., Gladden-Young,A., Lagerborg,K., Rudy,M., DeRuff,K., Carter,A., Normandin,E., Bauer,M., Reilly,S., Tomkins-Tinch,C., Loreth,C., Chaluvadi,S., Lemieux,J.E., Birren,B.W., Sabeti,P.C., Huard,R., King,E., Park,D.J., and MacInnis,B.L. |
| EPI_ISL_2133691 | Quest Diagnostics Incorporated | Centers for Disease Control and Prevention Division of Viral Diseases, Pathogen Discovery | Dakota Howard, Dhwani Batra, Peter W. Cook, Kara Moser, Adrian Paskey, Jason Caravas, Benjamin Rambo-Martin, Shatavia Morrison, Christopher Gulvick, Scott Sammons, Yvette Unoarumhi, Darlene Wagner, Matthew Schmerer, S. H. Rosenthal, A. Gerasimova, R. M. Kagan, B. Anderson, M. Hua, Y. Liu, L.E. Bernstein, K.E. Livingston, A. Perez, I. A. Shlyakhter, R. V. Rolando, R. Owen, P. Tanpaiboon, F. Lacbawan, Clinton R. Paden, Duncan MacCannell |
| EPI_ISL_2133692 | Rhode Island Department of Health | Infectious Disease Program, Broad Institute of Harvard and MIT | Siddle,K.J., Azevedo,K., Miller,A., Adams,G., Pearlman,L., Gladden-Young,A., Lagerborg,K., Rudy,M., DeRuff,K., Carter,A., Normandin,E., Bauer,M., Reilly,S., Tomkins-Tinch,C., Loreth,C., Chaluvadi,S., Lemieux,J.E., Birren,B.W., Sabeti,P.C., Huard,R., King,E., Park,D.J., and MacInnis,B.L. |
| EPI_ISL_2133693 | Quest Diagnostics Incorporated | Centers for Disease Control and Prevention Division of Viral Diseases, Pathogen Discovery | Dakota Howard, Dhwani Batra, Peter W. Cook, Kara Moser, Adrian Paskey, Jason Caravas, Benjamin Rambo-Martin, Shatavia Morrison, Christopher Gulvick, Scott Sammons, Yvette Unoarumhi, Darlene Wagner, Matthew Schmerer, S. H. Rosenthal, A. Gerasimova, R. M. Kagan, B. Anderson, M. Hua, Y. Liu, L.E. Bernstein, K.E. Livingston, A. Perez, I. A. Shlyakhter, R. V. Rolando, R. Owen, P. Tanpaiboon, F. Lacbawan, Clinton R. Paden, Duncan MacCannell |
| EPI_ISL_2133694 | Rhode Island Department of Health | Infectious Disease Program, Broad Institute of Harvard and MIT | Siddle,K.J., Azevedo,K., Miller,A., Adams,G., Pearlman,L., Gladden-Young,A., Lagerborg,K., Rudy,M., DeRuff,K., Carter,A., Normandin,E., Bauer,M., Reilly,S., Tomkins-Tinch,C., Loreth,C., Chaluvadi,S., Lemieux,J.E., Birren,B.W., Sabeti,P.C., Huard,R., King,E., Park,D.J., and MacInnis,B.L. |
| EPI_ISL_2133695 | Quest Diagnostics Incorporated | Centers for Disease Control and Prevention Division of Viral Diseases, Pathogen Discovery | Dakota Howard, Dhwani Batra, Peter W. Cook, Kara Moser, Adrian Paskey, Jason Caravas, Benjamin Rambo-Martin, Shatavia Morrison, Christopher Gulvick, Scott Sammons, Yvette Unoarumhi, Darlene Wagner, Matthew Schmerer, S. H. Rosenthal, A. Gerasimova, R. M. Kagan, B. Anderson, M. Hua, Y. Liu, L.E. Bernstein, K.E. Livingston, A. Perez, I. A. Shlyakhter, R. V. Rolando, R. Owen, P. Tanpaiboon, F. Lacbawan, Clinton R. Paden, Duncan MacCannell |
| EPI_ISL_2133696 | Rhode Island Department of Health | Infectious Disease Program, Broad Institute of Harvard and MIT | Siddle,K.J., Azevedo,K., Miller,A., Adams,G., Pearlman,L., Gladden-Young,A., Lagerborg,K., Rudy,M., DeRuff,K., Carter,A., Normandin,E., Bauer,M., Reilly,S., Tomkins-Tinch,C., Loreth,C., Chaluvadi,S., Lemieux,J.E., Birren,B.W., Sabeti,P.C., Huard,R., King,E., Park,D.J., and MacInnis,B.L. |
| EPI_ISL_2133697 | Quest Diagnostics Incorporated | Centers for Disease Control and Prevention Division of Viral Diseases, Pathogen Discovery | Dakota Howard, Dhwani Batra, Peter W. Cook, Kara Moser, Adrian Paskey, Jason Caravas, Benjamin Rambo-Martin, Shatavia Morrison, Christopher Gulvick, Scott Sammons, Yvette Unoarumhi, Darlene Wagner, Matthew Schmerer, S. H. Rosenthal, A. Gerasimova, R. M. Kagan, B. Anderson, M. Hua, Y. Liu, L.E. Bernstein, K.E. Livingston, A. Perez, I. A. Shlyakhter, R. V. Rolando, R. Owen, P. Tanpaiboon, F. Lacbawan, Clinton R. Paden, Duncan MacCannell |
| EPI_ISL_2133698 | Rhode Island Department of Health | Infectious Disease Program, Broad Institute of Harvard and MIT | Siddle,K.J., Azevedo,K., Miller,A., Adams,G., Pearlman,L., Gladden-Young,A., Lagerborg,K., Rudy,M., DeRuff,K., Carter,A., Normandin,E., Bauer,M., Reilly,S., Tomkins-Tinch,C., Loreth,C., Chaluvadi,S., Lemieux,J.E., Birren,B.W., Sabeti,P.C., Huard,R., King,E., Park,D.J., and MacInnis,B.L. |
| EPI_ISL_2133699 | Quest Diagnostics Incorporated | Centers for Disease Control and Prevention Division of Viral Diseases, Pathogen Discovery | Dakota Howard, Dhwani Batra, Peter W. Cook, Kara Moser, Adrian Paskey, Jason Caravas, Benjamin Rambo-Martin, Shatavia Morrison, Christopher Gulvick, Scott Sammons, Yvette Unoarumhi, Darlene Wagner, Matthew Schmerer, S. H. Rosenthal, A. Gerasimova, R. M. Kagan, B. Anderson, M. Hua, Y. Liu, L.E. Bernstein, K.E. Livingston, A. Perez, I. A. Shlyakhter, R. V. Rolando, R. Owen, P. Tanpaiboon, F. Lacbawan, Clinton R. Paden, Duncan MacCannell |
| EPI_ISL_2133700 | Rhode Island Department of Health | Infectious Disease Program, Broad Institute of Harvard and MIT | Siddle,K.J., Azevedo,K., Miller,A., Adams,G., Pearlman,L., Gladden-Young,A., Lagerborg,K., Rudy,M., DeRuff,K., Carter,A., Normandin,E., Bauer,M., Reilly,S., Tomkins-Tinch,C., Loreth,C., Chaluvadi,S., Lemieux,J.E., Birren,B.W., Sabeti,P.C., Huard,R., King,E., Park,D.J., and MacInnis,B.L. |
| EPI_ISL_2133701 | Quest Diagnostics Incorporated | Centers for Disease Control and Prevention Division of Viral Diseases, Pathogen Discovery | Dakota Howard, Dhwani Batra, Peter W. Cook, Kara Moser, Adrian Paskey, Jason Caravas, Benjamin Rambo-Martin, Shatavia Morrison, Christopher Gulvick, Scott Sammons, Yvette Unoarumhi, Darlene Wagner, Matthew Schmerer, S. H. Rosenthal, A. Gerasimova, R. M. Kagan, B. Anderson, M. Hua, Y. Liu, L.E. Bernstein, K.E. Livingston, A. Perez, I. A. Shlyakhter, R. V. Rolando, R. Owen, P. Tanpaiboon, F. Lacbawan, Clinton R. Paden, Duncan MacCannell |
| EPI_ISL_2133702 | Rhode Island Department of Health | Infectious Disease Program, Broad Institute of Harvard and MIT | Siddle,K.J., Azevedo,K., Miller,A., Adams,G., Pearlman,L., Gladden-Young,A., Lagerborg,K., Rudy,M., DeRuff,K., Carter,A., Normandin,E., Bauer,M., Reilly,S., Tomkins-Tinch,C., Loreth,C., Chaluvadi,S., Lemieux,J.E., Birren,B.W., Sabeti,P.C., Huard,R., King,E., Park,D.J., and MacInnis,B.L. |

| | | | |
|---|---|---|---|
| EPI_ISL_2133703 | Quest Diagnostics Incorporated | Centers for Disease Control and Prevention Division of Viral Diseases, Pathogen Discovery | Dakota Howard, Dhwani Batra, Peter W. Cook, Kara Moser, Adrian Paskey, Jason Caravas, Benjamin Rambo-Martin, Shatavia Morrison, Christopher Gulvick, Scott Sammons, Yvette Unoarumhi, Darlene Wagner, Matthew Schmerer, S. H. Rosenthal, A. Gerasimova, R. M. Kagan, B. Anderson, M. Hua, Y. Liu, L.E. Bernstein, K.E. Livingston, A. Perez, I. A. Shlyakhter, R. V. Rolando, R. Owen, P. Tanpaiboon, F. Lacbawan, Clinton R. Paden, Duncan MacCannell |
| EPI_ISL_2133704 | Rhode Island Department of Health | Infectious Disease Program, Broad Institute of Harvard and MIT | Siddle,K.J., Azevedo,K., Miller,A., Adams,G., Pearlman,L., Gladden-Young,A., Lagerborg,K., Rudy,M., DeRuff,K., Carter,A., Normandin,E., Bauer,M., Reilly,S., Tomkins-Tinch,C., Loreth,C., Chaluvadi,S., Lemieux,J.E., Birren,B.W., Sabeti,P.C., Huard,R., King,E., Park,D.J., and MacInnis,B.L. |
| EPI_ISL_2133705 | Quest Diagnostics Incorporated | Centers for Disease Control and Prevention Division of Viral Diseases, Pathogen Discovery | Dakota Howard, Dhwani Batra, Peter W. Cook, Kara Moser, Adrian Paskey, Jason Caravas, Benjamin Rambo-Martin, Shatavia Morrison, Christopher Gulvick, Scott Sammons, Yvette Unoarumhi, Darlene Wagner, Matthew Schmerer, S. H. Rosenthal, A. Gerasimova, R. M. Kagan, B. Anderson, M. Hua, Y. Liu, L.E. Bernstein, K.E. Livingston, A. Perez, I. A. Shlyakhter, R. V. Rolando, R. Owen, P. Tanpaiboon, F. Lacbawan, Clinton R. Paden, Duncan MacCannell |
| EPI_ISL_2133706 | Rhode Island Department of Health | Infectious Disease Program, Broad Institute of Harvard and MIT | Siddle,K.J., Azevedo,K., Miller,A., Adams,G., Pearlman,L., Gladden-Young,A., Lagerborg,K., Rudy,M., DeRuff,K., Carter,A., Normandin,E., Bauer,M., Reilly,S., Tomkins-Tinch,C., Loreth,C., Chaluvadi,S., Lemieux,J.E., Birren,B.W., Sabeti,P.C., Huard,R., King,E., Park,D.J., and MacInnis,B.L. |
| EPI_ISL_2133707 | Quest Diagnostics Incorporated | Centers for Disease Control and Prevention Division of Viral Diseases, Pathogen Discovery | Dakota Howard, Dhwani Batra, Peter W. Cook, Kara Moser, Adrian Paskey, Jason Caravas, Benjamin Rambo-Martin, Shatavia Morrison, Christopher Gulvick, Scott Sammons, Yvette Unoarumhi, Darlene Wagner, Matthew Schmerer, S. H. Rosenthal, A. Gerasimova, R. M. Kagan, B. Anderson, M. Hua, Y. Liu, L.E. Bernstein, K.E. Livingston, A. Perez, I. A. Shlyakhter, R. V. Rolando, R. Owen, P. Tanpaiboon, F. Lacbawan, Clinton R. Paden, Duncan MacCannell |
| EPI_ISL_2133708 | Rhode Island Department of Health | Infectious Disease Program, Broad Institute of Harvard and MIT | Siddle,K.J., Azevedo,K., Miller,A., Adams,G., Pearlman,L., Gladden-Young,A., Lagerborg,K., Rudy,M., DeRuff,K., Carter,A., Normandin,E., Bauer,M., Reilly,S., Tomkins-Tinch,C., Loreth,C., Chaluvadi,S., Lemieux,J.E., Birren,B.W., Sabeti,P.C., Huard,R., King,E., Park,D.J., and MacInnis,B.L. |
| EPI_ISL_2133710 | Quest Diagnostics Incorporated | Centers for Disease Control and Prevention Division of Viral Diseases, Pathogen Discovery | Dakota Howard, Dhwani Batra, Peter W. Cook, Kara Moser, Adrian Paskey, Jason Caravas, Benjamin Rambo-Martin, Shatavia Morrison, Christopher Gulvick, Scott Sammons, Yvette Unoarumhi, Darlene Wagner, Matthew Schmerer, S. H. Rosenthal, A. Gerasimova, R. M. Kagan, B. Anderson, M. Hua, Y. Liu, L.E. Bernstein, K.E. Livingston, A. Perez, I. A. Shlyakhter, R. V. Rolando, R. Owen, P. Tanpaiboon, F. Lacbawan, Clinton R. Paden, Duncan MacCannell |
| EPI_ISL_2133711 | Rhode Island Department of Health | Infectious Disease Program, Broad Institute of Harvard and MIT | Siddle,K.J., Azevedo,K., Miller,A., Adams,G., Pearlman,L., Gladden-Young,A., Lagerborg,K., Rudy,M., DeRuff,K., Carter,A., Normandin,E., Bauer,M., Reilly,S., Tomkins-Tinch,C., Loreth,C., Chaluvadi,S., Lemieux,J.E., Birren,B.W., Sabeti,P.C., Huard,R., King,E., Park,D.J., and MacInnis,B.L. |
| EPI_ISL_2133712 | Quest Diagnostics Incorporated | Centers for Disease Control and Prevention Division of Viral Diseases, Pathogen Discovery | Dakota Howard, Dhwani Batra, Peter W. Cook, Kara Moser, Adrian Paskey, Jason Caravas, Benjamin Rambo-Martin, Shatavia Morrison, Christopher Gulvick, Scott Sammons, Yvette Unoarumhi, Darlene Wagner, Matthew Schmerer, S. H. Rosenthal, A. Gerasimova, R. M. Kagan, B. Anderson, M. Hua, Y. Liu, L.E. Bernstein, K.E. Livingston, A. Perez, I. A. Shlyakhter, R. V. Rolando, R. Owen, P. Tanpaiboon, F. Lacbawan, Clinton R. Paden, Duncan MacCannell |
| EPI_ISL_2133713 | Rhode Island Department of Health | Infectious Disease Program, Broad Institute of Harvard and MIT | Siddle,K.J., Azevedo,K., Miller,A., Adams,G., Pearlman,L., Gladden-Young,A., Lagerborg,K., Rudy,M., DeRuff,K., Carter,A., Normandin,E., Bauer,M., Reilly,S., Tomkins-Tinch,C., Loreth,C., Chaluvadi,S., Lemieux,J.E., Birren,B.W., Sabeti,P.C., Huard,R., King,E., Park,D.J., and MacInnis,B.L. |
| EPI_ISL_2133714 | Quest Diagnostics Incorporated | Centers for Disease Control and Prevention Division of Viral Diseases, Pathogen Discovery | Dakota Howard, Dhwani Batra, Peter W. Cook, Kara Moser, Adrian Paskey, Jason Caravas, Benjamin Rambo-Martin, Shatavia Morrison, Christopher Gulvick, Scott Sammons, Yvette Unoarumhi, Darlene Wagner, Matthew Schmerer, S. H. Rosenthal, A. Gerasimova, R. M. Kagan, B. Anderson, M. Hua, Y. Liu, L.E. Bernstein, K.E. Livingston, A. Perez, I. A. Shlyakhter, R. V. Rolando, R. Owen, P. Tanpaiboon, F. Lacbawan, Clinton R. Paden, Duncan MacCannell |
| EPI_ISL_2133715 | Rhode Island Department of Health | Infectious Disease Program, Broad Institute of Harvard and MIT | Siddle,K.J., Azevedo,K., Miller,A., Adams,G., Pearlman,L., Gladden-Young,A., Lagerborg,K., Rudy,M., DeRuff,K., Carter,A., Normandin,E., Bauer,M., Reilly,S., Tomkins-Tinch,C., Loreth,C., Chaluvadi,S., Lemieux,J.E., Birren,B.W., Sabeti,P.C., Huard,R., King,E., Park,D.J., and MacInnis,B.L. |
| EPI_ISL_2133716 | Quest Diagnostics Incorporated | Centers for Disease Control and Prevention Division of Viral Diseases, Pathogen Discovery | Dakota Howard, Dhwani Batra, Peter W. Cook, Kara Moser, Adrian Paskey, Jason Caravas, Benjamin Rambo-Martin, Shatavia Morrison, Christopher Gulvick, Scott Sammons, Yvette Unoarumhi, Darlene Wagner, Matthew Schmerer, S. H. Rosenthal, A. Gerasimova, R. M. Kagan, B. Anderson, M. Hua, Y. Liu, L.E. Bernstein, K.E. Livingston, A. Perez, I. A. Shlyakhter, R. V. Rolando, R. Owen, P. Tanpaiboon, F. Lacbawan, Clinton R. Paden, Duncan MacCannell |
| EPI_ISL_2133717 | Rhode Island Department of Health | Infectious Disease Program, Broad Institute of Harvard and MIT | Siddle,K.J., Azevedo,K., Miller,A., Adams,G., Pearlman,L., Gladden-Young,A., Lagerborg,K., Rudy,M., DeRuff,K., Carter,A., Normandin,E., Bauer,M., Reilly,S., Tomkins-Tinch,C., Loreth,C., Chaluvadi,S., Lemieux,J.E., Birren,B.W., Sabeti,P.C., Huard,R., King,E., Park,D.J., and MacInnis,B.L. |
| EPI_ISL_2133718 | Quest Diagnostics Incorporated | Centers for Disease Control and Prevention Division of Viral Diseases, Pathogen Discovery | Dakota Howard, Dhwani Batra, Peter W. Cook, Kara Moser, Adrian Paskey, Jason Caravas, Benjamin Rambo-Martin, Shatavia Morrison, Christopher Gulvick, Scott Sammons, Yvette Unoarumhi, Darlene Wagner, Matthew Schmerer, S. H. Rosenthal, A. Gerasimova, R. M. Kagan, B. Anderson, M. Hua, Y. Liu, L.E. Bernstein, K.E. Livingston, A. Perez, I. A. Shlyakhter, R. V. Rolando, R. Owen, P. Tanpaiboon, F. Lacbawan, Clinton R. Paden, Duncan MacCannell |
| EPI_ISL_2133720 | Rhode Island Department of Health | Infectious Disease Program, Broad Institute of Harvard and MIT | Siddle,K.J., Azevedo,K., Miller,A., Adams,G., Pearlman,L., Gladden-Young,A., Lagerborg,K., Rudy,M., DeRuff,K., Carter,A., Normandin,E., Bauer,M., Reilly,S., Tomkins-Tinch,C., Loreth,C., Chaluvadi,S., Lemieux,J.E., Birren,B.W., Sabeti,P.C., Huard,R., King,E., Park,D.J., and MacInnis,B.L. |
| EPI_ISL_2133721 | Quest Diagnostics Incorporated | Centers for Disease Control and Prevention Division of Viral Diseases, Pathogen Discovery | Dakota Howard, Dhwani Batra, Peter W. Cook, Kara Moser, Adrian Paskey, Jason Caravas, Benjamin Rambo-Martin, Shatavia Morrison, Christopher Gulvick, Scott Sammons, Yvette Unoarumhi, Darlene Wagner, Matthew Schmerer, S. H. Rosenthal, A. Gerasimova, R. M. Kagan, B. Anderson, M. Hua, Y. Liu, L.E. Bernstein, K.E. Livingston, A. Perez, I. A. Shlyakhter, R. V. Rolando, R. Owen, P. Tanpaiboon, F. Lacbawan, Clinton R. Paden, Duncan MacCannell |
| EPI_ISL_2133722 | Rhode Island Department of Health | Infectious Disease Program, Broad Institute of Harvard and MIT | Siddle,K.J., Azevedo,K., Miller,A., Adams,G., Pearlman,L., Gladden-Young,A., Lagerborg,K., Rudy,M., DeRuff,K., Carter,A., Normandin,E., Bauer,M., Reilly,S., Tomkins-Tinch,C., Loreth,C., Chaluvadi,S., Lemieux,J.E., Birren,B.W., Sabeti,P.C., Huard,R., King,E., Park,D.J., and MacInnis,B.L. |
| EPI_ISL_2133723 | Quest Diagnostics Incorporated | Centers for Disease Control and Prevention Division of Viral Diseases, Pathogen Discovery | Dakota Howard, Dhwani Batra, Peter W. Cook, Kara Moser, Adrian Paskey, Jason Caravas, Benjamin Rambo-Martin, Shatavia Morrison, Christopher Gulvick, Scott Sammons, Yvette Unoarumhi, Darlene Wagner, Matthew Schmerer, S. H. Rosenthal, A. Gerasimova, R. M. Kagan, B. Anderson, M. Hua, Y. Liu, L.E. Bernstein, K.E. Livingston, A. Perez, I. A. Shlyakhter, R. V. Rolando, R. Owen, P. Tanpaiboon, F. Lacbawan, Clinton R. Paden, Duncan MacCannell |
| EPI_ISL_2133724 | Rhode Island Department of Health | Infectious Disease Program, Broad Institute of Harvard and MIT | Siddle,K.J., Azevedo,K., Miller,A., Adams,G., Pearlman,L., Gladden-Young,A., Lagerborg,K., Rudy,M., DeRuff,K., Carter,A., Normandin,E., Bauer,M., Reilly,S., Tomkins-Tinch,C., Loreth,C., Chaluvadi,S., Lemieux,J.E., Birren,B.W., Sabeti,P.C., Huard,R., King,E., Park,D.J., and MacInnis,B.L. |
| EPI_ISL_2133725, EPI_ISL_2133726 | Quest Diagnostics Incorporated | Centers for Disease Control and Prevention Division of Viral Diseases, Pathogen Discovery | Dakota Howard, Dhwani Batra, Peter W. Cook, Kara Moser, Adrian Paskey, Jason Caravas, Benjamin Rambo-Martin, Shatavia Morrison, Christopher Gulvick, Scott Sammons, Yvette Unoarumhi, Darlene Wagner, Matthew Schmerer, S. H. Rosenthal, A. Gerasimova, R. M. Kagan, B. Anderson, M. Hua, Y. Liu, L.E. Bernstein, K.E. Livingston, A. Perez, I. A. Shlyakhter, R. V. Rolando, R. Owen, P. Tanpaiboon, F. Lacbawan, Clinton R. Paden, Duncan MacCannell |
| EPI_ISL_2133727 | Rhode Island Department of Health | Infectious Disease Program, Broad Institute of Harvard and MIT | Siddle,K.J., Azevedo,K., Miller,A., Adams,G., Pearlman,L., Gladden-Young,A., Lagerborg,K., Rudy,M., DeRuff,K., Carter,A., Normandin,E., Bauer,M., Reilly,S., Tomkins-Tinch,C., Loreth,C., Chaluvadi,S., Lemieux,J.E., Birren,B.W., Sabeti,P.C., Huard,R., King,E., Park,D.J., and MacInnis,B.L. |
| EPI_ISL_2133728 | Quest Diagnostics Incorporated | Centers for Disease Control and Prevention Division of Viral Diseases, Pathogen Discovery | Dakota Howard, Dhwani Batra, Peter W. Cook, Kara Moser, Adrian Paskey, Jason Caravas, Benjamin Rambo-Martin, Shatavia Morrison, Christopher Gulvick, Scott Sammons, Yvette Unoarumhi, Darlene Wagner, Matthew Schmerer, S. H. Rosenthal, A. Gerasimova, R. M. Kagan, B. Anderson, M. Hua, Y. Liu, L.E. Bernstein, K.E. Livingston, A. Perez, I. A. Shlyakhter, R. V. Rolando, R. Owen, P. Tanpaiboon, F. Lacbawan, Clinton R. Paden, Duncan MacCannell |
| EPI_ISL_2133729 | Rhode Island Department of Health | Infectious Disease Program, Broad Institute of Harvard and MIT | Siddle,K.J., Azevedo,K., Miller,A., Adams,G., Pearlman,L., Gladden-Young,A., Lagerborg,K., Rudy,M., DeRuff,K., Carter,A., Normandin,E., Bauer,M., Reilly,S., Tomkins-Tinch,C., Loreth,C., Chaluvadi,S., Lemieux,J.E., Birren,B.W., Sabeti,P.C., Huard,R., King,E., Park,D.J., and MacInnis,B.L. |
| EPI_ISL_2133730 | Quest Diagnostics Incorporated | Centers for Disease Control and Prevention Division of Viral Diseases, Pathogen Discovery | Dakota Howard, Dhwani Batra, Peter W. Cook, Kara Moser, Adrian Paskey, Jason Caravas, Benjamin Rambo-Martin, Shatavia Morrison, Christopher Gulvick, Scott Sammons, Yvette Unoarumhi, Darlene Wagner, Matthew Schmerer, S. H. Rosenthal, A. Gerasimova, R. M. Kagan, B. Anderson, M. Hua, Y. Liu, L.E. Bernstein, K.E. Livingston, A. Perez, I. A. Shlyakhter, R. V. Rolando, R. Owen, P. Tanpaiboon, F. Lacbawan, Clinton R. Paden, Duncan MacCannell |
| EPI_ISL_2133731 | Rhode Island Department of Health | Infectious Disease Program, Broad Institute of Harvard and MIT | Siddle,K.J., Azevedo,K., Miller,A., Adams,G., Pearlman,L., Gladden-Young,A., Lagerborg,K., Rudy,M., DeRuff,K., Carter,A., Normandin,E., Bauer,M., Reilly,S., Tomkins-Tinch,C., Loreth,C., Chaluvadi,S., Lemieux,J.E., Birren,B.W., Sabeti,P.C., Huard,R., King,E., Park,D.J., and MacInnis,B.L. |
| EPI_ISL_2133732 | Quest Diagnostics Incorporated | Centers for Disease Control and Prevention Division of Viral Diseases, Pathogen Discovery | Dakota Howard, Dhwani Batra, Peter W. Cook, Kara Moser, Adrian Paskey, Jason Caravas, Benjamin Rambo-Martin, Shatavia Morrison, Christopher Gulvick, Scott Sammons, Yvette Unoarumhi, Darlene Wagner, Matthew Schmerer, S. H. Rosenthal, A. Gerasimova, R. M. Kagan, B. Anderson, M. Hua, Y. Liu, L.E. Bernstein, K.E. Livingston, A. Perez, I. A. Shlyakhter, R. V. Rolando, R. Owen, P. Tanpaiboon, F. Lacbawan, Clinton R. Paden, Duncan MacCannell |
| EPI_ISL_2133733 | Rhode Island Department of Health | Infectious Disease Program, Broad Institute of Harvard and MIT | Siddle,K.J., Azevedo,K., Miller,A., Adams,G., Pearlman,L., Gladden-Young,A., Lagerborg,K., Rudy,M., DeRuff,K., Carter,A., Normandin,E., Bauer,M., Reilly,S., Tomkins-Tinch,C., Loreth,C., Chaluvadi,S., Lemieux,J.E., Birren,B.W., Sabeti,P.C., Huard,R., King,E., Park,D.J., and MacInnis,B.L. |
| EPI_ISL_2133734 | Quest Diagnostics Incorporated | Centers for Disease Control and Prevention Division of Viral Diseases, Pathogen Discovery | Dakota Howard, Dhwani Batra, Peter W. Cook, Kara Moser, Adrian Paskey, Jason Caravas, Benjamin Rambo-Martin, Shatavia Morrison, Christopher Gulvick, Scott Sammons, Yvette Unoarumhi, Darlene Wagner, Matthew Schmerer, S. H. Rosenthal, A. Gerasimova, R. M. Kagan, B. Anderson, M. Hua, Y. Liu, L.E. Bernstein, K.E. Livingston, A. Perez, I. A. Shlyakhter, R. V. Rolando, R. Owen, P. Tanpaiboon, F. Lacbawan, Clinton R. Paden, Duncan MacCannell |
| EPI_ISL_2133735 | Rhode Island Department of Health | Infectious Disease Program, Broad Institute of Harvard and MIT | Siddle,K.J., Azevedo,K., Miller,A., Adams,G., Pearlman,L., Gladden-Young,A., Lagerborg,K., Rudy,M., DeRuff,K., Carter,A., Normandin,E., Bauer,M., Reilly,S., Tomkins-Tinch,C., Loreth,C., Chaluvadi,S., Lemieux,J.E., Birren,B.W., Sabeti,P.C., Huard,R., King,E., Park,D.J., and MacInnis,B.L. |
| EPI_ISL_2133736 | Quest Diagnostics Incorporated | Centers for Disease Control and Prevention Division of Viral Diseases, Pathogen Discovery | Dakota Howard, Dhwani Batra, Peter W. Cook, Kara Moser, Adrian Paskey, Jason Caravas, Benjamin Rambo-Martin, Shatavia Morrison, Christopher Gulvick, Scott Sammons, Yvette Unoarumhi, Darlene Wagner, Matthew Schmerer, S. H. Rosenthal, A. Gerasimova, R. M. Kagan, B. Anderson, M. Hua, Y. Liu, L.E. Bernstein, K.E. Livingston, A. Perez, I. A. Shlyakhter, R. V. Rolando, R. Owen, P. Tanpaiboon, F. Lacbawan, Clinton R. Paden, Duncan MacCannell |
| EPI_ISL_2133737, EPI_ISL_2133738 | Rhode Island Department of Health | Infectious Disease Program, Broad Institute of Harvard and MIT | Siddle,K.J., Azevedo,K., Miller,A., Adams,G., Pearlman,L., Gladden-Young,A., Lagerborg,K., Rudy,M., DeRuff,K., Carter,A., Normandin,E., Bauer,M., Reilly,S., Tomkins-Tinch,C., Loreth,C., Chaluvadi,S., Lemieux,J.E., Birren,B.W., Sabeti,P.C., Huard,R., King,E., Park,D.J., and MacInnis,B.L. |
| EPI_ISL_2133739 | Quest Diagnostics Incorporated | Centers for Disease Control and Prevention Division of Viral Diseases, Pathogen Discovery | Dakota Howard, Dhwani Batra, Peter W. Cook, Kara Moser, Adrian Paskey, Jason Caravas, Benjamin Rambo-Martin, Shatavia Morrison, Christopher Gulvick, Scott Sammons, Yvette Unoarumhi, Darlene Wagner, Matthew Schmerer, S. H. Rosenthal, A. Gerasimova, R. M. Kagan, B. Anderson, M. Hua, Y. Liu, L.E. Bernstein, K.E. Livingston, A. Perez, I. A. Shlyakhter, R. V. Rolando, R. Owen, P. Tanpaiboon, F. Lacbawan, Clinton R. Paden, Duncan MacCannell |
| EPI_ISL_2133740 | Rhode Island Department of Health | Infectious Disease Program, Broad Institute of Harvard and MIT | Siddle,K.J., Azevedo,K., Miller,A., Adams,G., Pearlman,L., Gladden-Young,A., Lagerborg,K., Rudy,M., DeRuff,K., Carter,A., Normandin,E., Bauer,M., Reilly,S., Tomkins-Tinch,C., Loreth,C., Chaluvadi,S., Lemieux,J.E., Birren,B.W., Sabeti,P.C., Huard,R., King,E., Park,D.J., and MacInnis,B.L. |
| EPI_ISL_2133741, EPI_ISL_2133743 | Quest Diagnostics Incorporated | Centers for Disease Control and Prevention Division of Viral Diseases, Pathogen Discovery | Dakota Howard, Dhwani Batra, Peter W. Cook, Kara Moser, Adrian Paskey, Jason Caravas, Benjamin Rambo-Martin, Shatavia Morrison, Christopher Gulvick, Scott Sammons, Yvette Unoarumhi, Darlene Wagner, Matthew Schmerer, S. H. Rosenthal, A. Gerasimova, R. M. Kagan, B. Anderson, M. Hua, Y. Liu, L.E. Bernstein, K.E. Livingston, A. Perez, I. A. Shlyakhter, R. V. Rolando, R. Owen, P. Tanpaiboon, F. Lacbawan, Clinton R. Paden, Duncan MacCannell |
| EPI_ISL_2133744 | Rhode Island Department of Health | Infectious Disease Program, Broad Institute of Harvard and MIT | Siddle,K.J., Azevedo,K., Miller,A., Adams,G., Pearlman,L., Gladden-Young,A., Lagerborg,K., Rudy,M., DeRuff,K., Carter,A., Normandin,E., Bauer,M., Reilly,S., Tomkins-Tinch,C., Loreth,C., Chaluvadi,S., Lemieux,J.E., Birren,B.W., Sabeti,P.C., Huard,R., King,E., Park,D.J., and MacInnis,B.L. |
| EPI_ISL_2133745 | Quest Diagnostics Incorporated | Centers for Disease Control and Prevention Division of Viral Diseases, Pathogen Discovery | Dakota Howard, Dhwani Batra, Peter W. Cook, Kara Moser, Adrian Paskey, Jason Caravas, Benjamin Rambo-Martin, Shatavia Morrison, Christopher Gulvick, Scott Sammons, Yvette Unoarumhi, Darlene Wagner, Matthew Schmerer, S. H. Rosenthal, A. Gerasimova, R. M. Kagan, B. Anderson, M. Hua, Y. Liu, L.E. Bernstein, K.E. Livingston, A. Perez, I. A. Shlyakhter, R. V. Rolando, R. Owen, P. Tanpaiboon, F. Lacbawan, Clinton R. Paden, Duncan MacCannell |
| EPI_ISL_2133746 | Rhode Island Department of Health | Infectious Disease Program, Broad Institute | Siddle,K.J., Azevedo,K., Miller,A., Adams,G., Pearlman,L., Gladden-Young,A., Lagerborg,K., Rudy,M., DeRuff,K., Carter,A., Normandin,E., Bauer,M., Reilly,S., Tomkins-Tinch,C., Loreth,C., Chaluvadi,S., Lemieux,J.E., Birren,B.W., Sabeti,P.C., Huard,R., King,E., Park,D.J., and MacInnis,B.L. |

| | | | |
|---|---|---|---|
| | | of Harvard and MIT | |
| EPI_ISL_2133747, EPI_ISL_2133749 | Quest Diagnostics Incorporated | Centers for Disease Control and Prevention Division of Viral Diseases, Pathogen Discovery | Dakota Howard, Dhwani Batra, Peter W. Cook, Kara Moser, Adrian Paskey, Jason Caravas, Benjamin Rambo-Martin, Shatavia Morrison, Christopher Gulvick, Scott Sammons, Yvette Unoarumhi, Darlene Wagner, Matthew Schmerer, S. H. Rosenthal, A. Gerasimova, R. M. Kagan, B. Anderson, M. Hua, Y. Liu, L.E. Bernstein, K.E. Livingston, A. Perez, I. A. Shlyakhter, R. V. Rolando, R. Owen, P. Tanpaiboon, F. Lacbawan, Clinton R. Paden, Duncan MacCannell |
| EPI_ISL_2133750 | Rhode Island Department of Health | Infectious Disease Program, Broad Institute of Harvard and MIT | Siddle,K.J., Azevedo,K., Miller,A., Adams,G., Pearlman,L., Gladden-Young,A., Lagerborg,K., Rudy,M., DeRuff,K., Carter,A., Normandin,E., Bauer,M., Reilly,S., Tomkins-Tinch,C., Loreth,C., Chaluvadi,S., Lemieux,J.E., Birren,B.W., Sabeti,P.C., Huard,R., King,E., Park,D.J., and MacInnis,B.L. |
| EPI_ISL_2133751 | Quest Diagnostics Incorporated | Centers for Disease Control and Prevention Division of Viral Diseases, Pathogen Discovery | Dakota Howard, Dhwani Batra, Peter W. Cook, Kara Moser, Adrian Paskey, Jason Caravas, Benjamin Rambo-Martin, Shatavia Morrison, Christopher Gulvick, Scott Sammons, Yvette Unoarumhi, Darlene Wagner, Matthew Schmerer, S. H. Rosenthal, A. Gerasimova, R. M. Kagan, B. Anderson, M. Hua, Y. Liu, L.E. Bernstein, K.E. Livingston, A. Perez, I. A. Shlyakhter, R. V. Rolando, R. Owen, P. Tanpaiboon, F. Lacbawan, Clinton R. Paden, Duncan MacCannell |
| EPI_ISL_2133752 | Rhode Island Department of Health | Infectious Disease Program, Broad Institute of Harvard and MIT | Siddle,K.J., Azevedo,K., Miller,A., Adams,G., Pearlman,L., Gladden-Young,A., Lagerborg,K., Rudy,M., DeRuff,K., Carter,A., Normandin,E., Bauer,M., Reilly,S., Tomkins-Tinch,C., Loreth,C., Chaluvadi,S., Lemieux,J.E., Birren,B.W., Sabeti,P.C., Huard,R., King,E., Park,D.J., and MacInnis,B.L. |
| EPI_ISL_2133753 | Quest Diagnostics Incorporated | Centers for Disease Control and Prevention Division of Viral Diseases, Pathogen Discovery | Dakota Howard, Dhwani Batra, Peter W. Cook, Kara Moser, Adrian Paskey, Jason Caravas, Benjamin Rambo-Martin, Shatavia Morrison, Christopher Gulvick, Scott Sammons, Yvette Unoarumhi, Darlene Wagner, Matthew Schmerer, S. H. Rosenthal, A. Gerasimova, R. M. Kagan, B. Anderson, M. Hua, Y. Liu, L.E. Bernstein, K.E. Livingston, A. Perez, I. A. Shlyakhter, R. V. Rolando, R. Owen, P. Tanpaiboon, F. Lacbawan, Clinton R. Paden, Duncan MacCannell |
| EPI_ISL_2133754 | Rhode Island Department of Health | Infectious Disease Program, Broad Institute of Harvard and MIT | Siddle,K.J., Azevedo,K., Miller,A., Adams,G., Pearlman,L., Gladden-Young,A., Lagerborg,K., Rudy,M., DeRuff,K., Carter,A., Normandin,E., Bauer,M., Reilly,S., Tomkins-Tinch,C., Loreth,C., Chaluvadi,S., Lemieux,J.E., Birren,B.W., Sabeti,P.C., Huard,R., King,E., Park,D.J., and MacInnis,B.L. |
| EPI_ISL_2133755 | Quest Diagnostics Incorporated | Centers for Disease Control and Prevention Division of Viral Diseases, Pathogen Discovery | Dakota Howard, Dhwani Batra, Peter W. Cook, Kara Moser, Adrian Paskey, Jason Caravas, Benjamin Rambo-Martin, Shatavia Morrison, Christopher Gulvick, Scott Sammons, Yvette Unoarumhi, Darlene Wagner, Matthew Schmerer, S. H. Rosenthal, A. Gerasimova, R. M. Kagan, B. Anderson, M. Hua, Y. Liu, L.E. Bernstein, K.E. Livingston, A. Perez, I. A. Shlyakhter, R. V. Rolando, R. Owen, P. Tanpaiboon, F. Lacbawan, Clinton R. Paden, Duncan MacCannell |
| EPI_ISL_2133756, EPI_ISL_2133757 | Rhode Island Department of Health | Infectious Disease Program, Broad Institute of Harvard and MIT | Siddle,K.J., Azevedo,K., Miller,A., Adams,G., Pearlman,L., Gladden-Young,A., Lagerborg,K., Rudy,M., DeRuff,K., Carter,A., Normandin,E., Bauer,M., Reilly,S., Tomkins-Tinch,C., Loreth,C., Chaluvadi,S., Lemieux,J.E., Birren,B.W., Sabeti,P.C., Huard,R., King,E., Park,D.J., and MacInnis,B.L. |
| EPI_ISL_2133758 | Quest Diagnostics Incorporated | Centers for Disease Control and Prevention Division of Viral Diseases, Pathogen Discovery | Dakota Howard, Dhwani Batra, Peter W. Cook, Kara Moser, Adrian Paskey, Jason Caravas, Benjamin Rambo-Martin, Shatavia Morrison, Christopher Gulvick, Scott Sammons, Yvette Unoarumhi, Darlene Wagner, Matthew Schmerer, S. H. Rosenthal, A. Gerasimova, R. M. Kagan, B. Anderson, M. Hua, Y. Liu, L.E. Bernstein, K.E. Livingston, A. Perez, I. A. Shlyakhter, R. V. Rolando, R. Owen, P. Tanpaiboon, F. Lacbawan, Clinton R. Paden, Duncan MacCannell |
| EPI_ISL_2133759 | Rhode Island Department of Health | Infectious Disease Program, Broad Institute of Harvard and MIT | Siddle,K.J., Azevedo,K., Miller,A., Adams,G., Pearlman,L., Gladden-Young,A., Lagerborg,K., Rudy,M., DeRuff,K., Carter,A., Normandin,E., Bauer,M., Reilly,S., Tomkins-Tinch,C., Loreth,C., Chaluvadi,S., Lemieux,J.E., Birren,B.W., Sabeti,P.C., Huard,R., King,E., Park,D.J., and MacInnis,B.L. |
| EPI_ISL_2133760 | Quest Diagnostics Incorporated | Centers for Disease Control and Prevention Division of Viral Diseases, Pathogen Discovery | Dakota Howard, Dhwani Batra, Peter W. Cook, Kara Moser, Adrian Paskey, Jason Caravas, Benjamin Rambo-Martin, Shatavia Morrison, Christopher Gulvick, Scott Sammons, Yvette Unoarumhi, Darlene Wagner, Matthew Schmerer, S. H. Rosenthal, A. Gerasimova, R. M. Kagan, B. Anderson, M. Hua, Y. Liu, L.E. Bernstein, K.E. Livingston, A. Perez, I. A. Shlyakhter, R. V. Rolando, R. Owen, P. Tanpaiboon, F. Lacbawan, Clinton R. Paden, Duncan MacCannell |
| EPI_ISL_2133761 | Rhode Island Department of Health | Infectious Disease Program, Broad Institute of Harvard and MIT | Siddle,K.J., Azevedo,K., Miller,A., Adams,G., Pearlman,L., Gladden-Young,A., Lagerborg,K., Rudy,M., DeRuff,K., Carter,A., Normandin,E., Bauer,M., Reilly,S., Tomkins-Tinch,C., Loreth,C., Chaluvadi,S., Lemieux,J.E., Birren,B.W., Sabeti,P.C., Huard,R., King,E., Park,D.J., and MacInnis,B.L. |
| EPI_ISL_2133762 | Quest Diagnostics Incorporated | Centers for Disease Control and Prevention Division of Viral Diseases, Pathogen Discovery | Dakota Howard, Dhwani Batra, Peter W. Cook, Kara Moser, Adrian Paskey, Jason Caravas, Benjamin Rambo-Martin, Shatavia Morrison, Christopher Gulvick, Scott Sammons, Yvette Unoarumhi, Darlene Wagner, Matthew Schmerer, S. H. Rosenthal, A. Gerasimova, R. M. Kagan, B. Anderson, M. Hua, Y. Liu, L.E. Bernstein, K.E. Livingston, A. Perez, I. A. Shlyakhter, R. V. Rolando, R. Owen, P. Tanpaiboon, F. Lacbawan, Clinton R. Paden, Duncan MacCannell |
| EPI_ISL_2133763 | Rhode Island Department of Health | Infectious Disease Program, Broad Institute of Harvard and MIT | Siddle,K.J., Azevedo,K., Miller,A., Adams,G., Pearlman,L., Gladden-Young,A., Lagerborg,K., Rudy,M., DeRuff,K., Carter,A., Normandin,E., Bauer,M., Reilly,S., Tomkins-Tinch,C., Loreth,C., Chaluvadi,S., Lemieux,J.E., Birren,B.W., Sabeti,P.C., Huard,R., King,E., Park,D.J., and MacInnis,B.L. |
| EPI_ISL_2133764 | Quest Diagnostics Incorporated | Centers for Disease Control and Prevention Division of Viral Diseases, Pathogen Discovery | Dakota Howard, Dhwani Batra, Peter W. Cook, Kara Moser, Adrian Paskey, Jason Caravas, Benjamin Rambo-Martin, Shatavia Morrison, Christopher Gulvick, Scott Sammons, Yvette Unoarumhi, Darlene Wagner, Matthew Schmerer, S. H. Rosenthal, A. Gerasimova, R. M. Kagan, B. Anderson, M. Hua, Y. Liu, L.E. Bernstein, K.E. Livingston, A. Perez, I. A. Shlyakhter, R. V. Rolando, R. Owen, P. Tanpaiboon, F. Lacbawan, Clinton R. Paden, Duncan MacCannell |
| EPI_ISL_2133765 | Rhode Island Department of Health | Infectious Disease Program, Broad Institute of Harvard and MIT | Siddle,K.J., Azevedo,K., Miller,A., Adams,G., Pearlman,L., Gladden-Young,A., Lagerborg,K., Rudy,M., DeRuff,K., Carter,A., Normandin,E., Bauer,M., Reilly,S., Tomkins-Tinch,C., Loreth,C., Chaluvadi,S., Lemieux,J.E., Birren,B.W., Sabeti,P.C., Huard,R., King,E., Park,D.J., and MacInnis,B.L. |
| EPI_ISL_2133766 | Quest Diagnostics Incorporated | Centers for Disease Control and Prevention Division of Viral Diseases, Pathogen Discovery | Dakota Howard, Dhwani Batra, Peter W. Cook, Kara Moser, Adrian Paskey, Jason Caravas, Benjamin Rambo-Martin, Shatavia Morrison, Christopher Gulvick, Scott Sammons, Yvette Unoarumhi, Darlene Wagner, Matthew Schmerer, S. H. Rosenthal, A. Gerasimova, R. M. Kagan, B. Anderson, M. Hua, Y. Liu, L.E. Bernstein, K.E. Livingston, A. Perez, I. A. Shlyakhter, R. V. Rolando, R. Owen, P. Tanpaiboon, F. Lacbawan, Clinton R. Paden, Duncan MacCannell |
| EPI_ISL_2133767 | Rhode Island Department of Health | Infectious Disease Program, Broad Institute of Harvard and MIT | Siddle,K.J., Azevedo,K., Miller,A., Adams,G., Pearlman,L., Gladden-Young,A., Lagerborg,K., Rudy,M., DeRuff,K., Carter,A., Normandin,E., Bauer,M., Reilly,S., Tomkins-Tinch,C., Loreth,C., Chaluvadi,S., Lemieux,J.E., Birren,B.W., Sabeti,P.C., Huard,R., King,E., Park,D.J., and MacInnis,B.L. |
| EPI_ISL_2133768 | Quest Diagnostics Incorporated | Centers for Disease Control and Prevention Division of Viral Diseases, Pathogen Discovery | Dakota Howard, Dhwani Batra, Peter W. Cook, Kara Moser, Adrian Paskey, Jason Caravas, Benjamin Rambo-Martin, Shatavia Morrison, Christopher Gulvick, Scott Sammons, Yvette Unoarumhi, Darlene Wagner, Matthew Schmerer, S. H. Rosenthal, A. Gerasimova, R. M. Kagan, B. Anderson, M. Hua, Y. Liu, L.E. Bernstein, K.E. Livingston, A. Perez, I. A. Shlyakhter, R. V. Rolando, R. Owen, P. Tanpaiboon, F. Lacbawan, Clinton R. Paden, Duncan MacCannell |
| EPI_ISL_2133769 | Rhode Island Department of Health | Infectious Disease Program, Broad Institute of Harvard and MIT | Siddle,K.J., Azevedo,K., Miller,A., Adams,G., Pearlman,L., Gladden-Young,A., Lagerborg,K., Rudy,M., DeRuff,K., Carter,A., Normandin,E., Bauer,M., Reilly,S., Tomkins-Tinch,C., Loreth,C., Chaluvadi,S., Lemieux,J.E., Birren,B.W., Sabeti,P.C., Huard,R., King,E., Park,D.J., and MacInnis,B.L. |
| EPI_ISL_2133770 | Quest Diagnostics Incorporated | Centers for Disease Control and Prevention Division of Viral Diseases, Pathogen Discovery | Dakota Howard, Dhwani Batra, Peter W. Cook, Kara Moser, Adrian Paskey, Jason Caravas, Benjamin Rambo-Martin, Shatavia Morrison, Christopher Gulvick, Scott Sammons, Yvette Unoarumhi, Darlene Wagner, Matthew Schmerer, S. H. Rosenthal, A. Gerasimova, R. M. Kagan, B. Anderson, M. Hua, Y. Liu, L.E. Bernstein, K.E. Livingston, A. Perez, I. A. Shlyakhter, R. V. Rolando, R. Owen, P. Tanpaiboon, F. Lacbawan, Clinton R. Paden, Duncan MacCannell |
| EPI_ISL_2133771 | Rhode Island Department of Health | Infectious Disease Program, Broad Institute of Harvard and MIT | Siddle,K.J., Azevedo,K., Miller,A., Adams,G., Pearlman,L., Gladden-Young,A., Lagerborg,K., Rudy,M., DeRuff,K., Carter,A., Normandin,E., Bauer,M., Reilly,S., Tomkins-Tinch,C., Loreth,C., Chaluvadi,S., Lemieux,J.E., Birren,B.W., Sabeti,P.C., Huard,R., King,E., Park,D.J., and MacInnis,B.L. |
| EPI_ISL_2133772 | Quest Diagnostics Incorporated | Centers for Disease Control and Prevention Division of Viral Diseases, Pathogen Discovery | Dakota Howard, Dhwani Batra, Peter W. Cook, Kara Moser, Adrian Paskey, Jason Caravas, Benjamin Rambo-Martin, Shatavia Morrison, Christopher Gulvick, Scott Sammons, Yvette Unoarumhi, Darlene Wagner, Matthew Schmerer, S. H. Rosenthal, A. Gerasimova, R. M. Kagan, B. Anderson, M. Hua, Y. Liu, L.E. Bernstein, K.E. Livingston, A. Perez, I. A. Shlyakhter, R. V. Rolando, R. Owen, P. Tanpaiboon, F. Lacbawan, Clinton R. Paden, Duncan MacCannell |
| EPI_ISL_2133773 | Rhode Island Department of Health | Infectious Disease Program, Broad Institute of Harvard and MIT | Siddle,K.J., Azevedo,K., Miller,A., Adams,G., Pearlman,L., Gladden-Young,A., Lagerborg,K., Rudy,M., DeRuff,K., Carter,A., Normandin,E., Bauer,M., Reilly,S., Tomkins-Tinch,C., Loreth,C., Chaluvadi,S., Lemieux,J.E., Birren,B.W., Sabeti,P.C., Huard,R., King,E., Park,D.J., and MacInnis,B.L. |
| EPI_ISL_2133774 | Quest Diagnostics Incorporated | Centers for Disease Control and Prevention Division of Viral Diseases, Pathogen Discovery | Dakota Howard, Dhwani Batra, Peter W. Cook, Kara Moser, Adrian Paskey, Jason Caravas, Benjamin Rambo-Martin, Shatavia Morrison, Christopher Gulvick, Scott Sammons, Yvette Unoarumhi, Darlene Wagner, Matthew Schmerer, S. H. Rosenthal, A. Gerasimova, R. M. Kagan, B. Anderson, M. Hua, Y. Liu, L.E. Bernstein, K.E. Livingston, A. Perez, I. A. Shlyakhter, R. V. Rolando, R. Owen, P. Tanpaiboon, F. Lacbawan, Clinton R. Paden, Duncan MacCannell |
| EPI_ISL_2133775 | Rhode Island Department of Health | Infectious Disease Program, Broad Institute of Harvard and MIT | Siddle,K.J., Azevedo,K., Miller,A., Adams,G., Pearlman,L., Gladden-Young,A., Lagerborg,K., Rudy,M., DeRuff,K., Carter,A., Normandin,E., Bauer,M., Reilly,S., Tomkins-Tinch,C., Loreth,C., Chaluvadi,S., Lemieux,J.E., Birren,B.W., Sabeti,P.C., Huard,R., King,E., Park,D.J., and MacInnis,B.L. |
| EPI_ISL_2133776 | Quest Diagnostics Incorporated | Centers for Disease Control and Prevention Division of Viral Diseases, Pathogen Discovery | Dakota Howard, Dhwani Batra, Peter W. Cook, Kara Moser, Adrian Paskey, Jason Caravas, Benjamin Rambo-Martin, Shatavia Morrison, Christopher Gulvick, Scott Sammons, Yvette Unoarumhi, Darlene Wagner, Matthew Schmerer, S. H. Rosenthal, A. Gerasimova, R. M. Kagan, B. Anderson, M. Hua, Y. Liu, L.E. Bernstein, K.E. Livingston, A. Perez, I. A. Shlyakhter, R. V. Rolando, R. Owen, P. Tanpaiboon, F. Lacbawan, Clinton R. Paden, Duncan MacCannell |
| EPI_ISL_2133777 | Rhode Island Department of Health | Infectious Disease Program, Broad Institute of Harvard and MIT | Siddle,K.J., Azevedo,K., Miller,A., Adams,G., Pearlman,L., Gladden-Young,A., Lagerborg,K., Rudy,M., DeRuff,K., Carter,A., Normandin,E., Bauer,M., Reilly,S., Tomkins-Tinch,C., Loreth,C., Chaluvadi,S., Lemieux,J.E., Birren,B.W., Sabeti,P.C., Huard,R., King,E., Park,D.J., and MacInnis,B.L. |
| EPI_ISL_2133778 | Quest Diagnostics Incorporated | Centers for Disease Control and Prevention Division of Viral Diseases, Pathogen Discovery | Dakota Howard, Dhwani Batra, Peter W. Cook, Kara Moser, Adrian Paskey, Jason Caravas, Benjamin Rambo-Martin, Shatavia Morrison, Christopher Gulvick, Scott Sammons, Yvette Unoarumhi, Darlene Wagner, Matthew Schmerer, S. H. Rosenthal, A. Gerasimova, R. M. Kagan, B. Anderson, M. Hua, Y. Liu, L.E. Bernstein, K.E. Livingston, A. Perez, I. A. Shlyakhter, R. V. Rolando, R. Owen, P. Tanpaiboon, F. Lacbawan, Clinton R. Paden, Duncan MacCannell |
| EPI_ISL_2133779, EPI_ISL_2133780 | Rhode Island Department of Health | Infectious Disease Program, Broad Institute of Harvard and MIT | Siddle,K.J., Azevedo,K., Miller,A., Adams,G., Pearlman,L., Gladden-Young,A., Lagerborg,K., Rudy,M., DeRuff,K., Carter,A., Normandin,E., Bauer,M., Reilly,S., Tomkins-Tinch,C., Loreth,C., Chaluvadi,S., Lemieux,J.E., Birren,B.W., Sabeti,P.C., Huard,R., King,E., Park,D.J., and MacInnis,B.L. |
| EPI_ISL_2133781 | Quest Diagnostics Incorporated | Centers for Disease Control and Prevention Division of Viral Diseases, Pathogen Discovery | Dakota Howard, Dhwani Batra, Peter W. Cook, Kara Moser, Adrian Paskey, Jason Caravas, Benjamin Rambo-Martin, Shatavia Morrison, Christopher Gulvick, Scott Sammons, Yvette Unoarumhi, Darlene Wagner, Matthew Schmerer, S. H. Rosenthal, A. Gerasimova, R. M. Kagan, B. Anderson, M. Hua, Y. Liu, L.E. Bernstein, K.E. Livingston, A. Perez, I. A. Shlyakhter, R. V. Rolando, R. Owen, P. Tanpaiboon, F. Lacbawan, Clinton R. Paden, Duncan MacCannell |
| EPI_ISL_2133782 | Rhode Island Department of Health | Infectious Disease Program, Broad Institute of Harvard and MIT | Siddle,K.J., Azevedo,K., Miller,A., Adams,G., Pearlman,L., Gladden-Young,A., Lagerborg,K., Rudy,M., DeRuff,K., Carter,A., Normandin,E., Bauer,M., Reilly,S., Tomkins-Tinch,C., Loreth,C., Chaluvadi,S., Lemieux,J.E., Birren,B.W., Sabeti,P.C., Huard,R., King,E., Park,D.J., and MacInnis,B.L. |
| EPI_ISL_2133783, EPI_ISL_2133784 | Quest Diagnostics Incorporated | Centers for Disease Control and Prevention Division of Viral Diseases, Pathogen Discovery | Dakota Howard, Dhwani Batra, Peter W. Cook, Kara Moser, Adrian Paskey, Jason Caravas, Benjamin Rambo-Martin, Shatavia Morrison, Christopher Gulvick, Scott Sammons, Yvette Unoarumhi, Darlene Wagner, Matthew Schmerer, S. H. Rosenthal, A. Gerasimova, R. M. Kagan, B. Anderson, M. Hua, Y. Liu, L.E. Bernstein, K.E. Livingston, A. Perez, I. A. Shlyakhter, R. V. Rolando, R. Owen, P. Tanpaiboon, F. Lacbawan, Clinton R. Paden, Duncan MacCannell |
| EPI_ISL_2133785 | Rhode Island Department of Health | Infectious Disease Program, Broad Institute of Harvard and MIT | Siddle,K.J., Azevedo,K., Miller,A., Adams,G., Pearlman,L., Gladden-Young,A., Lagerborg,K., Rudy,M., DeRuff,K., Carter,A., Normandin,E., Bauer,M., Reilly,S., Tomkins-Tinch,C., Loreth,C., Chaluvadi,S., Lemieux,J.E., Birren,B.W., Sabeti,P.C., Huard,R., King,E., Park,D.J., and MacInnis,B.L. |
| EPI_ISL_2133786 | Quest Diagnostics Incorporated | Centers for Disease Control and Prevention Division of Viral Diseases, Pathogen Discovery | Dakota Howard, Dhwani Batra, Peter W. Cook, Kara Moser, Adrian Paskey, Jason Caravas, Benjamin Rambo-Martin, Shatavia Morrison, Christopher Gulvick, Scott Sammons, Yvette Unoarumhi, Darlene Wagner, Matthew Schmerer, S. H. Rosenthal, A. Gerasimova, R. M. Kagan, B. Anderson, M. Hua, Y. Liu, L.E. Bernstein, K.E. Livingston, A. Perez, I. A. Shlyakhter, R. V. Rolando, R. Owen, P. Tanpaiboon, F. Lacbawan, Clinton R. Paden, Duncan MacCannell |
| EPI_ISL_2133787, EPI_ISL_2133788 | Rhode Island Department of Health | Infectious Disease Program, Broad Institute of Harvard and MIT | Siddle,K.J., Azevedo,K., Miller,A., Adams,G., Pearlman,L., Gladden-Young,A., Lagerborg,K., Rudy,M., DeRuff,K., Carter,A., Normandin,E., Bauer,M., Reilly,S., Tomkins-Tinch,C., Loreth,C., Chaluvadi,S., Lemieux,J.E., Birren,B.W., Sabeti,P.C., Huard,R., King,E., Park,D.J., and MacInnis,B.L. |
| EPI_ISL_2133789, EPI_ISL_2133790, EPI_ISL_2133791 | Quest Diagnostics Incorporated | Centers for Disease Control and Prevention Division of Viral Diseases, Pathogen Discovery | Dakota Howard, Dhwani Batra, Peter W. Cook, Kara Moser, Adrian Paskey, Jason Caravas, Benjamin Rambo-Martin, Shatavia Morrison, Christopher Gulvick, Scott Sammons, Yvette Unoarumhi, Darlene Wagner, Matthew Schmerer, S. H. Rosenthal, A. Gerasimova, R. M. Kagan, B. Anderson, M. Hua, Y. Liu, L.E. Bernstein, K.E. Livingston, A. Perez, I. A. Shlyakhter, R. V. Rolando, R. Owen, P. Tanpaiboon, F. Lacbawan, Clinton R. Paden, Duncan MacCannell |

| | | | |
|---|---|---|---|
| EPI_ISL_2133792 | Rhode Island Department of Health | Infectious Disease Program, Broad Institute of Harvard and MIT | Siddle,K.J., Azevedo,K., Miller,A., Adams,G., Pearlman,L., Gladden-Young,A., Lagerborg,K., Rudy,M., DeRuff,K., Carter,A., Normandin,E., Bauer,M., Reilly,S., Tomkins-Tinch,C., Loreth,C., Chaluvadi,S., Lemieux,J.E., Birren,B.W., Sabeti,P.C., Huard,R., King,E., Park,D.J., and MacInnis,B.L. |
| EPI_ISL_2133793 | Quest Diagnostics Incorporated | Centers for Disease Control and Prevention Division of Viral Diseases, Pathogen Discovery | Dakota Howard, Dhwani Batra, Peter W. Cook, Kara Moser, Adrian Paskey, Jason Caravas, Benjamin Rambo-Martin, Shatavia Morrison, Christopher Gulvick, Scott Sammons, Yvette Unoarumhi, Darlene Wagner, Matthew Schmerer, S. H. Rosenthal, A. Gerasimova, R. M. Kagan, B. Anderson, M. Hua, Y. Liu, L.E. Bernstein, K.E. Livingston, A. Perez, I. A. Shlyakhter, R. V. Rolando, R. Owen, P. Tanpaiboon, F. Lacbawan, Clinton R. Paden, Duncan MacCannell |
| EPI_ISL_2133794, EPI_ISL_2133795 | Rhode Island Department of Health | Infectious Disease Program, Broad Institute of Harvard and MIT | Siddle,K.J., Azevedo,K., Miller,A., Adams,G., Pearlman,L., Gladden-Young,A., Lagerborg,K., Rudy,M., DeRuff,K., Carter,A., Normandin,E., Bauer,M., Reilly,S., Tomkins-Tinch,C., Loreth,C., Chaluvadi,S., Lemieux,J.E., Birren,B.W., Sabeti,P.C., Huard,R., King,E., Park,D.J., and MacInnis,B.L. |
| EPI_ISL_2133796 | Quest Diagnostics Incorporated | Centers for Disease Control and Prevention Division of Viral Diseases, Pathogen Discovery | Dakota Howard, Dhwani Batra, Peter W. Cook, Kara Moser, Adrian Paskey, Jason Caravas, Benjamin Rambo-Martin, Shatavia Morrison, Christopher Gulvick, Scott Sammons, Yvette Unoarumhi, Darlene Wagner, Matthew Schmerer, S. H. Rosenthal, A. Gerasimova, R. M. Kagan, B. Anderson, M. Hua, Y. Liu, L.E. Bernstein, K.E. Livingston, A. Perez, I. A. Shlyakhter, R. V. Rolando, R. Owen, P. Tanpaiboon, F. Lacbawan, Clinton R. Paden, Duncan MacCannell |
| EPI_ISL_2133798 | Rhode Island Department of Health | Infectious Disease Program, Broad Institute of Harvard and MIT | Siddle,K.J., Azevedo,K., Miller,A., Adams,G., Pearlman,L., Gladden-Young,A., Lagerborg,K., Rudy,M., DeRuff,K., Carter,A., Normandin,E., Bauer,M., Reilly,S., Tomkins-Tinch,C., Loreth,C., Chaluvadi,S., Lemieux,J.E., Birren,B.W., Sabeti,P.C., Huard,R., King,E., Park,D.J., and MacInnis,B.L. |
| EPI_ISL_2133799 | Quest Diagnostics Incorporated | Centers for Disease Control and Prevention Division of Viral Diseases, Pathogen Discovery | Dakota Howard, Dhwani Batra, Peter W. Cook, Kara Moser, Adrian Paskey, Jason Caravas, Benjamin Rambo-Martin, Shatavia Morrison, Christopher Gulvick, Scott Sammons, Yvette Unoarumhi, Darlene Wagner, Matthew Schmerer, S. H. Rosenthal, A. Gerasimova, R. M. Kagan, B. Anderson, M. Hua, Y. Liu, L.E. Bernstein, K.E. Livingston, A. Perez, I. A. Shlyakhter, R. V. Rolando, R. Owen, P. Tanpaiboon, F. Lacbawan, Clinton R. Paden, Duncan MacCannell |
| EPI_ISL_2133800 | Rhode Island Department of Health | Infectious Disease Program, Broad Institute of Harvard and MIT | Siddle,K.J., Azevedo,K., Miller,A., Adams,G., Pearlman,L., Gladden-Young,A., Lagerborg,K., Rudy,M., DeRuff,K., Carter,A., Normandin,E., Bauer,M., Reilly,S., Tomkins-Tinch,C., Loreth,C., Chaluvadi,S., Lemieux,J.E., Birren,B.W., Sabeti,P.C., Huard,R., King,E., Park,D.J., and MacInnis,B.L. |
| EPI_ISL_2133801 | Quest Diagnostics Incorporated | Centers for Disease Control and Prevention Division of Viral Diseases, Pathogen Discovery | Dakota Howard, Dhwani Batra, Peter W. Cook, Kara Moser, Adrian Paskey, Jason Caravas, Benjamin Rambo-Martin, Shatavia Morrison, Christopher Gulvick, Scott Sammons, Yvette Unoarumhi, Darlene Wagner, Matthew Schmerer, S. H. Rosenthal, A. Gerasimova, R. M. Kagan, B. Anderson, M. Hua, Y. Liu, L.E. Bernstein, K.E. Livingston, A. Perez, I. A. Shlyakhter, R. V. Rolando, R. Owen, P. Tanpaiboon, F. Lacbawan, Clinton R. Paden, Duncan MacCannell |
| EPI_ISL_2133802, EPI_ISL_2133803 | Rhode Island Department of Health | Infectious Disease Program, Broad Institute of Harvard and MIT | Siddle,K.J., Azevedo,K., Miller,A., Adams,G., Pearlman,L., Gladden-Young,A., Lagerborg,K., Rudy,M., DeRuff,K., Carter,A., Normandin,E., Bauer,M., Reilly,S., Tomkins-Tinch,C., Loreth,C., Chaluvadi,S., Lemieux,J.E., Birren,B.W., Sabeti,P.C., Huard,R., King,E., Park,D.J., and MacInnis,B.L. |
| EPI_ISL_2133804 | Quest Diagnostics Incorporated | Centers for Disease Control and Prevention Division of Viral Diseases, Pathogen Discovery | Dakota Howard, Dhwani Batra, Peter W. Cook, Kara Moser, Adrian Paskey, Jason Caravas, Benjamin Rambo-Martin, Shatavia Morrison, Christopher Gulvick, Scott Sammons, Yvette Unoarumhi, Darlene Wagner, Matthew Schmerer, S. H. Rosenthal, A. Gerasimova, R. M. Kagan, B. Anderson, M. Hua, Y. Liu, L.E. Bernstein, K.E. Livingston, A. Perez, I. A. Shlyakhter, R. V. Rolando, R. Owen, P. Tanpaiboon, F. Lacbawan, Clinton R. Paden, Duncan MacCannell |
| EPI_ISL_2133805 | Rhode Island Department of Health | Infectious Disease Program, Broad Institute of Harvard and MIT | Siddle,K.J., Azevedo,K., Miller,A., Adams,G., Pearlman,L., Gladden-Young,A., Lagerborg,K., Rudy,M., DeRuff,K., Carter,A., Normandin,E., Bauer,M., Reilly,S., Tomkins-Tinch,C., Loreth,C., Chaluvadi,S., Lemieux,J.E., Birren,B.W., Sabeti,P.C., Huard,R., King,E., Park,D.J., and MacInnis,B.L. |
| EPI_ISL_2133806 | Quest Diagnostics Incorporated | Centers for Disease Control and Prevention Division of Viral Diseases, Pathogen Discovery | Dakota Howard, Dhwani Batra, Peter W. Cook, Kara Moser, Adrian Paskey, Jason Caravas, Benjamin Rambo-Martin, Shatavia Morrison, Christopher Gulvick, Scott Sammons, Yvette Unoarumhi, Darlene Wagner, Matthew Schmerer, S. H. Rosenthal, A. Gerasimova, R. M. Kagan, B. Anderson, M. Hua, Y. Liu, L.E. Bernstein, K.E. Livingston, A. Perez, I. A. Shlyakhter, R. V. Rolando, R. Owen, P. Tanpaiboon, F. Lacbawan, Clinton R. Paden, Duncan MacCannell |
| EPI_ISL_2133807, EPI_ISL_2133808 | Rhode Island Department of Health | Infectious Disease Program, Broad Institute of Harvard and MIT | Siddle,K.J., Azevedo,K., Miller,A., Adams,G., Pearlman,L., Gladden-Young,A., Lagerborg,K., Rudy,M., DeRuff,K., Carter,A., Normandin,E., Bauer,M., Reilly,S., Tomkins-Tinch,C., Loreth,C., Chaluvadi,S., Lemieux,J.E., Birren,B.W., Sabeti,P.C., Huard,R., King,E., Park,D.J., and MacInnis,B.L. |
| EPI_ISL_2133809 | Quest Diagnostics Incorporated | Centers for Disease Control and Prevention Division of Viral Diseases, Pathogen Discovery | Dakota Howard, Dhwani Batra, Peter W. Cook, Kara Moser, Adrian Paskey, Jason Caravas, Benjamin Rambo-Martin, Shatavia Morrison, Christopher Gulvick, Scott Sammons, Yvette Unoarumhi, Darlene Wagner, Matthew Schmerer, S. H. Rosenthal, A. Gerasimova, R. M. Kagan, B. Anderson, M. Hua, Y. Liu, L.E. Bernstein, K.E. Livingston, A. Perez, I. A. Shlyakhter, R. V. Rolando, R. Owen, P. Tanpaiboon, F. Lacbawan, Clinton R. Paden, Duncan MacCannell |
| EPI_ISL_2133810 | Rhode Island Department of Health | Infectious Disease Program, Broad Institute of Harvard and MIT | Siddle,K.J., Azevedo,K., Miller,A., Adams,G., Pearlman,L., Gladden-Young,A., Lagerborg,K., Rudy,M., DeRuff,K., Carter,A., Normandin,E., Bauer,M., Reilly,S., Tomkins-Tinch,C., Loreth,C., Chaluvadi,S., Lemieux,J.E., Birren,B.W., Sabeti,P.C., Huard,R., King,E., Park,D.J., and MacInnis,B.L. |
| EPI_ISL_2133811 | Quest Diagnostics Incorporated | Centers for Disease Control and Prevention Division of Viral Diseases, Pathogen Discovery | Dakota Howard, Dhwani Batra, Peter W. Cook, Kara Moser, Adrian Paskey, Jason Caravas, Benjamin Rambo-Martin, Shatavia Morrison, Christopher Gulvick, Scott Sammons, Yvette Unoarumhi, Darlene Wagner, Matthew Schmerer, S. H. Rosenthal, A. Gerasimova, R. M. Kagan, B. Anderson, M. Hua, Y. Liu, L.E. Bernstein, K.E. Livingston, A. Perez, I. A. Shlyakhter, R. V. Rolando, R. Owen, P. Tanpaiboon, F. Lacbawan, Clinton R. Paden, Duncan MacCannell |
| EPI_ISL_2133812 | Rhode Island Department of Health | Infectious Disease Program, Broad Institute of Harvard and MIT | Siddle,K.J., Azevedo,K., Miller,A., Adams,G., Pearlman,L., Gladden-Young,A., Lagerborg,K., Rudy,M., DeRuff,K., Carter,A., Normandin,E., Bauer,M., Reilly,S., Tomkins-Tinch,C., Loreth,C., Chaluvadi,S., Lemieux,J.E., Birren,B.W., Sabeti,P.C., Huard,R., King,E., Park,D.J., and MacInnis,B.L. |
| EPI_ISL_2133813 | Quest Diagnostics Incorporated | Centers for Disease Control and Prevention Division of Viral Diseases, Pathogen Discovery | Dakota Howard, Dhwani Batra, Peter W. Cook, Kara Moser, Adrian Paskey, Jason Caravas, Benjamin Rambo-Martin, Shatavia Morrison, Christopher Gulvick, Scott Sammons, Yvette Unoarumhi, Darlene Wagner, Matthew Schmerer, S. H. Rosenthal, A. Gerasimova, R. M. Kagan, B. Anderson, M. Hua, Y. Liu, L.E. Bernstein, K.E. Livingston, A. Perez, I. A. Shlyakhter, R. V. Rolando, R. Owen, P. Tanpaiboon, F. Lacbawan, Clinton R. Paden, Duncan MacCannell |
| EPI_ISL_2133814 | Rhode Island Department of Health | Infectious Disease Program, Broad Institute of Harvard and MIT | Siddle,K.J., Azevedo,K., Miller,A., Adams,G., Pearlman,L., Gladden-Young,A., Lagerborg,K., Rudy,M., DeRuff,K., Carter,A., Normandin,E., Bauer,M., Reilly,S., Tomkins-Tinch,C., Loreth,C., Chaluvadi,S., Lemieux,J.E., Birren,B.W., Sabeti,P.C., Huard,R., King,E., Park,D.J., and MacInnis,B.L. |
| EPI_ISL_2133815 | Quest Diagnostics Incorporated | Centers for Disease Control and Prevention Division of Viral Diseases, Pathogen Discovery | Dakota Howard, Dhwani Batra, Peter W. Cook, Kara Moser, Adrian Paskey, Jason Caravas, Benjamin Rambo-Martin, Shatavia Morrison, Christopher Gulvick, Scott Sammons, Yvette Unoarumhi, Darlene Wagner, Matthew Schmerer, S. H. Rosenthal, A. Gerasimova, R. M. Kagan, B. Anderson, M. Hua, Y. Liu, L.E. Bernstein, K.E. Livingston, A. Perez, I. A. Shlyakhter, R. V. Rolando, R. Owen, P. Tanpaiboon, F. Lacbawan, Clinton R. Paden, Duncan MacCannell |
| EPI_ISL_2133816 | Rhode Island Department of Health | Infectious Disease Program, Broad Institute of Harvard and MIT | Siddle,K.J., Azevedo,K., Miller,A., Adams,G., Pearlman,L., Gladden-Young,A., Lagerborg,K., Rudy,M., DeRuff,K., Carter,A., Normandin,E., Bauer,M., Reilly,S., Tomkins-Tinch,C., Loreth,C., Chaluvadi,S., Lemieux,J.E., Birren,B.W., Sabeti,P.C., Huard,R., King,E., Park,D.J., and MacInnis,B.L. |
| EPI_ISL_2133817, EPI_ISL_2133818 | Quest Diagnostics Incorporated | Centers for Disease Control and Prevention Division of Viral Diseases, Pathogen Discovery | Dakota Howard, Dhwani Batra, Peter W. Cook, Kara Moser, Adrian Paskey, Jason Caravas, Benjamin Rambo-Martin, Shatavia Morrison, Christopher Gulvick, Scott Sammons, Yvette Unoarumhi, Darlene Wagner, Matthew Schmerer, S. H. Rosenthal, A. Gerasimova, R. M. Kagan, B. Anderson, M. Hua, Y. Liu, L.E. Bernstein, K.E. Livingston, A. Perez, I. A. Shlyakhter, R. V. Rolando, R. Owen, P. Tanpaiboon, F. Lacbawan, Clinton R. Paden, Duncan MacCannell |
| EPI_ISL_2133819 | Rhode Island Department of Health | Infectious Disease Program, Broad Institute of Harvard and MIT | Siddle,K.J., Azevedo,K., Miller,A., Adams,G., Pearlman,L., Gladden-Young,A., Lagerborg,K., Rudy,M., DeRuff,K., Carter,A., Normandin,E., Bauer,M., Reilly,S., Tomkins-Tinch,C., Loreth,C., Chaluvadi,S., Lemieux,J.E., Birren,B.W., Sabeti,P.C., Huard,R., King,E., Park,D.J., and MacInnis,B.L. |
| EPI_ISL_2133820 | Quest Diagnostics Incorporated | Centers for Disease Control and Prevention Division of Viral Diseases, Pathogen Discovery | Dakota Howard, Dhwani Batra, Peter W. Cook, Kara Moser, Adrian Paskey, Jason Caravas, Benjamin Rambo-Martin, Shatavia Morrison, Christopher Gulvick, Scott Sammons, Yvette Unoarumhi, Darlene Wagner, Matthew Schmerer, S. H. Rosenthal, A. Gerasimova, R. M. Kagan, B. Anderson, M. Hua, Y. Liu, L.E. Bernstein, K.E. Livingston, A. Perez, I. A. Shlyakhter, R. V. Rolando, R. Owen, P. Tanpaiboon, F. Lacbawan, Clinton R. Paden, Duncan MacCannell |
| EPI_ISL_2133821 | Rhode Island Department of Health | Infectious Disease Program, Broad Institute of Harvard and MIT | Siddle,K.J., Azevedo,K., Miller,A., Adams,G., Pearlman,L., Gladden-Young,A., Lagerborg,K., Rudy,M., DeRuff,K., Carter,A., Normandin,E., Bauer,M., Reilly,S., Tomkins-Tinch,C., Loreth,C., Chaluvadi,S., Lemieux,J.E., Birren,B.W., Sabeti,P.C., Huard,R., King,E., Park,D.J., and MacInnis,B.L. |
| EPI_ISL_2133822 | Quest Diagnostics Incorporated | Centers for Disease Control and Prevention Division of Viral Diseases, Pathogen Discovery | Dakota Howard, Dhwani Batra, Peter W. Cook, Kara Moser, Adrian Paskey, Jason Caravas, Benjamin Rambo-Martin, Shatavia Morrison, Christopher Gulvick, Scott Sammons, Yvette Unoarumhi, Darlene Wagner, Matthew Schmerer, S. H. Rosenthal, A. Gerasimova, R. M. Kagan, B. Anderson, M. Hua, Y. Liu, L.E. Bernstein, K.E. Livingston, A. Perez, I. A. Shlyakhter, R. V. Rolando, R. Owen, P. Tanpaiboon, F. Lacbawan, Clinton R. Paden, Duncan MacCannell |
| EPI_ISL_2133823 | Rhode Island Department of Health | Infectious Disease Program, Broad Institute of Harvard and MIT | Siddle,K.J., Azevedo,K., Miller,A., Adams,G., Pearlman,L., Gladden-Young,A., Lagerborg,K., Rudy,M., DeRuff,K., Carter,A., Normandin,E., Bauer,M., Reilly,S., Tomkins-Tinch,C., Loreth,C., Chaluvadi,S., Lemieux,J.E., Birren,B.W., Sabeti,P.C., Huard,R., King,E., Park,D.J., and MacInnis,B.L. |
| EPI_ISL_2133824 | Quest Diagnostics Incorporated | Centers for Disease Control and Prevention Division of Viral Diseases, Pathogen Discovery | Dakota Howard, Dhwani Batra, Peter W. Cook, Kara Moser, Adrian Paskey, Jason Caravas, Benjamin Rambo-Martin, Shatavia Morrison, Christopher Gulvick, Scott Sammons, Yvette Unoarumhi, Darlene Wagner, Matthew Schmerer, S. H. Rosenthal, A. Gerasimova, R. M. Kagan, B. Anderson, M. Hua, Y. Liu, L.E. Bernstein, K.E. Livingston, A. Perez, I. A. Shlyakhter, R. V. Rolando, R. Owen, P. Tanpaiboon, F. Lacbawan, Clinton R. Paden, Duncan MacCannell |
| EPI_ISL_2133825 | Rhode Island Department of Health | Infectious Disease Program, Broad Institute of Harvard and MIT | Siddle,K.J., Azevedo,K., Miller,A., Adams,G., Pearlman,L., Gladden-Young,A., Lagerborg,K., Rudy,M., DeRuff,K., Carter,A., Normandin,E., Bauer,M., Reilly,S., Tomkins-Tinch,C., Loreth,C., Chaluvadi,S., Lemieux,J.E., Birren,B.W., Sabeti,P.C., Huard,R., King,E., Park,D.J., and MacInnis,B.L. |
| EPI_ISL_2133826 | Quest Diagnostics Incorporated | Centers for Disease Control and Prevention Division of Viral Diseases, Pathogen Discovery | Dakota Howard, Dhwani Batra, Peter W. Cook, Kara Moser, Adrian Paskey, Jason Caravas, Benjamin Rambo-Martin, Shatavia Morrison, Christopher Gulvick, Scott Sammons, Yvette Unoarumhi, Darlene Wagner, Matthew Schmerer, S. H. Rosenthal, A. Gerasimova, R. M. Kagan, B. Anderson, M. Hua, Y. Liu, L.E. Bernstein, K.E. Livingston, A. Perez, I. A. Shlyakhter, R. V. Rolando, R. Owen, P. Tanpaiboon, F. Lacbawan, Clinton R. Paden, Duncan MacCannell |
| EPI_ISL_2133827 | Rhode Island Department of Health | Infectious Disease Program, Broad Institute of Harvard and MIT | Siddle,K.J., Azevedo,K., Miller,A., Adams,G., Pearlman,L., Gladden-Young,A., Lagerborg,K., Rudy,M., DeRuff,K., Carter,A., Normandin,E., Bauer,M., Reilly,S., Tomkins-Tinch,C., Loreth,C., Chaluvadi,S., Lemieux,J.E., Birren,B.W., Sabeti,P.C., Huard,R., King,E., Park,D.J., and MacInnis,B.L. |
| EPI_ISL_2133828, EPI_ISL_2133830 | Quest Diagnostics Incorporated | Centers for Disease Control and Prevention Division of Viral Diseases, Pathogen Discovery | Dakota Howard, Dhwani Batra, Peter W. Cook, Kara Moser, Adrian Paskey, Jason Caravas, Benjamin Rambo-Martin, Shatavia Morrison, Christopher Gulvick, Scott Sammons, Yvette Unoarumhi, Darlene Wagner, Matthew Schmerer, S. H. Rosenthal, A. Gerasimova, R. M. Kagan, B. Anderson, M. Hua, Y. Liu, L.E. Bernstein, K.E. Livingston, A. Perez, I. A. Shlyakhter, R. V. Rolando, R. Owen, P. Tanpaiboon, F. Lacbawan, Clinton R. Paden, Duncan MacCannell |
| EPI_ISL_2133831 | Rhode Island Department of Health | Infectious Disease Program, Broad Institute of Harvard and MIT | Siddle,K.J., Azevedo,K., Miller,A., Adams,G., Pearlman,L., Gladden-Young,A., Lagerborg,K., Rudy,M., DeRuff,K., Carter,A., Normandin,E., Bauer,M., Reilly,S., Tomkins-Tinch,C., Loreth,C., Chaluvadi,S., Lemieux,J.E., Birren,B.W., Sabeti,P.C., Huard,R., King,E., Park,D.J., and MacInnis,B.L. |
| EPI_ISL_2133832 | Quest Diagnostics Incorporated | Centers for Disease Control and Prevention Division of Viral Diseases, Pathogen Discovery | Dakota Howard, Dhwani Batra, Peter W. Cook, Kara Moser, Adrian Paskey, Jason Caravas, Benjamin Rambo-Martin, Shatavia Morrison, Christopher Gulvick, Scott Sammons, Yvette Unoarumhi, Darlene Wagner, Matthew Schmerer, S. H. Rosenthal, A. Gerasimova, R. M. Kagan, B. Anderson, M. Hua, Y. Liu, L.E. Bernstein, K.E. Livingston, A. Perez, I. A. Shlyakhter, R. V. Rolando, R. Owen, P. Tanpaiboon, F. Lacbawan, Clinton R. Paden, Duncan MacCannell |
| EPI_ISL_2133833 | Rhode Island Department of Health | Infectious Disease Program, Broad Institute of Harvard and MIT | Siddle,K.J., Azevedo,K., Miller,A., Adams,G., Pearlman,L., Gladden-Young,A., Lagerborg,K., Rudy,M., DeRuff,K., Carter,A., Normandin,E., Bauer,M., Reilly,S., Tomkins-Tinch,C., Loreth,C., Chaluvadi,S., Lemieux,J.E., Birren,B.W., Sabeti,P.C., Huard,R., King,E., Park,D.J., and MacInnis,B.L. |
| EPI_ISL_2133834 | Quest Diagnostics Incorporated | Centers for Disease Control and Prevention Division of Viral Diseases, Pathogen Discovery | Dakota Howard, Dhwani Batra, Peter W. Cook, Kara Moser, Adrian Paskey, Jason Caravas, Benjamin Rambo-Martin, Shatavia Morrison, Christopher Gulvick, Scott Sammons, Yvette Unoarumhi, Darlene Wagner, Matthew Schmerer, S. H. Rosenthal, A. Gerasimova, R. M. Kagan, B. Anderson, M. Hua, Y. Liu, L.E. Bernstein, K.E. Livingston, A. Perez, I. A. Shlyakhter, R. V. Rolando, R. Owen, P. Tanpaiboon, F. Lacbawan, Clinton R. Paden, Duncan MacCannell |
| EPI_ISL_2133835 | Rhode Island Department of Health | Infectious Disease Program, Broad Institute of Harvard and MIT | Siddle,K.J., Azevedo,K., Miller,A., Adams,G., Pearlman,L., Gladden-Young,A., Lagerborg,K., Rudy,M., DeRuff,K., Carter,A., Normandin,E., Bauer,M., Reilly,S., Tomkins-Tinch,C., Loreth,C., Chaluvadi,S., Lemieux,J.E., Birren,B.W., Sabeti,P.C., Huard,R., King,E., Park,D.J., and MacInnis,B.L. |
| EPI_ISL_2133836 | Quest Diagnostics Incorporated | Centers for Disease Control and Prevention Division of Viral Diseases, Pathogen | Dakota Howard, Dhwani Batra, Peter W. Cook, Kara Moser, Adrian Paskey, Jason Caravas, Benjamin Rambo-Martin, Shatavia Morrison, Christopher Gulvick, Scott Sammons, Yvette Unoarumhi, Darlene Wagner, Matthew Schmerer, S. H. Rosenthal, A. Gerasimova, R. M. Kagan, B. Anderson, M. Hua, Y. Liu, L.E. Bernstein, K.E. Livingston, A. Perez, I. A. Shlyakhter, R. V. Rolando, R. Owen, P. Tanpaiboon, F. Lacbawan, Clinton R. Paden, Duncan MacCannell |

| | | | |
|---|---|---|---|
| | | Discovery | |
| EPI_ISL_2133837, EPI_ISL_2133838 | Rhode Island Department of Health | Infectious Disease Program, Broad Institute of Harvard and MIT | Siddle,K.J., Azevedo,K., Miller,A., Adams,G., Pearlman,L., Gladden-Young,A., Lagerborg,K., Rudy,M., DeRuff,K., Carter,A., Normandin,E., Bauer,M., Reilly,S., Tomkins-Tinch,C., Loreth,C., Chaluvadi,S., Lemieux,J.E., Birren,B.W., Sabeti,P.C., Huard,R., King,E., Park,D.J., and MacInnis,B.L. |
| EPI_ISL_2133839 | Quest Diagnostics Incorporated | Centers for Disease Control and Prevention Division of Viral Diseases, Pathogen Discovery | Dakota Howard, Dhwani Batra, Peter W. Cook, Kara Moser, Adrian Paskey, Jason Caravas, Benjamin Rambo-Martin, Shatavia Morrison, Christopher Gulvick, Scott Sammons, Yvette Unoarumhi, Darlene Wagner, Matthew Schmerer, S. H. Rosenthal, A. Gerasimova, R. M. Kagan, B. Anderson, M. Hua, Y. Liu, L.E. Bernstein, K.E. Livingston, A. Perez, I. A. Shlyakhter, R. V. Rolando, R. Owen, P. Tanpaiboon, F. Lacbawan, Clinton R. Paden, Duncan MacCannell |
| EPI_ISL_2133840, EPI_ISL_2133841 | Rhode Island Department of Health | Infectious Disease Program, Broad Institute of Harvard and MIT | Siddle,K.J., Azevedo,K., Miller,A., Adams,G., Pearlman,L., Gladden-Young,A., Lagerborg,K., Rudy,M., DeRuff,K., Carter,A., Normandin,E., Bauer,M., Reilly,S., Tomkins-Tinch,C., Loreth,C., Chaluvadi,S., Lemieux,J.E., Birren,B.W., Sabeti,P.C., Huard,R., King,E., Park,D.J., and MacInnis,B.L. |
| EPI_ISL_2133842 | Quest Diagnostics Incorporated | Centers for Disease Control and Prevention Division of Viral Diseases, Pathogen Discovery | Dakota Howard, Dhwani Batra, Peter W. Cook, Kara Moser, Adrian Paskey, Jason Caravas, Benjamin Rambo-Martin, Shatavia Morrison, Christopher Gulvick, Scott Sammons, Yvette Unoarumhi, Darlene Wagner, Matthew Schmerer, S. H. Rosenthal, A. Gerasimova, R. M. Kagan, B. Anderson, M. Hua, Y. Liu, L.E. Bernstein, K.E. Livingston, A. Perez, I. A. Shlyakhter, R. V. Rolando, R. Owen, P. Tanpaiboon, F. Lacbawan, Clinton R. Paden, Duncan MacCannell |
| EPI_ISL_2133843 | Rhode Island Department of Health | Infectious Disease Program, Broad Institute of Harvard and MIT | Siddle,K.J., Azevedo,K., Miller,A., Adams,G., Pearlman,L., Gladden-Young,A., Lagerborg,K., Rudy,M., DeRuff,K., Carter,A., Normandin,E., Bauer,M., Reilly,S., Tomkins-Tinch,C., Loreth,C., Chaluvadi,S., Lemieux,J.E., Birren,B.W., Sabeti,P.C., Huard,R., King,E., Park,D.J., and MacInnis,B.L. |
| EPI_ISL_2133844 | Quest Diagnostics Incorporated | Centers for Disease Control and Prevention Division of Viral Diseases, Pathogen Discovery | Dakota Howard, Dhwani Batra, Peter W. Cook, Kara Moser, Adrian Paskey, Jason Caravas, Benjamin Rambo-Martin, Shatavia Morrison, Christopher Gulvick, Scott Sammons, Yvette Unoarumhi, Darlene Wagner, Matthew Schmerer, S. H. Rosenthal, A. Gerasimova, R. M. Kagan, B. Anderson, M. Hua, Y. Liu, L.E. Bernstein, K.E. Livingston, A. Perez, I. A. Shlyakhter, R. V. Rolando, R. Owen, P. Tanpaiboon, F. Lacbawan, Clinton R. Paden, Duncan MacCannell |
| EPI_ISL_2133845 | Rhode Island Department of Health | Infectious Disease Program, Broad Institute of Harvard and MIT | Siddle,K.J., Azevedo,K., Miller,A., Adams,G., Pearlman,L., Gladden-Young,A., Lagerborg,K., Rudy,M., DeRuff,K., Carter,A., Normandin,E., Bauer,M., Reilly,S., Tomkins-Tinch,C., Loreth,C., Chaluvadi,S., Lemieux,J.E., Birren,B.W., Sabeti,P.C., Huard,R., King,E., Park,D.J., and MacInnis,B.L. |
| EPI_ISL_2133846 | Quest Diagnostics Incorporated | Centers for Disease Control and Prevention Division of Viral Diseases, Pathogen Discovery | Dakota Howard, Dhwani Batra, Peter W. Cook, Kara Moser, Adrian Paskey, Jason Caravas, Benjamin Rambo-Martin, Shatavia Morrison, Christopher Gulvick, Scott Sammons, Yvette Unoarumhi, Darlene Wagner, Matthew Schmerer, S. H. Rosenthal, A. Gerasimova, R. M. Kagan, B. Anderson, M. Hua, Y. Liu, L.E. Bernstein, K.E. Livingston, A. Perez, I. A. Shlyakhter, R. V. Rolando, R. Owen, P. Tanpaiboon, F. Lacbawan, Clinton R. Paden, Duncan MacCannell |
| EPI_ISL_2133847 | Rhode Island Department of Health | Infectious Disease Program, Broad Institute of Harvard and MIT | Siddle,K.J., Azevedo,K., Miller,A., Adams,G., Pearlman,L., Gladden-Young,A., Lagerborg,K., Rudy,M., DeRuff,K., Carter,A., Normandin,E., Bauer,M., Reilly,S., Tomkins-Tinch,C., Loreth,C., Chaluvadi,S., Lemieux,J.E., Birren,B.W., Sabeti,P.C., Huard,R., King,E., Park,D.J., and MacInnis,B.L. |
| EPI_ISL_2133848, EPI_ISL_2133850 | Quest Diagnostics Incorporated | Centers for Disease Control and Prevention Division of Viral Diseases, Pathogen Discovery | Dakota Howard, Dhwani Batra, Peter W. Cook, Kara Moser, Adrian Paskey, Jason Caravas, Benjamin Rambo-Martin, Shatavia Morrison, Christopher Gulvick, Scott Sammons, Yvette Unoarumhi, Darlene Wagner, Matthew Schmerer, S. H. Rosenthal, A. Gerasimova, R. M. Kagan, B. Anderson, M. Hua, Y. Liu, L.E. Bernstein, K.E. Livingston, A. Perez, I. A. Shlyakhter, R. V. Rolando, R. Owen, P. Tanpaiboon, F. Lacbawan, Clinton R. Paden, Duncan MacCannell |
| EPI_ISL_2133851, EPI_ISL_2133852 | Rhode Island Department of Health | Infectious Disease Program, Broad Institute of Harvard and MIT | Siddle,K.J., Azevedo,K., Miller,A., Adams,G., Pearlman,L., Gladden-Young,A., Lagerborg,K., Rudy,M., DeRuff,K., Carter,A., Normandin,E., Bauer,M., Reilly,S., Tomkins-Tinch,C., Loreth,C., Chaluvadi,S., Lemieux,J.E., Birren,B.W., Sabeti,P.C., Huard,R., King,E., Park,D.J., and MacInnis,B.L. |
| EPI_ISL_2133853 | Quest Diagnostics Incorporated | Centers for Disease Control and Prevention Division of Viral Diseases, Pathogen Discovery | Dakota Howard, Dhwani Batra, Peter W. Cook, Kara Moser, Adrian Paskey, Jason Caravas, Benjamin Rambo-Martin, Shatavia Morrison, Christopher Gulvick, Scott Sammons, Yvette Unoarumhi, Darlene Wagner, Matthew Schmerer, S. H. Rosenthal, A. Gerasimova, R. M. Kagan, B. Anderson, M. Hua, Y. Liu, L.E. Bernstein, K.E. Livingston, A. Perez, I. A. Shlyakhter, R. V. Rolando, R. Owen, P. Tanpaiboon, F. Lacbawan, Clinton R. Paden, Duncan MacCannell |
| EPI_ISL_2133854 | Rhode Island Department of Health | Infectious Disease Program, Broad Institute of Harvard and MIT | Siddle,K.J., Azevedo,K., Miller,A., Adams,G., Pearlman,L., Gladden-Young,A., Lagerborg,K., Rudy,M., DeRuff,K., Carter,A., Normandin,E., Bauer,M., Reilly,S., Tomkins-Tinch,C., Loreth,C., Chaluvadi,S., Lemieux,J.E., Birren,B.W., Sabeti,P.C., Huard,R., King,E., Park,D.J., and MacInnis,B.L. |
| EPI_ISL_2133855 | Quest Diagnostics Incorporated | Centers for Disease Control and Prevention Division of Viral Diseases, Pathogen Discovery | Dakota Howard, Dhwani Batra, Peter W. Cook, Kara Moser, Adrian Paskey, Jason Caravas, Benjamin Rambo-Martin, Shatavia Morrison, Christopher Gulvick, Scott Sammons, Yvette Unoarumhi, Darlene Wagner, Matthew Schmerer, S. H. Rosenthal, A. Gerasimova, R. M. Kagan, B. Anderson, M. Hua, Y. Liu, L.E. Bernstein, K.E. Livingston, A. Perez, I. A. Shlyakhter, R. V. Rolando, R. Owen, P. Tanpaiboon, F. Lacbawan, Clinton R. Paden, Duncan MacCannell |
| EPI_ISL_2133856, EPI_ISL_2133857 | Rhode Island Department of Health | Infectious Disease Program, Broad Institute of Harvard and MIT | Siddle,K.J., Azevedo,K., Miller,A., Adams,G., Pearlman,L., Gladden-Young,A., Lagerborg,K., Rudy,M., DeRuff,K., Carter,A., Normandin,E., Bauer,M., Reilly,S., Tomkins-Tinch,C., Loreth,C., Chaluvadi,S., Lemieux,J.E., Birren,B.W., Sabeti,P.C., Huard,R., King,E., Park,D.J., and MacInnis,B.L. |
| EPI_ISL_2133858 | Quest Diagnostics Incorporated | Centers for Disease Control and Prevention Division of Viral Diseases, Pathogen Discovery | Dakota Howard, Dhwani Batra, Peter W. Cook, Kara Moser, Adrian Paskey, Jason Caravas, Benjamin Rambo-Martin, Shatavia Morrison, Christopher Gulvick, Scott Sammons, Yvette Unoarumhi, Darlene Wagner, Matthew Schmerer, S. H. Rosenthal, A. Gerasimova, R. M. Kagan, B. Anderson, M. Hua, Y. Liu, L.E. Bernstein, K.E. Livingston, A. Perez, I. A. Shlyakhter, R. V. Rolando, R. Owen, P. Tanpaiboon, F. Lacbawan, Clinton R. Paden, Duncan MacCannell |
| EPI_ISL_2133859, EPI_ISL_2133860 | Rhode Island Department of Health | Infectious Disease Program, Broad Institute of Harvard and MIT | Siddle,K.J., Azevedo,K., Miller,A., Adams,G., Pearlman,L., Gladden-Young,A., Lagerborg,K., Rudy,M., DeRuff,K., Carter,A., Normandin,E., Bauer,M., Reilly,S., Tomkins-Tinch,C., Loreth,C., Chaluvadi,S., Lemieux,J.E., Birren,B.W., Sabeti,P.C., Huard,R., King,E., Park,D.J., and MacInnis,B.L. |
| EPI_ISL_2133861 | Quest Diagnostics Incorporated | Centers for Disease Control and Prevention Division of Viral Diseases, Pathogen Discovery | Dakota Howard, Dhwani Batra, Peter W. Cook, Kara Moser, Adrian Paskey, Jason Caravas, Benjamin Rambo-Martin, Shatavia Morrison, Christopher Gulvick, Scott Sammons, Yvette Unoarumhi, Darlene Wagner, Matthew Schmerer, S. H. Rosenthal, A. Gerasimova, R. M. Kagan, B. Anderson, M. Hua, Y. Liu, L.E. Bernstein, K.E. Livingston, A. Perez, I. A. Shlyakhter, R. V. Rolando, R. Owen, P. Tanpaiboon, F. Lacbawan, Clinton R. Paden, Duncan MacCannell |
| EPI_ISL_2133862 | Rhode Island Department of Health | Infectious Disease Program, Broad Institute of Harvard and MIT | Siddle,K.J., Azevedo,K., Miller,A., Adams,G., Pearlman,L., Gladden-Young,A., Lagerborg,K., Rudy,M., DeRuff,K., Carter,A., Normandin,E., Bauer,M., Reilly,S., Tomkins-Tinch,C., Loreth,C., Chaluvadi,S., Lemieux,J.E., Birren,B.W., Sabeti,P.C., Huard,R., King,E., Park,D.J., and MacInnis,B.L. |
| EPI_ISL_2133863 | Quest Diagnostics Incorporated | Centers for Disease Control and Prevention Division of Viral Diseases, Pathogen Discovery | Dakota Howard, Dhwani Batra, Peter W. Cook, Kara Moser, Adrian Paskey, Jason Caravas, Benjamin Rambo-Martin, Shatavia Morrison, Christopher Gulvick, Scott Sammons, Yvette Unoarumhi, Darlene Wagner, Matthew Schmerer, S. H. Rosenthal, A. Gerasimova, R. M. Kagan, B. Anderson, M. Hua, Y. Liu, L.E. Bernstein, K.E. Livingston, A. Perez, I. A. Shlyakhter, R. V. Rolando, R. Owen, P. Tanpaiboon, F. Lacbawan, Clinton R. Paden, Duncan MacCannell |
| EPI_ISL_2133864 | Rhode Island Department of Health | Infectious Disease Program, Broad Institute of Harvard and MIT | Siddle,K.J., Azevedo,K., Miller,A., Adams,G., Pearlman,L., Gladden-Young,A., Lagerborg,K., Rudy,M., DeRuff,K., Carter,A., Normandin,E., Bauer,M., Reilly,S., Tomkins-Tinch,C., Loreth,C., Chaluvadi,S., Lemieux,J.E., Birren,B.W., Sabeti,P.C., Huard,R., King,E., Park,D.J., and MacInnis,B.L. |
| EPI_ISL_2133865 | Quest Diagnostics Incorporated | Centers for Disease Control and Prevention Division of Viral Diseases, Pathogen Discovery | Dakota Howard, Dhwani Batra, Peter W. Cook, Kara Moser, Adrian Paskey, Jason Caravas, Benjamin Rambo-Martin, Shatavia Morrison, Christopher Gulvick, Scott Sammons, Yvette Unoarumhi, Darlene Wagner, Matthew Schmerer, S. H. Rosenthal, A. Gerasimova, R. M. Kagan, B. Anderson, M. Hua, Y. Liu, L.E. Bernstein, K.E. Livingston, A. Perez, I. A. Shlyakhter, R. V. Rolando, R. Owen, P. Tanpaiboon, F. Lacbawan, Clinton R. Paden, Duncan MacCannell |
| EPI_ISL_2133866, EPI_ISL_2133867 | Rhode Island Department of Health | Infectious Disease Program, Broad Institute of Harvard and MIT | Siddle,K.J., Azevedo,K., Miller,A., Adams,G., Pearlman,L., Gladden-Young,A., Lagerborg,K., Rudy,M., DeRuff,K., Carter,A., Normandin,E., Bauer,M., Reilly,S., Tomkins-Tinch,C., Loreth,C., Chaluvadi,S., Lemieux,J.E., Birren,B.W., Sabeti,P.C., Huard,R., King,E., Park,D.J., and MacInnis,B.L. |
| EPI_ISL_2133868 | Quest Diagnostics Incorporated | Centers for Disease Control and Prevention Division of Viral Diseases, Pathogen Discovery | Dakota Howard, Dhwani Batra, Peter W. Cook, Kara Moser, Adrian Paskey, Jason Caravas, Benjamin Rambo-Martin, Shatavia Morrison, Christopher Gulvick, Scott Sammons, Yvette Unoarumhi, Darlene Wagner, Matthew Schmerer, S. H. Rosenthal, A. Gerasimova, R. M. Kagan, B. Anderson, M. Hua, Y. Liu, L.E. Bernstein, K.E. Livingston, A. Perez, I. A. Shlyakhter, R. V. Rolando, R. Owen, P. Tanpaiboon, F. Lacbawan, Clinton R. Paden, Duncan MacCannell |
| EPI_ISL_2133869, EPI_ISL_2133870 | Rhode Island Department of Health | Infectious Disease Program, Broad Institute of Harvard and MIT | Siddle,K.J., Azevedo,K., Miller,A., Adams,G., Pearlman,L., Gladden-Young,A., Lagerborg,K., Rudy,M., DeRuff,K., Carter,A., Normandin,E., Bauer,M., Reilly,S., Tomkins-Tinch,C., Loreth,C., Chaluvadi,S., Lemieux,J.E., Birren,B.W., Sabeti,P.C., Huard,R., King,E., Park,D.J., and MacInnis,B.L. |
| EPI_ISL_2133871 | Quest Diagnostics Incorporated | Centers for Disease Control and Prevention Division of Viral Diseases, Pathogen Discovery | Dakota Howard, Dhwani Batra, Peter W. Cook, Kara Moser, Adrian Paskey, Jason Caravas, Benjamin Rambo-Martin, Shatavia Morrison, Christopher Gulvick, Scott Sammons, Yvette Unoarumhi, Darlene Wagner, Matthew Schmerer, S. H. Rosenthal, A. Gerasimova, R. M. Kagan, B. Anderson, M. Hua, Y. Liu, L.E. Bernstein, K.E. Livingston, A. Perez, I. A. Shlyakhter, R. V. Rolando, R. Owen, P. Tanpaiboon, F. Lacbawan, Clinton R. Paden, Duncan MacCannell |
| EPI_ISL_2133872, EPI_ISL_2133873, EPI_ISL_2133874, EPI_ISL_2133875, EPI_ISL_2133876, EPI_ISL_2133877, EPI_ISL_2133878, EPI_ISL_2133879, EPI_ISL_2133880, EPI_ISL_2133881, EPI_ISL_2133882, EPI_ISL_2133883, EPI_ISL_2133884, EPI_ISL_2133885, EPI_ISL_2133886, EPI_ISL_2133887, EPI_ISL_2133888, EPI_ISL_2133889, EPI_ISL_2133890, EPI_ISL_2133892, EPI_ISL_2133894, EPI_ISL_2133895, EPI_ISL_2133896, EPI_ISL_2133897, EPI_ISL_2133898, EPI_ISL_2133900, EPI_ISL_2133901, EPI_ISL_2133903, EPI_ISL_2133904, EPI_ISL_2133905, EPI_ISL_2133906, EPI_ISL_2133910, EPI_ISL_2133911, EPI_ISL_2133912, EPI_ISL_2133913, EPI_ISL_2133915, EPI_ISL_2133916, EPI_ISL_2133917, EPI_ISL_2133918, EPI_ISL_2133919, EPI_ISL_2133920, EPI_ISL_2133921, EPI_ISL_2133922, EPI_ISL_2133923, EPI_ISL_2133924, EPI_ISL_2133925, EPI_ISL_2133926 | | | |
| see above | Rhode Island Department of Health | Infectious Disease Program, Broad Institute of Harvard and MIT | Siddle,K.J., Azevedo,K., Miller,A., Adams,G., Pearlman,L., Gladden-Young,A., Lagerborg,K., Rudy,M., DeRuff,K., Carter,A., Normandin,E., Bauer,M., Reilly,S., Tomkins-Tinch,C., Loreth,C., Chaluvadi,S., Lemieux,J.E., Birren,B.W., Sabeti,P.C., Huard,R., King,E., Park,D.J., and MacInnis,B.L. |
| EPI_ISL_2133927 | Warrior Diagnostics Laboratory | Infectious Disease Program, Broad Institute of Harvard and MIT | Lemieux,J.E., Siddle,K.J., Marshall,J., ONeill,M., Bronson,A., Adams,G., Gladden-Young,A., Lagerborg,K., Rudy,M., DeRuff,K., Carter,A., Normandin,E., Bauer,M., Reilly,S., Tomkins-Tinch,C., Loreth,C., Chaluvadi,S., Birren,B.W., Gallagher,G., Smole,S., Park,D.J., MacInnis,B.L., and Sabeti,P.C. |
| EPI_ISL_2133929, EPI_ISL_2133930, EPI_ISL_2133932, EPI_ISL_2133933, EPI_ISL_2133934, EPI_ISL_2133935, EPI_ISL_2133936, EPI_ISL_2133937, EPI_ISL_2133938, EPI_ISL_2133939, EPI_ISL_2133940, EPI_ISL_2133941, EPI_ISL_2133942, EPI_ISL_2133943, EPI_ISL_2133944, EPI_ISL_2133945, EPI_ISL_2133946, EPI_ISL_2133948, EPI_ISL_2133949, EPI_ISL_2133951, EPI_ISL_2133952, EPI_ISL_2133953, EPI_ISL_2133954, EPI_ISL_2133955, EPI_ISL_2133956, EPI_ISL_2133957, EPI_ISL_2133958, EPI_ISL_2133959, EPI_ISL_2133960, EPI_ISL_2133961, EPI_ISL_2133962, EPI_ISL_2133963, EPI_ISL_2133964, EPI_ISL_2133966, EPI_ISL_2133967, EPI_ISL_2133968, EPI_ISL_2133969, EPI_ISL_2133970, EPI_ISL_2133972, EPI_ISL_2133973, EPI_ISL_2133974, EPI_ISL_2133975, EPI_ISL_2133976, EPI_ISL_2133977, EPI_ISL_2133978, EPI_ISL_2133979, EPI_ISL_2133980, EPI_ISL_2133981, EPI_ISL_2133982, EPI_ISL_2133983, EPI_ISL_2133984, EPI_ISL_2133985, EPI_ISL_2133986, EPI_ISL_2133987, EPI_ISL_2133988, EPI_ISL_2133989, EPI_ISL_2133990, EPI_ISL_2133992, EPI_ISL_2133993, EPI_ISL_2133994, EPI_ISL_2133995, EPI_ISL_2133996, EPI_ISL_2133997, EPI_ISL_2133998, EPI_ISL_2133999, EPI_ISL_2134000, EPI_ISL_2134001, EPI_ISL_2134002, EPI_ISL_2134003, EPI_ISL_2134004, EPI_ISL_2134005, EPI_ISL_2134006, EPI_ISL_2134007, EPI_ISL_2134008, EPI_ISL_2134009, EPI_ISL_2134010, EPI_ISL_2134011, EPI_ISL_2134012, EPI_ISL_2134013, EPI_ISL_2134014, EPI_ISL_2134015, EPI_ISL_2134016, EPI_ISL_2134017, EPI_ISL_2134018, EPI_ISL_2134019, EPI_ISL_2134020, EPI_ISL_2134021, EPI_ISL_2134023, EPI_ISL_2134024, EPI_ISL_2134025, EPI_ISL_2134026, EPI_ISL_2134027, EPI_ISL_2134029, EPI_ISL_2134030, EPI_ISL_2134031, EPI_ISL_2134032, EPI_ISL_2134033, EPI_ISL_2134034, EPI_ISL_2134035, EPI_ISL_2134036, EPI_ISL_2134037, EPI_ISL_2134038, EPI_ISL_2134039, EPI_ISL_2134040, EPI_ISL_2134041, EPI_ISL_2134042, EPI_ISL_2134043, EPI_ISL_2134044, EPI_ISL_2134047, EPI_ISL_2134048, EPI_ISL_2134049, EPI_ISL_2134050, EPI_ISL_2134051, EPI_ISL_2134052, EPI_ISL_2134053, EPI_ISL_2134055, EPI_ISL_2134056, EPI_ISL_2134057, EPI_ISL_2134058, EPI_ISL_2134059, EPI_ISL_2134060, EPI_ISL_2134061, EPI_ISL_2134062, EPI_ISL_2134063, EPI_ISL_2134064, EPI_ISL_2134065, EPI_ISL_2134066, EPI_ISL_2134067, EPI_ISL_2134068, EPI_ISL_2134069, EPI_ISL_2134070, EPI_ISL_2134071, EPI_ISL_2134073, EPI_ISL_2134074, EPI_ISL_2134075, EPI_ISL_2134076, EPI_ISL_2134077, EPI_ISL_2134078, EPI_ISL_2134079, EPI_ISL_2134080, EPI_ISL_2134082, EPI_ISL_2134083, EPI_ISL_2134084, EPI_ISL_2134085, EPI_ISL_2134086, EPI_ISL_2134087, EPI_ISL_2134089, EPI_ISL_2134090, EPI_ISL_2134091, EPI_ISL_2134092, EPI_ISL_2134093, EPI_ISL_2134095, EPI_ISL_2134096, EPI_ISL_2134097, EPI_ISL_2134098, EPI_ISL_2134099, EPI_ISL_2134100, EPI_ISL_2134101, EPI_ISL_2134102, EPI_ISL_2134103, EPI_ISL_2134104, EPI_ISL_2134105, EPI_ISL_2134106, EPI_ISL_2134107, EPI_ISL_2134108, EPI_ISL_2134109, EPI_ISL_2134110, EPI_ISL_2134111, EPI_ISL_2134112, EPI_ISL_2134113, EPI_ISL_2134114, EPI_ISL_2134115, EPI_ISL_2134117, EPI_ISL_2134118, EPI_ISL_2134119, EPI_ISL_2134120, EPI_ISL_2134121, EPI_ISL_2134122, EPI_ISL_2134123, EPI_ISL_2134124, EPI_ISL_2134125, EPI_ISL_2134126, EPI_ISL_2134127, EPI_ISL_2134128, EPI_ISL_2134129, EPI_ISL_2134130, EPI_ISL_2134131, EPI_ISL_2134132, EPI_ISL_2134133, EPI_ISL_2134134, EPI_ISL_2134135, EPI_ISL_2134137, EPI_ISL_2134138, EPI_ISL_2134139, EPI_ISL_2134140, EPI_ISL_2134142, EPI_ISL_2134143, EPI_ISL_2134144, EPI_ISL_2134145, EPI_ISL_2134146, EPI_ISL_2134147, EPI_ISL_2134148, EPI_ISL_2134149, EPI_ISL_2134150, EPI_ISL_2134151, EPI_ISL_2134152, EPI_ISL_2134153, EPI_ISL_2134154, EPI_ISL_2134155, EPI_ISL_2134156, EPI_ISL_2134157, EPI_ISL_2134158, EPI_ISL_2134159, EPI_ISL_2134160, EPI_ISL_2134161, EPI_ISL_2134162, EPI_ISL_2134163, EPI_ISL_2134164, EPI_ISL_2134165, EPI_ISL_2134166, EPI_ISL_2134167, EPI_ISL_2134168, EPI_ISL_2134169, EPI_ISL_2134170, EPI_ISL_2134171, EPI_ISL_2134172, EPI_ISL_2134173, EPI_ISL_2134174, EPI_ISL_2134175, EPI_ISL_2134176, EPI_ISL_2134177, EPI_ISL_2134178, EPI_ISL_2134179, EPI_ISL_2134180, EPI_ISL_2134181, EPI_ISL_2134182, EPI_ISL_2134183, EPI_ISL_2134184, EPI_ISL_2134186, EPI_ISL_2134187, EPI_ISL_2134188, EPI_ISL_2134189, EPI_ISL_2134190, EPI_ISL_2134191, EPI_ISL_2134192, EPI_ISL_2134193, EPI_ISL_2134194, EPI_ISL_2134195, EPI_ISL_2134196, EPI_ISL_2134197, EPI_ISL_2134198, EPI_ISL_2134199, EPI_ISL_2134200, EPI_ISL_2134201, EPI_ISL_2134202, EPI_ISL_2134203, EPI_ISL_2134204, EPI_ISL_2134205, EPI_ISL_2134206, EPI_ISL_2134207, EPI_ISL_2134208, EPI_ISL_2134209, EPI_ISL_2134210, EPI_ISL_2134211, EPI_ISL_2134212, EPI_ISL_2134213, EPI_ISL_2134214, EPI_ISL_2134215, EPI_ISL_2134216, EPI_ISL_2134218, EPI_ISL_2134219, EPI_ISL_2134220, EPI_ISL_2134221, EPI_ISL_2134222, EPI_ISL_2134223, EPI_ISL_2134224, EPI_ISL_2134225, EPI_ISL_2134226, EPI_ISL_2134227, EPI_ISL_2134228, EPI_ISL_2134229, EPI_ISL_2134232, EPI_ISL_2134233, EPI_ISL_2134234, EPI_ISL_2134235, EPI_ISL_2134236, EPI_ISL_2134237, EPI_ISL_2134238, EPI_ISL_2134240, EPI_ISL_2134241, EPI_ISL_2134242, EPI_ISL_2134243, EPI_ISL_2134244, EPI_ISL_2134245, EPI_ISL_2134246, EPI_ISL_2134248, EPI_ISL_2134249, EPI_ISL_2134250, EPI_ISL_2134251, EPI_ISL_2134253, EPI_ISL_2134254, EPI_ISL_2134255, EPI_ISL_2134256, EPI_ISL_2134257, EPI_ISL_2134258, EPI_ISL_2134259, EPI_ISL_2134260, EPI_ISL_2134261, EPI_ISL_2134262, EPI_ISL_2134263, EPI_ISL_2134264, EPI_ISL_2134265, EPI_ISL_2134267, EPI_ISL_2134268, EPI_ISL_2134269, EPI_ISL_2134270, EPI_ISL_2134271, EPI_ISL_2134273, EPI_ISL_2134274, EPI_ISL_2134275, EPI_ISL_2134276, EPI_ISL_2134278, EPI_ISL_2134279, EPI_ISL_2134280, EPI_ISL_2134281, EPI_ISL_2134283, EPI_ISL_2134284, EPI_ISL_2134285, EPI_ISL_2134288, EPI_ISL_2134289, EPI_ISL_2134290, EPI_ISL_2134291, EPI_ISL_2134292, EPI_ISL_2134294, EPI_ISL_2134295, EPI_ISL_2134296, EPI_ISL_2134297, EPI_ISL_2134298, EPI_ISL_2134299, EPI_ISL_2134300, EPI_ISL_2134301, EPI_ISL_2134302, EPI_ISL_2134303, EPI_ISL_2134304, EPI_ISL_2134306, EPI_ISL_2134307, EPI_ISL_2134309, EPI_ISL_2134310, EPI_ISL_2134311, EPI_ISL_2134312, EPI_ISL_2134313, EPI_ISL_2134314, EPI_ISL_2134315, EPI_ISL_2134316, EPI_ISL_2134318, EPI_ISL_2134319, EPI_ISL_2134320, EPI_ISL_2134321, EPI_ISL_2134322, EPI_ISL_2134323, EPI_ISL_2134324, EPI_ISL_2134325, EPI_ISL_2134326, EPI_ISL_2134327, EPI_ISL_2134328, EPI_ISL_2134329, EPI_ISL_2134331, EPI_ISL_2134332, EPI_ISL_2134333, EPI_ISL_2134334, EPI_ISL_2134335, EPI_ISL_2134337, EPI_ISL_2134338, EPI_ISL_2134339, EPI_ISL_2134340, EPI_ISL_2134341, EPI_ISL_2134342, EPI_ISL_2134343, EPI_ISL_2134344, EPI_ISL_2134345, EPI_ISL_2134346, EPI_ISL_2134347, EPI_ISL_2134348, EPI_ISL_2134349, EPI_ISL_2134350, EPI_ISL_2134351, EPI_ISL_2134352, EPI_ISL_2134353, EPI_ISL_2134354, EPI_ISL_2134355, EPI_ISL_2134356, EPI_ISL_2134357, EPI_ISL_2134358, EPI_ISL_2134359, EPI_ISL_2134360, EPI_ISL_2134361, EPI_ISL_2134362, EPI_ISL_2134364, EPI_ISL_2134365, EPI_ISL_2134366, EPI_ISL_2134367, EPI_ISL_2134368, EPI_ISL_2134369, EPI_ISL_2134370, EPI_ISL_2134371, EPI_ISL_2134372, EPI_ISL_2134373, EPI_ISL_2134374, EPI_ISL_2134375, EPI_ISL_2134376, EPI_ISL_2134377, EPI_ISL_2134378, EPI_ISL_2134379, EPI_ISL_2134380, EPI_ISL_2134381, EPI_ISL_2134382, EPI_ISL_2134383, EPI_ISL_2134384, EPI_ISL_2134385, EPI_ISL_2134386, EPI_ISL_2134387, EPI_ISL_2134388, EPI_ISL_2134389, EPI_ISL_2134390, EPI_ISL_2134391, EPI_ISL_2134392, EPI_ISL_2134393, EPI_ISL_2134394, EPI_ISL_2134396, EPI_ISL_2134397, EPI_ISL_2134400, EPI_ISL_2134401, EPI_ISL_2134402, EPI_ISL_2134403, EPI_ISL_2134404, EPI_ISL_2134405, EPI_ISL_2134406, EPI_ISL_2134407, EPI_ISL_2134408, EPI_ISL_2134409, EPI_ISL_2134410, EPI_ISL_2134411, EPI_ISL_2134412, EPI_ISL_2134413, EPI_ISL_2134414, EPI_ISL_2134415, EPI_ISL_2134416, EPI_ISL_2134417, EPI_ISL_2134418, EPI_ISL_2134419, EPI_ISL_2134420, EPI_ISL_2134421, EPI_ISL_2134422, EPI_ISL_2134423, EPI_ISL_2134424, EPI_ISL_2134425, EPI_ISL_2134426, EPI_ISL_2134427, EPI_ISL_2134428, EPI_ISL_2134429, EPI_ISL_2134430, EPI_ISL_2134431, EPI_ISL_2134432, EPI_ISL_2134433, EPI_ISL_2134434, EPI_ISL_2134435, EPI_ISL_2134436, EPI_ISL_2134438, EPI_ISL_2134439, EPI_ISL_2134440, EPI_ISL_2134441, EPI_ISL_2134442, EPI_ISL_2134443, EPI_ISL_2134444, EPI_ISL_2134445, EPI_ISL_2134446, EPI_ISL_2134447, EPI_ISL_2134448, EPI_ISL_2134449, EPI_ISL_2134451, EPI_ISL_2134452, EPI_ISL_2134453, EPI_ISL_2134454, EPI_ISL_2134455, EPI_ISL_2134456, EPI_ISL_2134457, EPI_ISL_2134458, EPI_ISL_2134459, EPI_ISL_2134460, EPI_ISL_2134461, EPI_ISL_2134462, EPI_ISL_2134463, EPI_ISL_2134464, EPI_ISL_2134465, EPI_ISL_2134466, EPI_ISL_2134467, EPI_ISL_2134468, EPI_ISL_2134469, EPI_ISL_2134470, EPI_ISL_2134471, EPI_ISL_2134472, EPI_ISL_2134473, EPI_ISL_2134474, EPI_ISL_2134475, EPI_ISL_2134476, EPI_ISL_2134477, | | | |

| | | | |
|---|---|---|---|
| EPI_ISL_2134478, EPI_ISL_2134479, EPI_ISL_2134480, EPI_ISL_2134481, EPI_ISL_2134482, EPI_ISL_2134483, EPI_ISL_2134484, EPI_ISL_2134487, EPI_ISL_2134488, EPI_ISL_2134489, EPI_ISL_2134490, EPI_ISL_2134491, EPI_ISL_2134492, EPI_ISL_2134493, EPI_ISL_2134494, EPI_ISL_2134495, EPI_ISL_2134496, EPI_ISL_2134497, EPI_ISL_2134498, EPI_ISL_2134499, EPI_ISL_2134500, EPI_ISL_2134501, EPI_ISL_2134502, EPI_ISL_2134503, EPI_ISL_2134504, EPI_ISL_2134505, EPI_ISL_2134506, EPI_ISL_2134507, EPI_ISL_2134508, EPI_ISL_2134510, EPI_ISL_2134512, EPI_ISL_2134513, EPI_ISL_2134514, EPI_ISL_2134517, EPI_ISL_2134519, EPI_ISL_2134520, EPI_ISL_2134522, EPI_ISL_2134523, EPI_ISL_2134525, EPI_ISL_2134526, EPI_ISL_2134527, EPI_ISL_2134528, EPI_ISL_2134529, EPI_ISL_2134530, EPI_ISL_2134531, EPI_ISL_2134532, EPI_ISL_2134533, EPI_ISL_2134535, EPI_ISL_2134536, EPI_ISL_2134537, EPI_ISL_2134538, EPI_ISL_2134539, EPI_ISL_2134540, EPI_ISL_2134541, EPI_ISL_2134542, EPI_ISL_2134543, EPI_ISL_2134544, EPI_ISL_2134545, EPI_ISL_2134546, EPI_ISL_2134547, EPI_ISL_2134548, EPI_ISL_2134549, EPI_ISL_2134550, EPI_ISL_2134551, EPI_ISL_2134552, EPI_ISL_2134554, EPI_ISL_2134555, EPI_ISL_2134556, EPI_ISL_2134557, EPI_ISL_2134558, EPI_ISL_2134559, EPI_ISL_2134560, EPI_ISL_2134561, EPI_ISL_2134562, EPI_ISL_2134563, EPI_ISL_2134564, EPI_ISL_2134565, EPI_ISL_2134567, EPI_ISL_2134568, EPI_ISL_2134569, EPI_ISL_2134571, EPI_ISL_2134572, EPI_ISL_2134573, EPI_ISL_2134574, EPI_ISL_2134575, EPI_ISL_2134576, EPI_ISL_2134577 | | | |
| see above | Fulgent Genetics | Centers for Disease Control and Prevention Division of Viral Diseases, Pathogen Discovery | Dakota Howard, Dhwani Batra, Peter W. Cook, Kara Moser, Adrian Paskey, Jason Caravas, Benjamin Rambo-Martin, Shatavia Morrison, Christopher Gulvick, Scott Sammons, Yvette Unoarumhi, Darlene Wagner, Matthew Schmerer, Harry Gao, Mickey Li, John Gao, Joseph Fierro, Benafsh Sapra, Becky Tsai, Yan Meng, Doreen Ng, James Xie, Clinton R. Paden, Duncan MacCannell |
| EPI_ISL_2134585, EPI_ISL_2134586, EPI_ISL_2134587, EPI_ISL_2134588, EPI_ISL_2134590, EPI_ISL_2134591, EPI_ISL_2134592, EPI_ISL_2134593, EPI_ISL_2134595, EPI_ISL_2134596, EPI_ISL_2134597, EPI_ISL_2134598, EPI_ISL_2134600, EPI_ISL_2134601, EPI_ISL_2134602, EPI_ISL_2134603, EPI_ISL_2134604, EPI_ISL_2134605, EPI_ISL_2134606, EPI_ISL_2134607, EPI_ISL_2134608, EPI_ISL_2134609, EPI_ISL_2134610, EPI_ISL_2134611, EPI_ISL_2134612, EPI_ISL_2134614, EPI_ISL_2134615, EPI_ISL_2134617, EPI_ISL_2134618, EPI_ISL_2134619, EPI_ISL_2134620, EPI_ISL_2134621, EPI_ISL_2134622, EPI_ISL_2134623, EPI_ISL_2134625, EPI_ISL_2134627, EPI_ISL_2134628 | | | |
| see above | IN State Department of Health Laboratory Services | IN State Department of Health Laboratory Services | Cassandra Campion, Jamie Yeadon, Brian Pope, Lixia Liu, Kyle Brownlee, Melissa Hindenlang, Mark Glazier |
| EPI_ISL_2134631 | Wyoming Public Health Laboratory | Wyoming Public Health Laboratory | Jim Mildenberger, Wanda Manley, Noah Hull, Taylor Fearing, Lynette Gumbleton, Channing Weber, Ashley Norberg, Chayse Rowley, Marley Goetz, Brian Dominguez, Elliot Thomasson, Sam Britz, Cari Sloma, and Rob Christensen |
| EPI_ISL_2134784 | BOSTON HEART DIAGNOSTICS CORP | Wadsworth Center, New York State Department of Health | Kirsten St. George, Daryl M. Lamson, Alexis Russell, Matthew Shudt, Melissa A Leisner, Jonathan Plitnick, Catharine Prussing, Navjot Singh, John Kelly, Erasmus Schneider, Erica Lasek-Nesselquist |
| EPI_ISL_2134785 | MONTEFIORE MEDICAL CENTER LABORATORIES | Wadsworth Center, New York State Department of Health | Kirsten St. George, Daryl M. Lamson, Alexis Russell, Matthew Shudt, Melissa A Leisner, Jonathan Plitnick, Catharine Prussing, Navjot Singh, John Kelly, Erasmus Schneider, Erica Lasek-Nesselquist |
| EPI_ISL_2134786, EPI_ISL_2134787 | NORTHWELL HEALTH LABORATORIES | Wadsworth Center, New York State Department of Health | Kirsten St. George, Daryl M. Lamson, Alexis Russell, Matthew Shudt, Melissa A Leisner, Jonathan Plitnick, Catharine Prussing, Navjot Singh, John Kelly, Erasmus Schneider, Erica Lasek-Nesselquist |
| EPI_ISL_2134788, EPI_ISL_2134789 | STONY BROOK UNIVERSITY HOSPITAL | Wadsworth Center, New York State Department of Health | Kirsten St. George, Daryl M. Lamson, Alexis Russell, Matthew Shudt, Melissa A Leisner, Jonathan Plitnick, Catharine Prussing, Navjot Singh, John Kelly, Erasmus Schneider, Erica Lasek-Nesselquist |
| EPI_ISL_2134790 | BOSTON HEART DIAGNOSTICS CORP | Wadsworth Center, New York State Department of Health | Kirsten St. George, Daryl M. Lamson, Alexis Russell, Matthew Shudt, Melissa A Leisner, Jonathan Plitnick, Catharine Prussing, Navjot Singh, John Kelly, Erasmus Schneider, Erica Lasek-Nesselquist |
| EPI_ISL_2134791 | MONTEFIORE MEDICAL CENTER LABORATORIES | Wadsworth Center, New York State Department of Health | Kirsten St. George, Daryl M. Lamson, Alexis Russell, Matthew Shudt, Melissa A Leisner, Jonathan Plitnick, Catharine Prussing, Navjot Singh, John Kelly, Erasmus Schneider, Erica Lasek-Nesselquist |
| EPI_ISL_2134792, EPI_ISL_2134793, EPI_ISL_2134794, EPI_ISL_2134795 | URMC LABS | Wadsworth Center, New York State Department of Health | Kirsten St. George, Daryl M. Lamson, Alexis Russell, Matthew Shudt, Melissa A Leisner, Jonathan Plitnick, Catharine Prussing, Navjot Singh, John Kelly, Erasmus Schneider, Erica Lasek-Nesselquist |
| EPI_ISL_2134796, EPI_ISL_2134797 | BOSTON HEART DIAGNOSTICS CORP | Wadsworth Center, New York State Department of Health | Kirsten St. George, Daryl M. Lamson, Alexis Russell, Matthew Shudt, Melissa A Leisner, Jonathan Plitnick, Catharine Prussing, Navjot Singh, John Kelly, Erasmus Schneider, Erica Lasek-Nesselquist |
| EPI_ISL_2134798 | SUNY UPSTATE MEDICAL UNIVERSITY | Wadsworth Center, New York State Department of Health | Kirsten St. George, Daryl M. Lamson, Alexis Russell, Matthew Shudt, Melissa A Leisner, Jonathan Plitnick, Catharine Prussing, Navjot Singh, John Kelly, Erasmus Schneider, Erica Lasek-Nesselquist |
| EPI_ISL_2134799, EPI_ISL_2134800, EPI_ISL_2134801, EPI_ISL_2134802, EPI_ISL_2134803, EPI_ISL_2134804, EPI_ISL_2134805, EPI_ISL_2134806, EPI_ISL_2134807, EPI_ISL_2134808, EPI_ISL_2134809, EPI_ISL_2134810, EPI_ISL_2134811, EPI_ISL_2134812, EPI_ISL_2134813, EPI_ISL_2134814, EPI_ISL_2134815, EPI_ISL_2134816, EPI_ISL_2134818, EPI_ISL_2134819 | | | |
| see above | DC Public Health Lab/ Dept. of Forensic Sciences | DC Public Health Lab/ Dept. of Forensic Sciences | Janis Doss, Scott Nguyen, Elizabeth Zelaya, Sarah Scott, Connie Maza, Monica Mann, Brittany Hamilton, David Payne, Jocelyn Hauser |
| EPI_ISL_2134820 | PrimBio Research institute | PrimBio Research institute | Yang Liu |
| EPI_ISL_2134972, EPI_ISL_2134974, EPI_ISL_2134978, EPI_ISL_2134979 | University of Mississippi Medical Center, Department of Pathology | University of Mississippi Medical Center, Molecular and Genomics Core Facility | Ashley C. Johnson, Ithiel J. Frame, Krishna K. Ayyalasomayajula, Michael R. Garrett, D. Ashley Robinson |
| EPI_ISL_2134985, EPI_ISL_2134987, EPI_ISL_2134988, EPI_ISL_2134989, EPI_ISL_2134990, EPI_ISL_2134992, EPI_ISL_2134993, EPI_ISL_2134994, EPI_ISL_2134995, EPI_ISL_2134996, EPI_ISL_2134997, EPI_ISL_2134999, EPI_ISL_2135000, EPI_ISL_2135001, EPI_ISL_2135002, EPI_ISL_2135003, EPI_ISL_2135004, EPI_ISL_2135005, EPI_ISL_2135006, EPI_ISL_2135007, EPI_ISL_2135008, EPI_ISL_2135010, EPI_ISL_2135011, EPI_ISL_2135012, EPI_ISL_2135013, EPI_ISL_2135014, EPI_ISL_2135015, EPI_ISL_2135016, EPI_ISL_2135017, EPI_ISL_2135018, EPI_ISL_2135019, EPI_ISL_2135020, EPI_ISL_2135021, EPI_ISL_2135022, EPI_ISL_2135023, EPI_ISL_2135024, EPI_ISL_2135025, EPI_ISL_2135026, EPI_ISL_2135027, EPI_ISL_2135028, EPI_ISL_2135029, EPI_ISL_2135030, EPI_ISL_2135031, EPI_ISL_2135032, EPI_ISL_2135033, EPI_ISL_2135034, EPI_ISL_2135035, EPI_ISL_2135036, EPI_ISL_2135037 | | | |
| see above | Texas Children's Hospital | Texas Children's Microbiome Center | Ruth Ann Luna, Jennifer K. Spinler, James Dunn, James Versalovic, Ila Singh |
| EPI_ISL_2135038, EPI_ISL_2135039, EPI_ISL_2135040, EPI_ISL_2135041, EPI_ISL_2135042, EPI_ISL_2135043, EPI_ISL_2135044, EPI_ISL_2135045, EPI_ISL_2135046, EPI_ISL_2135047, EPI_ISL_2135048, EPI_ISL_2135049, EPI_ISL_2135050, EPI_ISL_2135051, EPI_ISL_2135052, EPI_ISL_2135053, EPI_ISL_2135054, EPI_ISL_2135055, EPI_ISL_2135056, EPI_ISL_2135057, EPI_ISL_2135058, EPI_ISL_2135059, EPI_ISL_2135060, EPI_ISL_2135061, EPI_ISL_2135062, EPI_ISL_2135064, EPI_ISL_2135065, EPI_ISL_2135066, EPI_ISL_2135067, EPI_ISL_2135068, EPI_ISL_2135069, EPI_ISL_2135070, EPI_ISL_2135071, EPI_ISL_2135072, EPI_ISL_2135073, EPI_ISL_2135074, EPI_ISL_2135075, EPI_ISL_2135076, EPI_ISL_2135077, EPI_ISL_2135078, EPI_ISL_2135079, EPI_ISL_2135080, EPI_ISL_2135081, EPI_ISL_2135082, EPI_ISL_2135083, EPI_ISL_2135084, EPI_ISL_2135085, EPI_ISL_2135086, EPI_ISL_2135087, EPI_ISL_2135088, EPI_ISL_2135089, EPI_ISL_2135090, EPI_ISL_2135091, EPI_ISL_2135092, EPI_ISL_2135093, EPI_ISL_2135094, EPI_ISL_2135095, EPI_ISL_2135096, EPI_ISL_2135097, EPI_ISL_2135098, EPI_ISL_2135099, EPI_ISL_2135100, EPI_ISL_2135101, EPI_ISL_2135102, EPI_ISL_2135103, EPI_ISL_2135105, EPI_ISL_2135106, EPI_ISL_2135107, EPI_ISL_2135109, EPI_ISL_2135110, EPI_ISL_2135111, EPI_ISL_2135112, EPI_ISL_2135113, EPI_ISL_2135114, EPI_ISL_2135115, EPI_ISL_2135116, EPI_ISL_2135117, EPI_ISL_2135119, EPI_ISL_2135120, EPI_ISL_2135121, EPI_ISL_2135122, EPI_ISL_2135123, EPI_ISL_2135124, EPI_ISL_2135125, EPI_ISL_2135126, EPI_ISL_2135127, EPI_ISL_2135128, EPI_ISL_2135130, EPI_ISL_2135131 | | | |
| see above | Illinois Department of Public Health - Springfield Lab | Illinois Department of Public Health - Springfield Lab | Bryan Sim, Gordon McCall |
| EPI_ISL_2135132 | RI State Health Laboratories | Centers for Disease Control and Prevention Division of Viral Diseases, Pathogen Discovery | Mili Sheth, Sarah Nobles, Jasmine Padilla, Mark Burroughs, Shoshona Le, Katie Dillon, Peter Cook, Clinton R. Paden, Dhwani Batra, Krista Queen, Kristen Knipe, Dakota Howard, Yvette Unoarumhi, Darlene Wagner, Matthew Schmerer, Ben L. Rambo-Martin, Kristine Lacek, Sam Shepard, Alison Laufer Halpin, Dave Wentworth, Vivien Dugan, Suxiang Tong, Justin Lee |
| EPI_ISL_2135133 | SD Public Health Laboratory | Centers for Disease Control and Prevention Division of Viral Diseases, Pathogen Discovery | Mili Sheth, Sarah Nobles, Jasmine Padilla, Mark Burroughs, Shoshona Le, Katie Dillon, Peter Cook, Clinton R. Paden, Dhwani Batra, Krista Queen, Kristen Knipe, Dakota Howard, Yvette Unoarumhi, Darlene Wagner, Matthew Schmerer, Ben L. Rambo-Martin, Kristine Lacek, Sam Shepard, Alison Laufer Halpin, Dave Wentworth, Vivien Dugan, Suxiang Tong, Justin Lee |
| EPI_ISL_2135422, EPI_ISL_2135423, EPI_ISL_2135424, EPI_ISL_2135425, EPI_ISL_2135426, EPI_ISL_2135427, EPI_ISL_2135428, EPI_ISL_2135429, EPI_ISL_2135430, EPI_ISL_2135431, EPI_ISL_2135432, EPI_ISL_2135433, EPI_ISL_2135434, EPI_ISL_2135435, EPI_ISL_2135436, EPI_ISL_2135437, EPI_ISL_2135438, EPI_ISL_2135439, EPI_ISL_2135440, EPI_ISL_2135441, EPI_ISL_2135442, EPI_ISL_2135443, EPI_ISL_2135445, EPI_ISL_2135446, EPI_ISL_2135447, EPI_ISL_2135448, EPI_ISL_2135449, EPI_ISL_2135450, EPI_ISL_2135451, EPI_ISL_2135453, EPI_ISL_2135454, EPI_ISL_2135455, EPI_ISL_2135456, EPI_ISL_2135457, EPI_ISL_2135458, EPI_ISL_2135459, EPI_ISL_2135460, EPI_ISL_2135461, EPI_ISL_2135462, EPI_ISL_2135463, EPI_ISL_2135464, EPI_ISL_2135465, EPI_ISL_2135466, EPI_ISL_2135467, EPI_ISL_2135468, EPI_ISL_2135469, EPI_ISL_2135470, EPI_ISL_2135471, EPI_ISL_2135472, EPI_ISL_2135473, EPI_ISL_2135474, EPI_ISL_2135475, EPI_ISL_2135476, EPI_ISL_2135477, EPI_ISL_2135478, EPI_ISL_2135480, EPI_ISL_2135483, EPI_ISL_2135486, EPI_ISL_2135489, EPI_ISL_2135491, EPI_ISL_2135492, EPI_ISL_2135496, EPI_ISL_2135498, EPI_ISL_2135500, EPI_ISL_2135503, EPI_ISL_2135506, EPI_ISL_2135509, EPI_ISL_2135511, EPI_ISL_2135513, EPI_ISL_2135516, EPI_ISL_2135519, EPI_ISL_2135523, EPI_ISL_2135527, EPI_ISL_2135530, EPI_ISL_2135534, EPI_ISL_2135537, EPI_ISL_2135540, EPI_ISL_2135543, EPI_ISL_2135545, EPI_ISL_2135548, EPI_ISL_2135551, EPI_ISL_2135555, EPI_ISL_2135557, EPI_ISL_2135561, EPI_ISL_2135564, EPI_ISL_2135567, EPI_ISL_2135571 | | | |
| see above | State Laboratories Division, Hawaii State Department of Health | State Laboratories Division, Hawaii State Department of Health | Pamela O'Brien, Drew Kuwazaki, Ayana Garnet, Razvan Sultana, Edward Desmond |
| EPI_ISL_2135811, EPI_ISL_2135812, EPI_ISL_2135813, EPI_ISL_2135814, EPI_ISL_2135815, EPI_ISL_2135816, EPI_ISL_2135817, EPI_ISL_2135818, EPI_ISL_2135819, EPI_ISL_2135820, EPI_ISL_2135821, EPI_ISL_2135823, EPI_ISL_2135824, EPI_ISL_2135825, EPI_ISL_2135826, EPI_ISL_2135827, EPI_ISL_2135828, EPI_ISL_2135829, EPI_ISL_2135831, EPI_ISL_2135832, EPI_ISL_2135834, EPI_ISL_2135835, EPI_ISL_2135918, EPI_ISL_2135919, EPI_ISL_2135920, EPI_ISL_2135921, EPI_ISL_2135922, EPI_ISL_2135923, EPI_ISL_2135924, EPI_ISL_2135925, EPI_ISL_2135926, EPI_ISL_2135927, EPI_ISL_2135928, EPI_ISL_2135929, EPI_ISL_2135930, EPI_ISL_2135931, EPI_ISL_2135932, EPI_ISL_2135933, EPI_ISL_2135934, EPI_ISL_2135935, EPI_ISL_2135936, EPI_ISL_2135937, EPI_ISL_2135938, EPI_ISL_2135939, EPI_ISL_2135940, EPI_ISL_2135941, EPI_ISL_2135942, EPI_ISL_2135943, EPI_ISL_2135944, EPI_ISL_2135945, EPI_ISL_2135946, EPI_ISL_2135947, EPI_ISL_2135948, EPI_ISL_2135949, EPI_ISL_2135950, EPI_ISL_2135951, EPI_ISL_2135952, EPI_ISL_2135953, EPI_ISL_2135954, EPI_ISL_2135955, EPI_ISL_2135956, EPI_ISL_2135957, EPI_ISL_2135958, EPI_ISL_2135959, EPI_ISL_2135960, EPI_ISL_2135961, EPI_ISL_2135962, EPI_ISL_2135963, EPI_ISL_2135964, EPI_ISL_2135965, EPI_ISL_2135966, EPI_ISL_2135967, EPI_ISL_2135968, EPI_ISL_2135969, EPI_ISL_2135970, EPI_ISL_2135971, EPI_ISL_2135972, EPI_ISL_2135973, EPI_ISL_2135974, EPI_ISL_2135975, EPI_ISL_2135976, EPI_ISL_2135977, EPI_ISL_2135978, EPI_ISL_2135979, EPI_ISL_2135980, EPI_ISL_2135981, EPI_ISL_2135982, EPI_ISL_2135983, EPI_ISL_2135984 | | | |
| see above | Arizona State Public Health Laboratory | Arizona State Public Health Laboratory | Trung Huynh, Jessica Escobar, Katherine Fullerton, Nobuko Fukushima, Stacy White, Linda Getsinger, Victor Waddell |
| EPI_ISL_2136184, EPI_ISL_2136185, EPI_ISL_2136186, EPI_ISL_2136187, EPI_ISL_2136188, EPI_ISL_2136190, EPI_ISL_2136191, EPI_ISL_2136192, EPI_ISL_2136193 | Maryland Genomics, Institute for Genome Sciences, University of Maryland School of Medicine | Maryland Genomics, Institute for Genome Sciences, University of Maryland School of Medicine | Tallon, Luke J; Sadzewicz, Lisa D; Humphrys, Mike; Ott, Sandra; Roussey, Holly; Mehta, Aditya; Vavikolanu, Kranthi; Fraser, Claire M; Ravel, Jacques |
| EPI_ISL_2136215, EPI_ISL_2136216, EPI_ISL_2136217, EPI_ISL_2136218, EPI_ISL_2136219, EPI_ISL_2136220, EPI_ISL_2136221, EPI_ISL_2136222, EPI_ISL_2136223, EPI_ISL_2136224, EPI_ISL_2136225, EPI_ISL_2136226, EPI_ISL_2136227, EPI_ISL_2136228, EPI_ISL_2136229, EPI_ISL_2136230, EPI_ISL_2136231, EPI_ISL_2136232, EPI_ISL_2136233, EPI_ISL_2136234, EPI_ISL_2136235, EPI_ISL_2136236, EPI_ISL_2136237, EPI_ISL_2136238, EPI_ISL_2136239, EPI_ISL_2136240, EPI_ISL_2136242, EPI_ISL_2136243, EPI_ISL_2136244, EPI_ISL_2136245, EPI_ISL_2136246, EPI_ISL_2136247, EPI_ISL_2136248, EPI_ISL_2136249, EPI_ISL_2136250, EPI_ISL_2136251, EPI_ISL_2136252, EPI_ISL_2136253, EPI_ISL_2136254, EPI_ISL_2136255, EPI_ISL_2136256, EPI_ISL_2136257, EPI_ISL_2136259, EPI_ISL_2136260, EPI_ISL_2136261, EPI_ISL_2136263, EPI_ISL_2136265, EPI_ISL_2136266, EPI_ISL_2136267, EPI_ISL_2136268, EPI_ISL_2136269, EPI_ISL_2136270, EPI_ISL_2136271, EPI_ISL_2136272, EPI_ISL_2136273 | | | |
| see above | University of Michigan Clinical Microbiology Laboratory | Lauring Lab, University of Michigan, Department of Microbiology and Immunology | Valesano |
| EPI_ISL_2136337, EPI_ISL_2136338, EPI_ISL_2136339, EPI_ISL_2136340, EPI_ISL_2136341, EPI_ISL_2136342, EPI_ISL_2136343, EPI_ISL_2136344, EPI_ISL_2136345, EPI_ISL_2136346, EPI_ISL_2136347, EPI_ISL_2136348 | | | |
| see above | GA Department of Public Health | GA Department of Public Health | Stacy Reeves, Jonathan Edwards, Cynthia Dixey, Tonia Parrott, Aliyah Fields, Taylor Smith |
| EPI_ISL_2136372, EPI_ISL_2136373, EPI_ISL_2136374, EPI_ISL_2136375, EPI_ISL_2136376, EPI_ISL_2136378, EPI_ISL_2136379, EPI_ISL_2136380 | Minnesota Department of Health, Public Health Laboratory | Minnesota Department of Health, Public Health Laboratory | Alexandra Lorentz, Jacob Garfin, Matt Plumb, and Xiong Wang |
| EPI_ISL_2136381, EPI_ISL_2136382, | Centracare Laboratory Services | Minnesota Department of Health, Public Health Laboratory | Alexandra Lorentz, Jacob Garfin, Matt Plumb, and Xiong Wang |

| | | | |
|---|---|---|---|
| EPI_ISL_2136383 | | | |
| EPI_ISL_2136384, EPI_ISL_2136385, EPI_ISL_2136386, EPI_ISL_2136387, EPI_ISL_2136388, EPI_ISL_2136389, EPI_ISL_2136390, EPI_ISL_2136391, EPI_ISL_2136392, EPI_ISL_2136393, EPI_ISL_2136394, EPI_ISL_2136395, EPI_ISL_2136396, EPI_ISL_2136397, EPI_ISL_2136398, EPI_ISL_2136399, EPI_ISL_2136400, EPI_ISL_2136401, EPI_ISL_2136402 | | | |
| see above | Minnesota Department of Health, Public Health Laboratory | Minnesota Department of Health, Public Health Laboratory | Alexandra Lorentz, Jacob Garfin, Matt Plumb, and Xiong Wang |
| EPI_ISL_2136403, EPI_ISL_2136404 | Mayo Clinic & Mayo Clinic Laboratories | Minnesota Department of Health, Public Health Laboratory | Alexandra Lorentz, Jacob Garfin, Matt Plumb, and Xiong Wang |
| EPI_ISL_2136405, EPI_ISL_2136407, EPI_ISL_2136408, EPI_ISL_2136409, EPI_ISL_2136410, EPI_ISL_2136411 | Minnesota Department of Health, Public Health Laboratory | Minnesota Department of Health, Public Health Laboratory | Alexandra Lorentz, Jacob Garfin, Matt Plumb, and Xiong Wang |
| EPI_ISL_2136412 | Mayo Clinic & Mayo Clinic Laboratories | Minnesota Department of Health, Public Health Laboratory | Alexandra Lorentz, Jacob Garfin, Matt Plumb, and Xiong Wang |
| EPI_ISL_2136413, EPI_ISL_2136414, EPI_ISL_2136415, EPI_ISL_2136416, EPI_ISL_2136417, EPI_ISL_2136418, EPI_ISL_2136419, EPI_ISL_2136420, EPI_ISL_2136421, EPI_ISL_2136422, EPI_ISL_2136423, EPI_ISL_2136424, EPI_ISL_2136425, EPI_ISL_2136426, EPI_ISL_2136427, EPI_ISL_2136428 | | | |
| see above | Minnesota Department of Health, Public Health Laboratory | Minnesota Department of Health, Public Health Laboratory | Alexandra Lorentz, Jacob Garfin, Matt Plumb, and Xiong Wang |
| EPI_ISL_2136429, EPI_ISL_2136430, EPI_ISL_2136431, EPI_ISL_2136432, EPI_ISL_2136433, EPI_ISL_2136434, EPI_ISL_2136435, EPI_ISL_2136436, EPI_ISL_2136437, EPI_ISL_2136438, EPI_ISL_2136439, EPI_ISL_2136440, EPI_ISL_2136441, EPI_ISL_2136442, EPI_ISL_2136443, EPI_ISL_2136444, EPI_ISL_2136445, EPI_ISL_2136446, EPI_ISL_2136447, EPI_ISL_2136448, EPI_ISL_2136449, EPI_ISL_2136450, EPI_ISL_2136451, EPI_ISL_2136452, EPI_ISL_2136453, EPI_ISL_2136454, EPI_ISL_2136455, EPI_ISL_2136456, EPI_ISL_2136457, EPI_ISL_2136458, EPI_ISL_2136459, EPI_ISL_2136460, EPI_ISL_2136461, EPI_ISL_2136462, EPI_ISL_2136463, EPI_ISL_2136464, EPI_ISL_2136465, EPI_ISL_2136466, EPI_ISL_2136467, EPI_ISL_2136468, EPI_ISL_2136469 | | | |
| see above | Alaska State Virology Laboratory | Alaska State Virology Laboratory | Stephanie DeRonde, Elva House, Jacob Zidek, Lisa Smith, Ph.D., Jack Chen, Ph.D. |
| EPI_ISL_2136470, EPI_ISL_2136471, EPI_ISL_2136472, EPI_ISL_2136473, EPI_ISL_2136474, EPI_ISL_2136476, EPI_ISL_2136478, EPI_ISL_2136479, EPI_ISL_2136480, EPI_ISL_2136481, EPI_ISL_2136485, EPI_ISL_2136487, EPI_ISL_2136488, EPI_ISL_2136490, EPI_ISL_2136491, EPI_ISL_2136492, EPI_ISL_2136493, EPI_ISL_2136496, EPI_ISL_2136497, EPI_ISL_2136498, EPI_ISL_2136499, EPI_ISL_2136502, EPI_ISL_2136503, EPI_ISL_2136504, EPI_ISL_2136505, EPI_ISL_2136506, EPI_ISL_2136508, EPI_ISL_2136511, EPI_ISL_2136512, EPI_ISL_2136513, EPI_ISL_2136514, EPI_ISL_2136515, EPI_ISL_2136516, EPI_ISL_2136517, EPI_ISL_2136518, EPI_ISL_2136519, EPI_ISL_2136520, EPI_ISL_2136521, EPI_ISL_2136522 | | | |
| see above | Incyte Diagnostics Spokane | Seattle Flu Study | Deborah A. Nickerson, Chris D. Frazar, Jover Lee, Benjamin Pelle, Erica Ryke, Matthew Richardson, Amanda Adler, Elisabeth Brandstetter, Peter D. Han, Kairsten Fay, Misja Ilcisin, Kirsten Lacombe, Thomas R. Sibley, Melissa Truong, Caitlin R. Wolf, Romesh Gautom, Geoff Melly, Brian Hiatt, Philip Dykema, Scott Lindquist, Michael Boeckh, Janet A. Englund, Michael Famulare, Barry R. Lutz, Mark J. Rieder, Lea M. Starita, Matthew Thompson, Helen Y. Chu, Jay Shendure, Trevor Bedford |
| EPI_ISL_2136525, EPI_ISL_2136526, EPI_ISL_2136527, EPI_ISL_2136533, EPI_ISL_2136534, EPI_ISL_2136539, EPI_ISL_2136543, EPI_ISL_2136544, EPI_ISL_2136545, EPI_ISL_2136548, EPI_ISL_2136549, EPI_ISL_2136550, EPI_ISL_2136551, EPI_ISL_2136552, EPI_ISL_2136553, EPI_ISL_2136556, EPI_ISL_2136559, EPI_ISL_2136560, EPI_ISL_2136563, EPI_ISL_2136564, EPI_ISL_2136565, EPI_ISL_2136566, EPI_ISL_2136569, EPI_ISL_2136570, EPI_ISL_2136572, EPI_ISL_2136573, EPI_ISL_2136574, EPI_ISL_2136576, EPI_ISL_2136580, EPI_ISL_2136585, EPI_ISL_2136586, EPI_ISL_2136587, EPI_ISL_2136588, EPI_ISL_2136589, EPI_ISL_2136591, EPI_ISL_2136592, EPI_ISL_2136594, EPI_ISL_2136595, EPI_ISL_2136597, EPI_ISL_2136598, EPI_ISL_2136599, EPI_ISL_2136600, EPI_ISL_2136602, EPI_ISL_2136604, EPI_ISL_2136607, EPI_ISL_2136608, EPI_ISL_2136609, EPI_ISL_2136611, EPI_ISL_2136612, EPI_ISL_2136613, EPI_ISL_2136614, EPI_ISL_2136615, EPI_ISL_2136618, EPI_ISL_2136620, EPI_ISL_2136622, EPI_ISL_2136627, EPI_ISL_2136628, EPI_ISL_2136629, EPI_ISL_2136630, EPI_ISL_2136631, EPI_ISL_2136632, EPI_ISL_2136633, EPI_ISL_2136634, EPI_ISL_2136635, EPI_ISL_2136636, EPI_ISL_2136637, EPI_ISL_2136638, EPI_ISL_2136639, EPI_ISL_2136640, EPI_ISL_2136641, EPI_ISL_2136643, EPI_ISL_2136644, EPI_ISL_2136645, EPI_ISL_2136646, EPI_ISL_2136648, EPI_ISL_2136651, EPI_ISL_2136653, EPI_ISL_2136656, EPI_ISL_2136661, EPI_ISL_2136662 | | | |
| see above | Northwest Laboratory | Seattle Flu Study | Deborah A. Nickerson, Chris D. Frazar, Jover Lee, Benjamin Pelle, Erica Ryke, Matthew Richardson, Amanda Adler, Elisabeth Brandstetter, Peter D. Han, Kairsten Fay, Misja Ilcisin, Kirsten Lacombe, Thomas R. Sibley, Melissa Truong, Caitlin R. Wolf, Romesh Gautom, Geoff Melly, Brian Hiatt, Philip Dykema, Scott Lindquist, Michael Boeckh, Janet A. Englund, Michael Famulare, Barry R. Lutz, Mark J. Rieder, Lea M. Starita, Matthew Thompson, Helen Y. Chu, Jay Shendure, Trevor Bedford |
| EPI_ISL_2136665, EPI_ISL_2136667, EPI_ISL_2136668, EPI_ISL_2136669, EPI_ISL_2136670, EPI_ISL_2136671, EPI_ISL_2136672, EPI_ISL_2136673, EPI_ISL_2136674, EPI_ISL_2136675, EPI_ISL_2136676, EPI_ISL_2136677, EPI_ISL_2136678, EPI_ISL_2136679, EPI_ISL_2136681, EPI_ISL_2136682, EPI_ISL_2136684, EPI_ISL_2136685, EPI_ISL_2136686, EPI_ISL_2136687, EPI_ISL_2136688, EPI_ISL_2136689, EPI_ISL_2136690, EPI_ISL_2136691, EPI_ISL_2136693, EPI_ISL_2136694, EPI_ISL_2136695, EPI_ISL_2136696, EPI_ISL_2136697, EPI_ISL_2136698, EPI_ISL_2136699, EPI_ISL_2136700, EPI_ISL_2136701, EPI_ISL_2136703, EPI_ISL_2136705, EPI_ISL_2136706, EPI_ISL_2136708, EPI_ISL_2136710, EPI_ISL_2136711, EPI_ISL_2136712, EPI_ISL_2136713, EPI_ISL_2136714, EPI_ISL_2136716, EPI_ISL_2136718, EPI_ISL_2136721, EPI_ISL_2136723, EPI_ISL_2136724, EPI_ISL_2136725, EPI_ISL_2136727 | | | |
| see above | Atlas Genomics | Seattle Flu Study | Deborah A. Nickerson, Chris D. Frazar, Jover Lee, Benjamin Pelle, Erica Ryke, Matthew Richardson, Amanda Adler, Elisabeth Brandstetter, Peter D. Han, Kairsten Fay, Misja Ilcisin, Kirsten Lacombe, Thomas R. Sibley, Melissa Truong, Caitlin R. Wolf, Romesh Gautom, Geoff Melly, Brian Hiatt, Philip Dykema, Scott Lindquist, Michael Boeckh, Janet A. Englund, Michael Famulare, Barry R. Lutz, Mark J. Rieder, Lea M. Starita, Matthew Thompson, Helen Y. Chu, Jay Shendure, Trevor Bedford |
| EPI_ISL_2136730, EPI_ISL_2136732, EPI_ISL_2136733, EPI_ISL_2136734, EPI_ISL_2136735, EPI_ISL_2136736, EPI_ISL_2136737, EPI_ISL_2136739, EPI_ISL_2136740, EPI_ISL_2136741, EPI_ISL_2136742, EPI_ISL_2136744, EPI_ISL_2136745 | | | |
| see above | Incyte Diagnostics Spokane | Seattle Flu Study | Deborah A. Nickerson, Chris D. Frazar, Jover Lee, Benjamin Pelle, Erica Ryke, Matthew Richardson, Amanda Adler, Elisabeth Brandstetter, Peter D. Han, Kairsten Fay, Misja Ilcisin, Kirsten Lacombe, Thomas R. Sibley, Melissa Truong, Caitlin R. Wolf, Romesh Gautom, Geoff Melly, Brian Hiatt, Philip Dykema, Scott Lindquist, Michael Boeckh, Janet A. Englund, Michael Famulare, Barry R. Lutz, Mark J. Rieder, Lea M. Starita, Matthew Thompson, Helen Y. Chu, Jay Shendure, Trevor Bedford |
| EPI_ISL_2136747, EPI_ISL_2136748, EPI_ISL_2136749, EPI_ISL_2136750, EPI_ISL_2136751, EPI_ISL_2136752, EPI_ISL_2136753, EPI_ISL_2136754, EPI_ISL_2136755, EPI_ISL_2136756, EPI_ISL_2136757, EPI_ISL_2136758, EPI_ISL_2136759, EPI_ISL_2136760, EPI_ISL_2136761, EPI_ISL_2136762, EPI_ISL_2136763, EPI_ISL_2136764, EPI_ISL_2136765, EPI_ISL_2136766, EPI_ISL_2136767, EPI_ISL_2136768, EPI_ISL_2136769, EPI_ISL_2136770, EPI_ISL_2136771, EPI_ISL_2136772, EPI_ISL_2136773, EPI_ISL_2136774, EPI_ISL_2136775, EPI_ISL_2136776, EPI_ISL_2136777, EPI_ISL_2136778, EPI_ISL_2136779, EPI_ISL_2136780, EPI_ISL_2136781, EPI_ISL_2136782, EPI_ISL_2136783, EPI_ISL_2136784, EPI_ISL_2136785, EPI_ISL_2136786, EPI_ISL_2136787, EPI_ISL_2136788, EPI_ISL_2136789 | | | |
| see above | Alaska State Virology Laboratory | Alaska State Virology Laboratory | Stephanie DeRonde, Elva House, Jacob Zidek, Lisa Smith, Ph.D., Jack Chen, Ph.D. |
| EPI_ISL_2137381, EPI_ISL_2137382, EPI_ISL_2137383 | DC Public Health Lab/ Dept. of Forensic Sciences | DC Public Health Lab/ Dept. of Forensic Sciences | Janis Doss, Scott Nguyen, Elizabeth Zelaya, Sarah Scott, Connie Maza, Monica Mann, Brittany Hamilton, David Payne, Jocelyn Hauser |
| EPI_ISL_2137726, EPI_ISL_2137727, EPI_ISL_2137728, EPI_ISL_2137729, EPI_ISL_2137730, EPI_ISL_2137732 | Wyoming Public Health Laboratory | Wyoming Public Health Laboratory | Jim Mildenberger, Taylor Fearing, Channing Weber, Ashley Norberg, Chayse Rowley, Marley Goetz, Brian Dominguez, Elliot Thomasson, Brittany Oher, Sam Britz, Cari Sloma, and Rob Christensen |
| EPI_ISL_2137737, EPI_ISL_2137738, EPI_ISL_2137739, EPI_ISL_2137740, EPI_ISL_2137741, EPI_ISL_2137742, EPI_ISL_2137743, EPI_ISL_2137744, EPI_ISL_2137745, EPI_ISL_2137746, EPI_ISL_2137748, EPI_ISL_2137750, EPI_ISL_2137751, EPI_ISL_2137752, EPI_ISL_2137754 | | | |
| see above | Lafene Health Center | Kansas State Veterinary Diagnostic Laboratory | Tyler Doerksen, Andreea Lu, Lance Noll, Elizabeth Porter, Emily Cox, Jianfa Bai, Jamie Henningson, Rachel Palinski |
| EPI_ISL_2137759 | DC Public Health Lab/ Dept. of Forensic Sciences | DC Public Health Lab/ Dept. of Forensic Sciences | Janis Doss, Scott Nguyen, Elizabeth Zelaya, Sarah Scott, Connie Maza, Monica Mann, Brittany Hamilton, David Payne, Jocelyn Hauser |
| EPI_ISL_2137760, EPI_ISL_2137761, EPI_ISL_2137762, EPI_ISL_2137763, EPI_ISL_2137764, EPI_ISL_2137765, EPI_ISL_2137766 | North Carolina State Laboratory of Public Health (NCSLPH) | North Carolina State Laboratory of Public Health (NCSLPH) | Chase K, Miller MC, Greene S, Glover W |
| EPI_ISL_2137771, EPI_ISL_2137778, EPI_ISL_2137781, EPI_ISL_2137783, EPI_ISL_2137784, EPI_ISL_2137785, EPI_ISL_2137788 | Wyoming Public Health Laboratory | Wyoming Public Health Laboratory | Jim Mildenberger, Taylor Fearing, Channing Weber, Ashley Norberg, Chayse Rowley, Marley Goetz, Brian Dominguez, Elliot Thomasson, Brittany Oher, Sam Britz, Cari Sloma, and Rob Christensen |
| EPI_ISL_2138867, EPI_ISL_2138868, EPI_ISL_2138869, EPI_ISL_2138870, EPI_ISL_2138871, EPI_ISL_2138872, EPI_ISL_2138873, EPI_ISL_2138874, EPI_ISL_2138875, EPI_ISL_2138876, EPI_ISL_2138877, EPI_ISL_2138878, EPI_ISL_2138879, EPI_ISL_2138880, EPI_ISL_2138881, EPI_ISL_2138882, EPI_ISL_2138883, EPI_ISL_2138884, EPI_ISL_2138885, EPI_ISL_2138886, EPI_ISL_2138887, EPI_ISL_2138888, EPI_ISL_2138889, EPI_ISL_2138890, EPI_ISL_2138891, EPI_ISL_2138892, EPI_ISL_2138893, EPI_ISL_2138894, EPI_ISL_2138895, EPI_ISL_2138896, EPI_ISL_2138897, EPI_ISL_2138898, EPI_ISL_2138899, EPI_ISL_2138900, EPI_ISL_2138904, EPI_ISL_2138907, EPI_ISL_2139550, EPI_ISL_2139551, EPI_ISL_2139552, EPI_ISL_2139553, EPI_ISL_2139554, EPI_ISL_2139555, EPI_ISL_2139556, EPI_ISL_2139557, EPI_ISL_2139558, EPI_ISL_2139559, EPI_ISL_2139560, EPI_ISL_2139561, EPI_ISL_2139562, EPI_ISL_2139563, EPI_ISL_2139564, EPI_ISL_2139565, EPI_ISL_2139566, EPI_ISL_2139567, EPI_ISL_2139633, EPI_ISL_2139634, EPI_ISL_2139635, EPI_ISL_2139636, EPI_ISL_2139637, EPI_ISL_2139638, EPI_ISL_2139639, EPI_ISL_2139640, EPI_ISL_2139641, EPI_ISL_2139642, EPI_ISL_2139643, EPI_ISL_2139644, EPI_ISL_2139645, EPI_ISL_2139646, EPI_ISL_2139647, EPI_ISL_2139648 | | | |
| see above | Oregon State Public Health Laboratory | Oregon State Public Health Laboratory | Rafia Razzaque, Eugene Yeboah, Vanda Makris, Laura Tsaknaridis, John Fontana and Shane Sevey |
| EPI_ISL_2139649, EPI_ISL_2139650, EPI_ISL_2139651, EPI_ISL_2139652, EPI_ISL_2139653, EPI_ISL_2139654, EPI_ISL_2139655, EPI_ISL_2139656, EPI_ISL_2139657, EPI_ISL_2139658, EPI_ISL_2139659, EPI_ISL_2139660, EPI_ISL_2139661, EPI_ISL_2139662, EPI_ISL_2139663, EPI_ISL_2139664, EPI_ISL_2139665, EPI_ISL_2139666, EPI_ISL_2139667, EPI_ISL_2139668, EPI_ISL_2139669, EPI_ISL_2139670, EPI_ISL_2139671, EPI_ISL_2139672, EPI_ISL_2139673 | | | |
| see above | Wyoming Public Health Laboratory | Wyoming Public Health Laboratory | Jim Mildenberger, Taylor Fearing, Channing Weber, Ashley Norberg, Chayse Rowley, Marley Goetz, Brian Dominguez, Elliot Thomasson, Brittany Oher, Sam Britz, Cari Sloma, and Rob Christensen |
| EPI_ISL_2139674, EPI_ISL_2139675, EPI_ISL_2139676, EPI_ISL_2139677, EPI_ISL_2139678, EPI_ISL_2139679, EPI_ISL_2139680, EPI_ISL_2139681, EPI_ISL_2139682, EPI_ISL_2139683, EPI_ISL_2139684, EPI_ISL_2139685, EPI_ISL_2139686, EPI_ISL_2139687, EPI_ISL_2139688 | | | |
| see above | GA Department of Public Health | GA Department of Public Health | Stacy Reeves, Jonathan Edwards, Cynthia Dixey, Tonia Parrott, Aliyah Fields, Taylor Smith |
| EPI_ISL_2139908, EPI_ISL_2139909 | LESP Baja California | Instituto de Diagnostico y Referencia Epidemiologicos (INDRE) | Claudia Wong-Arambula, Abril Rodriguez-Maldonado, Vanessa Rivero-Arredondo, Ariadna Medina-Benitez, Joaquin Quiroz-Mercado, Sergio Rangel-Guerrero, Natividad Cruz-Ortiz, Tatiana Nunez-Garcia, Gisela Barrera-Badillo, Lucia Hernandez-Rivas, Irma Lopez-Martinez, Ernesto Ramirez-Gonzalez. |
| EPI_ISL_2139910 | LESP Nuevo Leon | Instituto de Diagnostico y Referencia Epidemiologicos (INDRE) | Claudia Wong-Arambula, Abril Rodriguez-Maldonado, Vanessa Rivero-Arredondo, Ariadna Medina-Benitez, Joaquin Quiroz-Mercado, Sergio Rangel-Guerrero, Natividad Cruz-Ortiz, Tatiana Nunez-Garcia, Gisela Barrera-Badillo, Lucia Hernandez-Rivas, Irma Lopez-Martinez, Ernesto Ramirez-Gonzalez. |
| EPI_ISL_2139911 | LESP Guanajuato | Instituto de Diagnostico y Referencia Epidemiologicos (INDRE) | Claudia Wong-Arambula, Abril Rodriguez-Maldonado, Vanessa Rivero-Arredondo, Ariadna Medina-Benitez, Joaquin Quiroz-Mercado, Sergio Rangel-Guerrero, Natividad Cruz-Ortiz, Tatiana Nunez-Garcia, Gisela Barrera-Badillo, Lucia Hernandez-Rivas, Irma Lopez-Martinez, Ernesto Ramirez-Gonzalez. |
| EPI_ISL_2139912 | LESP Colima | Instituto de Diagnostico y Referencia Epidemiologicos (INDRE) | Claudia Wong-Arambula, Abril Rodriguez-Maldonado, Vanessa Rivero-Arredondo, Ariadna Medina-Benitez, Joaquin Quiroz-Mercado, Sergio Rangel-Guerrero, Natividad Cruz-Ortiz, Tatiana Nunez-Garcia, Gisela Barrera-Badillo, Lucia Hernandez-Rivas, Irma Lopez-Martinez, Ernesto Ramirez-Gonzalez. |
| EPI_ISL_2139913 | LESP Baja California Sur | Instituto de Diagnostico y Referencia Epidemiologicos (INDRE) | Claudia Wong-Arambula, Abril Rodriguez-Maldonado, Vanessa Rivero-Arredondo, Ariadna Medina-Benitez, Joaquin Quiroz-Mercado, Sergio Rangel-Guerrero, Natividad Cruz-Ortiz, Tatiana Nunez-Garcia, Gisela Barrera-Badillo, Lucia Hernandez-Rivas, Irma Lopez-Martinez, Ernesto Ramirez-Gonzalez. |
| EPI_ISL_2139914 | LESP Baja California | Instituto de Diagnostico y Referencia Epidemiologicos (INDRE) | Claudia Wong-Arambula, Abril Rodriguez-Maldonado, Vanessa Rivero-Arredondo, Ariadna Medina-Benitez, Joaquin Quiroz-Mercado, Sergio Rangel-Guerrero, Natividad Cruz-Ortiz, Tatiana Nunez-Garcia, Gisela Barrera-Badillo, Lucia Hernandez-Rivas, Irma Lopez-Martinez, Ernesto Ramirez-Gonzalez. |
| EPI_ISL_2139915 | LESP Jalisco | Instituto de Diagnostico y Referencia Epidemiologicos (INDRE) | Claudia Wong-Arambula, Abril Rodriguez-Maldonado, Vanessa Rivero-Arredondo, Ariadna Medina-Benitez, Joaquin Quiroz-Mercado, Sergio Rangel-Guerrero, Natividad Cruz-Ortiz, Tatiana Nunez-Garcia, Gisela Barrera-Badillo, Lucia Hernandez-Rivas, Irma Lopez-Martinez, Ernesto Ramirez-Gonzalez. |

| | | | |
|---|---|---|---|
| EPI_ISL_2139916 | LESP Chihuahua | Instituto de Diagnostico y Referencia Epidemiologicos (INDRE) | Claudia Wong-Arambula, Abril Rodriguez-Maldonado, Vanessa Rivero-Arredondo, Ariadna Medina-Benitez, Joaquin Quiroz-Mercado, Sergio Rangel-Guerrero, Natividad Cruz-Ortiz, Tatiana Nunez-Garcia, Gisela Barrera-Badillo, Lucia Hernandez-Rivas, Irma Lopez-Martinez, Ernesto Ramirez-Gonzalez. |
| EPI_ISL_2139917 | LESP Michoacan | Instituto de Diagnostico y Referencia Epidemiologicos (INDRE) | Claudia Wong-Arambula, Abril Rodriguez-Maldonado, Vanessa Rivero-Arredondo, Ariadna Medina-Benitez, Joaquin Quiroz-Mercado, Sergio Rangel-Guerrero, Natividad Cruz-Ortiz, Tatiana Nunez-Garcia, Gisela Barrera-Badillo, Lucia Hernandez-Rivas, Irma Lopez-Martinez, Ernesto Ramirez-Gonzalez. |
| EPI_ISL_2139918 | LESP Sinaloa | Instituto de Diagnostico y Referencia Epidemiologicos (INDRE) | Claudia Wong-Arambula, Abril Rodriguez-Maldonado, Vanessa Rivero-Arredondo, Ariadna Medina-Benitez, Joaquin Quiroz-Mercado, Sergio Rangel-Guerrero, Natividad Cruz-Ortiz, Tatiana Nunez-Garcia, Gisela Barrera-Badillo, Lucia Hernandez-Rivas, Irma Lopez-Martinez, Ernesto Ramirez-Gonzalez. |
| EPI_ISL_2139919 | LESP Jalisco | Instituto de Diagnostico y Referencia Epidemiologicos (INDRE) | Claudia Wong-Arambula, Abril Rodriguez-Maldonado, Vanessa Rivero-Arredondo, Ariadna Medina-Benitez, Joaquin Quiroz-Mercado, Sergio Rangel-Guerrero, Natividad Cruz-Ortiz, Tatiana Nunez-Garcia, Gisela Barrera-Badillo, Lucia Hernandez-Rivas, Irma Lopez-Martinez, Ernesto Ramirez-Gonzalez. |
| EPI_ISL_2139920 | LESP Ciudad de Mexico | Instituto de Diagnostico y Referencia Epidemiologicos (INDRE) | Claudia Wong-Arambula, Abril Rodriguez-Maldonado, Vanessa Rivero-Arredondo, Ariadna Medina-Benitez, Joaquin Quiroz-Mercado, Sergio Rangel-Guerrero, Natividad Cruz-Ortiz, Tatiana Nunez-Garcia, Gisela Barrera-Badillo, Lucia Hernandez-Rivas, Irma Lopez-Martinez, Ernesto Ramirez-Gonzalez. |
| EPI_ISL_2139921 | LESP Campeche | Instituto de Diagnostico y Referencia Epidemiologicos (INDRE) | Claudia Wong-Arambula, Abril Rodriguez-Maldonado, Vanessa Rivero-Arredondo, Ariadna Medina-Benitez, Joaquin Quiroz-Mercado, Sergio Rangel-Guerrero, Natividad Cruz-Ortiz, Tatiana Nunez-Garcia, Gisela Barrera-Badillo, Lucia Hernandez-Rivas, Irma Lopez-Martinez, Ernesto Ramirez-Gonzalez. |
| EPI_ISL_2139922 | LESP Ciudad de Mexico | Instituto de Diagnostico y Referencia Epidemiologicos (INDRE) | Claudia Wong-Arambula, Abril Rodriguez-Maldonado, Vanessa Rivero-Arredondo, Ariadna Medina-Benitez, Joaquin Quiroz-Mercado, Sergio Rangel-Guerrero, Natividad Cruz-Ortiz, Tatiana Nunez-Garcia, Gisela Barrera-Badillo, Lucia Hernandez-Rivas, Irma Lopez-Martinez, Ernesto Ramirez-Gonzalez. |
| EPI_ISL_2139923, EPI_ISL_2139924, EPI_ISL_2139925, EPI_ISL_2139926, EPI_ISL_2139927, EPI_ISL_2139928, EPI_ISL_2139929, EPI_ISL_2139930, EPI_ISL_2139931, EPI_ISL_2139932, EPI_ISL_2139933, EPI_ISL_2139934, EPI_ISL_2139935, EPI_ISL_2139936, EPI_ISL_2139937, EPI_ISL_2139938, EPI_ISL_2139939, EPI_ISL_2139940, EPI_ISL_2139941, EPI_ISL_2139942, EPI_ISL_2139943, EPI_ISL_2139944, EPI_ISL_2139945, EPI_ISL_2139946, EPI_ISL_2139947, EPI_ISL_2139948, EPI_ISL_2139949, EPI_ISL_2139950, EPI_ISL_2139951, EPI_ISL_2139952, EPI_ISL_2139953, EPI_ISL_2139954, EPI_ISL_2139955, EPI_ISL_2139956, EPI_ISL_2139957, EPI_ISL_2139958, EPI_ISL_2139959, EPI_ISL_2139960, EPI_ISL_2139961, EPI_ISL_2139962, EPI_ISL_2139963, EPI_ISL_2139964, EPI_ISL_2139965, EPI_ISL_2139966, EPI_ISL_2139967, EPI_ISL_2139968, EPI_ISL_2139969, EPI_ISL_2139970, EPI_ISL_2139971, EPI_ISL_2139972, EPI_ISL_2139973, EPI_ISL_2139974, EPI_ISL_2139975, EPI_ISL_2139976, EPI_ISL_2139977, EPI_ISL_2139978, EPI_ISL_2139979, EPI_ISL_2139980, EPI_ISL_2139981, EPI_ISL_2139982, EPI_ISL_2139983, EPI_ISL_2139984, EPI_ISL_2139985, EPI_ISL_2139986, EPI_ISL_2139987, EPI_ISL_2139988, EPI_ISL_2139989, EPI_ISL_2139990, EPI_ISL_2139991, EPI_ISL_2139992, EPI_ISL_2139993, EPI_ISL_2139994, EPI_ISL_2139995, EPI_ISL_2139996, EPI_ISL_2139997, EPI_ISL_2139998, EPI_ISL_2139999, EPI_ISL_2140000, EPI_ISL_2140001, EPI_ISL_2140002, EPI_ISL_2140003, EPI_ISL_2140004, EPI_ISL_2140005, EPI_ISL_2140006, EPI_ISL_2140007, EPI_ISL_2140008, EPI_ISL_2140009, EPI_ISL_2140010, EPI_ISL_2140011, EPI_ISL_2140012, EPI_ISL_2140013, EPI_ISL_2140014, EPI_ISL_2140015, EPI_ISL_2140025, EPI_ISL_2140026, EPI_ISL_2140027, EPI_ISL_2140028, EPI_ISL_2140029, EPI_ISL_2140030, EPI_ISL_2140031, EPI_ISL_2140032, EPI_ISL_2140033, EPI_ISL_2140034, EPI_ISL_2140035, EPI_ISL_2140036, EPI_ISL_2140037 | | | |
| see above | Illinois Department of Public Health | Illinois Department of Public Health - Chicago Lab | Vineet K. Dhiman, Ira Heimler, Joel Price |
| EPI_ISL_2140147, EPI_ISL_2140148, EPI_ISL_2140149, EPI_ISL_2140150, EPI_ISL_2140151, EPI_ISL_2140152, EPI_ISL_2140153, EPI_ISL_2140154, EPI_ISL_2140155, EPI_ISL_2140156, EPI_ISL_2140157, EPI_ISL_2140158, EPI_ISL_2140159, EPI_ISL_2140160, EPI_ISL_2140161, EPI_ISL_2140162, EPI_ISL_2140163, EPI_ISL_2140164, EPI_ISL_2140165, EPI_ISL_2140166, EPI_ISL_2140167, EPI_ISL_2140168, EPI_ISL_2140169 | | | |
| see above | Idaho Bureau of Laboratories | Idaho Bureau of Laboratories | R. Beukelman, Matthew Charles Burns, Aimee Ceniseros, Robert L. Voermans, Christopher Ball |
| EPI_ISL_2140732, EPI_ISL_2140733, EPI_ISL_2140734, EPI_ISL_2140735, EPI_ISL_2140736, EPI_ISL_2140737, EPI_ISL_2140738, EPI_ISL_2140739, EPI_ISL_2140740, EPI_ISL_2140741, EPI_ISL_2140742, EPI_ISL_2140743, EPI_ISL_2140744, EPI_ISL_2140745, EPI_ISL_2140746, EPI_ISL_2140747, EPI_ISL_2140748, EPI_ISL_2140749, EPI_ISL_2140750, EPI_ISL_2140751, EPI_ISL_2140752, EPI_ISL_2140753, EPI_ISL_2140754, EPI_ISL_2140755, EPI_ISL_2140756, EPI_ISL_2140757, EPI_ISL_2140758, EPI_ISL_2140759, EPI_ISL_2140760, EPI_ISL_2140761, EPI_ISL_2140762, EPI_ISL_2140763, EPI_ISL_2140764, EPI_ISL_2140765, EPI_ISL_2140766, EPI_ISL_2140767, EPI_ISL_2140768, EPI_ISL_2140769, EPI_ISL_2140770, EPI_ISL_2140771, EPI_ISL_2140772, EPI_ISL_2140773, EPI_ISL_2140774, EPI_ISL_2140775, EPI_ISL_2140776, EPI_ISL_2140777, EPI_ISL_2140778, EPI_ISL_2140779, EPI_ISL_2140780, EPI_ISL_2140781, EPI_ISL_2140782, EPI_ISL_2140783, EPI_ISL_2140784, EPI_ISL_2140785, EPI_ISL_2140786, EPI_ISL_2140787, EPI_ISL_2140788, EPI_ISL_2140789, EPI_ISL_2140790, EPI_ISL_2140791, EPI_ISL_2140792, EPI_ISL_2140793, EPI_ISL_2140794, EPI_ISL_2140795, EPI_ISL_2140796, EPI_ISL_2140797, EPI_ISL_2140798 | | | |
| see above | Quest Diagnostics Incorporated | Centers for Disease Control and Prevention Division of Viral Diseases, Pathogen Discovery | Dakota Howard, Dhwani Batra, Peter W. Cook, Kara Moser, Adrian Paskey, Jason Caravas, Benjamin Rambo-Martin, Shatavia Morrison, Christopher Gulvick, Scott Sammons, Yvette Unoarumhi, Darlene Wagner, Matthew Schmerer, S. H. Rosenthal, A. Gerasimova, R. M. Kagan, B. Anderson, M. Hua, Y. Liu, L.E. Bernstein, K.E. Livingston, A. Perez, I. A. Shlyakhter, R. V. Rolando, R. Owen, P. Tanpaiboon, F. Lacbawan, Clinton R. Paden, Duncan MacCannell |
| EPI_ISL_2140799 | Fulgent Genetics | Centers for Disease Control and Prevention Division of Viral Diseases, Pathogen Discovery | Dakota Howard, Dhwani Batra, Peter W. Cook, Kara Moser, Adrian Paskey, Jason Caravas, Benjamin Rambo-Martin, Shatavia Morrison, Christopher Gulvick, Scott Sammons, Yvette Unoarumhi, Darlene Wagner, Matthew Schmerer, Harry Gao, Mickey Li, John Gao, Joseph Fierro, Benafsh Sapra, Becky Tsai, Yan Meng, Doreen Ng, James Xie, Clinton R. Paden, Duncan MacCannell |
| EPI_ISL_2140800, EPI_ISL_2140801, EPI_ISL_2140802, EPI_ISL_2140803, EPI_ISL_2140804, EPI_ISL_2140805, EPI_ISL_2140806, EPI_ISL_2140807, EPI_ISL_2140808, EPI_ISL_2140809, EPI_ISL_2140810, EPI_ISL_2140811, EPI_ISL_2140812, EPI_ISL_2140813, EPI_ISL_2140814, EPI_ISL_2140815, EPI_ISL_2140816, EPI_ISL_2140817, EPI_ISL_2140818, EPI_ISL_2140819, EPI_ISL_2140820, EPI_ISL_2140821, EPI_ISL_2140822, EPI_ISL_2140823, EPI_ISL_2140824, EPI_ISL_2140825, EPI_ISL_2140826, EPI_ISL_2140827, EPI_ISL_2140828, EPI_ISL_2140829, EPI_ISL_2140830, EPI_ISL_2140831, EPI_ISL_2140832, EPI_ISL_2140833, EPI_ISL_2140834, EPI_ISL_2140835, EPI_ISL_2140836, EPI_ISL_2140837, EPI_ISL_2140838, EPI_ISL_2140839, EPI_ISL_2140840, EPI_ISL_2140841, EPI_ISL_2140842, EPI_ISL_2140843, EPI_ISL_2140844, EPI_ISL_2140845, EPI_ISL_2140846, EPI_ISL_2140847, EPI_ISL_2140848, EPI_ISL_2140849, EPI_ISL_2140850, EPI_ISL_2140851, EPI_ISL_2140852, EPI_ISL_2140853, EPI_ISL_2140854, EPI_ISL_2140855, EPI_ISL_2140856, EPI_ISL_2140857, EPI_ISL_2140858, EPI_ISL_2140860, EPI_ISL_2140861, EPI_ISL_2140862, EPI_ISL_2140863, EPI_ISL_2140864, EPI_ISL_2140865, EPI_ISL_2140866, EPI_ISL_2140867, EPI_ISL_2140868, EPI_ISL_2140869, EPI_ISL_2140870, EPI_ISL_2140871, EPI_ISL_2140872, EPI_ISL_2140874, EPI_ISL_2140875, EPI_ISL_2140876, EPI_ISL_2140877, EPI_ISL_2140878, EPI_ISL_2140879, EPI_ISL_2140880, EPI_ISL_2140881, EPI_ISL_2140882, EPI_ISL_2140883, EPI_ISL_2140884 | | | |
| see above | The Ohio State University Applied Microbiology Services Laboratory | The Ohio State University Applied Microbiology Services Laboratory | Seth A. Faith PhD |
| EPI_ISL_2141421, EPI_ISL_2141422, EPI_ISL_2141423, EPI_ISL_2141424, EPI_ISL_2141425, EPI_ISL_2141426, EPI_ISL_2141427, EPI_ISL_2141428, EPI_ISL_2141429, EPI_ISL_2141430, EPI_ISL_2141431, EPI_ISL_2141432, EPI_ISL_2141433, EPI_ISL_2141434, EPI_ISL_2141435, EPI_ISL_2141436, EPI_ISL_2141437, EPI_ISL_2141438, EPI_ISL_2141441, EPI_ISL_2141442, EPI_ISL_2141443, EPI_ISL_2141444, EPI_ISL_2141445, EPI_ISL_2141446, EPI_ISL_2141447, EPI_ISL_2141448, EPI_ISL_2141449, EPI_ISL_2141450, EPI_ISL_2141451, EPI_ISL_2141452, EPI_ISL_2141453, EPI_ISL_2141454, EPI_ISL_2141455, EPI_ISL_2141456, EPI_ISL_2141457, EPI_ISL_2141458, EPI_ISL_2141459, EPI_ISL_2141460, EPI_ISL_2141461, EPI_ISL_2141462, EPI_ISL_2141463, EPI_ISL_2141464, EPI_ISL_2141465, EPI_ISL_2141466, EPI_ISL_2141467, EPI_ISL_2141468, EPI_ISL_2141469, EPI_ISL_2141470, EPI_ISL_2141471, EPI_ISL_2141472, EPI_ISL_2141473, EPI_ISL_2141474, EPI_ISL_2141475, EPI_ISL_2141476, EPI_ISL_2141481, EPI_ISL_2141482, EPI_ISL_2141484, EPI_ISL_2141492 | | | |
| see above | TXDSHS | TXDSHS | Rashmi Tuladhar, Bonnie Oh, Jenny Zhang, Maliha Rahman, Mayela Pedrueza, Anita Pokharel, Karen Bobier, Lorraine Rodriguez, Myong Koag, Chun Wang, Rachel Lee, Grace Kubin |
| EPI_ISL_2142878, EPI_ISL_2142879, EPI_ISL_2142880, EPI_ISL_2142881, EPI_ISL_2142882, EPI_ISL_2142883, EPI_ISL_2142884, EPI_ISL_2142885, EPI_ISL_2142886, EPI_ISL_2142887, EPI_ISL_2142888, EPI_ISL_2142889, EPI_ISL_2142890, EPI_ISL_2142891, EPI_ISL_2142892, EPI_ISL_2142893, EPI_ISL_2142894, EPI_ISL_2142895, EPI_ISL_2142896, EPI_ISL_2142897, EPI_ISL_2142898, EPI_ISL_2142899, EPI_ISL_2142900, EPI_ISL_2142901, EPI_ISL_2142902, EPI_ISL_2142903, EPI_ISL_2142904, EPI_ISL_2142905, EPI_ISL_2142906, EPI_ISL_2142907, EPI_ISL_2142908, EPI_ISL_2142909, EPI_ISL_2142910, EPI_ISL_2142911, EPI_ISL_2142912, EPI_ISL_2142913, EPI_ISL_2142914, EPI_ISL_2142915, EPI_ISL_2142916, EPI_ISL_2142917, EPI_ISL_2142918, EPI_ISL_2142919, EPI_ISL_2142920, EPI_ISL_2142921, EPI_ISL_2142922, EPI_ISL_2142923, EPI_ISL_2142924, EPI_ISL_2142925, EPI_ISL_2142926, EPI_ISL_2142927, EPI_ISL_2142928, EPI_ISL_2142929, EPI_ISL_2142930, EPI_ISL_2142931, EPI_ISL_2142932, EPI_ISL_2142933, EPI_ISL_2142934, EPI_ISL_2142935, EPI_ISL_2142936, EPI_ISL_2142937, EPI_ISL_2142938, EPI_ISL_2142939, EPI_ISL_2142940, EPI_ISL_2142941, EPI_ISL_2142942, EPI_ISL_2142943, EPI_ISL_2142944, EPI_ISL_2142945, EPI_ISL_2142946, EPI_ISL_2142947, EPI_ISL_2142948, EPI_ISL_2142949, EPI_ISL_2142950, EPI_ISL_2142951, EPI_ISL_2142952, EPI_ISL_2142953, EPI_ISL_2142954, EPI_ISL_2142955, EPI_ISL_2142956, EPI_ISL_2142957, EPI_ISL_2142958, EPI_ISL_2142959, EPI_ISL_2142960, EPI_ISL_2142961, EPI_ISL_2142962, EPI_ISL_2142963, EPI_ISL_2142964, EPI_ISL_2142965, EPI_ISL_2142966, EPI_ISL_2142967, EPI_ISL_2142968, EPI_ISL_2142969, EPI_ISL_2142970, EPI_ISL_2142971, EPI_ISL_2142972, EPI_ISL_2142973, EPI_ISL_2142974, EPI_ISL_2142975, EPI_ISL_2142976, EPI_ISL_2142977, EPI_ISL_2142978, EPI_ISL_2142979, EPI_ISL_2142980, EPI_ISL_2142981, EPI_ISL_2142982, EPI_ISL_2142983, EPI_ISL_2142984, EPI_ISL_2142985, EPI_ISL_2142986, EPI_ISL_2142987, EPI_ISL_2142988, EPI_ISL_2142989, EPI_ISL_2142990, EPI_ISL_2142991, EPI_ISL_2142992, EPI_ISL_2142993, EPI_ISL_2142994, EPI_ISL_2142995, EPI_ISL_2142996, EPI_ISL_2142997, EPI_ISL_2142998, EPI_ISL_2142999, EPI_ISL_2143000, EPI_ISL_2143001, EPI_ISL_2143002, EPI_ISL_2143003, EPI_ISL_2143004, EPI_ISL_2143005, EPI_ISL_2143006, EPI_ISL_2143007, EPI_ISL_2143008, EPI_ISL_2143009, EPI_ISL_2143010, EPI_ISL_2143011, EPI_ISL_2143012, EPI_ISL_2143013, EPI_ISL_2143014, EPI_ISL_2143015, EPI_ISL_2143016, EPI_ISL_2143017, EPI_ISL_2143018, EPI_ISL_2143019, EPI_ISL_2143020, EPI_ISL_2143021, EPI_ISL_2143022, EPI_ISL_2143023, EPI_ISL_2143024, EPI_ISL_2143025, EPI_ISL_2143026, EPI_ISL_2143027, EPI_ISL_2143028, EPI_ISL_2143029, EPI_ISL_2143030, EPI_ISL_2143031, EPI_ISL_2143032, EPI_ISL_2143033, EPI_ISL_2143034, EPI_ISL_2143035, EPI_ISL_2143036, EPI_ISL_2143037, EPI_ISL_2143038, EPI_ISL_2143039, EPI_ISL_2143040, EPI_ISL_2143041, EPI_ISL_2143042, EPI_ISL_2143043, EPI_ISL_2143044, EPI_ISL_2143045, EPI_ISL_2143046, EPI_ISL_2143047, EPI_ISL_2143048, EPI_ISL_2143049, EPI_ISL_2143050, EPI_ISL_2143051, EPI_ISL_2143052, EPI_ISL_2143053, EPI_ISL_2143054, EPI_ISL_2143055, EPI_ISL_2143056, EPI_ISL_2143057, EPI_ISL_2143058, EPI_ISL_2143059, EPI_ISL_2143060, EPI_ISL_2143061, EPI_ISL_2143062, EPI_ISL_2143063, EPI_ISL_2143064, EPI_ISL_2143065, EPI_ISL_2143066, EPI_ISL_2143067, EPI_ISL_2143068, EPI_ISL_2143069, EPI_ISL_2143070, EPI_ISL_2143071, EPI_ISL_2143072, EPI_ISL_2143073, EPI_ISL_2143074, EPI_ISL_2143075, EPI_ISL_2143076, EPI_ISL_2143077, EPI_ISL_2143078, EPI_ISL_2143079, EPI_ISL_2143080, EPI_ISL_2143081, EPI_ISL_2143082, EPI_ISL_2143083, EPI_ISL_2143084, EPI_ISL_2143085, EPI_ISL_2143086, EPI_ISL_2143087, EPI_ISL_2143088, EPI_ISL_2143089, EPI_ISL_2143090, EPI_ISL_2143091, EPI_ISL_2143092, EPI_ISL_2143093, EPI_ISL_2143094, EPI_ISL_2143095, EPI_ISL_2143096, EPI_ISL_2143097, EPI_ISL_2143098, EPI_ISL_2143099, EPI_ISL_2143100, EPI_ISL_2143101, EPI_ISL_2143102, EPI_ISL_2143103, EPI_ISL_2143104, EPI_ISL_2143105, EPI_ISL_2143106, EPI_ISL_2143107, EPI_ISL_2143108, EPI_ISL_2143109, EPI_ISL_2143110, EPI_ISL_2143111, EPI_ISL_2143112, EPI_ISL_2143113, EPI_ISL_2143114, EPI_ISL_2143115, EPI_ISL_2143116, EPI_ISL_2143117, EPI_ISL_2143118, EPI_ISL_2143119, EPI_ISL_2143120, EPI_ISL_2143121, EPI_ISL_2143122, EPI_ISL_2143123, EPI_ISL_2143124, EPI_ISL_2143125, EPI_ISL_2143126, EPI_ISL_2143127, EPI_ISL_2143128, EPI_ISL_2143129, EPI_ISL_2143130, EPI_ISL_2143131, EPI_ISL_2143132, EPI_ISL_2143133, EPI_ISL_2143134, EPI_ISL_2143135, EPI_ISL_2143136, EPI_ISL_2143137, EPI_ISL_2143138, EPI_ISL_2143139, EPI_ISL_2143140, EPI_ISL_2143141, EPI_ISL_2143142, EPI_ISL_2143143, EPI_ISL_2143144, EPI_ISL_2143145, EPI_ISL_2143146, EPI_ISL_2143147, EPI_ISL_2143148, EPI_ISL_2143149, EPI_ISL_2143150, EPI_ISL_2143151, EPI_ISL_2143152, EPI_ISL_2143153, EPI_ISL_2143154, EPI_ISL_2143155, EPI_ISL_2143156, EPI_ISL_2143157, EPI_ISL_2143158, EPI_ISL_2143159, EPI_ISL_2143160, EPI_ISL_2143161, EPI_ISL_2143162, EPI_ISL_2143163, EPI_ISL_2143164, EPI_ISL_2143165, EPI_ISL_2143166, EPI_ISL_2143167, EPI_ISL_2143168, EPI_ISL_2143169, EPI_ISL_2143170, EPI_ISL_2143171, EPI_ISL_2143172, EPI_ISL_2143173, EPI_ISL_2143174, EPI_ISL_2143175, EPI_ISL_2143176, EPI_ISL_2143177, EPI_ISL_2143178, EPI_ISL_2143179, EPI_ISL_2143180, EPI_ISL_2143181, EPI_ISL_2143182, EPI_ISL_2143183, EPI_ISL_2143184, EPI_ISL_2143185, EPI_ISL_2143186, EPI_ISL_2143187, EPI_ISL_2143188, EPI_ISL_2143189, EPI_ISL_2143190, EPI_ISL_2143191, EPI_ISL_2143192, EPI_ISL_2143193, EPI_ISL_2143194, EPI_ISL_2143195, EPI_ISL_2143196, EPI_ISL_2143197, EPI_ISL_2143198, EPI_ISL_2143199, EPI_ISL_2143200, EPI_ISL_2143201, EPI_ISL_2143202, EPI_ISL_2143203, EPI_ISL_2143204, EPI_ISL_2143205, EPI_ISL_2143206, EPI_ISL_2143207, EPI_ISL_2143208, EPI_ISL_2143209, EPI_ISL_2143210, EPI_ISL_2143211, EPI_ISL_2143212, EPI_ISL_2143213, EPI_ISL_2143214, EPI_ISL_2143215, EPI_ISL_2143216, EPI_ISL_2143217, EPI_ISL_2143218, EPI_ISL_2143219, EPI_ISL_2143220, EPI_ISL_2143221, EPI_ISL_2143222, EPI_ISL_2143223, EPI_ISL_2143224, EPI_ISL_2143225, EPI_ISL_2143226, EPI_ISL_2143227, EPI_ISL_2143228, EPI_ISL_2143229, EPI_ISL_2143230, EPI_ISL_2143231, EPI_ISL_2143232, EPI_ISL_2143233, EPI_ISL_2143234, EPI_ISL_2143235, EPI_ISL_2143236, EPI_ISL_2143237, EPI_ISL_2143238, EPI_ISL_2143239, EPI_ISL_2143240, EPI_ISL_2143241, EPI_ISL_2143242, EPI_ISL_2143243, EPI_ISL_2143244, EPI_ISL_2143245, EPI_ISL_2143246, EPI_ISL_2143247, EPI_ISL_2143248, EPI_ISL_2143249, EPI_ISL_2143250, EPI_ISL_2143251, EPI_ISL_2143252, EPI_ISL_2143253, EPI_ISL_2143254, EPI_ISL_2143255, EPI_ISL_2143256, EPI_ISL_2143257, EPI_ISL_2143258, EPI_ISL_2143259, EPI_ISL_2143260, EPI_ISL_2143261, EPI_ISL_2143262, EPI_ISL_2143263, EPI_ISL_2143264, EPI_ISL_2143265, EPI_ISL_2143266, EPI_ISL_2143267, EPI_ISL_2143268, EPI_ISL_2143269, EPI_ISL_2143270, EPI_ISL_2143271, EPI_ISL_2143272, EPI_ISL_2143273, EPI_ISL_2143274, EPI_ISL_2143275, EPI_ISL_2143276, EPI_ISL_2143277, EPI_ISL_2143278, EPI_ISL_2143279, EPI_ISL_2143280, EPI_ISL_2143281, EPI_ISL_2143282, EPI_ISL_2143283, EPI_ISL_2143284, EPI_ISL_2143285, EPI_ISL_2143286, EPI_ISL_2143287, EPI_ISL_2143288, EPI_ISL_2143289, EPI_ISL_2143290, EPI_ISL_2143291, EPI_ISL_2143292, EPI_ISL_2143293, EPI_ISL_2143294, EPI_ISL_2143295, EPI_ISL_2143296, EPI_ISL_2143297, EPI_ISL_2143298, EPI_ISL_2143299, EPI_ISL_2143300, EPI_ISL_2143301, EPI_ISL_2143302, EPI_ISL_2143303, EPI_ISL_2143304, EPI_ISL_2143305, EPI_ISL_2143306, EPI_ISL_2143307, EPI_ISL_2143308, EPI_ISL_2143309, EPI_ISL_2143310, EPI_ISL_2143311, EPI_ISL_2143312, EPI_ISL_2143313, EPI_ISL_2143314, EPI_ISL_2143315, EPI_ISL_2143316, EPI_ISL_2143317, EPI_ISL_2143318, EPI_ISL_2143319, EPI_ISL_2143320, EPI_ISL_2143321, EPI_ISL_2143322, EPI_ISL_2143323, EPI_ISL_2143324, EPI_ISL_2143325, EPI_ISL_2143326, EPI_ISL_2143327, EPI_ISL_2143328, EPI_ISL_2143329, EPI_ISL_2143330, EPI_ISL_2143331, EPI_ISL_2143332, EPI_ISL_2143333, EPI_ISL_2143334, EPI_ISL_2143335, EPI_ISL_2143336, EPI_ISL_2143337, EPI_ISL_2143338, EPI_ISL_2143339, EPI_ISL_2143340, EPI_ISL_2143341, EPI_ISL_2143342, EPI_ISL_2143343, EPI_ISL_2143344, EPI_ISL_2143345, EPI_ISL_2143346, EPI_ISL_2143347, EPI_ISL_2143348, EPI_ISL_2143349, EPI_ISL_2143350, EPI_ISL_2143351, EPI_ISL_2143352, EPI_ISL_2143353, EPI_ISL_2143354, EPI_ISL_2143355, EPI_ISL_2143356, EPI_ISL_2143357, EPI_ISL_2143358, EPI_ISL_2143359, EPI_ISL_2143360, EPI_ISL_2143361, EPI_ISL_2143362, EPI_ISL_2143363, EPI_ISL_2143364, EPI_ISL_2143365, EPI_ISL_2143366, EPI_ISL_2143367, EPI_ISL_2143368, EPI_ISL_2143369, EPI_ISL_2143370, EPI_ISL_2143371, EPI_ISL_2143372, EPI_ISL_2143373, EPI_ISL_2143374, EPI_ISL_2143375, EPI_ISL_2143376, EPI_ISL_2143377, EPI_ISL_2143378, EPI_ISL_2143379, EPI_ISL_2143380, EPI_ISL_2143381, EPI_ISL_2143382, EPI_ISL_2143383, EPI_ISL_2143384, EPI_ISL_2143387, EPI_ISL_2143389, EPI_ISL_2143392, EPI_ISL_2143394, EPI_ISL_2143397, EPI_ISL_2143399, EPI_ISL_2143402, EPI_ISL_2143405, EPI_ISL_2143407, EPI_ISL_2143410, EPI_ISL_2143412, EPI_ISL_2143414, EPI_ISL_2143415, EPI_ISL_2143418, EPI_ISL_2143420, EPI_ISL_2143423, EPI_ISL_2143425, EPI_ISL_2143428, EPI_ISL_2143431, EPI_ISL_2143434, EPI_ISL_2143436, EPI_ISL_2143438, EPI_ISL_2143441, EPI_ISL_2143444, EPI_ISL_2143447, EPI_ISL_2143449, EPI_ISL_2143452, EPI_ISL_2143455, EPI_ISL_2143458, EPI_ISL_2143461, EPI_ISL_2143463, EPI_ISL_2143465, EPI_ISL_2143468, EPI_ISL_2143470, EPI_ISL_2143473, EPI_ISL_2143476, EPI_ISL_2143478, EPI_ISL_2143481, EPI_ISL_2143484, EPI_ISL_2143488, EPI_ISL_2143491, EPI_ISL_2143496, EPI_ISL_2143500, EPI_ISL_2143505, EPI_ISL_2143509, EPI_ISL_2143514, EPI_ISL_2143518, EPI_ISL_2143522, EPI_ISL_2143527, EPI_ISL_2143530, EPI_ISL_2143535, EPI_ISL_2143537, EPI_ISL_2143542, EPI_ISL_2143545, EPI_ISL_2143549, EPI_ISL_2143552, EPI_ISL_2143557, EPI_ISL_2143562, EPI_ISL_2143574, EPI_ISL_2143627, EPI_ISL_2143635, EPI_ISL_2143658, EPI_ISL_2143665, EPI_ISL_2143667, EPI_ISL_2143670, EPI_ISL_2143673, EPI_ISL_2143675, EPI_ISL_2143677, EPI_ISL_2143679, EPI_ISL_2143682, EPI_ISL_2143684, EPI_ISL_2143686, EPI_ISL_2143689, EPI_ISL_2143691, EPI_ISL_2143695, EPI_ISL_2143697, EPI_ISL_2143699, EPI_ISL_2143700, EPI_ISL_2143702, EPI_ISL_2143705, EPI_ISL_2143708, EPI_ISL_2143710, EPI_ISL_2143713, EPI_ISL_2143716, EPI_ISL_2143718, EPI_ISL_2143721, EPI_ISL_2143723, EPI_ISL_2143726, EPI_ISL_2143728, EPI_ISL_2143730, EPI_ISL_2143733, EPI_ISL_2143736, EPI_ISL_2143738, EPI_ISL_2143741, EPI_ISL_2143743, EPI_ISL_2143745, EPI_ISL_2143748, EPI_ISL_2143750, EPI_ISL_2143753, EPI_ISL_2143755, EPI_ISL_2143758, EPI_ISL_2143760, EPI_ISL_2143763, EPI_ISL_2143765, EPI_ISL_2143768, EPI_ISL_2143770, EPI_ISL_2143772, EPI_ISL_2143775, EPI_ISL_2143778, EPI_ISL_2143780, EPI_ISL_2143783, EPI_ISL_2143785, EPI_ISL_2143786, EPI_ISL_2143789, EPI_ISL_2143791, EPI_ISL_2143794, EPI_ISL_2143797, EPI_ISL_2143800, EPI_ISL_2143803, EPI_ISL_2143805, EPI_ISL_2143807, EPI_ISL_2143809, EPI_ISL_2143811, EPI_ISL_2143814, EPI_ISL_2143817, EPI_ISL_2143819, EPI_ISL_2143822, EPI_ISL_2143824, EPI_ISL_2143826, EPI_ISL_2143828, EPI_ISL_2143832, EPI_ISL_2143834, EPI_ISL_2143837, EPI_ISL_2143839, EPI_ISL_2143841, EPI_ISL_2143843, EPI_ISL_2143845, EPI_ISL_2143848, EPI_ISL_2143850, EPI_ISL_2143853, EPI_ISL_2143856, EPI_ISL_2143858, EPI_ISL_2143861, EPI_ISL_2143862, EPI_ISL_2143863, EPI_ISL_2143865, EPI_ISL_2143866, EPI_ISL_2143869, EPI_ISL_2143871, EPI_ISL_2143873, EPI_ISL_2143875, EPI_ISL_2143878, EPI_ISL_2143880, EPI_ISL_2143883, EPI_ISL_2143885, EPI_ISL_2143887, EPI_ISL_2143889, EPI_ISL_2143892, EPI_ISL_2143894, EPI_ISL_2143897, EPI_ISL_2143899, EPI_ISL_2143901, EPI_ISL_2143904, EPI_ISL_2143906, EPI_ISL_2143908, EPI_ISL_2143911, EPI_ISL_2143914, EPI_ISL_2143916, EPI_ISL_2143919, EPI_ISL_2143921, EPI_ISL_2143923, EPI_ISL_2143926, EPI_ISL_2143929, EPI_ISL_2143931, EPI_ISL_2143934, EPI_ISL_2143936, EPI_ISL_2143938, EPI_ISL_2143941, EPI_ISL_2143944, EPI_ISL_2143946, EPI_ISL_2143948, EPI_ISL_2143951, EPI_ISL_2143953, EPI_ISL_2143956, EPI_ISL_2143958, EPI_ISL_2143961, EPI_ISL_2143963, EPI_ISL_2143966, EPI_ISL_2143968, EPI_ISL_2143971, EPI_ISL_2143973, EPI_ISL_2143976, EPI_ISL_2143978, EPI_ISL_2143980, EPI_ISL_2143983, EPI_ISL_2143986, EPI_ISL_2143988, EPI_ISL_2143990, EPI_ISL_2143993, EPI_ISL_2143995, EPI_ISL_2143997, EPI_ISL_2144000, EPI_ISL_2144002, EPI_ISL_2144004, EPI_ISL_2144006, EPI_ISL_2144009, EPI_ISL_2144011, EPI_ISL_2144014, EPI_ISL_2144016 | | | |
| see above | Quest Diagnostics Incorporated | Centers for Disease Control and Prevention Division of Viral Diseases, Pathogen Discovery | Dakota Howard, Dhwani Batra, Peter W. Cook, Kara Moser, Adrian Paskey, Jason Caravas, Benjamin Rambo-Martin, Shatavia Morrison, Christopher Gulvick, Scott Sammons, Yvette Unoarumhi, Darlene Wagner, Matthew Schmerer, S. H. Rosenthal, A. Gerasimova, R. M. Kagan, B. Anderson, M. Hua, Y. Liu, L.E. Bernstein, K.E. Livingston, A. Perez, I. A. Shlyakhter, R. V. Rolando, R. Owen, P. Tanpaiboon, F. Lacbawan, Clinton R. Paden, Duncan MacCannell |
| EPI_ISL_2144050, EPI_ISL_2144053, EPI_ISL_2144056, EPI_ISL_2144057, EPI_ISL_2144059, EPI_ISL_2144061, EPI_ISL_2144063, EPI_ISL_2144064, EPI_ISL_2144066, EPI_ISL_2144069, EPI_ISL_2144071, EPI_ISL_2144073, EPI_ISL_2144075, EPI_ISL_2144077, EPI_ISL_2144080, EPI_ISL_2144082, EPI_ISL_2144083, EPI_ISL_2144084, EPI_ISL_2144085, EPI_ISL_2144086, EPI_ISL_2144087, EPI_ISL_2144088, EPI_ISL_2144089, EPI_ISL_2144090, EPI_ISL_2144091, EPI_ISL_2144092, EPI_ISL_2144093, EPI_ISL_2144094, EPI_ISL_2144095, EPI_ISL_2144096, EPI_ISL_2144097, EPI_ISL_2144098, EPI_ISL_2144099, EPI_ISL_2144100, EPI_ISL_2144101, EPI_ISL_2144102, EPI_ISL_2144103, EPI_ISL_2144104, EPI_ISL_2144105, EPI_ISL_2144106, EPI_ISL_2144107, EPI_ISL_2144108, EPI_ISL_2144109, EPI_ISL_2144110, EPI_ISL_2144111, EPI_ISL_2144113, EPI_ISL_2144114, EPI_ISL_2144115, EPI_ISL_2144116, EPI_ISL_2144117, EPI_ISL_2144118, EPI_ISL_2144119, EPI_ISL_2144120, EPI_ISL_2144121, EPI_ISL_2144122, EPI_ISL_2144123, EPI_ISL_2144124, EPI_ISL_2144125, EPI_ISL_2144126, EPI_ISL_2144128, EPI_ISL_2144129, EPI_ISL_2144130, EPI_ISL_2144131, EPI_ISL_2144132, EPI_ISL_2144133, EPI_ISL_2144134, EPI_ISL_2144135, EPI_ISL_2144136, EPI_ISL_2144137, EPI_ISL_2144138, EPI_ISL_2144139, EPI_ISL_2144140, EPI_ISL_2144141, EPI_ISL_2144142, EPI_ISL_2144143, EPI_ISL_2144144, EPI_ISL_2144145, EPI_ISL_2144146, EPI_ISL_2144147, EPI_ISL_2144148, EPI_ISL_2144149, EPI_ISL_2144150, EPI_ISL_2144151, EPI_ISL_2144153, EPI_ISL_2144154, EPI_ISL_2144155, EPI_ISL_2144156, EPI_ISL_2144157, EPI_ISL_2144158, EPI_ISL_2144159, EPI_ISL_2144160, EPI_ISL_2144161, EPI_ISL_2144162, EPI_ISL_2144163, EPI_ISL_2144164, EPI_ISL_2144165, EPI_ISL_2144166, EPI_ISL_2144168, EPI_ISL_2144169, EPI_ISL_2144170, EPI_ISL_2144171, EPI_ISL_2144172, EPI_ISL_2144173, EPI_ISL_2144174, EPI_ISL_2144175, EPI_ISL_2144176, EPI_ISL_2144177, EPI_ISL_2144178, EPI_ISL_2144179, EPI_ISL_2144180, EPI_ISL_2144181, EPI_ISL_2144182, EPI_ISL_2144183, EPI_ISL_2144184, EPI_ISL_2144185, EPI_ISL_2144186, EPI_ISL_2144187, EPI_ISL_2144188, EPI_ISL_2144189, EPI_ISL_2144190, EPI_ISL_2144191, EPI_ISL_2144192, EPI_ISL_2144193, EPI_ISL_2144194, EPI_ISL_2144195, EPI_ISL_2144196, EPI_ISL_2144197, EPI_ISL_2144198, EPI_ISL_2144199, EPI_ISL_2144200, EPI_ISL_2144201, EPI_ISL_2144202, EPI_ISL_2144203, EPI_ISL_2144204, EPI_ISL_2144205, EPI_ISL_2144206, EPI_ISL_2144207, EPI_ISL_2144208, EPI_ISL_2144209, EPI_ISL_2144210, EPI_ISL_2144211, EPI_ISL_2144212, EPI_ISL_2144213, EPI_ISL_2144214, EPI_ISL_2144215, EPI_ISL_2144216, EPI_ISL_2144217, EPI_ISL_2144218, EPI_ISL_2144219, EPI_ISL_2144220, EPI_ISL_2144221, EPI_ISL_2144222, EPI_ISL_2144223, EPI_ISL_2144224, EPI_ISL_2144225, EPI_ISL_2144226, EPI_ISL_2144227, EPI_ISL_2144228, EPI_ISL_2144229, EPI_ISL_2144230, EPI_ISL_2144231, EPI_ISL_2144232, EPI_ISL_2144233, EPI_ISL_2144234, EPI_ISL_2144235, EPI_ISL_2144236, EPI_ISL_2144237, EPI_ISL_2144238, EPI_ISL_2144239, EPI_ISL_2144240, EPI_ISL_2144241, EPI_ISL_2144242, EPI_ISL_2144243, EPI_ISL_2144244, EPI_ISL_2144245, EPI_ISL_2144246, EPI_ISL_2144247, EPI_ISL_2144248, EPI_ISL_2144249, EPI_ISL_2144250, EPI_ISL_2144251, EPI_ISL_2144252, EPI_ISL_2144253, EPI_ISL_2144254, EPI_ISL_2144255, EPI_ISL_2144256, EPI_ISL_2144258, EPI_ISL_2144261, EPI_ISL_2144264, EPI_ISL_2144267, EPI_ISL_2144270, EPI_ISL_2144272, EPI_ISL_2144273, EPI_ISL_2144274, EPI_ISL_2144275, EPI_ISL_2144276, EPI_ISL_2144277, EPI_ISL_2144278, EPI_ISL_2144279, EPI_ISL_2144280, EPI_ISL_2144281, EPI_ISL_2144282, EPI_ISL_2144283, EPI_ISL_2144284, EPI_ISL_2144285, EPI_ISL_2144286, EPI_ISL_2144287, EPI_ISL_2144288, EPI_ISL_2144289, EPI_ISL_2144290, EPI_ISL_2144291, EPI_ISL_2144292, EPI_ISL_2144293, EPI_ISL_2144294, EPI_ISL_2144295, EPI_ISL_2144296, EPI_ISL_2144297, EPI_ISL_2144298, EPI_ISL_2144299, EPI_ISL_2144300, EPI_ISL_2144301, EPI_ISL_2144302, EPI_ISL_2144303, EPI_ISL_2144304, EPI_ISL_2144305, EPI_ISL_2144306, EPI_ISL_2144307, EPI_ISL_2144308, EPI_ISL_2144309, EPI_ISL_2144310, EPI_ISL_2144311, EPI_ISL_2144312, EPI_ISL_2144313, EPI_ISL_2144314, EPI_ISL_2144315, EPI_ISL_2144316, EPI_ISL_2144317, EPI_ISL_2144318, EPI_ISL_2144319, EPI_ISL_2144320, EPI_ISL_2144321, EPI_ISL_2144322, EPI_ISL_2144323, EPI_ISL_2144324, EPI_ISL_2144325, EPI_ISL_2144326, EPI_ISL_2144327, EPI_ISL_2144328, EPI_ISL_2144329, EPI_ISL_2144330, EPI_ISL_2144331, EPI_ISL_2144332, EPI_ISL_2144333, EPI_ISL_2144334, EPI_ISL_2144335, EPI_ISL_2144336, EPI_ISL_2144337, EPI_ISL_2144338, EPI_ISL_2144339, EPI_ISL_2144340, EPI_ISL_2144341, EPI_ISL_2144342, EPI_ISL_2144343, EPI_ISL_2144344, EPI_ISL_2144345, EPI_ISL_2144346, EPI_ISL_2144347, EPI_ISL_2144348, EPI_ISL_2144349, EPI_ISL_2144350, EPI_ISL_2144351, EPI_ISL_2144352, EPI_ISL_2144353, EPI_ISL_2144354, EPI_ISL_2144355, EPI_ISL_2144356, EPI_ISL_2144357, EPI_ISL_2144358, EPI_ISL_2144359, EPI_ISL_2144360, EPI_ISL_2144361, EPI_ISL_2144362, EPI_ISL_2144363, EPI_ISL_2144364, EPI_ISL_2144365, EPI_ISL_2144366, EPI_ISL_2144367, EPI_ISL_2144368, EPI_ISL_2144369, EPI_ISL_2144370, EPI_ISL_2144371, EPI_ISL_2144372, EPI_ISL_2144373, EPI_ISL_2144374, EPI_ISL_2144375, EPI_ISL_2144376, EPI_ISL_2144377, EPI_ISL_2144378, EPI_ISL_2144379, EPI_ISL_2144380, EPI_ISL_2144381, EPI_ISL_2144382, EPI_ISL_2144383, EPI_ISL_2144384, EPI_ISL_2144385, EPI_ISL_2144386, EPI_ISL_2144387, EPI_ISL_2144388, EPI_ISL_2144389, EPI_ISL_2144390, EPI_ISL_2144391, EPI_ISL_2144392, EPI_ISL_2144393, EPI_ISL_2144394, EPI_ISL_2144395, EPI_ISL_2144396, EPI_ISL_2144397, EPI_ISL_2144398, EPI_ISL_2144399, EPI_ISL_2144400, EPI_ISL_2144401, EPI_ISL_2144402, EPI_ISL_2144403, EPI_ISL_2144404, EPI_ISL_2144405, EPI_ISL_2144406, EPI_ISL_2144407, EPI_ISL_2144408, EPI_ISL_2144409, EPI_ISL_2144410, EPI_ISL_2144411, EPI_ISL_2144412, EPI_ISL_2144413, EPI_ISL_2144414, EPI_ISL_2144415, EPI_ISL_2144416, EPI_ISL_2144417, EPI_ISL_2144418, EPI_ISL_2144419, EPI_ISL_2144420, EPI_ISL_2144421, EPI_ISL_2144422, EPI_ISL_2144423, EPI_ISL_2144424, EPI_ISL_2144425, EPI_ISL_2144426, EPI_ISL_2144427, EPI_ISL_2144428, EPI_ISL_2144429, EPI_ISL_2144430, EPI_ISL_2144431, EPI_ISL_2144432, EPI_ISL_2144433, EPI_ISL_2144434, EPI_ISL_2144435, EPI_ISL_2144436, EPI_ISL_2144437, EPI_ISL_2144438, EPI_ISL_2144439, EPI_ISL_2144440, EPI_ISL_2144441, EPI_ISL_2144442, EPI_ISL_2144443, EPI_ISL_2144444, EPI_ISL_2144445, EPI_ISL_2144446, EPI_ISL_2144447, EPI_ISL_2144448, EPI_ISL_2144449, EPI_ISL_2144450, EPI_ISL_2144451, EPI_ISL_2144452, EPI_ISL_2144453, EPI_ISL_2144454, EPI_ISL_2144455, EPI_ISL_2144456, EPI_ISL_2144457, EPI_ISL_2144458, EPI_ISL_2144459, EPI_ISL_2144460, EPI_ISL_2144461, EPI_ISL_2144462, EPI_ISL_2144463, EPI_ISL_2144464, EPI_ISL_2144465, EPI_ISL_2144466, EPI_ISL_2144467, EPI_ISL_2144468, EPI_ISL_2144469, EPI_ISL_2144470, EPI_ISL_2144471, EPI_ISL_2144472, EPI_ISL_2144473, EPI_ISL_2144474, EPI_ISL_2144475, EPI_ISL_2144476, EPI_ISL_2144477, EPI_ISL_2144478, EPI_ISL_2144479, EPI_ISL_2144480, EPI_ISL_2144481, EPI_ISL_2144482, EPI_ISL_2144483, EPI_ISL_2144484, EPI_ISL_2144485, EPI_ISL_2144486, EPI_ISL_2144487, EPI_ISL_2144488, EPI_ISL_2144489, EPI_ISL_2144490, EPI_ISL_2144491, EPI_ISL_2144492, EPI_ISL_2144493, EPI_ISL_2144494, EPI_ISL_2144495, EPI_ISL_2144496, EPI_ISL_2144497, EPI_ISL_2144498, EPI_ISL_2144499, EPI_ISL_2144500, EPI_ISL_2144501, EPI_ISL_2144502, EPI_ISL_2144503, EPI_ISL_2144504, EPI_ISL_2144505, EPI_ISL_2144506, EPI_ISL_2144507, EPI_ISL_2144508, EPI_ISL_2144509, EPI_ISL_2144510, EPI_ISL_2144511, EPI_ISL_2144512, EPI_ISL_2144513, EPI_ISL_2144514, EPI_ISL_2144515, EPI_ISL_2144516, EPI_ISL_2144517, EPI_ISL_2144518, EPI_ISL_2144519, EPI_ISL_2144520, EPI_ISL_2144521, EPI_ISL_2144522, EPI_ISL_2144523, EPI_ISL_2144524, EPI_ISL_2144525, EPI_ISL_2144526, EPI_ISL_2144527, EPI_ISL_2144528, EPI_ISL_2144529, EPI_ISL_2144530, EPI_ISL_2144531, EPI_ISL_2144532, EPI_ISL_2144533, EPI_ISL_2144534, EPI_ISL_2144535, EPI_ISL_2144537, EPI_ISL_2144538, EPI_ISL_2144539, EPI_ISL_2144540, EPI_ISL_2144541, EPI_ISL_2144542, EPI_ISL_2144543, | | | |

| | | | |
|---|---|---|---|
| EPI_ISL_2144545, EPI_ISL_2144546, EPI_ISL_2144547, EPI_ISL_2144548, EPI_ISL_2144549, EPI_ISL_2144551, EPI_ISL_2144552, EPI_ISL_2144553, EPI_ISL_2144554, EPI_ISL_2144555, EPI_ISL_2144556, EPI_ISL_2144557, EPI_ISL_2144558, EPI_ISL_2144560, EPI_ISL_2144561, EPI_ISL_2144562, EPI_ISL_2144563, EPI_ISL_2144564, EPI_ISL_2144565, EPI_ISL_2144566, EPI_ISL_2144567, EPI_ISL_2144568, EPI_ISL_2144569, EPI_ISL_2144570, EPI_ISL_2144571, EPI_ISL_2144572, EPI_ISL_2144573, EPI_ISL_2144574, EPI_ISL_2144575, EPI_ISL_2144576, EPI_ISL_2144577, EPI_ISL_2144578, EPI_ISL_2144579, EPI_ISL_2144580, EPI_ISL_2144581, EPI_ISL_2144582, EPI_ISL_2144583, EPI_ISL_2144584, EPI_ISL_2144585, EPI_ISL_2144586, EPI_ISL_2144587, EPI_ISL_2144588, EPI_ISL_2144589, EPI_ISL_2144590, EPI_ISL_2144591, EPI_ISL_2144592, EPI_ISL_2144593, EPI_ISL_2144594, EPI_ISL_2144595, EPI_ISL_2144596, EPI_ISL_2144597, EPI_ISL_2144598, EPI_ISL_2144599, EPI_ISL_2144600, EPI_ISL_2144601, EPI_ISL_2144602, EPI_ISL_2144603, EPI_ISL_2144604, EPI_ISL_2144605, EPI_ISL_2144606, EPI_ISL_2144607, EPI_ISL_2144608, EPI_ISL_2144609, EPI_ISL_2144610, EPI_ISL_2144611, EPI_ISL_2144612, EPI_ISL_2144613, EPI_ISL_2144614, EPI_ISL_2144615, EPI_ISL_2144616, EPI_ISL_2144617, EPI_ISL_2144618, EPI_ISL_2144619, EPI_ISL_2144620, EPI_ISL_2144621, EPI_ISL_2144622, EPI_ISL_2144623, EPI_ISL_2144624, EPI_ISL_2144625, EPI_ISL_2144626, EPI_ISL_2144627, EPI_ISL_2144628, EPI_ISL_2144629, EPI_ISL_2144630, EPI_ISL_2144631, EPI_ISL_2144632, EPI_ISL_2144633, EPI_ISL_2144634, EPI_ISL_2144635, EPI_ISL_2144636, EPI_ISL_2144637, EPI_ISL_2144638, EPI_ISL_2144639, EPI_ISL_2144640, EPI_ISL_2144641, EPI_ISL_2144642, EPI_ISL_2144643, EPI_ISL_2144644, EPI_ISL_2144645, EPI_ISL_2144646, EPI_ISL_2144647, EPI_ISL_2144648, EPI_ISL_2144649, EPI_ISL_2144650, EPI_ISL_2144651, EPI_ISL_2144652, EPI_ISL_2144653, EPI_ISL_2144654, EPI_ISL_2144655, EPI_ISL_2144656, EPI_ISL_2144657, EPI_ISL_2144658, EPI_ISL_2144659, EPI_ISL_2144660, EPI_ISL_2144661, EPI_ISL_2144662, EPI_ISL_2144663, EPI_ISL_2144664, EPI_ISL_2144665, EPI_ISL_2144666, EPI_ISL_2144667, EPI_ISL_2144668, EPI_ISL_2144669, EPI_ISL_2144670, EPI_ISL_2144671, EPI_ISL_2144672, EPI_ISL_2144673, EPI_ISL_2144674, EPI_ISL_2144675, EPI_ISL_2144676, EPI_ISL_2144677, EPI_ISL_2144678, EPI_ISL_2144679, EPI_ISL_2144680, EPI_ISL_2144681, EPI_ISL_2144682, EPI_ISL_2144683, EPI_ISL_2144684, EPI_ISL_2144685, EPI_ISL_2144686, EPI_ISL_2144687, EPI_ISL_2144688, EPI_ISL_2144689, EPI_ISL_2144690, EPI_ISL_2144691, EPI_ISL_2144692, EPI_ISL_2144693, EPI_ISL_2144694, EPI_ISL_2144695, EPI_ISL_2144696, EPI_ISL_2144697, EPI_ISL_2144698, EPI_ISL_2144699, EPI_ISL_2144700, EPI_ISL_2144701, EPI_ISL_2144702, EPI_ISL_2144703, EPI_ISL_2144704, EPI_ISL_2144705, EPI_ISL_2144706, EPI_ISL_2144707, EPI_ISL_2144708, EPI_ISL_2144710, EPI_ISL_2144711, EPI_ISL_2144712, EPI_ISL_2144713, EPI_ISL_2144714, EPI_ISL_2144715, EPI_ISL_2144716, EPI_ISL_2144717, EPI_ISL_2144718, EPI_ISL_2144719, EPI_ISL_2144720, EPI_ISL_2144722, EPI_ISL_2144723, EPI_ISL_2144724, EPI_ISL_2144725, EPI_ISL_2144726, EPI_ISL_2144727, EPI_ISL_2144728, EPI_ISL_2144729, EPI_ISL_2144730, EPI_ISL_2144731, EPI_ISL_2144732, EPI_ISL_2144733, EPI_ISL_2144734, EPI_ISL_2144735, EPI_ISL_2144736, EPI_ISL_2144737, EPI_ISL_2144738, EPI_ISL_2144739, EPI_ISL_2144740, EPI_ISL_2144741, EPI_ISL_2144742, EPI_ISL_2144744, EPI_ISL_2144745, EPI_ISL_2144746, EPI_ISL_2144747, EPI_ISL_2144748, EPI_ISL_2144749, EPI_ISL_2144750, EPI_ISL_2144751, EPI_ISL_2144752, EPI_ISL_2144753, EPI_ISL_2144754, EPI_ISL_2144755, EPI_ISL_2144756, EPI_ISL_2144757, EPI_ISL_2144758, EPI_ISL_2144759, EPI_ISL_2144760, EPI_ISL_2144761, EPI_ISL_2144762, EPI_ISL_2144763, EPI_ISL_2144764, EPI_ISL_2144765, EPI_ISL_2144766, EPI_ISL_2144767, EPI_ISL_2144768, EPI_ISL_2144769, EPI_ISL_2144770, EPI_ISL_2144771, EPI_ISL_2144772, EPI_ISL_2144773, EPI_ISL_2144774, EPI_ISL_2144775, EPI_ISL_2144776, EPI_ISL_2144777, EPI_ISL_2144778, EPI_ISL_2144779, EPI_ISL_2144780, EPI_ISL_2144781, EPI_ISL_2144782, EPI_ISL_2144783, EPI_ISL_2144784, EPI_ISL_2144785, EPI_ISL_2144786, EPI_ISL_2144787, EPI_ISL_2144788, EPI_ISL_2144789, EPI_ISL_2144790, EPI_ISL_2144791, EPI_ISL_2144792, EPI_ISL_2144793, EPI_ISL_2144794, EPI_ISL_2144795, EPI_ISL_2144796, EPI_ISL_2144797, EPI_ISL_2144798, EPI_ISL_2144799, EPI_ISL_2144800, EPI_ISL_2144801, EPI_ISL_2144802, EPI_ISL_2144803, EPI_ISL_2144804, EPI_ISL_2144805, EPI_ISL_2144806, EPI_ISL_2144807, EPI_ISL_2144809, EPI_ISL_2144811, EPI_ISL_2144812, EPI_ISL_2144815, EPI_ISL_2144817, EPI_ISL_2144820, EPI_ISL_2144822, EPI_ISL_2144824, EPI_ISL_2144827, EPI_ISL_2144830, EPI_ISL_2144833, EPI_ISL_2144835, EPI_ISL_2144838, EPI_ISL_2144840, EPI_ISL_2144843, EPI_ISL_2144845, EPI_ISL_2144847, EPI_ISL_2144850, EPI_ISL_2144853, EPI_ISL_2144854, EPI_ISL_2144857, EPI_ISL_2144859, EPI_ISL_2144862, EPI_ISL_2144865, EPI_ISL_2144868, EPI_ISL_2144871, EPI_ISL_2144873, EPI_ISL_2144876, EPI_ISL_2144879, EPI_ISL_2144882, EPI_ISL_2144885, EPI_ISL_2144888, EPI_ISL_2144890, EPI_ISL_2144893, EPI_ISL_2144896, EPI_ISL_2144899, EPI_ISL_2144901, EPI_ISL_2144902, EPI_ISL_2144905, EPI_ISL_2144907, EPI_ISL_2144909, EPI_ISL_2144912, EPI_ISL_2144915, EPI_ISL_2144918, EPI_ISL_2144920, EPI_ISL_2144922, EPI_ISL_2144923, EPI_ISL_2144926, EPI_ISL_2144927, EPI_ISL_2144928, EPI_ISL_2144929, EPI_ISL_2144930, EPI_ISL_2144931, EPI_ISL_2144932, EPI_ISL_2144933, EPI_ISL_2144934, EPI_ISL_2144935, EPI_ISL_2144936, EPI_ISL_2144937, EPI_ISL_2144938, EPI_ISL_2144939, EPI_ISL_2144940, EPI_ISL_2144941, EPI_ISL_2144943, EPI_ISL_2144944, EPI_ISL_2144945, EPI_ISL_2144946, EPI_ISL_2144947, EPI_ISL_2144948, EPI_ISL_2144949, EPI_ISL_2144950, EPI_ISL_2144951, EPI_ISL_2144952, EPI_ISL_2144953, EPI_ISL_2144954, EPI_ISL_2144955, EPI_ISL_2144956, EPI_ISL_2144957, EPI_ISL_2144958, EPI_ISL_2144959, EPI_ISL_2144960, EPI_ISL_2144961, EPI_ISL_2144962, EPI_ISL_2144963, EPI_ISL_2144964, EPI_ISL_2144965, EPI_ISL_2144966, EPI_ISL_2144967, EPI_ISL_2144968, EPI_ISL_2144969, EPI_ISL_2144970, EPI_ISL_2144971, EPI_ISL_2144972, EPI_ISL_2144973, EPI_ISL_2144974, EPI_ISL_2144975, EPI_ISL_2144976, EPI_ISL_2144977, EPI_ISL_2144978, EPI_ISL_2144979, EPI_ISL_2144980, EPI_ISL_2144981, EPI_ISL_2144982, EPI_ISL_2144983, EPI_ISL_2144984, EPI_ISL_2144986, EPI_ISL_2144987, EPI_ISL_2144988, EPI_ISL_2144989, EPI_ISL_2144990, EPI_ISL_2144991, EPI_ISL_2144992, EPI_ISL_2144993, EPI_ISL_2144994, EPI_ISL_2144995, EPI_ISL_2144996, EPI_ISL_2144997, EPI_ISL_2144998, EPI_ISL_2144999, EPI_ISL_2145000, EPI_ISL_2145001, EPI_ISL_2145002, EPI_ISL_2145003, EPI_ISL_2145004, EPI_ISL_2145005, EPI_ISL_2145006, EPI_ISL_2145007, EPI_ISL_2145008, EPI_ISL_2145009, EPI_ISL_2145010, EPI_ISL_2145011, EPI_ISL_2145012, EPI_ISL_2145013, EPI_ISL_2145014, EPI_ISL_2145015, EPI_ISL_2145016, EPI_ISL_2145017, EPI_ISL_2145018, EPI_ISL_2145019, EPI_ISL_2145020, EPI_ISL_2145021, EPI_ISL_2145023, EPI_ISL_2145024, EPI_ISL_2145025, EPI_ISL_2145026, EPI_ISL_2145027, EPI_ISL_2145028, EPI_ISL_2145029, EPI_ISL_2145030, EPI_ISL_2145031, EPI_ISL_2145032, EPI_ISL_2145033, EPI_ISL_2145034, EPI_ISL_2145035, EPI_ISL_2145036, EPI_ISL_2145037, EPI_ISL_2145038, EPI_ISL_2145039, EPI_ISL_2145040, EPI_ISL_2145041, EPI_ISL_2145042, EPI_ISL_2145043, EPI_ISL_2145044, EPI_ISL_2145045, EPI_ISL_2145046, EPI_ISL_2145047, EPI_ISL_2145048, EPI_ISL_2145049, EPI_ISL_2145050, EPI_ISL_2145051, EPI_ISL_2145052, EPI_ISL_2145053, EPI_ISL_2145054, EPI_ISL_2145055, EPI_ISL_2145056, EPI_ISL_2145057, EPI_ISL_2145058, EPI_ISL_2145059, EPI_ISL_2145060, EPI_ISL_2145061, EPI_ISL_2145062, EPI_ISL_2145063, EPI_ISL_2145064, EPI_ISL_2145065, EPI_ISL_2145066, EPI_ISL_2145067, EPI_ISL_2145068, EPI_ISL_2145069, EPI_ISL_2145070, EPI_ISL_2145071, EPI_ISL_2145072, EPI_ISL_2145073, EPI_ISL_2145074, EPI_ISL_2145075, EPI_ISL_2145076, EPI_ISL_2145077, EPI_ISL_2145078, EPI_ISL_2145079, EPI_ISL_2145080, EPI_ISL_2145081, EPI_ISL_2145082, EPI_ISL_2145083, EPI_ISL_2145084, EPI_ISL_2145085, EPI_ISL_2145086, EPI_ISL_2145087, EPI_ISL_2145090, EPI_ISL_2145092, EPI_ISL_2145095, EPI_ISL_2145097, EPI_ISL_2145100, EPI_ISL_2145102, EPI_ISL_2145105, EPI_ISL_2145108, EPI_ISL_2145110, EPI_ISL_2145113, EPI_ISL_2145116, EPI_ISL_2145119, EPI_ISL_2145122, EPI_ISL_2145125, EPI_ISL_2145127, EPI_ISL_2145129, EPI_ISL_2145132, EPI_ISL_2145135, EPI_ISL_2145137, EPI_ISL_2145139, EPI_ISL_2145142, EPI_ISL_2145144, EPI_ISL_2145149, EPI_ISL_2145151, EPI_ISL_2145154, EPI_ISL_2145156, EPI_ISL_2145159, EPI_ISL_2145162, EPI_ISL_2145166, EPI_ISL_2145169, EPI_ISL_2145171, EPI_ISL_2145174, EPI_ISL_2145176, EPI_ISL_2145179, EPI_ISL_2145182, EPI_ISL_2145185, EPI_ISL_2145188, EPI_ISL_2145190, EPI_ISL_2145193, EPI_ISL_2145196, EPI_ISL_2145198, EPI_ISL_2145201, EPI_ISL_2145203, EPI_ISL_2145206, EPI_ISL_2145209, EPI_ISL_2145211, EPI_ISL_2145214, EPI_ISL_2145217, EPI_ISL_2145219, EPI_ISL_2145222, EPI_ISL_2145223, EPI_ISL_2145226, EPI_ISL_2145229, EPI_ISL_2145231, EPI_ISL_2145233, EPI_ISL_2145236, EPI_ISL_2145238, EPI_ISL_2145241, EPI_ISL_2145243, EPI_ISL_2145246, EPI_ISL_2145248, EPI_ISL_2145252, EPI_ISL_2145255, EPI_ISL_2145257, EPI_ISL_2145260, EPI_ISL_2145263, EPI_ISL_2145265, EPI_ISL_2145268, EPI_ISL_2145270, EPI_ISL_2145272, EPI_ISL_2145274, EPI_ISL_2145276, EPI_ISL_2145279, EPI_ISL_2145284, EPI_ISL_2145286, EPI_ISL_2145289, EPI_ISL_2145292, EPI_ISL_2145296, EPI_ISL_2145298, EPI_ISL_2145301, EPI_ISL_2145303, EPI_ISL_2145306, EPI_ISL_2145309, EPI_ISL_2145311, EPI_ISL_2145314, EPI_ISL_2145317, EPI_ISL_2145320, EPI_ISL_2145323, EPI_ISL_2145325, EPI_ISL_2145328, EPI_ISL_2145330, EPI_ISL_2145335, EPI_ISL_2145337, EPI_ISL_2145339, EPI_ISL_2145342, EPI_ISL_2145344, EPI_ISL_2145347, EPI_ISL_2145349, EPI_ISL_2145352, EPI_ISL_2145355, EPI_ISL_2145357, EPI_ISL_2145360, EPI_ISL_2145362, EPI_ISL_2145365, EPI_ISL_2145367, EPI_ISL_2145369, EPI_ISL_2145372, EPI_ISL_2145375, EPI_ISL_2145377, EPI_ISL_2145380, EPI_ISL_2145382, EPI_ISL_2145384, EPI_ISL_2145387, EPI_ISL_2145389, EPI_ISL_2145392, EPI_ISL_2145394, EPI_ISL_2145397, EPI_ISL_2145399, EPI_ISL_2145401, EPI_ISL_2145404, EPI_ISL_2145406, EPI_ISL_2145407, EPI_ISL_2145409, EPI_ISL_2145412, EPI_ISL_2145415, EPI_ISL_2145417, EPI_ISL_2145420, EPI_ISL_2145422, EPI_ISL_2145425, EPI_ISL_2145427, EPI_ISL_2145430, EPI_ISL_2145432, EPI_ISL_2145435, EPI_ISL_2145439, EPI_ISL_2145441, EPI_ISL_2145444, EPI_ISL_2145447, EPI_ISL_2145449, EPI_ISL_2145452, EPI_ISL_2145455, EPI_ISL_2145457, EPI_ISL_2145460, EPI_ISL_2145463, EPI_ISL_2145465, EPI_ISL_2145467, EPI_ISL_2145470, EPI_ISL_2145471, EPI_ISL_2145474, EPI_ISL_2145476, EPI_ISL_2145477, EPI_ISL_2145480, EPI_ISL_2145482, EPI_ISL_2145485, EPI_ISL_2145487, EPI_ISL_2145489, EPI_ISL_2145492, EPI_ISL_2145495, EPI_ISL_2145497, EPI_ISL_2145501, EPI_ISL_2145502, EPI_ISL_2145505, EPI_ISL_2145507, EPI_ISL_2145510, EPI_ISL_2145512, EPI_ISL_2145515, EPI_ISL_2145518, EPI_ISL_2145520, EPI_ISL_2145523, EPI_ISL_2145525, EPI_ISL_2145528, EPI_ISL_2145530, EPI_ISL_2145533, EPI_ISL_2145536, EPI_ISL_2145539, EPI_ISL_2145541, EPI_ISL_2145544, EPI_ISL_2145546, EPI_ISL_2145549, EPI_ISL_2145551, EPI_ISL_2145554, EPI_ISL_2145556, EPI_ISL_2145559, EPI_ISL_2145561, EPI_ISL_2145563, EPI_ISL_2145566, EPI_ISL_2145568, EPI_ISL_2145571, EPI_ISL_2145574, EPI_ISL_2145576, EPI_ISL_2145579, EPI_ISL_2145581, EPI_ISL_2145584, EPI_ISL_2145587, EPI_ISL_2145590, EPI_ISL_2145592, EPI_ISL_2145595, EPI_ISL_2145598, EPI_ISL_2145600, EPI_ISL_2145603, EPI_ISL_2145605, EPI_ISL_2145607, EPI_ISL_2145610, EPI_ISL_2145613, EPI_ISL_2145615, EPI_ISL_2145618, EPI_ISL_2145621, EPI_ISL_2145623, EPI_ISL_2145626, EPI_ISL_2145629, EPI_ISL_2145631, EPI_ISL_2145634, EPI_ISL_2145637, EPI_ISL_2145639, EPI_ISL_2145642, EPI_ISL_2145645, EPI_ISL_2145648, EPI_ISL_2145651, EPI_ISL_2145653, EPI_ISL_2145656, EPI_ISL_2145659, EPI_ISL_2145661, EPI_ISL_2145663, EPI_ISL_2145666, EPI_ISL_2145668, EPI_ISL_2145671, EPI_ISL_2145674, EPI_ISL_2145677, EPI_ISL_2145680, EPI_ISL_2145683, EPI_ISL_2145685, EPI_ISL_2145688, EPI_ISL_2145690, EPI_ISL_2145693, EPI_ISL_2145696, EPI_ISL_2145698, EPI_ISL_2145701, EPI_ISL_2145704, EPI_ISL_2145706, EPI_ISL_2145708, EPI_ISL_2145711, EPI_ISL_2145714, EPI_ISL_2145716, EPI_ISL_2145718, EPI_ISL_2145719, EPI_ISL_2145724, EPI_ISL_2145726, EPI_ISL_2145727, EPI_ISL_2145729, EPI_ISL_2145731, EPI_ISL_2145734, EPI_ISL_2145736, EPI_ISL_2145739, EPI_ISL_2145741, EPI_ISL_2145744, EPI_ISL_2145745, EPI_ISL_2145747, EPI_ISL_2145748, EPI_ISL_2145750, EPI_ISL_2145753, EPI_ISL_2145758, EPI_ISL_2145761, EPI_ISL_2145764, EPI_ISL_2145766, EPI_ISL_2145769, EPI_ISL_2145771, EPI_ISL_2145774, EPI_ISL_2145776, EPI_ISL_2145778, EPI_ISL_2145781 | | | |
| see above | Helix/Illumina | Centers for Disease Control and Prevention Division of Viral Diseases, Pathogen Discovery | Dakota Howard, Dhwani Batra, Peter W. Cook, Kara Moser, Adrian Paskey, Jason Caravas, Benjamin Rambo-Martin, Shatavia Morrison, Christopher Gulvick, Scott Sammons, Yvette Unoarumhi, Darlene Wagner, Matthew Schmerer, Eileen de Feo, Jan Antico, Christine Tran, Matthew Tolentino, Shannon Wickline, Kim Gietzen, Brad Sickler, Jingtao Liu, Eric Allen, Phil Febbo, Nicole L. Washington, Simon White, Geraint Levan, Kelly Schiabor Barrett, Elizabeth Cirulli, Alexandre Bolze, Ary Ascencio, Charlotte Rivera-Garcia, Ryan Cho, Jason Nguyen, Sherry Wang, Jimmy Ramirez, Tyler Cassens, Efren Sandoval, Magnus Isaksson, William Lee, David Becker, Marc Laurent, James Lu, Clinton R. Paden, Duncan MacCannell |
| EPI_ISL_2145795, EPI_ISL_2145796, EPI_ISL_2145797, EPI_ISL_2145798, EPI_ISL_2145799, EPI_ISL_2145800, EPI_ISL_2145801, EPI_ISL_2145802, EPI_ISL_2145803, EPI_ISL_2145804, EPI_ISL_2145805, EPI_ISL_2145806, EPI_ISL_2145807, EPI_ISL_2145808, EPI_ISL_2145809, EPI_ISL_2145810, EPI_ISL_2145811, EPI_ISL_2145812, EPI_ISL_2145813, EPI_ISL_2145814, EPI_ISL_2145815, EPI_ISL_2145816, EPI_ISL_2145817, EPI_ISL_2145818, EPI_ISL_2145819, EPI_ISL_2145820, EPI_ISL_2145821, EPI_ISL_2145822, EPI_ISL_2145823, EPI_ISL_2145824, EPI_ISL_2145825, EPI_ISL_2145826, EPI_ISL_2145827, EPI_ISL_2145828, EPI_ISL_2145829, EPI_ISL_2145830, EPI_ISL_2145831, EPI_ISL_2145832, EPI_ISL_2145833, EPI_ISL_2145834, EPI_ISL_2145835, EPI_ISL_2145836, EPI_ISL_2145837, EPI_ISL_2145838, EPI_ISL_2145839, EPI_ISL_2145840, EPI_ISL_2145841, EPI_ISL_2145842, EPI_ISL_2145843, EPI_ISL_2145844, EPI_ISL_2145845, EPI_ISL_2145846, EPI_ISL_2145847, EPI_ISL_2145849, EPI_ISL_2145850, EPI_ISL_2145851, EPI_ISL_2145852, EPI_ISL_2145853, EPI_ISL_2145854, EPI_ISL_2145855, EPI_ISL_2145856, EPI_ISL_2145857, EPI_ISL_2145858, EPI_ISL_2145859, EPI_ISL_2145860, EPI_ISL_2145861, EPI_ISL_2145862, EPI_ISL_2145863, EPI_ISL_2145864, EPI_ISL_2145865, EPI_ISL_2145866, EPI_ISL_2145867, EPI_ISL_2145868, EPI_ISL_2145869, EPI_ISL_2145870, EPI_ISL_2145871, EPI_ISL_2145872, EPI_ISL_2145873, EPI_ISL_2145874, EPI_ISL_2145875, EPI_ISL_2145876, EPI_ISL_2145877, EPI_ISL_2145878, EPI_ISL_2145879, EPI_ISL_2145880, EPI_ISL_2145881, EPI_ISL_2145882, EPI_ISL_2145883, EPI_ISL_2145884, EPI_ISL_2145885, EPI_ISL_2145886, EPI_ISL_2145887, EPI_ISL_2145888, EPI_ISL_2145889, EPI_ISL_2145890, EPI_ISL_2145891, EPI_ISL_2145892, EPI_ISL_2145893, EPI_ISL_2145894, EPI_ISL_2145895, EPI_ISL_2145896, EPI_ISL_2145897, EPI_ISL_2145898, EPI_ISL_2145899, EPI_ISL_2145900, EPI_ISL_2145901, EPI_ISL_2145902, EPI_ISL_2145903, EPI_ISL_2145904, EPI_ISL_2145905, EPI_ISL_2145906, EPI_ISL_2145907, EPI_ISL_2145908, EPI_ISL_2145909, EPI_ISL_2145910, EPI_ISL_2145911, EPI_ISL_2145912, EPI_ISL_2145913, EPI_ISL_2145914, EPI_ISL_2145915, EPI_ISL_2145916, EPI_ISL_2145917, EPI_ISL_2145918, EPI_ISL_2145919, EPI_ISL_2145920, EPI_ISL_2145922, EPI_ISL_2145923, EPI_ISL_2145924, EPI_ISL_2145925, EPI_ISL_2145926, EPI_ISL_2145928, EPI_ISL_2145929, EPI_ISL_2145930, EPI_ISL_2145931, EPI_ISL_2145932, EPI_ISL_2145933, EPI_ISL_2145934, EPI_ISL_2145935, EPI_ISL_2145936, EPI_ISL_2145937, EPI_ISL_2145938, EPI_ISL_2145939, EPI_ISL_2145940, EPI_ISL_2145941, EPI_ISL_2145942, EPI_ISL_2145944, EPI_ISL_2145945, EPI_ISL_2145946, EPI_ISL_2145947, EPI_ISL_2145948, EPI_ISL_2145949, EPI_ISL_2145950, EPI_ISL_2145951, EPI_ISL_2145952, EPI_ISL_2145953, EPI_ISL_2145954, EPI_ISL_2145955, EPI_ISL_2145956, EPI_ISL_2145957, EPI_ISL_2145958, EPI_ISL_2145959, EPI_ISL_2145960, EPI_ISL_2145961, EPI_ISL_2145962, EPI_ISL_2145963, EPI_ISL_2145964, EPI_ISL_2145965, EPI_ISL_2145966, EPI_ISL_2145967, EPI_ISL_2145968, EPI_ISL_2145969, EPI_ISL_2145970, EPI_ISL_2145971, EPI_ISL_2145972, EPI_ISL_2145973, EPI_ISL_2145974, EPI_ISL_2145975, EPI_ISL_2145976, EPI_ISL_2145977, EPI_ISL_2145978, EPI_ISL_2145980, EPI_ISL_2145981, EPI_ISL_2145982, EPI_ISL_2145983, EPI_ISL_2145984, EPI_ISL_2145985, EPI_ISL_2145986, EPI_ISL_2145987, EPI_ISL_2145988, EPI_ISL_2145989, EPI_ISL_2145990, EPI_ISL_2145991, EPI_ISL_2145992, EPI_ISL_2145993, EPI_ISL_2145994, EPI_ISL_2145995, EPI_ISL_2145996, EPI_ISL_2145997, EPI_ISL_2145998, EPI_ISL_2145999, EPI_ISL_2146000, EPI_ISL_2146001, EPI_ISL_2146002, EPI_ISL_2146003, EPI_ISL_2146004, EPI_ISL_2146005, EPI_ISL_2146006, EPI_ISL_2146007, EPI_ISL_2146008, EPI_ISL_2146009, EPI_ISL_2146010, EPI_ISL_2146011, EPI_ISL_2146012, EPI_ISL_2146013, EPI_ISL_2146014, EPI_ISL_2146015, EPI_ISL_2146016, EPI_ISL_2146017, EPI_ISL_2146018, EPI_ISL_2146019, EPI_ISL_2146021, EPI_ISL_2146024, EPI_ISL_2146026, EPI_ISL_2146029, EPI_ISL_2146032, EPI_ISL_2146033, EPI_ISL_2146034, EPI_ISL_2146035, EPI_ISL_2146036, EPI_ISL_2146037, EPI_ISL_2146038, EPI_ISL_2146039, EPI_ISL_2146040, EPI_ISL_2146041, EPI_ISL_2146042, EPI_ISL_2146043, EPI_ISL_2146044, EPI_ISL_2146045, EPI_ISL_2146046, EPI_ISL_2146047, EPI_ISL_2146048, EPI_ISL_2146049, EPI_ISL_2146050, EPI_ISL_2146051, EPI_ISL_2146052, EPI_ISL_2146053, EPI_ISL_2146054, EPI_ISL_2146055, EPI_ISL_2146056, EPI_ISL_2146057, EPI_ISL_2146058, EPI_ISL_2146059, EPI_ISL_2146060, EPI_ISL_2146061, EPI_ISL_2146062, EPI_ISL_2146063, EPI_ISL_2146064, EPI_ISL_2146065, EPI_ISL_2146066, EPI_ISL_2146067, EPI_ISL_2146068, EPI_ISL_2146069, EPI_ISL_2146070, EPI_ISL_2146071, EPI_ISL_2146072, EPI_ISL_2146073, EPI_ISL_2146074, EPI_ISL_2146075, EPI_ISL_2146076, EPI_ISL_2146077, EPI_ISL_2146078, EPI_ISL_2146079, EPI_ISL_2146080, EPI_ISL_2146081, EPI_ISL_2146082, EPI_ISL_2146083, EPI_ISL_2146084, EPI_ISL_2146085, EPI_ISL_2146086, EPI_ISL_2146087, EPI_ISL_2146088, EPI_ISL_2146089, EPI_ISL_2146090, EPI_ISL_2146091, EPI_ISL_2146092, EPI_ISL_2146093, EPI_ISL_2146094, EPI_ISL_2146095, EPI_ISL_2146096, EPI_ISL_2146097, EPI_ISL_2146098, EPI_ISL_2146100, EPI_ISL_2146101, EPI_ISL_2146102, EPI_ISL_2146103, EPI_ISL_2146104, EPI_ISL_2146105, EPI_ISL_2146106, EPI_ISL_2146107, EPI_ISL_2146108, EPI_ISL_2146109, EPI_ISL_2146110, EPI_ISL_2146111, EPI_ISL_2146112, EPI_ISL_2146113, EPI_ISL_2146114, EPI_ISL_2146116, EPI_ISL_2146117, EPI_ISL_2146118, EPI_ISL_2146119, EPI_ISL_2146120, EPI_ISL_2146121, EPI_ISL_2146122, EPI_ISL_2146123, EPI_ISL_2146124, EPI_ISL_2146125, EPI_ISL_2146126, EPI_ISL_2146127, EPI_ISL_2146129, EPI_ISL_2146131, EPI_ISL_2146132, EPI_ISL_2146133, EPI_ISL_2146134, EPI_ISL_2146135, EPI_ISL_2146136, EPI_ISL_2146137, EPI_ISL_2146138, EPI_ISL_2146139, EPI_ISL_2146140, EPI_ISL_2146141, EPI_ISL_2146142, EPI_ISL_2146143, EPI_ISL_2146144, EPI_ISL_2146145, EPI_ISL_2146146, EPI_ISL_2146147, EPI_ISL_2146148, EPI_ISL_2146150, EPI_ISL_2146151, EPI_ISL_2146152, EPI_ISL_2146153, EPI_ISL_2146154, EPI_ISL_2146155, EPI_ISL_2146156, EPI_ISL_2146157, EPI_ISL_2146158, EPI_ISL_2146160, EPI_ISL_2146161, EPI_ISL_2146162, EPI_ISL_2146163, EPI_ISL_2146165, EPI_ISL_2146166, EPI_ISL_2146167, EPI_ISL_2146168, EPI_ISL_2146169, EPI_ISL_2146170, EPI_ISL_2146171, EPI_ISL_2146172, EPI_ISL_2146173, EPI_ISL_2146174, EPI_ISL_2146175, EPI_ISL_2146176, EPI_ISL_2146177, EPI_ISL_2146178, EPI_ISL_2146179, EPI_ISL_2146180, EPI_ISL_2146181, EPI_ISL_2146182, EPI_ISL_2146183, EPI_ISL_2146184, EPI_ISL_2146185, EPI_ISL_2146186, EPI_ISL_2146187, EPI_ISL_2146188, EPI_ISL_2146189, EPI_ISL_2146190, EPI_ISL_2146191, EPI_ISL_2146192, EPI_ISL_2146193, EPI_ISL_2146194, EPI_ISL_2146195, EPI_ISL_2146196, EPI_ISL_2146197, EPI_ISL_2146198, EPI_ISL_2146199, EPI_ISL_2146200, EPI_ISL_2146201, EPI_ISL_2146202, EPI_ISL_2146203, EPI_ISL_2146204, EPI_ISL_2146205, EPI_ISL_2146206, EPI_ISL_2146207, EPI_ISL_2146208, EPI_ISL_2146209, EPI_ISL_2146210, EPI_ISL_2146211, EPI_ISL_2146212, EPI_ISL_2146213, EPI_ISL_2146214, EPI_ISL_2146215, EPI_ISL_2146216, EPI_ISL_2146217, EPI_ISL_2146218, EPI_ISL_2146219, EPI_ISL_2146220, EPI_ISL_2146221, EPI_ISL_2146222, EPI_ISL_2146223, EPI_ISL_2146225, EPI_ISL_2146226, EPI_ISL_2146227, EPI_ISL_2146228, EPI_ISL_2146229, EPI_ISL_2146230, EPI_ISL_2146231, EPI_ISL_2146232, EPI_ISL_2146234, EPI_ISL_2146235, EPI_ISL_2146236, EPI_ISL_2146237, EPI_ISL_2146238, EPI_ISL_2146239, EPI_ISL_2146240, EPI_ISL_2146241, EPI_ISL_2146242, EPI_ISL_2146243, EPI_ISL_2146244, EPI_ISL_2146245, EPI_ISL_2146246, EPI_ISL_2146247, EPI_ISL_2146248, EPI_ISL_2146249, EPI_ISL_2146250, EPI_ISL_2146251, EPI_ISL_2146252, EPI_ISL_2146253, EPI_ISL_2146254, EPI_ISL_2146255, EPI_ISL_2146256, EPI_ISL_2146257, EPI_ISL_2146258, EPI_ISL_2146259, EPI_ISL_2146260, EPI_ISL_2146261, EPI_ISL_2146262, EPI_ISL_2146263, EPI_ISL_2146264, EPI_ISL_2146265, EPI_ISL_2146266, EPI_ISL_2146267, EPI_ISL_2146268, EPI_ISL_2146269, EPI_ISL_2146270, EPI_ISL_2146271, EPI_ISL_2146272, EPI_ISL_2146273, EPI_ISL_2146274, EPI_ISL_2146275, EPI_ISL_2146276, EPI_ISL_2146277, EPI_ISL_2146278, EPI_ISL_2146279, EPI_ISL_2146280, EPI_ISL_2146281, EPI_ISL_2146282, EPI_ISL_2146283, EPI_ISL_2146284, EPI_ISL_2146285, EPI_ISL_2146286, EPI_ISL_2146287, EPI_ISL_2146288, EPI_ISL_2146289, EPI_ISL_2146290, EPI_ISL_2146291, EPI_ISL_2146292, EPI_ISL_2146293, EPI_ISL_2146294, EPI_ISL_2146295, EPI_ISL_2146296, EPI_ISL_2146297, EPI_ISL_2146298, EPI_ISL_2146299, EPI_ISL_2146300, EPI_ISL_2146301, EPI_ISL_2146302, EPI_ISL_2146303, EPI_ISL_2146304, EPI_ISL_2146305, EPI_ISL_2146306, EPI_ISL_2146307, EPI_ISL_2146308, EPI_ISL_2146309, EPI_ISL_2146310, EPI_ISL_2146311, EPI_ISL_2146312, EPI_ISL_2146313, EPI_ISL_2146314, EPI_ISL_2146315, EPI_ISL_2146316, EPI_ISL_2146317, EPI_ISL_2146318, EPI_ISL_2146319, EPI_ISL_2146320, EPI_ISL_2146321, EPI_ISL_2146322, EPI_ISL_2146323, EPI_ISL_2146324, EPI_ISL_2146325, EPI_ISL_2146326, EPI_ISL_2146327, EPI_ISL_2146328, EPI_ISL_2146329, EPI_ISL_2146330, EPI_ISL_2146331, EPI_ISL_2146332, EPI_ISL_2146333, EPI_ISL_2146334, EPI_ISL_2146335, EPI_ISL_2146336, EPI_ISL_2146337, EPI_ISL_2146338, EPI_ISL_2146339, EPI_ISL_2146340, EPI_ISL_2146341, EPI_ISL_2146342, EPI_ISL_2146343, EPI_ISL_2146344, EPI_ISL_2146345, EPI_ISL_2146346, EPI_ISL_2146348, EPI_ISL_2146349, EPI_ISL_2146350, EPI_ISL_2146351, EPI_ISL_2146352, EPI_ISL_2146353, EPI_ISL_2146354, EPI_ISL_2146355, EPI_ISL_2146356, EPI_ISL_2146357, EPI_ISL_2146358, EPI_ISL_2146359, EPI_ISL_2146361, EPI_ISL_2146363, EPI_ISL_2146364, EPI_ISL_2146365, EPI_ISL_2146366, EPI_ISL_2146367, EPI_ISL_2146368, EPI_ISL_2146369, EPI_ISL_2146370, EPI_ISL_2146371, EPI_ISL_2146372, EPI_ISL_2146373, EPI_ISL_2146374, EPI_ISL_2146375, EPI_ISL_2146376, EPI_ISL_2146377, EPI_ISL_2146378, EPI_ISL_2146379, EPI_ISL_2146381, EPI_ISL_2146382, EPI_ISL_2146383, EPI_ISL_2146384, EPI_ISL_2146385, EPI_ISL_2146386, EPI_ISL_2146387, EPI_ISL_2146388, EPI_ISL_2146390, EPI_ISL_2146391, EPI_ISL_2146392, EPI_ISL_2146393, EPI_ISL_2146394, EPI_ISL_2146395, EPI_ISL_2146396, EPI_ISL_2146397, EPI_ISL_2146398, EPI_ISL_2146399, EPI_ISL_2146400, EPI_ISL_2146401, EPI_ISL_2146403, EPI_ISL_2146404, EPI_ISL_2146405, EPI_ISL_2146406, EPI_ISL_2146407, EPI_ISL_2146408, EPI_ISL_2146409, EPI_ISL_2146410, EPI_ISL_2146411, EPI_ISL_2146412, EPI_ISL_2146413, EPI_ISL_2146414, EPI_ISL_2146415, EPI_ISL_2146416, EPI_ISL_2146417, EPI_ISL_2146418, EPI_ISL_2146419, EPI_ISL_2146420, EPI_ISL_2146421, EPI_ISL_2146422, EPI_ISL_2146423, EPI_ISL_2146424, EPI_ISL_2146425, EPI_ISL_2146426, EPI_ISL_2146427, EPI_ISL_2146428, EPI_ISL_2146430, EPI_ISL_2146431, EPI_ISL_2146432, EPI_ISL_2146433, EPI_ISL_2146434, EPI_ISL_2146435, EPI_ISL_2146436, EPI_ISL_2146437, EPI_ISL_2146439, EPI_ISL_2146440, EPI_ISL_2146441, EPI_ISL_2146442, EPI_ISL_2146443, EPI_ISL_2146444, EPI_ISL_2146445, EPI_ISL_2146446, EPI_ISL_2146447, EPI_ISL_2146448, EPI_ISL_2146449, EPI_ISL_2146450, EPI_ISL_2146451, EPI_ISL_2146452, EPI_ISL_2146453, EPI_ISL_2146454, EPI_ISL_2146455, EPI_ISL_2146456, EPI_ISL_2146457, EPI_ISL_2146458, EPI_ISL_2146459, EPI_ISL_2146460, EPI_ISL_2146461, EPI_ISL_2146462, EPI_ISL_2146463, EPI_ISL_2146464, EPI_ISL_2146465, EPI_ISL_2146466, EPI_ISL_2146467, EPI_ISL_2146468, EPI_ISL_2146469, EPI_ISL_2146470, EPI_ISL_2146471, EPI_ISL_2146472, EPI_ISL_2146473, EPI_ISL_2146474, EPI_ISL_2146475, EPI_ISL_2146476, EPI_ISL_2146477, EPI_ISL_2146478, EPI_ISL_2146479, EPI_ISL_2146480, EPI_ISL_2146481, EPI_ISL_2146482, EPI_ISL_2146483, EPI_ISL_2146484, EPI_ISL_2146485, EPI_ISL_2146486, EPI_ISL_2146487, EPI_ISL_2146488, EPI_ISL_2146489, EPI_ISL_2146490, EPI_ISL_2146491, EPI_ISL_2146492, EPI_ISL_2146493, EPI_ISL_2146494, EPI_ISL_2146495, EPI_ISL_2146496, EPI_ISL_2146497, EPI_ISL_2146498, EPI_ISL_2146499, EPI_ISL_2146500, EPI_ISL_2146501, EPI_ISL_2146502, EPI_ISL_2146503, EPI_ISL_2146504, EPI_ISL_2146505, EPI_ISL_2146506, EPI_ISL_2146507, EPI_ISL_2146508, EPI_ISL_2146509, EPI_ISL_2146510, EPI_ISL_2146511, EPI_ISL_2146512, EPI_ISL_2146513, EPI_ISL_2146514, EPI_ISL_2146515, EPI_ISL_2146516, EPI_ISL_2146517, EPI_ISL_2146518, EPI_ISL_2146519, EPI_ISL_2146520, EPI_ISL_2146521, EPI_ISL_2146522, EPI_ISL_2146523, EPI_ISL_2146524, EPI_ISL_2146525, EPI_ISL_2146526, EPI_ISL_2146527, EPI_ISL_2146528, EPI_ISL_2146529, EPI_ISL_2146530, EPI_ISL_2146531, EPI_ISL_2146532, EPI_ISL_2146533, EPI_ISL_2146534, EPI_ISL_2146535, EPI_ISL_2146536, EPI_ISL_2146537, EPI_ISL_2146538, EPI_ISL_2146539, EPI_ISL_2146540, EPI_ISL_2146541, EPI_ISL_2146542, EPI_ISL_2146543, EPI_ISL_2146545, EPI_ISL_2146546, EPI_ISL_2146547, EPI_ISL_2146548, EPI_ISL_2146549, EPI_ISL_2146550, EPI_ISL_2146551, EPI_ISL_2146552, EPI_ISL_2146553, EPI_ISL_2146554, EPI_ISL_2146555, EPI_ISL_2146556, EPI_ISL_2146557, EPI_ISL_2146558, EPI_ISL_2146559, EPI_ISL_2146560, EPI_ISL_2146561, EPI_ISL_2146562, EPI_ISL_2146563, EPI_ISL_2146564, EPI_ISL_2146565, EPI_ISL_2146566, EPI_ISL_2146568, EPI_ISL_2146569, EPI_ISL_2146570, EPI_ISL_2146571, EPI_ISL_2146572, EPI_ISL_2146573, EPI_ISL_2146574, EPI_ISL_2146575, EPI_ISL_2146576, EPI_ISL_2146577, EPI_ISL_2146578, EPI_ISL_2146579, EPI_ISL_2146580, EPI_ISL_2146581, EPI_ISL_2146582, EPI_ISL_2146583, EPI_ISL_2146584, EPI_ISL_2146585, EPI_ISL_2146586, EPI_ISL_2146587, EPI_ISL_2146588, EPI_ISL_2146589, EPI_ISL_2146590, EPI_ISL_2146591, EPI_ISL_2146592, EPI_ISL_2146593, EPI_ISL_2146594, EPI_ISL_2146596, EPI_ISL_2146597, EPI_ISL_2146598, EPI_ISL_2146599, EPI_ISL_2146600, EPI_ISL_2146601, EPI_ISL_2146602, EPI_ISL_2146603, EPI_ISL_2146604, EPI_ISL_2146605, EPI_ISL_2146606, EPI_ISL_2146607, EPI_ISL_2146608, EPI_ISL_2146609, EPI_ISL_2146610, EPI_ISL_2146611, EPI_ISL_2146612, EPI_ISL_2146613, EPI_ISL_2146614, EPI_ISL_2146615, EPI_ISL_2146616, EPI_ISL_2146617, EPI_ISL_2146618, EPI_ISL_2146620, EPI_ISL_2146621, EPI_ISL_2146622, EPI_ISL_2146623, EPI_ISL_2146624, EPI_ISL_2146625, EPI_ISL_2146626, EPI_ISL_2146627, EPI_ISL_2146628, EPI_ISL_2146629, EPI_ISL_2146630, EPI_ISL_2146631, EPI_ISL_2146632, EPI_ISL_2146633, EPI_ISL_2146634, EPI_ISL_2146635, EPI_ISL_2146636, EPI_ISL_2146637, EPI_ISL_2146638, EPI_ISL_2146639, EPI_ISL_2146640, EPI_ISL_2146641, EPI_ISL_2146642, EPI_ISL_2146643, EPI_ISL_2146644, EPI_ISL_2146645, EPI_ISL_2146646, EPI_ISL_2146647, EPI_ISL_2146648, EPI_ISL_2146649, EPI_ISL_2146650, EPI_ISL_2146651, EPI_ISL_2146652, EPI_ISL_2146653, EPI_ISL_2146655, EPI_ISL_2146656, EPI_ISL_2146657, EPI_ISL_2146658, EPI_ISL_2146659, EPI_ISL_2146660, EPI_ISL_2146661, EPI_ISL_2146662, EPI_ISL_2146663, EPI_ISL_2146664, EPI_ISL_2146665, EPI_ISL_2146666, EPI_ISL_2146667, EPI_ISL_2146668, EPI_ISL_2146669, EPI_ISL_2146670, EPI_ISL_2146671, EPI_ISL_2146672, EPI_ISL_2146673, EPI_ISL_2146674, EPI_ISL_2146675, EPI_ISL_2146676, EPI_ISL_2146677, EPI_ISL_2146678, EPI_ISL_2146679, EPI_ISL_2146680, EPI_ISL_2146681, EPI_ISL_2146682, EPI_ISL_2146683, EPI_ISL_2146684, EPI_ISL_2146685, EPI_ISL_2146686, EPI_ISL_2146687, EPI_ISL_2146688, EPI_ISL_2146689, EPI_ISL_2146690, EPI_ISL_2146691, EPI_ISL_2146692, EPI_ISL_2146693, EPI_ISL_2146694, EPI_ISL_2146695, EPI_ISL_2146696, EPI_ISL_2146697, EPI_ISL_2146699, EPI_ISL_2146700, EPI_ISL_2146701, EPI_ISL_2146702, EPI_ISL_2146703, EPI_ISL_2146704, EPI_ISL_2146705, EPI_ISL_2146706, EPI_ISL_2146707, EPI_ISL_2146708, EPI_ISL_2146709, EPI_ISL_2146710, EPI_ISL_2146711, EPI_ISL_2146712, EPI_ISL_2146713, EPI_ISL_2146714, EPI_ISL_2146716, EPI_ISL_2146717, EPI_ISL_2146718, EPI_ISL_2146719, EPI_ISL_2146720, EPI_ISL_2146721, EPI_ISL_2146722, EPI_ISL_2146723, EPI_ISL_2146724, EPI_ISL_2146725, EPI_ISL_2146726, EPI_ISL_2146727, EPI_ISL_2146729, EPI_ISL_2146730, EPI_ISL_2146731, EPI_ISL_2146732, EPI_ISL_2146733, EPI_ISL_2146734, EPI_ISL_2146735, EPI_ISL_2146736, EPI_ISL_2146737, EPI_ISL_2146738, EPI_ISL_2146739, EPI_ISL_2146740, EPI_ISL_2146741, EPI_ISL_2146742, EPI_ISL_2146743, EPI_ISL_2146744, EPI_ISL_2146745, EPI_ISL_2146746, EPI_ISL_2146747, EPI_ISL_2146748, EPI_ISL_2146749, EPI_ISL_2146750, EPI_ISL_2146751, EPI_ISL_2146752, EPI_ISL_2146753, EPI_ISL_2146754, EPI_ISL_2146755, EPI_ISL_2146756, EPI_ISL_2146757, EPI_ISL_2146758, EPI_ISL_2146759, EPI_ISL_2146760, EPI_ISL_2146761, EPI_ISL_2146762, EPI_ISL_2146763, EPI_ISL_2146764, EPI_ISL_2146765, EPI_ISL_2146766, EPI_ISL_2146767, EPI_ISL_2146768, EPI_ISL_2146769, EPI_ISL_2146770, EPI_ISL_2146771, EPI_ISL_2146772, EPI_ISL_2146773, EPI_ISL_2146774, EPI_ISL_2146775, EPI_ISL_2146776, EPI_ISL_2146777, EPI_ISL_2146778, EPI_ISL_2146779, EPI_ISL_2146780, EPI_ISL_2146781, EPI_ISL_2146782, EPI_ISL_2146783, EPI_ISL_2146784 | | | |
| see above | Aegis Sciences Corporation | Centers for Disease Control and Prevention Division of Viral Diseases, Pathogen Discovery | Dakota Howard, Dhwani Batra, Peter W. Cook, Kara Moser, Adrian Paskey, Jason Caravas, Benjamin Rambo-Martin, Shatavia Morrison, Christopher Gulvick, Scott Sammons, Yvette Unoarumhi, Darlene Wagner, Matthew Schmerer, Cyndi Clark, Patrick Campbell, Rob Case, Vikramsinha Ghorpade, Holly Houdeshell, Ola Kvalvaag, Dillon Nall, Ethan Sanders, Alec Vest, Shaun Westlund, Matthew Hardison, Clinton R. Paden, Duncan MacCannell |
| EPI_ISL_2146785 | The Ohio State University Applied Microbiology Services Laboratory | The Ohio State University Applied Microbiology Services Laboratory | Seth A. Faith PhD |
| EPI_ISL_2146786, EPI_ISL_2146787, EPI_ISL_2146788, EPI_ISL_2146789, EPI_ISL_2146790, EPI_ISL_2146791, EPI_ISL_2146792, EPI_ISL_2146793, EPI_ISL_2146794, EPI_ISL_2146795, EPI_ISL_2146796, EPI_ISL_2146797, EPI_ISL_2146798, EPI_ISL_2146799, EPI_ISL_2146800, EPI_ISL_2146801, EPI_ISL_2146802, EPI_ISL_2146803, EPI_ISL_2146804, EPI_ISL_2146805, EPI_ISL_2146806, EPI_ISL_2146807, EPI_ISL_2146808, EPI_ISL_2146809, EPI_ISL_2146810, EPI_ISL_2146811, EPI_ISL_2146812, EPI_ISL_2146813, EPI_ISL_2146814, EPI_ISL_2146815, EPI_ISL_2146816, EPI_ISL_2146817, EPI_ISL_2146818, EPI_ISL_2146819, EPI_ISL_2146820, EPI_ISL_2146821, EPI_ISL_2146822, EPI_ISL_2146823, EPI_ISL_2146824, EPI_ISL_2146825, EPI_ISL_2146826, EPI_ISL_2146827, EPI_ISL_2146828, EPI_ISL_2146829, EPI_ISL_2146830, EPI_ISL_2146831, EPI_ISL_2146832, EPI_ISL_2146833, EPI_ISL_2146834, EPI_ISL_2146835, EPI_ISL_2146836, EPI_ISL_2146837, EPI_ISL_2146838, EPI_ISL_2146839, EPI_ISL_2146840, EPI_ISL_2146841, EPI_ISL_2146842, EPI_ISL_2146843, EPI_ISL_2146844, EPI_ISL_2146845, EPI_ISL_2146846, EPI_ISL_2146847, EPI_ISL_2146848, EPI_ISL_2146849, EPI_ISL_2146850, EPI_ISL_2146851, EPI_ISL_2146852, EPI_ISL_2146853, EPI_ISL_2146854, EPI_ISL_2146855, EPI_ISL_2146856, EPI_ISL_2146857, EPI_ISL_2146858, EPI_ISL_2146859, EPI_ISL_2146860, EPI_ISL_2146861, EPI_ISL_2146862, EPI_ISL_2146863, EPI_ISL_2146864, EPI_ISL_2146866, EPI_ISL_2146867, EPI_ISL_2146868, EPI_ISL_2146869, EPI_ISL_2146870, EPI_ISL_2146871, EPI_ISL_2146872, EPI_ISL_2146873, EPI_ISL_2146874, EPI_ISL_2146875, EPI_ISL_2146876, EPI_ISL_2146877, EPI_ISL_2146878, EPI_ISL_2146879, EPI_ISL_2146880, EPI_ISL_2146881, EPI_ISL_2146882, EPI_ISL_2146883, EPI_ISL_2146884, EPI_ISL_2146885, EPI_ISL_2146886, EPI_ISL_2146887, EPI_ISL_2146888, EPI_ISL_2146889, EPI_ISL_2146890, EPI_ISL_2146891, EPI_ISL_2146892, EPI_ISL_2146893, EPI_ISL_2146894, EPI_ISL_2146895, EPI_ISL_2146896, EPI_ISL_2146897, EPI_ISL_2146898, EPI_ISL_2146899, EPI_ISL_2146900, EPI_ISL_2146901, EPI_ISL_2146902, EPI_ISL_2146903, EPI_ISL_2146904, EPI_ISL_2146905, EPI_ISL_2146906, EPI_ISL_2146907, EPI_ISL_2146908, EPI_ISL_2146909, EPI_ISL_2146910, EPI_ISL_2146911, EPI_ISL_2146912, EPI_ISL_2146913, EPI_ISL_2146914, EPI_ISL_2146915, EPI_ISL_2146916, EPI_ISL_2146917, EPI_ISL_2146918, EPI_ISL_2146919, EPI_ISL_2146920, EPI_ISL_2146921, EPI_ISL_2146922, EPI_ISL_2146923, EPI_ISL_2146924, EPI_ISL_2146925, EPI_ISL_2146926, EPI_ISL_2146927, EPI_ISL_2146928, EPI_ISL_2146929, EPI_ISL_2146930, EPI_ISL_2146931, EPI_ISL_2146932, EPI_ISL_2146933, EPI_ISL_2146934, EPI_ISL_2146935, EPI_ISL_2146936, EPI_ISL_2146937, EPI_ISL_2146938, EPI_ISL_2146939, EPI_ISL_2146940, EPI_ISL_2146941, EPI_ISL_2146942, EPI_ISL_2146943, EPI_ISL_2146944, EPI_ISL_2146945, EPI_ISL_2146946, EPI_ISL_2146947, EPI_ISL_2146948, EPI_ISL_2146949, EPI_ISL_2146950, EPI_ISL_2146951, EPI_ISL_2146952, EPI_ISL_2146953, EPI_ISL_2146954, EPI_ISL_2146955, EPI_ISL_2146956, EPI_ISL_2146957, EPI_ISL_2146958, EPI_ISL_2146959, EPI_ISL_2146960, EPI_ISL_2146961, EPI_ISL_2146962, EPI_ISL_2146963, EPI_ISL_2146964, EPI_ISL_2146965, EPI_ISL_2146966, EPI_ISL_2146967, EPI_ISL_2146968, EPI_ISL_2146969, EPI_ISL_2146970, EPI_ISL_2146971, EPI_ISL_2146972, EPI_ISL_2146973, EPI_ISL_2146974, EPI_ISL_2146975, EPI_ISL_2146976, EPI_ISL_2146977, EPI_ISL_2146978, EPI_ISL_2146979, EPI_ISL_2146980, EPI_ISL_2146983, EPI_ISL_2146985, EPI_ISL_2146988, EPI_ISL_2146990, EPI_ISL_2146991, EPI_ISL_2146992, EPI_ISL_2146993, EPI_ISL_2146994, EPI_ISL_2146995, EPI_ISL_2146996, EPI_ISL_2146998, EPI_ISL_2146999, EPI_ISL_2147000, EPI_ISL_2147001, EPI_ISL_2147002, EPI_ISL_2147003, EPI_ISL_2147004, EPI_ISL_2147005, EPI_ISL_2147006, EPI_ISL_2147007, EPI_ISL_2147008, EPI_ISL_2147009, EPI_ISL_2147010, EPI_ISL_2147011, EPI_ISL_2147012, EPI_ISL_2147013, EPI_ISL_2147014, EPI_ISL_2147016, EPI_ISL_2147017, EPI_ISL_2147018, EPI_ISL_2147019, EPI_ISL_2147020, EPI_ISL_2147021, EPI_ISL_2147022, EPI_ISL_2147023, EPI_ISL_2147024, EPI_ISL_2147025, EPI_ISL_2147026, EPI_ISL_2147027, EPI_ISL_2147028, EPI_ISL_2147029, EPI_ISL_2147030, EPI_ISL_2147031, EPI_ISL_2147032, EPI_ISL_2147033, EPI_ISL_2147034, EPI_ISL_2147035, EPI_ISL_2147036, EPI_ISL_2147037, EPI_ISL_2147038, EPI_ISL_2147039, EPI_ISL_2147040, EPI_ISL_2147041, EPI_ISL_2147042, EPI_ISL_2147043, EPI_ISL_2147044, EPI_ISL_2147045, EPI_ISL_2147046, EPI_ISL_2147047, EPI_ISL_2147048, EPI_ISL_2147049, EPI_ISL_2147050, EPI_ISL_2147051, EPI_ISL_2147052, EPI_ISL_2147053, EPI_ISL_2147054, EPI_ISL_2147055 | | | |
| see above | Aegis Sciences Corporation | Centers for Disease Control and Prevention Division of Viral Diseases, Pathogen Discovery | Dakota Howard, Dhwani Batra, Peter W. Cook, Kara Moser, Adrian Paskey, Jason Caravas, Benjamin Rambo-Martin, Shatavia Morrison, Christopher Gulvick, Scott Sammons, Yvette Unoarumhi, Darlene Wagner, Matthew Schmerer, Cyndi Clark, Patrick Campbell, Rob Case, Vikramsinha Ghorpade, Holly Houdeshell, Ola Kvalvaag, Dillon Nall, Ethan Sanders, Alec Vest, Shaun Westlund, Matthew Hardison, Clinton R. Paden, Duncan MacCannell |
| EPI_ISL_2147056 | The Ohio State University Applied Microbiology Services Laboratory | The Ohio State University Applied Microbiology Services Laboratory | Seth A. Faith PhD |
| EPI_ISL_2147057, EPI_ISL_2147058, EPI_ISL_2147059, EPI_ISL_2147060, EPI_ISL_2147061, EPI_ISL_2147062, EPI_ISL_2147063, EPI_ISL_2147064, EPI_ISL_2147065, EPI_ISL_2147066, EPI_ISL_2147067, EPI_ISL_2147068, EPI_ISL_2147069, EPI_ISL_2147070, EPI_ISL_2147071, EPI_ISL_2147072, EPI_ISL_2147074, EPI_ISL_2147075, EPI_ISL_2147076, EPI_ISL_2147077, EPI_ISL_2147078, EPI_ISL_2147079, EPI_ISL_2147080, EPI_ISL_2147081, EPI_ISL_2147082, EPI_ISL_2147083, EPI_ISL_2147084, EPI_ISL_2147085, EPI_ISL_2147086, EPI_ISL_2147087, EPI_ISL_2147088, EPI_ISL_2147089, EPI_ISL_2147090, EPI_ISL_2147091, EPI_ISL_2147092, EPI_ISL_2147094, EPI_ISL_2147095, EPI_ISL_2147096, EPI_ISL_2147097, EPI_ISL_2147098, EPI_ISL_2147099, EPI_ISL_2147100, EPI_ISL_2147101, EPI_ISL_2147102, EPI_ISL_2147103, EPI_ISL_2147104, EPI_ISL_2147105, EPI_ISL_2147106, EPI_ISL_2147107, EPI_ISL_2147108, EPI_ISL_2147109, EPI_ISL_2147110, EPI_ISL_2147111, EPI_ISL_2147112, EPI_ISL_2147113, EPI_ISL_2147114, EPI_ISL_2147115, EPI_ISL_2147116, EPI_ISL_2147117, EPI_ISL_2147118, EPI_ISL_2147119, EPI_ISL_2147120, EPI_ISL_2147121, EPI_ISL_2147122, EPI_ISL_2147123, EPI_ISL_2147124, | | | |

EPI_ISL_2147125, EPI_ISL_2147126, EPI_ISL_2147127, EPI_ISL_2147128, EPI_ISL_2147129, EPI_ISL_2147130, EPI_ISL_2147131, EPI_ISL_2147132, EPI_ISL_2147133, EPI_ISL_2147134, EPI_ISL_2147135, EPI_ISL_2147136, EPI_ISL_2147137, EPI_ISL_2147138, EPI_ISL_2147139, EPI_ISL_2147140, EPI_ISL_2147141, EPI_ISL_2147142, EPI_ISL_2147143, EPI_ISL_2147144, EPI_ISL_2147145, EPI_ISL_2147146,
EPI_ISL_2147147, EPI_ISL_2147148, EPI_ISL_2147151, EPI_ISL_2147152, EPI_ISL_2147153, EPI_ISL_2147154, EPI_ISL_2147155, EPI_ISL_2147156, EPI_ISL_2147157, EPI_ISL_2147158, EPI_ISL_2147159, EPI_ISL_2147160, EPI_ISL_2147161, EPI_ISL_2147162, EPI_ISL_2147163, EPI_ISL_2147164, EPI_ISL_2147165, EPI_ISL_2147166, EPI_ISL_2147167, EPI_ISL_2147168, EPI_ISL_2147170, EPI_ISL_2147171,
EPI_ISL_2147172, EPI_ISL_2147173, EPI_ISL_2147174, EPI_ISL_2147175, EPI_ISL_2147176, EPI_ISL_2147177, EPI_ISL_2147178, EPI_ISL_2147179, EPI_ISL_2147180, EPI_ISL_2147181, EPI_ISL_2147182, EPI_ISL_2147183, EPI_ISL_2147184, EPI_ISL_2147185, EPI_ISL_2147186, EPI_ISL_2147187, EPI_ISL_2147188, EPI_ISL_2147189, EPI_ISL_2147190, EPI_ISL_2147191, EPI_ISL_2147192, EPI_ISL_2147193,
EPI_ISL_2147194, EPI_ISL_2147195, EPI_ISL_2147196, EPI_ISL_2147198, EPI_ISL_2147199, EPI_ISL_2147200, EPI_ISL_2147201, EPI_ISL_2147202, EPI_ISL_2147203, EPI_ISL_2147204, EPI_ISL_2147205, EPI_ISL_2147206, EPI_ISL_2147207, EPI_ISL_2147208, EPI_ISL_2147209, EPI_ISL_2147210, EPI_ISL_2147211, EPI_ISL_2147212, EPI_ISL_2147213, EPI_ISL_2147214, EPI_ISL_2147215, EPI_ISL_2147216,
EPI_ISL_2147217, EPI_ISL_2147218, EPI_ISL_2147219, EPI_ISL_2147220, EPI_ISL_2147221, EPI_ISL_2147222, EPI_ISL_2147223, EPI_ISL_2147224, EPI_ISL_2147225, EPI_ISL_2147226, EPI_ISL_2147227, EPI_ISL_2147228, EPI_ISL_2147229, EPI_ISL_2147230, EPI_ISL_2147231, EPI_ISL_2147232, EPI_ISL_2147233, EPI_ISL_2147234, EPI_ISL_2147235, EPI_ISL_2147236, EPI_ISL_2147237, EPI_ISL_2147238,
EPI_ISL_2147239, EPI_ISL_2147240, EPI_ISL_2147241, EPI_ISL_2147242, EPI_ISL_2147243, EPI_ISL_2147244, EPI_ISL_2147245, EPI_ISL_2147246, EPI_ISL_2147248, EPI_ISL_2147249, EPI_ISL_2147250, EPI_ISL_2147251, EPI_ISL_2147252, EPI_ISL_2147253, EPI_ISL_2147255, EPI_ISL_2147258, EPI_ISL_2147260, EPI_ISL_2147262, EPI_ISL_2147265, EPI_ISL_2147268, EPI_ISL_2147270, EPI_ISL_2147273,
EPI_ISL_2147276, EPI_ISL_2147278, EPI_ISL_2147281, EPI_ISL_2147284, EPI_ISL_2147287, EPI_ISL_2147289, EPI_ISL_2147292, EPI_ISL_2147293, EPI_ISL_2147295, EPI_ISL_2147298, EPI_ISL_2147301, EPI_ISL_2147303, EPI_ISL_2147309, EPI_ISL_2147311, EPI_ISL_2147313, EPI_ISL_2147315, EPI_ISL_2147317, EPI_ISL_2147320, EPI_ISL_2147323, EPI_ISL_2147326, EPI_ISL_2147328, EPI_ISL_2147331,
EPI_ISL_2147333, EPI_ISL_2147336, EPI_ISL_2147339, EPI_ISL_2147342, EPI_ISL_2147344, EPI_ISL_2147346, EPI_ISL_2147348, EPI_ISL_2147350, EPI_ISL_2147352, EPI_ISL_2147354, EPI_ISL_2147356, EPI_ISL_2147358, EPI_ISL_2147360, EPI_ISL_2147362, EPI_ISL_2147365, EPI_ISL_2147367, EPI_ISL_2147369, EPI_ISL_2147372, EPI_ISL_2147374, EPI_ISL_2147376, EPI_ISL_2147378, EPI_ISL_2147380,
EPI_ISL_2147382, EPI_ISL_2147385, EPI_ISL_2147387, EPI_ISL_2147394, EPI_ISL_2147396, EPI_ISL_2147398, EPI_ISL_2147400, EPI_ISL_2147403, EPI_ISL_2147405, EPI_ISL_2147406, EPI_ISL_2147408, EPI_ISL_2147410, EPI_ISL_2147411, EPI_ISL_2147412, EPI_ISL_2147413, EPI_ISL_2147414, EPI_ISL_2147415, EPI_ISL_2147416, EPI_ISL_2147417, EPI_ISL_2147418, EPI_ISL_2147419, EPI_ISL_2147420,
EPI_ISL_2147421, EPI_ISL_2147422, EPI_ISL_2147423, EPI_ISL_2147424, EPI_ISL_2147425, EPI_ISL_2147426, EPI_ISL_2147427, EPI_ISL_2147428, EPI_ISL_2147429, EPI_ISL_2147430, EPI_ISL_2147431, EPI_ISL_2147432, EPI_ISL_2147433, EPI_ISL_2147434, EPI_ISL_2147436, EPI_ISL_2147438, EPI_ISL_2147439, EPI_ISL_2147440, EPI_ISL_2147441, EPI_ISL_2147442, EPI_ISL_2147443, EPI_ISL_2147444,
EPI_ISL_2147445, EPI_ISL_2147446, EPI_ISL_2147447, EPI_ISL_2147448, EPI_ISL_2147449, EPI_ISL_2147450, EPI_ISL_2147451, EPI_ISL_2147452, EPI_ISL_2147453, EPI_ISL_2147454, EPI_ISL_2147455, EPI_ISL_2147456, EPI_ISL_2147457, EPI_ISL_2147458, EPI_ISL_2147459, EPI_ISL_2147460, EPI_ISL_2147461, EPI_ISL_2147462, EPI_ISL_2147463, EPI_ISL_2147464, EPI_ISL_2147465, EPI_ISL_2147466,
EPI_ISL_2147467, EPI_ISL_2147468, EPI_ISL_2147469, EPI_ISL_2147470, EPI_ISL_2147471, EPI_ISL_2147472, EPI_ISL_2147473, EPI_ISL_2147474, EPI_ISL_2147475, EPI_ISL_2147476, EPI_ISL_2147477, EPI_ISL_2147478, EPI_ISL_2147479, EPI_ISL_2147480, EPI_ISL_2147481, EPI_ISL_2147482, EPI_ISL_2147483, EPI_ISL_2147484, EPI_ISL_2147485, EPI_ISL_2147486, EPI_ISL_2147487, EPI_ISL_2147488,
EPI_ISL_2147489, EPI_ISL_2147490, EPI_ISL_2147491, EPI_ISL_2147492, EPI_ISL_2147493, EPI_ISL_2147494, EPI_ISL_2147495, EPI_ISL_2147496, EPI_ISL_2147497, EPI_ISL_2147498, EPI_ISL_2147499, EPI_ISL_2147500, EPI_ISL_2147501, EPI_ISL_2147502, EPI_ISL_2147503, EPI_ISL_2147504, EPI_ISL_2147505, EPI_ISL_2147506, EPI_ISL_2147507, EPI_ISL_2147508, EPI_ISL_2147509, EPI_ISL_2147510,
EPI_ISL_2147511, EPI_ISL_2147512, EPI_ISL_2147513, EPI_ISL_2147514, EPI_ISL_2147515, EPI_ISL_2147516, EPI_ISL_2147517, EPI_ISL_2147518, EPI_ISL_2147519, EPI_ISL_2147520, EPI_ISL_2147521, EPI_ISL_2147522, EPI_ISL_2147523, EPI_ISL_2147524, EPI_ISL_2147525, EPI_ISL_2147526, EPI_ISL_2147527, EPI_ISL_2147528, EPI_ISL_2147529, EPI_ISL_2147530, EPI_ISL_2147531, EPI_ISL_2147532,
EPI_ISL_2147533, EPI_ISL_2147534, EPI_ISL_2147535, EPI_ISL_2147536, EPI_ISL_2147537, EPI_ISL_2147538, EPI_ISL_2147539, EPI_ISL_2147540, EPI_ISL_2147541, EPI_ISL_2147542, EPI_ISL_2147543, EPI_ISL_2147544, EPI_ISL_2147545, EPI_ISL_2147546, EPI_ISL_2147547, EPI_ISL_2147548, EPI_ISL_2147549, EPI_ISL_2147550, EPI_ISL_2147551, EPI_ISL_2147552, EPI_ISL_2147553, EPI_ISL_2147554,
EPI_ISL_2147555, EPI_ISL_2147556, EPI_ISL_2147557, EPI_ISL_2147559, EPI_ISL_2147560, EPI_ISL_2147561, EPI_ISL_2147562, EPI_ISL_2147563, EPI_ISL_2147564, EPI_ISL_2147565, EPI_ISL_2147566, EPI_ISL_2147567, EPI_ISL_2147568, EPI_ISL_2147569, EPI_ISL_2147570, EPI_ISL_2147571, EPI_ISL_2147572, EPI_ISL_2147573, EPI_ISL_2147574, EPI_ISL_2147575, EPI_ISL_2147576, EPI_ISL_2147577,
EPI_ISL_2147578, EPI_ISL_2147579, EPI_ISL_2147580, EPI_ISL_2147581, EPI_ISL_2147582, EPI_ISL_2147583, EPI_ISL_2147584, EPI_ISL_2147585, EPI_ISL_2147586, EPI_ISL_2147587, EPI_ISL_2147588, EPI_ISL_2147589, EPI_ISL_2147590, EPI_ISL_2147591, EPI_ISL_2147592, EPI_ISL_2147593, EPI_ISL_2147594, EPI_ISL_2147595, EPI_ISL_2147596, EPI_ISL_2147597, EPI_ISL_2147598, EPI_ISL_2147599,
EPI_ISL_2147600, EPI_ISL_2147601, EPI_ISL_2147602, EPI_ISL_2147604, EPI_ISL_2147605, EPI_ISL_2147606, EPI_ISL_2147607, EPI_ISL_2147608, EPI_ISL_2147609, EPI_ISL_2147610, EPI_ISL_2147611, EPI_ISL_2147612, EPI_ISL_2147613, EPI_ISL_2147614, EPI_ISL_2147615, EPI_ISL_2147616, EPI_ISL_2147617, EPI_ISL_2147619, EPI_ISL_2147620, EPI_ISL_2147621, EPI_ISL_2147622, EPI_ISL_2147623,
EPI_ISL_2147624, EPI_ISL_2147625, EPI_ISL_2147626, EPI_ISL_2147627, EPI_ISL_2147628, EPI_ISL_2147629, EPI_ISL_2147630, EPI_ISL_2147631, EPI_ISL_2147632, EPI_ISL_2147633, EPI_ISL_2147634, EPI_ISL_2147635, EPI_ISL_2147636, EPI_ISL_2147637, EPI_ISL_2147638, EPI_ISL_2147639, EPI_ISL_2147640, EPI_ISL_2147641, EPI_ISL_2147642, EPI_ISL_2147643, EPI_ISL_2147644, EPI_ISL_2147645,
EPI_ISL_2147646, EPI_ISL_2147647, EPI_ISL_2147648, EPI_ISL_2147649, EPI_ISL_2147650, EPI_ISL_2147651, EPI_ISL_2147652, EPI_ISL_2147653, EPI_ISL_2147654, EPI_ISL_2147655, EPI_ISL_2147656, EPI_ISL_2147657, EPI_ISL_2147658, EPI_ISL_2147660, EPI_ISL_2147661, EPI_ISL_2147662, EPI_ISL_2147663, EPI_ISL_2147665, EPI_ISL_2147666, EPI_ISL_2147667, EPI_ISL_2147669, EPI_ISL_2147670,
EPI_ISL_2147671, EPI_ISL_2147672, EPI_ISL_2147673, EPI_ISL_2147674, EPI_ISL_2147675, EPI_ISL_2147676, EPI_ISL_2147677, EPI_ISL_2147678, EPI_ISL_2147679, EPI_ISL_2147680, EPI_ISL_2147681, EPI_ISL_2147682, EPI_ISL_2147683, EPI_ISL_2147684, EPI_ISL_2147685, EPI_ISL_2147686, EPI_ISL_2147687, EPI_ISL_2147688, EPI_ISL_2147689, EPI_ISL_2147690, EPI_ISL_2147691, EPI_ISL_2147692,
EPI_ISL_2147693, EPI_ISL_2147695, EPI_ISL_2147697, EPI_ISL_2147699, EPI_ISL_2147701, EPI_ISL_2147703, EPI_ISL_2147705, EPI_ISL_2147707, EPI_ISL_2147709, EPI_ISL_2147711, EPI_ISL_2147713, EPI_ISL_2147715, EPI_ISL_2147717, EPI_ISL_2147718, EPI_ISL_2147720, EPI_ISL_2147722, EPI_ISL_2147724, EPI_ISL_2147726, EPI_ISL_2147728, EPI_ISL_2147730, EPI_ISL_2147733, EPI_ISL_2147735,
EPI_ISL_2147737, EPI_ISL_2147738, EPI_ISL_2147741, EPI_ISL_2147743, EPI_ISL_2147745, EPI_ISL_2147747, EPI_ISL_2147749, EPI_ISL_2147751, EPI_ISL_2147753, EPI_ISL_2147755, EPI_ISL_2147757, EPI_ISL_2147759, EPI_ISL_2147761, EPI_ISL_2147763, EPI_ISL_2147765, EPI_ISL_2147768, EPI_ISL_2147770, EPI_ISL_2147771, EPI_ISL_2147776, EPI_ISL_2147778, EPI_ISL_2147781, EPI_ISL_2147782,
EPI_ISL_2147784, EPI_ISL_2147786, EPI_ISL_2147788, EPI_ISL_2147789, EPI_ISL_2147791, EPI_ISL_2147793, EPI_ISL_2147795, EPI_ISL_2147797, EPI_ISL_2147798, EPI_ISL_2147800, EPI_ISL_2147801, EPI_ISL_2147803, EPI_ISL_2147804, EPI_ISL_2147806, EPI_ISL_2147807, EPI_ISL_2147809, EPI_ISL_2147811, EPI_ISL_2147813, EPI_ISL_2147815, EPI_ISL_2147817, EPI_ISL_2147819, EPI_ISL_2147821,
EPI_ISL_2147824, EPI_ISL_2147826, EPI_ISL_2147828, EPI_ISL_2147830, EPI_ISL_2147832, EPI_ISL_2147834, EPI_ISL_2147836, EPI_ISL_2147838, EPI_ISL_2147840, EPI_ISL_2147844, EPI_ISL_2147846, EPI_ISL_2147848, EPI_ISL_2147850, EPI_ISL_2147852, EPI_ISL_2147854, EPI_ISL_2147855, EPI_ISL_2147856, EPI_ISL_2147857, EPI_ISL_2147858, EPI_ISL_2147859, EPI_ISL_2147860, EPI_ISL_2147861,
EPI_ISL_2147862, EPI_ISL_2147863, EPI_ISL_2147864, EPI_ISL_2147865, EPI_ISL_2147866, EPI_ISL_2147867, EPI_ISL_2147868, EPI_ISL_2147869, EPI_ISL_2147870, EPI_ISL_2147871, EPI_ISL_2147872, EPI_ISL_2147873, EPI_ISL_2147874, EPI_ISL_2147875, EPI_ISL_2147876, EPI_ISL_2147877, EPI_ISL_2147878, EPI_ISL_2147879, EPI_ISL_2147880, EPI_ISL_2147881, EPI_ISL_2147882, EPI_ISL_2147883,
EPI_ISL_2147884, EPI_ISL_2147885, EPI_ISL_2147886, EPI_ISL_2147887, EPI_ISL_2147888, EPI_ISL_2147889, EPI_ISL_2147890, EPI_ISL_2147891, EPI_ISL_2147892, EPI_ISL_2147893, EPI_ISL_2147894, EPI_ISL_2147895, EPI_ISL_2147896, EPI_ISL_2147897, EPI_ISL_2147898, EPI_ISL_2147899, EPI_ISL_2147900, EPI_ISL_2147901, EPI_ISL_2147902, EPI_ISL_2147903, EPI_ISL_2147904, EPI_ISL_2147905,
EPI_ISL_2147906, EPI_ISL_2147907, EPI_ISL_2147908, EPI_ISL_2147909, EPI_ISL_2147910, EPI_ISL_2147911, EPI_ISL_2147912, EPI_ISL_2147913, EPI_ISL_2147914, EPI_ISL_2147915, EPI_ISL_2147916, EPI_ISL_2147917, EPI_ISL_2147918, EPI_ISL_2147919, EPI_ISL_2147920, EPI_ISL_2147921, EPI_ISL_2147922, EPI_ISL_2147923, EPI_ISL_2147924, EPI_ISL_2147925, EPI_ISL_2147926, EPI_ISL_2147927,
EPI_ISL_2147928, EPI_ISL_2147929, EPI_ISL_2147930, EPI_ISL_2147931, EPI_ISL_2147932, EPI_ISL_2147933, EPI_ISL_2147934, EPI_ISL_2147935, EPI_ISL_2147936, EPI_ISL_2147937, EPI_ISL_2147938, EPI_ISL_2147939, EPI_ISL_2147940, EPI_ISL_2147941, EPI_ISL_2147942, EPI_ISL_2147943, EPI_ISL_2147944, EPI_ISL_2147945, EPI_ISL_2147946, EPI_ISL_2147948, EPI_ISL_2147949, EPI_ISL_2147950,
EPI_ISL_2147951, EPI_ISL_2147952, EPI_ISL_2147954, EPI_ISL_2147955, EPI_ISL_2147956, EPI_ISL_2147957, EPI_ISL_2147958, EPI_ISL_2147959, EPI_ISL_2147960, EPI_ISL_2147961, EPI_ISL_2147962, EPI_ISL_2147963, EPI_ISL_2147964, EPI_ISL_2147965, EPI_ISL_2147966, EPI_ISL_2147967, EPI_ISL_2147968, EPI_ISL_2147969, EPI_ISL_2147971, EPI_ISL_2147972, EPI_ISL_2147973, EPI_ISL_2147974,
EPI_ISL_2147975, EPI_ISL_2147976, EPI_ISL_2147977, EPI_ISL_2147978, EPI_ISL_2147979, EPI_ISL_2147980, EPI_ISL_2147981, EPI_ISL_2147982, EPI_ISL_2147983, EPI_ISL_2147984, EPI_ISL_2147985, EPI_ISL_2147986, EPI_ISL_2147987, EPI_ISL_2147988, EPI_ISL_2147989, EPI_ISL_2147990, EPI_ISL_2147991, EPI_ISL_2147992, EPI_ISL_2147993, EPI_ISL_2147994, EPI_ISL_2147995, EPI_ISL_2147996,
EPI_ISL_2147997, EPI_ISL_2147998, EPI_ISL_2147999, EPI_ISL_2148000, EPI_ISL_2148001, EPI_ISL_2148002, EPI_ISL_2148003, EPI_ISL_2148004, EPI_ISL_2148005, EPI_ISL_2148006, EPI_ISL_2148007, EPI_ISL_2148008, EPI_ISL_2148009, EPI_ISL_2148010, EPI_ISL_2148011, EPI_ISL_2148012, EPI_ISL_2148013, EPI_ISL_2148014, EPI_ISL_2148015, EPI_ISL_2148016, EPI_ISL_2148017, EPI_ISL_2148018,
EPI_ISL_2148019, EPI_ISL_2148020, EPI_ISL_2148021, EPI_ISL_2148022, EPI_ISL_2148023, EPI_ISL_2148024, EPI_ISL_2148025, EPI_ISL_2148026, EPI_ISL_2148027, EPI_ISL_2148028, EPI_ISL_2148029, EPI_ISL_2148030, EPI_ISL_2148031, EPI_ISL_2148032, EPI_ISL_2148033, EPI_ISL_2148034, EPI_ISL_2148035, EPI_ISL_2148036, EPI_ISL_2148037, EPI_ISL_2148038, EPI_ISL_2148039, EPI_ISL_2148040,
EPI_ISL_2148041, EPI_ISL_2148042, EPI_ISL_2148043, EPI_ISL_2148044, EPI_ISL_2148045, EPI_ISL_2148047, EPI_ISL_2148048, EPI_ISL_2148049, EPI_ISL_2148050, EPI_ISL_2148051, EPI_ISL_2148052, EPI_ISL_2148053, EPI_ISL_2148054, EPI_ISL_2148055, EPI_ISL_2148056, EPI_ISL_2148057, EPI_ISL_2148058, EPI_ISL_2148059, EPI_ISL_2148060, EPI_ISL_2148061, EPI_ISL_2148062, EPI_ISL_2148063,
EPI_ISL_2148064, EPI_ISL_2148065, EPI_ISL_2148066, EPI_ISL_2148067, EPI_ISL_2148068, EPI_ISL_2148069, EPI_ISL_2148070, EPI_ISL_2148071, EPI_ISL_2148072, EPI_ISL_2148073, EPI_ISL_2148074, EPI_ISL_2148075, EPI_ISL_2148076, EPI_ISL_2148077, EPI_ISL_2148078, EPI_ISL_2148079, EPI_ISL_2148080, EPI_ISL_2148081, EPI_ISL_2148082, EPI_ISL_2148083, EPI_ISL_2148084, EPI_ISL_2148085,
EPI_ISL_2148086, EPI_ISL_2148087, EPI_ISL_2148088, EPI_ISL_2148089, EPI_ISL_2148090, EPI_ISL_2148091, EPI_ISL_2148092, EPI_ISL_2148093, EPI_ISL_2148094, EPI_ISL_2148095, EPI_ISL_2148096, EPI_ISL_2148097, EPI_ISL_2148098, EPI_ISL_2148099, EPI_ISL_2148100, EPI_ISL_2148101, EPI_ISL_2148102, EPI_ISL_2148103, EPI_ISL_2148104, EPI_ISL_2148105, EPI_ISL_2148106, EPI_ISL_2148107,
EPI_ISL_2148108, EPI_ISL_2148109, EPI_ISL_2148110, EPI_ISL_2148111, EPI_ISL_2148112, EPI_ISL_2148113, EPI_ISL_2148114, EPI_ISL_2148115, EPI_ISL_2148116, EPI_ISL_2148117, EPI_ISL_2148118, EPI_ISL_2148119, EPI_ISL_2148120, EPI_ISL_2148121, EPI_ISL_2148122, EPI_ISL_2148123, EPI_ISL_2148124, EPI_ISL_2148125, EPI_ISL_2148126, EPI_ISL_2148127, EPI_ISL_2148128, EPI_ISL_2148129,
EPI_ISL_2148130, EPI_ISL_2148131, EPI_ISL_2148132, EPI_ISL_2148133, EPI_ISL_2148134, EPI_ISL_2148135, EPI_ISL_2148136, EPI_ISL_2148137, EPI_ISL_2148138, EPI_ISL_2148139, EPI_ISL_2148140, EPI_ISL_2148141, EPI_ISL_2148143, EPI_ISL_2148144, EPI_ISL_2148145, EPI_ISL_2148147, EPI_ISL_2148148, EPI_ISL_2148149, EPI_ISL_2148150, EPI_ISL_2148151, EPI_ISL_2148152, EPI_ISL_2148153,
EPI_ISL_2148154, EPI_ISL_2148155, EPI_ISL_2148156, EPI_ISL_2148157, EPI_ISL_2148158, EPI_ISL_2148159, EPI_ISL_2148160, EPI_ISL_2148161, EPI_ISL_2148162, EPI_ISL_2148163, EPI_ISL_2148164, EPI_ISL_2148165, EPI_ISL_2148166, EPI_ISL_2148167, EPI_ISL_2148168, EPI_ISL_2148169, EPI_ISL_2148170, EPI_ISL_2148171, EPI_ISL_2148172, EPI_ISL_2148173, EPI_ISL_2148174, EPI_ISL_2148175,
EPI_ISL_2148176, EPI_ISL_2148177, EPI_ISL_2148178, EPI_ISL_2148179, EPI_ISL_2148180, EPI_ISL_2148181, EPI_ISL_2148182, EPI_ISL_2148183, EPI_ISL_2148184, EPI_ISL_2148185, EPI_ISL_2148186, EPI_ISL_2148187, EPI_ISL_2148188, EPI_ISL_2148189, EPI_ISL_2148190, EPI_ISL_2148191, EPI_ISL_2148192, EPI_ISL_2148193, EPI_ISL_2148195, EPI_ISL_2148196, EPI_ISL_2148197, EPI_ISL_2148198,
EPI_ISL_2148199, EPI_ISL_2148200, EPI_ISL_2148201, EPI_ISL_2148202, EPI_ISL_2148203, EPI_ISL_2148204, EPI_ISL_2148205, EPI_ISL_2148206, EPI_ISL_2148207, EPI_ISL_2148208, EPI_ISL_2148209, EPI_ISL_2148210, EPI_ISL_2148212, EPI_ISL_2148213, EPI_ISL_2148214, EPI_ISL_2148215, EPI_ISL_2148216, EPI_ISL_2148217, EPI_ISL_2148218, EPI_ISL_2148219, EPI_ISL_2148220, EPI_ISL_2148221,
EPI_ISL_2148223, EPI_ISL_2148224, EPI_ISL_2148225, EPI_ISL_2148226, EPI_ISL_2148227, EPI_ISL_2148228, EPI_ISL_2148229, EPI_ISL_2148230, EPI_ISL_2148231, EPI_ISL_2148232, EPI_ISL_2148233, EPI_ISL_2148234, EPI_ISL_2148235, EPI_ISL_2148236, EPI_ISL_2148237, EPI_ISL_2148238, EPI_ISL_2148239, EPI_ISL_2148240, EPI_ISL_2148241, EPI_ISL_2148242, EPI_ISL_2148243, EPI_ISL_2148245,
EPI_ISL_2148246, EPI_ISL_2148247, EPI_ISL_2148248, EPI_ISL_2148249, EPI_ISL_2148250, EPI_ISL_2148251, EPI_ISL_2148252, EPI_ISL_2148253, EPI_ISL_2148254, EPI_ISL_2148255, EPI_ISL_2148256, EPI_ISL_2148257, EPI_ISL_2148258, EPI_ISL_2148259, EPI_ISL_2148260, EPI_ISL_2148261, EPI_ISL_2148262, EPI_ISL_2148263, EPI_ISL_2148264, EPI_ISL_2148265, EPI_ISL_2148266, EPI_ISL_2148267,
EPI_ISL_2148268, EPI_ISL_2148269, EPI_ISL_2148270, EPI_ISL_2148271, EPI_ISL_2148272, EPI_ISL_2148273, EPI_ISL_2148274, EPI_ISL_2148275, EPI_ISL_2148276, EPI_ISL_2148277, EPI_ISL_2148278, EPI_ISL_2148279, EPI_ISL_2148280, EPI_ISL_2148281, EPI_ISL_2148282, EPI_ISL_2148283, EPI_ISL_2148284, EPI_ISL_2148285, EPI_ISL_2148286, EPI_ISL_2148287, EPI_ISL_2148288, EPI_ISL_2148290,
EPI_ISL_2148291, EPI_ISL_2148292, EPI_ISL_2148293, EPI_ISL_2148294, EPI_ISL_2148295, EPI_ISL_2148296, EPI_ISL_2148297, EPI_ISL_2148298, EPI_ISL_2148299, EPI_ISL_2148300, EPI_ISL_2148301, EPI_ISL_2148302, EPI_ISL_2148303, EPI_ISL_2148304, EPI_ISL_2148305, EPI_ISL_2148306, EPI_ISL_2148307, EPI_ISL_2148308, EPI_ISL_2148309, EPI_ISL_2148310, EPI_ISL_2148311, EPI_ISL_2148312,
EPI_ISL_2148313, EPI_ISL_2148314, EPI_ISL_2148315, EPI_ISL_2148316, EPI_ISL_2148317, EPI_ISL_2148318, EPI_ISL_2148319, EPI_ISL_2148320, EPI_ISL_2148321, EPI_ISL_2148322, EPI_ISL_2148323, EPI_ISL_2148324, EPI_ISL_2148325, EPI_ISL_2148326, EPI_ISL_2148327, EPI_ISL_2148328, EPI_ISL_2148329, EPI_ISL_2148330, EPI_ISL_2148331, EPI_ISL_2148332, EPI_ISL_2148334, EPI_ISL_2148335,
EPI_ISL_2148336, EPI_ISL_2148337, EPI_ISL_2148338, EPI_ISL_2148339, EPI_ISL_2148340, EPI_ISL_2148341, EPI_ISL_2148342, EPI_ISL_2148343, EPI_ISL_2148344, EPI_ISL_2148345, EPI_ISL_2148347, EPI_ISL_2148348, EPI_ISL_2148349, EPI_ISL_2148350, EPI_ISL_2148351, EPI_ISL_2148353, EPI_ISL_2148354, EPI_ISL_2148355, EPI_ISL_2148357, EPI_ISL_2148358, EPI_ISL_2148359, EPI_ISL_2148360,
EPI_ISL_2148361, EPI_ISL_2148362, EPI_ISL_2148363, EPI_ISL_2148365, EPI_ISL_2148366, EPI_ISL_2148367, EPI_ISL_2148368, EPI_ISL_2148369, EPI_ISL_2148370, EPI_ISL_2148371, EPI_ISL_2148372, EPI_ISL_2148373, EPI_ISL_2148374, EPI_ISL_2148375, EPI_ISL_2148376, EPI_ISL_2148377, EPI_ISL_2148378, EPI_ISL_2148379, EPI_ISL_2148380, EPI_ISL_2148381, EPI_ISL_2148382, EPI_ISL_2148383,
EPI_ISL_2148384, EPI_ISL_2148385, EPI_ISL_2148386, EPI_ISL_2148387, EPI_ISL_2148388, EPI_ISL_2148389, EPI_ISL_2148390, EPI_ISL_2148391, EPI_ISL_2148392, EPI_ISL_2148393, EPI_ISL_2148394, EPI_ISL_2148395, EPI_ISL_2148396, EPI_ISL_2148397, EPI_ISL_2148398, EPI_ISL_2148399, EPI_ISL_2148400, EPI_ISL_2148401, EPI_ISL_2148402, EPI_ISL_2148403, EPI_ISL_2148404, EPI_ISL_2148405,
EPI_ISL_2148406, EPI_ISL_2148407, EPI_ISL_2148408, EPI_ISL_2148409, EPI_ISL_2148410, EPI_ISL_2148411, EPI_ISL_2148412, EPI_ISL_2148413, EPI_ISL_2148414, EPI_ISL_2148415, EPI_ISL_2148416, EPI_ISL_2148417, EPI_ISL_2148418, EPI_ISL_2148419, EPI_ISL_2148420, EPI_ISL_2148421, EPI_ISL_2148422, EPI_ISL_2148423, EPI_ISL_2148424, EPI_ISL_2148425, EPI_ISL_2148426, EPI_ISL_2148427,
EPI_ISL_2148428, EPI_ISL_2148429, EPI_ISL_2148430, EPI_ISL_2148431, EPI_ISL_2148432, EPI_ISL_2148433, EPI_ISL_2148434, EPI_ISL_2148435, EPI_ISL_2148436, EPI_ISL_2148437, EPI_ISL_2148438, EPI_ISL_2148439, EPI_ISL_2148440, EPI_ISL_2148441, EPI_ISL_2148442, EPI_ISL_2148443, EPI_ISL_2148444, EPI_ISL_2148445, EPI_ISL_2148446, EPI_ISL_2148447, EPI_ISL_2148448, EPI_ISL_2148449,
EPI_ISL_2148450, EPI_ISL_2148451, EPI_ISL_2148452, EPI_ISL_2148453, EPI_ISL_2148454, EPI_ISL_2148455, EPI_ISL_2148456, EPI_ISL_2148457, EPI_ISL_2148458, EPI_ISL_2148459, EPI_ISL_2148460, EPI_ISL_2148461, EPI_ISL_2148462, EPI_ISL_2148463, EPI_ISL_2148464, EPI_ISL_2148465, EPI_ISL_2148466, EPI_ISL_2148467, EPI_ISL_2148468, EPI_ISL_2148469, EPI_ISL_2148470, EPI_ISL_2148471,
EPI_ISL_2148472, EPI_ISL_2148473, EPI_ISL_2148474, EPI_ISL_2148475, EPI_ISL_2148476, EPI_ISL_2148477, EPI_ISL_2148478, EPI_ISL_2148479, EPI_ISL_2148480, EPI_ISL_2148481, EPI_ISL_2148482, EPI_ISL_2148483, EPI_ISL_2148484, EPI_ISL_2148485, EPI_ISL_2148486, EPI_ISL_2148487, EPI_ISL_2148488, EPI_ISL_2148489, EPI_ISL_2148490, EPI_ISL_2148491, EPI_ISL_2148492, EPI_ISL_2148493,
EPI_ISL_2148494, EPI_ISL_2148495, EPI_ISL_2148496, EPI_ISL_2148497, EPI_ISL_2148498, EPI_ISL_2148499, EPI_ISL_2148500, EPI_ISL_2148501, EPI_ISL_2148502, EPI_ISL_2148504, EPI_ISL_2148505, EPI_ISL_2148506, EPI_ISL_2148507, EPI_ISL_2148508, EPI_ISL_2148509, EPI_ISL_2148510, EPI_ISL_2148511, EPI_ISL_2148512, EPI_ISL_2148513, EPI_ISL_2148514, EPI_ISL_2148515, EPI_ISL_2148516,
EPI_ISL_2148517, EPI_ISL_2148518, EPI_ISL_2148519, EPI_ISL_2148520, EPI_ISL_2148521, EPI_ISL_2148522, EPI_ISL_2148523, EPI_ISL_2148524, EPI_ISL_2148525, EPI_ISL_2148526, EPI_ISL_2148527, EPI_ISL_2148528, EPI_ISL_2148529, EPI_ISL_2148530, EPI_ISL_2148531, EPI_ISL_2148532, EPI_ISL_2148533, EPI_ISL_2148534, EPI_ISL_2148535, EPI_ISL_2148536, EPI_ISL_2148537, EPI_ISL_2148538,
EPI_ISL_2148539, EPI_ISL_2148540, EPI_ISL_2148541, EPI_ISL_2148542, EPI_ISL_2148543, EPI_ISL_2148544, EPI_ISL_2148545, EPI_ISL_2148546, EPI_ISL_2148547, EPI_ISL_2148548, EPI_ISL_2148549, EPI_ISL_2148550, EPI_ISL_2148551, EPI_ISL_2148552, EPI_ISL_2148553, EPI_ISL_2148554, EPI_ISL_2148555, EPI_ISL_2148556, EPI_ISL_2148557, EPI_ISL_2148558, EPI_ISL_2148559, EPI_ISL_2148560,
EPI_ISL_2148561, EPI_ISL_2148562, EPI_ISL_2148563, EPI_ISL_2148564, EPI_ISL_2148565, EPI_ISL_2148566, EPI_ISL_2148567, EPI_ISL_2148568, EPI_ISL_2148569, EPI_ISL_2148570, EPI_ISL_2148571, EPI_ISL_2148572, EPI_ISL_2148573, EPI_ISL_2148574, EPI_ISL_2148575, EPI_ISL_2148576, EPI_ISL_2148577, EPI_ISL_2148578, EPI_ISL_2148579, EPI_ISL_2148580, EPI_ISL_2148581, EPI_ISL_2148582,
EPI_ISL_2148583, EPI_ISL_2148584, EPI_ISL_2148585, EPI_ISL_2148586, EPI_ISL_2148587, EPI_ISL_2148588, EPI_ISL_2148589, EPI_ISL_2148590, EPI_ISL_2148591, EPI_ISL_2148592, EPI_ISL_2148593, EPI_ISL_2148594, EPI_ISL_2148595, EPI_ISL_2148596, EPI_ISL_2148597, EPI_ISL_2148598, EPI_ISL_2148599, EPI_ISL_2148600, EPI_ISL_2148601, EPI_ISL_2148603, EPI_ISL_2148605, EPI_ISL_2148606,
EPI_ISL_2148607, EPI_ISL_2148608, EPI_ISL_2148609, EPI_ISL_2148610, EPI_ISL_2148611, EPI_ISL_2148612, EPI_ISL_2148613, EPI_ISL_2148614, EPI_ISL_2148615, EPI_ISL_2148616, EPI_ISL_2148617, EPI_ISL_2148618, EPI_ISL_2148619, EPI_ISL_2148620, EPI_ISL_2148621, EPI_ISL_2148622, EPI_ISL_2148623, EPI_ISL_2148624, EPI_ISL_2148625, EPI_ISL_2148626, EPI_ISL_2148627, EPI_ISL_2148628,
EPI_ISL_2148629, EPI_ISL_2148630, EPI_ISL_2148631, EPI_ISL_2148632, EPI_ISL_2148633, EPI_ISL_2148634, EPI_ISL_2148635, EPI_ISL_2148636, EPI_ISL_2148637, EPI_ISL_2148638, EPI_ISL_2148639, EPI_ISL_2148640, EPI_ISL_2148641, EPI_ISL_2148642, EPI_ISL_2148643, EPI_ISL_2148645, EPI_ISL_2148646, EPI_ISL_2148647, EPI_ISL_2148648, EPI_ISL_2148649, EPI_ISL_2148650, EPI_ISL_2148651,
EPI_ISL_2148652, EPI_ISL_2148653, EPI_ISL_2148654, EPI_ISL_2148656, EPI_ISL_2148657, EPI_ISL_2148658, EPI_ISL_2148659, EPI_ISL_2148660, EPI_ISL_2148661, EPI_ISL_2148662, EPI_ISL_2148663, EPI_ISL_2148664, EPI_ISL_2148665, EPI_ISL_2148666, EPI_ISL_2148667, EPI_ISL_2148668, EPI_ISL_2148669, EPI_ISL_2148670, EPI_ISL_2148671, EPI_ISL_2148672, EPI_ISL_2148673, EPI_ISL_2148674,
EPI_ISL_2148675, EPI_ISL_2148676, EPI_ISL_2148678, EPI_ISL_2148679, EPI_ISL_2148680, EPI_ISL_2148681, EPI_ISL_2148682, EPI_ISL_2148683, EPI_ISL_2148684, EPI_ISL_2148685, EPI_ISL_2148686, EPI_ISL_2148687, EPI_ISL_2148688, EPI_ISL_2148689, EPI_ISL_2148690, EPI_ISL_2148691, EPI_ISL_2148692, EPI_ISL_2148693, EPI_ISL_2148694, EPI_ISL_2148695, EPI_ISL_2148696, EPI_ISL_2148697,
EPI_ISL_2148698, EPI_ISL_2148699, EPI_ISL_2148700, EPI_ISL_2148701, EPI_ISL_2148702, EPI_ISL_2148703, EPI_ISL_2148704, EPI_ISL_2148705, EPI_ISL_2148706, EPI_ISL_2148707, EPI_ISL_2148708, EPI_ISL_2148709, EPI_ISL_2148710, EPI_ISL_2148711, EPI_ISL_2148712, EPI_ISL_2148713, EPI_ISL_2148714, EPI_ISL_2148715, EPI_ISL_2148716, EPI_ISL_2148717, EPI_ISL_2148718, EPI_ISL_2148719,
EPI_ISL_2148720, EPI_ISL_2148721, EPI_ISL_2148722, EPI_ISL_2148723, EPI_ISL_2148724, EPI_ISL_2148725, EPI_ISL_2148726, EPI_ISL_2148727, EPI_ISL_2148728, EPI_ISL_2148729, EPI_ISL_2148730, EPI_ISL_2148731, EPI_ISL_2148732, EPI_ISL_2148733, EPI_ISL_2148734, EPI_ISL_2148735, EPI_ISL_2148736, EPI_ISL_2148737, EPI_ISL_2148738, EPI_ISL_2148739, EPI_ISL_2148740, EPI_ISL_2148741,
EPI_ISL_2148742, EPI_ISL_2148743, EPI_ISL_2148744, EPI_ISL_2148745, EPI_ISL_2148746, EPI_ISL_2148747, EPI_ISL_2148748, EPI_ISL_2148749, EPI_ISL_2148750, EPI_ISL_2148751, EPI_ISL_2148752, EPI_ISL_2148753, EPI_ISL_2148754, EPI_ISL_2148755, EPI_ISL_2148756, EPI_ISL_2148757, EPI_ISL_2148758, EPI_ISL_2148759, EPI_ISL_2148760, EPI_ISL_2148761, EPI_ISL_2148762, EPI_ISL_2148763,
EPI_ISL_2148764, EPI_ISL_2148765, EPI_ISL_2148766, EPI_ISL_2148767, EPI_ISL_2148768, EPI_ISL_2148769, EPI_ISL_2148770, EPI_ISL_2148772, EPI_ISL_2148773, EPI_ISL_2148774, EPI_ISL_2148775, EPI_ISL_2148776, EPI_ISL_2148777, EPI_ISL_2148778, EPI_ISL_2148779, EPI_ISL_2148780, EPI_ISL_2148781, EPI_ISL_2148782, EPI_ISL_2148783, EPI_ISL_2148784, EPI_ISL_2148785, EPI_ISL_2148786,
EPI_ISL_2148787, EPI_ISL_2148788, EPI_ISL_2148789, EPI_ISL_2148790, EPI_ISL_2148791, EPI_ISL_2148792, EPI_ISL_2148793, EPI_ISL_2148794, EPI_ISL_2148795, EPI_ISL_2148796, EPI_ISL_2148797, EPI_ISL_2148798, EPI_ISL_2148799, EPI_ISL_2148800, EPI_ISL_2148801, EPI_ISL_2148802, EPI_ISL_2148803, EPI_ISL_2148804, EPI_ISL_2148805, EPI_ISL_2148806, EPI_ISL_2148807, EPI_ISL_2148808,
EPI_ISL_2148809, EPI_ISL_2148810, EPI_ISL_2148811, EPI_ISL_2148812, EPI_ISL_2148813, EPI_ISL_2148814, EPI_ISL_2148815, EPI_ISL_2148816, EPI_ISL_2148817, EPI_ISL_2148818, EPI_ISL_2148819, EPI_ISL_2148820, EPI_ISL_2148821, EPI_ISL_2148822, EPI_ISL_2148823, EPI_ISL_2148824, EPI_ISL_2148825, EPI_ISL_2148826, EPI_ISL_2148827, EPI_ISL_2148828, EPI_ISL_2148829, EPI_ISL_2148830,
EPI_ISL_2148831, EPI_ISL_2148832, EPI_ISL_2148833, EPI_ISL_2148834, EPI_ISL_2148835, EPI_ISL_2148836, EPI_ISL_2148837, EPI_ISL_2148838, EPI_ISL_2148839, EPI_ISL_2148840, EPI_ISL_2148841, EPI_ISL_2148842, EPI_ISL_2148843, EPI_ISL_2148844, EPI_ISL_2148845, EPI_ISL_2148846, EPI_ISL_2148847, EPI_ISL_2148848, EPI_ISL_2148849, EPI_ISL_2148850, EPI_ISL_2148851, EPI_ISL_2148852,
EPI_ISL_2148853, EPI_ISL_2148854, EPI_ISL_2148856, EPI_ISL_2148857, EPI_ISL_2148858, EPI_ISL_2148859, EPI_ISL_2148860, EPI_ISL_2148861, EPI_ISL_2148862, EPI_ISL_2148864, EPI_ISL_2148865, EPI_ISL_2148866, EPI_ISL_2148867, EPI_ISL_2148868, EPI_ISL_2148869, EPI_ISL_2148870, EPI_ISL_2148871, EPI_ISL_2148872, EPI_ISL_2148873, EPI_ISL_2148875, EPI_ISL_2148876, EPI_ISL_2148877,
EPI_ISL_2148878, EPI_ISL_2148879, EPI_ISL_2148880, EPI_ISL_2148881, EPI_ISL_2148882, EPI_ISL_2148883, EPI_ISL_2148884, EPI_ISL_2148885, EPI_ISL_2148887, EPI_ISL_2148888, EPI_ISL_2148889, EPI_ISL_2148890, EPI_ISL_2148891, EPI_ISL_2148892, EPI_ISL_2148893, EPI_ISL_2148894, EPI_ISL_2148895, EPI_ISL_2148896, EPI_ISL_2148897, EPI_ISL_2148898, EPI_ISL_2148899, EPI_ISL_2148900,
EPI_ISL_2148901, EPI_ISL_2148902, EPI_ISL_2148903, EPI_ISL_2148904, EPI_ISL_2148905, EPI_ISL_2148906, EPI_ISL_2148907, EPI_ISL_2148908, EPI_ISL_2148909, EPI_ISL_2148910, EPI_ISL_2148911, EPI_ISL_2148912, EPI_ISL_2148913, EPI_ISL_2148914, EPI_ISL_2148915, EPI_ISL_2148916, EPI_ISL_2148917, EPI_ISL_2148918, EPI_ISL_2148919, EPI_ISL_2148920, EPI_ISL_2148921, EPI_ISL_2148922,
EPI_ISL_2148923, EPI_ISL_2148924, EPI_ISL_2148925, EPI_ISL_2148926, EPI_ISL_2148927, EPI_ISL_2148928, EPI_ISL_2148929, EPI_ISL_2148930, EPI_ISL_2148931, EPI_ISL_2148932, EPI_ISL_2148933, EPI_ISL_2148934, EPI_ISL_2148935, EPI_ISL_2148936, EPI_ISL_2148937, EPI_ISL_2148938, EPI_ISL_2148939, EPI_ISL_2148940, EPI_ISL_2148941, EPI_ISL_2148942, EPI_ISL_2148943, EPI_ISL_2148944,
EPI_ISL_2148945, EPI_ISL_2148946, EPI_ISL_2148947, EPI_ISL_2148948, EPI_ISL_2148949, EPI_ISL_2148950, EPI_ISL_2148951, EPI_ISL_2148952, EPI_ISL_2148953, EPI_ISL_2148954, EPI_ISL_2148955, EPI_ISL_2148956, EPI_ISL_2148957, EPI_ISL_2148958, EPI_ISL_2148959, EPI_ISL_2148960, EPI_ISL_2148961, EPI_ISL_2148962, EPI_ISL_2148963, EPI_ISL_2148964, EPI_ISL_2148965, EPI_ISL_2148966,
EPI_ISL_2148967, EPI_ISL_2148968, EPI_ISL_2148969, EPI_ISL_2148970, EPI_ISL_2148971, EPI_ISL_2148972, EPI_ISL_2148973, EPI_ISL_2148974, EPI_ISL_2148975, EPI_ISL_2149141, EPI_ISL_2149142, EPI_ISL_2149143, EPI_ISL_2149144, EPI_ISL_2149145, EPI_ISL_2149146, EPI_ISL_2149147, EPI_ISL_2149148, EPI_ISL_2149149, EPI_ISL_2149150, EPI_ISL_2149151, EPI_ISL_2149152, EPI_ISL_2149153,
EPI_ISL_2149155, EPI_ISL_2149156, EPI_ISL_2149157, EPI_ISL_2149158, EPI_ISL_2149159, EPI_ISL_2149160, EPI_ISL_2149161, EPI_ISL_2149162, EPI_ISL_2149163, EPI_ISL_2149164, EPI_ISL_2149165, EPI_ISL_2149166, EPI_ISL_2149168, EPI_ISL_2149169, EPI_ISL_2149170, EPI_ISL_2149171, EPI_ISL_2149172, EPI_ISL_2149173, EPI_ISL_2149174, EPI_ISL_2149175, EPI_ISL_2149176, EPI_ISL_2149177,
EPI_ISL_2149178, EPI_ISL_2149179, EPI_ISL_2149180, EPI_ISL_2149181, EPI_ISL_2149182, EPI_ISL_2149183, EPI_ISL_2149184, EPI_ISL_2149185, EPI_ISL_2149186, EPI_ISL_2149187, EPI_ISL_2149188, EPI_ISL_2149189, EPI_ISL_2149190, EPI_ISL_2149191, EPI_ISL_2149192, EPI_ISL_2149193, EPI_ISL_2149194, EPI_ISL_2149195, EPI_ISL_2149196, EPI_ISL_2149197, EPI_ISL_2149198, EPI_ISL_2149199,
EPI_ISL_2149200, EPI_ISL_2149201, EPI_ISL_2149202, EPI_ISL_2149203, EPI_ISL_2149204, EPI_ISL_2149205, EPI_ISL_2149206, EPI_ISL_2149207, EPI_ISL_2149208, EPI_ISL_2149209, EPI_ISL_2149210, EPI_ISL_2149211, EPI_ISL_2149212, EPI_ISL_2149213, EPI_ISL_2149215, EPI_ISL_2149216, EPI_ISL_2149217, EPI_ISL_2149218, EPI_ISL_2149219, EPI_ISL_2149220, EPI_ISL_2149221, EPI_ISL_2149222,
EPI_ISL_2149223, EPI_ISL_2149224, EPI_ISL_2149225, EPI_ISL_2149226, EPI_ISL_2149227, EPI_ISL_2149228, EPI_ISL_2149229, EPI_ISL_2149230, EPI_ISL_2149231, EPI_ISL_2149232, EPI_ISL_2149233, EPI_ISL_2149234, EPI_ISL_2149235, EPI_ISL_2149236, EPI_ISL_2149237, EPI_ISL_2149238, EPI_ISL_2149239, EPI_ISL_2149240, EPI_ISL_2149241, EPI_ISL_2149242, EPI_ISL_2149243, EPI_ISL_2149244,
EPI_ISL_2149245, EPI_ISL_2149246, EPI_ISL_2149247, EPI_ISL_2149248, EPI_ISL_2149249, EPI_ISL_2149250, EPI_ISL_2149251, EPI_ISL_2149252, EPI_ISL_2149253, EPI_ISL_2149254, EPI_ISL_2149255, EPI_ISL_2149256, EPI_ISL_2149257, EPI_ISL_2149258, EPI_ISL_2149259, EPI_ISL_2149260, EPI_ISL_2149261, EPI_ISL_2149262, EPI_ISL_2149263, EPI_ISL_2149264, EPI_ISL_2149265, EPI_ISL_2149266,
EPI_ISL_2149267, EPI_ISL_2149268, EPI_ISL_2149269, EPI_ISL_2149270, EPI_ISL_2149271, EPI_ISL_2149272, EPI_ISL_2149273, EPI_ISL_2149274, EPI_ISL_2149275, EPI_ISL_2149276, EPI_ISL_2149277, EPI_ISL_2149278, EPI_ISL_2149279, EPI_ISL_2149280, EPI_ISL_2149281, EPI_ISL_2149282, EPI_ISL_2149283, EPI_ISL_2149284, EPI_ISL_2149285, EPI_ISL_2149286, EPI_ISL_2149287, EPI_ISL_2149288,
EPI_ISL_2149289, EPI_ISL_2149290, EPI_ISL_2149291, EPI_ISL_2149292, EPI_ISL_2149293, EPI_ISL_2149294, EPI_ISL_2149295, EPI_ISL_2149296, EPI_ISL_2149297, EPI_ISL_2149298, EPI_ISL_2149299, EPI_ISL_2149300, EPI_ISL_2149301, EPI_ISL_2149302, EPI_ISL_2149303, EPI_ISL_2149304, EPI_ISL_2149305, EPI_ISL_2149306, EPI_ISL_2149307, EPI_ISL_2149308, EPI_ISL_2149309, EPI_ISL_2149310,
EPI_ISL_2149311, EPI_ISL_2149312, EPI_ISL_2149313, EPI_ISL_2149314, EPI_ISL_2149315, EPI_ISL_2149316, EPI_ISL_2149317, EPI_ISL_2149318, EPI_ISL_2149319, EPI_ISL_2149320, EPI_ISL_2149321, EPI_ISL_2149322, EPI_ISL_2149323, EPI_ISL_2149324, EPI_ISL_2149325, EPI_ISL_2149326, EPI_ISL_2149327, EPI_ISL_2149328, EPI_ISL_2149329, EPI_ISL_2149330, EPI_ISL_2149331, EPI_ISL_2149332,
EPI_ISL_2149333, EPI_ISL_2149334, EPI_ISL_2149335, EPI_ISL_2149336, EPI_ISL_2149337, EPI_ISL_2149338, EPI_ISL_2149339, EPI_ISL_2149340, EPI_ISL_2149341, EPI_ISL_2149342, EPI_ISL_2149343, EPI_ISL_2149344, EPI_ISL_2149345, EPI_ISL_2149347, EPI_ISL_2149349, EPI_ISL_2149350, EPI_ISL_2149351, EPI_ISL_2149353, EPI_ISL_2149354, EPI_ISL_2149355, EPI_ISL_2149356, EPI_ISL_2149357,
EPI_ISL_2149358, EPI_ISL_2149359, EPI_ISL_2149360, EPI_ISL_2149361, EPI_ISL_2149362, EPI_ISL_2149363, EPI_ISL_2149364, EPI_ISL_2149365, EPI_ISL_2149366, EPI_ISL_2149367, EPI_ISL_2149368, EPI_ISL_2149369, EPI_ISL_2149370, EPI_ISL_2149371, EPI_ISL_2149372, EPI_ISL_2149373, EPI_ISL_2149374, EPI_ISL_2149375, EPI_ISL_2149376, EPI_ISL_2149377, EPI_ISL_2149378, EPI_ISL_2149379,
EPI_ISL_2149380, EPI_ISL_2149381, EPI_ISL_2149382, EPI_ISL_2149383, EPI_ISL_2149384, EPI_ISL_2149385, EPI_ISL_2149386, EPI_ISL_2149388, EPI_ISL_2149389, EPI_ISL_2149390, EPI_ISL_2149391, EPI_ISL_2149392, EPI_ISL_2149393, EPI_ISL_2149394, EPI_ISL_2149395, EPI_ISL_2149396, EPI_ISL_2149397, EPI_ISL_2149398, EPI_ISL_2149399, EPI_ISL_2149400, EPI_ISL_2149401, EPI_ISL_2149402,
EPI_ISL_2149403, EPI_ISL_2149404, EPI_ISL_2149405, EPI_ISL_2149406, EPI_ISL_2149407, EPI_ISL_2149408, EPI_ISL_2149409, EPI_ISL_2149410, EPI_ISL_2149411, EPI_ISL_2149412, EPI_ISL_2149413, EPI_ISL_2149414, EPI_ISL_2149415, EPI_ISL_2149416, EPI_ISL_2149417, EPI_ISL_2149418, EPI_ISL_2149419, EPI_ISL_2149420, EPI_ISL_2149421, EPI_ISL_2149423, EPI_ISL_2149424, EPI_ISL_2149425,
EPI_ISL_2149426, EPI_ISL_2149427, EPI_ISL_2149428, EPI_ISL_2149429, EPI_ISL_2149430, EPI_ISL_2149431, EPI_ISL_2149432, EPI_ISL_2149434, EPI_ISL_2149435, EPI_ISL_2149437, EPI_ISL_2149438, EPI_ISL_2149439, EPI_ISL_2149440, EPI_ISL_2149441, EPI_ISL_2149442, EPI_ISL_2149443, EPI_ISL_2149444, EPI_ISL_2149446, EPI_ISL_2149447, EPI_ISL_2149448, EPI_ISL_2149449, EPI_ISL_2149450,
EPI_ISL_2149451, EPI_ISL_2149452, EPI_ISL_2149453, EPI_ISL_2149455, EPI_ISL_2149456, EPI_ISL_2149457, EPI_ISL_2149458, EPI_ISL_2149459, EPI_ISL_2149460, EPI_ISL_2149461, EPI_ISL_2149462, EPI_ISL_2149463, EPI_ISL_2149464, EPI_ISL_2149465, EPI_ISL_2149466, EPI_ISL_2149467, EPI_ISL_2149468, EPI_ISL_2149469, EPI_ISL_2149470, EPI_ISL_2149471, EPI_ISL_2149472, EPI_ISL_2149473,
EPI_ISL_2149474, EPI_ISL_2149475, EPI_ISL_2149476, EPI_ISL_2149477, EPI_ISL_2149478, EPI_ISL_2149479, EPI_ISL_2149480, EPI_ISL_2149481, EPI_ISL_2149482, EPI_ISL_2149483, EPI_ISL_2149484, EPI_ISL_2149485, EPI_ISL_2149486, EPI_ISL_2149487, EPI_ISL_2149488, EPI_ISL_2149489, EPI_ISL_2149490, EPI_ISL_2149491, EPI_ISL_2149492, EPI_ISL_2149493, EPI_ISL_2149494, EPI_ISL_2149495,
EPI_ISL_2149496, EPI_ISL_2149497, EPI_ISL_2149498, EPI_ISL_2149499, EPI_ISL_2149500, EPI_ISL_2149501, EPI_ISL_2149502, EPI_ISL_2149503, EPI_ISL_2149504, EPI_ISL_2149505, EPI_ISL_2149506, EPI_ISL_2149507, EPI_ISL_2149508, EPI_ISL_2149509, EPI_ISL_2149510, EPI_ISL_2149511, EPI_ISL_2149512, EPI_ISL_2149513, EPI_ISL_2149514, EPI_ISL_2149515, EPI_ISL_2149516, EPI_ISL_2149517,
EPI_ISL_2149518, EPI_ISL_2149519, EPI_ISL_2149520, EPI_ISL_2149521, EPI_ISL_2149522, EPI_ISL_2149523, EPI_ISL_2149524, EPI_ISL_2149525, EPI_ISL_2149526, EPI_ISL_2149527, EPI_ISL_2149528, EPI_ISL_2149529, EPI_ISL_2149530, EPI_ISL_2149531, EPI_ISL_2149532, EPI_ISL_2149533, EPI_ISL_2149534, EPI_ISL_2149535, EPI_ISL_2149536, EPI_ISL_2149537, EPI_ISL_2149538, EPI_ISL_2149539,
EPI_ISL_2149540, EPI_ISL_2149541, EPI_ISL_2149542, EPI_ISL_2149543, EPI_ISL_2149544, EPI_ISL_2149545, EPI_ISL_2149546, EPI_ISL_2149547, EPI_ISL_2149548, EPI_ISL_2149549, EPI_ISL_2149551, EPI_ISL_2149552, EPI_ISL_2149553, EPI_ISL_2149554, EPI_ISL_2149555, EPI_ISL_2149556, EPI_ISL_2149557, EPI_ISL_2149558, EPI_ISL_2149559, EPI_ISL_2149560, EPI_ISL_2149561, EPI_ISL_2149562,
EPI_ISL_2149563, EPI_ISL_2149564, EPI_ISL_2149565, EPI_ISL_2149566, EPI_ISL_2149567, EPI_ISL_2149568, EPI_ISL_2149569, EPI_ISL_2149570, EPI_ISL_2149571, EPI_ISL_2149572, EPI_ISL_2149573, EPI_ISL_2149574, EPI_ISL_2149575, EPI_ISL_2149576, EPI_ISL_2149577, EPI_ISL_2149578, EPI_ISL_2149579, EPI_ISL_2149580, EPI_ISL_2149581, EPI_ISL_2149582, EPI_ISL_2149583, EPI_ISL_2149584,
EPI_ISL_2149585, EPI_ISL_2149586, EPI_ISL_2149587, EPI_ISL_2149588, EPI_ISL_2149589, EPI_ISL_2149590, EPI_ISL_2149591, EPI_ISL_2149592, EPI_ISL_2149593, EPI_ISL_2149594, EPI_ISL_2149595, EPI_ISL_2149596, EPI_ISL_2149597, EPI_ISL_2149598, EPI_ISL_2149599, EPI_ISL_2149600, EPI_ISL_2149601, EPI_ISL_2149602, EPI_ISL_2149603, EPI_ISL_2149604, EPI_ISL_2149605, EPI_ISL_2149606,
EPI_ISL_2149607, EPI_ISL_2149608, EPI_ISL_2149609, EPI_ISL_2149610, EPI_ISL_2149611, EPI_ISL_2149612, EPI_ISL_2149613, EPI_ISL_2149614, EPI_ISL_2149615, EPI_ISL_2149616, EPI_ISL_2149617, EPI_ISL_2149618, EPI_ISL_2149619, EPI_ISL_2149620, EPI_ISL_2149621, EPI_ISL_2149622, EPI_ISL_2149623, EPI_ISL_2149624, EPI_ISL_2149625, EPI_ISL_2149626, EPI_ISL_2149627, EPI_ISL_2149628,
EPI_ISL_2149629, EPI_ISL_2149630, EPI_ISL_2149631, EPI_ISL_2149632, EPI_ISL_2149633, EPI_ISL_2149634, EPI_ISL_2149635, EPI_ISL_2149636, EPI_ISL_2149637, EPI_ISL_2149638, EPI_ISL_2149639, EPI_ISL_2149640, EPI_ISL_2149641, EPI_ISL_2149642, EPI_ISL_2149643, EPI_ISL_2149644, EPI_ISL_2149645, EPI_ISL_2149646, EPI_ISL_2149647, EPI_ISL_2149648, EPI_ISL_2149649, EPI_ISL_2149650,
EPI_ISL_2149651, EPI_ISL_2149652, EPI_ISL_2149653, EPI_ISL_2149654, EPI_ISL_2149655, EPI_ISL_2149656, EPI_ISL_2149657, EPI_ISL_2149658, EPI_ISL_2149659, EPI_ISL_2149660, EPI_ISL_2149661, EPI_ISL_2149662, EPI_ISL_2149663, EPI_ISL_2149664, EPI_ISL_2149665, EPI_ISL_2149666, EPI_ISL_2149667, EPI_ISL_2149668, EPI_ISL_2149669, EPI_ISL_2149670, EPI_ISL_2149671, EPI_ISL_2149672,
EPI_ISL_2149673, EPI_ISL_2149674, EPI_ISL_2149675, EPI_ISL_2149676, EPI_ISL_2149677, EPI_ISL_2149678, EPI_ISL_2149679, EPI_ISL_2149680, EPI_ISL_2149681, EPI_ISL_2149682, EPI_ISL_2149683, EPI_ISL_2149684, EPI_ISL_2149685, EPI_ISL_2149686, EPI_ISL_2149687, EPI_ISL_2149688, EPI_ISL_2149689, EPI_ISL_2149690, EPI_ISL_2149691, EPI_ISL_2149692, EPI_ISL_2149693, EPI_ISL_2149694,
EPI_ISL_2149695, EPI_ISL_2149696, EPI_ISL_2149697, EPI_ISL_2149698, EPI_ISL_2149699, EPI_ISL_2149700, EPI_ISL_2149701, EPI_ISL_2149702, EPI_ISL_2149703, EPI_ISL_2149704, EPI_ISL_2149705, EPI_ISL_2149706, EPI_ISL_2149707, EPI_ISL_2149708, EPI_ISL_2149709, EPI_ISL_2149710, EPI_ISL_2149711, EPI_ISL_2149712, EPI_ISL_2149713, EPI_ISL_2149714, EPI_ISL_2149715, EPI_ISL_2149716,
EPI_ISL_2149717, EPI_ISL_2149719, EPI_ISL_2149720, EPI_ISL_2149721, EPI_ISL_2149722, EPI_ISL_2149723, EPI_ISL_2149724, EPI_ISL_2149725, EPI_ISL_2149726, EPI_ISL_2149727, EPI_ISL_2149728, EPI_ISL_2149729, EPI_ISL_2149730, EPI_ISL_2149731, EPI_ISL_2149732, EPI_ISL_2149733, EPI_ISL_2149734, EPI_ISL_2149735, EPI_ISL_2149736, EPI_ISL_2149737, EPI_ISL_2149738, EPI_ISL_2149739,
EPI_ISL_2149740, EPI_ISL_2149741, EPI_ISL_2149742, EPI_ISL_2149743, EPI_ISL_2149744, EPI_ISL_2149745, EPI_ISL_2149747, EPI_ISL_2149748, EPI_ISL_2149749, EPI_ISL_2149750, EPI_ISL_2149751, EPI_ISL_2149752, EPI_ISL_2149753, EPI_ISL_2149754, EPI_ISL_2149755, EPI_ISL_2149756, EPI_ISL_2149757, EPI_ISL_2149758, EPI_ISL_2149759, EPI_ISL_2149760, EPI_ISL_2149761, EPI_ISL_2149762,
EPI_ISL_2149763, EPI_ISL_2149764, EPI_ISL_2149765, EPI_ISL_2149766, EPI_ISL_2149767, EPI_ISL_2149768, EPI_ISL_2149769, EPI_ISL_2149770, EPI_ISL_2149771, EPI_ISL_2149772, EPI_ISL_2149773, EPI_ISL_2149774, EPI_ISL_2149775, EPI_ISL_2149776, EPI_ISL_2149777, EPI_ISL_2149778, EPI_ISL_2149779, EPI_ISL_2149780, EPI_ISL_2149781, EPI_ISL_2149782, EPI_ISL_2149783, EPI_ISL_2149784,
EPI_ISL_2149785, EPI_ISL_2149786, EPI_ISL_2149787, EPI_ISL_2149788, EPI_ISL_2149789, EPI_ISL_2149790, EPI_ISL_2149791, EPI_ISL_2149792, EPI_ISL_2149793, EPI_ISL_2149794, EPI_ISL_2149795, EPI_ISL_2149796, EPI_ISL_2149797, EPI_ISL_2149798, EPI_ISL_2149799, EPI_ISL_2149800, EPI_ISL_2149801, EPI_ISL_2149802, EPI_ISL_2149803, EPI_ISL_2149804, EPI_ISL_2149805, EPI_ISL_2149806,
EPI_ISL_2149807, EPI_ISL_2149808, EPI_ISL_2149809, EPI_ISL_2149810, EPI_ISL_2149811, EPI_ISL_2149812, EPI_ISL_2149813, EPI_ISL_2149814, EPI_ISL_2149815, EPI_ISL_2149816, EPI_ISL_2149817, EPI_ISL_2149818, EPI_ISL_2149819, EPI_ISL_2149820, EPI_ISL_2149821, EPI_ISL_2149822, EPI_ISL_2149823, EPI_ISL_2149824, EPI_ISL_2149825, EPI_ISL_2149826, EPI_ISL_2149827, EPI_ISL_2149828,
EPI_ISL_2149829, EPI_ISL_2149830, EPI_ISL_2149831, EPI_ISL_2149832, EPI_ISL_2149833, EPI_ISL_2149834, EPI_ISL_2149835, EPI_ISL_2149836, EPI_ISL_2149837, EPI_ISL_2149838, EPI_ISL_2149839, EPI_ISL_2149840, EPI_ISL_2149841, EPI_ISL_2149842, EPI_ISL_2149843, EPI_ISL_2149844, EPI_ISL_2149845, EPI_ISL_2149846, EPI_ISL_2149847, EPI_ISL_2149848, EPI_ISL_2149849, EPI_ISL_2149850,
EPI_ISL_2149851, EPI_ISL_2149852, EPI_ISL_2149853, EPI_ISL_2149854, EPI_ISL_2149855, EPI_ISL_2149856, EPI_ISL_2149857, EPI_ISL_2149858, EPI_ISL_2149859, EPI_ISL_2149860, EPI_ISL_2149861, EPI_ISL_2149862, EPI_ISL_2149863, EPI_ISL_2149864, EPI_ISL_2149865, EPI_ISL_2149867, EPI_ISL_2149868, EPI_ISL_2149869, EPI_ISL_2149870, EPI_ISL_2149871, EPI_ISL_2149872, EPI_ISL_2149873,
EPI_ISL_2149874, EPI_ISL_2149875, EPI_ISL_2149876, EPI_ISL_2149877, EPI_ISL_2149878, EPI_ISL_2149879, EPI_ISL_2149880, EPI_ISL_2149881, EPI_ISL_2149882, EPI_ISL_2149883, EPI_ISL_2149884, EPI_ISL_2149885, EPI_ISL_2149886, EPI_ISL_2149887, EPI_ISL_2149888, EPI_ISL_2149889, EPI_ISL_2149890, EPI_ISL_2149891, EPI_ISL_2149892, EPI_ISL_2149893, EPI_ISL_2149894, EPI_ISL_2149895,
EPI_ISL_2149897, EPI_ISL_2149898, EPI_ISL_2149899, EPI_ISL_2149900, EPI_ISL_2149901, EPI_ISL_2149902, EPI_ISL_2149903, EPI_ISL_2149904, EPI_ISL_2149905, EPI_ISL_2149906, EPI_ISL_2149907, EPI_ISL_2149908, EPI_ISL_2149909, EPI_ISL_2149910, EPI_ISL_2149911, EPI_ISL_2149912, EPI_ISL_2149913, EPI_ISL_2149914, EPI_ISL_2149915, EPI_ISL_2149917, EPI_ISL_2149918, EPI_ISL_2149919,
EPI_ISL_2149920, EPI_ISL_2149921, EPI_ISL_2149922, EPI_ISL_2149923, EPI_ISL_2149924, EPI_ISL_2149925, EPI_ISL_2149926, EPI_ISL_2149927, EPI_ISL_2149928, EPI_ISL_2149929, EPI_ISL_2149930, EPI_ISL_2149931, EPI_ISL_2149932, EPI_ISL_2149933, EPI_ISL_2149934, EPI_ISL_2149935, EPI_ISL_2149936, EPI_ISL_2149937, EPI_ISL_2149938, EPI_ISL_2149939, EPI_ISL_2149940, EPI_ISL_2149941,

EPI_ISL_2149942, EPI_ISL_2149943, EPI_ISL_2149944, EPI_ISL_2149945, EPI_ISL_2149946, EPI_ISL_2149947, EPI_ISL_2149948, EPI_ISL_2149949, EPI_ISL_2149950, EPI_ISL_2149951, EPI_ISL_2149952, EPI_ISL_2149953, EPI_ISL_2149954, EPI_ISL_2149956, EPI_ISL_2149957, EPI_ISL_2149958, EPI_ISL_2149959, EPI_ISL_2149960, EPI_ISL_2149961, EPI_ISL_2149962, EPI_ISL_2149963, EPI_ISL_2149967, EPI_ISL_2149968, EPI_ISL_2149969, EPI_ISL_2149970, EPI_ISL_2149971, EPI_ISL_2149972, EPI_ISL_2149973, EPI_ISL_2149974, EPI_ISL_2149975, EPI_ISL_2149976, EPI_ISL_2149977, EPI_ISL_2149978, EPI_ISL_2149979, EPI_ISL_2149980, EPI_ISL_2149981, EPI_ISL_2149982, EPI_ISL_2149984, EPI_ISL_2149985, EPI_ISL_2149986, EPI_ISL_2149988, EPI_ISL_2149989, EPI_ISL_2149990, EPI_ISL_2149991, EPI_ISL_2149992, EPI_ISL_2149993, EPI_ISL_2149994, EPI_ISL_2149995, EPI_ISL_2149996, EPI_ISL_2149997, EPI_ISL_2149998, EPI_ISL_2149999, EPI_ISL_2150000, EPI_ISL_2150001, EPI_ISL_2150002, EPI_ISL_2150003, EPI_ISL_2150004, EPI_ISL_2150005, EPI_ISL_2150006, EPI_ISL_2150007, EPI_ISL_2150008, EPI_ISL_2150009, EPI_ISL_2150010, EPI_ISL_2150011, EPI_ISL_2150012, EPI_ISL_2150013, EPI_ISL_2150014, EPI_ISL_2150015, EPI_ISL_2150016, EPI_ISL_2150017, EPI_ISL_2150018, EPI_ISL_2150019, EPI_ISL_2150020, EPI_ISL_2150021, EPI_ISL_2150022, EPI_ISL_2150023, EPI_ISL_2150024, EPI_ISL_2150025, EPI_ISL_2150026, EPI_ISL_2150027, EPI_ISL_2150028, EPI_ISL_2150029, EPI_ISL_2150030, EPI_ISL_2150031, EPI_ISL_2150032, EPI_ISL_2150033, EPI_ISL_2150034, EPI_ISL_2150035, EPI_ISL_2150036, EPI_ISL_2150037, EPI_ISL_2150038, EPI_ISL_2150039, EPI_ISL_2150040, EPI_ISL_2150041, EPI_ISL_2150042, EPI_ISL_2150043, EPI_ISL_2150044, EPI_ISL_2150045, EPI_ISL_2150046, EPI_ISL_2150047, EPI_ISL_2150048, EPI_ISL_2150049, EPI_ISL_2150050, EPI_ISL_2150051, EPI_ISL_2150052, EPI_ISL_2150053, EPI_ISL_2150054, EPI_ISL_2150055, EPI_ISL_2150056, EPI_ISL_2150058, EPI_ISL_2150059, EPI_ISL_2150060, EPI_ISL_2150061, EPI_ISL_2150062, EPI_ISL_2150063, EPI_ISL_2150064, EPI_ISL_2150065, EPI_ISL_2150066, EPI_ISL_2150067, EPI_ISL_2150068, EPI_ISL_2150069, EPI_ISL_2150070, EPI_ISL_2150071, EPI_ISL_2150072, EPI_ISL_2150073, EPI_ISL_2150074, EPI_ISL_2150075, EPI_ISL_2150077, EPI_ISL_2150078, EPI_ISL_2150079, EPI_ISL_2150080, EPI_ISL_2150081, EPI_ISL_2150082, EPI_ISL_2150083, EPI_ISL_2150084, EPI_ISL_2150085, EPI_ISL_2150086, EPI_ISL_2150087, EPI_ISL_2150088, EPI_ISL_2150089, EPI_ISL_2150090, EPI_ISL_2150091, EPI_ISL_2150092, EPI_ISL_2150093, EPI_ISL_2150094, EPI_ISL_2150095, EPI_ISL_2150096, EPI_ISL_2150097, EPI_ISL_2150098, EPI_ISL_2150099, EPI_ISL_2150100, EPI_ISL_2150101, EPI_ISL_2150102, EPI_ISL_2150103, EPI_ISL_2150104, EPI_ISL_2150105, EPI_ISL_2150106, EPI_ISL_2150107, EPI_ISL_2150108, EPI_ISL_2150109, EPI_ISL_2150110, EPI_ISL_2150111, EPI_ISL_2150112, EPI_ISL_2150113, EPI_ISL_2150114, EPI_ISL_2150115, EPI_ISL_2150116, EPI_ISL_2150117, EPI_ISL_2150118, EPI_ISL_2150119, EPI_ISL_2150120, EPI_ISL_2150121, EPI_ISL_2150122, EPI_ISL_2150123, EPI_ISL_2150124, EPI_ISL_2150125, EPI_ISL_2150126, EPI_ISL_2150127, EPI_ISL_2150128, EPI_ISL_2150129, EPI_ISL_2150131, EPI_ISL_2150132, EPI_ISL_2150133, EPI_ISL_2150134, EPI_ISL_2150135, EPI_ISL_2150136, EPI_ISL_2150137, EPI_ISL_2150138, EPI_ISL_2150139, EPI_ISL_2150140, EPI_ISL_2150141, EPI_ISL_2150142, EPI_ISL_2150143, EPI_ISL_2150144, EPI_ISL_2150145, EPI_ISL_2150146, EPI_ISL_2150147, EPI_ISL_2150148, EPI_ISL_2150149, EPI_ISL_2150150, EPI_ISL_2150151, EPI_ISL_2150152, EPI_ISL_2150153, EPI_ISL_2150154, EPI_ISL_2150155, EPI_ISL_2150156, EPI_ISL_2150157, EPI_ISL_2150158, EPI_ISL_2150159, EPI_ISL_2150160, EPI_ISL_2150161, EPI_ISL_2150162, EPI_ISL_2150163, EPI_ISL_2150164, EPI_ISL_2150165, EPI_ISL_2150166, EPI_ISL_2150167, EPI_ISL_2150168, EPI_ISL_2150169, EPI_ISL_2150170, EPI_ISL_2150171, EPI_ISL_2150172, EPI_ISL_2150173, EPI_ISL_2150174, EPI_ISL_2150175, EPI_ISL_2150176, EPI_ISL_2150177, EPI_ISL_2150179, EPI_ISL_2150180, EPI_ISL_2150181, EPI_ISL_2150182, EPI_ISL_2150183, EPI_ISL_2150184, EPI_ISL_2150185, EPI_ISL_2150186, EPI_ISL_2150187, EPI_ISL_2150188, EPI_ISL_2150189, EPI_ISL_2150190, EPI_ISL_2150191, EPI_ISL_2150192, EPI_ISL_2150193, EPI_ISL_2150194, EPI_ISL_2150195, EPI_ISL_2150196, EPI_ISL_2150197, EPI_ISL_2150198, EPI_ISL_2150199, EPI_ISL_2150201, EPI_ISL_2150202, EPI_ISL_2150203, EPI_ISL_2150204, EPI_ISL_2150205, EPI_ISL_2150206, EPI_ISL_2150207, EPI_ISL_2150208, EPI_ISL_2150209, EPI_ISL_2150210, EPI_ISL_2150211, EPI_ISL_2150212, EPI_ISL_2150213, EPI_ISL_2150214, EPI_ISL_2150215, EPI_ISL_2150216, EPI_ISL_2150217, EPI_ISL_2150218, EPI_ISL_2150219, EPI_ISL_2150220, EPI_ISL_2150221, EPI_ISL_2150222, EPI_ISL_2150223, EPI_ISL_2150224, EPI_ISL_2150225, EPI_ISL_2150226, EPI_ISL_2150227, EPI_ISL_2150228, EPI_ISL_2150229, EPI_ISL_2150230, EPI_ISL_2150231, EPI_ISL_2150232, EPI_ISL_2150233, EPI_ISL_2150234, EPI_ISL_2150235, EPI_ISL_2150236, EPI_ISL_2150237, EPI_ISL_2150238, EPI_ISL_2150239, EPI_ISL_2150240, EPI_ISL_2150241, EPI_ISL_2150242, EPI_ISL_2150243, EPI_ISL_2150244, EPI_ISL_2150245, EPI_ISL_2150246, EPI_ISL_2150247, EPI_ISL_2150248, EPI_ISL_2150249, EPI_ISL_2150250, EPI_ISL_2150251, EPI_ISL_2150252, EPI_ISL_2150253, EPI_ISL_2150254, EPI_ISL_2150255, EPI_ISL_2150256, EPI_ISL_2150257, EPI_ISL_2150258, EPI_ISL_2150259, EPI_ISL_2150260, EPI_ISL_2150261, EPI_ISL_2150262, EPI_ISL_2150263, EPI_ISL_2150264, EPI_ISL_2150266, EPI_ISL_2150267, EPI_ISL_2150268, EPI_ISL_2150269, EPI_ISL_2150270, EPI_ISL_2150271, EPI_ISL_2150272, EPI_ISL_2150273, EPI_ISL_2150274, EPI_ISL_2150275, EPI_ISL_2150276, EPI_ISL_2150277, EPI_ISL_2150278, EPI_ISL_2150279, EPI_ISL_2150280, EPI_ISL_2150281, EPI_ISL_2150282, EPI_ISL_2150283, EPI_ISL_2150284, EPI_ISL_2150285, EPI_ISL_2150286, EPI_ISL_2150287, EPI_ISL_2150288, EPI_ISL_2150289, EPI_ISL_2150290, EPI_ISL_2150291, EPI_ISL_2150292, EPI_ISL_2150293, EPI_ISL_2150294, EPI_ISL_2150295, EPI_ISL_2150296, EPI_ISL_2150297, EPI_ISL_2150298, EPI_ISL_2150299, EPI_ISL_2150300, EPI_ISL_2150302, EPI_ISL_2150303, EPI_ISL_2150304, EPI_ISL_2150305, EPI_ISL_2150306, EPI_ISL_2150307, EPI_ISL_2150308, EPI_ISL_2150309, EPI_ISL_2150310, EPI_ISL_2150311, EPI_ISL_2150312, EPI_ISL_2150313, EPI_ISL_2150315, EPI_ISL_2150316, EPI_ISL_2150317, EPI_ISL_2150318, EPI_ISL_2150319, EPI_ISL_2150320, EPI_ISL_2150321, EPI_ISL_2150322, EPI_ISL_2150323, EPI_ISL_2150325, EPI_ISL_2150326, EPI_ISL_2150327, EPI_ISL_2150328, EPI_ISL_2150329, EPI_ISL_2150330, EPI_ISL_2150331, EPI_ISL_2150332, EPI_ISL_2150333, EPI_ISL_2150334, EPI_ISL_2150335, EPI_ISL_2150336, EPI_ISL_2150337, EPI_ISL_2150338, EPI_ISL_2150339, EPI_ISL_2150340, EPI_ISL_2150341, EPI_ISL_2150343, EPI_ISL_2150344, EPI_ISL_2150345, EPI_ISL_2150346, EPI_ISL_2150347, EPI_ISL_2150348, EPI_ISL_2150349, EPI_ISL_2150350, EPI_ISL_2150351, EPI_ISL_2150352, EPI_ISL_2150354, EPI_ISL_2150355, EPI_ISL_2150356, EPI_ISL_2150357, EPI_ISL_2150358, EPI_ISL_2150359, EPI_ISL_2150360, EPI_ISL_2150361, EPI_ISL_2150362, EPI_ISL_2150363, EPI_ISL_2150364, EPI_ISL_2150365, EPI_ISL_2150366, EPI_ISL_2150367, EPI_ISL_2150368, EPI_ISL_2150369, EPI_ISL_2150370, EPI_ISL_2150371, EPI_ISL_2150372, EPI_ISL_2150373, EPI_ISL_2150374, EPI_ISL_2150375, EPI_ISL_2150376, EPI_ISL_2150377, EPI_ISL_2150378, EPI_ISL_2150379, EPI_ISL_2150380, EPI_ISL_2150381, EPI_ISL_2150382, EPI_ISL_2150383, EPI_ISL_2150384, EPI_ISL_2150385, EPI_ISL_2150386, EPI_ISL_2150387, EPI_ISL_2150388, EPI_ISL_2150389, EPI_ISL_2150390, EPI_ISL_2150391, EPI_ISL_2150392, EPI_ISL_2150393, EPI_ISL_2150394, EPI_ISL_2150395, EPI_ISL_2150396, EPI_ISL_2150397, EPI_ISL_2150398, EPI_ISL_2150399, EPI_ISL_2150400, EPI_ISL_2150401, EPI_ISL_2150402, EPI_ISL_2150403, EPI_ISL_2150404, EPI_ISL_2150405, EPI_ISL_2150406, EPI_ISL_2150407, EPI_ISL_2150408, EPI_ISL_2150409, EPI_ISL_2150410, EPI_ISL_2150411, EPI_ISL_2150412, EPI_ISL_2150413, EPI_ISL_2150414, EPI_ISL_2150415, EPI_ISL_2150416, EPI_ISL_2150417, EPI_ISL_2150418, EPI_ISL_2150419, EPI_ISL_2150420, EPI_ISL_2150421, EPI_ISL_2150422, EPI_ISL_2150423, EPI_ISL_2150424, EPI_ISL_2150425, EPI_ISL_2150426, EPI_ISL_2150427, EPI_ISL_2150428, EPI_ISL_2150429, EPI_ISL_2150430, EPI_ISL_2150431, EPI_ISL_2150432, EPI_ISL_2150433, EPI_ISL_2150434, EPI_ISL_2150435, EPI_ISL_2150436, EPI_ISL_2150437, EPI_ISL_2150438, EPI_ISL_2150439, EPI_ISL_2150440, EPI_ISL_2150441, EPI_ISL_2150442, EPI_ISL_2150443, EPI_ISL_2150444, EPI_ISL_2150445, EPI_ISL_2150446, EPI_ISL_2150447, EPI_ISL_2150448, EPI_ISL_2150449, EPI_ISL_2150450, EPI_ISL_2150451, EPI_ISL_2150452, EPI_ISL_2150453, EPI_ISL_2150454, EPI_ISL_2150455, EPI_ISL_2150456, EPI_ISL_2150457, EPI_ISL_2150458, EPI_ISL_2150459, EPI_ISL_2150460, EPI_ISL_2150461, EPI_ISL_2150462, EPI_ISL_2150463, EPI_ISL_2150464, EPI_ISL_2150465, EPI_ISL_2150466, EPI_ISL_2150467, EPI_ISL_2150468, EPI_ISL_2150469, EPI_ISL_2150470, EPI_ISL_2150471, EPI_ISL_2150472, EPI_ISL_2150473, EPI_ISL_2150474, EPI_ISL_2150475, EPI_ISL_2150476, EPI_ISL_2150477, EPI_ISL_2150478, EPI_ISL_2150479, EPI_ISL_2150480, EPI_ISL_2150481, EPI_ISL_2150482, EPI_ISL_2150483, EPI_ISL_2150484, EPI_ISL_2150486, EPI_ISL_2150487, EPI_ISL_2150488, EPI_ISL_2150489, EPI_ISL_2150490, EPI_ISL_2150491, EPI_ISL_2150492, EPI_ISL_2150493, EPI_ISL_2150494, EPI_ISL_2150495, EPI_ISL_2150496, EPI_ISL_2150497, EPI_ISL_2150498, EPI_ISL_2150499, EPI_ISL_2150500, EPI_ISL_2150501, EPI_ISL_2150502, EPI_ISL_2150503, EPI_ISL_2150504, EPI_ISL_2150505, EPI_ISL_2150506, EPI_ISL_2150507, EPI_ISL_2150508, EPI_ISL_2150509, EPI_ISL_2150510, EPI_ISL_2150511, EPI_ISL_2150512, EPI_ISL_2150513, EPI_ISL_2150514, EPI_ISL_2150515, EPI_ISL_2150516, EPI_ISL_2150517, EPI_ISL_2150518, EPI_ISL_2150519, EPI_ISL_2150520, EPI_ISL_2150521, EPI_ISL_2150522, EPI_ISL_2150523, EPI_ISL_2150524, EPI_ISL_2150525, EPI_ISL_2150526, EPI_ISL_2150527, EPI_ISL_2150528, EPI_ISL_2150529, EPI_ISL_2150530, EPI_ISL_2150531, EPI_ISL_2150532, EPI_ISL_2150533, EPI_ISL_2150534, EPI_ISL_2150535, EPI_ISL_2150536, EPI_ISL_2150537, EPI_ISL_2150538, EPI_ISL_2150539, EPI_ISL_2150540, EPI_ISL_2150541, EPI_ISL_2150542, EPI_ISL_2150543, EPI_ISL_2150544, EPI_ISL_2150545, EPI_ISL_2150547, EPI_ISL_2150548, EPI_ISL_2150550, EPI_ISL_2150551, EPI_ISL_2150552, EPI_ISL_2150553, EPI_ISL_2150554, EPI_ISL_2150555, EPI_ISL_2150556, EPI_ISL_2150557, EPI_ISL_2150558, EPI_ISL_2150559, EPI_ISL_2150560, EPI_ISL_2150563, EPI_ISL_2150564, EPI_ISL_2150565, EPI_ISL_2150566, EPI_ISL_2150567, EPI_ISL_2150569, EPI_ISL_2150570, EPI_ISL_2150571, EPI_ISL_2150572, EPI_ISL_2150573, EPI_ISL_2150574, EPI_ISL_2150575, EPI_ISL_2150576, EPI_ISL_2150577, EPI_ISL_2150578, EPI_ISL_2150579, EPI_ISL_2150580, EPI_ISL_2150581, EPI_ISL_2150583, EPI_ISL_2150584, EPI_ISL_2150585, EPI_ISL_2150586, EPI_ISL_2150587, EPI_ISL_2150588, EPI_ISL_2150589, EPI_ISL_2150590, EPI_ISL_2150591, EPI_ISL_2150592, EPI_ISL_2150593, EPI_ISL_2150594, EPI_ISL_2150595, EPI_ISL_2150596, EPI_ISL_2150597, EPI_ISL_2150598, EPI_ISL_2150599, EPI_ISL_2150600, EPI_ISL_2150601, EPI_ISL_2150602, EPI_ISL_2150603, EPI_ISL_2150604, EPI_ISL_2150605, EPI_ISL_2150606, EPI_ISL_2150607, EPI_ISL_2150608, EPI_ISL_2150609, EPI_ISL_2150610, EPI_ISL_2150611, EPI_ISL_2150612, EPI_ISL_2150613, EPI_ISL_2150614, EPI_ISL_2150615, EPI_ISL_2150616, EPI_ISL_2150617, EPI_ISL_2150618, EPI_ISL_2150619, EPI_ISL_2150620, EPI_ISL_2150621, EPI_ISL_2150622, EPI_ISL_2150623, EPI_ISL_2150624, EPI_ISL_2150625, EPI_ISL_2150626, EPI_ISL_2150627, EPI_ISL_2150629, EPI_ISL_2150632, EPI_ISL_2150634, EPI_ISL_2150637, EPI_ISL_2150640, EPI_ISL_2150642, EPI_ISL_2150644, EPI_ISL_2150647, EPI_ISL_2150648, EPI_ISL_2150649, EPI_ISL_2150650, EPI_ISL_2150651, EPI_ISL_2150652, EPI_ISL_2150653, EPI_ISL_2150654, EPI_ISL_2150655, EPI_ISL_2150656, EPI_ISL_2150657, EPI_ISL_2150658, EPI_ISL_2150659, EPI_ISL_2150660, EPI_ISL_2150661, EPI_ISL_2150662, EPI_ISL_2150663, EPI_ISL_2150664, EPI_ISL_2150665, EPI_ISL_2150666, EPI_ISL_2150667, EPI_ISL_2150668, EPI_ISL_2150669, EPI_ISL_2150670, EPI_ISL_2150671, EPI_ISL_2150672, EPI_ISL_2150673, EPI_ISL_2150674, EPI_ISL_2150675, EPI_ISL_2150676, EPI_ISL_2150677, EPI_ISL_2150678, EPI_ISL_2150679, EPI_ISL_2150680, EPI_ISL_2150681, EPI_ISL_2150682, EPI_ISL_2150683, EPI_ISL_2150684, EPI_ISL_2150685, EPI_ISL_2150686, EPI_ISL_2150687, EPI_ISL_2150688, EPI_ISL_2150689, EPI_ISL_2150690, EPI_ISL_2150691, EPI_ISL_2150692, EPI_ISL_2150693, EPI_ISL_2150694, EPI_ISL_2150695, EPI_ISL_2150696, EPI_ISL_2150697, EPI_ISL_2150698, EPI_ISL_2150699, EPI_ISL_2150700, EPI_ISL_2150701, EPI_ISL_2150702, EPI_ISL_2150703, EPI_ISL_2150704, EPI_ISL_2150705, EPI_ISL_2150706, EPI_ISL_2150707, EPI_ISL_2150708, EPI_ISL_2150709, EPI_ISL_2150710, EPI_ISL_2150711, EPI_ISL_2150712, EPI_ISL_2150713, EPI_ISL_2150714, EPI_ISL_2150715, EPI_ISL_2150716, EPI_ISL_2150717, EPI_ISL_2150718, EPI_ISL_2150719, EPI_ISL_2150720, EPI_ISL_2150721, EPI_ISL_2150722, EPI_ISL_2150723, EPI_ISL_2150724, EPI_ISL_2150725, EPI_ISL_2150726, EPI_ISL_2150727, EPI_ISL_2150728, EPI_ISL_2150729, EPI_ISL_2150730, EPI_ISL_2150731, EPI_ISL_2150732, EPI_ISL_2150733, EPI_ISL_2150734, EPI_ISL_2150735, EPI_ISL_2150736, EPI_ISL_2150737, EPI_ISL_2150738, EPI_ISL_2150739, EPI_ISL_2150740, EPI_ISL_2150741, EPI_ISL_2150742, EPI_ISL_2150743, EPI_ISL_2150744, EPI_ISL_2150745, EPI_ISL_2150746, EPI_ISL_2150747, EPI_ISL_2150748, EPI_ISL_2150749, EPI_ISL_2150750, EPI_ISL_2150751, EPI_ISL_2150752, EPI_ISL_2150753, EPI_ISL_2150754, EPI_ISL_2150755, EPI_ISL_2150756, EPI_ISL_2150757, EPI_ISL_2150758, EPI_ISL_2150759, EPI_ISL_2150760, EPI_ISL_2150761, EPI_ISL_2150762, EPI_ISL_2150763, EPI_ISL_2150764, EPI_ISL_2150765, EPI_ISL_2150766, EPI_ISL_2150767, EPI_ISL_2150768, EPI_ISL_2150769, EPI_ISL_2150770, EPI_ISL_2150771, EPI_ISL_2150772, EPI_ISL_2150773, EPI_ISL_2150774, EPI_ISL_2150775, EPI_ISL_2150776, EPI_ISL_2150777, EPI_ISL_2150778, EPI_ISL_2150779, EPI_ISL_2150780, EPI_ISL_2150781, EPI_ISL_2150782, EPI_ISL_2150783, EPI_ISL_2150784, EPI_ISL_2150785, EPI_ISL_2150786, EPI_ISL_2150787, EPI_ISL_2150788, EPI_ISL_2150789, EPI_ISL_2150790, EPI_ISL_2150792, EPI_ISL_2150793, EPI_ISL_2150794, EPI_ISL_2150795, EPI_ISL_2150796, EPI_ISL_2150797, EPI_ISL_2150798, EPI_ISL_2150799, EPI_ISL_2150800, EPI_ISL_2150801, EPI_ISL_2150802, EPI_ISL_2150803, EPI_ISL_2150804, EPI_ISL_2150805, EPI_ISL_2150806, EPI_ISL_2150807, EPI_ISL_2150808, EPI_ISL_2150809, EPI_ISL_2150810, EPI_ISL_2150811, EPI_ISL_2150812, EPI_ISL_2150813, EPI_ISL_2150814, EPI_ISL_2150815, EPI_ISL_2150816, EPI_ISL_2150817, EPI_ISL_2150818, EPI_ISL_2150819, EPI_ISL_2150820, EPI_ISL_2150821, EPI_ISL_2150822, EPI_ISL_2150823, EPI_ISL_2150824, EPI_ISL_2150825, EPI_ISL_2150826, EPI_ISL_2150827, EPI_ISL_2150828, EPI_ISL_2150829, EPI_ISL_2150830, EPI_ISL_2150831, EPI_ISL_2150832, EPI_ISL_2150833, EPI_ISL_2150834, EPI_ISL_2150835, EPI_ISL_2150836, EPI_ISL_2150837, EPI_ISL_2150838, EPI_ISL_2150839, EPI_ISL_2150840, EPI_ISL_2150841, EPI_ISL_2150842, EPI_ISL_2150843, EPI_ISL_2150844, EPI_ISL_2150845, EPI_ISL_2150846, EPI_ISL_2150847, EPI_ISL_2150848, EPI_ISL_2150849, EPI_ISL_2150850, EPI_ISL_2150851, EPI_ISL_2150852, EPI_ISL_2150853, EPI_ISL_2150854, EPI_ISL_2150855, EPI_ISL_2150856, EPI_ISL_2150857, EPI_ISL_2150858, EPI_ISL_2150859, EPI_ISL_2150861, EPI_ISL_2150862, EPI_ISL_2150863, EPI_ISL_2150864, EPI_ISL_2150865, EPI_ISL_2150866, EPI_ISL_2150867, EPI_ISL_2150868, EPI_ISL_2150869, EPI_ISL_2150870, EPI_ISL_2150871, EPI_ISL_2150872, EPI_ISL_2150873, EPI_ISL_2150874, EPI_ISL_2150875, EPI_ISL_2150876, EPI_ISL_2150877, EPI_ISL_2150878, EPI_ISL_2150879, EPI_ISL_2150880, EPI_ISL_2150881, EPI_ISL_2150882, EPI_ISL_2150883, EPI_ISL_2150884, EPI_ISL_2150885, EPI_ISL_2150886, EPI_ISL_2150887, EPI_ISL_2150888, EPI_ISL_2150889, EPI_ISL_2150890, EPI_ISL_2150891, EPI_ISL_2150892, EPI_ISL_2150893, EPI_ISL_2150894, EPI_ISL_2150895, EPI_ISL_2150896, EPI_ISL_2150897, EPI_ISL_2150898, EPI_ISL_2150899, EPI_ISL_2150900, EPI_ISL_2150901, EPI_ISL_2150902, EPI_ISL_2150903, EPI_ISL_2150904, EPI_ISL_2150905, EPI_ISL_2150906, EPI_ISL_2150907, EPI_ISL_2150908, EPI_ISL_2150909, EPI_ISL_2150910, EPI_ISL_2150911, EPI_ISL_2150912, EPI_ISL_2150913, EPI_ISL_2150914, EPI_ISL_2150915, EPI_ISL_2150916, EPI_ISL_2150917, EPI_ISL_2150918, EPI_ISL_2150919, EPI_ISL_2150920, EPI_ISL_2150921, EPI_ISL_2150922, EPI_ISL_2150923, EPI_ISL_2150924, EPI_ISL_2150925, EPI_ISL_2150926, EPI_ISL_2150927, EPI_ISL_2150928, EPI_ISL_2150929, EPI_ISL_2150930, EPI_ISL_2150931, EPI_ISL_2150932, EPI_ISL_2150933, EPI_ISL_2150934, EPI_ISL_2150935, EPI_ISL_2150936, EPI_ISL_2150937, EPI_ISL_2150938, EPI_ISL_2150939, EPI_ISL_2150940, EPI_ISL_2150941, EPI_ISL_2150942, EPI_ISL_2150943, EPI_ISL_2150944, EPI_ISL_2150945, EPI_ISL_2150946, EPI_ISL_2150947, EPI_ISL_2150948, EPI_ISL_2150949, EPI_ISL_2150950, EPI_ISL_2150951, EPI_ISL_2150952, EPI_ISL_2150953, EPI_ISL_2150954, EPI_ISL_2150955, EPI_ISL_2150956, EPI_ISL_2150957, EPI_ISL_2150958, EPI_ISL_2150959, EPI_ISL_2150960, EPI_ISL_2150961, EPI_ISL_2150962, EPI_ISL_2150963, EPI_ISL_2150964, EPI_ISL_2150965, EPI_ISL_2150966, EPI_ISL_2150967, EPI_ISL_2150968, EPI_ISL_2150969, EPI_ISL_2150970, EPI_ISL_2150971, EPI_ISL_2150972, EPI_ISL_2150973, EPI_ISL_2150974

| | | | |
|---|---|---|---|
| see above | Aegis Sciences Corporation | Centers for Disease Control and Prevention Division of Viral Diseases, Pathogen Discovery | Dakota Howard, Dhwani Batra, Peter W. Cook, Kara Moser, Adrian Paskey, Jason Caravas, Benjamin Rambo-Martin, Shatavia Morrison, Christopher Gulvick, Scott Sammons, Yvette Unoarumhi, Darlene Wagner, Matthew Schmerer, Cyndi Clark, Patrick Campbell, Rob Case, Vikramsinha Ghorpade, Holly Houdeshell, Ola Kvalvaag, Dillon Nall, Ethan Sanders, Alec Vest, Shaun Westlund, Matthew Hardison, Clinton R. Paden, Duncan MacCannell |
| EPI_ISL_2151367 | DOHMH Crown Heights | New York City Public Health Laboratory | Jade Wang, et al. |
| EPI_ISL_2151368 | DOHMH Central Harlem | New York City Public Health Laboratory | Jade Wang, et al. |
| EPI_ISL_2151369, EPI_ISL_2151370 | DOHMH Jamaica | New York City Public Health Laboratory | Jade Wang, et al. |
| EPI_ISL_2151371, EPI_ISL_2151372, EPI_ISL_2151373, EPI_ISL_2151374 | DOHMH Riverside | New York City Public Health Laboratory | Jade Wang, et al. |
| EPI_ISL_2151375, EPI_ISL_2151376, EPI_ISL_2151377 | DOHMH Crown Heights | New York City Public Health Laboratory | Jade Wang, et al. |
| EPI_ISL_2151378 | Department of Homeless Services | New York City Public Health Laboratory | Jade Wang, et al. |
| EPI_ISL_2151379, EPI_ISL_2151380 | DOHMH Crown Heights | New York City Public Health Laboratory | Jade Wang, et al. |
| EPI_ISL_2151381 | DOHMH Morrisania | New York City Public Health Laboratory | Jade Wang, et al. |
| EPI_ISL_2151382 | DOHMH Central Harlem | New York City Public Health Laboratory | Jade Wang, et al. |
| EPI_ISL_2151383 | DOHMH Chelsea | New York City Public Health Laboratory | Jade Wang, et al. |
| EPI_ISL_2151384 | DOHMH Fort Greene | New York City Public Health Laboratory | Jade Wang, et al. |
| EPI_ISL_2151385 | DOHMH Crown Heights | New York City Public Health Laboratory | Jade Wang, et al. |
| EPI_ISL_2151386 | DOHMH PHL | New York City Public Health Laboratory | Jade Wang, et al. |
| EPI_ISL_2151387 | DOHMH Central Harlem | New York City Public Health Laboratory | Jade Wang, et al. |
| EPI_ISL_2151388 | DOHMH Riverside | New York City Public Health Laboratory | Jade Wang, et al. |
| EPI_ISL_2151389 | DOHMH Jamaica | New York City Public Health Laboratory | Jade Wang, et al. |
| EPI_ISL_2151390 | DOHMH Chelsea | New York City Public Health Laboratory | Jade Wang, et al. |
| EPI_ISL_2151391 | DOHMH Central Harlem | New York City Public Health Laboratory | Jade Wang, et al. |
| EPI_ISL_2151392 | DOHMH Jamaica | New York City Public Health Laboratory | Jade Wang, et al. |
| EPI_ISL_2151393, EPI_ISL_2151394 | DOHMH PHL | New York City Public Health Laboratory | Jade Wang, et al. |
| EPI_ISL_2151395, EPI_ISL_2151396, EPI_ISL_2151397, EPI_ISL_2151398, EPI_ISL_2151399, EPI_ISL_2151400, EPI_ISL_2151401 | HealthQuest Esoterics | New York City Public Health Laboratory | Jade Wang, et al. |
| EPI_ISL_2151402, EPI_ISL_2151403 | Department of Homeless Services | New York City Public Health Laboratory | Jade Wang, et al. |
| EPI_ISL_2151404, EPI_ISL_2151405, EPI_ISL_2151406, EPI_ISL_2151407, EPI_ISL_2151408, EPI_ISL_2151409, EPI_ISL_2151410, EPI_ISL_2151411 | OCME Office Of Chief Medical Examiner | New York City Public Health Laboratory | Jade Wang, et al. |
| EPI_ISL_2151412, EPI_ISL_2151413 | DOHMH Central Harlem | New York City Public Health Laboratory | Jade Wang, et al. |
| EPI_ISL_2151414 | DOHMH Crown Heights | New York City Public Health Laboratory | Jade Wang, et al. |
| EPI_ISL_2151415 | DOHMH Jamaica | New York City Public Health Laboratory | Jade Wang, et al. |

| | | | |
|---|---|---|---|
| EPI_ISL_2151416, EPI_ISL_2151417, EPI_ISL_2151418 | DOHMH Fort Greene | New York City Public Health Laboratory | Jade Wang, et al. |
| EPI_ISL_2151419 | DOHMH Jamaica | New York City Public Health Laboratory | Jade Wang, et al. |
| EPI_ISL_2151420 | SUNY Downstate Medical Center | New York City Public Health Laboratory | Jade Wang, et al. |
| EPI_ISL_2151421, EPI_ISL_2151422, EPI_ISL_2151423, EPI_ISL_2151424, EPI_ISL_2151425, EPI_ISL_2151426, EPI_ISL_2151427 | CDC Dengue Branch | Pathogen Discovery, Respiratory Viruses Branch, Division of Viral Diseases, Centers for Disease Control and Prevention | Yan Li, Jing Zhang, Anna Kelleher, Ying Tao, Brian Lynch, Krista Queen, Anna Uehara, Peter Cook, Han Jia Justin Ng, Rachel Marine, Clinton R. Paden, Dhwani Batra, Haibin Wang, Tara Coalter, Jasmine Padilla, Morgan Davis, Mili Sheth, Sarah Nobles, Mark Burroughs, Justin Lee, Adam Retchless, Suxiang Tong |
| EPI_ISL_2151428, EPI_ISL_2151429, EPI_ISL_2151430, EPI_ISL_2151431, EPI_ISL_2151432, EPI_ISL_2151433, EPI_ISL_2151434, EPI_ISL_2151435 | Johns Hopkins University | Pathogen Discovery, Respiratory Viruses Branch, Division of Viral Diseases, Centers for Disease Control and Prevention | Yan Li, Jing Zhang, Anna Kelleher, Ying Tao, Brian Lynch, Krista Queen, Anna Uehara, Peter Cook, Han Jia Justin Ng, Rachel Marine, Clinton R. Paden, Dhwani Batra, Haibin Wang, Tara Coalter, Jasmine Padilla, Morgan Davis, Mili Sheth, Sarah Nobles, Mark Burroughs, Justin Lee, Adam Retchless, Suxiang Tong |
| EPI_ISL_2151436 | Columbia University | Pathogen Discovery, Respiratory Viruses Branch, Division of Viral Diseases, Centers for Disease Control and Prevention | Yan Li, Jing Zhang, Anna Kelleher, Ying Tao, Brian Lynch, Krista Queen, Anna Uehara, Peter Cook, Han Jia Justin Ng, Rachel Marine, Clinton R. Paden, Dhwani Batra, Haibin Wang, Tara Coalter, Jasmine Padilla, Morgan Davis, Mili Sheth, Sarah Nobles, Mark Burroughs, Justin Lee, Adam Retchless, Suxiang Tong |
| EPI_ISL_2151437, EPI_ISL_2151438, EPI_ISL_2151439 | Johns Hopkins University | Pathogen Discovery, Respiratory Viruses Branch, Division of Viral Diseases, Centers for Disease Control and Prevention | Yan Li, Jing Zhang, Anna Kelleher, Ying Tao, Brian Lynch, Krista Queen, Anna Uehara, Peter Cook, Han Jia Justin Ng, Rachel Marine, Clinton R. Paden, Dhwani Batra, Haibin Wang, Tara Coalter, Jasmine Padilla, Morgan Davis, Mili Sheth, Sarah Nobles, Mark Burroughs, Justin Lee, Adam Retchless, Suxiang Tong |
| EPI_ISL_2151440 | Columbia University | Pathogen Discovery, Respiratory Viruses Branch, Division of Viral Diseases, Centers for Disease Control and Prevention | Yan Li, Jing Zhang, Anna Kelleher, Ying Tao, Brian Lynch, Krista Queen, Anna Uehara, Peter Cook, Han Jia Justin Ng, Rachel Marine, Clinton R. Paden, Dhwani Batra, Haibin Wang, Tara Coalter, Jasmine Padilla, Morgan Davis, Mili Sheth, Sarah Nobles, Mark Burroughs, Justin Lee, Adam Retchless, Suxiang Tong |
| EPI_ISL_2151441 | Johns Hopkins University | Pathogen Discovery, Respiratory Viruses Branch, Division of Viral Diseases, Centers for Disease Control and Prevention | Yan Li, Jing Zhang, Anna Kelleher, Ying Tao, Brian Lynch, Krista Queen, Anna Uehara, Peter Cook, Han Jia Justin Ng, Rachel Marine, Clinton R. Paden, Dhwani Batra, Haibin Wang, Tara Coalter, Jasmine Padilla, Morgan Davis, Mili Sheth, Sarah Nobles, Mark Burroughs, Justin Lee, Adam Retchless, Suxiang Tong |
| EPI_ISL_2151445, EPI_ISL_2151446, EPI_ISL_2151447, EPI_ISL_2151448, EPI_ISL_2151452, EPI_ISL_2151453, EPI_ISL_2151454, EPI_ISL_2151455, EPI_ISL_2151457, EPI_ISL_2151459, EPI_ISL_2151463, EPI_ISL_2151464, EPI_ISL_2151470, EPI_ISL_2151472, EPI_ISL_2151473, EPI_ISL_2151474, EPI_ISL_2151475, EPI_ISL_2151479, EPI_ISL_2151481, EPI_ISL_2151482, EPI_ISL_2151483, EPI_ISL_2151484, EPI_ISL_2151485, EPI_ISL_2151487, EPI_ISL_2151488, EPI_ISL_2151491, EPI_ISL_2151493, EPI_ISL_2151495, EPI_ISL_2151496, EPI_ISL_2151499, EPI_ISL_2151502, EPI_ISL_2151508 | | | |
| see above | North Dakota Department of Health, Public Health Laboratory | North Dakota Department of Health, Public Health Laboratory | Lisa Wingerter |
| EPI_ISL_2151800, EPI_ISL_2151801, EPI_ISL_2151802, EPI_ISL_2151803, EPI_ISL_2151804, EPI_ISL_2151805, EPI_ISL_2151806, EPI_ISL_2151807, EPI_ISL_2151808, EPI_ISL_2151809, EPI_ISL_2151811, EPI_ISL_2151813, EPI_ISL_2151814, EPI_ISL_2151815, EPI_ISL_2151816, EPI_ISL_2151817, EPI_ISL_2151818, EPI_ISL_2151819, EPI_ISL_2151820, EPI_ISL_2151821, EPI_ISL_2151822, EPI_ISL_2151823, EPI_ISL_2151824, EPI_ISL_2151825, EPI_ISL_2151826, EPI_ISL_2151827, EPI_ISL_2151828, EPI_ISL_2151829, EPI_ISL_2151830, EPI_ISL_2151831 | | | |
| see above | Wyoming Public Health Laboratory | Wyoming Public Health Laboratory | Jim Mildenberger, Taylor Fearing, Channing Weber, Ashley Norberg, Chayse Rowley, Marley Goetz, Brian Dominguez, Elliot Thomasson, Brittany Oher, Sam Britz, Cari Sloma, and Rob Christensen |
| EPI_ISL_2152597, EPI_ISL_2152598, EPI_ISL_2152599, EPI_ISL_2152600, EPI_ISL_2152601, EPI_ISL_2152602, EPI_ISL_2152603, EPI_ISL_2152604, EPI_ISL_2152605, EPI_ISL_2152606, EPI_ISL_2152607, EPI_ISL_2152608, EPI_ISL_2152609, EPI_ISL_2152610, EPI_ISL_2152611, EPI_ISL_2152612, EPI_ISL_2152613, EPI_ISL_2152614, EPI_ISL_2152615, EPI_ISL_2152616, EPI_ISL_2152617, EPI_ISL_2152618, EPI_ISL_2152619, EPI_ISL_2152620, EPI_ISL_2152621, EPI_ISL_2152622, EPI_ISL_2152623, EPI_ISL_2152624, EPI_ISL_2152625, EPI_ISL_2152626, EPI_ISL_2152627, EPI_ISL_2152628, EPI_ISL_2152629, EPI_ISL_2152630, EPI_ISL_2152631, EPI_ISL_2152632, EPI_ISL_2152633, EPI_ISL_2152634, EPI_ISL_2152635, EPI_ISL_2152636, EPI_ISL_2152637, EPI_ISL_2152638, EPI_ISL_2152639, EPI_ISL_2152640, EPI_ISL_2152641, EPI_ISL_2152642, EPI_ISL_2152643, EPI_ISL_2152644, EPI_ISL_2152645, EPI_ISL_2152646, EPI_ISL_2152647, EPI_ISL_2152648, EPI_ISL_2152649, EPI_ISL_2152650, EPI_ISL_2152651, EPI_ISL_2152652, EPI_ISL_2152653, EPI_ISL_2152654, EPI_ISL_2152655, EPI_ISL_2153096, EPI_ISL_2153097 | | | |
| see above | Hackensack Medical Center | New York Genome Center | Michael Zody, Andre Corvelo, Dayna M. Oschwald, Samantha Fennessey, Tom Maniatis, Liang Chen, Jose Mediavilla, Marcus Cunningham, Kaelea Composto, Kar Chow, David Perlin, Barry Kreiswirth |
| EPI_ISL_2153099, EPI_ISL_2153100, EPI_ISL_2153101 | Wyoming Public Health Laboratory | Wyoming Public Health Laboratory | Jim Mildenberger, Taylor Fearing, Channing Weber, Ashley Norberg, Chayse Rowley, Marley Goetz, Brian Dominguez, Elliot Thomasson, Brittany Oher, Sam Britz, Cari Sloma, and Rob Christensen |
| EPI_ISL_2153538, EPI_ISL_2153539, EPI_ISL_2153540, EPI_ISL_2153541, EPI_ISL_2153542, EPI_ISL_2153543, EPI_ISL_2153544, EPI_ISL_2153545, EPI_ISL_2153547, EPI_ISL_2153548, EPI_ISL_2153549, EPI_ISL_2153550, EPI_ISL_2153551, EPI_ISL_2153552, EPI_ISL_2153553 | | | |
| see above | Florida Bureau of Public Health Laboratories | Florida Bureau of Public Health Laboratories | Sarah Schmedes, Jason Blanton |
| EPI_ISL_2153554, EPI_ISL_2153555, EPI_ISL_2153556, EPI_ISL_2153557, EPI_ISL_2153558, EPI_ISL_2153559, EPI_ISL_2153560, EPI_ISL_2153561, EPI_ISL_2153562, EPI_ISL_2153563, EPI_ISL_2153564, EPI_ISL_2153565, EPI_ISL_2153566, EPI_ISL_2153567, EPI_ISL_2153568, EPI_ISL_2153569, EPI_ISL_2153570, EPI_ISL_2153571, EPI_ISL_2153572, EPI_ISL_2153573, EPI_ISL_2153574, EPI_ISL_2153575, EPI_ISL_2153576, EPI_ISL_2153577, EPI_ISL_2153578, EPI_ISL_2153579, EPI_ISL_2153580, EPI_ISL_2153581, EPI_ISL_2153582, EPI_ISL_2153583, EPI_ISL_2153584, EPI_ISL_2153585, EPI_ISL_2153586, EPI_ISL_2153587, EPI_ISL_2153588, EPI_ISL_2153589, EPI_ISL_2153590, EPI_ISL_2153591, EPI_ISL_2153592, EPI_ISL_2153593, EPI_ISL_2153594, EPI_ISL_2153595, EPI_ISL_2153596, EPI_ISL_2153597, EPI_ISL_2153598, EPI_ISL_2153599, EPI_ISL_2153600, EPI_ISL_2153601, EPI_ISL_2153602, EPI_ISL_2153603, EPI_ISL_2153604, EPI_ISL_2153605, EPI_ISL_2153606, EPI_ISL_2153607, EPI_ISL_2153608, EPI_ISL_2153609, EPI_ISL_2153610, EPI_ISL_2153611, EPI_ISL_2153612, EPI_ISL_2153613, EPI_ISL_2153614, EPI_ISL_2153659, EPI_ISL_2153660, EPI_ISL_2153661, EPI_ISL_2153662, EPI_ISL_2153663, EPI_ISL_2153664, EPI_ISL_2153665, EPI_ISL_2153666, EPI_ISL_2153667, EPI_ISL_2153668, EPI_ISL_2153669, EPI_ISL_2153670, EPI_ISL_2153671, EPI_ISL_2153672, EPI_ISL_2153673, EPI_ISL_2153674, EPI_ISL_2153675, EPI_ISL_2153676, EPI_ISL_2153677, EPI_ISL_2153678 | | | |
| see above | Arizona State University | Arizona State University | Peter T. Skidmore, LaRinda A. Holland, Matthew F. Smith, Rabia Maqsood, Nicholas J. Mellor, Joy M. Blain, Valerie Harris, Joshua LaBaer, Vel Murugan, Efrem S. Lim |
| EPI_ISL_2153679, EPI_ISL_2153680, EPI_ISL_2153681, EPI_ISL_2153683, EPI_ISL_2153684, EPI_ISL_2153685, EPI_ISL_2153686, EPI_ISL_2153687, EPI_ISL_2153688, EPI_ISL_2153689, EPI_ISL_2153690, EPI_ISL_2153691, EPI_ISL_2153692, EPI_ISL_2153693, EPI_ISL_2153696, EPI_ISL_2153697, EPI_ISL_2153698, EPI_ISL_2153699, EPI_ISL_2153700 | | | |
| see above | Kansas Health and Environmental Lab | Kansas Health and Environmental Lab | Mike Grose, Jonathan Barnell, Ben Olsen, and Phil Adam |
| EPI_ISL_2153703, EPI_ISL_2153704 | Incyte Diagnostics Spokane | Seattle Flu Study | Deborah A. Nickerson, Chris D. Frazar, Jover Lee, Benjamin Pelle, Erica Ryke, Matthew Richardson, Amanda Adler, Elisabeth Brandstetter, Peter D. Han, Kairsten Fay, Misja Ilcisin, Kirsten Lacombe, Thomas R. Sibley, Melissa Truong, Caitlin R. Wolf, Romesh Gautom, Geoff Melly, Brian Hiatt, Philip Dykema, Scott Lindquist, Michael Boeckh, Janet A. Englund, Michael Famulare, Barry R. Lutz, Mark J. Rieder, Lea M. Starita, Matthew Thompson, Helen Y. Chu, Jay Shendure, Trevor Bedford |
| EPI_ISL_2153706 | Northwest Laboratory | Seattle Flu Study | Deborah A. Nickerson, Chris D. Frazar, Jover Lee, Benjamin Pelle, Erica Ryke, Matthew Richardson, Amanda Adler, Elisabeth Brandstetter, Peter D. Han, Kairsten Fay, Misja Ilcisin, Kirsten Lacombe, Thomas R. Sibley, Melissa Truong, Caitlin R. Wolf, Romesh Gautom, Geoff Melly, Brian Hiatt, Philip Dykema, Scott Lindquist, Michael Boeckh, Janet A. Englund, Michael Famulare, Barry R. Lutz, Mark J. Rieder, Lea M. Starita, Matthew Thompson, Helen Y. Chu, Jay Shendure, Trevor Bedford |
| EPI_ISL_2153707 | Atlas Genomics | Seattle Flu Study | Deborah A. Nickerson, Chris D. Frazar, Jover Lee, Benjamin Pelle, Erica Ryke, Matthew Richardson, Amanda Adler, Elisabeth Brandstetter, Peter D. Han, Kairsten Fay, Misja Ilcisin, Kirsten Lacombe, Thomas R. Sibley, Melissa Truong, Caitlin R. Wolf, Romesh Gautom, Geoff Melly, Brian Hiatt, Philip Dykema, Scott Lindquist, Michael Boeckh, Janet A. Englund, Michael Famulare, Barry R. Lutz, Mark J. Rieder, Lea M. Starita, Matthew Thompson, Helen Y. Chu, Jay Shendure, Trevor Bedford |
| EPI_ISL_2153708, EPI_ISL_2153709, EPI_ISL_2153710, EPI_ISL_2153711 | Alaska State Virology Laboratory | Alaska State Virology Laboratory | Stephanie DeRonde, Elva House, Lisa Smith, Ph.D., Jack Chen, Ph.D. |
| EPI_ISL_2153712, EPI_ISL_2153713, EPI_ISL_2153714 | Alaska State Virology Laboratory | Alaska State Virology Laboratory | Stephanie DeRonde, Elva House, Jacob Zidek, Lisa Smith, Ph.D., Jack Chen, Ph.D. |
| EPI_ISL_2153715, EPI_ISL_2153725, EPI_ISL_2153727, EPI_ISL_2153730, EPI_ISL_2153732, EPI_ISL_2153733, EPI_ISL_2153735, EPI_ISL_2153736, EPI_ISL_2153737, EPI_ISL_2153741, EPI_ISL_2153743, EPI_ISL_2153744, EPI_ISL_2153745, EPI_ISL_2153747, EPI_ISL_2153748, EPI_ISL_2153749, EPI_ISL_2153752, EPI_ISL_2153754, EPI_ISL_2153756, EPI_ISL_2153758, EPI_ISL_2153759, EPI_ISL_2153760, EPI_ISL_2153761, EPI_ISL_2153763, EPI_ISL_2153764, EPI_ISL_2153765, EPI_ISL_2153766, EPI_ISL_2153767, EPI_ISL_2153768, EPI_ISL_2153769, EPI_ISL_2153770, EPI_ISL_2153771, EPI_ISL_2153772, EPI_ISL_2153773, EPI_ISL_2153774, EPI_ISL_2153775, EPI_ISL_2153776, EPI_ISL_2153777, EPI_ISL_2153778, EPI_ISL_2153779, EPI_ISL_2153780, EPI_ISL_2153781, EPI_ISL_2153782, EPI_ISL_2153783, EPI_ISL_2153784, EPI_ISL_2153785, EPI_ISL_2153786, EPI_ISL_2153787, EPI_ISL_2153788, EPI_ISL_2153789, EPI_ISL_2153790, EPI_ISL_2153791, EPI_ISL_2153792, EPI_ISL_2153793, EPI_ISL_2153794, EPI_ISL_2153795, EPI_ISL_2153796, EPI_ISL_2153797, EPI_ISL_2153798, EPI_ISL_2153799, EPI_ISL_2153800, EPI_ISL_2153801, EPI_ISL_2153802, EPI_ISL_2153803, EPI_ISL_2153804, EPI_ISL_2153805 | | | |
| see above | The Ohio State University Applied Microbiology Services Laboratory | The Ohio State University Applied Microbiology Services Laboratory | Seth A. Faith PhD |
| EPI_ISL_2154052, EPI_ISL_2154053, EPI_ISL_2154054, EPI_ISL_2154055, EPI_ISL_2154056, EPI_ISL_2154057, EPI_ISL_2154058, EPI_ISL_2154059, EPI_ISL_2154060, EPI_ISL_2154061, EPI_ISL_2154062, EPI_ISL_2154063, EPI_ISL_2154064, EPI_ISL_2154065, EPI_ISL_2154066, EPI_ISL_2154067, EPI_ISL_2154068, EPI_ISL_2154069, EPI_ISL_2154070, EPI_ISL_2154071, EPI_ISL_2154072, EPI_ISL_2154073, EPI_ISL_2154074, EPI_ISL_2154075, EPI_ISL_2154076, EPI_ISL_2154077, EPI_ISL_2154078, EPI_ISL_2154079, EPI_ISL_2154080, EPI_ISL_2154081, EPI_ISL_2154082, EPI_ISL_2154083, EPI_ISL_2154084, EPI_ISL_2154085, EPI_ISL_2154086, EPI_ISL_2154087 | | | |
| see above | Alaska State Virology Laboratory | Alaska State Virology Laboratory | Stephanie DeRonde, Elva House, Jacob Zidek, Lisa Smith, Ph.D., Jack Chen, Ph.D. |
| EPI_ISL_2155822, EPI_ISL_2155823 | LA Office of Public Health Laboratories | Pathogen Discovery, Respiratory Viruses Branch, Division of Viral Diseases, Centers for Disease Control and Prevention | Yan Li, Jing Zhang, Anna Kelleher, Ying Tao, Brian Lynch, Krista Queen, Anna Uehara, Peter Cook, Han Jia Justin Ng, Rachel Marine, Clinton R. Paden, Dhwani Batra, Haibin Wang, Tara Coalter, Jasmine Padilla, Morgan Davis, Mili Sheth, Sarah Nobles, Mark Burroughs, Justin Lee, Adam Retchless, Suxiang Tong |
| EPI_ISL_2155824, EPI_ISL_2155825 | AR Dept. of Health-PHL, Molecular Diagnostics | Pathogen Discovery, Respiratory Viruses Branch, Division of Viral Diseases, Centers for Disease Control and Prevention | Yan Li, Jing Zhang, Anna Kelleher, Ying Tao, Brian Lynch, Krista Queen, Anna Uehara, Peter Cook, Han Jia Justin Ng, Rachel Marine, Clinton R. Paden, Dhwani Batra, Haibin Wang, Tara Coalter, Jasmine Padilla, Morgan Davis, Mili Sheth, Sarah Nobles, Mark Burroughs, Justin Lee, Adam Retchless, Suxiang Tong |
| EPI_ISL_2155826, EPI_ISL_2155827, EPI_ISL_2155828 | WVDHHR - Office of Laboratory Services | Pathogen Discovery, Respiratory Viruses Branch, Division of Viral Diseases, Centers for Disease Control and Prevention | Yan Li, Jing Zhang, Anna Kelleher, Ying Tao, Brian Lynch, Krista Queen, Anna Uehara, Peter Cook, Han Jia Justin Ng, Rachel Marine, Clinton R. Paden, Dhwani Batra, Haibin Wang, Tara Coalter, Jasmine Padilla, Morgan Davis, Mili Sheth, Sarah Nobles, Mark Burroughs, Justin Lee, Adam Retchless, Suxiang Tong |
| EPI_ISL_2155829, EPI_ISL_2155830, EPI_ISL_2155831, EPI_ISL_2155832 | OK Public Health Laboratory, Oklahoma State DOH | Pathogen Discovery, Respiratory Viruses Branch, Division of Viral Diseases, Centers for Disease Control and Prevention | Yan Li, Jing Zhang, Anna Kelleher, Ying Tao, Brian Lynch, Krista Queen, Anna Uehara, Peter Cook, Han Jia Justin Ng, Rachel Marine, Clinton R. Paden, Dhwani Batra, Haibin Wang, Tara Coalter, Jasmine Padilla, Morgan Davis, Mili Sheth, Sarah Nobles, Mark Burroughs, Justin Lee, Adam Retchless, Suxiang Tong |
| EPI_ISL_2155833, EPI_ISL_2155834, EPI_ISL_2155835, EPI_ISL_2155836 | NC State Laboratory of Public Health | Pathogen Discovery, Respiratory Viruses Branch, Division of Viral Diseases, Centers for Disease Control and Prevention | Yan Li, Jing Zhang, Anna Kelleher, Ying Tao, Brian Lynch, Krista Queen, Anna Uehara, Peter Cook, Han Jia Justin Ng, Rachel Marine, Clinton R. Paden, Dhwani Batra, Haibin Wang, Tara Coalter, Jasmine Padilla, Morgan Davis, Mili Sheth, Sarah Nobles, Mark Burroughs, Justin Lee, Adam Retchless, Suxiang Tong |

| | | | |
|---|---|---|---|
| EPI_ISL_2155837, EPI_ISL_2155838, EPI_ISL_2155839, EPI_ISL_2155840 | TN Division of Laboratory Services | Pathogen Discovery, Respiratory Viruses Branch, Division of Viral Diseases, Centers for Disease Control and Prevention | Yan Li, Jing Zhang, Anna Kelleher, Ying Tao, Brian Lynch, Krista Queen, Anna Uehara, Peter Cook, Han Jia Justin Ng, Rachel Marine, Clinton R. Paden, Dhwani Batra, Haibin Wang, Tara Coalter, Jasmine Padilla, Morgan Davis, Mili Sheth, Sarah Nobles, Mark Burroughs, Justin Lee, Adam Retchless, Suxiang Tong |
| EPI_ISL_2156023, EPI_ISL_2156024, EPI_ISL_2156025, EPI_ISL_2156026, EPI_ISL_2156027, EPI_ISL_2156028, EPI_ISL_2156029, EPI_ISL_2156030, EPI_ISL_2156031, EPI_ISL_2156033, EPI_ISL_2156034 | | | |
| see above | Wichita State University - Molecular Diagnostics Lab | Kansas Health and Environmental Lab | Mike Grose, Katherine Wiggins, Jonathan Barnell, Ben Olsen, and Phil Adam |
| EPI_ISL_2156035, EPI_ISL_2156036, EPI_ISL_2156037, EPI_ISL_2156038, EPI_ISL_2156039, EPI_ISL_2156040, EPI_ISL_2156041, EPI_ISL_2156042, EPI_ISL_2156043 | Kansas Health and Environmental Lab | Kansas Health and Environmental Lab | Mike Grose, Katherine Wiggins, Jonathan Barnell, Ben Olsen, and Phil Adam |
| EPI_ISL_2156044, EPI_ISL_2156047 | Wichita State University - Molecular Diagnostics Lab | Kansas Health and Environmental Lab | Mike Grose, Katherine Wiggins, Jonathan Barnell, Ben Olsen, and Phil Adam |
| EPI_ISL_2156048, EPI_ISL_2156050 | Kansas Health and Environmental Lab | Kansas Health and Environmental Lab | Mike Grose, Katherine Wiggins, Jonathan Barnell, Ben Olsen, and Phil Adam |
| EPI_ISL_2156771, EPI_ISL_2156772, EPI_ISL_2156774, EPI_ISL_2156776, EPI_ISL_2156777, EPI_ISL_2156778, EPI_ISL_2156779, EPI_ISL_2156780, EPI_ISL_2156781, EPI_ISL_2156782, EPI_ISL_2156783, EPI_ISL_2156784, EPI_ISL_2156785, EPI_ISL_2156786, EPI_ISL_2156787, EPI_ISL_2156788, EPI_ISL_2156789, EPI_ISL_2156790, EPI_ISL_2156791, EPI_ISL_2156792, EPI_ISL_2156793, EPI_ISL_2156794 | | | |
| see above | Arkansas Children's Hospital | Center for Global Health, University of New Mexico Health Sciences Center | Daryl Domman, Kurt Schwalm, Valerie Morley, Catherine Kirkpatrick, Bobby Boyanton, Joshua L. Kennedy, Darrell Dinwiddie |
| EPI_ISL_2156797, EPI_ISL_2156798 | AR Dept. of Health-PHL, Molecular Diagnostics | Pathogen Discovery, Respiratory Viruses Branch, Division of Viral Diseases, Centers for Disease Control and Prevention | Yan Li, Jing Zhang, Anna Kelleher, Ying Tao, Brian Lynch, Krista Queen, Anna Uehara, Peter Cook, Han Jia Justin Ng, Rachel Marine, Clinton R. Paden, Dhwani Batra, Haibin Wang, Tara Coalter, Jasmine Padilla, Morgan Davis, Mili Sheth, Sarah Nobles, Mark Burroughs, Justin Lee, Adam Retchless, Suxiang Tong |
| EPI_ISL_2156799 | SD Public Health Laboratory | Pathogen Discovery, Respiratory Viruses Branch, Division of Viral Diseases, Centers for Disease Control and Prevention | Yan Li, Jing Zhang, Anna Kelleher, Ying Tao, Brian Lynch, Krista Queen, Anna Uehara, Peter Cook, Han Jia Justin Ng, Rachel Marine, Clinton R. Paden, Dhwani Batra, Haibin Wang, Tara Coalter, Jasmine Padilla, Morgan Davis, Mili Sheth, Sarah Nobles, Mark Burroughs, Justin Lee, Adam Retchless, Suxiang Tong |
| EPI_ISL_2156800, EPI_ISL_2156801 | NC State Laboratory of Public Health | Pathogen Discovery, Respiratory Viruses Branch, Division of Viral Diseases, Centers for Disease Control and Prevention | Yan Li, Jing Zhang, Anna Kelleher, Ying Tao, Brian Lynch, Krista Queen, Anna Uehara, Peter Cook, Han Jia Justin Ng, Rachel Marine, Clinton R. Paden, Dhwani Batra, Haibin Wang, Tara Coalter, Jasmine Padilla, Morgan Davis, Mili Sheth, Sarah Nobles, Mark Burroughs, Justin Lee, Adam Retchless, Suxiang Tong |
| EPI_ISL_2156802 | AR Dept. of Health-PHL, Molecular Diagnostics | Pathogen Discovery, Respiratory Viruses Branch, Division of Viral Diseases, Centers for Disease Control and Prevention | Yan Li, Jing Zhang, Anna Kelleher, Ying Tao, Brian Lynch, Krista Queen, Anna Uehara, Peter Cook, Han Jia Justin Ng, Rachel Marine, Clinton R. Paden, Dhwani Batra, Haibin Wang, Tara Coalter, Jasmine Padilla, Morgan Davis, Mili Sheth, Sarah Nobles, Mark Burroughs, Justin Lee, Adam Retchless, Suxiang Tong |
| EPI_ISL_2156803 | WVDHHR - Office of Laboratory Services | Pathogen Discovery, Respiratory Viruses Branch, Division of Viral Diseases, Centers for Disease Control and Prevention | Yan Li, Jing Zhang, Anna Kelleher, Ying Tao, Brian Lynch, Krista Queen, Anna Uehara, Peter Cook, Han Jia Justin Ng, Rachel Marine, Clinton R. Paden, Dhwani Batra, Haibin Wang, Tara Coalter, Jasmine Padilla, Morgan Davis, Mili Sheth, Sarah Nobles, Mark Burroughs, Justin Lee, Adam Retchless, Suxiang Tong |
| EPI_ISL_2156804, EPI_ISL_2156805 | TN Division of Laboratory Services | Pathogen Discovery, Respiratory Viruses Branch, Division of Viral Diseases, Centers for Disease Control and Prevention | Yan Li, Jing Zhang, Anna Kelleher, Ying Tao, Brian Lynch, Krista Queen, Anna Uehara, Peter Cook, Han Jia Justin Ng, Rachel Marine, Clinton R. Paden, Dhwani Batra, Haibin Wang, Tara Coalter, Jasmine Padilla, Morgan Davis, Mili Sheth, Sarah Nobles, Mark Burroughs, Justin Lee, Adam Retchless, Suxiang Tong |
| EPI_ISL_2156808, EPI_ISL_2156810, EPI_ISL_2156811, EPI_ISL_2156821, EPI_ISL_2156823, EPI_ISL_2156824, EPI_ISL_2156826, EPI_ISL_2156827 | Clinical Reference Laboratory | Diversigen | Aaron Garoutte, Tasha M. Santiago-Rodriguez, Heather L. Fehling , Rafal Iwasiow |
| EPI_ISL_2157214, EPI_ISL_2157215, EPI_ISL_2157216, EPI_ISL_2157218, EPI_ISL_2157219, EPI_ISL_2157220, EPI_ISL_2157221 | Kansas Health and Environmental Lab | Kansas Health and Environmental Lab | Mike Grose, Katherine Wiggins, Jonathan Barnell, Ben Olsen, and Phil Adam |
| EPI_ISL_2157222, EPI_ISL_2157223, EPI_ISL_2157224, EPI_ISL_2157225, EPI_ISL_2157226, EPI_ISL_2157227, EPI_ISL_2157228, EPI_ISL_2157229, EPI_ISL_2157230, EPI_ISL_2157231, EPI_ISL_2157232, EPI_ISL_2157233, EPI_ISL_2157234, EPI_ISL_2157236, EPI_ISL_2157237, EPI_ISL_2157238, EPI_ISL_2157239, EPI_ISL_2157240, EPI_ISL_2157241, EPI_ISL_2157242, EPI_ISL_2157243, EPI_ISL_2157244, EPI_ISL_2157245, EPI_ISL_2157246, EPI_ISL_2157247, EPI_ISL_2157248, EPI_ISL_2157249, EPI_ISL_2157250, EPI_ISL_2157251 | | | |
| see above | Gundersen Clinical Microbiology Laboratory | Kabara Cancer Research Institute | Craig S. Richmond, Paraic A. Kenny |
| EPI_ISL_2157253, EPI_ISL_2157254, EPI_ISL_2157255, EPI_ISL_2157256, EPI_ISL_2157257, EPI_ISL_2157258, EPI_ISL_2157259, EPI_ISL_2157260, EPI_ISL_2157261, EPI_ISL_2157262, EPI_ISL_2157263, EPI_ISL_2157264 | | | |
| see above | Arkansas Children's Hospital | Center for Global Health, University of New Mexico Health Sciences Center | Daryl Domman, Kurt Schwalm, Valerie Morley, Catherine Kirkpatrick, Bobby Boyanton, Joshua L. Kennedy, Darrell Dinwiddie |
| EPI_ISL_2157265, EPI_ISL_2157266, EPI_ISL_2157267, EPI_ISL_2157268, EPI_ISL_2157269, EPI_ISL_2157270, EPI_ISL_2157271, EPI_ISL_2157272, EPI_ISL_2157273, EPI_ISL_2157274, EPI_ISL_2157275 | | | |
| see above | Arkansas Children's Hospital | Center for Global Health, University of New Mexico Health Sciences Center | Daryl Domman, Kurt Schwalm, Valerie Morley, Amanda Novack, Dirk Haselow, Joshua L. Kennedy, Darrell Dinwiddie |
| EPI_ISL_2157276, EPI_ISL_2157277, EPI_ISL_2157278, EPI_ISL_2157279, EPI_ISL_2157280, EPI_ISL_2157281, EPI_ISL_2157282 | Arkansas Department of Health | Center for Global Health, University of New Mexico Health Sciences Center | Daryl Domman, Kurt Schwalm, Valerie Morley, Atul Kothari, Joshua L. Kennedy, Darrell Dinwiddie |
| EPI_ISL_2157283, EPI_ISL_2157284, EPI_ISL_2157285, EPI_ISL_2157286, EPI_ISL_2157287, EPI_ISL_2157288, EPI_ISL_2157289, EPI_ISL_2157290, EPI_ISL_2157291, EPI_ISL_2157292, EPI_ISL_2157293, EPI_ISL_2157294, EPI_ISL_2157295, EPI_ISL_2157296, EPI_ISL_2157297 | | | |
| see above | Kansas Health and Environmental Lab | Kansas Health and Environmental Lab | Mike Grose, Katherine Wiggins, Jonathan Barnell, Ben Olsen, and Phil Adam |
| EPI_ISL_2157298, EPI_ISL_2157299, EPI_ISL_2157300, EPI_ISL_2157301, EPI_ISL_2157302, EPI_ISL_2157303, EPI_ISL_2157304, EPI_ISL_2157305, EPI_ISL_2157306, EPI_ISL_2157307, EPI_ISL_2157308, EPI_ISL_2157309 | | | |
| see above | Arkansas Children's Hospital | Center for Global Health, University of New Mexico Health Sciences Center | Daryl Domman, Kurt Schwalm, Valerie Morley, Catherine Kirkpatrick, Bobby Boyanton, Joshua L. Kennedy, Darrell Dinwiddie |
| EPI_ISL_2157310 | LESP Nuevo Leon | Instituto de Diagnostico y Referencia Epidemiologicos (INDRE) | Claudia Wong-Arambula, Abril Rodriguez-Maldonado, Vanessa Rivero-Arredondo, Ariadna Medina-Benitez, Joaquin Quiroz-Mercado, Sergio Rangel-Guerrero, Natividad Cruz-Ortiz, Tatiana Nunez-Garcia, Gisela Barrera-Badillo, Lucia Hernandez-Rivas, Irma Lopez-Martinez, Ernesto Ramirez-Gonzalez. |
| EPI_ISL_2157311, EPI_ISL_2157312, EPI_ISL_2157313, EPI_ISL_2157314 | LESP Queretaro | Instituto de Diagnostico y Referencia Epidemiologicos (INDRE) | Claudia Wong-Arambula, Abril Rodriguez-Maldonado, Vanessa Rivero-Arredondo, Ariadna Medina-Benitez, Joaquin Quiroz-Mercado, Sergio Rangel-Guerrero, Natividad Cruz-Ortiz, Tatiana Nunez-Garcia, Gisela Barrera-Badillo, Lucia Hernandez-Rivas, Irma Lopez-Martinez, Ernesto Ramirez-Gonzalez. |
| EPI_ISL_2157315 | LESP Morelos | Instituto de Diagnostico y Referencia Epidemiologicos (INDRE) | Claudia Wong-Arambula, Abril Rodriguez-Maldonado, Vanessa Rivero-Arredondo, Ariadna Medina-Benitez, Joaquin Quiroz-Mercado, Sergio Rangel-Guerrero, Natividad Cruz-Ortiz, Tatiana Nunez-Garcia, Gisela Barrera-Badillo, Lucia Hernandez-Rivas, Irma Lopez-Martinez, Ernesto Ramirez-Gonzalez. |
| EPI_ISL_2157316, EPI_ISL_2157317, EPI_ISL_2157318 | LESP Chihuahua | Instituto de Diagnostico y Referencia Epidemiologicos (INDRE) | Claudia Wong-Arambula, Abril Rodriguez-Maldonado, Vanessa Rivero-Arredondo, Ariadna Medina-Benitez, Joaquin Quiroz-Mercado, Sergio Rangel-Guerrero, Natividad Cruz-Ortiz, Tatiana Nunez-Garcia, Gisela Barrera-Badillo, Lucia Hernandez-Rivas, Irma Lopez-Martinez, Ernesto Ramirez-Gonzalez. |
| EPI_ISL_2157319 | LESP Coahuila | Instituto de Diagnostico y Referencia Epidemiologicos (INDRE) | Claudia Wong-Arambula, Abril Rodriguez-Maldonado, Vanessa Rivero-Arredondo, Ariadna Medina-Benitez, Joaquin Quiroz-Mercado, Sergio Rangel-Guerrero, Natividad Cruz-Ortiz, Tatiana Nunez-Garcia, Gisela Barrera-Badillo, Lucia Hernandez-Rivas, Irma Lopez-Martinez, Ernesto Ramirez-Gonzalez. |
| EPI_ISL_2157320 | LESP Quintana Roo | Instituto de Diagnostico y Referencia Epidemiologicos (INDRE) | Claudia Wong-Arambula, Abril Rodriguez-Maldonado, Vanessa Rivero-Arredondo, Ariadna Medina-Benitez, Joaquin Quiroz-Mercado, Sergio Rangel-Guerrero, Natividad Cruz-Ortiz, Tatiana Nunez-Garcia, Gisela Barrera-Badillo, Lucia Hernandez-Rivas, Irma Lopez-Martinez, Ernesto Ramirez-Gonzalez. |
| EPI_ISL_2157321 | LESP Sonora | Instituto de Diagnostico y Referencia Epidemiologicos (INDRE) | Claudia Wong-Arambula, Abril Rodriguez-Maldonado, Vanessa Rivero-Arredondo, Ariadna Medina-Benitez, Joaquin Quiroz-Mercado, Sergio Rangel-Guerrero, Natividad Cruz-Ortiz, Tatiana Nunez-Garcia, Gisela Barrera-Badillo, Lucia Hernandez-Rivas, Irma Lopez-Martinez, Ernesto Ramirez-Gonzalez. |

| | | | |
|---|---|---|---|
| EPI_ISL_2157322 | LESP Chihuahua | Instituto de Diagnostico y Referencia Epidemiologicos (INDRE) | Claudia Wong-Arambula, Abril Rodriguez-Maldonado, Vanessa Rivero-Arredondo, Ariadna Medina-Benitez, Joaquin Quiroz-Mercado, Sergio Rangel-Guerrero, Natividad Cruz-Ortiz, Tatiana Nunez-Garcia, Gisela Barrera-Badillo, Lucia Hernandez-Rivas, Irma Lopez-Martinez, Ernesto Ramirez-Gonzalez. |
| EPI_ISL_2157323 | LESP Nuevo Leon | Instituto de Diagnostico y Referencia Epidemiologicos (INDRE) | Claudia Wong-Arambula, Abril Rodriguez-Maldonado, Vanessa Rivero-Arredondo, Ariadna Medina-Benitez, Joaquin Quiroz-Mercado, Sergio Rangel-Guerrero, Natividad Cruz-Ortiz, Tatiana Nunez-Garcia, Gisela Barrera-Badillo, Lucia Hernandez-Rivas, Irma Lopez-Martinez, Ernesto Ramirez-Gonzalez. |
| EPI_ISL_2157324, EPI_ISL_2157325, EPI_ISL_2157326 | LESP Guanajuato | Instituto de Diagnostico y Referencia Epidemiologicos (INDRE) | Claudia Wong-Arambula, Abril Rodriguez-Maldonado, Vanessa Rivero-Arredondo, Ariadna Medina-Benitez, Joaquin Quiroz-Mercado, Sergio Rangel-Guerrero, Natividad Cruz-Ortiz, Tatiana Nunez-Garcia, Gisela Barrera-Badillo, Lucia Hernandez-Rivas, Irma Lopez-Martinez, Ernesto Ramirez-Gonzalez. |
| EPI_ISL_2157327 | LESP Campeche | Instituto de Diagnostico y Referencia Epidemiologicos (INDRE) | Claudia Wong-Arambula, Abril Rodriguez-Maldonado, Vanessa Rivero-Arredondo, Ariadna Medina-Benitez, Joaquin Quiroz-Mercado, Sergio Rangel-Guerrero, Natividad Cruz-Ortiz, Tatiana Nunez-Garcia, Gisela Barrera-Badillo, Lucia Hernandez-Rivas, Irma Lopez-Martinez, Ernesto Ramirez-Gonzalez. |
| EPI_ISL_2157328 | LESP Baja California | Instituto de Diagnostico y Referencia Epidemiologicos (INDRE) | Claudia Wong-Arambula, Abril Rodriguez-Maldonado, Vanessa Rivero-Arredondo, Ariadna Medina-Benitez, Joaquin Quiroz-Mercado, Sergio Rangel-Guerrero, Natividad Cruz-Ortiz, Tatiana Nunez-Garcia, Gisela Barrera-Badillo, Lucia Hernandez-Rivas, Irma Lopez-Martinez, Ernesto Ramirez-Gonzalez. |
| EPI_ISL_2157329, EPI_ISL_2157330 | LESP Guerrero | Instituto de Diagnostico y Referencia Epidemiologicos (INDRE) | Claudia Wong-Arambula, Abril Rodriguez-Maldonado, Vanessa Rivero-Arredondo, Ariadna Medina-Benitez, Joaquin Quiroz-Mercado, Sergio Rangel-Guerrero, Natividad Cruz-Ortiz, Tatiana Nunez-Garcia, Gisela Barrera-Badillo, Lucia Hernandez-Rivas, Irma Lopez-Martinez, Ernesto Ramirez-Gonzalez. |
| EPI_ISL_2157331 | LESP Oaxaca | Instituto de Diagnostico y Referencia Epidemiologicos (INDRE) | Claudia Wong-Arambula, Abril Rodriguez-Maldonado, Vanessa Rivero-Arredondo, Ariadna Medina-Benitez, Joaquin Quiroz-Mercado, Sergio Rangel-Guerrero, Natividad Cruz-Ortiz, Tatiana Nunez-Garcia, Gisela Barrera-Badillo, Lucia Hernandez-Rivas, Irma Lopez-Martinez, Ernesto Ramirez-Gonzalez. |
| EPI_ISL_2157332 | LESP Puebla | Instituto de Diagnostico y Referencia Epidemiologicos (INDRE) | Claudia Wong-Arambula, Abril Rodriguez-Maldonado, Vanessa Rivero-Arredondo, Ariadna Medina-Benitez, Joaquin Quiroz-Mercado, Sergio Rangel-Guerrero, Natividad Cruz-Ortiz, Tatiana Nunez-Garcia, Gisela Barrera-Badillo, Lucia Hernandez-Rivas, Irma Lopez-Martinez, Ernesto Ramirez-Gonzalez. |
| EPI_ISL_2157333, EPI_ISL_2157334 | LESP Ciudad de Mexico | Instituto de Diagnostico y Referencia Epidemiologicos (INDRE) | Claudia Wong-Arambula, Abril Rodriguez-Maldonado, Vanessa Rivero-Arredondo, Ariadna Medina-Benitez, Joaquin Quiroz-Mercado, Sergio Rangel-Guerrero, Natividad Cruz-Ortiz, Tatiana Nunez-Garcia, Gisela Barrera-Badillo, Lucia Hernandez-Rivas, Irma Lopez-Martinez, Ernesto Ramirez-Gonzalez. |
| EPI_ISL_2157335, EPI_ISL_2157336, EPI_ISL_2157337, EPI_ISL_2157338, EPI_ISL_2157339, EPI_ISL_2157340, EPI_ISL_2157341, EPI_ISL_2157342 | Arkansas Children's Hospital | Center for Global Health, University of New Mexico Health Sciences Center | Daryl Domman, Kurt Schwalm, Valerie Morley, Catherine Kirkpatrick, Bobby Boyanton, Joshua L. Kennedy, Darrell Dinwiddie |
| EPI_ISL_2157600, EPI_ISL_2157601, EPI_ISL_2157602, EPI_ISL_2157603, EPI_ISL_2157604, EPI_ISL_2157605, EPI_ISL_2157606, EPI_ISL_2157607, EPI_ISL_2157609, EPI_ISL_2157610, EPI_ISL_2157611, EPI_ISL_2157612, EPI_ISL_2157613, EPI_ISL_2157614, EPI_ISL_2157615, EPI_ISL_2157616, EPI_ISL_2157617, EPI_ISL_2157618, EPI_ISL_2157619, EPI_ISL_2157620, EPI_ISL_2157621, EPI_ISL_2157622, EPI_ISL_2157623, EPI_ISL_2157624, EPI_ISL_2157625, EPI_ISL_2157626, EPI_ISL_2157627, EPI_ISL_2157628, EPI_ISL_2157629, EPI_ISL_2157630, EPI_ISL_2157631, EPI_ISL_2157632, EPI_ISL_2157634, EPI_ISL_2157635, EPI_ISL_2157637, EPI_ISL_2157638, EPI_ISL_2157639, EPI_ISL_2157640, EPI_ISL_2157641, EPI_ISL_2157642, EPI_ISL_2157643, EPI_ISL_2157644, EPI_ISL_2157645, EPI_ISL_2157646, EPI_ISL_2157647, EPI_ISL_2157648, EPI_ISL_2157649, EPI_ISL_2157650, EPI_ISL_2157651, EPI_ISL_2157652, EPI_ISL_2157653, EPI_ISL_2157654, EPI_ISL_2157655, EPI_ISL_2157656, EPI_ISL_2157658, EPI_ISL_2157659, EPI_ISL_2157660, EPI_ISL_2157661, EPI_ISL_2157662, EPI_ISL_2157663, EPI_ISL_2157664, EPI_ISL_2157665, EPI_ISL_2157666, EPI_ISL_2157667, EPI_ISL_2157668, EPI_ISL_2157669, EPI_ISL_2157670, EPI_ISL_2157671, EPI_ISL_2157672, EPI_ISL_2157673, EPI_ISL_2157674, EPI_ISL_2157675, EPI_ISL_2157676, EPI_ISL_2157677, EPI_ISL_2157678, EPI_ISL_2157679, EPI_ISL_2157680, EPI_ISL_2157682, EPI_ISL_2157683, EPI_ISL_2157684, EPI_ISL_2157685, EPI_ISL_2157686, EPI_ISL_2157687, EPI_ISL_2157688, EPI_ISL_2157689, EPI_ISL_2157690, EPI_ISL_2157691, EPI_ISL_2157692, EPI_ISL_2157693, EPI_ISL_2157694, EPI_ISL_2157695, EPI_ISL_2157696, EPI_ISL_2157697, EPI_ISL_2157698, EPI_ISL_2157699, EPI_ISL_2157700, EPI_ISL_2157701, EPI_ISL_2157703, EPI_ISL_2157704, EPI_ISL_2157705, EPI_ISL_2157706, EPI_ISL_2157707, EPI_ISL_2157708, EPI_ISL_2157709, EPI_ISL_2157710, EPI_ISL_2157711, EPI_ISL_2157712, EPI_ISL_2157713, EPI_ISL_2157714, EPI_ISL_2157715, EPI_ISL_2157716, EPI_ISL_2157719, EPI_ISL_2157720, EPI_ISL_2157722, EPI_ISL_2157723, EPI_ISL_2157724, EPI_ISL_2157725, EPI_ISL_2157726, EPI_ISL_2157727, EPI_ISL_2157728, EPI_ISL_2157729, EPI_ISL_2157730, EPI_ISL_2157731, EPI_ISL_2157732, EPI_ISL_2157733, EPI_ISL_2157734, EPI_ISL_2157735, EPI_ISL_2157736, EPI_ISL_2157737, EPI_ISL_2157738, EPI_ISL_2157739, EPI_ISL_2157740, EPI_ISL_2157741, EPI_ISL_2157742, EPI_ISL_2157743, EPI_ISL_2157744, EPI_ISL_2157745, EPI_ISL_2157746, EPI_ISL_2157747, EPI_ISL_2157748, EPI_ISL_2157749, EPI_ISL_2157750, EPI_ISL_2157751, EPI_ISL_2157752, EPI_ISL_2157753, EPI_ISL_2157754, EPI_ISL_2157755, EPI_ISL_2157756, EPI_ISL_2157757, EPI_ISL_2157758, EPI_ISL_2157759, EPI_ISL_2157760, EPI_ISL_2157761, EPI_ISL_2157762, EPI_ISL_2157763, EPI_ISL_2157764, EPI_ISL_2157765, EPI_ISL_2157766, EPI_ISL_2157767, EPI_ISL_2157768, EPI_ISL_2157769, EPI_ISL_2157770, EPI_ISL_2157771, EPI_ISL_2157772, EPI_ISL_2157773, EPI_ISL_2157774, EPI_ISL_2157775, EPI_ISL_2157776, EPI_ISL_2157777, EPI_ISL_2157778, EPI_ISL_2157779, EPI_ISL_2157780, EPI_ISL_2157781, EPI_ISL_2157782, EPI_ISL_2157783, EPI_ISL_2157784, EPI_ISL_2157785, EPI_ISL_2157787, EPI_ISL_2157789, EPI_ISL_2157790, EPI_ISL_2157791, EPI_ISL_2157792, EPI_ISL_2157793, EPI_ISL_2157794, EPI_ISL_2157795, EPI_ISL_2157796, EPI_ISL_2157797, EPI_ISL_2157798, EPI_ISL_2157799, EPI_ISL_2157800, EPI_ISL_2157801, EPI_ISL_2157802, EPI_ISL_2157803, EPI_ISL_2157805, EPI_ISL_2157806, EPI_ISL_2157807, EPI_ISL_2157808, EPI_ISL_2157809, EPI_ISL_2157810, EPI_ISL_2157811, EPI_ISL_2157812, EPI_ISL_2157813, EPI_ISL_2157814, EPI_ISL_2157815, EPI_ISL_2157816, EPI_ISL_2157817, EPI_ISL_2157818, EPI_ISL_2157819, EPI_ISL_2157820, EPI_ISL_2157821, EPI_ISL_2157822, EPI_ISL_2157823, EPI_ISL_2157824, EPI_ISL_2157825, EPI_ISL_2157826, EPI_ISL_2157827, EPI_ISL_2157828, EPI_ISL_2157829, EPI_ISL_2157830, EPI_ISL_2157831, EPI_ISL_2157832, EPI_ISL_2157833, EPI_ISL_2157834, EPI_ISL_2157835, EPI_ISL_2157836, EPI_ISL_2157837, EPI_ISL_2157838, EPI_ISL_2157839, EPI_ISL_2157840, EPI_ISL_2157841, EPI_ISL_2157842, EPI_ISL_2157843, EPI_ISL_2157844, EPI_ISL_2157845, EPI_ISL_2157846, EPI_ISL_2157847, EPI_ISL_2157848, EPI_ISL_2157849, EPI_ISL_2157850, EPI_ISL_2157851, EPI_ISL_2157852, EPI_ISL_2157853, EPI_ISL_2157854, EPI_ISL_2157855, EPI_ISL_2157856, EPI_ISL_2157857, EPI_ISL_2157858, EPI_ISL_2157859, EPI_ISL_2157860, EPI_ISL_2157861, EPI_ISL_2157862, EPI_ISL_2157863, EPI_ISL_2157864, EPI_ISL_2157865, EPI_ISL_2157868, EPI_ISL_2157869, EPI_ISL_2157870, EPI_ISL_2157871 | | | |
| see above | Reditus Laboratories | Reditus Laboratories | Joshua J. Geltz, Ph.D., Robert M. Sgambelluri, Ph.D., Cassy Philips, M.S., Alexa Eichelberger, M.S. |
| EPI_ISL_2158079, EPI_ISL_2158080, EPI_ISL_2158081, EPI_ISL_2158082, EPI_ISL_2158083, EPI_ISL_2158084, EPI_ISL_2158085, EPI_ISL_2158086, EPI_ISL_2158087, EPI_ISL_2158088, EPI_ISL_2158089, EPI_ISL_2158090, EPI_ISL_2158091, EPI_ISL_2158092, EPI_ISL_2158093, EPI_ISL_2158094, EPI_ISL_2158095, EPI_ISL_2158096, EPI_ISL_2158097, EPI_ISL_2158098, EPI_ISL_2158099, EPI_ISL_2158100, EPI_ISL_2158101, EPI_ISL_2158102, EPI_ISL_2158103 | | | |
| see above | DeRisi Lab, University of California, San Francisco | DeRisi Lab, University of California, San Francisco | Sabrina Mann, Anthea Mitchell, Jamin Liu, Sara Sunshine, Matthew Laurie, Genay Pilarowski, Amy Kistler, Manu Vanaerschot, Patrick Ayscue, Lucy Li, Eric Chow, IDseq Team, James Peng, Carina Marquez, Luis Rubio, Gabriel Chamie, Diane Jones, Jon Jacobo, Susana Rojas, Susy Rojas, Valerie Tulier-Laiwa, Douglas Black, Jackie Martinez, Maya Petersen, Diane Havlir, and Joseph DeRisi |
| EPI_ISL_2158108 | LESP Baja California | Instituto de Diagnostico y Referencia Epidemiologicos (INDRE) | Claudia Wong-Arambula, Abril Rodriguez-Maldonado, Vanessa Rivero-Arredondo, Ariadna Medina-Benitez, Joaquin Quiroz-Mercado, Sergio Rangel-Guerrero, Natividad Cruz-Ortiz, Tatiana Nunez-Garcia, Gisela Barrera-Badillo, Lucia Hernandez-Rivas, Irma Lopez-Martinez, Ernesto Ramirez-Gonzalez. |
| EPI_ISL_2158109, EPI_ISL_2158110, EPI_ISL_2158111, EPI_ISL_2158112, EPI_ISL_2158113 | LESP Nuevo Leon | Instituto de Diagnostico y Referencia Epidemiologicos (INDRE) | Claudia Wong-Arambula, Abril Rodriguez-Maldonado, Vanessa Rivero-Arredondo, Ariadna Medina-Benitez, Joaquin Quiroz-Mercado, Sergio Rangel-Guerrero, Natividad Cruz-Ortiz, Tatiana Nunez-Garcia, Gisela Barrera-Badillo, Lucia Hernandez-Rivas, Irma Lopez-Martinez, Ernesto Ramirez-Gonzalez. |
| EPI_ISL_2158114 | LESP Tamaulipas | Instituto de Diagnostico y Referencia Epidemiologicos (INDRE) | Claudia Wong-Arambula, Abril Rodriguez-Maldonado, Vanessa Rivero-Arredondo, Ariadna Medina-Benitez, Joaquin Quiroz-Mercado, Sergio Rangel-Guerrero, Natividad Cruz-Ortiz, Tatiana Nunez-Garcia, Gisela Barrera-Badillo, Lucia Hernandez-Rivas, Irma Lopez-Martinez, Ernesto Ramirez-Gonzalez. |
| EPI_ISL_2158115, EPI_ISL_2158116, EPI_ISL_2158117, EPI_ISL_2158118, EPI_ISL_2158119, EPI_ISL_2158120 | LESP Nuevo Leon | Instituto de Diagnostico y Referencia Epidemiologicos (INDRE) | Claudia Wong-Arambula, Abril Rodriguez-Maldonado, Vanessa Rivero-Arredondo, Ariadna Medina-Benitez, Joaquin Quiroz-Mercado, Sergio Rangel-Guerrero, Natividad Cruz-Ortiz, Tatiana Nunez-Garcia, Gisela Barrera-Badillo, Lucia Hernandez-Rivas, Irma Lopez-Martinez, Ernesto Ramirez-Gonzalez. |
| EPI_ISL_2158121, EPI_ISL_2158122, EPI_ISL_2158123, EPI_ISL_2158124, EPI_ISL_2158125, EPI_ISL_2158126, EPI_ISL_2158127 | LESP Hidalgo | Instituto de Diagnostico y Referencia Epidemiologicos (INDRE) | Claudia Wong-Arambula, Abril Rodriguez-Maldonado, Vanessa Rivero-Arredondo, Ariadna Medina-Benitez, Joaquin Quiroz-Mercado, Sergio Rangel-Guerrero, Natividad Cruz-Ortiz, Tatiana Nunez-Garcia, Gisela Barrera-Badillo, Lucia Hernandez-Rivas, Irma Lopez-Martinez, Ernesto Ramirez-Gonzalez. |
| EPI_ISL_2158128, EPI_ISL_2158129, EPI_ISL_2158130 | LESP Puebla | Instituto de Diagnostico y Referencia Epidemiologicos (INDRE) | Claudia Wong-Arambula, Abril Rodriguez-Maldonado, Vanessa Rivero-Arredondo, Ariadna Medina-Benitez, Joaquin Quiroz-Mercado, Sergio Rangel-Guerrero, Natividad Cruz-Ortiz, Tatiana Nunez-Garcia, Gisela Barrera-Badillo, Lucia Hernandez-Rivas, Irma Lopez-Martinez, Ernesto Ramirez-Gonzalez. |
| EPI_ISL_2158131 | LESP Tlaxcala | Instituto de Diagnostico y Referencia Epidemiologicos (INDRE) | Claudia Wong-Arambula, Abril Rodriguez-Maldonado, Vanessa Rivero-Arredondo, Ariadna Medina-Benitez, Joaquin Quiroz-Mercado, Sergio Rangel-Guerrero, Natividad Cruz-Ortiz, Tatiana Nunez-Garcia, Gisela Barrera-Badillo, Lucia Hernandez-Rivas, Irma Lopez-Martinez, Ernesto Ramirez-Gonzalez. |
| EPI_ISL_2158132, EPI_ISL_2158133, EPI_ISL_2158134 | LESP Guanajuato | Instituto de Diagnostico y Referencia Epidemiologicos (INDRE) | Claudia Wong-Arambula, Abril Rodriguez-Maldonado, Vanessa Rivero-Arredondo, Ariadna Medina-Benitez, Joaquin Quiroz-Mercado, Sergio Rangel-Guerrero, Natividad Cruz-Ortiz, Tatiana Nunez-Garcia, Gisela Barrera-Badillo, Lucia Hernandez-Rivas, Irma Lopez-Martinez, Ernesto Ramirez-Gonzalez. |
| EPI_ISL_2158135 | LESP Jalisco | Instituto de Diagnostico y Referencia Epidemiologicos (INDRE) | Claudia Wong-Arambula, Abril Rodriguez-Maldonado, Vanessa Rivero-Arredondo, Ariadna Medina-Benitez, Joaquin Quiroz-Mercado, Sergio Rangel-Guerrero, Natividad Cruz-Ortiz, Tatiana Nunez-Garcia, Gisela Barrera-Badillo, Lucia Hernandez-Rivas, Irma Lopez-Martinez, Ernesto Ramirez-Gonzalez. |
| EPI_ISL_2158136 | LESP Chihuahua | Instituto de Diagnostico y Referencia Epidemiologicos (INDRE) | Claudia Wong-Arambula, Abril Rodriguez-Maldonado, Vanessa Rivero-Arredondo, Ariadna Medina-Benitez, Joaquin Quiroz-Mercado, Sergio Rangel-Guerrero, Natividad Cruz-Ortiz, Tatiana Nunez-Garcia, Gisela Barrera-Badillo, Lucia Hernandez-Rivas, Irma Lopez-Martinez, Ernesto Ramirez-Gonzalez. |
| EPI_ISL_2158137 | LESP Nuevo Leon | Instituto de Diagnostico y Referencia Epidemiologicos (INDRE) | Claudia Wong-Arambula, Abril Rodriguez-Maldonado, Vanessa Rivero-Arredondo, Ariadna Medina-Benitez, Joaquin Quiroz-Mercado, Sergio Rangel-Guerrero, Natividad Cruz-Ortiz, Tatiana Nunez-Garcia, Gisela Barrera-Badillo, Lucia Hernandez-Rivas, Irma Lopez-Martinez, Ernesto Ramirez-Gonzalez. |
| EPI_ISL_2158138, EPI_ISL_2158139, EPI_ISL_2158140 | LESP Colima | Instituto de Diagnostico y Referencia Epidemiologicos (INDRE) | Claudia Wong-Arambula, Abril Rodriguez-Maldonado, Vanessa Rivero-Arredondo, Ariadna Medina-Benitez, Joaquin Quiroz-Mercado, Sergio Rangel-Guerrero, Natividad Cruz-Ortiz, Tatiana Nunez-Garcia, Gisela Barrera-Badillo, Lucia Hernandez-Rivas, Irma Lopez-Martinez, Ernesto Ramirez-Gonzalez. |
| EPI_ISL_2158141 | LESP Queretaro | Instituto de Diagnostico y Referencia Epidemiologicos (INDRE) | Claudia Wong-Arambula, Abril Rodriguez-Maldonado, Vanessa Rivero-Arredondo, Ariadna Medina-Benitez, Joaquin Quiroz-Mercado, Sergio Rangel-Guerrero, Natividad Cruz-Ortiz, Tatiana Nunez-Garcia, Gisela Barrera-Badillo, Lucia Hernandez-Rivas, Irma Lopez-Martinez, Ernesto Ramirez-Gonzalez. |
| EPI_ISL_2158142 | LESP Veracruz | Instituto de Diagnostico y Referencia Epidemiologicos (INDRE) | Claudia Wong-Arambula, Abril Rodriguez-Maldonado, Vanessa Rivero-Arredondo, Ariadna Medina-Benitez, Joaquin Quiroz-Mercado, Sergio Rangel-Guerrero, Natividad Cruz-Ortiz, Tatiana Nunez-Garcia, Gisela Barrera-Badillo, Lucia Hernandez-Rivas, Irma Lopez-Martinez, Ernesto Ramirez-Gonzalez. |
| EPI_ISL_2158143, EPI_ISL_2158144, EPI_ISL_2158145, EPI_ISL_2158146, EPI_ISL_2158147, EPI_ISL_2158148, EPI_ISL_2158149, EPI_ISL_2158150 | LESP Queretaro | Instituto de Diagnostico y Referencia Epidemiologicos (INDRE) | Claudia Wong-Arambula, Abril Rodriguez-Maldonado, Vanessa Rivero-Arredondo, Ariadna Medina-Benitez, Joaquin Quiroz-Mercado, Sergio Rangel-Guerrero, Natividad Cruz-Ortiz, Tatiana Nunez-Garcia, Gisela Barrera-Badillo, Lucia Hernandez-Rivas, Irma Lopez-Martinez, Ernesto Ramirez-Gonzalez. |
| EPI_ISL_2158151 | LESP Puebla | Instituto de Diagnostico y Referencia Epidemiologicos (INDRE) | Claudia Wong-Arambula, Abril Rodriguez-Maldonado, Vanessa Rivero-Arredondo, Ariadna Medina-Benitez, Joaquin Quiroz-Mercado, Sergio Rangel-Guerrero, Natividad Cruz-Ortiz, Tatiana Nunez-Garcia, Gisela Barrera-Badillo, Lucia Hernandez-Rivas, Irma Lopez-Martinez, Ernesto Ramirez-Gonzalez. |

| | | | |
|---|---|---|---|
| EPI_ISL_2158152, EPI_ISL_2158153, EPI_ISL_2158154, EPI_ISL_2158155, EPI_ISL_2158156, EPI_ISL_2158157, EPI_ISL_2158158, EPI_ISL_2158159, EPI_ISL_2158160, EPI_ISL_2158161 | LESP Morelos | Instituto de Diagnostico y Referencia Epidemiologicos (INDRE) | Claudia Wong-Arambula, Abril Rodriguez-Maldonado, Vanessa Rivero-Arredondo, Ariadna Medina-Benitez, Joaquin Quiroz-Mercado, Sergio Rangel-Guerrero, Natividad Cruz-Ortiz, Tatiana Nunez-Garcia, Gisela Barrera-Badillo, Lucia Hernandez-Rivas, Irma Lopez-Martinez, Ernesto Ramirez-Gonzalez. |
| EPI_ISL_2158162, EPI_ISL_2158163, EPI_ISL_2158164, EPI_ISL_2158165 | LESP Chihuahua | Instituto de Diagnostico y Referencia Epidemiologicos (INDRE) | Claudia Wong-Arambula, Abril Rodriguez-Maldonado, Vanessa Rivero-Arredondo, Ariadna Medina-Benitez, Joaquin Quiroz-Mercado, Sergio Rangel-Guerrero, Natividad Cruz-Ortiz, Tatiana Nunez-Garcia, Gisela Barrera-Badillo, Lucia Hernandez-Rivas, Irma Lopez-Martinez, Ernesto Ramirez-Gonzalez. |
| EPI_ISL_2158166 | LESP Ciudad de Mexico | Instituto de Diagnostico y Referencia Epidemiologicos (INDRE) | Claudia Wong-Arambula, Abril Rodriguez-Maldonado, Vanessa Rivero-Arredondo, Ariadna Medina-Benitez, Joaquin Quiroz-Mercado, Sergio Rangel-Guerrero, Natividad Cruz-Ortiz, Tatiana Nunez-Garcia, Gisela Barrera-Badillo, Lucia Hernandez-Rivas, Irma Lopez-Martinez, Ernesto Ramirez-Gonzalez. |
| EPI_ISL_2158167, EPI_ISL_2158168 | LESP Chihuahua | Instituto de Diagnostico y Referencia Epidemiologicos (INDRE) | Claudia Wong-Arambula, Abril Rodriguez-Maldonado, Vanessa Rivero-Arredondo, Ariadna Medina-Benitez, Joaquin Quiroz-Mercado, Sergio Rangel-Guerrero, Natividad Cruz-Ortiz, Tatiana Nunez-Garcia, Gisela Barrera-Badillo, Lucia Hernandez-Rivas, Irma Lopez-Martinez, Ernesto Ramirez-Gonzalez. |
| EPI_ISL_2158169 | LESP Coahuila | Instituto de Diagnostico y Referencia Epidemiologicos (INDRE) | Claudia Wong-Arambula, Abril Rodriguez-Maldonado, Vanessa Rivero-Arredondo, Ariadna Medina-Benitez, Joaquin Quiroz-Mercado, Sergio Rangel-Guerrero, Natividad Cruz-Ortiz, Tatiana Nunez-Garcia, Gisela Barrera-Badillo, Lucia Hernandez-Rivas, Irma Lopez-Martinez, Ernesto Ramirez-Gonzalez. |
| EPI_ISL_2158170 | LESP Guerrero | Instituto de Diagnostico y Referencia Epidemiologicos (INDRE) | Claudia Wong-Arambula, Abril Rodriguez-Maldonado, Vanessa Rivero-Arredondo, Ariadna Medina-Benitez, Joaquin Quiroz-Mercado, Sergio Rangel-Guerrero, Natividad Cruz-Ortiz, Tatiana Nunez-Garcia, Gisela Barrera-Badillo, Lucia Hernandez-Rivas, Irma Lopez-Martinez, Ernesto Ramirez-Gonzalez. |
| EPI_ISL_2158171, EPI_ISL_2158172 | LESP Jalisco | Instituto de Diagnostico y Referencia Epidemiologicos (INDRE) | Claudia Wong-Arambula, Abril Rodriguez-Maldonado, Vanessa Rivero-Arredondo, Ariadna Medina-Benitez, Joaquin Quiroz-Mercado, Sergio Rangel-Guerrero, Natividad Cruz-Ortiz, Tatiana Nunez-Garcia, Gisela Barrera-Badillo, Lucia Hernandez-Rivas, Irma Lopez-Martinez, Ernesto Ramirez-Gonzalez. |
| EPI_ISL_2158173 | LESP Sinaloa | Instituto de Diagnostico y Referencia Epidemiologicos (INDRE) | Claudia Wong-Arambula, Abril Rodriguez-Maldonado, Vanessa Rivero-Arredondo, Ariadna Medina-Benitez, Joaquin Quiroz-Mercado, Sergio Rangel-Guerrero, Natividad Cruz-Ortiz, Tatiana Nunez-Garcia, Gisela Barrera-Badillo, Lucia Hernandez-Rivas, Irma Lopez-Martinez, Ernesto Ramirez-Gonzalez. |
| EPI_ISL_2158174, EPI_ISL_2158175, EPI_ISL_2158176, EPI_ISL_2158177, EPI_ISL_2158178, EPI_ISL_2158179, EPI_ISL_2158180, EPI_ISL_2158181, EPI_ISL_2158182, EPI_ISL_2158183, EPI_ISL_2158184, EPI_ISL_2158185 | | | |
| see above | LESP Morelos | Instituto de Diagnostico y Referencia Epidemiologicos (INDRE) | Claudia Wong-Arambula, Abril Rodriguez-Maldonado, Vanessa Rivero-Arredondo, Ariadna Medina-Benitez, Joaquin Quiroz-Mercado, Sergio Rangel-Guerrero, Natividad Cruz-Ortiz, Tatiana Nunez-Garcia, Gisela Barrera-Badillo, Lucia Hernandez-Rivas, Irma Lopez-Martinez, Ernesto Ramirez-Gonzalez. |
| EPI_ISL_2158186, EPI_ISL_2158187, EPI_ISL_2158188, EPI_ISL_2158189, EPI_ISL_2158190, EPI_ISL_2158191, EPI_ISL_2158192, EPI_ISL_2158193, EPI_ISL_2158194, EPI_ISL_2158195, EPI_ISL_2158196 | | | |
| see above | LESP Chihuahua | Instituto de Diagnostico y Referencia Epidemiologicos (INDRE) | Claudia Wong-Arambula, Abril Rodriguez-Maldonado, Vanessa Rivero-Arredondo, Ariadna Medina-Benitez, Joaquin Quiroz-Mercado, Sergio Rangel-Guerrero, Natividad Cruz-Ortiz, Tatiana Nunez-Garcia, Gisela Barrera-Badillo, Lucia Hernandez-Rivas, Irma Lopez-Martinez, Ernesto Ramirez-Gonzalez. |
| EPI_ISL_2158197 | LESP Nuevo Leon | Instituto de Diagnostico y Referencia Epidemiologicos (INDRE) | Claudia Wong-Arambula, Abril Rodriguez-Maldonado, Vanessa Rivero-Arredondo, Ariadna Medina-Benitez, Joaquin Quiroz-Mercado, Sergio Rangel-Guerrero, Natividad Cruz-Ortiz, Tatiana Nunez-Garcia, Gisela Barrera-Badillo, Lucia Hernandez-Rivas, Irma Lopez-Martinez, Ernesto Ramirez-Gonzalez. |
| EPI_ISL_2158198 | LESP Chihuahua | Instituto de Diagnostico y Referencia Epidemiologicos (INDRE) | Claudia Wong-Arambula, Abril Rodriguez-Maldonado, Vanessa Rivero-Arredondo, Ariadna Medina-Benitez, Joaquin Quiroz-Mercado, Sergio Rangel-Guerrero, Natividad Cruz-Ortiz, Tatiana Nunez-Garcia, Gisela Barrera-Badillo, Lucia Hernandez-Rivas, Irma Lopez-Martinez, Ernesto Ramirez-Gonzalez. |
| EPI_ISL_2158199, EPI_ISL_2158200 | LESP Puebla | Instituto de Diagnostico y Referencia Epidemiologicos (INDRE) | Claudia Wong-Arambula, Abril Rodriguez-Maldonado, Vanessa Rivero-Arredondo, Ariadna Medina-Benitez, Joaquin Quiroz-Mercado, Sergio Rangel-Guerrero, Natividad Cruz-Ortiz, Tatiana Nunez-Garcia, Gisela Barrera-Badillo, Lucia Hernandez-Rivas, Irma Lopez-Martinez, Ernesto Ramirez-Gonzalez. |
| EPI_ISL_2158201, EPI_ISL_2158202 | LESP Jalisco | Instituto de Diagnostico y Referencia Epidemiologicos (INDRE) | Claudia Wong-Arambula, Abril Rodriguez-Maldonado, Vanessa Rivero-Arredondo, Ariadna Medina-Benitez, Joaquin Quiroz-Mercado, Sergio Rangel-Guerrero, Natividad Cruz-Ortiz, Tatiana Nunez-Garcia, Gisela Barrera-Badillo, Lucia Hernandez-Rivas, Irma Lopez-Martinez, Ernesto Ramirez-Gonzalez. |
| EPI_ISL_2158203 | LESP Hidalgo | Instituto de Diagnostico y Referencia Epidemiologicos (INDRE) | Claudia Wong-Arambula, Abril Rodriguez-Maldonado, Vanessa Rivero-Arredondo, Ariadna Medina-Benitez, Joaquin Quiroz-Mercado, Sergio Rangel-Guerrero, Natividad Cruz-Ortiz, Tatiana Nunez-Garcia, Gisela Barrera-Badillo, Lucia Hernandez-Rivas, Irma Lopez-Martinez, Ernesto Ramirez-Gonzalez. |
| EPI_ISL_2158204 | LESP Nayarit | Instituto de Diagnostico y Referencia Epidemiologicos (INDRE) | Claudia Wong-Arambula, Abril Rodriguez-Maldonado, Vanessa Rivero-Arredondo, Ariadna Medina-Benitez, Joaquin Quiroz-Mercado, Sergio Rangel-Guerrero, Natividad Cruz-Ortiz, Tatiana Nunez-Garcia, Gisela Barrera-Badillo, Lucia Hernandez-Rivas, Irma Lopez-Martinez, Ernesto Ramirez-Gonzalez. |
| EPI_ISL_2158205 | LESP Veracruz | Instituto de Diagnostico y Referencia Epidemiologicos (INDRE) | Claudia Wong-Arambula, Abril Rodriguez-Maldonado, Vanessa Rivero-Arredondo, Ariadna Medina-Benitez, Joaquin Quiroz-Mercado, Sergio Rangel-Guerrero, Natividad Cruz-Ortiz, Tatiana Nunez-Garcia, Gisela Barrera-Badillo, Lucia Hernandez-Rivas, Irma Lopez-Martinez, Ernesto Ramirez-Gonzalez. |
| EPI_ISL_2158206 | LESP Tamaulipas | Instituto de Diagnostico y Referencia Epidemiologicos (INDRE) | Claudia Wong-Arambula, Abril Rodriguez-Maldonado, Vanessa Rivero-Arredondo, Ariadna Medina-Benitez, Joaquin Quiroz-Mercado, Sergio Rangel-Guerrero, Natividad Cruz-Ortiz, Tatiana Nunez-Garcia, Gisela Barrera-Badillo, Lucia Hernandez-Rivas, Irma Lopez-Martinez, Ernesto Ramirez-Gonzalez. |
| EPI_ISL_2158207, EPI_ISL_2158208, EPI_ISL_2158209, EPI_ISL_2158210, EPI_ISL_2158211, EPI_ISL_2158212, EPI_ISL_2158213, EPI_ISL_2158214, EPI_ISL_2158215, EPI_ISL_2158216, EPI_ISL_2158217, EPI_ISL_2158218, EPI_ISL_2158219, EPI_ISL_2158220, EPI_ISL_2158221, EPI_ISL_2158222, EPI_ISL_2158223, EPI_ISL_2158224, EPI_ISL_2158225 | | | |
| see above | LESP Guanajuato | Instituto de Diagnostico y Referencia Epidemiologicos (INDRE) | Claudia Wong-Arambula, Abril Rodriguez-Maldonado, Vanessa Rivero-Arredondo, Ariadna Medina-Benitez, Joaquin Quiroz-Mercado, Sergio Rangel-Guerrero, Natividad Cruz-Ortiz, Tatiana Nunez-Garcia, Gisela Barrera-Badillo, Lucia Hernandez-Rivas, Irma Lopez-Martinez, Ernesto Ramirez-Gonzalez. |
| EPI_ISL_2158226 | LESP Puebla | Instituto de Diagnostico y Referencia Epidemiologicos (INDRE) | Claudia Wong-Arambula, Abril Rodriguez-Maldonado, Vanessa Rivero-Arredondo, Ariadna Medina-Benitez, Joaquin Quiroz-Mercado, Sergio Rangel-Guerrero, Natividad Cruz-Ortiz, Tatiana Nunez-Garcia, Gisela Barrera-Badillo, Lucia Hernandez-Rivas, Irma Lopez-Martinez, Ernesto Ramirez-Gonzalez. |
| EPI_ISL_2158227 | LESP Hidalgo | Instituto de Diagnostico y Referencia Epidemiologicos (INDRE) | Claudia Wong-Arambula, Abril Rodriguez-Maldonado, Vanessa Rivero-Arredondo, Ariadna Medina-Benitez, Joaquin Quiroz-Mercado, Sergio Rangel-Guerrero, Natividad Cruz-Ortiz, Tatiana Nunez-Garcia, Gisela Barrera-Badillo, Lucia Hernandez-Rivas, Irma Lopez-Martinez, Ernesto Ramirez-Gonzalez. |
| EPI_ISL_2158228 | LESP Veracruz | Instituto de Diagnostico y Referencia Epidemiologicos (INDRE) | Claudia Wong-Arambula, Abril Rodriguez-Maldonado, Vanessa Rivero-Arredondo, Ariadna Medina-Benitez, Joaquin Quiroz-Mercado, Sergio Rangel-Guerrero, Natividad Cruz-Ortiz, Tatiana Nunez-Garcia, Gisela Barrera-Badillo, Lucia Hernandez-Rivas, Irma Lopez-Martinez, Ernesto Ramirez-Gonzalez. |
| EPI_ISL_2158229 | LESP Tamaulipas | Instituto de Diagnostico y Referencia Epidemiologicos (INDRE) | Claudia Wong-Arambula, Abril Rodriguez-Maldonado, Vanessa Rivero-Arredondo, Ariadna Medina-Benitez, Joaquin Quiroz-Mercado, Sergio Rangel-Guerrero, Natividad Cruz-Ortiz, Tatiana Nunez-Garcia, Gisela Barrera-Badillo, Lucia Hernandez-Rivas, Irma Lopez-Martinez, Ernesto Ramirez-Gonzalez. |
| EPI_ISL_2158230 | LESP Chiapas | Instituto de Diagnostico y Referencia Epidemiologicos (INDRE) | Claudia Wong-Arambula, Abril Rodriguez-Maldonado, Vanessa Rivero-Arredondo, Ariadna Medina-Benitez, Joaquin Quiroz-Mercado, Sergio Rangel-Guerrero, Natividad Cruz-Ortiz, Tatiana Nunez-Garcia, Gisela Barrera-Badillo, Lucia Hernandez-Rivas, Irma Lopez-Martinez, Ernesto Ramirez-Gonzalez. |
| EPI_ISL_2158231 | LESP Baja California Sur | Instituto de Diagnostico y Referencia Epidemiologicos (INDRE) | Claudia Wong-Arambula, Abril Rodriguez-Maldonado, Vanessa Rivero-Arredondo, Ariadna Medina-Benitez, Joaquin Quiroz-Mercado, Sergio Rangel-Guerrero, Natividad Cruz-Ortiz, Tatiana Nunez-Garcia, Gisela Barrera-Badillo, Lucia Hernandez-Rivas, Irma Lopez-Martinez, Ernesto Ramirez-Gonzalez. |
| EPI_ISL_2158232 | LESP Guerrero | Instituto de Diagnostico y Referencia Epidemiologicos (INDRE) | Claudia Wong-Arambula, Abril Rodriguez-Maldonado, Vanessa Rivero-Arredondo, Ariadna Medina-Benitez, Joaquin Quiroz-Mercado, Sergio Rangel-Guerrero, Natividad Cruz-Ortiz, Tatiana Nunez-Garcia, Gisela Barrera-Badillo, Lucia Hernandez-Rivas, Irma Lopez-Martinez, Ernesto Ramirez-Gonzalez. |
| EPI_ISL_2158233 | LESP Jalisco | Instituto de Diagnostico y Referencia Epidemiologicos (INDRE) | Claudia Wong-Arambula, Abril Rodriguez-Maldonado, Vanessa Rivero-Arredondo, Ariadna Medina-Benitez, Joaquin Quiroz-Mercado, Sergio Rangel-Guerrero, Natividad Cruz-Ortiz, Tatiana Nunez-Garcia, Gisela Barrera-Badillo, Lucia Hernandez-Rivas, Irma Lopez-Martinez, Ernesto Ramirez-Gonzalez. |
| EPI_ISL_2158234 | LESP Coahuila | Instituto de Diagnostico y Referencia Epidemiologicos (INDRE) | Claudia Wong-Arambula, Abril Rodriguez-Maldonado, Vanessa Rivero-Arredondo, Ariadna Medina-Benitez, Joaquin Quiroz-Mercado, Sergio Rangel-Guerrero, Natividad Cruz-Ortiz, Tatiana Nunez-Garcia, Gisela Barrera-Badillo, Lucia Hernandez-Rivas, Irma Lopez-Martinez, Ernesto Ramirez-Gonzalez. |
| EPI_ISL_2158235, EPI_ISL_2158236 | LESP Queretaro | Instituto de Diagnostico y Referencia Epidemiologicos (INDRE) | Claudia Wong-Arambula, Abril Rodriguez-Maldonado, Vanessa Rivero-Arredondo, Ariadna Medina-Benitez, Joaquin Quiroz-Mercado, Sergio Rangel-Guerrero, Natividad Cruz-Ortiz, Tatiana Nunez-Garcia, Gisela Barrera-Badillo, Lucia Hernandez-Rivas, Irma Lopez-Martinez, Ernesto Ramirez-Gonzalez. |
| EPI_ISL_2158237 | LESP Quintana Roo | Instituto de Diagnostico y Referencia Epidemiologicos (INDRE) | Claudia Wong-Arambula, Abril Rodriguez-Maldonado, Vanessa Rivero-Arredondo, Ariadna Medina-Benitez, Joaquin Quiroz-Mercado, Sergio Rangel-Guerrero, Natividad Cruz-Ortiz, Tatiana Nunez-Garcia, Gisela Barrera-Badillo, Lucia Hernandez-Rivas, Irma Lopez-Martinez, Ernesto Ramirez-Gonzalez. |
| EPI_ISL_2158238, EPI_ISL_2158239, EPI_ISL_2158240, EPI_ISL_2158241 | LESP Ciudad de Mexico | Instituto de Diagnostico y Referencia Epidemiologicos (INDRE) | Claudia Wong-Arambula, Abril Rodriguez-Maldonado, Vanessa Rivero-Arredondo, Ariadna Medina-Benitez, Joaquin Quiroz-Mercado, Sergio Rangel-Guerrero, Natividad Cruz-Ortiz, Tatiana Nunez-Garcia, Gisela Barrera-Badillo, Lucia Hernandez-Rivas, Irma Lopez-Martinez, Ernesto Ramirez-Gonzalez. |
| EPI_ISL_2158242 | LESP Queretaro /BIMODI | Instituto de Diagnostico y Referencia Epidemiologicos (INDRE) | Claudia Wong-Arambula, Abril Rodriguez-Maldonado, Vanessa Rivero-Arredondo, Ariadna Medina-Benitez, Joaquin Quiroz-Mercado, Sergio Rangel-Guerrero, Natividad Cruz-Ortiz, Tatiana Nunez-Garcia, Gisela Barrera-Badillo, Lucia Hernandez-Rivas, Irma Lopez-Martinez, Ernesto Ramirez-Gonzalez. |
| EPI_ISL_2158243, EPI_ISL_2158244, EPI_ISL_2158245, EPI_ISL_2158246, EPI_ISL_2158247, EPI_ISL_2158248 | LESP Puebla | Instituto de Diagnostico y Referencia Epidemiologicos (INDRE) | Claudia Wong-Arambula, Abril Rodriguez-Maldonado, Vanessa Rivero-Arredondo, Ariadna Medina-Benitez, Joaquin Quiroz-Mercado, Sergio Rangel-Guerrero, Natividad Cruz-Ortiz, Tatiana Nunez-Garcia, Gisela Barrera-Badillo, Lucia Hernandez-Rivas, Irma Lopez-Martinez, Ernesto Ramirez-Gonzalez. |
| EPI_ISL_2158249 | LESP Guanajuato | Instituto de Diagnostico y Referencia Epidemiologicos (INDRE) | Claudia Wong-Arambula, Abril Rodriguez-Maldonado, Vanessa Rivero-Arredondo, Ariadna Medina-Benitez, Joaquin Quiroz-Mercado, Sergio Rangel-Guerrero, Natividad Cruz-Ortiz, Tatiana Nunez-Garcia, Gisela Barrera-Badillo, Lucia Hernandez-Rivas, Irma Lopez-Martinez, Ernesto Ramirez-Gonzalez. |
| EPI_ISL_2158250, EPI_ISL_2158251 | LESP Ciudad de Mexico | Instituto de Diagnostico y Referencia Epidemiologicos (INDRE) | Claudia Wong-Arambula, Abril Rodriguez-Maldonado, Vanessa Rivero-Arredondo, Ariadna Medina-Benitez, Joaquin Quiroz-Mercado, Sergio Rangel-Guerrero, Natividad Cruz-Ortiz, Tatiana Nunez-Garcia, Gisela Barrera-Badillo, Lucia Hernandez-Rivas, Irma Lopez-Martinez, Ernesto Ramirez-Gonzalez. |
| EPI_ISL_2158252, EPI_ISL_2158253 | LESP Ciudad de Mexico / Hospital Angeles | Instituto de Diagnostico y Referencia Epidemiologicos (INDRE) | Claudia Wong-Arambula, Abril Rodriguez-Maldonado, Vanessa Rivero-Arredondo, Ariadna Medina-Benitez, Joaquin Quiroz-Mercado, Sergio Rangel-Guerrero, Natividad Cruz-Ortiz, Tatiana Nunez-Garcia, Gisela Barrera-Badillo, Lucia Hernandez-Rivas, Irma Lopez-Martinez, Ernesto Ramirez-Gonzalez. |
| EPI_ISL_2158254 | LESP Sonora | Instituto de Diagnostico y Referencia Epidemiologicos (INDRE) | Claudia Wong-Arambula, Abril Rodriguez-Maldonado, Vanessa Rivero-Arredondo, Ariadna Medina-Benitez, Joaquin Quiroz-Mercado, Sergio Rangel-Guerrero, Natividad Cruz-Ortiz, Tatiana Nunez-Garcia, Gisela Barrera-Badillo, Lucia Hernandez-Rivas, Irma Lopez-Martinez, Ernesto Ramirez-Gonzalez. |

| | | | |
|---|---|---|---|
| EPI_ISL_2158255, EPI_ISL_2158256 | LESP Estado de Mexico | Instituto de Diagnostico y Referencia Epidemiologicos (INDRE) | Claudia Wong-Arambula, Abril Rodriguez-Maldonado, Vanessa Rivero-Arredondo, Ariadna Medina-Benitez, Joaquin Quiroz-Mercado, Sergio Rangel-Guerrero, Natividad Cruz-Ortiz, Tatiana Nunez-Garcia, Gisela Barrera-Badillo, Lucia Hernandez-Rivas, Irma Lopez-Martinez, Ernesto Ramirez-Gonzalez. |
| EPI_ISL_2158257, EPI_ISL_2158258, EPI_ISL_2158259, EPI_ISL_2158260, EPI_ISL_2158261, EPI_ISL_2158262, EPI_ISL_2158263 | LESP Ciudad de Mexico | Instituto de Diagnostico y Referencia Epidemiologicos (INDRE) | Claudia Wong-Arambula, Abril Rodriguez-Maldonado, Vanessa Rivero-Arredondo, Ariadna Medina-Benitez, Joaquin Quiroz-Mercado, Sergio Rangel-Guerrero, Natividad Cruz-Ortiz, Tatiana Nunez-Garcia, Gisela Barrera-Badillo, Lucia Hernandez-Rivas, Irma Lopez-Martinez, Ernesto Ramirez-Gonzalez. |
| EPI_ISL_2158264, EPI_ISL_2158265, EPI_ISL_2158266, EPI_ISL_2158267, EPI_ISL_2158268 | Alaska State Virology Laboratory | Alaska State Virology Laboratory | Stephanie DeRonde, Elva House, Jacob Zidek, Lisa Smith, Ph.D., Jack Chen, Ph.D. |
| EPI_ISL_2158308 | LESP Guanajuato | Instituto de Diagnostico y Referencia Epidemiologicos (INDRE) | Claudia Wong-Arambula, Abril Rodriguez-Maldonado, Vanessa Rivero-Arredondo, Ariadna Medina-Benitez, Joaquin Quiroz-Mercado, Sergio Rangel-Guerrero, Natividad Cruz-Ortiz, Tatiana Nunez-Garcia, Gisela Barrera-Badillo, Lucia Hernandez-Rivas, Irma Lopez-Martinez, Ernesto Ramirez-Gonzalez. |
| EPI_ISL_2158309 | LESP Chihuahua | Instituto de Diagnostico y Referencia Epidemiologicos (INDRE) | Claudia Wong-Arambula, Abril Rodriguez-Maldonado, Vanessa Rivero-Arredondo, Ariadna Medina-Benitez, Joaquin Quiroz-Mercado, Sergio Rangel-Guerrero, Natividad Cruz-Ortiz, Tatiana Nunez-Garcia, Gisela Barrera-Badillo, Lucia Hernandez-Rivas, Irma Lopez-Martinez, Ernesto Ramirez-Gonzalez. |
| EPI_ISL_2158310 | LESP Jalisco | Instituto de Diagnostico y Referencia Epidemiologicos (INDRE) | Claudia Wong-Arambula, Abril Rodriguez-Maldonado, Vanessa Rivero-Arredondo, Ariadna Medina-Benitez, Joaquin Quiroz-Mercado, Sergio Rangel-Guerrero, Natividad Cruz-Ortiz, Tatiana Nunez-Garcia, Gisela Barrera-Badillo, Lucia Hernandez-Rivas, Irma Lopez-Martinez, Ernesto Ramirez-Gonzalez. |
| EPI_ISL_2158311 | LESP Chihuahua | Instituto de Diagnostico y Referencia Epidemiologicos (INDRE) | Claudia Wong-Arambula, Abril Rodriguez-Maldonado, Vanessa Rivero-Arredondo, Ariadna Medina-Benitez, Joaquin Quiroz-Mercado, Sergio Rangel-Guerrero, Natividad Cruz-Ortiz, Tatiana Nunez-Garcia, Gisela Barrera-Badillo, Lucia Hernandez-Rivas, Irma Lopez-Martinez, Ernesto Ramirez-Gonzalez. |
| EPI_ISL_2158312, EPI_ISL_2158313 | LESP Nuevo Leon | Instituto de Diagnostico y Referencia Epidemiologicos (INDRE) | Claudia Wong-Arambula, Abril Rodriguez-Maldonado, Vanessa Rivero-Arredondo, Ariadna Medina-Benitez, Joaquin Quiroz-Mercado, Sergio Rangel-Guerrero, Natividad Cruz-Ortiz, Tatiana Nunez-Garcia, Gisela Barrera-Badillo, Lucia Hernandez-Rivas, Irma Lopez-Martinez, Ernesto Ramirez-Gonzalez. |
| EPI_ISL_2158314 | LESP Morelos | Instituto de Diagnostico y Referencia Epidemiologicos (INDRE) | Claudia Wong-Arambula, Abril Rodriguez-Maldonado, Vanessa Rivero-Arredondo, Ariadna Medina-Benitez, Joaquin Quiroz-Mercado, Sergio Rangel-Guerrero, Natividad Cruz-Ortiz, Tatiana Nunez-Garcia, Gisela Barrera-Badillo, Lucia Hernandez-Rivas, Irma Lopez-Martinez, Ernesto Ramirez-Gonzalez. |
| EPI_ISL_2158315, EPI_ISL_2158316, EPI_ISL_2158317, EPI_ISL_2158318 | LESP Ciudad de Mexico | Instituto de Diagnostico y Referencia Epidemiologicos (INDRE) | Claudia Wong-Arambula, Abril Rodriguez-Maldonado, Vanessa Rivero-Arredondo, Ariadna Medina-Benitez, Joaquin Quiroz-Mercado, Sergio Rangel-Guerrero, Natividad Cruz-Ortiz, Tatiana Nunez-Garcia, Gisela Barrera-Badillo, Lucia Hernandez-Rivas, Irma Lopez-Martinez, Ernesto Ramirez-Gonzalez. |
| EPI_ISL_2158319 | LESP Nuevo Leon | Instituto de Diagnostico y Referencia Epidemiologicos (INDRE) | Claudia Wong-Arambula, Abril Rodriguez-Maldonado, Vanessa Rivero-Arredondo, Ariadna Medina-Benitez, Joaquin Quiroz-Mercado, Sergio Rangel-Guerrero, Natividad Cruz-Ortiz, Tatiana Nunez-Garcia, Gisela Barrera-Badillo, Lucia Hernandez-Rivas, Irma Lopez-Martinez, Ernesto Ramirez-Gonzalez. |
| EPI_ISL_2158320 | LESP Chihuahua | Instituto de Diagnostico y Referencia Epidemiologicos (INDRE) | Claudia Wong-Arambula, Abril Rodriguez-Maldonado, Vanessa Rivero-Arredondo, Ariadna Medina-Benitez, Joaquin Quiroz-Mercado, Sergio Rangel-Guerrero, Natividad Cruz-Ortiz, Tatiana Nunez-Garcia, Gisela Barrera-Badillo, Lucia Hernandez-Rivas, Irma Lopez-Martinez, Ernesto Ramirez-Gonzalez. |
| EPI_ISL_2158321 | LESP Queretaro | Instituto de Diagnostico y Referencia Epidemiologicos (INDRE) | Claudia Wong-Arambula, Abril Rodriguez-Maldonado, Vanessa Rivero-Arredondo, Ariadna Medina-Benitez, Joaquin Quiroz-Mercado, Sergio Rangel-Guerrero, Natividad Cruz-Ortiz, Tatiana Nunez-Garcia, Gisela Barrera-Badillo, Lucia Hernandez-Rivas, Irma Lopez-Martinez, Ernesto Ramirez-Gonzalez. |
| EPI_ISL_2158546, EPI_ISL_2158547 | LSUHS Emerging Viral Threat Laboratory | LSUHS Emerging Viral Threat Laboratory | Alexander Mijalis, Jeremy P. Kamil, Jennifer L. Carroll, Gregory L. Ware, Maarten Van Diest, Rona S. Scott, Andrew D. Yurochko, Christopher G. Kevil, John A. Vanchiere |
| EPI_ISL_2158548 | LSUHS Emerging Viral Threat Laboratory | LSUHS Emerging Viral Threat Laboratory | Maarten Van Diest, Rona S. Scott, Gregory L. Ware, Alexander Mijalis, Jeremy P. Kamil, Jennifer L. Carroll, Andrew D. Yurochko, Christopher G. Kevil, John A. Vanchiere |
| EPI_ISL_2158549, EPI_ISL_2158550, EPI_ISL_2158551, EPI_ISL_2158552, EPI_ISL_2158553, EPI_ISL_2158554, EPI_ISL_2158555, EPI_ISL_2158556, EPI_ISL_2158557, EPI_ISL_2158558, EPI_ISL_2158559, EPI_ISL_2158560, EPI_ISL_2158561, EPI_ISL_2158562, EPI_ISL_2158563, EPI_ISL_2158564, EPI_ISL_2158565, EPI_ISL_2158566, EPI_ISL_2158567, EPI_ISL_2158568, EPI_ISL_2158569, EPI_ISL_2158570, EPI_ISL_2158571, EPI_ISL_2158572, EPI_ISL_2158573, EPI_ISL_2158574 | | | |
| see above | Oregon State Public Health Laboratory | Oregon State Public Health Laboratory | Rafia Razzaque, Eugene Yeboah, Vanda Makris, Laura Tsaknaridis, John Fontana and Shane Sevey |
| EPI_ISL_2158618, EPI_ISL_2158619, EPI_ISL_2158620, EPI_ISL_2158621, EPI_ISL_2158622, EPI_ISL_2158623, EPI_ISL_2158624, EPI_ISL_2158625, EPI_ISL_2158626, EPI_ISL_2158627, EPI_ISL_2158628, EPI_ISL_2158629, EPI_ISL_2158630, EPI_ISL_2158633, EPI_ISL_2158634, EPI_ISL_2158635, EPI_ISL_2158636, EPI_ISL_2158637, EPI_ISL_2158638, EPI_ISL_2158639, EPI_ISL_2158640, EPI_ISL_2158641, EPI_ISL_2158642, EPI_ISL_2158643, EPI_ISL_2158644, EPI_ISL_2158645, EPI_ISL_2158646, EPI_ISL_2158647, EPI_ISL_2158649, EPI_ISL_2158650, EPI_ISL_2158651, EPI_ISL_2158652, EPI_ISL_2158654, EPI_ISL_2158655, EPI_ISL_2158656, EPI_ISL_2158657, EPI_ISL_2158658, EPI_ISL_2158659, EPI_ISL_2158660, EPI_ISL_2158661, EPI_ISL_2158662, EPI_ISL_2158663, EPI_ISL_2158665, EPI_ISL_2158666, EPI_ISL_2158667, EPI_ISL_2158668, EPI_ISL_2158669, EPI_ISL_2158670, EPI_ISL_2158671, EPI_ISL_2158672, EPI_ISL_2158673, EPI_ISL_2158674, EPI_ISL_2158675, EPI_ISL_2158676, EPI_ISL_2158677, EPI_ISL_2158679, EPI_ISL_2158680, EPI_ISL_2158681, EPI_ISL_2158682, EPI_ISL_2158683, EPI_ISL_2158684, EPI_ISL_2158685, EPI_ISL_2158686, EPI_ISL_2158687, EPI_ISL_2158688, EPI_ISL_2158690, EPI_ISL_2158691, EPI_ISL_2158692 | | | |
| see above | Missouri State Public Health Laboratory | Missouri State Public Health Laboratory | Matthew Sinn, Joshua Barry, Ashley New |
| EPI_ISL_2158851, EPI_ISL_2158852, EPI_ISL_2158853, EPI_ISL_2158854, EPI_ISL_2158855, EPI_ISL_2158856, EPI_ISL_2158857, EPI_ISL_2158858, EPI_ISL_2158859, EPI_ISL_2158860, EPI_ISL_2158861, EPI_ISL_2158862, EPI_ISL_2158863, EPI_ISL_2158864, EPI_ISL_2158865, EPI_ISL_2158866, EPI_ISL_2158868, EPI_ISL_2158869, EPI_ISL_2158870, EPI_ISL_2158871, EPI_ISL_2158872, EPI_ISL_2158873, EPI_ISL_2158874, EPI_ISL_2158875, EPI_ISL_2158876, EPI_ISL_2158877, EPI_ISL_2158878, EPI_ISL_2158879, EPI_ISL_2158880, EPI_ISL_2158881, EPI_ISL_2158882, EPI_ISL_2158883, EPI_ISL_2158884, EPI_ISL_2158885, EPI_ISL_2158886, EPI_ISL_2158887, EPI_ISL_2158888, EPI_ISL_2158889, EPI_ISL_2158890, EPI_ISL_2158891, EPI_ISL_2158892, EPI_ISL_2158893, EPI_ISL_2158894, EPI_ISL_2158895, EPI_ISL_2158896, EPI_ISL_2158897, EPI_ISL_2158898, EPI_ISL_2158899, EPI_ISL_2158900, EPI_ISL_2158901, EPI_ISL_2158902, EPI_ISL_2158903, EPI_ISL_2158904, EPI_ISL_2158905, EPI_ISL_2158906, EPI_ISL_2158907, EPI_ISL_2158908, EPI_ISL_2158909, EPI_ISL_2158910, EPI_ISL_2158911, EPI_ISL_2158912, EPI_ISL_2158913, EPI_ISL_2158914, EPI_ISL_2158915, EPI_ISL_2158916, EPI_ISL_2158917, EPI_ISL_2158918, EPI_ISL_2158919, EPI_ISL_2158920, EPI_ISL_2158921, EPI_ISL_2158922, EPI_ISL_2158923, EPI_ISL_2158924, EPI_ISL_2158925, EPI_ISL_2158926, EPI_ISL_2158927, EPI_ISL_2158928, EPI_ISL_2158929, EPI_ISL_2158930, EPI_ISL_2158931, EPI_ISL_2158932, EPI_ISL_2158933, EPI_ISL_2158934, EPI_ISL_2158935, EPI_ISL_2158936, EPI_ISL_2158937, EPI_ISL_2158938, EPI_ISL_2158939, EPI_ISL_2158940, EPI_ISL_2158941, EPI_ISL_2158942, EPI_ISL_2158943, EPI_ISL_2158944, EPI_ISL_2158945, EPI_ISL_2158946, EPI_ISL_2158947, EPI_ISL_2158948, EPI_ISL_2158949, EPI_ISL_2158950, EPI_ISL_2158951, EPI_ISL_2158952, EPI_ISL_2158953, EPI_ISL_2158954, EPI_ISL_2158955, EPI_ISL_2158956, EPI_ISL_2158957, EPI_ISL_2158958, EPI_ISL_2158959, EPI_ISL_2158960, EPI_ISL_2158961, EPI_ISL_2158962, EPI_ISL_2158963, EPI_ISL_2158964, EPI_ISL_2158965, EPI_ISL_2158966, EPI_ISL_2158967, EPI_ISL_2158968, EPI_ISL_2158969, EPI_ISL_2158970, EPI_ISL_2158971, EPI_ISL_2158972, EPI_ISL_2158973, EPI_ISL_2158974 | | | |
| see above | Aegis Sciences Corporation | Centers for Disease Control and Prevention Division of Viral Diseases, Pathogen Discovery | Dakota Howard, Dhwani Batra, Peter W. Cook, Kara Moser, Adrian Paskey, Jason Caravas, Benjamin Rambo-Martin, Shatavia Morrison, Christopher Gulvick, Scott Sammons, Yvette Unoarumhi, Darlene Wagner, Matthew Schmerer, Cyndi Clark, Patrick Campbell, Rob Case, Vikramsinha Ghorpade, Holly Houdeshell, Ola Kvalvaag, Dillon Nall, Ethan Sanders, Alec Vest, Shaun Westlund, Matthew Hardison, Clinton R. Paden, Duncan MacCannell |
| EPI_ISL_2158975 | Yale Clinical Virology Lab | Yale Center for Genomic Analysis | Shrikant Mane, Kaya Bilguvar, Curt Scharfe, Irina Tikhonova, Brooke Sullivan |
| EPI_ISL_2158976 | Aegis Sciences Corporation | Centers for Disease Control and Prevention Division of Viral Diseases, Pathogen Discovery | Dakota Howard, Dhwani Batra, Peter W. Cook, Kara Moser, Adrian Paskey, Jason Caravas, Benjamin Rambo-Martin, Shatavia Morrison, Christopher Gulvick, Scott Sammons, Yvette Unoarumhi, Darlene Wagner, Matthew Schmerer, Cyndi Clark, Patrick Campbell, Rob Case, Vikramsinha Ghorpade, Holly Houdeshell, Ola Kvalvaag, Dillon Nall, Ethan Sanders, Alec Vest, Shaun Westlund, Matthew Hardison, Clinton R. Paden, Duncan MacCannell |
| EPI_ISL_2158977, EPI_ISL_2158978 | Yale Clinical Virology Lab | Yale Center for Genomic Analysis | Shrikant Mane, Kaya Bilguvar, Curt Scharfe, Irina Tikhonova, Brooke Sullivan |
| EPI_ISL_2158979 | Aegis Sciences Corporation | Centers for Disease Control and Prevention Division of Viral Diseases, Pathogen Discovery | Dakota Howard, Dhwani Batra, Peter W. Cook, Kara Moser, Adrian Paskey, Jason Caravas, Benjamin Rambo-Martin, Shatavia Morrison, Christopher Gulvick, Scott Sammons, Yvette Unoarumhi, Darlene Wagner, Matthew Schmerer, Cyndi Clark, Patrick Campbell, Rob Case, Vikramsinha Ghorpade, Holly Houdeshell, Ola Kvalvaag, Dillon Nall, Ethan Sanders, Alec Vest, Shaun Westlund, Matthew Hardison, Clinton R. Paden, Duncan MacCannell |
| EPI_ISL_2158980, EPI_ISL_2158981 | Yale Clinical Virology Lab | Yale Center for Genomic Analysis | Shrikant Mane, Kaya Bilguvar, Curt Scharfe, Irina Tikhonova, Brooke Sullivan |
| EPI_ISL_2158982 | Aegis Sciences Corporation | Centers for Disease Control and Prevention Division of Viral Diseases, Pathogen Discovery | Dakota Howard, Dhwani Batra, Peter W. Cook, Kara Moser, Adrian Paskey, Jason Caravas, Benjamin Rambo-Martin, Shatavia Morrison, Christopher Gulvick, Scott Sammons, Yvette Unoarumhi, Darlene Wagner, Matthew Schmerer, Cyndi Clark, Patrick Campbell, Rob Case, Vikramsinha Ghorpade, Holly Houdeshell, Ola Kvalvaag, Dillon Nall, Ethan Sanders, Alec Vest, Shaun Westlund, Matthew Hardison, Clinton R. Paden, Duncan MacCannell |
| EPI_ISL_2158983 | Yale Clinical Virology Lab | Yale Center for Genomic Analysis | Shrikant Mane, Kaya Bilguvar, Curt Scharfe, Irina Tikhonova, Brooke Sullivan |
| EPI_ISL_2158985 | Aegis Sciences Corporation | Centers for Disease Control and Prevention Division of Viral Diseases, Pathogen Discovery | Dakota Howard, Dhwani Batra, Peter W. Cook, Kara Moser, Adrian Paskey, Jason Caravas, Benjamin Rambo-Martin, Shatavia Morrison, Christopher Gulvick, Scott Sammons, Yvette Unoarumhi, Darlene Wagner, Matthew Schmerer, Cyndi Clark, Patrick Campbell, Rob Case, Vikramsinha Ghorpade, Holly Houdeshell, Ola Kvalvaag, Dillon Nall, Ethan Sanders, Alec Vest, Shaun Westlund, Matthew Hardison, Clinton R. Paden, Duncan MacCannell |
| EPI_ISL_2158986 | Yale Clinical Virology Lab | Yale Center for Genomic Analysis | Shrikant Mane, Kaya Bilguvar, Curt Scharfe, Irina Tikhonova, Brooke Sullivan |
| EPI_ISL_2158987 | Aegis Sciences Corporation | Centers for Disease Control and Prevention Division of Viral Diseases, Pathogen Discovery | Dakota Howard, Dhwani Batra, Peter W. Cook, Kara Moser, Adrian Paskey, Jason Caravas, Benjamin Rambo-Martin, Shatavia Morrison, Christopher Gulvick, Scott Sammons, Yvette Unoarumhi, Darlene Wagner, Matthew Schmerer, Cyndi Clark, Patrick Campbell, Rob Case, Vikramsinha Ghorpade, Holly Houdeshell, Ola Kvalvaag, Dillon Nall, Ethan Sanders, Alec Vest, Shaun Westlund, Matthew Hardison, Clinton R. Paden, Duncan MacCannell |
| EPI_ISL_2158988, EPI_ISL_2158989 | Yale Clinical Virology Lab | Yale Center for Genomic Analysis | Shrikant Mane, Kaya Bilguvar, Curt Scharfe, Irina Tikhonova, Brooke Sullivan |
| EPI_ISL_2158990 | Aegis Sciences Corporation | Centers for Disease Control and Prevention Division of Viral Diseases, Pathogen Discovery | Dakota Howard, Dhwani Batra, Peter W. Cook, Kara Moser, Adrian Paskey, Jason Caravas, Benjamin Rambo-Martin, Shatavia Morrison, Christopher Gulvick, Scott Sammons, Yvette Unoarumhi, Darlene Wagner, Matthew Schmerer, Cyndi Clark, Patrick Campbell, Rob Case, Vikramsinha Ghorpade, Holly Houdeshell, Ola Kvalvaag, Dillon Nall, Ethan Sanders, Alec Vest, Shaun Westlund, Matthew Hardison, Clinton R. Paden, Duncan MacCannell |
| EPI_ISL_2158991, EPI_ISL_2158992 | Yale Clinical Virology Lab | Yale Center for Genomic Analysis | Shrikant Mane, Kaya Bilguvar, Curt Scharfe, Irina Tikhonova, Brooke Sullivan |
| EPI_ISL_2158993 | Aegis Sciences Corporation | Centers for Disease Control and Prevention Division of Viral Diseases, Pathogen Discovery | Dakota Howard, Dhwani Batra, Peter W. Cook, Kara Moser, Adrian Paskey, Jason Caravas, Benjamin Rambo-Martin, Shatavia Morrison, Christopher Gulvick, Scott Sammons, Yvette Unoarumhi, Darlene Wagner, Matthew Schmerer, Cyndi Clark, Patrick Campbell, Rob Case, Vikramsinha Ghorpade, Holly Houdeshell, Ola Kvalvaag, Dillon Nall, Ethan Sanders, Alec Vest, Shaun Westlund, Matthew Hardison, Clinton R. Paden, Duncan MacCannell |
| EPI_ISL_2158994, EPI_ISL_2158995 | Yale Clinical Virology Lab | Yale Center for Genomic Analysis | Shrikant Mane, Kaya Bilguvar, Curt Scharfe, Irina Tikhonova, Brooke Sullivan |
| EPI_ISL_2158996 | Aegis Sciences Corporation | Centers for Disease Control and Prevention Division of Viral Diseases, Pathogen Discovery | Dakota Howard, Dhwani Batra, Peter W. Cook, Kara Moser, Adrian Paskey, Jason Caravas, Benjamin Rambo-Martin, Shatavia Morrison, Christopher Gulvick, Scott Sammons, Yvette Unoarumhi, Darlene Wagner, Matthew Schmerer, Cyndi Clark, Patrick Campbell, Rob Case, Vikramsinha Ghorpade, Holly Houdeshell, Ola Kvalvaag, Dillon Nall, Ethan Sanders, Alec Vest, Shaun Westlund, Matthew Hardison, Clinton R. Paden, Duncan MacCannell |

| | | | |
|---|---|---|---|
| EPI_ISL_2158997, EPI_ISL_2158998 | Yale Clinical Virology Lab | Yale Center for Genomic Analysis | Shrikant Mane, Kaya Bilguvar, Curt Scharfe, Irina Tikhonova, Brooke Sullivan |
| EPI_ISL_2158999, EPI_ISL_2159001 | Aegis Sciences Corporation | Centers for Disease Control and Prevention Division of Viral Diseases, Pathogen Discovery | Dakota Howard, Dhwani Batra, Peter W. Cook, Kara Moser, Adrian Paskey, Jason Caravas, Benjamin Rambo-Martin, Shatavia Morrison, Christopher Gulvick, Scott Sammons, Yvette Unoarumhi, Darlene Wagner, Matthew Schmerer, Cyndi Clark, Patrick Campbell, Rob Case, Vikramsinha Ghorpade, Holly Houdeshell, Ola Kvalvaag, Dillon Nall, Ethan Sanders, Alec Vest, Shaun Westlund, Matthew Hardison, Clinton R. Paden, Duncan MacCannell |
| EPI_ISL_2159002, EPI_ISL_2159003 | Yale Clinical Virology Lab | Yale Center for Genomic Analysis | Shrikant Mane, Kaya Bilguvar, Curt Scharfe, Irina Tikhonova, Brooke Sullivan |
| EPI_ISL_2159004 | Aegis Sciences Corporation | Centers for Disease Control and Prevention Division of Viral Diseases, Pathogen Discovery | Dakota Howard, Dhwani Batra, Peter W. Cook, Kara Moser, Adrian Paskey, Jason Caravas, Benjamin Rambo-Martin, Shatavia Morrison, Christopher Gulvick, Scott Sammons, Yvette Unoarumhi, Darlene Wagner, Matthew Schmerer, Cyndi Clark, Patrick Campbell, Rob Case, Vikramsinha Ghorpade, Holly Houdeshell, Ola Kvalvaag, Dillon Nall, Ethan Sanders, Alec Vest, Shaun Westlund, Matthew Hardison, Clinton R. Paden, Duncan MacCannell |
| EPI_ISL_2159005, EPI_ISL_2159006 | Yale Clinical Virology Lab | Yale Center for Genomic Analysis | Shrikant Mane, Kaya Bilguvar, Curt Scharfe, Irina Tikhonova, Brooke Sullivan |
| EPI_ISL_2159007 | Aegis Sciences Corporation | Centers for Disease Control and Prevention Division of Viral Diseases, Pathogen Discovery | Dakota Howard, Dhwani Batra, Peter W. Cook, Kara Moser, Adrian Paskey, Jason Caravas, Benjamin Rambo-Martin, Shatavia Morrison, Christopher Gulvick, Scott Sammons, Yvette Unoarumhi, Darlene Wagner, Matthew Schmerer, Cyndi Clark, Patrick Campbell, Rob Case, Vikramsinha Ghorpade, Holly Houdeshell, Ola Kvalvaag, Dillon Nall, Ethan Sanders, Alec Vest, Shaun Westlund, Matthew Hardison, Clinton R. Paden, Duncan MacCannell |
| EPI_ISL_2159008 | Yale Clinical Virology Lab | Yale Center for Genomic Analysis | Shrikant Mane, Kaya Bilguvar, Curt Scharfe, Irina Tikhonova, Brooke Sullivan |
| EPI_ISL_2159009 | Aegis Sciences Corporation | Centers for Disease Control and Prevention Division of Viral Diseases, Pathogen Discovery | Dakota Howard, Dhwani Batra, Peter W. Cook, Kara Moser, Adrian Paskey, Jason Caravas, Benjamin Rambo-Martin, Shatavia Morrison, Christopher Gulvick, Scott Sammons, Yvette Unoarumhi, Darlene Wagner, Matthew Schmerer, Cyndi Clark, Patrick Campbell, Rob Case, Vikramsinha Ghorpade, Holly Houdeshell, Ola Kvalvaag, Dillon Nall, Ethan Sanders, Alec Vest, Shaun Westlund, Matthew Hardison, Clinton R. Paden, Duncan MacCannell |
| EPI_ISL_2159010, EPI_ISL_2159011 | Yale Clinical Virology Lab | Yale Center for Genomic Analysis | Shrikant Mane, Kaya Bilguvar, Curt Scharfe, Irina Tikhonova, Brooke Sullivan |
| EPI_ISL_2159012 | Aegis Sciences Corporation | Centers for Disease Control and Prevention Division of Viral Diseases, Pathogen Discovery | Dakota Howard, Dhwani Batra, Peter W. Cook, Kara Moser, Adrian Paskey, Jason Caravas, Benjamin Rambo-Martin, Shatavia Morrison, Christopher Gulvick, Scott Sammons, Yvette Unoarumhi, Darlene Wagner, Matthew Schmerer, Cyndi Clark, Patrick Campbell, Rob Case, Vikramsinha Ghorpade, Holly Houdeshell, Ola Kvalvaag, Dillon Nall, Ethan Sanders, Alec Vest, Shaun Westlund, Matthew Hardison, Clinton R. Paden, Duncan MacCannell |
| EPI_ISL_2159013, EPI_ISL_2159014 | Yale Clinical Virology Lab | Yale Center for Genomic Analysis | Shrikant Mane, Kaya Bilguvar, Curt Scharfe, Irina Tikhonova, Brooke Sullivan |
| EPI_ISL_2159015 | Aegis Sciences Corporation | Centers for Disease Control and Prevention Division of Viral Diseases, Pathogen Discovery | Dakota Howard, Dhwani Batra, Peter W. Cook, Kara Moser, Adrian Paskey, Jason Caravas, Benjamin Rambo-Martin, Shatavia Morrison, Christopher Gulvick, Scott Sammons, Yvette Unoarumhi, Darlene Wagner, Matthew Schmerer, Cyndi Clark, Patrick Campbell, Rob Case, Vikramsinha Ghorpade, Holly Houdeshell, Ola Kvalvaag, Dillon Nall, Ethan Sanders, Alec Vest, Shaun Westlund, Matthew Hardison, Clinton R. Paden, Duncan MacCannell |
| EPI_ISL_2159016, EPI_ISL_2159017 | Yale Clinical Virology Lab | Yale Center for Genomic Analysis | Shrikant Mane, Kaya Bilguvar, Curt Scharfe, Irina Tikhonova, Brooke Sullivan |
| EPI_ISL_2159018 | Aegis Sciences Corporation | Centers for Disease Control and Prevention Division of Viral Diseases, Pathogen Discovery | Dakota Howard, Dhwani Batra, Peter W. Cook, Kara Moser, Adrian Paskey, Jason Caravas, Benjamin Rambo-Martin, Shatavia Morrison, Christopher Gulvick, Scott Sammons, Yvette Unoarumhi, Darlene Wagner, Matthew Schmerer, Cyndi Clark, Patrick Campbell, Rob Case, Vikramsinha Ghorpade, Holly Houdeshell, Ola Kvalvaag, Dillon Nall, Ethan Sanders, Alec Vest, Shaun Westlund, Matthew Hardison, Clinton R. Paden, Duncan MacCannell |
| EPI_ISL_2159019, EPI_ISL_2159020 | Yale Clinical Virology Lab | Yale Center for Genomic Analysis | Shrikant Mane, Kaya Bilguvar, Curt Scharfe, Irina Tikhonova, Brooke Sullivan |
| EPI_ISL_2159021 | Aegis Sciences Corporation | Centers for Disease Control and Prevention Division of Viral Diseases, Pathogen Discovery | Dakota Howard, Dhwani Batra, Peter W. Cook, Kara Moser, Adrian Paskey, Jason Caravas, Benjamin Rambo-Martin, Shatavia Morrison, Christopher Gulvick, Scott Sammons, Yvette Unoarumhi, Darlene Wagner, Matthew Schmerer, Cyndi Clark, Patrick Campbell, Rob Case, Vikramsinha Ghorpade, Holly Houdeshell, Ola Kvalvaag, Dillon Nall, Ethan Sanders, Alec Vest, Shaun Westlund, Matthew Hardison, Clinton R. Paden, Duncan MacCannell |
| EPI_ISL_2159022, EPI_ISL_2159023 | Yale Clinical Virology Lab | Yale Center for Genomic Analysis | Shrikant Mane, Kaya Bilguvar, Curt Scharfe, Irina Tikhonova, Brooke Sullivan |
| EPI_ISL_2159024 | Aegis Sciences Corporation | Centers for Disease Control and Prevention Division of Viral Diseases, Pathogen Discovery | Dakota Howard, Dhwani Batra, Peter W. Cook, Kara Moser, Adrian Paskey, Jason Caravas, Benjamin Rambo-Martin, Shatavia Morrison, Christopher Gulvick, Scott Sammons, Yvette Unoarumhi, Darlene Wagner, Matthew Schmerer, Cyndi Clark, Patrick Campbell, Rob Case, Vikramsinha Ghorpade, Holly Houdeshell, Ola Kvalvaag, Dillon Nall, Ethan Sanders, Alec Vest, Shaun Westlund, Matthew Hardison, Clinton R. Paden, Duncan MacCannell |
| EPI_ISL_2159025 | Yale Clinical Virology Lab | Yale Center for Genomic Analysis | Shrikant Mane, Kaya Bilguvar, Curt Scharfe, Irina Tikhonova, Brooke Sullivan |
| EPI_ISL_2159026 | Aegis Sciences Corporation | Centers for Disease Control and Prevention Division of Viral Diseases, Pathogen Discovery | Dakota Howard, Dhwani Batra, Peter W. Cook, Kara Moser, Adrian Paskey, Jason Caravas, Benjamin Rambo-Martin, Shatavia Morrison, Christopher Gulvick, Scott Sammons, Yvette Unoarumhi, Darlene Wagner, Matthew Schmerer, Cyndi Clark, Patrick Campbell, Rob Case, Vikramsinha Ghorpade, Holly Houdeshell, Ola Kvalvaag, Dillon Nall, Ethan Sanders, Alec Vest, Shaun Westlund, Matthew Hardison, Clinton R. Paden, Duncan MacCannell |
| EPI_ISL_2159027, EPI_ISL_2159028 | Yale Clinical Virology Lab | Yale Center for Genomic Analysis | Shrikant Mane, Kaya Bilguvar, Curt Scharfe, Irina Tikhonova, Brooke Sullivan |
| EPI_ISL_2159029 | Aegis Sciences Corporation | Centers for Disease Control and Prevention Division of Viral Diseases, Pathogen Discovery | Dakota Howard, Dhwani Batra, Peter W. Cook, Kara Moser, Adrian Paskey, Jason Caravas, Benjamin Rambo-Martin, Shatavia Morrison, Christopher Gulvick, Scott Sammons, Yvette Unoarumhi, Darlene Wagner, Matthew Schmerer, Cyndi Clark, Patrick Campbell, Rob Case, Vikramsinha Ghorpade, Holly Houdeshell, Ola Kvalvaag, Dillon Nall, Ethan Sanders, Alec Vest, Shaun Westlund, Matthew Hardison, Clinton R. Paden, Duncan MacCannell |
| EPI_ISL_2159030, EPI_ISL_2159031 | Yale Clinical Virology Lab | Yale Center for Genomic Analysis | Shrikant Mane, Kaya Bilguvar, Curt Scharfe, Irina Tikhonova, Brooke Sullivan |
| EPI_ISL_2159032 | Aegis Sciences Corporation | Centers for Disease Control and Prevention Division of Viral Diseases, Pathogen Discovery | Dakota Howard, Dhwani Batra, Peter W. Cook, Kara Moser, Adrian Paskey, Jason Caravas, Benjamin Rambo-Martin, Shatavia Morrison, Christopher Gulvick, Scott Sammons, Yvette Unoarumhi, Darlene Wagner, Matthew Schmerer, Cyndi Clark, Patrick Campbell, Rob Case, Vikramsinha Ghorpade, Holly Houdeshell, Ola Kvalvaag, Dillon Nall, Ethan Sanders, Alec Vest, Shaun Westlund, Matthew Hardison, Clinton R. Paden, Duncan MacCannell |
| EPI_ISL_2159033 | Yale Clinical Virology Lab | Yale Center for Genomic Analysis | Shrikant Mane, Kaya Bilguvar, Curt Scharfe, Irina Tikhonova, Brooke Sullivan |
| EPI_ISL_2159034 | Aegis Sciences Corporation | Centers for Disease Control and Prevention Division of Viral Diseases, Pathogen Discovery | Dakota Howard, Dhwani Batra, Peter W. Cook, Kara Moser, Adrian Paskey, Jason Caravas, Benjamin Rambo-Martin, Shatavia Morrison, Christopher Gulvick, Scott Sammons, Yvette Unoarumhi, Darlene Wagner, Matthew Schmerer, Cyndi Clark, Patrick Campbell, Rob Case, Vikramsinha Ghorpade, Holly Houdeshell, Ola Kvalvaag, Dillon Nall, Ethan Sanders, Alec Vest, Shaun Westlund, Matthew Hardison, Clinton R. Paden, Duncan MacCannell |
| EPI_ISL_2159035, EPI_ISL_2159036 | Yale Clinical Virology Lab | Yale Center for Genomic Analysis | Shrikant Mane, Kaya Bilguvar, Curt Scharfe, Irina Tikhonova, Brooke Sullivan |
| EPI_ISL_2159037 | Aegis Sciences Corporation | Centers for Disease Control and Prevention Division of Viral Diseases, Pathogen Discovery | Dakota Howard, Dhwani Batra, Peter W. Cook, Kara Moser, Adrian Paskey, Jason Caravas, Benjamin Rambo-Martin, Shatavia Morrison, Christopher Gulvick, Scott Sammons, Yvette Unoarumhi, Darlene Wagner, Matthew Schmerer, Cyndi Clark, Patrick Campbell, Rob Case, Vikramsinha Ghorpade, Holly Houdeshell, Ola Kvalvaag, Dillon Nall, Ethan Sanders, Alec Vest, Shaun Westlund, Matthew Hardison, Clinton R. Paden, Duncan MacCannell |
| EPI_ISL_2159038, EPI_ISL_2159039 | Yale Clinical Virology Lab | Yale Center for Genomic Analysis | Shrikant Mane, Kaya Bilguvar, Curt Scharfe, Irina Tikhonova, Brooke Sullivan |
| EPI_ISL_2159040 | Aegis Sciences Corporation | Centers for Disease Control and Prevention Division of Viral Diseases, Pathogen Discovery | Dakota Howard, Dhwani Batra, Peter W. Cook, Kara Moser, Adrian Paskey, Jason Caravas, Benjamin Rambo-Martin, Shatavia Morrison, Christopher Gulvick, Scott Sammons, Yvette Unoarumhi, Darlene Wagner, Matthew Schmerer, Cyndi Clark, Patrick Campbell, Rob Case, Vikramsinha Ghorpade, Holly Houdeshell, Ola Kvalvaag, Dillon Nall, Ethan Sanders, Alec Vest, Shaun Westlund, Matthew Hardison, Clinton R. Paden, Duncan MacCannell |
| EPI_ISL_2159041, EPI_ISL_2159042 | Yale Clinical Virology Lab | Yale Center for Genomic Analysis | Shrikant Mane, Kaya Bilguvar, Curt Scharfe, Irina Tikhonova, Brooke Sullivan |
| EPI_ISL_2159043 | Aegis Sciences Corporation | Centers for Disease Control and Prevention Division of Viral Diseases, Pathogen Discovery | Dakota Howard, Dhwani Batra, Peter W. Cook, Kara Moser, Adrian Paskey, Jason Caravas, Benjamin Rambo-Martin, Shatavia Morrison, Christopher Gulvick, Scott Sammons, Yvette Unoarumhi, Darlene Wagner, Matthew Schmerer, Cyndi Clark, Patrick Campbell, Rob Case, Vikramsinha Ghorpade, Holly Houdeshell, Ola Kvalvaag, Dillon Nall, Ethan Sanders, Alec Vest, Shaun Westlund, Matthew Hardison, Clinton R. Paden, Duncan MacCannell |
| EPI_ISL_2159044, EPI_ISL_2159045 | Yale Clinical Virology Lab | Yale Center for Genomic Analysis | Shrikant Mane, Kaya Bilguvar, Curt Scharfe, Irina Tikhonova, Brooke Sullivan |
| EPI_ISL_2159046 | Aegis Sciences Corporation | Centers for Disease Control and Prevention Division of Viral Diseases, Pathogen Discovery | Dakota Howard, Dhwani Batra, Peter W. Cook, Kara Moser, Adrian Paskey, Jason Caravas, Benjamin Rambo-Martin, Shatavia Morrison, Christopher Gulvick, Scott Sammons, Yvette Unoarumhi, Darlene Wagner, Matthew Schmerer, Cyndi Clark, Patrick Campbell, Rob Case, Vikramsinha Ghorpade, Holly Houdeshell, Ola Kvalvaag, Dillon Nall, Ethan Sanders, Alec Vest, Shaun Westlund, Matthew Hardison, Clinton R. Paden, Duncan MacCannell |
| EPI_ISL_2159047, EPI_ISL_2159048 | Yale Clinical Virology Lab | Yale Center for Genomic Analysis | Shrikant Mane, Kaya Bilguvar, Curt Scharfe, Irina Tikhonova, Brooke Sullivan |
| EPI_ISL_2159049 | Aegis Sciences Corporation | Centers for Disease Control and Prevention Division of Viral Diseases, Pathogen Discovery | Dakota Howard, Dhwani Batra, Peter W. Cook, Kara Moser, Adrian Paskey, Jason Caravas, Benjamin Rambo-Martin, Shatavia Morrison, Christopher Gulvick, Scott Sammons, Yvette Unoarumhi, Darlene Wagner, Matthew Schmerer, Cyndi Clark, Patrick Campbell, Rob Case, Vikramsinha Ghorpade, Holly Houdeshell, Ola Kvalvaag, Dillon Nall, Ethan Sanders, Alec Vest, Shaun Westlund, Matthew Hardison, Clinton R. Paden, Duncan MacCannell |
| EPI_ISL_2159050, EPI_ISL_2159051 | Yale Clinical Virology Lab | Yale Center for Genomic Analysis | Shrikant Mane, Kaya Bilguvar, Curt Scharfe, Irina Tikhonova, Brooke Sullivan |
| EPI_ISL_2159052 | Aegis Sciences Corporation | Centers for Disease Control and Prevention Division of Viral Diseases, Pathogen Discovery | Dakota Howard, Dhwani Batra, Peter W. Cook, Kara Moser, Adrian Paskey, Jason Caravas, Benjamin Rambo-Martin, Shatavia Morrison, Christopher Gulvick, Scott Sammons, Yvette Unoarumhi, Darlene Wagner, Matthew Schmerer, Cyndi Clark, Patrick Campbell, Rob Case, Vikramsinha Ghorpade, Holly Houdeshell, Ola Kvalvaag, Dillon Nall, Ethan Sanders, Alec Vest, Shaun Westlund, Matthew Hardison, Clinton R. Paden, Duncan MacCannell |
| EPI_ISL_2159053 | Yale Clinical Virology Lab | Yale Center for Genomic Analysis | Shrikant Mane, Kaya Bilguvar, Curt Scharfe, Irina Tikhonova, Brooke Sullivan |

| | | | |
|---|---|---|---|
| EPI_ISL_2159054 | Aegis Sciences Corporation | Centers for Disease Control and Prevention Division of Viral Diseases, Pathogen Discovery | Dakota Howard, Dhwani Batra, Peter W. Cook, Kara Moser, Adrian Paskey, Jason Caravas, Benjamin Rambo-Martin, Shatavia Morrison, Christopher Gulvick, Scott Sammons, Yvette Unoarumhi, Darlene Wagner, Matthew Schmerer, Cyndi Clark, Patrick Campbell, Rob Case, Vikramsinha Ghorpade, Holly Houdeshell, Ola Kvalvaag, Dillon Nall, Ethan Sanders, Alec Vest, Shaun Westlund, Matthew Hardison, Clinton R. Paden, Duncan MacCannell |
| EPI_ISL_2159055, EPI_ISL_2159056 | Yale Clinical Virology Lab | Yale Center for Genomic Analysis | Shrikant Mane, Kaya Bilguvar, Curt Scharfe, Irina Tikhonova, Brooke Sullivan |
| EPI_ISL_2159057 | Aegis Sciences Corporation | Centers for Disease Control and Prevention Division of Viral Diseases, Pathogen Discovery | Dakota Howard, Dhwani Batra, Peter W. Cook, Kara Moser, Adrian Paskey, Jason Caravas, Benjamin Rambo-Martin, Shatavia Morrison, Christopher Gulvick, Scott Sammons, Yvette Unoarumhi, Darlene Wagner, Matthew Schmerer, Cyndi Clark, Patrick Campbell, Rob Case, Vikramsinha Ghorpade, Holly Houdeshell, Ola Kvalvaag, Dillon Nall, Ethan Sanders, Alec Vest, Shaun Westlund, Matthew Hardison, Clinton R. Paden, Duncan MacCannell |
| EPI_ISL_2159058, EPI_ISL_2159059 | Yale Clinical Virology Lab | Yale Center for Genomic Analysis | Shrikant Mane, Kaya Bilguvar, Curt Scharfe, Irina Tikhonova, Brooke Sullivan |
| EPI_ISL_2159060 | Aegis Sciences Corporation | Centers for Disease Control and Prevention Division of Viral Diseases, Pathogen Discovery | Dakota Howard, Dhwani Batra, Peter W. Cook, Kara Moser, Adrian Paskey, Jason Caravas, Benjamin Rambo-Martin, Shatavia Morrison, Christopher Gulvick, Scott Sammons, Yvette Unoarumhi, Darlene Wagner, Matthew Schmerer, Cyndi Clark, Patrick Campbell, Rob Case, Vikramsinha Ghorpade, Holly Houdeshell, Ola Kvalvaag, Dillon Nall, Ethan Sanders, Alec Vest, Shaun Westlund, Matthew Hardison, Clinton R. Paden, Duncan MacCannell |
| EPI_ISL_2159061 | Yale Clinical Virology Lab | Yale Center for Genomic Analysis | Shrikant Mane, Kaya Bilguvar, Curt Scharfe, Irina Tikhonova, Brooke Sullivan |
| EPI_ISL_2159062 | Aegis Sciences Corporation | Centers for Disease Control and Prevention Division of Viral Diseases, Pathogen Discovery | Dakota Howard, Dhwani Batra, Peter W. Cook, Kara Moser, Adrian Paskey, Jason Caravas, Benjamin Rambo-Martin, Shatavia Morrison, Christopher Gulvick, Scott Sammons, Yvette Unoarumhi, Darlene Wagner, Matthew Schmerer, Cyndi Clark, Patrick Campbell, Rob Case, Vikramsinha Ghorpade, Holly Houdeshell, Ola Kvalvaag, Dillon Nall, Ethan Sanders, Alec Vest, Shaun Westlund, Matthew Hardison, Clinton R. Paden, Duncan MacCannell |
| EPI_ISL_2159063, EPI_ISL_2159064 | Yale Clinical Virology Lab | Yale Center for Genomic Analysis | Shrikant Mane, Kaya Bilguvar, Curt Scharfe, Irina Tikhonova, Brooke Sullivan |
| EPI_ISL_2159065 | Aegis Sciences Corporation | Centers for Disease Control and Prevention Division of Viral Diseases, Pathogen Discovery | Dakota Howard, Dhwani Batra, Peter W. Cook, Kara Moser, Adrian Paskey, Jason Caravas, Benjamin Rambo-Martin, Shatavia Morrison, Christopher Gulvick, Scott Sammons, Yvette Unoarumhi, Darlene Wagner, Matthew Schmerer, Cyndi Clark, Patrick Campbell, Rob Case, Vikramsinha Ghorpade, Holly Houdeshell, Ola Kvalvaag, Dillon Nall, Ethan Sanders, Alec Vest, Shaun Westlund, Matthew Hardison, Clinton R. Paden, Duncan MacCannell |
| EPI_ISL_2159066, EPI_ISL_2159067 | Yale Clinical Virology Lab | Yale Center for Genomic Analysis | Shrikant Mane, Kaya Bilguvar, Curt Scharfe, Irina Tikhonova, Brooke Sullivan |
| EPI_ISL_2159068 | Aegis Sciences Corporation | Centers for Disease Control and Prevention Division of Viral Diseases, Pathogen Discovery | Dakota Howard, Dhwani Batra, Peter W. Cook, Kara Moser, Adrian Paskey, Jason Caravas, Benjamin Rambo-Martin, Shatavia Morrison, Christopher Gulvick, Scott Sammons, Yvette Unoarumhi, Darlene Wagner, Matthew Schmerer, Cyndi Clark, Patrick Campbell, Rob Case, Vikramsinha Ghorpade, Holly Houdeshell, Ola Kvalvaag, Dillon Nall, Ethan Sanders, Alec Vest, Shaun Westlund, Matthew Hardison, Clinton R. Paden, Duncan MacCannell |
| EPI_ISL_2159069, EPI_ISL_2159070 | Yale Clinical Virology Lab | Yale Center for Genomic Analysis | Shrikant Mane, Kaya Bilguvar, Curt Scharfe, Irina Tikhonova, Brooke Sullivan |
| EPI_ISL_2159071 | Aegis Sciences Corporation | Centers for Disease Control and Prevention Division of Viral Diseases, Pathogen Discovery | Dakota Howard, Dhwani Batra, Peter W. Cook, Kara Moser, Adrian Paskey, Jason Caravas, Benjamin Rambo-Martin, Shatavia Morrison, Christopher Gulvick, Scott Sammons, Yvette Unoarumhi, Darlene Wagner, Matthew Schmerer, Cyndi Clark, Patrick Campbell, Rob Case, Vikramsinha Ghorpade, Holly Houdeshell, Ola Kvalvaag, Dillon Nall, Ethan Sanders, Alec Vest, Shaun Westlund, Matthew Hardison, Clinton R. Paden, Duncan MacCannell |
| EPI_ISL_2159072, EPI_ISL_2159073 | Yale Clinical Virology Lab | Yale Center for Genomic Analysis | Shrikant Mane, Kaya Bilguvar, Curt Scharfe, Irina Tikhonova, Brooke Sullivan |
| EPI_ISL_2159074 | Aegis Sciences Corporation | Centers for Disease Control and Prevention Division of Viral Diseases, Pathogen Discovery | Dakota Howard, Dhwani Batra, Peter W. Cook, Kara Moser, Adrian Paskey, Jason Caravas, Benjamin Rambo-Martin, Shatavia Morrison, Christopher Gulvick, Scott Sammons, Yvette Unoarumhi, Darlene Wagner, Matthew Schmerer, Cyndi Clark, Patrick Campbell, Rob Case, Vikramsinha Ghorpade, Holly Houdeshell, Ola Kvalvaag, Dillon Nall, Ethan Sanders, Alec Vest, Shaun Westlund, Matthew Hardison, Clinton R. Paden, Duncan MacCannell |
| EPI_ISL_2159075, EPI_ISL_2159076 | Yale Clinical Virology Lab | Yale Center for Genomic Analysis | Shrikant Mane, Kaya Bilguvar, Curt Scharfe, Irina Tikhonova, Brooke Sullivan |
| EPI_ISL_2159077 | Aegis Sciences Corporation | Centers for Disease Control and Prevention Division of Viral Diseases, Pathogen Discovery | Dakota Howard, Dhwani Batra, Peter W. Cook, Kara Moser, Adrian Paskey, Jason Caravas, Benjamin Rambo-Martin, Shatavia Morrison, Christopher Gulvick, Scott Sammons, Yvette Unoarumhi, Darlene Wagner, Matthew Schmerer, Cyndi Clark, Patrick Campbell, Rob Case, Vikramsinha Ghorpade, Holly Houdeshell, Ola Kvalvaag, Dillon Nall, Ethan Sanders, Alec Vest, Shaun Westlund, Matthew Hardison, Clinton R. Paden, Duncan MacCannell |
| EPI_ISL_2159078 | Yale Clinical Virology Lab | Yale Center for Genomic Analysis | Shrikant Mane, Kaya Bilguvar, Curt Scharfe, Irina Tikhonova, Brooke Sullivan |
| EPI_ISL_2159079 | Aegis Sciences Corporation | Centers for Disease Control and Prevention Division of Viral Diseases, Pathogen Discovery | Dakota Howard, Dhwani Batra, Peter W. Cook, Kara Moser, Adrian Paskey, Jason Caravas, Benjamin Rambo-Martin, Shatavia Morrison, Christopher Gulvick, Scott Sammons, Yvette Unoarumhi, Darlene Wagner, Matthew Schmerer, Cyndi Clark, Patrick Campbell, Rob Case, Vikramsinha Ghorpade, Holly Houdeshell, Ola Kvalvaag, Dillon Nall, Ethan Sanders, Alec Vest, Shaun Westlund, Matthew Hardison, Clinton R. Paden, Duncan MacCannell |
| EPI_ISL_2159080, EPI_ISL_2159081 | Yale Clinical Virology Lab | Yale Center for Genomic Analysis | Shrikant Mane, Kaya Bilguvar, Curt Scharfe, Irina Tikhonova, Brooke Sullivan |
| EPI_ISL_2159082 | Aegis Sciences Corporation | Centers for Disease Control and Prevention Division of Viral Diseases, Pathogen Discovery | Dakota Howard, Dhwani Batra, Peter W. Cook, Kara Moser, Adrian Paskey, Jason Caravas, Benjamin Rambo-Martin, Shatavia Morrison, Christopher Gulvick, Scott Sammons, Yvette Unoarumhi, Darlene Wagner, Matthew Schmerer, Cyndi Clark, Patrick Campbell, Rob Case, Vikramsinha Ghorpade, Holly Houdeshell, Ola Kvalvaag, Dillon Nall, Ethan Sanders, Alec Vest, Shaun Westlund, Matthew Hardison, Clinton R. Paden, Duncan MacCannell |
| EPI_ISL_2159083, EPI_ISL_2159084 | Yale Clinical Virology Lab | Yale Center for Genomic Analysis | Shrikant Mane, Kaya Bilguvar, Curt Scharfe, Irina Tikhonova, Brooke Sullivan |
| EPI_ISL_2159085 | Aegis Sciences Corporation | Centers for Disease Control and Prevention Division of Viral Diseases, Pathogen Discovery | Dakota Howard, Dhwani Batra, Peter W. Cook, Kara Moser, Adrian Paskey, Jason Caravas, Benjamin Rambo-Martin, Shatavia Morrison, Christopher Gulvick, Scott Sammons, Yvette Unoarumhi, Darlene Wagner, Matthew Schmerer, Cyndi Clark, Patrick Campbell, Rob Case, Vikramsinha Ghorpade, Holly Houdeshell, Ola Kvalvaag, Dillon Nall, Ethan Sanders, Alec Vest, Shaun Westlund, Matthew Hardison, Clinton R. Paden, Duncan MacCannell |
| EPI_ISL_2159086 | Yale Clinical Virology Lab | Yale Center for Genomic Analysis | Shrikant Mane, Kaya Bilguvar, Curt Scharfe, Irina Tikhonova, Brooke Sullivan |
| EPI_ISL_2159087 | Aegis Sciences Corporation | Centers for Disease Control and Prevention Division of Viral Diseases, Pathogen Discovery | Dakota Howard, Dhwani Batra, Peter W. Cook, Kara Moser, Adrian Paskey, Jason Caravas, Benjamin Rambo-Martin, Shatavia Morrison, Christopher Gulvick, Scott Sammons, Yvette Unoarumhi, Darlene Wagner, Matthew Schmerer, Cyndi Clark, Patrick Campbell, Rob Case, Vikramsinha Ghorpade, Holly Houdeshell, Ola Kvalvaag, Dillon Nall, Ethan Sanders, Alec Vest, Shaun Westlund, Matthew Hardison, Clinton R. Paden, Duncan MacCannell |
| EPI_ISL_2159088 | Yale Clinical Virology Lab | Yale Center for Genomic Analysis | Shrikant Mane, Kaya Bilguvar, Curt Scharfe, Irina Tikhonova, Brooke Sullivan |
| EPI_ISL_2159089 | Aegis Sciences Corporation | Centers for Disease Control and Prevention Division of Viral Diseases, Pathogen Discovery | Dakota Howard, Dhwani Batra, Peter W. Cook, Kara Moser, Adrian Paskey, Jason Caravas, Benjamin Rambo-Martin, Shatavia Morrison, Christopher Gulvick, Scott Sammons, Yvette Unoarumhi, Darlene Wagner, Matthew Schmerer, Cyndi Clark, Patrick Campbell, Rob Case, Vikramsinha Ghorpade, Holly Houdeshell, Ola Kvalvaag, Dillon Nall, Ethan Sanders, Alec Vest, Shaun Westlund, Matthew Hardison, Clinton R. Paden, Duncan MacCannell |
| EPI_ISL_2159090, EPI_ISL_2159091 | Yale Clinical Virology Lab | Yale Center for Genomic Analysis | Shrikant Mane, Kaya Bilguvar, Curt Scharfe, Irina Tikhonova, Brooke Sullivan |
| EPI_ISL_2159092 | Aegis Sciences Corporation | Centers for Disease Control and Prevention Division of Viral Diseases, Pathogen Discovery | Dakota Howard, Dhwani Batra, Peter W. Cook, Kara Moser, Adrian Paskey, Jason Caravas, Benjamin Rambo-Martin, Shatavia Morrison, Christopher Gulvick, Scott Sammons, Yvette Unoarumhi, Darlene Wagner, Matthew Schmerer, Cyndi Clark, Patrick Campbell, Rob Case, Vikramsinha Ghorpade, Holly Houdeshell, Ola Kvalvaag, Dillon Nall, Ethan Sanders, Alec Vest, Shaun Westlund, Matthew Hardison, Clinton R. Paden, Duncan MacCannell |
| EPI_ISL_2159093 | Yale Clinical Virology Lab | Yale Center for Genomic Analysis | Shrikant Mane, Kaya Bilguvar, Curt Scharfe, Irina Tikhonova, Brooke Sullivan |
| EPI_ISL_2159094 | Aegis Sciences Corporation | Centers for Disease Control and Prevention Division of Viral Diseases, Pathogen Discovery | Dakota Howard, Dhwani Batra, Peter W. Cook, Kara Moser, Adrian Paskey, Jason Caravas, Benjamin Rambo-Martin, Shatavia Morrison, Christopher Gulvick, Scott Sammons, Yvette Unoarumhi, Darlene Wagner, Matthew Schmerer, Cyndi Clark, Patrick Campbell, Rob Case, Vikramsinha Ghorpade, Holly Houdeshell, Ola Kvalvaag, Dillon Nall, Ethan Sanders, Alec Vest, Shaun Westlund, Matthew Hardison, Clinton R. Paden, Duncan MacCannell |
| EPI_ISL_2159095, EPI_ISL_2159096 | Yale Clinical Virology Lab | Yale Center for Genomic Analysis | Shrikant Mane, Kaya Bilguvar, Curt Scharfe, Irina Tikhonova, Brooke Sullivan |
| EPI_ISL_2159097 | Aegis Sciences Corporation | Centers for Disease Control and Prevention Division of Viral Diseases, Pathogen Discovery | Dakota Howard, Dhwani Batra, Peter W. Cook, Kara Moser, Adrian Paskey, Jason Caravas, Benjamin Rambo-Martin, Shatavia Morrison, Christopher Gulvick, Scott Sammons, Yvette Unoarumhi, Darlene Wagner, Matthew Schmerer, Cyndi Clark, Patrick Campbell, Rob Case, Vikramsinha Ghorpade, Holly Houdeshell, Ola Kvalvaag, Dillon Nall, Ethan Sanders, Alec Vest, Shaun Westlund, Matthew Hardison, Clinton R. Paden, Duncan MacCannell |
| EPI_ISL_2159098, EPI_ISL_2159099 | Yale Clinical Virology Lab | Yale Center for Genomic Analysis | Shrikant Mane, Kaya Bilguvar, Curt Scharfe, Irina Tikhonova, Brooke Sullivan |
| EPI_ISL_2159100 | Aegis Sciences Corporation | Centers for Disease Control and Prevention Division of Viral Diseases, Pathogen Discovery | Dakota Howard, Dhwani Batra, Peter W. Cook, Kara Moser, Adrian Paskey, Jason Caravas, Benjamin Rambo-Martin, Shatavia Morrison, Christopher Gulvick, Scott Sammons, Yvette Unoarumhi, Darlene Wagner, Matthew Schmerer, Cyndi Clark, Patrick Campbell, Rob Case, Vikramsinha Ghorpade, Holly Houdeshell, Ola Kvalvaag, Dillon Nall, Ethan Sanders, Alec Vest, Shaun Westlund, Matthew Hardison, Clinton R. Paden, Duncan MacCannell |
| EPI_ISL_2159101, EPI_ISL_2159102 | Yale Clinical Virology Lab | Yale Center for Genomic Analysis | Shrikant Mane, Kaya Bilguvar, Curt Scharfe, Irina Tikhonova, Brooke Sullivan |
| EPI_ISL_2159103 | Aegis Sciences Corporation | Centers for Disease Control and Prevention Division of Viral Diseases, Pathogen Discovery | Dakota Howard, Dhwani Batra, Peter W. Cook, Kara Moser, Adrian Paskey, Jason Caravas, Benjamin Rambo-Martin, Shatavia Morrison, Christopher Gulvick, Scott Sammons, Yvette Unoarumhi, Darlene Wagner, Matthew Schmerer, Cyndi Clark, Patrick Campbell, Rob Case, Vikramsinha Ghorpade, Holly Houdeshell, Ola Kvalvaag, Dillon Nall, Ethan Sanders, Alec Vest, Shaun Westlund, Matthew Hardison, Clinton R. Paden, Duncan MacCannell |
| EPI_ISL_2159104, EPI_ISL_2159105 | Yale Clinical Virology Lab | Yale Center for Genomic Analysis | Shrikant Mane, Kaya Bilguvar, Curt Scharfe, Irina Tikhonova, Brooke Sullivan |
| EPI_ISL_2159106 | Aegis Sciences Corporation | Centers for Disease Control and Prevention Division of Viral Diseases, Pathogen Discovery | Dakota Howard, Dhwani Batra, Peter W. Cook, Kara Moser, Adrian Paskey, Jason Caravas, Benjamin Rambo-Martin, Shatavia Morrison, Christopher Gulvick, Scott Sammons, Yvette Unoarumhi, Darlene Wagner, Matthew Schmerer, Cyndi Clark, Patrick Campbell, Rob Case, Vikramsinha Ghorpade, Holly Houdeshell, Ola Kvalvaag, Dillon Nall, Ethan Sanders, Alec Vest, Shaun Westlund, Matthew Hardison, Clinton R. Paden, Duncan MacCannell |

| | | | |
|---|---|---|---|
| EPI_ISL_2159107, EPI_ISL_2159108 | Yale Clinical Virology Lab | Yale Center for Genomic Analysis | Shrikant Mane, Kaya Bilguvar, Curt Scharfe, Irina Tikhonova, Brooke Sullivan |
| EPI_ISL_2159109 | Aegis Sciences Corporation | Centers for Disease Control and Prevention Division of Viral Diseases, Pathogen Discovery | Dakota Howard, Dhwani Batra, Peter W. Cook, Kara Moser, Adrian Paskey, Jason Caravas, Benjamin Rambo-Martin, Shatavia Morrison, Christopher Gulvick, Scott Sammons, Yvette Unoarumhi, Darlene Wagner, Matthew Schmerer, Cyndi Clark, Patrick Campbell, Rob Case, Vikramsinha Ghorpade, Holly Houdeshell, Ola Kvalvaag, Dillon Nall, Ethan Sanders, Alec Vest, Shaun Westlund, Matthew Hardison, Clinton R. Paden, Duncan MacCannell |
| EPI_ISL_2159110, EPI_ISL_2159111 | Yale Clinical Virology Lab | Yale Center for Genomic Analysis | Shrikant Mane, Kaya Bilguvar, Curt Scharfe, Irina Tikhonova, Brooke Sullivan |
| EPI_ISL_2159112 | Aegis Sciences Corporation | Centers for Disease Control and Prevention Division of Viral Diseases, Pathogen Discovery | Dakota Howard, Dhwani Batra, Peter W. Cook, Kara Moser, Adrian Paskey, Jason Caravas, Benjamin Rambo-Martin, Shatavia Morrison, Christopher Gulvick, Scott Sammons, Yvette Unoarumhi, Darlene Wagner, Matthew Schmerer, Cyndi Clark, Patrick Campbell, Rob Case, Vikramsinha Ghorpade, Holly Houdeshell, Ola Kvalvaag, Dillon Nall, Ethan Sanders, Alec Vest, Shaun Westlund, Matthew Hardison, Clinton R. Paden, Duncan MacCannell |
| EPI_ISL_2159113, EPI_ISL_2159114 | Yale Clinical Virology Lab | Yale Center for Genomic Analysis | Shrikant Mane, Kaya Bilguvar, Curt Scharfe, Irina Tikhonova, Brooke Sullivan |
| EPI_ISL_2159115, EPI_ISL_2159116, EPI_ISL_2159117, EPI_ISL_2159118, EPI_ISL_2159119, EPI_ISL_2159120, EPI_ISL_2159121, EPI_ISL_2159122, EPI_ISL_2159123, EPI_ISL_2159124, EPI_ISL_2159125, EPI_ISL_2159126, EPI_ISL_2159127, EPI_ISL_2159128, EPI_ISL_2159129, EPI_ISL_2159130, EPI_ISL_2159131, EPI_ISL_2159132, EPI_ISL_2159133, EPI_ISL_2159134, EPI_ISL_2159135, EPI_ISL_2159137, EPI_ISL_2159138, EPI_ISL_2159139, EPI_ISL_2159140, EPI_ISL_2159141, EPI_ISL_2159142, EPI_ISL_2159143, EPI_ISL_2159144, EPI_ISL_2159145, EPI_ISL_2159146, EPI_ISL_2159147, EPI_ISL_2159148, EPI_ISL_2159149, EPI_ISL_2159150, EPI_ISL_2159151, EPI_ISL_2159152, EPI_ISL_2159153, EPI_ISL_2159154, EPI_ISL_2159155, EPI_ISL_2159156, EPI_ISL_2159157, EPI_ISL_2159158, EPI_ISL_2159160, EPI_ISL_2159161, EPI_ISL_2159162, EPI_ISL_2159163, EPI_ISL_2159164, EPI_ISL_2159165, EPI_ISL_2159166, EPI_ISL_2159167, EPI_ISL_2159168, EPI_ISL_2159169, EPI_ISL_2159170, EPI_ISL_2159171, EPI_ISL_2159172, EPI_ISL_2159173, EPI_ISL_2159174, EPI_ISL_2159175, EPI_ISL_2159176, EPI_ISL_2159177, EPI_ISL_2159178, EPI_ISL_2159179, EPI_ISL_2159180, EPI_ISL_2159181, EPI_ISL_2159182, EPI_ISL_2159183, EPI_ISL_2159184, EPI_ISL_2159185, EPI_ISL_2159186, EPI_ISL_2159187, EPI_ISL_2159188, EPI_ISL_2159189, EPI_ISL_2159190, EPI_ISL_2159191, EPI_ISL_2159192, EPI_ISL_2159193, EPI_ISL_2159194, EPI_ISL_2159195, EPI_ISL_2159196, EPI_ISL_2159197, EPI_ISL_2159198, EPI_ISL_2159199, EPI_ISL_2159200, EPI_ISL_2159201, EPI_ISL_2159202, EPI_ISL_2159203, EPI_ISL_2159204, EPI_ISL_2159206, EPI_ISL_2159207, EPI_ISL_2159208, EPI_ISL_2159209, EPI_ISL_2159210, EPI_ISL_2159211, EPI_ISL_2159212, EPI_ISL_2159213, EPI_ISL_2159214, EPI_ISL_2159215, EPI_ISL_2159216, EPI_ISL_2159217, EPI_ISL_2159218, EPI_ISL_2159219, EPI_ISL_2159220, EPI_ISL_2159221, EPI_ISL_2159222, EPI_ISL_2159223, EPI_ISL_2159224, EPI_ISL_2159225, EPI_ISL_2159226, EPI_ISL_2159227, EPI_ISL_2159228, EPI_ISL_2159229, EPI_ISL_2159230, EPI_ISL_2159231, EPI_ISL_2159232, EPI_ISL_2159233, EPI_ISL_2159234, EPI_ISL_2159235, EPI_ISL_2159236, EPI_ISL_2159237, EPI_ISL_2159238, EPI_ISL_2159239, EPI_ISL_2159240, EPI_ISL_2159241, EPI_ISL_2159242, EPI_ISL_2159243, EPI_ISL_2159245, EPI_ISL_2159246, EPI_ISL_2159247, EPI_ISL_2159248, EPI_ISL_2159249, EPI_ISL_2159250, EPI_ISL_2159251, EPI_ISL_2159252, EPI_ISL_2159253, EPI_ISL_2159254, EPI_ISL_2159255, EPI_ISL_2159256, EPI_ISL_2159257, EPI_ISL_2159258, EPI_ISL_2159259, EPI_ISL_2159260, EPI_ISL_2159261, EPI_ISL_2159262, EPI_ISL_2159263, EPI_ISL_2159264, EPI_ISL_2159265, EPI_ISL_2159266, EPI_ISL_2159267, EPI_ISL_2159268, EPI_ISL_2159269, EPI_ISL_2159270, EPI_ISL_2159271, EPI_ISL_2159272, EPI_ISL_2159273, EPI_ISL_2159274, EPI_ISL_2159275, EPI_ISL_2159276, EPI_ISL_2159277, EPI_ISL_2159278, EPI_ISL_2159279, EPI_ISL_2159280, EPI_ISL_2159281, EPI_ISL_2159283, EPI_ISL_2159284, EPI_ISL_2159285, EPI_ISL_2159286, EPI_ISL_2159287, EPI_ISL_2159288, EPI_ISL_2159289, EPI_ISL_2159290, EPI_ISL_2159291, EPI_ISL_2159292, EPI_ISL_2159293, EPI_ISL_2159294, EPI_ISL_2159295, EPI_ISL_2159296, EPI_ISL_2159297, EPI_ISL_2159298, EPI_ISL_2159299, EPI_ISL_2159300, EPI_ISL_2159301, EPI_ISL_2159302, EPI_ISL_2159303, EPI_ISL_2159304, EPI_ISL_2159305, EPI_ISL_2159306, EPI_ISL_2159307, EPI_ISL_2159308, EPI_ISL_2159309, EPI_ISL_2159311, EPI_ISL_2159312, EPI_ISL_2159313, EPI_ISL_2159314, EPI_ISL_2159315, EPI_ISL_2159316, EPI_ISL_2159317, EPI_ISL_2159318, EPI_ISL_2159319, EPI_ISL_2159320, EPI_ISL_2159321, EPI_ISL_2159322, EPI_ISL_2159323, EPI_ISL_2159324, EPI_ISL_2159325, EPI_ISL_2159326, EPI_ISL_2159327, EPI_ISL_2159328, EPI_ISL_2159329, EPI_ISL_2159330, EPI_ISL_2159332, EPI_ISL_2159333, EPI_ISL_2159334, EPI_ISL_2159335, EPI_ISL_2159336, EPI_ISL_2159337, EPI_ISL_2159338, EPI_ISL_2159339, EPI_ISL_2159340, EPI_ISL_2159341, EPI_ISL_2159342, EPI_ISL_2159343, EPI_ISL_2159344, EPI_ISL_2159345, EPI_ISL_2159346, EPI_ISL_2159347, EPI_ISL_2159348, EPI_ISL_2159349, EPI_ISL_2159350, EPI_ISL_2159351, EPI_ISL_2159352, EPI_ISL_2159353, EPI_ISL_2159354, EPI_ISL_2159355, EPI_ISL_2159356, EPI_ISL_2159357, EPI_ISL_2159358, EPI_ISL_2159359, EPI_ISL_2159360, EPI_ISL_2159361, EPI_ISL_2159362, EPI_ISL_2159363, EPI_ISL_2159365, EPI_ISL_2159366, EPI_ISL_2159367, EPI_ISL_2159368, EPI_ISL_2159369, EPI_ISL_2159370, EPI_ISL_2159371, EPI_ISL_2159372, EPI_ISL_2159373, EPI_ISL_2159374, EPI_ISL_2159375, EPI_ISL_2159376, EPI_ISL_2159377, EPI_ISL_2159378, EPI_ISL_2159379, EPI_ISL_2159380, EPI_ISL_2159381, EPI_ISL_2159382, EPI_ISL_2159383, EPI_ISL_2159384, EPI_ISL_2159385, EPI_ISL_2159386, EPI_ISL_2159387, EPI_ISL_2159388, EPI_ISL_2159389, EPI_ISL_2159390, EPI_ISL_2159391, EPI_ISL_2159392, EPI_ISL_2159393, EPI_ISL_2159394, EPI_ISL_2159395, EPI_ISL_2159396, EPI_ISL_2159398, EPI_ISL_2159399, EPI_ISL_2159400, EPI_ISL_2159401, EPI_ISL_2159402, EPI_ISL_2159403, EPI_ISL_2159404, EPI_ISL_2159405, EPI_ISL_2159406, EPI_ISL_2159407, EPI_ISL_2159408, EPI_ISL_2159409, EPI_ISL_2159410, EPI_ISL_2159411, EPI_ISL_2159412, EPI_ISL_2159413, EPI_ISL_2159414, EPI_ISL_2159415, EPI_ISL_2159416, EPI_ISL_2159417, EPI_ISL_2159418, EPI_ISL_2159419, EPI_ISL_2159420, EPI_ISL_2159421, EPI_ISL_2159422, EPI_ISL_2159423, EPI_ISL_2159425, EPI_ISL_2159426, EPI_ISL_2159427, EPI_ISL_2159428, EPI_ISL_2159429, EPI_ISL_2159430, EPI_ISL_2159431, EPI_ISL_2159432, EPI_ISL_2159434, EPI_ISL_2159435, EPI_ISL_2159436, EPI_ISL_2159437, EPI_ISL_2159438, EPI_ISL_2159439, EPI_ISL_2159440, EPI_ISL_2159441, EPI_ISL_2159442, EPI_ISL_2159443, EPI_ISL_2159444, EPI_ISL_2159445, EPI_ISL_2159446, EPI_ISL_2159447, EPI_ISL_2159448, EPI_ISL_2159449, EPI_ISL_2159450, EPI_ISL_2159451, EPI_ISL_2159452, EPI_ISL_2159453, EPI_ISL_2159454, EPI_ISL_2159455, EPI_ISL_2159456, EPI_ISL_2159457, EPI_ISL_2159458, EPI_ISL_2159460, EPI_ISL_2159461, EPI_ISL_2159462, EPI_ISL_2159463, EPI_ISL_2159464, EPI_ISL_2159465, EPI_ISL_2159466, EPI_ISL_2159467, EPI_ISL_2159468, EPI_ISL_2159470 | | | |
| see above | Aegis Sciences Corporation | Centers for Disease Control and Prevention Division of Viral Diseases, Pathogen Discovery | Dakota Howard, Dhwani Batra, Peter W. Cook, Kara Moser, Adrian Paskey, Jason Caravas, Benjamin Rambo-Martin, Shatavia Morrison, Christopher Gulvick, Scott Sammons, Yvette Unoarumhi, Darlene Wagner, Matthew Schmerer, Cyndi Clark, Patrick Campbell, Rob Case, Vikramsinha Ghorpade, Holly Houdeshell, Ola Kvalvaag, Dillon Nall, Ethan Sanders, Alec Vest, Shaun Westlund, Matthew Hardison, Clinton R. Paden, Duncan MacCannell |
| EPI_ISL_2159471, EPI_ISL_2159472, EPI_ISL_2159473, EPI_ISL_2159474, EPI_ISL_2159475, EPI_ISL_2159476, EPI_ISL_2159477, EPI_ISL_2159478, EPI_ISL_2159479, EPI_ISL_2159480, EPI_ISL_2159482, EPI_ISL_2159483, EPI_ISL_2159485, EPI_ISL_2159486, EPI_ISL_2159487, EPI_ISL_2159488, EPI_ISL_2159489, EPI_ISL_2159490, EPI_ISL_2159491, EPI_ISL_2159492, EPI_ISL_2159493, EPI_ISL_2159494, EPI_ISL_2159495, EPI_ISL_2159496, EPI_ISL_2159497, EPI_ISL_2159498, EPI_ISL_2159499, EPI_ISL_2159500, EPI_ISL_2159501, EPI_ISL_2159502, EPI_ISL_2159503, EPI_ISL_2159504, EPI_ISL_2159505, EPI_ISL_2159506, EPI_ISL_2159507, EPI_ISL_2159508, EPI_ISL_2159509, EPI_ISL_2159510, EPI_ISL_2159511, EPI_ISL_2159512, EPI_ISL_2159513, EPI_ISL_2159514, EPI_ISL_2159515, EPI_ISL_2159516, EPI_ISL_2159517, EPI_ISL_2159518, EPI_ISL_2159519, EPI_ISL_2159520, EPI_ISL_2159521, EPI_ISL_2159522, EPI_ISL_2159523, EPI_ISL_2159524, EPI_ISL_2159525, EPI_ISL_2159526, EPI_ISL_2159527, EPI_ISL_2159528, EPI_ISL_2159529, EPI_ISL_2159530, EPI_ISL_2159531, EPI_ISL_2159532, EPI_ISL_2159533, EPI_ISL_2159534, EPI_ISL_2159535, EPI_ISL_2159536, EPI_ISL_2159537, EPI_ISL_2159538, EPI_ISL_2159539, EPI_ISL_2159540, EPI_ISL_2159541, EPI_ISL_2159542, EPI_ISL_2159543, EPI_ISL_2159544, EPI_ISL_2159545, EPI_ISL_2159546, EPI_ISL_2159547, EPI_ISL_2159548, EPI_ISL_2159549, EPI_ISL_2159550, EPI_ISL_2159551, EPI_ISL_2159552, EPI_ISL_2159553, EPI_ISL_2159554, EPI_ISL_2159555, EPI_ISL_2159556, EPI_ISL_2159557, EPI_ISL_2159558, EPI_ISL_2159559, EPI_ISL_2159560, EPI_ISL_2159561, EPI_ISL_2159562, EPI_ISL_2159563, EPI_ISL_2159564, EPI_ISL_2159565, EPI_ISL_2159566, EPI_ISL_2159567, EPI_ISL_2159568, EPI_ISL_2159569, EPI_ISL_2159570, EPI_ISL_2159571, EPI_ISL_2159572, EPI_ISL_2159573, EPI_ISL_2159574, EPI_ISL_2159575 | | | |
| see above | Helix/Illumina | Centers for Disease Control and Prevention Division of Viral Diseases, Pathogen Discovery | Dakota Howard, Dhwani Batra, Peter W. Cook, Kara Moser, Adrian Paskey, Jason Caravas, Benjamin Rambo-Martin, Shatavia Morrison, Christopher Gulvick, Scott Sammons, Yvette Unoarumhi, Darlene Wagner, Matthew Schmerer, Eileen de Feo, Jan Antico, Christine Tran, Matthew Tolentino, Shannon Wickline, Kim Gietzen, Brad Sickler, Jingtao Liu, Eric Allen, Phil Febbo, Nicole L. Washington, Simon White, Geraint Levan, Kelly Schiabor Barrett, Elizabeth Cirulli, Alexandre Bolze, Ary Ascencio, Charlotte Rivera-Garcia, Ryan Cho, Jason Nguyen, Sherry Wang, Jimmy Ramirez, Tyler Cassens, Efren Sandoval, Magnus Isaksson, William Lee, David Becker, Marc Laurent, James Lu, Clinton R. Paden, Duncan MacCannell |
| EPI_ISL_2159576 | Yale Clinical Virology Lab | Grubaugh Lab - Yale School of Public Health | Joseph Fauver, Mallery Breban, Isabel Ott, Tara Alpert, Mary Petrone, Anderson Brito, Chantal Vogels, Annie Watkins, Chaney Kalinich, Jessica Rothman, Marie L. Landry, Nathan Grubaugh |
| EPI_ISL_2159577 | Helix/Illumina | Centers for Disease Control and Prevention Division of Viral Diseases, Pathogen Discovery | Dakota Howard, Dhwani Batra, Peter W. Cook, Kara Moser, Adrian Paskey, Jason Caravas, Benjamin Rambo-Martin, Shatavia Morrison, Christopher Gulvick, Scott Sammons, Yvette Unoarumhi, Darlene Wagner, Matthew Schmerer, Eileen de Feo, Jan Antico, Christine Tran, Matthew Tolentino, Shannon Wickline, Kim Gietzen, Brad Sickler, Jingtao Liu, Eric Allen, Phil Febbo, Nicole L. Washington, Simon White, Geraint Levan, Kelly Schiabor Barrett, Elizabeth Cirulli, Alexandre Bolze, Ary Ascencio, Charlotte Rivera-Garcia, Ryan Cho, Jason Nguyen, Sherry Wang, Jimmy Ramirez, Tyler Cassens, Efren Sandoval, Magnus Isaksson, William Lee, David Becker, Marc Laurent, James Lu, Clinton R. Paden, Duncan MacCannell |
| EPI_ISL_2159578 | Yale Clinical Virology Lab | Grubaugh Lab - Yale School of Public Health | Joseph Fauver, Mallery Breban, Isabel Ott, Tara Alpert, Mary Petrone, Anderson Brito, Chantal Vogels, Annie Watkins, Chaney Kalinich, Jessica Rothman, Marie L. Landry, Nathan Grubaugh |
| EPI_ISL_2159579 | Helix/Illumina | Centers for Disease Control and Prevention Division of Viral Diseases, Pathogen Discovery | Dakota Howard, Dhwani Batra, Peter W. Cook, Kara Moser, Adrian Paskey, Jason Caravas, Benjamin Rambo-Martin, Shatavia Morrison, Christopher Gulvick, Scott Sammons, Yvette Unoarumhi, Darlene Wagner, Matthew Schmerer, Eileen de Feo, Jan Antico, Christine Tran, Matthew Tolentino, Shannon Wickline, Kim Gietzen, Brad Sickler, Jingtao Liu, Eric Allen, Phil Febbo, Nicole L. Washington, Simon White, Geraint Levan, Kelly Schiabor Barrett, Elizabeth Cirulli, Alexandre Bolze, Ary Ascencio, Charlotte Rivera-Garcia, Ryan Cho, Jason Nguyen, Sherry Wang, Jimmy Ramirez, Tyler Cassens, Efren Sandoval, Magnus Isaksson, William Lee, David Becker, Marc Laurent, James Lu, Clinton R. Paden, Duncan MacCannell |
| EPI_ISL_2159580, EPI_ISL_2159581 | Yale Clinical Virology Lab | Grubaugh Lab - Yale School of Public Health | Joseph Fauver, Mallery Breban, Isabel Ott, Tara Alpert, Mary Petrone, Anderson Brito, Chantal Vogels, Annie Watkins, Chaney Kalinich, Jessica Rothman, Marie L. Landry, Nathan Grubaugh |
| EPI_ISL_2159582 | Helix/Illumina | Centers for Disease Control and Prevention Division of Viral Diseases, Pathogen Discovery | Dakota Howard, Dhwani Batra, Peter W. Cook, Kara Moser, Adrian Paskey, Jason Caravas, Benjamin Rambo-Martin, Shatavia Morrison, Christopher Gulvick, Scott Sammons, Yvette Unoarumhi, Darlene Wagner, Matthew Schmerer, Eileen de Feo, Jan Antico, Christine Tran, Matthew Tolentino, Shannon Wickline, Kim Gietzen, Brad Sickler, Jingtao Liu, Eric Allen, Phil Febbo, Nicole L. Washington, Simon White, Geraint Levan, Kelly Schiabor Barrett, Elizabeth Cirulli, Alexandre Bolze, Ary Ascencio, Charlotte Rivera-Garcia, Ryan Cho, Jason Nguyen, Sherry Wang, Jimmy Ramirez, Tyler Cassens, Efren Sandoval, Magnus Isaksson, William Lee, David Becker, Marc Laurent, James Lu, Clinton R. Paden, Duncan MacCannell |
| EPI_ISL_2159583 | Yale Clinical Virology Lab | Grubaugh Lab - Yale School of Public Health | Joseph Fauver, Mallery Breban, Isabel Ott, Tara Alpert, Mary Petrone, Anderson Brito, Chantal Vogels, Annie Watkins, Chaney Kalinich, Jessica Rothman, Marie L. Landry, Nathan Grubaugh |
| EPI_ISL_2159584 | Helix/Illumina | Centers for Disease Control and Prevention Division of Viral Diseases, Pathogen Discovery | Dakota Howard, Dhwani Batra, Peter W. Cook, Kara Moser, Adrian Paskey, Jason Caravas, Benjamin Rambo-Martin, Shatavia Morrison, Christopher Gulvick, Scott Sammons, Yvette Unoarumhi, Darlene Wagner, Matthew Schmerer, Eileen de Feo, Jan Antico, Christine Tran, Matthew Tolentino, Shannon Wickline, Kim Gietzen, Brad Sickler, Jingtao Liu, Eric Allen, Phil Febbo, Nicole L. Washington, Simon White, Geraint Levan, Kelly Schiabor Barrett, Elizabeth Cirulli, Alexandre Bolze, Ary Ascencio, Charlotte Rivera-Garcia, Ryan Cho, Jason Nguyen, Sherry Wang, Jimmy Ramirez, Tyler Cassens, Efren Sandoval, Magnus Isaksson, William Lee, David Becker, Marc Laurent, James Lu, Clinton R. Paden, Duncan MacCannell |
| EPI_ISL_2159585, EPI_ISL_2159586 | Yale Clinical Virology Lab | Grubaugh Lab - Yale School of Public Health | Joseph Fauver, Mallery Breban, Isabel Ott, Tara Alpert, Mary Petrone, Anderson Brito, Chantal Vogels, Annie Watkins, Chaney Kalinich, Jessica Rothman, Marie L. Landry, Nathan Grubaugh |
| EPI_ISL_2159587 | Helix/Illumina | Centers for Disease Control and Prevention Division of Viral Diseases, Pathogen Discovery | Dakota Howard, Dhwani Batra, Peter W. Cook, Kara Moser, Adrian Paskey, Jason Caravas, Benjamin Rambo-Martin, Shatavia Morrison, Christopher Gulvick, Scott Sammons, Yvette Unoarumhi, Darlene Wagner, Matthew Schmerer, Eileen de Feo, Jan Antico, Christine Tran, Matthew Tolentino, Shannon Wickline, Kim Gietzen, Brad Sickler, Jingtao Liu, Eric Allen, Phil Febbo, Nicole L. Washington, Simon White, Geraint Levan, Kelly Schiabor Barrett, Elizabeth Cirulli, Alexandre Bolze, Ary Ascencio, Charlotte Rivera-Garcia, Ryan Cho, Jason Nguyen, Sherry Wang, Jimmy Ramirez, Tyler Cassens, Efren Sandoval, Magnus Isaksson, William Lee, David Becker, Marc Laurent, James Lu, Clinton R. Paden, Duncan MacCannell |
| EPI_ISL_2159588, EPI_ISL_2159589 | Yale Clinical Virology Lab | Grubaugh Lab - Yale School of Public Health | Joseph Fauver, Mallery Breban, Isabel Ott, Tara Alpert, Mary Petrone, Anderson Brito, Chantal Vogels, Annie Watkins, Chaney Kalinich, Jessica Rothman, Marie L. Landry, Nathan Grubaugh |
| EPI_ISL_2159590 | Helix/Illumina | Centers for Disease Control and Prevention Division of Viral Diseases, Pathogen Discovery | Dakota Howard, Dhwani Batra, Peter W. Cook, Kara Moser, Adrian Paskey, Jason Caravas, Benjamin Rambo-Martin, Shatavia Morrison, Christopher Gulvick, Scott Sammons, Yvette Unoarumhi, Darlene Wagner, Matthew Schmerer, Eileen de Feo, Jan Antico, Christine Tran, Matthew Tolentino, Shannon Wickline, Kim Gietzen, Brad Sickler, Jingtao Liu, Eric Allen, Phil Febbo, Nicole L. Washington, Simon White, Geraint Levan, Kelly Schiabor Barrett, Elizabeth Cirulli, Alexandre Bolze, Ary Ascencio, Charlotte Rivera-Garcia, Ryan Cho, Jason Nguyen, Sherry Wang, Jimmy Ramirez, Tyler Cassens, Efren Sandoval, Magnus Isaksson, William Lee, David Becker, Marc Laurent, James Lu, Clinton R. Paden, Duncan MacCannell |
| EPI_ISL_2159591 | Yale Clinical Virology Lab | Grubaugh Lab - Yale School of Public Health | Joseph Fauver, Mallery Breban, Isabel Ott, Tara Alpert, Mary Petrone, Anderson Brito, Chantal Vogels, Annie Watkins, Chaney Kalinich, Jessica Rothman, Marie L. Landry, Nathan Grubaugh |
| EPI_ISL_2159592 | Helix/Illumina | Centers for Disease Control and Prevention Division of Viral Diseases, Pathogen Discovery | Dakota Howard, Dhwani Batra, Peter W. Cook, Kara Moser, Adrian Paskey, Jason Caravas, Benjamin Rambo-Martin, Shatavia Morrison, Christopher Gulvick, Scott Sammons, Yvette Unoarumhi, Darlene Wagner, Matthew Schmerer, Eileen de Feo, Jan Antico, Christine Tran, Matthew Tolentino, Shannon Wickline, Kim Gietzen, Brad Sickler, Jingtao Liu, Eric Allen, Phil Febbo, Nicole L. Washington, Simon White, Geraint Levan, Kelly Schiabor Barrett, Elizabeth Cirulli, Alexandre Bolze, Ary Ascencio, Charlotte Rivera-Garcia, Ryan Cho, Jason Nguyen, Sherry Wang, Jimmy Ramirez, Tyler Cassens, Efren Sandoval, Magnus Isaksson, William Lee, David Becker, Marc Laurent, James Lu, Clinton R. Paden, Duncan MacCannell |
| EPI_ISL_2159593, EPI_ISL_2159594 | Yale Clinical Virology Lab | Grubaugh Lab - Yale School of Public Health | Joseph Fauver, Mallery Breban, Isabel Ott, Tara Alpert, Mary Petrone, Anderson Brito, Chantal Vogels, Annie Watkins, Chaney Kalinich, Jessica Rothman, Marie L. Landry, Nathan Grubaugh |
| EPI_ISL_2159595 | Helix/Illumina | Centers for Disease Control and Prevention Division of Viral Diseases, Pathogen Discovery | Dakota Howard, Dhwani Batra, Peter W. Cook, Kara Moser, Adrian Paskey, Jason Caravas, Benjamin Rambo-Martin, Shatavia Morrison, Christopher Gulvick, Scott Sammons, Yvette Unoarumhi, Darlene Wagner, Matthew Schmerer, Eileen de Feo, Jan Antico, Christine Tran, Matthew Tolentino, Shannon Wickline, Kim Gietzen, Brad Sickler, Jingtao Liu, Eric Allen, Phil Febbo, Nicole L. Washington, Simon White, Geraint Levan, Kelly Schiabor Barrett, Elizabeth Cirulli, Alexandre Bolze, Ary Ascencio, Charlotte Rivera-Garcia, Ryan Cho, Jason Nguyen, Sherry Wang, Jimmy Ramirez, Tyler Cassens, Efren Sandoval, Magnus Isaksson, William Lee, David Becker, Marc Laurent, James Lu, Clinton R. Paden, Duncan MacCannell |
| EPI_ISL_2159596, EPI_ISL_2159597 | Yale Clinical Virology Lab | Grubaugh Lab - Yale School of Public Health | Joseph Fauver, Mallery Breban, Isabel Ott, Tara Alpert, Mary Petrone, Anderson Brito, Chantal Vogels, Annie Watkins, Chaney Kalinich, Jessica Rothman, Marie L. Landry, Nathan Grubaugh |
| EPI_ISL_2159598 | Helix/Illumina | Centers for Disease Control and Prevention Division of Viral Diseases, Pathogen Discovery | Dakota Howard, Dhwani Batra, Peter W. Cook, Kara Moser, Adrian Paskey, Jason Caravas, Benjamin Rambo-Martin, Shatavia Morrison, Christopher Gulvick, Scott Sammons, Yvette Unoarumhi, Darlene Wagner, Matthew Schmerer, Eileen de Feo, Jan Antico, Christine Tran, Matthew Tolentino, Shannon Wickline, Kim Gietzen, Brad Sickler, Jingtao Liu, Eric Allen, Phil Febbo, Nicole L. Washington, Simon White, Geraint Levan, Kelly Schiabor Barrett, Elizabeth Cirulli, Alexandre Bolze, Ary Ascencio, Charlotte Rivera-Garcia, Ryan Cho, Jason Nguyen, Sherry Wang, Jimmy Ramirez, Tyler Cassens, Efren Sandoval, Magnus Isaksson, William Lee, David Becker, Marc Laurent, James Lu, Clinton R. Paden, Duncan MacCannell |
| EPI_ISL_2159599, EPI_ISL_2159600 | Yale Clinical Virology Lab | Grubaugh Lab - Yale School of Public Health | Joseph Fauver, Mallery Breban, Isabel Ott, Tara Alpert, Mary Petrone, Anderson Brito, Chantal Vogels, Annie Watkins, Chaney Kalinich, Jessica Rothman, Marie L. Landry, Nathan Grubaugh |
| EPI_ISL_2159601 | Helix/Illumina | Centers for Disease Control and Prevention Division of Viral Diseases, Pathogen Discovery | Dakota Howard, Dhwani Batra, Peter W. Cook, Kara Moser, Adrian Paskey, Jason Caravas, Benjamin Rambo-Martin, Shatavia Morrison, Christopher Gulvick, Scott Sammons, Yvette Unoarumhi, Darlene Wagner, Matthew Schmerer, Eileen de Feo, Jan Antico, Christine Tran, Matthew Tolentino, Shannon Wickline, Kim Gietzen, Brad Sickler, Jingtao Liu, Eric Allen, Phil Febbo, Nicole L. Washington, Simon White, Geraint Levan, Kelly Schiabor Barrett, Elizabeth Cirulli, Alexandre Bolze, Ary Ascencio, Charlotte Rivera-Garcia, Ryan Cho, Jason Nguyen, Sherry Wang, Jimmy Ramirez, Tyler Cassens, Efren Sandoval, Magnus Isaksson, William Lee, David Becker, Marc Laurent, James Lu, Clinton R. Paden, Duncan MacCannell |
| EPI_ISL_2159602, EPI_ISL_2159603 | Yale Clinical Virology Lab | Grubaugh Lab - Yale School of Public Health | Joseph Fauver, Mallery Breban, Isabel Ott, Tara Alpert, Mary Petrone, Anderson Brito, Chantal Vogels, Annie Watkins, Chaney Kalinich, Jessica Rothman, Marie L. Landry, Nathan Grubaugh |
| EPI_ISL_2159604 | Helix/Illumina | Centers for Disease Control and Prevention Division of Viral Diseases, Pathogen Discovery | Dakota Howard, Dhwani Batra, Peter W. Cook, Kara Moser, Adrian Paskey, Jason Caravas, Benjamin Rambo-Martin, Shatavia Morrison, Christopher Gulvick, Scott Sammons, Yvette Unoarumhi, Darlene Wagner, Matthew Schmerer, Eileen de Feo, Jan Antico, Christine Tran, Matthew Tolentino, Shannon Wickline, Kim Gietzen, Brad Sickler, Jingtao Liu, Eric Allen, Phil Febbo, Nicole L. Washington, Simon White, Geraint Levan, Kelly Schiabor Barrett, Elizabeth Cirulli, Alexandre Bolze, Ary Ascencio, Charlotte Rivera-Garcia, Ryan Cho, Jason Nguyen, Sherry Wang, Jimmy Ramirez, Tyler Cassens, Efren Sandoval, Magnus Isaksson, William Lee, David Becker, Marc Laurent, James Lu, Clinton R. Paden, Duncan MacCannell |
| EPI_ISL_2159605, EPI_ISL_2159606 | Yale Clinical Virology Lab | Grubaugh Lab - Yale School of Public Health | Joseph Fauver, Mallery Breban, Isabel Ott, Tara Alpert, Mary Petrone, Anderson Brito, Chantal Vogels, Annie Watkins, Chaney Kalinich, Jessica Rothman, Marie L. Landry, Nathan Grubaugh |
| EPI_ISL_2159607 | Helix/Illumina | Centers for Disease Control and Prevention Division of Viral Diseases, Pathogen Discovery | Dakota Howard, Dhwani Batra, Peter W. Cook, Kara Moser, Adrian Paskey, Jason Caravas, Benjamin Rambo-Martin, Shatavia Morrison, Christopher Gulvick, Scott Sammons, Yvette Unoarumhi, Darlene Wagner, Matthew Schmerer, Eileen de Feo, Jan Antico, Christine Tran, Matthew Tolentino, Shannon Wickline, Kim Gietzen, Brad Sickler, Jingtao Liu, Eric Allen, Phil Febbo, Nicole L. Washington, Simon White, Geraint Levan, Kelly Schiabor Barrett, Elizabeth Cirulli, Alexandre Bolze, Ary Ascencio, Charlotte Rivera-Garcia, Ryan Cho, Jason Nguyen, Sherry Wang, Jimmy Ramirez, Tyler Cassens, Efren Sandoval, Magnus Isaksson, William Lee, David Becker, Marc Laurent, James Lu, Clinton R. Paden, Duncan MacCannell |

| | | | |
|---|---|---|---|
| EPI_ISL_2159608, EPI_ISL_2159609 | Yale Clinical Virology Lab | Grubaugh Lab - Yale School of Public Health | Joseph Fauver, Mallery Breban, Isabel Ott, Tara Alpert, Mary Petrone, Anderson Brito, Chantal Vogels, Annie Watkins, Chaney Kalinich, Jessica Rothman, Marie L. Landry, Nathan Grubaugh |
| EPI_ISL_2159610 | Helix/Illumina | Centers for Disease Control and Prevention Division of Viral Diseases, Pathogen Discovery | Dakota Howard, Dhwani Batra, Peter W. Cook, Kara Moser, Adrian Paskey, Jason Caravas, Benjamin Rambo-Martin, Shatavia Morrison, Christopher Gulvick, Scott Sammons, Yvette Unoarumhi, Darlene Wagner, Matthew Schmerer, Eileen de Feo, Jan Antico, Christine Tran, Matthew Tolentino, Shannon Wickline, Kim Gietzen, Brad Sickler, Jingtao Liu, Eric Allen, Phil Febbo, Nicole L. Washington, Simon White, Geraint Levan, Kelly Schiabor Barrett, Elizabeth Cirulli, Alexandre Bolze, Ary Ascencio, Charlotte Rivera-Garcia, Ryan Cho, Jason Nguyen, Sherry Wang, Jimmy Ramirez, Tyler Cassens, Efren Sandoval, Magnus Isaksson, William Lee, David Becker, Marc Laurent, James Lu, Clinton R. Paden, Duncan MacCannell |
| EPI_ISL_2159611 | Yale Clinical Virology Lab | Grubaugh Lab - Yale School of Public Health | Joseph Fauver, Mallery Breban, Isabel Ott, Tara Alpert, Mary Petrone, Anderson Brito, Chantal Vogels, Annie Watkins, Chaney Kalinich, Jessica Rothman, Marie L. Landry, Nathan Grubaugh |
| EPI_ISL_2159612 | Helix/Illumina | Centers for Disease Control and Prevention Division of Viral Diseases, Pathogen Discovery | Dakota Howard, Dhwani Batra, Peter W. Cook, Kara Moser, Adrian Paskey, Jason Caravas, Benjamin Rambo-Martin, Shatavia Morrison, Christopher Gulvick, Scott Sammons, Yvette Unoarumhi, Darlene Wagner, Matthew Schmerer, Eileen de Feo, Jan Antico, Christine Tran, Matthew Tolentino, Shannon Wickline, Kim Gietzen, Brad Sickler, Jingtao Liu, Eric Allen, Phil Febbo, Nicole L. Washington, Simon White, Geraint Levan, Kelly Schiabor Barrett, Elizabeth Cirulli, Alexandre Bolze, Ary Ascencio, Charlotte Rivera-Garcia, Ryan Cho, Jason Nguyen, Sherry Wang, Jimmy Ramirez, Tyler Cassens, Efren Sandoval, Magnus Isaksson, William Lee, David Becker, Marc Laurent, James Lu, Clinton R. Paden, Duncan MacCannell |
| EPI_ISL_2159613 | Yale Clinical Virology Lab | Grubaugh Lab - Yale School of Public Health | Joseph Fauver, Mallery Breban, Isabel Ott, Tara Alpert, Mary Petrone, Anderson Brito, Chantal Vogels, Annie Watkins, Chaney Kalinich, Jessica Rothman, Marie L. Landry, Nathan Grubaugh |
| EPI_ISL_2159614 | Helix/Illumina | Centers for Disease Control and Prevention Division of Viral Diseases, Pathogen Discovery | Dakota Howard, Dhwani Batra, Peter W. Cook, Kara Moser, Adrian Paskey, Jason Caravas, Benjamin Rambo-Martin, Shatavia Morrison, Christopher Gulvick, Scott Sammons, Yvette Unoarumhi, Darlene Wagner, Matthew Schmerer, Eileen de Feo, Jan Antico, Christine Tran, Matthew Tolentino, Shannon Wickline, Kim Gietzen, Brad Sickler, Jingtao Liu, Eric Allen, Phil Febbo, Nicole L. Washington, Simon White, Geraint Levan, Kelly Schiabor Barrett, Elizabeth Cirulli, Alexandre Bolze, Ary Ascencio, Charlotte Rivera-Garcia, Ryan Cho, Jason Nguyen, Sherry Wang, Jimmy Ramirez, Tyler Cassens, Efren Sandoval, Magnus Isaksson, William Lee, David Becker, Marc Laurent, James Lu, Clinton R. Paden, Duncan MacCannell |
| EPI_ISL_2159615 | Yale Clinical Virology Lab | Grubaugh Lab - Yale School of Public Health | Joseph Fauver, Mallery Breban, Isabel Ott, Tara Alpert, Mary Petrone, Anderson Brito, Chantal Vogels, Annie Watkins, Chaney Kalinich, Jessica Rothman, Marie L. Landry, Nathan Grubaugh |
| EPI_ISL_2159616 | Helix/Illumina | Centers for Disease Control and Prevention Division of Viral Diseases, Pathogen Discovery | Dakota Howard, Dhwani Batra, Peter W. Cook, Kara Moser, Adrian Paskey, Jason Caravas, Benjamin Rambo-Martin, Shatavia Morrison, Christopher Gulvick, Scott Sammons, Yvette Unoarumhi, Darlene Wagner, Matthew Schmerer, Eileen de Feo, Jan Antico, Christine Tran, Matthew Tolentino, Shannon Wickline, Kim Gietzen, Brad Sickler, Jingtao Liu, Eric Allen, Phil Febbo, Nicole L. Washington, Simon White, Geraint Levan, Kelly Schiabor Barrett, Elizabeth Cirulli, Alexandre Bolze, Ary Ascencio, Charlotte Rivera-Garcia, Ryan Cho, Jason Nguyen, Sherry Wang, Jimmy Ramirez, Tyler Cassens, Efren Sandoval, Magnus Isaksson, William Lee, David Becker, Marc Laurent, James Lu, Clinton R. Paden, Duncan MacCannell |
| EPI_ISL_2159617, EPI_ISL_2159618 | Yale Clinical Virology Lab | Grubaugh Lab - Yale School of Public Health | Joseph Fauver, Mallery Breban, Isabel Ott, Tara Alpert, Mary Petrone, Anderson Brito, Chantal Vogels, Annie Watkins, Chaney Kalinich, Jessica Rothman, Marie L. Landry, Nathan Grubaugh |
| EPI_ISL_2159619, EPI_ISL_2159620 | Helix/Illumina | Centers for Disease Control and Prevention Division of Viral Diseases, Pathogen Discovery | Dakota Howard, Dhwani Batra, Peter W. Cook, Kara Moser, Adrian Paskey, Jason Caravas, Benjamin Rambo-Martin, Shatavia Morrison, Christopher Gulvick, Scott Sammons, Yvette Unoarumhi, Darlene Wagner, Matthew Schmerer, Eileen de Feo, Jan Antico, Christine Tran, Matthew Tolentino, Shannon Wickline, Kim Gietzen, Brad Sickler, Jingtao Liu, Eric Allen, Phil Febbo, Nicole L. Washington, Simon White, Geraint Levan, Kelly Schiabor Barrett, Elizabeth Cirulli, Alexandre Bolze, Ary Ascencio, Charlotte Rivera-Garcia, Ryan Cho, Jason Nguyen, Sherry Wang, Jimmy Ramirez, Tyler Cassens, Efren Sandoval, Magnus Isaksson, William Lee, David Becker, Marc Laurent, James Lu, Clinton R. Paden, Duncan MacCannell |
| EPI_ISL_2159621 | Yale Clinical Virology Lab | Grubaugh Lab - Yale School of Public Health | Joseph Fauver, Mallery Breban, Isabel Ott, Tara Alpert, Mary Petrone, Anderson Brito, Chantal Vogels, Annie Watkins, Chaney Kalinich, Jessica Rothman, Marie L. Landry, Nathan Grubaugh |
| EPI_ISL_2159622 | Helix/Illumina | Centers for Disease Control and Prevention Division of Viral Diseases, Pathogen Discovery | Dakota Howard, Dhwani Batra, Peter W. Cook, Kara Moser, Adrian Paskey, Jason Caravas, Benjamin Rambo-Martin, Shatavia Morrison, Christopher Gulvick, Scott Sammons, Yvette Unoarumhi, Darlene Wagner, Matthew Schmerer, Eileen de Feo, Jan Antico, Christine Tran, Matthew Tolentino, Shannon Wickline, Kim Gietzen, Brad Sickler, Jingtao Liu, Eric Allen, Phil Febbo, Nicole L. Washington, Simon White, Geraint Levan, Kelly Schiabor Barrett, Elizabeth Cirulli, Alexandre Bolze, Ary Ascencio, Charlotte Rivera-Garcia, Ryan Cho, Jason Nguyen, Sherry Wang, Jimmy Ramirez, Tyler Cassens, Efren Sandoval, Magnus Isaksson, William Lee, David Becker, Marc Laurent, James Lu, Clinton R. Paden, Duncan MacCannell |
| EPI_ISL_2159623 | Yale Clinical Virology Lab | Grubaugh Lab - Yale School of Public Health | Joseph Fauver, Mallery Breban, Isabel Ott, Tara Alpert, Mary Petrone, Anderson Brito, Chantal Vogels, Annie Watkins, Chaney Kalinich, Jessica Rothman, Marie L. Landry, Nathan Grubaugh |
| EPI_ISL_2159624 | Helix/Illumina | Centers for Disease Control and Prevention Division of Viral Diseases, Pathogen Discovery | Dakota Howard, Dhwani Batra, Peter W. Cook, Kara Moser, Adrian Paskey, Jason Caravas, Benjamin Rambo-Martin, Shatavia Morrison, Christopher Gulvick, Scott Sammons, Yvette Unoarumhi, Darlene Wagner, Matthew Schmerer, Eileen de Feo, Jan Antico, Christine Tran, Matthew Tolentino, Shannon Wickline, Kim Gietzen, Brad Sickler, Jingtao Liu, Eric Allen, Phil Febbo, Nicole L. Washington, Simon White, Geraint Levan, Kelly Schiabor Barrett, Elizabeth Cirulli, Alexandre Bolze, Ary Ascencio, Charlotte Rivera-Garcia, Ryan Cho, Jason Nguyen, Sherry Wang, Jimmy Ramirez, Tyler Cassens, Efren Sandoval, Magnus Isaksson, William Lee, David Becker, Marc Laurent, James Lu, Clinton R. Paden, Duncan MacCannell |
| EPI_ISL_2159625, EPI_ISL_2159626 | Yale Clinical Virology Lab | Grubaugh Lab - Yale School of Public Health | Joseph Fauver, Mallery Breban, Isabel Ott, Tara Alpert, Mary Petrone, Anderson Brito, Chantal Vogels, Annie Watkins, Chaney Kalinich, Jessica Rothman, Marie L. Landry, Nathan Grubaugh |
| EPI_ISL_2159627, EPI_ISL_2159628 | Helix/Illumina | Centers for Disease Control and Prevention Division of Viral Diseases, Pathogen Discovery | Dakota Howard, Dhwani Batra, Peter W. Cook, Kara Moser, Adrian Paskey, Jason Caravas, Benjamin Rambo-Martin, Shatavia Morrison, Christopher Gulvick, Scott Sammons, Yvette Unoarumhi, Darlene Wagner, Matthew Schmerer, Eileen de Feo, Jan Antico, Christine Tran, Matthew Tolentino, Shannon Wickline, Kim Gietzen, Brad Sickler, Jingtao Liu, Eric Allen, Phil Febbo, Nicole L. Washington, Simon White, Geraint Levan, Kelly Schiabor Barrett, Elizabeth Cirulli, Alexandre Bolze, Ary Ascencio, Charlotte Rivera-Garcia, Ryan Cho, Jason Nguyen, Sherry Wang, Jimmy Ramirez, Tyler Cassens, Efren Sandoval, Magnus Isaksson, William Lee, David Becker, Marc Laurent, James Lu, Clinton R. Paden, Duncan MacCannell |
| EPI_ISL_2159629 | Yale Clinical Virology Lab | Grubaugh Lab - Yale School of Public Health | Joseph Fauver, Mallery Breban, Isabel Ott, Tara Alpert, Mary Petrone, Anderson Brito, Chantal Vogels, Annie Watkins, Chaney Kalinich, Jessica Rothman, Marie L. Landry, Nathan Grubaugh |
| EPI_ISL_2159630 | Helix/Illumina | Centers for Disease Control and Prevention Division of Viral Diseases, Pathogen Discovery | Dakota Howard, Dhwani Batra, Peter W. Cook, Kara Moser, Adrian Paskey, Jason Caravas, Benjamin Rambo-Martin, Shatavia Morrison, Christopher Gulvick, Scott Sammons, Yvette Unoarumhi, Darlene Wagner, Matthew Schmerer, Eileen de Feo, Jan Antico, Christine Tran, Matthew Tolentino, Shannon Wickline, Kim Gietzen, Brad Sickler, Jingtao Liu, Eric Allen, Phil Febbo, Nicole L. Washington, Simon White, Geraint Levan, Kelly Schiabor Barrett, Elizabeth Cirulli, Alexandre Bolze, Ary Ascencio, Charlotte Rivera-Garcia, Ryan Cho, Jason Nguyen, Sherry Wang, Jimmy Ramirez, Tyler Cassens, Efren Sandoval, Magnus Isaksson, William Lee, David Becker, Marc Laurent, James Lu, Clinton R. Paden, Duncan MacCannell |
| EPI_ISL_2159631, EPI_ISL_2159632 | Yale Clinical Virology Lab | Grubaugh Lab - Yale School of Public Health | Joseph Fauver, Mallery Breban, Isabel Ott, Tara Alpert, Mary Petrone, Anderson Brito, Chantal Vogels, Annie Watkins, Chaney Kalinich, Jessica Rothman, Marie L. Landry, Nathan Grubaugh |
| EPI_ISL_2159633 | Helix/Illumina | Centers for Disease Control and Prevention Division of Viral Diseases, Pathogen Discovery | Dakota Howard, Dhwani Batra, Peter W. Cook, Kara Moser, Adrian Paskey, Jason Caravas, Benjamin Rambo-Martin, Shatavia Morrison, Christopher Gulvick, Scott Sammons, Yvette Unoarumhi, Darlene Wagner, Matthew Schmerer, Eileen de Feo, Jan Antico, Christine Tran, Matthew Tolentino, Shannon Wickline, Kim Gietzen, Brad Sickler, Jingtao Liu, Eric Allen, Phil Febbo, Nicole L. Washington, Simon White, Geraint Levan, Kelly Schiabor Barrett, Elizabeth Cirulli, Alexandre Bolze, Ary Ascencio, Charlotte Rivera-Garcia, Ryan Cho, Jason Nguyen, Sherry Wang, Jimmy Ramirez, Tyler Cassens, Efren Sandoval, Magnus Isaksson, William Lee, David Becker, Marc Laurent, James Lu, Clinton R. Paden, Duncan MacCannell |
| EPI_ISL_2159634 | Yale Clinical Virology Lab | Grubaugh Lab - Yale School of Public Health | Joseph Fauver, Mallery Breban, Isabel Ott, Tara Alpert, Mary Petrone, Anderson Brito, Chantal Vogels, Annie Watkins, Chaney Kalinich, Jessica Rothman, Marie L. Landry, Nathan Grubaugh |
| EPI_ISL_2159635 | Helix/Illumina | Centers for Disease Control and Prevention Division of Viral Diseases, Pathogen Discovery | Dakota Howard, Dhwani Batra, Peter W. Cook, Kara Moser, Adrian Paskey, Jason Caravas, Benjamin Rambo-Martin, Shatavia Morrison, Christopher Gulvick, Scott Sammons, Yvette Unoarumhi, Darlene Wagner, Matthew Schmerer, Eileen de Feo, Jan Antico, Christine Tran, Matthew Tolentino, Shannon Wickline, Kim Gietzen, Brad Sickler, Jingtao Liu, Eric Allen, Phil Febbo, Nicole L. Washington, Simon White, Geraint Levan, Kelly Schiabor Barrett, Elizabeth Cirulli, Alexandre Bolze, Ary Ascencio, Charlotte Rivera-Garcia, Ryan Cho, Jason Nguyen, Sherry Wang, Jimmy Ramirez, Tyler Cassens, Efren Sandoval, Magnus Isaksson, William Lee, David Becker, Marc Laurent, James Lu, Clinton R. Paden, Duncan MacCannell |
| EPI_ISL_2159636, EPI_ISL_2159637 | Yale Clinical Virology Lab | Grubaugh Lab - Yale School of Public Health | Joseph Fauver, Mallery Breban, Isabel Ott, Tara Alpert, Mary Petrone, Anderson Brito, Chantal Vogels, Annie Watkins, Chaney Kalinich, Jessica Rothman, Marie L. Landry, Nathan Grubaugh |
| EPI_ISL_2159638 | Helix/Illumina | Centers for Disease Control and Prevention Division of Viral Diseases, Pathogen Discovery | Dakota Howard, Dhwani Batra, Peter W. Cook, Kara Moser, Adrian Paskey, Jason Caravas, Benjamin Rambo-Martin, Shatavia Morrison, Christopher Gulvick, Scott Sammons, Yvette Unoarumhi, Darlene Wagner, Matthew Schmerer, Eileen de Feo, Jan Antico, Christine Tran, Matthew Tolentino, Shannon Wickline, Kim Gietzen, Brad Sickler, Jingtao Liu, Eric Allen, Phil Febbo, Nicole L. Washington, Simon White, Geraint Levan, Kelly Schiabor Barrett, Elizabeth Cirulli, Alexandre Bolze, Ary Ascencio, Charlotte Rivera-Garcia, Ryan Cho, Jason Nguyen, Sherry Wang, Jimmy Ramirez, Tyler Cassens, Efren Sandoval, Magnus Isaksson, William Lee, David Becker, Marc Laurent, James Lu, Clinton R. Paden, Duncan MacCannell |
| EPI_ISL_2159639, EPI_ISL_2159640 | Yale Clinical Virology Lab | Grubaugh Lab - Yale School of Public Health | Joseph Fauver, Mallery Breban, Isabel Ott, Tara Alpert, Mary Petrone, Anderson Brito, Chantal Vogels, Annie Watkins, Chaney Kalinich, Jessica Rothman, Marie L. Landry, Nathan Grubaugh |
| EPI_ISL_2159641 | Helix/Illumina | Centers for Disease Control and Prevention Division of Viral Diseases, Pathogen Discovery | Dakota Howard, Dhwani Batra, Peter W. Cook, Kara Moser, Adrian Paskey, Jason Caravas, Benjamin Rambo-Martin, Shatavia Morrison, Christopher Gulvick, Scott Sammons, Yvette Unoarumhi, Darlene Wagner, Matthew Schmerer, Eileen de Feo, Jan Antico, Christine Tran, Matthew Tolentino, Shannon Wickline, Kim Gietzen, Brad Sickler, Jingtao Liu, Eric Allen, Phil Febbo, Nicole L. Washington, Simon White, Geraint Levan, Kelly Schiabor Barrett, Elizabeth Cirulli, Alexandre Bolze, Ary Ascencio, Charlotte Rivera-Garcia, Ryan Cho, Jason Nguyen, Sherry Wang, Jimmy Ramirez, Tyler Cassens, Efren Sandoval, Magnus Isaksson, William Lee, David Becker, Marc Laurent, James Lu, Clinton R. Paden, Duncan MacCannell |
| EPI_ISL_2159643 | Yale Clinical Virology Lab | Grubaugh Lab - Yale School of Public Health | Joseph Fauver, Mallery Breban, Isabel Ott, Tara Alpert, Mary Petrone, Anderson Brito, Chantal Vogels, Annie Watkins, Chaney Kalinich, Jessica Rothman, Marie L. Landry, Nathan Grubaugh |
| EPI_ISL_2159644 | Helix/Illumina | Centers for Disease Control and Prevention Division of Viral Diseases, Pathogen Discovery | Dakota Howard, Dhwani Batra, Peter W. Cook, Kara Moser, Adrian Paskey, Jason Caravas, Benjamin Rambo-Martin, Shatavia Morrison, Christopher Gulvick, Scott Sammons, Yvette Unoarumhi, Darlene Wagner, Matthew Schmerer, Eileen de Feo, Jan Antico, Christine Tran, Matthew Tolentino, Shannon Wickline, Kim Gietzen, Brad Sickler, Jingtao Liu, Eric Allen, Phil Febbo, Nicole L. Washington, Simon White, Geraint Levan, Kelly Schiabor Barrett, Elizabeth Cirulli, Alexandre Bolze, Ary Ascencio, Charlotte Rivera-Garcia, Ryan Cho, Jason Nguyen, Sherry Wang, Jimmy Ramirez, Tyler Cassens, Efren Sandoval, Magnus Isaksson, William Lee, David Becker, Marc Laurent, James Lu, Clinton R. Paden, Duncan MacCannell |
| EPI_ISL_2159645, EPI_ISL_2159646 | Yale Clinical Virology Lab | Grubaugh Lab - Yale School of Public Health | Joseph Fauver, Mallery Breban, Isabel Ott, Tara Alpert, Mary Petrone, Anderson Brito, Chantal Vogels, Annie Watkins, Chaney Kalinich, Jessica Rothman, Marie L. Landry, Nathan Grubaugh |
| EPI_ISL_2159647 | Helix/Illumina | Centers for Disease Control and Prevention Division of Viral Diseases, Pathogen Discovery | Dakota Howard, Dhwani Batra, Peter W. Cook, Kara Moser, Adrian Paskey, Jason Caravas, Benjamin Rambo-Martin, Shatavia Morrison, Christopher Gulvick, Scott Sammons, Yvette Unoarumhi, Darlene Wagner, Matthew Schmerer, Eileen de Feo, Jan Antico, Christine Tran, Matthew Tolentino, Shannon Wickline, Kim Gietzen, Brad Sickler, Jingtao Liu, Eric Allen, Phil Febbo, Nicole L. Washington, Simon White, Geraint Levan, Kelly Schiabor Barrett, Elizabeth Cirulli, Alexandre Bolze, Ary Ascencio, Charlotte Rivera-Garcia, Ryan Cho, Jason Nguyen, Sherry Wang, Jimmy Ramirez, Tyler Cassens, Efren Sandoval, Magnus Isaksson, William Lee, David Becker, Marc Laurent, James Lu, Clinton R. Paden, Duncan MacCannell |
| EPI_ISL_2159648, EPI_ISL_2159649 | Yale Clinical Virology Lab | Grubaugh Lab - Yale School of Public Health | Joseph Fauver, Mallery Breban, Isabel Ott, Tara Alpert, Mary Petrone, Anderson Brito, Chantal Vogels, Annie Watkins, Chaney Kalinich, Jessica Rothman, Marie L. Landry, Nathan Grubaugh |
| EPI_ISL_2159650 | Helix/Illumina | Centers for Disease Control and Prevention Division of Viral Diseases, Pathogen Discovery | Dakota Howard, Dhwani Batra, Peter W. Cook, Kara Moser, Adrian Paskey, Jason Caravas, Benjamin Rambo-Martin, Shatavia Morrison, Christopher Gulvick, Scott Sammons, Yvette Unoarumhi, Darlene Wagner, Matthew Schmerer, Eileen de Feo, Jan Antico, Christine Tran, Matthew Tolentino, Shannon Wickline, Kim Gietzen, Brad Sickler, Jingtao Liu, Eric Allen, Phil Febbo, Nicole L. Washington, Simon White, Geraint Levan, Kelly Schiabor Barrett, Elizabeth Cirulli, Alexandre Bolze, Ary Ascencio, Charlotte Rivera-Garcia, Ryan Cho, Jason Nguyen, Sherry Wang, Jimmy Ramirez, Tyler Cassens, Efren Sandoval, Magnus Isaksson, William Lee, David Becker, Marc Laurent, James Lu, Clinton R. Paden, Duncan MacCannell |
| EPI_ISL_2159651 | Yale Clinical Virology Lab | Grubaugh Lab - Yale School of Public Health | Joseph Fauver, Mallery Breban, Isabel Ott, Tara Alpert, Mary Petrone, Anderson Brito, Chantal Vogels, Annie Watkins, Chaney Kalinich, Jessica Rothman, Marie L. Landry, Nathan Grubaugh |
| EPI_ISL_2159652 | Helix/Illumina | Centers for Disease Control and Prevention Division of Viral Diseases, Pathogen Discovery | Dakota Howard, Dhwani Batra, Peter W. Cook, Kara Moser, Adrian Paskey, Jason Caravas, Benjamin Rambo-Martin, Shatavia Morrison, Christopher Gulvick, Scott Sammons, Yvette Unoarumhi, Darlene Wagner, Matthew Schmerer, Eileen de Feo, Jan Antico, Christine Tran, Matthew Tolentino, Shannon Wickline, Kim Gietzen, Brad Sickler, Jingtao Liu, Eric Allen, Phil Febbo, Nicole L. Washington, Simon White, Geraint Levan, Kelly Schiabor Barrett, Elizabeth Cirulli, Alexandre Bolze, Ary Ascencio, Charlotte Rivera-Garcia, Ryan Cho, Jason Nguyen, Sherry Wang, Jimmy Ramirez, Tyler Cassens, Efren Sandoval, Magnus Isaksson, William Lee, David Becker, Marc Laurent, James Lu, Clinton R. Paden, Duncan MacCannell |
| EPI_ISL_2159653, EPI_ISL_2159654 | Yale Clinical Virology Lab | Grubaugh Lab - Yale School of Public Health | Joseph Fauver, Mallery Breban, Isabel Ott, Tara Alpert, Mary Petrone, Anderson Brito, Chantal Vogels, Annie Watkins, Chaney Kalinich, Jessica Rothman, Marie L. Landry, Nathan Grubaugh |
| EPI_ISL_2159655 | Helix/Illumina | Centers for Disease Control and Prevention Division of Viral Diseases, Pathogen Discovery | Dakota Howard, Dhwani Batra, Peter W. Cook, Kara Moser, Adrian Paskey, Jason Caravas, Benjamin Rambo-Martin, Shatavia Morrison, Christopher Gulvick, Scott Sammons, Yvette Unoarumhi, Darlene Wagner, Matthew Schmerer, Eileen de Feo, Jan Antico, Christine Tran, Matthew Tolentino, Shannon Wickline, Kim Gietzen, Brad Sickler, Jingtao Liu, Eric Allen, Phil Febbo, Nicole L. Washington, Simon White, Geraint Levan, Kelly Schiabor Barrett, Elizabeth Cirulli, Alexandre Bolze, Ary Ascencio, Charlotte Rivera-Garcia, Ryan Cho, Jason Nguyen, Sherry Wang, Jimmy Ramirez, Tyler Cassens, Efren Sandoval, Magnus Isaksson, William Lee, David Becker, Marc Laurent, James Lu, Clinton R. Paden, Duncan MacCannell |
| EPI_ISL_2159656, EPI_ISL_2159657 | Yale Clinical Virology Lab | Grubaugh Lab - Yale School of Public Health | Joseph Fauver, Mallery Breban, Isabel Ott, Tara Alpert, Mary Petrone, Anderson Brito, Chantal Vogels, Annie Watkins, Chaney Kalinich, Jessica Rothman, Marie L. Landry, Nathan Grubaugh |
| EPI_ISL_2159658 | Helix/Illumina | Centers for Disease Control and Prevention Division of Viral Diseases, Pathogen Discovery | Dakota Howard, Dhwani Batra, Peter W. Cook, Kara Moser, Adrian Paskey, Jason Caravas, Benjamin Rambo-Martin, Shatavia Morrison, Christopher Gulvick, Scott Sammons, Yvette Unoarumhi, Darlene Wagner, Matthew Schmerer, Eileen de Feo, Jan Antico, Christine Tran, Matthew Tolentino, Shannon Wickline, Kim Gietzen, Brad Sickler, Jingtao Liu, Eric Allen, Phil Febbo, Nicole L. Washington, Simon White, Geraint Levan, Kelly Schiabor Barrett, Elizabeth Cirulli, Alexandre Bolze, Ary Ascencio, Charlotte Rivera-Garcia, Ryan Cho, Jason Nguyen, Sherry Wang, Jimmy Ramirez, Tyler Cassens, Efren Sandoval, Magnus Isaksson, William Lee, David Becker, Marc Laurent, James Lu, Clinton R. Paden, Duncan MacCannell |
| EPI_ISL_2159659 | Yale Clinical Virology Lab | Grubaugh Lab - Yale School of Public Health | Joseph Fauver, Mallery Breban, Isabel Ott, Tara Alpert, Mary Petrone, Anderson Brito, Chantal Vogels, Annie Watkins, Chaney Kalinich, Jessica Rothman, Marie L. Landry, Nathan Grubaugh |
| EPI_ISL_2159660 | Helix/Illumina | Centers for Disease Control and Prevention Division of Viral Diseases, Pathogen Discovery | Dakota Howard, Dhwani Batra, Peter W. Cook, Kara Moser, Adrian Paskey, Jason Caravas, Benjamin Rambo-Martin, Shatavia Morrison, Christopher Gulvick, Scott Sammons, Yvette Unoarumhi, Darlene Wagner, Matthew Schmerer, Eileen de Feo, Jan Antico, Christine Tran, Matthew Tolentino, Shannon Wickline, Kim Gietzen, Brad Sickler, Jingtao Liu, Eric Allen, Phil Febbo, Nicole L. Washington, Simon White, Geraint Levan, Kelly Schiabor Barrett, Elizabeth Cirulli, Alexandre Bolze, Ary Ascencio, Charlotte Rivera-Garcia, Ryan Cho, Jason Nguyen, Sherry Wang, Jimmy Ramirez, Tyler Cassens, Efren Sandoval, Magnus Isaksson, William Lee, David Becker, Marc Laurent, James Lu, Clinton R. Paden, Duncan MacCannell |
| EPI_ISL_2159661, EPI_ISL_2159662 | Yale Clinical Virology Lab | Grubaugh Lab - Yale School of Public Health | Joseph Fauver, Mallery Breban, Isabel Ott, Tara Alpert, Mary Petrone, Anderson Brito, Chantal Vogels, Annie Watkins, Chaney Kalinich, Jessica Rothman, Marie L. Landry, Nathan Grubaugh |

| | | | |
|---|---|---|---|
| EPI_ISL_2159663 | Helix/Illumina | Centers for Disease Control and Prevention Division of Viral Diseases, Pathogen Discovery | Dakota Howard, Dhwani Batra, Peter W. Cook, Kara Moser, Adrian Paskey, Jason Caravas, Benjamin Rambo-Martin, Shatavia Morrison, Christopher Gulvick, Scott Sammons, Yvette Unoarumhi, Darlene Wagner, Matthew Schmerer, Eileen de Feo, Jan Antico, Christine Tran, Matthew Tolentino, Shannon Wickline, Kim Gietzen, Brad Sickler, Jingtao Liu, Eric Allen, Phil Febbo, Nicole L. Washington, Simon White, Geraint Levan, Kelly Schiabor Barrett, Elizabeth Cirulli, Alexandre Bolze, Ary Ascencio, Charlotte Rivera-Garcia, Ryan Cho, Jason Nguyen, Sherry Wang, Jimmy Ramirez, Tyler Cassens, Efren Sandoval, Magnus Isaksson, William Lee, David Becker, Marc Laurent, James Lu, Clinton R. Paden, Duncan MacCannell |
| EPI_ISL_2159664, EPI_ISL_2159665 | Yale Clinical Virology Lab | Grubaugh Lab - Yale School of Public Health | Joseph Fauver, Mallery Breban, Isabel Ott, Tara Alpert, Mary Petrone, Anderson Brito, Chantal Vogels, Annie Watkins, Chaney Kalinich, Jessica Rothman, Marie L. Landry, Nathan Grubaugh |
| EPI_ISL_2159666 | Helix/Illumina | Centers for Disease Control and Prevention Division of Viral Diseases, Pathogen Discovery | Dakota Howard, Dhwani Batra, Peter W. Cook, Kara Moser, Adrian Paskey, Jason Caravas, Benjamin Rambo-Martin, Shatavia Morrison, Christopher Gulvick, Scott Sammons, Yvette Unoarumhi, Darlene Wagner, Matthew Schmerer, Eileen de Feo, Jan Antico, Christine Tran, Matthew Tolentino, Shannon Wickline, Kim Gietzen, Brad Sickler, Jingtao Liu, Eric Allen, Phil Febbo, Nicole L. Washington, Simon White, Geraint Levan, Kelly Schiabor Barrett, Elizabeth Cirulli, Alexandre Bolze, Ary Ascencio, Charlotte Rivera-Garcia, Ryan Cho, Jason Nguyen, Sherry Wang, Jimmy Ramirez, Tyler Cassens, Efren Sandoval, Magnus Isaksson, William Lee, David Becker, Marc Laurent, James Lu, Clinton R. Paden, Duncan MacCannell |
| EPI_ISL_2159667 | Yale Clinical Virology Lab | Grubaugh Lab - Yale School of Public Health | Joseph Fauver, Mallery Breban, Isabel Ott, Tara Alpert, Mary Petrone, Anderson Brito, Chantal Vogels, Annie Watkins, Chaney Kalinich, Jessica Rothman, Marie L. Landry, Nathan Grubaugh |
| EPI_ISL_2159668 | Murphy Medical Association | Grubaugh Lab - Yale School of Public Health | Joseph Fauver, Mallery Breban, Isabel Ott, Tara Alpert, Mary Petrone, Anderson Brito, Chantal Vogels, Annie Watkins, Chaney Kalinich, Jessica Rothman, Caleb Neal, Eva Laszlo, Steven Murphy, Nathan Grubaugh |
| EPI_ISL_2159669 | Helix/Illumina | Centers for Disease Control and Prevention Division of Viral Diseases, Pathogen Discovery | Dakota Howard, Dhwani Batra, Peter W. Cook, Kara Moser, Adrian Paskey, Jason Caravas, Benjamin Rambo-Martin, Shatavia Morrison, Christopher Gulvick, Scott Sammons, Yvette Unoarumhi, Darlene Wagner, Matthew Schmerer, Eileen de Feo, Jan Antico, Christine Tran, Matthew Tolentino, Shannon Wickline, Kim Gietzen, Brad Sickler, Jingtao Liu, Eric Allen, Phil Febbo, Nicole L. Washington, Simon White, Geraint Levan, Kelly Schiabor Barrett, Elizabeth Cirulli, Alexandre Bolze, Ary Ascencio, Charlotte Rivera-Garcia, Ryan Cho, Jason Nguyen, Sherry Wang, Jimmy Ramirez, Tyler Cassens, Efren Sandoval, Magnus Isaksson, William Lee, David Becker, Marc Laurent, James Lu, Clinton R. Paden, Duncan MacCannell |
| EPI_ISL_2159670 | Murphy Medical Association | Grubaugh Lab - Yale School of Public Health | Joseph Fauver, Mallery Breban, Isabel Ott, Tara Alpert, Mary Petrone, Anderson Brito, Chantal Vogels, Annie Watkins, Chaney Kalinich, Jessica Rothman, Caleb Neal, Eva Laszlo, Steven Murphy, Nathan Grubaugh |
| EPI_ISL_2159671 | Helix/Illumina | Centers for Disease Control and Prevention Division of Viral Diseases, Pathogen Discovery | Dakota Howard, Dhwani Batra, Peter W. Cook, Kara Moser, Adrian Paskey, Jason Caravas, Benjamin Rambo-Martin, Shatavia Morrison, Christopher Gulvick, Scott Sammons, Yvette Unoarumhi, Darlene Wagner, Matthew Schmerer, Eileen de Feo, Jan Antico, Christine Tran, Matthew Tolentino, Shannon Wickline, Kim Gietzen, Brad Sickler, Jingtao Liu, Eric Allen, Phil Febbo, Nicole L. Washington, Simon White, Geraint Levan, Kelly Schiabor Barrett, Elizabeth Cirulli, Alexandre Bolze, Ary Ascencio, Charlotte Rivera-Garcia, Ryan Cho, Jason Nguyen, Sherry Wang, Jimmy Ramirez, Tyler Cassens, Efren Sandoval, Magnus Isaksson, William Lee, David Becker, Marc Laurent, James Lu, Clinton R. Paden, Duncan MacCannell |
| EPI_ISL_2159672 | Murphy Medical Association | Grubaugh Lab - Yale School of Public Health | Joseph Fauver, Mallery Breban, Isabel Ott, Tara Alpert, Mary Petrone, Anderson Brito, Chantal Vogels, Annie Watkins, Chaney Kalinich, Jessica Rothman, Caleb Neal, Eva Laszlo, Steven Murphy, Nathan Grubaugh |
| EPI_ISL_2159673 | Yale Pathology Lab | Grubaugh Lab - Yale School of Public Health | Joseph Fauver, Mallery Breban, Isabel Ott, Tara Alpert, Mary Petrone, Anderson Brito, Chantal Vogels, Annie Watkins, Chaney Kalinich, Jessica Rothman, Jianhui Wang, Chen Liu, Pei Hui, Nathan Grubaugh |
| EPI_ISL_2159674 | Helix/Illumina | Centers for Disease Control and Prevention Division of Viral Diseases, Pathogen Discovery | Dakota Howard, Dhwani Batra, Peter W. Cook, Kara Moser, Adrian Paskey, Jason Caravas, Benjamin Rambo-Martin, Shatavia Morrison, Christopher Gulvick, Scott Sammons, Yvette Unoarumhi, Darlene Wagner, Matthew Schmerer, Eileen de Feo, Jan Antico, Christine Tran, Matthew Tolentino, Shannon Wickline, Kim Gietzen, Brad Sickler, Jingtao Liu, Eric Allen, Phil Febbo, Nicole L. Washington, Simon White, Geraint Levan, Kelly Schiabor Barrett, Elizabeth Cirulli, Alexandre Bolze, Ary Ascencio, Charlotte Rivera-Garcia, Ryan Cho, Jason Nguyen, Sherry Wang, Jimmy Ramirez, Tyler Cassens, Efren Sandoval, Magnus Isaksson, William Lee, David Becker, Marc Laurent, James Lu, Clinton R. Paden, Duncan MacCannell |
| EPI_ISL_2159675 | Yale Pathology Lab | Grubaugh Lab - Yale School of Public Health | Joseph Fauver, Mallery Breban, Isabel Ott, Tara Alpert, Mary Petrone, Anderson Brito, Chantal Vogels, Annie Watkins, Chaney Kalinich, Jessica Rothman, Jianhui Wang, Chen Liu, Pei Hui, Nathan Grubaugh |
| EPI_ISL_2159676 | Grubaugh Lab - Yale School of Public Health | Grubaugh Lab - Yale School of Public Health | Joseph Fauver, Mallery Breban, Isabel Ott, Tara Alpert, Mary Petrone, Anderson Brito, Chantal Vogels, Annie Watkins, Chaney Kalinich, Jessica Rothman, Nathan Grubaugh |
| EPI_ISL_2159677, EPI_ISL_2159678, EPI_ISL_2159679, EPI_ISL_2159680, EPI_ISL_2159681, EPI_ISL_2159682, EPI_ISL_2159683, EPI_ISL_2159684, EPI_ISL_2159685, EPI_ISL_2159686, EPI_ISL_2159687, EPI_ISL_2159688, EPI_ISL_2159689, EPI_ISL_2159690, EPI_ISL_2159691, EPI_ISL_2159692, EPI_ISL_2159693, EPI_ISL_2159694, EPI_ISL_2159695, EPI_ISL_2159696, EPI_ISL_2159697, EPI_ISL_2159698, EPI_ISL_2159699, EPI_ISL_2159700, EPI_ISL_2159701, EPI_ISL_2159702, EPI_ISL_2159703, EPI_ISL_2159704, EPI_ISL_2159705, EPI_ISL_2159706, EPI_ISL_2159707, EPI_ISL_2159708, EPI_ISL_2159709, EPI_ISL_2159710, EPI_ISL_2159711, EPI_ISL_2159712, EPI_ISL_2159713, EPI_ISL_2159714, EPI_ISL_2159715, EPI_ISL_2159716, EPI_ISL_2159717, EPI_ISL_2159718, EPI_ISL_2159719, EPI_ISL_2159720, EPI_ISL_2159721, EPI_ISL_2159722, EPI_ISL_2159723, EPI_ISL_2159724, EPI_ISL_2159725, EPI_ISL_2159726, EPI_ISL_2159727, EPI_ISL_2159728, EPI_ISL_2159729, EPI_ISL_2159730, EPI_ISL_2159731, EPI_ISL_2159732, EPI_ISL_2159733, EPI_ISL_2159734, EPI_ISL_2159735, EPI_ISL_2159736, EPI_ISL_2159737, EPI_ISL_2159738, EPI_ISL_2159739, EPI_ISL_2159740, EPI_ISL_2159741, EPI_ISL_2159742, EPI_ISL_2159743, EPI_ISL_2159744, EPI_ISL_2159745, EPI_ISL_2159746, EPI_ISL_2159747, EPI_ISL_2159748, EPI_ISL_2159749, EPI_ISL_2159750, EPI_ISL_2159751, EPI_ISL_2159752, EPI_ISL_2159753, EPI_ISL_2159754, EPI_ISL_2159755, EPI_ISL_2159756, EPI_ISL_2159757, EPI_ISL_2159758, EPI_ISL_2159759, EPI_ISL_2159760, EPI_ISL_2159761, EPI_ISL_2159762, EPI_ISL_2159763, EPI_ISL_2159764, EPI_ISL_2159765, EPI_ISL_2159766, EPI_ISL_2159767, EPI_ISL_2159768, EPI_ISL_2159769, EPI_ISL_2159770, EPI_ISL_2159771, EPI_ISL_2159772, EPI_ISL_2159774, EPI_ISL_2159775, EPI_ISL_2159776, EPI_ISL_2159777, EPI_ISL_2159778, EPI_ISL_2159779, EPI_ISL_2159780, EPI_ISL_2159781, EPI_ISL_2159782, EPI_ISL_2159783, EPI_ISL_2159784, EPI_ISL_2159785, EPI_ISL_2159786, EPI_ISL_2159787, EPI_ISL_2159788, EPI_ISL_2159789, EPI_ISL_2159790, EPI_ISL_2159791, EPI_ISL_2159792, EPI_ISL_2159793, EPI_ISL_2159794, EPI_ISL_2159795, EPI_ISL_2159796, EPI_ISL_2159797, EPI_ISL_2159798, EPI_ISL_2159799, EPI_ISL_2159800, EPI_ISL_2159801, EPI_ISL_2159802, EPI_ISL_2159803, EPI_ISL_2159804, EPI_ISL_2159805, EPI_ISL_2159806, EPI_ISL_2159807, EPI_ISL_2159808, EPI_ISL_2159809, EPI_ISL_2159810, EPI_ISL_2159811, EPI_ISL_2159812, EPI_ISL_2159813, EPI_ISL_2159814, EPI_ISL_2159815, EPI_ISL_2159816, EPI_ISL_2159818, EPI_ISL_2159819, EPI_ISL_2159820, EPI_ISL_2159821, EPI_ISL_2159822, EPI_ISL_2159823, EPI_ISL_2159824, EPI_ISL_2159825, EPI_ISL_2159826, EPI_ISL_2159827, EPI_ISL_2159828, EPI_ISL_2159829, EPI_ISL_2159830, EPI_ISL_2159831, EPI_ISL_2159832, EPI_ISL_2159833, EPI_ISL_2159834, EPI_ISL_2159835, EPI_ISL_2159836, EPI_ISL_2159837, EPI_ISL_2159838, EPI_ISL_2159839, EPI_ISL_2159840, EPI_ISL_2159841, EPI_ISL_2159842, EPI_ISL_2159843, EPI_ISL_2159844, EPI_ISL_2159845, EPI_ISL_2159846, EPI_ISL_2159847, EPI_ISL_2159848, EPI_ISL_2159849, EPI_ISL_2159850, EPI_ISL_2159851, EPI_ISL_2159852, EPI_ISL_2159853, EPI_ISL_2159854, EPI_ISL_2159855, EPI_ISL_2159856, EPI_ISL_2159857, EPI_ISL_2159858, EPI_ISL_2159859, EPI_ISL_2159860, EPI_ISL_2159861, EPI_ISL_2159862, EPI_ISL_2159863, EPI_ISL_2159864, EPI_ISL_2159865, EPI_ISL_2159866, EPI_ISL_2159867, EPI_ISL_2159868, EPI_ISL_2159869, EPI_ISL_2159870, EPI_ISL_2159871, EPI_ISL_2159872, EPI_ISL_2159873, EPI_ISL_2159874, EPI_ISL_2159875, EPI_ISL_2159876, EPI_ISL_2159877, EPI_ISL_2159878, EPI_ISL_2159879, EPI_ISL_2159880, EPI_ISL_2159881, EPI_ISL_2159882, EPI_ISL_2159883, EPI_ISL_2159884, EPI_ISL_2159885, EPI_ISL_2159886, EPI_ISL_2159887, EPI_ISL_2159888, EPI_ISL_2159889, EPI_ISL_2159890, EPI_ISL_2159891, EPI_ISL_2159892, EPI_ISL_2159893, EPI_ISL_2159894, EPI_ISL_2159895, EPI_ISL_2159896, EPI_ISL_2159897, EPI_ISL_2159898, EPI_ISL_2159899, EPI_ISL_2159900, EPI_ISL_2159901, EPI_ISL_2159902, EPI_ISL_2159903, EPI_ISL_2159904, EPI_ISL_2159905, EPI_ISL_2159906, EPI_ISL_2159907, EPI_ISL_2159908, EPI_ISL_2159909, EPI_ISL_2159910, EPI_ISL_2159911, EPI_ISL_2159912, EPI_ISL_2159913, EPI_ISL_2159914, EPI_ISL_2159915, EPI_ISL_2159916, EPI_ISL_2159917, EPI_ISL_2159918, EPI_ISL_2159919, EPI_ISL_2159920, EPI_ISL_2159921, EPI_ISL_2159922, EPI_ISL_2159923, EPI_ISL_2159924, EPI_ISL_2159925, EPI_ISL_2159926, EPI_ISL_2159927, EPI_ISL_2159928, EPI_ISL_2159929, EPI_ISL_2159930, EPI_ISL_2159931, EPI_ISL_2159932, EPI_ISL_2159933, EPI_ISL_2159934, EPI_ISL_2159935, EPI_ISL_2159936, EPI_ISL_2159937, EPI_ISL_2159938, EPI_ISL_2159939, EPI_ISL_2159940, EPI_ISL_2159941, EPI_ISL_2159942, EPI_ISL_2159943, EPI_ISL_2159944, EPI_ISL_2159945, EPI_ISL_2159946, EPI_ISL_2159947, EPI_ISL_2159948, EPI_ISL_2159949, EPI_ISL_2159950, EPI_ISL_2159951, EPI_ISL_2159952, EPI_ISL_2159953, EPI_ISL_2159954, EPI_ISL_2159955, EPI_ISL_2159956, EPI_ISL_2159957, EPI_ISL_2159958, EPI_ISL_2159959, EPI_ISL_2159960, EPI_ISL_2159961, EPI_ISL_2159962, EPI_ISL_2159963, EPI_ISL_2159964, EPI_ISL_2159965, EPI_ISL_2159966, EPI_ISL_2159967, EPI_ISL_2159968, EPI_ISL_2159969, EPI_ISL_2159970, EPI_ISL_2159971, EPI_ISL_2159972, EPI_ISL_2159973, EPI_ISL_2159974, EPI_ISL_2159975, EPI_ISL_2159976, EPI_ISL_2159977, EPI_ISL_2159978, EPI_ISL_2159979, EPI_ISL_2159980, EPI_ISL_2159981, EPI_ISL_2159982, EPI_ISL_2159983, EPI_ISL_2159984, EPI_ISL_2159985, EPI_ISL_2159986, EPI_ISL_2159987, EPI_ISL_2159988, EPI_ISL_2159989, EPI_ISL_2159990, EPI_ISL_2159991, EPI_ISL_2159992, EPI_ISL_2159993, EPI_ISL_2159994, EPI_ISL_2159995, EPI_ISL_2159996, EPI_ISL_2159997, EPI_ISL_2159998, EPI_ISL_2159999, EPI_ISL_2160000, EPI_ISL_2160001, EPI_ISL_2160002, EPI_ISL_2160003, EPI_ISL_2160004, EPI_ISL_2160005, EPI_ISL_2160006, EPI_ISL_2160007, EPI_ISL_2160008, EPI_ISL_2160009, EPI_ISL_2160010, EPI_ISL_2160011, EPI_ISL_2160012, EPI_ISL_2160013, EPI_ISL_2160014, EPI_ISL_2160015, EPI_ISL_2160016, EPI_ISL_2160017, EPI_ISL_2160018, EPI_ISL_2160019, EPI_ISL_2160020, EPI_ISL_2160021, EPI_ISL_2160022, EPI_ISL_2160023, EPI_ISL_2160024, EPI_ISL_2160025, EPI_ISL_2160026, EPI_ISL_2160027, EPI_ISL_2160028, EPI_ISL_2160029, EPI_ISL_2160030, EPI_ISL_2160031, EPI_ISL_2160032, EPI_ISL_2160033, EPI_ISL_2160034, EPI_ISL_2160035, EPI_ISL_2160036, EPI_ISL_2160037, EPI_ISL_2160038, EPI_ISL_2160039, EPI_ISL_2160040, EPI_ISL_2160041, EPI_ISL_2160042, EPI_ISL_2160043, EPI_ISL_2160044, EPI_ISL_2160045, EPI_ISL_2160046, EPI_ISL_2160047, EPI_ISL_2160048, EPI_ISL_2160049, EPI_ISL_2160050, EPI_ISL_2160051, EPI_ISL_2160052, EPI_ISL_2160053, EPI_ISL_2160054, EPI_ISL_2160055, EPI_ISL_2160056, EPI_ISL_2160057, EPI_ISL_2160058, EPI_ISL_2160059, EPI_ISL_2160060, EPI_ISL_2160061, EPI_ISL_2160062, EPI_ISL_2160063, EPI_ISL_2160064, EPI_ISL_2160065, EPI_ISL_2160066, EPI_ISL_2160067, EPI_ISL_2160068, EPI_ISL_2160069, EPI_ISL_2160070, EPI_ISL_2160071, EPI_ISL_2160072, EPI_ISL_2160073, EPI_ISL_2160074, EPI_ISL_2160075, EPI_ISL_2160076, EPI_ISL_2160077, EPI_ISL_2160078, EPI_ISL_2160079, EPI_ISL_2160080, EPI_ISL_2160081, EPI_ISL_2160082, EPI_ISL_2160083, EPI_ISL_2160084, EPI_ISL_2160085, EPI_ISL_2160086, EPI_ISL_2160087, EPI_ISL_2160088, EPI_ISL_2160089, EPI_ISL_2160090, EPI_ISL_2160091, EPI_ISL_2160092, EPI_ISL_2160093, EPI_ISL_2160094, EPI_ISL_2160095, EPI_ISL_2160096, EPI_ISL_2160097, EPI_ISL_2160098, EPI_ISL_2160099, EPI_ISL_2160100, EPI_ISL_2160101, EPI_ISL_2160102, EPI_ISL_2160103, EPI_ISL_2160104, EPI_ISL_2160105, EPI_ISL_2160106, EPI_ISL_2160107, EPI_ISL_2160108, EPI_ISL_2160109, EPI_ISL_2160110, EPI_ISL_2160111, EPI_ISL_2160112, EPI_ISL_2160113, EPI_ISL_2160114, EPI_ISL_2160115, EPI_ISL_2160116, EPI_ISL_2160117, EPI_ISL_2160118, EPI_ISL_2160119, EPI_ISL_2160120, EPI_ISL_2160121, EPI_ISL_2160122, EPI_ISL_2160123, EPI_ISL_2160124, EPI_ISL_2160125, EPI_ISL_2160126, EPI_ISL_2160127, EPI_ISL_2160128, EPI_ISL_2160129, EPI_ISL_2160130, EPI_ISL_2160131, EPI_ISL_2160132, EPI_ISL_2160133, EPI_ISL_2160134, EPI_ISL_2160135, EPI_ISL_2160136, EPI_ISL_2160137, EPI_ISL_2160138, EPI_ISL_2160139, EPI_ISL_2160140, EPI_ISL_2160141, EPI_ISL_2160142, EPI_ISL_2160143, EPI_ISL_2160144, EPI_ISL_2160145, EPI_ISL_2160146, EPI_ISL_2160147, EPI_ISL_2160148, EPI_ISL_2160149, EPI_ISL_2160150, EPI_ISL_2160151, EPI_ISL_2160152, EPI_ISL_2160153, EPI_ISL_2160154, EPI_ISL_2160155, EPI_ISL_2160156, EPI_ISL_2160157, EPI_ISL_2160158, EPI_ISL_2160159, EPI_ISL_2160160, EPI_ISL_2160161, EPI_ISL_2160162, EPI_ISL_2160163, EPI_ISL_2160164, EPI_ISL_2160165, EPI_ISL_2160166, EPI_ISL_2160167, EPI_ISL_2160168, EPI_ISL_2160169, EPI_ISL_2160171, EPI_ISL_2160172, EPI_ISL_2160173, EPI_ISL_2160174, EPI_ISL_2160175, EPI_ISL_2160176, EPI_ISL_2160177, EPI_ISL_2160178, EPI_ISL_2160179, EPI_ISL_2160180, EPI_ISL_2160181, EPI_ISL_2160182, EPI_ISL_2160183, EPI_ISL_2160184, EPI_ISL_2160185, EPI_ISL_2160186, EPI_ISL_2160187, EPI_ISL_2160188, EPI_ISL_2160189, EPI_ISL_2160190, EPI_ISL_2160191, EPI_ISL_2160192, EPI_ISL_2160193, EPI_ISL_2160194, EPI_ISL_2160195, EPI_ISL_2160196, EPI_ISL_2160197, EPI_ISL_2160198, EPI_ISL_2160199, EPI_ISL_2160200, EPI_ISL_2160201, EPI_ISL_2160202, EPI_ISL_2160203, EPI_ISL_2160204, EPI_ISL_2160205, EPI_ISL_2160206, EPI_ISL_2160207, EPI_ISL_2160208, EPI_ISL_2160209, EPI_ISL_2160210, EPI_ISL_2160211, EPI_ISL_2160212, EPI_ISL_2160213, EPI_ISL_2160214, EPI_ISL_2160215, EPI_ISL_2160216, EPI_ISL_2160217, EPI_ISL_2160218, EPI_ISL_2160219, EPI_ISL_2160220, EPI_ISL_2160221, EPI_ISL_2160222, EPI_ISL_2160223, EPI_ISL_2160224, EPI_ISL_2160225, EPI_ISL_2160226, EPI_ISL_2160227, EPI_ISL_2160228, EPI_ISL_2160229, EPI_ISL_2160230, EPI_ISL_2160231, EPI_ISL_2160232, EPI_ISL_2160233, EPI_ISL_2160234, EPI_ISL_2160235, EPI_ISL_2160236, EPI_ISL_2160237, EPI_ISL_2160238, EPI_ISL_2160239, EPI_ISL_2160240, EPI_ISL_2160241, EPI_ISL_2160242, EPI_ISL_2160243, EPI_ISL_2160244, EPI_ISL_2160245, EPI_ISL_2160246, EPI_ISL_2160247, EPI_ISL_2160248, EPI_ISL_2160249, EPI_ISL_2160250, EPI_ISL_2160251, EPI_ISL_2160252, EPI_ISL_2160253, EPI_ISL_2160254, EPI_ISL_2160255, EPI_ISL_2160256, EPI_ISL_2160257, EPI_ISL_2160258, EPI_ISL_2160259, EPI_ISL_2160260, EPI_ISL_2160261, EPI_ISL_2160262, EPI_ISL_2160263, EPI_ISL_2160264, EPI_ISL_2160265, EPI_ISL_2160266, EPI_ISL_2160267, EPI_ISL_2160268, EPI_ISL_2160269, EPI_ISL_2160270, EPI_ISL_2160271, EPI_ISL_2160272, EPI_ISL_2160273, EPI_ISL_2160274, EPI_ISL_2160275, EPI_ISL_2160276, EPI_ISL_2160277, EPI_ISL_2160278, EPI_ISL_2160279, EPI_ISL_2160280, EPI_ISL_2160281, EPI_ISL_2160282, EPI_ISL_2160283, EPI_ISL_2160284, EPI_ISL_2160285, EPI_ISL_2160286, EPI_ISL_2160287, EPI_ISL_2160288, EPI_ISL_2160289, EPI_ISL_2160290, EPI_ISL_2160291, EPI_ISL_2160292, EPI_ISL_2160293, EPI_ISL_2160294, EPI_ISL_2160295, EPI_ISL_2160296, EPI_ISL_2160297, EPI_ISL_2160298, EPI_ISL_2160299, EPI_ISL_2160300, EPI_ISL_2160301, EPI_ISL_2160302, EPI_ISL_2160303, EPI_ISL_2160304, EPI_ISL_2160305, EPI_ISL_2160306, EPI_ISL_2160307, EPI_ISL_2160308, EPI_ISL_2160309, EPI_ISL_2160310, EPI_ISL_2160311, EPI_ISL_2160312, EPI_ISL_2160314, EPI_ISL_2160315, EPI_ISL_2160316, EPI_ISL_2160317, EPI_ISL_2160318, EPI_ISL_2160319, EPI_ISL_2160320, EPI_ISL_2160321, EPI_ISL_2160322, EPI_ISL_2160323, EPI_ISL_2160324, EPI_ISL_2160325, EPI_ISL_2160326, EPI_ISL_2160327, EPI_ISL_2160328, EPI_ISL_2160329, EPI_ISL_2160330, EPI_ISL_2160331, EPI_ISL_2160332, EPI_ISL_2160333, EPI_ISL_2160334, EPI_ISL_2160335, EPI_ISL_2160336, EPI_ISL_2160337, EPI_ISL_2160338, EPI_ISL_2160339, EPI_ISL_2160340, EPI_ISL_2160341, EPI_ISL_2160342, EPI_ISL_2160343, EPI_ISL_2160344, EPI_ISL_2160345, EPI_ISL_2160346, EPI_ISL_2160347, EPI_ISL_2160348, EPI_ISL_2160349, EPI_ISL_2160350, EPI_ISL_2160351, EPI_ISL_2160352, EPI_ISL_2160353, EPI_ISL_2160354, EPI_ISL_2160355, EPI_ISL_2160356, EPI_ISL_2160357, EPI_ISL_2160358, EPI_ISL_2160359, EPI_ISL_2160360, EPI_ISL_2160361, EPI_ISL_2160362, EPI_ISL_2160363, EPI_ISL_2160364, EPI_ISL_2160366, EPI_ISL_2160367, EPI_ISL_2160368, EPI_ISL_2160369, EPI_ISL_2160370, EPI_ISL_2160371, EPI_ISL_2160372, EPI_ISL_2160373, EPI_ISL_2160374, EPI_ISL_2160375, EPI_ISL_2160376, EPI_ISL_2160377, EPI_ISL_2160378, EPI_ISL_2160379, EPI_ISL_2160380, EPI_ISL_2160381, EPI_ISL_2160382, EPI_ISL_2160383, EPI_ISL_2160384, EPI_ISL_2160385, EPI_ISL_2160386, EPI_ISL_2160387, EPI_ISL_2160388, EPI_ISL_2160389, EPI_ISL_2160390, EPI_ISL_2160391, EPI_ISL_2160392, EPI_ISL_2160393, EPI_ISL_2160394, EPI_ISL_2160395, EPI_ISL_2160396, EPI_ISL_2160397, EPI_ISL_2160398, EPI_ISL_2160399, EPI_ISL_2160400, EPI_ISL_2160401, EPI_ISL_2160402, EPI_ISL_2160403, EPI_ISL_2160404, EPI_ISL_2160405, EPI_ISL_2160406, EPI_ISL_2160407, EPI_ISL_2160408, EPI_ISL_2160409, EPI_ISL_2160410, EPI_ISL_2160411, EPI_ISL_2160412, EPI_ISL_2160413, EPI_ISL_2160414, EPI_ISL_2160415, EPI_ISL_2160416, EPI_ISL_2160417, EPI_ISL_2160418, EPI_ISL_2160419, EPI_ISL_2160420, EPI_ISL_2160421, EPI_ISL_2160422, EPI_ISL_2160423, EPI_ISL_2160424, EPI_ISL_2160425, EPI_ISL_2160426, EPI_ISL_2160427, EPI_ISL_2160428, EPI_ISL_2160429, EPI_ISL_2160430, EPI_ISL_2160431, EPI_ISL_2160432, EPI_ISL_2160433, EPI_ISL_2160434, EPI_ISL_2160435, EPI_ISL_2160436, EPI_ISL_2160437, EPI_ISL_2160438, EPI_ISL_2160439, EPI_ISL_2160440, EPI_ISL_2160441, EPI_ISL_2160442, EPI_ISL_2160443, EPI_ISL_2160444, EPI_ISL_2160445, EPI_ISL_2160446, EPI_ISL_2160447, EPI_ISL_2160448, EPI_ISL_2160449, EPI_ISL_2160450, EPI_ISL_2160451, EPI_ISL_2160452, EPI_ISL_2160453, EPI_ISL_2160454, EPI_ISL_2160455, EPI_ISL_2160456, EPI_ISL_2160457, EPI_ISL_2160458, EPI_ISL_2160459, EPI_ISL_2160460, EPI_ISL_2160461, EPI_ISL_2160462, EPI_ISL_2160463, EPI_ISL_2160464, EPI_ISL_2160465, EPI_ISL_2160466, EPI_ISL_2160467, EPI_ISL_2160468, EPI_ISL_2160469, EPI_ISL_2160470, EPI_ISL_2160471, EPI_ISL_2160472, EPI_ISL_2160473, EPI_ISL_2160474, EPI_ISL_2160475, EPI_ISL_2160476, EPI_ISL_2160477, EPI_ISL_2160478, EPI_ISL_2160479, EPI_ISL_2160480, EPI_ISL_2160481, EPI_ISL_2160482, EPI_ISL_2160483, EPI_ISL_2160484, EPI_ISL_2160485, EPI_ISL_2160486, EPI_ISL_2160487, EPI_ISL_2160488, EPI_ISL_2160489, EPI_ISL_2160490, EPI_ISL_2160491, EPI_ISL_2160492, EPI_ISL_2160493, EPI_ISL_2160494, EPI_ISL_2160495, EPI_ISL_2160496, EPI_ISL_2160497, EPI_ISL_2160498, EPI_ISL_2160499, EPI_ISL_2160500, EPI_ISL_2160501, EPI_ISL_2160502, EPI_ISL_2160503, EPI_ISL_2160504, EPI_ISL_2160505, EPI_ISL_2160506, EPI_ISL_2160507, EPI_ISL_2160508, EPI_ISL_2160509, EPI_ISL_2160510, EPI_ISL_2160511, EPI_ISL_2160512, EPI_ISL_2160513, EPI_ISL_2160514, EPI_ISL_2160515, EPI_ISL_2160516, EPI_ISL_2160517, EPI_ISL_2160518, EPI_ISL_2160519, EPI_ISL_2160520, EPI_ISL_2160521, EPI_ISL_2160522, EPI_ISL_2160523, EPI_ISL_2160524, EPI_ISL_2160525, EPI_ISL_2160526, EPI_ISL_2160527, EPI_ISL_2160528, EPI_ISL_2160529, EPI_ISL_2160530, EPI_ISL_2160531, EPI_ISL_2160532, EPI_ISL_2160533, EPI_ISL_2160534, EPI_ISL_2160535, EPI_ISL_2160536, EPI_ISL_2160537, EPI_ISL_2160538, EPI_ISL_2160539, EPI_ISL_2160540, EPI_ISL_2160541, EPI_ISL_2160542, EPI_ISL_2160543, EPI_ISL_2160544, EPI_ISL_2160545, EPI_ISL_2160546, EPI_ISL_2160547, EPI_ISL_2160548, EPI_ISL_2160549, EPI_ISL_2160550, EPI_ISL_2160551, EPI_ISL_2160552, EPI_ISL_2160553, EPI_ISL_2160554, EPI_ISL_2160555, EPI_ISL_2160556, EPI_ISL_2160557, EPI_ISL_2160558, EPI_ISL_2160559, EPI_ISL_2160560, EPI_ISL_2160562, EPI_ISL_2160563, EPI_ISL_2160564, EPI_ISL_2160565, EPI_ISL_2160566, EPI_ISL_2160567, EPI_ISL_2160568, EPI_ISL_2160569, EPI_ISL_2160570, EPI_ISL_2160571, EPI_ISL_2160572, EPI_ISL_2160573, EPI_ISL_2160574, EPI_ISL_2160575, EPI_ISL_2160576, EPI_ISL_2160577, EPI_ISL_2160578, EPI_ISL_2160579, EPI_ISL_2160580, EPI_ISL_2160581, EPI_ISL_2160582, EPI_ISL_2160583, EPI_ISL_2160584, EPI_ISL_2160585, EPI_ISL_2160586, EPI_ISL_2160587, EPI_ISL_2160588, EPI_ISL_2160589, EPI_ISL_2160590, EPI_ISL_2160591, EPI_ISL_2160593, EPI_ISL_2160594, EPI_ISL_2160595, EPI_ISL_2160596, EPI_ISL_2160597, EPI_ISL_2160598, EPI_ISL_2160599, EPI_ISL_2160600, EPI_ISL_2160601, EPI_ISL_2160602, EPI_ISL_2160603, EPI_ISL_2160604, EPI_ISL_2160605, EPI_ISL_2160606, EPI_ISL_2160607, EPI_ISL_2160608, EPI_ISL_2160609, EPI_ISL_2160611, EPI_ISL_2160612, EPI_ISL_2160613, EPI_ISL_2160614, EPI_ISL_2160615, EPI_ISL_2160616, EPI_ISL_2160617, EPI_ISL_2160618, EPI_ISL_2160619, EPI_ISL_2160620, EPI_ISL_2160621, EPI_ISL_2160622, EPI_ISL_2160623, EPI_ISL_2160624, EPI_ISL_2160625, EPI_ISL_2160626, EPI_ISL_2160627, EPI_ISL_2160628, EPI_ISL_2160629, EPI_ISL_2160630, EPI_ISL_2160631, EPI_ISL_2160632, EPI_ISL_2160633, EPI_ISL_2160634, EPI_ISL_2160635, EPI_ISL_2160636, EPI_ISL_2160637, EPI_ISL_2160638, EPI_ISL_2160639, EPI_ISL_2160640, EPI_ISL_2160641 | | | |
| see above | Helix/Illumina | Centers for Disease Control and Prevention Division of Viral Diseases, Pathogen Discovery | Dakota Howard, Dhwani Batra, Peter W. Cook, Kara Moser, Adrian Paskey, Jason Caravas, Benjamin Rambo-Martin, Shatavia Morrison, Christopher Gulvick, Scott Sammons, Yvette Unoarumhi, Darlene Wagner, Matthew Schmerer, Eileen de Feo, Jan Antico, Christine Tran, Matthew Tolentino, Shannon Wickline, Kim Gietzen, Brad Sickler, Jingtao Liu, Eric Allen, Phil Febbo, Nicole L. Washington, Simon White, Geraint Levan, Kelly Schiabor Barrett, Elizabeth Cirulli, Alexandre Bolze, Ary Ascencio, Charlotte Rivera-Garcia, Ryan Cho, Jason Nguyen, Sherry Wang, Jimmy Ramirez, Tyler Cassens, Efren Sandoval, Magnus Isaksson, William Lee, David Becker, Marc Laurent, James Lu, Clinton R. Paden, Duncan MacCannell |
| EPI_ISL_2160642 | Instituto Nacional de Medicina Genomica | Centro de Investigación en Enfermedades Infecciosas (CIENI), Instituto Nacional de Enfermedades Respiratorias (INER) | Matias-Florentino M, Perez-Garcia M, Hidalgo-Miranda A, Mendoza-Vargas A, Reyes-Grajeda JP, Cedro-Tanda A, Arriaga-Canon C, Herrera-Montalvo LA, Reyes-Teran G, Avila-Rios S, Boukadida C |
| EPI_ISL_2160643 | Helix/Illumina | Centers for Disease Control and Prevention Division of Viral Diseases, Pathogen Discovery | Dakota Howard, Dhwani Batra, Peter W. Cook, Kara Moser, Adrian Paskey, Jason Caravas, Benjamin Rambo-Martin, Shatavia Morrison, Christopher Gulvick, Scott Sammons, Yvette Unoarumhi, Darlene Wagner, Matthew Schmerer, Eileen de Feo, Jan Antico, Christine Tran, Matthew Tolentino, Shannon Wickline, Kim Gietzen, Brad Sickler, Jingtao Liu, Eric Allen, Phil Febbo, Nicole L. Washington, Simon White, Geraint Levan, Kelly Schiabor Barrett, Elizabeth Cirulli, Alexandre Bolze, Ary Ascencio, Charlotte Rivera-Garcia, Ryan Cho, Jason Nguyen, Sherry Wang, Jimmy Ramirez, Tyler Cassens, Efren Sandoval, Magnus Isaksson, William Lee, David Becker, Marc Laurent, James Lu, Clinton R. Paden, Duncan MacCannell |
| EPI_ISL_2160644 | Instituto Nacional de Medicina Genomica | Centro de Investigación en Enfermedades Infecciosas (CIENI), Instituto Nacional de Enfermedades Respiratorias (INER) | Matias-Florentino M, Perez-Garcia M, Hidalgo-Miranda A, Mendoza-Vargas A, Reyes-Grajeda JP, Cedro-Tanda A, Arriaga-Canon C, Herrera-Montalvo LA, Reyes-Teran G, Avila-Rios S, Boukadida C |
| EPI_ISL_2160645 | Helix/Illumina | Centers for Disease Control and Prevention Division of Viral Diseases, Pathogen Discovery | Dakota Howard, Dhwani Batra, Peter W. Cook, Kara Moser, Adrian Paskey, Jason Caravas, Benjamin Rambo-Martin, Shatavia Morrison, Christopher Gulvick, Scott Sammons, Yvette Unoarumhi, Darlene Wagner, Matthew Schmerer, Eileen de Feo, Jan Antico, Christine Tran, Matthew Tolentino, Shannon Wickline, Kim Gietzen, Brad Sickler, Jingtao Liu, Eric Allen, Phil Febbo, Nicole L. Washington, Simon White, Geraint Levan, Kelly Schiabor Barrett, Elizabeth Cirulli, Alexandre Bolze, Ary Ascencio, Charlotte Rivera-Garcia, Ryan Cho, Jason Nguyen, Sherry Wang, Jimmy Ramirez, Tyler Cassens, Efren Sandoval, Magnus Isaksson, William Lee, David Becker, Marc Laurent, James Lu, Clinton R. Paden, Duncan MacCannell |
| EPI_ISL_2160646, EPI_ISL_2160647 | Instituto Nacional de Medicina Genomica | Centro de Investigación en Enfermedades Infecciosas (CIENI), Instituto Nacional de Enfermedades Respiratorias (INER) | Matias-Florentino M, Perez-Garcia M, Hidalgo-Miranda A, Mendoza-Vargas A, Reyes-Grajeda JP, Cedro-Tanda A, Arriaga-Canon C, Herrera-Montalvo LA, Reyes-Teran G, Avila-Rios S, Boukadida C |
| EPI_ISL_2160648 | Helix/Illumina | Centers for Disease Control and Prevention Division of Viral Diseases, Pathogen Discovery | Dakota Howard, Dhwani Batra, Peter W. Cook, Kara Moser, Adrian Paskey, Jason Caravas, Benjamin Rambo-Martin, Shatavia Morrison, Christopher Gulvick, Scott Sammons, Yvette Unoarumhi, Darlene Wagner, Matthew Schmerer, Eileen de Feo, Jan Antico, Christine Tran, Matthew Tolentino, Shannon Wickline, Kim Gietzen, Brad Sickler, Jingtao Liu, Eric Allen, Phil Febbo, Nicole L. Washington, Simon White, Geraint Levan, Kelly Schiabor Barrett, Elizabeth Cirulli, Alexandre Bolze, Ary Ascencio, Charlotte Rivera-Garcia, Ryan Cho, Jason Nguyen, Sherry Wang, Jimmy Ramirez, Tyler Cassens, Efren Sandoval, Magnus Isaksson, William Lee, David Becker, Marc Laurent, James Lu, Clinton R. Paden, Duncan MacCannell |
| EPI_ISL_2160649, EPI_ISL_2160650, EPI_ISL_2160651 | Instituto Nacional de Medicina Genomica | Centro de Investigación en Enfermedades Infecciosas (CIENI), Instituto Nacional de Enfermedades Respiratorias (INER) | Matias-Florentino M, Perez-Garcia M, Hidalgo-Miranda A, Mendoza-Vargas A, Reyes-Grajeda JP, Cedro-Tanda A, Arriaga-Canon C, Herrera-Montalvo LA, Reyes-Teran G, Avila-Rios S, Boukadida C |
| EPI_ISL_2160652 | Helix/Illumina | Centers for Disease Control and Prevention Division of Viral Diseases, Pathogen Discovery | Dakota Howard, Dhwani Batra, Peter W. Cook, Kara Moser, Adrian Paskey, Jason Caravas, Benjamin Rambo-Martin, Shatavia Morrison, Christopher Gulvick, Scott Sammons, Yvette Unoarumhi, Darlene Wagner, Matthew Schmerer, Eileen de Feo, Jan Antico, Christine Tran, Matthew Tolentino, Shannon Wickline, Kim Gietzen, Brad Sickler, Jingtao Liu, Eric Allen, Phil Febbo, Nicole L. Washington, Simon White, Geraint Levan, Kelly Schiabor Barrett, Elizabeth Cirulli, Alexandre Bolze, Ary Ascencio, Charlotte Rivera-Garcia, Ryan Cho, Jason Nguyen, Sherry Wang, Jimmy Ramirez, Tyler Cassens, Efren Sandoval, Magnus Isaksson, William Lee, David Becker, Marc Laurent, James Lu, Clinton R. Paden, Duncan MacCannell |
| EPI_ISL_2160653 | Instituto Nacional de Medicina Genomica | Centro de Investigación en Enfermedades Infecciosas (CIENI), Instituto Nacional de Enfermedades Respiratorias (INER) | Matias-Florentino M, Perez-Garcia M, Hidalgo-Miranda A, Mendoza-Vargas A, Reyes-Grajeda JP, Cedro-Tanda A, Arriaga-Canon C, Herrera-Montalvo LA, Reyes-Teran G, Avila-Rios S, Boukadida C |

| | | | |
|---|---|---|---|
| EPI_ISL_2160654 | Helix/Illumina | Centers for Disease Control and Prevention Division of Viral Diseases, Pathogen Discovery | Dakota Howard, Dhwani Batra, Peter W. Cook, Kara Moser, Adrian Paskey, Jason Caravas, Benjamin Rambo-Martin, Shatavia Morrison, Christopher Gulvick, Scott Sammons, Yvette Unoarumhi, Darlene Wagner, Matthew Schmerer, Eileen de Feo, Jan Antico, Christine Tran, Matthew Tolentino, Shannon Wickline, Kim Gietzen, Brad Sickler, Jingtao Liu, Eric Allen, Phil Febbo, Nicole L. Washington, Simon White, Geraint Levan, Kelly Schiabor Barrett, Elizabeth Cirulli, Alexandre Bolze, Ary Ascencio, Charlotte Rivera-Garcia, Ryan Cho, Jason Nguyen, Sherry Wang, Jimmy Ramirez, Tyler Cassens, Efren Sandoval, Magnus Isaksson, William Lee, David Becker, Marc Laurent, James Lu, Clinton R. Paden, Duncan MacCannell |
| EPI_ISL_2160655, EPI_ISL_2160656 | Instituto Nacional de Medicina Genomica | Centro de Investigación en Enfermedades Infecciosas (CIENI), Instituto Nacional de Enfermedades Respiratorias (INER) | Matias-Florentino M, Perez-Garcia M, Hidalgo-Miranda A, Mendoza-Vargas A, Reyes-Grajeda JP, Cedro-Tanda A, Arriaga-Canon C, Herrera-Montalvo LA, Reyes-Teran G, Avila-Rios S, Boukadida C |
| EPI_ISL_2160657 | Helix/Illumina | Centers for Disease Control and Prevention Division of Viral Diseases, Pathogen Discovery | Dakota Howard, Dhwani Batra, Peter W. Cook, Kara Moser, Adrian Paskey, Jason Caravas, Benjamin Rambo-Martin, Shatavia Morrison, Christopher Gulvick, Scott Sammons, Yvette Unoarumhi, Darlene Wagner, Matthew Schmerer, Eileen de Feo, Jan Antico, Christine Tran, Matthew Tolentino, Shannon Wickline, Kim Gietzen, Brad Sickler, Jingtao Liu, Eric Allen, Phil Febbo, Nicole L. Washington, Simon White, Geraint Levan, Kelly Schiabor Barrett, Elizabeth Cirulli, Alexandre Bolze, Ary Ascencio, Charlotte Rivera-Garcia, Ryan Cho, Jason Nguyen, Sherry Wang, Jimmy Ramirez, Tyler Cassens, Efren Sandoval, Magnus Isaksson, William Lee, David Becker, Marc Laurent, James Lu, Clinton R. Paden, Duncan MacCannell |
| EPI_ISL_2160658, EPI_ISL_2160659 | Instituto Nacional de Medicina Genomica | Centro de Investigación en Enfermedades Infecciosas (CIENI), Instituto Nacional de Enfermedades Respiratorias (INER) | Matias-Florentino M, Perez-Garcia M, Hidalgo-Miranda A, Mendoza-Vargas A, Reyes-Grajeda JP, Cedro-Tanda A, Arriaga-Canon C, Herrera-Montalvo LA, Reyes-Teran G, Avila-Rios S, Boukadida C |
| EPI_ISL_2160660 | Helix/Illumina | Centers for Disease Control and Prevention Division of Viral Diseases, Pathogen Discovery | Dakota Howard, Dhwani Batra, Peter W. Cook, Kara Moser, Adrian Paskey, Jason Caravas, Benjamin Rambo-Martin, Shatavia Morrison, Christopher Gulvick, Scott Sammons, Yvette Unoarumhi, Darlene Wagner, Matthew Schmerer, Eileen de Feo, Jan Antico, Christine Tran, Matthew Tolentino, Shannon Wickline, Kim Gietzen, Brad Sickler, Jingtao Liu, Eric Allen, Phil Febbo, Nicole L. Washington, Simon White, Geraint Levan, Kelly Schiabor Barrett, Elizabeth Cirulli, Alexandre Bolze, Ary Ascencio, Charlotte Rivera-Garcia, Ryan Cho, Jason Nguyen, Sherry Wang, Jimmy Ramirez, Tyler Cassens, Efren Sandoval, Magnus Isaksson, William Lee, David Becker, Marc Laurent, James Lu, Clinton R. Paden, Duncan MacCannell |
| EPI_ISL_2160661, EPI_ISL_2160662 | Instituto Nacional de Medicina Genomica | Centro de Investigación en Enfermedades Infecciosas (CIENI), Instituto Nacional de Enfermedades Respiratorias (INER) | Matias-Florentino M, Perez-Garcia M, Hidalgo-Miranda A, Mendoza-Vargas A, Reyes-Grajeda JP, Cedro-Tanda A, Arriaga-Canon C, Herrera-Montalvo LA, Reyes-Teran G, Avila-Rios S, Boukadida C |
| EPI_ISL_2160663, EPI_ISL_2160664 | Helix/Illumina | Centers for Disease Control and Prevention Division of Viral Diseases, Pathogen Discovery | Dakota Howard, Dhwani Batra, Peter W. Cook, Kara Moser, Adrian Paskey, Jason Caravas, Benjamin Rambo-Martin, Shatavia Morrison, Christopher Gulvick, Scott Sammons, Yvette Unoarumhi, Darlene Wagner, Matthew Schmerer, Eileen de Feo, Jan Antico, Christine Tran, Matthew Tolentino, Shannon Wickline, Kim Gietzen, Brad Sickler, Jingtao Liu, Eric Allen, Phil Febbo, Nicole L. Washington, Simon White, Geraint Levan, Kelly Schiabor Barrett, Elizabeth Cirulli, Alexandre Bolze, Ary Ascencio, Charlotte Rivera-Garcia, Ryan Cho, Jason Nguyen, Sherry Wang, Jimmy Ramirez, Tyler Cassens, Efren Sandoval, Magnus Isaksson, William Lee, David Becker, Marc Laurent, James Lu, Clinton R. Paden, Duncan MacCannell |
| EPI_ISL_2160665 | Instituto Nacional de Medicina Genomica | Centro de Investigación en Enfermedades Infecciosas (CIENI), Instituto Nacional de Enfermedades Respiratorias (INER) | Matias-Florentino M, Perez-Garcia M, Hidalgo-Miranda A, Mendoza-Vargas A, Reyes-Grajeda JP, Cedro-Tanda A, Arriaga-Canon C, Herrera-Montalvo LA, Reyes-Teran G, Avila-Rios S, Boukadida C |
| EPI_ISL_2160666 | Helix/Illumina | Centers for Disease Control and Prevention Division of Viral Diseases, Pathogen Discovery | Dakota Howard, Dhwani Batra, Peter W. Cook, Kara Moser, Adrian Paskey, Jason Caravas, Benjamin Rambo-Martin, Shatavia Morrison, Christopher Gulvick, Scott Sammons, Yvette Unoarumhi, Darlene Wagner, Matthew Schmerer, Eileen de Feo, Jan Antico, Christine Tran, Matthew Tolentino, Shannon Wickline, Kim Gietzen, Brad Sickler, Jingtao Liu, Eric Allen, Phil Febbo, Nicole L. Washington, Simon White, Geraint Levan, Kelly Schiabor Barrett, Elizabeth Cirulli, Alexandre Bolze, Ary Ascencio, Charlotte Rivera-Garcia, Ryan Cho, Jason Nguyen, Sherry Wang, Jimmy Ramirez, Tyler Cassens, Efren Sandoval, Magnus Isaksson, William Lee, David Becker, Marc Laurent, James Lu, Clinton R. Paden, Duncan MacCannell |
| EPI_ISL_2160667, EPI_ISL_2160668 | Instituto Nacional de Medicina Genomica | Centro de Investigación en Enfermedades Infecciosas (CIENI), Instituto Nacional de Enfermedades Respiratorias (INER) | Matias-Florentino M, Perez-Garcia M, Hidalgo-Miranda A, Mendoza-Vargas A, Reyes-Grajeda JP, Cedro-Tanda A, Arriaga-Canon C, Herrera-Montalvo LA, Reyes-Teran G, Avila-Rios S, Boukadida C |
| EPI_ISL_2160669 | Helix/Illumina | Centers for Disease Control and Prevention Division of Viral Diseases, Pathogen Discovery | Dakota Howard, Dhwani Batra, Peter W. Cook, Kara Moser, Adrian Paskey, Jason Caravas, Benjamin Rambo-Martin, Shatavia Morrison, Christopher Gulvick, Scott Sammons, Yvette Unoarumhi, Darlene Wagner, Matthew Schmerer, Eileen de Feo, Jan Antico, Christine Tran, Matthew Tolentino, Shannon Wickline, Kim Gietzen, Brad Sickler, Jingtao Liu, Eric Allen, Phil Febbo, Nicole L. Washington, Simon White, Geraint Levan, Kelly Schiabor Barrett, Elizabeth Cirulli, Alexandre Bolze, Ary Ascencio, Charlotte Rivera-Garcia, Ryan Cho, Jason Nguyen, Sherry Wang, Jimmy Ramirez, Tyler Cassens, Efren Sandoval, Magnus Isaksson, William Lee, David Becker, Marc Laurent, James Lu, Clinton R. Paden, Duncan MacCannell |
| EPI_ISL_2160670, EPI_ISL_2160671 | Instituto Nacional de Medicina Genomica | Centro de Investigación en Enfermedades Infecciosas (CIENI), Instituto Nacional de Enfermedades Respiratorias (INER) | Matias-Florentino M, Perez-Garcia M, Hidalgo-Miranda A, Mendoza-Vargas A, Reyes-Grajeda JP, Cedro-Tanda A, Arriaga-Canon C, Herrera-Montalvo LA, Reyes-Teran G, Avila-Rios S, Boukadida C |
| EPI_ISL_2160672 | Helix/Illumina | Centers for Disease Control and Prevention Division of Viral Diseases, Pathogen Discovery | Dakota Howard, Dhwani Batra, Peter W. Cook, Kara Moser, Adrian Paskey, Jason Caravas, Benjamin Rambo-Martin, Shatavia Morrison, Christopher Gulvick, Scott Sammons, Yvette Unoarumhi, Darlene Wagner, Matthew Schmerer, Eileen de Feo, Jan Antico, Christine Tran, Matthew Tolentino, Shannon Wickline, Kim Gietzen, Brad Sickler, Jingtao Liu, Eric Allen, Phil Febbo, Nicole L. Washington, Simon White, Geraint Levan, Kelly Schiabor Barrett, Elizabeth Cirulli, Alexandre Bolze, Ary Ascencio, Charlotte Rivera-Garcia, Ryan Cho, Jason Nguyen, Sherry Wang, Jimmy Ramirez, Tyler Cassens, Efren Sandoval, Magnus Isaksson, William Lee, David Becker, Marc Laurent, James Lu, Clinton R. Paden, Duncan MacCannell |
| EPI_ISL_2160673, EPI_ISL_2160674 | Instituto Nacional de Medicina Genomica | Centro de Investigación en Enfermedades Infecciosas (CIENI), Instituto Nacional de Enfermedades Respiratorias (INER) | Matias-Florentino M, Perez-Garcia M, Hidalgo-Miranda A, Mendoza-Vargas A, Reyes-Grajeda JP, Cedro-Tanda A, Arriaga-Canon C, Herrera-Montalvo LA, Reyes-Teran G, Avila-Rios S, Boukadida C |
| EPI_ISL_2160675 | Helix/Illumina | Centers for Disease Control and Prevention Division of Viral Diseases, Pathogen Discovery | Dakota Howard, Dhwani Batra, Peter W. Cook, Kara Moser, Adrian Paskey, Jason Caravas, Benjamin Rambo-Martin, Shatavia Morrison, Christopher Gulvick, Scott Sammons, Yvette Unoarumhi, Darlene Wagner, Matthew Schmerer, Eileen de Feo, Jan Antico, Christine Tran, Matthew Tolentino, Shannon Wickline, Kim Gietzen, Brad Sickler, Jingtao Liu, Eric Allen, Phil Febbo, Nicole L. Washington, Simon White, Geraint Levan, Kelly Schiabor Barrett, Elizabeth Cirulli, Alexandre Bolze, Ary Ascencio, Charlotte Rivera-Garcia, Ryan Cho, Jason Nguyen, Sherry Wang, Jimmy Ramirez, Tyler Cassens, Efren Sandoval, Magnus Isaksson, William Lee, David Becker, Marc Laurent, James Lu, Clinton R. Paden, Duncan MacCannell |
| EPI_ISL_2160676 | Instituto Nacional de Medicina Genomica | Centro de Investigación en Enfermedades Infecciosas (CIENI), Instituto Nacional de Enfermedades Respiratorias (INER) | Matias-Florentino M, Perez-Garcia M, Hidalgo-Miranda A, Mendoza-Vargas A, Reyes-Grajeda JP, Cedro-Tanda A, Arriaga-Canon C, Herrera-Montalvo LA, Reyes-Teran G, Avila-Rios S, Boukadida C |
| EPI_ISL_2160677 | Helix/Illumina | Centers for Disease Control and Prevention Division of Viral Diseases, Pathogen Discovery | Dakota Howard, Dhwani Batra, Peter W. Cook, Kara Moser, Adrian Paskey, Jason Caravas, Benjamin Rambo-Martin, Shatavia Morrison, Christopher Gulvick, Scott Sammons, Yvette Unoarumhi, Darlene Wagner, Matthew Schmerer, Eileen de Feo, Jan Antico, Christine Tran, Matthew Tolentino, Shannon Wickline, Kim Gietzen, Brad Sickler, Jingtao Liu, Eric Allen, Phil Febbo, Nicole L. Washington, Simon White, Geraint Levan, Kelly Schiabor Barrett, Elizabeth Cirulli, Alexandre Bolze, Ary Ascencio, Charlotte Rivera-Garcia, Ryan Cho, Jason Nguyen, Sherry Wang, Jimmy Ramirez, Tyler Cassens, Efren Sandoval, Magnus Isaksson, William Lee, David Becker, Marc Laurent, James Lu, Clinton R. Paden, Duncan MacCannell |
| EPI_ISL_2160678, EPI_ISL_2160679 | Instituto Nacional de Medicina Genomica | Centro de Investigación en Enfermedades Infecciosas (CIENI), Instituto Nacional de Enfermedades Respiratorias (INER) | Matias-Florentino M, Perez-Garcia M, Hidalgo-Miranda A, Mendoza-Vargas A, Reyes-Grajeda JP, Cedro-Tanda A, Arriaga-Canon C, Herrera-Montalvo LA, Reyes-Teran G, Avila-Rios S, Boukadida C |
| EPI_ISL_2160680 | Helix/Illumina | Centers for Disease Control and Prevention Division of Viral Diseases, Pathogen Discovery | Dakota Howard, Dhwani Batra, Peter W. Cook, Kara Moser, Adrian Paskey, Jason Caravas, Benjamin Rambo-Martin, Shatavia Morrison, Christopher Gulvick, Scott Sammons, Yvette Unoarumhi, Darlene Wagner, Matthew Schmerer, Eileen de Feo, Jan Antico, Christine Tran, Matthew Tolentino, Shannon Wickline, Kim Gietzen, Brad Sickler, Jingtao Liu, Eric Allen, Phil Febbo, Nicole L. Washington, Simon White, Geraint Levan, Kelly Schiabor Barrett, Elizabeth Cirulli, Alexandre Bolze, Ary Ascencio, Charlotte Rivera-Garcia, Ryan Cho, Jason Nguyen, Sherry Wang, Jimmy Ramirez, Tyler Cassens, Efren Sandoval, Magnus Isaksson, William Lee, David Becker, Marc Laurent, James Lu, Clinton R. Paden, Duncan MacCannell |
| EPI_ISL_2160681, EPI_ISL_2160682 | Instituto Nacional de Medicina Genomica | Centro de Investigación en Enfermedades Infecciosas (CIENI), Instituto Nacional de Enfermedades Respiratorias (INER) | Matias-Florentino M, Perez-Garcia M, Hidalgo-Miranda A, Mendoza-Vargas A, Reyes-Grajeda JP, Cedro-Tanda A, Arriaga-Canon C, Herrera-Montalvo LA, Reyes-Teran G, Avila-Rios S, Boukadida C |
| EPI_ISL_2160683 | Helix/Illumina | Centers for Disease Control and Prevention Division of Viral Diseases, Pathogen Discovery | Dakota Howard, Dhwani Batra, Peter W. Cook, Kara Moser, Adrian Paskey, Jason Caravas, Benjamin Rambo-Martin, Shatavia Morrison, Christopher Gulvick, Scott Sammons, Yvette Unoarumhi, Darlene Wagner, Matthew Schmerer, Eileen de Feo, Jan Antico, Christine Tran, Matthew Tolentino, Shannon Wickline, Kim Gietzen, Brad Sickler, Jingtao Liu, Eric Allen, Phil Febbo, Nicole L. Washington, Simon White, Geraint Levan, Kelly Schiabor Barrett, Elizabeth Cirulli, Alexandre Bolze, Ary Ascencio, Charlotte Rivera-Garcia, Ryan Cho, Jason Nguyen, Sherry Wang, Jimmy Ramirez, Tyler Cassens, Efren Sandoval, Magnus Isaksson, William Lee, David Becker, Marc Laurent, James Lu, Clinton R. Paden, Duncan MacCannell |
| EPI_ISL_2160684, EPI_ISL_2160685 | Instituto Nacional de Medicina Genomica | Centro de Investigación en Enfermedades Infecciosas (CIENI), Instituto Nacional de Enfermedades Respiratorias (INER) | Matias-Florentino M, Perez-Garcia M, Hidalgo-Miranda A, Mendoza-Vargas A, Reyes-Grajeda JP, Cedro-Tanda A, Arriaga-Canon C, Herrera-Montalvo LA, Reyes-Teran G, Avila-Rios S, Boukadida C |
| EPI_ISL_2160686 | Helix/Illumina | Centers for Disease Control and Prevention Division of Viral Diseases, Pathogen Discovery | Dakota Howard, Dhwani Batra, Peter W. Cook, Kara Moser, Adrian Paskey, Jason Caravas, Benjamin Rambo-Martin, Shatavia Morrison, Christopher Gulvick, Scott Sammons, Yvette Unoarumhi, Darlene Wagner, Matthew Schmerer, Eileen de Feo, Jan Antico, Christine Tran, Matthew Tolentino, Shannon Wickline, Kim Gietzen, Brad Sickler, Jingtao Liu, Eric Allen, Phil Febbo, Nicole L. Washington, Simon White, Geraint Levan, Kelly Schiabor Barrett, Elizabeth Cirulli, Alexandre Bolze, Ary Ascencio, Charlotte Rivera-Garcia, Ryan Cho, Jason Nguyen, Sherry Wang, Jimmy Ramirez, Tyler Cassens, Efren Sandoval, Magnus Isaksson, William Lee, David Becker, Marc Laurent, James Lu, Clinton R. Paden, Duncan MacCannell |
| EPI_ISL_2160687, EPI_ISL_2160689, EPI_ISL_2160690 | Instituto Nacional de Medicina Genomica | Centro de Investigación en Enfermedades Infecciosas (CIENI), Instituto Nacional de Enfermedades Respiratorias (INER) | Matias-Florentino M, Perez-Garcia M, Hidalgo-Miranda A, Mendoza-Vargas A, Reyes-Grajeda JP, Cedro-Tanda A, Arriaga-Canon C, Herrera-Montalvo LA, Reyes-Teran G, Avila-Rios S, Boukadida C |
| EPI_ISL_2160691 | Helix/Illumina | Centers for Disease Control and Prevention Division of Viral Diseases, Pathogen Discovery | Dakota Howard, Dhwani Batra, Peter W. Cook, Kara Moser, Adrian Paskey, Jason Caravas, Benjamin Rambo-Martin, Shatavia Morrison, Christopher Gulvick, Scott Sammons, Yvette Unoarumhi, Darlene Wagner, Matthew Schmerer, Eileen de Feo, Jan Antico, Christine Tran, Matthew Tolentino, Shannon Wickline, Kim Gietzen, Brad Sickler, Jingtao Liu, Eric Allen, Phil Febbo, Nicole L. Washington, Simon White, Geraint Levan, Kelly Schiabor Barrett, Elizabeth Cirulli, Alexandre Bolze, Ary Ascencio, Charlotte Rivera-Garcia, Ryan Cho, Jason Nguyen, Sherry Wang, Jimmy Ramirez, Tyler Cassens, Efren Sandoval, Magnus Isaksson, William Lee, David Becker, Marc Laurent, James Lu, Clinton R. Paden, Duncan MacCannell |
| EPI_ISL_2160692, EPI_ISL_2160693 | Instituto Nacional de Medicina Genomica | Centro de Investigación en Enfermedades Infecciosas (CIENI), Instituto Nacional de Enfermedades Respiratorias (INER) | Matias-Florentino M, Perez-Garcia M, Hidalgo-Miranda A, Mendoza-Vargas A, Reyes-Grajeda JP, Cedro-Tanda A, Arriaga-Canon C, Herrera-Montalvo LA, Reyes-Teran G, Avila-Rios S, Boukadida C |
| EPI_ISL_2160694 | Helix/Illumina | Centers for Disease Control and Prevention Division of Viral Diseases, Pathogen Discovery | Dakota Howard, Dhwani Batra, Peter W. Cook, Kara Moser, Adrian Paskey, Jason Caravas, Benjamin Rambo-Martin, Shatavia Morrison, Christopher Gulvick, Scott Sammons, Yvette Unoarumhi, Darlene Wagner, Matthew Schmerer, Eileen de Feo, Jan Antico, Christine Tran, Matthew Tolentino, Shannon Wickline, Kim Gietzen, Brad Sickler, Jingtao Liu, Eric Allen, Phil Febbo, Nicole L. Washington, Simon White, Geraint Levan, Kelly Schiabor Barrett, Elizabeth Cirulli, Alexandre Bolze, Ary Ascencio, Charlotte Rivera-Garcia, Ryan Cho, Jason Nguyen, Sherry Wang, Jimmy Ramirez, Tyler Cassens, Efren Sandoval, Magnus Isaksson, William Lee, David Becker, Marc Laurent, James Lu, Clinton R. Paden, Duncan MacCannell |
| EPI_ISL_2160695 | Instituto Nacional de Medicina Genomica | Centro de Investigación en Enfermedades Infecciosas (CIENI), Instituto Nacional de Enfermedades Respiratorias (INER) | Matias-Florentino M, Perez-Garcia M, Hidalgo-Miranda A, Mendoza-Vargas A, Reyes-Grajeda JP, Cedro-Tanda A, Arriaga-Canon C, Herrera-Montalvo LA, Reyes-Teran G, Avila-Rios S, Boukadida C |
| EPI_ISL_2160696, EPI_ISL_2160697, EPI_ISL_2160698, EPI_ISL_2160699, EPI_ISL_2160700, EPI_ISL_2160701, EPI_ISL_2160702, EPI_ISL_2160703, EPI_ISL_2160704, EPI_ISL_2160705, EPI_ISL_2160706, EPI_ISL_2160707, EPI_ISL_2160708, EPI_ISL_2160709, EPI_ISL_2160710, EPI_ISL_2160711, EPI_ISL_2160712, EPI_ISL_2160713 | | | |
| see above | Helix/Illumina | Centers for Disease Control and Prevention Division of Viral Diseases, Pathogen Discovery | Dakota Howard, Dhwani Batra, Peter W. Cook, Kara Moser, Adrian Paskey, Jason Caravas, Benjamin Rambo-Martin, Shatavia Morrison, Christopher Gulvick, Scott Sammons, Yvette Unoarumhi, Darlene Wagner, Matthew Schmerer, Eileen de Feo, Jan Antico, Christine Tran, Matthew Tolentino, Shannon Wickline, Kim Gietzen, Brad Sickler, Jingtao Liu, Eric Allen, Phil Febbo, Nicole L. Washington, Simon White, Geraint Levan, Kelly Schiabor Barrett, Elizabeth Cirulli, Alexandre Bolze, Ary Ascencio, Charlotte Rivera-Garcia, Ryan Cho, Jason Nguyen, Sherry Wang, Jimmy Ramirez, Tyler Cassens, Efren Sandoval, Magnus Isaksson, William Lee, David Becker, Marc Laurent, James Lu, Clinton R. Paden, Duncan MacCannell |
| EPI_ISL_2160714 | AR Dept. of Health-PHL, Molecular Diagnostics | Pathogen Discovery, Respiratory Viruses Branch, Division of Viral Diseases, Centers for Disease Control and Prevention | Yan Li, Jing Zhang, Anna Kelleher, Ying Tao, Brian Lynch, Krista Queen, Anna Uehara, Peter Cook, Han Jia Justin Ng, Rachel Marine, Clinton R. Paden, Dhwani Batra, Haibin Wang, Tara Coalter, Jasmine Padilla, Morgan Davis, Mili Sheth, Sarah Nobles, Mark Burroughs, Justin Lee, Adam Retchless, Suxiang Tong |
| EPI_ISL_2160715 | NC State Laboratory of Public Health | Pathogen Discovery, Respiratory Viruses Branch, Division of Viral Diseases, Centers for Disease Control and Prevention | Yan Li, Jing Zhang, Anna Kelleher, Ying Tao, Brian Lynch, Krista Queen, Anna Uehara, Peter Cook, Han Jia Justin Ng, Rachel Marine, Clinton R. Paden, Dhwani Batra, Haibin Wang, Tara Coalter, Jasmine Padilla, Morgan Davis, Mili Sheth, Sarah Nobles, Mark Burroughs, Justin Lee, Adam Retchless, Suxiang Tong |
| EPI_ISL_2160716, EPI_ISL_2160717, EPI_ISL_2160718, EPI_ISL_2160719, EPI_ISL_2160720, EPI_ISL_2160721, EPI_ISL_2160722, EPI_ISL_2160723, EPI_ISL_2160724, EPI_ISL_2160725, EPI_ISL_2160726, EPI_ISL_2160727, EPI_ISL_2160728, EPI_ISL_2160729, EPI_ISL_2160730, EPI_ISL_2160731, EPI_ISL_2160733, EPI_ISL_2160734, EPI_ISL_2160735, EPI_ISL_2160736, EPI_ISL_2160737, EPI_ISL_2160738, EPI_ISL_2160739, EPI_ISL_2160740, EPI_ISL_2160741, EPI_ISL_2160742 | | | |

| | | | |
|---|---|---|---|
| see above | Helix/Illumina | Centers for Disease Control and Prevention Division of Viral Diseases, Pathogen Discovery | Dakota Howard, Dhwani Batra, Peter W. Cook, Kara Moser, Adrian Paskey, Jason Caravas, Benjamin Rambo-Martin, Shatavia Morrison, Christopher Gulvick, Scott Sammons, Yvette Unoarumhi, Darlene Wagner, Matthew Schmerer, Eileen de Feo, Jan Antico, Christine Tran, Matthew Tolentino, Shannon Wickline, Kim Gietzen, Brad Sickler, Jingtao Liu, Eric Allen, Phil Febbo, Nicole L. Washington, Simon White, Geraint Levan, Kelly Schiabor Barrett, Elizabeth Cirulli, Alexandre Bolze, Ary Ascencio, Charlotte Rivera-Garcia, Ryan Cho, Jason Nguyen, Sherry Wang, Jimmy Ramirez, Tyler Cassens, Efren Sandoval, Magnus Isaksson, William Lee, David Becker, Marc Laurent, James Lu, Clinton R. Paden, Duncan MacCannell |
| EPI_ISL_2160743 | DC Public Health Lab/ Dept. of Forensic Sciences | DC Public Health Lab/ Dept. of Forensic Sciences | Janis Doss, Scott Nguyen, Elizabeth Zelaya, Sarah Scott, Connie Maza, Monica Mann, Brittany Hamilton, David Payne, Jocelyn Hauser |
| EPI_ISL_2160744, EPI_ISL_2160746, EPI_ISL_2160747, EPI_ISL_2160748, EPI_ISL_2160749, EPI_ISL_2160750, EPI_ISL_2160751, EPI_ISL_2160752, EPI_ISL_2160753, EPI_ISL_2160754, EPI_ISL_2160755, EPI_ISL_2160756, EPI_ISL_2160757, EPI_ISL_2160758, EPI_ISL_2160759, EPI_ISL_2160760, EPI_ISL_2160761, EPI_ISL_2160762, EPI_ISL_2160763, EPI_ISL_2160764, EPI_ISL_2160765, EPI_ISL_2160766, EPI_ISL_2160767, EPI_ISL_2160768, EPI_ISL_2160769, EPI_ISL_2160771, EPI_ISL_2160772, EPI_ISL_2160773, EPI_ISL_2160774, EPI_ISL_2160775, EPI_ISL_2160776, EPI_ISL_2160777, EPI_ISL_2160778, EPI_ISL_2160779, EPI_ISL_2160780, EPI_ISL_2160781, EPI_ISL_2160782, EPI_ISL_2160783, EPI_ISL_2160784, EPI_ISL_2160785, EPI_ISL_2160786, EPI_ISL_2160787, EPI_ISL_2160789, EPI_ISL_2160790, EPI_ISL_2160791, EPI_ISL_2160792, EPI_ISL_2160793, EPI_ISL_2160795, EPI_ISL_2160796, EPI_ISL_2160797, EPI_ISL_2160798, EPI_ISL_2160799, EPI_ISL_2160800, EPI_ISL_2160801, EPI_ISL_2160802, EPI_ISL_2160803, EPI_ISL_2160804, EPI_ISL_2160805, EPI_ISL_2160806, EPI_ISL_2160807, EPI_ISL_2160808, EPI_ISL_2160809 | | | |
| see above | Fulgent Genetics | Centers for Disease Control and Prevention Division of Viral Diseases, Pathogen Discovery | Dakota Howard, Dhwani Batra, Peter W. Cook, Kara Moser, Adrian Paskey, Jason Caravas, Benjamin Rambo-Martin, Shatavia Morrison, Christopher Gulvick, Scott Sammons, Yvette Unoarumhi, Darlene Wagner, Matthew Schmerer, Harry Gao, Mickey Li, John Gao, Joseph Fierro, Benafsh Sapra, Becky Tsai, Yan Meng, Doreen Ng, James Xie, Clinton R. Paden, Duncan MacCannell |
| EPI_ISL_2160810 | Wyoming Public Health Laboratory | Wyoming Public Health Laboratory | Jim Mildenberger, Wanda Manley, Noah Hull, Taylor Fearing, Lynette Gumbleton, Channing Weber, Ashley Norberg, Chayse Rowley, Marley Goetz, Brian Dominguez, Elliot Thomasson, Sam Britz, Cari Sloma, and Rob Christensen |
| EPI_ISL_2160811, EPI_ISL_2160812, EPI_ISL_2160813, EPI_ISL_2160814, EPI_ISL_2160815, EPI_ISL_2160816, EPI_ISL_2160817, EPI_ISL_2160818, EPI_ISL_2160819, EPI_ISL_2160820, EPI_ISL_2160821, EPI_ISL_2160822, EPI_ISL_2160823, EPI_ISL_2160824, EPI_ISL_2160825, EPI_ISL_2160826, EPI_ISL_2160827, EPI_ISL_2160828, EPI_ISL_2160829, EPI_ISL_2160830, EPI_ISL_2160831, EPI_ISL_2160832, EPI_ISL_2160833, EPI_ISL_2160834, EPI_ISL_2160835, EPI_ISL_2160836, EPI_ISL_2160837, EPI_ISL_2160838, EPI_ISL_2160839, EPI_ISL_2160840, EPI_ISL_2160841, EPI_ISL_2160842, EPI_ISL_2160843, EPI_ISL_2160844, EPI_ISL_2160845, EPI_ISL_2160846, EPI_ISL_2160847, EPI_ISL_2160848, EPI_ISL_2160849, EPI_ISL_2160850, EPI_ISL_2160851, EPI_ISL_2160852, EPI_ISL_2160853, EPI_ISL_2160854, EPI_ISL_2160855, EPI_ISL_2160856, EPI_ISL_2160857, EPI_ISL_2160859, EPI_ISL_2160860, EPI_ISL_2160861, EPI_ISL_2160862, EPI_ISL_2160863, EPI_ISL_2160864, EPI_ISL_2160865, EPI_ISL_2160866, EPI_ISL_2160868, EPI_ISL_2160869, EPI_ISL_2160870, EPI_ISL_2160871, EPI_ISL_2160872, EPI_ISL_2160873, EPI_ISL_2160874, EPI_ISL_2160875, EPI_ISL_2160877, EPI_ISL_2160880, EPI_ISL_2160881, EPI_ISL_2160882, EPI_ISL_2160883, EPI_ISL_2160884, EPI_ISL_2160885, EPI_ISL_2160887, EPI_ISL_2160889, EPI_ISL_2160891, EPI_ISL_2160892, EPI_ISL_2160894, EPI_ISL_2160895, EPI_ISL_2160896, EPI_ISL_2160898, EPI_ISL_2160900, EPI_ISL_2160901, EPI_ISL_2160902, EPI_ISL_2160903, EPI_ISL_2160904, EPI_ISL_2160905, EPI_ISL_2160908, EPI_ISL_2160909, EPI_ISL_2160912, EPI_ISL_2160913, EPI_ISL_2160914, EPI_ISL_2160915, EPI_ISL_2160916, EPI_ISL_2160918, EPI_ISL_2160919, EPI_ISL_2160920, EPI_ISL_2160921, EPI_ISL_2160922, EPI_ISL_2160923, EPI_ISL_2160924, EPI_ISL_2160925, EPI_ISL_2160926, EPI_ISL_2160927, EPI_ISL_2160928, EPI_ISL_2160929, EPI_ISL_2160931, EPI_ISL_2160932, EPI_ISL_2160933, EPI_ISL_2160934, EPI_ISL_2160935, EPI_ISL_2160936, EPI_ISL_2160937, EPI_ISL_2160938, EPI_ISL_2160939, EPI_ISL_2160940, EPI_ISL_2160941, EPI_ISL_2160942, EPI_ISL_2160943, EPI_ISL_2160944, EPI_ISL_2160945, EPI_ISL_2160946, EPI_ISL_2160947, EPI_ISL_2160948, EPI_ISL_2160949, EPI_ISL_2160950, EPI_ISL_2160951, EPI_ISL_2160952, EPI_ISL_2160953, EPI_ISL_2160954, EPI_ISL_2160955, EPI_ISL_2160956, EPI_ISL_2160957, EPI_ISL_2160958, EPI_ISL_2160959, EPI_ISL_2160960, EPI_ISL_2160961, EPI_ISL_2160962, EPI_ISL_2160963, EPI_ISL_2160964, EPI_ISL_2160965, EPI_ISL_2160966, EPI_ISL_2160967, EPI_ISL_2160968, EPI_ISL_2160969, EPI_ISL_2160970, EPI_ISL_2160972, EPI_ISL_2160973, EPI_ISL_2160974, EPI_ISL_2160975, EPI_ISL_2160976, EPI_ISL_2160977, EPI_ISL_2160978, EPI_ISL_2160979, EPI_ISL_2160980, EPI_ISL_2160981, EPI_ISL_2160982, EPI_ISL_2160983, EPI_ISL_2160984, EPI_ISL_2160985, EPI_ISL_2160986, EPI_ISL_2160988, EPI_ISL_2160989, EPI_ISL_2160990, EPI_ISL_2160991, EPI_ISL_2160992, EPI_ISL_2160993, EPI_ISL_2160994, EPI_ISL_2160996, EPI_ISL_2160997, EPI_ISL_2160999, EPI_ISL_2161000, EPI_ISL_2161001, EPI_ISL_2161002, EPI_ISL_2161003, EPI_ISL_2161004, EPI_ISL_2161005, EPI_ISL_2161006, EPI_ISL_2161007, EPI_ISL_2161008, EPI_ISL_2161009, EPI_ISL_2161010, EPI_ISL_2161012, EPI_ISL_2161013, EPI_ISL_2161014, EPI_ISL_2161015, EPI_ISL_2161016, EPI_ISL_2161017, EPI_ISL_2161018, EPI_ISL_2161019, EPI_ISL_2161020, EPI_ISL_2161021, EPI_ISL_2161022, EPI_ISL_2161023, EPI_ISL_2161024, EPI_ISL_2161025, EPI_ISL_2161026, EPI_ISL_2161027 | | | |
| see above | Fulgent Genetics | Centers for Disease Control and Prevention Division of Viral Diseases, Pathogen Discovery | Dakota Howard, Dhwani Batra, Peter W. Cook, Kara Moser, Adrian Paskey, Jason Caravas, Benjamin Rambo-Martin, Shatavia Morrison, Christopher Gulvick, Scott Sammons, Yvette Unoarumhi, Darlene Wagner, Matthew Schmerer, Harry Gao, Mickey Li, John Gao, Joseph Fierro, Benafsh Sapra, Becky Tsai, Yan Meng, Doreen Ng, James Xie, Clinton R. Paden, Duncan MacCannell |
| EPI_ISL_2161028 | University of Utah, School of Medicine | Genomics and Discovery, Respiratory Viruses Branch, Division of Viral Diseases, Centers for Disease Control and Prevention | Yan Li, Jing Zhang, Anna Kelleher, Ying Tao, Brian Lynch, Krista Queen, Anna Uehara, Peter Cook, Han Jia Justin Ng, Rachel Marine, Clinton R. Paden, Dhwani Batra, Haibin Wang, Tara Coalter, Jasmine Padilla, Morgan Davis, Mili Sheth, Sarah Nobles, Mark Burroughs, Justin Lee, Adam Retchless, Suxiang Tong |
| EPI_ISL_2161029 | Johns Hopkins University | Genomics and Discovery, Respiratory Viruses Branch, Division of Viral Diseases, Centers for Disease Control and Prevention | Yan Li, Jing Zhang, Anna Kelleher, Ying Tao, Brian Lynch, Krista Queen, Anna Uehara, Peter Cook, Han Jia Justin Ng, Rachel Marine, Clinton R. Paden, Dhwani Batra, Haibin Wang, Tara Coalter, Jasmine Padilla, Morgan Davis, Mili Sheth, Sarah Nobles, Mark Burroughs, Justin Lee, Adam Retchless, Suxiang Tong |
| EPI_ISL_2161438, EPI_ISL_2161439, EPI_ISL_2161440, EPI_ISL_2161441, EPI_ISL_2161442, EPI_ISL_2161443, EPI_ISL_2161444, EPI_ISL_2161445, EPI_ISL_2161446, EPI_ISL_2161447, EPI_ISL_2161448, EPI_ISL_2161449, EPI_ISL_2161450, EPI_ISL_2161451, EPI_ISL_2161452, EPI_ISL_2161453, EPI_ISL_2161454, EPI_ISL_2161455, EPI_ISL_2161456, EPI_ISL_2161457, EPI_ISL_2161458, EPI_ISL_2161459, EPI_ISL_2161460, EPI_ISL_2161461, EPI_ISL_2161462, EPI_ISL_2161463, EPI_ISL_2161464, EPI_ISL_2161465, EPI_ISL_2161466, EPI_ISL_2161467, EPI_ISL_2161468, EPI_ISL_2161469, EPI_ISL_2161470, EPI_ISL_2161471, EPI_ISL_2161472, EPI_ISL_2161473, EPI_ISL_2161474, EPI_ISL_2161475, EPI_ISL_2161476, EPI_ISL_2161477, EPI_ISL_2161478, EPI_ISL_2161479, EPI_ISL_2161480, EPI_ISL_2161481, EPI_ISL_2161482, EPI_ISL_2161483, EPI_ISL_2161484, EPI_ISL_2161485, EPI_ISL_2161486, EPI_ISL_2161487, EPI_ISL_2161489, EPI_ISL_2161490, EPI_ISL_2161491, EPI_ISL_2161492, EPI_ISL_2161493, EPI_ISL_2161494, EPI_ISL_2161495, EPI_ISL_2161496, EPI_ISL_2161497, EPI_ISL_2161498, EPI_ISL_2161499, EPI_ISL_2161500, EPI_ISL_2161501, EPI_ISL_2161502, EPI_ISL_2161503, EPI_ISL_2161504, EPI_ISL_2161506, EPI_ISL_2161507, EPI_ISL_2161508, EPI_ISL_2161509, EPI_ISL_2161510, EPI_ISL_2161511, EPI_ISL_2161512, EPI_ISL_2161513, EPI_ISL_2161514, EPI_ISL_2161516, EPI_ISL_2161517, EPI_ISL_2161518, EPI_ISL_2161519, EPI_ISL_2161520, EPI_ISL_2161521, EPI_ISL_2161522, EPI_ISL_2161523, EPI_ISL_2161524, EPI_ISL_2161525, EPI_ISL_2161526, EPI_ISL_2161527, EPI_ISL_2161528, EPI_ISL_2161529, EPI_ISL_2161530, EPI_ISL_2161532, EPI_ISL_2161533, EPI_ISL_2161534, EPI_ISL_2161535, EPI_ISL_2161536, EPI_ISL_2161537, EPI_ISL_2161538, EPI_ISL_2161539, EPI_ISL_2161540, EPI_ISL_2161541, EPI_ISL_2161542, EPI_ISL_2161543, EPI_ISL_2161544, EPI_ISL_2161545, EPI_ISL_2161546, EPI_ISL_2161547, EPI_ISL_2161548, EPI_ISL_2161549, EPI_ISL_2161550, EPI_ISL_2161551, EPI_ISL_2161552, EPI_ISL_2161553, EPI_ISL_2161554, EPI_ISL_2161555, EPI_ISL_2161556, EPI_ISL_2161557, EPI_ISL_2161558, EPI_ISL_2161559, EPI_ISL_2161560, EPI_ISL_2161561, EPI_ISL_2161562, EPI_ISL_2161563, EPI_ISL_2161564, EPI_ISL_2161565, EPI_ISL_2161566, EPI_ISL_2161567, EPI_ISL_2161568, EPI_ISL_2161569, EPI_ISL_2161570, EPI_ISL_2161571, EPI_ISL_2161572, EPI_ISL_2161573, EPI_ISL_2161574, EPI_ISL_2161576, EPI_ISL_2161577, EPI_ISL_2161578, EPI_ISL_2161580, EPI_ISL_2161582, EPI_ISL_2161583, EPI_ISL_2161584, EPI_ISL_2161585, EPI_ISL_2161586, EPI_ISL_2161587, EPI_ISL_2161588, EPI_ISL_2161589, EPI_ISL_2161590, EPI_ISL_2161591, EPI_ISL_2161592, EPI_ISL_2161593, EPI_ISL_2161594, EPI_ISL_2161595, EPI_ISL_2161596, EPI_ISL_2161597, EPI_ISL_2161598, EPI_ISL_2161599, EPI_ISL_2161600, EPI_ISL_2161601, EPI_ISL_2161602, EPI_ISL_2161603, EPI_ISL_2161604, EPI_ISL_2161605, EPI_ISL_2161606, EPI_ISL_2161607, EPI_ISL_2161608, EPI_ISL_2161609, EPI_ISL_2161610, EPI_ISL_2161611, EPI_ISL_2161612, EPI_ISL_2161613, EPI_ISL_2161614, EPI_ISL_2161615, EPI_ISL_2161616, EPI_ISL_2161617, EPI_ISL_2161618, EPI_ISL_2161620 | | | |
| see above | Maryland Genomics, Institute for Genome Sciences, University of Maryland School of Medicine | Maryland Genomics, Institute for Genome Sciences, University of Maryland School of Medicine | Tallon, Luke J; Sadzewicz, Lisa D; Humphrys, Mike; Ott, Sandra; Roussey, Holly; Mehta, Aditya; Vavikolanu, Kranthi; Fraser, Claire M; Ravel, Jacques |
| EPI_ISL_2161621, EPI_ISL_2161622, EPI_ISL_2161623, EPI_ISL_2161624, EPI_ISL_2161625, EPI_ISL_2161626, EPI_ISL_2161627, EPI_ISL_2161628, EPI_ISL_2161629, EPI_ISL_2161631, EPI_ISL_2161633, EPI_ISL_2161634, EPI_ISL_2161635, EPI_ISL_2161636, EPI_ISL_2161637, EPI_ISL_2161638, EPI_ISL_2161639, EPI_ISL_2161640, EPI_ISL_2161642, EPI_ISL_2161643, EPI_ISL_2161644, EPI_ISL_2161645, EPI_ISL_2161646, EPI_ISL_2161647, EPI_ISL_2161648, EPI_ISL_2161649, EPI_ISL_2161650, EPI_ISL_2161651, EPI_ISL_2161652, EPI_ISL_2161653, EPI_ISL_2161654, EPI_ISL_2161656, EPI_ISL_2161659, EPI_ISL_2161660, EPI_ISL_2161661, EPI_ISL_2161662, EPI_ISL_2161663, EPI_ISL_2161664, EPI_ISL_2161667, EPI_ISL_2161670, EPI_ISL_2161671, EPI_ISL_2161672, EPI_ISL_2161673, EPI_ISL_2161674, EPI_ISL_2161675, EPI_ISL_2161676, EPI_ISL_2161677, EPI_ISL_2161678, EPI_ISL_2161679, EPI_ISL_2161680, EPI_ISL_2161681, EPI_ISL_2161682, EPI_ISL_2161683, EPI_ISL_2161684, EPI_ISL_2161685, EPI_ISL_2161686, EPI_ISL_2161687, EPI_ISL_2161688, EPI_ISL_2161689, EPI_ISL_2161690, EPI_ISL_2161692, EPI_ISL_2161693, EPI_ISL_2161694, EPI_ISL_2161695, EPI_ISL_2161696, EPI_ISL_2161698, EPI_ISL_2161699, EPI_ISL_2161700, EPI_ISL_2161701, EPI_ISL_2161702, EPI_ISL_2161703, EPI_ISL_2161705, EPI_ISL_2161706, EPI_ISL_2161707, EPI_ISL_2161708, EPI_ISL_2161709, EPI_ISL_2161710, EPI_ISL_2161712, EPI_ISL_2161713, EPI_ISL_2161714, EPI_ISL_2161715, EPI_ISL_2161716, EPI_ISL_2161717, EPI_ISL_2161718, EPI_ISL_2161719, EPI_ISL_2161720, EPI_ISL_2161721, EPI_ISL_2161722, EPI_ISL_2161723, EPI_ISL_2161724, EPI_ISL_2161725, EPI_ISL_2161726, EPI_ISL_2161727, EPI_ISL_2161728, EPI_ISL_2161729, EPI_ISL_2161730, EPI_ISL_2161731, EPI_ISL_2161732, EPI_ISL_2161733, EPI_ISL_2161735, EPI_ISL_2161736, EPI_ISL_2161737, EPI_ISL_2161738, EPI_ISL_2161739, EPI_ISL_2161740, EPI_ISL_2161741, EPI_ISL_2161742, EPI_ISL_2161743, EPI_ISL_2161744, EPI_ISL_2161745, EPI_ISL_2161746, EPI_ISL_2161747, EPI_ISL_2161748, EPI_ISL_2161749, EPI_ISL_2161750, EPI_ISL_2161751, EPI_ISL_2161752, EPI_ISL_2161753, EPI_ISL_2161754, EPI_ISL_2161755, EPI_ISL_2161757, EPI_ISL_2161758, EPI_ISL_2161759, EPI_ISL_2161760, EPI_ISL_2161761, EPI_ISL_2161762, EPI_ISL_2161763, EPI_ISL_2161764, EPI_ISL_2161765, EPI_ISL_2161766, EPI_ISL_2161767, EPI_ISL_2161768, EPI_ISL_2161769, EPI_ISL_2161770, EPI_ISL_2161771, EPI_ISL_2161772, EPI_ISL_2161773, EPI_ISL_2161774, EPI_ISL_2161775, EPI_ISL_2161776, EPI_ISL_2161777, EPI_ISL_2161778, EPI_ISL_2161779, EPI_ISL_2161780 | | | |
| see above | NYU Langone Health | Departments of Pathology and Medicine, New York University School of Medicine | Adriana Heguy, Dacia Dimartino, Emily Guzman, Christian Marier, Peter Meyn, Sitharam Ramaswami, Gael Westby, Paul Zappile, Yutong Zhang, Paolo Cotzia, Guiqing Wang |
| EPI_ISL_2161812, EPI_ISL_2161813, EPI_ISL_2161814, EPI_ISL_2161815, EPI_ISL_2161816, EPI_ISL_2161817, EPI_ISL_2161818, EPI_ISL_2161819, EPI_ISL_2161820, EPI_ISL_2161821, EPI_ISL_2161822, EPI_ISL_2161823, EPI_ISL_2161824, EPI_ISL_2161825, EPI_ISL_2161826, EPI_ISL_2161827, EPI_ISL_2161828, EPI_ISL_2161829, EPI_ISL_2161830, EPI_ISL_2161831, EPI_ISL_2161832, EPI_ISL_2161833, EPI_ISL_2161834, EPI_ISL_2161835, EPI_ISL_2161836, EPI_ISL_2161837, EPI_ISL_2161838, EPI_ISL_2161839, EPI_ISL_2161840, EPI_ISL_2161841, EPI_ISL_2161842, EPI_ISL_2161843, EPI_ISL_2161844, EPI_ISL_2161845, EPI_ISL_2161846, EPI_ISL_2161847, EPI_ISL_2161848, EPI_ISL_2161850, EPI_ISL_2161851, EPI_ISL_2161852, EPI_ISL_2161853, EPI_ISL_2161854, EPI_ISL_2161855, EPI_ISL_2161856, EPI_ISL_2161857, EPI_ISL_2161858, EPI_ISL_2161859, EPI_ISL_2161860, EPI_ISL_2161861, EPI_ISL_2161862, EPI_ISL_2161863, EPI_ISL_2161864, EPI_ISL_2161865, EPI_ISL_2161866, EPI_ISL_2161867, EPI_ISL_2161868, EPI_ISL_2161869, EPI_ISL_2161870, EPI_ISL_2161871, EPI_ISL_2161872, EPI_ISL_2161873, EPI_ISL_2161874, EPI_ISL_2161875, EPI_ISL_2161876, EPI_ISL_2161877, EPI_ISL_2161878, EPI_ISL_2161879, EPI_ISL_2161880, EPI_ISL_2161881, EPI_ISL_2161882, EPI_ISL_2161883, EPI_ISL_2161884, EPI_ISL_2161886, EPI_ISL_2161887, EPI_ISL_2161888, EPI_ISL_2161889, EPI_ISL_2161890, EPI_ISL_2161891, EPI_ISL_2161892, EPI_ISL_2161893, EPI_ISL_2161894, EPI_ISL_2161895, EPI_ISL_2161896, EPI_ISL_2161897, EPI_ISL_2161898, EPI_ISL_2161899, EPI_ISL_2161900, EPI_ISL_2161901, EPI_ISL_2161902, EPI_ISL_2161903, EPI_ISL_2161904, EPI_ISL_2161905, EPI_ISL_2161906, EPI_ISL_2161907, EPI_ISL_2161908, EPI_ISL_2161909, EPI_ISL_2161910, EPI_ISL_2161911, EPI_ISL_2161912, EPI_ISL_2161913, EPI_ISL_2161914, EPI_ISL_2161916, EPI_ISL_2161917, EPI_ISL_2161918, EPI_ISL_2161919, EPI_ISL_2161920, EPI_ISL_2161921, EPI_ISL_2161922, EPI_ISL_2161923, EPI_ISL_2161924, EPI_ISL_2161925, EPI_ISL_2161926, EPI_ISL_2161927, EPI_ISL_2161928, EPI_ISL_2161929, EPI_ISL_2161930, EPI_ISL_2161931, EPI_ISL_2161932, EPI_ISL_2161933, EPI_ISL_2161934, EPI_ISL_2161935, EPI_ISL_2161936, EPI_ISL_2161937, EPI_ISL_2161938, EPI_ISL_2161939, EPI_ISL_2161940, EPI_ISL_2161941, EPI_ISL_2161942, EPI_ISL_2161943, EPI_ISL_2161944, EPI_ISL_2161945, EPI_ISL_2161946, EPI_ISL_2161947, EPI_ISL_2161948, EPI_ISL_2161949, EPI_ISL_2161950, EPI_ISL_2161951, EPI_ISL_2161952, EPI_ISL_2161953, EPI_ISL_2161954, EPI_ISL_2161955, EPI_ISL_2161956, EPI_ISL_2161957, EPI_ISL_2161958, EPI_ISL_2161959, EPI_ISL_2161960, EPI_ISL_2161961, EPI_ISL_2161962, EPI_ISL_2161963, EPI_ISL_2161964, EPI_ISL_2161965, EPI_ISL_2161966, EPI_ISL_2161967, EPI_ISL_2161968, EPI_ISL_2161969, EPI_ISL_2161970, EPI_ISL_2161971, EPI_ISL_2161972, EPI_ISL_2161973, EPI_ISL_2161974, EPI_ISL_2161975, EPI_ISL_2161976, EPI_ISL_2161977, EPI_ISL_2161978, EPI_ISL_2161979, EPI_ISL_2161980, EPI_ISL_2161981, EPI_ISL_2161982, EPI_ISL_2161983, EPI_ISL_2161984, EPI_ISL_2161985, EPI_ISL_2161986, EPI_ISL_2161987, EPI_ISL_2161988, EPI_ISL_2161989, EPI_ISL_2161990, EPI_ISL_2161991, EPI_ISL_2161992, EPI_ISL_2161993, EPI_ISL_2161994, EPI_ISL_2161995, EPI_ISL_2161996, EPI_ISL_2161997, EPI_ISL_2161998, EPI_ISL_2161999, EPI_ISL_2162000, EPI_ISL_2162001, EPI_ISL_2162002, EPI_ISL_2162003, EPI_ISL_2162004, EPI_ISL_2162005, EPI_ISL_2162006, EPI_ISL_2162007, EPI_ISL_2162008, EPI_ISL_2162009, EPI_ISL_2162010, EPI_ISL_2162011, EPI_ISL_2162012, EPI_ISL_2162013, EPI_ISL_2162014, EPI_ISL_2162015, EPI_ISL_2162016, EPI_ISL_2162017, EPI_ISL_2162018, EPI_ISL_2162019, EPI_ISL_2162020, EPI_ISL_2162021, EPI_ISL_2162022, EPI_ISL_2162023, EPI_ISL_2162024, EPI_ISL_2162025, EPI_ISL_2162026, EPI_ISL_2162027 | | | |
| see above | UW Virology Lab | UW Virology Lab | Pavitra Roychoudhury, Hong Xie, Lasata Shrestha, Tien V. Nguyen, Shah Mohamed Bakhash, Michelle Lin, Noah R. Baker, Ricardo Perez, Sean Ellis, Nathan Breit, Robert J. Livingston, Meei-Li Huang, Keith R Jerome, Patrick Mathias, Alexander Greninger |
| EPI_ISL_2162383 | Omega Diagnostics at Mounes | Omega Diagnostics at Mounes | Ashleigh MCGrail, Latira Haynes-Jacob, Cynthia Corley, Vivek Khare, MD |
| EPI_ISL_2162384, EPI_ISL_2162385, EPI_ISL_2162468, EPI_ISL_2162469 | Omega Diagnostics at Mounes | Omega Diagnostics at Mounes | Ashleigh McGrail, Latira Haynes-Jacob, Cynthia Corley, Vivek Khare, MD |
| EPI_ISL_2162472 | Omega Diagnostics at Mounes | Omega Diagnostics at Mounes | Ashleigh McGrail, Latira Haynes-Jacob, Cynthia Corley, Vivek Khare,MD |
| EPI_ISL_2162803, EPI_ISL_2162804, EPI_ISL_2162805, EPI_ISL_2162806 | Omega Diagnostics at Mounes | Omega Diagnostics at Mounes | Ashleigh McGrail, Latira Haynes-Jacob, Cynthia Corley, Vivek Khare, MD |
| EPI_ISL_2162807 | Omega Diagnostics at Mounes | Omega Diagnostics at Mounes | Ashleigh McGrail, Latira Haynes-JAcob, Cynthia Corley, Vivek Khare MD |
| EPI_ISL_2162808 | Omega Diagnostics at Mounes | Omega Diagnotics at Mounes | Ashleigh McGrail, Latira Haynes-Jacob, Cynthia Corley, Vivek Khare MD |
| EPI_ISL_2162809 | Omega Diagnostics | Omega Diagnostics at Mounes | Ashleigh McGrail, Latira Haynes-Jacob, Cynthia Corley, Vivek Khare MD |
| EPI_ISL_2162831 | Omega Diagnostics at Mounes | Omega Diagnostics at Mounes | Ashleigh McGrail, Latira Haynes-Jacob, Cynthia Corley, Vivek Khare MD |
| EPI_ISL_2163013 | Omega Diagnotics at Mounes | Omega Diagnostics | Ashleigh McGrail, Latira Haynes-Jacob, Cynthia Corley, Vivek Khare MD |
| EPI_ISL_2163980 | Omega Diagnostics at Mounes | Omega Diagnostics at Mounes | Ashleigh McGrail, Latira Haynes-Jacob, Cynthia Corley, Vivek Khare MD |
| EPI_ISL_2164289 | Omega Diagnostics at Mounes | Omega Diagnotics at Mounes | Ashleigh McGrail, Latira HAynes-Jacob, Cynthia Corley, Vivek Khare MD |
| EPI_ISL_2164546 | Omega Diagnostics at Mounes | Omega Diagnostics at Mounes | Ashleigh McGrail, Latira Haynes-Jacob, Cynthia Corley, Vivek Khare MD |
| EPI_ISL_2164823 | Omega Diagnostics at Mounes | Omega Diagnotics at Mounes | Ashleigh McGrail, Latira Haynes-Jacob, Cynthia Corley, vivek Khare MD |
| EPI_ISL_2165070 | Omega Diagnostics at Mounes | Omega Diagnostics at Mounes | Ashleigh McGrail, Latira Haynes-Jacob, Cynthia Corley, Vivek Khare MD |
| EPI_ISL_2165330 | Omega Diagnostics at Mounes | Omega Diagnostics at Mounes | Ashleigh McGrail, Latira Haynes-JAcob, Cynthia Corley, Vivek Khare MD |
| EPI_ISL_2165823, EPI_ISL_2166070 | Omega Diagnostics at Mounes | Omega Diagnostics at Mounes | Ashleigh McGrail, Latira Haynes-Jacob, Cynthia Corley, Vivek Khare MD |
| EPI_ISL_2166354 | Omega Diagnostics at Mounes | Omega Diagnostics at Mounes | Ashleigh McGrail, Latira HAynes-Jacob, Cynthia Corley, Vivek Khare MD |
| EPI_ISL_2171269, EPI_ISL_2171270, EPI_ISL_2171271, EPI_ISL_2171272, EPI_ISL_2171273, EPI_ISL_2171274, EPI_ISL_2171275, EPI_ISL_2171276, EPI_ISL_2171277, EPI_ISL_2171278, EPI_ISL_2171279, EPI_ISL_2171280 | | | |
| see above | COVID Outreach Wood River | UW Virology Lab | Pavitra Roychoudhury, Hong Xie, Shah Mohamed Bakhash, Tom Archie, Ryan Cole, Meei-Li Huang, Keith R. Jerome, Alexander Greninger |
| EPI_ISL_2171674 | Tulare County Public Health Laboratory | Tulare County Public Health Laboratory | Denise L Lopez, Joshua Kootstra |
| EPI_ISL_2172421, EPI_ISL_2172422 | Wyoming Public Health Laboratory | Wyoming Public Health Laboratory | Jim Mildenberger, Wanda Manley, Noah Hull, Taylor Fearing, Lynette Gumbleton, Channing Weber, Ashley Norberg, Chayse Rowley, Marley Goetz, Brian Dominguez, Elliot Thomasson, Sam Britz, Cari Sloma, and Rob Christensen |
| EPI_ISL_2172425, EPI_ISL_2172426, EPI_ISL_2172427, EPI_ISL_2172428, EPI_ISL_2172429, EPI_ISL_2172430, EPI_ISL_2172431, EPI_ISL_2172432, EPI_ISL_2172433, EPI_ISL_2172434, EPI_ISL_2172435, EPI_ISL_2172436, EPI_ISL_2172437, EPI_ISL_2172438, EPI_ISL_2172439, EPI_ISL_2172440, EPI_ISL_2172441, EPI_ISL_2172442, EPI_ISL_2172443, EPI_ISL_2172444, EPI_ISL_2172445, EPI_ISL_2172446, EPI_ISL_2172447, EPI_ISL_2172448, EPI_ISL_2172449, EPI_ISL_2172450, EPI_ISL_2172451, EPI_ISL_2172452, EPI_ISL_2172453, EPI_ISL_2172454, EPI_ISL_2172455, EPI_ISL_2172456, EPI_ISL_2172457, EPI_ISL_2172458, EPI_ISL_2172459, EPI_ISL_2172460, EPI_ISL_2172461, EPI_ISL_2172462, EPI_ISL_2172463, EPI_ISL_2172464, EPI_ISL_2172465, EPI_ISL_2172466, EPI_ISL_2172467, EPI_ISL_2172468, EPI_ISL_2172469, EPI_ISL_2172470, EPI_ISL_2172471, EPI_ISL_2172472, EPI_ISL_2172473, EPI_ISL_2172474, EPI_ISL_2172475, EPI_ISL_2172476, EPI_ISL_2172477, EPI_ISL_2172478, EPI_ISL_2172479, EPI_ISL_2172480, EPI_ISL_2172481, EPI_ISL_2172482, EPI_ISL_2172483, EPI_ISL_2172484, EPI_ISL_2172485, EPI_ISL_2172486, EPI_ISL_2172487, EPI_ISL_2172488, EPI_ISL_2172489, EPI_ISL_2172490, EPI_ISL_2172491, EPI_ISL_2172492, EPI_ISL_2172493, EPI_ISL_2172494, EPI_ISL_2172495, EPI_ISL_2172496, EPI_ISL_2172497, EPI_ISL_2172498, EPI_ISL_2172499, EPI_ISL_2172500, EPI_ISL_2172501, EPI_ISL_2172502, EPI_ISL_2172503, EPI_ISL_2172504, EPI_ISL_2172505, EPI_ISL_2172506, EPI_ISL_2172507, EPI_ISL_2172508, EPI_ISL_2172509, EPI_ISL_2172510, EPI_ISL_2172511, EPI_ISL_2172512, | | | |

| | | | |
|---|---|---|---|
| EPI_ISL_2172513, EPI_ISL_2172514, EPI_ISL_2172515, EPI_ISL_2172516 | | | |
| see above | UW Virology Lab | UW Virology Lab | Pavitra Roychoudhury, Hong Xie, Lasata Shrestha, Tien V. Nguyen, Shah Mohamed Bakhash, Michelle Lin, Noah R. Baker, Sean Ellis, Meei-Li Huang, Keith R Jerome, Alexander Greninger |
| EPI_ISL_2176237, EPI_ISL_2176238, EPI_ISL_2176240, EPI_ISL_2176242, EPI_ISL_2176244, EPI_ISL_2176248, EPI_ISL_2176250, EPI_ISL_2176253, EPI_ISL_2176257, EPI_ISL_2176261, EPI_ISL_2176263, EPI_ISL_2176266, EPI_ISL_2176270, EPI_ISL_2176276, EPI_ISL_2176279, EPI_ISL_2176284, EPI_ISL_2176287, EPI_ISL_2176290, EPI_ISL_2176294, EPI_ISL_2176297 | | | |
| see above | DC Public Health Lab/ Dept. of Forensic Sciences | DC Public Health Lab/ Dept. of Forensic Sciences | Janis Doss, Scott Nguyen, Elizabeth Zelaya, Sarah Scott, Connie Maza, Monica Mann, Brittany Hamilton, David Payne, Jocelyn Hauser |
| EPI_ISL_2177370, EPI_ISL_2177373, EPI_ISL_2177374, EPI_ISL_2177376, EPI_ISL_2177378, EPI_ISL_2177380, EPI_ISL_2177382, EPI_ISL_2177384, EPI_ISL_2177386, EPI_ISL_2177388, EPI_ISL_2177393, EPI_ISL_2177396, EPI_ISL_2177400, EPI_ISL_2177402, EPI_ISL_2177404, EPI_ISL_2177406, EPI_ISL_2177408, EPI_ISL_2177410, EPI_ISL_2177412, EPI_ISL_2177414, EPI_ISL_2177416, EPI_ISL_2177418, EPI_ISL_2177420, EPI_ISL_2177422, EPI_ISL_2177424, EPI_ISL_2177426, EPI_ISL_2177428, EPI_ISL_2177430, EPI_ISL_2177432, EPI_ISL_2177433, EPI_ISL_2177434, EPI_ISL_2177437, EPI_ISL_2177439, EPI_ISL_2177441, EPI_ISL_2177444, EPI_ISL_2177446, EPI_ISL_2177448, EPI_ISL_2177449, EPI_ISL_2177451, EPI_ISL_2177453, EPI_ISL_2177455, EPI_ISL_2177457, EPI_ISL_2177459, EPI_ISL_2177461, EPI_ISL_2177462, EPI_ISL_2177464, EPI_ISL_2177468, EPI_ISL_2177472, EPI_ISL_2177475, EPI_ISL_2177476, EPI_ISL_2177478, EPI_ISL_2177480, EPI_ISL_2177482, EPI_ISL_2177484, EPI_ISL_2177486, EPI_ISL_2177488, EPI_ISL_2177495, EPI_ISL_2177500, EPI_ISL_2177502, EPI_ISL_2177505, EPI_ISL_2177507, EPI_ISL_2177511, EPI_ISL_2177513, EPI_ISL_2177515, EPI_ISL_2177517, EPI_ISL_2177519 | | | |
| see above | TXDSHS | TXDSHS | Rashmi Tuladhar, Bonnie Oh, Jenny Zhang, Maliha Rahman, Mayela Pedrueza, Anita Pokharel, Karen Bobier, Lorraine Rodriguez, Myong Koag, Chun Wang, Rachel Lee, Grace Kubin |
| EPI_ISL_2178511, EPI_ISL_2178512, EPI_ISL_2178514, EPI_ISL_2178515, EPI_ISL_2178516, EPI_ISL_2178517, EPI_ISL_2178518, EPI_ISL_2178519, EPI_ISL_2178520, EPI_ISL_2178521, EPI_ISL_2178522, EPI_ISL_2178523, EPI_ISL_2178524, EPI_ISL_2178525, EPI_ISL_2178526, EPI_ISL_2178527 | | | |
| see above | INNO Diagnostics Reference Laboratory | RCMI-Center for Research Resources, Ponce Research Institute | Vanessa Rivera-Amill, Andrea Arias-García, Raphael Sánchez-Torres |
| EPI_ISL_2178528, EPI_ISL_2178531, EPI_ISL_2178532, EPI_ISL_2178533, EPI_ISL_2178534, EPI_ISL_2178535, EPI_ISL_2178536, EPI_ISL_2178539, EPI_ISL_2178540, EPI_ISL_2178541, EPI_ISL_2178542, EPI_ISL_2178543 | | | |
| see above | Wyoming Public Health Laboratory | Wyoming Public Health Laboratory | Jim Mildenberger, Taylor Fearing, Channing Weber, Ashley Norberg, Chayse Rowley, Marley Goetz, Brian Dominguez, Elliot Thomasson, Brittany Oher, Sam Britz, Cari Sloma, and Rob Christensen |
| EPI_ISL_2178544 | Mayo Clinic & Mayo Clinic Laboratories | Minnesota Department of Health, Public Health Laboratory | Alexandra Lorentz, Jacob Garfin, Matt Plumb, and Xiong Wang |
| EPI_ISL_2178545, EPI_ISL_2178546 | M Health Fairview | Minnesota Department of Health, Public Health Laboratory | Alexandra Lorentz, Jacob Garfin, Matt Plumb, and Xiong Wang |
| EPI_ISL_2178547, EPI_ISL_2178548 | Centracare Laboratory Services | Minnesota Department of Health, Public Health Laboratory | Alexandra Lorentz, Jacob Garfin, Matt Plumb, and Xiong Wang |
| EPI_ISL_2178549 | Mayo Clinic & Mayo Clinic Laboratories | Minnesota Department of Health, Public Health Laboratory | Alexandra Lorentz, Jacob Garfin, Matt Plumb, and Xiong Wang |
| EPI_ISL_2178550 | M Health Fairview | Minnesota Department of Health, Public Health Laboratory | Alexandra Lorentz, Jacob Garfin, Matt Plumb, and Xiong Wang |
| EPI_ISL_2178551 | Centracare Laboratory Services | Minnesota Department of Health, Public Health Laboratory | Alexandra Lorentz, Jacob Garfin, Matt Plumb, and Xiong Wang |
| EPI_ISL_2178552 | M Health Fairview | Minnesota Department of Health, Public Health Laboratory | Alexandra Lorentz, Jacob Garfin, Matt Plumb, and Xiong Wang |
| EPI_ISL_2178553, EPI_ISL_2178554 | Centracare Laboratory Services | Minnesota Department of Health, Public Health Laboratory | Alexandra Lorentz, Jacob Garfin, Matt Plumb, and Xiong Wang |
| EPI_ISL_2178555 | Mayo Clinic & Mayo Clinic Laboratories | Minnesota Department of Health, Public Health Laboratory | Alexandra Lorentz, Jacob Garfin, Matt Plumb, and Xiong Wang |
| EPI_ISL_2178556 | M Health Fairview | Minnesota Department of Health, Public Health Laboratory | Alexandra Lorentz, Jacob Garfin, Matt Plumb, and Xiong Wang |
| EPI_ISL_2178557 | Centracare Laboratory Services | Minnesota Department of Health, Public Health Laboratory | Alexandra Lorentz, Jacob Garfin, Matt Plumb, and Xiong Wang |
| EPI_ISL_2178558, EPI_ISL_2178559 | M Health Fairview | Minnesota Department of Health, Public Health Laboratory | Alexandra Lorentz, Jacob Garfin, Matt Plumb, and Xiong Wang |
| EPI_ISL_2178560, EPI_ISL_2178561, EPI_ISL_2178562 | Centracare Laboratory Services | Minnesota Department of Health, Public Health Laboratory | Alexandra Lorentz, Jacob Garfin, Matt Plumb, and Xiong Wang |
| EPI_ISL_2178563, EPI_ISL_2178564, EPI_ISL_2178565, EPI_ISL_2178566, EPI_ISL_2178567, EPI_ISL_2178568 | M Health Fairview | Minnesota Department of Health, Public Health Laboratory | Alexandra Lorentz, Jacob Garfin, Matt Plumb, and Xiong Wang |
| EPI_ISL_2178569, EPI_ISL_2178570, EPI_ISL_2178571, EPI_ISL_2178572, EPI_ISL_2178573, EPI_ISL_2178574, EPI_ISL_2178575, EPI_ISL_2178576, EPI_ISL_2178577, EPI_ISL_2178578, EPI_ISL_2178579, EPI_ISL_2178580, EPI_ISL_2178581, EPI_ISL_2178582 | | | |
| see above | Centracare Laboratory Services | Minnesota Department of Health, Public Health Laboratory | Alexandra Lorentz, Jacob Garfin, Matt Plumb, and Xiong Wang |
| EPI_ISL_2178583 | Mayo Clinic & Mayo Clinic Laboratories | Minnesota Department of Health, Public Health Laboratory | Alexandra Lorentz, Jacob Garfin, Matt Plumb, and Xiong Wang |
| EPI_ISL_2178584, EPI_ISL_2178585, EPI_ISL_2178586, EPI_ISL_2178587, EPI_ISL_2178588, EPI_ISL_2178589, EPI_ISL_2178590, EPI_ISL_2178591, EPI_ISL_2178592 | M Health Fairview | Minnesota Department of Health, Public Health Laboratory | Alexandra Lorentz, Jacob Garfin, Matt Plumb, and Xiong Wang |
| EPI_ISL_2178593, EPI_ISL_2178594, EPI_ISL_2178595, EPI_ISL_2178596, EPI_ISL_2178597, EPI_ISL_2178598, EPI_ISL_2178599 | Centracare Laboratory Services | Minnesota Department of Health, Public Health Laboratory | Alexandra Lorentz, Jacob Garfin, Matt Plumb, and Xiong Wang |
| EPI_ISL_2178600, EPI_ISL_2178601 | Mayo Clinic & Mayo Clinic Laboratories | Minnesota Department of Health, Public Health Laboratory | Alexandra Lorentz, Jacob Garfin, Matt Plumb, and Xiong Wang |
| EPI_ISL_2178602, EPI_ISL_2178603 | M Health Fairview | Minnesota Department of Health, Public Health Laboratory | Alexandra Lorentz, Jacob Garfin, Matt Plumb, and Xiong Wang |
| EPI_ISL_2178604, EPI_ISL_2178607, EPI_ISL_2178608, EPI_ISL_2178609 | Centracare Laboratory Services | Minnesota Department of Health, Public Health Laboratory | Alexandra Lorentz, Jacob Garfin, Matt Plumb, and Xiong Wang |
| EPI_ISL_2178610, EPI_ISL_2178611, EPI_ISL_2178612, EPI_ISL_2178613 | M Health Fairview | Minnesota Department of Health, Public Health Laboratory | Alexandra Lorentz, Jacob Garfin, Matt Plumb, and Xiong Wang |
| EPI_ISL_2178614, EPI_ISL_2178615, EPI_ISL_2178616, EPI_ISL_2178617, EPI_ISL_2178618, EPI_ISL_2178619 | Centracare Laboratory Services | Minnesota Department of Health, Public Health Laboratory | Alexandra Lorentz, Jacob Garfin, Matt Plumb, and Xiong Wang |
| EPI_ISL_2178620 | Mayo Clinic & Mayo Clinic Laboratories | Minnesota Department of Health, Public Health Laboratory | Alexandra Lorentz, Jacob Garfin, Matt Plumb, and Xiong Wang |
| EPI_ISL_2178621, EPI_ISL_2178622 | M Health Fairview | Minnesota Department of Health, Public Health Laboratory | Alexandra Lorentz, Jacob Garfin, Matt Plumb, and Xiong Wang |
| EPI_ISL_2178623, EPI_ISL_2178624, EPI_ISL_2178625, EPI_ISL_2178626, EPI_ISL_2178627, EPI_ISL_2178628, EPI_ISL_2178629, EPI_ISL_2178630, EPI_ISL_2178631, EPI_ISL_2178632, EPI_ISL_2178633, EPI_ISL_2178634, EPI_ISL_2178635, EPI_ISL_2178636, EPI_ISL_2178637, EPI_ISL_2178638, EPI_ISL_2178639, EPI_ISL_2178640, EPI_ISL_2178641, EPI_ISL_2178642, EPI_ISL_2178643, EPI_ISL_2178644, EPI_ISL_2178645, EPI_ISL_2178646, EPI_ISL_2178647, EPI_ISL_2178648, EPI_ISL_2178649, EPI_ISL_2178650, EPI_ISL_2178651, EPI_ISL_2178652, EPI_ISL_2178653, EPI_ISL_2178654, EPI_ISL_2178659, EPI_ISL_2178660, EPI_ISL_2178661, EPI_ISL_2178662, EPI_ISL_2178663, EPI_ISL_2178664, EPI_ISL_2178665 | | | |
| see above | Minnesota Department of Health, Public Health Laboratory | Minnesota Department of Health, Public Health Laboratory | Alexandra Lorentz, Jacob Garfin, Matt Plumb, and Xiong Wang |

| | | | |
|---|---|---|---|
| EPI_ISL_2178666 | Atlanta VA Medical Center | Pathogen Discovery, Respiratory Viruses Branch, Division of Viral Diseases, Centers for Disease Control and Prevention | Ying Tao, Yan Li, Jing Zhang, Anna Kelleher, Brian Lynch, Krista Queen, Anna Uehara, Peter Cook, Han Jia Justin Ng, Rachel Marine, Clinton R. Paden, Dhwani Batra, Haibin Wang, Tara Coalter, Jasmine Padilla, Morgan Davis, Mili Sheth, Sarah Nobles, Mark Burroughs, Justin Lee, Adam Retchless, Suxiang Tong |
| EPI_ISL_2178725, EPI_ISL_2178726 | Centro de Investigación Biomédica del Noreste (CIBIN) | Centro de Investigación en Enfermedades Infecciosas (CIENI), Instituto Nacional de Enfermedades Respiratorias (INER) | Consorcio Mexicano de Vigilancia Genómica (CoViGen-Mex). Authors (in alphabetical order): Julio Elias Alvarado-Yaah, Carlos F. Arias, Santiago Ávila-Ríos, Víctor Hugo Borja-Aburto, Celia Boukadida, Juan Bautista Chale-Dzul, Célida Duque Molina, José Antonio Enciso-Moreno, Gloria Elena Espinosa-Ayala, Fernando Fontove-Herrera, Victor Eduardo Garcia-Arias, Concepción Grajales-Muñiz, Ricardo Grande, Alfredo Herrera-Estrella, Carla Ivón Herrera-Najera, Pavel Isa, Brenda Irasema Maldonado-Meza, Bernardo Martínez-Miguel, Margarita Matías-Florentino, María Guadalupe de Jesús Mireles-Rivera, Gloria María Molina-Salinas, Hector Montoya-Fuentes, José Esteban Muñoz-Medina, José de Jesús Nuñez-Contreras, Alicia Ocaña-Mondragón, Luis Alberto Ochoa-Carrera, Hector Esteban Paz-Juárez, Francisco Pulido, Helen Haydee Fernanda Ramírez-Plascencia, Angel Gustavo Salas-Lais, Alejandro Sanchez-Flores, Clara Esperanza Santacruz-Tinoco, María Guadalupe Santiago-Mauricio, Nelly Sélem-Mojica, Blanca Taboada, Gloria Vazquez |
| EPI_ISL_2178727, EPI_ISL_2178728 | Unidad de Investigación Médica de Yucatán (UIMY) | Centro de Investigación en Enfermedades Infecciosas (CIENI), Instituto Nacional de Enfermedades Respiratorias (INER) | Consorcio Mexicano de Vigilancia Genómica (CoViGen-Mex). Authors (in alphabetical order): Julio Elias Alvarado-Yaah, Carlos F. Arias, Santiago Ávila-Ríos, Víctor Hugo Borja-Aburto, Celia Boukadida, Juan Bautista Chale-Dzul, Célida Duque Molina, José Antonio Enciso-Moreno, Gloria Elena Espinosa-Ayala, Fernando Fontove-Herrera, Victor Eduardo Garcia-Arias, Concepción Grajales-Muñiz, Ricardo Grande, Alfredo Herrera-Estrella, Carla Ivón Herrera-Najera, Pavel Isa, Brenda Irasema Maldonado-Meza, Bernardo Martínez-Miguel, Margarita Matías-Florentino, María Guadalupe de Jesús Mireles-Rivera, Gloria María Molina-Salinas, Hector Montoya-Fuentes, José Esteban Muñoz-Medina, José de Jesús Nuñez-Contreras, Alicia Ocaña-Mondragón, Luis Alberto Ochoa-Carrera, Hector Esteban Paz-Juárez, Francisco Pulido, Helen Haydee Fernanda Ramírez-Plascencia, Angel Gustavo Salas-Lais, Alejandro Sanchez-Flores, Clara Esperanza Santacruz-Tinoco, María Guadalupe Santiago-Mauricio, Nelly Sélem-Mojica, Blanca Taboada, Gloria Vazquez |
| EPI_ISL_2178729 | Centro de Investigación Biomédica de Occidente (CIBO) | Centro de Investigación en Enfermedades Infecciosas (CIENI), Instituto Nacional de Enfermedades Respiratorias (INER) | Consorcio Mexicano de Vigilancia Genómica (CoViGen-Mex). Authors (in alphabetical order): Julio Elias Alvarado-Yaah, Carlos F. Arias, Santiago Ávila-Ríos, Víctor Hugo Borja-Aburto, Celia Boukadida, Juan Bautista Chale-Dzul, Célida Duque Molina, José Antonio Enciso-Moreno, Gloria Elena Espinosa-Ayala, Fernando Fontove-Herrera, Victor Eduardo Garcia-Arias, Concepción Grajales-Muñiz, Ricardo Grande, Alfredo Herrera-Estrella, Carla Ivón Herrera-Najera, Pavel Isa, Brenda Irasema Maldonado-Meza, Bernardo Martínez-Miguel, Margarita Matías-Florentino, María Guadalupe de Jesús Mireles-Rivera, Gloria María Molina-Salinas, Hector Montoya-Fuentes, José Esteban Muñoz-Medina, José de Jesús Nuñez-Contreras, Alicia Ocaña-Mondragón, Luis Alberto Ochoa-Carrera, Hector Esteban Paz-Juárez, Francisco Pulido, Helen Haydee Fernanda Ramírez-Plascencia, Angel Gustavo Salas-Lais, Alejandro Sanchez-Flores, Clara Esperanza Santacruz-Tinoco, María Guadalupe Santiago-Mauricio, Nelly Sélem-Mojica, Blanca Taboada, Gloria Vazquez |
| EPI_ISL_2178730, EPI_ISL_2178731, EPI_ISL_2178732, EPI_ISL_2178733, EPI_ISL_2178734, EPI_ISL_2178735, EPI_ISL_2178736, EPI_ISL_2178737, EPI_ISL_2178738, EPI_ISL_2178739, EPI_ISL_2178740, EPI_ISL_2178741 | | | |
| see above | Laboratorio Central de Epidemiología (LCE) | Centro de Investigación en Enfermedades Infecciosas (CIENI), Instituto Nacional de Enfermedades Respiratorias (INER) | Consorcio Mexicano de Vigilancia Genómica (CoViGen-Mex). Authors (in alphabetical order): Julio Elias Alvarado-Yaah, Carlos F. Arias, Santiago Ávila-Ríos, Víctor Hugo Borja-Aburto, Celia Boukadida, Juan Bautista Chale-Dzul, Célida Duque Molina, José Antonio Enciso-Moreno, Gloria Elena Espinosa-Ayala, Fernando Fontove-Herrera, Victor Eduardo Garcia-Arias, Concepción Grajales-Muñiz, Ricardo Grande, Alfredo Herrera-Estrella, Carla Ivón Herrera-Najera, Pavel Isa, Brenda Irasema Maldonado-Meza, Bernardo Martínez-Miguel, Margarita Matías-Florentino, María Guadalupe de Jesús Mireles-Rivera, Gloria María Molina-Salinas, Hector Montoya-Fuentes, José Esteban Muñoz-Medina, José de Jesús Nuñez-Contreras, Alicia Ocaña-Mondragón, Luis Alberto Ochoa-Carrera, Hector Esteban Paz-Juárez, Francisco Pulido, Helen Haydee Fernanda Ramírez-Plascencia, Angel Gustavo Salas-Lais, Alejandro Sanchez-Flores, Clara Esperanza Santacruz-Tinoco, María Guadalupe Santiago-Mauricio, Nelly Sélem-Mojica, Blanca Taboada, Gloria Vazquez |
| EPI_ISL_2178742, EPI_ISL_2178743 | Instituto Nacional de Medicina Genomica | Centro de Investigación en Enfermedades Infecciosas (CIENI), Instituto Nacional de Enfermedades Respiratorias (INER) | Matias-Florentino M, Perez-Garcia M, Hidalgo-Miranda A, Mendoza-Vargas A, Reyes-Grajeda JP, Cedro-Tanda A, Arriaga-Canon C, Herrera-Montalvo LA, Reyes-Teran G, Avila-Rios S, Boukadida C |
| EPI_ISL_2178748 | Instituto Nacional de Medicina Genomica | Instituto Nacional de Medicina Genomica | Hidalgo-Miranda A, Cedro-Tanda A, Mendoza-Vargas A, Reyes-Grajeda JP, Cisneros-Villanueva M, Orjuela-Rodríguez M, Gonzalez-Barrera D, Rangel-DeLeon D, Munguia-Garza P, Ramirez-Vega O, Escobar-Arrazola, M, Herrera-Montalvo LA. |
| EPI_ISL_2178749, EPI_ISL_2178750, EPI_ISL_2178751, EPI_ISL_2178752, EPI_ISL_2178753, EPI_ISL_2178754, EPI_ISL_2178755, EPI_ISL_2178756, EPI_ISL_2178757, EPI_ISL_2178758, EPI_ISL_2178759 | | | |
| see above | Salud Digna | Instituto Nacional de Medicina Genomica | Hidalgo-Miranda A, Cedro-Tanda A, Mendoza-Vargas A, Reyes-Grajeda JP, Cisneros-Villanueva M, Orjuela-Rodríguez M, Abraham Campos-Romero, Moreno-Camacho José Luis, Rodriguez-Gallegos Jorge, Luna-Ruiz Marco, Gonzalez-Barrera D, Rangel-DeLeon D, Munguia-Garza P, Ramirez-Vega O, Escobar-Arrazola, M, Herrera-Montalvo LA. |
| EPI_ISL_2178765, EPI_ISL_2178766, EPI_ISL_2178767, EPI_ISL_2178768, EPI_ISL_2178769, EPI_ISL_2178770, EPI_ISL_2178771, EPI_ISL_2178772, EPI_ISL_2178773, EPI_ISL_2178774, EPI_ISL_2178775, EPI_ISL_2178776, EPI_ISL_2178777, EPI_ISL_2178778 | | | |
| see above | DeRisi Lab, University of California, San Francisco | DeRisi Lab, University of California, San Francisco | Sabrina Mann, Anthea Mitchell, Jamin Liu, Sara Sunshine, Matthew Laurie, Genay Pilarowski, Amy Kistler, Manu Vanaerschot, Patrick Ayscue, Lucy Li, Eric Chow, IDseq Team, James Peng, Carina Marquez, Luis Rubio, Gabriel Chamie, Diane Jones, Jon Jacobo, Susana Rojas, Susy Rojas, Valerie Tulier-Laiwa, Douglas Black, Jackie Martinez, Maya Petersen, Diane Havlir, and Joseph DeRisi |
| EPI_ISL_2179052, EPI_ISL_2179053, EPI_ISL_2179054, EPI_ISL_2179057, EPI_ISL_2179059, EPI_ISL_2179060, EPI_ISL_2179061, EPI_ISL_2179063, EPI_ISL_2179064, EPI_ISL_2179065, EPI_ISL_2179066, EPI_ISL_2179067, EPI_ISL_2179068, EPI_ISL_2179069, EPI_ISL_2179070, EPI_ISL_2179071, EPI_ISL_2179072, EPI_ISL_2179073, EPI_ISL_2179075, EPI_ISL_2179076, EPI_ISL_2179082, EPI_ISL_2179083, EPI_ISL_2179086 | | | |
| see above | City of Milwaukee Health Department Laboratory | City of Milwaukee Health Department Laboratory | Sanjib Bhattacharyya, Manjeet Khubbar, Amy Bauer, Jennifer Lentz, Samantha Scott |
| EPI_ISL_2179088 | AR Dept. of Health-PHL, Molecular Diagnostics | Genomics and Discovery, Respiratory Viruses Branch, Division of Viral Diseases, Centers for Disease Control and Prevention | Yan Li, Jing Zhang, Anna Kelleher, Ying Tao, Brian Lynch, Krista Queen, Anna Uehara, Peter Cook, Han Jia Justin Ng, Rachel Marine, Clinton R. Paden, Dhwani Batra, Haibin Wang, Tara Coalter, Jasmine Padilla, Morgan Davis, Mili Sheth, Sarah Nobles, Mark Burroughs, Justin Lee, Adam Retchless, Suxiang Tong |
| EPI_ISL_2179090 | WVDHHR - Office of Laboratory Services | Genomics and Discovery, Respiratory Viruses Branch, Division of Viral Diseases, Centers for Disease Control and Prevention | Yan Li, Jing Zhang, Anna Kelleher, Ying Tao, Brian Lynch, Krista Queen, Anna Uehara, Peter Cook, Han Jia Justin Ng, Rachel Marine, Clinton R. Paden, Dhwani Batra, Haibin Wang, Tara Coalter, Jasmine Padilla, Morgan Davis, Mili Sheth, Sarah Nobles, Mark Burroughs, Justin Lee, Adam Retchless, Suxiang Tong |
| EPI_ISL_2179091, EPI_ISL_2179092, EPI_ISL_2179093 | TN Division of Laboratory Services | Genomics and Discovery, Respiratory Viruses Branch, Division of Viral Diseases, Centers for Disease Control and Prevention | Yan Li, Jing Zhang, Anna Kelleher, Ying Tao, Brian Lynch, Krista Queen, Anna Uehara, Peter Cook, Han Jia Justin Ng, Rachel Marine, Clinton R. Paden, Dhwani Batra, Haibin Wang, Tara Coalter, Jasmine Padilla, Morgan Davis, Mili Sheth, Sarah Nobles, Mark Burroughs, Justin Lee, Adam Retchless, Suxiang Tong |
| EPI_ISL_2179094, EPI_ISL_2179095, EPI_ISL_2179097, EPI_ISL_2179098, EPI_ISL_2179099, EPI_ISL_2179101, EPI_ISL_2179102, EPI_ISL_2179103, EPI_ISL_2179104, EPI_ISL_2179105, EPI_ISL_2179106, EPI_ISL_2179107, EPI_ISL_2179108, EPI_ISL_2179109, EPI_ISL_2179110, EPI_ISL_2179111, EPI_ISL_2179112, EPI_ISL_2179113, EPI_ISL_2179114, EPI_ISL_2179115, EPI_ISL_2179116, EPI_ISL_2179117, EPI_ISL_2179118, EPI_ISL_2179119, EPI_ISL_2179120, EPI_ISL_2179121, EPI_ISL_2179122, EPI_ISL_2179125, EPI_ISL_2179126, EPI_ISL_2179127, EPI_ISL_2179128, EPI_ISL_2179129, EPI_ISL_2179130, EPI_ISL_2179131, EPI_ISL_2179132, EPI_ISL_2179133, EPI_ISL_2179134, EPI_ISL_2179135, EPI_ISL_2179136, EPI_ISL_2179137, EPI_ISL_2179138, EPI_ISL_2179139, EPI_ISL_2179140, EPI_ISL_2179141, EPI_ISL_2179142, EPI_ISL_2179144, EPI_ISL_2179145, EPI_ISL_2179146, EPI_ISL_2179148, EPI_ISL_2179149, EPI_ISL_2179150, EPI_ISL_2179152, EPI_ISL_2179153, EPI_ISL_2179154, EPI_ISL_2179155, EPI_ISL_2179156, EPI_ISL_2179158, EPI_ISL_2179161, EPI_ISL_2179162, EPI_ISL_2179163, EPI_ISL_2179164, EPI_ISL_2179165 | | | |
| see above | UTMG Pathology LLC | UTMG Pathology LLC | Parasakthy Kumaravelu, Timothy Hodge, Kelsey Matande, Tiara Hymon, Vickie Baselski, Abdallah Azouz |
| EPI_ISL_2179166 | HealthQuest Esoterics | New York City Public Health Laboratory | Jade Wang, et al. |
| EPI_ISL_2179167, EPI_ISL_2179168 | DOHMH Morrisania | New York City Public Health Laboratory | Jade Wang, et al. |
| EPI_ISL_2179169 | DOHMH Corona | New York City Public Health Laboratory | Jade Wang, et al. |
| EPI_ISL_2179170, EPI_ISL_2179171 | DOHMH PHL | New York City Public Health Laboratory | Jade Wang, et al. |
| EPI_ISL_2179172, EPI_ISL_2179173 | DOHMH Fort Greene | New York City Public Health Laboratory | Jade Wang, et al. |
| EPI_ISL_2179174 | DOHMH Crown Heights | New York City Public Health Laboratory | Jade Wang, et al. |
| EPI_ISL_2179175 | DOHMH Riverside | New York City Public Health Laboratory | Jade Wang, et al. |
| EPI_ISL_2179176 | DOHMH Morrisania | New York City Public Health Laboratory | Jade Wang, et al. |
| EPI_ISL_2179177, EPI_ISL_2179178 | DOHMH Central Harlem | New York City Public Health Laboratory | Jade Wang, et al. |
| EPI_ISL_2179179 | DOHMH PHL | New York City Public Health Laboratory | Jade Wang, et al. |
| EPI_ISL_2179180 | DOHMH Jamaica | New York City Public Health Laboratory | Jade Wang, et al. |
| EPI_ISL_2179181, EPI_ISL_2179182 | DOHMH Corona | New York City Public Health Laboratory | Jade Wang, et al. |
| EPI_ISL_2179183, EPI_ISL_2179184 | DOHMH Morrisania | New York City Public Health Laboratory | Jade Wang, et al. |
| EPI_ISL_2179185 | DOHMH Crown Heights | New York City Public Health Laboratory | Jade Wang, et al. |
| EPI_ISL_2179186, EPI_ISL_2179187 | DOHMH Jamaica | New York City Public Health Laboratory | Jade Wang, et al. |
| EPI_ISL_2179188, EPI_ISL_2179189 | Department of Homeless Services | New York City Public Health Laboratory | Jade Wang, et al. |
| EPI_ISL_2179190 | OCME Office Of Chief Medical Examiner | New York City Public Health Laboratory | Jade Wang, et al. |
| EPI_ISL_2179192, EPI_ISL_2179193, EPI_ISL_2179194, EPI_ISL_2179195, EPI_ISL_2179196, EPI_ISL_2179197, EPI_ISL_2179198, EPI_ISL_2179199, EPI_ISL_2179200, EPI_ISL_2179201, EPI_ISL_2179202, EPI_ISL_2179203, EPI_ISL_2179204, EPI_ISL_2179205, EPI_ISL_2179206, EPI_ISL_2179207, EPI_ISL_2179208, EPI_ISL_2179209, EPI_ISL_2179210, EPI_ISL_2179211, EPI_ISL_2179212, EPI_ISL_2179213, EPI_ISL_2179214, EPI_ISL_2179215, EPI_ISL_2179216, EPI_ISL_2179217, EPI_ISL_2179218, EPI_ISL_2179219, EPI_ISL_2179220, EPI_ISL_2179221, EPI_ISL_2179222, EPI_ISL_2179223, EPI_ISL_2179224, EPI_ISL_2179225, EPI_ISL_2179226, EPI_ISL_2179227, EPI_ISL_2179228, EPI_ISL_2179229, EPI_ISL_2179230, EPI_ISL_2179231, EPI_ISL_2179232, EPI_ISL_2179233, EPI_ISL_2179234, EPI_ISL_2179235, EPI_ISL_2179236, EPI_ISL_2179237, EPI_ISL_2179238, EPI_ISL_2179239, EPI_ISL_2179240, EPI_ISL_2179241, EPI_ISL_2179242, EPI_ISL_2179243, EPI_ISL_2179244, EPI_ISL_2179245, EPI_ISL_2179246, EPI_ISL_2179247, EPI_ISL_2179248, EPI_ISL_2179249, EPI_ISL_2179250, EPI_ISL_2179251, EPI_ISL_2179252, EPI_ISL_2179253, EPI_ISL_2179254, EPI_ISL_2179255, EPI_ISL_2179256, EPI_ISL_2179257, EPI_ISL_2179258, EPI_ISL_2179259, EPI_ISL_2179260, EPI_ISL_2179261, EPI_ISL_2179262, EPI_ISL_2179263, EPI_ISL_2179264, EPI_ISL_2179265, EPI_ISL_2179266, EPI_ISL_2179267, EPI_ISL_2179268, EPI_ISL_2179269, EPI_ISL_2179271, EPI_ISL_2179272, EPI_ISL_2179273, EPI_ISL_2179274 | | | |
| see above | UCSC Genomics Institute | UCSC Genomics Institute | Mark Akeson, Ikenna Anigbogu, Eric Beraut, Isabel Bjork, Molly Cassatt-Johnstone, Terren Chang, Russell Corbett-Detig, Namrita Dhillon, Maximilian Haeussler, David Haussler, Angie Hinrichs, Miten Jain, Joshua Kapp, A. Marm Kilpatrick, Jakob McBroome, Hugh Olsen, Jeremy Sanford, Beth Shapiro, Michael Stone, Bryan Thornlow, Yatish Turakhia, Ciara Wanket |
| EPI_ISL_2179648 | Oregon State Public Health Laboratory | Oregon State Public Health Laboratory | Rafia Razzaque, Eugene Yeboah, Vanda Makris, Laura Tsaknaridis, John Fontana and Shane Sevey |
| EPI_ISL_2179825, EPI_ISL_2179826, EPI_ISL_2179827, EPI_ISL_2179828, EPI_ISL_2179829, EPI_ISL_2179830, EPI_ISL_2179831, EPI_ISL_2179832, EPI_ISL_2179833, EPI_ISL_2179835, EPI_ISL_2179836, EPI_ISL_2179837, EPI_ISL_2179838, EPI_ISL_2179839, EPI_ISL_2179840, EPI_ISL_2179842, EPI_ISL_2179843, EPI_ISL_2179845, EPI_ISL_2179846, EPI_ISL_2179847, EPI_ISL_2179848, EPI_ISL_2179850, EPI_ISL_2179851, EPI_ISL_2179852, EPI_ISL_2179853, EPI_ISL_2179854, EPI_ISL_2179855, EPI_ISL_2179856, EPI_ISL_2179857, EPI_ISL_2179858, EPI_ISL_2179859, EPI_ISL_2179860, EPI_ISL_2179861, EPI_ISL_2179862, EPI_ISL_2179863, EPI_ISL_2179864, EPI_ISL_2179865, EPI_ISL_2179866, EPI_ISL_2179867, EPI_ISL_2179868, EPI_ISL_2179869, EPI_ISL_2179870, EPI_ISL_2179871, EPI_ISL_2179872, EPI_ISL_2179873, EPI_ISL_2179874, EPI_ISL_2179875, EPI_ISL_2179876, EPI_ISL_2179877, EPI_ISL_2179878, EPI_ISL_2179879, EPI_ISL_2179880, EPI_ISL_2179881, EPI_ISL_2179882, EPI_ISL_2179883, EPI_ISL_2179884, EPI_ISL_2179885, EPI_ISL_2179887, EPI_ISL_2179888, EPI_ISL_2179889, EPI_ISL_2179890, EPI_ISL_2179891, EPI_ISL_2179892, EPI_ISL_2179893, EPI_ISL_2179894, EPI_ISL_2179895, EPI_ISL_2179896, EPI_ISL_2179897, EPI_ISL_2179898, EPI_ISL_2179899, EPI_ISL_2179900, EPI_ISL_2179901, EPI_ISL_2179902, EPI_ISL_2179903, EPI_ISL_2179904, EPI_ISL_2179905, EPI_ISL_2179906, EPI_ISL_2179907, EPI_ISL_2179908, EPI_ISL_2179909, EPI_ISL_2179910, EPI_ISL_2179911, EPI_ISL_2179912, EPI_ISL_2179913, EPI_ISL_2179914, EPI_ISL_2179915, EPI_ISL_2179916, EPI_ISL_2179917, EPI_ISL_2179918, EPI_ISL_2179919, EPI_ISL_2179920 | | | |
| see above | Fulgent Genetics | Centers for Disease Control and Prevention Division of Viral Diseases, Pathogen Discovery | Dakota Howard, Dhwani Batra, Peter W. Cook, Kara Moser, Adrian Paskey, Jason Caravas, Benjamin Rambo-Martin, Shatavia Morrison, Christopher Gulvick, Scott Sammons, Yvette Unoarumhi, Darlene Wagner, Matthew Schmerer, Harry Gao, Mickey Li, John Gao, Joseph Fierro, Benafsh Sapra, Becky Tsai, Yan Meng, Doreen Ng, James Xie, Clinton R. Paden, Duncan MacCannell |
| EPI_ISL_2179921, EPI_ISL_2179922, EPI_ISL_2179923, EPI_ISL_2179924, EPI_ISL_2179925, EPI_ISL_2179926, EPI_ISL_2179927, EPI_ISL_2179928, EPI_ISL_2179929, EPI_ISL_2179930, EPI_ISL_2179931, EPI_ISL_2179932, EPI_ISL_2179933, EPI_ISL_2179934, EPI_ISL_2179935, EPI_ISL_2179936, EPI_ISL_2179937, EPI_ISL_2179938, EPI_ISL_2179939, EPI_ISL_2179940, EPI_ISL_2179941, EPI_ISL_2179942, EPI_ISL_2179943, EPI_ISL_2179944, EPI_ISL_2179945, EPI_ISL_2179946, EPI_ISL_2179947, EPI_ISL_2179948, EPI_ISL_2179949, EPI_ISL_2179950, EPI_ISL_2179951, EPI_ISL_2179952, EPI_ISL_2179953, EPI_ISL_2179954, EPI_ISL_2179955, EPI_ISL_2179956, EPI_ISL_2179957, EPI_ISL_2179958, EPI_ISL_2179959, EPI_ISL_2179960, EPI_ISL_2179961, EPI_ISL_2179962, EPI_ISL_2179963, EPI_ISL_2179964, EPI_ISL_2179965, EPI_ISL_2179966, EPI_ISL_2179967, EPI_ISL_2179968, EPI_ISL_2179969, EPI_ISL_2179970, EPI_ISL_2179971, EPI_ISL_2179972, EPI_ISL_2179973, EPI_ISL_2179974, EPI_ISL_2179975, EPI_ISL_2179976, EPI_ISL_2179977, EPI_ISL_2179978, EPI_ISL_2179979, EPI_ISL_2179980, EPI_ISL_2179981, EPI_ISL_2179982, EPI_ISL_2179983, EPI_ISL_2179984, EPI_ISL_2179985, EPI_ISL_2179986, EPI_ISL_2180458, EPI_ISL_2180459, EPI_ISL_2180460, EPI_ISL_2180461, EPI_ISL_2180462, EPI_ISL_2180463, EPI_ISL_2180464, EPI_ISL_2180465, EPI_ISL_2180466, EPI_ISL_2180467, EPI_ISL_2180468, EPI_ISL_2180469, EPI_ISL_2180470, EPI_ISL_2180471, EPI_ISL_2180472, EPI_ISL_2180473, EPI_ISL_2180474, EPI_ISL_2180475, EPI_ISL_2180476, EPI_ISL_2180477, EPI_ISL_2180478, EPI_ISL_2180479, | | | |

EPI_ISL_2180480, EPI_ISL_2180481, EPI_ISL_2180482, EPI_ISL_2180483, EPI_ISL_2180484, EPI_ISL_2180485, EPI_ISL_2180486, EPI_ISL_2180487, EPI_ISL_2180488, EPI_ISL_2180489, EPI_ISL_2180490, EPI_ISL_2180491, EPI_ISL_2180492, EPI_ISL_2180493, EPI_ISL_2180494, EPI_ISL_2180495, EPI_ISL_2180496, EPI_ISL_2180497, EPI_ISL_2180498, EPI_ISL_2180499, EPI_ISL_2180500, EPI_ISL_2180501, EPI_ISL_2180502, EPI_ISL_2180503, EPI_ISL_2180504, EPI_ISL_2180505, EPI_ISL_2180506, EPI_ISL_2180507, EPI_ISL_2180508, EPI_ISL_2180509, EPI_ISL_2180510, EPI_ISL_2180511, EPI_ISL_2180512, EPI_ISL_2180513, EPI_ISL_2180514, EPI_ISL_2180515, EPI_ISL_2180516, EPI_ISL_2180517, EPI_ISL_2180518, EPI_ISL_2180519, EPI_ISL_2180520, EPI_ISL_2180521, EPI_ISL_2180522, EPI_ISL_2180523, EPI_ISL_2180524, EPI_ISL_2180525, EPI_ISL_2180526, EPI_ISL_2180527, EPI_ISL_2180528, EPI_ISL_2180529, EPI_ISL_2180530, EPI_ISL_2180531, EPI_ISL_2180532, EPI_ISL_2180533

| | | | |
|---|---|---|---|
| see above | Quest Diagnostics Incorporated | Centers for Disease Control and Prevention Division of Viral Diseases, Pathogen Discovery | Dakota Howard, Dhwani Batra, Peter W. Cook, Kara Moser, Adrian Paskey, Jason Caravas, Benjamin Rambo-Martin, Shatavia Morrison, Christopher Gulvick, Scott Sammons, Yvette Unoarumhi, Darlene Wagner, Matthew Schmerer, S. H. Rosenthal, A. Gerasimova, R. M. Kagan, B. Anderson, M. Hua, Y. Liu, L.E. Bernstein, K.E. Livingston, A. Perez, I. A. Shlyakhter, R. V. Rolando, R. Owen, P. Tanpaiboon, F. Lacbawan, Clinton R. Paden, Duncan MacCannell |

EPI_ISL_2180534, EPI_ISL_2180536, EPI_ISL_2180537, EPI_ISL_2180538, EPI_ISL_2180539, EPI_ISL_2180540, EPI_ISL_2180541, EPI_ISL_2180542, EPI_ISL_2180543, EPI_ISL_2180544, EPI_ISL_2180545, EPI_ISL_2180546, EPI_ISL_2180547, EPI_ISL_2180548, EPI_ISL_2180549, EPI_ISL_2180550, EPI_ISL_2180551, EPI_ISL_2180552, EPI_ISL_2180553, EPI_ISL_2180554, EPI_ISL_2180555, EPI_ISL_2180556, EPI_ISL_2180557, EPI_ISL_2180558, EPI_ISL_2180559, EPI_ISL_2180560, EPI_ISL_2180561, EPI_ISL_2180562, EPI_ISL_2180563, EPI_ISL_2180564, EPI_ISL_2180565, EPI_ISL_2180566, EPI_ISL_2180567, EPI_ISL_2180568, EPI_ISL_2180569, EPI_ISL_2180570, EPI_ISL_2180571, EPI_ISL_2180572, EPI_ISL_2180573, EPI_ISL_2180574, EPI_ISL_2180575, EPI_ISL_2180576, EPI_ISL_2180577, EPI_ISL_2180578, EPI_ISL_2180579, EPI_ISL_2180580, EPI_ISL_2180581, EPI_ISL_2180582, EPI_ISL_2180583, EPI_ISL_2180584, EPI_ISL_2180586, EPI_ISL_2180587, EPI_ISL_2180588, EPI_ISL_2180589, EPI_ISL_2180590, EPI_ISL_2180591, EPI_ISL_2180592, EPI_ISL_2180593, EPI_ISL_2180594, EPI_ISL_2180595, EPI_ISL_2180596, EPI_ISL_2180597, EPI_ISL_2180598, EPI_ISL_2180599, EPI_ISL_2180600, EPI_ISL_2180601, EPI_ISL_2180602, EPI_ISL_2180603, EPI_ISL_2180604, EPI_ISL_2180605, EPI_ISL_2180606, EPI_ISL_2180607, EPI_ISL_2180608, EPI_ISL_2180609, EPI_ISL_2180610, EPI_ISL_2180611, EPI_ISL_2180612, EPI_ISL_2180613, EPI_ISL_2180614, EPI_ISL_2180615, EPI_ISL_2180616, EPI_ISL_2180617, EPI_ISL_2180618, EPI_ISL_2180619, EPI_ISL_2180620, EPI_ISL_2180621, EPI_ISL_2180622, EPI_ISL_2180623, EPI_ISL_2180624, EPI_ISL_2180625, EPI_ISL_2180626, EPI_ISL_2180627, EPI_ISL_2180628, EPI_ISL_2180629, EPI_ISL_2180630, EPI_ISL_2180631, EPI_ISL_2180632, EPI_ISL_2180633, EPI_ISL_2180634, EPI_ISL_2180635, EPI_ISL_2180636, EPI_ISL_2180637, EPI_ISL_2180638, EPI_ISL_2180639, EPI_ISL_2180640, EPI_ISL_2180641, EPI_ISL_2180642, EPI_ISL_2180643, EPI_ISL_2180644, EPI_ISL_2180645, EPI_ISL_2180646, EPI_ISL_2180647, EPI_ISL_2180648, EPI_ISL_2180649, EPI_ISL_2180650, EPI_ISL_2180651, EPI_ISL_2180652, EPI_ISL_2180653, EPI_ISL_2180654, EPI_ISL_2180655, EPI_ISL_2180656, EPI_ISL_2180657, EPI_ISL_2180658, EPI_ISL_2180659, EPI_ISL_2180660, EPI_ISL_2180661, EPI_ISL_2180662, EPI_ISL_2180663, EPI_ISL_2180664, EPI_ISL_2180665, EPI_ISL_2180666, EPI_ISL_2180667, EPI_ISL_2180668, EPI_ISL_2180669, EPI_ISL_2180670, EPI_ISL_2180671, EPI_ISL_2180672, EPI_ISL_2180673, EPI_ISL_2180674, EPI_ISL_2180675, EPI_ISL_2180676, EPI_ISL_2180677, EPI_ISL_2180678, EPI_ISL_2180680, EPI_ISL_2180681, EPI_ISL_2180682, EPI_ISL_2180684, EPI_ISL_2180685, EPI_ISL_2180686, EPI_ISL_2180687, EPI_ISL_2180688, EPI_ISL_2180689, EPI_ISL_2180690, EPI_ISL_2180691, EPI_ISL_2180692, EPI_ISL_2180693, EPI_ISL_2180694, EPI_ISL_2180695, EPI_ISL_2180696, EPI_ISL_2180697, EPI_ISL_2180698, EPI_ISL_2180699, EPI_ISL_2180700, EPI_ISL_2180701, EPI_ISL_2180702, EPI_ISL_2180703, EPI_ISL_2180704, EPI_ISL_2180705, EPI_ISL_2180706, EPI_ISL_2180707, EPI_ISL_2180708, EPI_ISL_2180711, EPI_ISL_2180712, EPI_ISL_2180713, EPI_ISL_2180714, EPI_ISL_2180715, EPI_ISL_2180716, EPI_ISL_2180717, EPI_ISL_2180718, EPI_ISL_2180719, EPI_ISL_2180720, EPI_ISL_2180721, EPI_ISL_2180722, EPI_ISL_2180723, EPI_ISL_2180724, EPI_ISL_2180725, EPI_ISL_2180726, EPI_ISL_2180727, EPI_ISL_2180728, EPI_ISL_2180729, EPI_ISL_2180731, EPI_ISL_2180732, EPI_ISL_2180733, EPI_ISL_2180734, EPI_ISL_2180735, EPI_ISL_2180736, EPI_ISL_2180737, EPI_ISL_2180738, EPI_ISL_2180739, EPI_ISL_2180740, EPI_ISL_2180741, EPI_ISL_2180742, EPI_ISL_2180743, EPI_ISL_2180744, EPI_ISL_2180745, EPI_ISL_2180746, EPI_ISL_2180747, EPI_ISL_2180748, EPI_ISL_2180749, EPI_ISL_2180750, EPI_ISL_2180751, EPI_ISL_2180752, EPI_ISL_2180753, EPI_ISL_2180755, EPI_ISL_2180758, EPI_ISL_2180761, EPI_ISL_2180764, EPI_ISL_2180767, EPI_ISL_2180772, EPI_ISL_2180774, EPI_ISL_2180777, EPI_ISL_2180780, EPI_ISL_2180781, EPI_ISL_2180783, EPI_ISL_2180786, EPI_ISL_2180791, EPI_ISL_2180794, EPI_ISL_2180797, EPI_ISL_2180799, EPI_ISL_2180804, EPI_ISL_2180807, EPI_ISL_2180809, EPI_ISL_2180813, EPI_ISL_2180815, EPI_ISL_2180818, EPI_ISL_2180821, EPI_ISL_2180824, EPI_ISL_2180826, EPI_ISL_2180827, EPI_ISL_2180828, EPI_ISL_2180829, EPI_ISL_2180830, EPI_ISL_2180831, EPI_ISL_2180832, EPI_ISL_2180833, EPI_ISL_2180834, EPI_ISL_2180835, EPI_ISL_2180836, EPI_ISL_2180837, EPI_ISL_2180838, EPI_ISL_2180839, EPI_ISL_2180840, EPI_ISL_2180841, EPI_ISL_2180842, EPI_ISL_2180843, EPI_ISL_2180844, EPI_ISL_2180845, EPI_ISL_2180848, EPI_ISL_2180849, EPI_ISL_2180850, EPI_ISL_2180851, EPI_ISL_2180852, EPI_ISL_2180853, EPI_ISL_2180854, EPI_ISL_2180856, EPI_ISL_2180857, EPI_ISL_2180858, EPI_ISL_2180859, EPI_ISL_2180860, EPI_ISL_2180861, EPI_ISL_2180862, EPI_ISL_2180863, EPI_ISL_2180864, EPI_ISL_2180865, EPI_ISL_2180866, EPI_ISL_2180867, EPI_ISL_2180868, EPI_ISL_2180869, EPI_ISL_2180870, EPI_ISL_2180871, EPI_ISL_2180872, EPI_ISL_2180873, EPI_ISL_2180874, EPI_ISL_2180875, EPI_ISL_2180876, EPI_ISL_2180877, EPI_ISL_2180879, EPI_ISL_2180880, EPI_ISL_2180881, EPI_ISL_2180882, EPI_ISL_2180883, EPI_ISL_2180884, EPI_ISL_2180885, EPI_ISL_2180886, EPI_ISL_2180887, EPI_ISL_2180888, EPI_ISL_2180889, EPI_ISL_2180890, EPI_ISL_2180891, EPI_ISL_2180892, EPI_ISL_2180893, EPI_ISL_2180894, EPI_ISL_2180895, EPI_ISL_2180896, EPI_ISL_2180897, EPI_ISL_2180898, EPI_ISL_2180899, EPI_ISL_2180900, EPI_ISL_2180901, EPI_ISL_2180902, EPI_ISL_2180903, EPI_ISL_2180904, EPI_ISL_2180905, EPI_ISL_2180907, EPI_ISL_2180909, EPI_ISL_2180910, EPI_ISL_2180911, EPI_ISL_2180912, EPI_ISL_2180913, EPI_ISL_2180914, EPI_ISL_2180915, EPI_ISL_2180916, EPI_ISL_2180917, EPI_ISL_2180918, EPI_ISL_2180919, EPI_ISL_2180920, EPI_ISL_2180921, EPI_ISL_2180922, EPI_ISL_2180923, EPI_ISL_2180926, EPI_ISL_2180928, EPI_ISL_2180930, EPI_ISL_2180933, EPI_ISL_2180936, EPI_ISL_2180938, EPI_ISL_2180941, EPI_ISL_2180943, EPI_ISL_2180945, EPI_ISL_2180948, EPI_ISL_2180951, EPI_ISL_2180953, EPI_ISL_2180956, EPI_ISL_2180958, EPI_ISL_2180961, EPI_ISL_2180964, EPI_ISL_2180966, EPI_ISL_2180969, EPI_ISL_2180971, EPI_ISL_2180974, EPI_ISL_2180976, EPI_ISL_2180979, EPI_ISL_2180982, EPI_ISL_2180984, EPI_ISL_2180987, EPI_ISL_2180991, EPI_ISL_2180993, EPI_ISL_2180996, EPI_ISL_2180998, EPI_ISL_2181001, EPI_ISL_2181003, EPI_ISL_2181006, EPI_ISL_2181007, EPI_ISL_2181008, EPI_ISL_2181009, EPI_ISL_2181010, EPI_ISL_2181011, EPI_ISL_2181012, EPI_ISL_2181013, EPI_ISL_2181014, EPI_ISL_2181015, EPI_ISL_2181016, EPI_ISL_2181017, EPI_ISL_2181018, EPI_ISL_2181019, EPI_ISL_2181020, EPI_ISL_2181021, EPI_ISL_2181022, EPI_ISL_2181023, EPI_ISL_2181024, EPI_ISL_2181025, EPI_ISL_2181026, EPI_ISL_2181027, EPI_ISL_2181028, EPI_ISL_2181029, EPI_ISL_2181031, EPI_ISL_2181032, EPI_ISL_2181033, EPI_ISL_2181034, EPI_ISL_2181035, EPI_ISL_2181036, EPI_ISL_2181037, EPI_ISL_2181038, EPI_ISL_2181039, EPI_ISL_2181040, EPI_ISL_2181041, EPI_ISL_2181042, EPI_ISL_2181043, EPI_ISL_2181044, EPI_ISL_2181045, EPI_ISL_2181046, EPI_ISL_2181047, EPI_ISL_2181048, EPI_ISL_2181050, EPI_ISL_2181051, EPI_ISL_2181053, EPI_ISL_2181054, EPI_ISL_2181056, EPI_ISL_2181057, EPI_ISL_2181058, EPI_ISL_2181059, EPI_ISL_2181060, EPI_ISL_2181061, EPI_ISL_2181062, EPI_ISL_2181063, EPI_ISL_2181065, EPI_ISL_2181066, EPI_ISL_2181067, EPI_ISL_2181068, EPI_ISL_2181069, EPI_ISL_2181070, EPI_ISL_2181071, EPI_ISL_2181072, EPI_ISL_2181073, EPI_ISL_2181074, EPI_ISL_2181075, EPI_ISL_2181076, EPI_ISL_2181077, EPI_ISL_2181078, EPI_ISL_2181079, EPI_ISL_2181080, EPI_ISL_2181081, EPI_ISL_2181082, EPI_ISL_2181083, EPI_ISL_2181084, EPI_ISL_2181085, EPI_ISL_2181086, EPI_ISL_2181087, EPI_ISL_2181088, EPI_ISL_2181089, EPI_ISL_2181090, EPI_ISL_2181091, EPI_ISL_2181092, EPI_ISL_2181093, EPI_ISL_2181094, EPI_ISL_2181095, EPI_ISL_2181096, EPI_ISL_2181097, EPI_ISL_2181098, EPI_ISL_2181099, EPI_ISL_2181100, EPI_ISL_2181101, EPI_ISL_2181102, EPI_ISL_2181103, EPI_ISL_2181104, EPI_ISL_2181105, EPI_ISL_2181106, EPI_ISL_2181107, EPI_ISL_2181108, EPI_ISL_2181109, EPI_ISL_2181110, EPI_ISL_2181111, EPI_ISL_2181113, EPI_ISL_2181114, EPI_ISL_2181115, EPI_ISL_2181116, EPI_ISL_2181117, EPI_ISL_2181118, EPI_ISL_2181119, EPI_ISL_2181120, EPI_ISL_2181121, EPI_ISL_2181122, EPI_ISL_2181123, EPI_ISL_2181124, EPI_ISL_2181125, EPI_ISL_2181126, EPI_ISL_2181127, EPI_ISL_2181128, EPI_ISL_2181129, EPI_ISL_2181130, EPI_ISL_2181131, EPI_ISL_2181133, EPI_ISL_2181134, EPI_ISL_2181135, EPI_ISL_2181136, EPI_ISL_2181137, EPI_ISL_2181138, EPI_ISL_2181139, EPI_ISL_2181140, EPI_ISL_2181141, EPI_ISL_2181142, EPI_ISL_2181143, EPI_ISL_2181144, EPI_ISL_2181145, EPI_ISL_2181146, EPI_ISL_2181147, EPI_ISL_2181148, EPI_ISL_2181149, EPI_ISL_2181150, EPI_ISL_2181151, EPI_ISL_2181152, EPI_ISL_2181153, EPI_ISL_2181155, EPI_ISL_2181156, EPI_ISL_2181157, EPI_ISL_2181158, EPI_ISL_2181159, EPI_ISL_2181160, EPI_ISL_2181161, EPI_ISL_2181162, EPI_ISL_2181163, EPI_ISL_2181164, EPI_ISL_2181165, EPI_ISL_2181166, EPI_ISL_2181167, EPI_ISL_2181168, EPI_ISL_2181169, EPI_ISL_2181170, EPI_ISL_2181171, EPI_ISL_2181172, EPI_ISL_2181173, EPI_ISL_2181174, EPI_ISL_2181175, EPI_ISL_2181176, EPI_ISL_2181177, EPI_ISL_2181178, EPI_ISL_2181179, EPI_ISL_2181180, EPI_ISL_2181181, EPI_ISL_2181182, EPI_ISL_2181183, EPI_ISL_2181184, EPI_ISL_2181185, EPI_ISL_2181186, EPI_ISL_2181187, EPI_ISL_2181188, EPI_ISL_2181189, EPI_ISL_2181190, EPI_ISL_2181191, EPI_ISL_2181192, EPI_ISL_2181193, EPI_ISL_2181194, EPI_ISL_2181195, EPI_ISL_2181196, EPI_ISL_2181197, EPI_ISL_2181198, EPI_ISL_2181199, EPI_ISL_2181200, EPI_ISL_2181201, EPI_ISL_2181202, EPI_ISL_2181203, EPI_ISL_2181204, EPI_ISL_2181205, EPI_ISL_2181206, EPI_ISL_2181207, EPI_ISL_2181208, EPI_ISL_2181209, EPI_ISL_2181210, EPI_ISL_2181211, EPI_ISL_2181212, EPI_ISL_2181213, EPI_ISL_2181214, EPI_ISL_2181215, EPI_ISL_2181216, EPI_ISL_2181217, EPI_ISL_2181218, EPI_ISL_2181219, EPI_ISL_2181220, EPI_ISL_2181221, EPI_ISL_2181222, EPI_ISL_2181223, EPI_ISL_2181224, EPI_ISL_2181225, EPI_ISL_2181226, EPI_ISL_2181227, EPI_ISL_2181228, EPI_ISL_2181229, EPI_ISL_2181230, EPI_ISL_2181231, EPI_ISL_2181232, EPI_ISL_2181233, EPI_ISL_2181234, EPI_ISL_2181235, EPI_ISL_2181236, EPI_ISL_2181237, EPI_ISL_2181238, EPI_ISL_2181239, EPI_ISL_2181240, EPI_ISL_2181241, EPI_ISL_2181242, EPI_ISL_2181243, EPI_ISL_2181244, EPI_ISL_2181245, EPI_ISL_2181246, EPI_ISL_2181247, EPI_ISL_2181248, EPI_ISL_2181249, EPI_ISL_2181250, EPI_ISL_2181251, EPI_ISL_2181252, EPI_ISL_2181253, EPI_ISL_2181254, EPI_ISL_2181255, EPI_ISL_2181256, EPI_ISL_2181257, EPI_ISL_2181258, EPI_ISL_2181259, EPI_ISL_2181260, EPI_ISL_2181261, EPI_ISL_2181262, EPI_ISL_2181264, EPI_ISL_2181265, EPI_ISL_2181266, EPI_ISL_2181267, EPI_ISL_2181268, EPI_ISL_2181269, EPI_ISL_2181270, EPI_ISL_2181271, EPI_ISL_2181272, EPI_ISL_2181273, EPI_ISL_2181274, EPI_ISL_2181275, EPI_ISL_2181276, EPI_ISL_2181277, EPI_ISL_2181278, EPI_ISL_2181279, EPI_ISL_2181280, EPI_ISL_2181281, EPI_ISL_2181282, EPI_ISL_2181283, EPI_ISL_2181284, EPI_ISL_2181285, EPI_ISL_2181287, EPI_ISL_2181288, EPI_ISL_2181289, EPI_ISL_2181291, EPI_ISL_2181292, EPI_ISL_2181293, EPI_ISL_2181294, EPI_ISL_2181295, EPI_ISL_2181296, EPI_ISL_2181298, EPI_ISL_2181299, EPI_ISL_2181300, EPI_ISL_2181301, EPI_ISL_2181302, EPI_ISL_2181303, EPI_ISL_2181304, EPI_ISL_2181305, EPI_ISL_2181306, EPI_ISL_2181307, EPI_ISL_2181308, EPI_ISL_2181309, EPI_ISL_2181310, EPI_ISL_2181311, EPI_ISL_2181312, EPI_ISL_2181313, EPI_ISL_2181314, EPI_ISL_2181315, EPI_ISL_2181316, EPI_ISL_2181317, EPI_ISL_2181318, EPI_ISL_2181319, EPI_ISL_2181320, EPI_ISL_2181321, EPI_ISL_2181322, EPI_ISL_2181323, EPI_ISL_2181324, EPI_ISL_2181325, EPI_ISL_2181326, EPI_ISL_2181327, EPI_ISL_2181328, EPI_ISL_2181329, EPI_ISL_2181330, EPI_ISL_2181331, EPI_ISL_2181332, EPI_ISL_2181333, EPI_ISL_2181334, EPI_ISL_2181335, EPI_ISL_2181336, EPI_ISL_2181337, EPI_ISL_2181338, EPI_ISL_2181339, EPI_ISL_2181340, EPI_ISL_2181341, EPI_ISL_2181342, EPI_ISL_2181343, EPI_ISL_2181344, EPI_ISL_2181345, EPI_ISL_2181346, EPI_ISL_2181347, EPI_ISL_2181348, EPI_ISL_2181349, EPI_ISL_2181350, EPI_ISL_2181351, EPI_ISL_2181352, EPI_ISL_2181353, EPI_ISL_2181354, EPI_ISL_2181355, EPI_ISL_2181356, EPI_ISL_2181357, EPI_ISL_2181358, EPI_ISL_2181359, EPI_ISL_2181360, EPI_ISL_2181361, EPI_ISL_2181362, EPI_ISL_2181363, EPI_ISL_2181364, EPI_ISL_2181365, EPI_ISL_2181366, EPI_ISL_2181367, EPI_ISL_2181368, EPI_ISL_2181369, EPI_ISL_2181370, EPI_ISL_2181371, EPI_ISL_2181372, EPI_ISL_2181373, EPI_ISL_2181374, EPI_ISL_2181375, EPI_ISL_2181376, EPI_ISL_2181377, EPI_ISL_2181378, EPI_ISL_2181379, EPI_ISL_2181380, EPI_ISL_2181381, EPI_ISL_2181382, EPI_ISL_2181383, EPI_ISL_2181384, EPI_ISL_2181385, EPI_ISL_2181386, EPI_ISL_2181387, EPI_ISL_2181388, EPI_ISL_2181389, EPI_ISL_2181390, EPI_ISL_2181391, EPI_ISL_2181392, EPI_ISL_2181393, EPI_ISL_2181394, EPI_ISL_2181395, EPI_ISL_2181396, EPI_ISL_2181397, EPI_ISL_2181399, EPI_ISL_2181400, EPI_ISL_2181401, EPI_ISL_2181402, EPI_ISL_2181403, EPI_ISL_2181404, EPI_ISL_2181405, EPI_ISL_2181406, EPI_ISL_2181407, EPI_ISL_2181408, EPI_ISL_2181409, EPI_ISL_2181410, EPI_ISL_2181411, EPI_ISL_2181412, EPI_ISL_2181413, EPI_ISL_2181414, EPI_ISL_2181415, EPI_ISL_2181416, EPI_ISL_2181417, EPI_ISL_2181418, EPI_ISL_2181419, EPI_ISL_2181420, EPI_ISL_2181421, EPI_ISL_2181422, EPI_ISL_2181423, EPI_ISL_2181424, EPI_ISL_2181425, EPI_ISL_2181426, EPI_ISL_2181427, EPI_ISL_2181428, EPI_ISL_2181429, EPI_ISL_2181430, EPI_ISL_2181431, EPI_ISL_2181432, EPI_ISL_2181433, EPI_ISL_2181434, EPI_ISL_2181435, EPI_ISL_2181436, EPI_ISL_2181437, EPI_ISL_2181438, EPI_ISL_2181439, EPI_ISL_2181440, EPI_ISL_2181441, EPI_ISL_2181442, EPI_ISL_2181443, EPI_ISL_2181444, EPI_ISL_2181445, EPI_ISL_2181446, EPI_ISL_2181447, EPI_ISL_2181448, EPI_ISL_2181449, EPI_ISL_2181450, EPI_ISL_2181451, EPI_ISL_2181452, EPI_ISL_2181453, EPI_ISL_2181454, EPI_ISL_2181455, EPI_ISL_2181456, EPI_ISL_2181457, EPI_ISL_2181458, EPI_ISL_2181459, EPI_ISL_2181460, EPI_ISL_2181461, EPI_ISL_2181462, EPI_ISL_2181463, EPI_ISL_2181464, EPI_ISL_2181465, EPI_ISL_2181466, EPI_ISL_2181467, EPI_ISL_2181468, EPI_ISL_2181469, EPI_ISL_2181470, EPI_ISL_2181472, EPI_ISL_2181473, EPI_ISL_2181474, EPI_ISL_2181475, EPI_ISL_2181476, EPI_ISL_2181477, EPI_ISL_2181478, EPI_ISL_2181479, EPI_ISL_2181480, EPI_ISL_2181481, EPI_ISL_2181482, EPI_ISL_2181483, EPI_ISL_2181484, EPI_ISL_2181485, EPI_ISL_2181486, EPI_ISL_2181487, EPI_ISL_2181488, EPI_ISL_2181489, EPI_ISL_2181490, EPI_ISL_2181491, EPI_ISL_2181492, EPI_ISL_2181493, EPI_ISL_2181494, EPI_ISL_2181495, EPI_ISL_2181496, EPI_ISL_2181497, EPI_ISL_2181498, EPI_ISL_2181499, EPI_ISL_2181500, EPI_ISL_2181501, EPI_ISL_2181502, EPI_ISL_2181503, EPI_ISL_2181504, EPI_ISL_2181505, EPI_ISL_2181506, EPI_ISL_2181507, EPI_ISL_2181508, EPI_ISL_2181509, EPI_ISL_2181510, EPI_ISL_2181511, EPI_ISL_2181512, EPI_ISL_2181513, EPI_ISL_2181514, EPI_ISL_2181515, EPI_ISL_2181516, EPI_ISL_2181517, EPI_ISL_2181518, EPI_ISL_2181519, EPI_ISL_2181520, EPI_ISL_2181521, EPI_ISL_2181522, EPI_ISL_2181523, EPI_ISL_2181524, EPI_ISL_2181525, EPI_ISL_2181526, EPI_ISL_2181527, EPI_ISL_2181528, EPI_ISL_2181529, EPI_ISL_2181530, EPI_ISL_2181531, EPI_ISL_2181532, EPI_ISL_2181533, EPI_ISL_2181534, EPI_ISL_2181535, EPI_ISL_2181536, EPI_ISL_2181537, EPI_ISL_2181538, EPI_ISL_2181539, EPI_ISL_2181540, EPI_ISL_2181541, EPI_ISL_2181542, EPI_ISL_2181543, EPI_ISL_2181544, EPI_ISL_2181545, EPI_ISL_2181546, EPI_ISL_2181547, EPI_ISL_2181548, EPI_ISL_2181549, EPI_ISL_2181550, EPI_ISL_2181552, EPI_ISL_2181553, EPI_ISL_2181554, EPI_ISL_2181555, EPI_ISL_2181556, EPI_ISL_2181557, EPI_ISL_2181558, EPI_ISL_2181559, EPI_ISL_2181560, EPI_ISL_2181561, EPI_ISL_2181562, EPI_ISL_2181563, EPI_ISL_2181564, EPI_ISL_2181565, EPI_ISL_2181566, EPI_ISL_2181567, EPI_ISL_2181568, EPI_ISL_2181569, EPI_ISL_2181570, EPI_ISL_2181571, EPI_ISL_2181572, EPI_ISL_2181573, EPI_ISL_2181574, EPI_ISL_2181575, EPI_ISL_2181576, EPI_ISL_2181577, EPI_ISL_2181578, EPI_ISL_2181579, EPI_ISL_2181580, EPI_ISL_2181581, EPI_ISL_2181582, EPI_ISL_2181583, EPI_ISL_2181584, EPI_ISL_2181585, EPI_ISL_2181586, EPI_ISL_2181587, EPI_ISL_2181588, EPI_ISL_2181589, EPI_ISL_2181590, EPI_ISL_2181591, EPI_ISL_2181592, EPI_ISL_2181593, EPI_ISL_2181594, EPI_ISL_2181595, EPI_ISL_2181596, EPI_ISL_2181598, EPI_ISL_2181599, EPI_ISL_2181600, EPI_ISL_2181601, EPI_ISL_2181602, EPI_ISL_2181603, EPI_ISL_2181604, EPI_ISL_2181605, EPI_ISL_2181606, EPI_ISL_2181607, EPI_ISL_2181608, EPI_ISL_2181609, EPI_ISL_2181610, EPI_ISL_2181612, EPI_ISL_2181613, EPI_ISL_2181614, EPI_ISL_2181615, EPI_ISL_2181616, EPI_ISL_2181617, EPI_ISL_2181618, EPI_ISL_2181619, EPI_ISL_2181620, EPI_ISL_2181621, EPI_ISL_2181622, EPI_ISL_2181623, EPI_ISL_2181624, EPI_ISL_2181625, EPI_ISL_2181626, EPI_ISL_2181627, EPI_ISL_2181628, EPI_ISL_2181629, EPI_ISL_2181630, EPI_ISL_2181631, EPI_ISL_2181632, EPI_ISL_2181633, EPI_ISL_2181634, EPI_ISL_2181635, EPI_ISL_2181636, EPI_ISL_2181637, EPI_ISL_2181638, EPI_ISL_2181639, EPI_ISL_2181640, EPI_ISL_2181641, EPI_ISL_2181642, EPI_ISL_2181643, EPI_ISL_2181644, EPI_ISL_2181645, EPI_ISL_2181646, EPI_ISL_2181647, EPI_ISL_2181648, EPI_ISL_2181649, EPI_ISL_2181650, EPI_ISL_2181651, EPI_ISL_2181652, EPI_ISL_2181653, EPI_ISL_2181654, EPI_ISL_2181655, EPI_ISL_2181656, EPI_ISL_2181657

| | | | |
|---|---|---|---|
| see above | Aegis Sciences Corporation | Centers for Disease Control and Prevention Division of Viral Diseases, Pathogen Discovery | Dakota Howard, Dhwani Batra, Peter W. Cook, Kara Moser, Adrian Paskey, Jason Caravas, Benjamin Rambo-Martin, Shatavia Morrison, Christopher Gulvick, Scott Sammons, Yvette Unoarumhi, Darlene Wagner, Matthew Schmerer, Cyndi Clark, Patrick Campbell, Rob Case, Vikramsinha Ghorpade, Holly Houdeshell, Ola Kvalvaag, Dillon Nall, Ethan Sanders, Alec Vest, Shaun Westlund, Matthew Hardison, Clinton R. Paden, Duncan MacCannell |

EPI_ISL_2181658, EPI_ISL_2181660, EPI_ISL_2181661, EPI_ISL_2181662, EPI_ISL_2181663, EPI_ISL_2181664, EPI_ISL_2181665, EPI_ISL_2181666, EPI_ISL_2181667, EPI_ISL_2181670, EPI_ISL_2181671, EPI_ISL_2181672, EPI_ISL_2181673, EPI_ISL_2181675, EPI_ISL_2181676, EPI_ISL_2181677, EPI_ISL_2181678, EPI_ISL_2181679, EPI_ISL_2181680, EPI_ISL_2181681, EPI_ISL_2181683, EPI_ISL_2181685, EPI_ISL_2181686, EPI_ISL_2181687, EPI_ISL_2181688, EPI_ISL_2181689, EPI_ISL_2181692, EPI_ISL_2181693, EPI_ISL_2181694, EPI_ISL_2181695, EPI_ISL_2181696, EPI_ISL_2181697, EPI_ISL_2181698, EPI_ISL_2181699, EPI_ISL_2181700, EPI_ISL_2181701, EPI_ISL_2181702, EPI_ISL_2181703, EPI_ISL_2181704, EPI_ISL_2181705, EPI_ISL_2181706, EPI_ISL_2181708, EPI_ISL_2181709, EPI_ISL_2181710, EPI_ISL_2181711, EPI_ISL_2181712, EPI_ISL_2181713, EPI_ISL_2181715, EPI_ISL_2181716, EPI_ISL_2181717, EPI_ISL_2181719, EPI_ISL_2181720, EPI_ISL_2181721, EPI_ISL_2181722, EPI_ISL_2181723, EPI_ISL_2181724, EPI_ISL_2181725, EPI_ISL_2181726, EPI_ISL_2181728, EPI_ISL_2181729, EPI_ISL_2181730, EPI_ISL_2181731, EPI_ISL_2181732, EPI_ISL_2181733, EPI_ISL_2181734, EPI_ISL_2181735, EPI_ISL_2181736, EPI_ISL_2181737, EPI_ISL_2181738, EPI_ISL_2181739, EPI_ISL_2181740, EPI_ISL_2181742, EPI_ISL_2181743, EPI_ISL_2181744, EPI_ISL_2181745, EPI_ISL_2181746, EPI_ISL_2181747, EPI_ISL_2181748, EPI_ISL_2181749, EPI_ISL_2181750, EPI_ISL_2181751, EPI_ISL_2181752, EPI_ISL_2181753, EPI_ISL_2181754, EPI_ISL_2181755, EPI_ISL_2181756, EPI_ISL_2181759, EPI_ISL_2181761, EPI_ISL_2181764, EPI_ISL_2181767, EPI_ISL_2181770, EPI_ISL_2181773, EPI_ISL_2181775, EPI_ISL_2181778, EPI_ISL_2181781, EPI_ISL_2181786, EPI_ISL_2181789, EPI_ISL_2181793, EPI_ISL_2181796, EPI_ISL_2181799, EPI_ISL_2181802, EPI_ISL_2181808, EPI_ISL_2181809, EPI_ISL_2181811, EPI_ISL_2181814, EPI_ISL_2181816, EPI_ISL_2181822, EPI_ISL_2181825, EPI_ISL_2181828, EPI_ISL_2181831, EPI_ISL_2181836, EPI_ISL_2181842, EPI_ISL_2181845, EPI_ISL_2181847, EPI_ISL_2181850, EPI_ISL_2181853, EPI_ISL_2181857, EPI_ISL_2181859, EPI_ISL_2181862, EPI_ISL_2181865, EPI_ISL_2181869, EPI_ISL_2181877, EPI_ISL_2181883, EPI_ISL_2181886, EPI_ISL_2181889, EPI_ISL_2181892, EPI_ISL_2181895, EPI_ISL_2181898, EPI_ISL_2181901, EPI_ISL_2181903, EPI_ISL_2181906, EPI_ISL_2181909, EPI_ISL_2181912, EPI_ISL_2181917, EPI_ISL_2181920, EPI_ISL_2181923, EPI_ISL_2181927, EPI_ISL_2181932, EPI_ISL_2181935, EPI_ISL_2181938, EPI_ISL_2181941, EPI_ISL_2181944, EPI_ISL_2181947, EPI_ISL_2181949, EPI_ISL_2181952, EPI_ISL_2181955, EPI_ISL_2181958, EPI_ISL_2181961, EPI_ISL_2181964, EPI_ISL_2181969, EPI_ISL_2181972, EPI_ISL_2181975, EPI_ISL_2181978, EPI_ISL_2181981, EPI_ISL_2181983, EPI_ISL_2181986, EPI_ISL_2181989, EPI_ISL_2181992, EPI_ISL_2181995, EPI_ISL_2181997, EPI_ISL_2182000, EPI_ISL_2182003, EPI_ISL_2182006, EPI_ISL_2182009, EPI_ISL_2182015, EPI_ISL_2182021, EPI_ISL_2182024, EPI_ISL_2182027, EPI_ISL_2182029, EPI_ISL_2182038, EPI_ISL_2182041, EPI_ISL_2182044, EPI_ISL_2182046, EPI_ISL_2182049, EPI_ISL_2182052, EPI_ISL_2182055, EPI_ISL_2182058, EPI_ISL_2182060, EPI_ISL_2182063, EPI_ISL_2182066, EPI_ISL_2182069, EPI_ISL_2182071, EPI_ISL_2182074, EPI_ISL_2182077, EPI_ISL_2182078, EPI_ISL_2182080, EPI_ISL_2182083, EPI_ISL_2182086, EPI_ISL_2182088, EPI_ISL_2182092, EPI_ISL_2182094, EPI_ISL_2182097, EPI_ISL_2182100, EPI_ISL_2182103, EPI_ISL_2182105, EPI_ISL_2182108, EPI_ISL_2182110, EPI_ISL_2182113, EPI_ISL_2182115, EPI_ISL_2182118, EPI_ISL_2182121, EPI_ISL_2182123, EPI_ISL_2182126, EPI_ISL_2182132, EPI_ISL_2182135, EPI_ISL_2182137, EPI_ISL_2182140, EPI_ISL_2182142, EPI_ISL_2182145, EPI_ISL_2182150, EPI_ISL_2182153, EPI_ISL_2182155, EPI_ISL_2182158, EPI_ISL_2182162, EPI_ISL_2182167, EPI_ISL_2182169, EPI_ISL_2182171, EPI_ISL_2182174, EPI_ISL_2182176, EPI_ISL_2182182, EPI_ISL_2182185, EPI_ISL_2182188, EPI_ISL_2182190, EPI_ISL_2182193, EPI_ISL_2182196, EPI_ISL_2182198, EPI_ISL_2182200, EPI_ISL_2182202, EPI_ISL_2182205, EPI_ISL_2182208, EPI_ISL_2182211, EPI_ISL_2182214, EPI_ISL_2182217, EPI_ISL_2182220, EPI_ISL_2182222, EPI_ISL_2182225, EPI_ISL_2182228, EPI_ISL_2182231, EPI_ISL_2182233, EPI_ISL_2182236, EPI_ISL_2182239, EPI_ISL_2182242, EPI_ISL_2182245, EPI_ISL_2182248, EPI_ISL_2182250, EPI_ISL_2182254, EPI_ISL_2182256, EPI_ISL_2182259, EPI_ISL_2182262, EPI_ISL_2182265, EPI_ISL_2182267, EPI_ISL_2182270, EPI_ISL_2182276, EPI_ISL_2182279, EPI_ISL_2182281, EPI_ISL_2182284, EPI_ISL_2182286, EPI_ISL_2182289, EPI_ISL_2182292, EPI_ISL_2182294, EPI_ISL_2182297, EPI_ISL_2182299, EPI_ISL_2182302, EPI_ISL_2182305, EPI_ISL_2182308, EPI_ISL_2182310, EPI_ISL_2182313, EPI_ISL_2182316, EPI_ISL_2182319, EPI_ISL_2182322, EPI_ISL_2182325, EPI_ISL_2182327, EPI_ISL_2182330, EPI_ISL_2182333, EPI_ISL_2182338, EPI_ISL_2182341, EPI_ISL_2182344, EPI_ISL_2182349, EPI_ISL_2182352, EPI_ISL_2182355, EPI_ISL_2182357, EPI_ISL_2182360, EPI_ISL_2182363, EPI_ISL_2182365, EPI_ISL_2182371, EPI_ISL_2182375, EPI_ISL_2182378, EPI_ISL_2182381, EPI_ISL_2182384, EPI_ISL_2182386, EPI_ISL_2182389, EPI_ISL_2182392, EPI_ISL_2182395, EPI_ISL_2182399, EPI_ISL_2182402, EPI_ISL_2182405, EPI_ISL_2182407, EPI_ISL_2182410, EPI_ISL_2182413, EPI_ISL_2182416, EPI_ISL_2182419, EPI_ISL_2182421, EPI_ISL_2182424, EPI_ISL_2182426, EPI_ISL_2182429, EPI_ISL_2182432, EPI_ISL_2182434, EPI_ISL_2182437, EPI_ISL_2182439, EPI_ISL_2182442, EPI_ISL_2182444, EPI_ISL_2182447, EPI_ISL_2182449, EPI_ISL_2182452, EPI_ISL_2182455, EPI_ISL_2182457, EPI_ISL_2182460, EPI_ISL_2182463, EPI_ISL_2182465, EPI_ISL_2182468, EPI_ISL_2182471, EPI_ISL_2182474, EPI_ISL_2182479, EPI_ISL_2182482, EPI_ISL_2182485, EPI_ISL_2182487, EPI_ISL_2182490, EPI_ISL_2182493, EPI_ISL_2182496, EPI_ISL_2182499, EPI_ISL_2182501, EPI_ISL_2182504, EPI_ISL_2182506, EPI_ISL_2182509, EPI_ISL_2182511, EPI_ISL_2182514, EPI_ISL_2182517, EPI_ISL_2182520, EPI_ISL_2182522, EPI_ISL_2182525, EPI_ISL_2182527, EPI_ISL_2182530, EPI_ISL_2182532, EPI_ISL_2182535, EPI_ISL_2182538, EPI_ISL_2182541, EPI_ISL_2182544, EPI_ISL_2182546, EPI_ISL_2182549, EPI_ISL_2182552, EPI_ISL_2182555, EPI_ISL_2182557, EPI_ISL_2182560, EPI_ISL_2182563, EPI_ISL_2182565, EPI_ISL_2182568, EPI_ISL_2182572, EPI_ISL_2182574, EPI_ISL_2182577, EPI_ISL_2182580, EPI_ISL_2182583, EPI_ISL_2182586, EPI_ISL_2182588, EPI_ISL_2182590, EPI_ISL_2182593, EPI_ISL_2182595, EPI_ISL_2182598, EPI_ISL_2182601, EPI_ISL_2182604, EPI_ISL_2182606, EPI_ISL_2182608, EPI_ISL_2182611, EPI_ISL_2182614, EPI_ISL_2182617, EPI_ISL_2182619, EPI_ISL_2182622, EPI_ISL_2182625, EPI_ISL_2182628, EPI_ISL_2182631, EPI_ISL_2182634, EPI_ISL_2182636, EPI_ISL_2182639, EPI_ISL_2182642, EPI_ISL_2182645, EPI_ISL_2182647, EPI_ISL_2182649, EPI_ISL_2182652, EPI_ISL_2182655, EPI_ISL_2182658, EPI_ISL_2182660, EPI_ISL_2182663, EPI_ISL_2182666, EPI_ISL_2182668, EPI_ISL_2182671, EPI_ISL_2182674, EPI_ISL_2182677, EPI_ISL_2182679, EPI_ISL_2182682, EPI_ISL_2182685, EPI_ISL_2182688, EPI_ISL_2182691, EPI_ISL_2182694, EPI_ISL_2182697, EPI_ISL_2182699, EPI_ISL_2182702, EPI_ISL_2182705, EPI_ISL_2182708, EPI_ISL_2182710, EPI_ISL_2182713, EPI_ISL_2182715, EPI_ISL_2182717, EPI_ISL_2182720, EPI_ISL_2182725, EPI_ISL_2182728, EPI_ISL_2182731, EPI_ISL_2182733, EPI_ISL_2182736, EPI_ISL_2182739, EPI_ISL_2182741, EPI_ISL_2182744, EPI_ISL_2182745, EPI_ISL_2182747, EPI_ISL_2182749, EPI_ISL_2182752, EPI_ISL_2182755, EPI_ISL_2182759, EPI_ISL_2182761, EPI_ISL_2182764, EPI_ISL_2182767, EPI_ISL_2182770, EPI_ISL_2182772, EPI_ISL_2182775, EPI_ISL_2182778, EPI_ISL_2182780, EPI_ISL_2182783, EPI_ISL_2182786, EPI_ISL_2182789, EPI_ISL_2182792, EPI_ISL_2182795, EPI_ISL_2182797, EPI_ISL_2182800, EPI_ISL_2182803, EPI_ISL_2182805, EPI_ISL_2182808, EPI_ISL_2182815, EPI_ISL_2182817, EPI_ISL_2182820, EPI_ISL_2182823, EPI_ISL_2182825, EPI_ISL_2182828, EPI_ISL_2182831, EPI_ISL_2182834, EPI_ISL_2182837, EPI_ISL_2182840, EPI_ISL_2182842, EPI_ISL_2182845, EPI_ISL_2182848, EPI_ISL_2182850, EPI_ISL_2182853, EPI_ISL_2182861, EPI_ISL_2182863, EPI_ISL_2182867, EPI_ISL_2182869, EPI_ISL_2182872, EPI_ISL_2182875, EPI_ISL_2182877, EPI_ISL_2182878, EPI_ISL_2182881, EPI_ISL_2182883, EPI_ISL_2182886, EPI_ISL_2182889, EPI_ISL_2182891, EPI_ISL_2182894, EPI_ISL_2182896, EPI_ISL_2182899, EPI_ISL_2182902, EPI_ISL_2182905, EPI_ISL_2182907, EPI_ISL_2182910, EPI_ISL_2182913, EPI_ISL_2182916, EPI_ISL_2182919, EPI_ISL_2182922, EPI_ISL_2182924, EPI_ISL_2182927, EPI_ISL_2182930, EPI_ISL_2182933, EPI_ISL_2182935, EPI_ISL_2182938, EPI_ISL_2182941, EPI_ISL_2182944, EPI_ISL_2182946, EPI_ISL_2182949, EPI_ISL_2182952, EPI_ISL_2182954, EPI_ISL_2182957, EPI_ISL_2182960, EPI_ISL_2182965, EPI_ISL_2182968, EPI_ISL_2182971, EPI_ISL_2182973, EPI_ISL_2182976, EPI_ISL_2182979, EPI_ISL_2182985, EPI_ISL_2182987, EPI_ISL_2182990, EPI_ISL_2182993, EPI_ISL_2182995, EPI_ISL_2182998, EPI_ISL_2183000, EPI_ISL_2183003, EPI_ISL_2183006, EPI_ISL_2183008, EPI_ISL_2183011, EPI_ISL_2183014, EPI_ISL_2183017, EPI_ISL_2183018, EPI_ISL_2183021, EPI_ISL_2183024, EPI_ISL_2183027, EPI_ISL_2183030, EPI_ISL_2183033, EPI_ISL_2183036, EPI_ISL_2183039, EPI_ISL_2183042, EPI_ISL_2183044, EPI_ISL_2183047, EPI_ISL_2183050, EPI_ISL_2183053, EPI_ISL_2183055, EPI_ISL_2183058, EPI_ISL_2183061, EPI_ISL_2183064, EPI_ISL_2183066, EPI_ISL_2183068, EPI_ISL_2183071, EPI_ISL_2183074, EPI_ISL_2183077, EPI_ISL_2183080, EPI_ISL_2183083, EPI_ISL_2183085, EPI_ISL_2183088, EPI_ISL_2183091, EPI_ISL_2183094, EPI_ISL_2183097, EPI_ISL_2183100, EPI_ISL_2183103, EPI_ISL_2183105, EPI_ISL_2183108, EPI_ISL_2183110, EPI_ISL_2183113, EPI_ISL_2183115, EPI_ISL_2183118, EPI_ISL_2183121, EPI_ISL_2183124, EPI_ISL_2183127, EPI_ISL_2183129, EPI_ISL_2183132, EPI_ISL_2183134, EPI_ISL_2183137, EPI_ISL_2183140, EPI_ISL_2183142, EPI_ISL_2183145, EPI_ISL_2183147, EPI_ISL_2183150, EPI_ISL_2183153, EPI_ISL_2183156, EPI_ISL_2183159, EPI_ISL_2183162, EPI_ISL_2183165, EPI_ISL_2183167, EPI_ISL_2183169, EPI_ISL_2183172, EPI_ISL_2183175, EPI_ISL_2183177, EPI_ISL_2183178, EPI_ISL_2183181, EPI_ISL_2183183, EPI_ISL_2183184, EPI_ISL_2183187, EPI_ISL_2183190, EPI_ISL_2183195, EPI_ISL_2183198, EPI_ISL_2183201, EPI_ISL_2183206, EPI_ISL_2183208, EPI_ISL_2183211, EPI_ISL_2183213, EPI_ISL_2183216, EPI_ISL_2183219, EPI_ISL_2183221, EPI_ISL_2183224, EPI_ISL_2183227, EPI_ISL_2183229, EPI_ISL_2183232, EPI_ISL_2183235, EPI_ISL_2183238, EPI_ISL_2183241, EPI_ISL_2183243, EPI_ISL_2183246, EPI_ISL_2183249, EPI_ISL_2183251, EPI_ISL_2183254, EPI_ISL_2183256, EPI_ISL_2183259, EPI_ISL_2183261, EPI_ISL_2183264, EPI_ISL_2183268, EPI_ISL_2183270, EPI_ISL_2183273, EPI_ISL_2183275, EPI_ISL_2183278, EPI_ISL_2183280, EPI_ISL_2183283, EPI_ISL_2183286, EPI_ISL_2183288, EPI_ISL_2183291, EPI_ISL_2183294, EPI_ISL_2183297, EPI_ISL_2183299, EPI_ISL_2183302, EPI_ISL_2183305, EPI_ISL_2183308, EPI_ISL_2183311, EPI_ISL_2183313, EPI_ISL_2183316, EPI_ISL_2183319, EPI_ISL_2183322, EPI_ISL_2183324, EPI_ISL_2183327, EPI_ISL_2183329, EPI_ISL_2183332, EPI_ISL_2183334, EPI_ISL_2183337, EPI_ISL_2183339, EPI_ISL_2183342, EPI_ISL_2183345, EPI_ISL_2183348, EPI_ISL_2183350, EPI_ISL_2183352, EPI_ISL_2183355, EPI_ISL_2183358, EPI_ISL_2183361, EPI_ISL_2183363, EPI_ISL_2183369, EPI_ISL_2183371, EPI_ISL_2183374, EPI_ISL_2183377, EPI_ISL_2183380, EPI_ISL_2183382, EPI_ISL_2183385, EPI_ISL_2183388, EPI_ISL_2183391, EPI_ISL_2183393, EPI_ISL_2183395, EPI_ISL_2183397, EPI_ISL_2183400, EPI_ISL_2183402, EPI_ISL_2183405, EPI_ISL_2183408, EPI_ISL_2183411, EPI_ISL_2183413, EPI_ISL_2183415, EPI_ISL_2183418, EPI_ISL_2183419, EPI_ISL_2183421, EPI_ISL_2183423, EPI_ISL_2183426, EPI_ISL_2183428, EPI_ISL_2183430, EPI_ISL_2183432, EPI_ISL_2183434, EPI_ISL_2183437, EPI_ISL_2183438, EPI_ISL_2183440, EPI_ISL_2183443, EPI_ISL_2183445, EPI_ISL_2183447, EPI_ISL_2183450, EPI_ISL_2183451, EPI_ISL_2183454, EPI_ISL_2183456, EPI_ISL_2183458, EPI_ISL_2183460, EPI_ISL_2183461, EPI_ISL_2183464, EPI_ISL_2183467, EPI_ISL_2183470, EPI_ISL_2183472, EPI_ISL_2183475, EPI_ISL_2183478, EPI_ISL_2183480, EPI_ISL_2183482, EPI_ISL_2183485, EPI_ISL_2183486, EPI_ISL_2183489, EPI_ISL_2183491, EPI_ISL_2183493, EPI_ISL_2183495, EPI_ISL_2183497, EPI_ISL_2183499, EPI_ISL_2183501, EPI_ISL_2183503, EPI_ISL_2183505, EPI_ISL_2183507, EPI_ISL_2183509, EPI_ISL_2183512, EPI_ISL_2183515, EPI_ISL_2183516, EPI_ISL_2183518, EPI_ISL_2183521, EPI_ISL_2183524, EPI_ISL_2183526, EPI_ISL_2183529, EPI_ISL_2183534, EPI_ISL_2183537, EPI_ISL_2183540, EPI_ISL_2183543, EPI_ISL_2183545, EPI_ISL_2183548, EPI_ISL_2183553, EPI_ISL_2183555, EPI_ISL_2183558, EPI_ISL_2183561, EPI_ISL_2183564, EPI_ISL_2183569, EPI_ISL_2183572, EPI_ISL_2183574, EPI_ISL_2183579, EPI_ISL_2183582, EPI_ISL_2183584, EPI_ISL_2183587, EPI_ISL_2183594, EPI_ISL_2183597, EPI_ISL_2183600, EPI_ISL_2183603, EPI_ISL_2183606, EPI_ISL_2183608, EPI_ISL_2183610, EPI_ISL_2183612, EPI_ISL_2183614, EPI_ISL_2183617, EPI_ISL_2183620, EPI_ISL_2183623, EPI_ISL_2183624, EPI_ISL_2183626, EPI_ISL_2183629, EPI_ISL_2183632, EPI_ISL_2183634, EPI_ISL_2183638, EPI_ISL_2183639, EPI_ISL_2183642, EPI_ISL_2183645, EPI_ISL_2183648, EPI_ISL_2183653, EPI_ISL_2183656, EPI_ISL_2183658, EPI_ISL_2183661, EPI_ISL_2183664, EPI_ISL_2183667, EPI_ISL_2183669, EPI_ISL_2183672, EPI_ISL_2183674, EPI_ISL_2183677, EPI_ISL_2183680, EPI_ISL_2183682, EPI_ISL_2183684, EPI_ISL_2183690, EPI_ISL_2183695, EPI_ISL_2183698, EPI_ISL_2183700, EPI_ISL_2183705, EPI_ISL_2183710, EPI_ISL_2183712, EPI_ISL_2183714, EPI_ISL_2183716, EPI_ISL_2183720, EPI_ISL_2183722, EPI_ISL_2183725, EPI_ISL_2183729, EPI_ISL_2183731, EPI_ISL_2183734, EPI_ISL_2183735, EPI_ISL_2183737, EPI_ISL_2183739, EPI_ISL_2183745, EPI_ISL_2183748, EPI_ISL_2183751, EPI_ISL_2183753, EPI_ISL_2183757, EPI_ISL_2183761, EPI_ISL_2183764, EPI_ISL_2183766, EPI_ISL_2183768, EPI_ISL_2183778, EPI_ISL_2183781, EPI_ISL_2183783, EPI_ISL_2183785, EPI_ISL_2183788, EPI_ISL_2183790, EPI_ISL_2183792, EPI_ISL_2183794, EPI_ISL_2183796, EPI_ISL_2183798, EPI_ISL_2183799, EPI_ISL_2183801, EPI_ISL_2183804, EPI_ISL_2183807, EPI_ISL_2183812, EPI_ISL_2183814, EPI_ISL_2183816, EPI_ISL_2183821, EPI_ISL_2183822, EPI_ISL_2183825, EPI_ISL_2183826, EPI_ISL_2183828, EPI_ISL_2183830, EPI_ISL_2183833, EPI_ISL_2183835, EPI_ISL_2183840, EPI_ISL_2183843, EPI_ISL_2183845, EPI_ISL_2183847, EPI_ISL_2183851, EPI_ISL_2183853, EPI_ISL_2183854, EPI_ISL_2183855, EPI_ISL_2183856, EPI_ISL_2183857, EPI_ISL_2183858, EPI_ISL_2183859, EPI_ISL_2183860, EPI_ISL_2183861, EPI_ISL_2183862, EPI_ISL_2183863, EPI_ISL_2183865, EPI_ISL_2183866, EPI_ISL_2183867, EPI_ISL_2183868, EPI_ISL_2183870, EPI_ISL_2183871, EPI_ISL_2183872, EPI_ISL_2183873, EPI_ISL_2183874, EPI_ISL_2183875, EPI_ISL_2183876, EPI_ISL_2183878, EPI_ISL_2183879, EPI_ISL_2183880, EPI_ISL_2183882, EPI_ISL_2183883, EPI_ISL_2183884, EPI_ISL_2183885, EPI_ISL_2183886, EPI_ISL_2183887, EPI_ISL_2183888, EPI_ISL_2183889, EPI_ISL_2183890, EPI_ISL_2183892, EPI_ISL_2183894, EPI_ISL_2183895, EPI_ISL_2183896, EPI_ISL_2183897, EPI_ISL_2183898, EPI_ISL_2183899, EPI_ISL_2183900, EPI_ISL_2183901, EPI_ISL_2183902, EPI_ISL_2183903, EPI_ISL_2183904, EPI_ISL_2183905, EPI_ISL_2183907, EPI_ISL_2183908, EPI_ISL_2183909, EPI_ISL_2183910, EPI_ISL_2183911, EPI_ISL_2183912, EPI_ISL_2183913, EPI_ISL_2183915, EPI_ISL_2183916, EPI_ISL_2183917, EPI_ISL_2183918, EPI_ISL_2183920, EPI_ISL_2183921, EPI_ISL_2183922, EPI_ISL_2183923, EPI_ISL_2183924, EPI_ISL_2183925, EPI_ISL_2183926, EPI_ISL_2183927, EPI_ISL_2183928, EPI_ISL_2183929, EPI_ISL_2183930, EPI_ISL_2183931, EPI_ISL_2183932, EPI_ISL_2183933, EPI_ISL_2183934, EPI_ISL_2183935, EPI_ISL_2183936, EPI_ISL_2183937, EPI_ISL_2183938, EPI_ISL_2183939, EPI_ISL_2183940, EPI_ISL_2183941, EPI_ISL_2183942, EPI_ISL_2183943, EPI_ISL_2183944, EPI_ISL_2183945, EPI_ISL_2183947, EPI_ISL_2183948, EPI_ISL_2183950, EPI_ISL_2183951, EPI_ISL_2183952, EPI_ISL_2183953, EPI_ISL_2183954, EPI_ISL_2183955, EPI_ISL_2183956, EPI_ISL_2183957, EPI_ISL_2183958, EPI_ISL_2183959, EPI_ISL_2183960, EPI_ISL_2183961, EPI_ISL_2183962, EPI_ISL_2183963, EPI_ISL_2183964, EPI_ISL_2183965, EPI_ISL_2183966, EPI_ISL_2183967, EPI_ISL_2183968, EPI_ISL_2183969, EPI_ISL_2183970, EPI_ISL_2183971, EPI_ISL_2183972, EPI_ISL_2183973, EPI_ISL_2183974, EPI_ISL_2183975, EPI_ISL_2183976, EPI_ISL_2183978, EPI_ISL_2183979, EPI_ISL_2183980, EPI_ISL_2183981, EPI_ISL_2183982, EPI_ISL_2183983, EPI_ISL_2183985, EPI_ISL_2183986, EPI_ISL_2183987, EPI_ISL_2183988, EPI_ISL_2183989, EPI_ISL_2183992, EPI_ISL_2183993, EPI_ISL_2183994, EPI_ISL_2183995, EPI_ISL_2183996, EPI_ISL_2183997, EPI_ISL_2183998, EPI_ISL_2183999, EPI_ISL_2184000, EPI_ISL_2184003, EPI_ISL_2184004, EPI_ISL_2184005, EPI_ISL_2184007, EPI_ISL_2184008, EPI_ISL_2184009, EPI_ISL_2184010, EPI_ISL_2184011, EPI_ISL_2184012, EPI_ISL_2184013, EPI_ISL_2184014, EPI_ISL_2184016, EPI_ISL_2184017, EPI_ISL_2184019, EPI_ISL_2184020, EPI_ISL_2184021, EPI_ISL_2184022, EPI_ISL_2184023, EPI_ISL_2184024, EPI_ISL_2184025, EPI_ISL_2184026, EPI_ISL_2184027, EPI_ISL_2184028, EPI_ISL_2184029, EPI_ISL_2184032, EPI_ISL_2184033, EPI_ISL_2184034, EPI_ISL_2184035, EPI_ISL_2184036, EPI_ISL_2184038, EPI_ISL_2184039, EPI_ISL_2184040, EPI_ISL_2184041, EPI_ISL_2184042, EPI_ISL_2184044, EPI_ISL_2184045, EPI_ISL_2184046, EPI_ISL_2184047, EPI_ISL_2184048, EPI_ISL_2184049, EPI_ISL_2184050, EPI_ISL_2184051, EPI_ISL_2184052, EPI_ISL_2184054, EPI_ISL_2184055, EPI_ISL_2184056, EPI_ISL_2184058, EPI_ISL_2184059, EPI_ISL_2184061, EPI_ISL_2184063, EPI_ISL_2184064, EPI_ISL_2184065, EPI_ISL_2184066, EPI_ISL_2184067, EPI_ISL_2184068, EPI_ISL_2184069, EPI_ISL_2184071, EPI_ISL_2184072, EPI_ISL_2184074, EPI_ISL_2184075, EPI_ISL_2184076, EPI_ISL_2184077, EPI_ISL_2184078, EPI_ISL_2184079, EPI_ISL_2184080, EPI_ISL_2184081, EPI_ISL_2184082, EPI_ISL_2184083, EPI_ISL_2184085, EPI_ISL_2184086, EPI_ISL_2184088, EPI_ISL_2184089, EPI_ISL_2184090, EPI_ISL_2184091, EPI_ISL_2184092, EPI_ISL_2184093, EPI_ISL_2184094, EPI_ISL_2184095, EPI_ISL_2184096, EPI_ISL_2184097, EPI_ISL_2184099, EPI_ISL_2184100, EPI_ISL_2184102, EPI_ISL_2184103, EPI_ISL_2184104, EPI_ISL_2184105, EPI_ISL_2184106, EPI_ISL_2184107, EPI_ISL_2184108, EPI_ISL_2184109, EPI_ISL_2184111, EPI_ISL_2184112, EPI_ISL_2184113, EPI_ISL_2184114, EPI_ISL_2184115, EPI_ISL_2184117, EPI_ISL_2184119, EPI_ISL_2184120, EPI_ISL_2184121, EPI_ISL_2184122, EPI_ISL_2184123, EPI_ISL_2184124, EPI_ISL_2184125, EPI_ISL_2184126, EPI_ISL_2184127, EPI_ISL_2184128, EPI_ISL_2184129, EPI_ISL_2184130, EPI_ISL_2184131, EPI_ISL_2184133, EPI_ISL_2184134, EPI_ISL_2184135, EPI_ISL_2184136, EPI_ISL_2184137, EPI_ISL_2184139, EPI_ISL_2184141, EPI_ISL_2184142, EPI_ISL_2184143, EPI_ISL_2184144, EPI_ISL_2184145, EPI_ISL_2184146, EPI_ISL_2184147, EPI_ISL_2184150, EPI_ISL_2184151, EPI_ISL_2184152, EPI_ISL_2184153, EPI_ISL_2184154, EPI_ISL_2184155, EPI_ISL_2184156, EPI_ISL_2184157, EPI_ISL_2184158, EPI_ISL_2184160, EPI_ISL_2184161, EPI_ISL_2184162, EPI_ISL_2184163, EPI_ISL_2184165, EPI_ISL_2184166, EPI_ISL_2184167, EPI_ISL_2184168, EPI_ISL_2184169, EPI_ISL_2184170, EPI_ISL_2184171, EPI_ISL_2184172, EPI_ISL_2184173, EPI_ISL_2184174, EPI_ISL_2184175, EPI_ISL_2184176, EPI_ISL_2184177, EPI_ISL_2184178, EPI_ISL_2184179, EPI_ISL_2184180, EPI_ISL_2184181, EPI_ISL_2184182, EPI_ISL_2184183, EPI_ISL_2184184, EPI_ISL_2184185, EPI_ISL_2184186, EPI_ISL_2184187, EPI_ISL_2184188, EPI_ISL_2184189, EPI_ISL_2184190, EPI_ISL_2184192, EPI_ISL_2184193, EPI_ISL_2184194, EPI_ISL_2184195, EPI_ISL_2184196, EPI_ISL_2184197, EPI_ISL_2184198, EPI_ISL_2184199, EPI_ISL_2184200, EPI_ISL_2184202, EPI_ISL_2184203, EPI_ISL_2184204, EPI_ISL_2184206, EPI_ISL_2184207, EPI_ISL_2184208, EPI_ISL_2184209, EPI_ISL_2184210, EPI_ISL_2184211, EPI_ISL_2184212, EPI_ISL_2184213, EPI_ISL_2184214, EPI_ISL_2184216, EPI_ISL_2184217, EPI_ISL_2184218, EPI_ISL_2184220, EPI_ISL_2184221, EPI_ISL_2184222, EPI_ISL_2184223, EPI_ISL_2184225, EPI_ISL_2184226, EPI_ISL_2184227, EPI_ISL_2184229, EPI_ISL_2184230, EPI_ISL_2184231, EPI_ISL_2184232, EPI_ISL_2184233, EPI_ISL_2184234, EPI_ISL_2184235, EPI_ISL_2184236, EPI_ISL_2184237, EPI_ISL_2184238, EPI_ISL_2184239, EPI_ISL_2184240, EPI_ISL_2184241, EPI_ISL_2184242, EPI_ISL_2184243, EPI_ISL_2184244, EPI_ISL_2184246,

EPI_ISL_2184247, EPI_ISL_2184248, EPI_ISL_2184249, EPI_ISL_2184250, EPI_ISL_2184251, EPI_ISL_2184252, EPI_ISL_2184253, EPI_ISL_2184254, EPI_ISL_2184255, EPI_ISL_2184256, EPI_ISL_2184257, EPI_ISL_2184258, EPI_ISL_2184259, EPI_ISL_2184260, EPI_ISL_2184262, EPI_ISL_2184263, EPI_ISL_2184264, EPI_ISL_2184266, EPI_ISL_2184267, EPI_ISL_2184268, EPI_ISL_2184269, EPI_ISL_2184270, EPI_ISL_2184271, EPI_ISL_2184274, EPI_ISL_2184275, EPI_ISL_2184276, EPI_ISL_2184277, EPI_ISL_2184278, EPI_ISL_2184279, EPI_ISL_2184280, EPI_ISL_2184281, EPI_ISL_2184282, EPI_ISL_2184283, EPI_ISL_2184285, EPI_ISL_2184286, EPI_ISL_2184287, EPI_ISL_2184288, EPI_ISL_2184290, EPI_ISL_2184292, EPI_ISL_2184293, EPI_ISL_2184294, EPI_ISL_2184296, EPI_ISL_2184297, EPI_ISL_2184300, EPI_ISL_2184301, EPI_ISL_2184302, EPI_ISL_2184303, EPI_ISL_2184304, EPI_ISL_2184305, EPI_ISL_2184306, EPI_ISL_2184308, EPI_ISL_2184312, EPI_ISL_2184313, EPI_ISL_2184314, EPI_ISL_2184315, EPI_ISL_2184316, EPI_ISL_2184317, EPI_ISL_2184318, EPI_ISL_2184319, EPI_ISL_2184321, EPI_ISL_2184322, EPI_ISL_2184323, EPI_ISL_2184324, EPI_ISL_2184325, EPI_ISL_2184326, EPI_ISL_2184327, EPI_ISL_2184328, EPI_ISL_2184329, EPI_ISL_2184330, EPI_ISL_2184331, EPI_ISL_2184332, EPI_ISL_2184333, EPI_ISL_2184337, EPI_ISL_2184338, EPI_ISL_2184339, EPI_ISL_2184340, EPI_ISL_2184341, EPI_ISL_2184342, EPI_ISL_2184343, EPI_ISL_2184344, EPI_ISL_2184345, EPI_ISL_2184346, EPI_ISL_2184348, EPI_ISL_2184349, EPI_ISL_2184350, EPI_ISL_2184353, EPI_ISL_2184354, EPI_ISL_2184355, EPI_ISL_2184356, EPI_ISL_2184359, EPI_ISL_2184361, EPI_ISL_2184362, EPI_ISL_2184363, EPI_ISL_2184364, EPI_ISL_2184365, EPI_ISL_2184366, EPI_ISL_2184367, EPI_ISL_2184368, EPI_ISL_2184369, EPI_ISL_2184370, EPI_ISL_2184371, EPI_ISL_2184372, EPI_ISL_2184374, EPI_ISL_2184375, EPI_ISL_2184376, EPI_ISL_2184377, EPI_ISL_2184378, EPI_ISL_2184380, EPI_ISL_2184381, EPI_ISL_2184383, EPI_ISL_2184385, EPI_ISL_2184386, EPI_ISL_2184387, EPI_ISL_2184392, EPI_ISL_2184394, EPI_ISL_2184395, EPI_ISL_2184396, EPI_ISL_2184397, EPI_ISL_2184398, EPI_ISL_2184399, EPI_ISL_2184401, EPI_ISL_2184404, EPI_ISL_2184405, EPI_ISL_2184407, EPI_ISL_2184409, EPI_ISL_2184410, EPI_ISL_2184412, EPI_ISL_2184413, EPI_ISL_2184414, EPI_ISL_2184416, EPI_ISL_2184418, EPI_ISL_2184419, EPI_ISL_2184421, EPI_ISL_2184422, EPI_ISL_2184423, EPI_ISL_2184424, EPI_ISL_2184425, EPI_ISL_2184426, EPI_ISL_2184428, EPI_ISL_2184429, EPI_ISL_2184430, EPI_ISL_2184431, EPI_ISL_2184432, EPI_ISL_2184433, EPI_ISL_2184434, EPI_ISL_2184435, EPI_ISL_2184437, EPI_ISL_2184438, EPI_ISL_2184439, EPI_ISL_2184440, EPI_ISL_2184442, EPI_ISL_2184444, EPI_ISL_2184445, EPI_ISL_2184446, EPI_ISL_2184447, EPI_ISL_2184449, EPI_ISL_2184450, EPI_ISL_2184452, EPI_ISL_2184454, EPI_ISL_2184455, EPI_ISL_2184456, EPI_ISL_2184457, EPI_ISL_2184461, EPI_ISL_2184463, EPI_ISL_2184464, EPI_ISL_2184465, EPI_ISL_2184466, EPI_ISL_2184469, EPI_ISL_2184470, EPI_ISL_2184471, EPI_ISL_2184472, EPI_ISL_2184473, EPI_ISL_2184474, EPI_ISL_2184475, EPI_ISL_2184476, EPI_ISL_2184477, EPI_ISL_2184480, EPI_ISL_2184481, EPI_ISL_2184484, EPI_ISL_2184485, EPI_ISL_2184486, EPI_ISL_2184488, EPI_ISL_2184489, EPI_ISL_2184490, EPI_ISL_2184491, EPI_ISL_2184492, EPI_ISL_2184493, EPI_ISL_2184494, EPI_ISL_2184495, EPI_ISL_2184498, EPI_ISL_2184499, EPI_ISL_2184500, EPI_ISL_2184501, EPI_ISL_2184503, EPI_ISL_2184505, EPI_ISL_2184507, EPI_ISL_2184508, EPI_ISL_2184509, EPI_ISL_2184510, EPI_ISL_2184511, EPI_ISL_2184513, EPI_ISL_2184514, EPI_ISL_2184516, EPI_ISL_2184517, EPI_ISL_2184518, EPI_ISL_2184520, EPI_ISL_2184521, EPI_ISL_2184522, EPI_ISL_2184523, EPI_ISL_2184525, EPI_ISL_2184526, EPI_ISL_2184528, EPI_ISL_2184529, EPI_ISL_2184530, EPI_ISL_2184532, EPI_ISL_2184533, EPI_ISL_2184534, EPI_ISL_2184536, EPI_ISL_2184537, EPI_ISL_2184538, EPI_ISL_2184539, EPI_ISL_2184540, EPI_ISL_2184541, EPI_ISL_2184542, EPI_ISL_2184543, EPI_ISL_2184544, EPI_ISL_2184545, EPI_ISL_2184546, EPI_ISL_2184547, EPI_ISL_2184548, EPI_ISL_2184549, EPI_ISL_2184550, EPI_ISL_2184551, EPI_ISL_2184552, EPI_ISL_2184553, EPI_ISL_2184555, EPI_ISL_2184556, EPI_ISL_2184557, EPI_ISL_2184559, EPI_ISL_2184560, EPI_ISL_2184561, EPI_ISL_2184563, EPI_ISL_2184564, EPI_ISL_2184565, EPI_ISL_2184566, EPI_ISL_2184567, EPI_ISL_2184568, EPI_ISL_2184571, EPI_ISL_2184572, EPI_ISL_2184573, EPI_ISL_2184574, EPI_ISL_2184575, EPI_ISL_2184576, EPI_ISL_2184577, EPI_ISL_2184578, EPI_ISL_2184579, EPI_ISL_2184580, EPI_ISL_2184582, EPI_ISL_2184584, EPI_ISL_2184587, EPI_ISL_2184588, EPI_ISL_2184589, EPI_ISL_2184590, EPI_ISL_2184591, EPI_ISL_2184592, EPI_ISL_2184593, EPI_ISL_2184594, EPI_ISL_2184595, EPI_ISL_2184596, EPI_ISL_2184599, EPI_ISL_2184600, EPI_ISL_2184602, EPI_ISL_2184603, EPI_ISL_2184604, EPI_ISL_2184606, EPI_ISL_2184607, EPI_ISL_2184608, EPI_ISL_2184609, EPI_ISL_2184610, EPI_ISL_2184611, EPI_ISL_2184612, EPI_ISL_2184613, EPI_ISL_2184614, EPI_ISL_2184615, EPI_ISL_2184616, EPI_ISL_2184617, EPI_ISL_2184618, EPI_ISL_2184620, EPI_ISL_2184621, EPI_ISL_2184622, EPI_ISL_2184623, EPI_ISL_2184624, EPI_ISL_2184625, EPI_ISL_2184626, EPI_ISL_2184628, EPI_ISL_2184629, EPI_ISL_2184632, EPI_ISL_2184633, EPI_ISL_2184634, EPI_ISL_2184635, EPI_ISL_2184637, EPI_ISL_2184638, EPI_ISL_2184639, EPI_ISL_2184641, EPI_ISL_2184642, EPI_ISL_2184643, EPI_ISL_2184644, EPI_ISL_2184645, EPI_ISL_2184646, EPI_ISL_2184647, EPI_ISL_2184648, EPI_ISL_2184649, EPI_ISL_2184650, EPI_ISL_2184651, EPI_ISL_2184652, EPI_ISL_2184653, EPI_ISL_2184655, EPI_ISL_2184658, EPI_ISL_2184659, EPI_ISL_2184660, EPI_ISL_2184661, EPI_ISL_2184662, EPI_ISL_2184663, EPI_ISL_2184666, EPI_ISL_2184667, EPI_ISL_2184668, EPI_ISL_2184671, EPI_ISL_2184672, EPI_ISL_2184675, EPI_ISL_2184676, EPI_ISL_2184677, EPI_ISL_2184678, EPI_ISL_2184679, EPI_ISL_2184680, EPI_ISL_2184682, EPI_ISL_2184683, EPI_ISL_2184685, EPI_ISL_2184686, EPI_ISL_2184687, EPI_ISL_2184688, EPI_ISL_2184689, EPI_ISL_2184691, EPI_ISL_2184692, EPI_ISL_2184694, EPI_ISL_2184697, EPI_ISL_2184698, EPI_ISL_2184699, EPI_ISL_2184701, EPI_ISL_2184704, EPI_ISL_2184705, EPI_ISL_2184706, EPI_ISL_2184707, EPI_ISL_2184708, EPI_ISL_2184709, EPI_ISL_2184710, EPI_ISL_2184711, EPI_ISL_2184712, EPI_ISL_2184714, EPI_ISL_2184715, EPI_ISL_2184718, EPI_ISL_2184719, EPI_ISL_2184720, EPI_ISL_2184722, EPI_ISL_2184723, EPI_ISL_2184724, EPI_ISL_2184725, EPI_ISL_2184726, EPI_ISL_2184727, EPI_ISL_2184728, EPI_ISL_2184729, EPI_ISL_2184730, EPI_ISL_2184731, EPI_ISL_2184732, EPI_ISL_2184733, EPI_ISL_2184734, EPI_ISL_2184735, EPI_ISL_2184736, EPI_ISL_2184737, EPI_ISL_2184738, EPI_ISL_2184739, EPI_ISL_2184740, EPI_ISL_2184742, EPI_ISL_2184743, EPI_ISL_2184744, EPI_ISL_2184745, EPI_ISL_2184746, EPI_ISL_2184747, EPI_ISL_2184748, EPI_ISL_2184749, EPI_ISL_2184750, EPI_ISL_2184751, EPI_ISL_2184752, EPI_ISL_2184753, EPI_ISL_2184754, EPI_ISL_2184757, EPI_ISL_2184758, EPI_ISL_2184759, EPI_ISL_2184760, EPI_ISL_2184762, EPI_ISL_2184765, EPI_ISL_2184766, EPI_ISL_2184768, EPI_ISL_2184769, EPI_ISL_2184770, EPI_ISL_2184771, EPI_ISL_2184772, EPI_ISL_2184773, EPI_ISL_2184775, EPI_ISL_2184776, EPI_ISL_2184778, EPI_ISL_2184779, EPI_ISL_2184780, EPI_ISL_2184781, EPI_ISL_2184782, EPI_ISL_2184783, EPI_ISL_2184784, EPI_ISL_2184786, EPI_ISL_2184788, EPI_ISL_2184789, EPI_ISL_2184790, EPI_ISL_2184791, EPI_ISL_2184792, EPI_ISL_2184794, EPI_ISL_2184795, EPI_ISL_2184796, EPI_ISL_2184797, EPI_ISL_2184799, EPI_ISL_2184800, EPI_ISL_2184801, EPI_ISL_2184802, EPI_ISL_2184803, EPI_ISL_2184804, EPI_ISL_2184805, EPI_ISL_2184806, EPI_ISL_2184807, EPI_ISL_2184808, EPI_ISL_2184809, EPI_ISL_2184811, EPI_ISL_2184814, EPI_ISL_2184815, EPI_ISL_2184816, EPI_ISL_2184817, EPI_ISL_2184818, EPI_ISL_2184819, EPI_ISL_2184820, EPI_ISL_2184821, EPI_ISL_2184825, EPI_ISL_2184826, EPI_ISL_2184827, EPI_ISL_2184828, EPI_ISL_2184829, EPI_ISL_2184831, EPI_ISL_2184833, EPI_ISL_2184834, EPI_ISL_2184835, EPI_ISL_2184837, EPI_ISL_2184838, EPI_ISL_2184839, EPI_ISL_2184840, EPI_ISL_2184841, EPI_ISL_2184842, EPI_ISL_2184843, EPI_ISL_2184844, EPI_ISL_2184846, EPI_ISL_2184847, EPI_ISL_2184848, EPI_ISL_2184849, EPI_ISL_2184851, EPI_ISL_2184852, EPI_ISL_2184854, EPI_ISL_2184855, EPI_ISL_2184856, EPI_ISL_2184857, EPI_ISL_2184858, EPI_ISL_2184859, EPI_ISL_2184860, EPI_ISL_2184862, EPI_ISL_2184863, EPI_ISL_2184864, EPI_ISL_2184865, EPI_ISL_2184867, EPI_ISL_2184868, EPI_ISL_2184869, EPI_ISL_2184870, EPI_ISL_2184871, EPI_ISL_2184872, EPI_ISL_2184873, EPI_ISL_2184874, EPI_ISL_2184876, EPI_ISL_2184877, EPI_ISL_2184878, EPI_ISL_2184879, EPI_ISL_2184880, EPI_ISL_2184882, EPI_ISL_2184883, EPI_ISL_2184885, EPI_ISL_2184886, EPI_ISL_2184887, EPI_ISL_2184888, EPI_ISL_2184889, EPI_ISL_2184890, EPI_ISL_2184891, EPI_ISL_2184894, EPI_ISL_2184895, EPI_ISL_2184897, EPI_ISL_2184898, EPI_ISL_2184899, EPI_ISL_2184900, EPI_ISL_2184901, EPI_ISL_2184902, EPI_ISL_2184903, EPI_ISL_2184904, EPI_ISL_2184906, EPI_ISL_2184907, EPI_ISL_2184908, EPI_ISL_2184909, EPI_ISL_2184910, EPI_ISL_2184912, EPI_ISL_2184915, EPI_ISL_2184916, EPI_ISL_2184917, EPI_ISL_2184918, EPI_ISL_2184919, EPI_ISL_2184920, EPI_ISL_2184921, EPI_ISL_2184922, EPI_ISL_2184924, EPI_ISL_2184925, EPI_ISL_2184926, EPI_ISL_2184927, EPI_ISL_2184928, EPI_ISL_2184929, EPI_ISL_2184931, EPI_ISL_2184932, EPI_ISL_2184934, EPI_ISL_2184936, EPI_ISL_2184937, EPI_ISL_2184938, EPI_ISL_2184939, EPI_ISL_2184940, EPI_ISL_2184941, EPI_ISL_2184942, EPI_ISL_2184943, EPI_ISL_2184944, EPI_ISL_2184945, EPI_ISL_2184947, EPI_ISL_2184948, EPI_ISL_2184949, EPI_ISL_2184950, EPI_ISL_2184951, EPI_ISL_2184952, EPI_ISL_2184953, EPI_ISL_2184954, EPI_ISL_2184955, EPI_ISL_2184956, EPI_ISL_2184958, EPI_ISL_2184959, EPI_ISL_2184960, EPI_ISL_2184961, EPI_ISL_2184962, EPI_ISL_2184963, EPI_ISL_2184964, EPI_ISL_2184965, EPI_ISL_2184967, EPI_ISL_2184968, EPI_ISL_2184971, EPI_ISL_2184972, EPI_ISL_2184973, EPI_ISL_2184974, EPI_ISL_2184975, EPI_ISL_2184976, EPI_ISL_2184977, EPI_ISL_2184978, EPI_ISL_2184979, EPI_ISL_2184980, EPI_ISL_2184982, EPI_ISL_2184983, EPI_ISL_2184984, EPI_ISL_2184985, EPI_ISL_2184986, EPI_ISL_2184987, EPI_ISL_2184988, EPI_ISL_2184989, EPI_ISL_2184990, EPI_ISL_2184991, EPI_ISL_2184992, EPI_ISL_2184993, EPI_ISL_2184994, EPI_ISL_2184995, EPI_ISL_2184996, EPI_ISL_2184998, EPI_ISL_2185000, EPI_ISL_2185001, EPI_ISL_2185002, EPI_ISL_2185006, EPI_ISL_2185008, EPI_ISL_2185010, EPI_ISL_2185011, EPI_ISL_2185012, EPI_ISL_2185013, EPI_ISL_2185014, EPI_ISL_2185015, EPI_ISL_2185017, EPI_ISL_2185020, EPI_ISL_2185022, EPI_ISL_2185024, EPI_ISL_2185025, EPI_ISL_2185026, EPI_ISL_2185027, EPI_ISL_2185028, EPI_ISL_2185029, EPI_ISL_2185030, EPI_ISL_2185031, EPI_ISL_2185032, EPI_ISL_2185033, EPI_ISL_2185037, EPI_ISL_2185038, EPI_ISL_2185039, EPI_ISL_2185040, EPI_ISL_2185041, EPI_ISL_2185042, EPI_ISL_2185043, EPI_ISL_2185044, EPI_ISL_2185045, EPI_ISL_2185047, EPI_ISL_2185048, EPI_ISL_2185050, EPI_ISL_2185051, EPI_ISL_2185052, EPI_ISL_2185053, EPI_ISL_2185054, EPI_ISL_2185055, EPI_ISL_2185056, EPI_ISL_2185057, EPI_ISL_2185058, EPI_ISL_2185059, EPI_ISL_2185060, EPI_ISL_2185061, EPI_ISL_2185062, EPI_ISL_2185066, EPI_ISL_2185068, EPI_ISL_2185069, EPI_ISL_2185070, EPI_ISL_2185071, EPI_ISL_2185072, EPI_ISL_2185073, EPI_ISL_2185074, EPI_ISL_2185075, EPI_ISL_2185076, EPI_ISL_2185078, EPI_ISL_2185079, EPI_ISL_2185080, EPI_ISL_2185081, EPI_ISL_2185082, EPI_ISL_2185084, EPI_ISL_2185085, EPI_ISL_2185086, EPI_ISL_2185088, EPI_ISL_2185089, EPI_ISL_2185090, EPI_ISL_2185091, EPI_ISL_2185092, EPI_ISL_2185093, EPI_ISL_2185094, EPI_ISL_2185095, EPI_ISL_2185096, EPI_ISL_2185097, EPI_ISL_2185099, EPI_ISL_2185101, EPI_ISL_2185103, EPI_ISL_2185106, EPI_ISL_2185108, EPI_ISL_2185110, EPI_ISL_2185113, EPI_ISL_2185114, EPI_ISL_2185115, EPI_ISL_2185116, EPI_ISL_2185117, EPI_ISL_2185119, EPI_ISL_2185120, EPI_ISL_2185121, EPI_ISL_2185123, EPI_ISL_2185124, EPI_ISL_2185126, EPI_ISL_2185127, EPI_ISL_2185128, EPI_ISL_2185129, EPI_ISL_2185130, EPI_ISL_2185131, EPI_ISL_2185132, EPI_ISL_2185133, EPI_ISL_2185134, EPI_ISL_2185137, EPI_ISL_2185138, EPI_ISL_2185139, EPI_ISL_2185140, EPI_ISL_2185141, EPI_ISL_2185142, EPI_ISL_2185143, EPI_ISL_2185145, EPI_ISL_2185146, EPI_ISL_2185147, EPI_ISL_2185150, EPI_ISL_2185151, EPI_ISL_2185152, EPI_ISL_2185153, EPI_ISL_2185154, EPI_ISL_2185155, EPI_ISL_2185157, EPI_ISL_2185159, EPI_ISL_2185160, EPI_ISL_2185161, EPI_ISL_2185162, EPI_ISL_2185164, EPI_ISL_2185167, EPI_ISL_2185168, EPI_ISL_2185169, EPI_ISL_2185171, EPI_ISL_2185172, EPI_ISL_2185173, EPI_ISL_2185175, EPI_ISL_2185176, EPI_ISL_2185177, EPI_ISL_2185179, EPI_ISL_2185180, EPI_ISL_2185183, EPI_ISL_2185184, EPI_ISL_2185185, EPI_ISL_2185186, EPI_ISL_2185187, EPI_ISL_2185188, EPI_ISL_2185190, EPI_ISL_2185191, EPI_ISL_2185192, EPI_ISL_2185194, EPI_ISL_2185195, EPI_ISL_2185196, EPI_ISL_2185197, EPI_ISL_2185199, EPI_ISL_2185200, EPI_ISL_2185201, EPI_ISL_2185202, EPI_ISL_2185203, EPI_ISL_2185204, EPI_ISL_2185205, EPI_ISL_2185206, EPI_ISL_2185207, EPI_ISL_2185208, EPI_ISL_2185209, EPI_ISL_2185211, EPI_ISL_2185212, EPI_ISL_2185213, EPI_ISL_2185215, EPI_ISL_2185217, EPI_ISL_2185218, EPI_ISL_2185219, EPI_ISL_2185220, EPI_ISL_2185221, EPI_ISL_2185225, EPI_ISL_2185226, EPI_ISL_2185227, EPI_ISL_2185230, EPI_ISL_2185231, EPI_ISL_2185233, EPI_ISL_2185234, EPI_ISL_2185235, EPI_ISL_2185236, EPI_ISL_2185237, EPI_ISL_2185238, EPI_ISL_2185240, EPI_ISL_2185241, EPI_ISL_2185242, EPI_ISL_2185243, EPI_ISL_2185244, EPI_ISL_2185245, EPI_ISL_2185246, EPI_ISL_2185247, EPI_ISL_2185248, EPI_ISL_2185250, EPI_ISL_2185251, EPI_ISL_2185252, EPI_ISL_2185253, EPI_ISL_2185255, EPI_ISL_2185256, EPI_ISL_2185258, EPI_ISL_2185260, EPI_ISL_2185261, EPI_ISL_2185262, EPI_ISL_2185263, EPI_ISL_2185264, EPI_ISL_2185265, EPI_ISL_2185266, EPI_ISL_2185267, EPI_ISL_2185268, EPI_ISL_2185270, EPI_ISL_2185271, EPI_ISL_2185272, EPI_ISL_2185273, EPI_ISL_2185275, EPI_ISL_2185277, EPI_ISL_2185278, EPI_ISL_2185279, EPI_ISL_2185281, EPI_ISL_2185282, EPI_ISL_2185284, EPI_ISL_2185285, EPI_ISL_2185286, EPI_ISL_2185288, EPI_ISL_2185289, EPI_ISL_2185290, EPI_ISL_2185291, EPI_ISL_2185292, EPI_ISL_2185293, EPI_ISL_2185294, EPI_ISL_2185295, EPI_ISL_2185296, EPI_ISL_2185298, EPI_ISL_2185299, EPI_ISL_2185301, EPI_ISL_2185303, EPI_ISL_2185304, EPI_ISL_2185305, EPI_ISL_2185307, EPI_ISL_2185308, EPI_ISL_2185309, EPI_ISL_2185310, EPI_ISL_2185311, EPI_ISL_2185312, EPI_ISL_2185313, EPI_ISL_2185315, EPI_ISL_2185316, EPI_ISL_2185317, EPI_ISL_2185318, EPI_ISL_2185319, EPI_ISL_2185320, EPI_ISL_2185321, EPI_ISL_2185324, EPI_ISL_2185327, EPI_ISL_2185328, EPI_ISL_2185329, EPI_ISL_2185330, EPI_ISL_2185331, EPI_ISL_2185332, EPI_ISL_2185333, EPI_ISL_2185334, EPI_ISL_2185335, EPI_ISL_2185337, EPI_ISL_2185338, EPI_ISL_2185340, EPI_ISL_2185341, EPI_ISL_2185342, EPI_ISL_2185343, EPI_ISL_2185346, EPI_ISL_2185347, EPI_ISL_2185348, EPI_ISL_2185349, EPI_ISL_2185351, EPI_ISL_2185352, EPI_ISL_2185353, EPI_ISL_2185354, EPI_ISL_2185355, EPI_ISL_2185356, EPI_ISL_2185357, EPI_ISL_2185358, EPI_ISL_2185359, EPI_ISL_2185360, EPI_ISL_2185362, EPI_ISL_2185363, EPI_ISL_2185365, EPI_ISL_2185366, EPI_ISL_2185367, EPI_ISL_2185368, EPI_ISL_2185369, EPI_ISL_2185370, EPI_ISL_2185371, EPI_ISL_2185372, EPI_ISL_2185373, EPI_ISL_2185375, EPI_ISL_2185376, EPI_ISL_2185377, EPI_ISL_2185380, EPI_ISL_2185381, EPI_ISL_2185382, EPI_ISL_2185383, EPI_ISL_2185384, EPI_ISL_2185385, EPI_ISL_2185386, EPI_ISL_2185387, EPI_ISL_2185388, EPI_ISL_2185389, EPI_ISL_2185390, EPI_ISL_2185391, EPI_ISL_2185392, EPI_ISL_2185393, EPI_ISL_2185394, EPI_ISL_2185395, EPI_ISL_2185397, EPI_ISL_2185398, EPI_ISL_2185399, EPI_ISL_2185400, EPI_ISL_2185401, EPI_ISL_2185403, EPI_ISL_2185404, EPI_ISL_2185406, EPI_ISL_2185408, EPI_ISL_2185409, EPI_ISL_2185410, EPI_ISL_2185411, EPI_ISL_2185413, EPI_ISL_2185414, EPI_ISL_2185415, EPI_ISL_2185417, EPI_ISL_2185418, EPI_ISL_2185419, EPI_ISL_2185420, EPI_ISL_2185421, EPI_ISL_2185422, EPI_ISL_2185423, EPI_ISL_2185425, EPI_ISL_2185426, EPI_ISL_2185427, EPI_ISL_2185429, EPI_ISL_2185430, EPI_ISL_2185431, EPI_ISL_2185432, EPI_ISL_2185433, EPI_ISL_2185434, EPI_ISL_2185435, EPI_ISL_2185436, EPI_ISL_2185437, EPI_ISL_2185438, EPI_ISL_2185440, EPI_ISL_2185441, EPI_ISL_2185442, EPI_ISL_2185444, EPI_ISL_2185445, EPI_ISL_2185449, EPI_ISL_2185451, EPI_ISL_2185452, EPI_ISL_2185453, EPI_ISL_2185454, EPI_ISL_2185455, EPI_ISL_2185456, EPI_ISL_2185457, EPI_ISL_2185458, EPI_ISL_2185459, EPI_ISL_2185460, EPI_ISL_2185461, EPI_ISL_2185462, EPI_ISL_2185463, EPI_ISL_2185465, EPI_ISL_2185466, EPI_ISL_2185467, EPI_ISL_2185468, EPI_ISL_2185469, EPI_ISL_2185470, EPI_ISL_2185471, EPI_ISL_2185472, EPI_ISL_2185473, EPI_ISL_2185475, EPI_ISL_2185476, EPI_ISL_2185477, EPI_ISL_2185479, EPI_ISL_2185480, EPI_ISL_2185481, EPI_ISL_2185482, EPI_ISL_2185483, EPI_ISL_2185484, EPI_ISL_2185485, EPI_ISL_2185486, EPI_ISL_2185487, EPI_ISL_2185489, EPI_ISL_2185490, EPI_ISL_2185491, EPI_ISL_2185492, EPI_ISL_2185494, EPI_ISL_2185495, EPI_ISL_2185497, EPI_ISL_2185499, EPI_ISL_2185500, EPI_ISL_2185502, EPI_ISL_2185503, EPI_ISL_2185504, EPI_ISL_2185506, EPI_ISL_2185507, EPI_ISL_2185509, EPI_ISL_2185510, EPI_ISL_2185511, EPI_ISL_2185512, EPI_ISL_2185513, EPI_ISL_2185514, EPI_ISL_2185516, EPI_ISL_2185517, EPI_ISL_2185518, EPI_ISL_2185520, EPI_ISL_2185521, EPI_ISL_2185522, EPI_ISL_2185523, EPI_ISL_2185524, EPI_ISL_2185525, EPI_ISL_2185526, EPI_ISL_2185527, EPI_ISL_2185528, EPI_ISL_2185529

see above | Laboratory Corporation of America | Centers for Disease Control and Prevention Division of Viral Diseases, Pathogen Discovery | Dakota Howard, Dhwani Batra, Peter W. Cook, Kara Moser, Adrian Paskey, Jason Caravas, Benjamin Rambo-Martin, Shatavia Morrison, Christopher Gulvick, Scott Sammons, Yvette Unoarumhi, Darlene Wagner, Matthew Schmerer, Minoo Agarwal, Eyad Almasri, Debbie Boles, Ayla Burns, Nuthawin Charoensri, Oren Cohen, Susan Countryman, Mary Ann Cristobal, Bobbi Croy, Suzanne Dale, Hrushikesh Deshmukh, Amanda Douglas, Vincent Drouillon, Marcia Eisenberg, Howard Engler, Rama Ghatti, Prashant Gupta, Susan Hicks, Jake Humphrey, Lax Iyer, Manoj Jain, Mohan Kolli, Brian Krueger, Tim Kuphal, Stanley Letovsky, Michael Levandoski, Craig Lukasik, Jonathan Meltzer, Brian Norvell, Mindy Nye, Scott Parker, Christos Petropoulos, John Pruitt, Steven Ragan, Scott Ryan, Mike Sapeta, Jana Schroth, Suresh Babu Selvaraju, Goran Stevovic, Amanda Suchanek, Andrea Throop, Lyndon Tilson, Thomas Urban, Joe Voshell, Kimberly Wagner, Jonathan Williams, Mary Williamson, Qian Zeng, Tricia Zwiefelhofer, Clinton R. Paden, Duncan MacCannell

EPI_ISL_2185530, EPI_ISL_2185531, EPI_ISL_2185532, EPI_ISL_2185533, EPI_ISL_2185534, EPI_ISL_2185536, EPI_ISL_2185537, EPI_ISL_2185538, EPI_ISL_2185539, EPI_ISL_2185540, EPI_ISL_2185541, EPI_ISL_2185542, EPI_ISL_2185543, EPI_ISL_2185544, EPI_ISL_2185545, EPI_ISL_2185546, EPI_ISL_2185548, EPI_ISL_2185550, EPI_ISL_2185551, EPI_ISL_2185552, EPI_ISL_2185553, EPI_ISL_2185556, EPI_ISL_2185557, EPI_ISL_2185558, EPI_ISL_2185559, EPI_ISL_2185560, EPI_ISL_2185562, EPI_ISL_2185563, EPI_ISL_2185564, EPI_ISL_2185565, EPI_ISL_2185566, EPI_ISL_2185568, EPI_ISL_2185569, EPI_ISL_2185570, EPI_ISL_2185571

see above | Curative | New Mexico Department of Health Scientific Laboratory | Ellie Johnson, D'eldra Malone, Jennifer Benoit, Ratheesh Rajan, Linda Salazar, Anastacia Griego-Fisher

EPI_ISL_2185575, EPI_ISL_2185576, EPI_ISL_2185577, EPI_ISL_2185578, EPI_ISL_2185579, EPI_ISL_2185580, EPI_ISL_2185581, EPI_ISL_2185582, EPI_ISL_2185583, EPI_ISL_2185584, EPI_ISL_2185585, EPI_ISL_2185586, EPI_ISL_2185587, EPI_ISL_2185588, EPI_ISL_2185589, EPI_ISL_2185590, EPI_ISL_2185591, EPI_ISL_2185592, EPI_ISL_2185593, EPI_ISL_2185594, EPI_ISL_2185595, EPI_ISL_2185596, EPI_ISL_2185597, EPI_ISL_2185598, EPI_ISL_2185599, EPI_ISL_2185600, EPI_ISL_2185601, EPI_ISL_2185602, EPI_ISL_2185603, EPI_ISL_2185604, EPI_ISL_2185605, EPI_ISL_2185606, EPI_ISL_2185607, EPI_ISL_2185608, EPI_ISL_2185609, EPI_ISL_2185610, EPI_ISL_2185611, EPI_ISL_2185612, EPI_ISL_2185613, EPI_ISL_2185614, EPI_ISL_2185616, EPI_ISL_2185617, EPI_ISL_2185618, EPI_ISL_2185619, EPI_ISL_2185620, EPI_ISL_2185621, EPI_ISL_2185622, EPI_ISL_2185623, EPI_ISL_2185624, EPI_ISL_2185625, EPI_ISL_2185626, EPI_ISL_2185627, EPI_ISL_2185629

see above | NJDOH, Public Health and Environmental Laboratories | NJ_PHEL | Lindsey Bodnar, Shiv K. Verma, Jacquelyn Deverell, Matthew Scarnati, Dana Woell, Byeong Jeong

EPI_ISL_2185630, EPI_ISL_2185631, EPI_ISL_2185632, EPI_ISL_2185633, EPI_ISL_2185634, EPI_ISL_2185635, EPI_ISL_2185636, EPI_ISL_2185637, EPI_ISL_2185638, EPI_ISL_2185639, EPI_ISL_2185640, EPI_ISL_2185641, EPI_ISL_2185642, EPI_ISL_2185643, EPI_ISL_2185644, EPI_ISL_2185645, EPI_ISL_2185646, EPI_ISL_2185647, EPI_ISL_2185648, EPI_ISL_2185649, EPI_ISL_2185650, EPI_ISL_2185651, EPI_ISL_2185652, EPI_ISL_2185653, EPI_ISL_2185654, EPI_ISL_2185656, EPI_ISL_2185657, EPI_ISL_2185658, EPI_ISL_2185659, EPI_ISL_2185660, EPI_ISL_2185661, EPI_ISL_2185662, EPI_ISL_2185663, EPI_ISL_2185664, EPI_ISL_2185665, EPI_ISL_2185666, EPI_ISL_2185667, EPI_ISL_2185668, EPI_ISL_2185669, EPI_ISL_2185670, EPI_ISL_2185672, EPI_ISL_2185673, EPI_ISL_2185674, EPI_ISL_2185675, EPI_ISL_2185676, EPI_ISL_2185678, EPI_ISL_2185679, EPI_ISL_2185680, EPI_ISL_2185681, EPI_ISL_2185682, EPI_ISL_2185684, EPI_ISL_2185685, EPI_ISL_2185686, EPI_ISL_2185687, EPI_ISL_2185688, EPI_ISL_2185690, EPI_ISL_2185691, EPI_ISL_2185692, EPI_ISL_2185693, EPI_ISL_2185694, EPI_ISL_2185695, EPI_ISL_2185696, EPI_ISL_2185697, EPI_ISL_2185698, EPI_ISL_2185699, EPI_ISL_2185701, EPI_ISL_2185702, EPI_ISL_2185703, EPI_ISL_2185704, EPI_ISL_2185705, EPI_ISL_2185706, EPI_ISL_2185707, EPI_ISL_2185708, EPI_ISL_2185710, EPI_ISL_2185711, EPI_ISL_2185712, EPI_ISL_2185713, EPI_ISL_2185714, EPI_ISL_2185715, EPI_ISL_2185716, EPI_ISL_2185717, EPI_ISL_2185718, EPI_ISL_2185719, EPI_ISL_2185720, EPI_ISL_2185721, EPI_ISL_2185722, EPI_ISL_2185723, EPI_ISL_2185725, EPI_ISL_2185726, EPI_ISL_2185727, EPI_ISL_2185728, EPI_ISL_2185730, EPI_ISL_2185731, EPI_ISL_2185732, EPI_ISL_2185733, EPI_ISL_2185734, EPI_ISL_2185735, EPI_ISL_2185736, EPI_ISL_2185737, EPI_ISL_2185738, EPI_ISL_2185739, EPI_ISL_2185740, EPI_ISL_2185741, EPI_ISL_2185742, EPI_ISL_2185743, EPI_ISL_2185744, EPI_ISL_2185745, EPI_ISL_2185747, EPI_ISL_2185748, EPI_ISL_2185749, EPI_ISL_2185750, EPI_ISL_2185751, EPI_ISL_2185752, EPI_ISL_2185753, EPI_ISL_2185754, EPI_ISL_2185755, EPI_ISL_2185756, EPI_ISL_2185757, EPI_ISL_2185758, EPI_ISL_2185759, EPI_ISL_2185760, EPI_ISL_2185761, EPI_ISL_2185762, EPI_ISL_2185763, EPI_ISL_2185764, EPI_ISL_2185765, EPI_ISL_2185766, EPI_ISL_2185767, EPI_ISL_2185768, EPI_ISL_2185769, EPI_ISL_2185770, EPI_ISL_2185771, EPI_ISL_2185772, EPI_ISL_2185773, EPI_ISL_2185775, EPI_ISL_2185776, EPI_ISL_2185777, EPI_ISL_2185778, EPI_ISL_2185779, EPI_ISL_2185780, EPI_ISL_2185781, EPI_ISL_2185782, EPI_ISL_2185783, EPI_ISL_2185784, EPI_ISL_2185785, EPI_ISL_2185786, EPI_ISL_2185787, EPI_ISL_2185788, EPI_ISL_2185789, EPI_ISL_2185790, EPI_ISL_2185791, EPI_ISL_2185792, EPI_ISL_2185793, EPI_ISL_2185796, EPI_ISL_2185797, EPI_ISL_2185798, EPI_ISL_2185799, EPI_ISL_2185800, EPI_ISL_2185801, EPI_ISL_2185802, EPI_ISL_2185803, EPI_ISL_2185804, EPI_ISL_2185805, EPI_ISL_2185806, EPI_ISL_2185807, EPI_ISL_2185808, EPI_ISL_2185809, EPI_ISL_2185810, EPI_ISL_2185811, EPI_ISL_2185812, EPI_ISL_2185813, EPI_ISL_2185814, EPI_ISL_2185815, EPI_ISL_2185816, EPI_ISL_2185817, EPI_ISL_2185818, EPI_ISL_2185820, EPI_ISL_2185821, EPI_ISL_2185822, EPI_ISL_2185823, EPI_ISL_2185824, EPI_ISL_2185825, EPI_ISL_2185826, EPI_ISL_2185827, EPI_ISL_2185828, EPI_ISL_2185829, EPI_ISL_2185830, EPI_ISL_2185831, EPI_ISL_2185832, EPI_ISL_2185833, EPI_ISL_2185834, EPI_ISL_2185836, EPI_ISL_2185837, EPI_ISL_2185838, EPI_ISL_2185839, EPI_ISL_2185840, EPI_ISL_2185841, EPI_ISL_2185843, EPI_ISL_2185844, EPI_ISL_2185845, EPI_ISL_2185846, EPI_ISL_2185847, EPI_ISL_2185848, EPI_ISL_2185849, EPI_ISL_2185850, EPI_ISL_2185851, EPI_ISL_2185852, EPI_ISL_2185853, EPI_ISL_2185854, EPI_ISL_2185855, EPI_ISL_2185856, EPI_ISL_2185857, EPI_ISL_2185858, EPI_ISL_2185859, EPI_ISL_2185860, EPI_ISL_2185861, EPI_ISL_2185862, EPI_ISL_2185863, EPI_ISL_2185864, EPI_ISL_2185865, EPI_ISL_2185866, EPI_ISL_2185867, EPI_ISL_2185868, EPI_ISL_2185869, EPI_ISL_2185871, EPI_ISL_2185872, EPI_ISL_2185873, EPI_ISL_2185874, EPI_ISL_2185875, EPI_ISL_2185876, EPI_ISL_2185877, EPI_ISL_2185878, EPI_ISL_2185879, EPI_ISL_2185880, EPI_ISL_2185881, EPI_ISL_2185882, EPI_ISL_2185883, EPI_ISL_2185884, EPI_ISL_2185885, EPI_ISL_2185886, EPI_ISL_2185887, EPI_ISL_2185888, EPI_ISL_2185889, EPI_ISL_2185890, EPI_ISL_2185893, EPI_ISL_2185894, EPI_ISL_2185895, EPI_ISL_2185896, EPI_ISL_2185897, EPI_ISL_2185898, EPI_ISL_2185899, EPI_ISL_2185900, EPI_ISL_2185901, EPI_ISL_2185902, EPI_ISL_2185903, EPI_ISL_2185904, EPI_ISL_2185905, EPI_ISL_2185906, EPI_ISL_2185907, EPI_ISL_2185908, EPI_ISL_2185909, EPI_ISL_2185910, EPI_ISL_2185911, EPI_ISL_2185912, EPI_ISL_2185913, EPI_ISL_2185914, EPI_ISL_2185915, EPI_ISL_2185916, EPI_ISL_2185917, EPI_ISL_2185918, EPI_ISL_2185919, EPI_ISL_2185920, EPI_ISL_2185921, EPI_ISL_2185922, EPI_ISL_2185923, EPI_ISL_2185924, EPI_ISL_2185925, EPI_ISL_2185926, EPI_ISL_2185927, EPI_ISL_2185928, EPI_ISL_2185929, EPI_ISL_2185930, EPI_ISL_2185931, EPI_ISL_2185932, EPI_ISL_2185933, EPI_ISL_2185934, EPI_ISL_2185936, EPI_ISL_2185937, EPI_ISL_2185938, EPI_ISL_2185939, EPI_ISL_2185940, EPI_ISL_2185941, EPI_ISL_2185942, EPI_ISL_2185943, EPI_ISL_2185944, EPI_ISL_2185945, EPI_ISL_2185946, EPI_ISL_2185947, EPI_ISL_2185948, EPI_ISL_2185950, EPI_ISL_2185951, EPI_ISL_2185952, EPI_ISL_2185953, EPI_ISL_2185954, EPI_ISL_2185955, EPI_ISL_2185956, EPI_ISL_2185957, EPI_ISL_2185958, EPI_ISL_2185959, EPI_ISL_2185961, EPI_ISL_2185962, EPI_ISL_2185963, EPI_ISL_2185964, EPI_ISL_2185965, EPI_ISL_2185966, EPI_ISL_2185967, EPI_ISL_2185968, EPI_ISL_2185969, EPI_ISL_2185970, EPI_ISL_2185971, EPI_ISL_2185972, EPI_ISL_2185973, EPI_ISL_2185974, EPI_ISL_2185975, EPI_ISL_2185976, EPI_ISL_2185977, EPI_ISL_2185978, EPI_ISL_2185979, EPI_ISL_2185980, EPI_ISL_2185981, EPI_ISL_2185982, EPI_ISL_2185983, EPI_ISL_2185984, EPI_ISL_2185985, EPI_ISL_2185986, EPI_ISL_2185987, EPI_ISL_2185988, EPI_ISL_2185989, EPI_ISL_2185990, EPI_ISL_2185991, EPI_ISL_2185992, EPI_ISL_2185993, EPI_ISL_2185994, EPI_ISL_2185995, EPI_ISL_2185996, EPI_ISL_2185998, EPI_ISL_2185999, EPI_ISL_2186000, EPI_ISL_2186001, EPI_ISL_2186002, EPI_ISL_2186003, EPI_ISL_2186005, EPI_ISL_2186006, EPI_ISL_2186007, EPI_ISL_2186008, EPI_ISL_2186009, EPI_ISL_2186010, EPI_ISL_2186011, EPI_ISL_2186012, EPI_ISL_2186013, EPI_ISL_2186014, EPI_ISL_2186015, EPI_ISL_2186016, EPI_ISL_2186017, EPI_ISL_2186018, EPI_ISL_2186019, EPI_ISL_2186020, EPI_ISL_2186021, EPI_ISL_2186023, EPI_ISL_2186024, EPI_ISL_2186025, EPI_ISL_2186026, EPI_ISL_2186027, EPI_ISL_2186028, EPI_ISL_2186029, EPI_ISL_2186030, EPI_ISL_2186031, EPI_ISL_2186032, EPI_ISL_2186033, EPI_ISL_2186034, EPI_ISL_2186035, EPI_ISL_2186036, EPI_ISL_2186037, EPI_ISL_2186038, EPI_ISL_2186039, EPI_ISL_2186040, EPI_ISL_2186041, EPI_ISL_2186042, EPI_ISL_2186043, EPI_ISL_2186044, EPI_ISL_2186045, EPI_ISL_2186047, EPI_ISL_2186048, EPI_ISL_2186049, EPI_ISL_2186050, EPI_ISL_2186051, EPI_ISL_2186052, EPI_ISL_2186053, EPI_ISL_2186054, EPI_ISL_2186055, EPI_ISL_2186056, EPI_ISL_2186057, EPI_ISL_2186058, EPI_ISL_2186059, EPI_ISL_2186060, EPI_ISL_2186061, EPI_ISL_2186062, EPI_ISL_2186063, EPI_ISL_2186064, EPI_ISL_2186065, EPI_ISL_2186066, EPI_ISL_2186067, EPI_ISL_2186068, EPI_ISL_2186069, EPI_ISL_2186071, EPI_ISL_2186072, EPI_ISL_2186073, EPI_ISL_2186074, EPI_ISL_2186075, EPI_ISL_2186076, EPI_ISL_2186077, EPI_ISL_2186078, EPI_ISL_2186079, EPI_ISL_2186080, EPI_ISL_2186081, EPI_ISL_2186082, EPI_ISL_2186083, EPI_ISL_2186084, EPI_ISL_2186085, EPI_ISL_2186086, EPI_ISL_2186088, EPI_ISL_2186089, EPI_ISL_2186090, EPI_ISL_2186091, EPI_ISL_2186092, EPI_ISL_2186093, EPI_ISL_2186094, EPI_ISL_2186095, EPI_ISL_2186096, EPI_ISL_2186097, EPI_ISL_2186098, EPI_ISL_2186100, EPI_ISL_2186101, EPI_ISL_2186102, EPI_ISL_2186103, EPI_ISL_2186105, EPI_ISL_2186106, EPI_ISL_2186107, EPI_ISL_2186108, EPI_ISL_2186110, EPI_ISL_2186111, EPI_ISL_2186112, EPI_ISL_2186113, EPI_ISL_2186114, EPI_ISL_2186115, EPI_ISL_2186116, EPI_ISL_2186117, EPI_ISL_2186119, EPI_ISL_2186120, EPI_ISL_2186121, EPI_ISL_2186122, EPI_ISL_2186123, EPI_ISL_2186124, EPI_ISL_2186125, EPI_ISL_2186126, EPI_ISL_2186127, EPI_ISL_2186128, EPI_ISL_2186129, EPI_ISL_2186130, EPI_ISL_2186131, EPI_ISL_2186132, EPI_ISL_2186133, EPI_ISL_2186135, EPI_ISL_2186136, EPI_ISL_2186137, EPI_ISL_2186138, EPI_ISL_2186139, EPI_ISL_2186141, EPI_ISL_2186142, EPI_ISL_2186143, EPI_ISL_2186144, EPI_ISL_2186145, EPI_ISL_2186146, EPI_ISL_2186149, EPI_ISL_2186150, EPI_ISL_2186151, EPI_ISL_2186152, EPI_ISL_2186153, EPI_ISL_2186154, EPI_ISL_2186155, EPI_ISL_2186156, EPI_ISL_2186157, EPI_ISL_2186158, EPI_ISL_2186159, EPI_ISL_2186160, EPI_ISL_2186161, EPI_ISL_2186162, EPI_ISL_2186163, EPI_ISL_2186165, EPI_ISL_2186166, EPI_ISL_2186167, EPI_ISL_2186168, EPI_ISL_2186169, EPI_ISL_2186170, EPI_ISL_2186171, EPI_ISL_2186172, EPI_ISL_2186173, EPI_ISL_2186174, EPI_ISL_2186175, EPI_ISL_2186176, EPI_ISL_2186177, EPI_ISL_2186178, EPI_ISL_2186179, EPI_ISL_2186180, EPI_ISL_2186181, EPI_ISL_2186182, EPI_ISL_2186183, EPI_ISL_2186184, EPI_ISL_2186185, EPI_ISL_2186186, EPI_ISL_2186187, EPI_ISL_2186188, EPI_ISL_2186189, EPI_ISL_2186190, EPI_ISL_2186191, EPI_ISL_2186192, EPI_ISL_2186193, EPI_ISL_2186194, EPI_ISL_2186195, EPI_ISL_2186196, EPI_ISL_2186197, EPI_ISL_2186198, EPI_ISL_2186199, EPI_ISL_2186200, EPI_ISL_2186201, EPI_ISL_2186203, EPI_ISL_2186204, EPI_ISL_2186206, EPI_ISL_2186207, EPI_ISL_2186208, EPI_ISL_2186210, EPI_ISL_2186211, EPI_ISL_2186212, EPI_ISL_2186214, EPI_ISL_2186216, EPI_ISL_2186217, EPI_ISL_2186218, EPI_ISL_2186219, EPI_ISL_2186220, EPI_ISL_2186221, EPI_ISL_2186222, EPI_ISL_2186223, EPI_ISL_2186224, EPI_ISL_2186225, EPI_ISL_2186226, EPI_ISL_2186227, EPI_ISL_2186228, EPI_ISL_2186229, EPI_ISL_2186230, EPI_ISL_2186231, EPI_ISL_2186232, EPI_ISL_2186233, EPI_ISL_2186234, EPI_ISL_2186235, EPI_ISL_2186236, EPI_ISL_2186237, EPI_ISL_2186238, EPI_ISL_2186239, EPI_ISL_2186240, EPI_ISL_2186241, EPI_ISL_2186242, EPI_ISL_2186244, EPI_ISL_2186245, EPI_ISL_2186246, EPI_ISL_2186247, EPI_ISL_2186248, EPI_ISL_2186249, EPI_ISL_2186250, EPI_ISL_2186251, EPI_ISL_2186252, EPI_ISL_2186253, EPI_ISL_2186254, EPI_ISL_2186255, EPI_ISL_2186256, EPI_ISL_2186257, EPI_ISL_2186258, EPI_ISL_2186259, EPI_ISL_2186260, EPI_ISL_2186261, EPI_ISL_2186262, EPI_ISL_2186263, EPI_ISL_2186264, EPI_ISL_2186265

see above | Aegis Sciences Corporation | Centers for Disease Control and Prevention Division of Viral Diseases, Pathogen Discovery | Dakota Howard, Dhwani Batra, Peter W. Cook, Kara Moser, Adrian Paskey, Jason Caravas, Benjamin Rambo-Martin, Shatavia Morrison, Christopher Gulvick, Scott Sammons, Yvette Unoarumhi, Darlene Wagner, Matthew Schmerer, Cyndi Clark, Patrick Campbell, Rob Case, Vikramsinha Ghorpade, Holly Houdeshell, Ola Kvalvaag, Dillon Nall, Ethan Sanders, Alec Vest, Shaun Westlund, Matthew Hardison, Clinton R. Paden, Duncan MacCannell

EPI_ISL_2186270, EPI_ISL_2186271, EPI_ISL_2186272, EPI_ISL_2186275, EPI_ISL_2186276, EPI_ISL_2186278, EPI_ISL_2186279, EPI_ISL_2186280, EPI_ISL_2186281, EPI_ISL_2186285, EPI_ISL_2186289, EPI_ISL_2186291, EPI_ISL_2186294, EPI_ISL_2186299, EPI_ISL_2186304, EPI_ISL_2186305, EPI_ISL_2186309, EPI_ISL_2186313, EPI_ISL_2186314, EPI_ISL_2186316, EPI_ISL_2186322, EPI_ISL_2186324, EPI_ISL_2186334, EPI_ISL_2186340, EPI_ISL_2186341, EPI_ISL_2186346, EPI_ISL_2186352, EPI_ISL_2186356, EPI_ISL_2186357, EPI_ISL_2186362, EPI_ISL_2186363, EPI_ISL_2186368, EPI_ISL_2186372, EPI_ISL_2186376, EPI_ISL_2186387, EPI_ISL_2186390, EPI_ISL_2186392, EPI_ISL_2186393, EPI_ISL_2186398, EPI_ISL_2186399, EPI_ISL_2186401, EPI_ISL_2186406, EPI_ISL_2186407, EPI_ISL_2186408, EPI_ISL_2186411, EPI_ISL_2186413, EPI_ISL_2186415, EPI_ISL_2186416, EPI_ISL_2186417, EPI_ISL_2186418, EPI_ISL_2186420, EPI_ISL_2186421, EPI_ISL_2186422, EPI_ISL_2186423, EPI_ISL_2186424, EPI_ISL_2186425, EPI_ISL_2186426, EPI_ISL_2186427, EPI_ISL_2186428, EPI_ISL_2186429, EPI_ISL_2186430, EPI_ISL_2186431, EPI_ISL_2186432, EPI_ISL_2186433, EPI_ISL_2186434, EPI_ISL_2186435, EPI_ISL_2186436, EPI_ISL_2186437, EPI_ISL_2186438, EPI_ISL_2186439, EPI_ISL_2186440, EPI_ISL_2186441, EPI_ISL_2186442, EPI_ISL_2186443, EPI_ISL_2186444, EPI_ISL_2186445, EPI_ISL_2186446, EPI_ISL_2186447, EPI_ISL_2186448, EPI_ISL_2186449, EPI_ISL_2186450, EPI_ISL_2186451, EPI_ISL_2186452, EPI_ISL_2186453, EPI_ISL_2186455, EPI_ISL_2186456, EPI_ISL_2186458, EPI_ISL_2186459, EPI_ISL_2186460, EPI_ISL_2186461, EPI_ISL_2186462, EPI_ISL_2186463, EPI_ISL_2186464, EPI_ISL_2186465, EPI_ISL_2186467, EPI_ISL_2186468, EPI_ISL_2186469, EPI_ISL_2186470, EPI_ISL_2186471, EPI_ISL_2186472, EPI_ISL_2186473, EPI_ISL_2186474, EPI_ISL_2186475, EPI_ISL_2186476, EPI_ISL_2186477, EPI_ISL_2186478, EPI_ISL_2186479, EPI_ISL_2186480, EPI_ISL_2186481, EPI_ISL_2186482, EPI_ISL_2186483, EPI_ISL_2186484, EPI_ISL_2186485, EPI_ISL_2186486, EPI_ISL_2186487, EPI_ISL_2186488, EPI_ISL_2186489, EPI_ISL_2186490, EPI_ISL_2186491, EPI_ISL_2186493, EPI_ISL_2186494, EPI_ISL_2186495, EPI_ISL_2186496, EPI_ISL_2186497, EPI_ISL_2186498, EPI_ISL_2186499, EPI_ISL_2186500, EPI_ISL_2186501, EPI_ISL_2186502, EPI_ISL_2186503, EPI_ISL_2186504, EPI_ISL_2186505, EPI_ISL_2186506, EPI_ISL_2186507, EPI_ISL_2186508, EPI_ISL_2186509, EPI_ISL_2186510, EPI_ISL_2186511, EPI_ISL_2186512, EPI_ISL_2186513, EPI_ISL_2186514, EPI_ISL_2186515, EPI_ISL_2186516, EPI_ISL_2186517, EPI_ISL_2186518, EPI_ISL_2186519, EPI_ISL_2186520, EPI_ISL_2186521, EPI_ISL_2186522, EPI_ISL_2186523, EPI_ISL_2186524, EPI_ISL_2186525, EPI_ISL_2186526, EPI_ISL_2186527, EPI_ISL_2186528, EPI_ISL_2186529, EPI_ISL_2186530, EPI_ISL_2186531, EPI_ISL_2186532, EPI_ISL_2186533, EPI_ISL_2186534, EPI_ISL_2186535, EPI_ISL_2186536, EPI_ISL_2186537, EPI_ISL_2186538, EPI_ISL_2186539, EPI_ISL_2186540, EPI_ISL_2186541, EPI_ISL_2186542, EPI_ISL_2186543, EPI_ISL_2186544, EPI_ISL_2186545, EPI_ISL_2186546, EPI_ISL_2186547, EPI_ISL_2186548, EPI_ISL_2186549, EPI_ISL_2186550, EPI_ISL_2186551, EPI_ISL_2186552, EPI_ISL_2186553, EPI_ISL_2186554, EPI_ISL_2186555, EPI_ISL_2186556, EPI_ISL_2186557, EPI_ISL_2186558, EPI_ISL_2186559, EPI_ISL_2186560, EPI_ISL_2186561, EPI_ISL_2186562, EPI_ISL_2186563, EPI_ISL_2186564, EPI_ISL_2186565, EPI_ISL_2186566, EPI_ISL_2186567, EPI_ISL_2186568, EPI_ISL_2186569, EPI_ISL_2186570, EPI_ISL_2186571, EPI_ISL_2186572, EPI_ISL_2186573, EPI_ISL_2186574, EPI_ISL_2186575, EPI_ISL_2186576, EPI_ISL_2186578, EPI_ISL_2186579, EPI_ISL_2186580, EPI_ISL_2186581, EPI_ISL_2186583, EPI_ISL_2186584, EPI_ISL_2186585, EPI_ISL_2186586, EPI_ISL_2186587, EPI_ISL_2186588, EPI_ISL_2186589, EPI_ISL_2186590, EPI_ISL_2186591, EPI_ISL_2186592, EPI_ISL_2186593, EPI_ISL_2186594, EPI_ISL_2186595, EPI_ISL_2186596, EPI_ISL_2186597, EPI_ISL_2186598, EPI_ISL_2186599, EPI_ISL_2186600, EPI_ISL_2186602, EPI_ISL_2186603, EPI_ISL_2186604, EPI_ISL_2186605, EPI_ISL_2186606, EPI_ISL_2186607, EPI_ISL_2186608, EPI_ISL_2186609, EPI_ISL_2186610, EPI_ISL_2186611, EPI_ISL_2186612, EPI_ISL_2186613, EPI_ISL_2186614, EPI_ISL_2186615, EPI_ISL_2186616, EPI_ISL_2186617, EPI_ISL_2186618, EPI_ISL_2186619, EPI_ISL_2186620, EPI_ISL_2186621, EPI_ISL_2186622, EPI_ISL_2186623, EPI_ISL_2186624, EPI_ISL_2186625, EPI_ISL_2186626, EPI_ISL_2186627, EPI_ISL_2186628, EPI_ISL_2186629, EPI_ISL_2186630, EPI_ISL_2186631, EPI_ISL_2186632, EPI_ISL_2186633, EPI_ISL_2186634, EPI_ISL_2186635, EPI_ISL_2186636, EPI_ISL_2186637, EPI_ISL_2186638, EPI_ISL_2186639, EPI_ISL_2186640, EPI_ISL_2186641, EPI_ISL_2186642, EPI_ISL_2186643, EPI_ISL_2186644, EPI_ISL_2186645, EPI_ISL_2186646, EPI_ISL_2186647, EPI_ISL_2186649, EPI_ISL_2186650, EPI_ISL_2186651, EPI_ISL_2186653, EPI_ISL_2186654, EPI_ISL_2186655, EPI_ISL_2186656, EPI_ISL_2186657, EPI_ISL_2186658, EPI_ISL_2186659, EPI_ISL_2186660, EPI_ISL_2186661, EPI_ISL_2186662, EPI_ISL_2186663, EPI_ISL_2186664, EPI_ISL_2186665, EPI_ISL_2186666, EPI_ISL_2186667, EPI_ISL_2186668, EPI_ISL_2186669, EPI_ISL_2186670, EPI_ISL_2186671, EPI_ISL_2186672, EPI_ISL_2186673, EPI_ISL_2186674, EPI_ISL_2186675, EPI_ISL_2186676, EPI_ISL_2186677, EPI_ISL_2186678, EPI_ISL_2186679, EPI_ISL_2186680, EPI_ISL_2186681, EPI_ISL_2186683, EPI_ISL_2186684, EPI_ISL_2186685, EPI_ISL_2186686, EPI_ISL_2186687, EPI_ISL_2186688, EPI_ISL_2186689, EPI_ISL_2186690, EPI_ISL_2186691, EPI_ISL_2186693, EPI_ISL_2186694, EPI_ISL_2186695, EPI_ISL_2186696, EPI_ISL_2186697, EPI_ISL_2186698, EPI_ISL_2186699, EPI_ISL_2186700, EPI_ISL_2186701, EPI_ISL_2186702, EPI_ISL_2186703, EPI_ISL_2186704, EPI_ISL_2186705, EPI_ISL_2186706, EPI_ISL_2186707, EPI_ISL_2186708, EPI_ISL_2186709, EPI_ISL_2186711, EPI_ISL_2186712, EPI_ISL_2186714, EPI_ISL_2186715, EPI_ISL_2186716, EPI_ISL_2186718, EPI_ISL_2186719, EPI_ISL_2186720, EPI_ISL_2186721, EPI_ISL_2186722, EPI_ISL_2186723, EPI_ISL_2186724, EPI_ISL_2186725, EPI_ISL_2186726, EPI_ISL_2186727, EPI_ISL_2186728, EPI_ISL_2186729, EPI_ISL_2186730, EPI_ISL_2186733, EPI_ISL_2186735, EPI_ISL_2186736, EPI_ISL_2186738, EPI_ISL_2186739, EPI_ISL_2186740, EPI_ISL_2186741, EPI_ISL_2186742, EPI_ISL_2186743, EPI_ISL_2186744, EPI_ISL_2186745, EPI_ISL_2186746, EPI_ISL_2186748, EPI_ISL_2186749, EPI_ISL_2186750, EPI_ISL_2186751, EPI_ISL_2186752, EPI_ISL_2186753, EPI_ISL_2186755, EPI_ISL_2186756, EPI_ISL_2186757, EPI_ISL_2186758, EPI_ISL_2186759, EPI_ISL_2186760,

| | | | |
|---|---|---|---|
| EPI_ISL_2186761, EPI_ISL_2186763, EPI_ISL_2186765, EPI_ISL_2186767, EPI_ISL_2186768, EPI_ISL_2186769, EPI_ISL_2186771, EPI_ISL_2186772, EPI_ISL_2186773, EPI_ISL_2186774, EPI_ISL_2186775, EPI_ISL_2186776, EPI_ISL_2186777, EPI_ISL_2186778, EPI_ISL_2186779, EPI_ISL_2186780, EPI_ISL_2186781, EPI_ISL_2186782, EPI_ISL_2186783, EPI_ISL_2186784, EPI_ISL_2186785, EPI_ISL_2186786, EPI_ISL_2186787, EPI_ISL_2186788, EPI_ISL_2186789, EPI_ISL_2186790, EPI_ISL_2186791, EPI_ISL_2186793, EPI_ISL_2186794, EPI_ISL_2186795, EPI_ISL_2186796, EPI_ISL_2186797, EPI_ISL_2186798, EPI_ISL_2186799, EPI_ISL_2186800, EPI_ISL_2186801, EPI_ISL_2186802, EPI_ISL_2186803, EPI_ISL_2186805, EPI_ISL_2186807, EPI_ISL_2186808, EPI_ISL_2186809, EPI_ISL_2186810, EPI_ISL_2186811, EPI_ISL_2186812, EPI_ISL_2186813, EPI_ISL_2186815, EPI_ISL_2186816, EPI_ISL_2186817, EPI_ISL_2186818, EPI_ISL_2186819, EPI_ISL_2186820, EPI_ISL_2186821, EPI_ISL_2186822, EPI_ISL_2186823, EPI_ISL_2186824, EPI_ISL_2186825, EPI_ISL_2186826, EPI_ISL_2186827, EPI_ISL_2186828, EPI_ISL_2186829, EPI_ISL_2186830, EPI_ISL_2186831, EPI_ISL_2186832, EPI_ISL_2186833, EPI_ISL_2186834, EPI_ISL_2186835, EPI_ISL_2186836, EPI_ISL_2186837, EPI_ISL_2186838, EPI_ISL_2186839, EPI_ISL_2186840, EPI_ISL_2186841, EPI_ISL_2186842, EPI_ISL_2186843, EPI_ISL_2186844, EPI_ISL_2186845, EPI_ISL_2186846, EPI_ISL_2186847, EPI_ISL_2186848, EPI_ISL_2186849, EPI_ISL_2186851, EPI_ISL_2186852, EPI_ISL_2186853, EPI_ISL_2186855, EPI_ISL_2186856, EPI_ISL_2186857, EPI_ISL_2186858, EPI_ISL_2186859, EPI_ISL_2186860, EPI_ISL_2186861, EPI_ISL_2186862, EPI_ISL_2186863, EPI_ISL_2186864, EPI_ISL_2186865, EPI_ISL_2186866, EPI_ISL_2186867, EPI_ISL_2186868, EPI_ISL_2186870, EPI_ISL_2186871, EPI_ISL_2186873, EPI_ISL_2186874, EPI_ISL_2186875, EPI_ISL_2186876, EPI_ISL_2186877, EPI_ISL_2186878, EPI_ISL_2186879, EPI_ISL_2186880, EPI_ISL_2186881, EPI_ISL_2186882, EPI_ISL_2186883, EPI_ISL_2186885, EPI_ISL_2186886, EPI_ISL_2186887, EPI_ISL_2186888, EPI_ISL_2186889, EPI_ISL_2186890, EPI_ISL_2186891, EPI_ISL_2186892, EPI_ISL_2186893, EPI_ISL_2186894, EPI_ISL_2186895, EPI_ISL_2186896, EPI_ISL_2186897, EPI_ISL_2186898, EPI_ISL_2186899, EPI_ISL_2186900, EPI_ISL_2186901, EPI_ISL_2186902, EPI_ISL_2186903, EPI_ISL_2186904, EPI_ISL_2186905, EPI_ISL_2186906, EPI_ISL_2186907, EPI_ISL_2186908, EPI_ISL_2186909, EPI_ISL_2186910, EPI_ISL_2186911, EPI_ISL_2186912, EPI_ISL_2186913, EPI_ISL_2186914, EPI_ISL_2186915, EPI_ISL_2186916, EPI_ISL_2186917, EPI_ISL_2186918, EPI_ISL_2186919, EPI_ISL_2186920, EPI_ISL_2186921, EPI_ISL_2186922, EPI_ISL_2186923, EPI_ISL_2186924, EPI_ISL_2186927, EPI_ISL_2186928, EPI_ISL_2186929, EPI_ISL_2186930, EPI_ISL_2186931, EPI_ISL_2186932, EPI_ISL_2186933, EPI_ISL_2186934, EPI_ISL_2186935, EPI_ISL_2186936, EPI_ISL_2186937, EPI_ISL_2186938, EPI_ISL_2186939, EPI_ISL_2186940, EPI_ISL_2186941, EPI_ISL_2186942, EPI_ISL_2186943, EPI_ISL_2186944, EPI_ISL_2186946, EPI_ISL_2186947, EPI_ISL_2186949, EPI_ISL_2186951, EPI_ISL_2186952, EPI_ISL_2186953, EPI_ISL_2186954, EPI_ISL_2186955, EPI_ISL_2186956, EPI_ISL_2186957, EPI_ISL_2186958, EPI_ISL_2186959, EPI_ISL_2186960, EPI_ISL_2186961, EPI_ISL_2186962, EPI_ISL_2186963, EPI_ISL_2186964, EPI_ISL_2186965, EPI_ISL_2186966, EPI_ISL_2186967, EPI_ISL_2186968, EPI_ISL_2186969, EPI_ISL_2186970, EPI_ISL_2186971, EPI_ISL_2186972, EPI_ISL_2186973, EPI_ISL_2186974, EPI_ISL_2186975, EPI_ISL_2186976, EPI_ISL_2186977, EPI_ISL_2186978, EPI_ISL_2186979, EPI_ISL_2186980, EPI_ISL_2186981, EPI_ISL_2186982, EPI_ISL_2186983, EPI_ISL_2186984, EPI_ISL_2186985, EPI_ISL_2186986, EPI_ISL_2186988, EPI_ISL_2186989, EPI_ISL_2186990, EPI_ISL_2186992, EPI_ISL_2186993, EPI_ISL_2186994, EPI_ISL_2186995, EPI_ISL_2186996, EPI_ISL_2186997, EPI_ISL_2186998, EPI_ISL_2186999, EPI_ISL_2187000, EPI_ISL_2187001, EPI_ISL_2187002, EPI_ISL_2187003, EPI_ISL_2187004, EPI_ISL_2187005, EPI_ISL_2187006, EPI_ISL_2187007, EPI_ISL_2187008, EPI_ISL_2187010, EPI_ISL_2187011, EPI_ISL_2187012, EPI_ISL_2187014, EPI_ISL_2187015, EPI_ISL_2187016, EPI_ISL_2187017, EPI_ISL_2187018, EPI_ISL_2187019, EPI_ISL_2187020, EPI_ISL_2187022, EPI_ISL_2187023, EPI_ISL_2187024, EPI_ISL_2187025, EPI_ISL_2187026, EPI_ISL_2187027, EPI_ISL_2187028, EPI_ISL_2187029, EPI_ISL_2187030, EPI_ISL_2187031, EPI_ISL_2187032, EPI_ISL_2187033, EPI_ISL_2187035, EPI_ISL_2187036, EPI_ISL_2187037, EPI_ISL_2187038, EPI_ISL_2187039, EPI_ISL_2187040, EPI_ISL_2187041, EPI_ISL_2187042, EPI_ISL_2187043, EPI_ISL_2187044, EPI_ISL_2187045, EPI_ISL_2187046, EPI_ISL_2187047, EPI_ISL_2187048, EPI_ISL_2187049, EPI_ISL_2187050, EPI_ISL_2187051, EPI_ISL_2187052, EPI_ISL_2187053, EPI_ISL_2187054, EPI_ISL_2187055, EPI_ISL_2187056, EPI_ISL_2187057, EPI_ISL_2187058, EPI_ISL_2187059, EPI_ISL_2187060, EPI_ISL_2187061, EPI_ISL_2187062, EPI_ISL_2187063, EPI_ISL_2187064, EPI_ISL_2187065, EPI_ISL_2187066, EPI_ISL_2187067, EPI_ISL_2187068, EPI_ISL_2187069, EPI_ISL_2187070, EPI_ISL_2187071, EPI_ISL_2187072, EPI_ISL_2187073, EPI_ISL_2187074, EPI_ISL_2187075, EPI_ISL_2187076, EPI_ISL_2187077, EPI_ISL_2187078, EPI_ISL_2187079, EPI_ISL_2187080, EPI_ISL_2187081, EPI_ISL_2187082, EPI_ISL_2187083, EPI_ISL_2187084, EPI_ISL_2187085, EPI_ISL_2187086, EPI_ISL_2187087, EPI_ISL_2187088, EPI_ISL_2187089, EPI_ISL_2187090, EPI_ISL_2187091, EPI_ISL_2187093, EPI_ISL_2187094, EPI_ISL_2187095, EPI_ISL_2187096, EPI_ISL_2187097, EPI_ISL_2187098, EPI_ISL_2187099, EPI_ISL_2187100, EPI_ISL_2187101, EPI_ISL_2187102, EPI_ISL_2187103, EPI_ISL_2187104, EPI_ISL_2187105, EPI_ISL_2187106, EPI_ISL_2187107, EPI_ISL_2187109, EPI_ISL_2187110, EPI_ISL_2187111, EPI_ISL_2187112, EPI_ISL_2187113, EPI_ISL_2187114, EPI_ISL_2187115, EPI_ISL_2187116, EPI_ISL_2187119, EPI_ISL_2187120, EPI_ISL_2187122, EPI_ISL_2187123, EPI_ISL_2187124, EPI_ISL_2187125, EPI_ISL_2187126, EPI_ISL_2187128, EPI_ISL_2187129, EPI_ISL_2187130, EPI_ISL_2187131, EPI_ISL_2187132, EPI_ISL_2187133, EPI_ISL_2187134, EPI_ISL_2187135, EPI_ISL_2187136, EPI_ISL_2187137, EPI_ISL_2187138, EPI_ISL_2187139, EPI_ISL_2187140, EPI_ISL_2187141, EPI_ISL_2187142, EPI_ISL_2187143, EPI_ISL_2187144, EPI_ISL_2187145, EPI_ISL_2187146, EPI_ISL_2187147, EPI_ISL_2187148, EPI_ISL_2187149, EPI_ISL_2187150, EPI_ISL_2187151, EPI_ISL_2187152, EPI_ISL_2187153, EPI_ISL_2187154, EPI_ISL_2187156, EPI_ISL_2187157, EPI_ISL_2187158, EPI_ISL_2187159, EPI_ISL_2187160, EPI_ISL_2187161, EPI_ISL_2187162, EPI_ISL_2187163, EPI_ISL_2187164, EPI_ISL_2187165, EPI_ISL_2187166, EPI_ISL_2187167, EPI_ISL_2187168, EPI_ISL_2187169, EPI_ISL_2187170, EPI_ISL_2187171, EPI_ISL_2187172, EPI_ISL_2187174, EPI_ISL_2187175, EPI_ISL_2187176, EPI_ISL_2187177, EPI_ISL_2187178, EPI_ISL_2187179, EPI_ISL_2187181, EPI_ISL_2187182, EPI_ISL_2187183, EPI_ISL_2187184, EPI_ISL_2187185, EPI_ISL_2187186, EPI_ISL_2187187, EPI_ISL_2187189, EPI_ISL_2187190, EPI_ISL_2187191, EPI_ISL_2187192, EPI_ISL_2187193, EPI_ISL_2187194, EPI_ISL_2187195, EPI_ISL_2187196, EPI_ISL_2187197, EPI_ISL_2187198, EPI_ISL_2187199, EPI_ISL_2187200, EPI_ISL_2187201, EPI_ISL_2187202, EPI_ISL_2187203, EPI_ISL_2187204, EPI_ISL_2187205, EPI_ISL_2187206, EPI_ISL_2187208, EPI_ISL_2187209, EPI_ISL_2187210, EPI_ISL_2187212, EPI_ISL_2187213, EPI_ISL_2187214, EPI_ISL_2187216, EPI_ISL_2187217, EPI_ISL_2187218, EPI_ISL_2187219, EPI_ISL_2187220, EPI_ISL_2187222, EPI_ISL_2187223, EPI_ISL_2187224, EPI_ISL_2187225, EPI_ISL_2187226, EPI_ISL_2187227, EPI_ISL_2187228, EPI_ISL_2187229 | | | |
| see above | Infinity Biologix | Centers for Disease Control and Prevention Division of Viral Diseases, Pathogen Discovery | Dakota Howard, Dhwani Batra, Peter W. Cook, Kara Moser, Adrian Paskey, Jason Caravas, Benjamin Rambo-Martin, Shatavia Morrison, Christopher Gulvick, Scott Sammons, Yvette Unoarumhi, Darlene Wagner, Matthew Schmerer, Christian Bixby, Yihe Wang, Jonathan Schultz, Chirayu Goswami, Russ Hager, Robin Grimwood, Clinton R. Paden, Duncan MacCannell |
| EPI_ISL_2187230, EPI_ISL_2187231, EPI_ISL_2187232, EPI_ISL_2187233, EPI_ISL_2187234, EPI_ISL_2187235, EPI_ISL_2187236, EPI_ISL_2187237, EPI_ISL_2187238, EPI_ISL_2187239, EPI_ISL_2187240, EPI_ISL_2187241, EPI_ISL_2187242, EPI_ISL_2187243, EPI_ISL_2187244, EPI_ISL_2187245, EPI_ISL_2187246, EPI_ISL_2187247, EPI_ISL_2187248, EPI_ISL_2187249, EPI_ISL_2187250, EPI_ISL_2187251, EPI_ISL_2187252, EPI_ISL_2187253, EPI_ISL_2187254, EPI_ISL_2187255, EPI_ISL_2187256, EPI_ISL_2187257, EPI_ISL_2187258, EPI_ISL_2187259, EPI_ISL_2187260, EPI_ISL_2187261, EPI_ISL_2187262, EPI_ISL_2187263, EPI_ISL_2187264, EPI_ISL_2187265, EPI_ISL_2187266, EPI_ISL_2187267, EPI_ISL_2187268, EPI_ISL_2187269, EPI_ISL_2187270, EPI_ISL_2187271, EPI_ISL_2187272, EPI_ISL_2187273, EPI_ISL_2187274, EPI_ISL_2187275, EPI_ISL_2187276, EPI_ISL_2187277, EPI_ISL_2187278, EPI_ISL_2187279, EPI_ISL_2187280, EPI_ISL_2187281, EPI_ISL_2187282, EPI_ISL_2187283, EPI_ISL_2187284, EPI_ISL_2187285, EPI_ISL_2187286, EPI_ISL_2187287, EPI_ISL_2187288, EPI_ISL_2187289, EPI_ISL_2187290, EPI_ISL_2187291, EPI_ISL_2187292, EPI_ISL_2187293, EPI_ISL_2187294, EPI_ISL_2187295, EPI_ISL_2187296, EPI_ISL_2187297, EPI_ISL_2187298, EPI_ISL_2187299, EPI_ISL_2187300, EPI_ISL_2187301, EPI_ISL_2187302, EPI_ISL_2187303, EPI_ISL_2187304, EPI_ISL_2187305, EPI_ISL_2187306, EPI_ISL_2187307, EPI_ISL_2187308, EPI_ISL_2187309, EPI_ISL_2187310, EPI_ISL_2187311, EPI_ISL_2187312, EPI_ISL_2187313, EPI_ISL_2187314, EPI_ISL_2187315, EPI_ISL_2187316, EPI_ISL_2187317 | | | |
| see above | Quest Diagnostics Incorporated | Centers for Disease Control and Prevention Division of Viral Diseases, Pathogen Discovery | Dakota Howard, Dhwani Batra, Peter W. Cook, Kara Moser, Adrian Paskey, Jason Caravas, Benjamin Rambo-Martin, Shatavia Morrison, Christopher Gulvick, Scott Sammons, Yvette Unoarumhi, Darlene Wagner, Matthew Schmerer, S. H. Rosenthal, A. Gerasimova, R. M. Kagan, B. Anderson, M. Hua, Y. Liu, L.E. Bernstein, K.E. Livingston, A. Perez, I. A. Shlyakhter, R. V. Rolando, R. Owen, P. Tanpaiboon, F. Lacbawan, Clinton R. Paden, Duncan MacCannell |
| EPI_ISL_2187318, EPI_ISL_2187319, EPI_ISL_2187320, EPI_ISL_2187321, EPI_ISL_2187322, EPI_ISL_2187323, EPI_ISL_2187324, EPI_ISL_2187325, EPI_ISL_2187326, EPI_ISL_2187327, EPI_ISL_2187328, EPI_ISL_2187329, EPI_ISL_2187330, EPI_ISL_2187331, EPI_ISL_2187332, EPI_ISL_2187333, EPI_ISL_2187334, EPI_ISL_2187335, EPI_ISL_2187336, EPI_ISL_2187337, EPI_ISL_2187338, EPI_ISL_2187339, EPI_ISL_2187340, EPI_ISL_2187341, EPI_ISL_2187342, EPI_ISL_2187343, EPI_ISL_2187344, EPI_ISL_2187345, EPI_ISL_2187346, EPI_ISL_2187347, EPI_ISL_2187348, EPI_ISL_2187349, EPI_ISL_2187351, EPI_ISL_2187352, EPI_ISL_2187353, EPI_ISL_2187354 | | | |
| see above | Helix/Illumina | Centers for Disease Control and Prevention Division of Viral Diseases, Pathogen Discovery | Dakota Howard, Dhwani Batra, Peter W. Cook, Kara Moser, Adrian Paskey, Jason Caravas, Benjamin Rambo-Martin, Shatavia Morrison, Christopher Gulvick, Scott Sammons, Yvette Unoarumhi, Darlene Wagner, Matthew Schmerer, Eileen de Feo, Jan Antico, Christine Tran, Matthew Tolentino, Shannon Wickline, Kim Gietzen, Brad Sickler, Jingtao Liu, Eric Allen, Phil Febbo, Nicole L. Washington, Simon White, Geraint Levan, Kelly Schiabor Barrett, Elizabeth Cirulli, Alexandre Bolze, Ary Ascencio, Charlotte Rivera-Garcia, Ryan Cho, Jason Nguyen, Sherry Wang, Jimmy Ramirez, Tyler Cassens, Efren Sandoval, Magnus Isaksson, William Lee, David Becker, Marc Laurent, James Lu, Clinton R. Paden, Duncan MacCannell |
| EPI_ISL_2187355, EPI_ISL_2187356, EPI_ISL_2187357, EPI_ISL_2187358, EPI_ISL_2187359, EPI_ISL_2187360, EPI_ISL_2187361, EPI_ISL_2187362, EPI_ISL_2187363, EPI_ISL_2187364, EPI_ISL_2187365, EPI_ISL_2187366, EPI_ISL_2187367, EPI_ISL_2187368, EPI_ISL_2187369, EPI_ISL_2187370, EPI_ISL_2187371, EPI_ISL_2187372, EPI_ISL_2187373, EPI_ISL_2187374, EPI_ISL_2187375, EPI_ISL_2187376, EPI_ISL_2187377, EPI_ISL_2187378, EPI_ISL_2187379, EPI_ISL_2187380, EPI_ISL_2187381, EPI_ISL_2187382, EPI_ISL_2187383, EPI_ISL_2187384, EPI_ISL_2187385, EPI_ISL_2187386, EPI_ISL_2187387, EPI_ISL_2187388, EPI_ISL_2187389, EPI_ISL_2187390, EPI_ISL_2187391, EPI_ISL_2187392, EPI_ISL_2187394, EPI_ISL_2187395, EPI_ISL_2187396, EPI_ISL_2187397, EPI_ISL_2187398, EPI_ISL_2187399, EPI_ISL_2187401, EPI_ISL_2187403, EPI_ISL_2187404, EPI_ISL_2187405, EPI_ISL_2187406, EPI_ISL_2187407, EPI_ISL_2187408, EPI_ISL_2187409, EPI_ISL_2187410, EPI_ISL_2187411, EPI_ISL_2187412, EPI_ISL_2187413, EPI_ISL_2187414, EPI_ISL_2187415, EPI_ISL_2187416, EPI_ISL_2187417, EPI_ISL_2187419, EPI_ISL_2187420, EPI_ISL_2187421, EPI_ISL_2187422, EPI_ISL_2187423, EPI_ISL_2187424, EPI_ISL_2187425, EPI_ISL_2187426, EPI_ISL_2187427, EPI_ISL_2187428, EPI_ISL_2187429, EPI_ISL_2187430, EPI_ISL_2187431, EPI_ISL_2187432, EPI_ISL_2187433, EPI_ISL_2187434, EPI_ISL_2187435, EPI_ISL_2187436, EPI_ISL_2187437, EPI_ISL_2187438, EPI_ISL_2187439, EPI_ISL_2187440, EPI_ISL_2187442, EPI_ISL_2187443, EPI_ISL_2187444, EPI_ISL_2187445, EPI_ISL_2187446, EPI_ISL_2187448, EPI_ISL_2187449, EPI_ISL_2187450, EPI_ISL_2187451, EPI_ISL_2187452, EPI_ISL_2187453, EPI_ISL_2187454, EPI_ISL_2187455, EPI_ISL_2187456, EPI_ISL_2187457, EPI_ISL_2187458, EPI_ISL_2187459, EPI_ISL_2187460, EPI_ISL_2187461, EPI_ISL_2187462, EPI_ISL_2187463, EPI_ISL_2187464, EPI_ISL_2187466, EPI_ISL_2187467, EPI_ISL_2187468, EPI_ISL_2187469, EPI_ISL_2187470, EPI_ISL_2187471, EPI_ISL_2187472, EPI_ISL_2187473, EPI_ISL_2187474, EPI_ISL_2187475, EPI_ISL_2187476, EPI_ISL_2187477, EPI_ISL_2187478, EPI_ISL_2187479, EPI_ISL_2187480, EPI_ISL_2187481, EPI_ISL_2187482, EPI_ISL_2187483, EPI_ISL_2187484, EPI_ISL_2187485, EPI_ISL_2187486, EPI_ISL_2187487, EPI_ISL_2187488, EPI_ISL_2187489, EPI_ISL_2187490, EPI_ISL_2187491, EPI_ISL_2187492, EPI_ISL_2187493, EPI_ISL_2187494, EPI_ISL_2187495, EPI_ISL_2187496, EPI_ISL_2187497, EPI_ISL_2187498, EPI_ISL_2187499, EPI_ISL_2187500, EPI_ISL_2187501, EPI_ISL_2187502, EPI_ISL_2187503, EPI_ISL_2187504, EPI_ISL_2187505, EPI_ISL_2187506, EPI_ISL_2187507, EPI_ISL_2187508, EPI_ISL_2187509, EPI_ISL_2187510, EPI_ISL_2187511, EPI_ISL_2187512, EPI_ISL_2187513, EPI_ISL_2187515, EPI_ISL_2187516, EPI_ISL_2187517, EPI_ISL_2187518, EPI_ISL_2187519, EPI_ISL_2187520, EPI_ISL_2187521, EPI_ISL_2187522, EPI_ISL_2187523, EPI_ISL_2187524, EPI_ISL_2187525, EPI_ISL_2187526, EPI_ISL_2187528, EPI_ISL_2187529, EPI_ISL_2187530, EPI_ISL_2187531, EPI_ISL_2187532, EPI_ISL_2187533, EPI_ISL_2187534, EPI_ISL_2187535, EPI_ISL_2187536, EPI_ISL_2187537, EPI_ISL_2187538, EPI_ISL_2187539, EPI_ISL_2187540, EPI_ISL_2187541, EPI_ISL_2187542, EPI_ISL_2187543, EPI_ISL_2187544, EPI_ISL_2187545, EPI_ISL_2187546, EPI_ISL_2187549, EPI_ISL_2187550, EPI_ISL_2187551, EPI_ISL_2187553, EPI_ISL_2187554, EPI_ISL_2187555, EPI_ISL_2187556, EPI_ISL_2187557, EPI_ISL_2187560, EPI_ISL_2187561, EPI_ISL_2187562, EPI_ISL_2187563, EPI_ISL_2187564, EPI_ISL_2187565, EPI_ISL_2187566, EPI_ISL_2187567, EPI_ISL_2187568, EPI_ISL_2187569, EPI_ISL_2187570, EPI_ISL_2187571, EPI_ISL_2187572, EPI_ISL_2187573, EPI_ISL_2187574, EPI_ISL_2187575, EPI_ISL_2187576, EPI_ISL_2187577, EPI_ISL_2187578, EPI_ISL_2187579, EPI_ISL_2187580, EPI_ISL_2187581, EPI_ISL_2187582, EPI_ISL_2187583, EPI_ISL_2187584, EPI_ISL_2187585, EPI_ISL_2187586, EPI_ISL_2187587, EPI_ISL_2187588, EPI_ISL_2187590, EPI_ISL_2187591, EPI_ISL_2187592, EPI_ISL_2187593, EPI_ISL_2187594, EPI_ISL_2187595, EPI_ISL_2187596, EPI_ISL_2187597, EPI_ISL_2187598, EPI_ISL_2187599, EPI_ISL_2187600, EPI_ISL_2187601, EPI_ISL_2187602, EPI_ISL_2187603, EPI_ISL_2187604, EPI_ISL_2187605, EPI_ISL_2187606, EPI_ISL_2187607, EPI_ISL_2187608, EPI_ISL_2187609, EPI_ISL_2187610, EPI_ISL_2187611, EPI_ISL_2187612, EPI_ISL_2187613, EPI_ISL_2187614, EPI_ISL_2187615, EPI_ISL_2187616 | | | |
| see above | Aegis Sciences Corporation | Centers for Disease Control and Prevention Division of Viral Diseases, Pathogen Discovery | Dakota Howard, Dhwani Batra, Peter W. Cook, Kara Moser, Adrian Paskey, Jason Caravas, Benjamin Rambo-Martin, Shatavia Morrison, Christopher Gulvick, Scott Sammons, Yvette Unoarumhi, Darlene Wagner, Matthew Schmerer, Cyndi Clark, Patrick Campbell, Rob Case, Vikramsinha Ghorpade, Holly Houdeshell, Ola Kvalvaag, Dillon Nall, Ethan Sanders, Alec Vest, Shaun Westlund, Matthew Hardison, Clinton R. Paden, Duncan MacCannell |
| EPI_ISL_2187617, EPI_ISL_2187618, EPI_ISL_2187619, EPI_ISL_2187620, EPI_ISL_2187621, EPI_ISL_2187622, EPI_ISL_2187623, EPI_ISL_2187624, EPI_ISL_2187625, EPI_ISL_2187626, EPI_ISL_2187627, EPI_ISL_2187628, EPI_ISL_2187629, EPI_ISL_2187630, EPI_ISL_2187631, EPI_ISL_2187632, EPI_ISL_2187633, EPI_ISL_2187634, EPI_ISL_2187635, EPI_ISL_2187636, EPI_ISL_2187637, EPI_ISL_2187638, EPI_ISL_2187639, EPI_ISL_2187640, EPI_ISL_2187641, EPI_ISL_2187642 | | | |
| see above | Helix/Illumina | Centers for Disease Control and Prevention Division of Viral Diseases, Pathogen Discovery | Dakota Howard, Dhwani Batra, Peter W. Cook, Kara Moser, Adrian Paskey, Jason Caravas, Benjamin Rambo-Martin, Shatavia Morrison, Christopher Gulvick, Scott Sammons, Yvette Unoarumhi, Darlene Wagner, Matthew Schmerer, Eileen de Feo, Jan Antico, Christine Tran, Matthew Tolentino, Shannon Wickline, Kim Gietzen, Brad Sickler, Jingtao Liu, Eric Allen, Phil Febbo, Nicole L. Washington, Simon White, Geraint Levan, Kelly Schiabor Barrett, Elizabeth Cirulli, Alexandre Bolze, Ary Ascencio, Charlotte Rivera-Garcia, Ryan Cho, Jason Nguyen, Sherry Wang, Jimmy Ramirez, Tyler Cassens, Efren Sandoval, Magnus Isaksson, William Lee, David Becker, Marc Laurent, James Lu, Clinton R. Paden, Duncan MacCannell |
| EPI_ISL_2187643, EPI_ISL_2187644 | Fulgent Genetics | Centers for Disease Control and Prevention Division of Viral Diseases, Pathogen Discovery | Dakota Howard, Dhwani Batra, Peter W. Cook, Kara Moser, Adrian Paskey, Jason Caravas, Benjamin Rambo-Martin, Shatavia Morrison, Christopher Gulvick, Scott Sammons, Yvette Unoarumhi, Darlene Wagner, Matthew Schmerer, Harry Gao, Mickey Li, John Gao, Joseph Fierro, Benafsh Sapra, Becky Tsai, Yan Meng, Doreen Ng, James Xie, Clinton R. Paden, Duncan MacCannell |
| EPI_ISL_2187645, EPI_ISL_2187646, EPI_ISL_2187647, EPI_ISL_2187648, EPI_ISL_2187649, EPI_ISL_2187650, EPI_ISL_2187651, EPI_ISL_2187652, EPI_ISL_2187653, EPI_ISL_2187654, EPI_ISL_2187655, EPI_ISL_2187656, EPI_ISL_2187657, EPI_ISL_2187658, EPI_ISL_2187659, EPI_ISL_2187660, EPI_ISL_2187661, EPI_ISL_2187662, EPI_ISL_2187663, EPI_ISL_2187664, EPI_ISL_2187665, EPI_ISL_2187666, EPI_ISL_2187667, EPI_ISL_2187668, EPI_ISL_2187669, EPI_ISL_2187670, EPI_ISL_2187671, EPI_ISL_2187672, EPI_ISL_2187673, EPI_ISL_2187674, EPI_ISL_2187675, EPI_ISL_2187676, EPI_ISL_2187677, EPI_ISL_2187678, EPI_ISL_2187679, EPI_ISL_2187680, EPI_ISL_2187681, EPI_ISL_2187682, EPI_ISL_2187683 | | | |
| see above | Alaska State Virology Laboratory | Alaska State Virology Laboratory | Stephanie DeRonde, Elva House, Jacob Zidek, Lisa Smith, Ph.D., Jack Chen, Ph.D. |
| EPI_ISL_2188490, EPI_ISL_2188491, EPI_ISL_2188492, EPI_ISL_2188493, EPI_ISL_2188494, EPI_ISL_2188495, EPI_ISL_2188496, EPI_ISL_2188497, EPI_ISL_2188498, EPI_ISL_2188499, EPI_ISL_2188500, EPI_ISL_2188501, EPI_ISL_2188502, EPI_ISL_2188503, EPI_ISL_2188504, EPI_ISL_2188505, EPI_ISL_2188506, EPI_ISL_2188507, EPI_ISL_2188508, EPI_ISL_2188509, EPI_ISL_2188510, EPI_ISL_2188511, EPI_ISL_2188512, EPI_ISL_2188513, EPI_ISL_2188514, EPI_ISL_2188515, EPI_ISL_2188516, EPI_ISL_2188517, EPI_ISL_2188518, EPI_ISL_2188519, EPI_ISL_2188520, EPI_ISL_2188521, EPI_ISL_2188522, EPI_ISL_2188523, EPI_ISL_2188524, EPI_ISL_2188525, EPI_ISL_2188526, EPI_ISL_2188527, EPI_ISL_2188528, EPI_ISL_2188529, EPI_ISL_2188530, EPI_ISL_2188531, EPI_ISL_2188532, EPI_ISL_2188533, EPI_ISL_2188534, EPI_ISL_2188535, EPI_ISL_2188536, EPI_ISL_2188537, EPI_ISL_2188538, EPI_ISL_2188539, EPI_ISL_2188540, EPI_ISL_2188541, EPI_ISL_2188542, EPI_ISL_2188543, EPI_ISL_2188544, EPI_ISL_2188545, EPI_ISL_2188546, EPI_ISL_2188547, EPI_ISL_2188548, EPI_ISL_2188549, EPI_ISL_2188550, EPI_ISL_2188551, EPI_ISL_2188552, EPI_ISL_2188553, EPI_ISL_2188554, EPI_ISL_2188555, EPI_ISL_2188556, EPI_ISL_2188557, EPI_ISL_2188558, EPI_ISL_2188559, EPI_ISL_2188560, EPI_ISL_2188561, EPI_ISL_2188562, EPI_ISL_2188563, EPI_ISL_2188564, EPI_ISL_2188565, EPI_ISL_2188566, EPI_ISL_2188567, EPI_ISL_2188568, EPI_ISL_2188569, EPI_ISL_2188570, EPI_ISL_2188571, EPI_ISL_2188572, EPI_ISL_2188573, EPI_ISL_2188574, EPI_ISL_2188575, EPI_ISL_2188576, EPI_ISL_2188577, EPI_ISL_2188578, EPI_ISL_2188579, EPI_ISL_2188580, EPI_ISL_2188581, EPI_ISL_2188582, EPI_ISL_2188583, EPI_ISL_2188584, EPI_ISL_2188585, EPI_ISL_2188586, EPI_ISL_2188587, EPI_ISL_2188588, EPI_ISL_2188589, EPI_ISL_2188590, EPI_ISL_2188591, EPI_ISL_2188592, EPI_ISL_2188593, EPI_ISL_2188594, EPI_ISL_2188595, EPI_ISL_2188596, EPI_ISL_2188597, EPI_ISL_2188598, EPI_ISL_2188599, EPI_ISL_2188600, EPI_ISL_2188601, EPI_ISL_2188602, EPI_ISL_2188603, EPI_ISL_2188604, EPI_ISL_2188605, EPI_ISL_2188606, EPI_ISL_2188607, EPI_ISL_2188608, EPI_ISL_2188609, EPI_ISL_2188610, EPI_ISL_2188611, EPI_ISL_2188612, EPI_ISL_2188613, EPI_ISL_2188614, EPI_ISL_2188615, EPI_ISL_2188616, EPI_ISL_2188617, EPI_ISL_2188618, EPI_ISL_2188619, EPI_ISL_2188620, EPI_ISL_2188622, EPI_ISL_2188623, EPI_ISL_2188624, EPI_ISL_2188625, EPI_ISL_2188626, EPI_ISL_2188627, EPI_ISL_2188628, EPI_ISL_2188629, EPI_ISL_2188630, EPI_ISL_2188631, EPI_ISL_2188632, EPI_ISL_2188633, EPI_ISL_2188634, EPI_ISL_2188635, EPI_ISL_2188636, EPI_ISL_2188638, EPI_ISL_2188639, EPI_ISL_2188640, EPI_ISL_2188641, EPI_ISL_2188642, EPI_ISL_2188643, EPI_ISL_2188644, EPI_ISL_2188645, EPI_ISL_2188648, EPI_ISL_2188649, EPI_ISL_2188650, EPI_ISL_2188651, EPI_ISL_2188652, EPI_ISL_2188653, EPI_ISL_2188654, EPI_ISL_2188655, EPI_ISL_2188657, EPI_ISL_2188659, EPI_ISL_2188660, EPI_ISL_2188661, EPI_ISL_2188663, EPI_ISL_2188665, EPI_ISL_2188666, EPI_ISL_2188667, EPI_ISL_2188668, EPI_ISL_2188669, EPI_ISL_2188672 | | | |
| see above | Mako Medical | Centers for Disease Control and Prevention Division of Viral Diseases, Pathogen Discovery | Dakota Howard, Dhwani Batra, Peter W. Cook, Kara Moser, Adrian Paskey, Jason Caravas, Benjamin Rambo-Martin, Shatavia Morrison, Christopher Gulvick, Scott Sammons, Yvette Unoarumhi, Darlene Wagner, Matthew Schmerer,Matthew Tugwell, Lauren Moon, Clinton R. Paden, Duncan MacCannell |
| EPI_ISL_2189528 | Aegis Sciences Corporation | Centers for Disease Control and Prevention Division of Viral Diseases, Pathogen Discovery | Dakota Howard, Dhwani Batra, Peter W. Cook, Kara Moser, Adrian Paskey, Jason Caravas, Benjamin Rambo-Martin, Shatavia Morrison, Christopher Gulvick, Scott Sammons, Yvette Unoarumhi, Darlene Wagner, Matthew Schmerer, Cyndi Clark, Patrick Campbell, Rob Case, Vikramsinha Ghorpade, Holly Houdeshell, Ola Kvalvaag, Dillon Nall, Ethan Sanders, Alec Vest, Shaun Westlund, Matthew Hardison, Clinton R. Paden, Duncan MacCannell |
| EPI_ISL_2189891, EPI_ISL_2189892, EPI_ISL_2189893, EPI_ISL_2189894, EPI_ISL_2189895, EPI_ISL_2189896, EPI_ISL_2189897, EPI_ISL_2189898, EPI_ISL_2189899, EPI_ISL_2189900, EPI_ISL_2189901, EPI_ISL_2189902, EPI_ISL_2189903, EPI_ISL_2189904, EPI_ISL_2189905, EPI_ISL_2189906, EPI_ISL_2189907, EPI_ISL_2189908, EPI_ISL_2189909, EPI_ISL_2189910, EPI_ISL_2189911, EPI_ISL_2189912, EPI_ISL_2189913, EPI_ISL_2189914, EPI_ISL_2189915, EPI_ISL_2189916, EPI_ISL_2189917, EPI_ISL_2189918, EPI_ISL_2189919, EPI_ISL_2189920, EPI_ISL_2189921, EPI_ISL_2189922, EPI_ISL_2189923, EPI_ISL_2189924, EPI_ISL_2189925, EPI_ISL_2189926, EPI_ISL_2189927, EPI_ISL_2189928, EPI_ISL_2189929, EPI_ISL_2189930, EPI_ISL_2189931, EPI_ISL_2189932, EPI_ISL_2189933, EPI_ISL_2189934, EPI_ISL_2189935, EPI_ISL_2189936, EPI_ISL_2189937, EPI_ISL_2189938, EPI_ISL_2189939, EPI_ISL_2189940, EPI_ISL_2189941, EPI_ISL_2189943, EPI_ISL_2189944, EPI_ISL_2189945, EPI_ISL_2189946, EPI_ISL_2189947, EPI_ISL_2189948, EPI_ISL_2189949, EPI_ISL_2189950, EPI_ISL_2189951, EPI_ISL_2189952, EPI_ISL_2189953, EPI_ISL_2189954, EPI_ISL_2189955, EPI_ISL_2189956, EPI_ISL_2189957, EPI_ISL_2189958, EPI_ISL_2189959, EPI_ISL_2189960, EPI_ISL_2189961, EPI_ISL_2189962, EPI_ISL_2189963, EPI_ISL_2189964, EPI_ISL_2189965, EPI_ISL_2189966, EPI_ISL_2189967, EPI_ISL_2189968, EPI_ISL_2189969, EPI_ISL_2189970, EPI_ISL_2189971, EPI_ISL_2189972, EPI_ISL_2189973, EPI_ISL_2189974, EPI_ISL_2189975, EPI_ISL_2189976, EPI_ISL_2189977, EPI_ISL_2189978, EPI_ISL_2189979, EPI_ISL_2189980, EPI_ISL_2189981, EPI_ISL_2189982, EPI_ISL_2189983, EPI_ISL_2189984, EPI_ISL_2189985, EPI_ISL_2189986, EPI_ISL_2189987, EPI_ISL_2189988, EPI_ISL_2189989, EPI_ISL_2189990, EPI_ISL_2189993, EPI_ISL_2189994, EPI_ISL_2189995, EPI_ISL_2189996, EPI_ISL_2189997, EPI_ISL_2189998, EPI_ISL_2189999, EPI_ISL_2190000, EPI_ISL_2190001, EPI_ISL_2190002, EPI_ISL_2190003, EPI_ISL_2190004, EPI_ISL_2190005, EPI_ISL_2190006, EPI_ISL_2190007, EPI_ISL_2190008, EPI_ISL_2190009, EPI_ISL_2190010, EPI_ISL_2190011, EPI_ISL_2190012, EPI_ISL_2190013, EPI_ISL_2190014, EPI_ISL_2190015, EPI_ISL_2190016, EPI_ISL_2190017, EPI_ISL_2190018, EPI_ISL_2190019, EPI_ISL_2190020, EPI_ISL_2190021, EPI_ISL_2190022, EPI_ISL_2190023, EPI_ISL_2190024, EPI_ISL_2190025, EPI_ISL_2190026, EPI_ISL_2190027, EPI_ISL_2190028, EPI_ISL_2190029, EPI_ISL_2190030, EPI_ISL_2190031, EPI_ISL_2190032, EPI_ISL_2190033, EPI_ISL_2190034, EPI_ISL_2190035, EPI_ISL_2190036, EPI_ISL_2190037, EPI_ISL_2190038, EPI_ISL_2190039, EPI_ISL_2190040, EPI_ISL_2190041, EPI_ISL_2190042, EPI_ISL_2190043, EPI_ISL_2190044, EPI_ISL_2190045, EPI_ISL_2190046, EPI_ISL_2190047, EPI_ISL_2190048, EPI_ISL_2190049, EPI_ISL_2190050, EPI_ISL_2190051, EPI_ISL_2190052, EPI_ISL_2190053, EPI_ISL_2190054, EPI_ISL_2190055, EPI_ISL_2190056, EPI_ISL_2190057, EPI_ISL_2190058, EPI_ISL_2190059, EPI_ISL_2190060, EPI_ISL_2190061, EPI_ISL_2190062, EPI_ISL_2190063, EPI_ISL_2190064, EPI_ISL_2190065, EPI_ISL_2190066, EPI_ISL_2190067, EPI_ISL_2190068, EPI_ISL_2190069, EPI_ISL_2190070, EPI_ISL_2190071, EPI_ISL_2190072, EPI_ISL_2190073, EPI_ISL_2190074, EPI_ISL_2190075, EPI_ISL_2190076, EPI_ISL_2190077, EPI_ISL_2190078, EPI_ISL_2190079, EPI_ISL_2190080, EPI_ISL_2190081, EPI_ISL_2190082, EPI_ISL_2190083, EPI_ISL_2190084, EPI_ISL_2190085, EPI_ISL_2190086, EPI_ISL_2190087, EPI_ISL_2190088, EPI_ISL_2190089, EPI_ISL_2190090, EPI_ISL_2190091, EPI_ISL_2190092, EPI_ISL_2190093, EPI_ISL_2190094, EPI_ISL_2190095, EPI_ISL_2190096, EPI_ISL_2190097, EPI_ISL_2190099, EPI_ISL_2190102, EPI_ISL_2190105, EPI_ISL_2190109, EPI_ISL_2190112, EPI_ISL_2190115, EPI_ISL_2190118, EPI_ISL_2190124, EPI_ISL_2190128, EPI_ISL_2190131, EPI_ISL_2190135, EPI_ISL_2190139, EPI_ISL_2190142, EPI_ISL_2190143, EPI_ISL_2190144, EPI_ISL_2190145, EPI_ISL_2190146, EPI_ISL_2190147, EPI_ISL_2190148, EPI_ISL_2190149, EPI_ISL_2190150, EPI_ISL_2190151, EPI_ISL_2190152, EPI_ISL_2190153, EPI_ISL_2190154, EPI_ISL_2190155, EPI_ISL_2190156, EPI_ISL_2190157, EPI_ISL_2190158, EPI_ISL_2190159, EPI_ISL_2190160, EPI_ISL_2190161, EPI_ISL_2190162, EPI_ISL_2190163, EPI_ISL_2190164, EPI_ISL_2190165, EPI_ISL_2190166, EPI_ISL_2190167, EPI_ISL_2190168, EPI_ISL_2190169, EPI_ISL_2190170, EPI_ISL_2190171, EPI_ISL_2190172, EPI_ISL_2190173, EPI_ISL_2190174, EPI_ISL_2190175, EPI_ISL_2190176, EPI_ISL_2190177, EPI_ISL_2190178, EPI_ISL_2190179, EPI_ISL_2190180, EPI_ISL_2190181, EPI_ISL_2190182, EPI_ISL_2190183, EPI_ISL_2190184, EPI_ISL_2190185, EPI_ISL_2190186, EPI_ISL_2190187, EPI_ISL_2190188, EPI_ISL_2190189, EPI_ISL_2190190, EPI_ISL_2190191, EPI_ISL_2190192, EPI_ISL_2190193, EPI_ISL_2190194, EPI_ISL_2190195, EPI_ISL_2190196, EPI_ISL_2190197, EPI_ISL_2190198, EPI_ISL_2190199, EPI_ISL_2190200, EPI_ISL_2190201, EPI_ISL_2190202, EPI_ISL_2190203, EPI_ISL_2190204, EPI_ISL_2190205, EPI_ISL_2190206, EPI_ISL_2190207, EPI_ISL_2190208, EPI_ISL_2190209, EPI_ISL_2190210, EPI_ISL_2190211, EPI_ISL_2190212, EPI_ISL_2190213, EPI_ISL_2190214, EPI_ISL_2190215, EPI_ISL_2190216, EPI_ISL_2190217, EPI_ISL_2190218, EPI_ISL_2190219, EPI_ISL_2190220, EPI_ISL_2190221, EPI_ISL_2190222, EPI_ISL_2190223, EPI_ISL_2190224, EPI_ISL_2190225, EPI_ISL_2190226, EPI_ISL_2190227, EPI_ISL_2190228, EPI_ISL_2190229, EPI_ISL_2190230, EPI_ISL_2190231, EPI_ISL_2190232, EPI_ISL_2190233, EPI_ISL_2190234, EPI_ISL_2190235, EPI_ISL_2190236, EPI_ISL_2190237, EPI_ISL_2190238, EPI_ISL_2190239, EPI_ISL_2190240, EPI_ISL_2190241, EPI_ISL_2190242, EPI_ISL_2190243, EPI_ISL_2190244, EPI_ISL_2190245, EPI_ISL_2190246, EPI_ISL_2190247, EPI_ISL_2190248, EPI_ISL_2190249, EPI_ISL_2190250, EPI_ISL_2190251, EPI_ISL_2190252, EPI_ISL_2190253, EPI_ISL_2190254, EPI_ISL_2190255, EPI_ISL_2190256, EPI_ISL_2190257, EPI_ISL_2190258, EPI_ISL_2190259, EPI_ISL_2190260, EPI_ISL_2190261, EPI_ISL_2190262, EPI_ISL_2190263, EPI_ISL_2190264, EPI_ISL_2190265, EPI_ISL_2190266, EPI_ISL_2190267, EPI_ISL_2190268, EPI_ISL_2190269, EPI_ISL_2190270, EPI_ISL_2190271, EPI_ISL_2190272, EPI_ISL_2190273, EPI_ISL_2190274, EPI_ISL_2190275, EPI_ISL_2190276, EPI_ISL_2190277, EPI_ISL_2190278, EPI_ISL_2190279, EPI_ISL_2190280, EPI_ISL_2190281, EPI_ISL_2190282, EPI_ISL_2190283, EPI_ISL_2190284, EPI_ISL_2190285, EPI_ISL_2190286, EPI_ISL_2190287, EPI_ISL_2190288, EPI_ISL_2190289, EPI_ISL_2190290, EPI_ISL_2190291, EPI_ISL_2190292, EPI_ISL_2190293, EPI_ISL_2190294, EPI_ISL_2190295, EPI_ISL_2190296, EPI_ISL_2190297, EPI_ISL_2190298, EPI_ISL_2190299, EPI_ISL_2190300, EPI_ISL_2190301, EPI_ISL_2190302, EPI_ISL_2190303, EPI_ISL_2190304, EPI_ISL_2190305, EPI_ISL_2190306, EPI_ISL_2190307, EPI_ISL_2190309, EPI_ISL_2190310, EPI_ISL_2190311, EPI_ISL_2190312, EPI_ISL_2190314, EPI_ISL_2190315, EPI_ISL_2190316, EPI_ISL_2190317, EPI_ISL_2190319, EPI_ISL_2190320, EPI_ISL_2190321, EPI_ISL_2190322, EPI_ISL_2190323, EPI_ISL_2190324, EPI_ISL_2190325, EPI_ISL_2190326, EPI_ISL_2190327, EPI_ISL_2190328, EPI_ISL_2190329, EPI_ISL_2190330, EPI_ISL_2190331, EPI_ISL_2190332, EPI_ISL_2190333, EPI_ISL_2190334, EPI_ISL_2190336, EPI_ISL_2190337, EPI_ISL_2190338, EPI_ISL_2190339, EPI_ISL_2190340, EPI_ISL_2190343, EPI_ISL_2190344, EPI_ISL_2190345, EPI_ISL_2190347, EPI_ISL_2190349, EPI_ISL_2190350, EPI_ISL_2190351, EPI_ISL_2190352, EPI_ISL_2190353, EPI_ISL_2190355, EPI_ISL_2190356, EPI_ISL_2190357, EPI_ISL_2190359, EPI_ISL_2190360, EPI_ISL_2190362, EPI_ISL_2190363, EPI_ISL_2190364, EPI_ISL_2190367, EPI_ISL_2190368, EPI_ISL_2190369, EPI_ISL_2190370, EPI_ISL_2190371, EPI_ISL_2190372, EPI_ISL_2190373, EPI_ISL_2190374, EPI_ISL_2190375, EPI_ISL_2190376, EPI_ISL_2190377, EPI_ISL_2190378, EPI_ISL_2190379, EPI_ISL_2190380, EPI_ISL_2190381, EPI_ISL_2190382, EPI_ISL_2190383, EPI_ISL_2190384, EPI_ISL_2190385, EPI_ISL_2190386, EPI_ISL_2190387, EPI_ISL_2190388, EPI_ISL_2190389, EPI_ISL_2190390, EPI_ISL_2190391, EPI_ISL_2190392, EPI_ISL_2190393, EPI_ISL_2190394, EPI_ISL_2190395, EPI_ISL_2190396, EPI_ISL_2190397, EPI_ISL_2190399, EPI_ISL_2190401, EPI_ISL_2190402, EPI_ISL_2190403, EPI_ISL_2190404, EPI_ISL_2190405, EPI_ISL_2190406, EPI_ISL_2190407, EPI_ISL_2190409, EPI_ISL_2190410, EPI_ISL_2190411, EPI_ISL_2190415, EPI_ISL_2190416, EPI_ISL_2190417, EPI_ISL_2190418, EPI_ISL_2190419, EPI_ISL_2190420, EPI_ISL_2190421, EPI_ISL_2190422, EPI_ISL_2190423, EPI_ISL_2190424, EPI_ISL_2190425, EPI_ISL_2190426, EPI_ISL_2190427, EPI_ISL_2190428, EPI_ISL_2190429, EPI_ISL_2190431, EPI_ISL_2190432, EPI_ISL_2190434, EPI_ISL_2190435, EPI_ISL_2190436, EPI_ISL_2190437, EPI_ISL_2190438, EPI_ISL_2190439, EPI_ISL_2190440, EPI_ISL_2190441, EPI_ISL_2190444, EPI_ISL_2190445, EPI_ISL_2190447, EPI_ISL_2190449, EPI_ISL_2190450, EPI_ISL_2190451, EPI_ISL_2190452, EPI_ISL_2190456, EPI_ISL_2190457, EPI_ISL_2190459, EPI_ISL_2190461, EPI_ISL_2190462, EPI_ISL_2190463, EPI_ISL_2190465, EPI_ISL_2190466, EPI_ISL_2190467, EPI_ISL_2190468, EPI_ISL_2190469, EPI_ISL_2190470, EPI_ISL_2190471, EPI_ISL_2190472, EPI_ISL_2190473, EPI_ISL_2190474, EPI_ISL_2190475, EPI_ISL_2190476, EPI_ISL_2190477, EPI_ISL_2190478, EPI_ISL_2190479, EPI_ISL_2190480, EPI_ISL_2190481, EPI_ISL_2190482, EPI_ISL_2190483, EPI_ISL_2190484, EPI_ISL_2190485, EPI_ISL_2190486, EPI_ISL_2190487, EPI_ISL_2190489, EPI_ISL_2190490, EPI_ISL_2190491, EPI_ISL_2190493, EPI_ISL_2190494, EPI_ISL_2190495, EPI_ISL_2190496, EPI_ISL_2190497, EPI_ISL_2190498, EPI_ISL_2190499, EPI_ISL_2190500, EPI_ISL_2190501, EPI_ISL_2190502, EPI_ISL_2190503, EPI_ISL_2190504, EPI_ISL_2190505, EPI_ISL_2190506, EPI_ISL_2190507, EPI_ISL_2190508, EPI_ISL_2190509, EPI_ISL_2190510, EPI_ISL_2190511, EPI_ISL_2190513, EPI_ISL_2190514, EPI_ISL_2190515, EPI_ISL_2190516, EPI_ISL_2190517, EPI_ISL_2190518, EPI_ISL_2190519, EPI_ISL_2190520, EPI_ISL_2190521, EPI_ISL_2190522, EPI_ISL_2190523, EPI_ISL_2190524, EPI_ISL_2190525, EPI_ISL_2190526, EPI_ISL_2190527, EPI_ISL_2190528, EPI_ISL_2190529, EPI_ISL_2190530, EPI_ISL_2190531, EPI_ISL_2190532, EPI_ISL_2190533, EPI_ISL_2190534, EPI_ISL_2190535, EPI_ISL_2190536, EPI_ISL_2190537, EPI_ISL_2190539, EPI_ISL_2190540, EPI_ISL_2190541, EPI_ISL_2190542, EPI_ISL_2190543, EPI_ISL_2190544, EPI_ISL_2190545, EPI_ISL_2190546, EPI_ISL_2190547, EPI_ISL_2190548, EPI_ISL_2190549, EPI_ISL_2190550, EPI_ISL_2190551, EPI_ISL_2190552, EPI_ISL_2190553, EPI_ISL_2190555, EPI_ISL_2190556, EPI_ISL_2190557, EPI_ISL_2190558, EPI_ISL_2190559, EPI_ISL_2190560, EPI_ISL_2190561, EPI_ISL_2190562, EPI_ISL_2190563, EPI_ISL_2190564, EPI_ISL_2190565, EPI_ISL_2190566, EPI_ISL_2190567, EPI_ISL_2190568, EPI_ISL_2190569, EPI_ISL_2190570, EPI_ISL_2190571, EPI_ISL_2190572, EPI_ISL_2190573, EPI_ISL_2190574, EPI_ISL_2190575, EPI_ISL_2190576, EPI_ISL_2190577, | | | |

EPI_ISL_2190578, EPI_ISL_2190579, EPI_ISL_2190580, EPI_ISL_2190581, EPI_ISL_2190583, EPI_ISL_2190584, EPI_ISL_2190585, EPI_ISL_2190586, EPI_ISL_2190587, EPI_ISL_2190588, EPI_ISL_2190589, EPI_ISL_2190591, EPI_ISL_2190592, EPI_ISL_2190593, EPI_ISL_2190595, EPI_ISL_2190596, EPI_ISL_2190597, EPI_ISL_2190598, EPI_ISL_2190599, EPI_ISL_2190600, EPI_ISL_2190601, EPI_ISL_2190602, EPI_ISL_2190603, EPI_ISL_2190604, EPI_ISL_2190605, EPI_ISL_2190606, EPI_ISL_2190607, EPI_ISL_2190608, EPI_ISL_2190609, EPI_ISL_2190610, EPI_ISL_2190612, EPI_ISL_2190614, EPI_ISL_2190615, EPI_ISL_2190616, EPI_ISL_2190617, EPI_ISL_2190618, EPI_ISL_2190619, EPI_ISL_2190620, EPI_ISL_2190621, EPI_ISL_2190622, EPI_ISL_2190623, EPI_ISL_2190624, EPI_ISL_2190625, EPI_ISL_2190626, EPI_ISL_2190627, EPI_ISL_2190628, EPI_ISL_2190629, EPI_ISL_2190630, EPI_ISL_2190631, EPI_ISL_2190632, EPI_ISL_2190633, EPI_ISL_2190634, EPI_ISL_2190636, EPI_ISL_2190637, EPI_ISL_2190638, EPI_ISL_2190639, EPI_ISL_2190640, EPI_ISL_2190641, EPI_ISL_2190642, EPI_ISL_2190643, EPI_ISL_2190644, EPI_ISL_2190645, EPI_ISL_2190648, EPI_ISL_2190649, EPI_ISL_2190650, EPI_ISL_2190652, EPI_ISL_2190653, EPI_ISL_2190654, EPI_ISL_2190655, EPI_ISL_2190656, EPI_ISL_2190657, EPI_ISL_2190658, EPI_ISL_2190659, EPI_ISL_2190660, EPI_ISL_2190661, EPI_ISL_2190662, EPI_ISL_2190663, EPI_ISL_2190664, EPI_ISL_2190665, EPI_ISL_2190666, EPI_ISL_2190667, EPI_ISL_2190668, EPI_ISL_2190670, EPI_ISL_2190671, EPI_ISL_2190673, EPI_ISL_2190674, EPI_ISL_2190675, EPI_ISL_2190676, EPI_ISL_2190677, EPI_ISL_2190678, EPI_ISL_2190679, EPI_ISL_2190680, EPI_ISL_2190681, EPI_ISL_2190682, EPI_ISL_2190683, EPI_ISL_2190684, EPI_ISL_2190685, EPI_ISL_2190686, EPI_ISL_2190687, EPI_ISL_2190688, EPI_ISL_2190689, EPI_ISL_2190690, EPI_ISL_2190691, EPI_ISL_2190692, EPI_ISL_2190693, EPI_ISL_2190694, EPI_ISL_2190695, EPI_ISL_2190696, EPI_ISL_2190697, EPI_ISL_2190698, EPI_ISL_2190699, EPI_ISL_2190700, EPI_ISL_2190701, EPI_ISL_2190702, EPI_ISL_2190704, EPI_ISL_2190705, EPI_ISL_2190706, EPI_ISL_2190707, EPI_ISL_2190708, EPI_ISL_2190709, EPI_ISL_2190710, EPI_ISL_2190711, EPI_ISL_2190713, EPI_ISL_2190714, EPI_ISL_2190716, EPI_ISL_2190717, EPI_ISL_2190718, EPI_ISL_2190719, EPI_ISL_2190722, EPI_ISL_2190723, EPI_ISL_2190724, EPI_ISL_2190725, EPI_ISL_2190726, EPI_ISL_2190728, EPI_ISL_2190729, EPI_ISL_2190730, EPI_ISL_2190731, EPI_ISL_2190733, EPI_ISL_2190734, EPI_ISL_2190735, EPI_ISL_2190736, EPI_ISL_2190737, EPI_ISL_2190738, EPI_ISL_2190739, EPI_ISL_2190740, EPI_ISL_2190743, EPI_ISL_2190744, EPI_ISL_2190745, EPI_ISL_2190747, EPI_ISL_2190748, EPI_ISL_2190749, EPI_ISL_2190751, EPI_ISL_2190752, EPI_ISL_2190753, EPI_ISL_2190754, EPI_ISL_2190755, EPI_ISL_2190756, EPI_ISL_2190757, EPI_ISL_2190758, EPI_ISL_2190759, EPI_ISL_2190760, EPI_ISL_2190761, EPI_ISL_2190762, EPI_ISL_2190763, EPI_ISL_2190764, EPI_ISL_2190765, EPI_ISL_2190766, EPI_ISL_2190767, EPI_ISL_2190768, EPI_ISL_2190770, EPI_ISL_2190771, EPI_ISL_2190772, EPI_ISL_2190773, EPI_ISL_2190774, EPI_ISL_2190775, EPI_ISL_2190776, EPI_ISL_2190777, EPI_ISL_2190780, EPI_ISL_2190781, EPI_ISL_2190782, EPI_ISL_2190785, EPI_ISL_2190786, EPI_ISL_2190787, EPI_ISL_2190788, EPI_ISL_2190789, EPI_ISL_2190790, EPI_ISL_2190791, EPI_ISL_2190792, EPI_ISL_2190793, EPI_ISL_2190794, EPI_ISL_2190795, EPI_ISL_2190796, EPI_ISL_2190797, EPI_ISL_2190798, EPI_ISL_2190799, EPI_ISL_2190800, EPI_ISL_2190801, EPI_ISL_2190802, EPI_ISL_2190803, EPI_ISL_2190804, EPI_ISL_2190805, EPI_ISL_2190806, EPI_ISL_2190808, EPI_ISL_2190809, EPI_ISL_2190810, EPI_ISL_2190812, EPI_ISL_2190813, EPI_ISL_2190814, EPI_ISL_2190815, EPI_ISL_2190816, EPI_ISL_2190820, EPI_ISL_2190821, EPI_ISL_2190823, EPI_ISL_2190824, EPI_ISL_2190825, EPI_ISL_2190826, EPI_ISL_2190828, EPI_ISL_2190829, EPI_ISL_2190831, EPI_ISL_2190832, EPI_ISL_2190833, EPI_ISL_2190834, EPI_ISL_2190835, EPI_ISL_2190836, EPI_ISL_2190838, EPI_ISL_2190839, EPI_ISL_2190841, EPI_ISL_2190842, EPI_ISL_2190843, EPI_ISL_2190845, EPI_ISL_2190846, EPI_ISL_2190849, EPI_ISL_2190850, EPI_ISL_2190851, EPI_ISL_2190852, EPI_ISL_2190854, EPI_ISL_2190855, EPI_ISL_2190856, EPI_ISL_2190857, EPI_ISL_2190858, EPI_ISL_2190860, EPI_ISL_2190863, EPI_ISL_2190864, EPI_ISL_2190865, EPI_ISL_2190866, EPI_ISL_2190868, EPI_ISL_2190870, EPI_ISL_2190872, EPI_ISL_2190873, EPI_ISL_2190874, EPI_ISL_2190877, EPI_ISL_2190880, EPI_ISL_2190881, EPI_ISL_2190883, EPI_ISL_2190884, EPI_ISL_2190885, EPI_ISL_2190886, EPI_ISL_2190887, EPI_ISL_2190888, EPI_ISL_2190889, EPI_ISL_2190890, EPI_ISL_2190891, EPI_ISL_2190892, EPI_ISL_2190893, EPI_ISL_2190896, EPI_ISL_2190897, EPI_ISL_2190898, EPI_ISL_2190899, EPI_ISL_2190900, EPI_ISL_2190901, EPI_ISL_2190902, EPI_ISL_2190903, EPI_ISL_2190904, EPI_ISL_2190905, EPI_ISL_2190907, EPI_ISL_2190909, EPI_ISL_2190910, EPI_ISL_2190911, EPI_ISL_2190912, EPI_ISL_2190913, EPI_ISL_2190914, EPI_ISL_2190915, EPI_ISL_2190917, EPI_ISL_2190919, EPI_ISL_2190920, EPI_ISL_2190921, EPI_ISL_2190923, EPI_ISL_2190924, EPI_ISL_2190926, EPI_ISL_2190927, EPI_ISL_2190928, EPI_ISL_2190929, EPI_ISL_2190930, EPI_ISL_2190931, EPI_ISL_2190932, EPI_ISL_2190933, EPI_ISL_2190935, EPI_ISL_2190936, EPI_ISL_2190937, EPI_ISL_2190938, EPI_ISL_2190939, EPI_ISL_2190940, EPI_ISL_2190941, EPI_ISL_2190943, EPI_ISL_2190944, EPI_ISL_2190945, EPI_ISL_2190946, EPI_ISL_2190947, EPI_ISL_2190948, EPI_ISL_2190949, EPI_ISL_2190950, EPI_ISL_2190951, EPI_ISL_2190952, EPI_ISL_2190953, EPI_ISL_2190954, EPI_ISL_2190955, EPI_ISL_2190956, EPI_ISL_2190957, EPI_ISL_2190958, EPI_ISL_2190959, EPI_ISL_2190960, EPI_ISL_2190961, EPI_ISL_2190962, EPI_ISL_2190963, EPI_ISL_2190964, EPI_ISL_2190967, EPI_ISL_2190968, EPI_ISL_2190969, EPI_ISL_2190970, EPI_ISL_2190971, EPI_ISL_2190972, EPI_ISL_2190973, EPI_ISL_2190974, EPI_ISL_2190975, EPI_ISL_2190976, EPI_ISL_2190977, EPI_ISL_2190978, EPI_ISL_2190979, EPI_ISL_2190980, EPI_ISL_2190981, EPI_ISL_2190982, EPI_ISL_2190983, EPI_ISL_2190985, EPI_ISL_2190986, EPI_ISL_2190988, EPI_ISL_2190989, EPI_ISL_2190990, EPI_ISL_2190991, EPI_ISL_2190992, EPI_ISL_2190993, EPI_ISL_2190994, EPI_ISL_2190995, EPI_ISL_2190997, EPI_ISL_2190998, EPI_ISL_2190999, EPI_ISL_2191000, EPI_ISL_2191001, EPI_ISL_2191002, EPI_ISL_2191003, EPI_ISL_2191004, EPI_ISL_2191005, EPI_ISL_2191006, EPI_ISL_2191007, EPI_ISL_2191009, EPI_ISL_2191010, EPI_ISL_2191011, EPI_ISL_2191012, EPI_ISL_2191013, EPI_ISL_2191014, EPI_ISL_2191016, EPI_ISL_2191017, EPI_ISL_2191018, EPI_ISL_2191019, EPI_ISL_2191020, EPI_ISL_2191021, EPI_ISL_2191023, EPI_ISL_2191024, EPI_ISL_2191025, EPI_ISL_2191026, EPI_ISL_2191028, EPI_ISL_2191029, EPI_ISL_2191030, EPI_ISL_2191032, EPI_ISL_2191033, EPI_ISL_2191035, EPI_ISL_2191036, EPI_ISL_2191037, EPI_ISL_2191038, EPI_ISL_2191039, EPI_ISL_2191040, EPI_ISL_2191041, EPI_ISL_2191042, EPI_ISL_2191044, EPI_ISL_2191045, EPI_ISL_2191046, EPI_ISL_2191047, EPI_ISL_2191048, EPI_ISL_2191049, EPI_ISL_2191050, EPI_ISL_2191051, EPI_ISL_2191052, EPI_ISL_2191053, EPI_ISL_2191054, EPI_ISL_2191055, EPI_ISL_2191056, EPI_ISL_2191057, EPI_ISL_2191058, EPI_ISL_2191059, EPI_ISL_2191060, EPI_ISL_2191061, EPI_ISL_2191062, EPI_ISL_2191063, EPI_ISL_2191064, EPI_ISL_2191066, EPI_ISL_2191067, EPI_ISL_2191069, EPI_ISL_2191070, EPI_ISL_2191071, EPI_ISL_2191072, EPI_ISL_2191073, EPI_ISL_2191074, EPI_ISL_2191075, EPI_ISL_2191076, EPI_ISL_2191077, EPI_ISL_2191078, EPI_ISL_2191079, EPI_ISL_2191081, EPI_ISL_2191082, EPI_ISL_2191083, EPI_ISL_2191084, EPI_ISL_2191085, EPI_ISL_2191086, EPI_ISL_2191087, EPI_ISL_2191088, EPI_ISL_2191090, EPI_ISL_2191091, EPI_ISL_2191092, EPI_ISL_2191093, EPI_ISL_2191094, EPI_ISL_2191095, EPI_ISL_2191096, EPI_ISL_2191097, EPI_ISL_2191098, EPI_ISL_2191100, EPI_ISL_2191101, EPI_ISL_2191102, EPI_ISL_2191103, EPI_ISL_2191104, EPI_ISL_2191105, EPI_ISL_2191106, EPI_ISL_2191107, EPI_ISL_2191108, EPI_ISL_2191109, EPI_ISL_2191110, EPI_ISL_2191111, EPI_ISL_2191112, EPI_ISL_2191113, EPI_ISL_2191114, EPI_ISL_2191117, EPI_ISL_2191118, EPI_ISL_2191119, EPI_ISL_2191120, EPI_ISL_2191121, EPI_ISL_2191122, EPI_ISL_2191123, EPI_ISL_2191124, EPI_ISL_2191126, EPI_ISL_2191127, EPI_ISL_2191128, EPI_ISL_2191129, EPI_ISL_2191130, EPI_ISL_2191131, EPI_ISL_2191133, EPI_ISL_2191135, EPI_ISL_2191136, EPI_ISL_2191137, EPI_ISL_2191138, EPI_ISL_2191139, EPI_ISL_2191140, EPI_ISL_2191141, EPI_ISL_2191143, EPI_ISL_2191144, EPI_ISL_2191145, EPI_ISL_2191146, EPI_ISL_2191147, EPI_ISL_2191149, EPI_ISL_2191150, EPI_ISL_2191151, EPI_ISL_2191152, EPI_ISL_2191153, EPI_ISL_2191154, EPI_ISL_2191155, EPI_ISL_2191156, EPI_ISL_2191157, EPI_ISL_2191159, EPI_ISL_2191160, EPI_ISL_2191161, EPI_ISL_2191162, EPI_ISL_2191163, EPI_ISL_2191164, EPI_ISL_2191165, EPI_ISL_2191166, EPI_ISL_2191167, EPI_ISL_2191169, EPI_ISL_2191170, EPI_ISL_2191171, EPI_ISL_2191172, EPI_ISL_2191173, EPI_ISL_2191174, EPI_ISL_2191175, EPI_ISL_2191176, EPI_ISL_2191177, EPI_ISL_2191178, EPI_ISL_2191179, EPI_ISL_2191180, EPI_ISL_2191181, EPI_ISL_2191182, EPI_ISL_2191183, EPI_ISL_2191184, EPI_ISL_2191185, EPI_ISL_2191186, EPI_ISL_2191187, EPI_ISL_2191188, EPI_ISL_2191189, EPI_ISL_2191190, EPI_ISL_2191191, EPI_ISL_2191192, EPI_ISL_2191193, EPI_ISL_2191194, EPI_ISL_2191195, EPI_ISL_2191196, EPI_ISL_2191197, EPI_ISL_2191199, EPI_ISL_2191200, EPI_ISL_2191201, EPI_ISL_2191202, EPI_ISL_2191203, EPI_ISL_2191204, EPI_ISL_2191205, EPI_ISL_2191206, EPI_ISL_2191207, EPI_ISL_2191208, EPI_ISL_2191209, EPI_ISL_2191210, EPI_ISL_2191211, EPI_ISL_2191212, EPI_ISL_2191213, EPI_ISL_2191214, EPI_ISL_2191215, EPI_ISL_2191216, EPI_ISL_2191217, EPI_ISL_2191218, EPI_ISL_2191219, EPI_ISL_2191220, EPI_ISL_2191221, EPI_ISL_2191222, EPI_ISL_2191223, EPI_ISL_2191224, EPI_ISL_2191225, EPI_ISL_2191226, EPI_ISL_2191227, EPI_ISL_2191228, EPI_ISL_2191229, EPI_ISL_2191231, EPI_ISL_2191232, EPI_ISL_2191233, EPI_ISL_2191234, EPI_ISL_2191235, EPI_ISL_2191236, EPI_ISL_2191237, EPI_ISL_2191238, EPI_ISL_2191239, EPI_ISL_2191240, EPI_ISL_2191241, EPI_ISL_2191242, EPI_ISL_2191243, EPI_ISL_2191244, EPI_ISL_2191245, EPI_ISL_2191246, EPI_ISL_2191247, EPI_ISL_2191248, EPI_ISL_2191249, EPI_ISL_2191250, EPI_ISL_2191251, EPI_ISL_2191252, EPI_ISL_2191253, EPI_ISL_2191255, EPI_ISL_2191256, EPI_ISL_2191257, EPI_ISL_2191258, EPI_ISL_2191260, EPI_ISL_2191261, EPI_ISL_2191263, EPI_ISL_2191264, EPI_ISL_2191265, EPI_ISL_2191268, EPI_ISL_2191270, EPI_ISL_2191271, EPI_ISL_2191272, EPI_ISL_2191273, EPI_ISL_2191274, EPI_ISL_2191276, EPI_ISL_2191277, EPI_ISL_2191278, EPI_ISL_2191279, EPI_ISL_2191280, EPI_ISL_2191281, EPI_ISL_2191282, EPI_ISL_2191283, EPI_ISL_2191284, EPI_ISL_2191285, EPI_ISL_2191286, EPI_ISL_2191287, EPI_ISL_2191288, EPI_ISL_2191289, EPI_ISL_2191290, EPI_ISL_2191291, EPI_ISL_2191292, EPI_ISL_2191293, EPI_ISL_2191294, EPI_ISL_2191295, EPI_ISL_2191296, EPI_ISL_2191297, EPI_ISL_2191298, EPI_ISL_2191299, EPI_ISL_2191300, EPI_ISL_2191301, EPI_ISL_2191303, EPI_ISL_2191304, EPI_ISL_2191305, EPI_ISL_2191307, EPI_ISL_2191308, EPI_ISL_2191309, EPI_ISL_2191310, EPI_ISL_2191312, EPI_ISL_2191313, EPI_ISL_2191314, EPI_ISL_2191315, EPI_ISL_2191316, EPI_ISL_2191317, EPI_ISL_2191318, EPI_ISL_2191319, EPI_ISL_2191320, EPI_ISL_2191321, EPI_ISL_2191322, EPI_ISL_2191323, EPI_ISL_2191324, EPI_ISL_2191325, EPI_ISL_2191326, EPI_ISL_2191327, EPI_ISL_2191328, EPI_ISL_2191329, EPI_ISL_2191330, EPI_ISL_2191331, EPI_ISL_2191332, EPI_ISL_2191333, EPI_ISL_2191334, EPI_ISL_2191335, EPI_ISL_2191336, EPI_ISL_2191337, EPI_ISL_2191338, EPI_ISL_2191339, EPI_ISL_2191340, EPI_ISL_2191341, EPI_ISL_2191342, EPI_ISL_2191343, EPI_ISL_2191344, EPI_ISL_2191345, EPI_ISL_2191346, EPI_ISL_2191347, EPI_ISL_2191348, EPI_ISL_2191349, EPI_ISL_2191350, EPI_ISL_2191351, EPI_ISL_2191352, EPI_ISL_2191353, EPI_ISL_2191354, EPI_ISL_2191355, EPI_ISL_2191356, EPI_ISL_2191357, EPI_ISL_2191358, EPI_ISL_2191359, EPI_ISL_2191360, EPI_ISL_2191361, EPI_ISL_2191362, EPI_ISL_2191363, EPI_ISL_2191364, EPI_ISL_2191365, EPI_ISL_2191366, EPI_ISL_2191367, EPI_ISL_2191368, EPI_ISL_2191369, EPI_ISL_2191371, EPI_ISL_2191372, EPI_ISL_2191373, EPI_ISL_2191375, EPI_ISL_2191377, EPI_ISL_2191378, EPI_ISL_2191379, EPI_ISL_2191380, EPI_ISL_2191381, EPI_ISL_2191382, EPI_ISL_2191383, EPI_ISL_2191384, EPI_ISL_2191385, EPI_ISL_2191386, EPI_ISL_2191387, EPI_ISL_2191388, EPI_ISL_2191389, EPI_ISL_2191391, EPI_ISL_2191392, EPI_ISL_2191393, EPI_ISL_2191394, EPI_ISL_2191395, EPI_ISL_2191396, EPI_ISL_2191397, EPI_ISL_2191398, EPI_ISL_2191399, EPI_ISL_2191400, EPI_ISL_2191401, EPI_ISL_2191402, EPI_ISL_2191403, EPI_ISL_2191404, EPI_ISL_2191405, EPI_ISL_2191406, EPI_ISL_2191407, EPI_ISL_2191408, EPI_ISL_2191409, EPI_ISL_2191410, EPI_ISL_2191411, EPI_ISL_2191412, EPI_ISL_2191413, EPI_ISL_2191414, EPI_ISL_2191415, EPI_ISL_2191416, EPI_ISL_2191417, EPI_ISL_2191418, EPI_ISL_2191419, EPI_ISL_2191420, EPI_ISL_2191421, EPI_ISL_2191422, EPI_ISL_2191423, EPI_ISL_2191424, EPI_ISL_2191425, EPI_ISL_2191426, EPI_ISL_2191427, EPI_ISL_2191428, EPI_ISL_2191429, EPI_ISL_2191430, EPI_ISL_2191431, EPI_ISL_2191432, EPI_ISL_2191433, EPI_ISL_2191434, EPI_ISL_2191435, EPI_ISL_2191436, EPI_ISL_2191437, EPI_ISL_2191438, EPI_ISL_2191439, EPI_ISL_2191440, EPI_ISL_2191441, EPI_ISL_2191442, EPI_ISL_2191443, EPI_ISL_2191444, EPI_ISL_2191445, EPI_ISL_2191446, EPI_ISL_2191447, EPI_ISL_2191448, EPI_ISL_2191449, EPI_ISL_2191450, EPI_ISL_2191451, EPI_ISL_2191452, EPI_ISL_2191453, EPI_ISL_2191454, EPI_ISL_2191455, EPI_ISL_2191456, EPI_ISL_2191457, EPI_ISL_2191458, EPI_ISL_2191459, EPI_ISL_2191460, EPI_ISL_2191461, EPI_ISL_2191462, EPI_ISL_2191463, EPI_ISL_2191464, EPI_ISL_2191465, EPI_ISL_2191466, EPI_ISL_2191467, EPI_ISL_2191468, EPI_ISL_2191469, EPI_ISL_2191470, EPI_ISL_2191471, EPI_ISL_2191472, EPI_ISL_2191473, EPI_ISL_2191474, EPI_ISL_2191475, EPI_ISL_2191476, EPI_ISL_2191477, EPI_ISL_2191478, EPI_ISL_2191479, EPI_ISL_2191480, EPI_ISL_2191481, EPI_ISL_2191482, EPI_ISL_2191483, EPI_ISL_2191484, EPI_ISL_2191485, EPI_ISL_2191486, EPI_ISL_2191487, EPI_ISL_2191488, EPI_ISL_2191489, EPI_ISL_2191490, EPI_ISL_2191491, EPI_ISL_2191492, EPI_ISL_2191493, EPI_ISL_2191494, EPI_ISL_2191495, EPI_ISL_2191496, EPI_ISL_2191497, EPI_ISL_2191498, EPI_ISL_2191499, EPI_ISL_2191500, EPI_ISL_2191501, EPI_ISL_2191502, EPI_ISL_2191503, EPI_ISL_2191504, EPI_ISL_2191505, EPI_ISL_2191506, EPI_ISL_2191507, EPI_ISL_2191508, EPI_ISL_2191509, EPI_ISL_2191510, EPI_ISL_2191511, EPI_ISL_2191512, EPI_ISL_2191513, EPI_ISL_2191514, EPI_ISL_2191515, EPI_ISL_2191516, EPI_ISL_2191517, EPI_ISL_2191518, EPI_ISL_2191519, EPI_ISL_2191520, EPI_ISL_2191521, EPI_ISL_2191522, EPI_ISL_2191523, EPI_ISL_2191525, EPI_ISL_2191527, EPI_ISL_2191529, EPI_ISL_2191531, EPI_ISL_2191534, EPI_ISL_2191537, EPI_ISL_2191540, EPI_ISL_2191542, EPI_ISL_2191543, EPI_ISL_2191544, EPI_ISL_2191545, EPI_ISL_2191546, EPI_ISL_2191547, EPI_ISL_2191548, EPI_ISL_2191549, EPI_ISL_2191550, EPI_ISL_2191551, EPI_ISL_2191552, EPI_ISL_2191553, EPI_ISL_2191554, EPI_ISL_2191555, EPI_ISL_2191556, EPI_ISL_2191557, EPI_ISL_2191558, EPI_ISL_2191559, EPI_ISL_2191560, EPI_ISL_2191561, EPI_ISL_2191562, EPI_ISL_2191563, EPI_ISL_2191564, EPI_ISL_2191565, EPI_ISL_2191566, EPI_ISL_2191567, EPI_ISL_2191568, EPI_ISL_2191569, EPI_ISL_2191570, EPI_ISL_2191571, EPI_ISL_2191572, EPI_ISL_2191573, EPI_ISL_2191574, EPI_ISL_2191575, EPI_ISL_2191576, EPI_ISL_2191577, EPI_ISL_2191578, EPI_ISL_2191579, EPI_ISL_2191580, EPI_ISL_2191581, EPI_ISL_2191582, EPI_ISL_2191583, EPI_ISL_2191584, EPI_ISL_2191585, EPI_ISL_2191586, EPI_ISL_2191587, EPI_ISL_2191588, EPI_ISL_2191589, EPI_ISL_2191590, EPI_ISL_2191591, EPI_ISL_2191592, EPI_ISL_2191593, EPI_ISL_2191594, EPI_ISL_2191595, EPI_ISL_2191596, EPI_ISL_2191597, EPI_ISL_2191598, EPI_ISL_2191599, EPI_ISL_2191600, EPI_ISL_2191601, EPI_ISL_2191602, EPI_ISL_2191603, EPI_ISL_2191604, EPI_ISL_2191605, EPI_ISL_2191606, EPI_ISL_2191607, EPI_ISL_2191608, EPI_ISL_2191609, EPI_ISL_2191610, EPI_ISL_2191611, EPI_ISL_2191612, EPI_ISL_2191613, EPI_ISL_2191614, EPI_ISL_2191615, EPI_ISL_2191616, EPI_ISL_2191617, EPI_ISL_2191618, EPI_ISL_2191619, EPI_ISL_2191620, EPI_ISL_2191621, EPI_ISL_2191622, EPI_ISL_2191623, EPI_ISL_2191624, EPI_ISL_2191625, EPI_ISL_2191626, EPI_ISL_2191627, EPI_ISL_2191628, EPI_ISL_2191629, EPI_ISL_2191630, EPI_ISL_2191631, EPI_ISL_2191632, EPI_ISL_2191633, EPI_ISL_2191634, EPI_ISL_2191635, EPI_ISL_2191636, EPI_ISL_2191637, EPI_ISL_2191638, EPI_ISL_2191639, EPI_ISL_2191640, EPI_ISL_2191641, EPI_ISL_2191642, EPI_ISL_2191643, EPI_ISL_2191644, EPI_ISL_2191645, EPI_ISL_2191646, EPI_ISL_2191647, EPI_ISL_2191648, EPI_ISL_2191649, EPI_ISL_2191650, EPI_ISL_2191651, EPI_ISL_2191652, EPI_ISL_2191653, EPI_ISL_2191654, EPI_ISL_2191655, EPI_ISL_2191656, EPI_ISL_2191657, EPI_ISL_2191658, EPI_ISL_2191659, EPI_ISL_2191660, EPI_ISL_2191661, EPI_ISL_2191662, EPI_ISL_2191663, EPI_ISL_2191664, EPI_ISL_2191665, EPI_ISL_2191666, EPI_ISL_2191667, EPI_ISL_2191668, EPI_ISL_2191669, EPI_ISL_2191670, EPI_ISL_2191671, EPI_ISL_2191672, EPI_ISL_2191673, EPI_ISL_2191674, EPI_ISL_2191675, EPI_ISL_2191676, EPI_ISL_2191677, EPI_ISL_2191678, EPI_ISL_2191679, EPI_ISL_2191680, EPI_ISL_2191681, EPI_ISL_2191682, EPI_ISL_2191683, EPI_ISL_2191684, EPI_ISL_2191685, EPI_ISL_2191686, EPI_ISL_2191687, EPI_ISL_2191688, EPI_ISL_2191689, EPI_ISL_2191690, EPI_ISL_2191691, EPI_ISL_2191692, EPI_ISL_2191693, EPI_ISL_2191694, EPI_ISL_2191695, EPI_ISL_2191696, EPI_ISL_2191697, EPI_ISL_2191698, EPI_ISL_2191699, EPI_ISL_2191700, EPI_ISL_2191701, EPI_ISL_2191702, EPI_ISL_2191703, EPI_ISL_2191704, EPI_ISL_2191705, EPI_ISL_2191706, EPI_ISL_2191707, EPI_ISL_2191708, EPI_ISL_2191709, EPI_ISL_2191710, EPI_ISL_2191711, EPI_ISL_2191712, EPI_ISL_2191713, EPI_ISL_2191714, EPI_ISL_2191715, EPI_ISL_2191716, EPI_ISL_2191717, EPI_ISL_2191718, EPI_ISL_2191719, EPI_ISL_2191720, EPI_ISL_2191721, EPI_ISL_2191722, EPI_ISL_2191723, EPI_ISL_2191724, EPI_ISL_2191725, EPI_ISL_2191726, EPI_ISL_2191727, EPI_ISL_2191728, EPI_ISL_2191729, EPI_ISL_2191730, EPI_ISL_2191731, EPI_ISL_2191732, EPI_ISL_2191733, EPI_ISL_2191734, EPI_ISL_2191735, EPI_ISL_2191736, EPI_ISL_2191737, EPI_ISL_2191738, EPI_ISL_2191739, EPI_ISL_2191740, EPI_ISL_2191741, EPI_ISL_2191742, EPI_ISL_2191743, EPI_ISL_2191744, EPI_ISL_2191745, EPI_ISL_2191746, EPI_ISL_2191747, EPI_ISL_2191748, EPI_ISL_2191749, EPI_ISL_2191750, EPI_ISL_2191751, EPI_ISL_2191752, EPI_ISL_2191753, EPI_ISL_2191754, EPI_ISL_2191755, EPI_ISL_2191756, EPI_ISL_2191757, EPI_ISL_2191758, EPI_ISL_2191759, EPI_ISL_2191760, EPI_ISL_2191761, EPI_ISL_2191762, EPI_ISL_2191763, EPI_ISL_2191764, EPI_ISL_2191765, EPI_ISL_2191766, EPI_ISL_2191767, EPI_ISL_2191768, EPI_ISL_2191769, EPI_ISL_2191770, EPI_ISL_2191771, EPI_ISL_2191772, EPI_ISL_2191773, EPI_ISL_2191774, EPI_ISL_2191775, EPI_ISL_2191776, EPI_ISL_2191777, EPI_ISL_2191778, EPI_ISL_2191779, EPI_ISL_2191780, EPI_ISL_2191781, EPI_ISL_2191782, EPI_ISL_2191783, EPI_ISL_2191784, EPI_ISL_2191785, EPI_ISL_2191786, EPI_ISL_2191787, EPI_ISL_2191788, EPI_ISL_2191789, EPI_ISL_2191790, EPI_ISL_2191791, EPI_ISL_2191792, EPI_ISL_2191793, EPI_ISL_2191794, EPI_ISL_2191795, EPI_ISL_2191796, EPI_ISL_2191797, EPI_ISL_2191798, EPI_ISL_2191799, EPI_ISL_2191800, EPI_ISL_2191801, EPI_ISL_2191802, EPI_ISL_2191803, EPI_ISL_2191804, EPI_ISL_2191805, EPI_ISL_2191806, EPI_ISL_2191807, EPI_ISL_2191808, EPI_ISL_2191809, EPI_ISL_2191810, EPI_ISL_2191811, EPI_ISL_2191812, EPI_ISL_2191813, EPI_ISL_2191814, EPI_ISL_2191815, EPI_ISL_2191816, EPI_ISL_2191817, EPI_ISL_2191818, EPI_ISL_2191819, EPI_ISL_2191820, EPI_ISL_2191821, EPI_ISL_2191822, EPI_ISL_2191823, EPI_ISL_2191824, EPI_ISL_2191825, EPI_ISL_2191826, EPI_ISL_2191827, EPI_ISL_2191828, EPI_ISL_2191829, EPI_ISL_2191830, EPI_ISL_2191831, EPI_ISL_2191832, EPI_ISL_2191833, EPI_ISL_2191834, EPI_ISL_2191835, EPI_ISL_2191836, EPI_ISL_2191837, EPI_ISL_2191838, EPI_ISL_2191839, EPI_ISL_2191840, EPI_ISL_2191841, EPI_ISL_2191842, EPI_ISL_2191843, EPI_ISL_2191844, EPI_ISL_2191845, EPI_ISL_2191846, EPI_ISL_2191847, EPI_ISL_2191848

see above | Houston Methodist Hospital | Houston Methodist Hospital | Randall J. Olsen, Paul A. Christensen, S. Wesley Long, Sishir Subedi, Robert Olson, Marcus Nguyen, James J. Davis, Matthew Ojeda Saavedra, Prasanti Yerramilli, Layne Pruitt, Kristina Reppond, Madison N. Shyer, Jessica Cambric, Ryan Gadd, Ilya J. Finkelstein, Jimmy Gollihar, and James M. Musser

EPI_ISL_2192589, EPI_ISL_2192590, EPI_ISL_2192591, EPI_ISL_2192592, EPI_ISL_2192593, EPI_ISL_2192594, EPI_ISL_2192595, EPI_ISL_2192596, EPI_ISL_2192597, EPI_ISL_2192598, EPI_ISL_2192599, EPI_ISL_2192600, EPI_ISL_2192602, EPI_ISL_2192603, EPI_ISL_2192604, EPI_ISL_2192605, EPI_ISL_2192606, EPI_ISL_2192607, EPI_ISL_2192608, EPI_ISL_2192609, EPI_ISL_2192610, EPI_ISL_2192611, EPI_ISL_2192612, EPI_ISL_2192613, EPI_ISL_2192614, EPI_ISL_2192615, EPI_ISL_2192616, EPI_ISL_2192617, EPI_ISL_2192618, EPI_ISL_2192619, EPI_ISL_2192620, EPI_ISL_2192621, EPI_ISL_2192622, EPI_ISL_2192623, EPI_ISL_2192624, EPI_ISL_2192625, EPI_ISL_2192626, EPI_ISL_2192627, EPI_ISL_2192628, EPI_ISL_2192629, EPI_ISL_2192630, EPI_ISL_2192631, EPI_ISL_2192632, EPI_ISL_2192633, EPI_ISL_2192634, EPI_ISL_2192635, EPI_ISL_2192636, EPI_ISL_2192637, EPI_ISL_2192638, EPI_ISL_2192639, EPI_ISL_2192640, EPI_ISL_2192641, EPI_ISL_2192642, EPI_ISL_2192643, EPI_ISL_2192644, EPI_ISL_2192645, EPI_ISL_2192646, EPI_ISL_2192647, EPI_ISL_2192648, EPI_ISL_2192649, EPI_ISL_2192650, EPI_ISL_2192651, EPI_ISL_2192652, EPI_ISL_2192653, EPI_ISL_2192654, EPI_ISL_2192655, EPI_ISL_2192656, EPI_ISL_2192657, EPI_ISL_2192658, EPI_ISL_2192659, EPI_ISL_2192660, EPI_ISL_2192661, EPI_ISL_2192662, EPI_ISL_2192663, EPI_ISL_2192664, EPI_ISL_2192665, EPI_ISL_2192666, EPI_ISL_2192667, EPI_ISL_2192668, EPI_ISL_2192669, EPI_ISL_2192670, EPI_ISL_2192671, EPI_ISL_2192672, EPI_ISL_2192673, EPI_ISL_2192674, EPI_ISL_2192675, EPI_ISL_2192676, EPI_ISL_2192677, EPI_ISL_2192678, EPI_ISL_2192679, EPI_ISL_2192680, EPI_ISL_2192681, EPI_ISL_2192682, EPI_ISL_2192683, EPI_ISL_2192684, EPI_ISL_2192685, EPI_ISL_2192686, EPI_ISL_2192687, EPI_ISL_2192688, EPI_ISL_2192689, EPI_ISL_2192690, EPI_ISL_2192691, EPI_ISL_2192692, EPI_ISL_2192693, EPI_ISL_2192694, EPI_ISL_2192695, EPI_ISL_2192696, EPI_ISL_2192697, EPI_ISL_2192698, EPI_ISL_2192699, EPI_ISL_2192700, EPI_ISL_2192701, EPI_ISL_2192702, EPI_ISL_2192703, EPI_ISL_2192704, EPI_ISL_2192705, EPI_ISL_2192706, EPI_ISL_2192707, EPI_ISL_2192708, EPI_ISL_2192709, EPI_ISL_2192710, EPI_ISL_2192711, EPI_ISL_2192712, EPI_ISL_2192713, EPI_ISL_2192714, EPI_ISL_2192715, EPI_ISL_2192716, EPI_ISL_2192717, EPI_ISL_2192718, EPI_ISL_2192719, EPI_ISL_2192720, EPI_ISL_2192721, EPI_ISL_2192722, EPI_ISL_2192723, EPI_ISL_2192724, EPI_ISL_2192725, EPI_ISL_2192726, EPI_ISL_2192727, EPI_ISL_2192728, EPI_ISL_2192729, EPI_ISL_2192730, EPI_ISL_2192731, EPI_ISL_2192732, EPI_ISL_2192733, EPI_ISL_2192735, EPI_ISL_2192736, EPI_ISL_2192737, EPI_ISL_2192738, EPI_ISL_2192739, EPI_ISL_2192740, EPI_ISL_2192741, EPI_ISL_2192742, EPI_ISL_2192743, EPI_ISL_2192744, EPI_ISL_2192745, EPI_ISL_2192746, EPI_ISL_2192747, EPI_ISL_2192748, EPI_ISL_2192749, EPI_ISL_2192750, EPI_ISL_2192751, EPI_ISL_2192752, EPI_ISL_2192753, EPI_ISL_2192754, EPI_ISL_2192755, EPI_ISL_2192756, EPI_ISL_2192757, EPI_ISL_2192758, EPI_ISL_2192759, EPI_ISL_2192760, EPI_ISL_2192761, EPI_ISL_2192762, EPI_ISL_2192763, EPI_ISL_2192764, EPI_ISL_2192765, EPI_ISL_2192766, EPI_ISL_2192767, EPI_ISL_2192768, EPI_ISL_2192769, EPI_ISL_2192770, EPI_ISL_2192771, EPI_ISL_2192772, EPI_ISL_2192773, EPI_ISL_2192774, EPI_ISL_2192775, EPI_ISL_2192776, EPI_ISL_2192777, EPI_ISL_2192778, EPI_ISL_2192779, EPI_ISL_2192780, EPI_ISL_2192781, EPI_ISL_2192782, EPI_ISL_2192783, EPI_ISL_2192784, EPI_ISL_2192785, EPI_ISL_2192786, EPI_ISL_2192787, EPI_ISL_2192788, EPI_ISL_2192789, EPI_ISL_2192790, EPI_ISL_2192791, EPI_ISL_2192792, EPI_ISL_2192793, EPI_ISL_2192794, EPI_ISL_2192796, EPI_ISL_2192797, EPI_ISL_2192798, EPI_ISL_2192799, EPI_ISL_2192800, EPI_ISL_2192801, EPI_ISL_2192802, EPI_ISL_2192803, EPI_ISL_2192804, EPI_ISL_2192805, EPI_ISL_2192806, EPI_ISL_2192807, EPI_ISL_2192808, EPI_ISL_2192809, EPI_ISL_2192810, EPI_ISL_2192811, EPI_ISL_2192812, EPI_ISL_2192813, EPI_ISL_2192814, EPI_ISL_2192815, EPI_ISL_2192816, EPI_ISL_2192817, EPI_ISL_2192818, EPI_ISL_2192819, EPI_ISL_2192820, EPI_ISL_2192821, EPI_ISL_2192822, EPI_ISL_2192823, EPI_ISL_2192824, EPI_ISL_2192825, EPI_ISL_2192826, EPI_ISL_2192827, EPI_ISL_2192828, EPI_ISL_2192829, EPI_ISL_2192830, EPI_ISL_2192831, EPI_ISL_2192832, EPI_ISL_2192833, EPI_ISL_2192834, EPI_ISL_2192835, EPI_ISL_2192836, EPI_ISL_2192837, EPI_ISL_2192838, EPI_ISL_2192839, EPI_ISL_2192841, EPI_ISL_2192842, EPI_ISL_2192843, EPI_ISL_2192845, EPI_ISL_2192846, EPI_ISL_2192847, EPI_ISL_2192848, EPI_ISL_2192849, EPI_ISL_2192850, EPI_ISL_2192851, EPI_ISL_2192852, EPI_ISL_2192853, EPI_ISL_2192854, EPI_ISL_2192855, EPI_ISL_2192856, EPI_ISL_2192857, EPI_ISL_2192858, EPI_ISL_2192860, EPI_ISL_2192861, EPI_ISL_2192862, EPI_ISL_2192863, EPI_ISL_2192864, EPI_ISL_2192865, EPI_ISL_2192866, EPI_ISL_2192867, EPI_ISL_2192868, EPI_ISL_2192869, EPI_ISL_2192870, EPI_ISL_2192871, EPI_ISL_2192873, EPI_ISL_2192874, EPI_ISL_2192875, EPI_ISL_2192876, EPI_ISL_2192878, EPI_ISL_2192879, EPI_ISL_2192880, EPI_ISL_2192881, EPI_ISL_2192882, EPI_ISL_2192883, EPI_ISL_2192884, EPI_ISL_2192885, EPI_ISL_2192886, EPI_ISL_2192887, EPI_ISL_2192888, EPI_ISL_2192889, EPI_ISL_2192890, EPI_ISL_2192891, EPI_ISL_2192892, EPI_ISL_2192893, EPI_ISL_2192894, EPI_ISL_2192895, EPI_ISL_2192896, EPI_ISL_2192897, EPI_ISL_2192898, EPI_ISL_2192899, EPI_ISL_2192900, EPI_ISL_2192901, EPI_ISL_2192902, EPI_ISL_2192903, EPI_ISL_2192904, EPI_ISL_2192905, EPI_ISL_2192906, EPI_ISL_2192908, EPI_ISL_2192909, EPI_ISL_2192910, EPI_ISL_2192911, EPI_ISL_2192912, EPI_ISL_2192913, EPI_ISL_2192914, EPI_ISL_2192916, EPI_ISL_2192917, EPI_ISL_2192918, EPI_ISL_2192919, EPI_ISL_2192920, EPI_ISL_2192921, EPI_ISL_2192922, EPI_ISL_2192923, EPI_ISL_2192924, EPI_ISL_2192925, EPI_ISL_2192926, EPI_ISL_2192927, EPI_ISL_2192928, EPI_ISL_2192929, EPI_ISL_2192930, EPI_ISL_2192931, EPI_ISL_2192932, EPI_ISL_2192933, EPI_ISL_2192934, EPI_ISL_2192935, EPI_ISL_2192936, EPI_ISL_2192937, EPI_ISL_2192938, EPI_ISL_2192939, EPI_ISL_2192940, EPI_ISL_2192941, EPI_ISL_2192942, EPI_ISL_2192943, EPI_ISL_2192944, EPI_ISL_2192945, EPI_ISL_2192946, EPI_ISL_2192947, EPI_ISL_2192948, EPI_ISL_2192949, EPI_ISL_2192950, EPI_ISL_2192951, EPI_ISL_2192952, EPI_ISL_2192953, EPI_ISL_2192954, EPI_ISL_2192955, EPI_ISL_2192956, EPI_ISL_2192957, EPI_ISL_2192958, EPI_ISL_2192960, EPI_ISL_2192961, EPI_ISL_2192962, EPI_ISL_2192963, EPI_ISL_2192964, EPI_ISL_2192965, EPI_ISL_2192966, EPI_ISL_2192967, EPI_ISL_2192968, EPI_ISL_2192969, EPI_ISL_2192970, EPI_ISL_2192971, EPI_ISL_2192972, EPI_ISL_2192973, EPI_ISL_2192974, EPI_ISL_2192975, EPI_ISL_2192976, EPI_ISL_2192977, EPI_ISL_2192978, EPI_ISL_2192979, EPI_ISL_2192980, EPI_ISL_2192981, EPI_ISL_2192982, EPI_ISL_2192983, EPI_ISL_2192984, EPI_ISL_2192985, EPI_ISL_2192986, EPI_ISL_2192987, EPI_ISL_2192988, EPI_ISL_2192989, EPI_ISL_2192990, EPI_ISL_2192991, EPI_ISL_2192992, EPI_ISL_2192993, EPI_ISL_2192994, EPI_ISL_2192995, EPI_ISL_2192996, EPI_ISL_2192997, EPI_ISL_2192998, EPI_ISL_2192999, EPI_ISL_2193000, EPI_ISL_2193001, EPI_ISL_2193002, EPI_ISL_2193003, EPI_ISL_2193004, EPI_ISL_2193005, EPI_ISL_2193006, EPI_ISL_2193007, EPI_ISL_2193008, EPI_ISL_2193009, EPI_ISL_2193010, EPI_ISL_2193011, EPI_ISL_2193012, EPI_ISL_2193013, EPI_ISL_2193014, EPI_ISL_2193015, EPI_ISL_2193016, EPI_ISL_2193017, EPI_ISL_2193018, EPI_ISL_2193019, EPI_ISL_2193020, EPI_ISL_2193021, EPI_ISL_2193022, EPI_ISL_2193023, EPI_ISL_2193024, EPI_ISL_2193025, EPI_ISL_2193026, EPI_ISL_2193027, EPI_ISL_2193028, EPI_ISL_2193029, EPI_ISL_2193030, EPI_ISL_2193031, EPI_ISL_2193032, EPI_ISL_2193033, EPI_ISL_2193034, EPI_ISL_2193035, EPI_ISL_2193036, EPI_ISL_2193037, EPI_ISL_2193038, EPI_ISL_2193039, EPI_ISL_2193040, EPI_ISL_2193041, EPI_ISL_2193042, EPI_ISL_2193043, EPI_ISL_2193044, EPI_ISL_2193045, EPI_ISL_2193046, EPI_ISL_2193047, EPI_ISL_2193048, EPI_ISL_2193049, EPI_ISL_2193050, EPI_ISL_2193051, EPI_ISL_2193052, EPI_ISL_2193053, EPI_ISL_2193054, EPI_ISL_2193055, EPI_ISL_2193056, EPI_ISL_2193057, EPI_ISL_2193058, EPI_ISL_2193059, EPI_ISL_2193060, EPI_ISL_2193061, EPI_ISL_2193062, EPI_ISL_2193063, EPI_ISL_2193064, EPI_ISL_2193065, EPI_ISL_2193066, EPI_ISL_2193067, EPI_ISL_2193068, EPI_ISL_2193069, EPI_ISL_2193070, EPI_ISL_2193071, EPI_ISL_2193072, EPI_ISL_2193073, EPI_ISL_2193074, EPI_ISL_2193075, EPI_ISL_2193076, EPI_ISL_2193077, EPI_ISL_2193078, EPI_ISL_2193079, EPI_ISL_2193080, EPI_ISL_2193081, EPI_ISL_2193082, EPI_ISL_2193083, EPI_ISL_2193084, EPI_ISL_2193085, EPI_ISL_2193087, EPI_ISL_2193088, EPI_ISL_2193089, EPI_ISL_2193090, EPI_ISL_2193091, EPI_ISL_2193092, EPI_ISL_2193093, EPI_ISL_2193094, EPI_ISL_2193095, EPI_ISL_2193096, EPI_ISL_2193097, EPI_ISL_2193098, EPI_ISL_2193099, EPI_ISL_2193100, EPI_ISL_2193101, EPI_ISL_2193102, EPI_ISL_2193103, EPI_ISL_2193104, EPI_ISL_2193105, EPI_ISL_2193106, EPI_ISL_2193107, EPI_ISL_2193108, EPI_ISL_2193109, EPI_ISL_2193110, EPI_ISL_2193111, EPI_ISL_2193112, EPI_ISL_2193113, EPI_ISL_2193114, EPI_ISL_2193115, EPI_ISL_2193116, EPI_ISL_2193117, EPI_ISL_2193118, EPI_ISL_2193119, EPI_ISL_2193120, EPI_ISL_2193121, EPI_ISL_2193122, EPI_ISL_2193123, EPI_ISL_2193124, EPI_ISL_2193125, EPI_ISL_2193126, EPI_ISL_2193127, EPI_ISL_2193128, EPI_ISL_2193129, EPI_ISL_2193130, EPI_ISL_2193131, EPI_ISL_2193132, EPI_ISL_2193133, EPI_ISL_2193134, EPI_ISL_2193135, EPI_ISL_2193136, EPI_ISL_2193137, EPI_ISL_2193138, EPI_ISL_2193139, EPI_ISL_2193140, EPI_ISL_2193141, EPI_ISL_2193142, EPI_ISL_2193143, EPI_ISL_2193144, EPI_ISL_2193145, EPI_ISL_2193146, EPI_ISL_2193147, EPI_ISL_2193148, EPI_ISL_2193149, EPI_ISL_2193150, EPI_ISL_2193151, EPI_ISL_2193152, EPI_ISL_2193153, EPI_ISL_2193154, EPI_ISL_2193155, EPI_ISL_2193156, EPI_ISL_2193157, EPI_ISL_2193158, EPI_ISL_2193159, EPI_ISL_2193160, EPI_ISL_2193161, EPI_ISL_2193162, EPI_ISL_2193163, EPI_ISL_2193165, EPI_ISL_2193166, EPI_ISL_2193167, EPI_ISL_2193168, EPI_ISL_2193169, EPI_ISL_2193170, EPI_ISL_2193171, EPI_ISL_2193172, EPI_ISL_2193173, EPI_ISL_2193174, EPI_ISL_2193175, EPI_ISL_2193176, EPI_ISL_2193177, EPI_ISL_2193178, EPI_ISL_2193179, EPI_ISL_2193180, EPI_ISL_2193181, EPI_ISL_2193182, EPI_ISL_2193183, EPI_ISL_2193184, EPI_ISL_2193185, EPI_ISL_2193186, EPI_ISL_2193187, EPI_ISL_2193188, EPI_ISL_2193189, EPI_ISL_2193190, EPI_ISL_2193191, EPI_ISL_2193192, EPI_ISL_2193193, EPI_ISL_2193194, EPI_ISL_2193195, EPI_ISL_2193196, EPI_ISL_2193197, EPI_ISL_2193198, EPI_ISL_2193199, EPI_ISL_2193200, EPI_ISL_2193201, EPI_ISL_2193202, EPI_ISL_2193203, EPI_ISL_2193204, EPI_ISL_2193205, EPI_ISL_2193206, EPI_ISL_2193207, EPI_ISL_2193209, EPI_ISL_2193210, EPI_ISL_2193211, EPI_ISL_2193212, EPI_ISL_2193213, EPI_ISL_2193214, EPI_ISL_2193215, EPI_ISL_2193216, EPI_ISL_2193217

see above | Broad Institute Clinical Research Sequencing Platform | Infectious Disease Program, Broad Institute of Harvard and MIT | Siddle,K.J., Adams,G., Pearlman,L., Gladden-Young,A., Vicente,G., Blumenstiel,B., DeFelice,M., Lee,M., McGovern,S., Lagerborg,K., Rudy,M., DeRuff,K., Carter,A., Normandin,E., Bauer,M., Reilly,S., Tomkins-Tinch,C., Loreth,C., Chaluvadi,S., Meldrim,J., Granger,B., Lemieux,J.E., Birren,B.W., Sabeti,P.C., Larkin,K., Dodge,S., Lennon,N., Madoff,L., Brown,C., Gallagher,G., Smole,S., Park,D.J., Gabriel,S., and MacInnis,B.L.

EPI_ISL_2193229, EPI_ISL_2193230, EPI_ISL_2193231, EPI_ISL_2193232, EPI_ISL_2193233, EPI_ISL_2193234, EPI_ISL_2193235, EPI_ISL_2193236, EPI_ISL_2193237, EPI_ISL_2193238, EPI_ISL_2193239, EPI_ISL_2193240, EPI_ISL_2193241, EPI_ISL_2193242, EPI_ISL_2193243, EPI_ISL_2193244, EPI_ISL_2193245, EPI_ISL_2193246, EPI_ISL_2193247, EPI_ISL_2193248, EPI_ISL_2193249, EPI_ISL_2193250, EPI_ISL_2193251, EPI_ISL_2193252, EPI_ISL_2193253, EPI_ISL_2193254, EPI_ISL_2193255, EPI_ISL_2193256, EPI_ISL_2193257, EPI_ISL_2193258, EPI_ISL_2193259, EPI_ISL_2193260, EPI_ISL_2193261, EPI_ISL_2193262, EPI_ISL_2193263, EPI_ISL_2193264, EPI_ISL_2193265, EPI_ISL_2193266, EPI_ISL_2193267, EPI_ISL_2193268, EPI_ISL_2193269, EPI_ISL_2193270, EPI_ISL_2193271, EPI_ISL_2193272, EPI_ISL_2193273, EPI_ISL_2193274, EPI_ISL_2193275, EPI_ISL_2193276, EPI_ISL_2193277, EPI_ISL_2193278, EPI_ISL_2193279, EPI_ISL_2193280, EPI_ISL_2193281, EPI_ISL_2193282, EPI_ISL_2193283, EPI_ISL_2193284, EPI_ISL_2193285

see above | Baylor Scott & White-Temple | Baylor Scott & White-Temple | Ari Rao, Linden Morales, Kimberly Walker, Marcus Volz, Shelby Hendrickson

EPI_ISL_2193305, EPI_ISL_2193306, EPI_ISL_2193308, EPI_ISL_2193309, EPI_ISL_2193311, EPI_ISL_2193315, EPI_ISL_2193319, EPI_ISL_2193321, EPI_ISL_2193327, EPI_ISL_2193333, EPI_ISL_2193334, EPI_ISL_2193335, EPI_ISL_2193341

see above | Wyoming Public Health Laboratory | Wyoming Public Health Laboratory | Jim Mildenberger, Wanda Manley, Noah Hull, Taylor Fearing, Lynette Gumbleton, Channing Weber, Ashley Norberg, Chayse Rowley, Marley Goetz, Brian Dominguez, Elliot Thomasson, Sam Britz, Cari Sloma, and Rob Christensen

EPI_ISL_2193365, EPI_ISL_2193366, EPI_ISL_2193367, EPI_ISL_2193368, EPI_ISL_2193369, EPI_ISL_2193370, EPI_ISL_2193371, EPI_ISL_2193372, EPI_ISL_2193373, EPI_ISL_2193374, EPI_ISL_2193375, EPI_ISL_2193376, EPI_ISL_2193377, EPI_ISL_2193378, EPI_ISL_2193379, EPI_ISL_2193380, EPI_ISL_2193381

see above | University of Wisconsin-Madison AIDS Vaccine Research Laboratories | University of Wisconsin-Madison AIDS Vaccine Research Laboratories | Gage Moreno, Katarina Braun, et al. AIDS Vaccine Research Laboratories

EPI_ISL_2193384, EPI_ISL_2193385, EPI_ISL_2193386, EPI_ISL_2193387, EPI_ISL_2193388, EPI_ISL_2193389, EPI_ISL_2193390, EPI_ISL_2193391, EPI_ISL_2193392, EPI_ISL_2193393, EPI_ISL_2193394, EPI_ISL_2193395, EPI_ISL_2193396, EPI_ISL_2193397, EPI_ISL_2193398, EPI_ISL_2193399, EPI_ISL_2193400, EPI_ISL_2193401, EPI_ISL_2193402, EPI_ISL_2193403, EPI_ISL_2193404, EPI_ISL_2193405, EPI_ISL_2193406, EPI_ISL_2193407, EPI_ISL_2193408, EPI_ISL_2193409, EPI_ISL_2193410, EPI_ISL_2193411, EPI_ISL_2193412, EPI_ISL_2193413, EPI_ISL_2193414, EPI_ISL_2193415, EPI_ISL_2193416, EPI_ISL_2193417, EPI_ISL_2193418, EPI_ISL_2193419, EPI_ISL_2193420, EPI_ISL_2193421, EPI_ISL_2193422, EPI_ISL_2193423, EPI_ISL_2193424, EPI_ISL_2193425, EPI_ISL_2193426, EPI_ISL_2193427, EPI_ISL_2193428, EPI_ISL_2193429, EPI_ISL_2193430, EPI_ISL_2193431, EPI_ISL_2193432, EPI_ISL_2193433, EPI_ISL_2193434, EPI_ISL_2193435, EPI_ISL_2193436, EPI_ISL_2193437, EPI_ISL_2193438, EPI_ISL_2193439, EPI_ISL_2193440, EPI_ISL_2193441, EPI_ISL_2193442, EPI_ISL_2193443, EPI_ISL_2193444, EPI_ISL_2193445, EPI_ISL_2193446, EPI_ISL_2193447, EPI_ISL_2193448, EPI_ISL_2193449, EPI_ISL_2193450, EPI_ISL_2193451

see above | University Hospitals Translational Laboratory (UHTL), University Hospitals | University Hospitals Translational Laboratory (UHTL), University Hospitals | Sadri,N., Alouani,D., Song,X.

EPI_ISL_2193453, EPI_ISL_2193454, EPI_ISL_2193455, EPI_ISL_2193456, EPI_ISL_2193457, EPI_ISL_2193458, EPI_ISL_2193459, EPI_ISL_2193460, EPI_ISL_2193461, EPI_ISL_2193462, EPI_ISL_2193463, EPI_ISL_2193464, EPI_ISL_2193465, EPI_ISL_2193466, EPI_ISL_2193467, EPI_ISL_2193468, EPI_ISL_2193469, EPI_ISL_2193470, EPI_ISL_2193471, EPI_ISL_2193472, EPI_ISL_2193473, EPI_ISL_2193474, EPI_ISL_2193475, EPI_ISL_2193476, EPI_ISL_2193477, EPI_ISL_2193478, EPI_ISL_2193479, EPI_ISL_2193480, EPI_ISL_2193481, EPI_ISL_2193482, EPI_ISL_2193483, EPI_ISL_2193484, EPI_ISL_2193485, EPI_ISL_2193486, EPI_ISL_2193487, EPI_ISL_2193488, EPI_ISL_2193489, EPI_ISL_2193490, EPI_ISL_2193491, EPI_ISL_2193492, EPI_ISL_2193493, EPI_ISL_2193494, EPI_ISL_2193495, EPI_ISL_2193496, EPI_ISL_2193497, EPI_ISL_2193498, EPI_ISL_2193499, EPI_ISL_2193500, EPI_ISL_2193501, EPI_ISL_2193502, EPI_ISL_2193503, EPI_ISL_2193504, EPI_ISL_2193505, EPI_ISL_2193506, EPI_ISL_2193507, EPI_ISL_2193508, EPI_ISL_2193509, EPI_ISL_2193510, EPI_ISL_2193511, EPI_ISL_2193512, EPI_ISL_2193513, EPI_ISL_2193514, EPI_ISL_2193515, EPI_ISL_2193516, EPI_ISL_2193517, EPI_ISL_2193518, EPI_ISL_2193519, EPI_ISL_2193520, EPI_ISL_2193521, EPI_ISL_2193522, EPI_ISL_2193523, EPI_ISL_2193524, EPI_ISL_2193525, EPI_ISL_2193526, EPI_ISL_2193527, EPI_ISL_2193528, EPI_ISL_2193529, EPI_ISL_2193530, EPI_ISL_2193531, EPI_ISL_2193532, EPI_ISL_2193533, EPI_ISL_2193534, EPI_ISL_2193535, EPI_ISL_2193536, EPI_ISL_2193537, EPI_ISL_2193538, EPI_ISL_2193539, EPI_ISL_2193540, EPI_ISL_2193541, EPI_ISL_2193542, EPI_ISL_2193543, EPI_ISL_2193544, EPI_ISL_2193545, EPI_ISL_2193546, EPI_ISL_2193547, EPI_ISL_2193548, EPI_ISL_2193549, EPI_ISL_2193550, EPI_ISL_2193551, EPI_ISL_2193552, EPI_ISL_2193553, EPI_ISL_2193554, EPI_ISL_2193555, EPI_ISL_2193556, EPI_ISL_2193557, EPI_ISL_2193558, EPI_ISL_2193559, EPI_ISL_2193560, EPI_ISL_2193561, EPI_ISL_2193562,

| | | | |
|---|---|---|---|
| EPI_ISL_2193563, EPI_ISL_2193564, EPI_ISL_2193565, EPI_ISL_2193566, EPI_ISL_2193567, EPI_ISL_2193568, EPI_ISL_2193569, EPI_ISL_2193570, EPI_ISL_2193571, EPI_ISL_2193572, EPI_ISL_2193573, EPI_ISL_2193574, EPI_ISL_2193575, EPI_ISL_2193576, EPI_ISL_2193577, EPI_ISL_2193578, EPI_ISL_2193579, EPI_ISL_2193580, EPI_ISL_2193581, EPI_ISL_2193582, EPI_ISL_2193583, EPI_ISL_2193584, EPI_ISL_2193585, EPI_ISL_2193586, EPI_ISL_2193587, EPI_ISL_2193588, EPI_ISL_2193589, EPI_ISL_2193590, EPI_ISL_2193591, EPI_ISL_2193592, EPI_ISL_2193593, EPI_ISL_2193594, EPI_ISL_2193595, EPI_ISL_2193596, EPI_ISL_2193597, EPI_ISL_2193598, EPI_ISL_2193599, EPI_ISL_2193600, EPI_ISL_2193601, EPI_ISL_2193602, EPI_ISL_2193603, EPI_ISL_2193604, EPI_ISL_2193605, EPI_ISL_2193606, EPI_ISL_2193607, EPI_ISL_2193608, EPI_ISL_2193609, EPI_ISL_2193610, EPI_ISL_2193611, EPI_ISL_2193612, EPI_ISL_2193613, EPI_ISL_2193614, EPI_ISL_2193615, EPI_ISL_2193616, EPI_ISL_2193617, EPI_ISL_2193618, EPI_ISL_2193619, EPI_ISL_2193620, EPI_ISL_2193621, EPI_ISL_2193622, EPI_ISL_2193623, EPI_ISL_2193624, EPI_ISL_2193625, EPI_ISL_2193626, EPI_ISL_2193627, EPI_ISL_2193628, EPI_ISL_2193629, EPI_ISL_2193630, EPI_ISL_2193631, EPI_ISL_2193632, EPI_ISL_2193633, EPI_ISL_2193634, EPI_ISL_2193635, EPI_ISL_2193636, EPI_ISL_2193637, EPI_ISL_2193638, EPI_ISL_2193639, EPI_ISL_2193640, EPI_ISL_2193641, EPI_ISL_2193642, EPI_ISL_2193643, EPI_ISL_2193644, EPI_ISL_2193645, EPI_ISL_2193646, EPI_ISL_2193647, EPI_ISL_2193648, EPI_ISL_2193649, EPI_ISL_2193650, EPI_ISL_2193651, EPI_ISL_2193652, EPI_ISL_2193653, EPI_ISL_2193654, EPI_ISL_2193655, EPI_ISL_2193656, EPI_ISL_2193657, EPI_ISL_2193658, EPI_ISL_2193659, EPI_ISL_2193660, EPI_ISL_2193661, EPI_ISL_2193662, EPI_ISL_2193663, EPI_ISL_2193664, EPI_ISL_2193665, EPI_ISL_2193666, EPI_ISL_2193667, EPI_ISL_2193668, EPI_ISL_2193669, EPI_ISL_2193670, EPI_ISL_2193671, EPI_ISL_2193672, EPI_ISL_2193673, EPI_ISL_2193674, EPI_ISL_2193675, EPI_ISL_2193676, EPI_ISL_2193677, EPI_ISL_2193678, EPI_ISL_2193679, EPI_ISL_2193680, EPI_ISL_2193681, EPI_ISL_2193682, EPI_ISL_2193683, EPI_ISL_2193684, EPI_ISL_2193685, EPI_ISL_2193686, EPI_ISL_2193687, EPI_ISL_2193688, EPI_ISL_2193689, EPI_ISL_2193690, EPI_ISL_2193691, EPI_ISL_2193692, EPI_ISL_2193693, EPI_ISL_2193694, EPI_ISL_2193695, EPI_ISL_2193696, EPI_ISL_2193697, EPI_ISL_2193698, EPI_ISL_2193699, EPI_ISL_2193700, EPI_ISL_2193701, EPI_ISL_2193702, EPI_ISL_2193703, EPI_ISL_2193705, EPI_ISL_2193706, EPI_ISL_2193707, EPI_ISL_2193708, EPI_ISL_2193709, EPI_ISL_2193710, EPI_ISL_2193711, EPI_ISL_2193712, EPI_ISL_2193713, EPI_ISL_2193714, EPI_ISL_2193715, EPI_ISL_2193716, EPI_ISL_2193717, EPI_ISL_2193718, EPI_ISL_2193719, EPI_ISL_2193720, EPI_ISL_2193721, EPI_ISL_2193722, EPI_ISL_2193723, EPI_ISL_2193724, EPI_ISL_2193725, EPI_ISL_2193726, EPI_ISL_2193727, EPI_ISL_2193728, EPI_ISL_2193729, EPI_ISL_2193730, EPI_ISL_2193731, EPI_ISL_2193732, EPI_ISL_2193733, EPI_ISL_2193734, EPI_ISL_2193735, EPI_ISL_2193736, EPI_ISL_2193737, EPI_ISL_2193738, EPI_ISL_2193739, EPI_ISL_2193740, EPI_ISL_2193741, EPI_ISL_2193742, EPI_ISL_2193743, EPI_ISL_2193744, EPI_ISL_2193745, EPI_ISL_2193746, EPI_ISL_2193747, EPI_ISL_2193748, EPI_ISL_2193749, EPI_ISL_2193750, EPI_ISL_2193751, EPI_ISL_2193752, EPI_ISL_2193753, EPI_ISL_2193754, EPI_ISL_2193755, EPI_ISL_2193756, EPI_ISL_2193757, EPI_ISL_2193758, EPI_ISL_2193759, EPI_ISL_2193760, EPI_ISL_2193761 | | | |
| see above | Hackensack Medical Center | New York Genome Center | Michael Zody, Andre Corvelo, Dayna M. Oschwald, Samantha Fennessey, Tom Maniatis, Liang Chen, Jose Mediavilla, Marcus Cunningham, Kaelea Composto, Kar Chow, David Perlin, Barry Kreiswirth |
| EPI_ISL_2193762, EPI_ISL_2193763, EPI_ISL_2193764, EPI_ISL_2193765, EPI_ISL_2193766 | Weill Cornell Medicine | New York Genome Center | Michael Zody, Andre Corvelo, Dayna M. Oschwald, Samantha Fennessey, Tom Maniatis, Melissa Cushing, Olivier Elemento, Margaret Elizabeth Ross, Chris Mason, Priya Velu, Hanna Rennert, Arryn Craney, Lars F Westblade |
| EPI_ISL_2193781, EPI_ISL_2193782, EPI_ISL_2193783, EPI_ISL_2193784, EPI_ISL_2193785, EPI_ISL_2193786, EPI_ISL_2193787, EPI_ISL_2193788, EPI_ISL_2193789, EPI_ISL_2193790, EPI_ISL_2193791, EPI_ISL_2193792, EPI_ISL_2193793, EPI_ISL_2193794, EPI_ISL_2193795, EPI_ISL_2193796, EPI_ISL_2193797, EPI_ISL_2193798, EPI_ISL_2193799, EPI_ISL_2193800, EPI_ISL_2193801, EPI_ISL_2193802, EPI_ISL_2193803, EPI_ISL_2193804, EPI_ISL_2193805, EPI_ISL_2193806, EPI_ISL_2193807, EPI_ISL_2193808, EPI_ISL_2193809, EPI_ISL_2193810, EPI_ISL_2193811 | | | |
| see above | Hackensack Medical Center | New York Genome Center | Michael Zody, Andre Corvelo, Dayna M. Oschwald, Samantha Fennessey, Tom Maniatis, Liang Chen, Jose Mediavilla, Marcus Cunningham, Kaelea Composto, Kar Chow, David Perlin, Barry Kreiswirth |
| EPI_ISL_2193968, EPI_ISL_2193969, EPI_ISL_2193970, EPI_ISL_2193971, EPI_ISL_2193972, EPI_ISL_2193973, EPI_ISL_2193974, EPI_ISL_2193975, EPI_ISL_2193976, EPI_ISL_2193977, EPI_ISL_2193979, EPI_ISL_2193980, EPI_ISL_2193981, EPI_ISL_2193982 | | | |
| see above | Wyoming Public Health Laboratory | Wyoming Public Health Laboratory | Jim Mildenberger, Wanda Manley, Noah Hull, Taylor Fearing, Lynette Gumbleton, Channing Weber, Ashley Norberg, Chayse Rowley, Marley Goetz, Brian Dominguez, Elliot Thomasson, Sam Britz, Cari Sloma, and Rob Christensen |
| EPI_ISL_2194037, EPI_ISL_2194038, EPI_ISL_2194039, EPI_ISL_2194040, EPI_ISL_2194042, EPI_ISL_2194043, EPI_ISL_2194044, EPI_ISL_2194045, EPI_ISL_2194046, EPI_ISL_2194047, EPI_ISL_2194048, EPI_ISL_2194049, EPI_ISL_2194050, EPI_ISL_2194051, EPI_ISL_2194053, EPI_ISL_2194054, EPI_ISL_2194055, EPI_ISL_2194056, EPI_ISL_2194057, EPI_ISL_2194058, EPI_ISL_2194059, EPI_ISL_2194060, EPI_ISL_2194061, EPI_ISL_2194062, EPI_ISL_2194063, EPI_ISL_2194064, EPI_ISL_2194065, EPI_ISL_2194066, EPI_ISL_2194067, EPI_ISL_2194068, EPI_ISL_2194070, EPI_ISL_2194071, EPI_ISL_2194072, EPI_ISL_2194073, EPI_ISL_2194074, EPI_ISL_2194076, EPI_ISL_2194077, EPI_ISL_2194078, EPI_ISL_2194079, EPI_ISL_2194080, EPI_ISL_2194081, EPI_ISL_2194082, EPI_ISL_2194083, EPI_ISL_2194084, EPI_ISL_2194085, EPI_ISL_2194086, EPI_ISL_2194087, EPI_ISL_2194088, EPI_ISL_2194089, EPI_ISL_2194090, EPI_ISL_2194091, EPI_ISL_2194092, EPI_ISL_2194093, EPI_ISL_2194094, EPI_ISL_2194095, EPI_ISL_2194096, EPI_ISL_2194097, EPI_ISL_2194098, EPI_ISL_2194099, EPI_ISL_2194101, EPI_ISL_2194102, EPI_ISL_2194103, EPI_ISL_2194104, EPI_ISL_2194105, EPI_ISL_2194106, EPI_ISL_2194107, EPI_ISL_2194108, EPI_ISL_2194110, EPI_ISL_2194111, EPI_ISL_2194112, EPI_ISL_2194113, EPI_ISL_2194115, EPI_ISL_2194116, EPI_ISL_2194117, EPI_ISL_2194118, EPI_ISL_2194119, EPI_ISL_2194120, EPI_ISL_2194121, EPI_ISL_2194122, EPI_ISL_2194123, EPI_ISL_2194124, EPI_ISL_2194125, EPI_ISL_2194126, EPI_ISL_2194127, EPI_ISL_2194128, EPI_ISL_2194129, EPI_ISL_2194130, EPI_ISL_2194131, EPI_ISL_2194133, EPI_ISL_2194134, EPI_ISL_2194136, EPI_ISL_2194137, EPI_ISL_2194138, EPI_ISL_2194139, EPI_ISL_2194140, EPI_ISL_2194141, EPI_ISL_2194142, EPI_ISL_2194143, EPI_ISL_2194144, EPI_ISL_2194145, EPI_ISL_2194146, EPI_ISL_2194147, EPI_ISL_2194148, EPI_ISL_2194149, EPI_ISL_2194150, EPI_ISL_2194151, EPI_ISL_2194152, EPI_ISL_2194153, EPI_ISL_2194154, EPI_ISL_2194155, EPI_ISL_2194156, EPI_ISL_2194158, EPI_ISL_2194160, EPI_ISL_2194161, EPI_ISL_2194162, EPI_ISL_2194163, EPI_ISL_2194164, EPI_ISL_2194165, EPI_ISL_2194166, EPI_ISL_2194167, EPI_ISL_2194168, EPI_ISL_2194169, EPI_ISL_2194170, EPI_ISL_2194171, EPI_ISL_2194172, EPI_ISL_2194173, EPI_ISL_2194174, EPI_ISL_2194175, EPI_ISL_2194176, EPI_ISL_2194177, EPI_ISL_2194178, EPI_ISL_2194179, EPI_ISL_2194180, EPI_ISL_2194181, EPI_ISL_2194182, EPI_ISL_2194183, EPI_ISL_2194184, EPI_ISL_2194186, EPI_ISL_2194187, EPI_ISL_2194188, EPI_ISL_2194189, EPI_ISL_2194190, EPI_ISL_2194191, EPI_ISL_2194192, EPI_ISL_2194193, EPI_ISL_2194194, EPI_ISL_2194195, EPI_ISL_2194196, EPI_ISL_2194197, EPI_ISL_2194198, EPI_ISL_2194199, EPI_ISL_2194200, EPI_ISL_2194201, EPI_ISL_2194202, EPI_ISL_2194203, EPI_ISL_2194204, EPI_ISL_2194205, EPI_ISL_2194206, EPI_ISL_2194207, EPI_ISL_2194208, EPI_ISL_2194209, EPI_ISL_2194210, EPI_ISL_2194211, EPI_ISL_2194212, EPI_ISL_2194213, EPI_ISL_2194214, EPI_ISL_2194215, EPI_ISL_2194216 | | | |
| see above | UW Virology Lab | UW Virology Lab | Pavitra Roychoudhury, Hong Xie, Lasata Shrestha, Tien V. Nguyen, Shah Mohamed Bakhash, Michelle Lin, Noah R. Baker, Ricardo Perez, Sean Ellis, Nathan Breit, Robert J. Livingston, Meei-Li Huang, Keith R Jerome, Patrick Mathias, Alexander Greninger |
| EPI_ISL_2194546, EPI_ISL_2194547, EPI_ISL_2194548, EPI_ISL_2194549, EPI_ISL_2194550, EPI_ISL_2194551, EPI_ISL_2194552, EPI_ISL_2194553, EPI_ISL_2194555, EPI_ISL_2194557, EPI_ISL_2194558, EPI_ISL_2194559, EPI_ISL_2194561, EPI_ISL_2194562, EPI_ISL_2194563, EPI_ISL_2194565, EPI_ISL_2194567, EPI_ISL_2194568, EPI_ISL_2194572, EPI_ISL_2194574, EPI_ISL_2194575, EPI_ISL_2194576, EPI_ISL_2194577, EPI_ISL_2194578, EPI_ISL_2194580, EPI_ISL_2194581, EPI_ISL_2194582, EPI_ISL_2194583, EPI_ISL_2194585, EPI_ISL_2194586, EPI_ISL_2194587, EPI_ISL_2194588, EPI_ISL_2194589, EPI_ISL_2194592, EPI_ISL_2194593, EPI_ISL_2194594, EPI_ISL_2194596, EPI_ISL_2194597, EPI_ISL_2194598, EPI_ISL_2194599, EPI_ISL_2194601, EPI_ISL_2194604, EPI_ISL_2194605, EPI_ISL_2194606, EPI_ISL_2194607, EPI_ISL_2194608, EPI_ISL_2194611, EPI_ISL_2194613, EPI_ISL_2194636, EPI_ISL_2194655, EPI_ISL_2194664, EPI_ISL_2194676, EPI_ISL_2194678, EPI_ISL_2194679, EPI_ISL_2194681, EPI_ISL_2194682, EPI_ISL_2194713, EPI_ISL_2194715, EPI_ISL_2194716, EPI_ISL_2194719, EPI_ISL_2194723, EPI_ISL_2194726, EPI_ISL_2194727, EPI_ISL_2194729, EPI_ISL_2194732, EPI_ISL_2194733, EPI_ISL_2194734, EPI_ISL_2194735, EPI_ISL_2194739, EPI_ISL_2194755, EPI_ISL_2194756, EPI_ISL_2194757, EPI_ISL_2194758, EPI_ISL_2194759, EPI_ISL_2194760, EPI_ISL_2194761, EPI_ISL_2194762, EPI_ISL_2194768, EPI_ISL_2194769, EPI_ISL_2194771, EPI_ISL_2194780, EPI_ISL_2194781, EPI_ISL_2194782, EPI_ISL_2194783, EPI_ISL_2194784, EPI_ISL_2194785, EPI_ISL_2194787, EPI_ISL_2194792, EPI_ISL_2194796, EPI_ISL_2194797, EPI_ISL_2194805, EPI_ISL_2194806, EPI_ISL_2194810, EPI_ISL_2194812, EPI_ISL_2194813, EPI_ISL_2194815, EPI_ISL_2194816, EPI_ISL_2194817, EPI_ISL_2194818, EPI_ISL_2194819, EPI_ISL_2194820, EPI_ISL_2194821, EPI_ISL_2194822, EPI_ISL_2194823, EPI_ISL_2194824, EPI_ISL_2194825, EPI_ISL_2194827, EPI_ISL_2194828, EPI_ISL_2194829, EPI_ISL_2194830, EPI_ISL_2194832, EPI_ISL_2194833, EPI_ISL_2194834, EPI_ISL_2194835, EPI_ISL_2194836, EPI_ISL_2194837, EPI_ISL_2194838, EPI_ISL_2194839, EPI_ISL_2194841, EPI_ISL_2194843, EPI_ISL_2194844, EPI_ISL_2194845, EPI_ISL_2194846, EPI_ISL_2194848, EPI_ISL_2194849, EPI_ISL_2194850, EPI_ISL_2194852, EPI_ISL_2194853, EPI_ISL_2194854, EPI_ISL_2194855, EPI_ISL_2194857, EPI_ISL_2194858, EPI_ISL_2194860, EPI_ISL_2194861, EPI_ISL_2194862, EPI_ISL_2194863, EPI_ISL_2194865, EPI_ISL_2194866, EPI_ISL_2194867, EPI_ISL_2194868, EPI_ISL_2194869, EPI_ISL_2194870, EPI_ISL_2194871, EPI_ISL_2194872, EPI_ISL_2194875, EPI_ISL_2194876, EPI_ISL_2194877, EPI_ISL_2194878, EPI_ISL_2194879, EPI_ISL_2194880, EPI_ISL_2194882, EPI_ISL_2194883, EPI_ISL_2194884, EPI_ISL_2194885, EPI_ISL_2194887, EPI_ISL_2194888, EPI_ISL_2194889, EPI_ISL_2194891, EPI_ISL_2194892, EPI_ISL_2194894, EPI_ISL_2194897, EPI_ISL_2194898, EPI_ISL_2194900, EPI_ISL_2194901, EPI_ISL_2194902, EPI_ISL_2194904, EPI_ISL_2194906, EPI_ISL_2194907, EPI_ISL_2194909, EPI_ISL_2194910, EPI_ISL_2194912, EPI_ISL_2194913, EPI_ISL_2194914, EPI_ISL_2194915, EPI_ISL_2194916, EPI_ISL_2194919, EPI_ISL_2194922, EPI_ISL_2194923, EPI_ISL_2194924, EPI_ISL_2194925, EPI_ISL_2194927, EPI_ISL_2194928, EPI_ISL_2194929, EPI_ISL_2194931, EPI_ISL_2194932, EPI_ISL_2194933, EPI_ISL_2194938, EPI_ISL_2194939, EPI_ISL_2194940, EPI_ISL_2194941, EPI_ISL_2194943, EPI_ISL_2194944, EPI_ISL_2194945, EPI_ISL_2194946, EPI_ISL_2194948, EPI_ISL_2194950, EPI_ISL_2194951, EPI_ISL_2194952, EPI_ISL_2194953, EPI_ISL_2194956, EPI_ISL_2194957, EPI_ISL_2194960, EPI_ISL_2194961, EPI_ISL_2194964, EPI_ISL_2194965, EPI_ISL_2194966, EPI_ISL_2194969, EPI_ISL_2194971, EPI_ISL_2194972, EPI_ISL_2194973, EPI_ISL_2194975, EPI_ISL_2194976, EPI_ISL_2194979, EPI_ISL_2194980, EPI_ISL_2194981, EPI_ISL_2194987, EPI_ISL_2194989, EPI_ISL_2194990, EPI_ISL_2194991, EPI_ISL_2194995, EPI_ISL_2194996, EPI_ISL_2194999, EPI_ISL_2195000, EPI_ISL_2195001, EPI_ISL_2195003, EPI_ISL_2195004, EPI_ISL_2195005, EPI_ISL_2195006, EPI_ISL_2195007, EPI_ISL_2195008, EPI_ISL_2195010, EPI_ISL_2195011, EPI_ISL_2195012, EPI_ISL_2195013, EPI_ISL_2195014, EPI_ISL_2195016, EPI_ISL_2195018, EPI_ISL_2195020, EPI_ISL_2195021, EPI_ISL_2195023, EPI_ISL_2195024, EPI_ISL_2195025, EPI_ISL_2195030, EPI_ISL_2195031, EPI_ISL_2195032, EPI_ISL_2195034, EPI_ISL_2195035, EPI_ISL_2195037, EPI_ISL_2195039, EPI_ISL_2195040, EPI_ISL_2195044, EPI_ISL_2195045, EPI_ISL_2195046, EPI_ISL_2195048, EPI_ISL_2195050, EPI_ISL_2195051, EPI_ISL_2195052, EPI_ISL_2195053, EPI_ISL_2195054, EPI_ISL_2195055, EPI_ISL_2195056, EPI_ISL_2195057, EPI_ISL_2195058, EPI_ISL_2195059, EPI_ISL_2195060, EPI_ISL_2195064, EPI_ISL_2195065, EPI_ISL_2195068, EPI_ISL_2195069, EPI_ISL_2195070, EPI_ISL_2195071, EPI_ISL_2195073, EPI_ISL_2195075, EPI_ISL_2195077, EPI_ISL_2195078, EPI_ISL_2195079, EPI_ISL_2195080, EPI_ISL_2195081, EPI_ISL_2195082, EPI_ISL_2195083, EPI_ISL_2195084, EPI_ISL_2195085, EPI_ISL_2195086, EPI_ISL_2195087, EPI_ISL_2195088, EPI_ISL_2195091, EPI_ISL_2195093, EPI_ISL_2195094, EPI_ISL_2195098, EPI_ISL_2195099, EPI_ISL_2195101, EPI_ISL_2195102, EPI_ISL_2195103, EPI_ISL_2195106, EPI_ISL_2195107, EPI_ISL_2195108, EPI_ISL_2195111, EPI_ISL_2195112, EPI_ISL_2195113, EPI_ISL_2195115, EPI_ISL_2195116, EPI_ISL_2195117, EPI_ISL_2195134, EPI_ISL_2195135, EPI_ISL_2195136, EPI_ISL_2195137, EPI_ISL_2195140, EPI_ISL_2195141, EPI_ISL_2195142, EPI_ISL_2195146, EPI_ISL_2195153, EPI_ISL_2195157, EPI_ISL_2195158, EPI_ISL_2195159, EPI_ISL_2195160, EPI_ISL_2195161, EPI_ISL_2195162, EPI_ISL_2195163, EPI_ISL_2195164, EPI_ISL_2195165, EPI_ISL_2195166, EPI_ISL_2195167, EPI_ISL_2195168, EPI_ISL_2195169, EPI_ISL_2195170, EPI_ISL_2195171, EPI_ISL_2195173, EPI_ISL_2195176, EPI_ISL_2195177, EPI_ISL_2195178, EPI_ISL_2195179, EPI_ISL_2195180, EPI_ISL_2195181, EPI_ISL_2195182, EPI_ISL_2195183, EPI_ISL_2195187, EPI_ISL_2195188, EPI_ISL_2195189, EPI_ISL_2195191, EPI_ISL_2195193, EPI_ISL_2195197, EPI_ISL_2195198, EPI_ISL_2195199, EPI_ISL_2195200, EPI_ISL_2195201, EPI_ISL_2195203, EPI_ISL_2195204, EPI_ISL_2195205, EPI_ISL_2195206, EPI_ISL_2195207, EPI_ISL_2195208, EPI_ISL_2195209, EPI_ISL_2195210, EPI_ISL_2195211, EPI_ISL_2195212, EPI_ISL_2195213, EPI_ISL_2195214, EPI_ISL_2195217, EPI_ISL_2195218, EPI_ISL_2195219, EPI_ISL_2195220, EPI_ISL_2195221, EPI_ISL_2195222, EPI_ISL_2195223, EPI_ISL_2195224, EPI_ISL_2195227, EPI_ISL_2195228, EPI_ISL_2195230, EPI_ISL_2195231, EPI_ISL_2195232, EPI_ISL_2195233, EPI_ISL_2195235, EPI_ISL_2195238, EPI_ISL_2195243, EPI_ISL_2195244, EPI_ISL_2195247, EPI_ISL_2195248, EPI_ISL_2195251, EPI_ISL_2195253, EPI_ISL_2195254, EPI_ISL_2195255, EPI_ISL_2195256, EPI_ISL_2195257, EPI_ISL_2195258, EPI_ISL_2195259, EPI_ISL_2195260, EPI_ISL_2195261, EPI_ISL_2195262, EPI_ISL_2195263, EPI_ISL_2195267, EPI_ISL_2195268, EPI_ISL_2195269, EPI_ISL_2195270, EPI_ISL_2195271, EPI_ISL_2195272, EPI_ISL_2195273, EPI_ISL_2195274, EPI_ISL_2195275, EPI_ISL_2195276, EPI_ISL_2195277, EPI_ISL_2195278, EPI_ISL_2195279, EPI_ISL_2195280, EPI_ISL_2195281, EPI_ISL_2195282, EPI_ISL_2195283, EPI_ISL_2195284, EPI_ISL_2195285, EPI_ISL_2195286, EPI_ISL_2195287, EPI_ISL_2195288, EPI_ISL_2195289, EPI_ISL_2195290, EPI_ISL_2195291, EPI_ISL_2195292, EPI_ISL_2195293, EPI_ISL_2195294, EPI_ISL_2195295, EPI_ISL_2195296, EPI_ISL_2195297, EPI_ISL_2195298, EPI_ISL_2195299, EPI_ISL_2195301, EPI_ISL_2195302, EPI_ISL_2195303, EPI_ISL_2195304, EPI_ISL_2195305, EPI_ISL_2195306, EPI_ISL_2195307, EPI_ISL_2195308, EPI_ISL_2195309, EPI_ISL_2195311, EPI_ISL_2195312, EPI_ISL_2195313, EPI_ISL_2195314, EPI_ISL_2195315, EPI_ISL_2195316, EPI_ISL_2195317, EPI_ISL_2195318, EPI_ISL_2195319, EPI_ISL_2195320, EPI_ISL_2195322, EPI_ISL_2195323, EPI_ISL_2195325, EPI_ISL_2195326, EPI_ISL_2195327, EPI_ISL_2195328, EPI_ISL_2195329, EPI_ISL_2195330, EPI_ISL_2195331, EPI_ISL_2195333, EPI_ISL_2195335, EPI_ISL_2195336, EPI_ISL_2195337, EPI_ISL_2195338, EPI_ISL_2195340, EPI_ISL_2195341, EPI_ISL_2195342, EPI_ISL_2195343, EPI_ISL_2195344, EPI_ISL_2195345, EPI_ISL_2195346, EPI_ISL_2195347, EPI_ISL_2195350, EPI_ISL_2195351, EPI_ISL_2195352, EPI_ISL_2195355, EPI_ISL_2195358, EPI_ISL_2195359, EPI_ISL_2195360, EPI_ISL_2195362, EPI_ISL_2195364, EPI_ISL_2195365, EPI_ISL_2195366, EPI_ISL_2195367, EPI_ISL_2195368, EPI_ISL_2195370, EPI_ISL_2195371, EPI_ISL_2195372 | | | |
| see above | Colorado Department of Public Health and Environment | Colorado Department of Public Health and Environment | Laura Bankers, Molly C. Hetherington-Rauth, Diana Ir, Alexandria Rossheim, Shannon R. Matzinger, Sarah Elizabeth Totten, Emily A. Travanty |
| EPI_ISL_2195373, EPI_ISL_2195374, EPI_ISL_2195375, EPI_ISL_2195376, EPI_ISL_2195377, EPI_ISL_2195378, EPI_ISL_2195379, EPI_ISL_2195380 | J.W. Ruby Memorial Hospital | WVU and Marshall University Combined Genomics Core Facilities | James Denvir, Peter Stoilov, Peter Perrotta, Wesley Kimble, Ryan Percifield |
| EPI_ISL_2195381, EPI_ISL_2195382, EPI_ISL_2195383, EPI_ISL_2195384, EPI_ISL_2195385, EPI_ISL_2195386, EPI_ISL_2195387, EPI_ISL_2195388, EPI_ISL_2195389, EPI_ISL_2195390, EPI_ISL_2195391, EPI_ISL_2195392, EPI_ISL_2195393, EPI_ISL_2195394, EPI_ISL_2195395, EPI_ISL_2195396, EPI_ISL_2195397, EPI_ISL_2195398, EPI_ISL_2195399, EPI_ISL_2195400, EPI_ISL_2195401, EPI_ISL_2195402, EPI_ISL_2195403, EPI_ISL_2195404, EPI_ISL_2195405, EPI_ISL_2195406, EPI_ISL_2195407, EPI_ISL_2195408, EPI_ISL_2195409, EPI_ISL_2195410, EPI_ISL_2195411, EPI_ISL_2195412, EPI_ISL_2195413, EPI_ISL_2195414, EPI_ISL_2195415, EPI_ISL_2195416, EPI_ISL_2195417, EPI_ISL_2195418, EPI_ISL_2195419, EPI_ISL_2195420, EPI_ISL_2195421, EPI_ISL_2195422, EPI_ISL_2195423, EPI_ISL_2195424, EPI_ISL_2195425, EPI_ISL_2195426, EPI_ISL_2195427, EPI_ISL_2195428, EPI_ISL_2195429, EPI_ISL_2195430, EPI_ISL_2195431, EPI_ISL_2195432, EPI_ISL_2195433, EPI_ISL_2195434, EPI_ISL_2195435, EPI_ISL_2195436, EPI_ISL_2195437, EPI_ISL_2195438, EPI_ISL_2195439, EPI_ISL_2195440, EPI_ISL_2195441, EPI_ISL_2195442, EPI_ISL_2195443, EPI_ISL_2195444, EPI_ISL_2195445, EPI_ISL_2195446, EPI_ISL_2195447, EPI_ISL_2195448, EPI_ISL_2195449, EPI_ISL_2195450, EPI_ISL_2195451, EPI_ISL_2195452, EPI_ISL_2195453, EPI_ISL_2195454, EPI_ISL_2195455, EPI_ISL_2195456, EPI_ISL_2195457, EPI_ISL_2195458, EPI_ISL_2195459, EPI_ISL_2195460, EPI_ISL_2195461, EPI_ISL_2195462, EPI_ISL_2195463, EPI_ISL_2195464, EPI_ISL_2195465, EPI_ISL_2195466, EPI_ISL_2195467, EPI_ISL_2195468, EPI_ISL_2195469, EPI_ISL_2195470, EPI_ISL_2195471, EPI_ISL_2195472, EPI_ISL_2195473, EPI_ISL_2195474, EPI_ISL_2195475, EPI_ISL_2195476, EPI_ISL_2195477, EPI_ISL_2195479, EPI_ISL_2195480, EPI_ISL_2195481, EPI_ISL_2195482, EPI_ISL_2195483, EPI_ISL_2195484, EPI_ISL_2195485, EPI_ISL_2195486, EPI_ISL_2195487, EPI_ISL_2195489, EPI_ISL_2195490, EPI_ISL_2195491, EPI_ISL_2195492, EPI_ISL_2195493, EPI_ISL_2195494, EPI_ISL_2195495, EPI_ISL_2195496, EPI_ISL_2195497, EPI_ISL_2195498, EPI_ISL_2195499, EPI_ISL_2195500, EPI_ISL_2195501, EPI_ISL_2195502, EPI_ISL_2195503, EPI_ISL_2195504, EPI_ISL_2195505, EPI_ISL_2195506, EPI_ISL_2195507, EPI_ISL_2195508, EPI_ISL_2195510, EPI_ISL_2195512, EPI_ISL_2195513, EPI_ISL_2195514, EPI_ISL_2195515, EPI_ISL_2195516, EPI_ISL_2195517, EPI_ISL_2195518, EPI_ISL_2195519, EPI_ISL_2195520, EPI_ISL_2195521, EPI_ISL_2195522, EPI_ISL_2195523, EPI_ISL_2195524, EPI_ISL_2195525, EPI_ISL_2195526, EPI_ISL_2195527, EPI_ISL_2195528, EPI_ISL_2195529, EPI_ISL_2195530, EPI_ISL_2195531, EPI_ISL_2195532, EPI_ISL_2195533, EPI_ISL_2195534, EPI_ISL_2195535, EPI_ISL_2195536, EPI_ISL_2195537, EPI_ISL_2195538, EPI_ISL_2195539, EPI_ISL_2195540, EPI_ISL_2195541, EPI_ISL_2195542, EPI_ISL_2195543, EPI_ISL_2195544, EPI_ISL_2195545, EPI_ISL_2195547, EPI_ISL_2195548, EPI_ISL_2195549, EPI_ISL_2195550, EPI_ISL_2195551, EPI_ISL_2195552, EPI_ISL_2195553, EPI_ISL_2195554, EPI_ISL_2195557, EPI_ISL_2195558, EPI_ISL_2195559, EPI_ISL_2195560, EPI_ISL_2195561, EPI_ISL_2195564, EPI_ISL_2195565, EPI_ISL_2195566, EPI_ISL_2195567, EPI_ISL_2195568, EPI_ISL_2195569, EPI_ISL_2195570, EPI_ISL_2195571, EPI_ISL_2195572, EPI_ISL_2195573, EPI_ISL_2195574, EPI_ISL_2195575, EPI_ISL_2195576, EPI_ISL_2195577, EPI_ISL_2195578, EPI_ISL_2195579, EPI_ISL_2195580, EPI_ISL_2195581, EPI_ISL_2195583, EPI_ISL_2195584, EPI_ISL_2195585, EPI_ISL_2195586, EPI_ISL_2195587 | | | |
| see above | WVU Rapid Development Lab | WVU and Marshall University Combined Genomics Core Facilities | James Denvir, Peter Stoilov, Peter Perrotta, Wesley Kimble, Ryan Percifield |
| EPI_ISL_2195588, EPI_ISL_2195589, EPI_ISL_2195590, EPI_ISL_2195591, EPI_ISL_2195592, EPI_ISL_2195593, EPI_ISL_2195594, EPI_ISL_2195596, EPI_ISL_2195598, EPI_ISL_2195599, EPI_ISL_2195600, EPI_ISL_2195601, EPI_ISL_2195602, EPI_ISL_2195603, EPI_ISL_2195604, EPI_ISL_2195605, EPI_ISL_2195606, EPI_ISL_2195607, EPI_ISL_2195609, EPI_ISL_2195610, EPI_ISL_2195611, EPI_ISL_2195612, EPI_ISL_2195613, EPI_ISL_2195615, EPI_ISL_2195616 | | | |
| see above | J.W. Ruby Memorial Hospital | WVU and Marshall University Combined Genomics Core Facilities | James Denvir, Peter Stoilov, Peter Perrotta, Wesley Kimble, Ryan Percifield |
| EPI_ISL_2195617, EPI_ISL_2195618, EPI_ISL_2195619, EPI_ISL_2195620 | WVU Rapid Development Lab | WVU and Marshall University Combined Genomics Core Facilities | James Denvir, Peter Stoilov, Peter Perrotta, Wesley Kimble, Ryan Percifield |
| EPI_ISL_2195621, EPI_ISL_2195622, EPI_ISL_2195623, EPI_ISL_2195625, EPI_ISL_2195626, EPI_ISL_2195627, EPI_ISL_2195628, EPI_ISL_2195629, EPI_ISL_2195630, EPI_ISL_2195631, EPI_ISL_2195632, EPI_ISL_2195633, EPI_ISL_2195634, EPI_ISL_2195635, EPI_ISL_2195636, EPI_ISL_2195637, EPI_ISL_2195638, EPI_ISL_2195639, EPI_ISL_2195640, EPI_ISL_2195641, EPI_ISL_2195643, EPI_ISL_2195644, EPI_ISL_2195645, EPI_ISL_2195646, EPI_ISL_2195647, EPI_ISL_2195648, EPI_ISL_2195649, EPI_ISL_2195650, EPI_ISL_2195651, EPI_ISL_2195652, EPI_ISL_2195653, EPI_ISL_2195654, EPI_ISL_2195655, EPI_ISL_2195656, EPI_ISL_2195657, EPI_ISL_2195658, EPI_ISL_2195659, EPI_ISL_2195660, EPI_ISL_2195661 | | | |
| see above | J.W. Ruby Memorial Hospital | WVU and Marshall University Combined Genomics Core Facilities | James Denvir, Peter Stoilov, Peter Perrotta, Wesley Kimble, Ryan Percifield |
| EPI_ISL_2195662, EPI_ISL_2195664, EPI_ISL_2195665, EPI_ISL_2195666, EPI_ISL_2195667, EPI_ISL_2195668, EPI_ISL_2195669, EPI_ISL_2195670, EPI_ISL_2195671, EPI_ISL_2195672, EPI_ISL_2195673, EPI_ISL_2195674, EPI_ISL_2195675, EPI_ISL_2195676, EPI_ISL_2195677, EPI_ISL_2195678, EPI_ISL_2195679, EPI_ISL_2195680, EPI_ISL_2195681, EPI_ISL_2195682, EPI_ISL_2195683, EPI_ISL_2195684, EPI_ISL_2195685, EPI_ISL_2195686, EPI_ISL_2195687, EPI_ISL_2195688, EPI_ISL_2195689, EPI_ISL_2195690, EPI_ISL_2195691, EPI_ISL_2195693, EPI_ISL_2195694, EPI_ISL_2195695, EPI_ISL_2195696, EPI_ISL_2195697, EPI_ISL_2195698, EPI_ISL_2195699, EPI_ISL_2195700, EPI_ISL_2195701, EPI_ISL_2195702, EPI_ISL_2195703, EPI_ISL_2195704, EPI_ISL_2195705, EPI_ISL_2195706, EPI_ISL_2195707, EPI_ISL_2195708, EPI_ISL_2195709, EPI_ISL_2195710, EPI_ISL_2195711, EPI_ISL_2195713, EPI_ISL_2195714, EPI_ISL_2195715, EPI_ISL_2195716 | | | |
| see above | QLabs | WVU and Marshall University Combined Genomics Core Facilities | James Denvir, Peter Stoilov, Peter Perrotta, Wesley Kimble, Ryan Percifield |
| EPI_ISL_2195717, EPI_ISL_2195719 | Wheeling Hospital | WVU and Marshall University Combined Genomics Core Facilities | James Denvir, Peter Stoilov, Peter Perrotta, Wesley Kimble, Ryan Percifield |
| EPI_ISL_2195721, EPI_ISL_2195722, EPI_ISL_2195723, EPI_ISL_2195724 | J.W. Ruby Memorial Hospital | WVU and Marshall University Combined Genomics Core Facilities | James Denvir, Peter Stoilov, Peter Perrotta, Wesley Kimble, Ryan Percifield |
| EPI_ISL_2195725, EPI_ISL_2195726, EPI_ISL_2195727, EPI_ISL_2195728, EPI_ISL_2195729, EPI_ISL_2195730, EPI_ISL_2195731, EPI_ISL_2195732, EPI_ISL_2195733, EPI_ISL_2195734, EPI_ISL_2195735, EPI_ISL_2195736, EPI_ISL_2195738, EPI_ISL_2195739, EPI_ISL_2195740, EPI_ISL_2195741, EPI_ISL_2195742, EPI_ISL_2195743, EPI_ISL_2195744, EPI_ISL_2195745 | | | |
| see above | Wheeling Hospital | WVU and Marshall University Combined Genomics Core Facilities | James Denvir, Peter Stoilov, Peter Perrotta, Wesley Kimble, Ryan Percifield |
| EPI_ISL_2195746, | Berkeley Medical Center | WVU and Marshall University Combined | James Denvir, Peter Stoilov, Peter Perrotta, Wesley Kimble, Ryan Percifield |

| | | | |
|---|---|---|---|
| EPI_ISL_2195747 | | Genomics Core Facilities | |
| EPI_ISL_2195748, EPI_ISL_2195749, EPI_ISL_2195750, EPI_ISL_2195751, EPI_ISL_2195752, EPI_ISL_2195753, EPI_ISL_2195754, EPI_ISL_2195755, EPI_ISL_2195756, EPI_ISL_2195757, EPI_ISL_2195758, EPI_ISL_2195759, EPI_ISL_2195760, EPI_ISL_2195761 | | | |
| see above | Wheeling Hospital | WVU and Marshall University Combined Genomics Core Facilities | James Denvir, Peter Stoilov, Peter Perrotta, Wesley Kimble, Ryan Percifield |
| EPI_ISL_2195762, EPI_ISL_2195763, EPI_ISL_2195764, EPI_ISL_2195765, EPI_ISL_2195766, EPI_ISL_2195767, EPI_ISL_2195768, EPI_ISL_2195769, EPI_ISL_2195770, EPI_ISL_2195771, EPI_ISL_2195772, EPI_ISL_2195773, EPI_ISL_2195774, EPI_ISL_2195775, EPI_ISL_2195777, EPI_ISL_2195778, EPI_ISL_2195779, EPI_ISL_2195781, EPI_ISL_2195782 | | | |
| see above | J.W. Ruby Memorial Hospital | WVU and Marshall University Combined Genomics Core Facilities | James Denvir, Peter Stoilov, Peter Perrotta, Wesley Kimble, Ryan Percifield |
| EPI_ISL_2195783 | UHC | WVU and Marshall University Combined Genomics Core Facilities | James Denvir, Peter Stoilov, Peter Perrotta, Wesley Kimble, Ryan Percifield |
| EPI_ISL_2195784 | J.W. Ruby Memorial Hospital | WVU and Marshall University Combined Genomics Core Facilities | James Denvir, Peter Stoilov, Peter Perrotta, Wesley Kimble, Ryan Percifield |
| EPI_ISL_2195785, EPI_ISL_2195786, EPI_ISL_2195787, EPI_ISL_2195788, EPI_ISL_2195789, EPI_ISL_2195790, EPI_ISL_2195791, EPI_ISL_2195792, EPI_ISL_2195793, EPI_ISL_2195794, EPI_ISL_2195795, EPI_ISL_2195796, EPI_ISL_2195797, EPI_ISL_2195798, EPI_ISL_2195799, EPI_ISL_2195800, EPI_ISL_2195801, EPI_ISL_2195803, EPI_ISL_2195804, EPI_ISL_2195805, EPI_ISL_2195806, EPI_ISL_2195807, EPI_ISL_2195808, EPI_ISL_2195809, EPI_ISL_2195810, EPI_ISL_2195811, EPI_ISL_2195812, EPI_ISL_2195813 | | | |
| see above | Berkeley Medical Center | WVU and Marshall University Combined Genomics Core Facilities | James Denvir, Peter Stoilov, Peter Perrotta, Wesley Kimble, Ryan Percifield |
| EPI_ISL_2195814, EPI_ISL_2195815, EPI_ISL_2195816, EPI_ISL_2195817 | Wheeling Hospital | WVU and Marshall University Combined Genomics Core Facilities | James Denvir, Peter Stoilov, Peter Perrotta, Wesley Kimble, Ryan Percifield |
| EPI_ISL_2195818, EPI_ISL_2195819, EPI_ISL_2195820, EPI_ISL_2195821, EPI_ISL_2195822, EPI_ISL_2195823, EPI_ISL_2195824, EPI_ISL_2195825, EPI_ISL_2195826, EPI_ISL_2195827, EPI_ISL_2195828, EPI_ISL_2195830, EPI_ISL_2195831, EPI_ISL_2195832, EPI_ISL_2195834, EPI_ISL_2195835, EPI_ISL_2195836, EPI_ISL_2195837, EPI_ISL_2195838, EPI_ISL_2195839, EPI_ISL_2195840, EPI_ISL_2195841, EPI_ISL_2195842, EPI_ISL_2195843, EPI_ISL_2195844, EPI_ISL_2195845, EPI_ISL_2195846, EPI_ISL_2195848, EPI_ISL_2195849, EPI_ISL_2195850, EPI_ISL_2195851 | | | |
| see above | J.W. Ruby Memorial Hospital | WVU and Marshall University Combined Genomics Core Facilities | James Denvir, Peter Stoilov, Peter Perrotta, Wesley Kimble, Ryan Percifield |
| EPI_ISL_2195852, EPI_ISL_2195853, EPI_ISL_2195854 | Raleigh General Hospital Microbiology Dept. | WVU and Marshall University Combined Genomics Core Facilities | James Denvir, Peter Stoilov, Peter Perrotta, Wesley Kimble, Ryan Percifield |
| EPI_ISL_2195855 | J.W. Ruby Memorial Hospital | WVU and Marshall University Combined Genomics Core Facilities | James Denvir, Peter Stoilov, Peter Perrotta, Wesley Kimble, Ryan Percifield |
| EPI_ISL_2195856, EPI_ISL_2195857, EPI_ISL_2195858, EPI_ISL_2195859, EPI_ISL_2195860, EPI_ISL_2195861, EPI_ISL_2195862, EPI_ISL_2195863 | Raleigh General Hospital Microbiology Dept. | WVU and Marshall University Combined Genomics Core Facilities | James Denvir, Peter Stoilov, Peter Perrotta, Wesley Kimble, Ryan Percifield |
| EPI_ISL_2195864, EPI_ISL_2195865, EPI_ISL_2195866, EPI_ISL_2195867, EPI_ISL_2195868, EPI_ISL_2195869, EPI_ISL_2195870, EPI_ISL_2195871, EPI_ISL_2195872, EPI_ISL_2195873, EPI_ISL_2195874, EPI_ISL_2195875, EPI_ISL_2195876, EPI_ISL_2195877, EPI_ISL_2195878, EPI_ISL_2195879, EPI_ISL_2195880, EPI_ISL_2195881, EPI_ISL_2195882, EPI_ISL_2195883, EPI_ISL_2195884, EPI_ISL_2195885, EPI_ISL_2195886, EPI_ISL_2195887, EPI_ISL_2195888, EPI_ISL_2195889, EPI_ISL_2195890, EPI_ISL_2195891 | | | |
| see above | Berkeley Medical Center | WVU and Marshall University Combined Genomics Core Facilities | James Denvir, Peter Stoilov, Peter Perrotta, Wesley Kimble, Ryan Percifield |
| EPI_ISL_2195892, EPI_ISL_2195893, EPI_ISL_2195894, EPI_ISL_2195895, EPI_ISL_2195896, EPI_ISL_2195897, EPI_ISL_2195898, EPI_ISL_2195899, EPI_ISL_2195900, EPI_ISL_2195902, EPI_ISL_2195903 | | | |
| see above | WVU Rapid Development Lab | WVU and Marshall University Combined Genomics Core Facilities | James Denvir, Peter Stoilov, Peter Perrotta, Wesley Kimble, Ryan Percifield |
| EPI_ISL_2195904, EPI_ISL_2195905, EPI_ISL_2195906, EPI_ISL_2195908, EPI_ISL_2195909, EPI_ISL_2195910, EPI_ISL_2195911, EPI_ISL_2195912, EPI_ISL_2195913, EPI_ISL_2195914, EPI_ISL_2195915, EPI_ISL_2195916, EPI_ISL_2195917, EPI_ISL_2195918, EPI_ISL_2195919, EPI_ISL_2195920, EPI_ISL_2195921, EPI_ISL_2195922, EPI_ISL_2195925, EPI_ISL_2195926 | | | |
| see above | J.W. Ruby Memorial Hospital | WVU and Marshall University Combined Genomics Core Facilities | James Denvir, Peter Stoilov, Peter Perrotta, Wesley Kimble, Ryan Percifield |
| EPI_ISL_2195928, EPI_ISL_2195929, EPI_ISL_2195930, EPI_ISL_2195931, EPI_ISL_2195932, EPI_ISL_2195933, EPI_ISL_2195934, EPI_ISL_2195936, EPI_ISL_2195937, EPI_ISL_2195938, EPI_ISL_2195939, EPI_ISL_2195940, EPI_ISL_2195941, EPI_ISL_2195942, EPI_ISL_2195943, EPI_ISL_2195944, EPI_ISL_2195945, EPI_ISL_2195946, EPI_ISL_2195947, EPI_ISL_2195948, EPI_ISL_2195949, EPI_ISL_2195950, EPI_ISL_2195951, EPI_ISL_2195952, EPI_ISL_2195953, EPI_ISL_2195954, EPI_ISL_2195955, EPI_ISL_2195956, EPI_ISL_2195957, EPI_ISL_2195958, EPI_ISL_2195959, EPI_ISL_2195960, EPI_ISL_2195961, EPI_ISL_2195962, EPI_ISL_2195963, EPI_ISL_2195964, EPI_ISL_2195965, EPI_ISL_2195966, EPI_ISL_2195967, EPI_ISL_2195968, EPI_ISL_2195969, EPI_ISL_2195970, EPI_ISL_2195971 | | | |
| see above | QLabs | WVU and Marshall University Combined Genomics Core Facilities | James Denvir, Peter Stoilov, Peter Perrotta, Wesley Kimble, Ryan Percifield |
| EPI_ISL_2195972, EPI_ISL_2195973 | Preston Taylor Community Health Center | WVU and Marshall University Combined Genomics Core Facilities | James Denvir, Peter Stoilov, Peter Perrotta, Wesley Kimble, Ryan Percifield |
| EPI_ISL_2195974, EPI_ISL_2195975 | J.W. Ruby Memorial Hospital | WVU and Marshall University Combined Genomics Core Facilities | James Denvir, Peter Stoilov, Peter Perrotta, Wesley Kimble, Ryan Percifield |
| EPI_ISL_2195976 | UHC | WVU and Marshall University Combined Genomics Core Facilities | James Denvir, Peter Stoilov, Peter Perrotta, Wesley Kimble, Ryan Percifield |
| EPI_ISL_2195977 | Summersville Regional Medical Center | WVU and Marshall University Combined Genomics Core Facilities | James Denvir, Peter Stoilov, Peter Perrotta, Wesley Kimble, Ryan Percifield |
| EPI_ISL_2195978, EPI_ISL_2195979, EPI_ISL_2195980, EPI_ISL_2195981, EPI_ISL_2195982, EPI_ISL_2195983, EPI_ISL_2195984, EPI_ISL_2195985, EPI_ISL_2195986, EPI_ISL_2195987, EPI_ISL_2195988, EPI_ISL_2195989, EPI_ISL_2195990, EPI_ISL_2195991, EPI_ISL_2195992, EPI_ISL_2195993, EPI_ISL_2195994, EPI_ISL_2195995, EPI_ISL_2195996, EPI_ISL_2195997, EPI_ISL_2195998, EPI_ISL_2195999, EPI_ISL_2196001, EPI_ISL_2196002, EPI_ISL_2196003, EPI_ISL_2196004, EPI_ISL_2196005, EPI_ISL_2196006, EPI_ISL_2196007, EPI_ISL_2196008, EPI_ISL_2196009, EPI_ISL_2196010, EPI_ISL_2196011, EPI_ISL_2196012, EPI_ISL_2196013, EPI_ISL_2196014, EPI_ISL_2196015, EPI_ISL_2196016, EPI_ISL_2196017, EPI_ISL_2196018, EPI_ISL_2196019, EPI_ISL_2196020, EPI_ISL_2196021, EPI_ISL_2196022, EPI_ISL_2196023, EPI_ISL_2196024, EPI_ISL_2196025, EPI_ISL_2196026, EPI_ISL_2196027, EPI_ISL_2196028, EPI_ISL_2196029, EPI_ISL_2196030, EPI_ISL_2196031, EPI_ISL_2196032, EPI_ISL_2196033, EPI_ISL_2196034, EPI_ISL_2196035, EPI_ISL_2196036, EPI_ISL_2196037, EPI_ISL_2196038, EPI_ISL_2196039, EPI_ISL_2196040, EPI_ISL_2196041, EPI_ISL_2196042, EPI_ISL_2196043, EPI_ISL_2196044, EPI_ISL_2196045, EPI_ISL_2196046 | | | |
| see above | J.W. Ruby Memorial Hospital | WVU and Marshall University Combined Genomics Core Facilities | James Denvir, Peter Stoilov, Peter Perrotta, Wesley Kimble, Ryan Percifield |
| EPI_ISL_2196047, EPI_ISL_2196048, EPI_ISL_2196049, EPI_ISL_2196050, EPI_ISL_2196051, EPI_ISL_2196052, EPI_ISL_2196053, EPI_ISL_2196054 | Raleigh General Hospital Microbiology Dept. | WVU and Marshall University Combined Genomics Core Facilities | James Denvir, Peter Stoilov, Peter Perrotta, Wesley Kimble, Ryan Percifield |
| EPI_ISL_2196055, EPI_ISL_2196056, EPI_ISL_2196057, EPI_ISL_2196058, EPI_ISL_2196059, EPI_ISL_2196060, EPI_ISL_2196061, EPI_ISL_2196062, EPI_ISL_2196063, EPI_ISL_2196064, EPI_ISL_2196065, EPI_ISL_2196066, EPI_ISL_2196067, EPI_ISL_2196068, EPI_ISL_2196069, EPI_ISL_2196070, EPI_ISL_2196071, EPI_ISL_2196072, EPI_ISL_2196073, EPI_ISL_2196074, EPI_ISL_2196075, EPI_ISL_2196076 | | | |
| see above | BMC LAB | WVU and Marshall University Combined Genomics Core Facilities | James Denvir, Peter Stoilov, Peter Perrotta, Wesley Kimble, Ryan Percifield |
| EPI_ISL_2196077 | UHC | WVU and Marshall University Combined Genomics Core Facilities | James Denvir, Peter Stoilov, Peter Perrotta, Wesley Kimble, Ryan Percifield |
| EPI_ISL_2196078 | J.W. Ruby Memorial Hospital | WVU and Marshall University Combined Genomics Core Facilities | James Denvir, Peter Stoilov, Peter Perrotta, Wesley Kimble, Ryan Percifield |
| EPI_ISL_2196079, EPI_ISL_2196080, EPI_ISL_2196081, EPI_ISL_2196082, EPI_ISL_2196083, EPI_ISL_2196084, EPI_ISL_2196085, EPI_ISL_2196086, EPI_ISL_2196087, EPI_ISL_2196088, EPI_ISL_2196089, EPI_ISL_2196090, EPI_ISL_2196091, EPI_ISL_2196092, EPI_ISL_2196093, EPI_ISL_2196094, EPI_ISL_2196095, EPI_ISL_2196096, EPI_ISL_2196097, EPI_ISL_2196098, EPI_ISL_2196099, EPI_ISL_2196100, EPI_ISL_2196101, EPI_ISL_2196102, EPI_ISL_2196103, EPI_ISL_2196104, EPI_ISL_2196105, EPI_ISL_2196106, EPI_ISL_2196107, EPI_ISL_2196108, EPI_ISL_2196109 | | | |
| see above | QLabs | WVU and Marshall University Combined Genomics Core Facilities | James Denvir, Peter Stoilov, Peter Perrotta, Wesley Kimble, Ryan Percifield |
| EPI_ISL_2196111 | West Virigina Labs | WVU and Marshall University Combined Genomics Core Facilities | James Denvir, Peter Stoilov, Peter Perrotta, Wesley Kimble, Ryan Percifield |
| EPI_ISL_2196112, EPI_ISL_2196113, EPI_ISL_2196114, EPI_ISL_2196115, EPI_ISL_2196116, EPI_ISL_2196117 | WV Labs | WVU and Marshall University Combined Genomics Core Facilities | James Denvir, Peter Stoilov, Peter Perrotta, Wesley Kimble, Ryan Percifield |
| EPI_ISL_2196118, EPI_ISL_2196119, EPI_ISL_2196120, EPI_ISL_2196121, EPI_ISL_2196122, EPI_ISL_2196123, EPI_ISL_2196124, EPI_ISL_2196126, EPI_ISL_2196127, EPI_ISL_2196128, EPI_ISL_2196129, EPI_ISL_2196130, EPI_ISL_2196131, EPI_ISL_2196132, EPI_ISL_2196133, EPI_ISL_2196134, EPI_ISL_2196135, EPI_ISL_2196136, EPI_ISL_2196137, EPI_ISL_2196138, EPI_ISL_2196139, EPI_ISL_2196140, EPI_ISL_2196141, EPI_ISL_2196142, EPI_ISL_2196143, EPI_ISL_2196144, EPI_ISL_2196145, EPI_ISL_2196146, EPI_ISL_2196147, EPI_ISL_2196148, EPI_ISL_2196149, EPI_ISL_2196150, EPI_ISL_2196151, EPI_ISL_2196152, EPI_ISL_2196153, EPI_ISL_2196154, EPI_ISL_2196155, EPI_ISL_2196157, EPI_ISL_2196158, EPI_ISL_2196159, EPI_ISL_2196160, EPI_ISL_2196161, EPI_ISL_2196162, EPI_ISL_2196163, | | | |

| | | | |
|---|---|---|---|
| EPI_ISL_2196164, EPI_ISL_2196165, EPI_ISL_2196166, EPI_ISL_2196167, EPI_ISL_2196168, EPI_ISL_2196169, EPI_ISL_2196170, EPI_ISL_2196171, EPI_ISL_2196173, EPI_ISL_2196174, EPI_ISL_2196175, EPI_ISL_2196176, EPI_ISL_2196177 | | | |
| see above | QLabs | WVU and Marshall University Combined Genomics Core Facilities | James Denvir, Peter Stoilov, Peter Perrotta, Wesley Kimble, Ryan Percifield |
| EPI_ISL_2196178, EPI_ISL_2196179, EPI_ISL_2196180, EPI_ISL_2196181, EPI_ISL_2196182, EPI_ISL_2196183 | J.W. Ruby Memorial Hospital | WVU and Marshall University Combined Genomics Core Facilities | James Denvir, Peter Stoilov, Peter Perrotta, Wesley Kimble, Ryan Percifield |
| EPI_ISL_2196702 | Baylor Scott & White-Waxahachie | Baylor Scott & White-Temple | Ari Rao, Linden Morales, Kimberly Walker, Marcus Volz, Shelby Hendrickson |
| EPI_ISL_2198343, EPI_ISL_2199998, EPI_ISL_2200062, EPI_ISL_2200063 | Ohio Department of Health Laboratory | Ohio Department of Health Laboratory | Holmes, Jennifer; Eric Brandt, Keoni Omura, Glen McGillivary, Caitlin McDonnell, Jade Mowery, Stephanie Mccracken, Tyler Payne, Kirtana Ramadugu, Erica Leasure, Brent Lee, Kelsey Florek, Heather Blankenship, Allison Black, Quanta Brown, and Tammy Bannerman |
| EPI_ISL_2200071, EPI_ISL_2200078, EPI_ISL_2200079, EPI_ISL_2200081, EPI_ISL_2200082, EPI_ISL_2200083, EPI_ISL_2200084, EPI_ISL_2200085, EPI_ISL_2200086, EPI_ISL_2200087, EPI_ISL_2200088, EPI_ISL_2200089, EPI_ISL_2200091, EPI_ISL_2200092, EPI_ISL_2200093, EPI_ISL_2200094, EPI_ISL_2200096, EPI_ISL_2200097, EPI_ISL_2200099, EPI_ISL_2200100, EPI_ISL_2200101, EPI_ISL_2200102, EPI_ISL_2200103, EPI_ISL_2200104, EPI_ISL_2200105, EPI_ISL_2200106, EPI_ISL_2200107, EPI_ISL_2200108, EPI_ISL_2200109, EPI_ISL_2200110, EPI_ISL_2200111, EPI_ISL_2200112, EPI_ISL_2200113, EPI_ISL_2200114, EPI_ISL_2200115, EPI_ISL_2200116, EPI_ISL_2200117, EPI_ISL_2200118, EPI_ISL_2200119, EPI_ISL_2200120, EPI_ISL_2200121, EPI_ISL_2200122, EPI_ISL_2200123, EPI_ISL_2200124, EPI_ISL_2200125, EPI_ISL_2200126, EPI_ISL_2200127, EPI_ISL_2200128, EPI_ISL_2200129, EPI_ISL_2200130, EPI_ISL_2200131, EPI_ISL_2200132, EPI_ISL_2200133, EPI_ISL_2200134, EPI_ISL_2200135, EPI_ISL_2200136, EPI_ISL_2200137, EPI_ISL_2200138, EPI_ISL_2200139, EPI_ISL_2200140, EPI_ISL_2200141, EPI_ISL_2200142, EPI_ISL_2200143, EPI_ISL_2200144, EPI_ISL_2200145, EPI_ISL_2200146, EPI_ISL_2200147, EPI_ISL_2200148, EPI_ISL_2200149, EPI_ISL_2200150, EPI_ISL_2200151, EPI_ISL_2200152, EPI_ISL_2200153, EPI_ISL_2200154 | | | |
| see above | The Ohio State University Applied Microbiology Services Laboratory | The Ohio State University Applied Microbiology Services Laboratory | Seth A. Faith PhD |
| EPI_ISL_2200158, EPI_ISL_2200159, EPI_ISL_2200160, EPI_ISL_2200161, EPI_ISL_2200162, EPI_ISL_2200163, EPI_ISL_2200164, EPI_ISL_2200165, EPI_ISL_2200166, EPI_ISL_2200167, EPI_ISL_2200168, EPI_ISL_2200169, EPI_ISL_2200170, EPI_ISL_2200171, EPI_ISL_2200172, EPI_ISL_2200173, EPI_ISL_2200174, EPI_ISL_2200175, EPI_ISL_2200176, EPI_ISL_2200177, EPI_ISL_2200178, EPI_ISL_2200179, EPI_ISL_2200180, EPI_ISL_2200181, EPI_ISL_2200182, EPI_ISL_2200183, EPI_ISL_2200184, EPI_ISL_2200185, EPI_ISL_2200186, EPI_ISL_2200187, EPI_ISL_2200188, EPI_ISL_2200189, EPI_ISL_2200190, EPI_ISL_2200191, EPI_ISL_2200192, EPI_ISL_2200193, EPI_ISL_2200194, EPI_ISL_2200195, EPI_ISL_2200196, EPI_ISL_2200197, EPI_ISL_2200198, EPI_ISL_2200199, EPI_ISL_2200200, EPI_ISL_2200201, EPI_ISL_2200202, EPI_ISL_2200203, EPI_ISL_2200204, EPI_ISL_2200205, EPI_ISL_2200206, EPI_ISL_2200207, EPI_ISL_2200208, EPI_ISL_2200209, EPI_ISL_2200210, EPI_ISL_2200211, EPI_ISL_2200212, EPI_ISL_2200213, EPI_ISL_2200214, EPI_ISL_2200215, EPI_ISL_2200216, EPI_ISL_2200217, EPI_ISL_2200218, EPI_ISL_2200219, EPI_ISL_2200220, EPI_ISL_2200221, EPI_ISL_2200222, EPI_ISL_2200223, EPI_ISL_2200224, EPI_ISL_2200225, EPI_ISL_2200226, EPI_ISL_2200227, EPI_ISL_2200228, EPI_ISL_2200229, EPI_ISL_2200230, EPI_ISL_2200231, EPI_ISL_2200232, EPI_ISL_2200233, EPI_ISL_2200234, EPI_ISL_2200235, EPI_ISL_2200236, EPI_ISL_2200237, EPI_ISL_2200238, EPI_ISL_2200239, EPI_ISL_2200240, EPI_ISL_2200241, EPI_ISL_2200242, EPI_ISL_2200243, EPI_ISL_2200244, EPI_ISL_2200245, EPI_ISL_2200246, EPI_ISL_2200247, EPI_ISL_2200248, EPI_ISL_2200249, EPI_ISL_2200250, EPI_ISL_2200251, EPI_ISL_2200252, EPI_ISL_2200253, EPI_ISL_2200254, EPI_ISL_2200255, EPI_ISL_2200256, EPI_ISL_2200257, EPI_ISL_2200258, EPI_ISL_2200259, EPI_ISL_2200260, EPI_ISL_2200261, EPI_ISL_2200262, EPI_ISL_2200263, EPI_ISL_2200264, EPI_ISL_2200265, EPI_ISL_2200267, EPI_ISL_2200268, EPI_ISL_2200269, EPI_ISL_2200270, EPI_ISL_2200271, EPI_ISL_2200272, EPI_ISL_2200273, EPI_ISL_2200274, EPI_ISL_2200275, EPI_ISL_2200276, EPI_ISL_2200277, EPI_ISL_2200278, EPI_ISL_2200279, EPI_ISL_2200280, EPI_ISL_2200281, EPI_ISL_2200282, EPI_ISL_2200283, EPI_ISL_2200284, EPI_ISL_2200285, EPI_ISL_2200286, EPI_ISL_2200287, EPI_ISL_2200288, EPI_ISL_2200289, EPI_ISL_2200290, EPI_ISL_2200291, EPI_ISL_2200292, EPI_ISL_2200293, EPI_ISL_2200294, EPI_ISL_2200295, EPI_ISL_2200296, EPI_ISL_2200297, EPI_ISL_2200298, EPI_ISL_2200299, EPI_ISL_2200300, EPI_ISL_2200301, EPI_ISL_2200302, EPI_ISL_2200303, EPI_ISL_2200304, EPI_ISL_2200305, EPI_ISL_2200306, EPI_ISL_2200307, EPI_ISL_2200308, EPI_ISL_2200309, EPI_ISL_2200311, EPI_ISL_2200312, EPI_ISL_2200313, EPI_ISL_2200314, EPI_ISL_2200315, EPI_ISL_2200316, EPI_ISL_2200317, EPI_ISL_2200318, EPI_ISL_2200319, EPI_ISL_2200320, EPI_ISL_2200321, EPI_ISL_2200322, EPI_ISL_2200323, EPI_ISL_2200324, EPI_ISL_2200325, EPI_ISL_2200326, EPI_ISL_2200327, EPI_ISL_2200328, EPI_ISL_2200329, EPI_ISL_2200330, EPI_ISL_2200331, EPI_ISL_2200332, EPI_ISL_2200333, EPI_ISL_2200334, EPI_ISL_2200335, EPI_ISL_2200336, EPI_ISL_2200337, EPI_ISL_2200338, EPI_ISL_2200339, EPI_ISL_2200340, EPI_ISL_2200341, EPI_ISL_2200342, EPI_ISL_2200343, EPI_ISL_2200344, EPI_ISL_2200345, EPI_ISL_2200346, EPI_ISL_2200347, EPI_ISL_2200348, EPI_ISL_2200349, EPI_ISL_2200350, EPI_ISL_2200351, EPI_ISL_2200352, EPI_ISL_2200353, EPI_ISL_2200354, EPI_ISL_2200355, EPI_ISL_2200356, EPI_ISL_2200357, EPI_ISL_2200358, EPI_ISL_2200359, EPI_ISL_2200360, EPI_ISL_2200361, EPI_ISL_2200362, EPI_ISL_2200363, EPI_ISL_2200364, EPI_ISL_2200365, EPI_ISL_2200366, EPI_ISL_2200367, EPI_ISL_2200368, EPI_ISL_2200369, EPI_ISL_2200370, EPI_ISL_2200371, EPI_ISL_2200372, EPI_ISL_2200373, EPI_ISL_2200374, EPI_ISL_2200375, EPI_ISL_2200376, EPI_ISL_2200377, EPI_ISL_2200378, EPI_ISL_2200379, EPI_ISL_2200380, EPI_ISL_2200381, EPI_ISL_2200382, EPI_ISL_2200383, EPI_ISL_2200384, EPI_ISL_2200385, EPI_ISL_2200386, EPI_ISL_2200387, EPI_ISL_2200388, EPI_ISL_2200389, EPI_ISL_2200390, EPI_ISL_2200391, EPI_ISL_2200392, EPI_ISL_2200393, EPI_ISL_2200394, EPI_ISL_2200395, EPI_ISL_2200396, EPI_ISL_2200397, EPI_ISL_2200398, EPI_ISL_2200399, EPI_ISL_2200400, EPI_ISL_2200401, EPI_ISL_2200402, EPI_ISL_2200403, EPI_ISL_2200404, EPI_ISL_2200405, EPI_ISL_2200407, EPI_ISL_2200408, EPI_ISL_2200409, EPI_ISL_2200410, EPI_ISL_2200411, EPI_ISL_2200412, EPI_ISL_2200413, EPI_ISL_2200414, EPI_ISL_2200415, EPI_ISL_2200416, EPI_ISL_2200417, EPI_ISL_2200418, EPI_ISL_2200419, EPI_ISL_2200420, EPI_ISL_2200421, EPI_ISL_2200422, EPI_ISL_2200423, EPI_ISL_2200424, EPI_ISL_2200425, EPI_ISL_2200426, EPI_ISL_2200427, EPI_ISL_2200428, EPI_ISL_2200429, EPI_ISL_2200430, EPI_ISL_2200431, EPI_ISL_2200432, EPI_ISL_2200433, EPI_ISL_2200434, EPI_ISL_2200435, EPI_ISL_2200436, EPI_ISL_2200437, EPI_ISL_2200438, EPI_ISL_2200439, EPI_ISL_2200440, EPI_ISL_2200441, EPI_ISL_2200442, EPI_ISL_2200443, EPI_ISL_2200444, EPI_ISL_2200445, EPI_ISL_2200446, EPI_ISL_2200447, EPI_ISL_2200448, EPI_ISL_2200449, EPI_ISL_2200450, EPI_ISL_2200451, EPI_ISL_2200452, EPI_ISL_2200453, EPI_ISL_2200454, EPI_ISL_2200455, EPI_ISL_2200456, EPI_ISL_2200457, EPI_ISL_2200458, EPI_ISL_2200459, EPI_ISL_2200460, EPI_ISL_2200461, EPI_ISL_2200462, EPI_ISL_2200463, EPI_ISL_2200464, EPI_ISL_2200465, EPI_ISL_2200466, EPI_ISL_2200467 | | | |
| see above | Houston Methodist Hospital | Houston Methodist Hospital | Randall J. Olsen, Paul A. Christensen, S. Wesley Long, Sishir Subedi, Robert Olson, Marcus Nguyen, James J. Davis, Matthew Ojeda Saavedra, Prasanti Yerramilli, Layne Pruitt, Kristina Reppond, Madison N. Shyer, Jessica Cambric, Ryan Gadd, Ilya J. Finkelstein, Jimmy Gollihar, and James M. Musser |
| EPI_ISL_2200468 | Ohio Department of Health Laboratory | Ohio Department of Health Laboratory | Holmes, Jennifer; Eric Brandt, Keoni Omura, Glen McGillivary, Caitlin McDonnell, Jade Mowery, Stephanie Mccracken, Tyler Payne, Kirtana Ramadugu, Erica Leasure, Brent Lee, Kelsey Florek, Heather Blankenship, Allison Black, Quanta Brown, and Tammy Bannerman |
| EPI_ISL_2200469, EPI_ISL_2200470, EPI_ISL_2200471, EPI_ISL_2200472, EPI_ISL_2200473, EPI_ISL_2200474, EPI_ISL_2200475, EPI_ISL_2200476 | Houston Methodist Hospital | Houston Methodist Hospital | Randall J. Olsen, Paul A. Christensen, S. Wesley Long, Sishir Subedi, Robert Olson, Marcus Nguyen, James J. Davis, Matthew Ojeda Saavedra, Prasanti Yerramilli, Layne Pruitt, Kristina Reppond, Madison N. Shyer, Jessica Cambric, Ryan Gadd, Ilya J. Finkelstein, Jimmy Gollihar, and James M. Musser |
| EPI_ISL_2200477 | Kentucky State Public Health Lab | Kentucky State Public Health Lab | Anjayya Tummala, Karim George, Joshua Tobias, William Grooms, Vaneet Arora, Matthew Johnson, Rachel Zinner, Rhonda Lucas |
| EPI_ISL_2200478 | Houston Methodist Hospital | Houston Methodist Hospital | Randall J. Olsen, Paul A. Christensen, S. Wesley Long, Sishir Subedi, Robert Olson, Marcus Nguyen, James J. Davis, Matthew Ojeda Saavedra, Prasanti Yerramilli, Layne Pruitt, Kristina Reppond, Madison N. Shyer, Jessica Cambric, Ryan Gadd, Ilya J. Finkelstein, Jimmy Gollihar, and James M. Musser |
| EPI_ISL_2200479, EPI_ISL_2200480, EPI_ISL_2200481 | Kentucky State Public Health Lab | Kentucky State Public Health Lab | Anjayya Tummala, Karim George, Joshua Tobias, William Grooms, Vaneet Arora, Matthew Johnson, Rachel Zinner, Rhonda Lucas |
| EPI_ISL_2200482 | Houston Methodist Hospital | Houston Methodist Hospital | Randall J. Olsen, Paul A. Christensen, S. Wesley Long, Sishir Subedi, Robert Olson, Marcus Nguyen, James J. Davis, Matthew Ojeda Saavedra, Prasanti Yerramilli, Layne Pruitt, Kristina Reppond, Madison N. Shyer, Jessica Cambric, Ryan Gadd, Ilya J. Finkelstein, Jimmy Gollihar, and James M. Musser |
| EPI_ISL_2200483 | Kentucky State Public Health Lab | Kentucky State Public Health Lab | Anjayya Tummala, Karim George, Joshua Tobias, William Grooms, Vaneet Arora, Matthew Johnson, Rachel Zinner, Rhonda Lucas |
| EPI_ISL_2200484 | Houston Methodist Hospital | Houston Methodist Hospital | Randall J. Olsen, Paul A. Christensen, S. Wesley Long, Sishir Subedi, Robert Olson, Marcus Nguyen, James J. Davis, Matthew Ojeda Saavedra, Prasanti Yerramilli, Layne Pruitt, Kristina Reppond, Madison N. Shyer, Jessica Cambric, Ryan Gadd, Ilya J. Finkelstein, Jimmy Gollihar, and James M. Musser |
| EPI_ISL_2200485, EPI_ISL_2200486 | Kentucky State Public Health Lab | Kentucky State Public Health Lab | Anjayya Tummala, Karim George, Joshua Tobias, William Grooms, Vaneet Arora, Matthew Johnson, Rachel Zinner, Rhonda Lucas |
| EPI_ISL_2200487 | Houston Methodist Hospital | Houston Methodist Hospital | Randall J. Olsen, Paul A. Christensen, S. Wesley Long, Sishir Subedi, Robert Olson, Marcus Nguyen, James J. Davis, Matthew Ojeda Saavedra, Prasanti Yerramilli, Layne Pruitt, Kristina Reppond, Madison N. Shyer, Jessica Cambric, Ryan Gadd, Ilya J. Finkelstein, Jimmy Gollihar, and James M. Musser |
| EPI_ISL_2200488, EPI_ISL_2200489 | Kentucky State Public Health Lab | Kentucky State Public Health Lab | Anjayya Tummala, Karim George, Joshua Tobias, William Grooms, Vaneet Arora, Matthew Johnson, Rachel Zinner, Rhonda Lucas |
| EPI_ISL_2200490 | Houston Methodist Hospital | Houston Methodist Hospital | Randall J. Olsen, Paul A. Christensen, S. Wesley Long, Sishir Subedi, Robert Olson, Marcus Nguyen, James J. Davis, Matthew Ojeda Saavedra, Prasanti Yerramilli, Layne Pruitt, Kristina Reppond, Madison N. Shyer, Jessica Cambric, Ryan Gadd, Ilya J. Finkelstein, Jimmy Gollihar, and James M. Musser |
| EPI_ISL_2200492 | Kentucky State Public Health Lab | Kentucky State Public Health Lab | Anjayya Tummala, Karim George, Joshua Tobias, William Grooms, Vaneet Arora, Matthew Johnson, Rachel Zinner, Rhonda Lucas |
| EPI_ISL_2200493 | Houston Methodist Hospital | Houston Methodist Hospital | Randall J. Olsen, Paul A. Christensen, S. Wesley Long, Sishir Subedi, Robert Olson, Marcus Nguyen, James J. Davis, Matthew Ojeda Saavedra, Prasanti Yerramilli, Layne Pruitt, Kristina Reppond, Madison N. Shyer, Jessica Cambric, Ryan Gadd, Ilya J. Finkelstein, Jimmy Gollihar, and James M. Musser |
| EPI_ISL_2200494, EPI_ISL_2200495, EPI_ISL_2200496 | Kentucky State Public Health Lab | Kentucky State Public Health Lab | Anjayya Tummala, Karim George, Joshua Tobias, William Grooms, Vaneet Arora, Matthew Johnson, Rachel Zinner, Rhonda Lucas |
| EPI_ISL_2200497 | Houston Methodist Hospital | Houston Methodist Hospital | Randall J. Olsen, Paul A. Christensen, S. Wesley Long, Sishir Subedi, Robert Olson, Marcus Nguyen, James J. Davis, Matthew Ojeda Saavedra, Prasanti Yerramilli, Layne Pruitt, Kristina Reppond, Madison N. Shyer, Jessica Cambric, Ryan Gadd, Ilya J. Finkelstein, Jimmy Gollihar, and James M. Musser |
| EPI_ISL_2200498, EPI_ISL_2200499 | Kentucky State Public Health Lab | Kentucky State Public Health Lab | Anjayya Tummala, Karim George, Joshua Tobias, William Grooms, Vaneet Arora, Matthew Johnson, Rachel Zinner, Rhonda Lucas |
| EPI_ISL_2200500 | Houston Methodist Hospital | Houston Methodist Hospital | Randall J. Olsen, Paul A. Christensen, S. Wesley Long, Sishir Subedi, Robert Olson, Marcus Nguyen, James J. Davis, Matthew Ojeda Saavedra, Prasanti Yerramilli, Layne Pruitt, Kristina Reppond, Madison N. Shyer, Jessica Cambric, Ryan Gadd, Ilya J. Finkelstein, Jimmy Gollihar, and James M. Musser |
| EPI_ISL_2200501 | Kentucky State Public Health Lab | Kentucky State Public Health Lab | Anjayya Tummala, Karim George, Joshua Tobias, William Grooms, Vaneet Arora, Matthew Johnson, Rachel Zinner, Rhonda Lucas |
| EPI_ISL_2200503 | Houston Methodist Hospital | Houston Methodist Hospital | Randall J. Olsen, Paul A. Christensen, S. Wesley Long, Sishir Subedi, Robert Olson, Marcus Nguyen, James J. Davis, Matthew Ojeda Saavedra, Prasanti Yerramilli, Layne Pruitt, Kristina Reppond, Madison N. Shyer, Jessica Cambric, Ryan Gadd, Ilya J. Finkelstein, Jimmy Gollihar, and James M. Musser |
| EPI_ISL_2200504 | Kentucky State Public Health Lab | Kentucky State Public Health Lab | Anjayya Tummala, Karim George, Joshua Tobias, William Grooms, Vaneet Arora, Matthew Johnson, Rachel Zinner, Rhonda Lucas |
| EPI_ISL_2200505, EPI_ISL_2200506, EPI_ISL_2200507, EPI_ISL_2200508, EPI_ISL_2200509, EPI_ISL_2200510, EPI_ISL_2200511, EPI_ISL_2200512, EPI_ISL_2200513, EPI_ISL_2200514, EPI_ISL_2200515, EPI_ISL_2200516, EPI_ISL_2200517, EPI_ISL_2200518, EPI_ISL_2200519, EPI_ISL_2200520, EPI_ISL_2200521, EPI_ISL_2200522, EPI_ISL_2200523, EPI_ISL_2200524, EPI_ISL_2200525, EPI_ISL_2200526, EPI_ISL_2200527, EPI_ISL_2200528, EPI_ISL_2200529, EPI_ISL_2200530, EPI_ISL_2200531, EPI_ISL_2200532, EPI_ISL_2200533, EPI_ISL_2200534, EPI_ISL_2200535, EPI_ISL_2200536, EPI_ISL_2200537, EPI_ISL_2200538, EPI_ISL_2200539, EPI_ISL_2200540, EPI_ISL_2200541, EPI_ISL_2200542, EPI_ISL_2200543, EPI_ISL_2200544, EPI_ISL_2200545, EPI_ISL_2200546, EPI_ISL_2200547, EPI_ISL_2200548, EPI_ISL_2200549, EPI_ISL_2200550, EPI_ISL_2200551, EPI_ISL_2200552, EPI_ISL_2200553, EPI_ISL_2200554, EPI_ISL_2200555, EPI_ISL_2200556, EPI_ISL_2200557, EPI_ISL_2200558, EPI_ISL_2200559, EPI_ISL_2200560, EPI_ISL_2200561, EPI_ISL_2200562, EPI_ISL_2200563, EPI_ISL_2200564, EPI_ISL_2200565, EPI_ISL_2200566, EPI_ISL_2200567, EPI_ISL_2200568, EPI_ISL_2200569, EPI_ISL_2200570, EPI_ISL_2200571, EPI_ISL_2200572, EPI_ISL_2200573, EPI_ISL_2200574, EPI_ISL_2200575, EPI_ISL_2200576, EPI_ISL_2200577, EPI_ISL_2200578, EPI_ISL_2200579, EPI_ISL_2200580, EPI_ISL_2200581, EPI_ISL_2200582, EPI_ISL_2200583, EPI_ISL_2200584, EPI_ISL_2200585, EPI_ISL_2200586, EPI_ISL_2200587, EPI_ISL_2200588, EPI_ISL_2200589, EPI_ISL_2200590, EPI_ISL_2200591, EPI_ISL_2200592, EPI_ISL_2200593, EPI_ISL_2200594, EPI_ISL_2200595, EPI_ISL_2200596, EPI_ISL_2200597, EPI_ISL_2200598, EPI_ISL_2200599, EPI_ISL_2200600, EPI_ISL_2200601, EPI_ISL_2200602, EPI_ISL_2200603, EPI_ISL_2200604, EPI_ISL_2200605, EPI_ISL_2200606, EPI_ISL_2200607, EPI_ISL_2200608, EPI_ISL_2200609, EPI_ISL_2200611, EPI_ISL_2200612, EPI_ISL_2200613, EPI_ISL_2200614, EPI_ISL_2200615, EPI_ISL_2200617, EPI_ISL_2200619, EPI_ISL_2200620, EPI_ISL_2200621, EPI_ISL_2200622, EPI_ISL_2200623, EPI_ISL_2200624, EPI_ISL_2200625, EPI_ISL_2200626, EPI_ISL_2200627, EPI_ISL_2200628, EPI_ISL_2200629, EPI_ISL_2200630, EPI_ISL_2200631, EPI_ISL_2200632, EPI_ISL_2200634, EPI_ISL_2200637, EPI_ISL_2200640, EPI_ISL_2200644, EPI_ISL_2200647, EPI_ISL_2200650, EPI_ISL_2200653, EPI_ISL_2200657, EPI_ISL_2200661, EPI_ISL_2200665, EPI_ISL_2200668, EPI_ISL_2200671, EPI_ISL_2200674, EPI_ISL_2200677, EPI_ISL_2200680, EPI_ISL_2200683, EPI_ISL_2200686, EPI_ISL_2200689, EPI_ISL_2200691, EPI_ISL_2200692, EPI_ISL_2200693, EPI_ISL_2200694, EPI_ISL_2200695, EPI_ISL_2200696, EPI_ISL_2200697, EPI_ISL_2200698, EPI_ISL_2200699, EPI_ISL_2200700, EPI_ISL_2200701, EPI_ISL_2200702, EPI_ISL_2200703, EPI_ISL_2200704, EPI_ISL_2200705, EPI_ISL_2200706, EPI_ISL_2200707, EPI_ISL_2200708, EPI_ISL_2200709, EPI_ISL_2200710, EPI_ISL_2200711, EPI_ISL_2200712, EPI_ISL_2200713, EPI_ISL_2200714, EPI_ISL_2200715, EPI_ISL_2200716, EPI_ISL_2200717, EPI_ISL_2200719, EPI_ISL_2200720, EPI_ISL_2200721, EPI_ISL_2200722, EPI_ISL_2200724, EPI_ISL_2200725, EPI_ISL_2200726, EPI_ISL_2200727, EPI_ISL_2200728, EPI_ISL_2200729, EPI_ISL_2200730, EPI_ISL_2200732, EPI_ISL_2200733, EPI_ISL_2200734, EPI_ISL_2200735, EPI_ISL_2200736, EPI_ISL_2200737, EPI_ISL_2200738, EPI_ISL_2200739, EPI_ISL_2200740, EPI_ISL_2200741, EPI_ISL_2200742, EPI_ISL_2200743, EPI_ISL_2200744, EPI_ISL_2200745, EPI_ISL_2200746, EPI_ISL_2200747, EPI_ISL_2200748, EPI_ISL_2200749, EPI_ISL_2200750, EPI_ISL_2200751 | | | |
| see above | Houston Methodist Hospital | Houston Methodist Hospital | Randall J. Olsen, Paul A. Christensen, S. Wesley Long, Sishir Subedi, Robert Olson, Marcus Nguyen, James J. Davis, Matthew Ojeda Saavedra, Prasanti Yerramilli, Layne Pruitt, Kristina Reppond, Madison N. Shyer, Jessica Cambric, Ryan Gadd, Ilya J. Finkelstein, Jimmy Gollihar, and James M. Musser |
| EPI_ISL_2200752 | Ohio Department of Health Laboratory | Ohio Department of Health Laboratory | Holmes, Jennifer; Eric Brandt, Keoni Omura, Glen McGillivary, Caitlin McDonnell, Jade Mowery, Stephanie Mccracken, Tyler Payne, Kirtana Ramadugu, Erica Leasure, Brent Lee, Kelsey Florek, Heather Blankenship, Allison Black, Quanta Brown, and Tammy Bannerman |
| EPI_ISL_2200753, EPI_ISL_2200754, EPI_ISL_2200755, EPI_ISL_2200756, EPI_ISL_2200757, EPI_ISL_2200758, EPI_ISL_2200759, EPI_ISL_2200760, EPI_ISL_2200761, EPI_ISL_2200762, EPI_ISL_2200763, EPI_ISL_2200764, EPI_ISL_2200765, EPI_ISL_2200766, EPI_ISL_2200767, EPI_ISL_2200768, EPI_ISL_2200769, EPI_ISL_2200770, EPI_ISL_2200771, EPI_ISL_2200772, EPI_ISL_2200773, EPI_ISL_2200774, EPI_ISL_2200775, EPI_ISL_2200776, EPI_ISL_2200777, EPI_ISL_2200778, EPI_ISL_2200779, EPI_ISL_2200780, EPI_ISL_2200781, EPI_ISL_2200782, EPI_ISL_2200783, EPI_ISL_2200784, EPI_ISL_2200785, EPI_ISL_2200786, EPI_ISL_2200787, EPI_ISL_2200788, EPI_ISL_2200789, EPI_ISL_2200790, EPI_ISL_2200791, EPI_ISL_2200792, EPI_ISL_2200793, EPI_ISL_2200794, EPI_ISL_2200795, EPI_ISL_2200796, | | | |

EPI_ISL_2200797, EPI_ISL_2200798, EPI_ISL_2200799, EPI_ISL_2200800, EPI_ISL_2200801, EPI_ISL_2200802, EPI_ISL_2200803, EPI_ISL_2200804, EPI_ISL_2200805, EPI_ISL_2200806, EPI_ISL_2200807, EPI_ISL_2200808, EPI_ISL_2200809, EPI_ISL_2200810, EPI_ISL_2200811, EPI_ISL_2200812, EPI_ISL_2200813, EPI_ISL_2200814, EPI_ISL_2200815, EPI_ISL_2200816, EPI_ISL_2200818, EPI_ISL_2200819, EPI_ISL_2200820, EPI_ISL_2200821, EPI_ISL_2200822, EPI_ISL_2200823, EPI_ISL_2200824, EPI_ISL_2200825, EPI_ISL_2200826, EPI_ISL_2200827, EPI_ISL_2200828, EPI_ISL_2200829, EPI_ISL_2200830, EPI_ISL_2200831, EPI_ISL_2200832, EPI_ISL_2200833, EPI_ISL_2200834, EPI_ISL_2200835, EPI_ISL_2200836, EPI_ISL_2200837, EPI_ISL_2200838, EPI_ISL_2200840, EPI_ISL_2200841, EPI_ISL_2200842, EPI_ISL_2200843, EPI_ISL_2200844, EPI_ISL_2200846, EPI_ISL_2200847, EPI_ISL_2200848, EPI_ISL_2200849, EPI_ISL_2200850, EPI_ISL_2200851, EPI_ISL_2200852, EPI_ISL_2200853, EPI_ISL_2200854, EPI_ISL_2200855, EPI_ISL_2200856, EPI_ISL_2200857, EPI_ISL_2200858, EPI_ISL_2200859, EPI_ISL_2200860, EPI_ISL_2200861, EPI_ISL_2200862, EPI_ISL_2200863, EPI_ISL_2200864, EPI_ISL_2200865, EPI_ISL_2200866, EPI_ISL_2200867, EPI_ISL_2200869, EPI_ISL_2200870, EPI_ISL_2200871, EPI_ISL_2200872, EPI_ISL_2200873, EPI_ISL_2200874, EPI_ISL_2200875, EPI_ISL_2200876, EPI_ISL_2200877, EPI_ISL_2200878, EPI_ISL_2200879, EPI_ISL_2200880, EPI_ISL_2200881, EPI_ISL_2200882, EPI_ISL_2200883, EPI_ISL_2200884, EPI_ISL_2200885, EPI_ISL_2200886, EPI_ISL_2200887, EPI_ISL_2200888, EPI_ISL_2200889, EPI_ISL_2200890, EPI_ISL_2200891, EPI_ISL_2200892, EPI_ISL_2200893, EPI_ISL_2200894, EPI_ISL_2200895, EPI_ISL_2200896, EPI_ISL_2200897, EPI_ISL_2200898, EPI_ISL_2200899, EPI_ISL_2200900, EPI_ISL_2200901, EPI_ISL_2200902, EPI_ISL_2200903, EPI_ISL_2200904, EPI_ISL_2200905, EPI_ISL_2200906, EPI_ISL_2200907, EPI_ISL_2200908, EPI_ISL_2200909, EPI_ISL_2200910, EPI_ISL_2200911, EPI_ISL_2200912, EPI_ISL_2200913, EPI_ISL_2200914, EPI_ISL_2200915, EPI_ISL_2200916, EPI_ISL_2200917, EPI_ISL_2200918, EPI_ISL_2200919, EPI_ISL_2200920, EPI_ISL_2200921, EPI_ISL_2200922, EPI_ISL_2200923, EPI_ISL_2200924, EPI_ISL_2200925, EPI_ISL_2200926, EPI_ISL_2200927, EPI_ISL_2200928, EPI_ISL_2200929, EPI_ISL_2200930, EPI_ISL_2200931, EPI_ISL_2200932, EPI_ISL_2200933, EPI_ISL_2200934, EPI_ISL_2200935, EPI_ISL_2200936, EPI_ISL_2200937, EPI_ISL_2200938, EPI_ISL_2200939, EPI_ISL_2200941, EPI_ISL_2200942, EPI_ISL_2200943, EPI_ISL_2200944, EPI_ISL_2200945, EPI_ISL_2200946, EPI_ISL_2200947, EPI_ISL_2200948, EPI_ISL_2200949, EPI_ISL_2200950, EPI_ISL_2200951, EPI_ISL_2200953, EPI_ISL_2200955, EPI_ISL_2200956, EPI_ISL_2200957, EPI_ISL_2200958, EPI_ISL_2200959, EPI_ISL_2200960, EPI_ISL_2200961, EPI_ISL_2200962, EPI_ISL_2200963, EPI_ISL_2200964, EPI_ISL_2200965, EPI_ISL_2200966, EPI_ISL_2200967, EPI_ISL_2200968, EPI_ISL_2200969, EPI_ISL_2200970, EPI_ISL_2200971, EPI_ISL_2200972, EPI_ISL_2200973, EPI_ISL_2200974, EPI_ISL_2200975, EPI_ISL_2200976, EPI_ISL_2200977, EPI_ISL_2200978, EPI_ISL_2200979, EPI_ISL_2200980, EPI_ISL_2200981, EPI_ISL_2200982, EPI_ISL_2200983, EPI_ISL_2200984, EPI_ISL_2200985, EPI_ISL_2200986, EPI_ISL_2200987, EPI_ISL_2200988, EPI_ISL_2200989, EPI_ISL_2200992, EPI_ISL_2200993, EPI_ISL_2200994, EPI_ISL_2200995, EPI_ISL_2200997, EPI_ISL_2200998, EPI_ISL_2200999, EPI_ISL_2201000, EPI_ISL_2201001, EPI_ISL_2201002, EPI_ISL_2201003, EPI_ISL_2201004, EPI_ISL_2201005, EPI_ISL_2201006, EPI_ISL_2201007, EPI_ISL_2201008, EPI_ISL_2201009, EPI_ISL_2201011, EPI_ISL_2201012, EPI_ISL_2201013, EPI_ISL_2201014, EPI_ISL_2201015, EPI_ISL_2201016, EPI_ISL_2201017, EPI_ISL_2201018, EPI_ISL_2201019, EPI_ISL_2201020, EPI_ISL_2201021, EPI_ISL_2201022, EPI_ISL_2201023, EPI_ISL_2201024, EPI_ISL_2201025, EPI_ISL_2201026, EPI_ISL_2201027, EPI_ISL_2201028, EPI_ISL_2201029, EPI_ISL_2201030, EPI_ISL_2201031, EPI_ISL_2201032, EPI_ISL_2201033, EPI_ISL_2201034, EPI_ISL_2201035, EPI_ISL_2201036, EPI_ISL_2201037, EPI_ISL_2201039, EPI_ISL_2201040, EPI_ISL_2201041, EPI_ISL_2201042, EPI_ISL_2201043, EPI_ISL_2201044, EPI_ISL_2201045, EPI_ISL_2201046, EPI_ISL_2201047, EPI_ISL_2201048, EPI_ISL_2201049, EPI_ISL_2201050, EPI_ISL_2201051, EPI_ISL_2201052, EPI_ISL_2201053, EPI_ISL_2201054, EPI_ISL_2201055, EPI_ISL_2201057, EPI_ISL_2201058, EPI_ISL_2201059, EPI_ISL_2201060, EPI_ISL_2201061, EPI_ISL_2201062, EPI_ISL_2201063, EPI_ISL_2201064, EPI_ISL_2201065, EPI_ISL_2201066, EPI_ISL_2201067, EPI_ISL_2201068, EPI_ISL_2201069, EPI_ISL_2201070, EPI_ISL_2201071, EPI_ISL_2201072, EPI_ISL_2201073, EPI_ISL_2201074, EPI_ISL_2201075, EPI_ISL_2201076, EPI_ISL_2201077, EPI_ISL_2201078, EPI_ISL_2201079, EPI_ISL_2201080, EPI_ISL_2201082, EPI_ISL_2201083, EPI_ISL_2201084, EPI_ISL_2201085, EPI_ISL_2201086, EPI_ISL_2201087, EPI_ISL_2201088, EPI_ISL_2201089, EPI_ISL_2201090, EPI_ISL_2201091, EPI_ISL_2201092, EPI_ISL_2201093, EPI_ISL_2201094, EPI_ISL_2201095, EPI_ISL_2201096, EPI_ISL_2201097, EPI_ISL_2201098, EPI_ISL_2201099, EPI_ISL_2201100, EPI_ISL_2201101, EPI_ISL_2201102, EPI_ISL_2201103, EPI_ISL_2201104, EPI_ISL_2201105, EPI_ISL_2201106, EPI_ISL_2201107, EPI_ISL_2201108, EPI_ISL_2201109, EPI_ISL_2201110, EPI_ISL_2201111, EPI_ISL_2201112, EPI_ISL_2201113, EPI_ISL_2201114, EPI_ISL_2201115, EPI_ISL_2201116, EPI_ISL_2201117, EPI_ISL_2201118, EPI_ISL_2201119

| | | | |
|---|---|---|---|
| see above | Houston Methodist Hospital | Houston Methodist Hospital | Randall J. Olsen, Paul A. Christensen, S. Wesley Long, Sishir Subedi, Robert Olson, Marcus Nguyen, James J. Davis, Matthew Ojeda Saavedra, Prasanti Yerramilli, Layne Pruitt, Kristina Reppond, Madison N. Shyer, Jessica Cambric, Ryan Gadd, Ilya J. Finkelstein, Jimmy Gollihar, and James M. Musser |
| EPI_ISL_2201120, EPI_ISL_2201121 | Aegis Sciences Corporation | Centers for Disease Control and Prevention Division of Viral Diseases, Pathogen Discovery | Dakota Howard, Dhwani Batra, Peter W. Cook, Kara Moser, Adrian Paskey, Jason Caravas, Benjamin Rambo-Martin, Shatavia Morrison, Christopher Gulvick, Scott Sammons, Yvette Unoarumhi, Darlene Wagner, Matthew Schmerer, Cyndi Clark, Patrick Campbell, Rob Case, Vikramsinha Ghorpade, Holly Houdeshell, Ola Kvalvaag, Dillon Nall, Ethan Sanders, Alec Vest, Shaun Westlund, Matthew Hardison, Clinton R. Paden, Duncan MacCannell |
| EPI_ISL_2201122 | Houston Methodist Hospital | Houston Methodist Hospital | Randall J. Olsen, Paul A. Christensen, S. Wesley Long, Sishir Subedi, Robert Olson, Marcus Nguyen, James J. Davis, Matthew Ojeda Saavedra, Prasanti Yerramilli, Layne Pruitt, Kristina Reppond, Madison N. Shyer, Jessica Cambric, Ryan Gadd, Ilya J. Finkelstein, Jimmy Gollihar, and James M. Musser |
| EPI_ISL_2201123 | Aegis Sciences Corporation | Centers for Disease Control and Prevention Division of Viral Diseases, Pathogen Discovery | Dakota Howard, Dhwani Batra, Peter W. Cook, Kara Moser, Adrian Paskey, Jason Caravas, Benjamin Rambo-Martin, Shatavia Morrison, Christopher Gulvick, Scott Sammons, Yvette Unoarumhi, Darlene Wagner, Matthew Schmerer, Cyndi Clark, Patrick Campbell, Rob Case, Vikramsinha Ghorpade, Holly Houdeshell, Ola Kvalvaag, Dillon Nall, Ethan Sanders, Alec Vest, Shaun Westlund, Matthew Hardison, Clinton R. Paden, Duncan MacCannell |
| EPI_ISL_2201124 | Houston Methodist Hospital | Houston Methodist Hospital | Randall J. Olsen, Paul A. Christensen, S. Wesley Long, Sishir Subedi, Robert Olson, Marcus Nguyen, James J. Davis, Matthew Ojeda Saavedra, Prasanti Yerramilli, Layne Pruitt, Kristina Reppond, Madison N. Shyer, Jessica Cambric, Ryan Gadd, Ilya J. Finkelstein, Jimmy Gollihar, and James M. Musser |
| EPI_ISL_2201125 | Aegis Sciences Corporation | Centers for Disease Control and Prevention Division of Viral Diseases, Pathogen Discovery | Dakota Howard, Dhwani Batra, Peter W. Cook, Kara Moser, Adrian Paskey, Jason Caravas, Benjamin Rambo-Martin, Shatavia Morrison, Christopher Gulvick, Scott Sammons, Yvette Unoarumhi, Darlene Wagner, Matthew Schmerer, Cyndi Clark, Patrick Campbell, Rob Case, Vikramsinha Ghorpade, Holly Houdeshell, Ola Kvalvaag, Dillon Nall, Ethan Sanders, Alec Vest, Shaun Westlund, Matthew Hardison, Clinton R. Paden, Duncan MacCannell |
| EPI_ISL_2201126 | Houston Methodist Hospital | Houston Methodist Hospital | Randall J. Olsen, Paul A. Christensen, S. Wesley Long, Sishir Subedi, Robert Olson, Marcus Nguyen, James J. Davis, Matthew Ojeda Saavedra, Prasanti Yerramilli, Layne Pruitt, Kristina Reppond, Madison N. Shyer, Jessica Cambric, Ryan Gadd, Ilya J. Finkelstein, Jimmy Gollihar, and James M. Musser |
| EPI_ISL_2201127 | Aegis Sciences Corporation | Centers for Disease Control and Prevention Division of Viral Diseases, Pathogen Discovery | Dakota Howard, Dhwani Batra, Peter W. Cook, Kara Moser, Adrian Paskey, Jason Caravas, Benjamin Rambo-Martin, Shatavia Morrison, Christopher Gulvick, Scott Sammons, Yvette Unoarumhi, Darlene Wagner, Matthew Schmerer, Cyndi Clark, Patrick Campbell, Rob Case, Vikramsinha Ghorpade, Holly Houdeshell, Ola Kvalvaag, Dillon Nall, Ethan Sanders, Alec Vest, Shaun Westlund, Matthew Hardison, Clinton R. Paden, Duncan MacCannell |
| EPI_ISL_2201128 | Houston Methodist Hospital | Houston Methodist Hospital | Randall J. Olsen, Paul A. Christensen, S. Wesley Long, Sishir Subedi, Robert Olson, Marcus Nguyen, James J. Davis, Matthew Ojeda Saavedra, Prasanti Yerramilli, Layne Pruitt, Kristina Reppond, Madison N. Shyer, Jessica Cambric, Ryan Gadd, Ilya J. Finkelstein, Jimmy Gollihar, and James M. Musser |
| EPI_ISL_2201129 | Aegis Sciences Corporation | Centers for Disease Control and Prevention Division of Viral Diseases, Pathogen Discovery | Dakota Howard, Dhwani Batra, Peter W. Cook, Kara Moser, Adrian Paskey, Jason Caravas, Benjamin Rambo-Martin, Shatavia Morrison, Christopher Gulvick, Scott Sammons, Yvette Unoarumhi, Darlene Wagner, Matthew Schmerer, Cyndi Clark, Patrick Campbell, Rob Case, Vikramsinha Ghorpade, Holly Houdeshell, Ola Kvalvaag, Dillon Nall, Ethan Sanders, Alec Vest, Shaun Westlund, Matthew Hardison, Clinton R. Paden, Duncan MacCannell |
| EPI_ISL_2201130 | Houston Methodist Hospital | Houston Methodist Hospital | Randall J. Olsen, Paul A. Christensen, S. Wesley Long, Sishir Subedi, Robert Olson, Marcus Nguyen, James J. Davis, Matthew Ojeda Saavedra, Prasanti Yerramilli, Layne Pruitt, Kristina Reppond, Madison N. Shyer, Jessica Cambric, Ryan Gadd, Ilya J. Finkelstein, Jimmy Gollihar, and James M. Musser |
| EPI_ISL_2201131 | Aegis Sciences Corporation | Centers for Disease Control and Prevention Division of Viral Diseases, Pathogen Discovery | Dakota Howard, Dhwani Batra, Peter W. Cook, Kara Moser, Adrian Paskey, Jason Caravas, Benjamin Rambo-Martin, Shatavia Morrison, Christopher Gulvick, Scott Sammons, Yvette Unoarumhi, Darlene Wagner, Matthew Schmerer, Cyndi Clark, Patrick Campbell, Rob Case, Vikramsinha Ghorpade, Holly Houdeshell, Ola Kvalvaag, Dillon Nall, Ethan Sanders, Alec Vest, Shaun Westlund, Matthew Hardison, Clinton R. Paden, Duncan MacCannell |
| EPI_ISL_2201132 | Houston Methodist Hospital | Houston Methodist Hospital | Randall J. Olsen, Paul A. Christensen, S. Wesley Long, Sishir Subedi, Robert Olson, Marcus Nguyen, James J. Davis, Matthew Ojeda Saavedra, Prasanti Yerramilli, Layne Pruitt, Kristina Reppond, Madison N. Shyer, Jessica Cambric, Ryan Gadd, Ilya J. Finkelstein, Jimmy Gollihar, and James M. Musser |
| EPI_ISL_2201133 | Aegis Sciences Corporation | Centers for Disease Control and Prevention Division of Viral Diseases, Pathogen Discovery | Dakota Howard, Dhwani Batra, Peter W. Cook, Kara Moser, Adrian Paskey, Jason Caravas, Benjamin Rambo-Martin, Shatavia Morrison, Christopher Gulvick, Scott Sammons, Yvette Unoarumhi, Darlene Wagner, Matthew Schmerer, Cyndi Clark, Patrick Campbell, Rob Case, Vikramsinha Ghorpade, Holly Houdeshell, Ola Kvalvaag, Dillon Nall, Ethan Sanders, Alec Vest, Shaun Westlund, Matthew Hardison, Clinton R. Paden, Duncan MacCannell |
| EPI_ISL_2201134 | Houston Methodist Hospital | Houston Methodist Hospital | Randall J. Olsen, Paul A. Christensen, S. Wesley Long, Sishir Subedi, Robert Olson, Marcus Nguyen, James J. Davis, Matthew Ojeda Saavedra, Prasanti Yerramilli, Layne Pruitt, Kristina Reppond, Madison N. Shyer, Jessica Cambric, Ryan Gadd, Ilya J. Finkelstein, Jimmy Gollihar, and James M. Musser |
| EPI_ISL_2201135 | Aegis Sciences Corporation | Centers for Disease Control and Prevention Division of Viral Diseases, Pathogen Discovery | Dakota Howard, Dhwani Batra, Peter W. Cook, Kara Moser, Adrian Paskey, Jason Caravas, Benjamin Rambo-Martin, Shatavia Morrison, Christopher Gulvick, Scott Sammons, Yvette Unoarumhi, Darlene Wagner, Matthew Schmerer, Cyndi Clark, Patrick Campbell, Rob Case, Vikramsinha Ghorpade, Holly Houdeshell, Ola Kvalvaag, Dillon Nall, Ethan Sanders, Alec Vest, Shaun Westlund, Matthew Hardison, Clinton R. Paden, Duncan MacCannell |
| EPI_ISL_2201136 | Houston Methodist Hospital | Houston Methodist Hospital | Randall J. Olsen, Paul A. Christensen, S. Wesley Long, Sishir Subedi, Robert Olson, Marcus Nguyen, James J. Davis, Matthew Ojeda Saavedra, Prasanti Yerramilli, Layne Pruitt, Kristina Reppond, Madison N. Shyer, Jessica Cambric, Ryan Gadd, Ilya J. Finkelstein, Jimmy Gollihar, and James M. Musser |
| EPI_ISL_2201137 | Aegis Sciences Corporation | Centers for Disease Control and Prevention Division of Viral Diseases, Pathogen Discovery | Dakota Howard, Dhwani Batra, Peter W. Cook, Kara Moser, Adrian Paskey, Jason Caravas, Benjamin Rambo-Martin, Shatavia Morrison, Christopher Gulvick, Scott Sammons, Yvette Unoarumhi, Darlene Wagner, Matthew Schmerer, Cyndi Clark, Patrick Campbell, Rob Case, Vikramsinha Ghorpade, Holly Houdeshell, Ola Kvalvaag, Dillon Nall, Ethan Sanders, Alec Vest, Shaun Westlund, Matthew Hardison, Clinton R. Paden, Duncan MacCannell |
| EPI_ISL_2201138 | Houston Methodist Hospital | Houston Methodist Hospital | Randall J. Olsen, Paul A. Christensen, S. Wesley Long, Sishir Subedi, Robert Olson, Marcus Nguyen, James J. Davis, Matthew Ojeda Saavedra, Prasanti Yerramilli, Layne Pruitt, Kristina Reppond, Madison N. Shyer, Jessica Cambric, Ryan Gadd, Ilya J. Finkelstein, Jimmy Gollihar, and James M. Musser |
| EPI_ISL_2201139 | Aegis Sciences Corporation | Centers for Disease Control and Prevention Division of Viral Diseases, Pathogen Discovery | Dakota Howard, Dhwani Batra, Peter W. Cook, Kara Moser, Adrian Paskey, Jason Caravas, Benjamin Rambo-Martin, Shatavia Morrison, Christopher Gulvick, Scott Sammons, Yvette Unoarumhi, Darlene Wagner, Matthew Schmerer, Cyndi Clark, Patrick Campbell, Rob Case, Vikramsinha Ghorpade, Holly Houdeshell, Ola Kvalvaag, Dillon Nall, Ethan Sanders, Alec Vest, Shaun Westlund, Matthew Hardison, Clinton R. Paden, Duncan MacCannell |
| EPI_ISL_2201140 | Houston Methodist Hospital | Houston Methodist Hospital | Randall J. Olsen, Paul A. Christensen, S. Wesley Long, Sishir Subedi, Robert Olson, Marcus Nguyen, James J. Davis, Matthew Ojeda Saavedra, Prasanti Yerramilli, Layne Pruitt, Kristina Reppond, Madison N. Shyer, Jessica Cambric, Ryan Gadd, Ilya J. Finkelstein, Jimmy Gollihar, and James M. Musser |
| EPI_ISL_2201141 | Aegis Sciences Corporation | Centers for Disease Control and Prevention Division of Viral Diseases, Pathogen Discovery | Dakota Howard, Dhwani Batra, Peter W. Cook, Kara Moser, Adrian Paskey, Jason Caravas, Benjamin Rambo-Martin, Shatavia Morrison, Christopher Gulvick, Scott Sammons, Yvette Unoarumhi, Darlene Wagner, Matthew Schmerer, Cyndi Clark, Patrick Campbell, Rob Case, Vikramsinha Ghorpade, Holly Houdeshell, Ola Kvalvaag, Dillon Nall, Ethan Sanders, Alec Vest, Shaun Westlund, Matthew Hardison, Clinton R. Paden, Duncan MacCannell |
| EPI_ISL_2201142 | Houston Methodist Hospital | Houston Methodist Hospital | Randall J. Olsen, Paul A. Christensen, S. Wesley Long, Sishir Subedi, Robert Olson, Marcus Nguyen, James J. Davis, Matthew Ojeda Saavedra, Prasanti Yerramilli, Layne Pruitt, Kristina Reppond, Madison N. Shyer, Jessica Cambric, Ryan Gadd, Ilya J. Finkelstein, Jimmy Gollihar, and James M. Musser |
| EPI_ISL_2201143 | Aegis Sciences Corporation | Centers for Disease Control and Prevention Division of Viral Diseases, Pathogen Discovery | Dakota Howard, Dhwani Batra, Peter W. Cook, Kara Moser, Adrian Paskey, Jason Caravas, Benjamin Rambo-Martin, Shatavia Morrison, Christopher Gulvick, Scott Sammons, Yvette Unoarumhi, Darlene Wagner, Matthew Schmerer, Cyndi Clark, Patrick Campbell, Rob Case, Vikramsinha Ghorpade, Holly Houdeshell, Ola Kvalvaag, Dillon Nall, Ethan Sanders, Alec Vest, Shaun Westlund, Matthew Hardison, Clinton R. Paden, Duncan MacCannell |
| EPI_ISL_2201144 | Houston Methodist Hospital | Houston Methodist Hospital | Randall J. Olsen, Paul A. Christensen, S. Wesley Long, Sishir Subedi, Robert Olson, Marcus Nguyen, James J. Davis, Matthew Ojeda Saavedra, Prasanti Yerramilli, Layne Pruitt, Kristina Reppond, Madison N. Shyer, Jessica Cambric, Ryan Gadd, Ilya J. Finkelstein, Jimmy Gollihar, and James M. Musser |
| EPI_ISL_2201145 | Aegis Sciences Corporation | Centers for Disease Control and Prevention Division of Viral Diseases, Pathogen Discovery | Dakota Howard, Dhwani Batra, Peter W. Cook, Kara Moser, Adrian Paskey, Jason Caravas, Benjamin Rambo-Martin, Shatavia Morrison, Christopher Gulvick, Scott Sammons, Yvette Unoarumhi, Darlene Wagner, Matthew Schmerer, Cyndi Clark, Patrick Campbell, Rob Case, Vikramsinha Ghorpade, Holly Houdeshell, Ola Kvalvaag, Dillon Nall, Ethan Sanders, Alec Vest, Shaun Westlund, Matthew Hardison, Clinton R. Paden, Duncan MacCannell |
| EPI_ISL_2201146, EPI_ISL_2201147 | Houston Methodist Hospital | Houston Methodist Hospital | Randall J. Olsen, Paul A. Christensen, S. Wesley Long, Sishir Subedi, Robert Olson, Marcus Nguyen, James J. Davis, Matthew Ojeda Saavedra, Prasanti Yerramilli, Layne Pruitt, Kristina Reppond, Madison N. Shyer, Jessica Cambric, Ryan Gadd, Ilya J. Finkelstein, Jimmy Gollihar, and James M. Musser |
| EPI_ISL_2201148, EPI_ISL_2201149 | Aegis Sciences Corporation | Centers for Disease Control and Prevention Division of Viral Diseases, Pathogen Discovery | Dakota Howard, Dhwani Batra, Peter W. Cook, Kara Moser, Adrian Paskey, Jason Caravas, Benjamin Rambo-Martin, Shatavia Morrison, Christopher Gulvick, Scott Sammons, Yvette Unoarumhi, Darlene Wagner, Matthew Schmerer, Cyndi Clark, Patrick Campbell, Rob Case, Vikramsinha Ghorpade, Holly Houdeshell, Ola Kvalvaag, Dillon Nall, Ethan Sanders, Alec Vest, Shaun Westlund, Matthew Hardison, Clinton R. Paden, Duncan MacCannell |
| EPI_ISL_2201150 | Houston Methodist Hospital | Houston Methodist Hospital | Randall J. Olsen, Paul A. Christensen, S. Wesley Long, Sishir Subedi, Robert Olson, Marcus Nguyen, James J. Davis, Matthew Ojeda Saavedra, Prasanti Yerramilli, Layne Pruitt, Kristina Reppond, Madison N. Shyer, Jessica Cambric, Ryan Gadd, Ilya J. Finkelstein, Jimmy Gollihar, and James M. Musser |
| EPI_ISL_2201151 | Aegis Sciences Corporation | Centers for Disease Control and Prevention Division of Viral Diseases, Pathogen Discovery | Dakota Howard, Dhwani Batra, Peter W. Cook, Kara Moser, Adrian Paskey, Jason Caravas, Benjamin Rambo-Martin, Shatavia Morrison, Christopher Gulvick, Scott Sammons, Yvette Unoarumhi, Darlene Wagner, Matthew Schmerer, Cyndi Clark, Patrick Campbell, Rob Case, Vikramsinha Ghorpade, Holly Houdeshell, Ola Kvalvaag, Dillon Nall, Ethan Sanders, Alec Vest, Shaun Westlund, Matthew Hardison, Clinton R. Paden, Duncan MacCannell |
| EPI_ISL_2201152, EPI_ISL_2201153 | Houston Methodist Hospital | Houston Methodist Hospital | Randall J. Olsen, Paul A. Christensen, S. Wesley Long, Sishir Subedi, Robert Olson, Marcus Nguyen, James J. Davis, Matthew Ojeda Saavedra, Prasanti Yerramilli, Layne Pruitt, Kristina Reppond, Madison N. Shyer, Jessica Cambric, Ryan Gadd, Ilya J. Finkelstein, Jimmy Gollihar, and James M. Musser |
| EPI_ISL_2201154, EPI_ISL_2201155 | Aegis Sciences Corporation | Centers for Disease Control and Prevention Division of Viral Diseases, Pathogen Discovery | Dakota Howard, Dhwani Batra, Peter W. Cook, Kara Moser, Adrian Paskey, Jason Caravas, Benjamin Rambo-Martin, Shatavia Morrison, Christopher Gulvick, Scott Sammons, Yvette Unoarumhi, Darlene Wagner, Matthew Schmerer, Cyndi Clark, Patrick Campbell, Rob Case, Vikramsinha Ghorpade, Holly Houdeshell, Ola Kvalvaag, Dillon Nall, Ethan Sanders, Alec Vest, Shaun Westlund, Matthew Hardison, Clinton R. Paden, Duncan MacCannell |

| | | | |
|---|---|---|---|
| EPI_ISL_2201156 | Houston Methodist Hospital | Houston Methodist Hospital | Randall J. Olsen, Paul A. Christensen, S. Wesley Long, Sishir Subedi, Robert Olson, Marcus Nguyen, James J. Davis, Matthew Ojeda Saavedra, Prasanti Yerramilli, Layne Pruitt, Kristina Reppond, Madison N. Shyer, Jessica Cambric, Ryan Gadd, Ilya J. Finkelstein, Jimmy Gollihar, and James M. Musser |
| EPI_ISL_2201157 | Aegis Sciences Corporation | Centers for Disease Control and Prevention Division of Viral Diseases, Pathogen Discovery | Dakota Howard, Dhwani Batra, Peter W. Cook, Kara Moser, Adrian Paskey, Jason Caravas, Benjamin Rambo-Martin, Shatavia Morrison, Christopher Gulvick, Scott Sammons, Yvette Unoarumhi, Darlene Wagner, Matthew Schmerer, Cyndi Clark, Patrick Campbell, Rob Case, Vikramsinha Ghorpade, Holly Houdeshell, Ola Kvalvaag, Dillon Nall, Ethan Sanders, Alec Vest, Shaun Westlund, Matthew Hardison, Clinton R. Paden, Duncan MacCannell |
| EPI_ISL_2201158, EPI_ISL_2201159 | Houston Methodist Hospital | Houston Methodist Hospital | Randall J. Olsen, Paul A. Christensen, S. Wesley Long, Sishir Subedi, Robert Olson, Marcus Nguyen, James J. Davis, Matthew Ojeda Saavedra, Prasanti Yerramilli, Layne Pruitt, Kristina Reppond, Madison N. Shyer, Jessica Cambric, Ryan Gadd, Ilya J. Finkelstein, Jimmy Gollihar, and James M. Musser |
| EPI_ISL_2201160, EPI_ISL_2201161 | Aegis Sciences Corporation | Centers for Disease Control and Prevention Division of Viral Diseases, Pathogen Discovery | Dakota Howard, Dhwani Batra, Peter W. Cook, Kara Moser, Adrian Paskey, Jason Caravas, Benjamin Rambo-Martin, Shatavia Morrison, Christopher Gulvick, Scott Sammons, Yvette Unoarumhi, Darlene Wagner, Matthew Schmerer, Cyndi Clark, Patrick Campbell, Rob Case, Vikramsinha Ghorpade, Holly Houdeshell, Ola Kvalvaag, Dillon Nall, Ethan Sanders, Alec Vest, Shaun Westlund, Matthew Hardison, Clinton R. Paden, Duncan MacCannell |
| EPI_ISL_2201162 | Houston Methodist Hospital | Houston Methodist Hospital | Randall J. Olsen, Paul A. Christensen, S. Wesley Long, Sishir Subedi, Robert Olson, Marcus Nguyen, James J. Davis, Matthew Ojeda Saavedra, Prasanti Yerramilli, Layne Pruitt, Kristina Reppond, Madison N. Shyer, Jessica Cambric, Ryan Gadd, Ilya J. Finkelstein, Jimmy Gollihar, and James M. Musser |
| EPI_ISL_2201163, EPI_ISL_2201164 | Aegis Sciences Corporation | Centers for Disease Control and Prevention Division of Viral Diseases, Pathogen Discovery | Dakota Howard, Dhwani Batra, Peter W. Cook, Kara Moser, Adrian Paskey, Jason Caravas, Benjamin Rambo-Martin, Shatavia Morrison, Christopher Gulvick, Scott Sammons, Yvette Unoarumhi, Darlene Wagner, Matthew Schmerer, Cyndi Clark, Patrick Campbell, Rob Case, Vikramsinha Ghorpade, Holly Houdeshell, Ola Kvalvaag, Dillon Nall, Ethan Sanders, Alec Vest, Shaun Westlund, Matthew Hardison, Clinton R. Paden, Duncan MacCannell |
| EPI_ISL_2201165 | Houston Methodist Hospital | Houston Methodist Hospital | Randall J. Olsen, Paul A. Christensen, S. Wesley Long, Sishir Subedi, Robert Olson, Marcus Nguyen, James J. Davis, Matthew Ojeda Saavedra, Prasanti Yerramilli, Layne Pruitt, Kristina Reppond, Madison N. Shyer, Jessica Cambric, Ryan Gadd, Ilya J. Finkelstein, Jimmy Gollihar, and James M. Musser |
| EPI_ISL_2201166, EPI_ISL_2201168 | Aegis Sciences Corporation | Centers for Disease Control and Prevention Division of Viral Diseases, Pathogen Discovery | Dakota Howard, Dhwani Batra, Peter W. Cook, Kara Moser, Adrian Paskey, Jason Caravas, Benjamin Rambo-Martin, Shatavia Morrison, Christopher Gulvick, Scott Sammons, Yvette Unoarumhi, Darlene Wagner, Matthew Schmerer, Cyndi Clark, Patrick Campbell, Rob Case, Vikramsinha Ghorpade, Holly Houdeshell, Ola Kvalvaag, Dillon Nall, Ethan Sanders, Alec Vest, Shaun Westlund, Matthew Hardison, Clinton R. Paden, Duncan MacCannell |
| EPI_ISL_2201169 | Houston Methodist Hospital | Houston Methodist Hospital | Randall J. Olsen, Paul A. Christensen, S. Wesley Long, Sishir Subedi, Robert Olson, Marcus Nguyen, James J. Davis, Matthew Ojeda Saavedra, Prasanti Yerramilli, Layne Pruitt, Kristina Reppond, Madison N. Shyer, Jessica Cambric, Ryan Gadd, Ilya J. Finkelstein, Jimmy Gollihar, and James M. Musser |
| EPI_ISL_2201170, EPI_ISL_2201171 | Aegis Sciences Corporation | Centers for Disease Control and Prevention Division of Viral Diseases, Pathogen Discovery | Dakota Howard, Dhwani Batra, Peter W. Cook, Kara Moser, Adrian Paskey, Jason Caravas, Benjamin Rambo-Martin, Shatavia Morrison, Christopher Gulvick, Scott Sammons, Yvette Unoarumhi, Darlene Wagner, Matthew Schmerer, Cyndi Clark, Patrick Campbell, Rob Case, Vikramsinha Ghorpade, Holly Houdeshell, Ola Kvalvaag, Dillon Nall, Ethan Sanders, Alec Vest, Shaun Westlund, Matthew Hardison, Clinton R. Paden, Duncan MacCannell |
| EPI_ISL_2201172 | Houston Methodist Hospital | Houston Methodist Hospital | Randall J. Olsen, Paul A. Christensen, S. Wesley Long, Sishir Subedi, Robert Olson, Marcus Nguyen, James J. Davis, Matthew Ojeda Saavedra, Prasanti Yerramilli, Layne Pruitt, Kristina Reppond, Madison N. Shyer, Jessica Cambric, Ryan Gadd, Ilya J. Finkelstein, Jimmy Gollihar, and James M. Musser |
| EPI_ISL_2201173 | Aegis Sciences Corporation | Centers for Disease Control and Prevention Division of Viral Diseases, Pathogen Discovery | Dakota Howard, Dhwani Batra, Peter W. Cook, Kara Moser, Adrian Paskey, Jason Caravas, Benjamin Rambo-Martin, Shatavia Morrison, Christopher Gulvick, Scott Sammons, Yvette Unoarumhi, Darlene Wagner, Matthew Schmerer, Cyndi Clark, Patrick Campbell, Rob Case, Vikramsinha Ghorpade, Holly Houdeshell, Ola Kvalvaag, Dillon Nall, Ethan Sanders, Alec Vest, Shaun Westlund, Matthew Hardison, Clinton R. Paden, Duncan MacCannell |
| EPI_ISL_2201174 | Houston Methodist Hospital | Houston Methodist Hospital | Randall J. Olsen, Paul A. Christensen, S. Wesley Long, Sishir Subedi, Robert Olson, Marcus Nguyen, James J. Davis, Matthew Ojeda Saavedra, Prasanti Yerramilli, Layne Pruitt, Kristina Reppond, Madison N. Shyer, Jessica Cambric, Ryan Gadd, Ilya J. Finkelstein, Jimmy Gollihar, and James M. Musser |
| EPI_ISL_2201175 | Aegis Sciences Corporation | Centers for Disease Control and Prevention Division of Viral Diseases, Pathogen Discovery | Dakota Howard, Dhwani Batra, Peter W. Cook, Kara Moser, Adrian Paskey, Jason Caravas, Benjamin Rambo-Martin, Shatavia Morrison, Christopher Gulvick, Scott Sammons, Yvette Unoarumhi, Darlene Wagner, Matthew Schmerer, Cyndi Clark, Patrick Campbell, Rob Case, Vikramsinha Ghorpade, Holly Houdeshell, Ola Kvalvaag, Dillon Nall, Ethan Sanders, Alec Vest, Shaun Westlund, Matthew Hardison, Clinton R. Paden, Duncan MacCannell |
| EPI_ISL_2201176 | Houston Methodist Hospital | Houston Methodist Hospital | Randall J. Olsen, Paul A. Christensen, S. Wesley Long, Sishir Subedi, Robert Olson, Marcus Nguyen, James J. Davis, Matthew Ojeda Saavedra, Prasanti Yerramilli, Layne Pruitt, Kristina Reppond, Madison N. Shyer, Jessica Cambric, Ryan Gadd, Ilya J. Finkelstein, Jimmy Gollihar, and James M. Musser |
| EPI_ISL_2201177 | Aegis Sciences Corporation | Centers for Disease Control and Prevention Division of Viral Diseases, Pathogen Discovery | Dakota Howard, Dhwani Batra, Peter W. Cook, Kara Moser, Adrian Paskey, Jason Caravas, Benjamin Rambo-Martin, Shatavia Morrison, Christopher Gulvick, Scott Sammons, Yvette Unoarumhi, Darlene Wagner, Matthew Schmerer, Cyndi Clark, Patrick Campbell, Rob Case, Vikramsinha Ghorpade, Holly Houdeshell, Ola Kvalvaag, Dillon Nall, Ethan Sanders, Alec Vest, Shaun Westlund, Matthew Hardison, Clinton R. Paden, Duncan MacCannell |
| EPI_ISL_2201178 | Houston Methodist Hospital | Houston Methodist Hospital | Randall J. Olsen, Paul A. Christensen, S. Wesley Long, Sishir Subedi, Robert Olson, Marcus Nguyen, James J. Davis, Matthew Ojeda Saavedra, Prasanti Yerramilli, Layne Pruitt, Kristina Reppond, Madison N. Shyer, Jessica Cambric, Ryan Gadd, Ilya J. Finkelstein, Jimmy Gollihar, and James M. Musser |
| EPI_ISL_2201179 | Aegis Sciences Corporation | Centers for Disease Control and Prevention Division of Viral Diseases, Pathogen Discovery | Dakota Howard, Dhwani Batra, Peter W. Cook, Kara Moser, Adrian Paskey, Jason Caravas, Benjamin Rambo-Martin, Shatavia Morrison, Christopher Gulvick, Scott Sammons, Yvette Unoarumhi, Darlene Wagner, Matthew Schmerer, Cyndi Clark, Patrick Campbell, Rob Case, Vikramsinha Ghorpade, Holly Houdeshell, Ola Kvalvaag, Dillon Nall, Ethan Sanders, Alec Vest, Shaun Westlund, Matthew Hardison, Clinton R. Paden, Duncan MacCannell |
| EPI_ISL_2201180 | Houston Methodist Hospital | Houston Methodist Hospital | Randall J. Olsen, Paul A. Christensen, S. Wesley Long, Sishir Subedi, Robert Olson, Marcus Nguyen, James J. Davis, Matthew Ojeda Saavedra, Prasanti Yerramilli, Layne Pruitt, Kristina Reppond, Madison N. Shyer, Jessica Cambric, Ryan Gadd, Ilya J. Finkelstein, Jimmy Gollihar, and James M. Musser |
| EPI_ISL_2201181 | Aegis Sciences Corporation | Centers for Disease Control and Prevention Division of Viral Diseases, Pathogen Discovery | Dakota Howard, Dhwani Batra, Peter W. Cook, Kara Moser, Adrian Paskey, Jason Caravas, Benjamin Rambo-Martin, Shatavia Morrison, Christopher Gulvick, Scott Sammons, Yvette Unoarumhi, Darlene Wagner, Matthew Schmerer, Cyndi Clark, Patrick Campbell, Rob Case, Vikramsinha Ghorpade, Holly Houdeshell, Ola Kvalvaag, Dillon Nall, Ethan Sanders, Alec Vest, Shaun Westlund, Matthew Hardison, Clinton R. Paden, Duncan MacCannell |
| EPI_ISL_2201182 | Houston Methodist Hospital | Houston Methodist Hospital | Randall J. Olsen, Paul A. Christensen, S. Wesley Long, Sishir Subedi, Robert Olson, Marcus Nguyen, James J. Davis, Matthew Ojeda Saavedra, Prasanti Yerramilli, Layne Pruitt, Kristina Reppond, Madison N. Shyer, Jessica Cambric, Ryan Gadd, Ilya J. Finkelstein, Jimmy Gollihar, and James M. Musser |
| EPI_ISL_2201183 | Aegis Sciences Corporation | Centers for Disease Control and Prevention Division of Viral Diseases, Pathogen Discovery | Dakota Howard, Dhwani Batra, Peter W. Cook, Kara Moser, Adrian Paskey, Jason Caravas, Benjamin Rambo-Martin, Shatavia Morrison, Christopher Gulvick, Scott Sammons, Yvette Unoarumhi, Darlene Wagner, Matthew Schmerer, Cyndi Clark, Patrick Campbell, Rob Case, Vikramsinha Ghorpade, Holly Houdeshell, Ola Kvalvaag, Dillon Nall, Ethan Sanders, Alec Vest, Shaun Westlund, Matthew Hardison, Clinton R. Paden, Duncan MacCannell |
| EPI_ISL_2201184 | Houston Methodist Hospital | Houston Methodist Hospital | Randall J. Olsen, Paul A. Christensen, S. Wesley Long, Sishir Subedi, Robert Olson, Marcus Nguyen, James J. Davis, Matthew Ojeda Saavedra, Prasanti Yerramilli, Layne Pruitt, Kristina Reppond, Madison N. Shyer, Jessica Cambric, Ryan Gadd, Ilya J. Finkelstein, Jimmy Gollihar, and James M. Musser |
| EPI_ISL_2201185 | Aegis Sciences Corporation | Centers for Disease Control and Prevention Division of Viral Diseases, Pathogen Discovery | Dakota Howard, Dhwani Batra, Peter W. Cook, Kara Moser, Adrian Paskey, Jason Caravas, Benjamin Rambo-Martin, Shatavia Morrison, Christopher Gulvick, Scott Sammons, Yvette Unoarumhi, Darlene Wagner, Matthew Schmerer, Cyndi Clark, Patrick Campbell, Rob Case, Vikramsinha Ghorpade, Holly Houdeshell, Ola Kvalvaag, Dillon Nall, Ethan Sanders, Alec Vest, Shaun Westlund, Matthew Hardison, Clinton R. Paden, Duncan MacCannell |
| EPI_ISL_2201186 | Houston Methodist Hospital | Houston Methodist Hospital | Randall J. Olsen, Paul A. Christensen, S. Wesley Long, Sishir Subedi, Robert Olson, Marcus Nguyen, James J. Davis, Matthew Ojeda Saavedra, Prasanti Yerramilli, Layne Pruitt, Kristina Reppond, Madison N. Shyer, Jessica Cambric, Ryan Gadd, Ilya J. Finkelstein, Jimmy Gollihar, and James M. Musser |
| EPI_ISL_2201187 | Aegis Sciences Corporation | Centers for Disease Control and Prevention Division of Viral Diseases, Pathogen Discovery | Dakota Howard, Dhwani Batra, Peter W. Cook, Kara Moser, Adrian Paskey, Jason Caravas, Benjamin Rambo-Martin, Shatavia Morrison, Christopher Gulvick, Scott Sammons, Yvette Unoarumhi, Darlene Wagner, Matthew Schmerer, Cyndi Clark, Patrick Campbell, Rob Case, Vikramsinha Ghorpade, Holly Houdeshell, Ola Kvalvaag, Dillon Nall, Ethan Sanders, Alec Vest, Shaun Westlund, Matthew Hardison, Clinton R. Paden, Duncan MacCannell |
| EPI_ISL_2201188 | Houston Methodist Hospital | Houston Methodist Hospital | Randall J. Olsen, Paul A. Christensen, S. Wesley Long, Sishir Subedi, Robert Olson, Marcus Nguyen, James J. Davis, Matthew Ojeda Saavedra, Prasanti Yerramilli, Layne Pruitt, Kristina Reppond, Madison N. Shyer, Jessica Cambric, Ryan Gadd, Ilya J. Finkelstein, Jimmy Gollihar, and James M. Musser |
| EPI_ISL_2201189 | Aegis Sciences Corporation | Centers for Disease Control and Prevention Division of Viral Diseases, Pathogen Discovery | Dakota Howard, Dhwani Batra, Peter W. Cook, Kara Moser, Adrian Paskey, Jason Caravas, Benjamin Rambo-Martin, Shatavia Morrison, Christopher Gulvick, Scott Sammons, Yvette Unoarumhi, Darlene Wagner, Matthew Schmerer, Cyndi Clark, Patrick Campbell, Rob Case, Vikramsinha Ghorpade, Holly Houdeshell, Ola Kvalvaag, Dillon Nall, Ethan Sanders, Alec Vest, Shaun Westlund, Matthew Hardison, Clinton R. Paden, Duncan MacCannell |
| EPI_ISL_2201190 | Houston Methodist Hospital | Houston Methodist Hospital | Randall J. Olsen, Paul A. Christensen, S. Wesley Long, Sishir Subedi, Robert Olson, Marcus Nguyen, James J. Davis, Matthew Ojeda Saavedra, Prasanti Yerramilli, Layne Pruitt, Kristina Reppond, Madison N. Shyer, Jessica Cambric, Ryan Gadd, Ilya J. Finkelstein, Jimmy Gollihar, and James M. Musser |
| EPI_ISL_2201191 | Aegis Sciences Corporation | Centers for Disease Control and Prevention Division of Viral Diseases, Pathogen Discovery | Dakota Howard, Dhwani Batra, Peter W. Cook, Kara Moser, Adrian Paskey, Jason Caravas, Benjamin Rambo-Martin, Shatavia Morrison, Christopher Gulvick, Scott Sammons, Yvette Unoarumhi, Darlene Wagner, Matthew Schmerer, Cyndi Clark, Patrick Campbell, Rob Case, Vikramsinha Ghorpade, Holly Houdeshell, Ola Kvalvaag, Dillon Nall, Ethan Sanders, Alec Vest, Shaun Westlund, Matthew Hardison, Clinton R. Paden, Duncan MacCannell |
| EPI_ISL_2201192 | Houston Methodist Hospital | Houston Methodist Hospital | Randall J. Olsen, Paul A. Christensen, S. Wesley Long, Sishir Subedi, Robert Olson, Marcus Nguyen, James J. Davis, Matthew Ojeda Saavedra, Prasanti Yerramilli, Layne Pruitt, Kristina Reppond, Madison N. Shyer, Jessica Cambric, Ryan Gadd, Ilya J. Finkelstein, Jimmy Gollihar, and James M. Musser |
| EPI_ISL_2201193 | Aegis Sciences Corporation | Centers for Disease Control and Prevention Division of Viral Diseases, Pathogen Discovery | Dakota Howard, Dhwani Batra, Peter W. Cook, Kara Moser, Adrian Paskey, Jason Caravas, Benjamin Rambo-Martin, Shatavia Morrison, Christopher Gulvick, Scott Sammons, Yvette Unoarumhi, Darlene Wagner, Matthew Schmerer, Cyndi Clark, Patrick Campbell, Rob Case, Vikramsinha Ghorpade, Holly Houdeshell, Ola Kvalvaag, Dillon Nall, Ethan Sanders, Alec Vest, Shaun Westlund, Matthew Hardison, Clinton R. Paden, Duncan MacCannell |
| EPI_ISL_2201194 | Houston Methodist Hospital | Houston Methodist Hospital | Randall J. Olsen, Paul A. Christensen, S. Wesley Long, Sishir Subedi, Robert Olson, Marcus Nguyen, James J. Davis, Matthew Ojeda Saavedra, Prasanti Yerramilli, Layne Pruitt, Kristina Reppond, Madison N. Shyer, Jessica Cambric, Ryan Gadd, Ilya J. Finkelstein, Jimmy Gollihar, and James M. Musser |
| EPI_ISL_2201195, EPI_ISL_2201196 | Aegis Sciences Corporation | Centers for Disease Control and Prevention Division of Viral Diseases, Pathogen Discovery | Dakota Howard, Dhwani Batra, Peter W. Cook, Kara Moser, Adrian Paskey, Jason Caravas, Benjamin Rambo-Martin, Shatavia Morrison, Christopher Gulvick, Scott Sammons, Yvette Unoarumhi, Darlene Wagner, Matthew Schmerer, Cyndi Clark, Patrick Campbell, Rob Case, Vikramsinha Ghorpade, Holly Houdeshell, Ola Kvalvaag, Dillon Nall, Ethan Sanders, Alec Vest, Shaun Westlund, Matthew Hardison, Clinton R. Paden, Duncan MacCannell |
| EPI_ISL_2201197 | Houston Methodist Hospital | Houston Methodist Hospital | Randall J. Olsen, Paul A. Christensen, S. Wesley Long, Sishir Subedi, Robert Olson, Marcus Nguyen, James J. Davis, Matthew Ojeda Saavedra, Prasanti Yerramilli, Layne Pruitt, Kristina Reppond, Madison N. Shyer, Jessica Cambric, Ryan Gadd, Ilya J. Finkelstein, Jimmy Gollihar, and James M. Musser |
| EPI_ISL_2201198, EPI_ISL_2201199 | Aegis Sciences Corporation | Centers for Disease Control and Prevention Division of Viral Diseases, Pathogen Discovery | Dakota Howard, Dhwani Batra, Peter W. Cook, Kara Moser, Adrian Paskey, Jason Caravas, Benjamin Rambo-Martin, Shatavia Morrison, Christopher Gulvick, Scott Sammons, Yvette Unoarumhi, Darlene Wagner, Matthew Schmerer, Cyndi Clark, Patrick Campbell, Rob Case, Vikramsinha Ghorpade, Holly Houdeshell, Ola Kvalvaag, Dillon Nall, Ethan Sanders, Alec Vest, Shaun Westlund, Matthew Hardison, Clinton R. Paden, Duncan MacCannell |
| EPI_ISL_2201200 | Houston Methodist Hospital | Houston Methodist Hospital | Randall J. Olsen, Paul A. Christensen, S. Wesley Long, Sishir Subedi, Robert Olson, Marcus Nguyen, James J. Davis, Matthew Ojeda Saavedra, Prasanti Yerramilli, Layne Pruitt, Kristina Reppond, Madison N. Shyer, Jessica Cambric, Ryan Gadd, Ilya J. Finkelstein, Jimmy Gollihar, and James M. Musser |
| EPI_ISL_2201201 | Aegis Sciences Corporation | Centers for Disease Control and Prevention | Dakota Howard, Dhwani Batra, Peter W. Cook, Kara Moser, Adrian Paskey, Jason Caravas, Benjamin Rambo-Martin, Shatavia Morrison, Christopher Gulvick, Scott Sammons, Yvette Unoarumhi, Darlene Wagner, Matthew Schmerer, Cyndi Clark, Patrick Campbell, Rob Case, Vikramsinha |

| | | | |
|---|---|---|---|
| | | Division of Viral Diseases, Pathogen Discovery | Ghorpade, Holly Houdeshell, Ola Kvalvaag, Dillon Nall, Ethan Sanders, Alec Vest, Shaun Westlund, Matthew Hardison, Clinton R. Paden, Duncan MacCannell |
| EPI_ISL_2201202 | Houston Methodist Hospital | Houston Methodist Hospital | Randall J. Olsen, Paul A. Christensen, S. Wesley Long, Sishir Subedi, Robert Olson, Marcus Nguyen, James J. Davis, Matthew Ojeda Saavedra, Prasanti Yerramilli, Layne Pruitt, Kristina Reppond, Madison N. Shyer, Jessica Cambric, Ryan Gadd, Ilya J. Finkelstein, Jimmy Gollihar, and James M. Musser |
| EPI_ISL_2201203 | Aegis Sciences Corporation | Centers for Disease Control and Prevention Division of Viral Diseases, Pathogen Discovery | Dakota Howard, Dhwani Batra, Peter W. Cook, Kara Moser, Adrian Paskey, Jason Caravas, Benjamin Rambo-Martin, Shatavia Morrison, Christopher Gulvick, Scott Sammons, Yvette Unoarumhi, Darlene Wagner, Matthew Schmerer, Cyndi Clark, Patrick Campbell, Rob Case, Vikramsinha Ghorpade, Holly Houdeshell, Ola Kvalvaag, Dillon Nall, Ethan Sanders, Alec Vest, Shaun Westlund, Matthew Hardison, Clinton R. Paden, Duncan MacCannell |
| EPI_ISL_2201204 | Houston Methodist Hospital | Houston Methodist Hospital | Randall J. Olsen, Paul A. Christensen, S. Wesley Long, Sishir Subedi, Robert Olson, Marcus Nguyen, James J. Davis, Matthew Ojeda Saavedra, Prasanti Yerramilli, Layne Pruitt, Kristina Reppond, Madison N. Shyer, Jessica Cambric, Ryan Gadd, Ilya J. Finkelstein, Jimmy Gollihar, and James M. Musser |
| EPI_ISL_2201205 | Aegis Sciences Corporation | Centers for Disease Control and Prevention Division of Viral Diseases, Pathogen Discovery | Dakota Howard, Dhwani Batra, Peter W. Cook, Kara Moser, Adrian Paskey, Jason Caravas, Benjamin Rambo-Martin, Shatavia Morrison, Christopher Gulvick, Scott Sammons, Yvette Unoarumhi, Darlene Wagner, Matthew Schmerer, Cyndi Clark, Patrick Campbell, Rob Case, Vikramsinha Ghorpade, Holly Houdeshell, Ola Kvalvaag, Dillon Nall, Ethan Sanders, Alec Vest, Shaun Westlund, Matthew Hardison, Clinton R. Paden, Duncan MacCannell |
| EPI_ISL_2201206 | Houston Methodist Hospital | Houston Methodist Hospital | Randall J. Olsen, Paul A. Christensen, S. Wesley Long, Sishir Subedi, Robert Olson, Marcus Nguyen, James J. Davis, Matthew Ojeda Saavedra, Prasanti Yerramilli, Layne Pruitt, Kristina Reppond, Madison N. Shyer, Jessica Cambric, Ryan Gadd, Ilya J. Finkelstein, Jimmy Gollihar, and James M. Musser |
| EPI_ISL_2201207, EPI_ISL_2201208 | Aegis Sciences Corporation | Centers for Disease Control and Prevention Division of Viral Diseases, Pathogen Discovery | Dakota Howard, Dhwani Batra, Peter W. Cook, Kara Moser, Adrian Paskey, Jason Caravas, Benjamin Rambo-Martin, Shatavia Morrison, Christopher Gulvick, Scott Sammons, Yvette Unoarumhi, Darlene Wagner, Matthew Schmerer, Cyndi Clark, Patrick Campbell, Rob Case, Vikramsinha Ghorpade, Holly Houdeshell, Ola Kvalvaag, Dillon Nall, Ethan Sanders, Alec Vest, Shaun Westlund, Matthew Hardison, Clinton R. Paden, Duncan MacCannell |
| EPI_ISL_2201209 | Houston Methodist Hospital | Houston Methodist Hospital | Randall J. Olsen, Paul A. Christensen, S. Wesley Long, Sishir Subedi, Robert Olson, Marcus Nguyen, James J. Davis, Matthew Ojeda Saavedra, Prasanti Yerramilli, Layne Pruitt, Kristina Reppond, Madison N. Shyer, Jessica Cambric, Ryan Gadd, Ilya J. Finkelstein, Jimmy Gollihar, and James M. Musser |
| EPI_ISL_2201210, EPI_ISL_2201211 | Aegis Sciences Corporation | Centers for Disease Control and Prevention Division of Viral Diseases, Pathogen Discovery | Dakota Howard, Dhwani Batra, Peter W. Cook, Kara Moser, Adrian Paskey, Jason Caravas, Benjamin Rambo-Martin, Shatavia Morrison, Christopher Gulvick, Scott Sammons, Yvette Unoarumhi, Darlene Wagner, Matthew Schmerer, Cyndi Clark, Patrick Campbell, Rob Case, Vikramsinha Ghorpade, Holly Houdeshell, Ola Kvalvaag, Dillon Nall, Ethan Sanders, Alec Vest, Shaun Westlund, Matthew Hardison, Clinton R. Paden, Duncan MacCannell |
| EPI_ISL_2201212 | Houston Methodist Hospital | Houston Methodist Hospital | Randall J. Olsen, Paul A. Christensen, S. Wesley Long, Sishir Subedi, Robert Olson, Marcus Nguyen, James J. Davis, Matthew Ojeda Saavedra, Prasanti Yerramilli, Layne Pruitt, Kristina Reppond, Madison N. Shyer, Jessica Cambric, Ryan Gadd, Ilya J. Finkelstein, Jimmy Gollihar, and James M. Musser |
| EPI_ISL_2201213 | Aegis Sciences Corporation | Centers for Disease Control and Prevention Division of Viral Diseases, Pathogen Discovery | Dakota Howard, Dhwani Batra, Peter W. Cook, Kara Moser, Adrian Paskey, Jason Caravas, Benjamin Rambo-Martin, Shatavia Morrison, Christopher Gulvick, Scott Sammons, Yvette Unoarumhi, Darlene Wagner, Matthew Schmerer, Cyndi Clark, Patrick Campbell, Rob Case, Vikramsinha Ghorpade, Holly Houdeshell, Ola Kvalvaag, Dillon Nall, Ethan Sanders, Alec Vest, Shaun Westlund, Matthew Hardison, Clinton R. Paden, Duncan MacCannell |
| EPI_ISL_2201214 | Houston Methodist Hospital | Houston Methodist Hospital | Randall J. Olsen, Paul A. Christensen, S. Wesley Long, Sishir Subedi, Robert Olson, Marcus Nguyen, James J. Davis, Matthew Ojeda Saavedra, Prasanti Yerramilli, Layne Pruitt, Kristina Reppond, Madison N. Shyer, Jessica Cambric, Ryan Gadd, Ilya J. Finkelstein, Jimmy Gollihar, and James M. Musser |
| EPI_ISL_2201215 | Aegis Sciences Corporation | Centers for Disease Control and Prevention Division of Viral Diseases, Pathogen Discovery | Dakota Howard, Dhwani Batra, Peter W. Cook, Kara Moser, Adrian Paskey, Jason Caravas, Benjamin Rambo-Martin, Shatavia Morrison, Christopher Gulvick, Scott Sammons, Yvette Unoarumhi, Darlene Wagner, Matthew Schmerer, Cyndi Clark, Patrick Campbell, Rob Case, Vikramsinha Ghorpade, Holly Houdeshell, Ola Kvalvaag, Dillon Nall, Ethan Sanders, Alec Vest, Shaun Westlund, Matthew Hardison, Clinton R. Paden, Duncan MacCannell |
| EPI_ISL_2201216 | Houston Methodist Hospital | Houston Methodist Hospital | Randall J. Olsen, Paul A. Christensen, S. Wesley Long, Sishir Subedi, Robert Olson, Marcus Nguyen, James J. Davis, Matthew Ojeda Saavedra, Prasanti Yerramilli, Layne Pruitt, Kristina Reppond, Madison N. Shyer, Jessica Cambric, Ryan Gadd, Ilya J. Finkelstein, Jimmy Gollihar, and James M. Musser |
| EPI_ISL_2201217, EPI_ISL_2201218 | Aegis Sciences Corporation | Centers for Disease Control and Prevention Division of Viral Diseases, Pathogen Discovery | Dakota Howard, Dhwani Batra, Peter W. Cook, Kara Moser, Adrian Paskey, Jason Caravas, Benjamin Rambo-Martin, Shatavia Morrison, Christopher Gulvick, Scott Sammons, Yvette Unoarumhi, Darlene Wagner, Matthew Schmerer, Cyndi Clark, Patrick Campbell, Rob Case, Vikramsinha Ghorpade, Holly Houdeshell, Ola Kvalvaag, Dillon Nall, Ethan Sanders, Alec Vest, Shaun Westlund, Matthew Hardison, Clinton R. Paden, Duncan MacCannell |
| EPI_ISL_2201219 | Houston Methodist Hospital | Houston Methodist Hospital | Randall J. Olsen, Paul A. Christensen, S. Wesley Long, Sishir Subedi, Robert Olson, Marcus Nguyen, James J. Davis, Matthew Ojeda Saavedra, Prasanti Yerramilli, Layne Pruitt, Kristina Reppond, Madison N. Shyer, Jessica Cambric, Ryan Gadd, Ilya J. Finkelstein, Jimmy Gollihar, and James M. Musser |
| EPI_ISL_2201220 | Aegis Sciences Corporation | Centers for Disease Control and Prevention Division of Viral Diseases, Pathogen Discovery | Dakota Howard, Dhwani Batra, Peter W. Cook, Kara Moser, Adrian Paskey, Jason Caravas, Benjamin Rambo-Martin, Shatavia Morrison, Christopher Gulvick, Scott Sammons, Yvette Unoarumhi, Darlene Wagner, Matthew Schmerer, Cyndi Clark, Patrick Campbell, Rob Case, Vikramsinha Ghorpade, Holly Houdeshell, Ola Kvalvaag, Dillon Nall, Ethan Sanders, Alec Vest, Shaun Westlund, Matthew Hardison, Clinton R. Paden, Duncan MacCannell |
| EPI_ISL_2201221 | Houston Methodist Hospital | Houston Methodist Hospital | Randall J. Olsen, Paul A. Christensen, S. Wesley Long, Sishir Subedi, Robert Olson, Marcus Nguyen, James J. Davis, Matthew Ojeda Saavedra, Prasanti Yerramilli, Layne Pruitt, Kristina Reppond, Madison N. Shyer, Jessica Cambric, Ryan Gadd, Ilya J. Finkelstein, Jimmy Gollihar, and James M. Musser |
| EPI_ISL_2201222 | Aegis Sciences Corporation | Centers for Disease Control and Prevention Division of Viral Diseases, Pathogen Discovery | Dakota Howard, Dhwani Batra, Peter W. Cook, Kara Moser, Adrian Paskey, Jason Caravas, Benjamin Rambo-Martin, Shatavia Morrison, Christopher Gulvick, Scott Sammons, Yvette Unoarumhi, Darlene Wagner, Matthew Schmerer, Cyndi Clark, Patrick Campbell, Rob Case, Vikramsinha Ghorpade, Holly Houdeshell, Ola Kvalvaag, Dillon Nall, Ethan Sanders, Alec Vest, Shaun Westlund, Matthew Hardison, Clinton R. Paden, Duncan MacCannell |
| EPI_ISL_2201223 | Houston Methodist Hospital | Houston Methodist Hospital | Randall J. Olsen, Paul A. Christensen, S. Wesley Long, Sishir Subedi, Robert Olson, Marcus Nguyen, James J. Davis, Matthew Ojeda Saavedra, Prasanti Yerramilli, Layne Pruitt, Kristina Reppond, Madison N. Shyer, Jessica Cambric, Ryan Gadd, Ilya J. Finkelstein, Jimmy Gollihar, and James M. Musser |
| EPI_ISL_2201224, EPI_ISL_2201225 | Aegis Sciences Corporation | Centers for Disease Control and Prevention Division of Viral Diseases, Pathogen Discovery | Dakota Howard, Dhwani Batra, Peter W. Cook, Kara Moser, Adrian Paskey, Jason Caravas, Benjamin Rambo-Martin, Shatavia Morrison, Christopher Gulvick, Scott Sammons, Yvette Unoarumhi, Darlene Wagner, Matthew Schmerer, Cyndi Clark, Patrick Campbell, Rob Case, Vikramsinha Ghorpade, Holly Houdeshell, Ola Kvalvaag, Dillon Nall, Ethan Sanders, Alec Vest, Shaun Westlund, Matthew Hardison, Clinton R. Paden, Duncan MacCannell |
| EPI_ISL_2201226 | Houston Methodist Hospital | Houston Methodist Hospital | Randall J. Olsen, Paul A. Christensen, S. Wesley Long, Sishir Subedi, Robert Olson, Marcus Nguyen, James J. Davis, Matthew Ojeda Saavedra, Prasanti Yerramilli, Layne Pruitt, Kristina Reppond, Madison N. Shyer, Jessica Cambric, Ryan Gadd, Ilya J. Finkelstein, Jimmy Gollihar, and James M. Musser |
| EPI_ISL_2201227 | Aegis Sciences Corporation | Centers for Disease Control and Prevention Division of Viral Diseases, Pathogen Discovery | Dakota Howard, Dhwani Batra, Peter W. Cook, Kara Moser, Adrian Paskey, Jason Caravas, Benjamin Rambo-Martin, Shatavia Morrison, Christopher Gulvick, Scott Sammons, Yvette Unoarumhi, Darlene Wagner, Matthew Schmerer, Cyndi Clark, Patrick Campbell, Rob Case, Vikramsinha Ghorpade, Holly Houdeshell, Ola Kvalvaag, Dillon Nall, Ethan Sanders, Alec Vest, Shaun Westlund, Matthew Hardison, Clinton R. Paden, Duncan MacCannell |
| EPI_ISL_2201228 | Houston Methodist Hospital | Houston Methodist Hospital | Randall J. Olsen, Paul A. Christensen, S. Wesley Long, Sishir Subedi, Robert Olson, Marcus Nguyen, James J. Davis, Matthew Ojeda Saavedra, Prasanti Yerramilli, Layne Pruitt, Kristina Reppond, Madison N. Shyer, Jessica Cambric, Ryan Gadd, Ilya J. Finkelstein, Jimmy Gollihar, and James M. Musser |
| EPI_ISL_2201229 | Aegis Sciences Corporation | Centers for Disease Control and Prevention Division of Viral Diseases, Pathogen Discovery | Dakota Howard, Dhwani Batra, Peter W. Cook, Kara Moser, Adrian Paskey, Jason Caravas, Benjamin Rambo-Martin, Shatavia Morrison, Christopher Gulvick, Scott Sammons, Yvette Unoarumhi, Darlene Wagner, Matthew Schmerer, Cyndi Clark, Patrick Campbell, Rob Case, Vikramsinha Ghorpade, Holly Houdeshell, Ola Kvalvaag, Dillon Nall, Ethan Sanders, Alec Vest, Shaun Westlund, Matthew Hardison, Clinton R. Paden, Duncan MacCannell |
| EPI_ISL_2201230 | Houston Methodist Hospital | Houston Methodist Hospital | Randall J. Olsen, Paul A. Christensen, S. Wesley Long, Sishir Subedi, Robert Olson, Marcus Nguyen, James J. Davis, Matthew Ojeda Saavedra, Prasanti Yerramilli, Layne Pruitt, Kristina Reppond, Madison N. Shyer, Jessica Cambric, Ryan Gadd, Ilya J. Finkelstein, Jimmy Gollihar, and James M. Musser |
| EPI_ISL_2201231 | Aegis Sciences Corporation | Centers for Disease Control and Prevention Division of Viral Diseases, Pathogen Discovery | Dakota Howard, Dhwani Batra, Peter W. Cook, Kara Moser, Adrian Paskey, Jason Caravas, Benjamin Rambo-Martin, Shatavia Morrison, Christopher Gulvick, Scott Sammons, Yvette Unoarumhi, Darlene Wagner, Matthew Schmerer, Cyndi Clark, Patrick Campbell, Rob Case, Vikramsinha Ghorpade, Holly Houdeshell, Ola Kvalvaag, Dillon Nall, Ethan Sanders, Alec Vest, Shaun Westlund, Matthew Hardison, Clinton R. Paden, Duncan MacCannell |
| EPI_ISL_2201232, EPI_ISL_2201233 | Houston Methodist Hospital | Houston Methodist Hospital | Randall J. Olsen, Paul A. Christensen, S. Wesley Long, Sishir Subedi, Robert Olson, Marcus Nguyen, James J. Davis, Matthew Ojeda Saavedra, Prasanti Yerramilli, Layne Pruitt, Kristina Reppond, Madison N. Shyer, Jessica Cambric, Ryan Gadd, Ilya J. Finkelstein, Jimmy Gollihar, and James M. Musser |
| EPI_ISL_2201234, EPI_ISL_2201235 | Aegis Sciences Corporation | Centers for Disease Control and Prevention Division of Viral Diseases, Pathogen Discovery | Dakota Howard, Dhwani Batra, Peter W. Cook, Kara Moser, Adrian Paskey, Jason Caravas, Benjamin Rambo-Martin, Shatavia Morrison, Christopher Gulvick, Scott Sammons, Yvette Unoarumhi, Darlene Wagner, Matthew Schmerer, Cyndi Clark, Patrick Campbell, Rob Case, Vikramsinha Ghorpade, Holly Houdeshell, Ola Kvalvaag, Dillon Nall, Ethan Sanders, Alec Vest, Shaun Westlund, Matthew Hardison, Clinton R. Paden, Duncan MacCannell |
| EPI_ISL_2201236 | Houston Methodist Hospital | Houston Methodist Hospital | Randall J. Olsen, Paul A. Christensen, S. Wesley Long, Sishir Subedi, Robert Olson, Marcus Nguyen, James J. Davis, Matthew Ojeda Saavedra, Prasanti Yerramilli, Layne Pruitt, Kristina Reppond, Madison N. Shyer, Jessica Cambric, Ryan Gadd, Ilya J. Finkelstein, Jimmy Gollihar, and James M. Musser |
| EPI_ISL_2201237 | Aegis Sciences Corporation | Centers for Disease Control and Prevention Division of Viral Diseases, Pathogen Discovery | Dakota Howard, Dhwani Batra, Peter W. Cook, Kara Moser, Adrian Paskey, Jason Caravas, Benjamin Rambo-Martin, Shatavia Morrison, Christopher Gulvick, Scott Sammons, Yvette Unoarumhi, Darlene Wagner, Matthew Schmerer, Cyndi Clark, Patrick Campbell, Rob Case, Vikramsinha Ghorpade, Holly Houdeshell, Ola Kvalvaag, Dillon Nall, Ethan Sanders, Alec Vest, Shaun Westlund, Matthew Hardison, Clinton R. Paden, Duncan MacCannell |
| EPI_ISL_2201238 | Houston Methodist Hospital | Houston Methodist Hospital | Randall J. Olsen, Paul A. Christensen, S. Wesley Long, Sishir Subedi, Robert Olson, Marcus Nguyen, James J. Davis, Matthew Ojeda Saavedra, Prasanti Yerramilli, Layne Pruitt, Kristina Reppond, Madison N. Shyer, Jessica Cambric, Ryan Gadd, Ilya J. Finkelstein, Jimmy Gollihar, and James M. Musser |
| EPI_ISL_2201239 | Aegis Sciences Corporation | Centers for Disease Control and Prevention Division of Viral Diseases, Pathogen Discovery | Dakota Howard, Dhwani Batra, Peter W. Cook, Kara Moser, Adrian Paskey, Jason Caravas, Benjamin Rambo-Martin, Shatavia Morrison, Christopher Gulvick, Scott Sammons, Yvette Unoarumhi, Darlene Wagner, Matthew Schmerer, Cyndi Clark, Patrick Campbell, Rob Case, Vikramsinha Ghorpade, Holly Houdeshell, Ola Kvalvaag, Dillon Nall, Ethan Sanders, Alec Vest, Shaun Westlund, Matthew Hardison, Clinton R. Paden, Duncan MacCannell |
| EPI_ISL_2201240 | Houston Methodist Hospital | Houston Methodist Hospital | Randall J. Olsen, Paul A. Christensen, S. Wesley Long, Sishir Subedi, Robert Olson, Marcus Nguyen, James J. Davis, Matthew Ojeda Saavedra, Prasanti Yerramilli, Layne Pruitt, Kristina Reppond, Madison N. Shyer, Jessica Cambric, Ryan Gadd, Ilya J. Finkelstein, Jimmy Gollihar, and James M. Musser |
| EPI_ISL_2201241, EPI_ISL_2201242 | Aegis Sciences Corporation | Centers for Disease Control and Prevention Division of Viral Diseases, Pathogen Discovery | Dakota Howard, Dhwani Batra, Peter W. Cook, Kara Moser, Adrian Paskey, Jason Caravas, Benjamin Rambo-Martin, Shatavia Morrison, Christopher Gulvick, Scott Sammons, Yvette Unoarumhi, Darlene Wagner, Matthew Schmerer, Cyndi Clark, Patrick Campbell, Rob Case, Vikramsinha Ghorpade, Holly Houdeshell, Ola Kvalvaag, Dillon Nall, Ethan Sanders, Alec Vest, Shaun Westlund, Matthew Hardison, Clinton R. Paden, Duncan MacCannell |
| EPI_ISL_2201243 | Houston Methodist Hospital | Houston Methodist Hospital | Randall J. Olsen, Paul A. Christensen, S. Wesley Long, Sishir Subedi, Robert Olson, Marcus Nguyen, James J. Davis, Matthew Ojeda Saavedra, Prasanti Yerramilli, Layne Pruitt, Kristina Reppond, Madison N. Shyer, Jessica Cambric, Ryan Gadd, Ilya J. Finkelstein, Jimmy Gollihar, and James M. Musser |
| EPI_ISL_2201244 | Aegis Sciences Corporation | Centers for Disease Control and Prevention Division of Viral Diseases, Pathogen Discovery | Dakota Howard, Dhwani Batra, Peter W. Cook, Kara Moser, Adrian Paskey, Jason Caravas, Benjamin Rambo-Martin, Shatavia Morrison, Christopher Gulvick, Scott Sammons, Yvette Unoarumhi, Darlene Wagner, Matthew Schmerer, Cyndi Clark, Patrick Campbell, Rob Case, Vikramsinha Ghorpade, Holly Houdeshell, Ola Kvalvaag, Dillon Nall, Ethan Sanders, Alec Vest, Shaun Westlund, Matthew Hardison, Clinton R. Paden, Duncan MacCannell |
| EPI_ISL_2201245 | Houston Methodist Hospital | Houston Methodist Hospital | Randall J. Olsen, Paul A. Christensen, S. Wesley Long, Sishir Subedi, Robert Olson, Marcus Nguyen, James J. Davis, Matthew Ojeda Saavedra, Prasanti Yerramilli, Layne Pruitt, Kristina Reppond, Madison N. Shyer, Jessica Cambric, Ryan Gadd, Ilya J. Finkelstein, Jimmy Gollihar, and James M. Musser |

| | | | |
|---|---|---|---|
| EPI_ISL_2201246 | Aegis Sciences Corporation | Centers for Disease Control and Prevention Division of Viral Diseases, Pathogen Discovery | Dakota Howard, Dhwani Batra, Peter W. Cook, Kara Moser, Adrian Paskey, Jason Caravas, Benjamin Rambo-Martin, Shatavia Morrison, Christopher Gulvick, Scott Sammons, Yvette Unoarumhi, Darlene Wagner, Matthew Schmerer, Cyndi Clark, Patrick Campbell, Rob Case, Vikramsinha Ghorpade, Holly Houdeshell, Ola Kvalvaag, Dillon Nall, Ethan Sanders, Alec Vest, Shaun Westlund, Matthew Hardison, Clinton R. Paden, Duncan MacCannell |
| EPI_ISL_2201247 | Houston Methodist Hospital | Houston Methodist Hospital | Randall J. Olsen, Paul A. Christensen, S. Wesley Long, Sishir Subedi, Robert Olson, Marcus Nguyen, James J. Davis, Matthew Ojeda Saavedra, Prasanti Yerramilli, Layne Pruitt, Kristina Reppond, Madison N. Shyer, Jessica Cambric, Ryan Gadd, Ilya J. Finkelstein, Jimmy Gollihar, and James M. Musser |
| EPI_ISL_2201248, EPI_ISL_2201250, EPI_ISL_2201251 | Aegis Sciences Corporation | Centers for Disease Control and Prevention Division of Viral Diseases, Pathogen Discovery | Dakota Howard, Dhwani Batra, Peter W. Cook, Kara Moser, Adrian Paskey, Jason Caravas, Benjamin Rambo-Martin, Shatavia Morrison, Christopher Gulvick, Scott Sammons, Yvette Unoarumhi, Darlene Wagner, Matthew Schmerer, Cyndi Clark, Patrick Campbell, Rob Case, Vikramsinha Ghorpade, Holly Houdeshell, Ola Kvalvaag, Dillon Nall, Ethan Sanders, Alec Vest, Shaun Westlund, Matthew Hardison, Clinton R. Paden, Duncan MacCannell |
| EPI_ISL_2201252 | Houston Methodist Hospital | Houston Methodist Hospital | Randall J. Olsen, Paul A. Christensen, S. Wesley Long, Sishir Subedi, Robert Olson, Marcus Nguyen, James J. Davis, Matthew Ojeda Saavedra, Prasanti Yerramilli, Layne Pruitt, Kristina Reppond, Madison N. Shyer, Jessica Cambric, Ryan Gadd, Ilya J. Finkelstein, Jimmy Gollihar, and James M. Musser |
| EPI_ISL_2201253 | Aegis Sciences Corporation | Centers for Disease Control and Prevention Division of Viral Diseases, Pathogen Discovery | Dakota Howard, Dhwani Batra, Peter W. Cook, Kara Moser, Adrian Paskey, Jason Caravas, Benjamin Rambo-Martin, Shatavia Morrison, Christopher Gulvick, Scott Sammons, Yvette Unoarumhi, Darlene Wagner, Matthew Schmerer, Cyndi Clark, Patrick Campbell, Rob Case, Vikramsinha Ghorpade, Holly Houdeshell, Ola Kvalvaag, Dillon Nall, Ethan Sanders, Alec Vest, Shaun Westlund, Matthew Hardison, Clinton R. Paden, Duncan MacCannell |
| EPI_ISL_2201254 | Houston Methodist Hospital | Houston Methodist Hospital | Randall J. Olsen, Paul A. Christensen, S. Wesley Long, Sishir Subedi, Robert Olson, Marcus Nguyen, James J. Davis, Matthew Ojeda Saavedra, Prasanti Yerramilli, Layne Pruitt, Kristina Reppond, Madison N. Shyer, Jessica Cambric, Ryan Gadd, Ilya J. Finkelstein, Jimmy Gollihar, and James M. Musser |
| EPI_ISL_2201255, EPI_ISL_2201256 | Aegis Sciences Corporation | Centers for Disease Control and Prevention Division of Viral Diseases, Pathogen Discovery | Dakota Howard, Dhwani Batra, Peter W. Cook, Kara Moser, Adrian Paskey, Jason Caravas, Benjamin Rambo-Martin, Shatavia Morrison, Christopher Gulvick, Scott Sammons, Yvette Unoarumhi, Darlene Wagner, Matthew Schmerer, Cyndi Clark, Patrick Campbell, Rob Case, Vikramsinha Ghorpade, Holly Houdeshell, Ola Kvalvaag, Dillon Nall, Ethan Sanders, Alec Vest, Shaun Westlund, Matthew Hardison, Clinton R. Paden, Duncan MacCannell |
| EPI_ISL_2201257 | Houston Methodist Hospital | Houston Methodist Hospital | Randall J. Olsen, Paul A. Christensen, S. Wesley Long, Sishir Subedi, Robert Olson, Marcus Nguyen, James J. Davis, Matthew Ojeda Saavedra, Prasanti Yerramilli, Layne Pruitt, Kristina Reppond, Madison N. Shyer, Jessica Cambric, Ryan Gadd, Ilya J. Finkelstein, Jimmy Gollihar, and James M. Musser |
| EPI_ISL_2201258 | Aegis Sciences Corporation | Centers for Disease Control and Prevention Division of Viral Diseases, Pathogen Discovery | Dakota Howard, Dhwani Batra, Peter W. Cook, Kara Moser, Adrian Paskey, Jason Caravas, Benjamin Rambo-Martin, Shatavia Morrison, Christopher Gulvick, Scott Sammons, Yvette Unoarumhi, Darlene Wagner, Matthew Schmerer, Cyndi Clark, Patrick Campbell, Rob Case, Vikramsinha Ghorpade, Holly Houdeshell, Ola Kvalvaag, Dillon Nall, Ethan Sanders, Alec Vest, Shaun Westlund, Matthew Hardison, Clinton R. Paden, Duncan MacCannell |
| EPI_ISL_2201259 | Houston Methodist Hospital | Houston Methodist Hospital | Randall J. Olsen, Paul A. Christensen, S. Wesley Long, Sishir Subedi, Robert Olson, Marcus Nguyen, James J. Davis, Matthew Ojeda Saavedra, Prasanti Yerramilli, Layne Pruitt, Kristina Reppond, Madison N. Shyer, Jessica Cambric, Ryan Gadd, Ilya J. Finkelstein, Jimmy Gollihar, and James M. Musser |
| EPI_ISL_2201260 | Aegis Sciences Corporation | Centers for Disease Control and Prevention Division of Viral Diseases, Pathogen Discovery | Dakota Howard, Dhwani Batra, Peter W. Cook, Kara Moser, Adrian Paskey, Jason Caravas, Benjamin Rambo-Martin, Shatavia Morrison, Christopher Gulvick, Scott Sammons, Yvette Unoarumhi, Darlene Wagner, Matthew Schmerer, Cyndi Clark, Patrick Campbell, Rob Case, Vikramsinha Ghorpade, Holly Houdeshell, Ola Kvalvaag, Dillon Nall, Ethan Sanders, Alec Vest, Shaun Westlund, Matthew Hardison, Clinton R. Paden, Duncan MacCannell |
| EPI_ISL_2201261 | Houston Methodist Hospital | Houston Methodist Hospital | Randall J. Olsen, Paul A. Christensen, S. Wesley Long, Sishir Subedi, Robert Olson, Marcus Nguyen, James J. Davis, Matthew Ojeda Saavedra, Prasanti Yerramilli, Layne Pruitt, Kristina Reppond, Madison N. Shyer, Jessica Cambric, Ryan Gadd, Ilya J. Finkelstein, Jimmy Gollihar, and James M. Musser |
| EPI_ISL_2201262 | Aegis Sciences Corporation | Centers for Disease Control and Prevention Division of Viral Diseases, Pathogen Discovery | Dakota Howard, Dhwani Batra, Peter W. Cook, Kara Moser, Adrian Paskey, Jason Caravas, Benjamin Rambo-Martin, Shatavia Morrison, Christopher Gulvick, Scott Sammons, Yvette Unoarumhi, Darlene Wagner, Matthew Schmerer, Cyndi Clark, Patrick Campbell, Rob Case, Vikramsinha Ghorpade, Holly Houdeshell, Ola Kvalvaag, Dillon Nall, Ethan Sanders, Alec Vest, Shaun Westlund, Matthew Hardison, Clinton R. Paden, Duncan MacCannell |
| EPI_ISL_2201263 | Houston Methodist Hospital | Houston Methodist Hospital | Randall J. Olsen, Paul A. Christensen, S. Wesley Long, Sishir Subedi, Robert Olson, Marcus Nguyen, James J. Davis, Matthew Ojeda Saavedra, Prasanti Yerramilli, Layne Pruitt, Kristina Reppond, Madison N. Shyer, Jessica Cambric, Ryan Gadd, Ilya J. Finkelstein, Jimmy Gollihar, and James M. Musser |
| EPI_ISL_2201264, EPI_ISL_2201265 | Aegis Sciences Corporation | Centers for Disease Control and Prevention Division of Viral Diseases, Pathogen Discovery | Dakota Howard, Dhwani Batra, Peter W. Cook, Kara Moser, Adrian Paskey, Jason Caravas, Benjamin Rambo-Martin, Shatavia Morrison, Christopher Gulvick, Scott Sammons, Yvette Unoarumhi, Darlene Wagner, Matthew Schmerer, Cyndi Clark, Patrick Campbell, Rob Case, Vikramsinha Ghorpade, Holly Houdeshell, Ola Kvalvaag, Dillon Nall, Ethan Sanders, Alec Vest, Shaun Westlund, Matthew Hardison, Clinton R. Paden, Duncan MacCannell |
| EPI_ISL_2201266 | Houston Methodist Hospital | Houston Methodist Hospital | Randall J. Olsen, Paul A. Christensen, S. Wesley Long, Sishir Subedi, Robert Olson, Marcus Nguyen, James J. Davis, Matthew Ojeda Saavedra, Prasanti Yerramilli, Layne Pruitt, Kristina Reppond, Madison N. Shyer, Jessica Cambric, Ryan Gadd, Ilya J. Finkelstein, Jimmy Gollihar, and James M. Musser |
| EPI_ISL_2201267, EPI_ISL_2201268 | Aegis Sciences Corporation | Centers for Disease Control and Prevention Division of Viral Diseases, Pathogen Discovery | Dakota Howard, Dhwani Batra, Peter W. Cook, Kara Moser, Adrian Paskey, Jason Caravas, Benjamin Rambo-Martin, Shatavia Morrison, Christopher Gulvick, Scott Sammons, Yvette Unoarumhi, Darlene Wagner, Matthew Schmerer, Cyndi Clark, Patrick Campbell, Rob Case, Vikramsinha Ghorpade, Holly Houdeshell, Ola Kvalvaag, Dillon Nall, Ethan Sanders, Alec Vest, Shaun Westlund, Matthew Hardison, Clinton R. Paden, Duncan MacCannell |
| EPI_ISL_2201269 | Houston Methodist Hospital | Houston Methodist Hospital | Randall J. Olsen, Paul A. Christensen, S. Wesley Long, Sishir Subedi, Robert Olson, Marcus Nguyen, James J. Davis, Matthew Ojeda Saavedra, Prasanti Yerramilli, Layne Pruitt, Kristina Reppond, Madison N. Shyer, Jessica Cambric, Ryan Gadd, Ilya J. Finkelstein, Jimmy Gollihar, and James M. Musser |
| EPI_ISL_2201270, EPI_ISL_2201272 | Aegis Sciences Corporation | Centers for Disease Control and Prevention Division of Viral Diseases, Pathogen Discovery | Dakota Howard, Dhwani Batra, Peter W. Cook, Kara Moser, Adrian Paskey, Jason Caravas, Benjamin Rambo-Martin, Shatavia Morrison, Christopher Gulvick, Scott Sammons, Yvette Unoarumhi, Darlene Wagner, Matthew Schmerer, Cyndi Clark, Patrick Campbell, Rob Case, Vikramsinha Ghorpade, Holly Houdeshell, Ola Kvalvaag, Dillon Nall, Ethan Sanders, Alec Vest, Shaun Westlund, Matthew Hardison, Clinton R. Paden, Duncan MacCannell |
| EPI_ISL_2201273 | Houston Methodist Hospital | Houston Methodist Hospital | Randall J. Olsen, Paul A. Christensen, S. Wesley Long, Sishir Subedi, Robert Olson, Marcus Nguyen, James J. Davis, Matthew Ojeda Saavedra, Prasanti Yerramilli, Layne Pruitt, Kristina Reppond, Madison N. Shyer, Jessica Cambric, Ryan Gadd, Ilya J. Finkelstein, Jimmy Gollihar, and James M. Musser |
| EPI_ISL_2201274, EPI_ISL_2201275 | Aegis Sciences Corporation | Centers for Disease Control and Prevention Division of Viral Diseases, Pathogen Discovery | Dakota Howard, Dhwani Batra, Peter W. Cook, Kara Moser, Adrian Paskey, Jason Caravas, Benjamin Rambo-Martin, Shatavia Morrison, Christopher Gulvick, Scott Sammons, Yvette Unoarumhi, Darlene Wagner, Matthew Schmerer, Cyndi Clark, Patrick Campbell, Rob Case, Vikramsinha Ghorpade, Holly Houdeshell, Ola Kvalvaag, Dillon Nall, Ethan Sanders, Alec Vest, Shaun Westlund, Matthew Hardison, Clinton R. Paden, Duncan MacCannell |
| EPI_ISL_2201276 | Houston Methodist Hospital | Houston Methodist Hospital | Randall J. Olsen, Paul A. Christensen, S. Wesley Long, Sishir Subedi, Robert Olson, Marcus Nguyen, James J. Davis, Matthew Ojeda Saavedra, Prasanti Yerramilli, Layne Pruitt, Kristina Reppond, Madison N. Shyer, Jessica Cambric, Ryan Gadd, Ilya J. Finkelstein, Jimmy Gollihar, and James M. Musser |
| EPI_ISL_2201277, EPI_ISL_2201278 | Aegis Sciences Corporation | Centers for Disease Control and Prevention Division of Viral Diseases, Pathogen Discovery | Dakota Howard, Dhwani Batra, Peter W. Cook, Kara Moser, Adrian Paskey, Jason Caravas, Benjamin Rambo-Martin, Shatavia Morrison, Christopher Gulvick, Scott Sammons, Yvette Unoarumhi, Darlene Wagner, Matthew Schmerer, Cyndi Clark, Patrick Campbell, Rob Case, Vikramsinha Ghorpade, Holly Houdeshell, Ola Kvalvaag, Dillon Nall, Ethan Sanders, Alec Vest, Shaun Westlund, Matthew Hardison, Clinton R. Paden, Duncan MacCannell |
| EPI_ISL_2201279 | Houston Methodist Hospital | Houston Methodist Hospital | Randall J. Olsen, Paul A. Christensen, S. Wesley Long, Sishir Subedi, Robert Olson, Marcus Nguyen, James J. Davis, Matthew Ojeda Saavedra, Prasanti Yerramilli, Layne Pruitt, Kristina Reppond, Madison N. Shyer, Jessica Cambric, Ryan Gadd, Ilya J. Finkelstein, Jimmy Gollihar, and James M. Musser |
| EPI_ISL_2201280, EPI_ISL_2201281 | Aegis Sciences Corporation | Centers for Disease Control and Prevention Division of Viral Diseases, Pathogen Discovery | Dakota Howard, Dhwani Batra, Peter W. Cook, Kara Moser, Adrian Paskey, Jason Caravas, Benjamin Rambo-Martin, Shatavia Morrison, Christopher Gulvick, Scott Sammons, Yvette Unoarumhi, Darlene Wagner, Matthew Schmerer, Cyndi Clark, Patrick Campbell, Rob Case, Vikramsinha Ghorpade, Holly Houdeshell, Ola Kvalvaag, Dillon Nall, Ethan Sanders, Alec Vest, Shaun Westlund, Matthew Hardison, Clinton R. Paden, Duncan MacCannell |
| EPI_ISL_2201282 | Houston Methodist Hospital | Houston Methodist Hospital | Randall J. Olsen, Paul A. Christensen, S. Wesley Long, Sishir Subedi, Robert Olson, Marcus Nguyen, James J. Davis, Matthew Ojeda Saavedra, Prasanti Yerramilli, Layne Pruitt, Kristina Reppond, Madison N. Shyer, Jessica Cambric, Ryan Gadd, Ilya J. Finkelstein, Jimmy Gollihar, and James M. Musser |
| EPI_ISL_2201283 | Aegis Sciences Corporation | Centers for Disease Control and Prevention Division of Viral Diseases, Pathogen Discovery | Dakota Howard, Dhwani Batra, Peter W. Cook, Kara Moser, Adrian Paskey, Jason Caravas, Benjamin Rambo-Martin, Shatavia Morrison, Christopher Gulvick, Scott Sammons, Yvette Unoarumhi, Darlene Wagner, Matthew Schmerer, Cyndi Clark, Patrick Campbell, Rob Case, Vikramsinha Ghorpade, Holly Houdeshell, Ola Kvalvaag, Dillon Nall, Ethan Sanders, Alec Vest, Shaun Westlund, Matthew Hardison, Clinton R. Paden, Duncan MacCannell |
| EPI_ISL_2201284, EPI_ISL_2201285 | Houston Methodist Hospital | Houston Methodist Hospital | Randall J. Olsen, Paul A. Christensen, S. Wesley Long, Sishir Subedi, Robert Olson, Marcus Nguyen, James J. Davis, Matthew Ojeda Saavedra, Prasanti Yerramilli, Layne Pruitt, Kristina Reppond, Madison N. Shyer, Jessica Cambric, Ryan Gadd, Ilya J. Finkelstein, Jimmy Gollihar, and James M. Musser |
| EPI_ISL_2201286 | Aegis Sciences Corporation | Centers for Disease Control and Prevention Division of Viral Diseases, Pathogen Discovery | Dakota Howard, Dhwani Batra, Peter W. Cook, Kara Moser, Adrian Paskey, Jason Caravas, Benjamin Rambo-Martin, Shatavia Morrison, Christopher Gulvick, Scott Sammons, Yvette Unoarumhi, Darlene Wagner, Matthew Schmerer, Cyndi Clark, Patrick Campbell, Rob Case, Vikramsinha Ghorpade, Holly Houdeshell, Ola Kvalvaag, Dillon Nall, Ethan Sanders, Alec Vest, Shaun Westlund, Matthew Hardison, Clinton R. Paden, Duncan MacCannell |
| EPI_ISL_2201287 | Houston Methodist Hospital | Houston Methodist Hospital | Randall J. Olsen, Paul A. Christensen, S. Wesley Long, Sishir Subedi, Robert Olson, Marcus Nguyen, James J. Davis, Matthew Ojeda Saavedra, Prasanti Yerramilli, Layne Pruitt, Kristina Reppond, Madison N. Shyer, Jessica Cambric, Ryan Gadd, Ilya J. Finkelstein, Jimmy Gollihar, and James M. Musser |
| EPI_ISL_2201288 | Aegis Sciences Corporation | Centers for Disease Control and Prevention Division of Viral Diseases, Pathogen Discovery | Dakota Howard, Dhwani Batra, Peter W. Cook, Kara Moser, Adrian Paskey, Jason Caravas, Benjamin Rambo-Martin, Shatavia Morrison, Christopher Gulvick, Scott Sammons, Yvette Unoarumhi, Darlene Wagner, Matthew Schmerer, Cyndi Clark, Patrick Campbell, Rob Case, Vikramsinha Ghorpade, Holly Houdeshell, Ola Kvalvaag, Dillon Nall, Ethan Sanders, Alec Vest, Shaun Westlund, Matthew Hardison, Clinton R. Paden, Duncan MacCannell |
| EPI_ISL_2201290 | Houston Methodist Hospital | Houston Methodist Hospital | Randall J. Olsen, Paul A. Christensen, S. Wesley Long, Sishir Subedi, Robert Olson, Marcus Nguyen, James J. Davis, Matthew Ojeda Saavedra, Prasanti Yerramilli, Layne Pruitt, Kristina Reppond, Madison N. Shyer, Jessica Cambric, Ryan Gadd, Ilya J. Finkelstein, Jimmy Gollihar, and James M. Musser |
| EPI_ISL_2201291 | Aegis Sciences Corporation | Centers for Disease Control and Prevention Division of Viral Diseases, Pathogen Discovery | Dakota Howard, Dhwani Batra, Peter W. Cook, Kara Moser, Adrian Paskey, Jason Caravas, Benjamin Rambo-Martin, Shatavia Morrison, Christopher Gulvick, Scott Sammons, Yvette Unoarumhi, Darlene Wagner, Matthew Schmerer, Cyndi Clark, Patrick Campbell, Rob Case, Vikramsinha Ghorpade, Holly Houdeshell, Ola Kvalvaag, Dillon Nall, Ethan Sanders, Alec Vest, Shaun Westlund, Matthew Hardison, Clinton R. Paden, Duncan MacCannell |
| EPI_ISL_2201292 | Houston Methodist Hospital | Houston Methodist Hospital | Randall J. Olsen, Paul A. Christensen, S. Wesley Long, Sishir Subedi, Robert Olson, Marcus Nguyen, James J. Davis, Matthew Ojeda Saavedra, Prasanti Yerramilli, Layne Pruitt, Kristina Reppond, Madison N. Shyer, Jessica Cambric, Ryan Gadd, Ilya J. Finkelstein, Jimmy Gollihar, and James M. Musser |
| EPI_ISL_2201293 | Aegis Sciences Corporation | Centers for Disease Control and Prevention Division of Viral Diseases, Pathogen Discovery | Dakota Howard, Dhwani Batra, Peter W. Cook, Kara Moser, Adrian Paskey, Jason Caravas, Benjamin Rambo-Martin, Shatavia Morrison, Christopher Gulvick, Scott Sammons, Yvette Unoarumhi, Darlene Wagner, Matthew Schmerer, Cyndi Clark, Patrick Campbell, Rob Case, Vikramsinha Ghorpade, Holly Houdeshell, Ola Kvalvaag, Dillon Nall, Ethan Sanders, Alec Vest, Shaun Westlund, Matthew Hardison, Clinton R. Paden, Duncan MacCannell |
| EPI_ISL_2201294 | Houston Methodist Hospital | Houston Methodist Hospital | Randall J. Olsen, Paul A. Christensen, S. Wesley Long, Sishir Subedi, Robert Olson, Marcus Nguyen, James J. Davis, Matthew Ojeda Saavedra, Prasanti Yerramilli, Layne Pruitt, Kristina Reppond, Madison N. Shyer, Jessica Cambric, Ryan Gadd, Ilya J. Finkelstein, Jimmy Gollihar, and James M. Musser |
| EPI_ISL_2201295, EPI_ISL_2201296 | Aegis Sciences Corporation | Centers for Disease Control and Prevention Division of Viral Diseases, Pathogen Discovery | Dakota Howard, Dhwani Batra, Peter W. Cook, Kara Moser, Adrian Paskey, Jason Caravas, Benjamin Rambo-Martin, Shatavia Morrison, Christopher Gulvick, Scott Sammons, Yvette Unoarumhi, Darlene Wagner, Matthew Schmerer, Cyndi Clark, Patrick Campbell, Rob Case, Vikramsinha Ghorpade, Holly Houdeshell, Ola Kvalvaag, Dillon Nall, Ethan Sanders, Alec Vest, Shaun Westlund, Matthew Hardison, Clinton R. Paden, Duncan MacCannell |

| | | | |
|---|---|---|---|
| EPI_ISL_2201297 | Houston Methodist Hospital | Houston Methodist Hospital | Randall J. Olsen, Paul A. Christensen, S. Wesley Long, Sishir Subedi, Robert Olson, Marcus Nguyen, James J. Davis, Matthew Ojeda Saavedra, Prasanti Yerramilli, Layne Pruitt, Kristina Reppond, Madison N. Shyer, Jessica Cambric, Ryan Gadd, Ilya J. Finkelstein, Jimmy Gollihar, and James M. Musser |
| EPI_ISL_2201298 | Aegis Sciences Corporation | Centers for Disease Control and Prevention Division of Viral Diseases, Pathogen Discovery | Dakota Howard, Dhwani Batra, Peter W. Cook, Kara Moser, Adrian Paskey, Jason Caravas, Benjamin Rambo-Martin, Shatavia Morrison, Christopher Gulvick, Scott Sammons, Yvette Unoarumhi, Darlene Wagner, Matthew Schmerer, Cyndi Clark, Patrick Campbell, Rob Case, Vikramsinha Ghorpade, Holly Houdeshell, Ola Kvalvaag, Dillon Nall, Ethan Sanders, Alec Vest, Shaun Westlund, Matthew Hardison, Clinton R. Paden, Duncan MacCannell |
| EPI_ISL_2201299 | Houston Methodist Hospital | Houston Methodist Hospital | Randall J. Olsen, Paul A. Christensen, S. Wesley Long, Sishir Subedi, Robert Olson, Marcus Nguyen, James J. Davis, Matthew Ojeda Saavedra, Prasanti Yerramilli, Layne Pruitt, Kristina Reppond, Madison N. Shyer, Jessica Cambric, Ryan Gadd, Ilya J. Finkelstein, Jimmy Gollihar, and James M. Musser |
| EPI_ISL_2201300, EPI_ISL_2201301 | Aegis Sciences Corporation | Centers for Disease Control and Prevention Division of Viral Diseases, Pathogen Discovery | Dakota Howard, Dhwani Batra, Peter W. Cook, Kara Moser, Adrian Paskey, Jason Caravas, Benjamin Rambo-Martin, Shatavia Morrison, Christopher Gulvick, Scott Sammons, Yvette Unoarumhi, Darlene Wagner, Matthew Schmerer, Cyndi Clark, Patrick Campbell, Rob Case, Vikramsinha Ghorpade, Holly Houdeshell, Ola Kvalvaag, Dillon Nall, Ethan Sanders, Alec Vest, Shaun Westlund, Matthew Hardison, Clinton R. Paden, Duncan MacCannell |
| EPI_ISL_2201302 | Houston Methodist Hospital | Houston Methodist Hospital | Randall J. Olsen, Paul A. Christensen, S. Wesley Long, Sishir Subedi, Robert Olson, Marcus Nguyen, James J. Davis, Matthew Ojeda Saavedra, Prasanti Yerramilli, Layne Pruitt, Kristina Reppond, Madison N. Shyer, Jessica Cambric, Ryan Gadd, Ilya J. Finkelstein, Jimmy Gollihar, and James M. Musser |
| EPI_ISL_2201303 | Aegis Sciences Corporation | Centers for Disease Control and Prevention Division of Viral Diseases, Pathogen Discovery | Dakota Howard, Dhwani Batra, Peter W. Cook, Kara Moser, Adrian Paskey, Jason Caravas, Benjamin Rambo-Martin, Shatavia Morrison, Christopher Gulvick, Scott Sammons, Yvette Unoarumhi, Darlene Wagner, Matthew Schmerer, Cyndi Clark, Patrick Campbell, Rob Case, Vikramsinha Ghorpade, Holly Houdeshell, Ola Kvalvaag, Dillon Nall, Ethan Sanders, Alec Vest, Shaun Westlund, Matthew Hardison, Clinton R. Paden, Duncan MacCannell |
| EPI_ISL_2201304 | Houston Methodist Hospital | Houston Methodist Hospital | Randall J. Olsen, Paul A. Christensen, S. Wesley Long, Sishir Subedi, Robert Olson, Marcus Nguyen, James J. Davis, Matthew Ojeda Saavedra, Prasanti Yerramilli, Layne Pruitt, Kristina Reppond, Madison N. Shyer, Jessica Cambric, Ryan Gadd, Ilya J. Finkelstein, Jimmy Gollihar, and James M. Musser |
| EPI_ISL_2201305 | Aegis Sciences Corporation | Centers for Disease Control and Prevention Division of Viral Diseases, Pathogen Discovery | Dakota Howard, Dhwani Batra, Peter W. Cook, Kara Moser, Adrian Paskey, Jason Caravas, Benjamin Rambo-Martin, Shatavia Morrison, Christopher Gulvick, Scott Sammons, Yvette Unoarumhi, Darlene Wagner, Matthew Schmerer, Cyndi Clark, Patrick Campbell, Rob Case, Vikramsinha Ghorpade, Holly Houdeshell, Ola Kvalvaag, Dillon Nall, Ethan Sanders, Alec Vest, Shaun Westlund, Matthew Hardison, Clinton R. Paden, Duncan MacCannell |
| EPI_ISL_2201306 | Houston Methodist Hospital | Houston Methodist Hospital | Randall J. Olsen, Paul A. Christensen, S. Wesley Long, Sishir Subedi, Robert Olson, Marcus Nguyen, James J. Davis, Matthew Ojeda Saavedra, Prasanti Yerramilli, Layne Pruitt, Kristina Reppond, Madison N. Shyer, Jessica Cambric, Ryan Gadd, Ilya J. Finkelstein, Jimmy Gollihar, and James M. Musser |
| EPI_ISL_2201307 | Aegis Sciences Corporation | Centers for Disease Control and Prevention Division of Viral Diseases, Pathogen Discovery | Dakota Howard, Dhwani Batra, Peter W. Cook, Kara Moser, Adrian Paskey, Jason Caravas, Benjamin Rambo-Martin, Shatavia Morrison, Christopher Gulvick, Scott Sammons, Yvette Unoarumhi, Darlene Wagner, Matthew Schmerer, Cyndi Clark, Patrick Campbell, Rob Case, Vikramsinha Ghorpade, Holly Houdeshell, Ola Kvalvaag, Dillon Nall, Ethan Sanders, Alec Vest, Shaun Westlund, Matthew Hardison, Clinton R. Paden, Duncan MacCannell |
| EPI_ISL_2201308 | Houston Methodist Hospital | Houston Methodist Hospital | Randall J. Olsen, Paul A. Christensen, S. Wesley Long, Sishir Subedi, Robert Olson, Marcus Nguyen, James J. Davis, Matthew Ojeda Saavedra, Prasanti Yerramilli, Layne Pruitt, Kristina Reppond, Madison N. Shyer, Jessica Cambric, Ryan Gadd, Ilya J. Finkelstein, Jimmy Gollihar, and James M. Musser |
| EPI_ISL_2201309, EPI_ISL_2201310 | Aegis Sciences Corporation | Centers for Disease Control and Prevention Division of Viral Diseases, Pathogen Discovery | Dakota Howard, Dhwani Batra, Peter W. Cook, Kara Moser, Adrian Paskey, Jason Caravas, Benjamin Rambo-Martin, Shatavia Morrison, Christopher Gulvick, Scott Sammons, Yvette Unoarumhi, Darlene Wagner, Matthew Schmerer, Cyndi Clark, Patrick Campbell, Rob Case, Vikramsinha Ghorpade, Holly Houdeshell, Ola Kvalvaag, Dillon Nall, Ethan Sanders, Alec Vest, Shaun Westlund, Matthew Hardison, Clinton R. Paden, Duncan MacCannell |
| EPI_ISL_2201311 | Houston Methodist Hospital | Houston Methodist Hospital | Randall J. Olsen, Paul A. Christensen, S. Wesley Long, Sishir Subedi, Robert Olson, Marcus Nguyen, James J. Davis, Matthew Ojeda Saavedra, Prasanti Yerramilli, Layne Pruitt, Kristina Reppond, Madison N. Shyer, Jessica Cambric, Ryan Gadd, Ilya J. Finkelstein, Jimmy Gollihar, and James M. Musser |
| EPI_ISL_2201312 | Aegis Sciences Corporation | Centers for Disease Control and Prevention Division of Viral Diseases, Pathogen Discovery | Dakota Howard, Dhwani Batra, Peter W. Cook, Kara Moser, Adrian Paskey, Jason Caravas, Benjamin Rambo-Martin, Shatavia Morrison, Christopher Gulvick, Scott Sammons, Yvette Unoarumhi, Darlene Wagner, Matthew Schmerer, Cyndi Clark, Patrick Campbell, Rob Case, Vikramsinha Ghorpade, Holly Houdeshell, Ola Kvalvaag, Dillon Nall, Ethan Sanders, Alec Vest, Shaun Westlund, Matthew Hardison, Clinton R. Paden, Duncan MacCannell |
| EPI_ISL_2201313 | Houston Methodist Hospital | Houston Methodist Hospital | Randall J. Olsen, Paul A. Christensen, S. Wesley Long, Sishir Subedi, Robert Olson, Marcus Nguyen, James J. Davis, Matthew Ojeda Saavedra, Prasanti Yerramilli, Layne Pruitt, Kristina Reppond, Madison N. Shyer, Jessica Cambric, Ryan Gadd, Ilya J. Finkelstein, Jimmy Gollihar, and James M. Musser |
| EPI_ISL_2201314 | Aegis Sciences Corporation | Centers for Disease Control and Prevention Division of Viral Diseases, Pathogen Discovery | Dakota Howard, Dhwani Batra, Peter W. Cook, Kara Moser, Adrian Paskey, Jason Caravas, Benjamin Rambo-Martin, Shatavia Morrison, Christopher Gulvick, Scott Sammons, Yvette Unoarumhi, Darlene Wagner, Matthew Schmerer, Cyndi Clark, Patrick Campbell, Rob Case, Vikramsinha Ghorpade, Holly Houdeshell, Ola Kvalvaag, Dillon Nall, Ethan Sanders, Alec Vest, Shaun Westlund, Matthew Hardison, Clinton R. Paden, Duncan MacCannell |
| EPI_ISL_2201315 | Houston Methodist Hospital | Houston Methodist Hospital | Randall J. Olsen, Paul A. Christensen, S. Wesley Long, Sishir Subedi, Robert Olson, Marcus Nguyen, James J. Davis, Matthew Ojeda Saavedra, Prasanti Yerramilli, Layne Pruitt, Kristina Reppond, Madison N. Shyer, Jessica Cambric, Ryan Gadd, Ilya J. Finkelstein, Jimmy Gollihar, and James M. Musser |
| EPI_ISL_2201316, EPI_ISL_2201317 | Aegis Sciences Corporation | Centers for Disease Control and Prevention Division of Viral Diseases, Pathogen Discovery | Dakota Howard, Dhwani Batra, Peter W. Cook, Kara Moser, Adrian Paskey, Jason Caravas, Benjamin Rambo-Martin, Shatavia Morrison, Christopher Gulvick, Scott Sammons, Yvette Unoarumhi, Darlene Wagner, Matthew Schmerer, Cyndi Clark, Patrick Campbell, Rob Case, Vikramsinha Ghorpade, Holly Houdeshell, Ola Kvalvaag, Dillon Nall, Ethan Sanders, Alec Vest, Shaun Westlund, Matthew Hardison, Clinton R. Paden, Duncan MacCannell |
| EPI_ISL_2201318 | Houston Methodist Hospital | Houston Methodist Hospital | Randall J. Olsen, Paul A. Christensen, S. Wesley Long, Sishir Subedi, Robert Olson, Marcus Nguyen, James J. Davis, Matthew Ojeda Saavedra, Prasanti Yerramilli, Layne Pruitt, Kristina Reppond, Madison N. Shyer, Jessica Cambric, Ryan Gadd, Ilya J. Finkelstein, Jimmy Gollihar, and James M. Musser |
| EPI_ISL_2201319 | Aegis Sciences Corporation | Centers for Disease Control and Prevention Division of Viral Diseases, Pathogen Discovery | Dakota Howard, Dhwani Batra, Peter W. Cook, Kara Moser, Adrian Paskey, Jason Caravas, Benjamin Rambo-Martin, Shatavia Morrison, Christopher Gulvick, Scott Sammons, Yvette Unoarumhi, Darlene Wagner, Matthew Schmerer, Cyndi Clark, Patrick Campbell, Rob Case, Vikramsinha Ghorpade, Holly Houdeshell, Ola Kvalvaag, Dillon Nall, Ethan Sanders, Alec Vest, Shaun Westlund, Matthew Hardison, Clinton R. Paden, Duncan MacCannell |
| EPI_ISL_2201321 | Houston Methodist Hospital | Houston Methodist Hospital | Randall J. Olsen, Paul A. Christensen, S. Wesley Long, Sishir Subedi, Robert Olson, Marcus Nguyen, James J. Davis, Matthew Ojeda Saavedra, Prasanti Yerramilli, Layne Pruitt, Kristina Reppond, Madison N. Shyer, Jessica Cambric, Ryan Gadd, Ilya J. Finkelstein, Jimmy Gollihar, and James M. Musser |
| EPI_ISL_2201322, EPI_ISL_2201323 | Aegis Sciences Corporation | Centers for Disease Control and Prevention Division of Viral Diseases, Pathogen Discovery | Dakota Howard, Dhwani Batra, Peter W. Cook, Kara Moser, Adrian Paskey, Jason Caravas, Benjamin Rambo-Martin, Shatavia Morrison, Christopher Gulvick, Scott Sammons, Yvette Unoarumhi, Darlene Wagner, Matthew Schmerer, Cyndi Clark, Patrick Campbell, Rob Case, Vikramsinha Ghorpade, Holly Houdeshell, Ola Kvalvaag, Dillon Nall, Ethan Sanders, Alec Vest, Shaun Westlund, Matthew Hardison, Clinton R. Paden, Duncan MacCannell |
| EPI_ISL_2201324 | Houston Methodist Hospital | Houston Methodist Hospital | Randall J. Olsen, Paul A. Christensen, S. Wesley Long, Sishir Subedi, Robert Olson, Marcus Nguyen, James J. Davis, Matthew Ojeda Saavedra, Prasanti Yerramilli, Layne Pruitt, Kristina Reppond, Madison N. Shyer, Jessica Cambric, Ryan Gadd, Ilya J. Finkelstein, Jimmy Gollihar, and James M. Musser |
| EPI_ISL_2201325 | Aegis Sciences Corporation | Centers for Disease Control and Prevention Division of Viral Diseases, Pathogen Discovery | Dakota Howard, Dhwani Batra, Peter W. Cook, Kara Moser, Adrian Paskey, Jason Caravas, Benjamin Rambo-Martin, Shatavia Morrison, Christopher Gulvick, Scott Sammons, Yvette Unoarumhi, Darlene Wagner, Matthew Schmerer, Cyndi Clark, Patrick Campbell, Rob Case, Vikramsinha Ghorpade, Holly Houdeshell, Ola Kvalvaag, Dillon Nall, Ethan Sanders, Alec Vest, Shaun Westlund, Matthew Hardison, Clinton R. Paden, Duncan MacCannell |
| EPI_ISL_2201326 | Houston Methodist Hospital | Houston Methodist Hospital | Randall J. Olsen, Paul A. Christensen, S. Wesley Long, Sishir Subedi, Robert Olson, Marcus Nguyen, James J. Davis, Matthew Ojeda Saavedra, Prasanti Yerramilli, Layne Pruitt, Kristina Reppond, Madison N. Shyer, Jessica Cambric, Ryan Gadd, Ilya J. Finkelstein, Jimmy Gollihar, and James M. Musser |
| EPI_ISL_2201327, EPI_ISL_2201328 | Aegis Sciences Corporation | Centers for Disease Control and Prevention Division of Viral Diseases, Pathogen Discovery | Dakota Howard, Dhwani Batra, Peter W. Cook, Kara Moser, Adrian Paskey, Jason Caravas, Benjamin Rambo-Martin, Shatavia Morrison, Christopher Gulvick, Scott Sammons, Yvette Unoarumhi, Darlene Wagner, Matthew Schmerer, Cyndi Clark, Patrick Campbell, Rob Case, Vikramsinha Ghorpade, Holly Houdeshell, Ola Kvalvaag, Dillon Nall, Ethan Sanders, Alec Vest, Shaun Westlund, Matthew Hardison, Clinton R. Paden, Duncan MacCannell |
| EPI_ISL_2201329 | Houston Methodist Hospital | Houston Methodist Hospital | Randall J. Olsen, Paul A. Christensen, S. Wesley Long, Sishir Subedi, Robert Olson, Marcus Nguyen, James J. Davis, Matthew Ojeda Saavedra, Prasanti Yerramilli, Layne Pruitt, Kristina Reppond, Madison N. Shyer, Jessica Cambric, Ryan Gadd, Ilya J. Finkelstein, Jimmy Gollihar, and James M. Musser |
| EPI_ISL_2201330, EPI_ISL_2201331 | Aegis Sciences Corporation | Centers for Disease Control and Prevention Division of Viral Diseases, Pathogen Discovery | Dakota Howard, Dhwani Batra, Peter W. Cook, Kara Moser, Adrian Paskey, Jason Caravas, Benjamin Rambo-Martin, Shatavia Morrison, Christopher Gulvick, Scott Sammons, Yvette Unoarumhi, Darlene Wagner, Matthew Schmerer, Cyndi Clark, Patrick Campbell, Rob Case, Vikramsinha Ghorpade, Holly Houdeshell, Ola Kvalvaag, Dillon Nall, Ethan Sanders, Alec Vest, Shaun Westlund, Matthew Hardison, Clinton R. Paden, Duncan MacCannell |
| EPI_ISL_2201333 | Houston Methodist Hospital | Houston Methodist Hospital | Randall J. Olsen, Paul A. Christensen, S. Wesley Long, Sishir Subedi, Robert Olson, Marcus Nguyen, James J. Davis, Matthew Ojeda Saavedra, Prasanti Yerramilli, Layne Pruitt, Kristina Reppond, Madison N. Shyer, Jessica Cambric, Ryan Gadd, Ilya J. Finkelstein, Jimmy Gollihar, and James M. Musser |
| EPI_ISL_2201334 | Aegis Sciences Corporation | Centers for Disease Control and Prevention Division of Viral Diseases, Pathogen Discovery | Dakota Howard, Dhwani Batra, Peter W. Cook, Kara Moser, Adrian Paskey, Jason Caravas, Benjamin Rambo-Martin, Shatavia Morrison, Christopher Gulvick, Scott Sammons, Yvette Unoarumhi, Darlene Wagner, Matthew Schmerer, Cyndi Clark, Patrick Campbell, Rob Case, Vikramsinha Ghorpade, Holly Houdeshell, Ola Kvalvaag, Dillon Nall, Ethan Sanders, Alec Vest, Shaun Westlund, Matthew Hardison, Clinton R. Paden, Duncan MacCannell |
| EPI_ISL_2201335 | Houston Methodist Hospital | Houston Methodist Hospital | Randall J. Olsen, Paul A. Christensen, S. Wesley Long, Sishir Subedi, Robert Olson, Marcus Nguyen, James J. Davis, Matthew Ojeda Saavedra, Prasanti Yerramilli, Layne Pruitt, Kristina Reppond, Madison N. Shyer, Jessica Cambric, Ryan Gadd, Ilya J. Finkelstein, Jimmy Gollihar, and James M. Musser |
| EPI_ISL_2201336 | Aegis Sciences Corporation | Centers for Disease Control and Prevention Division of Viral Diseases, Pathogen Discovery | Dakota Howard, Dhwani Batra, Peter W. Cook, Kara Moser, Adrian Paskey, Jason Caravas, Benjamin Rambo-Martin, Shatavia Morrison, Christopher Gulvick, Scott Sammons, Yvette Unoarumhi, Darlene Wagner, Matthew Schmerer, Cyndi Clark, Patrick Campbell, Rob Case, Vikramsinha Ghorpade, Holly Houdeshell, Ola Kvalvaag, Dillon Nall, Ethan Sanders, Alec Vest, Shaun Westlund, Matthew Hardison, Clinton R. Paden, Duncan MacCannell |
| EPI_ISL_2201337 | Houston Methodist Hospital | Houston Methodist Hospital | Randall J. Olsen, Paul A. Christensen, S. Wesley Long, Sishir Subedi, Robert Olson, Marcus Nguyen, James J. Davis, Matthew Ojeda Saavedra, Prasanti Yerramilli, Layne Pruitt, Kristina Reppond, Madison N. Shyer, Jessica Cambric, Ryan Gadd, Ilya J. Finkelstein, Jimmy Gollihar, and James M. Musser |
| EPI_ISL_2201338, EPI_ISL_2201339 | Aegis Sciences Corporation | Centers for Disease Control and Prevention Division of Viral Diseases, Pathogen Discovery | Dakota Howard, Dhwani Batra, Peter W. Cook, Kara Moser, Adrian Paskey, Jason Caravas, Benjamin Rambo-Martin, Shatavia Morrison, Christopher Gulvick, Scott Sammons, Yvette Unoarumhi, Darlene Wagner, Matthew Schmerer, Cyndi Clark, Patrick Campbell, Rob Case, Vikramsinha Ghorpade, Holly Houdeshell, Ola Kvalvaag, Dillon Nall, Ethan Sanders, Alec Vest, Shaun Westlund, Matthew Hardison, Clinton R. Paden, Duncan MacCannell |
| EPI_ISL_2201340 | Houston Methodist Hospital | Houston Methodist Hospital | Randall J. Olsen, Paul A. Christensen, S. Wesley Long, Sishir Subedi, Robert Olson, Marcus Nguyen, James J. Davis, Matthew Ojeda Saavedra, Prasanti Yerramilli, Layne Pruitt, Kristina Reppond, Madison N. Shyer, Jessica Cambric, Ryan Gadd, Ilya J. Finkelstein, Jimmy Gollihar, and James M. Musser |
| EPI_ISL_2201341 | Aegis Sciences Corporation | Centers for Disease Control and Prevention Division of Viral Diseases, Pathogen Discovery | Dakota Howard, Dhwani Batra, Peter W. Cook, Kara Moser, Adrian Paskey, Jason Caravas, Benjamin Rambo-Martin, Shatavia Morrison, Christopher Gulvick, Scott Sammons, Yvette Unoarumhi, Darlene Wagner, Matthew Schmerer, Cyndi Clark, Patrick Campbell, Rob Case, Vikramsinha Ghorpade, Holly Houdeshell, Ola Kvalvaag, Dillon Nall, Ethan Sanders, Alec Vest, Shaun Westlund, Matthew Hardison, Clinton R. Paden, Duncan MacCannell |
| EPI_ISL_2201342 | Houston Methodist Hospital | Houston Methodist Hospital | Randall J. Olsen, Paul A. Christensen, S. Wesley Long, Sishir Subedi, Robert Olson, Marcus Nguyen, James J. Davis, Matthew Ojeda Saavedra, Prasanti Yerramilli, Layne Pruitt, Kristina Reppond, Madison N. Shyer, Jessica Cambric, Ryan Gadd, Ilya J. Finkelstein, Jimmy Gollihar, and James M. Musser |
| EPI_ISL_2201343, | Aegis Sciences Corporation | Centers for Disease Control and Prevention | Dakota Howard, Dhwani Batra, Peter W. Cook, Kara Moser, Adrian Paskey, Jason Caravas, Benjamin Rambo-Martin, Shatavia Morrison, Christopher Gulvick, Scott Sammons, Yvette Unoarumhi, Darlene Wagner, Matthew Schmerer, Cyndi Clark, Patrick Campbell, Rob Case, Vikramsinha |

| | | | |
|---|---|---|---|
| EPI_ISL_2201344 | | Division of Viral Diseases, Pathogen Discovery | Ghorpade, Holly Houdeshell, Ola Kvalvaag, Dillon Nall, Ethan Sanders, Alec Vest, Shaun Westlund, Matthew Hardison, Clinton R. Paden, Duncan MacCannell |
| EPI_ISL_2201345 | Houston Methodist Hospital | Houston Methodist Hospital | Randall J. Olsen, Paul A. Christensen, S. Wesley Long, Sishir Subedi, Robert Olson, Marcus Nguyen, James J. Davis, Matthew Ojeda Saavedra, Prasanti Yerramilli, Layne Pruitt, Kristina Reppond, Madison N. Shyer, Jessica Cambric, Ryan Gadd, Ilya J. Finkelstein, Jimmy Gollihar, and James M. Musser |
| EPI_ISL_2201346 | Aegis Sciences Corporation | Centers for Disease Control and Prevention Division of Viral Diseases, Pathogen Discovery | Dakota Howard, Dhwani Batra, Peter W. Cook, Kara Moser, Adrian Paskey, Jason Caravas, Benjamin Rambo-Martin, Shatavia Morrison, Christopher Gulvick, Scott Sammons, Yvette Unoarumhi, Darlene Wagner, Matthew Schmerer, Cyndi Clark, Patrick Campbell, Rob Case, Vikramsinha Ghorpade, Holly Houdeshell, Ola Kvalvaag, Dillon Nall, Ethan Sanders, Alec Vest, Shaun Westlund, Matthew Hardison, Clinton R. Paden, Duncan MacCannell |
| EPI_ISL_2201347 | Houston Methodist Hospital | Houston Methodist Hospital | Randall J. Olsen, Paul A. Christensen, S. Wesley Long, Sishir Subedi, Robert Olson, Marcus Nguyen, James J. Davis, Matthew Ojeda Saavedra, Prasanti Yerramilli, Layne Pruitt, Kristina Reppond, Madison N. Shyer, Jessica Cambric, Ryan Gadd, Ilya J. Finkelstein, Jimmy Gollihar, and James M. Musser |
| EPI_ISL_2201348, EPI_ISL_2201349 | Aegis Sciences Corporation | Centers for Disease Control and Prevention Division of Viral Diseases, Pathogen Discovery | Dakota Howard, Dhwani Batra, Peter W. Cook, Kara Moser, Adrian Paskey, Jason Caravas, Benjamin Rambo-Martin, Shatavia Morrison, Christopher Gulvick, Scott Sammons, Yvette Unoarumhi, Darlene Wagner, Matthew Schmerer, Cyndi Clark, Patrick Campbell, Rob Case, Vikramsinha Ghorpade, Holly Houdeshell, Ola Kvalvaag, Dillon Nall, Ethan Sanders, Alec Vest, Shaun Westlund, Matthew Hardison, Clinton R. Paden, Duncan MacCannell |
| EPI_ISL_2201350 | Houston Methodist Hospital | Houston Methodist Hospital | Randall J. Olsen, Paul A. Christensen, S. Wesley Long, Sishir Subedi, Robert Olson, Marcus Nguyen, James J. Davis, Matthew Ojeda Saavedra, Prasanti Yerramilli, Layne Pruitt, Kristina Reppond, Madison N. Shyer, Jessica Cambric, Ryan Gadd, Ilya J. Finkelstein, Jimmy Gollihar, and James M. Musser |
| EPI_ISL_2201351 | Aegis Sciences Corporation | Centers for Disease Control and Prevention Division of Viral Diseases, Pathogen Discovery | Dakota Howard, Dhwani Batra, Peter W. Cook, Kara Moser, Adrian Paskey, Jason Caravas, Benjamin Rambo-Martin, Shatavia Morrison, Christopher Gulvick, Scott Sammons, Yvette Unoarumhi, Darlene Wagner, Matthew Schmerer, Cyndi Clark, Patrick Campbell, Rob Case, Vikramsinha Ghorpade, Holly Houdeshell, Ola Kvalvaag, Dillon Nall, Ethan Sanders, Alec Vest, Shaun Westlund, Matthew Hardison, Clinton R. Paden, Duncan MacCannell |
| EPI_ISL_2201352 | Houston Methodist Hospital | Houston Methodist Hospital | Randall J. Olsen, Paul A. Christensen, S. Wesley Long, Sishir Subedi, Robert Olson, Marcus Nguyen, James J. Davis, Matthew Ojeda Saavedra, Prasanti Yerramilli, Layne Pruitt, Kristina Reppond, Madison N. Shyer, Jessica Cambric, Ryan Gadd, Ilya J. Finkelstein, Jimmy Gollihar, and James M. Musser |
| EPI_ISL_2201353, EPI_ISL_2201354, EPI_ISL_2201355, EPI_ISL_2201356, EPI_ISL_2201357 | Aegis Sciences Corporation | Centers for Disease Control and Prevention Division of Viral Diseases, Pathogen Discovery | Dakota Howard, Dhwani Batra, Peter W. Cook, Kara Moser, Adrian Paskey, Jason Caravas, Benjamin Rambo-Martin, Shatavia Morrison, Christopher Gulvick, Scott Sammons, Yvette Unoarumhi, Darlene Wagner, Matthew Schmerer, Cyndi Clark, Patrick Campbell, Rob Case, Vikramsinha Ghorpade, Holly Houdeshell, Ola Kvalvaag, Dillon Nall, Ethan Sanders, Alec Vest, Shaun Westlund, Matthew Hardison, Clinton R. Paden, Duncan MacCannell |
| EPI_ISL_2201358 | Houston Methodist Hospital | Houston Methodist Hospital | Randall J. Olsen, Paul A. Christensen, S. Wesley Long, Sishir Subedi, Robert Olson, Marcus Nguyen, James J. Davis, Matthew Ojeda Saavedra, Prasanti Yerramilli, Layne Pruitt, Kristina Reppond, Madison N. Shyer, Jessica Cambric, Ryan Gadd, Ilya J. Finkelstein, Jimmy Gollihar, and James M. Musser |
| EPI_ISL_2201359 | Aegis Sciences Corporation | Centers for Disease Control and Prevention Division of Viral Diseases, Pathogen Discovery | Dakota Howard, Dhwani Batra, Peter W. Cook, Kara Moser, Adrian Paskey, Jason Caravas, Benjamin Rambo-Martin, Shatavia Morrison, Christopher Gulvick, Scott Sammons, Yvette Unoarumhi, Darlene Wagner, Matthew Schmerer, Cyndi Clark, Patrick Campbell, Rob Case, Vikramsinha Ghorpade, Holly Houdeshell, Ola Kvalvaag, Dillon Nall, Ethan Sanders, Alec Vest, Shaun Westlund, Matthew Hardison, Clinton R. Paden, Duncan MacCannell |
| EPI_ISL_2201361 | Houston Methodist Hospital | Houston Methodist Hospital | Randall J. Olsen, Paul A. Christensen, S. Wesley Long, Sishir Subedi, Robert Olson, Marcus Nguyen, James J. Davis, Matthew Ojeda Saavedra, Prasanti Yerramilli, Layne Pruitt, Kristina Reppond, Madison N. Shyer, Jessica Cambric, Ryan Gadd, Ilya J. Finkelstein, Jimmy Gollihar, and James M. Musser |
| EPI_ISL_2201362 | Aegis Sciences Corporation | Centers for Disease Control and Prevention Division of Viral Diseases, Pathogen Discovery | Dakota Howard, Dhwani Batra, Peter W. Cook, Kara Moser, Adrian Paskey, Jason Caravas, Benjamin Rambo-Martin, Shatavia Morrison, Christopher Gulvick, Scott Sammons, Yvette Unoarumhi, Darlene Wagner, Matthew Schmerer, Cyndi Clark, Patrick Campbell, Rob Case, Vikramsinha Ghorpade, Holly Houdeshell, Ola Kvalvaag, Dillon Nall, Ethan Sanders, Alec Vest, Shaun Westlund, Matthew Hardison, Clinton R. Paden, Duncan MacCannell |
| EPI_ISL_2201363 | Houston Methodist Hospital | Houston Methodist Hospital | Randall J. Olsen, Paul A. Christensen, S. Wesley Long, Sishir Subedi, Robert Olson, Marcus Nguyen, James J. Davis, Matthew Ojeda Saavedra, Prasanti Yerramilli, Layne Pruitt, Kristina Reppond, Madison N. Shyer, Jessica Cambric, Ryan Gadd, Ilya J. Finkelstein, Jimmy Gollihar, and James M. Musser |
| EPI_ISL_2201364, EPI_ISL_2201365 | Aegis Sciences Corporation | Centers for Disease Control and Prevention Division of Viral Diseases, Pathogen Discovery | Dakota Howard, Dhwani Batra, Peter W. Cook, Kara Moser, Adrian Paskey, Jason Caravas, Benjamin Rambo-Martin, Shatavia Morrison, Christopher Gulvick, Scott Sammons, Yvette Unoarumhi, Darlene Wagner, Matthew Schmerer, Cyndi Clark, Patrick Campbell, Rob Case, Vikramsinha Ghorpade, Holly Houdeshell, Ola Kvalvaag, Dillon Nall, Ethan Sanders, Alec Vest, Shaun Westlund, Matthew Hardison, Clinton R. Paden, Duncan MacCannell |
| EPI_ISL_2201366 | Houston Methodist Hospital | Houston Methodist Hospital | Randall J. Olsen, Paul A. Christensen, S. Wesley Long, Sishir Subedi, Robert Olson, Marcus Nguyen, James J. Davis, Matthew Ojeda Saavedra, Prasanti Yerramilli, Layne Pruitt, Kristina Reppond, Madison N. Shyer, Jessica Cambric, Ryan Gadd, Ilya J. Finkelstein, Jimmy Gollihar, and James M. Musser |
| EPI_ISL_2201367 | Aegis Sciences Corporation | Centers for Disease Control and Prevention Division of Viral Diseases, Pathogen Discovery | Dakota Howard, Dhwani Batra, Peter W. Cook, Kara Moser, Adrian Paskey, Jason Caravas, Benjamin Rambo-Martin, Shatavia Morrison, Christopher Gulvick, Scott Sammons, Yvette Unoarumhi, Darlene Wagner, Matthew Schmerer, Cyndi Clark, Patrick Campbell, Rob Case, Vikramsinha Ghorpade, Holly Houdeshell, Ola Kvalvaag, Dillon Nall, Ethan Sanders, Alec Vest, Shaun Westlund, Matthew Hardison, Clinton R. Paden, Duncan MacCannell |
| EPI_ISL_2201368 | Houston Methodist Hospital | Houston Methodist Hospital | Randall J. Olsen, Paul A. Christensen, S. Wesley Long, Sishir Subedi, Robert Olson, Marcus Nguyen, James J. Davis, Matthew Ojeda Saavedra, Prasanti Yerramilli, Layne Pruitt, Kristina Reppond, Madison N. Shyer, Jessica Cambric, Ryan Gadd, Ilya J. Finkelstein, Jimmy Gollihar, and James M. Musser |
| EPI_ISL_2201369, EPI_ISL_2201370 | Aegis Sciences Corporation | Centers for Disease Control and Prevention Division of Viral Diseases, Pathogen Discovery | Dakota Howard, Dhwani Batra, Peter W. Cook, Kara Moser, Adrian Paskey, Jason Caravas, Benjamin Rambo-Martin, Shatavia Morrison, Christopher Gulvick, Scott Sammons, Yvette Unoarumhi, Darlene Wagner, Matthew Schmerer, Cyndi Clark, Patrick Campbell, Rob Case, Vikramsinha Ghorpade, Holly Houdeshell, Ola Kvalvaag, Dillon Nall, Ethan Sanders, Alec Vest, Shaun Westlund, Matthew Hardison, Clinton R. Paden, Duncan MacCannell |
| EPI_ISL_2201371 | Houston Methodist Hospital | Houston Methodist Hospital | Randall J. Olsen, Paul A. Christensen, S. Wesley Long, Sishir Subedi, Robert Olson, Marcus Nguyen, James J. Davis, Matthew Ojeda Saavedra, Prasanti Yerramilli, Layne Pruitt, Kristina Reppond, Madison N. Shyer, Jessica Cambric, Ryan Gadd, Ilya J. Finkelstein, Jimmy Gollihar, and James M. Musser |
| EPI_ISL_2201372 | Aegis Sciences Corporation | Centers for Disease Control and Prevention Division of Viral Diseases, Pathogen Discovery | Dakota Howard, Dhwani Batra, Peter W. Cook, Kara Moser, Adrian Paskey, Jason Caravas, Benjamin Rambo-Martin, Shatavia Morrison, Christopher Gulvick, Scott Sammons, Yvette Unoarumhi, Darlene Wagner, Matthew Schmerer, Cyndi Clark, Patrick Campbell, Rob Case, Vikramsinha Ghorpade, Holly Houdeshell, Ola Kvalvaag, Dillon Nall, Ethan Sanders, Alec Vest, Shaun Westlund, Matthew Hardison, Clinton R. Paden, Duncan MacCannell |
| EPI_ISL_2201373 | Houston Methodist Hospital | Houston Methodist Hospital | Randall J. Olsen, Paul A. Christensen, S. Wesley Long, Sishir Subedi, Robert Olson, Marcus Nguyen, James J. Davis, Matthew Ojeda Saavedra, Prasanti Yerramilli, Layne Pruitt, Kristina Reppond, Madison N. Shyer, Jessica Cambric, Ryan Gadd, Ilya J. Finkelstein, Jimmy Gollihar, and James M. Musser |
| EPI_ISL_2201374 | Aegis Sciences Corporation | Centers for Disease Control and Prevention Division of Viral Diseases, Pathogen Discovery | Dakota Howard, Dhwani Batra, Peter W. Cook, Kara Moser, Adrian Paskey, Jason Caravas, Benjamin Rambo-Martin, Shatavia Morrison, Christopher Gulvick, Scott Sammons, Yvette Unoarumhi, Darlene Wagner, Matthew Schmerer, Cyndi Clark, Patrick Campbell, Rob Case, Vikramsinha Ghorpade, Holly Houdeshell, Ola Kvalvaag, Dillon Nall, Ethan Sanders, Alec Vest, Shaun Westlund, Matthew Hardison, Clinton R. Paden, Duncan MacCannell |
| EPI_ISL_2201375 | Houston Methodist Hospital | Houston Methodist Hospital | Randall J. Olsen, Paul A. Christensen, S. Wesley Long, Sishir Subedi, Robert Olson, Marcus Nguyen, James J. Davis, Matthew Ojeda Saavedra, Prasanti Yerramilli, Layne Pruitt, Kristina Reppond, Madison N. Shyer, Jessica Cambric, Ryan Gadd, Ilya J. Finkelstein, Jimmy Gollihar, and James M. Musser |
| EPI_ISL_2201376 | Aegis Sciences Corporation | Centers for Disease Control and Prevention Division of Viral Diseases, Pathogen Discovery | Dakota Howard, Dhwani Batra, Peter W. Cook, Kara Moser, Adrian Paskey, Jason Caravas, Benjamin Rambo-Martin, Shatavia Morrison, Christopher Gulvick, Scott Sammons, Yvette Unoarumhi, Darlene Wagner, Matthew Schmerer, Cyndi Clark, Patrick Campbell, Rob Case, Vikramsinha Ghorpade, Holly Houdeshell, Ola Kvalvaag, Dillon Nall, Ethan Sanders, Alec Vest, Shaun Westlund, Matthew Hardison, Clinton R. Paden, Duncan MacCannell |
| EPI_ISL_2201377 | Houston Methodist Hospital | Houston Methodist Hospital | Randall J. Olsen, Paul A. Christensen, S. Wesley Long, Sishir Subedi, Robert Olson, Marcus Nguyen, James J. Davis, Matthew Ojeda Saavedra, Prasanti Yerramilli, Layne Pruitt, Kristina Reppond, Madison N. Shyer, Jessica Cambric, Ryan Gadd, Ilya J. Finkelstein, Jimmy Gollihar, and James M. Musser |
| EPI_ISL_2201378 | Aegis Sciences Corporation | Centers for Disease Control and Prevention Division of Viral Diseases, Pathogen Discovery | Dakota Howard, Dhwani Batra, Peter W. Cook, Kara Moser, Adrian Paskey, Jason Caravas, Benjamin Rambo-Martin, Shatavia Morrison, Christopher Gulvick, Scott Sammons, Yvette Unoarumhi, Darlene Wagner, Matthew Schmerer, Cyndi Clark, Patrick Campbell, Rob Case, Vikramsinha Ghorpade, Holly Houdeshell, Ola Kvalvaag, Dillon Nall, Ethan Sanders, Alec Vest, Shaun Westlund, Matthew Hardison, Clinton R. Paden, Duncan MacCannell |
| EPI_ISL_2201379 | Houston Methodist Hospital | Houston Methodist Hospital | Randall J. Olsen, Paul A. Christensen, S. Wesley Long, Sishir Subedi, Robert Olson, Marcus Nguyen, James J. Davis, Matthew Ojeda Saavedra, Prasanti Yerramilli, Layne Pruitt, Kristina Reppond, Madison N. Shyer, Jessica Cambric, Ryan Gadd, Ilya J. Finkelstein, Jimmy Gollihar, and James M. Musser |
| EPI_ISL_2201380 | Aegis Sciences Corporation | Centers for Disease Control and Prevention Division of Viral Diseases, Pathogen Discovery | Dakota Howard, Dhwani Batra, Peter W. Cook, Kara Moser, Adrian Paskey, Jason Caravas, Benjamin Rambo-Martin, Shatavia Morrison, Christopher Gulvick, Scott Sammons, Yvette Unoarumhi, Darlene Wagner, Matthew Schmerer, Cyndi Clark, Patrick Campbell, Rob Case, Vikramsinha Ghorpade, Holly Houdeshell, Ola Kvalvaag, Dillon Nall, Ethan Sanders, Alec Vest, Shaun Westlund, Matthew Hardison, Clinton R. Paden, Duncan MacCannell |
| EPI_ISL_2201381 | Houston Methodist Hospital | Houston Methodist Hospital | Randall J. Olsen, Paul A. Christensen, S. Wesley Long, Sishir Subedi, Robert Olson, Marcus Nguyen, James J. Davis, Matthew Ojeda Saavedra, Prasanti Yerramilli, Layne Pruitt, Kristina Reppond, Madison N. Shyer, Jessica Cambric, Ryan Gadd, Ilya J. Finkelstein, Jimmy Gollihar, and James M. Musser |
| EPI_ISL_2201382, EPI_ISL_2201383 | Aegis Sciences Corporation | Centers for Disease Control and Prevention Division of Viral Diseases, Pathogen Discovery | Dakota Howard, Dhwani Batra, Peter W. Cook, Kara Moser, Adrian Paskey, Jason Caravas, Benjamin Rambo-Martin, Shatavia Morrison, Christopher Gulvick, Scott Sammons, Yvette Unoarumhi, Darlene Wagner, Matthew Schmerer, Cyndi Clark, Patrick Campbell, Rob Case, Vikramsinha Ghorpade, Holly Houdeshell, Ola Kvalvaag, Dillon Nall, Ethan Sanders, Alec Vest, Shaun Westlund, Matthew Hardison, Clinton R. Paden, Duncan MacCannell |
| EPI_ISL_2201384 | Houston Methodist Hospital | Houston Methodist Hospital | Randall J. Olsen, Paul A. Christensen, S. Wesley Long, Sishir Subedi, Robert Olson, Marcus Nguyen, James J. Davis, Matthew Ojeda Saavedra, Prasanti Yerramilli, Layne Pruitt, Kristina Reppond, Madison N. Shyer, Jessica Cambric, Ryan Gadd, Ilya J. Finkelstein, Jimmy Gollihar, and James M. Musser |
| EPI_ISL_2201385 | Aegis Sciences Corporation | Centers for Disease Control and Prevention Division of Viral Diseases, Pathogen Discovery | Dakota Howard, Dhwani Batra, Peter W. Cook, Kara Moser, Adrian Paskey, Jason Caravas, Benjamin Rambo-Martin, Shatavia Morrison, Christopher Gulvick, Scott Sammons, Yvette Unoarumhi, Darlene Wagner, Matthew Schmerer, Cyndi Clark, Patrick Campbell, Rob Case, Vikramsinha Ghorpade, Holly Houdeshell, Ola Kvalvaag, Dillon Nall, Ethan Sanders, Alec Vest, Shaun Westlund, Matthew Hardison, Clinton R. Paden, Duncan MacCannell |
| EPI_ISL_2201386 | Houston Methodist Hospital | Houston Methodist Hospital | Randall J. Olsen, Paul A. Christensen, S. Wesley Long, Sishir Subedi, Robert Olson, Marcus Nguyen, James J. Davis, Matthew Ojeda Saavedra, Prasanti Yerramilli, Layne Pruitt, Kristina Reppond, Madison N. Shyer, Jessica Cambric, Ryan Gadd, Ilya J. Finkelstein, Jimmy Gollihar, and James M. Musser |
| EPI_ISL_2201387, EPI_ISL_2201388 | Aegis Sciences Corporation | Centers for Disease Control and Prevention Division of Viral Diseases, Pathogen Discovery | Dakota Howard, Dhwani Batra, Peter W. Cook, Kara Moser, Adrian Paskey, Jason Caravas, Benjamin Rambo-Martin, Shatavia Morrison, Christopher Gulvick, Scott Sammons, Yvette Unoarumhi, Darlene Wagner, Matthew Schmerer, Cyndi Clark, Patrick Campbell, Rob Case, Vikramsinha Ghorpade, Holly Houdeshell, Ola Kvalvaag, Dillon Nall, Ethan Sanders, Alec Vest, Shaun Westlund, Matthew Hardison, Clinton R. Paden, Duncan MacCannell |
| EPI_ISL_2201389 | Houston Methodist Hospital | Houston Methodist Hospital | Randall J. Olsen, Paul A. Christensen, S. Wesley Long, Sishir Subedi, Robert Olson, Marcus Nguyen, James J. Davis, Matthew Ojeda Saavedra, Prasanti Yerramilli, Layne Pruitt, Kristina Reppond, Madison N. Shyer, Jessica Cambric, Ryan Gadd, Ilya J. Finkelstein, Jimmy Gollihar, and James M. Musser |
| EPI_ISL_2201390 | Aegis Sciences Corporation | Centers for Disease Control and Prevention Division of Viral Diseases, Pathogen Discovery | Dakota Howard, Dhwani Batra, Peter W. Cook, Kara Moser, Adrian Paskey, Jason Caravas, Benjamin Rambo-Martin, Shatavia Morrison, Christopher Gulvick, Scott Sammons, Yvette Unoarumhi, Darlene Wagner, Matthew Schmerer, Cyndi Clark, Patrick Campbell, Rob Case, Vikramsinha Ghorpade, Holly Houdeshell, Ola Kvalvaag, Dillon Nall, Ethan Sanders, Alec Vest, Shaun Westlund, Matthew Hardison, Clinton R. Paden, Duncan MacCannell |

| | | | |
|---|---|---|---|
| EPI_ISL_2201391 | Houston Methodist Hospital | Houston Methodist Hospital | Randall J. Olsen, Paul A. Christensen, S. Wesley Long, Sishir Subedi, Robert Olson, Marcus Nguyen, James J. Davis, Matthew Ojeda Saavedra, Prasanti Yerramilli, Layne Pruitt, Kristina Reppond, Madison N. Shyer, Jessica Cambric, Ryan Gadd, Ilya J. Finkelstein, Jimmy Gollihar, and James M. Musser |
| EPI_ISL_2201392 | Aegis Sciences Corporation | Centers for Disease Control and Prevention Division of Viral Diseases, Pathogen Discovery | Dakota Howard, Dhwani Batra, Peter W. Cook, Kara Moser, Adrian Paskey, Jason Caravas, Benjamin Rambo-Martin, Shatavia Morrison, Christopher Gulvick, Scott Sammons, Yvette Unoarumhi, Darlene Wagner, Matthew Schmerer, Cyndi Clark, Patrick Campbell, Rob Case, Vikramsinha Ghorpade, Holly Houdeshell, Ola Kvalvaag, Dillon Nall, Ethan Sanders, Alec Vest, Shaun Westlund, Matthew Hardison, Clinton R. Paden, Duncan MacCannell |
| EPI_ISL_2201393 | Houston Methodist Hospital | Houston Methodist Hospital | Randall J. Olsen, Paul A. Christensen, S. Wesley Long, Sishir Subedi, Robert Olson, Marcus Nguyen, James J. Davis, Matthew Ojeda Saavedra, Prasanti Yerramilli, Layne Pruitt, Kristina Reppond, Madison N. Shyer, Jessica Cambric, Ryan Gadd, Ilya J. Finkelstein, Jimmy Gollihar, and James M. Musser |
| EPI_ISL_2201394, EPI_ISL_2201395 | Aegis Sciences Corporation | Centers for Disease Control and Prevention Division of Viral Diseases, Pathogen Discovery | Dakota Howard, Dhwani Batra, Peter W. Cook, Kara Moser, Adrian Paskey, Jason Caravas, Benjamin Rambo-Martin, Shatavia Morrison, Christopher Gulvick, Scott Sammons, Yvette Unoarumhi, Darlene Wagner, Matthew Schmerer, Cyndi Clark, Patrick Campbell, Rob Case, Vikramsinha Ghorpade, Holly Houdeshell, Ola Kvalvaag, Dillon Nall, Ethan Sanders, Alec Vest, Shaun Westlund, Matthew Hardison, Clinton R. Paden, Duncan MacCannell |
| EPI_ISL_2201396 | Houston Methodist Hospital | Houston Methodist Hospital | Randall J. Olsen, Paul A. Christensen, S. Wesley Long, Sishir Subedi, Robert Olson, Marcus Nguyen, James J. Davis, Matthew Ojeda Saavedra, Prasanti Yerramilli, Layne Pruitt, Kristina Reppond, Madison N. Shyer, Jessica Cambric, Ryan Gadd, Ilya J. Finkelstein, Jimmy Gollihar, and James M. Musser |
| EPI_ISL_2201397 | Aegis Sciences Corporation | Centers for Disease Control and Prevention Division of Viral Diseases, Pathogen Discovery | Dakota Howard, Dhwani Batra, Peter W. Cook, Kara Moser, Adrian Paskey, Jason Caravas, Benjamin Rambo-Martin, Shatavia Morrison, Christopher Gulvick, Scott Sammons, Yvette Unoarumhi, Darlene Wagner, Matthew Schmerer, Cyndi Clark, Patrick Campbell, Rob Case, Vikramsinha Ghorpade, Holly Houdeshell, Ola Kvalvaag, Dillon Nall, Ethan Sanders, Alec Vest, Shaun Westlund, Matthew Hardison, Clinton R. Paden, Duncan MacCannell |
| EPI_ISL_2201398 | Houston Methodist Hospital | Houston Methodist Hospital | Randall J. Olsen, Paul A. Christensen, S. Wesley Long, Sishir Subedi, Robert Olson, Marcus Nguyen, James J. Davis, Matthew Ojeda Saavedra, Prasanti Yerramilli, Layne Pruitt, Kristina Reppond, Madison N. Shyer, Jessica Cambric, Ryan Gadd, Ilya J. Finkelstein, Jimmy Gollihar, and James M. Musser |
| EPI_ISL_2201399, EPI_ISL_2201400 | Aegis Sciences Corporation | Centers for Disease Control and Prevention Division of Viral Diseases, Pathogen Discovery | Dakota Howard, Dhwani Batra, Peter W. Cook, Kara Moser, Adrian Paskey, Jason Caravas, Benjamin Rambo-Martin, Shatavia Morrison, Christopher Gulvick, Scott Sammons, Yvette Unoarumhi, Darlene Wagner, Matthew Schmerer, Cyndi Clark, Patrick Campbell, Rob Case, Vikramsinha Ghorpade, Holly Houdeshell, Ola Kvalvaag, Dillon Nall, Ethan Sanders, Alec Vest, Shaun Westlund, Matthew Hardison, Clinton R. Paden, Duncan MacCannell |
| EPI_ISL_2201401 | Houston Methodist Hospital | Houston Methodist Hospital | Randall J. Olsen, Paul A. Christensen, S. Wesley Long, Sishir Subedi, Robert Olson, Marcus Nguyen, James J. Davis, Matthew Ojeda Saavedra, Prasanti Yerramilli, Layne Pruitt, Kristina Reppond, Madison N. Shyer, Jessica Cambric, Ryan Gadd, Ilya J. Finkelstein, Jimmy Gollihar, and James M. Musser |
| EPI_ISL_2201402 | Aegis Sciences Corporation | Centers for Disease Control and Prevention Division of Viral Diseases, Pathogen Discovery | Dakota Howard, Dhwani Batra, Peter W. Cook, Kara Moser, Adrian Paskey, Jason Caravas, Benjamin Rambo-Martin, Shatavia Morrison, Christopher Gulvick, Scott Sammons, Yvette Unoarumhi, Darlene Wagner, Matthew Schmerer, Cyndi Clark, Patrick Campbell, Rob Case, Vikramsinha Ghorpade, Holly Houdeshell, Ola Kvalvaag, Dillon Nall, Ethan Sanders, Alec Vest, Shaun Westlund, Matthew Hardison, Clinton R. Paden, Duncan MacCannell |
| EPI_ISL_2201403 | Houston Methodist Hospital | Houston Methodist Hospital | Randall J. Olsen, Paul A. Christensen, S. Wesley Long, Sishir Subedi, Robert Olson, Marcus Nguyen, James J. Davis, Matthew Ojeda Saavedra, Prasanti Yerramilli, Layne Pruitt, Kristina Reppond, Madison N. Shyer, Jessica Cambric, Ryan Gadd, Ilya J. Finkelstein, Jimmy Gollihar, and James M. Musser |
| EPI_ISL_2201404, EPI_ISL_2201405 | Aegis Sciences Corporation | Centers for Disease Control and Prevention Division of Viral Diseases, Pathogen Discovery | Dakota Howard, Dhwani Batra, Peter W. Cook, Kara Moser, Adrian Paskey, Jason Caravas, Benjamin Rambo-Martin, Shatavia Morrison, Christopher Gulvick, Scott Sammons, Yvette Unoarumhi, Darlene Wagner, Matthew Schmerer, Cyndi Clark, Patrick Campbell, Rob Case, Vikramsinha Ghorpade, Holly Houdeshell, Ola Kvalvaag, Dillon Nall, Ethan Sanders, Alec Vest, Shaun Westlund, Matthew Hardison, Clinton R. Paden, Duncan MacCannell |
| EPI_ISL_2201406 | Houston Methodist Hospital | Houston Methodist Hospital | Randall J. Olsen, Paul A. Christensen, S. Wesley Long, Sishir Subedi, Robert Olson, Marcus Nguyen, James J. Davis, Matthew Ojeda Saavedra, Prasanti Yerramilli, Layne Pruitt, Kristina Reppond, Madison N. Shyer, Jessica Cambric, Ryan Gadd, Ilya J. Finkelstein, Jimmy Gollihar, and James M. Musser |
| EPI_ISL_2201407 | Aegis Sciences Corporation | Centers for Disease Control and Prevention Division of Viral Diseases, Pathogen Discovery | Dakota Howard, Dhwani Batra, Peter W. Cook, Kara Moser, Adrian Paskey, Jason Caravas, Benjamin Rambo-Martin, Shatavia Morrison, Christopher Gulvick, Scott Sammons, Yvette Unoarumhi, Darlene Wagner, Matthew Schmerer, Cyndi Clark, Patrick Campbell, Rob Case, Vikramsinha Ghorpade, Holly Houdeshell, Ola Kvalvaag, Dillon Nall, Ethan Sanders, Alec Vest, Shaun Westlund, Matthew Hardison, Clinton R. Paden, Duncan MacCannell |
| EPI_ISL_2201408 | Houston Methodist Hospital | Houston Methodist Hospital | Randall J. Olsen, Paul A. Christensen, S. Wesley Long, Sishir Subedi, Robert Olson, Marcus Nguyen, James J. Davis, Matthew Ojeda Saavedra, Prasanti Yerramilli, Layne Pruitt, Kristina Reppond, Madison N. Shyer, Jessica Cambric, Ryan Gadd, Ilya J. Finkelstein, Jimmy Gollihar, and James M. Musser |
| EPI_ISL_2201409 | Aegis Sciences Corporation | Centers for Disease Control and Prevention Division of Viral Diseases, Pathogen Discovery | Dakota Howard, Dhwani Batra, Peter W. Cook, Kara Moser, Adrian Paskey, Jason Caravas, Benjamin Rambo-Martin, Shatavia Morrison, Christopher Gulvick, Scott Sammons, Yvette Unoarumhi, Darlene Wagner, Matthew Schmerer, Cyndi Clark, Patrick Campbell, Rob Case, Vikramsinha Ghorpade, Holly Houdeshell, Ola Kvalvaag, Dillon Nall, Ethan Sanders, Alec Vest, Shaun Westlund, Matthew Hardison, Clinton R. Paden, Duncan MacCannell |
| EPI_ISL_2201410 | Houston Methodist Hospital | Houston Methodist Hospital | Randall J. Olsen, Paul A. Christensen, S. Wesley Long, Sishir Subedi, Robert Olson, Marcus Nguyen, James J. Davis, Matthew Ojeda Saavedra, Prasanti Yerramilli, Layne Pruitt, Kristina Reppond, Madison N. Shyer, Jessica Cambric, Ryan Gadd, Ilya J. Finkelstein, Jimmy Gollihar, and James M. Musser |
| EPI_ISL_2201411 | Aegis Sciences Corporation | Centers for Disease Control and Prevention Division of Viral Diseases, Pathogen Discovery | Dakota Howard, Dhwani Batra, Peter W. Cook, Kara Moser, Adrian Paskey, Jason Caravas, Benjamin Rambo-Martin, Shatavia Morrison, Christopher Gulvick, Scott Sammons, Yvette Unoarumhi, Darlene Wagner, Matthew Schmerer, Cyndi Clark, Patrick Campbell, Rob Case, Vikramsinha Ghorpade, Holly Houdeshell, Ola Kvalvaag, Dillon Nall, Ethan Sanders, Alec Vest, Shaun Westlund, Matthew Hardison, Clinton R. Paden, Duncan MacCannell |
| EPI_ISL_2201412 | Houston Methodist Hospital | Houston Methodist Hospital | Randall J. Olsen, Paul A. Christensen, S. Wesley Long, Sishir Subedi, Robert Olson, Marcus Nguyen, James J. Davis, Matthew Ojeda Saavedra, Prasanti Yerramilli, Layne Pruitt, Kristina Reppond, Madison N. Shyer, Jessica Cambric, Ryan Gadd, Ilya J. Finkelstein, Jimmy Gollihar, and James M. Musser |
| EPI_ISL_2201413, EPI_ISL_2201414 | Aegis Sciences Corporation | Centers for Disease Control and Prevention Division of Viral Diseases, Pathogen Discovery | Dakota Howard, Dhwani Batra, Peter W. Cook, Kara Moser, Adrian Paskey, Jason Caravas, Benjamin Rambo-Martin, Shatavia Morrison, Christopher Gulvick, Scott Sammons, Yvette Unoarumhi, Darlene Wagner, Matthew Schmerer, Cyndi Clark, Patrick Campbell, Rob Case, Vikramsinha Ghorpade, Holly Houdeshell, Ola Kvalvaag, Dillon Nall, Ethan Sanders, Alec Vest, Shaun Westlund, Matthew Hardison, Clinton R. Paden, Duncan MacCannell |
| EPI_ISL_2201415 | Houston Methodist Hospital | Houston Methodist Hospital | Randall J. Olsen, Paul A. Christensen, S. Wesley Long, Sishir Subedi, Robert Olson, Marcus Nguyen, James J. Davis, Matthew Ojeda Saavedra, Prasanti Yerramilli, Layne Pruitt, Kristina Reppond, Madison N. Shyer, Jessica Cambric, Ryan Gadd, Ilya J. Finkelstein, Jimmy Gollihar, and James M. Musser |
| EPI_ISL_2201416 | Aegis Sciences Corporation | Centers for Disease Control and Prevention Division of Viral Diseases, Pathogen Discovery | Dakota Howard, Dhwani Batra, Peter W. Cook, Kara Moser, Adrian Paskey, Jason Caravas, Benjamin Rambo-Martin, Shatavia Morrison, Christopher Gulvick, Scott Sammons, Yvette Unoarumhi, Darlene Wagner, Matthew Schmerer, Cyndi Clark, Patrick Campbell, Rob Case, Vikramsinha Ghorpade, Holly Houdeshell, Ola Kvalvaag, Dillon Nall, Ethan Sanders, Alec Vest, Shaun Westlund, Matthew Hardison, Clinton R. Paden, Duncan MacCannell |
| EPI_ISL_2201417 | Houston Methodist Hospital | Houston Methodist Hospital | Randall J. Olsen, Paul A. Christensen, S. Wesley Long, Sishir Subedi, Robert Olson, Marcus Nguyen, James J. Davis, Matthew Ojeda Saavedra, Prasanti Yerramilli, Layne Pruitt, Kristina Reppond, Madison N. Shyer, Jessica Cambric, Ryan Gadd, Ilya J. Finkelstein, Jimmy Gollihar, and James M. Musser |
| EPI_ISL_2201418 | Aegis Sciences Corporation | Centers for Disease Control and Prevention Division of Viral Diseases, Pathogen Discovery | Dakota Howard, Dhwani Batra, Peter W. Cook, Kara Moser, Adrian Paskey, Jason Caravas, Benjamin Rambo-Martin, Shatavia Morrison, Christopher Gulvick, Scott Sammons, Yvette Unoarumhi, Darlene Wagner, Matthew Schmerer, Cyndi Clark, Patrick Campbell, Rob Case, Vikramsinha Ghorpade, Holly Houdeshell, Ola Kvalvaag, Dillon Nall, Ethan Sanders, Alec Vest, Shaun Westlund, Matthew Hardison, Clinton R. Paden, Duncan MacCannell |
| EPI_ISL_2201419 | Houston Methodist Hospital | Houston Methodist Hospital | Randall J. Olsen, Paul A. Christensen, S. Wesley Long, Sishir Subedi, Robert Olson, Marcus Nguyen, James J. Davis, Matthew Ojeda Saavedra, Prasanti Yerramilli, Layne Pruitt, Kristina Reppond, Madison N. Shyer, Jessica Cambric, Ryan Gadd, Ilya J. Finkelstein, Jimmy Gollihar, and James M. Musser |
| EPI_ISL_2201420 | Aegis Sciences Corporation | Centers for Disease Control and Prevention Division of Viral Diseases, Pathogen Discovery | Dakota Howard, Dhwani Batra, Peter W. Cook, Kara Moser, Adrian Paskey, Jason Caravas, Benjamin Rambo-Martin, Shatavia Morrison, Christopher Gulvick, Scott Sammons, Yvette Unoarumhi, Darlene Wagner, Matthew Schmerer, Cyndi Clark, Patrick Campbell, Rob Case, Vikramsinha Ghorpade, Holly Houdeshell, Ola Kvalvaag, Dillon Nall, Ethan Sanders, Alec Vest, Shaun Westlund, Matthew Hardison, Clinton R. Paden, Duncan MacCannell |
| EPI_ISL_2201421 | Houston Methodist Hospital | Houston Methodist Hospital | Randall J. Olsen, Paul A. Christensen, S. Wesley Long, Sishir Subedi, Robert Olson, Marcus Nguyen, James J. Davis, Matthew Ojeda Saavedra, Prasanti Yerramilli, Layne Pruitt, Kristina Reppond, Madison N. Shyer, Jessica Cambric, Ryan Gadd, Ilya J. Finkelstein, Jimmy Gollihar, and James M. Musser |
| EPI_ISL_2201422 | Aegis Sciences Corporation | Centers for Disease Control and Prevention Division of Viral Diseases, Pathogen Discovery | Dakota Howard, Dhwani Batra, Peter W. Cook, Kara Moser, Adrian Paskey, Jason Caravas, Benjamin Rambo-Martin, Shatavia Morrison, Christopher Gulvick, Scott Sammons, Yvette Unoarumhi, Darlene Wagner, Matthew Schmerer, Cyndi Clark, Patrick Campbell, Rob Case, Vikramsinha Ghorpade, Holly Houdeshell, Ola Kvalvaag, Dillon Nall, Ethan Sanders, Alec Vest, Shaun Westlund, Matthew Hardison, Clinton R. Paden, Duncan MacCannell |
| EPI_ISL_2201423 | Houston Methodist Hospital | Houston Methodist Hospital | Randall J. Olsen, Paul A. Christensen, S. Wesley Long, Sishir Subedi, Robert Olson, Marcus Nguyen, James J. Davis, Matthew Ojeda Saavedra, Prasanti Yerramilli, Layne Pruitt, Kristina Reppond, Madison N. Shyer, Jessica Cambric, Ryan Gadd, Ilya J. Finkelstein, Jimmy Gollihar, and James M. Musser |
| EPI_ISL_2201424 | Aegis Sciences Corporation | Centers for Disease Control and Prevention Division of Viral Diseases, Pathogen Discovery | Dakota Howard, Dhwani Batra, Peter W. Cook, Kara Moser, Adrian Paskey, Jason Caravas, Benjamin Rambo-Martin, Shatavia Morrison, Christopher Gulvick, Scott Sammons, Yvette Unoarumhi, Darlene Wagner, Matthew Schmerer, Cyndi Clark, Patrick Campbell, Rob Case, Vikramsinha Ghorpade, Holly Houdeshell, Ola Kvalvaag, Dillon Nall, Ethan Sanders, Alec Vest, Shaun Westlund, Matthew Hardison, Clinton R. Paden, Duncan MacCannell |
| EPI_ISL_2201425 | Houston Methodist Hospital | Houston Methodist Hospital | Randall J. Olsen, Paul A. Christensen, S. Wesley Long, Sishir Subedi, Robert Olson, Marcus Nguyen, James J. Davis, Matthew Ojeda Saavedra, Prasanti Yerramilli, Layne Pruitt, Kristina Reppond, Madison N. Shyer, Jessica Cambric, Ryan Gadd, Ilya J. Finkelstein, Jimmy Gollihar, and James M. Musser |
| EPI_ISL_2201426 | Aegis Sciences Corporation | Centers for Disease Control and Prevention Division of Viral Diseases, Pathogen Discovery | Dakota Howard, Dhwani Batra, Peter W. Cook, Kara Moser, Adrian Paskey, Jason Caravas, Benjamin Rambo-Martin, Shatavia Morrison, Christopher Gulvick, Scott Sammons, Yvette Unoarumhi, Darlene Wagner, Matthew Schmerer, Cyndi Clark, Patrick Campbell, Rob Case, Vikramsinha Ghorpade, Holly Houdeshell, Ola Kvalvaag, Dillon Nall, Ethan Sanders, Alec Vest, Shaun Westlund, Matthew Hardison, Clinton R. Paden, Duncan MacCannell |
| EPI_ISL_2201427 | Houston Methodist Hospital | Houston Methodist Hospital | Randall J. Olsen, Paul A. Christensen, S. Wesley Long, Sishir Subedi, Robert Olson, Marcus Nguyen, James J. Davis, Matthew Ojeda Saavedra, Prasanti Yerramilli, Layne Pruitt, Kristina Reppond, Madison N. Shyer, Jessica Cambric, Ryan Gadd, Ilya J. Finkelstein, Jimmy Gollihar, and James M. Musser |
| EPI_ISL_2201428 | Aegis Sciences Corporation | Centers for Disease Control and Prevention Division of Viral Diseases, Pathogen Discovery | Dakota Howard, Dhwani Batra, Peter W. Cook, Kara Moser, Adrian Paskey, Jason Caravas, Benjamin Rambo-Martin, Shatavia Morrison, Christopher Gulvick, Scott Sammons, Yvette Unoarumhi, Darlene Wagner, Matthew Schmerer, Cyndi Clark, Patrick Campbell, Rob Case, Vikramsinha Ghorpade, Holly Houdeshell, Ola Kvalvaag, Dillon Nall, Ethan Sanders, Alec Vest, Shaun Westlund, Matthew Hardison, Clinton R. Paden, Duncan MacCannell |
| EPI_ISL_2201429 | Houston Methodist Hospital | Houston Methodist Hospital | Randall J. Olsen, Paul A. Christensen, S. Wesley Long, Sishir Subedi, Robert Olson, Marcus Nguyen, James J. Davis, Matthew Ojeda Saavedra, Prasanti Yerramilli, Layne Pruitt, Kristina Reppond, Madison N. Shyer, Jessica Cambric, Ryan Gadd, Ilya J. Finkelstein, Jimmy Gollihar, and James M. Musser |
| EPI_ISL_2201430, EPI_ISL_2201431 | Aegis Sciences Corporation | Centers for Disease Control and Prevention Division of Viral Diseases, Pathogen Discovery | Dakota Howard, Dhwani Batra, Peter W. Cook, Kara Moser, Adrian Paskey, Jason Caravas, Benjamin Rambo-Martin, Shatavia Morrison, Christopher Gulvick, Scott Sammons, Yvette Unoarumhi, Darlene Wagner, Matthew Schmerer, Cyndi Clark, Patrick Campbell, Rob Case, Vikramsinha Ghorpade, Holly Houdeshell, Ola Kvalvaag, Dillon Nall, Ethan Sanders, Alec Vest, Shaun Westlund, Matthew Hardison, Clinton R. Paden, Duncan MacCannell |
| EPI_ISL_2201432 | Houston Methodist Hospital | Houston Methodist Hospital | Randall J. Olsen, Paul A. Christensen, S. Wesley Long, Sishir Subedi, Robert Olson, Marcus Nguyen, James J. Davis, Matthew Ojeda Saavedra, Prasanti Yerramilli, Layne Pruitt, Kristina Reppond, Madison N. Shyer, Jessica Cambric, Ryan Gadd, Ilya J. Finkelstein, Jimmy Gollihar, and James M. Musser |
| EPI_ISL_2201433 | Aegis Sciences Corporation | Centers for Disease Control and Prevention | Dakota Howard, Dhwani Batra, Peter W. Cook, Kara Moser, Adrian Paskey, Jason Caravas, Benjamin Rambo-Martin, Shatavia Morrison, Christopher Gulvick, Scott Sammons, Yvette Unoarumhi, Darlene Wagner, Matthew Schmerer, Cyndi Clark, Patrick Campbell, Rob Case, Vikramsinha |

| | | | |
|---|---|---|---|
| | | Division of Viral Diseases, Pathogen Discovery | Ghorpade, Holly Houdeshell, Ola Kvalvaag, Dillon Nall, Ethan Sanders, Alec Vest, Shaun Westlund, Matthew Hardison, Clinton R. Paden, Duncan MacCannell |
| EPI_ISL_2201434 | Houston Methodist Hospital | Houston Methodist Hospital | Randall J. Olsen, Paul A. Christensen, S. Wesley Long, Sishir Subedi, Robert Olson, Marcus Nguyen, James J. Davis, Matthew Ojeda Saavedra, Prasanti Yerramilli, Layne Pruitt, Kristina Reppond, Madison N. Shyer, Jessica Cambric, Ryan Gadd, Ilya J. Finkelstein, Jimmy Gollihar, and James M. Musser |
| EPI_ISL_2201435, EPI_ISL_2201436 | Aegis Sciences Corporation | Centers for Disease Control and Prevention Division of Viral Diseases, Pathogen Discovery | Dakota Howard, Dhwani Batra, Peter W. Cook, Kara Moser, Adrian Paskey, Jason Caravas, Benjamin Rambo-Martin, Shatavia Morrison, Christopher Gulvick, Scott Sammons, Yvette Unoarumhi, Darlene Wagner, Matthew Schmerer, Cyndi Clark, Patrick Campbell, Rob Case, Vikramsinha Ghorpade, Holly Houdeshell, Ola Kvalvaag, Dillon Nall, Ethan Sanders, Alec Vest, Shaun Westlund, Matthew Hardison, Clinton R. Paden, Duncan MacCannell |
| EPI_ISL_2201437 | Houston Methodist Hospital | Houston Methodist Hospital | Randall J. Olsen, Paul A. Christensen, S. Wesley Long, Sishir Subedi, Robert Olson, Marcus Nguyen, James J. Davis, Matthew Ojeda Saavedra, Prasanti Yerramilli, Layne Pruitt, Kristina Reppond, Madison N. Shyer, Jessica Cambric, Ryan Gadd, Ilya J. Finkelstein, Jimmy Gollihar, and James M. Musser |
| EPI_ISL_2201438 | Aegis Sciences Corporation | Centers for Disease Control and Prevention Division of Viral Diseases, Pathogen Discovery | Dakota Howard, Dhwani Batra, Peter W. Cook, Kara Moser, Adrian Paskey, Jason Caravas, Benjamin Rambo-Martin, Shatavia Morrison, Christopher Gulvick, Scott Sammons, Yvette Unoarumhi, Darlene Wagner, Matthew Schmerer, Cyndi Clark, Patrick Campbell, Rob Case, Vikramsinha Ghorpade, Holly Houdeshell, Ola Kvalvaag, Dillon Nall, Ethan Sanders, Alec Vest, Shaun Westlund, Matthew Hardison, Clinton R. Paden, Duncan MacCannell |
| EPI_ISL_2201439 | Houston Methodist Hospital | Houston Methodist Hospital | Randall J. Olsen, Paul A. Christensen, S. Wesley Long, Sishir Subedi, Robert Olson, Marcus Nguyen, James J. Davis, Matthew Ojeda Saavedra, Prasanti Yerramilli, Layne Pruitt, Kristina Reppond, Madison N. Shyer, Jessica Cambric, Ryan Gadd, Ilya J. Finkelstein, Jimmy Gollihar, and James M. Musser |
| EPI_ISL_2201440 | Aegis Sciences Corporation | Centers for Disease Control and Prevention Division of Viral Diseases, Pathogen Discovery | Dakota Howard, Dhwani Batra, Peter W. Cook, Kara Moser, Adrian Paskey, Jason Caravas, Benjamin Rambo-Martin, Shatavia Morrison, Christopher Gulvick, Scott Sammons, Yvette Unoarumhi, Darlene Wagner, Matthew Schmerer, Cyndi Clark, Patrick Campbell, Rob Case, Vikramsinha Ghorpade, Holly Houdeshell, Ola Kvalvaag, Dillon Nall, Ethan Sanders, Alec Vest, Shaun Westlund, Matthew Hardison, Clinton R. Paden, Duncan MacCannell |
| EPI_ISL_2201441 | Houston Methodist Hospital | Houston Methodist Hospital | Randall J. Olsen, Paul A. Christensen, S. Wesley Long, Sishir Subedi, Robert Olson, Marcus Nguyen, James J. Davis, Matthew Ojeda Saavedra, Prasanti Yerramilli, Layne Pruitt, Kristina Reppond, Madison N. Shyer, Jessica Cambric, Ryan Gadd, Ilya J. Finkelstein, Jimmy Gollihar, and James M. Musser |
| EPI_ISL_2201442 | Aegis Sciences Corporation | Centers for Disease Control and Prevention Division of Viral Diseases, Pathogen Discovery | Dakota Howard, Dhwani Batra, Peter W. Cook, Kara Moser, Adrian Paskey, Jason Caravas, Benjamin Rambo-Martin, Shatavia Morrison, Christopher Gulvick, Scott Sammons, Yvette Unoarumhi, Darlene Wagner, Matthew Schmerer, Cyndi Clark, Patrick Campbell, Rob Case, Vikramsinha Ghorpade, Holly Houdeshell, Ola Kvalvaag, Dillon Nall, Ethan Sanders, Alec Vest, Shaun Westlund, Matthew Hardison, Clinton R. Paden, Duncan MacCannell |
| EPI_ISL_2201443 | Houston Methodist Hospital | Houston Methodist Hospital | Randall J. Olsen, Paul A. Christensen, S. Wesley Long, Sishir Subedi, Robert Olson, Marcus Nguyen, James J. Davis, Matthew Ojeda Saavedra, Prasanti Yerramilli, Layne Pruitt, Kristina Reppond, Madison N. Shyer, Jessica Cambric, Ryan Gadd, Ilya J. Finkelstein, Jimmy Gollihar, and James M. Musser |
| EPI_ISL_2201444, EPI_ISL_2201445 | Aegis Sciences Corporation | Centers for Disease Control and Prevention Division of Viral Diseases, Pathogen Discovery | Dakota Howard, Dhwani Batra, Peter W. Cook, Kara Moser, Adrian Paskey, Jason Caravas, Benjamin Rambo-Martin, Shatavia Morrison, Christopher Gulvick, Scott Sammons, Yvette Unoarumhi, Darlene Wagner, Matthew Schmerer, Cyndi Clark, Patrick Campbell, Rob Case, Vikramsinha Ghorpade, Holly Houdeshell, Ola Kvalvaag, Dillon Nall, Ethan Sanders, Alec Vest, Shaun Westlund, Matthew Hardison, Clinton R. Paden, Duncan MacCannell |
| EPI_ISL_2201446 | Houston Methodist Hospital | Houston Methodist Hospital | Randall J. Olsen, Paul A. Christensen, S. Wesley Long, Sishir Subedi, Robert Olson, Marcus Nguyen, James J. Davis, Matthew Ojeda Saavedra, Prasanti Yerramilli, Layne Pruitt, Kristina Reppond, Madison N. Shyer, Jessica Cambric, Ryan Gadd, Ilya J. Finkelstein, Jimmy Gollihar, and James M. Musser |
| EPI_ISL_2201447 | Aegis Sciences Corporation | Centers for Disease Control and Prevention Division of Viral Diseases, Pathogen Discovery | Dakota Howard, Dhwani Batra, Peter W. Cook, Kara Moser, Adrian Paskey, Jason Caravas, Benjamin Rambo-Martin, Shatavia Morrison, Christopher Gulvick, Scott Sammons, Yvette Unoarumhi, Darlene Wagner, Matthew Schmerer, Cyndi Clark, Patrick Campbell, Rob Case, Vikramsinha Ghorpade, Holly Houdeshell, Ola Kvalvaag, Dillon Nall, Ethan Sanders, Alec Vest, Shaun Westlund, Matthew Hardison, Clinton R. Paden, Duncan MacCannell |
| EPI_ISL_2201448 | Houston Methodist Hospital | Houston Methodist Hospital | Randall J. Olsen, Paul A. Christensen, S. Wesley Long, Sishir Subedi, Robert Olson, Marcus Nguyen, James J. Davis, Matthew Ojeda Saavedra, Prasanti Yerramilli, Layne Pruitt, Kristina Reppond, Madison N. Shyer, Jessica Cambric, Ryan Gadd, Ilya J. Finkelstein, Jimmy Gollihar, and James M. Musser |
| EPI_ISL_2201449 | Aegis Sciences Corporation | Centers for Disease Control and Prevention Division of Viral Diseases, Pathogen Discovery | Dakota Howard, Dhwani Batra, Peter W. Cook, Kara Moser, Adrian Paskey, Jason Caravas, Benjamin Rambo-Martin, Shatavia Morrison, Christopher Gulvick, Scott Sammons, Yvette Unoarumhi, Darlene Wagner, Matthew Schmerer, Cyndi Clark, Patrick Campbell, Rob Case, Vikramsinha Ghorpade, Holly Houdeshell, Ola Kvalvaag, Dillon Nall, Ethan Sanders, Alec Vest, Shaun Westlund, Matthew Hardison, Clinton R. Paden, Duncan MacCannell |
| EPI_ISL_2201450 | Houston Methodist Hospital | Houston Methodist Hospital | Randall J. Olsen, Paul A. Christensen, S. Wesley Long, Sishir Subedi, Robert Olson, Marcus Nguyen, James J. Davis, Matthew Ojeda Saavedra, Prasanti Yerramilli, Layne Pruitt, Kristina Reppond, Madison N. Shyer, Jessica Cambric, Ryan Gadd, Ilya J. Finkelstein, Jimmy Gollihar, and James M. Musser |
| EPI_ISL_2201451, EPI_ISL_2201452 | Aegis Sciences Corporation | Centers for Disease Control and Prevention Division of Viral Diseases, Pathogen Discovery | Dakota Howard, Dhwani Batra, Peter W. Cook, Kara Moser, Adrian Paskey, Jason Caravas, Benjamin Rambo-Martin, Shatavia Morrison, Christopher Gulvick, Scott Sammons, Yvette Unoarumhi, Darlene Wagner, Matthew Schmerer, Cyndi Clark, Patrick Campbell, Rob Case, Vikramsinha Ghorpade, Holly Houdeshell, Ola Kvalvaag, Dillon Nall, Ethan Sanders, Alec Vest, Shaun Westlund, Matthew Hardison, Clinton R. Paden, Duncan MacCannell |
| EPI_ISL_2201453 | Houston Methodist Hospital | Houston Methodist Hospital | Randall J. Olsen, Paul A. Christensen, S. Wesley Long, Sishir Subedi, Robert Olson, Marcus Nguyen, James J. Davis, Matthew Ojeda Saavedra, Prasanti Yerramilli, Layne Pruitt, Kristina Reppond, Madison N. Shyer, Jessica Cambric, Ryan Gadd, Ilya J. Finkelstein, Jimmy Gollihar, and James M. Musser |
| EPI_ISL_2201454, EPI_ISL_2201455 | Aegis Sciences Corporation | Centers for Disease Control and Prevention Division of Viral Diseases, Pathogen Discovery | Dakota Howard, Dhwani Batra, Peter W. Cook, Kara Moser, Adrian Paskey, Jason Caravas, Benjamin Rambo-Martin, Shatavia Morrison, Christopher Gulvick, Scott Sammons, Yvette Unoarumhi, Darlene Wagner, Matthew Schmerer, Cyndi Clark, Patrick Campbell, Rob Case, Vikramsinha Ghorpade, Holly Houdeshell, Ola Kvalvaag, Dillon Nall, Ethan Sanders, Alec Vest, Shaun Westlund, Matthew Hardison, Clinton R. Paden, Duncan MacCannell |
| EPI_ISL_2201456 | Houston Methodist Hospital | Houston Methodist Hospital | Randall J. Olsen, Paul A. Christensen, S. Wesley Long, Sishir Subedi, Robert Olson, Marcus Nguyen, James J. Davis, Matthew Ojeda Saavedra, Prasanti Yerramilli, Layne Pruitt, Kristina Reppond, Madison N. Shyer, Jessica Cambric, Ryan Gadd, Ilya J. Finkelstein, Jimmy Gollihar, and James M. Musser |
| EPI_ISL_2201457 | Aegis Sciences Corporation | Centers for Disease Control and Prevention Division of Viral Diseases, Pathogen Discovery | Dakota Howard, Dhwani Batra, Peter W. Cook, Kara Moser, Adrian Paskey, Jason Caravas, Benjamin Rambo-Martin, Shatavia Morrison, Christopher Gulvick, Scott Sammons, Yvette Unoarumhi, Darlene Wagner, Matthew Schmerer, Cyndi Clark, Patrick Campbell, Rob Case, Vikramsinha Ghorpade, Holly Houdeshell, Ola Kvalvaag, Dillon Nall, Ethan Sanders, Alec Vest, Shaun Westlund, Matthew Hardison, Clinton R. Paden, Duncan MacCannell |
| EPI_ISL_2201458 | Houston Methodist Hospital | Houston Methodist Hospital | Randall J. Olsen, Paul A. Christensen, S. Wesley Long, Sishir Subedi, Robert Olson, Marcus Nguyen, James J. Davis, Matthew Ojeda Saavedra, Prasanti Yerramilli, Layne Pruitt, Kristina Reppond, Madison N. Shyer, Jessica Cambric, Ryan Gadd, Ilya J. Finkelstein, Jimmy Gollihar, and James M. Musser |
| EPI_ISL_2201459 | Aegis Sciences Corporation | Centers for Disease Control and Prevention Division of Viral Diseases, Pathogen Discovery | Dakota Howard, Dhwani Batra, Peter W. Cook, Kara Moser, Adrian Paskey, Jason Caravas, Benjamin Rambo-Martin, Shatavia Morrison, Christopher Gulvick, Scott Sammons, Yvette Unoarumhi, Darlene Wagner, Matthew Schmerer, Cyndi Clark, Patrick Campbell, Rob Case, Vikramsinha Ghorpade, Holly Houdeshell, Ola Kvalvaag, Dillon Nall, Ethan Sanders, Alec Vest, Shaun Westlund, Matthew Hardison, Clinton R. Paden, Duncan MacCannell |
| EPI_ISL_2201460 | Houston Methodist Hospital | Houston Methodist Hospital | Randall J. Olsen, Paul A. Christensen, S. Wesley Long, Sishir Subedi, Robert Olson, Marcus Nguyen, James J. Davis, Matthew Ojeda Saavedra, Prasanti Yerramilli, Layne Pruitt, Kristina Reppond, Madison N. Shyer, Jessica Cambric, Ryan Gadd, Ilya J. Finkelstein, Jimmy Gollihar, and James M. Musser |
| EPI_ISL_2201461 | Aegis Sciences Corporation | Centers for Disease Control and Prevention Division of Viral Diseases, Pathogen Discovery | Dakota Howard, Dhwani Batra, Peter W. Cook, Kara Moser, Adrian Paskey, Jason Caravas, Benjamin Rambo-Martin, Shatavia Morrison, Christopher Gulvick, Scott Sammons, Yvette Unoarumhi, Darlene Wagner, Matthew Schmerer, Cyndi Clark, Patrick Campbell, Rob Case, Vikramsinha Ghorpade, Holly Houdeshell, Ola Kvalvaag, Dillon Nall, Ethan Sanders, Alec Vest, Shaun Westlund, Matthew Hardison, Clinton R. Paden, Duncan MacCannell |
| EPI_ISL_2201463 | Houston Methodist Hospital | Houston Methodist Hospital | Randall J. Olsen, Paul A. Christensen, S. Wesley Long, Sishir Subedi, Robert Olson, Marcus Nguyen, James J. Davis, Matthew Ojeda Saavedra, Prasanti Yerramilli, Layne Pruitt, Kristina Reppond, Madison N. Shyer, Jessica Cambric, Ryan Gadd, Ilya J. Finkelstein, Jimmy Gollihar, and James M. Musser |
| EPI_ISL_2201464 | Aegis Sciences Corporation | Centers for Disease Control and Prevention Division of Viral Diseases, Pathogen Discovery | Dakota Howard, Dhwani Batra, Peter W. Cook, Kara Moser, Adrian Paskey, Jason Caravas, Benjamin Rambo-Martin, Shatavia Morrison, Christopher Gulvick, Scott Sammons, Yvette Unoarumhi, Darlene Wagner, Matthew Schmerer, Cyndi Clark, Patrick Campbell, Rob Case, Vikramsinha Ghorpade, Holly Houdeshell, Ola Kvalvaag, Dillon Nall, Ethan Sanders, Alec Vest, Shaun Westlund, Matthew Hardison, Clinton R. Paden, Duncan MacCannell |
| EPI_ISL_2201465 | Houston Methodist Hospital | Houston Methodist Hospital | Randall J. Olsen, Paul A. Christensen, S. Wesley Long, Sishir Subedi, Robert Olson, Marcus Nguyen, James J. Davis, Matthew Ojeda Saavedra, Prasanti Yerramilli, Layne Pruitt, Kristina Reppond, Madison N. Shyer, Jessica Cambric, Ryan Gadd, Ilya J. Finkelstein, Jimmy Gollihar, and James M. Musser |
| EPI_ISL_2201466 | Aegis Sciences Corporation | Centers for Disease Control and Prevention Division of Viral Diseases, Pathogen Discovery | Dakota Howard, Dhwani Batra, Peter W. Cook, Kara Moser, Adrian Paskey, Jason Caravas, Benjamin Rambo-Martin, Shatavia Morrison, Christopher Gulvick, Scott Sammons, Yvette Unoarumhi, Darlene Wagner, Matthew Schmerer, Cyndi Clark, Patrick Campbell, Rob Case, Vikramsinha Ghorpade, Holly Houdeshell, Ola Kvalvaag, Dillon Nall, Ethan Sanders, Alec Vest, Shaun Westlund, Matthew Hardison, Clinton R. Paden, Duncan MacCannell |
| EPI_ISL_2201467, EPI_ISL_2201468 | Houston Methodist Hospital | Houston Methodist Hospital | Randall J. Olsen, Paul A. Christensen, S. Wesley Long, Sishir Subedi, Robert Olson, Marcus Nguyen, James J. Davis, Matthew Ojeda Saavedra, Prasanti Yerramilli, Layne Pruitt, Kristina Reppond, Madison N. Shyer, Jessica Cambric, Ryan Gadd, Ilya J. Finkelstein, Jimmy Gollihar, and James M. Musser |
| EPI_ISL_2201469 | Aegis Sciences Corporation | Centers for Disease Control and Prevention Division of Viral Diseases, Pathogen Discovery | Dakota Howard, Dhwani Batra, Peter W. Cook, Kara Moser, Adrian Paskey, Jason Caravas, Benjamin Rambo-Martin, Shatavia Morrison, Christopher Gulvick, Scott Sammons, Yvette Unoarumhi, Darlene Wagner, Matthew Schmerer, Cyndi Clark, Patrick Campbell, Rob Case, Vikramsinha Ghorpade, Holly Houdeshell, Ola Kvalvaag, Dillon Nall, Ethan Sanders, Alec Vest, Shaun Westlund, Matthew Hardison, Clinton R. Paden, Duncan MacCannell |
| EPI_ISL_2201470 | Houston Methodist Hospital | Houston Methodist Hospital | Randall J. Olsen, Paul A. Christensen, S. Wesley Long, Sishir Subedi, Robert Olson, Marcus Nguyen, James J. Davis, Matthew Ojeda Saavedra, Prasanti Yerramilli, Layne Pruitt, Kristina Reppond, Madison N. Shyer, Jessica Cambric, Ryan Gadd, Ilya J. Finkelstein, Jimmy Gollihar, and James M. Musser |
| EPI_ISL_2201471 | Aegis Sciences Corporation | Centers for Disease Control and Prevention Division of Viral Diseases, Pathogen Discovery | Dakota Howard, Dhwani Batra, Peter W. Cook, Kara Moser, Adrian Paskey, Jason Caravas, Benjamin Rambo-Martin, Shatavia Morrison, Christopher Gulvick, Scott Sammons, Yvette Unoarumhi, Darlene Wagner, Matthew Schmerer, Cyndi Clark, Patrick Campbell, Rob Case, Vikramsinha Ghorpade, Holly Houdeshell, Ola Kvalvaag, Dillon Nall, Ethan Sanders, Alec Vest, Shaun Westlund, Matthew Hardison, Clinton R. Paden, Duncan MacCannell |
| EPI_ISL_2201472 | Houston Methodist Hospital | Houston Methodist Hospital | Randall J. Olsen, Paul A. Christensen, S. Wesley Long, Sishir Subedi, Robert Olson, Marcus Nguyen, James J. Davis, Matthew Ojeda Saavedra, Prasanti Yerramilli, Layne Pruitt, Kristina Reppond, Madison N. Shyer, Jessica Cambric, Ryan Gadd, Ilya J. Finkelstein, Jimmy Gollihar, and James M. Musser |
| EPI_ISL_2201473 | Aegis Sciences Corporation | Centers for Disease Control and Prevention Division of Viral Diseases, Pathogen Discovery | Dakota Howard, Dhwani Batra, Peter W. Cook, Kara Moser, Adrian Paskey, Jason Caravas, Benjamin Rambo-Martin, Shatavia Morrison, Christopher Gulvick, Scott Sammons, Yvette Unoarumhi, Darlene Wagner, Matthew Schmerer, Cyndi Clark, Patrick Campbell, Rob Case, Vikramsinha Ghorpade, Holly Houdeshell, Ola Kvalvaag, Dillon Nall, Ethan Sanders, Alec Vest, Shaun Westlund, Matthew Hardison, Clinton R. Paden, Duncan MacCannell |
| EPI_ISL_2201474 | Houston Methodist Hospital | Houston Methodist Hospital | Randall J. Olsen, Paul A. Christensen, S. Wesley Long, Sishir Subedi, Robert Olson, Marcus Nguyen, James J. Davis, Matthew Ojeda Saavedra, Prasanti Yerramilli, Layne Pruitt, Kristina Reppond, Madison N. Shyer, Jessica Cambric, Ryan Gadd, Ilya J. Finkelstein, Jimmy Gollihar, and James M. Musser |
| EPI_ISL_2201475 | Aegis Sciences Corporation | Centers for Disease Control and Prevention Division of Viral Diseases, Pathogen Discovery | Dakota Howard, Dhwani Batra, Peter W. Cook, Kara Moser, Adrian Paskey, Jason Caravas, Benjamin Rambo-Martin, Shatavia Morrison, Christopher Gulvick, Scott Sammons, Yvette Unoarumhi, Darlene Wagner, Matthew Schmerer, Cyndi Clark, Patrick Campbell, Rob Case, Vikramsinha Ghorpade, Holly Houdeshell, Ola Kvalvaag, Dillon Nall, Ethan Sanders, Alec Vest, Shaun Westlund, Matthew Hardison, Clinton R. Paden, Duncan MacCannell |
| EPI_ISL_2201476, | Houston Methodist Hospital | Houston Methodist Hospital | Randall J. Olsen, Paul A. Christensen, S. Wesley Long, Sishir Subedi, Robert Olson, Marcus Nguyen, James J. Davis, Matthew Ojeda Saavedra, Prasanti Yerramilli, Layne Pruitt, Kristina Reppond, Madison N. Shyer, Jessica Cambric, Ryan Gadd, Ilya J. Finkelstein, Jimmy Gollihar, and James |

| | | | |
|---|---|---|---|
| EPI_ISL_2201477 | | | M. Musser |
| EPI_ISL_2201478 | Aegis Sciences Corporation | Centers for Disease Control and Prevention Division of Viral Diseases, Pathogen Discovery | Dakota Howard, Dhwani Batra, Peter W. Cook, Kara Moser, Adrian Paskey, Jason Caravas, Benjamin Rambo-Martin, Shatavia Morrison, Christopher Gulvick, Scott Sammons, Yvette Unoarumhi, Darlene Wagner, Matthew Schmerer, Cyndi Clark, Patrick Campbell, Rob Case, Vikramsinha Ghorpade, Holly Houdeshell, Ola Kvalvaag, Dillon Nall, Ethan Sanders, Alec Vest, Shaun Westlund, Matthew Hardison, Clinton R. Paden, Duncan MacCannell |
| EPI_ISL_2201479 | Houston Methodist Hospital | Houston Methodist Hospital | Randall J. Olsen, Paul A. Christensen, S. Wesley Long, Sishir Subedi, Robert Olson, Marcus Nguyen, James J. Davis, Matthew Ojeda Saavedra, Prasanti Yerramilli, Layne Pruitt, Kristina Reppond, Madison N. Shyer, Jessica Cambric, Ryan Gadd, Ilya J. Finkelstein, Jimmy Gollihar, and James M. Musser |
| EPI_ISL_2201480, EPI_ISL_2201481 | Aegis Sciences Corporation | Centers for Disease Control and Prevention Division of Viral Diseases, Pathogen Discovery | Dakota Howard, Dhwani Batra, Peter W. Cook, Kara Moser, Adrian Paskey, Jason Caravas, Benjamin Rambo-Martin, Shatavia Morrison, Christopher Gulvick, Scott Sammons, Yvette Unoarumhi, Darlene Wagner, Matthew Schmerer, Cyndi Clark, Patrick Campbell, Rob Case, Vikramsinha Ghorpade, Holly Houdeshell, Ola Kvalvaag, Dillon Nall, Ethan Sanders, Alec Vest, Shaun Westlund, Matthew Hardison, Clinton R. Paden, Duncan MacCannell |
| EPI_ISL_2201482, EPI_ISL_2201483 | Houston Methodist Hospital | Houston Methodist Hospital | Randall J. Olsen, Paul A. Christensen, S. Wesley Long, Sishir Subedi, Robert Olson, Marcus Nguyen, James J. Davis, Matthew Ojeda Saavedra, Prasanti Yerramilli, Layne Pruitt, Kristina Reppond, Madison N. Shyer, Jessica Cambric, Ryan Gadd, Ilya J. Finkelstein, Jimmy Gollihar, and James M. Musser |
| EPI_ISL_2201484 | Aegis Sciences Corporation | Centers for Disease Control and Prevention Division of Viral Diseases, Pathogen Discovery | Dakota Howard, Dhwani Batra, Peter W. Cook, Kara Moser, Adrian Paskey, Jason Caravas, Benjamin Rambo-Martin, Shatavia Morrison, Christopher Gulvick, Scott Sammons, Yvette Unoarumhi, Darlene Wagner, Matthew Schmerer, Cyndi Clark, Patrick Campbell, Rob Case, Vikramsinha Ghorpade, Holly Houdeshell, Ola Kvalvaag, Dillon Nall, Ethan Sanders, Alec Vest, Shaun Westlund, Matthew Hardison, Clinton R. Paden, Duncan MacCannell |
| EPI_ISL_2201485 | Houston Methodist Hospital | Houston Methodist Hospital | Randall J. Olsen, Paul A. Christensen, S. Wesley Long, Sishir Subedi, Robert Olson, Marcus Nguyen, James J. Davis, Matthew Ojeda Saavedra, Prasanti Yerramilli, Layne Pruitt, Kristina Reppond, Madison N. Shyer, Jessica Cambric, Ryan Gadd, Ilya J. Finkelstein, Jimmy Gollihar, and James M. Musser |
| EPI_ISL_2201486 | Aegis Sciences Corporation | Centers for Disease Control and Prevention Division of Viral Diseases, Pathogen Discovery | Dakota Howard, Dhwani Batra, Peter W. Cook, Kara Moser, Adrian Paskey, Jason Caravas, Benjamin Rambo-Martin, Shatavia Morrison, Christopher Gulvick, Scott Sammons, Yvette Unoarumhi, Darlene Wagner, Matthew Schmerer, Cyndi Clark, Patrick Campbell, Rob Case, Vikramsinha Ghorpade, Holly Houdeshell, Ola Kvalvaag, Dillon Nall, Ethan Sanders, Alec Vest, Shaun Westlund, Matthew Hardison, Clinton R. Paden, Duncan MacCannell |
| EPI_ISL_2201487 | Houston Methodist Hospital | Houston Methodist Hospital | Randall J. Olsen, Paul A. Christensen, S. Wesley Long, Sishir Subedi, Robert Olson, Marcus Nguyen, James J. Davis, Matthew Ojeda Saavedra, Prasanti Yerramilli, Layne Pruitt, Kristina Reppond, Madison N. Shyer, Jessica Cambric, Ryan Gadd, Ilya J. Finkelstein, Jimmy Gollihar, and James M. Musser |
| EPI_ISL_2201488, EPI_ISL_2201489 | Aegis Sciences Corporation | Centers for Disease Control and Prevention Division of Viral Diseases, Pathogen Discovery | Dakota Howard, Dhwani Batra, Peter W. Cook, Kara Moser, Adrian Paskey, Jason Caravas, Benjamin Rambo-Martin, Shatavia Morrison, Christopher Gulvick, Scott Sammons, Yvette Unoarumhi, Darlene Wagner, Matthew Schmerer, Cyndi Clark, Patrick Campbell, Rob Case, Vikramsinha Ghorpade, Holly Houdeshell, Ola Kvalvaag, Dillon Nall, Ethan Sanders, Alec Vest, Shaun Westlund, Matthew Hardison, Clinton R. Paden, Duncan MacCannell |
| EPI_ISL_2201490 | Houston Methodist Hospital | Houston Methodist Hospital | Randall J. Olsen, Paul A. Christensen, S. Wesley Long, Sishir Subedi, Robert Olson, Marcus Nguyen, James J. Davis, Matthew Ojeda Saavedra, Prasanti Yerramilli, Layne Pruitt, Kristina Reppond, Madison N. Shyer, Jessica Cambric, Ryan Gadd, Ilya J. Finkelstein, Jimmy Gollihar, and James M. Musser |
| EPI_ISL_2201491 | Aegis Sciences Corporation | Centers for Disease Control and Prevention Division of Viral Diseases, Pathogen Discovery | Dakota Howard, Dhwani Batra, Peter W. Cook, Kara Moser, Adrian Paskey, Jason Caravas, Benjamin Rambo-Martin, Shatavia Morrison, Christopher Gulvick, Scott Sammons, Yvette Unoarumhi, Darlene Wagner, Matthew Schmerer, Cyndi Clark, Patrick Campbell, Rob Case, Vikramsinha Ghorpade, Holly Houdeshell, Ola Kvalvaag, Dillon Nall, Ethan Sanders, Alec Vest, Shaun Westlund, Matthew Hardison, Clinton R. Paden, Duncan MacCannell |
| EPI_ISL_2201492 | Houston Methodist Hospital | Houston Methodist Hospital | Randall J. Olsen, Paul A. Christensen, S. Wesley Long, Sishir Subedi, Robert Olson, Marcus Nguyen, James J. Davis, Matthew Ojeda Saavedra, Prasanti Yerramilli, Layne Pruitt, Kristina Reppond, Madison N. Shyer, Jessica Cambric, Ryan Gadd, Ilya J. Finkelstein, Jimmy Gollihar, and James M. Musser |
| EPI_ISL_2201493 | Aegis Sciences Corporation | Centers for Disease Control and Prevention Division of Viral Diseases, Pathogen Discovery | Dakota Howard, Dhwani Batra, Peter W. Cook, Kara Moser, Adrian Paskey, Jason Caravas, Benjamin Rambo-Martin, Shatavia Morrison, Christopher Gulvick, Scott Sammons, Yvette Unoarumhi, Darlene Wagner, Matthew Schmerer, Cyndi Clark, Patrick Campbell, Rob Case, Vikramsinha Ghorpade, Holly Houdeshell, Ola Kvalvaag, Dillon Nall, Ethan Sanders, Alec Vest, Shaun Westlund, Matthew Hardison, Clinton R. Paden, Duncan MacCannell |
| EPI_ISL_2201494 | Houston Methodist Hospital | Houston Methodist Hospital | Randall J. Olsen, Paul A. Christensen, S. Wesley Long, Sishir Subedi, Robert Olson, Marcus Nguyen, James J. Davis, Matthew Ojeda Saavedra, Prasanti Yerramilli, Layne Pruitt, Kristina Reppond, Madison N. Shyer, Jessica Cambric, Ryan Gadd, Ilya J. Finkelstein, Jimmy Gollihar, and James M. Musser |
| EPI_ISL_2201495 | Aegis Sciences Corporation | Centers for Disease Control and Prevention Division of Viral Diseases, Pathogen Discovery | Dakota Howard, Dhwani Batra, Peter W. Cook, Kara Moser, Adrian Paskey, Jason Caravas, Benjamin Rambo-Martin, Shatavia Morrison, Christopher Gulvick, Scott Sammons, Yvette Unoarumhi, Darlene Wagner, Matthew Schmerer, Cyndi Clark, Patrick Campbell, Rob Case, Vikramsinha Ghorpade, Holly Houdeshell, Ola Kvalvaag, Dillon Nall, Ethan Sanders, Alec Vest, Shaun Westlund, Matthew Hardison, Clinton R. Paden, Duncan MacCannell |
| EPI_ISL_2201496 | Houston Methodist Hospital | Houston Methodist Hospital | Randall J. Olsen, Paul A. Christensen, S. Wesley Long, Sishir Subedi, Robert Olson, Marcus Nguyen, James J. Davis, Matthew Ojeda Saavedra, Prasanti Yerramilli, Layne Pruitt, Kristina Reppond, Madison N. Shyer, Jessica Cambric, Ryan Gadd, Ilya J. Finkelstein, Jimmy Gollihar, and James M. Musser |
| EPI_ISL_2201497, EPI_ISL_2201498 | Aegis Sciences Corporation | Centers for Disease Control and Prevention Division of Viral Diseases, Pathogen Discovery | Dakota Howard, Dhwani Batra, Peter W. Cook, Kara Moser, Adrian Paskey, Jason Caravas, Benjamin Rambo-Martin, Shatavia Morrison, Christopher Gulvick, Scott Sammons, Yvette Unoarumhi, Darlene Wagner, Matthew Schmerer, Cyndi Clark, Patrick Campbell, Rob Case, Vikramsinha Ghorpade, Holly Houdeshell, Ola Kvalvaag, Dillon Nall, Ethan Sanders, Alec Vest, Shaun Westlund, Matthew Hardison, Clinton R. Paden, Duncan MacCannell |
| EPI_ISL_2201499 | Houston Methodist Hospital | Houston Methodist Hospital | Randall J. Olsen, Paul A. Christensen, S. Wesley Long, Sishir Subedi, Robert Olson, Marcus Nguyen, James J. Davis, Matthew Ojeda Saavedra, Prasanti Yerramilli, Layne Pruitt, Kristina Reppond, Madison N. Shyer, Jessica Cambric, Ryan Gadd, Ilya J. Finkelstein, Jimmy Gollihar, and James M. Musser |
| EPI_ISL_2201500 | Aegis Sciences Corporation | Centers for Disease Control and Prevention Division of Viral Diseases, Pathogen Discovery | Dakota Howard, Dhwani Batra, Peter W. Cook, Kara Moser, Adrian Paskey, Jason Caravas, Benjamin Rambo-Martin, Shatavia Morrison, Christopher Gulvick, Scott Sammons, Yvette Unoarumhi, Darlene Wagner, Matthew Schmerer, Cyndi Clark, Patrick Campbell, Rob Case, Vikramsinha Ghorpade, Holly Houdeshell, Ola Kvalvaag, Dillon Nall, Ethan Sanders, Alec Vest, Shaun Westlund, Matthew Hardison, Clinton R. Paden, Duncan MacCannell |
| EPI_ISL_2201501 | Houston Methodist Hospital | Houston Methodist Hospital | Randall J. Olsen, Paul A. Christensen, S. Wesley Long, Sishir Subedi, Robert Olson, Marcus Nguyen, James J. Davis, Matthew Ojeda Saavedra, Prasanti Yerramilli, Layne Pruitt, Kristina Reppond, Madison N. Shyer, Jessica Cambric, Ryan Gadd, Ilya J. Finkelstein, Jimmy Gollihar, and James M. Musser |
| EPI_ISL_2201502, EPI_ISL_2201503 | Aegis Sciences Corporation | Centers for Disease Control and Prevention Division of Viral Diseases, Pathogen Discovery | Dakota Howard, Dhwani Batra, Peter W. Cook, Kara Moser, Adrian Paskey, Jason Caravas, Benjamin Rambo-Martin, Shatavia Morrison, Christopher Gulvick, Scott Sammons, Yvette Unoarumhi, Darlene Wagner, Matthew Schmerer, Cyndi Clark, Patrick Campbell, Rob Case, Vikramsinha Ghorpade, Holly Houdeshell, Ola Kvalvaag, Dillon Nall, Ethan Sanders, Alec Vest, Shaun Westlund, Matthew Hardison, Clinton R. Paden, Duncan MacCannell |
| EPI_ISL_2201504 | Houston Methodist Hospital | Houston Methodist Hospital | Randall J. Olsen, Paul A. Christensen, S. Wesley Long, Sishir Subedi, Robert Olson, Marcus Nguyen, James J. Davis, Matthew Ojeda Saavedra, Prasanti Yerramilli, Layne Pruitt, Kristina Reppond, Madison N. Shyer, Jessica Cambric, Ryan Gadd, Ilya J. Finkelstein, Jimmy Gollihar, and James M. Musser |
| EPI_ISL_2201505 | Aegis Sciences Corporation | Centers for Disease Control and Prevention Division of Viral Diseases, Pathogen Discovery | Dakota Howard, Dhwani Batra, Peter W. Cook, Kara Moser, Adrian Paskey, Jason Caravas, Benjamin Rambo-Martin, Shatavia Morrison, Christopher Gulvick, Scott Sammons, Yvette Unoarumhi, Darlene Wagner, Matthew Schmerer, Cyndi Clark, Patrick Campbell, Rob Case, Vikramsinha Ghorpade, Holly Houdeshell, Ola Kvalvaag, Dillon Nall, Ethan Sanders, Alec Vest, Shaun Westlund, Matthew Hardison, Clinton R. Paden, Duncan MacCannell |
| EPI_ISL_2201506 | Houston Methodist Hospital | Houston Methodist Hospital | Randall J. Olsen, Paul A. Christensen, S. Wesley Long, Sishir Subedi, Robert Olson, Marcus Nguyen, James J. Davis, Matthew Ojeda Saavedra, Prasanti Yerramilli, Layne Pruitt, Kristina Reppond, Madison N. Shyer, Jessica Cambric, Ryan Gadd, Ilya J. Finkelstein, Jimmy Gollihar, and James M. Musser |
| EPI_ISL_2201507 | Aegis Sciences Corporation | Centers for Disease Control and Prevention Division of Viral Diseases, Pathogen Discovery | Dakota Howard, Dhwani Batra, Peter W. Cook, Kara Moser, Adrian Paskey, Jason Caravas, Benjamin Rambo-Martin, Shatavia Morrison, Christopher Gulvick, Scott Sammons, Yvette Unoarumhi, Darlene Wagner, Matthew Schmerer, Cyndi Clark, Patrick Campbell, Rob Case, Vikramsinha Ghorpade, Holly Houdeshell, Ola Kvalvaag, Dillon Nall, Ethan Sanders, Alec Vest, Shaun Westlund, Matthew Hardison, Clinton R. Paden, Duncan MacCannell |
| EPI_ISL_2201508 | Houston Methodist Hospital | Houston Methodist Hospital | Randall J. Olsen, Paul A. Christensen, S. Wesley Long, Sishir Subedi, Robert Olson, Marcus Nguyen, James J. Davis, Matthew Ojeda Saavedra, Prasanti Yerramilli, Layne Pruitt, Kristina Reppond, Madison N. Shyer, Jessica Cambric, Ryan Gadd, Ilya J. Finkelstein, Jimmy Gollihar, and James M. Musser |
| EPI_ISL_2201509 | Aegis Sciences Corporation | Centers for Disease Control and Prevention Division of Viral Diseases, Pathogen Discovery | Dakota Howard, Dhwani Batra, Peter W. Cook, Kara Moser, Adrian Paskey, Jason Caravas, Benjamin Rambo-Martin, Shatavia Morrison, Christopher Gulvick, Scott Sammons, Yvette Unoarumhi, Darlene Wagner, Matthew Schmerer, Cyndi Clark, Patrick Campbell, Rob Case, Vikramsinha Ghorpade, Holly Houdeshell, Ola Kvalvaag, Dillon Nall, Ethan Sanders, Alec Vest, Shaun Westlund, Matthew Hardison, Clinton R. Paden, Duncan MacCannell |
| EPI_ISL_2201510, EPI_ISL_2201513 | Houston Methodist Hospital | Houston Methodist Hospital | Randall J. Olsen, Paul A. Christensen, S. Wesley Long, Sishir Subedi, Robert Olson, Marcus Nguyen, James J. Davis, Matthew Ojeda Saavedra, Prasanti Yerramilli, Layne Pruitt, Kristina Reppond, Madison N. Shyer, Jessica Cambric, Ryan Gadd, Ilya J. Finkelstein, Jimmy Gollihar, and James M. Musser |
| EPI_ISL_2201514, EPI_ISL_2201515 | Aegis Sciences Corporation | Centers for Disease Control and Prevention Division of Viral Diseases, Pathogen Discovery | Dakota Howard, Dhwani Batra, Peter W. Cook, Kara Moser, Adrian Paskey, Jason Caravas, Benjamin Rambo-Martin, Shatavia Morrison, Christopher Gulvick, Scott Sammons, Yvette Unoarumhi, Darlene Wagner, Matthew Schmerer, Cyndi Clark, Patrick Campbell, Rob Case, Vikramsinha Ghorpade, Holly Houdeshell, Ola Kvalvaag, Dillon Nall, Ethan Sanders, Alec Vest, Shaun Westlund, Matthew Hardison, Clinton R. Paden, Duncan MacCannell |
| EPI_ISL_2201516 | Houston Methodist Hospital | Houston Methodist Hospital | Randall J. Olsen, Paul A. Christensen, S. Wesley Long, Sishir Subedi, Robert Olson, Marcus Nguyen, James J. Davis, Matthew Ojeda Saavedra, Prasanti Yerramilli, Layne Pruitt, Kristina Reppond, Madison N. Shyer, Jessica Cambric, Ryan Gadd, Ilya J. Finkelstein, Jimmy Gollihar, and James M. Musser |
| EPI_ISL_2201517 | Aegis Sciences Corporation | Centers for Disease Control and Prevention Division of Viral Diseases, Pathogen Discovery | Dakota Howard, Dhwani Batra, Peter W. Cook, Kara Moser, Adrian Paskey, Jason Caravas, Benjamin Rambo-Martin, Shatavia Morrison, Christopher Gulvick, Scott Sammons, Yvette Unoarumhi, Darlene Wagner, Matthew Schmerer, Cyndi Clark, Patrick Campbell, Rob Case, Vikramsinha Ghorpade, Holly Houdeshell, Ola Kvalvaag, Dillon Nall, Ethan Sanders, Alec Vest, Shaun Westlund, Matthew Hardison, Clinton R. Paden, Duncan MacCannell |
| EPI_ISL_2201518, EPI_ISL_2201520 | Houston Methodist Hospital | Houston Methodist Hospital | Randall J. Olsen, Paul A. Christensen, S. Wesley Long, Sishir Subedi, Robert Olson, Marcus Nguyen, James J. Davis, Matthew Ojeda Saavedra, Prasanti Yerramilli, Layne Pruitt, Kristina Reppond, Madison N. Shyer, Jessica Cambric, Ryan Gadd, Ilya J. Finkelstein, Jimmy Gollihar, and James M. Musser |
| EPI_ISL_2201521 | Aegis Sciences Corporation | Centers for Disease Control and Prevention Division of Viral Diseases, Pathogen Discovery | Dakota Howard, Dhwani Batra, Peter W. Cook, Kara Moser, Adrian Paskey, Jason Caravas, Benjamin Rambo-Martin, Shatavia Morrison, Christopher Gulvick, Scott Sammons, Yvette Unoarumhi, Darlene Wagner, Matthew Schmerer, Cyndi Clark, Patrick Campbell, Rob Case, Vikramsinha Ghorpade, Holly Houdeshell, Ola Kvalvaag, Dillon Nall, Ethan Sanders, Alec Vest, Shaun Westlund, Matthew Hardison, Clinton R. Paden, Duncan MacCannell |
| EPI_ISL_2201522 | Houston Methodist Hospital | Houston Methodist Hospital | Randall J. Olsen, Paul A. Christensen, S. Wesley Long, Sishir Subedi, Robert Olson, Marcus Nguyen, James J. Davis, Matthew Ojeda Saavedra, Prasanti Yerramilli, Layne Pruitt, Kristina Reppond, Madison N. Shyer, Jessica Cambric, Ryan Gadd, Ilya J. Finkelstein, Jimmy Gollihar, and James M. Musser |
| EPI_ISL_2201523 | Aegis Sciences Corporation | Centers for Disease Control and Prevention Division of Viral Diseases, Pathogen Discovery | Dakota Howard, Dhwani Batra, Peter W. Cook, Kara Moser, Adrian Paskey, Jason Caravas, Benjamin Rambo-Martin, Shatavia Morrison, Christopher Gulvick, Scott Sammons, Yvette Unoarumhi, Darlene Wagner, Matthew Schmerer, Cyndi Clark, Patrick Campbell, Rob Case, Vikramsinha Ghorpade, Holly Houdeshell, Ola Kvalvaag, Dillon Nall, Ethan Sanders, Alec Vest, Shaun Westlund, Matthew Hardison, Clinton R. Paden, Duncan MacCannell |
| EPI_ISL_2201524 | Houston Methodist Hospital | Houston Methodist Hospital | Randall J. Olsen, Paul A. Christensen, S. Wesley Long, Sishir Subedi, Robert Olson, Marcus Nguyen, James J. Davis, Matthew Ojeda Saavedra, Prasanti Yerramilli, Layne Pruitt, Kristina Reppond, Madison N. Shyer, Jessica Cambric, Ryan Gadd, Ilya J. Finkelstein, Jimmy Gollihar, and James M. Musser |
| EPI_ISL_2201525, EPI_ISL_2201526 | Aegis Sciences Corporation | Centers for Disease Control and Prevention Division of Viral Diseases, Pathogen | Dakota Howard, Dhwani Batra, Peter W. Cook, Kara Moser, Adrian Paskey, Jason Caravas, Benjamin Rambo-Martin, Shatavia Morrison, Christopher Gulvick, Scott Sammons, Yvette Unoarumhi, Darlene Wagner, Matthew Schmerer, Cyndi Clark, Patrick Campbell, Rob Case, Vikramsinha Ghorpade, Holly Houdeshell, Ola Kvalvaag, Dillon Nall, Ethan Sanders, Alec Vest, Shaun Westlund, Matthew Hardison, Clinton R. Paden, Duncan MacCannell |

| | | | |
|---|---|---|---|
| | | Discovery | |
| EPI_ISL_2201527 | Houston Methodist Hospital | Houston Methodist Hospital | Randall J. Olsen, Paul A. Christensen, S. Wesley Long, Sishir Subedi, Robert Olson, Marcus Nguyen, James J. Davis, Matthew Ojeda Saavedra, Prasanti Yerramilli, Layne Pruitt, Kristina Reppond, Madison N. Shyer, Jessica Cambric, Ryan Gadd, Ilya J. Finkelstein, Jimmy Gollihar, and James M. Musser |
| EPI_ISL_2201528 | Aegis Sciences Corporation | Centers for Disease Control and Prevention Division of Viral Diseases, Pathogen Discovery | Dakota Howard, Dhwani Batra, Peter W. Cook, Kara Moser, Adrian Paskey, Jason Caravas, Benjamin Rambo-Martin, Shatavia Morrison, Christopher Gulvick, Scott Sammons, Yvette Unoarumhi, Darlene Wagner, Matthew Schmerer, Cyndi Clark, Patrick Campbell, Rob Case, Vikramsinha Ghorpade, Holly Houdeshell, Ola Kvalvaag, Dillon Nall, Ethan Sanders, Alec Vest, Shaun Westlund, Matthew Hardison, Clinton R. Paden, Duncan MacCannell |
| EPI_ISL_2201529 | Houston Methodist Hospital | Houston Methodist Hospital | Randall J. Olsen, Paul A. Christensen, S. Wesley Long, Sishir Subedi, Robert Olson, Marcus Nguyen, James J. Davis, Matthew Ojeda Saavedra, Prasanti Yerramilli, Layne Pruitt, Kristina Reppond, Madison N. Shyer, Jessica Cambric, Ryan Gadd, Ilya J. Finkelstein, Jimmy Gollihar, and James M. Musser |
| EPI_ISL_2201530 | Aegis Sciences Corporation | Centers for Disease Control and Prevention Division of Viral Diseases, Pathogen Discovery | Dakota Howard, Dhwani Batra, Peter W. Cook, Kara Moser, Adrian Paskey, Jason Caravas, Benjamin Rambo-Martin, Shatavia Morrison, Christopher Gulvick, Scott Sammons, Yvette Unoarumhi, Darlene Wagner, Matthew Schmerer, Cyndi Clark, Patrick Campbell, Rob Case, Vikramsinha Ghorpade, Holly Houdeshell, Ola Kvalvaag, Dillon Nall, Ethan Sanders, Alec Vest, Shaun Westlund, Matthew Hardison, Clinton R. Paden, Duncan MacCannell |
| EPI_ISL_2201531 | Houston Methodist Hospital | Houston Methodist Hospital | Randall J. Olsen, Paul A. Christensen, S. Wesley Long, Sishir Subedi, Robert Olson, Marcus Nguyen, James J. Davis, Matthew Ojeda Saavedra, Prasanti Yerramilli, Layne Pruitt, Kristina Reppond, Madison N. Shyer, Jessica Cambric, Ryan Gadd, Ilya J. Finkelstein, Jimmy Gollihar, and James M. Musser |
| EPI_ISL_2201532 | Aegis Sciences Corporation | Centers for Disease Control and Prevention Division of Viral Diseases, Pathogen Discovery | Dakota Howard, Dhwani Batra, Peter W. Cook, Kara Moser, Adrian Paskey, Jason Caravas, Benjamin Rambo-Martin, Shatavia Morrison, Christopher Gulvick, Scott Sammons, Yvette Unoarumhi, Darlene Wagner, Matthew Schmerer, Cyndi Clark, Patrick Campbell, Rob Case, Vikramsinha Ghorpade, Holly Houdeshell, Ola Kvalvaag, Dillon Nall, Ethan Sanders, Alec Vest, Shaun Westlund, Matthew Hardison, Clinton R. Paden, Duncan MacCannell |
| EPI_ISL_2201533 | Houston Methodist Hospital | Houston Methodist Hospital | Randall J. Olsen, Paul A. Christensen, S. Wesley Long, Sishir Subedi, Robert Olson, Marcus Nguyen, James J. Davis, Matthew Ojeda Saavedra, Prasanti Yerramilli, Layne Pruitt, Kristina Reppond, Madison N. Shyer, Jessica Cambric, Ryan Gadd, Ilya J. Finkelstein, Jimmy Gollihar, and James M. Musser |
| EPI_ISL_2201534 | Aegis Sciences Corporation | Centers for Disease Control and Prevention Division of Viral Diseases, Pathogen Discovery | Dakota Howard, Dhwani Batra, Peter W. Cook, Kara Moser, Adrian Paskey, Jason Caravas, Benjamin Rambo-Martin, Shatavia Morrison, Christopher Gulvick, Scott Sammons, Yvette Unoarumhi, Darlene Wagner, Matthew Schmerer, Cyndi Clark, Patrick Campbell, Rob Case, Vikramsinha Ghorpade, Holly Houdeshell, Ola Kvalvaag, Dillon Nall, Ethan Sanders, Alec Vest, Shaun Westlund, Matthew Hardison, Clinton R. Paden, Duncan MacCannell |
| EPI_ISL_2201535 | Houston Methodist Hospital | Houston Methodist Hospital | Randall J. Olsen, Paul A. Christensen, S. Wesley Long, Sishir Subedi, Robert Olson, Marcus Nguyen, James J. Davis, Matthew Ojeda Saavedra, Prasanti Yerramilli, Layne Pruitt, Kristina Reppond, Madison N. Shyer, Jessica Cambric, Ryan Gadd, Ilya J. Finkelstein, Jimmy Gollihar, and James M. Musser |
| EPI_ISL_2201536 | Aegis Sciences Corporation | Centers for Disease Control and Prevention Division of Viral Diseases, Pathogen Discovery | Dakota Howard, Dhwani Batra, Peter W. Cook, Kara Moser, Adrian Paskey, Jason Caravas, Benjamin Rambo-Martin, Shatavia Morrison, Christopher Gulvick, Scott Sammons, Yvette Unoarumhi, Darlene Wagner, Matthew Schmerer, Cyndi Clark, Patrick Campbell, Rob Case, Vikramsinha Ghorpade, Holly Houdeshell, Ola Kvalvaag, Dillon Nall, Ethan Sanders, Alec Vest, Shaun Westlund, Matthew Hardison, Clinton R. Paden, Duncan MacCannell |
| EPI_ISL_2201537 | Houston Methodist Hospital | Houston Methodist Hospital | Randall J. Olsen, Paul A. Christensen, S. Wesley Long, Sishir Subedi, Robert Olson, Marcus Nguyen, James J. Davis, Matthew Ojeda Saavedra, Prasanti Yerramilli, Layne Pruitt, Kristina Reppond, Madison N. Shyer, Jessica Cambric, Ryan Gadd, Ilya J. Finkelstein, Jimmy Gollihar, and James M. Musser |
| EPI_ISL_2201538 | Aegis Sciences Corporation | Centers for Disease Control and Prevention Division of Viral Diseases, Pathogen Discovery | Dakota Howard, Dhwani Batra, Peter W. Cook, Kara Moser, Adrian Paskey, Jason Caravas, Benjamin Rambo-Martin, Shatavia Morrison, Christopher Gulvick, Scott Sammons, Yvette Unoarumhi, Darlene Wagner, Matthew Schmerer, Cyndi Clark, Patrick Campbell, Rob Case, Vikramsinha Ghorpade, Holly Houdeshell, Ola Kvalvaag, Dillon Nall, Ethan Sanders, Alec Vest, Shaun Westlund, Matthew Hardison, Clinton R. Paden, Duncan MacCannell |
| EPI_ISL_2201540 | Houston Methodist Hospital | Houston Methodist Hospital | Randall J. Olsen, Paul A. Christensen, S. Wesley Long, Sishir Subedi, Robert Olson, Marcus Nguyen, James J. Davis, Matthew Ojeda Saavedra, Prasanti Yerramilli, Layne Pruitt, Kristina Reppond, Madison N. Shyer, Jessica Cambric, Ryan Gadd, Ilya J. Finkelstein, Jimmy Gollihar, and James M. Musser |
| EPI_ISL_2201541 | Aegis Sciences Corporation | Centers for Disease Control and Prevention Division of Viral Diseases, Pathogen Discovery | Dakota Howard, Dhwani Batra, Peter W. Cook, Kara Moser, Adrian Paskey, Jason Caravas, Benjamin Rambo-Martin, Shatavia Morrison, Christopher Gulvick, Scott Sammons, Yvette Unoarumhi, Darlene Wagner, Matthew Schmerer, Cyndi Clark, Patrick Campbell, Rob Case, Vikramsinha Ghorpade, Holly Houdeshell, Ola Kvalvaag, Dillon Nall, Ethan Sanders, Alec Vest, Shaun Westlund, Matthew Hardison, Clinton R. Paden, Duncan MacCannell |
| EPI_ISL_2201542 | Houston Methodist Hospital | Houston Methodist Hospital | Randall J. Olsen, Paul A. Christensen, S. Wesley Long, Sishir Subedi, Robert Olson, Marcus Nguyen, James J. Davis, Matthew Ojeda Saavedra, Prasanti Yerramilli, Layne Pruitt, Kristina Reppond, Madison N. Shyer, Jessica Cambric, Ryan Gadd, Ilya J. Finkelstein, Jimmy Gollihar, and James M. Musser |
| EPI_ISL_2201543 | Aegis Sciences Corporation | Centers for Disease Control and Prevention Division of Viral Diseases, Pathogen Discovery | Dakota Howard, Dhwani Batra, Peter W. Cook, Kara Moser, Adrian Paskey, Jason Caravas, Benjamin Rambo-Martin, Shatavia Morrison, Christopher Gulvick, Scott Sammons, Yvette Unoarumhi, Darlene Wagner, Matthew Schmerer, Cyndi Clark, Patrick Campbell, Rob Case, Vikramsinha Ghorpade, Holly Houdeshell, Ola Kvalvaag, Dillon Nall, Ethan Sanders, Alec Vest, Shaun Westlund, Matthew Hardison, Clinton R. Paden, Duncan MacCannell |
| EPI_ISL_2201544 | Houston Methodist Hospital | Houston Methodist Hospital | Randall J. Olsen, Paul A. Christensen, S. Wesley Long, Sishir Subedi, Robert Olson, Marcus Nguyen, James J. Davis, Matthew Ojeda Saavedra, Prasanti Yerramilli, Layne Pruitt, Kristina Reppond, Madison N. Shyer, Jessica Cambric, Ryan Gadd, Ilya J. Finkelstein, Jimmy Gollihar, and James M. Musser |
| EPI_ISL_2201545, EPI_ISL_2201546, EPI_ISL_2201547 | Aegis Sciences Corporation | Centers for Disease Control and Prevention Division of Viral Diseases, Pathogen Discovery | Dakota Howard, Dhwani Batra, Peter W. Cook, Kara Moser, Adrian Paskey, Jason Caravas, Benjamin Rambo-Martin, Shatavia Morrison, Christopher Gulvick, Scott Sammons, Yvette Unoarumhi, Darlene Wagner, Matthew Schmerer, Cyndi Clark, Patrick Campbell, Rob Case, Vikramsinha Ghorpade, Holly Houdeshell, Ola Kvalvaag, Dillon Nall, Ethan Sanders, Alec Vest, Shaun Westlund, Matthew Hardison, Clinton R. Paden, Duncan MacCannell |
| EPI_ISL_2201548 | Houston Methodist Hospital | Houston Methodist Hospital | Randall J. Olsen, Paul A. Christensen, S. Wesley Long, Sishir Subedi, Robert Olson, Marcus Nguyen, James J. Davis, Matthew Ojeda Saavedra, Prasanti Yerramilli, Layne Pruitt, Kristina Reppond, Madison N. Shyer, Jessica Cambric, Ryan Gadd, Ilya J. Finkelstein, Jimmy Gollihar, and James M. Musser |
| EPI_ISL_2201549 | Aegis Sciences Corporation | Centers for Disease Control and Prevention Division of Viral Diseases, Pathogen Discovery | Dakota Howard, Dhwani Batra, Peter W. Cook, Kara Moser, Adrian Paskey, Jason Caravas, Benjamin Rambo-Martin, Shatavia Morrison, Christopher Gulvick, Scott Sammons, Yvette Unoarumhi, Darlene Wagner, Matthew Schmerer, Cyndi Clark, Patrick Campbell, Rob Case, Vikramsinha Ghorpade, Holly Houdeshell, Ola Kvalvaag, Dillon Nall, Ethan Sanders, Alec Vest, Shaun Westlund, Matthew Hardison, Clinton R. Paden, Duncan MacCannell |
| EPI_ISL_2201550 | Houston Methodist Hospital | Houston Methodist Hospital | Randall J. Olsen, Paul A. Christensen, S. Wesley Long, Sishir Subedi, Robert Olson, Marcus Nguyen, James J. Davis, Matthew Ojeda Saavedra, Prasanti Yerramilli, Layne Pruitt, Kristina Reppond, Madison N. Shyer, Jessica Cambric, Ryan Gadd, Ilya J. Finkelstein, Jimmy Gollihar, and James M. Musser |
| EPI_ISL_2201551 | Aegis Sciences Corporation | Centers for Disease Control and Prevention Division of Viral Diseases, Pathogen Discovery | Dakota Howard, Dhwani Batra, Peter W. Cook, Kara Moser, Adrian Paskey, Jason Caravas, Benjamin Rambo-Martin, Shatavia Morrison, Christopher Gulvick, Scott Sammons, Yvette Unoarumhi, Darlene Wagner, Matthew Schmerer, Cyndi Clark, Patrick Campbell, Rob Case, Vikramsinha Ghorpade, Holly Houdeshell, Ola Kvalvaag, Dillon Nall, Ethan Sanders, Alec Vest, Shaun Westlund, Matthew Hardison, Clinton R. Paden, Duncan MacCannell |
| EPI_ISL_2201552 | Houston Methodist Hospital | Houston Methodist Hospital | Randall J. Olsen, Paul A. Christensen, S. Wesley Long, Sishir Subedi, Robert Olson, Marcus Nguyen, James J. Davis, Matthew Ojeda Saavedra, Prasanti Yerramilli, Layne Pruitt, Kristina Reppond, Madison N. Shyer, Jessica Cambric, Ryan Gadd, Ilya J. Finkelstein, Jimmy Gollihar, and James M. Musser |
| EPI_ISL_2201553 | Aegis Sciences Corporation | Centers for Disease Control and Prevention Division of Viral Diseases, Pathogen Discovery | Dakota Howard, Dhwani Batra, Peter W. Cook, Kara Moser, Adrian Paskey, Jason Caravas, Benjamin Rambo-Martin, Shatavia Morrison, Christopher Gulvick, Scott Sammons, Yvette Unoarumhi, Darlene Wagner, Matthew Schmerer, Cyndi Clark, Patrick Campbell, Rob Case, Vikramsinha Ghorpade, Holly Houdeshell, Ola Kvalvaag, Dillon Nall, Ethan Sanders, Alec Vest, Shaun Westlund, Matthew Hardison, Clinton R. Paden, Duncan MacCannell |
| EPI_ISL_2201554 | Houston Methodist Hospital | Houston Methodist Hospital | Randall J. Olsen, Paul A. Christensen, S. Wesley Long, Sishir Subedi, Robert Olson, Marcus Nguyen, James J. Davis, Matthew Ojeda Saavedra, Prasanti Yerramilli, Layne Pruitt, Kristina Reppond, Madison N. Shyer, Jessica Cambric, Ryan Gadd, Ilya J. Finkelstein, Jimmy Gollihar, and James M. Musser |
| EPI_ISL_2201555 | Aegis Sciences Corporation | Centers for Disease Control and Prevention Division of Viral Diseases, Pathogen Discovery | Dakota Howard, Dhwani Batra, Peter W. Cook, Kara Moser, Adrian Paskey, Jason Caravas, Benjamin Rambo-Martin, Shatavia Morrison, Christopher Gulvick, Scott Sammons, Yvette Unoarumhi, Darlene Wagner, Matthew Schmerer, Cyndi Clark, Patrick Campbell, Rob Case, Vikramsinha Ghorpade, Holly Houdeshell, Ola Kvalvaag, Dillon Nall, Ethan Sanders, Alec Vest, Shaun Westlund, Matthew Hardison, Clinton R. Paden, Duncan MacCannell |
| EPI_ISL_2201556 | Houston Methodist Hospital | Houston Methodist Hospital | Randall J. Olsen, Paul A. Christensen, S. Wesley Long, Sishir Subedi, Robert Olson, Marcus Nguyen, James J. Davis, Matthew Ojeda Saavedra, Prasanti Yerramilli, Layne Pruitt, Kristina Reppond, Madison N. Shyer, Jessica Cambric, Ryan Gadd, Ilya J. Finkelstein, Jimmy Gollihar, and James M. Musser |
| EPI_ISL_2201557 | Aegis Sciences Corporation | Centers for Disease Control and Prevention Division of Viral Diseases, Pathogen Discovery | Dakota Howard, Dhwani Batra, Peter W. Cook, Kara Moser, Adrian Paskey, Jason Caravas, Benjamin Rambo-Martin, Shatavia Morrison, Christopher Gulvick, Scott Sammons, Yvette Unoarumhi, Darlene Wagner, Matthew Schmerer, Cyndi Clark, Patrick Campbell, Rob Case, Vikramsinha Ghorpade, Holly Houdeshell, Ola Kvalvaag, Dillon Nall, Ethan Sanders, Alec Vest, Shaun Westlund, Matthew Hardison, Clinton R. Paden, Duncan MacCannell |
| EPI_ISL_2201558 | Houston Methodist Hospital | Houston Methodist Hospital | Randall J. Olsen, Paul A. Christensen, S. Wesley Long, Sishir Subedi, Robert Olson, Marcus Nguyen, James J. Davis, Matthew Ojeda Saavedra, Prasanti Yerramilli, Layne Pruitt, Kristina Reppond, Madison N. Shyer, Jessica Cambric, Ryan Gadd, Ilya J. Finkelstein, Jimmy Gollihar, and James M. Musser |
| EPI_ISL_2201559, EPI_ISL_2201560 | Aegis Sciences Corporation | Centers for Disease Control and Prevention Division of Viral Diseases, Pathogen Discovery | Dakota Howard, Dhwani Batra, Peter W. Cook, Kara Moser, Adrian Paskey, Jason Caravas, Benjamin Rambo-Martin, Shatavia Morrison, Christopher Gulvick, Scott Sammons, Yvette Unoarumhi, Darlene Wagner, Matthew Schmerer, Cyndi Clark, Patrick Campbell, Rob Case, Vikramsinha Ghorpade, Holly Houdeshell, Ola Kvalvaag, Dillon Nall, Ethan Sanders, Alec Vest, Shaun Westlund, Matthew Hardison, Clinton R. Paden, Duncan MacCannell |
| EPI_ISL_2201561 | Houston Methodist Hospital | Houston Methodist Hospital | Randall J. Olsen, Paul A. Christensen, S. Wesley Long, Sishir Subedi, Robert Olson, Marcus Nguyen, James J. Davis, Matthew Ojeda Saavedra, Prasanti Yerramilli, Layne Pruitt, Kristina Reppond, Madison N. Shyer, Jessica Cambric, Ryan Gadd, Ilya J. Finkelstein, Jimmy Gollihar, and James M. Musser |
| EPI_ISL_2201562 | Aegis Sciences Corporation | Centers for Disease Control and Prevention Division of Viral Diseases, Pathogen Discovery | Dakota Howard, Dhwani Batra, Peter W. Cook, Kara Moser, Adrian Paskey, Jason Caravas, Benjamin Rambo-Martin, Shatavia Morrison, Christopher Gulvick, Scott Sammons, Yvette Unoarumhi, Darlene Wagner, Matthew Schmerer, Cyndi Clark, Patrick Campbell, Rob Case, Vikramsinha Ghorpade, Holly Houdeshell, Ola Kvalvaag, Dillon Nall, Ethan Sanders, Alec Vest, Shaun Westlund, Matthew Hardison, Clinton R. Paden, Duncan MacCannell |
| EPI_ISL_2201563 | Houston Methodist Hospital | Houston Methodist Hospital | Randall J. Olsen, Paul A. Christensen, S. Wesley Long, Sishir Subedi, Robert Olson, Marcus Nguyen, James J. Davis, Matthew Ojeda Saavedra, Prasanti Yerramilli, Layne Pruitt, Kristina Reppond, Madison N. Shyer, Jessica Cambric, Ryan Gadd, Ilya J. Finkelstein, Jimmy Gollihar, and James M. Musser |
| EPI_ISL_2201564 | Aegis Sciences Corporation | Centers for Disease Control and Prevention Division of Viral Diseases, Pathogen Discovery | Dakota Howard, Dhwani Batra, Peter W. Cook, Kara Moser, Adrian Paskey, Jason Caravas, Benjamin Rambo-Martin, Shatavia Morrison, Christopher Gulvick, Scott Sammons, Yvette Unoarumhi, Darlene Wagner, Matthew Schmerer, Cyndi Clark, Patrick Campbell, Rob Case, Vikramsinha Ghorpade, Holly Houdeshell, Ola Kvalvaag, Dillon Nall, Ethan Sanders, Alec Vest, Shaun Westlund, Matthew Hardison, Clinton R. Paden, Duncan MacCannell |
| EPI_ISL_2201565 | Houston Methodist Hospital | Houston Methodist Hospital | Randall J. Olsen, Paul A. Christensen, S. Wesley Long, Sishir Subedi, Robert Olson, Marcus Nguyen, James J. Davis, Matthew Ojeda Saavedra, Prasanti Yerramilli, Layne Pruitt, Kristina Reppond, Madison N. Shyer, Jessica Cambric, Ryan Gadd, Ilya J. Finkelstein, Jimmy Gollihar, and James M. Musser |
| EPI_ISL_2201566, EPI_ISL_2201567 | Aegis Sciences Corporation | Centers for Disease Control and Prevention Division of Viral Diseases, Pathogen Discovery | Dakota Howard, Dhwani Batra, Peter W. Cook, Kara Moser, Adrian Paskey, Jason Caravas, Benjamin Rambo-Martin, Shatavia Morrison, Christopher Gulvick, Scott Sammons, Yvette Unoarumhi, Darlene Wagner, Matthew Schmerer, Cyndi Clark, Patrick Campbell, Rob Case, Vikramsinha Ghorpade, Holly Houdeshell, Ola Kvalvaag, Dillon Nall, Ethan Sanders, Alec Vest, Shaun Westlund, Matthew Hardison, Clinton R. Paden, Duncan MacCannell |
| EPI_ISL_2201568 | Houston Methodist Hospital | Houston Methodist Hospital | Randall J. Olsen, Paul A. Christensen, S. Wesley Long, Sishir Subedi, Robert Olson, Marcus Nguyen, James J. Davis, Matthew Ojeda Saavedra, Prasanti Yerramilli, Layne Pruitt, Kristina Reppond, Madison N. Shyer, Jessica Cambric, Ryan Gadd, Ilya J. Finkelstein, Jimmy Gollihar, and James M. Musser |

| | | | |
|---|---|---|---|
| EPI_ISL_2201569 | Aegis Sciences Corporation | Centers for Disease Control and Prevention Division of Viral Diseases, Pathogen Discovery | Dakota Howard, Dhwani Batra, Peter W. Cook, Kara Moser, Adrian Paskey, Jason Caravas, Benjamin Rambo-Martin, Shatavia Morrison, Christopher Gulvick, Scott Sammons, Yvette Unoarumhi, Darlene Wagner, Matthew Schmerer, Cyndi Clark, Patrick Campbell, Rob Case, Vikramsinha Ghorpade, Holly Houdeshell, Ola Kvalvaag, Dillon Nall, Ethan Sanders, Alec Vest, Shaun Westlund, Matthew Hardison, Clinton R. Paden, Duncan MacCannell |
| EPI_ISL_2201570 | Houston Methodist Hospital | Houston Methodist Hospital | Randall J. Olsen, Paul A. Christensen, S. Wesley Long, Sishir Subedi, Robert Olson, Marcus Nguyen, James J. Davis, Matthew Ojeda Saavedra, Prasanti Yerramilli, Layne Pruitt, Kristina Reppond, Madison N. Shyer, Jessica Cambric, Ryan Gadd, Ilya J. Finkelstein, Jimmy Gollihar, and James M. Musser |
| EPI_ISL_2201571 | Aegis Sciences Corporation | Centers for Disease Control and Prevention Division of Viral Diseases, Pathogen Discovery | Dakota Howard, Dhwani Batra, Peter W. Cook, Kara Moser, Adrian Paskey, Jason Caravas, Benjamin Rambo-Martin, Shatavia Morrison, Christopher Gulvick, Scott Sammons, Yvette Unoarumhi, Darlene Wagner, Matthew Schmerer, Cyndi Clark, Patrick Campbell, Rob Case, Vikramsinha Ghorpade, Holly Houdeshell, Ola Kvalvaag, Dillon Nall, Ethan Sanders, Alec Vest, Shaun Westlund, Matthew Hardison, Clinton R. Paden, Duncan MacCannell |
| EPI_ISL_2201572 | Houston Methodist Hospital | Houston Methodist Hospital | Randall J. Olsen, Paul A. Christensen, S. Wesley Long, Sishir Subedi, Robert Olson, Marcus Nguyen, James J. Davis, Matthew Ojeda Saavedra, Prasanti Yerramilli, Layne Pruitt, Kristina Reppond, Madison N. Shyer, Jessica Cambric, Ryan Gadd, Ilya J. Finkelstein, Jimmy Gollihar, and James M. Musser |
| EPI_ISL_2201573, EPI_ISL_2201574 | Aegis Sciences Corporation | Centers for Disease Control and Prevention Division of Viral Diseases, Pathogen Discovery | Dakota Howard, Dhwani Batra, Peter W. Cook, Kara Moser, Adrian Paskey, Jason Caravas, Benjamin Rambo-Martin, Shatavia Morrison, Christopher Gulvick, Scott Sammons, Yvette Unoarumhi, Darlene Wagner, Matthew Schmerer, Cyndi Clark, Patrick Campbell, Rob Case, Vikramsinha Ghorpade, Holly Houdeshell, Ola Kvalvaag, Dillon Nall, Ethan Sanders, Alec Vest, Shaun Westlund, Matthew Hardison, Clinton R. Paden, Duncan MacCannell |
| EPI_ISL_2201575 | Houston Methodist Hospital | Houston Methodist Hospital | Randall J. Olsen, Paul A. Christensen, S. Wesley Long, Sishir Subedi, Robert Olson, Marcus Nguyen, James J. Davis, Matthew Ojeda Saavedra, Prasanti Yerramilli, Layne Pruitt, Kristina Reppond, Madison N. Shyer, Jessica Cambric, Ryan Gadd, Ilya J. Finkelstein, Jimmy Gollihar, and James M. Musser |
| EPI_ISL_2201576 | Aegis Sciences Corporation | Centers for Disease Control and Prevention Division of Viral Diseases, Pathogen Discovery | Dakota Howard, Dhwani Batra, Peter W. Cook, Kara Moser, Adrian Paskey, Jason Caravas, Benjamin Rambo-Martin, Shatavia Morrison, Christopher Gulvick, Scott Sammons, Yvette Unoarumhi, Darlene Wagner, Matthew Schmerer, Cyndi Clark, Patrick Campbell, Rob Case, Vikramsinha Ghorpade, Holly Houdeshell, Ola Kvalvaag, Dillon Nall, Ethan Sanders, Alec Vest, Shaun Westlund, Matthew Hardison, Clinton R. Paden, Duncan MacCannell |
| EPI_ISL_2201577 | Houston Methodist Hospital | Houston Methodist Hospital | Randall J. Olsen, Paul A. Christensen, S. Wesley Long, Sishir Subedi, Robert Olson, Marcus Nguyen, James J. Davis, Matthew Ojeda Saavedra, Prasanti Yerramilli, Layne Pruitt, Kristina Reppond, Madison N. Shyer, Jessica Cambric, Ryan Gadd, Ilya J. Finkelstein, Jimmy Gollihar, and James M. Musser |
| EPI_ISL_2201578, EPI_ISL_2201579 | Aegis Sciences Corporation | Centers for Disease Control and Prevention Division of Viral Diseases, Pathogen Discovery | Dakota Howard, Dhwani Batra, Peter W. Cook, Kara Moser, Adrian Paskey, Jason Caravas, Benjamin Rambo-Martin, Shatavia Morrison, Christopher Gulvick, Scott Sammons, Yvette Unoarumhi, Darlene Wagner, Matthew Schmerer, Cyndi Clark, Patrick Campbell, Rob Case, Vikramsinha Ghorpade, Holly Houdeshell, Ola Kvalvaag, Dillon Nall, Ethan Sanders, Alec Vest, Shaun Westlund, Matthew Hardison, Clinton R. Paden, Duncan MacCannell |
| EPI_ISL_2201580 | Houston Methodist Hospital | Houston Methodist Hospital | Randall J. Olsen, Paul A. Christensen, S. Wesley Long, Sishir Subedi, Robert Olson, Marcus Nguyen, James J. Davis, Matthew Ojeda Saavedra, Prasanti Yerramilli, Layne Pruitt, Kristina Reppond, Madison N. Shyer, Jessica Cambric, Ryan Gadd, Ilya J. Finkelstein, Jimmy Gollihar, and James M. Musser |
| EPI_ISL_2201581, EPI_ISL_2201582 | Aegis Sciences Corporation | Centers for Disease Control and Prevention Division of Viral Diseases, Pathogen Discovery | Dakota Howard, Dhwani Batra, Peter W. Cook, Kara Moser, Adrian Paskey, Jason Caravas, Benjamin Rambo-Martin, Shatavia Morrison, Christopher Gulvick, Scott Sammons, Yvette Unoarumhi, Darlene Wagner, Matthew Schmerer, Cyndi Clark, Patrick Campbell, Rob Case, Vikramsinha Ghorpade, Holly Houdeshell, Ola Kvalvaag, Dillon Nall, Ethan Sanders, Alec Vest, Shaun Westlund, Matthew Hardison, Clinton R. Paden, Duncan MacCannell |
| EPI_ISL_2201583 | Houston Methodist Hospital | Houston Methodist Hospital | Randall J. Olsen, Paul A. Christensen, S. Wesley Long, Sishir Subedi, Robert Olson, Marcus Nguyen, James J. Davis, Matthew Ojeda Saavedra, Prasanti Yerramilli, Layne Pruitt, Kristina Reppond, Madison N. Shyer, Jessica Cambric, Ryan Gadd, Ilya J. Finkelstein, Jimmy Gollihar, and James M. Musser |
| EPI_ISL_2201584 | Utah Public Health Laboratory | Utah Public Health Laboratory | Erin L. Young, Kelly F. Oakeson, Tara Gallagher |
| EPI_ISL_2201585 | Aegis Sciences Corporation | Centers for Disease Control and Prevention Division of Viral Diseases, Pathogen Discovery | Dakota Howard, Dhwani Batra, Peter W. Cook, Kara Moser, Adrian Paskey, Jason Caravas, Benjamin Rambo-Martin, Shatavia Morrison, Christopher Gulvick, Scott Sammons, Yvette Unoarumhi, Darlene Wagner, Matthew Schmerer, Cyndi Clark, Patrick Campbell, Rob Case, Vikramsinha Ghorpade, Holly Houdeshell, Ola Kvalvaag, Dillon Nall, Ethan Sanders, Alec Vest, Shaun Westlund, Matthew Hardison, Clinton R. Paden, Duncan MacCannell |
| EPI_ISL_2201587 | Houston Methodist Hospital | Houston Methodist Hospital | Randall J. Olsen, Paul A. Christensen, S. Wesley Long, Sishir Subedi, Robert Olson, Marcus Nguyen, James J. Davis, Matthew Ojeda Saavedra, Prasanti Yerramilli, Layne Pruitt, Kristina Reppond, Madison N. Shyer, Jessica Cambric, Ryan Gadd, Ilya J. Finkelstein, Jimmy Gollihar, and James M. Musser |
| EPI_ISL_2201588 | Aegis Sciences Corporation | Centers for Disease Control and Prevention Division of Viral Diseases, Pathogen Discovery | Dakota Howard, Dhwani Batra, Peter W. Cook, Kara Moser, Adrian Paskey, Jason Caravas, Benjamin Rambo-Martin, Shatavia Morrison, Christopher Gulvick, Scott Sammons, Yvette Unoarumhi, Darlene Wagner, Matthew Schmerer, Cyndi Clark, Patrick Campbell, Rob Case, Vikramsinha Ghorpade, Holly Houdeshell, Ola Kvalvaag, Dillon Nall, Ethan Sanders, Alec Vest, Shaun Westlund, Matthew Hardison, Clinton R. Paden, Duncan MacCannell |
| EPI_ISL_2201589, EPI_ISL_2201590 | Utah Public Health Laboratory | Utah Public Health Laboratory | Erin L. Young, Kelly F. Oakeson, Tara Gallagher |
| EPI_ISL_2201591 | Houston Methodist Hospital | Houston Methodist Hospital | Randall J. Olsen, Paul A. Christensen, S. Wesley Long, Sishir Subedi, Robert Olson, Marcus Nguyen, James J. Davis, Matthew Ojeda Saavedra, Prasanti Yerramilli, Layne Pruitt, Kristina Reppond, Madison N. Shyer, Jessica Cambric, Ryan Gadd, Ilya J. Finkelstein, Jimmy Gollihar, and James M. Musser |
| EPI_ISL_2201592 | Aegis Sciences Corporation | Centers for Disease Control and Prevention Division of Viral Diseases, Pathogen Discovery | Dakota Howard, Dhwani Batra, Peter W. Cook, Kara Moser, Adrian Paskey, Jason Caravas, Benjamin Rambo-Martin, Shatavia Morrison, Christopher Gulvick, Scott Sammons, Yvette Unoarumhi, Darlene Wagner, Matthew Schmerer, Cyndi Clark, Patrick Campbell, Rob Case, Vikramsinha Ghorpade, Holly Houdeshell, Ola Kvalvaag, Dillon Nall, Ethan Sanders, Alec Vest, Shaun Westlund, Matthew Hardison, Clinton R. Paden, Duncan MacCannell |
| EPI_ISL_2201593, EPI_ISL_2201594 | Utah Public Health Laboratory | Utah Public Health Laboratory | Erin L. Young, Kelly F. Oakeson, Tara Gallagher |
| EPI_ISL_2201595 | Aegis Sciences Corporation | Centers for Disease Control and Prevention Division of Viral Diseases, Pathogen Discovery | Dakota Howard, Dhwani Batra, Peter W. Cook, Kara Moser, Adrian Paskey, Jason Caravas, Benjamin Rambo-Martin, Shatavia Morrison, Christopher Gulvick, Scott Sammons, Yvette Unoarumhi, Darlene Wagner, Matthew Schmerer, Cyndi Clark, Patrick Campbell, Rob Case, Vikramsinha Ghorpade, Holly Houdeshell, Ola Kvalvaag, Dillon Nall, Ethan Sanders, Alec Vest, Shaun Westlund, Matthew Hardison, Clinton R. Paden, Duncan MacCannell |
| EPI_ISL_2201596 | Utah Public Health Laboratory | Utah Public Health Laboratory | Erin L. Young, Kelly F. Oakeson, Tara Gallagher |
| EPI_ISL_2201597 | Houston Methodist Hospital | Houston Methodist Hospital | Randall J. Olsen, Paul A. Christensen, S. Wesley Long, Sishir Subedi, Robert Olson, Marcus Nguyen, James J. Davis, Matthew Ojeda Saavedra, Prasanti Yerramilli, Layne Pruitt, Kristina Reppond, Madison N. Shyer, Jessica Cambric, Ryan Gadd, Ilya J. Finkelstein, Jimmy Gollihar, and James M. Musser |
| EPI_ISL_2201599 | Aegis Sciences Corporation | Centers for Disease Control and Prevention Division of Viral Diseases, Pathogen Discovery | Dakota Howard, Dhwani Batra, Peter W. Cook, Kara Moser, Adrian Paskey, Jason Caravas, Benjamin Rambo-Martin, Shatavia Morrison, Christopher Gulvick, Scott Sammons, Yvette Unoarumhi, Darlene Wagner, Matthew Schmerer, Cyndi Clark, Patrick Campbell, Rob Case, Vikramsinha Ghorpade, Holly Houdeshell, Ola Kvalvaag, Dillon Nall, Ethan Sanders, Alec Vest, Shaun Westlund, Matthew Hardison, Clinton R. Paden, Duncan MacCannell |
| EPI_ISL_2201600 | Utah Public Health Laboratory | Utah Public Health Laboratory | Erin L. Young, Kelly F. Oakeson, Tara Gallagher |
| EPI_ISL_2201601 | Aegis Sciences Corporation | Centers for Disease Control and Prevention Division of Viral Diseases, Pathogen Discovery | Dakota Howard, Dhwani Batra, Peter W. Cook, Kara Moser, Adrian Paskey, Jason Caravas, Benjamin Rambo-Martin, Shatavia Morrison, Christopher Gulvick, Scott Sammons, Yvette Unoarumhi, Darlene Wagner, Matthew Schmerer, Cyndi Clark, Patrick Campbell, Rob Case, Vikramsinha Ghorpade, Holly Houdeshell, Ola Kvalvaag, Dillon Nall, Ethan Sanders, Alec Vest, Shaun Westlund, Matthew Hardison, Clinton R. Paden, Duncan MacCannell |
| EPI_ISL_2201602, EPI_ISL_2201603 | Utah Public Health Laboratory | Utah Public Health Laboratory | Erin L. Young, Kelly F. Oakeson, Tara Gallagher |
| EPI_ISL_2201604 | Houston Methodist Hospital | Houston Methodist Hospital | Randall J. Olsen, Paul A. Christensen, S. Wesley Long, Sishir Subedi, Robert Olson, Marcus Nguyen, James J. Davis, Matthew Ojeda Saavedra, Prasanti Yerramilli, Layne Pruitt, Kristina Reppond, Madison N. Shyer, Jessica Cambric, Ryan Gadd, Ilya J. Finkelstein, Jimmy Gollihar, and James M. Musser |
| EPI_ISL_2201605 | Aegis Sciences Corporation | Centers for Disease Control and Prevention Division of Viral Diseases, Pathogen Discovery | Dakota Howard, Dhwani Batra, Peter W. Cook, Kara Moser, Adrian Paskey, Jason Caravas, Benjamin Rambo-Martin, Shatavia Morrison, Christopher Gulvick, Scott Sammons, Yvette Unoarumhi, Darlene Wagner, Matthew Schmerer, Cyndi Clark, Patrick Campbell, Rob Case, Vikramsinha Ghorpade, Holly Houdeshell, Ola Kvalvaag, Dillon Nall, Ethan Sanders, Alec Vest, Shaun Westlund, Matthew Hardison, Clinton R. Paden, Duncan MacCannell |
| EPI_ISL_2201606, EPI_ISL_2201607 | Utah Public Health Laboratory | Utah Public Health Laboratory | Erin L. Young, Kelly F. Oakeson, Tara Gallagher |
| EPI_ISL_2201608 | Houston Methodist Hospital | Houston Methodist Hospital | Randall J. Olsen, Paul A. Christensen, S. Wesley Long, Sishir Subedi, Robert Olson, Marcus Nguyen, James J. Davis, Matthew Ojeda Saavedra, Prasanti Yerramilli, Layne Pruitt, Kristina Reppond, Madison N. Shyer, Jessica Cambric, Ryan Gadd, Ilya J. Finkelstein, Jimmy Gollihar, and James M. Musser |
| EPI_ISL_2201609 | Aegis Sciences Corporation | Centers for Disease Control and Prevention Division of Viral Diseases, Pathogen Discovery | Dakota Howard, Dhwani Batra, Peter W. Cook, Kara Moser, Adrian Paskey, Jason Caravas, Benjamin Rambo-Martin, Shatavia Morrison, Christopher Gulvick, Scott Sammons, Yvette Unoarumhi, Darlene Wagner, Matthew Schmerer, Cyndi Clark, Patrick Campbell, Rob Case, Vikramsinha Ghorpade, Holly Houdeshell, Ola Kvalvaag, Dillon Nall, Ethan Sanders, Alec Vest, Shaun Westlund, Matthew Hardison, Clinton R. Paden, Duncan MacCannell |
| EPI_ISL_2201610, EPI_ISL_2201611 | Utah Public Health Laboratory | Utah Public Health Laboratory | Erin L. Young, Kelly F. Oakeson, Tara Gallagher |
| EPI_ISL_2201612 | Aegis Sciences Corporation | Centers for Disease Control and Prevention Division of Viral Diseases, Pathogen Discovery | Dakota Howard, Dhwani Batra, Peter W. Cook, Kara Moser, Adrian Paskey, Jason Caravas, Benjamin Rambo-Martin, Shatavia Morrison, Christopher Gulvick, Scott Sammons, Yvette Unoarumhi, Darlene Wagner, Matthew Schmerer, Cyndi Clark, Patrick Campbell, Rob Case, Vikramsinha Ghorpade, Holly Houdeshell, Ola Kvalvaag, Dillon Nall, Ethan Sanders, Alec Vest, Shaun Westlund, Matthew Hardison, Clinton R. Paden, Duncan MacCannell |
| EPI_ISL_2201613 | Houston Methodist Hospital | Houston Methodist Hospital | Randall J. Olsen, Paul A. Christensen, S. Wesley Long, Sishir Subedi, Robert Olson, Marcus Nguyen, James J. Davis, Matthew Ojeda Saavedra, Prasanti Yerramilli, Layne Pruitt, Kristina Reppond, Madison N. Shyer, Jessica Cambric, Ryan Gadd, Ilya J. Finkelstein, Jimmy Gollihar, and James M. Musser |
| EPI_ISL_2201614, EPI_ISL_2201615 | Utah Public Health Laboratory | Utah Public Health Laboratory | Erin L. Young, Kelly F. Oakeson, Tara Gallagher |
| EPI_ISL_2201616 | Aegis Sciences Corporation | Centers for Disease Control and Prevention Division of Viral Diseases, Pathogen Discovery | Dakota Howard, Dhwani Batra, Peter W. Cook, Kara Moser, Adrian Paskey, Jason Caravas, Benjamin Rambo-Martin, Shatavia Morrison, Christopher Gulvick, Scott Sammons, Yvette Unoarumhi, Darlene Wagner, Matthew Schmerer, Cyndi Clark, Patrick Campbell, Rob Case, Vikramsinha Ghorpade, Holly Houdeshell, Ola Kvalvaag, Dillon Nall, Ethan Sanders, Alec Vest, Shaun Westlund, Matthew Hardison, Clinton R. Paden, Duncan MacCannell |
| EPI_ISL_2201617 | Utah Public Health Laboratory | Utah Public Health Laboratory | Erin L. Young, Kelly F. Oakeson, Tara Gallagher |
| EPI_ISL_2201618 | Houston Methodist Hospital | Houston Methodist Hospital | Randall J. Olsen, Paul A. Christensen, S. Wesley Long, Sishir Subedi, Robert Olson, Marcus Nguyen, James J. Davis, Matthew Ojeda Saavedra, Prasanti Yerramilli, Layne Pruitt, Kristina Reppond, Madison N. Shyer, Jessica Cambric, Ryan Gadd, Ilya J. Finkelstein, Jimmy Gollihar, and James M. Musser |
| EPI_ISL_2201619 | Utah Public Health Laboratory | Utah Public Health Laboratory | Erin L. Young, Kelly F. Oakeson, Tara Gallagher |
| EPI_ISL_2201620 | Aegis Sciences Corporation | Centers for Disease Control and Prevention Division of Viral Diseases, Pathogen | Dakota Howard, Dhwani Batra, Peter W. Cook, Kara Moser, Adrian Paskey, Jason Caravas, Benjamin Rambo-Martin, Shatavia Morrison, Christopher Gulvick, Scott Sammons, Yvette Unoarumhi, Darlene Wagner, Matthew Schmerer, Cyndi Clark, Patrick Campbell, Rob Case, Vikramsinha Ghorpade, Holly Houdeshell, Ola Kvalvaag, Dillon Nall, Ethan Sanders, Alec Vest, Shaun Westlund, Matthew Hardison, Clinton R. Paden, Duncan MacCannell |

| | | | |
|---|---|---|---|
| | | Discovery | |
| EPI_ISL_2201621 | Utah Public Health Laboratory | Utah Public Health Laboratory | Erin L. Young, Kelly F. Oakeson, Tara Gallagher |
| EPI_ISL_2201622 | Houston Methodist Hospital | Houston Methodist Hospital | Randall J. Olsen, Paul A. Christensen, S. Wesley Long, Sishir Subedi, Robert Olson, Marcus Nguyen, James J. Davis, Matthew Ojeda Saavedra, Prasanti Yerramilli, Layne Pruitt, Kristina Reppond, Madison N. Shyer, Jessica Cambric, Ryan Gadd, Ilya J. Finkelstein, Jimmy Gollihar, and James M. Musser |
| EPI_ISL_2201624 | Aegis Sciences Corporation | Centers for Disease Control and Prevention Division of Viral Diseases, Pathogen Discovery | Dakota Howard, Dhwani Batra, Peter W. Cook, Kara Moser, Adrian Paskey, Jason Caravas, Benjamin Rambo-Martin, Shatavia Morrison, Christopher Gulvick, Scott Sammons, Yvette Unoarumhi, Darlene Wagner, Matthew Schmerer, Cyndi Clark, Patrick Campbell, Rob Case, Vikramsinha Ghorpade, Holly Houdeshell, Ola Kvalvaag, Dillon Nall, Ethan Sanders, Alec Vest, Shaun Westlund, Matthew Hardison, Clinton R. Paden, Duncan MacCannell |
| EPI_ISL_2201626 | Houston Methodist Hospital | Houston Methodist Hospital | Randall J. Olsen, Paul A. Christensen, S. Wesley Long, Sishir Subedi, Robert Olson, Marcus Nguyen, James J. Davis, Matthew Ojeda Saavedra, Prasanti Yerramilli, Layne Pruitt, Kristina Reppond, Madison N. Shyer, Jessica Cambric, Ryan Gadd, Ilya J. Finkelstein, Jimmy Gollihar, and James M. Musser |
| EPI_ISL_2201627 | Utah Public Health Laboratory | Utah Public Health Laboratory | Erin L. Young, Kelly F. Oakeson, Tara Gallagher |
| EPI_ISL_2201628 | Aegis Sciences Corporation | Centers for Disease Control and Prevention Division of Viral Diseases, Pathogen Discovery | Dakota Howard, Dhwani Batra, Peter W. Cook, Kara Moser, Adrian Paskey, Jason Caravas, Benjamin Rambo-Martin, Shatavia Morrison, Christopher Gulvick, Scott Sammons, Yvette Unoarumhi, Darlene Wagner, Matthew Schmerer, Cyndi Clark, Patrick Campbell, Rob Case, Vikramsinha Ghorpade, Holly Houdeshell, Ola Kvalvaag, Dillon Nall, Ethan Sanders, Alec Vest, Shaun Westlund, Matthew Hardison, Clinton R. Paden, Duncan MacCannell |
| EPI_ISL_2201629 | Utah Public Health Laboratory | Utah Public Health Laboratory | Erin L. Young, Kelly F. Oakeson, Tara Gallagher |
| EPI_ISL_2201630 | Houston Methodist Hospital | Houston Methodist Hospital | Randall J. Olsen, Paul A. Christensen, S. Wesley Long, Sishir Subedi, Robert Olson, Marcus Nguyen, James J. Davis, Matthew Ojeda Saavedra, Prasanti Yerramilli, Layne Pruitt, Kristina Reppond, Madison N. Shyer, Jessica Cambric, Ryan Gadd, Ilya J. Finkelstein, Jimmy Gollihar, and James M. Musser |
| EPI_ISL_2201631 | Aegis Sciences Corporation | Centers for Disease Control and Prevention Division of Viral Diseases, Pathogen Discovery | Dakota Howard, Dhwani Batra, Peter W. Cook, Kara Moser, Adrian Paskey, Jason Caravas, Benjamin Rambo-Martin, Shatavia Morrison, Christopher Gulvick, Scott Sammons, Yvette Unoarumhi, Darlene Wagner, Matthew Schmerer, Cyndi Clark, Patrick Campbell, Rob Case, Vikramsinha Ghorpade, Holly Houdeshell, Ola Kvalvaag, Dillon Nall, Ethan Sanders, Alec Vest, Shaun Westlund, Matthew Hardison, Clinton R. Paden, Duncan MacCannell |
| EPI_ISL_2201632, EPI_ISL_2201633 | Utah Public Health Laboratory | Utah Public Health Laboratory | Erin L. Young, Kelly F. Oakeson, Tara Gallagher |
| EPI_ISL_2201634 | Aegis Sciences Corporation | Centers for Disease Control and Prevention Division of Viral Diseases, Pathogen Discovery | Dakota Howard, Dhwani Batra, Peter W. Cook, Kara Moser, Adrian Paskey, Jason Caravas, Benjamin Rambo-Martin, Shatavia Morrison, Christopher Gulvick, Scott Sammons, Yvette Unoarumhi, Darlene Wagner, Matthew Schmerer, Cyndi Clark, Patrick Campbell, Rob Case, Vikramsinha Ghorpade, Holly Houdeshell, Ola Kvalvaag, Dillon Nall, Ethan Sanders, Alec Vest, Shaun Westlund, Matthew Hardison, Clinton R. Paden, Duncan MacCannell |
| EPI_ISL_2201635 | Utah Public Health Laboratory | Utah Public Health Laboratory | Erin L. Young, Kelly F. Oakeson, Tara Gallagher |
| EPI_ISL_2201636 | Houston Methodist Hospital | Houston Methodist Hospital | Randall J. Olsen, Paul A. Christensen, S. Wesley Long, Sishir Subedi, Robert Olson, Marcus Nguyen, James J. Davis, Matthew Ojeda Saavedra, Prasanti Yerramilli, Layne Pruitt, Kristina Reppond, Madison N. Shyer, Jessica Cambric, Ryan Gadd, Ilya J. Finkelstein, Jimmy Gollihar, and James M. Musser |
| EPI_ISL_2201637 | Utah Public Health Laboratory | Utah Public Health Laboratory | Erin L. Young, Kelly F. Oakeson, Tara Gallagher |
| EPI_ISL_2201638 | Aegis Sciences Corporation | Centers for Disease Control and Prevention Division of Viral Diseases, Pathogen Discovery | Dakota Howard, Dhwani Batra, Peter W. Cook, Kara Moser, Adrian Paskey, Jason Caravas, Benjamin Rambo-Martin, Shatavia Morrison, Christopher Gulvick, Scott Sammons, Yvette Unoarumhi, Darlene Wagner, Matthew Schmerer, Cyndi Clark, Patrick Campbell, Rob Case, Vikramsinha Ghorpade, Holly Houdeshell, Ola Kvalvaag, Dillon Nall, Ethan Sanders, Alec Vest, Shaun Westlund, Matthew Hardison, Clinton R. Paden, Duncan MacCannell |
| EPI_ISL_2201639 | Utah Public Health Laboratory | Utah Public Health Laboratory | Erin L. Young, Kelly F. Oakeson, Tara Gallagher |
| EPI_ISL_2201640 | Houston Methodist Hospital | Houston Methodist Hospital | Randall J. Olsen, Paul A. Christensen, S. Wesley Long, Sishir Subedi, Robert Olson, Marcus Nguyen, James J. Davis, Matthew Ojeda Saavedra, Prasanti Yerramilli, Layne Pruitt, Kristina Reppond, Madison N. Shyer, Jessica Cambric, Ryan Gadd, Ilya J. Finkelstein, Jimmy Gollihar, and James M. Musser |
| EPI_ISL_2201641 | Utah Public Health Laboratory | Utah Public Health Laboratory | Erin L. Young, Kelly F. Oakeson, Tara Gallagher |
| EPI_ISL_2201642 | Aegis Sciences Corporation | Centers for Disease Control and Prevention Division of Viral Diseases, Pathogen Discovery | Dakota Howard, Dhwani Batra, Peter W. Cook, Kara Moser, Adrian Paskey, Jason Caravas, Benjamin Rambo-Martin, Shatavia Morrison, Christopher Gulvick, Scott Sammons, Yvette Unoarumhi, Darlene Wagner, Matthew Schmerer, Cyndi Clark, Patrick Campbell, Rob Case, Vikramsinha Ghorpade, Holly Houdeshell, Ola Kvalvaag, Dillon Nall, Ethan Sanders, Alec Vest, Shaun Westlund, Matthew Hardison, Clinton R. Paden, Duncan MacCannell |
| EPI_ISL_2201644 | Utah Public Health Laboratory | Utah Public Health Laboratory | Erin L. Young, Kelly F. Oakeson, Tara Gallagher |
| EPI_ISL_2201645 | Aegis Sciences Corporation | Centers for Disease Control and Prevention Division of Viral Diseases, Pathogen Discovery | Dakota Howard, Dhwani Batra, Peter W. Cook, Kara Moser, Adrian Paskey, Jason Caravas, Benjamin Rambo-Martin, Shatavia Morrison, Christopher Gulvick, Scott Sammons, Yvette Unoarumhi, Darlene Wagner, Matthew Schmerer, Cyndi Clark, Patrick Campbell, Rob Case, Vikramsinha Ghorpade, Holly Houdeshell, Ola Kvalvaag, Dillon Nall, Ethan Sanders, Alec Vest, Shaun Westlund, Matthew Hardison, Clinton R. Paden, Duncan MacCannell |
| EPI_ISL_2201646 | Houston Methodist Hospital | Houston Methodist Hospital | Randall J. Olsen, Paul A. Christensen, S. Wesley Long, Sishir Subedi, Robert Olson, Marcus Nguyen, James J. Davis, Matthew Ojeda Saavedra, Prasanti Yerramilli, Layne Pruitt, Kristina Reppond, Madison N. Shyer, Jessica Cambric, Ryan Gadd, Ilya J. Finkelstein, Jimmy Gollihar, and James M. Musser |
| EPI_ISL_2201649 | Utah Public Health Laboratory | Utah Public Health Laboratory | Erin L. Young, Kelly F. Oakeson, Tara Gallagher |
| EPI_ISL_2201650 | Houston Methodist Hospital | Houston Methodist Hospital | Randall J. Olsen, Paul A. Christensen, S. Wesley Long, Sishir Subedi, Robert Olson, Marcus Nguyen, James J. Davis, Matthew Ojeda Saavedra, Prasanti Yerramilli, Layne Pruitt, Kristina Reppond, Madison N. Shyer, Jessica Cambric, Ryan Gadd, Ilya J. Finkelstein, Jimmy Gollihar, and James M. Musser |
| EPI_ISL_2201651 | Aegis Sciences Corporation | Centers for Disease Control and Prevention Division of Viral Diseases, Pathogen Discovery | Dakota Howard, Dhwani Batra, Peter W. Cook, Kara Moser, Adrian Paskey, Jason Caravas, Benjamin Rambo-Martin, Shatavia Morrison, Christopher Gulvick, Scott Sammons, Yvette Unoarumhi, Darlene Wagner, Matthew Schmerer, Cyndi Clark, Patrick Campbell, Rob Case, Vikramsinha Ghorpade, Holly Houdeshell, Ola Kvalvaag, Dillon Nall, Ethan Sanders, Alec Vest, Shaun Westlund, Matthew Hardison, Clinton R. Paden, Duncan MacCannell |
| EPI_ISL_2201652 | Utah Public Health Laboratory | Utah Public Health Laboratory | Erin L. Young, Kelly F. Oakeson, Tara Gallagher |
| EPI_ISL_2201654 | Houston Methodist Hospital | Houston Methodist Hospital | Randall J. Olsen, Paul A. Christensen, S. Wesley Long, Sishir Subedi, Robert Olson, Marcus Nguyen, James J. Davis, Matthew Ojeda Saavedra, Prasanti Yerramilli, Layne Pruitt, Kristina Reppond, Madison N. Shyer, Jessica Cambric, Ryan Gadd, Ilya J. Finkelstein, Jimmy Gollihar, and James M. Musser |
| EPI_ISL_2201655 | Aegis Sciences Corporation | Centers for Disease Control and Prevention Division of Viral Diseases, Pathogen Discovery | Dakota Howard, Dhwani Batra, Peter W. Cook, Kara Moser, Adrian Paskey, Jason Caravas, Benjamin Rambo-Martin, Shatavia Morrison, Christopher Gulvick, Scott Sammons, Yvette Unoarumhi, Darlene Wagner, Matthew Schmerer, Cyndi Clark, Patrick Campbell, Rob Case, Vikramsinha Ghorpade, Holly Houdeshell, Ola Kvalvaag, Dillon Nall, Ethan Sanders, Alec Vest, Shaun Westlund, Matthew Hardison, Clinton R. Paden, Duncan MacCannell |
| EPI_ISL_2201656, EPI_ISL_2201657 | Utah Public Health Laboratory | Utah Public Health Laboratory | Erin L. Young, Kelly F. Oakeson, Tara Gallagher |
| EPI_ISL_2201658 | Aegis Sciences Corporation | Centers for Disease Control and Prevention Division of Viral Diseases, Pathogen Discovery | Dakota Howard, Dhwani Batra, Peter W. Cook, Kara Moser, Adrian Paskey, Jason Caravas, Benjamin Rambo-Martin, Shatavia Morrison, Christopher Gulvick, Scott Sammons, Yvette Unoarumhi, Darlene Wagner, Matthew Schmerer, Cyndi Clark, Patrick Campbell, Rob Case, Vikramsinha Ghorpade, Holly Houdeshell, Ola Kvalvaag, Dillon Nall, Ethan Sanders, Alec Vest, Shaun Westlund, Matthew Hardison, Clinton R. Paden, Duncan MacCannell |
| EPI_ISL_2201659 | Houston Methodist Hospital | Houston Methodist Hospital | Randall J. Olsen, Paul A. Christensen, S. Wesley Long, Sishir Subedi, Robert Olson, Marcus Nguyen, James J. Davis, Matthew Ojeda Saavedra, Prasanti Yerramilli, Layne Pruitt, Kristina Reppond, Madison N. Shyer, Jessica Cambric, Ryan Gadd, Ilya J. Finkelstein, Jimmy Gollihar, and James M. Musser |
| EPI_ISL_2201660 | Utah Public Health Laboratory | Utah Public Health Laboratory | Erin L. Young, Kelly F. Oakeson, Tara Gallagher |
| EPI_ISL_2201661 | Aegis Sciences Corporation | Centers for Disease Control and Prevention Division of Viral Diseases, Pathogen Discovery | Dakota Howard, Dhwani Batra, Peter W. Cook, Kara Moser, Adrian Paskey, Jason Caravas, Benjamin Rambo-Martin, Shatavia Morrison, Christopher Gulvick, Scott Sammons, Yvette Unoarumhi, Darlene Wagner, Matthew Schmerer, Cyndi Clark, Patrick Campbell, Rob Case, Vikramsinha Ghorpade, Holly Houdeshell, Ola Kvalvaag, Dillon Nall, Ethan Sanders, Alec Vest, Shaun Westlund, Matthew Hardison, Clinton R. Paden, Duncan MacCannell |
| EPI_ISL_2201662, EPI_ISL_2201664 | Utah Public Health Laboratory | Utah Public Health Laboratory | Erin L. Young, Kelly F. Oakeson, Tara Gallagher |
| EPI_ISL_2201665 | Aegis Sciences Corporation | Centers for Disease Control and Prevention Division of Viral Diseases, Pathogen Discovery | Dakota Howard, Dhwani Batra, Peter W. Cook, Kara Moser, Adrian Paskey, Jason Caravas, Benjamin Rambo-Martin, Shatavia Morrison, Christopher Gulvick, Scott Sammons, Yvette Unoarumhi, Darlene Wagner, Matthew Schmerer, Cyndi Clark, Patrick Campbell, Rob Case, Vikramsinha Ghorpade, Holly Houdeshell, Ola Kvalvaag, Dillon Nall, Ethan Sanders, Alec Vest, Shaun Westlund, Matthew Hardison, Clinton R. Paden, Duncan MacCannell |
| EPI_ISL_2201666 | Utah Public Health Laboratory | Utah Public Health Laboratory | Erin L. Young, Kelly F. Oakeson, Tara Gallagher |
| EPI_ISL_2201667 | Houston Methodist Hospital | Houston Methodist Hospital | Randall J. Olsen, Paul A. Christensen, S. Wesley Long, Sishir Subedi, Robert Olson, Marcus Nguyen, James J. Davis, Matthew Ojeda Saavedra, Prasanti Yerramilli, Layne Pruitt, Kristina Reppond, Madison N. Shyer, Jessica Cambric, Ryan Gadd, Ilya J. Finkelstein, Jimmy Gollihar, and James M. Musser |
| EPI_ISL_2201668 | Aegis Sciences Corporation | Centers for Disease Control and Prevention Division of Viral Diseases, Pathogen Discovery | Dakota Howard, Dhwani Batra, Peter W. Cook, Kara Moser, Adrian Paskey, Jason Caravas, Benjamin Rambo-Martin, Shatavia Morrison, Christopher Gulvick, Scott Sammons, Yvette Unoarumhi, Darlene Wagner, Matthew Schmerer, Cyndi Clark, Patrick Campbell, Rob Case, Vikramsinha Ghorpade, Holly Houdeshell, Ola Kvalvaag, Dillon Nall, Ethan Sanders, Alec Vest, Shaun Westlund, Matthew Hardison, Clinton R. Paden, Duncan MacCannell |
| EPI_ISL_2201669, EPI_ISL_2201670 | Utah Public Health Laboratory | Utah Public Health Laboratory | Erin L. Young, Kelly F. Oakeson, Tara Gallagher |
| EPI_ISL_2201671 | Houston Methodist Hospital | Houston Methodist Hospital | Randall J. Olsen, Paul A. Christensen, S. Wesley Long, Sishir Subedi, Robert Olson, Marcus Nguyen, James J. Davis, Matthew Ojeda Saavedra, Prasanti Yerramilli, Layne Pruitt, Kristina Reppond, Madison N. Shyer, Jessica Cambric, Ryan Gadd, Ilya J. Finkelstein, Jimmy Gollihar, and James M. Musser |
| EPI_ISL_2201672 | Aegis Sciences Corporation | Centers for Disease Control and Prevention Division of Viral Diseases, Pathogen Discovery | Dakota Howard, Dhwani Batra, Peter W. Cook, Kara Moser, Adrian Paskey, Jason Caravas, Benjamin Rambo-Martin, Shatavia Morrison, Christopher Gulvick, Scott Sammons, Yvette Unoarumhi, Darlene Wagner, Matthew Schmerer, Cyndi Clark, Patrick Campbell, Rob Case, Vikramsinha Ghorpade, Holly Houdeshell, Ola Kvalvaag, Dillon Nall, Ethan Sanders, Alec Vest, Shaun Westlund, Matthew Hardison, Clinton R. Paden, Duncan MacCannell |
| EPI_ISL_2201673, EPI_ISL_2201674 | Utah Public Health Laboratory | Utah Public Health Laboratory | Erin L. Young, Kelly F. Oakeson, Tara Gallagher |
| EPI_ISL_2201675 | Houston Methodist Hospital | Houston Methodist Hospital | Randall J. Olsen, Paul A. Christensen, S. Wesley Long, Sishir Subedi, Robert Olson, Marcus Nguyen, James J. Davis, Matthew Ojeda Saavedra, Prasanti Yerramilli, Layne Pruitt, Kristina Reppond, Madison N. Shyer, Jessica Cambric, Ryan Gadd, Ilya J. Finkelstein, Jimmy Gollihar, and James M. Musser |
| EPI_ISL_2201676 | Aegis Sciences Corporation | Centers for Disease Control and Prevention Division of Viral Diseases, Pathogen | Dakota Howard, Dhwani Batra, Peter W. Cook, Kara Moser, Adrian Paskey, Jason Caravas, Benjamin Rambo-Martin, Shatavia Morrison, Christopher Gulvick, Scott Sammons, Yvette Unoarumhi, Darlene Wagner, Matthew Schmerer, Cyndi Clark, Patrick Campbell, Rob Case, Vikramsinha Ghorpade, Holly Houdeshell, Ola Kvalvaag, Dillon Nall, Ethan Sanders, Alec Vest, Shaun Westlund, Matthew Hardison, Clinton R. Paden, Duncan MacCannell |

| | | | |
|---|---|---|---|
| | | Discovery | |
| EPI_ISL_2201677, EPI_ISL_2201679 | Utah Public Health Laboratory | Utah Public Health Laboratory | Erin L. Young, Kelly F. Oakeson, Tara Gallagher |
| EPI_ISL_2201680 | Houston Methodist Hospital | Houston Methodist Hospital | Randall J. Olsen, Paul A. Christensen, S. Wesley Long, Sishir Subedi, Robert Olson, Marcus Nguyen, James J. Davis, Matthew Ojeda Saavedra, Prasanti Yerramilli, Layne Pruitt, Kristina Reppond, Madison N. Shyer, Jessica Cambric, Ryan Gadd, Ilya J. Finkelstein, Jimmy Gollihar, and James M. Musser |
| EPI_ISL_2201681 | Utah Public Health Laboratory | Utah Public Health Laboratory | Erin L. Young, Kelly F. Oakeson, Tara Gallagher |
| EPI_ISL_2201682 | Aegis Sciences Corporation | Centers for Disease Control and Prevention Division of Viral Diseases, Pathogen Discovery | Dakota Howard, Dhwani Batra, Peter W. Cook, Kara Moser, Adrian Paskey, Jason Caravas, Benjamin Rambo-Martin, Shatavia Morrison, Christopher Gulvick, Scott Sammons, Yvette Unoarumhi, Darlene Wagner, Matthew Schmerer, Cyndi Clark, Patrick Campbell, Rob Case, Vikramsinha Ghorpade, Holly Houdeshell, Ola Kvalvaag, Dillon Nall, Ethan Sanders, Alec Vest, Shaun Westlund, Matthew Hardison, Clinton R. Paden, Duncan MacCannell |
| EPI_ISL_2201683 | Utah Public Health Laboratory | Utah Public Health Laboratory | Erin L. Young, Kelly F. Oakeson, Tara Gallagher |
| EPI_ISL_2201684 | Houston Methodist Hospital | Houston Methodist Hospital | Randall J. Olsen, Paul A. Christensen, S. Wesley Long, Sishir Subedi, Robert Olson, Marcus Nguyen, James J. Davis, Matthew Ojeda Saavedra, Prasanti Yerramilli, Layne Pruitt, Kristina Reppond, Madison N. Shyer, Jessica Cambric, Ryan Gadd, Ilya J. Finkelstein, Jimmy Gollihar, and James M. Musser |
| EPI_ISL_2201685 | Utah Public Health Laboratory | Utah Public Health Laboratory | Erin L. Young, Kelly F. Oakeson, Tara Gallagher |
| EPI_ISL_2201686 | Aegis Sciences Corporation | Centers for Disease Control and Prevention Division of Viral Diseases, Pathogen Discovery | Dakota Howard, Dhwani Batra, Peter W. Cook, Kara Moser, Adrian Paskey, Jason Caravas, Benjamin Rambo-Martin, Shatavia Morrison, Christopher Gulvick, Scott Sammons, Yvette Unoarumhi, Darlene Wagner, Matthew Schmerer, Cyndi Clark, Patrick Campbell, Rob Case, Vikramsinha Ghorpade, Holly Houdeshell, Ola Kvalvaag, Dillon Nall, Ethan Sanders, Alec Vest, Shaun Westlund, Matthew Hardison, Clinton R. Paden, Duncan MacCannell |
| EPI_ISL_2201687, EPI_ISL_2201688 | Utah Public Health Laboratory | Utah Public Health Laboratory | Erin L. Young, Kelly F. Oakeson, Tara Gallagher |
| EPI_ISL_2201689 | Aegis Sciences Corporation | Centers for Disease Control and Prevention Division of Viral Diseases, Pathogen Discovery | Dakota Howard, Dhwani Batra, Peter W. Cook, Kara Moser, Adrian Paskey, Jason Caravas, Benjamin Rambo-Martin, Shatavia Morrison, Christopher Gulvick, Scott Sammons, Yvette Unoarumhi, Darlene Wagner, Matthew Schmerer, Cyndi Clark, Patrick Campbell, Rob Case, Vikramsinha Ghorpade, Holly Houdeshell, Ola Kvalvaag, Dillon Nall, Ethan Sanders, Alec Vest, Shaun Westlund, Matthew Hardison, Clinton R. Paden, Duncan MacCannell |
| EPI_ISL_2201690 | Houston Methodist Hospital | Houston Methodist Hospital | Randall J. Olsen, Paul A. Christensen, S. Wesley Long, Sishir Subedi, Robert Olson, Marcus Nguyen, James J. Davis, Matthew Ojeda Saavedra, Prasanti Yerramilli, Layne Pruitt, Kristina Reppond, Madison N. Shyer, Jessica Cambric, Ryan Gadd, Ilya J. Finkelstein, Jimmy Gollihar, and James M. Musser |
| EPI_ISL_2201691, EPI_ISL_2201692 | Utah Public Health Laboratory | Utah Public Health Laboratory | Erin L. Young, Kelly F. Oakeson, Tara Gallagher |
| EPI_ISL_2201693 | Aegis Sciences Corporation | Centers for Disease Control and Prevention Division of Viral Diseases, Pathogen Discovery | Dakota Howard, Dhwani Batra, Peter W. Cook, Kara Moser, Adrian Paskey, Jason Caravas, Benjamin Rambo-Martin, Shatavia Morrison, Christopher Gulvick, Scott Sammons, Yvette Unoarumhi, Darlene Wagner, Matthew Schmerer, Cyndi Clark, Patrick Campbell, Rob Case, Vikramsinha Ghorpade, Holly Houdeshell, Ola Kvalvaag, Dillon Nall, Ethan Sanders, Alec Vest, Shaun Westlund, Matthew Hardison, Clinton R. Paden, Duncan MacCannell |
| EPI_ISL_2201695 | Houston Methodist Hospital | Houston Methodist Hospital | Randall J. Olsen, Paul A. Christensen, S. Wesley Long, Sishir Subedi, Robert Olson, Marcus Nguyen, James J. Davis, Matthew Ojeda Saavedra, Prasanti Yerramilli, Layne Pruitt, Kristina Reppond, Madison N. Shyer, Jessica Cambric, Ryan Gadd, Ilya J. Finkelstein, Jimmy Gollihar, and James M. Musser |
| EPI_ISL_2201696 | Utah Public Health Laboratory | Utah Public Health Laboratory | Erin L. Young, Kelly F. Oakeson, Tara Gallagher |
| EPI_ISL_2201697 | Aegis Sciences Corporation | Centers for Disease Control and Prevention Division of Viral Diseases, Pathogen Discovery | Dakota Howard, Dhwani Batra, Peter W. Cook, Kara Moser, Adrian Paskey, Jason Caravas, Benjamin Rambo-Martin, Shatavia Morrison, Christopher Gulvick, Scott Sammons, Yvette Unoarumhi, Darlene Wagner, Matthew Schmerer, Cyndi Clark, Patrick Campbell, Rob Case, Vikramsinha Ghorpade, Holly Houdeshell, Ola Kvalvaag, Dillon Nall, Ethan Sanders, Alec Vest, Shaun Westlund, Matthew Hardison, Clinton R. Paden, Duncan MacCannell |
| EPI_ISL_2201698 | Utah Public Health Laboratory | Utah Public Health Laboratory | Erin L. Young, Kelly F. Oakeson, Tara Gallagher |
| EPI_ISL_2201699 | Aegis Sciences Corporation | Centers for Disease Control and Prevention Division of Viral Diseases, Pathogen Discovery | Dakota Howard, Dhwani Batra, Peter W. Cook, Kara Moser, Adrian Paskey, Jason Caravas, Benjamin Rambo-Martin, Shatavia Morrison, Christopher Gulvick, Scott Sammons, Yvette Unoarumhi, Darlene Wagner, Matthew Schmerer, Cyndi Clark, Patrick Campbell, Rob Case, Vikramsinha Ghorpade, Holly Houdeshell, Ola Kvalvaag, Dillon Nall, Ethan Sanders, Alec Vest, Shaun Westlund, Matthew Hardison, Clinton R. Paden, Duncan MacCannell |
| EPI_ISL_2201700 | Houston Methodist Hospital | Houston Methodist Hospital | Randall J. Olsen, Paul A. Christensen, S. Wesley Long, Sishir Subedi, Robert Olson, Marcus Nguyen, James J. Davis, Matthew Ojeda Saavedra, Prasanti Yerramilli, Layne Pruitt, Kristina Reppond, Madison N. Shyer, Jessica Cambric, Ryan Gadd, Ilya J. Finkelstein, Jimmy Gollihar, and James M. Musser |
| EPI_ISL_2201701, EPI_ISL_2201702 | Utah Public Health Laboratory | Utah Public Health Laboratory | Erin L. Young, Kelly F. Oakeson, Tara Gallagher |
| EPI_ISL_2201703 | Houston Methodist Hospital | Houston Methodist Hospital | Randall J. Olsen, Paul A. Christensen, S. Wesley Long, Sishir Subedi, Robert Olson, Marcus Nguyen, James J. Davis, Matthew Ojeda Saavedra, Prasanti Yerramilli, Layne Pruitt, Kristina Reppond, Madison N. Shyer, Jessica Cambric, Ryan Gadd, Ilya J. Finkelstein, Jimmy Gollihar, and James M. Musser |
| EPI_ISL_2201704, EPI_ISL_2201705, EPI_ISL_2201706 | Utah Public Health Laboratory | Utah Public Health Laboratory | Erin L. Young, Kelly F. Oakeson, Tara Gallagher |
| EPI_ISL_2201707 | Aegis Sciences Corporation | Centers for Disease Control and Prevention Division of Viral Diseases, Pathogen Discovery | Dakota Howard, Dhwani Batra, Peter W. Cook, Kara Moser, Adrian Paskey, Jason Caravas, Benjamin Rambo-Martin, Shatavia Morrison, Christopher Gulvick, Scott Sammons, Yvette Unoarumhi, Darlene Wagner, Matthew Schmerer, Cyndi Clark, Patrick Campbell, Rob Case, Vikramsinha Ghorpade, Holly Houdeshell, Ola Kvalvaag, Dillon Nall, Ethan Sanders, Alec Vest, Shaun Westlund, Matthew Hardison, Clinton R. Paden, Duncan MacCannell |
| EPI_ISL_2201708 | Houston Methodist Hospital | Houston Methodist Hospital | Randall J. Olsen, Paul A. Christensen, S. Wesley Long, Sishir Subedi, Robert Olson, Marcus Nguyen, James J. Davis, Matthew Ojeda Saavedra, Prasanti Yerramilli, Layne Pruitt, Kristina Reppond, Madison N. Shyer, Jessica Cambric, Ryan Gadd, Ilya J. Finkelstein, Jimmy Gollihar, and James M. Musser |
| EPI_ISL_2201709, EPI_ISL_2201710 | Utah Public Health Laboratory | Utah Public Health Laboratory | Erin L. Young, Kelly F. Oakeson, Tara Gallagher |
| EPI_ISL_2201711 | Aegis Sciences Corporation | Centers for Disease Control and Prevention Division of Viral Diseases, Pathogen Discovery | Dakota Howard, Dhwani Batra, Peter W. Cook, Kara Moser, Adrian Paskey, Jason Caravas, Benjamin Rambo-Martin, Shatavia Morrison, Christopher Gulvick, Scott Sammons, Yvette Unoarumhi, Darlene Wagner, Matthew Schmerer, Cyndi Clark, Patrick Campbell, Rob Case, Vikramsinha Ghorpade, Holly Houdeshell, Ola Kvalvaag, Dillon Nall, Ethan Sanders, Alec Vest, Shaun Westlund, Matthew Hardison, Clinton R. Paden, Duncan MacCannell |
| EPI_ISL_2201712 | Utah Public Health Laboratory | Utah Public Health Laboratory | Erin L. Young, Kelly F. Oakeson, Tara Gallagher |
| EPI_ISL_2201713 | Houston Methodist Hospital | Houston Methodist Hospital | Randall J. Olsen, Paul A. Christensen, S. Wesley Long, Sishir Subedi, Robert Olson, Marcus Nguyen, James J. Davis, Matthew Ojeda Saavedra, Prasanti Yerramilli, Layne Pruitt, Kristina Reppond, Madison N. Shyer, Jessica Cambric, Ryan Gadd, Ilya J. Finkelstein, Jimmy Gollihar, and James M. Musser |
| EPI_ISL_2201714 | Utah Public Health Laboratory | Utah Public Health Laboratory | Erin L. Young, Kelly F. Oakeson, Tara Gallagher |
| EPI_ISL_2201715 | Aegis Sciences Corporation | Centers for Disease Control and Prevention Division of Viral Diseases, Pathogen Discovery | Dakota Howard, Dhwani Batra, Peter W. Cook, Kara Moser, Adrian Paskey, Jason Caravas, Benjamin Rambo-Martin, Shatavia Morrison, Christopher Gulvick, Scott Sammons, Yvette Unoarumhi, Darlene Wagner, Matthew Schmerer, Cyndi Clark, Patrick Campbell, Rob Case, Vikramsinha Ghorpade, Holly Houdeshell, Ola Kvalvaag, Dillon Nall, Ethan Sanders, Alec Vest, Shaun Westlund, Matthew Hardison, Clinton R. Paden, Duncan MacCannell |
| EPI_ISL_2201716 | Utah Public Health Laboratory | Utah Public Health Laboratory | Erin L. Young, Kelly F. Oakeson, Tara Gallagher |
| EPI_ISL_2201717 | Houston Methodist Hospital | Houston Methodist Hospital | Randall J. Olsen, Paul A. Christensen, S. Wesley Long, Sishir Subedi, Robert Olson, Marcus Nguyen, James J. Davis, Matthew Ojeda Saavedra, Prasanti Yerramilli, Layne Pruitt, Kristina Reppond, Madison N. Shyer, Jessica Cambric, Ryan Gadd, Ilya J. Finkelstein, Jimmy Gollihar, and James M. Musser |
| EPI_ISL_2201718 | Aegis Sciences Corporation | Centers for Disease Control and Prevention Division of Viral Diseases, Pathogen Discovery | Dakota Howard, Dhwani Batra, Peter W. Cook, Kara Moser, Adrian Paskey, Jason Caravas, Benjamin Rambo-Martin, Shatavia Morrison, Christopher Gulvick, Scott Sammons, Yvette Unoarumhi, Darlene Wagner, Matthew Schmerer, Cyndi Clark, Patrick Campbell, Rob Case, Vikramsinha Ghorpade, Holly Houdeshell, Ola Kvalvaag, Dillon Nall, Ethan Sanders, Alec Vest, Shaun Westlund, Matthew Hardison, Clinton R. Paden, Duncan MacCannell |
| EPI_ISL_2201719 | Utah Public Health Laboratory | Utah Public Health Laboratory | Erin L. Young, Kelly F. Oakeson, Tara Gallagher |
| EPI_ISL_2201720 | Houston Methodist Hospital | Houston Methodist Hospital | Randall J. Olsen, Paul A. Christensen, S. Wesley Long, Sishir Subedi, Robert Olson, Marcus Nguyen, James J. Davis, Matthew Ojeda Saavedra, Prasanti Yerramilli, Layne Pruitt, Kristina Reppond, Madison N. Shyer, Jessica Cambric, Ryan Gadd, Ilya J. Finkelstein, Jimmy Gollihar, and James M. Musser |
| EPI_ISL_2201721 | Aegis Sciences Corporation | Centers for Disease Control and Prevention Division of Viral Diseases, Pathogen Discovery | Dakota Howard, Dhwani Batra, Peter W. Cook, Kara Moser, Adrian Paskey, Jason Caravas, Benjamin Rambo-Martin, Shatavia Morrison, Christopher Gulvick, Scott Sammons, Yvette Unoarumhi, Darlene Wagner, Matthew Schmerer, Cyndi Clark, Patrick Campbell, Rob Case, Vikramsinha Ghorpade, Holly Houdeshell, Ola Kvalvaag, Dillon Nall, Ethan Sanders, Alec Vest, Shaun Westlund, Matthew Hardison, Clinton R. Paden, Duncan MacCannell |
| EPI_ISL_2201722 | Utah Public Health Laboratory | Utah Public Health Laboratory | Erin L. Young, Kelly F. Oakeson, Tara Gallagher |
| EPI_ISL_2201723 | Aegis Sciences Corporation | Centers for Disease Control and Prevention Division of Viral Diseases, Pathogen Discovery | Dakota Howard, Dhwani Batra, Peter W. Cook, Kara Moser, Adrian Paskey, Jason Caravas, Benjamin Rambo-Martin, Shatavia Morrison, Christopher Gulvick, Scott Sammons, Yvette Unoarumhi, Darlene Wagner, Matthew Schmerer, Cyndi Clark, Patrick Campbell, Rob Case, Vikramsinha Ghorpade, Holly Houdeshell, Ola Kvalvaag, Dillon Nall, Ethan Sanders, Alec Vest, Shaun Westlund, Matthew Hardison, Clinton R. Paden, Duncan MacCannell |
| EPI_ISL_2201724 | Houston Methodist Hospital | Houston Methodist Hospital | Randall J. Olsen, Paul A. Christensen, S. Wesley Long, Sishir Subedi, Robert Olson, Marcus Nguyen, James J. Davis, Matthew Ojeda Saavedra, Prasanti Yerramilli, Layne Pruitt, Kristina Reppond, Madison N. Shyer, Jessica Cambric, Ryan Gadd, Ilya J. Finkelstein, Jimmy Gollihar, and James M. Musser |
| EPI_ISL_2201725, EPI_ISL_2201726 | Utah Public Health Laboratory | Utah Public Health Laboratory | Erin L. Young, Kelly F. Oakeson, Tara Gallagher |
| EPI_ISL_2201727 | Aegis Sciences Corporation | Centers for Disease Control and Prevention Division of Viral Diseases, Pathogen Discovery | Dakota Howard, Dhwani Batra, Peter W. Cook, Kara Moser, Adrian Paskey, Jason Caravas, Benjamin Rambo-Martin, Shatavia Morrison, Christopher Gulvick, Scott Sammons, Yvette Unoarumhi, Darlene Wagner, Matthew Schmerer, Cyndi Clark, Patrick Campbell, Rob Case, Vikramsinha Ghorpade, Holly Houdeshell, Ola Kvalvaag, Dillon Nall, Ethan Sanders, Alec Vest, Shaun Westlund, Matthew Hardison, Clinton R. Paden, Duncan MacCannell |
| EPI_ISL_2201728 | Utah Public Health Laboratory | Utah Public Health Laboratory | Erin L. Young, Kelly F. Oakeson, Tara Gallagher |
| EPI_ISL_2201729 | Houston Methodist Hospital | Houston Methodist Hospital | Randall J. Olsen, Paul A. Christensen, S. Wesley Long, Sishir Subedi, Robert Olson, Marcus Nguyen, James J. Davis, Matthew Ojeda Saavedra, Prasanti Yerramilli, Layne Pruitt, Kristina Reppond, Madison N. Shyer, Jessica Cambric, Ryan Gadd, Ilya J. Finkelstein, Jimmy Gollihar, and James M. Musser |
| EPI_ISL_2201730 | Utah Public Health Laboratory | Utah Public Health Laboratory | Erin L. Young, Kelly F. Oakeson, Tara Gallagher |

| | | | |
|---|---|---|---|
| EPI_ISL_2201731 | Aegis Sciences Corporation | Centers for Disease Control and Prevention Division of Viral Diseases, Pathogen Discovery | Dakota Howard, Dhwani Batra, Peter W. Cook, Kara Moser, Adrian Paskey, Jason Caravas, Benjamin Rambo-Martin, Shatavia Morrison, Christopher Gulvick, Scott Sammons, Yvette Unoarumhi, Darlene Wagner, Matthew Schmerer, Cyndi Clark, Patrick Campbell, Rob Case, Vikramsinha Ghorpade, Holly Houdeshell, Ola Kvalvaag, Dillon Nall, Ethan Sanders, Alec Vest, Shaun Westlund, Matthew Hardison, Clinton R. Paden, Duncan MacCannell |
| EPI_ISL_2201732 | Houston Methodist Hospital | Houston Methodist Hospital | Randall J. Olsen, Paul A. Christensen, S. Wesley Long, Sishir Subedi, Robert Olson, Marcus Nguyen, James J. Davis, Matthew Ojeda Saavedra, Prasanti Yerramilli, Layne Pruitt, Kristina Reppond, Madison N. Shyer, Jessica Cambric, Ryan Gadd, Ilya J. Finkelstein, Jimmy Gollihar, and James M. Musser |
| EPI_ISL_2201733 | Aegis Sciences Corporation | Centers for Disease Control and Prevention Division of Viral Diseases, Pathogen Discovery | Dakota Howard, Dhwani Batra, Peter W. Cook, Kara Moser, Adrian Paskey, Jason Caravas, Benjamin Rambo-Martin, Shatavia Morrison, Christopher Gulvick, Scott Sammons, Yvette Unoarumhi, Darlene Wagner, Matthew Schmerer, Cyndi Clark, Patrick Campbell, Rob Case, Vikramsinha Ghorpade, Holly Houdeshell, Ola Kvalvaag, Dillon Nall, Ethan Sanders, Alec Vest, Shaun Westlund, Matthew Hardison, Clinton R. Paden, Duncan MacCannell |
| EPI_ISL_2201734 | Houston Methodist Hospital | Houston Methodist Hospital | Randall J. Olsen, Paul A. Christensen, S. Wesley Long, Sishir Subedi, Robert Olson, Marcus Nguyen, James J. Davis, Matthew Ojeda Saavedra, Prasanti Yerramilli, Layne Pruitt, Kristina Reppond, Madison N. Shyer, Jessica Cambric, Ryan Gadd, Ilya J. Finkelstein, Jimmy Gollihar, and James M. Musser |
| EPI_ISL_2201735 | Utah Public Health Laboratory | Utah Public Health Laboratory | Erin L. Young, Kelly F. Oakeson, Tara Gallagher |
| EPI_ISL_2201736 | Aegis Sciences Corporation | Centers for Disease Control and Prevention Division of Viral Diseases, Pathogen Discovery | Dakota Howard, Dhwani Batra, Peter W. Cook, Kara Moser, Adrian Paskey, Jason Caravas, Benjamin Rambo-Martin, Shatavia Morrison, Christopher Gulvick, Scott Sammons, Yvette Unoarumhi, Darlene Wagner, Matthew Schmerer, Cyndi Clark, Patrick Campbell, Rob Case, Vikramsinha Ghorpade, Holly Houdeshell, Ola Kvalvaag, Dillon Nall, Ethan Sanders, Alec Vest, Shaun Westlund, Matthew Hardison, Clinton R. Paden, Duncan MacCannell |
| EPI_ISL_2201737 | Utah Public Health Laboratory | Utah Public Health Laboratory | Erin L. Young, Kelly F. Oakeson, Tara Gallagher |
| EPI_ISL_2201738 | Houston Methodist Hospital | Houston Methodist Hospital | Randall J. Olsen, Paul A. Christensen, S. Wesley Long, Sishir Subedi, Robert Olson, Marcus Nguyen, James J. Davis, Matthew Ojeda Saavedra, Prasanti Yerramilli, Layne Pruitt, Kristina Reppond, Madison N. Shyer, Jessica Cambric, Ryan Gadd, Ilya J. Finkelstein, Jimmy Gollihar, and James M. Musser |
| EPI_ISL_2201739, EPI_ISL_2201740 | Aegis Sciences Corporation | Centers for Disease Control and Prevention Division of Viral Diseases, Pathogen Discovery | Dakota Howard, Dhwani Batra, Peter W. Cook, Kara Moser, Adrian Paskey, Jason Caravas, Benjamin Rambo-Martin, Shatavia Morrison, Christopher Gulvick, Scott Sammons, Yvette Unoarumhi, Darlene Wagner, Matthew Schmerer, Cyndi Clark, Patrick Campbell, Rob Case, Vikramsinha Ghorpade, Holly Houdeshell, Ola Kvalvaag, Dillon Nall, Ethan Sanders, Alec Vest, Shaun Westlund, Matthew Hardison, Clinton R. Paden, Duncan MacCannell |
| EPI_ISL_2201741 | Houston Methodist Hospital | Houston Methodist Hospital | Randall J. Olsen, Paul A. Christensen, S. Wesley Long, Sishir Subedi, Robert Olson, Marcus Nguyen, James J. Davis, Matthew Ojeda Saavedra, Prasanti Yerramilli, Layne Pruitt, Kristina Reppond, Madison N. Shyer, Jessica Cambric, Ryan Gadd, Ilya J. Finkelstein, Jimmy Gollihar, and James M. Musser |
| EPI_ISL_2201742 | Aegis Sciences Corporation | Centers for Disease Control and Prevention Division of Viral Diseases, Pathogen Discovery | Dakota Howard, Dhwani Batra, Peter W. Cook, Kara Moser, Adrian Paskey, Jason Caravas, Benjamin Rambo-Martin, Shatavia Morrison, Christopher Gulvick, Scott Sammons, Yvette Unoarumhi, Darlene Wagner, Matthew Schmerer, Cyndi Clark, Patrick Campbell, Rob Case, Vikramsinha Ghorpade, Holly Houdeshell, Ola Kvalvaag, Dillon Nall, Ethan Sanders, Alec Vest, Shaun Westlund, Matthew Hardison, Clinton R. Paden, Duncan MacCannell |
| EPI_ISL_2201743 | Houston Methodist Hospital | Houston Methodist Hospital | Randall J. Olsen, Paul A. Christensen, S. Wesley Long, Sishir Subedi, Robert Olson, Marcus Nguyen, James J. Davis, Matthew Ojeda Saavedra, Prasanti Yerramilli, Layne Pruitt, Kristina Reppond, Madison N. Shyer, Jessica Cambric, Ryan Gadd, Ilya J. Finkelstein, Jimmy Gollihar, and James M. Musser |
| EPI_ISL_2201744 | Aegis Sciences Corporation | Centers for Disease Control and Prevention Division of Viral Diseases, Pathogen Discovery | Dakota Howard, Dhwani Batra, Peter W. Cook, Kara Moser, Adrian Paskey, Jason Caravas, Benjamin Rambo-Martin, Shatavia Morrison, Christopher Gulvick, Scott Sammons, Yvette Unoarumhi, Darlene Wagner, Matthew Schmerer, Cyndi Clark, Patrick Campbell, Rob Case, Vikramsinha Ghorpade, Holly Houdeshell, Ola Kvalvaag, Dillon Nall, Ethan Sanders, Alec Vest, Shaun Westlund, Matthew Hardison, Clinton R. Paden, Duncan MacCannell |
| EPI_ISL_2201745, EPI_ISL_2201746 | Houston Methodist Hospital | Houston Methodist Hospital | Randall J. Olsen, Paul A. Christensen, S. Wesley Long, Sishir Subedi, Robert Olson, Marcus Nguyen, James J. Davis, Matthew Ojeda Saavedra, Prasanti Yerramilli, Layne Pruitt, Kristina Reppond, Madison N. Shyer, Jessica Cambric, Ryan Gadd, Ilya J. Finkelstein, Jimmy Gollihar, and James M. Musser |
| EPI_ISL_2201747 | Aegis Sciences Corporation | Centers for Disease Control and Prevention Division of Viral Diseases, Pathogen Discovery | Dakota Howard, Dhwani Batra, Peter W. Cook, Kara Moser, Adrian Paskey, Jason Caravas, Benjamin Rambo-Martin, Shatavia Morrison, Christopher Gulvick, Scott Sammons, Yvette Unoarumhi, Darlene Wagner, Matthew Schmerer, Cyndi Clark, Patrick Campbell, Rob Case, Vikramsinha Ghorpade, Holly Houdeshell, Ola Kvalvaag, Dillon Nall, Ethan Sanders, Alec Vest, Shaun Westlund, Matthew Hardison, Clinton R. Paden, Duncan MacCannell |
| EPI_ISL_2201748 | Houston Methodist Hospital | Houston Methodist Hospital | Randall J. Olsen, Paul A. Christensen, S. Wesley Long, Sishir Subedi, Robert Olson, Marcus Nguyen, James J. Davis, Matthew Ojeda Saavedra, Prasanti Yerramilli, Layne Pruitt, Kristina Reppond, Madison N. Shyer, Jessica Cambric, Ryan Gadd, Ilya J. Finkelstein, Jimmy Gollihar, and James M. Musser |
| EPI_ISL_2201749 | Aegis Sciences Corporation | Centers for Disease Control and Prevention Division of Viral Diseases, Pathogen Discovery | Dakota Howard, Dhwani Batra, Peter W. Cook, Kara Moser, Adrian Paskey, Jason Caravas, Benjamin Rambo-Martin, Shatavia Morrison, Christopher Gulvick, Scott Sammons, Yvette Unoarumhi, Darlene Wagner, Matthew Schmerer, Cyndi Clark, Patrick Campbell, Rob Case, Vikramsinha Ghorpade, Holly Houdeshell, Ola Kvalvaag, Dillon Nall, Ethan Sanders, Alec Vest, Shaun Westlund, Matthew Hardison, Clinton R. Paden, Duncan MacCannell |
| EPI_ISL_2201750 | Houston Methodist Hospital | Houston Methodist Hospital | Randall J. Olsen, Paul A. Christensen, S. Wesley Long, Sishir Subedi, Robert Olson, Marcus Nguyen, James J. Davis, Matthew Ojeda Saavedra, Prasanti Yerramilli, Layne Pruitt, Kristina Reppond, Madison N. Shyer, Jessica Cambric, Ryan Gadd, Ilya J. Finkelstein, Jimmy Gollihar, and James M. Musser |
| EPI_ISL_2201751 | Aegis Sciences Corporation | Centers for Disease Control and Prevention Division of Viral Diseases, Pathogen Discovery | Dakota Howard, Dhwani Batra, Peter W. Cook, Kara Moser, Adrian Paskey, Jason Caravas, Benjamin Rambo-Martin, Shatavia Morrison, Christopher Gulvick, Scott Sammons, Yvette Unoarumhi, Darlene Wagner, Matthew Schmerer, Cyndi Clark, Patrick Campbell, Rob Case, Vikramsinha Ghorpade, Holly Houdeshell, Ola Kvalvaag, Dillon Nall, Ethan Sanders, Alec Vest, Shaun Westlund, Matthew Hardison, Clinton R. Paden, Duncan MacCannell |
| EPI_ISL_2201752 | Houston Methodist Hospital | Houston Methodist Hospital | Randall J. Olsen, Paul A. Christensen, S. Wesley Long, Sishir Subedi, Robert Olson, Marcus Nguyen, James J. Davis, Matthew Ojeda Saavedra, Prasanti Yerramilli, Layne Pruitt, Kristina Reppond, Madison N. Shyer, Jessica Cambric, Ryan Gadd, Ilya J. Finkelstein, Jimmy Gollihar, and James M. Musser |
| EPI_ISL_2201753, EPI_ISL_2201754 | Aegis Sciences Corporation | Centers for Disease Control and Prevention Division of Viral Diseases, Pathogen Discovery | Dakota Howard, Dhwani Batra, Peter W. Cook, Kara Moser, Adrian Paskey, Jason Caravas, Benjamin Rambo-Martin, Shatavia Morrison, Christopher Gulvick, Scott Sammons, Yvette Unoarumhi, Darlene Wagner, Matthew Schmerer, Cyndi Clark, Patrick Campbell, Rob Case, Vikramsinha Ghorpade, Holly Houdeshell, Ola Kvalvaag, Dillon Nall, Ethan Sanders, Alec Vest, Shaun Westlund, Matthew Hardison, Clinton R. Paden, Duncan MacCannell |
| EPI_ISL_2201755 | Houston Methodist Hospital | Houston Methodist Hospital | Randall J. Olsen, Paul A. Christensen, S. Wesley Long, Sishir Subedi, Robert Olson, Marcus Nguyen, James J. Davis, Matthew Ojeda Saavedra, Prasanti Yerramilli, Layne Pruitt, Kristina Reppond, Madison N. Shyer, Jessica Cambric, Ryan Gadd, Ilya J. Finkelstein, Jimmy Gollihar, and James M. Musser |
| EPI_ISL_2201756 | Aegis Sciences Corporation | Centers for Disease Control and Prevention Division of Viral Diseases, Pathogen Discovery | Dakota Howard, Dhwani Batra, Peter W. Cook, Kara Moser, Adrian Paskey, Jason Caravas, Benjamin Rambo-Martin, Shatavia Morrison, Christopher Gulvick, Scott Sammons, Yvette Unoarumhi, Darlene Wagner, Matthew Schmerer, Cyndi Clark, Patrick Campbell, Rob Case, Vikramsinha Ghorpade, Holly Houdeshell, Ola Kvalvaag, Dillon Nall, Ethan Sanders, Alec Vest, Shaun Westlund, Matthew Hardison, Clinton R. Paden, Duncan MacCannell |
| EPI_ISL_2201757 | Houston Methodist Hospital | Houston Methodist Hospital | Randall J. Olsen, Paul A. Christensen, S. Wesley Long, Sishir Subedi, Robert Olson, Marcus Nguyen, James J. Davis, Matthew Ojeda Saavedra, Prasanti Yerramilli, Layne Pruitt, Kristina Reppond, Madison N. Shyer, Jessica Cambric, Ryan Gadd, Ilya J. Finkelstein, Jimmy Gollihar, and James M. Musser |
| EPI_ISL_2201758 | Aegis Sciences Corporation | Centers for Disease Control and Prevention Division of Viral Diseases, Pathogen Discovery | Dakota Howard, Dhwani Batra, Peter W. Cook, Kara Moser, Adrian Paskey, Jason Caravas, Benjamin Rambo-Martin, Shatavia Morrison, Christopher Gulvick, Scott Sammons, Yvette Unoarumhi, Darlene Wagner, Matthew Schmerer, Cyndi Clark, Patrick Campbell, Rob Case, Vikramsinha Ghorpade, Holly Houdeshell, Ola Kvalvaag, Dillon Nall, Ethan Sanders, Alec Vest, Shaun Westlund, Matthew Hardison, Clinton R. Paden, Duncan MacCannell |
| EPI_ISL_2201759, EPI_ISL_2201762 | Houston Methodist Hospital | Houston Methodist Hospital | Randall J. Olsen, Paul A. Christensen, S. Wesley Long, Sishir Subedi, Robert Olson, Marcus Nguyen, James J. Davis, Matthew Ojeda Saavedra, Prasanti Yerramilli, Layne Pruitt, Kristina Reppond, Madison N. Shyer, Jessica Cambric, Ryan Gadd, Ilya J. Finkelstein, Jimmy Gollihar, and James M. Musser |
| EPI_ISL_2201763, EPI_ISL_2201765, EPI_ISL_2201766, EPI_ISL_2201768 | Aegis Sciences Corporation | Centers for Disease Control and Prevention Division of Viral Diseases, Pathogen Discovery | Dakota Howard, Dhwani Batra, Peter W. Cook, Kara Moser, Adrian Paskey, Jason Caravas, Benjamin Rambo-Martin, Shatavia Morrison, Christopher Gulvick, Scott Sammons, Yvette Unoarumhi, Darlene Wagner, Matthew Schmerer, Cyndi Clark, Patrick Campbell, Rob Case, Vikramsinha Ghorpade, Holly Houdeshell, Ola Kvalvaag, Dillon Nall, Ethan Sanders, Alec Vest, Shaun Westlund, Matthew Hardison, Clinton R. Paden, Duncan MacCannell |
| EPI_ISL_2201769 | Houston Methodist Hospital | Houston Methodist Hospital | Randall J. Olsen, Paul A. Christensen, S. Wesley Long, Sishir Subedi, Robert Olson, Marcus Nguyen, James J. Davis, Matthew Ojeda Saavedra, Prasanti Yerramilli, Layne Pruitt, Kristina Reppond, Madison N. Shyer, Jessica Cambric, Ryan Gadd, Ilya J. Finkelstein, Jimmy Gollihar, and James M. Musser |
| EPI_ISL_2201770 | Aegis Sciences Corporation | Centers for Disease Control and Prevention Division of Viral Diseases, Pathogen Discovery | Dakota Howard, Dhwani Batra, Peter W. Cook, Kara Moser, Adrian Paskey, Jason Caravas, Benjamin Rambo-Martin, Shatavia Morrison, Christopher Gulvick, Scott Sammons, Yvette Unoarumhi, Darlene Wagner, Matthew Schmerer, Cyndi Clark, Patrick Campbell, Rob Case, Vikramsinha Ghorpade, Holly Houdeshell, Ola Kvalvaag, Dillon Nall, Ethan Sanders, Alec Vest, Shaun Westlund, Matthew Hardison, Clinton R. Paden, Duncan MacCannell |
| EPI_ISL_2201771 | Houston Methodist Hospital | Houston Methodist Hospital | Randall J. Olsen, Paul A. Christensen, S. Wesley Long, Sishir Subedi, Robert Olson, Marcus Nguyen, James J. Davis, Matthew Ojeda Saavedra, Prasanti Yerramilli, Layne Pruitt, Kristina Reppond, Madison N. Shyer, Jessica Cambric, Ryan Gadd, Ilya J. Finkelstein, Jimmy Gollihar, and James M. Musser |
| EPI_ISL_2201772, EPI_ISL_2201773, EPI_ISL_2201774 | Aegis Sciences Corporation | Centers for Disease Control and Prevention Division of Viral Diseases, Pathogen Discovery | Dakota Howard, Dhwani Batra, Peter W. Cook, Kara Moser, Adrian Paskey, Jason Caravas, Benjamin Rambo-Martin, Shatavia Morrison, Christopher Gulvick, Scott Sammons, Yvette Unoarumhi, Darlene Wagner, Matthew Schmerer, Cyndi Clark, Patrick Campbell, Rob Case, Vikramsinha Ghorpade, Holly Houdeshell, Ola Kvalvaag, Dillon Nall, Ethan Sanders, Alec Vest, Shaun Westlund, Matthew Hardison, Clinton R. Paden, Duncan MacCannell |
| EPI_ISL_2201777 | Houston Methodist Hospital | Houston Methodist Hospital | Randall J. Olsen, Paul A. Christensen, S. Wesley Long, Sishir Subedi, Robert Olson, Marcus Nguyen, James J. Davis, Matthew Ojeda Saavedra, Prasanti Yerramilli, Layne Pruitt, Kristina Reppond, Madison N. Shyer, Jessica Cambric, Ryan Gadd, Ilya J. Finkelstein, Jimmy Gollihar, and James M. Musser |
| EPI_ISL_2201778 | Aegis Sciences Corporation | Centers for Disease Control and Prevention Division of Viral Diseases, Pathogen Discovery | Dakota Howard, Dhwani Batra, Peter W. Cook, Kara Moser, Adrian Paskey, Jason Caravas, Benjamin Rambo-Martin, Shatavia Morrison, Christopher Gulvick, Scott Sammons, Yvette Unoarumhi, Darlene Wagner, Matthew Schmerer, Cyndi Clark, Patrick Campbell, Rob Case, Vikramsinha Ghorpade, Holly Houdeshell, Ola Kvalvaag, Dillon Nall, Ethan Sanders, Alec Vest, Shaun Westlund, Matthew Hardison, Clinton R. Paden, Duncan MacCannell |
| EPI_ISL_2201779 | Houston Methodist Hospital | Houston Methodist Hospital | Randall J. Olsen, Paul A. Christensen, S. Wesley Long, Sishir Subedi, Robert Olson, Marcus Nguyen, James J. Davis, Matthew Ojeda Saavedra, Prasanti Yerramilli, Layne Pruitt, Kristina Reppond, Madison N. Shyer, Jessica Cambric, Ryan Gadd, Ilya J. Finkelstein, Jimmy Gollihar, and James M. Musser |
| EPI_ISL_2201780, EPI_ISL_2201782 | Aegis Sciences Corporation | Centers for Disease Control and Prevention Division of Viral Diseases, Pathogen Discovery | Dakota Howard, Dhwani Batra, Peter W. Cook, Kara Moser, Adrian Paskey, Jason Caravas, Benjamin Rambo-Martin, Shatavia Morrison, Christopher Gulvick, Scott Sammons, Yvette Unoarumhi, Darlene Wagner, Matthew Schmerer, Cyndi Clark, Patrick Campbell, Rob Case, Vikramsinha Ghorpade, Holly Houdeshell, Ola Kvalvaag, Dillon Nall, Ethan Sanders, Alec Vest, Shaun Westlund, Matthew Hardison, Clinton R. Paden, Duncan MacCannell |
| EPI_ISL_2201783 | Houston Methodist Hospital | Houston Methodist Hospital | Randall J. Olsen, Paul A. Christensen, S. Wesley Long, Sishir Subedi, Robert Olson, Marcus Nguyen, James J. Davis, Matthew Ojeda Saavedra, Prasanti Yerramilli, Layne Pruitt, Kristina Reppond, Madison N. Shyer, Jessica Cambric, Ryan Gadd, Ilya J. Finkelstein, Jimmy Gollihar, and James M. Musser |
| EPI_ISL_2201784, EPI_ISL_2201785 | Aegis Sciences Corporation | Centers for Disease Control and Prevention Division of Viral Diseases, Pathogen Discovery | Dakota Howard, Dhwani Batra, Peter W. Cook, Kara Moser, Adrian Paskey, Jason Caravas, Benjamin Rambo-Martin, Shatavia Morrison, Christopher Gulvick, Scott Sammons, Yvette Unoarumhi, Darlene Wagner, Matthew Schmerer, Cyndi Clark, Patrick Campbell, Rob Case, Vikramsinha Ghorpade, Holly Houdeshell, Ola Kvalvaag, Dillon Nall, Ethan Sanders, Alec Vest, Shaun Westlund, Matthew Hardison, Clinton R. Paden, Duncan MacCannell |
| EPI_ISL_2201786 | Houston Methodist Hospital | Houston Methodist Hospital | Randall J. Olsen, Paul A. Christensen, S. Wesley Long, Sishir Subedi, Robert Olson, Marcus Nguyen, James J. Davis, Matthew Ojeda Saavedra, Prasanti Yerramilli, Layne Pruitt, Kristina Reppond, Madison N. Shyer, Jessica Cambric, Ryan Gadd, Ilya J. Finkelstein, Jimmy Gollihar, and James |

| | | | |
|---|---|---|---|
| | | | M. Musser |
| EPI_ISL_2201787, EPI_ISL_2201789 | Aegis Sciences Corporation | Centers for Disease Control and Prevention Division of Viral Diseases, Pathogen Discovery | Dakota Howard, Dhwani Batra, Peter W. Cook, Kara Moser, Adrian Paskey, Jason Caravas, Benjamin Rambo-Martin, Shatavia Morrison, Christopher Gulvick, Scott Sammons, Yvette Unoarumhi, Darlene Wagner, Matthew Schmerer, Cyndi Clark, Patrick Campbell, Rob Case, Vikramsinha Ghorpade, Holly Houdeshell, Ola Kvalvaag, Dillon Nall, Ethan Sanders, Alec Vest, Shaun Westlund, Matthew Hardison, Clinton R. Paden, Duncan MacCannell |
| EPI_ISL_2201790 | Houston Methodist Hospital | Houston Methodist Hospital | Randall J. Olsen, Paul A. Christensen, S. Wesley Long, Sishir Subedi, Robert Olson, Marcus Nguyen, James J. Davis, Matthew Ojeda Saavedra, Prasanti Yerramilli, Layne Pruitt, Kristina Reppond, Madison N. Shyer, Jessica Cambric, Ryan Gadd, Ilya J. Finkelstein, Jimmy Gollihar, and James M. Musser |
| EPI_ISL_2201791 | Aegis Sciences Corporation | Centers for Disease Control and Prevention Division of Viral Diseases, Pathogen Discovery | Dakota Howard, Dhwani Batra, Peter W. Cook, Kara Moser, Adrian Paskey, Jason Caravas, Benjamin Rambo-Martin, Shatavia Morrison, Christopher Gulvick, Scott Sammons, Yvette Unoarumhi, Darlene Wagner, Matthew Schmerer, Cyndi Clark, Patrick Campbell, Rob Case, Vikramsinha Ghorpade, Holly Houdeshell, Ola Kvalvaag, Dillon Nall, Ethan Sanders, Alec Vest, Shaun Westlund, Matthew Hardison, Clinton R. Paden, Duncan MacCannell |
| EPI_ISL_2201792 | Houston Methodist Hospital | Houston Methodist Hospital | Randall J. Olsen, Paul A. Christensen, S. Wesley Long, Sishir Subedi, Robert Olson, Marcus Nguyen, James J. Davis, Matthew Ojeda Saavedra, Prasanti Yerramilli, Layne Pruitt, Kristina Reppond, Madison N. Shyer, Jessica Cambric, Ryan Gadd, Ilya J. Finkelstein, Jimmy Gollihar, and James M. Musser |
| EPI_ISL_2201793, EPI_ISL_2201794 | Aegis Sciences Corporation | Centers for Disease Control and Prevention Division of Viral Diseases, Pathogen Discovery | Dakota Howard, Dhwani Batra, Peter W. Cook, Kara Moser, Adrian Paskey, Jason Caravas, Benjamin Rambo-Martin, Shatavia Morrison, Christopher Gulvick, Scott Sammons, Yvette Unoarumhi, Darlene Wagner, Matthew Schmerer, Cyndi Clark, Patrick Campbell, Rob Case, Vikramsinha Ghorpade, Holly Houdeshell, Ola Kvalvaag, Dillon Nall, Ethan Sanders, Alec Vest, Shaun Westlund, Matthew Hardison, Clinton R. Paden, Duncan MacCannell |
| EPI_ISL_2201795 | Houston Methodist Hospital | Houston Methodist Hospital | Randall J. Olsen, Paul A. Christensen, S. Wesley Long, Sishir Subedi, Robert Olson, Marcus Nguyen, James J. Davis, Matthew Ojeda Saavedra, Prasanti Yerramilli, Layne Pruitt, Kristina Reppond, Madison N. Shyer, Jessica Cambric, Ryan Gadd, Ilya J. Finkelstein, Jimmy Gollihar, and James M. Musser |
| EPI_ISL_2201796 | Aegis Sciences Corporation | Centers for Disease Control and Prevention Division of Viral Diseases, Pathogen Discovery | Dakota Howard, Dhwani Batra, Peter W. Cook, Kara Moser, Adrian Paskey, Jason Caravas, Benjamin Rambo-Martin, Shatavia Morrison, Christopher Gulvick, Scott Sammons, Yvette Unoarumhi, Darlene Wagner, Matthew Schmerer, Cyndi Clark, Patrick Campbell, Rob Case, Vikramsinha Ghorpade, Holly Houdeshell, Ola Kvalvaag, Dillon Nall, Ethan Sanders, Alec Vest, Shaun Westlund, Matthew Hardison, Clinton R. Paden, Duncan MacCannell |
| EPI_ISL_2201797 | Houston Methodist Hospital | Houston Methodist Hospital | Randall J. Olsen, Paul A. Christensen, S. Wesley Long, Sishir Subedi, Robert Olson, Marcus Nguyen, James J. Davis, Matthew Ojeda Saavedra, Prasanti Yerramilli, Layne Pruitt, Kristina Reppond, Madison N. Shyer, Jessica Cambric, Ryan Gadd, Ilya J. Finkelstein, Jimmy Gollihar, and James M. Musser |
| EPI_ISL_2201798, EPI_ISL_2201799 | Aegis Sciences Corporation | Centers for Disease Control and Prevention Division of Viral Diseases, Pathogen Discovery | Dakota Howard, Dhwani Batra, Peter W. Cook, Kara Moser, Adrian Paskey, Jason Caravas, Benjamin Rambo-Martin, Shatavia Morrison, Christopher Gulvick, Scott Sammons, Yvette Unoarumhi, Darlene Wagner, Matthew Schmerer, Cyndi Clark, Patrick Campbell, Rob Case, Vikramsinha Ghorpade, Holly Houdeshell, Ola Kvalvaag, Dillon Nall, Ethan Sanders, Alec Vest, Shaun Westlund, Matthew Hardison, Clinton R. Paden, Duncan MacCannell |
| EPI_ISL_2201800 | Houston Methodist Hospital | Houston Methodist Hospital | Randall J. Olsen, Paul A. Christensen, S. Wesley Long, Sishir Subedi, Robert Olson, Marcus Nguyen, James J. Davis, Matthew Ojeda Saavedra, Prasanti Yerramilli, Layne Pruitt, Kristina Reppond, Madison N. Shyer, Jessica Cambric, Ryan Gadd, Ilya J. Finkelstein, Jimmy Gollihar, and James M. Musser |
| EPI_ISL_2201801 | Aegis Sciences Corporation | Centers for Disease Control and Prevention Division of Viral Diseases, Pathogen Discovery | Dakota Howard, Dhwani Batra, Peter W. Cook, Kara Moser, Adrian Paskey, Jason Caravas, Benjamin Rambo-Martin, Shatavia Morrison, Christopher Gulvick, Scott Sammons, Yvette Unoarumhi, Darlene Wagner, Matthew Schmerer, Cyndi Clark, Patrick Campbell, Rob Case, Vikramsinha Ghorpade, Holly Houdeshell, Ola Kvalvaag, Dillon Nall, Ethan Sanders, Alec Vest, Shaun Westlund, Matthew Hardison, Clinton R. Paden, Duncan MacCannell |
| EPI_ISL_2201802 | Houston Methodist Hospital | Houston Methodist Hospital | Randall J. Olsen, Paul A. Christensen, S. Wesley Long, Sishir Subedi, Robert Olson, Marcus Nguyen, James J. Davis, Matthew Ojeda Saavedra, Prasanti Yerramilli, Layne Pruitt, Kristina Reppond, Madison N. Shyer, Jessica Cambric, Ryan Gadd, Ilya J. Finkelstein, Jimmy Gollihar, and James M. Musser |
| EPI_ISL_2201803 | Aegis Sciences Corporation | Centers for Disease Control and Prevention Division of Viral Diseases, Pathogen Discovery | Dakota Howard, Dhwani Batra, Peter W. Cook, Kara Moser, Adrian Paskey, Jason Caravas, Benjamin Rambo-Martin, Shatavia Morrison, Christopher Gulvick, Scott Sammons, Yvette Unoarumhi, Darlene Wagner, Matthew Schmerer, Cyndi Clark, Patrick Campbell, Rob Case, Vikramsinha Ghorpade, Holly Houdeshell, Ola Kvalvaag, Dillon Nall, Ethan Sanders, Alec Vest, Shaun Westlund, Matthew Hardison, Clinton R. Paden, Duncan MacCannell |
| EPI_ISL_2201804 | Houston Methodist Hospital | Houston Methodist Hospital | Randall J. Olsen, Paul A. Christensen, S. Wesley Long, Sishir Subedi, Robert Olson, Marcus Nguyen, James J. Davis, Matthew Ojeda Saavedra, Prasanti Yerramilli, Layne Pruitt, Kristina Reppond, Madison N. Shyer, Jessica Cambric, Ryan Gadd, Ilya J. Finkelstein, Jimmy Gollihar, and James M. Musser |
| EPI_ISL_2201805, EPI_ISL_2201806 | Aegis Sciences Corporation | Centers for Disease Control and Prevention Division of Viral Diseases, Pathogen Discovery | Dakota Howard, Dhwani Batra, Peter W. Cook, Kara Moser, Adrian Paskey, Jason Caravas, Benjamin Rambo-Martin, Shatavia Morrison, Christopher Gulvick, Scott Sammons, Yvette Unoarumhi, Darlene Wagner, Matthew Schmerer, Cyndi Clark, Patrick Campbell, Rob Case, Vikramsinha Ghorpade, Holly Houdeshell, Ola Kvalvaag, Dillon Nall, Ethan Sanders, Alec Vest, Shaun Westlund, Matthew Hardison, Clinton R. Paden, Duncan MacCannell |
| EPI_ISL_2201807 | Houston Methodist Hospital | Houston Methodist Hospital | Randall J. Olsen, Paul A. Christensen, S. Wesley Long, Sishir Subedi, Robert Olson, Marcus Nguyen, James J. Davis, Matthew Ojeda Saavedra, Prasanti Yerramilli, Layne Pruitt, Kristina Reppond, Madison N. Shyer, Jessica Cambric, Ryan Gadd, Ilya J. Finkelstein, Jimmy Gollihar, and James M. Musser |
| EPI_ISL_2201808 | Aegis Sciences Corporation | Centers for Disease Control and Prevention Division of Viral Diseases, Pathogen Discovery | Dakota Howard, Dhwani Batra, Peter W. Cook, Kara Moser, Adrian Paskey, Jason Caravas, Benjamin Rambo-Martin, Shatavia Morrison, Christopher Gulvick, Scott Sammons, Yvette Unoarumhi, Darlene Wagner, Matthew Schmerer, Cyndi Clark, Patrick Campbell, Rob Case, Vikramsinha Ghorpade, Holly Houdeshell, Ola Kvalvaag, Dillon Nall, Ethan Sanders, Alec Vest, Shaun Westlund, Matthew Hardison, Clinton R. Paden, Duncan MacCannell |
| EPI_ISL_2201809 | Houston Methodist Hospital | Houston Methodist Hospital | Randall J. Olsen, Paul A. Christensen, S. Wesley Long, Sishir Subedi, Robert Olson, Marcus Nguyen, James J. Davis, Matthew Ojeda Saavedra, Prasanti Yerramilli, Layne Pruitt, Kristina Reppond, Madison N. Shyer, Jessica Cambric, Ryan Gadd, Ilya J. Finkelstein, Jimmy Gollihar, and James M. Musser |
| EPI_ISL_2201810 | Aegis Sciences Corporation | Centers for Disease Control and Prevention Division of Viral Diseases, Pathogen Discovery | Dakota Howard, Dhwani Batra, Peter W. Cook, Kara Moser, Adrian Paskey, Jason Caravas, Benjamin Rambo-Martin, Shatavia Morrison, Christopher Gulvick, Scott Sammons, Yvette Unoarumhi, Darlene Wagner, Matthew Schmerer, Cyndi Clark, Patrick Campbell, Rob Case, Vikramsinha Ghorpade, Holly Houdeshell, Ola Kvalvaag, Dillon Nall, Ethan Sanders, Alec Vest, Shaun Westlund, Matthew Hardison, Clinton R. Paden, Duncan MacCannell |
| EPI_ISL_2201811 | Houston Methodist Hospital | Houston Methodist Hospital | Randall J. Olsen, Paul A. Christensen, S. Wesley Long, Sishir Subedi, Robert Olson, Marcus Nguyen, James J. Davis, Matthew Ojeda Saavedra, Prasanti Yerramilli, Layne Pruitt, Kristina Reppond, Madison N. Shyer, Jessica Cambric, Ryan Gadd, Ilya J. Finkelstein, Jimmy Gollihar, and James M. Musser |
| EPI_ISL_2201812 | Aegis Sciences Corporation | Centers for Disease Control and Prevention Division of Viral Diseases, Pathogen Discovery | Dakota Howard, Dhwani Batra, Peter W. Cook, Kara Moser, Adrian Paskey, Jason Caravas, Benjamin Rambo-Martin, Shatavia Morrison, Christopher Gulvick, Scott Sammons, Yvette Unoarumhi, Darlene Wagner, Matthew Schmerer, Cyndi Clark, Patrick Campbell, Rob Case, Vikramsinha Ghorpade, Holly Houdeshell, Ola Kvalvaag, Dillon Nall, Ethan Sanders, Alec Vest, Shaun Westlund, Matthew Hardison, Clinton R. Paden, Duncan MacCannell |
| EPI_ISL_2201813 | Houston Methodist Hospital | Houston Methodist Hospital | Randall J. Olsen, Paul A. Christensen, S. Wesley Long, Sishir Subedi, Robert Olson, Marcus Nguyen, James J. Davis, Matthew Ojeda Saavedra, Prasanti Yerramilli, Layne Pruitt, Kristina Reppond, Madison N. Shyer, Jessica Cambric, Ryan Gadd, Ilya J. Finkelstein, Jimmy Gollihar, and James M. Musser |
| EPI_ISL_2201814, EPI_ISL_2201815 | Aegis Sciences Corporation | Centers for Disease Control and Prevention Division of Viral Diseases, Pathogen Discovery | Dakota Howard, Dhwani Batra, Peter W. Cook, Kara Moser, Adrian Paskey, Jason Caravas, Benjamin Rambo-Martin, Shatavia Morrison, Christopher Gulvick, Scott Sammons, Yvette Unoarumhi, Darlene Wagner, Matthew Schmerer, Cyndi Clark, Patrick Campbell, Rob Case, Vikramsinha Ghorpade, Holly Houdeshell, Ola Kvalvaag, Dillon Nall, Ethan Sanders, Alec Vest, Shaun Westlund, Matthew Hardison, Clinton R. Paden, Duncan MacCannell |
| EPI_ISL_2201816 | Houston Methodist Hospital | Houston Methodist Hospital | Randall J. Olsen, Paul A. Christensen, S. Wesley Long, Sishir Subedi, Robert Olson, Marcus Nguyen, James J. Davis, Matthew Ojeda Saavedra, Prasanti Yerramilli, Layne Pruitt, Kristina Reppond, Madison N. Shyer, Jessica Cambric, Ryan Gadd, Ilya J. Finkelstein, Jimmy Gollihar, and James M. Musser |
| EPI_ISL_2201817 | Aegis Sciences Corporation | Centers for Disease Control and Prevention Division of Viral Diseases, Pathogen Discovery | Dakota Howard, Dhwani Batra, Peter W. Cook, Kara Moser, Adrian Paskey, Jason Caravas, Benjamin Rambo-Martin, Shatavia Morrison, Christopher Gulvick, Scott Sammons, Yvette Unoarumhi, Darlene Wagner, Matthew Schmerer, Cyndi Clark, Patrick Campbell, Rob Case, Vikramsinha Ghorpade, Holly Houdeshell, Ola Kvalvaag, Dillon Nall, Ethan Sanders, Alec Vest, Shaun Westlund, Matthew Hardison, Clinton R. Paden, Duncan MacCannell |
| EPI_ISL_2201818 | Houston Methodist Hospital | Houston Methodist Hospital | Randall J. Olsen, Paul A. Christensen, S. Wesley Long, Sishir Subedi, Robert Olson, Marcus Nguyen, James J. Davis, Matthew Ojeda Saavedra, Prasanti Yerramilli, Layne Pruitt, Kristina Reppond, Madison N. Shyer, Jessica Cambric, Ryan Gadd, Ilya J. Finkelstein, Jimmy Gollihar, and James M. Musser |
| EPI_ISL_2201819 | Aegis Sciences Corporation | Centers for Disease Control and Prevention Division of Viral Diseases, Pathogen Discovery | Dakota Howard, Dhwani Batra, Peter W. Cook, Kara Moser, Adrian Paskey, Jason Caravas, Benjamin Rambo-Martin, Shatavia Morrison, Christopher Gulvick, Scott Sammons, Yvette Unoarumhi, Darlene Wagner, Matthew Schmerer, Cyndi Clark, Patrick Campbell, Rob Case, Vikramsinha Ghorpade, Holly Houdeshell, Ola Kvalvaag, Dillon Nall, Ethan Sanders, Alec Vest, Shaun Westlund, Matthew Hardison, Clinton R. Paden, Duncan MacCannell |
| EPI_ISL_2201820 | Houston Methodist Hospital | Houston Methodist Hospital | Randall J. Olsen, Paul A. Christensen, S. Wesley Long, Sishir Subedi, Robert Olson, Marcus Nguyen, James J. Davis, Matthew Ojeda Saavedra, Prasanti Yerramilli, Layne Pruitt, Kristina Reppond, Madison N. Shyer, Jessica Cambric, Ryan Gadd, Ilya J. Finkelstein, Jimmy Gollihar, and James M. Musser |
| EPI_ISL_2201821 | Aegis Sciences Corporation | Centers for Disease Control and Prevention Division of Viral Diseases, Pathogen Discovery | Dakota Howard, Dhwani Batra, Peter W. Cook, Kara Moser, Adrian Paskey, Jason Caravas, Benjamin Rambo-Martin, Shatavia Morrison, Christopher Gulvick, Scott Sammons, Yvette Unoarumhi, Darlene Wagner, Matthew Schmerer, Cyndi Clark, Patrick Campbell, Rob Case, Vikramsinha Ghorpade, Holly Houdeshell, Ola Kvalvaag, Dillon Nall, Ethan Sanders, Alec Vest, Shaun Westlund, Matthew Hardison, Clinton R. Paden, Duncan MacCannell |
| EPI_ISL_2201822 | Houston Methodist Hospital | Houston Methodist Hospital | Randall J. Olsen, Paul A. Christensen, S. Wesley Long, Sishir Subedi, Robert Olson, Marcus Nguyen, James J. Davis, Matthew Ojeda Saavedra, Prasanti Yerramilli, Layne Pruitt, Kristina Reppond, Madison N. Shyer, Jessica Cambric, Ryan Gadd, Ilya J. Finkelstein, Jimmy Gollihar, and James M. Musser |
| EPI_ISL_2201823 | Aegis Sciences Corporation | Centers for Disease Control and Prevention Division of Viral Diseases, Pathogen Discovery | Dakota Howard, Dhwani Batra, Peter W. Cook, Kara Moser, Adrian Paskey, Jason Caravas, Benjamin Rambo-Martin, Shatavia Morrison, Christopher Gulvick, Scott Sammons, Yvette Unoarumhi, Darlene Wagner, Matthew Schmerer, Cyndi Clark, Patrick Campbell, Rob Case, Vikramsinha Ghorpade, Holly Houdeshell, Ola Kvalvaag, Dillon Nall, Ethan Sanders, Alec Vest, Shaun Westlund, Matthew Hardison, Clinton R. Paden, Duncan MacCannell |
| EPI_ISL_2201824 | Houston Methodist Hospital | Houston Methodist Hospital | Randall J. Olsen, Paul A. Christensen, S. Wesley Long, Sishir Subedi, Robert Olson, Marcus Nguyen, James J. Davis, Matthew Ojeda Saavedra, Prasanti Yerramilli, Layne Pruitt, Kristina Reppond, Madison N. Shyer, Jessica Cambric, Ryan Gadd, Ilya J. Finkelstein, Jimmy Gollihar, and James M. Musser |
| EPI_ISL_2201825, EPI_ISL_2201826 | Aegis Sciences Corporation | Centers for Disease Control and Prevention Division of Viral Diseases, Pathogen Discovery | Dakota Howard, Dhwani Batra, Peter W. Cook, Kara Moser, Adrian Paskey, Jason Caravas, Benjamin Rambo-Martin, Shatavia Morrison, Christopher Gulvick, Scott Sammons, Yvette Unoarumhi, Darlene Wagner, Matthew Schmerer, Cyndi Clark, Patrick Campbell, Rob Case, Vikramsinha Ghorpade, Holly Houdeshell, Ola Kvalvaag, Dillon Nall, Ethan Sanders, Alec Vest, Shaun Westlund, Matthew Hardison, Clinton R. Paden, Duncan MacCannell |
| EPI_ISL_2201827 | Houston Methodist Hospital | Houston Methodist Hospital | Randall J. Olsen, Paul A. Christensen, S. Wesley Long, Sishir Subedi, Robert Olson, Marcus Nguyen, James J. Davis, Matthew Ojeda Saavedra, Prasanti Yerramilli, Layne Pruitt, Kristina Reppond, Madison N. Shyer, Jessica Cambric, Ryan Gadd, Ilya J. Finkelstein, Jimmy Gollihar, and James M. Musser |
| EPI_ISL_2201828 | Aegis Sciences Corporation | Centers for Disease Control and Prevention Division of Viral Diseases, Pathogen Discovery | Dakota Howard, Dhwani Batra, Peter W. Cook, Kara Moser, Adrian Paskey, Jason Caravas, Benjamin Rambo-Martin, Shatavia Morrison, Christopher Gulvick, Scott Sammons, Yvette Unoarumhi, Darlene Wagner, Matthew Schmerer, Cyndi Clark, Patrick Campbell, Rob Case, Vikramsinha Ghorpade, Holly Houdeshell, Ola Kvalvaag, Dillon Nall, Ethan Sanders, Alec Vest, Shaun Westlund, Matthew Hardison, Clinton R. Paden, Duncan MacCannell |
| EPI_ISL_2201829 | Houston Methodist Hospital | Houston Methodist Hospital | Randall J. Olsen, Paul A. Christensen, S. Wesley Long, Sishir Subedi, Robert Olson, Marcus Nguyen, James J. Davis, Matthew Ojeda Saavedra, Prasanti Yerramilli, Layne Pruitt, Kristina Reppond, Madison N. Shyer, Jessica Cambric, Ryan Gadd, Ilya J. Finkelstein, Jimmy Gollihar, and James M. Musser |
| EPI_ISL_2201830, EPI_ISL_2201831 | Aegis Sciences Corporation | Centers for Disease Control and Prevention Division of Viral Diseases, Pathogen Discovery | Dakota Howard, Dhwani Batra, Peter W. Cook, Kara Moser, Adrian Paskey, Jason Caravas, Benjamin Rambo-Martin, Shatavia Morrison, Christopher Gulvick, Scott Sammons, Yvette Unoarumhi, Darlene Wagner, Matthew Schmerer, Cyndi Clark, Patrick Campbell, Rob Case, Vikramsinha Ghorpade, Holly Houdeshell, Ola Kvalvaag, Dillon Nall, Ethan Sanders, Alec Vest, Shaun Westlund, Matthew Hardison, Clinton R. Paden, Duncan MacCannell |

| | | | |
|---|---|---|---|
| EPI_ISL_2201832 | Houston Methodist Hospital | Houston Methodist Hospital | Randall J. Olsen, Paul A. Christensen, S. Wesley Long, Sishir Subedi, Robert Olson, Marcus Nguyen, James J. Davis, Matthew Ojeda Saavedra, Prasanti Yerramilli, Layne Pruitt, Kristina Reppond, Madison N. Shyer, Jessica Cambric, Ryan Gadd, Ilya J. Finkelstein, Jimmy Gollihar, and James M. Musser |
| EPI_ISL_2201833 | Aegis Sciences Corporation | Centers for Disease Control and Prevention Division of Viral Diseases, Pathogen Discovery | Dakota Howard, Dhwani Batra, Peter W. Cook, Kara Moser, Adrian Paskey, Jason Caravas, Benjamin Rambo-Martin, Shatavia Morrison, Christopher Gulvick, Scott Sammons, Yvette Unoarumhi, Darlene Wagner, Matthew Schmerer, Cyndi Clark, Patrick Campbell, Rob Case, Vikramsinha Ghorpade, Holly Houdeshell, Ola Kvalvaag, Dillon Nall, Ethan Sanders, Alec Vest, Shaun Westlund, Matthew Hardison, Clinton R. Paden, Duncan MacCannell |
| EPI_ISL_2201834 | Houston Methodist Hospital | Houston Methodist Hospital | Randall J. Olsen, Paul A. Christensen, S. Wesley Long, Sishir Subedi, Robert Olson, Marcus Nguyen, James J. Davis, Matthew Ojeda Saavedra, Prasanti Yerramilli, Layne Pruitt, Kristina Reppond, Madison N. Shyer, Jessica Cambric, Ryan Gadd, Ilya J. Finkelstein, Jimmy Gollihar, and James M. Musser |
| EPI_ISL_2201835 | Aegis Sciences Corporation | Centers for Disease Control and Prevention Division of Viral Diseases, Pathogen Discovery | Dakota Howard, Dhwani Batra, Peter W. Cook, Kara Moser, Adrian Paskey, Jason Caravas, Benjamin Rambo-Martin, Shatavia Morrison, Christopher Gulvick, Scott Sammons, Yvette Unoarumhi, Darlene Wagner, Matthew Schmerer, Cyndi Clark, Patrick Campbell, Rob Case, Vikramsinha Ghorpade, Holly Houdeshell, Ola Kvalvaag, Dillon Nall, Ethan Sanders, Alec Vest, Shaun Westlund, Matthew Hardison, Clinton R. Paden, Duncan MacCannell |
| EPI_ISL_2201836 | Houston Methodist Hospital | Houston Methodist Hospital | Randall J. Olsen, Paul A. Christensen, S. Wesley Long, Sishir Subedi, Robert Olson, Marcus Nguyen, James J. Davis, Matthew Ojeda Saavedra, Prasanti Yerramilli, Layne Pruitt, Kristina Reppond, Madison N. Shyer, Jessica Cambric, Ryan Gadd, Ilya J. Finkelstein, Jimmy Gollihar, and James M. Musser |
| EPI_ISL_2201837, EPI_ISL_2201838 | Aegis Sciences Corporation | Centers for Disease Control and Prevention Division of Viral Diseases, Pathogen Discovery | Dakota Howard, Dhwani Batra, Peter W. Cook, Kara Moser, Adrian Paskey, Jason Caravas, Benjamin Rambo-Martin, Shatavia Morrison, Christopher Gulvick, Scott Sammons, Yvette Unoarumhi, Darlene Wagner, Matthew Schmerer, Cyndi Clark, Patrick Campbell, Rob Case, Vikramsinha Ghorpade, Holly Houdeshell, Ola Kvalvaag, Dillon Nall, Ethan Sanders, Alec Vest, Shaun Westlund, Matthew Hardison, Clinton R. Paden, Duncan MacCannell |
| EPI_ISL_2201839 | Houston Methodist Hospital | Houston Methodist Hospital | Randall J. Olsen, Paul A. Christensen, S. Wesley Long, Sishir Subedi, Robert Olson, Marcus Nguyen, James J. Davis, Matthew Ojeda Saavedra, Prasanti Yerramilli, Layne Pruitt, Kristina Reppond, Madison N. Shyer, Jessica Cambric, Ryan Gadd, Ilya J. Finkelstein, Jimmy Gollihar, and James M. Musser |
| EPI_ISL_2201840 | Aegis Sciences Corporation | Centers for Disease Control and Prevention Division of Viral Diseases, Pathogen Discovery | Dakota Howard, Dhwani Batra, Peter W. Cook, Kara Moser, Adrian Paskey, Jason Caravas, Benjamin Rambo-Martin, Shatavia Morrison, Christopher Gulvick, Scott Sammons, Yvette Unoarumhi, Darlene Wagner, Matthew Schmerer, Cyndi Clark, Patrick Campbell, Rob Case, Vikramsinha Ghorpade, Holly Houdeshell, Ola Kvalvaag, Dillon Nall, Ethan Sanders, Alec Vest, Shaun Westlund, Matthew Hardison, Clinton R. Paden, Duncan MacCannell |
| EPI_ISL_2201841 | Houston Methodist Hospital | Houston Methodist Hospital | Randall J. Olsen, Paul A. Christensen, S. Wesley Long, Sishir Subedi, Robert Olson, Marcus Nguyen, James J. Davis, Matthew Ojeda Saavedra, Prasanti Yerramilli, Layne Pruitt, Kristina Reppond, Madison N. Shyer, Jessica Cambric, Ryan Gadd, Ilya J. Finkelstein, Jimmy Gollihar, and James M. Musser |
| EPI_ISL_2201842, EPI_ISL_2201843 | Aegis Sciences Corporation | Centers for Disease Control and Prevention Division of Viral Diseases, Pathogen Discovery | Dakota Howard, Dhwani Batra, Peter W. Cook, Kara Moser, Adrian Paskey, Jason Caravas, Benjamin Rambo-Martin, Shatavia Morrison, Christopher Gulvick, Scott Sammons, Yvette Unoarumhi, Darlene Wagner, Matthew Schmerer, Cyndi Clark, Patrick Campbell, Rob Case, Vikramsinha Ghorpade, Holly Houdeshell, Ola Kvalvaag, Dillon Nall, Ethan Sanders, Alec Vest, Shaun Westlund, Matthew Hardison, Clinton R. Paden, Duncan MacCannell |
| EPI_ISL_2201844 | Houston Methodist Hospital | Houston Methodist Hospital | Randall J. Olsen, Paul A. Christensen, S. Wesley Long, Sishir Subedi, Robert Olson, Marcus Nguyen, James J. Davis, Matthew Ojeda Saavedra, Prasanti Yerramilli, Layne Pruitt, Kristina Reppond, Madison N. Shyer, Jessica Cambric, Ryan Gadd, Ilya J. Finkelstein, Jimmy Gollihar, and James M. Musser |
| EPI_ISL_2201845 | Aegis Sciences Corporation | Centers for Disease Control and Prevention Division of Viral Diseases, Pathogen Discovery | Dakota Howard, Dhwani Batra, Peter W. Cook, Kara Moser, Adrian Paskey, Jason Caravas, Benjamin Rambo-Martin, Shatavia Morrison, Christopher Gulvick, Scott Sammons, Yvette Unoarumhi, Darlene Wagner, Matthew Schmerer, Cyndi Clark, Patrick Campbell, Rob Case, Vikramsinha Ghorpade, Holly Houdeshell, Ola Kvalvaag, Dillon Nall, Ethan Sanders, Alec Vest, Shaun Westlund, Matthew Hardison, Clinton R. Paden, Duncan MacCannell |
| EPI_ISL_2201846 | Houston Methodist Hospital | Houston Methodist Hospital | Randall J. Olsen, Paul A. Christensen, S. Wesley Long, Sishir Subedi, Robert Olson, Marcus Nguyen, James J. Davis, Matthew Ojeda Saavedra, Prasanti Yerramilli, Layne Pruitt, Kristina Reppond, Madison N. Shyer, Jessica Cambric, Ryan Gadd, Ilya J. Finkelstein, Jimmy Gollihar, and James M. Musser |
| EPI_ISL_2201847 | Aegis Sciences Corporation | Centers for Disease Control and Prevention Division of Viral Diseases, Pathogen Discovery | Dakota Howard, Dhwani Batra, Peter W. Cook, Kara Moser, Adrian Paskey, Jason Caravas, Benjamin Rambo-Martin, Shatavia Morrison, Christopher Gulvick, Scott Sammons, Yvette Unoarumhi, Darlene Wagner, Matthew Schmerer, Cyndi Clark, Patrick Campbell, Rob Case, Vikramsinha Ghorpade, Holly Houdeshell, Ola Kvalvaag, Dillon Nall, Ethan Sanders, Alec Vest, Shaun Westlund, Matthew Hardison, Clinton R. Paden, Duncan MacCannell |
| EPI_ISL_2201848 | Houston Methodist Hospital | Houston Methodist Hospital | Randall J. Olsen, Paul A. Christensen, S. Wesley Long, Sishir Subedi, Robert Olson, Marcus Nguyen, James J. Davis, Matthew Ojeda Saavedra, Prasanti Yerramilli, Layne Pruitt, Kristina Reppond, Madison N. Shyer, Jessica Cambric, Ryan Gadd, Ilya J. Finkelstein, Jimmy Gollihar, and James M. Musser |
| EPI_ISL_2201849 | Aegis Sciences Corporation | Centers for Disease Control and Prevention Division of Viral Diseases, Pathogen Discovery | Dakota Howard, Dhwani Batra, Peter W. Cook, Kara Moser, Adrian Paskey, Jason Caravas, Benjamin Rambo-Martin, Shatavia Morrison, Christopher Gulvick, Scott Sammons, Yvette Unoarumhi, Darlene Wagner, Matthew Schmerer, Cyndi Clark, Patrick Campbell, Rob Case, Vikramsinha Ghorpade, Holly Houdeshell, Ola Kvalvaag, Dillon Nall, Ethan Sanders, Alec Vest, Shaun Westlund, Matthew Hardison, Clinton R. Paden, Duncan MacCannell |
| EPI_ISL_2201850 | Houston Methodist Hospital | Houston Methodist Hospital | Randall J. Olsen, Paul A. Christensen, S. Wesley Long, Sishir Subedi, Robert Olson, Marcus Nguyen, James J. Davis, Matthew Ojeda Saavedra, Prasanti Yerramilli, Layne Pruitt, Kristina Reppond, Madison N. Shyer, Jessica Cambric, Ryan Gadd, Ilya J. Finkelstein, Jimmy Gollihar, and James M. Musser |
| EPI_ISL_2201851, EPI_ISL_2201852 | Aegis Sciences Corporation | Centers for Disease Control and Prevention Division of Viral Diseases, Pathogen Discovery | Dakota Howard, Dhwani Batra, Peter W. Cook, Kara Moser, Adrian Paskey, Jason Caravas, Benjamin Rambo-Martin, Shatavia Morrison, Christopher Gulvick, Scott Sammons, Yvette Unoarumhi, Darlene Wagner, Matthew Schmerer, Cyndi Clark, Patrick Campbell, Rob Case, Vikramsinha Ghorpade, Holly Houdeshell, Ola Kvalvaag, Dillon Nall, Ethan Sanders, Alec Vest, Shaun Westlund, Matthew Hardison, Clinton R. Paden, Duncan MacCannell |
| EPI_ISL_2201853 | Houston Methodist Hospital | Houston Methodist Hospital | Randall J. Olsen, Paul A. Christensen, S. Wesley Long, Sishir Subedi, Robert Olson, Marcus Nguyen, James J. Davis, Matthew Ojeda Saavedra, Prasanti Yerramilli, Layne Pruitt, Kristina Reppond, Madison N. Shyer, Jessica Cambric, Ryan Gadd, Ilya J. Finkelstein, Jimmy Gollihar, and James M. Musser |
| EPI_ISL_2201854 | Aegis Sciences Corporation | Centers for Disease Control and Prevention Division of Viral Diseases, Pathogen Discovery | Dakota Howard, Dhwani Batra, Peter W. Cook, Kara Moser, Adrian Paskey, Jason Caravas, Benjamin Rambo-Martin, Shatavia Morrison, Christopher Gulvick, Scott Sammons, Yvette Unoarumhi, Darlene Wagner, Matthew Schmerer, Cyndi Clark, Patrick Campbell, Rob Case, Vikramsinha Ghorpade, Holly Houdeshell, Ola Kvalvaag, Dillon Nall, Ethan Sanders, Alec Vest, Shaun Westlund, Matthew Hardison, Clinton R. Paden, Duncan MacCannell |
| EPI_ISL_2201855 | Houston Methodist Hospital | Houston Methodist Hospital | Randall J. Olsen, Paul A. Christensen, S. Wesley Long, Sishir Subedi, Robert Olson, Marcus Nguyen, James J. Davis, Matthew Ojeda Saavedra, Prasanti Yerramilli, Layne Pruitt, Kristina Reppond, Madison N. Shyer, Jessica Cambric, Ryan Gadd, Ilya J. Finkelstein, Jimmy Gollihar, and James M. Musser |
| EPI_ISL_2201856 | Aegis Sciences Corporation | Centers for Disease Control and Prevention Division of Viral Diseases, Pathogen Discovery | Dakota Howard, Dhwani Batra, Peter W. Cook, Kara Moser, Adrian Paskey, Jason Caravas, Benjamin Rambo-Martin, Shatavia Morrison, Christopher Gulvick, Scott Sammons, Yvette Unoarumhi, Darlene Wagner, Matthew Schmerer, Cyndi Clark, Patrick Campbell, Rob Case, Vikramsinha Ghorpade, Holly Houdeshell, Ola Kvalvaag, Dillon Nall, Ethan Sanders, Alec Vest, Shaun Westlund, Matthew Hardison, Clinton R. Paden, Duncan MacCannell |
| EPI_ISL_2201857 | Houston Methodist Hospital | Houston Methodist Hospital | Randall J. Olsen, Paul A. Christensen, S. Wesley Long, Sishir Subedi, Robert Olson, Marcus Nguyen, James J. Davis, Matthew Ojeda Saavedra, Prasanti Yerramilli, Layne Pruitt, Kristina Reppond, Madison N. Shyer, Jessica Cambric, Ryan Gadd, Ilya J. Finkelstein, Jimmy Gollihar, and James M. Musser |
| EPI_ISL_2201858 | Aegis Sciences Corporation | Centers for Disease Control and Prevention Division of Viral Diseases, Pathogen Discovery | Dakota Howard, Dhwani Batra, Peter W. Cook, Kara Moser, Adrian Paskey, Jason Caravas, Benjamin Rambo-Martin, Shatavia Morrison, Christopher Gulvick, Scott Sammons, Yvette Unoarumhi, Darlene Wagner, Matthew Schmerer, Cyndi Clark, Patrick Campbell, Rob Case, Vikramsinha Ghorpade, Holly Houdeshell, Ola Kvalvaag, Dillon Nall, Ethan Sanders, Alec Vest, Shaun Westlund, Matthew Hardison, Clinton R. Paden, Duncan MacCannell |
| EPI_ISL_2201859 | Houston Methodist Hospital | Houston Methodist Hospital | Randall J. Olsen, Paul A. Christensen, S. Wesley Long, Sishir Subedi, Robert Olson, Marcus Nguyen, James J. Davis, Matthew Ojeda Saavedra, Prasanti Yerramilli, Layne Pruitt, Kristina Reppond, Madison N. Shyer, Jessica Cambric, Ryan Gadd, Ilya J. Finkelstein, Jimmy Gollihar, and James M. Musser |
| EPI_ISL_2201860 | Aegis Sciences Corporation | Centers for Disease Control and Prevention Division of Viral Diseases, Pathogen Discovery | Dakota Howard, Dhwani Batra, Peter W. Cook, Kara Moser, Adrian Paskey, Jason Caravas, Benjamin Rambo-Martin, Shatavia Morrison, Christopher Gulvick, Scott Sammons, Yvette Unoarumhi, Darlene Wagner, Matthew Schmerer, Cyndi Clark, Patrick Campbell, Rob Case, Vikramsinha Ghorpade, Holly Houdeshell, Ola Kvalvaag, Dillon Nall, Ethan Sanders, Alec Vest, Shaun Westlund, Matthew Hardison, Clinton R. Paden, Duncan MacCannell |
| EPI_ISL_2201861 | Houston Methodist Hospital | Houston Methodist Hospital | Randall J. Olsen, Paul A. Christensen, S. Wesley Long, Sishir Subedi, Robert Olson, Marcus Nguyen, James J. Davis, Matthew Ojeda Saavedra, Prasanti Yerramilli, Layne Pruitt, Kristina Reppond, Madison N. Shyer, Jessica Cambric, Ryan Gadd, Ilya J. Finkelstein, Jimmy Gollihar, and James M. Musser |
| EPI_ISL_2201862 | Aegis Sciences Corporation | Centers for Disease Control and Prevention Division of Viral Diseases, Pathogen Discovery | Dakota Howard, Dhwani Batra, Peter W. Cook, Kara Moser, Adrian Paskey, Jason Caravas, Benjamin Rambo-Martin, Shatavia Morrison, Christopher Gulvick, Scott Sammons, Yvette Unoarumhi, Darlene Wagner, Matthew Schmerer, Cyndi Clark, Patrick Campbell, Rob Case, Vikramsinha Ghorpade, Holly Houdeshell, Ola Kvalvaag, Dillon Nall, Ethan Sanders, Alec Vest, Shaun Westlund, Matthew Hardison, Clinton R. Paden, Duncan MacCannell |
| EPI_ISL_2201863 | Houston Methodist Hospital | Houston Methodist Hospital | Randall J. Olsen, Paul A. Christensen, S. Wesley Long, Sishir Subedi, Robert Olson, Marcus Nguyen, James J. Davis, Matthew Ojeda Saavedra, Prasanti Yerramilli, Layne Pruitt, Kristina Reppond, Madison N. Shyer, Jessica Cambric, Ryan Gadd, Ilya J. Finkelstein, Jimmy Gollihar, and James M. Musser |
| EPI_ISL_2201864 | Aegis Sciences Corporation | Centers for Disease Control and Prevention Division of Viral Diseases, Pathogen Discovery | Dakota Howard, Dhwani Batra, Peter W. Cook, Kara Moser, Adrian Paskey, Jason Caravas, Benjamin Rambo-Martin, Shatavia Morrison, Christopher Gulvick, Scott Sammons, Yvette Unoarumhi, Darlene Wagner, Matthew Schmerer, Cyndi Clark, Patrick Campbell, Rob Case, Vikramsinha Ghorpade, Holly Houdeshell, Ola Kvalvaag, Dillon Nall, Ethan Sanders, Alec Vest, Shaun Westlund, Matthew Hardison, Clinton R. Paden, Duncan MacCannell |
| EPI_ISL_2201865 | Houston Methodist Hospital | Houston Methodist Hospital | Randall J. Olsen, Paul A. Christensen, S. Wesley Long, Sishir Subedi, Robert Olson, Marcus Nguyen, James J. Davis, Matthew Ojeda Saavedra, Prasanti Yerramilli, Layne Pruitt, Kristina Reppond, Madison N. Shyer, Jessica Cambric, Ryan Gadd, Ilya J. Finkelstein, Jimmy Gollihar, and James M. Musser |
| EPI_ISL_2201866, EPI_ISL_2201867 | Aegis Sciences Corporation | Centers for Disease Control and Prevention Division of Viral Diseases, Pathogen Discovery | Dakota Howard, Dhwani Batra, Peter W. Cook, Kara Moser, Adrian Paskey, Jason Caravas, Benjamin Rambo-Martin, Shatavia Morrison, Christopher Gulvick, Scott Sammons, Yvette Unoarumhi, Darlene Wagner, Matthew Schmerer, Cyndi Clark, Patrick Campbell, Rob Case, Vikramsinha Ghorpade, Holly Houdeshell, Ola Kvalvaag, Dillon Nall, Ethan Sanders, Alec Vest, Shaun Westlund, Matthew Hardison, Clinton R. Paden, Duncan MacCannell |
| EPI_ISL_2201868, EPI_ISL_2201869 | Houston Methodist Hospital | Houston Methodist Hospital | Randall J. Olsen, Paul A. Christensen, S. Wesley Long, Sishir Subedi, Robert Olson, Marcus Nguyen, James J. Davis, Matthew Ojeda Saavedra, Prasanti Yerramilli, Layne Pruitt, Kristina Reppond, Madison N. Shyer, Jessica Cambric, Ryan Gadd, Ilya J. Finkelstein, Jimmy Gollihar, and James M. Musser |
| EPI_ISL_2201870, EPI_ISL_2201871 | Aegis Sciences Corporation | Centers for Disease Control and Prevention Division of Viral Diseases, Pathogen Discovery | Dakota Howard, Dhwani Batra, Peter W. Cook, Kara Moser, Adrian Paskey, Jason Caravas, Benjamin Rambo-Martin, Shatavia Morrison, Christopher Gulvick, Scott Sammons, Yvette Unoarumhi, Darlene Wagner, Matthew Schmerer, Cyndi Clark, Patrick Campbell, Rob Case, Vikramsinha Ghorpade, Holly Houdeshell, Ola Kvalvaag, Dillon Nall, Ethan Sanders, Alec Vest, Shaun Westlund, Matthew Hardison, Clinton R. Paden, Duncan MacCannell |
| EPI_ISL_2201872 | Houston Methodist Hospital | Houston Methodist Hospital | Randall J. Olsen, Paul A. Christensen, S. Wesley Long, Sishir Subedi, Robert Olson, Marcus Nguyen, James J. Davis, Matthew Ojeda Saavedra, Prasanti Yerramilli, Layne Pruitt, Kristina Reppond, Madison N. Shyer, Jessica Cambric, Ryan Gadd, Ilya J. Finkelstein, Jimmy Gollihar, and James M. Musser |
| EPI_ISL_2201873 | Aegis Sciences Corporation | Centers for Disease Control and Prevention Division of Viral Diseases, Pathogen Discovery | Dakota Howard, Dhwani Batra, Peter W. Cook, Kara Moser, Adrian Paskey, Jason Caravas, Benjamin Rambo-Martin, Shatavia Morrison, Christopher Gulvick, Scott Sammons, Yvette Unoarumhi, Darlene Wagner, Matthew Schmerer, Cyndi Clark, Patrick Campbell, Rob Case, Vikramsinha Ghorpade, Holly Houdeshell, Ola Kvalvaag, Dillon Nall, Ethan Sanders, Alec Vest, Shaun Westlund, Matthew Hardison, Clinton R. Paden, Duncan MacCannell |
| EPI_ISL_2201874 | Houston Methodist Hospital | Houston Methodist Hospital | Randall J. Olsen, Paul A. Christensen, S. Wesley Long, Sishir Subedi, Robert Olson, Marcus Nguyen, James J. Davis, Matthew Ojeda Saavedra, Prasanti Yerramilli, Layne Pruitt, Kristina Reppond, Madison N. Shyer, Jessica Cambric, Ryan Gadd, Ilya J. Finkelstein, Jimmy Gollihar, and James M. Musser |
| EPI_ISL_2201875 | Aegis Sciences Corporation | Centers for Disease Control and Prevention | Dakota Howard, Dhwani Batra, Peter W. Cook, Kara Moser, Adrian Paskey, Jason Caravas, Benjamin Rambo-Martin, Shatavia Morrison, Christopher Gulvick, Scott Sammons, Yvette Unoarumhi, Darlene Wagner, Matthew Schmerer, Cyndi Clark, Patrick Campbell, Rob Case, Vikramsinha |

| | | | |
|---|---|---|---|
| | | Division of Viral Diseases, Pathogen Discovery | Ghorpade, Holly Houdeshell, Ola Kvalvaag, Dillon Nall, Ethan Sanders, Alec Vest, Shaun Westlund, Matthew Hardison, Clinton R. Paden, Duncan MacCannell |
| EPI_ISL_2201876 | Houston Methodist Hospital | Houston Methodist Hospital | Randall J. Olsen, Paul A. Christensen, S. Wesley Long, Sishir Subedi, Robert Olson, Marcus Nguyen, James J. Davis, Matthew Ojeda Saavedra, Prasanti Yerramilli, Layne Pruitt, Kristina Reppond, Madison N. Shyer, Jessica Cambric, Ryan Gadd, Ilya J. Finkelstein, Jimmy Gollihar, and James M. Musser |
| EPI_ISL_2201877, EPI_ISL_2201878 | Aegis Sciences Corporation | Centers for Disease Control and Prevention Division of Viral Diseases, Pathogen Discovery | Dakota Howard, Dhwani Batra, Peter W. Cook, Kara Moser, Adrian Paskey, Jason Caravas, Benjamin Rambo-Martin, Shatavia Morrison, Christopher Gulvick, Scott Sammons, Yvette Unoarumhi, Darlene Wagner, Matthew Schmerer, Cyndi Clark, Patrick Campbell, Rob Case, Vikramsinha Ghorpade, Holly Houdeshell, Ola Kvalvaag, Dillon Nall, Ethan Sanders, Alec Vest, Shaun Westlund, Matthew Hardison, Clinton R. Paden, Duncan MacCannell |
| EPI_ISL_2201879 | Houston Methodist Hospital | Houston Methodist Hospital | Randall J. Olsen, Paul A. Christensen, S. Wesley Long, Sishir Subedi, Robert Olson, Marcus Nguyen, James J. Davis, Matthew Ojeda Saavedra, Prasanti Yerramilli, Layne Pruitt, Kristina Reppond, Madison N. Shyer, Jessica Cambric, Ryan Gadd, Ilya J. Finkelstein, Jimmy Gollihar, and James M. Musser |
| EPI_ISL_2201880 | Aegis Sciences Corporation | Centers for Disease Control and Prevention Division of Viral Diseases, Pathogen Discovery | Dakota Howard, Dhwani Batra, Peter W. Cook, Kara Moser, Adrian Paskey, Jason Caravas, Benjamin Rambo-Martin, Shatavia Morrison, Christopher Gulvick, Scott Sammons, Yvette Unoarumhi, Darlene Wagner, Matthew Schmerer, Cyndi Clark, Patrick Campbell, Rob Case, Vikramsinha Ghorpade, Holly Houdeshell, Ola Kvalvaag, Dillon Nall, Ethan Sanders, Alec Vest, Shaun Westlund, Matthew Hardison, Clinton R. Paden, Duncan MacCannell |
| EPI_ISL_2201881 | Houston Methodist Hospital | Houston Methodist Hospital | Randall J. Olsen, Paul A. Christensen, S. Wesley Long, Sishir Subedi, Robert Olson, Marcus Nguyen, James J. Davis, Matthew Ojeda Saavedra, Prasanti Yerramilli, Layne Pruitt, Kristina Reppond, Madison N. Shyer, Jessica Cambric, Ryan Gadd, Ilya J. Finkelstein, Jimmy Gollihar, and James M. Musser |
| EPI_ISL_2201882 | Aegis Sciences Corporation | Centers for Disease Control and Prevention Division of Viral Diseases, Pathogen Discovery | Dakota Howard, Dhwani Batra, Peter W. Cook, Kara Moser, Adrian Paskey, Jason Caravas, Benjamin Rambo-Martin, Shatavia Morrison, Christopher Gulvick, Scott Sammons, Yvette Unoarumhi, Darlene Wagner, Matthew Schmerer, Cyndi Clark, Patrick Campbell, Rob Case, Vikramsinha Ghorpade, Holly Houdeshell, Ola Kvalvaag, Dillon Nall, Ethan Sanders, Alec Vest, Shaun Westlund, Matthew Hardison, Clinton R. Paden, Duncan MacCannell |
| EPI_ISL_2201883 | Houston Methodist Hospital | Houston Methodist Hospital | Randall J. Olsen, Paul A. Christensen, S. Wesley Long, Sishir Subedi, Robert Olson, Marcus Nguyen, James J. Davis, Matthew Ojeda Saavedra, Prasanti Yerramilli, Layne Pruitt, Kristina Reppond, Madison N. Shyer, Jessica Cambric, Ryan Gadd, Ilya J. Finkelstein, Jimmy Gollihar, and James M. Musser |
| EPI_ISL_2201884 | Aegis Sciences Corporation | Centers for Disease Control and Prevention Division of Viral Diseases, Pathogen Discovery | Dakota Howard, Dhwani Batra, Peter W. Cook, Kara Moser, Adrian Paskey, Jason Caravas, Benjamin Rambo-Martin, Shatavia Morrison, Christopher Gulvick, Scott Sammons, Yvette Unoarumhi, Darlene Wagner, Matthew Schmerer, Cyndi Clark, Patrick Campbell, Rob Case, Vikramsinha Ghorpade, Holly Houdeshell, Ola Kvalvaag, Dillon Nall, Ethan Sanders, Alec Vest, Shaun Westlund, Matthew Hardison, Clinton R. Paden, Duncan MacCannell |
| EPI_ISL_2201885 | Houston Methodist Hospital | Houston Methodist Hospital | Randall J. Olsen, Paul A. Christensen, S. Wesley Long, Sishir Subedi, Robert Olson, Marcus Nguyen, James J. Davis, Matthew Ojeda Saavedra, Prasanti Yerramilli, Layne Pruitt, Kristina Reppond, Madison N. Shyer, Jessica Cambric, Ryan Gadd, Ilya J. Finkelstein, Jimmy Gollihar, and James M. Musser |
| EPI_ISL_2201886 | Aegis Sciences Corporation | Centers for Disease Control and Prevention Division of Viral Diseases, Pathogen Discovery | Dakota Howard, Dhwani Batra, Peter W. Cook, Kara Moser, Adrian Paskey, Jason Caravas, Benjamin Rambo-Martin, Shatavia Morrison, Christopher Gulvick, Scott Sammons, Yvette Unoarumhi, Darlene Wagner, Matthew Schmerer, Cyndi Clark, Patrick Campbell, Rob Case, Vikramsinha Ghorpade, Holly Houdeshell, Ola Kvalvaag, Dillon Nall, Ethan Sanders, Alec Vest, Shaun Westlund, Matthew Hardison, Clinton R. Paden, Duncan MacCannell |
| EPI_ISL_2201887 | Houston Methodist Hospital | Houston Methodist Hospital | Randall J. Olsen, Paul A. Christensen, S. Wesley Long, Sishir Subedi, Robert Olson, Marcus Nguyen, James J. Davis, Matthew Ojeda Saavedra, Prasanti Yerramilli, Layne Pruitt, Kristina Reppond, Madison N. Shyer, Jessica Cambric, Ryan Gadd, Ilya J. Finkelstein, Jimmy Gollihar, and James M. Musser |
| EPI_ISL_2201888, EPI_ISL_2201889 | Aegis Sciences Corporation | Centers for Disease Control and Prevention Division of Viral Diseases, Pathogen Discovery | Dakota Howard, Dhwani Batra, Peter W. Cook, Kara Moser, Adrian Paskey, Jason Caravas, Benjamin Rambo-Martin, Shatavia Morrison, Christopher Gulvick, Scott Sammons, Yvette Unoarumhi, Darlene Wagner, Matthew Schmerer, Cyndi Clark, Patrick Campbell, Rob Case, Vikramsinha Ghorpade, Holly Houdeshell, Ola Kvalvaag, Dillon Nall, Ethan Sanders, Alec Vest, Shaun Westlund, Matthew Hardison, Clinton R. Paden, Duncan MacCannell |
| EPI_ISL_2201890 | Houston Methodist Hospital | Houston Methodist Hospital | Randall J. Olsen, Paul A. Christensen, S. Wesley Long, Sishir Subedi, Robert Olson, Marcus Nguyen, James J. Davis, Matthew Ojeda Saavedra, Prasanti Yerramilli, Layne Pruitt, Kristina Reppond, Madison N. Shyer, Jessica Cambric, Ryan Gadd, Ilya J. Finkelstein, Jimmy Gollihar, and James M. Musser |
| EPI_ISL_2201891 | Aegis Sciences Corporation | Centers for Disease Control and Prevention Division of Viral Diseases, Pathogen Discovery | Dakota Howard, Dhwani Batra, Peter W. Cook, Kara Moser, Adrian Paskey, Jason Caravas, Benjamin Rambo-Martin, Shatavia Morrison, Christopher Gulvick, Scott Sammons, Yvette Unoarumhi, Darlene Wagner, Matthew Schmerer, Cyndi Clark, Patrick Campbell, Rob Case, Vikramsinha Ghorpade, Holly Houdeshell, Ola Kvalvaag, Dillon Nall, Ethan Sanders, Alec Vest, Shaun Westlund, Matthew Hardison, Clinton R. Paden, Duncan MacCannell |
| EPI_ISL_2201892 | Houston Methodist Hospital | Houston Methodist Hospital | Randall J. Olsen, Paul A. Christensen, S. Wesley Long, Sishir Subedi, Robert Olson, Marcus Nguyen, James J. Davis, Matthew Ojeda Saavedra, Prasanti Yerramilli, Layne Pruitt, Kristina Reppond, Madison N. Shyer, Jessica Cambric, Ryan Gadd, Ilya J. Finkelstein, Jimmy Gollihar, and James M. Musser |
| EPI_ISL_2201893 | Aegis Sciences Corporation | Centers for Disease Control and Prevention Division of Viral Diseases, Pathogen Discovery | Dakota Howard, Dhwani Batra, Peter W. Cook, Kara Moser, Adrian Paskey, Jason Caravas, Benjamin Rambo-Martin, Shatavia Morrison, Christopher Gulvick, Scott Sammons, Yvette Unoarumhi, Darlene Wagner, Matthew Schmerer, Cyndi Clark, Patrick Campbell, Rob Case, Vikramsinha Ghorpade, Holly Houdeshell, Ola Kvalvaag, Dillon Nall, Ethan Sanders, Alec Vest, Shaun Westlund, Matthew Hardison, Clinton R. Paden, Duncan MacCannell |
| EPI_ISL_2201894 | Houston Methodist Hospital | Houston Methodist Hospital | Randall J. Olsen, Paul A. Christensen, S. Wesley Long, Sishir Subedi, Robert Olson, Marcus Nguyen, James J. Davis, Matthew Ojeda Saavedra, Prasanti Yerramilli, Layne Pruitt, Kristina Reppond, Madison N. Shyer, Jessica Cambric, Ryan Gadd, Ilya J. Finkelstein, Jimmy Gollihar, and James M. Musser |
| EPI_ISL_2201895 | Aegis Sciences Corporation | Centers for Disease Control and Prevention Division of Viral Diseases, Pathogen Discovery | Dakota Howard, Dhwani Batra, Peter W. Cook, Kara Moser, Adrian Paskey, Jason Caravas, Benjamin Rambo-Martin, Shatavia Morrison, Christopher Gulvick, Scott Sammons, Yvette Unoarumhi, Darlene Wagner, Matthew Schmerer, Cyndi Clark, Patrick Campbell, Rob Case, Vikramsinha Ghorpade, Holly Houdeshell, Ola Kvalvaag, Dillon Nall, Ethan Sanders, Alec Vest, Shaun Westlund, Matthew Hardison, Clinton R. Paden, Duncan MacCannell |
| EPI_ISL_2201896 | Houston Methodist Hospital | Houston Methodist Hospital | Randall J. Olsen, Paul A. Christensen, S. Wesley Long, Sishir Subedi, Robert Olson, Marcus Nguyen, James J. Davis, Matthew Ojeda Saavedra, Prasanti Yerramilli, Layne Pruitt, Kristina Reppond, Madison N. Shyer, Jessica Cambric, Ryan Gadd, Ilya J. Finkelstein, Jimmy Gollihar, and James M. Musser |
| EPI_ISL_2201897 | Aegis Sciences Corporation | Centers for Disease Control and Prevention Division of Viral Diseases, Pathogen Discovery | Dakota Howard, Dhwani Batra, Peter W. Cook, Kara Moser, Adrian Paskey, Jason Caravas, Benjamin Rambo-Martin, Shatavia Morrison, Christopher Gulvick, Scott Sammons, Yvette Unoarumhi, Darlene Wagner, Matthew Schmerer, Cyndi Clark, Patrick Campbell, Rob Case, Vikramsinha Ghorpade, Holly Houdeshell, Ola Kvalvaag, Dillon Nall, Ethan Sanders, Alec Vest, Shaun Westlund, Matthew Hardison, Clinton R. Paden, Duncan MacCannell |
| EPI_ISL_2201898 | Houston Methodist Hospital | Houston Methodist Hospital | Randall J. Olsen, Paul A. Christensen, S. Wesley Long, Sishir Subedi, Robert Olson, Marcus Nguyen, James J. Davis, Matthew Ojeda Saavedra, Prasanti Yerramilli, Layne Pruitt, Kristina Reppond, Madison N. Shyer, Jessica Cambric, Ryan Gadd, Ilya J. Finkelstein, Jimmy Gollihar, and James M. Musser |
| EPI_ISL_2201899, EPI_ISL_2201900 | Aegis Sciences Corporation | Centers for Disease Control and Prevention Division of Viral Diseases, Pathogen Discovery | Dakota Howard, Dhwani Batra, Peter W. Cook, Kara Moser, Adrian Paskey, Jason Caravas, Benjamin Rambo-Martin, Shatavia Morrison, Christopher Gulvick, Scott Sammons, Yvette Unoarumhi, Darlene Wagner, Matthew Schmerer, Cyndi Clark, Patrick Campbell, Rob Case, Vikramsinha Ghorpade, Holly Houdeshell, Ola Kvalvaag, Dillon Nall, Ethan Sanders, Alec Vest, Shaun Westlund, Matthew Hardison, Clinton R. Paden, Duncan MacCannell |
| EPI_ISL_2201901 | Houston Methodist Hospital | Houston Methodist Hospital | Randall J. Olsen, Paul A. Christensen, S. Wesley Long, Sishir Subedi, Robert Olson, Marcus Nguyen, James J. Davis, Matthew Ojeda Saavedra, Prasanti Yerramilli, Layne Pruitt, Kristina Reppond, Madison N. Shyer, Jessica Cambric, Ryan Gadd, Ilya J. Finkelstein, Jimmy Gollihar, and James M. Musser |
| EPI_ISL_2201902 | Aegis Sciences Corporation | Centers for Disease Control and Prevention Division of Viral Diseases, Pathogen Discovery | Dakota Howard, Dhwani Batra, Peter W. Cook, Kara Moser, Adrian Paskey, Jason Caravas, Benjamin Rambo-Martin, Shatavia Morrison, Christopher Gulvick, Scott Sammons, Yvette Unoarumhi, Darlene Wagner, Matthew Schmerer, Cyndi Clark, Patrick Campbell, Rob Case, Vikramsinha Ghorpade, Holly Houdeshell, Ola Kvalvaag, Dillon Nall, Ethan Sanders, Alec Vest, Shaun Westlund, Matthew Hardison, Clinton R. Paden, Duncan MacCannell |
| EPI_ISL_2201903 | Houston Methodist Hospital | Houston Methodist Hospital | Randall J. Olsen, Paul A. Christensen, S. Wesley Long, Sishir Subedi, Robert Olson, Marcus Nguyen, James J. Davis, Matthew Ojeda Saavedra, Prasanti Yerramilli, Layne Pruitt, Kristina Reppond, Madison N. Shyer, Jessica Cambric, Ryan Gadd, Ilya J. Finkelstein, Jimmy Gollihar, and James M. Musser |
| EPI_ISL_2201904, EPI_ISL_2201905 | Aegis Sciences Corporation | Centers for Disease Control and Prevention Division of Viral Diseases, Pathogen Discovery | Dakota Howard, Dhwani Batra, Peter W. Cook, Kara Moser, Adrian Paskey, Jason Caravas, Benjamin Rambo-Martin, Shatavia Morrison, Christopher Gulvick, Scott Sammons, Yvette Unoarumhi, Darlene Wagner, Matthew Schmerer, Cyndi Clark, Patrick Campbell, Rob Case, Vikramsinha Ghorpade, Holly Houdeshell, Ola Kvalvaag, Dillon Nall, Ethan Sanders, Alec Vest, Shaun Westlund, Matthew Hardison, Clinton R. Paden, Duncan MacCannell |
| EPI_ISL_2201906 | Houston Methodist Hospital | Houston Methodist Hospital | Randall J. Olsen, Paul A. Christensen, S. Wesley Long, Sishir Subedi, Robert Olson, Marcus Nguyen, James J. Davis, Matthew Ojeda Saavedra, Prasanti Yerramilli, Layne Pruitt, Kristina Reppond, Madison N. Shyer, Jessica Cambric, Ryan Gadd, Ilya J. Finkelstein, Jimmy Gollihar, and James M. Musser |
| EPI_ISL_2201907 | Aegis Sciences Corporation | Centers for Disease Control and Prevention Division of Viral Diseases, Pathogen Discovery | Dakota Howard, Dhwani Batra, Peter W. Cook, Kara Moser, Adrian Paskey, Jason Caravas, Benjamin Rambo-Martin, Shatavia Morrison, Christopher Gulvick, Scott Sammons, Yvette Unoarumhi, Darlene Wagner, Matthew Schmerer, Cyndi Clark, Patrick Campbell, Rob Case, Vikramsinha Ghorpade, Holly Houdeshell, Ola Kvalvaag, Dillon Nall, Ethan Sanders, Alec Vest, Shaun Westlund, Matthew Hardison, Clinton R. Paden, Duncan MacCannell |
| EPI_ISL_2201908 | Houston Methodist Hospital | Houston Methodist Hospital | Randall J. Olsen, Paul A. Christensen, S. Wesley Long, Sishir Subedi, Robert Olson, Marcus Nguyen, James J. Davis, Matthew Ojeda Saavedra, Prasanti Yerramilli, Layne Pruitt, Kristina Reppond, Madison N. Shyer, Jessica Cambric, Ryan Gadd, Ilya J. Finkelstein, Jimmy Gollihar, and James M. Musser |
| EPI_ISL_2201909 | Aegis Sciences Corporation | Centers for Disease Control and Prevention Division of Viral Diseases, Pathogen Discovery | Dakota Howard, Dhwani Batra, Peter W. Cook, Kara Moser, Adrian Paskey, Jason Caravas, Benjamin Rambo-Martin, Shatavia Morrison, Christopher Gulvick, Scott Sammons, Yvette Unoarumhi, Darlene Wagner, Matthew Schmerer, Cyndi Clark, Patrick Campbell, Rob Case, Vikramsinha Ghorpade, Holly Houdeshell, Ola Kvalvaag, Dillon Nall, Ethan Sanders, Alec Vest, Shaun Westlund, Matthew Hardison, Clinton R. Paden, Duncan MacCannell |
| EPI_ISL_2201910 | Houston Methodist Hospital | Houston Methodist Hospital | Randall J. Olsen, Paul A. Christensen, S. Wesley Long, Sishir Subedi, Robert Olson, Marcus Nguyen, James J. Davis, Matthew Ojeda Saavedra, Prasanti Yerramilli, Layne Pruitt, Kristina Reppond, Madison N. Shyer, Jessica Cambric, Ryan Gadd, Ilya J. Finkelstein, Jimmy Gollihar, and James M. Musser |
| EPI_ISL_2201911, EPI_ISL_2201912 | Aegis Sciences Corporation | Centers for Disease Control and Prevention Division of Viral Diseases, Pathogen Discovery | Dakota Howard, Dhwani Batra, Peter W. Cook, Kara Moser, Adrian Paskey, Jason Caravas, Benjamin Rambo-Martin, Shatavia Morrison, Christopher Gulvick, Scott Sammons, Yvette Unoarumhi, Darlene Wagner, Matthew Schmerer, Cyndi Clark, Patrick Campbell, Rob Case, Vikramsinha Ghorpade, Holly Houdeshell, Ola Kvalvaag, Dillon Nall, Ethan Sanders, Alec Vest, Shaun Westlund, Matthew Hardison, Clinton R. Paden, Duncan MacCannell |
| EPI_ISL_2201913 | Houston Methodist Hospital | Houston Methodist Hospital | Randall J. Olsen, Paul A. Christensen, S. Wesley Long, Sishir Subedi, Robert Olson, Marcus Nguyen, James J. Davis, Matthew Ojeda Saavedra, Prasanti Yerramilli, Layne Pruitt, Kristina Reppond, Madison N. Shyer, Jessica Cambric, Ryan Gadd, Ilya J. Finkelstein, Jimmy Gollihar, and James M. Musser |
| EPI_ISL_2201914 | Aegis Sciences Corporation | Centers for Disease Control and Prevention Division of Viral Diseases, Pathogen Discovery | Dakota Howard, Dhwani Batra, Peter W. Cook, Kara Moser, Adrian Paskey, Jason Caravas, Benjamin Rambo-Martin, Shatavia Morrison, Christopher Gulvick, Scott Sammons, Yvette Unoarumhi, Darlene Wagner, Matthew Schmerer, Cyndi Clark, Patrick Campbell, Rob Case, Vikramsinha Ghorpade, Holly Houdeshell, Ola Kvalvaag, Dillon Nall, Ethan Sanders, Alec Vest, Shaun Westlund, Matthew Hardison, Clinton R. Paden, Duncan MacCannell |
| EPI_ISL_2201915 | Houston Methodist Hospital | Houston Methodist Hospital | Randall J. Olsen, Paul A. Christensen, S. Wesley Long, Sishir Subedi, Robert Olson, Marcus Nguyen, James J. Davis, Matthew Ojeda Saavedra, Prasanti Yerramilli, Layne Pruitt, Kristina Reppond, Madison N. Shyer, Jessica Cambric, Ryan Gadd, Ilya J. Finkelstein, Jimmy Gollihar, and James M. Musser |
| EPI_ISL_2201916 | Aegis Sciences Corporation | Centers for Disease Control and Prevention Division of Viral Diseases, Pathogen Discovery | Dakota Howard, Dhwani Batra, Peter W. Cook, Kara Moser, Adrian Paskey, Jason Caravas, Benjamin Rambo-Martin, Shatavia Morrison, Christopher Gulvick, Scott Sammons, Yvette Unoarumhi, Darlene Wagner, Matthew Schmerer, Cyndi Clark, Patrick Campbell, Rob Case, Vikramsinha Ghorpade, Holly Houdeshell, Ola Kvalvaag, Dillon Nall, Ethan Sanders, Alec Vest, Shaun Westlund, Matthew Hardison, Clinton R. Paden, Duncan MacCannell |

| | | | |
|---|---|---|---|
| EPI_ISL_2201917 | Houston Methodist Hospital | Houston Methodist Hospital | Randall J. Olsen, Paul A. Christensen, S. Wesley Long, Sishir Subedi, Robert Olson, Marcus Nguyen, James J. Davis, Matthew Ojeda Saavedra, Prasanti Yerramilli, Layne Pruitt, Kristina Reppond, Madison N. Shyer, Jessica Cambric, Ryan Gadd, Ilya J. Finkelstein, Jimmy Gollihar, and James M. Musser |
| EPI_ISL_2201918 | Aegis Sciences Corporation | Centers for Disease Control and Prevention Division of Viral Diseases, Pathogen Discovery | Dakota Howard, Dhwani Batra, Peter W. Cook, Kara Moser, Adrian Paskey, Jason Caravas, Benjamin Rambo-Martin, Shatavia Morrison, Christopher Gulvick, Scott Sammons, Yvette Unoarumhi, Darlene Wagner, Matthew Schmerer, Cyndi Clark, Patrick Campbell, Rob Case, Vikramsinha Ghorpade, Holly Houdeshell, Ola Kvalvaag, Dillon Nall, Ethan Sanders, Alec Vest, Shaun Westlund, Matthew Hardison, Clinton R. Paden, Duncan MacCannell |
| EPI_ISL_2201919 | Houston Methodist Hospital | Houston Methodist Hospital | Randall J. Olsen, Paul A. Christensen, S. Wesley Long, Sishir Subedi, Robert Olson, Marcus Nguyen, James J. Davis, Matthew Ojeda Saavedra, Prasanti Yerramilli, Layne Pruitt, Kristina Reppond, Madison N. Shyer, Jessica Cambric, Ryan Gadd, Ilya J. Finkelstein, Jimmy Gollihar, and James M. Musser |
| EPI_ISL_2201920, EPI_ISL_2201921 | Aegis Sciences Corporation | Centers for Disease Control and Prevention Division of Viral Diseases, Pathogen Discovery | Dakota Howard, Dhwani Batra, Peter W. Cook, Kara Moser, Adrian Paskey, Jason Caravas, Benjamin Rambo-Martin, Shatavia Morrison, Christopher Gulvick, Scott Sammons, Yvette Unoarumhi, Darlene Wagner, Matthew Schmerer, Cyndi Clark, Patrick Campbell, Rob Case, Vikramsinha Ghorpade, Holly Houdeshell, Ola Kvalvaag, Dillon Nall, Ethan Sanders, Alec Vest, Shaun Westlund, Matthew Hardison, Clinton R. Paden, Duncan MacCannell |
| EPI_ISL_2201922 | Houston Methodist Hospital | Houston Methodist Hospital | Randall J. Olsen, Paul A. Christensen, S. Wesley Long, Sishir Subedi, Robert Olson, Marcus Nguyen, James J. Davis, Matthew Ojeda Saavedra, Prasanti Yerramilli, Layne Pruitt, Kristina Reppond, Madison N. Shyer, Jessica Cambric, Ryan Gadd, Ilya J. Finkelstein, Jimmy Gollihar, and James M. Musser |
| EPI_ISL_2201923 | Aegis Sciences Corporation | Centers for Disease Control and Prevention Division of Viral Diseases, Pathogen Discovery | Dakota Howard, Dhwani Batra, Peter W. Cook, Kara Moser, Adrian Paskey, Jason Caravas, Benjamin Rambo-Martin, Shatavia Morrison, Christopher Gulvick, Scott Sammons, Yvette Unoarumhi, Darlene Wagner, Matthew Schmerer, Cyndi Clark, Patrick Campbell, Rob Case, Vikramsinha Ghorpade, Holly Houdeshell, Ola Kvalvaag, Dillon Nall, Ethan Sanders, Alec Vest, Shaun Westlund, Matthew Hardison, Clinton R. Paden, Duncan MacCannell |
| EPI_ISL_2201924 | Houston Methodist Hospital | Houston Methodist Hospital | Randall J. Olsen, Paul A. Christensen, S. Wesley Long, Sishir Subedi, Robert Olson, Marcus Nguyen, James J. Davis, Matthew Ojeda Saavedra, Prasanti Yerramilli, Layne Pruitt, Kristina Reppond, Madison N. Shyer, Jessica Cambric, Ryan Gadd, Ilya J. Finkelstein, Jimmy Gollihar, and James M. Musser |
| EPI_ISL_2201925, EPI_ISL_2201926 | Aegis Sciences Corporation | Centers for Disease Control and Prevention Division of Viral Diseases, Pathogen Discovery | Dakota Howard, Dhwani Batra, Peter W. Cook, Kara Moser, Adrian Paskey, Jason Caravas, Benjamin Rambo-Martin, Shatavia Morrison, Christopher Gulvick, Scott Sammons, Yvette Unoarumhi, Darlene Wagner, Matthew Schmerer, Cyndi Clark, Patrick Campbell, Rob Case, Vikramsinha Ghorpade, Holly Houdeshell, Ola Kvalvaag, Dillon Nall, Ethan Sanders, Alec Vest, Shaun Westlund, Matthew Hardison, Clinton R. Paden, Duncan MacCannell |
| EPI_ISL_2201927 | Houston Methodist Hospital | Houston Methodist Hospital | Randall J. Olsen, Paul A. Christensen, S. Wesley Long, Sishir Subedi, Robert Olson, Marcus Nguyen, James J. Davis, Matthew Ojeda Saavedra, Prasanti Yerramilli, Layne Pruitt, Kristina Reppond, Madison N. Shyer, Jessica Cambric, Ryan Gadd, Ilya J. Finkelstein, Jimmy Gollihar, and James M. Musser |
| EPI_ISL_2201928 | Aegis Sciences Corporation | Centers for Disease Control and Prevention Division of Viral Diseases, Pathogen Discovery | Dakota Howard, Dhwani Batra, Peter W. Cook, Kara Moser, Adrian Paskey, Jason Caravas, Benjamin Rambo-Martin, Shatavia Morrison, Christopher Gulvick, Scott Sammons, Yvette Unoarumhi, Darlene Wagner, Matthew Schmerer, Cyndi Clark, Patrick Campbell, Rob Case, Vikramsinha Ghorpade, Holly Houdeshell, Ola Kvalvaag, Dillon Nall, Ethan Sanders, Alec Vest, Shaun Westlund, Matthew Hardison, Clinton R. Paden, Duncan MacCannell |
| EPI_ISL_2201929 | Houston Methodist Hospital | Houston Methodist Hospital | Randall J. Olsen, Paul A. Christensen, S. Wesley Long, Sishir Subedi, Robert Olson, Marcus Nguyen, James J. Davis, Matthew Ojeda Saavedra, Prasanti Yerramilli, Layne Pruitt, Kristina Reppond, Madison N. Shyer, Jessica Cambric, Ryan Gadd, Ilya J. Finkelstein, Jimmy Gollihar, and James M. Musser |
| EPI_ISL_2201930, EPI_ISL_2201931, EPI_ISL_2201932 | Aegis Sciences Corporation | Centers for Disease Control and Prevention Division of Viral Diseases, Pathogen Discovery | Dakota Howard, Dhwani Batra, Peter W. Cook, Kara Moser, Adrian Paskey, Jason Caravas, Benjamin Rambo-Martin, Shatavia Morrison, Christopher Gulvick, Scott Sammons, Yvette Unoarumhi, Darlene Wagner, Matthew Schmerer, Cyndi Clark, Patrick Campbell, Rob Case, Vikramsinha Ghorpade, Holly Houdeshell, Ola Kvalvaag, Dillon Nall, Ethan Sanders, Alec Vest, Shaun Westlund, Matthew Hardison, Clinton R. Paden, Duncan MacCannell |
| EPI_ISL_2201933 | Houston Methodist Hospital | Houston Methodist Hospital | Randall J. Olsen, Paul A. Christensen, S. Wesley Long, Sishir Subedi, Robert Olson, Marcus Nguyen, James J. Davis, Matthew Ojeda Saavedra, Prasanti Yerramilli, Layne Pruitt, Kristina Reppond, Madison N. Shyer, Jessica Cambric, Ryan Gadd, Ilya J. Finkelstein, Jimmy Gollihar, and James M. Musser |
| EPI_ISL_2201934 | Aegis Sciences Corporation | Centers for Disease Control and Prevention Division of Viral Diseases, Pathogen Discovery | Dakota Howard, Dhwani Batra, Peter W. Cook, Kara Moser, Adrian Paskey, Jason Caravas, Benjamin Rambo-Martin, Shatavia Morrison, Christopher Gulvick, Scott Sammons, Yvette Unoarumhi, Darlene Wagner, Matthew Schmerer, Cyndi Clark, Patrick Campbell, Rob Case, Vikramsinha Ghorpade, Holly Houdeshell, Ola Kvalvaag, Dillon Nall, Ethan Sanders, Alec Vest, Shaun Westlund, Matthew Hardison, Clinton R. Paden, Duncan MacCannell |
| EPI_ISL_2201935 | Houston Methodist Hospital | Houston Methodist Hospital | Randall J. Olsen, Paul A. Christensen, S. Wesley Long, Sishir Subedi, Robert Olson, Marcus Nguyen, James J. Davis, Matthew Ojeda Saavedra, Prasanti Yerramilli, Layne Pruitt, Kristina Reppond, Madison N. Shyer, Jessica Cambric, Ryan Gadd, Ilya J. Finkelstein, Jimmy Gollihar, and James M. Musser |
| EPI_ISL_2201936 | Aegis Sciences Corporation | Centers for Disease Control and Prevention Division of Viral Diseases, Pathogen Discovery | Dakota Howard, Dhwani Batra, Peter W. Cook, Kara Moser, Adrian Paskey, Jason Caravas, Benjamin Rambo-Martin, Shatavia Morrison, Christopher Gulvick, Scott Sammons, Yvette Unoarumhi, Darlene Wagner, Matthew Schmerer, Cyndi Clark, Patrick Campbell, Rob Case, Vikramsinha Ghorpade, Holly Houdeshell, Ola Kvalvaag, Dillon Nall, Ethan Sanders, Alec Vest, Shaun Westlund, Matthew Hardison, Clinton R. Paden, Duncan MacCannell |
| EPI_ISL_2201938 | Houston Methodist Hospital | Houston Methodist Hospital | Randall J. Olsen, Paul A. Christensen, S. Wesley Long, Sishir Subedi, Robert Olson, Marcus Nguyen, James J. Davis, Matthew Ojeda Saavedra, Prasanti Yerramilli, Layne Pruitt, Kristina Reppond, Madison N. Shyer, Jessica Cambric, Ryan Gadd, Ilya J. Finkelstein, Jimmy Gollihar, and James M. Musser |
| EPI_ISL_2201939, EPI_ISL_2201940 | Aegis Sciences Corporation | Centers for Disease Control and Prevention Division of Viral Diseases, Pathogen Discovery | Dakota Howard, Dhwani Batra, Peter W. Cook, Kara Moser, Adrian Paskey, Jason Caravas, Benjamin Rambo-Martin, Shatavia Morrison, Christopher Gulvick, Scott Sammons, Yvette Unoarumhi, Darlene Wagner, Matthew Schmerer, Cyndi Clark, Patrick Campbell, Rob Case, Vikramsinha Ghorpade, Holly Houdeshell, Ola Kvalvaag, Dillon Nall, Ethan Sanders, Alec Vest, Shaun Westlund, Matthew Hardison, Clinton R. Paden, Duncan MacCannell |
| EPI_ISL_2201941 | Houston Methodist Hospital | Houston Methodist Hospital | Randall J. Olsen, Paul A. Christensen, S. Wesley Long, Sishir Subedi, Robert Olson, Marcus Nguyen, James J. Davis, Matthew Ojeda Saavedra, Prasanti Yerramilli, Layne Pruitt, Kristina Reppond, Madison N. Shyer, Jessica Cambric, Ryan Gadd, Ilya J. Finkelstein, Jimmy Gollihar, and James M. Musser |
| EPI_ISL_2201942, EPI_ISL_2201943, EPI_ISL_2201944 | Aegis Sciences Corporation | Centers for Disease Control and Prevention Division of Viral Diseases, Pathogen Discovery | Dakota Howard, Dhwani Batra, Peter W. Cook, Kara Moser, Adrian Paskey, Jason Caravas, Benjamin Rambo-Martin, Shatavia Morrison, Christopher Gulvick, Scott Sammons, Yvette Unoarumhi, Darlene Wagner, Matthew Schmerer, Cyndi Clark, Patrick Campbell, Rob Case, Vikramsinha Ghorpade, Holly Houdeshell, Ola Kvalvaag, Dillon Nall, Ethan Sanders, Alec Vest, Shaun Westlund, Matthew Hardison, Clinton R. Paden, Duncan MacCannell |
| EPI_ISL_2201945 | Houston Methodist Hospital | Houston Methodist Hospital | Randall J. Olsen, Paul A. Christensen, S. Wesley Long, Sishir Subedi, Robert Olson, Marcus Nguyen, James J. Davis, Matthew Ojeda Saavedra, Prasanti Yerramilli, Layne Pruitt, Kristina Reppond, Madison N. Shyer, Jessica Cambric, Ryan Gadd, Ilya J. Finkelstein, Jimmy Gollihar, and James M. Musser |
| EPI_ISL_2201946 | Aegis Sciences Corporation | Centers for Disease Control and Prevention Division of Viral Diseases, Pathogen Discovery | Dakota Howard, Dhwani Batra, Peter W. Cook, Kara Moser, Adrian Paskey, Jason Caravas, Benjamin Rambo-Martin, Shatavia Morrison, Christopher Gulvick, Scott Sammons, Yvette Unoarumhi, Darlene Wagner, Matthew Schmerer, Cyndi Clark, Patrick Campbell, Rob Case, Vikramsinha Ghorpade, Holly Houdeshell, Ola Kvalvaag, Dillon Nall, Ethan Sanders, Alec Vest, Shaun Westlund, Matthew Hardison, Clinton R. Paden, Duncan MacCannell |
| EPI_ISL_2201947, EPI_ISL_2201948 | Houston Methodist Hospital | Houston Methodist Hospital | Randall J. Olsen, Paul A. Christensen, S. Wesley Long, Sishir Subedi, Robert Olson, Marcus Nguyen, James J. Davis, Matthew Ojeda Saavedra, Prasanti Yerramilli, Layne Pruitt, Kristina Reppond, Madison N. Shyer, Jessica Cambric, Ryan Gadd, Ilya J. Finkelstein, Jimmy Gollihar, and James M. Musser |
| EPI_ISL_2201949, EPI_ISL_2201950 | Aegis Sciences Corporation | Centers for Disease Control and Prevention Division of Viral Diseases, Pathogen Discovery | Dakota Howard, Dhwani Batra, Peter W. Cook, Kara Moser, Adrian Paskey, Jason Caravas, Benjamin Rambo-Martin, Shatavia Morrison, Christopher Gulvick, Scott Sammons, Yvette Unoarumhi, Darlene Wagner, Matthew Schmerer, Cyndi Clark, Patrick Campbell, Rob Case, Vikramsinha Ghorpade, Holly Houdeshell, Ola Kvalvaag, Dillon Nall, Ethan Sanders, Alec Vest, Shaun Westlund, Matthew Hardison, Clinton R. Paden, Duncan MacCannell |
| EPI_ISL_2201951 | Houston Methodist Hospital | Houston Methodist Hospital | Randall J. Olsen, Paul A. Christensen, S. Wesley Long, Sishir Subedi, Robert Olson, Marcus Nguyen, James J. Davis, Matthew Ojeda Saavedra, Prasanti Yerramilli, Layne Pruitt, Kristina Reppond, Madison N. Shyer, Jessica Cambric, Ryan Gadd, Ilya J. Finkelstein, Jimmy Gollihar, and James M. Musser |
| EPI_ISL_2201952 | Aegis Sciences Corporation | Centers for Disease Control and Prevention Division of Viral Diseases, Pathogen Discovery | Dakota Howard, Dhwani Batra, Peter W. Cook, Kara Moser, Adrian Paskey, Jason Caravas, Benjamin Rambo-Martin, Shatavia Morrison, Christopher Gulvick, Scott Sammons, Yvette Unoarumhi, Darlene Wagner, Matthew Schmerer, Cyndi Clark, Patrick Campbell, Rob Case, Vikramsinha Ghorpade, Holly Houdeshell, Ola Kvalvaag, Dillon Nall, Ethan Sanders, Alec Vest, Shaun Westlund, Matthew Hardison, Clinton R. Paden, Duncan MacCannell |
| EPI_ISL_2201953 | Marshfield Labs | Genomics and Discovery, Respiratory Viruses Branch, Division of Viral Diseases, Centers for Disease Control and Prevention | Yan Li, Jing Zhang, Anna Kelleher, Ying Tao, Brian Lynch, Krista Queen, Anna Uehara, Peter Cook, Han Jia Justin Ng, Rachel Marine, Clinton R. Paden, Dhwani Batra, Haibin Wang, Tara Coalter, Jasmine Padilla, Morgan Davis, Mili Sheth, Sarah Nobles, Mark Burroughs, Justin Lee, Adam Retchless, Suxiang Tong |
| EPI_ISL_2201954 | Houston Methodist Hospital | Houston Methodist Hospital | Randall J. Olsen, Paul A. Christensen, S. Wesley Long, Sishir Subedi, Robert Olson, Marcus Nguyen, James J. Davis, Matthew Ojeda Saavedra, Prasanti Yerramilli, Layne Pruitt, Kristina Reppond, Madison N. Shyer, Jessica Cambric, Ryan Gadd, Ilya J. Finkelstein, Jimmy Gollihar, and James M. Musser |
| EPI_ISL_2201955 | Aegis Sciences Corporation | Centers for Disease Control and Prevention Division of Viral Diseases, Pathogen Discovery | Dakota Howard, Dhwani Batra, Peter W. Cook, Kara Moser, Adrian Paskey, Jason Caravas, Benjamin Rambo-Martin, Shatavia Morrison, Christopher Gulvick, Scott Sammons, Yvette Unoarumhi, Darlene Wagner, Matthew Schmerer, Cyndi Clark, Patrick Campbell, Rob Case, Vikramsinha Ghorpade, Holly Houdeshell, Ola Kvalvaag, Dillon Nall, Ethan Sanders, Alec Vest, Shaun Westlund, Matthew Hardison, Clinton R. Paden, Duncan MacCannell |
| EPI_ISL_2201956, EPI_ISL_2201957 | Marshfield Labs | Genomics and Discovery, Respiratory Viruses Branch, Division of Viral Diseases, Centers for Disease Control and Prevention | Yan Li, Jing Zhang, Anna Kelleher, Ying Tao, Brian Lynch, Krista Queen, Anna Uehara, Peter Cook, Han Jia Justin Ng, Rachel Marine, Clinton R. Paden, Dhwani Batra, Haibin Wang, Tara Coalter, Jasmine Padilla, Morgan Davis, Mili Sheth, Sarah Nobles, Mark Burroughs, Justin Lee, Adam Retchless, Suxiang Tong |
| EPI_ISL_2201958 | Aegis Sciences Corporation | Centers for Disease Control and Prevention Division of Viral Diseases, Pathogen Discovery | Dakota Howard, Dhwani Batra, Peter W. Cook, Kara Moser, Adrian Paskey, Jason Caravas, Benjamin Rambo-Martin, Shatavia Morrison, Christopher Gulvick, Scott Sammons, Yvette Unoarumhi, Darlene Wagner, Matthew Schmerer, Cyndi Clark, Patrick Campbell, Rob Case, Vikramsinha Ghorpade, Holly Houdeshell, Ola Kvalvaag, Dillon Nall, Ethan Sanders, Alec Vest, Shaun Westlund, Matthew Hardison, Clinton R. Paden, Duncan MacCannell |
| EPI_ISL_2201959 | Houston Methodist Hospital | Houston Methodist Hospital | Randall J. Olsen, Paul A. Christensen, S. Wesley Long, Sishir Subedi, Robert Olson, Marcus Nguyen, James J. Davis, Matthew Ojeda Saavedra, Prasanti Yerramilli, Layne Pruitt, Kristina Reppond, Madison N. Shyer, Jessica Cambric, Ryan Gadd, Ilya J. Finkelstein, Jimmy Gollihar, and James M. Musser |
| EPI_ISL_2201960 | Marshfield Labs | Genomics and Discovery, Respiratory Viruses Branch, Division of Viral Diseases, Centers for Disease Control and Prevention | Yan Li, Jing Zhang, Anna Kelleher, Ying Tao, Brian Lynch, Krista Queen, Anna Uehara, Peter Cook, Han Jia Justin Ng, Rachel Marine, Clinton R. Paden, Dhwani Batra, Haibin Wang, Tara Coalter, Jasmine Padilla, Morgan Davis, Mili Sheth, Sarah Nobles, Mark Burroughs, Justin Lee, Adam Retchless, Suxiang Tong |
| EPI_ISL_2201961 | Aegis Sciences Corporation | Centers for Disease Control and Prevention Division of Viral Diseases, Pathogen Discovery | Dakota Howard, Dhwani Batra, Peter W. Cook, Kara Moser, Adrian Paskey, Jason Caravas, Benjamin Rambo-Martin, Shatavia Morrison, Christopher Gulvick, Scott Sammons, Yvette Unoarumhi, Darlene Wagner, Matthew Schmerer, Cyndi Clark, Patrick Campbell, Rob Case, Vikramsinha Ghorpade, Holly Houdeshell, Ola Kvalvaag, Dillon Nall, Ethan Sanders, Alec Vest, Shaun Westlund, Matthew Hardison, Clinton R. Paden, Duncan MacCannell |
| EPI_ISL_2201962 | Marshfield Labs | Genomics and Discovery, Respiratory Viruses Branch, Division of Viral Diseases, Centers for Disease Control and Prevention | Yan Li, Jing Zhang, Anna Kelleher, Ying Tao, Brian Lynch, Krista Queen, Anna Uehara, Peter Cook, Han Jia Justin Ng, Rachel Marine, Clinton R. Paden, Dhwani Batra, Haibin Wang, Tara Coalter, Jasmine Padilla, Morgan Davis, Mili Sheth, Sarah Nobles, Mark Burroughs, Justin Lee, Adam Retchless, Suxiang Tong |
| EPI_ISL_2201963 | Houston Methodist Hospital | Houston Methodist Hospital | Randall J. Olsen, Paul A. Christensen, S. Wesley Long, Sishir Subedi, Robert Olson, Marcus Nguyen, James J. Davis, Matthew Ojeda Saavedra, Prasanti Yerramilli, Layne Pruitt, Kristina Reppond, Madison N. Shyer, Jessica Cambric, Ryan Gadd, Ilya J. Finkelstein, Jimmy Gollihar, and James M. Musser |
| EPI_ISL_2201964 | Marshfield Labs | Genomics and Discovery, Respiratory Viruses Branch, Division of Viral Diseases, | Yan Li, Jing Zhang, Anna Kelleher, Ying Tao, Brian Lynch, Krista Queen, Anna Uehara, Peter Cook, Han Jia Justin Ng, Rachel Marine, Clinton R. Paden, Dhwani Batra, Haibin Wang, Tara Coalter, Jasmine Padilla, Morgan Davis, Mili Sheth, Sarah Nobles, Mark Burroughs, Justin Lee, Adam Retchless, Suxiang Tong |

| | | | |
|---|---|---|---|
| | | Centers for Disease Control and Prevention | |
| EPI_ISL_2201965 | Aegis Sciences Corporation | Centers for Disease Control and Prevention Division of Viral Diseases, Pathogen Discovery | Dakota Howard, Dhwani Batra, Peter W. Cook, Kara Moser, Adrian Paskey, Jason Caravas, Benjamin Rambo-Martin, Shatavia Morrison, Christopher Gulvick, Scott Sammons, Yvette Unoarumhi, Darlene Wagner, Matthew Schmerer, Cyndi Clark, Patrick Campbell, Rob Case, Vikramsinha Ghorpade, Holly Houdeshell, Ola Kvalvaag, Dillon Nall, Ethan Sanders, Alec Vest, Shaun Westlund, Matthew Hardison, Clinton R. Paden, Duncan MacCannell |
| EPI_ISL_2201966, EPI_ISL_2201967 | Marshfield Labs | Genomics and Discovery, Respiratory Viruses Branch, Division of Viral Diseases, Centers for Disease Control and Prevention | Yan Li, Jing Zhang, Anna Kelleher, Ying Tao, Brian Lynch, Krista Queen, Anna Uehara, Peter Cook, Han Jia Justin Ng, Rachel Marine, Clinton R. Paden, Dhwani Batra, Haibin Wang, Tara Coalter, Jasmine Padilla, Morgan Davis, Mili Sheth, Sarah Nobles, Mark Burroughs, Justin Lee, Adam Retchless, Suxiang Tong |
| EPI_ISL_2201968 | Aegis Sciences Corporation | Centers for Disease Control and Prevention Division of Viral Diseases, Pathogen Discovery | Dakota Howard, Dhwani Batra, Peter W. Cook, Kara Moser, Adrian Paskey, Jason Caravas, Benjamin Rambo-Martin, Shatavia Morrison, Christopher Gulvick, Scott Sammons, Yvette Unoarumhi, Darlene Wagner, Matthew Schmerer, Cyndi Clark, Patrick Campbell, Rob Case, Vikramsinha Ghorpade, Holly Houdeshell, Ola Kvalvaag, Dillon Nall, Ethan Sanders, Alec Vest, Shaun Westlund, Matthew Hardison, Clinton R. Paden, Duncan MacCannell |
| EPI_ISL_2201969 | Houston Methodist Hospital | Houston Methodist Hospital | Randall J. Olsen, Paul A. Christensen, S. Wesley Long, Sishir Subedi, Robert Olson, Marcus Nguyen, James J. Davis, Matthew Ojeda Saavedra, Prasanti Yerramilli, Layne Pruitt, Kristina Reppond, Madison N. Shyer, Jessica Cambric, Ryan Gadd, Ilya J. Finkelstein, Jimmy Gollihar, and James M. Musser |
| EPI_ISL_2201970 | Marshfield Labs | Genomics and Discovery, Respiratory Viruses Branch, Division of Viral Diseases, Centers for Disease Control and Prevention | Yan Li, Jing Zhang, Anna Kelleher, Ying Tao, Brian Lynch, Krista Queen, Anna Uehara, Peter Cook, Han Jia Justin Ng, Rachel Marine, Clinton R. Paden, Dhwani Batra, Haibin Wang, Tara Coalter, Jasmine Padilla, Morgan Davis, Mili Sheth, Sarah Nobles, Mark Burroughs, Justin Lee, Adam Retchless, Suxiang Tong |
| EPI_ISL_2201971 | Aegis Sciences Corporation | Centers for Disease Control and Prevention Division of Viral Diseases, Pathogen Discovery | Dakota Howard, Dhwani Batra, Peter W. Cook, Kara Moser, Adrian Paskey, Jason Caravas, Benjamin Rambo-Martin, Shatavia Morrison, Christopher Gulvick, Scott Sammons, Yvette Unoarumhi, Darlene Wagner, Matthew Schmerer, Cyndi Clark, Patrick Campbell, Rob Case, Vikramsinha Ghorpade, Holly Houdeshell, Ola Kvalvaag, Dillon Nall, Ethan Sanders, Alec Vest, Shaun Westlund, Matthew Hardison, Clinton R. Paden, Duncan MacCannell |
| EPI_ISL_2201972 | Houston Methodist Hospital | Houston Methodist Hospital | Randall J. Olsen, Paul A. Christensen, S. Wesley Long, Sishir Subedi, Robert Olson, Marcus Nguyen, James J. Davis, Matthew Ojeda Saavedra, Prasanti Yerramilli, Layne Pruitt, Kristina Reppond, Madison N. Shyer, Jessica Cambric, Ryan Gadd, Ilya J. Finkelstein, Jimmy Gollihar, and James M. Musser |
| EPI_ISL_2201973 | Aegis Sciences Corporation | Centers for Disease Control and Prevention Division of Viral Diseases, Pathogen Discovery | Dakota Howard, Dhwani Batra, Peter W. Cook, Kara Moser, Adrian Paskey, Jason Caravas, Benjamin Rambo-Martin, Shatavia Morrison, Christopher Gulvick, Scott Sammons, Yvette Unoarumhi, Darlene Wagner, Matthew Schmerer, Cyndi Clark, Patrick Campbell, Rob Case, Vikramsinha Ghorpade, Holly Houdeshell, Ola Kvalvaag, Dillon Nall, Ethan Sanders, Alec Vest, Shaun Westlund, Matthew Hardison, Clinton R. Paden, Duncan MacCannell |
| EPI_ISL_2201974 | Marshfield Labs | Genomics and Discovery, Respiratory Viruses Branch, Division of Viral Diseases, Centers for Disease Control and Prevention | Yan Li, Jing Zhang, Anna Kelleher, Ying Tao, Brian Lynch, Krista Queen, Anna Uehara, Peter Cook, Han Jia Justin Ng, Rachel Marine, Clinton R. Paden, Dhwani Batra, Haibin Wang, Tara Coalter, Jasmine Padilla, Morgan Davis, Mili Sheth, Sarah Nobles, Mark Burroughs, Justin Lee, Adam Retchless, Suxiang Tong |
| EPI_ISL_2201975 | Aegis Sciences Corporation | Centers for Disease Control and Prevention Division of Viral Diseases, Pathogen Discovery | Dakota Howard, Dhwani Batra, Peter W. Cook, Kara Moser, Adrian Paskey, Jason Caravas, Benjamin Rambo-Martin, Shatavia Morrison, Christopher Gulvick, Scott Sammons, Yvette Unoarumhi, Darlene Wagner, Matthew Schmerer, Cyndi Clark, Patrick Campbell, Rob Case, Vikramsinha Ghorpade, Holly Houdeshell, Ola Kvalvaag, Dillon Nall, Ethan Sanders, Alec Vest, Shaun Westlund, Matthew Hardison, Clinton R. Paden, Duncan MacCannell |
| EPI_ISL_2201976 | Houston Methodist Hospital | Houston Methodist Hospital | Randall J. Olsen, Paul A. Christensen, S. Wesley Long, Sishir Subedi, Robert Olson, Marcus Nguyen, James J. Davis, Matthew Ojeda Saavedra, Prasanti Yerramilli, Layne Pruitt, Kristina Reppond, Madison N. Shyer, Jessica Cambric, Ryan Gadd, Ilya J. Finkelstein, Jimmy Gollihar, and James M. Musser |
| EPI_ISL_2201977, EPI_ISL_2201978, EPI_ISL_2201979, EPI_ISL_2201980 | Aegis Sciences Corporation | Centers for Disease Control and Prevention Division of Viral Diseases, Pathogen Discovery | Dakota Howard, Dhwani Batra, Peter W. Cook, Kara Moser, Adrian Paskey, Jason Caravas, Benjamin Rambo-Martin, Shatavia Morrison, Christopher Gulvick, Scott Sammons, Yvette Unoarumhi, Darlene Wagner, Matthew Schmerer, Cyndi Clark, Patrick Campbell, Rob Case, Vikramsinha Ghorpade, Holly Houdeshell, Ola Kvalvaag, Dillon Nall, Ethan Sanders, Alec Vest, Shaun Westlund, Matthew Hardison, Clinton R. Paden, Duncan MacCannell |
| EPI_ISL_2201981 | Houston Methodist Hospital | Houston Methodist Hospital | Randall J. Olsen, Paul A. Christensen, S. Wesley Long, Sishir Subedi, Robert Olson, Marcus Nguyen, James J. Davis, Matthew Ojeda Saavedra, Prasanti Yerramilli, Layne Pruitt, Kristina Reppond, Madison N. Shyer, Jessica Cambric, Ryan Gadd, Ilya J. Finkelstein, Jimmy Gollihar, and James M. Musser |
| EPI_ISL_2201982 | Aegis Sciences Corporation | Centers for Disease Control and Prevention Division of Viral Diseases, Pathogen Discovery | Dakota Howard, Dhwani Batra, Peter W. Cook, Kara Moser, Adrian Paskey, Jason Caravas, Benjamin Rambo-Martin, Shatavia Morrison, Christopher Gulvick, Scott Sammons, Yvette Unoarumhi, Darlene Wagner, Matthew Schmerer, Cyndi Clark, Patrick Campbell, Rob Case, Vikramsinha Ghorpade, Holly Houdeshell, Ola Kvalvaag, Dillon Nall, Ethan Sanders, Alec Vest, Shaun Westlund, Matthew Hardison, Clinton R. Paden, Duncan MacCannell |
| EPI_ISL_2201983 | Houston Methodist Hospital | Houston Methodist Hospital | Randall J. Olsen, Paul A. Christensen, S. Wesley Long, Sishir Subedi, Robert Olson, Marcus Nguyen, James J. Davis, Matthew Ojeda Saavedra, Prasanti Yerramilli, Layne Pruitt, Kristina Reppond, Madison N. Shyer, Jessica Cambric, Ryan Gadd, Ilya J. Finkelstein, Jimmy Gollihar, and James M. Musser |
| EPI_ISL_2201984, EPI_ISL_2201985 | Aegis Sciences Corporation | Centers for Disease Control and Prevention Division of Viral Diseases, Pathogen Discovery | Dakota Howard, Dhwani Batra, Peter W. Cook, Kara Moser, Adrian Paskey, Jason Caravas, Benjamin Rambo-Martin, Shatavia Morrison, Christopher Gulvick, Scott Sammons, Yvette Unoarumhi, Darlene Wagner, Matthew Schmerer, Cyndi Clark, Patrick Campbell, Rob Case, Vikramsinha Ghorpade, Holly Houdeshell, Ola Kvalvaag, Dillon Nall, Ethan Sanders, Alec Vest, Shaun Westlund, Matthew Hardison, Clinton R. Paden, Duncan MacCannell |
| EPI_ISL_2201986, EPI_ISL_2201988 | Houston Methodist Hospital | Houston Methodist Hospital | Randall J. Olsen, Paul A. Christensen, S. Wesley Long, Sishir Subedi, Robert Olson, Marcus Nguyen, James J. Davis, Matthew Ojeda Saavedra, Prasanti Yerramilli, Layne Pruitt, Kristina Reppond, Madison N. Shyer, Jessica Cambric, Ryan Gadd, Ilya J. Finkelstein, Jimmy Gollihar, and James M. Musser |
| EPI_ISL_2201989, EPI_ISL_2201990 | Aegis Sciences Corporation | Centers for Disease Control and Prevention Division of Viral Diseases, Pathogen Discovery | Dakota Howard, Dhwani Batra, Peter W. Cook, Kara Moser, Adrian Paskey, Jason Caravas, Benjamin Rambo-Martin, Shatavia Morrison, Christopher Gulvick, Scott Sammons, Yvette Unoarumhi, Darlene Wagner, Matthew Schmerer, Cyndi Clark, Patrick Campbell, Rob Case, Vikramsinha Ghorpade, Holly Houdeshell, Ola Kvalvaag, Dillon Nall, Ethan Sanders, Alec Vest, Shaun Westlund, Matthew Hardison, Clinton R. Paden, Duncan MacCannell |
| EPI_ISL_2201991 | Houston Methodist Hospital | Houston Methodist Hospital | Randall J. Olsen, Paul A. Christensen, S. Wesley Long, Sishir Subedi, Robert Olson, Marcus Nguyen, James J. Davis, Matthew Ojeda Saavedra, Prasanti Yerramilli, Layne Pruitt, Kristina Reppond, Madison N. Shyer, Jessica Cambric, Ryan Gadd, Ilya J. Finkelstein, Jimmy Gollihar, and James M. Musser |
| EPI_ISL_2201992, EPI_ISL_2201993 | Aegis Sciences Corporation | Centers for Disease Control and Prevention Division of Viral Diseases, Pathogen Discovery | Dakota Howard, Dhwani Batra, Peter W. Cook, Kara Moser, Adrian Paskey, Jason Caravas, Benjamin Rambo-Martin, Shatavia Morrison, Christopher Gulvick, Scott Sammons, Yvette Unoarumhi, Darlene Wagner, Matthew Schmerer, Cyndi Clark, Patrick Campbell, Rob Case, Vikramsinha Ghorpade, Holly Houdeshell, Ola Kvalvaag, Dillon Nall, Ethan Sanders, Alec Vest, Shaun Westlund, Matthew Hardison, Clinton R. Paden, Duncan MacCannell |
| EPI_ISL_2201994 | Houston Methodist Hospital | Houston Methodist Hospital | Randall J. Olsen, Paul A. Christensen, S. Wesley Long, Sishir Subedi, Robert Olson, Marcus Nguyen, James J. Davis, Matthew Ojeda Saavedra, Prasanti Yerramilli, Layne Pruitt, Kristina Reppond, Madison N. Shyer, Jessica Cambric, Ryan Gadd, Ilya J. Finkelstein, Jimmy Gollihar, and James M. Musser |
| EPI_ISL_2201995 | Aegis Sciences Corporation | Centers for Disease Control and Prevention Division of Viral Diseases, Pathogen Discovery | Dakota Howard, Dhwani Batra, Peter W. Cook, Kara Moser, Adrian Paskey, Jason Caravas, Benjamin Rambo-Martin, Shatavia Morrison, Christopher Gulvick, Scott Sammons, Yvette Unoarumhi, Darlene Wagner, Matthew Schmerer, Cyndi Clark, Patrick Campbell, Rob Case, Vikramsinha Ghorpade, Holly Houdeshell, Ola Kvalvaag, Dillon Nall, Ethan Sanders, Alec Vest, Shaun Westlund, Matthew Hardison, Clinton R. Paden, Duncan MacCannell |
| EPI_ISL_2201996 | Houston Methodist Hospital | Houston Methodist Hospital | Randall J. Olsen, Paul A. Christensen, S. Wesley Long, Sishir Subedi, Robert Olson, Marcus Nguyen, James J. Davis, Matthew Ojeda Saavedra, Prasanti Yerramilli, Layne Pruitt, Kristina Reppond, Madison N. Shyer, Jessica Cambric, Ryan Gadd, Ilya J. Finkelstein, Jimmy Gollihar, and James M. Musser |
| EPI_ISL_2201997 | Aegis Sciences Corporation | Centers for Disease Control and Prevention Division of Viral Diseases, Pathogen Discovery | Dakota Howard, Dhwani Batra, Peter W. Cook, Kara Moser, Adrian Paskey, Jason Caravas, Benjamin Rambo-Martin, Shatavia Morrison, Christopher Gulvick, Scott Sammons, Yvette Unoarumhi, Darlene Wagner, Matthew Schmerer, Cyndi Clark, Patrick Campbell, Rob Case, Vikramsinha Ghorpade, Holly Houdeshell, Ola Kvalvaag, Dillon Nall, Ethan Sanders, Alec Vest, Shaun Westlund, Matthew Hardison, Clinton R. Paden, Duncan MacCannell |
| EPI_ISL_2201998 | Houston Methodist Hospital | Houston Methodist Hospital | Randall J. Olsen, Paul A. Christensen, S. Wesley Long, Sishir Subedi, Robert Olson, Marcus Nguyen, James J. Davis, Matthew Ojeda Saavedra, Prasanti Yerramilli, Layne Pruitt, Kristina Reppond, Madison N. Shyer, Jessica Cambric, Ryan Gadd, Ilya J. Finkelstein, Jimmy Gollihar, and James M. Musser |
| EPI_ISL_2201999, EPI_ISL_2202000 | Aegis Sciences Corporation | Centers for Disease Control and Prevention Division of Viral Diseases, Pathogen Discovery | Dakota Howard, Dhwani Batra, Peter W. Cook, Kara Moser, Adrian Paskey, Jason Caravas, Benjamin Rambo-Martin, Shatavia Morrison, Christopher Gulvick, Scott Sammons, Yvette Unoarumhi, Darlene Wagner, Matthew Schmerer, Cyndi Clark, Patrick Campbell, Rob Case, Vikramsinha Ghorpade, Holly Houdeshell, Ola Kvalvaag, Dillon Nall, Ethan Sanders, Alec Vest, Shaun Westlund, Matthew Hardison, Clinton R. Paden, Duncan MacCannell |
| EPI_ISL_2202001 | Houston Methodist Hospital | Houston Methodist Hospital | Randall J. Olsen, Paul A. Christensen, S. Wesley Long, Sishir Subedi, Robert Olson, Marcus Nguyen, James J. Davis, Matthew Ojeda Saavedra, Prasanti Yerramilli, Layne Pruitt, Kristina Reppond, Madison N. Shyer, Jessica Cambric, Ryan Gadd, Ilya J. Finkelstein, Jimmy Gollihar, and James M. Musser |
| EPI_ISL_2202002 | Aegis Sciences Corporation | Centers for Disease Control and Prevention Division of Viral Diseases, Pathogen Discovery | Dakota Howard, Dhwani Batra, Peter W. Cook, Kara Moser, Adrian Paskey, Jason Caravas, Benjamin Rambo-Martin, Shatavia Morrison, Christopher Gulvick, Scott Sammons, Yvette Unoarumhi, Darlene Wagner, Matthew Schmerer, Cyndi Clark, Patrick Campbell, Rob Case, Vikramsinha Ghorpade, Holly Houdeshell, Ola Kvalvaag, Dillon Nall, Ethan Sanders, Alec Vest, Shaun Westlund, Matthew Hardison, Clinton R. Paden, Duncan MacCannell |
| EPI_ISL_2202003 | Houston Methodist Hospital | Houston Methodist Hospital | Randall J. Olsen, Paul A. Christensen, S. Wesley Long, Sishir Subedi, Robert Olson, Marcus Nguyen, James J. Davis, Matthew Ojeda Saavedra, Prasanti Yerramilli, Layne Pruitt, Kristina Reppond, Madison N. Shyer, Jessica Cambric, Ryan Gadd, Ilya J. Finkelstein, Jimmy Gollihar, and James M. Musser |
| EPI_ISL_2202004, EPI_ISL_2202005 | Aegis Sciences Corporation | Centers for Disease Control and Prevention Division of Viral Diseases, Pathogen Discovery | Dakota Howard, Dhwani Batra, Peter W. Cook, Kara Moser, Adrian Paskey, Jason Caravas, Benjamin Rambo-Martin, Shatavia Morrison, Christopher Gulvick, Scott Sammons, Yvette Unoarumhi, Darlene Wagner, Matthew Schmerer, Cyndi Clark, Patrick Campbell, Rob Case, Vikramsinha Ghorpade, Holly Houdeshell, Ola Kvalvaag, Dillon Nall, Ethan Sanders, Alec Vest, Shaun Westlund, Matthew Hardison, Clinton R. Paden, Duncan MacCannell |
| EPI_ISL_2202006 | Houston Methodist Hospital | Houston Methodist Hospital | Randall J. Olsen, Paul A. Christensen, S. Wesley Long, Sishir Subedi, Robert Olson, Marcus Nguyen, James J. Davis, Matthew Ojeda Saavedra, Prasanti Yerramilli, Layne Pruitt, Kristina Reppond, Madison N. Shyer, Jessica Cambric, Ryan Gadd, Ilya J. Finkelstein, Jimmy Gollihar, and James M. Musser |
| EPI_ISL_2202007 | Aegis Sciences Corporation | Centers for Disease Control and Prevention Division of Viral Diseases, Pathogen Discovery | Dakota Howard, Dhwani Batra, Peter W. Cook, Kara Moser, Adrian Paskey, Jason Caravas, Benjamin Rambo-Martin, Shatavia Morrison, Christopher Gulvick, Scott Sammons, Yvette Unoarumhi, Darlene Wagner, Matthew Schmerer, Cyndi Clark, Patrick Campbell, Rob Case, Vikramsinha Ghorpade, Holly Houdeshell, Ola Kvalvaag, Dillon Nall, Ethan Sanders, Alec Vest, Shaun Westlund, Matthew Hardison, Clinton R. Paden, Duncan MacCannell |
| EPI_ISL_2202008 | Houston Methodist Hospital | Houston Methodist Hospital | Randall J. Olsen, Paul A. Christensen, S. Wesley Long, Sishir Subedi, Robert Olson, Marcus Nguyen, James J. Davis, Matthew Ojeda Saavedra, Prasanti Yerramilli, Layne Pruitt, Kristina Reppond, Madison N. Shyer, Jessica Cambric, Ryan Gadd, Ilya J. Finkelstein, Jimmy Gollihar, and James M. Musser |
| EPI_ISL_2202009 | Aegis Sciences Corporation | Centers for Disease Control and Prevention Division of Viral Diseases, Pathogen Discovery | Dakota Howard, Dhwani Batra, Peter W. Cook, Kara Moser, Adrian Paskey, Jason Caravas, Benjamin Rambo-Martin, Shatavia Morrison, Christopher Gulvick, Scott Sammons, Yvette Unoarumhi, Darlene Wagner, Matthew Schmerer, Cyndi Clark, Patrick Campbell, Rob Case, Vikramsinha Ghorpade, Holly Houdeshell, Ola Kvalvaag, Dillon Nall, Ethan Sanders, Alec Vest, Shaun Westlund, Matthew Hardison, Clinton R. Paden, Duncan MacCannell |
| EPI_ISL_2202010 | Houston Methodist Hospital | Houston Methodist Hospital | Randall J. Olsen, Paul A. Christensen, S. Wesley Long, Sishir Subedi, Robert Olson, Marcus Nguyen, James J. Davis, Matthew Ojeda Saavedra, Prasanti Yerramilli, Layne Pruitt, Kristina Reppond, Madison N. Shyer, Jessica Cambric, Ryan Gadd, Ilya J. Finkelstein, Jimmy Gollihar, and James M. Musser |
| EPI_ISL_2202011 | Aegis Sciences Corporation | Centers for Disease Control and Prevention Division of Viral Diseases, Pathogen | Dakota Howard, Dhwani Batra, Peter W. Cook, Kara Moser, Adrian Paskey, Jason Caravas, Benjamin Rambo-Martin, Shatavia Morrison, Christopher Gulvick, Scott Sammons, Yvette Unoarumhi, Darlene Wagner, Matthew Schmerer, Cyndi Clark, Patrick Campbell, Rob Case, Vikramsinha Ghorpade, Holly Houdeshell, Ola Kvalvaag, Dillon Nall, Ethan Sanders, Alec Vest, Shaun Westlund, Matthew Hardison, Clinton R. Paden, Duncan MacCannell |

| | | | |
|---|---|---|---|
| | | Discovery | |
| EPI_ISL_2202012 | Houston Methodist Hospital | Houston Methodist Hospital | Randall J. Olsen, Paul A. Christensen, S. Wesley Long, Sishir Subedi, Robert Olson, Marcus Nguyen, James J. Davis, Matthew Ojeda Saavedra, Prasanti Yerramilli, Layne Pruitt, Kristina Reppond, Madison N. Shyer, Jessica Cambric, Ryan Gadd, Ilya J. Finkelstein, Jimmy Gollihar, and James M. Musser |
| EPI_ISL_2202013, EPI_ISL_2202014 | Aegis Sciences Corporation | Centers for Disease Control and Prevention Division of Viral Diseases, Pathogen Discovery | Dakota Howard, Dhwani Batra, Peter W. Cook, Kara Moser, Adrian Paskey, Jason Caravas, Benjamin Rambo-Martin, Shatavia Morrison, Christopher Gulvick, Scott Sammons, Yvette Unoarumhi, Darlene Wagner, Matthew Schmerer, Cyndi Clark, Patrick Campbell, Rob Case, Vikramsinha Ghorpade, Holly Houdeshell, Ola Kvalvaag, Dillon Nall, Ethan Sanders, Alec Vest, Shaun Westlund, Matthew Hardison, Clinton R. Paden, Duncan MacCannell |
| EPI_ISL_2202015 | Houston Methodist Hospital | Houston Methodist Hospital | Randall J. Olsen, Paul A. Christensen, S. Wesley Long, Sishir Subedi, Robert Olson, Marcus Nguyen, James J. Davis, Matthew Ojeda Saavedra, Prasanti Yerramilli, Layne Pruitt, Kristina Reppond, Madison N. Shyer, Jessica Cambric, Ryan Gadd, Ilya J. Finkelstein, Jimmy Gollihar, and James M. Musser |
| EPI_ISL_2202016 | Aegis Sciences Corporation | Centers for Disease Control and Prevention Division of Viral Diseases, Pathogen Discovery | Dakota Howard, Dhwani Batra, Peter W. Cook, Kara Moser, Adrian Paskey, Jason Caravas, Benjamin Rambo-Martin, Shatavia Morrison, Christopher Gulvick, Scott Sammons, Yvette Unoarumhi, Darlene Wagner, Matthew Schmerer, Cyndi Clark, Patrick Campbell, Rob Case, Vikramsinha Ghorpade, Holly Houdeshell, Ola Kvalvaag, Dillon Nall, Ethan Sanders, Alec Vest, Shaun Westlund, Matthew Hardison, Clinton R. Paden, Duncan MacCannell |
| EPI_ISL_2202017 | Houston Methodist Hospital | Houston Methodist Hospital | Randall J. Olsen, Paul A. Christensen, S. Wesley Long, Sishir Subedi, Robert Olson, Marcus Nguyen, James J. Davis, Matthew Ojeda Saavedra, Prasanti Yerramilli, Layne Pruitt, Kristina Reppond, Madison N. Shyer, Jessica Cambric, Ryan Gadd, Ilya J. Finkelstein, Jimmy Gollihar, and James M. Musser |
| EPI_ISL_2202018 | Aegis Sciences Corporation | Centers for Disease Control and Prevention Division of Viral Diseases, Pathogen Discovery | Dakota Howard, Dhwani Batra, Peter W. Cook, Kara Moser, Adrian Paskey, Jason Caravas, Benjamin Rambo-Martin, Shatavia Morrison, Christopher Gulvick, Scott Sammons, Yvette Unoarumhi, Darlene Wagner, Matthew Schmerer, Cyndi Clark, Patrick Campbell, Rob Case, Vikramsinha Ghorpade, Holly Houdeshell, Ola Kvalvaag, Dillon Nall, Ethan Sanders, Alec Vest, Shaun Westlund, Matthew Hardison, Clinton R. Paden, Duncan MacCannell |
| EPI_ISL_2202019 | Houston Methodist Hospital | Houston Methodist Hospital | Randall J. Olsen, Paul A. Christensen, S. Wesley Long, Sishir Subedi, Robert Olson, Marcus Nguyen, James J. Davis, Matthew Ojeda Saavedra, Prasanti Yerramilli, Layne Pruitt, Kristina Reppond, Madison N. Shyer, Jessica Cambric, Ryan Gadd, Ilya J. Finkelstein, Jimmy Gollihar, and James M. Musser |
| EPI_ISL_2202020, EPI_ISL_2202021 | Aegis Sciences Corporation | Centers for Disease Control and Prevention Division of Viral Diseases, Pathogen Discovery | Dakota Howard, Dhwani Batra, Peter W. Cook, Kara Moser, Adrian Paskey, Jason Caravas, Benjamin Rambo-Martin, Shatavia Morrison, Christopher Gulvick, Scott Sammons, Yvette Unoarumhi, Darlene Wagner, Matthew Schmerer, Cyndi Clark, Patrick Campbell, Rob Case, Vikramsinha Ghorpade, Holly Houdeshell, Ola Kvalvaag, Dillon Nall, Ethan Sanders, Alec Vest, Shaun Westlund, Matthew Hardison, Clinton R. Paden, Duncan MacCannell |
| EPI_ISL_2202022 | Houston Methodist Hospital | Houston Methodist Hospital | Randall J. Olsen, Paul A. Christensen, S. Wesley Long, Sishir Subedi, Robert Olson, Marcus Nguyen, James J. Davis, Matthew Ojeda Saavedra, Prasanti Yerramilli, Layne Pruitt, Kristina Reppond, Madison N. Shyer, Jessica Cambric, Ryan Gadd, Ilya J. Finkelstein, Jimmy Gollihar, and James M. Musser |
| EPI_ISL_2202023 | Aegis Sciences Corporation | Centers for Disease Control and Prevention Division of Viral Diseases, Pathogen Discovery | Dakota Howard, Dhwani Batra, Peter W. Cook, Kara Moser, Adrian Paskey, Jason Caravas, Benjamin Rambo-Martin, Shatavia Morrison, Christopher Gulvick, Scott Sammons, Yvette Unoarumhi, Darlene Wagner, Matthew Schmerer, Cyndi Clark, Patrick Campbell, Rob Case, Vikramsinha Ghorpade, Holly Houdeshell, Ola Kvalvaag, Dillon Nall, Ethan Sanders, Alec Vest, Shaun Westlund, Matthew Hardison, Clinton R. Paden, Duncan MacCannell |
| EPI_ISL_2202024 | Houston Methodist Hospital | Houston Methodist Hospital | Randall J. Olsen, Paul A. Christensen, S. Wesley Long, Sishir Subedi, Robert Olson, Marcus Nguyen, James J. Davis, Matthew Ojeda Saavedra, Prasanti Yerramilli, Layne Pruitt, Kristina Reppond, Madison N. Shyer, Jessica Cambric, Ryan Gadd, Ilya J. Finkelstein, Jimmy Gollihar, and James M. Musser |
| EPI_ISL_2202025 | Aegis Sciences Corporation | Centers for Disease Control and Prevention Division of Viral Diseases, Pathogen Discovery | Dakota Howard, Dhwani Batra, Peter W. Cook, Kara Moser, Adrian Paskey, Jason Caravas, Benjamin Rambo-Martin, Shatavia Morrison, Christopher Gulvick, Scott Sammons, Yvette Unoarumhi, Darlene Wagner, Matthew Schmerer, Cyndi Clark, Patrick Campbell, Rob Case, Vikramsinha Ghorpade, Holly Houdeshell, Ola Kvalvaag, Dillon Nall, Ethan Sanders, Alec Vest, Shaun Westlund, Matthew Hardison, Clinton R. Paden, Duncan MacCannell |
| EPI_ISL_2202026 | Houston Methodist Hospital | Houston Methodist Hospital | Randall J. Olsen, Paul A. Christensen, S. Wesley Long, Sishir Subedi, Robert Olson, Marcus Nguyen, James J. Davis, Matthew Ojeda Saavedra, Prasanti Yerramilli, Layne Pruitt, Kristina Reppond, Madison N. Shyer, Jessica Cambric, Ryan Gadd, Ilya J. Finkelstein, Jimmy Gollihar, and James M. Musser |
| EPI_ISL_2202027, EPI_ISL_2202028 | Aegis Sciences Corporation | Centers for Disease Control and Prevention Division of Viral Diseases, Pathogen Discovery | Dakota Howard, Dhwani Batra, Peter W. Cook, Kara Moser, Adrian Paskey, Jason Caravas, Benjamin Rambo-Martin, Shatavia Morrison, Christopher Gulvick, Scott Sammons, Yvette Unoarumhi, Darlene Wagner, Matthew Schmerer, Cyndi Clark, Patrick Campbell, Rob Case, Vikramsinha Ghorpade, Holly Houdeshell, Ola Kvalvaag, Dillon Nall, Ethan Sanders, Alec Vest, Shaun Westlund, Matthew Hardison, Clinton R. Paden, Duncan MacCannell |
| EPI_ISL_2202029 | Houston Methodist Hospital | Houston Methodist Hospital | Randall J. Olsen, Paul A. Christensen, S. Wesley Long, Sishir Subedi, Robert Olson, Marcus Nguyen, James J. Davis, Matthew Ojeda Saavedra, Prasanti Yerramilli, Layne Pruitt, Kristina Reppond, Madison N. Shyer, Jessica Cambric, Ryan Gadd, Ilya J. Finkelstein, Jimmy Gollihar, and James M. Musser |
| EPI_ISL_2202030, EPI_ISL_2202031 | Aegis Sciences Corporation | Centers for Disease Control and Prevention Division of Viral Diseases, Pathogen Discovery | Dakota Howard, Dhwani Batra, Peter W. Cook, Kara Moser, Adrian Paskey, Jason Caravas, Benjamin Rambo-Martin, Shatavia Morrison, Christopher Gulvick, Scott Sammons, Yvette Unoarumhi, Darlene Wagner, Matthew Schmerer, Cyndi Clark, Patrick Campbell, Rob Case, Vikramsinha Ghorpade, Holly Houdeshell, Ola Kvalvaag, Dillon Nall, Ethan Sanders, Alec Vest, Shaun Westlund, Matthew Hardison, Clinton R. Paden, Duncan MacCannell |
| EPI_ISL_2202032 | Houston Methodist Hospital | Houston Methodist Hospital | Randall J. Olsen, Paul A. Christensen, S. Wesley Long, Sishir Subedi, Robert Olson, Marcus Nguyen, James J. Davis, Matthew Ojeda Saavedra, Prasanti Yerramilli, Layne Pruitt, Kristina Reppond, Madison N. Shyer, Jessica Cambric, Ryan Gadd, Ilya J. Finkelstein, Jimmy Gollihar, and James M. Musser |
| EPI_ISL_2202033 | Aegis Sciences Corporation | Centers for Disease Control and Prevention Division of Viral Diseases, Pathogen Discovery | Dakota Howard, Dhwani Batra, Peter W. Cook, Kara Moser, Adrian Paskey, Jason Caravas, Benjamin Rambo-Martin, Shatavia Morrison, Christopher Gulvick, Scott Sammons, Yvette Unoarumhi, Darlene Wagner, Matthew Schmerer, Cyndi Clark, Patrick Campbell, Rob Case, Vikramsinha Ghorpade, Holly Houdeshell, Ola Kvalvaag, Dillon Nall, Ethan Sanders, Alec Vest, Shaun Westlund, Matthew Hardison, Clinton R. Paden, Duncan MacCannell |
| EPI_ISL_2202034 | Houston Methodist Hospital | Houston Methodist Hospital | Randall J. Olsen, Paul A. Christensen, S. Wesley Long, Sishir Subedi, Robert Olson, Marcus Nguyen, James J. Davis, Matthew Ojeda Saavedra, Prasanti Yerramilli, Layne Pruitt, Kristina Reppond, Madison N. Shyer, Jessica Cambric, Ryan Gadd, Ilya J. Finkelstein, Jimmy Gollihar, and James M. Musser |
| EPI_ISL_2202035 | Aegis Sciences Corporation | Centers for Disease Control and Prevention Division of Viral Diseases, Pathogen Discovery | Dakota Howard, Dhwani Batra, Peter W. Cook, Kara Moser, Adrian Paskey, Jason Caravas, Benjamin Rambo-Martin, Shatavia Morrison, Christopher Gulvick, Scott Sammons, Yvette Unoarumhi, Darlene Wagner, Matthew Schmerer, Cyndi Clark, Patrick Campbell, Rob Case, Vikramsinha Ghorpade, Holly Houdeshell, Ola Kvalvaag, Dillon Nall, Ethan Sanders, Alec Vest, Shaun Westlund, Matthew Hardison, Clinton R. Paden, Duncan MacCannell |
| EPI_ISL_2202036 | Houston Methodist Hospital | Houston Methodist Hospital | Randall J. Olsen, Paul A. Christensen, S. Wesley Long, Sishir Subedi, Robert Olson, Marcus Nguyen, James J. Davis, Matthew Ojeda Saavedra, Prasanti Yerramilli, Layne Pruitt, Kristina Reppond, Madison N. Shyer, Jessica Cambric, Ryan Gadd, Ilya J. Finkelstein, Jimmy Gollihar, and James M. Musser |
| EPI_ISL_2202037, EPI_ISL_2202038 | Aegis Sciences Corporation | Centers for Disease Control and Prevention Division of Viral Diseases, Pathogen Discovery | Dakota Howard, Dhwani Batra, Peter W. Cook, Kara Moser, Adrian Paskey, Jason Caravas, Benjamin Rambo-Martin, Shatavia Morrison, Christopher Gulvick, Scott Sammons, Yvette Unoarumhi, Darlene Wagner, Matthew Schmerer, Cyndi Clark, Patrick Campbell, Rob Case, Vikramsinha Ghorpade, Holly Houdeshell, Ola Kvalvaag, Dillon Nall, Ethan Sanders, Alec Vest, Shaun Westlund, Matthew Hardison, Clinton R. Paden, Duncan MacCannell |
| EPI_ISL_2202039, EPI_ISL_2202040 | Houston Methodist Hospital | Houston Methodist Hospital | Randall J. Olsen, Paul A. Christensen, S. Wesley Long, Sishir Subedi, Robert Olson, Marcus Nguyen, James J. Davis, Matthew Ojeda Saavedra, Prasanti Yerramilli, Layne Pruitt, Kristina Reppond, Madison N. Shyer, Jessica Cambric, Ryan Gadd, Ilya J. Finkelstein, Jimmy Gollihar, and James M. Musser |
| EPI_ISL_2202041, EPI_ISL_2202042 | Aegis Sciences Corporation | Centers for Disease Control and Prevention Division of Viral Diseases, Pathogen Discovery | Dakota Howard, Dhwani Batra, Peter W. Cook, Kara Moser, Adrian Paskey, Jason Caravas, Benjamin Rambo-Martin, Shatavia Morrison, Christopher Gulvick, Scott Sammons, Yvette Unoarumhi, Darlene Wagner, Matthew Schmerer, Cyndi Clark, Patrick Campbell, Rob Case, Vikramsinha Ghorpade, Holly Houdeshell, Ola Kvalvaag, Dillon Nall, Ethan Sanders, Alec Vest, Shaun Westlund, Matthew Hardison, Clinton R. Paden, Duncan MacCannell |
| EPI_ISL_2202043 | Houston Methodist Hospital | Houston Methodist Hospital | Randall J. Olsen, Paul A. Christensen, S. Wesley Long, Sishir Subedi, Robert Olson, Marcus Nguyen, James J. Davis, Matthew Ojeda Saavedra, Prasanti Yerramilli, Layne Pruitt, Kristina Reppond, Madison N. Shyer, Jessica Cambric, Ryan Gadd, Ilya J. Finkelstein, Jimmy Gollihar, and James M. Musser |
| EPI_ISL_2202044 | Aegis Sciences Corporation | Centers for Disease Control and Prevention Division of Viral Diseases, Pathogen Discovery | Dakota Howard, Dhwani Batra, Peter W. Cook, Kara Moser, Adrian Paskey, Jason Caravas, Benjamin Rambo-Martin, Shatavia Morrison, Christopher Gulvick, Scott Sammons, Yvette Unoarumhi, Darlene Wagner, Matthew Schmerer, Cyndi Clark, Patrick Campbell, Rob Case, Vikramsinha Ghorpade, Holly Houdeshell, Ola Kvalvaag, Dillon Nall, Ethan Sanders, Alec Vest, Shaun Westlund, Matthew Hardison, Clinton R. Paden, Duncan MacCannell |
| EPI_ISL_2202045 | Houston Methodist Hospital | Houston Methodist Hospital | Randall J. Olsen, Paul A. Christensen, S. Wesley Long, Sishir Subedi, Robert Olson, Marcus Nguyen, James J. Davis, Matthew Ojeda Saavedra, Prasanti Yerramilli, Layne Pruitt, Kristina Reppond, Madison N. Shyer, Jessica Cambric, Ryan Gadd, Ilya J. Finkelstein, Jimmy Gollihar, and James M. Musser |
| EPI_ISL_2202046 | Aegis Sciences Corporation | Centers for Disease Control and Prevention Division of Viral Diseases, Pathogen Discovery | Dakota Howard, Dhwani Batra, Peter W. Cook, Kara Moser, Adrian Paskey, Jason Caravas, Benjamin Rambo-Martin, Shatavia Morrison, Christopher Gulvick, Scott Sammons, Yvette Unoarumhi, Darlene Wagner, Matthew Schmerer, Cyndi Clark, Patrick Campbell, Rob Case, Vikramsinha Ghorpade, Holly Houdeshell, Ola Kvalvaag, Dillon Nall, Ethan Sanders, Alec Vest, Shaun Westlund, Matthew Hardison, Clinton R. Paden, Duncan MacCannell |
| EPI_ISL_2202047 | Houston Methodist Hospital | Houston Methodist Hospital | Randall J. Olsen, Paul A. Christensen, S. Wesley Long, Sishir Subedi, Robert Olson, Marcus Nguyen, James J. Davis, Matthew Ojeda Saavedra, Prasanti Yerramilli, Layne Pruitt, Kristina Reppond, Madison N. Shyer, Jessica Cambric, Ryan Gadd, Ilya J. Finkelstein, Jimmy Gollihar, and James M. Musser |
| EPI_ISL_2202048 | Aegis Sciences Corporation | Centers for Disease Control and Prevention Division of Viral Diseases, Pathogen Discovery | Dakota Howard, Dhwani Batra, Peter W. Cook, Kara Moser, Adrian Paskey, Jason Caravas, Benjamin Rambo-Martin, Shatavia Morrison, Christopher Gulvick, Scott Sammons, Yvette Unoarumhi, Darlene Wagner, Matthew Schmerer, Cyndi Clark, Patrick Campbell, Rob Case, Vikramsinha Ghorpade, Holly Houdeshell, Ola Kvalvaag, Dillon Nall, Ethan Sanders, Alec Vest, Shaun Westlund, Matthew Hardison, Clinton R. Paden, Duncan MacCannell |
| EPI_ISL_2202049 | Houston Methodist Hospital | Houston Methodist Hospital | Randall J. Olsen, Paul A. Christensen, S. Wesley Long, Sishir Subedi, Robert Olson, Marcus Nguyen, James J. Davis, Matthew Ojeda Saavedra, Prasanti Yerramilli, Layne Pruitt, Kristina Reppond, Madison N. Shyer, Jessica Cambric, Ryan Gadd, Ilya J. Finkelstein, Jimmy Gollihar, and James M. Musser |
| EPI_ISL_2202050, EPI_ISL_2202051 | Aegis Sciences Corporation | Centers for Disease Control and Prevention Division of Viral Diseases, Pathogen Discovery | Dakota Howard, Dhwani Batra, Peter W. Cook, Kara Moser, Adrian Paskey, Jason Caravas, Benjamin Rambo-Martin, Shatavia Morrison, Christopher Gulvick, Scott Sammons, Yvette Unoarumhi, Darlene Wagner, Matthew Schmerer, Cyndi Clark, Patrick Campbell, Rob Case, Vikramsinha Ghorpade, Holly Houdeshell, Ola Kvalvaag, Dillon Nall, Ethan Sanders, Alec Vest, Shaun Westlund, Matthew Hardison, Clinton R. Paden, Duncan MacCannell |
| EPI_ISL_2202052 | Houston Methodist Hospital | Houston Methodist Hospital | Randall J. Olsen, Paul A. Christensen, S. Wesley Long, Sishir Subedi, Robert Olson, Marcus Nguyen, James J. Davis, Matthew Ojeda Saavedra, Prasanti Yerramilli, Layne Pruitt, Kristina Reppond, Madison N. Shyer, Jessica Cambric, Ryan Gadd, Ilya J. Finkelstein, Jimmy Gollihar, and James M. Musser |
| EPI_ISL_2202053, EPI_ISL_2202054 | Aegis Sciences Corporation | Centers for Disease Control and Prevention Division of Viral Diseases, Pathogen Discovery | Dakota Howard, Dhwani Batra, Peter W. Cook, Kara Moser, Adrian Paskey, Jason Caravas, Benjamin Rambo-Martin, Shatavia Morrison, Christopher Gulvick, Scott Sammons, Yvette Unoarumhi, Darlene Wagner, Matthew Schmerer, Cyndi Clark, Patrick Campbell, Rob Case, Vikramsinha Ghorpade, Holly Houdeshell, Ola Kvalvaag, Dillon Nall, Ethan Sanders, Alec Vest, Shaun Westlund, Matthew Hardison, Clinton R. Paden, Duncan MacCannell |
| EPI_ISL_2202055 | Houston Methodist Hospital | Houston Methodist Hospital | Randall J. Olsen, Paul A. Christensen, S. Wesley Long, Sishir Subedi, Robert Olson, Marcus Nguyen, James J. Davis, Matthew Ojeda Saavedra, Prasanti Yerramilli, Layne Pruitt, Kristina Reppond, Madison N. Shyer, Jessica Cambric, Ryan Gadd, Ilya J. Finkelstein, Jimmy Gollihar, and James M. Musser |
| EPI_ISL_2202056 | Aegis Sciences Corporation | Centers for Disease Control and Prevention Division of Viral Diseases, Pathogen Discovery | Dakota Howard, Dhwani Batra, Peter W. Cook, Kara Moser, Adrian Paskey, Jason Caravas, Benjamin Rambo-Martin, Shatavia Morrison, Christopher Gulvick, Scott Sammons, Yvette Unoarumhi, Darlene Wagner, Matthew Schmerer, Cyndi Clark, Patrick Campbell, Rob Case, Vikramsinha Ghorpade, Holly Houdeshell, Ola Kvalvaag, Dillon Nall, Ethan Sanders, Alec Vest, Shaun Westlund, Matthew Hardison, Clinton R. Paden, Duncan MacCannell |
| EPI_ISL_2202057 | Houston Methodist Hospital | Houston Methodist Hospital | Randall J. Olsen, Paul A. Christensen, S. Wesley Long, Sishir Subedi, Robert Olson, Marcus Nguyen, James J. Davis, Matthew Ojeda Saavedra, Prasanti Yerramilli, Layne Pruitt, Kristina Reppond, Madison N. Shyer, Jessica Cambric, Ryan Gadd, Ilya J. Finkelstein, Jimmy Gollihar, and James M. Musser |

| | | | |
|---|---|---|---|
| EPI_ISL_2202058, EPI_ISL_2202059, EPI_ISL_2202060, EPI_ISL_2202061 | Aegis Sciences Corporation | Centers for Disease Control and Prevention Division of Viral Diseases, Pathogen Discovery | Dakota Howard, Dhwani Batra, Peter W. Cook, Kara Moser, Adrian Paskey, Jason Caravas, Benjamin Rambo-Martin, Shatavia Morrison, Christopher Gulvick, Scott Sammons, Yvette Unoarumhi, Darlene Wagner, Matthew Schmerer, Cyndi Clark, Patrick Campbell, Rob Case, Vikramsinha Ghorpade, Holly Houdeshell, Ola Kvalvaag, Dillon Nall, Ethan Sanders, Alec Vest, Shaun Westlund, Matthew Hardison, Clinton R. Paden, Duncan MacCannell |
| EPI_ISL_2202062 | Houston Methodist Hospital | Houston Methodist Hospital | Randall J. Olsen, Paul A. Christensen, S. Wesley Long, Sishir Subedi, Robert Olson, Marcus Nguyen, James J. Davis, Matthew Ojeda Saavedra, Prasanti Yerramilli, Layne Pruitt, Kristina Reppond, Madison N. Shyer, Jessica Cambric, Ryan Gadd, Ilya J. Finkelstein, Jimmy Gollihar, and James M. Musser |
| EPI_ISL_2202063 | Aegis Sciences Corporation | Centers for Disease Control and Prevention Division of Viral Diseases, Pathogen Discovery | Dakota Howard, Dhwani Batra, Peter W. Cook, Kara Moser, Adrian Paskey, Jason Caravas, Benjamin Rambo-Martin, Shatavia Morrison, Christopher Gulvick, Scott Sammons, Yvette Unoarumhi, Darlene Wagner, Matthew Schmerer, Cyndi Clark, Patrick Campbell, Rob Case, Vikramsinha Ghorpade, Holly Houdeshell, Ola Kvalvaag, Dillon Nall, Ethan Sanders, Alec Vest, Shaun Westlund, Matthew Hardison, Clinton R. Paden, Duncan MacCannell |
| EPI_ISL_2202064 | Houston Methodist Hospital | Houston Methodist Hospital | Randall J. Olsen, Paul A. Christensen, S. Wesley Long, Sishir Subedi, Robert Olson, Marcus Nguyen, James J. Davis, Matthew Ojeda Saavedra, Prasanti Yerramilli, Layne Pruitt, Kristina Reppond, Madison N. Shyer, Jessica Cambric, Ryan Gadd, Ilya J. Finkelstein, Jimmy Gollihar, and James M. Musser |
| EPI_ISL_2202065, EPI_ISL_2202066 | Aegis Sciences Corporation | Centers for Disease Control and Prevention Division of Viral Diseases, Pathogen Discovery | Dakota Howard, Dhwani Batra, Peter W. Cook, Kara Moser, Adrian Paskey, Jason Caravas, Benjamin Rambo-Martin, Shatavia Morrison, Christopher Gulvick, Scott Sammons, Yvette Unoarumhi, Darlene Wagner, Matthew Schmerer, Cyndi Clark, Patrick Campbell, Rob Case, Vikramsinha Ghorpade, Holly Houdeshell, Ola Kvalvaag, Dillon Nall, Ethan Sanders, Alec Vest, Shaun Westlund, Matthew Hardison, Clinton R. Paden, Duncan MacCannell |
| EPI_ISL_2202067 | Houston Methodist Hospital | Houston Methodist Hospital | Randall J. Olsen, Paul A. Christensen, S. Wesley Long, Sishir Subedi, Robert Olson, Marcus Nguyen, James J. Davis, Matthew Ojeda Saavedra, Prasanti Yerramilli, Layne Pruitt, Kristina Reppond, Madison N. Shyer, Jessica Cambric, Ryan Gadd, Ilya J. Finkelstein, Jimmy Gollihar, and James M. Musser |
| EPI_ISL_2202068 | Aegis Sciences Corporation | Centers for Disease Control and Prevention Division of Viral Diseases, Pathogen Discovery | Dakota Howard, Dhwani Batra, Peter W. Cook, Kara Moser, Adrian Paskey, Jason Caravas, Benjamin Rambo-Martin, Shatavia Morrison, Christopher Gulvick, Scott Sammons, Yvette Unoarumhi, Darlene Wagner, Matthew Schmerer, Cyndi Clark, Patrick Campbell, Rob Case, Vikramsinha Ghorpade, Holly Houdeshell, Ola Kvalvaag, Dillon Nall, Ethan Sanders, Alec Vest, Shaun Westlund, Matthew Hardison, Clinton R. Paden, Duncan MacCannell |
| EPI_ISL_2202069 | Houston Methodist Hospital | Houston Methodist Hospital | Randall J. Olsen, Paul A. Christensen, S. Wesley Long, Sishir Subedi, Robert Olson, Marcus Nguyen, James J. Davis, Matthew Ojeda Saavedra, Prasanti Yerramilli, Layne Pruitt, Kristina Reppond, Madison N. Shyer, Jessica Cambric, Ryan Gadd, Ilya J. Finkelstein, Jimmy Gollihar, and James M. Musser |
| EPI_ISL_2202070 | Aegis Sciences Corporation | Centers for Disease Control and Prevention Division of Viral Diseases, Pathogen Discovery | Dakota Howard, Dhwani Batra, Peter W. Cook, Kara Moser, Adrian Paskey, Jason Caravas, Benjamin Rambo-Martin, Shatavia Morrison, Christopher Gulvick, Scott Sammons, Yvette Unoarumhi, Darlene Wagner, Matthew Schmerer, Cyndi Clark, Patrick Campbell, Rob Case, Vikramsinha Ghorpade, Holly Houdeshell, Ola Kvalvaag, Dillon Nall, Ethan Sanders, Alec Vest, Shaun Westlund, Matthew Hardison, Clinton R. Paden, Duncan MacCannell |
| EPI_ISL_2202071 | Houston Methodist Hospital | Houston Methodist Hospital | Randall J. Olsen, Paul A. Christensen, S. Wesley Long, Sishir Subedi, Robert Olson, Marcus Nguyen, James J. Davis, Matthew Ojeda Saavedra, Prasanti Yerramilli, Layne Pruitt, Kristina Reppond, Madison N. Shyer, Jessica Cambric, Ryan Gadd, Ilya J. Finkelstein, Jimmy Gollihar, and James M. Musser |
| EPI_ISL_2202072 | Aegis Sciences Corporation | Centers for Disease Control and Prevention Division of Viral Diseases, Pathogen Discovery | Dakota Howard, Dhwani Batra, Peter W. Cook, Kara Moser, Adrian Paskey, Jason Caravas, Benjamin Rambo-Martin, Shatavia Morrison, Christopher Gulvick, Scott Sammons, Yvette Unoarumhi, Darlene Wagner, Matthew Schmerer, Cyndi Clark, Patrick Campbell, Rob Case, Vikramsinha Ghorpade, Holly Houdeshell, Ola Kvalvaag, Dillon Nall, Ethan Sanders, Alec Vest, Shaun Westlund, Matthew Hardison, Clinton R. Paden, Duncan MacCannell |
| EPI_ISL_2202073 | Houston Methodist Hospital | Houston Methodist Hospital | Randall J. Olsen, Paul A. Christensen, S. Wesley Long, Sishir Subedi, Robert Olson, Marcus Nguyen, James J. Davis, Matthew Ojeda Saavedra, Prasanti Yerramilli, Layne Pruitt, Kristina Reppond, Madison N. Shyer, Jessica Cambric, Ryan Gadd, Ilya J. Finkelstein, Jimmy Gollihar, and James M. Musser |
| EPI_ISL_2202074, EPI_ISL_2202075 | Aegis Sciences Corporation | Centers for Disease Control and Prevention Division of Viral Diseases, Pathogen Discovery | Dakota Howard, Dhwani Batra, Peter W. Cook, Kara Moser, Adrian Paskey, Jason Caravas, Benjamin Rambo-Martin, Shatavia Morrison, Christopher Gulvick, Scott Sammons, Yvette Unoarumhi, Darlene Wagner, Matthew Schmerer, Cyndi Clark, Patrick Campbell, Rob Case, Vikramsinha Ghorpade, Holly Houdeshell, Ola Kvalvaag, Dillon Nall, Ethan Sanders, Alec Vest, Shaun Westlund, Matthew Hardison, Clinton R. Paden, Duncan MacCannell |
| EPI_ISL_2202076 | Houston Methodist Hospital | Houston Methodist Hospital | Randall J. Olsen, Paul A. Christensen, S. Wesley Long, Sishir Subedi, Robert Olson, Marcus Nguyen, James J. Davis, Matthew Ojeda Saavedra, Prasanti Yerramilli, Layne Pruitt, Kristina Reppond, Madison N. Shyer, Jessica Cambric, Ryan Gadd, Ilya J. Finkelstein, Jimmy Gollihar, and James M. Musser |
| EPI_ISL_2202077 | Aegis Sciences Corporation | Centers for Disease Control and Prevention Division of Viral Diseases, Pathogen Discovery | Dakota Howard, Dhwani Batra, Peter W. Cook, Kara Moser, Adrian Paskey, Jason Caravas, Benjamin Rambo-Martin, Shatavia Morrison, Christopher Gulvick, Scott Sammons, Yvette Unoarumhi, Darlene Wagner, Matthew Schmerer, Cyndi Clark, Patrick Campbell, Rob Case, Vikramsinha Ghorpade, Holly Houdeshell, Ola Kvalvaag, Dillon Nall, Ethan Sanders, Alec Vest, Shaun Westlund, Matthew Hardison, Clinton R. Paden, Duncan MacCannell |
| EPI_ISL_2202078 | Houston Methodist Hospital | Houston Methodist Hospital | Randall J. Olsen, Paul A. Christensen, S. Wesley Long, Sishir Subedi, Robert Olson, Marcus Nguyen, James J. Davis, Matthew Ojeda Saavedra, Prasanti Yerramilli, Layne Pruitt, Kristina Reppond, Madison N. Shyer, Jessica Cambric, Ryan Gadd, Ilya J. Finkelstein, Jimmy Gollihar, and James M. Musser |
| EPI_ISL_2202079 | Aegis Sciences Corporation | Centers for Disease Control and Prevention Division of Viral Diseases, Pathogen Discovery | Dakota Howard, Dhwani Batra, Peter W. Cook, Kara Moser, Adrian Paskey, Jason Caravas, Benjamin Rambo-Martin, Shatavia Morrison, Christopher Gulvick, Scott Sammons, Yvette Unoarumhi, Darlene Wagner, Matthew Schmerer, Cyndi Clark, Patrick Campbell, Rob Case, Vikramsinha Ghorpade, Holly Houdeshell, Ola Kvalvaag, Dillon Nall, Ethan Sanders, Alec Vest, Shaun Westlund, Matthew Hardison, Clinton R. Paden, Duncan MacCannell |
| EPI_ISL_2202080 | Houston Methodist Hospital | Houston Methodist Hospital | Randall J. Olsen, Paul A. Christensen, S. Wesley Long, Sishir Subedi, Robert Olson, Marcus Nguyen, James J. Davis, Matthew Ojeda Saavedra, Prasanti Yerramilli, Layne Pruitt, Kristina Reppond, Madison N. Shyer, Jessica Cambric, Ryan Gadd, Ilya J. Finkelstein, Jimmy Gollihar, and James M. Musser |
| EPI_ISL_2202081, EPI_ISL_2202082 | Aegis Sciences Corporation | Centers for Disease Control and Prevention Division of Viral Diseases, Pathogen Discovery | Dakota Howard, Dhwani Batra, Peter W. Cook, Kara Moser, Adrian Paskey, Jason Caravas, Benjamin Rambo-Martin, Shatavia Morrison, Christopher Gulvick, Scott Sammons, Yvette Unoarumhi, Darlene Wagner, Matthew Schmerer, Cyndi Clark, Patrick Campbell, Rob Case, Vikramsinha Ghorpade, Holly Houdeshell, Ola Kvalvaag, Dillon Nall, Ethan Sanders, Alec Vest, Shaun Westlund, Matthew Hardison, Clinton R. Paden, Duncan MacCannell |
| EPI_ISL_2202083 | Houston Methodist Hospital | Houston Methodist Hospital | Randall J. Olsen, Paul A. Christensen, S. Wesley Long, Sishir Subedi, Robert Olson, Marcus Nguyen, James J. Davis, Matthew Ojeda Saavedra, Prasanti Yerramilli, Layne Pruitt, Kristina Reppond, Madison N. Shyer, Jessica Cambric, Ryan Gadd, Ilya J. Finkelstein, Jimmy Gollihar, and James M. Musser |
| EPI_ISL_2202084 | Aegis Sciences Corporation | Centers for Disease Control and Prevention Division of Viral Diseases, Pathogen Discovery | Dakota Howard, Dhwani Batra, Peter W. Cook, Kara Moser, Adrian Paskey, Jason Caravas, Benjamin Rambo-Martin, Shatavia Morrison, Christopher Gulvick, Scott Sammons, Yvette Unoarumhi, Darlene Wagner, Matthew Schmerer, Cyndi Clark, Patrick Campbell, Rob Case, Vikramsinha Ghorpade, Holly Houdeshell, Ola Kvalvaag, Dillon Nall, Ethan Sanders, Alec Vest, Shaun Westlund, Matthew Hardison, Clinton R. Paden, Duncan MacCannell |
| EPI_ISL_2202085, EPI_ISL_2202086, EPI_ISL_2202087 | Houston Methodist Hospital | Houston Methodist Hospital | Randall J. Olsen, Paul A. Christensen, S. Wesley Long, Sishir Subedi, Robert Olson, Marcus Nguyen, James J. Davis, Matthew Ojeda Saavedra, Prasanti Yerramilli, Layne Pruitt, Kristina Reppond, Madison N. Shyer, Jessica Cambric, Ryan Gadd, Ilya J. Finkelstein, Jimmy Gollihar, and James M. Musser |
| EPI_ISL_2202088 | Aegis Sciences Corporation | Centers for Disease Control and Prevention Division of Viral Diseases, Pathogen Discovery | Dakota Howard, Dhwani Batra, Peter W. Cook, Kara Moser, Adrian Paskey, Jason Caravas, Benjamin Rambo-Martin, Shatavia Morrison, Christopher Gulvick, Scott Sammons, Yvette Unoarumhi, Darlene Wagner, Matthew Schmerer, Cyndi Clark, Patrick Campbell, Rob Case, Vikramsinha Ghorpade, Holly Houdeshell, Ola Kvalvaag, Dillon Nall, Ethan Sanders, Alec Vest, Shaun Westlund, Matthew Hardison, Clinton R. Paden, Duncan MacCannell |
| EPI_ISL_2202089 | Houston Methodist Hospital | Houston Methodist Hospital | Randall J. Olsen, Paul A. Christensen, S. Wesley Long, Sishir Subedi, Robert Olson, Marcus Nguyen, James J. Davis, Matthew Ojeda Saavedra, Prasanti Yerramilli, Layne Pruitt, Kristina Reppond, Madison N. Shyer, Jessica Cambric, Ryan Gadd, Ilya J. Finkelstein, Jimmy Gollihar, and James M. Musser |
| EPI_ISL_2202090, EPI_ISL_2202091 | Aegis Sciences Corporation | Centers for Disease Control and Prevention Division of Viral Diseases, Pathogen Discovery | Dakota Howard, Dhwani Batra, Peter W. Cook, Kara Moser, Adrian Paskey, Jason Caravas, Benjamin Rambo-Martin, Shatavia Morrison, Christopher Gulvick, Scott Sammons, Yvette Unoarumhi, Darlene Wagner, Matthew Schmerer, Cyndi Clark, Patrick Campbell, Rob Case, Vikramsinha Ghorpade, Holly Houdeshell, Ola Kvalvaag, Dillon Nall, Ethan Sanders, Alec Vest, Shaun Westlund, Matthew Hardison, Clinton R. Paden, Duncan MacCannell |
| EPI_ISL_2202092 | Houston Methodist Hospital | Houston Methodist Hospital | Randall J. Olsen, Paul A. Christensen, S. Wesley Long, Sishir Subedi, Robert Olson, Marcus Nguyen, James J. Davis, Matthew Ojeda Saavedra, Prasanti Yerramilli, Layne Pruitt, Kristina Reppond, Madison N. Shyer, Jessica Cambric, Ryan Gadd, Ilya J. Finkelstein, Jimmy Gollihar, and James M. Musser |
| EPI_ISL_2202093 | Aegis Sciences Corporation | Centers for Disease Control and Prevention Division of Viral Diseases, Pathogen Discovery | Dakota Howard, Dhwani Batra, Peter W. Cook, Kara Moser, Adrian Paskey, Jason Caravas, Benjamin Rambo-Martin, Shatavia Morrison, Christopher Gulvick, Scott Sammons, Yvette Unoarumhi, Darlene Wagner, Matthew Schmerer, Cyndi Clark, Patrick Campbell, Rob Case, Vikramsinha Ghorpade, Holly Houdeshell, Ola Kvalvaag, Dillon Nall, Ethan Sanders, Alec Vest, Shaun Westlund, Matthew Hardison, Clinton R. Paden, Duncan MacCannell |
| EPI_ISL_2202094 | Houston Methodist Hospital | Houston Methodist Hospital | Randall J. Olsen, Paul A. Christensen, S. Wesley Long, Sishir Subedi, Robert Olson, Marcus Nguyen, James J. Davis, Matthew Ojeda Saavedra, Prasanti Yerramilli, Layne Pruitt, Kristina Reppond, Madison N. Shyer, Jessica Cambric, Ryan Gadd, Ilya J. Finkelstein, Jimmy Gollihar, and James M. Musser |
| EPI_ISL_2202095 | Aegis Sciences Corporation | Centers for Disease Control and Prevention Division of Viral Diseases, Pathogen Discovery | Dakota Howard, Dhwani Batra, Peter W. Cook, Kara Moser, Adrian Paskey, Jason Caravas, Benjamin Rambo-Martin, Shatavia Morrison, Christopher Gulvick, Scott Sammons, Yvette Unoarumhi, Darlene Wagner, Matthew Schmerer, Cyndi Clark, Patrick Campbell, Rob Case, Vikramsinha Ghorpade, Holly Houdeshell, Ola Kvalvaag, Dillon Nall, Ethan Sanders, Alec Vest, Shaun Westlund, Matthew Hardison, Clinton R. Paden, Duncan MacCannell |
| EPI_ISL_2202096 | Houston Methodist Hospital | Houston Methodist Hospital | Randall J. Olsen, Paul A. Christensen, S. Wesley Long, Sishir Subedi, Robert Olson, Marcus Nguyen, James J. Davis, Matthew Ojeda Saavedra, Prasanti Yerramilli, Layne Pruitt, Kristina Reppond, Madison N. Shyer, Jessica Cambric, Ryan Gadd, Ilya J. Finkelstein, Jimmy Gollihar, and James M. Musser |
| EPI_ISL_2202097, EPI_ISL_2202098 | Aegis Sciences Corporation | Centers for Disease Control and Prevention Division of Viral Diseases, Pathogen Discovery | Dakota Howard, Dhwani Batra, Peter W. Cook, Kara Moser, Adrian Paskey, Jason Caravas, Benjamin Rambo-Martin, Shatavia Morrison, Christopher Gulvick, Scott Sammons, Yvette Unoarumhi, Darlene Wagner, Matthew Schmerer, Cyndi Clark, Patrick Campbell, Rob Case, Vikramsinha Ghorpade, Holly Houdeshell, Ola Kvalvaag, Dillon Nall, Ethan Sanders, Alec Vest, Shaun Westlund, Matthew Hardison, Clinton R. Paden, Duncan MacCannell |
| EPI_ISL_2202099 | Houston Methodist Hospital | Houston Methodist Hospital | Randall J. Olsen, Paul A. Christensen, S. Wesley Long, Sishir Subedi, Robert Olson, Marcus Nguyen, James J. Davis, Matthew Ojeda Saavedra, Prasanti Yerramilli, Layne Pruitt, Kristina Reppond, Madison N. Shyer, Jessica Cambric, Ryan Gadd, Ilya J. Finkelstein, Jimmy Gollihar, and James M. Musser |
| EPI_ISL_2202100 | Aegis Sciences Corporation | Centers for Disease Control and Prevention Division of Viral Diseases, Pathogen Discovery | Dakota Howard, Dhwani Batra, Peter W. Cook, Kara Moser, Adrian Paskey, Jason Caravas, Benjamin Rambo-Martin, Shatavia Morrison, Christopher Gulvick, Scott Sammons, Yvette Unoarumhi, Darlene Wagner, Matthew Schmerer, Cyndi Clark, Patrick Campbell, Rob Case, Vikramsinha Ghorpade, Holly Houdeshell, Ola Kvalvaag, Dillon Nall, Ethan Sanders, Alec Vest, Shaun Westlund, Matthew Hardison, Clinton R. Paden, Duncan MacCannell |
| EPI_ISL_2202101 | Houston Methodist Hospital | Houston Methodist Hospital | Randall J. Olsen, Paul A. Christensen, S. Wesley Long, Sishir Subedi, Robert Olson, Marcus Nguyen, James J. Davis, Matthew Ojeda Saavedra, Prasanti Yerramilli, Layne Pruitt, Kristina Reppond, Madison N. Shyer, Jessica Cambric, Ryan Gadd, Ilya J. Finkelstein, Jimmy Gollihar, and James M. Musser |
| EPI_ISL_2202102 | Aegis Sciences Corporation | Centers for Disease Control and Prevention Division of Viral Diseases, Pathogen Discovery | Dakota Howard, Dhwani Batra, Peter W. Cook, Kara Moser, Adrian Paskey, Jason Caravas, Benjamin Rambo-Martin, Shatavia Morrison, Christopher Gulvick, Scott Sammons, Yvette Unoarumhi, Darlene Wagner, Matthew Schmerer, Cyndi Clark, Patrick Campbell, Rob Case, Vikramsinha Ghorpade, Holly Houdeshell, Ola Kvalvaag, Dillon Nall, Ethan Sanders, Alec Vest, Shaun Westlund, Matthew Hardison, Clinton R. Paden, Duncan MacCannell |
| EPI_ISL_2202103 | Houston Methodist Hospital | Houston Methodist Hospital | Randall J. Olsen, Paul A. Christensen, S. Wesley Long, Sishir Subedi, Robert Olson, Marcus Nguyen, James J. Davis, Matthew Ojeda Saavedra, Prasanti Yerramilli, Layne Pruitt, Kristina Reppond, Madison N. Shyer, Jessica Cambric, Ryan Gadd, Ilya J. Finkelstein, Jimmy Gollihar, and James M. Musser |
| EPI_ISL_2202104, EPI_ISL_2202105 | Aegis Sciences Corporation | Centers for Disease Control and Prevention Division of Viral Diseases, Pathogen | Dakota Howard, Dhwani Batra, Peter W. Cook, Kara Moser, Adrian Paskey, Jason Caravas, Benjamin Rambo-Martin, Shatavia Morrison, Christopher Gulvick, Scott Sammons, Yvette Unoarumhi, Darlene Wagner, Matthew Schmerer, Cyndi Clark, Patrick Campbell, Rob Case, Vikramsinha Ghorpade, Holly Houdeshell, Ola Kvalvaag, Dillon Nall, Ethan Sanders, Alec Vest, Shaun Westlund, Matthew Hardison, Clinton R. Paden, Duncan MacCannell |

| | | | |
|---|---|---|---|
| | | Discovery | |
| EPI_ISL_2202106 | Houston Methodist Hospital | Houston Methodist Hospital | Randall J. Olsen, Paul A. Christensen, S. Wesley Long, Sishir Subedi, Robert Olson, Marcus Nguyen, James J. Davis, Matthew Ojeda Saavedra, Prasanti Yerramilli, Layne Pruitt, Kristina Reppond, Madison N. Shyer, Jessica Cambric, Ryan Gadd, Ilya J. Finkelstein, Jimmy Gollihar, and James M. Musser |
| EPI_ISL_2202107 | Aegis Sciences Corporation | Centers for Disease Control and Prevention Division of Viral Diseases, Pathogen Discovery | Dakota Howard, Dhwani Batra, Peter W. Cook, Kara Moser, Adrian Paskey, Jason Caravas, Benjamin Rambo-Martin, Shatavia Morrison, Christopher Gulvick, Scott Sammons, Yvette Unoarumhi, Darlene Wagner, Matthew Schmerer, Cyndi Clark, Patrick Campbell, Rob Case, Vikramsinha Ghorpade, Holly Houdeshell, Ola Kvalvaag, Dillon Nall, Ethan Sanders, Alec Vest, Shaun Westlund, Matthew Hardison, Clinton R. Paden, Duncan MacCannell |
| EPI_ISL_2202108 | Houston Methodist Hospital | Houston Methodist Hospital | Randall J. Olsen, Paul A. Christensen, S. Wesley Long, Sishir Subedi, Robert Olson, Marcus Nguyen, James J. Davis, Matthew Ojeda Saavedra, Prasanti Yerramilli, Layne Pruitt, Kristina Reppond, Madison N. Shyer, Jessica Cambric, Ryan Gadd, Ilya J. Finkelstein, Jimmy Gollihar, and James M. Musser |
| EPI_ISL_2202109, EPI_ISL_2202110 | Aegis Sciences Corporation | Centers for Disease Control and Prevention Division of Viral Diseases, Pathogen Discovery | Dakota Howard, Dhwani Batra, Peter W. Cook, Kara Moser, Adrian Paskey, Jason Caravas, Benjamin Rambo-Martin, Shatavia Morrison, Christopher Gulvick, Scott Sammons, Yvette Unoarumhi, Darlene Wagner, Matthew Schmerer, Cyndi Clark, Patrick Campbell, Rob Case, Vikramsinha Ghorpade, Holly Houdeshell, Ola Kvalvaag, Dillon Nall, Ethan Sanders, Alec Vest, Shaun Westlund, Matthew Hardison, Clinton R. Paden, Duncan MacCannell |
| EPI_ISL_2202111 | Houston Methodist Hospital | Houston Methodist Hospital | Randall J. Olsen, Paul A. Christensen, S. Wesley Long, Sishir Subedi, Robert Olson, Marcus Nguyen, James J. Davis, Matthew Ojeda Saavedra, Prasanti Yerramilli, Layne Pruitt, Kristina Reppond, Madison N. Shyer, Jessica Cambric, Ryan Gadd, Ilya J. Finkelstein, Jimmy Gollihar, and James M. Musser |
| EPI_ISL_2202112 | Aegis Sciences Corporation | Centers for Disease Control and Prevention Division of Viral Diseases, Pathogen Discovery | Dakota Howard, Dhwani Batra, Peter W. Cook, Kara Moser, Adrian Paskey, Jason Caravas, Benjamin Rambo-Martin, Shatavia Morrison, Christopher Gulvick, Scott Sammons, Yvette Unoarumhi, Darlene Wagner, Matthew Schmerer, Cyndi Clark, Patrick Campbell, Rob Case, Vikramsinha Ghorpade, Holly Houdeshell, Ola Kvalvaag, Dillon Nall, Ethan Sanders, Alec Vest, Shaun Westlund, Matthew Hardison, Clinton R. Paden, Duncan MacCannell |
| EPI_ISL_2202113 | Houston Methodist Hospital | Houston Methodist Hospital | Randall J. Olsen, Paul A. Christensen, S. Wesley Long, Sishir Subedi, Robert Olson, Marcus Nguyen, James J. Davis, Matthew Ojeda Saavedra, Prasanti Yerramilli, Layne Pruitt, Kristina Reppond, Madison N. Shyer, Jessica Cambric, Ryan Gadd, Ilya J. Finkelstein, Jimmy Gollihar, and James M. Musser |
| EPI_ISL_2202114, EPI_ISL_2202115 | Aegis Sciences Corporation | Centers for Disease Control and Prevention Division of Viral Diseases, Pathogen Discovery | Dakota Howard, Dhwani Batra, Peter W. Cook, Kara Moser, Adrian Paskey, Jason Caravas, Benjamin Rambo-Martin, Shatavia Morrison, Christopher Gulvick, Scott Sammons, Yvette Unoarumhi, Darlene Wagner, Matthew Schmerer, Cyndi Clark, Patrick Campbell, Rob Case, Vikramsinha Ghorpade, Holly Houdeshell, Ola Kvalvaag, Dillon Nall, Ethan Sanders, Alec Vest, Shaun Westlund, Matthew Hardison, Clinton R. Paden, Duncan MacCannell |
| EPI_ISL_2202116 | Houston Methodist Hospital | Houston Methodist Hospital | Randall J. Olsen, Paul A. Christensen, S. Wesley Long, Sishir Subedi, Robert Olson, Marcus Nguyen, James J. Davis, Matthew Ojeda Saavedra, Prasanti Yerramilli, Layne Pruitt, Kristina Reppond, Madison N. Shyer, Jessica Cambric, Ryan Gadd, Ilya J. Finkelstein, Jimmy Gollihar, and James M. Musser |
| EPI_ISL_2202117 | Aegis Sciences Corporation | Centers for Disease Control and Prevention Division of Viral Diseases, Pathogen Discovery | Dakota Howard, Dhwani Batra, Peter W. Cook, Kara Moser, Adrian Paskey, Jason Caravas, Benjamin Rambo-Martin, Shatavia Morrison, Christopher Gulvick, Scott Sammons, Yvette Unoarumhi, Darlene Wagner, Matthew Schmerer, Cyndi Clark, Patrick Campbell, Rob Case, Vikramsinha Ghorpade, Holly Houdeshell, Ola Kvalvaag, Dillon Nall, Ethan Sanders, Alec Vest, Shaun Westlund, Matthew Hardison, Clinton R. Paden, Duncan MacCannell |
| EPI_ISL_2202118 | Houston Methodist Hospital | Houston Methodist Hospital | Randall J. Olsen, Paul A. Christensen, S. Wesley Long, Sishir Subedi, Robert Olson, Marcus Nguyen, James J. Davis, Matthew Ojeda Saavedra, Prasanti Yerramilli, Layne Pruitt, Kristina Reppond, Madison N. Shyer, Jessica Cambric, Ryan Gadd, Ilya J. Finkelstein, Jimmy Gollihar, and James M. Musser |
| EPI_ISL_2202119 | Aegis Sciences Corporation | Centers for Disease Control and Prevention Division of Viral Diseases, Pathogen Discovery | Dakota Howard, Dhwani Batra, Peter W. Cook, Kara Moser, Adrian Paskey, Jason Caravas, Benjamin Rambo-Martin, Shatavia Morrison, Christopher Gulvick, Scott Sammons, Yvette Unoarumhi, Darlene Wagner, Matthew Schmerer, Cyndi Clark, Patrick Campbell, Rob Case, Vikramsinha Ghorpade, Holly Houdeshell, Ola Kvalvaag, Dillon Nall, Ethan Sanders, Alec Vest, Shaun Westlund, Matthew Hardison, Clinton R. Paden, Duncan MacCannell |
| EPI_ISL_2202120 | Houston Methodist Hospital | Houston Methodist Hospital | Randall J. Olsen, Paul A. Christensen, S. Wesley Long, Sishir Subedi, Robert Olson, Marcus Nguyen, James J. Davis, Matthew Ojeda Saavedra, Prasanti Yerramilli, Layne Pruitt, Kristina Reppond, Madison N. Shyer, Jessica Cambric, Ryan Gadd, Ilya J. Finkelstein, Jimmy Gollihar, and James M. Musser |
| EPI_ISL_2202121 | Aegis Sciences Corporation | Centers for Disease Control and Prevention Division of Viral Diseases, Pathogen Discovery | Dakota Howard, Dhwani Batra, Peter W. Cook, Kara Moser, Adrian Paskey, Jason Caravas, Benjamin Rambo-Martin, Shatavia Morrison, Christopher Gulvick, Scott Sammons, Yvette Unoarumhi, Darlene Wagner, Matthew Schmerer, Cyndi Clark, Patrick Campbell, Rob Case, Vikramsinha Ghorpade, Holly Houdeshell, Ola Kvalvaag, Dillon Nall, Ethan Sanders, Alec Vest, Shaun Westlund, Matthew Hardison, Clinton R. Paden, Duncan MacCannell |
| EPI_ISL_2202122 | Houston Methodist Hospital | Houston Methodist Hospital | Randall J. Olsen, Paul A. Christensen, S. Wesley Long, Sishir Subedi, Robert Olson, Marcus Nguyen, James J. Davis, Matthew Ojeda Saavedra, Prasanti Yerramilli, Layne Pruitt, Kristina Reppond, Madison N. Shyer, Jessica Cambric, Ryan Gadd, Ilya J. Finkelstein, Jimmy Gollihar, and James M. Musser |
| EPI_ISL_2202123, EPI_ISL_2202124 | Aegis Sciences Corporation | Centers for Disease Control and Prevention Division of Viral Diseases, Pathogen Discovery | Dakota Howard, Dhwani Batra, Peter W. Cook, Kara Moser, Adrian Paskey, Jason Caravas, Benjamin Rambo-Martin, Shatavia Morrison, Christopher Gulvick, Scott Sammons, Yvette Unoarumhi, Darlene Wagner, Matthew Schmerer, Cyndi Clark, Patrick Campbell, Rob Case, Vikramsinha Ghorpade, Holly Houdeshell, Ola Kvalvaag, Dillon Nall, Ethan Sanders, Alec Vest, Shaun Westlund, Matthew Hardison, Clinton R. Paden, Duncan MacCannell |
| EPI_ISL_2202125 | Houston Methodist Hospital | Houston Methodist Hospital | Randall J. Olsen, Paul A. Christensen, S. Wesley Long, Sishir Subedi, Robert Olson, Marcus Nguyen, James J. Davis, Matthew Ojeda Saavedra, Prasanti Yerramilli, Layne Pruitt, Kristina Reppond, Madison N. Shyer, Jessica Cambric, Ryan Gadd, Ilya J. Finkelstein, Jimmy Gollihar, and James M. Musser |
| EPI_ISL_2202126 | Aegis Sciences Corporation | Centers for Disease Control and Prevention Division of Viral Diseases, Pathogen Discovery | Dakota Howard, Dhwani Batra, Peter W. Cook, Kara Moser, Adrian Paskey, Jason Caravas, Benjamin Rambo-Martin, Shatavia Morrison, Christopher Gulvick, Scott Sammons, Yvette Unoarumhi, Darlene Wagner, Matthew Schmerer, Cyndi Clark, Patrick Campbell, Rob Case, Vikramsinha Ghorpade, Holly Houdeshell, Ola Kvalvaag, Dillon Nall, Ethan Sanders, Alec Vest, Shaun Westlund, Matthew Hardison, Clinton R. Paden, Duncan MacCannell |
| EPI_ISL_2202127 | Houston Methodist Hospital | Houston Methodist Hospital | Randall J. Olsen, Paul A. Christensen, S. Wesley Long, Sishir Subedi, Robert Olson, Marcus Nguyen, James J. Davis, Matthew Ojeda Saavedra, Prasanti Yerramilli, Layne Pruitt, Kristina Reppond, Madison N. Shyer, Jessica Cambric, Ryan Gadd, Ilya J. Finkelstein, Jimmy Gollihar, and James M. Musser |
| EPI_ISL_2202129 | Aegis Sciences Corporation | Centers for Disease Control and Prevention Division of Viral Diseases, Pathogen Discovery | Dakota Howard, Dhwani Batra, Peter W. Cook, Kara Moser, Adrian Paskey, Jason Caravas, Benjamin Rambo-Martin, Shatavia Morrison, Christopher Gulvick, Scott Sammons, Yvette Unoarumhi, Darlene Wagner, Matthew Schmerer, Cyndi Clark, Patrick Campbell, Rob Case, Vikramsinha Ghorpade, Holly Houdeshell, Ola Kvalvaag, Dillon Nall, Ethan Sanders, Alec Vest, Shaun Westlund, Matthew Hardison, Clinton R. Paden, Duncan MacCannell |
| EPI_ISL_2202130 | Houston Methodist Hospital | Houston Methodist Hospital | Randall J. Olsen, Paul A. Christensen, S. Wesley Long, Sishir Subedi, Robert Olson, Marcus Nguyen, James J. Davis, Matthew Ojeda Saavedra, Prasanti Yerramilli, Layne Pruitt, Kristina Reppond, Madison N. Shyer, Jessica Cambric, Ryan Gadd, Ilya J. Finkelstein, Jimmy Gollihar, and James M. Musser |
| EPI_ISL_2202131 | Aegis Sciences Corporation | Centers for Disease Control and Prevention Division of Viral Diseases, Pathogen Discovery | Dakota Howard, Dhwani Batra, Peter W. Cook, Kara Moser, Adrian Paskey, Jason Caravas, Benjamin Rambo-Martin, Shatavia Morrison, Christopher Gulvick, Scott Sammons, Yvette Unoarumhi, Darlene Wagner, Matthew Schmerer, Cyndi Clark, Patrick Campbell, Rob Case, Vikramsinha Ghorpade, Holly Houdeshell, Ola Kvalvaag, Dillon Nall, Ethan Sanders, Alec Vest, Shaun Westlund, Matthew Hardison, Clinton R. Paden, Duncan MacCannell |
| EPI_ISL_2202132 | Houston Methodist Hospital | Houston Methodist Hospital | Randall J. Olsen, Paul A. Christensen, S. Wesley Long, Sishir Subedi, Robert Olson, Marcus Nguyen, James J. Davis, Matthew Ojeda Saavedra, Prasanti Yerramilli, Layne Pruitt, Kristina Reppond, Madison N. Shyer, Jessica Cambric, Ryan Gadd, Ilya J. Finkelstein, Jimmy Gollihar, and James M. Musser |
| EPI_ISL_2202133, EPI_ISL_2202134 | Aegis Sciences Corporation | Centers for Disease Control and Prevention Division of Viral Diseases, Pathogen Discovery | Dakota Howard, Dhwani Batra, Peter W. Cook, Kara Moser, Adrian Paskey, Jason Caravas, Benjamin Rambo-Martin, Shatavia Morrison, Christopher Gulvick, Scott Sammons, Yvette Unoarumhi, Darlene Wagner, Matthew Schmerer, Cyndi Clark, Patrick Campbell, Rob Case, Vikramsinha Ghorpade, Holly Houdeshell, Ola Kvalvaag, Dillon Nall, Ethan Sanders, Alec Vest, Shaun Westlund, Matthew Hardison, Clinton R. Paden, Duncan MacCannell |
| EPI_ISL_2202135 | Houston Methodist Hospital | Houston Methodist Hospital | Randall J. Olsen, Paul A. Christensen, S. Wesley Long, Sishir Subedi, Robert Olson, Marcus Nguyen, James J. Davis, Matthew Ojeda Saavedra, Prasanti Yerramilli, Layne Pruitt, Kristina Reppond, Madison N. Shyer, Jessica Cambric, Ryan Gadd, Ilya J. Finkelstein, Jimmy Gollihar, and James M. Musser |
| EPI_ISL_2202136 | Aegis Sciences Corporation | Centers for Disease Control and Prevention Division of Viral Diseases, Pathogen Discovery | Dakota Howard, Dhwani Batra, Peter W. Cook, Kara Moser, Adrian Paskey, Jason Caravas, Benjamin Rambo-Martin, Shatavia Morrison, Christopher Gulvick, Scott Sammons, Yvette Unoarumhi, Darlene Wagner, Matthew Schmerer, Cyndi Clark, Patrick Campbell, Rob Case, Vikramsinha Ghorpade, Holly Houdeshell, Ola Kvalvaag, Dillon Nall, Ethan Sanders, Alec Vest, Shaun Westlund, Matthew Hardison, Clinton R. Paden, Duncan MacCannell |
| EPI_ISL_2202137 | Houston Methodist Hospital | Houston Methodist Hospital | Randall J. Olsen, Paul A. Christensen, S. Wesley Long, Sishir Subedi, Robert Olson, Marcus Nguyen, James J. Davis, Matthew Ojeda Saavedra, Prasanti Yerramilli, Layne Pruitt, Kristina Reppond, Madison N. Shyer, Jessica Cambric, Ryan Gadd, Ilya J. Finkelstein, Jimmy Gollihar, and James M. Musser |
| EPI_ISL_2202138 | Aegis Sciences Corporation | Centers for Disease Control and Prevention Division of Viral Diseases, Pathogen Discovery | Dakota Howard, Dhwani Batra, Peter W. Cook, Kara Moser, Adrian Paskey, Jason Caravas, Benjamin Rambo-Martin, Shatavia Morrison, Christopher Gulvick, Scott Sammons, Yvette Unoarumhi, Darlene Wagner, Matthew Schmerer, Cyndi Clark, Patrick Campbell, Rob Case, Vikramsinha Ghorpade, Holly Houdeshell, Ola Kvalvaag, Dillon Nall, Ethan Sanders, Alec Vest, Shaun Westlund, Matthew Hardison, Clinton R. Paden, Duncan MacCannell |
| EPI_ISL_2202139 | Houston Methodist Hospital | Houston Methodist Hospital | Randall J. Olsen, Paul A. Christensen, S. Wesley Long, Sishir Subedi, Robert Olson, Marcus Nguyen, James J. Davis, Matthew Ojeda Saavedra, Prasanti Yerramilli, Layne Pruitt, Kristina Reppond, Madison N. Shyer, Jessica Cambric, Ryan Gadd, Ilya J. Finkelstein, Jimmy Gollihar, and James M. Musser |
| EPI_ISL_2202140, EPI_ISL_2202141 | Aegis Sciences Corporation | Centers for Disease Control and Prevention Division of Viral Diseases, Pathogen Discovery | Dakota Howard, Dhwani Batra, Peter W. Cook, Kara Moser, Adrian Paskey, Jason Caravas, Benjamin Rambo-Martin, Shatavia Morrison, Christopher Gulvick, Scott Sammons, Yvette Unoarumhi, Darlene Wagner, Matthew Schmerer, Cyndi Clark, Patrick Campbell, Rob Case, Vikramsinha Ghorpade, Holly Houdeshell, Ola Kvalvaag, Dillon Nall, Ethan Sanders, Alec Vest, Shaun Westlund, Matthew Hardison, Clinton R. Paden, Duncan MacCannell |
| EPI_ISL_2202142 | Houston Methodist Hospital | Houston Methodist Hospital | Randall J. Olsen, Paul A. Christensen, S. Wesley Long, Sishir Subedi, Robert Olson, Marcus Nguyen, James J. Davis, Matthew Ojeda Saavedra, Prasanti Yerramilli, Layne Pruitt, Kristina Reppond, Madison N. Shyer, Jessica Cambric, Ryan Gadd, Ilya J. Finkelstein, Jimmy Gollihar, and James M. Musser |
| EPI_ISL_2202143 | Aegis Sciences Corporation | Centers for Disease Control and Prevention Division of Viral Diseases, Pathogen Discovery | Dakota Howard, Dhwani Batra, Peter W. Cook, Kara Moser, Adrian Paskey, Jason Caravas, Benjamin Rambo-Martin, Shatavia Morrison, Christopher Gulvick, Scott Sammons, Yvette Unoarumhi, Darlene Wagner, Matthew Schmerer, Cyndi Clark, Patrick Campbell, Rob Case, Vikramsinha Ghorpade, Holly Houdeshell, Ola Kvalvaag, Dillon Nall, Ethan Sanders, Alec Vest, Shaun Westlund, Matthew Hardison, Clinton R. Paden, Duncan MacCannell |
| EPI_ISL_2202144, EPI_ISL_2202146 | Houston Methodist Hospital | Houston Methodist Hospital | Randall J. Olsen, Paul A. Christensen, S. Wesley Long, Sishir Subedi, Robert Olson, Marcus Nguyen, James J. Davis, Matthew Ojeda Saavedra, Prasanti Yerramilli, Layne Pruitt, Kristina Reppond, Madison N. Shyer, Jessica Cambric, Ryan Gadd, Ilya J. Finkelstein, Jimmy Gollihar, and James M. Musser |
| EPI_ISL_2202147 | Aegis Sciences Corporation | Centers for Disease Control and Prevention Division of Viral Diseases, Pathogen Discovery | Dakota Howard, Dhwani Batra, Peter W. Cook, Kara Moser, Adrian Paskey, Jason Caravas, Benjamin Rambo-Martin, Shatavia Morrison, Christopher Gulvick, Scott Sammons, Yvette Unoarumhi, Darlene Wagner, Matthew Schmerer, Cyndi Clark, Patrick Campbell, Rob Case, Vikramsinha Ghorpade, Holly Houdeshell, Ola Kvalvaag, Dillon Nall, Ethan Sanders, Alec Vest, Shaun Westlund, Matthew Hardison, Clinton R. Paden, Duncan MacCannell |
| EPI_ISL_2202148 | Houston Methodist Hospital | Houston Methodist Hospital | Randall J. Olsen, Paul A. Christensen, S. Wesley Long, Sishir Subedi, Robert Olson, Marcus Nguyen, James J. Davis, Matthew Ojeda Saavedra, Prasanti Yerramilli, Layne Pruitt, Kristina Reppond, Madison N. Shyer, Jessica Cambric, Ryan Gadd, Ilya J. Finkelstein, Jimmy Gollihar, and James M. Musser |
| EPI_ISL_2202149 | Aegis Sciences Corporation | Centers for Disease Control and Prevention Division of Viral Diseases, Pathogen Discovery | Dakota Howard, Dhwani Batra, Peter W. Cook, Kara Moser, Adrian Paskey, Jason Caravas, Benjamin Rambo-Martin, Shatavia Morrison, Christopher Gulvick, Scott Sammons, Yvette Unoarumhi, Darlene Wagner, Matthew Schmerer, Cyndi Clark, Patrick Campbell, Rob Case, Vikramsinha Ghorpade, Holly Houdeshell, Ola Kvalvaag, Dillon Nall, Ethan Sanders, Alec Vest, Shaun Westlund, Matthew Hardison, Clinton R. Paden, Duncan MacCannell |
| EPI_ISL_2202150 | Houston Methodist Hospital | Houston Methodist Hospital | Randall J. Olsen, Paul A. Christensen, S. Wesley Long, Sishir Subedi, Robert Olson, Marcus Nguyen, James J. Davis, Matthew Ojeda Saavedra, Prasanti Yerramilli, Layne Pruitt, Kristina Reppond, Madison N. Shyer, Jessica Cambric, Ryan Gadd, Ilya J. Finkelstein, Jimmy Gollihar, and James M. Musser |

| | | | |
|---|---|---|---|
| EPI_ISL_2202151 | Aegis Sciences Corporation | Centers for Disease Control and Prevention Division of Viral Diseases, Pathogen Discovery | Dakota Howard, Dhwani Batra, Peter W. Cook, Kara Moser, Adrian Paskey, Jason Caravas, Benjamin Rambo-Martin, Shatavia Morrison, Christopher Gulvick, Scott Sammons, Yvette Unoarumhi, Darlene Wagner, Matthew Schmerer, Cyndi Clark, Patrick Campbell, Rob Case, Vikramsinha Ghorpade, Holly Houdeshell, Ola Kvalvaag, Dillon Nall, Ethan Sanders, Alec Vest, Shaun Westlund, Matthew Hardison, Clinton R. Paden, Duncan MacCannell |
| EPI_ISL_2202152 | Houston Methodist Hospital | Houston Methodist Hospital | Randall J. Olsen, Paul A. Christensen, S. Wesley Long, Sishir Subedi, Robert Olson, Marcus Nguyen, James J. Davis, Matthew Ojeda Saavedra, Prasanti Yerramilli, Layne Pruitt, Kristina Reppond, Madison N. Shyer, Jessica Cambric, Ryan Gadd, Ilya J. Finkelstein, Jimmy Gollihar, and James M. Musser |
| EPI_ISL_2202153 | Aegis Sciences Corporation | Centers for Disease Control and Prevention Division of Viral Diseases, Pathogen Discovery | Dakota Howard, Dhwani Batra, Peter W. Cook, Kara Moser, Adrian Paskey, Jason Caravas, Benjamin Rambo-Martin, Shatavia Morrison, Christopher Gulvick, Scott Sammons, Yvette Unoarumhi, Darlene Wagner, Matthew Schmerer, Cyndi Clark, Patrick Campbell, Rob Case, Vikramsinha Ghorpade, Holly Houdeshell, Ola Kvalvaag, Dillon Nall, Ethan Sanders, Alec Vest, Shaun Westlund, Matthew Hardison, Clinton R. Paden, Duncan MacCannell |
| EPI_ISL_2202154 | Houston Methodist Hospital | Houston Methodist Hospital | Randall J. Olsen, Paul A. Christensen, S. Wesley Long, Sishir Subedi, Robert Olson, Marcus Nguyen, James J. Davis, Matthew Ojeda Saavedra, Prasanti Yerramilli, Layne Pruitt, Kristina Reppond, Madison N. Shyer, Jessica Cambric, Ryan Gadd, Ilya J. Finkelstein, Jimmy Gollihar, and James M. Musser |
| EPI_ISL_2202155, EPI_ISL_2202156 | Aegis Sciences Corporation | Centers for Disease Control and Prevention Division of Viral Diseases, Pathogen Discovery | Dakota Howard, Dhwani Batra, Peter W. Cook, Kara Moser, Adrian Paskey, Jason Caravas, Benjamin Rambo-Martin, Shatavia Morrison, Christopher Gulvick, Scott Sammons, Yvette Unoarumhi, Darlene Wagner, Matthew Schmerer, Cyndi Clark, Patrick Campbell, Rob Case, Vikramsinha Ghorpade, Holly Houdeshell, Ola Kvalvaag, Dillon Nall, Ethan Sanders, Alec Vest, Shaun Westlund, Matthew Hardison, Clinton R. Paden, Duncan MacCannell |
| EPI_ISL_2202157 | Houston Methodist Hospital | Houston Methodist Hospital | Randall J. Olsen, Paul A. Christensen, S. Wesley Long, Sishir Subedi, Robert Olson, Marcus Nguyen, James J. Davis, Matthew Ojeda Saavedra, Prasanti Yerramilli, Layne Pruitt, Kristina Reppond, Madison N. Shyer, Jessica Cambric, Ryan Gadd, Ilya J. Finkelstein, Jimmy Gollihar, and James M. Musser |
| EPI_ISL_2202158 | Aegis Sciences Corporation | Centers for Disease Control and Prevention Division of Viral Diseases, Pathogen Discovery | Dakota Howard, Dhwani Batra, Peter W. Cook, Kara Moser, Adrian Paskey, Jason Caravas, Benjamin Rambo-Martin, Shatavia Morrison, Christopher Gulvick, Scott Sammons, Yvette Unoarumhi, Darlene Wagner, Matthew Schmerer, Cyndi Clark, Patrick Campbell, Rob Case, Vikramsinha Ghorpade, Holly Houdeshell, Ola Kvalvaag, Dillon Nall, Ethan Sanders, Alec Vest, Shaun Westlund, Matthew Hardison, Clinton R. Paden, Duncan MacCannell |
| EPI_ISL_2202159 | Houston Methodist Hospital | Houston Methodist Hospital | Randall J. Olsen, Paul A. Christensen, S. Wesley Long, Sishir Subedi, Robert Olson, Marcus Nguyen, James J. Davis, Matthew Ojeda Saavedra, Prasanti Yerramilli, Layne Pruitt, Kristina Reppond, Madison N. Shyer, Jessica Cambric, Ryan Gadd, Ilya J. Finkelstein, Jimmy Gollihar, and James M. Musser |
| EPI_ISL_2202160, EPI_ISL_2202161 | Aegis Sciences Corporation | Centers for Disease Control and Prevention Division of Viral Diseases, Pathogen Discovery | Dakota Howard, Dhwani Batra, Peter W. Cook, Kara Moser, Adrian Paskey, Jason Caravas, Benjamin Rambo-Martin, Shatavia Morrison, Christopher Gulvick, Scott Sammons, Yvette Unoarumhi, Darlene Wagner, Matthew Schmerer, Cyndi Clark, Patrick Campbell, Rob Case, Vikramsinha Ghorpade, Holly Houdeshell, Ola Kvalvaag, Dillon Nall, Ethan Sanders, Alec Vest, Shaun Westlund, Matthew Hardison, Clinton R. Paden, Duncan MacCannell |
| EPI_ISL_2202162 | Houston Methodist Hospital | Houston Methodist Hospital | Randall J. Olsen, Paul A. Christensen, S. Wesley Long, Sishir Subedi, Robert Olson, Marcus Nguyen, James J. Davis, Matthew Ojeda Saavedra, Prasanti Yerramilli, Layne Pruitt, Kristina Reppond, Madison N. Shyer, Jessica Cambric, Ryan Gadd, Ilya J. Finkelstein, Jimmy Gollihar, and James M. Musser |
| EPI_ISL_2202163 | Aegis Sciences Corporation | Centers for Disease Control and Prevention Division of Viral Diseases, Pathogen Discovery | Dakota Howard, Dhwani Batra, Peter W. Cook, Kara Moser, Adrian Paskey, Jason Caravas, Benjamin Rambo-Martin, Shatavia Morrison, Christopher Gulvick, Scott Sammons, Yvette Unoarumhi, Darlene Wagner, Matthew Schmerer, Cyndi Clark, Patrick Campbell, Rob Case, Vikramsinha Ghorpade, Holly Houdeshell, Ola Kvalvaag, Dillon Nall, Ethan Sanders, Alec Vest, Shaun Westlund, Matthew Hardison, Clinton R. Paden, Duncan MacCannell |
| EPI_ISL_2202164 | Houston Methodist Hospital | Houston Methodist Hospital | Randall J. Olsen, Paul A. Christensen, S. Wesley Long, Sishir Subedi, Robert Olson, Marcus Nguyen, James J. Davis, Matthew Ojeda Saavedra, Prasanti Yerramilli, Layne Pruitt, Kristina Reppond, Madison N. Shyer, Jessica Cambric, Ryan Gadd, Ilya J. Finkelstein, Jimmy Gollihar, and James M. Musser |
| EPI_ISL_2202165 | Aegis Sciences Corporation | Centers for Disease Control and Prevention Division of Viral Diseases, Pathogen Discovery | Dakota Howard, Dhwani Batra, Peter W. Cook, Kara Moser, Adrian Paskey, Jason Caravas, Benjamin Rambo-Martin, Shatavia Morrison, Christopher Gulvick, Scott Sammons, Yvette Unoarumhi, Darlene Wagner, Matthew Schmerer, Cyndi Clark, Patrick Campbell, Rob Case, Vikramsinha Ghorpade, Holly Houdeshell, Ola Kvalvaag, Dillon Nall, Ethan Sanders, Alec Vest, Shaun Westlund, Matthew Hardison, Clinton R. Paden, Duncan MacCannell |
| EPI_ISL_2202166 | Houston Methodist Hospital | Houston Methodist Hospital | Randall J. Olsen, Paul A. Christensen, S. Wesley Long, Sishir Subedi, Robert Olson, Marcus Nguyen, James J. Davis, Matthew Ojeda Saavedra, Prasanti Yerramilli, Layne Pruitt, Kristina Reppond, Madison N. Shyer, Jessica Cambric, Ryan Gadd, Ilya J. Finkelstein, Jimmy Gollihar, and James M. Musser |
| EPI_ISL_2202167 | Aegis Sciences Corporation | Centers for Disease Control and Prevention Division of Viral Diseases, Pathogen Discovery | Dakota Howard, Dhwani Batra, Peter W. Cook, Kara Moser, Adrian Paskey, Jason Caravas, Benjamin Rambo-Martin, Shatavia Morrison, Christopher Gulvick, Scott Sammons, Yvette Unoarumhi, Darlene Wagner, Matthew Schmerer, Cyndi Clark, Patrick Campbell, Rob Case, Vikramsinha Ghorpade, Holly Houdeshell, Ola Kvalvaag, Dillon Nall, Ethan Sanders, Alec Vest, Shaun Westlund, Matthew Hardison, Clinton R. Paden, Duncan MacCannell |
| EPI_ISL_2202168 | Houston Methodist Hospital | Houston Methodist Hospital | Randall J. Olsen, Paul A. Christensen, S. Wesley Long, Sishir Subedi, Robert Olson, Marcus Nguyen, James J. Davis, Matthew Ojeda Saavedra, Prasanti Yerramilli, Layne Pruitt, Kristina Reppond, Madison N. Shyer, Jessica Cambric, Ryan Gadd, Ilya J. Finkelstein, Jimmy Gollihar, and James M. Musser |
| EPI_ISL_2202169 | Aegis Sciences Corporation | Centers for Disease Control and Prevention Division of Viral Diseases, Pathogen Discovery | Dakota Howard, Dhwani Batra, Peter W. Cook, Kara Moser, Adrian Paskey, Jason Caravas, Benjamin Rambo-Martin, Shatavia Morrison, Christopher Gulvick, Scott Sammons, Yvette Unoarumhi, Darlene Wagner, Matthew Schmerer, Cyndi Clark, Patrick Campbell, Rob Case, Vikramsinha Ghorpade, Holly Houdeshell, Ola Kvalvaag, Dillon Nall, Ethan Sanders, Alec Vest, Shaun Westlund, Matthew Hardison, Clinton R. Paden, Duncan MacCannell |
| EPI_ISL_2202170 | Houston Methodist Hospital | Houston Methodist Hospital | Randall J. Olsen, Paul A. Christensen, S. Wesley Long, Sishir Subedi, Robert Olson, Marcus Nguyen, James J. Davis, Matthew Ojeda Saavedra, Prasanti Yerramilli, Layne Pruitt, Kristina Reppond, Madison N. Shyer, Jessica Cambric, Ryan Gadd, Ilya J. Finkelstein, Jimmy Gollihar, and James M. Musser |
| EPI_ISL_2202171, EPI_ISL_2202172 | Aegis Sciences Corporation | Centers for Disease Control and Prevention Division of Viral Diseases, Pathogen Discovery | Dakota Howard, Dhwani Batra, Peter W. Cook, Kara Moser, Adrian Paskey, Jason Caravas, Benjamin Rambo-Martin, Shatavia Morrison, Christopher Gulvick, Scott Sammons, Yvette Unoarumhi, Darlene Wagner, Matthew Schmerer, Cyndi Clark, Patrick Campbell, Rob Case, Vikramsinha Ghorpade, Holly Houdeshell, Ola Kvalvaag, Dillon Nall, Ethan Sanders, Alec Vest, Shaun Westlund, Matthew Hardison, Clinton R. Paden, Duncan MacCannell |
| EPI_ISL_2202173 | Houston Methodist Hospital | Houston Methodist Hospital | Randall J. Olsen, Paul A. Christensen, S. Wesley Long, Sishir Subedi, Robert Olson, Marcus Nguyen, James J. Davis, Matthew Ojeda Saavedra, Prasanti Yerramilli, Layne Pruitt, Kristina Reppond, Madison N. Shyer, Jessica Cambric, Ryan Gadd, Ilya J. Finkelstein, Jimmy Gollihar, and James M. Musser |
| EPI_ISL_2202174 | Aegis Sciences Corporation | Centers for Disease Control and Prevention Division of Viral Diseases, Pathogen Discovery | Dakota Howard, Dhwani Batra, Peter W. Cook, Kara Moser, Adrian Paskey, Jason Caravas, Benjamin Rambo-Martin, Shatavia Morrison, Christopher Gulvick, Scott Sammons, Yvette Unoarumhi, Darlene Wagner, Matthew Schmerer, Cyndi Clark, Patrick Campbell, Rob Case, Vikramsinha Ghorpade, Holly Houdeshell, Ola Kvalvaag, Dillon Nall, Ethan Sanders, Alec Vest, Shaun Westlund, Matthew Hardison, Clinton R. Paden, Duncan MacCannell |
| EPI_ISL_2202175 | Houston Methodist Hospital | Houston Methodist Hospital | Randall J. Olsen, Paul A. Christensen, S. Wesley Long, Sishir Subedi, Robert Olson, Marcus Nguyen, James J. Davis, Matthew Ojeda Saavedra, Prasanti Yerramilli, Layne Pruitt, Kristina Reppond, Madison N. Shyer, Jessica Cambric, Ryan Gadd, Ilya J. Finkelstein, Jimmy Gollihar, and James M. Musser |
| EPI_ISL_2202176 | Aegis Sciences Corporation | Centers for Disease Control and Prevention Division of Viral Diseases, Pathogen Discovery | Dakota Howard, Dhwani Batra, Peter W. Cook, Kara Moser, Adrian Paskey, Jason Caravas, Benjamin Rambo-Martin, Shatavia Morrison, Christopher Gulvick, Scott Sammons, Yvette Unoarumhi, Darlene Wagner, Matthew Schmerer, Cyndi Clark, Patrick Campbell, Rob Case, Vikramsinha Ghorpade, Holly Houdeshell, Ola Kvalvaag, Dillon Nall, Ethan Sanders, Alec Vest, Shaun Westlund, Matthew Hardison, Clinton R. Paden, Duncan MacCannell |
| EPI_ISL_2202177 | Houston Methodist Hospital | Houston Methodist Hospital | Randall J. Olsen, Paul A. Christensen, S. Wesley Long, Sishir Subedi, Robert Olson, Marcus Nguyen, James J. Davis, Matthew Ojeda Saavedra, Prasanti Yerramilli, Layne Pruitt, Kristina Reppond, Madison N. Shyer, Jessica Cambric, Ryan Gadd, Ilya J. Finkelstein, Jimmy Gollihar, and James M. Musser |
| EPI_ISL_2202179 | Aegis Sciences Corporation | Centers for Disease Control and Prevention Division of Viral Diseases, Pathogen Discovery | Dakota Howard, Dhwani Batra, Peter W. Cook, Kara Moser, Adrian Paskey, Jason Caravas, Benjamin Rambo-Martin, Shatavia Morrison, Christopher Gulvick, Scott Sammons, Yvette Unoarumhi, Darlene Wagner, Matthew Schmerer, Cyndi Clark, Patrick Campbell, Rob Case, Vikramsinha Ghorpade, Holly Houdeshell, Ola Kvalvaag, Dillon Nall, Ethan Sanders, Alec Vest, Shaun Westlund, Matthew Hardison, Clinton R. Paden, Duncan MacCannell |
| EPI_ISL_2202180 | Houston Methodist Hospital | Houston Methodist Hospital | Randall J. Olsen, Paul A. Christensen, S. Wesley Long, Sishir Subedi, Robert Olson, Marcus Nguyen, James J. Davis, Matthew Ojeda Saavedra, Prasanti Yerramilli, Layne Pruitt, Kristina Reppond, Madison N. Shyer, Jessica Cambric, Ryan Gadd, Ilya J. Finkelstein, Jimmy Gollihar, and James M. Musser |
| EPI_ISL_2202181 | Aegis Sciences Corporation | Centers for Disease Control and Prevention Division of Viral Diseases, Pathogen Discovery | Dakota Howard, Dhwani Batra, Peter W. Cook, Kara Moser, Adrian Paskey, Jason Caravas, Benjamin Rambo-Martin, Shatavia Morrison, Christopher Gulvick, Scott Sammons, Yvette Unoarumhi, Darlene Wagner, Matthew Schmerer, Cyndi Clark, Patrick Campbell, Rob Case, Vikramsinha Ghorpade, Holly Houdeshell, Ola Kvalvaag, Dillon Nall, Ethan Sanders, Alec Vest, Shaun Westlund, Matthew Hardison, Clinton R. Paden, Duncan MacCannell |
| EPI_ISL_2202182 | Houston Methodist Hospital | Houston Methodist Hospital | Randall J. Olsen, Paul A. Christensen, S. Wesley Long, Sishir Subedi, Robert Olson, Marcus Nguyen, James J. Davis, Matthew Ojeda Saavedra, Prasanti Yerramilli, Layne Pruitt, Kristina Reppond, Madison N. Shyer, Jessica Cambric, Ryan Gadd, Ilya J. Finkelstein, Jimmy Gollihar, and James M. Musser |
| EPI_ISL_2202183 | Aegis Sciences Corporation | Centers for Disease Control and Prevention Division of Viral Diseases, Pathogen Discovery | Dakota Howard, Dhwani Batra, Peter W. Cook, Kara Moser, Adrian Paskey, Jason Caravas, Benjamin Rambo-Martin, Shatavia Morrison, Christopher Gulvick, Scott Sammons, Yvette Unoarumhi, Darlene Wagner, Matthew Schmerer, Cyndi Clark, Patrick Campbell, Rob Case, Vikramsinha Ghorpade, Holly Houdeshell, Ola Kvalvaag, Dillon Nall, Ethan Sanders, Alec Vest, Shaun Westlund, Matthew Hardison, Clinton R. Paden, Duncan MacCannell |
| EPI_ISL_2202184 | Houston Methodist Hospital | Houston Methodist Hospital | Randall J. Olsen, Paul A. Christensen, S. Wesley Long, Sishir Subedi, Robert Olson, Marcus Nguyen, James J. Davis, Matthew Ojeda Saavedra, Prasanti Yerramilli, Layne Pruitt, Kristina Reppond, Madison N. Shyer, Jessica Cambric, Ryan Gadd, Ilya J. Finkelstein, Jimmy Gollihar, and James M. Musser |
| EPI_ISL_2202185 | Aegis Sciences Corporation | Centers for Disease Control and Prevention Division of Viral Diseases, Pathogen Discovery | Dakota Howard, Dhwani Batra, Peter W. Cook, Kara Moser, Adrian Paskey, Jason Caravas, Benjamin Rambo-Martin, Shatavia Morrison, Christopher Gulvick, Scott Sammons, Yvette Unoarumhi, Darlene Wagner, Matthew Schmerer, Cyndi Clark, Patrick Campbell, Rob Case, Vikramsinha Ghorpade, Holly Houdeshell, Ola Kvalvaag, Dillon Nall, Ethan Sanders, Alec Vest, Shaun Westlund, Matthew Hardison, Clinton R. Paden, Duncan MacCannell |
| EPI_ISL_2202187 | Houston Methodist Hospital | Houston Methodist Hospital | Randall J. Olsen, Paul A. Christensen, S. Wesley Long, Sishir Subedi, Robert Olson, Marcus Nguyen, James J. Davis, Matthew Ojeda Saavedra, Prasanti Yerramilli, Layne Pruitt, Kristina Reppond, Madison N. Shyer, Jessica Cambric, Ryan Gadd, Ilya J. Finkelstein, Jimmy Gollihar, and James M. Musser |
| EPI_ISL_2202188 | Aegis Sciences Corporation | Centers for Disease Control and Prevention Division of Viral Diseases, Pathogen Discovery | Dakota Howard, Dhwani Batra, Peter W. Cook, Kara Moser, Adrian Paskey, Jason Caravas, Benjamin Rambo-Martin, Shatavia Morrison, Christopher Gulvick, Scott Sammons, Yvette Unoarumhi, Darlene Wagner, Matthew Schmerer, Cyndi Clark, Patrick Campbell, Rob Case, Vikramsinha Ghorpade, Holly Houdeshell, Ola Kvalvaag, Dillon Nall, Ethan Sanders, Alec Vest, Shaun Westlund, Matthew Hardison, Clinton R. Paden, Duncan MacCannell |
| EPI_ISL_2202189 | Houston Methodist Hospital | Houston Methodist Hospital | Randall J. Olsen, Paul A. Christensen, S. Wesley Long, Sishir Subedi, Robert Olson, Marcus Nguyen, James J. Davis, Matthew Ojeda Saavedra, Prasanti Yerramilli, Layne Pruitt, Kristina Reppond, Madison N. Shyer, Jessica Cambric, Ryan Gadd, Ilya J. Finkelstein, Jimmy Gollihar, and James M. Musser |
| EPI_ISL_2202190 | Aegis Sciences Corporation | Centers for Disease Control and Prevention Division of Viral Diseases, Pathogen Discovery | Dakota Howard, Dhwani Batra, Peter W. Cook, Kara Moser, Adrian Paskey, Jason Caravas, Benjamin Rambo-Martin, Shatavia Morrison, Christopher Gulvick, Scott Sammons, Yvette Unoarumhi, Darlene Wagner, Matthew Schmerer, Cyndi Clark, Patrick Campbell, Rob Case, Vikramsinha Ghorpade, Holly Houdeshell, Ola Kvalvaag, Dillon Nall, Ethan Sanders, Alec Vest, Shaun Westlund, Matthew Hardison, Clinton R. Paden, Duncan MacCannell |
| EPI_ISL_2202191 | Houston Methodist Hospital | Houston Methodist Hospital | Randall J. Olsen, Paul A. Christensen, S. Wesley Long, Sishir Subedi, Robert Olson, Marcus Nguyen, James J. Davis, Matthew Ojeda Saavedra, Prasanti Yerramilli, Layne Pruitt, Kristina Reppond, Madison N. Shyer, Jessica Cambric, Ryan Gadd, Ilya J. Finkelstein, Jimmy Gollihar, and James M. Musser |
| EPI_ISL_2202192 | Aegis Sciences Corporation | Centers for Disease Control and Prevention Division of Viral Diseases, Pathogen Discovery | Dakota Howard, Dhwani Batra, Peter W. Cook, Kara Moser, Adrian Paskey, Jason Caravas, Benjamin Rambo-Martin, Shatavia Morrison, Christopher Gulvick, Scott Sammons, Yvette Unoarumhi, Darlene Wagner, Matthew Schmerer, Cyndi Clark, Patrick Campbell, Rob Case, Vikramsinha Ghorpade, Holly Houdeshell, Ola Kvalvaag, Dillon Nall, Ethan Sanders, Alec Vest, Shaun Westlund, Matthew Hardison, Clinton R. Paden, Duncan MacCannell |
| EPI_ISL_2202193 | Houston Methodist Hospital | Houston Methodist Hospital | Randall J. Olsen, Paul A. Christensen, S. Wesley Long, Sishir Subedi, Robert Olson, Marcus Nguyen, James J. Davis, Matthew Ojeda Saavedra, Prasanti Yerramilli, Layne Pruitt, Kristina Reppond, Madison N. Shyer, Jessica Cambric, Ryan Gadd, Ilya J. Finkelstein, Jimmy Gollihar, and James |

| | | | |
|---|---|---|---|
| | | | M. Musser |
| EPI_ISL_2202194, EPI_ISL_2202195 | Aegis Sciences Corporation | Centers for Disease Control and Prevention Division of Viral Diseases, Pathogen Discovery | Dakota Howard, Dhwani Batra, Peter W. Cook, Kara Moser, Adrian Paskey, Jason Caravas, Benjamin Rambo-Martin, Shatavia Morrison, Christopher Gulvick, Scott Sammons, Yvette Unoarumhi, Darlene Wagner, Matthew Schmerer, Cyndi Clark, Patrick Campbell, Rob Case, Vikramsinha Ghorpade, Holly Houdeshell, Ola Kvalvaag, Dillon Nall, Ethan Sanders, Alec Vest, Shaun Westlund, Matthew Hardison, Clinton R. Paden, Duncan MacCannell |
| EPI_ISL_2202196 | Houston Methodist Hospital | Houston Methodist Hospital | Randall J. Olsen, Paul A. Christensen, S. Wesley Long, Sishir Subedi, Robert Olson, Marcus Nguyen, James J. Davis, Matthew Ojeda Saavedra, Prasanti Yerramilli, Layne Pruitt, Kristina Reppond, Madison N. Shyer, Jessica Cambric, Ryan Gadd, Ilya J. Finkelstein, Jimmy Gollihar, and James M. Musser |
| EPI_ISL_2202197 | Aegis Sciences Corporation | Centers for Disease Control and Prevention Division of Viral Diseases, Pathogen Discovery | Dakota Howard, Dhwani Batra, Peter W. Cook, Kara Moser, Adrian Paskey, Jason Caravas, Benjamin Rambo-Martin, Shatavia Morrison, Christopher Gulvick, Scott Sammons, Yvette Unoarumhi, Darlene Wagner, Matthew Schmerer, Cyndi Clark, Patrick Campbell, Rob Case, Vikramsinha Ghorpade, Holly Houdeshell, Ola Kvalvaag, Dillon Nall, Ethan Sanders, Alec Vest, Shaun Westlund, Matthew Hardison, Clinton R. Paden, Duncan MacCannell |
| EPI_ISL_2202198 | Houston Methodist Hospital | Houston Methodist Hospital | Randall J. Olsen, Paul A. Christensen, S. Wesley Long, Sishir Subedi, Robert Olson, Marcus Nguyen, James J. Davis, Matthew Ojeda Saavedra, Prasanti Yerramilli, Layne Pruitt, Kristina Reppond, Madison N. Shyer, Jessica Cambric, Ryan Gadd, Ilya J. Finkelstein, Jimmy Gollihar, and James M. Musser |
| EPI_ISL_2202199 | Aegis Sciences Corporation | Centers for Disease Control and Prevention Division of Viral Diseases, Pathogen Discovery | Dakota Howard, Dhwani Batra, Peter W. Cook, Kara Moser, Adrian Paskey, Jason Caravas, Benjamin Rambo-Martin, Shatavia Morrison, Christopher Gulvick, Scott Sammons, Yvette Unoarumhi, Darlene Wagner, Matthew Schmerer, Cyndi Clark, Patrick Campbell, Rob Case, Vikramsinha Ghorpade, Holly Houdeshell, Ola Kvalvaag, Dillon Nall, Ethan Sanders, Alec Vest, Shaun Westlund, Matthew Hardison, Clinton R. Paden, Duncan MacCannell |
| EPI_ISL_2202200 | Houston Methodist Hospital | Houston Methodist Hospital | Randall J. Olsen, Paul A. Christensen, S. Wesley Long, Sishir Subedi, Robert Olson, Marcus Nguyen, James J. Davis, Matthew Ojeda Saavedra, Prasanti Yerramilli, Layne Pruitt, Kristina Reppond, Madison N. Shyer, Jessica Cambric, Ryan Gadd, Ilya J. Finkelstein, Jimmy Gollihar, and James M. Musser |
| EPI_ISL_2202201, EPI_ISL_2202202 | Aegis Sciences Corporation | Centers for Disease Control and Prevention Division of Viral Diseases, Pathogen Discovery | Dakota Howard, Dhwani Batra, Peter W. Cook, Kara Moser, Adrian Paskey, Jason Caravas, Benjamin Rambo-Martin, Shatavia Morrison, Christopher Gulvick, Scott Sammons, Yvette Unoarumhi, Darlene Wagner, Matthew Schmerer, Cyndi Clark, Patrick Campbell, Rob Case, Vikramsinha Ghorpade, Holly Houdeshell, Ola Kvalvaag, Dillon Nall, Ethan Sanders, Alec Vest, Shaun Westlund, Matthew Hardison, Clinton R. Paden, Duncan MacCannell |
| EPI_ISL_2202203 | Houston Methodist Hospital | Houston Methodist Hospital | Randall J. Olsen, Paul A. Christensen, S. Wesley Long, Sishir Subedi, Robert Olson, Marcus Nguyen, James J. Davis, Matthew Ojeda Saavedra, Prasanti Yerramilli, Layne Pruitt, Kristina Reppond, Madison N. Shyer, Jessica Cambric, Ryan Gadd, Ilya J. Finkelstein, Jimmy Gollihar, and James M. Musser |
| EPI_ISL_2202204 | Aegis Sciences Corporation | Centers for Disease Control and Prevention Division of Viral Diseases, Pathogen Discovery | Dakota Howard, Dhwani Batra, Peter W. Cook, Kara Moser, Adrian Paskey, Jason Caravas, Benjamin Rambo-Martin, Shatavia Morrison, Christopher Gulvick, Scott Sammons, Yvette Unoarumhi, Darlene Wagner, Matthew Schmerer, Cyndi Clark, Patrick Campbell, Rob Case, Vikramsinha Ghorpade, Holly Houdeshell, Ola Kvalvaag, Dillon Nall, Ethan Sanders, Alec Vest, Shaun Westlund, Matthew Hardison, Clinton R. Paden, Duncan MacCannell |
| EPI_ISL_2202205 | Houston Methodist Hospital | Houston Methodist Hospital | Randall J. Olsen, Paul A. Christensen, S. Wesley Long, Sishir Subedi, Robert Olson, Marcus Nguyen, James J. Davis, Matthew Ojeda Saavedra, Prasanti Yerramilli, Layne Pruitt, Kristina Reppond, Madison N. Shyer, Jessica Cambric, Ryan Gadd, Ilya J. Finkelstein, Jimmy Gollihar, and James M. Musser |
| EPI_ISL_2202206 | Aegis Sciences Corporation | Centers for Disease Control and Prevention Division of Viral Diseases, Pathogen Discovery | Dakota Howard, Dhwani Batra, Peter W. Cook, Kara Moser, Adrian Paskey, Jason Caravas, Benjamin Rambo-Martin, Shatavia Morrison, Christopher Gulvick, Scott Sammons, Yvette Unoarumhi, Darlene Wagner, Matthew Schmerer, Cyndi Clark, Patrick Campbell, Rob Case, Vikramsinha Ghorpade, Holly Houdeshell, Ola Kvalvaag, Dillon Nall, Ethan Sanders, Alec Vest, Shaun Westlund, Matthew Hardison, Clinton R. Paden, Duncan MacCannell |
| EPI_ISL_2202207 | Houston Methodist Hospital | Houston Methodist Hospital | Randall J. Olsen, Paul A. Christensen, S. Wesley Long, Sishir Subedi, Robert Olson, Marcus Nguyen, James J. Davis, Matthew Ojeda Saavedra, Prasanti Yerramilli, Layne Pruitt, Kristina Reppond, Madison N. Shyer, Jessica Cambric, Ryan Gadd, Ilya J. Finkelstein, Jimmy Gollihar, and James M. Musser |
| EPI_ISL_2202208, EPI_ISL_2202209 | Aegis Sciences Corporation | Centers for Disease Control and Prevention Division of Viral Diseases, Pathogen Discovery | Dakota Howard, Dhwani Batra, Peter W. Cook, Kara Moser, Adrian Paskey, Jason Caravas, Benjamin Rambo-Martin, Shatavia Morrison, Christopher Gulvick, Scott Sammons, Yvette Unoarumhi, Darlene Wagner, Matthew Schmerer, Cyndi Clark, Patrick Campbell, Rob Case, Vikramsinha Ghorpade, Holly Houdeshell, Ola Kvalvaag, Dillon Nall, Ethan Sanders, Alec Vest, Shaun Westlund, Matthew Hardison, Clinton R. Paden, Duncan MacCannell |
| EPI_ISL_2202210 | Houston Methodist Hospital | Houston Methodist Hospital | Randall J. Olsen, Paul A. Christensen, S. Wesley Long, Sishir Subedi, Robert Olson, Marcus Nguyen, James J. Davis, Matthew Ojeda Saavedra, Prasanti Yerramilli, Layne Pruitt, Kristina Reppond, Madison N. Shyer, Jessica Cambric, Ryan Gadd, Ilya J. Finkelstein, Jimmy Gollihar, and James M. Musser |
| EPI_ISL_2202211 | Aegis Sciences Corporation | Centers for Disease Control and Prevention Division of Viral Diseases, Pathogen Discovery | Dakota Howard, Dhwani Batra, Peter W. Cook, Kara Moser, Adrian Paskey, Jason Caravas, Benjamin Rambo-Martin, Shatavia Morrison, Christopher Gulvick, Scott Sammons, Yvette Unoarumhi, Darlene Wagner, Matthew Schmerer, Cyndi Clark, Patrick Campbell, Rob Case, Vikramsinha Ghorpade, Holly Houdeshell, Ola Kvalvaag, Dillon Nall, Ethan Sanders, Alec Vest, Shaun Westlund, Matthew Hardison, Clinton R. Paden, Duncan MacCannell |
| EPI_ISL_2202212 | Houston Methodist Hospital | Houston Methodist Hospital | Randall J. Olsen, Paul A. Christensen, S. Wesley Long, Sishir Subedi, Robert Olson, Marcus Nguyen, James J. Davis, Matthew Ojeda Saavedra, Prasanti Yerramilli, Layne Pruitt, Kristina Reppond, Madison N. Shyer, Jessica Cambric, Ryan Gadd, Ilya J. Finkelstein, Jimmy Gollihar, and James M. Musser |
| EPI_ISL_2202213 | Aegis Sciences Corporation | Centers for Disease Control and Prevention Division of Viral Diseases, Pathogen Discovery | Dakota Howard, Dhwani Batra, Peter W. Cook, Kara Moser, Adrian Paskey, Jason Caravas, Benjamin Rambo-Martin, Shatavia Morrison, Christopher Gulvick, Scott Sammons, Yvette Unoarumhi, Darlene Wagner, Matthew Schmerer, Cyndi Clark, Patrick Campbell, Rob Case, Vikramsinha Ghorpade, Holly Houdeshell, Ola Kvalvaag, Dillon Nall, Ethan Sanders, Alec Vest, Shaun Westlund, Matthew Hardison, Clinton R. Paden, Duncan MacCannell |
| EPI_ISL_2202214 | Houston Methodist Hospital | Houston Methodist Hospital | Randall J. Olsen, Paul A. Christensen, S. Wesley Long, Sishir Subedi, Robert Olson, Marcus Nguyen, James J. Davis, Matthew Ojeda Saavedra, Prasanti Yerramilli, Layne Pruitt, Kristina Reppond, Madison N. Shyer, Jessica Cambric, Ryan Gadd, Ilya J. Finkelstein, Jimmy Gollihar, and James M. Musser |
| EPI_ISL_2202215 | Aegis Sciences Corporation | Centers for Disease Control and Prevention Division of Viral Diseases, Pathogen Discovery | Dakota Howard, Dhwani Batra, Peter W. Cook, Kara Moser, Adrian Paskey, Jason Caravas, Benjamin Rambo-Martin, Shatavia Morrison, Christopher Gulvick, Scott Sammons, Yvette Unoarumhi, Darlene Wagner, Matthew Schmerer, Cyndi Clark, Patrick Campbell, Rob Case, Vikramsinha Ghorpade, Holly Houdeshell, Ola Kvalvaag, Dillon Nall, Ethan Sanders, Alec Vest, Shaun Westlund, Matthew Hardison, Clinton R. Paden, Duncan MacCannell |
| EPI_ISL_2202216 | Houston Methodist Hospital | Houston Methodist Hospital | Randall J. Olsen, Paul A. Christensen, S. Wesley Long, Sishir Subedi, Robert Olson, Marcus Nguyen, James J. Davis, Matthew Ojeda Saavedra, Prasanti Yerramilli, Layne Pruitt, Kristina Reppond, Madison N. Shyer, Jessica Cambric, Ryan Gadd, Ilya J. Finkelstein, Jimmy Gollihar, and James M. Musser |
| EPI_ISL_2202217 | Aegis Sciences Corporation | Centers for Disease Control and Prevention Division of Viral Diseases, Pathogen Discovery | Dakota Howard, Dhwani Batra, Peter W. Cook, Kara Moser, Adrian Paskey, Jason Caravas, Benjamin Rambo-Martin, Shatavia Morrison, Christopher Gulvick, Scott Sammons, Yvette Unoarumhi, Darlene Wagner, Matthew Schmerer, Cyndi Clark, Patrick Campbell, Rob Case, Vikramsinha Ghorpade, Holly Houdeshell, Ola Kvalvaag, Dillon Nall, Ethan Sanders, Alec Vest, Shaun Westlund, Matthew Hardison, Clinton R. Paden, Duncan MacCannell |
| EPI_ISL_2202218 | Houston Methodist Hospital | Houston Methodist Hospital | Randall J. Olsen, Paul A. Christensen, S. Wesley Long, Sishir Subedi, Robert Olson, Marcus Nguyen, James J. Davis, Matthew Ojeda Saavedra, Prasanti Yerramilli, Layne Pruitt, Kristina Reppond, Madison N. Shyer, Jessica Cambric, Ryan Gadd, Ilya J. Finkelstein, Jimmy Gollihar, and James M. Musser |
| EPI_ISL_2202219 | Aegis Sciences Corporation | Centers for Disease Control and Prevention Division of Viral Diseases, Pathogen Discovery | Dakota Howard, Dhwani Batra, Peter W. Cook, Kara Moser, Adrian Paskey, Jason Caravas, Benjamin Rambo-Martin, Shatavia Morrison, Christopher Gulvick, Scott Sammons, Yvette Unoarumhi, Darlene Wagner, Matthew Schmerer, Cyndi Clark, Patrick Campbell, Rob Case, Vikramsinha Ghorpade, Holly Houdeshell, Ola Kvalvaag, Dillon Nall, Ethan Sanders, Alec Vest, Shaun Westlund, Matthew Hardison, Clinton R. Paden, Duncan MacCannell |
| EPI_ISL_2202220 | Houston Methodist Hospital | Houston Methodist Hospital | Randall J. Olsen, Paul A. Christensen, S. Wesley Long, Sishir Subedi, Robert Olson, Marcus Nguyen, James J. Davis, Matthew Ojeda Saavedra, Prasanti Yerramilli, Layne Pruitt, Kristina Reppond, Madison N. Shyer, Jessica Cambric, Ryan Gadd, Ilya J. Finkelstein, Jimmy Gollihar, and James M. Musser |
| EPI_ISL_2202221, EPI_ISL_2202222 | Aegis Sciences Corporation | Centers for Disease Control and Prevention Division of Viral Diseases, Pathogen Discovery | Dakota Howard, Dhwani Batra, Peter W. Cook, Kara Moser, Adrian Paskey, Jason Caravas, Benjamin Rambo-Martin, Shatavia Morrison, Christopher Gulvick, Scott Sammons, Yvette Unoarumhi, Darlene Wagner, Matthew Schmerer, Cyndi Clark, Patrick Campbell, Rob Case, Vikramsinha Ghorpade, Holly Houdeshell, Ola Kvalvaag, Dillon Nall, Ethan Sanders, Alec Vest, Shaun Westlund, Matthew Hardison, Clinton R. Paden, Duncan MacCannell |
| EPI_ISL_2202223 | Houston Methodist Hospital | Houston Methodist Hospital | Randall J. Olsen, Paul A. Christensen, S. Wesley Long, Sishir Subedi, Robert Olson, Marcus Nguyen, James J. Davis, Matthew Ojeda Saavedra, Prasanti Yerramilli, Layne Pruitt, Kristina Reppond, Madison N. Shyer, Jessica Cambric, Ryan Gadd, Ilya J. Finkelstein, Jimmy Gollihar, and James M. Musser |
| EPI_ISL_2202224, EPI_ISL_2202225 | Aegis Sciences Corporation | Centers for Disease Control and Prevention Division of Viral Diseases, Pathogen Discovery | Dakota Howard, Dhwani Batra, Peter W. Cook, Kara Moser, Adrian Paskey, Jason Caravas, Benjamin Rambo-Martin, Shatavia Morrison, Christopher Gulvick, Scott Sammons, Yvette Unoarumhi, Darlene Wagner, Matthew Schmerer, Cyndi Clark, Patrick Campbell, Rob Case, Vikramsinha Ghorpade, Holly Houdeshell, Ola Kvalvaag, Dillon Nall, Ethan Sanders, Alec Vest, Shaun Westlund, Matthew Hardison, Clinton R. Paden, Duncan MacCannell |
| EPI_ISL_2202226 | Houston Methodist Hospital | Houston Methodist Hospital | Randall J. Olsen, Paul A. Christensen, S. Wesley Long, Sishir Subedi, Robert Olson, Marcus Nguyen, James J. Davis, Matthew Ojeda Saavedra, Prasanti Yerramilli, Layne Pruitt, Kristina Reppond, Madison N. Shyer, Jessica Cambric, Ryan Gadd, Ilya J. Finkelstein, Jimmy Gollihar, and James M. Musser |
| EPI_ISL_2202227 | Aegis Sciences Corporation | Centers for Disease Control and Prevention Division of Viral Diseases, Pathogen Discovery | Dakota Howard, Dhwani Batra, Peter W. Cook, Kara Moser, Adrian Paskey, Jason Caravas, Benjamin Rambo-Martin, Shatavia Morrison, Christopher Gulvick, Scott Sammons, Yvette Unoarumhi, Darlene Wagner, Matthew Schmerer, Cyndi Clark, Patrick Campbell, Rob Case, Vikramsinha Ghorpade, Holly Houdeshell, Ola Kvalvaag, Dillon Nall, Ethan Sanders, Alec Vest, Shaun Westlund, Matthew Hardison, Clinton R. Paden, Duncan MacCannell |
| EPI_ISL_2202228 | Houston Methodist Hospital | Houston Methodist Hospital | Randall J. Olsen, Paul A. Christensen, S. Wesley Long, Sishir Subedi, Robert Olson, Marcus Nguyen, James J. Davis, Matthew Ojeda Saavedra, Prasanti Yerramilli, Layne Pruitt, Kristina Reppond, Madison N. Shyer, Jessica Cambric, Ryan Gadd, Ilya J. Finkelstein, Jimmy Gollihar, and James M. Musser |
| EPI_ISL_2202230 | Aegis Sciences Corporation | Centers for Disease Control and Prevention Division of Viral Diseases, Pathogen Discovery | Dakota Howard, Dhwani Batra, Peter W. Cook, Kara Moser, Adrian Paskey, Jason Caravas, Benjamin Rambo-Martin, Shatavia Morrison, Christopher Gulvick, Scott Sammons, Yvette Unoarumhi, Darlene Wagner, Matthew Schmerer, Cyndi Clark, Patrick Campbell, Rob Case, Vikramsinha Ghorpade, Holly Houdeshell, Ola Kvalvaag, Dillon Nall, Ethan Sanders, Alec Vest, Shaun Westlund, Matthew Hardison, Clinton R. Paden, Duncan MacCannell |
| EPI_ISL_2202231 | Houston Methodist Hospital | Houston Methodist Hospital | Randall J. Olsen, Paul A. Christensen, S. Wesley Long, Sishir Subedi, Robert Olson, Marcus Nguyen, James J. Davis, Matthew Ojeda Saavedra, Prasanti Yerramilli, Layne Pruitt, Kristina Reppond, Madison N. Shyer, Jessica Cambric, Ryan Gadd, Ilya J. Finkelstein, Jimmy Gollihar, and James M. Musser |
| EPI_ISL_2202232 | Aegis Sciences Corporation | Centers for Disease Control and Prevention Division of Viral Diseases, Pathogen Discovery | Dakota Howard, Dhwani Batra, Peter W. Cook, Kara Moser, Adrian Paskey, Jason Caravas, Benjamin Rambo-Martin, Shatavia Morrison, Christopher Gulvick, Scott Sammons, Yvette Unoarumhi, Darlene Wagner, Matthew Schmerer, Cyndi Clark, Patrick Campbell, Rob Case, Vikramsinha Ghorpade, Holly Houdeshell, Ola Kvalvaag, Dillon Nall, Ethan Sanders, Alec Vest, Shaun Westlund, Matthew Hardison, Clinton R. Paden, Duncan MacCannell |
| EPI_ISL_2202233 | Houston Methodist Hospital | Houston Methodist Hospital | Randall J. Olsen, Paul A. Christensen, S. Wesley Long, Sishir Subedi, Robert Olson, Marcus Nguyen, James J. Davis, Matthew Ojeda Saavedra, Prasanti Yerramilli, Layne Pruitt, Kristina Reppond, Madison N. Shyer, Jessica Cambric, Ryan Gadd, Ilya J. Finkelstein, Jimmy Gollihar, and James M. Musser |
| EPI_ISL_2202234 | Aegis Sciences Corporation | Centers for Disease Control and Prevention Division of Viral Diseases, Pathogen Discovery | Dakota Howard, Dhwani Batra, Peter W. Cook, Kara Moser, Adrian Paskey, Jason Caravas, Benjamin Rambo-Martin, Shatavia Morrison, Christopher Gulvick, Scott Sammons, Yvette Unoarumhi, Darlene Wagner, Matthew Schmerer, Cyndi Clark, Patrick Campbell, Rob Case, Vikramsinha Ghorpade, Holly Houdeshell, Ola Kvalvaag, Dillon Nall, Ethan Sanders, Alec Vest, Shaun Westlund, Matthew Hardison, Clinton R. Paden, Duncan MacCannell |
| EPI_ISL_2202236 | Houston Methodist Hospital | Houston Methodist Hospital | Randall J. Olsen, Paul A. Christensen, S. Wesley Long, Sishir Subedi, Robert Olson, Marcus Nguyen, James J. Davis, Matthew Ojeda Saavedra, Prasanti Yerramilli, Layne Pruitt, Kristina Reppond, Madison N. Shyer, Jessica Cambric, Ryan Gadd, Ilya J. Finkelstein, Jimmy Gollihar, and James M. Musser |
| EPI_ISL_2202237 | Aegis Sciences Corporation | Centers for Disease Control and Prevention Division of Viral Diseases, Pathogen Discovery | Dakota Howard, Dhwani Batra, Peter W. Cook, Kara Moser, Adrian Paskey, Jason Caravas, Benjamin Rambo-Martin, Shatavia Morrison, Christopher Gulvick, Scott Sammons, Yvette Unoarumhi, Darlene Wagner, Matthew Schmerer, Cyndi Clark, Patrick Campbell, Rob Case, Vikramsinha Ghorpade, Holly Houdeshell, Ola Kvalvaag, Dillon Nall, Ethan Sanders, Alec Vest, Shaun Westlund, Matthew Hardison, Clinton R. Paden, Duncan MacCannell |

| | | | |
|---|---|---|---|
| EPI_ISL_2202238 | Houston Methodist Hospital | Houston Methodist Hospital | Randall J. Olsen, Paul A. Christensen, S. Wesley Long, Sishir Subedi, Robert Olson, Marcus Nguyen, James J. Davis, Matthew Ojeda Saavedra, Prasanti Yerramilli, Layne Pruitt, Kristina Reppond, Madison N. Shyer, Jessica Cambric, Ryan Gadd, Ilya J. Finkelstein, Jimmy Gollihar, and James M. Musser |
| EPI_ISL_2202239 | Aegis Sciences Corporation | Centers for Disease Control and Prevention Division of Viral Diseases, Pathogen Discovery | Dakota Howard, Dhwani Batra, Peter W. Cook, Kara Moser, Adrian Paskey, Jason Caravas, Benjamin Rambo-Martin, Shatavia Morrison, Christopher Gulvick, Scott Sammons, Yvette Unoarumhi, Darlene Wagner, Matthew Schmerer, Cyndi Clark, Patrick Campbell, Rob Case, Vikramsinha Ghorpade, Holly Houdeshell, Ola Kvalvaag, Dillon Nall, Ethan Sanders, Alec Vest, Shaun Westlund, Matthew Hardison, Clinton R. Paden, Duncan MacCannell |
| EPI_ISL_2202240 | Houston Methodist Hospital | Houston Methodist Hospital | Randall J. Olsen, Paul A. Christensen, S. Wesley Long, Sishir Subedi, Robert Olson, Marcus Nguyen, James J. Davis, Matthew Ojeda Saavedra, Prasanti Yerramilli, Layne Pruitt, Kristina Reppond, Madison N. Shyer, Jessica Cambric, Ryan Gadd, Ilya J. Finkelstein, Jimmy Gollihar, and James M. Musser |
| EPI_ISL_2202241 | Aegis Sciences Corporation | Centers for Disease Control and Prevention Division of Viral Diseases, Pathogen Discovery | Dakota Howard, Dhwani Batra, Peter W. Cook, Kara Moser, Adrian Paskey, Jason Caravas, Benjamin Rambo-Martin, Shatavia Morrison, Christopher Gulvick, Scott Sammons, Yvette Unoarumhi, Darlene Wagner, Matthew Schmerer, Cyndi Clark, Patrick Campbell, Rob Case, Vikramsinha Ghorpade, Holly Houdeshell, Ola Kvalvaag, Dillon Nall, Ethan Sanders, Alec Vest, Shaun Westlund, Matthew Hardison, Clinton R. Paden, Duncan MacCannell |
| EPI_ISL_2202242 | Houston Methodist Hospital | Houston Methodist Hospital | Randall J. Olsen, Paul A. Christensen, S. Wesley Long, Sishir Subedi, Robert Olson, Marcus Nguyen, James J. Davis, Matthew Ojeda Saavedra, Prasanti Yerramilli, Layne Pruitt, Kristina Reppond, Madison N. Shyer, Jessica Cambric, Ryan Gadd, Ilya J. Finkelstein, Jimmy Gollihar, and James M. Musser |
| EPI_ISL_2202243, EPI_ISL_2202244 | Aegis Sciences Corporation | Centers for Disease Control and Prevention Division of Viral Diseases, Pathogen Discovery | Dakota Howard, Dhwani Batra, Peter W. Cook, Kara Moser, Adrian Paskey, Jason Caravas, Benjamin Rambo-Martin, Shatavia Morrison, Christopher Gulvick, Scott Sammons, Yvette Unoarumhi, Darlene Wagner, Matthew Schmerer, Cyndi Clark, Patrick Campbell, Rob Case, Vikramsinha Ghorpade, Holly Houdeshell, Ola Kvalvaag, Dillon Nall, Ethan Sanders, Alec Vest, Shaun Westlund, Matthew Hardison, Clinton R. Paden, Duncan MacCannell |
| EPI_ISL_2202245 | Houston Methodist Hospital | Houston Methodist Hospital | Randall J. Olsen, Paul A. Christensen, S. Wesley Long, Sishir Subedi, Robert Olson, Marcus Nguyen, James J. Davis, Matthew Ojeda Saavedra, Prasanti Yerramilli, Layne Pruitt, Kristina Reppond, Madison N. Shyer, Jessica Cambric, Ryan Gadd, Ilya J. Finkelstein, Jimmy Gollihar, and James M. Musser |
| EPI_ISL_2202246 | Aegis Sciences Corporation | Centers for Disease Control and Prevention Division of Viral Diseases, Pathogen Discovery | Dakota Howard, Dhwani Batra, Peter W. Cook, Kara Moser, Adrian Paskey, Jason Caravas, Benjamin Rambo-Martin, Shatavia Morrison, Christopher Gulvick, Scott Sammons, Yvette Unoarumhi, Darlene Wagner, Matthew Schmerer, Cyndi Clark, Patrick Campbell, Rob Case, Vikramsinha Ghorpade, Holly Houdeshell, Ola Kvalvaag, Dillon Nall, Ethan Sanders, Alec Vest, Shaun Westlund, Matthew Hardison, Clinton R. Paden, Duncan MacCannell |
| EPI_ISL_2202247 | Houston Methodist Hospital | Houston Methodist Hospital | Randall J. Olsen, Paul A. Christensen, S. Wesley Long, Sishir Subedi, Robert Olson, Marcus Nguyen, James J. Davis, Matthew Ojeda Saavedra, Prasanti Yerramilli, Layne Pruitt, Kristina Reppond, Madison N. Shyer, Jessica Cambric, Ryan Gadd, Ilya J. Finkelstein, Jimmy Gollihar, and James M. Musser |
| EPI_ISL_2202248, EPI_ISL_2202249 | Aegis Sciences Corporation | Centers for Disease Control and Prevention Division of Viral Diseases, Pathogen Discovery | Dakota Howard, Dhwani Batra, Peter W. Cook, Kara Moser, Adrian Paskey, Jason Caravas, Benjamin Rambo-Martin, Shatavia Morrison, Christopher Gulvick, Scott Sammons, Yvette Unoarumhi, Darlene Wagner, Matthew Schmerer, Cyndi Clark, Patrick Campbell, Rob Case, Vikramsinha Ghorpade, Holly Houdeshell, Ola Kvalvaag, Dillon Nall, Ethan Sanders, Alec Vest, Shaun Westlund, Matthew Hardison, Clinton R. Paden, Duncan MacCannell |
| EPI_ISL_2202250 | Houston Methodist Hospital | Houston Methodist Hospital | Randall J. Olsen, Paul A. Christensen, S. Wesley Long, Sishir Subedi, Robert Olson, Marcus Nguyen, James J. Davis, Matthew Ojeda Saavedra, Prasanti Yerramilli, Layne Pruitt, Kristina Reppond, Madison N. Shyer, Jessica Cambric, Ryan Gadd, Ilya J. Finkelstein, Jimmy Gollihar, and James M. Musser |
| EPI_ISL_2202251, EPI_ISL_2202252 | Aegis Sciences Corporation | Centers for Disease Control and Prevention Division of Viral Diseases, Pathogen Discovery | Dakota Howard, Dhwani Batra, Peter W. Cook, Kara Moser, Adrian Paskey, Jason Caravas, Benjamin Rambo-Martin, Shatavia Morrison, Christopher Gulvick, Scott Sammons, Yvette Unoarumhi, Darlene Wagner, Matthew Schmerer, Cyndi Clark, Patrick Campbell, Rob Case, Vikramsinha Ghorpade, Holly Houdeshell, Ola Kvalvaag, Dillon Nall, Ethan Sanders, Alec Vest, Shaun Westlund, Matthew Hardison, Clinton R. Paden, Duncan MacCannell |
| EPI_ISL_2202253 | Houston Methodist Hospital | Houston Methodist Hospital | Randall J. Olsen, Paul A. Christensen, S. Wesley Long, Sishir Subedi, Robert Olson, Marcus Nguyen, James J. Davis, Matthew Ojeda Saavedra, Prasanti Yerramilli, Layne Pruitt, Kristina Reppond, Madison N. Shyer, Jessica Cambric, Ryan Gadd, Ilya J. Finkelstein, Jimmy Gollihar, and James M. Musser |
| EPI_ISL_2202254 | Aegis Sciences Corporation | Centers for Disease Control and Prevention Division of Viral Diseases, Pathogen Discovery | Dakota Howard, Dhwani Batra, Peter W. Cook, Kara Moser, Adrian Paskey, Jason Caravas, Benjamin Rambo-Martin, Shatavia Morrison, Christopher Gulvick, Scott Sammons, Yvette Unoarumhi, Darlene Wagner, Matthew Schmerer, Cyndi Clark, Patrick Campbell, Rob Case, Vikramsinha Ghorpade, Holly Houdeshell, Ola Kvalvaag, Dillon Nall, Ethan Sanders, Alec Vest, Shaun Westlund, Matthew Hardison, Clinton R. Paden, Duncan MacCannell |
| EPI_ISL_2202255 | Houston Methodist Hospital | Houston Methodist Hospital | Randall J. Olsen, Paul A. Christensen, S. Wesley Long, Sishir Subedi, Robert Olson, Marcus Nguyen, James J. Davis, Matthew Ojeda Saavedra, Prasanti Yerramilli, Layne Pruitt, Kristina Reppond, Madison N. Shyer, Jessica Cambric, Ryan Gadd, Ilya J. Finkelstein, Jimmy Gollihar, and James M. Musser |
| EPI_ISL_2202256 | Aegis Sciences Corporation | Centers for Disease Control and Prevention Division of Viral Diseases, Pathogen Discovery | Dakota Howard, Dhwani Batra, Peter W. Cook, Kara Moser, Adrian Paskey, Jason Caravas, Benjamin Rambo-Martin, Shatavia Morrison, Christopher Gulvick, Scott Sammons, Yvette Unoarumhi, Darlene Wagner, Matthew Schmerer, Cyndi Clark, Patrick Campbell, Rob Case, Vikramsinha Ghorpade, Holly Houdeshell, Ola Kvalvaag, Dillon Nall, Ethan Sanders, Alec Vest, Shaun Westlund, Matthew Hardison, Clinton R. Paden, Duncan MacCannell |
| EPI_ISL_2202257 | Houston Methodist Hospital | Houston Methodist Hospital | Randall J. Olsen, Paul A. Christensen, S. Wesley Long, Sishir Subedi, Robert Olson, Marcus Nguyen, James J. Davis, Matthew Ojeda Saavedra, Prasanti Yerramilli, Layne Pruitt, Kristina Reppond, Madison N. Shyer, Jessica Cambric, Ryan Gadd, Ilya J. Finkelstein, Jimmy Gollihar, and James M. Musser |
| EPI_ISL_2202258, EPI_ISL_2202259 | Aegis Sciences Corporation | Centers for Disease Control and Prevention Division of Viral Diseases, Pathogen Discovery | Dakota Howard, Dhwani Batra, Peter W. Cook, Kara Moser, Adrian Paskey, Jason Caravas, Benjamin Rambo-Martin, Shatavia Morrison, Christopher Gulvick, Scott Sammons, Yvette Unoarumhi, Darlene Wagner, Matthew Schmerer, Cyndi Clark, Patrick Campbell, Rob Case, Vikramsinha Ghorpade, Holly Houdeshell, Ola Kvalvaag, Dillon Nall, Ethan Sanders, Alec Vest, Shaun Westlund, Matthew Hardison, Clinton R. Paden, Duncan MacCannell |
| EPI_ISL_2202260 | Houston Methodist Hospital | Houston Methodist Hospital | Randall J. Olsen, Paul A. Christensen, S. Wesley Long, Sishir Subedi, Robert Olson, Marcus Nguyen, James J. Davis, Matthew Ojeda Saavedra, Prasanti Yerramilli, Layne Pruitt, Kristina Reppond, Madison N. Shyer, Jessica Cambric, Ryan Gadd, Ilya J. Finkelstein, Jimmy Gollihar, and James M. Musser |
| EPI_ISL_2202261 | Aegis Sciences Corporation | Centers for Disease Control and Prevention Division of Viral Diseases, Pathogen Discovery | Dakota Howard, Dhwani Batra, Peter W. Cook, Kara Moser, Adrian Paskey, Jason Caravas, Benjamin Rambo-Martin, Shatavia Morrison, Christopher Gulvick, Scott Sammons, Yvette Unoarumhi, Darlene Wagner, Matthew Schmerer, Cyndi Clark, Patrick Campbell, Rob Case, Vikramsinha Ghorpade, Holly Houdeshell, Ola Kvalvaag, Dillon Nall, Ethan Sanders, Alec Vest, Shaun Westlund, Matthew Hardison, Clinton R. Paden, Duncan MacCannell |
| EPI_ISL_2202262 | Houston Methodist Hospital | Houston Methodist Hospital | Randall J. Olsen, Paul A. Christensen, S. Wesley Long, Sishir Subedi, Robert Olson, Marcus Nguyen, James J. Davis, Matthew Ojeda Saavedra, Prasanti Yerramilli, Layne Pruitt, Kristina Reppond, Madison N. Shyer, Jessica Cambric, Ryan Gadd, Ilya J. Finkelstein, Jimmy Gollihar, and James M. Musser |
| EPI_ISL_2202263 | Aegis Sciences Corporation | Centers for Disease Control and Prevention Division of Viral Diseases, Pathogen Discovery | Dakota Howard, Dhwani Batra, Peter W. Cook, Kara Moser, Adrian Paskey, Jason Caravas, Benjamin Rambo-Martin, Shatavia Morrison, Christopher Gulvick, Scott Sammons, Yvette Unoarumhi, Darlene Wagner, Matthew Schmerer, Cyndi Clark, Patrick Campbell, Rob Case, Vikramsinha Ghorpade, Holly Houdeshell, Ola Kvalvaag, Dillon Nall, Ethan Sanders, Alec Vest, Shaun Westlund, Matthew Hardison, Clinton R. Paden, Duncan MacCannell |
| EPI_ISL_2202264 | Houston Methodist Hospital | Houston Methodist Hospital | Randall J. Olsen, Paul A. Christensen, S. Wesley Long, Sishir Subedi, Robert Olson, Marcus Nguyen, James J. Davis, Matthew Ojeda Saavedra, Prasanti Yerramilli, Layne Pruitt, Kristina Reppond, Madison N. Shyer, Jessica Cambric, Ryan Gadd, Ilya J. Finkelstein, Jimmy Gollihar, and James M. Musser |
| EPI_ISL_2202265 | Aegis Sciences Corporation | Centers for Disease Control and Prevention Division of Viral Diseases, Pathogen Discovery | Dakota Howard, Dhwani Batra, Peter W. Cook, Kara Moser, Adrian Paskey, Jason Caravas, Benjamin Rambo-Martin, Shatavia Morrison, Christopher Gulvick, Scott Sammons, Yvette Unoarumhi, Darlene Wagner, Matthew Schmerer, Cyndi Clark, Patrick Campbell, Rob Case, Vikramsinha Ghorpade, Holly Houdeshell, Ola Kvalvaag, Dillon Nall, Ethan Sanders, Alec Vest, Shaun Westlund, Matthew Hardison, Clinton R. Paden, Duncan MacCannell |
| EPI_ISL_2202266 | Houston Methodist Hospital | Houston Methodist Hospital | Randall J. Olsen, Paul A. Christensen, S. Wesley Long, Sishir Subedi, Robert Olson, Marcus Nguyen, James J. Davis, Matthew Ojeda Saavedra, Prasanti Yerramilli, Layne Pruitt, Kristina Reppond, Madison N. Shyer, Jessica Cambric, Ryan Gadd, Ilya J. Finkelstein, Jimmy Gollihar, and James M. Musser |
| EPI_ISL_2202267, EPI_ISL_2202268 | Aegis Sciences Corporation | Centers for Disease Control and Prevention Division of Viral Diseases, Pathogen Discovery | Dakota Howard, Dhwani Batra, Peter W. Cook, Kara Moser, Adrian Paskey, Jason Caravas, Benjamin Rambo-Martin, Shatavia Morrison, Christopher Gulvick, Scott Sammons, Yvette Unoarumhi, Darlene Wagner, Matthew Schmerer, Cyndi Clark, Patrick Campbell, Rob Case, Vikramsinha Ghorpade, Holly Houdeshell, Ola Kvalvaag, Dillon Nall, Ethan Sanders, Alec Vest, Shaun Westlund, Matthew Hardison, Clinton R. Paden, Duncan MacCannell |
| EPI_ISL_2202269 | Houston Methodist Hospital | Houston Methodist Hospital | Randall J. Olsen, Paul A. Christensen, S. Wesley Long, Sishir Subedi, Robert Olson, Marcus Nguyen, James J. Davis, Matthew Ojeda Saavedra, Prasanti Yerramilli, Layne Pruitt, Kristina Reppond, Madison N. Shyer, Jessica Cambric, Ryan Gadd, Ilya J. Finkelstein, Jimmy Gollihar, and James M. Musser |
| EPI_ISL_2202270 | Aegis Sciences Corporation | Centers for Disease Control and Prevention Division of Viral Diseases, Pathogen Discovery | Dakota Howard, Dhwani Batra, Peter W. Cook, Kara Moser, Adrian Paskey, Jason Caravas, Benjamin Rambo-Martin, Shatavia Morrison, Christopher Gulvick, Scott Sammons, Yvette Unoarumhi, Darlene Wagner, Matthew Schmerer, Cyndi Clark, Patrick Campbell, Rob Case, Vikramsinha Ghorpade, Holly Houdeshell, Ola Kvalvaag, Dillon Nall, Ethan Sanders, Alec Vest, Shaun Westlund, Matthew Hardison, Clinton R. Paden, Duncan MacCannell |
| EPI_ISL_2202271 | Houston Methodist Hospital | Houston Methodist Hospital | Randall J. Olsen, Paul A. Christensen, S. Wesley Long, Sishir Subedi, Robert Olson, Marcus Nguyen, James J. Davis, Matthew Ojeda Saavedra, Prasanti Yerramilli, Layne Pruitt, Kristina Reppond, Madison N. Shyer, Jessica Cambric, Ryan Gadd, Ilya J. Finkelstein, Jimmy Gollihar, and James M. Musser |
| EPI_ISL_2202272 | Aegis Sciences Corporation | Centers for Disease Control and Prevention Division of Viral Diseases, Pathogen Discovery | Dakota Howard, Dhwani Batra, Peter W. Cook, Kara Moser, Adrian Paskey, Jason Caravas, Benjamin Rambo-Martin, Shatavia Morrison, Christopher Gulvick, Scott Sammons, Yvette Unoarumhi, Darlene Wagner, Matthew Schmerer, Cyndi Clark, Patrick Campbell, Rob Case, Vikramsinha Ghorpade, Holly Houdeshell, Ola Kvalvaag, Dillon Nall, Ethan Sanders, Alec Vest, Shaun Westlund, Matthew Hardison, Clinton R. Paden, Duncan MacCannell |
| EPI_ISL_2202273 | Houston Methodist Hospital | Houston Methodist Hospital | Randall J. Olsen, Paul A. Christensen, S. Wesley Long, Sishir Subedi, Robert Olson, Marcus Nguyen, James J. Davis, Matthew Ojeda Saavedra, Prasanti Yerramilli, Layne Pruitt, Kristina Reppond, Madison N. Shyer, Jessica Cambric, Ryan Gadd, Ilya J. Finkelstein, Jimmy Gollihar, and James M. Musser |
| EPI_ISL_2202274, EPI_ISL_2202275 | Aegis Sciences Corporation | Centers for Disease Control and Prevention Division of Viral Diseases, Pathogen Discovery | Dakota Howard, Dhwani Batra, Peter W. Cook, Kara Moser, Adrian Paskey, Jason Caravas, Benjamin Rambo-Martin, Shatavia Morrison, Christopher Gulvick, Scott Sammons, Yvette Unoarumhi, Darlene Wagner, Matthew Schmerer, Cyndi Clark, Patrick Campbell, Rob Case, Vikramsinha Ghorpade, Holly Houdeshell, Ola Kvalvaag, Dillon Nall, Ethan Sanders, Alec Vest, Shaun Westlund, Matthew Hardison, Clinton R. Paden, Duncan MacCannell |
| EPI_ISL_2202276 | Houston Methodist Hospital | Houston Methodist Hospital | Randall J. Olsen, Paul A. Christensen, S. Wesley Long, Sishir Subedi, Robert Olson, Marcus Nguyen, James J. Davis, Matthew Ojeda Saavedra, Prasanti Yerramilli, Layne Pruitt, Kristina Reppond, Madison N. Shyer, Jessica Cambric, Ryan Gadd, Ilya J. Finkelstein, Jimmy Gollihar, and James M. Musser |
| EPI_ISL_2202277 | Aegis Sciences Corporation | Centers for Disease Control and Prevention Division of Viral Diseases, Pathogen Discovery | Dakota Howard, Dhwani Batra, Peter W. Cook, Kara Moser, Adrian Paskey, Jason Caravas, Benjamin Rambo-Martin, Shatavia Morrison, Christopher Gulvick, Scott Sammons, Yvette Unoarumhi, Darlene Wagner, Matthew Schmerer, Cyndi Clark, Patrick Campbell, Rob Case, Vikramsinha Ghorpade, Holly Houdeshell, Ola Kvalvaag, Dillon Nall, Ethan Sanders, Alec Vest, Shaun Westlund, Matthew Hardison, Clinton R. Paden, Duncan MacCannell |
| EPI_ISL_2202278 | Houston Methodist Hospital | Houston Methodist Hospital | Randall J. Olsen, Paul A. Christensen, S. Wesley Long, Sishir Subedi, Robert Olson, Marcus Nguyen, James J. Davis, Matthew Ojeda Saavedra, Prasanti Yerramilli, Layne Pruitt, Kristina Reppond, Madison N. Shyer, Jessica Cambric, Ryan Gadd, Ilya J. Finkelstein, Jimmy Gollihar, and James M. Musser |
| EPI_ISL_2202279 | Aegis Sciences Corporation | Centers for Disease Control and Prevention Division of Viral Diseases, Pathogen Discovery | Dakota Howard, Dhwani Batra, Peter W. Cook, Kara Moser, Adrian Paskey, Jason Caravas, Benjamin Rambo-Martin, Shatavia Morrison, Christopher Gulvick, Scott Sammons, Yvette Unoarumhi, Darlene Wagner, Matthew Schmerer, Cyndi Clark, Patrick Campbell, Rob Case, Vikramsinha Ghorpade, Holly Houdeshell, Ola Kvalvaag, Dillon Nall, Ethan Sanders, Alec Vest, Shaun Westlund, Matthew Hardison, Clinton R. Paden, Duncan MacCannell |
| EPI_ISL_2202280 | Houston Methodist Hospital | Houston Methodist Hospital | Randall J. Olsen, Paul A. Christensen, S. Wesley Long, Sishir Subedi, Robert Olson, Marcus Nguyen, James J. Davis, Matthew Ojeda Saavedra, Prasanti Yerramilli, Layne Pruitt, Kristina Reppond, Madison N. Shyer, Jessica Cambric, Ryan Gadd, Ilya J. Finkelstein, Jimmy Gollihar, and James M. Musser |
| EPI_ISL_2202281 | Aegis Sciences Corporation | Centers for Disease Control and Prevention | Dakota Howard, Dhwani Batra, Peter W. Cook, Kara Moser, Adrian Paskey, Jason Caravas, Benjamin Rambo-Martin, Shatavia Morrison, Christopher Gulvick, Scott Sammons, Yvette Unoarumhi, Darlene Wagner, Matthew Schmerer, Cyndi Clark, Patrick Campbell, Rob Case, Vikramsinha |

| | | | |
|---|---|---|---|
| | | Division of Viral Diseases, Pathogen Discovery | Ghorpade, Holly Houdeshell, Ola Kvalvaag, Dillon Nall, Ethan Sanders, Alec Vest, Shaun Westlund, Matthew Hardison, Clinton R. Paden, Duncan MacCannell |
| EPI_ISL_2202282 | Houston Methodist Hospital | Houston Methodist Hospital | Randall J. Olsen, Paul A. Christensen, S. Wesley Long, Sishir Subedi, Robert Olson, Marcus Nguyen, James J. Davis, Matthew Ojeda Saavedra, Prasanti Yerramilli, Layne Pruitt, Kristina Reppond, Madison N. Shyer, Jessica Cambric, Ryan Gadd, Ilya J. Finkelstein, Jimmy Gollihar, and James M. Musser |
| EPI_ISL_2202283, EPI_ISL_2202284 | Aegis Sciences Corporation | Centers for Disease Control and Prevention Division of Viral Diseases, Pathogen Discovery | Dakota Howard, Dhwani Batra, Peter W. Cook, Kara Moser, Adrian Paskey, Jason Caravas, Benjamin Rambo-Martin, Shatavia Morrison, Christopher Gulvick, Scott Sammons, Yvette Unoarumhi, Darlene Wagner, Matthew Schmerer, Cyndi Clark, Patrick Campbell, Rob Case, Vikramsinha Ghorpade, Holly Houdeshell, Ola Kvalvaag, Dillon Nall, Ethan Sanders, Alec Vest, Shaun Westlund, Matthew Hardison, Clinton R. Paden, Duncan MacCannell |
| EPI_ISL_2202285, EPI_ISL_2202287 | Houston Methodist Hospital | Houston Methodist Hospital | Randall J. Olsen, Paul A. Christensen, S. Wesley Long, Sishir Subedi, Robert Olson, Marcus Nguyen, James J. Davis, Matthew Ojeda Saavedra, Prasanti Yerramilli, Layne Pruitt, Kristina Reppond, Madison N. Shyer, Jessica Cambric, Ryan Gadd, Ilya J. Finkelstein, Jimmy Gollihar, and James M. Musser |
| EPI_ISL_2202289, EPI_ISL_2202290 | Aegis Sciences Corporation | Centers for Disease Control and Prevention Division of Viral Diseases, Pathogen Discovery | Dakota Howard, Dhwani Batra, Peter W. Cook, Kara Moser, Adrian Paskey, Jason Caravas, Benjamin Rambo-Martin, Shatavia Morrison, Christopher Gulvick, Scott Sammons, Yvette Unoarumhi, Darlene Wagner, Matthew Schmerer, Cyndi Clark, Patrick Campbell, Rob Case, Vikramsinha Ghorpade, Holly Houdeshell, Ola Kvalvaag, Dillon Nall, Ethan Sanders, Alec Vest, Shaun Westlund, Matthew Hardison, Clinton R. Paden, Duncan MacCannell |
| EPI_ISL_2202291 | Houston Methodist Hospital | Houston Methodist Hospital | Randall J. Olsen, Paul A. Christensen, S. Wesley Long, Sishir Subedi, Robert Olson, Marcus Nguyen, James J. Davis, Matthew Ojeda Saavedra, Prasanti Yerramilli, Layne Pruitt, Kristina Reppond, Madison N. Shyer, Jessica Cambric, Ryan Gadd, Ilya J. Finkelstein, Jimmy Gollihar, and James M. Musser |
| EPI_ISL_2202292 | Aegis Sciences Corporation | Centers for Disease Control and Prevention Division of Viral Diseases, Pathogen Discovery | Dakota Howard, Dhwani Batra, Peter W. Cook, Kara Moser, Adrian Paskey, Jason Caravas, Benjamin Rambo-Martin, Shatavia Morrison, Christopher Gulvick, Scott Sammons, Yvette Unoarumhi, Darlene Wagner, Matthew Schmerer, Cyndi Clark, Patrick Campbell, Rob Case, Vikramsinha Ghorpade, Holly Houdeshell, Ola Kvalvaag, Dillon Nall, Ethan Sanders, Alec Vest, Shaun Westlund, Matthew Hardison, Clinton R. Paden, Duncan MacCannell |
| EPI_ISL_2202293 | Houston Methodist Hospital | Houston Methodist Hospital | Randall J. Olsen, Paul A. Christensen, S. Wesley Long, Sishir Subedi, Robert Olson, Marcus Nguyen, James J. Davis, Matthew Ojeda Saavedra, Prasanti Yerramilli, Layne Pruitt, Kristina Reppond, Madison N. Shyer, Jessica Cambric, Ryan Gadd, Ilya J. Finkelstein, Jimmy Gollihar, and James M. Musser |
| EPI_ISL_2202294, EPI_ISL_2202295 | Aegis Sciences Corporation | Centers for Disease Control and Prevention Division of Viral Diseases, Pathogen Discovery | Dakota Howard, Dhwani Batra, Peter W. Cook, Kara Moser, Adrian Paskey, Jason Caravas, Benjamin Rambo-Martin, Shatavia Morrison, Christopher Gulvick, Scott Sammons, Yvette Unoarumhi, Darlene Wagner, Matthew Schmerer, Cyndi Clark, Patrick Campbell, Rob Case, Vikramsinha Ghorpade, Holly Houdeshell, Ola Kvalvaag, Dillon Nall, Ethan Sanders, Alec Vest, Shaun Westlund, Matthew Hardison, Clinton R. Paden, Duncan MacCannell |
| EPI_ISL_2202296 | Houston Methodist Hospital | Houston Methodist Hospital | Randall J. Olsen, Paul A. Christensen, S. Wesley Long, Sishir Subedi, Robert Olson, Marcus Nguyen, James J. Davis, Matthew Ojeda Saavedra, Prasanti Yerramilli, Layne Pruitt, Kristina Reppond, Madison N. Shyer, Jessica Cambric, Ryan Gadd, Ilya J. Finkelstein, Jimmy Gollihar, and James M. Musser |
| EPI_ISL_2202297 | Aegis Sciences Corporation | Centers for Disease Control and Prevention Division of Viral Diseases, Pathogen Discovery | Dakota Howard, Dhwani Batra, Peter W. Cook, Kara Moser, Adrian Paskey, Jason Caravas, Benjamin Rambo-Martin, Shatavia Morrison, Christopher Gulvick, Scott Sammons, Yvette Unoarumhi, Darlene Wagner, Matthew Schmerer, Cyndi Clark, Patrick Campbell, Rob Case, Vikramsinha Ghorpade, Holly Houdeshell, Ola Kvalvaag, Dillon Nall, Ethan Sanders, Alec Vest, Shaun Westlund, Matthew Hardison, Clinton R. Paden, Duncan MacCannell |
| EPI_ISL_2202298 | Houston Methodist Hospital | Houston Methodist Hospital | Randall J. Olsen, Paul A. Christensen, S. Wesley Long, Sishir Subedi, Robert Olson, Marcus Nguyen, James J. Davis, Matthew Ojeda Saavedra, Prasanti Yerramilli, Layne Pruitt, Kristina Reppond, Madison N. Shyer, Jessica Cambric, Ryan Gadd, Ilya J. Finkelstein, Jimmy Gollihar, and James M. Musser |
| EPI_ISL_2202299 | Aegis Sciences Corporation | Centers for Disease Control and Prevention Division of Viral Diseases, Pathogen Discovery | Dakota Howard, Dhwani Batra, Peter W. Cook, Kara Moser, Adrian Paskey, Jason Caravas, Benjamin Rambo-Martin, Shatavia Morrison, Christopher Gulvick, Scott Sammons, Yvette Unoarumhi, Darlene Wagner, Matthew Schmerer, Cyndi Clark, Patrick Campbell, Rob Case, Vikramsinha Ghorpade, Holly Houdeshell, Ola Kvalvaag, Dillon Nall, Ethan Sanders, Alec Vest, Shaun Westlund, Matthew Hardison, Clinton R. Paden, Duncan MacCannell |
| EPI_ISL_2202300 | Houston Methodist Hospital | Houston Methodist Hospital | Randall J. Olsen, Paul A. Christensen, S. Wesley Long, Sishir Subedi, Robert Olson, Marcus Nguyen, James J. Davis, Matthew Ojeda Saavedra, Prasanti Yerramilli, Layne Pruitt, Kristina Reppond, Madison N. Shyer, Jessica Cambric, Ryan Gadd, Ilya J. Finkelstein, Jimmy Gollihar, and James M. Musser |
| EPI_ISL_2202301, EPI_ISL_2202302 | Aegis Sciences Corporation | Centers for Disease Control and Prevention Division of Viral Diseases, Pathogen Discovery | Dakota Howard, Dhwani Batra, Peter W. Cook, Kara Moser, Adrian Paskey, Jason Caravas, Benjamin Rambo-Martin, Shatavia Morrison, Christopher Gulvick, Scott Sammons, Yvette Unoarumhi, Darlene Wagner, Matthew Schmerer, Cyndi Clark, Patrick Campbell, Rob Case, Vikramsinha Ghorpade, Holly Houdeshell, Ola Kvalvaag, Dillon Nall, Ethan Sanders, Alec Vest, Shaun Westlund, Matthew Hardison, Clinton R. Paden, Duncan MacCannell |
| EPI_ISL_2202303 | Houston Methodist Hospital | Houston Methodist Hospital | Randall J. Olsen, Paul A. Christensen, S. Wesley Long, Sishir Subedi, Robert Olson, Marcus Nguyen, James J. Davis, Matthew Ojeda Saavedra, Prasanti Yerramilli, Layne Pruitt, Kristina Reppond, Madison N. Shyer, Jessica Cambric, Ryan Gadd, Ilya J. Finkelstein, Jimmy Gollihar, and James M. Musser |
| EPI_ISL_2202304 | Aegis Sciences Corporation | Centers for Disease Control and Prevention Division of Viral Diseases, Pathogen Discovery | Dakota Howard, Dhwani Batra, Peter W. Cook, Kara Moser, Adrian Paskey, Jason Caravas, Benjamin Rambo-Martin, Shatavia Morrison, Christopher Gulvick, Scott Sammons, Yvette Unoarumhi, Darlene Wagner, Matthew Schmerer, Cyndi Clark, Patrick Campbell, Rob Case, Vikramsinha Ghorpade, Holly Houdeshell, Ola Kvalvaag, Dillon Nall, Ethan Sanders, Alec Vest, Shaun Westlund, Matthew Hardison, Clinton R. Paden, Duncan MacCannell |
| EPI_ISL_2202305 | Houston Methodist Hospital | Houston Methodist Hospital | Randall J. Olsen, Paul A. Christensen, S. Wesley Long, Sishir Subedi, Robert Olson, Marcus Nguyen, James J. Davis, Matthew Ojeda Saavedra, Prasanti Yerramilli, Layne Pruitt, Kristina Reppond, Madison N. Shyer, Jessica Cambric, Ryan Gadd, Ilya J. Finkelstein, Jimmy Gollihar, and James M. Musser |
| EPI_ISL_2202306 | Aegis Sciences Corporation | Centers for Disease Control and Prevention Division of Viral Diseases, Pathogen Discovery | Dakota Howard, Dhwani Batra, Peter W. Cook, Kara Moser, Adrian Paskey, Jason Caravas, Benjamin Rambo-Martin, Shatavia Morrison, Christopher Gulvick, Scott Sammons, Yvette Unoarumhi, Darlene Wagner, Matthew Schmerer, Cyndi Clark, Patrick Campbell, Rob Case, Vikramsinha Ghorpade, Holly Houdeshell, Ola Kvalvaag, Dillon Nall, Ethan Sanders, Alec Vest, Shaun Westlund, Matthew Hardison, Clinton R. Paden, Duncan MacCannell |
| EPI_ISL_2202307 | Houston Methodist Hospital | Houston Methodist Hospital | Randall J. Olsen, Paul A. Christensen, S. Wesley Long, Sishir Subedi, Robert Olson, Marcus Nguyen, James J. Davis, Matthew Ojeda Saavedra, Prasanti Yerramilli, Layne Pruitt, Kristina Reppond, Madison N. Shyer, Jessica Cambric, Ryan Gadd, Ilya J. Finkelstein, Jimmy Gollihar, and James M. Musser |
| EPI_ISL_2202308, EPI_ISL_2202309 | Aegis Sciences Corporation | Centers for Disease Control and Prevention Division of Viral Diseases, Pathogen Discovery | Dakota Howard, Dhwani Batra, Peter W. Cook, Kara Moser, Adrian Paskey, Jason Caravas, Benjamin Rambo-Martin, Shatavia Morrison, Christopher Gulvick, Scott Sammons, Yvette Unoarumhi, Darlene Wagner, Matthew Schmerer, Cyndi Clark, Patrick Campbell, Rob Case, Vikramsinha Ghorpade, Holly Houdeshell, Ola Kvalvaag, Dillon Nall, Ethan Sanders, Alec Vest, Shaun Westlund, Matthew Hardison, Clinton R. Paden, Duncan MacCannell |
| EPI_ISL_2202310 | Houston Methodist Hospital | Houston Methodist Hospital | Randall J. Olsen, Paul A. Christensen, S. Wesley Long, Sishir Subedi, Robert Olson, Marcus Nguyen, James J. Davis, Matthew Ojeda Saavedra, Prasanti Yerramilli, Layne Pruitt, Kristina Reppond, Madison N. Shyer, Jessica Cambric, Ryan Gadd, Ilya J. Finkelstein, Jimmy Gollihar, and James M. Musser |
| EPI_ISL_2202311 | Aegis Sciences Corporation | Centers for Disease Control and Prevention Division of Viral Diseases, Pathogen Discovery | Dakota Howard, Dhwani Batra, Peter W. Cook, Kara Moser, Adrian Paskey, Jason Caravas, Benjamin Rambo-Martin, Shatavia Morrison, Christopher Gulvick, Scott Sammons, Yvette Unoarumhi, Darlene Wagner, Matthew Schmerer, Cyndi Clark, Patrick Campbell, Rob Case, Vikramsinha Ghorpade, Holly Houdeshell, Ola Kvalvaag, Dillon Nall, Ethan Sanders, Alec Vest, Shaun Westlund, Matthew Hardison, Clinton R. Paden, Duncan MacCannell |
| EPI_ISL_2202312, EPI_ISL_2202313 | Houston Methodist Hospital | Houston Methodist Hospital | Randall J. Olsen, Paul A. Christensen, S. Wesley Long, Sishir Subedi, Robert Olson, Marcus Nguyen, James J. Davis, Matthew Ojeda Saavedra, Prasanti Yerramilli, Layne Pruitt, Kristina Reppond, Madison N. Shyer, Jessica Cambric, Ryan Gadd, Ilya J. Finkelstein, Jimmy Gollihar, and James M. Musser |
| EPI_ISL_2202314 | Aegis Sciences Corporation | Centers for Disease Control and Prevention Division of Viral Diseases, Pathogen Discovery | Dakota Howard, Dhwani Batra, Peter W. Cook, Kara Moser, Adrian Paskey, Jason Caravas, Benjamin Rambo-Martin, Shatavia Morrison, Christopher Gulvick, Scott Sammons, Yvette Unoarumhi, Darlene Wagner, Matthew Schmerer, Cyndi Clark, Patrick Campbell, Rob Case, Vikramsinha Ghorpade, Holly Houdeshell, Ola Kvalvaag, Dillon Nall, Ethan Sanders, Alec Vest, Shaun Westlund, Matthew Hardison, Clinton R. Paden, Duncan MacCannell |
| EPI_ISL_2202315 | Houston Methodist Hospital | Houston Methodist Hospital | Randall J. Olsen, Paul A. Christensen, S. Wesley Long, Sishir Subedi, Robert Olson, Marcus Nguyen, James J. Davis, Matthew Ojeda Saavedra, Prasanti Yerramilli, Layne Pruitt, Kristina Reppond, Madison N. Shyer, Jessica Cambric, Ryan Gadd, Ilya J. Finkelstein, Jimmy Gollihar, and James M. Musser |
| EPI_ISL_2202316, EPI_ISL_2202317 | Aegis Sciences Corporation | Centers for Disease Control and Prevention Division of Viral Diseases, Pathogen Discovery | Dakota Howard, Dhwani Batra, Peter W. Cook, Kara Moser, Adrian Paskey, Jason Caravas, Benjamin Rambo-Martin, Shatavia Morrison, Christopher Gulvick, Scott Sammons, Yvette Unoarumhi, Darlene Wagner, Matthew Schmerer, Cyndi Clark, Patrick Campbell, Rob Case, Vikramsinha Ghorpade, Holly Houdeshell, Ola Kvalvaag, Dillon Nall, Ethan Sanders, Alec Vest, Shaun Westlund, Matthew Hardison, Clinton R. Paden, Duncan MacCannell |
| EPI_ISL_2202318 | Houston Methodist Hospital | Houston Methodist Hospital | Randall J. Olsen, Paul A. Christensen, S. Wesley Long, Sishir Subedi, Robert Olson, Marcus Nguyen, James J. Davis, Matthew Ojeda Saavedra, Prasanti Yerramilli, Layne Pruitt, Kristina Reppond, Madison N. Shyer, Jessica Cambric, Ryan Gadd, Ilya J. Finkelstein, Jimmy Gollihar, and James M. Musser |
| EPI_ISL_2202319 | Aegis Sciences Corporation | Centers for Disease Control and Prevention Division of Viral Diseases, Pathogen Discovery | Dakota Howard, Dhwani Batra, Peter W. Cook, Kara Moser, Adrian Paskey, Jason Caravas, Benjamin Rambo-Martin, Shatavia Morrison, Christopher Gulvick, Scott Sammons, Yvette Unoarumhi, Darlene Wagner, Matthew Schmerer, Cyndi Clark, Patrick Campbell, Rob Case, Vikramsinha Ghorpade, Holly Houdeshell, Ola Kvalvaag, Dillon Nall, Ethan Sanders, Alec Vest, Shaun Westlund, Matthew Hardison, Clinton R. Paden, Duncan MacCannell |
| EPI_ISL_2202320 | Houston Methodist Hospital | Houston Methodist Hospital | Randall J. Olsen, Paul A. Christensen, S. Wesley Long, Sishir Subedi, Robert Olson, Marcus Nguyen, James J. Davis, Matthew Ojeda Saavedra, Prasanti Yerramilli, Layne Pruitt, Kristina Reppond, Madison N. Shyer, Jessica Cambric, Ryan Gadd, Ilya J. Finkelstein, Jimmy Gollihar, and James M. Musser |
| EPI_ISL_2202321, EPI_ISL_2202322 | Aegis Sciences Corporation | Centers for Disease Control and Prevention Division of Viral Diseases, Pathogen Discovery | Dakota Howard, Dhwani Batra, Peter W. Cook, Kara Moser, Adrian Paskey, Jason Caravas, Benjamin Rambo-Martin, Shatavia Morrison, Christopher Gulvick, Scott Sammons, Yvette Unoarumhi, Darlene Wagner, Matthew Schmerer, Cyndi Clark, Patrick Campbell, Rob Case, Vikramsinha Ghorpade, Holly Houdeshell, Ola Kvalvaag, Dillon Nall, Ethan Sanders, Alec Vest, Shaun Westlund, Matthew Hardison, Clinton R. Paden, Duncan MacCannell |
| EPI_ISL_2202323 | Houston Methodist Hospital | Houston Methodist Hospital | Randall J. Olsen, Paul A. Christensen, S. Wesley Long, Sishir Subedi, Robert Olson, Marcus Nguyen, James J. Davis, Matthew Ojeda Saavedra, Prasanti Yerramilli, Layne Pruitt, Kristina Reppond, Madison N. Shyer, Jessica Cambric, Ryan Gadd, Ilya J. Finkelstein, Jimmy Gollihar, and James M. Musser |
| EPI_ISL_2202324 | Aegis Sciences Corporation | Centers for Disease Control and Prevention Division of Viral Diseases, Pathogen Discovery | Dakota Howard, Dhwani Batra, Peter W. Cook, Kara Moser, Adrian Paskey, Jason Caravas, Benjamin Rambo-Martin, Shatavia Morrison, Christopher Gulvick, Scott Sammons, Yvette Unoarumhi, Darlene Wagner, Matthew Schmerer, Cyndi Clark, Patrick Campbell, Rob Case, Vikramsinha Ghorpade, Holly Houdeshell, Ola Kvalvaag, Dillon Nall, Ethan Sanders, Alec Vest, Shaun Westlund, Matthew Hardison, Clinton R. Paden, Duncan MacCannell |
| EPI_ISL_2202325 | Houston Methodist Hospital | Houston Methodist Hospital | Randall J. Olsen, Paul A. Christensen, S. Wesley Long, Sishir Subedi, Robert Olson, Marcus Nguyen, James J. Davis, Matthew Ojeda Saavedra, Prasanti Yerramilli, Layne Pruitt, Kristina Reppond, Madison N. Shyer, Jessica Cambric, Ryan Gadd, Ilya J. Finkelstein, Jimmy Gollihar, and James M. Musser |
| EPI_ISL_2202326 | Aegis Sciences Corporation | Centers for Disease Control and Prevention Division of Viral Diseases, Pathogen Discovery | Dakota Howard, Dhwani Batra, Peter W. Cook, Kara Moser, Adrian Paskey, Jason Caravas, Benjamin Rambo-Martin, Shatavia Morrison, Christopher Gulvick, Scott Sammons, Yvette Unoarumhi, Darlene Wagner, Matthew Schmerer, Cyndi Clark, Patrick Campbell, Rob Case, Vikramsinha Ghorpade, Holly Houdeshell, Ola Kvalvaag, Dillon Nall, Ethan Sanders, Alec Vest, Shaun Westlund, Matthew Hardison, Clinton R. Paden, Duncan MacCannell |
| EPI_ISL_2202327 | Houston Methodist Hospital | Houston Methodist Hospital | Randall J. Olsen, Paul A. Christensen, S. Wesley Long, Sishir Subedi, Robert Olson, Marcus Nguyen, James J. Davis, Matthew Ojeda Saavedra, Prasanti Yerramilli, Layne Pruitt, Kristina Reppond, Madison N. Shyer, Jessica Cambric, Ryan Gadd, Ilya J. Finkelstein, Jimmy Gollihar, and James M. Musser |
| EPI_ISL_2202328, EPI_ISL_2202329 | Aegis Sciences Corporation | Centers for Disease Control and Prevention Division of Viral Diseases, Pathogen Discovery | Dakota Howard, Dhwani Batra, Peter W. Cook, Kara Moser, Adrian Paskey, Jason Caravas, Benjamin Rambo-Martin, Shatavia Morrison, Christopher Gulvick, Scott Sammons, Yvette Unoarumhi, Darlene Wagner, Matthew Schmerer, Cyndi Clark, Patrick Campbell, Rob Case, Vikramsinha Ghorpade, Holly Houdeshell, Ola Kvalvaag, Dillon Nall, Ethan Sanders, Alec Vest, Shaun Westlund, Matthew Hardison, Clinton R. Paden, Duncan MacCannell |
| EPI_ISL_2202330 | Houston Methodist Hospital | Houston Methodist Hospital | Randall J. Olsen, Paul A. Christensen, S. Wesley Long, Sishir Subedi, Robert Olson, Marcus Nguyen, James J. Davis, Matthew Ojeda Saavedra, Prasanti Yerramilli, Layne Pruitt, Kristina Reppond, Madison N. Shyer, Jessica Cambric, Ryan Gadd, Ilya J. Finkelstein, Jimmy Gollihar, and James M. Musser |

| | | | |
|---|---|---|---|
| EPI_ISL_2202331 | Aegis Sciences Corporation | Centers for Disease Control and Prevention Division of Viral Diseases, Pathogen Discovery | Dakota Howard, Dhwani Batra, Peter W. Cook, Kara Moser, Adrian Paskey, Jason Caravas, Benjamin Rambo-Martin, Shatavia Morrison, Christopher Gulvick, Scott Sammons, Yvette Unoarumhi, Darlene Wagner, Matthew Schmerer, Cyndi Clark, Patrick Campbell, Rob Case, Vikramsinha Ghorpade, Holly Houdeshell, Ola Kvalvaag, Dillon Nall, Ethan Sanders, Alec Vest, Shaun Westlund, Matthew Hardison, Clinton R. Paden, Duncan MacCannell |
| EPI_ISL_2202332 | Houston Methodist Hospital | Houston Methodist Hospital | Randall J. Olsen, Paul A. Christensen, S. Wesley Long, Sishir Subedi, Robert Olson, Marcus Nguyen, James J. Davis, Matthew Ojeda Saavedra, Prasanti Yerramilli, Layne Pruitt, Kristina Reppond, Madison N. Shyer, Jessica Cambric, Ryan Gadd, Ilya J. Finkelstein, Jimmy Gollihar, and James M. Musser |
| EPI_ISL_2202333 | Aegis Sciences Corporation | Centers for Disease Control and Prevention Division of Viral Diseases, Pathogen Discovery | Dakota Howard, Dhwani Batra, Peter W. Cook, Kara Moser, Adrian Paskey, Jason Caravas, Benjamin Rambo-Martin, Shatavia Morrison, Christopher Gulvick, Scott Sammons, Yvette Unoarumhi, Darlene Wagner, Matthew Schmerer, Cyndi Clark, Patrick Campbell, Rob Case, Vikramsinha Ghorpade, Holly Houdeshell, Ola Kvalvaag, Dillon Nall, Ethan Sanders, Alec Vest, Shaun Westlund, Matthew Hardison, Clinton R. Paden, Duncan MacCannell |
| EPI_ISL_2202334 | Houston Methodist Hospital | Houston Methodist Hospital | Randall J. Olsen, Paul A. Christensen, S. Wesley Long, Sishir Subedi, Robert Olson, Marcus Nguyen, James J. Davis, Matthew Ojeda Saavedra, Prasanti Yerramilli, Layne Pruitt, Kristina Reppond, Madison N. Shyer, Jessica Cambric, Ryan Gadd, Ilya J. Finkelstein, Jimmy Gollihar, and James M. Musser |
| EPI_ISL_2202335 | Aegis Sciences Corporation | Centers for Disease Control and Prevention Division of Viral Diseases, Pathogen Discovery | Dakota Howard, Dhwani Batra, Peter W. Cook, Kara Moser, Adrian Paskey, Jason Caravas, Benjamin Rambo-Martin, Shatavia Morrison, Christopher Gulvick, Scott Sammons, Yvette Unoarumhi, Darlene Wagner, Matthew Schmerer, Cyndi Clark, Patrick Campbell, Rob Case, Vikramsinha Ghorpade, Holly Houdeshell, Ola Kvalvaag, Dillon Nall, Ethan Sanders, Alec Vest, Shaun Westlund, Matthew Hardison, Clinton R. Paden, Duncan MacCannell |
| EPI_ISL_2202336 | Houston Methodist Hospital | Houston Methodist Hospital | Randall J. Olsen, Paul A. Christensen, S. Wesley Long, Sishir Subedi, Robert Olson, Marcus Nguyen, James J. Davis, Matthew Ojeda Saavedra, Prasanti Yerramilli, Layne Pruitt, Kristina Reppond, Madison N. Shyer, Jessica Cambric, Ryan Gadd, Ilya J. Finkelstein, Jimmy Gollihar, and James M. Musser |
| EPI_ISL_2202337, EPI_ISL_2202338 | Aegis Sciences Corporation | Centers for Disease Control and Prevention Division of Viral Diseases, Pathogen Discovery | Dakota Howard, Dhwani Batra, Peter W. Cook, Kara Moser, Adrian Paskey, Jason Caravas, Benjamin Rambo-Martin, Shatavia Morrison, Christopher Gulvick, Scott Sammons, Yvette Unoarumhi, Darlene Wagner, Matthew Schmerer, Cyndi Clark, Patrick Campbell, Rob Case, Vikramsinha Ghorpade, Holly Houdeshell, Ola Kvalvaag, Dillon Nall, Ethan Sanders, Alec Vest, Shaun Westlund, Matthew Hardison, Clinton R. Paden, Duncan MacCannell |
| EPI_ISL_2202339 | Houston Methodist Hospital | Houston Methodist Hospital | Randall J. Olsen, Paul A. Christensen, S. Wesley Long, Sishir Subedi, Robert Olson, Marcus Nguyen, James J. Davis, Matthew Ojeda Saavedra, Prasanti Yerramilli, Layne Pruitt, Kristina Reppond, Madison N. Shyer, Jessica Cambric, Ryan Gadd, Ilya J. Finkelstein, Jimmy Gollihar, and James M. Musser |
| EPI_ISL_2202340 | Aegis Sciences Corporation | Centers for Disease Control and Prevention Division of Viral Diseases, Pathogen Discovery | Dakota Howard, Dhwani Batra, Peter W. Cook, Kara Moser, Adrian Paskey, Jason Caravas, Benjamin Rambo-Martin, Shatavia Morrison, Christopher Gulvick, Scott Sammons, Yvette Unoarumhi, Darlene Wagner, Matthew Schmerer, Cyndi Clark, Patrick Campbell, Rob Case, Vikramsinha Ghorpade, Holly Houdeshell, Ola Kvalvaag, Dillon Nall, Ethan Sanders, Alec Vest, Shaun Westlund, Matthew Hardison, Clinton R. Paden, Duncan MacCannell |
| EPI_ISL_2202341 | Houston Methodist Hospital | Houston Methodist Hospital | Randall J. Olsen, Paul A. Christensen, S. Wesley Long, Sishir Subedi, Robert Olson, Marcus Nguyen, James J. Davis, Matthew Ojeda Saavedra, Prasanti Yerramilli, Layne Pruitt, Kristina Reppond, Madison N. Shyer, Jessica Cambric, Ryan Gadd, Ilya J. Finkelstein, Jimmy Gollihar, and James M. Musser |
| EPI_ISL_2202342 | Aegis Sciences Corporation | Centers for Disease Control and Prevention Division of Viral Diseases, Pathogen Discovery | Dakota Howard, Dhwani Batra, Peter W. Cook, Kara Moser, Adrian Paskey, Jason Caravas, Benjamin Rambo-Martin, Shatavia Morrison, Christopher Gulvick, Scott Sammons, Yvette Unoarumhi, Darlene Wagner, Matthew Schmerer, Cyndi Clark, Patrick Campbell, Rob Case, Vikramsinha Ghorpade, Holly Houdeshell, Ola Kvalvaag, Dillon Nall, Ethan Sanders, Alec Vest, Shaun Westlund, Matthew Hardison, Clinton R. Paden, Duncan MacCannell |
| EPI_ISL_2202343 | Houston Methodist Hospital | Houston Methodist Hospital | Randall J. Olsen, Paul A. Christensen, S. Wesley Long, Sishir Subedi, Robert Olson, Marcus Nguyen, James J. Davis, Matthew Ojeda Saavedra, Prasanti Yerramilli, Layne Pruitt, Kristina Reppond, Madison N. Shyer, Jessica Cambric, Ryan Gadd, Ilya J. Finkelstein, Jimmy Gollihar, and James M. Musser |
| EPI_ISL_2202344 | Aegis Sciences Corporation | Centers for Disease Control and Prevention Division of Viral Diseases, Pathogen Discovery | Dakota Howard, Dhwani Batra, Peter W. Cook, Kara Moser, Adrian Paskey, Jason Caravas, Benjamin Rambo-Martin, Shatavia Morrison, Christopher Gulvick, Scott Sammons, Yvette Unoarumhi, Darlene Wagner, Matthew Schmerer, Cyndi Clark, Patrick Campbell, Rob Case, Vikramsinha Ghorpade, Holly Houdeshell, Ola Kvalvaag, Dillon Nall, Ethan Sanders, Alec Vest, Shaun Westlund, Matthew Hardison, Clinton R. Paden, Duncan MacCannell |
| EPI_ISL_2202346 | Houston Methodist Hospital | Houston Methodist Hospital | Randall J. Olsen, Paul A. Christensen, S. Wesley Long, Sishir Subedi, Robert Olson, Marcus Nguyen, James J. Davis, Matthew Ojeda Saavedra, Prasanti Yerramilli, Layne Pruitt, Kristina Reppond, Madison N. Shyer, Jessica Cambric, Ryan Gadd, Ilya J. Finkelstein, Jimmy Gollihar, and James M. Musser |
| EPI_ISL_2202347, EPI_ISL_2202348 | Aegis Sciences Corporation | Centers for Disease Control and Prevention Division of Viral Diseases, Pathogen Discovery | Dakota Howard, Dhwani Batra, Peter W. Cook, Kara Moser, Adrian Paskey, Jason Caravas, Benjamin Rambo-Martin, Shatavia Morrison, Christopher Gulvick, Scott Sammons, Yvette Unoarumhi, Darlene Wagner, Matthew Schmerer, Cyndi Clark, Patrick Campbell, Rob Case, Vikramsinha Ghorpade, Holly Houdeshell, Ola Kvalvaag, Dillon Nall, Ethan Sanders, Alec Vest, Shaun Westlund, Matthew Hardison, Clinton R. Paden, Duncan MacCannell |
| EPI_ISL_2202349 | Houston Methodist Hospital | Houston Methodist Hospital | Randall J. Olsen, Paul A. Christensen, S. Wesley Long, Sishir Subedi, Robert Olson, Marcus Nguyen, James J. Davis, Matthew Ojeda Saavedra, Prasanti Yerramilli, Layne Pruitt, Kristina Reppond, Madison N. Shyer, Jessica Cambric, Ryan Gadd, Ilya J. Finkelstein, Jimmy Gollihar, and James M. Musser |
| EPI_ISL_2202350, EPI_ISL_2202351 | Aegis Sciences Corporation | Centers for Disease Control and Prevention Division of Viral Diseases, Pathogen Discovery | Dakota Howard, Dhwani Batra, Peter W. Cook, Kara Moser, Adrian Paskey, Jason Caravas, Benjamin Rambo-Martin, Shatavia Morrison, Christopher Gulvick, Scott Sammons, Yvette Unoarumhi, Darlene Wagner, Matthew Schmerer, Cyndi Clark, Patrick Campbell, Rob Case, Vikramsinha Ghorpade, Holly Houdeshell, Ola Kvalvaag, Dillon Nall, Ethan Sanders, Alec Vest, Shaun Westlund, Matthew Hardison, Clinton R. Paden, Duncan MacCannell |
| EPI_ISL_2202352 | Houston Methodist Hospital | Houston Methodist Hospital | Randall J. Olsen, Paul A. Christensen, S. Wesley Long, Sishir Subedi, Robert Olson, Marcus Nguyen, James J. Davis, Matthew Ojeda Saavedra, Prasanti Yerramilli, Layne Pruitt, Kristina Reppond, Madison N. Shyer, Jessica Cambric, Ryan Gadd, Ilya J. Finkelstein, Jimmy Gollihar, and James M. Musser |
| EPI_ISL_2202353 | Aegis Sciences Corporation | Centers for Disease Control and Prevention Division of Viral Diseases, Pathogen Discovery | Dakota Howard, Dhwani Batra, Peter W. Cook, Kara Moser, Adrian Paskey, Jason Caravas, Benjamin Rambo-Martin, Shatavia Morrison, Christopher Gulvick, Scott Sammons, Yvette Unoarumhi, Darlene Wagner, Matthew Schmerer, Cyndi Clark, Patrick Campbell, Rob Case, Vikramsinha Ghorpade, Holly Houdeshell, Ola Kvalvaag, Dillon Nall, Ethan Sanders, Alec Vest, Shaun Westlund, Matthew Hardison, Clinton R. Paden, Duncan MacCannell |
| EPI_ISL_2202354 | Houston Methodist Hospital | Houston Methodist Hospital | Randall J. Olsen, Paul A. Christensen, S. Wesley Long, Sishir Subedi, Robert Olson, Marcus Nguyen, James J. Davis, Matthew Ojeda Saavedra, Prasanti Yerramilli, Layne Pruitt, Kristina Reppond, Madison N. Shyer, Jessica Cambric, Ryan Gadd, Ilya J. Finkelstein, Jimmy Gollihar, and James M. Musser |
| EPI_ISL_2202355 | Aegis Sciences Corporation | Centers for Disease Control and Prevention Division of Viral Diseases, Pathogen Discovery | Dakota Howard, Dhwani Batra, Peter W. Cook, Kara Moser, Adrian Paskey, Jason Caravas, Benjamin Rambo-Martin, Shatavia Morrison, Christopher Gulvick, Scott Sammons, Yvette Unoarumhi, Darlene Wagner, Matthew Schmerer, Cyndi Clark, Patrick Campbell, Rob Case, Vikramsinha Ghorpade, Holly Houdeshell, Ola Kvalvaag, Dillon Nall, Ethan Sanders, Alec Vest, Shaun Westlund, Matthew Hardison, Clinton R. Paden, Duncan MacCannell |
| EPI_ISL_2202356 | Houston Methodist Hospital | Houston Methodist Hospital | Randall J. Olsen, Paul A. Christensen, S. Wesley Long, Sishir Subedi, Robert Olson, Marcus Nguyen, James J. Davis, Matthew Ojeda Saavedra, Prasanti Yerramilli, Layne Pruitt, Kristina Reppond, Madison N. Shyer, Jessica Cambric, Ryan Gadd, Ilya J. Finkelstein, Jimmy Gollihar, and James M. Musser |
| EPI_ISL_2202357, EPI_ISL_2202358 | Aegis Sciences Corporation | Centers for Disease Control and Prevention Division of Viral Diseases, Pathogen Discovery | Dakota Howard, Dhwani Batra, Peter W. Cook, Kara Moser, Adrian Paskey, Jason Caravas, Benjamin Rambo-Martin, Shatavia Morrison, Christopher Gulvick, Scott Sammons, Yvette Unoarumhi, Darlene Wagner, Matthew Schmerer, Cyndi Clark, Patrick Campbell, Rob Case, Vikramsinha Ghorpade, Holly Houdeshell, Ola Kvalvaag, Dillon Nall, Ethan Sanders, Alec Vest, Shaun Westlund, Matthew Hardison, Clinton R. Paden, Duncan MacCannell |
| EPI_ISL_2202359 | Houston Methodist Hospital | Houston Methodist Hospital | Randall J. Olsen, Paul A. Christensen, S. Wesley Long, Sishir Subedi, Robert Olson, Marcus Nguyen, James J. Davis, Matthew Ojeda Saavedra, Prasanti Yerramilli, Layne Pruitt, Kristina Reppond, Madison N. Shyer, Jessica Cambric, Ryan Gadd, Ilya J. Finkelstein, Jimmy Gollihar, and James M. Musser |
| EPI_ISL_2202360 | Aegis Sciences Corporation | Centers for Disease Control and Prevention Division of Viral Diseases, Pathogen Discovery | Dakota Howard, Dhwani Batra, Peter W. Cook, Kara Moser, Adrian Paskey, Jason Caravas, Benjamin Rambo-Martin, Shatavia Morrison, Christopher Gulvick, Scott Sammons, Yvette Unoarumhi, Darlene Wagner, Matthew Schmerer, Cyndi Clark, Patrick Campbell, Rob Case, Vikramsinha Ghorpade, Holly Houdeshell, Ola Kvalvaag, Dillon Nall, Ethan Sanders, Alec Vest, Shaun Westlund, Matthew Hardison, Clinton R. Paden, Duncan MacCannell |
| EPI_ISL_2202361 | Houston Methodist Hospital | Houston Methodist Hospital | Randall J. Olsen, Paul A. Christensen, S. Wesley Long, Sishir Subedi, Robert Olson, Marcus Nguyen, James J. Davis, Matthew Ojeda Saavedra, Prasanti Yerramilli, Layne Pruitt, Kristina Reppond, Madison N. Shyer, Jessica Cambric, Ryan Gadd, Ilya J. Finkelstein, Jimmy Gollihar, and James M. Musser |
| EPI_ISL_2202362, EPI_ISL_2202363 | Aegis Sciences Corporation | Centers for Disease Control and Prevention Division of Viral Diseases, Pathogen Discovery | Dakota Howard, Dhwani Batra, Peter W. Cook, Kara Moser, Adrian Paskey, Jason Caravas, Benjamin Rambo-Martin, Shatavia Morrison, Christopher Gulvick, Scott Sammons, Yvette Unoarumhi, Darlene Wagner, Matthew Schmerer, Cyndi Clark, Patrick Campbell, Rob Case, Vikramsinha Ghorpade, Holly Houdeshell, Ola Kvalvaag, Dillon Nall, Ethan Sanders, Alec Vest, Shaun Westlund, Matthew Hardison, Clinton R. Paden, Duncan MacCannell |
| EPI_ISL_2202364 | Houston Methodist Hospital | Houston Methodist Hospital | Randall J. Olsen, Paul A. Christensen, S. Wesley Long, Sishir Subedi, Robert Olson, Marcus Nguyen, James J. Davis, Matthew Ojeda Saavedra, Prasanti Yerramilli, Layne Pruitt, Kristina Reppond, Madison N. Shyer, Jessica Cambric, Ryan Gadd, Ilya J. Finkelstein, Jimmy Gollihar, and James M. Musser |
| EPI_ISL_2202365, EPI_ISL_2202366, EPI_ISL_2202367 | Aegis Sciences Corporation | Centers for Disease Control and Prevention Division of Viral Diseases, Pathogen Discovery | Dakota Howard, Dhwani Batra, Peter W. Cook, Kara Moser, Adrian Paskey, Jason Caravas, Benjamin Rambo-Martin, Shatavia Morrison, Christopher Gulvick, Scott Sammons, Yvette Unoarumhi, Darlene Wagner, Matthew Schmerer, Cyndi Clark, Patrick Campbell, Rob Case, Vikramsinha Ghorpade, Holly Houdeshell, Ola Kvalvaag, Dillon Nall, Ethan Sanders, Alec Vest, Shaun Westlund, Matthew Hardison, Clinton R. Paden, Duncan MacCannell |
| EPI_ISL_2202368 | Houston Methodist Hospital | Houston Methodist Hospital | Randall J. Olsen, Paul A. Christensen, S. Wesley Long, Sishir Subedi, Robert Olson, Marcus Nguyen, James J. Davis, Matthew Ojeda Saavedra, Prasanti Yerramilli, Layne Pruitt, Kristina Reppond, Madison N. Shyer, Jessica Cambric, Ryan Gadd, Ilya J. Finkelstein, Jimmy Gollihar, and James M. Musser |
| EPI_ISL_2202369 | Aegis Sciences Corporation | Centers for Disease Control and Prevention Division of Viral Diseases, Pathogen Discovery | Dakota Howard, Dhwani Batra, Peter W. Cook, Kara Moser, Adrian Paskey, Jason Caravas, Benjamin Rambo-Martin, Shatavia Morrison, Christopher Gulvick, Scott Sammons, Yvette Unoarumhi, Darlene Wagner, Matthew Schmerer, Cyndi Clark, Patrick Campbell, Rob Case, Vikramsinha Ghorpade, Holly Houdeshell, Ola Kvalvaag, Dillon Nall, Ethan Sanders, Alec Vest, Shaun Westlund, Matthew Hardison, Clinton R. Paden, Duncan MacCannell |
| EPI_ISL_2202370 | Houston Methodist Hospital | Houston Methodist Hospital | Randall J. Olsen, Paul A. Christensen, S. Wesley Long, Sishir Subedi, Robert Olson, Marcus Nguyen, James J. Davis, Matthew Ojeda Saavedra, Prasanti Yerramilli, Layne Pruitt, Kristina Reppond, Madison N. Shyer, Jessica Cambric, Ryan Gadd, Ilya J. Finkelstein, Jimmy Gollihar, and James M. Musser |
| EPI_ISL_2202371 | Aegis Sciences Corporation | Centers for Disease Control and Prevention Division of Viral Diseases, Pathogen Discovery | Dakota Howard, Dhwani Batra, Peter W. Cook, Kara Moser, Adrian Paskey, Jason Caravas, Benjamin Rambo-Martin, Shatavia Morrison, Christopher Gulvick, Scott Sammons, Yvette Unoarumhi, Darlene Wagner, Matthew Schmerer, Cyndi Clark, Patrick Campbell, Rob Case, Vikramsinha Ghorpade, Holly Houdeshell, Ola Kvalvaag, Dillon Nall, Ethan Sanders, Alec Vest, Shaun Westlund, Matthew Hardison, Clinton R. Paden, Duncan MacCannell |
| EPI_ISL_2202372 | Houston Methodist Hospital | Houston Methodist Hospital | Randall J. Olsen, Paul A. Christensen, S. Wesley Long, Sishir Subedi, Robert Olson, Marcus Nguyen, James J. Davis, Matthew Ojeda Saavedra, Prasanti Yerramilli, Layne Pruitt, Kristina Reppond, Madison N. Shyer, Jessica Cambric, Ryan Gadd, Ilya J. Finkelstein, Jimmy Gollihar, and James M. Musser |
| EPI_ISL_2202373 | Aegis Sciences Corporation | Centers for Disease Control and Prevention Division of Viral Diseases, Pathogen Discovery | Dakota Howard, Dhwani Batra, Peter W. Cook, Kara Moser, Adrian Paskey, Jason Caravas, Benjamin Rambo-Martin, Shatavia Morrison, Christopher Gulvick, Scott Sammons, Yvette Unoarumhi, Darlene Wagner, Matthew Schmerer, Cyndi Clark, Patrick Campbell, Rob Case, Vikramsinha Ghorpade, Holly Houdeshell, Ola Kvalvaag, Dillon Nall, Ethan Sanders, Alec Vest, Shaun Westlund, Matthew Hardison, Clinton R. Paden, Duncan MacCannell |
| EPI_ISL_2202374 | Houston Methodist Hospital | Houston Methodist Hospital | Randall J. Olsen, Paul A. Christensen, S. Wesley Long, Sishir Subedi, Robert Olson, Marcus Nguyen, James J. Davis, Matthew Ojeda Saavedra, Prasanti Yerramilli, Layne Pruitt, Kristina Reppond, Madison N. Shyer, Jessica Cambric, Ryan Gadd, Ilya J. Finkelstein, Jimmy Gollihar, and James M. Musser |
| EPI_ISL_2202375, EPI_ISL_2202376, EPI_ISL_2202377 | Aegis Sciences Corporation | Centers for Disease Control and Prevention Division of Viral Diseases, Pathogen Discovery | Dakota Howard, Dhwani Batra, Peter W. Cook, Kara Moser, Adrian Paskey, Jason Caravas, Benjamin Rambo-Martin, Shatavia Morrison, Christopher Gulvick, Scott Sammons, Yvette Unoarumhi, Darlene Wagner, Matthew Schmerer, Cyndi Clark, Patrick Campbell, Rob Case, Vikramsinha Ghorpade, Holly Houdeshell, Ola Kvalvaag, Dillon Nall, Ethan Sanders, Alec Vest, Shaun Westlund, Matthew Hardison, Clinton R. Paden, Duncan MacCannell |
| EPI_ISL_2202378 | Houston Methodist Hospital | Houston Methodist Hospital | Randall J. Olsen, Paul A. Christensen, S. Wesley Long, Sishir Subedi, Robert Olson, Marcus Nguyen, James J. Davis, Matthew Ojeda Saavedra, Prasanti Yerramilli, Layne Pruitt, Kristina Reppond, Madison N. Shyer, Jessica Cambric, Ryan Gadd, Ilya J. Finkelstein, Jimmy Gollihar, and James |

| | | | |
|---|---|---|---|
| | | | M. Musser |
| EPI_ISL_2202379 | Aegis Sciences Corporation | Centers for Disease Control and Prevention Division of Viral Diseases, Pathogen Discovery | Dakota Howard, Dhwani Batra, Peter W. Cook, Kara Moser, Adrian Paskey, Jason Caravas, Benjamin Rambo-Martin, Shatavia Morrison, Christopher Gulvick, Scott Sammons, Yvette Unoarumhi, Darlene Wagner, Matthew Schmerer, Cyndi Clark, Patrick Campbell, Rob Case, Vikramsinha Ghorpade, Holly Houdeshell, Ola Kvalvaag, Dillon Nall, Ethan Sanders, Alec Vest, Shaun Westlund, Matthew Hardison, Clinton R. Paden, Duncan MacCannell |
| EPI_ISL_2202380 | Houston Methodist Hospital | Houston Methodist Hospital | Randall J. Olsen, Paul A. Christensen, S. Wesley Long, Sishir Subedi, Robert Olson, Marcus Nguyen, James J. Davis, Matthew Ojeda Saavedra, Prasanti Yerramilli, Layne Pruitt, Kristina Reppond, Madison N. Shyer, Jessica Cambric, Ryan Gadd, Ilya J. Finkelstein, Jimmy Gollihar, and James M. Musser |
| EPI_ISL_2202381, EPI_ISL_2202382 | Aegis Sciences Corporation | Centers for Disease Control and Prevention Division of Viral Diseases, Pathogen Discovery | Dakota Howard, Dhwani Batra, Peter W. Cook, Kara Moser, Adrian Paskey, Jason Caravas, Benjamin Rambo-Martin, Shatavia Morrison, Christopher Gulvick, Scott Sammons, Yvette Unoarumhi, Darlene Wagner, Matthew Schmerer, Cyndi Clark, Patrick Campbell, Rob Case, Vikramsinha Ghorpade, Holly Houdeshell, Ola Kvalvaag, Dillon Nall, Ethan Sanders, Alec Vest, Shaun Westlund, Matthew Hardison, Clinton R. Paden, Duncan MacCannell |
| EPI_ISL_2202383, EPI_ISL_2202384 | Houston Methodist Hospital | Houston Methodist Hospital | Randall J. Olsen, Paul A. Christensen, S. Wesley Long, Sishir Subedi, Robert Olson, Marcus Nguyen, James J. Davis, Matthew Ojeda Saavedra, Prasanti Yerramilli, Layne Pruitt, Kristina Reppond, Madison N. Shyer, Jessica Cambric, Ryan Gadd, Ilya J. Finkelstein, Jimmy Gollihar, and James M. Musser |
| EPI_ISL_2202385, EPI_ISL_2202388 | Aegis Sciences Corporation | Centers for Disease Control and Prevention Division of Viral Diseases, Pathogen Discovery | Dakota Howard, Dhwani Batra, Peter W. Cook, Kara Moser, Adrian Paskey, Jason Caravas, Benjamin Rambo-Martin, Shatavia Morrison, Christopher Gulvick, Scott Sammons, Yvette Unoarumhi, Darlene Wagner, Matthew Schmerer, Cyndi Clark, Patrick Campbell, Rob Case, Vikramsinha Ghorpade, Holly Houdeshell, Ola Kvalvaag, Dillon Nall, Ethan Sanders, Alec Vest, Shaun Westlund, Matthew Hardison, Clinton R. Paden, Duncan MacCannell |
| EPI_ISL_2202390 | Houston Methodist Hospital | Houston Methodist Hospital | Randall J. Olsen, Paul A. Christensen, S. Wesley Long, Sishir Subedi, Robert Olson, Marcus Nguyen, James J. Davis, Matthew Ojeda Saavedra, Prasanti Yerramilli, Layne Pruitt, Kristina Reppond, Madison N. Shyer, Jessica Cambric, Ryan Gadd, Ilya J. Finkelstein, Jimmy Gollihar, and James M. Musser |
| EPI_ISL_2202392, EPI_ISL_2202395 | Aegis Sciences Corporation | Centers for Disease Control and Prevention Division of Viral Diseases, Pathogen Discovery | Dakota Howard, Dhwani Batra, Peter W. Cook, Kara Moser, Adrian Paskey, Jason Caravas, Benjamin Rambo-Martin, Shatavia Morrison, Christopher Gulvick, Scott Sammons, Yvette Unoarumhi, Darlene Wagner, Matthew Schmerer, Cyndi Clark, Patrick Campbell, Rob Case, Vikramsinha Ghorpade, Holly Houdeshell, Ola Kvalvaag, Dillon Nall, Ethan Sanders, Alec Vest, Shaun Westlund, Matthew Hardison, Clinton R. Paden, Duncan MacCannell |
| EPI_ISL_2202397 | Houston Methodist Hospital | Houston Methodist Hospital | Randall J. Olsen, Paul A. Christensen, S. Wesley Long, Sishir Subedi, Robert Olson, Marcus Nguyen, James J. Davis, Matthew Ojeda Saavedra, Prasanti Yerramilli, Layne Pruitt, Kristina Reppond, Madison N. Shyer, Jessica Cambric, Ryan Gadd, Ilya J. Finkelstein, Jimmy Gollihar, and James M. Musser |
| EPI_ISL_2202399 | Aegis Sciences Corporation | Centers for Disease Control and Prevention Division of Viral Diseases, Pathogen Discovery | Dakota Howard, Dhwani Batra, Peter W. Cook, Kara Moser, Adrian Paskey, Jason Caravas, Benjamin Rambo-Martin, Shatavia Morrison, Christopher Gulvick, Scott Sammons, Yvette Unoarumhi, Darlene Wagner, Matthew Schmerer, Cyndi Clark, Patrick Campbell, Rob Case, Vikramsinha Ghorpade, Holly Houdeshell, Ola Kvalvaag, Dillon Nall, Ethan Sanders, Alec Vest, Shaun Westlund, Matthew Hardison, Clinton R. Paden, Duncan MacCannell |
| EPI_ISL_2202401 | Houston Methodist Hospital | Houston Methodist Hospital | Randall J. Olsen, Paul A. Christensen, S. Wesley Long, Sishir Subedi, Robert Olson, Marcus Nguyen, James J. Davis, Matthew Ojeda Saavedra, Prasanti Yerramilli, Layne Pruitt, Kristina Reppond, Madison N. Shyer, Jessica Cambric, Ryan Gadd, Ilya J. Finkelstein, Jimmy Gollihar, and James M. Musser |
| EPI_ISL_2202403 | Aegis Sciences Corporation | Centers for Disease Control and Prevention Division of Viral Diseases, Pathogen Discovery | Dakota Howard, Dhwani Batra, Peter W. Cook, Kara Moser, Adrian Paskey, Jason Caravas, Benjamin Rambo-Martin, Shatavia Morrison, Christopher Gulvick, Scott Sammons, Yvette Unoarumhi, Darlene Wagner, Matthew Schmerer, Cyndi Clark, Patrick Campbell, Rob Case, Vikramsinha Ghorpade, Holly Houdeshell, Ola Kvalvaag, Dillon Nall, Ethan Sanders, Alec Vest, Shaun Westlund, Matthew Hardison, Clinton R. Paden, Duncan MacCannell |
| EPI_ISL_2202405 | Houston Methodist Hospital | Houston Methodist Hospital | Randall J. Olsen, Paul A. Christensen, S. Wesley Long, Sishir Subedi, Robert Olson, Marcus Nguyen, James J. Davis, Matthew Ojeda Saavedra, Prasanti Yerramilli, Layne Pruitt, Kristina Reppond, Madison N. Shyer, Jessica Cambric, Ryan Gadd, Ilya J. Finkelstein, Jimmy Gollihar, and James M. Musser |
| EPI_ISL_2202407 | Aegis Sciences Corporation | Centers for Disease Control and Prevention Division of Viral Diseases, Pathogen Discovery | Dakota Howard, Dhwani Batra, Peter W. Cook, Kara Moser, Adrian Paskey, Jason Caravas, Benjamin Rambo-Martin, Shatavia Morrison, Christopher Gulvick, Scott Sammons, Yvette Unoarumhi, Darlene Wagner, Matthew Schmerer, Cyndi Clark, Patrick Campbell, Rob Case, Vikramsinha Ghorpade, Holly Houdeshell, Ola Kvalvaag, Dillon Nall, Ethan Sanders, Alec Vest, Shaun Westlund, Matthew Hardison, Clinton R. Paden, Duncan MacCannell |
| EPI_ISL_2202409 | Houston Methodist Hospital | Houston Methodist Hospital | Randall J. Olsen, Paul A. Christensen, S. Wesley Long, Sishir Subedi, Robert Olson, Marcus Nguyen, James J. Davis, Matthew Ojeda Saavedra, Prasanti Yerramilli, Layne Pruitt, Kristina Reppond, Madison N. Shyer, Jessica Cambric, Ryan Gadd, Ilya J. Finkelstein, Jimmy Gollihar, and James M. Musser |
| EPI_ISL_2202410, EPI_ISL_2202413 | Aegis Sciences Corporation | Centers for Disease Control and Prevention Division of Viral Diseases, Pathogen Discovery | Dakota Howard, Dhwani Batra, Peter W. Cook, Kara Moser, Adrian Paskey, Jason Caravas, Benjamin Rambo-Martin, Shatavia Morrison, Christopher Gulvick, Scott Sammons, Yvette Unoarumhi, Darlene Wagner, Matthew Schmerer, Cyndi Clark, Patrick Campbell, Rob Case, Vikramsinha Ghorpade, Holly Houdeshell, Ola Kvalvaag, Dillon Nall, Ethan Sanders, Alec Vest, Shaun Westlund, Matthew Hardison, Clinton R. Paden, Duncan MacCannell |
| EPI_ISL_2202414 | Houston Methodist Hospital | Houston Methodist Hospital | Randall J. Olsen, Paul A. Christensen, S. Wesley Long, Sishir Subedi, Robert Olson, Marcus Nguyen, James J. Davis, Matthew Ojeda Saavedra, Prasanti Yerramilli, Layne Pruitt, Kristina Reppond, Madison N. Shyer, Jessica Cambric, Ryan Gadd, Ilya J. Finkelstein, Jimmy Gollihar, and James M. Musser |
| EPI_ISL_2202417 | Aegis Sciences Corporation | Centers for Disease Control and Prevention Division of Viral Diseases, Pathogen Discovery | Dakota Howard, Dhwani Batra, Peter W. Cook, Kara Moser, Adrian Paskey, Jason Caravas, Benjamin Rambo-Martin, Shatavia Morrison, Christopher Gulvick, Scott Sammons, Yvette Unoarumhi, Darlene Wagner, Matthew Schmerer, Cyndi Clark, Patrick Campbell, Rob Case, Vikramsinha Ghorpade, Holly Houdeshell, Ola Kvalvaag, Dillon Nall, Ethan Sanders, Alec Vest, Shaun Westlund, Matthew Hardison, Clinton R. Paden, Duncan MacCannell |
| EPI_ISL_2202419 | Houston Methodist Hospital | Houston Methodist Hospital | Randall J. Olsen, Paul A. Christensen, S. Wesley Long, Sishir Subedi, Robert Olson, Marcus Nguyen, James J. Davis, Matthew Ojeda Saavedra, Prasanti Yerramilli, Layne Pruitt, Kristina Reppond, Madison N. Shyer, Jessica Cambric, Ryan Gadd, Ilya J. Finkelstein, Jimmy Gollihar, and James M. Musser |
| EPI_ISL_2202421, EPI_ISL_2202424 | Aegis Sciences Corporation | Centers for Disease Control and Prevention Division of Viral Diseases, Pathogen Discovery | Dakota Howard, Dhwani Batra, Peter W. Cook, Kara Moser, Adrian Paskey, Jason Caravas, Benjamin Rambo-Martin, Shatavia Morrison, Christopher Gulvick, Scott Sammons, Yvette Unoarumhi, Darlene Wagner, Matthew Schmerer, Cyndi Clark, Patrick Campbell, Rob Case, Vikramsinha Ghorpade, Holly Houdeshell, Ola Kvalvaag, Dillon Nall, Ethan Sanders, Alec Vest, Shaun Westlund, Matthew Hardison, Clinton R. Paden, Duncan MacCannell |
| EPI_ISL_2202425 | Houston Methodist Hospital | Houston Methodist Hospital | Randall J. Olsen, Paul A. Christensen, S. Wesley Long, Sishir Subedi, Robert Olson, Marcus Nguyen, James J. Davis, Matthew Ojeda Saavedra, Prasanti Yerramilli, Layne Pruitt, Kristina Reppond, Madison N. Shyer, Jessica Cambric, Ryan Gadd, Ilya J. Finkelstein, Jimmy Gollihar, and James M. Musser |
| EPI_ISL_2202428, EPI_ISL_2202430 | Aegis Sciences Corporation | Centers for Disease Control and Prevention Division of Viral Diseases, Pathogen Discovery | Dakota Howard, Dhwani Batra, Peter W. Cook, Kara Moser, Adrian Paskey, Jason Caravas, Benjamin Rambo-Martin, Shatavia Morrison, Christopher Gulvick, Scott Sammons, Yvette Unoarumhi, Darlene Wagner, Matthew Schmerer, Cyndi Clark, Patrick Campbell, Rob Case, Vikramsinha Ghorpade, Holly Houdeshell, Ola Kvalvaag, Dillon Nall, Ethan Sanders, Alec Vest, Shaun Westlund, Matthew Hardison, Clinton R. Paden, Duncan MacCannell |
| EPI_ISL_2202431 | Houston Methodist Hospital | Houston Methodist Hospital | Randall J. Olsen, Paul A. Christensen, S. Wesley Long, Sishir Subedi, Robert Olson, Marcus Nguyen, James J. Davis, Matthew Ojeda Saavedra, Prasanti Yerramilli, Layne Pruitt, Kristina Reppond, Madison N. Shyer, Jessica Cambric, Ryan Gadd, Ilya J. Finkelstein, Jimmy Gollihar, and James M. Musser |
| EPI_ISL_2202433 | Aegis Sciences Corporation | Centers for Disease Control and Prevention Division of Viral Diseases, Pathogen Discovery | Dakota Howard, Dhwani Batra, Peter W. Cook, Kara Moser, Adrian Paskey, Jason Caravas, Benjamin Rambo-Martin, Shatavia Morrison, Christopher Gulvick, Scott Sammons, Yvette Unoarumhi, Darlene Wagner, Matthew Schmerer, Cyndi Clark, Patrick Campbell, Rob Case, Vikramsinha Ghorpade, Holly Houdeshell, Ola Kvalvaag, Dillon Nall, Ethan Sanders, Alec Vest, Shaun Westlund, Matthew Hardison, Clinton R. Paden, Duncan MacCannell |
| EPI_ISL_2202436 | Houston Methodist Hospital | Houston Methodist Hospital | Randall J. Olsen, Paul A. Christensen, S. Wesley Long, Sishir Subedi, Robert Olson, Marcus Nguyen, James J. Davis, Matthew Ojeda Saavedra, Prasanti Yerramilli, Layne Pruitt, Kristina Reppond, Madison N. Shyer, Jessica Cambric, Ryan Gadd, Ilya J. Finkelstein, Jimmy Gollihar, and James M. Musser |
| EPI_ISL_2202438, EPI_ISL_2202441 | Aegis Sciences Corporation | Centers for Disease Control and Prevention Division of Viral Diseases, Pathogen Discovery | Dakota Howard, Dhwani Batra, Peter W. Cook, Kara Moser, Adrian Paskey, Jason Caravas, Benjamin Rambo-Martin, Shatavia Morrison, Christopher Gulvick, Scott Sammons, Yvette Unoarumhi, Darlene Wagner, Matthew Schmerer, Cyndi Clark, Patrick Campbell, Rob Case, Vikramsinha Ghorpade, Holly Houdeshell, Ola Kvalvaag, Dillon Nall, Ethan Sanders, Alec Vest, Shaun Westlund, Matthew Hardison, Clinton R. Paden, Duncan MacCannell |
| EPI_ISL_2202442 | Houston Methodist Hospital | Houston Methodist Hospital | Randall J. Olsen, Paul A. Christensen, S. Wesley Long, Sishir Subedi, Robert Olson, Marcus Nguyen, James J. Davis, Matthew Ojeda Saavedra, Prasanti Yerramilli, Layne Pruitt, Kristina Reppond, Madison N. Shyer, Jessica Cambric, Ryan Gadd, Ilya J. Finkelstein, Jimmy Gollihar, and James M. Musser |
| EPI_ISL_2202444 | Aegis Sciences Corporation | Centers for Disease Control and Prevention Division of Viral Diseases, Pathogen Discovery | Dakota Howard, Dhwani Batra, Peter W. Cook, Kara Moser, Adrian Paskey, Jason Caravas, Benjamin Rambo-Martin, Shatavia Morrison, Christopher Gulvick, Scott Sammons, Yvette Unoarumhi, Darlene Wagner, Matthew Schmerer, Cyndi Clark, Patrick Campbell, Rob Case, Vikramsinha Ghorpade, Holly Houdeshell, Ola Kvalvaag, Dillon Nall, Ethan Sanders, Alec Vest, Shaun Westlund, Matthew Hardison, Clinton R. Paden, Duncan MacCannell |
| EPI_ISL_2202446 | Houston Methodist Hospital | Houston Methodist Hospital | Randall J. Olsen, Paul A. Christensen, S. Wesley Long, Sishir Subedi, Robert Olson, Marcus Nguyen, James J. Davis, Matthew Ojeda Saavedra, Prasanti Yerramilli, Layne Pruitt, Kristina Reppond, Madison N. Shyer, Jessica Cambric, Ryan Gadd, Ilya J. Finkelstein, Jimmy Gollihar, and James M. Musser |
| EPI_ISL_2202448 | Aegis Sciences Corporation | Centers for Disease Control and Prevention Division of Viral Diseases, Pathogen Discovery | Dakota Howard, Dhwani Batra, Peter W. Cook, Kara Moser, Adrian Paskey, Jason Caravas, Benjamin Rambo-Martin, Shatavia Morrison, Christopher Gulvick, Scott Sammons, Yvette Unoarumhi, Darlene Wagner, Matthew Schmerer, Cyndi Clark, Patrick Campbell, Rob Case, Vikramsinha Ghorpade, Holly Houdeshell, Ola Kvalvaag, Dillon Nall, Ethan Sanders, Alec Vest, Shaun Westlund, Matthew Hardison, Clinton R. Paden, Duncan MacCannell |
| EPI_ISL_2202451 | Houston Methodist Hospital | Houston Methodist Hospital | Randall J. Olsen, Paul A. Christensen, S. Wesley Long, Sishir Subedi, Robert Olson, Marcus Nguyen, James J. Davis, Matthew Ojeda Saavedra, Prasanti Yerramilli, Layne Pruitt, Kristina Reppond, Madison N. Shyer, Jessica Cambric, Ryan Gadd, Ilya J. Finkelstein, Jimmy Gollihar, and James M. Musser |
| EPI_ISL_2202452, EPI_ISL_2202455 | Aegis Sciences Corporation | Centers for Disease Control and Prevention Division of Viral Diseases, Pathogen Discovery | Dakota Howard, Dhwani Batra, Peter W. Cook, Kara Moser, Adrian Paskey, Jason Caravas, Benjamin Rambo-Martin, Shatavia Morrison, Christopher Gulvick, Scott Sammons, Yvette Unoarumhi, Darlene Wagner, Matthew Schmerer, Cyndi Clark, Patrick Campbell, Rob Case, Vikramsinha Ghorpade, Holly Houdeshell, Ola Kvalvaag, Dillon Nall, Ethan Sanders, Alec Vest, Shaun Westlund, Matthew Hardison, Clinton R. Paden, Duncan MacCannell |
| EPI_ISL_2202456 | Houston Methodist Hospital | Houston Methodist Hospital | Randall J. Olsen, Paul A. Christensen, S. Wesley Long, Sishir Subedi, Robert Olson, Marcus Nguyen, James J. Davis, Matthew Ojeda Saavedra, Prasanti Yerramilli, Layne Pruitt, Kristina Reppond, Madison N. Shyer, Jessica Cambric, Ryan Gadd, Ilya J. Finkelstein, Jimmy Gollihar, and James M. Musser |
| EPI_ISL_2202459 | Aegis Sciences Corporation | Centers for Disease Control and Prevention Division of Viral Diseases, Pathogen Discovery | Dakota Howard, Dhwani Batra, Peter W. Cook, Kara Moser, Adrian Paskey, Jason Caravas, Benjamin Rambo-Martin, Shatavia Morrison, Christopher Gulvick, Scott Sammons, Yvette Unoarumhi, Darlene Wagner, Matthew Schmerer, Cyndi Clark, Patrick Campbell, Rob Case, Vikramsinha Ghorpade, Holly Houdeshell, Ola Kvalvaag, Dillon Nall, Ethan Sanders, Alec Vest, Shaun Westlund, Matthew Hardison, Clinton R. Paden, Duncan MacCannell |
| EPI_ISL_2202461 | Houston Methodist Hospital | Houston Methodist Hospital | Randall J. Olsen, Paul A. Christensen, S. Wesley Long, Sishir Subedi, Robert Olson, Marcus Nguyen, James J. Davis, Matthew Ojeda Saavedra, Prasanti Yerramilli, Layne Pruitt, Kristina Reppond, Madison N. Shyer, Jessica Cambric, Ryan Gadd, Ilya J. Finkelstein, Jimmy Gollihar, and James M. Musser |
| EPI_ISL_2202463 | Aegis Sciences Corporation | Centers for Disease Control and Prevention Division of Viral Diseases, Pathogen Discovery | Dakota Howard, Dhwani Batra, Peter W. Cook, Kara Moser, Adrian Paskey, Jason Caravas, Benjamin Rambo-Martin, Shatavia Morrison, Christopher Gulvick, Scott Sammons, Yvette Unoarumhi, Darlene Wagner, Matthew Schmerer, Cyndi Clark, Patrick Campbell, Rob Case, Vikramsinha Ghorpade, Holly Houdeshell, Ola Kvalvaag, Dillon Nall, Ethan Sanders, Alec Vest, Shaun Westlund, Matthew Hardison, Clinton R. Paden, Duncan MacCannell |
| EPI_ISL_2202465 | Houston Methodist Hospital | Houston Methodist Hospital | Randall J. Olsen, Paul A. Christensen, S. Wesley Long, Sishir Subedi, Robert Olson, Marcus Nguyen, James J. Davis, Matthew Ojeda Saavedra, Prasanti Yerramilli, Layne Pruitt, Kristina Reppond, Madison N. Shyer, Jessica Cambric, Ryan Gadd, Ilya J. Finkelstein, Jimmy Gollihar, and James M. Musser |
| EPI_ISL_2202467, EPI_ISL_2202469 | Aegis Sciences Corporation | Centers for Disease Control and Prevention Division of Viral Diseases, Pathogen | Dakota Howard, Dhwani Batra, Peter W. Cook, Kara Moser, Adrian Paskey, Jason Caravas, Benjamin Rambo-Martin, Shatavia Morrison, Christopher Gulvick, Scott Sammons, Yvette Unoarumhi, Darlene Wagner, Matthew Schmerer, Cyndi Clark, Patrick Campbell, Rob Case, Vikramsinha Ghorpade, Holly Houdeshell, Ola Kvalvaag, Dillon Nall, Ethan Sanders, Alec Vest, Shaun Westlund, Matthew Hardison, Clinton R. Paden, Duncan MacCannell |

| | | | |
|---|---|---|---|
| | | Discovery | |
| EPI_ISL_2202471 | Houston Methodist Hospital | Houston Methodist Hospital | Randall J. Olsen, Paul A. Christensen, S. Wesley Long, Sishir Subedi, Robert Olson, Marcus Nguyen, James J. Davis, Matthew Ojeda Saavedra, Prasanti Yerramilli, Layne Pruitt, Kristina Reppond, Madison N. Shyer, Jessica Cambric, Ryan Gadd, Ilya J. Finkelstein, Jimmy Gollihar, and James M. Musser |
| EPI_ISL_2202473 | Aegis Sciences Corporation | Centers for Disease Control and Prevention Division of Viral Diseases, Pathogen Discovery | Dakota Howard, Dhwani Batra, Peter W. Cook, Kara Moser, Adrian Paskey, Jason Caravas, Benjamin Rambo-Martin, Shatavia Morrison, Christopher Gulvick, Scott Sammons, Yvette Unoarumhi, Darlene Wagner, Matthew Schmerer, Cyndi Clark, Patrick Campbell, Rob Case, Vikramsinha Ghorpade, Holly Houdeshell, Ola Kvalvaag, Dillon Nall, Ethan Sanders, Alec Vest, Shaun Westlund, Matthew Hardison, Clinton R. Paden, Duncan MacCannell |
| EPI_ISL_2202475 | Houston Methodist Hospital | Houston Methodist Hospital | Randall J. Olsen, Paul A. Christensen, S. Wesley Long, Sishir Subedi, Robert Olson, Marcus Nguyen, James J. Davis, Matthew Ojeda Saavedra, Prasanti Yerramilli, Layne Pruitt, Kristina Reppond, Madison N. Shyer, Jessica Cambric, Ryan Gadd, Ilya J. Finkelstein, Jimmy Gollihar, and James M. Musser |
| EPI_ISL_2202477 | Aegis Sciences Corporation | Centers for Disease Control and Prevention Division of Viral Diseases, Pathogen Discovery | Dakota Howard, Dhwani Batra, Peter W. Cook, Kara Moser, Adrian Paskey, Jason Caravas, Benjamin Rambo-Martin, Shatavia Morrison, Christopher Gulvick, Scott Sammons, Yvette Unoarumhi, Darlene Wagner, Matthew Schmerer, Cyndi Clark, Patrick Campbell, Rob Case, Vikramsinha Ghorpade, Holly Houdeshell, Ola Kvalvaag, Dillon Nall, Ethan Sanders, Alec Vest, Shaun Westlund, Matthew Hardison, Clinton R. Paden, Duncan MacCannell |
| EPI_ISL_2202479 | Houston Methodist Hospital | Houston Methodist Hospital | Randall J. Olsen, Paul A. Christensen, S. Wesley Long, Sishir Subedi, Robert Olson, Marcus Nguyen, James J. Davis, Matthew Ojeda Saavedra, Prasanti Yerramilli, Layne Pruitt, Kristina Reppond, Madison N. Shyer, Jessica Cambric, Ryan Gadd, Ilya J. Finkelstein, Jimmy Gollihar, and James M. Musser |
| EPI_ISL_2202481, EPI_ISL_2202484 | Aegis Sciences Corporation | Centers for Disease Control and Prevention Division of Viral Diseases, Pathogen Discovery | Dakota Howard, Dhwani Batra, Peter W. Cook, Kara Moser, Adrian Paskey, Jason Caravas, Benjamin Rambo-Martin, Shatavia Morrison, Christopher Gulvick, Scott Sammons, Yvette Unoarumhi, Darlene Wagner, Matthew Schmerer, Cyndi Clark, Patrick Campbell, Rob Case, Vikramsinha Ghorpade, Holly Houdeshell, Ola Kvalvaag, Dillon Nall, Ethan Sanders, Alec Vest, Shaun Westlund, Matthew Hardison, Clinton R. Paden, Duncan MacCannell |
| EPI_ISL_2202485 | Houston Methodist Hospital | Houston Methodist Hospital | Randall J. Olsen, Paul A. Christensen, S. Wesley Long, Sishir Subedi, Robert Olson, Marcus Nguyen, James J. Davis, Matthew Ojeda Saavedra, Prasanti Yerramilli, Layne Pruitt, Kristina Reppond, Madison N. Shyer, Jessica Cambric, Ryan Gadd, Ilya J. Finkelstein, Jimmy Gollihar, and James M. Musser |
| EPI_ISL_2202488 | Aegis Sciences Corporation | Centers for Disease Control and Prevention Division of Viral Diseases, Pathogen Discovery | Dakota Howard, Dhwani Batra, Peter W. Cook, Kara Moser, Adrian Paskey, Jason Caravas, Benjamin Rambo-Martin, Shatavia Morrison, Christopher Gulvick, Scott Sammons, Yvette Unoarumhi, Darlene Wagner, Matthew Schmerer, Cyndi Clark, Patrick Campbell, Rob Case, Vikramsinha Ghorpade, Holly Houdeshell, Ola Kvalvaag, Dillon Nall, Ethan Sanders, Alec Vest, Shaun Westlund, Matthew Hardison, Clinton R. Paden, Duncan MacCannell |
| EPI_ISL_2202490 | Houston Methodist Hospital | Houston Methodist Hospital | Randall J. Olsen, Paul A. Christensen, S. Wesley Long, Sishir Subedi, Robert Olson, Marcus Nguyen, James J. Davis, Matthew Ojeda Saavedra, Prasanti Yerramilli, Layne Pruitt, Kristina Reppond, Madison N. Shyer, Jessica Cambric, Ryan Gadd, Ilya J. Finkelstein, Jimmy Gollihar, and James M. Musser |
| EPI_ISL_2202492, EPI_ISL_2202493, EPI_ISL_2202495 | Aegis Sciences Corporation | Centers for Disease Control and Prevention Division of Viral Diseases, Pathogen Discovery | Dakota Howard, Dhwani Batra, Peter W. Cook, Kara Moser, Adrian Paskey, Jason Caravas, Benjamin Rambo-Martin, Shatavia Morrison, Christopher Gulvick, Scott Sammons, Yvette Unoarumhi, Darlene Wagner, Matthew Schmerer, Cyndi Clark, Patrick Campbell, Rob Case, Vikramsinha Ghorpade, Holly Houdeshell, Ola Kvalvaag, Dillon Nall, Ethan Sanders, Alec Vest, Shaun Westlund, Matthew Hardison, Clinton R. Paden, Duncan MacCannell |
| EPI_ISL_2202497 | Houston Methodist Hospital | Houston Methodist Hospital | Randall J. Olsen, Paul A. Christensen, S. Wesley Long, Sishir Subedi, Robert Olson, Marcus Nguyen, James J. Davis, Matthew Ojeda Saavedra, Prasanti Yerramilli, Layne Pruitt, Kristina Reppond, Madison N. Shyer, Jessica Cambric, Ryan Gadd, Ilya J. Finkelstein, Jimmy Gollihar, and James M. Musser |
| EPI_ISL_2202499 | Aegis Sciences Corporation | Centers for Disease Control and Prevention Division of Viral Diseases, Pathogen Discovery | Dakota Howard, Dhwani Batra, Peter W. Cook, Kara Moser, Adrian Paskey, Jason Caravas, Benjamin Rambo-Martin, Shatavia Morrison, Christopher Gulvick, Scott Sammons, Yvette Unoarumhi, Darlene Wagner, Matthew Schmerer, Cyndi Clark, Patrick Campbell, Rob Case, Vikramsinha Ghorpade, Holly Houdeshell, Ola Kvalvaag, Dillon Nall, Ethan Sanders, Alec Vest, Shaun Westlund, Matthew Hardison, Clinton R. Paden, Duncan MacCannell |
| EPI_ISL_2202501 | Houston Methodist Hospital | Houston Methodist Hospital | Randall J. Olsen, Paul A. Christensen, S. Wesley Long, Sishir Subedi, Robert Olson, Marcus Nguyen, James J. Davis, Matthew Ojeda Saavedra, Prasanti Yerramilli, Layne Pruitt, Kristina Reppond, Madison N. Shyer, Jessica Cambric, Ryan Gadd, Ilya J. Finkelstein, Jimmy Gollihar, and James M. Musser |
| EPI_ISL_2202502 | Aegis Sciences Corporation | Centers for Disease Control and Prevention Division of Viral Diseases, Pathogen Discovery | Dakota Howard, Dhwani Batra, Peter W. Cook, Kara Moser, Adrian Paskey, Jason Caravas, Benjamin Rambo-Martin, Shatavia Morrison, Christopher Gulvick, Scott Sammons, Yvette Unoarumhi, Darlene Wagner, Matthew Schmerer, Cyndi Clark, Patrick Campbell, Rob Case, Vikramsinha Ghorpade, Holly Houdeshell, Ola Kvalvaag, Dillon Nall, Ethan Sanders, Alec Vest, Shaun Westlund, Matthew Hardison, Clinton R. Paden, Duncan MacCannell |
| EPI_ISL_2202505 | Houston Methodist Hospital | Houston Methodist Hospital | Randall J. Olsen, Paul A. Christensen, S. Wesley Long, Sishir Subedi, Robert Olson, Marcus Nguyen, James J. Davis, Matthew Ojeda Saavedra, Prasanti Yerramilli, Layne Pruitt, Kristina Reppond, Madison N. Shyer, Jessica Cambric, Ryan Gadd, Ilya J. Finkelstein, Jimmy Gollihar, and James M. Musser |
| EPI_ISL_2202506 | Aegis Sciences Corporation | Centers for Disease Control and Prevention Division of Viral Diseases, Pathogen Discovery | Dakota Howard, Dhwani Batra, Peter W. Cook, Kara Moser, Adrian Paskey, Jason Caravas, Benjamin Rambo-Martin, Shatavia Morrison, Christopher Gulvick, Scott Sammons, Yvette Unoarumhi, Darlene Wagner, Matthew Schmerer, Cyndi Clark, Patrick Campbell, Rob Case, Vikramsinha Ghorpade, Holly Houdeshell, Ola Kvalvaag, Dillon Nall, Ethan Sanders, Alec Vest, Shaun Westlund, Matthew Hardison, Clinton R. Paden, Duncan MacCannell |
| EPI_ISL_2202509 | Houston Methodist Hospital | Houston Methodist Hospital | Randall J. Olsen, Paul A. Christensen, S. Wesley Long, Sishir Subedi, Robert Olson, Marcus Nguyen, James J. Davis, Matthew Ojeda Saavedra, Prasanti Yerramilli, Layne Pruitt, Kristina Reppond, Madison N. Shyer, Jessica Cambric, Ryan Gadd, Ilya J. Finkelstein, Jimmy Gollihar, and James M. Musser |
| EPI_ISL_2202510, EPI_ISL_2202513 | Aegis Sciences Corporation | Centers for Disease Control and Prevention Division of Viral Diseases, Pathogen Discovery | Dakota Howard, Dhwani Batra, Peter W. Cook, Kara Moser, Adrian Paskey, Jason Caravas, Benjamin Rambo-Martin, Shatavia Morrison, Christopher Gulvick, Scott Sammons, Yvette Unoarumhi, Darlene Wagner, Matthew Schmerer, Cyndi Clark, Patrick Campbell, Rob Case, Vikramsinha Ghorpade, Holly Houdeshell, Ola Kvalvaag, Dillon Nall, Ethan Sanders, Alec Vest, Shaun Westlund, Matthew Hardison, Clinton R. Paden, Duncan MacCannell |
| EPI_ISL_2202515, EPI_ISL_2202517 | Houston Methodist Hospital | Houston Methodist Hospital | Randall J. Olsen, Paul A. Christensen, S. Wesley Long, Sishir Subedi, Robert Olson, Marcus Nguyen, James J. Davis, Matthew Ojeda Saavedra, Prasanti Yerramilli, Layne Pruitt, Kristina Reppond, Madison N. Shyer, Jessica Cambric, Ryan Gadd, Ilya J. Finkelstein, Jimmy Gollihar, and James M. Musser |
| EPI_ISL_2202518, EPI_ISL_2202522 | Aegis Sciences Corporation | Centers for Disease Control and Prevention Division of Viral Diseases, Pathogen Discovery | Dakota Howard, Dhwani Batra, Peter W. Cook, Kara Moser, Adrian Paskey, Jason Caravas, Benjamin Rambo-Martin, Shatavia Morrison, Christopher Gulvick, Scott Sammons, Yvette Unoarumhi, Darlene Wagner, Matthew Schmerer, Cyndi Clark, Patrick Campbell, Rob Case, Vikramsinha Ghorpade, Holly Houdeshell, Ola Kvalvaag, Dillon Nall, Ethan Sanders, Alec Vest, Shaun Westlund, Matthew Hardison, Clinton R. Paden, Duncan MacCannell |
| EPI_ISL_2202523 | Houston Methodist Hospital | Houston Methodist Hospital | Randall J. Olsen, Paul A. Christensen, S. Wesley Long, Sishir Subedi, Robert Olson, Marcus Nguyen, James J. Davis, Matthew Ojeda Saavedra, Prasanti Yerramilli, Layne Pruitt, Kristina Reppond, Madison N. Shyer, Jessica Cambric, Ryan Gadd, Ilya J. Finkelstein, Jimmy Gollihar, and James M. Musser |
| EPI_ISL_2202525 | Aegis Sciences Corporation | Centers for Disease Control and Prevention Division of Viral Diseases, Pathogen Discovery | Dakota Howard, Dhwani Batra, Peter W. Cook, Kara Moser, Adrian Paskey, Jason Caravas, Benjamin Rambo-Martin, Shatavia Morrison, Christopher Gulvick, Scott Sammons, Yvette Unoarumhi, Darlene Wagner, Matthew Schmerer, Cyndi Clark, Patrick Campbell, Rob Case, Vikramsinha Ghorpade, Holly Houdeshell, Ola Kvalvaag, Dillon Nall, Ethan Sanders, Alec Vest, Shaun Westlund, Matthew Hardison, Clinton R. Paden, Duncan MacCannell |
| EPI_ISL_2202528 | Houston Methodist Hospital | Houston Methodist Hospital | Randall J. Olsen, Paul A. Christensen, S. Wesley Long, Sishir Subedi, Robert Olson, Marcus Nguyen, James J. Davis, Matthew Ojeda Saavedra, Prasanti Yerramilli, Layne Pruitt, Kristina Reppond, Madison N. Shyer, Jessica Cambric, Ryan Gadd, Ilya J. Finkelstein, Jimmy Gollihar, and James M. Musser |
| EPI_ISL_2202529, EPI_ISL_2202532 | Aegis Sciences Corporation | Centers for Disease Control and Prevention Division of Viral Diseases, Pathogen Discovery | Dakota Howard, Dhwani Batra, Peter W. Cook, Kara Moser, Adrian Paskey, Jason Caravas, Benjamin Rambo-Martin, Shatavia Morrison, Christopher Gulvick, Scott Sammons, Yvette Unoarumhi, Darlene Wagner, Matthew Schmerer, Cyndi Clark, Patrick Campbell, Rob Case, Vikramsinha Ghorpade, Holly Houdeshell, Ola Kvalvaag, Dillon Nall, Ethan Sanders, Alec Vest, Shaun Westlund, Matthew Hardison, Clinton R. Paden, Duncan MacCannell |
| EPI_ISL_2202533 | Houston Methodist Hospital | Houston Methodist Hospital | Randall J. Olsen, Paul A. Christensen, S. Wesley Long, Sishir Subedi, Robert Olson, Marcus Nguyen, James J. Davis, Matthew Ojeda Saavedra, Prasanti Yerramilli, Layne Pruitt, Kristina Reppond, Madison N. Shyer, Jessica Cambric, Ryan Gadd, Ilya J. Finkelstein, Jimmy Gollihar, and James M. Musser |
| EPI_ISL_2202535 | Aegis Sciences Corporation | Centers for Disease Control and Prevention Division of Viral Diseases, Pathogen Discovery | Dakota Howard, Dhwani Batra, Peter W. Cook, Kara Moser, Adrian Paskey, Jason Caravas, Benjamin Rambo-Martin, Shatavia Morrison, Christopher Gulvick, Scott Sammons, Yvette Unoarumhi, Darlene Wagner, Matthew Schmerer, Cyndi Clark, Patrick Campbell, Rob Case, Vikramsinha Ghorpade, Holly Houdeshell, Ola Kvalvaag, Dillon Nall, Ethan Sanders, Alec Vest, Shaun Westlund, Matthew Hardison, Clinton R. Paden, Duncan MacCannell |
| EPI_ISL_2202537 | Houston Methodist Hospital | Houston Methodist Hospital | Randall J. Olsen, Paul A. Christensen, S. Wesley Long, Sishir Subedi, Robert Olson, Marcus Nguyen, James J. Davis, Matthew Ojeda Saavedra, Prasanti Yerramilli, Layne Pruitt, Kristina Reppond, Madison N. Shyer, Jessica Cambric, Ryan Gadd, Ilya J. Finkelstein, Jimmy Gollihar, and James M. Musser |
| EPI_ISL_2202540 | Aegis Sciences Corporation | Centers for Disease Control and Prevention Division of Viral Diseases, Pathogen Discovery | Dakota Howard, Dhwani Batra, Peter W. Cook, Kara Moser, Adrian Paskey, Jason Caravas, Benjamin Rambo-Martin, Shatavia Morrison, Christopher Gulvick, Scott Sammons, Yvette Unoarumhi, Darlene Wagner, Matthew Schmerer, Cyndi Clark, Patrick Campbell, Rob Case, Vikramsinha Ghorpade, Holly Houdeshell, Ola Kvalvaag, Dillon Nall, Ethan Sanders, Alec Vest, Shaun Westlund, Matthew Hardison, Clinton R. Paden, Duncan MacCannell |
| EPI_ISL_2202542 | Houston Methodist Hospital | Houston Methodist Hospital | Randall J. Olsen, Paul A. Christensen, S. Wesley Long, Sishir Subedi, Robert Olson, Marcus Nguyen, James J. Davis, Matthew Ojeda Saavedra, Prasanti Yerramilli, Layne Pruitt, Kristina Reppond, Madison N. Shyer, Jessica Cambric, Ryan Gadd, Ilya J. Finkelstein, Jimmy Gollihar, and James M. Musser |
| EPI_ISL_2202544 | Aegis Sciences Corporation | Centers for Disease Control and Prevention Division of Viral Diseases, Pathogen Discovery | Dakota Howard, Dhwani Batra, Peter W. Cook, Kara Moser, Adrian Paskey, Jason Caravas, Benjamin Rambo-Martin, Shatavia Morrison, Christopher Gulvick, Scott Sammons, Yvette Unoarumhi, Darlene Wagner, Matthew Schmerer, Cyndi Clark, Patrick Campbell, Rob Case, Vikramsinha Ghorpade, Holly Houdeshell, Ola Kvalvaag, Dillon Nall, Ethan Sanders, Alec Vest, Shaun Westlund, Matthew Hardison, Clinton R. Paden, Duncan MacCannell |
| EPI_ISL_2202546 | Houston Methodist Hospital | Houston Methodist Hospital | Randall J. Olsen, Paul A. Christensen, S. Wesley Long, Sishir Subedi, Robert Olson, Marcus Nguyen, James J. Davis, Matthew Ojeda Saavedra, Prasanti Yerramilli, Layne Pruitt, Kristina Reppond, Madison N. Shyer, Jessica Cambric, Ryan Gadd, Ilya J. Finkelstein, Jimmy Gollihar, and James M. Musser |
| EPI_ISL_2202547, EPI_ISL_2202550 | Aegis Sciences Corporation | Centers for Disease Control and Prevention Division of Viral Diseases, Pathogen Discovery | Dakota Howard, Dhwani Batra, Peter W. Cook, Kara Moser, Adrian Paskey, Jason Caravas, Benjamin Rambo-Martin, Shatavia Morrison, Christopher Gulvick, Scott Sammons, Yvette Unoarumhi, Darlene Wagner, Matthew Schmerer, Cyndi Clark, Patrick Campbell, Rob Case, Vikramsinha Ghorpade, Holly Houdeshell, Ola Kvalvaag, Dillon Nall, Ethan Sanders, Alec Vest, Shaun Westlund, Matthew Hardison, Clinton R. Paden, Duncan MacCannell |
| EPI_ISL_2202552 | Houston Methodist Hospital | Houston Methodist Hospital | Randall J. Olsen, Paul A. Christensen, S. Wesley Long, Sishir Subedi, Robert Olson, Marcus Nguyen, James J. Davis, Matthew Ojeda Saavedra, Prasanti Yerramilli, Layne Pruitt, Kristina Reppond, Madison N. Shyer, Jessica Cambric, Ryan Gadd, Ilya J. Finkelstein, Jimmy Gollihar, and James M. Musser |
| EPI_ISL_2202554 | Aegis Sciences Corporation | Centers for Disease Control and Prevention Division of Viral Diseases, Pathogen Discovery | Dakota Howard, Dhwani Batra, Peter W. Cook, Kara Moser, Adrian Paskey, Jason Caravas, Benjamin Rambo-Martin, Shatavia Morrison, Christopher Gulvick, Scott Sammons, Yvette Unoarumhi, Darlene Wagner, Matthew Schmerer, Cyndi Clark, Patrick Campbell, Rob Case, Vikramsinha Ghorpade, Holly Houdeshell, Ola Kvalvaag, Dillon Nall, Ethan Sanders, Alec Vest, Shaun Westlund, Matthew Hardison, Clinton R. Paden, Duncan MacCannell |
| EPI_ISL_2202557 | Houston Methodist Hospital | Houston Methodist Hospital | Randall J. Olsen, Paul A. Christensen, S. Wesley Long, Sishir Subedi, Robert Olson, Marcus Nguyen, James J. Davis, Matthew Ojeda Saavedra, Prasanti Yerramilli, Layne Pruitt, Kristina Reppond, Madison N. Shyer, Jessica Cambric, Ryan Gadd, Ilya J. Finkelstein, Jimmy Gollihar, and James M. Musser |
| EPI_ISL_2202558, EPI_ISL_2202561 | Aegis Sciences Corporation | Centers for Disease Control and Prevention Division of Viral Diseases, Pathogen Discovery | Dakota Howard, Dhwani Batra, Peter W. Cook, Kara Moser, Adrian Paskey, Jason Caravas, Benjamin Rambo-Martin, Shatavia Morrison, Christopher Gulvick, Scott Sammons, Yvette Unoarumhi, Darlene Wagner, Matthew Schmerer, Cyndi Clark, Patrick Campbell, Rob Case, Vikramsinha Ghorpade, Holly Houdeshell, Ola Kvalvaag, Dillon Nall, Ethan Sanders, Alec Vest, Shaun Westlund, Matthew Hardison, Clinton R. Paden, Duncan MacCannell |
| EPI_ISL_2202562 | Houston Methodist Hospital | Houston Methodist Hospital | Randall J. Olsen, Paul A. Christensen, S. Wesley Long, Sishir Subedi, Robert Olson, Marcus Nguyen, James J. Davis, Matthew Ojeda Saavedra, Prasanti Yerramilli, Layne Pruitt, Kristina Reppond, Madison N. Shyer, Jessica Cambric, Ryan Gadd, Ilya J. Finkelstein, Jimmy Gollihar, and James M. Musser |

| | | | |
|---|---|---|---|
| EPI_ISL_2202565 | Aegis Sciences Corporation | Centers for Disease Control and Prevention Division of Viral Diseases, Pathogen Discovery | Dakota Howard, Dhwani Batra, Peter W. Cook, Kara Moser, Adrian Paskey, Jason Caravas, Benjamin Rambo-Martin, Shatavia Morrison, Christopher Gulvick, Scott Sammons, Yvette Unoarumhi, Darlene Wagner, Matthew Schmerer, Cyndi Clark, Patrick Campbell, Rob Case, Vikramsinha Ghorpade, Holly Houdeshell, Ola Kvalvaag, Dillon Nall, Ethan Sanders, Alec Vest, Shaun Westlund, Matthew Hardison, Clinton R. Paden, Duncan MacCannell |
| EPI_ISL_2202567 | Houston Methodist Hospital | Houston Methodist Hospital | Randall J. Olsen, Paul A. Christensen, S. Wesley Long, Sishir Subedi, Robert Olson, Marcus Nguyen, James J. Davis, Matthew Ojeda Saavedra, Prasanti Yerramilli, Layne Pruitt, Kristina Reppond, Madison N. Shyer, Jessica Cambric, Ryan Gadd, Ilya J. Finkelstein, Jimmy Gollihar, and James M. Musser |
| EPI_ISL_2202569 | Aegis Sciences Corporation | Centers for Disease Control and Prevention Division of Viral Diseases, Pathogen Discovery | Dakota Howard, Dhwani Batra, Peter W. Cook, Kara Moser, Adrian Paskey, Jason Caravas, Benjamin Rambo-Martin, Shatavia Morrison, Christopher Gulvick, Scott Sammons, Yvette Unoarumhi, Darlene Wagner, Matthew Schmerer, Cyndi Clark, Patrick Campbell, Rob Case, Vikramsinha Ghorpade, Holly Houdeshell, Ola Kvalvaag, Dillon Nall, Ethan Sanders, Alec Vest, Shaun Westlund, Matthew Hardison, Clinton R. Paden, Duncan MacCannell |
| EPI_ISL_2202571 | Houston Methodist Hospital | Houston Methodist Hospital | Randall J. Olsen, Paul A. Christensen, S. Wesley Long, Sishir Subedi, Robert Olson, Marcus Nguyen, James J. Davis, Matthew Ojeda Saavedra, Prasanti Yerramilli, Layne Pruitt, Kristina Reppond, Madison N. Shyer, Jessica Cambric, Ryan Gadd, Ilya J. Finkelstein, Jimmy Gollihar, and James M. Musser |
| EPI_ISL_2202572, EPI_ISL_2202573 | Aegis Sciences Corporation | Centers for Disease Control and Prevention Division of Viral Diseases, Pathogen Discovery | Dakota Howard, Dhwani Batra, Peter W. Cook, Kara Moser, Adrian Paskey, Jason Caravas, Benjamin Rambo-Martin, Shatavia Morrison, Christopher Gulvick, Scott Sammons, Yvette Unoarumhi, Darlene Wagner, Matthew Schmerer, Cyndi Clark, Patrick Campbell, Rob Case, Vikramsinha Ghorpade, Holly Houdeshell, Ola Kvalvaag, Dillon Nall, Ethan Sanders, Alec Vest, Shaun Westlund, Matthew Hardison, Clinton R. Paden, Duncan MacCannell |
| EPI_ISL_2202574 | Houston Methodist Hospital | Houston Methodist Hospital | Randall J. Olsen, Paul A. Christensen, S. Wesley Long, Sishir Subedi, Robert Olson, Marcus Nguyen, James J. Davis, Matthew Ojeda Saavedra, Prasanti Yerramilli, Layne Pruitt, Kristina Reppond, Madison N. Shyer, Jessica Cambric, Ryan Gadd, Ilya J. Finkelstein, Jimmy Gollihar, and James M. Musser |
| EPI_ISL_2202575 | Aegis Sciences Corporation | Centers for Disease Control and Prevention Division of Viral Diseases, Pathogen Discovery | Dakota Howard, Dhwani Batra, Peter W. Cook, Kara Moser, Adrian Paskey, Jason Caravas, Benjamin Rambo-Martin, Shatavia Morrison, Christopher Gulvick, Scott Sammons, Yvette Unoarumhi, Darlene Wagner, Matthew Schmerer, Cyndi Clark, Patrick Campbell, Rob Case, Vikramsinha Ghorpade, Holly Houdeshell, Ola Kvalvaag, Dillon Nall, Ethan Sanders, Alec Vest, Shaun Westlund, Matthew Hardison, Clinton R. Paden, Duncan MacCannell |
| EPI_ISL_2202576 | Houston Methodist Hospital | Houston Methodist Hospital | Randall J. Olsen, Paul A. Christensen, S. Wesley Long, Sishir Subedi, Robert Olson, Marcus Nguyen, James J. Davis, Matthew Ojeda Saavedra, Prasanti Yerramilli, Layne Pruitt, Kristina Reppond, Madison N. Shyer, Jessica Cambric, Ryan Gadd, Ilya J. Finkelstein, Jimmy Gollihar, and James M. Musser |
| EPI_ISL_2202577 | Aegis Sciences Corporation | Centers for Disease Control and Prevention Division of Viral Diseases, Pathogen Discovery | Dakota Howard, Dhwani Batra, Peter W. Cook, Kara Moser, Adrian Paskey, Jason Caravas, Benjamin Rambo-Martin, Shatavia Morrison, Christopher Gulvick, Scott Sammons, Yvette Unoarumhi, Darlene Wagner, Matthew Schmerer, Cyndi Clark, Patrick Campbell, Rob Case, Vikramsinha Ghorpade, Holly Houdeshell, Ola Kvalvaag, Dillon Nall, Ethan Sanders, Alec Vest, Shaun Westlund, Matthew Hardison, Clinton R. Paden, Duncan MacCannell |
| EPI_ISL_2202578 | Houston Methodist Hospital | Houston Methodist Hospital | Randall J. Olsen, Paul A. Christensen, S. Wesley Long, Sishir Subedi, Robert Olson, Marcus Nguyen, James J. Davis, Matthew Ojeda Saavedra, Prasanti Yerramilli, Layne Pruitt, Kristina Reppond, Madison N. Shyer, Jessica Cambric, Ryan Gadd, Ilya J. Finkelstein, Jimmy Gollihar, and James M. Musser |
| EPI_ISL_2202579 | Aegis Sciences Corporation | Centers for Disease Control and Prevention Division of Viral Diseases, Pathogen Discovery | Dakota Howard, Dhwani Batra, Peter W. Cook, Kara Moser, Adrian Paskey, Jason Caravas, Benjamin Rambo-Martin, Shatavia Morrison, Christopher Gulvick, Scott Sammons, Yvette Unoarumhi, Darlene Wagner, Matthew Schmerer, Cyndi Clark, Patrick Campbell, Rob Case, Vikramsinha Ghorpade, Holly Houdeshell, Ola Kvalvaag, Dillon Nall, Ethan Sanders, Alec Vest, Shaun Westlund, Matthew Hardison, Clinton R. Paden, Duncan MacCannell |
| EPI_ISL_2202580 | Houston Methodist Hospital | Houston Methodist Hospital | Randall J. Olsen, Paul A. Christensen, S. Wesley Long, Sishir Subedi, Robert Olson, Marcus Nguyen, James J. Davis, Matthew Ojeda Saavedra, Prasanti Yerramilli, Layne Pruitt, Kristina Reppond, Madison N. Shyer, Jessica Cambric, Ryan Gadd, Ilya J. Finkelstein, Jimmy Gollihar, and James M. Musser |
| EPI_ISL_2202581 | Aegis Sciences Corporation | Centers for Disease Control and Prevention Division of Viral Diseases, Pathogen Discovery | Dakota Howard, Dhwani Batra, Peter W. Cook, Kara Moser, Adrian Paskey, Jason Caravas, Benjamin Rambo-Martin, Shatavia Morrison, Christopher Gulvick, Scott Sammons, Yvette Unoarumhi, Darlene Wagner, Matthew Schmerer, Cyndi Clark, Patrick Campbell, Rob Case, Vikramsinha Ghorpade, Holly Houdeshell, Ola Kvalvaag, Dillon Nall, Ethan Sanders, Alec Vest, Shaun Westlund, Matthew Hardison, Clinton R. Paden, Duncan MacCannell |
| EPI_ISL_2202582 | Houston Methodist Hospital | Houston Methodist Hospital | Randall J. Olsen, Paul A. Christensen, S. Wesley Long, Sishir Subedi, Robert Olson, Marcus Nguyen, James J. Davis, Matthew Ojeda Saavedra, Prasanti Yerramilli, Layne Pruitt, Kristina Reppond, Madison N. Shyer, Jessica Cambric, Ryan Gadd, Ilya J. Finkelstein, Jimmy Gollihar, and James M. Musser |
| EPI_ISL_2202583 | Aegis Sciences Corporation | Centers for Disease Control and Prevention Division of Viral Diseases, Pathogen Discovery | Dakota Howard, Dhwani Batra, Peter W. Cook, Kara Moser, Adrian Paskey, Jason Caravas, Benjamin Rambo-Martin, Shatavia Morrison, Christopher Gulvick, Scott Sammons, Yvette Unoarumhi, Darlene Wagner, Matthew Schmerer, Cyndi Clark, Patrick Campbell, Rob Case, Vikramsinha Ghorpade, Holly Houdeshell, Ola Kvalvaag, Dillon Nall, Ethan Sanders, Alec Vest, Shaun Westlund, Matthew Hardison, Clinton R. Paden, Duncan MacCannell |
| EPI_ISL_2202584 | Houston Methodist Hospital | Houston Methodist Hospital | Randall J. Olsen, Paul A. Christensen, S. Wesley Long, Sishir Subedi, Robert Olson, Marcus Nguyen, James J. Davis, Matthew Ojeda Saavedra, Prasanti Yerramilli, Layne Pruitt, Kristina Reppond, Madison N. Shyer, Jessica Cambric, Ryan Gadd, Ilya J. Finkelstein, Jimmy Gollihar, and James M. Musser |
| EPI_ISL_2202585, EPI_ISL_2202586 | Aegis Sciences Corporation | Centers for Disease Control and Prevention Division of Viral Diseases, Pathogen Discovery | Dakota Howard, Dhwani Batra, Peter W. Cook, Kara Moser, Adrian Paskey, Jason Caravas, Benjamin Rambo-Martin, Shatavia Morrison, Christopher Gulvick, Scott Sammons, Yvette Unoarumhi, Darlene Wagner, Matthew Schmerer, Cyndi Clark, Patrick Campbell, Rob Case, Vikramsinha Ghorpade, Holly Houdeshell, Ola Kvalvaag, Dillon Nall, Ethan Sanders, Alec Vest, Shaun Westlund, Matthew Hardison, Clinton R. Paden, Duncan MacCannell |
| EPI_ISL_2202587 | Houston Methodist Hospital | Houston Methodist Hospital | Randall J. Olsen, Paul A. Christensen, S. Wesley Long, Sishir Subedi, Robert Olson, Marcus Nguyen, James J. Davis, Matthew Ojeda Saavedra, Prasanti Yerramilli, Layne Pruitt, Kristina Reppond, Madison N. Shyer, Jessica Cambric, Ryan Gadd, Ilya J. Finkelstein, Jimmy Gollihar, and James M. Musser |
| EPI_ISL_2202588 | Aegis Sciences Corporation | Centers for Disease Control and Prevention Division of Viral Diseases, Pathogen Discovery | Dakota Howard, Dhwani Batra, Peter W. Cook, Kara Moser, Adrian Paskey, Jason Caravas, Benjamin Rambo-Martin, Shatavia Morrison, Christopher Gulvick, Scott Sammons, Yvette Unoarumhi, Darlene Wagner, Matthew Schmerer, Cyndi Clark, Patrick Campbell, Rob Case, Vikramsinha Ghorpade, Holly Houdeshell, Ola Kvalvaag, Dillon Nall, Ethan Sanders, Alec Vest, Shaun Westlund, Matthew Hardison, Clinton R. Paden, Duncan MacCannell |
| EPI_ISL_2202589 | Houston Methodist Hospital | Houston Methodist Hospital | Randall J. Olsen, Paul A. Christensen, S. Wesley Long, Sishir Subedi, Robert Olson, Marcus Nguyen, James J. Davis, Matthew Ojeda Saavedra, Prasanti Yerramilli, Layne Pruitt, Kristina Reppond, Madison N. Shyer, Jessica Cambric, Ryan Gadd, Ilya J. Finkelstein, Jimmy Gollihar, and James M. Musser |
| EPI_ISL_2202590 | Aegis Sciences Corporation | Centers for Disease Control and Prevention Division of Viral Diseases, Pathogen Discovery | Dakota Howard, Dhwani Batra, Peter W. Cook, Kara Moser, Adrian Paskey, Jason Caravas, Benjamin Rambo-Martin, Shatavia Morrison, Christopher Gulvick, Scott Sammons, Yvette Unoarumhi, Darlene Wagner, Matthew Schmerer, Cyndi Clark, Patrick Campbell, Rob Case, Vikramsinha Ghorpade, Holly Houdeshell, Ola Kvalvaag, Dillon Nall, Ethan Sanders, Alec Vest, Shaun Westlund, Matthew Hardison, Clinton R. Paden, Duncan MacCannell |
| EPI_ISL_2202591, EPI_ISL_2202594 | Houston Methodist Hospital | Houston Methodist Hospital | Randall J. Olsen, Paul A. Christensen, S. Wesley Long, Sishir Subedi, Robert Olson, Marcus Nguyen, James J. Davis, Matthew Ojeda Saavedra, Prasanti Yerramilli, Layne Pruitt, Kristina Reppond, Madison N. Shyer, Jessica Cambric, Ryan Gadd, Ilya J. Finkelstein, Jimmy Gollihar, and James M. Musser |
| EPI_ISL_2202595 | Aegis Sciences Corporation | Centers for Disease Control and Prevention Division of Viral Diseases, Pathogen Discovery | Dakota Howard, Dhwani Batra, Peter W. Cook, Kara Moser, Adrian Paskey, Jason Caravas, Benjamin Rambo-Martin, Shatavia Morrison, Christopher Gulvick, Scott Sammons, Yvette Unoarumhi, Darlene Wagner, Matthew Schmerer, Cyndi Clark, Patrick Campbell, Rob Case, Vikramsinha Ghorpade, Holly Houdeshell, Ola Kvalvaag, Dillon Nall, Ethan Sanders, Alec Vest, Shaun Westlund, Matthew Hardison, Clinton R. Paden, Duncan MacCannell |
| EPI_ISL_2202596 | Houston Methodist Hospital | Houston Methodist Hospital | Randall J. Olsen, Paul A. Christensen, S. Wesley Long, Sishir Subedi, Robert Olson, Marcus Nguyen, James J. Davis, Matthew Ojeda Saavedra, Prasanti Yerramilli, Layne Pruitt, Kristina Reppond, Madison N. Shyer, Jessica Cambric, Ryan Gadd, Ilya J. Finkelstein, Jimmy Gollihar, and James M. Musser |
| EPI_ISL_2202597 | Aegis Sciences Corporation | Centers for Disease Control and Prevention Division of Viral Diseases, Pathogen Discovery | Dakota Howard, Dhwani Batra, Peter W. Cook, Kara Moser, Adrian Paskey, Jason Caravas, Benjamin Rambo-Martin, Shatavia Morrison, Christopher Gulvick, Scott Sammons, Yvette Unoarumhi, Darlene Wagner, Matthew Schmerer, Cyndi Clark, Patrick Campbell, Rob Case, Vikramsinha Ghorpade, Holly Houdeshell, Ola Kvalvaag, Dillon Nall, Ethan Sanders, Alec Vest, Shaun Westlund, Matthew Hardison, Clinton R. Paden, Duncan MacCannell |
| EPI_ISL_2202598 | Houston Methodist Hospital | Houston Methodist Hospital | Randall J. Olsen, Paul A. Christensen, S. Wesley Long, Sishir Subedi, Robert Olson, Marcus Nguyen, James J. Davis, Matthew Ojeda Saavedra, Prasanti Yerramilli, Layne Pruitt, Kristina Reppond, Madison N. Shyer, Jessica Cambric, Ryan Gadd, Ilya J. Finkelstein, Jimmy Gollihar, and James M. Musser |
| EPI_ISL_2202599 | Aegis Sciences Corporation | Centers for Disease Control and Prevention Division of Viral Diseases, Pathogen Discovery | Dakota Howard, Dhwani Batra, Peter W. Cook, Kara Moser, Adrian Paskey, Jason Caravas, Benjamin Rambo-Martin, Shatavia Morrison, Christopher Gulvick, Scott Sammons, Yvette Unoarumhi, Darlene Wagner, Matthew Schmerer, Cyndi Clark, Patrick Campbell, Rob Case, Vikramsinha Ghorpade, Holly Houdeshell, Ola Kvalvaag, Dillon Nall, Ethan Sanders, Alec Vest, Shaun Westlund, Matthew Hardison, Clinton R. Paden, Duncan MacCannell |
| EPI_ISL_2202600 | Houston Methodist Hospital | Houston Methodist Hospital | Randall J. Olsen, Paul A. Christensen, S. Wesley Long, Sishir Subedi, Robert Olson, Marcus Nguyen, James J. Davis, Matthew Ojeda Saavedra, Prasanti Yerramilli, Layne Pruitt, Kristina Reppond, Madison N. Shyer, Jessica Cambric, Ryan Gadd, Ilya J. Finkelstein, Jimmy Gollihar, and James M. Musser |
| EPI_ISL_2202602 | Aegis Sciences Corporation | Centers for Disease Control and Prevention Division of Viral Diseases, Pathogen Discovery | Dakota Howard, Dhwani Batra, Peter W. Cook, Kara Moser, Adrian Paskey, Jason Caravas, Benjamin Rambo-Martin, Shatavia Morrison, Christopher Gulvick, Scott Sammons, Yvette Unoarumhi, Darlene Wagner, Matthew Schmerer, Cyndi Clark, Patrick Campbell, Rob Case, Vikramsinha Ghorpade, Holly Houdeshell, Ola Kvalvaag, Dillon Nall, Ethan Sanders, Alec Vest, Shaun Westlund, Matthew Hardison, Clinton R. Paden, Duncan MacCannell |
| EPI_ISL_2202603 | Houston Methodist Hospital | Houston Methodist Hospital | Randall J. Olsen, Paul A. Christensen, S. Wesley Long, Sishir Subedi, Robert Olson, Marcus Nguyen, James J. Davis, Matthew Ojeda Saavedra, Prasanti Yerramilli, Layne Pruitt, Kristina Reppond, Madison N. Shyer, Jessica Cambric, Ryan Gadd, Ilya J. Finkelstein, Jimmy Gollihar, and James M. Musser |
| EPI_ISL_2202604 | Aegis Sciences Corporation | Centers for Disease Control and Prevention Division of Viral Diseases, Pathogen Discovery | Dakota Howard, Dhwani Batra, Peter W. Cook, Kara Moser, Adrian Paskey, Jason Caravas, Benjamin Rambo-Martin, Shatavia Morrison, Christopher Gulvick, Scott Sammons, Yvette Unoarumhi, Darlene Wagner, Matthew Schmerer, Cyndi Clark, Patrick Campbell, Rob Case, Vikramsinha Ghorpade, Holly Houdeshell, Ola Kvalvaag, Dillon Nall, Ethan Sanders, Alec Vest, Shaun Westlund, Matthew Hardison, Clinton R. Paden, Duncan MacCannell |
| EPI_ISL_2202605 | Houston Methodist Hospital | Houston Methodist Hospital | Randall J. Olsen, Paul A. Christensen, S. Wesley Long, Sishir Subedi, Robert Olson, Marcus Nguyen, James J. Davis, Matthew Ojeda Saavedra, Prasanti Yerramilli, Layne Pruitt, Kristina Reppond, Madison N. Shyer, Jessica Cambric, Ryan Gadd, Ilya J. Finkelstein, Jimmy Gollihar, and James M. Musser |
| EPI_ISL_2202606 | Aegis Sciences Corporation | Centers for Disease Control and Prevention Division of Viral Diseases, Pathogen Discovery | Dakota Howard, Dhwani Batra, Peter W. Cook, Kara Moser, Adrian Paskey, Jason Caravas, Benjamin Rambo-Martin, Shatavia Morrison, Christopher Gulvick, Scott Sammons, Yvette Unoarumhi, Darlene Wagner, Matthew Schmerer, Cyndi Clark, Patrick Campbell, Rob Case, Vikramsinha Ghorpade, Holly Houdeshell, Ola Kvalvaag, Dillon Nall, Ethan Sanders, Alec Vest, Shaun Westlund, Matthew Hardison, Clinton R. Paden, Duncan MacCannell |
| EPI_ISL_2202607 | Houston Methodist Hospital | Houston Methodist Hospital | Randall J. Olsen, Paul A. Christensen, S. Wesley Long, Sishir Subedi, Robert Olson, Marcus Nguyen, James J. Davis, Matthew Ojeda Saavedra, Prasanti Yerramilli, Layne Pruitt, Kristina Reppond, Madison N. Shyer, Jessica Cambric, Ryan Gadd, Ilya J. Finkelstein, Jimmy Gollihar, and James M. Musser |
| EPI_ISL_2202608, EPI_ISL_2202609 | Aegis Sciences Corporation | Centers for Disease Control and Prevention Division of Viral Diseases, Pathogen Discovery | Dakota Howard, Dhwani Batra, Peter W. Cook, Kara Moser, Adrian Paskey, Jason Caravas, Benjamin Rambo-Martin, Shatavia Morrison, Christopher Gulvick, Scott Sammons, Yvette Unoarumhi, Darlene Wagner, Matthew Schmerer, Cyndi Clark, Patrick Campbell, Rob Case, Vikramsinha Ghorpade, Holly Houdeshell, Ola Kvalvaag, Dillon Nall, Ethan Sanders, Alec Vest, Shaun Westlund, Matthew Hardison, Clinton R. Paden, Duncan MacCannell |
| EPI_ISL_2202610 | Houston Methodist Hospital | Houston Methodist Hospital | Randall J. Olsen, Paul A. Christensen, S. Wesley Long, Sishir Subedi, Robert Olson, Marcus Nguyen, James J. Davis, Matthew Ojeda Saavedra, Prasanti Yerramilli, Layne Pruitt, Kristina Reppond, Madison N. Shyer, Jessica Cambric, Ryan Gadd, Ilya J. Finkelstein, Jimmy Gollihar, and James M. Musser |
| EPI_ISL_2202611 | Aegis Sciences Corporation | Centers for Disease Control and Prevention Division of Viral Diseases, Pathogen Discovery | Dakota Howard, Dhwani Batra, Peter W. Cook, Kara Moser, Adrian Paskey, Jason Caravas, Benjamin Rambo-Martin, Shatavia Morrison, Christopher Gulvick, Scott Sammons, Yvette Unoarumhi, Darlene Wagner, Matthew Schmerer, Cyndi Clark, Patrick Campbell, Rob Case, Vikramsinha Ghorpade, Holly Houdeshell, Ola Kvalvaag, Dillon Nall, Ethan Sanders, Alec Vest, Shaun Westlund, Matthew Hardison, Clinton R. Paden, Duncan MacCannell |
| EPI_ISL_2202612 | Houston Methodist Hospital | Houston Methodist Hospital | Randall J. Olsen, Paul A. Christensen, S. Wesley Long, Sishir Subedi, Robert Olson, Marcus Nguyen, James J. Davis, Matthew Ojeda Saavedra, Prasanti Yerramilli, Layne Pruitt, Kristina Reppond, Madison N. Shyer, Jessica Cambric, Ryan Gadd, Ilya J. Finkelstein, Jimmy Gollihar, and James |

| | | | |
|---|---|---|---|
| | | | M. Musser |
| EPI_ISL_2202613 | Aegis Sciences Corporation | Centers for Disease Control and Prevention Division of Viral Diseases, Pathogen Discovery | Dakota Howard, Dhwani Batra, Peter W. Cook, Kara Moser, Adrian Paskey, Jason Caravas, Benjamin Rambo-Martin, Shatavia Morrison, Christopher Gulvick, Scott Sammons, Yvette Unoarumhi, Darlene Wagner, Matthew Schmerer, Cyndi Clark, Patrick Campbell, Rob Case, Vikramsinha Ghorpade, Holly Houdeshell, Ola Kvalvaag, Dillon Nall, Ethan Sanders, Alec Vest, Shaun Westlund, Matthew Hardison, Clinton R. Paden, Duncan MacCannell |
| EPI_ISL_2202614 | Houston Methodist Hospital | Houston Methodist Hospital | Randall J. Olsen, Paul A. Christensen, S. Wesley Long, Sishir Subedi, Robert Olson, Marcus Nguyen, James J. Davis, Matthew Ojeda Saavedra, Prasanti Yerramilli, Layne Pruitt, Kristina Reppond, Madison N. Shyer, Jessica Cambric, Ryan Gadd, Ilya J. Finkelstein, Jimmy Gollihar, and James M. Musser |
| EPI_ISL_2202615 | Aegis Sciences Corporation | Centers for Disease Control and Prevention Division of Viral Diseases, Pathogen Discovery | Dakota Howard, Dhwani Batra, Peter W. Cook, Kara Moser, Adrian Paskey, Jason Caravas, Benjamin Rambo-Martin, Shatavia Morrison, Christopher Gulvick, Scott Sammons, Yvette Unoarumhi, Darlene Wagner, Matthew Schmerer, Cyndi Clark, Patrick Campbell, Rob Case, Vikramsinha Ghorpade, Holly Houdeshell, Ola Kvalvaag, Dillon Nall, Ethan Sanders, Alec Vest, Shaun Westlund, Matthew Hardison, Clinton R. Paden, Duncan MacCannell |
| EPI_ISL_2202616 | Houston Methodist Hospital | Houston Methodist Hospital | Randall J. Olsen, Paul A. Christensen, S. Wesley Long, Sishir Subedi, Robert Olson, Marcus Nguyen, James J. Davis, Matthew Ojeda Saavedra, Prasanti Yerramilli, Layne Pruitt, Kristina Reppond, Madison N. Shyer, Jessica Cambric, Ryan Gadd, Ilya J. Finkelstein, Jimmy Gollihar, and James M. Musser |
| EPI_ISL_2202617, EPI_ISL_2202618, EPI_ISL_2202619 | Aegis Sciences Corporation | Centers for Disease Control and Prevention Division of Viral Diseases, Pathogen Discovery | Dakota Howard, Dhwani Batra, Peter W. Cook, Kara Moser, Adrian Paskey, Jason Caravas, Benjamin Rambo-Martin, Shatavia Morrison, Christopher Gulvick, Scott Sammons, Yvette Unoarumhi, Darlene Wagner, Matthew Schmerer, Cyndi Clark, Patrick Campbell, Rob Case, Vikramsinha Ghorpade, Holly Houdeshell, Ola Kvalvaag, Dillon Nall, Ethan Sanders, Alec Vest, Shaun Westlund, Matthew Hardison, Clinton R. Paden, Duncan MacCannell |
| EPI_ISL_2202620 | Houston Methodist Hospital | Houston Methodist Hospital | Randall J. Olsen, Paul A. Christensen, S. Wesley Long, Sishir Subedi, Robert Olson, Marcus Nguyen, James J. Davis, Matthew Ojeda Saavedra, Prasanti Yerramilli, Layne Pruitt, Kristina Reppond, Madison N. Shyer, Jessica Cambric, Ryan Gadd, Ilya J. Finkelstein, Jimmy Gollihar, and James M. Musser |
| EPI_ISL_2202621 | Aegis Sciences Corporation | Centers for Disease Control and Prevention Division of Viral Diseases, Pathogen Discovery | Dakota Howard, Dhwani Batra, Peter W. Cook, Kara Moser, Adrian Paskey, Jason Caravas, Benjamin Rambo-Martin, Shatavia Morrison, Christopher Gulvick, Scott Sammons, Yvette Unoarumhi, Darlene Wagner, Matthew Schmerer, Cyndi Clark, Patrick Campbell, Rob Case, Vikramsinha Ghorpade, Holly Houdeshell, Ola Kvalvaag, Dillon Nall, Ethan Sanders, Alec Vest, Shaun Westlund, Matthew Hardison, Clinton R. Paden, Duncan MacCannell |
| EPI_ISL_2202622 | Houston Methodist Hospital | Houston Methodist Hospital | Randall J. Olsen, Paul A. Christensen, S. Wesley Long, Sishir Subedi, Robert Olson, Marcus Nguyen, James J. Davis, Matthew Ojeda Saavedra, Prasanti Yerramilli, Layne Pruitt, Kristina Reppond, Madison N. Shyer, Jessica Cambric, Ryan Gadd, Ilya J. Finkelstein, Jimmy Gollihar, and James M. Musser |
| EPI_ISL_2202623, EPI_ISL_2202624 | Aegis Sciences Corporation | Centers for Disease Control and Prevention Division of Viral Diseases, Pathogen Discovery | Dakota Howard, Dhwani Batra, Peter W. Cook, Kara Moser, Adrian Paskey, Jason Caravas, Benjamin Rambo-Martin, Shatavia Morrison, Christopher Gulvick, Scott Sammons, Yvette Unoarumhi, Darlene Wagner, Matthew Schmerer, Cyndi Clark, Patrick Campbell, Rob Case, Vikramsinha Ghorpade, Holly Houdeshell, Ola Kvalvaag, Dillon Nall, Ethan Sanders, Alec Vest, Shaun Westlund, Matthew Hardison, Clinton R. Paden, Duncan MacCannell |
| EPI_ISL_2202625, EPI_ISL_2202627 | Houston Methodist Hospital | Houston Methodist Hospital | Randall J. Olsen, Paul A. Christensen, S. Wesley Long, Sishir Subedi, Robert Olson, Marcus Nguyen, James J. Davis, Matthew Ojeda Saavedra, Prasanti Yerramilli, Layne Pruitt, Kristina Reppond, Madison N. Shyer, Jessica Cambric, Ryan Gadd, Ilya J. Finkelstein, Jimmy Gollihar, and James M. Musser |
| EPI_ISL_2202628 | Aegis Sciences Corporation | Centers for Disease Control and Prevention Division of Viral Diseases, Pathogen Discovery | Dakota Howard, Dhwani Batra, Peter W. Cook, Kara Moser, Adrian Paskey, Jason Caravas, Benjamin Rambo-Martin, Shatavia Morrison, Christopher Gulvick, Scott Sammons, Yvette Unoarumhi, Darlene Wagner, Matthew Schmerer, Cyndi Clark, Patrick Campbell, Rob Case, Vikramsinha Ghorpade, Holly Houdeshell, Ola Kvalvaag, Dillon Nall, Ethan Sanders, Alec Vest, Shaun Westlund, Matthew Hardison, Clinton R. Paden, Duncan MacCannell |
| EPI_ISL_2202629 | Houston Methodist Hospital | Houston Methodist Hospital | Randall J. Olsen, Paul A. Christensen, S. Wesley Long, Sishir Subedi, Robert Olson, Marcus Nguyen, James J. Davis, Matthew Ojeda Saavedra, Prasanti Yerramilli, Layne Pruitt, Kristina Reppond, Madison N. Shyer, Jessica Cambric, Ryan Gadd, Ilya J. Finkelstein, Jimmy Gollihar, and James M. Musser |
| EPI_ISL_2202630 | Aegis Sciences Corporation | Centers for Disease Control and Prevention Division of Viral Diseases, Pathogen Discovery | Dakota Howard, Dhwani Batra, Peter W. Cook, Kara Moser, Adrian Paskey, Jason Caravas, Benjamin Rambo-Martin, Shatavia Morrison, Christopher Gulvick, Scott Sammons, Yvette Unoarumhi, Darlene Wagner, Matthew Schmerer, Cyndi Clark, Patrick Campbell, Rob Case, Vikramsinha Ghorpade, Holly Houdeshell, Ola Kvalvaag, Dillon Nall, Ethan Sanders, Alec Vest, Shaun Westlund, Matthew Hardison, Clinton R. Paden, Duncan MacCannell |
| EPI_ISL_2202631 | Houston Methodist Hospital | Houston Methodist Hospital | Randall J. Olsen, Paul A. Christensen, S. Wesley Long, Sishir Subedi, Robert Olson, Marcus Nguyen, James J. Davis, Matthew Ojeda Saavedra, Prasanti Yerramilli, Layne Pruitt, Kristina Reppond, Madison N. Shyer, Jessica Cambric, Ryan Gadd, Ilya J. Finkelstein, Jimmy Gollihar, and James M. Musser |
| EPI_ISL_2202632, EPI_ISL_2202633 | Aegis Sciences Corporation | Centers for Disease Control and Prevention Division of Viral Diseases, Pathogen Discovery | Dakota Howard, Dhwani Batra, Peter W. Cook, Kara Moser, Adrian Paskey, Jason Caravas, Benjamin Rambo-Martin, Shatavia Morrison, Christopher Gulvick, Scott Sammons, Yvette Unoarumhi, Darlene Wagner, Matthew Schmerer, Cyndi Clark, Patrick Campbell, Rob Case, Vikramsinha Ghorpade, Holly Houdeshell, Ola Kvalvaag, Dillon Nall, Ethan Sanders, Alec Vest, Shaun Westlund, Matthew Hardison, Clinton R. Paden, Duncan MacCannell |
| EPI_ISL_2202634 | Houston Methodist Hospital | Houston Methodist Hospital | Randall J. Olsen, Paul A. Christensen, S. Wesley Long, Sishir Subedi, Robert Olson, Marcus Nguyen, James J. Davis, Matthew Ojeda Saavedra, Prasanti Yerramilli, Layne Pruitt, Kristina Reppond, Madison N. Shyer, Jessica Cambric, Ryan Gadd, Ilya J. Finkelstein, Jimmy Gollihar, and James M. Musser |
| EPI_ISL_2202635 | Aegis Sciences Corporation | Centers for Disease Control and Prevention Division of Viral Diseases, Pathogen Discovery | Dakota Howard, Dhwani Batra, Peter W. Cook, Kara Moser, Adrian Paskey, Jason Caravas, Benjamin Rambo-Martin, Shatavia Morrison, Christopher Gulvick, Scott Sammons, Yvette Unoarumhi, Darlene Wagner, Matthew Schmerer, Cyndi Clark, Patrick Campbell, Rob Case, Vikramsinha Ghorpade, Holly Houdeshell, Ola Kvalvaag, Dillon Nall, Ethan Sanders, Alec Vest, Shaun Westlund, Matthew Hardison, Clinton R. Paden, Duncan MacCannell |
| EPI_ISL_2202636 | Houston Methodist Hospital | Houston Methodist Hospital | Randall J. Olsen, Paul A. Christensen, S. Wesley Long, Sishir Subedi, Robert Olson, Marcus Nguyen, James J. Davis, Matthew Ojeda Saavedra, Prasanti Yerramilli, Layne Pruitt, Kristina Reppond, Madison N. Shyer, Jessica Cambric, Ryan Gadd, Ilya J. Finkelstein, Jimmy Gollihar, and James M. Musser |
| EPI_ISL_2202637 | Aegis Sciences Corporation | Centers for Disease Control and Prevention Division of Viral Diseases, Pathogen Discovery | Dakota Howard, Dhwani Batra, Peter W. Cook, Kara Moser, Adrian Paskey, Jason Caravas, Benjamin Rambo-Martin, Shatavia Morrison, Christopher Gulvick, Scott Sammons, Yvette Unoarumhi, Darlene Wagner, Matthew Schmerer, Cyndi Clark, Patrick Campbell, Rob Case, Vikramsinha Ghorpade, Holly Houdeshell, Ola Kvalvaag, Dillon Nall, Ethan Sanders, Alec Vest, Shaun Westlund, Matthew Hardison, Clinton R. Paden, Duncan MacCannell |
| EPI_ISL_2202638 | Houston Methodist Hospital | Houston Methodist Hospital | Randall J. Olsen, Paul A. Christensen, S. Wesley Long, Sishir Subedi, Robert Olson, Marcus Nguyen, James J. Davis, Matthew Ojeda Saavedra, Prasanti Yerramilli, Layne Pruitt, Kristina Reppond, Madison N. Shyer, Jessica Cambric, Ryan Gadd, Ilya J. Finkelstein, Jimmy Gollihar, and James M. Musser |
| EPI_ISL_2202639, EPI_ISL_2202640 | Aegis Sciences Corporation | Centers for Disease Control and Prevention Division of Viral Diseases, Pathogen Discovery | Dakota Howard, Dhwani Batra, Peter W. Cook, Kara Moser, Adrian Paskey, Jason Caravas, Benjamin Rambo-Martin, Shatavia Morrison, Christopher Gulvick, Scott Sammons, Yvette Unoarumhi, Darlene Wagner, Matthew Schmerer, Cyndi Clark, Patrick Campbell, Rob Case, Vikramsinha Ghorpade, Holly Houdeshell, Ola Kvalvaag, Dillon Nall, Ethan Sanders, Alec Vest, Shaun Westlund, Matthew Hardison, Clinton R. Paden, Duncan MacCannell |
| EPI_ISL_2202641 | Houston Methodist Hospital | Houston Methodist Hospital | Randall J. Olsen, Paul A. Christensen, S. Wesley Long, Sishir Subedi, Robert Olson, Marcus Nguyen, James J. Davis, Matthew Ojeda Saavedra, Prasanti Yerramilli, Layne Pruitt, Kristina Reppond, Madison N. Shyer, Jessica Cambric, Ryan Gadd, Ilya J. Finkelstein, Jimmy Gollihar, and James M. Musser |
| EPI_ISL_2202642 | Aegis Sciences Corporation | Centers for Disease Control and Prevention Division of Viral Diseases, Pathogen Discovery | Dakota Howard, Dhwani Batra, Peter W. Cook, Kara Moser, Adrian Paskey, Jason Caravas, Benjamin Rambo-Martin, Shatavia Morrison, Christopher Gulvick, Scott Sammons, Yvette Unoarumhi, Darlene Wagner, Matthew Schmerer, Cyndi Clark, Patrick Campbell, Rob Case, Vikramsinha Ghorpade, Holly Houdeshell, Ola Kvalvaag, Dillon Nall, Ethan Sanders, Alec Vest, Shaun Westlund, Matthew Hardison, Clinton R. Paden, Duncan MacCannell |
| EPI_ISL_2202643 | Houston Methodist Hospital | Houston Methodist Hospital | Randall J. Olsen, Paul A. Christensen, S. Wesley Long, Sishir Subedi, Robert Olson, Marcus Nguyen, James J. Davis, Matthew Ojeda Saavedra, Prasanti Yerramilli, Layne Pruitt, Kristina Reppond, Madison N. Shyer, Jessica Cambric, Ryan Gadd, Ilya J. Finkelstein, Jimmy Gollihar, and James M. Musser |
| EPI_ISL_2202644 | Aegis Sciences Corporation | Centers for Disease Control and Prevention Division of Viral Diseases, Pathogen Discovery | Dakota Howard, Dhwani Batra, Peter W. Cook, Kara Moser, Adrian Paskey, Jason Caravas, Benjamin Rambo-Martin, Shatavia Morrison, Christopher Gulvick, Scott Sammons, Yvette Unoarumhi, Darlene Wagner, Matthew Schmerer, Cyndi Clark, Patrick Campbell, Rob Case, Vikramsinha Ghorpade, Holly Houdeshell, Ola Kvalvaag, Dillon Nall, Ethan Sanders, Alec Vest, Shaun Westlund, Matthew Hardison, Clinton R. Paden, Duncan MacCannell |
| EPI_ISL_2202645 | Houston Methodist Hospital | Houston Methodist Hospital | Randall J. Olsen, Paul A. Christensen, S. Wesley Long, Sishir Subedi, Robert Olson, Marcus Nguyen, James J. Davis, Matthew Ojeda Saavedra, Prasanti Yerramilli, Layne Pruitt, Kristina Reppond, Madison N. Shyer, Jessica Cambric, Ryan Gadd, Ilya J. Finkelstein, Jimmy Gollihar, and James M. Musser |
| EPI_ISL_2202646 | Aegis Sciences Corporation | Centers for Disease Control and Prevention Division of Viral Diseases, Pathogen Discovery | Dakota Howard, Dhwani Batra, Peter W. Cook, Kara Moser, Adrian Paskey, Jason Caravas, Benjamin Rambo-Martin, Shatavia Morrison, Christopher Gulvick, Scott Sammons, Yvette Unoarumhi, Darlene Wagner, Matthew Schmerer, Cyndi Clark, Patrick Campbell, Rob Case, Vikramsinha Ghorpade, Holly Houdeshell, Ola Kvalvaag, Dillon Nall, Ethan Sanders, Alec Vest, Shaun Westlund, Matthew Hardison, Clinton R. Paden, Duncan MacCannell |
| EPI_ISL_2202647 | Houston Methodist Hospital | Houston Methodist Hospital | Randall J. Olsen, Paul A. Christensen, S. Wesley Long, Sishir Subedi, Robert Olson, Marcus Nguyen, James J. Davis, Matthew Ojeda Saavedra, Prasanti Yerramilli, Layne Pruitt, Kristina Reppond, Madison N. Shyer, Jessica Cambric, Ryan Gadd, Ilya J. Finkelstein, Jimmy Gollihar, and James M. Musser |
| EPI_ISL_2202648, EPI_ISL_2202649 | Aegis Sciences Corporation | Centers for Disease Control and Prevention Division of Viral Diseases, Pathogen Discovery | Dakota Howard, Dhwani Batra, Peter W. Cook, Kara Moser, Adrian Paskey, Jason Caravas, Benjamin Rambo-Martin, Shatavia Morrison, Christopher Gulvick, Scott Sammons, Yvette Unoarumhi, Darlene Wagner, Matthew Schmerer, Cyndi Clark, Patrick Campbell, Rob Case, Vikramsinha Ghorpade, Holly Houdeshell, Ola Kvalvaag, Dillon Nall, Ethan Sanders, Alec Vest, Shaun Westlund, Matthew Hardison, Clinton R. Paden, Duncan MacCannell |
| EPI_ISL_2202650 | Houston Methodist Hospital | Houston Methodist Hospital | Randall J. Olsen, Paul A. Christensen, S. Wesley Long, Sishir Subedi, Robert Olson, Marcus Nguyen, James J. Davis, Matthew Ojeda Saavedra, Prasanti Yerramilli, Layne Pruitt, Kristina Reppond, Madison N. Shyer, Jessica Cambric, Ryan Gadd, Ilya J. Finkelstein, Jimmy Gollihar, and James M. Musser |
| EPI_ISL_2202651 | Aegis Sciences Corporation | Centers for Disease Control and Prevention Division of Viral Diseases, Pathogen Discovery | Dakota Howard, Dhwani Batra, Peter W. Cook, Kara Moser, Adrian Paskey, Jason Caravas, Benjamin Rambo-Martin, Shatavia Morrison, Christopher Gulvick, Scott Sammons, Yvette Unoarumhi, Darlene Wagner, Matthew Schmerer, Cyndi Clark, Patrick Campbell, Rob Case, Vikramsinha Ghorpade, Holly Houdeshell, Ola Kvalvaag, Dillon Nall, Ethan Sanders, Alec Vest, Shaun Westlund, Matthew Hardison, Clinton R. Paden, Duncan MacCannell |
| EPI_ISL_2202652 | Houston Methodist Hospital | Houston Methodist Hospital | Randall J. Olsen, Paul A. Christensen, S. Wesley Long, Sishir Subedi, Robert Olson, Marcus Nguyen, James J. Davis, Matthew Ojeda Saavedra, Prasanti Yerramilli, Layne Pruitt, Kristina Reppond, Madison N. Shyer, Jessica Cambric, Ryan Gadd, Ilya J. Finkelstein, Jimmy Gollihar, and James M. Musser |
| EPI_ISL_2202653, EPI_ISL_2202654 | Aegis Sciences Corporation | Centers for Disease Control and Prevention Division of Viral Diseases, Pathogen Discovery | Dakota Howard, Dhwani Batra, Peter W. Cook, Kara Moser, Adrian Paskey, Jason Caravas, Benjamin Rambo-Martin, Shatavia Morrison, Christopher Gulvick, Scott Sammons, Yvette Unoarumhi, Darlene Wagner, Matthew Schmerer, Cyndi Clark, Patrick Campbell, Rob Case, Vikramsinha Ghorpade, Holly Houdeshell, Ola Kvalvaag, Dillon Nall, Ethan Sanders, Alec Vest, Shaun Westlund, Matthew Hardison, Clinton R. Paden, Duncan MacCannell |
| EPI_ISL_2202655 | Houston Methodist Hospital | Houston Methodist Hospital | Randall J. Olsen, Paul A. Christensen, S. Wesley Long, Sishir Subedi, Robert Olson, Marcus Nguyen, James J. Davis, Matthew Ojeda Saavedra, Prasanti Yerramilli, Layne Pruitt, Kristina Reppond, Madison N. Shyer, Jessica Cambric, Ryan Gadd, Ilya J. Finkelstein, Jimmy Gollihar, and James M. Musser |
| EPI_ISL_2202656 | Aegis Sciences Corporation | Centers for Disease Control and Prevention Division of Viral Diseases, Pathogen Discovery | Dakota Howard, Dhwani Batra, Peter W. Cook, Kara Moser, Adrian Paskey, Jason Caravas, Benjamin Rambo-Martin, Shatavia Morrison, Christopher Gulvick, Scott Sammons, Yvette Unoarumhi, Darlene Wagner, Matthew Schmerer, Cyndi Clark, Patrick Campbell, Rob Case, Vikramsinha Ghorpade, Holly Houdeshell, Ola Kvalvaag, Dillon Nall, Ethan Sanders, Alec Vest, Shaun Westlund, Matthew Hardison, Clinton R. Paden, Duncan MacCannell |
| EPI_ISL_2202657 | Houston Methodist Hospital | Houston Methodist Hospital | Randall J. Olsen, Paul A. Christensen, S. Wesley Long, Sishir Subedi, Robert Olson, Marcus Nguyen, James J. Davis, Matthew Ojeda Saavedra, Prasanti Yerramilli, Layne Pruitt, Kristina Reppond, Madison N. Shyer, Jessica Cambric, Ryan Gadd, Ilya J. Finkelstein, Jimmy Gollihar, and James M. Musser |
| EPI_ISL_2202658, EPI_ISL_2202659 | Aegis Sciences Corporation | Centers for Disease Control and Prevention Division of Viral Diseases, Pathogen | Dakota Howard, Dhwani Batra, Peter W. Cook, Kara Moser, Adrian Paskey, Jason Caravas, Benjamin Rambo-Martin, Shatavia Morrison, Christopher Gulvick, Scott Sammons, Yvette Unoarumhi, Darlene Wagner, Matthew Schmerer, Cyndi Clark, Patrick Campbell, Rob Case, Vikramsinha Ghorpade, Holly Houdeshell, Ola Kvalvaag, Dillon Nall, Ethan Sanders, Alec Vest, Shaun Westlund, Matthew Hardison, Clinton R. Paden, Duncan MacCannell |

| | | | |
|---|---|---|---|
| | | Discovery | |
| EPI_ISL_2202660 | Houston Methodist Hospital | Houston Methodist Hospital | Randall J. Olsen, Paul A. Christensen, S. Wesley Long, Sishir Subedi, Robert Olson, Marcus Nguyen, James J. Davis, Matthew Ojeda Saavedra, Prasanti Yerramilli, Layne Pruitt, Kristina Reppond, Madison N. Shyer, Jessica Cambric, Ryan Gadd, Ilya J. Finkelstein, Jimmy Gollihar, and James M. Musser |
| EPI_ISL_2202661 | Aegis Sciences Corporation | Centers for Disease Control and Prevention Division of Viral Diseases, Pathogen Discovery | Dakota Howard, Dhwani Batra, Peter W. Cook, Kara Moser, Adrian Paskey, Jason Caravas, Benjamin Rambo-Martin, Shatavia Morrison, Christopher Gulvick, Scott Sammons, Yvette Unoarumhi, Darlene Wagner, Matthew Schmerer, Cyndi Clark, Patrick Campbell, Rob Case, Vikramsinha Ghorpade, Holly Houdeshell, Ola Kvalvaag, Dillon Nall, Ethan Sanders, Alec Vest, Shaun Westlund, Matthew Hardison, Clinton R. Paden, Duncan MacCannell |
| EPI_ISL_2202662 | Houston Methodist Hospital | Houston Methodist Hospital | Randall J. Olsen, Paul A. Christensen, S. Wesley Long, Sishir Subedi, Robert Olson, Marcus Nguyen, James J. Davis, Matthew Ojeda Saavedra, Prasanti Yerramilli, Layne Pruitt, Kristina Reppond, Madison N. Shyer, Jessica Cambric, Ryan Gadd, Ilya J. Finkelstein, Jimmy Gollihar, and James M. Musser |
| EPI_ISL_2202663 | Aegis Sciences Corporation | Centers for Disease Control and Prevention Division of Viral Diseases, Pathogen Discovery | Dakota Howard, Dhwani Batra, Peter W. Cook, Kara Moser, Adrian Paskey, Jason Caravas, Benjamin Rambo-Martin, Shatavia Morrison, Christopher Gulvick, Scott Sammons, Yvette Unoarumhi, Darlene Wagner, Matthew Schmerer, Cyndi Clark, Patrick Campbell, Rob Case, Vikramsinha Ghorpade, Holly Houdeshell, Ola Kvalvaag, Dillon Nall, Ethan Sanders, Alec Vest, Shaun Westlund, Matthew Hardison, Clinton R. Paden, Duncan MacCannell |
| EPI_ISL_2202664 | Houston Methodist Hospital | Houston Methodist Hospital | Randall J. Olsen, Paul A. Christensen, S. Wesley Long, Sishir Subedi, Robert Olson, Marcus Nguyen, James J. Davis, Matthew Ojeda Saavedra, Prasanti Yerramilli, Layne Pruitt, Kristina Reppond, Madison N. Shyer, Jessica Cambric, Ryan Gadd, Ilya J. Finkelstein, Jimmy Gollihar, and James M. Musser |
| EPI_ISL_2202665 | Aegis Sciences Corporation | Centers for Disease Control and Prevention Division of Viral Diseases, Pathogen Discovery | Dakota Howard, Dhwani Batra, Peter W. Cook, Kara Moser, Adrian Paskey, Jason Caravas, Benjamin Rambo-Martin, Shatavia Morrison, Christopher Gulvick, Scott Sammons, Yvette Unoarumhi, Darlene Wagner, Matthew Schmerer, Cyndi Clark, Patrick Campbell, Rob Case, Vikramsinha Ghorpade, Holly Houdeshell, Ola Kvalvaag, Dillon Nall, Ethan Sanders, Alec Vest, Shaun Westlund, Matthew Hardison, Clinton R. Paden, Duncan MacCannell |
| EPI_ISL_2202666 | Houston Methodist Hospital | Houston Methodist Hospital | Randall J. Olsen, Paul A. Christensen, S. Wesley Long, Sishir Subedi, Robert Olson, Marcus Nguyen, James J. Davis, Matthew Ojeda Saavedra, Prasanti Yerramilli, Layne Pruitt, Kristina Reppond, Madison N. Shyer, Jessica Cambric, Ryan Gadd, Ilya J. Finkelstein, Jimmy Gollihar, and James M. Musser |
| EPI_ISL_2202667, EPI_ISL_2202668 | Aegis Sciences Corporation | Centers for Disease Control and Prevention Division of Viral Diseases, Pathogen Discovery | Dakota Howard, Dhwani Batra, Peter W. Cook, Kara Moser, Adrian Paskey, Jason Caravas, Benjamin Rambo-Martin, Shatavia Morrison, Christopher Gulvick, Scott Sammons, Yvette Unoarumhi, Darlene Wagner, Matthew Schmerer, Cyndi Clark, Patrick Campbell, Rob Case, Vikramsinha Ghorpade, Holly Houdeshell, Ola Kvalvaag, Dillon Nall, Ethan Sanders, Alec Vest, Shaun Westlund, Matthew Hardison, Clinton R. Paden, Duncan MacCannell |
| EPI_ISL_2202669 | Houston Methodist Hospital | Houston Methodist Hospital | Randall J. Olsen, Paul A. Christensen, S. Wesley Long, Sishir Subedi, Robert Olson, Marcus Nguyen, James J. Davis, Matthew Ojeda Saavedra, Prasanti Yerramilli, Layne Pruitt, Kristina Reppond, Madison N. Shyer, Jessica Cambric, Ryan Gadd, Ilya J. Finkelstein, Jimmy Gollihar, and James M. Musser |
| EPI_ISL_2202670, EPI_ISL_2202671 | Aegis Sciences Corporation | Centers for Disease Control and Prevention Division of Viral Diseases, Pathogen Discovery | Dakota Howard, Dhwani Batra, Peter W. Cook, Kara Moser, Adrian Paskey, Jason Caravas, Benjamin Rambo-Martin, Shatavia Morrison, Christopher Gulvick, Scott Sammons, Yvette Unoarumhi, Darlene Wagner, Matthew Schmerer, Cyndi Clark, Patrick Campbell, Rob Case, Vikramsinha Ghorpade, Holly Houdeshell, Ola Kvalvaag, Dillon Nall, Ethan Sanders, Alec Vest, Shaun Westlund, Matthew Hardison, Clinton R. Paden, Duncan MacCannell |
| EPI_ISL_2202672 | Houston Methodist Hospital | Houston Methodist Hospital | Randall J. Olsen, Paul A. Christensen, S. Wesley Long, Sishir Subedi, Robert Olson, Marcus Nguyen, James J. Davis, Matthew Ojeda Saavedra, Prasanti Yerramilli, Layne Pruitt, Kristina Reppond, Madison N. Shyer, Jessica Cambric, Ryan Gadd, Ilya J. Finkelstein, Jimmy Gollihar, and James M. Musser |
| EPI_ISL_2202673 | The Ohio State University Applied Microbiology Services Laboratory | The Ohio State University Applied Microbiology Services Laboratory | Seth A. Faith PhD |
| EPI_ISL_2202674 | Aegis Sciences Corporation | Centers for Disease Control and Prevention Division of Viral Diseases, Pathogen Discovery | Dakota Howard, Dhwani Batra, Peter W. Cook, Kara Moser, Adrian Paskey, Jason Caravas, Benjamin Rambo-Martin, Shatavia Morrison, Christopher Gulvick, Scott Sammons, Yvette Unoarumhi, Darlene Wagner, Matthew Schmerer, Cyndi Clark, Patrick Campbell, Rob Case, Vikramsinha Ghorpade, Holly Houdeshell, Ola Kvalvaag, Dillon Nall, Ethan Sanders, Alec Vest, Shaun Westlund, Matthew Hardison, Clinton R. Paden, Duncan MacCannell |
| EPI_ISL_2202675 | The Ohio State University Applied Microbiology Services Laboratory | The Ohio State University Applied Microbiology Services Laboratory | Seth A. Faith PhD |
| EPI_ISL_2202676 | Houston Methodist Hospital | Houston Methodist Hospital | Randall J. Olsen, Paul A. Christensen, S. Wesley Long, Sishir Subedi, Robert Olson, Marcus Nguyen, James J. Davis, Matthew Ojeda Saavedra, Prasanti Yerramilli, Layne Pruitt, Kristina Reppond, Madison N. Shyer, Jessica Cambric, Ryan Gadd, Ilya J. Finkelstein, Jimmy Gollihar, and James M. Musser |
| EPI_ISL_2202677 | Aegis Sciences Corporation | Centers for Disease Control and Prevention Division of Viral Diseases, Pathogen Discovery | Dakota Howard, Dhwani Batra, Peter W. Cook, Kara Moser, Adrian Paskey, Jason Caravas, Benjamin Rambo-Martin, Shatavia Morrison, Christopher Gulvick, Scott Sammons, Yvette Unoarumhi, Darlene Wagner, Matthew Schmerer, Cyndi Clark, Patrick Campbell, Rob Case, Vikramsinha Ghorpade, Holly Houdeshell, Ola Kvalvaag, Dillon Nall, Ethan Sanders, Alec Vest, Shaun Westlund, Matthew Hardison, Clinton R. Paden, Duncan MacCannell |
| EPI_ISL_2202678 | The Ohio State University Applied Microbiology Services Laboratory | The Ohio State University Applied Microbiology Services Laboratory | Seth A. Faith PhD |
| EPI_ISL_2202679 | Houston Methodist Hospital | Houston Methodist Hospital | Randall J. Olsen, Paul A. Christensen, S. Wesley Long, Sishir Subedi, Robert Olson, Marcus Nguyen, James J. Davis, Matthew Ojeda Saavedra, Prasanti Yerramilli, Layne Pruitt, Kristina Reppond, Madison N. Shyer, Jessica Cambric, Ryan Gadd, Ilya J. Finkelstein, Jimmy Gollihar, and James M. Musser |
| EPI_ISL_2202680, EPI_ISL_2202681 | Aegis Sciences Corporation | Centers for Disease Control and Prevention Division of Viral Diseases, Pathogen Discovery | Dakota Howard, Dhwani Batra, Peter W. Cook, Kara Moser, Adrian Paskey, Jason Caravas, Benjamin Rambo-Martin, Shatavia Morrison, Christopher Gulvick, Scott Sammons, Yvette Unoarumhi, Darlene Wagner, Matthew Schmerer, Cyndi Clark, Patrick Campbell, Rob Case, Vikramsinha Ghorpade, Holly Houdeshell, Ola Kvalvaag, Dillon Nall, Ethan Sanders, Alec Vest, Shaun Westlund, Matthew Hardison, Clinton R. Paden, Duncan MacCannell |
| EPI_ISL_2202682 | Houston Methodist Hospital | Houston Methodist Hospital | Randall J. Olsen, Paul A. Christensen, S. Wesley Long, Sishir Subedi, Robert Olson, Marcus Nguyen, James J. Davis, Matthew Ojeda Saavedra, Prasanti Yerramilli, Layne Pruitt, Kristina Reppond, Madison N. Shyer, Jessica Cambric, Ryan Gadd, Ilya J. Finkelstein, Jimmy Gollihar, and James M. Musser |
| EPI_ISL_2202683 | Aegis Sciences Corporation | Centers for Disease Control and Prevention Division of Viral Diseases, Pathogen Discovery | Dakota Howard, Dhwani Batra, Peter W. Cook, Kara Moser, Adrian Paskey, Jason Caravas, Benjamin Rambo-Martin, Shatavia Morrison, Christopher Gulvick, Scott Sammons, Yvette Unoarumhi, Darlene Wagner, Matthew Schmerer, Cyndi Clark, Patrick Campbell, Rob Case, Vikramsinha Ghorpade, Holly Houdeshell, Ola Kvalvaag, Dillon Nall, Ethan Sanders, Alec Vest, Shaun Westlund, Matthew Hardison, Clinton R. Paden, Duncan MacCannell |
| EPI_ISL_2202684 | Houston Methodist Hospital | Houston Methodist Hospital | Randall J. Olsen, Paul A. Christensen, S. Wesley Long, Sishir Subedi, Robert Olson, Marcus Nguyen, James J. Davis, Matthew Ojeda Saavedra, Prasanti Yerramilli, Layne Pruitt, Kristina Reppond, Madison N. Shyer, Jessica Cambric, Ryan Gadd, Ilya J. Finkelstein, Jimmy Gollihar, and James M. Musser |
| EPI_ISL_2202685 | Aegis Sciences Corporation | Centers for Disease Control and Prevention Division of Viral Diseases, Pathogen Discovery | Dakota Howard, Dhwani Batra, Peter W. Cook, Kara Moser, Adrian Paskey, Jason Caravas, Benjamin Rambo-Martin, Shatavia Morrison, Christopher Gulvick, Scott Sammons, Yvette Unoarumhi, Darlene Wagner, Matthew Schmerer, Cyndi Clark, Patrick Campbell, Rob Case, Vikramsinha Ghorpade, Holly Houdeshell, Ola Kvalvaag, Dillon Nall, Ethan Sanders, Alec Vest, Shaun Westlund, Matthew Hardison, Clinton R. Paden, Duncan MacCannell |
| EPI_ISL_2202686 | Houston Methodist Hospital | Houston Methodist Hospital | Randall J. Olsen, Paul A. Christensen, S. Wesley Long, Sishir Subedi, Robert Olson, Marcus Nguyen, James J. Davis, Matthew Ojeda Saavedra, Prasanti Yerramilli, Layne Pruitt, Kristina Reppond, Madison N. Shyer, Jessica Cambric, Ryan Gadd, Ilya J. Finkelstein, Jimmy Gollihar, and James M. Musser |
| EPI_ISL_2202687 | Aegis Sciences Corporation | Centers for Disease Control and Prevention Division of Viral Diseases, Pathogen Discovery | Dakota Howard, Dhwani Batra, Peter W. Cook, Kara Moser, Adrian Paskey, Jason Caravas, Benjamin Rambo-Martin, Shatavia Morrison, Christopher Gulvick, Scott Sammons, Yvette Unoarumhi, Darlene Wagner, Matthew Schmerer, Cyndi Clark, Patrick Campbell, Rob Case, Vikramsinha Ghorpade, Holly Houdeshell, Ola Kvalvaag, Dillon Nall, Ethan Sanders, Alec Vest, Shaun Westlund, Matthew Hardison, Clinton R. Paden, Duncan MacCannell |
| EPI_ISL_2202688 | Houston Methodist Hospital | Houston Methodist Hospital | Randall J. Olsen, Paul A. Christensen, S. Wesley Long, Sishir Subedi, Robert Olson, Marcus Nguyen, James J. Davis, Matthew Ojeda Saavedra, Prasanti Yerramilli, Layne Pruitt, Kristina Reppond, Madison N. Shyer, Jessica Cambric, Ryan Gadd, Ilya J. Finkelstein, Jimmy Gollihar, and James M. Musser |
| EPI_ISL_2202689 | Aegis Sciences Corporation | Centers for Disease Control and Prevention Division of Viral Diseases, Pathogen Discovery | Dakota Howard, Dhwani Batra, Peter W. Cook, Kara Moser, Adrian Paskey, Jason Caravas, Benjamin Rambo-Martin, Shatavia Morrison, Christopher Gulvick, Scott Sammons, Yvette Unoarumhi, Darlene Wagner, Matthew Schmerer, Cyndi Clark, Patrick Campbell, Rob Case, Vikramsinha Ghorpade, Holly Houdeshell, Ola Kvalvaag, Dillon Nall, Ethan Sanders, Alec Vest, Shaun Westlund, Matthew Hardison, Clinton R. Paden, Duncan MacCannell |
| EPI_ISL_2202690 | Houston Methodist Hospital | Houston Methodist Hospital | Randall J. Olsen, Paul A. Christensen, S. Wesley Long, Sishir Subedi, Robert Olson, Marcus Nguyen, James J. Davis, Matthew Ojeda Saavedra, Prasanti Yerramilli, Layne Pruitt, Kristina Reppond, Madison N. Shyer, Jessica Cambric, Ryan Gadd, Ilya J. Finkelstein, Jimmy Gollihar, and James M. Musser |
| EPI_ISL_2202691, EPI_ISL_2202692 | Aegis Sciences Corporation | Centers for Disease Control and Prevention Division of Viral Diseases, Pathogen Discovery | Dakota Howard, Dhwani Batra, Peter W. Cook, Kara Moser, Adrian Paskey, Jason Caravas, Benjamin Rambo-Martin, Shatavia Morrison, Christopher Gulvick, Scott Sammons, Yvette Unoarumhi, Darlene Wagner, Matthew Schmerer, Cyndi Clark, Patrick Campbell, Rob Case, Vikramsinha Ghorpade, Holly Houdeshell, Ola Kvalvaag, Dillon Nall, Ethan Sanders, Alec Vest, Shaun Westlund, Matthew Hardison, Clinton R. Paden, Duncan MacCannell |
| EPI_ISL_2202693 | Houston Methodist Hospital | Houston Methodist Hospital | Randall J. Olsen, Paul A. Christensen, S. Wesley Long, Sishir Subedi, Robert Olson, Marcus Nguyen, James J. Davis, Matthew Ojeda Saavedra, Prasanti Yerramilli, Layne Pruitt, Kristina Reppond, Madison N. Shyer, Jessica Cambric, Ryan Gadd, Ilya J. Finkelstein, Jimmy Gollihar, and James M. Musser |
| EPI_ISL_2202694 | Aegis Sciences Corporation | Centers for Disease Control and Prevention Division of Viral Diseases, Pathogen Discovery | Dakota Howard, Dhwani Batra, Peter W. Cook, Kara Moser, Adrian Paskey, Jason Caravas, Benjamin Rambo-Martin, Shatavia Morrison, Christopher Gulvick, Scott Sammons, Yvette Unoarumhi, Darlene Wagner, Matthew Schmerer, Cyndi Clark, Patrick Campbell, Rob Case, Vikramsinha Ghorpade, Holly Houdeshell, Ola Kvalvaag, Dillon Nall, Ethan Sanders, Alec Vest, Shaun Westlund, Matthew Hardison, Clinton R. Paden, Duncan MacCannell |
| EPI_ISL_2202695 | Houston Methodist Hospital | Houston Methodist Hospital | Randall J. Olsen, Paul A. Christensen, S. Wesley Long, Sishir Subedi, Robert Olson, Marcus Nguyen, James J. Davis, Matthew Ojeda Saavedra, Prasanti Yerramilli, Layne Pruitt, Kristina Reppond, Madison N. Shyer, Jessica Cambric, Ryan Gadd, Ilya J. Finkelstein, Jimmy Gollihar, and James M. Musser |
| EPI_ISL_2202696 | Aegis Sciences Corporation | Centers for Disease Control and Prevention Division of Viral Diseases, Pathogen Discovery | Dakota Howard, Dhwani Batra, Peter W. Cook, Kara Moser, Adrian Paskey, Jason Caravas, Benjamin Rambo-Martin, Shatavia Morrison, Christopher Gulvick, Scott Sammons, Yvette Unoarumhi, Darlene Wagner, Matthew Schmerer, Cyndi Clark, Patrick Campbell, Rob Case, Vikramsinha Ghorpade, Holly Houdeshell, Ola Kvalvaag, Dillon Nall, Ethan Sanders, Alec Vest, Shaun Westlund, Matthew Hardison, Clinton R. Paden, Duncan MacCannell |
| EPI_ISL_2202697 | Houston Methodist Hospital | Houston Methodist Hospital | Randall J. Olsen, Paul A. Christensen, S. Wesley Long, Sishir Subedi, Robert Olson, Marcus Nguyen, James J. Davis, Matthew Ojeda Saavedra, Prasanti Yerramilli, Layne Pruitt, Kristina Reppond, Madison N. Shyer, Jessica Cambric, Ryan Gadd, Ilya J. Finkelstein, Jimmy Gollihar, and James M. Musser |
| EPI_ISL_2202698, EPI_ISL_2202699, EPI_ISL_2202700 | Aegis Sciences Corporation | Centers for Disease Control and Prevention Division of Viral Diseases, Pathogen Discovery | Dakota Howard, Dhwani Batra, Peter W. Cook, Kara Moser, Adrian Paskey, Jason Caravas, Benjamin Rambo-Martin, Shatavia Morrison, Christopher Gulvick, Scott Sammons, Yvette Unoarumhi, Darlene Wagner, Matthew Schmerer, Cyndi Clark, Patrick Campbell, Rob Case, Vikramsinha Ghorpade, Holly Houdeshell, Ola Kvalvaag, Dillon Nall, Ethan Sanders, Alec Vest, Shaun Westlund, Matthew Hardison, Clinton R. Paden, Duncan MacCannell |
| EPI_ISL_2202701 | Houston Methodist Hospital | Houston Methodist Hospital | Randall J. Olsen, Paul A. Christensen, S. Wesley Long, Sishir Subedi, Robert Olson, Marcus Nguyen, James J. Davis, Matthew Ojeda Saavedra, Prasanti Yerramilli, Layne Pruitt, Kristina Reppond, Madison N. Shyer, Jessica Cambric, Ryan Gadd, Ilya J. Finkelstein, Jimmy Gollihar, and James M. Musser |
| EPI_ISL_2202702 | Aegis Sciences Corporation | Centers for Disease Control and Prevention Division of Viral Diseases, Pathogen Discovery | Dakota Howard, Dhwani Batra, Peter W. Cook, Kara Moser, Adrian Paskey, Jason Caravas, Benjamin Rambo-Martin, Shatavia Morrison, Christopher Gulvick, Scott Sammons, Yvette Unoarumhi, Darlene Wagner, Matthew Schmerer, Cyndi Clark, Patrick Campbell, Rob Case, Vikramsinha Ghorpade, Holly Houdeshell, Ola Kvalvaag, Dillon Nall, Ethan Sanders, Alec Vest, Shaun Westlund, Matthew Hardison, Clinton R. Paden, Duncan MacCannell |
| EPI_ISL_2202703 | Houston Methodist Hospital | Houston Methodist Hospital | Randall J. Olsen, Paul A. Christensen, S. Wesley Long, Sishir Subedi, Robert Olson, Marcus Nguyen, James J. Davis, Matthew Ojeda Saavedra, Prasanti Yerramilli, Layne Pruitt, Kristina Reppond, Madison N. Shyer, Jessica Cambric, Ryan Gadd, Ilya J. Finkelstein, Jimmy Gollihar, and James |

| | | | |
|---|---|---|---|
| | | | M. Musser |
| EPI_ISL_2202704 | Aegis Sciences Corporation | Centers for Disease Control and Prevention Division of Viral Diseases, Pathogen Discovery | Dakota Howard, Dhwani Batra, Peter W. Cook, Kara Moser, Adrian Paskey, Jason Caravas, Benjamin Rambo-Martin, Shatavia Morrison, Christopher Gulvick, Scott Sammons, Yvette Unoarumhi, Darlene Wagner, Matthew Schmerer, Cyndi Clark, Patrick Campbell, Rob Case, Vikramsinha Ghorpade, Holly Houdeshell, Ola Kvalvaag, Dillon Nall, Ethan Sanders, Alec Vest, Shaun Westlund, Matthew Hardison, Clinton R. Paden, Duncan MacCannell |
| EPI_ISL_2202705 | Houston Methodist Hospital | Houston Methodist Hospital | Randall J. Olsen, Paul A. Christensen, S. Wesley Long, Sishir Subedi, Robert Olson, Marcus Nguyen, James J. Davis, Matthew Ojeda Saavedra, Prasanti Yerramilli, Layne Pruitt, Kristina Reppond, Madison N. Shyer, Jessica Cambric, Ryan Gadd, Ilya J. Finkelstein, Jimmy Gollihar, and James M. Musser |
| EPI_ISL_2202706 | Aegis Sciences Corporation | Centers for Disease Control and Prevention Division of Viral Diseases, Pathogen Discovery | Dakota Howard, Dhwani Batra, Peter W. Cook, Kara Moser, Adrian Paskey, Jason Caravas, Benjamin Rambo-Martin, Shatavia Morrison, Christopher Gulvick, Scott Sammons, Yvette Unoarumhi, Darlene Wagner, Matthew Schmerer, Cyndi Clark, Patrick Campbell, Rob Case, Vikramsinha Ghorpade, Holly Houdeshell, Ola Kvalvaag, Dillon Nall, Ethan Sanders, Alec Vest, Shaun Westlund, Matthew Hardison, Clinton R. Paden, Duncan MacCannell |
| EPI_ISL_2202707 | Houston Methodist Hospital | Houston Methodist Hospital | Randall J. Olsen, Paul A. Christensen, S. Wesley Long, Sishir Subedi, Robert Olson, Marcus Nguyen, James J. Davis, Matthew Ojeda Saavedra, Prasanti Yerramilli, Layne Pruitt, Kristina Reppond, Madison N. Shyer, Jessica Cambric, Ryan Gadd, Ilya J. Finkelstein, Jimmy Gollihar, and James M. Musser |
| EPI_ISL_2202708, EPI_ISL_2202709, EPI_ISL_2202710 | Aegis Sciences Corporation | Centers for Disease Control and Prevention Division of Viral Diseases, Pathogen Discovery | Dakota Howard, Dhwani Batra, Peter W. Cook, Kara Moser, Adrian Paskey, Jason Caravas, Benjamin Rambo-Martin, Shatavia Morrison, Christopher Gulvick, Scott Sammons, Yvette Unoarumhi, Darlene Wagner, Matthew Schmerer, Cyndi Clark, Patrick Campbell, Rob Case, Vikramsinha Ghorpade, Holly Houdeshell, Ola Kvalvaag, Dillon Nall, Ethan Sanders, Alec Vest, Shaun Westlund, Matthew Hardison, Clinton R. Paden, Duncan MacCannell |
| EPI_ISL_2202713 | Houston Methodist Hospital | Houston Methodist Hospital | Randall J. Olsen, Paul A. Christensen, S. Wesley Long, Sishir Subedi, Robert Olson, Marcus Nguyen, James J. Davis, Matthew Ojeda Saavedra, Prasanti Yerramilli, Layne Pruitt, Kristina Reppond, Madison N. Shyer, Jessica Cambric, Ryan Gadd, Ilya J. Finkelstein, Jimmy Gollihar, and James M. Musser |
| EPI_ISL_2202714 | Aegis Sciences Corporation | Centers for Disease Control and Prevention Division of Viral Diseases, Pathogen Discovery | Dakota Howard, Dhwani Batra, Peter W. Cook, Kara Moser, Adrian Paskey, Jason Caravas, Benjamin Rambo-Martin, Shatavia Morrison, Christopher Gulvick, Scott Sammons, Yvette Unoarumhi, Darlene Wagner, Matthew Schmerer, Cyndi Clark, Patrick Campbell, Rob Case, Vikramsinha Ghorpade, Holly Houdeshell, Ola Kvalvaag, Dillon Nall, Ethan Sanders, Alec Vest, Shaun Westlund, Matthew Hardison, Clinton R. Paden, Duncan MacCannell |
| EPI_ISL_2202715 | Houston Methodist Hospital | Houston Methodist Hospital | Randall J. Olsen, Paul A. Christensen, S. Wesley Long, Sishir Subedi, Robert Olson, Marcus Nguyen, James J. Davis, Matthew Ojeda Saavedra, Prasanti Yerramilli, Layne Pruitt, Kristina Reppond, Madison N. Shyer, Jessica Cambric, Ryan Gadd, Ilya J. Finkelstein, Jimmy Gollihar, and James M. Musser |
| EPI_ISL_2202716 | Aegis Sciences Corporation | Centers for Disease Control and Prevention Division of Viral Diseases, Pathogen Discovery | Dakota Howard, Dhwani Batra, Peter W. Cook, Kara Moser, Adrian Paskey, Jason Caravas, Benjamin Rambo-Martin, Shatavia Morrison, Christopher Gulvick, Scott Sammons, Yvette Unoarumhi, Darlene Wagner, Matthew Schmerer, Cyndi Clark, Patrick Campbell, Rob Case, Vikramsinha Ghorpade, Holly Houdeshell, Ola Kvalvaag, Dillon Nall, Ethan Sanders, Alec Vest, Shaun Westlund, Matthew Hardison, Clinton R. Paden, Duncan MacCannell |
| EPI_ISL_2202717 | NYU Langone Health | Departments of Pathology and Medicine, New York University School of Medicine | Adriana Heguy, Dacia Dimartino, Emily Guzman, Christian Marier, Peter Meyn, Sitharam Ramaswami, Gael Westby, Paul Zappile, Yutong Zhang, Paolo Cotzia, Guiqing Wang |
| EPI_ISL_2202718 | Aegis Sciences Corporation | Centers for Disease Control and Prevention Division of Viral Diseases, Pathogen Discovery | Dakota Howard, Dhwani Batra, Peter W. Cook, Kara Moser, Adrian Paskey, Jason Caravas, Benjamin Rambo-Martin, Shatavia Morrison, Christopher Gulvick, Scott Sammons, Yvette Unoarumhi, Darlene Wagner, Matthew Schmerer, Cyndi Clark, Patrick Campbell, Rob Case, Vikramsinha Ghorpade, Holly Houdeshell, Ola Kvalvaag, Dillon Nall, Ethan Sanders, Alec Vest, Shaun Westlund, Matthew Hardison, Clinton R. Paden, Duncan MacCannell |
| EPI_ISL_2202719 | Houston Methodist Hospital | Houston Methodist Hospital | Randall J. Olsen, Paul A. Christensen, S. Wesley Long, Sishir Subedi, Robert Olson, Marcus Nguyen, James J. Davis, Matthew Ojeda Saavedra, Prasanti Yerramilli, Layne Pruitt, Kristina Reppond, Madison N. Shyer, Jessica Cambric, Ryan Gadd, Ilya J. Finkelstein, Jimmy Gollihar, and James M. Musser |
| EPI_ISL_2202720 | NYU Langone Health | Departments of Pathology and Medicine, New York University School of Medicine | Adriana Heguy, Dacia Dimartino, Emily Guzman, Christian Marier, Peter Meyn, Sitharam Ramaswami, Gael Westby, Paul Zappile, Yutong Zhang, Paolo Cotzia, Guiqing Wang |
| EPI_ISL_2202721 | Aegis Sciences Corporation | Centers for Disease Control and Prevention Division of Viral Diseases, Pathogen Discovery | Dakota Howard, Dhwani Batra, Peter W. Cook, Kara Moser, Adrian Paskey, Jason Caravas, Benjamin Rambo-Martin, Shatavia Morrison, Christopher Gulvick, Scott Sammons, Yvette Unoarumhi, Darlene Wagner, Matthew Schmerer, Cyndi Clark, Patrick Campbell, Rob Case, Vikramsinha Ghorpade, Holly Houdeshell, Ola Kvalvaag, Dillon Nall, Ethan Sanders, Alec Vest, Shaun Westlund, Matthew Hardison, Clinton R. Paden, Duncan MacCannell |
| EPI_ISL_2202722 | NYU Langone Health | Departments of Pathology and Medicine, New York University School of Medicine | Adriana Heguy, Dacia Dimartino, Emily Guzman, Christian Marier, Peter Meyn, Sitharam Ramaswami, Gael Westby, Paul Zappile, Yutong Zhang, Paolo Cotzia, Guiqing Wang |
| EPI_ISL_2202723 | Houston Methodist Hospital | Houston Methodist Hospital | Randall J. Olsen, Paul A. Christensen, S. Wesley Long, Sishir Subedi, Robert Olson, Marcus Nguyen, James J. Davis, Matthew Ojeda Saavedra, Prasanti Yerramilli, Layne Pruitt, Kristina Reppond, Madison N. Shyer, Jessica Cambric, Ryan Gadd, Ilya J. Finkelstein, Jimmy Gollihar, and James M. Musser |
| EPI_ISL_2202724 | NYU Langone Health | Departments of Pathology and Medicine, New York University School of Medicine | Adriana Heguy, Dacia Dimartino, Emily Guzman, Christian Marier, Peter Meyn, Sitharam Ramaswami, Gael Westby, Paul Zappile, Yutong Zhang, Paolo Cotzia, Guiqing Wang |
| EPI_ISL_2202725 | Aegis Sciences Corporation | Centers for Disease Control and Prevention Division of Viral Diseases, Pathogen Discovery | Dakota Howard, Dhwani Batra, Peter W. Cook, Kara Moser, Adrian Paskey, Jason Caravas, Benjamin Rambo-Martin, Shatavia Morrison, Christopher Gulvick, Scott Sammons, Yvette Unoarumhi, Darlene Wagner, Matthew Schmerer, Cyndi Clark, Patrick Campbell, Rob Case, Vikramsinha Ghorpade, Holly Houdeshell, Ola Kvalvaag, Dillon Nall, Ethan Sanders, Alec Vest, Shaun Westlund, Matthew Hardison, Clinton R. Paden, Duncan MacCannell |
| EPI_ISL_2202726 | NYU Langone Health | Departments of Pathology and Medicine, New York University School of Medicine | Adriana Heguy, Dacia Dimartino, Emily Guzman, Christian Marier, Peter Meyn, Sitharam Ramaswami, Gael Westby, Paul Zappile, Yutong Zhang, Paolo Cotzia, Guiqing Wang |
| EPI_ISL_2202727 | Houston Methodist Hospital | Houston Methodist Hospital | Randall J. Olsen, Paul A. Christensen, S. Wesley Long, Sishir Subedi, Robert Olson, Marcus Nguyen, James J. Davis, Matthew Ojeda Saavedra, Prasanti Yerramilli, Layne Pruitt, Kristina Reppond, Madison N. Shyer, Jessica Cambric, Ryan Gadd, Ilya J. Finkelstein, Jimmy Gollihar, and James M. Musser |
| EPI_ISL_2202728 | NYU Langone Health | Departments of Pathology and Medicine, New York University School of Medicine | Adriana Heguy, Dacia Dimartino, Emily Guzman, Christian Marier, Peter Meyn, Sitharam Ramaswami, Gael Westby, Paul Zappile, Yutong Zhang, Paolo Cotzia, Guiqing Wang |
| EPI_ISL_2202729 | Aegis Sciences Corporation | Centers for Disease Control and Prevention Division of Viral Diseases, Pathogen Discovery | Dakota Howard, Dhwani Batra, Peter W. Cook, Kara Moser, Adrian Paskey, Jason Caravas, Benjamin Rambo-Martin, Shatavia Morrison, Christopher Gulvick, Scott Sammons, Yvette Unoarumhi, Darlene Wagner, Matthew Schmerer, Cyndi Clark, Patrick Campbell, Rob Case, Vikramsinha Ghorpade, Holly Houdeshell, Ola Kvalvaag, Dillon Nall, Ethan Sanders, Alec Vest, Shaun Westlund, Matthew Hardison, Clinton R. Paden, Duncan MacCannell |
| EPI_ISL_2202730, EPI_ISL_2202731 | NYU Langone Health | Departments of Pathology and Medicine, New York University School of Medicine | Adriana Heguy, Dacia Dimartino, Emily Guzman, Christian Marier, Peter Meyn, Sitharam Ramaswami, Gael Westby, Paul Zappile, Yutong Zhang, Paolo Cotzia, Guiqing Wang |
| EPI_ISL_2202732 | Houston Methodist Hospital | Houston Methodist Hospital | Randall J. Olsen, Paul A. Christensen, S. Wesley Long, Sishir Subedi, Robert Olson, Marcus Nguyen, James J. Davis, Matthew Ojeda Saavedra, Prasanti Yerramilli, Layne Pruitt, Kristina Reppond, Madison N. Shyer, Jessica Cambric, Ryan Gadd, Ilya J. Finkelstein, Jimmy Gollihar, and James M. Musser |
| EPI_ISL_2202733 | Aegis Sciences Corporation | Centers for Disease Control and Prevention Division of Viral Diseases, Pathogen Discovery | Dakota Howard, Dhwani Batra, Peter W. Cook, Kara Moser, Adrian Paskey, Jason Caravas, Benjamin Rambo-Martin, Shatavia Morrison, Christopher Gulvick, Scott Sammons, Yvette Unoarumhi, Darlene Wagner, Matthew Schmerer, Cyndi Clark, Patrick Campbell, Rob Case, Vikramsinha Ghorpade, Holly Houdeshell, Ola Kvalvaag, Dillon Nall, Ethan Sanders, Alec Vest, Shaun Westlund, Matthew Hardison, Clinton R. Paden, Duncan MacCannell |
| EPI_ISL_2202734 | NYU Langone Health | Departments of Pathology and Medicine, New York University School of Medicine | Adriana Heguy, Dacia Dimartino, Emily Guzman, Christian Marier, Peter Meyn, Sitharam Ramaswami, Gael Westby, Paul Zappile, Yutong Zhang, Paolo Cotzia, Guiqing Wang |
| EPI_ISL_2202735 | Aegis Sciences Corporation | Centers for Disease Control and Prevention Division of Viral Diseases, Pathogen Discovery | Dakota Howard, Dhwani Batra, Peter W. Cook, Kara Moser, Adrian Paskey, Jason Caravas, Benjamin Rambo-Martin, Shatavia Morrison, Christopher Gulvick, Scott Sammons, Yvette Unoarumhi, Darlene Wagner, Matthew Schmerer, Cyndi Clark, Patrick Campbell, Rob Case, Vikramsinha Ghorpade, Holly Houdeshell, Ola Kvalvaag, Dillon Nall, Ethan Sanders, Alec Vest, Shaun Westlund, Matthew Hardison, Clinton R. Paden, Duncan MacCannell |
| EPI_ISL_2202736 | NYU Langone Health | Departments of Pathology and Medicine, New York University School of Medicine | Adriana Heguy, Dacia Dimartino, Emily Guzman, Christian Marier, Peter Meyn, Sitharam Ramaswami, Gael Westby, Paul Zappile, Yutong Zhang, Paolo Cotzia, Guiqing Wang |
| EPI_ISL_2202737 | Houston Methodist Hospital | Houston Methodist Hospital | Randall J. Olsen, Paul A. Christensen, S. Wesley Long, Sishir Subedi, Robert Olson, Marcus Nguyen, James J. Davis, Matthew Ojeda Saavedra, Prasanti Yerramilli, Layne Pruitt, Kristina Reppond, Madison N. Shyer, Jessica Cambric, Ryan Gadd, Ilya J. Finkelstein, Jimmy Gollihar, and James M. Musser |
| EPI_ISL_2202738 | NYU Langone Health | Departments of Pathology and Medicine, New York University School of Medicine | Adriana Heguy, Dacia Dimartino, Emily Guzman, Christian Marier, Peter Meyn, Sitharam Ramaswami, Gael Westby, Paul Zappile, Yutong Zhang, Paolo Cotzia, Guiqing Wang |
| EPI_ISL_2202739 | Aegis Sciences Corporation | Centers for Disease Control and Prevention Division of Viral Diseases, Pathogen Discovery | Dakota Howard, Dhwani Batra, Peter W. Cook, Kara Moser, Adrian Paskey, Jason Caravas, Benjamin Rambo-Martin, Shatavia Morrison, Christopher Gulvick, Scott Sammons, Yvette Unoarumhi, Darlene Wagner, Matthew Schmerer, Cyndi Clark, Patrick Campbell, Rob Case, Vikramsinha Ghorpade, Holly Houdeshell, Ola Kvalvaag, Dillon Nall, Ethan Sanders, Alec Vest, Shaun Westlund, Matthew Hardison, Clinton R. Paden, Duncan MacCannell |
| EPI_ISL_2202740 | NYU Langone Health | Departments of Pathology and Medicine, New York University School of Medicine | Adriana Heguy, Dacia Dimartino, Emily Guzman, Christian Marier, Peter Meyn, Sitharam Ramaswami, Gael Westby, Paul Zappile, Yutong Zhang, Paolo Cotzia, Guiqing Wang |
| EPI_ISL_2202741 | Houston Methodist Hospital | Houston Methodist Hospital | Randall J. Olsen, Paul A. Christensen, S. Wesley Long, Sishir Subedi, Robert Olson, Marcus Nguyen, James J. Davis, Matthew Ojeda Saavedra, Prasanti Yerramilli, Layne Pruitt, Kristina Reppond, Madison N. Shyer, Jessica Cambric, Ryan Gadd, Ilya J. Finkelstein, Jimmy Gollihar, and James M. Musser |
| EPI_ISL_2202742 | NYU Langone Health | Departments of Pathology and Medicine, New York University School of Medicine | Adriana Heguy, Dacia Dimartino, Emily Guzman, Christian Marier, Peter Meyn, Sitharam Ramaswami, Gael Westby, Paul Zappile, Yutong Zhang, Paolo Cotzia, Guiqing Wang |
| EPI_ISL_2202743 | Aegis Sciences Corporation | Centers for Disease Control and Prevention Division of Viral Diseases, Pathogen Discovery | Dakota Howard, Dhwani Batra, Peter W. Cook, Kara Moser, Adrian Paskey, Jason Caravas, Benjamin Rambo-Martin, Shatavia Morrison, Christopher Gulvick, Scott Sammons, Yvette Unoarumhi, Darlene Wagner, Matthew Schmerer, Cyndi Clark, Patrick Campbell, Rob Case, Vikramsinha Ghorpade, Holly Houdeshell, Ola Kvalvaag, Dillon Nall, Ethan Sanders, Alec Vest, Shaun Westlund, Matthew Hardison, Clinton R. Paden, Duncan MacCannell |
| EPI_ISL_2202745 | Houston Methodist Hospital | Houston Methodist Hospital | Randall J. Olsen, Paul A. Christensen, S. Wesley Long, Sishir Subedi, Robert Olson, Marcus Nguyen, James J. Davis, Matthew Ojeda Saavedra, Prasanti Yerramilli, Layne Pruitt, Kristina Reppond, Madison N. Shyer, Jessica Cambric, Ryan Gadd, Ilya J. Finkelstein, Jimmy Gollihar, and James M. Musser |
| EPI_ISL_2202747, EPI_ISL_2202749 | Aegis Sciences Corporation | Centers for Disease Control and Prevention Division of Viral Diseases, Pathogen Discovery | Dakota Howard, Dhwani Batra, Peter W. Cook, Kara Moser, Adrian Paskey, Jason Caravas, Benjamin Rambo-Martin, Shatavia Morrison, Christopher Gulvick, Scott Sammons, Yvette Unoarumhi, Darlene Wagner, Matthew Schmerer, Cyndi Clark, Patrick Campbell, Rob Case, Vikramsinha Ghorpade, Holly Houdeshell, Ola Kvalvaag, Dillon Nall, Ethan Sanders, Alec Vest, Shaun Westlund, Matthew Hardison, Clinton R. Paden, Duncan MacCannell |
| EPI_ISL_2202750 | Houston Methodist Hospital | Houston Methodist Hospital | Randall J. Olsen, Paul A. Christensen, S. Wesley Long, Sishir Subedi, Robert Olson, Marcus Nguyen, James J. Davis, Matthew Ojeda Saavedra, Prasanti Yerramilli, Layne Pruitt, Kristina Reppond, Madison N. Shyer, Jessica Cambric, Ryan Gadd, Ilya J. Finkelstein, Jimmy Gollihar, and James M. Musser |
| EPI_ISL_2202752 | NYU Langone Health | Departments of Pathology and Medicine, New York University School of Medicine | Adriana Heguy, Dacia Dimartino, Emily Guzman, Christian Marier, Peter Meyn, Sitharam Ramaswami, Gael Westby, Paul Zappile, Yutong Zhang, Paolo Cotzia, Guiqing Wang |

| | | | |
|---|---|---|---|
| EPI_ISL_2202753 | Aegis Sciences Corporation | Centers for Disease Control and Prevention Division of Viral Diseases, Pathogen Discovery | Dakota Howard, Dhwani Batra, Peter W. Cook, Kara Moser, Adrian Paskey, Jason Caravas, Benjamin Rambo-Martin, Shatavia Morrison, Christopher Gulvick, Scott Sammons, Yvette Unoarumhi, Darlene Wagner, Matthew Schmerer, Cyndi Clark, Patrick Campbell, Rob Case, Vikramsinha Ghorpade, Holly Houdeshell, Ola Kvalvaag, Dillon Nall, Ethan Sanders, Alec Vest, Shaun Westlund, Matthew Hardison, Clinton R. Paden, Duncan MacCannell |
| EPI_ISL_2202754 | NYU Langone Health | Departments of Pathology and Medicine, New York University School of Medicine | Adriana Heguy, Dacia Dimartino, Emily Guzman, Christian Marier, Peter Meyn, Sitharam Ramaswami, Gael Westby, Paul Zappile, Yutong Zhang, Paolo Cotzia, Guiqing Wang |
| EPI_ISL_2202755 | Houston Methodist Hospital | Houston Methodist Hospital | Randall J. Olsen, Paul A. Christensen, S. Wesley Long, Sishir Subedi, Robert Olson, Marcus Nguyen, James J. Davis, Matthew Ojeda Saavedra, Prasanti Yerramilli, Layne Pruitt, Kristina Reppond, Madison N. Shyer, Jessica Cambric, Ryan Gadd, Ilya J. Finkelstein, Jimmy Gollihar, and James M. Musser |
| EPI_ISL_2202756 | Aegis Sciences Corporation | Centers for Disease Control and Prevention Division of Viral Diseases, Pathogen Discovery | Dakota Howard, Dhwani Batra, Peter W. Cook, Kara Moser, Adrian Paskey, Jason Caravas, Benjamin Rambo-Martin, Shatavia Morrison, Christopher Gulvick, Scott Sammons, Yvette Unoarumhi, Darlene Wagner, Matthew Schmerer, Cyndi Clark, Patrick Campbell, Rob Case, Vikramsinha Ghorpade, Holly Houdeshell, Ola Kvalvaag, Dillon Nall, Ethan Sanders, Alec Vest, Shaun Westlund, Matthew Hardison, Clinton R. Paden, Duncan MacCannell |
| EPI_ISL_2202757 | NYU Langone Health | Departments of Pathology and Medicine, New York University School of Medicine | Adriana Heguy, Dacia Dimartino, Emily Guzman, Christian Marier, Peter Meyn, Sitharam Ramaswami, Gael Westby, Paul Zappile, Yutong Zhang, Paolo Cotzia, Guiqing Wang |
| EPI_ISL_2202758 | Houston Methodist Hospital | Houston Methodist Hospital | Randall J. Olsen, Paul A. Christensen, S. Wesley Long, Sishir Subedi, Robert Olson, Marcus Nguyen, James J. Davis, Matthew Ojeda Saavedra, Prasanti Yerramilli, Layne Pruitt, Kristina Reppond, Madison N. Shyer, Jessica Cambric, Ryan Gadd, Ilya J. Finkelstein, Jimmy Gollihar, and James M. Musser |
| EPI_ISL_2202759 | NYU Langone Health | Departments of Pathology and Medicine, New York University School of Medicine | Adriana Heguy, Dacia Dimartino, Emily Guzman, Christian Marier, Peter Meyn, Sitharam Ramaswami, Gael Westby, Paul Zappile, Yutong Zhang, Paolo Cotzia, Guiqing Wang |
| EPI_ISL_2202760, EPI_ISL_2202761 | Aegis Sciences Corporation | Centers for Disease Control and Prevention Division of Viral Diseases, Pathogen Discovery | Dakota Howard, Dhwani Batra, Peter W. Cook, Kara Moser, Adrian Paskey, Jason Caravas, Benjamin Rambo-Martin, Shatavia Morrison, Christopher Gulvick, Scott Sammons, Yvette Unoarumhi, Darlene Wagner, Matthew Schmerer, Cyndi Clark, Patrick Campbell, Rob Case, Vikramsinha Ghorpade, Holly Houdeshell, Ola Kvalvaag, Dillon Nall, Ethan Sanders, Alec Vest, Shaun Westlund, Matthew Hardison, Clinton R. Paden, Duncan MacCannell |
| EPI_ISL_2202762 | NYU Langone Health | Departments of Pathology and Medicine, New York University School of Medicine | Adriana Heguy, Dacia Dimartino, Emily Guzman, Christian Marier, Peter Meyn, Sitharam Ramaswami, Gael Westby, Paul Zappile, Yutong Zhang, Paolo Cotzia, Guiqing Wang |
| EPI_ISL_2202763 | Houston Methodist Hospital | Houston Methodist Hospital | Randall J. Olsen, Paul A. Christensen, S. Wesley Long, Sishir Subedi, Robert Olson, Marcus Nguyen, James J. Davis, Matthew Ojeda Saavedra, Prasanti Yerramilli, Layne Pruitt, Kristina Reppond, Madison N. Shyer, Jessica Cambric, Ryan Gadd, Ilya J. Finkelstein, Jimmy Gollihar, and James M. Musser |
| EPI_ISL_2202764 | NYU Langone Health | Departments of Pathology and Medicine, New York University School of Medicine | Adriana Heguy, Dacia Dimartino, Emily Guzman, Christian Marier, Peter Meyn, Sitharam Ramaswami, Gael Westby, Paul Zappile, Yutong Zhang, Paolo Cotzia, Guiqing Wang |
| EPI_ISL_2202765 | Aegis Sciences Corporation | Centers for Disease Control and Prevention Division of Viral Diseases, Pathogen Discovery | Dakota Howard, Dhwani Batra, Peter W. Cook, Kara Moser, Adrian Paskey, Jason Caravas, Benjamin Rambo-Martin, Shatavia Morrison, Christopher Gulvick, Scott Sammons, Yvette Unoarumhi, Darlene Wagner, Matthew Schmerer, Cyndi Clark, Patrick Campbell, Rob Case, Vikramsinha Ghorpade, Holly Houdeshell, Ola Kvalvaag, Dillon Nall, Ethan Sanders, Alec Vest, Shaun Westlund, Matthew Hardison, Clinton R. Paden, Duncan MacCannell |
| EPI_ISL_2202766 | NYU Langone Health | Departments of Pathology and Medicine, New York University School of Medicine | Adriana Heguy, Dacia Dimartino, Emily Guzman, Christian Marier, Peter Meyn, Sitharam Ramaswami, Gael Westby, Paul Zappile, Yutong Zhang, Paolo Cotzia, Guiqing Wang |
| EPI_ISL_2202767 | Houston Methodist Hospital | Houston Methodist Hospital | Randall J. Olsen, Paul A. Christensen, S. Wesley Long, Sishir Subedi, Robert Olson, Marcus Nguyen, James J. Davis, Matthew Ojeda Saavedra, Prasanti Yerramilli, Layne Pruitt, Kristina Reppond, Madison N. Shyer, Jessica Cambric, Ryan Gadd, Ilya J. Finkelstein, Jimmy Gollihar, and James M. Musser |
| EPI_ISL_2202768 | NYU Langone Health | Departments of Pathology and Medicine, New York University School of Medicine | Adriana Heguy, Dacia Dimartino, Emily Guzman, Christian Marier, Peter Meyn, Sitharam Ramaswami, Gael Westby, Paul Zappile, Yutong Zhang, Paolo Cotzia, Guiqing Wang |
| EPI_ISL_2202769 | Aegis Sciences Corporation | Centers for Disease Control and Prevention Division of Viral Diseases, Pathogen Discovery | Dakota Howard, Dhwani Batra, Peter W. Cook, Kara Moser, Adrian Paskey, Jason Caravas, Benjamin Rambo-Martin, Shatavia Morrison, Christopher Gulvick, Scott Sammons, Yvette Unoarumhi, Darlene Wagner, Matthew Schmerer, Cyndi Clark, Patrick Campbell, Rob Case, Vikramsinha Ghorpade, Holly Houdeshell, Ola Kvalvaag, Dillon Nall, Ethan Sanders, Alec Vest, Shaun Westlund, Matthew Hardison, Clinton R. Paden, Duncan MacCannell |
| EPI_ISL_2202770, EPI_ISL_2202771 | NYU Langone Health | Departments of Pathology and Medicine, New York University School of Medicine | Adriana Heguy, Dacia Dimartino, Emily Guzman, Christian Marier, Peter Meyn, Sitharam Ramaswami, Gael Westby, Paul Zappile, Yutong Zhang, Paolo Cotzia, Guiqing Wang |
| EPI_ISL_2202772 | Houston Methodist Hospital | Houston Methodist Hospital | Randall J. Olsen, Paul A. Christensen, S. Wesley Long, Sishir Subedi, Robert Olson, Marcus Nguyen, James J. Davis, Matthew Ojeda Saavedra, Prasanti Yerramilli, Layne Pruitt, Kristina Reppond, Madison N. Shyer, Jessica Cambric, Ryan Gadd, Ilya J. Finkelstein, Jimmy Gollihar, and James M. Musser |
| EPI_ISL_2202773 | Aegis Sciences Corporation | Centers for Disease Control and Prevention Division of Viral Diseases, Pathogen Discovery | Dakota Howard, Dhwani Batra, Peter W. Cook, Kara Moser, Adrian Paskey, Jason Caravas, Benjamin Rambo-Martin, Shatavia Morrison, Christopher Gulvick, Scott Sammons, Yvette Unoarumhi, Darlene Wagner, Matthew Schmerer, Cyndi Clark, Patrick Campbell, Rob Case, Vikramsinha Ghorpade, Holly Houdeshell, Ola Kvalvaag, Dillon Nall, Ethan Sanders, Alec Vest, Shaun Westlund, Matthew Hardison, Clinton R. Paden, Duncan MacCannell |
| EPI_ISL_2202774, EPI_ISL_2202775 | NYU Langone Health | Departments of Pathology and Medicine, New York University School of Medicine | Adriana Heguy, Dacia Dimartino, Emily Guzman, Christian Marier, Peter Meyn, Sitharam Ramaswami, Gael Westby, Paul Zappile, Yutong Zhang, Paolo Cotzia, Guiqing Wang |
| EPI_ISL_2202776 | Aegis Sciences Corporation | Centers for Disease Control and Prevention Division of Viral Diseases, Pathogen Discovery | Dakota Howard, Dhwani Batra, Peter W. Cook, Kara Moser, Adrian Paskey, Jason Caravas, Benjamin Rambo-Martin, Shatavia Morrison, Christopher Gulvick, Scott Sammons, Yvette Unoarumhi, Darlene Wagner, Matthew Schmerer, Cyndi Clark, Patrick Campbell, Rob Case, Vikramsinha Ghorpade, Holly Houdeshell, Ola Kvalvaag, Dillon Nall, Ethan Sanders, Alec Vest, Shaun Westlund, Matthew Hardison, Clinton R. Paden, Duncan MacCannell |
| EPI_ISL_2202777 | NYU Langone Health | Departments of Pathology and Medicine, New York University School of Medicine | Adriana Heguy, Dacia Dimartino, Emily Guzman, Christian Marier, Peter Meyn, Sitharam Ramaswami, Gael Westby, Paul Zappile, Yutong Zhang, Paolo Cotzia, Guiqing Wang |
| EPI_ISL_2202778 | Houston Methodist Hospital | Houston Methodist Hospital | Randall J. Olsen, Paul A. Christensen, S. Wesley Long, Sishir Subedi, Robert Olson, Marcus Nguyen, James J. Davis, Matthew Ojeda Saavedra, Prasanti Yerramilli, Layne Pruitt, Kristina Reppond, Madison N. Shyer, Jessica Cambric, Ryan Gadd, Ilya J. Finkelstein, Jimmy Gollihar, and James M. Musser |
| EPI_ISL_2202779 | Aegis Sciences Corporation | Centers for Disease Control and Prevention Division of Viral Diseases, Pathogen Discovery | Dakota Howard, Dhwani Batra, Peter W. Cook, Kara Moser, Adrian Paskey, Jason Caravas, Benjamin Rambo-Martin, Shatavia Morrison, Christopher Gulvick, Scott Sammons, Yvette Unoarumhi, Darlene Wagner, Matthew Schmerer, Cyndi Clark, Patrick Campbell, Rob Case, Vikramsinha Ghorpade, Holly Houdeshell, Ola Kvalvaag, Dillon Nall, Ethan Sanders, Alec Vest, Shaun Westlund, Matthew Hardison, Clinton R. Paden, Duncan MacCannell |
| EPI_ISL_2202780, EPI_ISL_2202781 | NYU Langone Health | Departments of Pathology and Medicine, New York University School of Medicine | Adriana Heguy, Dacia Dimartino, Emily Guzman, Christian Marier, Peter Meyn, Sitharam Ramaswami, Gael Westby, Paul Zappile, Yutong Zhang, Paolo Cotzia, Guiqing Wang |
| EPI_ISL_2202782 | Aegis Sciences Corporation | Centers for Disease Control and Prevention Division of Viral Diseases, Pathogen Discovery | Dakota Howard, Dhwani Batra, Peter W. Cook, Kara Moser, Adrian Paskey, Jason Caravas, Benjamin Rambo-Martin, Shatavia Morrison, Christopher Gulvick, Scott Sammons, Yvette Unoarumhi, Darlene Wagner, Matthew Schmerer, Cyndi Clark, Patrick Campbell, Rob Case, Vikramsinha Ghorpade, Holly Houdeshell, Ola Kvalvaag, Dillon Nall, Ethan Sanders, Alec Vest, Shaun Westlund, Matthew Hardison, Clinton R. Paden, Duncan MacCannell |
| EPI_ISL_2202783 | NYU Langone Health | Departments of Pathology and Medicine, New York University School of Medicine | Adriana Heguy, Dacia Dimartino, Emily Guzman, Christian Marier, Peter Meyn, Sitharam Ramaswami, Gael Westby, Paul Zappile, Yutong Zhang, Paolo Cotzia, Guiqing Wang |
| EPI_ISL_2202784 | Houston Methodist Hospital | Houston Methodist Hospital | Randall J. Olsen, Paul A. Christensen, S. Wesley Long, Sishir Subedi, Robert Olson, Marcus Nguyen, James J. Davis, Matthew Ojeda Saavedra, Prasanti Yerramilli, Layne Pruitt, Kristina Reppond, Madison N. Shyer, Jessica Cambric, Ryan Gadd, Ilya J. Finkelstein, Jimmy Gollihar, and James M. Musser |
| EPI_ISL_2202785 | NYU Langone Health | Departments of Pathology and Medicine, New York University School of Medicine | Adriana Heguy, Dacia Dimartino, Emily Guzman, Christian Marier, Peter Meyn, Sitharam Ramaswami, Gael Westby, Paul Zappile, Yutong Zhang, Paolo Cotzia, Guiqing Wang |
| EPI_ISL_2202786 | Aegis Sciences Corporation | Centers for Disease Control and Prevention Division of Viral Diseases, Pathogen Discovery | Dakota Howard, Dhwani Batra, Peter W. Cook, Kara Moser, Adrian Paskey, Jason Caravas, Benjamin Rambo-Martin, Shatavia Morrison, Christopher Gulvick, Scott Sammons, Yvette Unoarumhi, Darlene Wagner, Matthew Schmerer, Cyndi Clark, Patrick Campbell, Rob Case, Vikramsinha Ghorpade, Holly Houdeshell, Ola Kvalvaag, Dillon Nall, Ethan Sanders, Alec Vest, Shaun Westlund, Matthew Hardison, Clinton R. Paden, Duncan MacCannell |
| EPI_ISL_2202787 | NYU Langone Health | Departments of Pathology and Medicine, New York University School of Medicine | Adriana Heguy, Dacia Dimartino, Emily Guzman, Christian Marier, Peter Meyn, Sitharam Ramaswami, Gael Westby, Paul Zappile, Yutong Zhang, Paolo Cotzia, Guiqing Wang |
| EPI_ISL_2202788 | Houston Methodist Hospital | Houston Methodist Hospital | Randall J. Olsen, Paul A. Christensen, S. Wesley Long, Sishir Subedi, Robert Olson, Marcus Nguyen, James J. Davis, Matthew Ojeda Saavedra, Prasanti Yerramilli, Layne Pruitt, Kristina Reppond, Madison N. Shyer, Jessica Cambric, Ryan Gadd, Ilya J. Finkelstein, Jimmy Gollihar, and James M. Musser |
| EPI_ISL_2202789 | Aegis Sciences Corporation | Centers for Disease Control and Prevention Division of Viral Diseases, Pathogen Discovery | Dakota Howard, Dhwani Batra, Peter W. Cook, Kara Moser, Adrian Paskey, Jason Caravas, Benjamin Rambo-Martin, Shatavia Morrison, Christopher Gulvick, Scott Sammons, Yvette Unoarumhi, Darlene Wagner, Matthew Schmerer, Cyndi Clark, Patrick Campbell, Rob Case, Vikramsinha Ghorpade, Holly Houdeshell, Ola Kvalvaag, Dillon Nall, Ethan Sanders, Alec Vest, Shaun Westlund, Matthew Hardison, Clinton R. Paden, Duncan MacCannell |
| EPI_ISL_2202790, EPI_ISL_2202791 | NYU Langone Health | Departments of Pathology and Medicine, New York University School of Medicine | Adriana Heguy, Dacia Dimartino, Emily Guzman, Christian Marier, Peter Meyn, Sitharam Ramaswami, Gael Westby, Paul Zappile, Yutong Zhang, Paolo Cotzia, Guiqing Wang |
| EPI_ISL_2202792 | Aegis Sciences Corporation | Centers for Disease Control and Prevention Division of Viral Diseases, Pathogen Discovery | Dakota Howard, Dhwani Batra, Peter W. Cook, Kara Moser, Adrian Paskey, Jason Caravas, Benjamin Rambo-Martin, Shatavia Morrison, Christopher Gulvick, Scott Sammons, Yvette Unoarumhi, Darlene Wagner, Matthew Schmerer, Cyndi Clark, Patrick Campbell, Rob Case, Vikramsinha Ghorpade, Holly Houdeshell, Ola Kvalvaag, Dillon Nall, Ethan Sanders, Alec Vest, Shaun Westlund, Matthew Hardison, Clinton R. Paden, Duncan MacCannell |
| EPI_ISL_2202793 | Houston Methodist Hospital | Houston Methodist Hospital | Randall J. Olsen, Paul A. Christensen, S. Wesley Long, Sishir Subedi, Robert Olson, Marcus Nguyen, James J. Davis, Matthew Ojeda Saavedra, Prasanti Yerramilli, Layne Pruitt, Kristina Reppond, Madison N. Shyer, Jessica Cambric, Ryan Gadd, Ilya J. Finkelstein, Jimmy Gollihar, and James M. Musser |
| EPI_ISL_2202794, EPI_ISL_2202795 | NYU Langone Health | Departments of Pathology and Medicine, New York University School of Medicine | Adriana Heguy, Dacia Dimartino, Emily Guzman, Christian Marier, Peter Meyn, Sitharam Ramaswami, Gael Westby, Paul Zappile, Yutong Zhang, Paolo Cotzia, Guiqing Wang |
| EPI_ISL_2202796 | Aegis Sciences Corporation | Centers for Disease Control and Prevention Division of Viral Diseases, Pathogen Discovery | Dakota Howard, Dhwani Batra, Peter W. Cook, Kara Moser, Adrian Paskey, Jason Caravas, Benjamin Rambo-Martin, Shatavia Morrison, Christopher Gulvick, Scott Sammons, Yvette Unoarumhi, Darlene Wagner, Matthew Schmerer, Cyndi Clark, Patrick Campbell, Rob Case, Vikramsinha Ghorpade, Holly Houdeshell, Ola Kvalvaag, Dillon Nall, Ethan Sanders, Alec Vest, Shaun Westlund, Matthew Hardison, Clinton R. Paden, Duncan MacCannell |
| EPI_ISL_2202797 | NYU Langone Health | Departments of Pathology and Medicine, New York University School of Medicine | Adriana Heguy, Dacia Dimartino, Emily Guzman, Christian Marier, Peter Meyn, Sitharam Ramaswami, Gael Westby, Paul Zappile, Yutong Zhang, Paolo Cotzia, Guiqing Wang |
| EPI_ISL_2202798 | Houston Methodist Hospital | Houston Methodist Hospital | Randall J. Olsen, Paul A. Christensen, S. Wesley Long, Sishir Subedi, Robert Olson, Marcus Nguyen, James J. Davis, Matthew Ojeda Saavedra, Prasanti Yerramilli, Layne Pruitt, Kristina Reppond, Madison N. Shyer, Jessica Cambric, Ryan Gadd, Ilya J. Finkelstein, Jimmy Gollihar, and James M. Musser |

| | | | |
|---|---|---|---|
| EPI_ISL_2202799 | Aegis Sciences Corporation | Centers for Disease Control and Prevention Division of Viral Diseases, Pathogen Discovery | Dakota Howard, Dhwani Batra, Peter W. Cook, Kara Moser, Adrian Paskey, Jason Caravas, Benjamin Rambo-Martin, Shatavia Morrison, Christopher Gulvick, Scott Sammons, Yvette Unoarumhi, Darlene Wagner, Matthew Schmerer, Cyndi Clark, Patrick Campbell, Rob Case, Vikramsinha Ghorpade, Holly Houdeshell, Ola Kvalvaag, Dillon Nall, Ethan Sanders, Alec Vest, Shaun Westlund, Matthew Hardison, Clinton R. Paden, Duncan MacCannell |
| EPI_ISL_2202800, EPI_ISL_2202801 | NYU Langone Health | Departments of Pathology and Medicine, New York University School of Medicine | Adriana Heguy, Dacia Dimartino, Emily Guzman, Christian Marier, Peter Meyn, Sitharam Ramaswami, Gael Westby, Paul Zappile, Yutong Zhang, Paolo Cotzia, Guiqing Wang |
| EPI_ISL_2202802 | Aegis Sciences Corporation | Centers for Disease Control and Prevention Division of Viral Diseases, Pathogen Discovery | Dakota Howard, Dhwani Batra, Peter W. Cook, Kara Moser, Adrian Paskey, Jason Caravas, Benjamin Rambo-Martin, Shatavia Morrison, Christopher Gulvick, Scott Sammons, Yvette Unoarumhi, Darlene Wagner, Matthew Schmerer, Cyndi Clark, Patrick Campbell, Rob Case, Vikramsinha Ghorpade, Holly Houdeshell, Ola Kvalvaag, Dillon Nall, Ethan Sanders, Alec Vest, Shaun Westlund, Matthew Hardison, Clinton R. Paden, Duncan MacCannell |
| EPI_ISL_2202803, EPI_ISL_2202804 | NYU Langone Health | Departments of Pathology and Medicine, New York University School of Medicine | Adriana Heguy, Dacia Dimartino, Emily Guzman, Christian Marier, Peter Meyn, Sitharam Ramaswami, Gael Westby, Paul Zappile, Yutong Zhang, Paolo Cotzia, Guiqing Wang |
| EPI_ISL_2202805 | Houston Methodist Hospital | Houston Methodist Hospital | Randall J. Olsen, Paul A. Christensen, S. Wesley Long, Sishir Subedi, Robert Olson, Marcus Nguyen, James J. Davis, Matthew Ojeda Saavedra, Prasanti Yerramilli, Layne Pruitt, Kristina Reppond, Madison N. Shyer, Jessica Cambric, Ryan Gadd, Ilya J. Finkelstein, Jimmy Gollihar, and James M. Musser |
| EPI_ISL_2202806 | Aegis Sciences Corporation | Centers for Disease Control and Prevention Division of Viral Diseases, Pathogen Discovery | Dakota Howard, Dhwani Batra, Peter W. Cook, Kara Moser, Adrian Paskey, Jason Caravas, Benjamin Rambo-Martin, Shatavia Morrison, Christopher Gulvick, Scott Sammons, Yvette Unoarumhi, Darlene Wagner, Matthew Schmerer, Cyndi Clark, Patrick Campbell, Rob Case, Vikramsinha Ghorpade, Holly Houdeshell, Ola Kvalvaag, Dillon Nall, Ethan Sanders, Alec Vest, Shaun Westlund, Matthew Hardison, Clinton R. Paden, Duncan MacCannell |
| EPI_ISL_2202807, EPI_ISL_2202808 | NYU Langone Health | Departments of Pathology and Medicine, New York University School of Medicine | Adriana Heguy, Dacia Dimartino, Emily Guzman, Christian Marier, Peter Meyn, Sitharam Ramaswami, Gael Westby, Paul Zappile, Yutong Zhang, Paolo Cotzia, Guiqing Wang |
| EPI_ISL_2202809 | Aegis Sciences Corporation | Centers for Disease Control and Prevention Division of Viral Diseases, Pathogen Discovery | Dakota Howard, Dhwani Batra, Peter W. Cook, Kara Moser, Adrian Paskey, Jason Caravas, Benjamin Rambo-Martin, Shatavia Morrison, Christopher Gulvick, Scott Sammons, Yvette Unoarumhi, Darlene Wagner, Matthew Schmerer, Cyndi Clark, Patrick Campbell, Rob Case, Vikramsinha Ghorpade, Holly Houdeshell, Ola Kvalvaag, Dillon Nall, Ethan Sanders, Alec Vest, Shaun Westlund, Matthew Hardison, Clinton R. Paden, Duncan MacCannell |
| EPI_ISL_2202810 | Houston Methodist Hospital | Houston Methodist Hospital | Randall J. Olsen, Paul A. Christensen, S. Wesley Long, Sishir Subedi, Robert Olson, Marcus Nguyen, James J. Davis, Matthew Ojeda Saavedra, Prasanti Yerramilli, Layne Pruitt, Kristina Reppond, Madison N. Shyer, Jessica Cambric, Ryan Gadd, Ilya J. Finkelstein, Jimmy Gollihar, and James M. Musser |
| EPI_ISL_2202811 | NYU Langone Health | Departments of Pathology and Medicine, New York University School of Medicine | Adriana Heguy, Dacia Dimartino, Emily Guzman, Christian Marier, Peter Meyn, Sitharam Ramaswami, Gael Westby, Paul Zappile, Yutong Zhang, Paolo Cotzia, Guiqing Wang |
| EPI_ISL_2202812 | Aegis Sciences Corporation | Centers for Disease Control and Prevention Division of Viral Diseases, Pathogen Discovery | Dakota Howard, Dhwani Batra, Peter W. Cook, Kara Moser, Adrian Paskey, Jason Caravas, Benjamin Rambo-Martin, Shatavia Morrison, Christopher Gulvick, Scott Sammons, Yvette Unoarumhi, Darlene Wagner, Matthew Schmerer, Cyndi Clark, Patrick Campbell, Rob Case, Vikramsinha Ghorpade, Holly Houdeshell, Ola Kvalvaag, Dillon Nall, Ethan Sanders, Alec Vest, Shaun Westlund, Matthew Hardison, Clinton R. Paden, Duncan MacCannell |
| EPI_ISL_2202813 | Houston Methodist Hospital | Houston Methodist Hospital | Randall J. Olsen, Paul A. Christensen, S. Wesley Long, Sishir Subedi, Robert Olson, Marcus Nguyen, James J. Davis, Matthew Ojeda Saavedra, Prasanti Yerramilli, Layne Pruitt, Kristina Reppond, Madison N. Shyer, Jessica Cambric, Ryan Gadd, Ilya J. Finkelstein, Jimmy Gollihar, and James M. Musser |
| EPI_ISL_2202814, EPI_ISL_2202815 | Aegis Sciences Corporation | Centers for Disease Control and Prevention Division of Viral Diseases, Pathogen Discovery | Dakota Howard, Dhwani Batra, Peter W. Cook, Kara Moser, Adrian Paskey, Jason Caravas, Benjamin Rambo-Martin, Shatavia Morrison, Christopher Gulvick, Scott Sammons, Yvette Unoarumhi, Darlene Wagner, Matthew Schmerer, Cyndi Clark, Patrick Campbell, Rob Case, Vikramsinha Ghorpade, Holly Houdeshell, Ola Kvalvaag, Dillon Nall, Ethan Sanders, Alec Vest, Shaun Westlund, Matthew Hardison, Clinton R. Paden, Duncan MacCannell |
| EPI_ISL_2202816 | Houston Methodist Hospital | Houston Methodist Hospital | Randall J. Olsen, Paul A. Christensen, S. Wesley Long, Sishir Subedi, Robert Olson, Marcus Nguyen, James J. Davis, Matthew Ojeda Saavedra, Prasanti Yerramilli, Layne Pruitt, Kristina Reppond, Madison N. Shyer, Jessica Cambric, Ryan Gadd, Ilya J. Finkelstein, Jimmy Gollihar, and James M. Musser |
| EPI_ISL_2202817 | Aegis Sciences Corporation | Centers for Disease Control and Prevention Division of Viral Diseases, Pathogen Discovery | Dakota Howard, Dhwani Batra, Peter W. Cook, Kara Moser, Adrian Paskey, Jason Caravas, Benjamin Rambo-Martin, Shatavia Morrison, Christopher Gulvick, Scott Sammons, Yvette Unoarumhi, Darlene Wagner, Matthew Schmerer, Cyndi Clark, Patrick Campbell, Rob Case, Vikramsinha Ghorpade, Holly Houdeshell, Ola Kvalvaag, Dillon Nall, Ethan Sanders, Alec Vest, Shaun Westlund, Matthew Hardison, Clinton R. Paden, Duncan MacCannell |
| EPI_ISL_2202818 | Houston Methodist Hospital | Houston Methodist Hospital | Randall J. Olsen, Paul A. Christensen, S. Wesley Long, Sishir Subedi, Robert Olson, Marcus Nguyen, James J. Davis, Matthew Ojeda Saavedra, Prasanti Yerramilli, Layne Pruitt, Kristina Reppond, Madison N. Shyer, Jessica Cambric, Ryan Gadd, Ilya J. Finkelstein, Jimmy Gollihar, and James M. Musser |
| EPI_ISL_2202819 | Aegis Sciences Corporation | Centers for Disease Control and Prevention Division of Viral Diseases, Pathogen Discovery | Dakota Howard, Dhwani Batra, Peter W. Cook, Kara Moser, Adrian Paskey, Jason Caravas, Benjamin Rambo-Martin, Shatavia Morrison, Christopher Gulvick, Scott Sammons, Yvette Unoarumhi, Darlene Wagner, Matthew Schmerer, Cyndi Clark, Patrick Campbell, Rob Case, Vikramsinha Ghorpade, Holly Houdeshell, Ola Kvalvaag, Dillon Nall, Ethan Sanders, Alec Vest, Shaun Westlund, Matthew Hardison, Clinton R. Paden, Duncan MacCannell |
| EPI_ISL_2202820 | Houston Methodist Hospital | Houston Methodist Hospital | Randall J. Olsen, Paul A. Christensen, S. Wesley Long, Sishir Subedi, Robert Olson, Marcus Nguyen, James J. Davis, Matthew Ojeda Saavedra, Prasanti Yerramilli, Layne Pruitt, Kristina Reppond, Madison N. Shyer, Jessica Cambric, Ryan Gadd, Ilya J. Finkelstein, Jimmy Gollihar, and James M. Musser |
| EPI_ISL_2202821 | Aegis Sciences Corporation | Centers for Disease Control and Prevention Division of Viral Diseases, Pathogen Discovery | Dakota Howard, Dhwani Batra, Peter W. Cook, Kara Moser, Adrian Paskey, Jason Caravas, Benjamin Rambo-Martin, Shatavia Morrison, Christopher Gulvick, Scott Sammons, Yvette Unoarumhi, Darlene Wagner, Matthew Schmerer, Cyndi Clark, Patrick Campbell, Rob Case, Vikramsinha Ghorpade, Holly Houdeshell, Ola Kvalvaag, Dillon Nall, Ethan Sanders, Alec Vest, Shaun Westlund, Matthew Hardison, Clinton R. Paden, Duncan MacCannell |
| EPI_ISL_2202822 | Houston Methodist Hospital | Houston Methodist Hospital | Randall J. Olsen, Paul A. Christensen, S. Wesley Long, Sishir Subedi, Robert Olson, Marcus Nguyen, James J. Davis, Matthew Ojeda Saavedra, Prasanti Yerramilli, Layne Pruitt, Kristina Reppond, Madison N. Shyer, Jessica Cambric, Ryan Gadd, Ilya J. Finkelstein, Jimmy Gollihar, and James M. Musser |
| EPI_ISL_2202823, EPI_ISL_2202824 | Aegis Sciences Corporation | Centers for Disease Control and Prevention Division of Viral Diseases, Pathogen Discovery | Dakota Howard, Dhwani Batra, Peter W. Cook, Kara Moser, Adrian Paskey, Jason Caravas, Benjamin Rambo-Martin, Shatavia Morrison, Christopher Gulvick, Scott Sammons, Yvette Unoarumhi, Darlene Wagner, Matthew Schmerer, Cyndi Clark, Patrick Campbell, Rob Case, Vikramsinha Ghorpade, Holly Houdeshell, Ola Kvalvaag, Dillon Nall, Ethan Sanders, Alec Vest, Shaun Westlund, Matthew Hardison, Clinton R. Paden, Duncan MacCannell |
| EPI_ISL_2202825 | Houston Methodist Hospital | Houston Methodist Hospital | Randall J. Olsen, Paul A. Christensen, S. Wesley Long, Sishir Subedi, Robert Olson, Marcus Nguyen, James J. Davis, Matthew Ojeda Saavedra, Prasanti Yerramilli, Layne Pruitt, Kristina Reppond, Madison N. Shyer, Jessica Cambric, Ryan Gadd, Ilya J. Finkelstein, Jimmy Gollihar, and James M. Musser |
| EPI_ISL_2202826 | Aegis Sciences Corporation | Centers for Disease Control and Prevention Division of Viral Diseases, Pathogen Discovery | Dakota Howard, Dhwani Batra, Peter W. Cook, Kara Moser, Adrian Paskey, Jason Caravas, Benjamin Rambo-Martin, Shatavia Morrison, Christopher Gulvick, Scott Sammons, Yvette Unoarumhi, Darlene Wagner, Matthew Schmerer, Cyndi Clark, Patrick Campbell, Rob Case, Vikramsinha Ghorpade, Holly Houdeshell, Ola Kvalvaag, Dillon Nall, Ethan Sanders, Alec Vest, Shaun Westlund, Matthew Hardison, Clinton R. Paden, Duncan MacCannell |
| EPI_ISL_2202827 | Houston Methodist Hospital | Houston Methodist Hospital | Randall J. Olsen, Paul A. Christensen, S. Wesley Long, Sishir Subedi, Robert Olson, Marcus Nguyen, James J. Davis, Matthew Ojeda Saavedra, Prasanti Yerramilli, Layne Pruitt, Kristina Reppond, Madison N. Shyer, Jessica Cambric, Ryan Gadd, Ilya J. Finkelstein, Jimmy Gollihar, and James M. Musser |
| EPI_ISL_2202828 | Aegis Sciences Corporation | Centers for Disease Control and Prevention Division of Viral Diseases, Pathogen Discovery | Dakota Howard, Dhwani Batra, Peter W. Cook, Kara Moser, Adrian Paskey, Jason Caravas, Benjamin Rambo-Martin, Shatavia Morrison, Christopher Gulvick, Scott Sammons, Yvette Unoarumhi, Darlene Wagner, Matthew Schmerer, Cyndi Clark, Patrick Campbell, Rob Case, Vikramsinha Ghorpade, Holly Houdeshell, Ola Kvalvaag, Dillon Nall, Ethan Sanders, Alec Vest, Shaun Westlund, Matthew Hardison, Clinton R. Paden, Duncan MacCannell |
| EPI_ISL_2202829 | Houston Methodist Hospital | Houston Methodist Hospital | Randall J. Olsen, Paul A. Christensen, S. Wesley Long, Sishir Subedi, Robert Olson, Marcus Nguyen, James J. Davis, Matthew Ojeda Saavedra, Prasanti Yerramilli, Layne Pruitt, Kristina Reppond, Madison N. Shyer, Jessica Cambric, Ryan Gadd, Ilya J. Finkelstein, Jimmy Gollihar, and James M. Musser |
| EPI_ISL_2202830 | Aegis Sciences Corporation | Centers for Disease Control and Prevention Division of Viral Diseases, Pathogen Discovery | Dakota Howard, Dhwani Batra, Peter W. Cook, Kara Moser, Adrian Paskey, Jason Caravas, Benjamin Rambo-Martin, Shatavia Morrison, Christopher Gulvick, Scott Sammons, Yvette Unoarumhi, Darlene Wagner, Matthew Schmerer, Cyndi Clark, Patrick Campbell, Rob Case, Vikramsinha Ghorpade, Holly Houdeshell, Ola Kvalvaag, Dillon Nall, Ethan Sanders, Alec Vest, Shaun Westlund, Matthew Hardison, Clinton R. Paden, Duncan MacCannell |
| EPI_ISL_2202831 | Houston Methodist Hospital | Houston Methodist Hospital | Randall J. Olsen, Paul A. Christensen, S. Wesley Long, Sishir Subedi, Robert Olson, Marcus Nguyen, James J. Davis, Matthew Ojeda Saavedra, Prasanti Yerramilli, Layne Pruitt, Kristina Reppond, Madison N. Shyer, Jessica Cambric, Ryan Gadd, Ilya J. Finkelstein, Jimmy Gollihar, and James M. Musser |
| EPI_ISL_2202832, EPI_ISL_2202833 | Aegis Sciences Corporation | Centers for Disease Control and Prevention Division of Viral Diseases, Pathogen Discovery | Dakota Howard, Dhwani Batra, Peter W. Cook, Kara Moser, Adrian Paskey, Jason Caravas, Benjamin Rambo-Martin, Shatavia Morrison, Christopher Gulvick, Scott Sammons, Yvette Unoarumhi, Darlene Wagner, Matthew Schmerer, Cyndi Clark, Patrick Campbell, Rob Case, Vikramsinha Ghorpade, Holly Houdeshell, Ola Kvalvaag, Dillon Nall, Ethan Sanders, Alec Vest, Shaun Westlund, Matthew Hardison, Clinton R. Paden, Duncan MacCannell |
| EPI_ISL_2202834 | Houston Methodist Hospital | Houston Methodist Hospital | Randall J. Olsen, Paul A. Christensen, S. Wesley Long, Sishir Subedi, Robert Olson, Marcus Nguyen, James J. Davis, Matthew Ojeda Saavedra, Prasanti Yerramilli, Layne Pruitt, Kristina Reppond, Madison N. Shyer, Jessica Cambric, Ryan Gadd, Ilya J. Finkelstein, Jimmy Gollihar, and James M. Musser |
| EPI_ISL_2202835, EPI_ISL_2202836 | Aegis Sciences Corporation | Centers for Disease Control and Prevention Division of Viral Diseases, Pathogen Discovery | Dakota Howard, Dhwani Batra, Peter W. Cook, Kara Moser, Adrian Paskey, Jason Caravas, Benjamin Rambo-Martin, Shatavia Morrison, Christopher Gulvick, Scott Sammons, Yvette Unoarumhi, Darlene Wagner, Matthew Schmerer, Cyndi Clark, Patrick Campbell, Rob Case, Vikramsinha Ghorpade, Holly Houdeshell, Ola Kvalvaag, Dillon Nall, Ethan Sanders, Alec Vest, Shaun Westlund, Matthew Hardison, Clinton R. Paden, Duncan MacCannell |
| EPI_ISL_2202837 | Houston Methodist Hospital | Houston Methodist Hospital | Randall J. Olsen, Paul A. Christensen, S. Wesley Long, Sishir Subedi, Robert Olson, Marcus Nguyen, James J. Davis, Matthew Ojeda Saavedra, Prasanti Yerramilli, Layne Pruitt, Kristina Reppond, Madison N. Shyer, Jessica Cambric, Ryan Gadd, Ilya J. Finkelstein, Jimmy Gollihar, and James M. Musser |
| EPI_ISL_2202838 | Aegis Sciences Corporation | Centers for Disease Control and Prevention Division of Viral Diseases, Pathogen Discovery | Dakota Howard, Dhwani Batra, Peter W. Cook, Kara Moser, Adrian Paskey, Jason Caravas, Benjamin Rambo-Martin, Shatavia Morrison, Christopher Gulvick, Scott Sammons, Yvette Unoarumhi, Darlene Wagner, Matthew Schmerer, Cyndi Clark, Patrick Campbell, Rob Case, Vikramsinha Ghorpade, Holly Houdeshell, Ola Kvalvaag, Dillon Nall, Ethan Sanders, Alec Vest, Shaun Westlund, Matthew Hardison, Clinton R. Paden, Duncan MacCannell |
| EPI_ISL_2202839 | Houston Methodist Hospital | Houston Methodist Hospital | Randall J. Olsen, Paul A. Christensen, S. Wesley Long, Sishir Subedi, Robert Olson, Marcus Nguyen, James J. Davis, Matthew Ojeda Saavedra, Prasanti Yerramilli, Layne Pruitt, Kristina Reppond, Madison N. Shyer, Jessica Cambric, Ryan Gadd, Ilya J. Finkelstein, Jimmy Gollihar, and James M. Musser |
| EPI_ISL_2202840 | Aegis Sciences Corporation | Centers for Disease Control and Prevention Division of Viral Diseases, Pathogen Discovery | Dakota Howard, Dhwani Batra, Peter W. Cook, Kara Moser, Adrian Paskey, Jason Caravas, Benjamin Rambo-Martin, Shatavia Morrison, Christopher Gulvick, Scott Sammons, Yvette Unoarumhi, Darlene Wagner, Matthew Schmerer, Cyndi Clark, Patrick Campbell, Rob Case, Vikramsinha Ghorpade, Holly Houdeshell, Ola Kvalvaag, Dillon Nall, Ethan Sanders, Alec Vest, Shaun Westlund, Matthew Hardison, Clinton R. Paden, Duncan MacCannell |
| EPI_ISL_2202841 | Houston Methodist Hospital | Houston Methodist Hospital | Randall J. Olsen, Paul A. Christensen, S. Wesley Long, Sishir Subedi, Robert Olson, Marcus Nguyen, James J. Davis, Matthew Ojeda Saavedra, Prasanti Yerramilli, Layne Pruitt, Kristina Reppond, Madison N. Shyer, Jessica Cambric, Ryan Gadd, Ilya J. Finkelstein, Jimmy Gollihar, and James M. Musser |
| EPI_ISL_2202842, EPI_ISL_2202843 | Aegis Sciences Corporation | Centers for Disease Control and Prevention Division of Viral Diseases, Pathogen Discovery | Dakota Howard, Dhwani Batra, Peter W. Cook, Kara Moser, Adrian Paskey, Jason Caravas, Benjamin Rambo-Martin, Shatavia Morrison, Christopher Gulvick, Scott Sammons, Yvette Unoarumhi, Darlene Wagner, Matthew Schmerer, Cyndi Clark, Patrick Campbell, Rob Case, Vikramsinha Ghorpade, Holly Houdeshell, Ola Kvalvaag, Dillon Nall, Ethan Sanders, Alec Vest, Shaun Westlund, Matthew Hardison, Clinton R. Paden, Duncan MacCannell |
| EPI_ISL_2202844 | Houston Methodist Hospital | Houston Methodist Hospital | Randall J. Olsen, Paul A. Christensen, S. Wesley Long, Sishir Subedi, Robert Olson, Marcus Nguyen, James J. Davis, Matthew Ojeda Saavedra, Prasanti Yerramilli, Layne Pruitt, Kristina Reppond, Madison N. Shyer, Jessica Cambric, Ryan Gadd, Ilya J. Finkelstein, Jimmy Gollihar, and James |

| | | | |
|---|---|---|---|
| | | | M. Musser |
| EPI_ISL_2202845 | Aegis Sciences Corporation | Centers for Disease Control and Prevention Division of Viral Diseases, Pathogen Discovery | Dakota Howard, Dhwani Batra, Peter W. Cook, Kara Moser, Adrian Paskey, Jason Caravas, Benjamin Rambo-Martin, Shatavia Morrison, Christopher Gulvick, Scott Sammons, Yvette Unoarumhi, Darlene Wagner, Matthew Schmerer, Cyndi Clark, Patrick Campbell, Rob Case, Vikramsinha Ghorpade, Holly Houdeshell, Ola Kvalvaag, Dillon Nall, Ethan Sanders, Alec Vest, Shaun Westlund, Matthew Hardison, Clinton R. Paden, Duncan MacCannell |
| EPI_ISL_2202846 | Houston Methodist Hospital | Houston Methodist Hospital | Randall J. Olsen, Paul A. Christensen, S. Wesley Long, Sishir Subedi, Robert Olson, Marcus Nguyen, James J. Davis, Matthew Ojeda Saavedra, Prasanti Yerramilli, Layne Pruitt, Kristina Reppond, Madison N. Shyer, Jessica Cambric, Ryan Gadd, Ilya J. Finkelstein, Jimmy Gollihar, and James M. Musser |
| EPI_ISL_2202847, EPI_ISL_2202848 | Aegis Sciences Corporation | Centers for Disease Control and Prevention Division of Viral Diseases, Pathogen Discovery | Dakota Howard, Dhwani Batra, Peter W. Cook, Kara Moser, Adrian Paskey, Jason Caravas, Benjamin Rambo-Martin, Shatavia Morrison, Christopher Gulvick, Scott Sammons, Yvette Unoarumhi, Darlene Wagner, Matthew Schmerer, Cyndi Clark, Patrick Campbell, Rob Case, Vikramsinha Ghorpade, Holly Houdeshell, Ola Kvalvaag, Dillon Nall, Ethan Sanders, Alec Vest, Shaun Westlund, Matthew Hardison, Clinton R. Paden, Duncan MacCannell |
| EPI_ISL_2202849 | Houston Methodist Hospital | Houston Methodist Hospital | Randall J. Olsen, Paul A. Christensen, S. Wesley Long, Sishir Subedi, Robert Olson, Marcus Nguyen, James J. Davis, Matthew Ojeda Saavedra, Prasanti Yerramilli, Layne Pruitt, Kristina Reppond, Madison N. Shyer, Jessica Cambric, Ryan Gadd, Ilya J. Finkelstein, Jimmy Gollihar, and James M. Musser |
| EPI_ISL_2202850 | Aegis Sciences Corporation | Centers for Disease Control and Prevention Division of Viral Diseases, Pathogen Discovery | Dakota Howard, Dhwani Batra, Peter W. Cook, Kara Moser, Adrian Paskey, Jason Caravas, Benjamin Rambo-Martin, Shatavia Morrison, Christopher Gulvick, Scott Sammons, Yvette Unoarumhi, Darlene Wagner, Matthew Schmerer, Cyndi Clark, Patrick Campbell, Rob Case, Vikramsinha Ghorpade, Holly Houdeshell, Ola Kvalvaag, Dillon Nall, Ethan Sanders, Alec Vest, Shaun Westlund, Matthew Hardison, Clinton R. Paden, Duncan MacCannell |
| EPI_ISL_2202851 | Houston Methodist Hospital | Houston Methodist Hospital | Randall J. Olsen, Paul A. Christensen, S. Wesley Long, Sishir Subedi, Robert Olson, Marcus Nguyen, James J. Davis, Matthew Ojeda Saavedra, Prasanti Yerramilli, Layne Pruitt, Kristina Reppond, Madison N. Shyer, Jessica Cambric, Ryan Gadd, Ilya J. Finkelstein, Jimmy Gollihar, and James M. Musser |
| EPI_ISL_2202852, EPI_ISL_2202853 | Aegis Sciences Corporation | Centers for Disease Control and Prevention Division of Viral Diseases, Pathogen Discovery | Dakota Howard, Dhwani Batra, Peter W. Cook, Kara Moser, Adrian Paskey, Jason Caravas, Benjamin Rambo-Martin, Shatavia Morrison, Christopher Gulvick, Scott Sammons, Yvette Unoarumhi, Darlene Wagner, Matthew Schmerer, Cyndi Clark, Patrick Campbell, Rob Case, Vikramsinha Ghorpade, Holly Houdeshell, Ola Kvalvaag, Dillon Nall, Ethan Sanders, Alec Vest, Shaun Westlund, Matthew Hardison, Clinton R. Paden, Duncan MacCannell |
| EPI_ISL_2202854 | Houston Methodist Hospital | Houston Methodist Hospital | Randall J. Olsen, Paul A. Christensen, S. Wesley Long, Sishir Subedi, Robert Olson, Marcus Nguyen, James J. Davis, Matthew Ojeda Saavedra, Prasanti Yerramilli, Layne Pruitt, Kristina Reppond, Madison N. Shyer, Jessica Cambric, Ryan Gadd, Ilya J. Finkelstein, Jimmy Gollihar, and James M. Musser |
| EPI_ISL_2202855 | Aegis Sciences Corporation | Centers for Disease Control and Prevention Division of Viral Diseases, Pathogen Discovery | Dakota Howard, Dhwani Batra, Peter W. Cook, Kara Moser, Adrian Paskey, Jason Caravas, Benjamin Rambo-Martin, Shatavia Morrison, Christopher Gulvick, Scott Sammons, Yvette Unoarumhi, Darlene Wagner, Matthew Schmerer, Cyndi Clark, Patrick Campbell, Rob Case, Vikramsinha Ghorpade, Holly Houdeshell, Ola Kvalvaag, Dillon Nall, Ethan Sanders, Alec Vest, Shaun Westlund, Matthew Hardison, Clinton R. Paden, Duncan MacCannell |
| EPI_ISL_2202856 | Houston Methodist Hospital | Houston Methodist Hospital | Randall J. Olsen, Paul A. Christensen, S. Wesley Long, Sishir Subedi, Robert Olson, Marcus Nguyen, James J. Davis, Matthew Ojeda Saavedra, Prasanti Yerramilli, Layne Pruitt, Kristina Reppond, Madison N. Shyer, Jessica Cambric, Ryan Gadd, Ilya J. Finkelstein, Jimmy Gollihar, and James M. Musser |
| EPI_ISL_2202857, EPI_ISL_2202858 | Aegis Sciences Corporation | Centers for Disease Control and Prevention Division of Viral Diseases, Pathogen Discovery | Dakota Howard, Dhwani Batra, Peter W. Cook, Kara Moser, Adrian Paskey, Jason Caravas, Benjamin Rambo-Martin, Shatavia Morrison, Christopher Gulvick, Scott Sammons, Yvette Unoarumhi, Darlene Wagner, Matthew Schmerer, Cyndi Clark, Patrick Campbell, Rob Case, Vikramsinha Ghorpade, Holly Houdeshell, Ola Kvalvaag, Dillon Nall, Ethan Sanders, Alec Vest, Shaun Westlund, Matthew Hardison, Clinton R. Paden, Duncan MacCannell |
| EPI_ISL_2202859 | Houston Methodist Hospital | Houston Methodist Hospital | Randall J. Olsen, Paul A. Christensen, S. Wesley Long, Sishir Subedi, Robert Olson, Marcus Nguyen, James J. Davis, Matthew Ojeda Saavedra, Prasanti Yerramilli, Layne Pruitt, Kristina Reppond, Madison N. Shyer, Jessica Cambric, Ryan Gadd, Ilya J. Finkelstein, Jimmy Gollihar, and James M. Musser |
| EPI_ISL_2202860 | Aegis Sciences Corporation | Centers for Disease Control and Prevention Division of Viral Diseases, Pathogen Discovery | Dakota Howard, Dhwani Batra, Peter W. Cook, Kara Moser, Adrian Paskey, Jason Caravas, Benjamin Rambo-Martin, Shatavia Morrison, Christopher Gulvick, Scott Sammons, Yvette Unoarumhi, Darlene Wagner, Matthew Schmerer, Cyndi Clark, Patrick Campbell, Rob Case, Vikramsinha Ghorpade, Holly Houdeshell, Ola Kvalvaag, Dillon Nall, Ethan Sanders, Alec Vest, Shaun Westlund, Matthew Hardison, Clinton R. Paden, Duncan MacCannell |
| EPI_ISL_2202861 | Houston Methodist Hospital | Houston Methodist Hospital | Randall J. Olsen, Paul A. Christensen, S. Wesley Long, Sishir Subedi, Robert Olson, Marcus Nguyen, James J. Davis, Matthew Ojeda Saavedra, Prasanti Yerramilli, Layne Pruitt, Kristina Reppond, Madison N. Shyer, Jessica Cambric, Ryan Gadd, Ilya J. Finkelstein, Jimmy Gollihar, and James M. Musser |
| EPI_ISL_2202862, EPI_ISL_2202863 | Aegis Sciences Corporation | Centers for Disease Control and Prevention Division of Viral Diseases, Pathogen Discovery | Dakota Howard, Dhwani Batra, Peter W. Cook, Kara Moser, Adrian Paskey, Jason Caravas, Benjamin Rambo-Martin, Shatavia Morrison, Christopher Gulvick, Scott Sammons, Yvette Unoarumhi, Darlene Wagner, Matthew Schmerer, Cyndi Clark, Patrick Campbell, Rob Case, Vikramsinha Ghorpade, Holly Houdeshell, Ola Kvalvaag, Dillon Nall, Ethan Sanders, Alec Vest, Shaun Westlund, Matthew Hardison, Clinton R. Paden, Duncan MacCannell |
| EPI_ISL_2202864 | Houston Methodist Hospital | Houston Methodist Hospital | Randall J. Olsen, Paul A. Christensen, S. Wesley Long, Sishir Subedi, Robert Olson, Marcus Nguyen, James J. Davis, Matthew Ojeda Saavedra, Prasanti Yerramilli, Layne Pruitt, Kristina Reppond, Madison N. Shyer, Jessica Cambric, Ryan Gadd, Ilya J. Finkelstein, Jimmy Gollihar, and James M. Musser |
| EPI_ISL_2202865 | Aegis Sciences Corporation | Centers for Disease Control and Prevention Division of Viral Diseases, Pathogen Discovery | Dakota Howard, Dhwani Batra, Peter W. Cook, Kara Moser, Adrian Paskey, Jason Caravas, Benjamin Rambo-Martin, Shatavia Morrison, Christopher Gulvick, Scott Sammons, Yvette Unoarumhi, Darlene Wagner, Matthew Schmerer, Cyndi Clark, Patrick Campbell, Rob Case, Vikramsinha Ghorpade, Holly Houdeshell, Ola Kvalvaag, Dillon Nall, Ethan Sanders, Alec Vest, Shaun Westlund, Matthew Hardison, Clinton R. Paden, Duncan MacCannell |
| EPI_ISL_2202866 | Houston Methodist Hospital | Houston Methodist Hospital | Randall J. Olsen, Paul A. Christensen, S. Wesley Long, Sishir Subedi, Robert Olson, Marcus Nguyen, James J. Davis, Matthew Ojeda Saavedra, Prasanti Yerramilli, Layne Pruitt, Kristina Reppond, Madison N. Shyer, Jessica Cambric, Ryan Gadd, Ilya J. Finkelstein, Jimmy Gollihar, and James M. Musser |
| EPI_ISL_2202867 | Aegis Sciences Corporation | Centers for Disease Control and Prevention Division of Viral Diseases, Pathogen Discovery | Dakota Howard, Dhwani Batra, Peter W. Cook, Kara Moser, Adrian Paskey, Jason Caravas, Benjamin Rambo-Martin, Shatavia Morrison, Christopher Gulvick, Scott Sammons, Yvette Unoarumhi, Darlene Wagner, Matthew Schmerer, Cyndi Clark, Patrick Campbell, Rob Case, Vikramsinha Ghorpade, Holly Houdeshell, Ola Kvalvaag, Dillon Nall, Ethan Sanders, Alec Vest, Shaun Westlund, Matthew Hardison, Clinton R. Paden, Duncan MacCannell |
| EPI_ISL_2202868 | Houston Methodist Hospital | Houston Methodist Hospital | Randall J. Olsen, Paul A. Christensen, S. Wesley Long, Sishir Subedi, Robert Olson, Marcus Nguyen, James J. Davis, Matthew Ojeda Saavedra, Prasanti Yerramilli, Layne Pruitt, Kristina Reppond, Madison N. Shyer, Jessica Cambric, Ryan Gadd, Ilya J. Finkelstein, Jimmy Gollihar, and James M. Musser |
| EPI_ISL_2202869 | Aegis Sciences Corporation | Centers for Disease Control and Prevention Division of Viral Diseases, Pathogen Discovery | Dakota Howard, Dhwani Batra, Peter W. Cook, Kara Moser, Adrian Paskey, Jason Caravas, Benjamin Rambo-Martin, Shatavia Morrison, Christopher Gulvick, Scott Sammons, Yvette Unoarumhi, Darlene Wagner, Matthew Schmerer, Cyndi Clark, Patrick Campbell, Rob Case, Vikramsinha Ghorpade, Holly Houdeshell, Ola Kvalvaag, Dillon Nall, Ethan Sanders, Alec Vest, Shaun Westlund, Matthew Hardison, Clinton R. Paden, Duncan MacCannell |
| EPI_ISL_2202870 | Houston Methodist Hospital | Houston Methodist Hospital | Randall J. Olsen, Paul A. Christensen, S. Wesley Long, Sishir Subedi, Robert Olson, Marcus Nguyen, James J. Davis, Matthew Ojeda Saavedra, Prasanti Yerramilli, Layne Pruitt, Kristina Reppond, Madison N. Shyer, Jessica Cambric, Ryan Gadd, Ilya J. Finkelstein, Jimmy Gollihar, and James M. Musser |
| EPI_ISL_2202871 | Aegis Sciences Corporation | Centers for Disease Control and Prevention Division of Viral Diseases, Pathogen Discovery | Dakota Howard, Dhwani Batra, Peter W. Cook, Kara Moser, Adrian Paskey, Jason Caravas, Benjamin Rambo-Martin, Shatavia Morrison, Christopher Gulvick, Scott Sammons, Yvette Unoarumhi, Darlene Wagner, Matthew Schmerer, Cyndi Clark, Patrick Campbell, Rob Case, Vikramsinha Ghorpade, Holly Houdeshell, Ola Kvalvaag, Dillon Nall, Ethan Sanders, Alec Vest, Shaun Westlund, Matthew Hardison, Clinton R. Paden, Duncan MacCannell |
| EPI_ISL_2202872 | New Mexico Department of Health Scientific Laboratory Division | Los Alamos National Laboratory Bioscience Division | Chien-Chi Lo, Migun Shakya, Cheryl Gleasner, Kim McMurry, Alina Deshpande, Twila Kunde, Joseph Hicks, Michael Edwards, Patrick Chain |
| EPI_ISL_2202873 | Aegis Sciences Corporation | Centers for Disease Control and Prevention Division of Viral Diseases, Pathogen Discovery | Dakota Howard, Dhwani Batra, Peter W. Cook, Kara Moser, Adrian Paskey, Jason Caravas, Benjamin Rambo-Martin, Shatavia Morrison, Christopher Gulvick, Scott Sammons, Yvette Unoarumhi, Darlene Wagner, Matthew Schmerer, Cyndi Clark, Patrick Campbell, Rob Case, Vikramsinha Ghorpade, Holly Houdeshell, Ola Kvalvaag, Dillon Nall, Ethan Sanders, Alec Vest, Shaun Westlund, Matthew Hardison, Clinton R. Paden, Duncan MacCannell |
| EPI_ISL_2202874, EPI_ISL_2202875 | New Mexico Department of Health Scientific Laboratory Division | Los Alamos National Laboratory Bioscience Division | Chien-Chi Lo, Migun Shakya, Cheryl Gleasner, Kim McMurry, Alina Deshpande, Twila Kunde, Joseph Hicks, Michael Edwards, Patrick Chain |
| EPI_ISL_2202876 | Aegis Sciences Corporation | Centers for Disease Control and Prevention Division of Viral Diseases, Pathogen Discovery | Dakota Howard, Dhwani Batra, Peter W. Cook, Kara Moser, Adrian Paskey, Jason Caravas, Benjamin Rambo-Martin, Shatavia Morrison, Christopher Gulvick, Scott Sammons, Yvette Unoarumhi, Darlene Wagner, Matthew Schmerer, Cyndi Clark, Patrick Campbell, Rob Case, Vikramsinha Ghorpade, Holly Houdeshell, Ola Kvalvaag, Dillon Nall, Ethan Sanders, Alec Vest, Shaun Westlund, Matthew Hardison, Clinton R. Paden, Duncan MacCannell |
| EPI_ISL_2202877 | New Mexico Department of Health Scientific Laboratory Division | Los Alamos National Laboratory Bioscience Division | Chien-Chi Lo, Migun Shakya, Cheryl Gleasner, Kim McMurry, Alina Deshpande, Twila Kunde, Joseph Hicks, Michael Edwards, Patrick Chain |
| EPI_ISL_2202878 | Houston Methodist Hospital | Houston Methodist Hospital | Randall J. Olsen, Paul A. Christensen, S. Wesley Long, Sishir Subedi, Robert Olson, Marcus Nguyen, James J. Davis, Matthew Ojeda Saavedra, Prasanti Yerramilli, Layne Pruitt, Kristina Reppond, Madison N. Shyer, Jessica Cambric, Ryan Gadd, Ilya J. Finkelstein, Jimmy Gollihar, and James M. Musser |
| EPI_ISL_2202879 | New Mexico Department of Health Scientific Laboratory Division | Los Alamos National Laboratory Bioscience Division | Chien-Chi Lo, Migun Shakya, Cheryl Gleasner, Kim McMurry, Alina Deshpande, Twila Kunde, Joseph Hicks, Michael Edwards, Patrick Chain |
| EPI_ISL_2202880 | Aegis Sciences Corporation | Centers for Disease Control and Prevention Division of Viral Diseases, Pathogen Discovery | Dakota Howard, Dhwani Batra, Peter W. Cook, Kara Moser, Adrian Paskey, Jason Caravas, Benjamin Rambo-Martin, Shatavia Morrison, Christopher Gulvick, Scott Sammons, Yvette Unoarumhi, Darlene Wagner, Matthew Schmerer, Cyndi Clark, Patrick Campbell, Rob Case, Vikramsinha Ghorpade, Holly Houdeshell, Ola Kvalvaag, Dillon Nall, Ethan Sanders, Alec Vest, Shaun Westlund, Matthew Hardison, Clinton R. Paden, Duncan MacCannell |
| EPI_ISL_2202881 | New Mexico Department of Health Scientific Laboratory Division | Los Alamos National Laboratory Bioscience Division | Chien-Chi Lo, Migun Shakya, Cheryl Gleasner, Kim McMurry, Alina Deshpande, Twila Kunde, Joseph Hicks, Michael Edwards, Patrick Chain |
| EPI_ISL_2202882 | Houston Methodist Hospital | Houston Methodist Hospital | Randall J. Olsen, Paul A. Christensen, S. Wesley Long, Sishir Subedi, Robert Olson, Marcus Nguyen, James J. Davis, Matthew Ojeda Saavedra, Prasanti Yerramilli, Layne Pruitt, Kristina Reppond, Madison N. Shyer, Jessica Cambric, Ryan Gadd, Ilya J. Finkelstein, Jimmy Gollihar, and James M. Musser |
| EPI_ISL_2202883 | New Mexico Department of Health Scientific Laboratory Division | Los Alamos National Laboratory Bioscience Division | Chien-Chi Lo, Migun Shakya, Cheryl Gleasner, Kim McMurry, Alina Deshpande, Twila Kunde, Joseph Hicks, Michael Edwards, Patrick Chain |
| EPI_ISL_2202884 | Aegis Sciences Corporation | Centers for Disease Control and Prevention Division of Viral Diseases, Pathogen Discovery | Dakota Howard, Dhwani Batra, Peter W. Cook, Kara Moser, Adrian Paskey, Jason Caravas, Benjamin Rambo-Martin, Shatavia Morrison, Christopher Gulvick, Scott Sammons, Yvette Unoarumhi, Darlene Wagner, Matthew Schmerer, Cyndi Clark, Patrick Campbell, Rob Case, Vikramsinha Ghorpade, Holly Houdeshell, Ola Kvalvaag, Dillon Nall, Ethan Sanders, Alec Vest, Shaun Westlund, Matthew Hardison, Clinton R. Paden, Duncan MacCannell |
| EPI_ISL_2202885 | New Mexico Department of Health Scientific Laboratory Division | Los Alamos National Laboratory Bioscience Division | Chien-Chi Lo, Migun Shakya, Cheryl Gleasner, Kim McMurry, Alina Deshpande, Twila Kunde, Joseph Hicks, Michael Edwards, Patrick Chain |
| EPI_ISL_2202886 | Houston Methodist Hospital | Houston Methodist Hospital | Randall J. Olsen, Paul A. Christensen, S. Wesley Long, Sishir Subedi, Robert Olson, Marcus Nguyen, James J. Davis, Matthew Ojeda Saavedra, Prasanti Yerramilli, Layne Pruitt, Kristina Reppond, Madison N. Shyer, Jessica Cambric, Ryan Gadd, Ilya J. Finkelstein, Jimmy Gollihar, and James M. Musser |
| EPI_ISL_2202887 | New Mexico Department of Health Scientific Laboratory Division | Los Alamos National Laboratory Bioscience Division | Chien-Chi Lo, Migun Shakya, Cheryl Gleasner, Kim McMurry, Alina Deshpande, Twila Kunde, Joseph Hicks, Michael Edwards, Patrick Chain |

| | | | |
|---|---|---|---|
| EPI_ISL_2202888 | Aegis Sciences Corporation | Centers for Disease Control and Prevention Division of Viral Diseases, Pathogen Discovery | Dakota Howard, Dhwani Batra, Peter W. Cook, Kara Moser, Adrian Paskey, Jason Caravas, Benjamin Rambo-Martin, Shatavia Morrison, Christopher Gulvick, Scott Sammons, Yvette Unoarumhi, Darlene Wagner, Matthew Schmerer, Cyndi Clark, Patrick Campbell, Rob Case, Vikramsinha Ghorpade, Holly Houdeshell, Ola Kvalvaag, Dillon Nall, Ethan Sanders, Alec Vest, Shaun Westlund, Matthew Hardison, Clinton R. Paden, Duncan MacCannell |
| EPI_ISL_2202889 | New Mexico Department of Health Scientific Laboratory Division | Los Alamos National Laboratory Bioscience Division | Chien-Chi Lo, Migun Shakya, Cheryl Gleasner, Kim McMurry, Alina Deshpande, Twila Kunde, Joseph Hicks, Michael Edwards, Patrick Chain |
| EPI_ISL_2202890 | Houston Methodist Hospital | Houston Methodist Hospital | Randall J. Olsen, Paul A. Christensen, S. Wesley Long, Sishir Subedi, Robert Olson, Marcus Nguyen, James J. Davis, Matthew Ojeda Saavedra, Prasanti Yerramilli, Layne Pruitt, Kristina Reppond, Madison N. Shyer, Jessica Cambric, Ryan Gadd, Ilya J. Finkelstein, Jimmy Gollihar, and James M. Musser |
| EPI_ISL_2202891 | New Mexico Department of Health Scientific Laboratory Division | Los Alamos National Laboratory Bioscience Division | Chien-Chi Lo, Migun Shakya, Cheryl Gleasner, Kim McMurry, Alina Deshpande, Twila Kunde, Joseph Hicks, Michael Edwards, Patrick Chain |
| EPI_ISL_2202892 | Aegis Sciences Corporation | Centers for Disease Control and Prevention Division of Viral Diseases, Pathogen Discovery | Dakota Howard, Dhwani Batra, Peter W. Cook, Kara Moser, Adrian Paskey, Jason Caravas, Benjamin Rambo-Martin, Shatavia Morrison, Christopher Gulvick, Scott Sammons, Yvette Unoarumhi, Darlene Wagner, Matthew Schmerer, Cyndi Clark, Patrick Campbell, Rob Case, Vikramsinha Ghorpade, Holly Houdeshell, Ola Kvalvaag, Dillon Nall, Ethan Sanders, Alec Vest, Shaun Westlund, Matthew Hardison, Clinton R. Paden, Duncan MacCannell |
| EPI_ISL_2202893 | New Mexico Department of Health Scientific Laboratory Division | Los Alamos National Laboratory Bioscience Division | Chien-Chi Lo, Migun Shakya, Cheryl Gleasner, Kim McMurry, Alina Deshpande, Twila Kunde, Joseph Hicks, Michael Edwards, Patrick Chain |
| EPI_ISL_2202894 | Houston Methodist Hospital | Houston Methodist Hospital | Randall J. Olsen, Paul A. Christensen, S. Wesley Long, Sishir Subedi, Robert Olson, Marcus Nguyen, James J. Davis, Matthew Ojeda Saavedra, Prasanti Yerramilli, Layne Pruitt, Kristina Reppond, Madison N. Shyer, Jessica Cambric, Ryan Gadd, Ilya J. Finkelstein, Jimmy Gollihar, and James M. Musser |
| EPI_ISL_2202895 | New Mexico Department of Health Scientific Laboratory Division | Los Alamos National Laboratory Bioscience Division | Chien-Chi Lo, Migun Shakya, Cheryl Gleasner, Kim McMurry, Alina Deshpande, Twila Kunde, Joseph Hicks, Michael Edwards, Patrick Chain |
| EPI_ISL_2202896 | Aegis Sciences Corporation | Centers for Disease Control and Prevention Division of Viral Diseases, Pathogen Discovery | Dakota Howard, Dhwani Batra, Peter W. Cook, Kara Moser, Adrian Paskey, Jason Caravas, Benjamin Rambo-Martin, Shatavia Morrison, Christopher Gulvick, Scott Sammons, Yvette Unoarumhi, Darlene Wagner, Matthew Schmerer, Cyndi Clark, Patrick Campbell, Rob Case, Vikramsinha Ghorpade, Holly Houdeshell, Ola Kvalvaag, Dillon Nall, Ethan Sanders, Alec Vest, Shaun Westlund, Matthew Hardison, Clinton R. Paden, Duncan MacCannell |
| EPI_ISL_2202897, EPI_ISL_2202898 | New Mexico Department of Health Scientific Laboratory Division | Los Alamos National Laboratory Bioscience Division | Chien-Chi Lo, Migun Shakya, Cheryl Gleasner, Kim McMurry, Alina Deshpande, Twila Kunde, Joseph Hicks, Michael Edwards, Patrick Chain |
| EPI_ISL_2202899 | Houston Methodist Hospital | Houston Methodist Hospital | Randall J. Olsen, Paul A. Christensen, S. Wesley Long, Sishir Subedi, Robert Olson, Marcus Nguyen, James J. Davis, Matthew Ojeda Saavedra, Prasanti Yerramilli, Layne Pruitt, Kristina Reppond, Madison N. Shyer, Jessica Cambric, Ryan Gadd, Ilya J. Finkelstein, Jimmy Gollihar, and James M. Musser |
| EPI_ISL_2202900 | Aegis Sciences Corporation | Centers for Disease Control and Prevention Division of Viral Diseases, Pathogen Discovery | Dakota Howard, Dhwani Batra, Peter W. Cook, Kara Moser, Adrian Paskey, Jason Caravas, Benjamin Rambo-Martin, Shatavia Morrison, Christopher Gulvick, Scott Sammons, Yvette Unoarumhi, Darlene Wagner, Matthew Schmerer, Cyndi Clark, Patrick Campbell, Rob Case, Vikramsinha Ghorpade, Holly Houdeshell, Ola Kvalvaag, Dillon Nall, Ethan Sanders, Alec Vest, Shaun Westlund, Matthew Hardison, Clinton R. Paden, Duncan MacCannell |
| EPI_ISL_2202901 | New Mexico Department of Health Scientific Laboratory Division | Los Alamos National Laboratory Bioscience Division | Chien-Chi Lo, Migun Shakya, Cheryl Gleasner, Kim McMurry, Alina Deshpande, Twila Kunde, Joseph Hicks, Michael Edwards, Patrick Chain |
| EPI_ISL_2202902 | Aegis Sciences Corporation | Centers for Disease Control and Prevention Division of Viral Diseases, Pathogen Discovery | Dakota Howard, Dhwani Batra, Peter W. Cook, Kara Moser, Adrian Paskey, Jason Caravas, Benjamin Rambo-Martin, Shatavia Morrison, Christopher Gulvick, Scott Sammons, Yvette Unoarumhi, Darlene Wagner, Matthew Schmerer, Cyndi Clark, Patrick Campbell, Rob Case, Vikramsinha Ghorpade, Holly Houdeshell, Ola Kvalvaag, Dillon Nall, Ethan Sanders, Alec Vest, Shaun Westlund, Matthew Hardison, Clinton R. Paden, Duncan MacCannell |
| EPI_ISL_2202903 | New Mexico Department of Health Scientific Laboratory Division | Los Alamos National Laboratory Bioscience Division | Chien-Chi Lo, Migun Shakya, Cheryl Gleasner, Kim McMurry, Alina Deshpande, Twila Kunde, Joseph Hicks, Michael Edwards, Patrick Chain |
| EPI_ISL_2202904 | Houston Methodist Hospital | Houston Methodist Hospital | Randall J. Olsen, Paul A. Christensen, S. Wesley Long, Sishir Subedi, Robert Olson, Marcus Nguyen, James J. Davis, Matthew Ojeda Saavedra, Prasanti Yerramilli, Layne Pruitt, Kristina Reppond, Madison N. Shyer, Jessica Cambric, Ryan Gadd, Ilya J. Finkelstein, Jimmy Gollihar, and James M. Musser |
| EPI_ISL_2202905 | New Mexico Department of Health Scientific Laboratory Division | Los Alamos National Laboratory Bioscience Division | Chien-Chi Lo, Migun Shakya, Cheryl Gleasner, Kim McMurry, Alina Deshpande, Twila Kunde, Joseph Hicks, Michael Edwards, Patrick Chain |
| EPI_ISL_2202906 | Aegis Sciences Corporation | Centers for Disease Control and Prevention Division of Viral Diseases, Pathogen Discovery | Dakota Howard, Dhwani Batra, Peter W. Cook, Kara Moser, Adrian Paskey, Jason Caravas, Benjamin Rambo-Martin, Shatavia Morrison, Christopher Gulvick, Scott Sammons, Yvette Unoarumhi, Darlene Wagner, Matthew Schmerer, Cyndi Clark, Patrick Campbell, Rob Case, Vikramsinha Ghorpade, Holly Houdeshell, Ola Kvalvaag, Dillon Nall, Ethan Sanders, Alec Vest, Shaun Westlund, Matthew Hardison, Clinton R. Paden, Duncan MacCannell |
| EPI_ISL_2202907, EPI_ISL_2202908 | New Mexico Department of Health Scientific Laboratory Division | Los Alamos National Laboratory Bioscience Division | Chien-Chi Lo, Migun Shakya, Cheryl Gleasner, Kim McMurry, Alina Deshpande, Twila Kunde, Joseph Hicks, Michael Edwards, Patrick Chain |
| EPI_ISL_2202909 | Houston Methodist Hospital | Houston Methodist Hospital | Randall J. Olsen, Paul A. Christensen, S. Wesley Long, Sishir Subedi, Robert Olson, Marcus Nguyen, James J. Davis, Matthew Ojeda Saavedra, Prasanti Yerramilli, Layne Pruitt, Kristina Reppond, Madison N. Shyer, Jessica Cambric, Ryan Gadd, Ilya J. Finkelstein, Jimmy Gollihar, and James M. Musser |
| EPI_ISL_2202910 | Aegis Sciences Corporation | Centers for Disease Control and Prevention Division of Viral Diseases, Pathogen Discovery | Dakota Howard, Dhwani Batra, Peter W. Cook, Kara Moser, Adrian Paskey, Jason Caravas, Benjamin Rambo-Martin, Shatavia Morrison, Christopher Gulvick, Scott Sammons, Yvette Unoarumhi, Darlene Wagner, Matthew Schmerer, Cyndi Clark, Patrick Campbell, Rob Case, Vikramsinha Ghorpade, Holly Houdeshell, Ola Kvalvaag, Dillon Nall, Ethan Sanders, Alec Vest, Shaun Westlund, Matthew Hardison, Clinton R. Paden, Duncan MacCannell |
| EPI_ISL_2202911, EPI_ISL_2202912 | New Mexico Department of Health Scientific Laboratory Division | Los Alamos National Laboratory Bioscience Division | Chien-Chi Lo, Migun Shakya, Cheryl Gleasner, Kim McMurry, Alina Deshpande, Twila Kunde, Joseph Hicks, Michael Edwards, Patrick Chain |
| EPI_ISL_2202913 | Houston Methodist Hospital | Houston Methodist Hospital | Randall J. Olsen, Paul A. Christensen, S. Wesley Long, Sishir Subedi, Robert Olson, Marcus Nguyen, James J. Davis, Matthew Ojeda Saavedra, Prasanti Yerramilli, Layne Pruitt, Kristina Reppond, Madison N. Shyer, Jessica Cambric, Ryan Gadd, Ilya J. Finkelstein, Jimmy Gollihar, and James M. Musser |
| EPI_ISL_2202914 | Aegis Sciences Corporation | Centers for Disease Control and Prevention Division of Viral Diseases, Pathogen Discovery | Dakota Howard, Dhwani Batra, Peter W. Cook, Kara Moser, Adrian Paskey, Jason Caravas, Benjamin Rambo-Martin, Shatavia Morrison, Christopher Gulvick, Scott Sammons, Yvette Unoarumhi, Darlene Wagner, Matthew Schmerer, Cyndi Clark, Patrick Campbell, Rob Case, Vikramsinha Ghorpade, Holly Houdeshell, Ola Kvalvaag, Dillon Nall, Ethan Sanders, Alec Vest, Shaun Westlund, Matthew Hardison, Clinton R. Paden, Duncan MacCannell |
| EPI_ISL_2202915, EPI_ISL_2202916 | New Mexico Department of Health Scientific Laboratory Division | Los Alamos National Laboratory Bioscience Division | Chien-Chi Lo, Migun Shakya, Cheryl Gleasner, Kim McMurry, Alina Deshpande, Twila Kunde, Joseph Hicks, Michael Edwards, Patrick Chain |
| EPI_ISL_2202917 | Aegis Sciences Corporation | Centers for Disease Control and Prevention Division of Viral Diseases, Pathogen Discovery | Dakota Howard, Dhwani Batra, Peter W. Cook, Kara Moser, Adrian Paskey, Jason Caravas, Benjamin Rambo-Martin, Shatavia Morrison, Christopher Gulvick, Scott Sammons, Yvette Unoarumhi, Darlene Wagner, Matthew Schmerer, Cyndi Clark, Patrick Campbell, Rob Case, Vikramsinha Ghorpade, Holly Houdeshell, Ola Kvalvaag, Dillon Nall, Ethan Sanders, Alec Vest, Shaun Westlund, Matthew Hardison, Clinton R. Paden, Duncan MacCannell |
| EPI_ISL_2202918 | Houston Methodist Hospital | Houston Methodist Hospital | Randall J. Olsen, Paul A. Christensen, S. Wesley Long, Sishir Subedi, Robert Olson, Marcus Nguyen, James J. Davis, Matthew Ojeda Saavedra, Prasanti Yerramilli, Layne Pruitt, Kristina Reppond, Madison N. Shyer, Jessica Cambric, Ryan Gadd, Ilya J. Finkelstein, Jimmy Gollihar, and James M. Musser |
| EPI_ISL_2202919, EPI_ISL_2202920 | New Mexico Department of Health Scientific Laboratory Division | Los Alamos National Laboratory Bioscience Division | Chien-Chi Lo, Migun Shakya, Cheryl Gleasner, Kim McMurry, Alina Deshpande, Twila Kunde, Joseph Hicks, Michael Edwards, Patrick Chain |
| EPI_ISL_2202921 | Aegis Sciences Corporation | Centers for Disease Control and Prevention Division of Viral Diseases, Pathogen Discovery | Dakota Howard, Dhwani Batra, Peter W. Cook, Kara Moser, Adrian Paskey, Jason Caravas, Benjamin Rambo-Martin, Shatavia Morrison, Christopher Gulvick, Scott Sammons, Yvette Unoarumhi, Darlene Wagner, Matthew Schmerer, Cyndi Clark, Patrick Campbell, Rob Case, Vikramsinha Ghorpade, Holly Houdeshell, Ola Kvalvaag, Dillon Nall, Ethan Sanders, Alec Vest, Shaun Westlund, Matthew Hardison, Clinton R. Paden, Duncan MacCannell |
| EPI_ISL_2202922 | New Mexico Department of Health Scientific Laboratory Division | Los Alamos National Laboratory Bioscience Division | Chien-Chi Lo, Migun Shakya, Cheryl Gleasner, Kim McMurry, Alina Deshpande, Twila Kunde, Joseph Hicks, Michael Edwards, Patrick Chain |
| EPI_ISL_2202923 | Houston Methodist Hospital | Houston Methodist Hospital | Randall J. Olsen, Paul A. Christensen, S. Wesley Long, Sishir Subedi, Robert Olson, Marcus Nguyen, James J. Davis, Matthew Ojeda Saavedra, Prasanti Yerramilli, Layne Pruitt, Kristina Reppond, Madison N. Shyer, Jessica Cambric, Ryan Gadd, Ilya J. Finkelstein, Jimmy Gollihar, and James M. Musser |
| EPI_ISL_2202924 | New Mexico Department of Health Scientific Laboratory Division | Los Alamos National Laboratory Bioscience Division | Chien-Chi Lo, Migun Shakya, Cheryl Gleasner, Kim McMurry, Alina Deshpande, Twila Kunde, Joseph Hicks, Michael Edwards, Patrick Chain |
| EPI_ISL_2202925 | Aegis Sciences Corporation | Centers for Disease Control and Prevention Division of Viral Diseases, Pathogen Discovery | Dakota Howard, Dhwani Batra, Peter W. Cook, Kara Moser, Adrian Paskey, Jason Caravas, Benjamin Rambo-Martin, Shatavia Morrison, Christopher Gulvick, Scott Sammons, Yvette Unoarumhi, Darlene Wagner, Matthew Schmerer, Cyndi Clark, Patrick Campbell, Rob Case, Vikramsinha Ghorpade, Holly Houdeshell, Ola Kvalvaag, Dillon Nall, Ethan Sanders, Alec Vest, Shaun Westlund, Matthew Hardison, Clinton R. Paden, Duncan MacCannell |
| EPI_ISL_2202926 | New Mexico Department of Health Scientific Laboratory Division | Los Alamos National Laboratory Bioscience Division | Chien-Chi Lo, Migun Shakya, Cheryl Gleasner, Kim McMurry, Alina Deshpande, Twila Kunde, Joseph Hicks, Michael Edwards, Patrick Chain |
| EPI_ISL_2202927 | Houston Methodist Hospital | Houston Methodist Hospital | Randall J. Olsen, Paul A. Christensen, S. Wesley Long, Sishir Subedi, Robert Olson, Marcus Nguyen, James J. Davis, Matthew Ojeda Saavedra, Prasanti Yerramilli, Layne Pruitt, Kristina Reppond, Madison N. Shyer, Jessica Cambric, Ryan Gadd, Ilya J. Finkelstein, Jimmy Gollihar, and James M. Musser |
| EPI_ISL_2202928 | New Mexico Department of Health Scientific Laboratory Division | Los Alamos National Laboratory Bioscience Division | Chien-Chi Lo, Migun Shakya, Cheryl Gleasner, Kim McMurry, Alina Deshpande, Twila Kunde, Joseph Hicks, Michael Edwards, Patrick Chain |
| EPI_ISL_2202929 | Aegis Sciences Corporation | Centers for Disease Control and Prevention Division of Viral Diseases, Pathogen Discovery | Dakota Howard, Dhwani Batra, Peter W. Cook, Kara Moser, Adrian Paskey, Jason Caravas, Benjamin Rambo-Martin, Shatavia Morrison, Christopher Gulvick, Scott Sammons, Yvette Unoarumhi, Darlene Wagner, Matthew Schmerer, Cyndi Clark, Patrick Campbell, Rob Case, Vikramsinha Ghorpade, Holly Houdeshell, Ola Kvalvaag, Dillon Nall, Ethan Sanders, Alec Vest, Shaun Westlund, Matthew Hardison, Clinton R. Paden, Duncan MacCannell |
| EPI_ISL_2202930 | New Mexico Department of Health Scientific Laboratory Division | Los Alamos National Laboratory Bioscience Division | Chien-Chi Lo, Migun Shakya, Cheryl Gleasner, Kim McMurry, Alina Deshpande, Twila Kunde, Joseph Hicks, Michael Edwards, Patrick Chain |
| EPI_ISL_2202931 | Aegis Sciences Corporation | Centers for Disease Control and Prevention Division of Viral Diseases, Pathogen Discovery | Dakota Howard, Dhwani Batra, Peter W. Cook, Kara Moser, Adrian Paskey, Jason Caravas, Benjamin Rambo-Martin, Shatavia Morrison, Christopher Gulvick, Scott Sammons, Yvette Unoarumhi, Darlene Wagner, Matthew Schmerer, Cyndi Clark, Patrick Campbell, Rob Case, Vikramsinha Ghorpade, Holly Houdeshell, Ola Kvalvaag, Dillon Nall, Ethan Sanders, Alec Vest, Shaun Westlund, Matthew Hardison, Clinton R. Paden, Duncan MacCannell |
| EPI_ISL_2202932 | New Mexico Department of Health | Los Alamos National Laboratory Bioscience | Chien-Chi Lo, Migun Shakya, Cheryl Gleasner, Kim McMurry, Alina Deshpande, Twila Kunde, Joseph Hicks, Michael Edwards, Patrick Chain |

| | | | |
|---|---|---|---|
| | Scientific Laboratory Division | Division | |
| EPI_ISL_2202933 | Houston Methodist Hospital | Houston Methodist Hospital | Randall J. Olsen, Paul A. Christensen, S. Wesley Long, Sishir Subedi, Robert Olson, Marcus Nguyen, James J. Davis, Matthew Ojeda Saavedra, Prasanti Yerramilli, Layne Pruitt, Kristina Reppond, Madison N. Shyer, Jessica Cambric, Ryan Gadd, Ilya J. Finkelstein, Jimmy Gollihar, and James M. Musser |
| EPI_ISL_2202934 | New Mexico Department of Health Scientific Laboratory Division | Los Alamos National Laboratory Bioscience Division | Chien-Chi Lo, Migun Shakya, Cheryl Gleasner, Kim McMurry, Alina Deshpande, Twila Kunde, Joseph Hicks, Michael Edwards, Patrick Chain |
| EPI_ISL_2202935 | Aegis Sciences Corporation | Centers for Disease Control and Prevention Division of Viral Diseases, Pathogen Discovery | Dakota Howard, Dhwani Batra, Peter W. Cook, Kara Moser, Adrian Paskey, Jason Caravas, Benjamin Rambo-Martin, Shatavia Morrison, Christopher Gulvick, Scott Sammons, Yvette Unoarumhi, Darlene Wagner, Matthew Schmerer, Cyndi Clark, Patrick Campbell, Rob Case, Vikramsinha Ghorpade, Holly Houdeshell, Ola Kvalvaag, Dillon Nall, Ethan Sanders, Alec Vest, Shaun Westlund, Matthew Hardison, Clinton R. Paden, Duncan MacCannell |
| EPI_ISL_2202936, EPI_ISL_2202937 | New Mexico Department of Health Scientific Laboratory Division | Los Alamos National Laboratory Bioscience Division | Chien-Chi Lo, Migun Shakya, Cheryl Gleasner, Kim McMurry, Alina Deshpande, Twila Kunde, Joseph Hicks, Michael Edwards, Patrick Chain |
| EPI_ISL_2202938 | Houston Methodist Hospital | Houston Methodist Hospital | Randall J. Olsen, Paul A. Christensen, S. Wesley Long, Sishir Subedi, Robert Olson, Marcus Nguyen, James J. Davis, Matthew Ojeda Saavedra, Prasanti Yerramilli, Layne Pruitt, Kristina Reppond, Madison N. Shyer, Jessica Cambric, Ryan Gadd, Ilya J. Finkelstein, Jimmy Gollihar, and James M. Musser |
| EPI_ISL_2202939 | Aegis Sciences Corporation | Centers for Disease Control and Prevention Division of Viral Diseases, Pathogen Discovery | Dakota Howard, Dhwani Batra, Peter W. Cook, Kara Moser, Adrian Paskey, Jason Caravas, Benjamin Rambo-Martin, Shatavia Morrison, Christopher Gulvick, Scott Sammons, Yvette Unoarumhi, Darlene Wagner, Matthew Schmerer, Cyndi Clark, Patrick Campbell, Rob Case, Vikramsinha Ghorpade, Holly Houdeshell, Ola Kvalvaag, Dillon Nall, Ethan Sanders, Alec Vest, Shaun Westlund, Matthew Hardison, Clinton R. Paden, Duncan MacCannell |
| EPI_ISL_2202940 | New Mexico Department of Health Scientific Laboratory Division | Los Alamos National Laboratory Bioscience Division | Chien-Chi Lo, Migun Shakya, Cheryl Gleasner, Kim McMurry, Alina Deshpande, Twila Kunde, Joseph Hicks, Michael Edwards, Patrick Chain |
| EPI_ISL_2202941 | Aegis Sciences Corporation | Centers for Disease Control and Prevention Division of Viral Diseases, Pathogen Discovery | Dakota Howard, Dhwani Batra, Peter W. Cook, Kara Moser, Adrian Paskey, Jason Caravas, Benjamin Rambo-Martin, Shatavia Morrison, Christopher Gulvick, Scott Sammons, Yvette Unoarumhi, Darlene Wagner, Matthew Schmerer, Cyndi Clark, Patrick Campbell, Rob Case, Vikramsinha Ghorpade, Holly Houdeshell, Ola Kvalvaag, Dillon Nall, Ethan Sanders, Alec Vest, Shaun Westlund, Matthew Hardison, Clinton R. Paden, Duncan MacCannell |
| EPI_ISL_2202942 | Houston Methodist Hospital | Houston Methodist Hospital | Randall J. Olsen, Paul A. Christensen, S. Wesley Long, Sishir Subedi, Robert Olson, Marcus Nguyen, James J. Davis, Matthew Ojeda Saavedra, Prasanti Yerramilli, Layne Pruitt, Kristina Reppond, Madison N. Shyer, Jessica Cambric, Ryan Gadd, Ilya J. Finkelstein, Jimmy Gollihar, and James M. Musser |
| EPI_ISL_2202943 | Aegis Sciences Corporation | Centers for Disease Control and Prevention Division of Viral Diseases, Pathogen Discovery | Dakota Howard, Dhwani Batra, Peter W. Cook, Kara Moser, Adrian Paskey, Jason Caravas, Benjamin Rambo-Martin, Shatavia Morrison, Christopher Gulvick, Scott Sammons, Yvette Unoarumhi, Darlene Wagner, Matthew Schmerer, Cyndi Clark, Patrick Campbell, Rob Case, Vikramsinha Ghorpade, Holly Houdeshell, Ola Kvalvaag, Dillon Nall, Ethan Sanders, Alec Vest, Shaun Westlund, Matthew Hardison, Clinton R. Paden, Duncan MacCannell |
| EPI_ISL_2202944 | Houston Methodist Hospital | Houston Methodist Hospital | Randall J. Olsen, Paul A. Christensen, S. Wesley Long, Sishir Subedi, Robert Olson, Marcus Nguyen, James J. Davis, Matthew Ojeda Saavedra, Prasanti Yerramilli, Layne Pruitt, Kristina Reppond, Madison N. Shyer, Jessica Cambric, Ryan Gadd, Ilya J. Finkelstein, Jimmy Gollihar, and James M. Musser |
| EPI_ISL_2202945, EPI_ISL_2202946, EPI_ISL_2202947, EPI_ISL_2202948, EPI_ISL_2202949 | Aegis Sciences Corporation | Centers for Disease Control and Prevention Division of Viral Diseases, Pathogen Discovery | Dakota Howard, Dhwani Batra, Peter W. Cook, Kara Moser, Adrian Paskey, Jason Caravas, Benjamin Rambo-Martin, Shatavia Morrison, Christopher Gulvick, Scott Sammons, Yvette Unoarumhi, Darlene Wagner, Matthew Schmerer, Cyndi Clark, Patrick Campbell, Rob Case, Vikramsinha Ghorpade, Holly Houdeshell, Ola Kvalvaag, Dillon Nall, Ethan Sanders, Alec Vest, Shaun Westlund, Matthew Hardison, Clinton R. Paden, Duncan MacCannell |
| EPI_ISL_2202950 | Houston Methodist Hospital | Houston Methodist Hospital | Randall J. Olsen, Paul A. Christensen, S. Wesley Long, Sishir Subedi, Robert Olson, Marcus Nguyen, James J. Davis, Matthew Ojeda Saavedra, Prasanti Yerramilli, Layne Pruitt, Kristina Reppond, Madison N. Shyer, Jessica Cambric, Ryan Gadd, Ilya J. Finkelstein, Jimmy Gollihar, and James M. Musser |
| EPI_ISL_2202951, EPI_ISL_2202952 | Aegis Sciences Corporation | Centers for Disease Control and Prevention Division of Viral Diseases, Pathogen Discovery | Dakota Howard, Dhwani Batra, Peter W. Cook, Kara Moser, Adrian Paskey, Jason Caravas, Benjamin Rambo-Martin, Shatavia Morrison, Christopher Gulvick, Scott Sammons, Yvette Unoarumhi, Darlene Wagner, Matthew Schmerer, Cyndi Clark, Patrick Campbell, Rob Case, Vikramsinha Ghorpade, Holly Houdeshell, Ola Kvalvaag, Dillon Nall, Ethan Sanders, Alec Vest, Shaun Westlund, Matthew Hardison, Clinton R. Paden, Duncan MacCannell |
| EPI_ISL_2202953 | Houston Methodist Hospital | Houston Methodist Hospital | Randall J. Olsen, Paul A. Christensen, S. Wesley Long, Sishir Subedi, Robert Olson, Marcus Nguyen, James J. Davis, Matthew Ojeda Saavedra, Prasanti Yerramilli, Layne Pruitt, Kristina Reppond, Madison N. Shyer, Jessica Cambric, Ryan Gadd, Ilya J. Finkelstein, Jimmy Gollihar, and James M. Musser |
| EPI_ISL_2202954, EPI_ISL_2202955, EPI_ISL_2202956 | Aegis Sciences Corporation | Centers for Disease Control and Prevention Division of Viral Diseases, Pathogen Discovery | Dakota Howard, Dhwani Batra, Peter W. Cook, Kara Moser, Adrian Paskey, Jason Caravas, Benjamin Rambo-Martin, Shatavia Morrison, Christopher Gulvick, Scott Sammons, Yvette Unoarumhi, Darlene Wagner, Matthew Schmerer, Cyndi Clark, Patrick Campbell, Rob Case, Vikramsinha Ghorpade, Holly Houdeshell, Ola Kvalvaag, Dillon Nall, Ethan Sanders, Alec Vest, Shaun Westlund, Matthew Hardison, Clinton R. Paden, Duncan MacCannell |
| EPI_ISL_2202957 | Houston Methodist Hospital | Houston Methodist Hospital | Randall J. Olsen, Paul A. Christensen, S. Wesley Long, Sishir Subedi, Robert Olson, Marcus Nguyen, James J. Davis, Matthew Ojeda Saavedra, Prasanti Yerramilli, Layne Pruitt, Kristina Reppond, Madison N. Shyer, Jessica Cambric, Ryan Gadd, Ilya J. Finkelstein, Jimmy Gollihar, and James M. Musser |
| EPI_ISL_2202958, EPI_ISL_2202959 | Aegis Sciences Corporation | Centers for Disease Control and Prevention Division of Viral Diseases, Pathogen Discovery | Dakota Howard, Dhwani Batra, Peter W. Cook, Kara Moser, Adrian Paskey, Jason Caravas, Benjamin Rambo-Martin, Shatavia Morrison, Christopher Gulvick, Scott Sammons, Yvette Unoarumhi, Darlene Wagner, Matthew Schmerer, Cyndi Clark, Patrick Campbell, Rob Case, Vikramsinha Ghorpade, Holly Houdeshell, Ola Kvalvaag, Dillon Nall, Ethan Sanders, Alec Vest, Shaun Westlund, Matthew Hardison, Clinton R. Paden, Duncan MacCannell |
| EPI_ISL_2202960 | Houston Methodist Hospital | Houston Methodist Hospital | Randall J. Olsen, Paul A. Christensen, S. Wesley Long, Sishir Subedi, Robert Olson, Marcus Nguyen, James J. Davis, Matthew Ojeda Saavedra, Prasanti Yerramilli, Layne Pruitt, Kristina Reppond, Madison N. Shyer, Jessica Cambric, Ryan Gadd, Ilya J. Finkelstein, Jimmy Gollihar, and James M. Musser |
| EPI_ISL_2202961, EPI_ISL_2202962, EPI_ISL_2202963 | Aegis Sciences Corporation | Centers for Disease Control and Prevention Division of Viral Diseases, Pathogen Discovery | Dakota Howard, Dhwani Batra, Peter W. Cook, Kara Moser, Adrian Paskey, Jason Caravas, Benjamin Rambo-Martin, Shatavia Morrison, Christopher Gulvick, Scott Sammons, Yvette Unoarumhi, Darlene Wagner, Matthew Schmerer, Cyndi Clark, Patrick Campbell, Rob Case, Vikramsinha Ghorpade, Holly Houdeshell, Ola Kvalvaag, Dillon Nall, Ethan Sanders, Alec Vest, Shaun Westlund, Matthew Hardison, Clinton R. Paden, Duncan MacCannell |
| EPI_ISL_2202964 | Houston Methodist Hospital | Houston Methodist Hospital | Randall J. Olsen, Paul A. Christensen, S. Wesley Long, Sishir Subedi, Robert Olson, Marcus Nguyen, James J. Davis, Matthew Ojeda Saavedra, Prasanti Yerramilli, Layne Pruitt, Kristina Reppond, Madison N. Shyer, Jessica Cambric, Ryan Gadd, Ilya J. Finkelstein, Jimmy Gollihar, and James M. Musser |
| EPI_ISL_2202965, EPI_ISL_2202966 | Aegis Sciences Corporation | Centers for Disease Control and Prevention Division of Viral Diseases, Pathogen Discovery | Dakota Howard, Dhwani Batra, Peter W. Cook, Kara Moser, Adrian Paskey, Jason Caravas, Benjamin Rambo-Martin, Shatavia Morrison, Christopher Gulvick, Scott Sammons, Yvette Unoarumhi, Darlene Wagner, Matthew Schmerer, Cyndi Clark, Patrick Campbell, Rob Case, Vikramsinha Ghorpade, Holly Houdeshell, Ola Kvalvaag, Dillon Nall, Ethan Sanders, Alec Vest, Shaun Westlund, Matthew Hardison, Clinton R. Paden, Duncan MacCannell |
| EPI_ISL_2202967 | Houston Methodist Hospital | Houston Methodist Hospital | Randall J. Olsen, Paul A. Christensen, S. Wesley Long, Sishir Subedi, Robert Olson, Marcus Nguyen, James J. Davis, Matthew Ojeda Saavedra, Prasanti Yerramilli, Layne Pruitt, Kristina Reppond, Madison N. Shyer, Jessica Cambric, Ryan Gadd, Ilya J. Finkelstein, Jimmy Gollihar, and James M. Musser |
| EPI_ISL_2202968 | Aegis Sciences Corporation | Centers for Disease Control and Prevention Division of Viral Diseases, Pathogen Discovery | Dakota Howard, Dhwani Batra, Peter W. Cook, Kara Moser, Adrian Paskey, Jason Caravas, Benjamin Rambo-Martin, Shatavia Morrison, Christopher Gulvick, Scott Sammons, Yvette Unoarumhi, Darlene Wagner, Matthew Schmerer, Cyndi Clark, Patrick Campbell, Rob Case, Vikramsinha Ghorpade, Holly Houdeshell, Ola Kvalvaag, Dillon Nall, Ethan Sanders, Alec Vest, Shaun Westlund, Matthew Hardison, Clinton R. Paden, Duncan MacCannell |
| EPI_ISL_2202969 | Houston Methodist Hospital | Houston Methodist Hospital | Randall J. Olsen, Paul A. Christensen, S. Wesley Long, Sishir Subedi, Robert Olson, Marcus Nguyen, James J. Davis, Matthew Ojeda Saavedra, Prasanti Yerramilli, Layne Pruitt, Kristina Reppond, Madison N. Shyer, Jessica Cambric, Ryan Gadd, Ilya J. Finkelstein, Jimmy Gollihar, and James M. Musser |
| EPI_ISL_2202970, EPI_ISL_2202971 | Aegis Sciences Corporation | Centers for Disease Control and Prevention Division of Viral Diseases, Pathogen Discovery | Dakota Howard, Dhwani Batra, Peter W. Cook, Kara Moser, Adrian Paskey, Jason Caravas, Benjamin Rambo-Martin, Shatavia Morrison, Christopher Gulvick, Scott Sammons, Yvette Unoarumhi, Darlene Wagner, Matthew Schmerer, Cyndi Clark, Patrick Campbell, Rob Case, Vikramsinha Ghorpade, Holly Houdeshell, Ola Kvalvaag, Dillon Nall, Ethan Sanders, Alec Vest, Shaun Westlund, Matthew Hardison, Clinton R. Paden, Duncan MacCannell |
| EPI_ISL_2202972 | Houston Methodist Hospital | Houston Methodist Hospital | Randall J. Olsen, Paul A. Christensen, S. Wesley Long, Sishir Subedi, Robert Olson, Marcus Nguyen, James J. Davis, Matthew Ojeda Saavedra, Prasanti Yerramilli, Layne Pruitt, Kristina Reppond, Madison N. Shyer, Jessica Cambric, Ryan Gadd, Ilya J. Finkelstein, Jimmy Gollihar, and James M. Musser |
| EPI_ISL_2202973 | Aegis Sciences Corporation | Centers for Disease Control and Prevention Division of Viral Diseases, Pathogen Discovery | Dakota Howard, Dhwani Batra, Peter W. Cook, Kara Moser, Adrian Paskey, Jason Caravas, Benjamin Rambo-Martin, Shatavia Morrison, Christopher Gulvick, Scott Sammons, Yvette Unoarumhi, Darlene Wagner, Matthew Schmerer, Cyndi Clark, Patrick Campbell, Rob Case, Vikramsinha Ghorpade, Holly Houdeshell, Ola Kvalvaag, Dillon Nall, Ethan Sanders, Alec Vest, Shaun Westlund, Matthew Hardison, Clinton R. Paden, Duncan MacCannell |
| EPI_ISL_2202974 | Houston Methodist Hospital | Houston Methodist Hospital | Randall J. Olsen, Paul A. Christensen, S. Wesley Long, Sishir Subedi, Robert Olson, Marcus Nguyen, James J. Davis, Matthew Ojeda Saavedra, Prasanti Yerramilli, Layne Pruitt, Kristina Reppond, Madison N. Shyer, Jessica Cambric, Ryan Gadd, Ilya J. Finkelstein, Jimmy Gollihar, and James M. Musser |
| EPI_ISL_2202975 | Aegis Sciences Corporation | Centers for Disease Control and Prevention Division of Viral Diseases, Pathogen Discovery | Dakota Howard, Dhwani Batra, Peter W. Cook, Kara Moser, Adrian Paskey, Jason Caravas, Benjamin Rambo-Martin, Shatavia Morrison, Christopher Gulvick, Scott Sammons, Yvette Unoarumhi, Darlene Wagner, Matthew Schmerer, Cyndi Clark, Patrick Campbell, Rob Case, Vikramsinha Ghorpade, Holly Houdeshell, Ola Kvalvaag, Dillon Nall, Ethan Sanders, Alec Vest, Shaun Westlund, Matthew Hardison, Clinton R. Paden, Duncan MacCannell |
| EPI_ISL_2202976 | Houston Methodist Hospital | Houston Methodist Hospital | Randall J. Olsen, Paul A. Christensen, S. Wesley Long, Sishir Subedi, Robert Olson, Marcus Nguyen, James J. Davis, Matthew Ojeda Saavedra, Prasanti Yerramilli, Layne Pruitt, Kristina Reppond, Madison N. Shyer, Jessica Cambric, Ryan Gadd, Ilya J. Finkelstein, Jimmy Gollihar, and James M. Musser |
| EPI_ISL_2202977, EPI_ISL_2202978 | Aegis Sciences Corporation | Centers for Disease Control and Prevention Division of Viral Diseases, Pathogen Discovery | Dakota Howard, Dhwani Batra, Peter W. Cook, Kara Moser, Adrian Paskey, Jason Caravas, Benjamin Rambo-Martin, Shatavia Morrison, Christopher Gulvick, Scott Sammons, Yvette Unoarumhi, Darlene Wagner, Matthew Schmerer, Cyndi Clark, Patrick Campbell, Rob Case, Vikramsinha Ghorpade, Holly Houdeshell, Ola Kvalvaag, Dillon Nall, Ethan Sanders, Alec Vest, Shaun Westlund, Matthew Hardison, Clinton R. Paden, Duncan MacCannell |
| EPI_ISL_2202979 | Houston Methodist Hospital | Houston Methodist Hospital | Randall J. Olsen, Paul A. Christensen, S. Wesley Long, Sishir Subedi, Robert Olson, Marcus Nguyen, James J. Davis, Matthew Ojeda Saavedra, Prasanti Yerramilli, Layne Pruitt, Kristina Reppond, Madison N. Shyer, Jessica Cambric, Ryan Gadd, Ilya J. Finkelstein, Jimmy Gollihar, and James M. Musser |
| EPI_ISL_2202980 | Aegis Sciences Corporation | Centers for Disease Control and Prevention Division of Viral Diseases, Pathogen Discovery | Dakota Howard, Dhwani Batra, Peter W. Cook, Kara Moser, Adrian Paskey, Jason Caravas, Benjamin Rambo-Martin, Shatavia Morrison, Christopher Gulvick, Scott Sammons, Yvette Unoarumhi, Darlene Wagner, Matthew Schmerer, Cyndi Clark, Patrick Campbell, Rob Case, Vikramsinha Ghorpade, Holly Houdeshell, Ola Kvalvaag, Dillon Nall, Ethan Sanders, Alec Vest, Shaun Westlund, Matthew Hardison, Clinton R. Paden, Duncan MacCannell |
| EPI_ISL_2202981 | Houston Methodist Hospital | Houston Methodist Hospital | Randall J. Olsen, Paul A. Christensen, S. Wesley Long, Sishir Subedi, Robert Olson, Marcus Nguyen, James J. Davis, Matthew Ojeda Saavedra, Prasanti Yerramilli, Layne Pruitt, Kristina Reppond, Madison N. Shyer, Jessica Cambric, Ryan Gadd, Ilya J. Finkelstein, Jimmy Gollihar, and James M. Musser |
| EPI_ISL_2202982, EPI_ISL_2202983, EPI_ISL_2202984 | Aegis Sciences Corporation | Centers for Disease Control and Prevention Division of Viral Diseases, Pathogen Discovery | Dakota Howard, Dhwani Batra, Peter W. Cook, Kara Moser, Adrian Paskey, Jason Caravas, Benjamin Rambo-Martin, Shatavia Morrison, Christopher Gulvick, Scott Sammons, Yvette Unoarumhi, Darlene Wagner, Matthew Schmerer, Cyndi Clark, Patrick Campbell, Rob Case, Vikramsinha Ghorpade, Holly Houdeshell, Ola Kvalvaag, Dillon Nall, Ethan Sanders, Alec Vest, Shaun Westlund, Matthew Hardison, Clinton R. Paden, Duncan MacCannell |
| EPI_ISL_2202985 | Houston Methodist Hospital | Houston Methodist Hospital | Randall J. Olsen, Paul A. Christensen, S. Wesley Long, Sishir Subedi, Robert Olson, Marcus Nguyen, James J. Davis, Matthew Ojeda Saavedra, Prasanti Yerramilli, Layne Pruitt, Kristina Reppond, Madison N. Shyer, Jessica Cambric, Ryan Gadd, Ilya J. Finkelstein, Jimmy Gollihar, and James M. Musser |

| | | | |
|---|---|---|---|
| EPI_ISL_2202986 | Aegis Sciences Corporation | Centers for Disease Control and Prevention Division of Viral Diseases, Pathogen Discovery | Dakota Howard, Dhwani Batra, Peter W. Cook, Kara Moser, Adrian Paskey, Jason Caravas, Benjamin Rambo-Martin, Shatavia Morrison, Christopher Gulvick, Scott Sammons, Yvette Unoarumhi, Darlene Wagner, Matthew Schmerer, Cyndi Clark, Patrick Campbell, Rob Case, Vikramsinha Ghorpade, Holly Houdeshell, Ola Kvalvaag, Dillon Nall, Ethan Sanders, Alec Vest, Shaun Westlund, Matthew Hardison, Clinton R. Paden, Duncan MacCannell |
| EPI_ISL_2202987 | Houston Methodist Hospital | Houston Methodist Hospital | Randall J. Olsen, Paul A. Christensen, S. Wesley Long, Sishir Subedi, Robert Olson, Marcus Nguyen, James J. Davis, Matthew Ojeda Saavedra, Prasanti Yerramilli, Layne Pruitt, Kristina Reppond, Madison N. Shyer, Jessica Cambric, Ryan Gadd, Ilya J. Finkelstein, Jimmy Gollihar, and James M. Musser |
| EPI_ISL_2202988, EPI_ISL_2202989 | Aegis Sciences Corporation | Centers for Disease Control and Prevention Division of Viral Diseases, Pathogen Discovery | Dakota Howard, Dhwani Batra, Peter W. Cook, Kara Moser, Adrian Paskey, Jason Caravas, Benjamin Rambo-Martin, Shatavia Morrison, Christopher Gulvick, Scott Sammons, Yvette Unoarumhi, Darlene Wagner, Matthew Schmerer, Cyndi Clark, Patrick Campbell, Rob Case, Vikramsinha Ghorpade, Holly Houdeshell, Ola Kvalvaag, Dillon Nall, Ethan Sanders, Alec Vest, Shaun Westlund, Matthew Hardison, Clinton R. Paden, Duncan MacCannell |
| EPI_ISL_2202990 | Houston Methodist Hospital | Houston Methodist Hospital | Randall J. Olsen, Paul A. Christensen, S. Wesley Long, Sishir Subedi, Robert Olson, Marcus Nguyen, James J. Davis, Matthew Ojeda Saavedra, Prasanti Yerramilli, Layne Pruitt, Kristina Reppond, Madison N. Shyer, Jessica Cambric, Ryan Gadd, Ilya J. Finkelstein, Jimmy Gollihar, and James M. Musser |
| EPI_ISL_2202991 | Aegis Sciences Corporation | Centers for Disease Control and Prevention Division of Viral Diseases, Pathogen Discovery | Dakota Howard, Dhwani Batra, Peter W. Cook, Kara Moser, Adrian Paskey, Jason Caravas, Benjamin Rambo-Martin, Shatavia Morrison, Christopher Gulvick, Scott Sammons, Yvette Unoarumhi, Darlene Wagner, Matthew Schmerer, Cyndi Clark, Patrick Campbell, Rob Case, Vikramsinha Ghorpade, Holly Houdeshell, Ola Kvalvaag, Dillon Nall, Ethan Sanders, Alec Vest, Shaun Westlund, Matthew Hardison, Clinton R. Paden, Duncan MacCannell |
| EPI_ISL_2202992 | Houston Methodist Hospital | Houston Methodist Hospital | Randall J. Olsen, Paul A. Christensen, S. Wesley Long, Sishir Subedi, Robert Olson, Marcus Nguyen, James J. Davis, Matthew Ojeda Saavedra, Prasanti Yerramilli, Layne Pruitt, Kristina Reppond, Madison N. Shyer, Jessica Cambric, Ryan Gadd, Ilya J. Finkelstein, Jimmy Gollihar, and James M. Musser |
| EPI_ISL_2202993 | Aegis Sciences Corporation | Centers for Disease Control and Prevention Division of Viral Diseases, Pathogen Discovery | Dakota Howard, Dhwani Batra, Peter W. Cook, Kara Moser, Adrian Paskey, Jason Caravas, Benjamin Rambo-Martin, Shatavia Morrison, Christopher Gulvick, Scott Sammons, Yvette Unoarumhi, Darlene Wagner, Matthew Schmerer, Cyndi Clark, Patrick Campbell, Rob Case, Vikramsinha Ghorpade, Holly Houdeshell, Ola Kvalvaag, Dillon Nall, Ethan Sanders, Alec Vest, Shaun Westlund, Matthew Hardison, Clinton R. Paden, Duncan MacCannell |
| EPI_ISL_2202994 | Houston Methodist Hospital | Houston Methodist Hospital | Randall J. Olsen, Paul A. Christensen, S. Wesley Long, Sishir Subedi, Robert Olson, Marcus Nguyen, James J. Davis, Matthew Ojeda Saavedra, Prasanti Yerramilli, Layne Pruitt, Kristina Reppond, Madison N. Shyer, Jessica Cambric, Ryan Gadd, Ilya J. Finkelstein, Jimmy Gollihar, and James M. Musser |
| EPI_ISL_2202995, EPI_ISL_2202996, EPI_ISL_2202997 | Aegis Sciences Corporation | Centers for Disease Control and Prevention Division of Viral Diseases, Pathogen Discovery | Dakota Howard, Dhwani Batra, Peter W. Cook, Kara Moser, Adrian Paskey, Jason Caravas, Benjamin Rambo-Martin, Shatavia Morrison, Christopher Gulvick, Scott Sammons, Yvette Unoarumhi, Darlene Wagner, Matthew Schmerer, Cyndi Clark, Patrick Campbell, Rob Case, Vikramsinha Ghorpade, Holly Houdeshell, Ola Kvalvaag, Dillon Nall, Ethan Sanders, Alec Vest, Shaun Westlund, Matthew Hardison, Clinton R. Paden, Duncan MacCannell |
| EPI_ISL_2202998 | Houston Methodist Hospital | Houston Methodist Hospital | Randall J. Olsen, Paul A. Christensen, S. Wesley Long, Sishir Subedi, Robert Olson, Marcus Nguyen, James J. Davis, Matthew Ojeda Saavedra, Prasanti Yerramilli, Layne Pruitt, Kristina Reppond, Madison N. Shyer, Jessica Cambric, Ryan Gadd, Ilya J. Finkelstein, Jimmy Gollihar, and James M. Musser |
| EPI_ISL_2202999 | Aegis Sciences Corporation | Centers for Disease Control and Prevention Division of Viral Diseases, Pathogen Discovery | Dakota Howard, Dhwani Batra, Peter W. Cook, Kara Moser, Adrian Paskey, Jason Caravas, Benjamin Rambo-Martin, Shatavia Morrison, Christopher Gulvick, Scott Sammons, Yvette Unoarumhi, Darlene Wagner, Matthew Schmerer, Cyndi Clark, Patrick Campbell, Rob Case, Vikramsinha Ghorpade, Holly Houdeshell, Ola Kvalvaag, Dillon Nall, Ethan Sanders, Alec Vest, Shaun Westlund, Matthew Hardison, Clinton R. Paden, Duncan MacCannell |
| EPI_ISL_2203000 | Houston Methodist Hospital | Houston Methodist Hospital | Randall J. Olsen, Paul A. Christensen, S. Wesley Long, Sishir Subedi, Robert Olson, Marcus Nguyen, James J. Davis, Matthew Ojeda Saavedra, Prasanti Yerramilli, Layne Pruitt, Kristina Reppond, Madison N. Shyer, Jessica Cambric, Ryan Gadd, Ilya J. Finkelstein, Jimmy Gollihar, and James M. Musser |
| EPI_ISL_2203001 | Aegis Sciences Corporation | Centers for Disease Control and Prevention Division of Viral Diseases, Pathogen Discovery | Dakota Howard, Dhwani Batra, Peter W. Cook, Kara Moser, Adrian Paskey, Jason Caravas, Benjamin Rambo-Martin, Shatavia Morrison, Christopher Gulvick, Scott Sammons, Yvette Unoarumhi, Darlene Wagner, Matthew Schmerer, Cyndi Clark, Patrick Campbell, Rob Case, Vikramsinha Ghorpade, Holly Houdeshell, Ola Kvalvaag, Dillon Nall, Ethan Sanders, Alec Vest, Shaun Westlund, Matthew Hardison, Clinton R. Paden, Duncan MacCannell |
| EPI_ISL_2203002 | Houston Methodist Hospital | Houston Methodist Hospital | Randall J. Olsen, Paul A. Christensen, S. Wesley Long, Sishir Subedi, Robert Olson, Marcus Nguyen, James J. Davis, Matthew Ojeda Saavedra, Prasanti Yerramilli, Layne Pruitt, Kristina Reppond, Madison N. Shyer, Jessica Cambric, Ryan Gadd, Ilya J. Finkelstein, Jimmy Gollihar, and James M. Musser |
| EPI_ISL_2203003 | Aegis Sciences Corporation | Centers for Disease Control and Prevention Division of Viral Diseases, Pathogen Discovery | Dakota Howard, Dhwani Batra, Peter W. Cook, Kara Moser, Adrian Paskey, Jason Caravas, Benjamin Rambo-Martin, Shatavia Morrison, Christopher Gulvick, Scott Sammons, Yvette Unoarumhi, Darlene Wagner, Matthew Schmerer, Cyndi Clark, Patrick Campbell, Rob Case, Vikramsinha Ghorpade, Holly Houdeshell, Ola Kvalvaag, Dillon Nall, Ethan Sanders, Alec Vest, Shaun Westlund, Matthew Hardison, Clinton R. Paden, Duncan MacCannell |
| EPI_ISL_2203004 | Houston Methodist Hospital | Houston Methodist Hospital | Randall J. Olsen, Paul A. Christensen, S. Wesley Long, Sishir Subedi, Robert Olson, Marcus Nguyen, James J. Davis, Matthew Ojeda Saavedra, Prasanti Yerramilli, Layne Pruitt, Kristina Reppond, Madison N. Shyer, Jessica Cambric, Ryan Gadd, Ilya J. Finkelstein, Jimmy Gollihar, and James M. Musser |
| EPI_ISL_2203005, EPI_ISL_2203006 | Aegis Sciences Corporation | Centers for Disease Control and Prevention Division of Viral Diseases, Pathogen Discovery | Dakota Howard, Dhwani Batra, Peter W. Cook, Kara Moser, Adrian Paskey, Jason Caravas, Benjamin Rambo-Martin, Shatavia Morrison, Christopher Gulvick, Scott Sammons, Yvette Unoarumhi, Darlene Wagner, Matthew Schmerer, Cyndi Clark, Patrick Campbell, Rob Case, Vikramsinha Ghorpade, Holly Houdeshell, Ola Kvalvaag, Dillon Nall, Ethan Sanders, Alec Vest, Shaun Westlund, Matthew Hardison, Clinton R. Paden, Duncan MacCannell |
| EPI_ISL_2203007 | Houston Methodist Hospital | Houston Methodist Hospital | Randall J. Olsen, Paul A. Christensen, S. Wesley Long, Sishir Subedi, Robert Olson, Marcus Nguyen, James J. Davis, Matthew Ojeda Saavedra, Prasanti Yerramilli, Layne Pruitt, Kristina Reppond, Madison N. Shyer, Jessica Cambric, Ryan Gadd, Ilya J. Finkelstein, Jimmy Gollihar, and James M. Musser |
| EPI_ISL_2203008, EPI_ISL_2203009 | Aegis Sciences Corporation | Centers for Disease Control and Prevention Division of Viral Diseases, Pathogen Discovery | Dakota Howard, Dhwani Batra, Peter W. Cook, Kara Moser, Adrian Paskey, Jason Caravas, Benjamin Rambo-Martin, Shatavia Morrison, Christopher Gulvick, Scott Sammons, Yvette Unoarumhi, Darlene Wagner, Matthew Schmerer, Cyndi Clark, Patrick Campbell, Rob Case, Vikramsinha Ghorpade, Holly Houdeshell, Ola Kvalvaag, Dillon Nall, Ethan Sanders, Alec Vest, Shaun Westlund, Matthew Hardison, Clinton R. Paden, Duncan MacCannell |
| EPI_ISL_2203010 | Houston Methodist Hospital | Houston Methodist Hospital | Randall J. Olsen, Paul A. Christensen, S. Wesley Long, Sishir Subedi, Robert Olson, Marcus Nguyen, James J. Davis, Matthew Ojeda Saavedra, Prasanti Yerramilli, Layne Pruitt, Kristina Reppond, Madison N. Shyer, Jessica Cambric, Ryan Gadd, Ilya J. Finkelstein, Jimmy Gollihar, and James M. Musser |
| EPI_ISL_2203012 | Aegis Sciences Corporation | Centers for Disease Control and Prevention Division of Viral Diseases, Pathogen Discovery | Dakota Howard, Dhwani Batra, Peter W. Cook, Kara Moser, Adrian Paskey, Jason Caravas, Benjamin Rambo-Martin, Shatavia Morrison, Christopher Gulvick, Scott Sammons, Yvette Unoarumhi, Darlene Wagner, Matthew Schmerer, Cyndi Clark, Patrick Campbell, Rob Case, Vikramsinha Ghorpade, Holly Houdeshell, Ola Kvalvaag, Dillon Nall, Ethan Sanders, Alec Vest, Shaun Westlund, Matthew Hardison, Clinton R. Paden, Duncan MacCannell |
| EPI_ISL_2203013 | Houston Methodist Hospital | Houston Methodist Hospital | Randall J. Olsen, Paul A. Christensen, S. Wesley Long, Sishir Subedi, Robert Olson, Marcus Nguyen, James J. Davis, Matthew Ojeda Saavedra, Prasanti Yerramilli, Layne Pruitt, Kristina Reppond, Madison N. Shyer, Jessica Cambric, Ryan Gadd, Ilya J. Finkelstein, Jimmy Gollihar, and James M. Musser |
| EPI_ISL_2203014, EPI_ISL_2203015 | Aegis Sciences Corporation | Centers for Disease Control and Prevention Division of Viral Diseases, Pathogen Discovery | Dakota Howard, Dhwani Batra, Peter W. Cook, Kara Moser, Adrian Paskey, Jason Caravas, Benjamin Rambo-Martin, Shatavia Morrison, Christopher Gulvick, Scott Sammons, Yvette Unoarumhi, Darlene Wagner, Matthew Schmerer, Cyndi Clark, Patrick Campbell, Rob Case, Vikramsinha Ghorpade, Holly Houdeshell, Ola Kvalvaag, Dillon Nall, Ethan Sanders, Alec Vest, Shaun Westlund, Matthew Hardison, Clinton R. Paden, Duncan MacCannell |
| EPI_ISL_2203016 | Houston Methodist Hospital | Houston Methodist Hospital | Randall J. Olsen, Paul A. Christensen, S. Wesley Long, Sishir Subedi, Robert Olson, Marcus Nguyen, James J. Davis, Matthew Ojeda Saavedra, Prasanti Yerramilli, Layne Pruitt, Kristina Reppond, Madison N. Shyer, Jessica Cambric, Ryan Gadd, Ilya J. Finkelstein, Jimmy Gollihar, and James M. Musser |
| EPI_ISL_2203017, EPI_ISL_2203018, EPI_ISL_2203019 | Aegis Sciences Corporation | Centers for Disease Control and Prevention Division of Viral Diseases, Pathogen Discovery | Dakota Howard, Dhwani Batra, Peter W. Cook, Kara Moser, Adrian Paskey, Jason Caravas, Benjamin Rambo-Martin, Shatavia Morrison, Christopher Gulvick, Scott Sammons, Yvette Unoarumhi, Darlene Wagner, Matthew Schmerer, Cyndi Clark, Patrick Campbell, Rob Case, Vikramsinha Ghorpade, Holly Houdeshell, Ola Kvalvaag, Dillon Nall, Ethan Sanders, Alec Vest, Shaun Westlund, Matthew Hardison, Clinton R. Paden, Duncan MacCannell |
| EPI_ISL_2203020 | Houston Methodist Hospital | Houston Methodist Hospital | Randall J. Olsen, Paul A. Christensen, S. Wesley Long, Sishir Subedi, Robert Olson, Marcus Nguyen, James J. Davis, Matthew Ojeda Saavedra, Prasanti Yerramilli, Layne Pruitt, Kristina Reppond, Madison N. Shyer, Jessica Cambric, Ryan Gadd, Ilya J. Finkelstein, Jimmy Gollihar, and James M. Musser |
| EPI_ISL_2203021 | Aegis Sciences Corporation | Centers for Disease Control and Prevention Division of Viral Diseases, Pathogen Discovery | Dakota Howard, Dhwani Batra, Peter W. Cook, Kara Moser, Adrian Paskey, Jason Caravas, Benjamin Rambo-Martin, Shatavia Morrison, Christopher Gulvick, Scott Sammons, Yvette Unoarumhi, Darlene Wagner, Matthew Schmerer, Cyndi Clark, Patrick Campbell, Rob Case, Vikramsinha Ghorpade, Holly Houdeshell, Ola Kvalvaag, Dillon Nall, Ethan Sanders, Alec Vest, Shaun Westlund, Matthew Hardison, Clinton R. Paden, Duncan MacCannell |
| EPI_ISL_2203022 | Houston Methodist Hospital | Houston Methodist Hospital | Randall J. Olsen, Paul A. Christensen, S. Wesley Long, Sishir Subedi, Robert Olson, Marcus Nguyen, James J. Davis, Matthew Ojeda Saavedra, Prasanti Yerramilli, Layne Pruitt, Kristina Reppond, Madison N. Shyer, Jessica Cambric, Ryan Gadd, Ilya J. Finkelstein, Jimmy Gollihar, and James M. Musser |
| EPI_ISL_2203023 | Aegis Sciences Corporation | Centers for Disease Control and Prevention Division of Viral Diseases, Pathogen Discovery | Dakota Howard, Dhwani Batra, Peter W. Cook, Kara Moser, Adrian Paskey, Jason Caravas, Benjamin Rambo-Martin, Shatavia Morrison, Christopher Gulvick, Scott Sammons, Yvette Unoarumhi, Darlene Wagner, Matthew Schmerer, Cyndi Clark, Patrick Campbell, Rob Case, Vikramsinha Ghorpade, Holly Houdeshell, Ola Kvalvaag, Dillon Nall, Ethan Sanders, Alec Vest, Shaun Westlund, Matthew Hardison, Clinton R. Paden, Duncan MacCannell |
| EPI_ISL_2203024, EPI_ISL_2203025 | Oregon State Public Health Laboratory | Oregon State Public Health Laboratory | Rafia Razzaque, Eugene Yeboah, Vanda Makris, Laura Tsaknaridis, John Fontana and Shane Sevey |
| EPI_ISL_2203026 | Aegis Sciences Corporation | Centers for Disease Control and Prevention Division of Viral Diseases, Pathogen Discovery | Dakota Howard, Dhwani Batra, Peter W. Cook, Kara Moser, Adrian Paskey, Jason Caravas, Benjamin Rambo-Martin, Shatavia Morrison, Christopher Gulvick, Scott Sammons, Yvette Unoarumhi, Darlene Wagner, Matthew Schmerer, Cyndi Clark, Patrick Campbell, Rob Case, Vikramsinha Ghorpade, Holly Houdeshell, Ola Kvalvaag, Dillon Nall, Ethan Sanders, Alec Vest, Shaun Westlund, Matthew Hardison, Clinton R. Paden, Duncan MacCannell |
| EPI_ISL_2203027 | Oregon State Public Health Laboratory | Oregon State Public Health Laboratory | Rafia Razzaque, Eugene Yeboah, Vanda Makris, Laura Tsaknaridis, John Fontana and Shane Sevey |
| EPI_ISL_2203028 | Aegis Sciences Corporation | Centers for Disease Control and Prevention Division of Viral Diseases, Pathogen Discovery | Dakota Howard, Dhwani Batra, Peter W. Cook, Kara Moser, Adrian Paskey, Jason Caravas, Benjamin Rambo-Martin, Shatavia Morrison, Christopher Gulvick, Scott Sammons, Yvette Unoarumhi, Darlene Wagner, Matthew Schmerer, Cyndi Clark, Patrick Campbell, Rob Case, Vikramsinha Ghorpade, Holly Houdeshell, Ola Kvalvaag, Dillon Nall, Ethan Sanders, Alec Vest, Shaun Westlund, Matthew Hardison, Clinton R. Paden, Duncan MacCannell |
| EPI_ISL_2203029 | Oregon State Public Health Laboratory | Oregon State Public Health Laboratory | Rafia Razzaque, Eugene Yeboah, Vanda Makris, Laura Tsaknaridis, John Fontana and Shane Sevey |
| EPI_ISL_2203030 | Houston Methodist Hospital | Houston Methodist Hospital | Randall J. Olsen, Paul A. Christensen, S. Wesley Long, Sishir Subedi, Robert Olson, Marcus Nguyen, James J. Davis, Matthew Ojeda Saavedra, Prasanti Yerramilli, Layne Pruitt, Kristina Reppond, Madison N. Shyer, Jessica Cambric, Ryan Gadd, Ilya J. Finkelstein, Jimmy Gollihar, and James M. Musser |
| EPI_ISL_2203031 | Aegis Sciences Corporation | Centers for Disease Control and Prevention Division of Viral Diseases, Pathogen Discovery | Dakota Howard, Dhwani Batra, Peter W. Cook, Kara Moser, Adrian Paskey, Jason Caravas, Benjamin Rambo-Martin, Shatavia Morrison, Christopher Gulvick, Scott Sammons, Yvette Unoarumhi, Darlene Wagner, Matthew Schmerer, Cyndi Clark, Patrick Campbell, Rob Case, Vikramsinha Ghorpade, Holly Houdeshell, Ola Kvalvaag, Dillon Nall, Ethan Sanders, Alec Vest, Shaun Westlund, Matthew Hardison, Clinton R. Paden, Duncan MacCannell |
| EPI_ISL_2203032 | Oregon State Public Health Laboratory | Oregon State Public Health Laboratory | Rafia Razzaque, Eugene Yeboah, Vanda Makris, Laura Tsaknaridis, John Fontana and Shane Sevey |
| EPI_ISL_2203033 | Aegis Sciences Corporation | Centers for Disease Control and Prevention Division of Viral Diseases, Pathogen Discovery | Dakota Howard, Dhwani Batra, Peter W. Cook, Kara Moser, Adrian Paskey, Jason Caravas, Benjamin Rambo-Martin, Shatavia Morrison, Christopher Gulvick, Scott Sammons, Yvette Unoarumhi, Darlene Wagner, Matthew Schmerer, Cyndi Clark, Patrick Campbell, Rob Case, Vikramsinha Ghorpade, Holly Houdeshell, Ola Kvalvaag, Dillon Nall, Ethan Sanders, Alec Vest, Shaun Westlund, Matthew Hardison, Clinton R. Paden, Duncan MacCannell |
| EPI_ISL_2203034, | Oregon State Public Health Laboratory | Oregon State Public Health Laboratory | Rafia Razzaque, Eugene Yeboah, Vanda Makris, Laura Tsaknaridis, John Fontana and Shane Sevey |

| | | | |
|---|---|---|---|
| EPI_ISL_2203035 | | | |
| EPI_ISL_2203036 | Aegis Sciences Corporation | Centers for Disease Control and Prevention Division of Viral Diseases, Pathogen Discovery | Dakota Howard, Dhwani Batra, Peter W. Cook, Kara Moser, Adrian Paskey, Jason Caravas, Benjamin Rambo-Martin, Shatavia Morrison, Christopher Gulvick, Scott Sammons, Yvette Unoarumhi, Darlene Wagner, Matthew Schmerer, Cyndi Clark, Patrick Campbell, Rob Case, Vikramsinha Ghorpade, Holly Houdeshell, Ola Kvalvaag, Dillon Nall, Ethan Sanders, Alec Vest, Shaun Westlund, Matthew Hardison, Clinton R. Paden, Duncan MacCannell |
| EPI_ISL_2203037 | Oregon State Public Health Laboratory | Oregon State Public Health Laboratory | Rafia Razzaque, Eugene Yeboah, Vanda Makris, Laura Tsaknaridis, John Fontana and Shane Sevey |
| EPI_ISL_2203038 | Houston Methodist Hospital | Houston Methodist Hospital | Randall J. Olsen, Paul A. Christensen, S. Wesley Long, Sishir Subedi, Robert Olson, Marcus Nguyen, James J. Davis, Matthew Ojeda Saavedra, Prasanti Yerramilli, Layne Pruitt, Kristina Reppond, Madison N. Shyer, Jessica Cambric, Ryan Gadd, Ilya J. Finkelstein, Jimmy Gollihar, and James M. Musser |
| EPI_ISL_2203039 | Oregon State Public Health Laboratory | Oregon State Public Health Laboratory | Rafia Razzaque, Eugene Yeboah, Vanda Makris, Laura Tsaknaridis, John Fontana and Shane Sevey |
| EPI_ISL_2203040 | Aegis Sciences Corporation | Centers for Disease Control and Prevention Division of Viral Diseases, Pathogen Discovery | Dakota Howard, Dhwani Batra, Peter W. Cook, Kara Moser, Adrian Paskey, Jason Caravas, Benjamin Rambo-Martin, Shatavia Morrison, Christopher Gulvick, Scott Sammons, Yvette Unoarumhi, Darlene Wagner, Matthew Schmerer, Cyndi Clark, Patrick Campbell, Rob Case, Vikramsinha Ghorpade, Holly Houdeshell, Ola Kvalvaag, Dillon Nall, Ethan Sanders, Alec Vest, Shaun Westlund, Matthew Hardison, Clinton R. Paden, Duncan MacCannell |
| EPI_ISL_2203041 | Houston Methodist Hospital | Houston Methodist Hospital | Randall J. Olsen, Paul A. Christensen, S. Wesley Long, Sishir Subedi, Robert Olson, Marcus Nguyen, James J. Davis, Matthew Ojeda Saavedra, Prasanti Yerramilli, Layne Pruitt, Kristina Reppond, Madison N. Shyer, Jessica Cambric, Ryan Gadd, Ilya J. Finkelstein, Jimmy Gollihar, and James M. Musser |
| EPI_ISL_2203042 | Oregon State Public Health Laboratory | Oregon State Public Health Laboratory | Rafia Razzaque, Eugene Yeboah, Vanda Makris, Laura Tsaknaridis, John Fontana and Shane Sevey |
| EPI_ISL_2203043 | Aegis Sciences Corporation | Centers for Disease Control and Prevention Division of Viral Diseases, Pathogen Discovery | Dakota Howard, Dhwani Batra, Peter W. Cook, Kara Moser, Adrian Paskey, Jason Caravas, Benjamin Rambo-Martin, Shatavia Morrison, Christopher Gulvick, Scott Sammons, Yvette Unoarumhi, Darlene Wagner, Matthew Schmerer, Cyndi Clark, Patrick Campbell, Rob Case, Vikramsinha Ghorpade, Holly Houdeshell, Ola Kvalvaag, Dillon Nall, Ethan Sanders, Alec Vest, Shaun Westlund, Matthew Hardison, Clinton R. Paden, Duncan MacCannell |
| EPI_ISL_2203044 | Oregon State Public Health Laboratory | Oregon State Public Health Laboratory | Rafia Razzaque, Eugene Yeboah, Vanda Makris, Laura Tsaknaridis, John Fontana and Shane Sevey |
| EPI_ISL_2203045 | Houston Methodist Hospital | Houston Methodist Hospital | Randall J. Olsen, Paul A. Christensen, S. Wesley Long, Sishir Subedi, Robert Olson, Marcus Nguyen, James J. Davis, Matthew Ojeda Saavedra, Prasanti Yerramilli, Layne Pruitt, Kristina Reppond, Madison N. Shyer, Jessica Cambric, Ryan Gadd, Ilya J. Finkelstein, Jimmy Gollihar, and James M. Musser |
| EPI_ISL_2203046 | Aegis Sciences Corporation | Centers for Disease Control and Prevention Division of Viral Diseases, Pathogen Discovery | Dakota Howard, Dhwani Batra, Peter W. Cook, Kara Moser, Adrian Paskey, Jason Caravas, Benjamin Rambo-Martin, Shatavia Morrison, Christopher Gulvick, Scott Sammons, Yvette Unoarumhi, Darlene Wagner, Matthew Schmerer, Cyndi Clark, Patrick Campbell, Rob Case, Vikramsinha Ghorpade, Holly Houdeshell, Ola Kvalvaag, Dillon Nall, Ethan Sanders, Alec Vest, Shaun Westlund, Matthew Hardison, Clinton R. Paden, Duncan MacCannell |
| EPI_ISL_2203047, EPI_ISL_2203048 | Oregon State Public Health Laboratory | Oregon State Public Health Laboratory | Rafia Razzaque, Eugene Yeboah, Vanda Makris, Laura Tsaknaridis, John Fontana and Shane Sevey |
| EPI_ISL_2203049 | Aegis Sciences Corporation | Centers for Disease Control and Prevention Division of Viral Diseases, Pathogen Discovery | Dakota Howard, Dhwani Batra, Peter W. Cook, Kara Moser, Adrian Paskey, Jason Caravas, Benjamin Rambo-Martin, Shatavia Morrison, Christopher Gulvick, Scott Sammons, Yvette Unoarumhi, Darlene Wagner, Matthew Schmerer, Cyndi Clark, Patrick Campbell, Rob Case, Vikramsinha Ghorpade, Holly Houdeshell, Ola Kvalvaag, Dillon Nall, Ethan Sanders, Alec Vest, Shaun Westlund, Matthew Hardison, Clinton R. Paden, Duncan MacCannell |
| EPI_ISL_2203050 | Houston Methodist Hospital | Houston Methodist Hospital | Randall J. Olsen, Paul A. Christensen, S. Wesley Long, Sishir Subedi, Robert Olson, Marcus Nguyen, James J. Davis, Matthew Ojeda Saavedra, Prasanti Yerramilli, Layne Pruitt, Kristina Reppond, Madison N. Shyer, Jessica Cambric, Ryan Gadd, Ilya J. Finkelstein, Jimmy Gollihar, and James M. Musser |
| EPI_ISL_2203051 | Aegis Sciences Corporation | Centers for Disease Control and Prevention Division of Viral Diseases, Pathogen Discovery | Dakota Howard, Dhwani Batra, Peter W. Cook, Kara Moser, Adrian Paskey, Jason Caravas, Benjamin Rambo-Martin, Shatavia Morrison, Christopher Gulvick, Scott Sammons, Yvette Unoarumhi, Darlene Wagner, Matthew Schmerer, Cyndi Clark, Patrick Campbell, Rob Case, Vikramsinha Ghorpade, Holly Houdeshell, Ola Kvalvaag, Dillon Nall, Ethan Sanders, Alec Vest, Shaun Westlund, Matthew Hardison, Clinton R. Paden, Duncan MacCannell |
| EPI_ISL_2203052 | Oregon State Public Health Laboratory | Oregon State Public Health Laboratory | Rafia Razzaque, Eugene Yeboah, Vanda Makris, Laura Tsaknaridis, John Fontana and Shane Sevey |
| EPI_ISL_2203053 | Aegis Sciences Corporation | Centers for Disease Control and Prevention Division of Viral Diseases, Pathogen Discovery | Dakota Howard, Dhwani Batra, Peter W. Cook, Kara Moser, Adrian Paskey, Jason Caravas, Benjamin Rambo-Martin, Shatavia Morrison, Christopher Gulvick, Scott Sammons, Yvette Unoarumhi, Darlene Wagner, Matthew Schmerer, Cyndi Clark, Patrick Campbell, Rob Case, Vikramsinha Ghorpade, Holly Houdeshell, Ola Kvalvaag, Dillon Nall, Ethan Sanders, Alec Vest, Shaun Westlund, Matthew Hardison, Clinton R. Paden, Duncan MacCannell |
| EPI_ISL_2203054 | Houston Methodist Hospital | Houston Methodist Hospital | Randall J. Olsen, Paul A. Christensen, S. Wesley Long, Sishir Subedi, Robert Olson, Marcus Nguyen, James J. Davis, Matthew Ojeda Saavedra, Prasanti Yerramilli, Layne Pruitt, Kristina Reppond, Madison N. Shyer, Jessica Cambric, Ryan Gadd, Ilya J. Finkelstein, Jimmy Gollihar, and James M. Musser |
| EPI_ISL_2203055 | Aegis Sciences Corporation | Centers for Disease Control and Prevention Division of Viral Diseases, Pathogen Discovery | Dakota Howard, Dhwani Batra, Peter W. Cook, Kara Moser, Adrian Paskey, Jason Caravas, Benjamin Rambo-Martin, Shatavia Morrison, Christopher Gulvick, Scott Sammons, Yvette Unoarumhi, Darlene Wagner, Matthew Schmerer, Cyndi Clark, Patrick Campbell, Rob Case, Vikramsinha Ghorpade, Holly Houdeshell, Ola Kvalvaag, Dillon Nall, Ethan Sanders, Alec Vest, Shaun Westlund, Matthew Hardison, Clinton R. Paden, Duncan MacCannell |
| EPI_ISL_2203056, EPI_ISL_2203057 | Oregon State Public Health Laboratory | Oregon State Public Health Laboratory | Rafia Razzaque, Eugene Yeboah, Vanda Makris, Laura Tsaknaridis, John Fontana and Shane Sevey |
| EPI_ISL_2203058 | Aegis Sciences Corporation | Centers for Disease Control and Prevention Division of Viral Diseases, Pathogen Discovery | Dakota Howard, Dhwani Batra, Peter W. Cook, Kara Moser, Adrian Paskey, Jason Caravas, Benjamin Rambo-Martin, Shatavia Morrison, Christopher Gulvick, Scott Sammons, Yvette Unoarumhi, Darlene Wagner, Matthew Schmerer, Cyndi Clark, Patrick Campbell, Rob Case, Vikramsinha Ghorpade, Holly Houdeshell, Ola Kvalvaag, Dillon Nall, Ethan Sanders, Alec Vest, Shaun Westlund, Matthew Hardison, Clinton R. Paden, Duncan MacCannell |
| EPI_ISL_2203059 | Houston Methodist Hospital | Houston Methodist Hospital | Randall J. Olsen, Paul A. Christensen, S. Wesley Long, Sishir Subedi, Robert Olson, Marcus Nguyen, James J. Davis, Matthew Ojeda Saavedra, Prasanti Yerramilli, Layne Pruitt, Kristina Reppond, Madison N. Shyer, Jessica Cambric, Ryan Gadd, Ilya J. Finkelstein, Jimmy Gollihar, and James M. Musser |
| EPI_ISL_2203060 | Oregon State Public Health Laboratory | Oregon State Public Health Laboratory | Rafia Razzaque, Eugene Yeboah, Vanda Makris, Laura Tsaknaridis, John Fontana and Shane Sevey |
| EPI_ISL_2203061 | Aegis Sciences Corporation | Centers for Disease Control and Prevention Division of Viral Diseases, Pathogen Discovery | Dakota Howard, Dhwani Batra, Peter W. Cook, Kara Moser, Adrian Paskey, Jason Caravas, Benjamin Rambo-Martin, Shatavia Morrison, Christopher Gulvick, Scott Sammons, Yvette Unoarumhi, Darlene Wagner, Matthew Schmerer, Cyndi Clark, Patrick Campbell, Rob Case, Vikramsinha Ghorpade, Holly Houdeshell, Ola Kvalvaag, Dillon Nall, Ethan Sanders, Alec Vest, Shaun Westlund, Matthew Hardison, Clinton R. Paden, Duncan MacCannell |
| EPI_ISL_2203062 | Oregon State Public Health Laboratory | Oregon State Public Health Laboratory | Rafia Razzaque, Eugene Yeboah, Vanda Makris, Laura Tsaknaridis, John Fontana and Shane Sevey |
| EPI_ISL_2203063 | Houston Methodist Hospital | Houston Methodist Hospital | Randall J. Olsen, Paul A. Christensen, S. Wesley Long, Sishir Subedi, Robert Olson, Marcus Nguyen, James J. Davis, Matthew Ojeda Saavedra, Prasanti Yerramilli, Layne Pruitt, Kristina Reppond, Madison N. Shyer, Jessica Cambric, Ryan Gadd, Ilya J. Finkelstein, Jimmy Gollihar, and James M. Musser |
| EPI_ISL_2203064 | Oregon State Public Health Laboratory | Oregon State Public Health Laboratory | Rafia Razzaque, Eugene Yeboah, Vanda Makris, Laura Tsaknaridis, John Fontana and Shane Sevey |
| EPI_ISL_2203065 | Aegis Sciences Corporation | Centers for Disease Control and Prevention Division of Viral Diseases, Pathogen Discovery | Dakota Howard, Dhwani Batra, Peter W. Cook, Kara Moser, Adrian Paskey, Jason Caravas, Benjamin Rambo-Martin, Shatavia Morrison, Christopher Gulvick, Scott Sammons, Yvette Unoarumhi, Darlene Wagner, Matthew Schmerer, Cyndi Clark, Patrick Campbell, Rob Case, Vikramsinha Ghorpade, Holly Houdeshell, Ola Kvalvaag, Dillon Nall, Ethan Sanders, Alec Vest, Shaun Westlund, Matthew Hardison, Clinton R. Paden, Duncan MacCannell |
| EPI_ISL_2203066, EPI_ISL_2203067 | Oregon State Public Health Laboratory | Oregon State Public Health Laboratory | Rafia Razzaque, Eugene Yeboah, Vanda Makris, Laura Tsaknaridis, John Fontana and Shane Sevey |
| EPI_ISL_2203068 | Houston Methodist Hospital | Houston Methodist Hospital | Randall J. Olsen, Paul A. Christensen, S. Wesley Long, Sishir Subedi, Robert Olson, Marcus Nguyen, James J. Davis, Matthew Ojeda Saavedra, Prasanti Yerramilli, Layne Pruitt, Kristina Reppond, Madison N. Shyer, Jessica Cambric, Ryan Gadd, Ilya J. Finkelstein, Jimmy Gollihar, and James M. Musser |
| EPI_ISL_2203069 | Aegis Sciences Corporation | Centers for Disease Control and Prevention Division of Viral Diseases, Pathogen Discovery | Dakota Howard, Dhwani Batra, Peter W. Cook, Kara Moser, Adrian Paskey, Jason Caravas, Benjamin Rambo-Martin, Shatavia Morrison, Christopher Gulvick, Scott Sammons, Yvette Unoarumhi, Darlene Wagner, Matthew Schmerer, Cyndi Clark, Patrick Campbell, Rob Case, Vikramsinha Ghorpade, Holly Houdeshell, Ola Kvalvaag, Dillon Nall, Ethan Sanders, Alec Vest, Shaun Westlund, Matthew Hardison, Clinton R. Paden, Duncan MacCannell |
| EPI_ISL_2203070, EPI_ISL_2203071 | Oregon State Public Health Laboratory | Oregon State Public Health Laboratory | Rafia Razzaque, Eugene Yeboah, Vanda Makris, Laura Tsaknaridis, John Fontana and Shane Sevey |
| EPI_ISL_2203072 | Houston Methodist Hospital | Houston Methodist Hospital | Randall J. Olsen, Paul A. Christensen, S. Wesley Long, Sishir Subedi, Robert Olson, Marcus Nguyen, James J. Davis, Matthew Ojeda Saavedra, Prasanti Yerramilli, Layne Pruitt, Kristina Reppond, Madison N. Shyer, Jessica Cambric, Ryan Gadd, Ilya J. Finkelstein, Jimmy Gollihar, and James M. Musser |
| EPI_ISL_2203073 | Aegis Sciences Corporation | Centers for Disease Control and Prevention Division of Viral Diseases, Pathogen Discovery | Dakota Howard, Dhwani Batra, Peter W. Cook, Kara Moser, Adrian Paskey, Jason Caravas, Benjamin Rambo-Martin, Shatavia Morrison, Christopher Gulvick, Scott Sammons, Yvette Unoarumhi, Darlene Wagner, Matthew Schmerer, Cyndi Clark, Patrick Campbell, Rob Case, Vikramsinha Ghorpade, Holly Houdeshell, Ola Kvalvaag, Dillon Nall, Ethan Sanders, Alec Vest, Shaun Westlund, Matthew Hardison, Clinton R. Paden, Duncan MacCannell |
| EPI_ISL_2203074 | Oregon State Public Health Laboratory | Oregon State Public Health Laboratory | Rafia Razzaque, Eugene Yeboah, Vanda Makris, Laura Tsaknaridis, John Fontana and Shane Sevey |
| EPI_ISL_2203075 | Houston Methodist Hospital | Houston Methodist Hospital | Randall J. Olsen, Paul A. Christensen, S. Wesley Long, Sishir Subedi, Robert Olson, Marcus Nguyen, James J. Davis, Matthew Ojeda Saavedra, Prasanti Yerramilli, Layne Pruitt, Kristina Reppond, Madison N. Shyer, Jessica Cambric, Ryan Gadd, Ilya J. Finkelstein, Jimmy Gollihar, and James M. Musser |
| EPI_ISL_2203076 | Oregon State Public Health Laboratory | Oregon State Public Health Laboratory | Rafia Razzaque, Eugene Yeboah, Vanda Makris, Laura Tsaknaridis, John Fontana and Shane Sevey |
| EPI_ISL_2203077 | Aegis Sciences Corporation | Centers for Disease Control and Prevention Division of Viral Diseases, Pathogen Discovery | Dakota Howard, Dhwani Batra, Peter W. Cook, Kara Moser, Adrian Paskey, Jason Caravas, Benjamin Rambo-Martin, Shatavia Morrison, Christopher Gulvick, Scott Sammons, Yvette Unoarumhi, Darlene Wagner, Matthew Schmerer, Cyndi Clark, Patrick Campbell, Rob Case, Vikramsinha Ghorpade, Holly Houdeshell, Ola Kvalvaag, Dillon Nall, Ethan Sanders, Alec Vest, Shaun Westlund, Matthew Hardison, Clinton R. Paden, Duncan MacCannell |
| EPI_ISL_2203078 | Oregon State Public Health Laboratory | Oregon State Public Health Laboratory | Rafia Razzaque, Eugene Yeboah, Vanda Makris, Laura Tsaknaridis, John Fontana and Shane Sevey |
| EPI_ISL_2203079 | Houston Methodist Hospital | Houston Methodist Hospital | Randall J. Olsen, Paul A. Christensen, S. Wesley Long, Sishir Subedi, Robert Olson, Marcus Nguyen, James J. Davis, Matthew Ojeda Saavedra, Prasanti Yerramilli, Layne Pruitt, Kristina Reppond, Madison N. Shyer, Jessica Cambric, Ryan Gadd, Ilya J. Finkelstein, Jimmy Gollihar, and James M. Musser |
| EPI_ISL_2203080, EPI_ISL_2203081 | Oregon State Public Health Laboratory | Oregon State Public Health Laboratory | Rafia Razzaque, Eugene Yeboah, Vanda Makris, Laura Tsaknaridis, John Fontana and Shane Sevey |
| EPI_ISL_2203082 | Aegis Sciences Corporation | Centers for Disease Control and Prevention Division of Viral Diseases, Pathogen Discovery | Dakota Howard, Dhwani Batra, Peter W. Cook, Kara Moser, Adrian Paskey, Jason Caravas, Benjamin Rambo-Martin, Shatavia Morrison, Christopher Gulvick, Scott Sammons, Yvette Unoarumhi, Darlene Wagner, Matthew Schmerer, Cyndi Clark, Patrick Campbell, Rob Case, Vikramsinha Ghorpade, Holly Houdeshell, Ola Kvalvaag, Dillon Nall, Ethan Sanders, Alec Vest, Shaun Westlund, Matthew Hardison, Clinton R. Paden, Duncan MacCannell |
| EPI_ISL_2203083 | Oregon State Public Health Laboratory | Oregon State Public Health Laboratory | Rafia Razzaque, Eugene Yeboah, Vanda Makris, Laura Tsaknaridis, John Fontana and Shane Sevey |

| | | | |
|---|---|---|---|
| EPI_ISL_2203084 | Houston Methodist Hospital | Houston Methodist Hospital | Randall J. Olsen, Paul A. Christensen, S. Wesley Long, Sishir Subedi, Robert Olson, Marcus Nguyen, James J. Davis, Matthew Ojeda Saavedra, Prasanti Yerramilli, Layne Pruitt, Kristina Reppond, Madison N. Shyer, Jessica Cambric, Ryan Gadd, Ilya J. Finkelstein, Jimmy Gollihar, and James M. Musser |
| EPI_ISL_2203085 | Oregon State Public Health Laboratory | Oregon State Public Health Laboratory | Rafia Razzaque, Eugene Yeboah, Vanda Makris, Laura Tsaknaridis, John Fontana and Shane Sevey |
| EPI_ISL_2203086 | Aegis Sciences Corporation | Centers for Disease Control and Prevention Division of Viral Diseases, Pathogen Discovery | Dakota Howard, Dhwani Batra, Peter W. Cook, Kara Moser, Adrian Paskey, Jason Caravas, Benjamin Rambo-Martin, Shatavia Morrison, Christopher Gulvick, Scott Sammons, Yvette Unoarumhi, Darlene Wagner, Matthew Schmerer, Cyndi Clark, Patrick Campbell, Rob Case, Vikramsinha Ghorpade, Holly Houdeshell, Ola Kvalvaag, Dillon Nall, Ethan Sanders, Alec Vest, Shaun Westlund, Matthew Hardison, Clinton R. Paden, Duncan MacCannell |
| EPI_ISL_2203087 | Oregon State Public Health Laboratory | Oregon State Public Health Laboratory | Rafia Razzaque, Eugene Yeboah, Vanda Makris, Laura Tsaknaridis, John Fontana and Shane Sevey |
| EPI_ISL_2203088 | Houston Methodist Hospital | Houston Methodist Hospital | Randall J. Olsen, Paul A. Christensen, S. Wesley Long, Sishir Subedi, Robert Olson, Marcus Nguyen, James J. Davis, Matthew Ojeda Saavedra, Prasanti Yerramilli, Layne Pruitt, Kristina Reppond, Madison N. Shyer, Jessica Cambric, Ryan Gadd, Ilya J. Finkelstein, Jimmy Gollihar, and James M. Musser |
| EPI_ISL_2203089 | Oregon State Public Health Laboratory | Oregon State Public Health Laboratory | Rafia Razzaque, Eugene Yeboah, Vanda Makris, Laura Tsaknaridis, John Fontana and Shane Sevey |
| EPI_ISL_2203090 | Aegis Sciences Corporation | Centers for Disease Control and Prevention Division of Viral Diseases, Pathogen Discovery | Dakota Howard, Dhwani Batra, Peter W. Cook, Kara Moser, Adrian Paskey, Jason Caravas, Benjamin Rambo-Martin, Shatavia Morrison, Christopher Gulvick, Scott Sammons, Yvette Unoarumhi, Darlene Wagner, Matthew Schmerer, Cyndi Clark, Patrick Campbell, Rob Case, Vikramsinha Ghorpade, Holly Houdeshell, Ola Kvalvaag, Dillon Nall, Ethan Sanders, Alec Vest, Shaun Westlund, Matthew Hardison, Clinton R. Paden, Duncan MacCannell |
| EPI_ISL_2203091, EPI_ISL_2203092 | Oregon State Public Health Laboratory | Oregon State Public Health Laboratory | Rafia Razzaque, Eugene Yeboah, Vanda Makris, Laura Tsaknaridis, John Fontana and Shane Sevey |
| EPI_ISL_2203093 | Houston Methodist Hospital | Houston Methodist Hospital | Randall J. Olsen, Paul A. Christensen, S. Wesley Long, Sishir Subedi, Robert Olson, Marcus Nguyen, James J. Davis, Matthew Ojeda Saavedra, Prasanti Yerramilli, Layne Pruitt, Kristina Reppond, Madison N. Shyer, Jessica Cambric, Ryan Gadd, Ilya J. Finkelstein, Jimmy Gollihar, and James M. Musser |
| EPI_ISL_2203094 | Aegis Sciences Corporation | Centers for Disease Control and Prevention Division of Viral Diseases, Pathogen Discovery | Dakota Howard, Dhwani Batra, Peter W. Cook, Kara Moser, Adrian Paskey, Jason Caravas, Benjamin Rambo-Martin, Shatavia Morrison, Christopher Gulvick, Scott Sammons, Yvette Unoarumhi, Darlene Wagner, Matthew Schmerer, Cyndi Clark, Patrick Campbell, Rob Case, Vikramsinha Ghorpade, Holly Houdeshell, Ola Kvalvaag, Dillon Nall, Ethan Sanders, Alec Vest, Shaun Westlund, Matthew Hardison, Clinton R. Paden, Duncan MacCannell |
| EPI_ISL_2203095 | Oregon State Public Health Laboratory | Oregon State Public Health Laboratory | Rafia Razzaque, Eugene Yeboah, Vanda Makris, Laura Tsaknaridis, John Fontana and Shane Sevey |
| EPI_ISL_2203096 | Aegis Sciences Corporation | Centers for Disease Control and Prevention Division of Viral Diseases, Pathogen Discovery | Dakota Howard, Dhwani Batra, Peter W. Cook, Kara Moser, Adrian Paskey, Jason Caravas, Benjamin Rambo-Martin, Shatavia Morrison, Christopher Gulvick, Scott Sammons, Yvette Unoarumhi, Darlene Wagner, Matthew Schmerer, Cyndi Clark, Patrick Campbell, Rob Case, Vikramsinha Ghorpade, Holly Houdeshell, Ola Kvalvaag, Dillon Nall, Ethan Sanders, Alec Vest, Shaun Westlund, Matthew Hardison, Clinton R. Paden, Duncan MacCannell |
| EPI_ISL_2203097 | Oregon State Public Health Laboratory | Oregon State Public Health Laboratory | Rafia Razzaque, Eugene Yeboah, Vanda Makris, Laura Tsaknaridis, John Fontana and Shane Sevey |
| EPI_ISL_2203098 | Houston Methodist Hospital | Houston Methodist Hospital | Randall J. Olsen, Paul A. Christensen, S. Wesley Long, Sishir Subedi, Robert Olson, Marcus Nguyen, James J. Davis, Matthew Ojeda Saavedra, Prasanti Yerramilli, Layne Pruitt, Kristina Reppond, Madison N. Shyer, Jessica Cambric, Ryan Gadd, Ilya J. Finkelstein, Jimmy Gollihar, and James M. Musser |
| EPI_ISL_2203100 | Oregon State Public Health Laboratory | Oregon State Public Health Laboratory | Rafia Razzaque, Eugene Yeboah, Vanda Makris, Laura Tsaknaridis, John Fontana and Shane Sevey |
| EPI_ISL_2203101 | Houston Methodist Hospital | Houston Methodist Hospital | Randall J. Olsen, Paul A. Christensen, S. Wesley Long, Sishir Subedi, Robert Olson, Marcus Nguyen, James J. Davis, Matthew Ojeda Saavedra, Prasanti Yerramilli, Layne Pruitt, Kristina Reppond, Madison N. Shyer, Jessica Cambric, Ryan Gadd, Ilya J. Finkelstein, Jimmy Gollihar, and James M. Musser |
| EPI_ISL_2203102 | Oregon State Public Health Laboratory | Oregon State Public Health Laboratory | Rafia Razzaque, Eugene Yeboah, Vanda Makris, Laura Tsaknaridis, John Fontana and Shane Sevey |
| EPI_ISL_2203103 | Aegis Sciences Corporation | Centers for Disease Control and Prevention Division of Viral Diseases, Pathogen Discovery | Dakota Howard, Dhwani Batra, Peter W. Cook, Kara Moser, Adrian Paskey, Jason Caravas, Benjamin Rambo-Martin, Shatavia Morrison, Christopher Gulvick, Scott Sammons, Yvette Unoarumhi, Darlene Wagner, Matthew Schmerer, Cyndi Clark, Patrick Campbell, Rob Case, Vikramsinha Ghorpade, Holly Houdeshell, Ola Kvalvaag, Dillon Nall, Ethan Sanders, Alec Vest, Shaun Westlund, Matthew Hardison, Clinton R. Paden, Duncan MacCannell |
| EPI_ISL_2203104 | Houston Methodist Hospital | Houston Methodist Hospital | Randall J. Olsen, Paul A. Christensen, S. Wesley Long, Sishir Subedi, Robert Olson, Marcus Nguyen, James J. Davis, Matthew Ojeda Saavedra, Prasanti Yerramilli, Layne Pruitt, Kristina Reppond, Madison N. Shyer, Jessica Cambric, Ryan Gadd, Ilya J. Finkelstein, Jimmy Gollihar, and James M. Musser |
| EPI_ISL_2203105 | Aegis Sciences Corporation | Centers for Disease Control and Prevention Division of Viral Diseases, Pathogen Discovery | Dakota Howard, Dhwani Batra, Peter W. Cook, Kara Moser, Adrian Paskey, Jason Caravas, Benjamin Rambo-Martin, Shatavia Morrison, Christopher Gulvick, Scott Sammons, Yvette Unoarumhi, Darlene Wagner, Matthew Schmerer, Cyndi Clark, Patrick Campbell, Rob Case, Vikramsinha Ghorpade, Holly Houdeshell, Ola Kvalvaag, Dillon Nall, Ethan Sanders, Alec Vest, Shaun Westlund, Matthew Hardison, Clinton R. Paden, Duncan MacCannell |
| EPI_ISL_2203106 | Houston Methodist Hospital | Houston Methodist Hospital | Randall J. Olsen, Paul A. Christensen, S. Wesley Long, Sishir Subedi, Robert Olson, Marcus Nguyen, James J. Davis, Matthew Ojeda Saavedra, Prasanti Yerramilli, Layne Pruitt, Kristina Reppond, Madison N. Shyer, Jessica Cambric, Ryan Gadd, Ilya J. Finkelstein, Jimmy Gollihar, and James M. Musser |
| EPI_ISL_2203107 | Aegis Sciences Corporation | Centers for Disease Control and Prevention Division of Viral Diseases, Pathogen Discovery | Dakota Howard, Dhwani Batra, Peter W. Cook, Kara Moser, Adrian Paskey, Jason Caravas, Benjamin Rambo-Martin, Shatavia Morrison, Christopher Gulvick, Scott Sammons, Yvette Unoarumhi, Darlene Wagner, Matthew Schmerer, Cyndi Clark, Patrick Campbell, Rob Case, Vikramsinha Ghorpade, Holly Houdeshell, Ola Kvalvaag, Dillon Nall, Ethan Sanders, Alec Vest, Shaun Westlund, Matthew Hardison, Clinton R. Paden, Duncan MacCannell |
| EPI_ISL_2203108 | Houston Methodist Hospital | Houston Methodist Hospital | Randall J. Olsen, Paul A. Christensen, S. Wesley Long, Sishir Subedi, Robert Olson, Marcus Nguyen, James J. Davis, Matthew Ojeda Saavedra, Prasanti Yerramilli, Layne Pruitt, Kristina Reppond, Madison N. Shyer, Jessica Cambric, Ryan Gadd, Ilya J. Finkelstein, Jimmy Gollihar, and James M. Musser |
| EPI_ISL_2203109 | Aegis Sciences Corporation | Centers for Disease Control and Prevention Division of Viral Diseases, Pathogen Discovery | Dakota Howard, Dhwani Batra, Peter W. Cook, Kara Moser, Adrian Paskey, Jason Caravas, Benjamin Rambo-Martin, Shatavia Morrison, Christopher Gulvick, Scott Sammons, Yvette Unoarumhi, Darlene Wagner, Matthew Schmerer, Cyndi Clark, Patrick Campbell, Rob Case, Vikramsinha Ghorpade, Holly Houdeshell, Ola Kvalvaag, Dillon Nall, Ethan Sanders, Alec Vest, Shaun Westlund, Matthew Hardison, Clinton R. Paden, Duncan MacCannell |
| EPI_ISL_2203110 | Houston Methodist Hospital | Houston Methodist Hospital | Randall J. Olsen, Paul A. Christensen, S. Wesley Long, Sishir Subedi, Robert Olson, Marcus Nguyen, James J. Davis, Matthew Ojeda Saavedra, Prasanti Yerramilli, Layne Pruitt, Kristina Reppond, Madison N. Shyer, Jessica Cambric, Ryan Gadd, Ilya J. Finkelstein, Jimmy Gollihar, and James M. Musser |
| EPI_ISL_2203111 | Aegis Sciences Corporation | Centers for Disease Control and Prevention Division of Viral Diseases, Pathogen Discovery | Dakota Howard, Dhwani Batra, Peter W. Cook, Kara Moser, Adrian Paskey, Jason Caravas, Benjamin Rambo-Martin, Shatavia Morrison, Christopher Gulvick, Scott Sammons, Yvette Unoarumhi, Darlene Wagner, Matthew Schmerer, Cyndi Clark, Patrick Campbell, Rob Case, Vikramsinha Ghorpade, Holly Houdeshell, Ola Kvalvaag, Dillon Nall, Ethan Sanders, Alec Vest, Shaun Westlund, Matthew Hardison, Clinton R. Paden, Duncan MacCannell |
| EPI_ISL_2203112 | Houston Methodist Hospital | Houston Methodist Hospital | Randall J. Olsen, Paul A. Christensen, S. Wesley Long, Sishir Subedi, Robert Olson, Marcus Nguyen, James J. Davis, Matthew Ojeda Saavedra, Prasanti Yerramilli, Layne Pruitt, Kristina Reppond, Madison N. Shyer, Jessica Cambric, Ryan Gadd, Ilya J. Finkelstein, Jimmy Gollihar, and James M. Musser |
| EPI_ISL_2203113, EPI_ISL_2203114 | Aegis Sciences Corporation | Centers for Disease Control and Prevention Division of Viral Diseases, Pathogen Discovery | Dakota Howard, Dhwani Batra, Peter W. Cook, Kara Moser, Adrian Paskey, Jason Caravas, Benjamin Rambo-Martin, Shatavia Morrison, Christopher Gulvick, Scott Sammons, Yvette Unoarumhi, Darlene Wagner, Matthew Schmerer, Cyndi Clark, Patrick Campbell, Rob Case, Vikramsinha Ghorpade, Holly Houdeshell, Ola Kvalvaag, Dillon Nall, Ethan Sanders, Alec Vest, Shaun Westlund, Matthew Hardison, Clinton R. Paden, Duncan MacCannell |
| EPI_ISL_2203115 | Houston Methodist Hospital | Houston Methodist Hospital | Randall J. Olsen, Paul A. Christensen, S. Wesley Long, Sishir Subedi, Robert Olson, Marcus Nguyen, James J. Davis, Matthew Ojeda Saavedra, Prasanti Yerramilli, Layne Pruitt, Kristina Reppond, Madison N. Shyer, Jessica Cambric, Ryan Gadd, Ilya J. Finkelstein, Jimmy Gollihar, and James M. Musser |
| EPI_ISL_2203116 | Aegis Sciences Corporation | Centers for Disease Control and Prevention Division of Viral Diseases, Pathogen Discovery | Dakota Howard, Dhwani Batra, Peter W. Cook, Kara Moser, Adrian Paskey, Jason Caravas, Benjamin Rambo-Martin, Shatavia Morrison, Christopher Gulvick, Scott Sammons, Yvette Unoarumhi, Darlene Wagner, Matthew Schmerer, Cyndi Clark, Patrick Campbell, Rob Case, Vikramsinha Ghorpade, Holly Houdeshell, Ola Kvalvaag, Dillon Nall, Ethan Sanders, Alec Vest, Shaun Westlund, Matthew Hardison, Clinton R. Paden, Duncan MacCannell |
| EPI_ISL_2203117 | Houston Methodist Hospital | Houston Methodist Hospital | Randall J. Olsen, Paul A. Christensen, S. Wesley Long, Sishir Subedi, Robert Olson, Marcus Nguyen, James J. Davis, Matthew Ojeda Saavedra, Prasanti Yerramilli, Layne Pruitt, Kristina Reppond, Madison N. Shyer, Jessica Cambric, Ryan Gadd, Ilya J. Finkelstein, Jimmy Gollihar, and James M. Musser |
| EPI_ISL_2203118 | Aegis Sciences Corporation | Centers for Disease Control and Prevention Division of Viral Diseases, Pathogen Discovery | Dakota Howard, Dhwani Batra, Peter W. Cook, Kara Moser, Adrian Paskey, Jason Caravas, Benjamin Rambo-Martin, Shatavia Morrison, Christopher Gulvick, Scott Sammons, Yvette Unoarumhi, Darlene Wagner, Matthew Schmerer, Cyndi Clark, Patrick Campbell, Rob Case, Vikramsinha Ghorpade, Holly Houdeshell, Ola Kvalvaag, Dillon Nall, Ethan Sanders, Alec Vest, Shaun Westlund, Matthew Hardison, Clinton R. Paden, Duncan MacCannell |
| EPI_ISL_2203119 | Houston Methodist Hospital | Houston Methodist Hospital | Randall J. Olsen, Paul A. Christensen, S. Wesley Long, Sishir Subedi, Robert Olson, Marcus Nguyen, James J. Davis, Matthew Ojeda Saavedra, Prasanti Yerramilli, Layne Pruitt, Kristina Reppond, Madison N. Shyer, Jessica Cambric, Ryan Gadd, Ilya J. Finkelstein, Jimmy Gollihar, and James M. Musser |
| EPI_ISL_2203120, EPI_ISL_2203121 | Aegis Sciences Corporation | Centers for Disease Control and Prevention Division of Viral Diseases, Pathogen Discovery | Dakota Howard, Dhwani Batra, Peter W. Cook, Kara Moser, Adrian Paskey, Jason Caravas, Benjamin Rambo-Martin, Shatavia Morrison, Christopher Gulvick, Scott Sammons, Yvette Unoarumhi, Darlene Wagner, Matthew Schmerer, Cyndi Clark, Patrick Campbell, Rob Case, Vikramsinha Ghorpade, Holly Houdeshell, Ola Kvalvaag, Dillon Nall, Ethan Sanders, Alec Vest, Shaun Westlund, Matthew Hardison, Clinton R. Paden, Duncan MacCannell |
| EPI_ISL_2203122 | Houston Methodist Hospital | Houston Methodist Hospital | Randall J. Olsen, Paul A. Christensen, S. Wesley Long, Sishir Subedi, Robert Olson, Marcus Nguyen, James J. Davis, Matthew Ojeda Saavedra, Prasanti Yerramilli, Layne Pruitt, Kristina Reppond, Madison N. Shyer, Jessica Cambric, Ryan Gadd, Ilya J. Finkelstein, Jimmy Gollihar, and James M. Musser |
| EPI_ISL_2203123, EPI_ISL_2203124 | Aegis Sciences Corporation | Centers for Disease Control and Prevention Division of Viral Diseases, Pathogen Discovery | Dakota Howard, Dhwani Batra, Peter W. Cook, Kara Moser, Adrian Paskey, Jason Caravas, Benjamin Rambo-Martin, Shatavia Morrison, Christopher Gulvick, Scott Sammons, Yvette Unoarumhi, Darlene Wagner, Matthew Schmerer, Cyndi Clark, Patrick Campbell, Rob Case, Vikramsinha Ghorpade, Holly Houdeshell, Ola Kvalvaag, Dillon Nall, Ethan Sanders, Alec Vest, Shaun Westlund, Matthew Hardison, Clinton R. Paden, Duncan MacCannell |
| EPI_ISL_2203125 | Houston Methodist Hospital | Houston Methodist Hospital | Randall J. Olsen, Paul A. Christensen, S. Wesley Long, Sishir Subedi, Robert Olson, Marcus Nguyen, James J. Davis, Matthew Ojeda Saavedra, Prasanti Yerramilli, Layne Pruitt, Kristina Reppond, Madison N. Shyer, Jessica Cambric, Ryan Gadd, Ilya J. Finkelstein, Jimmy Gollihar, and James M. Musser |
| EPI_ISL_2203126, EPI_ISL_2203127 | Aegis Sciences Corporation | Centers for Disease Control and Prevention Division of Viral Diseases, Pathogen Discovery | Dakota Howard, Dhwani Batra, Peter W. Cook, Kara Moser, Adrian Paskey, Jason Caravas, Benjamin Rambo-Martin, Shatavia Morrison, Christopher Gulvick, Scott Sammons, Yvette Unoarumhi, Darlene Wagner, Matthew Schmerer, Cyndi Clark, Patrick Campbell, Rob Case, Vikramsinha Ghorpade, Holly Houdeshell, Ola Kvalvaag, Dillon Nall, Ethan Sanders, Alec Vest, Shaun Westlund, Matthew Hardison, Clinton R. Paden, Duncan MacCannell |
| EPI_ISL_2203128 | Houston Methodist Hospital | Houston Methodist Hospital | Randall J. Olsen, Paul A. Christensen, S. Wesley Long, Sishir Subedi, Robert Olson, Marcus Nguyen, James J. Davis, Matthew Ojeda Saavedra, Prasanti Yerramilli, Layne Pruitt, Kristina Reppond, Madison N. Shyer, Jessica Cambric, Ryan Gadd, Ilya J. Finkelstein, Jimmy Gollihar, and James M. Musser |
| EPI_ISL_2203129 | Aegis Sciences Corporation | Centers for Disease Control and Prevention Division of Viral Diseases, Pathogen Discovery | Dakota Howard, Dhwani Batra, Peter W. Cook, Kara Moser, Adrian Paskey, Jason Caravas, Benjamin Rambo-Martin, Shatavia Morrison, Christopher Gulvick, Scott Sammons, Yvette Unoarumhi, Darlene Wagner, Matthew Schmerer, Cyndi Clark, Patrick Campbell, Rob Case, Vikramsinha Ghorpade, Holly Houdeshell, Ola Kvalvaag, Dillon Nall, Ethan Sanders, Alec Vest, Shaun Westlund, Matthew Hardison, Clinton R. Paden, Duncan MacCannell |
| EPI_ISL_2203130 | Houston Methodist Hospital | Houston Methodist Hospital | Randall J. Olsen, Paul A. Christensen, S. Wesley Long, Sishir Subedi, Robert Olson, Marcus Nguyen, James J. Davis, Matthew Ojeda Saavedra, Prasanti Yerramilli, Layne Pruitt, Kristina Reppond, Madison N. Shyer, Jessica Cambric, Ryan Gadd, Ilya J. Finkelstein, Jimmy Gollihar, and James M. Musser |
| EPI_ISL_2203131 | Aegis Sciences Corporation | Centers for Disease Control and Prevention | Dakota Howard, Dhwani Batra, Peter W. Cook, Kara Moser, Adrian Paskey, Jason Caravas, Benjamin Rambo-Martin, Shatavia Morrison, Christopher Gulvick, Scott Sammons, Yvette Unoarumhi, Darlene Wagner, Matthew Schmerer, Cyndi Clark, Patrick Campbell, Rob Case, Vikramsinha |

| | | | |
|---|---|---|---|
| | | Division of Viral Diseases, Pathogen Discovery | Ghorpade, Holly Houdeshell, Ola Kvalvaag, Dillon Nall, Ethan Sanders, Alec Vest, Shaun Westlund, Matthew Hardison, Clinton R. Paden, Duncan MacCannell |
| EPI_ISL_2203132 | Houston Methodist Hospital | Houston Methodist Hospital | Randall J. Olsen, Paul A. Christensen, S. Wesley Long, Sishir Subedi, Robert Olson, Marcus Nguyen, James J. Davis, Matthew Ojeda Saavedra, Prasanti Yerramilli, Layne Pruitt, Kristina Reppond, Madison N. Shyer, Jessica Cambric, Ryan Gadd, Ilya J. Finkelstein, Jimmy Gollihar, and James M. Musser |
| EPI_ISL_2203133, EPI_ISL_2203134, EPI_ISL_2203135 | Aegis Sciences Corporation | Centers for Disease Control and Prevention Division of Viral Diseases, Pathogen Discovery | Dakota Howard, Dhwani Batra, Peter W. Cook, Kara Moser, Adrian Paskey, Jason Caravas, Benjamin Rambo-Martin, Shatavia Morrison, Christopher Gulvick, Scott Sammons, Yvette Unoarumhi, Darlene Wagner, Matthew Schmerer, Cyndi Clark, Patrick Campbell, Rob Case, Vikramsinha Ghorpade, Holly Houdeshell, Ola Kvalvaag, Dillon Nall, Ethan Sanders, Alec Vest, Shaun Westlund, Matthew Hardison, Clinton R. Paden, Duncan MacCannell |
| EPI_ISL_2203136 | Houston Methodist Hospital | Houston Methodist Hospital | Randall J. Olsen, Paul A. Christensen, S. Wesley Long, Sishir Subedi, Robert Olson, Marcus Nguyen, James J. Davis, Matthew Ojeda Saavedra, Prasanti Yerramilli, Layne Pruitt, Kristina Reppond, Madison N. Shyer, Jessica Cambric, Ryan Gadd, Ilya J. Finkelstein, Jimmy Gollihar, and James M. Musser |
| EPI_ISL_2203137, EPI_ISL_2203138 | Aegis Sciences Corporation | Centers for Disease Control and Prevention Division of Viral Diseases, Pathogen Discovery | Dakota Howard, Dhwani Batra, Peter W. Cook, Kara Moser, Adrian Paskey, Jason Caravas, Benjamin Rambo-Martin, Shatavia Morrison, Christopher Gulvick, Scott Sammons, Yvette Unoarumhi, Darlene Wagner, Matthew Schmerer, Cyndi Clark, Patrick Campbell, Rob Case, Vikramsinha Ghorpade, Holly Houdeshell, Ola Kvalvaag, Dillon Nall, Ethan Sanders, Alec Vest, Shaun Westlund, Matthew Hardison, Clinton R. Paden, Duncan MacCannell |
| EPI_ISL_2203139 | Houston Methodist Hospital | Houston Methodist Hospital | Randall J. Olsen, Paul A. Christensen, S. Wesley Long, Sishir Subedi, Robert Olson, Marcus Nguyen, James J. Davis, Matthew Ojeda Saavedra, Prasanti Yerramilli, Layne Pruitt, Kristina Reppond, Madison N. Shyer, Jessica Cambric, Ryan Gadd, Ilya J. Finkelstein, Jimmy Gollihar, and James M. Musser |
| EPI_ISL_2203140 | Aegis Sciences Corporation | Centers for Disease Control and Prevention Division of Viral Diseases, Pathogen Discovery | Dakota Howard, Dhwani Batra, Peter W. Cook, Kara Moser, Adrian Paskey, Jason Caravas, Benjamin Rambo-Martin, Shatavia Morrison, Christopher Gulvick, Scott Sammons, Yvette Unoarumhi, Darlene Wagner, Matthew Schmerer, Cyndi Clark, Patrick Campbell, Rob Case, Vikramsinha Ghorpade, Holly Houdeshell, Ola Kvalvaag, Dillon Nall, Ethan Sanders, Alec Vest, Shaun Westlund, Matthew Hardison, Clinton R. Paden, Duncan MacCannell |
| EPI_ISL_2203141 | Houston Methodist Hospital | Houston Methodist Hospital | Randall J. Olsen, Paul A. Christensen, S. Wesley Long, Sishir Subedi, Robert Olson, Marcus Nguyen, James J. Davis, Matthew Ojeda Saavedra, Prasanti Yerramilli, Layne Pruitt, Kristina Reppond, Madison N. Shyer, Jessica Cambric, Ryan Gadd, Ilya J. Finkelstein, Jimmy Gollihar, and James M. Musser |
| EPI_ISL_2203142 | Aegis Sciences Corporation | Centers for Disease Control and Prevention Division of Viral Diseases, Pathogen Discovery | Dakota Howard, Dhwani Batra, Peter W. Cook, Kara Moser, Adrian Paskey, Jason Caravas, Benjamin Rambo-Martin, Shatavia Morrison, Christopher Gulvick, Scott Sammons, Yvette Unoarumhi, Darlene Wagner, Matthew Schmerer, Cyndi Clark, Patrick Campbell, Rob Case, Vikramsinha Ghorpade, Holly Houdeshell, Ola Kvalvaag, Dillon Nall, Ethan Sanders, Alec Vest, Shaun Westlund, Matthew Hardison, Clinton R. Paden, Duncan MacCannell |
| EPI_ISL_2203143 | Houston Methodist Hospital | Houston Methodist Hospital | Randall J. Olsen, Paul A. Christensen, S. Wesley Long, Sishir Subedi, Robert Olson, Marcus Nguyen, James J. Davis, Matthew Ojeda Saavedra, Prasanti Yerramilli, Layne Pruitt, Kristina Reppond, Madison N. Shyer, Jessica Cambric, Ryan Gadd, Ilya J. Finkelstein, Jimmy Gollihar, and James M. Musser |
| EPI_ISL_2203144, EPI_ISL_2203145 | Aegis Sciences Corporation | Centers for Disease Control and Prevention Division of Viral Diseases, Pathogen Discovery | Dakota Howard, Dhwani Batra, Peter W. Cook, Kara Moser, Adrian Paskey, Jason Caravas, Benjamin Rambo-Martin, Shatavia Morrison, Christopher Gulvick, Scott Sammons, Yvette Unoarumhi, Darlene Wagner, Matthew Schmerer, Cyndi Clark, Patrick Campbell, Rob Case, Vikramsinha Ghorpade, Holly Houdeshell, Ola Kvalvaag, Dillon Nall, Ethan Sanders, Alec Vest, Shaun Westlund, Matthew Hardison, Clinton R. Paden, Duncan MacCannell |
| EPI_ISL_2203146 | Houston Methodist Hospital | Houston Methodist Hospital | Randall J. Olsen, Paul A. Christensen, S. Wesley Long, Sishir Subedi, Robert Olson, Marcus Nguyen, James J. Davis, Matthew Ojeda Saavedra, Prasanti Yerramilli, Layne Pruitt, Kristina Reppond, Madison N. Shyer, Jessica Cambric, Ryan Gadd, Ilya J. Finkelstein, Jimmy Gollihar, and James M. Musser |
| EPI_ISL_2203147, EPI_ISL_2203148 | Aegis Sciences Corporation | Centers for Disease Control and Prevention Division of Viral Diseases, Pathogen Discovery | Dakota Howard, Dhwani Batra, Peter W. Cook, Kara Moser, Adrian Paskey, Jason Caravas, Benjamin Rambo-Martin, Shatavia Morrison, Christopher Gulvick, Scott Sammons, Yvette Unoarumhi, Darlene Wagner, Matthew Schmerer, Cyndi Clark, Patrick Campbell, Rob Case, Vikramsinha Ghorpade, Holly Houdeshell, Ola Kvalvaag, Dillon Nall, Ethan Sanders, Alec Vest, Shaun Westlund, Matthew Hardison, Clinton R. Paden, Duncan MacCannell |
| EPI_ISL_2203149 | Houston Methodist Hospital | Houston Methodist Hospital | Randall J. Olsen, Paul A. Christensen, S. Wesley Long, Sishir Subedi, Robert Olson, Marcus Nguyen, James J. Davis, Matthew Ojeda Saavedra, Prasanti Yerramilli, Layne Pruitt, Kristina Reppond, Madison N. Shyer, Jessica Cambric, Ryan Gadd, Ilya J. Finkelstein, Jimmy Gollihar, and James M. Musser |
| EPI_ISL_2203150 | Aegis Sciences Corporation | Centers for Disease Control and Prevention Division of Viral Diseases, Pathogen Discovery | Dakota Howard, Dhwani Batra, Peter W. Cook, Kara Moser, Adrian Paskey, Jason Caravas, Benjamin Rambo-Martin, Shatavia Morrison, Christopher Gulvick, Scott Sammons, Yvette Unoarumhi, Darlene Wagner, Matthew Schmerer, Cyndi Clark, Patrick Campbell, Rob Case, Vikramsinha Ghorpade, Holly Houdeshell, Ola Kvalvaag, Dillon Nall, Ethan Sanders, Alec Vest, Shaun Westlund, Matthew Hardison, Clinton R. Paden, Duncan MacCannell |
| EPI_ISL_2203151 | Houston Methodist Hospital | Houston Methodist Hospital | Randall J. Olsen, Paul A. Christensen, S. Wesley Long, Sishir Subedi, Robert Olson, Marcus Nguyen, James J. Davis, Matthew Ojeda Saavedra, Prasanti Yerramilli, Layne Pruitt, Kristina Reppond, Madison N. Shyer, Jessica Cambric, Ryan Gadd, Ilya J. Finkelstein, Jimmy Gollihar, and James M. Musser |
| EPI_ISL_2203152 | Aegis Sciences Corporation | Centers for Disease Control and Prevention Division of Viral Diseases, Pathogen Discovery | Dakota Howard, Dhwani Batra, Peter W. Cook, Kara Moser, Adrian Paskey, Jason Caravas, Benjamin Rambo-Martin, Shatavia Morrison, Christopher Gulvick, Scott Sammons, Yvette Unoarumhi, Darlene Wagner, Matthew Schmerer, Cyndi Clark, Patrick Campbell, Rob Case, Vikramsinha Ghorpade, Holly Houdeshell, Ola Kvalvaag, Dillon Nall, Ethan Sanders, Alec Vest, Shaun Westlund, Matthew Hardison, Clinton R. Paden, Duncan MacCannell |
| EPI_ISL_2203153 | Houston Methodist Hospital | Houston Methodist Hospital | Randall J. Olsen, Paul A. Christensen, S. Wesley Long, Sishir Subedi, Robert Olson, Marcus Nguyen, James J. Davis, Matthew Ojeda Saavedra, Prasanti Yerramilli, Layne Pruitt, Kristina Reppond, Madison N. Shyer, Jessica Cambric, Ryan Gadd, Ilya J. Finkelstein, Jimmy Gollihar, and James M. Musser |
| EPI_ISL_2203154, EPI_ISL_2203155 | Aegis Sciences Corporation | Centers for Disease Control and Prevention Division of Viral Diseases, Pathogen Discovery | Dakota Howard, Dhwani Batra, Peter W. Cook, Kara Moser, Adrian Paskey, Jason Caravas, Benjamin Rambo-Martin, Shatavia Morrison, Christopher Gulvick, Scott Sammons, Yvette Unoarumhi, Darlene Wagner, Matthew Schmerer, Cyndi Clark, Patrick Campbell, Rob Case, Vikramsinha Ghorpade, Holly Houdeshell, Ola Kvalvaag, Dillon Nall, Ethan Sanders, Alec Vest, Shaun Westlund, Matthew Hardison, Clinton R. Paden, Duncan MacCannell |
| EPI_ISL_2203156 | Houston Methodist Hospital | Houston Methodist Hospital | Randall J. Olsen, Paul A. Christensen, S. Wesley Long, Sishir Subedi, Robert Olson, Marcus Nguyen, James J. Davis, Matthew Ojeda Saavedra, Prasanti Yerramilli, Layne Pruitt, Kristina Reppond, Madison N. Shyer, Jessica Cambric, Ryan Gadd, Ilya J. Finkelstein, Jimmy Gollihar, and James M. Musser |
| EPI_ISL_2203157 | Aegis Sciences Corporation | Centers for Disease Control and Prevention Division of Viral Diseases, Pathogen Discovery | Dakota Howard, Dhwani Batra, Peter W. Cook, Kara Moser, Adrian Paskey, Jason Caravas, Benjamin Rambo-Martin, Shatavia Morrison, Christopher Gulvick, Scott Sammons, Yvette Unoarumhi, Darlene Wagner, Matthew Schmerer, Cyndi Clark, Patrick Campbell, Rob Case, Vikramsinha Ghorpade, Holly Houdeshell, Ola Kvalvaag, Dillon Nall, Ethan Sanders, Alec Vest, Shaun Westlund, Matthew Hardison, Clinton R. Paden, Duncan MacCannell |
| EPI_ISL_2203158 | Houston Methodist Hospital | Houston Methodist Hospital | Randall J. Olsen, Paul A. Christensen, S. Wesley Long, Sishir Subedi, Robert Olson, Marcus Nguyen, James J. Davis, Matthew Ojeda Saavedra, Prasanti Yerramilli, Layne Pruitt, Kristina Reppond, Madison N. Shyer, Jessica Cambric, Ryan Gadd, Ilya J. Finkelstein, Jimmy Gollihar, and James M. Musser |
| EPI_ISL_2203159 | Aegis Sciences Corporation | Centers for Disease Control and Prevention Division of Viral Diseases, Pathogen Discovery | Dakota Howard, Dhwani Batra, Peter W. Cook, Kara Moser, Adrian Paskey, Jason Caravas, Benjamin Rambo-Martin, Shatavia Morrison, Christopher Gulvick, Scott Sammons, Yvette Unoarumhi, Darlene Wagner, Matthew Schmerer, Cyndi Clark, Patrick Campbell, Rob Case, Vikramsinha Ghorpade, Holly Houdeshell, Ola Kvalvaag, Dillon Nall, Ethan Sanders, Alec Vest, Shaun Westlund, Matthew Hardison, Clinton R. Paden, Duncan MacCannell |
| EPI_ISL_2203160, EPI_ISL_2203162, EPI_ISL_2203163 | Houston Methodist Hospital | Houston Methodist Hospital | Randall J. Olsen, Paul A. Christensen, S. Wesley Long, Sishir Subedi, Robert Olson, Marcus Nguyen, James J. Davis, Matthew Ojeda Saavedra, Prasanti Yerramilli, Layne Pruitt, Kristina Reppond, Madison N. Shyer, Jessica Cambric, Ryan Gadd, Ilya J. Finkelstein, Jimmy Gollihar, and James M. Musser |
| EPI_ISL_2203164, EPI_ISL_2203165 | Aegis Sciences Corporation | Centers for Disease Control and Prevention Division of Viral Diseases, Pathogen Discovery | Dakota Howard, Dhwani Batra, Peter W. Cook, Kara Moser, Adrian Paskey, Jason Caravas, Benjamin Rambo-Martin, Shatavia Morrison, Christopher Gulvick, Scott Sammons, Yvette Unoarumhi, Darlene Wagner, Matthew Schmerer, Cyndi Clark, Patrick Campbell, Rob Case, Vikramsinha Ghorpade, Holly Houdeshell, Ola Kvalvaag, Dillon Nall, Ethan Sanders, Alec Vest, Shaun Westlund, Matthew Hardison, Clinton R. Paden, Duncan MacCannell |
| EPI_ISL_2203166 | Houston Methodist Hospital | Houston Methodist Hospital | Randall J. Olsen, Paul A. Christensen, S. Wesley Long, Sishir Subedi, Robert Olson, Marcus Nguyen, James J. Davis, Matthew Ojeda Saavedra, Prasanti Yerramilli, Layne Pruitt, Kristina Reppond, Madison N. Shyer, Jessica Cambric, Ryan Gadd, Ilya J. Finkelstein, Jimmy Gollihar, and James M. Musser |
| EPI_ISL_2203167 | Aegis Sciences Corporation | Centers for Disease Control and Prevention Division of Viral Diseases, Pathogen Discovery | Dakota Howard, Dhwani Batra, Peter W. Cook, Kara Moser, Adrian Paskey, Jason Caravas, Benjamin Rambo-Martin, Shatavia Morrison, Christopher Gulvick, Scott Sammons, Yvette Unoarumhi, Darlene Wagner, Matthew Schmerer, Cyndi Clark, Patrick Campbell, Rob Case, Vikramsinha Ghorpade, Holly Houdeshell, Ola Kvalvaag, Dillon Nall, Ethan Sanders, Alec Vest, Shaun Westlund, Matthew Hardison, Clinton R. Paden, Duncan MacCannell |
| EPI_ISL_2203168 | Houston Methodist Hospital | Houston Methodist Hospital | Randall J. Olsen, Paul A. Christensen, S. Wesley Long, Sishir Subedi, Robert Olson, Marcus Nguyen, James J. Davis, Matthew Ojeda Saavedra, Prasanti Yerramilli, Layne Pruitt, Kristina Reppond, Madison N. Shyer, Jessica Cambric, Ryan Gadd, Ilya J. Finkelstein, Jimmy Gollihar, and James M. Musser |
| EPI_ISL_2203169, EPI_ISL_2203170 | Aegis Sciences Corporation | Centers for Disease Control and Prevention Division of Viral Diseases, Pathogen Discovery | Dakota Howard, Dhwani Batra, Peter W. Cook, Kara Moser, Adrian Paskey, Jason Caravas, Benjamin Rambo-Martin, Shatavia Morrison, Christopher Gulvick, Scott Sammons, Yvette Unoarumhi, Darlene Wagner, Matthew Schmerer, Cyndi Clark, Patrick Campbell, Rob Case, Vikramsinha Ghorpade, Holly Houdeshell, Ola Kvalvaag, Dillon Nall, Ethan Sanders, Alec Vest, Shaun Westlund, Matthew Hardison, Clinton R. Paden, Duncan MacCannell |
| EPI_ISL_2203171 | Houston Methodist Hospital | Houston Methodist Hospital | Randall J. Olsen, Paul A. Christensen, S. Wesley Long, Sishir Subedi, Robert Olson, Marcus Nguyen, James J. Davis, Matthew Ojeda Saavedra, Prasanti Yerramilli, Layne Pruitt, Kristina Reppond, Madison N. Shyer, Jessica Cambric, Ryan Gadd, Ilya J. Finkelstein, Jimmy Gollihar, and James M. Musser |
| EPI_ISL_2203172 | Aegis Sciences Corporation | Centers for Disease Control and Prevention Division of Viral Diseases, Pathogen Discovery | Dakota Howard, Dhwani Batra, Peter W. Cook, Kara Moser, Adrian Paskey, Jason Caravas, Benjamin Rambo-Martin, Shatavia Morrison, Christopher Gulvick, Scott Sammons, Yvette Unoarumhi, Darlene Wagner, Matthew Schmerer, Cyndi Clark, Patrick Campbell, Rob Case, Vikramsinha Ghorpade, Holly Houdeshell, Ola Kvalvaag, Dillon Nall, Ethan Sanders, Alec Vest, Shaun Westlund, Matthew Hardison, Clinton R. Paden, Duncan MacCannell |
| EPI_ISL_2203173 | Houston Methodist Hospital | Houston Methodist Hospital | Randall J. Olsen, Paul A. Christensen, S. Wesley Long, Sishir Subedi, Robert Olson, Marcus Nguyen, James J. Davis, Matthew Ojeda Saavedra, Prasanti Yerramilli, Layne Pruitt, Kristina Reppond, Madison N. Shyer, Jessica Cambric, Ryan Gadd, Ilya J. Finkelstein, Jimmy Gollihar, and James M. Musser |
| EPI_ISL_2203174 | Aegis Sciences Corporation | Centers for Disease Control and Prevention Division of Viral Diseases, Pathogen Discovery | Dakota Howard, Dhwani Batra, Peter W. Cook, Kara Moser, Adrian Paskey, Jason Caravas, Benjamin Rambo-Martin, Shatavia Morrison, Christopher Gulvick, Scott Sammons, Yvette Unoarumhi, Darlene Wagner, Matthew Schmerer, Cyndi Clark, Patrick Campbell, Rob Case, Vikramsinha Ghorpade, Holly Houdeshell, Ola Kvalvaag, Dillon Nall, Ethan Sanders, Alec Vest, Shaun Westlund, Matthew Hardison, Clinton R. Paden, Duncan MacCannell |
| EPI_ISL_2203175 | Houston Methodist Hospital | Houston Methodist Hospital | Randall J. Olsen, Paul A. Christensen, S. Wesley Long, Sishir Subedi, Robert Olson, Marcus Nguyen, James J. Davis, Matthew Ojeda Saavedra, Prasanti Yerramilli, Layne Pruitt, Kristina Reppond, Madison N. Shyer, Jessica Cambric, Ryan Gadd, Ilya J. Finkelstein, Jimmy Gollihar, and James M. Musser |
| EPI_ISL_2203176, EPI_ISL_2203177 | Aegis Sciences Corporation | Centers for Disease Control and Prevention Division of Viral Diseases, Pathogen Discovery | Dakota Howard, Dhwani Batra, Peter W. Cook, Kara Moser, Adrian Paskey, Jason Caravas, Benjamin Rambo-Martin, Shatavia Morrison, Christopher Gulvick, Scott Sammons, Yvette Unoarumhi, Darlene Wagner, Matthew Schmerer, Cyndi Clark, Patrick Campbell, Rob Case, Vikramsinha Ghorpade, Holly Houdeshell, Ola Kvalvaag, Dillon Nall, Ethan Sanders, Alec Vest, Shaun Westlund, Matthew Hardison, Clinton R. Paden, Duncan MacCannell |
| EPI_ISL_2203178 | Houston Methodist Hospital | Houston Methodist Hospital | Randall J. Olsen, Paul A. Christensen, S. Wesley Long, Sishir Subedi, Robert Olson, Marcus Nguyen, James J. Davis, Matthew Ojeda Saavedra, Prasanti Yerramilli, Layne Pruitt, Kristina Reppond, Madison N. Shyer, Jessica Cambric, Ryan Gadd, Ilya J. Finkelstein, Jimmy Gollihar, and James M. Musser |
| EPI_ISL_2203179 | Aegis Sciences Corporation | Centers for Disease Control and Prevention Division of Viral Diseases, Pathogen Discovery | Dakota Howard, Dhwani Batra, Peter W. Cook, Kara Moser, Adrian Paskey, Jason Caravas, Benjamin Rambo-Martin, Shatavia Morrison, Christopher Gulvick, Scott Sammons, Yvette Unoarumhi, Darlene Wagner, Matthew Schmerer, Cyndi Clark, Patrick Campbell, Rob Case, Vikramsinha Ghorpade, Holly Houdeshell, Ola Kvalvaag, Dillon Nall, Ethan Sanders, Alec Vest, Shaun Westlund, Matthew Hardison, Clinton R. Paden, Duncan MacCannell |

| | | | |
|---|---|---|---|
| EPI_ISL_2203180 | Houston Methodist Hospital | Houston Methodist Hospital | Randall J. Olsen, Paul A. Christensen, S. Wesley Long, Sishir Subedi, Robert Olson, Marcus Nguyen, James J. Davis, Matthew Ojeda Saavedra, Prasanti Yerramilli, Layne Pruitt, Kristina Reppond, Madison N. Shyer, Jessica Cambric, Ryan Gadd, Ilya J. Finkelstein, Jimmy Gollihar, and James M. Musser |
| EPI_ISL_2203181, EPI_ISL_2203182 | Aegis Sciences Corporation | Centers for Disease Control and Prevention Division of Viral Diseases, Pathogen Discovery | Dakota Howard, Dhwani Batra, Peter W. Cook, Kara Moser, Adrian Paskey, Jason Caravas, Benjamin Rambo-Martin, Shatavia Morrison, Christopher Gulvick, Scott Sammons, Yvette Unoarumhi, Darlene Wagner, Matthew Schmerer, Cyndi Clark, Patrick Campbell, Rob Case, Vikramsinha Ghorpade, Holly Houdeshell, Ola Kvalvaag, Dillon Nall, Ethan Sanders, Alec Vest, Shaun Westlund, Matthew Hardison, Clinton R. Paden, Duncan MacCannell |
| EPI_ISL_2203183 | Houston Methodist Hospital | Houston Methodist Hospital | Randall J. Olsen, Paul A. Christensen, S. Wesley Long, Sishir Subedi, Robert Olson, Marcus Nguyen, James J. Davis, Matthew Ojeda Saavedra, Prasanti Yerramilli, Layne Pruitt, Kristina Reppond, Madison N. Shyer, Jessica Cambric, Ryan Gadd, Ilya J. Finkelstein, Jimmy Gollihar, and James M. Musser |
| EPI_ISL_2203184 | Aegis Sciences Corporation | Centers for Disease Control and Prevention Division of Viral Diseases, Pathogen Discovery | Dakota Howard, Dhwani Batra, Peter W. Cook, Kara Moser, Adrian Paskey, Jason Caravas, Benjamin Rambo-Martin, Shatavia Morrison, Christopher Gulvick, Scott Sammons, Yvette Unoarumhi, Darlene Wagner, Matthew Schmerer, Cyndi Clark, Patrick Campbell, Rob Case, Vikramsinha Ghorpade, Holly Houdeshell, Ola Kvalvaag, Dillon Nall, Ethan Sanders, Alec Vest, Shaun Westlund, Matthew Hardison, Clinton R. Paden, Duncan MacCannell |
| EPI_ISL_2203185, EPI_ISL_2203187 | Houston Methodist Hospital | Houston Methodist Hospital | Randall J. Olsen, Paul A. Christensen, S. Wesley Long, Sishir Subedi, Robert Olson, Marcus Nguyen, James J. Davis, Matthew Ojeda Saavedra, Prasanti Yerramilli, Layne Pruitt, Kristina Reppond, Madison N. Shyer, Jessica Cambric, Ryan Gadd, Ilya J. Finkelstein, Jimmy Gollihar, and James M. Musser |
| EPI_ISL_2203188 | Aegis Sciences Corporation | Centers for Disease Control and Prevention Division of Viral Diseases, Pathogen Discovery | Dakota Howard, Dhwani Batra, Peter W. Cook, Kara Moser, Adrian Paskey, Jason Caravas, Benjamin Rambo-Martin, Shatavia Morrison, Christopher Gulvick, Scott Sammons, Yvette Unoarumhi, Darlene Wagner, Matthew Schmerer, Cyndi Clark, Patrick Campbell, Rob Case, Vikramsinha Ghorpade, Holly Houdeshell, Ola Kvalvaag, Dillon Nall, Ethan Sanders, Alec Vest, Shaun Westlund, Matthew Hardison, Clinton R. Paden, Duncan MacCannell |
| EPI_ISL_2203189 | Houston Methodist Hospital | Houston Methodist Hospital | Randall J. Olsen, Paul A. Christensen, S. Wesley Long, Sishir Subedi, Robert Olson, Marcus Nguyen, James J. Davis, Matthew Ojeda Saavedra, Prasanti Yerramilli, Layne Pruitt, Kristina Reppond, Madison N. Shyer, Jessica Cambric, Ryan Gadd, Ilya J. Finkelstein, Jimmy Gollihar, and James M. Musser |
| EPI_ISL_2203190, EPI_ISL_2203191 | Aegis Sciences Corporation | Centers for Disease Control and Prevention Division of Viral Diseases, Pathogen Discovery | Dakota Howard, Dhwani Batra, Peter W. Cook, Kara Moser, Adrian Paskey, Jason Caravas, Benjamin Rambo-Martin, Shatavia Morrison, Christopher Gulvick, Scott Sammons, Yvette Unoarumhi, Darlene Wagner, Matthew Schmerer, Cyndi Clark, Patrick Campbell, Rob Case, Vikramsinha Ghorpade, Holly Houdeshell, Ola Kvalvaag, Dillon Nall, Ethan Sanders, Alec Vest, Shaun Westlund, Matthew Hardison, Clinton R. Paden, Duncan MacCannell |
| EPI_ISL_2203192 | Houston Methodist Hospital | Houston Methodist Hospital | Randall J. Olsen, Paul A. Christensen, S. Wesley Long, Sishir Subedi, Robert Olson, Marcus Nguyen, James J. Davis, Matthew Ojeda Saavedra, Prasanti Yerramilli, Layne Pruitt, Kristina Reppond, Madison N. Shyer, Jessica Cambric, Ryan Gadd, Ilya J. Finkelstein, Jimmy Gollihar, and James M. Musser |
| EPI_ISL_2203193, EPI_ISL_2203194, EPI_ISL_2203195 | Aegis Sciences Corporation | Centers for Disease Control and Prevention Division of Viral Diseases, Pathogen Discovery | Dakota Howard, Dhwani Batra, Peter W. Cook, Kara Moser, Adrian Paskey, Jason Caravas, Benjamin Rambo-Martin, Shatavia Morrison, Christopher Gulvick, Scott Sammons, Yvette Unoarumhi, Darlene Wagner, Matthew Schmerer, Cyndi Clark, Patrick Campbell, Rob Case, Vikramsinha Ghorpade, Holly Houdeshell, Ola Kvalvaag, Dillon Nall, Ethan Sanders, Alec Vest, Shaun Westlund, Matthew Hardison, Clinton R. Paden, Duncan MacCannell |
| EPI_ISL_2203196 | Houston Methodist Hospital | Houston Methodist Hospital | Randall J. Olsen, Paul A. Christensen, S. Wesley Long, Sishir Subedi, Robert Olson, Marcus Nguyen, James J. Davis, Matthew Ojeda Saavedra, Prasanti Yerramilli, Layne Pruitt, Kristina Reppond, Madison N. Shyer, Jessica Cambric, Ryan Gadd, Ilya J. Finkelstein, Jimmy Gollihar, and James M. Musser |
| EPI_ISL_2203197, EPI_ISL_2203198 | Aegis Sciences Corporation | Centers for Disease Control and Prevention Division of Viral Diseases, Pathogen Discovery | Dakota Howard, Dhwani Batra, Peter W. Cook, Kara Moser, Adrian Paskey, Jason Caravas, Benjamin Rambo-Martin, Shatavia Morrison, Christopher Gulvick, Scott Sammons, Yvette Unoarumhi, Darlene Wagner, Matthew Schmerer, Cyndi Clark, Patrick Campbell, Rob Case, Vikramsinha Ghorpade, Holly Houdeshell, Ola Kvalvaag, Dillon Nall, Ethan Sanders, Alec Vest, Shaun Westlund, Matthew Hardison, Clinton R. Paden, Duncan MacCannell |
| EPI_ISL_2203199 | Houston Methodist Hospital | Houston Methodist Hospital | Randall J. Olsen, Paul A. Christensen, S. Wesley Long, Sishir Subedi, Robert Olson, Marcus Nguyen, James J. Davis, Matthew Ojeda Saavedra, Prasanti Yerramilli, Layne Pruitt, Kristina Reppond, Madison N. Shyer, Jessica Cambric, Ryan Gadd, Ilya J. Finkelstein, Jimmy Gollihar, and James M. Musser |
| EPI_ISL_2203200 | Aegis Sciences Corporation | Centers for Disease Control and Prevention Division of Viral Diseases, Pathogen Discovery | Dakota Howard, Dhwani Batra, Peter W. Cook, Kara Moser, Adrian Paskey, Jason Caravas, Benjamin Rambo-Martin, Shatavia Morrison, Christopher Gulvick, Scott Sammons, Yvette Unoarumhi, Darlene Wagner, Matthew Schmerer, Cyndi Clark, Patrick Campbell, Rob Case, Vikramsinha Ghorpade, Holly Houdeshell, Ola Kvalvaag, Dillon Nall, Ethan Sanders, Alec Vest, Shaun Westlund, Matthew Hardison, Clinton R. Paden, Duncan MacCannell |
| EPI_ISL_2203201, EPI_ISL_2203203 | Houston Methodist Hospital | Houston Methodist Hospital | Randall J. Olsen, Paul A. Christensen, S. Wesley Long, Sishir Subedi, Robert Olson, Marcus Nguyen, James J. Davis, Matthew Ojeda Saavedra, Prasanti Yerramilli, Layne Pruitt, Kristina Reppond, Madison N. Shyer, Jessica Cambric, Ryan Gadd, Ilya J. Finkelstein, Jimmy Gollihar, and James M. Musser |
| EPI_ISL_2203204, EPI_ISL_2203205, EPI_ISL_2203206 | Aegis Sciences Corporation | Centers for Disease Control and Prevention Division of Viral Diseases, Pathogen Discovery | Dakota Howard, Dhwani Batra, Peter W. Cook, Kara Moser, Adrian Paskey, Jason Caravas, Benjamin Rambo-Martin, Shatavia Morrison, Christopher Gulvick, Scott Sammons, Yvette Unoarumhi, Darlene Wagner, Matthew Schmerer, Cyndi Clark, Patrick Campbell, Rob Case, Vikramsinha Ghorpade, Holly Houdeshell, Ola Kvalvaag, Dillon Nall, Ethan Sanders, Alec Vest, Shaun Westlund, Matthew Hardison, Clinton R. Paden, Duncan MacCannell |
| EPI_ISL_2203207 | Houston Methodist Hospital | Houston Methodist Hospital | Randall J. Olsen, Paul A. Christensen, S. Wesley Long, Sishir Subedi, Robert Olson, Marcus Nguyen, James J. Davis, Matthew Ojeda Saavedra, Prasanti Yerramilli, Layne Pruitt, Kristina Reppond, Madison N. Shyer, Jessica Cambric, Ryan Gadd, Ilya J. Finkelstein, Jimmy Gollihar, and James M. Musser |
| EPI_ISL_2203208 | Aegis Sciences Corporation | Centers for Disease Control and Prevention Division of Viral Diseases, Pathogen Discovery | Dakota Howard, Dhwani Batra, Peter W. Cook, Kara Moser, Adrian Paskey, Jason Caravas, Benjamin Rambo-Martin, Shatavia Morrison, Christopher Gulvick, Scott Sammons, Yvette Unoarumhi, Darlene Wagner, Matthew Schmerer, Cyndi Clark, Patrick Campbell, Rob Case, Vikramsinha Ghorpade, Holly Houdeshell, Ola Kvalvaag, Dillon Nall, Ethan Sanders, Alec Vest, Shaun Westlund, Matthew Hardison, Clinton R. Paden, Duncan MacCannell |
| EPI_ISL_2203209 | Houston Methodist Hospital | Houston Methodist Hospital | Randall J. Olsen, Paul A. Christensen, S. Wesley Long, Sishir Subedi, Robert Olson, Marcus Nguyen, James J. Davis, Matthew Ojeda Saavedra, Prasanti Yerramilli, Layne Pruitt, Kristina Reppond, Madison N. Shyer, Jessica Cambric, Ryan Gadd, Ilya J. Finkelstein, Jimmy Gollihar, and James M. Musser |
| EPI_ISL_2203210, EPI_ISL_2203211 | Aegis Sciences Corporation | Centers for Disease Control and Prevention Division of Viral Diseases, Pathogen Discovery | Dakota Howard, Dhwani Batra, Peter W. Cook, Kara Moser, Adrian Paskey, Jason Caravas, Benjamin Rambo-Martin, Shatavia Morrison, Christopher Gulvick, Scott Sammons, Yvette Unoarumhi, Darlene Wagner, Matthew Schmerer, Cyndi Clark, Patrick Campbell, Rob Case, Vikramsinha Ghorpade, Holly Houdeshell, Ola Kvalvaag, Dillon Nall, Ethan Sanders, Alec Vest, Shaun Westlund, Matthew Hardison, Clinton R. Paden, Duncan MacCannell |
| EPI_ISL_2203212 | Houston Methodist Hospital | Houston Methodist Hospital | Randall J. Olsen, Paul A. Christensen, S. Wesley Long, Sishir Subedi, Robert Olson, Marcus Nguyen, James J. Davis, Matthew Ojeda Saavedra, Prasanti Yerramilli, Layne Pruitt, Kristina Reppond, Madison N. Shyer, Jessica Cambric, Ryan Gadd, Ilya J. Finkelstein, Jimmy Gollihar, and James M. Musser |
| EPI_ISL_2203213 | Aegis Sciences Corporation | Centers for Disease Control and Prevention Division of Viral Diseases, Pathogen Discovery | Dakota Howard, Dhwani Batra, Peter W. Cook, Kara Moser, Adrian Paskey, Jason Caravas, Benjamin Rambo-Martin, Shatavia Morrison, Christopher Gulvick, Scott Sammons, Yvette Unoarumhi, Darlene Wagner, Matthew Schmerer, Cyndi Clark, Patrick Campbell, Rob Case, Vikramsinha Ghorpade, Holly Houdeshell, Ola Kvalvaag, Dillon Nall, Ethan Sanders, Alec Vest, Shaun Westlund, Matthew Hardison, Clinton R. Paden, Duncan MacCannell |
| EPI_ISL_2203214 | Houston Methodist Hospital | Houston Methodist Hospital | Randall J. Olsen, Paul A. Christensen, S. Wesley Long, Sishir Subedi, Robert Olson, Marcus Nguyen, James J. Davis, Matthew Ojeda Saavedra, Prasanti Yerramilli, Layne Pruitt, Kristina Reppond, Madison N. Shyer, Jessica Cambric, Ryan Gadd, Ilya J. Finkelstein, Jimmy Gollihar, and James M. Musser |
| EPI_ISL_2203215 | Aegis Sciences Corporation | Centers for Disease Control and Prevention Division of Viral Diseases, Pathogen Discovery | Dakota Howard, Dhwani Batra, Peter W. Cook, Kara Moser, Adrian Paskey, Jason Caravas, Benjamin Rambo-Martin, Shatavia Morrison, Christopher Gulvick, Scott Sammons, Yvette Unoarumhi, Darlene Wagner, Matthew Schmerer, Cyndi Clark, Patrick Campbell, Rob Case, Vikramsinha Ghorpade, Holly Houdeshell, Ola Kvalvaag, Dillon Nall, Ethan Sanders, Alec Vest, Shaun Westlund, Matthew Hardison, Clinton R. Paden, Duncan MacCannell |
| EPI_ISL_2203216 | Houston Methodist Hospital | Houston Methodist Hospital | Randall J. Olsen, Paul A. Christensen, S. Wesley Long, Sishir Subedi, Robert Olson, Marcus Nguyen, James J. Davis, Matthew Ojeda Saavedra, Prasanti Yerramilli, Layne Pruitt, Kristina Reppond, Madison N. Shyer, Jessica Cambric, Ryan Gadd, Ilya J. Finkelstein, Jimmy Gollihar, and James M. Musser |
| EPI_ISL_2203217 | Aegis Sciences Corporation | Centers for Disease Control and Prevention Division of Viral Diseases, Pathogen Discovery | Dakota Howard, Dhwani Batra, Peter W. Cook, Kara Moser, Adrian Paskey, Jason Caravas, Benjamin Rambo-Martin, Shatavia Morrison, Christopher Gulvick, Scott Sammons, Yvette Unoarumhi, Darlene Wagner, Matthew Schmerer, Cyndi Clark, Patrick Campbell, Rob Case, Vikramsinha Ghorpade, Holly Houdeshell, Ola Kvalvaag, Dillon Nall, Ethan Sanders, Alec Vest, Shaun Westlund, Matthew Hardison, Clinton R. Paden, Duncan MacCannell |
| EPI_ISL_2203218 | Houston Methodist Hospital | Houston Methodist Hospital | Randall J. Olsen, Paul A. Christensen, S. Wesley Long, Sishir Subedi, Robert Olson, Marcus Nguyen, James J. Davis, Matthew Ojeda Saavedra, Prasanti Yerramilli, Layne Pruitt, Kristina Reppond, Madison N. Shyer, Jessica Cambric, Ryan Gadd, Ilya J. Finkelstein, Jimmy Gollihar, and James M. Musser |
| EPI_ISL_2203219 | Aegis Sciences Corporation | Centers for Disease Control and Prevention Division of Viral Diseases, Pathogen Discovery | Dakota Howard, Dhwani Batra, Peter W. Cook, Kara Moser, Adrian Paskey, Jason Caravas, Benjamin Rambo-Martin, Shatavia Morrison, Christopher Gulvick, Scott Sammons, Yvette Unoarumhi, Darlene Wagner, Matthew Schmerer, Cyndi Clark, Patrick Campbell, Rob Case, Vikramsinha Ghorpade, Holly Houdeshell, Ola Kvalvaag, Dillon Nall, Ethan Sanders, Alec Vest, Shaun Westlund, Matthew Hardison, Clinton R. Paden, Duncan MacCannell |
| EPI_ISL_2203220 | Houston Methodist Hospital | Houston Methodist Hospital | Randall J. Olsen, Paul A. Christensen, S. Wesley Long, Sishir Subedi, Robert Olson, Marcus Nguyen, James J. Davis, Matthew Ojeda Saavedra, Prasanti Yerramilli, Layne Pruitt, Kristina Reppond, Madison N. Shyer, Jessica Cambric, Ryan Gadd, Ilya J. Finkelstein, Jimmy Gollihar, and James M. Musser |
| EPI_ISL_2203221, EPI_ISL_2203222, EPI_ISL_2203223 | Aegis Sciences Corporation | Centers for Disease Control and Prevention Division of Viral Diseases, Pathogen Discovery | Dakota Howard, Dhwani Batra, Peter W. Cook, Kara Moser, Adrian Paskey, Jason Caravas, Benjamin Rambo-Martin, Shatavia Morrison, Christopher Gulvick, Scott Sammons, Yvette Unoarumhi, Darlene Wagner, Matthew Schmerer, Cyndi Clark, Patrick Campbell, Rob Case, Vikramsinha Ghorpade, Holly Houdeshell, Ola Kvalvaag, Dillon Nall, Ethan Sanders, Alec Vest, Shaun Westlund, Matthew Hardison, Clinton R. Paden, Duncan MacCannell |
| EPI_ISL_2203224 | Houston Methodist Hospital | Houston Methodist Hospital | Randall J. Olsen, Paul A. Christensen, S. Wesley Long, Sishir Subedi, Robert Olson, Marcus Nguyen, James J. Davis, Matthew Ojeda Saavedra, Prasanti Yerramilli, Layne Pruitt, Kristina Reppond, Madison N. Shyer, Jessica Cambric, Ryan Gadd, Ilya J. Finkelstein, Jimmy Gollihar, and James M. Musser |
| EPI_ISL_2203225 | Aegis Sciences Corporation | Centers for Disease Control and Prevention Division of Viral Diseases, Pathogen Discovery | Dakota Howard, Dhwani Batra, Peter W. Cook, Kara Moser, Adrian Paskey, Jason Caravas, Benjamin Rambo-Martin, Shatavia Morrison, Christopher Gulvick, Scott Sammons, Yvette Unoarumhi, Darlene Wagner, Matthew Schmerer, Cyndi Clark, Patrick Campbell, Rob Case, Vikramsinha Ghorpade, Holly Houdeshell, Ola Kvalvaag, Dillon Nall, Ethan Sanders, Alec Vest, Shaun Westlund, Matthew Hardison, Clinton R. Paden, Duncan MacCannell |
| EPI_ISL_2203226 | Houston Methodist Hospital | Houston Methodist Hospital | Randall J. Olsen, Paul A. Christensen, S. Wesley Long, Sishir Subedi, Robert Olson, Marcus Nguyen, James J. Davis, Matthew Ojeda Saavedra, Prasanti Yerramilli, Layne Pruitt, Kristina Reppond, Madison N. Shyer, Jessica Cambric, Ryan Gadd, Ilya J. Finkelstein, Jimmy Gollihar, and James M. Musser |
| EPI_ISL_2203227 | Aegis Sciences Corporation | Centers for Disease Control and Prevention Division of Viral Diseases, Pathogen Discovery | Dakota Howard, Dhwani Batra, Peter W. Cook, Kara Moser, Adrian Paskey, Jason Caravas, Benjamin Rambo-Martin, Shatavia Morrison, Christopher Gulvick, Scott Sammons, Yvette Unoarumhi, Darlene Wagner, Matthew Schmerer, Cyndi Clark, Patrick Campbell, Rob Case, Vikramsinha Ghorpade, Holly Houdeshell, Ola Kvalvaag, Dillon Nall, Ethan Sanders, Alec Vest, Shaun Westlund, Matthew Hardison, Clinton R. Paden, Duncan MacCannell |
| EPI_ISL_2203228 | Houston Methodist Hospital | Houston Methodist Hospital | Randall J. Olsen, Paul A. Christensen, S. Wesley Long, Sishir Subedi, Robert Olson, Marcus Nguyen, James J. Davis, Matthew Ojeda Saavedra, Prasanti Yerramilli, Layne Pruitt, Kristina Reppond, Madison N. Shyer, Jessica Cambric, Ryan Gadd, Ilya J. Finkelstein, Jimmy Gollihar, and James M. Musser |
| EPI_ISL_2203229, EPI_ISL_2203230 | Aegis Sciences Corporation | Centers for Disease Control and Prevention Division of Viral Diseases, Pathogen Discovery | Dakota Howard, Dhwani Batra, Peter W. Cook, Kara Moser, Adrian Paskey, Jason Caravas, Benjamin Rambo-Martin, Shatavia Morrison, Christopher Gulvick, Scott Sammons, Yvette Unoarumhi, Darlene Wagner, Matthew Schmerer, Cyndi Clark, Patrick Campbell, Rob Case, Vikramsinha Ghorpade, Holly Houdeshell, Ola Kvalvaag, Dillon Nall, Ethan Sanders, Alec Vest, Shaun Westlund, Matthew Hardison, Clinton R. Paden, Duncan MacCannell |
| EPI_ISL_2203231 | Houston Methodist Hospital | Houston Methodist Hospital | Randall J. Olsen, Paul A. Christensen, S. Wesley Long, Sishir Subedi, Robert Olson, Marcus Nguyen, James J. Davis, Matthew Ojeda Saavedra, Prasanti Yerramilli, Layne Pruitt, Kristina Reppond, Madison N. Shyer, Jessica Cambric, Ryan Gadd, Ilya J. Finkelstein, Jimmy Gollihar, and James M. Musser |
| EPI_ISL_2203232 | Aegis Sciences Corporation | Centers for Disease Control and Prevention | Dakota Howard, Dhwani Batra, Peter W. Cook, Kara Moser, Adrian Paskey, Jason Caravas, Benjamin Rambo-Martin, Shatavia Morrison, Christopher Gulvick, Scott Sammons, Yvette Unoarumhi, Darlene Wagner, Matthew Schmerer, Cyndi Clark, Patrick Campbell, Rob Case, Vikramsinha |

| | | | |
|---|---|---|---|
| | | Division of Viral Diseases, Pathogen Discovery | Ghorpade, Holly Houdeshell, Ola Kvalvaag, Dillon Nall, Ethan Sanders, Alec Vest, Shaun Westlund, Matthew Hardison, Clinton R. Paden, Duncan MacCannell |
| EPI_ISL_2203233 | Houston Methodist Hospital | Houston Methodist Hospital | Randall J. Olsen, Paul A. Christensen, S. Wesley Long, Sishir Subedi, Robert Olson, Marcus Nguyen, James J. Davis, Matthew Ojeda Saavedra, Prasanti Yerramilli, Layne Pruitt, Kristina Reppond, Madison N. Shyer, Jessica Cambric, Ryan Gadd, Ilya J. Finkelstein, Jimmy Gollihar, and James M. Musser |
| EPI_ISL_2203234 | Aegis Sciences Corporation | Centers for Disease Control and Prevention Division of Viral Diseases, Pathogen Discovery | Dakota Howard, Dhwani Batra, Peter W. Cook, Kara Moser, Adrian Paskey, Jason Caravas, Benjamin Rambo-Martin, Shatavia Morrison, Christopher Gulvick, Scott Sammons, Yvette Unoarumhi, Darlene Wagner, Matthew Schmerer, Cyndi Clark, Patrick Campbell, Rob Case, Vikramsinha Ghorpade, Holly Houdeshell, Ola Kvalvaag, Dillon Nall, Ethan Sanders, Alec Vest, Shaun Westlund, Matthew Hardison, Clinton R. Paden, Duncan MacCannell |
| EPI_ISL_2203235 | Maryland Genomics, Institute for Genome Sciences, University of Maryland School of Medicine | Maryland Genomics, Institute for Genome Sciences, University of Maryland School of Medicine | Tallon, Luke J; Sadzewicz, Lisa D; Humphrys, Mike; Ott, Sandra; Roussey, Holly; Mehta, Aditya; Vavikolanu, Kranthi; Fraser, Claire M; Ravel, Jacques |
| EPI_ISL_2203236 | Aegis Sciences Corporation | Centers for Disease Control and Prevention Division of Viral Diseases, Pathogen Discovery | Dakota Howard, Dhwani Batra, Peter W. Cook, Kara Moser, Adrian Paskey, Jason Caravas, Benjamin Rambo-Martin, Shatavia Morrison, Christopher Gulvick, Scott Sammons, Yvette Unoarumhi, Darlene Wagner, Matthew Schmerer, Cyndi Clark, Patrick Campbell, Rob Case, Vikramsinha Ghorpade, Holly Houdeshell, Ola Kvalvaag, Dillon Nall, Ethan Sanders, Alec Vest, Shaun Westlund, Matthew Hardison, Clinton R. Paden, Duncan MacCannell |
| EPI_ISL_2203238 | Maryland Genomics, Institute for Genome Sciences, University of Maryland School of Medicine | Maryland Genomics, Institute for Genome Sciences, University of Maryland School of Medicine | Tallon, Luke J; Sadzewicz, Lisa D; Humphrys, Mike; Ott, Sandra; Roussey, Holly; Mehta, Aditya; Vavikolanu, Kranthi; Fraser, Claire M; Ravel, Jacques |
| EPI_ISL_2203239 | Houston Methodist Hospital | Houston Methodist Hospital | Randall J. Olsen, Paul A. Christensen, S. Wesley Long, Sishir Subedi, Robert Olson, Marcus Nguyen, James J. Davis, Matthew Ojeda Saavedra, Prasanti Yerramilli, Layne Pruitt, Kristina Reppond, Madison N. Shyer, Jessica Cambric, Ryan Gadd, Ilya J. Finkelstein, Jimmy Gollihar, and James M. Musser |
| EPI_ISL_2203240 | Aegis Sciences Corporation | Centers for Disease Control and Prevention Division of Viral Diseases, Pathogen Discovery | Dakota Howard, Dhwani Batra, Peter W. Cook, Kara Moser, Adrian Paskey, Jason Caravas, Benjamin Rambo-Martin, Shatavia Morrison, Christopher Gulvick, Scott Sammons, Yvette Unoarumhi, Darlene Wagner, Matthew Schmerer, Cyndi Clark, Patrick Campbell, Rob Case, Vikramsinha Ghorpade, Holly Houdeshell, Ola Kvalvaag, Dillon Nall, Ethan Sanders, Alec Vest, Shaun Westlund, Matthew Hardison, Clinton R. Paden, Duncan MacCannell |
| EPI_ISL_2203241, EPI_ISL_2203242 | Maryland Genomics, Institute for Genome Sciences, University of Maryland School of Medicine | Maryland Genomics, Institute for Genome Sciences, University of Maryland School of Medicine | Tallon, Luke J; Sadzewicz, Lisa D; Humphrys, Mike; Ott, Sandra; Roussey, Holly; Mehta, Aditya; Vavikolanu, Kranthi; Fraser, Claire M; Ravel, Jacques |
| EPI_ISL_2203243 | Aegis Sciences Corporation | Centers for Disease Control and Prevention Division of Viral Diseases, Pathogen Discovery | Dakota Howard, Dhwani Batra, Peter W. Cook, Kara Moser, Adrian Paskey, Jason Caravas, Benjamin Rambo-Martin, Shatavia Morrison, Christopher Gulvick, Scott Sammons, Yvette Unoarumhi, Darlene Wagner, Matthew Schmerer, Cyndi Clark, Patrick Campbell, Rob Case, Vikramsinha Ghorpade, Holly Houdeshell, Ola Kvalvaag, Dillon Nall, Ethan Sanders, Alec Vest, Shaun Westlund, Matthew Hardison, Clinton R. Paden, Duncan MacCannell |
| EPI_ISL_2203244 | Houston Methodist Hospital | Houston Methodist Hospital | Randall J. Olsen, Paul A. Christensen, S. Wesley Long, Sishir Subedi, Robert Olson, Marcus Nguyen, James J. Davis, Matthew Ojeda Saavedra, Prasanti Yerramilli, Layne Pruitt, Kristina Reppond, Madison N. Shyer, Jessica Cambric, Ryan Gadd, Ilya J. Finkelstein, Jimmy Gollihar, and James M. Musser |
| EPI_ISL_2203245 | Maryland Genomics, Institute for Genome Sciences, University of Maryland School of Medicine | Maryland Genomics, Institute for Genome Sciences, University of Maryland School of Medicine | Tallon, Luke J; Sadzewicz, Lisa D; Humphrys, Mike; Ott, Sandra; Roussey, Holly; Mehta, Aditya; Vavikolanu, Kranthi; Fraser, Claire M; Ravel, Jacques |
| EPI_ISL_2203246 | Aegis Sciences Corporation | Centers for Disease Control and Prevention Division of Viral Diseases, Pathogen Discovery | Dakota Howard, Dhwani Batra, Peter W. Cook, Kara Moser, Adrian Paskey, Jason Caravas, Benjamin Rambo-Martin, Shatavia Morrison, Christopher Gulvick, Scott Sammons, Yvette Unoarumhi, Darlene Wagner, Matthew Schmerer, Cyndi Clark, Patrick Campbell, Rob Case, Vikramsinha Ghorpade, Holly Houdeshell, Ola Kvalvaag, Dillon Nall, Ethan Sanders, Alec Vest, Shaun Westlund, Matthew Hardison, Clinton R. Paden, Duncan MacCannell |
| EPI_ISL_2203247, EPI_ISL_2203248 | Maryland Genomics, Institute for Genome Sciences, University of Maryland School of Medicine | Maryland Genomics, Institute for Genome Sciences, University of Maryland School of Medicine | Tallon, Luke J; Sadzewicz, Lisa D; Humphrys, Mike; Ott, Sandra; Roussey, Holly; Mehta, Aditya; Vavikolanu, Kranthi; Fraser, Claire M; Ravel, Jacques |
| EPI_ISL_2203249 | Houston Methodist Hospital | Houston Methodist Hospital | Randall J. Olsen, Paul A. Christensen, S. Wesley Long, Sishir Subedi, Robert Olson, Marcus Nguyen, James J. Davis, Matthew Ojeda Saavedra, Prasanti Yerramilli, Layne Pruitt, Kristina Reppond, Madison N. Shyer, Jessica Cambric, Ryan Gadd, Ilya J. Finkelstein, Jimmy Gollihar, and James M. Musser |
| EPI_ISL_2203250 | Aegis Sciences Corporation | Centers for Disease Control and Prevention Division of Viral Diseases, Pathogen Discovery | Dakota Howard, Dhwani Batra, Peter W. Cook, Kara Moser, Adrian Paskey, Jason Caravas, Benjamin Rambo-Martin, Shatavia Morrison, Christopher Gulvick, Scott Sammons, Yvette Unoarumhi, Darlene Wagner, Matthew Schmerer, Cyndi Clark, Patrick Campbell, Rob Case, Vikramsinha Ghorpade, Holly Houdeshell, Ola Kvalvaag, Dillon Nall, Ethan Sanders, Alec Vest, Shaun Westlund, Matthew Hardison, Clinton R. Paden, Duncan MacCannell |
| EPI_ISL_2203251 | Maryland Genomics, Institute for Genome Sciences, University of Maryland School of Medicine | Maryland Genomics, Institute for Genome Sciences, University of Maryland School of Medicine | Tallon, Luke J; Sadzewicz, Lisa D; Humphrys, Mike; Ott, Sandra; Roussey, Holly; Mehta, Aditya; Vavikolanu, Kranthi; Fraser, Claire M; Ravel, Jacques |
| EPI_ISL_2203252 | Aegis Sciences Corporation | Centers for Disease Control and Prevention Division of Viral Diseases, Pathogen Discovery | Dakota Howard, Dhwani Batra, Peter W. Cook, Kara Moser, Adrian Paskey, Jason Caravas, Benjamin Rambo-Martin, Shatavia Morrison, Christopher Gulvick, Scott Sammons, Yvette Unoarumhi, Darlene Wagner, Matthew Schmerer, Cyndi Clark, Patrick Campbell, Rob Case, Vikramsinha Ghorpade, Holly Houdeshell, Ola Kvalvaag, Dillon Nall, Ethan Sanders, Alec Vest, Shaun Westlund, Matthew Hardison, Clinton R. Paden, Duncan MacCannell |
| EPI_ISL_2203253 | Houston Methodist Hospital | Houston Methodist Hospital | Randall J. Olsen, Paul A. Christensen, S. Wesley Long, Sishir Subedi, Robert Olson, Marcus Nguyen, James J. Davis, Matthew Ojeda Saavedra, Prasanti Yerramilli, Layne Pruitt, Kristina Reppond, Madison N. Shyer, Jessica Cambric, Ryan Gadd, Ilya J. Finkelstein, Jimmy Gollihar, and James M. Musser |
| EPI_ISL_2203254 | Aegis Sciences Corporation | Centers for Disease Control and Prevention Division of Viral Diseases, Pathogen Discovery | Dakota Howard, Dhwani Batra, Peter W. Cook, Kara Moser, Adrian Paskey, Jason Caravas, Benjamin Rambo-Martin, Shatavia Morrison, Christopher Gulvick, Scott Sammons, Yvette Unoarumhi, Darlene Wagner, Matthew Schmerer, Cyndi Clark, Patrick Campbell, Rob Case, Vikramsinha Ghorpade, Holly Houdeshell, Ola Kvalvaag, Dillon Nall, Ethan Sanders, Alec Vest, Shaun Westlund, Matthew Hardison, Clinton R. Paden, Duncan MacCannell |
| EPI_ISL_2203255 | Houston Methodist Hospital | Houston Methodist Hospital | Randall J. Olsen, Paul A. Christensen, S. Wesley Long, Sishir Subedi, Robert Olson, Marcus Nguyen, James J. Davis, Matthew Ojeda Saavedra, Prasanti Yerramilli, Layne Pruitt, Kristina Reppond, Madison N. Shyer, Jessica Cambric, Ryan Gadd, Ilya J. Finkelstein, Jimmy Gollihar, and James M. Musser |
| EPI_ISL_2203256, EPI_ISL_2203257 | Aegis Sciences Corporation | Centers for Disease Control and Prevention Division of Viral Diseases, Pathogen Discovery | Dakota Howard, Dhwani Batra, Peter W. Cook, Kara Moser, Adrian Paskey, Jason Caravas, Benjamin Rambo-Martin, Shatavia Morrison, Christopher Gulvick, Scott Sammons, Yvette Unoarumhi, Darlene Wagner, Matthew Schmerer, Cyndi Clark, Patrick Campbell, Rob Case, Vikramsinha Ghorpade, Holly Houdeshell, Ola Kvalvaag, Dillon Nall, Ethan Sanders, Alec Vest, Shaun Westlund, Matthew Hardison, Clinton R. Paden, Duncan MacCannell |
| EPI_ISL_2203258 | Houston Methodist Hospital | Houston Methodist Hospital | Randall J. Olsen, Paul A. Christensen, S. Wesley Long, Sishir Subedi, Robert Olson, Marcus Nguyen, James J. Davis, Matthew Ojeda Saavedra, Prasanti Yerramilli, Layne Pruitt, Kristina Reppond, Madison N. Shyer, Jessica Cambric, Ryan Gadd, Ilya J. Finkelstein, Jimmy Gollihar, and James M. Musser |
| EPI_ISL_2203259 | Aegis Sciences Corporation | Centers for Disease Control and Prevention Division of Viral Diseases, Pathogen Discovery | Dakota Howard, Dhwani Batra, Peter W. Cook, Kara Moser, Adrian Paskey, Jason Caravas, Benjamin Rambo-Martin, Shatavia Morrison, Christopher Gulvick, Scott Sammons, Yvette Unoarumhi, Darlene Wagner, Matthew Schmerer, Cyndi Clark, Patrick Campbell, Rob Case, Vikramsinha Ghorpade, Holly Houdeshell, Ola Kvalvaag, Dillon Nall, Ethan Sanders, Alec Vest, Shaun Westlund, Matthew Hardison, Clinton R. Paden, Duncan MacCannell |
| EPI_ISL_2203260 | Houston Methodist Hospital | Houston Methodist Hospital | Randall J. Olsen, Paul A. Christensen, S. Wesley Long, Sishir Subedi, Robert Olson, Marcus Nguyen, James J. Davis, Matthew Ojeda Saavedra, Prasanti Yerramilli, Layne Pruitt, Kristina Reppond, Madison N. Shyer, Jessica Cambric, Ryan Gadd, Ilya J. Finkelstein, Jimmy Gollihar, and James M. Musser |
| EPI_ISL_2203261, EPI_ISL_2203262 | Aegis Sciences Corporation | Centers for Disease Control and Prevention Division of Viral Diseases, Pathogen Discovery | Dakota Howard, Dhwani Batra, Peter W. Cook, Kara Moser, Adrian Paskey, Jason Caravas, Benjamin Rambo-Martin, Shatavia Morrison, Christopher Gulvick, Scott Sammons, Yvette Unoarumhi, Darlene Wagner, Matthew Schmerer, Cyndi Clark, Patrick Campbell, Rob Case, Vikramsinha Ghorpade, Holly Houdeshell, Ola Kvalvaag, Dillon Nall, Ethan Sanders, Alec Vest, Shaun Westlund, Matthew Hardison, Clinton R. Paden, Duncan MacCannell |
| EPI_ISL_2203263 | Houston Methodist Hospital | Houston Methodist Hospital | Randall J. Olsen, Paul A. Christensen, S. Wesley Long, Sishir Subedi, Robert Olson, Marcus Nguyen, James J. Davis, Matthew Ojeda Saavedra, Prasanti Yerramilli, Layne Pruitt, Kristina Reppond, Madison N. Shyer, Jessica Cambric, Ryan Gadd, Ilya J. Finkelstein, Jimmy Gollihar, and James M. Musser |
| EPI_ISL_2203264 | Aegis Sciences Corporation | Centers for Disease Control and Prevention Division of Viral Diseases, Pathogen Discovery | Dakota Howard, Dhwani Batra, Peter W. Cook, Kara Moser, Adrian Paskey, Jason Caravas, Benjamin Rambo-Martin, Shatavia Morrison, Christopher Gulvick, Scott Sammons, Yvette Unoarumhi, Darlene Wagner, Matthew Schmerer, Cyndi Clark, Patrick Campbell, Rob Case, Vikramsinha Ghorpade, Holly Houdeshell, Ola Kvalvaag, Dillon Nall, Ethan Sanders, Alec Vest, Shaun Westlund, Matthew Hardison, Clinton R. Paden, Duncan MacCannell |
| EPI_ISL_2203266 | Houston Methodist Hospital | Houston Methodist Hospital | Randall J. Olsen, Paul A. Christensen, S. Wesley Long, Sishir Subedi, Robert Olson, Marcus Nguyen, James J. Davis, Matthew Ojeda Saavedra, Prasanti Yerramilli, Layne Pruitt, Kristina Reppond, Madison N. Shyer, Jessica Cambric, Ryan Gadd, Ilya J. Finkelstein, Jimmy Gollihar, and James M. Musser |
| EPI_ISL_2203267 | Aegis Sciences Corporation | Centers for Disease Control and Prevention Division of Viral Diseases, Pathogen Discovery | Dakota Howard, Dhwani Batra, Peter W. Cook, Kara Moser, Adrian Paskey, Jason Caravas, Benjamin Rambo-Martin, Shatavia Morrison, Christopher Gulvick, Scott Sammons, Yvette Unoarumhi, Darlene Wagner, Matthew Schmerer, Cyndi Clark, Patrick Campbell, Rob Case, Vikramsinha Ghorpade, Holly Houdeshell, Ola Kvalvaag, Dillon Nall, Ethan Sanders, Alec Vest, Shaun Westlund, Matthew Hardison, Clinton R. Paden, Duncan MacCannell |
| EPI_ISL_2203268, EPI_ISL_2203270 | Houston Methodist Hospital | Houston Methodist Hospital | Randall J. Olsen, Paul A. Christensen, S. Wesley Long, Sishir Subedi, Robert Olson, Marcus Nguyen, James J. Davis, Matthew Ojeda Saavedra, Prasanti Yerramilli, Layne Pruitt, Kristina Reppond, Madison N. Shyer, Jessica Cambric, Ryan Gadd, Ilya J. Finkelstein, Jimmy Gollihar, and James M. Musser |
| EPI_ISL_2203271 | Aegis Sciences Corporation | Centers for Disease Control and Prevention Division of Viral Diseases, Pathogen Discovery | Dakota Howard, Dhwani Batra, Peter W. Cook, Kara Moser, Adrian Paskey, Jason Caravas, Benjamin Rambo-Martin, Shatavia Morrison, Christopher Gulvick, Scott Sammons, Yvette Unoarumhi, Darlene Wagner, Matthew Schmerer, Cyndi Clark, Patrick Campbell, Rob Case, Vikramsinha Ghorpade, Holly Houdeshell, Ola Kvalvaag, Dillon Nall, Ethan Sanders, Alec Vest, Shaun Westlund, Matthew Hardison, Clinton R. Paden, Duncan MacCannell |
| EPI_ISL_2203272 | Houston Methodist Hospital | Houston Methodist Hospital | Randall J. Olsen, Paul A. Christensen, S. Wesley Long, Sishir Subedi, Robert Olson, Marcus Nguyen, James J. Davis, Matthew Ojeda Saavedra, Prasanti Yerramilli, Layne Pruitt, Kristina Reppond, Madison N. Shyer, Jessica Cambric, Ryan Gadd, Ilya J. Finkelstein, Jimmy Gollihar, and James M. Musser |
| EPI_ISL_2203273 | Aegis Sciences Corporation | Centers for Disease Control and Prevention Division of Viral Diseases, Pathogen Discovery | Dakota Howard, Dhwani Batra, Peter W. Cook, Kara Moser, Adrian Paskey, Jason Caravas, Benjamin Rambo-Martin, Shatavia Morrison, Christopher Gulvick, Scott Sammons, Yvette Unoarumhi, Darlene Wagner, Matthew Schmerer, Cyndi Clark, Patrick Campbell, Rob Case, Vikramsinha Ghorpade, Holly Houdeshell, Ola Kvalvaag, Dillon Nall, Ethan Sanders, Alec Vest, Shaun Westlund, Matthew Hardison, Clinton R. Paden, Duncan MacCannell |
| EPI_ISL_2203274 | Houston Methodist Hospital | Houston Methodist Hospital | Randall J. Olsen, Paul A. Christensen, S. Wesley Long, Sishir Subedi, Robert Olson, Marcus Nguyen, James J. Davis, Matthew Ojeda Saavedra, Prasanti Yerramilli, Layne Pruitt, Kristina Reppond, Madison N. Shyer, Jessica Cambric, Ryan Gadd, Ilya J. Finkelstein, Jimmy Gollihar, and James M. Musser |
| EPI_ISL_2203275, EPI_ISL_2203276 | Aegis Sciences Corporation | Centers for Disease Control and Prevention Division of Viral Diseases, Pathogen Discovery | Dakota Howard, Dhwani Batra, Peter W. Cook, Kara Moser, Adrian Paskey, Jason Caravas, Benjamin Rambo-Martin, Shatavia Morrison, Christopher Gulvick, Scott Sammons, Yvette Unoarumhi, Darlene Wagner, Matthew Schmerer, Cyndi Clark, Patrick Campbell, Rob Case, Vikramsinha Ghorpade, Holly Houdeshell, Ola Kvalvaag, Dillon Nall, Ethan Sanders, Alec Vest, Shaun Westlund, Matthew Hardison, Clinton R. Paden, Duncan MacCannell |

| | | | |
|---|---|---|---|
| EPI_ISL_2203277 | Houston Methodist Hospital | Houston Methodist Hospital | Randall J. Olsen, Paul A. Christensen, S. Wesley Long, Sishir Subedi, Robert Olson, Marcus Nguyen, James J. Davis, Matthew Ojeda Saavedra, Prasanti Yerramilli, Layne Pruitt, Kristina Reppond, Madison N. Shyer, Jessica Cambric, Ryan Gadd, Ilya J. Finkelstein, Jimmy Gollihar, and James M. Musser |
| EPI_ISL_2203278 | Aegis Sciences Corporation | Centers for Disease Control and Prevention Division of Viral Diseases, Pathogen Discovery | Dakota Howard, Dhwani Batra, Peter W. Cook, Kara Moser, Adrian Paskey, Jason Caravas, Benjamin Rambo-Martin, Shatavia Morrison, Christopher Gulvick, Scott Sammons, Yvette Unoarumhi, Darlene Wagner, Matthew Schmerer, Cyndi Clark, Patrick Campbell, Rob Case, Vikramsinha Ghorpade, Holly Houdeshell, Ola Kvalvaag, Dillon Nall, Ethan Sanders, Alec Vest, Shaun Westlund, Matthew Hardison, Clinton R. Paden, Duncan MacCannell |
| EPI_ISL_2203279, EPI_ISL_2203281 | Houston Methodist Hospital | Houston Methodist Hospital | Randall J. Olsen, Paul A. Christensen, S. Wesley Long, Sishir Subedi, Robert Olson, Marcus Nguyen, James J. Davis, Matthew Ojeda Saavedra, Prasanti Yerramilli, Layne Pruitt, Kristina Reppond, Madison N. Shyer, Jessica Cambric, Ryan Gadd, Ilya J. Finkelstein, Jimmy Gollihar, and James M. Musser |
| EPI_ISL_2203282, EPI_ISL_2203283 | Aegis Sciences Corporation | Centers for Disease Control and Prevention Division of Viral Diseases, Pathogen Discovery | Dakota Howard, Dhwani Batra, Peter W. Cook, Kara Moser, Adrian Paskey, Jason Caravas, Benjamin Rambo-Martin, Shatavia Morrison, Christopher Gulvick, Scott Sammons, Yvette Unoarumhi, Darlene Wagner, Matthew Schmerer, Cyndi Clark, Patrick Campbell, Rob Case, Vikramsinha Ghorpade, Holly Houdeshell, Ola Kvalvaag, Dillon Nall, Ethan Sanders, Alec Vest, Shaun Westlund, Matthew Hardison, Clinton R. Paden, Duncan MacCannell |
| EPI_ISL_2203284 | Houston Methodist Hospital | Houston Methodist Hospital | Randall J. Olsen, Paul A. Christensen, S. Wesley Long, Sishir Subedi, Robert Olson, Marcus Nguyen, James J. Davis, Matthew Ojeda Saavedra, Prasanti Yerramilli, Layne Pruitt, Kristina Reppond, Madison N. Shyer, Jessica Cambric, Ryan Gadd, Ilya J. Finkelstein, Jimmy Gollihar, and James M. Musser |
| EPI_ISL_2203285 | Aegis Sciences Corporation | Centers for Disease Control and Prevention Division of Viral Diseases, Pathogen Discovery | Dakota Howard, Dhwani Batra, Peter W. Cook, Kara Moser, Adrian Paskey, Jason Caravas, Benjamin Rambo-Martin, Shatavia Morrison, Christopher Gulvick, Scott Sammons, Yvette Unoarumhi, Darlene Wagner, Matthew Schmerer, Cyndi Clark, Patrick Campbell, Rob Case, Vikramsinha Ghorpade, Holly Houdeshell, Ola Kvalvaag, Dillon Nall, Ethan Sanders, Alec Vest, Shaun Westlund, Matthew Hardison, Clinton R. Paden, Duncan MacCannell |
| EPI_ISL_2203286 | Houston Methodist Hospital | Houston Methodist Hospital | Randall J. Olsen, Paul A. Christensen, S. Wesley Long, Sishir Subedi, Robert Olson, Marcus Nguyen, James J. Davis, Matthew Ojeda Saavedra, Prasanti Yerramilli, Layne Pruitt, Kristina Reppond, Madison N. Shyer, Jessica Cambric, Ryan Gadd, Ilya J. Finkelstein, Jimmy Gollihar, and James M. Musser |
| EPI_ISL_2203287 | Aegis Sciences Corporation | Centers for Disease Control and Prevention Division of Viral Diseases, Pathogen Discovery | Dakota Howard, Dhwani Batra, Peter W. Cook, Kara Moser, Adrian Paskey, Jason Caravas, Benjamin Rambo-Martin, Shatavia Morrison, Christopher Gulvick, Scott Sammons, Yvette Unoarumhi, Darlene Wagner, Matthew Schmerer, Cyndi Clark, Patrick Campbell, Rob Case, Vikramsinha Ghorpade, Holly Houdeshell, Ola Kvalvaag, Dillon Nall, Ethan Sanders, Alec Vest, Shaun Westlund, Matthew Hardison, Clinton R. Paden, Duncan MacCannell |
| EPI_ISL_2203288 | Houston Methodist Hospital | Houston Methodist Hospital | Randall J. Olsen, Paul A. Christensen, S. Wesley Long, Sishir Subedi, Robert Olson, Marcus Nguyen, James J. Davis, Matthew Ojeda Saavedra, Prasanti Yerramilli, Layne Pruitt, Kristina Reppond, Madison N. Shyer, Jessica Cambric, Ryan Gadd, Ilya J. Finkelstein, Jimmy Gollihar, and James M. Musser |
| EPI_ISL_2203289, EPI_ISL_2203290 | Aegis Sciences Corporation | Centers for Disease Control and Prevention Division of Viral Diseases, Pathogen Discovery | Dakota Howard, Dhwani Batra, Peter W. Cook, Kara Moser, Adrian Paskey, Jason Caravas, Benjamin Rambo-Martin, Shatavia Morrison, Christopher Gulvick, Scott Sammons, Yvette Unoarumhi, Darlene Wagner, Matthew Schmerer, Cyndi Clark, Patrick Campbell, Rob Case, Vikramsinha Ghorpade, Holly Houdeshell, Ola Kvalvaag, Dillon Nall, Ethan Sanders, Alec Vest, Shaun Westlund, Matthew Hardison, Clinton R. Paden, Duncan MacCannell |
| EPI_ISL_2203291 | Houston Methodist Hospital | Houston Methodist Hospital | Randall J. Olsen, Paul A. Christensen, S. Wesley Long, Sishir Subedi, Robert Olson, Marcus Nguyen, James J. Davis, Matthew Ojeda Saavedra, Prasanti Yerramilli, Layne Pruitt, Kristina Reppond, Madison N. Shyer, Jessica Cambric, Ryan Gadd, Ilya J. Finkelstein, Jimmy Gollihar, and James M. Musser |
| EPI_ISL_2203292 | Aegis Sciences Corporation | Centers for Disease Control and Prevention Division of Viral Diseases, Pathogen Discovery | Dakota Howard, Dhwani Batra, Peter W. Cook, Kara Moser, Adrian Paskey, Jason Caravas, Benjamin Rambo-Martin, Shatavia Morrison, Christopher Gulvick, Scott Sammons, Yvette Unoarumhi, Darlene Wagner, Matthew Schmerer, Cyndi Clark, Patrick Campbell, Rob Case, Vikramsinha Ghorpade, Holly Houdeshell, Ola Kvalvaag, Dillon Nall, Ethan Sanders, Alec Vest, Shaun Westlund, Matthew Hardison, Clinton R. Paden, Duncan MacCannell |
| EPI_ISL_2203293, EPI_ISL_2203294, EPI_ISL_2203295, EPI_ISL_2203296, EPI_ISL_2203297, EPI_ISL_2203298, EPI_ISL_2203299, EPI_ISL_2203300, EPI_ISL_2203301, EPI_ISL_2203302, EPI_ISL_2203303, EPI_ISL_2203304, EPI_ISL_2203305, EPI_ISL_2203306, EPI_ISL_2203307, EPI_ISL_2203308, EPI_ISL_2203309, EPI_ISL_2203310, EPI_ISL_2203311, EPI_ISL_2203312, EPI_ISL_2203313, EPI_ISL_2203314, EPI_ISL_2203315, EPI_ISL_2203316, EPI_ISL_2203317, EPI_ISL_2203318, EPI_ISL_2203319, EPI_ISL_2203320, EPI_ISL_2203321, EPI_ISL_2203322, EPI_ISL_2203323, EPI_ISL_2203324, EPI_ISL_2203325, EPI_ISL_2203326, EPI_ISL_2203327, EPI_ISL_2203328, EPI_ISL_2203329, EPI_ISL_2203330, EPI_ISL_2203331, EPI_ISL_2203332, EPI_ISL_2203333, EPI_ISL_2203334, EPI_ISL_2203335, EPI_ISL_2203336, EPI_ISL_2203337, EPI_ISL_2203338, EPI_ISL_2203339, EPI_ISL_2203340, EPI_ISL_2203341, EPI_ISL_2203342, EPI_ISL_2203343, EPI_ISL_2203344, EPI_ISL_2203345, EPI_ISL_2203346, EPI_ISL_2203347, EPI_ISL_2203348, EPI_ISL_2203349, EPI_ISL_2203350, EPI_ISL_2203351, EPI_ISL_2203352, EPI_ISL_2203353, EPI_ISL_2203354, EPI_ISL_2203355, EPI_ISL_2203356, EPI_ISL_2203357, EPI_ISL_2203358, EPI_ISL_2203359, EPI_ISL_2203360, EPI_ISL_2203361, EPI_ISL_2203362, EPI_ISL_2203363, EPI_ISL_2203364, EPI_ISL_2203365, EPI_ISL_2203366, EPI_ISL_2203367, EPI_ISL_2203368, EPI_ISL_2203369, EPI_ISL_2203370, EPI_ISL_2203371, EPI_ISL_2203372, EPI_ISL_2203373, EPI_ISL_2203374, EPI_ISL_2203375, EPI_ISL_2203376, EPI_ISL_2203377, EPI_ISL_2203378, EPI_ISL_2203379, EPI_ISL_2203380, EPI_ISL_2203381, EPI_ISL_2203382, EPI_ISL_2203383, EPI_ISL_2203384, EPI_ISL_2203385, EPI_ISL_2203386, EPI_ISL_2203387, EPI_ISL_2203388, EPI_ISL_2203389, EPI_ISL_2203390, EPI_ISL_2203391, EPI_ISL_2203392, EPI_ISL_2203393, EPI_ISL_2203394, EPI_ISL_2203395, EPI_ISL_2203396, EPI_ISL_2203397, EPI_ISL_2203398, EPI_ISL_2203399, EPI_ISL_2203400, EPI_ISL_2203401, EPI_ISL_2203402, EPI_ISL_2203403, EPI_ISL_2203404, EPI_ISL_2203405, EPI_ISL_2203406, EPI_ISL_2203407, EPI_ISL_2203408, EPI_ISL_2203409, EPI_ISL_2203410, EPI_ISL_2203411, EPI_ISL_2203412, EPI_ISL_2203413, EPI_ISL_2203414, EPI_ISL_2203415, EPI_ISL_2203416, EPI_ISL_2203417, EPI_ISL_2203418, EPI_ISL_2203419, EPI_ISL_2203420, EPI_ISL_2203421, EPI_ISL_2203422, EPI_ISL_2203423, EPI_ISL_2203424, EPI_ISL_2203425, EPI_ISL_2203426, EPI_ISL_2203427, EPI_ISL_2203428, EPI_ISL_2203429, EPI_ISL_2203430, EPI_ISL_2203431, EPI_ISL_2203432, EPI_ISL_2203433, EPI_ISL_2203434, EPI_ISL_2203435, EPI_ISL_2203436, EPI_ISL_2203437, EPI_ISL_2203438, EPI_ISL_2203439, EPI_ISL_2203440, EPI_ISL_2203441, EPI_ISL_2203442, EPI_ISL_2203443, EPI_ISL_2203444, EPI_ISL_2203445, EPI_ISL_2203446, EPI_ISL_2203447, EPI_ISL_2203448, EPI_ISL_2203449, EPI_ISL_2203450, EPI_ISL_2203451, EPI_ISL_2203452, EPI_ISL_2203453, EPI_ISL_2203454, EPI_ISL_2203455, EPI_ISL_2203457, EPI_ISL_2203458, EPI_ISL_2203459, EPI_ISL_2203460, EPI_ISL_2203461, EPI_ISL_2203462, EPI_ISL_2203463, EPI_ISL_2203464, EPI_ISL_2203465, EPI_ISL_2203466, EPI_ISL_2203468, EPI_ISL_2203469, EPI_ISL_2203470, EPI_ISL_2203471, EPI_ISL_2203472, EPI_ISL_2203473, EPI_ISL_2203474, EPI_ISL_2203475, EPI_ISL_2203476, EPI_ISL_2203477, EPI_ISL_2203478, EPI_ISL_2203479, EPI_ISL_2203481, EPI_ISL_2203482, EPI_ISL_2203483, EPI_ISL_2203484, EPI_ISL_2203485, EPI_ISL_2203486, EPI_ISL_2203487, EPI_ISL_2203488, EPI_ISL_2203489 | | | |
| see above | Houston Methodist Hospital | Houston Methodist Hospital | Randall J. Olsen, Paul A. Christensen, S. Wesley Long, Sishir Subedi, Robert Olson, Marcus Nguyen, James J. Davis, Matthew Ojeda Saavedra, Prasanti Yerramilli, Layne Pruitt, Kristina Reppond, Madison N. Shyer, Jessica Cambric, Ryan Gadd, Ilya J. Finkelstein, Jimmy Gollihar, and James M. Musser |
| EPI_ISL_2203490, EPI_ISL_2203492, EPI_ISL_2203493 | Illinois Department of Public Health - Springfield Lab | Illinois Department of Public Health - Springfield Lab | Bryan Sim, Gordon McCall |
| EPI_ISL_2203494 | Houston Methodist Hospital | Houston Methodist Hospital | Randall J. Olsen, Paul A. Christensen, S. Wesley Long, Sishir Subedi, Robert Olson, Marcus Nguyen, James J. Davis, Matthew Ojeda Saavedra, Prasanti Yerramilli, Layne Pruitt, Kristina Reppond, Madison N. Shyer, Jessica Cambric, Ryan Gadd, Ilya J. Finkelstein, Jimmy Gollihar, and James M. Musser |
| EPI_ISL_2203495, EPI_ISL_2203496 | Illinois Department of Public Health - Springfield Lab | Illinois Department of Public Health - Springfield Lab | Bryan Sim, Gordon McCall |
| EPI_ISL_2203497 | Houston Methodist Hospital | Houston Methodist Hospital | Randall J. Olsen, Paul A. Christensen, S. Wesley Long, Sishir Subedi, Robert Olson, Marcus Nguyen, James J. Davis, Matthew Ojeda Saavedra, Prasanti Yerramilli, Layne Pruitt, Kristina Reppond, Madison N. Shyer, Jessica Cambric, Ryan Gadd, Ilya J. Finkelstein, Jimmy Gollihar, and James M. Musser |
| EPI_ISL_2203498, EPI_ISL_2203499, EPI_ISL_2203500, EPI_ISL_2203502, EPI_ISL_2203503, EPI_ISL_2203505, EPI_ISL_2203506 | Illinois Department of Public Health - Springfield Lab | Illinois Department of Public Health - Springfield Lab | Bryan Sim, Gordon McCall |
| EPI_ISL_2203507 | Houston Methodist Hospital | Houston Methodist Hospital | Randall J. Olsen, Paul A. Christensen, S. Wesley Long, Sishir Subedi, Robert Olson, Marcus Nguyen, James J. Davis, Matthew Ojeda Saavedra, Prasanti Yerramilli, Layne Pruitt, Kristina Reppond, Madison N. Shyer, Jessica Cambric, Ryan Gadd, Ilya J. Finkelstein, Jimmy Gollihar, and James M. Musser |
| EPI_ISL_2203508, EPI_ISL_2203511, EPI_ISL_2203512, EPI_ISL_2203513 | Illinois Department of Public Health - Springfield Lab | Illinois Department of Public Health - Springfield Lab | Bryan Sim, Gordon McCall |
| EPI_ISL_2203514 | Houston Methodist Hospital | Houston Methodist Hospital | Randall J. Olsen, Paul A. Christensen, S. Wesley Long, Sishir Subedi, Robert Olson, Marcus Nguyen, James J. Davis, Matthew Ojeda Saavedra, Prasanti Yerramilli, Layne Pruitt, Kristina Reppond, Madison N. Shyer, Jessica Cambric, Ryan Gadd, Ilya J. Finkelstein, Jimmy Gollihar, and James M. Musser |
| EPI_ISL_2203515, EPI_ISL_2203516 | Illinois Department of Public Health - Springfield Lab | Illinois Department of Public Health - Springfield Lab | Bryan Sim, Gordon McCall |
| EPI_ISL_2203517 | Houston Methodist Hospital | Houston Methodist Hospital | Randall J. Olsen, Paul A. Christensen, S. Wesley Long, Sishir Subedi, Robert Olson, Marcus Nguyen, James J. Davis, Matthew Ojeda Saavedra, Prasanti Yerramilli, Layne Pruitt, Kristina Reppond, Madison N. Shyer, Jessica Cambric, Ryan Gadd, Ilya J. Finkelstein, Jimmy Gollihar, and James M. Musser |
| EPI_ISL_2203518, EPI_ISL_2203519 | Illinois Department of Public Health - Springfield Lab | Illinois Department of Public Health - Springfield Lab | Bryan Sim, Gordon McCall |
| EPI_ISL_2203520 | Houston Methodist Hospital | Houston Methodist Hospital | Randall J. Olsen, Paul A. Christensen, S. Wesley Long, Sishir Subedi, Robert Olson, Marcus Nguyen, James J. Davis, Matthew Ojeda Saavedra, Prasanti Yerramilli, Layne Pruitt, Kristina Reppond, Madison N. Shyer, Jessica Cambric, Ryan Gadd, Ilya J. Finkelstein, Jimmy Gollihar, and James M. Musser |
| EPI_ISL_2203521, EPI_ISL_2203522 | Illinois Department of Public Health - Springfield Lab | Illinois Department of Public Health - Springfield Lab | Bryan Sim, Gordon McCall |
| EPI_ISL_2203523 | Houston Methodist Hospital | Houston Methodist Hospital | Randall J. Olsen, Paul A. Christensen, S. Wesley Long, Sishir Subedi, Robert Olson, Marcus Nguyen, James J. Davis, Matthew Ojeda Saavedra, Prasanti Yerramilli, Layne Pruitt, Kristina Reppond, Madison N. Shyer, Jessica Cambric, Ryan Gadd, Ilya J. Finkelstein, Jimmy Gollihar, and James M. Musser |
| EPI_ISL_2203524, EPI_ISL_2203525 | Illinois Department of Public Health - Springfield Lab | Illinois Department of Public Health - Springfield Lab | Bryan Sim, Gordon McCall |
| EPI_ISL_2203526 | Houston Methodist Hospital | Houston Methodist Hospital | Randall J. Olsen, Paul A. Christensen, S. Wesley Long, Sishir Subedi, Robert Olson, Marcus Nguyen, James J. Davis, Matthew Ojeda Saavedra, Prasanti Yerramilli, Layne Pruitt, Kristina Reppond, Madison N. Shyer, Jessica Cambric, Ryan Gadd, Ilya J. Finkelstein, Jimmy Gollihar, and James M. Musser |
| EPI_ISL_2203527, EPI_ISL_2203528 | Illinois Department of Public Health - Springfield Lab | Illinois Department of Public Health - Springfield Lab | Bryan Sim, Gordon McCall |
| EPI_ISL_2203529 | Houston Methodist Hospital | Houston Methodist Hospital | Randall J. Olsen, Paul A. Christensen, S. Wesley Long, Sishir Subedi, Robert Olson, Marcus Nguyen, James J. Davis, Matthew Ojeda Saavedra, Prasanti Yerramilli, Layne Pruitt, Kristina Reppond, Madison N. Shyer, Jessica Cambric, Ryan Gadd, Ilya J. Finkelstein, Jimmy Gollihar, and James M. Musser |
| EPI_ISL_2203530, EPI_ISL_2203531 | Illinois Department of Public Health - Springfield Lab | Illinois Department of Public Health - Springfield Lab | Bryan Sim, Gordon McCall |
| EPI_ISL_2203532 | Houston Methodist Hospital | Houston Methodist Hospital | Randall J. Olsen, Paul A. Christensen, S. Wesley Long, Sishir Subedi, Robert Olson, Marcus Nguyen, James J. Davis, Matthew Ojeda Saavedra, Prasanti Yerramilli, Layne Pruitt, Kristina Reppond, Madison N. Shyer, Jessica Cambric, Ryan Gadd, Ilya J. Finkelstein, Jimmy Gollihar, and James M. Musser |
| EPI_ISL_2203533, EPI_ISL_2203534, EPI_ISL_2203535 | Illinois Department of Public Health - Springfield Lab | Illinois Department of Public Health - Springfield Lab | Bryan Sim, Gordon McCall |
| EPI_ISL_2203536 | Houston Methodist Hospital | Houston Methodist Hospital | Randall J. Olsen, Paul A. Christensen, S. Wesley Long, Sishir Subedi, Robert Olson, Marcus Nguyen, James J. Davis, Matthew Ojeda Saavedra, Prasanti Yerramilli, Layne Pruitt, Kristina Reppond, Madison N. Shyer, Jessica Cambric, Ryan Gadd, Ilya J. Finkelstein, Jimmy Gollihar, and James |

| | | | |
|---|---|---|---|
| | | | M. Musser |
| EPI_ISL_2203537, EPI_ISL_2203538 | Illinois Department of Public Health - Springfield Lab | Illinois Department of Public Health - Springfield Lab | Bryan Sim, Gordon McCall |
| EPI_ISL_2203539 | Houston Methodist Hospital | Houston Methodist Hospital | Randall J. Olsen, Paul A. Christensen, S. Wesley Long, Sishir Subedi, Robert Olson, Marcus Nguyen, James J. Davis, Matthew Ojeda Saavedra, Prasanti Yerramilli, Layne Pruitt, Kristina Reppond, Madison N. Shyer, Jessica Cambric, Ryan Gadd, Ilya J. Finkelstein, Jimmy Gollihar, and James M. Musser |
| EPI_ISL_2203540 | Illinois Department of Public Health - Springfield Lab | Illinois Department of Public Health - Springfield Lab | Bryan Sim, Gordon McCall |
| EPI_ISL_2203541 | Houston Methodist Hospital | Houston Methodist Hospital | Randall J. Olsen, Paul A. Christensen, S. Wesley Long, Sishir Subedi, Robert Olson, Marcus Nguyen, James J. Davis, Matthew Ojeda Saavedra, Prasanti Yerramilli, Layne Pruitt, Kristina Reppond, Madison N. Shyer, Jessica Cambric, Ryan Gadd, Ilya J. Finkelstein, Jimmy Gollihar, and James M. Musser |
| EPI_ISL_2203542, EPI_ISL_2203543 | Illinois Department of Public Health - Springfield Lab | Illinois Department of Public Health - Springfield Lab | Bryan Sim, Gordon McCall |
| EPI_ISL_2203544 | Houston Methodist Hospital | Houston Methodist Hospital | Randall J. Olsen, Paul A. Christensen, S. Wesley Long, Sishir Subedi, Robert Olson, Marcus Nguyen, James J. Davis, Matthew Ojeda Saavedra, Prasanti Yerramilli, Layne Pruitt, Kristina Reppond, Madison N. Shyer, Jessica Cambric, Ryan Gadd, Ilya J. Finkelstein, Jimmy Gollihar, and James M. Musser |
| EPI_ISL_2203545, EPI_ISL_2203546 | Illinois Department of Public Health - Springfield Lab | Illinois Department of Public Health - Springfield Lab | Bryan Sim, Gordon McCall |
| EPI_ISL_2203547 | Houston Methodist Hospital | Houston Methodist Hospital | Randall J. Olsen, Paul A. Christensen, S. Wesley Long, Sishir Subedi, Robert Olson, Marcus Nguyen, James J. Davis, Matthew Ojeda Saavedra, Prasanti Yerramilli, Layne Pruitt, Kristina Reppond, Madison N. Shyer, Jessica Cambric, Ryan Gadd, Ilya J. Finkelstein, Jimmy Gollihar, and James M. Musser |
| EPI_ISL_2203548, EPI_ISL_2203549, EPI_ISL_2203550 | Illinois Department of Public Health - Springfield Lab | Illinois Department of Public Health - Springfield Lab | Bryan Sim, Gordon McCall |
| EPI_ISL_2203551 | Houston Methodist Hospital | Houston Methodist Hospital | Randall J. Olsen, Paul A. Christensen, S. Wesley Long, Sishir Subedi, Robert Olson, Marcus Nguyen, James J. Davis, Matthew Ojeda Saavedra, Prasanti Yerramilli, Layne Pruitt, Kristina Reppond, Madison N. Shyer, Jessica Cambric, Ryan Gadd, Ilya J. Finkelstein, Jimmy Gollihar, and James M. Musser |
| EPI_ISL_2203552, EPI_ISL_2203553 | Illinois Department of Public Health - Springfield Lab | Illinois Department of Public Health - Springfield Lab | Bryan Sim, Gordon McCall |
| EPI_ISL_2203554 | Houston Methodist Hospital | Houston Methodist Hospital | Randall J. Olsen, Paul A. Christensen, S. Wesley Long, Sishir Subedi, Robert Olson, Marcus Nguyen, James J. Davis, Matthew Ojeda Saavedra, Prasanti Yerramilli, Layne Pruitt, Kristina Reppond, Madison N. Shyer, Jessica Cambric, Ryan Gadd, Ilya J. Finkelstein, Jimmy Gollihar, and James M. Musser |
| EPI_ISL_2203555, EPI_ISL_2203556 | Illinois Department of Public Health - Springfield Lab | Illinois Department of Public Health - Springfield Lab | Bryan Sim, Gordon McCall |
| EPI_ISL_2203557 | Houston Methodist Hospital | Houston Methodist Hospital | Randall J. Olsen, Paul A. Christensen, S. Wesley Long, Sishir Subedi, Robert Olson, Marcus Nguyen, James J. Davis, Matthew Ojeda Saavedra, Prasanti Yerramilli, Layne Pruitt, Kristina Reppond, Madison N. Shyer, Jessica Cambric, Ryan Gadd, Ilya J. Finkelstein, Jimmy Gollihar, and James M. Musser |
| EPI_ISL_2203558, EPI_ISL_2203559 | Illinois Department of Public Health - Springfield Lab | Illinois Department of Public Health - Springfield Lab | Bryan Sim, Gordon McCall |
| EPI_ISL_2203560 | Houston Methodist Hospital | Houston Methodist Hospital | Randall J. Olsen, Paul A. Christensen, S. Wesley Long, Sishir Subedi, Robert Olson, Marcus Nguyen, James J. Davis, Matthew Ojeda Saavedra, Prasanti Yerramilli, Layne Pruitt, Kristina Reppond, Madison N. Shyer, Jessica Cambric, Ryan Gadd, Ilya J. Finkelstein, Jimmy Gollihar, and James M. Musser |
| EPI_ISL_2203561, EPI_ISL_2203562 | Illinois Department of Public Health - Springfield Lab | Illinois Department of Public Health - Springfield Lab | Bryan Sim, Gordon McCall |
| EPI_ISL_2203563 | Houston Methodist Hospital | Houston Methodist Hospital | Randall J. Olsen, Paul A. Christensen, S. Wesley Long, Sishir Subedi, Robert Olson, Marcus Nguyen, James J. Davis, Matthew Ojeda Saavedra, Prasanti Yerramilli, Layne Pruitt, Kristina Reppond, Madison N. Shyer, Jessica Cambric, Ryan Gadd, Ilya J. Finkelstein, Jimmy Gollihar, and James M. Musser |
| EPI_ISL_2203564, EPI_ISL_2203565 | Illinois Department of Public Health - Springfield Lab | Illinois Department of Public Health - Springfield Lab | Bryan Sim, Gordon McCall |
| EPI_ISL_2203566 | Houston Methodist Hospital | Houston Methodist Hospital | Randall J. Olsen, Paul A. Christensen, S. Wesley Long, Sishir Subedi, Robert Olson, Marcus Nguyen, James J. Davis, Matthew Ojeda Saavedra, Prasanti Yerramilli, Layne Pruitt, Kristina Reppond, Madison N. Shyer, Jessica Cambric, Ryan Gadd, Ilya J. Finkelstein, Jimmy Gollihar, and James M. Musser |
| EPI_ISL_2203567, EPI_ISL_2203568, EPI_ISL_2203570, EPI_ISL_2203571, EPI_ISL_2203573, EPI_ISL_2203574, EPI_ISL_2203576, EPI_ISL_2203577 | Illinois Department of Public Health - Springfield Lab | Illinois Department of Public Health - Springfield Lab | Bryan Sim, Gordon McCall |
| EPI_ISL_2203578 | Houston Methodist Hospital | Houston Methodist Hospital | Randall J. Olsen, Paul A. Christensen, S. Wesley Long, Sishir Subedi, Robert Olson, Marcus Nguyen, James J. Davis, Matthew Ojeda Saavedra, Prasanti Yerramilli, Layne Pruitt, Kristina Reppond, Madison N. Shyer, Jessica Cambric, Ryan Gadd, Ilya J. Finkelstein, Jimmy Gollihar, and James M. Musser |
| EPI_ISL_2203579, EPI_ISL_2203580, EPI_ISL_2203581 | Illinois Department of Public Health - Springfield Lab | Illinois Department of Public Health - Springfield Lab | Bryan Sim, Gordon McCall |
| EPI_ISL_2203582 | Houston Methodist Hospital | Houston Methodist Hospital | Randall J. Olsen, Paul A. Christensen, S. Wesley Long, Sishir Subedi, Robert Olson, Marcus Nguyen, James J. Davis, Matthew Ojeda Saavedra, Prasanti Yerramilli, Layne Pruitt, Kristina Reppond, Madison N. Shyer, Jessica Cambric, Ryan Gadd, Ilya J. Finkelstein, Jimmy Gollihar, and James M. Musser |
| EPI_ISL_2203583, EPI_ISL_2203584 | Illinois Department of Public Health - Springfield Lab | Illinois Department of Public Health - Springfield Lab | Bryan Sim, Gordon McCall |
| EPI_ISL_2203585 | Houston Methodist Hospital | Houston Methodist Hospital | Randall J. Olsen, Paul A. Christensen, S. Wesley Long, Sishir Subedi, Robert Olson, Marcus Nguyen, James J. Davis, Matthew Ojeda Saavedra, Prasanti Yerramilli, Layne Pruitt, Kristina Reppond, Madison N. Shyer, Jessica Cambric, Ryan Gadd, Ilya J. Finkelstein, Jimmy Gollihar, and James M. Musser |
| EPI_ISL_2203586, EPI_ISL_2203587, EPI_ISL_2203588 | Illinois Department of Public Health - Springfield Lab | Illinois Department of Public Health - Springfield Lab | Bryan Sim, Gordon McCall |
| EPI_ISL_2203589 | Houston Methodist Hospital | Houston Methodist Hospital | Randall J. Olsen, Paul A. Christensen, S. Wesley Long, Sishir Subedi, Robert Olson, Marcus Nguyen, James J. Davis, Matthew Ojeda Saavedra, Prasanti Yerramilli, Layne Pruitt, Kristina Reppond, Madison N. Shyer, Jessica Cambric, Ryan Gadd, Ilya J. Finkelstein, Jimmy Gollihar, and James M. Musser |
| EPI_ISL_2203590, EPI_ISL_2203591, EPI_ISL_2203593, EPI_ISL_2203594 | Illinois Department of Public Health - Springfield Lab | Illinois Department of Public Health - Springfield Lab | Bryan Sim, Gordon McCall |
| EPI_ISL_2203595 | Houston Methodist Hospital | Houston Methodist Hospital | Randall J. Olsen, Paul A. Christensen, S. Wesley Long, Sishir Subedi, Robert Olson, Marcus Nguyen, James J. Davis, Matthew Ojeda Saavedra, Prasanti Yerramilli, Layne Pruitt, Kristina Reppond, Madison N. Shyer, Jessica Cambric, Ryan Gadd, Ilya J. Finkelstein, Jimmy Gollihar, and James M. Musser |
| EPI_ISL_2203596, EPI_ISL_2203598, EPI_ISL_2203599 | Illinois Department of Public Health - Springfield Lab | Illinois Department of Public Health - Springfield Lab | Bryan Sim, Gordon McCall |
| EPI_ISL_2203600 | Houston Methodist Hospital | Houston Methodist Hospital | Randall J. Olsen, Paul A. Christensen, S. Wesley Long, Sishir Subedi, Robert Olson, Marcus Nguyen, James J. Davis, Matthew Ojeda Saavedra, Prasanti Yerramilli, Layne Pruitt, Kristina Reppond, Madison N. Shyer, Jessica Cambric, Ryan Gadd, Ilya J. Finkelstein, Jimmy Gollihar, and James M. Musser |
| EPI_ISL_2203601, EPI_ISL_2203602 | Illinois Department of Public Health - Springfield Lab | Illinois Department of Public Health - Springfield Lab | Bryan Sim, Gordon McCall |
| EPI_ISL_2203603 | Houston Methodist Hospital | Houston Methodist Hospital | Randall J. Olsen, Paul A. Christensen, S. Wesley Long, Sishir Subedi, Robert Olson, Marcus Nguyen, James J. Davis, Matthew Ojeda Saavedra, Prasanti Yerramilli, Layne Pruitt, Kristina Reppond, Madison N. Shyer, Jessica Cambric, Ryan Gadd, Ilya J. Finkelstein, Jimmy Gollihar, and James M. Musser |
| EPI_ISL_2203604, EPI_ISL_2203605, EPI_ISL_2203607, EPI_ISL_2203608 | Illinois Department of Public Health - Springfield Lab | Illinois Department of Public Health - Springfield Lab | Bryan Sim, Gordon McCall |
| EPI_ISL_2203609 | Houston Methodist Hospital | Houston Methodist Hospital | Randall J. Olsen, Paul A. Christensen, S. Wesley Long, Sishir Subedi, Robert Olson, Marcus Nguyen, James J. Davis, Matthew Ojeda Saavedra, Prasanti Yerramilli, Layne Pruitt, Kristina Reppond, Madison N. Shyer, Jessica Cambric, Ryan Gadd, Ilya J. Finkelstein, Jimmy Gollihar, and James M. Musser |
| EPI_ISL_2203610, EPI_ISL_2203611, EPI_ISL_2203613, EPI_ISL_2203614 | Illinois Department of Public Health - Springfield Lab | Illinois Department of Public Health - Springfield Lab | Bryan Sim, Gordon McCall |
| EPI_ISL_2203615 | Houston Methodist Hospital | Houston Methodist Hospital | Randall J. Olsen, Paul A. Christensen, S. Wesley Long, Sishir Subedi, Robert Olson, Marcus Nguyen, James J. Davis, Matthew Ojeda Saavedra, Prasanti Yerramilli, Layne Pruitt, Kristina Reppond, Madison N. Shyer, Jessica Cambric, Ryan Gadd, Ilya J. Finkelstein, Jimmy Gollihar, and James M. Musser |

| | | | |
|---|---|---|---|
| EPI_ISL_2203616 | Illinois Department of Public Health - Springfield Lab | Illinois Department of Public Health - Springfield Lab | Bryan Sim, Gordon McCall |
| EPI_ISL_2203618 | Houston Methodist Hospital | Houston Methodist Hospital | Randall J. Olsen, Paul A. Christensen, S. Wesley Long, Sishir Subedi, Robert Olson, Marcus Nguyen, James J. Davis, Matthew Ojeda Saavedra, Prasanti Yerramilli, Layne Pruitt, Kristina Reppond, Madison N. Shyer, Jessica Cambric, Ryan Gadd, Ilya J. Finkelstein, Jimmy Gollihar, and James M. Musser |
| EPI_ISL_2203619, EPI_ISL_2203620 | Illinois Department of Public Health - Springfield Lab | Illinois Department of Public Health - Springfield Lab | Bryan Sim, Gordon McCall |
| EPI_ISL_2203621 | Houston Methodist Hospital | Houston Methodist Hospital | Randall J. Olsen, Paul A. Christensen, S. Wesley Long, Sishir Subedi, Robert Olson, Marcus Nguyen, James J. Davis, Matthew Ojeda Saavedra, Prasanti Yerramilli, Layne Pruitt, Kristina Reppond, Madison N. Shyer, Jessica Cambric, Ryan Gadd, Ilya J. Finkelstein, Jimmy Gollihar, and James M. Musser |
| EPI_ISL_2203622, EPI_ISL_2203623, EPI_ISL_2203624 | Illinois Department of Public Health - Springfield Lab | Illinois Department of Public Health - Springfield Lab | Bryan Sim, Gordon McCall |
| EPI_ISL_2203625 | Houston Methodist Hospital | Houston Methodist Hospital | Randall J. Olsen, Paul A. Christensen, S. Wesley Long, Sishir Subedi, Robert Olson, Marcus Nguyen, James J. Davis, Matthew Ojeda Saavedra, Prasanti Yerramilli, Layne Pruitt, Kristina Reppond, Madison N. Shyer, Jessica Cambric, Ryan Gadd, Ilya J. Finkelstein, Jimmy Gollihar, and James M. Musser |
| EPI_ISL_2203626, EPI_ISL_2203627 | Illinois Department of Public Health - Springfield Lab | Illinois Department of Public Health - Springfield Lab | Bryan Sim, Gordon McCall |
| EPI_ISL_2203628 | Houston Methodist Hospital | Houston Methodist Hospital | Randall J. Olsen, Paul A. Christensen, S. Wesley Long, Sishir Subedi, Robert Olson, Marcus Nguyen, James J. Davis, Matthew Ojeda Saavedra, Prasanti Yerramilli, Layne Pruitt, Kristina Reppond, Madison N. Shyer, Jessica Cambric, Ryan Gadd, Ilya J. Finkelstein, Jimmy Gollihar, and James M. Musser |
| EPI_ISL_2203629, EPI_ISL_2203630 | Illinois Department of Public Health - Springfield Lab | Illinois Department of Public Health - Springfield Lab | Bryan Sim, Gordon McCall |
| EPI_ISL_2203631 | Houston Methodist Hospital | Houston Methodist Hospital | Randall J. Olsen, Paul A. Christensen, S. Wesley Long, Sishir Subedi, Robert Olson, Marcus Nguyen, James J. Davis, Matthew Ojeda Saavedra, Prasanti Yerramilli, Layne Pruitt, Kristina Reppond, Madison N. Shyer, Jessica Cambric, Ryan Gadd, Ilya J. Finkelstein, Jimmy Gollihar, and James M. Musser |
| EPI_ISL_2203632, EPI_ISL_2203633 | Illinois Department of Public Health - Springfield Lab | Illinois Department of Public Health - Springfield Lab | Bryan Sim, Gordon McCall |
| EPI_ISL_2203634 | Houston Methodist Hospital | Houston Methodist Hospital | Randall J. Olsen, Paul A. Christensen, S. Wesley Long, Sishir Subedi, Robert Olson, Marcus Nguyen, James J. Davis, Matthew Ojeda Saavedra, Prasanti Yerramilli, Layne Pruitt, Kristina Reppond, Madison N. Shyer, Jessica Cambric, Ryan Gadd, Ilya J. Finkelstein, Jimmy Gollihar, and James M. Musser |
| EPI_ISL_2203635 | Illinois Department of Public Health - Springfield Lab | Illinois Department of Public Health - Springfield Lab | Bryan Sim, Gordon McCall |
| EPI_ISL_2203636 | Quest Diagnostics Incorporated | Centers for Disease Control and Prevention Division of Viral Diseases, Pathogen Discovery | Dakota Howard, Dhwani Batra, Peter W. Cook, Kara Moser, Adrian Paskey, Jason Caravas, Benjamin Rambo-Martin, Shatavia Morrison, Christopher Gulvick, Scott Sammons, Yvette Unoarumhi, Darlene Wagner, Matthew Schmerer, S. H. Rosenthal, A. Gerasimova, R. M. Kagan, B. Anderson, M. Hua, Y. Liu, L.E. Bernstein, K.E. Livingston, A. Perez, I. A. Shlyakhter, R. V. Rolando, R. Owen, P. Tanpaiboon, F. Lacbawan, Clinton R. Paden, Duncan MacCannell |
| EPI_ISL_2203637 | Houston Methodist Hospital | Houston Methodist Hospital | Randall J. Olsen, Paul A. Christensen, S. Wesley Long, Sishir Subedi, Robert Olson, Marcus Nguyen, James J. Davis, Matthew Ojeda Saavedra, Prasanti Yerramilli, Layne Pruitt, Kristina Reppond, Madison N. Shyer, Jessica Cambric, Ryan Gadd, Ilya J. Finkelstein, Jimmy Gollihar, and James M. Musser |
| EPI_ISL_2203638, EPI_ISL_2203639 | Illinois Department of Public Health - Springfield Lab | Illinois Department of Public Health - Springfield Lab | Bryan Sim, Gordon McCall |
| EPI_ISL_2203640 | Quest Diagnostics Incorporated | Centers for Disease Control and Prevention Division of Viral Diseases, Pathogen Discovery | Dakota Howard, Dhwani Batra, Peter W. Cook, Kara Moser, Adrian Paskey, Jason Caravas, Benjamin Rambo-Martin, Shatavia Morrison, Christopher Gulvick, Scott Sammons, Yvette Unoarumhi, Darlene Wagner, Matthew Schmerer, S. H. Rosenthal, A. Gerasimova, R. M. Kagan, B. Anderson, M. Hua, Y. Liu, L.E. Bernstein, K.E. Livingston, A. Perez, I. A. Shlyakhter, R. V. Rolando, R. Owen, P. Tanpaiboon, F. Lacbawan, Clinton R. Paden, Duncan MacCannell |
| EPI_ISL_2203641 | Houston Methodist Hospital | Houston Methodist Hospital | Randall J. Olsen, Paul A. Christensen, S. Wesley Long, Sishir Subedi, Robert Olson, Marcus Nguyen, James J. Davis, Matthew Ojeda Saavedra, Prasanti Yerramilli, Layne Pruitt, Kristina Reppond, Madison N. Shyer, Jessica Cambric, Ryan Gadd, Ilya J. Finkelstein, Jimmy Gollihar, and James M. Musser |
| EPI_ISL_2203642 | Quest Diagnostics Incorporated | Centers for Disease Control and Prevention Division of Viral Diseases, Pathogen Discovery | Dakota Howard, Dhwani Batra, Peter W. Cook, Kara Moser, Adrian Paskey, Jason Caravas, Benjamin Rambo-Martin, Shatavia Morrison, Christopher Gulvick, Scott Sammons, Yvette Unoarumhi, Darlene Wagner, Matthew Schmerer, S. H. Rosenthal, A. Gerasimova, R. M. Kagan, B. Anderson, M. Hua, Y. Liu, L.E. Bernstein, K.E. Livingston, A. Perez, I. A. Shlyakhter, R. V. Rolando, R. Owen, P. Tanpaiboon, F. Lacbawan, Clinton R. Paden, Duncan MacCannell |
| EPI_ISL_2203643 | Illinois Department of Public Health - Springfield Lab | Illinois Department of Public Health - Springfield Lab | Bryan Sim, Gordon McCall |
| EPI_ISL_2203644 | Quest Diagnostics Incorporated | Centers for Disease Control and Prevention Division of Viral Diseases, Pathogen Discovery | Dakota Howard, Dhwani Batra, Peter W. Cook, Kara Moser, Adrian Paskey, Jason Caravas, Benjamin Rambo-Martin, Shatavia Morrison, Christopher Gulvick, Scott Sammons, Yvette Unoarumhi, Darlene Wagner, Matthew Schmerer, S. H. Rosenthal, A. Gerasimova, R. M. Kagan, B. Anderson, M. Hua, Y. Liu, L.E. Bernstein, K.E. Livingston, A. Perez, I. A. Shlyakhter, R. V. Rolando, R. Owen, P. Tanpaiboon, F. Lacbawan, Clinton R. Paden, Duncan MacCannell |
| EPI_ISL_2203645 | Houston Methodist Hospital | Houston Methodist Hospital | Randall J. Olsen, Paul A. Christensen, S. Wesley Long, Sishir Subedi, Robert Olson, Marcus Nguyen, James J. Davis, Matthew Ojeda Saavedra, Prasanti Yerramilli, Layne Pruitt, Kristina Reppond, Madison N. Shyer, Jessica Cambric, Ryan Gadd, Ilya J. Finkelstein, Jimmy Gollihar, and James M. Musser |
| EPI_ISL_2203646 | Quest Diagnostics Incorporated | Centers for Disease Control and Prevention Division of Viral Diseases, Pathogen Discovery | Dakota Howard, Dhwani Batra, Peter W. Cook, Kara Moser, Adrian Paskey, Jason Caravas, Benjamin Rambo-Martin, Shatavia Morrison, Christopher Gulvick, Scott Sammons, Yvette Unoarumhi, Darlene Wagner, Matthew Schmerer, S. H. Rosenthal, A. Gerasimova, R. M. Kagan, B. Anderson, M. Hua, Y. Liu, L.E. Bernstein, K.E. Livingston, A. Perez, I. A. Shlyakhter, R. V. Rolando, R. Owen, P. Tanpaiboon, F. Lacbawan, Clinton R. Paden, Duncan MacCannell |
| EPI_ISL_2203647 | Houston Methodist Hospital | Houston Methodist Hospital | Randall J. Olsen, Paul A. Christensen, S. Wesley Long, Sishir Subedi, Robert Olson, Marcus Nguyen, James J. Davis, Matthew Ojeda Saavedra, Prasanti Yerramilli, Layne Pruitt, Kristina Reppond, Madison N. Shyer, Jessica Cambric, Ryan Gadd, Ilya J. Finkelstein, Jimmy Gollihar, and James M. Musser |
| EPI_ISL_2203648 | Illinois Department of Public Health - Springfield Lab | Illinois Department of Public Health - Springfield Lab | Bryan Sim, Gordon McCall |
| EPI_ISL_2203649 | Quest Diagnostics Incorporated | Centers for Disease Control and Prevention Division of Viral Diseases, Pathogen Discovery | Dakota Howard, Dhwani Batra, Peter W. Cook, Kara Moser, Adrian Paskey, Jason Caravas, Benjamin Rambo-Martin, Shatavia Morrison, Christopher Gulvick, Scott Sammons, Yvette Unoarumhi, Darlene Wagner, Matthew Schmerer, S. H. Rosenthal, A. Gerasimova, R. M. Kagan, B. Anderson, M. Hua, Y. Liu, L.E. Bernstein, K.E. Livingston, A. Perez, I. A. Shlyakhter, R. V. Rolando, R. Owen, P. Tanpaiboon, F. Lacbawan, Clinton R. Paden, Duncan MacCannell |
| EPI_ISL_2203650 | Illinois Department of Public Health - Springfield Lab | Illinois Department of Public Health - Springfield Lab | Bryan Sim, Gordon McCall |
| EPI_ISL_2203651 | Houston Methodist Hospital | Houston Methodist Hospital | Randall J. Olsen, Paul A. Christensen, S. Wesley Long, Sishir Subedi, Robert Olson, Marcus Nguyen, James J. Davis, Matthew Ojeda Saavedra, Prasanti Yerramilli, Layne Pruitt, Kristina Reppond, Madison N. Shyer, Jessica Cambric, Ryan Gadd, Ilya J. Finkelstein, Jimmy Gollihar, and James M. Musser |
| EPI_ISL_2203652 | Quest Diagnostics Incorporated | Centers for Disease Control and Prevention Division of Viral Diseases, Pathogen Discovery | Dakota Howard, Dhwani Batra, Peter W. Cook, Kara Moser, Adrian Paskey, Jason Caravas, Benjamin Rambo-Martin, Shatavia Morrison, Christopher Gulvick, Scott Sammons, Yvette Unoarumhi, Darlene Wagner, Matthew Schmerer, S. H. Rosenthal, A. Gerasimova, R. M. Kagan, B. Anderson, M. Hua, Y. Liu, L.E. Bernstein, K.E. Livingston, A. Perez, I. A. Shlyakhter, R. V. Rolando, R. Owen, P. Tanpaiboon, F. Lacbawan, Clinton R. Paden, Duncan MacCannell |
| EPI_ISL_2203653 | Illinois Department of Public Health - Springfield Lab | Illinois Department of Public Health - Springfield Lab | Bryan Sim, Gordon McCall |
| EPI_ISL_2203654 | Quest Diagnostics Incorporated | Centers for Disease Control and Prevention Division of Viral Diseases, Pathogen Discovery | Dakota Howard, Dhwani Batra, Peter W. Cook, Kara Moser, Adrian Paskey, Jason Caravas, Benjamin Rambo-Martin, Shatavia Morrison, Christopher Gulvick, Scott Sammons, Yvette Unoarumhi, Darlene Wagner, Matthew Schmerer, S. H. Rosenthal, A. Gerasimova, R. M. Kagan, B. Anderson, M. Hua, Y. Liu, L.E. Bernstein, K.E. Livingston, A. Perez, I. A. Shlyakhter, R. V. Rolando, R. Owen, P. Tanpaiboon, F. Lacbawan, Clinton R. Paden, Duncan MacCannell |
| EPI_ISL_2203655 | Houston Methodist Hospital | Houston Methodist Hospital | Randall J. Olsen, Paul A. Christensen, S. Wesley Long, Sishir Subedi, Robert Olson, Marcus Nguyen, James J. Davis, Matthew Ojeda Saavedra, Prasanti Yerramilli, Layne Pruitt, Kristina Reppond, Madison N. Shyer, Jessica Cambric, Ryan Gadd, Ilya J. Finkelstein, Jimmy Gollihar, and James M. Musser |
| EPI_ISL_2203656 | Illinois Department of Public Health - Springfield Lab | Illinois Department of Public Health - Springfield Lab | Bryan Sim, Gordon McCall |
| EPI_ISL_2203657 | Quest Diagnostics Incorporated | Centers for Disease Control and Prevention Division of Viral Diseases, Pathogen Discovery | Dakota Howard, Dhwani Batra, Peter W. Cook, Kara Moser, Adrian Paskey, Jason Caravas, Benjamin Rambo-Martin, Shatavia Morrison, Christopher Gulvick, Scott Sammons, Yvette Unoarumhi, Darlene Wagner, Matthew Schmerer, S. H. Rosenthal, A. Gerasimova, R. M. Kagan, B. Anderson, M. Hua, Y. Liu, L.E. Bernstein, K.E. Livingston, A. Perez, I. A. Shlyakhter, R. V. Rolando, R. Owen, P. Tanpaiboon, F. Lacbawan, Clinton R. Paden, Duncan MacCannell |
| EPI_ISL_2203658 | Houston Methodist Hospital | Houston Methodist Hospital | Randall J. Olsen, Paul A. Christensen, S. Wesley Long, Sishir Subedi, Robert Olson, Marcus Nguyen, James J. Davis, Matthew Ojeda Saavedra, Prasanti Yerramilli, Layne Pruitt, Kristina Reppond, Madison N. Shyer, Jessica Cambric, Ryan Gadd, Ilya J. Finkelstein, Jimmy Gollihar, and James M. Musser |
| EPI_ISL_2203659 | Quest Diagnostics Incorporated | Centers for Disease Control and Prevention Division of Viral Diseases, Pathogen Discovery | Dakota Howard, Dhwani Batra, Peter W. Cook, Kara Moser, Adrian Paskey, Jason Caravas, Benjamin Rambo-Martin, Shatavia Morrison, Christopher Gulvick, Scott Sammons, Yvette Unoarumhi, Darlene Wagner, Matthew Schmerer, S. H. Rosenthal, A. Gerasimova, R. M. Kagan, B. Anderson, M. Hua, Y. Liu, L.E. Bernstein, K.E. Livingston, A. Perez, I. A. Shlyakhter, R. V. Rolando, R. Owen, P. Tanpaiboon, F. Lacbawan, Clinton R. Paden, Duncan MacCannell |
| EPI_ISL_2203660 | Illinois Department of Public Health - Springfield Lab | Illinois Department of Public Health - Springfield Lab | Bryan Sim, Gordon McCall |
| EPI_ISL_2203661 | Houston Methodist Hospital | Houston Methodist Hospital | Randall J. Olsen, Paul A. Christensen, S. Wesley Long, Sishir Subedi, Robert Olson, Marcus Nguyen, James J. Davis, Matthew Ojeda Saavedra, Prasanti Yerramilli, Layne Pruitt, Kristina Reppond, Madison N. Shyer, Jessica Cambric, Ryan Gadd, Ilya J. Finkelstein, Jimmy Gollihar, and James M. Musser |
| EPI_ISL_2203662 | Quest Diagnostics Incorporated | Centers for Disease Control and Prevention Division of Viral Diseases, Pathogen Discovery | Dakota Howard, Dhwani Batra, Peter W. Cook, Kara Moser, Adrian Paskey, Jason Caravas, Benjamin Rambo-Martin, Shatavia Morrison, Christopher Gulvick, Scott Sammons, Yvette Unoarumhi, Darlene Wagner, Matthew Schmerer, S. H. Rosenthal, A. Gerasimova, R. M. Kagan, B. Anderson, M. Hua, Y. Liu, L.E. Bernstein, K.E. Livingston, A. Perez, I. A. Shlyakhter, R. V. Rolando, R. Owen, P. Tanpaiboon, F. Lacbawan, Clinton R. Paden, Duncan MacCannell |
| EPI_ISL_2203663 | Illinois Department of Public Health - Springfield Lab | Illinois Department of Public Health - Springfield Lab | Bryan Sim, Gordon McCall |

| | | | |
|---|---|---|---|
| EPI_ISL_2203664 | Houston Methodist Hospital | Houston Methodist Hospital | Randall J. Olsen, Paul A. Christensen, S. Wesley Long, Sishir Subedi, Robert Olson, Marcus Nguyen, James J. Davis, Matthew Ojeda Saavedra, Prasanti Yerramilli, Layne Pruitt, Kristina Reppond, Madison N. Shyer, Jessica Cambric, Ryan Gadd, Ilya J. Finkelstein, Jimmy Gollihar, and James M. Musser |
| EPI_ISL_2203665 | Quest Diagnostics Incorporated | Centers for Disease Control and Prevention Division of Viral Diseases, Pathogen Discovery | Dakota Howard, Dhwani Batra, Peter W. Cook, Kara Moser, Adrian Paskey, Jason Caravas, Benjamin Rambo-Martin, Shatavia Morrison, Christopher Gulvick, Scott Sammons, Yvette Unoarumhi, Darlene Wagner, Matthew Schmerer, S. H. Rosenthal, A. Gerasimova, R. M. Kagan, B. Anderson, M. Hua, Y. Liu, L.E. Bernstein, K.E. Livingston, A. Perez, I. A. Shlyakhter, R. V. Rolando, R. Owen, P. Tanpaiboon, F. Lacbawan, Clinton R. Paden, Duncan MacCannell |
| EPI_ISL_2203666 | Illinois Department of Public Health - Springfield Lab | Illinois Department of Public Health - Springfield Lab | Bryan Sim, Gordon McCall |
| EPI_ISL_2203667 | Quest Diagnostics Incorporated | Centers for Disease Control and Prevention Division of Viral Diseases, Pathogen Discovery | Dakota Howard, Dhwani Batra, Peter W. Cook, Kara Moser, Adrian Paskey, Jason Caravas, Benjamin Rambo-Martin, Shatavia Morrison, Christopher Gulvick, Scott Sammons, Yvette Unoarumhi, Darlene Wagner, Matthew Schmerer, S. H. Rosenthal, A. Gerasimova, R. M. Kagan, B. Anderson, M. Hua, Y. Liu, L.E. Bernstein, K.E. Livingston, A. Perez, I. A. Shlyakhter, R. V. Rolando, R. Owen, P. Tanpaiboon, F. Lacbawan, Clinton R. Paden, Duncan MacCannell |
| EPI_ISL_2203668 | Houston Methodist Hospital | Houston Methodist Hospital | Randall J. Olsen, Paul A. Christensen, S. Wesley Long, Sishir Subedi, Robert Olson, Marcus Nguyen, James J. Davis, Matthew Ojeda Saavedra, Prasanti Yerramilli, Layne Pruitt, Kristina Reppond, Madison N. Shyer, Jessica Cambric, Ryan Gadd, Ilya J. Finkelstein, Jimmy Gollihar, and James M. Musser |
| EPI_ISL_2203669 | Illinois Department of Public Health - Springfield Lab | Illinois Department of Public Health - Springfield Lab | Bryan Sim, Gordon McCall |
| EPI_ISL_2203670 | Houston Methodist Hospital | Houston Methodist Hospital | Randall J. Olsen, Paul A. Christensen, S. Wesley Long, Sishir Subedi, Robert Olson, Marcus Nguyen, James J. Davis, Matthew Ojeda Saavedra, Prasanti Yerramilli, Layne Pruitt, Kristina Reppond, Madison N. Shyer, Jessica Cambric, Ryan Gadd, Ilya J. Finkelstein, Jimmy Gollihar, and James M. Musser |
| EPI_ISL_2203671 | Illinois Department of Public Health - Springfield Lab | Illinois Department of Public Health - Springfield Lab | Bryan Sim, Gordon McCall |
| EPI_ISL_2203672 | Quest Diagnostics Incorporated | Centers for Disease Control and Prevention Division of Viral Diseases, Pathogen Discovery | Dakota Howard, Dhwani Batra, Peter W. Cook, Kara Moser, Adrian Paskey, Jason Caravas, Benjamin Rambo-Martin, Shatavia Morrison, Christopher Gulvick, Scott Sammons, Yvette Unoarumhi, Darlene Wagner, Matthew Schmerer, S. H. Rosenthal, A. Gerasimova, R. M. Kagan, B. Anderson, M. Hua, Y. Liu, L.E. Bernstein, K.E. Livingston, A. Perez, I. A. Shlyakhter, R. V. Rolando, R. Owen, P. Tanpaiboon, F. Lacbawan, Clinton R. Paden, Duncan MacCannell |
| EPI_ISL_2203673, EPI_ISL_2203674 | Houston Methodist Hospital | Houston Methodist Hospital | Randall J. Olsen, Paul A. Christensen, S. Wesley Long, Sishir Subedi, Robert Olson, Marcus Nguyen, James J. Davis, Matthew Ojeda Saavedra, Prasanti Yerramilli, Layne Pruitt, Kristina Reppond, Madison N. Shyer, Jessica Cambric, Ryan Gadd, Ilya J. Finkelstein, Jimmy Gollihar, and James M. Musser |
| EPI_ISL_2203675 | Quest Diagnostics Incorporated | Centers for Disease Control and Prevention Division of Viral Diseases, Pathogen Discovery | Dakota Howard, Dhwani Batra, Peter W. Cook, Kara Moser, Adrian Paskey, Jason Caravas, Benjamin Rambo-Martin, Shatavia Morrison, Christopher Gulvick, Scott Sammons, Yvette Unoarumhi, Darlene Wagner, Matthew Schmerer, S. H. Rosenthal, A. Gerasimova, R. M. Kagan, B. Anderson, M. Hua, Y. Liu, L.E. Bernstein, K.E. Livingston, A. Perez, I. A. Shlyakhter, R. V. Rolando, R. Owen, P. Tanpaiboon, F. Lacbawan, Clinton R. Paden, Duncan MacCannell |
| EPI_ISL_2203676 | Illinois Department of Public Health - Springfield Lab | Illinois Department of Public Health - Springfield Lab | Bryan Sim, Gordon McCall |
| EPI_ISL_2203677 | Quest Diagnostics Incorporated | Centers for Disease Control and Prevention Division of Viral Diseases, Pathogen Discovery | Dakota Howard, Dhwani Batra, Peter W. Cook, Kara Moser, Adrian Paskey, Jason Caravas, Benjamin Rambo-Martin, Shatavia Morrison, Christopher Gulvick, Scott Sammons, Yvette Unoarumhi, Darlene Wagner, Matthew Schmerer, S. H. Rosenthal, A. Gerasimova, R. M. Kagan, B. Anderson, M. Hua, Y. Liu, L.E. Bernstein, K.E. Livingston, A. Perez, I. A. Shlyakhter, R. V. Rolando, R. Owen, P. Tanpaiboon, F. Lacbawan, Clinton R. Paden, Duncan MacCannell |
| EPI_ISL_2203678 | Houston Methodist Hospital | Houston Methodist Hospital | Randall J. Olsen, Paul A. Christensen, S. Wesley Long, Sishir Subedi, Robert Olson, Marcus Nguyen, James J. Davis, Matthew Ojeda Saavedra, Prasanti Yerramilli, Layne Pruitt, Kristina Reppond, Madison N. Shyer, Jessica Cambric, Ryan Gadd, Ilya J. Finkelstein, Jimmy Gollihar, and James M. Musser |
| EPI_ISL_2203679 | Illinois Department of Public Health - Springfield Lab | Illinois Department of Public Health - Springfield Lab | Bryan Sim, Gordon McCall |
| EPI_ISL_2203680 | Quest Diagnostics Incorporated | Centers for Disease Control and Prevention Division of Viral Diseases, Pathogen Discovery | Dakota Howard, Dhwani Batra, Peter W. Cook, Kara Moser, Adrian Paskey, Jason Caravas, Benjamin Rambo-Martin, Shatavia Morrison, Christopher Gulvick, Scott Sammons, Yvette Unoarumhi, Darlene Wagner, Matthew Schmerer, S. H. Rosenthal, A. Gerasimova, R. M. Kagan, B. Anderson, M. Hua, Y. Liu, L.E. Bernstein, K.E. Livingston, A. Perez, I. A. Shlyakhter, R. V. Rolando, R. Owen, P. Tanpaiboon, F. Lacbawan, Clinton R. Paden, Duncan MacCannell |
| EPI_ISL_2203681 | Houston Methodist Hospital | Houston Methodist Hospital | Randall J. Olsen, Paul A. Christensen, S. Wesley Long, Sishir Subedi, Robert Olson, Marcus Nguyen, James J. Davis, Matthew Ojeda Saavedra, Prasanti Yerramilli, Layne Pruitt, Kristina Reppond, Madison N. Shyer, Jessica Cambric, Ryan Gadd, Ilya J. Finkelstein, Jimmy Gollihar, and James M. Musser |
| EPI_ISL_2203682 | Quest Diagnostics Incorporated | Centers for Disease Control and Prevention Division of Viral Diseases, Pathogen Discovery | Dakota Howard, Dhwani Batra, Peter W. Cook, Kara Moser, Adrian Paskey, Jason Caravas, Benjamin Rambo-Martin, Shatavia Morrison, Christopher Gulvick, Scott Sammons, Yvette Unoarumhi, Darlene Wagner, Matthew Schmerer, S. H. Rosenthal, A. Gerasimova, R. M. Kagan, B. Anderson, M. Hua, Y. Liu, L.E. Bernstein, K.E. Livingston, A. Perez, I. A. Shlyakhter, R. V. Rolando, R. Owen, P. Tanpaiboon, F. Lacbawan, Clinton R. Paden, Duncan MacCannell |
| EPI_ISL_2203683 | Illinois Department of Public Health - Springfield Lab | Illinois Department of Public Health - Springfield Lab | Bryan Sim, Gordon McCall |
| EPI_ISL_2203684 | Quest Diagnostics Incorporated | Centers for Disease Control and Prevention Division of Viral Diseases, Pathogen Discovery | Dakota Howard, Dhwani Batra, Peter W. Cook, Kara Moser, Adrian Paskey, Jason Caravas, Benjamin Rambo-Martin, Shatavia Morrison, Christopher Gulvick, Scott Sammons, Yvette Unoarumhi, Darlene Wagner, Matthew Schmerer, S. H. Rosenthal, A. Gerasimova, R. M. Kagan, B. Anderson, M. Hua, Y. Liu, L.E. Bernstein, K.E. Livingston, A. Perez, I. A. Shlyakhter, R. V. Rolando, R. Owen, P. Tanpaiboon, F. Lacbawan, Clinton R. Paden, Duncan MacCannell |
| EPI_ISL_2203685 | Illinois Department of Public Health - Springfield Lab | Illinois Department of Public Health - Springfield Lab | Bryan Sim, Gordon McCall |
| EPI_ISL_2203687 | Quest Diagnostics Incorporated | Centers for Disease Control and Prevention Division of Viral Diseases, Pathogen Discovery | Dakota Howard, Dhwani Batra, Peter W. Cook, Kara Moser, Adrian Paskey, Jason Caravas, Benjamin Rambo-Martin, Shatavia Morrison, Christopher Gulvick, Scott Sammons, Yvette Unoarumhi, Darlene Wagner, Matthew Schmerer, S. H. Rosenthal, A. Gerasimova, R. M. Kagan, B. Anderson, M. Hua, Y. Liu, L.E. Bernstein, K.E. Livingston, A. Perez, I. A. Shlyakhter, R. V. Rolando, R. Owen, P. Tanpaiboon, F. Lacbawan, Clinton R. Paden, Duncan MacCannell |
| EPI_ISL_2203688, EPI_ISL_2203689 | Illinois Department of Public Health - Springfield Lab | Illinois Department of Public Health - Springfield Lab | Bryan Sim, Gordon McCall |
| EPI_ISL_2203691 | Quest Diagnostics Incorporated | Centers for Disease Control and Prevention Division of Viral Diseases, Pathogen Discovery | Dakota Howard, Dhwani Batra, Peter W. Cook, Kara Moser, Adrian Paskey, Jason Caravas, Benjamin Rambo-Martin, Shatavia Morrison, Christopher Gulvick, Scott Sammons, Yvette Unoarumhi, Darlene Wagner, Matthew Schmerer, S. H. Rosenthal, A. Gerasimova, R. M. Kagan, B. Anderson, M. Hua, Y. Liu, L.E. Bernstein, K.E. Livingston, A. Perez, I. A. Shlyakhter, R. V. Rolando, R. Owen, P. Tanpaiboon, F. Lacbawan, Clinton R. Paden, Duncan MacCannell |
| EPI_ISL_2203692 | Illinois Department of Public Health - Springfield Lab | Illinois Department of Public Health - Springfield Lab | Bryan Sim, Gordon McCall |
| EPI_ISL_2203693 | Quest Diagnostics Incorporated | Centers for Disease Control and Prevention Division of Viral Diseases, Pathogen Discovery | Dakota Howard, Dhwani Batra, Peter W. Cook, Kara Moser, Adrian Paskey, Jason Caravas, Benjamin Rambo-Martin, Shatavia Morrison, Christopher Gulvick, Scott Sammons, Yvette Unoarumhi, Darlene Wagner, Matthew Schmerer, S. H. Rosenthal, A. Gerasimova, R. M. Kagan, B. Anderson, M. Hua, Y. Liu, L.E. Bernstein, K.E. Livingston, A. Perez, I. A. Shlyakhter, R. V. Rolando, R. Owen, P. Tanpaiboon, F. Lacbawan, Clinton R. Paden, Duncan MacCannell |
| EPI_ISL_2203694 | Houston Methodist Hospital | Houston Methodist Hospital | Randall J. Olsen, Paul A. Christensen, S. Wesley Long, Sishir Subedi, Robert Olson, Marcus Nguyen, James J. Davis, Matthew Ojeda Saavedra, Prasanti Yerramilli, Layne Pruitt, Kristina Reppond, Madison N. Shyer, Jessica Cambric, Ryan Gadd, Ilya J. Finkelstein, Jimmy Gollihar, and James M. Musser |
| EPI_ISL_2203695 | Quest Diagnostics Incorporated | Centers for Disease Control and Prevention Division of Viral Diseases, Pathogen Discovery | Dakota Howard, Dhwani Batra, Peter W. Cook, Kara Moser, Adrian Paskey, Jason Caravas, Benjamin Rambo-Martin, Shatavia Morrison, Christopher Gulvick, Scott Sammons, Yvette Unoarumhi, Darlene Wagner, Matthew Schmerer, S. H. Rosenthal, A. Gerasimova, R. M. Kagan, B. Anderson, M. Hua, Y. Liu, L.E. Bernstein, K.E. Livingston, A. Perez, I. A. Shlyakhter, R. V. Rolando, R. Owen, P. Tanpaiboon, F. Lacbawan, Clinton R. Paden, Duncan MacCannell |
| EPI_ISL_2203696, EPI_ISL_2203698 | Illinois Department of Public Health - Springfield Lab | Illinois Department of Public Health - Springfield Lab | Bryan Sim, Gordon McCall |
| EPI_ISL_2203699 | Quest Diagnostics Incorporated | Centers for Disease Control and Prevention Division of Viral Diseases, Pathogen Discovery | Dakota Howard, Dhwani Batra, Peter W. Cook, Kara Moser, Adrian Paskey, Jason Caravas, Benjamin Rambo-Martin, Shatavia Morrison, Christopher Gulvick, Scott Sammons, Yvette Unoarumhi, Darlene Wagner, Matthew Schmerer, S. H. Rosenthal, A. Gerasimova, R. M. Kagan, B. Anderson, M. Hua, Y. Liu, L.E. Bernstein, K.E. Livingston, A. Perez, I. A. Shlyakhter, R. V. Rolando, R. Owen, P. Tanpaiboon, F. Lacbawan, Clinton R. Paden, Duncan MacCannell |
| EPI_ISL_2203700 | Illinois Department of Public Health - Springfield Lab | Illinois Department of Public Health - Springfield Lab | Bryan Sim, Gordon McCall |
| EPI_ISL_2203701 | Houston Methodist Hospital | Houston Methodist Hospital | Randall J. Olsen, Paul A. Christensen, S. Wesley Long, Sishir Subedi, Robert Olson, Marcus Nguyen, James J. Davis, Matthew Ojeda Saavedra, Prasanti Yerramilli, Layne Pruitt, Kristina Reppond, Madison N. Shyer, Jessica Cambric, Ryan Gadd, Ilya J. Finkelstein, Jimmy Gollihar, and James M. Musser |
| EPI_ISL_2203702, EPI_ISL_2203703 | Quest Diagnostics Incorporated | Centers for Disease Control and Prevention Division of Viral Diseases, Pathogen Discovery | Dakota Howard, Dhwani Batra, Peter W. Cook, Kara Moser, Adrian Paskey, Jason Caravas, Benjamin Rambo-Martin, Shatavia Morrison, Christopher Gulvick, Scott Sammons, Yvette Unoarumhi, Darlene Wagner, Matthew Schmerer, S. H. Rosenthal, A. Gerasimova, R. M. Kagan, B. Anderson, M. Hua, Y. Liu, L.E. Bernstein, K.E. Livingston, A. Perez, I. A. Shlyakhter, R. V. Rolando, R. Owen, P. Tanpaiboon, F. Lacbawan, Clinton R. Paden, Duncan MacCannell |
| EPI_ISL_2203704 | Houston Methodist Hospital | Houston Methodist Hospital | Randall J. Olsen, Paul A. Christensen, S. Wesley Long, Sishir Subedi, Robert Olson, Marcus Nguyen, James J. Davis, Matthew Ojeda Saavedra, Prasanti Yerramilli, Layne Pruitt, Kristina Reppond, Madison N. Shyer, Jessica Cambric, Ryan Gadd, Ilya J. Finkelstein, Jimmy Gollihar, and James M. Musser |
| EPI_ISL_2203705 | Illinois Department of Public Health - Springfield Lab | Illinois Department of Public Health - Springfield Lab | Bryan Sim, Gordon McCall |
| EPI_ISL_2203706 | Quest Diagnostics Incorporated | Centers for Disease Control and Prevention Division of Viral Diseases, Pathogen Discovery | Dakota Howard, Dhwani Batra, Peter W. Cook, Kara Moser, Adrian Paskey, Jason Caravas, Benjamin Rambo-Martin, Shatavia Morrison, Christopher Gulvick, Scott Sammons, Yvette Unoarumhi, Darlene Wagner, Matthew Schmerer, S. H. Rosenthal, A. Gerasimova, R. M. Kagan, B. Anderson, M. Hua, Y. Liu, L.E. Bernstein, K.E. Livingston, A. Perez, I. A. Shlyakhter, R. V. Rolando, R. Owen, P. Tanpaiboon, F. Lacbawan, Clinton R. Paden, Duncan MacCannell |
| EPI_ISL_2203707 | Houston Methodist Hospital | Houston Methodist Hospital | Randall J. Olsen, Paul A. Christensen, S. Wesley Long, Sishir Subedi, Robert Olson, Marcus Nguyen, James J. Davis, Matthew Ojeda Saavedra, Prasanti Yerramilli, Layne Pruitt, Kristina Reppond, Madison N. Shyer, Jessica Cambric, Ryan Gadd, Ilya J. Finkelstein, Jimmy Gollihar, and James M. Musser |
| EPI_ISL_2203708 | Quest Diagnostics Incorporated | Centers for Disease Control and Prevention Division of Viral Diseases, Pathogen Discovery | Dakota Howard, Dhwani Batra, Peter W. Cook, Kara Moser, Adrian Paskey, Jason Caravas, Benjamin Rambo-Martin, Shatavia Morrison, Christopher Gulvick, Scott Sammons, Yvette Unoarumhi, Darlene Wagner, Matthew Schmerer, S. H. Rosenthal, A. Gerasimova, R. M. Kagan, B. Anderson, M. Hua, Y. Liu, L.E. Bernstein, K.E. Livingston, A. Perez, I. A. Shlyakhter, R. V. Rolando, R. Owen, P. Tanpaiboon, F. Lacbawan, Clinton R. Paden, Duncan MacCannell |
| EPI_ISL_2203709 | Illinois Department of Public Health - | Illinois Department of Public Health - | Bryan Sim, Gordon McCall |

| | | | |
|---|---|---|---|
| | Springfield Lab | Springfield Lab | |
| EPI_ISL_2203710 | Houston Methodist Hospital | Houston Methodist Hospital | Randall J. Olsen, Paul A. Christensen, S. Wesley Long, Sishir Subedi, Robert Olson, Marcus Nguyen, James J. Davis, Matthew Ojeda Saavedra, Prasanti Yerramilli, Layne Pruitt, Kristina Reppond, Madison N. Shyer, Jessica Cambric, Ryan Gadd, Ilya J. Finkelstein, Jimmy Gollihar, and James M. Musser |
| EPI_ISL_2203711 | Quest Diagnostics Incorporated | Centers for Disease Control and Prevention Division of Viral Diseases, Pathogen Discovery | Dakota Howard, Dhwani Batra, Peter W. Cook, Kara Moser, Adrian Paskey, Jason Caravas, Benjamin Rambo-Martin, Shatavia Morrison, Christopher Gulvick, Scott Sammons, Yvette Unoarumhi, Darlene Wagner, Matthew Schmerer, S. H. Rosenthal, A. Gerasimova, R. M. Kagan, B. Anderson, M. Hua, Y. Liu, L.E. Bernstein, K.E. Livingston, A. Perez, I. A. Shlyakhter, R. V. Rolando, R. Owen, P. Tanpaiboon, F. Lacbawan, Clinton R. Paden, Duncan MacCannell |
| EPI_ISL_2203712 | Illinois Department of Public Health - Springfield Lab | Illinois Department of Public Health - Springfield Lab | Bryan Sim, Gordon McCall |
| EPI_ISL_2203713 | Quest Diagnostics Incorporated | Centers for Disease Control and Prevention Division of Viral Diseases, Pathogen Discovery | Dakota Howard, Dhwani Batra, Peter W. Cook, Kara Moser, Adrian Paskey, Jason Caravas, Benjamin Rambo-Martin, Shatavia Morrison, Christopher Gulvick, Scott Sammons, Yvette Unoarumhi, Darlene Wagner, Matthew Schmerer, S. H. Rosenthal, A. Gerasimova, R. M. Kagan, B. Anderson, M. Hua, Y. Liu, L.E. Bernstein, K.E. Livingston, A. Perez, I. A. Shlyakhter, R. V. Rolando, R. Owen, P. Tanpaiboon, F. Lacbawan, Clinton R. Paden, Duncan MacCannell |
| EPI_ISL_2203714 | Houston Methodist Hospital | Houston Methodist Hospital | Randall J. Olsen, Paul A. Christensen, S. Wesley Long, Sishir Subedi, Robert Olson, Marcus Nguyen, James J. Davis, Matthew Ojeda Saavedra, Prasanti Yerramilli, Layne Pruitt, Kristina Reppond, Madison N. Shyer, Jessica Cambric, Ryan Gadd, Ilya J. Finkelstein, Jimmy Gollihar, and James M. Musser |
| EPI_ISL_2203715 | Illinois Department of Public Health - Springfield Lab | Illinois Department of Public Health - Springfield Lab | Bryan Sim, Gordon McCall |
| EPI_ISL_2203716 | Quest Diagnostics Incorporated | Centers for Disease Control and Prevention Division of Viral Diseases, Pathogen Discovery | Dakota Howard, Dhwani Batra, Peter W. Cook, Kara Moser, Adrian Paskey, Jason Caravas, Benjamin Rambo-Martin, Shatavia Morrison, Christopher Gulvick, Scott Sammons, Yvette Unoarumhi, Darlene Wagner, Matthew Schmerer, S. H. Rosenthal, A. Gerasimova, R. M. Kagan, B. Anderson, M. Hua, Y. Liu, L.E. Bernstein, K.E. Livingston, A. Perez, I. A. Shlyakhter, R. V. Rolando, R. Owen, P. Tanpaiboon, F. Lacbawan, Clinton R. Paden, Duncan MacCannell |
| EPI_ISL_2203717 | Houston Methodist Hospital | Houston Methodist Hospital | Randall J. Olsen, Paul A. Christensen, S. Wesley Long, Sishir Subedi, Robert Olson, Marcus Nguyen, James J. Davis, Matthew Ojeda Saavedra, Prasanti Yerramilli, Layne Pruitt, Kristina Reppond, Madison N. Shyer, Jessica Cambric, Ryan Gadd, Ilya J. Finkelstein, Jimmy Gollihar, and James M. Musser |
| EPI_ISL_2203718 | Quest Diagnostics Incorporated | Centers for Disease Control and Prevention Division of Viral Diseases, Pathogen Discovery | Dakota Howard, Dhwani Batra, Peter W. Cook, Kara Moser, Adrian Paskey, Jason Caravas, Benjamin Rambo-Martin, Shatavia Morrison, Christopher Gulvick, Scott Sammons, Yvette Unoarumhi, Darlene Wagner, Matthew Schmerer, S. H. Rosenthal, A. Gerasimova, R. M. Kagan, B. Anderson, M. Hua, Y. Liu, L.E. Bernstein, K.E. Livingston, A. Perez, I. A. Shlyakhter, R. V. Rolando, R. Owen, P. Tanpaiboon, F. Lacbawan, Clinton R. Paden, Duncan MacCannell |
| EPI_ISL_2203719 | Illinois Department of Public Health - Springfield Lab | Illinois Department of Public Health - Springfield Lab | Bryan Sim, Gordon McCall |
| EPI_ISL_2203720 | Houston Methodist Hospital | Houston Methodist Hospital | Randall J. Olsen, Paul A. Christensen, S. Wesley Long, Sishir Subedi, Robert Olson, Marcus Nguyen, James J. Davis, Matthew Ojeda Saavedra, Prasanti Yerramilli, Layne Pruitt, Kristina Reppond, Madison N. Shyer, Jessica Cambric, Ryan Gadd, Ilya J. Finkelstein, Jimmy Gollihar, and James M. Musser |
| EPI_ISL_2203721 | Quest Diagnostics Incorporated | Centers for Disease Control and Prevention Division of Viral Diseases, Pathogen Discovery | Dakota Howard, Dhwani Batra, Peter W. Cook, Kara Moser, Adrian Paskey, Jason Caravas, Benjamin Rambo-Martin, Shatavia Morrison, Christopher Gulvick, Scott Sammons, Yvette Unoarumhi, Darlene Wagner, Matthew Schmerer, S. H. Rosenthal, A. Gerasimova, R. M. Kagan, B. Anderson, M. Hua, Y. Liu, L.E. Bernstein, K.E. Livingston, A. Perez, I. A. Shlyakhter, R. V. Rolando, R. Owen, P. Tanpaiboon, F. Lacbawan, Clinton R. Paden, Duncan MacCannell |
| EPI_ISL_2203722 | Illinois Department of Public Health - Springfield Lab | Illinois Department of Public Health - Springfield Lab | Bryan Sim, Gordon McCall |
| EPI_ISL_2203723 | Houston Methodist Hospital | Houston Methodist Hospital | Randall J. Olsen, Paul A. Christensen, S. Wesley Long, Sishir Subedi, Robert Olson, Marcus Nguyen, James J. Davis, Matthew Ojeda Saavedra, Prasanti Yerramilli, Layne Pruitt, Kristina Reppond, Madison N. Shyer, Jessica Cambric, Ryan Gadd, Ilya J. Finkelstein, Jimmy Gollihar, and James M. Musser |
| EPI_ISL_2203724, EPI_ISL_2203725 | Quest Diagnostics Incorporated | Centers for Disease Control and Prevention Division of Viral Diseases, Pathogen Discovery | Dakota Howard, Dhwani Batra, Peter W. Cook, Kara Moser, Adrian Paskey, Jason Caravas, Benjamin Rambo-Martin, Shatavia Morrison, Christopher Gulvick, Scott Sammons, Yvette Unoarumhi, Darlene Wagner, Matthew Schmerer, S. H. Rosenthal, A. Gerasimova, R. M. Kagan, B. Anderson, M. Hua, Y. Liu, L.E. Bernstein, K.E. Livingston, A. Perez, I. A. Shlyakhter, R. V. Rolando, R. Owen, P. Tanpaiboon, F. Lacbawan, Clinton R. Paden, Duncan MacCannell |
| EPI_ISL_2203726 | Illinois Department of Public Health - Springfield Lab | Illinois Department of Public Health - Springfield Lab | Bryan Sim, Gordon McCall |
| EPI_ISL_2203727 | Houston Methodist Hospital | Houston Methodist Hospital | Randall J. Olsen, Paul A. Christensen, S. Wesley Long, Sishir Subedi, Robert Olson, Marcus Nguyen, James J. Davis, Matthew Ojeda Saavedra, Prasanti Yerramilli, Layne Pruitt, Kristina Reppond, Madison N. Shyer, Jessica Cambric, Ryan Gadd, Ilya J. Finkelstein, Jimmy Gollihar, and James M. Musser |
| EPI_ISL_2203728 | Quest Diagnostics Incorporated | Centers for Disease Control and Prevention Division of Viral Diseases, Pathogen Discovery | Dakota Howard, Dhwani Batra, Peter W. Cook, Kara Moser, Adrian Paskey, Jason Caravas, Benjamin Rambo-Martin, Shatavia Morrison, Christopher Gulvick, Scott Sammons, Yvette Unoarumhi, Darlene Wagner, Matthew Schmerer, S. H. Rosenthal, A. Gerasimova, R. M. Kagan, B. Anderson, M. Hua, Y. Liu, L.E. Bernstein, K.E. Livingston, A. Perez, I. A. Shlyakhter, R. V. Rolando, R. Owen, P. Tanpaiboon, F. Lacbawan, Clinton R. Paden, Duncan MacCannell |
| EPI_ISL_2203729 | Illinois Department of Public Health - Springfield Lab | Illinois Department of Public Health - Springfield Lab | Bryan Sim, Gordon McCall |
| EPI_ISL_2203730 | Houston Methodist Hospital | Houston Methodist Hospital | Randall J. Olsen, Paul A. Christensen, S. Wesley Long, Sishir Subedi, Robert Olson, Marcus Nguyen, James J. Davis, Matthew Ojeda Saavedra, Prasanti Yerramilli, Layne Pruitt, Kristina Reppond, Madison N. Shyer, Jessica Cambric, Ryan Gadd, Ilya J. Finkelstein, Jimmy Gollihar, and James M. Musser |
| EPI_ISL_2203731, EPI_ISL_2203732 | Quest Diagnostics Incorporated | Centers for Disease Control and Prevention Division of Viral Diseases, Pathogen Discovery | Dakota Howard, Dhwani Batra, Peter W. Cook, Kara Moser, Adrian Paskey, Jason Caravas, Benjamin Rambo-Martin, Shatavia Morrison, Christopher Gulvick, Scott Sammons, Yvette Unoarumhi, Darlene Wagner, Matthew Schmerer, S. H. Rosenthal, A. Gerasimova, R. M. Kagan, B. Anderson, M. Hua, Y. Liu, L.E. Bernstein, K.E. Livingston, A. Perez, I. A. Shlyakhter, R. V. Rolando, R. Owen, P. Tanpaiboon, F. Lacbawan, Clinton R. Paden, Duncan MacCannell |
| EPI_ISL_2203733 | Illinois Department of Public Health - Springfield Lab | Illinois Department of Public Health - Springfield Lab | Bryan Sim, Gordon McCall |
| EPI_ISL_2203734 | Houston Methodist Hospital | Houston Methodist Hospital | Randall J. Olsen, Paul A. Christensen, S. Wesley Long, Sishir Subedi, Robert Olson, Marcus Nguyen, James J. Davis, Matthew Ojeda Saavedra, Prasanti Yerramilli, Layne Pruitt, Kristina Reppond, Madison N. Shyer, Jessica Cambric, Ryan Gadd, Ilya J. Finkelstein, Jimmy Gollihar, and James M. Musser |
| EPI_ISL_2203735 | Quest Diagnostics Incorporated | Centers for Disease Control and Prevention Division of Viral Diseases, Pathogen Discovery | Dakota Howard, Dhwani Batra, Peter W. Cook, Kara Moser, Adrian Paskey, Jason Caravas, Benjamin Rambo-Martin, Shatavia Morrison, Christopher Gulvick, Scott Sammons, Yvette Unoarumhi, Darlene Wagner, Matthew Schmerer, S. H. Rosenthal, A. Gerasimova, R. M. Kagan, B. Anderson, M. Hua, Y. Liu, L.E. Bernstein, K.E. Livingston, A. Perez, I. A. Shlyakhter, R. V. Rolando, R. Owen, P. Tanpaiboon, F. Lacbawan, Clinton R. Paden, Duncan MacCannell |
| EPI_ISL_2203736 | Illinois Department of Public Health - Springfield Lab | Illinois Department of Public Health - Springfield Lab | Bryan Sim, Gordon McCall |
| EPI_ISL_2203737 | Houston Methodist Hospital | Houston Methodist Hospital | Randall J. Olsen, Paul A. Christensen, S. Wesley Long, Sishir Subedi, Robert Olson, Marcus Nguyen, James J. Davis, Matthew Ojeda Saavedra, Prasanti Yerramilli, Layne Pruitt, Kristina Reppond, Madison N. Shyer, Jessica Cambric, Ryan Gadd, Ilya J. Finkelstein, Jimmy Gollihar, and James M. Musser |
| EPI_ISL_2203738, EPI_ISL_2203739 | Quest Diagnostics Incorporated | Centers for Disease Control and Prevention Division of Viral Diseases, Pathogen Discovery | Dakota Howard, Dhwani Batra, Peter W. Cook, Kara Moser, Adrian Paskey, Jason Caravas, Benjamin Rambo-Martin, Shatavia Morrison, Christopher Gulvick, Scott Sammons, Yvette Unoarumhi, Darlene Wagner, Matthew Schmerer, S. H. Rosenthal, A. Gerasimova, R. M. Kagan, B. Anderson, M. Hua, Y. Liu, L.E. Bernstein, K.E. Livingston, A. Perez, I. A. Shlyakhter, R. V. Rolando, R. Owen, P. Tanpaiboon, F. Lacbawan, Clinton R. Paden, Duncan MacCannell |
| EPI_ISL_2203740 | Houston Methodist Hospital | Houston Methodist Hospital | Randall J. Olsen, Paul A. Christensen, S. Wesley Long, Sishir Subedi, Robert Olson, Marcus Nguyen, James J. Davis, Matthew Ojeda Saavedra, Prasanti Yerramilli, Layne Pruitt, Kristina Reppond, Madison N. Shyer, Jessica Cambric, Ryan Gadd, Ilya J. Finkelstein, Jimmy Gollihar, and James M. Musser |
| EPI_ISL_2203741 | Illinois Department of Public Health - Springfield Lab | Illinois Department of Public Health - Springfield Lab | Bryan Sim, Gordon McCall |
| EPI_ISL_2203742 | Quest Diagnostics Incorporated | Centers for Disease Control and Prevention Division of Viral Diseases, Pathogen Discovery | Dakota Howard, Dhwani Batra, Peter W. Cook, Kara Moser, Adrian Paskey, Jason Caravas, Benjamin Rambo-Martin, Shatavia Morrison, Christopher Gulvick, Scott Sammons, Yvette Unoarumhi, Darlene Wagner, Matthew Schmerer, S. H. Rosenthal, A. Gerasimova, R. M. Kagan, B. Anderson, M. Hua, Y. Liu, L.E. Bernstein, K.E. Livingston, A. Perez, I. A. Shlyakhter, R. V. Rolando, R. Owen, P. Tanpaiboon, F. Lacbawan, Clinton R. Paden, Duncan MacCannell |
| EPI_ISL_2203743 | Houston Methodist Hospital | Houston Methodist Hospital | Randall J. Olsen, Paul A. Christensen, S. Wesley Long, Sishir Subedi, Robert Olson, Marcus Nguyen, James J. Davis, Matthew Ojeda Saavedra, Prasanti Yerramilli, Layne Pruitt, Kristina Reppond, Madison N. Shyer, Jessica Cambric, Ryan Gadd, Ilya J. Finkelstein, Jimmy Gollihar, and James M. Musser |
| EPI_ISL_2203744 | Illinois Department of Public Health - Springfield Lab | Illinois Department of Public Health - Springfield Lab | Bryan Sim, Gordon McCall |
| EPI_ISL_2203745 | Quest Diagnostics Incorporated | Centers for Disease Control and Prevention Division of Viral Diseases, Pathogen Discovery | Dakota Howard, Dhwani Batra, Peter W. Cook, Kara Moser, Adrian Paskey, Jason Caravas, Benjamin Rambo-Martin, Shatavia Morrison, Christopher Gulvick, Scott Sammons, Yvette Unoarumhi, Darlene Wagner, Matthew Schmerer, S. H. Rosenthal, A. Gerasimova, R. M. Kagan, B. Anderson, M. Hua, Y. Liu, L.E. Bernstein, K.E. Livingston, A. Perez, I. A. Shlyakhter, R. V. Rolando, R. Owen, P. Tanpaiboon, F. Lacbawan, Clinton R. Paden, Duncan MacCannell |
| EPI_ISL_2203746 | Houston Methodist Hospital | Houston Methodist Hospital | Randall J. Olsen, Paul A. Christensen, S. Wesley Long, Sishir Subedi, Robert Olson, Marcus Nguyen, James J. Davis, Matthew Ojeda Saavedra, Prasanti Yerramilli, Layne Pruitt, Kristina Reppond, Madison N. Shyer, Jessica Cambric, Ryan Gadd, Ilya J. Finkelstein, Jimmy Gollihar, and James M. Musser |
| EPI_ISL_2203747 | Quest Diagnostics Incorporated | Centers for Disease Control and Prevention Division of Viral Diseases, Pathogen Discovery | Dakota Howard, Dhwani Batra, Peter W. Cook, Kara Moser, Adrian Paskey, Jason Caravas, Benjamin Rambo-Martin, Shatavia Morrison, Christopher Gulvick, Scott Sammons, Yvette Unoarumhi, Darlene Wagner, Matthew Schmerer, S. H. Rosenthal, A. Gerasimova, R. M. Kagan, B. Anderson, M. Hua, Y. Liu, L.E. Bernstein, K.E. Livingston, A. Perez, I. A. Shlyakhter, R. V. Rolando, R. Owen, P. Tanpaiboon, F. Lacbawan, Clinton R. Paden, Duncan MacCannell |
| EPI_ISL_2203748 | Illinois Department of Public Health - Springfield Lab | Illinois Department of Public Health - Springfield Lab | Bryan Sim, Gordon McCall |
| EPI_ISL_2203749 | Quest Diagnostics Incorporated | Centers for Disease Control and Prevention Division of Viral Diseases, Pathogen Discovery | Dakota Howard, Dhwani Batra, Peter W. Cook, Kara Moser, Adrian Paskey, Jason Caravas, Benjamin Rambo-Martin, Shatavia Morrison, Christopher Gulvick, Scott Sammons, Yvette Unoarumhi, Darlene Wagner, Matthew Schmerer, S. H. Rosenthal, A. Gerasimova, R. M. Kagan, B. Anderson, M. Hua, Y. Liu, L.E. Bernstein, K.E. Livingston, A. Perez, I. A. Shlyakhter, R. V. Rolando, R. Owen, P. Tanpaiboon, F. Lacbawan, Clinton R. Paden, Duncan MacCannell |
| EPI_ISL_2203750 | Houston Methodist Hospital | Houston Methodist Hospital | Randall J. Olsen, Paul A. Christensen, S. Wesley Long, Sishir Subedi, Robert Olson, Marcus Nguyen, James J. Davis, Matthew Ojeda Saavedra, Prasanti Yerramilli, Layne Pruitt, Kristina Reppond, Madison N. Shyer, Jessica Cambric, Ryan Gadd, Ilya J. Finkelstein, Jimmy Gollihar, and James M. Musser |
| EPI_ISL_2203751 | Quest Diagnostics Incorporated | Centers for Disease Control and Prevention Division of Viral Diseases, Pathogen | Dakota Howard, Dhwani Batra, Peter W. Cook, Kara Moser, Adrian Paskey, Jason Caravas, Benjamin Rambo-Martin, Shatavia Morrison, Christopher Gulvick, Scott Sammons, Yvette Unoarumhi, Darlene Wagner, Matthew Schmerer, S. H. Rosenthal, A. Gerasimova, R. M. Kagan, B. Anderson, M. Hua, Y. Liu, L.E. Bernstein, K.E. Livingston, A. Perez, I. A. Shlyakhter, R. V. Rolando, R. Owen, P. Tanpaiboon, F. Lacbawan, Clinton R. Paden, Duncan MacCannell |

| | | | |
|---|---|---|---|
| | | Discovery | |
| EPI_ISL_2203752 | Illinois Department of Public Health - Springfield Lab | Illinois Department of Public Health - Springfield Lab | Bryan Sim, Gordon McCall |
| EPI_ISL_2203753 | Houston Methodist Hospital | Houston Methodist Hospital | Randall J. Olsen, Paul A. Christensen, S. Wesley Long, Sishir Subedi, Robert Olson, Marcus Nguyen, James J. Davis, Matthew Ojeda Saavedra, Prasanti Yerramilli, Layne Pruitt, Kristina Reppond, Madison N. Shyer, Jessica Cambric, Ryan Gadd, Ilya J. Finkelstein, Jimmy Gollihar, and James M. Musser |
| EPI_ISL_2203754 | Quest Diagnostics Incorporated | Centers for Disease Control and Prevention Division of Viral Diseases, Pathogen Discovery | Dakota Howard, Dhwani Batra, Peter W. Cook, Kara Moser, Adrian Paskey, Jason Caravas, Benjamin Rambo-Martin, Shatavia Morrison, Christopher Gulvick, Scott Sammons, Yvette Unoarumhi, Darlene Wagner, Matthew Schmerer, S. H. Rosenthal, A. Gerasimova, R. M. Kagan, B. Anderson, M. Hua, Y. Liu, L.E. Bernstein, K.E. Livingston, A. Perez, I. A. Shlyakhter, R. V. Rolando, R. Owen, P. Tanpaiboon, F. Lacbawan, Clinton R. Paden, Duncan MacCannell |
| EPI_ISL_2203755 | Illinois Department of Public Health - Springfield Lab | Illinois Department of Public Health - Springfield Lab | Bryan Sim, Gordon McCall |
| EPI_ISL_2203756 | Quest Diagnostics Incorporated | Centers for Disease Control and Prevention Division of Viral Diseases, Pathogen Discovery | Dakota Howard, Dhwani Batra, Peter W. Cook, Kara Moser, Adrian Paskey, Jason Caravas, Benjamin Rambo-Martin, Shatavia Morrison, Christopher Gulvick, Scott Sammons, Yvette Unoarumhi, Darlene Wagner, Matthew Schmerer, S. H. Rosenthal, A. Gerasimova, R. M. Kagan, B. Anderson, M. Hua, Y. Liu, L.E. Bernstein, K.E. Livingston, A. Perez, I. A. Shlyakhter, R. V. Rolando, R. Owen, P. Tanpaiboon, F. Lacbawan, Clinton R. Paden, Duncan MacCannell |
| EPI_ISL_2203757 | Houston Methodist Hospital | Houston Methodist Hospital | Randall J. Olsen, Paul A. Christensen, S. Wesley Long, Sishir Subedi, Robert Olson, Marcus Nguyen, James J. Davis, Matthew Ojeda Saavedra, Prasanti Yerramilli, Layne Pruitt, Kristina Reppond, Madison N. Shyer, Jessica Cambric, Ryan Gadd, Ilya J. Finkelstein, Jimmy Gollihar, and James M. Musser |
| EPI_ISL_2203758 | Quest Diagnostics Incorporated | Centers for Disease Control and Prevention Division of Viral Diseases, Pathogen Discovery | Dakota Howard, Dhwani Batra, Peter W. Cook, Kara Moser, Adrian Paskey, Jason Caravas, Benjamin Rambo-Martin, Shatavia Morrison, Christopher Gulvick, Scott Sammons, Yvette Unoarumhi, Darlene Wagner, Matthew Schmerer, S. H. Rosenthal, A. Gerasimova, R. M. Kagan, B. Anderson, M. Hua, Y. Liu, L.E. Bernstein, K.E. Livingston, A. Perez, I. A. Shlyakhter, R. V. Rolando, R. Owen, P. Tanpaiboon, F. Lacbawan, Clinton R. Paden, Duncan MacCannell |
| EPI_ISL_2203759 | Illinois Department of Public Health - Springfield Lab | Illinois Department of Public Health - Springfield Lab | Bryan Sim, Gordon McCall |
| EPI_ISL_2203760 | Houston Methodist Hospital | Houston Methodist Hospital | Randall J. Olsen, Paul A. Christensen, S. Wesley Long, Sishir Subedi, Robert Olson, Marcus Nguyen, James J. Davis, Matthew Ojeda Saavedra, Prasanti Yerramilli, Layne Pruitt, Kristina Reppond, Madison N. Shyer, Jessica Cambric, Ryan Gadd, Ilya J. Finkelstein, Jimmy Gollihar, and James M. Musser |
| EPI_ISL_2203761 | Quest Diagnostics Incorporated | Centers for Disease Control and Prevention Division of Viral Diseases, Pathogen Discovery | Dakota Howard, Dhwani Batra, Peter W. Cook, Kara Moser, Adrian Paskey, Jason Caravas, Benjamin Rambo-Martin, Shatavia Morrison, Christopher Gulvick, Scott Sammons, Yvette Unoarumhi, Darlene Wagner, Matthew Schmerer, S. H. Rosenthal, A. Gerasimova, R. M. Kagan, B. Anderson, M. Hua, Y. Liu, L.E. Bernstein, K.E. Livingston, A. Perez, I. A. Shlyakhter, R. V. Rolando, R. Owen, P. Tanpaiboon, F. Lacbawan, Clinton R. Paden, Duncan MacCannell |
| EPI_ISL_2203762 | Houston Methodist Hospital | Houston Methodist Hospital | Randall J. Olsen, Paul A. Christensen, S. Wesley Long, Sishir Subedi, Robert Olson, Marcus Nguyen, James J. Davis, Matthew Ojeda Saavedra, Prasanti Yerramilli, Layne Pruitt, Kristina Reppond, Madison N. Shyer, Jessica Cambric, Ryan Gadd, Ilya J. Finkelstein, Jimmy Gollihar, and James M. Musser |
| EPI_ISL_2203763 | Illinois Department of Public Health - Springfield Lab | Illinois Department of Public Health - Springfield Lab | Bryan Sim, Gordon McCall |
| EPI_ISL_2203764 | Quest Diagnostics Incorporated | Centers for Disease Control and Prevention Division of Viral Diseases, Pathogen Discovery | Dakota Howard, Dhwani Batra, Peter W. Cook, Kara Moser, Adrian Paskey, Jason Caravas, Benjamin Rambo-Martin, Shatavia Morrison, Christopher Gulvick, Scott Sammons, Yvette Unoarumhi, Darlene Wagner, Matthew Schmerer, S. H. Rosenthal, A. Gerasimova, R. M. Kagan, B. Anderson, M. Hua, Y. Liu, L.E. Bernstein, K.E. Livingston, A. Perez, I. A. Shlyakhter, R. V. Rolando, R. Owen, P. Tanpaiboon, F. Lacbawan, Clinton R. Paden, Duncan MacCannell |
| EPI_ISL_2203765 | Houston Methodist Hospital | Houston Methodist Hospital | Randall J. Olsen, Paul A. Christensen, S. Wesley Long, Sishir Subedi, Robert Olson, Marcus Nguyen, James J. Davis, Matthew Ojeda Saavedra, Prasanti Yerramilli, Layne Pruitt, Kristina Reppond, Madison N. Shyer, Jessica Cambric, Ryan Gadd, Ilya J. Finkelstein, Jimmy Gollihar, and James M. Musser |
| EPI_ISL_2203766, EPI_ISL_2203767 | Quest Diagnostics Incorporated | Centers for Disease Control and Prevention Division of Viral Diseases, Pathogen Discovery | Dakota Howard, Dhwani Batra, Peter W. Cook, Kara Moser, Adrian Paskey, Jason Caravas, Benjamin Rambo-Martin, Shatavia Morrison, Christopher Gulvick, Scott Sammons, Yvette Unoarumhi, Darlene Wagner, Matthew Schmerer, S. H. Rosenthal, A. Gerasimova, R. M. Kagan, B. Anderson, M. Hua, Y. Liu, L.E. Bernstein, K.E. Livingston, A. Perez, I. A. Shlyakhter, R. V. Rolando, R. Owen, P. Tanpaiboon, F. Lacbawan, Clinton R. Paden, Duncan MacCannell |
| EPI_ISL_2203768 | Houston Methodist Hospital | Houston Methodist Hospital | Randall J. Olsen, Paul A. Christensen, S. Wesley Long, Sishir Subedi, Robert Olson, Marcus Nguyen, James J. Davis, Matthew Ojeda Saavedra, Prasanti Yerramilli, Layne Pruitt, Kristina Reppond, Madison N. Shyer, Jessica Cambric, Ryan Gadd, Ilya J. Finkelstein, Jimmy Gollihar, and James M. Musser |
| EPI_ISL_2203769 | Quest Diagnostics Incorporated | Centers for Disease Control and Prevention Division of Viral Diseases, Pathogen Discovery | Dakota Howard, Dhwani Batra, Peter W. Cook, Kara Moser, Adrian Paskey, Jason Caravas, Benjamin Rambo-Martin, Shatavia Morrison, Christopher Gulvick, Scott Sammons, Yvette Unoarumhi, Darlene Wagner, Matthew Schmerer, S. H. Rosenthal, A. Gerasimova, R. M. Kagan, B. Anderson, M. Hua, Y. Liu, L.E. Bernstein, K.E. Livingston, A. Perez, I. A. Shlyakhter, R. V. Rolando, R. Owen, P. Tanpaiboon, F. Lacbawan, Clinton R. Paden, Duncan MacCannell |
| EPI_ISL_2203770, EPI_ISL_2203771 | Houston Methodist Hospital | Houston Methodist Hospital | Randall J. Olsen, Paul A. Christensen, S. Wesley Long, Sishir Subedi, Robert Olson, Marcus Nguyen, James J. Davis, Matthew Ojeda Saavedra, Prasanti Yerramilli, Layne Pruitt, Kristina Reppond, Madison N. Shyer, Jessica Cambric, Ryan Gadd, Ilya J. Finkelstein, Jimmy Gollihar, and James M. Musser |
| EPI_ISL_2203772, EPI_ISL_2203773 | Quest Diagnostics Incorporated | Centers for Disease Control and Prevention Division of Viral Diseases, Pathogen Discovery | Dakota Howard, Dhwani Batra, Peter W. Cook, Kara Moser, Adrian Paskey, Jason Caravas, Benjamin Rambo-Martin, Shatavia Morrison, Christopher Gulvick, Scott Sammons, Yvette Unoarumhi, Darlene Wagner, Matthew Schmerer, S. H. Rosenthal, A. Gerasimova, R. M. Kagan, B. Anderson, M. Hua, Y. Liu, L.E. Bernstein, K.E. Livingston, A. Perez, I. A. Shlyakhter, R. V. Rolando, R. Owen, P. Tanpaiboon, F. Lacbawan, Clinton R. Paden, Duncan MacCannell |
| EPI_ISL_2203774 | Houston Methodist Hospital | Houston Methodist Hospital | Randall J. Olsen, Paul A. Christensen, S. Wesley Long, Sishir Subedi, Robert Olson, Marcus Nguyen, James J. Davis, Matthew Ojeda Saavedra, Prasanti Yerramilli, Layne Pruitt, Kristina Reppond, Madison N. Shyer, Jessica Cambric, Ryan Gadd, Ilya J. Finkelstein, Jimmy Gollihar, and James M. Musser |
| EPI_ISL_2203775 | Quest Diagnostics Incorporated | Centers for Disease Control and Prevention Division of Viral Diseases, Pathogen Discovery | Dakota Howard, Dhwani Batra, Peter W. Cook, Kara Moser, Adrian Paskey, Jason Caravas, Benjamin Rambo-Martin, Shatavia Morrison, Christopher Gulvick, Scott Sammons, Yvette Unoarumhi, Darlene Wagner, Matthew Schmerer, S. H. Rosenthal, A. Gerasimova, R. M. Kagan, B. Anderson, M. Hua, Y. Liu, L.E. Bernstein, K.E. Livingston, A. Perez, I. A. Shlyakhter, R. V. Rolando, R. Owen, P. Tanpaiboon, F. Lacbawan, Clinton R. Paden, Duncan MacCannell |
| EPI_ISL_2203776 | Houston Methodist Hospital | Houston Methodist Hospital | Randall J. Olsen, Paul A. Christensen, S. Wesley Long, Sishir Subedi, Robert Olson, Marcus Nguyen, James J. Davis, Matthew Ojeda Saavedra, Prasanti Yerramilli, Layne Pruitt, Kristina Reppond, Madison N. Shyer, Jessica Cambric, Ryan Gadd, Ilya J. Finkelstein, Jimmy Gollihar, and James M. Musser |
| EPI_ISL_2203777 | Quest Diagnostics Incorporated | Centers for Disease Control and Prevention Division of Viral Diseases, Pathogen Discovery | Dakota Howard, Dhwani Batra, Peter W. Cook, Kara Moser, Adrian Paskey, Jason Caravas, Benjamin Rambo-Martin, Shatavia Morrison, Christopher Gulvick, Scott Sammons, Yvette Unoarumhi, Darlene Wagner, Matthew Schmerer, S. H. Rosenthal, A. Gerasimova, R. M. Kagan, B. Anderson, M. Hua, Y. Liu, L.E. Bernstein, K.E. Livingston, A. Perez, I. A. Shlyakhter, R. V. Rolando, R. Owen, P. Tanpaiboon, F. Lacbawan, Clinton R. Paden, Duncan MacCannell |
| EPI_ISL_2203778 | Houston Methodist Hospital | Houston Methodist Hospital | Randall J. Olsen, Paul A. Christensen, S. Wesley Long, Sishir Subedi, Robert Olson, Marcus Nguyen, James J. Davis, Matthew Ojeda Saavedra, Prasanti Yerramilli, Layne Pruitt, Kristina Reppond, Madison N. Shyer, Jessica Cambric, Ryan Gadd, Ilya J. Finkelstein, Jimmy Gollihar, and James M. Musser |
| EPI_ISL_2203779 | Quest Diagnostics Incorporated | Centers for Disease Control and Prevention Division of Viral Diseases, Pathogen Discovery | Dakota Howard, Dhwani Batra, Peter W. Cook, Kara Moser, Adrian Paskey, Jason Caravas, Benjamin Rambo-Martin, Shatavia Morrison, Christopher Gulvick, Scott Sammons, Yvette Unoarumhi, Darlene Wagner, Matthew Schmerer, S. H. Rosenthal, A. Gerasimova, R. M. Kagan, B. Anderson, M. Hua, Y. Liu, L.E. Bernstein, K.E. Livingston, A. Perez, I. A. Shlyakhter, R. V. Rolando, R. Owen, P. Tanpaiboon, F. Lacbawan, Clinton R. Paden, Duncan MacCannell |
| EPI_ISL_2203780 | Houston Methodist Hospital | Houston Methodist Hospital | Randall J. Olsen, Paul A. Christensen, S. Wesley Long, Sishir Subedi, Robert Olson, Marcus Nguyen, James J. Davis, Matthew Ojeda Saavedra, Prasanti Yerramilli, Layne Pruitt, Kristina Reppond, Madison N. Shyer, Jessica Cambric, Ryan Gadd, Ilya J. Finkelstein, Jimmy Gollihar, and James M. Musser |
| EPI_ISL_2203781 | Quest Diagnostics Incorporated | Centers for Disease Control and Prevention Division of Viral Diseases, Pathogen Discovery | Dakota Howard, Dhwani Batra, Peter W. Cook, Kara Moser, Adrian Paskey, Jason Caravas, Benjamin Rambo-Martin, Shatavia Morrison, Christopher Gulvick, Scott Sammons, Yvette Unoarumhi, Darlene Wagner, Matthew Schmerer, S. H. Rosenthal, A. Gerasimova, R. M. Kagan, B. Anderson, M. Hua, Y. Liu, L.E. Bernstein, K.E. Livingston, A. Perez, I. A. Shlyakhter, R. V. Rolando, R. Owen, P. Tanpaiboon, F. Lacbawan, Clinton R. Paden, Duncan MacCannell |
| EPI_ISL_2203782 | Houston Methodist Hospital | Houston Methodist Hospital | Randall J. Olsen, Paul A. Christensen, S. Wesley Long, Sishir Subedi, Robert Olson, Marcus Nguyen, James J. Davis, Matthew Ojeda Saavedra, Prasanti Yerramilli, Layne Pruitt, Kristina Reppond, Madison N. Shyer, Jessica Cambric, Ryan Gadd, Ilya J. Finkelstein, Jimmy Gollihar, and James M. Musser |
| EPI_ISL_2203783 | Quest Diagnostics Incorporated | Centers for Disease Control and Prevention Division of Viral Diseases, Pathogen Discovery | Dakota Howard, Dhwani Batra, Peter W. Cook, Kara Moser, Adrian Paskey, Jason Caravas, Benjamin Rambo-Martin, Shatavia Morrison, Christopher Gulvick, Scott Sammons, Yvette Unoarumhi, Darlene Wagner, Matthew Schmerer, S. H. Rosenthal, A. Gerasimova, R. M. Kagan, B. Anderson, M. Hua, Y. Liu, L.E. Bernstein, K.E. Livingston, A. Perez, I. A. Shlyakhter, R. V. Rolando, R. Owen, P. Tanpaiboon, F. Lacbawan, Clinton R. Paden, Duncan MacCannell |
| EPI_ISL_2203784 | Houston Methodist Hospital | Houston Methodist Hospital | Randall J. Olsen, Paul A. Christensen, S. Wesley Long, Sishir Subedi, Robert Olson, Marcus Nguyen, James J. Davis, Matthew Ojeda Saavedra, Prasanti Yerramilli, Layne Pruitt, Kristina Reppond, Madison N. Shyer, Jessica Cambric, Ryan Gadd, Ilya J. Finkelstein, Jimmy Gollihar, and James M. Musser |
| EPI_ISL_2203785, EPI_ISL_2203786 | Quest Diagnostics Incorporated | Centers for Disease Control and Prevention Division of Viral Diseases, Pathogen Discovery | Dakota Howard, Dhwani Batra, Peter W. Cook, Kara Moser, Adrian Paskey, Jason Caravas, Benjamin Rambo-Martin, Shatavia Morrison, Christopher Gulvick, Scott Sammons, Yvette Unoarumhi, Darlene Wagner, Matthew Schmerer, S. H. Rosenthal, A. Gerasimova, R. M. Kagan, B. Anderson, M. Hua, Y. Liu, L.E. Bernstein, K.E. Livingston, A. Perez, I. A. Shlyakhter, R. V. Rolando, R. Owen, P. Tanpaiboon, F. Lacbawan, Clinton R. Paden, Duncan MacCannell |
| EPI_ISL_2203787 | Houston Methodist Hospital | Houston Methodist Hospital | Randall J. Olsen, Paul A. Christensen, S. Wesley Long, Sishir Subedi, Robert Olson, Marcus Nguyen, James J. Davis, Matthew Ojeda Saavedra, Prasanti Yerramilli, Layne Pruitt, Kristina Reppond, Madison N. Shyer, Jessica Cambric, Ryan Gadd, Ilya J. Finkelstein, Jimmy Gollihar, and James M. Musser |
| EPI_ISL_2203788 | Quest Diagnostics Incorporated | Centers for Disease Control and Prevention Division of Viral Diseases, Pathogen Discovery | Dakota Howard, Dhwani Batra, Peter W. Cook, Kara Moser, Adrian Paskey, Jason Caravas, Benjamin Rambo-Martin, Shatavia Morrison, Christopher Gulvick, Scott Sammons, Yvette Unoarumhi, Darlene Wagner, Matthew Schmerer, S. H. Rosenthal, A. Gerasimova, R. M. Kagan, B. Anderson, M. Hua, Y. Liu, L.E. Bernstein, K.E. Livingston, A. Perez, I. A. Shlyakhter, R. V. Rolando, R. Owen, P. Tanpaiboon, F. Lacbawan, Clinton R. Paden, Duncan MacCannell |
| EPI_ISL_2203789 | Houston Methodist Hospital | Houston Methodist Hospital | Randall J. Olsen, Paul A. Christensen, S. Wesley Long, Sishir Subedi, Robert Olson, Marcus Nguyen, James J. Davis, Matthew Ojeda Saavedra, Prasanti Yerramilli, Layne Pruitt, Kristina Reppond, Madison N. Shyer, Jessica Cambric, Ryan Gadd, Ilya J. Finkelstein, Jimmy Gollihar, and James M. Musser |
| EPI_ISL_2203790 | Quest Diagnostics Incorporated | Centers for Disease Control and Prevention Division of Viral Diseases, Pathogen Discovery | Dakota Howard, Dhwani Batra, Peter W. Cook, Kara Moser, Adrian Paskey, Jason Caravas, Benjamin Rambo-Martin, Shatavia Morrison, Christopher Gulvick, Scott Sammons, Yvette Unoarumhi, Darlene Wagner, Matthew Schmerer, S. H. Rosenthal, A. Gerasimova, R. M. Kagan, B. Anderson, M. Hua, Y. Liu, L.E. Bernstein, K.E. Livingston, A. Perez, I. A. Shlyakhter, R. V. Rolando, R. Owen, P. Tanpaiboon, F. Lacbawan, Clinton R. Paden, Duncan MacCannell |
| EPI_ISL_2203791 | Houston Methodist Hospital | Houston Methodist Hospital | Randall J. Olsen, Paul A. Christensen, S. Wesley Long, Sishir Subedi, Robert Olson, Marcus Nguyen, James J. Davis, Matthew Ojeda Saavedra, Prasanti Yerramilli, Layne Pruitt, Kristina Reppond, Madison N. Shyer, Jessica Cambric, Ryan Gadd, Ilya J. Finkelstein, Jimmy Gollihar, and James M. Musser |
| EPI_ISL_2203792 | Quest Diagnostics Incorporated | Centers for Disease Control and Prevention Division of Viral Diseases, Pathogen Discovery | Dakota Howard, Dhwani Batra, Peter W. Cook, Kara Moser, Adrian Paskey, Jason Caravas, Benjamin Rambo-Martin, Shatavia Morrison, Christopher Gulvick, Scott Sammons, Yvette Unoarumhi, Darlene Wagner, Matthew Schmerer, S. H. Rosenthal, A. Gerasimova, R. M. Kagan, B. Anderson, M. Hua, Y. Liu, L.E. Bernstein, K.E. Livingston, A. Perez, I. A. Shlyakhter, R. V. Rolando, R. Owen, P. Tanpaiboon, F. Lacbawan, Clinton R. Paden, Duncan MacCannell |
| EPI_ISL_2203793 | Houston Methodist Hospital | Houston Methodist Hospital | Randall J. Olsen, Paul A. Christensen, S. Wesley Long, Sishir Subedi, Robert Olson, Marcus Nguyen, James J. Davis, Matthew Ojeda Saavedra, Prasanti Yerramilli, Layne Pruitt, Kristina Reppond, Madison N. Shyer, Jessica Cambric, Ryan Gadd, Ilya J. Finkelstein, Jimmy Gollihar, and James |

| | | | |
|---|---|---|---|
| | | | M. Musser |
| EPI_ISL_2203794 | Quest Diagnostics Incorporated | Centers for Disease Control and Prevention Division of Viral Diseases, Pathogen Discovery | Dakota Howard, Dhwani Batra, Peter W. Cook, Kara Moser, Adrian Paskey, Jason Caravas, Benjamin Rambo-Martin, Shatavia Morrison, Christopher Gulvick, Scott Sammons, Yvette Unoarumhi, Darlene Wagner, Matthew Schmerer, S. H. Rosenthal, A. Gerasimova, R. M. Kagan, B. Anderson, M. Hua, Y. Liu, L.E. Bernstein, K.E. Livingston, A. Perez, I. A. Shlyakhter, R. V. Rolando, R. Owen, P. Tanpaiboon, F. Lacbawan, Clinton R. Paden, Duncan MacCannell |
| EPI_ISL_2203795 | Houston Methodist Hospital | Houston Methodist Hospital | Randall J. Olsen, Paul A. Christensen, S. Wesley Long, Sishir Subedi, Robert Olson, Marcus Nguyen, James J. Davis, Matthew Ojeda Saavedra, Prasanti Yerramilli, Layne Pruitt, Kristina Reppond, Madison N. Shyer, Jessica Cambric, Ryan Gadd, Ilya J. Finkelstein, Jimmy Gollihar, and James M. Musser |
| EPI_ISL_2203796 | Quest Diagnostics Incorporated | Centers for Disease Control and Prevention Division of Viral Diseases, Pathogen Discovery | Dakota Howard, Dhwani Batra, Peter W. Cook, Kara Moser, Adrian Paskey, Jason Caravas, Benjamin Rambo-Martin, Shatavia Morrison, Christopher Gulvick, Scott Sammons, Yvette Unoarumhi, Darlene Wagner, Matthew Schmerer, S. H. Rosenthal, A. Gerasimova, R. M. Kagan, B. Anderson, M. Hua, Y. Liu, L.E. Bernstein, K.E. Livingston, A. Perez, I. A. Shlyakhter, R. V. Rolando, R. Owen, P. Tanpaiboon, F. Lacbawan, Clinton R. Paden, Duncan MacCannell |
| EPI_ISL_2203797 | Houston Methodist Hospital | Houston Methodist Hospital | Randall J. Olsen, Paul A. Christensen, S. Wesley Long, Sishir Subedi, Robert Olson, Marcus Nguyen, James J. Davis, Matthew Ojeda Saavedra, Prasanti Yerramilli, Layne Pruitt, Kristina Reppond, Madison N. Shyer, Jessica Cambric, Ryan Gadd, Ilya J. Finkelstein, Jimmy Gollihar, and James M. Musser |
| EPI_ISL_2203798, EPI_ISL_2203799 | Quest Diagnostics Incorporated | Centers for Disease Control and Prevention Division of Viral Diseases, Pathogen Discovery | Dakota Howard, Dhwani Batra, Peter W. Cook, Kara Moser, Adrian Paskey, Jason Caravas, Benjamin Rambo-Martin, Shatavia Morrison, Christopher Gulvick, Scott Sammons, Yvette Unoarumhi, Darlene Wagner, Matthew Schmerer, S. H. Rosenthal, A. Gerasimova, R. M. Kagan, B. Anderson, M. Hua, Y. Liu, L.E. Bernstein, K.E. Livingston, A. Perez, I. A. Shlyakhter, R. V. Rolando, R. Owen, P. Tanpaiboon, F. Lacbawan, Clinton R. Paden, Duncan MacCannell |
| EPI_ISL_2203800 | Houston Methodist Hospital | Houston Methodist Hospital | Randall J. Olsen, Paul A. Christensen, S. Wesley Long, Sishir Subedi, Robert Olson, Marcus Nguyen, James J. Davis, Matthew Ojeda Saavedra, Prasanti Yerramilli, Layne Pruitt, Kristina Reppond, Madison N. Shyer, Jessica Cambric, Ryan Gadd, Ilya J. Finkelstein, Jimmy Gollihar, and James M. Musser |
| EPI_ISL_2203801 | Quest Diagnostics Incorporated | Centers for Disease Control and Prevention Division of Viral Diseases, Pathogen Discovery | Dakota Howard, Dhwani Batra, Peter W. Cook, Kara Moser, Adrian Paskey, Jason Caravas, Benjamin Rambo-Martin, Shatavia Morrison, Christopher Gulvick, Scott Sammons, Yvette Unoarumhi, Darlene Wagner, Matthew Schmerer, S. H. Rosenthal, A. Gerasimova, R. M. Kagan, B. Anderson, M. Hua, Y. Liu, L.E. Bernstein, K.E. Livingston, A. Perez, I. A. Shlyakhter, R. V. Rolando, R. Owen, P. Tanpaiboon, F. Lacbawan, Clinton R. Paden, Duncan MacCannell |
| EPI_ISL_2203802 | Houston Methodist Hospital | Houston Methodist Hospital | Randall J. Olsen, Paul A. Christensen, S. Wesley Long, Sishir Subedi, Robert Olson, Marcus Nguyen, James J. Davis, Matthew Ojeda Saavedra, Prasanti Yerramilli, Layne Pruitt, Kristina Reppond, Madison N. Shyer, Jessica Cambric, Ryan Gadd, Ilya J. Finkelstein, Jimmy Gollihar, and James M. Musser |
| EPI_ISL_2203803 | Quest Diagnostics Incorporated | Centers for Disease Control and Prevention Division of Viral Diseases, Pathogen Discovery | Dakota Howard, Dhwani Batra, Peter W. Cook, Kara Moser, Adrian Paskey, Jason Caravas, Benjamin Rambo-Martin, Shatavia Morrison, Christopher Gulvick, Scott Sammons, Yvette Unoarumhi, Darlene Wagner, Matthew Schmerer, S. H. Rosenthal, A. Gerasimova, R. M. Kagan, B. Anderson, M. Hua, Y. Liu, L.E. Bernstein, K.E. Livingston, A. Perez, I. A. Shlyakhter, R. V. Rolando, R. Owen, P. Tanpaiboon, F. Lacbawan, Clinton R. Paden, Duncan MacCannell |
| EPI_ISL_2203804 | Houston Methodist Hospital | Houston Methodist Hospital | Randall J. Olsen, Paul A. Christensen, S. Wesley Long, Sishir Subedi, Robert Olson, Marcus Nguyen, James J. Davis, Matthew Ojeda Saavedra, Prasanti Yerramilli, Layne Pruitt, Kristina Reppond, Madison N. Shyer, Jessica Cambric, Ryan Gadd, Ilya J. Finkelstein, Jimmy Gollihar, and James M. Musser |
| EPI_ISL_2203805 | Quest Diagnostics Incorporated | Centers for Disease Control and Prevention Division of Viral Diseases, Pathogen Discovery | Dakota Howard, Dhwani Batra, Peter W. Cook, Kara Moser, Adrian Paskey, Jason Caravas, Benjamin Rambo-Martin, Shatavia Morrison, Christopher Gulvick, Scott Sammons, Yvette Unoarumhi, Darlene Wagner, Matthew Schmerer, S. H. Rosenthal, A. Gerasimova, R. M. Kagan, B. Anderson, M. Hua, Y. Liu, L.E. Bernstein, K.E. Livingston, A. Perez, I. A. Shlyakhter, R. V. Rolando, R. Owen, P. Tanpaiboon, F. Lacbawan, Clinton R. Paden, Duncan MacCannell |
| EPI_ISL_2203806 | Houston Methodist Hospital | Houston Methodist Hospital | Randall J. Olsen, Paul A. Christensen, S. Wesley Long, Sishir Subedi, Robert Olson, Marcus Nguyen, James J. Davis, Matthew Ojeda Saavedra, Prasanti Yerramilli, Layne Pruitt, Kristina Reppond, Madison N. Shyer, Jessica Cambric, Ryan Gadd, Ilya J. Finkelstein, Jimmy Gollihar, and James M. Musser |
| EPI_ISL_2203807, EPI_ISL_2203808 | Quest Diagnostics Incorporated | Centers for Disease Control and Prevention Division of Viral Diseases, Pathogen Discovery | Dakota Howard, Dhwani Batra, Peter W. Cook, Kara Moser, Adrian Paskey, Jason Caravas, Benjamin Rambo-Martin, Shatavia Morrison, Christopher Gulvick, Scott Sammons, Yvette Unoarumhi, Darlene Wagner, Matthew Schmerer, S. H. Rosenthal, A. Gerasimova, R. M. Kagan, B. Anderson, M. Hua, Y. Liu, L.E. Bernstein, K.E. Livingston, A. Perez, I. A. Shlyakhter, R. V. Rolando, R. Owen, P. Tanpaiboon, F. Lacbawan, Clinton R. Paden, Duncan MacCannell |
| EPI_ISL_2203809 | Houston Methodist Hospital | Houston Methodist Hospital | Randall J. Olsen, Paul A. Christensen, S. Wesley Long, Sishir Subedi, Robert Olson, Marcus Nguyen, James J. Davis, Matthew Ojeda Saavedra, Prasanti Yerramilli, Layne Pruitt, Kristina Reppond, Madison N. Shyer, Jessica Cambric, Ryan Gadd, Ilya J. Finkelstein, Jimmy Gollihar, and James M. Musser |
| EPI_ISL_2203810 | Quest Diagnostics Incorporated | Centers for Disease Control and Prevention Division of Viral Diseases, Pathogen Discovery | Dakota Howard, Dhwani Batra, Peter W. Cook, Kara Moser, Adrian Paskey, Jason Caravas, Benjamin Rambo-Martin, Shatavia Morrison, Christopher Gulvick, Scott Sammons, Yvette Unoarumhi, Darlene Wagner, Matthew Schmerer, S. H. Rosenthal, A. Gerasimova, R. M. Kagan, B. Anderson, M. Hua, Y. Liu, L.E. Bernstein, K.E. Livingston, A. Perez, I. A. Shlyakhter, R. V. Rolando, R. Owen, P. Tanpaiboon, F. Lacbawan, Clinton R. Paden, Duncan MacCannell |
| EPI_ISL_2203811 | Houston Methodist Hospital | Houston Methodist Hospital | Randall J. Olsen, Paul A. Christensen, S. Wesley Long, Sishir Subedi, Robert Olson, Marcus Nguyen, James J. Davis, Matthew Ojeda Saavedra, Prasanti Yerramilli, Layne Pruitt, Kristina Reppond, Madison N. Shyer, Jessica Cambric, Ryan Gadd, Ilya J. Finkelstein, Jimmy Gollihar, and James M. Musser |
| EPI_ISL_2203812 | Quest Diagnostics Incorporated | Centers for Disease Control and Prevention Division of Viral Diseases, Pathogen Discovery | Dakota Howard, Dhwani Batra, Peter W. Cook, Kara Moser, Adrian Paskey, Jason Caravas, Benjamin Rambo-Martin, Shatavia Morrison, Christopher Gulvick, Scott Sammons, Yvette Unoarumhi, Darlene Wagner, Matthew Schmerer, S. H. Rosenthal, A. Gerasimova, R. M. Kagan, B. Anderson, M. Hua, Y. Liu, L.E. Bernstein, K.E. Livingston, A. Perez, I. A. Shlyakhter, R. V. Rolando, R. Owen, P. Tanpaiboon, F. Lacbawan, Clinton R. Paden, Duncan MacCannell |
| EPI_ISL_2203813 | Houston Methodist Hospital | Houston Methodist Hospital | Randall J. Olsen, Paul A. Christensen, S. Wesley Long, Sishir Subedi, Robert Olson, Marcus Nguyen, James J. Davis, Matthew Ojeda Saavedra, Prasanti Yerramilli, Layne Pruitt, Kristina Reppond, Madison N. Shyer, Jessica Cambric, Ryan Gadd, Ilya J. Finkelstein, Jimmy Gollihar, and James M. Musser |
| EPI_ISL_2203814 | Quest Diagnostics Incorporated | Centers for Disease Control and Prevention Division of Viral Diseases, Pathogen Discovery | Dakota Howard, Dhwani Batra, Peter W. Cook, Kara Moser, Adrian Paskey, Jason Caravas, Benjamin Rambo-Martin, Shatavia Morrison, Christopher Gulvick, Scott Sammons, Yvette Unoarumhi, Darlene Wagner, Matthew Schmerer, S. H. Rosenthal, A. Gerasimova, R. M. Kagan, B. Anderson, M. Hua, Y. Liu, L.E. Bernstein, K.E. Livingston, A. Perez, I. A. Shlyakhter, R. V. Rolando, R. Owen, P. Tanpaiboon, F. Lacbawan, Clinton R. Paden, Duncan MacCannell |
| EPI_ISL_2203815 | Houston Methodist Hospital | Houston Methodist Hospital | Randall J. Olsen, Paul A. Christensen, S. Wesley Long, Sishir Subedi, Robert Olson, Marcus Nguyen, James J. Davis, Matthew Ojeda Saavedra, Prasanti Yerramilli, Layne Pruitt, Kristina Reppond, Madison N. Shyer, Jessica Cambric, Ryan Gadd, Ilya J. Finkelstein, Jimmy Gollihar, and James M. Musser |
| EPI_ISL_2203816, EPI_ISL_2203817 | Quest Diagnostics Incorporated | Centers for Disease Control and Prevention Division of Viral Diseases, Pathogen Discovery | Dakota Howard, Dhwani Batra, Peter W. Cook, Kara Moser, Adrian Paskey, Jason Caravas, Benjamin Rambo-Martin, Shatavia Morrison, Christopher Gulvick, Scott Sammons, Yvette Unoarumhi, Darlene Wagner, Matthew Schmerer, S. H. Rosenthal, A. Gerasimova, R. M. Kagan, B. Anderson, M. Hua, Y. Liu, L.E. Bernstein, K.E. Livingston, A. Perez, I. A. Shlyakhter, R. V. Rolando, R. Owen, P. Tanpaiboon, F. Lacbawan, Clinton R. Paden, Duncan MacCannell |
| EPI_ISL_2203818 | Houston Methodist Hospital | Houston Methodist Hospital | Randall J. Olsen, Paul A. Christensen, S. Wesley Long, Sishir Subedi, Robert Olson, Marcus Nguyen, James J. Davis, Matthew Ojeda Saavedra, Prasanti Yerramilli, Layne Pruitt, Kristina Reppond, Madison N. Shyer, Jessica Cambric, Ryan Gadd, Ilya J. Finkelstein, Jimmy Gollihar, and James M. Musser |
| EPI_ISL_2203819 | Quest Diagnostics Incorporated | Centers for Disease Control and Prevention Division of Viral Diseases, Pathogen Discovery | Dakota Howard, Dhwani Batra, Peter W. Cook, Kara Moser, Adrian Paskey, Jason Caravas, Benjamin Rambo-Martin, Shatavia Morrison, Christopher Gulvick, Scott Sammons, Yvette Unoarumhi, Darlene Wagner, Matthew Schmerer, S. H. Rosenthal, A. Gerasimova, R. M. Kagan, B. Anderson, M. Hua, Y. Liu, L.E. Bernstein, K.E. Livingston, A. Perez, I. A. Shlyakhter, R. V. Rolando, R. Owen, P. Tanpaiboon, F. Lacbawan, Clinton R. Paden, Duncan MacCannell |
| EPI_ISL_2203820 | Houston Methodist Hospital | Houston Methodist Hospital | Randall J. Olsen, Paul A. Christensen, S. Wesley Long, Sishir Subedi, Robert Olson, Marcus Nguyen, James J. Davis, Matthew Ojeda Saavedra, Prasanti Yerramilli, Layne Pruitt, Kristina Reppond, Madison N. Shyer, Jessica Cambric, Ryan Gadd, Ilya J. Finkelstein, Jimmy Gollihar, and James M. Musser |
| EPI_ISL_2203821, EPI_ISL_2203822 | Quest Diagnostics Incorporated | Centers for Disease Control and Prevention Division of Viral Diseases, Pathogen Discovery | Dakota Howard, Dhwani Batra, Peter W. Cook, Kara Moser, Adrian Paskey, Jason Caravas, Benjamin Rambo-Martin, Shatavia Morrison, Christopher Gulvick, Scott Sammons, Yvette Unoarumhi, Darlene Wagner, Matthew Schmerer, S. H. Rosenthal, A. Gerasimova, R. M. Kagan, B. Anderson, M. Hua, Y. Liu, L.E. Bernstein, K.E. Livingston, A. Perez, I. A. Shlyakhter, R. V. Rolando, R. Owen, P. Tanpaiboon, F. Lacbawan, Clinton R. Paden, Duncan MacCannell |
| EPI_ISL_2203823 | Houston Methodist Hospital | Houston Methodist Hospital | Randall J. Olsen, Paul A. Christensen, S. Wesley Long, Sishir Subedi, Robert Olson, Marcus Nguyen, James J. Davis, Matthew Ojeda Saavedra, Prasanti Yerramilli, Layne Pruitt, Kristina Reppond, Madison N. Shyer, Jessica Cambric, Ryan Gadd, Ilya J. Finkelstein, Jimmy Gollihar, and James M. Musser |
| EPI_ISL_2203824 | Quest Diagnostics Incorporated | Centers for Disease Control and Prevention Division of Viral Diseases, Pathogen Discovery | Dakota Howard, Dhwani Batra, Peter W. Cook, Kara Moser, Adrian Paskey, Jason Caravas, Benjamin Rambo-Martin, Shatavia Morrison, Christopher Gulvick, Scott Sammons, Yvette Unoarumhi, Darlene Wagner, Matthew Schmerer, S. H. Rosenthal, A. Gerasimova, R. M. Kagan, B. Anderson, M. Hua, Y. Liu, L.E. Bernstein, K.E. Livingston, A. Perez, I. A. Shlyakhter, R. V. Rolando, R. Owen, P. Tanpaiboon, F. Lacbawan, Clinton R. Paden, Duncan MacCannell |
| EPI_ISL_2203825 | Houston Methodist Hospital | Houston Methodist Hospital | Randall J. Olsen, Paul A. Christensen, S. Wesley Long, Sishir Subedi, Robert Olson, Marcus Nguyen, James J. Davis, Matthew Ojeda Saavedra, Prasanti Yerramilli, Layne Pruitt, Kristina Reppond, Madison N. Shyer, Jessica Cambric, Ryan Gadd, Ilya J. Finkelstein, Jimmy Gollihar, and James M. Musser |
| EPI_ISL_2203826 | Quest Diagnostics Incorporated | Centers for Disease Control and Prevention Division of Viral Diseases, Pathogen Discovery | Dakota Howard, Dhwani Batra, Peter W. Cook, Kara Moser, Adrian Paskey, Jason Caravas, Benjamin Rambo-Martin, Shatavia Morrison, Christopher Gulvick, Scott Sammons, Yvette Unoarumhi, Darlene Wagner, Matthew Schmerer, S. H. Rosenthal, A. Gerasimova, R. M. Kagan, B. Anderson, M. Hua, Y. Liu, L.E. Bernstein, K.E. Livingston, A. Perez, I. A. Shlyakhter, R. V. Rolando, R. Owen, P. Tanpaiboon, F. Lacbawan, Clinton R. Paden, Duncan MacCannell |
| EPI_ISL_2203827 | Houston Methodist Hospital | Houston Methodist Hospital | Randall J. Olsen, Paul A. Christensen, S. Wesley Long, Sishir Subedi, Robert Olson, Marcus Nguyen, James J. Davis, Matthew Ojeda Saavedra, Prasanti Yerramilli, Layne Pruitt, Kristina Reppond, Madison N. Shyer, Jessica Cambric, Ryan Gadd, Ilya J. Finkelstein, Jimmy Gollihar, and James M. Musser |
| EPI_ISL_2203828 | Quest Diagnostics Incorporated | Centers for Disease Control and Prevention Division of Viral Diseases, Pathogen Discovery | Dakota Howard, Dhwani Batra, Peter W. Cook, Kara Moser, Adrian Paskey, Jason Caravas, Benjamin Rambo-Martin, Shatavia Morrison, Christopher Gulvick, Scott Sammons, Yvette Unoarumhi, Darlene Wagner, Matthew Schmerer, S. H. Rosenthal, A. Gerasimova, R. M. Kagan, B. Anderson, M. Hua, Y. Liu, L.E. Bernstein, K.E. Livingston, A. Perez, I. A. Shlyakhter, R. V. Rolando, R. Owen, P. Tanpaiboon, F. Lacbawan, Clinton R. Paden, Duncan MacCannell |
| EPI_ISL_2203829 | Houston Methodist Hospital | Houston Methodist Hospital | Randall J. Olsen, Paul A. Christensen, S. Wesley Long, Sishir Subedi, Robert Olson, Marcus Nguyen, James J. Davis, Matthew Ojeda Saavedra, Prasanti Yerramilli, Layne Pruitt, Kristina Reppond, Madison N. Shyer, Jessica Cambric, Ryan Gadd, Ilya J. Finkelstein, Jimmy Gollihar, and James M. Musser |
| EPI_ISL_2203830, EPI_ISL_2203831 | Quest Diagnostics Incorporated | Centers for Disease Control and Prevention Division of Viral Diseases, Pathogen Discovery | Dakota Howard, Dhwani Batra, Peter W. Cook, Kara Moser, Adrian Paskey, Jason Caravas, Benjamin Rambo-Martin, Shatavia Morrison, Christopher Gulvick, Scott Sammons, Yvette Unoarumhi, Darlene Wagner, Matthew Schmerer, S. H. Rosenthal, A. Gerasimova, R. M. Kagan, B. Anderson, M. Hua, Y. Liu, L.E. Bernstein, K.E. Livingston, A. Perez, I. A. Shlyakhter, R. V. Rolando, R. Owen, P. Tanpaiboon, F. Lacbawan, Clinton R. Paden, Duncan MacCannell |
| EPI_ISL_2203832 | Houston Methodist Hospital | Houston Methodist Hospital | Randall J. Olsen, Paul A. Christensen, S. Wesley Long, Sishir Subedi, Robert Olson, Marcus Nguyen, James J. Davis, Matthew Ojeda Saavedra, Prasanti Yerramilli, Layne Pruitt, Kristina Reppond, Madison N. Shyer, Jessica Cambric, Ryan Gadd, Ilya J. Finkelstein, Jimmy Gollihar, and James M. Musser |
| EPI_ISL_2203833, EPI_ISL_2203834 | Quest Diagnostics Incorporated | Centers for Disease Control and Prevention Division of Viral Diseases, Pathogen Discovery | Dakota Howard, Dhwani Batra, Peter W. Cook, Kara Moser, Adrian Paskey, Jason Caravas, Benjamin Rambo-Martin, Shatavia Morrison, Christopher Gulvick, Scott Sammons, Yvette Unoarumhi, Darlene Wagner, Matthew Schmerer, S. H. Rosenthal, A. Gerasimova, R. M. Kagan, B. Anderson, M. Hua, Y. Liu, L.E. Bernstein, K.E. Livingston, A. Perez, I. A. Shlyakhter, R. V. Rolando, R. Owen, P. Tanpaiboon, F. Lacbawan, Clinton R. Paden, Duncan MacCannell |
| EPI_ISL_2203835 | Houston Methodist Hospital | Houston Methodist Hospital | Randall J. Olsen, Paul A. Christensen, S. Wesley Long, Sishir Subedi, Robert Olson, Marcus Nguyen, James J. Davis, Matthew Ojeda Saavedra, Prasanti Yerramilli, Layne Pruitt, Kristina Reppond, Madison N. Shyer, Jessica Cambric, Ryan Gadd, Ilya J. Finkelstein, Jimmy Gollihar, and James M. Musser |
| EPI_ISL_2203836, EPI_ISL_2203837 | Quest Diagnostics Incorporated | Centers for Disease Control and Prevention Division of Viral Diseases, Pathogen | Dakota Howard, Dhwani Batra, Peter W. Cook, Kara Moser, Adrian Paskey, Jason Caravas, Benjamin Rambo-Martin, Shatavia Morrison, Christopher Gulvick, Scott Sammons, Yvette Unoarumhi, Darlene Wagner, Matthew Schmerer, S. H. Rosenthal, A. Gerasimova, R. M. Kagan, B. Anderson, M. Hua, Y. Liu, L.E. Bernstein, K.E. Livingston, A. Perez, I. A. Shlyakhter, R. V. Rolando, R. Owen, P. Tanpaiboon, F. Lacbawan, Clinton R. Paden, Duncan MacCannell |

| | | | |
|---|---|---|---|
| | | Discovery | |
| EPI_ISL_2203838 | Houston Methodist Hospital | Houston Methodist Hospital | Randall J. Olsen, Paul A. Christensen, S. Wesley Long, Sishir Subedi, Robert Olson, Marcus Nguyen, James J. Davis, Matthew Ojeda Saavedra, Prasanti Yerramilli, Layne Pruitt, Kristina Reppond, Madison N. Shyer, Jessica Cambric, Ryan Gadd, Ilya J. Finkelstein, Jimmy Gollihar, and James M. Musser |
| EPI_ISL_2203839, EPI_ISL_2203840 | Quest Diagnostics Incorporated | Centers for Disease Control and Prevention Division of Viral Diseases, Pathogen Discovery | Dakota Howard, Dhwani Batra, Peter W. Cook, Kara Moser, Adrian Paskey, Jason Caravas, Benjamin Rambo-Martin, Shatavia Morrison, Christopher Gulvick, Scott Sammons, Yvette Unoarumhi, Darlene Wagner, Matthew Schmerer, S. H. Rosenthal, A. Gerasimova, R. M. Kagan, B. Anderson, M. Hua, Y. Liu, L.E. Bernstein, K.E. Livingston, A. Perez, I. A. Shlyakhter, R. V. Rolando, R. Owen, P. Tanpaiboon, F. Lacbawan, Clinton R. Paden, Duncan MacCannell |
| EPI_ISL_2203841 | Houston Methodist Hospital | Houston Methodist Hospital | Randall J. Olsen, Paul A. Christensen, S. Wesley Long, Sishir Subedi, Robert Olson, Marcus Nguyen, James J. Davis, Matthew Ojeda Saavedra, Prasanti Yerramilli, Layne Pruitt, Kristina Reppond, Madison N. Shyer, Jessica Cambric, Ryan Gadd, Ilya J. Finkelstein, Jimmy Gollihar, and James M. Musser |
| EPI_ISL_2203842, EPI_ISL_2203843 | Quest Diagnostics Incorporated | Centers for Disease Control and Prevention Division of Viral Diseases, Pathogen Discovery | Dakota Howard, Dhwani Batra, Peter W. Cook, Kara Moser, Adrian Paskey, Jason Caravas, Benjamin Rambo-Martin, Shatavia Morrison, Christopher Gulvick, Scott Sammons, Yvette Unoarumhi, Darlene Wagner, Matthew Schmerer, S. H. Rosenthal, A. Gerasimova, R. M. Kagan, B. Anderson, M. Hua, Y. Liu, L.E. Bernstein, K.E. Livingston, A. Perez, I. A. Shlyakhter, R. V. Rolando, R. Owen, P. Tanpaiboon, F. Lacbawan, Clinton R. Paden, Duncan MacCannell |
| EPI_ISL_2203844 | Houston Methodist Hospital | Houston Methodist Hospital | Randall J. Olsen, Paul A. Christensen, S. Wesley Long, Sishir Subedi, Robert Olson, Marcus Nguyen, James J. Davis, Matthew Ojeda Saavedra, Prasanti Yerramilli, Layne Pruitt, Kristina Reppond, Madison N. Shyer, Jessica Cambric, Ryan Gadd, Ilya J. Finkelstein, Jimmy Gollihar, and James M. Musser |
| EPI_ISL_2203845, EPI_ISL_2203846, EPI_ISL_2203847, EPI_ISL_2203848 | Quest Diagnostics Incorporated | Centers for Disease Control and Prevention Division of Viral Diseases, Pathogen Discovery | Dakota Howard, Dhwani Batra, Peter W. Cook, Kara Moser, Adrian Paskey, Jason Caravas, Benjamin Rambo-Martin, Shatavia Morrison, Christopher Gulvick, Scott Sammons, Yvette Unoarumhi, Darlene Wagner, Matthew Schmerer, S. H. Rosenthal, A. Gerasimova, R. M. Kagan, B. Anderson, M. Hua, Y. Liu, L.E. Bernstein, K.E. Livingston, A. Perez, I. A. Shlyakhter, R. V. Rolando, R. Owen, P. Tanpaiboon, F. Lacbawan, Clinton R. Paden, Duncan MacCannell |
| EPI_ISL_2203849 | Houston Methodist Hospital | Houston Methodist Hospital | Randall J. Olsen, Paul A. Christensen, S. Wesley Long, Sishir Subedi, Robert Olson, Marcus Nguyen, James J. Davis, Matthew Ojeda Saavedra, Prasanti Yerramilli, Layne Pruitt, Kristina Reppond, Madison N. Shyer, Jessica Cambric, Ryan Gadd, Ilya J. Finkelstein, Jimmy Gollihar, and James M. Musser |
| EPI_ISL_2203850, EPI_ISL_2203851, EPI_ISL_2203852 | Quest Diagnostics Incorporated | Centers for Disease Control and Prevention Division of Viral Diseases, Pathogen Discovery | Dakota Howard, Dhwani Batra, Peter W. Cook, Kara Moser, Adrian Paskey, Jason Caravas, Benjamin Rambo-Martin, Shatavia Morrison, Christopher Gulvick, Scott Sammons, Yvette Unoarumhi, Darlene Wagner, Matthew Schmerer, S. H. Rosenthal, A. Gerasimova, R. M. Kagan, B. Anderson, M. Hua, Y. Liu, L.E. Bernstein, K.E. Livingston, A. Perez, I. A. Shlyakhter, R. V. Rolando, R. Owen, P. Tanpaiboon, F. Lacbawan, Clinton R. Paden, Duncan MacCannell |
| EPI_ISL_2203853 | Houston Methodist Hospital | Houston Methodist Hospital | Randall J. Olsen, Paul A. Christensen, S. Wesley Long, Sishir Subedi, Robert Olson, Marcus Nguyen, James J. Davis, Matthew Ojeda Saavedra, Prasanti Yerramilli, Layne Pruitt, Kristina Reppond, Madison N. Shyer, Jessica Cambric, Ryan Gadd, Ilya J. Finkelstein, Jimmy Gollihar, and James M. Musser |
| EPI_ISL_2203854 | Quest Diagnostics Incorporated | Centers for Disease Control and Prevention Division of Viral Diseases, Pathogen Discovery | Dakota Howard, Dhwani Batra, Peter W. Cook, Kara Moser, Adrian Paskey, Jason Caravas, Benjamin Rambo-Martin, Shatavia Morrison, Christopher Gulvick, Scott Sammons, Yvette Unoarumhi, Darlene Wagner, Matthew Schmerer, S. H. Rosenthal, A. Gerasimova, R. M. Kagan, B. Anderson, M. Hua, Y. Liu, L.E. Bernstein, K.E. Livingston, A. Perez, I. A. Shlyakhter, R. V. Rolando, R. Owen, P. Tanpaiboon, F. Lacbawan, Clinton R. Paden, Duncan MacCannell |
| EPI_ISL_2203855 | Houston Methodist Hospital | Houston Methodist Hospital | Randall J. Olsen, Paul A. Christensen, S. Wesley Long, Sishir Subedi, Robert Olson, Marcus Nguyen, James J. Davis, Matthew Ojeda Saavedra, Prasanti Yerramilli, Layne Pruitt, Kristina Reppond, Madison N. Shyer, Jessica Cambric, Ryan Gadd, Ilya J. Finkelstein, Jimmy Gollihar, and James M. Musser |
| EPI_ISL_2203856 | Quest Diagnostics Incorporated | Centers for Disease Control and Prevention Division of Viral Diseases, Pathogen Discovery | Dakota Howard, Dhwani Batra, Peter W. Cook, Kara Moser, Adrian Paskey, Jason Caravas, Benjamin Rambo-Martin, Shatavia Morrison, Christopher Gulvick, Scott Sammons, Yvette Unoarumhi, Darlene Wagner, Matthew Schmerer, S. H. Rosenthal, A. Gerasimova, R. M. Kagan, B. Anderson, M. Hua, Y. Liu, L.E. Bernstein, K.E. Livingston, A. Perez, I. A. Shlyakhter, R. V. Rolando, R. Owen, P. Tanpaiboon, F. Lacbawan, Clinton R. Paden, Duncan MacCannell |
| EPI_ISL_2203857 | Houston Methodist Hospital | Houston Methodist Hospital | Randall J. Olsen, Paul A. Christensen, S. Wesley Long, Sishir Subedi, Robert Olson, Marcus Nguyen, James J. Davis, Matthew Ojeda Saavedra, Prasanti Yerramilli, Layne Pruitt, Kristina Reppond, Madison N. Shyer, Jessica Cambric, Ryan Gadd, Ilya J. Finkelstein, Jimmy Gollihar, and James M. Musser |
| EPI_ISL_2203858 | Quest Diagnostics Incorporated | Centers for Disease Control and Prevention Division of Viral Diseases, Pathogen Discovery | Dakota Howard, Dhwani Batra, Peter W. Cook, Kara Moser, Adrian Paskey, Jason Caravas, Benjamin Rambo-Martin, Shatavia Morrison, Christopher Gulvick, Scott Sammons, Yvette Unoarumhi, Darlene Wagner, Matthew Schmerer, S. H. Rosenthal, A. Gerasimova, R. M. Kagan, B. Anderson, M. Hua, Y. Liu, L.E. Bernstein, K.E. Livingston, A. Perez, I. A. Shlyakhter, R. V. Rolando, R. Owen, P. Tanpaiboon, F. Lacbawan, Clinton R. Paden, Duncan MacCannell |
| EPI_ISL_2203859 | Houston Methodist Hospital | Houston Methodist Hospital | Randall J. Olsen, Paul A. Christensen, S. Wesley Long, Sishir Subedi, Robert Olson, Marcus Nguyen, James J. Davis, Matthew Ojeda Saavedra, Prasanti Yerramilli, Layne Pruitt, Kristina Reppond, Madison N. Shyer, Jessica Cambric, Ryan Gadd, Ilya J. Finkelstein, Jimmy Gollihar, and James M. Musser |
| EPI_ISL_2203860, EPI_ISL_2203861 | Quest Diagnostics Incorporated | Centers for Disease Control and Prevention Division of Viral Diseases, Pathogen Discovery | Dakota Howard, Dhwani Batra, Peter W. Cook, Kara Moser, Adrian Paskey, Jason Caravas, Benjamin Rambo-Martin, Shatavia Morrison, Christopher Gulvick, Scott Sammons, Yvette Unoarumhi, Darlene Wagner, Matthew Schmerer, S. H. Rosenthal, A. Gerasimova, R. M. Kagan, B. Anderson, M. Hua, Y. Liu, L.E. Bernstein, K.E. Livingston, A. Perez, I. A. Shlyakhter, R. V. Rolando, R. Owen, P. Tanpaiboon, F. Lacbawan, Clinton R. Paden, Duncan MacCannell |
| EPI_ISL_2203862 | Houston Methodist Hospital | Houston Methodist Hospital | Randall J. Olsen, Paul A. Christensen, S. Wesley Long, Sishir Subedi, Robert Olson, Marcus Nguyen, James J. Davis, Matthew Ojeda Saavedra, Prasanti Yerramilli, Layne Pruitt, Kristina Reppond, Madison N. Shyer, Jessica Cambric, Ryan Gadd, Ilya J. Finkelstein, Jimmy Gollihar, and James M. Musser |
| EPI_ISL_2203863, EPI_ISL_2203864, EPI_ISL_2203865 | Quest Diagnostics Incorporated | Centers for Disease Control and Prevention Division of Viral Diseases, Pathogen Discovery | Dakota Howard, Dhwani Batra, Peter W. Cook, Kara Moser, Adrian Paskey, Jason Caravas, Benjamin Rambo-Martin, Shatavia Morrison, Christopher Gulvick, Scott Sammons, Yvette Unoarumhi, Darlene Wagner, Matthew Schmerer, S. H. Rosenthal, A. Gerasimova, R. M. Kagan, B. Anderson, M. Hua, Y. Liu, L.E. Bernstein, K.E. Livingston, A. Perez, I. A. Shlyakhter, R. V. Rolando, R. Owen, P. Tanpaiboon, F. Lacbawan, Clinton R. Paden, Duncan MacCannell |
| EPI_ISL_2203866 | Houston Methodist Hospital | Houston Methodist Hospital | Randall J. Olsen, Paul A. Christensen, S. Wesley Long, Sishir Subedi, Robert Olson, Marcus Nguyen, James J. Davis, Matthew Ojeda Saavedra, Prasanti Yerramilli, Layne Pruitt, Kristina Reppond, Madison N. Shyer, Jessica Cambric, Ryan Gadd, Ilya J. Finkelstein, Jimmy Gollihar, and James M. Musser |
| EPI_ISL_2203867, EPI_ISL_2203868 | Quest Diagnostics Incorporated | Centers for Disease Control and Prevention Division of Viral Diseases, Pathogen Discovery | Dakota Howard, Dhwani Batra, Peter W. Cook, Kara Moser, Adrian Paskey, Jason Caravas, Benjamin Rambo-Martin, Shatavia Morrison, Christopher Gulvick, Scott Sammons, Yvette Unoarumhi, Darlene Wagner, Matthew Schmerer, S. H. Rosenthal, A. Gerasimova, R. M. Kagan, B. Anderson, M. Hua, Y. Liu, L.E. Bernstein, K.E. Livingston, A. Perez, I. A. Shlyakhter, R. V. Rolando, R. Owen, P. Tanpaiboon, F. Lacbawan, Clinton R. Paden, Duncan MacCannell |
| EPI_ISL_2203869 | Houston Methodist Hospital | Houston Methodist Hospital | Randall J. Olsen, Paul A. Christensen, S. Wesley Long, Sishir Subedi, Robert Olson, Marcus Nguyen, James J. Davis, Matthew Ojeda Saavedra, Prasanti Yerramilli, Layne Pruitt, Kristina Reppond, Madison N. Shyer, Jessica Cambric, Ryan Gadd, Ilya J. Finkelstein, Jimmy Gollihar, and James M. Musser |
| EPI_ISL_2203870, EPI_ISL_2203872 | Quest Diagnostics Incorporated | Centers for Disease Control and Prevention Division of Viral Diseases, Pathogen Discovery | Dakota Howard, Dhwani Batra, Peter W. Cook, Kara Moser, Adrian Paskey, Jason Caravas, Benjamin Rambo-Martin, Shatavia Morrison, Christopher Gulvick, Scott Sammons, Yvette Unoarumhi, Darlene Wagner, Matthew Schmerer, S. H. Rosenthal, A. Gerasimova, R. M. Kagan, B. Anderson, M. Hua, Y. Liu, L.E. Bernstein, K.E. Livingston, A. Perez, I. A. Shlyakhter, R. V. Rolando, R. Owen, P. Tanpaiboon, F. Lacbawan, Clinton R. Paden, Duncan MacCannell |
| EPI_ISL_2203873 | Houston Methodist Hospital | Houston Methodist Hospital | Randall J. Olsen, Paul A. Christensen, S. Wesley Long, Sishir Subedi, Robert Olson, Marcus Nguyen, James J. Davis, Matthew Ojeda Saavedra, Prasanti Yerramilli, Layne Pruitt, Kristina Reppond, Madison N. Shyer, Jessica Cambric, Ryan Gadd, Ilya J. Finkelstein, Jimmy Gollihar, and James M. Musser |
| EPI_ISL_2203874, EPI_ISL_2203875 | Quest Diagnostics Incorporated | Centers for Disease Control and Prevention Division of Viral Diseases, Pathogen Discovery | Dakota Howard, Dhwani Batra, Peter W. Cook, Kara Moser, Adrian Paskey, Jason Caravas, Benjamin Rambo-Martin, Shatavia Morrison, Christopher Gulvick, Scott Sammons, Yvette Unoarumhi, Darlene Wagner, Matthew Schmerer, S. H. Rosenthal, A. Gerasimova, R. M. Kagan, B. Anderson, M. Hua, Y. Liu, L.E. Bernstein, K.E. Livingston, A. Perez, I. A. Shlyakhter, R. V. Rolando, R. Owen, P. Tanpaiboon, F. Lacbawan, Clinton R. Paden, Duncan MacCannell |
| EPI_ISL_2203876 | Houston Methodist Hospital | Houston Methodist Hospital | Randall J. Olsen, Paul A. Christensen, S. Wesley Long, Sishir Subedi, Robert Olson, Marcus Nguyen, James J. Davis, Matthew Ojeda Saavedra, Prasanti Yerramilli, Layne Pruitt, Kristina Reppond, Madison N. Shyer, Jessica Cambric, Ryan Gadd, Ilya J. Finkelstein, Jimmy Gollihar, and James M. Musser |
| EPI_ISL_2203877 | Quest Diagnostics Incorporated | Centers for Disease Control and Prevention Division of Viral Diseases, Pathogen Discovery | Dakota Howard, Dhwani Batra, Peter W. Cook, Kara Moser, Adrian Paskey, Jason Caravas, Benjamin Rambo-Martin, Shatavia Morrison, Christopher Gulvick, Scott Sammons, Yvette Unoarumhi, Darlene Wagner, Matthew Schmerer, S. H. Rosenthal, A. Gerasimova, R. M. Kagan, B. Anderson, M. Hua, Y. Liu, L.E. Bernstein, K.E. Livingston, A. Perez, I. A. Shlyakhter, R. V. Rolando, R. Owen, P. Tanpaiboon, F. Lacbawan, Clinton R. Paden, Duncan MacCannell |
| EPI_ISL_2203878 | Houston Methodist Hospital | Houston Methodist Hospital | Randall J. Olsen, Paul A. Christensen, S. Wesley Long, Sishir Subedi, Robert Olson, Marcus Nguyen, James J. Davis, Matthew Ojeda Saavedra, Prasanti Yerramilli, Layne Pruitt, Kristina Reppond, Madison N. Shyer, Jessica Cambric, Ryan Gadd, Ilya J. Finkelstein, Jimmy Gollihar, and James M. Musser |
| EPI_ISL_2203879 | Quest Diagnostics Incorporated | Centers for Disease Control and Prevention Division of Viral Diseases, Pathogen Discovery | Dakota Howard, Dhwani Batra, Peter W. Cook, Kara Moser, Adrian Paskey, Jason Caravas, Benjamin Rambo-Martin, Shatavia Morrison, Christopher Gulvick, Scott Sammons, Yvette Unoarumhi, Darlene Wagner, Matthew Schmerer, S. H. Rosenthal, A. Gerasimova, R. M. Kagan, B. Anderson, M. Hua, Y. Liu, L.E. Bernstein, K.E. Livingston, A. Perez, I. A. Shlyakhter, R. V. Rolando, R. Owen, P. Tanpaiboon, F. Lacbawan, Clinton R. Paden, Duncan MacCannell |
| EPI_ISL_2203880 | Houston Methodist Hospital | Houston Methodist Hospital | Randall J. Olsen, Paul A. Christensen, S. Wesley Long, Sishir Subedi, Robert Olson, Marcus Nguyen, James J. Davis, Matthew Ojeda Saavedra, Prasanti Yerramilli, Layne Pruitt, Kristina Reppond, Madison N. Shyer, Jessica Cambric, Ryan Gadd, Ilya J. Finkelstein, Jimmy Gollihar, and James M. Musser |
| EPI_ISL_2203881, EPI_ISL_2203882 | Quest Diagnostics Incorporated | Centers for Disease Control and Prevention Division of Viral Diseases, Pathogen Discovery | Dakota Howard, Dhwani Batra, Peter W. Cook, Kara Moser, Adrian Paskey, Jason Caravas, Benjamin Rambo-Martin, Shatavia Morrison, Christopher Gulvick, Scott Sammons, Yvette Unoarumhi, Darlene Wagner, Matthew Schmerer, S. H. Rosenthal, A. Gerasimova, R. M. Kagan, B. Anderson, M. Hua, Y. Liu, L.E. Bernstein, K.E. Livingston, A. Perez, I. A. Shlyakhter, R. V. Rolando, R. Owen, P. Tanpaiboon, F. Lacbawan, Clinton R. Paden, Duncan MacCannell |
| EPI_ISL_2203883 | Houston Methodist Hospital | Houston Methodist Hospital | Randall J. Olsen, Paul A. Christensen, S. Wesley Long, Sishir Subedi, Robert Olson, Marcus Nguyen, James J. Davis, Matthew Ojeda Saavedra, Prasanti Yerramilli, Layne Pruitt, Kristina Reppond, Madison N. Shyer, Jessica Cambric, Ryan Gadd, Ilya J. Finkelstein, Jimmy Gollihar, and James M. Musser |
| EPI_ISL_2203884, EPI_ISL_2203885 | Quest Diagnostics Incorporated | Centers for Disease Control and Prevention Division of Viral Diseases, Pathogen Discovery | Dakota Howard, Dhwani Batra, Peter W. Cook, Kara Moser, Adrian Paskey, Jason Caravas, Benjamin Rambo-Martin, Shatavia Morrison, Christopher Gulvick, Scott Sammons, Yvette Unoarumhi, Darlene Wagner, Matthew Schmerer, S. H. Rosenthal, A. Gerasimova, R. M. Kagan, B. Anderson, M. Hua, Y. Liu, L.E. Bernstein, K.E. Livingston, A. Perez, I. A. Shlyakhter, R. V. Rolando, R. Owen, P. Tanpaiboon, F. Lacbawan, Clinton R. Paden, Duncan MacCannell |
| EPI_ISL_2203886 | Houston Methodist Hospital | Houston Methodist Hospital | Randall J. Olsen, Paul A. Christensen, S. Wesley Long, Sishir Subedi, Robert Olson, Marcus Nguyen, James J. Davis, Matthew Ojeda Saavedra, Prasanti Yerramilli, Layne Pruitt, Kristina Reppond, Madison N. Shyer, Jessica Cambric, Ryan Gadd, Ilya J. Finkelstein, Jimmy Gollihar, and James M. Musser |
| EPI_ISL_2203887 | Quest Diagnostics Incorporated | Centers for Disease Control and Prevention Division of Viral Diseases, Pathogen Discovery | Dakota Howard, Dhwani Batra, Peter W. Cook, Kara Moser, Adrian Paskey, Jason Caravas, Benjamin Rambo-Martin, Shatavia Morrison, Christopher Gulvick, Scott Sammons, Yvette Unoarumhi, Darlene Wagner, Matthew Schmerer, S. H. Rosenthal, A. Gerasimova, R. M. Kagan, B. Anderson, M. Hua, Y. Liu, L.E. Bernstein, K.E. Livingston, A. Perez, I. A. Shlyakhter, R. V. Rolando, R. Owen, P. Tanpaiboon, F. Lacbawan, Clinton R. Paden, Duncan MacCannell |
| EPI_ISL_2203888 | Houston Methodist Hospital | Houston Methodist Hospital | Randall J. Olsen, Paul A. Christensen, S. Wesley Long, Sishir Subedi, Robert Olson, Marcus Nguyen, James J. Davis, Matthew Ojeda Saavedra, Prasanti Yerramilli, Layne Pruitt, Kristina Reppond, Madison N. Shyer, Jessica Cambric, Ryan Gadd, Ilya J. Finkelstein, Jimmy Gollihar, and James M. Musser |
| EPI_ISL_2203889, EPI_ISL_2203890 | Quest Diagnostics Incorporated | Centers for Disease Control and Prevention Division of Viral Diseases, Pathogen Discovery | Dakota Howard, Dhwani Batra, Peter W. Cook, Kara Moser, Adrian Paskey, Jason Caravas, Benjamin Rambo-Martin, Shatavia Morrison, Christopher Gulvick, Scott Sammons, Yvette Unoarumhi, Darlene Wagner, Matthew Schmerer, S. H. Rosenthal, A. Gerasimova, R. M. Kagan, B. Anderson, M. Hua, Y. Liu, L.E. Bernstein, K.E. Livingston, A. Perez, I. A. Shlyakhter, R. V. Rolando, R. Owen, P. Tanpaiboon, F. Lacbawan, Clinton R. Paden, Duncan MacCannell |
| EPI_ISL_2203891 | Houston Methodist Hospital | Houston Methodist Hospital | Randall J. Olsen, Paul A. Christensen, S. Wesley Long, Sishir Subedi, Robert Olson, Marcus Nguyen, James J. Davis, Matthew Ojeda Saavedra, Prasanti Yerramilli, Layne Pruitt, Kristina Reppond, Madison N. Shyer, Jessica Cambric, Ryan Gadd, Ilya J. Finkelstein, Jimmy Gollihar, and James |

| | | | |
|---|---|---|---|
| | | | M. Musser |
| EPI_ISL_2203892 | Quest Diagnostics Incorporated | Centers for Disease Control and Prevention Division of Viral Diseases, Pathogen Discovery | Dakota Howard, Dhwani Batra, Peter W. Cook, Kara Moser, Adrian Paskey, Jason Caravas, Benjamin Rambo-Martin, Shatavia Morrison, Christopher Gulvick, Scott Sammons, Yvette Unoarumhi, Darlene Wagner, Matthew Schmerer, S. H. Rosenthal, A. Gerasimova, R. M. Kagan, B. Anderson, M. Hua, Y. Liu, L.E. Bernstein, K.E. Livingston, A. Perez, I. A. Shlyakhter, R. V. Rolando, R. Owen, P. Tanpaiboon, F. Lacbawan, Clinton R. Paden, Duncan MacCannell |
| EPI_ISL_2203893 | Houston Methodist Hospital | Houston Methodist Hospital | Randall J. Olsen, Paul A. Christensen, S. Wesley Long, Sishir Subedi, Robert Olson, Marcus Nguyen, James J. Davis, Matthew Ojeda Saavedra, Prasanti Yerramilli, Layne Pruitt, Kristina Reppond, Madison N. Shyer, Jessica Cambric, Ryan Gadd, Ilya J. Finkelstein, Jimmy Gollihar, and James M. Musser |
| EPI_ISL_2203894 | Quest Diagnostics Incorporated | Centers for Disease Control and Prevention Division of Viral Diseases, Pathogen Discovery | Dakota Howard, Dhwani Batra, Peter W. Cook, Kara Moser, Adrian Paskey, Jason Caravas, Benjamin Rambo-Martin, Shatavia Morrison, Christopher Gulvick, Scott Sammons, Yvette Unoarumhi, Darlene Wagner, Matthew Schmerer, S. H. Rosenthal, A. Gerasimova, R. M. Kagan, B. Anderson, M. Hua, Y. Liu, L.E. Bernstein, K.E. Livingston, A. Perez, I. A. Shlyakhter, R. V. Rolando, R. Owen, P. Tanpaiboon, F. Lacbawan, Clinton R. Paden, Duncan MacCannell |
| EPI_ISL_2203895 | Houston Methodist Hospital | Houston Methodist Hospital | Randall J. Olsen, Paul A. Christensen, S. Wesley Long, Sishir Subedi, Robert Olson, Marcus Nguyen, James J. Davis, Matthew Ojeda Saavedra, Prasanti Yerramilli, Layne Pruitt, Kristina Reppond, Madison N. Shyer, Jessica Cambric, Ryan Gadd, Ilya J. Finkelstein, Jimmy Gollihar, and James M. Musser |
| EPI_ISL_2203896 | Quest Diagnostics Incorporated | Centers for Disease Control and Prevention Division of Viral Diseases, Pathogen Discovery | Dakota Howard, Dhwani Batra, Peter W. Cook, Kara Moser, Adrian Paskey, Jason Caravas, Benjamin Rambo-Martin, Shatavia Morrison, Christopher Gulvick, Scott Sammons, Yvette Unoarumhi, Darlene Wagner, Matthew Schmerer, S. H. Rosenthal, A. Gerasimova, R. M. Kagan, B. Anderson, M. Hua, Y. Liu, L.E. Bernstein, K.E. Livingston, A. Perez, I. A. Shlyakhter, R. V. Rolando, R. Owen, P. Tanpaiboon, F. Lacbawan, Clinton R. Paden, Duncan MacCannell |
| EPI_ISL_2203897 | Houston Methodist Hospital | Houston Methodist Hospital | Randall J. Olsen, Paul A. Christensen, S. Wesley Long, Sishir Subedi, Robert Olson, Marcus Nguyen, James J. Davis, Matthew Ojeda Saavedra, Prasanti Yerramilli, Layne Pruitt, Kristina Reppond, Madison N. Shyer, Jessica Cambric, Ryan Gadd, Ilya J. Finkelstein, Jimmy Gollihar, and James M. Musser |
| EPI_ISL_2203898 | Quest Diagnostics Incorporated | Centers for Disease Control and Prevention Division of Viral Diseases, Pathogen Discovery | Dakota Howard, Dhwani Batra, Peter W. Cook, Kara Moser, Adrian Paskey, Jason Caravas, Benjamin Rambo-Martin, Shatavia Morrison, Christopher Gulvick, Scott Sammons, Yvette Unoarumhi, Darlene Wagner, Matthew Schmerer, S. H. Rosenthal, A. Gerasimova, R. M. Kagan, B. Anderson, M. Hua, Y. Liu, L.E. Bernstein, K.E. Livingston, A. Perez, I. A. Shlyakhter, R. V. Rolando, R. Owen, P. Tanpaiboon, F. Lacbawan, Clinton R. Paden, Duncan MacCannell |
| EPI_ISL_2203899, EPI_ISL_2203900, EPI_ISL_2203901 | Houston Methodist Hospital | Houston Methodist Hospital | Randall J. Olsen, Paul A. Christensen, S. Wesley Long, Sishir Subedi, Robert Olson, Marcus Nguyen, James J. Davis, Matthew Ojeda Saavedra, Prasanti Yerramilli, Layne Pruitt, Kristina Reppond, Madison N. Shyer, Jessica Cambric, Ryan Gadd, Ilya J. Finkelstein, Jimmy Gollihar, and James M. Musser |
| EPI_ISL_2203902 | Quest Diagnostics Incorporated | Centers for Disease Control and Prevention Division of Viral Diseases, Pathogen Discovery | Dakota Howard, Dhwani Batra, Peter W. Cook, Kara Moser, Adrian Paskey, Jason Caravas, Benjamin Rambo-Martin, Shatavia Morrison, Christopher Gulvick, Scott Sammons, Yvette Unoarumhi, Darlene Wagner, Matthew Schmerer, S. H. Rosenthal, A. Gerasimova, R. M. Kagan, B. Anderson, M. Hua, Y. Liu, L.E. Bernstein, K.E. Livingston, A. Perez, I. A. Shlyakhter, R. V. Rolando, R. Owen, P. Tanpaiboon, F. Lacbawan, Clinton R. Paden, Duncan MacCannell |
| EPI_ISL_2203903 | Houston Methodist Hospital | Houston Methodist Hospital | Randall J. Olsen, Paul A. Christensen, S. Wesley Long, Sishir Subedi, Robert Olson, Marcus Nguyen, James J. Davis, Matthew Ojeda Saavedra, Prasanti Yerramilli, Layne Pruitt, Kristina Reppond, Madison N. Shyer, Jessica Cambric, Ryan Gadd, Ilya J. Finkelstein, Jimmy Gollihar, and James M. Musser |
| EPI_ISL_2203904 | Quest Diagnostics Incorporated | Centers for Disease Control and Prevention Division of Viral Diseases, Pathogen Discovery | Dakota Howard, Dhwani Batra, Peter W. Cook, Kara Moser, Adrian Paskey, Jason Caravas, Benjamin Rambo-Martin, Shatavia Morrison, Christopher Gulvick, Scott Sammons, Yvette Unoarumhi, Darlene Wagner, Matthew Schmerer, S. H. Rosenthal, A. Gerasimova, R. M. Kagan, B. Anderson, M. Hua, Y. Liu, L.E. Bernstein, K.E. Livingston, A. Perez, I. A. Shlyakhter, R. V. Rolando, R. Owen, P. Tanpaiboon, F. Lacbawan, Clinton R. Paden, Duncan MacCannell |
| EPI_ISL_2203905 | Houston Methodist Hospital | Houston Methodist Hospital | Randall J. Olsen, Paul A. Christensen, S. Wesley Long, Sishir Subedi, Robert Olson, Marcus Nguyen, James J. Davis, Matthew Ojeda Saavedra, Prasanti Yerramilli, Layne Pruitt, Kristina Reppond, Madison N. Shyer, Jessica Cambric, Ryan Gadd, Ilya J. Finkelstein, Jimmy Gollihar, and James M. Musser |
| EPI_ISL_2203906 | Quest Diagnostics Incorporated | Centers for Disease Control and Prevention Division of Viral Diseases, Pathogen Discovery | Dakota Howard, Dhwani Batra, Peter W. Cook, Kara Moser, Adrian Paskey, Jason Caravas, Benjamin Rambo-Martin, Shatavia Morrison, Christopher Gulvick, Scott Sammons, Yvette Unoarumhi, Darlene Wagner, Matthew Schmerer, S. H. Rosenthal, A. Gerasimova, R. M. Kagan, B. Anderson, M. Hua, Y. Liu, L.E. Bernstein, K.E. Livingston, A. Perez, I. A. Shlyakhter, R. V. Rolando, R. Owen, P. Tanpaiboon, F. Lacbawan, Clinton R. Paden, Duncan MacCannell |
| EPI_ISL_2203907 | Houston Methodist Hospital | Houston Methodist Hospital | Randall J. Olsen, Paul A. Christensen, S. Wesley Long, Sishir Subedi, Robert Olson, Marcus Nguyen, James J. Davis, Matthew Ojeda Saavedra, Prasanti Yerramilli, Layne Pruitt, Kristina Reppond, Madison N. Shyer, Jessica Cambric, Ryan Gadd, Ilya J. Finkelstein, Jimmy Gollihar, and James M. Musser |
| EPI_ISL_2203908, EPI_ISL_2203909 | Quest Diagnostics Incorporated | Centers for Disease Control and Prevention Division of Viral Diseases, Pathogen Discovery | Dakota Howard, Dhwani Batra, Peter W. Cook, Kara Moser, Adrian Paskey, Jason Caravas, Benjamin Rambo-Martin, Shatavia Morrison, Christopher Gulvick, Scott Sammons, Yvette Unoarumhi, Darlene Wagner, Matthew Schmerer, S. H. Rosenthal, A. Gerasimova, R. M. Kagan, B. Anderson, M. Hua, Y. Liu, L.E. Bernstein, K.E. Livingston, A. Perez, I. A. Shlyakhter, R. V. Rolando, R. Owen, P. Tanpaiboon, F. Lacbawan, Clinton R. Paden, Duncan MacCannell |
| EPI_ISL_2203910 | Houston Methodist Hospital | Houston Methodist Hospital | Randall J. Olsen, Paul A. Christensen, S. Wesley Long, Sishir Subedi, Robert Olson, Marcus Nguyen, James J. Davis, Matthew Ojeda Saavedra, Prasanti Yerramilli, Layne Pruitt, Kristina Reppond, Madison N. Shyer, Jessica Cambric, Ryan Gadd, Ilya J. Finkelstein, Jimmy Gollihar, and James M. Musser |
| EPI_ISL_2203911 | Quest Diagnostics Incorporated | Centers for Disease Control and Prevention Division of Viral Diseases, Pathogen Discovery | Dakota Howard, Dhwani Batra, Peter W. Cook, Kara Moser, Adrian Paskey, Jason Caravas, Benjamin Rambo-Martin, Shatavia Morrison, Christopher Gulvick, Scott Sammons, Yvette Unoarumhi, Darlene Wagner, Matthew Schmerer, S. H. Rosenthal, A. Gerasimova, R. M. Kagan, B. Anderson, M. Hua, Y. Liu, L.E. Bernstein, K.E. Livingston, A. Perez, I. A. Shlyakhter, R. V. Rolando, R. Owen, P. Tanpaiboon, F. Lacbawan, Clinton R. Paden, Duncan MacCannell |
| EPI_ISL_2203912 | Houston Methodist Hospital | Houston Methodist Hospital | Randall J. Olsen, Paul A. Christensen, S. Wesley Long, Sishir Subedi, Robert Olson, Marcus Nguyen, James J. Davis, Matthew Ojeda Saavedra, Prasanti Yerramilli, Layne Pruitt, Kristina Reppond, Madison N. Shyer, Jessica Cambric, Ryan Gadd, Ilya J. Finkelstein, Jimmy Gollihar, and James M. Musser |
| EPI_ISL_2203913 | Quest Diagnostics Incorporated | Centers for Disease Control and Prevention Division of Viral Diseases, Pathogen Discovery | Dakota Howard, Dhwani Batra, Peter W. Cook, Kara Moser, Adrian Paskey, Jason Caravas, Benjamin Rambo-Martin, Shatavia Morrison, Christopher Gulvick, Scott Sammons, Yvette Unoarumhi, Darlene Wagner, Matthew Schmerer, S. H. Rosenthal, A. Gerasimova, R. M. Kagan, B. Anderson, M. Hua, Y. Liu, L.E. Bernstein, K.E. Livingston, A. Perez, I. A. Shlyakhter, R. V. Rolando, R. Owen, P. Tanpaiboon, F. Lacbawan, Clinton R. Paden, Duncan MacCannell |
| EPI_ISL_2203914 | Houston Methodist Hospital | Houston Methodist Hospital | Randall J. Olsen, Paul A. Christensen, S. Wesley Long, Sishir Subedi, Robert Olson, Marcus Nguyen, James J. Davis, Matthew Ojeda Saavedra, Prasanti Yerramilli, Layne Pruitt, Kristina Reppond, Madison N. Shyer, Jessica Cambric, Ryan Gadd, Ilya J. Finkelstein, Jimmy Gollihar, and James M. Musser |
| EPI_ISL_2203915 | Quest Diagnostics Incorporated | Centers for Disease Control and Prevention Division of Viral Diseases, Pathogen Discovery | Dakota Howard, Dhwani Batra, Peter W. Cook, Kara Moser, Adrian Paskey, Jason Caravas, Benjamin Rambo-Martin, Shatavia Morrison, Christopher Gulvick, Scott Sammons, Yvette Unoarumhi, Darlene Wagner, Matthew Schmerer, S. H. Rosenthal, A. Gerasimova, R. M. Kagan, B. Anderson, M. Hua, Y. Liu, L.E. Bernstein, K.E. Livingston, A. Perez, I. A. Shlyakhter, R. V. Rolando, R. Owen, P. Tanpaiboon, F. Lacbawan, Clinton R. Paden, Duncan MacCannell |
| EPI_ISL_2203916 | Houston Methodist Hospital | Houston Methodist Hospital | Randall J. Olsen, Paul A. Christensen, S. Wesley Long, Sishir Subedi, Robert Olson, Marcus Nguyen, James J. Davis, Matthew Ojeda Saavedra, Prasanti Yerramilli, Layne Pruitt, Kristina Reppond, Madison N. Shyer, Jessica Cambric, Ryan Gadd, Ilya J. Finkelstein, Jimmy Gollihar, and James M. Musser |
| EPI_ISL_2203917 | Quest Diagnostics Incorporated | Centers for Disease Control and Prevention Division of Viral Diseases, Pathogen Discovery | Dakota Howard, Dhwani Batra, Peter W. Cook, Kara Moser, Adrian Paskey, Jason Caravas, Benjamin Rambo-Martin, Shatavia Morrison, Christopher Gulvick, Scott Sammons, Yvette Unoarumhi, Darlene Wagner, Matthew Schmerer, S. H. Rosenthal, A. Gerasimova, R. M. Kagan, B. Anderson, M. Hua, Y. Liu, L.E. Bernstein, K.E. Livingston, A. Perez, I. A. Shlyakhter, R. V. Rolando, R. Owen, P. Tanpaiboon, F. Lacbawan, Clinton R. Paden, Duncan MacCannell |
| EPI_ISL_2203918 | Houston Methodist Hospital | Houston Methodist Hospital | Randall J. Olsen, Paul A. Christensen, S. Wesley Long, Sishir Subedi, Robert Olson, Marcus Nguyen, James J. Davis, Matthew Ojeda Saavedra, Prasanti Yerramilli, Layne Pruitt, Kristina Reppond, Madison N. Shyer, Jessica Cambric, Ryan Gadd, Ilya J. Finkelstein, Jimmy Gollihar, and James M. Musser |
| EPI_ISL_2203919 | Quest Diagnostics Incorporated | Centers for Disease Control and Prevention Division of Viral Diseases, Pathogen Discovery | Dakota Howard, Dhwani Batra, Peter W. Cook, Kara Moser, Adrian Paskey, Jason Caravas, Benjamin Rambo-Martin, Shatavia Morrison, Christopher Gulvick, Scott Sammons, Yvette Unoarumhi, Darlene Wagner, Matthew Schmerer, S. H. Rosenthal, A. Gerasimova, R. M. Kagan, B. Anderson, M. Hua, Y. Liu, L.E. Bernstein, K.E. Livingston, A. Perez, I. A. Shlyakhter, R. V. Rolando, R. Owen, P. Tanpaiboon, F. Lacbawan, Clinton R. Paden, Duncan MacCannell |
| EPI_ISL_2203920 | Houston Methodist Hospital | Houston Methodist Hospital | Randall J. Olsen, Paul A. Christensen, S. Wesley Long, Sishir Subedi, Robert Olson, Marcus Nguyen, James J. Davis, Matthew Ojeda Saavedra, Prasanti Yerramilli, Layne Pruitt, Kristina Reppond, Madison N. Shyer, Jessica Cambric, Ryan Gadd, Ilya J. Finkelstein, Jimmy Gollihar, and James M. Musser |
| EPI_ISL_2203921, EPI_ISL_2203922 | Quest Diagnostics Incorporated | Centers for Disease Control and Prevention Division of Viral Diseases, Pathogen Discovery | Dakota Howard, Dhwani Batra, Peter W. Cook, Kara Moser, Adrian Paskey, Jason Caravas, Benjamin Rambo-Martin, Shatavia Morrison, Christopher Gulvick, Scott Sammons, Yvette Unoarumhi, Darlene Wagner, Matthew Schmerer, S. H. Rosenthal, A. Gerasimova, R. M. Kagan, B. Anderson, M. Hua, Y. Liu, L.E. Bernstein, K.E. Livingston, A. Perez, I. A. Shlyakhter, R. V. Rolando, R. Owen, P. Tanpaiboon, F. Lacbawan, Clinton R. Paden, Duncan MacCannell |
| EPI_ISL_2203923 | Houston Methodist Hospital | Houston Methodist Hospital | Randall J. Olsen, Paul A. Christensen, S. Wesley Long, Sishir Subedi, Robert Olson, Marcus Nguyen, James J. Davis, Matthew Ojeda Saavedra, Prasanti Yerramilli, Layne Pruitt, Kristina Reppond, Madison N. Shyer, Jessica Cambric, Ryan Gadd, Ilya J. Finkelstein, Jimmy Gollihar, and James M. Musser |
| EPI_ISL_2203924 | Quest Diagnostics Incorporated | Centers for Disease Control and Prevention Division of Viral Diseases, Pathogen Discovery | Dakota Howard, Dhwani Batra, Peter W. Cook, Kara Moser, Adrian Paskey, Jason Caravas, Benjamin Rambo-Martin, Shatavia Morrison, Christopher Gulvick, Scott Sammons, Yvette Unoarumhi, Darlene Wagner, Matthew Schmerer, S. H. Rosenthal, A. Gerasimova, R. M. Kagan, B. Anderson, M. Hua, Y. Liu, L.E. Bernstein, K.E. Livingston, A. Perez, I. A. Shlyakhter, R. V. Rolando, R. Owen, P. Tanpaiboon, F. Lacbawan, Clinton R. Paden, Duncan MacCannell |
| EPI_ISL_2203925 | Houston Methodist Hospital | Houston Methodist Hospital | Randall J. Olsen, Paul A. Christensen, S. Wesley Long, Sishir Subedi, Robert Olson, Marcus Nguyen, James J. Davis, Matthew Ojeda Saavedra, Prasanti Yerramilli, Layne Pruitt, Kristina Reppond, Madison N. Shyer, Jessica Cambric, Ryan Gadd, Ilya J. Finkelstein, Jimmy Gollihar, and James M. Musser |
| EPI_ISL_2203926 | Quest Diagnostics Incorporated | Centers for Disease Control and Prevention Division of Viral Diseases, Pathogen Discovery | Dakota Howard, Dhwani Batra, Peter W. Cook, Kara Moser, Adrian Paskey, Jason Caravas, Benjamin Rambo-Martin, Shatavia Morrison, Christopher Gulvick, Scott Sammons, Yvette Unoarumhi, Darlene Wagner, Matthew Schmerer, S. H. Rosenthal, A. Gerasimova, R. M. Kagan, B. Anderson, M. Hua, Y. Liu, L.E. Bernstein, K.E. Livingston, A. Perez, I. A. Shlyakhter, R. V. Rolando, R. Owen, P. Tanpaiboon, F. Lacbawan, Clinton R. Paden, Duncan MacCannell |
| EPI_ISL_2203927 | Houston Methodist Hospital | Houston Methodist Hospital | Randall J. Olsen, Paul A. Christensen, S. Wesley Long, Sishir Subedi, Robert Olson, Marcus Nguyen, James J. Davis, Matthew Ojeda Saavedra, Prasanti Yerramilli, Layne Pruitt, Kristina Reppond, Madison N. Shyer, Jessica Cambric, Ryan Gadd, Ilya J. Finkelstein, Jimmy Gollihar, and James M. Musser |
| EPI_ISL_2203928 | Quest Diagnostics Incorporated | Centers for Disease Control and Prevention Division of Viral Diseases, Pathogen Discovery | Dakota Howard, Dhwani Batra, Peter W. Cook, Kara Moser, Adrian Paskey, Jason Caravas, Benjamin Rambo-Martin, Shatavia Morrison, Christopher Gulvick, Scott Sammons, Yvette Unoarumhi, Darlene Wagner, Matthew Schmerer, S. H. Rosenthal, A. Gerasimova, R. M. Kagan, B. Anderson, M. Hua, Y. Liu, L.E. Bernstein, K.E. Livingston, A. Perez, I. A. Shlyakhter, R. V. Rolando, R. Owen, P. Tanpaiboon, F. Lacbawan, Clinton R. Paden, Duncan MacCannell |
| EPI_ISL_2203929 | Houston Methodist Hospital | Houston Methodist Hospital | Randall J. Olsen, Paul A. Christensen, S. Wesley Long, Sishir Subedi, Robert Olson, Marcus Nguyen, James J. Davis, Matthew Ojeda Saavedra, Prasanti Yerramilli, Layne Pruitt, Kristina Reppond, Madison N. Shyer, Jessica Cambric, Ryan Gadd, Ilya J. Finkelstein, Jimmy Gollihar, and James M. Musser |
| EPI_ISL_2203930 | Quest Diagnostics Incorporated | Centers for Disease Control and Prevention Division of Viral Diseases, Pathogen Discovery | Dakota Howard, Dhwani Batra, Peter W. Cook, Kara Moser, Adrian Paskey, Jason Caravas, Benjamin Rambo-Martin, Shatavia Morrison, Christopher Gulvick, Scott Sammons, Yvette Unoarumhi, Darlene Wagner, Matthew Schmerer, S. H. Rosenthal, A. Gerasimova, R. M. Kagan, B. Anderson, M. Hua, Y. Liu, L.E. Bernstein, K.E. Livingston, A. Perez, I. A. Shlyakhter, R. V. Rolando, R. Owen, P. Tanpaiboon, F. Lacbawan, Clinton R. Paden, Duncan MacCannell |
| EPI_ISL_2203931 | Houston Methodist Hospital | Houston Methodist Hospital | Randall J. Olsen, Paul A. Christensen, S. Wesley Long, Sishir Subedi, Robert Olson, Marcus Nguyen, James J. Davis, Matthew Ojeda Saavedra, Prasanti Yerramilli, Layne Pruitt, Kristina Reppond, Madison N. Shyer, Jessica Cambric, Ryan Gadd, Ilya J. Finkelstein, Jimmy Gollihar, and James M. Musser |
| EPI_ISL_2203932, EPI_ISL_2203933, | Quest Diagnostics Incorporated | Centers for Disease Control and Prevention Division of Viral Diseases, Pathogen | Dakota Howard, Dhwani Batra, Peter W. Cook, Kara Moser, Adrian Paskey, Jason Caravas, Benjamin Rambo-Martin, Shatavia Morrison, Christopher Gulvick, Scott Sammons, Yvette Unoarumhi, Darlene Wagner, Matthew Schmerer, S. H. Rosenthal, A. Gerasimova, R. M. Kagan, B. Anderson, M. Hua, Y. Liu, L.E. Bernstein, K.E. Livingston, A. Perez, I. A. Shlyakhter, R. V. Rolando, R. Owen, P. Tanpaiboon, F. Lacbawan, Clinton R. Paden, Duncan MacCannell |

| | | | |
|---|---|---|---|
| EPI_ISL_2203935 | | Discovery | |
| EPI_ISL_2203936 | Houston Methodist Hospital | Houston Methodist Hospital | Randall J. Olsen, Paul A. Christensen, S. Wesley Long, Sishir Subedi, Robert Olson, Marcus Nguyen, James J. Davis, Matthew Ojeda Saavedra, Prasanti Yerramilli, Layne Pruitt, Kristina Reppond, Madison N. Shyer, Jessica Cambric, Ryan Gadd, Ilya J. Finkelstein, Jimmy Gollihar, and James M. Musser |
| EPI_ISL_2203937, EPI_ISL_2203938 | Quest Diagnostics Incorporated | Centers for Disease Control and Prevention Division of Viral Diseases, Pathogen Discovery | Dakota Howard, Dhwani Batra, Peter W. Cook, Kara Moser, Adrian Paskey, Jason Caravas, Benjamin Rambo-Martin, Shatavia Morrison, Christopher Gulvick, Scott Sammons, Yvette Unoarumhi, Darlene Wagner, Matthew Schmerer, S. H. Rosenthal, A. Gerasimova, R. M. Kagan, B. Anderson, M. Hua, Y. Liu, L.E. Bernstein, K.E. Livingston, A. Perez, I. A. Shlyakhter, R. V. Rolando, R. Owen, P. Tanpaiboon, F. Lacbawan, Clinton R. Paden, Duncan MacCannell |
| EPI_ISL_2203939 | Houston Methodist Hospital | Houston Methodist Hospital | Randall J. Olsen, Paul A. Christensen, S. Wesley Long, Sishir Subedi, Robert Olson, Marcus Nguyen, James J. Davis, Matthew Ojeda Saavedra, Prasanti Yerramilli, Layne Pruitt, Kristina Reppond, Madison N. Shyer, Jessica Cambric, Ryan Gadd, Ilya J. Finkelstein, Jimmy Gollihar, and James M. Musser |
| EPI_ISL_2203940, EPI_ISL_2203941 | Quest Diagnostics Incorporated | Centers for Disease Control and Prevention Division of Viral Diseases, Pathogen Discovery | Dakota Howard, Dhwani Batra, Peter W. Cook, Kara Moser, Adrian Paskey, Jason Caravas, Benjamin Rambo-Martin, Shatavia Morrison, Christopher Gulvick, Scott Sammons, Yvette Unoarumhi, Darlene Wagner, Matthew Schmerer, S. H. Rosenthal, A. Gerasimova, R. M. Kagan, B. Anderson, M. Hua, Y. Liu, L.E. Bernstein, K.E. Livingston, A. Perez, I. A. Shlyakhter, R. V. Rolando, R. Owen, P. Tanpaiboon, F. Lacbawan, Clinton R. Paden, Duncan MacCannell |
| EPI_ISL_2203942 | Houston Methodist Hospital | Houston Methodist Hospital | Randall J. Olsen, Paul A. Christensen, S. Wesley Long, Sishir Subedi, Robert Olson, Marcus Nguyen, James J. Davis, Matthew Ojeda Saavedra, Prasanti Yerramilli, Layne Pruitt, Kristina Reppond, Madison N. Shyer, Jessica Cambric, Ryan Gadd, Ilya J. Finkelstein, Jimmy Gollihar, and James M. Musser |
| EPI_ISL_2203943 | Quest Diagnostics Incorporated | Centers for Disease Control and Prevention Division of Viral Diseases, Pathogen Discovery | Dakota Howard, Dhwani Batra, Peter W. Cook, Kara Moser, Adrian Paskey, Jason Caravas, Benjamin Rambo-Martin, Shatavia Morrison, Christopher Gulvick, Scott Sammons, Yvette Unoarumhi, Darlene Wagner, Matthew Schmerer, S. H. Rosenthal, A. Gerasimova, R. M. Kagan, B. Anderson, M. Hua, Y. Liu, L.E. Bernstein, K.E. Livingston, A. Perez, I. A. Shlyakhter, R. V. Rolando, R. Owen, P. Tanpaiboon, F. Lacbawan, Clinton R. Paden, Duncan MacCannell |
| EPI_ISL_2203944 | Houston Methodist Hospital | Houston Methodist Hospital | Randall J. Olsen, Paul A. Christensen, S. Wesley Long, Sishir Subedi, Robert Olson, Marcus Nguyen, James J. Davis, Matthew Ojeda Saavedra, Prasanti Yerramilli, Layne Pruitt, Kristina Reppond, Madison N. Shyer, Jessica Cambric, Ryan Gadd, Ilya J. Finkelstein, Jimmy Gollihar, and James M. Musser |
| EPI_ISL_2203945 | Quest Diagnostics Incorporated | Centers for Disease Control and Prevention Division of Viral Diseases, Pathogen Discovery | Dakota Howard, Dhwani Batra, Peter W. Cook, Kara Moser, Adrian Paskey, Jason Caravas, Benjamin Rambo-Martin, Shatavia Morrison, Christopher Gulvick, Scott Sammons, Yvette Unoarumhi, Darlene Wagner, Matthew Schmerer, S. H. Rosenthal, A. Gerasimova, R. M. Kagan, B. Anderson, M. Hua, Y. Liu, L.E. Bernstein, K.E. Livingston, A. Perez, I. A. Shlyakhter, R. V. Rolando, R. Owen, P. Tanpaiboon, F. Lacbawan, Clinton R. Paden, Duncan MacCannell |
| EPI_ISL_2203946 | Houston Methodist Hospital | Houston Methodist Hospital | Randall J. Olsen, Paul A. Christensen, S. Wesley Long, Sishir Subedi, Robert Olson, Marcus Nguyen, James J. Davis, Matthew Ojeda Saavedra, Prasanti Yerramilli, Layne Pruitt, Kristina Reppond, Madison N. Shyer, Jessica Cambric, Ryan Gadd, Ilya J. Finkelstein, Jimmy Gollihar, and James M. Musser |
| EPI_ISL_2203947 | Quest Diagnostics Incorporated | Centers for Disease Control and Prevention Division of Viral Diseases, Pathogen Discovery | Dakota Howard, Dhwani Batra, Peter W. Cook, Kara Moser, Adrian Paskey, Jason Caravas, Benjamin Rambo-Martin, Shatavia Morrison, Christopher Gulvick, Scott Sammons, Yvette Unoarumhi, Darlene Wagner, Matthew Schmerer, S. H. Rosenthal, A. Gerasimova, R. M. Kagan, B. Anderson, M. Hua, Y. Liu, L.E. Bernstein, K.E. Livingston, A. Perez, I. A. Shlyakhter, R. V. Rolando, R. Owen, P. Tanpaiboon, F. Lacbawan, Clinton R. Paden, Duncan MacCannell |
| EPI_ISL_2203948 | Houston Methodist Hospital | Houston Methodist Hospital | Randall J. Olsen, Paul A. Christensen, S. Wesley Long, Sishir Subedi, Robert Olson, Marcus Nguyen, James J. Davis, Matthew Ojeda Saavedra, Prasanti Yerramilli, Layne Pruitt, Kristina Reppond, Madison N. Shyer, Jessica Cambric, Ryan Gadd, Ilya J. Finkelstein, Jimmy Gollihar, and James M. Musser |
| EPI_ISL_2203949, EPI_ISL_2203950 | Quest Diagnostics Incorporated | Centers for Disease Control and Prevention Division of Viral Diseases, Pathogen Discovery | Dakota Howard, Dhwani Batra, Peter W. Cook, Kara Moser, Adrian Paskey, Jason Caravas, Benjamin Rambo-Martin, Shatavia Morrison, Christopher Gulvick, Scott Sammons, Yvette Unoarumhi, Darlene Wagner, Matthew Schmerer, S. H. Rosenthal, A. Gerasimova, R. M. Kagan, B. Anderson, M. Hua, Y. Liu, L.E. Bernstein, K.E. Livingston, A. Perez, I. A. Shlyakhter, R. V. Rolando, R. Owen, P. Tanpaiboon, F. Lacbawan, Clinton R. Paden, Duncan MacCannell |
| EPI_ISL_2203951 | Houston Methodist Hospital | Houston Methodist Hospital | Randall J. Olsen, Paul A. Christensen, S. Wesley Long, Sishir Subedi, Robert Olson, Marcus Nguyen, James J. Davis, Matthew Ojeda Saavedra, Prasanti Yerramilli, Layne Pruitt, Kristina Reppond, Madison N. Shyer, Jessica Cambric, Ryan Gadd, Ilya J. Finkelstein, Jimmy Gollihar, and James M. Musser |
| EPI_ISL_2203952, EPI_ISL_2203953 | Quest Diagnostics Incorporated | Centers for Disease Control and Prevention Division of Viral Diseases, Pathogen Discovery | Dakota Howard, Dhwani Batra, Peter W. Cook, Kara Moser, Adrian Paskey, Jason Caravas, Benjamin Rambo-Martin, Shatavia Morrison, Christopher Gulvick, Scott Sammons, Yvette Unoarumhi, Darlene Wagner, Matthew Schmerer, S. H. Rosenthal, A. Gerasimova, R. M. Kagan, B. Anderson, M. Hua, Y. Liu, L.E. Bernstein, K.E. Livingston, A. Perez, I. A. Shlyakhter, R. V. Rolando, R. Owen, P. Tanpaiboon, F. Lacbawan, Clinton R. Paden, Duncan MacCannell |
| EPI_ISL_2203954 | Houston Methodist Hospital | Houston Methodist Hospital | Randall J. Olsen, Paul A. Christensen, S. Wesley Long, Sishir Subedi, Robert Olson, Marcus Nguyen, James J. Davis, Matthew Ojeda Saavedra, Prasanti Yerramilli, Layne Pruitt, Kristina Reppond, Madison N. Shyer, Jessica Cambric, Ryan Gadd, Ilya J. Finkelstein, Jimmy Gollihar, and James M. Musser |
| EPI_ISL_2203955, EPI_ISL_2203956 | Quest Diagnostics Incorporated | Centers for Disease Control and Prevention Division of Viral Diseases, Pathogen Discovery | Dakota Howard, Dhwani Batra, Peter W. Cook, Kara Moser, Adrian Paskey, Jason Caravas, Benjamin Rambo-Martin, Shatavia Morrison, Christopher Gulvick, Scott Sammons, Yvette Unoarumhi, Darlene Wagner, Matthew Schmerer, S. H. Rosenthal, A. Gerasimova, R. M. Kagan, B. Anderson, M. Hua, Y. Liu, L.E. Bernstein, K.E. Livingston, A. Perez, I. A. Shlyakhter, R. V. Rolando, R. Owen, P. Tanpaiboon, F. Lacbawan, Clinton R. Paden, Duncan MacCannell |
| EPI_ISL_2203957 | Houston Methodist Hospital | Houston Methodist Hospital | Randall J. Olsen, Paul A. Christensen, S. Wesley Long, Sishir Subedi, Robert Olson, Marcus Nguyen, James J. Davis, Matthew Ojeda Saavedra, Prasanti Yerramilli, Layne Pruitt, Kristina Reppond, Madison N. Shyer, Jessica Cambric, Ryan Gadd, Ilya J. Finkelstein, Jimmy Gollihar, and James M. Musser |
| EPI_ISL_2203958, EPI_ISL_2203959 | Quest Diagnostics Incorporated | Centers for Disease Control and Prevention Division of Viral Diseases, Pathogen Discovery | Dakota Howard, Dhwani Batra, Peter W. Cook, Kara Moser, Adrian Paskey, Jason Caravas, Benjamin Rambo-Martin, Shatavia Morrison, Christopher Gulvick, Scott Sammons, Yvette Unoarumhi, Darlene Wagner, Matthew Schmerer, S. H. Rosenthal, A. Gerasimova, R. M. Kagan, B. Anderson, M. Hua, Y. Liu, L.E. Bernstein, K.E. Livingston, A. Perez, I. A. Shlyakhter, R. V. Rolando, R. Owen, P. Tanpaiboon, F. Lacbawan, Clinton R. Paden, Duncan MacCannell |
| EPI_ISL_2203960 | Houston Methodist Hospital | Houston Methodist Hospital | Randall J. Olsen, Paul A. Christensen, S. Wesley Long, Sishir Subedi, Robert Olson, Marcus Nguyen, James J. Davis, Matthew Ojeda Saavedra, Prasanti Yerramilli, Layne Pruitt, Kristina Reppond, Madison N. Shyer, Jessica Cambric, Ryan Gadd, Ilya J. Finkelstein, Jimmy Gollihar, and James M. Musser |
| EPI_ISL_2203961, EPI_ISL_2203963 | Quest Diagnostics Incorporated | Centers for Disease Control and Prevention Division of Viral Diseases, Pathogen Discovery | Dakota Howard, Dhwani Batra, Peter W. Cook, Kara Moser, Adrian Paskey, Jason Caravas, Benjamin Rambo-Martin, Shatavia Morrison, Christopher Gulvick, Scott Sammons, Yvette Unoarumhi, Darlene Wagner, Matthew Schmerer, S. H. Rosenthal, A. Gerasimova, R. M. Kagan, B. Anderson, M. Hua, Y. Liu, L.E. Bernstein, K.E. Livingston, A. Perez, I. A. Shlyakhter, R. V. Rolando, R. Owen, P. Tanpaiboon, F. Lacbawan, Clinton R. Paden, Duncan MacCannell |
| EPI_ISL_2203965 | Houston Methodist Hospital | Houston Methodist Hospital | Randall J. Olsen, Paul A. Christensen, S. Wesley Long, Sishir Subedi, Robert Olson, Marcus Nguyen, James J. Davis, Matthew Ojeda Saavedra, Prasanti Yerramilli, Layne Pruitt, Kristina Reppond, Madison N. Shyer, Jessica Cambric, Ryan Gadd, Ilya J. Finkelstein, Jimmy Gollihar, and James M. Musser |
| EPI_ISL_2203966, EPI_ISL_2203967, EPI_ISL_2203968 | Quest Diagnostics Incorporated | Centers for Disease Control and Prevention Division of Viral Diseases, Pathogen Discovery | Dakota Howard, Dhwani Batra, Peter W. Cook, Kara Moser, Adrian Paskey, Jason Caravas, Benjamin Rambo-Martin, Shatavia Morrison, Christopher Gulvick, Scott Sammons, Yvette Unoarumhi, Darlene Wagner, Matthew Schmerer, S. H. Rosenthal, A. Gerasimova, R. M. Kagan, B. Anderson, M. Hua, Y. Liu, L.E. Bernstein, K.E. Livingston, A. Perez, I. A. Shlyakhter, R. V. Rolando, R. Owen, P. Tanpaiboon, F. Lacbawan, Clinton R. Paden, Duncan MacCannell |
| EPI_ISL_2203969 | Houston Methodist Hospital | Houston Methodist Hospital | Randall J. Olsen, Paul A. Christensen, S. Wesley Long, Sishir Subedi, Robert Olson, Marcus Nguyen, James J. Davis, Matthew Ojeda Saavedra, Prasanti Yerramilli, Layne Pruitt, Kristina Reppond, Madison N. Shyer, Jessica Cambric, Ryan Gadd, Ilya J. Finkelstein, Jimmy Gollihar, and James M. Musser |
| EPI_ISL_2203970, EPI_ISL_2203972 | Quest Diagnostics Incorporated | Centers for Disease Control and Prevention Division of Viral Diseases, Pathogen Discovery | Dakota Howard, Dhwani Batra, Peter W. Cook, Kara Moser, Adrian Paskey, Jason Caravas, Benjamin Rambo-Martin, Shatavia Morrison, Christopher Gulvick, Scott Sammons, Yvette Unoarumhi, Darlene Wagner, Matthew Schmerer, S. H. Rosenthal, A. Gerasimova, R. M. Kagan, B. Anderson, M. Hua, Y. Liu, L.E. Bernstein, K.E. Livingston, A. Perez, I. A. Shlyakhter, R. V. Rolando, R. Owen, P. Tanpaiboon, F. Lacbawan, Clinton R. Paden, Duncan MacCannell |
| EPI_ISL_2203973 | Houston Methodist Hospital | Houston Methodist Hospital | Randall J. Olsen, Paul A. Christensen, S. Wesley Long, Sishir Subedi, Robert Olson, Marcus Nguyen, James J. Davis, Matthew Ojeda Saavedra, Prasanti Yerramilli, Layne Pruitt, Kristina Reppond, Madison N. Shyer, Jessica Cambric, Ryan Gadd, Ilya J. Finkelstein, Jimmy Gollihar, and James M. Musser |
| EPI_ISL_2203974, EPI_ISL_2203975 | Quest Diagnostics Incorporated | Centers for Disease Control and Prevention Division of Viral Diseases, Pathogen Discovery | Dakota Howard, Dhwani Batra, Peter W. Cook, Kara Moser, Adrian Paskey, Jason Caravas, Benjamin Rambo-Martin, Shatavia Morrison, Christopher Gulvick, Scott Sammons, Yvette Unoarumhi, Darlene Wagner, Matthew Schmerer, S. H. Rosenthal, A. Gerasimova, R. M. Kagan, B. Anderson, M. Hua, Y. Liu, L.E. Bernstein, K.E. Livingston, A. Perez, I. A. Shlyakhter, R. V. Rolando, R. Owen, P. Tanpaiboon, F. Lacbawan, Clinton R. Paden, Duncan MacCannell |
| EPI_ISL_2203976 | Houston Methodist Hospital | Houston Methodist Hospital | Randall J. Olsen, Paul A. Christensen, S. Wesley Long, Sishir Subedi, Robert Olson, Marcus Nguyen, James J. Davis, Matthew Ojeda Saavedra, Prasanti Yerramilli, Layne Pruitt, Kristina Reppond, Madison N. Shyer, Jessica Cambric, Ryan Gadd, Ilya J. Finkelstein, Jimmy Gollihar, and James M. Musser |
| EPI_ISL_2203977, EPI_ISL_2203978, EPI_ISL_2203980, EPI_ISL_2203982 | Quest Diagnostics Incorporated | Centers for Disease Control and Prevention Division of Viral Diseases, Pathogen Discovery | Dakota Howard, Dhwani Batra, Peter W. Cook, Kara Moser, Adrian Paskey, Jason Caravas, Benjamin Rambo-Martin, Shatavia Morrison, Christopher Gulvick, Scott Sammons, Yvette Unoarumhi, Darlene Wagner, Matthew Schmerer, S. H. Rosenthal, A. Gerasimova, R. M. Kagan, B. Anderson, M. Hua, Y. Liu, L.E. Bernstein, K.E. Livingston, A. Perez, I. A. Shlyakhter, R. V. Rolando, R. Owen, P. Tanpaiboon, F. Lacbawan, Clinton R. Paden, Duncan MacCannell |
| EPI_ISL_2203983 | Houston Methodist Hospital | Houston Methodist Hospital | Randall J. Olsen, Paul A. Christensen, S. Wesley Long, Sishir Subedi, Robert Olson, Marcus Nguyen, James J. Davis, Matthew Ojeda Saavedra, Prasanti Yerramilli, Layne Pruitt, Kristina Reppond, Madison N. Shyer, Jessica Cambric, Ryan Gadd, Ilya J. Finkelstein, Jimmy Gollihar, and James M. Musser |
| EPI_ISL_2203984 | Quest Diagnostics Incorporated | Centers for Disease Control and Prevention Division of Viral Diseases, Pathogen Discovery | Dakota Howard, Dhwani Batra, Peter W. Cook, Kara Moser, Adrian Paskey, Jason Caravas, Benjamin Rambo-Martin, Shatavia Morrison, Christopher Gulvick, Scott Sammons, Yvette Unoarumhi, Darlene Wagner, Matthew Schmerer, S. H. Rosenthal, A. Gerasimova, R. M. Kagan, B. Anderson, M. Hua, Y. Liu, L.E. Bernstein, K.E. Livingston, A. Perez, I. A. Shlyakhter, R. V. Rolando, R. Owen, P. Tanpaiboon, F. Lacbawan, Clinton R. Paden, Duncan MacCannell |
| EPI_ISL_2203985 | Houston Methodist Hospital | Houston Methodist Hospital | Randall J. Olsen, Paul A. Christensen, S. Wesley Long, Sishir Subedi, Robert Olson, Marcus Nguyen, James J. Davis, Matthew Ojeda Saavedra, Prasanti Yerramilli, Layne Pruitt, Kristina Reppond, Madison N. Shyer, Jessica Cambric, Ryan Gadd, Ilya J. Finkelstein, Jimmy Gollihar, and James M. Musser |
| EPI_ISL_2203986, EPI_ISL_2203987 | Quest Diagnostics Incorporated | Centers for Disease Control and Prevention Division of Viral Diseases, Pathogen Discovery | Dakota Howard, Dhwani Batra, Peter W. Cook, Kara Moser, Adrian Paskey, Jason Caravas, Benjamin Rambo-Martin, Shatavia Morrison, Christopher Gulvick, Scott Sammons, Yvette Unoarumhi, Darlene Wagner, Matthew Schmerer, S. H. Rosenthal, A. Gerasimova, R. M. Kagan, B. Anderson, M. Hua, Y. Liu, L.E. Bernstein, K.E. Livingston, A. Perez, I. A. Shlyakhter, R. V. Rolando, R. Owen, P. Tanpaiboon, F. Lacbawan, Clinton R. Paden, Duncan MacCannell |
| EPI_ISL_2203988 | Houston Methodist Hospital | Houston Methodist Hospital | Randall J. Olsen, Paul A. Christensen, S. Wesley Long, Sishir Subedi, Robert Olson, Marcus Nguyen, James J. Davis, Matthew Ojeda Saavedra, Prasanti Yerramilli, Layne Pruitt, Kristina Reppond, Madison N. Shyer, Jessica Cambric, Ryan Gadd, Ilya J. Finkelstein, Jimmy Gollihar, and James M. Musser |
| EPI_ISL_2203989, EPI_ISL_2203990, EPI_ISL_2203992 | Quest Diagnostics Incorporated | Centers for Disease Control and Prevention Division of Viral Diseases, Pathogen Discovery | Dakota Howard, Dhwani Batra, Peter W. Cook, Kara Moser, Adrian Paskey, Jason Caravas, Benjamin Rambo-Martin, Shatavia Morrison, Christopher Gulvick, Scott Sammons, Yvette Unoarumhi, Darlene Wagner, Matthew Schmerer, S. H. Rosenthal, A. Gerasimova, R. M. Kagan, B. Anderson, M. Hua, Y. Liu, L.E. Bernstein, K.E. Livingston, A. Perez, I. A. Shlyakhter, R. V. Rolando, R. Owen, P. Tanpaiboon, F. Lacbawan, Clinton R. Paden, Duncan MacCannell |
| EPI_ISL_2203993 | Houston Methodist Hospital | Houston Methodist Hospital | Randall J. Olsen, Paul A. Christensen, S. Wesley Long, Sishir Subedi, Robert Olson, Marcus Nguyen, James J. Davis, Matthew Ojeda Saavedra, Prasanti Yerramilli, Layne Pruitt, Kristina Reppond, Madison N. Shyer, Jessica Cambric, Ryan Gadd, Ilya J. Finkelstein, Jimmy Gollihar, and James M. Musser |
| EPI_ISL_2203994, EPI_ISL_2203996, EPI_ISL_2203998, EPI_ISL_2203999 | Quest Diagnostics Incorporated | Centers for Disease Control and Prevention Division of Viral Diseases, Pathogen Discovery | Dakota Howard, Dhwani Batra, Peter W. Cook, Kara Moser, Adrian Paskey, Jason Caravas, Benjamin Rambo-Martin, Shatavia Morrison, Christopher Gulvick, Scott Sammons, Yvette Unoarumhi, Darlene Wagner, Matthew Schmerer, S. H. Rosenthal, A. Gerasimova, R. M. Kagan, B. Anderson, M. Hua, Y. Liu, L.E. Bernstein, K.E. Livingston, A. Perez, I. A. Shlyakhter, R. V. Rolando, R. Owen, P. Tanpaiboon, F. Lacbawan, Clinton R. Paden, Duncan MacCannell |
| EPI_ISL_2204000 | Houston Methodist Hospital | Houston Methodist Hospital | Randall J. Olsen, Paul A. Christensen, S. Wesley Long, Sishir Subedi, Robert Olson, Marcus Nguyen, James J. Davis, Matthew Ojeda Saavedra, Prasanti Yerramilli, Layne Pruitt, Kristina Reppond, Madison N. Shyer, Jessica Cambric, Ryan Gadd, Ilya J. Finkelstein, Jimmy Gollihar, and James |

| | | | |
|---|---|---|---|
| | | | M. Musser |
| EPI_ISL_2204001 | Quest Diagnostics Incorporated | Centers for Disease Control and Prevention Division of Viral Diseases, Pathogen Discovery | Dakota Howard, Dhwani Batra, Peter W. Cook, Kara Moser, Adrian Paskey, Jason Caravas, Benjamin Rambo-Martin, Shatavia Morrison, Christopher Gulvick, Scott Sammons, Yvette Unoarumhi, Darlene Wagner, Matthew Schmerer, S. H. Rosenthal, A. Gerasimova, R. M. Kagan, B. Anderson, M. Hua, Y. Liu, L.E. Bernstein, K.E. Livingston, A. Perez, I. A. Shlyakhter, R. V. Rolando, R. Owen, P. Tanpaiboon, F. Lacbawan, Clinton R. Paden, Duncan MacCannell |
| EPI_ISL_2204002 | Houston Methodist Hospital | Houston Methodist Hospital | Randall J. Olsen, Paul A. Christensen, S. Wesley Long, Sishir Subedi, Robert Olson, Marcus Nguyen, James J. Davis, Matthew Ojeda Saavedra, Prasanti Yerramilli, Layne Pruitt, Kristina Reppond, Madison N. Shyer, Jessica Cambric, Ryan Gadd, Ilya J. Finkelstein, Jimmy Gollihar, and James M. Musser |
| EPI_ISL_2204003 | Quest Diagnostics Incorporated | Centers for Disease Control and Prevention Division of Viral Diseases, Pathogen Discovery | Dakota Howard, Dhwani Batra, Peter W. Cook, Kara Moser, Adrian Paskey, Jason Caravas, Benjamin Rambo-Martin, Shatavia Morrison, Christopher Gulvick, Scott Sammons, Yvette Unoarumhi, Darlene Wagner, Matthew Schmerer, S. H. Rosenthal, A. Gerasimova, R. M. Kagan, B. Anderson, M. Hua, Y. Liu, L.E. Bernstein, K.E. Livingston, A. Perez, I. A. Shlyakhter, R. V. Rolando, R. Owen, P. Tanpaiboon, F. Lacbawan, Clinton R. Paden, Duncan MacCannell |
| EPI_ISL_2204004 | Houston Methodist Hospital | Houston Methodist Hospital | Randall J. Olsen, Paul A. Christensen, S. Wesley Long, Sishir Subedi, Robert Olson, Marcus Nguyen, James J. Davis, Matthew Ojeda Saavedra, Prasanti Yerramilli, Layne Pruitt, Kristina Reppond, Madison N. Shyer, Jessica Cambric, Ryan Gadd, Ilya J. Finkelstein, Jimmy Gollihar, and James M. Musser |
| EPI_ISL_2204005 | Quest Diagnostics Incorporated | Centers for Disease Control and Prevention Division of Viral Diseases, Pathogen Discovery | Dakota Howard, Dhwani Batra, Peter W. Cook, Kara Moser, Adrian Paskey, Jason Caravas, Benjamin Rambo-Martin, Shatavia Morrison, Christopher Gulvick, Scott Sammons, Yvette Unoarumhi, Darlene Wagner, Matthew Schmerer, S. H. Rosenthal, A. Gerasimova, R. M. Kagan, B. Anderson, M. Hua, Y. Liu, L.E. Bernstein, K.E. Livingston, A. Perez, I. A. Shlyakhter, R. V. Rolando, R. Owen, P. Tanpaiboon, F. Lacbawan, Clinton R. Paden, Duncan MacCannell |
| EPI_ISL_2204006 | Houston Methodist Hospital | Houston Methodist Hospital | Randall J. Olsen, Paul A. Christensen, S. Wesley Long, Sishir Subedi, Robert Olson, Marcus Nguyen, James J. Davis, Matthew Ojeda Saavedra, Prasanti Yerramilli, Layne Pruitt, Kristina Reppond, Madison N. Shyer, Jessica Cambric, Ryan Gadd, Ilya J. Finkelstein, Jimmy Gollihar, and James M. Musser |
| EPI_ISL_2204007, EPI_ISL_2204008 | Quest Diagnostics Incorporated | Centers for Disease Control and Prevention Division of Viral Diseases, Pathogen Discovery | Dakota Howard, Dhwani Batra, Peter W. Cook, Kara Moser, Adrian Paskey, Jason Caravas, Benjamin Rambo-Martin, Shatavia Morrison, Christopher Gulvick, Scott Sammons, Yvette Unoarumhi, Darlene Wagner, Matthew Schmerer, S. H. Rosenthal, A. Gerasimova, R. M. Kagan, B. Anderson, M. Hua, Y. Liu, L.E. Bernstein, K.E. Livingston, A. Perez, I. A. Shlyakhter, R. V. Rolando, R. Owen, P. Tanpaiboon, F. Lacbawan, Clinton R. Paden, Duncan MacCannell |
| EPI_ISL_2204009 | Houston Methodist Hospital | Houston Methodist Hospital | Randall J. Olsen, Paul A. Christensen, S. Wesley Long, Sishir Subedi, Robert Olson, Marcus Nguyen, James J. Davis, Matthew Ojeda Saavedra, Prasanti Yerramilli, Layne Pruitt, Kristina Reppond, Madison N. Shyer, Jessica Cambric, Ryan Gadd, Ilya J. Finkelstein, Jimmy Gollihar, and James M. Musser |
| EPI_ISL_2204010, EPI_ISL_2204011 | Quest Diagnostics Incorporated | Centers for Disease Control and Prevention Division of Viral Diseases, Pathogen Discovery | Dakota Howard, Dhwani Batra, Peter W. Cook, Kara Moser, Adrian Paskey, Jason Caravas, Benjamin Rambo-Martin, Shatavia Morrison, Christopher Gulvick, Scott Sammons, Yvette Unoarumhi, Darlene Wagner, Matthew Schmerer, S. H. Rosenthal, A. Gerasimova, R. M. Kagan, B. Anderson, M. Hua, Y. Liu, L.E. Bernstein, K.E. Livingston, A. Perez, I. A. Shlyakhter, R. V. Rolando, R. Owen, P. Tanpaiboon, F. Lacbawan, Clinton R. Paden, Duncan MacCannell |
| EPI_ISL_2204012 | Houston Methodist Hospital | Houston Methodist Hospital | Randall J. Olsen, Paul A. Christensen, S. Wesley Long, Sishir Subedi, Robert Olson, Marcus Nguyen, James J. Davis, Matthew Ojeda Saavedra, Prasanti Yerramilli, Layne Pruitt, Kristina Reppond, Madison N. Shyer, Jessica Cambric, Ryan Gadd, Ilya J. Finkelstein, Jimmy Gollihar, and James M. Musser |
| EPI_ISL_2204013 | Quest Diagnostics Incorporated | Centers for Disease Control and Prevention Division of Viral Diseases, Pathogen Discovery | Dakota Howard, Dhwani Batra, Peter W. Cook, Kara Moser, Adrian Paskey, Jason Caravas, Benjamin Rambo-Martin, Shatavia Morrison, Christopher Gulvick, Scott Sammons, Yvette Unoarumhi, Darlene Wagner, Matthew Schmerer, S. H. Rosenthal, A. Gerasimova, R. M. Kagan, B. Anderson, M. Hua, Y. Liu, L.E. Bernstein, K.E. Livingston, A. Perez, I. A. Shlyakhter, R. V. Rolando, R. Owen, P. Tanpaiboon, F. Lacbawan, Clinton R. Paden, Duncan MacCannell |
| EPI_ISL_2204014 | Houston Methodist Hospital | Houston Methodist Hospital | Randall J. Olsen, Paul A. Christensen, S. Wesley Long, Sishir Subedi, Robert Olson, Marcus Nguyen, James J. Davis, Matthew Ojeda Saavedra, Prasanti Yerramilli, Layne Pruitt, Kristina Reppond, Madison N. Shyer, Jessica Cambric, Ryan Gadd, Ilya J. Finkelstein, Jimmy Gollihar, and James M. Musser |
| EPI_ISL_2204015, EPI_ISL_2204017 | Quest Diagnostics Incorporated | Centers for Disease Control and Prevention Division of Viral Diseases, Pathogen Discovery | Dakota Howard, Dhwani Batra, Peter W. Cook, Kara Moser, Adrian Paskey, Jason Caravas, Benjamin Rambo-Martin, Shatavia Morrison, Christopher Gulvick, Scott Sammons, Yvette Unoarumhi, Darlene Wagner, Matthew Schmerer, S. H. Rosenthal, A. Gerasimova, R. M. Kagan, B. Anderson, M. Hua, Y. Liu, L.E. Bernstein, K.E. Livingston, A. Perez, I. A. Shlyakhter, R. V. Rolando, R. Owen, P. Tanpaiboon, F. Lacbawan, Clinton R. Paden, Duncan MacCannell |
| EPI_ISL_2204018 | Houston Methodist Hospital | Houston Methodist Hospital | Randall J. Olsen, Paul A. Christensen, S. Wesley Long, Sishir Subedi, Robert Olson, Marcus Nguyen, James J. Davis, Matthew Ojeda Saavedra, Prasanti Yerramilli, Layne Pruitt, Kristina Reppond, Madison N. Shyer, Jessica Cambric, Ryan Gadd, Ilya J. Finkelstein, Jimmy Gollihar, and James M. Musser |
| EPI_ISL_2204019, EPI_ISL_2204020 | Quest Diagnostics Incorporated | Centers for Disease Control and Prevention Division of Viral Diseases, Pathogen Discovery | Dakota Howard, Dhwani Batra, Peter W. Cook, Kara Moser, Adrian Paskey, Jason Caravas, Benjamin Rambo-Martin, Shatavia Morrison, Christopher Gulvick, Scott Sammons, Yvette Unoarumhi, Darlene Wagner, Matthew Schmerer, S. H. Rosenthal, A. Gerasimova, R. M. Kagan, B. Anderson, M. Hua, Y. Liu, L.E. Bernstein, K.E. Livingston, A. Perez, I. A. Shlyakhter, R. V. Rolando, R. Owen, P. Tanpaiboon, F. Lacbawan, Clinton R. Paden, Duncan MacCannell |
| EPI_ISL_2204021 | Houston Methodist Hospital | Houston Methodist Hospital | Randall J. Olsen, Paul A. Christensen, S. Wesley Long, Sishir Subedi, Robert Olson, Marcus Nguyen, James J. Davis, Matthew Ojeda Saavedra, Prasanti Yerramilli, Layne Pruitt, Kristina Reppond, Madison N. Shyer, Jessica Cambric, Ryan Gadd, Ilya J. Finkelstein, Jimmy Gollihar, and James M. Musser |
| EPI_ISL_2204022 | Quest Diagnostics Incorporated | Centers for Disease Control and Prevention Division of Viral Diseases, Pathogen Discovery | Dakota Howard, Dhwani Batra, Peter W. Cook, Kara Moser, Adrian Paskey, Jason Caravas, Benjamin Rambo-Martin, Shatavia Morrison, Christopher Gulvick, Scott Sammons, Yvette Unoarumhi, Darlene Wagner, Matthew Schmerer, S. H. Rosenthal, A. Gerasimova, R. M. Kagan, B. Anderson, M. Hua, Y. Liu, L.E. Bernstein, K.E. Livingston, A. Perez, I. A. Shlyakhter, R. V. Rolando, R. Owen, P. Tanpaiboon, F. Lacbawan, Clinton R. Paden, Duncan MacCannell |
| EPI_ISL_2204023 | Houston Methodist Hospital | Houston Methodist Hospital | Randall J. Olsen, Paul A. Christensen, S. Wesley Long, Sishir Subedi, Robert Olson, Marcus Nguyen, James J. Davis, Matthew Ojeda Saavedra, Prasanti Yerramilli, Layne Pruitt, Kristina Reppond, Madison N. Shyer, Jessica Cambric, Ryan Gadd, Ilya J. Finkelstein, Jimmy Gollihar, and James M. Musser |
| EPI_ISL_2204024 | Quest Diagnostics Incorporated | Centers for Disease Control and Prevention Division of Viral Diseases, Pathogen Discovery | Dakota Howard, Dhwani Batra, Peter W. Cook, Kara Moser, Adrian Paskey, Jason Caravas, Benjamin Rambo-Martin, Shatavia Morrison, Christopher Gulvick, Scott Sammons, Yvette Unoarumhi, Darlene Wagner, Matthew Schmerer, S. H. Rosenthal, A. Gerasimova, R. M. Kagan, B. Anderson, M. Hua, Y. Liu, L.E. Bernstein, K.E. Livingston, A. Perez, I. A. Shlyakhter, R. V. Rolando, R. Owen, P. Tanpaiboon, F. Lacbawan, Clinton R. Paden, Duncan MacCannell |
| EPI_ISL_2204025 | Houston Methodist Hospital | Houston Methodist Hospital | Randall J. Olsen, Paul A. Christensen, S. Wesley Long, Sishir Subedi, Robert Olson, Marcus Nguyen, James J. Davis, Matthew Ojeda Saavedra, Prasanti Yerramilli, Layne Pruitt, Kristina Reppond, Madison N. Shyer, Jessica Cambric, Ryan Gadd, Ilya J. Finkelstein, Jimmy Gollihar, and James M. Musser |
| EPI_ISL_2204026, EPI_ISL_2204027 | Quest Diagnostics Incorporated | Centers for Disease Control and Prevention Division of Viral Diseases, Pathogen Discovery | Dakota Howard, Dhwani Batra, Peter W. Cook, Kara Moser, Adrian Paskey, Jason Caravas, Benjamin Rambo-Martin, Shatavia Morrison, Christopher Gulvick, Scott Sammons, Yvette Unoarumhi, Darlene Wagner, Matthew Schmerer, S. H. Rosenthal, A. Gerasimova, R. M. Kagan, B. Anderson, M. Hua, Y. Liu, L.E. Bernstein, K.E. Livingston, A. Perez, I. A. Shlyakhter, R. V. Rolando, R. Owen, P. Tanpaiboon, F. Lacbawan, Clinton R. Paden, Duncan MacCannell |
| EPI_ISL_2204028 | Houston Methodist Hospital | Houston Methodist Hospital | Randall J. Olsen, Paul A. Christensen, S. Wesley Long, Sishir Subedi, Robert Olson, Marcus Nguyen, James J. Davis, Matthew Ojeda Saavedra, Prasanti Yerramilli, Layne Pruitt, Kristina Reppond, Madison N. Shyer, Jessica Cambric, Ryan Gadd, Ilya J. Finkelstein, Jimmy Gollihar, and James M. Musser |
| EPI_ISL_2204029, EPI_ISL_2204030, EPI_ISL_2204031, EPI_ISL_2204033 | Quest Diagnostics Incorporated | Centers for Disease Control and Prevention Division of Viral Diseases, Pathogen Discovery | Dakota Howard, Dhwani Batra, Peter W. Cook, Kara Moser, Adrian Paskey, Jason Caravas, Benjamin Rambo-Martin, Shatavia Morrison, Christopher Gulvick, Scott Sammons, Yvette Unoarumhi, Darlene Wagner, Matthew Schmerer, S. H. Rosenthal, A. Gerasimova, R. M. Kagan, B. Anderson, M. Hua, Y. Liu, L.E. Bernstein, K.E. Livingston, A. Perez, I. A. Shlyakhter, R. V. Rolando, R. Owen, P. Tanpaiboon, F. Lacbawan, Clinton R. Paden, Duncan MacCannell |
| EPI_ISL_2204034 | Houston Methodist Hospital | Houston Methodist Hospital | Randall J. Olsen, Paul A. Christensen, S. Wesley Long, Sishir Subedi, Robert Olson, Marcus Nguyen, James J. Davis, Matthew Ojeda Saavedra, Prasanti Yerramilli, Layne Pruitt, Kristina Reppond, Madison N. Shyer, Jessica Cambric, Ryan Gadd, Ilya J. Finkelstein, Jimmy Gollihar, and James M. Musser |
| EPI_ISL_2204035, EPI_ISL_2204036 | Quest Diagnostics Incorporated | Centers for Disease Control and Prevention Division of Viral Diseases, Pathogen Discovery | Dakota Howard, Dhwani Batra, Peter W. Cook, Kara Moser, Adrian Paskey, Jason Caravas, Benjamin Rambo-Martin, Shatavia Morrison, Christopher Gulvick, Scott Sammons, Yvette Unoarumhi, Darlene Wagner, Matthew Schmerer, S. H. Rosenthal, A. Gerasimova, R. M. Kagan, B. Anderson, M. Hua, Y. Liu, L.E. Bernstein, K.E. Livingston, A. Perez, I. A. Shlyakhter, R. V. Rolando, R. Owen, P. Tanpaiboon, F. Lacbawan, Clinton R. Paden, Duncan MacCannell |
| EPI_ISL_2204037 | Houston Methodist Hospital | Houston Methodist Hospital | Randall J. Olsen, Paul A. Christensen, S. Wesley Long, Sishir Subedi, Robert Olson, Marcus Nguyen, James J. Davis, Matthew Ojeda Saavedra, Prasanti Yerramilli, Layne Pruitt, Kristina Reppond, Madison N. Shyer, Jessica Cambric, Ryan Gadd, Ilya J. Finkelstein, Jimmy Gollihar, and James M. Musser |
| EPI_ISL_2204038, EPI_ISL_2204039 | Quest Diagnostics Incorporated | Centers for Disease Control and Prevention Division of Viral Diseases, Pathogen Discovery | Dakota Howard, Dhwani Batra, Peter W. Cook, Kara Moser, Adrian Paskey, Jason Caravas, Benjamin Rambo-Martin, Shatavia Morrison, Christopher Gulvick, Scott Sammons, Yvette Unoarumhi, Darlene Wagner, Matthew Schmerer, S. H. Rosenthal, A. Gerasimova, R. M. Kagan, B. Anderson, M. Hua, Y. Liu, L.E. Bernstein, K.E. Livingston, A. Perez, I. A. Shlyakhter, R. V. Rolando, R. Owen, P. Tanpaiboon, F. Lacbawan, Clinton R. Paden, Duncan MacCannell |
| EPI_ISL_2204040 | Houston Methodist Hospital | Houston Methodist Hospital | Randall J. Olsen, Paul A. Christensen, S. Wesley Long, Sishir Subedi, Robert Olson, Marcus Nguyen, James J. Davis, Matthew Ojeda Saavedra, Prasanti Yerramilli, Layne Pruitt, Kristina Reppond, Madison N. Shyer, Jessica Cambric, Ryan Gadd, Ilya J. Finkelstein, Jimmy Gollihar, and James M. Musser |
| EPI_ISL_2204041 | Quest Diagnostics Incorporated | Centers for Disease Control and Prevention Division of Viral Diseases, Pathogen Discovery | Dakota Howard, Dhwani Batra, Peter W. Cook, Kara Moser, Adrian Paskey, Jason Caravas, Benjamin Rambo-Martin, Shatavia Morrison, Christopher Gulvick, Scott Sammons, Yvette Unoarumhi, Darlene Wagner, Matthew Schmerer, S. H. Rosenthal, A. Gerasimova, R. M. Kagan, B. Anderson, M. Hua, Y. Liu, L.E. Bernstein, K.E. Livingston, A. Perez, I. A. Shlyakhter, R. V. Rolando, R. Owen, P. Tanpaiboon, F. Lacbawan, Clinton R. Paden, Duncan MacCannell |
| EPI_ISL_2204042 | Houston Methodist Hospital | Houston Methodist Hospital | Randall J. Olsen, Paul A. Christensen, S. Wesley Long, Sishir Subedi, Robert Olson, Marcus Nguyen, James J. Davis, Matthew Ojeda Saavedra, Prasanti Yerramilli, Layne Pruitt, Kristina Reppond, Madison N. Shyer, Jessica Cambric, Ryan Gadd, Ilya J. Finkelstein, Jimmy Gollihar, and James M. Musser |
| EPI_ISL_2204043, EPI_ISL_2204044 | Quest Diagnostics Incorporated | Centers for Disease Control and Prevention Division of Viral Diseases, Pathogen Discovery | Dakota Howard, Dhwani Batra, Peter W. Cook, Kara Moser, Adrian Paskey, Jason Caravas, Benjamin Rambo-Martin, Shatavia Morrison, Christopher Gulvick, Scott Sammons, Yvette Unoarumhi, Darlene Wagner, Matthew Schmerer, S. H. Rosenthal, A. Gerasimova, R. M. Kagan, B. Anderson, M. Hua, Y. Liu, L.E. Bernstein, K.E. Livingston, A. Perez, I. A. Shlyakhter, R. V. Rolando, R. Owen, P. Tanpaiboon, F. Lacbawan, Clinton R. Paden, Duncan MacCannell |
| EPI_ISL_2204045 | Houston Methodist Hospital | Houston Methodist Hospital | Randall J. Olsen, Paul A. Christensen, S. Wesley Long, Sishir Subedi, Robert Olson, Marcus Nguyen, James J. Davis, Matthew Ojeda Saavedra, Prasanti Yerramilli, Layne Pruitt, Kristina Reppond, Madison N. Shyer, Jessica Cambric, Ryan Gadd, Ilya J. Finkelstein, Jimmy Gollihar, and James M. Musser |
| EPI_ISL_2204046, EPI_ISL_2204047, EPI_ISL_2204048 | Quest Diagnostics Incorporated | Centers for Disease Control and Prevention Division of Viral Diseases, Pathogen Discovery | Dakota Howard, Dhwani Batra, Peter W. Cook, Kara Moser, Adrian Paskey, Jason Caravas, Benjamin Rambo-Martin, Shatavia Morrison, Christopher Gulvick, Scott Sammons, Yvette Unoarumhi, Darlene Wagner, Matthew Schmerer, S. H. Rosenthal, A. Gerasimova, R. M. Kagan, B. Anderson, M. Hua, Y. Liu, L.E. Bernstein, K.E. Livingston, A. Perez, I. A. Shlyakhter, R. V. Rolando, R. Owen, P. Tanpaiboon, F. Lacbawan, Clinton R. Paden, Duncan MacCannell |
| EPI_ISL_2204049 | Houston Methodist Hospital | Houston Methodist Hospital | Randall J. Olsen, Paul A. Christensen, S. Wesley Long, Sishir Subedi, Robert Olson, Marcus Nguyen, James J. Davis, Matthew Ojeda Saavedra, Prasanti Yerramilli, Layne Pruitt, Kristina Reppond, Madison N. Shyer, Jessica Cambric, Ryan Gadd, Ilya J. Finkelstein, Jimmy Gollihar, and James M. Musser |
| EPI_ISL_2204050, EPI_ISL_2204051 | Quest Diagnostics Incorporated | Centers for Disease Control and Prevention Division of Viral Diseases, Pathogen Discovery | Dakota Howard, Dhwani Batra, Peter W. Cook, Kara Moser, Adrian Paskey, Jason Caravas, Benjamin Rambo-Martin, Shatavia Morrison, Christopher Gulvick, Scott Sammons, Yvette Unoarumhi, Darlene Wagner, Matthew Schmerer, S. H. Rosenthal, A. Gerasimova, R. M. Kagan, B. Anderson, M. Hua, Y. Liu, L.E. Bernstein, K.E. Livingston, A. Perez, I. A. Shlyakhter, R. V. Rolando, R. Owen, P. Tanpaiboon, F. Lacbawan, Clinton R. Paden, Duncan MacCannell |
| EPI_ISL_2204052 | Houston Methodist Hospital | Houston Methodist Hospital | Randall J. Olsen, Paul A. Christensen, S. Wesley Long, Sishir Subedi, Robert Olson, Marcus Nguyen, James J. Davis, Matthew Ojeda Saavedra, Prasanti Yerramilli, Layne Pruitt, Kristina Reppond, Madison N. Shyer, Jessica Cambric, Ryan Gadd, Ilya J. Finkelstein, Jimmy Gollihar, and James M. Musser |
| EPI_ISL_2204053, | Quest Diagnostics Incorporated | Centers for Disease Control and Prevention | Dakota Howard, Dhwani Batra, Peter W. Cook, Kara Moser, Adrian Paskey, Jason Caravas, Benjamin Rambo-Martin, Shatavia Morrison, Christopher Gulvick, Scott Sammons, Yvette Unoarumhi, Darlene Wagner, Matthew Schmerer, S. H. Rosenthal, A. Gerasimova, R. M. Kagan, B. |

| | | | |
|---|---|---|---|
| EPI_ISL_2204054, EPI_ISL_2204055, EPI_ISL_2204056 | | Division of Viral Diseases, Pathogen Discovery | Anderson, M. Hua, Y. Liu, L.E. Bernstein, K.E. Livingston, A. Perez, I. A. Shlyakhter, R. V. Rolando, R. Owen, P. Tanpaiboon, F. Lacbawan, Clinton R. Paden, Duncan MacCannell |
| EPI_ISL_2204057 | Houston Methodist Hospital | Houston Methodist Hospital | Randall J. Olsen, Paul A. Christensen, S. Wesley Long, Sishir Subedi, Robert Olson, Marcus Nguyen, James J. Davis, Matthew Ojeda Saavedra, Prasanti Yerramilli, Layne Pruitt, Kristina Reppond, Madison N. Shyer, Jessica Cambric, Ryan Gadd, Ilya J. Finkelstein, Jimmy Gollihar, and James M. Musser |
| EPI_ISL_2204058, EPI_ISL_2204059 | Quest Diagnostics Incorporated | Centers for Disease Control and Prevention Division of Viral Diseases, Pathogen Discovery | Dakota Howard, Dhwani Batra, Peter W. Cook, Kara Moser, Adrian Paskey, Jason Caravas, Benjamin Rambo-Martin, Shatavia Morrison, Christopher Gulvick, Scott Sammons, Yvette Unoarumhi, Darlene Wagner, Matthew Schmerer, S. H. Rosenthal, A. Gerasimova, R. M. Kagan, B. Anderson, M. Hua, Y. Liu, L.E. Bernstein, K.E. Livingston, A. Perez, I. A. Shlyakhter, R. V. Rolando, R. Owen, P. Tanpaiboon, F. Lacbawan, Clinton R. Paden, Duncan MacCannell |
| EPI_ISL_2204060 | Houston Methodist Hospital | Houston Methodist Hospital | Randall J. Olsen, Paul A. Christensen, S. Wesley Long, Sishir Subedi, Robert Olson, Marcus Nguyen, James J. Davis, Matthew Ojeda Saavedra, Prasanti Yerramilli, Layne Pruitt, Kristina Reppond, Madison N. Shyer, Jessica Cambric, Ryan Gadd, Ilya J. Finkelstein, Jimmy Gollihar, and James M. Musser |
| EPI_ISL_2204061, EPI_ISL_2204062, EPI_ISL_2204064 | Quest Diagnostics Incorporated | Centers for Disease Control and Prevention Division of Viral Diseases, Pathogen Discovery | Dakota Howard, Dhwani Batra, Peter W. Cook, Kara Moser, Adrian Paskey, Jason Caravas, Benjamin Rambo-Martin, Shatavia Morrison, Christopher Gulvick, Scott Sammons, Yvette Unoarumhi, Darlene Wagner, Matthew Schmerer, S. H. Rosenthal, A. Gerasimova, R. M. Kagan, B. Anderson, M. Hua, Y. Liu, L.E. Bernstein, K.E. Livingston, A. Perez, I. A. Shlyakhter, R. V. Rolando, R. Owen, P. Tanpaiboon, F. Lacbawan, Clinton R. Paden, Duncan MacCannell |
| EPI_ISL_2204065 | Houston Methodist Hospital | Houston Methodist Hospital | Randall J. Olsen, Paul A. Christensen, S. Wesley Long, Sishir Subedi, Robert Olson, Marcus Nguyen, James J. Davis, Matthew Ojeda Saavedra, Prasanti Yerramilli, Layne Pruitt, Kristina Reppond, Madison N. Shyer, Jessica Cambric, Ryan Gadd, Ilya J. Finkelstein, Jimmy Gollihar, and James M. Musser |
| EPI_ISL_2204066, EPI_ISL_2204067 | Quest Diagnostics Incorporated | Centers for Disease Control and Prevention Division of Viral Diseases, Pathogen Discovery | Dakota Howard, Dhwani Batra, Peter W. Cook, Kara Moser, Adrian Paskey, Jason Caravas, Benjamin Rambo-Martin, Shatavia Morrison, Christopher Gulvick, Scott Sammons, Yvette Unoarumhi, Darlene Wagner, Matthew Schmerer, S. H. Rosenthal, A. Gerasimova, R. M. Kagan, B. Anderson, M. Hua, Y. Liu, L.E. Bernstein, K.E. Livingston, A. Perez, I. A. Shlyakhter, R. V. Rolando, R. Owen, P. Tanpaiboon, F. Lacbawan, Clinton R. Paden, Duncan MacCannell |
| EPI_ISL_2204068 | Houston Methodist Hospital | Houston Methodist Hospital | Randall J. Olsen, Paul A. Christensen, S. Wesley Long, Sishir Subedi, Robert Olson, Marcus Nguyen, James J. Davis, Matthew Ojeda Saavedra, Prasanti Yerramilli, Layne Pruitt, Kristina Reppond, Madison N. Shyer, Jessica Cambric, Ryan Gadd, Ilya J. Finkelstein, Jimmy Gollihar, and James M. Musser |
| EPI_ISL_2204069, EPI_ISL_2204070, EPI_ISL_2204072, EPI_ISL_2204073, EPI_ISL_2204075, EPI_ISL_2204076 | Quest Diagnostics Incorporated | Centers for Disease Control and Prevention Division of Viral Diseases, Pathogen Discovery | Dakota Howard, Dhwani Batra, Peter W. Cook, Kara Moser, Adrian Paskey, Jason Caravas, Benjamin Rambo-Martin, Shatavia Morrison, Christopher Gulvick, Scott Sammons, Yvette Unoarumhi, Darlene Wagner, Matthew Schmerer, S. H. Rosenthal, A. Gerasimova, R. M. Kagan, B. Anderson, M. Hua, Y. Liu, L.E. Bernstein, K.E. Livingston, A. Perez, I. A. Shlyakhter, R. V. Rolando, R. Owen, P. Tanpaiboon, F. Lacbawan, Clinton R. Paden, Duncan MacCannell |
| EPI_ISL_2204077 | Houston Methodist Hospital | Houston Methodist Hospital | Randall J. Olsen, Paul A. Christensen, S. Wesley Long, Sishir Subedi, Robert Olson, Marcus Nguyen, James J. Davis, Matthew Ojeda Saavedra, Prasanti Yerramilli, Layne Pruitt, Kristina Reppond, Madison N. Shyer, Jessica Cambric, Ryan Gadd, Ilya J. Finkelstein, Jimmy Gollihar, and James M. Musser |
| EPI_ISL_2204078, EPI_ISL_2204079 | Quest Diagnostics Incorporated | Centers for Disease Control and Prevention Division of Viral Diseases, Pathogen Discovery | Dakota Howard, Dhwani Batra, Peter W. Cook, Kara Moser, Adrian Paskey, Jason Caravas, Benjamin Rambo-Martin, Shatavia Morrison, Christopher Gulvick, Scott Sammons, Yvette Unoarumhi, Darlene Wagner, Matthew Schmerer, S. H. Rosenthal, A. Gerasimova, R. M. Kagan, B. Anderson, M. Hua, Y. Liu, L.E. Bernstein, K.E. Livingston, A. Perez, I. A. Shlyakhter, R. V. Rolando, R. Owen, P. Tanpaiboon, F. Lacbawan, Clinton R. Paden, Duncan MacCannell |
| EPI_ISL_2204080 | Houston Methodist Hospital | Houston Methodist Hospital | Randall J. Olsen, Paul A. Christensen, S. Wesley Long, Sishir Subedi, Robert Olson, Marcus Nguyen, James J. Davis, Matthew Ojeda Saavedra, Prasanti Yerramilli, Layne Pruitt, Kristina Reppond, Madison N. Shyer, Jessica Cambric, Ryan Gadd, Ilya J. Finkelstein, Jimmy Gollihar, and James M. Musser |
| EPI_ISL_2204081, EPI_ISL_2204082 | Quest Diagnostics Incorporated | Centers for Disease Control and Prevention Division of Viral Diseases, Pathogen Discovery | Dakota Howard, Dhwani Batra, Peter W. Cook, Kara Moser, Adrian Paskey, Jason Caravas, Benjamin Rambo-Martin, Shatavia Morrison, Christopher Gulvick, Scott Sammons, Yvette Unoarumhi, Darlene Wagner, Matthew Schmerer, S. H. Rosenthal, A. Gerasimova, R. M. Kagan, B. Anderson, M. Hua, Y. Liu, L.E. Bernstein, K.E. Livingston, A. Perez, I. A. Shlyakhter, R. V. Rolando, R. Owen, P. Tanpaiboon, F. Lacbawan, Clinton R. Paden, Duncan MacCannell |
| EPI_ISL_2204083 | J.W. Ruby Memorial Hospital | WVU and Marshall University Combined Genomics Core Facilities | James Denvir, Peter Stoilov, Peter Perrotta, Wesley Kimble, Ryan Percifield |
| EPI_ISL_2204084 | Quest Diagnostics Incorporated | Centers for Disease Control and Prevention Division of Viral Diseases, Pathogen Discovery | Dakota Howard, Dhwani Batra, Peter W. Cook, Kara Moser, Adrian Paskey, Jason Caravas, Benjamin Rambo-Martin, Shatavia Morrison, Christopher Gulvick, Scott Sammons, Yvette Unoarumhi, Darlene Wagner, Matthew Schmerer, S. H. Rosenthal, A. Gerasimova, R. M. Kagan, B. Anderson, M. Hua, Y. Liu, L.E. Bernstein, K.E. Livingston, A. Perez, I. A. Shlyakhter, R. V. Rolando, R. Owen, P. Tanpaiboon, F. Lacbawan, Clinton R. Paden, Duncan MacCannell |
| EPI_ISL_2204085 | J.W. Ruby Memorial Hospital | WVU and Marshall University Combined Genomics Core Facilities | James Denvir, Peter Stoilov, Peter Perrotta, Wesley Kimble, Ryan Percifield |
| EPI_ISL_2204086 | Houston Methodist Hospital | Houston Methodist Hospital | Randall J. Olsen, Paul A. Christensen, S. Wesley Long, Sishir Subedi, Robert Olson, Marcus Nguyen, James J. Davis, Matthew Ojeda Saavedra, Prasanti Yerramilli, Layne Pruitt, Kristina Reppond, Madison N. Shyer, Jessica Cambric, Ryan Gadd, Ilya J. Finkelstein, Jimmy Gollihar, and James M. Musser |
| EPI_ISL_2204087, EPI_ISL_2204088 | Quest Diagnostics Incorporated | Centers for Disease Control and Prevention Division of Viral Diseases, Pathogen Discovery | Dakota Howard, Dhwani Batra, Peter W. Cook, Kara Moser, Adrian Paskey, Jason Caravas, Benjamin Rambo-Martin, Shatavia Morrison, Christopher Gulvick, Scott Sammons, Yvette Unoarumhi, Darlene Wagner, Matthew Schmerer, S. H. Rosenthal, A. Gerasimova, R. M. Kagan, B. Anderson, M. Hua, Y. Liu, L.E. Bernstein, K.E. Livingston, A. Perez, I. A. Shlyakhter, R. V. Rolando, R. Owen, P. Tanpaiboon, F. Lacbawan, Clinton R. Paden, Duncan MacCannell |
| EPI_ISL_2204089 | J.W. Ruby Memorial Hospital | WVU and Marshall University Combined Genomics Core Facilities | James Denvir, Peter Stoilov, Peter Perrotta, Wesley Kimble, Ryan Percifield |
| EPI_ISL_2204090 | Quest Diagnostics Incorporated | Centers for Disease Control and Prevention Division of Viral Diseases, Pathogen Discovery | Dakota Howard, Dhwani Batra, Peter W. Cook, Kara Moser, Adrian Paskey, Jason Caravas, Benjamin Rambo-Martin, Shatavia Morrison, Christopher Gulvick, Scott Sammons, Yvette Unoarumhi, Darlene Wagner, Matthew Schmerer, S. H. Rosenthal, A. Gerasimova, R. M. Kagan, B. Anderson, M. Hua, Y. Liu, L.E. Bernstein, K.E. Livingston, A. Perez, I. A. Shlyakhter, R. V. Rolando, R. Owen, P. Tanpaiboon, F. Lacbawan, Clinton R. Paden, Duncan MacCannell |
| EPI_ISL_2204091 | J.W. Ruby Memorial Hospital | WVU and Marshall University Combined Genomics Core Facilities | James Denvir, Peter Stoilov, Peter Perrotta, Wesley Kimble, Ryan Percifield |
| EPI_ISL_2204092 | Quest Diagnostics Incorporated | Centers for Disease Control and Prevention Division of Viral Diseases, Pathogen Discovery | Dakota Howard, Dhwani Batra, Peter W. Cook, Kara Moser, Adrian Paskey, Jason Caravas, Benjamin Rambo-Martin, Shatavia Morrison, Christopher Gulvick, Scott Sammons, Yvette Unoarumhi, Darlene Wagner, Matthew Schmerer, S. H. Rosenthal, A. Gerasimova, R. M. Kagan, B. Anderson, M. Hua, Y. Liu, L.E. Bernstein, K.E. Livingston, A. Perez, I. A. Shlyakhter, R. V. Rolando, R. Owen, P. Tanpaiboon, F. Lacbawan, Clinton R. Paden, Duncan MacCannell |
| EPI_ISL_2204093 | J.W. Ruby Memorial Hospital | WVU and Marshall University Combined Genomics Core Facilities | James Denvir, Peter Stoilov, Peter Perrotta, Wesley Kimble, Ryan Percifield |
| EPI_ISL_2204094 | Quest Diagnostics Incorporated | Centers for Disease Control and Prevention Division of Viral Diseases, Pathogen Discovery | Dakota Howard, Dhwani Batra, Peter W. Cook, Kara Moser, Adrian Paskey, Jason Caravas, Benjamin Rambo-Martin, Shatavia Morrison, Christopher Gulvick, Scott Sammons, Yvette Unoarumhi, Darlene Wagner, Matthew Schmerer, S. H. Rosenthal, A. Gerasimova, R. M. Kagan, B. Anderson, M. Hua, Y. Liu, L.E. Bernstein, K.E. Livingston, A. Perez, I. A. Shlyakhter, R. V. Rolando, R. Owen, P. Tanpaiboon, F. Lacbawan, Clinton R. Paden, Duncan MacCannell |
| EPI_ISL_2204095 | J.W. Ruby Memorial Hospital | WVU and Marshall University Combined Genomics Core Facilities | James Denvir, Peter Stoilov, Peter Perrotta, Wesley Kimble, Ryan Percifield |
| EPI_ISL_2204097 | Quest Diagnostics Incorporated | Centers for Disease Control and Prevention Division of Viral Diseases, Pathogen Discovery | Dakota Howard, Dhwani Batra, Peter W. Cook, Kara Moser, Adrian Paskey, Jason Caravas, Benjamin Rambo-Martin, Shatavia Morrison, Christopher Gulvick, Scott Sammons, Yvette Unoarumhi, Darlene Wagner, Matthew Schmerer, S. H. Rosenthal, A. Gerasimova, R. M. Kagan, B. Anderson, M. Hua, Y. Liu, L.E. Bernstein, K.E. Livingston, A. Perez, I. A. Shlyakhter, R. V. Rolando, R. Owen, P. Tanpaiboon, F. Lacbawan, Clinton R. Paden, Duncan MacCannell |
| EPI_ISL_2204098, EPI_ISL_2204099 | J.W. Ruby Memorial Hospital | WVU and Marshall University Combined Genomics Core Facilities | James Denvir, Peter Stoilov, Peter Perrotta, Wesley Kimble, Ryan Percifield |
| EPI_ISL_2204100 | Quest Diagnostics Incorporated | Centers for Disease Control and Prevention Division of Viral Diseases, Pathogen Discovery | Dakota Howard, Dhwani Batra, Peter W. Cook, Kara Moser, Adrian Paskey, Jason Caravas, Benjamin Rambo-Martin, Shatavia Morrison, Christopher Gulvick, Scott Sammons, Yvette Unoarumhi, Darlene Wagner, Matthew Schmerer, S. H. Rosenthal, A. Gerasimova, R. M. Kagan, B. Anderson, M. Hua, Y. Liu, L.E. Bernstein, K.E. Livingston, A. Perez, I. A. Shlyakhter, R. V. Rolando, R. Owen, P. Tanpaiboon, F. Lacbawan, Clinton R. Paden, Duncan MacCannell |
| EPI_ISL_2204101 | Houston Methodist Hospital | Houston Methodist Hospital | Randall J. Olsen, Paul A. Christensen, S. Wesley Long, Sishir Subedi, Robert Olson, Marcus Nguyen, James J. Davis, Matthew Ojeda Saavedra, Prasanti Yerramilli, Layne Pruitt, Kristina Reppond, Madison N. Shyer, Jessica Cambric, Ryan Gadd, Ilya J. Finkelstein, Jimmy Gollihar, and James M. Musser |
| EPI_ISL_2204102 | Quest Diagnostics Incorporated | Centers for Disease Control and Prevention Division of Viral Diseases, Pathogen Discovery | Dakota Howard, Dhwani Batra, Peter W. Cook, Kara Moser, Adrian Paskey, Jason Caravas, Benjamin Rambo-Martin, Shatavia Morrison, Christopher Gulvick, Scott Sammons, Yvette Unoarumhi, Darlene Wagner, Matthew Schmerer, S. H. Rosenthal, A. Gerasimova, R. M. Kagan, B. Anderson, M. Hua, Y. Liu, L.E. Bernstein, K.E. Livingston, A. Perez, I. A. Shlyakhter, R. V. Rolando, R. Owen, P. Tanpaiboon, F. Lacbawan, Clinton R. Paden, Duncan MacCannell |
| EPI_ISL_2204103 | J.W. Ruby Memorial Hospital | WVU and Marshall University Combined Genomics Core Facilities | James Denvir, Peter Stoilov, Peter Perrotta, Wesley Kimble, Ryan Percifield |
| EPI_ISL_2204104 | Quest Diagnostics Incorporated | Centers for Disease Control and Prevention Division of Viral Diseases, Pathogen Discovery | Dakota Howard, Dhwani Batra, Peter W. Cook, Kara Moser, Adrian Paskey, Jason Caravas, Benjamin Rambo-Martin, Shatavia Morrison, Christopher Gulvick, Scott Sammons, Yvette Unoarumhi, Darlene Wagner, Matthew Schmerer, S. H. Rosenthal, A. Gerasimova, R. M. Kagan, B. Anderson, M. Hua, Y. Liu, L.E. Bernstein, K.E. Livingston, A. Perez, I. A. Shlyakhter, R. V. Rolando, R. Owen, P. Tanpaiboon, F. Lacbawan, Clinton R. Paden, Duncan MacCannell |
| EPI_ISL_2204105 | J.W. Ruby Memorial Hospital | WVU and Marshall University Combined Genomics Core Facilities | James Denvir, Peter Stoilov, Peter Perrotta, Wesley Kimble, Ryan Percifield |
| EPI_ISL_2204106 | Quest Diagnostics Incorporated | Centers for Disease Control and Prevention Division of Viral Diseases, Pathogen Discovery | Dakota Howard, Dhwani Batra, Peter W. Cook, Kara Moser, Adrian Paskey, Jason Caravas, Benjamin Rambo-Martin, Shatavia Morrison, Christopher Gulvick, Scott Sammons, Yvette Unoarumhi, Darlene Wagner, Matthew Schmerer, S. H. Rosenthal, A. Gerasimova, R. M. Kagan, B. Anderson, M. Hua, Y. Liu, L.E. Bernstein, K.E. Livingston, A. Perez, I. A. Shlyakhter, R. V. Rolando, R. Owen, P. Tanpaiboon, F. Lacbawan, Clinton R. Paden, Duncan MacCannell |
| EPI_ISL_2204107, EPI_ISL_2204109 | J.W. Ruby Memorial Hospital | WVU and Marshall University Combined Genomics Core Facilities | James Denvir, Peter Stoilov, Peter Perrotta, Wesley Kimble, Ryan Percifield |
| EPI_ISL_2204110 | Quest Diagnostics Incorporated | Centers for Disease Control and Prevention Division of Viral Diseases, Pathogen Discovery | Dakota Howard, Dhwani Batra, Peter W. Cook, Kara Moser, Adrian Paskey, Jason Caravas, Benjamin Rambo-Martin, Shatavia Morrison, Christopher Gulvick, Scott Sammons, Yvette Unoarumhi, Darlene Wagner, Matthew Schmerer, S. H. Rosenthal, A. Gerasimova, R. M. Kagan, B. Anderson, M. Hua, Y. Liu, L.E. Bernstein, K.E. Livingston, A. Perez, I. A. Shlyakhter, R. V. Rolando, R. Owen, P. Tanpaiboon, F. Lacbawan, Clinton R. Paden, Duncan MacCannell |
| EPI_ISL_2204111 | J.W. Ruby Memorial Hospital | WVU and Marshall University Combined | James Denvir, Peter Stoilov, Peter Perrotta, Wesley Kimble, Ryan Percifield |

| | | | |
|---|---|---|---|
| | | Genomics Core Facilities | |
| EPI_ISL_2204112 | Houston Methodist Hospital | Houston Methodist Hospital | Randall J. Olsen, Paul A. Christensen, S. Wesley Long, Sishir Subedi, Robert Olson, Marcus Nguyen, James J. Davis, Matthew Ojeda Saavedra, Prasanti Yerramilli, Layne Pruitt, Kristina Reppond, Madison N. Shyer, Jessica Cambric, Ryan Gadd, Ilya J. Finkelstein, Jimmy Gollihar, and James M. Musser |
| EPI_ISL_2204113 | Quest Diagnostics Incorporated | Centers for Disease Control and Prevention Division of Viral Diseases, Pathogen Discovery | Dakota Howard, Dhwani Batra, Peter W. Cook, Kara Moser, Adrian Paskey, Jason Caravas, Benjamin Rambo-Martin, Shatavia Morrison, Christopher Gulvick, Scott Sammons, Yvette Unoarumhi, Darlene Wagner, Matthew Schmerer, S. H. Rosenthal, A. Gerasimova, R. M. Kagan, B. Anderson, M. Hua, Y. Liu, L.E. Bernstein, K.E. Livingston, A. Perez, I. A. Shlyakhter, R. V. Rolando, R. Owen, P. Tanpaiboon, F. Lacbawan, Clinton R. Paden, Duncan MacCannell |
| EPI_ISL_2204114 | J.W. Ruby Memorial Hospital | WVU and Marshall University Combined Genomics Core Facilities | James Denvir, Peter Stoilov, Peter Perrotta, Wesley Kimble, Ryan Percifield |
| EPI_ISL_2204116 | Houston Methodist Hospital | Houston Methodist Hospital | Randall J. Olsen, Paul A. Christensen, S. Wesley Long, Sishir Subedi, Robert Olson, Marcus Nguyen, James J. Davis, Matthew Ojeda Saavedra, Prasanti Yerramilli, Layne Pruitt, Kristina Reppond, Madison N. Shyer, Jessica Cambric, Ryan Gadd, Ilya J. Finkelstein, Jimmy Gollihar, and James M. Musser |
| EPI_ISL_2204117 | Quest Diagnostics Incorporated | Centers for Disease Control and Prevention Division of Viral Diseases, Pathogen Discovery | Dakota Howard, Dhwani Batra, Peter W. Cook, Kara Moser, Adrian Paskey, Jason Caravas, Benjamin Rambo-Martin, Shatavia Morrison, Christopher Gulvick, Scott Sammons, Yvette Unoarumhi, Darlene Wagner, Matthew Schmerer, S. H. Rosenthal, A. Gerasimova, R. M. Kagan, B. Anderson, M. Hua, Y. Liu, L.E. Bernstein, K.E. Livingston, A. Perez, I. A. Shlyakhter, R. V. Rolando, R. Owen, P. Tanpaiboon, F. Lacbawan, Clinton R. Paden, Duncan MacCannell |
| EPI_ISL_2204118 | J.W. Ruby Memorial Hospital | WVU and Marshall University Combined Genomics Core Facilities | James Denvir, Peter Stoilov, Peter Perrotta, Wesley Kimble, Ryan Percifield |
| EPI_ISL_2204119 | Quest Diagnostics Incorporated | Centers for Disease Control and Prevention Division of Viral Diseases, Pathogen Discovery | Dakota Howard, Dhwani Batra, Peter W. Cook, Kara Moser, Adrian Paskey, Jason Caravas, Benjamin Rambo-Martin, Shatavia Morrison, Christopher Gulvick, Scott Sammons, Yvette Unoarumhi, Darlene Wagner, Matthew Schmerer, S. H. Rosenthal, A. Gerasimova, R. M. Kagan, B. Anderson, M. Hua, Y. Liu, L.E. Bernstein, K.E. Livingston, A. Perez, I. A. Shlyakhter, R. V. Rolando, R. Owen, P. Tanpaiboon, F. Lacbawan, Clinton R. Paden, Duncan MacCannell |
| EPI_ISL_2204120 | J.W. Ruby Memorial Hospital | WVU and Marshall University Combined Genomics Core Facilities | James Denvir, Peter Stoilov, Peter Perrotta, Wesley Kimble, Ryan Percifield |
| EPI_ISL_2204121 | Quest Diagnostics Incorporated | Centers for Disease Control and Prevention Division of Viral Diseases, Pathogen Discovery | Dakota Howard, Dhwani Batra, Peter W. Cook, Kara Moser, Adrian Paskey, Jason Caravas, Benjamin Rambo-Martin, Shatavia Morrison, Christopher Gulvick, Scott Sammons, Yvette Unoarumhi, Darlene Wagner, Matthew Schmerer, S. H. Rosenthal, A. Gerasimova, R. M. Kagan, B. Anderson, M. Hua, Y. Liu, L.E. Bernstein, K.E. Livingston, A. Perez, I. A. Shlyakhter, R. V. Rolando, R. Owen, P. Tanpaiboon, F. Lacbawan, Clinton R. Paden, Duncan MacCannell |
| EPI_ISL_2204122, EPI_ISL_2204124 | J.W. Ruby Memorial Hospital | WVU and Marshall University Combined Genomics Core Facilities | James Denvir, Peter Stoilov, Peter Perrotta, Wesley Kimble, Ryan Percifield |
| EPI_ISL_2204125 | Quest Diagnostics Incorporated | Centers for Disease Control and Prevention Division of Viral Diseases, Pathogen Discovery | Dakota Howard, Dhwani Batra, Peter W. Cook, Kara Moser, Adrian Paskey, Jason Caravas, Benjamin Rambo-Martin, Shatavia Morrison, Christopher Gulvick, Scott Sammons, Yvette Unoarumhi, Darlene Wagner, Matthew Schmerer, S. H. Rosenthal, A. Gerasimova, R. M. Kagan, B. Anderson, M. Hua, Y. Liu, L.E. Bernstein, K.E. Livingston, A. Perez, I. A. Shlyakhter, R. V. Rolando, R. Owen, P. Tanpaiboon, F. Lacbawan, Clinton R. Paden, Duncan MacCannell |
| EPI_ISL_2204126 | J.W. Ruby Memorial Hospital | WVU and Marshall University Combined Genomics Core Facilities | James Denvir, Peter Stoilov, Peter Perrotta, Wesley Kimble, Ryan Percifield |
| EPI_ISL_2204127 | Houston Methodist Hospital | Houston Methodist Hospital | Randall J. Olsen, Paul A. Christensen, S. Wesley Long, Sishir Subedi, Robert Olson, Marcus Nguyen, James J. Davis, Matthew Ojeda Saavedra, Prasanti Yerramilli, Layne Pruitt, Kristina Reppond, Madison N. Shyer, Jessica Cambric, Ryan Gadd, Ilya J. Finkelstein, Jimmy Gollihar, and James M. Musser |
| EPI_ISL_2204128 | J.W. Ruby Memorial Hospital | WVU and Marshall University Combined Genomics Core Facilities | James Denvir, Peter Stoilov, Peter Perrotta, Wesley Kimble, Ryan Percifield |
| EPI_ISL_2204129 | Quest Diagnostics Incorporated | Centers for Disease Control and Prevention Division of Viral Diseases, Pathogen Discovery | Dakota Howard, Dhwani Batra, Peter W. Cook, Kara Moser, Adrian Paskey, Jason Caravas, Benjamin Rambo-Martin, Shatavia Morrison, Christopher Gulvick, Scott Sammons, Yvette Unoarumhi, Darlene Wagner, Matthew Schmerer, S. H. Rosenthal, A. Gerasimova, R. M. Kagan, B. Anderson, M. Hua, Y. Liu, L.E. Bernstein, K.E. Livingston, A. Perez, I. A. Shlyakhter, R. V. Rolando, R. Owen, P. Tanpaiboon, F. Lacbawan, Clinton R. Paden, Duncan MacCannell |
| EPI_ISL_2204130 | J.W. Ruby Memorial Hospital | WVU and Marshall University Combined Genomics Core Facilities | James Denvir, Peter Stoilov, Peter Perrotta, Wesley Kimble, Ryan Percifield |
| EPI_ISL_2204131 | Quest Diagnostics Incorporated | Centers for Disease Control and Prevention Division of Viral Diseases, Pathogen Discovery | Dakota Howard, Dhwani Batra, Peter W. Cook, Kara Moser, Adrian Paskey, Jason Caravas, Benjamin Rambo-Martin, Shatavia Morrison, Christopher Gulvick, Scott Sammons, Yvette Unoarumhi, Darlene Wagner, Matthew Schmerer, S. H. Rosenthal, A. Gerasimova, R. M. Kagan, B. Anderson, M. Hua, Y. Liu, L.E. Bernstein, K.E. Livingston, A. Perez, I. A. Shlyakhter, R. V. Rolando, R. Owen, P. Tanpaiboon, F. Lacbawan, Clinton R. Paden, Duncan MacCannell |
| EPI_ISL_2204132 | Houston Methodist Hospital | Houston Methodist Hospital | Randall J. Olsen, Paul A. Christensen, S. Wesley Long, Sishir Subedi, Robert Olson, Marcus Nguyen, James J. Davis, Matthew Ojeda Saavedra, Prasanti Yerramilli, Layne Pruitt, Kristina Reppond, Madison N. Shyer, Jessica Cambric, Ryan Gadd, Ilya J. Finkelstein, Jimmy Gollihar, and James M. Musser |
| EPI_ISL_2204134 | Quest Diagnostics Incorporated | Centers for Disease Control and Prevention Division of Viral Diseases, Pathogen Discovery | Dakota Howard, Dhwani Batra, Peter W. Cook, Kara Moser, Adrian Paskey, Jason Caravas, Benjamin Rambo-Martin, Shatavia Morrison, Christopher Gulvick, Scott Sammons, Yvette Unoarumhi, Darlene Wagner, Matthew Schmerer, S. H. Rosenthal, A. Gerasimova, R. M. Kagan, B. Anderson, M. Hua, Y. Liu, L.E. Bernstein, K.E. Livingston, A. Perez, I. A. Shlyakhter, R. V. Rolando, R. Owen, P. Tanpaiboon, F. Lacbawan, Clinton R. Paden, Duncan MacCannell |
| EPI_ISL_2204135 | Houston Methodist Hospital | Houston Methodist Hospital | Randall J. Olsen, Paul A. Christensen, S. Wesley Long, Sishir Subedi, Robert Olson, Marcus Nguyen, James J. Davis, Matthew Ojeda Saavedra, Prasanti Yerramilli, Layne Pruitt, Kristina Reppond, Madison N. Shyer, Jessica Cambric, Ryan Gadd, Ilya J. Finkelstein, Jimmy Gollihar, and James M. Musser |
| EPI_ISL_2204136 | Quest Diagnostics Incorporated | Centers for Disease Control and Prevention Division of Viral Diseases, Pathogen Discovery | Dakota Howard, Dhwani Batra, Peter W. Cook, Kara Moser, Adrian Paskey, Jason Caravas, Benjamin Rambo-Martin, Shatavia Morrison, Christopher Gulvick, Scott Sammons, Yvette Unoarumhi, Darlene Wagner, Matthew Schmerer, S. H. Rosenthal, A. Gerasimova, R. M. Kagan, B. Anderson, M. Hua, Y. Liu, L.E. Bernstein, K.E. Livingston, A. Perez, I. A. Shlyakhter, R. V. Rolando, R. Owen, P. Tanpaiboon, F. Lacbawan, Clinton R. Paden, Duncan MacCannell |
| EPI_ISL_2204137 | J.W. Ruby Memorial Hospital | WVU and Marshall University Combined Genomics Core Facilities | James Denvir, Peter Stoilov, Peter Perrotta, Wesley Kimble, Ryan Percifield |
| EPI_ISL_2204138 | Houston Methodist Hospital | Houston Methodist Hospital | Randall J. Olsen, Paul A. Christensen, S. Wesley Long, Sishir Subedi, Robert Olson, Marcus Nguyen, James J. Davis, Matthew Ojeda Saavedra, Prasanti Yerramilli, Layne Pruitt, Kristina Reppond, Madison N. Shyer, Jessica Cambric, Ryan Gadd, Ilya J. Finkelstein, Jimmy Gollihar, and James M. Musser |
| EPI_ISL_2204139, EPI_ISL_2204141 | Quest Diagnostics Incorporated | Centers for Disease Control and Prevention Division of Viral Diseases, Pathogen Discovery | Dakota Howard, Dhwani Batra, Peter W. Cook, Kara Moser, Adrian Paskey, Jason Caravas, Benjamin Rambo-Martin, Shatavia Morrison, Christopher Gulvick, Scott Sammons, Yvette Unoarumhi, Darlene Wagner, Matthew Schmerer, S. H. Rosenthal, A. Gerasimova, R. M. Kagan, B. Anderson, M. Hua, Y. Liu, L.E. Bernstein, K.E. Livingston, A. Perez, I. A. Shlyakhter, R. V. Rolando, R. Owen, P. Tanpaiboon, F. Lacbawan, Clinton R. Paden, Duncan MacCannell |
| EPI_ISL_2204142 | J.W. Ruby Memorial Hospital | WVU and Marshall University Combined Genomics Core Facilities | James Denvir, Peter Stoilov, Peter Perrotta, Wesley Kimble, Ryan Percifield |
| EPI_ISL_2204143 | Houston Methodist Hospital | Houston Methodist Hospital | Randall J. Olsen, Paul A. Christensen, S. Wesley Long, Sishir Subedi, Robert Olson, Marcus Nguyen, James J. Davis, Matthew Ojeda Saavedra, Prasanti Yerramilli, Layne Pruitt, Kristina Reppond, Madison N. Shyer, Jessica Cambric, Ryan Gadd, Ilya J. Finkelstein, Jimmy Gollihar, and James M. Musser |
| EPI_ISL_2204144 | Quest Diagnostics Incorporated | Centers for Disease Control and Prevention Division of Viral Diseases, Pathogen Discovery | Dakota Howard, Dhwani Batra, Peter W. Cook, Kara Moser, Adrian Paskey, Jason Caravas, Benjamin Rambo-Martin, Shatavia Morrison, Christopher Gulvick, Scott Sammons, Yvette Unoarumhi, Darlene Wagner, Matthew Schmerer, S. H. Rosenthal, A. Gerasimova, R. M. Kagan, B. Anderson, M. Hua, Y. Liu, L.E. Bernstein, K.E. Livingston, A. Perez, I. A. Shlyakhter, R. V. Rolando, R. Owen, P. Tanpaiboon, F. Lacbawan, Clinton R. Paden, Duncan MacCannell |
| EPI_ISL_2204145 | J.W. Ruby Memorial Hospital | WVU and Marshall University Combined Genomics Core Facilities | James Denvir, Peter Stoilov, Peter Perrotta, Wesley Kimble, Ryan Percifield |
| EPI_ISL_2204146 | Quest Diagnostics Incorporated | Centers for Disease Control and Prevention Division of Viral Diseases, Pathogen Discovery | Dakota Howard, Dhwani Batra, Peter W. Cook, Kara Moser, Adrian Paskey, Jason Caravas, Benjamin Rambo-Martin, Shatavia Morrison, Christopher Gulvick, Scott Sammons, Yvette Unoarumhi, Darlene Wagner, Matthew Schmerer, S. H. Rosenthal, A. Gerasimova, R. M. Kagan, B. Anderson, M. Hua, Y. Liu, L.E. Bernstein, K.E. Livingston, A. Perez, I. A. Shlyakhter, R. V. Rolando, R. Owen, P. Tanpaiboon, F. Lacbawan, Clinton R. Paden, Duncan MacCannell |
| EPI_ISL_2204147 | Houston Methodist Hospital | Houston Methodist Hospital | Randall J. Olsen, Paul A. Christensen, S. Wesley Long, Sishir Subedi, Robert Olson, Marcus Nguyen, James J. Davis, Matthew Ojeda Saavedra, Prasanti Yerramilli, Layne Pruitt, Kristina Reppond, Madison N. Shyer, Jessica Cambric, Ryan Gadd, Ilya J. Finkelstein, Jimmy Gollihar, and James M. Musser |
| EPI_ISL_2204148 | Quest Diagnostics Incorporated | Centers for Disease Control and Prevention Division of Viral Diseases, Pathogen Discovery | Dakota Howard, Dhwani Batra, Peter W. Cook, Kara Moser, Adrian Paskey, Jason Caravas, Benjamin Rambo-Martin, Shatavia Morrison, Christopher Gulvick, Scott Sammons, Yvette Unoarumhi, Darlene Wagner, Matthew Schmerer, S. H. Rosenthal, A. Gerasimova, R. M. Kagan, B. Anderson, M. Hua, Y. Liu, L.E. Bernstein, K.E. Livingston, A. Perez, I. A. Shlyakhter, R. V. Rolando, R. Owen, P. Tanpaiboon, F. Lacbawan, Clinton R. Paden, Duncan MacCannell |
| EPI_ISL_2204149 | Houston Methodist Hospital | Houston Methodist Hospital | Randall J. Olsen, Paul A. Christensen, S. Wesley Long, Sishir Subedi, Robert Olson, Marcus Nguyen, James J. Davis, Matthew Ojeda Saavedra, Prasanti Yerramilli, Layne Pruitt, Kristina Reppond, Madison N. Shyer, Jessica Cambric, Ryan Gadd, Ilya J. Finkelstein, Jimmy Gollihar, and James M. Musser |
| EPI_ISL_2204150 | J.W. Ruby Memorial Hospital | WVU and Marshall University Combined Genomics Core Facilities | James Denvir, Peter Stoilov, Peter Perrotta, Wesley Kimble, Ryan Percifield |
| EPI_ISL_2204151 | Quest Diagnostics Incorporated | Centers for Disease Control and Prevention Division of Viral Diseases, Pathogen Discovery | Dakota Howard, Dhwani Batra, Peter W. Cook, Kara Moser, Adrian Paskey, Jason Caravas, Benjamin Rambo-Martin, Shatavia Morrison, Christopher Gulvick, Scott Sammons, Yvette Unoarumhi, Darlene Wagner, Matthew Schmerer, S. H. Rosenthal, A. Gerasimova, R. M. Kagan, B. Anderson, M. Hua, Y. Liu, L.E. Bernstein, K.E. Livingston, A. Perez, I. A. Shlyakhter, R. V. Rolando, R. Owen, P. Tanpaiboon, F. Lacbawan, Clinton R. Paden, Duncan MacCannell |
| EPI_ISL_2204152 | J.W. Ruby Memorial Hospital | WVU and Marshall University Combined Genomics Core Facilities | James Denvir, Peter Stoilov, Peter Perrotta, Wesley Kimble, Ryan Percifield |
| EPI_ISL_2204153 | Houston Methodist Hospital | Houston Methodist Hospital | Randall J. Olsen, Paul A. Christensen, S. Wesley Long, Sishir Subedi, Robert Olson, Marcus Nguyen, James J. Davis, Matthew Ojeda Saavedra, Prasanti Yerramilli, Layne Pruitt, Kristina Reppond, Madison N. Shyer, Jessica Cambric, Ryan Gadd, Ilya J. Finkelstein, Jimmy Gollihar, and James M. Musser |
| EPI_ISL_2204154 | Quest Diagnostics Incorporated | Centers for Disease Control and Prevention Division of Viral Diseases, Pathogen Discovery | Dakota Howard, Dhwani Batra, Peter W. Cook, Kara Moser, Adrian Paskey, Jason Caravas, Benjamin Rambo-Martin, Shatavia Morrison, Christopher Gulvick, Scott Sammons, Yvette Unoarumhi, Darlene Wagner, Matthew Schmerer, S. H. Rosenthal, A. Gerasimova, R. M. Kagan, B. Anderson, M. Hua, Y. Liu, L.E. Bernstein, K.E. Livingston, A. Perez, I. A. Shlyakhter, R. V. Rolando, R. Owen, P. Tanpaiboon, F. Lacbawan, Clinton R. Paden, Duncan MacCannell |
| EPI_ISL_2204155, EPI_ISL_2204156 | J.W. Ruby Memorial Hospital | WVU and Marshall University Combined Genomics Core Facilities | James Denvir, Peter Stoilov, Peter Perrotta, Wesley Kimble, Ryan Percifield |
| EPI_ISL_2204157 | Quest Diagnostics Incorporated | Centers for Disease Control and Prevention | Dakota Howard, Dhwani Batra, Peter W. Cook, Kara Moser, Adrian Paskey, Jason Caravas, Benjamin Rambo-Martin, Shatavia Morrison, Christopher Gulvick, Scott Sammons, Yvette Unoarumhi, Darlene Wagner, Matthew Schmerer, S. H. Rosenthal, A. Gerasimova, R. M. Kagan, B. |

| | | | |
|---|---|---|---|
| | | Division of Viral Diseases, Pathogen Discovery | Anderson, M. Hua, Y. Liu, L.E. Bernstein, K.E. Livingston, A. Perez, I. A. Shlyakhter, R. V. Rolando, R. Owen, P. Tanpaiboon, F. Lacbawan, Clinton R. Paden, Duncan MacCannell |
| EPI_ISL_2204158 | Houston Methodist Hospital | Houston Methodist Hospital | Randall J. Olsen, Paul A. Christensen, S. Wesley Long, Sishir Subedi, Robert Olson, Marcus Nguyen, James J. Davis, Matthew Ojeda Saavedra, Prasanti Yerramilli, Layne Pruitt, Kristina Reppond, Madison N. Shyer, Jessica Cambric, Ryan Gadd, Ilya J. Finkelstein, Jimmy Gollihar, and James M. Musser |
| EPI_ISL_2204159 | Quest Diagnostics Incorporated | Centers for Disease Control and Prevention Division of Viral Diseases, Pathogen Discovery | Dakota Howard, Dhwani Batra, Peter W. Cook, Kara Moser, Adrian Paskey, Jason Caravas, Benjamin Rambo-Martin, Shatavia Morrison, Christopher Gulvick, Scott Sammons, Yvette Unoarumhi, Darlene Wagner, Matthew Schmerer, S. H. Rosenthal, A. Gerasimova, R. M. Kagan, B. Anderson, M. Hua, Y. Liu, L.E. Bernstein, K.E. Livingston, A. Perez, I. A. Shlyakhter, R. V. Rolando, R. Owen, P. Tanpaiboon, F. Lacbawan, Clinton R. Paden, Duncan MacCannell |
| EPI_ISL_2204160 | Houston Methodist Hospital | Houston Methodist Hospital | Randall J. Olsen, Paul A. Christensen, S. Wesley Long, Sishir Subedi, Robert Olson, Marcus Nguyen, James J. Davis, Matthew Ojeda Saavedra, Prasanti Yerramilli, Layne Pruitt, Kristina Reppond, Madison N. Shyer, Jessica Cambric, Ryan Gadd, Ilya J. Finkelstein, Jimmy Gollihar, and James M. Musser |
| EPI_ISL_2204161 | J.W. Ruby Memorial Hospital | WVU and Marshall University Combined Genomics Core Facilities | James Denvir, Peter Stoilov, Peter Perrotta, Wesley Kimble, Ryan Percifield |
| EPI_ISL_2204162 | Quest Diagnostics Incorporated | Centers for Disease Control and Prevention Division of Viral Diseases, Pathogen Discovery | Dakota Howard, Dhwani Batra, Peter W. Cook, Kara Moser, Adrian Paskey, Jason Caravas, Benjamin Rambo-Martin, Shatavia Morrison, Christopher Gulvick, Scott Sammons, Yvette Unoarumhi, Darlene Wagner, Matthew Schmerer, S. H. Rosenthal, A. Gerasimova, R. M. Kagan, B. Anderson, M. Hua, Y. Liu, L.E. Bernstein, K.E. Livingston, A. Perez, I. A. Shlyakhter, R. V. Rolando, R. Owen, P. Tanpaiboon, F. Lacbawan, Clinton R. Paden, Duncan MacCannell |
| EPI_ISL_2204164 | J.W. Ruby Memorial Hospital | WVU and Marshall University Combined Genomics Core Facilities | James Denvir, Peter Stoilov, Peter Perrotta, Wesley Kimble, Ryan Percifield |
| EPI_ISL_2204165 | Quest Diagnostics Incorporated | Centers for Disease Control and Prevention Division of Viral Diseases, Pathogen Discovery | Dakota Howard, Dhwani Batra, Peter W. Cook, Kara Moser, Adrian Paskey, Jason Caravas, Benjamin Rambo-Martin, Shatavia Morrison, Christopher Gulvick, Scott Sammons, Yvette Unoarumhi, Darlene Wagner, Matthew Schmerer, S. H. Rosenthal, A. Gerasimova, R. M. Kagan, B. Anderson, M. Hua, Y. Liu, L.E. Bernstein, K.E. Livingston, A. Perez, I. A. Shlyakhter, R. V. Rolando, R. Owen, P. Tanpaiboon, F. Lacbawan, Clinton R. Paden, Duncan MacCannell |
| EPI_ISL_2204166 | J.W. Ruby Memorial Hospital | WVU and Marshall University Combined Genomics Core Facilities | James Denvir, Peter Stoilov, Peter Perrotta, Wesley Kimble, Ryan Percifield |
| EPI_ISL_2204167 | Houston Methodist Hospital | Houston Methodist Hospital | Randall J. Olsen, Paul A. Christensen, S. Wesley Long, Sishir Subedi, Robert Olson, Marcus Nguyen, James J. Davis, Matthew Ojeda Saavedra, Prasanti Yerramilli, Layne Pruitt, Kristina Reppond, Madison N. Shyer, Jessica Cambric, Ryan Gadd, Ilya J. Finkelstein, Jimmy Gollihar, and James M. Musser |
| EPI_ISL_2204168 | Quest Diagnostics Incorporated | Centers for Disease Control and Prevention Division of Viral Diseases, Pathogen Discovery | Dakota Howard, Dhwani Batra, Peter W. Cook, Kara Moser, Adrian Paskey, Jason Caravas, Benjamin Rambo-Martin, Shatavia Morrison, Christopher Gulvick, Scott Sammons, Yvette Unoarumhi, Darlene Wagner, Matthew Schmerer, S. H. Rosenthal, A. Gerasimova, R. M. Kagan, B. Anderson, M. Hua, Y. Liu, L.E. Bernstein, K.E. Livingston, A. Perez, I. A. Shlyakhter, R. V. Rolando, R. Owen, P. Tanpaiboon, F. Lacbawan, Clinton R. Paden, Duncan MacCannell |
| EPI_ISL_2204169 | J.W. Ruby Memorial Hospital | WVU and Marshall University Combined Genomics Core Facilities | James Denvir, Peter Stoilov, Peter Perrotta, Wesley Kimble, Ryan Percifield |
| EPI_ISL_2204170 | Quest Diagnostics Incorporated | Centers for Disease Control and Prevention Division of Viral Diseases, Pathogen Discovery | Dakota Howard, Dhwani Batra, Peter W. Cook, Kara Moser, Adrian Paskey, Jason Caravas, Benjamin Rambo-Martin, Shatavia Morrison, Christopher Gulvick, Scott Sammons, Yvette Unoarumhi, Darlene Wagner, Matthew Schmerer, S. H. Rosenthal, A. Gerasimova, R. M. Kagan, B. Anderson, M. Hua, Y. Liu, L.E. Bernstein, K.E. Livingston, A. Perez, I. A. Shlyakhter, R. V. Rolando, R. Owen, P. Tanpaiboon, F. Lacbawan, Clinton R. Paden, Duncan MacCannell |
| EPI_ISL_2204171 | Houston Methodist Hospital | Houston Methodist Hospital | Randall J. Olsen, Paul A. Christensen, S. Wesley Long, Sishir Subedi, Robert Olson, Marcus Nguyen, James J. Davis, Matthew Ojeda Saavedra, Prasanti Yerramilli, Layne Pruitt, Kristina Reppond, Madison N. Shyer, Jessica Cambric, Ryan Gadd, Ilya J. Finkelstein, Jimmy Gollihar, and James M. Musser |
| EPI_ISL_2204172 | J.W. Ruby Memorial Hospital | WVU and Marshall University Combined Genomics Core Facilities | James Denvir, Peter Stoilov, Peter Perrotta, Wesley Kimble, Ryan Percifield |
| EPI_ISL_2204173 | Quest Diagnostics Incorporated | Centers for Disease Control and Prevention Division of Viral Diseases, Pathogen Discovery | Dakota Howard, Dhwani Batra, Peter W. Cook, Kara Moser, Adrian Paskey, Jason Caravas, Benjamin Rambo-Martin, Shatavia Morrison, Christopher Gulvick, Scott Sammons, Yvette Unoarumhi, Darlene Wagner, Matthew Schmerer, S. H. Rosenthal, A. Gerasimova, R. M. Kagan, B. Anderson, M. Hua, Y. Liu, L.E. Bernstein, K.E. Livingston, A. Perez, I. A. Shlyakhter, R. V. Rolando, R. Owen, P. Tanpaiboon, F. Lacbawan, Clinton R. Paden, Duncan MacCannell |
| EPI_ISL_2204174 | Houston Methodist Hospital | Houston Methodist Hospital | Randall J. Olsen, Paul A. Christensen, S. Wesley Long, Sishir Subedi, Robert Olson, Marcus Nguyen, James J. Davis, Matthew Ojeda Saavedra, Prasanti Yerramilli, Layne Pruitt, Kristina Reppond, Madison N. Shyer, Jessica Cambric, Ryan Gadd, Ilya J. Finkelstein, Jimmy Gollihar, and James M. Musser |
| EPI_ISL_2204175 | Quest Diagnostics Incorporated | Centers for Disease Control and Prevention Division of Viral Diseases, Pathogen Discovery | Dakota Howard, Dhwani Batra, Peter W. Cook, Kara Moser, Adrian Paskey, Jason Caravas, Benjamin Rambo-Martin, Shatavia Morrison, Christopher Gulvick, Scott Sammons, Yvette Unoarumhi, Darlene Wagner, Matthew Schmerer, S. H. Rosenthal, A. Gerasimova, R. M. Kagan, B. Anderson, M. Hua, Y. Liu, L.E. Bernstein, K.E. Livingston, A. Perez, I. A. Shlyakhter, R. V. Rolando, R. Owen, P. Tanpaiboon, F. Lacbawan, Clinton R. Paden, Duncan MacCannell |
| EPI_ISL_2204176 | J.W. Ruby Memorial Hospital | WVU and Marshall University Combined Genomics Core Facilities | James Denvir, Peter Stoilov, Peter Perrotta, Wesley Kimble, Ryan Percifield |
| EPI_ISL_2204177 | Quest Diagnostics Incorporated | Centers for Disease Control and Prevention Division of Viral Diseases, Pathogen Discovery | Dakota Howard, Dhwani Batra, Peter W. Cook, Kara Moser, Adrian Paskey, Jason Caravas, Benjamin Rambo-Martin, Shatavia Morrison, Christopher Gulvick, Scott Sammons, Yvette Unoarumhi, Darlene Wagner, Matthew Schmerer, S. H. Rosenthal, A. Gerasimova, R. M. Kagan, B. Anderson, M. Hua, Y. Liu, L.E. Bernstein, K.E. Livingston, A. Perez, I. A. Shlyakhter, R. V. Rolando, R. Owen, P. Tanpaiboon, F. Lacbawan, Clinton R. Paden, Duncan MacCannell |
| EPI_ISL_2204178 | Houston Methodist Hospital | Houston Methodist Hospital | Randall J. Olsen, Paul A. Christensen, S. Wesley Long, Sishir Subedi, Robert Olson, Marcus Nguyen, James J. Davis, Matthew Ojeda Saavedra, Prasanti Yerramilli, Layne Pruitt, Kristina Reppond, Madison N. Shyer, Jessica Cambric, Ryan Gadd, Ilya J. Finkelstein, Jimmy Gollihar, and James M. Musser |
| EPI_ISL_2204179 | J.W. Ruby Memorial Hospital | WVU and Marshall University Combined Genomics Core Facilities | James Denvir, Peter Stoilov, Peter Perrotta, Wesley Kimble, Ryan Percifield |
| EPI_ISL_2204180 | Quest Diagnostics Incorporated | Centers for Disease Control and Prevention Division of Viral Diseases, Pathogen Discovery | Dakota Howard, Dhwani Batra, Peter W. Cook, Kara Moser, Adrian Paskey, Jason Caravas, Benjamin Rambo-Martin, Shatavia Morrison, Christopher Gulvick, Scott Sammons, Yvette Unoarumhi, Darlene Wagner, Matthew Schmerer, S. H. Rosenthal, A. Gerasimova, R. M. Kagan, B. Anderson, M. Hua, Y. Liu, L.E. Bernstein, K.E. Livingston, A. Perez, I. A. Shlyakhter, R. V. Rolando, R. Owen, P. Tanpaiboon, F. Lacbawan, Clinton R. Paden, Duncan MacCannell |
| EPI_ISL_2204181 | Houston Methodist Hospital | Houston Methodist Hospital | Randall J. Olsen, Paul A. Christensen, S. Wesley Long, Sishir Subedi, Robert Olson, Marcus Nguyen, James J. Davis, Matthew Ojeda Saavedra, Prasanti Yerramilli, Layne Pruitt, Kristina Reppond, Madison N. Shyer, Jessica Cambric, Ryan Gadd, Ilya J. Finkelstein, Jimmy Gollihar, and James M. Musser |
| EPI_ISL_2204182 | Quest Diagnostics Incorporated | Centers for Disease Control and Prevention Division of Viral Diseases, Pathogen Discovery | Dakota Howard, Dhwani Batra, Peter W. Cook, Kara Moser, Adrian Paskey, Jason Caravas, Benjamin Rambo-Martin, Shatavia Morrison, Christopher Gulvick, Scott Sammons, Yvette Unoarumhi, Darlene Wagner, Matthew Schmerer, S. H. Rosenthal, A. Gerasimova, R. M. Kagan, B. Anderson, M. Hua, Y. Liu, L.E. Bernstein, K.E. Livingston, A. Perez, I. A. Shlyakhter, R. V. Rolando, R. Owen, P. Tanpaiboon, F. Lacbawan, Clinton R. Paden, Duncan MacCannell |
| EPI_ISL_2204183 | Houston Methodist Hospital | Houston Methodist Hospital | Randall J. Olsen, Paul A. Christensen, S. Wesley Long, Sishir Subedi, Robert Olson, Marcus Nguyen, James J. Davis, Matthew Ojeda Saavedra, Prasanti Yerramilli, Layne Pruitt, Kristina Reppond, Madison N. Shyer, Jessica Cambric, Ryan Gadd, Ilya J. Finkelstein, Jimmy Gollihar, and James M. Musser |
| EPI_ISL_2204184 | J.W. Ruby Memorial Hospital | WVU and Marshall University Combined Genomics Core Facilities | James Denvir, Peter Stoilov, Peter Perrotta, Wesley Kimble, Ryan Percifield |
| EPI_ISL_2204185 | Quest Diagnostics Incorporated | Centers for Disease Control and Prevention Division of Viral Diseases, Pathogen Discovery | Dakota Howard, Dhwani Batra, Peter W. Cook, Kara Moser, Adrian Paskey, Jason Caravas, Benjamin Rambo-Martin, Shatavia Morrison, Christopher Gulvick, Scott Sammons, Yvette Unoarumhi, Darlene Wagner, Matthew Schmerer, S. H. Rosenthal, A. Gerasimova, R. M. Kagan, B. Anderson, M. Hua, Y. Liu, L.E. Bernstein, K.E. Livingston, A. Perez, I. A. Shlyakhter, R. V. Rolando, R. Owen, P. Tanpaiboon, F. Lacbawan, Clinton R. Paden, Duncan MacCannell |
| EPI_ISL_2204186 | Houston Methodist Hospital | Houston Methodist Hospital | Randall J. Olsen, Paul A. Christensen, S. Wesley Long, Sishir Subedi, Robert Olson, Marcus Nguyen, James J. Davis, Matthew Ojeda Saavedra, Prasanti Yerramilli, Layne Pruitt, Kristina Reppond, Madison N. Shyer, Jessica Cambric, Ryan Gadd, Ilya J. Finkelstein, Jimmy Gollihar, and James M. Musser |
| EPI_ISL_2204187 | Quest Diagnostics Incorporated | Centers for Disease Control and Prevention Division of Viral Diseases, Pathogen Discovery | Dakota Howard, Dhwani Batra, Peter W. Cook, Kara Moser, Adrian Paskey, Jason Caravas, Benjamin Rambo-Martin, Shatavia Morrison, Christopher Gulvick, Scott Sammons, Yvette Unoarumhi, Darlene Wagner, Matthew Schmerer, S. H. Rosenthal, A. Gerasimova, R. M. Kagan, B. Anderson, M. Hua, Y. Liu, L.E. Bernstein, K.E. Livingston, A. Perez, I. A. Shlyakhter, R. V. Rolando, R. Owen, P. Tanpaiboon, F. Lacbawan, Clinton R. Paden, Duncan MacCannell |
| EPI_ISL_2204188 | J.W. Ruby Memorial Hospital | WVU and Marshall University Combined Genomics Core Facilities | James Denvir, Peter Stoilov, Peter Perrotta, Wesley Kimble, Ryan Percifield |
| EPI_ISL_2204189 | Quest Diagnostics Incorporated | Centers for Disease Control and Prevention Division of Viral Diseases, Pathogen Discovery | Dakota Howard, Dhwani Batra, Peter W. Cook, Kara Moser, Adrian Paskey, Jason Caravas, Benjamin Rambo-Martin, Shatavia Morrison, Christopher Gulvick, Scott Sammons, Yvette Unoarumhi, Darlene Wagner, Matthew Schmerer, S. H. Rosenthal, A. Gerasimova, R. M. Kagan, B. Anderson, M. Hua, Y. Liu, L.E. Bernstein, K.E. Livingston, A. Perez, I. A. Shlyakhter, R. V. Rolando, R. Owen, P. Tanpaiboon, F. Lacbawan, Clinton R. Paden, Duncan MacCannell |
| EPI_ISL_2204190 | Houston Methodist Hospital | Houston Methodist Hospital | Randall J. Olsen, Paul A. Christensen, S. Wesley Long, Sishir Subedi, Robert Olson, Marcus Nguyen, James J. Davis, Matthew Ojeda Saavedra, Prasanti Yerramilli, Layne Pruitt, Kristina Reppond, Madison N. Shyer, Jessica Cambric, Ryan Gadd, Ilya J. Finkelstein, Jimmy Gollihar, and James M. Musser |
| EPI_ISL_2204191 | J.W. Ruby Memorial Hospital | WVU and Marshall University Combined Genomics Core Facilities | James Denvir, Peter Stoilov, Peter Perrotta, Wesley Kimble, Ryan Percifield |
| EPI_ISL_2204192 | Quest Diagnostics Incorporated | Centers for Disease Control and Prevention Division of Viral Diseases, Pathogen Discovery | Dakota Howard, Dhwani Batra, Peter W. Cook, Kara Moser, Adrian Paskey, Jason Caravas, Benjamin Rambo-Martin, Shatavia Morrison, Christopher Gulvick, Scott Sammons, Yvette Unoarumhi, Darlene Wagner, Matthew Schmerer, S. H. Rosenthal, A. Gerasimova, R. M. Kagan, B. Anderson, M. Hua, Y. Liu, L.E. Bernstein, K.E. Livingston, A. Perez, I. A. Shlyakhter, R. V. Rolando, R. Owen, P. Tanpaiboon, F. Lacbawan, Clinton R. Paden, Duncan MacCannell |
| EPI_ISL_2204193 | Houston Methodist Hospital | Houston Methodist Hospital | Randall J. Olsen, Paul A. Christensen, S. Wesley Long, Sishir Subedi, Robert Olson, Marcus Nguyen, James J. Davis, Matthew Ojeda Saavedra, Prasanti Yerramilli, Layne Pruitt, Kristina Reppond, Madison N. Shyer, Jessica Cambric, Ryan Gadd, Ilya J. Finkelstein, Jimmy Gollihar, and James M. Musser |
| EPI_ISL_2204194 | Quest Diagnostics Incorporated | Centers for Disease Control and Prevention Division of Viral Diseases, Pathogen Discovery | Dakota Howard, Dhwani Batra, Peter W. Cook, Kara Moser, Adrian Paskey, Jason Caravas, Benjamin Rambo-Martin, Shatavia Morrison, Christopher Gulvick, Scott Sammons, Yvette Unoarumhi, Darlene Wagner, Matthew Schmerer, S. H. Rosenthal, A. Gerasimova, R. M. Kagan, B. Anderson, M. Hua, Y. Liu, L.E. Bernstein, K.E. Livingston, A. Perez, I. A. Shlyakhter, R. V. Rolando, R. Owen, P. Tanpaiboon, F. Lacbawan, Clinton R. Paden, Duncan MacCannell |
| EPI_ISL_2204195 | J.W. Ruby Memorial Hospital | WVU and Marshall University Combined Genomics Core Facilities | James Denvir, Peter Stoilov, Peter Perrotta, Wesley Kimble, Ryan Percifield |
| EPI_ISL_2204196 | Houston Methodist Hospital | Houston Methodist Hospital | Randall J. Olsen, Paul A. Christensen, S. Wesley Long, Sishir Subedi, Robert Olson, Marcus Nguyen, James J. Davis, Matthew Ojeda Saavedra, Prasanti Yerramilli, Layne Pruitt, Kristina Reppond, Madison N. Shyer, Jessica Cambric, Ryan Gadd, Ilya J. Finkelstein, Jimmy Gollihar, and James M. Musser |

| | | | |
|---|---|---|---|
| EPI_ISL_2204197 | Quest Diagnostics Incorporated | Centers for Disease Control and Prevention Division of Viral Diseases, Pathogen Discovery | Dakota Howard, Dhwani Batra, Peter W. Cook, Kara Moser, Adrian Paskey, Jason Caravas, Benjamin Rambo-Martin, Shatavia Morrison, Christopher Gulvick, Scott Sammons, Yvette Unoarumhi, Darlene Wagner, Matthew Schmerer, S. H. Rosenthal, A. Gerasimova, R. M. Kagan, B. Anderson, M. Hua, Y. Liu, L.E. Bernstein, K.E. Livingston, A. Perez, I. A. Shlyakhter, R. V. Rolando, R. Owen, P. Tanpaiboon, F. Lacbawan, Clinton R. Paden, Duncan MacCannell |
| EPI_ISL_2204198 | J.W. Ruby Memorial Hospital | WVU and Marshall University Combined Genomics Core Facilities | James Denvir, Peter Stoilov, Peter Perrotta, Wesley Kimble, Ryan Percifield |
| EPI_ISL_2204199 | Quest Diagnostics Incorporated | Centers for Disease Control and Prevention Division of Viral Diseases, Pathogen Discovery | Dakota Howard, Dhwani Batra, Peter W. Cook, Kara Moser, Adrian Paskey, Jason Caravas, Benjamin Rambo-Martin, Shatavia Morrison, Christopher Gulvick, Scott Sammons, Yvette Unoarumhi, Darlene Wagner, Matthew Schmerer, S. H. Rosenthal, A. Gerasimova, R. M. Kagan, B. Anderson, M. Hua, Y. Liu, L.E. Bernstein, K.E. Livingston, A. Perez, I. A. Shlyakhter, R. V. Rolando, R. Owen, P. Tanpaiboon, F. Lacbawan, Clinton R. Paden, Duncan MacCannell |
| EPI_ISL_2204200 | Houston Methodist Hospital | Houston Methodist Hospital | Randall J. Olsen, Paul A. Christensen, S. Wesley Long, Sishir Subedi, Robert Olson, Marcus Nguyen, James J. Davis, Matthew Ojeda Saavedra, Prasanti Yerramilli, Layne Pruitt, Kristina Reppond, Madison N. Shyer, Jessica Cambric, Ryan Gadd, Ilya J. Finkelstein, Jimmy Gollihar, and James M. Musser |
| EPI_ISL_2204201 | J.W. Ruby Memorial Hospital | WVU and Marshall University Combined Genomics Core Facilities | James Denvir, Peter Stoilov, Peter Perrotta, Wesley Kimble, Ryan Percifield |
| EPI_ISL_2204202 | Quest Diagnostics Incorporated | Centers for Disease Control and Prevention Division of Viral Diseases, Pathogen Discovery | Dakota Howard, Dhwani Batra, Peter W. Cook, Kara Moser, Adrian Paskey, Jason Caravas, Benjamin Rambo-Martin, Shatavia Morrison, Christopher Gulvick, Scott Sammons, Yvette Unoarumhi, Darlene Wagner, Matthew Schmerer, S. H. Rosenthal, A. Gerasimova, R. M. Kagan, B. Anderson, M. Hua, Y. Liu, L.E. Bernstein, K.E. Livingston, A. Perez, I. A. Shlyakhter, R. V. Rolando, R. Owen, P. Tanpaiboon, F. Lacbawan, Clinton R. Paden, Duncan MacCannell |
| EPI_ISL_2204203 | J.W. Ruby Memorial Hospital | WVU and Marshall University Combined Genomics Core Facilities | James Denvir, Peter Stoilov, Peter Perrotta, Wesley Kimble, Ryan Percifield |
| EPI_ISL_2204204 | Houston Methodist Hospital | Houston Methodist Hospital | Randall J. Olsen, Paul A. Christensen, S. Wesley Long, Sishir Subedi, Robert Olson, Marcus Nguyen, James J. Davis, Matthew Ojeda Saavedra, Prasanti Yerramilli, Layne Pruitt, Kristina Reppond, Madison N. Shyer, Jessica Cambric, Ryan Gadd, Ilya J. Finkelstein, Jimmy Gollihar, and James M. Musser |
| EPI_ISL_2204205 | Quest Diagnostics Incorporated | Centers for Disease Control and Prevention Division of Viral Diseases, Pathogen Discovery | Dakota Howard, Dhwani Batra, Peter W. Cook, Kara Moser, Adrian Paskey, Jason Caravas, Benjamin Rambo-Martin, Shatavia Morrison, Christopher Gulvick, Scott Sammons, Yvette Unoarumhi, Darlene Wagner, Matthew Schmerer, S. H. Rosenthal, A. Gerasimova, R. M. Kagan, B. Anderson, M. Hua, Y. Liu, L.E. Bernstein, K.E. Livingston, A. Perez, I. A. Shlyakhter, R. V. Rolando, R. Owen, P. Tanpaiboon, F. Lacbawan, Clinton R. Paden, Duncan MacCannell |
| EPI_ISL_2204206 | J.W. Ruby Memorial Hospital | WVU and Marshall University Combined Genomics Core Facilities | James Denvir, Peter Stoilov, Peter Perrotta, Wesley Kimble, Ryan Percifield |
| EPI_ISL_2204207 | Houston Methodist Hospital | Houston Methodist Hospital | Randall J. Olsen, Paul A. Christensen, S. Wesley Long, Sishir Subedi, Robert Olson, Marcus Nguyen, James J. Davis, Matthew Ojeda Saavedra, Prasanti Yerramilli, Layne Pruitt, Kristina Reppond, Madison N. Shyer, Jessica Cambric, Ryan Gadd, Ilya J. Finkelstein, Jimmy Gollihar, and James M. Musser |
| EPI_ISL_2204208 | Quest Diagnostics Incorporated | Centers for Disease Control and Prevention Division of Viral Diseases, Pathogen Discovery | Dakota Howard, Dhwani Batra, Peter W. Cook, Kara Moser, Adrian Paskey, Jason Caravas, Benjamin Rambo-Martin, Shatavia Morrison, Christopher Gulvick, Scott Sammons, Yvette Unoarumhi, Darlene Wagner, Matthew Schmerer, S. H. Rosenthal, A. Gerasimova, R. M. Kagan, B. Anderson, M. Hua, Y. Liu, L.E. Bernstein, K.E. Livingston, A. Perez, I. A. Shlyakhter, R. V. Rolando, R. Owen, P. Tanpaiboon, F. Lacbawan, Clinton R. Paden, Duncan MacCannell |
| EPI_ISL_2204209 | J.W. Ruby Memorial Hospital | WVU and Marshall University Combined Genomics Core Facilities | James Denvir, Peter Stoilov, Peter Perrotta, Wesley Kimble, Ryan Percifield |
| EPI_ISL_2204210 | Quest Diagnostics Incorporated | Centers for Disease Control and Prevention Division of Viral Diseases, Pathogen Discovery | Dakota Howard, Dhwani Batra, Peter W. Cook, Kara Moser, Adrian Paskey, Jason Caravas, Benjamin Rambo-Martin, Shatavia Morrison, Christopher Gulvick, Scott Sammons, Yvette Unoarumhi, Darlene Wagner, Matthew Schmerer, S. H. Rosenthal, A. Gerasimova, R. M. Kagan, B. Anderson, M. Hua, Y. Liu, L.E. Bernstein, K.E. Livingston, A. Perez, I. A. Shlyakhter, R. V. Rolando, R. Owen, P. Tanpaiboon, F. Lacbawan, Clinton R. Paden, Duncan MacCannell |
| EPI_ISL_2204211 | Houston Methodist Hospital | Houston Methodist Hospital | Randall J. Olsen, Paul A. Christensen, S. Wesley Long, Sishir Subedi, Robert Olson, Marcus Nguyen, James J. Davis, Matthew Ojeda Saavedra, Prasanti Yerramilli, Layne Pruitt, Kristina Reppond, Madison N. Shyer, Jessica Cambric, Ryan Gadd, Ilya J. Finkelstein, Jimmy Gollihar, and James M. Musser |
| EPI_ISL_2204212 | Quest Diagnostics Incorporated | Centers for Disease Control and Prevention Division of Viral Diseases, Pathogen Discovery | Dakota Howard, Dhwani Batra, Peter W. Cook, Kara Moser, Adrian Paskey, Jason Caravas, Benjamin Rambo-Martin, Shatavia Morrison, Christopher Gulvick, Scott Sammons, Yvette Unoarumhi, Darlene Wagner, Matthew Schmerer, S. H. Rosenthal, A. Gerasimova, R. M. Kagan, B. Anderson, M. Hua, Y. Liu, L.E. Bernstein, K.E. Livingston, A. Perez, I. A. Shlyakhter, R. V. Rolando, R. Owen, P. Tanpaiboon, F. Lacbawan, Clinton R. Paden, Duncan MacCannell |
| EPI_ISL_2204213 | J.W. Ruby Memorial Hospital | WVU and Marshall University Combined Genomics Core Facilities | James Denvir, Peter Stoilov, Peter Perrotta, Wesley Kimble, Ryan Percifield |
| EPI_ISL_2204214 | Houston Methodist Hospital | Houston Methodist Hospital | Randall J. Olsen, Paul A. Christensen, S. Wesley Long, Sishir Subedi, Robert Olson, Marcus Nguyen, James J. Davis, Matthew Ojeda Saavedra, Prasanti Yerramilli, Layne Pruitt, Kristina Reppond, Madison N. Shyer, Jessica Cambric, Ryan Gadd, Ilya J. Finkelstein, Jimmy Gollihar, and James M. Musser |
| EPI_ISL_2204215 | Quest Diagnostics Incorporated | Centers for Disease Control and Prevention Division of Viral Diseases, Pathogen Discovery | Dakota Howard, Dhwani Batra, Peter W. Cook, Kara Moser, Adrian Paskey, Jason Caravas, Benjamin Rambo-Martin, Shatavia Morrison, Christopher Gulvick, Scott Sammons, Yvette Unoarumhi, Darlene Wagner, Matthew Schmerer, S. H. Rosenthal, A. Gerasimova, R. M. Kagan, B. Anderson, M. Hua, Y. Liu, L.E. Bernstein, K.E. Livingston, A. Perez, I. A. Shlyakhter, R. V. Rolando, R. Owen, P. Tanpaiboon, F. Lacbawan, Clinton R. Paden, Duncan MacCannell |
| EPI_ISL_2204216 | Houston Methodist Hospital | Houston Methodist Hospital | Randall J. Olsen, Paul A. Christensen, S. Wesley Long, Sishir Subedi, Robert Olson, Marcus Nguyen, James J. Davis, Matthew Ojeda Saavedra, Prasanti Yerramilli, Layne Pruitt, Kristina Reppond, Madison N. Shyer, Jessica Cambric, Ryan Gadd, Ilya J. Finkelstein, Jimmy Gollihar, and James M. Musser |
| EPI_ISL_2204217 | J.W. Ruby Memorial Hospital | WVU and Marshall University Combined Genomics Core Facilities | James Denvir, Peter Stoilov, Peter Perrotta, Wesley Kimble, Ryan Percifield |
| EPI_ISL_2204218, EPI_ISL_2204219 | Quest Diagnostics Incorporated | Centers for Disease Control and Prevention Division of Viral Diseases, Pathogen Discovery | Dakota Howard, Dhwani Batra, Peter W. Cook, Kara Moser, Adrian Paskey, Jason Caravas, Benjamin Rambo-Martin, Shatavia Morrison, Christopher Gulvick, Scott Sammons, Yvette Unoarumhi, Darlene Wagner, Matthew Schmerer, S. H. Rosenthal, A. Gerasimova, R. M. Kagan, B. Anderson, M. Hua, Y. Liu, L.E. Bernstein, K.E. Livingston, A. Perez, I. A. Shlyakhter, R. V. Rolando, R. Owen, P. Tanpaiboon, F. Lacbawan, Clinton R. Paden, Duncan MacCannell |
| EPI_ISL_2204220 | Houston Methodist Hospital | Houston Methodist Hospital | Randall J. Olsen, Paul A. Christensen, S. Wesley Long, Sishir Subedi, Robert Olson, Marcus Nguyen, James J. Davis, Matthew Ojeda Saavedra, Prasanti Yerramilli, Layne Pruitt, Kristina Reppond, Madison N. Shyer, Jessica Cambric, Ryan Gadd, Ilya J. Finkelstein, Jimmy Gollihar, and James M. Musser |
| EPI_ISL_2204221 | J.W. Ruby Memorial Hospital | WVU and Marshall University Combined Genomics Core Facilities | James Denvir, Peter Stoilov, Peter Perrotta, Wesley Kimble, Ryan Percifield |
| EPI_ISL_2204222 | Quest Diagnostics Incorporated | Centers for Disease Control and Prevention Division of Viral Diseases, Pathogen Discovery | Dakota Howard, Dhwani Batra, Peter W. Cook, Kara Moser, Adrian Paskey, Jason Caravas, Benjamin Rambo-Martin, Shatavia Morrison, Christopher Gulvick, Scott Sammons, Yvette Unoarumhi, Darlene Wagner, Matthew Schmerer, S. H. Rosenthal, A. Gerasimova, R. M. Kagan, B. Anderson, M. Hua, Y. Liu, L.E. Bernstein, K.E. Livingston, A. Perez, I. A. Shlyakhter, R. V. Rolando, R. Owen, P. Tanpaiboon, F. Lacbawan, Clinton R. Paden, Duncan MacCannell |
| EPI_ISL_2204223 | Houston Methodist Hospital | Houston Methodist Hospital | Randall J. Olsen, Paul A. Christensen, S. Wesley Long, Sishir Subedi, Robert Olson, Marcus Nguyen, James J. Davis, Matthew Ojeda Saavedra, Prasanti Yerramilli, Layne Pruitt, Kristina Reppond, Madison N. Shyer, Jessica Cambric, Ryan Gadd, Ilya J. Finkelstein, Jimmy Gollihar, and James M. Musser |
| EPI_ISL_2204224 | J.W. Ruby Memorial Hospital | WVU and Marshall University Combined Genomics Core Facilities | James Denvir, Peter Stoilov, Peter Perrotta, Wesley Kimble, Ryan Percifield |
| EPI_ISL_2204225 | Quest Diagnostics Incorporated | Centers for Disease Control and Prevention Division of Viral Diseases, Pathogen Discovery | Dakota Howard, Dhwani Batra, Peter W. Cook, Kara Moser, Adrian Paskey, Jason Caravas, Benjamin Rambo-Martin, Shatavia Morrison, Christopher Gulvick, Scott Sammons, Yvette Unoarumhi, Darlene Wagner, Matthew Schmerer, S. H. Rosenthal, A. Gerasimova, R. M. Kagan, B. Anderson, M. Hua, Y. Liu, L.E. Bernstein, K.E. Livingston, A. Perez, I. A. Shlyakhter, R. V. Rolando, R. Owen, P. Tanpaiboon, F. Lacbawan, Clinton R. Paden, Duncan MacCannell |
| EPI_ISL_2204226 | Houston Methodist Hospital | Houston Methodist Hospital | Randall J. Olsen, Paul A. Christensen, S. Wesley Long, Sishir Subedi, Robert Olson, Marcus Nguyen, James J. Davis, Matthew Ojeda Saavedra, Prasanti Yerramilli, Layne Pruitt, Kristina Reppond, Madison N. Shyer, Jessica Cambric, Ryan Gadd, Ilya J. Finkelstein, Jimmy Gollihar, and James M. Musser |
| EPI_ISL_2204227 | Quest Diagnostics Incorporated | Centers for Disease Control and Prevention Division of Viral Diseases, Pathogen Discovery | Dakota Howard, Dhwani Batra, Peter W. Cook, Kara Moser, Adrian Paskey, Jason Caravas, Benjamin Rambo-Martin, Shatavia Morrison, Christopher Gulvick, Scott Sammons, Yvette Unoarumhi, Darlene Wagner, Matthew Schmerer, S. H. Rosenthal, A. Gerasimova, R. M. Kagan, B. Anderson, M. Hua, Y. Liu, L.E. Bernstein, K.E. Livingston, A. Perez, I. A. Shlyakhter, R. V. Rolando, R. Owen, P. Tanpaiboon, F. Lacbawan, Clinton R. Paden, Duncan MacCannell |
| EPI_ISL_2204228 | J.W. Ruby Memorial Hospital | WVU and Marshall University Combined Genomics Core Facilities | James Denvir, Peter Stoilov, Peter Perrotta, Wesley Kimble, Ryan Percifield |
| EPI_ISL_2204229, EPI_ISL_2204230 | Quest Diagnostics Incorporated | Centers for Disease Control and Prevention Division of Viral Diseases, Pathogen Discovery | Dakota Howard, Dhwani Batra, Peter W. Cook, Kara Moser, Adrian Paskey, Jason Caravas, Benjamin Rambo-Martin, Shatavia Morrison, Christopher Gulvick, Scott Sammons, Yvette Unoarumhi, Darlene Wagner, Matthew Schmerer, S. H. Rosenthal, A. Gerasimova, R. M. Kagan, B. Anderson, M. Hua, Y. Liu, L.E. Bernstein, K.E. Livingston, A. Perez, I. A. Shlyakhter, R. V. Rolando, R. Owen, P. Tanpaiboon, F. Lacbawan, Clinton R. Paden, Duncan MacCannell |
| EPI_ISL_2204231 | J.W. Ruby Memorial Hospital | WVU and Marshall University Combined Genomics Core Facilities | James Denvir, Peter Stoilov, Peter Perrotta, Wesley Kimble, Ryan Percifield |
| EPI_ISL_2204232 | Quest Diagnostics Incorporated | Centers for Disease Control and Prevention Division of Viral Diseases, Pathogen Discovery | Dakota Howard, Dhwani Batra, Peter W. Cook, Kara Moser, Adrian Paskey, Jason Caravas, Benjamin Rambo-Martin, Shatavia Morrison, Christopher Gulvick, Scott Sammons, Yvette Unoarumhi, Darlene Wagner, Matthew Schmerer, S. H. Rosenthal, A. Gerasimova, R. M. Kagan, B. Anderson, M. Hua, Y. Liu, L.E. Bernstein, K.E. Livingston, A. Perez, I. A. Shlyakhter, R. V. Rolando, R. Owen, P. Tanpaiboon, F. Lacbawan, Clinton R. Paden, Duncan MacCannell |
| EPI_ISL_2204233 | Houston Methodist Hospital | Houston Methodist Hospital | Randall J. Olsen, Paul A. Christensen, S. Wesley Long, Sishir Subedi, Robert Olson, Marcus Nguyen, James J. Davis, Matthew Ojeda Saavedra, Prasanti Yerramilli, Layne Pruitt, Kristina Reppond, Madison N. Shyer, Jessica Cambric, Ryan Gadd, Ilya J. Finkelstein, Jimmy Gollihar, and James M. Musser |
| EPI_ISL_2204234 | J.W. Ruby Memorial Hospital | WVU and Marshall University Combined Genomics Core Facilities | James Denvir, Peter Stoilov, Peter Perrotta, Wesley Kimble, Ryan Percifield |
| EPI_ISL_2204235, EPI_ISL_2204236 | Quest Diagnostics Incorporated | Centers for Disease Control and Prevention Division of Viral Diseases, Pathogen Discovery | Dakota Howard, Dhwani Batra, Peter W. Cook, Kara Moser, Adrian Paskey, Jason Caravas, Benjamin Rambo-Martin, Shatavia Morrison, Christopher Gulvick, Scott Sammons, Yvette Unoarumhi, Darlene Wagner, Matthew Schmerer, S. H. Rosenthal, A. Gerasimova, R. M. Kagan, B. Anderson, M. Hua, Y. Liu, L.E. Bernstein, K.E. Livingston, A. Perez, I. A. Shlyakhter, R. V. Rolando, R. Owen, P. Tanpaiboon, F. Lacbawan, Clinton R. Paden, Duncan MacCannell |
| EPI_ISL_2204237 | J.W. Ruby Memorial Hospital | WVU and Marshall University Combined Genomics Core Facilities | James Denvir, Peter Stoilov, Peter Perrotta, Wesley Kimble, Ryan Percifield |
| EPI_ISL_2204238 | Houston Methodist Hospital | Houston Methodist Hospital | Randall J. Olsen, Paul A. Christensen, S. Wesley Long, Sishir Subedi, Robert Olson, Marcus Nguyen, James J. Davis, Matthew Ojeda Saavedra, Prasanti Yerramilli, Layne Pruitt, Kristina Reppond, Madison N. Shyer, Jessica Cambric, Ryan Gadd, Ilya J. Finkelstein, Jimmy Gollihar, and James M. Musser |

| | | | |
|---|---|---|---|
| EPI_ISL_2204239 | Quest Diagnostics Incorporated | Centers for Disease Control and Prevention Division of Viral Diseases, Pathogen Discovery | Dakota Howard, Dhwani Batra, Peter W. Cook, Kara Moser, Adrian Paskey, Jason Caravas, Benjamin Rambo-Martin, Shatavia Morrison, Christopher Gulvick, Scott Sammons, Yvette Unoarumhi, Darlene Wagner, Matthew Schmerer, S. H. Rosenthal, A. Gerasimova, R. M. Kagan, B. Anderson, M. Hua, Y. Liu, L.E. Bernstein, K.E. Livingston, A. Perez, I. A. Shlyakhter, R. V. Rolando, R. Owen, P. Tanpaiboon, F. Lacbawan, Clinton R. Paden, Duncan MacCannell |
| EPI_ISL_2204240 | Houston Methodist Hospital | Houston Methodist Hospital | Randall J. Olsen, Paul A. Christensen, S. Wesley Long, Sishir Subedi, Robert Olson, Marcus Nguyen, James J. Davis, Matthew Ojeda Saavedra, Prasanti Yerramilli, Layne Pruitt, Kristina Reppond, Madison N. Shyer, Jessica Cambric, Ryan Gadd, Ilya J. Finkelstein, Jimmy Gollihar, and James M. Musser |
| EPI_ISL_2204241 | J.W. Ruby Memorial Hospital | WVU and Marshall University Combined Genomics Core Facilities | James Denvir, Peter Stoilov, Peter Perrotta, Wesley Kimble, Ryan Percifield |
| EPI_ISL_2204242 | Quest Diagnostics Incorporated | Centers for Disease Control and Prevention Division of Viral Diseases, Pathogen Discovery | Dakota Howard, Dhwani Batra, Peter W. Cook, Kara Moser, Adrian Paskey, Jason Caravas, Benjamin Rambo-Martin, Shatavia Morrison, Christopher Gulvick, Scott Sammons, Yvette Unoarumhi, Darlene Wagner, Matthew Schmerer, S. H. Rosenthal, A. Gerasimova, R. M. Kagan, B. Anderson, M. Hua, Y. Liu, L.E. Bernstein, K.E. Livingston, A. Perez, I. A. Shlyakhter, R. V. Rolando, R. Owen, P. Tanpaiboon, F. Lacbawan, Clinton R. Paden, Duncan MacCannell |
| EPI_ISL_2204243 | Houston Methodist Hospital | Houston Methodist Hospital | Randall J. Olsen, Paul A. Christensen, S. Wesley Long, Sishir Subedi, Robert Olson, Marcus Nguyen, James J. Davis, Matthew Ojeda Saavedra, Prasanti Yerramilli, Layne Pruitt, Kristina Reppond, Madison N. Shyer, Jessica Cambric, Ryan Gadd, Ilya J. Finkelstein, Jimmy Gollihar, and James M. Musser |
| EPI_ISL_2204244 | Quest Diagnostics Incorporated | Centers for Disease Control and Prevention Division of Viral Diseases, Pathogen Discovery | Dakota Howard, Dhwani Batra, Peter W. Cook, Kara Moser, Adrian Paskey, Jason Caravas, Benjamin Rambo-Martin, Shatavia Morrison, Christopher Gulvick, Scott Sammons, Yvette Unoarumhi, Darlene Wagner, Matthew Schmerer, S. H. Rosenthal, A. Gerasimova, R. M. Kagan, B. Anderson, M. Hua, Y. Liu, L.E. Bernstein, K.E. Livingston, A. Perez, I. A. Shlyakhter, R. V. Rolando, R. Owen, P. Tanpaiboon, F. Lacbawan, Clinton R. Paden, Duncan MacCannell |
| EPI_ISL_2204245 | J.W. Ruby Memorial Hospital | WVU and Marshall University Combined Genomics Core Facilities | James Denvir, Peter Stoilov, Peter Perrotta, Wesley Kimble, Ryan Percifield |
| EPI_ISL_2204246 | Houston Methodist Hospital | Houston Methodist Hospital | Randall J. Olsen, Paul A. Christensen, S. Wesley Long, Sishir Subedi, Robert Olson, Marcus Nguyen, James J. Davis, Matthew Ojeda Saavedra, Prasanti Yerramilli, Layne Pruitt, Kristina Reppond, Madison N. Shyer, Jessica Cambric, Ryan Gadd, Ilya J. Finkelstein, Jimmy Gollihar, and James M. Musser |
| EPI_ISL_2204247 | Quest Diagnostics Incorporated | Centers for Disease Control and Prevention Division of Viral Diseases, Pathogen Discovery | Dakota Howard, Dhwani Batra, Peter W. Cook, Kara Moser, Adrian Paskey, Jason Caravas, Benjamin Rambo-Martin, Shatavia Morrison, Christopher Gulvick, Scott Sammons, Yvette Unoarumhi, Darlene Wagner, Matthew Schmerer, S. H. Rosenthal, A. Gerasimova, R. M. Kagan, B. Anderson, M. Hua, Y. Liu, L.E. Bernstein, K.E. Livingston, A. Perez, I. A. Shlyakhter, R. V. Rolando, R. Owen, P. Tanpaiboon, F. Lacbawan, Clinton R. Paden, Duncan MacCannell |
| EPI_ISL_2204248 | J.W. Ruby Memorial Hospital | WVU and Marshall University Combined Genomics Core Facilities | James Denvir, Peter Stoilov, Peter Perrotta, Wesley Kimble, Ryan Percifield |
| EPI_ISL_2204249 | Quest Diagnostics Incorporated | Centers for Disease Control and Prevention Division of Viral Diseases, Pathogen Discovery | Dakota Howard, Dhwani Batra, Peter W. Cook, Kara Moser, Adrian Paskey, Jason Caravas, Benjamin Rambo-Martin, Shatavia Morrison, Christopher Gulvick, Scott Sammons, Yvette Unoarumhi, Darlene Wagner, Matthew Schmerer, S. H. Rosenthal, A. Gerasimova, R. M. Kagan, B. Anderson, M. Hua, Y. Liu, L.E. Bernstein, K.E. Livingston, A. Perez, I. A. Shlyakhter, R. V. Rolando, R. Owen, P. Tanpaiboon, F. Lacbawan, Clinton R. Paden, Duncan MacCannell |
| EPI_ISL_2204250 | Houston Methodist Hospital | Houston Methodist Hospital | Randall J. Olsen, Paul A. Christensen, S. Wesley Long, Sishir Subedi, Robert Olson, Marcus Nguyen, James J. Davis, Matthew Ojeda Saavedra, Prasanti Yerramilli, Layne Pruitt, Kristina Reppond, Madison N. Shyer, Jessica Cambric, Ryan Gadd, Ilya J. Finkelstein, Jimmy Gollihar, and James M. Musser |
| EPI_ISL_2204251 | Quest Diagnostics Incorporated | Centers for Disease Control and Prevention Division of Viral Diseases, Pathogen Discovery | Dakota Howard, Dhwani Batra, Peter W. Cook, Kara Moser, Adrian Paskey, Jason Caravas, Benjamin Rambo-Martin, Shatavia Morrison, Christopher Gulvick, Scott Sammons, Yvette Unoarumhi, Darlene Wagner, Matthew Schmerer, S. H. Rosenthal, A. Gerasimova, R. M. Kagan, B. Anderson, M. Hua, Y. Liu, L.E. Bernstein, K.E. Livingston, A. Perez, I. A. Shlyakhter, R. V. Rolando, R. Owen, P. Tanpaiboon, F. Lacbawan, Clinton R. Paden, Duncan MacCannell |
| EPI_ISL_2204252 | J.W. Ruby Memorial Hospital | WVU and Marshall University Combined Genomics Core Facilities | James Denvir, Peter Stoilov, Peter Perrotta, Wesley Kimble, Ryan Percifield |
| EPI_ISL_2204253 | Houston Methodist Hospital | Houston Methodist Hospital | Randall J. Olsen, Paul A. Christensen, S. Wesley Long, Sishir Subedi, Robert Olson, Marcus Nguyen, James J. Davis, Matthew Ojeda Saavedra, Prasanti Yerramilli, Layne Pruitt, Kristina Reppond, Madison N. Shyer, Jessica Cambric, Ryan Gadd, Ilya J. Finkelstein, Jimmy Gollihar, and James M. Musser |
| EPI_ISL_2204254 | Quest Diagnostics Incorporated | Centers for Disease Control and Prevention Division of Viral Diseases, Pathogen Discovery | Dakota Howard, Dhwani Batra, Peter W. Cook, Kara Moser, Adrian Paskey, Jason Caravas, Benjamin Rambo-Martin, Shatavia Morrison, Christopher Gulvick, Scott Sammons, Yvette Unoarumhi, Darlene Wagner, Matthew Schmerer, S. H. Rosenthal, A. Gerasimova, R. M. Kagan, B. Anderson, M. Hua, Y. Liu, L.E. Bernstein, K.E. Livingston, A. Perez, I. A. Shlyakhter, R. V. Rolando, R. Owen, P. Tanpaiboon, F. Lacbawan, Clinton R. Paden, Duncan MacCannell |
| EPI_ISL_2204255 | J.W. Ruby Memorial Hospital | WVU and Marshall University Combined Genomics Core Facilities | James Denvir, Peter Stoilov, Peter Perrotta, Wesley Kimble, Ryan Percifield |
| EPI_ISL_2204256 | Quest Diagnostics Incorporated | Centers for Disease Control and Prevention Division of Viral Diseases, Pathogen Discovery | Dakota Howard, Dhwani Batra, Peter W. Cook, Kara Moser, Adrian Paskey, Jason Caravas, Benjamin Rambo-Martin, Shatavia Morrison, Christopher Gulvick, Scott Sammons, Yvette Unoarumhi, Darlene Wagner, Matthew Schmerer, S. H. Rosenthal, A. Gerasimova, R. M. Kagan, B. Anderson, M. Hua, Y. Liu, L.E. Bernstein, K.E. Livingston, A. Perez, I. A. Shlyakhter, R. V. Rolando, R. Owen, P. Tanpaiboon, F. Lacbawan, Clinton R. Paden, Duncan MacCannell |
| EPI_ISL_2204257 | Houston Methodist Hospital | Houston Methodist Hospital | Randall J. Olsen, Paul A. Christensen, S. Wesley Long, Sishir Subedi, Robert Olson, Marcus Nguyen, James J. Davis, Matthew Ojeda Saavedra, Prasanti Yerramilli, Layne Pruitt, Kristina Reppond, Madison N. Shyer, Jessica Cambric, Ryan Gadd, Ilya J. Finkelstein, Jimmy Gollihar, and James M. Musser |
| EPI_ISL_2204258 | Quest Diagnostics Incorporated | Centers for Disease Control and Prevention Division of Viral Diseases, Pathogen Discovery | Dakota Howard, Dhwani Batra, Peter W. Cook, Kara Moser, Adrian Paskey, Jason Caravas, Benjamin Rambo-Martin, Shatavia Morrison, Christopher Gulvick, Scott Sammons, Yvette Unoarumhi, Darlene Wagner, Matthew Schmerer, S. H. Rosenthal, A. Gerasimova, R. M. Kagan, B. Anderson, M. Hua, Y. Liu, L.E. Bernstein, K.E. Livingston, A. Perez, I. A. Shlyakhter, R. V. Rolando, R. Owen, P. Tanpaiboon, F. Lacbawan, Clinton R. Paden, Duncan MacCannell |
| EPI_ISL_2204259 | J.W. Ruby Memorial Hospital | WVU and Marshall University Combined Genomics Core Facilities | James Denvir, Peter Stoilov, Peter Perrotta, Wesley Kimble, Ryan Percifield |
| EPI_ISL_2204260 | Quest Diagnostics Incorporated | Centers for Disease Control and Prevention Division of Viral Diseases, Pathogen Discovery | Dakota Howard, Dhwani Batra, Peter W. Cook, Kara Moser, Adrian Paskey, Jason Caravas, Benjamin Rambo-Martin, Shatavia Morrison, Christopher Gulvick, Scott Sammons, Yvette Unoarumhi, Darlene Wagner, Matthew Schmerer, S. H. Rosenthal, A. Gerasimova, R. M. Kagan, B. Anderson, M. Hua, Y. Liu, L.E. Bernstein, K.E. Livingston, A. Perez, I. A. Shlyakhter, R. V. Rolando, R. Owen, P. Tanpaiboon, F. Lacbawan, Clinton R. Paden, Duncan MacCannell |
| EPI_ISL_2204261 | J.W. Ruby Memorial Hospital | WVU and Marshall University Combined Genomics Core Facilities | James Denvir, Peter Stoilov, Peter Perrotta, Wesley Kimble, Ryan Percifield |
| EPI_ISL_2204262 | Quest Diagnostics Incorporated | Centers for Disease Control and Prevention Division of Viral Diseases, Pathogen Discovery | Dakota Howard, Dhwani Batra, Peter W. Cook, Kara Moser, Adrian Paskey, Jason Caravas, Benjamin Rambo-Martin, Shatavia Morrison, Christopher Gulvick, Scott Sammons, Yvette Unoarumhi, Darlene Wagner, Matthew Schmerer, S. H. Rosenthal, A. Gerasimova, R. M. Kagan, B. Anderson, M. Hua, Y. Liu, L.E. Bernstein, K.E. Livingston, A. Perez, I. A. Shlyakhter, R. V. Rolando, R. Owen, P. Tanpaiboon, F. Lacbawan, Clinton R. Paden, Duncan MacCannell |
| EPI_ISL_2204263 | Houston Methodist Hospital | Houston Methodist Hospital | Randall J. Olsen, Paul A. Christensen, S. Wesley Long, Sishir Subedi, Robert Olson, Marcus Nguyen, James J. Davis, Matthew Ojeda Saavedra, Prasanti Yerramilli, Layne Pruitt, Kristina Reppond, Madison N. Shyer, Jessica Cambric, Ryan Gadd, Ilya J. Finkelstein, Jimmy Gollihar, and James M. Musser |
| EPI_ISL_2204264, EPI_ISL_2204265 | J.W. Ruby Memorial Hospital | WVU and Marshall University Combined Genomics Core Facilities | James Denvir, Peter Stoilov, Peter Perrotta, Wesley Kimble, Ryan Percifield |
| EPI_ISL_2204266 | Houston Methodist Hospital | Houston Methodist Hospital | Randall J. Olsen, Paul A. Christensen, S. Wesley Long, Sishir Subedi, Robert Olson, Marcus Nguyen, James J. Davis, Matthew Ojeda Saavedra, Prasanti Yerramilli, Layne Pruitt, Kristina Reppond, Madison N. Shyer, Jessica Cambric, Ryan Gadd, Ilya J. Finkelstein, Jimmy Gollihar, and James M. Musser |
| EPI_ISL_2204267, EPI_ISL_2204268 | J.W. Ruby Memorial Hospital | WVU and Marshall University Combined Genomics Core Facilities | James Denvir, Peter Stoilov, Peter Perrotta, Wesley Kimble, Ryan Percifield |
| EPI_ISL_2204269 | Houston Methodist Hospital | Houston Methodist Hospital | Randall J. Olsen, Paul A. Christensen, S. Wesley Long, Sishir Subedi, Robert Olson, Marcus Nguyen, James J. Davis, Matthew Ojeda Saavedra, Prasanti Yerramilli, Layne Pruitt, Kristina Reppond, Madison N. Shyer, Jessica Cambric, Ryan Gadd, Ilya J. Finkelstein, Jimmy Gollihar, and James M. Musser |
| EPI_ISL_2204270 | J.W. Ruby Memorial Hospital | WVU and Marshall University Combined Genomics Core Facilities | James Denvir, Peter Stoilov, Peter Perrotta, Wesley Kimble, Ryan Percifield |
| EPI_ISL_2204271 | Houston Methodist Hospital | Houston Methodist Hospital | Randall J. Olsen, Paul A. Christensen, S. Wesley Long, Sishir Subedi, Robert Olson, Marcus Nguyen, James J. Davis, Matthew Ojeda Saavedra, Prasanti Yerramilli, Layne Pruitt, Kristina Reppond, Madison N. Shyer, Jessica Cambric, Ryan Gadd, Ilya J. Finkelstein, Jimmy Gollihar, and James M. Musser |
| EPI_ISL_2204272 | J.W. Ruby Memorial Hospital | WVU and Marshall University Combined Genomics Core Facilities | James Denvir, Peter Stoilov, Peter Perrotta, Wesley Kimble, Ryan Percifield |
| EPI_ISL_2204273, EPI_ISL_2204274, EPI_ISL_2204276 | Houston Methodist Hospital | Houston Methodist Hospital | Randall J. Olsen, Paul A. Christensen, S. Wesley Long, Sishir Subedi, Robert Olson, Marcus Nguyen, James J. Davis, Matthew Ojeda Saavedra, Prasanti Yerramilli, Layne Pruitt, Kristina Reppond, Madison N. Shyer, Jessica Cambric, Ryan Gadd, Ilya J. Finkelstein, Jimmy Gollihar, and James M. Musser |
| EPI_ISL_2204277 | J.W. Ruby Memorial Hospital | WVU and Marshall University Combined Genomics Core Facilities | James Denvir, Peter Stoilov, Peter Perrotta, Wesley Kimble, Ryan Percifield |
| EPI_ISL_2204278 | Houston Methodist Hospital | Houston Methodist Hospital | Randall J. Olsen, Paul A. Christensen, S. Wesley Long, Sishir Subedi, Robert Olson, Marcus Nguyen, James J. Davis, Matthew Ojeda Saavedra, Prasanti Yerramilli, Layne Pruitt, Kristina Reppond, Madison N. Shyer, Jessica Cambric, Ryan Gadd, Ilya J. Finkelstein, Jimmy Gollihar, and James M. Musser |
| EPI_ISL_2204279 | J.W. Ruby Memorial Hospital | WVU and Marshall University Combined Genomics Core Facilities | James Denvir, Peter Stoilov, Peter Perrotta, Wesley Kimble, Ryan Percifield |
| EPI_ISL_2204280 | Houston Methodist Hospital | Houston Methodist Hospital | Randall J. Olsen, Paul A. Christensen, S. Wesley Long, Sishir Subedi, Robert Olson, Marcus Nguyen, James J. Davis, Matthew Ojeda Saavedra, Prasanti Yerramilli, Layne Pruitt, Kristina Reppond, Madison N. Shyer, Jessica Cambric, Ryan Gadd, Ilya J. Finkelstein, Jimmy Gollihar, and James M. Musser |
| EPI_ISL_2204281 | J.W. Ruby Memorial Hospital | WVU and Marshall University Combined Genomics Core Facilities | James Denvir, Peter Stoilov, Peter Perrotta, Wesley Kimble, Ryan Percifield |
| EPI_ISL_2204282 | Houston Methodist Hospital | Houston Methodist Hospital | Randall J. Olsen, Paul A. Christensen, S. Wesley Long, Sishir Subedi, Robert Olson, Marcus Nguyen, James J. Davis, Matthew Ojeda Saavedra, Prasanti Yerramilli, Layne Pruitt, Kristina Reppond, Madison N. Shyer, Jessica Cambric, Ryan Gadd, Ilya J. Finkelstein, Jimmy Gollihar, and James M. Musser |
| EPI_ISL_2204283 | J.W. Ruby Memorial Hospital | WVU and Marshall University Combined Genomics Core Facilities | James Denvir, Peter Stoilov, Peter Perrotta, Wesley Kimble, Ryan Percifield |

| | | | |
|---|---|---|---|
| EPI_ISL_2204284, EPI_ISL_2204285 | Houston Methodist Hospital | Houston Methodist Hospital | Randall J. Olsen, Paul A. Christensen, S. Wesley Long, Sishir Subedi, Robert Olson, Marcus Nguyen, James J. Davis, Matthew Ojeda Saavedra, Prasanti Yerramilli, Layne Pruitt, Kristina Reppond, Madison N. Shyer, Jessica Cambric, Ryan Gadd, Ilya J. Finkelstein, Jimmy Gollihar, and James M. Musser |
| EPI_ISL_2204286 | J.W. Ruby Memorial Hospital | WVU and Marshall University Combined Genomics Core Facilities | James Denvir, Peter Stoilov, Peter Perrotta, Wesley Kimble, Ryan Percifield |
| EPI_ISL_2204287 | Houston Methodist Hospital | Houston Methodist Hospital | Randall J. Olsen, Paul A. Christensen, S. Wesley Long, Sishir Subedi, Robert Olson, Marcus Nguyen, James J. Davis, Matthew Ojeda Saavedra, Prasanti Yerramilli, Layne Pruitt, Kristina Reppond, Madison N. Shyer, Jessica Cambric, Ryan Gadd, Ilya J. Finkelstein, Jimmy Gollihar, and James M. Musser |
| EPI_ISL_2204288 | Wheeling Hospital | WVU and Marshall University Combined Genomics Core Facilities | James Denvir, Peter Stoilov, Peter Perrotta, Wesley Kimble, Ryan Percifield |
| EPI_ISL_2204289, EPI_ISL_2204290 | Houston Methodist Hospital | Houston Methodist Hospital | Randall J. Olsen, Paul A. Christensen, S. Wesley Long, Sishir Subedi, Robert Olson, Marcus Nguyen, James J. Davis, Matthew Ojeda Saavedra, Prasanti Yerramilli, Layne Pruitt, Kristina Reppond, Madison N. Shyer, Jessica Cambric, Ryan Gadd, Ilya J. Finkelstein, Jimmy Gollihar, and James M. Musser |
| EPI_ISL_2204291 | J.W. Ruby Memorial Hospital | WVU and Marshall University Combined Genomics Core Facilities | James Denvir, Peter Stoilov, Peter Perrotta, Wesley Kimble, Ryan Percifield |
| EPI_ISL_2204292 | Houston Methodist Hospital | Houston Methodist Hospital | Randall J. Olsen, Paul A. Christensen, S. Wesley Long, Sishir Subedi, Robert Olson, Marcus Nguyen, James J. Davis, Matthew Ojeda Saavedra, Prasanti Yerramilli, Layne Pruitt, Kristina Reppond, Madison N. Shyer, Jessica Cambric, Ryan Gadd, Ilya J. Finkelstein, Jimmy Gollihar, and James M. Musser |
| EPI_ISL_2204293 | J.W. Ruby Memorial Hospital | WVU and Marshall University Combined Genomics Core Facilities | James Denvir, Peter Stoilov, Peter Perrotta, Wesley Kimble, Ryan Percifield |
| EPI_ISL_2204294 | Houston Methodist Hospital | Houston Methodist Hospital | Randall J. Olsen, Paul A. Christensen, S. Wesley Long, Sishir Subedi, Robert Olson, Marcus Nguyen, James J. Davis, Matthew Ojeda Saavedra, Prasanti Yerramilli, Layne Pruitt, Kristina Reppond, Madison N. Shyer, Jessica Cambric, Ryan Gadd, Ilya J. Finkelstein, Jimmy Gollihar, and James M. Musser |
| EPI_ISL_2204295 | J.W. Ruby Memorial Hospital | WVU and Marshall University Combined Genomics Core Facilities | James Denvir, Peter Stoilov, Peter Perrotta, Wesley Kimble, Ryan Percifield |
| EPI_ISL_2204296, EPI_ISL_2204297 | Houston Methodist Hospital | Houston Methodist Hospital | Randall J. Olsen, Paul A. Christensen, S. Wesley Long, Sishir Subedi, Robert Olson, Marcus Nguyen, James J. Davis, Matthew Ojeda Saavedra, Prasanti Yerramilli, Layne Pruitt, Kristina Reppond, Madison N. Shyer, Jessica Cambric, Ryan Gadd, Ilya J. Finkelstein, Jimmy Gollihar, and James M. Musser |
| EPI_ISL_2204298 | J.W. Ruby Memorial Hospital | WVU and Marshall University Combined Genomics Core Facilities | James Denvir, Peter Stoilov, Peter Perrotta, Wesley Kimble, Ryan Percifield |
| EPI_ISL_2204299 | Houston Methodist Hospital | Houston Methodist Hospital | Randall J. Olsen, Paul A. Christensen, S. Wesley Long, Sishir Subedi, Robert Olson, Marcus Nguyen, James J. Davis, Matthew Ojeda Saavedra, Prasanti Yerramilli, Layne Pruitt, Kristina Reppond, Madison N. Shyer, Jessica Cambric, Ryan Gadd, Ilya J. Finkelstein, Jimmy Gollihar, and James M. Musser |
| EPI_ISL_2204300, EPI_ISL_2204302 | J.W. Ruby Memorial Hospital | WVU and Marshall University Combined Genomics Core Facilities | James Denvir, Peter Stoilov, Peter Perrotta, Wesley Kimble, Ryan Percifield |
| EPI_ISL_2204303 | Houston Methodist Hospital | Houston Methodist Hospital | Randall J. Olsen, Paul A. Christensen, S. Wesley Long, Sishir Subedi, Robert Olson, Marcus Nguyen, James J. Davis, Matthew Ojeda Saavedra, Prasanti Yerramilli, Layne Pruitt, Kristina Reppond, Madison N. Shyer, Jessica Cambric, Ryan Gadd, Ilya J. Finkelstein, Jimmy Gollihar, and James M. Musser |
| EPI_ISL_2204304 | Wheeling Hospital | WVU and Marshall University Combined Genomics Core Facilities | James Denvir, Peter Stoilov, Peter Perrotta, Wesley Kimble, Ryan Percifield |
| EPI_ISL_2204305 | Houston Methodist Hospital | Houston Methodist Hospital | Randall J. Olsen, Paul A. Christensen, S. Wesley Long, Sishir Subedi, Robert Olson, Marcus Nguyen, James J. Davis, Matthew Ojeda Saavedra, Prasanti Yerramilli, Layne Pruitt, Kristina Reppond, Madison N. Shyer, Jessica Cambric, Ryan Gadd, Ilya J. Finkelstein, Jimmy Gollihar, and James M. Musser |
| EPI_ISL_2204306 | Wheeling Hospital | WVU and Marshall University Combined Genomics Core Facilities | James Denvir, Peter Stoilov, Peter Perrotta, Wesley Kimble, Ryan Percifield |
| EPI_ISL_2204307 | Houston Methodist Hospital | Houston Methodist Hospital | Randall J. Olsen, Paul A. Christensen, S. Wesley Long, Sishir Subedi, Robert Olson, Marcus Nguyen, James J. Davis, Matthew Ojeda Saavedra, Prasanti Yerramilli, Layne Pruitt, Kristina Reppond, Madison N. Shyer, Jessica Cambric, Ryan Gadd, Ilya J. Finkelstein, Jimmy Gollihar, and James M. Musser |
| EPI_ISL_2204308, EPI_ISL_2204310, EPI_ISL_2204311 | Wheeling Hospital | WVU and Marshall University Combined Genomics Core Facilities | James Denvir, Peter Stoilov, Peter Perrotta, Wesley Kimble, Ryan Percifield |
| EPI_ISL_2204312, EPI_ISL_2204313 | Houston Methodist Hospital | Houston Methodist Hospital | Randall J. Olsen, Paul A. Christensen, S. Wesley Long, Sishir Subedi, Robert Olson, Marcus Nguyen, James J. Davis, Matthew Ojeda Saavedra, Prasanti Yerramilli, Layne Pruitt, Kristina Reppond, Madison N. Shyer, Jessica Cambric, Ryan Gadd, Ilya J. Finkelstein, Jimmy Gollihar, and James M. Musser |
| EPI_ISL_2204314, EPI_ISL_2204315 | Wheeling Hospital | WVU and Marshall University Combined Genomics Core Facilities | James Denvir, Peter Stoilov, Peter Perrotta, Wesley Kimble, Ryan Percifield |
| EPI_ISL_2204316, EPI_ISL_2204317 | Houston Methodist Hospital | Houston Methodist Hospital | Randall J. Olsen, Paul A. Christensen, S. Wesley Long, Sishir Subedi, Robert Olson, Marcus Nguyen, James J. Davis, Matthew Ojeda Saavedra, Prasanti Yerramilli, Layne Pruitt, Kristina Reppond, Madison N. Shyer, Jessica Cambric, Ryan Gadd, Ilya J. Finkelstein, Jimmy Gollihar, and James M. Musser |
| EPI_ISL_2204318, EPI_ISL_2204319 | Wheeling Hospital | WVU and Marshall University Combined Genomics Core Facilities | James Denvir, Peter Stoilov, Peter Perrotta, Wesley Kimble, Ryan Percifield |
| EPI_ISL_2204320 | Houston Methodist Hospital | Houston Methodist Hospital | Randall J. Olsen, Paul A. Christensen, S. Wesley Long, Sishir Subedi, Robert Olson, Marcus Nguyen, James J. Davis, Matthew Ojeda Saavedra, Prasanti Yerramilli, Layne Pruitt, Kristina Reppond, Madison N. Shyer, Jessica Cambric, Ryan Gadd, Ilya J. Finkelstein, Jimmy Gollihar, and James M. Musser |
| EPI_ISL_2204321, EPI_ISL_2204323 | Wheeling Hospital | WVU and Marshall University Combined Genomics Core Facilities | James Denvir, Peter Stoilov, Peter Perrotta, Wesley Kimble, Ryan Percifield |
| EPI_ISL_2204324 | Houston Methodist Hospital | Houston Methodist Hospital | Randall J. Olsen, Paul A. Christensen, S. Wesley Long, Sishir Subedi, Robert Olson, Marcus Nguyen, James J. Davis, Matthew Ojeda Saavedra, Prasanti Yerramilli, Layne Pruitt, Kristina Reppond, Madison N. Shyer, Jessica Cambric, Ryan Gadd, Ilya J. Finkelstein, Jimmy Gollihar, and James M. Musser |
| EPI_ISL_2204325 | Wheeling Hospital | WVU and Marshall University Combined Genomics Core Facilities | James Denvir, Peter Stoilov, Peter Perrotta, Wesley Kimble, Ryan Percifield |
| EPI_ISL_2204326 | Houston Methodist Hospital | Houston Methodist Hospital | Randall J. Olsen, Paul A. Christensen, S. Wesley Long, Sishir Subedi, Robert Olson, Marcus Nguyen, James J. Davis, Matthew Ojeda Saavedra, Prasanti Yerramilli, Layne Pruitt, Kristina Reppond, Madison N. Shyer, Jessica Cambric, Ryan Gadd, Ilya J. Finkelstein, Jimmy Gollihar, and James M. Musser |
| EPI_ISL_2204328 | Wheeling Hospital | WVU and Marshall University Combined Genomics Core Facilities | James Denvir, Peter Stoilov, Peter Perrotta, Wesley Kimble, Ryan Percifield |
| EPI_ISL_2204329, EPI_ISL_2204331 | Houston Methodist Hospital | Houston Methodist Hospital | Randall J. Olsen, Paul A. Christensen, S. Wesley Long, Sishir Subedi, Robert Olson, Marcus Nguyen, James J. Davis, Matthew Ojeda Saavedra, Prasanti Yerramilli, Layne Pruitt, Kristina Reppond, Madison N. Shyer, Jessica Cambric, Ryan Gadd, Ilya J. Finkelstein, Jimmy Gollihar, and James M. Musser |
| EPI_ISL_2204332 | BMC LAB | WVU and Marshall University Combined Genomics Core Facilities | James Denvir, Peter Stoilov, Peter Perrotta, Wesley Kimble, Ryan Percifield |
| EPI_ISL_2204333 | Houston Methodist Hospital | Houston Methodist Hospital | Randall J. Olsen, Paul A. Christensen, S. Wesley Long, Sishir Subedi, Robert Olson, Marcus Nguyen, James J. Davis, Matthew Ojeda Saavedra, Prasanti Yerramilli, Layne Pruitt, Kristina Reppond, Madison N. Shyer, Jessica Cambric, Ryan Gadd, Ilya J. Finkelstein, Jimmy Gollihar, and James M. Musser |
| EPI_ISL_2204334 | BMC LAB | WVU and Marshall University Combined Genomics Core Facilities | James Denvir, Peter Stoilov, Peter Perrotta, Wesley Kimble, Ryan Percifield |
| EPI_ISL_2204335, EPI_ISL_2204336 | Houston Methodist Hospital | Houston Methodist Hospital | Randall J. Olsen, Paul A. Christensen, S. Wesley Long, Sishir Subedi, Robert Olson, Marcus Nguyen, James J. Davis, Matthew Ojeda Saavedra, Prasanti Yerramilli, Layne Pruitt, Kristina Reppond, Madison N. Shyer, Jessica Cambric, Ryan Gadd, Ilya J. Finkelstein, Jimmy Gollihar, and James M. Musser |
| EPI_ISL_2204337 | BMC LAB | WVU and Marshall University Combined Genomics Core Facilities | James Denvir, Peter Stoilov, Peter Perrotta, Wesley Kimble, Ryan Percifield |
| EPI_ISL_2204338 | Houston Methodist Hospital | Houston Methodist Hospital | Randall J. Olsen, Paul A. Christensen, S. Wesley Long, Sishir Subedi, Robert Olson, Marcus Nguyen, James J. Davis, Matthew Ojeda Saavedra, Prasanti Yerramilli, Layne Pruitt, Kristina Reppond, Madison N. Shyer, Jessica Cambric, Ryan Gadd, Ilya J. Finkelstein, Jimmy Gollihar, and James M. Musser |
| EPI_ISL_2204339 | BMC LAB | WVU and Marshall University Combined Genomics Core Facilities | James Denvir, Peter Stoilov, Peter Perrotta, Wesley Kimble, Ryan Percifield |
| EPI_ISL_2204340 | Houston Methodist Hospital | Houston Methodist Hospital | Randall J. Olsen, Paul A. Christensen, S. Wesley Long, Sishir Subedi, Robert Olson, Marcus Nguyen, James J. Davis, Matthew Ojeda Saavedra, Prasanti Yerramilli, Layne Pruitt, Kristina Reppond, Madison N. Shyer, Jessica Cambric, Ryan Gadd, Ilya J. Finkelstein, Jimmy Gollihar, and James M. Musser |
| EPI_ISL_2204341 | BMC LAB | WVU and Marshall University Combined Genomics Core Facilities | James Denvir, Peter Stoilov, Peter Perrotta, Wesley Kimble, Ryan Percifield |
| EPI_ISL_2204342 | Wheeling Hospital | WVU and Marshall University Combined Genomics Core Facilities | James Denvir, Peter Stoilov, Peter Perrotta, Wesley Kimble, Ryan Percifield |
| EPI_ISL_2204343 | Houston Methodist Hospital | Houston Methodist Hospital | Randall J. Olsen, Paul A. Christensen, S. Wesley Long, Sishir Subedi, Robert Olson, Marcus Nguyen, James J. Davis, Matthew Ojeda Saavedra, Prasanti Yerramilli, Layne Pruitt, Kristina Reppond, Madison N. Shyer, Jessica Cambric, Ryan Gadd, Ilya J. Finkelstein, Jimmy Gollihar, and James M. Musser |
| EPI_ISL_2204344 | BMC LAB | WVU and Marshall University Combined Genomics Core Facilities | James Denvir, Peter Stoilov, Peter Perrotta, Wesley Kimble, Ryan Percifield |
| EPI_ISL_2204345 | Houston Methodist Hospital | Houston Methodist Hospital | Randall J. Olsen, Paul A. Christensen, S. Wesley Long, Sishir Subedi, Robert Olson, Marcus Nguyen, James J. Davis, Matthew Ojeda Saavedra, Prasanti Yerramilli, Layne Pruitt, Kristina Reppond, Madison N. Shyer, Jessica Cambric, Ryan Gadd, Ilya J. Finkelstein, Jimmy Gollihar, and James M. Musser |
| EPI_ISL_2204347 | BMC LAB | WVU and Marshall University Combined | James Denvir, Peter Stoilov, Peter Perrotta, Wesley Kimble, Ryan Percifield |

| | | | |
|---|---|---|---|
| | | Genomics Core Facilities | |
| EPI_ISL_2204348 | Houston Methodist Hospital | Houston Methodist Hospital | Randall J. Olsen, Paul A. Christensen, S. Wesley Long, Sishir Subedi, Robert Olson, Marcus Nguyen, James J. Davis, Matthew Ojeda Saavedra, Prasanti Yerramilli, Layne Pruitt, Kristina Reppond, Madison N. Shyer, Jessica Cambric, Ryan Gadd, Ilya J. Finkelstein, Jimmy Gollihar, and James M. Musser |
| EPI_ISL_2204349 | BMC LAB | WVU and Marshall University Combined Genomics Core Facilities | James Denvir, Peter Stoilov, Peter Perrotta, Wesley Kimble, Ryan Percifield |
| EPI_ISL_2204350 | Houston Methodist Hospital | Houston Methodist Hospital | Randall J. Olsen, Paul A. Christensen, S. Wesley Long, Sishir Subedi, Robert Olson, Marcus Nguyen, James J. Davis, Matthew Ojeda Saavedra, Prasanti Yerramilli, Layne Pruitt, Kristina Reppond, Madison N. Shyer, Jessica Cambric, Ryan Gadd, Ilya J. Finkelstein, Jimmy Gollihar, and James M. Musser |
| EPI_ISL_2204351 | BMC LAB | WVU and Marshall University Combined Genomics Core Facilities | James Denvir, Peter Stoilov, Peter Perrotta, Wesley Kimble, Ryan Percifield |
| EPI_ISL_2204352 | Houston Methodist Hospital | Houston Methodist Hospital | Randall J. Olsen, Paul A. Christensen, S. Wesley Long, Sishir Subedi, Robert Olson, Marcus Nguyen, James J. Davis, Matthew Ojeda Saavedra, Prasanti Yerramilli, Layne Pruitt, Kristina Reppond, Madison N. Shyer, Jessica Cambric, Ryan Gadd, Ilya J. Finkelstein, Jimmy Gollihar, and James M. Musser |
| EPI_ISL_2204353 | BMC LAB | WVU and Marshall University Combined Genomics Core Facilities | James Denvir, Peter Stoilov, Peter Perrotta, Wesley Kimble, Ryan Percifield |
| EPI_ISL_2204354 | Houston Methodist Hospital | Houston Methodist Hospital | Randall J. Olsen, Paul A. Christensen, S. Wesley Long, Sishir Subedi, Robert Olson, Marcus Nguyen, James J. Davis, Matthew Ojeda Saavedra, Prasanti Yerramilli, Layne Pruitt, Kristina Reppond, Madison N. Shyer, Jessica Cambric, Ryan Gadd, Ilya J. Finkelstein, Jimmy Gollihar, and James M. Musser |
| EPI_ISL_2204355, EPI_ISL_2204356 | BMC LAB | WVU and Marshall University Combined Genomics Core Facilities | James Denvir, Peter Stoilov, Peter Perrotta, Wesley Kimble, Ryan Percifield |
| EPI_ISL_2204357 | Houston Methodist Hospital | Houston Methodist Hospital | Randall J. Olsen, Paul A. Christensen, S. Wesley Long, Sishir Subedi, Robert Olson, Marcus Nguyen, James J. Davis, Matthew Ojeda Saavedra, Prasanti Yerramilli, Layne Pruitt, Kristina Reppond, Madison N. Shyer, Jessica Cambric, Ryan Gadd, Ilya J. Finkelstein, Jimmy Gollihar, and James M. Musser |
| EPI_ISL_2204358 | BMC LAB | WVU and Marshall University Combined Genomics Core Facilities | James Denvir, Peter Stoilov, Peter Perrotta, Wesley Kimble, Ryan Percifield |
| EPI_ISL_2204359 | Wheeling Hospital | WVU and Marshall University Combined Genomics Core Facilities | James Denvir, Peter Stoilov, Peter Perrotta, Wesley Kimble, Ryan Percifield |
| EPI_ISL_2204360 | Houston Methodist Hospital | Houston Methodist Hospital | Randall J. Olsen, Paul A. Christensen, S. Wesley Long, Sishir Subedi, Robert Olson, Marcus Nguyen, James J. Davis, Matthew Ojeda Saavedra, Prasanti Yerramilli, Layne Pruitt, Kristina Reppond, Madison N. Shyer, Jessica Cambric, Ryan Gadd, Ilya J. Finkelstein, Jimmy Gollihar, and James M. Musser |
| EPI_ISL_2204361, EPI_ISL_2204362, EPI_ISL_2204363 | Wheeling Hospital | WVU and Marshall University Combined Genomics Core Facilities | James Denvir, Peter Stoilov, Peter Perrotta, Wesley Kimble, Ryan Percifield |
| EPI_ISL_2204364 | Houston Methodist Hospital | Houston Methodist Hospital | Randall J. Olsen, Paul A. Christensen, S. Wesley Long, Sishir Subedi, Robert Olson, Marcus Nguyen, James J. Davis, Matthew Ojeda Saavedra, Prasanti Yerramilli, Layne Pruitt, Kristina Reppond, Madison N. Shyer, Jessica Cambric, Ryan Gadd, Ilya J. Finkelstein, Jimmy Gollihar, and James M. Musser |
| EPI_ISL_2204365 | Wheeling Hospital | WVU and Marshall University Combined Genomics Core Facilities | James Denvir, Peter Stoilov, Peter Perrotta, Wesley Kimble, Ryan Percifield |
| EPI_ISL_2204366, EPI_ISL_2204367 | BMC LAB | WVU and Marshall University Combined Genomics Core Facilities | James Denvir, Peter Stoilov, Peter Perrotta, Wesley Kimble, Ryan Percifield |
| EPI_ISL_2204368 | Houston Methodist Hospital | Houston Methodist Hospital | Randall J. Olsen, Paul A. Christensen, S. Wesley Long, Sishir Subedi, Robert Olson, Marcus Nguyen, James J. Davis, Matthew Ojeda Saavedra, Prasanti Yerramilli, Layne Pruitt, Kristina Reppond, Madison N. Shyer, Jessica Cambric, Ryan Gadd, Ilya J. Finkelstein, Jimmy Gollihar, and James M. Musser |
| EPI_ISL_2204369 | BMC LAB | WVU and Marshall University Combined Genomics Core Facilities | James Denvir, Peter Stoilov, Peter Perrotta, Wesley Kimble, Ryan Percifield |
| EPI_ISL_2204370 | J.W. Ruby Memorial Hospital | WVU and Marshall University Combined Genomics Core Facilities | James Denvir, Peter Stoilov, Peter Perrotta, Wesley Kimble, Ryan Percifield |
| EPI_ISL_2204371 | Houston Methodist Hospital | Houston Methodist Hospital | Randall J. Olsen, Paul A. Christensen, S. Wesley Long, Sishir Subedi, Robert Olson, Marcus Nguyen, James J. Davis, Matthew Ojeda Saavedra, Prasanti Yerramilli, Layne Pruitt, Kristina Reppond, Madison N. Shyer, Jessica Cambric, Ryan Gadd, Ilya J. Finkelstein, Jimmy Gollihar, and James M. Musser |
| EPI_ISL_2204372 | Wheeling Hospital | WVU and Marshall University Combined Genomics Core Facilities | James Denvir, Peter Stoilov, Peter Perrotta, Wesley Kimble, Ryan Percifield |
| EPI_ISL_2204373 | BMC LAB | WVU and Marshall University Combined Genomics Core Facilities | James Denvir, Peter Stoilov, Peter Perrotta, Wesley Kimble, Ryan Percifield |
| EPI_ISL_2204374 | Wheeling Hospital | WVU and Marshall University Combined Genomics Core Facilities | James Denvir, Peter Stoilov, Peter Perrotta, Wesley Kimble, Ryan Percifield |
| EPI_ISL_2204375 | Houston Methodist Hospital | Houston Methodist Hospital | Randall J. Olsen, Paul A. Christensen, S. Wesley Long, Sishir Subedi, Robert Olson, Marcus Nguyen, James J. Davis, Matthew Ojeda Saavedra, Prasanti Yerramilli, Layne Pruitt, Kristina Reppond, Madison N. Shyer, Jessica Cambric, Ryan Gadd, Ilya J. Finkelstein, Jimmy Gollihar, and James M. Musser |
| EPI_ISL_2204376 | Wheeling Hospital | WVU and Marshall University Combined Genomics Core Facilities | James Denvir, Peter Stoilov, Peter Perrotta, Wesley Kimble, Ryan Percifield |
| EPI_ISL_2204378, EPI_ISL_2204380, EPI_ISL_2204381 | BMC LAB | WVU and Marshall University Combined Genomics Core Facilities | James Denvir, Peter Stoilov, Peter Perrotta, Wesley Kimble, Ryan Percifield |
| EPI_ISL_2204382 | Houston Methodist Hospital | Houston Methodist Hospital | Randall J. Olsen, Paul A. Christensen, S. Wesley Long, Sishir Subedi, Robert Olson, Marcus Nguyen, James J. Davis, Matthew Ojeda Saavedra, Prasanti Yerramilli, Layne Pruitt, Kristina Reppond, Madison N. Shyer, Jessica Cambric, Ryan Gadd, Ilya J. Finkelstein, Jimmy Gollihar, and James M. Musser |
| EPI_ISL_2204383, EPI_ISL_2204384 | BMC LAB | WVU and Marshall University Combined Genomics Core Facilities | James Denvir, Peter Stoilov, Peter Perrotta, Wesley Kimble, Ryan Percifield |
| EPI_ISL_2204385 | Houston Methodist Hospital | Houston Methodist Hospital | Randall J. Olsen, Paul A. Christensen, S. Wesley Long, Sishir Subedi, Robert Olson, Marcus Nguyen, James J. Davis, Matthew Ojeda Saavedra, Prasanti Yerramilli, Layne Pruitt, Kristina Reppond, Madison N. Shyer, Jessica Cambric, Ryan Gadd, Ilya J. Finkelstein, Jimmy Gollihar, and James M. Musser |
| EPI_ISL_2204386, EPI_ISL_2204387 | BMC LAB | WVU and Marshall University Combined Genomics Core Facilities | James Denvir, Peter Stoilov, Peter Perrotta, Wesley Kimble, Ryan Percifield |
| EPI_ISL_2204388 | Houston Methodist Hospital | Houston Methodist Hospital | Randall J. Olsen, Paul A. Christensen, S. Wesley Long, Sishir Subedi, Robert Olson, Marcus Nguyen, James J. Davis, Matthew Ojeda Saavedra, Prasanti Yerramilli, Layne Pruitt, Kristina Reppond, Madison N. Shyer, Jessica Cambric, Ryan Gadd, Ilya J. Finkelstein, Jimmy Gollihar, and James M. Musser |
| EPI_ISL_2204389, EPI_ISL_2204390 | BMC LAB | WVU and Marshall University Combined Genomics Core Facilities | James Denvir, Peter Stoilov, Peter Perrotta, Wesley Kimble, Ryan Percifield |
| EPI_ISL_2204391 | Houston Methodist Hospital | Houston Methodist Hospital | Randall J. Olsen, Paul A. Christensen, S. Wesley Long, Sishir Subedi, Robert Olson, Marcus Nguyen, James J. Davis, Matthew Ojeda Saavedra, Prasanti Yerramilli, Layne Pruitt, Kristina Reppond, Madison N. Shyer, Jessica Cambric, Ryan Gadd, Ilya J. Finkelstein, Jimmy Gollihar, and James M. Musser |
| EPI_ISL_2204392, EPI_ISL_2204393, EPI_ISL_2204395 | BMC LAB | WVU and Marshall University Combined Genomics Core Facilities | James Denvir, Peter Stoilov, Peter Perrotta, Wesley Kimble, Ryan Percifield |
| EPI_ISL_2204396 | Raleigh General Hospital Microbiology Dept. | WVU and Marshall University Combined Genomics Core Facilities | James Denvir, Peter Stoilov, Peter Perrotta, Wesley Kimble, Ryan Percifield |
| EPI_ISL_2204397 | Houston Methodist Hospital | Houston Methodist Hospital | Randall J. Olsen, Paul A. Christensen, S. Wesley Long, Sishir Subedi, Robert Olson, Marcus Nguyen, James J. Davis, Matthew Ojeda Saavedra, Prasanti Yerramilli, Layne Pruitt, Kristina Reppond, Madison N. Shyer, Jessica Cambric, Ryan Gadd, Ilya J. Finkelstein, Jimmy Gollihar, and James M. Musser |
| EPI_ISL_2204398, EPI_ISL_2204399 | Wheeling Hospital | WVU and Marshall University Combined Genomics Core Facilities | James Denvir, Peter Stoilov, Peter Perrotta, Wesley Kimble, Ryan Percifield |
| EPI_ISL_2204400, EPI_ISL_2204401 | QLabs | WVU and Marshall University Combined Genomics Core Facilities | James Denvir, Peter Stoilov, Peter Perrotta, Wesley Kimble, Ryan Percifield |
| EPI_ISL_2204402 | Berkeley Medical Center | WVU and Marshall University Combined Genomics Core Facilities | James Denvir, Peter Stoilov, Peter Perrotta, Wesley Kimble, Ryan Percifield |
| EPI_ISL_2204403 | Houston Methodist Hospital | Houston Methodist Hospital | Randall J. Olsen, Paul A. Christensen, S. Wesley Long, Sishir Subedi, Robert Olson, Marcus Nguyen, James J. Davis, Matthew Ojeda Saavedra, Prasanti Yerramilli, Layne Pruitt, Kristina Reppond, Madison N. Shyer, Jessica Cambric, Ryan Gadd, Ilya J. Finkelstein, Jimmy Gollihar, and James M. Musser |
| EPI_ISL_2204404 | QLabs | WVU and Marshall University Combined Genomics Core Facilities | James Denvir, Peter Stoilov, Peter Perrotta, Wesley Kimble, Ryan Percifield |
| EPI_ISL_2204405 | Raleigh General Hospital Microbiology Dept. | WVU and Marshall University Combined Genomics Core Facilities | James Denvir, Peter Stoilov, Peter Perrotta, Wesley Kimble, Ryan Percifield |
| EPI_ISL_2204406 | Summersville Regiol Medical Center | WVU and Marshall University Combined Genomics Core Facilities | James Denvir, Peter Stoilov, Peter Perrotta, Wesley Kimble, Ryan Percifield |
| EPI_ISL_2204407 | Houston Methodist Hospital | Houston Methodist Hospital | Randall J. Olsen, Paul A. Christensen, S. Wesley Long, Sishir Subedi, Robert Olson, Marcus Nguyen, James J. Davis, Matthew Ojeda Saavedra, Prasanti Yerramilli, Layne Pruitt, Kristina Reppond, Madison N. Shyer, Jessica Cambric, Ryan Gadd, Ilya J. Finkelstein, Jimmy Gollihar, and James |

| | | | |
|---|---|---|---|
| | | | M. Musser |
| EPI_ISL_2204408, EPI_ISL_2204409 | Berkeley Medical Center | WVU and Marshall University Combined Genomics Core Facilities | James Denvir, Peter Stoilov, Peter Perrotta, Wesley Kimble, Ryan Percifield |
| EPI_ISL_2204410 | Houston Methodist Hospital | Houston Methodist Hospital | Randall J. Olsen, Paul A. Christensen, S. Wesley Long, Sishir Subedi, Robert Olson, Marcus Nguyen, James J. Davis, Matthew Ojeda Saavedra, Prasanti Yerramilli, Layne Pruitt, Kristina Reppond, Madison N. Shyer, Jessica Cambric, Ryan Gadd, Ilya J. Finkelstein, Jimmy Gollihar, and James M. Musser |
| EPI_ISL_2204411, EPI_ISL_2204412, EPI_ISL_2204413, EPI_ISL_2204414 | Berkeley Medical Center | WVU and Marshall University Combined Genomics Core Facilities | James Denvir, Peter Stoilov, Peter Perrotta, Wesley Kimble, Ryan Percifield |
| EPI_ISL_2204415 | Houston Methodist Hospital | Houston Methodist Hospital | Randall J. Olsen, Paul A. Christensen, S. Wesley Long, Sishir Subedi, Robert Olson, Marcus Nguyen, James J. Davis, Matthew Ojeda Saavedra, Prasanti Yerramilli, Layne Pruitt, Kristina Reppond, Madison N. Shyer, Jessica Cambric, Ryan Gadd, Ilya J. Finkelstein, Jimmy Gollihar, and James M. Musser |
| EPI_ISL_2204416 | Berkeley Medical Center | WVU and Marshall University Combined Genomics Core Facilities | James Denvir, Peter Stoilov, Peter Perrotta, Wesley Kimble, Ryan Percifield |
| EPI_ISL_2204417 | Houston Methodist Hospital | Houston Methodist Hospital | Randall J. Olsen, Paul A. Christensen, S. Wesley Long, Sishir Subedi, Robert Olson, Marcus Nguyen, James J. Davis, Matthew Ojeda Saavedra, Prasanti Yerramilli, Layne Pruitt, Kristina Reppond, Madison N. Shyer, Jessica Cambric, Ryan Gadd, Ilya J. Finkelstein, Jimmy Gollihar, and James M. Musser |
| EPI_ISL_2204418, EPI_ISL_2204419 | Berkeley Medical Center | WVU and Marshall University Combined Genomics Core Facilities | James Denvir, Peter Stoilov, Peter Perrotta, Wesley Kimble, Ryan Percifield |
| EPI_ISL_2204420 | Houston Methodist Hospital | Houston Methodist Hospital | Randall J. Olsen, Paul A. Christensen, S. Wesley Long, Sishir Subedi, Robert Olson, Marcus Nguyen, James J. Davis, Matthew Ojeda Saavedra, Prasanti Yerramilli, Layne Pruitt, Kristina Reppond, Madison N. Shyer, Jessica Cambric, Ryan Gadd, Ilya J. Finkelstein, Jimmy Gollihar, and James M. Musser |
| EPI_ISL_2204421, EPI_ISL_2204423, EPI_ISL_2204424, EPI_ISL_2204425 | Berkeley Medical Center | WVU and Marshall University Combined Genomics Core Facilities | James Denvir, Peter Stoilov, Peter Perrotta, Wesley Kimble, Ryan Percifield |
| EPI_ISL_2204426 | Houston Methodist Hospital | Houston Methodist Hospital | Randall J. Olsen, Paul A. Christensen, S. Wesley Long, Sishir Subedi, Robert Olson, Marcus Nguyen, James J. Davis, Matthew Ojeda Saavedra, Prasanti Yerramilli, Layne Pruitt, Kristina Reppond, Madison N. Shyer, Jessica Cambric, Ryan Gadd, Ilya J. Finkelstein, Jimmy Gollihar, and James M. Musser |
| EPI_ISL_2204427 | Berkeley Medical Center | WVU and Marshall University Combined Genomics Core Facilities | James Denvir, Peter Stoilov, Peter Perrotta, Wesley Kimble, Ryan Percifield |
| EPI_ISL_2204428 | Houston Methodist Hospital | Houston Methodist Hospital | Randall J. Olsen, Paul A. Christensen, S. Wesley Long, Sishir Subedi, Robert Olson, Marcus Nguyen, James J. Davis, Matthew Ojeda Saavedra, Prasanti Yerramilli, Layne Pruitt, Kristina Reppond, Madison N. Shyer, Jessica Cambric, Ryan Gadd, Ilya J. Finkelstein, Jimmy Gollihar, and James M. Musser |
| EPI_ISL_2204429 | Berkeley Medical Center | WVU and Marshall University Combined Genomics Core Facilities | James Denvir, Peter Stoilov, Peter Perrotta, Wesley Kimble, Ryan Percifield |
| EPI_ISL_2204430 | Houston Methodist Hospital | Houston Methodist Hospital | Randall J. Olsen, Paul A. Christensen, S. Wesley Long, Sishir Subedi, Robert Olson, Marcus Nguyen, James J. Davis, Matthew Ojeda Saavedra, Prasanti Yerramilli, Layne Pruitt, Kristina Reppond, Madison N. Shyer, Jessica Cambric, Ryan Gadd, Ilya J. Finkelstein, Jimmy Gollihar, and James M. Musser |
| EPI_ISL_2204431 | Berkeley Medical Center | WVU and Marshall University Combined Genomics Core Facilities | James Denvir, Peter Stoilov, Peter Perrotta, Wesley Kimble, Ryan Percifield |
| EPI_ISL_2204432 | Houston Methodist Hospital | Houston Methodist Hospital | Randall J. Olsen, Paul A. Christensen, S. Wesley Long, Sishir Subedi, Robert Olson, Marcus Nguyen, James J. Davis, Matthew Ojeda Saavedra, Prasanti Yerramilli, Layne Pruitt, Kristina Reppond, Madison N. Shyer, Jessica Cambric, Ryan Gadd, Ilya J. Finkelstein, Jimmy Gollihar, and James M. Musser |
| EPI_ISL_2204433 | Raleigh General Hospital Microbiology Dept. | WVU and Marshall University Combined Genomics Core Facilities | James Denvir, Peter Stoilov, Peter Perrotta, Wesley Kimble, Ryan Percifield |
| EPI_ISL_2204434, EPI_ISL_2204435 | Houston Methodist Hospital | Houston Methodist Hospital | Randall J. Olsen, Paul A. Christensen, S. Wesley Long, Sishir Subedi, Robert Olson, Marcus Nguyen, James J. Davis, Matthew Ojeda Saavedra, Prasanti Yerramilli, Layne Pruitt, Kristina Reppond, Madison N. Shyer, Jessica Cambric, Ryan Gadd, Ilya J. Finkelstein, Jimmy Gollihar, and James M. Musser |
| EPI_ISL_2204436 | Summersville Regiol Medical Center | WVU and Marshall University Combined Genomics Core Facilities | James Denvir, Peter Stoilov, Peter Perrotta, Wesley Kimble, Ryan Percifield |
| EPI_ISL_2204437 | Houston Methodist Hospital | Houston Methodist Hospital | Randall J. Olsen, Paul A. Christensen, S. Wesley Long, Sishir Subedi, Robert Olson, Marcus Nguyen, James J. Davis, Matthew Ojeda Saavedra, Prasanti Yerramilli, Layne Pruitt, Kristina Reppond, Madison N. Shyer, Jessica Cambric, Ryan Gadd, Ilya J. Finkelstein, Jimmy Gollihar, and James M. Musser |
| EPI_ISL_2204438 | Berkeley Medical Center | WVU and Marshall University Combined Genomics Core Facilities | James Denvir, Peter Stoilov, Peter Perrotta, Wesley Kimble, Ryan Percifield |
| EPI_ISL_2204439 | Houston Methodist Hospital | Houston Methodist Hospital | Randall J. Olsen, Paul A. Christensen, S. Wesley Long, Sishir Subedi, Robert Olson, Marcus Nguyen, James J. Davis, Matthew Ojeda Saavedra, Prasanti Yerramilli, Layne Pruitt, Kristina Reppond, Madison N. Shyer, Jessica Cambric, Ryan Gadd, Ilya J. Finkelstein, Jimmy Gollihar, and James M. Musser |
| EPI_ISL_2204440 | Berkeley Medical Center | WVU and Marshall University Combined Genomics Core Facilities | James Denvir, Peter Stoilov, Peter Perrotta, Wesley Kimble, Ryan Percifield |
| EPI_ISL_2204441 | Houston Methodist Hospital | Houston Methodist Hospital | Randall J. Olsen, Paul A. Christensen, S. Wesley Long, Sishir Subedi, Robert Olson, Marcus Nguyen, James J. Davis, Matthew Ojeda Saavedra, Prasanti Yerramilli, Layne Pruitt, Kristina Reppond, Madison N. Shyer, Jessica Cambric, Ryan Gadd, Ilya J. Finkelstein, Jimmy Gollihar, and James M. Musser |
| EPI_ISL_2204442 | Berkeley Medical Center | WVU and Marshall University Combined Genomics Core Facilities | James Denvir, Peter Stoilov, Peter Perrotta, Wesley Kimble, Ryan Percifield |
| EPI_ISL_2204443 | Houston Methodist Hospital | Houston Methodist Hospital | Randall J. Olsen, Paul A. Christensen, S. Wesley Long, Sishir Subedi, Robert Olson, Marcus Nguyen, James J. Davis, Matthew Ojeda Saavedra, Prasanti Yerramilli, Layne Pruitt, Kristina Reppond, Madison N. Shyer, Jessica Cambric, Ryan Gadd, Ilya J. Finkelstein, Jimmy Gollihar, and James M. Musser |
| EPI_ISL_2204444 | Berkeley Medical Center | WVU and Marshall University Combined Genomics Core Facilities | James Denvir, Peter Stoilov, Peter Perrotta, Wesley Kimble, Ryan Percifield |
| EPI_ISL_2204445 | Houston Methodist Hospital | Houston Methodist Hospital | Randall J. Olsen, Paul A. Christensen, S. Wesley Long, Sishir Subedi, Robert Olson, Marcus Nguyen, James J. Davis, Matthew Ojeda Saavedra, Prasanti Yerramilli, Layne Pruitt, Kristina Reppond, Madison N. Shyer, Jessica Cambric, Ryan Gadd, Ilya J. Finkelstein, Jimmy Gollihar, and James M. Musser |
| EPI_ISL_2204446 | Berkeley Medical Center | WVU and Marshall University Combined Genomics Core Facilities | James Denvir, Peter Stoilov, Peter Perrotta, Wesley Kimble, Ryan Percifield |
| EPI_ISL_2204447 | Houston Methodist Hospital | Houston Methodist Hospital | Randall J. Olsen, Paul A. Christensen, S. Wesley Long, Sishir Subedi, Robert Olson, Marcus Nguyen, James J. Davis, Matthew Ojeda Saavedra, Prasanti Yerramilli, Layne Pruitt, Kristina Reppond, Madison N. Shyer, Jessica Cambric, Ryan Gadd, Ilya J. Finkelstein, Jimmy Gollihar, and James M. Musser |
| EPI_ISL_2204448, EPI_ISL_2204449 | Berkeley Medical Center | WVU and Marshall University Combined Genomics Core Facilities | James Denvir, Peter Stoilov, Peter Perrotta, Wesley Kimble, Ryan Percifield |
| EPI_ISL_2204450 | Houston Methodist Hospital | Houston Methodist Hospital | Randall J. Olsen, Paul A. Christensen, S. Wesley Long, Sishir Subedi, Robert Olson, Marcus Nguyen, James J. Davis, Matthew Ojeda Saavedra, Prasanti Yerramilli, Layne Pruitt, Kristina Reppond, Madison N. Shyer, Jessica Cambric, Ryan Gadd, Ilya J. Finkelstein, Jimmy Gollihar, and James M. Musser |
| EPI_ISL_2204451 | Berkeley Medical Center | WVU and Marshall University Combined Genomics Core Facilities | James Denvir, Peter Stoilov, Peter Perrotta, Wesley Kimble, Ryan Percifield |
| EPI_ISL_2204452, EPI_ISL_2204453 | Houston Methodist Hospital | Houston Methodist Hospital | Randall J. Olsen, Paul A. Christensen, S. Wesley Long, Sishir Subedi, Robert Olson, Marcus Nguyen, James J. Davis, Matthew Ojeda Saavedra, Prasanti Yerramilli, Layne Pruitt, Kristina Reppond, Madison N. Shyer, Jessica Cambric, Ryan Gadd, Ilya J. Finkelstein, Jimmy Gollihar, and James M. Musser |
| EPI_ISL_2204454 | WV Labs | WVU and Marshall University Combined Genomics Core Facilities | James Denvir, Peter Stoilov, Peter Perrotta, Wesley Kimble, Ryan Percifield |
| EPI_ISL_2204455 | Houston Methodist Hospital | Houston Methodist Hospital | Randall J. Olsen, Paul A. Christensen, S. Wesley Long, Sishir Subedi, Robert Olson, Marcus Nguyen, James J. Davis, Matthew Ojeda Saavedra, Prasanti Yerramilli, Layne Pruitt, Kristina Reppond, Madison N. Shyer, Jessica Cambric, Ryan Gadd, Ilya J. Finkelstein, Jimmy Gollihar, and James M. Musser |
| EPI_ISL_2204456 | WV Labs | WVU and Marshall University Combined Genomics Core Facilities | James Denvir, Peter Stoilov, Peter Perrotta, Wesley Kimble, Ryan Percifield |
| EPI_ISL_2204457 | QLabs | WVU and Marshall University Combined Genomics Core Facilities | James Denvir, Peter Stoilov, Peter Perrotta, Wesley Kimble, Ryan Percifield |
| EPI_ISL_2204458 | Houston Methodist Hospital | Houston Methodist Hospital | Randall J. Olsen, Paul A. Christensen, S. Wesley Long, Sishir Subedi, Robert Olson, Marcus Nguyen, James J. Davis, Matthew Ojeda Saavedra, Prasanti Yerramilli, Layne Pruitt, Kristina Reppond, Madison N. Shyer, Jessica Cambric, Ryan Gadd, Ilya J. Finkelstein, Jimmy Gollihar, and James M. Musser |
| EPI_ISL_2204459, EPI_ISL_2204460 | Berkeley Medical Center | WVU and Marshall University Combined Genomics Core Facilities | James Denvir, Peter Stoilov, Peter Perrotta, Wesley Kimble, Ryan Percifield |
| EPI_ISL_2204461 | Houston Methodist Hospital | Houston Methodist Hospital | Randall J. Olsen, Paul A. Christensen, S. Wesley Long, Sishir Subedi, Robert Olson, Marcus Nguyen, James J. Davis, Matthew Ojeda Saavedra, Prasanti Yerramilli, Layne Pruitt, Kristina Reppond, Madison N. Shyer, Jessica Cambric, Ryan Gadd, Ilya J. Finkelstein, Jimmy Gollihar, and James M. Musser |
| EPI_ISL_2204462, EPI_ISL_2204463 | Berkeley Medical Center | WVU and Marshall University Combined Genomics Core Facilities | James Denvir, Peter Stoilov, Peter Perrotta, Wesley Kimble, Ryan Percifield |
| EPI_ISL_2204464 | Houston Methodist Hospital | Houston Methodist Hospital | Randall J. Olsen, Paul A. Christensen, S. Wesley Long, Sishir Subedi, Robert Olson, Marcus Nguyen, James J. Davis, Matthew Ojeda Saavedra, Prasanti Yerramilli, Layne Pruitt, Kristina Reppond, Madison N. Shyer, Jessica Cambric, Ryan Gadd, Ilya J. Finkelstein, Jimmy Gollihar, and James M. Musser |

| | | | |
|---|---|---|---|
| EPI_ISL_2204465 | Berkeley Medical Center | WVU and Marshall University Combined Genomics Core Facilities | James Denvir, Peter Stoilov, Peter Perrotta, Wesley Kimble, Ryan Percifield |
| EPI_ISL_2204466 | Houston Methodist Hospital | Houston Methodist Hospital | Randall J. Olsen, Paul A. Christensen, S. Wesley Long, Sishir Subedi, Robert Olson, Marcus Nguyen, James J. Davis, Matthew Ojeda Saavedra, Prasanti Yerramilli, Layne Pruitt, Kristina Reppond, Madison N. Shyer, Jessica Cambric, Ryan Gadd, Ilya J. Finkelstein, Jimmy Gollihar, and James M. Musser |
| EPI_ISL_2204467 | Berkeley Medical Center | WVU and Marshall University Combined Genomics Core Facilities | James Denvir, Peter Stoilov, Peter Perrotta, Wesley Kimble, Ryan Percifield |
| EPI_ISL_2204468 | Houston Methodist Hospital | Houston Methodist Hospital | Randall J. Olsen, Paul A. Christensen, S. Wesley Long, Sishir Subedi, Robert Olson, Marcus Nguyen, James J. Davis, Matthew Ojeda Saavedra, Prasanti Yerramilli, Layne Pruitt, Kristina Reppond, Madison N. Shyer, Jessica Cambric, Ryan Gadd, Ilya J. Finkelstein, Jimmy Gollihar, and James M. Musser |
| EPI_ISL_2204469 | Berkeley Medical Center | WVU and Marshall University Combined Genomics Core Facilities | James Denvir, Peter Stoilov, Peter Perrotta, Wesley Kimble, Ryan Percifield |
| EPI_ISL_2204470 | Houston Methodist Hospital | Houston Methodist Hospital | Randall J. Olsen, Paul A. Christensen, S. Wesley Long, Sishir Subedi, Robert Olson, Marcus Nguyen, James J. Davis, Matthew Ojeda Saavedra, Prasanti Yerramilli, Layne Pruitt, Kristina Reppond, Madison N. Shyer, Jessica Cambric, Ryan Gadd, Ilya J. Finkelstein, Jimmy Gollihar, and James M. Musser |
| EPI_ISL_2204471, EPI_ISL_2204472 | Berkeley Medical Center | WVU and Marshall University Combined Genomics Core Facilities | James Denvir, Peter Stoilov, Peter Perrotta, Wesley Kimble, Ryan Percifield |
| EPI_ISL_2204473 | Houston Methodist Hospital | Houston Methodist Hospital | Randall J. Olsen, Paul A. Christensen, S. Wesley Long, Sishir Subedi, Robert Olson, Marcus Nguyen, James J. Davis, Matthew Ojeda Saavedra, Prasanti Yerramilli, Layne Pruitt, Kristina Reppond, Madison N. Shyer, Jessica Cambric, Ryan Gadd, Ilya J. Finkelstein, Jimmy Gollihar, and James M. Musser |
| EPI_ISL_2204474 | Berkeley Medical Center | WVU and Marshall University Combined Genomics Core Facilities | James Denvir, Peter Stoilov, Peter Perrotta, Wesley Kimble, Ryan Percifield |
| EPI_ISL_2204475 | Houston Methodist Hospital | Houston Methodist Hospital | Randall J. Olsen, Paul A. Christensen, S. Wesley Long, Sishir Subedi, Robert Olson, Marcus Nguyen, James J. Davis, Matthew Ojeda Saavedra, Prasanti Yerramilli, Layne Pruitt, Kristina Reppond, Madison N. Shyer, Jessica Cambric, Ryan Gadd, Ilya J. Finkelstein, Jimmy Gollihar, and James M. Musser |
| EPI_ISL_2204476 | Berkeley Medical Center | WVU and Marshall University Combined Genomics Core Facilities | James Denvir, Peter Stoilov, Peter Perrotta, Wesley Kimble, Ryan Percifield |
| EPI_ISL_2204477 | Houston Methodist Hospital | Houston Methodist Hospital | Randall J. Olsen, Paul A. Christensen, S. Wesley Long, Sishir Subedi, Robert Olson, Marcus Nguyen, James J. Davis, Matthew Ojeda Saavedra, Prasanti Yerramilli, Layne Pruitt, Kristina Reppond, Madison N. Shyer, Jessica Cambric, Ryan Gadd, Ilya J. Finkelstein, Jimmy Gollihar, and James M. Musser |
| EPI_ISL_2204478, EPI_ISL_2204479 | Berkeley Medical Center | WVU and Marshall University Combined Genomics Core Facilities | James Denvir, Peter Stoilov, Peter Perrotta, Wesley Kimble, Ryan Percifield |
| EPI_ISL_2204480, EPI_ISL_2204481 | WV Labs | WVU and Marshall University Combined Genomics Core Facilities | James Denvir, Peter Stoilov, Peter Perrotta, Wesley Kimble, Ryan Percifield |
| EPI_ISL_2204482 | Houston Methodist Hospital | Houston Methodist Hospital | Randall J. Olsen, Paul A. Christensen, S. Wesley Long, Sishir Subedi, Robert Olson, Marcus Nguyen, James J. Davis, Matthew Ojeda Saavedra, Prasanti Yerramilli, Layne Pruitt, Kristina Reppond, Madison N. Shyer, Jessica Cambric, Ryan Gadd, Ilya J. Finkelstein, Jimmy Gollihar, and James M. Musser |
| EPI_ISL_2204483 | WV Labs | WVU and Marshall University Combined Genomics Core Facilities | James Denvir, Peter Stoilov, Peter Perrotta, Wesley Kimble, Ryan Percifield |
| EPI_ISL_2204484 | Raleigh General Hospital Microbiology Dept. | WVU and Marshall University Combined Genomics Core Facilities | James Denvir, Peter Stoilov, Peter Perrotta, Wesley Kimble, Ryan Percifield |
| EPI_ISL_2204485 | Houston Methodist Hospital | Houston Methodist Hospital | Randall J. Olsen, Paul A. Christensen, S. Wesley Long, Sishir Subedi, Robert Olson, Marcus Nguyen, James J. Davis, Matthew Ojeda Saavedra, Prasanti Yerramilli, Layne Pruitt, Kristina Reppond, Madison N. Shyer, Jessica Cambric, Ryan Gadd, Ilya J. Finkelstein, Jimmy Gollihar, and James M. Musser |
| EPI_ISL_2204486 | Raleigh General Hospital Microbiology Dept. | WVU and Marshall University Combined Genomics Core Facilities | James Denvir, Peter Stoilov, Peter Perrotta, Wesley Kimble, Ryan Percifield |
| EPI_ISL_2204487, EPI_ISL_2204489 | Berkeley Medical Center | WVU and Marshall University Combined Genomics Core Facilities | James Denvir, Peter Stoilov, Peter Perrotta, Wesley Kimble, Ryan Percifield |
| EPI_ISL_2204490 | Houston Methodist Hospital | Houston Methodist Hospital | Randall J. Olsen, Paul A. Christensen, S. Wesley Long, Sishir Subedi, Robert Olson, Marcus Nguyen, James J. Davis, Matthew Ojeda Saavedra, Prasanti Yerramilli, Layne Pruitt, Kristina Reppond, Madison N. Shyer, Jessica Cambric, Ryan Gadd, Ilya J. Finkelstein, Jimmy Gollihar, and James M. Musser |
| EPI_ISL_2204491 | Berkeley Medical Center | WVU and Marshall University Combined Genomics Core Facilities | James Denvir, Peter Stoilov, Peter Perrotta, Wesley Kimble, Ryan Percifield |
| EPI_ISL_2204492 | Houston Methodist Hospital | Houston Methodist Hospital | Randall J. Olsen, Paul A. Christensen, S. Wesley Long, Sishir Subedi, Robert Olson, Marcus Nguyen, James J. Davis, Matthew Ojeda Saavedra, Prasanti Yerramilli, Layne Pruitt, Kristina Reppond, Madison N. Shyer, Jessica Cambric, Ryan Gadd, Ilya J. Finkelstein, Jimmy Gollihar, and James M. Musser |
| EPI_ISL_2204493 | Berkeley Medical Center | WVU and Marshall University Combined Genomics Core Facilities | James Denvir, Peter Stoilov, Peter Perrotta, Wesley Kimble, Ryan Percifield |
| EPI_ISL_2204494 | J.W. Ruby Memorial Hospital | WVU and Marshall University Combined Genomics Core Facilities | James Denvir, Peter Stoilov, Peter Perrotta, Wesley Kimble, Ryan Percifield |
| EPI_ISL_2204495 | Houston Methodist Hospital | Houston Methodist Hospital | Randall J. Olsen, Paul A. Christensen, S. Wesley Long, Sishir Subedi, Robert Olson, Marcus Nguyen, James J. Davis, Matthew Ojeda Saavedra, Prasanti Yerramilli, Layne Pruitt, Kristina Reppond, Madison N. Shyer, Jessica Cambric, Ryan Gadd, Ilya J. Finkelstein, Jimmy Gollihar, and James M. Musser |
| EPI_ISL_2204496, EPI_ISL_2204498, EPI_ISL_2204500 | QLabs | WVU and Marshall University Combined Genomics Core Facilities | James Denvir, Peter Stoilov, Peter Perrotta, Wesley Kimble, Ryan Percifield |
| EPI_ISL_2204501 | Houston Methodist Hospital | Houston Methodist Hospital | Randall J. Olsen, Paul A. Christensen, S. Wesley Long, Sishir Subedi, Robert Olson, Marcus Nguyen, James J. Davis, Matthew Ojeda Saavedra, Prasanti Yerramilli, Layne Pruitt, Kristina Reppond, Madison N. Shyer, Jessica Cambric, Ryan Gadd, Ilya J. Finkelstein, Jimmy Gollihar, and James M. Musser |
| EPI_ISL_2204502 | QLabs | WVU and Marshall University Combined Genomics Core Facilities | James Denvir, Peter Stoilov, Peter Perrotta, Wesley Kimble, Ryan Percifield |
| EPI_ISL_2204503 | Houston Methodist Hospital | Houston Methodist Hospital | Randall J. Olsen, Paul A. Christensen, S. Wesley Long, Sishir Subedi, Robert Olson, Marcus Nguyen, James J. Davis, Matthew Ojeda Saavedra, Prasanti Yerramilli, Layne Pruitt, Kristina Reppond, Madison N. Shyer, Jessica Cambric, Ryan Gadd, Ilya J. Finkelstein, Jimmy Gollihar, and James M. Musser |
| EPI_ISL_2204504 | QLabs | WVU and Marshall University Combined Genomics Core Facilities | James Denvir, Peter Stoilov, Peter Perrotta, Wesley Kimble, Ryan Percifield |
| EPI_ISL_2204505 | Raleigh General Hospital Microbiology Dept. | WVU and Marshall University Combined Genomics Core Facilities | James Denvir, Peter Stoilov, Peter Perrotta, Wesley Kimble, Ryan Percifield |
| EPI_ISL_2204506 | Aegis Sciences Corporation | Centers for Disease Control and Prevention Division of Viral Diseases, Pathogen Discovery | Dakota Howard, Dhwani Batra, Peter W. Cook, Kara Moser, Adrian Paskey, Jason Caravas, Benjamin Rambo-Martin, Shatavia Morrison, Christopher Gulvick, Scott Sammons, Yvette Unoarumhi, Darlene Wagner, Matthew Schmerer, Cyndi Clark, Patrick Campbell, Rob Case, Vikramsinha Ghorpade, Holly Houdeshell, Ola Kvalvaag, Dillon Nall, Ethan Sanders, Alec Vest, Shaun Westlund, Matthew Hardison, Clinton R. Paden, Duncan MacCannell |
| EPI_ISL_2204507 | Houston Methodist Hospital | Houston Methodist Hospital | Randall J. Olsen, Paul A. Christensen, S. Wesley Long, Sishir Subedi, Robert Olson, Marcus Nguyen, James J. Davis, Matthew Ojeda Saavedra, Prasanti Yerramilli, Layne Pruitt, Kristina Reppond, Madison N. Shyer, Jessica Cambric, Ryan Gadd, Ilya J. Finkelstein, Jimmy Gollihar, and James M. Musser |
| EPI_ISL_2204508 | Berkeley Medical Center | WVU and Marshall University Combined Genomics Core Facilities | James Denvir, Peter Stoilov, Peter Perrotta, Wesley Kimble, Ryan Percifield |
| EPI_ISL_2204509 | Aegis Sciences Corporation | Centers for Disease Control and Prevention Division of Viral Diseases, Pathogen Discovery | Dakota Howard, Dhwani Batra, Peter W. Cook, Kara Moser, Adrian Paskey, Jason Caravas, Benjamin Rambo-Martin, Shatavia Morrison, Christopher Gulvick, Scott Sammons, Yvette Unoarumhi, Darlene Wagner, Matthew Schmerer, Cyndi Clark, Patrick Campbell, Rob Case, Vikramsinha Ghorpade, Holly Houdeshell, Ola Kvalvaag, Dillon Nall, Ethan Sanders, Alec Vest, Shaun Westlund, Matthew Hardison, Clinton R. Paden, Duncan MacCannell |
| EPI_ISL_2204510 | Berkeley Medical Center | WVU and Marshall University Combined Genomics Core Facilities | James Denvir, Peter Stoilov, Peter Perrotta, Wesley Kimble, Ryan Percifield |
| EPI_ISL_2204511 | Houston Methodist Hospital | Houston Methodist Hospital | Randall J. Olsen, Paul A. Christensen, S. Wesley Long, Sishir Subedi, Robert Olson, Marcus Nguyen, James J. Davis, Matthew Ojeda Saavedra, Prasanti Yerramilli, Layne Pruitt, Kristina Reppond, Madison N. Shyer, Jessica Cambric, Ryan Gadd, Ilya J. Finkelstein, Jimmy Gollihar, and James M. Musser |
| EPI_ISL_2204512 | Berkeley Medical Center | WVU and Marshall University Combined Genomics Core Facilities | James Denvir, Peter Stoilov, Peter Perrotta, Wesley Kimble, Ryan Percifield |
| EPI_ISL_2204513 | Aegis Sciences Corporation | Centers for Disease Control and Prevention Division of Viral Diseases, Pathogen Discovery | Dakota Howard, Dhwani Batra, Peter W. Cook, Kara Moser, Adrian Paskey, Jason Caravas, Benjamin Rambo-Martin, Shatavia Morrison, Christopher Gulvick, Scott Sammons, Yvette Unoarumhi, Darlene Wagner, Matthew Schmerer, Cyndi Clark, Patrick Campbell, Rob Case, Vikramsinha Ghorpade, Holly Houdeshell, Ola Kvalvaag, Dillon Nall, Ethan Sanders, Alec Vest, Shaun Westlund, Matthew Hardison, Clinton R. Paden, Duncan MacCannell |
| EPI_ISL_2204514 | Berkeley Medical Center | WVU and Marshall University Combined Genomics Core Facilities | James Denvir, Peter Stoilov, Peter Perrotta, Wesley Kimble, Ryan Percifield |
| EPI_ISL_2204515 | Houston Methodist Hospital | Houston Methodist Hospital | Randall J. Olsen, Paul A. Christensen, S. Wesley Long, Sishir Subedi, Robert Olson, Marcus Nguyen, James J. Davis, Matthew Ojeda Saavedra, Prasanti Yerramilli, Layne Pruitt, Kristina Reppond, Madison N. Shyer, Jessica Cambric, Ryan Gadd, Ilya J. Finkelstein, Jimmy Gollihar, and James M. Musser |
| EPI_ISL_2204516 | Aegis Sciences Corporation | Centers for Disease Control and Prevention Division of Viral Diseases, Pathogen Discovery | Dakota Howard, Dhwani Batra, Peter W. Cook, Kara Moser, Adrian Paskey, Jason Caravas, Benjamin Rambo-Martin, Shatavia Morrison, Christopher Gulvick, Scott Sammons, Yvette Unoarumhi, Darlene Wagner, Matthew Schmerer, Cyndi Clark, Patrick Campbell, Rob Case, Vikramsinha Ghorpade, Holly Houdeshell, Ola Kvalvaag, Dillon Nall, Ethan Sanders, Alec Vest, Shaun Westlund, Matthew Hardison, Clinton R. Paden, Duncan MacCannell |

| | | | |
|---|---|---|---|
| EPI_ISL_2204517, EPI_ISL_2204518 | Berkeley Medical Center | WVU and Marshall University Combined Genomics Core Facilities | James Denvir, Peter Stoilov, Peter Perrotta, Wesley Kimble, Ryan Percifield |
| EPI_ISL_2204519 | Aegis Sciences Corporation | Centers for Disease Control and Prevention Division of Viral Diseases, Pathogen Discovery | Dakota Howard, Dhwani Batra, Peter W. Cook, Kara Moser, Adrian Paskey, Jason Caravas, Benjamin Rambo-Martin, Shatavia Morrison, Christopher Gulvick, Scott Sammons, Yvette Unoarumhi, Darlene Wagner, Matthew Schmerer, Cyndi Clark, Patrick Campbell, Rob Case, Vikramsinha Ghorpade, Holly Houdeshell, Ola Kvalvaag, Dillon Nall, Ethan Sanders, Alec Vest, Shaun Westlund, Matthew Hardison, Clinton R. Paden, Duncan MacCannell |
| EPI_ISL_2204521 | Berkeley Medical Center | WVU and Marshall University Combined Genomics Core Facilities | James Denvir, Peter Stoilov, Peter Perrotta, Wesley Kimble, Ryan Percifield |
| EPI_ISL_2204522 | Aegis Sciences Corporation | Centers for Disease Control and Prevention Division of Viral Diseases, Pathogen Discovery | Dakota Howard, Dhwani Batra, Peter W. Cook, Kara Moser, Adrian Paskey, Jason Caravas, Benjamin Rambo-Martin, Shatavia Morrison, Christopher Gulvick, Scott Sammons, Yvette Unoarumhi, Darlene Wagner, Matthew Schmerer, Cyndi Clark, Patrick Campbell, Rob Case, Vikramsinha Ghorpade, Holly Houdeshell, Ola Kvalvaag, Dillon Nall, Ethan Sanders, Alec Vest, Shaun Westlund, Matthew Hardison, Clinton R. Paden, Duncan MacCannell |
| EPI_ISL_2204523 | Houston Methodist Hospital | Houston Methodist Hospital | Randall J. Olsen, Paul A. Christensen, S. Wesley Long, Sishir Subedi, Robert Olson, Marcus Nguyen, James J. Davis, Matthew Ojeda Saavedra, Prasanti Yerramilli, Layne Pruitt, Kristina Reppond, Madison N. Shyer, Jessica Cambric, Ryan Gadd, Ilya J. Finkelstein, Jimmy Gollihar, and James M. Musser |
| EPI_ISL_2204524 | Aegis Sciences Corporation | Centers for Disease Control and Prevention Division of Viral Diseases, Pathogen Discovery | Dakota Howard, Dhwani Batra, Peter W. Cook, Kara Moser, Adrian Paskey, Jason Caravas, Benjamin Rambo-Martin, Shatavia Morrison, Christopher Gulvick, Scott Sammons, Yvette Unoarumhi, Darlene Wagner, Matthew Schmerer, Cyndi Clark, Patrick Campbell, Rob Case, Vikramsinha Ghorpade, Holly Houdeshell, Ola Kvalvaag, Dillon Nall, Ethan Sanders, Alec Vest, Shaun Westlund, Matthew Hardison, Clinton R. Paden, Duncan MacCannell |
| EPI_ISL_2204525 | Berkeley Medical Center | WVU and Marshall University Combined Genomics Core Facilities | James Denvir, Peter Stoilov, Peter Perrotta, Wesley Kimble, Ryan Percifield |
| EPI_ISL_2204526 | Houston Methodist Hospital | Houston Methodist Hospital | Randall J. Olsen, Paul A. Christensen, S. Wesley Long, Sishir Subedi, Robert Olson, Marcus Nguyen, James J. Davis, Matthew Ojeda Saavedra, Prasanti Yerramilli, Layne Pruitt, Kristina Reppond, Madison N. Shyer, Jessica Cambric, Ryan Gadd, Ilya J. Finkelstein, Jimmy Gollihar, and James M. Musser |
| EPI_ISL_2204527, EPI_ISL_2204529 | Berkeley Medical Center | WVU and Marshall University Combined Genomics Core Facilities | James Denvir, Peter Stoilov, Peter Perrotta, Wesley Kimble, Ryan Percifield |
| EPI_ISL_2204530 | Houston Methodist Hospital | Houston Methodist Hospital | Randall J. Olsen, Paul A. Christensen, S. Wesley Long, Sishir Subedi, Robert Olson, Marcus Nguyen, James J. Davis, Matthew Ojeda Saavedra, Prasanti Yerramilli, Layne Pruitt, Kristina Reppond, Madison N. Shyer, Jessica Cambric, Ryan Gadd, Ilya J. Finkelstein, Jimmy Gollihar, and James M. Musser |
| EPI_ISL_2204531 | Berkeley Medical Center | WVU and Marshall University Combined Genomics Core Facilities | James Denvir, Peter Stoilov, Peter Perrotta, Wesley Kimble, Ryan Percifield |
| EPI_ISL_2204533 | WV Labs | WVU and Marshall University Combined Genomics Core Facilities | James Denvir, Peter Stoilov, Peter Perrotta, Wesley Kimble, Ryan Percifield |
| EPI_ISL_2204534 | Houston Methodist Hospital | Houston Methodist Hospital | Randall J. Olsen, Paul A. Christensen, S. Wesley Long, Sishir Subedi, Robert Olson, Marcus Nguyen, James J. Davis, Matthew Ojeda Saavedra, Prasanti Yerramilli, Layne Pruitt, Kristina Reppond, Madison N. Shyer, Jessica Cambric, Ryan Gadd, Ilya J. Finkelstein, Jimmy Gollihar, and James M. Musser |
| EPI_ISL_2204535 | Aegis Sciences Corporation | Centers for Disease Control and Prevention Division of Viral Diseases, Pathogen Discovery | Dakota Howard, Dhwani Batra, Peter W. Cook, Kara Moser, Adrian Paskey, Jason Caravas, Benjamin Rambo-Martin, Shatavia Morrison, Christopher Gulvick, Scott Sammons, Yvette Unoarumhi, Darlene Wagner, Matthew Schmerer, Cyndi Clark, Patrick Campbell, Rob Case, Vikramsinha Ghorpade, Holly Houdeshell, Ola Kvalvaag, Dillon Nall, Ethan Sanders, Alec Vest, Shaun Westlund, Matthew Hardison, Clinton R. Paden, Duncan MacCannell |
| EPI_ISL_2204536, EPI_ISL_2204537 | WV Labs | WVU and Marshall University Combined Genomics Core Facilities | James Denvir, Peter Stoilov, Peter Perrotta, Wesley Kimble, Ryan Percifield |
| EPI_ISL_2204538 | Aegis Sciences Corporation | Centers for Disease Control and Prevention Division of Viral Diseases, Pathogen Discovery | Dakota Howard, Dhwani Batra, Peter W. Cook, Kara Moser, Adrian Paskey, Jason Caravas, Benjamin Rambo-Martin, Shatavia Morrison, Christopher Gulvick, Scott Sammons, Yvette Unoarumhi, Darlene Wagner, Matthew Schmerer, Cyndi Clark, Patrick Campbell, Rob Case, Vikramsinha Ghorpade, Holly Houdeshell, Ola Kvalvaag, Dillon Nall, Ethan Sanders, Alec Vest, Shaun Westlund, Matthew Hardison, Clinton R. Paden, Duncan MacCannell |
| EPI_ISL_2204539 | Houston Methodist Hospital | Houston Methodist Hospital | Randall J. Olsen, Paul A. Christensen, S. Wesley Long, Sishir Subedi, Robert Olson, Marcus Nguyen, James J. Davis, Matthew Ojeda Saavedra, Prasanti Yerramilli, Layne Pruitt, Kristina Reppond, Madison N. Shyer, Jessica Cambric, Ryan Gadd, Ilya J. Finkelstein, Jimmy Gollihar, and James M. Musser |
| EPI_ISL_2204540, EPI_ISL_2204541 | WV Labs | WVU and Marshall University Combined Genomics Core Facilities | James Denvir, Peter Stoilov, Peter Perrotta, Wesley Kimble, Ryan Percifield |
| EPI_ISL_2204542 | Aegis Sciences Corporation | Centers for Disease Control and Prevention Division of Viral Diseases, Pathogen Discovery | Dakota Howard, Dhwani Batra, Peter W. Cook, Kara Moser, Adrian Paskey, Jason Caravas, Benjamin Rambo-Martin, Shatavia Morrison, Christopher Gulvick, Scott Sammons, Yvette Unoarumhi, Darlene Wagner, Matthew Schmerer, Cyndi Clark, Patrick Campbell, Rob Case, Vikramsinha Ghorpade, Holly Houdeshell, Ola Kvalvaag, Dillon Nall, Ethan Sanders, Alec Vest, Shaun Westlund, Matthew Hardison, Clinton R. Paden, Duncan MacCannell |
| EPI_ISL_2204543 | Houston Methodist Hospital | Houston Methodist Hospital | Randall J. Olsen, Paul A. Christensen, S. Wesley Long, Sishir Subedi, Robert Olson, Marcus Nguyen, James J. Davis, Matthew Ojeda Saavedra, Prasanti Yerramilli, Layne Pruitt, Kristina Reppond, Madison N. Shyer, Jessica Cambric, Ryan Gadd, Ilya J. Finkelstein, Jimmy Gollihar, and James M. Musser |
| EPI_ISL_2204544 | WV Labs | WVU and Marshall University Combined Genomics Core Facilities | James Denvir, Peter Stoilov, Peter Perrotta, Wesley Kimble, Ryan Percifield |
| EPI_ISL_2204545 | QLabs | WVU and Marshall University Combined Genomics Core Facilities | James Denvir, Peter Stoilov, Peter Perrotta, Wesley Kimble, Ryan Percifield |
| EPI_ISL_2204546 | Aegis Sciences Corporation | Centers for Disease Control and Prevention Division of Viral Diseases, Pathogen Discovery | Dakota Howard, Dhwani Batra, Peter W. Cook, Kara Moser, Adrian Paskey, Jason Caravas, Benjamin Rambo-Martin, Shatavia Morrison, Christopher Gulvick, Scott Sammons, Yvette Unoarumhi, Darlene Wagner, Matthew Schmerer, Cyndi Clark, Patrick Campbell, Rob Case, Vikramsinha Ghorpade, Holly Houdeshell, Ola Kvalvaag, Dillon Nall, Ethan Sanders, Alec Vest, Shaun Westlund, Matthew Hardison, Clinton R. Paden, Duncan MacCannell |
| EPI_ISL_2204547 | Houston Methodist Hospital | Houston Methodist Hospital | Randall J. Olsen, Paul A. Christensen, S. Wesley Long, Sishir Subedi, Robert Olson, Marcus Nguyen, James J. Davis, Matthew Ojeda Saavedra, Prasanti Yerramilli, Layne Pruitt, Kristina Reppond, Madison N. Shyer, Jessica Cambric, Ryan Gadd, Ilya J. Finkelstein, Jimmy Gollihar, and James M. Musser |
| EPI_ISL_2204548, EPI_ISL_2204550, EPI_ISL_2204551 | QLabs | WVU and Marshall University Combined Genomics Core Facilities | James Denvir, Peter Stoilov, Peter Perrotta, Wesley Kimble, Ryan Percifield |
| EPI_ISL_2204552 | Aegis Sciences Corporation | Centers for Disease Control and Prevention Division of Viral Diseases, Pathogen Discovery | Dakota Howard, Dhwani Batra, Peter W. Cook, Kara Moser, Adrian Paskey, Jason Caravas, Benjamin Rambo-Martin, Shatavia Morrison, Christopher Gulvick, Scott Sammons, Yvette Unoarumhi, Darlene Wagner, Matthew Schmerer, Cyndi Clark, Patrick Campbell, Rob Case, Vikramsinha Ghorpade, Holly Houdeshell, Ola Kvalvaag, Dillon Nall, Ethan Sanders, Alec Vest, Shaun Westlund, Matthew Hardison, Clinton R. Paden, Duncan MacCannell |
| EPI_ISL_2204553 | QLabs | WVU and Marshall University Combined Genomics Core Facilities | James Denvir, Peter Stoilov, Peter Perrotta, Wesley Kimble, Ryan Percifield |
| EPI_ISL_2204555 | Aegis Sciences Corporation | Centers for Disease Control and Prevention Division of Viral Diseases, Pathogen Discovery | Dakota Howard, Dhwani Batra, Peter W. Cook, Kara Moser, Adrian Paskey, Jason Caravas, Benjamin Rambo-Martin, Shatavia Morrison, Christopher Gulvick, Scott Sammons, Yvette Unoarumhi, Darlene Wagner, Matthew Schmerer, Cyndi Clark, Patrick Campbell, Rob Case, Vikramsinha Ghorpade, Holly Houdeshell, Ola Kvalvaag, Dillon Nall, Ethan Sanders, Alec Vest, Shaun Westlund, Matthew Hardison, Clinton R. Paden, Duncan MacCannell |
| EPI_ISL_2204556, EPI_ISL_2204557 | QLabs | WVU and Marshall University Combined Genomics Core Facilities | James Denvir, Peter Stoilov, Peter Perrotta, Wesley Kimble, Ryan Percifield |
| EPI_ISL_2204558 | Aegis Sciences Corporation | Centers for Disease Control and Prevention Division of Viral Diseases, Pathogen Discovery | Dakota Howard, Dhwani Batra, Peter W. Cook, Kara Moser, Adrian Paskey, Jason Caravas, Benjamin Rambo-Martin, Shatavia Morrison, Christopher Gulvick, Scott Sammons, Yvette Unoarumhi, Darlene Wagner, Matthew Schmerer, Cyndi Clark, Patrick Campbell, Rob Case, Vikramsinha Ghorpade, Holly Houdeshell, Ola Kvalvaag, Dillon Nall, Ethan Sanders, Alec Vest, Shaun Westlund, Matthew Hardison, Clinton R. Paden, Duncan MacCannell |
| EPI_ISL_2204559 | Houston Methodist Hospital | Houston Methodist Hospital | Randall J. Olsen, Paul A. Christensen, S. Wesley Long, Sishir Subedi, Robert Olson, Marcus Nguyen, James J. Davis, Matthew Ojeda Saavedra, Prasanti Yerramilli, Layne Pruitt, Kristina Reppond, Madison N. Shyer, Jessica Cambric, Ryan Gadd, Ilya J. Finkelstein, Jimmy Gollihar, and James M. Musser |
| EPI_ISL_2204560, EPI_ISL_2204561 | QLabs | WVU and Marshall University Combined Genomics Core Facilities | James Denvir, Peter Stoilov, Peter Perrotta, Wesley Kimble, Ryan Percifield |
| EPI_ISL_2204562 | Aegis Sciences Corporation | Centers for Disease Control and Prevention Division of Viral Diseases, Pathogen Discovery | Dakota Howard, Dhwani Batra, Peter W. Cook, Kara Moser, Adrian Paskey, Jason Caravas, Benjamin Rambo-Martin, Shatavia Morrison, Christopher Gulvick, Scott Sammons, Yvette Unoarumhi, Darlene Wagner, Matthew Schmerer, Cyndi Clark, Patrick Campbell, Rob Case, Vikramsinha Ghorpade, Holly Houdeshell, Ola Kvalvaag, Dillon Nall, Ethan Sanders, Alec Vest, Shaun Westlund, Matthew Hardison, Clinton R. Paden, Duncan MacCannell |
| EPI_ISL_2204564 | QLabs | WVU and Marshall University Combined Genomics Core Facilities | James Denvir, Peter Stoilov, Peter Perrotta, Wesley Kimble, Ryan Percifield |
| EPI_ISL_2204565 | Aegis Sciences Corporation | Centers for Disease Control and Prevention Division of Viral Diseases, Pathogen Discovery | Dakota Howard, Dhwani Batra, Peter W. Cook, Kara Moser, Adrian Paskey, Jason Caravas, Benjamin Rambo-Martin, Shatavia Morrison, Christopher Gulvick, Scott Sammons, Yvette Unoarumhi, Darlene Wagner, Matthew Schmerer, Cyndi Clark, Patrick Campbell, Rob Case, Vikramsinha Ghorpade, Holly Houdeshell, Ola Kvalvaag, Dillon Nall, Ethan Sanders, Alec Vest, Shaun Westlund, Matthew Hardison, Clinton R. Paden, Duncan MacCannell |
| EPI_ISL_2204566 | QLabs | WVU and Marshall University Combined Genomics Core Facilities | James Denvir, Peter Stoilov, Peter Perrotta, Wesley Kimble, Ryan Percifield |
| EPI_ISL_2204568 | Aegis Sciences Corporation | Centers for Disease Control and Prevention Division of Viral Diseases, Pathogen Discovery | Dakota Howard, Dhwani Batra, Peter W. Cook, Kara Moser, Adrian Paskey, Jason Caravas, Benjamin Rambo-Martin, Shatavia Morrison, Christopher Gulvick, Scott Sammons, Yvette Unoarumhi, Darlene Wagner, Matthew Schmerer, Cyndi Clark, Patrick Campbell, Rob Case, Vikramsinha Ghorpade, Holly Houdeshell, Ola Kvalvaag, Dillon Nall, Ethan Sanders, Alec Vest, Shaun Westlund, Matthew Hardison, Clinton R. Paden, Duncan MacCannell |
| EPI_ISL_2204569 | QLabs | WVU and Marshall University Combined Genomics Core Facilities | James Denvir, Peter Stoilov, Peter Perrotta, Wesley Kimble, Ryan Percifield |
| EPI_ISL_2204570 | Houston Methodist Hospital | Houston Methodist Hospital | Randall J. Olsen, Paul A. Christensen, S. Wesley Long, Sishir Subedi, Robert Olson, Marcus Nguyen, James J. Davis, Matthew Ojeda Saavedra, Prasanti Yerramilli, Layne Pruitt, Kristina Reppond, Madison N. Shyer, Jessica Cambric, Ryan Gadd, Ilya J. Finkelstein, Jimmy Gollihar, and James M. Musser |

| | | | |
|---|---|---|---|
| EPI_ISL_2204571 | Aegis Sciences Corporation | Centers for Disease Control and Prevention Division of Viral Diseases, Pathogen Discovery | Dakota Howard, Dhwani Batra, Peter W. Cook, Kara Moser, Adrian Paskey, Jason Caravas, Benjamin Rambo-Martin, Shatavia Morrison, Christopher Gulvick, Scott Sammons, Yvette Unoarumhi, Darlene Wagner, Matthew Schmerer, Cyndi Clark, Patrick Campbell, Rob Case, Vikramsinha Ghorpade, Holly Houdeshell, Ola Kvalvaag, Dillon Nall, Ethan Sanders, Alec Vest, Shaun Westlund, Matthew Hardison, Clinton R. Paden, Duncan MacCannell |
| EPI_ISL_2204572 | QLabs | WVU and Marshall University Combined Genomics Core Facilities | James Denvir, Peter Stoilov, Peter Perrotta, Wesley Kimble, Ryan Percifield |
| EPI_ISL_2204573 | Houston Methodist Hospital | Houston Methodist Hospital | Randall J. Olsen, Paul A. Christensen, S. Wesley Long, Sishir Subedi, Robert Olson, Marcus Nguyen, James J. Davis, Matthew Ojeda Saavedra, Prasanti Yerramilli, Layne Pruitt, Kristina Reppond, Madison N. Shyer, Jessica Cambric, Ryan Gadd, Ilya J. Finkelstein, Jimmy Gollihar, and James M. Musser |
| EPI_ISL_2204574 | Aegis Sciences Corporation | Centers for Disease Control and Prevention Division of Viral Diseases, Pathogen Discovery | Dakota Howard, Dhwani Batra, Peter W. Cook, Kara Moser, Adrian Paskey, Jason Caravas, Benjamin Rambo-Martin, Shatavia Morrison, Christopher Gulvick, Scott Sammons, Yvette Unoarumhi, Darlene Wagner, Matthew Schmerer, Cyndi Clark, Patrick Campbell, Rob Case, Vikramsinha Ghorpade, Holly Houdeshell, Ola Kvalvaag, Dillon Nall, Ethan Sanders, Alec Vest, Shaun Westlund, Matthew Hardison, Clinton R. Paden, Duncan MacCannell |
| EPI_ISL_2204576 | QLabs | WVU and Marshall University Combined Genomics Core Facilities | James Denvir, Peter Stoilov, Peter Perrotta, Wesley Kimble, Ryan Percifield |
| EPI_ISL_2204577 | Houston Methodist Hospital | Houston Methodist Hospital | Randall J. Olsen, Paul A. Christensen, S. Wesley Long, Sishir Subedi, Robert Olson, Marcus Nguyen, James J. Davis, Matthew Ojeda Saavedra, Prasanti Yerramilli, Layne Pruitt, Kristina Reppond, Madison N. Shyer, Jessica Cambric, Ryan Gadd, Ilya J. Finkelstein, Jimmy Gollihar, and James M. Musser |
| EPI_ISL_2204578, EPI_ISL_2204580 | QLabs | WVU and Marshall University Combined Genomics Core Facilities | James Denvir, Peter Stoilov, Peter Perrotta, Wesley Kimble, Ryan Percifield |
| EPI_ISL_2204581 | Houston Methodist Hospital | Houston Methodist Hospital | Randall J. Olsen, Paul A. Christensen, S. Wesley Long, Sishir Subedi, Robert Olson, Marcus Nguyen, James J. Davis, Matthew Ojeda Saavedra, Prasanti Yerramilli, Layne Pruitt, Kristina Reppond, Madison N. Shyer, Jessica Cambric, Ryan Gadd, Ilya J. Finkelstein, Jimmy Gollihar, and James M. Musser |
| EPI_ISL_2204582 | QLabs | WVU and Marshall University Combined Genomics Core Facilities | James Denvir, Peter Stoilov, Peter Perrotta, Wesley Kimble, Ryan Percifield |
| EPI_ISL_2204583 | Aegis Sciences Corporation | Centers for Disease Control and Prevention Division of Viral Diseases, Pathogen Discovery | Dakota Howard, Dhwani Batra, Peter W. Cook, Kara Moser, Adrian Paskey, Jason Caravas, Benjamin Rambo-Martin, Shatavia Morrison, Christopher Gulvick, Scott Sammons, Yvette Unoarumhi, Darlene Wagner, Matthew Schmerer, Cyndi Clark, Patrick Campbell, Rob Case, Vikramsinha Ghorpade, Holly Houdeshell, Ola Kvalvaag, Dillon Nall, Ethan Sanders, Alec Vest, Shaun Westlund, Matthew Hardison, Clinton R. Paden, Duncan MacCannell |
| EPI_ISL_2204584 | QLabs | WVU and Marshall University Combined Genomics Core Facilities | James Denvir, Peter Stoilov, Peter Perrotta, Wesley Kimble, Ryan Percifield |
| EPI_ISL_2204585 | Houston Methodist Hospital | Houston Methodist Hospital | Randall J. Olsen, Paul A. Christensen, S. Wesley Long, Sishir Subedi, Robert Olson, Marcus Nguyen, James J. Davis, Matthew Ojeda Saavedra, Prasanti Yerramilli, Layne Pruitt, Kristina Reppond, Madison N. Shyer, Jessica Cambric, Ryan Gadd, Ilya J. Finkelstein, Jimmy Gollihar, and James M. Musser |
| EPI_ISL_2204586 | QLabs | WVU and Marshall University Combined Genomics Core Facilities | James Denvir, Peter Stoilov, Peter Perrotta, Wesley Kimble, Ryan Percifield |
| EPI_ISL_2204587 | Aegis Sciences Corporation | Centers for Disease Control and Prevention Division of Viral Diseases, Pathogen Discovery | Dakota Howard, Dhwani Batra, Peter W. Cook, Kara Moser, Adrian Paskey, Jason Caravas, Benjamin Rambo-Martin, Shatavia Morrison, Christopher Gulvick, Scott Sammons, Yvette Unoarumhi, Darlene Wagner, Matthew Schmerer, Cyndi Clark, Patrick Campbell, Rob Case, Vikramsinha Ghorpade, Holly Houdeshell, Ola Kvalvaag, Dillon Nall, Ethan Sanders, Alec Vest, Shaun Westlund, Matthew Hardison, Clinton R. Paden, Duncan MacCannell |
| EPI_ISL_2204588 | QLabs | WVU and Marshall University Combined Genomics Core Facilities | James Denvir, Peter Stoilov, Peter Perrotta, Wesley Kimble, Ryan Percifield |
| EPI_ISL_2204589 | Houston Methodist Hospital | Houston Methodist Hospital | Randall J. Olsen, Paul A. Christensen, S. Wesley Long, Sishir Subedi, Robert Olson, Marcus Nguyen, James J. Davis, Matthew Ojeda Saavedra, Prasanti Yerramilli, Layne Pruitt, Kristina Reppond, Madison N. Shyer, Jessica Cambric, Ryan Gadd, Ilya J. Finkelstein, Jimmy Gollihar, and James M. Musser |
| EPI_ISL_2204590 | QLabs | WVU and Marshall University Combined Genomics Core Facilities | James Denvir, Peter Stoilov, Peter Perrotta, Wesley Kimble, Ryan Percifield |
| EPI_ISL_2204591 | Aegis Sciences Corporation | Centers for Disease Control and Prevention Division of Viral Diseases, Pathogen Discovery | Dakota Howard, Dhwani Batra, Peter W. Cook, Kara Moser, Adrian Paskey, Jason Caravas, Benjamin Rambo-Martin, Shatavia Morrison, Christopher Gulvick, Scott Sammons, Yvette Unoarumhi, Darlene Wagner, Matthew Schmerer, Cyndi Clark, Patrick Campbell, Rob Case, Vikramsinha Ghorpade, Holly Houdeshell, Ola Kvalvaag, Dillon Nall, Ethan Sanders, Alec Vest, Shaun Westlund, Matthew Hardison, Clinton R. Paden, Duncan MacCannell |
| EPI_ISL_2204592, EPI_ISL_2204594 | QLabs | WVU and Marshall University Combined Genomics Core Facilities | James Denvir, Peter Stoilov, Peter Perrotta, Wesley Kimble, Ryan Percifield |
| EPI_ISL_2204595 | Aegis Sciences Corporation | Centers for Disease Control and Prevention Division of Viral Diseases, Pathogen Discovery | Dakota Howard, Dhwani Batra, Peter W. Cook, Kara Moser, Adrian Paskey, Jason Caravas, Benjamin Rambo-Martin, Shatavia Morrison, Christopher Gulvick, Scott Sammons, Yvette Unoarumhi, Darlene Wagner, Matthew Schmerer, Cyndi Clark, Patrick Campbell, Rob Case, Vikramsinha Ghorpade, Holly Houdeshell, Ola Kvalvaag, Dillon Nall, Ethan Sanders, Alec Vest, Shaun Westlund, Matthew Hardison, Clinton R. Paden, Duncan MacCannell |
| EPI_ISL_2204598 | QLabs | WVU and Marshall University Combined Genomics Core Facilities | James Denvir, Peter Stoilov, Peter Perrotta, Wesley Kimble, Ryan Percifield |
| EPI_ISL_2204599 | Aegis Sciences Corporation | Centers for Disease Control and Prevention Division of Viral Diseases, Pathogen Discovery | Dakota Howard, Dhwani Batra, Peter W. Cook, Kara Moser, Adrian Paskey, Jason Caravas, Benjamin Rambo-Martin, Shatavia Morrison, Christopher Gulvick, Scott Sammons, Yvette Unoarumhi, Darlene Wagner, Matthew Schmerer, Cyndi Clark, Patrick Campbell, Rob Case, Vikramsinha Ghorpade, Holly Houdeshell, Ola Kvalvaag, Dillon Nall, Ethan Sanders, Alec Vest, Shaun Westlund, Matthew Hardison, Clinton R. Paden, Duncan MacCannell |
| EPI_ISL_2204600 | QLabs | WVU and Marshall University Combined Genomics Core Facilities | James Denvir, Peter Stoilov, Peter Perrotta, Wesley Kimble, Ryan Percifield |
| EPI_ISL_2204601 | Houston Methodist Hospital | Houston Methodist Hospital | Randall J. Olsen, Paul A. Christensen, S. Wesley Long, Sishir Subedi, Robert Olson, Marcus Nguyen, James J. Davis, Matthew Ojeda Saavedra, Prasanti Yerramilli, Layne Pruitt, Kristina Reppond, Madison N. Shyer, Jessica Cambric, Ryan Gadd, Ilya J. Finkelstein, Jimmy Gollihar, and James M. Musser |
| EPI_ISL_2204602 | Aegis Sciences Corporation | Centers for Disease Control and Prevention Division of Viral Diseases, Pathogen Discovery | Dakota Howard, Dhwani Batra, Peter W. Cook, Kara Moser, Adrian Paskey, Jason Caravas, Benjamin Rambo-Martin, Shatavia Morrison, Christopher Gulvick, Scott Sammons, Yvette Unoarumhi, Darlene Wagner, Matthew Schmerer, Cyndi Clark, Patrick Campbell, Rob Case, Vikramsinha Ghorpade, Holly Houdeshell, Ola Kvalvaag, Dillon Nall, Ethan Sanders, Alec Vest, Shaun Westlund, Matthew Hardison, Clinton R. Paden, Duncan MacCannell |
| EPI_ISL_2204603 | QLabs | WVU and Marshall University Combined Genomics Core Facilities | James Denvir, Peter Stoilov, Peter Perrotta, Wesley Kimble, Ryan Percifield |
| EPI_ISL_2204604 | Aegis Sciences Corporation | Centers for Disease Control and Prevention Division of Viral Diseases, Pathogen Discovery | Dakota Howard, Dhwani Batra, Peter W. Cook, Kara Moser, Adrian Paskey, Jason Caravas, Benjamin Rambo-Martin, Shatavia Morrison, Christopher Gulvick, Scott Sammons, Yvette Unoarumhi, Darlene Wagner, Matthew Schmerer, Cyndi Clark, Patrick Campbell, Rob Case, Vikramsinha Ghorpade, Holly Houdeshell, Ola Kvalvaag, Dillon Nall, Ethan Sanders, Alec Vest, Shaun Westlund, Matthew Hardison, Clinton R. Paden, Duncan MacCannell |
| EPI_ISL_2204605 | Houston Methodist Hospital | Houston Methodist Hospital | Randall J. Olsen, Paul A. Christensen, S. Wesley Long, Sishir Subedi, Robert Olson, Marcus Nguyen, James J. Davis, Matthew Ojeda Saavedra, Prasanti Yerramilli, Layne Pruitt, Kristina Reppond, Madison N. Shyer, Jessica Cambric, Ryan Gadd, Ilya J. Finkelstein, Jimmy Gollihar, and James M. Musser |
| EPI_ISL_2204606 | Raleigh General Hospital Microbiology Dept. | WVU and Marshall University Combined Genomics Core Facilities | James Denvir, Peter Stoilov, Peter Perrotta, Wesley Kimble, Ryan Percifield |
| EPI_ISL_2204609 | Aegis Sciences Corporation | Centers for Disease Control and Prevention Division of Viral Diseases, Pathogen Discovery | Dakota Howard, Dhwani Batra, Peter W. Cook, Kara Moser, Adrian Paskey, Jason Caravas, Benjamin Rambo-Martin, Shatavia Morrison, Christopher Gulvick, Scott Sammons, Yvette Unoarumhi, Darlene Wagner, Matthew Schmerer, Cyndi Clark, Patrick Campbell, Rob Case, Vikramsinha Ghorpade, Holly Houdeshell, Ola Kvalvaag, Dillon Nall, Ethan Sanders, Alec Vest, Shaun Westlund, Matthew Hardison, Clinton R. Paden, Duncan MacCannell |
| EPI_ISL_2204610 | BMC LAB | WVU and Marshall University Combined Genomics Core Facilities | James Denvir, Peter Stoilov, Peter Perrotta, Wesley Kimble, Ryan Percifield |
| EPI_ISL_2204611 | Houston Methodist Hospital | Houston Methodist Hospital | Randall J. Olsen, Paul A. Christensen, S. Wesley Long, Sishir Subedi, Robert Olson, Marcus Nguyen, James J. Davis, Matthew Ojeda Saavedra, Prasanti Yerramilli, Layne Pruitt, Kristina Reppond, Madison N. Shyer, Jessica Cambric, Ryan Gadd, Ilya J. Finkelstein, Jimmy Gollihar, and James M. Musser |
| EPI_ISL_2204612 | Aegis Sciences Corporation | Centers for Disease Control and Prevention Division of Viral Diseases, Pathogen Discovery | Dakota Howard, Dhwani Batra, Peter W. Cook, Kara Moser, Adrian Paskey, Jason Caravas, Benjamin Rambo-Martin, Shatavia Morrison, Christopher Gulvick, Scott Sammons, Yvette Unoarumhi, Darlene Wagner, Matthew Schmerer, Cyndi Clark, Patrick Campbell, Rob Case, Vikramsinha Ghorpade, Holly Houdeshell, Ola Kvalvaag, Dillon Nall, Ethan Sanders, Alec Vest, Shaun Westlund, Matthew Hardison, Clinton R. Paden, Duncan MacCannell |
| EPI_ISL_2204613 | Houston Methodist Hospital | Houston Methodist Hospital | Randall J. Olsen, Paul A. Christensen, S. Wesley Long, Sishir Subedi, Robert Olson, Marcus Nguyen, James J. Davis, Matthew Ojeda Saavedra, Prasanti Yerramilli, Layne Pruitt, Kristina Reppond, Madison N. Shyer, Jessica Cambric, Ryan Gadd, Ilya J. Finkelstein, Jimmy Gollihar, and James M. Musser |
| EPI_ISL_2204614, EPI_ISL_2204616 | BMC LAB | WVU and Marshall University Combined Genomics Core Facilities | James Denvir, Peter Stoilov, Peter Perrotta, Wesley Kimble, Ryan Percifield |
| EPI_ISL_2204617 | Houston Methodist Hospital | Houston Methodist Hospital | Randall J. Olsen, Paul A. Christensen, S. Wesley Long, Sishir Subedi, Robert Olson, Marcus Nguyen, James J. Davis, Matthew Ojeda Saavedra, Prasanti Yerramilli, Layne Pruitt, Kristina Reppond, Madison N. Shyer, Jessica Cambric, Ryan Gadd, Ilya J. Finkelstein, Jimmy Gollihar, and James M. Musser |
| EPI_ISL_2204618 | Aegis Sciences Corporation | Centers for Disease Control and Prevention Division of Viral Diseases, Pathogen Discovery | Dakota Howard, Dhwani Batra, Peter W. Cook, Kara Moser, Adrian Paskey, Jason Caravas, Benjamin Rambo-Martin, Shatavia Morrison, Christopher Gulvick, Scott Sammons, Yvette Unoarumhi, Darlene Wagner, Matthew Schmerer, Cyndi Clark, Patrick Campbell, Rob Case, Vikramsinha Ghorpade, Holly Houdeshell, Ola Kvalvaag, Dillon Nall, Ethan Sanders, Alec Vest, Shaun Westlund, Matthew Hardison, Clinton R. Paden, Duncan MacCannell |
| EPI_ISL_2204619 | BMC LAB | WVU and Marshall University Combined Genomics Core Facilities | James Denvir, Peter Stoilov, Peter Perrotta, Wesley Kimble, Ryan Percifield |
| EPI_ISL_2204620 | Houston Methodist Hospital | Houston Methodist Hospital | Randall J. Olsen, Paul A. Christensen, S. Wesley Long, Sishir Subedi, Robert Olson, Marcus Nguyen, James J. Davis, Matthew Ojeda Saavedra, Prasanti Yerramilli, Layne Pruitt, Kristina Reppond, Madison N. Shyer, Jessica Cambric, Ryan Gadd, Ilya J. Finkelstein, Jimmy Gollihar, and James M. Musser |
| EPI_ISL_2204621 | BMC LAB | WVU and Marshall University Combined Genomics Core Facilities | James Denvir, Peter Stoilov, Peter Perrotta, Wesley Kimble, Ryan Percifield |

| | | | |
|---|---|---|---|
| EPI_ISL_2204622 | Aegis Sciences Corporation | Centers for Disease Control and Prevention Division of Viral Diseases, Pathogen Discovery | Dakota Howard, Dhwani Batra, Peter W. Cook, Kara Moser, Adrian Paskey, Jason Caravas, Benjamin Rambo-Martin, Shatavia Morrison, Christopher Gulvick, Scott Sammons, Yvette Unoarumhi, Darlene Wagner, Matthew Schmerer, Cyndi Clark, Patrick Campbell, Rob Case, Vikramsinha Ghorpade, Holly Houdeshell, Ola Kvalvaag, Dillon Nall, Ethan Sanders, Alec Vest, Shaun Westlund, Matthew Hardison, Clinton R. Paden, Duncan MacCannell |
| EPI_ISL_2204623 | BMC LAB | WVU and Marshall University Combined Genomics Core Facilities | James Denvir, Peter Stoilov, Peter Perrotta, Wesley Kimble, Ryan Percifield |
| EPI_ISL_2204624 | Houston Methodist Hospital | Houston Methodist Hospital | Randall J. Olsen, Paul A. Christensen, S. Wesley Long, Sishir Subedi, Robert Olson, Marcus Nguyen, James J. Davis, Matthew Ojeda Saavedra, Prasanti Yerramilli, Layne Pruitt, Kristina Reppond, Madison N. Shyer, Jessica Cambric, Ryan Gadd, Ilya J. Finkelstein, Jimmy Gollihar, and James M. Musser |
| EPI_ISL_2204625 | Aegis Sciences Corporation | Centers for Disease Control and Prevention Division of Viral Diseases, Pathogen Discovery | Dakota Howard, Dhwani Batra, Peter W. Cook, Kara Moser, Adrian Paskey, Jason Caravas, Benjamin Rambo-Martin, Shatavia Morrison, Christopher Gulvick, Scott Sammons, Yvette Unoarumhi, Darlene Wagner, Matthew Schmerer, Cyndi Clark, Patrick Campbell, Rob Case, Vikramsinha Ghorpade, Holly Houdeshell, Ola Kvalvaag, Dillon Nall, Ethan Sanders, Alec Vest, Shaun Westlund, Matthew Hardison, Clinton R. Paden, Duncan MacCannell |
| EPI_ISL_2204626, EPI_ISL_2204627 | WV Labs | WVU and Marshall University Combined Genomics Core Facilities | James Denvir, Peter Stoilov, Peter Perrotta, Wesley Kimble, Ryan Percifield |
| EPI_ISL_2204628 | Aegis Sciences Corporation | Centers for Disease Control and Prevention Division of Viral Diseases, Pathogen Discovery | Dakota Howard, Dhwani Batra, Peter W. Cook, Kara Moser, Adrian Paskey, Jason Caravas, Benjamin Rambo-Martin, Shatavia Morrison, Christopher Gulvick, Scott Sammons, Yvette Unoarumhi, Darlene Wagner, Matthew Schmerer, Cyndi Clark, Patrick Campbell, Rob Case, Vikramsinha Ghorpade, Holly Houdeshell, Ola Kvalvaag, Dillon Nall, Ethan Sanders, Alec Vest, Shaun Westlund, Matthew Hardison, Clinton R. Paden, Duncan MacCannell |
| EPI_ISL_2204629 | Houston Methodist Hospital | Houston Methodist Hospital | Randall J. Olsen, Paul A. Christensen, S. Wesley Long, Sishir Subedi, Robert Olson, Marcus Nguyen, James J. Davis, Matthew Ojeda Saavedra, Prasanti Yerramilli, Layne Pruitt, Kristina Reppond, Madison N. Shyer, Jessica Cambric, Ryan Gadd, Ilya J. Finkelstein, Jimmy Gollihar, and James M. Musser |
| EPI_ISL_2204630 | Aegis Sciences Corporation | Centers for Disease Control and Prevention Division of Viral Diseases, Pathogen Discovery | Dakota Howard, Dhwani Batra, Peter W. Cook, Kara Moser, Adrian Paskey, Jason Caravas, Benjamin Rambo-Martin, Shatavia Morrison, Christopher Gulvick, Scott Sammons, Yvette Unoarumhi, Darlene Wagner, Matthew Schmerer, Cyndi Clark, Patrick Campbell, Rob Case, Vikramsinha Ghorpade, Holly Houdeshell, Ola Kvalvaag, Dillon Nall, Ethan Sanders, Alec Vest, Shaun Westlund, Matthew Hardison, Clinton R. Paden, Duncan MacCannell |
| EPI_ISL_2204631 | Houston Methodist Hospital | Houston Methodist Hospital | Randall J. Olsen, Paul A. Christensen, S. Wesley Long, Sishir Subedi, Robert Olson, Marcus Nguyen, James J. Davis, Matthew Ojeda Saavedra, Prasanti Yerramilli, Layne Pruitt, Kristina Reppond, Madison N. Shyer, Jessica Cambric, Ryan Gadd, Ilya J. Finkelstein, Jimmy Gollihar, and James M. Musser |
| EPI_ISL_2204633 | WV Labs | WVU and Marshall University Combined Genomics Core Facilities | James Denvir, Peter Stoilov, Peter Perrotta, Wesley Kimble, Ryan Percifield |
| EPI_ISL_2204634 | Houston Methodist Hospital | Houston Methodist Hospital | Randall J. Olsen, Paul A. Christensen, S. Wesley Long, Sishir Subedi, Robert Olson, Marcus Nguyen, James J. Davis, Matthew Ojeda Saavedra, Prasanti Yerramilli, Layne Pruitt, Kristina Reppond, Madison N. Shyer, Jessica Cambric, Ryan Gadd, Ilya J. Finkelstein, Jimmy Gollihar, and James M. Musser |
| EPI_ISL_2204635, EPI_ISL_2204636 | Aegis Sciences Corporation | Centers for Disease Control and Prevention Division of Viral Diseases, Pathogen Discovery | Dakota Howard, Dhwani Batra, Peter W. Cook, Kara Moser, Adrian Paskey, Jason Caravas, Benjamin Rambo-Martin, Shatavia Morrison, Christopher Gulvick, Scott Sammons, Yvette Unoarumhi, Darlene Wagner, Matthew Schmerer, Cyndi Clark, Patrick Campbell, Rob Case, Vikramsinha Ghorpade, Holly Houdeshell, Ola Kvalvaag, Dillon Nall, Ethan Sanders, Alec Vest, Shaun Westlund, Matthew Hardison, Clinton R. Paden, Duncan MacCannell |
| EPI_ISL_2204637 | Houston Methodist Hospital | Houston Methodist Hospital | Randall J. Olsen, Paul A. Christensen, S. Wesley Long, Sishir Subedi, Robert Olson, Marcus Nguyen, James J. Davis, Matthew Ojeda Saavedra, Prasanti Yerramilli, Layne Pruitt, Kristina Reppond, Madison N. Shyer, Jessica Cambric, Ryan Gadd, Ilya J. Finkelstein, Jimmy Gollihar, and James M. Musser |
| EPI_ISL_2204638 | QLabs | WVU and Marshall University Combined Genomics Core Facilities | James Denvir, Peter Stoilov, Peter Perrotta, Wesley Kimble, Ryan Percifield |
| EPI_ISL_2204639, EPI_ISL_2204641 | Houston Methodist Hospital | Houston Methodist Hospital | Randall J. Olsen, Paul A. Christensen, S. Wesley Long, Sishir Subedi, Robert Olson, Marcus Nguyen, James J. Davis, Matthew Ojeda Saavedra, Prasanti Yerramilli, Layne Pruitt, Kristina Reppond, Madison N. Shyer, Jessica Cambric, Ryan Gadd, Ilya J. Finkelstein, Jimmy Gollihar, and James M. Musser |
| EPI_ISL_2204642 | Aegis Sciences Corporation | Centers for Disease Control and Prevention Division of Viral Diseases, Pathogen Discovery | Dakota Howard, Dhwani Batra, Peter W. Cook, Kara Moser, Adrian Paskey, Jason Caravas, Benjamin Rambo-Martin, Shatavia Morrison, Christopher Gulvick, Scott Sammons, Yvette Unoarumhi, Darlene Wagner, Matthew Schmerer, Cyndi Clark, Patrick Campbell, Rob Case, Vikramsinha Ghorpade, Holly Houdeshell, Ola Kvalvaag, Dillon Nall, Ethan Sanders, Alec Vest, Shaun Westlund, Matthew Hardison, Clinton R. Paden, Duncan MacCannell |
| EPI_ISL_2204643 | QLabs | WVU and Marshall University Combined Genomics Core Facilities | James Denvir, Peter Stoilov, Peter Perrotta, Wesley Kimble, Ryan Percifield |
| EPI_ISL_2204645 | Aegis Sciences Corporation | Centers for Disease Control and Prevention Division of Viral Diseases, Pathogen Discovery | Dakota Howard, Dhwani Batra, Peter W. Cook, Kara Moser, Adrian Paskey, Jason Caravas, Benjamin Rambo-Martin, Shatavia Morrison, Christopher Gulvick, Scott Sammons, Yvette Unoarumhi, Darlene Wagner, Matthew Schmerer, Cyndi Clark, Patrick Campbell, Rob Case, Vikramsinha Ghorpade, Holly Houdeshell, Ola Kvalvaag, Dillon Nall, Ethan Sanders, Alec Vest, Shaun Westlund, Matthew Hardison, Clinton R. Paden, Duncan MacCannell |
| EPI_ISL_2204646 | QLabs | WVU and Marshall University Combined Genomics Core Facilities | James Denvir, Peter Stoilov, Peter Perrotta, Wesley Kimble, Ryan Percifield |
| EPI_ISL_2204647 | Houston Methodist Hospital | Houston Methodist Hospital | Randall J. Olsen, Paul A. Christensen, S. Wesley Long, Sishir Subedi, Robert Olson, Marcus Nguyen, James J. Davis, Matthew Ojeda Saavedra, Prasanti Yerramilli, Layne Pruitt, Kristina Reppond, Madison N. Shyer, Jessica Cambric, Ryan Gadd, Ilya J. Finkelstein, Jimmy Gollihar, and James M. Musser |
| EPI_ISL_2204648 | Aegis Sciences Corporation | Centers for Disease Control and Prevention Division of Viral Diseases, Pathogen Discovery | Dakota Howard, Dhwani Batra, Peter W. Cook, Kara Moser, Adrian Paskey, Jason Caravas, Benjamin Rambo-Martin, Shatavia Morrison, Christopher Gulvick, Scott Sammons, Yvette Unoarumhi, Darlene Wagner, Matthew Schmerer, Cyndi Clark, Patrick Campbell, Rob Case, Vikramsinha Ghorpade, Holly Houdeshell, Ola Kvalvaag, Dillon Nall, Ethan Sanders, Alec Vest, Shaun Westlund, Matthew Hardison, Clinton R. Paden, Duncan MacCannell |
| EPI_ISL_2204650 | QLabs | WVU and Marshall University Combined Genomics Core Facilities | James Denvir, Peter Stoilov, Peter Perrotta, Wesley Kimble, Ryan Percifield |
| EPI_ISL_2204651, EPI_ISL_2204653 | Houston Methodist Hospital | Houston Methodist Hospital | Randall J. Olsen, Paul A. Christensen, S. Wesley Long, Sishir Subedi, Robert Olson, Marcus Nguyen, James J. Davis, Matthew Ojeda Saavedra, Prasanti Yerramilli, Layne Pruitt, Kristina Reppond, Madison N. Shyer, Jessica Cambric, Ryan Gadd, Ilya J. Finkelstein, Jimmy Gollihar, and James M. Musser |
| EPI_ISL_2204654 | QLabs | WVU and Marshall University Combined Genomics Core Facilities | James Denvir, Peter Stoilov, Peter Perrotta, Wesley Kimble, Ryan Percifield |
| EPI_ISL_2204655 | Aegis Sciences Corporation | Centers for Disease Control and Prevention Division of Viral Diseases, Pathogen Discovery | Dakota Howard, Dhwani Batra, Peter W. Cook, Kara Moser, Adrian Paskey, Jason Caravas, Benjamin Rambo-Martin, Shatavia Morrison, Christopher Gulvick, Scott Sammons, Yvette Unoarumhi, Darlene Wagner, Matthew Schmerer, Cyndi Clark, Patrick Campbell, Rob Case, Vikramsinha Ghorpade, Holly Houdeshell, Ola Kvalvaag, Dillon Nall, Ethan Sanders, Alec Vest, Shaun Westlund, Matthew Hardison, Clinton R. Paden, Duncan MacCannell |
| EPI_ISL_2204656 | Houston Methodist Hospital | Houston Methodist Hospital | Randall J. Olsen, Paul A. Christensen, S. Wesley Long, Sishir Subedi, Robert Olson, Marcus Nguyen, James J. Davis, Matthew Ojeda Saavedra, Prasanti Yerramilli, Layne Pruitt, Kristina Reppond, Madison N. Shyer, Jessica Cambric, Ryan Gadd, Ilya J. Finkelstein, Jimmy Gollihar, and James M. Musser |
| EPI_ISL_2204657 | QLabs | WVU and Marshall University Combined Genomics Core Facilities | James Denvir, Peter Stoilov, Peter Perrotta, Wesley Kimble, Ryan Percifield |
| EPI_ISL_2204659 | Houston Methodist Hospital | Houston Methodist Hospital | Randall J. Olsen, Paul A. Christensen, S. Wesley Long, Sishir Subedi, Robert Olson, Marcus Nguyen, James J. Davis, Matthew Ojeda Saavedra, Prasanti Yerramilli, Layne Pruitt, Kristina Reppond, Madison N. Shyer, Jessica Cambric, Ryan Gadd, Ilya J. Finkelstein, Jimmy Gollihar, and James M. Musser |
| EPI_ISL_2204660 | QLabs | WVU and Marshall University Combined Genomics Core Facilities | James Denvir, Peter Stoilov, Peter Perrotta, Wesley Kimble, Ryan Percifield |
| EPI_ISL_2204662 | Houston Methodist Hospital | Houston Methodist Hospital | Randall J. Olsen, Paul A. Christensen, S. Wesley Long, Sishir Subedi, Robert Olson, Marcus Nguyen, James J. Davis, Matthew Ojeda Saavedra, Prasanti Yerramilli, Layne Pruitt, Kristina Reppond, Madison N. Shyer, Jessica Cambric, Ryan Gadd, Ilya J. Finkelstein, Jimmy Gollihar, and James M. Musser |
| EPI_ISL_2204663 | Aegis Sciences Corporation | Centers for Disease Control and Prevention Division of Viral Diseases, Pathogen Discovery | Dakota Howard, Dhwani Batra, Peter W. Cook, Kara Moser, Adrian Paskey, Jason Caravas, Benjamin Rambo-Martin, Shatavia Morrison, Christopher Gulvick, Scott Sammons, Yvette Unoarumhi, Darlene Wagner, Matthew Schmerer, Cyndi Clark, Patrick Campbell, Rob Case, Vikramsinha Ghorpade, Holly Houdeshell, Ola Kvalvaag, Dillon Nall, Ethan Sanders, Alec Vest, Shaun Westlund, Matthew Hardison, Clinton R. Paden, Duncan MacCannell |
| EPI_ISL_2204664 | QLabs | WVU and Marshall University Combined Genomics Core Facilities | James Denvir, Peter Stoilov, Peter Perrotta, Wesley Kimble, Ryan Percifield |
| EPI_ISL_2204665 | Aegis Sciences Corporation | Centers for Disease Control and Prevention Division of Viral Diseases, Pathogen Discovery | Dakota Howard, Dhwani Batra, Peter W. Cook, Kara Moser, Adrian Paskey, Jason Caravas, Benjamin Rambo-Martin, Shatavia Morrison, Christopher Gulvick, Scott Sammons, Yvette Unoarumhi, Darlene Wagner, Matthew Schmerer, Cyndi Clark, Patrick Campbell, Rob Case, Vikramsinha Ghorpade, Holly Houdeshell, Ola Kvalvaag, Dillon Nall, Ethan Sanders, Alec Vest, Shaun Westlund, Matthew Hardison, Clinton R. Paden, Duncan MacCannell |
| EPI_ISL_2204666 | QLabs | WVU and Marshall University Combined Genomics Core Facilities | James Denvir, Peter Stoilov, Peter Perrotta, Wesley Kimble, Ryan Percifield |
| EPI_ISL_2204667 | Houston Methodist Hospital | Houston Methodist Hospital | Randall J. Olsen, Paul A. Christensen, S. Wesley Long, Sishir Subedi, Robert Olson, Marcus Nguyen, James J. Davis, Matthew Ojeda Saavedra, Prasanti Yerramilli, Layne Pruitt, Kristina Reppond, Madison N. Shyer, Jessica Cambric, Ryan Gadd, Ilya J. Finkelstein, Jimmy Gollihar, and James M. Musser |
| EPI_ISL_2204668, EPI_ISL_2204669 | Aegis Sciences Corporation | Centers for Disease Control and Prevention Division of Viral Diseases, Pathogen Discovery | Dakota Howard, Dhwani Batra, Peter W. Cook, Kara Moser, Adrian Paskey, Jason Caravas, Benjamin Rambo-Martin, Shatavia Morrison, Christopher Gulvick, Scott Sammons, Yvette Unoarumhi, Darlene Wagner, Matthew Schmerer, Cyndi Clark, Patrick Campbell, Rob Case, Vikramsinha Ghorpade, Holly Houdeshell, Ola Kvalvaag, Dillon Nall, Ethan Sanders, Alec Vest, Shaun Westlund, Matthew Hardison, Clinton R. Paden, Duncan MacCannell |
| EPI_ISL_2204670 | QLabs | WVU and Marshall University Combined Genomics Core Facilities | James Denvir, Peter Stoilov, Peter Perrotta, Wesley Kimble, Ryan Percifield |
| EPI_ISL_2204672 | Aegis Sciences Corporation | Centers for Disease Control and Prevention Division of Viral Diseases, Pathogen Discovery | Dakota Howard, Dhwani Batra, Peter W. Cook, Kara Moser, Adrian Paskey, Jason Caravas, Benjamin Rambo-Martin, Shatavia Morrison, Christopher Gulvick, Scott Sammons, Yvette Unoarumhi, Darlene Wagner, Matthew Schmerer, Cyndi Clark, Patrick Campbell, Rob Case, Vikramsinha Ghorpade, Holly Houdeshell, Ola Kvalvaag, Dillon Nall, Ethan Sanders, Alec Vest, Shaun Westlund, Matthew Hardison, Clinton R. Paden, Duncan MacCannell |
| EPI_ISL_2204673, EPI_ISL_2204674 | QLabs | WVU and Marshall University Combined Genomics Core Facilities | James Denvir, Peter Stoilov, Peter Perrotta, Wesley Kimble, Ryan Percifield |
| EPI_ISL_2204675 | Aegis Sciences Corporation | Centers for Disease Control and Prevention Division of Viral Diseases, Pathogen | Dakota Howard, Dhwani Batra, Peter W. Cook, Kara Moser, Adrian Paskey, Jason Caravas, Benjamin Rambo-Martin, Shatavia Morrison, Christopher Gulvick, Scott Sammons, Yvette Unoarumhi, Darlene Wagner, Matthew Schmerer, Cyndi Clark, Patrick Campbell, Rob Case, Vikramsinha Ghorpade, Holly Houdeshell, Ola Kvalvaag, Dillon Nall, Ethan Sanders, Alec Vest, Shaun Westlund, Matthew Hardison, Clinton R. Paden, Duncan MacCannell |

| | | | |
|---|---|---|---|
| | | Discovery | |
| EPI_ISL_2204676 | Houston Methodist Hospital | Houston Methodist Hospital | Randall J. Olsen, Paul A. Christensen, S. Wesley Long, Sishir Subedi, Robert Olson, Marcus Nguyen, James J. Davis, Matthew Ojeda Saavedra, Prasanti Yerramilli, Layne Pruitt, Kristina Reppond, Madison N. Shyer, Jessica Cambric, Ryan Gadd, Ilya J. Finkelstein, Jimmy Gollihar, and James M. Musser |
| EPI_ISL_2204677 | QLabs | WVU and Marshall University Combined Genomics Core Facilities | James Denvir, Peter Stoilov, Peter Perrotta, Wesley Kimble, Ryan Percifield |
| EPI_ISL_2204678 | Aegis Sciences Corporation | Centers for Disease Control and Prevention Division of Viral Diseases, Pathogen Discovery | Dakota Howard, Dhwani Batra, Peter W. Cook, Kara Moser, Adrian Paskey, Jason Caravas, Benjamin Rambo-Martin, Shatavia Morrison, Christopher Gulvick, Scott Sammons, Yvette Unoarumhi, Darlene Wagner, Matthew Schmerer, Cyndi Clark, Patrick Campbell, Rob Case, Vikramsinha Ghorpade, Holly Houdeshell, Ola Kvalvaag, Dillon Nall, Ethan Sanders, Alec Vest, Shaun Westlund, Matthew Hardison, Clinton R. Paden, Duncan MacCannell |
| EPI_ISL_2204679 | QLabs | WVU and Marshall University Combined Genomics Core Facilities | James Denvir, Peter Stoilov, Peter Perrotta, Wesley Kimble, Ryan Percifield |
| EPI_ISL_2204680 | Houston Methodist Hospital | Houston Methodist Hospital | Randall J. Olsen, Paul A. Christensen, S. Wesley Long, Sishir Subedi, Robert Olson, Marcus Nguyen, James J. Davis, Matthew Ojeda Saavedra, Prasanti Yerramilli, Layne Pruitt, Kristina Reppond, Madison N. Shyer, Jessica Cambric, Ryan Gadd, Ilya J. Finkelstein, Jimmy Gollihar, and James M. Musser |
| EPI_ISL_2204681 | Aegis Sciences Corporation | Centers for Disease Control and Prevention Division of Viral Diseases, Pathogen Discovery | Dakota Howard, Dhwani Batra, Peter W. Cook, Kara Moser, Adrian Paskey, Jason Caravas, Benjamin Rambo-Martin, Shatavia Morrison, Christopher Gulvick, Scott Sammons, Yvette Unoarumhi, Darlene Wagner, Matthew Schmerer, Cyndi Clark, Patrick Campbell, Rob Case, Vikramsinha Ghorpade, Holly Houdeshell, Ola Kvalvaag, Dillon Nall, Ethan Sanders, Alec Vest, Shaun Westlund, Matthew Hardison, Clinton R. Paden, Duncan MacCannell |
| EPI_ISL_2204682 | QLabs | WVU and Marshall University Combined Genomics Core Facilities | James Denvir, Peter Stoilov, Peter Perrotta, Wesley Kimble, Ryan Percifield |
| EPI_ISL_2204683 | Aegis Sciences Corporation | Centers for Disease Control and Prevention Division of Viral Diseases, Pathogen Discovery | Dakota Howard, Dhwani Batra, Peter W. Cook, Kara Moser, Adrian Paskey, Jason Caravas, Benjamin Rambo-Martin, Shatavia Morrison, Christopher Gulvick, Scott Sammons, Yvette Unoarumhi, Darlene Wagner, Matthew Schmerer, Cyndi Clark, Patrick Campbell, Rob Case, Vikramsinha Ghorpade, Holly Houdeshell, Ola Kvalvaag, Dillon Nall, Ethan Sanders, Alec Vest, Shaun Westlund, Matthew Hardison, Clinton R. Paden, Duncan MacCannell |
| EPI_ISL_2204684 | Houston Methodist Hospital | Houston Methodist Hospital | Randall J. Olsen, Paul A. Christensen, S. Wesley Long, Sishir Subedi, Robert Olson, Marcus Nguyen, James J. Davis, Matthew Ojeda Saavedra, Prasanti Yerramilli, Layne Pruitt, Kristina Reppond, Madison N. Shyer, Jessica Cambric, Ryan Gadd, Ilya J. Finkelstein, Jimmy Gollihar, and James M. Musser |
| EPI_ISL_2204685 | QLabs | WVU and Marshall University Combined Genomics Core Facilities | James Denvir, Peter Stoilov, Peter Perrotta, Wesley Kimble, Ryan Percifield |
| EPI_ISL_2204688 | Houston Methodist Hospital | Houston Methodist Hospital | Randall J. Olsen, Paul A. Christensen, S. Wesley Long, Sishir Subedi, Robert Olson, Marcus Nguyen, James J. Davis, Matthew Ojeda Saavedra, Prasanti Yerramilli, Layne Pruitt, Kristina Reppond, Madison N. Shyer, Jessica Cambric, Ryan Gadd, Ilya J. Finkelstein, Jimmy Gollihar, and James M. Musser |
| EPI_ISL_2204690 | Aegis Sciences Corporation | Centers for Disease Control and Prevention Division of Viral Diseases, Pathogen Discovery | Dakota Howard, Dhwani Batra, Peter W. Cook, Kara Moser, Adrian Paskey, Jason Caravas, Benjamin Rambo-Martin, Shatavia Morrison, Christopher Gulvick, Scott Sammons, Yvette Unoarumhi, Darlene Wagner, Matthew Schmerer, Cyndi Clark, Patrick Campbell, Rob Case, Vikramsinha Ghorpade, Holly Houdeshell, Ola Kvalvaag, Dillon Nall, Ethan Sanders, Alec Vest, Shaun Westlund, Matthew Hardison, Clinton R. Paden, Duncan MacCannell |
| EPI_ISL_2204691 | QLabs | WVU and Marshall University Combined Genomics Core Facilities | James Denvir, Peter Stoilov, Peter Perrotta, Wesley Kimble, Ryan Percifield |
| EPI_ISL_2204692 | Houston Methodist Hospital | Houston Methodist Hospital | Randall J. Olsen, Paul A. Christensen, S. Wesley Long, Sishir Subedi, Robert Olson, Marcus Nguyen, James J. Davis, Matthew Ojeda Saavedra, Prasanti Yerramilli, Layne Pruitt, Kristina Reppond, Madison N. Shyer, Jessica Cambric, Ryan Gadd, Ilya J. Finkelstein, Jimmy Gollihar, and James M. Musser |
| EPI_ISL_2204693 | Aegis Sciences Corporation | Centers for Disease Control and Prevention Division of Viral Diseases, Pathogen Discovery | Dakota Howard, Dhwani Batra, Peter W. Cook, Kara Moser, Adrian Paskey, Jason Caravas, Benjamin Rambo-Martin, Shatavia Morrison, Christopher Gulvick, Scott Sammons, Yvette Unoarumhi, Darlene Wagner, Matthew Schmerer, Cyndi Clark, Patrick Campbell, Rob Case, Vikramsinha Ghorpade, Holly Houdeshell, Ola Kvalvaag, Dillon Nall, Ethan Sanders, Alec Vest, Shaun Westlund, Matthew Hardison, Clinton R. Paden, Duncan MacCannell |
| EPI_ISL_2204694, EPI_ISL_2204696 | QLabs | WVU and Marshall University Combined Genomics Core Facilities | James Denvir, Peter Stoilov, Peter Perrotta, Wesley Kimble, Ryan Percifield |
| EPI_ISL_2204697 | Aegis Sciences Corporation | Centers for Disease Control and Prevention Division of Viral Diseases, Pathogen Discovery | Dakota Howard, Dhwani Batra, Peter W. Cook, Kara Moser, Adrian Paskey, Jason Caravas, Benjamin Rambo-Martin, Shatavia Morrison, Christopher Gulvick, Scott Sammons, Yvette Unoarumhi, Darlene Wagner, Matthew Schmerer, Cyndi Clark, Patrick Campbell, Rob Case, Vikramsinha Ghorpade, Holly Houdeshell, Ola Kvalvaag, Dillon Nall, Ethan Sanders, Alec Vest, Shaun Westlund, Matthew Hardison, Clinton R. Paden, Duncan MacCannell |
| EPI_ISL_2204698 | QLabs | WVU and Marshall University Combined Genomics Core Facilities | James Denvir, Peter Stoilov, Peter Perrotta, Wesley Kimble, Ryan Percifield |
| EPI_ISL_2204699 | Houston Methodist Hospital | Houston Methodist Hospital | Randall J. Olsen, Paul A. Christensen, S. Wesley Long, Sishir Subedi, Robert Olson, Marcus Nguyen, James J. Davis, Matthew Ojeda Saavedra, Prasanti Yerramilli, Layne Pruitt, Kristina Reppond, Madison N. Shyer, Jessica Cambric, Ryan Gadd, Ilya J. Finkelstein, Jimmy Gollihar, and James M. Musser |
| EPI_ISL_2204700 | Aegis Sciences Corporation | Centers for Disease Control and Prevention Division of Viral Diseases, Pathogen Discovery | Dakota Howard, Dhwani Batra, Peter W. Cook, Kara Moser, Adrian Paskey, Jason Caravas, Benjamin Rambo-Martin, Shatavia Morrison, Christopher Gulvick, Scott Sammons, Yvette Unoarumhi, Darlene Wagner, Matthew Schmerer, Cyndi Clark, Patrick Campbell, Rob Case, Vikramsinha Ghorpade, Holly Houdeshell, Ola Kvalvaag, Dillon Nall, Ethan Sanders, Alec Vest, Shaun Westlund, Matthew Hardison, Clinton R. Paden, Duncan MacCannell |
| EPI_ISL_2204701, EPI_ISL_2204702 | QLabs | WVU and Marshall University Combined Genomics Core Facilities | James Denvir, Peter Stoilov, Peter Perrotta, Wesley Kimble, Ryan Percifield |
| EPI_ISL_2204703 | Aegis Sciences Corporation | Centers for Disease Control and Prevention Division of Viral Diseases, Pathogen Discovery | Dakota Howard, Dhwani Batra, Peter W. Cook, Kara Moser, Adrian Paskey, Jason Caravas, Benjamin Rambo-Martin, Shatavia Morrison, Christopher Gulvick, Scott Sammons, Yvette Unoarumhi, Darlene Wagner, Matthew Schmerer, Cyndi Clark, Patrick Campbell, Rob Case, Vikramsinha Ghorpade, Holly Houdeshell, Ola Kvalvaag, Dillon Nall, Ethan Sanders, Alec Vest, Shaun Westlund, Matthew Hardison, Clinton R. Paden, Duncan MacCannell |
| EPI_ISL_2204704 | Houston Methodist Hospital | Houston Methodist Hospital | Randall J. Olsen, Paul A. Christensen, S. Wesley Long, Sishir Subedi, Robert Olson, Marcus Nguyen, James J. Davis, Matthew Ojeda Saavedra, Prasanti Yerramilli, Layne Pruitt, Kristina Reppond, Madison N. Shyer, Jessica Cambric, Ryan Gadd, Ilya J. Finkelstein, Jimmy Gollihar, and James M. Musser |
| EPI_ISL_2204705 | Aegis Sciences Corporation | Centers for Disease Control and Prevention Division of Viral Diseases, Pathogen Discovery | Dakota Howard, Dhwani Batra, Peter W. Cook, Kara Moser, Adrian Paskey, Jason Caravas, Benjamin Rambo-Martin, Shatavia Morrison, Christopher Gulvick, Scott Sammons, Yvette Unoarumhi, Darlene Wagner, Matthew Schmerer, Cyndi Clark, Patrick Campbell, Rob Case, Vikramsinha Ghorpade, Holly Houdeshell, Ola Kvalvaag, Dillon Nall, Ethan Sanders, Alec Vest, Shaun Westlund, Matthew Hardison, Clinton R. Paden, Duncan MacCannell |
| EPI_ISL_2204706 | Houston Methodist Hospital | Houston Methodist Hospital | Randall J. Olsen, Paul A. Christensen, S. Wesley Long, Sishir Subedi, Robert Olson, Marcus Nguyen, James J. Davis, Matthew Ojeda Saavedra, Prasanti Yerramilli, Layne Pruitt, Kristina Reppond, Madison N. Shyer, Jessica Cambric, Ryan Gadd, Ilya J. Finkelstein, Jimmy Gollihar, and James M. Musser |
| EPI_ISL_2204707 | QLabs | WVU and Marshall University Combined Genomics Core Facilities | James Denvir, Peter Stoilov, Peter Perrotta, Wesley Kimble, Ryan Percifield |
| EPI_ISL_2204708 | Aegis Sciences Corporation | Centers for Disease Control and Prevention Division of Viral Diseases, Pathogen Discovery | Dakota Howard, Dhwani Batra, Peter W. Cook, Kara Moser, Adrian Paskey, Jason Caravas, Benjamin Rambo-Martin, Shatavia Morrison, Christopher Gulvick, Scott Sammons, Yvette Unoarumhi, Darlene Wagner, Matthew Schmerer, Cyndi Clark, Patrick Campbell, Rob Case, Vikramsinha Ghorpade, Holly Houdeshell, Ola Kvalvaag, Dillon Nall, Ethan Sanders, Alec Vest, Shaun Westlund, Matthew Hardison, Clinton R. Paden, Duncan MacCannell |
| EPI_ISL_2204709 | Houston Methodist Hospital | Houston Methodist Hospital | Randall J. Olsen, Paul A. Christensen, S. Wesley Long, Sishir Subedi, Robert Olson, Marcus Nguyen, James J. Davis, Matthew Ojeda Saavedra, Prasanti Yerramilli, Layne Pruitt, Kristina Reppond, Madison N. Shyer, Jessica Cambric, Ryan Gadd, Ilya J. Finkelstein, Jimmy Gollihar, and James M. Musser |
| EPI_ISL_2204710 | Summersville Regiol Medical Center | WVU and Marshall University Combined Genomics Core Facilities | James Denvir, Peter Stoilov, Peter Perrotta, Wesley Kimble, Ryan Percifield |
| EPI_ISL_2204711 | Aegis Sciences Corporation | Centers for Disease Control and Prevention Division of Viral Diseases, Pathogen Discovery | Dakota Howard, Dhwani Batra, Peter W. Cook, Kara Moser, Adrian Paskey, Jason Caravas, Benjamin Rambo-Martin, Shatavia Morrison, Christopher Gulvick, Scott Sammons, Yvette Unoarumhi, Darlene Wagner, Matthew Schmerer, Cyndi Clark, Patrick Campbell, Rob Case, Vikramsinha Ghorpade, Holly Houdeshell, Ola Kvalvaag, Dillon Nall, Ethan Sanders, Alec Vest, Shaun Westlund, Matthew Hardison, Clinton R. Paden, Duncan MacCannell |
| EPI_ISL_2204712 | Summersville Regiol Medical Center | WVU and Marshall University Combined Genomics Core Facilities | James Denvir, Peter Stoilov, Peter Perrotta, Wesley Kimble, Ryan Percifield |
| EPI_ISL_2204713 | Houston Methodist Hospital | Houston Methodist Hospital | Randall J. Olsen, Paul A. Christensen, S. Wesley Long, Sishir Subedi, Robert Olson, Marcus Nguyen, James J. Davis, Matthew Ojeda Saavedra, Prasanti Yerramilli, Layne Pruitt, Kristina Reppond, Madison N. Shyer, Jessica Cambric, Ryan Gadd, Ilya J. Finkelstein, Jimmy Gollihar, and James M. Musser |
| EPI_ISL_2204714 | Summersville Regiol Medical Center | WVU and Marshall University Combined Genomics Core Facilities | James Denvir, Peter Stoilov, Peter Perrotta, Wesley Kimble, Ryan Percifield |
| EPI_ISL_2204715, EPI_ISL_2204716 | Aegis Sciences Corporation | Centers for Disease Control and Prevention Division of Viral Diseases, Pathogen Discovery | Dakota Howard, Dhwani Batra, Peter W. Cook, Kara Moser, Adrian Paskey, Jason Caravas, Benjamin Rambo-Martin, Shatavia Morrison, Christopher Gulvick, Scott Sammons, Yvette Unoarumhi, Darlene Wagner, Matthew Schmerer, Cyndi Clark, Patrick Campbell, Rob Case, Vikramsinha Ghorpade, Holly Houdeshell, Ola Kvalvaag, Dillon Nall, Ethan Sanders, Alec Vest, Shaun Westlund, Matthew Hardison, Clinton R. Paden, Duncan MacCannell |
| EPI_ISL_2204717, EPI_ISL_2204718 | Houston Methodist Hospital | Houston Methodist Hospital | Randall J. Olsen, Paul A. Christensen, S. Wesley Long, Sishir Subedi, Robert Olson, Marcus Nguyen, James J. Davis, Matthew Ojeda Saavedra, Prasanti Yerramilli, Layne Pruitt, Kristina Reppond, Madison N. Shyer, Jessica Cambric, Ryan Gadd, Ilya J. Finkelstein, Jimmy Gollihar, and James M. Musser |
| EPI_ISL_2204719, EPI_ISL_2204721 | Summersville Regiol Medical Center | WVU and Marshall University Combined Genomics Core Facilities | James Denvir, Peter Stoilov, Peter Perrotta, Wesley Kimble, Ryan Percifield |
| EPI_ISL_2204722 | Aegis Sciences Corporation | Centers for Disease Control and Prevention Division of Viral Diseases, Pathogen Discovery | Dakota Howard, Dhwani Batra, Peter W. Cook, Kara Moser, Adrian Paskey, Jason Caravas, Benjamin Rambo-Martin, Shatavia Morrison, Christopher Gulvick, Scott Sammons, Yvette Unoarumhi, Darlene Wagner, Matthew Schmerer, Cyndi Clark, Patrick Campbell, Rob Case, Vikramsinha Ghorpade, Holly Houdeshell, Ola Kvalvaag, Dillon Nall, Ethan Sanders, Alec Vest, Shaun Westlund, Matthew Hardison, Clinton R. Paden, Duncan MacCannell |
| EPI_ISL_2204723 | Houston Methodist Hospital | Houston Methodist Hospital | Randall J. Olsen, Paul A. Christensen, S. Wesley Long, Sishir Subedi, Robert Olson, Marcus Nguyen, James J. Davis, Matthew Ojeda Saavedra, Prasanti Yerramilli, Layne Pruitt, Kristina Reppond, Madison N. Shyer, Jessica Cambric, Ryan Gadd, Ilya J. Finkelstein, Jimmy Gollihar, and James M. Musser |
| EPI_ISL_2204724 | Summersville Regiol Medical Center | WVU and Marshall University Combined Genomics Core Facilities | James Denvir, Peter Stoilov, Peter Perrotta, Wesley Kimble, Ryan Percifield |

| | | | |
|---|---|---|---|
| EPI_ISL_2204725 | Aegis Sciences Corporation | Centers for Disease Control and Prevention Division of Viral Diseases, Pathogen Discovery | Dakota Howard, Dhwani Batra, Peter W. Cook, Kara Moser, Adrian Paskey, Jason Caravas, Benjamin Rambo-Martin, Shatavia Morrison, Christopher Gulvick, Scott Sammons, Yvette Unoarumhi, Darlene Wagner, Matthew Schmerer, Cyndi Clark, Patrick Campbell, Rob Case, Vikramsinha Ghorpade, Holly Houdeshell, Ola Kvalvaag, Dillon Nall, Ethan Sanders, Alec Vest, Shaun Westlund, Matthew Hardison, Clinton R. Paden, Duncan MacCannell |
| EPI_ISL_2204726 | United Hospital Center | WVU and Marshall University Combined Genomics Core Facilities | James Denvir, Peter Stoilov, Peter Perrotta, Wesley Kimble, Ryan Percifield |
| EPI_ISL_2204727 | Houston Methodist Hospital | Houston Methodist Hospital | Randall J. Olsen, Paul A. Christensen, S. Wesley Long, Sishir Subedi, Robert Olson, Marcus Nguyen, James J. Davis, Matthew Ojeda Saavedra, Prasanti Yerramilli, Layne Pruitt, Kristina Reppond, Madison N. Shyer, Jessica Cambric, Ryan Gadd, Ilya J. Finkelstein, Jimmy Gollihar, and James M. Musser |
| EPI_ISL_2204728 | BMC LAB | WVU and Marshall University Combined Genomics Core Facilities | James Denvir, Peter Stoilov, Peter Perrotta, Wesley Kimble, Ryan Percifield |
| EPI_ISL_2204729 | Aegis Sciences Corporation | Centers for Disease Control and Prevention Division of Viral Diseases, Pathogen Discovery | Dakota Howard, Dhwani Batra, Peter W. Cook, Kara Moser, Adrian Paskey, Jason Caravas, Benjamin Rambo-Martin, Shatavia Morrison, Christopher Gulvick, Scott Sammons, Yvette Unoarumhi, Darlene Wagner, Matthew Schmerer, Cyndi Clark, Patrick Campbell, Rob Case, Vikramsinha Ghorpade, Holly Houdeshell, Ola Kvalvaag, Dillon Nall, Ethan Sanders, Alec Vest, Shaun Westlund, Matthew Hardison, Clinton R. Paden, Duncan MacCannell |
| EPI_ISL_2204730 | Houston Methodist Hospital | Houston Methodist Hospital | Randall J. Olsen, Paul A. Christensen, S. Wesley Long, Sishir Subedi, Robert Olson, Marcus Nguyen, James J. Davis, Matthew Ojeda Saavedra, Prasanti Yerramilli, Layne Pruitt, Kristina Reppond, Madison N. Shyer, Jessica Cambric, Ryan Gadd, Ilya J. Finkelstein, Jimmy Gollihar, and James M. Musser |
| EPI_ISL_2204731 | Aegis Sciences Corporation | Centers for Disease Control and Prevention Division of Viral Diseases, Pathogen Discovery | Dakota Howard, Dhwani Batra, Peter W. Cook, Kara Moser, Adrian Paskey, Jason Caravas, Benjamin Rambo-Martin, Shatavia Morrison, Christopher Gulvick, Scott Sammons, Yvette Unoarumhi, Darlene Wagner, Matthew Schmerer, Cyndi Clark, Patrick Campbell, Rob Case, Vikramsinha Ghorpade, Holly Houdeshell, Ola Kvalvaag, Dillon Nall, Ethan Sanders, Alec Vest, Shaun Westlund, Matthew Hardison, Clinton R. Paden, Duncan MacCannell |
| EPI_ISL_2204732 | BMC LAB | WVU and Marshall University Combined Genomics Core Facilities | James Denvir, Peter Stoilov, Peter Perrotta, Wesley Kimble, Ryan Percifield |
| EPI_ISL_2204733 | Houston Methodist Hospital | Houston Methodist Hospital | Randall J. Olsen, Paul A. Christensen, S. Wesley Long, Sishir Subedi, Robert Olson, Marcus Nguyen, James J. Davis, Matthew Ojeda Saavedra, Prasanti Yerramilli, Layne Pruitt, Kristina Reppond, Madison N. Shyer, Jessica Cambric, Ryan Gadd, Ilya J. Finkelstein, Jimmy Gollihar, and James M. Musser |
| EPI_ISL_2204734, EPI_ISL_2204736 | Aegis Sciences Corporation | Centers for Disease Control and Prevention Division of Viral Diseases, Pathogen Discovery | Dakota Howard, Dhwani Batra, Peter W. Cook, Kara Moser, Adrian Paskey, Jason Caravas, Benjamin Rambo-Martin, Shatavia Morrison, Christopher Gulvick, Scott Sammons, Yvette Unoarumhi, Darlene Wagner, Matthew Schmerer, Cyndi Clark, Patrick Campbell, Rob Case, Vikramsinha Ghorpade, Holly Houdeshell, Ola Kvalvaag, Dillon Nall, Ethan Sanders, Alec Vest, Shaun Westlund, Matthew Hardison, Clinton R. Paden, Duncan MacCannell |
| EPI_ISL_2204737 | Houston Methodist Hospital | Houston Methodist Hospital | Randall J. Olsen, Paul A. Christensen, S. Wesley Long, Sishir Subedi, Robert Olson, Marcus Nguyen, James J. Davis, Matthew Ojeda Saavedra, Prasanti Yerramilli, Layne Pruitt, Kristina Reppond, Madison N. Shyer, Jessica Cambric, Ryan Gadd, Ilya J. Finkelstein, Jimmy Gollihar, and James M. Musser |
| EPI_ISL_2204738 | BMC LAB | WVU and Marshall University Combined Genomics Core Facilities | James Denvir, Peter Stoilov, Peter Perrotta, Wesley Kimble, Ryan Percifield |
| EPI_ISL_2204739 | Aegis Sciences Corporation | Centers for Disease Control and Prevention Division of Viral Diseases, Pathogen Discovery | Dakota Howard, Dhwani Batra, Peter W. Cook, Kara Moser, Adrian Paskey, Jason Caravas, Benjamin Rambo-Martin, Shatavia Morrison, Christopher Gulvick, Scott Sammons, Yvette Unoarumhi, Darlene Wagner, Matthew Schmerer, Cyndi Clark, Patrick Campbell, Rob Case, Vikramsinha Ghorpade, Holly Houdeshell, Ola Kvalvaag, Dillon Nall, Ethan Sanders, Alec Vest, Shaun Westlund, Matthew Hardison, Clinton R. Paden, Duncan MacCannell |
| EPI_ISL_2204740 | Houston Methodist Hospital | Houston Methodist Hospital | Randall J. Olsen, Paul A. Christensen, S. Wesley Long, Sishir Subedi, Robert Olson, Marcus Nguyen, James J. Davis, Matthew Ojeda Saavedra, Prasanti Yerramilli, Layne Pruitt, Kristina Reppond, Madison N. Shyer, Jessica Cambric, Ryan Gadd, Ilya J. Finkelstein, Jimmy Gollihar, and James M. Musser |
| EPI_ISL_2204741 | BMC LAB | WVU and Marshall University Combined Genomics Core Facilities | James Denvir, Peter Stoilov, Peter Perrotta, Wesley Kimble, Ryan Percifield |
| EPI_ISL_2204742 | Aegis Sciences Corporation | Centers for Disease Control and Prevention Division of Viral Diseases, Pathogen Discovery | Dakota Howard, Dhwani Batra, Peter W. Cook, Kara Moser, Adrian Paskey, Jason Caravas, Benjamin Rambo-Martin, Shatavia Morrison, Christopher Gulvick, Scott Sammons, Yvette Unoarumhi, Darlene Wagner, Matthew Schmerer, Cyndi Clark, Patrick Campbell, Rob Case, Vikramsinha Ghorpade, Holly Houdeshell, Ola Kvalvaag, Dillon Nall, Ethan Sanders, Alec Vest, Shaun Westlund, Matthew Hardison, Clinton R. Paden, Duncan MacCannell |
| EPI_ISL_2204743 | Houston Methodist Hospital | Houston Methodist Hospital | Randall J. Olsen, Paul A. Christensen, S. Wesley Long, Sishir Subedi, Robert Olson, Marcus Nguyen, James J. Davis, Matthew Ojeda Saavedra, Prasanti Yerramilli, Layne Pruitt, Kristina Reppond, Madison N. Shyer, Jessica Cambric, Ryan Gadd, Ilya J. Finkelstein, Jimmy Gollihar, and James M. Musser |
| EPI_ISL_2204744, EPI_ISL_2204745 | BMC LAB | WVU and Marshall University Combined Genomics Core Facilities | James Denvir, Peter Stoilov, Peter Perrotta, Wesley Kimble, Ryan Percifield |
| EPI_ISL_2204746 | Aegis Sciences Corporation | Centers for Disease Control and Prevention Division of Viral Diseases, Pathogen Discovery | Dakota Howard, Dhwani Batra, Peter W. Cook, Kara Moser, Adrian Paskey, Jason Caravas, Benjamin Rambo-Martin, Shatavia Morrison, Christopher Gulvick, Scott Sammons, Yvette Unoarumhi, Darlene Wagner, Matthew Schmerer, Cyndi Clark, Patrick Campbell, Rob Case, Vikramsinha Ghorpade, Holly Houdeshell, Ola Kvalvaag, Dillon Nall, Ethan Sanders, Alec Vest, Shaun Westlund, Matthew Hardison, Clinton R. Paden, Duncan MacCannell |
| EPI_ISL_2204747 | Houston Methodist Hospital | Houston Methodist Hospital | Randall J. Olsen, Paul A. Christensen, S. Wesley Long, Sishir Subedi, Robert Olson, Marcus Nguyen, James J. Davis, Matthew Ojeda Saavedra, Prasanti Yerramilli, Layne Pruitt, Kristina Reppond, Madison N. Shyer, Jessica Cambric, Ryan Gadd, Ilya J. Finkelstein, Jimmy Gollihar, and James M. Musser |
| EPI_ISL_2204748 | Berkeley Medical Center | WVU and Marshall University Combined Genomics Core Facilities | James Denvir, Peter Stoilov, Peter Perrotta, Wesley Kimble, Ryan Percifield |
| EPI_ISL_2204749 | Aegis Sciences Corporation | Centers for Disease Control and Prevention Division of Viral Diseases, Pathogen Discovery | Dakota Howard, Dhwani Batra, Peter W. Cook, Kara Moser, Adrian Paskey, Jason Caravas, Benjamin Rambo-Martin, Shatavia Morrison, Christopher Gulvick, Scott Sammons, Yvette Unoarumhi, Darlene Wagner, Matthew Schmerer, Cyndi Clark, Patrick Campbell, Rob Case, Vikramsinha Ghorpade, Holly Houdeshell, Ola Kvalvaag, Dillon Nall, Ethan Sanders, Alec Vest, Shaun Westlund, Matthew Hardison, Clinton R. Paden, Duncan MacCannell |
| EPI_ISL_2204750 | Berkeley Medical Center | WVU and Marshall University Combined Genomics Core Facilities | James Denvir, Peter Stoilov, Peter Perrotta, Wesley Kimble, Ryan Percifield |
| EPI_ISL_2204751 | Houston Methodist Hospital | Houston Methodist Hospital | Randall J. Olsen, Paul A. Christensen, S. Wesley Long, Sishir Subedi, Robert Olson, Marcus Nguyen, James J. Davis, Matthew Ojeda Saavedra, Prasanti Yerramilli, Layne Pruitt, Kristina Reppond, Madison N. Shyer, Jessica Cambric, Ryan Gadd, Ilya J. Finkelstein, Jimmy Gollihar, and James M. Musser |
| EPI_ISL_2204752 | Berkeley Medical Center | WVU and Marshall University Combined Genomics Core Facilities | James Denvir, Peter Stoilov, Peter Perrotta, Wesley Kimble, Ryan Percifield |
| EPI_ISL_2204753 | Aegis Sciences Corporation | Centers for Disease Control and Prevention Division of Viral Diseases, Pathogen Discovery | Dakota Howard, Dhwani Batra, Peter W. Cook, Kara Moser, Adrian Paskey, Jason Caravas, Benjamin Rambo-Martin, Shatavia Morrison, Christopher Gulvick, Scott Sammons, Yvette Unoarumhi, Darlene Wagner, Matthew Schmerer, Cyndi Clark, Patrick Campbell, Rob Case, Vikramsinha Ghorpade, Holly Houdeshell, Ola Kvalvaag, Dillon Nall, Ethan Sanders, Alec Vest, Shaun Westlund, Matthew Hardison, Clinton R. Paden, Duncan MacCannell |
| EPI_ISL_2204754 | Berkeley Medical Center | WVU and Marshall University Combined Genomics Core Facilities | James Denvir, Peter Stoilov, Peter Perrotta, Wesley Kimble, Ryan Percifield |
| EPI_ISL_2204755 | Houston Methodist Hospital | Houston Methodist Hospital | Randall J. Olsen, Paul A. Christensen, S. Wesley Long, Sishir Subedi, Robert Olson, Marcus Nguyen, James J. Davis, Matthew Ojeda Saavedra, Prasanti Yerramilli, Layne Pruitt, Kristina Reppond, Madison N. Shyer, Jessica Cambric, Ryan Gadd, Ilya J. Finkelstein, Jimmy Gollihar, and James M. Musser |
| EPI_ISL_2204756, EPI_ISL_2204758 | Berkeley Medical Center | WVU and Marshall University Combined Genomics Core Facilities | James Denvir, Peter Stoilov, Peter Perrotta, Wesley Kimble, Ryan Percifield |
| EPI_ISL_2204759 | Aegis Sciences Corporation | Centers for Disease Control and Prevention Division of Viral Diseases, Pathogen Discovery | Dakota Howard, Dhwani Batra, Peter W. Cook, Kara Moser, Adrian Paskey, Jason Caravas, Benjamin Rambo-Martin, Shatavia Morrison, Christopher Gulvick, Scott Sammons, Yvette Unoarumhi, Darlene Wagner, Matthew Schmerer, Cyndi Clark, Patrick Campbell, Rob Case, Vikramsinha Ghorpade, Holly Houdeshell, Ola Kvalvaag, Dillon Nall, Ethan Sanders, Alec Vest, Shaun Westlund, Matthew Hardison, Clinton R. Paden, Duncan MacCannell |
| EPI_ISL_2204760 | Houston Methodist Hospital | Houston Methodist Hospital | Randall J. Olsen, Paul A. Christensen, S. Wesley Long, Sishir Subedi, Robert Olson, Marcus Nguyen, James J. Davis, Matthew Ojeda Saavedra, Prasanti Yerramilli, Layne Pruitt, Kristina Reppond, Madison N. Shyer, Jessica Cambric, Ryan Gadd, Ilya J. Finkelstein, Jimmy Gollihar, and James M. Musser |
| EPI_ISL_2204761, EPI_ISL_2204762 | Berkeley Medical Center | WVU and Marshall University Combined Genomics Core Facilities | James Denvir, Peter Stoilov, Peter Perrotta, Wesley Kimble, Ryan Percifield |
| EPI_ISL_2204763, EPI_ISL_2204764 | Aegis Sciences Corporation | Centers for Disease Control and Prevention Division of Viral Diseases, Pathogen Discovery | Dakota Howard, Dhwani Batra, Peter W. Cook, Kara Moser, Adrian Paskey, Jason Caravas, Benjamin Rambo-Martin, Shatavia Morrison, Christopher Gulvick, Scott Sammons, Yvette Unoarumhi, Darlene Wagner, Matthew Schmerer, Cyndi Clark, Patrick Campbell, Rob Case, Vikramsinha Ghorpade, Holly Houdeshell, Ola Kvalvaag, Dillon Nall, Ethan Sanders, Alec Vest, Shaun Westlund, Matthew Hardison, Clinton R. Paden, Duncan MacCannell |
| EPI_ISL_2204765 | Houston Methodist Hospital | Houston Methodist Hospital | Randall J. Olsen, Paul A. Christensen, S. Wesley Long, Sishir Subedi, Robert Olson, Marcus Nguyen, James J. Davis, Matthew Ojeda Saavedra, Prasanti Yerramilli, Layne Pruitt, Kristina Reppond, Madison N. Shyer, Jessica Cambric, Ryan Gadd, Ilya J. Finkelstein, Jimmy Gollihar, and James M. Musser |
| EPI_ISL_2204766 | Berkeley Medical Center | WVU and Marshall University Combined Genomics Core Facilities | James Denvir, Peter Stoilov, Peter Perrotta, Wesley Kimble, Ryan Percifield |
| EPI_ISL_2204767 | Aegis Sciences Corporation | Centers for Disease Control and Prevention Division of Viral Diseases, Pathogen Discovery | Dakota Howard, Dhwani Batra, Peter W. Cook, Kara Moser, Adrian Paskey, Jason Caravas, Benjamin Rambo-Martin, Shatavia Morrison, Christopher Gulvick, Scott Sammons, Yvette Unoarumhi, Darlene Wagner, Matthew Schmerer, Cyndi Clark, Patrick Campbell, Rob Case, Vikramsinha Ghorpade, Holly Houdeshell, Ola Kvalvaag, Dillon Nall, Ethan Sanders, Alec Vest, Shaun Westlund, Matthew Hardison, Clinton R. Paden, Duncan MacCannell |
| EPI_ISL_2204768 | Houston Methodist Hospital | Houston Methodist Hospital | Randall J. Olsen, Paul A. Christensen, S. Wesley Long, Sishir Subedi, Robert Olson, Marcus Nguyen, James J. Davis, Matthew Ojeda Saavedra, Prasanti Yerramilli, Layne Pruitt, Kristina Reppond, Madison N. Shyer, Jessica Cambric, Ryan Gadd, Ilya J. Finkelstein, Jimmy Gollihar, and James M. Musser |
| EPI_ISL_2204769 | Berkeley Medical Center | WVU and Marshall University Combined Genomics Core Facilities | James Denvir, Peter Stoilov, Peter Perrotta, Wesley Kimble, Ryan Percifield |
| EPI_ISL_2204770 | Summersville Regiol Medical Center | WVU and Marshall University Combined Genomics Core Facilities | James Denvir, Peter Stoilov, Peter Perrotta, Wesley Kimble, Ryan Percifield |
| EPI_ISL_2204771 | Aegis Sciences Corporation | Centers for Disease Control and Prevention Division of Viral Diseases, Pathogen Discovery | Dakota Howard, Dhwani Batra, Peter W. Cook, Kara Moser, Adrian Paskey, Jason Caravas, Benjamin Rambo-Martin, Shatavia Morrison, Christopher Gulvick, Scott Sammons, Yvette Unoarumhi, Darlene Wagner, Matthew Schmerer, Cyndi Clark, Patrick Campbell, Rob Case, Vikramsinha Ghorpade, Holly Houdeshell, Ola Kvalvaag, Dillon Nall, Ethan Sanders, Alec Vest, Shaun Westlund, Matthew Hardison, Clinton R. Paden, Duncan MacCannell |

| | | | |
|---|---|---|---|
| EPI_ISL_2204772 | Berkeley Medical Center | WVU and Marshall University Combined Genomics Core Facilities | James Denvir, Peter Stoilov, Peter Perrotta, Wesley Kimble, Ryan Percifield |
| EPI_ISL_2204773 | Houston Methodist Hospital | Houston Methodist Hospital | Randall J. Olsen, Paul A. Christensen, S. Wesley Long, Sishir Subedi, Robert Olson, Marcus Nguyen, James J. Davis, Matthew Ojeda Saavedra, Prasanti Yerramilli, Layne Pruitt, Kristina Reppond, Madison N. Shyer, Jessica Cambric, Ryan Gadd, Ilya J. Finkelstein, Jimmy Gollihar, and James M. Musser |
| EPI_ISL_2204774 | Berkeley Medical Center | WVU and Marshall University Combined Genomics Core Facilities | James Denvir, Peter Stoilov, Peter Perrotta, Wesley Kimble, Ryan Percifield |
| EPI_ISL_2204775 | Aegis Sciences Corporation | Centers for Disease Control and Prevention Division of Viral Diseases, Pathogen Discovery | Dakota Howard, Dhwani Batra, Peter W. Cook, Kara Moser, Adrian Paskey, Jason Caravas, Benjamin Rambo-Martin, Shatavia Morrison, Christopher Gulvick, Scott Sammons, Yvette Unoarumhi, Darlene Wagner, Matthew Schmerer, Cyndi Clark, Patrick Campbell, Rob Case, Vikramsinha Ghorpade, Holly Houdeshell, Ola Kvalvaag, Dillon Nall, Ethan Sanders, Alec Vest, Shaun Westlund, Matthew Hardison, Clinton R. Paden, Duncan MacCannell |
| EPI_ISL_2204776 | Berkeley Medical Center | WVU and Marshall University Combined Genomics Core Facilities | James Denvir, Peter Stoilov, Peter Perrotta, Wesley Kimble, Ryan Percifield |
| EPI_ISL_2204778 | Aegis Sciences Corporation | Centers for Disease Control and Prevention Division of Viral Diseases, Pathogen Discovery | Dakota Howard, Dhwani Batra, Peter W. Cook, Kara Moser, Adrian Paskey, Jason Caravas, Benjamin Rambo-Martin, Shatavia Morrison, Christopher Gulvick, Scott Sammons, Yvette Unoarumhi, Darlene Wagner, Matthew Schmerer, Cyndi Clark, Patrick Campbell, Rob Case, Vikramsinha Ghorpade, Holly Houdeshell, Ola Kvalvaag, Dillon Nall, Ethan Sanders, Alec Vest, Shaun Westlund, Matthew Hardison, Clinton R. Paden, Duncan MacCannell |
| EPI_ISL_2204779 | Berkeley Medical Center | WVU and Marshall University Combined Genomics Core Facilities | James Denvir, Peter Stoilov, Peter Perrotta, Wesley Kimble, Ryan Percifield |
| EPI_ISL_2204780 | Houston Methodist Hospital | Houston Methodist Hospital | Randall J. Olsen, Paul A. Christensen, S. Wesley Long, Sishir Subedi, Robert Olson, Marcus Nguyen, James J. Davis, Matthew Ojeda Saavedra, Prasanti Yerramilli, Layne Pruitt, Kristina Reppond, Madison N. Shyer, Jessica Cambric, Ryan Gadd, Ilya J. Finkelstein, Jimmy Gollihar, and James M. Musser |
| EPI_ISL_2204781, EPI_ISL_2204783 | Berkeley Medical Center | WVU and Marshall University Combined Genomics Core Facilities | James Denvir, Peter Stoilov, Peter Perrotta, Wesley Kimble, Ryan Percifield |
| EPI_ISL_2204785 | Aegis Sciences Corporation | Centers for Disease Control and Prevention Division of Viral Diseases, Pathogen Discovery | Dakota Howard, Dhwani Batra, Peter W. Cook, Kara Moser, Adrian Paskey, Jason Caravas, Benjamin Rambo-Martin, Shatavia Morrison, Christopher Gulvick, Scott Sammons, Yvette Unoarumhi, Darlene Wagner, Matthew Schmerer, Cyndi Clark, Patrick Campbell, Rob Case, Vikramsinha Ghorpade, Holly Houdeshell, Ola Kvalvaag, Dillon Nall, Ethan Sanders, Alec Vest, Shaun Westlund, Matthew Hardison, Clinton R. Paden, Duncan MacCannell |
| EPI_ISL_2204786 | Berkeley Medical Center | WVU and Marshall University Combined Genomics Core Facilities | James Denvir, Peter Stoilov, Peter Perrotta, Wesley Kimble, Ryan Percifield |
| EPI_ISL_2204787 | Houston Methodist Hospital | Houston Methodist Hospital | Randall J. Olsen, Paul A. Christensen, S. Wesley Long, Sishir Subedi, Robert Olson, Marcus Nguyen, James J. Davis, Matthew Ojeda Saavedra, Prasanti Yerramilli, Layne Pruitt, Kristina Reppond, Madison N. Shyer, Jessica Cambric, Ryan Gadd, Ilya J. Finkelstein, Jimmy Gollihar, and James M. Musser |
| EPI_ISL_2204788, EPI_ISL_2204789 | Aegis Sciences Corporation | Centers for Disease Control and Prevention Division of Viral Diseases, Pathogen Discovery | Dakota Howard, Dhwani Batra, Peter W. Cook, Kara Moser, Adrian Paskey, Jason Caravas, Benjamin Rambo-Martin, Shatavia Morrison, Christopher Gulvick, Scott Sammons, Yvette Unoarumhi, Darlene Wagner, Matthew Schmerer, Cyndi Clark, Patrick Campbell, Rob Case, Vikramsinha Ghorpade, Holly Houdeshell, Ola Kvalvaag, Dillon Nall, Ethan Sanders, Alec Vest, Shaun Westlund, Matthew Hardison, Clinton R. Paden, Duncan MacCannell |
| EPI_ISL_2204790, EPI_ISL_2204793 | Berkeley Medical Center | WVU and Marshall University Combined Genomics Core Facilities | James Denvir, Peter Stoilov, Peter Perrotta, Wesley Kimble, Ryan Percifield |
| EPI_ISL_2204794 | Houston Methodist Hospital | Houston Methodist Hospital | Randall J. Olsen, Paul A. Christensen, S. Wesley Long, Sishir Subedi, Robert Olson, Marcus Nguyen, James J. Davis, Matthew Ojeda Saavedra, Prasanti Yerramilli, Layne Pruitt, Kristina Reppond, Madison N. Shyer, Jessica Cambric, Ryan Gadd, Ilya J. Finkelstein, Jimmy Gollihar, and James M. Musser |
| EPI_ISL_2204795 | Aegis Sciences Corporation | Centers for Disease Control and Prevention Division of Viral Diseases, Pathogen Discovery | Dakota Howard, Dhwani Batra, Peter W. Cook, Kara Moser, Adrian Paskey, Jason Caravas, Benjamin Rambo-Martin, Shatavia Morrison, Christopher Gulvick, Scott Sammons, Yvette Unoarumhi, Darlene Wagner, Matthew Schmerer, Cyndi Clark, Patrick Campbell, Rob Case, Vikramsinha Ghorpade, Holly Houdeshell, Ola Kvalvaag, Dillon Nall, Ethan Sanders, Alec Vest, Shaun Westlund, Matthew Hardison, Clinton R. Paden, Duncan MacCannell |
| EPI_ISL_2204796 | WV Labs | WVU and Marshall University Combined Genomics Core Facilities | James Denvir, Peter Stoilov, Peter Perrotta, Wesley Kimble, Ryan Percifield |
| EPI_ISL_2204797 | Aegis Sciences Corporation | Centers for Disease Control and Prevention Division of Viral Diseases, Pathogen Discovery | Dakota Howard, Dhwani Batra, Peter W. Cook, Kara Moser, Adrian Paskey, Jason Caravas, Benjamin Rambo-Martin, Shatavia Morrison, Christopher Gulvick, Scott Sammons, Yvette Unoarumhi, Darlene Wagner, Matthew Schmerer, Cyndi Clark, Patrick Campbell, Rob Case, Vikramsinha Ghorpade, Holly Houdeshell, Ola Kvalvaag, Dillon Nall, Ethan Sanders, Alec Vest, Shaun Westlund, Matthew Hardison, Clinton R. Paden, Duncan MacCannell |
| EPI_ISL_2204798 | Houston Methodist Hospital | Houston Methodist Hospital | Randall J. Olsen, Paul A. Christensen, S. Wesley Long, Sishir Subedi, Robert Olson, Marcus Nguyen, James J. Davis, Matthew Ojeda Saavedra, Prasanti Yerramilli, Layne Pruitt, Kristina Reppond, Madison N. Shyer, Jessica Cambric, Ryan Gadd, Ilya J. Finkelstein, Jimmy Gollihar, and James M. Musser |
| EPI_ISL_2204799 | Aegis Sciences Corporation | Centers for Disease Control and Prevention Division of Viral Diseases, Pathogen Discovery | Dakota Howard, Dhwani Batra, Peter W. Cook, Kara Moser, Adrian Paskey, Jason Caravas, Benjamin Rambo-Martin, Shatavia Morrison, Christopher Gulvick, Scott Sammons, Yvette Unoarumhi, Darlene Wagner, Matthew Schmerer, Cyndi Clark, Patrick Campbell, Rob Case, Vikramsinha Ghorpade, Holly Houdeshell, Ola Kvalvaag, Dillon Nall, Ethan Sanders, Alec Vest, Shaun Westlund, Matthew Hardison, Clinton R. Paden, Duncan MacCannell |
| EPI_ISL_2204800 | WV Labs | WVU and Marshall University Combined Genomics Core Facilities | James Denvir, Peter Stoilov, Peter Perrotta, Wesley Kimble, Ryan Percifield |
| EPI_ISL_2204801 | Houston Methodist Hospital | Houston Methodist Hospital | Randall J. Olsen, Paul A. Christensen, S. Wesley Long, Sishir Subedi, Robert Olson, Marcus Nguyen, James J. Davis, Matthew Ojeda Saavedra, Prasanti Yerramilli, Layne Pruitt, Kristina Reppond, Madison N. Shyer, Jessica Cambric, Ryan Gadd, Ilya J. Finkelstein, Jimmy Gollihar, and James M. Musser |
| EPI_ISL_2204802 | WV Labs | WVU and Marshall University Combined Genomics Core Facilities | James Denvir, Peter Stoilov, Peter Perrotta, Wesley Kimble, Ryan Percifield |
| EPI_ISL_2204803, EPI_ISL_2204805 | Aegis Sciences Corporation | Centers for Disease Control and Prevention Division of Viral Diseases, Pathogen Discovery | Dakota Howard, Dhwani Batra, Peter W. Cook, Kara Moser, Adrian Paskey, Jason Caravas, Benjamin Rambo-Martin, Shatavia Morrison, Christopher Gulvick, Scott Sammons, Yvette Unoarumhi, Darlene Wagner, Matthew Schmerer, Cyndi Clark, Patrick Campbell, Rob Case, Vikramsinha Ghorpade, Holly Houdeshell, Ola Kvalvaag, Dillon Nall, Ethan Sanders, Alec Vest, Shaun Westlund, Matthew Hardison, Clinton R. Paden, Duncan MacCannell |
| EPI_ISL_2204806 | Houston Methodist Hospital | Houston Methodist Hospital | Randall J. Olsen, Paul A. Christensen, S. Wesley Long, Sishir Subedi, Robert Olson, Marcus Nguyen, James J. Davis, Matthew Ojeda Saavedra, Prasanti Yerramilli, Layne Pruitt, Kristina Reppond, Madison N. Shyer, Jessica Cambric, Ryan Gadd, Ilya J. Finkelstein, Jimmy Gollihar, and James M. Musser |
| EPI_ISL_2204807 | WV Labs | WVU and Marshall University Combined Genomics Core Facilities | James Denvir, Peter Stoilov, Peter Perrotta, Wesley Kimble, Ryan Percifield |
| EPI_ISL_2204808 | Aegis Sciences Corporation | Centers for Disease Control and Prevention Division of Viral Diseases, Pathogen Discovery | Dakota Howard, Dhwani Batra, Peter W. Cook, Kara Moser, Adrian Paskey, Jason Caravas, Benjamin Rambo-Martin, Shatavia Morrison, Christopher Gulvick, Scott Sammons, Yvette Unoarumhi, Darlene Wagner, Matthew Schmerer, Cyndi Clark, Patrick Campbell, Rob Case, Vikramsinha Ghorpade, Holly Houdeshell, Ola Kvalvaag, Dillon Nall, Ethan Sanders, Alec Vest, Shaun Westlund, Matthew Hardison, Clinton R. Paden, Duncan MacCannell |
| EPI_ISL_2204809 | Houston Methodist Hospital | Houston Methodist Hospital | Randall J. Olsen, Paul A. Christensen, S. Wesley Long, Sishir Subedi, Robert Olson, Marcus Nguyen, James J. Davis, Matthew Ojeda Saavedra, Prasanti Yerramilli, Layne Pruitt, Kristina Reppond, Madison N. Shyer, Jessica Cambric, Ryan Gadd, Ilya J. Finkelstein, Jimmy Gollihar, and James M. Musser |
| EPI_ISL_2204810 | WV Labs | WVU and Marshall University Combined Genomics Core Facilities | James Denvir, Peter Stoilov, Peter Perrotta, Wesley Kimble, Ryan Percifield |
| EPI_ISL_2204811 | Aegis Sciences Corporation | Centers for Disease Control and Prevention Division of Viral Diseases, Pathogen Discovery | Dakota Howard, Dhwani Batra, Peter W. Cook, Kara Moser, Adrian Paskey, Jason Caravas, Benjamin Rambo-Martin, Shatavia Morrison, Christopher Gulvick, Scott Sammons, Yvette Unoarumhi, Darlene Wagner, Matthew Schmerer, Cyndi Clark, Patrick Campbell, Rob Case, Vikramsinha Ghorpade, Holly Houdeshell, Ola Kvalvaag, Dillon Nall, Ethan Sanders, Alec Vest, Shaun Westlund, Matthew Hardison, Clinton R. Paden, Duncan MacCannell |
| EPI_ISL_2204812 | Berkeley Medical Center | WVU and Marshall University Combined Genomics Core Facilities | James Denvir, Peter Stoilov, Peter Perrotta, Wesley Kimble, Ryan Percifield |
| EPI_ISL_2204813 | Houston Methodist Hospital | Houston Methodist Hospital | Randall J. Olsen, Paul A. Christensen, S. Wesley Long, Sishir Subedi, Robert Olson, Marcus Nguyen, James J. Davis, Matthew Ojeda Saavedra, Prasanti Yerramilli, Layne Pruitt, Kristina Reppond, Madison N. Shyer, Jessica Cambric, Ryan Gadd, Ilya J. Finkelstein, Jimmy Gollihar, and James M. Musser |
| EPI_ISL_2204814 | Aegis Sciences Corporation | Centers for Disease Control and Prevention Division of Viral Diseases, Pathogen Discovery | Dakota Howard, Dhwani Batra, Peter W. Cook, Kara Moser, Adrian Paskey, Jason Caravas, Benjamin Rambo-Martin, Shatavia Morrison, Christopher Gulvick, Scott Sammons, Yvette Unoarumhi, Darlene Wagner, Matthew Schmerer, Cyndi Clark, Patrick Campbell, Rob Case, Vikramsinha Ghorpade, Holly Houdeshell, Ola Kvalvaag, Dillon Nall, Ethan Sanders, Alec Vest, Shaun Westlund, Matthew Hardison, Clinton R. Paden, Duncan MacCannell |
| EPI_ISL_2204815, EPI_ISL_2204816 | Berkeley Medical Center | WVU and Marshall University Combined Genomics Core Facilities | James Denvir, Peter Stoilov, Peter Perrotta, Wesley Kimble, Ryan Percifield |
| EPI_ISL_2204817 | Aegis Sciences Corporation | Centers for Disease Control and Prevention Division of Viral Diseases, Pathogen Discovery | Dakota Howard, Dhwani Batra, Peter W. Cook, Kara Moser, Adrian Paskey, Jason Caravas, Benjamin Rambo-Martin, Shatavia Morrison, Christopher Gulvick, Scott Sammons, Yvette Unoarumhi, Darlene Wagner, Matthew Schmerer, Cyndi Clark, Patrick Campbell, Rob Case, Vikramsinha Ghorpade, Holly Houdeshell, Ola Kvalvaag, Dillon Nall, Ethan Sanders, Alec Vest, Shaun Westlund, Matthew Hardison, Clinton R. Paden, Duncan MacCannell |
| EPI_ISL_2204818 | Houston Methodist Hospital | Houston Methodist Hospital | Randall J. Olsen, Paul A. Christensen, S. Wesley Long, Sishir Subedi, Robert Olson, Marcus Nguyen, James J. Davis, Matthew Ojeda Saavedra, Prasanti Yerramilli, Layne Pruitt, Kristina Reppond, Madison N. Shyer, Jessica Cambric, Ryan Gadd, Ilya J. Finkelstein, Jimmy Gollihar, and James M. Musser |
| EPI_ISL_2204819, EPI_ISL_2204820 | Berkeley Medical Center | WVU and Marshall University Combined Genomics Core Facilities | James Denvir, Peter Stoilov, Peter Perrotta, Wesley Kimble, Ryan Percifield |
| EPI_ISL_2204821 | Aegis Sciences Corporation | Centers for Disease Control and Prevention Division of Viral Diseases, Pathogen Discovery | Dakota Howard, Dhwani Batra, Peter W. Cook, Kara Moser, Adrian Paskey, Jason Caravas, Benjamin Rambo-Martin, Shatavia Morrison, Christopher Gulvick, Scott Sammons, Yvette Unoarumhi, Darlene Wagner, Matthew Schmerer, Cyndi Clark, Patrick Campbell, Rob Case, Vikramsinha Ghorpade, Holly Houdeshell, Ola Kvalvaag, Dillon Nall, Ethan Sanders, Alec Vest, Shaun Westlund, Matthew Hardison, Clinton R. Paden, Duncan MacCannell |
| EPI_ISL_2204823 | Berkeley Medical Center | WVU and Marshall University Combined Genomics Core Facilities | James Denvir, Peter Stoilov, Peter Perrotta, Wesley Kimble, Ryan Percifield |
| EPI_ISL_2204824 | Aegis Sciences Corporation | Centers for Disease Control and Prevention | Dakota Howard, Dhwani Batra, Peter W. Cook, Kara Moser, Adrian Paskey, Jason Caravas, Benjamin Rambo-Martin, Shatavia Morrison, Christopher Gulvick, Scott Sammons, Yvette Unoarumhi, Darlene Wagner, Matthew Schmerer, Cyndi Clark, Patrick Campbell, Rob Case, Vikramsinha |

| | | | |
|---|---|---|---|
| | | Division of Viral Diseases, Pathogen Discovery | Ghorpade, Holly Houdeshell, Ola Kvalvaag, Dillon Nall, Ethan Sanders, Alec Vest, Shaun Westlund, Matthew Hardison, Clinton R. Paden, Duncan MacCannell |
| EPI_ISL_2204825 | Berkeley Medical Center | WVU and Marshall University Combined Genomics Core Facilities | James Denvir, Peter Stoilov, Peter Perrotta, Wesley Kimble, Ryan Percifield |
| EPI_ISL_2204826 | Houston Methodist Hospital | Houston Methodist Hospital | Randall J. Olsen, Paul A. Christensen, S. Wesley Long, Sishir Subedi, Robert Olson, Marcus Nguyen, James J. Davis, Matthew Ojeda Saavedra, Prasanti Yerramilli, Layne Pruitt, Kristina Reppond, Madison N. Shyer, Jessica Cambric, Ryan Gadd, Ilya J. Finkelstein, Jimmy Gollihar, and James M. Musser |
| EPI_ISL_2204827 | Berkeley Medical Center | WVU and Marshall University Combined Genomics Core Facilities | James Denvir, Peter Stoilov, Peter Perrotta, Wesley Kimble, Ryan Percifield |
| EPI_ISL_2204828 | Aegis Sciences Corporation | Centers for Disease Control and Prevention Division of Viral Diseases, Pathogen Discovery | Dakota Howard, Dhwani Batra, Peter W. Cook, Kara Moser, Adrian Paskey, Jason Caravas, Benjamin Rambo-Martin, Shatavia Morrison, Christopher Gulvick, Scott Sammons, Yvette Unoarumhi, Darlene Wagner, Matthew Schmerer, Cyndi Clark, Patrick Campbell, Rob Case, Vikramsinha Ghorpade, Holly Houdeshell, Ola Kvalvaag, Dillon Nall, Ethan Sanders, Alec Vest, Shaun Westlund, Matthew Hardison, Clinton R. Paden, Duncan MacCannell |
| EPI_ISL_2204829, EPI_ISL_2204830 | Berkeley Medical Center | WVU and Marshall University Combined Genomics Core Facilities | James Denvir, Peter Stoilov, Peter Perrotta, Wesley Kimble, Ryan Percifield |
| EPI_ISL_2204831 | Houston Methodist Hospital | Houston Methodist Hospital | Randall J. Olsen, Paul A. Christensen, S. Wesley Long, Sishir Subedi, Robert Olson, Marcus Nguyen, James J. Davis, Matthew Ojeda Saavedra, Prasanti Yerramilli, Layne Pruitt, Kristina Reppond, Madison N. Shyer, Jessica Cambric, Ryan Gadd, Ilya J. Finkelstein, Jimmy Gollihar, and James M. Musser |
| EPI_ISL_2204832, EPI_ISL_2204833 | Aegis Sciences Corporation | Centers for Disease Control and Prevention Division of Viral Diseases, Pathogen Discovery | Dakota Howard, Dhwani Batra, Peter W. Cook, Kara Moser, Adrian Paskey, Jason Caravas, Benjamin Rambo-Martin, Shatavia Morrison, Christopher Gulvick, Scott Sammons, Yvette Unoarumhi, Darlene Wagner, Matthew Schmerer, Cyndi Clark, Patrick Campbell, Rob Case, Vikramsinha Ghorpade, Holly Houdeshell, Ola Kvalvaag, Dillon Nall, Ethan Sanders, Alec Vest, Shaun Westlund, Matthew Hardison, Clinton R. Paden, Duncan MacCannell |
| EPI_ISL_2204834 | Berkeley Medical Center | WVU and Marshall University Combined Genomics Core Facilities | James Denvir, Peter Stoilov, Peter Perrotta, Wesley Kimble, Ryan Percifield |
| EPI_ISL_2204835 | Houston Methodist Hospital | Houston Methodist Hospital | Randall J. Olsen, Paul A. Christensen, S. Wesley Long, Sishir Subedi, Robert Olson, Marcus Nguyen, James J. Davis, Matthew Ojeda Saavedra, Prasanti Yerramilli, Layne Pruitt, Kristina Reppond, Madison N. Shyer, Jessica Cambric, Ryan Gadd, Ilya J. Finkelstein, Jimmy Gollihar, and James M. Musser |
| EPI_ISL_2204837 | Aegis Sciences Corporation | Centers for Disease Control and Prevention Division of Viral Diseases, Pathogen Discovery | Dakota Howard, Dhwani Batra, Peter W. Cook, Kara Moser, Adrian Paskey, Jason Caravas, Benjamin Rambo-Martin, Shatavia Morrison, Christopher Gulvick, Scott Sammons, Yvette Unoarumhi, Darlene Wagner, Matthew Schmerer, Cyndi Clark, Patrick Campbell, Rob Case, Vikramsinha Ghorpade, Holly Houdeshell, Ola Kvalvaag, Dillon Nall, Ethan Sanders, Alec Vest, Shaun Westlund, Matthew Hardison, Clinton R. Paden, Duncan MacCannell |
| EPI_ISL_2204838 | Houston Methodist Hospital | Houston Methodist Hospital | Randall J. Olsen, Paul A. Christensen, S. Wesley Long, Sishir Subedi, Robert Olson, Marcus Nguyen, James J. Davis, Matthew Ojeda Saavedra, Prasanti Yerramilli, Layne Pruitt, Kristina Reppond, Madison N. Shyer, Jessica Cambric, Ryan Gadd, Ilya J. Finkelstein, Jimmy Gollihar, and James M. Musser |
| EPI_ISL_2204839 | Aegis Sciences Corporation | Centers for Disease Control and Prevention Division of Viral Diseases, Pathogen Discovery | Dakota Howard, Dhwani Batra, Peter W. Cook, Kara Moser, Adrian Paskey, Jason Caravas, Benjamin Rambo-Martin, Shatavia Morrison, Christopher Gulvick, Scott Sammons, Yvette Unoarumhi, Darlene Wagner, Matthew Schmerer, Cyndi Clark, Patrick Campbell, Rob Case, Vikramsinha Ghorpade, Holly Houdeshell, Ola Kvalvaag, Dillon Nall, Ethan Sanders, Alec Vest, Shaun Westlund, Matthew Hardison, Clinton R. Paden, Duncan MacCannell |
| EPI_ISL_2204840, EPI_ISL_2204842 | Berkeley Medical Center | WVU and Marshall University Combined Genomics Core Facilities | James Denvir, Peter Stoilov, Peter Perrotta, Wesley Kimble, Ryan Percifield |
| EPI_ISL_2204843 | Aegis Sciences Corporation | Centers for Disease Control and Prevention Division of Viral Diseases, Pathogen Discovery | Dakota Howard, Dhwani Batra, Peter W. Cook, Kara Moser, Adrian Paskey, Jason Caravas, Benjamin Rambo-Martin, Shatavia Morrison, Christopher Gulvick, Scott Sammons, Yvette Unoarumhi, Darlene Wagner, Matthew Schmerer, Cyndi Clark, Patrick Campbell, Rob Case, Vikramsinha Ghorpade, Holly Houdeshell, Ola Kvalvaag, Dillon Nall, Ethan Sanders, Alec Vest, Shaun Westlund, Matthew Hardison, Clinton R. Paden, Duncan MacCannell |
| EPI_ISL_2204844 | Berkeley Medical Center | WVU and Marshall University Combined Genomics Core Facilities | James Denvir, Peter Stoilov, Peter Perrotta, Wesley Kimble, Ryan Percifield |
| EPI_ISL_2204845 | Aegis Sciences Corporation | Centers for Disease Control and Prevention Division of Viral Diseases, Pathogen Discovery | Dakota Howard, Dhwani Batra, Peter W. Cook, Kara Moser, Adrian Paskey, Jason Caravas, Benjamin Rambo-Martin, Shatavia Morrison, Christopher Gulvick, Scott Sammons, Yvette Unoarumhi, Darlene Wagner, Matthew Schmerer, Cyndi Clark, Patrick Campbell, Rob Case, Vikramsinha Ghorpade, Holly Houdeshell, Ola Kvalvaag, Dillon Nall, Ethan Sanders, Alec Vest, Shaun Westlund, Matthew Hardison, Clinton R. Paden, Duncan MacCannell |
| EPI_ISL_2204846 | Houston Methodist Hospital | Houston Methodist Hospital | Randall J. Olsen, Paul A. Christensen, S. Wesley Long, Sishir Subedi, Robert Olson, Marcus Nguyen, James J. Davis, Matthew Ojeda Saavedra, Prasanti Yerramilli, Layne Pruitt, Kristina Reppond, Madison N. Shyer, Jessica Cambric, Ryan Gadd, Ilya J. Finkelstein, Jimmy Gollihar, and James M. Musser |
| EPI_ISL_2204847, EPI_ISL_2204848 | Berkeley Medical Center | WVU and Marshall University Combined Genomics Core Facilities | James Denvir, Peter Stoilov, Peter Perrotta, Wesley Kimble, Ryan Percifield |
| EPI_ISL_2204849 | Aegis Sciences Corporation | Centers for Disease Control and Prevention Division of Viral Diseases, Pathogen Discovery | Dakota Howard, Dhwani Batra, Peter W. Cook, Kara Moser, Adrian Paskey, Jason Caravas, Benjamin Rambo-Martin, Shatavia Morrison, Christopher Gulvick, Scott Sammons, Yvette Unoarumhi, Darlene Wagner, Matthew Schmerer, Cyndi Clark, Patrick Campbell, Rob Case, Vikramsinha Ghorpade, Holly Houdeshell, Ola Kvalvaag, Dillon Nall, Ethan Sanders, Alec Vest, Shaun Westlund, Matthew Hardison, Clinton R. Paden, Duncan MacCannell |
| EPI_ISL_2204850 | Summersville Regiol Medical Center | WVU and Marshall University Combined Genomics Core Facilities | James Denvir, Peter Stoilov, Peter Perrotta, Wesley Kimble, Ryan Percifield |
| EPI_ISL_2204851 | Houston Methodist Hospital | Houston Methodist Hospital | Randall J. Olsen, Paul A. Christensen, S. Wesley Long, Sishir Subedi, Robert Olson, Marcus Nguyen, James J. Davis, Matthew Ojeda Saavedra, Prasanti Yerramilli, Layne Pruitt, Kristina Reppond, Madison N. Shyer, Jessica Cambric, Ryan Gadd, Ilya J. Finkelstein, Jimmy Gollihar, and James M. Musser |
| EPI_ISL_2204852 | Summersville Regiol Medical Center | WVU and Marshall University Combined Genomics Core Facilities | James Denvir, Peter Stoilov, Peter Perrotta, Wesley Kimble, Ryan Percifield |
| EPI_ISL_2204854 | Aegis Sciences Corporation | Centers for Disease Control and Prevention Division of Viral Diseases, Pathogen Discovery | Dakota Howard, Dhwani Batra, Peter W. Cook, Kara Moser, Adrian Paskey, Jason Caravas, Benjamin Rambo-Martin, Shatavia Morrison, Christopher Gulvick, Scott Sammons, Yvette Unoarumhi, Darlene Wagner, Matthew Schmerer, Cyndi Clark, Patrick Campbell, Rob Case, Vikramsinha Ghorpade, Holly Houdeshell, Ola Kvalvaag, Dillon Nall, Ethan Sanders, Alec Vest, Shaun Westlund, Matthew Hardison, Clinton R. Paden, Duncan MacCannell |
| EPI_ISL_2204855 | Houston Methodist Hospital | Houston Methodist Hospital | Randall J. Olsen, Paul A. Christensen, S. Wesley Long, Sishir Subedi, Robert Olson, Marcus Nguyen, James J. Davis, Matthew Ojeda Saavedra, Prasanti Yerramilli, Layne Pruitt, Kristina Reppond, Madison N. Shyer, Jessica Cambric, Ryan Gadd, Ilya J. Finkelstein, Jimmy Gollihar, and James M. Musser |
| EPI_ISL_2204856 | Summersville Regiol Medical Center | WVU and Marshall University Combined Genomics Core Facilities | James Denvir, Peter Stoilov, Peter Perrotta, Wesley Kimble, Ryan Percifield |
| EPI_ISL_2204857 | Aegis Sciences Corporation | Centers for Disease Control and Prevention Division of Viral Diseases, Pathogen Discovery | Dakota Howard, Dhwani Batra, Peter W. Cook, Kara Moser, Adrian Paskey, Jason Caravas, Benjamin Rambo-Martin, Shatavia Morrison, Christopher Gulvick, Scott Sammons, Yvette Unoarumhi, Darlene Wagner, Matthew Schmerer, Cyndi Clark, Patrick Campbell, Rob Case, Vikramsinha Ghorpade, Holly Houdeshell, Ola Kvalvaag, Dillon Nall, Ethan Sanders, Alec Vest, Shaun Westlund, Matthew Hardison, Clinton R. Paden, Duncan MacCannell |
| EPI_ISL_2204858 | Houston Methodist Hospital | Houston Methodist Hospital | Randall J. Olsen, Paul A. Christensen, S. Wesley Long, Sishir Subedi, Robert Olson, Marcus Nguyen, James J. Davis, Matthew Ojeda Saavedra, Prasanti Yerramilli, Layne Pruitt, Kristina Reppond, Madison N. Shyer, Jessica Cambric, Ryan Gadd, Ilya J. Finkelstein, Jimmy Gollihar, and James M. Musser |
| EPI_ISL_2204859 | Aegis Sciences Corporation | Centers for Disease Control and Prevention Division of Viral Diseases, Pathogen Discovery | Dakota Howard, Dhwani Batra, Peter W. Cook, Kara Moser, Adrian Paskey, Jason Caravas, Benjamin Rambo-Martin, Shatavia Morrison, Christopher Gulvick, Scott Sammons, Yvette Unoarumhi, Darlene Wagner, Matthew Schmerer, Cyndi Clark, Patrick Campbell, Rob Case, Vikramsinha Ghorpade, Holly Houdeshell, Ola Kvalvaag, Dillon Nall, Ethan Sanders, Alec Vest, Shaun Westlund, Matthew Hardison, Clinton R. Paden, Duncan MacCannell |
| EPI_ISL_2204860, EPI_ISL_2204861 | Summersville Regiol Medical Center | WVU and Marshall University Combined Genomics Core Facilities | James Denvir, Peter Stoilov, Peter Perrotta, Wesley Kimble, Ryan Percifield |
| EPI_ISL_2204863 | Houston Methodist Hospital | Houston Methodist Hospital | Randall J. Olsen, Paul A. Christensen, S. Wesley Long, Sishir Subedi, Robert Olson, Marcus Nguyen, James J. Davis, Matthew Ojeda Saavedra, Prasanti Yerramilli, Layne Pruitt, Kristina Reppond, Madison N. Shyer, Jessica Cambric, Ryan Gadd, Ilya J. Finkelstein, Jimmy Gollihar, and James M. Musser |
| EPI_ISL_2204864 | Summersville Regiol Medical Center | WVU and Marshall University Combined Genomics Core Facilities | James Denvir, Peter Stoilov, Peter Perrotta, Wesley Kimble, Ryan Percifield |
| EPI_ISL_2204865, EPI_ISL_2204866 | Aegis Sciences Corporation | Centers for Disease Control and Prevention Division of Viral Diseases, Pathogen Discovery | Dakota Howard, Dhwani Batra, Peter W. Cook, Kara Moser, Adrian Paskey, Jason Caravas, Benjamin Rambo-Martin, Shatavia Morrison, Christopher Gulvick, Scott Sammons, Yvette Unoarumhi, Darlene Wagner, Matthew Schmerer, Cyndi Clark, Patrick Campbell, Rob Case, Vikramsinha Ghorpade, Holly Houdeshell, Ola Kvalvaag, Dillon Nall, Ethan Sanders, Alec Vest, Shaun Westlund, Matthew Hardison, Clinton R. Paden, Duncan MacCannell |
| EPI_ISL_2204867 | WV Labs | WVU and Marshall University Combined Genomics Core Facilities | James Denvir, Peter Stoilov, Peter Perrotta, Wesley Kimble, Ryan Percifield |
| EPI_ISL_2204868 | Houston Methodist Hospital | Houston Methodist Hospital | Randall J. Olsen, Paul A. Christensen, S. Wesley Long, Sishir Subedi, Robert Olson, Marcus Nguyen, James J. Davis, Matthew Ojeda Saavedra, Prasanti Yerramilli, Layne Pruitt, Kristina Reppond, Madison N. Shyer, Jessica Cambric, Ryan Gadd, Ilya J. Finkelstein, Jimmy Gollihar, and James M. Musser |
| EPI_ISL_2204869 | Aegis Sciences Corporation | Centers for Disease Control and Prevention Division of Viral Diseases, Pathogen Discovery | Dakota Howard, Dhwani Batra, Peter W. Cook, Kara Moser, Adrian Paskey, Jason Caravas, Benjamin Rambo-Martin, Shatavia Morrison, Christopher Gulvick, Scott Sammons, Yvette Unoarumhi, Darlene Wagner, Matthew Schmerer, Cyndi Clark, Patrick Campbell, Rob Case, Vikramsinha Ghorpade, Holly Houdeshell, Ola Kvalvaag, Dillon Nall, Ethan Sanders, Alec Vest, Shaun Westlund, Matthew Hardison, Clinton R. Paden, Duncan MacCannell |
| EPI_ISL_2204870 | WV Labs | WVU and Marshall University Combined Genomics Core Facilities | James Denvir, Peter Stoilov, Peter Perrotta, Wesley Kimble, Ryan Percifield |
| EPI_ISL_2204871 | Aegis Sciences Corporation | Centers for Disease Control and Prevention Division of Viral Diseases, Pathogen Discovery | Dakota Howard, Dhwani Batra, Peter W. Cook, Kara Moser, Adrian Paskey, Jason Caravas, Benjamin Rambo-Martin, Shatavia Morrison, Christopher Gulvick, Scott Sammons, Yvette Unoarumhi, Darlene Wagner, Matthew Schmerer, Cyndi Clark, Patrick Campbell, Rob Case, Vikramsinha Ghorpade, Holly Houdeshell, Ola Kvalvaag, Dillon Nall, Ethan Sanders, Alec Vest, Shaun Westlund, Matthew Hardison, Clinton R. Paden, Duncan MacCannell |
| EPI_ISL_2204872 | WV Labs | WVU and Marshall University Combined Genomics Core Facilities | James Denvir, Peter Stoilov, Peter Perrotta, Wesley Kimble, Ryan Percifield |
| EPI_ISL_2204873 | Aegis Sciences Corporation | Centers for Disease Control and Prevention | Dakota Howard, Dhwani Batra, Peter W. Cook, Kara Moser, Adrian Paskey, Jason Caravas, Benjamin Rambo-Martin, Shatavia Morrison, Christopher Gulvick, Scott Sammons, Yvette Unoarumhi, Darlene Wagner, Matthew Schmerer, Cyndi Clark, Patrick Campbell, Rob Case, Vikramsinha |

| | | | |
|---|---|---|---|
| | | Division of Viral Diseases, Pathogen Discovery | Ghorpade, Holly Houdeshell, Ola Kvalvaag, Dillon Nall, Ethan Sanders, Alec Vest, Shaun Westlund, Matthew Hardison, Clinton R. Paden, Duncan MacCannell |
| EPI_ISL_2204874 | Houston Methodist Hospital | Houston Methodist Hospital | Randall J. Olsen, Paul A. Christensen, S. Wesley Long, Sishir Subedi, Robert Olson, Marcus Nguyen, James J. Davis, Matthew Ojeda Saavedra, Prasanti Yerramilli, Layne Pruitt, Kristina Reppond, Madison N. Shyer, Jessica Cambric, Ryan Gadd, Ilya J. Finkelstein, Jimmy Gollihar, and James M. Musser |
| EPI_ISL_2204876 | Aegis Sciences Corporation | Centers for Disease Control and Prevention Division of Viral Diseases, Pathogen Discovery | Dakota Howard, Dhwani Batra, Peter W. Cook, Kara Moser, Adrian Paskey, Jason Caravas, Benjamin Rambo-Martin, Shatavia Morrison, Christopher Gulvick, Scott Sammons, Yvette Unoarumhi, Darlene Wagner, Matthew Schmerer, Cyndi Clark, Patrick Campbell, Rob Case, Vikramsinha Ghorpade, Holly Houdeshell, Ola Kvalvaag, Dillon Nall, Ethan Sanders, Alec Vest, Shaun Westlund, Matthew Hardison, Clinton R. Paden, Duncan MacCannell |
| EPI_ISL_2204877 | WV Labs | WVU and Marshall University Combined Genomics Core Facilities | James Denvir, Peter Stoilov, Peter Perrotta, Wesley Kimble, Ryan Percifield |
| EPI_ISL_2204878 | Houston Methodist Hospital | Houston Methodist Hospital | Randall J. Olsen, Paul A. Christensen, S. Wesley Long, Sishir Subedi, Robert Olson, Marcus Nguyen, James J. Davis, Matthew Ojeda Saavedra, Prasanti Yerramilli, Layne Pruitt, Kristina Reppond, Madison N. Shyer, Jessica Cambric, Ryan Gadd, Ilya J. Finkelstein, Jimmy Gollihar, and James M. Musser |
| EPI_ISL_2204879 | Aegis Sciences Corporation | Centers for Disease Control and Prevention Division of Viral Diseases, Pathogen Discovery | Dakota Howard, Dhwani Batra, Peter W. Cook, Kara Moser, Adrian Paskey, Jason Caravas, Benjamin Rambo-Martin, Shatavia Morrison, Christopher Gulvick, Scott Sammons, Yvette Unoarumhi, Darlene Wagner, Matthew Schmerer, Cyndi Clark, Patrick Campbell, Rob Case, Vikramsinha Ghorpade, Holly Houdeshell, Ola Kvalvaag, Dillon Nall, Ethan Sanders, Alec Vest, Shaun Westlund, Matthew Hardison, Clinton R. Paden, Duncan MacCannell |
| EPI_ISL_2204880 | WV Labs | WVU and Marshall University Combined Genomics Core Facilities | James Denvir, Peter Stoilov, Peter Perrotta, Wesley Kimble, Ryan Percifield |
| EPI_ISL_2204881 | Aegis Sciences Corporation | Centers for Disease Control and Prevention Division of Viral Diseases, Pathogen Discovery | Dakota Howard, Dhwani Batra, Peter W. Cook, Kara Moser, Adrian Paskey, Jason Caravas, Benjamin Rambo-Martin, Shatavia Morrison, Christopher Gulvick, Scott Sammons, Yvette Unoarumhi, Darlene Wagner, Matthew Schmerer, Cyndi Clark, Patrick Campbell, Rob Case, Vikramsinha Ghorpade, Holly Houdeshell, Ola Kvalvaag, Dillon Nall, Ethan Sanders, Alec Vest, Shaun Westlund, Matthew Hardison, Clinton R. Paden, Duncan MacCannell |
| EPI_ISL_2204882 | Houston Methodist Hospital | Houston Methodist Hospital | Randall J. Olsen, Paul A. Christensen, S. Wesley Long, Sishir Subedi, Robert Olson, Marcus Nguyen, James J. Davis, Matthew Ojeda Saavedra, Prasanti Yerramilli, Layne Pruitt, Kristina Reppond, Madison N. Shyer, Jessica Cambric, Ryan Gadd, Ilya J. Finkelstein, Jimmy Gollihar, and James M. Musser |
| EPI_ISL_2204883 | Aegis Sciences Corporation | Centers for Disease Control and Prevention Division of Viral Diseases, Pathogen Discovery | Dakota Howard, Dhwani Batra, Peter W. Cook, Kara Moser, Adrian Paskey, Jason Caravas, Benjamin Rambo-Martin, Shatavia Morrison, Christopher Gulvick, Scott Sammons, Yvette Unoarumhi, Darlene Wagner, Matthew Schmerer, Cyndi Clark, Patrick Campbell, Rob Case, Vikramsinha Ghorpade, Holly Houdeshell, Ola Kvalvaag, Dillon Nall, Ethan Sanders, Alec Vest, Shaun Westlund, Matthew Hardison, Clinton R. Paden, Duncan MacCannell |
| EPI_ISL_2204884 | WV Labs | WVU and Marshall University Combined Genomics Core Facilities | James Denvir, Peter Stoilov, Peter Perrotta, Wesley Kimble, Ryan Percifield |
| EPI_ISL_2204885 | Houston Methodist Hospital | Houston Methodist Hospital | Randall J. Olsen, Paul A. Christensen, S. Wesley Long, Sishir Subedi, Robert Olson, Marcus Nguyen, James J. Davis, Matthew Ojeda Saavedra, Prasanti Yerramilli, Layne Pruitt, Kristina Reppond, Madison N. Shyer, Jessica Cambric, Ryan Gadd, Ilya J. Finkelstein, Jimmy Gollihar, and James M. Musser |
| EPI_ISL_2204886 | Aegis Sciences Corporation | Centers for Disease Control and Prevention Division of Viral Diseases, Pathogen Discovery | Dakota Howard, Dhwani Batra, Peter W. Cook, Kara Moser, Adrian Paskey, Jason Caravas, Benjamin Rambo-Martin, Shatavia Morrison, Christopher Gulvick, Scott Sammons, Yvette Unoarumhi, Darlene Wagner, Matthew Schmerer, Cyndi Clark, Patrick Campbell, Rob Case, Vikramsinha Ghorpade, Holly Houdeshell, Ola Kvalvaag, Dillon Nall, Ethan Sanders, Alec Vest, Shaun Westlund, Matthew Hardison, Clinton R. Paden, Duncan MacCannell |
| EPI_ISL_2204887 | WV Labs | WVU and Marshall University Combined Genomics Core Facilities | James Denvir, Peter Stoilov, Peter Perrotta, Wesley Kimble, Ryan Percifield |
| EPI_ISL_2204889 | Aegis Sciences Corporation | Centers for Disease Control and Prevention Division of Viral Diseases, Pathogen Discovery | Dakota Howard, Dhwani Batra, Peter W. Cook, Kara Moser, Adrian Paskey, Jason Caravas, Benjamin Rambo-Martin, Shatavia Morrison, Christopher Gulvick, Scott Sammons, Yvette Unoarumhi, Darlene Wagner, Matthew Schmerer, Cyndi Clark, Patrick Campbell, Rob Case, Vikramsinha Ghorpade, Holly Houdeshell, Ola Kvalvaag, Dillon Nall, Ethan Sanders, Alec Vest, Shaun Westlund, Matthew Hardison, Clinton R. Paden, Duncan MacCannell |
| EPI_ISL_2204890 | Houston Methodist Hospital | Houston Methodist Hospital | Randall J. Olsen, Paul A. Christensen, S. Wesley Long, Sishir Subedi, Robert Olson, Marcus Nguyen, James J. Davis, Matthew Ojeda Saavedra, Prasanti Yerramilli, Layne Pruitt, Kristina Reppond, Madison N. Shyer, Jessica Cambric, Ryan Gadd, Ilya J. Finkelstein, Jimmy Gollihar, and James M. Musser |
| EPI_ISL_2204891 | WV Labs | WVU and Marshall University Combined Genomics Core Facilities | James Denvir, Peter Stoilov, Peter Perrotta, Wesley Kimble, Ryan Percifield |
| EPI_ISL_2204892 | Houston Methodist Hospital | Houston Methodist Hospital | Randall J. Olsen, Paul A. Christensen, S. Wesley Long, Sishir Subedi, Robert Olson, Marcus Nguyen, James J. Davis, Matthew Ojeda Saavedra, Prasanti Yerramilli, Layne Pruitt, Kristina Reppond, Madison N. Shyer, Jessica Cambric, Ryan Gadd, Ilya J. Finkelstein, Jimmy Gollihar, and James M. Musser |
| EPI_ISL_2204893 | WV Labs | WVU and Marshall University Combined Genomics Core Facilities | James Denvir, Peter Stoilov, Peter Perrotta, Wesley Kimble, Ryan Percifield |
| EPI_ISL_2204894 | Aegis Sciences Corporation | Centers for Disease Control and Prevention Division of Viral Diseases, Pathogen Discovery | Dakota Howard, Dhwani Batra, Peter W. Cook, Kara Moser, Adrian Paskey, Jason Caravas, Benjamin Rambo-Martin, Shatavia Morrison, Christopher Gulvick, Scott Sammons, Yvette Unoarumhi, Darlene Wagner, Matthew Schmerer, Cyndi Clark, Patrick Campbell, Rob Case, Vikramsinha Ghorpade, Holly Houdeshell, Ola Kvalvaag, Dillon Nall, Ethan Sanders, Alec Vest, Shaun Westlund, Matthew Hardison, Clinton R. Paden, Duncan MacCannell |
| EPI_ISL_2204895 | Houston Methodist Hospital | Houston Methodist Hospital | Randall J. Olsen, Paul A. Christensen, S. Wesley Long, Sishir Subedi, Robert Olson, Marcus Nguyen, James J. Davis, Matthew Ojeda Saavedra, Prasanti Yerramilli, Layne Pruitt, Kristina Reppond, Madison N. Shyer, Jessica Cambric, Ryan Gadd, Ilya J. Finkelstein, Jimmy Gollihar, and James M. Musser |
| EPI_ISL_2204896 | WV Labs | WVU and Marshall University Combined Genomics Core Facilities | James Denvir, Peter Stoilov, Peter Perrotta, Wesley Kimble, Ryan Percifield |
| EPI_ISL_2204897 | Aegis Sciences Corporation | Centers for Disease Control and Prevention Division of Viral Diseases, Pathogen Discovery | Dakota Howard, Dhwani Batra, Peter W. Cook, Kara Moser, Adrian Paskey, Jason Caravas, Benjamin Rambo-Martin, Shatavia Morrison, Christopher Gulvick, Scott Sammons, Yvette Unoarumhi, Darlene Wagner, Matthew Schmerer, Cyndi Clark, Patrick Campbell, Rob Case, Vikramsinha Ghorpade, Holly Houdeshell, Ola Kvalvaag, Dillon Nall, Ethan Sanders, Alec Vest, Shaun Westlund, Matthew Hardison, Clinton R. Paden, Duncan MacCannell |
| EPI_ISL_2204898 | WV Labs | WVU and Marshall University Combined Genomics Core Facilities | James Denvir, Peter Stoilov, Peter Perrotta, Wesley Kimble, Ryan Percifield |
| EPI_ISL_2204899 | Houston Methodist Hospital | Houston Methodist Hospital | Randall J. Olsen, Paul A. Christensen, S. Wesley Long, Sishir Subedi, Robert Olson, Marcus Nguyen, James J. Davis, Matthew Ojeda Saavedra, Prasanti Yerramilli, Layne Pruitt, Kristina Reppond, Madison N. Shyer, Jessica Cambric, Ryan Gadd, Ilya J. Finkelstein, Jimmy Gollihar, and James M. Musser |
| EPI_ISL_2204901 | Aegis Sciences Corporation | Centers for Disease Control and Prevention Division of Viral Diseases, Pathogen Discovery | Dakota Howard, Dhwani Batra, Peter W. Cook, Kara Moser, Adrian Paskey, Jason Caravas, Benjamin Rambo-Martin, Shatavia Morrison, Christopher Gulvick, Scott Sammons, Yvette Unoarumhi, Darlene Wagner, Matthew Schmerer, Cyndi Clark, Patrick Campbell, Rob Case, Vikramsinha Ghorpade, Holly Houdeshell, Ola Kvalvaag, Dillon Nall, Ethan Sanders, Alec Vest, Shaun Westlund, Matthew Hardison, Clinton R. Paden, Duncan MacCannell |
| EPI_ISL_2204902 | WV Labs | WVU and Marshall University Combined Genomics Core Facilities | James Denvir, Peter Stoilov, Peter Perrotta, Wesley Kimble, Ryan Percifield |
| EPI_ISL_2204903 | Houston Methodist Hospital | Houston Methodist Hospital | Randall J. Olsen, Paul A. Christensen, S. Wesley Long, Sishir Subedi, Robert Olson, Marcus Nguyen, James J. Davis, Matthew Ojeda Saavedra, Prasanti Yerramilli, Layne Pruitt, Kristina Reppond, Madison N. Shyer, Jessica Cambric, Ryan Gadd, Ilya J. Finkelstein, Jimmy Gollihar, and James M. Musser |
| EPI_ISL_2204904 | Aegis Sciences Corporation | Centers for Disease Control and Prevention Division of Viral Diseases, Pathogen Discovery | Dakota Howard, Dhwani Batra, Peter W. Cook, Kara Moser, Adrian Paskey, Jason Caravas, Benjamin Rambo-Martin, Shatavia Morrison, Christopher Gulvick, Scott Sammons, Yvette Unoarumhi, Darlene Wagner, Matthew Schmerer, Cyndi Clark, Patrick Campbell, Rob Case, Vikramsinha Ghorpade, Holly Houdeshell, Ola Kvalvaag, Dillon Nall, Ethan Sanders, Alec Vest, Shaun Westlund, Matthew Hardison, Clinton R. Paden, Duncan MacCannell |
| EPI_ISL_2204905 | United Hospital Center | WVU and Marshall University Combined Genomics Core Facilities | James Denvir, Peter Stoilov, Peter Perrotta, Wesley Kimble, Ryan Percifield |
| EPI_ISL_2204907, EPI_ISL_2204908 | Aegis Sciences Corporation | Centers for Disease Control and Prevention Division of Viral Diseases, Pathogen Discovery | Dakota Howard, Dhwani Batra, Peter W. Cook, Kara Moser, Adrian Paskey, Jason Caravas, Benjamin Rambo-Martin, Shatavia Morrison, Christopher Gulvick, Scott Sammons, Yvette Unoarumhi, Darlene Wagner, Matthew Schmerer, Cyndi Clark, Patrick Campbell, Rob Case, Vikramsinha Ghorpade, Holly Houdeshell, Ola Kvalvaag, Dillon Nall, Ethan Sanders, Alec Vest, Shaun Westlund, Matthew Hardison, Clinton R. Paden, Duncan MacCannell |
| EPI_ISL_2204909 | WVU Rapid Development Lab | WVU and Marshall University Combined Genomics Core Facilities | James Denvir, Peter Stoilov, Peter Perrotta, Wesley Kimble, Ryan Percifield |
| EPI_ISL_2204910 | Houston Methodist Hospital | Houston Methodist Hospital | Randall J. Olsen, Paul A. Christensen, S. Wesley Long, Sishir Subedi, Robert Olson, Marcus Nguyen, James J. Davis, Matthew Ojeda Saavedra, Prasanti Yerramilli, Layne Pruitt, Kristina Reppond, Madison N. Shyer, Jessica Cambric, Ryan Gadd, Ilya J. Finkelstein, Jimmy Gollihar, and James M. Musser |
| EPI_ISL_2204911 | WVU Rapid Development Lab | WVU and Marshall University Combined Genomics Core Facilities | James Denvir, Peter Stoilov, Peter Perrotta, Wesley Kimble, Ryan Percifield |
| EPI_ISL_2204912 | Aegis Sciences Corporation | Centers for Disease Control and Prevention Division of Viral Diseases, Pathogen Discovery | Dakota Howard, Dhwani Batra, Peter W. Cook, Kara Moser, Adrian Paskey, Jason Caravas, Benjamin Rambo-Martin, Shatavia Morrison, Christopher Gulvick, Scott Sammons, Yvette Unoarumhi, Darlene Wagner, Matthew Schmerer, Cyndi Clark, Patrick Campbell, Rob Case, Vikramsinha Ghorpade, Holly Houdeshell, Ola Kvalvaag, Dillon Nall, Ethan Sanders, Alec Vest, Shaun Westlund, Matthew Hardison, Clinton R. Paden, Duncan MacCannell |
| EPI_ISL_2204913 | WVU Rapid Development Lab | WVU and Marshall University Combined Genomics Core Facilities | James Denvir, Peter Stoilov, Peter Perrotta, Wesley Kimble, Ryan Percifield |
| EPI_ISL_2204914 | Houston Methodist Hospital | Houston Methodist Hospital | Randall J. Olsen, Paul A. Christensen, S. Wesley Long, Sishir Subedi, Robert Olson, Marcus Nguyen, James J. Davis, Matthew Ojeda Saavedra, Prasanti Yerramilli, Layne Pruitt, Kristina Reppond, Madison N. Shyer, Jessica Cambric, Ryan Gadd, Ilya J. Finkelstein, Jimmy Gollihar, and James M. Musser |
| EPI_ISL_2204915 | Aegis Sciences Corporation | Centers for Disease Control and Prevention Division of Viral Diseases, Pathogen Discovery | Dakota Howard, Dhwani Batra, Peter W. Cook, Kara Moser, Adrian Paskey, Jason Caravas, Benjamin Rambo-Martin, Shatavia Morrison, Christopher Gulvick, Scott Sammons, Yvette Unoarumhi, Darlene Wagner, Matthew Schmerer, Cyndi Clark, Patrick Campbell, Rob Case, Vikramsinha Ghorpade, Holly Houdeshell, Ola Kvalvaag, Dillon Nall, Ethan Sanders, Alec Vest, Shaun Westlund, Matthew Hardison, Clinton R. Paden, Duncan MacCannell |
| EPI_ISL_2204916, EPI_ISL_2204917 | WVU Rapid Development Lab | WVU and Marshall University Combined Genomics Core Facilities | James Denvir, Peter Stoilov, Peter Perrotta, Wesley Kimble, Ryan Percifield |
| EPI_ISL_2204918 | Houston Methodist Hospital | Houston Methodist Hospital | Randall J. Olsen, Paul A. Christensen, S. Wesley Long, Sishir Subedi, Robert Olson, Marcus Nguyen, James J. Davis, Matthew Ojeda Saavedra, Prasanti Yerramilli, Layne Pruitt, Kristina Reppond, Madison N. Shyer, Jessica Cambric, Ryan Gadd, Ilya J. Finkelstein, Jimmy Gollihar, and James |

| | | | |
|---|---|---|---|
| | | | M. Musser |
| EPI_ISL_2204919, EPI_ISL_2204920 | Aegis Sciences Corporation | Centers for Disease Control and Prevention Division of Viral Diseases, Pathogen Discovery | Dakota Howard, Dhwani Batra, Peter W. Cook, Kara Moser, Adrian Paskey, Jason Caravas, Benjamin Rambo-Martin, Shatavia Morrison, Christopher Gulvick, Scott Sammons, Yvette Unoarumhi, Darlene Wagner, Matthew Schmerer, Cyndi Clark, Patrick Campbell, Rob Case, Vikramsinha Ghorpade, Holly Houdeshell, Ola Kvalvaag, Dillon Nall, Ethan Sanders, Alec Vest, Shaun Westlund, Matthew Hardison, Clinton R. Paden, Duncan MacCannell |
| EPI_ISL_2204921 | Houston Methodist Hospital | Houston Methodist Hospital | Randall J. Olsen, Paul A. Christensen, S. Wesley Long, Sishir Subedi, Robert Olson, Marcus Nguyen, James J. Davis, Matthew Ojeda Saavedra, Prasanti Yerramilli, Layne Pruitt, Kristina Reppond, Madison N. Shyer, Jessica Cambric, Ryan Gadd, Ilya J. Finkelstein, Jimmy Gollihar, and James M. Musser |
| EPI_ISL_2204922 | J.W. Ruby Memorial Hospital | WVU and Marshall University Combined Genomics Core Facilities | James Denvir, Peter Stoilov, Peter Perrotta, Wesley Kimble, Ryan Percifield |
| EPI_ISL_2204924 | Houston Methodist Hospital | Houston Methodist Hospital | Randall J. Olsen, Paul A. Christensen, S. Wesley Long, Sishir Subedi, Robert Olson, Marcus Nguyen, James J. Davis, Matthew Ojeda Saavedra, Prasanti Yerramilli, Layne Pruitt, Kristina Reppond, Madison N. Shyer, Jessica Cambric, Ryan Gadd, Ilya J. Finkelstein, Jimmy Gollihar, and James M. Musser |
| EPI_ISL_2204925 | Aegis Sciences Corporation | Centers for Disease Control and Prevention Division of Viral Diseases, Pathogen Discovery | Dakota Howard, Dhwani Batra, Peter W. Cook, Kara Moser, Adrian Paskey, Jason Caravas, Benjamin Rambo-Martin, Shatavia Morrison, Christopher Gulvick, Scott Sammons, Yvette Unoarumhi, Darlene Wagner, Matthew Schmerer, Cyndi Clark, Patrick Campbell, Rob Case, Vikramsinha Ghorpade, Holly Houdeshell, Ola Kvalvaag, Dillon Nall, Ethan Sanders, Alec Vest, Shaun Westlund, Matthew Hardison, Clinton R. Paden, Duncan MacCannell |
| EPI_ISL_2204926 | Camden Clark Medical Center | WVU and Marshall University Combined Genomics Core Facilities | James Denvir, Peter Stoilov, Peter Perrotta, Wesley Kimble, Ryan Percifield |
| EPI_ISL_2204927 | Aegis Sciences Corporation | Centers for Disease Control and Prevention Division of Viral Diseases, Pathogen Discovery | Dakota Howard, Dhwani Batra, Peter W. Cook, Kara Moser, Adrian Paskey, Jason Caravas, Benjamin Rambo-Martin, Shatavia Morrison, Christopher Gulvick, Scott Sammons, Yvette Unoarumhi, Darlene Wagner, Matthew Schmerer, Cyndi Clark, Patrick Campbell, Rob Case, Vikramsinha Ghorpade, Holly Houdeshell, Ola Kvalvaag, Dillon Nall, Ethan Sanders, Alec Vest, Shaun Westlund, Matthew Hardison, Clinton R. Paden, Duncan MacCannell |
| EPI_ISL_2204929 | Wetzel County Hospital | WVU and Marshall University Combined Genomics Core Facilities | James Denvir, Peter Stoilov, Peter Perrotta, Wesley Kimble, Ryan Percifield |
| EPI_ISL_2204931 | Houston Methodist Hospital | Houston Methodist Hospital | Randall J. Olsen, Paul A. Christensen, S. Wesley Long, Sishir Subedi, Robert Olson, Marcus Nguyen, James J. Davis, Matthew Ojeda Saavedra, Prasanti Yerramilli, Layne Pruitt, Kristina Reppond, Madison N. Shyer, Jessica Cambric, Ryan Gadd, Ilya J. Finkelstein, Jimmy Gollihar, and James M. Musser |
| EPI_ISL_2204932 | Aegis Sciences Corporation | Centers for Disease Control and Prevention Division of Viral Diseases, Pathogen Discovery | Dakota Howard, Dhwani Batra, Peter W. Cook, Kara Moser, Adrian Paskey, Jason Caravas, Benjamin Rambo-Martin, Shatavia Morrison, Christopher Gulvick, Scott Sammons, Yvette Unoarumhi, Darlene Wagner, Matthew Schmerer, Cyndi Clark, Patrick Campbell, Rob Case, Vikramsinha Ghorpade, Holly Houdeshell, Ola Kvalvaag, Dillon Nall, Ethan Sanders, Alec Vest, Shaun Westlund, Matthew Hardison, Clinton R. Paden, Duncan MacCannell |
| EPI_ISL_2204933 | WVU Rapid Development Lab | WVU and Marshall University Combined Genomics Core Facilities | James Denvir, Peter Stoilov, Peter Perrotta, Wesley Kimble, Ryan Percifield |
| EPI_ISL_2204934 | Aegis Sciences Corporation | Centers for Disease Control and Prevention Division of Viral Diseases, Pathogen Discovery | Dakota Howard, Dhwani Batra, Peter W. Cook, Kara Moser, Adrian Paskey, Jason Caravas, Benjamin Rambo-Martin, Shatavia Morrison, Christopher Gulvick, Scott Sammons, Yvette Unoarumhi, Darlene Wagner, Matthew Schmerer, Cyndi Clark, Patrick Campbell, Rob Case, Vikramsinha Ghorpade, Holly Houdeshell, Ola Kvalvaag, Dillon Nall, Ethan Sanders, Alec Vest, Shaun Westlund, Matthew Hardison, Clinton R. Paden, Duncan MacCannell |
| EPI_ISL_2204935 | Houston Methodist Hospital | Houston Methodist Hospital | Randall J. Olsen, Paul A. Christensen, S. Wesley Long, Sishir Subedi, Robert Olson, Marcus Nguyen, James J. Davis, Matthew Ojeda Saavedra, Prasanti Yerramilli, Layne Pruitt, Kristina Reppond, Madison N. Shyer, Jessica Cambric, Ryan Gadd, Ilya J. Finkelstein, Jimmy Gollihar, and James M. Musser |
| EPI_ISL_2204936 | WVU Rapid Development Lab | WVU and Marshall University Combined Genomics Core Facilities | James Denvir, Peter Stoilov, Peter Perrotta, Wesley Kimble, Ryan Percifield |
| EPI_ISL_2204937 | Aegis Sciences Corporation | Centers for Disease Control and Prevention Division of Viral Diseases, Pathogen Discovery | Dakota Howard, Dhwani Batra, Peter W. Cook, Kara Moser, Adrian Paskey, Jason Caravas, Benjamin Rambo-Martin, Shatavia Morrison, Christopher Gulvick, Scott Sammons, Yvette Unoarumhi, Darlene Wagner, Matthew Schmerer, Cyndi Clark, Patrick Campbell, Rob Case, Vikramsinha Ghorpade, Holly Houdeshell, Ola Kvalvaag, Dillon Nall, Ethan Sanders, Alec Vest, Shaun Westlund, Matthew Hardison, Clinton R. Paden, Duncan MacCannell |
| EPI_ISL_2204938 | Houston Methodist Hospital | Houston Methodist Hospital | Randall J. Olsen, Paul A. Christensen, S. Wesley Long, Sishir Subedi, Robert Olson, Marcus Nguyen, James J. Davis, Matthew Ojeda Saavedra, Prasanti Yerramilli, Layne Pruitt, Kristina Reppond, Madison N. Shyer, Jessica Cambric, Ryan Gadd, Ilya J. Finkelstein, Jimmy Gollihar, and James M. Musser |
| EPI_ISL_2204939 | Aegis Sciences Corporation | Centers for Disease Control and Prevention Division of Viral Diseases, Pathogen Discovery | Dakota Howard, Dhwani Batra, Peter W. Cook, Kara Moser, Adrian Paskey, Jason Caravas, Benjamin Rambo-Martin, Shatavia Morrison, Christopher Gulvick, Scott Sammons, Yvette Unoarumhi, Darlene Wagner, Matthew Schmerer, Cyndi Clark, Patrick Campbell, Rob Case, Vikramsinha Ghorpade, Holly Houdeshell, Ola Kvalvaag, Dillon Nall, Ethan Sanders, Alec Vest, Shaun Westlund, Matthew Hardison, Clinton R. Paden, Duncan MacCannell |
| EPI_ISL_2204940 | WVU Rapid Development Lab | WVU and Marshall University Combined Genomics Core Facilities | James Denvir, Peter Stoilov, Peter Perrotta, Wesley Kimble, Ryan Percifield |
| EPI_ISL_2204941 | Aegis Sciences Corporation | Centers for Disease Control and Prevention Division of Viral Diseases, Pathogen Discovery | Dakota Howard, Dhwani Batra, Peter W. Cook, Kara Moser, Adrian Paskey, Jason Caravas, Benjamin Rambo-Martin, Shatavia Morrison, Christopher Gulvick, Scott Sammons, Yvette Unoarumhi, Darlene Wagner, Matthew Schmerer, Cyndi Clark, Patrick Campbell, Rob Case, Vikramsinha Ghorpade, Holly Houdeshell, Ola Kvalvaag, Dillon Nall, Ethan Sanders, Alec Vest, Shaun Westlund, Matthew Hardison, Clinton R. Paden, Duncan MacCannell |
| EPI_ISL_2204942 | Houston Methodist Hospital | Houston Methodist Hospital | Randall J. Olsen, Paul A. Christensen, S. Wesley Long, Sishir Subedi, Robert Olson, Marcus Nguyen, James J. Davis, Matthew Ojeda Saavedra, Prasanti Yerramilli, Layne Pruitt, Kristina Reppond, Madison N. Shyer, Jessica Cambric, Ryan Gadd, Ilya J. Finkelstein, Jimmy Gollihar, and James M. Musser |
| EPI_ISL_2204943 | WVU Rapid Development Lab | WVU and Marshall University Combined Genomics Core Facilities | James Denvir, Peter Stoilov, Peter Perrotta, Wesley Kimble, Ryan Percifield |
| EPI_ISL_2204944 | Aegis Sciences Corporation | Centers for Disease Control and Prevention Division of Viral Diseases, Pathogen Discovery | Dakota Howard, Dhwani Batra, Peter W. Cook, Kara Moser, Adrian Paskey, Jason Caravas, Benjamin Rambo-Martin, Shatavia Morrison, Christopher Gulvick, Scott Sammons, Yvette Unoarumhi, Darlene Wagner, Matthew Schmerer, Cyndi Clark, Patrick Campbell, Rob Case, Vikramsinha Ghorpade, Holly Houdeshell, Ola Kvalvaag, Dillon Nall, Ethan Sanders, Alec Vest, Shaun Westlund, Matthew Hardison, Clinton R. Paden, Duncan MacCannell |
| EPI_ISL_2204945 | Houston Methodist Hospital | Houston Methodist Hospital | Randall J. Olsen, Paul A. Christensen, S. Wesley Long, Sishir Subedi, Robert Olson, Marcus Nguyen, James J. Davis, Matthew Ojeda Saavedra, Prasanti Yerramilli, Layne Pruitt, Kristina Reppond, Madison N. Shyer, Jessica Cambric, Ryan Gadd, Ilya J. Finkelstein, Jimmy Gollihar, and James M. Musser |
| EPI_ISL_2204946 | WVU Rapid Development Lab | WVU and Marshall University Combined Genomics Core Facilities | James Denvir, Peter Stoilov, Peter Perrotta, Wesley Kimble, Ryan Percifield |
| EPI_ISL_2204947 | Aegis Sciences Corporation | Centers for Disease Control and Prevention Division of Viral Diseases, Pathogen Discovery | Dakota Howard, Dhwani Batra, Peter W. Cook, Kara Moser, Adrian Paskey, Jason Caravas, Benjamin Rambo-Martin, Shatavia Morrison, Christopher Gulvick, Scott Sammons, Yvette Unoarumhi, Darlene Wagner, Matthew Schmerer, Cyndi Clark, Patrick Campbell, Rob Case, Vikramsinha Ghorpade, Holly Houdeshell, Ola Kvalvaag, Dillon Nall, Ethan Sanders, Alec Vest, Shaun Westlund, Matthew Hardison, Clinton R. Paden, Duncan MacCannell |
| EPI_ISL_2204948 | Houston Methodist Hospital | Houston Methodist Hospital | Randall J. Olsen, Paul A. Christensen, S. Wesley Long, Sishir Subedi, Robert Olson, Marcus Nguyen, James J. Davis, Matthew Ojeda Saavedra, Prasanti Yerramilli, Layne Pruitt, Kristina Reppond, Madison N. Shyer, Jessica Cambric, Ryan Gadd, Ilya J. Finkelstein, Jimmy Gollihar, and James M. Musser |
| EPI_ISL_2204949 | WVU Rapid Development Lab | WVU and Marshall University Combined Genomics Core Facilities | James Denvir, Peter Stoilov, Peter Perrotta, Wesley Kimble, Ryan Percifield |
| EPI_ISL_2204950 | Aegis Sciences Corporation | Centers for Disease Control and Prevention Division of Viral Diseases, Pathogen Discovery | Dakota Howard, Dhwani Batra, Peter W. Cook, Kara Moser, Adrian Paskey, Jason Caravas, Benjamin Rambo-Martin, Shatavia Morrison, Christopher Gulvick, Scott Sammons, Yvette Unoarumhi, Darlene Wagner, Matthew Schmerer, Cyndi Clark, Patrick Campbell, Rob Case, Vikramsinha Ghorpade, Holly Houdeshell, Ola Kvalvaag, Dillon Nall, Ethan Sanders, Alec Vest, Shaun Westlund, Matthew Hardison, Clinton R. Paden, Duncan MacCannell |
| EPI_ISL_2204951 | WVU Rapid Development Lab | WVU and Marshall University Combined Genomics Core Facilities | James Denvir, Peter Stoilov, Peter Perrotta, Wesley Kimble, Ryan Percifield |
| EPI_ISL_2204952 | Houston Methodist Hospital | Houston Methodist Hospital | Randall J. Olsen, Paul A. Christensen, S. Wesley Long, Sishir Subedi, Robert Olson, Marcus Nguyen, James J. Davis, Matthew Ojeda Saavedra, Prasanti Yerramilli, Layne Pruitt, Kristina Reppond, Madison N. Shyer, Jessica Cambric, Ryan Gadd, Ilya J. Finkelstein, Jimmy Gollihar, and James M. Musser |
| EPI_ISL_2204953 | Aegis Sciences Corporation | Centers for Disease Control and Prevention Division of Viral Diseases, Pathogen Discovery | Dakota Howard, Dhwani Batra, Peter W. Cook, Kara Moser, Adrian Paskey, Jason Caravas, Benjamin Rambo-Martin, Shatavia Morrison, Christopher Gulvick, Scott Sammons, Yvette Unoarumhi, Darlene Wagner, Matthew Schmerer, Cyndi Clark, Patrick Campbell, Rob Case, Vikramsinha Ghorpade, Holly Houdeshell, Ola Kvalvaag, Dillon Nall, Ethan Sanders, Alec Vest, Shaun Westlund, Matthew Hardison, Clinton R. Paden, Duncan MacCannell |
| EPI_ISL_2204954 | WVU Rapid Development Lab | WVU and Marshall University Combined Genomics Core Facilities | James Denvir, Peter Stoilov, Peter Perrotta, Wesley Kimble, Ryan Percifield |
| EPI_ISL_2204955 | Aegis Sciences Corporation | Centers for Disease Control and Prevention Division of Viral Diseases, Pathogen Discovery | Dakota Howard, Dhwani Batra, Peter W. Cook, Kara Moser, Adrian Paskey, Jason Caravas, Benjamin Rambo-Martin, Shatavia Morrison, Christopher Gulvick, Scott Sammons, Yvette Unoarumhi, Darlene Wagner, Matthew Schmerer, Cyndi Clark, Patrick Campbell, Rob Case, Vikramsinha Ghorpade, Holly Houdeshell, Ola Kvalvaag, Dillon Nall, Ethan Sanders, Alec Vest, Shaun Westlund, Matthew Hardison, Clinton R. Paden, Duncan MacCannell |
| EPI_ISL_2204956 | Houston Methodist Hospital | Houston Methodist Hospital | Randall J. Olsen, Paul A. Christensen, S. Wesley Long, Sishir Subedi, Robert Olson, Marcus Nguyen, James J. Davis, Matthew Ojeda Saavedra, Prasanti Yerramilli, Layne Pruitt, Kristina Reppond, Madison N. Shyer, Jessica Cambric, Ryan Gadd, Ilya J. Finkelstein, Jimmy Gollihar, and James M. Musser |
| EPI_ISL_2204957 | Aegis Sciences Corporation | Centers for Disease Control and Prevention Division of Viral Diseases, Pathogen Discovery | Dakota Howard, Dhwani Batra, Peter W. Cook, Kara Moser, Adrian Paskey, Jason Caravas, Benjamin Rambo-Martin, Shatavia Morrison, Christopher Gulvick, Scott Sammons, Yvette Unoarumhi, Darlene Wagner, Matthew Schmerer, Cyndi Clark, Patrick Campbell, Rob Case, Vikramsinha Ghorpade, Holly Houdeshell, Ola Kvalvaag, Dillon Nall, Ethan Sanders, Alec Vest, Shaun Westlund, Matthew Hardison, Clinton R. Paden, Duncan MacCannell |
| EPI_ISL_2204958 | Houston Methodist Hospital | Houston Methodist Hospital | Randall J. Olsen, Paul A. Christensen, S. Wesley Long, Sishir Subedi, Robert Olson, Marcus Nguyen, James J. Davis, Matthew Ojeda Saavedra, Prasanti Yerramilli, Layne Pruitt, Kristina Reppond, Madison N. Shyer, Jessica Cambric, Ryan Gadd, Ilya J. Finkelstein, Jimmy Gollihar, and James M. Musser |
| EPI_ISL_2204959, EPI_ISL_2204960 | Aegis Sciences Corporation | Centers for Disease Control and Prevention Division of Viral Diseases, Pathogen Discovery | Dakota Howard, Dhwani Batra, Peter W. Cook, Kara Moser, Adrian Paskey, Jason Caravas, Benjamin Rambo-Martin, Shatavia Morrison, Christopher Gulvick, Scott Sammons, Yvette Unoarumhi, Darlene Wagner, Matthew Schmerer, Cyndi Clark, Patrick Campbell, Rob Case, Vikramsinha Ghorpade, Holly Houdeshell, Ola Kvalvaag, Dillon Nall, Ethan Sanders, Alec Vest, Shaun Westlund, Matthew Hardison, Clinton R. Paden, Duncan MacCannell |
| EPI_ISL_2204961 | Houston Methodist Hospital | Houston Methodist Hospital | Randall J. Olsen, Paul A. Christensen, S. Wesley Long, Sishir Subedi, Robert Olson, Marcus Nguyen, James J. Davis, Matthew Ojeda Saavedra, Prasanti Yerramilli, Layne Pruitt, Kristina Reppond, Madison N. Shyer, Jessica Cambric, Ryan Gadd, Ilya J. Finkelstein, Jimmy Gollihar, and James M. Musser |
| EPI_ISL_2204962 | Aegis Sciences Corporation | Centers for Disease Control and Prevention | Dakota Howard, Dhwani Batra, Peter W. Cook, Kara Moser, Adrian Paskey, Jason Caravas, Benjamin Rambo-Martin, Shatavia Morrison, Christopher Gulvick, Scott Sammons, Yvette Unoarumhi, Darlene Wagner, Matthew Schmerer, Cyndi Clark, Patrick Campbell, Rob Case, Vikramsinha |

| | | | |
|---|---|---|---|
| | | Division of Viral Diseases, Pathogen Discovery | Ghorpade, Holly Houdeshell, Ola Kvalvaag, Dillon Nall, Ethan Sanders, Alec Vest, Shaun Westlund, Matthew Hardison, Clinton R. Paden, Duncan MacCannell |
| EPI_ISL_2204963 | Houston Methodist Hospital | Houston Methodist Hospital | Randall J. Olsen, Paul A. Christensen, S. Wesley Long, Sishir Subedi, Robert Olson, Marcus Nguyen, James J. Davis, Matthew Ojeda Saavedra, Prasanti Yerramilli, Layne Pruitt, Kristina Reppond, Madison N. Shyer, Jessica Cambric, Ryan Gadd, Ilya J. Finkelstein, Jimmy Gollihar, and James M. Musser |
| EPI_ISL_2204964 | Aegis Sciences Corporation | Centers for Disease Control and Prevention Division of Viral Diseases, Pathogen Discovery | Dakota Howard, Dhwani Batra, Peter W. Cook, Kara Moser, Adrian Paskey, Jason Caravas, Benjamin Rambo-Martin, Shatavia Morrison, Christopher Gulvick, Scott Sammons, Yvette Unoarumhi, Darlene Wagner, Matthew Schmerer, Cyndi Clark, Patrick Campbell, Rob Case, Vikramsinha Ghorpade, Holly Houdeshell, Ola Kvalvaag, Dillon Nall, Ethan Sanders, Alec Vest, Shaun Westlund, Matthew Hardison, Clinton R. Paden, Duncan MacCannell |
| EPI_ISL_2204965 | Houston Methodist Hospital | Houston Methodist Hospital | Randall J. Olsen, Paul A. Christensen, S. Wesley Long, Sishir Subedi, Robert Olson, Marcus Nguyen, James J. Davis, Matthew Ojeda Saavedra, Prasanti Yerramilli, Layne Pruitt, Kristina Reppond, Madison N. Shyer, Jessica Cambric, Ryan Gadd, Ilya J. Finkelstein, Jimmy Gollihar, and James M. Musser |
| EPI_ISL_2204966 | Aegis Sciences Corporation | Centers for Disease Control and Prevention Division of Viral Diseases, Pathogen Discovery | Dakota Howard, Dhwani Batra, Peter W. Cook, Kara Moser, Adrian Paskey, Jason Caravas, Benjamin Rambo-Martin, Shatavia Morrison, Christopher Gulvick, Scott Sammons, Yvette Unoarumhi, Darlene Wagner, Matthew Schmerer, Cyndi Clark, Patrick Campbell, Rob Case, Vikramsinha Ghorpade, Holly Houdeshell, Ola Kvalvaag, Dillon Nall, Ethan Sanders, Alec Vest, Shaun Westlund, Matthew Hardison, Clinton R. Paden, Duncan MacCannell |
| EPI_ISL_2204967 | Houston Methodist Hospital | Houston Methodist Hospital | Randall J. Olsen, Paul A. Christensen, S. Wesley Long, Sishir Subedi, Robert Olson, Marcus Nguyen, James J. Davis, Matthew Ojeda Saavedra, Prasanti Yerramilli, Layne Pruitt, Kristina Reppond, Madison N. Shyer, Jessica Cambric, Ryan Gadd, Ilya J. Finkelstein, Jimmy Gollihar, and James M. Musser |
| EPI_ISL_2204968 | Aegis Sciences Corporation | Centers for Disease Control and Prevention Division of Viral Diseases, Pathogen Discovery | Dakota Howard, Dhwani Batra, Peter W. Cook, Kara Moser, Adrian Paskey, Jason Caravas, Benjamin Rambo-Martin, Shatavia Morrison, Christopher Gulvick, Scott Sammons, Yvette Unoarumhi, Darlene Wagner, Matthew Schmerer, Cyndi Clark, Patrick Campbell, Rob Case, Vikramsinha Ghorpade, Holly Houdeshell, Ola Kvalvaag, Dillon Nall, Ethan Sanders, Alec Vest, Shaun Westlund, Matthew Hardison, Clinton R. Paden, Duncan MacCannell |
| EPI_ISL_2204969 | Houston Methodist Hospital | Houston Methodist Hospital | Randall J. Olsen, Paul A. Christensen, S. Wesley Long, Sishir Subedi, Robert Olson, Marcus Nguyen, James J. Davis, Matthew Ojeda Saavedra, Prasanti Yerramilli, Layne Pruitt, Kristina Reppond, Madison N. Shyer, Jessica Cambric, Ryan Gadd, Ilya J. Finkelstein, Jimmy Gollihar, and James M. Musser |
| EPI_ISL_2204970 | Aegis Sciences Corporation | Centers for Disease Control and Prevention Division of Viral Diseases, Pathogen Discovery | Dakota Howard, Dhwani Batra, Peter W. Cook, Kara Moser, Adrian Paskey, Jason Caravas, Benjamin Rambo-Martin, Shatavia Morrison, Christopher Gulvick, Scott Sammons, Yvette Unoarumhi, Darlene Wagner, Matthew Schmerer, Cyndi Clark, Patrick Campbell, Rob Case, Vikramsinha Ghorpade, Holly Houdeshell, Ola Kvalvaag, Dillon Nall, Ethan Sanders, Alec Vest, Shaun Westlund, Matthew Hardison, Clinton R. Paden, Duncan MacCannell |
| EPI_ISL_2204971 | Houston Methodist Hospital | Houston Methodist Hospital | Randall J. Olsen, Paul A. Christensen, S. Wesley Long, Sishir Subedi, Robert Olson, Marcus Nguyen, James J. Davis, Matthew Ojeda Saavedra, Prasanti Yerramilli, Layne Pruitt, Kristina Reppond, Madison N. Shyer, Jessica Cambric, Ryan Gadd, Ilya J. Finkelstein, Jimmy Gollihar, and James M. Musser |
| EPI_ISL_2204973 | Aegis Sciences Corporation | Centers for Disease Control and Prevention Division of Viral Diseases, Pathogen Discovery | Dakota Howard, Dhwani Batra, Peter W. Cook, Kara Moser, Adrian Paskey, Jason Caravas, Benjamin Rambo-Martin, Shatavia Morrison, Christopher Gulvick, Scott Sammons, Yvette Unoarumhi, Darlene Wagner, Matthew Schmerer, Cyndi Clark, Patrick Campbell, Rob Case, Vikramsinha Ghorpade, Holly Houdeshell, Ola Kvalvaag, Dillon Nall, Ethan Sanders, Alec Vest, Shaun Westlund, Matthew Hardison, Clinton R. Paden, Duncan MacCannell |
| EPI_ISL_2204974, EPI_ISL_2204976 | Houston Methodist Hospital | Houston Methodist Hospital | Randall J. Olsen, Paul A. Christensen, S. Wesley Long, Sishir Subedi, Robert Olson, Marcus Nguyen, James J. Davis, Matthew Ojeda Saavedra, Prasanti Yerramilli, Layne Pruitt, Kristina Reppond, Madison N. Shyer, Jessica Cambric, Ryan Gadd, Ilya J. Finkelstein, Jimmy Gollihar, and James M. Musser |
| EPI_ISL_2204977 | Aegis Sciences Corporation | Centers for Disease Control and Prevention Division of Viral Diseases, Pathogen Discovery | Dakota Howard, Dhwani Batra, Peter W. Cook, Kara Moser, Adrian Paskey, Jason Caravas, Benjamin Rambo-Martin, Shatavia Morrison, Christopher Gulvick, Scott Sammons, Yvette Unoarumhi, Darlene Wagner, Matthew Schmerer, Cyndi Clark, Patrick Campbell, Rob Case, Vikramsinha Ghorpade, Holly Houdeshell, Ola Kvalvaag, Dillon Nall, Ethan Sanders, Alec Vest, Shaun Westlund, Matthew Hardison, Clinton R. Paden, Duncan MacCannell |
| EPI_ISL_2204978 | Houston Methodist Hospital | Houston Methodist Hospital | Randall J. Olsen, Paul A. Christensen, S. Wesley Long, Sishir Subedi, Robert Olson, Marcus Nguyen, James J. Davis, Matthew Ojeda Saavedra, Prasanti Yerramilli, Layne Pruitt, Kristina Reppond, Madison N. Shyer, Jessica Cambric, Ryan Gadd, Ilya J. Finkelstein, Jimmy Gollihar, and James M. Musser |
| EPI_ISL_2204979, EPI_ISL_2204980 | Aegis Sciences Corporation | Centers for Disease Control and Prevention Division of Viral Diseases, Pathogen Discovery | Dakota Howard, Dhwani Batra, Peter W. Cook, Kara Moser, Adrian Paskey, Jason Caravas, Benjamin Rambo-Martin, Shatavia Morrison, Christopher Gulvick, Scott Sammons, Yvette Unoarumhi, Darlene Wagner, Matthew Schmerer, Cyndi Clark, Patrick Campbell, Rob Case, Vikramsinha Ghorpade, Holly Houdeshell, Ola Kvalvaag, Dillon Nall, Ethan Sanders, Alec Vest, Shaun Westlund, Matthew Hardison, Clinton R. Paden, Duncan MacCannell |
| EPI_ISL_2204981 | Houston Methodist Hospital | Houston Methodist Hospital | Randall J. Olsen, Paul A. Christensen, S. Wesley Long, Sishir Subedi, Robert Olson, Marcus Nguyen, James J. Davis, Matthew Ojeda Saavedra, Prasanti Yerramilli, Layne Pruitt, Kristina Reppond, Madison N. Shyer, Jessica Cambric, Ryan Gadd, Ilya J. Finkelstein, Jimmy Gollihar, and James M. Musser |
| EPI_ISL_2204982, EPI_ISL_2204984 | Aegis Sciences Corporation | Centers for Disease Control and Prevention Division of Viral Diseases, Pathogen Discovery | Dakota Howard, Dhwani Batra, Peter W. Cook, Kara Moser, Adrian Paskey, Jason Caravas, Benjamin Rambo-Martin, Shatavia Morrison, Christopher Gulvick, Scott Sammons, Yvette Unoarumhi, Darlene Wagner, Matthew Schmerer, Cyndi Clark, Patrick Campbell, Rob Case, Vikramsinha Ghorpade, Holly Houdeshell, Ola Kvalvaag, Dillon Nall, Ethan Sanders, Alec Vest, Shaun Westlund, Matthew Hardison, Clinton R. Paden, Duncan MacCannell |
| EPI_ISL_2204985 | Houston Methodist Hospital | Houston Methodist Hospital | Randall J. Olsen, Paul A. Christensen, S. Wesley Long, Sishir Subedi, Robert Olson, Marcus Nguyen, James J. Davis, Matthew Ojeda Saavedra, Prasanti Yerramilli, Layne Pruitt, Kristina Reppond, Madison N. Shyer, Jessica Cambric, Ryan Gadd, Ilya J. Finkelstein, Jimmy Gollihar, and James M. Musser |
| EPI_ISL_2204986 | Aegis Sciences Corporation | Centers for Disease Control and Prevention Division of Viral Diseases, Pathogen Discovery | Dakota Howard, Dhwani Batra, Peter W. Cook, Kara Moser, Adrian Paskey, Jason Caravas, Benjamin Rambo-Martin, Shatavia Morrison, Christopher Gulvick, Scott Sammons, Yvette Unoarumhi, Darlene Wagner, Matthew Schmerer, Cyndi Clark, Patrick Campbell, Rob Case, Vikramsinha Ghorpade, Holly Houdeshell, Ola Kvalvaag, Dillon Nall, Ethan Sanders, Alec Vest, Shaun Westlund, Matthew Hardison, Clinton R. Paden, Duncan MacCannell |
| EPI_ISL_2204987 | Nebraska Public Health Laboratory | NPHL COVID-19 Response Team | NPHL COVID-19 Response Team |
| EPI_ISL_2204988 | Houston Methodist Hospital | Houston Methodist Hospital | Randall J. Olsen, Paul A. Christensen, S. Wesley Long, Sishir Subedi, Robert Olson, Marcus Nguyen, James J. Davis, Matthew Ojeda Saavedra, Prasanti Yerramilli, Layne Pruitt, Kristina Reppond, Madison N. Shyer, Jessica Cambric, Ryan Gadd, Ilya J. Finkelstein, Jimmy Gollihar, and James M. Musser |
| EPI_ISL_2204989 | Aegis Sciences Corporation | Centers for Disease Control and Prevention Division of Viral Diseases, Pathogen Discovery | Dakota Howard, Dhwani Batra, Peter W. Cook, Kara Moser, Adrian Paskey, Jason Caravas, Benjamin Rambo-Martin, Shatavia Morrison, Christopher Gulvick, Scott Sammons, Yvette Unoarumhi, Darlene Wagner, Matthew Schmerer, Cyndi Clark, Patrick Campbell, Rob Case, Vikramsinha Ghorpade, Holly Houdeshell, Ola Kvalvaag, Dillon Nall, Ethan Sanders, Alec Vest, Shaun Westlund, Matthew Hardison, Clinton R. Paden, Duncan MacCannell |
| EPI_ISL_2204990, EPI_ISL_2204991 | Nebraska Public Health Laboratory | NPHL COVID-19 Response Team | NPHL COVID-19 Response Team |
| EPI_ISL_2204992 | Aegis Sciences Corporation | Centers for Disease Control and Prevention Division of Viral Diseases, Pathogen Discovery | Dakota Howard, Dhwani Batra, Peter W. Cook, Kara Moser, Adrian Paskey, Jason Caravas, Benjamin Rambo-Martin, Shatavia Morrison, Christopher Gulvick, Scott Sammons, Yvette Unoarumhi, Darlene Wagner, Matthew Schmerer, Cyndi Clark, Patrick Campbell, Rob Case, Vikramsinha Ghorpade, Holly Houdeshell, Ola Kvalvaag, Dillon Nall, Ethan Sanders, Alec Vest, Shaun Westlund, Matthew Hardison, Clinton R. Paden, Duncan MacCannell |
| EPI_ISL_2204993 | Houston Methodist Hospital | Houston Methodist Hospital | Randall J. Olsen, Paul A. Christensen, S. Wesley Long, Sishir Subedi, Robert Olson, Marcus Nguyen, James J. Davis, Matthew Ojeda Saavedra, Prasanti Yerramilli, Layne Pruitt, Kristina Reppond, Madison N. Shyer, Jessica Cambric, Ryan Gadd, Ilya J. Finkelstein, Jimmy Gollihar, and James M. Musser |
| EPI_ISL_2204994 | Nebraska Public Health Laboratory | NPHL COVID-19 Response Team | NPHL COVID-19 Response Team |
| EPI_ISL_2204995 | Houston Methodist Hospital | Houston Methodist Hospital | Randall J. Olsen, Paul A. Christensen, S. Wesley Long, Sishir Subedi, Robert Olson, Marcus Nguyen, James J. Davis, Matthew Ojeda Saavedra, Prasanti Yerramilli, Layne Pruitt, Kristina Reppond, Madison N. Shyer, Jessica Cambric, Ryan Gadd, Ilya J. Finkelstein, Jimmy Gollihar, and James M. Musser |
| EPI_ISL_2204996 | Aegis Sciences Corporation | Centers for Disease Control and Prevention Division of Viral Diseases, Pathogen Discovery | Dakota Howard, Dhwani Batra, Peter W. Cook, Kara Moser, Adrian Paskey, Jason Caravas, Benjamin Rambo-Martin, Shatavia Morrison, Christopher Gulvick, Scott Sammons, Yvette Unoarumhi, Darlene Wagner, Matthew Schmerer, Cyndi Clark, Patrick Campbell, Rob Case, Vikramsinha Ghorpade, Holly Houdeshell, Ola Kvalvaag, Dillon Nall, Ethan Sanders, Alec Vest, Shaun Westlund, Matthew Hardison, Clinton R. Paden, Duncan MacCannell |
| EPI_ISL_2204997, EPI_ISL_2204998 | Nebraska Public Health Laboratory | NPHL COVID-19 Response Team | NPHL COVID-19 Response Team |
| EPI_ISL_2204999 | Aegis Sciences Corporation | Centers for Disease Control and Prevention Division of Viral Diseases, Pathogen Discovery | Dakota Howard, Dhwani Batra, Peter W. Cook, Kara Moser, Adrian Paskey, Jason Caravas, Benjamin Rambo-Martin, Shatavia Morrison, Christopher Gulvick, Scott Sammons, Yvette Unoarumhi, Darlene Wagner, Matthew Schmerer, Cyndi Clark, Patrick Campbell, Rob Case, Vikramsinha Ghorpade, Holly Houdeshell, Ola Kvalvaag, Dillon Nall, Ethan Sanders, Alec Vest, Shaun Westlund, Matthew Hardison, Clinton R. Paden, Duncan MacCannell |
| EPI_ISL_2205000 | Houston Methodist Hospital | Houston Methodist Hospital | Randall J. Olsen, Paul A. Christensen, S. Wesley Long, Sishir Subedi, Robert Olson, Marcus Nguyen, James J. Davis, Matthew Ojeda Saavedra, Prasanti Yerramilli, Layne Pruitt, Kristina Reppond, Madison N. Shyer, Jessica Cambric, Ryan Gadd, Ilya J. Finkelstein, Jimmy Gollihar, and James M. Musser |
| EPI_ISL_2205001 | Nebraska Public Health Laboratory | NPHL COVID-19 Response Team | NPHL COVID-19 Response Team |
| EPI_ISL_2205002 | Aegis Sciences Corporation | Centers for Disease Control and Prevention Division of Viral Diseases, Pathogen Discovery | Dakota Howard, Dhwani Batra, Peter W. Cook, Kara Moser, Adrian Paskey, Jason Caravas, Benjamin Rambo-Martin, Shatavia Morrison, Christopher Gulvick, Scott Sammons, Yvette Unoarumhi, Darlene Wagner, Matthew Schmerer, Cyndi Clark, Patrick Campbell, Rob Case, Vikramsinha Ghorpade, Holly Houdeshell, Ola Kvalvaag, Dillon Nall, Ethan Sanders, Alec Vest, Shaun Westlund, Matthew Hardison, Clinton R. Paden, Duncan MacCannell |
| EPI_ISL_2205003 | Nebraska Public Health Laboratory | NPHL COVID-19 Response Team | NPHL COVID-19 Response Team |
| EPI_ISL_2205004 | Houston Methodist Hospital | Houston Methodist Hospital | Randall J. Olsen, Paul A. Christensen, S. Wesley Long, Sishir Subedi, Robert Olson, Marcus Nguyen, James J. Davis, Matthew Ojeda Saavedra, Prasanti Yerramilli, Layne Pruitt, Kristina Reppond, Madison N. Shyer, Jessica Cambric, Ryan Gadd, Ilya J. Finkelstein, Jimmy Gollihar, and James M. Musser |
| EPI_ISL_2205005, EPI_ISL_2205007 | Nebraska Public Health Laboratory | NPHL COVID-19 Response Team | NPHL COVID-19 Response Team |
| EPI_ISL_2205008 | Houston Methodist Hospital | Houston Methodist Hospital | Randall J. Olsen, Paul A. Christensen, S. Wesley Long, Sishir Subedi, Robert Olson, Marcus Nguyen, James J. Davis, Matthew Ojeda Saavedra, Prasanti Yerramilli, Layne Pruitt, Kristina Reppond, Madison N. Shyer, Jessica Cambric, Ryan Gadd, Ilya J. Finkelstein, Jimmy Gollihar, and James M. Musser |
| EPI_ISL_2205009 | Nebraska Public Health Laboratory | NPHL COVID-19 Response Team | NPHL COVID-19 Response Team |
| EPI_ISL_2205010 | Aegis Sciences Corporation | Centers for Disease Control and Prevention Division of Viral Diseases, Pathogen Discovery | Dakota Howard, Dhwani Batra, Peter W. Cook, Kara Moser, Adrian Paskey, Jason Caravas, Benjamin Rambo-Martin, Shatavia Morrison, Christopher Gulvick, Scott Sammons, Yvette Unoarumhi, Darlene Wagner, Matthew Schmerer, Cyndi Clark, Patrick Campbell, Rob Case, Vikramsinha Ghorpade, Holly Houdeshell, Ola Kvalvaag, Dillon Nall, Ethan Sanders, Alec Vest, Shaun Westlund, Matthew Hardison, Clinton R. Paden, Duncan MacCannell |
| EPI_ISL_2205011 | Nebraska Public Health Laboratory | NPHL COVID-19 Response Team | NPHL COVID-19 Response Team |
| EPI_ISL_2205012 | Aegis Sciences Corporation | Centers for Disease Control and Prevention Division of Viral Diseases, Pathogen Discovery | Dakota Howard, Dhwani Batra, Peter W. Cook, Kara Moser, Adrian Paskey, Jason Caravas, Benjamin Rambo-Martin, Shatavia Morrison, Christopher Gulvick, Scott Sammons, Yvette Unoarumhi, Darlene Wagner, Matthew Schmerer, Cyndi Clark, Patrick Campbell, Rob Case, Vikramsinha Ghorpade, Holly Houdeshell, Ola Kvalvaag, Dillon Nall, Ethan Sanders, Alec Vest, Shaun Westlund, Matthew Hardison, Clinton R. Paden, Duncan MacCannell |

| | | | |
|---|---|---|---|
| EPI_ISL_2205013 | Houston Methodist Hospital | Houston Methodist Hospital | Randall J. Olsen, Paul A. Christensen, S. Wesley Long, Sishir Subedi, Robert Olson, Marcus Nguyen, James J. Davis, Matthew Ojeda Saavedra, Prasanti Yerramilli, Layne Pruitt, Kristina Reppond, Madison N. Shyer, Jessica Cambric, Ryan Gadd, Ilya J. Finkelstein, Jimmy Gollihar, and James M. Musser |
| EPI_ISL_2205014, EPI_ISL_2205015 | Nebraska Public Health Laboratory | NPHL COVID-19 Response Team | NPHL COVID-19 Response Team |
| EPI_ISL_2205016 | Aegis Sciences Corporation | Centers for Disease Control and Prevention Division of Viral Diseases, Pathogen Discovery | Dakota Howard, Dhwani Batra, Peter W. Cook, Kara Moser, Adrian Paskey, Jason Caravas, Benjamin Rambo-Martin, Shatavia Morrison, Christopher Gulvick, Scott Sammons, Yvette Unoarumhi, Darlene Wagner, Matthew Schmerer, Cyndi Clark, Patrick Campbell, Rob Case, Vikramsinha Ghorpade, Holly Houdeshell, Ola Kvalvaag, Dillon Nall, Ethan Sanders, Alec Vest, Shaun Westlund, Matthew Hardison, Clinton R. Paden, Duncan MacCannell |
| EPI_ISL_2205017, EPI_ISL_2205019 | Nebraska Public Health Laboratory | NPHL COVID-19 Response Team | NPHL COVID-19 Response Team |
| EPI_ISL_2205020 | Aegis Sciences Corporation | Centers for Disease Control and Prevention Division of Viral Diseases, Pathogen Discovery | Dakota Howard, Dhwani Batra, Peter W. Cook, Kara Moser, Adrian Paskey, Jason Caravas, Benjamin Rambo-Martin, Shatavia Morrison, Christopher Gulvick, Scott Sammons, Yvette Unoarumhi, Darlene Wagner, Matthew Schmerer, Cyndi Clark, Patrick Campbell, Rob Case, Vikramsinha Ghorpade, Holly Houdeshell, Ola Kvalvaag, Dillon Nall, Ethan Sanders, Alec Vest, Shaun Westlund, Matthew Hardison, Clinton R. Paden, Duncan MacCannell |
| EPI_ISL_2205021 | Nebraska Public Health Laboratory | NPHL COVID-19 Response Team | NPHL COVID-19 Response Team |
| EPI_ISL_2205022 | Houston Methodist Hospital | Houston Methodist Hospital | Randall J. Olsen, Paul A. Christensen, S. Wesley Long, Sishir Subedi, Robert Olson, Marcus Nguyen, James J. Davis, Matthew Ojeda Saavedra, Prasanti Yerramilli, Layne Pruitt, Kristina Reppond, Madison N. Shyer, Jessica Cambric, Ryan Gadd, Ilya J. Finkelstein, Jimmy Gollihar, and James M. Musser |
| EPI_ISL_2205023, EPI_ISL_2205024, EPI_ISL_2205026, EPI_ISL_2205027 | Nebraska Public Health Laboratory | NPHL COVID-19 Response Team | NPHL COVID-19 Response Team |
| EPI_ISL_2205028 | Houston Methodist Hospital | Houston Methodist Hospital | Randall J. Olsen, Paul A. Christensen, S. Wesley Long, Sishir Subedi, Robert Olson, Marcus Nguyen, James J. Davis, Matthew Ojeda Saavedra, Prasanti Yerramilli, Layne Pruitt, Kristina Reppond, Madison N. Shyer, Jessica Cambric, Ryan Gadd, Ilya J. Finkelstein, Jimmy Gollihar, and James M. Musser |
| EPI_ISL_2205029 | Nebraska Public Health Laboratory | NPHL COVID-19 Response Team | NPHL COVID-19 Response Team |
| EPI_ISL_2205030 | Houston Methodist Hospital | Houston Methodist Hospital | Randall J. Olsen, Paul A. Christensen, S. Wesley Long, Sishir Subedi, Robert Olson, Marcus Nguyen, James J. Davis, Matthew Ojeda Saavedra, Prasanti Yerramilli, Layne Pruitt, Kristina Reppond, Madison N. Shyer, Jessica Cambric, Ryan Gadd, Ilya J. Finkelstein, Jimmy Gollihar, and James M. Musser |
| EPI_ISL_2205031 | Nebraska Public Health Laboratory | NPHL COVID-19 Response Team | NPHL COVID-19 Response Team |
| EPI_ISL_2205032, EPI_ISL_2205033, EPI_ISL_2205034, EPI_ISL_2205035, EPI_ISL_2205037, EPI_ISL_2205044, EPI_ISL_2205046, EPI_ISL_2205053, EPI_ISL_2205064, EPI_ISL_2205067, EPI_ISL_2205068, EPI_ISL_2205071, EPI_ISL_2205074, EPI_ISL_2205077, EPI_ISL_2205082, EPI_ISL_2205086, EPI_ISL_2205089, EPI_ISL_2205092, EPI_ISL_2205094, EPI_ISL_2205097, EPI_ISL_2205101, EPI_ISL_2205110, EPI_ISL_2205112, EPI_ISL_2205116, EPI_ISL_2205118, EPI_ISL_2205121, EPI_ISL_2205127, EPI_ISL_2205130, EPI_ISL_2205134, EPI_ISL_2205137, EPI_ISL_2205143, EPI_ISL_2205146, EPI_ISL_2205149, EPI_ISL_2205154, EPI_ISL_2205156, EPI_ISL_2205158, EPI_ISL_2205161, EPI_ISL_2205164, EPI_ISL_2205167, EPI_ISL_2205174, EPI_ISL_2205177, EPI_ISL_2205180, EPI_ISL_2205186, EPI_ISL_2205189, EPI_ISL_2205192, EPI_ISL_2205195, EPI_ISL_2205198, EPI_ISL_2205201, EPI_ISL_2205204, EPI_ISL_2205206, EPI_ISL_2205209, EPI_ISL_2205212, EPI_ISL_2205216, EPI_ISL_2205219, EPI_ISL_2205222, EPI_ISL_2205226, EPI_ISL_2205229, EPI_ISL_2205232, EPI_ISL_2205235, EPI_ISL_2205238, EPI_ISL_2205241, EPI_ISL_2205243, EPI_ISL_2205246, EPI_ISL_2205249, EPI_ISL_2205252, EPI_ISL_2205254, EPI_ISL_2205257, EPI_ISL_2205260, EPI_ISL_2205263, EPI_ISL_2205266, EPI_ISL_2205269, EPI_ISL_2205272, EPI_ISL_2205275, EPI_ISL_2205277, EPI_ISL_2205280, EPI_ISL_2205284, EPI_ISL_2205290, EPI_ISL_2205296, EPI_ISL_2205299, EPI_ISL_2205303, EPI_ISL_2205306, EPI_ISL_2205308, EPI_ISL_2205311, EPI_ISL_2205315, EPI_ISL_2205317, EPI_ISL_2205320, EPI_ISL_2205331, EPI_ISL_2205334, EPI_ISL_2205337, EPI_ISL_2205340, EPI_ISL_2205345, EPI_ISL_2205348, EPI_ISL_2205360, EPI_ISL_2205366, EPI_ISL_2205369, EPI_ISL_2205376, EPI_ISL_2205380, EPI_ISL_2205383, EPI_ISL_2205386, EPI_ISL_2205389, EPI_ISL_2205392, EPI_ISL_2205395, EPI_ISL_2205398, EPI_ISL_2205401, EPI_ISL_2205405, EPI_ISL_2205408, EPI_ISL_2205411, EPI_ISL_2205414, EPI_ISL_2205417, EPI_ISL_2205420, EPI_ISL_2205425, EPI_ISL_2205428, EPI_ISL_2205433, EPI_ISL_2205434, EPI_ISL_2205440, EPI_ISL_2205443, EPI_ISL_2205449, EPI_ISL_2205451, EPI_ISL_2205457, EPI_ISL_2205460, EPI_ISL_2205463, EPI_ISL_2205466, EPI_ISL_2205469, EPI_ISL_2205473, EPI_ISL_2205475, EPI_ISL_2205478, EPI_ISL_2205482, EPI_ISL_2205485, EPI_ISL_2205488, EPI_ISL_2205490, EPI_ISL_2205496, EPI_ISL_2205502, EPI_ISL_2205506, EPI_ISL_2205509, EPI_ISL_2205515, EPI_ISL_2205518, EPI_ISL_2205520, EPI_ISL_2205523, EPI_ISL_2205525, EPI_ISL_2205528, EPI_ISL_2205532, EPI_ISL_2205535, EPI_ISL_2205538, EPI_ISL_2205542, EPI_ISL_2205545, EPI_ISL_2205548, EPI_ISL_2205551, EPI_ISL_2205554, EPI_ISL_2205557, EPI_ISL_2205560, EPI_ISL_2205563, EPI_ISL_2205566, EPI_ISL_2205570, EPI_ISL_2205573 | | | |
| see above | Houston Methodist Hospital | Houston Methodist Hospital | Randall J. Olsen, Paul A. Christensen, S. Wesley Long, Sishir Subedi, Robert Olson, Marcus Nguyen, James J. Davis, Matthew Ojeda Saavedra, Prasanti Yerramilli, Layne Pruitt, Kristina Reppond, Madison N. Shyer, Jessica Cambric, Ryan Gadd, Ilya J. Finkelstein, Jimmy Gollihar, and James M. Musser |
| EPI_ISL_2205574 | Nebraska Public Health Laboratory | NPHL COVID-19 Response Team | NPHL COVID-19 Response Team |
| EPI_ISL_2205576 | Houston Methodist Hospital | Houston Methodist Hospital | Randall J. Olsen, Paul A. Christensen, S. Wesley Long, Sishir Subedi, Robert Olson, Marcus Nguyen, James J. Davis, Matthew Ojeda Saavedra, Prasanti Yerramilli, Layne Pruitt, Kristina Reppond, Madison N. Shyer, Jessica Cambric, Ryan Gadd, Ilya J. Finkelstein, Jimmy Gollihar, and James M. Musser |
| EPI_ISL_2205578 | Nebraska Public Health Laboratory | NPHL COVID-19 Response Team | NPHL COVID-19 Response Team |
| EPI_ISL_2205581 | Houston Methodist Hospital | Houston Methodist Hospital | Randall J. Olsen, Paul A. Christensen, S. Wesley Long, Sishir Subedi, Robert Olson, Marcus Nguyen, James J. Davis, Matthew Ojeda Saavedra, Prasanti Yerramilli, Layne Pruitt, Kristina Reppond, Madison N. Shyer, Jessica Cambric, Ryan Gadd, Ilya J. Finkelstein, Jimmy Gollihar, and James M. Musser |
| EPI_ISL_2205582 | Nebraska Public Health Laboratory | NPHL COVID-19 Response Team | NPHL COVID-19 Response Team |
| EPI_ISL_2205585 | Houston Methodist Hospital | Houston Methodist Hospital | Randall J. Olsen, Paul A. Christensen, S. Wesley Long, Sishir Subedi, Robert Olson, Marcus Nguyen, James J. Davis, Matthew Ojeda Saavedra, Prasanti Yerramilli, Layne Pruitt, Kristina Reppond, Madison N. Shyer, Jessica Cambric, Ryan Gadd, Ilya J. Finkelstein, Jimmy Gollihar, and James M. Musser |
| EPI_ISL_2205586, EPI_ISL_2205588, EPI_ISL_2205591, EPI_ISL_2205593 | Nebraska Public Health Laboratory | NPHL COVID-19 Response Team | NPHL COVID-19 Response Team |
| EPI_ISL_2205594, EPI_ISL_2205597 | Houston Methodist Hospital | Houston Methodist Hospital | Randall J. Olsen, Paul A. Christensen, S. Wesley Long, Sishir Subedi, Robert Olson, Marcus Nguyen, James J. Davis, Matthew Ojeda Saavedra, Prasanti Yerramilli, Layne Pruitt, Kristina Reppond, Madison N. Shyer, Jessica Cambric, Ryan Gadd, Ilya J. Finkelstein, Jimmy Gollihar, and James M. Musser |
| EPI_ISL_2205599, EPI_ISL_2205604 | Nebraska Public Health Laboratory | NPHL COVID-19 Response Team | NPHL COVID-19 Response Team |
| EPI_ISL_2205606 | Houston Methodist Hospital | Houston Methodist Hospital | Randall J. Olsen, Paul A. Christensen, S. Wesley Long, Sishir Subedi, Robert Olson, Marcus Nguyen, James J. Davis, Matthew Ojeda Saavedra, Prasanti Yerramilli, Layne Pruitt, Kristina Reppond, Madison N. Shyer, Jessica Cambric, Ryan Gadd, Ilya J. Finkelstein, Jimmy Gollihar, and James M. Musser |
| EPI_ISL_2205608 | Nebraska Public Health Laboratory | NPHL COVID-19 Response Team | NPHL COVID-19 Response Team |
| EPI_ISL_2205610 | Houston Methodist Hospital | Houston Methodist Hospital | Randall J. Olsen, Paul A. Christensen, S. Wesley Long, Sishir Subedi, Robert Olson, Marcus Nguyen, James J. Davis, Matthew Ojeda Saavedra, Prasanti Yerramilli, Layne Pruitt, Kristina Reppond, Madison N. Shyer, Jessica Cambric, Ryan Gadd, Ilya J. Finkelstein, Jimmy Gollihar, and James M. Musser |
| EPI_ISL_2205612 | Nebraska Public Health Laboratory | NPHL COVID-19 Response Team | NPHL COVID-19 Response Team |
| EPI_ISL_2205613, EPI_ISL_2205616 | Houston Methodist Hospital | Houston Methodist Hospital | Randall J. Olsen, Paul A. Christensen, S. Wesley Long, Sishir Subedi, Robert Olson, Marcus Nguyen, James J. Davis, Matthew Ojeda Saavedra, Prasanti Yerramilli, Layne Pruitt, Kristina Reppond, Madison N. Shyer, Jessica Cambric, Ryan Gadd, Ilya J. Finkelstein, Jimmy Gollihar, and James M. Musser |
| EPI_ISL_2205618, EPI_ISL_2205620 | Nebraska Public Health Laboratory | NPHL COVID-19 Response Team | NPHL COVID-19 Response Team |
| EPI_ISL_2205621 | Houston Methodist Hospital | Houston Methodist Hospital | Randall J. Olsen, Paul A. Christensen, S. Wesley Long, Sishir Subedi, Robert Olson, Marcus Nguyen, James J. Davis, Matthew Ojeda Saavedra, Prasanti Yerramilli, Layne Pruitt, Kristina Reppond, Madison N. Shyer, Jessica Cambric, Ryan Gadd, Ilya J. Finkelstein, Jimmy Gollihar, and James M. Musser |
| EPI_ISL_2205624 | Nebraska Public Health Laboratory | NPHL COVID-19 Response Team | NPHL COVID-19 Response Team |
| EPI_ISL_2205627 | Houston Methodist Hospital | Houston Methodist Hospital | Randall J. Olsen, Paul A. Christensen, S. Wesley Long, Sishir Subedi, Robert Olson, Marcus Nguyen, James J. Davis, Matthew Ojeda Saavedra, Prasanti Yerramilli, Layne Pruitt, Kristina Reppond, Madison N. Shyer, Jessica Cambric, Ryan Gadd, Ilya J. Finkelstein, Jimmy Gollihar, and James M. Musser |
| EPI_ISL_2205628, EPI_ISL_2205632 | Nebraska Public Health Laboratory | NPHL COVID-19 Response Team | NPHL COVID-19 Response Team |
| EPI_ISL_2205633 | Houston Methodist Hospital | Houston Methodist Hospital | Randall J. Olsen, Paul A. Christensen, S. Wesley Long, Sishir Subedi, Robert Olson, Marcus Nguyen, James J. Davis, Matthew Ojeda Saavedra, Prasanti Yerramilli, Layne Pruitt, Kristina Reppond, Madison N. Shyer, Jessica Cambric, Ryan Gadd, Ilya J. Finkelstein, Jimmy Gollihar, and James M. Musser |
| EPI_ISL_2205636 | Nebraska Public Health Laboratory | NPHL COVID-19 Response Team | NPHL COVID-19 Response Team |
| EPI_ISL_2205639 | Houston Methodist Hospital | Houston Methodist Hospital | Randall J. Olsen, Paul A. Christensen, S. Wesley Long, Sishir Subedi, Robert Olson, Marcus Nguyen, James J. Davis, Matthew Ojeda Saavedra, Prasanti Yerramilli, Layne Pruitt, Kristina Reppond, Madison N. Shyer, Jessica Cambric, Ryan Gadd, Ilya J. Finkelstein, Jimmy Gollihar, and James M. Musser |
| EPI_ISL_2205640, EPI_ISL_2205643, EPI_ISL_2205652 | Nebraska Public Health Laboratory | NPHL COVID-19 Response Team | NPHL COVID-19 Response Team |
| EPI_ISL_2205653, EPI_ISL_2205655 | Houston Methodist Hospital | Houston Methodist Hospital | Randall J. Olsen, Paul A. Christensen, S. Wesley Long, Sishir Subedi, Robert Olson, Marcus Nguyen, James J. Davis, Matthew Ojeda Saavedra, Prasanti Yerramilli, Layne Pruitt, Kristina Reppond, Madison N. Shyer, Jessica Cambric, Ryan Gadd, Ilya J. Finkelstein, Jimmy Gollihar, and James M. Musser |
| EPI_ISL_2205657 | Nebraska Public Health Laboratory | NPHL COVID-19 Response Team | NPHL COVID-19 Response Team |
| EPI_ISL_2205659, EPI_ISL_2205662, EPI_ISL_2205666, EPI_ISL_2205669, EPI_ISL_2205672, EPI_ISL_2205677, EPI_ISL_2205681, EPI_ISL_2205685, EPI_ISL_2205689, EPI_ISL_2205692, EPI_ISL_2205695, EPI_ISL_2205700, EPI_ISL_2205706, EPI_ISL_2205709, EPI_ISL_2205711, EPI_ISL_2205715, EPI_ISL_2205718, EPI_ISL_2205721, EPI_ISL_2205724, EPI_ISL_2205726, EPI_ISL_2205729, EPI_ISL_2205732, EPI_ISL_2205734, EPI_ISL_2205737, EPI_ISL_2205740, EPI_ISL_2205742, EPI_ISL_2205746, EPI_ISL_2205750, EPI_ISL_2205753, EPI_ISL_2205757, EPI_ISL_2205761, EPI_ISL_2205763, EPI_ISL_2205766, EPI_ISL_2205769, EPI_ISL_2205772, EPI_ISL_2205775, EPI_ISL_2205778, EPI_ISL_2205781, EPI_ISL_2205785, EPI_ISL_2205788, EPI_ISL_2205791, EPI_ISL_2205795, EPI_ISL_2205797, EPI_ISL_2205798, EPI_ISL_2205800, EPI_ISL_2205803, EPI_ISL_2205806, EPI_ISL_2205809, EPI_ISL_2205812, EPI_ISL_2205815, EPI_ISL_2205818, EPI_ISL_2205822, EPI_ISL_2205825, EPI_ISL_2205828, EPI_ISL_2205831, EPI_ISL_2205833, EPI_ISL_2205835, EPI_ISL_2205837, EPI_ISL_2205840, EPI_ISL_2205842, EPI_ISL_2205845, EPI_ISL_2205848, EPI_ISL_2205850, EPI_ISL_2205852, EPI_ISL_2205855, EPI_ISL_2205858, EPI_ISL_2205861, EPI_ISL_2205864, EPI_ISL_2205867, EPI_ISL_2205869, EPI_ISL_2205872, EPI_ISL_2205875, EPI_ISL_2205878, EPI_ISL_2205881, EPI_ISL_2205884, EPI_ISL_2205887, EPI_ISL_2205891, EPI_ISL_2205893, EPI_ISL_2205896, EPI_ISL_2205899, EPI_ISL_2205902, EPI_ISL_2205905, EPI_ISL_2205908, EPI_ISL_2205910, EPI_ISL_2205913, EPI_ISL_2205916, EPI_ISL_2205917, EPI_ISL_2205920, EPI_ISL_2205923, EPI_ISL_2205926, EPI_ISL_2205929, EPI_ISL_2205932, EPI_ISL_2205935, EPI_ISL_2205938, EPI_ISL_2205941, EPI_ISL_2205944, EPI_ISL_2205947, EPI_ISL_2205950, EPI_ISL_2205953, EPI_ISL_2205955, EPI_ISL_2205958, EPI_ISL_2205960, EPI_ISL_2205964, EPI_ISL_2205967, EPI_ISL_2205970, EPI_ISL_2205973, EPI_ISL_2205976, EPI_ISL_2205979, EPI_ISL_2205983, EPI_ISL_2205988, EPI_ISL_2205991, EPI_ISL_2205995, EPI_ISL_2205999, EPI_ISL_2206003, EPI_ISL_2206006, EPI_ISL_2206009, EPI_ISL_2206012, EPI_ISL_2206015, EPI_ISL_2206018, EPI_ISL_2206021, EPI_ISL_2206024, EPI_ISL_2206026, EPI_ISL_2206029, EPI_ISL_2206032, EPI_ISL_2206035, EPI_ISL_2206038, EPI_ISL_2206041, EPI_ISL_2206044, EPI_ISL_2206048, EPI_ISL_2206053, EPI_ISL_2206057, EPI_ISL_2206060, EPI_ISL_2206063, EPI_ISL_2206067, EPI_ISL_2206069, EPI_ISL_2206072, EPI_ISL_2206074, EPI_ISL_2206077, EPI_ISL_2206083, EPI_ISL_2206086, EPI_ISL_2206088, EPI_ISL_2206093, EPI_ISL_2206095, EPI_ISL_2206099, EPI_ISL_2206101, EPI_ISL_2206104, EPI_ISL_2206107, EPI_ISL_2206110, EPI_ISL_2206113, EPI_ISL_2206116, EPI_ISL_2206119, EPI_ISL_2206122, EPI_ISL_2206125, EPI_ISL_2206128, EPI_ISL_2206131, EPI_ISL_2206134, EPI_ISL_2206136, EPI_ISL_2206140, EPI_ISL_2206142, EPI_ISL_2206146, EPI_ISL_2206149, EPI_ISL_2206152, EPI_ISL_2206155, EPI_ISL_2206156, EPI_ISL_2206159, EPI_ISL_2206161, EPI_ISL_2206166, EPI_ISL_2206169, EPI_ISL_2206172, EPI_ISL_2206175, EPI_ISL_2206178, EPI_ISL_2206181, EPI_ISL_2206184, EPI_ISL_2206187, EPI_ISL_2206190, EPI_ISL_2206193, EPI_ISL_2206196, EPI_ISL_2206199, EPI_ISL_2206201, EPI_ISL_2206203, EPI_ISL_2206206, EPI_ISL_2206209, EPI_ISL_2206212, EPI_ISL_2206215, EPI_ISL_2206219, EPI_ISL_2206222, EPI_ISL_2206227, EPI_ISL_2206229, EPI_ISL_2206231, EPI_ISL_2206234, EPI_ISL_2206237, EPI_ISL_2206241, EPI_ISL_2206245, EPI_ISL_2206249, EPI_ISL_2206253, EPI_ISL_2206257, EPI_ISL_2206261, EPI_ISL_2206263, EPI_ISL_2206268 | | | |
| see above | Houston Methodist Hospital | Houston Methodist Hospital | Randall J. Olsen, Paul A. Christensen, S. Wesley Long, Sishir Subedi, Robert Olson, Marcus Nguyen, James J. Davis, Matthew Ojeda Saavedra, Prasanti Yerramilli, Layne Pruitt, Kristina Reppond, Madison N. Shyer, Jessica Cambric, Ryan Gadd, Ilya J. Finkelstein, Jimmy Gollihar, and James M. Musser |
| EPI_ISL_2206271, | Nebraska Public Health Laboratory | NPHL COVID-19 Response Team | NPHL COVID-19 Response Team |

| | | | |
|---|---|---|---|
| EPI_ISL_2206273, EPI_ISL_2206275 | | | |
| EPI_ISL_2206276 | Houston Methodist Hospital | Houston Methodist Hospital | Randall J. Olsen, Paul A. Christensen, S. Wesley Long, Sishir Subedi, Robert Olson, Marcus Nguyen, James J. Davis, Matthew Ojeda Saavedra, Prasanti Yerramilli, Layne Pruitt, Kristina Reppond, Madison N. Shyer, Jessica Cambric, Ryan Gadd, Ilya J. Finkelstein, Jimmy Gollihar, and James M. Musser |
| EPI_ISL_2206278, EPI_ISL_2206280 | Nebraska Public Health Laboratory | NPHL COVID-19 Response Team | NPHL COVID-19 Response Team |
| EPI_ISL_2206281 | Houston Methodist Hospital | Houston Methodist Hospital | Randall J. Olsen, Paul A. Christensen, S. Wesley Long, Sishir Subedi, Robert Olson, Marcus Nguyen, James J. Davis, Matthew Ojeda Saavedra, Prasanti Yerramilli, Layne Pruitt, Kristina Reppond, Madison N. Shyer, Jessica Cambric, Ryan Gadd, Ilya J. Finkelstein, Jimmy Gollihar, and James M. Musser |
| EPI_ISL_2206283, EPI_ISL_2206285 | Nebraska Public Health Laboratory | NPHL COVID-19 Response Team | NPHL COVID-19 Response Team |
| EPI_ISL_2206286 | Houston Methodist Hospital | Houston Methodist Hospital | Randall J. Olsen, Paul A. Christensen, S. Wesley Long, Sishir Subedi, Robert Olson, Marcus Nguyen, James J. Davis, Matthew Ojeda Saavedra, Prasanti Yerramilli, Layne Pruitt, Kristina Reppond, Madison N. Shyer, Jessica Cambric, Ryan Gadd, Ilya J. Finkelstein, Jimmy Gollihar, and James M. Musser |
| EPI_ISL_2206288, EPI_ISL_2206290 | Nebraska Public Health Laboratory | NPHL COVID-19 Response Team | NPHL COVID-19 Response Team |
| EPI_ISL_2206291 | Houston Methodist Hospital | Houston Methodist Hospital | Randall J. Olsen, Paul A. Christensen, S. Wesley Long, Sishir Subedi, Robert Olson, Marcus Nguyen, James J. Davis, Matthew Ojeda Saavedra, Prasanti Yerramilli, Layne Pruitt, Kristina Reppond, Madison N. Shyer, Jessica Cambric, Ryan Gadd, Ilya J. Finkelstein, Jimmy Gollihar, and James M. Musser |
| EPI_ISL_2206293 | Nebraska Public Health Laboratory | NPHL COVID-19 Response Team | NPHL COVID-19 Response Team |
| EPI_ISL_2206295, EPI_ISL_2206300, EPI_ISL_2206309, EPI_ISL_2206315, EPI_ISL_2206322, EPI_ISL_2206328, EPI_ISL_2206331, EPI_ISL_2206333, EPI_ISL_2206337, EPI_ISL_2206340, EPI_ISL_2206343, EPI_ISL_2206345, EPI_ISL_2206348, EPI_ISL_2206351, EPI_ISL_2206355, EPI_ISL_2206361, EPI_ISL_2206365, EPI_ISL_2206369, EPI_ISL_2206372, EPI_ISL_2206375, EPI_ISL_2206378, EPI_ISL_2206382, EPI_ISL_2206385, EPI_ISL_2206386, EPI_ISL_2206389, EPI_ISL_2206393, EPI_ISL_2206395, EPI_ISL_2206399, EPI_ISL_2206401, EPI_ISL_2206405, EPI_ISL_2206408, EPI_ISL_2206411, EPI_ISL_2206414, EPI_ISL_2206417, EPI_ISL_2206420, EPI_ISL_2206423, EPI_ISL_2206426, EPI_ISL_2206429, EPI_ISL_2206433, EPI_ISL_2206436, EPI_ISL_2206439, EPI_ISL_2206443, EPI_ISL_2206445, EPI_ISL_2206448, EPI_ISL_2206451, EPI_ISL_2206454, EPI_ISL_2206458, EPI_ISL_2206461, EPI_ISL_2206464, EPI_ISL_2206467, EPI_ISL_2206470, EPI_ISL_2206472, EPI_ISL_2206478, EPI_ISL_2206481, EPI_ISL_2206484, EPI_ISL_2206486, EPI_ISL_2206490, EPI_ISL_2206492, EPI_ISL_2206496, EPI_ISL_2206497, EPI_ISL_2206501, EPI_ISL_2206504, EPI_ISL_2206507, EPI_ISL_2206510, EPI_ISL_2206513, EPI_ISL_2206516, EPI_ISL_2206523, EPI_ISL_2206528, EPI_ISL_2206535, EPI_ISL_2206538, EPI_ISL_2206539, EPI_ISL_2206540, EPI_ISL_2206543, EPI_ISL_2206546, EPI_ISL_2206550, EPI_ISL_2206553, EPI_ISL_2206558, EPI_ISL_2206562, EPI_ISL_2206565, EPI_ISL_2206569, EPI_ISL_2206572, EPI_ISL_2206576, EPI_ISL_2206579, EPI_ISL_2206582, EPI_ISL_2206586, EPI_ISL_2206590, EPI_ISL_2206594, EPI_ISL_2206597, EPI_ISL_2206600, EPI_ISL_2206604, EPI_ISL_2206609, EPI_ISL_2206614, EPI_ISL_2206617, EPI_ISL_2206619, EPI_ISL_2206622, EPI_ISL_2206626, EPI_ISL_2206629, EPI_ISL_2206631, EPI_ISL_2206634, EPI_ISL_2206638, EPI_ISL_2206641, EPI_ISL_2206644, EPI_ISL_2206647, EPI_ISL_2206650, EPI_ISL_2206652, EPI_ISL_2206656, EPI_ISL_2206659, EPI_ISL_2206662, EPI_ISL_2206666, EPI_ISL_2206669, EPI_ISL_2206673, EPI_ISL_2206675, EPI_ISL_2206678, EPI_ISL_2206680, EPI_ISL_2206683, EPI_ISL_2206685, EPI_ISL_2206688, EPI_ISL_2206691, EPI_ISL_2206695, EPI_ISL_2206698, EPI_ISL_2206701, EPI_ISL_2206704, EPI_ISL_2206707, EPI_ISL_2206711, EPI_ISL_2206714, EPI_ISL_2206716, EPI_ISL_2206718, EPI_ISL_2206722, EPI_ISL_2206726, EPI_ISL_2206729, EPI_ISL_2206734, EPI_ISL_2206738, EPI_ISL_2206742, EPI_ISL_2206745, EPI_ISL_2206748, EPI_ISL_2206752, EPI_ISL_2206755, EPI_ISL_2206758, EPI_ISL_2206761, EPI_ISL_2206764, EPI_ISL_2206768, EPI_ISL_2206773, EPI_ISL_2206776, EPI_ISL_2206779, EPI_ISL_2206782, EPI_ISL_2206785, EPI_ISL_2206787, EPI_ISL_2206792, EPI_ISL_2206797, EPI_ISL_2206800, EPI_ISL_2206806, EPI_ISL_2206812, EPI_ISL_2206817, EPI_ISL_2206821, EPI_ISL_2206823, EPI_ISL_2206826, EPI_ISL_2206829, EPI_ISL_2206832, EPI_ISL_2206834, EPI_ISL_2206837, EPI_ISL_2206840, EPI_ISL_2206843, EPI_ISL_2206849, EPI_ISL_2206852, EPI_ISL_2206855, EPI_ISL_2206858, EPI_ISL_2206861, EPI_ISL_2206865, EPI_ISL_2206869, EPI_ISL_2206872, EPI_ISL_2206875, EPI_ISL_2206878, EPI_ISL_2206881, EPI_ISL_2206882, EPI_ISL_2206886, EPI_ISL_2206889, EPI_ISL_2206893, EPI_ISL_2206897, EPI_ISL_2206900, EPI_ISL_2206903, EPI_ISL_2206906, EPI_ISL_2206909, EPI_ISL_2206912, EPI_ISL_2206915, EPI_ISL_2206919, EPI_ISL_2206922, EPI_ISL_2206925, EPI_ISL_2206929, EPI_ISL_2206932, EPI_ISL_2206936, EPI_ISL_2206939, EPI_ISL_2206942, EPI_ISL_2206945, EPI_ISL_2206947, EPI_ISL_2206950, EPI_ISL_2206953, EPI_ISL_2206956, EPI_ISL_2206959, EPI_ISL_2206962, EPI_ISL_2206965, EPI_ISL_2206969, EPI_ISL_2206972, EPI_ISL_2206975, EPI_ISL_2206978, EPI_ISL_2206982 | | | |
| see above | Houston Methodist Hospital | Houston Methodist Hospital | Randall J. Olsen, Paul A. Christensen, S. Wesley Long, Sishir Subedi, Robert Olson, Marcus Nguyen, James J. Davis, Matthew Ojeda Saavedra, Prasanti Yerramilli, Layne Pruitt, Kristina Reppond, Madison N. Shyer, Jessica Cambric, Ryan Gadd, Ilya J. Finkelstein, Jimmy Gollihar, and James M. Musser |
| EPI_ISL_2206984, EPI_ISL_2206986 | Nebraska Public Health Laboratory | NPHL COVID-19 Response Team | NPHL COVID-19 Response Team |
| EPI_ISL_2206987 | Houston Methodist Hospital | Houston Methodist Hospital | Randall J. Olsen, Paul A. Christensen, S. Wesley Long, Sishir Subedi, Robert Olson, Marcus Nguyen, James J. Davis, Matthew Ojeda Saavedra, Prasanti Yerramilli, Layne Pruitt, Kristina Reppond, Madison N. Shyer, Jessica Cambric, Ryan Gadd, Ilya J. Finkelstein, Jimmy Gollihar, and James M. Musser |
| EPI_ISL_2206989, EPI_ISL_2206991 | Nebraska Public Health Laboratory | NPHL COVID-19 Response Team | NPHL COVID-19 Response Team |
| EPI_ISL_2206992, EPI_ISL_2206996 | Houston Methodist Hospital | Houston Methodist Hospital | Randall J. Olsen, Paul A. Christensen, S. Wesley Long, Sishir Subedi, Robert Olson, Marcus Nguyen, James J. Davis, Matthew Ojeda Saavedra, Prasanti Yerramilli, Layne Pruitt, Kristina Reppond, Madison N. Shyer, Jessica Cambric, Ryan Gadd, Ilya J. Finkelstein, Jimmy Gollihar, and James M. Musser |
| EPI_ISL_2206997, EPI_ISL_2206999, EPI_ISL_2207001, EPI_ISL_2207003, EPI_ISL_2207005, EPI_ISL_2207007, EPI_ISL_2207009, EPI_ISL_2207011 | Nebraska Public Health Laboratory | NPHL COVID-19 Response Team | NPHL COVID-19 Response Team |
| EPI_ISL_2207013, EPI_ISL_2207015 | Houston Methodist Hospital | Houston Methodist Hospital | Randall J. Olsen, Paul A. Christensen, S. Wesley Long, Sishir Subedi, Robert Olson, Marcus Nguyen, James J. Davis, Matthew Ojeda Saavedra, Prasanti Yerramilli, Layne Pruitt, Kristina Reppond, Madison N. Shyer, Jessica Cambric, Ryan Gadd, Ilya J. Finkelstein, Jimmy Gollihar, and James M. Musser |
| EPI_ISL_2207016, EPI_ISL_2207018 | Nebraska Public Health Laboratory | NPHL COVID-19 Response Team | NPHL COVID-19 Response Team |
| EPI_ISL_2207020, EPI_ISL_2207023, EPI_ISL_2207026, EPI_ISL_2207029, EPI_ISL_2207032, EPI_ISL_2207035, EPI_ISL_2207039, EPI_ISL_2207043, EPI_ISL_2207046, EPI_ISL_2207049, EPI_ISL_2207056, EPI_ISL_2207061, EPI_ISL_2207064, EPI_ISL_2207069, EPI_ISL_2207073, EPI_ISL_2207078, EPI_ISL_2207082, EPI_ISL_2207088, EPI_ISL_2207092, EPI_ISL_2207094, EPI_ISL_2207097, EPI_ISL_2207099, EPI_ISL_2207101, EPI_ISL_2207102, EPI_ISL_2207107, EPI_ISL_2207112, EPI_ISL_2207116, EPI_ISL_2207120, EPI_ISL_2207124, EPI_ISL_2207129, EPI_ISL_2207135, EPI_ISL_2207140, EPI_ISL_2207144, EPI_ISL_2207148, EPI_ISL_2207151, EPI_ISL_2207154, EPI_ISL_2207157, EPI_ISL_2207160, EPI_ISL_2207162, EPI_ISL_2207165, EPI_ISL_2207168, EPI_ISL_2207170, EPI_ISL_2207174, EPI_ISL_2207176, EPI_ISL_2207179, EPI_ISL_2207182, EPI_ISL_2207185, EPI_ISL_2207187, EPI_ISL_2207190, EPI_ISL_2207192, EPI_ISL_2207195, EPI_ISL_2207198, EPI_ISL_2207200, EPI_ISL_2207203, EPI_ISL_2207206, EPI_ISL_2207209, EPI_ISL_2207212, EPI_ISL_2207215, EPI_ISL_2207219, EPI_ISL_2207223, EPI_ISL_2207225, EPI_ISL_2207228, EPI_ISL_2207231, EPI_ISL_2207234, EPI_ISL_2207237, EPI_ISL_2207240, EPI_ISL_2207243, EPI_ISL_2207246 | | | |
| see above | Houston Methodist Hospital | Houston Methodist Hospital | Randall J. Olsen, Paul A. Christensen, S. Wesley Long, Sishir Subedi, Robert Olson, Marcus Nguyen, James J. Davis, Matthew Ojeda Saavedra, Prasanti Yerramilli, Layne Pruitt, Kristina Reppond, Madison N. Shyer, Jessica Cambric, Ryan Gadd, Ilya J. Finkelstein, Jimmy Gollihar, and James M. Musser |
| EPI_ISL_2207247, EPI_ISL_2207249, EPI_ISL_2207251 | TriCore Reference Laboratories | Center for Global Health, University of New Mexico Health Sciences Center | Daryl Domman, Kurt Schwalm, Valerie Morley, Cecilia Thompson, Kendra Pesko, Karissa Culbreath, Darrell Dinwiddie |
| EPI_ISL_2207252 | Houston Methodist Hospital | Houston Methodist Hospital | Randall J. Olsen, Paul A. Christensen, S. Wesley Long, Sishir Subedi, Robert Olson, Marcus Nguyen, James J. Davis, Matthew Ojeda Saavedra, Prasanti Yerramilli, Layne Pruitt, Kristina Reppond, Madison N. Shyer, Jessica Cambric, Ryan Gadd, Ilya J. Finkelstein, Jimmy Gollihar, and James M. Musser |
| EPI_ISL_2207253, EPI_ISL_2207255, EPI_ISL_2207257 | TriCore Reference Laboratories | Center for Global Health, University of New Mexico Health Sciences Center | Daryl Domman, Kurt Schwalm, Valerie Morley, Cecilia Thompson, Kendra Pesko, Karissa Culbreath, Darrell Dinwiddie |
| EPI_ISL_2207258 | Houston Methodist Hospital | Houston Methodist Hospital | Randall J. Olsen, Paul A. Christensen, S. Wesley Long, Sishir Subedi, Robert Olson, Marcus Nguyen, James J. Davis, Matthew Ojeda Saavedra, Prasanti Yerramilli, Layne Pruitt, Kristina Reppond, Madison N. Shyer, Jessica Cambric, Ryan Gadd, Ilya J. Finkelstein, Jimmy Gollihar, and James M. Musser |
| EPI_ISL_2207259, EPI_ISL_2207261 | TriCore Reference Laboratories | Center for Global Health, University of New Mexico Health Sciences Center | Daryl Domman, Kurt Schwalm, Valerie Morley, Cecilia Thompson, Kendra Pesko, Karissa Culbreath, Darrell Dinwiddie |
| EPI_ISL_2207262 | Houston Methodist Hospital | Houston Methodist Hospital | Randall J. Olsen, Paul A. Christensen, S. Wesley Long, Sishir Subedi, Robert Olson, Marcus Nguyen, James J. Davis, Matthew Ojeda Saavedra, Prasanti Yerramilli, Layne Pruitt, Kristina Reppond, Madison N. Shyer, Jessica Cambric, Ryan Gadd, Ilya J. Finkelstein, Jimmy Gollihar, and James M. Musser |
| EPI_ISL_2207264 | TriCore Reference Laboratories | Center for Global Health, University of New Mexico Health Sciences Center | Daryl Domman, Kurt Schwalm, Valerie Morley, Cecilia Thompson, Kendra Pesko, Karissa Culbreath, Darrell Dinwiddie |
| EPI_ISL_2207266, EPI_ISL_2207267, EPI_ISL_2207269 | Univeristy of New Mexico Hospital | Center for Global Health, University of New Mexico Health Sciences Center | Daryl Domman, Kurt Schwalm, Valerie Morley, Justin Bacca, Jon Femling, Darrell Dinwiddie |
| EPI_ISL_2207270 | Houston Methodist Hospital | Houston Methodist Hospital | Randall J. Olsen, Paul A. Christensen, S. Wesley Long, Sishir Subedi, Robert Olson, Marcus Nguyen, James J. Davis, Matthew Ojeda Saavedra, Prasanti Yerramilli, Layne Pruitt, Kristina Reppond, Madison N. Shyer, Jessica Cambric, Ryan Gadd, Ilya J. Finkelstein, Jimmy Gollihar, and James M. Musser |
| EPI_ISL_2207272, EPI_ISL_2207274 | TriCore Reference Laboratories | Center for Global Health, University of New Mexico Health Sciences Center | Daryl Domman, Kurt Schwalm, Valerie Morley, Cecilia Thompson, Kendra Pesko, Karissa Culbreath, Darrell Dinwiddie |
| EPI_ISL_2207275, EPI_ISL_2207277 | Houston Methodist Hospital | Houston Methodist Hospital | Randall J. Olsen, Paul A. Christensen, S. Wesley Long, Sishir Subedi, Robert Olson, Marcus Nguyen, James J. Davis, Matthew Ojeda Saavedra, Prasanti Yerramilli, Layne Pruitt, Kristina Reppond, Madison N. Shyer, Jessica Cambric, Ryan Gadd, Ilya J. Finkelstein, Jimmy Gollihar, and James M. Musser |
| EPI_ISL_2207279, EPI_ISL_2207282 | TriCore Reference Laboratories | Center for Global Health, University of New Mexico Health Sciences Center | Daryl Domman, Kurt Schwalm, Valerie Morley, Cecilia Thompson, Kendra Pesko, Karissa Culbreath, Darrell Dinwiddie |
| EPI_ISL_2207284 | Houston Methodist Hospital | Houston Methodist Hospital | Randall J. Olsen, Paul A. Christensen, S. Wesley Long, Sishir Subedi, Robert Olson, Marcus Nguyen, James J. Davis, Matthew Ojeda Saavedra, Prasanti Yerramilli, Layne Pruitt, Kristina Reppond, Madison N. Shyer, Jessica Cambric, Ryan Gadd, Ilya J. Finkelstein, Jimmy Gollihar, and James M. Musser |
| EPI_ISL_2207285 | TriCore Reference Laboratories | Center for Global Health, University of New Mexico Health Sciences Center | Daryl Domman, Kurt Schwalm, Valerie Morley, Cecilia Thompson, Kendra Pesko, Karissa Culbreath, Darrell Dinwiddie |
| EPI_ISL_2207286 | Univeristy of New Mexico Hospital | Center for Global Health, University of New Mexico Health Sciences Center | Daryl Domman, Kurt Schwalm, Valerie Morley, Justin Bacca, Jon Femling, Darrell Dinwiddie |
| EPI_ISL_2207287 | Houston Methodist Hospital | Houston Methodist Hospital | Randall J. Olsen, Paul A. Christensen, S. Wesley Long, Sishir Subedi, Robert Olson, Marcus Nguyen, James J. Davis, Matthew Ojeda Saavedra, Prasanti Yerramilli, Layne Pruitt, Kristina Reppond, Madison N. Shyer, Jessica Cambric, Ryan Gadd, Ilya J. Finkelstein, Jimmy Gollihar, and James M. Musser |
| EPI_ISL_2207288 | Univeristy of New Mexico Hospital | Center for Global Health, University of New Mexico Health Sciences Center | Daryl Domman, Kurt Schwalm, Valerie Morley, Justin Bacca, Jon Femling, Darrell Dinwiddie |
| EPI_ISL_2207289 | TriCore Reference Laboratories | Center for Global Health, University of New Mexico Health Sciences Center | Daryl Domman, Kurt Schwalm, Valerie Morley, Cecilia Thompson, Kendra Pesko, Karissa Culbreath, Darrell Dinwiddie |

| | | | |
|---|---|---|---|
| EPI_ISL_2207290 | Houston Methodist Hospital | Houston Methodist Hospital | Randall J. Olsen, Paul A. Christensen, S. Wesley Long, Sishir Subedi, Robert Olson, Marcus Nguyen, James J. Davis, Matthew Ojeda Saavedra, Prasanti Yerramilli, Layne Pruitt, Kristina Reppond, Madison N. Shyer, Jessica Cambric, Ryan Gadd, Ilya J. Finkelstein, Jimmy Gollihar, and James M. Musser |
| EPI_ISL_2207291, EPI_ISL_2207293, EPI_ISL_2207295 | TriCore Reference Laboratories | Center for Global Health, University of New Mexico Health Sciences Center | Daryl Domman, Kurt Schwalm, Valerie Morley, Cecilia Thompson, Kendra Pesko, Karissa Culbreath, Darrell Dinwiddie |
| EPI_ISL_2207296 | Houston Methodist Hospital | Houston Methodist Hospital | Randall J. Olsen, Paul A. Christensen, S. Wesley Long, Sishir Subedi, Robert Olson, Marcus Nguyen, James J. Davis, Matthew Ojeda Saavedra, Prasanti Yerramilli, Layne Pruitt, Kristina Reppond, Madison N. Shyer, Jessica Cambric, Ryan Gadd, Ilya J. Finkelstein, Jimmy Gollihar, and James M. Musser |
| EPI_ISL_2207297, EPI_ISL_2207299 | TriCore Reference Laboratories | Center for Global Health, University of New Mexico Health Sciences Center | Daryl Domman, Kurt Schwalm, Valerie Morley, Cecilia Thompson, Kendra Pesko, Karissa Culbreath, Darrell Dinwiddie |
| EPI_ISL_2207301 | Houston Methodist Hospital | Houston Methodist Hospital | Randall J. Olsen, Paul A. Christensen, S. Wesley Long, Sishir Subedi, Robert Olson, Marcus Nguyen, James J. Davis, Matthew Ojeda Saavedra, Prasanti Yerramilli, Layne Pruitt, Kristina Reppond, Madison N. Shyer, Jessica Cambric, Ryan Gadd, Ilya J. Finkelstein, Jimmy Gollihar, and James M. Musser |
| EPI_ISL_2207302 | TriCore Reference Laboratories | Center for Global Health, University of New Mexico Health Sciences Center | Daryl Domman, Kurt Schwalm, Valerie Morley, Cecilia Thompson, Kendra Pesko, Karissa Culbreath, Darrell Dinwiddie |
| EPI_ISL_2207305 | Houston Methodist Hospital | Houston Methodist Hospital | Randall J. Olsen, Paul A. Christensen, S. Wesley Long, Sishir Subedi, Robert Olson, Marcus Nguyen, James J. Davis, Matthew Ojeda Saavedra, Prasanti Yerramilli, Layne Pruitt, Kristina Reppond, Madison N. Shyer, Jessica Cambric, Ryan Gadd, Ilya J. Finkelstein, Jimmy Gollihar, and James M. Musser |
| EPI_ISL_2207308 | TriCore Reference Laboratories | Center for Global Health, University of New Mexico Health Sciences Center | Daryl Domman, Kurt Schwalm, Valerie Morley, Cecilia Thompson, Kendra Pesko, Karissa Culbreath, Darrell Dinwiddie |
| EPI_ISL_2207310 | Houston Methodist Hospital | Houston Methodist Hospital | Randall J. Olsen, Paul A. Christensen, S. Wesley Long, Sishir Subedi, Robert Olson, Marcus Nguyen, James J. Davis, Matthew Ojeda Saavedra, Prasanti Yerramilli, Layne Pruitt, Kristina Reppond, Madison N. Shyer, Jessica Cambric, Ryan Gadd, Ilya J. Finkelstein, Jimmy Gollihar, and James M. Musser |
| EPI_ISL_2207311 | TriCore Reference Laboratories | Center for Global Health, University of New Mexico Health Sciences Center | Daryl Domman, Kurt Schwalm, Valerie Morley, Cecilia Thompson, Kendra Pesko, Karissa Culbreath, Darrell Dinwiddie |
| EPI_ISL_2207314 | Houston Methodist Hospital | Houston Methodist Hospital | Randall J. Olsen, Paul A. Christensen, S. Wesley Long, Sishir Subedi, Robert Olson, Marcus Nguyen, James J. Davis, Matthew Ojeda Saavedra, Prasanti Yerramilli, Layne Pruitt, Kristina Reppond, Madison N. Shyer, Jessica Cambric, Ryan Gadd, Ilya J. Finkelstein, Jimmy Gollihar, and James M. Musser |
| EPI_ISL_2207315, EPI_ISL_2207316 | TriCore Reference Laboratories | Center for Global Health, University of New Mexico Health Sciences Center | Daryl Domman, Kurt Schwalm, Valerie Morley, Cecilia Thompson, Kendra Pesko, Karissa Culbreath, Darrell Dinwiddie |
| EPI_ISL_2207318 | Houston Methodist Hospital | Houston Methodist Hospital | Randall J. Olsen, Paul A. Christensen, S. Wesley Long, Sishir Subedi, Robert Olson, Marcus Nguyen, James J. Davis, Matthew Ojeda Saavedra, Prasanti Yerramilli, Layne Pruitt, Kristina Reppond, Madison N. Shyer, Jessica Cambric, Ryan Gadd, Ilya J. Finkelstein, Jimmy Gollihar, and James M. Musser |
| EPI_ISL_2207320, EPI_ISL_2207322 | Univeristy of New Mexico Hospital | Center for Global Health, University of New Mexico Health Sciences Center | Daryl Domman, Kurt Schwalm, Valerie Morley, Justin Bacca, Jon Femling, Darrell Dinwiddie |
| EPI_ISL_2207323 | Houston Methodist Hospital | Houston Methodist Hospital | Randall J. Olsen, Paul A. Christensen, S. Wesley Long, Sishir Subedi, Robert Olson, Marcus Nguyen, James J. Davis, Matthew Ojeda Saavedra, Prasanti Yerramilli, Layne Pruitt, Kristina Reppond, Madison N. Shyer, Jessica Cambric, Ryan Gadd, Ilya J. Finkelstein, Jimmy Gollihar, and James M. Musser |
| EPI_ISL_2207324, EPI_ISL_2207326 | TriCore Reference Laboratories | Center for Global Health, University of New Mexico Health Sciences Center | Daryl Domman, Kurt Schwalm, Valerie Morley, Cecilia Thompson, Kendra Pesko, Karissa Culbreath, Darrell Dinwiddie |
| EPI_ISL_2207328 | Univeristy of New Mexico Hospital | Center for Global Health, University of New Mexico Health Sciences Center | Daryl Domman, Kurt Schwalm, Valerie Morley, Justin Bacca, Jon Femling, Darrell Dinwiddie |
| EPI_ISL_2207329 | Houston Methodist Hospital | Houston Methodist Hospital | Randall J. Olsen, Paul A. Christensen, S. Wesley Long, Sishir Subedi, Robert Olson, Marcus Nguyen, James J. Davis, Matthew Ojeda Saavedra, Prasanti Yerramilli, Layne Pruitt, Kristina Reppond, Madison N. Shyer, Jessica Cambric, Ryan Gadd, Ilya J. Finkelstein, Jimmy Gollihar, and James M. Musser |
| EPI_ISL_2207331 | Univeristy of New Mexico Hospital | Center for Global Health, University of New Mexico Health Sciences Center | Daryl Domman, Kurt Schwalm, Valerie Morley, Justin Bacca, Jon Femling, Darrell Dinwiddie |
| EPI_ISL_2207333 | TriCore Reference Laboratories | Center for Global Health, University of New Mexico Health Sciences Center | Daryl Domman, Kurt Schwalm, Valerie Morley, Cecilia Thompson, Kendra Pesko, Karissa Culbreath, Darrell Dinwiddie |
| EPI_ISL_2207334 | Houston Methodist Hospital | Houston Methodist Hospital | Randall J. Olsen, Paul A. Christensen, S. Wesley Long, Sishir Subedi, Robert Olson, Marcus Nguyen, James J. Davis, Matthew Ojeda Saavedra, Prasanti Yerramilli, Layne Pruitt, Kristina Reppond, Madison N. Shyer, Jessica Cambric, Ryan Gadd, Ilya J. Finkelstein, Jimmy Gollihar, and James M. Musser |
| EPI_ISL_2207335, EPI_ISL_2207337 | TriCore Reference Laboratories | Center for Global Health, University of New Mexico Health Sciences Center | Daryl Domman, Kurt Schwalm, Valerie Morley, Cecilia Thompson, Kendra Pesko, Karissa Culbreath, Darrell Dinwiddie |
| EPI_ISL_2207339 | Houston Methodist Hospital | Houston Methodist Hospital | Randall J. Olsen, Paul A. Christensen, S. Wesley Long, Sishir Subedi, Robert Olson, Marcus Nguyen, James J. Davis, Matthew Ojeda Saavedra, Prasanti Yerramilli, Layne Pruitt, Kristina Reppond, Madison N. Shyer, Jessica Cambric, Ryan Gadd, Ilya J. Finkelstein, Jimmy Gollihar, and James M. Musser |
| EPI_ISL_2207341 | TriCore Reference Laboratories | Center for Global Health, University of New Mexico Health Sciences Center | Daryl Domman, Kurt Schwalm, Valerie Morley, Cecilia Thompson, Kendra Pesko, Karissa Culbreath, Darrell Dinwiddie |
| EPI_ISL_2207342 | Houston Methodist Hospital | Houston Methodist Hospital | Randall J. Olsen, Paul A. Christensen, S. Wesley Long, Sishir Subedi, Robert Olson, Marcus Nguyen, James J. Davis, Matthew Ojeda Saavedra, Prasanti Yerramilli, Layne Pruitt, Kristina Reppond, Madison N. Shyer, Jessica Cambric, Ryan Gadd, Ilya J. Finkelstein, Jimmy Gollihar, and James M. Musser |
| EPI_ISL_2207344 | TriCore Reference Laboratories | Center for Global Health, University of New Mexico Health Sciences Center | Daryl Domman, Kurt Schwalm, Valerie Morley, Cecilia Thompson, Kendra Pesko, Karissa Culbreath, Darrell Dinwiddie |
| EPI_ISL_2207346 | Houston Methodist Hospital | Houston Methodist Hospital | Randall J. Olsen, Paul A. Christensen, S. Wesley Long, Sishir Subedi, Robert Olson, Marcus Nguyen, James J. Davis, Matthew Ojeda Saavedra, Prasanti Yerramilli, Layne Pruitt, Kristina Reppond, Madison N. Shyer, Jessica Cambric, Ryan Gadd, Ilya J. Finkelstein, Jimmy Gollihar, and James M. Musser |
| EPI_ISL_2207348 | TriCore Reference Laboratories | Center for Global Health, University of New Mexico Health Sciences Center | Daryl Domman, Kurt Schwalm, Valerie Morley, Cecilia Thompson, Kendra Pesko, Karissa Culbreath, Darrell Dinwiddie |
| EPI_ISL_2207350 | Houston Methodist Hospital | Houston Methodist Hospital | Randall J. Olsen, Paul A. Christensen, S. Wesley Long, Sishir Subedi, Robert Olson, Marcus Nguyen, James J. Davis, Matthew Ojeda Saavedra, Prasanti Yerramilli, Layne Pruitt, Kristina Reppond, Madison N. Shyer, Jessica Cambric, Ryan Gadd, Ilya J. Finkelstein, Jimmy Gollihar, and James M. Musser |
| EPI_ISL_2207351 | TriCore Reference Laboratories | Center for Global Health, University of New Mexico Health Sciences Center | Daryl Domman, Kurt Schwalm, Valerie Morley, Cecilia Thompson, Kendra Pesko, Karissa Culbreath, Darrell Dinwiddie |
| EPI_ISL_2207354, EPI_ISL_2207357, EPI_ISL_2207361, EPI_ISL_2207364, EPI_ISL_2207366, EPI_ISL_2207369, EPI_ISL_2207372, EPI_ISL_2207375, EPI_ISL_2207379, EPI_ISL_2207382, EPI_ISL_2207385, EPI_ISL_2207390, EPI_ISL_2207394, EPI_ISL_2207397, EPI_ISL_2207400, EPI_ISL_2207403, EPI_ISL_2207406, EPI_ISL_2207410, EPI_ISL_2207413, EPI_ISL_2207416, EPI_ISL_2207421, EPI_ISL_2207424, EPI_ISL_2207428, EPI_ISL_2207432, EPI_ISL_2207438, EPI_ISL_2207442, EPI_ISL_2207445, EPI_ISL_2207454, EPI_ISL_2207457, EPI_ISL_2207460, EPI_ISL_2207461, EPI_ISL_2207464, EPI_ISL_2207469, EPI_ISL_2207476, EPI_ISL_2207481, EPI_ISL_2207484, EPI_ISL_2207487, EPI_ISL_2207489, EPI_ISL_2207491, EPI_ISL_2207494, EPI_ISL_2207498, EPI_ISL_2207502, EPI_ISL_2207505, EPI_ISL_2207508, EPI_ISL_2207511, EPI_ISL_2207514, EPI_ISL_2207516, EPI_ISL_2207522, EPI_ISL_2207524, EPI_ISL_2207528, EPI_ISL_2207531, EPI_ISL_2207534, EPI_ISL_2207539, EPI_ISL_2207542, EPI_ISL_2207546, EPI_ISL_2207550, EPI_ISL_2207554, EPI_ISL_2207556, EPI_ISL_2207559, EPI_ISL_2207565, EPI_ISL_2207570, EPI_ISL_2207573, EPI_ISL_2207577, EPI_ISL_2207581, EPI_ISL_2207583, EPI_ISL_2207587, EPI_ISL_2207589, EPI_ISL_2207592, EPI_ISL_2207595, EPI_ISL_2207598, EPI_ISL_2207601, EPI_ISL_2207604, EPI_ISL_2207608, EPI_ISL_2207610, EPI_ISL_2207612, EPI_ISL_2207615, EPI_ISL_2207618, EPI_ISL_2207621, EPI_ISL_2207623, EPI_ISL_2207627, EPI_ISL_2207630, EPI_ISL_2207633, EPI_ISL_2207636, EPI_ISL_2207638, EPI_ISL_2207641, EPI_ISL_2207644, EPI_ISL_2207648, EPI_ISL_2207651, EPI_ISL_2207654, EPI_ISL_2207658, EPI_ISL_2207660, EPI_ISL_2207663, EPI_ISL_2207666, EPI_ISL_2207669, EPI_ISL_2207672, EPI_ISL_2207675, EPI_ISL_2207678, EPI_ISL_2207680, EPI_ISL_2207683, EPI_ISL_2207685, EPI_ISL_2207687, EPI_ISL_2207691, EPI_ISL_2207694, EPI_ISL_2207697, EPI_ISL_2207700, EPI_ISL_2207702, EPI_ISL_2207705, EPI_ISL_2207708, EPI_ISL_2207712, EPI_ISL_2207716, EPI_ISL_2207720, EPI_ISL_2207723, EPI_ISL_2207726, EPI_ISL_2207730, EPI_ISL_2207733, EPI_ISL_2207735, EPI_ISL_2207738, EPI_ISL_2207741, EPI_ISL_2207744, EPI_ISL_2207747, EPI_ISL_2207751, EPI_ISL_2207754, EPI_ISL_2207758, EPI_ISL_2207761, EPI_ISL_2207765, EPI_ISL_2207768, EPI_ISL_2207771, EPI_ISL_2207775, EPI_ISL_2207781, EPI_ISL_2207784, EPI_ISL_2207788, EPI_ISL_2207791, EPI_ISL_2207794, EPI_ISL_2207797, EPI_ISL_2207799, EPI_ISL_2207802, EPI_ISL_2207806, EPI_ISL_2207809, EPI_ISL_2207811, EPI_ISL_2207814, EPI_ISL_2207817, EPI_ISL_2207821, EPI_ISL_2207825, EPI_ISL_2207827, EPI_ISL_2207831, EPI_ISL_2207834, EPI_ISL_2207837, EPI_ISL_2207840, EPI_ISL_2207843, EPI_ISL_2207846, EPI_ISL_2207849, EPI_ISL_2207852, EPI_ISL_2207856, EPI_ISL_2207858, EPI_ISL_2207861, EPI_ISL_2207865, EPI_ISL_2207867, EPI_ISL_2207870, EPI_ISL_2207873, EPI_ISL_2207876, EPI_ISL_2207878, EPI_ISL_2207881, EPI_ISL_2207887, EPI_ISL_2207890, EPI_ISL_2207893, EPI_ISL_2207896, EPI_ISL_2207900, EPI_ISL_2207902, EPI_ISL_2207903, EPI_ISL_2207905, EPI_ISL_2207908, EPI_ISL_2207911, EPI_ISL_2207913, EPI_ISL_2207916, EPI_ISL_2207918, EPI_ISL_2207921, EPI_ISL_2207924, EPI_ISL_2207927, EPI_ISL_2207930, EPI_ISL_2207932, EPI_ISL_2207935, EPI_ISL_2207938, EPI_ISL_2207940, EPI_ISL_2207943, EPI_ISL_2207946, EPI_ISL_2207949, EPI_ISL_2207951, EPI_ISL_2207954, EPI_ISL_2207958, EPI_ISL_2207961, EPI_ISL_2207963, EPI_ISL_2207966, EPI_ISL_2207968, EPI_ISL_2207970, EPI_ISL_2207973, EPI_ISL_2207976, EPI_ISL_2207979, EPI_ISL_2207982, EPI_ISL_2207985, EPI_ISL_2207988, EPI_ISL_2207992, EPI_ISL_2207995, EPI_ISL_2207998, EPI_ISL_2208002, EPI_ISL_2208006, EPI_ISL_2208011, EPI_ISL_2208014, EPI_ISL_2208019, EPI_ISL_2208022, EPI_ISL_2208026, EPI_ISL_2208030, EPI_ISL_2208037, EPI_ISL_2208044, EPI_ISL_2208047, EPI_ISL_2208050, EPI_ISL_2208053, EPI_ISL_2208054, EPI_ISL_2208056, EPI_ISL_2208059, EPI_ISL_2208062, EPI_ISL_2208065, EPI_ISL_2208068, EPI_ISL_2208071, EPI_ISL_2208074, EPI_ISL_2208077, EPI_ISL_2208078, EPI_ISL_2208081, EPI_ISL_2208084, EPI_ISL_2208087, EPI_ISL_2208090, EPI_ISL_2208093, EPI_ISL_2208098, EPI_ISL_2208100, EPI_ISL_2208104, EPI_ISL_2208109, EPI_ISL_2208111, EPI_ISL_2208115, EPI_ISL_2208119, EPI_ISL_2208122, EPI_ISL_2208126, EPI_ISL_2208129, EPI_ISL_2208132, EPI_ISL_2208135, EPI_ISL_2208138, EPI_ISL_2208141, EPI_ISL_2208145, EPI_ISL_2208148, EPI_ISL_2208154, EPI_ISL_2208157, EPI_ISL_2208161, EPI_ISL_2208165, EPI_ISL_2208169, EPI_ISL_2208172, EPI_ISL_2208174, EPI_ISL_2208177, EPI_ISL_2208180, EPI_ISL_2208182, EPI_ISL_2208185, EPI_ISL_2208188, EPI_ISL_2208191, EPI_ISL_2208194, EPI_ISL_2208197, EPI_ISL_2208201, EPI_ISL_2208206, EPI_ISL_2208211, EPI_ISL_2208216, EPI_ISL_2208221, EPI_ISL_2208226, EPI_ISL_2208231, EPI_ISL_2208236, EPI_ISL_2208242, EPI_ISL_2208246, EPI_ISL_2208249, EPI_ISL_2208253, EPI_ISL_2208257, EPI_ISL_2208262, EPI_ISL_2208267, EPI_ISL_2208272, EPI_ISL_2208277, EPI_ISL_2208282, EPI_ISL_2208287, EPI_ISL_2208292, EPI_ISL_2208296, EPI_ISL_2208301, EPI_ISL_2208305, EPI_ISL_2208308, EPI_ISL_2208313, EPI_ISL_2208317, EPI_ISL_2208324, EPI_ISL_2208328, EPI_ISL_2208333, EPI_ISL_2208337, EPI_ISL_2208343, EPI_ISL_2208348, EPI_ISL_2208353, EPI_ISL_2208357, EPI_ISL_2208364, EPI_ISL_2208369, EPI_ISL_2208374, EPI_ISL_2208383, EPI_ISL_2208388, EPI_ISL_2208392, EPI_ISL_2208396, EPI_ISL_2208401, EPI_ISL_2208404, EPI_ISL_2208409, EPI_ISL_2208413, EPI_ISL_2208419, EPI_ISL_2208425, EPI_ISL_2208430, EPI_ISL_2208435, EPI_ISL_2208440, EPI_ISL_2208445, EPI_ISL_2208450, EPI_ISL_2208456, EPI_ISL_2208462, EPI_ISL_2208467, EPI_ISL_2208472, EPI_ISL_2208477, EPI_ISL_2208484, EPI_ISL_2208488, EPI_ISL_2208494, EPI_ISL_2208498, EPI_ISL_2208503, EPI_ISL_2208514, EPI_ISL_2208520, EPI_ISL_2208525, EPI_ISL_2208531, EPI_ISL_2208537, EPI_ISL_2208541, EPI_ISL_2208548, EPI_ISL_2208551, EPI_ISL_2208557, EPI_ISL_2208565, EPI_ISL_2208569, EPI_ISL_2208574, EPI_ISL_2208580, EPI_ISL_2208584, EPI_ISL_2208592, EPI_ISL_2208597, EPI_ISL_2208603, EPI_ISL_2208606, EPI_ISL_2208612, EPI_ISL_2208617, EPI_ISL_2208622, EPI_ISL_2208626, EPI_ISL_2208633, EPI_ISL_2208638, EPI_ISL_2208644, EPI_ISL_2208649, EPI_ISL_2208654, EPI_ISL_2208657, EPI_ISL_2208668, EPI_ISL_2208677, EPI_ISL_2208681, EPI_ISL_2208687, EPI_ISL_2208691, EPI_ISL_2208696, EPI_ISL_2208700, EPI_ISL_2208704, EPI_ISL_2208709, EPI_ISL_2208721, EPI_ISL_2208726, EPI_ISL_2208731, EPI_ISL_2208738, EPI_ISL_2208744, EPI_ISL_2208749, EPI_ISL_2208756, EPI_ISL_2208760, EPI_ISL_2208767, EPI_ISL_2208774, EPI_ISL_2208781, EPI_ISL_2208786, EPI_ISL_2208791, EPI_ISL_2208796, EPI_ISL_2208801, EPI_ISL_2208807, EPI_ISL_2208811, EPI_ISL_2208816, EPI_ISL_2208819, EPI_ISL_2208825, EPI_ISL_2208830, EPI_ISL_2208835, EPI_ISL_2208838, EPI_ISL_2208843, EPI_ISL_2208847, EPI_ISL_2208852, EPI_ISL_2208855, EPI_ISL_2208859, EPI_ISL_2208867, EPI_ISL_2208875, EPI_ISL_2208880, EPI_ISL_2208886, EPI_ISL_2208893, EPI_ISL_2208901, EPI_ISL_2208909, EPI_ISL_2208919, EPI_ISL_2208925, EPI_ISL_2208932, EPI_ISL_2208936, EPI_ISL_2208941, EPI_ISL_2208947, EPI_ISL_2208953, EPI_ISL_2208959, EPI_ISL_2208964, EPI_ISL_2208969, EPI_ISL_2208976, EPI_ISL_2209005, EPI_ISL_2209010, EPI_ISL_2209015, EPI_ISL_2209019, EPI_ISL_2209026, EPI_ISL_2209039, EPI_ISL_2209048, EPI_ISL_2209053, EPI_ISL_2209058, EPI_ISL_2209062, EPI_ISL_2209067, EPI_ISL_2209071, EPI_ISL_2209079, EPI_ISL_2209085, EPI_ISL_2209090, EPI_ISL_2209096, EPI_ISL_2209102, EPI_ISL_2209105, EPI_ISL_2209113, EPI_ISL_2209120, EPI_ISL_2209126, EPI_ISL_2209131, EPI_ISL_2209136, EPI_ISL_2209144, EPI_ISL_2209150, EPI_ISL_2209155, EPI_ISL_2209160, EPI_ISL_2209166, EPI_ISL_2209174, EPI_ISL_2209181, EPI_ISL_2209186, EPI_ISL_2209194, EPI_ISL_2209198, EPI_ISL_2209203, EPI_ISL_2209209, EPI_ISL_2209219, EPI_ISL_2209225, EPI_ISL_2209230, EPI_ISL_2209239, EPI_ISL_2209245, EPI_ISL_2209251, EPI_ISL_2209256, EPI_ISL_2209262, EPI_ISL_2209268, EPI_ISL_2209274, EPI_ISL_2209280, EPI_ISL_2209286, EPI_ISL_2209291, EPI_ISL_2209299, EPI_ISL_2209306, EPI_ISL_2209311, EPI_ISL_2209318, EPI_ISL_2209328, EPI_ISL_2209336, EPI_ISL_2209343, EPI_ISL_2209358, EPI_ISL_2209372, EPI_ISL_2209381, EPI_ISL_2209390, EPI_ISL_2209400, EPI_ISL_2209411, EPI_ISL_2209418, EPI_ISL_2209426, EPI_ISL_2209440, EPI_ISL_2209460, EPI_ISL_2209470, EPI_ISL_2209482, EPI_ISL_2209491, EPI_ISL_2209501, EPI_ISL_2209506, EPI_ISL_2209518, EPI_ISL_2209525, EPI_ISL_2209539, EPI_ISL_2209547, EPI_ISL_2209554, EPI_ISL_2209562, EPI_ISL_2209568, EPI_ISL_2209574, EPI_ISL_2209580, EPI_ISL_2209587, EPI_ISL_2209594, EPI_ISL_2209599, EPI_ISL_2209606, EPI_ISL_2209613, EPI_ISL_2209619, EPI_ISL_2209625, EPI_ISL_2209633, EPI_ISL_2209640, EPI_ISL_2209648, EPI_ISL_2209654, EPI_ISL_2209660, EPI_ISL_2209667, EPI_ISL_2209673, EPI_ISL_2209677, EPI_ISL_2209683, EPI_ISL_2209689, EPI_ISL_2209694, EPI_ISL_2209700, EPI_ISL_2209708, EPI_ISL_2209713, EPI_ISL_2209720, EPI_ISL_2209727, EPI_ISL_2209740, EPI_ISL_2209748, EPI_ISL_2209752, EPI_ISL_2209758, EPI_ISL_2209762, EPI_ISL_2209767, EPI_ISL_2209771, EPI_ISL_2209776, EPI_ISL_2209782, EPI_ISL_2209789, EPI_ISL_2209794, EPI_ISL_2209800, EPI_ISL_2209805, EPI_ISL_2209812, EPI_ISL_2209817, EPI_ISL_2209823, EPI_ISL_2209829, EPI_ISL_2209833, EPI_ISL_2209837, EPI_ISL_2209842, EPI_ISL_2209847, EPI_ISL_2209853, EPI_ISL_2209860, EPI_ISL_2209866, EPI_ISL_2209873, EPI_ISL_2209877, EPI_ISL_2209881, EPI_ISL_2209885, EPI_ISL_2209892, EPI_ISL_2209897, EPI_ISL_2209903, EPI_ISL_2209909, EPI_ISL_2209915, EPI_ISL_2209926, EPI_ISL_2209932, EPI_ISL_2209940, EPI_ISL_2209944, EPI_ISL_2209950, EPI_ISL_2209956, EPI_ISL_2209959, EPI_ISL_2209965, EPI_ISL_2209971, EPI_ISL_2209981, EPI_ISL_2209988, EPI_ISL_2209993, EPI_ISL_2209999, EPI_ISL_2210006, EPI_ISL_2210013, EPI_ISL_2210017, EPI_ISL_2210023, EPI_ISL_2210027, EPI_ISL_2210031, EPI_ISL_2210038, EPI_ISL_2210044, EPI_ISL_2210053, EPI_ISL_2210059, EPI_ISL_2210065, EPI_ISL_2210068, EPI_ISL_2210075, EPI_ISL_2210080, EPI_ISL_2210089, EPI_ISL_2210095, EPI_ISL_2210099, EPI_ISL_2210107, EPI_ISL_2210115, EPI_ISL_2210122, EPI_ISL_2210128, EPI_ISL_2210135, EPI_ISL_2210140, EPI_ISL_2210145, EPI_ISL_2210150, EPI_ISL_2210155, EPI_ISL_2210161, EPI_ISL_2210167, EPI_ISL_2210172, EPI_ISL_2210178, EPI_ISL_2210184, EPI_ISL_2210191, EPI_ISL_2210197, EPI_ISL_2210201, EPI_ISL_2210206, EPI_ISL_2210212, EPI_ISL_2210216, EPI_ISL_2210220, EPI_ISL_2210227, EPI_ISL_2210234, EPI_ISL_2210241, EPI_ISL_2210248, EPI_ISL_2210255, EPI_ISL_2210264, EPI_ISL_2210267, EPI_ISL_2210277, EPI_ISL_2210284, EPI_ISL_2210291, EPI_ISL_2210300, EPI_ISL_2210306, EPI_ISL_2210311, EPI_ISL_2210316, EPI_ISL_2210320, EPI_ISL_2210326 | | | |
| see above | Houston Methodist Hospital | Houston Methodist Hospital | Randall J. Olsen, Paul A. Christensen, S. Wesley Long, Sishir Subedi, Robert Olson, Marcus Nguyen, James J. Davis, Matthew Ojeda Saavedra, Prasanti Yerramilli, Layne Pruitt, Kristina Reppond, Madison N. Shyer, Jessica Cambric, Ryan Gadd, Ilya J. Finkelstein, Jimmy Gollihar, and James M. Musser |
| EPI_ISL_2210327 | UTMG Pathology LLC | UTMG Pathology LLC | Parasakthy Kumaravelu, Timothy Hodge, Kelsey Matande, Tiara Hymon, Vickie Baselski, Abdallah Azouz |
| EPI_ISL_2210330 | Houston Methodist Hospital | Houston Methodist Hospital | Randall J. Olsen, Paul A. Christensen, S. Wesley Long, Sishir Subedi, Robert Olson, Marcus Nguyen, James J. Davis, Matthew Ojeda Saavedra, Prasanti Yerramilli, Layne Pruitt, Kristina Reppond, Madison N. Shyer, Jessica Cambric, Ryan Gadd, Ilya J. Finkelstein, Jimmy Gollihar, and James M. Musser |
| EPI_ISL_2210332, | UTMG Pathology LLC | UTMG Pathology LLC | Parasakthy Kumaravelu, Timothy Hodge, Kelsey Matande, Tiara Hymon, Vickie Baselski, Abdallah Azouz |

| | | | |
|---|---|---|---|
| EPI_ISL_2210336 | | | |
| EPI_ISL_2210337 | Houston Methodist Hospital | Houston Methodist Hospital | Randall J. Olsen, Paul A. Christensen, S. Wesley Long, Sishir Subedi, Robert Olson, Marcus Nguyen, James J. Davis, Matthew Ojeda Saavedra, Prasanti Yerramilli, Layne Pruitt, Kristina Reppond, Madison N. Shyer, Jessica Cambric, Ryan Gadd, Ilya J. Finkelstein, Jimmy Gollihar, and James M. Musser |
| EPI_ISL_2210340, EPI_ISL_2210341 | UTMG Pathology LLC | UTMG Pathology LLC | Parasakthy Kumaravelu, Timothy Hodge, Kelsey Matande, Tiara Hymon, Vickie Baselski, Abdallah Azouz |
| EPI_ISL_2210344, EPI_ISL_2210349, EPI_ISL_2210354, EPI_ISL_2210360, EPI_ISL_2210365, EPI_ISL_2210368, EPI_ISL_2210373, EPI_ISL_2210377, EPI_ISL_2210382, EPI_ISL_2210387, EPI_ISL_2210393, EPI_ISL_2210396, EPI_ISL_2210401, EPI_ISL_2210406, EPI_ISL_2210411, EPI_ISL_2210414, EPI_ISL_2210419, EPI_ISL_2210424 | | | |
| see above | Houston Methodist Hospital | Houston Methodist Hospital | Randall J. Olsen, Paul A. Christensen, S. Wesley Long, Sishir Subedi, Robert Olson, Marcus Nguyen, James J. Davis, Matthew Ojeda Saavedra, Prasanti Yerramilli, Layne Pruitt, Kristina Reppond, Madison N. Shyer, Jessica Cambric, Ryan Gadd, Ilya J. Finkelstein, Jimmy Gollihar, and James M. Musser |
| EPI_ISL_2210429 | Houston Health Dept. | Houston Health Dept. | Ryker Penn, Pamela Brown, Adolpho Lara, Yanlai Lai |
| EPI_ISL_2210431 | Houston Methodist Hospital | Houston Methodist Hospital | Randall J. Olsen, Paul A. Christensen, S. Wesley Long, Sishir Subedi, Robert Olson, Marcus Nguyen, James J. Davis, Matthew Ojeda Saavedra, Prasanti Yerramilli, Layne Pruitt, Kristina Reppond, Madison N. Shyer, Jessica Cambric, Ryan Gadd, Ilya J. Finkelstein, Jimmy Gollihar, and James M. Musser |
| EPI_ISL_2210433 | Houston Health Dept. | Houston Health Dept. | Ryker Penn, Pamela Brown, Adolpho Lara, Yanlai Lai |
| EPI_ISL_2210438 | Houston Methodist Hospital | Houston Methodist Hospital | Randall J. Olsen, Paul A. Christensen, S. Wesley Long, Sishir Subedi, Robert Olson, Marcus Nguyen, James J. Davis, Matthew Ojeda Saavedra, Prasanti Yerramilli, Layne Pruitt, Kristina Reppond, Madison N. Shyer, Jessica Cambric, Ryan Gadd, Ilya J. Finkelstein, Jimmy Gollihar, and James M. Musser |
| EPI_ISL_2210440 | Houston Health Dept. | Houston Health Dept. | Ryker Penn, Pamela Brown, Adolpho Lara, Yanlai Lai |
| EPI_ISL_2210443 | Houston Methodist Hospital | Houston Methodist Hospital | Randall J. Olsen, Paul A. Christensen, S. Wesley Long, Sishir Subedi, Robert Olson, Marcus Nguyen, James J. Davis, Matthew Ojeda Saavedra, Prasanti Yerramilli, Layne Pruitt, Kristina Reppond, Madison N. Shyer, Jessica Cambric, Ryan Gadd, Ilya J. Finkelstein, Jimmy Gollihar, and James M. Musser |
| EPI_ISL_2210446 | Houston Health Dept. | Houston Health Dept. | Ryker Penn, Pamela Brown, Adolpho Lara, Yanlai Lai |
| EPI_ISL_2210448, EPI_ISL_2210453 | Houston Methodist Hospital | Houston Methodist Hospital | Randall J. Olsen, Paul A. Christensen, S. Wesley Long, Sishir Subedi, Robert Olson, Marcus Nguyen, James J. Davis, Matthew Ojeda Saavedra, Prasanti Yerramilli, Layne Pruitt, Kristina Reppond, Madison N. Shyer, Jessica Cambric, Ryan Gadd, Ilya J. Finkelstein, Jimmy Gollihar, and James M. Musser |
| EPI_ISL_2210454, EPI_ISL_2210458 | Houston Health Dept. | Houston Health Dept. | Ryker Penn, Pamela Brown, Adolpho Lara, Yanlai Lai |
| EPI_ISL_2210460 | Houston Methodist Hospital | Houston Methodist Hospital | Randall J. Olsen, Paul A. Christensen, S. Wesley Long, Sishir Subedi, Robert Olson, Marcus Nguyen, James J. Davis, Matthew Ojeda Saavedra, Prasanti Yerramilli, Layne Pruitt, Kristina Reppond, Madison N. Shyer, Jessica Cambric, Ryan Gadd, Ilya J. Finkelstein, Jimmy Gollihar, and James M. Musser |
| EPI_ISL_2210465 | Houston Health Dept. | Houston Health Dept. | Ryker Penn, Pamela Brown, Adolpho Lara, Yanlai Lai |
| EPI_ISL_2210466, EPI_ISL_2210470 | Houston Methodist Hospital | Houston Methodist Hospital | Randall J. Olsen, Paul A. Christensen, S. Wesley Long, Sishir Subedi, Robert Olson, Marcus Nguyen, James J. Davis, Matthew Ojeda Saavedra, Prasanti Yerramilli, Layne Pruitt, Kristina Reppond, Madison N. Shyer, Jessica Cambric, Ryan Gadd, Ilya J. Finkelstein, Jimmy Gollihar, and James M. Musser |
| EPI_ISL_2210472 | Houston Health Dept. | Houston Health Dept. | Ryker Penn, Pamela Brown, Adolpho Lara, Yanlai Lai |
| EPI_ISL_2210476 | Houston Methodist Hospital | Houston Methodist Hospital | Randall J. Olsen, Paul A. Christensen, S. Wesley Long, Sishir Subedi, Robert Olson, Marcus Nguyen, James J. Davis, Matthew Ojeda Saavedra, Prasanti Yerramilli, Layne Pruitt, Kristina Reppond, Madison N. Shyer, Jessica Cambric, Ryan Gadd, Ilya J. Finkelstein, Jimmy Gollihar, and James M. Musser |
| EPI_ISL_2210479 | Houston Health Dept. | Houston Health Dept. | Ryker Penn, Pamela Brown, Adolpho Lara, Yanlai Lai |
| EPI_ISL_2210483 | Houston Methodist Hospital | Houston Methodist Hospital | Randall J. Olsen, Paul A. Christensen, S. Wesley Long, Sishir Subedi, Robert Olson, Marcus Nguyen, James J. Davis, Matthew Ojeda Saavedra, Prasanti Yerramilli, Layne Pruitt, Kristina Reppond, Madison N. Shyer, Jessica Cambric, Ryan Gadd, Ilya J. Finkelstein, Jimmy Gollihar, and James M. Musser |
| EPI_ISL_2210484, EPI_ISL_2210487, EPI_ISL_2210490, EPI_ISL_2210493 | Houston Health Dept. | Houston Health Dept. | Ryker Penn, Pamela Brown, Adolpho Lara, Yanlai Lai |
| EPI_ISL_2210495 | Houston Methodist Hospital | Houston Methodist Hospital | Randall J. Olsen, Paul A. Christensen, S. Wesley Long, Sishir Subedi, Robert Olson, Marcus Nguyen, James J. Davis, Matthew Ojeda Saavedra, Prasanti Yerramilli, Layne Pruitt, Kristina Reppond, Madison N. Shyer, Jessica Cambric, Ryan Gadd, Ilya J. Finkelstein, Jimmy Gollihar, and James M. Musser |
| EPI_ISL_2210498, EPI_ISL_2210500, EPI_ISL_2210504 | Houston Health Dept. | Houston Health Dept. | Ryker Penn, Pamela Brown, Adolpho Lara, Yanlai Lai |
| EPI_ISL_2210505 | Houston Methodist Hospital | Houston Methodist Hospital | Randall J. Olsen, Paul A. Christensen, S. Wesley Long, Sishir Subedi, Robert Olson, Marcus Nguyen, James J. Davis, Matthew Ojeda Saavedra, Prasanti Yerramilli, Layne Pruitt, Kristina Reppond, Madison N. Shyer, Jessica Cambric, Ryan Gadd, Ilya J. Finkelstein, Jimmy Gollihar, and James M. Musser |
| EPI_ISL_2210507, EPI_ISL_2210510 | Houston Health Dept. | Houston Health Dept. | Ryker Penn, Pamela Brown, Adolpho Lara, Yanlai Lai |
| EPI_ISL_2210513 | Houston Methodist Hospital | Houston Methodist Hospital | Randall J. Olsen, Paul A. Christensen, S. Wesley Long, Sishir Subedi, Robert Olson, Marcus Nguyen, James J. Davis, Matthew Ojeda Saavedra, Prasanti Yerramilli, Layne Pruitt, Kristina Reppond, Madison N. Shyer, Jessica Cambric, Ryan Gadd, Ilya J. Finkelstein, Jimmy Gollihar, and James M. Musser |
| EPI_ISL_2210515 | Houston Health Dept. | Houston Health Dept. | Ryker Penn, Pamela Brown, Adolpho Lara, Yanlai Lai |
| EPI_ISL_2210521, EPI_ISL_2210526, EPI_ISL_2210531, EPI_ISL_2210535, EPI_ISL_2210539, EPI_ISL_2210544, EPI_ISL_2210549, EPI_ISL_2210556, EPI_ISL_2210562, EPI_ISL_2210567, EPI_ISL_2210572, EPI_ISL_2210577, EPI_ISL_2210583, EPI_ISL_2210589, EPI_ISL_2210594, EPI_ISL_2210600, EPI_ISL_2210608, EPI_ISL_2210621, EPI_ISL_2210626, EPI_ISL_2210631, EPI_ISL_2210636, EPI_ISL_2210643, EPI_ISL_2210648, EPI_ISL_2210655, EPI_ISL_2210662, EPI_ISL_2210667, EPI_ISL_2210674, EPI_ISL_2210679, EPI_ISL_2210686, EPI_ISL_2210691, EPI_ISL_2210695, EPI_ISL_2210705, EPI_ISL_2210710, EPI_ISL_2210716, EPI_ISL_2210720, EPI_ISL_2210726, EPI_ISL_2210732, EPI_ISL_2210739, EPI_ISL_2210744, EPI_ISL_2210750, EPI_ISL_2210756, EPI_ISL_2210761, EPI_ISL_2210765, EPI_ISL_2210770, EPI_ISL_2210788, EPI_ISL_2210793, EPI_ISL_2210800 | | | |
| see above | Houston Methodist Hospital | Houston Methodist Hospital | Randall J. Olsen, Paul A. Christensen, S. Wesley Long, Sishir Subedi, Robert Olson, Marcus Nguyen, James J. Davis, Matthew Ojeda Saavedra, Prasanti Yerramilli, Layne Pruitt, Kristina Reppond, Madison N. Shyer, Jessica Cambric, Ryan Gadd, Ilya J. Finkelstein, Jimmy Gollihar, and James M. Musser |
| EPI_ISL_2210805 | Wyoming Public Health Laboratory | Wyoming Public Health Laboratory | Jim Mildenberger, Wanda Manley, Noah Hull, Taylor Fearing, Lynette Gumbleton, Channing Weber, Ashley Norberg, Chayse Rowley, Marley Goetz, Brian Dominguez, Elliot Thomasson, Sam Britz, Cari Sloma, and Rob Christensen |
| EPI_ISL_2210806, EPI_ISL_2210813, EPI_ISL_2210825, EPI_ISL_2210830, EPI_ISL_2210835, EPI_ISL_2210841, EPI_ISL_2210854, EPI_ISL_2210862, EPI_ISL_2210867, EPI_ISL_2210876, EPI_ISL_2210885, EPI_ISL_2210889, EPI_ISL_2210894, EPI_ISL_2210897, EPI_ISL_2210901, EPI_ISL_2210905, EPI_ISL_2210910, EPI_ISL_2210917, EPI_ISL_2210921, EPI_ISL_2210927, EPI_ISL_2210932, EPI_ISL_2210938, EPI_ISL_2210947, EPI_ISL_2210955, EPI_ISL_2210962, EPI_ISL_2210968, EPI_ISL_2210980, EPI_ISL_2210986, EPI_ISL_2210989, EPI_ISL_2210995, EPI_ISL_2211008, EPI_ISL_2211011, EPI_ISL_2211014, EPI_ISL_2211030, EPI_ISL_2211034, EPI_ISL_2211037, EPI_ISL_2211042, EPI_ISL_2211045, EPI_ISL_2211046, EPI_ISL_2211049, EPI_ISL_2211053, EPI_ISL_2211057, EPI_ISL_2211060, EPI_ISL_2211063, EPI_ISL_2211068, EPI_ISL_2211072, EPI_ISL_2211076 | | | |
| see above | Houston Methodist Hospital | Houston Methodist Hospital | Randall J. Olsen, Paul A. Christensen, S. Wesley Long, Sishir Subedi, Robert Olson, Marcus Nguyen, James J. Davis, Matthew Ojeda Saavedra, Prasanti Yerramilli, Layne Pruitt, Kristina Reppond, Madison N. Shyer, Jessica Cambric, Ryan Gadd, Ilya J. Finkelstein, Jimmy Gollihar, and James M. Musser |
| EPI_ISL_2211077 | Nevada State Public Health Laboratory | Nevada State Public Health Laboratory | Andrew Gorzalski, Mark Pandori |
| EPI_ISL_2211084, EPI_ISL_2211089 | Houston Methodist Hospital | Houston Methodist Hospital | Randall J. Olsen, Paul A. Christensen, S. Wesley Long, Sishir Subedi, Robert Olson, Marcus Nguyen, James J. Davis, Matthew Ojeda Saavedra, Prasanti Yerramilli, Layne Pruitt, Kristina Reppond, Madison N. Shyer, Jessica Cambric, Ryan Gadd, Ilya J. Finkelstein, Jimmy Gollihar, and James M. Musser |
| EPI_ISL_2211091 | Nevada State Public Health Laboratory | Nevada State Public Health Laboratory | Andrew Gorzalski, Mark Pandori |
| EPI_ISL_2211094 | Houston Methodist Hospital | Houston Methodist Hospital | Randall J. Olsen, Paul A. Christensen, S. Wesley Long, Sishir Subedi, Robert Olson, Marcus Nguyen, James J. Davis, Matthew Ojeda Saavedra, Prasanti Yerramilli, Layne Pruitt, Kristina Reppond, Madison N. Shyer, Jessica Cambric, Ryan Gadd, Ilya J. Finkelstein, Jimmy Gollihar, and James M. Musser |
| EPI_ISL_2211096 | Nevada State Public Health Laboratory | Nevada State Public Health Laboratory | Andrew Gorzalski, Mark Pandori |
| EPI_ISL_2211100 | Houston Methodist Hospital | Houston Methodist Hospital | Randall J. Olsen, Paul A. Christensen, S. Wesley Long, Sishir Subedi, Robert Olson, Marcus Nguyen, James J. Davis, Matthew Ojeda Saavedra, Prasanti Yerramilli, Layne Pruitt, Kristina Reppond, Madison N. Shyer, Jessica Cambric, Ryan Gadd, Ilya J. Finkelstein, Jimmy Gollihar, and James M. Musser |
| EPI_ISL_2211103 | Nevada State Public Health Laboratory | Nevada State Public Health Laboratory | Andrew Gorzalski, Mark Pandori |
| EPI_ISL_2211104, EPI_ISL_2211109 | Houston Methodist Hospital | Houston Methodist Hospital | Randall J. Olsen, Paul A. Christensen, S. Wesley Long, Sishir Subedi, Robert Olson, Marcus Nguyen, James J. Davis, Matthew Ojeda Saavedra, Prasanti Yerramilli, Layne Pruitt, Kristina Reppond, Madison N. Shyer, Jessica Cambric, Ryan Gadd, Ilya J. Finkelstein, Jimmy Gollihar, and James M. Musser |
| EPI_ISL_2211110 | Nevada State Public Health Laboratory | Nevada State Public Health Laboratory | Andrew Gorzalski, Mark Pandori |
| EPI_ISL_2211113 | Houston Methodist Hospital | Houston Methodist Hospital | Randall J. Olsen, Paul A. Christensen, S. Wesley Long, Sishir Subedi, Robert Olson, Marcus Nguyen, James J. Davis, Matthew Ojeda Saavedra, Prasanti Yerramilli, Layne Pruitt, Kristina Reppond, Madison N. Shyer, Jessica Cambric, Ryan Gadd, Ilya J. Finkelstein, Jimmy Gollihar, and James M. Musser |
| EPI_ISL_2211116, EPI_ISL_2211118 | Nevada State Public Health Laboratory | Nevada State Public Health Laboratory | Andrew Gorzalski, Mark Pandori |
| EPI_ISL_2211121 | Houston Methodist Hospital | Houston Methodist Hospital | Randall J. Olsen, Paul A. Christensen, S. Wesley Long, Sishir Subedi, Robert Olson, Marcus Nguyen, James J. Davis, Matthew Ojeda Saavedra, Prasanti Yerramilli, Layne Pruitt, Kristina Reppond, Madison N. Shyer, Jessica Cambric, Ryan Gadd, Ilya J. Finkelstein, Jimmy Gollihar, and James M. Musser |
| EPI_ISL_2211123, EPI_ISL_2211125 | Nevada State Public Health Laboratory | Nevada State Public Health Laboratory | Andrew Gorzalski, Mark Pandori |
| EPI_ISL_2211127, EPI_ISL_2211130, EPI_ISL_2211134, EPI_ISL_2211140, EPI_ISL_2211144, EPI_ISL_2211147, EPI_ISL_2211151, EPI_ISL_2211155, EPI_ISL_2211158, EPI_ISL_2211161, EPI_ISL_2211166, EPI_ISL_2211169, EPI_ISL_2211171, EPI_ISL_2211174, EPI_ISL_2211177 | | | |
| see above | Houston Methodist Hospital | Houston Methodist Hospital | Randall J. Olsen, Paul A. Christensen, S. Wesley Long, Sishir Subedi, Robert Olson, Marcus Nguyen, James J. Davis, Matthew Ojeda Saavedra, Prasanti Yerramilli, Layne Pruitt, Kristina Reppond, Madison N. Shyer, Jessica Cambric, Ryan Gadd, Ilya J. Finkelstein, Jimmy Gollihar, and James M. Musser |
| EPI_ISL_2211181 | New Mexico Department of Health Scientific Laboratory | New Mexico Department of Health Scientific Laboratory | Ellie Johnson, D'eldra Malone, Jennifer Benoit, Ratheesh Rajan, Linda Salazar, Anastacia Griego-Fisher |
| EPI_ISL_2211182, EPI_ISL_2211185 | Houston Methodist Hospital | Houston Methodist Hospital | Randall J. Olsen, Paul A. Christensen, S. Wesley Long, Sishir Subedi, Robert Olson, Marcus Nguyen, James J. Davis, Matthew Ojeda Saavedra, Prasanti Yerramilli, Layne Pruitt, Kristina Reppond, Madison N. Shyer, Jessica Cambric, Ryan Gadd, Ilya J. Finkelstein, Jimmy Gollihar, and James M. Musser |

| | | | |
|---|---|---|---|
| EPI_ISL_2211187 | New Mexico Department of Health Scientific Laboratory | New Mexico Department of Health Scientific Laboratory | Ellie Johnson, D'eldra Malone, Jennifer Benoit, Ratheesh Rajan, Linda Salazar, Anastacia Griego-Fisher |
| EPI_ISL_2211189 | Houston Methodist Hospital | Houston Methodist Hospital | Randall J. Olsen, Paul A. Christensen, S. Wesley Long, Sishir Subedi, Robert Olson, Marcus Nguyen, James J. Davis, Matthew Ojeda Saavedra, Prasanti Yerramilli, Layne Pruitt, Kristina Reppond, Madison N. Shyer, Jessica Cambric, Ryan Gadd, Ilya J. Finkelstein, Jimmy Gollihar, and James M. Musser |
| EPI_ISL_2211191, EPI_ISL_2211193 | New Mexico Department of Health Scientific Laboratory | New Mexico Department of Health Scientific Laboratory | Ellie Johnson, D'eldra Malone, Jennifer Benoit, Ratheesh Rajan, Linda Salazar, Anastacia Griego-Fisher |
| EPI_ISL_2211195 | Houston Methodist Hospital | Houston Methodist Hospital | Randall J. Olsen, Paul A. Christensen, S. Wesley Long, Sishir Subedi, Robert Olson, Marcus Nguyen, James J. Davis, Matthew Ojeda Saavedra, Prasanti Yerramilli, Layne Pruitt, Kristina Reppond, Madison N. Shyer, Jessica Cambric, Ryan Gadd, Ilya J. Finkelstein, Jimmy Gollihar, and James M. Musser |
| EPI_ISL_2211196 | New Mexico Department of Health Scientific Laboratory | New Mexico Department of Health Scientific Laboratory | Ellie Johnson, D'eldra Malone, Jennifer Benoit, Ratheesh Rajan, Linda Salazar, Anastacia Griego-Fisher |
| EPI_ISL_2211199 | Houston Methodist Hospital | Houston Methodist Hospital | Randall J. Olsen, Paul A. Christensen, S. Wesley Long, Sishir Subedi, Robert Olson, Marcus Nguyen, James J. Davis, Matthew Ojeda Saavedra, Prasanti Yerramilli, Layne Pruitt, Kristina Reppond, Madison N. Shyer, Jessica Cambric, Ryan Gadd, Ilya J. Finkelstein, Jimmy Gollihar, and James M. Musser |
| EPI_ISL_2211201 | New Mexico Department of Health Scientific Laboratory | New Mexico Department of Health Scientific Laboratory | Ellie Johnson, D'eldra Malone, Jennifer Benoit, Ratheesh Rajan, Linda Salazar, Anastacia Griego-Fisher |
| EPI_ISL_2211203 | Houston Methodist Hospital | Houston Methodist Hospital | Randall J. Olsen, Paul A. Christensen, S. Wesley Long, Sishir Subedi, Robert Olson, Marcus Nguyen, James J. Davis, Matthew Ojeda Saavedra, Prasanti Yerramilli, Layne Pruitt, Kristina Reppond, Madison N. Shyer, Jessica Cambric, Ryan Gadd, Ilya J. Finkelstein, Jimmy Gollihar, and James M. Musser |
| EPI_ISL_2211206 | New Mexico Department of Health Scientific Laboratory | New Mexico Department of Health Scientific Laboratory | Ellie Johnson, D'eldra Malone, Jennifer Benoit, Ratheesh Rajan, Linda Salazar, Anastacia Griego-Fisher |
| EPI_ISL_2211207 | Houston Methodist Hospital | Houston Methodist Hospital | Randall J. Olsen, Paul A. Christensen, S. Wesley Long, Sishir Subedi, Robert Olson, Marcus Nguyen, James J. Davis, Matthew Ojeda Saavedra, Prasanti Yerramilli, Layne Pruitt, Kristina Reppond, Madison N. Shyer, Jessica Cambric, Ryan Gadd, Ilya J. Finkelstein, Jimmy Gollihar, and James M. Musser |
| EPI_ISL_2211210 | TriCore Reference Laboratory Inc. | New Mexico Department of Health Scientific Laboratory | Ellie Johnson, D'eldra Malone, Jennifer Benoit, Ratheesh Rajan, Linda Salazar, Anastacia Griego-Fisher |
| EPI_ISL_2211211 | Houston Methodist Hospital | Houston Methodist Hospital | Randall J. Olsen, Paul A. Christensen, S. Wesley Long, Sishir Subedi, Robert Olson, Marcus Nguyen, James J. Davis, Matthew Ojeda Saavedra, Prasanti Yerramilli, Layne Pruitt, Kristina Reppond, Madison N. Shyer, Jessica Cambric, Ryan Gadd, Ilya J. Finkelstein, Jimmy Gollihar, and James M. Musser |
| EPI_ISL_2211215, EPI_ISL_2211219 | New Mexico Department of Health Scientific Laboratory | New Mexico Department of Health Scientific Laboratory | Ellie Johnson, D'eldra Malone, Jennifer Benoit, Ratheesh Rajan, Linda Salazar, Anastacia Griego-Fisher |
| EPI_ISL_2211220 | Houston Methodist Hospital | Houston Methodist Hospital | Randall J. Olsen, Paul A. Christensen, S. Wesley Long, Sishir Subedi, Robert Olson, Marcus Nguyen, James J. Davis, Matthew Ojeda Saavedra, Prasanti Yerramilli, Layne Pruitt, Kristina Reppond, Madison N. Shyer, Jessica Cambric, Ryan Gadd, Ilya J. Finkelstein, Jimmy Gollihar, and James M. Musser |
| EPI_ISL_2211222 | New Mexico Department of Health Scientific Laboratory | New Mexico Department of Health Scientific Laboratory | Ellie Johnson, D'eldra Malone, Jennifer Benoit, Ratheesh Rajan, Linda Salazar, Anastacia Griego-Fisher |
| EPI_ISL_2211223 | Houston Methodist Hospital | Houston Methodist Hospital | Randall J. Olsen, Paul A. Christensen, S. Wesley Long, Sishir Subedi, Robert Olson, Marcus Nguyen, James J. Davis, Matthew Ojeda Saavedra, Prasanti Yerramilli, Layne Pruitt, Kristina Reppond, Madison N. Shyer, Jessica Cambric, Ryan Gadd, Ilya J. Finkelstein, Jimmy Gollihar, and James M. Musser |
| EPI_ISL_2211226 | New Mexico Department of Health Scientific Laboratory | New Mexico Department of Health Scientific Laboratory | Ellie Johnson, D'eldra Malone, Jennifer Benoit, Ratheesh Rajan, Linda Salazar, Anastacia Griego-Fisher |
| EPI_ISL_2211229 | Houston Methodist Hospital | Houston Methodist Hospital | Randall J. Olsen, Paul A. Christensen, S. Wesley Long, Sishir Subedi, Robert Olson, Marcus Nguyen, James J. Davis, Matthew Ojeda Saavedra, Prasanti Yerramilli, Layne Pruitt, Kristina Reppond, Madison N. Shyer, Jessica Cambric, Ryan Gadd, Ilya J. Finkelstein, Jimmy Gollihar, and James M. Musser |
| EPI_ISL_2211231 | New Mexico Department of Health Scientific Laboratory | New Mexico Department of Health Scientific Laboratory | Ellie Johnson, D'eldra Malone, Jennifer Benoit, Ratheesh Rajan, Linda Salazar, Anastacia Griego-Fisher |
| EPI_ISL_2211234 | Houston Methodist Hospital | Houston Methodist Hospital | Randall J. Olsen, Paul A. Christensen, S. Wesley Long, Sishir Subedi, Robert Olson, Marcus Nguyen, James J. Davis, Matthew Ojeda Saavedra, Prasanti Yerramilli, Layne Pruitt, Kristina Reppond, Madison N. Shyer, Jessica Cambric, Ryan Gadd, Ilya J. Finkelstein, Jimmy Gollihar, and James M. Musser |
| EPI_ISL_2211235 | New Mexico Department of Health Scientific Laboratory | New Mexico Department of Health Scientific Laboratory | Ellie Johnson, D'eldra Malone, Jennifer Benoit, Ratheesh Rajan, Linda Salazar, Anastacia Griego-Fisher |
| EPI_ISL_2211238 | Houston Methodist Hospital | Houston Methodist Hospital | Randall J. Olsen, Paul A. Christensen, S. Wesley Long, Sishir Subedi, Robert Olson, Marcus Nguyen, James J. Davis, Matthew Ojeda Saavedra, Prasanti Yerramilli, Layne Pruitt, Kristina Reppond, Madison N. Shyer, Jessica Cambric, Ryan Gadd, Ilya J. Finkelstein, Jimmy Gollihar, and James M. Musser |
| EPI_ISL_2211239, EPI_ISL_2211242 | New Mexico Department of Health Scientific Laboratory | New Mexico Department of Health Scientific Laboratory | Ellie Johnson, D'eldra Malone, Jennifer Benoit, Ratheesh Rajan, Linda Salazar, Anastacia Griego-Fisher |
| EPI_ISL_2211245 | Houston Methodist Hospital | Houston Methodist Hospital | Randall J. Olsen, Paul A. Christensen, S. Wesley Long, Sishir Subedi, Robert Olson, Marcus Nguyen, James J. Davis, Matthew Ojeda Saavedra, Prasanti Yerramilli, Layne Pruitt, Kristina Reppond, Madison N. Shyer, Jessica Cambric, Ryan Gadd, Ilya J. Finkelstein, Jimmy Gollihar, and James M. Musser |
| EPI_ISL_2211247 | San Juan Regional Medical Center | New Mexico Department of Health Scientific Laboratory | Ellie Johnson, D'eldra Malone, Jennifer Benoit, Ratheesh Rajan, Linda Salazar, Anastacia Griego-Fisher |
| EPI_ISL_2211249 | Houston Methodist Hospital | Houston Methodist Hospital | Randall J. Olsen, Paul A. Christensen, S. Wesley Long, Sishir Subedi, Robert Olson, Marcus Nguyen, James J. Davis, Matthew Ojeda Saavedra, Prasanti Yerramilli, Layne Pruitt, Kristina Reppond, Madison N. Shyer, Jessica Cambric, Ryan Gadd, Ilya J. Finkelstein, Jimmy Gollihar, and James M. Musser |
| EPI_ISL_2211251 | San Juan Regional Medical Center | New Mexico Department of Health Scientific Laboratory | Ellie Johnson, D'eldra Malone, Jennifer Benoit, Ratheesh Rajan, Linda Salazar, Anastacia Griego-Fisher |
| EPI_ISL_2211253 | Houston Methodist Hospital | Houston Methodist Hospital | Randall J. Olsen, Paul A. Christensen, S. Wesley Long, Sishir Subedi, Robert Olson, Marcus Nguyen, James J. Davis, Matthew Ojeda Saavedra, Prasanti Yerramilli, Layne Pruitt, Kristina Reppond, Madison N. Shyer, Jessica Cambric, Ryan Gadd, Ilya J. Finkelstein, Jimmy Gollihar, and James M. Musser |
| EPI_ISL_2211256 | San Juan Regional Medical Center | New Mexico Department of Health Scientific Laboratory | Ellie Johnson, D'eldra Malone, Jennifer Benoit, Ratheesh Rajan, Linda Salazar, Anastacia Griego-Fisher |
| EPI_ISL_2211257, EPI_ISL_2211260 | Houston Methodist Hospital | Houston Methodist Hospital | Randall J. Olsen, Paul A. Christensen, S. Wesley Long, Sishir Subedi, Robert Olson, Marcus Nguyen, James J. Davis, Matthew Ojeda Saavedra, Prasanti Yerramilli, Layne Pruitt, Kristina Reppond, Madison N. Shyer, Jessica Cambric, Ryan Gadd, Ilya J. Finkelstein, Jimmy Gollihar, and James M. Musser |
| EPI_ISL_2211262 | San Juan Regional Medical Center | New Mexico Department of Health Scientific Laboratory | Ellie Johnson, D'eldra Malone, Jennifer Benoit, Ratheesh Rajan, Linda Salazar, Anastacia Griego-Fisher |
| EPI_ISL_2211265 | Houston Methodist Hospital | Houston Methodist Hospital | Randall J. Olsen, Paul A. Christensen, S. Wesley Long, Sishir Subedi, Robert Olson, Marcus Nguyen, James J. Davis, Matthew Ojeda Saavedra, Prasanti Yerramilli, Layne Pruitt, Kristina Reppond, Madison N. Shyer, Jessica Cambric, Ryan Gadd, Ilya J. Finkelstein, Jimmy Gollihar, and James M. Musser |
| EPI_ISL_2211267 | San Juan Regional Medical Center | New Mexico Department of Health Scientific Laboratory | Ellie Johnson, D'eldra Malone, Jennifer Benoit, Ratheesh Rajan, Linda Salazar, Anastacia Griego-Fisher |
| EPI_ISL_2211269, EPI_ISL_2211272 | Houston Methodist Hospital | Houston Methodist Hospital | Randall J. Olsen, Paul A. Christensen, S. Wesley Long, Sishir Subedi, Robert Olson, Marcus Nguyen, James J. Davis, Matthew Ojeda Saavedra, Prasanti Yerramilli, Layne Pruitt, Kristina Reppond, Madison N. Shyer, Jessica Cambric, Ryan Gadd, Ilya J. Finkelstein, Jimmy Gollihar, and James M. Musser |
| EPI_ISL_2211273 | San Juan Regional Medical Center | New Mexico Department of Health Scientific Laboratory | Ellie Johnson, D'eldra Malone, Jennifer Benoit, Ratheesh Rajan, Linda Salazar, Anastacia Griego-Fisher |
| EPI_ISL_2211275 | Houston Methodist Hospital | Houston Methodist Hospital | Randall J. Olsen, Paul A. Christensen, S. Wesley Long, Sishir Subedi, Robert Olson, Marcus Nguyen, James J. Davis, Matthew Ojeda Saavedra, Prasanti Yerramilli, Layne Pruitt, Kristina Reppond, Madison N. Shyer, Jessica Cambric, Ryan Gadd, Ilya J. Finkelstein, Jimmy Gollihar, and James M. Musser |
| EPI_ISL_2211277 | San Juan Regional Medical Center | New Mexico Department of Health Scientific Laboratory | Ellie Johnson, D'eldra Malone, Jennifer Benoit, Ratheesh Rajan, Linda Salazar, Anastacia Griego-Fisher |
| EPI_ISL_2211279, EPI_ISL_2211282, EPI_ISL_2211286 | Houston Methodist Hospital | Houston Methodist Hospital | Randall J. Olsen, Paul A. Christensen, S. Wesley Long, Sishir Subedi, Robert Olson, Marcus Nguyen, James J. Davis, Matthew Ojeda Saavedra, Prasanti Yerramilli, Layne Pruitt, Kristina Reppond, Madison N. Shyer, Jessica Cambric, Ryan Gadd, Ilya J. Finkelstein, Jimmy Gollihar, and James M. Musser |
| EPI_ISL_2211287 | San Juan Regional Medical Center | New Mexico Department of Health Scientific Laboratory | Ellie Johnson, D'eldra Malone, Jennifer Benoit, Ratheesh Rajan, Linda Salazar, Anastacia Griego-Fisher |
| EPI_ISL_2211291, EPI_ISL_2211295, EPI_ISL_2211298 | Houston Methodist Hospital | Houston Methodist Hospital | Randall J. Olsen, Paul A. Christensen, S. Wesley Long, Sishir Subedi, Robert Olson, Marcus Nguyen, James J. Davis, Matthew Ojeda Saavedra, Prasanti Yerramilli, Layne Pruitt, Kristina Reppond, Madison N. Shyer, Jessica Cambric, Ryan Gadd, Ilya J. Finkelstein, Jimmy Gollihar, and James M. Musser |
| EPI_ISL_2211300 | San Juan Regional Medical Center | New Mexico Department of Health Scientific Laboratory | Ellie Johnson, D'eldra Malone, Jennifer Benoit, Ratheesh Rajan, Linda Salazar, Anastacia Griego-Fisher |
| EPI_ISL_2211301 | Houston Methodist Hospital | Houston Methodist Hospital | Randall J. Olsen, Paul A. Christensen, S. Wesley Long, Sishir Subedi, Robert Olson, Marcus Nguyen, James J. Davis, Matthew Ojeda Saavedra, Prasanti Yerramilli, Layne Pruitt, Kristina Reppond, Madison N. Shyer, Jessica Cambric, Ryan Gadd, Ilya J. Finkelstein, Jimmy Gollihar, and James M. Musser |
| EPI_ISL_2211304 | San Juan Regional Medical Center | New Mexico Department of Health Scientific Laboratory | Ellie Johnson, D'eldra Malone, Jennifer Benoit, Ratheesh Rajan, Linda Salazar, Anastacia Griego-Fisher |
| EPI_ISL_2211305, EPI_ISL_2211309 | Houston Methodist Hospital | Houston Methodist Hospital | Randall J. Olsen, Paul A. Christensen, S. Wesley Long, Sishir Subedi, Robert Olson, Marcus Nguyen, James J. Davis, Matthew Ojeda Saavedra, Prasanti Yerramilli, Layne Pruitt, Kristina Reppond, Madison N. Shyer, Jessica Cambric, Ryan Gadd, Ilya J. Finkelstein, Jimmy Gollihar, and James M. Musser |

| | | | |
|---|---|---|---|
| EPI_ISL_2211310 | San Juan Regional Medical Center | New Mexico Department of Health Scientific Laboratory | Ellie Johnson, D'eldra Malone, Jennifer Benoit, Ratheesh Rajan, Linda Salazar, Anastacia Griego-Fisher |
| EPI_ISL_2211313 | Houston Methodist Hospital | Houston Methodist Hospital | Randall J. Olsen, Paul A. Christensen, S. Wesley Long, Sishir Subedi, Robert Olson, Marcus Nguyen, James J. Davis, Matthew Ojeda Saavedra, Prasanti Yerramilli, Layne Pruitt, Kristina Reppond, Madison N. Shyer, Jessica Cambric, Ryan Gadd, Ilya J. Finkelstein, Jimmy Gollihar, and James M. Musser |
| EPI_ISL_2211315 | San Juan Regional Medical Center | New Mexico Department of Health Scientific Laboratory | Ellie Johnson, D'eldra Malone, Jennifer Benoit, Ratheesh Rajan, Linda Salazar, Anastacia Griego-Fisher |
| EPI_ISL_2211317, EPI_ISL_2211319 | Houston Methodist Hospital | Houston Methodist Hospital | Randall J. Olsen, Paul A. Christensen, S. Wesley Long, Sishir Subedi, Robert Olson, Marcus Nguyen, James J. Davis, Matthew Ojeda Saavedra, Prasanti Yerramilli, Layne Pruitt, Kristina Reppond, Madison N. Shyer, Jessica Cambric, Ryan Gadd, Ilya J. Finkelstein, Jimmy Gollihar, and James M. Musser |
| EPI_ISL_2211321 | San Juan Regional Medical Center | New Mexico Department of Health Scientific Laboratory | Ellie Johnson, D'eldra Malone, Jennifer Benoit, Ratheesh Rajan, Linda Salazar, Anastacia Griego-Fisher |
| EPI_ISL_2211324 | Houston Methodist Hospital | Houston Methodist Hospital | Randall J. Olsen, Paul A. Christensen, S. Wesley Long, Sishir Subedi, Robert Olson, Marcus Nguyen, James J. Davis, Matthew Ojeda Saavedra, Prasanti Yerramilli, Layne Pruitt, Kristina Reppond, Madison N. Shyer, Jessica Cambric, Ryan Gadd, Ilya J. Finkelstein, Jimmy Gollihar, and James M. Musser |
| EPI_ISL_2211326 | San Juan Regional Medical Center | New Mexico Department of Health Scientific Laboratory | Ellie Johnson, D'eldra Malone, Jennifer Benoit, Ratheesh Rajan, Linda Salazar, Anastacia Griego-Fisher |
| EPI_ISL_2211328 | Houston Methodist Hospital | Houston Methodist Hospital | Randall J. Olsen, Paul A. Christensen, S. Wesley Long, Sishir Subedi, Robert Olson, Marcus Nguyen, James J. Davis, Matthew Ojeda Saavedra, Prasanti Yerramilli, Layne Pruitt, Kristina Reppond, Madison N. Shyer, Jessica Cambric, Ryan Gadd, Ilya J. Finkelstein, Jimmy Gollihar, and James M. Musser |
| EPI_ISL_2211330 | San Juan Regional Medical Center | New Mexico Department of Health Scientific Laboratory | Ellie Johnson, D'eldra Malone, Jennifer Benoit, Ratheesh Rajan, Linda Salazar, Anastacia Griego-Fisher |
| EPI_ISL_2211332 | Houston Methodist Hospital | Houston Methodist Hospital | Randall J. Olsen, Paul A. Christensen, S. Wesley Long, Sishir Subedi, Robert Olson, Marcus Nguyen, James J. Davis, Matthew Ojeda Saavedra, Prasanti Yerramilli, Layne Pruitt, Kristina Reppond, Madison N. Shyer, Jessica Cambric, Ryan Gadd, Ilya J. Finkelstein, Jimmy Gollihar, and James M. Musser |
| EPI_ISL_2211335 | San Juan Regional Medical Center | New Mexico Department of Health Scientific Laboratory | Ellie Johnson, D'eldra Malone, Jennifer Benoit, Ratheesh Rajan, Linda Salazar, Anastacia Griego-Fisher |
| EPI_ISL_2211337, EPI_ISL_2211339 | Houston Methodist Hospital | Houston Methodist Hospital | Randall J. Olsen, Paul A. Christensen, S. Wesley Long, Sishir Subedi, Robert Olson, Marcus Nguyen, James J. Davis, Matthew Ojeda Saavedra, Prasanti Yerramilli, Layne Pruitt, Kristina Reppond, Madison N. Shyer, Jessica Cambric, Ryan Gadd, Ilya J. Finkelstein, Jimmy Gollihar, and James M. Musser |
| EPI_ISL_2211341 | San Juan Regional Medical Center | New Mexico Department of Health Scientific Laboratory | Ellie Johnson, D'eldra Malone, Jennifer Benoit, Ratheesh Rajan, Linda Salazar, Anastacia Griego-Fisher |
| EPI_ISL_2211344 | Houston Methodist Hospital | Houston Methodist Hospital | Randall J. Olsen, Paul A. Christensen, S. Wesley Long, Sishir Subedi, Robert Olson, Marcus Nguyen, James J. Davis, Matthew Ojeda Saavedra, Prasanti Yerramilli, Layne Pruitt, Kristina Reppond, Madison N. Shyer, Jessica Cambric, Ryan Gadd, Ilya J. Finkelstein, Jimmy Gollihar, and James M. Musser |
| EPI_ISL_2211346 | San Juan Regional Medical Center | New Mexico Department of Health Scientific Laboratory | Ellie Johnson, D'eldra Malone, Jennifer Benoit, Ratheesh Rajan, Linda Salazar, Anastacia Griego-Fisher |
| EPI_ISL_2211348 | Houston Methodist Hospital | Houston Methodist Hospital | Randall J. Olsen, Paul A. Christensen, S. Wesley Long, Sishir Subedi, Robert Olson, Marcus Nguyen, James J. Davis, Matthew Ojeda Saavedra, Prasanti Yerramilli, Layne Pruitt, Kristina Reppond, Madison N. Shyer, Jessica Cambric, Ryan Gadd, Ilya J. Finkelstein, Jimmy Gollihar, and James M. Musser |
| EPI_ISL_2211351 | San Juan Regional Medical Center | New Mexico Department of Health Scientific Laboratory | Ellie Johnson, D'eldra Malone, Jennifer Benoit, Ratheesh Rajan, Linda Salazar, Anastacia Griego-Fisher |
| EPI_ISL_2211352, EPI_ISL_2211355 | Houston Methodist Hospital | Houston Methodist Hospital | Randall J. Olsen, Paul A. Christensen, S. Wesley Long, Sishir Subedi, Robert Olson, Marcus Nguyen, James J. Davis, Matthew Ojeda Saavedra, Prasanti Yerramilli, Layne Pruitt, Kristina Reppond, Madison N. Shyer, Jessica Cambric, Ryan Gadd, Ilya J. Finkelstein, Jimmy Gollihar, and James M. Musser |
| EPI_ISL_2211356 | San Juan Regional Medical Center | New Mexico Department of Health Scientific Laboratory | Ellie Johnson, D'eldra Malone, Jennifer Benoit, Ratheesh Rajan, Linda Salazar, Anastacia Griego-Fisher |
| EPI_ISL_2211360 | New Mexico Department of Health Scientific Laboratory | New Mexico Department of Health Scientific Laboratory | Ellie Johnson, D'eldra Malone, Jennifer Benoit, Ratheesh Rajan, Linda Salazar, Anastacia Griego-Fisher |
| EPI_ISL_2211362 | Houston Methodist Hospital | Houston Methodist Hospital | Randall J. Olsen, Paul A. Christensen, S. Wesley Long, Sishir Subedi, Robert Olson, Marcus Nguyen, James J. Davis, Matthew Ojeda Saavedra, Prasanti Yerramilli, Layne Pruitt, Kristina Reppond, Madison N. Shyer, Jessica Cambric, Ryan Gadd, Ilya J. Finkelstein, Jimmy Gollihar, and James M. Musser |
| EPI_ISL_2211365 | New Mexico Department of Health Scientific Laboratory | New Mexico Department of Health Scientific Laboratory | Ellie Johnson, D'eldra Malone, Jennifer Benoit, Ratheesh Rajan, Linda Salazar, Anastacia Griego-Fisher |
| EPI_ISL_2211367 | Houston Methodist Hospital | Houston Methodist Hospital | Randall J. Olsen, Paul A. Christensen, S. Wesley Long, Sishir Subedi, Robert Olson, Marcus Nguyen, James J. Davis, Matthew Ojeda Saavedra, Prasanti Yerramilli, Layne Pruitt, Kristina Reppond, Madison N. Shyer, Jessica Cambric, Ryan Gadd, Ilya J. Finkelstein, Jimmy Gollihar, and James M. Musser |
| EPI_ISL_2211370 | New Mexico Department of Health Scientific Laboratory | New Mexico Department of Health Scientific Laboratory | Ellie Johnson, D'eldra Malone, Jennifer Benoit, Ratheesh Rajan, Linda Salazar, Anastacia Griego-Fisher |
| EPI_ISL_2211371, EPI_ISL_2211374 | Houston Methodist Hospital | Houston Methodist Hospital | Randall J. Olsen, Paul A. Christensen, S. Wesley Long, Sishir Subedi, Robert Olson, Marcus Nguyen, James J. Davis, Matthew Ojeda Saavedra, Prasanti Yerramilli, Layne Pruitt, Kristina Reppond, Madison N. Shyer, Jessica Cambric, Ryan Gadd, Ilya J. Finkelstein, Jimmy Gollihar, and James M. Musser |
| EPI_ISL_2211376 | New Mexico Department of Health Scientific Laboratory | New Mexico Department of Health Scientific Laboratory | Ellie Johnson, D'eldra Malone, Jennifer Benoit, Ratheesh Rajan, Linda Salazar, Anastacia Griego-Fisher |
| EPI_ISL_2211378 | Houston Methodist Hospital | Houston Methodist Hospital | Randall J. Olsen, Paul A. Christensen, S. Wesley Long, Sishir Subedi, Robert Olson, Marcus Nguyen, James J. Davis, Matthew Ojeda Saavedra, Prasanti Yerramilli, Layne Pruitt, Kristina Reppond, Madison N. Shyer, Jessica Cambric, Ryan Gadd, Ilya J. Finkelstein, Jimmy Gollihar, and James M. Musser |
| EPI_ISL_2211380 | New Mexico Department of Health Scientific Laboratory | New Mexico Department of Health Scientific Laboratory | Ellie Johnson, D'eldra Malone, Jennifer Benoit, Ratheesh Rajan, Linda Salazar, Anastacia Griego-Fisher |
| EPI_ISL_2211382, EPI_ISL_2211385 | Houston Methodist Hospital | Houston Methodist Hospital | Randall J. Olsen, Paul A. Christensen, S. Wesley Long, Sishir Subedi, Robert Olson, Marcus Nguyen, James J. Davis, Matthew Ojeda Saavedra, Prasanti Yerramilli, Layne Pruitt, Kristina Reppond, Madison N. Shyer, Jessica Cambric, Ryan Gadd, Ilya J. Finkelstein, Jimmy Gollihar, and James M. Musser |
| EPI_ISL_2211386 | New Mexico Department of Health Scientific Laboratory | New Mexico Department of Health Scientific Laboratory | Ellie Johnson, D'eldra Malone, Jennifer Benoit, Ratheesh Rajan, Linda Salazar, Anastacia Griego-Fisher |
| EPI_ISL_2211390, EPI_ISL_2211393, EPI_ISL_2211395, EPI_ISL_2211398, EPI_ISL_2211401, EPI_ISL_2211405, EPI_ISL_2211409, EPI_ISL_2211413, EPI_ISL_2211416, EPI_ISL_2211420, EPI_ISL_2211424, EPI_ISL_2211427, EPI_ISL_2211430, EPI_ISL_2211433, EPI_ISL_2211436, EPI_ISL_2211439, EPI_ISL_2211448, EPI_ISL_2211452, EPI_ISL_2211455, EPI_ISL_2211458, EPI_ISL_2211461, EPI_ISL_2211466, EPI_ISL_2211469, EPI_ISL_2211472, EPI_ISL_2211475, EPI_ISL_2211478, EPI_ISL_2211482, EPI_ISL_2211485, EPI_ISL_2211489, EPI_ISL_2211491, EPI_ISL_2211495, EPI_ISL_2211498, EPI_ISL_2211502, EPI_ISL_2211505, EPI_ISL_2211508, EPI_ISL_2211511, EPI_ISL_2211515, EPI_ISL_2211518, EPI_ISL_2211521, EPI_ISL_2211525, EPI_ISL_2211528, EPI_ISL_2211531, EPI_ISL_2211534, EPI_ISL_2211537, EPI_ISL_2211541, EPI_ISL_2211546, EPI_ISL_2211549, EPI_ISL_2211552, EPI_ISL_2211554, EPI_ISL_2211558, EPI_ISL_2211563, EPI_ISL_2211566, EPI_ISL_2211570, EPI_ISL_2211574, EPI_ISL_2211577, EPI_ISL_2211580, EPI_ISL_2211585, EPI_ISL_2211589, EPI_ISL_2211593, EPI_ISL_2211597, EPI_ISL_2211601, EPI_ISL_2211607, EPI_ISL_2211610, EPI_ISL_2211615, EPI_ISL_2211620, EPI_ISL_2211624, EPI_ISL_2211628, EPI_ISL_2211638, EPI_ISL_2211642, EPI_ISL_2211648, EPI_ISL_2211650, EPI_ISL_2211655, EPI_ISL_2211662, EPI_ISL_2211668, EPI_ISL_2211673, EPI_ISL_2211678, EPI_ISL_2211683, EPI_ISL_2211690, EPI_ISL_2211697, EPI_ISL_2211702, EPI_ISL_2211707, EPI_ISL_2211713, EPI_ISL_2211718, EPI_ISL_2211723, EPI_ISL_2211729, EPI_ISL_2211736, EPI_ISL_2211741, EPI_ISL_2211746, EPI_ISL_2211751, EPI_ISL_2211758, EPI_ISL_2211764, EPI_ISL_2211770, EPI_ISL_2211775, EPI_ISL_2211781, EPI_ISL_2211795, EPI_ISL_2211802, EPI_ISL_2211807, EPI_ISL_2211812, EPI_ISL_2211817, EPI_ISL_2211824, EPI_ISL_2211829, EPI_ISL_2211834, EPI_ISL_2211838, EPI_ISL_2211842, EPI_ISL_2211850, EPI_ISL_2211853, EPI_ISL_2211856, EPI_ISL_2211861, EPI_ISL_2211865, EPI_ISL_2211869, EPI_ISL_2211873, EPI_ISL_2211878, EPI_ISL_2211882, EPI_ISL_2211885, EPI_ISL_2211891, EPI_ISL_2211896, EPI_ISL_2211901, EPI_ISL_2211906, EPI_ISL_2211911, EPI_ISL_2211918, EPI_ISL_2211924, EPI_ISL_2211929, EPI_ISL_2211932, EPI_ISL_2211936, EPI_ISL_2211940, EPI_ISL_2211944, EPI_ISL_2211947, EPI_ISL_2211950, EPI_ISL_2211954, EPI_ISL_2211958, EPI_ISL_2211959, EPI_ISL_2211961, EPI_ISL_2211965, EPI_ISL_2211970, EPI_ISL_2211974, EPI_ISL_2211977, EPI_ISL_2211982, EPI_ISL_2211985, EPI_ISL_2211991, EPI_ISL_2211995, EPI_ISL_2211998, EPI_ISL_2212003, EPI_ISL_2212007, EPI_ISL_2212010, EPI_ISL_2212015, EPI_ISL_2212018, EPI_ISL_2212021, EPI_ISL_2212025, EPI_ISL_2212029, EPI_ISL_2212032, EPI_ISL_2212037, EPI_ISL_2212042, EPI_ISL_2212045, EPI_ISL_2212047, EPI_ISL_2212051, EPI_ISL_2212055, EPI_ISL_2212059, EPI_ISL_2212063, EPI_ISL_2212067, EPI_ISL_2212070, EPI_ISL_2212073, EPI_ISL_2212078, EPI_ISL_2212082, EPI_ISL_2212086, EPI_ISL_2212090, EPI_ISL_2212093, EPI_ISL_2212100, EPI_ISL_2212103, EPI_ISL_2212105, EPI_ISL_2212110, EPI_ISL_2212114, EPI_ISL_2212119, EPI_ISL_2212123, EPI_ISL_2212126, EPI_ISL_2212130, EPI_ISL_2212132, EPI_ISL_2212136, EPI_ISL_2212141, EPI_ISL_2212145, EPI_ISL_2212148, EPI_ISL_2212152, EPI_ISL_2212157, EPI_ISL_2212161, EPI_ISL_2212165, EPI_ISL_2212168, EPI_ISL_2212172, EPI_ISL_2212177, EPI_ISL_2212181, EPI_ISL_2212187, EPI_ISL_2212190, EPI_ISL_2212195, EPI_ISL_2212197, EPI_ISL_2212202, EPI_ISL_2212211, EPI_ISL_2212215, EPI_ISL_2212219, EPI_ISL_2212222, EPI_ISL_2212227, EPI_ISL_2212231, EPI_ISL_2212235, EPI_ISL_2212238, EPI_ISL_2212243, EPI_ISL_2212248, EPI_ISL_2212255, EPI_ISL_2212259, EPI_ISL_2212264, EPI_ISL_2212268, EPI_ISL_2212272, EPI_ISL_2212274, EPI_ISL_2212278, EPI_ISL_2212283, EPI_ISL_2212287, EPI_ISL_2212291, EPI_ISL_2212296, EPI_ISL_2212298, EPI_ISL_2212303, EPI_ISL_2212308, EPI_ISL_2212313, EPI_ISL_2212317, EPI_ISL_2212322, EPI_ISL_2212327, EPI_ISL_2212331, EPI_ISL_2212335, EPI_ISL_2212338, EPI_ISL_2212343, EPI_ISL_2212348, EPI_ISL_2212353, EPI_ISL_2212359, EPI_ISL_2212363, EPI_ISL_2212367, EPI_ISL_2212372, EPI_ISL_2212384, EPI_ISL_2212388, EPI_ISL_2212392, EPI_ISL_2212397, EPI_ISL_2212403, EPI_ISL_2212407, EPI_ISL_2212413, EPI_ISL_2212416, EPI_ISL_2212420, EPI_ISL_2212425, EPI_ISL_2212429, EPI_ISL_2212433, EPI_ISL_2212437, EPI_ISL_2212440, EPI_ISL_2212447, EPI_ISL_2212451, EPI_ISL_2212456, EPI_ISL_2212459, EPI_ISL_2212465, EPI_ISL_2212468, EPI_ISL_2212472, EPI_ISL_2212475, EPI_ISL_2212478, EPI_ISL_2212484, EPI_ISL_2212489, EPI_ISL_2212496, EPI_ISL_2212500, EPI_ISL_2212505, EPI_ISL_2212508, EPI_ISL_2212515, EPI_ISL_2212520, EPI_ISL_2212524, EPI_ISL_2212530, EPI_ISL_2212539, EPI_ISL_2212545, EPI_ISL_2212549, EPI_ISL_2212557, EPI_ISL_2212561, EPI_ISL_2212566, EPI_ISL_2212571, EPI_ISL_2212580, EPI_ISL_2212583, EPI_ISL_2212587, EPI_ISL_2212591, EPI_ISL_2212595, EPI_ISL_2212598, EPI_ISL_2212603, EPI_ISL_2212606, EPI_ISL_2212610, EPI_ISL_2212616, EPI_ISL_2212620, EPI_ISL_2212625, EPI_ISL_2212630, EPI_ISL_2212634, EPI_ISL_2212640, EPI_ISL_2212645, EPI_ISL_2212649, EPI_ISL_2212653, EPI_ISL_2212656, EPI_ISL_2212660, EPI_ISL_2212664, EPI_ISL_2212668, EPI_ISL_2212672, EPI_ISL_2212676, EPI_ISL_2212681 | | | |
| see above | Houston Methodist Hospital | Houston Methodist Hospital | Randall J. Olsen, Paul A. Christensen, S. Wesley Long, Sishir Subedi, Robert Olson, Marcus Nguyen, James J. Davis, Matthew Ojeda Saavedra, Prasanti Yerramilli, Layne Pruitt, Kristina Reppond, Madison N. Shyer, Jessica Cambric, Ryan Gadd, Ilya J. Finkelstein, Jimmy Gollihar, and James M. Musser |
| EPI_ISL_2212684 | Ohio Department of Health Laboratory | Ohio Department of Health Laboratory | Holmes, Jennifer; Eric Brandt, Keoni Omura, Glen McGillivary, Caitlin McDonnell, Jade Mowery, Stephanie Mccracken, Tyler Payne, Kirtana Ramadugu, Erica Leasure, Brent Lee, Kelsey Florek, Heather Blankenship, Allison Black, Quanta Brown, and Tammy Bannerman |
| EPI_ISL_2212686, EPI_ISL_2212690, EPI_ISL_2212695, EPI_ISL_2212698, EPI_ISL_2212701, EPI_ISL_2212706, EPI_ISL_2212710, EPI_ISL_2212713, EPI_ISL_2212717, EPI_ISL_2212721, EPI_ISL_2212724, EPI_ISL_2212730, EPI_ISL_2212732, EPI_ISL_2212738, EPI_ISL_2212744, EPI_ISL_2212750, EPI_ISL_2212754, EPI_ISL_2212757, EPI_ISL_2212761, EPI_ISL_2212765, EPI_ISL_2212769, EPI_ISL_2212772, EPI_ISL_2212777, EPI_ISL_2212782, EPI_ISL_2212785, EPI_ISL_2212789, EPI_ISL_2212793, EPI_ISL_2212798, EPI_ISL_2212803, EPI_ISL_2212809, EPI_ISL_2212815, EPI_ISL_2212819, EPI_ISL_2212823, EPI_ISL_2212827, EPI_ISL_2212831, EPI_ISL_2212834, EPI_ISL_2212837, EPI_ISL_2212840, EPI_ISL_2212843, EPI_ISL_2212845, EPI_ISL_2212849, EPI_ISL_2212852, EPI_ISL_2212855, EPI_ISL_2212860, EPI_ISL_2212863, EPI_ISL_2212866, EPI_ISL_2212869, EPI_ISL_2212873, EPI_ISL_2212877, EPI_ISL_2212880, EPI_ISL_2212883, EPI_ISL_2212886, EPI_ISL_2212889, EPI_ISL_2212893, EPI_ISL_2212896, EPI_ISL_2212899, EPI_ISL_2212901, EPI_ISL_2212902, EPI_ISL_2212905, EPI_ISL_2212907, EPI_ISL_2212911, EPI_ISL_2212913, EPI_ISL_2212916, EPI_ISL_2212919, EPI_ISL_2212922, EPI_ISL_2212925, EPI_ISL_2212928, EPI_ISL_2212931, EPI_ISL_2212934, EPI_ISL_2212937, EPI_ISL_2212943, EPI_ISL_2212947, EPI_ISL_2212950, EPI_ISL_2212952, EPI_ISL_2212955, EPI_ISL_2212959, EPI_ISL_2212962, EPI_ISL_2212965, EPI_ISL_2212969, EPI_ISL_2212971, EPI_ISL_2212974, EPI_ISL_2212977, EPI_ISL_2212981, EPI_ISL_2212986, EPI_ISL_2212991, EPI_ISL_2212998, EPI_ISL_2213003, EPI_ISL_2213009, EPI_ISL_2213015, EPI_ISL_2213018, EPI_ISL_2213021, EPI_ISL_2213026, EPI_ISL_2213031, EPI_ISL_2213036, EPI_ISL_2213041, EPI_ISL_2213047, EPI_ISL_2213053, EPI_ISL_2213057, EPI_ISL_2213061, EPI_ISL_2213065, EPI_ISL_2213068, EPI_ISL_2213071, EPI_ISL_2213073, EPI_ISL_2213076, EPI_ISL_2213080, EPI_ISL_2213083, EPI_ISL_2213086, EPI_ISL_2213089, EPI_ISL_2213092, EPI_ISL_2213095, EPI_ISL_2213098, EPI_ISL_2213101, EPI_ISL_2213104, EPI_ISL_2213108, EPI_ISL_2213111, EPI_ISL_2213114, EPI_ISL_2213117, EPI_ISL_2213122, EPI_ISL_2213125, EPI_ISL_2213128, EPI_ISL_2213130, EPI_ISL_2213132, EPI_ISL_2213135, EPI_ISL_2213139, EPI_ISL_2213142, EPI_ISL_2213146, EPI_ISL_2213150 | | | |
| see above | Houston Methodist Hospital | Houston Methodist Hospital | Randall J. Olsen, Paul A. Christensen, S. Wesley Long, Sishir Subedi, Robert Olson, Marcus Nguyen, James J. Davis, Matthew Ojeda Saavedra, Prasanti Yerramilli, Layne Pruitt, Kristina Reppond, Madison N. Shyer, Jessica Cambric, Ryan Gadd, Ilya J. Finkelstein, Jimmy Gollihar, and James M. Musser |
| EPI_ISL_2213152, EPI_ISL_2213154 | Alaska State Virology Laboratory | Alaska State Virology Laboratory | Stephanie DeRonde, Elva House, Jacob Zidek, Lisa Smith, Ph.D., Jack Chen, Ph.D. |
| EPI_ISL_2213155 | Houston Methodist Hospital | Houston Methodist Hospital | Randall J. Olsen, Paul A. Christensen, S. Wesley Long, Sishir Subedi, Robert Olson, Marcus Nguyen, James J. Davis, Matthew Ojeda Saavedra, Prasanti Yerramilli, Layne Pruitt, Kristina Reppond, Madison N. Shyer, Jessica Cambric, Ryan Gadd, Ilya J. Finkelstein, Jimmy Gollihar, and James M. Musser |
| EPI_ISL_2213157, EPI_ISL_2213159 | Alaska State Virology Laboratory | Alaska State Virology Laboratory | Stephanie DeRonde, Elva House, Jacob Zidek, Lisa Smith, Ph.D., Jack Chen, Ph.D. |
| EPI_ISL_2213160 | Houston Methodist Hospital | Houston Methodist Hospital | Randall J. Olsen, Paul A. Christensen, S. Wesley Long, Sishir Subedi, Robert Olson, Marcus Nguyen, James J. Davis, Matthew Ojeda Saavedra, Prasanti Yerramilli, Layne Pruitt, Kristina Reppond, Madison N. Shyer, Jessica Cambric, Ryan Gadd, Ilya J. Finkelstein, Jimmy Gollihar, and James M. Musser |

| | | | |
|---|---|---|---|
| EPI_ISL_2213162 | Alaska State Virology Laboratory | Alaska State Virology Laboratory | Stephanie DeRonde, Elva House, Jacob Zidek, Lisa Smith, Ph.D., Jack Chen, Ph.D. |
| EPI_ISL_2213164 | Houston Methodist Hospital | Houston Methodist Hospital | Randall J. Olsen, Paul A. Christensen, S. Wesley Long, Sishir Subedi, Robert Olson, Marcus Nguyen, James J. Davis, Matthew Ojeda Saavedra, Prasanti Yerramilli, Layne Pruitt, Kristina Reppond, Madison N. Shyer, Jessica Cambric, Ryan Gadd, Ilya J. Finkelstein, Jimmy Gollihar, and James M. Musser |
| EPI_ISL_2213165, EPI_ISL_2213167, EPI_ISL_2213169 | Alaska State Virology Laboratory | Alaska State Virology Laboratory | Stephanie DeRonde, Elva House, Jacob Zidek, Lisa Smith, Ph.D., Jack Chen, Ph.D. |
| EPI_ISL_2213171 | Houston Methodist Hospital | Houston Methodist Hospital | Randall J. Olsen, Paul A. Christensen, S. Wesley Long, Sishir Subedi, Robert Olson, Marcus Nguyen, James J. Davis, Matthew Ojeda Saavedra, Prasanti Yerramilli, Layne Pruitt, Kristina Reppond, Madison N. Shyer, Jessica Cambric, Ryan Gadd, Ilya J. Finkelstein, Jimmy Gollihar, and James M. Musser |
| EPI_ISL_2213172, EPI_ISL_2213174, EPI_ISL_2213176 | Alaska State Virology Laboratory | Alaska State Virology Laboratory | Stephanie DeRonde, Elva House, Jacob Zidek, Lisa Smith, Ph.D., Jack Chen, Ph.D. |
| EPI_ISL_2213178 | Houston Methodist Hospital | Houston Methodist Hospital | Randall J. Olsen, Paul A. Christensen, S. Wesley Long, Sishir Subedi, Robert Olson, Marcus Nguyen, James J. Davis, Matthew Ojeda Saavedra, Prasanti Yerramilli, Layne Pruitt, Kristina Reppond, Madison N. Shyer, Jessica Cambric, Ryan Gadd, Ilya J. Finkelstein, Jimmy Gollihar, and James M. Musser |
| EPI_ISL_2213179, EPI_ISL_2213181 | Alaska State Virology Laboratory | Alaska State Virology Laboratory | Stephanie DeRonde, Elva House, Jacob Zidek, Lisa Smith, Ph.D., Jack Chen, Ph.D. |
| EPI_ISL_2213183 | Houston Methodist Hospital | Houston Methodist Hospital | Randall J. Olsen, Paul A. Christensen, S. Wesley Long, Sishir Subedi, Robert Olson, Marcus Nguyen, James J. Davis, Matthew Ojeda Saavedra, Prasanti Yerramilli, Layne Pruitt, Kristina Reppond, Madison N. Shyer, Jessica Cambric, Ryan Gadd, Ilya J. Finkelstein, Jimmy Gollihar, and James M. Musser |
| EPI_ISL_2213184, EPI_ISL_2213186 | Alaska State Virology Laboratory | Alaska State Virology Laboratory | Stephanie DeRonde, Elva House, Jacob Zidek, Lisa Smith, Ph.D., Jack Chen, Ph.D. |
| EPI_ISL_2213187 | Houston Methodist Hospital | Houston Methodist Hospital | Randall J. Olsen, Paul A. Christensen, S. Wesley Long, Sishir Subedi, Robert Olson, Marcus Nguyen, James J. Davis, Matthew Ojeda Saavedra, Prasanti Yerramilli, Layne Pruitt, Kristina Reppond, Madison N. Shyer, Jessica Cambric, Ryan Gadd, Ilya J. Finkelstein, Jimmy Gollihar, and James M. Musser |
| EPI_ISL_2213189, EPI_ISL_2213191 | Alaska State Virology Laboratory | Alaska State Virology Laboratory | Stephanie DeRonde, Elva House, Jacob Zidek, Lisa Smith, Ph.D., Jack Chen, Ph.D. |
| EPI_ISL_2213193 | Houston Methodist Hospital | Houston Methodist Hospital | Randall J. Olsen, Paul A. Christensen, S. Wesley Long, Sishir Subedi, Robert Olson, Marcus Nguyen, James J. Davis, Matthew Ojeda Saavedra, Prasanti Yerramilli, Layne Pruitt, Kristina Reppond, Madison N. Shyer, Jessica Cambric, Ryan Gadd, Ilya J. Finkelstein, Jimmy Gollihar, and James M. Musser |
| EPI_ISL_2213194, EPI_ISL_2213196 | Alaska State Virology Laboratory | Alaska State Virology Laboratory | Stephanie DeRonde, Elva House, Jacob Zidek, Lisa Smith, Ph.D., Jack Chen, Ph.D. |
| EPI_ISL_2213198 | Houston Methodist Hospital | Houston Methodist Hospital | Randall J. Olsen, Paul A. Christensen, S. Wesley Long, Sishir Subedi, Robert Olson, Marcus Nguyen, James J. Davis, Matthew Ojeda Saavedra, Prasanti Yerramilli, Layne Pruitt, Kristina Reppond, Madison N. Shyer, Jessica Cambric, Ryan Gadd, Ilya J. Finkelstein, Jimmy Gollihar, and James M. Musser |
| EPI_ISL_2213199, EPI_ISL_2213201 | Alaska State Virology Laboratory | Alaska State Virology Laboratory | Stephanie DeRonde, Elva House, Jacob Zidek, Lisa Smith, Ph.D., Jack Chen, Ph.D. |
| EPI_ISL_2213202 | Houston Methodist Hospital | Houston Methodist Hospital | Randall J. Olsen, Paul A. Christensen, S. Wesley Long, Sishir Subedi, Robert Olson, Marcus Nguyen, James J. Davis, Matthew Ojeda Saavedra, Prasanti Yerramilli, Layne Pruitt, Kristina Reppond, Madison N. Shyer, Jessica Cambric, Ryan Gadd, Ilya J. Finkelstein, Jimmy Gollihar, and James M. Musser |
| EPI_ISL_2213203, EPI_ISL_2213204, EPI_ISL_2213207 | Alaska State Virology Laboratory | Alaska State Virology Laboratory | Stephanie DeRonde, Elva House, Jacob Zidek, Lisa Smith, Ph.D., Jack Chen, Ph.D. |
| EPI_ISL_2213208 | Houston Methodist Hospital | Houston Methodist Hospital | Randall J. Olsen, Paul A. Christensen, S. Wesley Long, Sishir Subedi, Robert Olson, Marcus Nguyen, James J. Davis, Matthew Ojeda Saavedra, Prasanti Yerramilli, Layne Pruitt, Kristina Reppond, Madison N. Shyer, Jessica Cambric, Ryan Gadd, Ilya J. Finkelstein, Jimmy Gollihar, and James M. Musser |
| EPI_ISL_2213210, EPI_ISL_2213212 | Alaska State Virology Laboratory | Alaska State Virology Laboratory | Stephanie DeRonde, Elva House, Jacob Zidek, Lisa Smith, Ph.D., Jack Chen, Ph.D. |
| EPI_ISL_2213213 | Houston Methodist Hospital | Houston Methodist Hospital | Randall J. Olsen, Paul A. Christensen, S. Wesley Long, Sishir Subedi, Robert Olson, Marcus Nguyen, James J. Davis, Matthew Ojeda Saavedra, Prasanti Yerramilli, Layne Pruitt, Kristina Reppond, Madison N. Shyer, Jessica Cambric, Ryan Gadd, Ilya J. Finkelstein, Jimmy Gollihar, and James M. Musser |
| EPI_ISL_2213215, EPI_ISL_2213216 | Alaska State Virology Laboratory | Alaska State Virology Laboratory | Stephanie DeRonde, Elva House, Jacob Zidek, Lisa Smith, Ph.D., Jack Chen, Ph.D. |
| EPI_ISL_2213218 | Houston Methodist Hospital | Houston Methodist Hospital | Randall J. Olsen, Paul A. Christensen, S. Wesley Long, Sishir Subedi, Robert Olson, Marcus Nguyen, James J. Davis, Matthew Ojeda Saavedra, Prasanti Yerramilli, Layne Pruitt, Kristina Reppond, Madison N. Shyer, Jessica Cambric, Ryan Gadd, Ilya J. Finkelstein, Jimmy Gollihar, and James M. Musser |
| EPI_ISL_2213219, EPI_ISL_2213221 | Alaska State Virology Laboratory | Alaska State Virology Laboratory | Stephanie DeRonde, Elva House, Jacob Zidek, Lisa Smith, Ph.D., Jack Chen, Ph.D. |
| EPI_ISL_2213222 | Houston Methodist Hospital | Houston Methodist Hospital | Randall J. Olsen, Paul A. Christensen, S. Wesley Long, Sishir Subedi, Robert Olson, Marcus Nguyen, James J. Davis, Matthew Ojeda Saavedra, Prasanti Yerramilli, Layne Pruitt, Kristina Reppond, Madison N. Shyer, Jessica Cambric, Ryan Gadd, Ilya J. Finkelstein, Jimmy Gollihar, and James M. Musser |
| EPI_ISL_2213224, EPI_ISL_2213225 | Alaska State Virology Laboratory | Alaska State Virology Laboratory | Stephanie DeRonde, Elva House, Jacob Zidek, Lisa Smith, Ph.D., Jack Chen, Ph.D. |
| EPI_ISL_2213227 | Houston Methodist Hospital | Houston Methodist Hospital | Randall J. Olsen, Paul A. Christensen, S. Wesley Long, Sishir Subedi, Robert Olson, Marcus Nguyen, James J. Davis, Matthew Ojeda Saavedra, Prasanti Yerramilli, Layne Pruitt, Kristina Reppond, Madison N. Shyer, Jessica Cambric, Ryan Gadd, Ilya J. Finkelstein, Jimmy Gollihar, and James M. Musser |
| EPI_ISL_2213228, EPI_ISL_2213231, EPI_ISL_2213232 | Alaska State Virology Laboratory | Alaska State Virology Laboratory | Stephanie DeRonde, Elva House, Jacob Zidek, Lisa Smith, Ph.D., Jack Chen, Ph.D. |
| EPI_ISL_2213233 | Houston Methodist Hospital | Houston Methodist Hospital | Randall J. Olsen, Paul A. Christensen, S. Wesley Long, Sishir Subedi, Robert Olson, Marcus Nguyen, James J. Davis, Matthew Ojeda Saavedra, Prasanti Yerramilli, Layne Pruitt, Kristina Reppond, Madison N. Shyer, Jessica Cambric, Ryan Gadd, Ilya J. Finkelstein, Jimmy Gollihar, and James M. Musser |
| EPI_ISL_2213235 | Alaska State Virology Laboratory | Alaska State Virology Laboratory | Stephanie DeRonde, Elva House, Jacob Zidek, Lisa Smith, Ph.D., Jack Chen, Ph.D. |
| EPI_ISL_2213239, EPI_ISL_2213243, EPI_ISL_2213246, EPI_ISL_2213250, EPI_ISL_2213253, EPI_ISL_2213256, EPI_ISL_2213261, EPI_ISL_2213264, EPI_ISL_2213267, EPI_ISL_2213272, EPI_ISL_2213276, EPI_ISL_2213278, EPI_ISL_2213282, EPI_ISL_2213285, EPI_ISL_2213288, EPI_ISL_2213291, EPI_ISL_2213294, EPI_ISL_2213298, EPI_ISL_2213301, EPI_ISL_2213304, EPI_ISL_2213309, EPI_ISL_2213312, EPI_ISL_2213315, EPI_ISL_2213318, EPI_ISL_2213321, EPI_ISL_2213324, EPI_ISL_2213327, EPI_ISL_2213329, EPI_ISL_2213332, EPI_ISL_2213337, EPI_ISL_2213340, EPI_ISL_2213343, EPI_ISL_2213347, EPI_ISL_2213350, EPI_ISL_2213353, EPI_ISL_2213355, EPI_ISL_2213358, EPI_ISL_2213361, EPI_ISL_2213364, EPI_ISL_2213367, EPI_ISL_2213370, EPI_ISL_2213374, EPI_ISL_2213376, EPI_ISL_2213379, EPI_ISL_2213382, EPI_ISL_2213386, EPI_ISL_2213390, EPI_ISL_2213393, EPI_ISL_2213396, EPI_ISL_2213399, EPI_ISL_2213403, EPI_ISL_2213406, EPI_ISL_2213408, EPI_ISL_2213411, EPI_ISL_2213414, EPI_ISL_2213417, EPI_ISL_2213422, EPI_ISL_2213425, EPI_ISL_2213428, EPI_ISL_2213432, EPI_ISL_2213434, EPI_ISL_2213438, EPI_ISL_2213441, EPI_ISL_2213444, EPI_ISL_2213447, EPI_ISL_2213450, EPI_ISL_2213453, EPI_ISL_2213456, EPI_ISL_2213458, EPI_ISL_2213461, EPI_ISL_2213465, EPI_ISL_2213468, EPI_ISL_2213471, EPI_ISL_2213474, EPI_ISL_2213477, EPI_ISL_2213479, EPI_ISL_2213482, EPI_ISL_2213489, EPI_ISL_2213494, EPI_ISL_2213499, EPI_ISL_2213502, EPI_ISL_2213508, EPI_ISL_2213512, EPI_ISL_2213516, EPI_ISL_2213520, EPI_ISL_2213524, EPI_ISL_2213530, EPI_ISL_2213533, EPI_ISL_2213539, EPI_ISL_2213542, EPI_ISL_2213545, EPI_ISL_2213550, EPI_ISL_2213554, EPI_ISL_2213556, EPI_ISL_2213561, EPI_ISL_2213565, EPI_ISL_2213570, EPI_ISL_2213574, EPI_ISL_2213576, EPI_ISL_2213581 | | | |
| see above | Houston Methodist Hospital | Houston Methodist Hospital | Randall J. Olsen, Paul A. Christensen, S. Wesley Long, Sishir Subedi, Robert Olson, Marcus Nguyen, James J. Davis, Matthew Ojeda Saavedra, Prasanti Yerramilli, Layne Pruitt, Kristina Reppond, Madison N. Shyer, Jessica Cambric, Ryan Gadd, Ilya J. Finkelstein, Jimmy Gollihar, and James M. Musser |
| EPI_ISL_2213586 | Ohio Department of Health Laboratory | Ohio Department of Health Laboratory | Holmes, Jennifer; Eric Brandt, Keoni Omura, Glen McGillivary, Caitlin McDonnell, Jade Mowery, Stephanie Mccracken, Tyler Payne, Kirtana Ramadugu, Erica Leasure, Brent Lee, Kelsey Florek, Heather Blankenship, Allison Black, Quanta Brown, and Tammy Bannerman |
| EPI_ISL_2213588, EPI_ISL_2213594, EPI_ISL_2213600, EPI_ISL_2213602, EPI_ISL_2213606, EPI_ISL_2213610, EPI_ISL_2213613, EPI_ISL_2213620, EPI_ISL_2213626, EPI_ISL_2213633, EPI_ISL_2213640, EPI_ISL_2213643, EPI_ISL_2213647, EPI_ISL_2213652, EPI_ISL_2213661, EPI_ISL_2213665, EPI_ISL_2213669, EPI_ISL_2213674, EPI_ISL_2213678, EPI_ISL_2213681, EPI_ISL_2213685, EPI_ISL_2213689, EPI_ISL_2213693, EPI_ISL_2213697, EPI_ISL_2213701, EPI_ISL_2213704, EPI_ISL_2213708, EPI_ISL_2213713, EPI_ISL_2213717, EPI_ISL_2213721, EPI_ISL_2213726, EPI_ISL_2213730, EPI_ISL_2213734, EPI_ISL_2213738, EPI_ISL_2213741, EPI_ISL_2213745, EPI_ISL_2213750, EPI_ISL_2213754, EPI_ISL_2213760, EPI_ISL_2213765, EPI_ISL_2213770, EPI_ISL_2213773, EPI_ISL_2213777, EPI_ISL_2213781, EPI_ISL_2213786, EPI_ISL_2213791, EPI_ISL_2213797, EPI_ISL_2213802, EPI_ISL_2213805, EPI_ISL_2213807, EPI_ISL_2213811, EPI_ISL_2213817, EPI_ISL_2213820, EPI_ISL_2213823, EPI_ISL_2213828, EPI_ISL_2213840, EPI_ISL_2213843, EPI_ISL_2213847, EPI_ISL_2213852, EPI_ISL_2213857, EPI_ISL_2213862, EPI_ISL_2213868, EPI_ISL_2213873, EPI_ISL_2213878, EPI_ISL_2213882, EPI_ISL_2213886, EPI_ISL_2213891, EPI_ISL_2213895, EPI_ISL_2213898, EPI_ISL_2213902, EPI_ISL_2213907, EPI_ISL_2213914, EPI_ISL_2213921, EPI_ISL_2213926, EPI_ISL_2213930, EPI_ISL_2213933, EPI_ISL_2213936, EPI_ISL_2213939, EPI_ISL_2213944, EPI_ISL_2213950, EPI_ISL_2213956, EPI_ISL_2213962, EPI_ISL_2213967, EPI_ISL_2213970, EPI_ISL_2213974, EPI_ISL_2213978, EPI_ISL_2213983, EPI_ISL_2213986, EPI_ISL_2213990, EPI_ISL_2213995, EPI_ISL_2213999, EPI_ISL_2214009, EPI_ISL_2214012, EPI_ISL_2214019, EPI_ISL_2214021, EPI_ISL_2214030, EPI_ISL_2214045, EPI_ISL_2214049, EPI_ISL_2214054, EPI_ISL_2214060, EPI_ISL_2214064, EPI_ISL_2214069, EPI_ISL_2214073, EPI_ISL_2214077, EPI_ISL_2214080, EPI_ISL_2214091, EPI_ISL_2214095, EPI_ISL_2214112, EPI_ISL_2214115, EPI_ISL_2214119, EPI_ISL_2214122, EPI_ISL_2214126, EPI_ISL_2214130, EPI_ISL_2214135, EPI_ISL_2214138, EPI_ISL_2214142, EPI_ISL_2214146, EPI_ISL_2214149, EPI_ISL_2214153, EPI_ISL_2214157, EPI_ISL_2214162, EPI_ISL_2214167, EPI_ISL_2214171, EPI_ISL_2214174, EPI_ISL_2214178, EPI_ISL_2214183, EPI_ISL_2214189, EPI_ISL_2214192, EPI_ISL_2214195, EPI_ISL_2214199, EPI_ISL_2214202, EPI_ISL_2214206, EPI_ISL_2214211, EPI_ISL_2214213, EPI_ISL_2214218, EPI_ISL_2214223, EPI_ISL_2214228, EPI_ISL_2214233, EPI_ISL_2214240, EPI_ISL_2214243, EPI_ISL_2214245, EPI_ISL_2214253, EPI_ISL_2214256, EPI_ISL_2214259, EPI_ISL_2214262, EPI_ISL_2214266, EPI_ISL_2214270, EPI_ISL_2214279, EPI_ISL_2214282, EPI_ISL_2214287, EPI_ISL_2214289, EPI_ISL_2214299, EPI_ISL_2214303, EPI_ISL_2214308, EPI_ISL_2214310, EPI_ISL_2214315, EPI_ISL_2214319, EPI_ISL_2214323, EPI_ISL_2214326, EPI_ISL_2214330, EPI_ISL_2214334, EPI_ISL_2214337, EPI_ISL_2214340, EPI_ISL_2214345, EPI_ISL_2214348, EPI_ISL_2214351, EPI_ISL_2214355, EPI_ISL_2214358, EPI_ISL_2214367, EPI_ISL_2214370, EPI_ISL_2214374, EPI_ISL_2214380, EPI_ISL_2214383, EPI_ISL_2214386, EPI_ISL_2214390, EPI_ISL_2214394, EPI_ISL_2214398, EPI_ISL_2214401, EPI_ISL_2214404, EPI_ISL_2214408, EPI_ISL_2214410, EPI_ISL_2214413, EPI_ISL_2214419, EPI_ISL_2214422, EPI_ISL_2214425, EPI_ISL_2214428, EPI_ISL_2214430, EPI_ISL_2214433, EPI_ISL_2214436, EPI_ISL_2214439, EPI_ISL_2214442, EPI_ISL_2214445, EPI_ISL_2214448, EPI_ISL_2214450, EPI_ISL_2214452, EPI_ISL_2214456, EPI_ISL_2214459, EPI_ISL_2214462, EPI_ISL_2214465, EPI_ISL_2214468, EPI_ISL_2214471, EPI_ISL_2214474, EPI_ISL_2214476, EPI_ISL_2214479, EPI_ISL_2214482, EPI_ISL_2214485, EPI_ISL_2214488, EPI_ISL_2214491, EPI_ISL_2214494, EPI_ISL_2214498, EPI_ISL_2214500, EPI_ISL_2214503, EPI_ISL_2214506, EPI_ISL_2214509, EPI_ISL_2214512, EPI_ISL_2214514, EPI_ISL_2214517, EPI_ISL_2214520, EPI_ISL_2214523, EPI_ISL_2214526, EPI_ISL_2214529, EPI_ISL_2214533, EPI_ISL_2214539, EPI_ISL_2214543, EPI_ISL_2214546, EPI_ISL_2214549, EPI_ISL_2214552, EPI_ISL_2214555, EPI_ISL_2214557, EPI_ISL_2214560, EPI_ISL_2214563, EPI_ISL_2214566, EPI_ISL_2214570, EPI_ISL_2214573, EPI_ISL_2214575, EPI_ISL_2214577, EPI_ISL_2214579, EPI_ISL_2214582, EPI_ISL_2214585, EPI_ISL_2214588, EPI_ISL_2214591, EPI_ISL_2214594, EPI_ISL_2214598, EPI_ISL_2214601, EPI_ISL_2214603, EPI_ISL_2214605, EPI_ISL_2214610, EPI_ISL_2214613, EPI_ISL_2214616, EPI_ISL_2214619, EPI_ISL_2214622, EPI_ISL_2214625, EPI_ISL_2214627, EPI_ISL_2214631, EPI_ISL_2214634, EPI_ISL_2214637, EPI_ISL_2214641, EPI_ISL_2214644, EPI_ISL_2214647, EPI_ISL_2214652, EPI_ISL_2214656, EPI_ISL_2214660, EPI_ISL_2214664, EPI_ISL_2214667, EPI_ISL_2214670, EPI_ISL_2214674, EPI_ISL_2214678, EPI_ISL_2214682, EPI_ISL_2214686, EPI_ISL_2214689, EPI_ISL_2214698, EPI_ISL_2214708, EPI_ISL_2214711, EPI_ISL_2214715, EPI_ISL_2214719, EPI_ISL_2214723, EPI_ISL_2214726, EPI_ISL_2214729, EPI_ISL_2214734, EPI_ISL_2214738, EPI_ISL_2214740, EPI_ISL_2214741, EPI_ISL_2214744, EPI_ISL_2214748, EPI_ISL_2214751, EPI_ISL_2214755, EPI_ISL_2214759, EPI_ISL_2214764, EPI_ISL_2214767, EPI_ISL_2214771, EPI_ISL_2214775, EPI_ISL_2214778, EPI_ISL_2214781, EPI_ISL_2214782, EPI_ISL_2214785, EPI_ISL_2214789, EPI_ISL_2214792, EPI_ISL_2214796, EPI_ISL_2214800, EPI_ISL_2214803, EPI_ISL_2214807, EPI_ISL_2214810, EPI_ISL_2214813, EPI_ISL_2214817, EPI_ISL_2214821, EPI_ISL_2214824, EPI_ISL_2214828, EPI_ISL_2214832, EPI_ISL_2214835, EPI_ISL_2214838, EPI_ISL_2214842, EPI_ISL_2214847, EPI_ISL_2214850, EPI_ISL_2214855, EPI_ISL_2214858, EPI_ISL_2214862, EPI_ISL_2214867, EPI_ISL_2214871, EPI_ISL_2214874, EPI_ISL_2214877, EPI_ISL_2214880, EPI_ISL_2214882, EPI_ISL_2214886, EPI_ISL_2214890, EPI_ISL_2214893, EPI_ISL_2214897, EPI_ISL_2214901, EPI_ISL_2214904, EPI_ISL_2214908, EPI_ISL_2214912, EPI_ISL_2214915, EPI_ISL_2214919, EPI_ISL_2214923, EPI_ISL_2214928, EPI_ISL_2214930, EPI_ISL_2214935, EPI_ISL_2214938, EPI_ISL_2214943, EPI_ISL_2214947, EPI_ISL_2214951, EPI_ISL_2214956, EPI_ISL_2214959, EPI_ISL_2214962, EPI_ISL_2214966, EPI_ISL_2214970, EPI_ISL_2214974, EPI_ISL_2214977, EPI_ISL_2214981, EPI_ISL_2214984, EPI_ISL_2214987, EPI_ISL_2214990, EPI_ISL_2214995, EPI_ISL_2214998, EPI_ISL_2215001, EPI_ISL_2215004, EPI_ISL_2215007, EPI_ISL_2215009, EPI_ISL_2215012, EPI_ISL_2215015, EPI_ISL_2215018, EPI_ISL_2215021, EPI_ISL_2215026, EPI_ISL_2215028, EPI_ISL_2215030, EPI_ISL_2215033, EPI_ISL_2215035, EPI_ISL_2215039, EPI_ISL_2215042, EPI_ISL_2215045, EPI_ISL_2215048, EPI_ISL_2215050, EPI_ISL_2215054, EPI_ISL_2215057, EPI_ISL_2215060, EPI_ISL_2215064, EPI_ISL_2215067, EPI_ISL_2215070, EPI_ISL_2215073, EPI_ISL_2215076, EPI_ISL_2215079, EPI_ISL_2215082, EPI_ISL_2215085, EPI_ISL_2215088, EPI_ISL_2215092, EPI_ISL_2215095, EPI_ISL_2215097, EPI_ISL_2215100, EPI_ISL_2215105, EPI_ISL_2215108, EPI_ISL_2215111, EPI_ISL_2215112, EPI_ISL_2215114, EPI_ISL_2215117, EPI_ISL_2215120, EPI_ISL_2215125, EPI_ISL_2215128, EPI_ISL_2215131, EPI_ISL_2215134, EPI_ISL_2215137, EPI_ISL_2215140, EPI_ISL_2215145, EPI_ISL_2215148, EPI_ISL_2215151, EPI_ISL_2215154, EPI_ISL_2215157, EPI_ISL_2215160, EPI_ISL_2215163, EPI_ISL_2215166, EPI_ISL_2215169, EPI_ISL_2215171, EPI_ISL_2215173, EPI_ISL_2215176, EPI_ISL_2215179, EPI_ISL_2215182, EPI_ISL_2215186, EPI_ISL_2215189, EPI_ISL_2215191, EPI_ISL_2215195, EPI_ISL_2215198, EPI_ISL_2215201, EPI_ISL_2215204, EPI_ISL_2215207, EPI_ISL_2215210, EPI_ISL_2215213, EPI_ISL_2215216, EPI_ISL_2215219, EPI_ISL_2215222, EPI_ISL_2215224, EPI_ISL_2215227, EPI_ISL_2215230, EPI_ISL_2215232, EPI_ISL_2215235, EPI_ISL_2215237, EPI_ISL_2215239, EPI_ISL_2215242, EPI_ISL_2215245, EPI_ISL_2215248, EPI_ISL_2215252, EPI_ISL_2215255, EPI_ISL_2215259, EPI_ISL_2215262, EPI_ISL_2215265, EPI_ISL_2215267, EPI_ISL_2215270, EPI_ISL_2215273, EPI_ISL_2215276, EPI_ISL_2215279, EPI_ISL_2215280, EPI_ISL_2215283, EPI_ISL_2215286, EPI_ISL_2215289, EPI_ISL_2215292, EPI_ISL_2215295, EPI_ISL_2215298, EPI_ISL_2215302, EPI_ISL_2215304, EPI_ISL_2215307, EPI_ISL_2215310, EPI_ISL_2215313, EPI_ISL_2215316, EPI_ISL_2215319, EPI_ISL_2215322, EPI_ISL_2215325, EPI_ISL_2215328, EPI_ISL_2215332, EPI_ISL_2215334, EPI_ISL_2215337, EPI_ISL_2215340, EPI_ISL_2215343, EPI_ISL_2215346, EPI_ISL_2215349, EPI_ISL_2215352, EPI_ISL_2215355, EPI_ISL_2215357, EPI_ISL_2215359, EPI_ISL_2215362, EPI_ISL_2215365, EPI_ISL_2215368, EPI_ISL_2215369, EPI_ISL_2215371, EPI_ISL_2215375, EPI_ISL_2215378, EPI_ISL_2215381, EPI_ISL_2215383, EPI_ISL_2215386, EPI_ISL_2215388, EPI_ISL_2215390, EPI_ISL_2215392, EPI_ISL_2215395, EPI_ISL_2215398, EPI_ISL_2215401, EPI_ISL_2215405, EPI_ISL_2215408, EPI_ISL_2215411, EPI_ISL_2215414, EPI_ISL_2215415, EPI_ISL_2215418, EPI_ISL_2215421, EPI_ISL_2215425, EPI_ISL_2215428, EPI_ISL_2215432, EPI_ISL_2215435, EPI_ISL_2215439, EPI_ISL_2215442, EPI_ISL_2215446, EPI_ISL_2215448, EPI_ISL_2215453, EPI_ISL_2215456, EPI_ISL_2215459, EPI_ISL_2215463, EPI_ISL_2215466, EPI_ISL_2215471, EPI_ISL_2215476, EPI_ISL_2215479, EPI_ISL_2215483, EPI_ISL_2215485, EPI_ISL_2215489, EPI_ISL_2215492, EPI_ISL_2215496, EPI_ISL_2215499, EPI_ISL_2215502, EPI_ISL_2215506, EPI_ISL_2215510, EPI_ISL_2215515, EPI_ISL_2215518, EPI_ISL_2215522, EPI_ISL_2215524, EPI_ISL_2215529, EPI_ISL_2215533, EPI_ISL_2215536, EPI_ISL_2215540, EPI_ISL_2215543, EPI_ISL_2215547, EPI_ISL_2215551, EPI_ISL_2215554, EPI_ISL_2215557, EPI_ISL_2215561, EPI_ISL_2215566, EPI_ISL_2215569, EPI_ISL_2215576, EPI_ISL_2215579, EPI_ISL_2215586, EPI_ISL_2215589, EPI_ISL_2215593, EPI_ISL_2215596, EPI_ISL_2215598, EPI_ISL_2215600, EPI_ISL_2215604, EPI_ISL_2215608, EPI_ISL_2215614, EPI_ISL_2215617, EPI_ISL_2215620, EPI_ISL_2215624, EPI_ISL_2215627, EPI_ISL_2215632, EPI_ISL_2215634, EPI_ISL_2215638, EPI_ISL_2215642, EPI_ISL_2215645, EPI_ISL_2215648, EPI_ISL_2215651, EPI_ISL_2215656, EPI_ISL_2215660, EPI_ISL_2215664, EPI_ISL_2215667, EPI_ISL_2215670, EPI_ISL_2215673, EPI_ISL_2215677, EPI_ISL_2215682, EPI_ISL_2215685, EPI_ISL_2215690, EPI_ISL_2215694, EPI_ISL_2215697, EPI_ISL_2215701, EPI_ISL_2215703, EPI_ISL_2215707, EPI_ISL_2215710, EPI_ISL_2215714, EPI_ISL_2215718, EPI_ISL_2215721, EPI_ISL_2215724, EPI_ISL_2215728, EPI_ISL_2215731, EPI_ISL_2215736, EPI_ISL_2215740, EPI_ISL_2215742, EPI_ISL_2215746, EPI_ISL_2215751, EPI_ISL_2215754, EPI_ISL_2215758, EPI_ISL_2215761, EPI_ISL_2215764, EPI_ISL_2215768, EPI_ISL_2215772, EPI_ISL_2215775, EPI_ISL_2215779, EPI_ISL_2215783, EPI_ISL_2215788, EPI_ISL_2215794, EPI_ISL_2215799, EPI_ISL_2215803, EPI_ISL_2215808, EPI_ISL_2215812, EPI_ISL_2215819, EPI_ISL_2215822, EPI_ISL_2215827, EPI_ISL_2215831, EPI_ISL_2215834, EPI_ISL_2215839, EPI_ISL_2215843, EPI_ISL_2215847, EPI_ISL_2215850, EPI_ISL_2215854, EPI_ISL_2215858, EPI_ISL_2215861, EPI_ISL_2215865, | | | |

EPI_ISL_2215869, EPI_ISL_2215872, EPI_ISL_2215877, EPI_ISL_2215880, EPI_ISL_2215885, EPI_ISL_2215891, EPI_ISL_2215896, EPI_ISL_2215900, EPI_ISL_2215905, EPI_ISL_2215910, EPI_ISL_2215915, EPI_ISL_2215922, EPI_ISL_2215927, EPI_ISL_2215935, EPI_ISL_2215942, EPI_ISL_2215948, EPI_ISL_2215952, EPI_ISL_2215957, EPI_ISL_2215962, EPI_ISL_2215967, EPI_ISL_2215972, EPI_ISL_2215978, EPI_ISL_2215984, EPI_ISL_2215988, EPI_ISL_2215992, EPI_ISL_2215995, EPI_ISL_2215999, EPI_ISL_2216004, EPI_ISL_2216009, EPI_ISL_2216015, EPI_ISL_2216018, EPI_ISL_2216021, EPI_ISL_2216025, EPI_ISL_2216030, EPI_ISL_2216035, EPI_ISL_2216039, EPI_ISL_2216045, EPI_ISL_2216048, EPI_ISL_2216051, EPI_ISL_2216055, EPI_ISL_2216060, EPI_ISL_2216064, EPI_ISL_2216067, EPI_ISL_2216073, EPI_ISL_2216078, EPI_ISL_2216081, EPI_ISL_2216084, EPI_ISL_2216088, EPI_ISL_2216092, EPI_ISL_2216097, EPI_ISL_2216100, EPI_ISL_2216106, EPI_ISL_2216110, EPI_ISL_2216113, EPI_ISL_2216117, EPI_ISL_2216121, EPI_ISL_2216125, EPI_ISL_2216129, EPI_ISL_2216133, EPI_ISL_2216136, EPI_ISL_2216140, EPI_ISL_2216147, EPI_ISL_2216151, EPI_ISL_2216154, EPI_ISL_2216158, EPI_ISL_2216161, EPI_ISL_2216166, EPI_ISL_2216171, EPI_ISL_2216175, EPI_ISL_2216185, EPI_ISL_2216189, EPI_ISL_2216193, EPI_ISL_2216196, EPI_ISL_2216198, EPI_ISL_2216205, EPI_ISL_2216210, EPI_ISL_2216215, EPI_ISL_2216219, EPI_ISL_2216222, EPI_ISL_2216226, EPI_ISL_2216230, EPI_ISL_2216233, EPI_ISL_2216235, EPI_ISL_2216241, EPI_ISL_2216247, EPI_ISL_2216250, EPI_ISL_2216257, EPI_ISL_2216260, EPI_ISL_2216267, EPI_ISL_2216274, EPI_ISL_2216279, EPI_ISL_2216283, EPI_ISL_2216287, EPI_ISL_2216295, EPI_ISL_2216298, EPI_ISL_2216303, EPI_ISL_2216306, EPI_ISL_2216310, EPI_ISL_2216313, EPI_ISL_2216318, EPI_ISL_2216321, EPI_ISL_2216325, EPI_ISL_2216328, EPI_ISL_2216334, EPI_ISL_2216338, EPI_ISL_2216341, EPI_ISL_2216346, EPI_ISL_2216351, EPI_ISL_2216354, EPI_ISL_2216358, EPI_ISL_2216362, EPI_ISL_2216367, EPI_ISL_2216372, EPI_ISL_2216375, EPI_ISL_2216380, EPI_ISL_2216385, EPI_ISL_2216390, EPI_ISL_2216395, EPI_ISL_2216401, EPI_ISL_2216406, EPI_ISL_2216411, EPI_ISL_2216416, EPI_ISL_2216420, EPI_ISL_2216425, EPI_ISL_2216430, EPI_ISL_2216437, EPI_ISL_2216442, EPI_ISL_2216446, EPI_ISL_2216450, EPI_ISL_2216455, EPI_ISL_2216460, EPI_ISL_2216466, EPI_ISL_2216472, EPI_ISL_2216476, EPI_ISL_2216482, EPI_ISL_2216487, EPI_ISL_2216492, EPI_ISL_2216497, EPI_ISL_2216502, EPI_ISL_2216507, EPI_ISL_2216512, EPI_ISL_2216517, EPI_ISL_2216522, EPI_ISL_2216526, EPI_ISL_2216530, EPI_ISL_2216535, EPI_ISL_2216544, EPI_ISL_2216549, EPI_ISL_2216554, EPI_ISL_2216558, EPI_ISL_2216564, EPI_ISL_2216570, EPI_ISL_2216575, EPI_ISL_2216580, EPI_ISL_2216584, EPI_ISL_2216588, EPI_ISL_2216591, EPI_ISL_2216595, EPI_ISL_2216599, EPI_ISL_2216602, EPI_ISL_2216606, EPI_ISL_2216611, EPI_ISL_2216615, EPI_ISL_2216621, EPI_ISL_2216626, EPI_ISL_2216631, EPI_ISL_2216637, EPI_ISL_2216640, EPI_ISL_2216641, EPI_ISL_2216645, EPI_ISL_2216648, EPI_ISL_2216652, EPI_ISL_2216657, EPI_ISL_2216662, EPI_ISL_2216667, EPI_ISL_2216672, EPI_ISL_2216678, EPI_ISL_2216683, EPI_ISL_2216689, EPI_ISL_2216693, EPI_ISL_2216697, EPI_ISL_2216702, EPI_ISL_2216707, EPI_ISL_2216712, EPI_ISL_2216717, EPI_ISL_2216722, EPI_ISL_2216727, EPI_ISL_2216732, EPI_ISL_2216738, EPI_ISL_2216742, EPI_ISL_2216747, EPI_ISL_2216753, EPI_ISL_2216758, EPI_ISL_2216763, EPI_ISL_2216769, EPI_ISL_2216773, EPI_ISL_2216777, EPI_ISL_2216782, EPI_ISL_2216789, EPI_ISL_2216794, EPI_ISL_2216799, EPI_ISL_2216802, EPI_ISL_2216806, EPI_ISL_2216809, EPI_ISL_2216813, EPI_ISL_2216818, EPI_ISL_2216822, EPI_ISL_2216825, EPI_ISL_2216829, EPI_ISL_2216833, EPI_ISL_2216838, EPI_ISL_2216840, EPI_ISL_2216844, EPI_ISL_2216848, EPI_ISL_2216852, EPI_ISL_2216855, EPI_ISL_2216859, EPI_ISL_2216863, EPI_ISL_2216867, EPI_ISL_2216873, EPI_ISL_2216878, EPI_ISL_2216882, EPI_ISL_2216887, EPI_ISL_2216891, EPI_ISL_2216896, EPI_ISL_2216899, EPI_ISL_2216902, EPI_ISL_2216906, EPI_ISL_2216909, EPI_ISL_2216913, EPI_ISL_2216917, EPI_ISL_2216920, EPI_ISL_2216926, EPI_ISL_2216929, EPI_ISL_2216933, EPI_ISL_2216937, EPI_ISL_2216941, EPI_ISL_2216945, EPI_ISL_2216950, EPI_ISL_2216955, EPI_ISL_2216958, EPI_ISL_2216962, EPI_ISL_2216966, EPI_ISL_2216974, EPI_ISL_2216976, EPI_ISL_2216980, EPI_ISL_2216983, EPI_ISL_2216988, EPI_ISL_2216992, EPI_ISL_2216995, EPI_ISL_2216998, EPI_ISL_2217001, EPI_ISL_2217005, EPI_ISL_2217009, EPI_ISL_2217012, EPI_ISL_2217015, EPI_ISL_2217020, EPI_ISL_2217025, EPI_ISL_2217030, EPI_ISL_2217035, EPI_ISL_2217040, EPI_ISL_2217045, EPI_ISL_2217049, EPI_ISL_2217052, EPI_ISL_2217056, EPI_ISL_2217060, EPI_ISL_2217064, EPI_ISL_2217067, EPI_ISL_2217071, EPI_ISL_2217073, EPI_ISL_2217077, EPI_ISL_2217082, EPI_ISL_2217085, EPI_ISL_2217092, EPI_ISL_2217096, EPI_ISL_2217100, EPI_ISL_2217103, EPI_ISL_2217106, EPI_ISL_2217110, EPI_ISL_2217113, EPI_ISL_2217118, EPI_ISL_2217122, EPI_ISL_2217125, EPI_ISL_2217131, EPI_ISL_2217137, EPI_ISL_2217144, EPI_ISL_2217147, EPI_ISL_2217150, EPI_ISL_2217154, EPI_ISL_2217158, EPI_ISL_2217159, EPI_ISL_2217163, EPI_ISL_2217166, EPI_ISL_2217170, EPI_ISL_2217173, EPI_ISL_2217176, EPI_ISL_2217180, EPI_ISL_2217184, EPI_ISL_2217187, EPI_ISL_2217191, EPI_ISL_2217195, EPI_ISL_2217200, EPI_ISL_2217205, EPI_ISL_2217210, EPI_ISL_2217213, EPI_ISL_2217217, EPI_ISL_2217221, EPI_ISL_2217223, EPI_ISL_2217228, EPI_ISL_2217232, EPI_ISL_2217236, EPI_ISL_2217241, EPI_ISL_2217246, EPI_ISL_2217250, EPI_ISL_2217254, EPI_ISL_2217257, EPI_ISL_2217261, EPI_ISL_2217265, EPI_ISL_2217269, EPI_ISL_2217272, EPI_ISL_2217276, EPI_ISL_2217280, EPI_ISL_2217284, EPI_ISL_2217288, EPI_ISL_2217292, EPI_ISL_2217297, EPI_ISL_2217302, EPI_ISL_2217307, EPI_ISL_2217311, EPI_ISL_2217316, EPI_ISL_2217321, EPI_ISL_2217326, EPI_ISL_2217331, EPI_ISL_2217337, EPI_ISL_2217341, EPI_ISL_2217344, EPI_ISL_2217348, EPI_ISL_2217353, EPI_ISL_2217356, EPI_ISL_2217359, EPI_ISL_2217364, EPI_ISL_2217370, EPI_ISL_2217375, EPI_ISL_2217379, EPI_ISL_2217384, EPI_ISL_2217390, EPI_ISL_2217394, EPI_ISL_2217396, EPI_ISL_2217399, EPI_ISL_2217402, EPI_ISL_2217406, EPI_ISL_2217410, EPI_ISL_2217413, EPI_ISL_2217417, EPI_ISL_2217420, EPI_ISL_2217424, EPI_ISL_2217428, EPI_ISL_2217433, EPI_ISL_2217437, EPI_ISL_2217441, EPI_ISL_2217444, EPI_ISL_2217448, EPI_ISL_2217451, EPI_ISL_2217454, EPI_ISL_2217459, EPI_ISL_2217470, EPI_ISL_2217476, EPI_ISL_2217481, EPI_ISL_2217486, EPI_ISL_2217490, EPI_ISL_2217493, EPI_ISL_2217497, EPI_ISL_2217501, EPI_ISL_2217505, EPI_ISL_2217511, EPI_ISL_2217518, EPI_ISL_2217527, EPI_ISL_2217532, EPI_ISL_2217536, EPI_ISL_2217539, EPI_ISL_2217546, EPI_ISL_2217549, EPI_ISL_2217553, EPI_ISL_2217557, EPI_ISL_2217562, EPI_ISL_2217569, EPI_ISL_2217573, EPI_ISL_2217579, EPI_ISL_2217583, EPI_ISL_2217585, EPI_ISL_2217589, EPI_ISL_2217594, EPI_ISL_2217598, EPI_ISL_2217601, EPI_ISL_2217605, EPI_ISL_2217609, EPI_ISL_2217614, EPI_ISL_2217617, EPI_ISL_2217620, EPI_ISL_2217623, EPI_ISL_2217627, EPI_ISL_2217631, EPI_ISL_2217635, EPI_ISL_2217639, EPI_ISL_2217642, EPI_ISL_2217646, EPI_ISL_2217649, EPI_ISL_2217656, EPI_ISL_2217661, EPI_ISL_2217664, EPI_ISL_2217670, EPI_ISL_2217675, EPI_ISL_2217678, EPI_ISL_2217681, EPI_ISL_2217686, EPI_ISL_2217690, EPI_ISL_2217706, EPI_ISL_2217712, EPI_ISL_2217716, EPI_ISL_2217724, EPI_ISL_2217728, EPI_ISL_2217733, EPI_ISL_2217739, EPI_ISL_2217743, EPI_ISL_2217747, EPI_ISL_2217750, EPI_ISL_2217755, EPI_ISL_2217759, EPI_ISL_2217762, EPI_ISL_2217766, EPI_ISL_2217769, EPI_ISL_2217773, EPI_ISL_2217775, EPI_ISL_2217778, EPI_ISL_2217781, EPI_ISL_2217786, EPI_ISL_2217790, EPI_ISL_2217792, EPI_ISL_2217794, EPI_ISL_2217799, EPI_ISL_2217802, EPI_ISL_2217806, EPI_ISL_2217810, EPI_ISL_2217815, EPI_ISL_2217819, EPI_ISL_2217822, EPI_ISL_2217826, EPI_ISL_2217829, EPI_ISL_2217833, EPI_ISL_2217837, EPI_ISL_2217839, EPI_ISL_2217844, EPI_ISL_2217847, EPI_ISL_2217851, EPI_ISL_2217854, EPI_ISL_2217858, EPI_ISL_2217863, EPI_ISL_2217868, EPI_ISL_2217873, EPI_ISL_2217878, EPI_ISL_2217884, EPI_ISL_2217888, EPI_ISL_2217892, EPI_ISL_2217899, EPI_ISL_2217903, EPI_ISL_2217909, EPI_ISL_2217913, EPI_ISL_2217915, EPI_ISL_2217919, EPI_ISL_2217921, EPI_ISL_2217924, EPI_ISL_2217927, EPI_ISL_2217931, EPI_ISL_2217934, EPI_ISL_2217937, EPI_ISL_2217941, EPI_ISL_2217944, EPI_ISL_2217948, EPI_ISL_2217949, EPI_ISL_2217953, EPI_ISL_2217959, EPI_ISL_2217963, EPI_ISL_2217966, EPI_ISL_2217970, EPI_ISL_2217972, EPI_ISL_2217976, EPI_ISL_2217979, EPI_ISL_2217986, EPI_ISL_2217989, EPI_ISL_2217994, EPI_ISL_2217998, EPI_ISL_2218001, EPI_ISL_2218006, EPI_ISL_2218009, EPI_ISL_2218013, EPI_ISL_2218016, EPI_ISL_2218020, EPI_ISL_2218024, EPI_ISL_2218027, EPI_ISL_2218032, EPI_ISL_2218035, EPI_ISL_2218040, EPI_ISL_2218045, EPI_ISL_2218050, EPI_ISL_2218055, EPI_ISL_2218063, EPI_ISL_2218069, EPI_ISL_2218074, EPI_ISL_2218079, EPI_ISL_2218084, EPI_ISL_2218088, EPI_ISL_2218093, EPI_ISL_2218099, EPI_ISL_2218105, EPI_ISL_2218110, EPI_ISL_2218114, EPI_ISL_2218119, EPI_ISL_2218128, EPI_ISL_2218135, EPI_ISL_2218140, EPI_ISL_2218143, EPI_ISL_2218147, EPI_ISL_2218152, EPI_ISL_2218157, EPI_ISL_2218162, EPI_ISL_2218167, EPI_ISL_2218173, EPI_ISL_2218178, EPI_ISL_2218184, EPI_ISL_2218189, EPI_ISL_2218194, EPI_ISL_2218199, EPI_ISL_2218204, EPI_ISL_2218209, EPI_ISL_2218215, EPI_ISL_2218220, EPI_ISL_2218225, EPI_ISL_2218230, EPI_ISL_2218234, EPI_ISL_2218240, EPI_ISL_2218244, EPI_ISL_2218250, EPI_ISL_2218255, EPI_ISL_2218260, EPI_ISL_2218265, EPI_ISL_2218270, EPI_ISL_2218275, EPI_ISL_2218279, EPI_ISL_2218286, EPI_ISL_2218291, EPI_ISL_2218298, EPI_ISL_2218303, EPI_ISL_2218310, EPI_ISL_2218315, EPI_ISL_2218322, EPI_ISL_2218327, EPI_ISL_2218334, EPI_ISL_2218341, EPI_ISL_2218346, EPI_ISL_2218353, EPI_ISL_2218358, EPI_ISL_2218369, EPI_ISL_2218373, EPI_ISL_2218378, EPI_ISL_2218383, EPI_ISL_2218392, EPI_ISL_2218396, EPI_ISL_2218403, EPI_ISL_2218408, EPI_ISL_2218412, EPI_ISL_2218420, EPI_ISL_2218425, EPI_ISL_2218430, EPI_ISL_2218434, EPI_ISL_2218439, EPI_ISL_2218443, EPI_ISL_2218447, EPI_ISL_2218452, EPI_ISL_2218459, EPI_ISL_2218465, EPI_ISL_2218471, EPI_ISL_2218474, EPI_ISL_2218480, EPI_ISL_2218488, EPI_ISL_2218494, EPI_ISL_2218499, EPI_ISL_2218504, EPI_ISL_2218509, EPI_ISL_2218514, EPI_ISL_2218523, EPI_ISL_2218527, EPI_ISL_2218538, EPI_ISL_2218543, EPI_ISL_2218548, EPI_ISL_2218553, EPI_ISL_2218557, EPI_ISL_2218560, EPI_ISL_2218566, EPI_ISL_2218572, EPI_ISL_2218577, EPI_ISL_2218582, EPI_ISL_2218585, EPI_ISL_2218590, EPI_ISL_2218593, EPI_ISL_2218598, EPI_ISL_2218603, EPI_ISL_2218609, EPI_ISL_2218615, EPI_ISL_2218619, EPI_ISL_2218624, EPI_ISL_2218629, EPI_ISL_2218634, EPI_ISL_2218638, EPI_ISL_2218645, EPI_ISL_2218653, EPI_ISL_2218658, EPI_ISL_2218663, EPI_ISL_2218668, EPI_ISL_2218673, EPI_ISL_2218678, EPI_ISL_2218683, EPI_ISL_2218689, EPI_ISL_2218694, EPI_ISL_2218698, EPI_ISL_2218702, EPI_ISL_2218708, EPI_ISL_2218712, EPI_ISL_2218718, EPI_ISL_2218722, EPI_ISL_2218727, EPI_ISL_2218732, EPI_ISL_2218751, EPI_ISL_2218755, EPI_ISL_2218760, EPI_ISL_2218775, EPI_ISL_2218778, EPI_ISL_2218788, EPI_ISL_2218793, EPI_ISL_2218798, EPI_ISL_2218803, EPI_ISL_2218808, EPI_ISL_2218812, EPI_ISL_2218814, EPI_ISL_2218817, EPI_ISL_2218820, EPI_ISL_2218823, EPI_ISL_2218827, EPI_ISL_2218830, EPI_ISL_2218834, EPI_ISL_2218837, EPI_ISL_2218840, EPI_ISL_2218841, EPI_ISL_2218844, EPI_ISL_2218847, EPI_ISL_2218851, EPI_ISL_2218854, EPI_ISL_2218857, EPI_ISL_2218862, EPI_ISL_2218866, EPI_ISL_2218869, EPI_ISL_2218873, EPI_ISL_2218879, EPI_ISL_2218882, EPI_ISL_2218885, EPI_ISL_2218891, EPI_ISL_2218896, EPI_ISL_2218901, EPI_ISL_2218904, EPI_ISL_2218909, EPI_ISL_2218915, EPI_ISL_2218920, EPI_ISL_2218925, EPI_ISL_2218930, EPI_ISL_2218935, EPI_ISL_2218940, EPI_ISL_2218945, EPI_ISL_2218949, EPI_ISL_2218954, EPI_ISL_2218959, EPI_ISL_2218964, EPI_ISL_2218972, EPI_ISL_2218977, EPI_ISL_2218982, EPI_ISL_2218987, EPI_ISL_2218998, EPI_ISL_2219005, EPI_ISL_2219008, EPI_ISL_2219011, EPI_ISL_2219016, EPI_ISL_2219021, EPI_ISL_2219026, EPI_ISL_2219031, EPI_ISL_2219036, EPI_ISL_2219040, EPI_ISL_2219044, EPI_ISL_2219056, EPI_ISL_2219061, EPI_ISL_2219066, EPI_ISL_2219071, EPI_ISL_2219074, EPI_ISL_2219079, EPI_ISL_2219082, EPI_ISL_2219089, EPI_ISL_2219093, EPI_ISL_2219097, EPI_ISL_2219103, EPI_ISL_2219108, EPI_ISL_2219113, EPI_ISL_2219117, EPI_ISL_2219120, EPI_ISL_2219125, EPI_ISL_2219129, EPI_ISL_2219134, EPI_ISL_2219139, EPI_ISL_2219144, EPI_ISL_2219149, EPI_ISL_2219154, EPI_ISL_2219159, EPI_ISL_2219162, EPI_ISL_2219167, EPI_ISL_2219177, EPI_ISL_2219181, EPI_ISL_2219193, EPI_ISL_2219219, EPI_ISL_2219231, EPI_ISL_2219243, EPI_ISL_2219253, EPI_ISL_2219265, EPI_ISL_2219273, EPI_ISL_2219291, EPI_ISL_2219300, EPI_ISL_2219311, EPI_ISL_2219324, EPI_ISL_2219332, EPI_ISL_2219339, EPI_ISL_2219344, EPI_ISL_2219348, EPI_ISL_2219353, EPI_ISL_2219358, EPI_ISL_2219363, EPI_ISL_2219368, EPI_ISL_2219374, EPI_ISL_2219380, EPI_ISL_2219384, EPI_ISL_2219388, EPI_ISL_2219393, EPI_ISL_2219398, EPI_ISL_2219403, EPI_ISL_2219409, EPI_ISL_2219415, EPI_ISL_2219420, EPI_ISL_2219423, EPI_ISL_2219426, EPI_ISL_2219430, EPI_ISL_2219433, EPI_ISL_2219437, EPI_ISL_2219440, EPI_ISL_2219444, EPI_ISL_2219446, EPI_ISL_2219449, EPI_ISL_2219453, EPI_ISL_2219457, EPI_ISL_2219460, EPI_ISL_2219464, EPI_ISL_2219467, EPI_ISL_2219471, EPI_ISL_2219477, EPI_ISL_2219480, EPI_ISL_2219484, EPI_ISL_2219491, EPI_ISL_2219497, EPI_ISL_2219501, EPI_ISL_2219521, EPI_ISL_2219527, EPI_ISL_2219535, EPI_ISL_2219540, EPI_ISL_2219550, EPI_ISL_2219559, EPI_ISL_2219563, EPI_ISL_2219569, EPI_ISL_2219572, EPI_ISL_2219577, EPI_ISL_2219582, EPI_ISL_2219587, EPI_ISL_2219593, EPI_ISL_2219597, EPI_ISL_2219609, EPI_ISL_2219612, EPI_ISL_2219616, EPI_ISL_2219621, EPI_ISL_2219625, EPI_ISL_2219630, EPI_ISL_2219635, EPI_ISL_2219640, EPI_ISL_2219644, EPI_ISL_2219649, EPI_ISL_2219654, EPI_ISL_2219659, EPI_ISL_2219664, EPI_ISL_2219669, EPI_ISL_2219674, EPI_ISL_2219683, EPI_ISL_2219687, EPI_ISL_2219692, EPI_ISL_2219697, EPI_ISL_2219702, EPI_ISL_2219712, EPI_ISL_2219715, EPI_ISL_2219720, EPI_ISL_2219725, EPI_ISL_2219730, EPI_ISL_2219735, EPI_ISL_2219739, EPI_ISL_2219745, EPI_ISL_2219751, EPI_ISL_2219756, EPI_ISL_2219761, EPI_ISL_2219765, EPI_ISL_2219770, EPI_ISL_2219773, EPI_ISL_2219778, EPI_ISL_2219783, EPI_ISL_2219795, EPI_ISL_2219798, EPI_ISL_2219803, EPI_ISL_2219808, EPI_ISL_2219813, EPI_ISL_2219818, EPI_ISL_2219821, EPI_ISL_2219825, EPI_ISL_2219829, EPI_ISL_2219835, EPI_ISL_2219839, EPI_ISL_2219848, EPI_ISL_2219854, EPI_ISL_2219857, EPI_ISL_2219864, EPI_ISL_2219869, EPI_ISL_2219872, EPI_ISL_2219887, EPI_ISL_2219892, EPI_ISL_2219901, EPI_ISL_2219906, EPI_ISL_2219911, EPI_ISL_2219916, EPI_ISL_2219919, EPI_ISL_2219930, EPI_ISL_2219938, EPI_ISL_2219953, EPI_ISL_2219970, EPI_ISL_2219983, EPI_ISL_2220002, EPI_ISL_2220006, EPI_ISL_2220009, EPI_ISL_2220013, EPI_ISL_2220016, EPI_ISL_2220021, EPI_ISL_2220032, EPI_ISL_2220037, EPI_ISL_2220045, EPI_ISL_2220064, EPI_ISL_2220075, EPI_ISL_2220080, EPI_ISL_2220094, EPI_ISL_2220099, EPI_ISL_2220104, EPI_ISL_2220112, EPI_ISL_2220127, EPI_ISL_2220133, EPI_ISL_2220137, EPI_ISL_2220142, EPI_ISL_2220146, EPI_ISL_2220151, EPI_ISL_2220155, EPI_ISL_2220170, EPI_ISL_2220174, EPI_ISL_2220179, EPI_ISL_2220184, EPI_ISL_2220188, EPI_ISL_2220194, EPI_ISL_2220199, EPI_ISL_2220204, EPI_ISL_2220207, EPI_ISL_2220212, EPI_ISL_2220217, EPI_ISL_2220221, EPI_ISL_2220223, EPI_ISL_2220232, EPI_ISL_2220237, EPI_ISL_2220247, EPI_ISL_2220251, EPI_ISL_2220261, EPI_ISL_2220265, EPI_ISL_2220272, EPI_ISL_2220277, EPI_ISL_2220283, EPI_ISL_2220292, EPI_ISL_2220302, EPI_ISL_2220306, EPI_ISL_2220312, EPI_ISL_2220316, EPI_ISL_2220321, EPI_ISL_2220326, EPI_ISL_2220329, EPI_ISL_2220334, EPI_ISL_2220339, EPI_ISL_2220343, EPI_ISL_2220348, EPI_ISL_2220352, EPI_ISL_2220360, EPI_ISL_2220365, EPI_ISL_2220370, EPI_ISL_2220374, EPI_ISL_2220381, EPI_ISL_2220386, EPI_ISL_2220391, EPI_ISL_2220396, EPI_ISL_2220400, EPI_ISL_2220406, EPI_ISL_2220410, EPI_ISL_2220414, EPI_ISL_2220417, EPI_ISL_2220422, EPI_ISL_2220427, EPI_ISL_2220434, EPI_ISL_2220436, EPI_ISL_2220441, EPI_ISL_2220446, EPI_ISL_2220451, EPI_ISL_2220455, EPI_ISL_2220463, EPI_ISL_2220468, EPI_ISL_2220473, EPI_ISL_2220477, EPI_ISL_2220482, EPI_ISL_2220487, EPI_ISL_2220492, EPI_ISL_2220497, EPI_ISL_2220500, EPI_ISL_2220504, EPI_ISL_2220508, EPI_ISL_2220512, EPI_ISL_2220516, EPI_ISL_2220521, EPI_ISL_2220527, EPI_ISL_2220531, EPI_ISL_2220535, EPI_ISL_2220541, EPI_ISL_2220546, EPI_ISL_2220550, EPI_ISL_2220555, EPI_ISL_2220560, EPI_ISL_2220566, EPI_ISL_2220571, EPI_ISL_2220575, EPI_ISL_2220578, EPI_ISL_2220581, EPI_ISL_2220585, EPI_ISL_2220588, EPI_ISL_2220590, EPI_ISL_2220593, EPI_ISL_2220596, EPI_ISL_2220599, EPI_ISL_2220602, EPI_ISL_2220604, EPI_ISL_2220607, EPI_ISL_2220610, EPI_ISL_2220613, EPI_ISL_2220616, EPI_ISL_2220619, EPI_ISL_2220622, EPI_ISL_2220625, EPI_ISL_2220628, EPI_ISL_2220631, EPI_ISL_2220634, EPI_ISL_2220637, EPI_ISL_2220640, EPI_ISL_2220643, EPI_ISL_2220646, EPI_ISL_2220649, EPI_ISL_2220652, EPI_ISL_2220656, EPI_ISL_2220659, EPI_ISL_2220662, EPI_ISL_2220665, EPI_ISL_2220668, EPI_ISL_2220672, EPI_ISL_2220674, EPI_ISL_2220677, EPI_ISL_2220680, EPI_ISL_2220684, EPI_ISL_2220686, EPI_ISL_2220689, EPI_ISL_2220692, EPI_ISL_2220695, EPI_ISL_2220700, EPI_ISL_2220706, EPI_ISL_2220708, EPI_ISL_2220712, EPI_ISL_2220715, EPI_ISL_2220717, EPI_ISL_2220721, EPI_ISL_2220724, EPI_ISL_2220727, EPI_ISL_2220730, EPI_ISL_2220734, EPI_ISL_2220737, EPI_ISL_2220740, EPI_ISL_2220741, EPI_ISL_2220744, EPI_ISL_2220747, EPI_ISL_2220750, EPI_ISL_2220753, EPI_ISL_2220756, EPI_ISL_2220759, EPI_ISL_2220762, EPI_ISL_2220764, EPI_ISL_2220766, EPI_ISL_2220770, EPI_ISL_2220774, EPI_ISL_2220776, EPI_ISL_2220778, EPI_ISL_2220782, EPI_ISL_2220785, EPI_ISL_2220788, EPI_ISL_2220791, EPI_ISL_2220794, EPI_ISL_2220796, EPI_ISL_2220800, EPI_ISL_2220803, EPI_ISL_2220805, EPI_ISL_2220808, EPI_ISL_2220811, EPI_ISL_2220814, EPI_ISL_2220818, EPI_ISL_2220820, EPI_ISL_2220823, EPI_ISL_2220825, EPI_ISL_2220828, EPI_ISL_2220831, EPI_ISL_2220834, EPI_ISL_2220837, EPI_ISL_2220840, EPI_ISL_2220843, EPI_ISL_2220845, EPI_ISL_2220847, EPI_ISL_2220850, EPI_ISL_2220853, EPI_ISL_2220855, EPI_ISL_2220858, EPI_ISL_2220861, EPI_ISL_2220866, EPI_ISL_2220869, EPI_ISL_2220872, EPI_ISL_2220876, EPI_ISL_2220879, EPI_ISL_2220881, EPI_ISL_2220884, EPI_ISL_2220886, EPI_ISL_2220889, EPI_ISL_2220892, EPI_ISL_2220895, EPI_ISL_2220898, EPI_ISL_2220901, EPI_ISL_2220904, EPI_ISL_2220907, EPI_ISL_2220910, EPI_ISL_2220913, EPI_ISL_2220915, EPI_ISL_2220918, EPI_ISL_2220919, EPI_ISL_2220922, EPI_ISL_2220926, EPI_ISL_2220928, EPI_ISL_2220930, EPI_ISL_2220932, EPI_ISL_2220935, EPI_ISL_2220938, EPI_ISL_2220942, EPI_ISL_2220944, EPI_ISL_2220947, EPI_ISL_2220949, EPI_ISL_2220952, EPI_ISL_2220955, EPI_ISL_2220958, EPI_ISL_2220960, EPI_ISL_2220963, EPI_ISL_2220965, EPI_ISL_2220968, EPI_ISL_2220970, EPI_ISL_2220973, EPI_ISL_2220975, EPI_ISL_2220978, EPI_ISL_2220981, EPI_ISL_2220985, EPI_ISL_2220988, EPI_ISL_2220990, EPI_ISL_2220993, EPI_ISL_2220997, EPI_ISL_2221000, EPI_ISL_2221003, EPI_ISL_2221005, EPI_ISL_2221009, EPI_ISL_2221012, EPI_ISL_2221013, EPI_ISL_2221015, EPI_ISL_2221017, EPI_ISL_2221020, EPI_ISL_2221023, EPI_ISL_2221026, EPI_ISL_2221030, EPI_ISL_2221034, EPI_ISL_2221037, EPI_ISL_2221040, EPI_ISL_2221043, EPI_ISL_2221044, EPI_ISL_2221046, EPI_ISL_2221050, EPI_ISL_2221054, EPI_ISL_2221056, EPI_ISL_2221059, EPI_ISL_2221063, EPI_ISL_2221065, EPI_ISL_2221068, EPI_ISL_2221072, EPI_ISL_2221075, EPI_ISL_2221078, EPI_ISL_2221081, EPI_ISL_2221084, EPI_ISL_2221086, EPI_ISL_2221089, EPI_ISL_2221092, EPI_ISL_2221095, EPI_ISL_2221098, EPI_ISL_2221100, EPI_ISL_2221106, EPI_ISL_2221109, EPI_ISL_2221113, EPI_ISL_2221115, EPI_ISL_2221118, EPI_ISL_2221121, EPI_ISL_2221124, EPI_ISL_2221127, EPI_ISL_2221129, EPI_ISL_2221132, EPI_ISL_2221135, EPI_ISL_2221139, EPI_ISL_2221142, EPI_ISL_2221145, EPI_ISL_2221148, EPI_ISL_2221151, EPI_ISL_2221156, EPI_ISL_2221159, EPI_ISL_2221162, EPI_ISL_2221164, EPI_ISL_2221167, EPI_ISL_2221170, EPI_ISL_2221175, EPI_ISL_2221178, EPI_ISL_2221182, EPI_ISL_2221203, EPI_ISL_2221206, EPI_ISL_2221209, EPI_ISL_2221212, EPI_ISL_2221215, EPI_ISL_2221217, EPI_ISL_2221219, EPI_ISL_2221222, EPI_ISL_2221224, EPI_ISL_2221227, EPI_ISL_2221230, EPI_ISL_2221233, EPI_ISL_2221236, EPI_ISL_2221239, EPI_ISL_2221242, EPI_ISL_2221245, EPI_ISL_2221247, EPI_ISL_2221250, EPI_ISL_2221252, EPI_ISL_2221255, EPI_ISL_2221260, EPI_ISL_2221262, EPI_ISL_2221265, EPI_ISL_2221268, EPI_ISL_2221271, EPI_ISL_2221273, EPI_ISL_2221276, EPI_ISL_2221279, EPI_ISL_2221282, EPI_ISL_2221285, EPI_ISL_2221288, EPI_ISL_2221289, EPI_ISL_2221297, EPI_ISL_2221302, EPI_ISL_2221304, EPI_ISL_2221306, EPI_ISL_2221309, EPI_ISL_2221312, EPI_ISL_2221315, EPI_ISL_2221318, EPI_ISL_2221321, EPI_ISL_2221324, EPI_ISL_2221327, EPI_ISL_2221330, EPI_ISL_2221333, EPI_ISL_2221336, EPI_ISL_2221339, EPI_ISL_2221342, EPI_ISL_2221345, EPI_ISL_2221348, EPI_ISL_2221351, EPI_ISL_2221354, EPI_ISL_2221356, EPI_ISL_2221359, EPI_ISL_2221362, EPI_ISL_2221365, EPI_ISL_2221367, EPI_ISL_2221370, EPI_ISL_2221373, EPI_ISL_2221375, EPI_ISL_2221378, EPI_ISL_2221382, EPI_ISL_2221385, EPI_ISL_2221387, EPI_ISL_2221391, EPI_ISL_2221394, EPI_ISL_2221397, EPI_ISL_2221399, EPI_ISL_2221403, EPI_ISL_2221406, EPI_ISL_2221409, EPI_ISL_2221412, EPI_ISL_2221416, EPI_ISL_2221419, EPI_ISL_2221422, EPI_ISL_2221423, EPI_ISL_2221425, EPI_ISL_2221428, EPI_ISL_2221431, EPI_ISL_2221435, EPI_ISL_2221439, EPI_ISL_2221441, EPI_ISL_2221445, EPI_ISL_2221447, EPI_ISL_2221451, EPI_ISL_2221454, EPI_ISL_2221457, EPI_ISL_2221460, EPI_ISL_2221462, EPI_ISL_2221465, EPI_ISL_2221466, EPI_ISL_2221469, EPI_ISL_2221472, EPI_ISL_2221474, EPI_ISL_2221477, EPI_ISL_2221481, EPI_ISL_2221486, EPI_ISL_2221489, EPI_ISL_2221492, EPI_ISL_2221495, EPI_ISL_2221497, EPI_ISL_2221499, EPI_ISL_2221502, EPI_ISL_2221504, EPI_ISL_2221507, EPI_ISL_2221509, EPI_ISL_2221511, EPI_ISL_2221514, EPI_ISL_2221517, EPI_ISL_2221519, EPI_ISL_2221522, EPI_ISL_2221525, EPI_ISL_2221528, EPI_ISL_2221531, EPI_ISL_2221534, EPI_ISL_2221536, EPI_ISL_2221538, EPI_ISL_2221542, EPI_ISL_2221547, EPI_ISL_2221553, EPI_ISL_2221556, EPI_ISL_2221560, EPI_ISL_2221563, EPI_ISL_2221568, EPI_ISL_2221571, EPI_ISL_2221574, EPI_ISL_2221577, EPI_ISL_2221580, EPI_ISL_2221583, EPI_ISL_2221586, EPI_ISL_2221589, EPI_ISL_2221592, EPI_ISL_2221594, EPI_ISL_2221599, EPI_ISL_2221602, EPI_ISL_2221604, EPI_ISL_2221607, EPI_ISL_2221610, EPI_ISL_2221613, EPI_ISL_2221617, EPI_ISL_2221620, EPI_ISL_2221624, EPI_ISL_2221626, EPI_ISL_2221629, EPI_ISL_2221632, EPI_ISL_2221635, EPI_ISL_2221642, EPI_ISL_2221644, EPI_ISL_2221647, EPI_ISL_2221651, EPI_ISL_2221654, EPI_ISL_2221656, EPI_ISL_2221659, EPI_ISL_2221662, EPI_ISL_2221665, EPI_ISL_2221668, EPI_ISL_2221671, EPI_ISL_2221674, EPI_ISL_2221677, EPI_ISL_2221680, EPI_ISL_2221682, EPI_ISL_2221684, EPI_ISL_2221687, EPI_ISL_2221690, EPI_ISL_2221693, EPI_ISL_2221695, EPI_ISL_2221696, EPI_ISL_2221699, EPI_ISL_2221700, EPI_ISL_2221703, EPI_ISL_2221707, EPI_ISL_2221709, EPI_ISL_2221712, EPI_ISL_2221715, EPI_ISL_2221717, EPI_ISL_2221720, EPI_ISL_2221725, EPI_ISL_2221730, EPI_ISL_2221733, EPI_ISL_2221736, EPI_ISL_2221740, EPI_ISL_2221743, EPI_ISL_2221746, EPI_ISL_2221749, EPI_ISL_2221752, EPI_ISL_2221755, EPI_ISL_2221757, EPI_ISL_2221762, EPI_ISL_2221768, EPI_ISL_2221770, EPI_ISL_2221772, EPI_ISL_2221775, EPI_ISL_2221778, EPI_ISL_2221781, EPI_ISL_2221784, EPI_ISL_2221787, EPI_ISL_2221790, EPI_ISL_2221793, EPI_ISL_2221796, EPI_ISL_2221799, EPI_ISL_2221802, EPI_ISL_2221805, EPI_ISL_2221808, EPI_ISL_2221811, EPI_ISL_2221814, EPI_ISL_2221817, EPI_ISL_2221820, EPI_ISL_2221823, EPI_ISL_2221826, EPI_ISL_2221829, EPI_ISL_2221833, EPI_ISL_2221836, EPI_ISL_2221839, EPI_ISL_2221842, EPI_ISL_2221846, EPI_ISL_2221849, EPI_ISL_2221852, EPI_ISL_2221854, EPI_ISL_2221857, EPI_ISL_2221862, EPI_ISL_2221867, EPI_ISL_2221871, EPI_ISL_2221875, EPI_ISL_2221878, EPI_ISL_2221881, EPI_ISL_2221884, EPI_ISL_2221887, EPI_ISL_2221891, EPI_ISL_2221894, EPI_ISL_2221897, EPI_ISL_2221900, EPI_ISL_2221904, EPI_ISL_2221907, EPI_ISL_2221910, EPI_ISL_2221913, EPI_ISL_2221916, EPI_ISL_2221919, EPI_ISL_2221922, EPI_ISL_2221925, EPI_ISL_2221928, EPI_ISL_2221931, EPI_ISL_2221934, EPI_ISL_2221936, EPI_ISL_2221939, EPI_ISL_2221942, EPI_ISL_2221945, EPI_ISL_2221948, EPI_ISL_2221951, EPI_ISL_2221954, EPI_ISL_2221957, EPI_ISL_2221960, EPI_ISL_2221963, EPI_ISL_2221966, EPI_ISL_2221969, EPI_ISL_2221971, EPI_ISL_2221974, EPI_ISL_2221977, EPI_ISL_2221979, EPI_ISL_2221984, EPI_ISL_2221988, EPI_ISL_2221991, EPI_ISL_2221994, EPI_ISL_2221997, EPI_ISL_2222000, EPI_ISL_2222003, EPI_ISL_2222006, EPI_ISL_2222008, EPI_ISL_2222011, EPI_ISL_2222015, EPI_ISL_2222018, EPI_ISL_2222021, EPI_ISL_2222023, EPI_ISL_2222028, EPI_ISL_2222032, EPI_ISL_2222034, EPI_ISL_2222036, EPI_ISL_2222040, EPI_ISL_2222043, EPI_ISL_2222046, EPI_ISL_2222048, EPI_ISL_2222051, EPI_ISL_2222056, EPI_ISL_2222059, EPI_ISL_2222062, EPI_ISL_2222065, EPI_ISL_2222067, EPI_ISL_2222070, EPI_ISL_2222074, EPI_ISL_2222077, EPI_ISL_2222079, EPI_ISL_2222085, EPI_ISL_2222090, EPI_ISL_2222093, EPI_ISL_2222096, EPI_ISL_2222099, EPI_ISL_2222103, EPI_ISL_2222106, EPI_ISL_2222108, EPI_ISL_2222112, EPI_ISL_2222115, EPI_ISL_2222118, EPI_ISL_2222121, EPI_ISL_2222124, EPI_ISL_2222128, EPI_ISL_2222131, EPI_ISL_2222134, EPI_ISL_2222137, EPI_ISL_2222141, EPI_ISL_2222143, EPI_ISL_2222146, EPI_ISL_2222149, EPI_ISL_2222153, EPI_ISL_2222154, EPI_ISL_2222157, EPI_ISL_2222160, EPI_ISL_2222162, EPI_ISL_2222165, EPI_ISL_2222168, EPI_ISL_2222171, EPI_ISL_2222174, EPI_ISL_2222177, EPI_ISL_2222180, EPI_ISL_2222184, EPI_ISL_2222187, EPI_ISL_2222190, EPI_ISL_2222193, EPI_ISL_2222195, EPI_ISL_2222198, EPI_ISL_2222200, EPI_ISL_2222203, EPI_ISL_2222206, EPI_ISL_2222209, EPI_ISL_2222212, EPI_ISL_2222216, EPI_ISL_2222219, EPI_ISL_2222222, EPI_ISL_2222225, EPI_ISL_2222227, EPI_ISL_2222231, EPI_ISL_2222234, EPI_ISL_2222236, EPI_ISL_2222239, EPI_ISL_2222242, EPI_ISL_2222245, EPI_ISL_2222247, EPI_ISL_2222250, EPI_ISL_2222253, EPI_ISL_2222256, EPI_ISL_2222259, EPI_ISL_2222262, EPI_ISL_2222265, EPI_ISL_2222269, EPI_ISL_2222272, EPI_ISL_2222275, EPI_ISL_2222278, EPI_ISL_2222281, EPI_ISL_2222284, EPI_ISL_2222287, EPI_ISL_2222290, EPI_ISL_2222292, EPI_ISL_2222296, EPI_ISL_2222299, EPI_ISL_2222302, EPI_ISL_2222305, EPI_ISL_2222308, EPI_ISL_2222311, EPI_ISL_2222315, EPI_ISL_2222318, EPI_ISL_2222321, EPI_ISL_2222324, EPI_ISL_2222327, EPI_ISL_2222330, EPI_ISL_2222332, EPI_ISL_2222335, EPI_ISL_2222338, EPI_ISL_2222341, EPI_ISL_2222343, EPI_ISL_2222347, EPI_ISL_2222350, EPI_ISL_2222353, EPI_ISL_2222356, EPI_ISL_2222359, EPI_ISL_2222361, EPI_ISL_2222364, EPI_ISL_2222367, EPI_ISL_2222370, EPI_ISL_2222373, EPI_ISL_2222376, EPI_ISL_2222380, EPI_ISL_2222383, EPI_ISL_2222386, EPI_ISL_2222390, EPI_ISL_2222392, EPI_ISL_2222396, EPI_ISL_2222399, EPI_ISL_2222402, EPI_ISL_2222404, EPI_ISL_2222407, EPI_ISL_2222410, EPI_ISL_2222413, EPI_ISL_2222415, EPI_ISL_2222418, EPI_ISL_2222421, EPI_ISL_2222424, EPI_ISL_2222427, EPI_ISL_2222430, EPI_ISL_2222433, EPI_ISL_2222436, EPI_ISL_2222439, EPI_ISL_2222442, EPI_ISL_2222445, EPI_ISL_2222448, EPI_ISL_2222450, EPI_ISL_2222453, EPI_ISL_2222455, EPI_ISL_2222458, EPI_ISL_2222461, EPI_ISL_2222464, EPI_ISL_2222467, EPI_ISL_2222470, EPI_ISL_2222473, EPI_ISL_2222476, EPI_ISL_2222479, EPI_ISL_2222482, EPI_ISL_2222485, EPI_ISL_2222487, EPI_ISL_2222490, EPI_ISL_2222493, EPI_ISL_2222496, EPI_ISL_2222499, EPI_ISL_2222502, EPI_ISL_2222505, EPI_ISL_2222508, EPI_ISL_2222511, EPI_ISL_2222515, EPI_ISL_2222518, EPI_ISL_2222520, EPI_ISL_2222523, EPI_ISL_2222526, EPI_ISL_2222530, EPI_ISL_2222533, EPI_ISL_2222538, EPI_ISL_2222541, EPI_ISL_2222544, EPI_ISL_2222547, EPI_ISL_2222550, EPI_ISL_2222552, EPI_ISL_2222554, EPI_ISL_2222557, EPI_ISL_2222560, EPI_ISL_2222563, EPI_ISL_2222566, EPI_ISL_2222568, EPI_ISL_2222571, EPI_ISL_2222574, EPI_ISL_2222577, EPI_ISL_2222580, EPI_ISL_2222582, EPI_ISL_2222585, EPI_ISL_2222588, EPI_ISL_2222591, EPI_ISL_2222594, EPI_ISL_2222597, EPI_ISL_2222600, EPI_ISL_2222603, EPI_ISL_2222606, EPI_ISL_2222609, EPI_ISL_2222612, EPI_ISL_2222615, EPI_ISL_2222618, EPI_ISL_2222621, EPI_ISL_2222624, EPI_ISL_2222627, EPI_ISL_2222630, EPI_ISL_2222632, EPI_ISL_2222635, EPI_ISL_2222638, EPI_ISL_2222641, EPI_ISL_2222646, EPI_ISL_2222649, EPI_ISL_2222652, EPI_ISL_2222655, EPI_ISL_2222658, EPI_ISL_2222660, EPI_ISL_2222663, EPI_ISL_2222666, EPI_ISL_2222670, EPI_ISL_2222674, EPI_ISL_2222678, EPI_ISL_2222681, EPI_ISL_2222686, EPI_ISL_2222691, EPI_ISL_2222696, EPI_ISL_2222703, EPI_ISL_2222711, EPI_ISL_2222714, EPI_ISL_2222715, EPI_ISL_2222716, EPI_ISL_2222717, EPI_ISL_2222718, EPI_ISL_2222719, EPI_ISL_2222720, EPI_ISL_2222721, EPI_ISL_2222722, EPI_ISL_2222723, EPI_ISL_2222724, EPI_ISL_2222725, EPI_ISL_2222726, EPI_ISL_2222727, EPI_ISL_2222728, EPI_ISL_2222729, EPI_ISL_2222730, EPI_ISL_2222731, EPI_ISL_2222732, EPI_ISL_2222733, EPI_ISL_2222734, EPI_ISL_2222735, EPI_ISL_2222736, EPI_ISL_2222737, EPI_ISL_2222738, EPI_ISL_2222739, EPI_ISL_2222740, EPI_ISL_2222741, EPI_ISL_2222742, EPI_ISL_2222743, EPI_ISL_2222744, EPI_ISL_2222745, EPI_ISL_2222746, EPI_ISL_2222747, EPI_ISL_2222748, EPI_ISL_2222749, EPI_ISL_2222750, EPI_ISL_2222751, EPI_ISL_2222752, EPI_ISL_2222753, EPI_ISL_2222756, EPI_ISL_2222757, EPI_ISL_2222758, EPI_ISL_2222759, EPI_ISL_2222760, EPI_ISL_2222761, EPI_ISL_2222762, EPI_ISL_2222763, EPI_ISL_2222764, EPI_ISL_2222765, EPI_ISL_2222766, EPI_ISL_2222767, EPI_ISL_2222768, EPI_ISL_2222769, EPI_ISL_2222770, EPI_ISL_2222771, EPI_ISL_2222772, EPI_ISL_2222773, EPI_ISL_2222774, EPI_ISL_2222775, EPI_ISL_2222776, EPI_ISL_2222777, EPI_ISL_2222778, EPI_ISL_2222779, EPI_ISL_2222780, EPI_ISL_2222781, EPI_ISL_2222782, EPI_ISL_2222783, EPI_ISL_2222784, EPI_ISL_2222785, EPI_ISL_2222786, EPI_ISL_2222787, EPI_ISL_2222788, EPI_ISL_2222789, EPI_ISL_2222790, EPI_ISL_2222791, EPI_ISL_2222792, EPI_ISL_2222793, EPI_ISL_2222794, EPI_ISL_2222795, EPI_ISL_2222796, EPI_ISL_2222797, EPI_ISL_2222798, EPI_ISL_2222799, EPI_ISL_2222800, EPI_ISL_2222801, EPI_ISL_2222802, EPI_ISL_2222803, EPI_ISL_2222804, EPI_ISL_2222805, EPI_ISL_2222806, EPI_ISL_2222807, EPI_ISL_2222808, EPI_ISL_2222809, EPI_ISL_2222810, EPI_ISL_2222811, EPI_ISL_2222812, EPI_ISL_2222813, EPI_ISL_2222814, EPI_ISL_2222815, EPI_ISL_2222816, EPI_ISL_2222817, EPI_ISL_2222818, EPI_ISL_2222819, EPI_ISL_2222820, EPI_ISL_2222821, EPI_ISL_2222822, EPI_ISL_2222823, EPI_ISL_2222824, EPI_ISL_2222825, EPI_ISL_2222826, EPI_ISL_2222827, EPI_ISL_2222828, EPI_ISL_2222829, EPI_ISL_2222830, EPI_ISL_2222831, EPI_ISL_2222832, EPI_ISL_2222833, EPI_ISL_2222834, EPI_ISL_2222835, EPI_ISL_2222836, EPI_ISL_2222837, EPI_ISL_2222838, EPI_ISL_2222839, EPI_ISL_2222840, EPI_ISL_2222841, EPI_ISL_2222842, EPI_ISL_2222843, EPI_ISL_2222844, EPI_ISL_2222845, EPI_ISL_2222846, EPI_ISL_2222847, EPI_ISL_2222848, EPI_ISL_2222849, EPI_ISL_2222850, EPI_ISL_2222851, EPI_ISL_2222852, EPI_ISL_2222853, EPI_ISL_2222854, EPI_ISL_2222855, EPI_ISL_2222856, EPI_ISL_2222857, EPI_ISL_2222858, EPI_ISL_2222859, EPI_ISL_2222860, EPI_ISL_2222861, EPI_ISL_2222862, EPI_ISL_2222863, EPI_ISL_2222864, EPI_ISL_2222865, EPI_ISL_2222866, EPI_ISL_2222867, EPI_ISL_2222868, EPI_ISL_2222869, EPI_ISL_2222871, EPI_ISL_2222872, EPI_ISL_2222873, EPI_ISL_2222874, EPI_ISL_2222875, EPI_ISL_2222876, EPI_ISL_2222877, EPI_ISL_2222879, EPI_ISL_2222880, EPI_ISL_2222882, EPI_ISL_2222884, EPI_ISL_2222886, EPI_ISL_2222888, EPI_ISL_2222889, EPI_ISL_2222891, EPI_ISL_2222893, EPI_ISL_2222895, EPI_ISL_2222896, EPI_ISL_2222897, EPI_ISL_2222899, EPI_ISL_2222901, EPI_ISL_2222903, EPI_ISL_2222904, EPI_ISL_2222906, EPI_ISL_2222907, EPI_ISL_2222909, EPI_ISL_2222910, EPI_ISL_2222912, EPI_ISL_2222914, EPI_ISL_2222915, EPI_ISL_2222919, EPI_ISL_2222920, EPI_ISL_2222921, EPI_ISL_2222922, EPI_ISL_2222923, EPI_ISL_2222924, EPI_ISL_2222925, EPI_ISL_2222926, EPI_ISL_2222927, EPI_ISL_2222928, EPI_ISL_2222929, EPI_ISL_2222930, EPI_ISL_2222931, EPI_ISL_2222932, EPI_ISL_2222933, EPI_ISL_2222934, EPI_ISL_2222935, EPI_ISL_2222936, EPI_ISL_2222937, EPI_ISL_2222938, EPI_ISL_2222939, EPI_ISL_2222940, EPI_ISL_2222941, EPI_ISL_2222942, EPI_ISL_2222943, EPI_ISL_2222944, EPI_ISL_2222945, EPI_ISL_2222946, EPI_ISL_2222947, EPI_ISL_2222948, EPI_ISL_2222949, EPI_ISL_2222950, EPI_ISL_2222951, EPI_ISL_2222952, EPI_ISL_2222953, EPI_ISL_2222954, EPI_ISL_2222955, EPI_ISL_2222956, EPI_ISL_2222957, EPI_ISL_2222958, EPI_ISL_2222959, EPI_ISL_2222960, EPI_ISL_2222961, EPI_ISL_2222962, EPI_ISL_2222963, EPI_ISL_2222964, EPI_ISL_2222965, EPI_ISL_2222966, EPI_ISL_2222967, EPI_ISL_2222968, EPI_ISL_2222969, EPI_ISL_2222970, EPI_ISL_2222971, EPI_ISL_2222972, EPI_ISL_2222974, EPI_ISL_2222975, EPI_ISL_2222976, EPI_ISL_2222977, EPI_ISL_2222978, EPI_ISL_2222979, EPI_ISL_2222980, EPI_ISL_2222981, EPI_ISL_2222982, EPI_ISL_2222983, EPI_ISL_2222984, EPI_ISL_2222985, EPI_ISL_2222986, EPI_ISL_2222988, EPI_ISL_2222989, EPI_ISL_2222990, EPI_ISL_2222991, EPI_ISL_2222992, EPI_ISL_2222993, EPI_ISL_2222995, EPI_ISL_2222996, EPI_ISL_2222997, EPI_ISL_2222998, EPI_ISL_2222999, EPI_ISL_2223000, EPI_ISL_2223001, EPI_ISL_2223002, EPI_ISL_2223003, EPI_ISL_2223004, EPI_ISL_2223005, EPI_ISL_2223006, EPI_ISL_2223007, EPI_ISL_2223008, EPI_ISL_2223009, EPI_ISL_2223010, EPI_ISL_2223011, EPI_ISL_2223012, EPI_ISL_2223013, EPI_ISL_2223014, EPI_ISL_2223015, EPI_ISL_2223016, EPI_ISL_2223017, EPI_ISL_2223018, EPI_ISL_2223019, EPI_ISL_2223020, EPI_ISL_2223021, EPI_ISL_2223022, EPI_ISL_2223023, EPI_ISL_2223024, EPI_ISL_2223025, EPI_ISL_2223026, EPI_ISL_2223027, EPI_ISL_2223028, EPI_ISL_2223029, EPI_ISL_2223030, EPI_ISL_2223031, EPI_ISL_2223032, EPI_ISL_2223033, EPI_ISL_2223034, EPI_ISL_2223035, EPI_ISL_2223036, EPI_ISL_2223037, EPI_ISL_2223038, EPI_ISL_2223039, EPI_ISL_2223040, EPI_ISL_2223041, EPI_ISL_2223042, EPI_ISL_2223043, EPI_ISL_2223044, EPI_ISL_2223045, EPI_ISL_2223046, EPI_ISL_2223047, EPI_ISL_2223048, EPI_ISL_2223049, EPI_ISL_2223050, EPI_ISL_2223051, EPI_ISL_2223052, EPI_ISL_2223053, EPI_ISL_2223054, EPI_ISL_2223055, EPI_ISL_2223056, EPI_ISL_2223057, EPI_ISL_2223058, EPI_ISL_2223059, EPI_ISL_2223060, EPI_ISL_2223061, EPI_ISL_2223062, EPI_ISL_2223063, EPI_ISL_2223064, EPI_ISL_2223065, EPI_ISL_2223066, EPI_ISL_2223067, EPI_ISL_2223068, EPI_ISL_2223069, EPI_ISL_2223070, EPI_ISL_2223071, EPI_ISL_2223072, EPI_ISL_2223073, EPI_ISL_2223074, EPI_ISL_2223075, EPI_ISL_2223076, EPI_ISL_2223077, EPI_ISL_2223078, EPI_ISL_2223079, EPI_ISL_2223080, EPI_ISL_2223081, EPI_ISL_2223082, EPI_ISL_2223083, EPI_ISL_2223084, EPI_ISL_2223085, EPI_ISL_2223086, EPI_ISL_2223087, EPI_ISL_2223088, EPI_ISL_2223089, EPI_ISL_2223091, EPI_ISL_2223093, EPI_ISL_2223098, EPI_ISL_2223099, EPI_ISL_2223100, EPI_ISL_2223101, EPI_ISL_2223102, EPI_ISL_2223104, EPI_ISL_2223105, EPI_ISL_2223106, EPI_ISL_2223107, EPI_ISL_2223108, EPI_ISL_2223110, EPI_ISL_2223111, EPI_ISL_2223112, EPI_ISL_2223114, EPI_ISL_2223117, EPI_ISL_2223124, EPI_ISL_2223125, EPI_ISL_2223126, EPI_ISL_2223127, EPI_ISL_2223128, EPI_ISL_2223129, EPI_ISL_2223130, EPI_ISL_2223131, EPI_ISL_2223132, EPI_ISL_2223133, EPI_ISL_2223134, EPI_ISL_2223135, EPI_ISL_2223136, EPI_ISL_2223137, EPI_ISL_2223138, EPI_ISL_2223139, EPI_ISL_2223140, EPI_ISL_2223141, EPI_ISL_2223144, EPI_ISL_2223145, EPI_ISL_2223146, EPI_ISL_2223148, EPI_ISL_2223149, EPI_ISL_2223151, EPI_ISL_2223152, EPI_ISL_2223154, EPI_ISL_2223156, EPI_ISL_2223157, EPI_ISL_2223158, EPI_ISL_2223160, EPI_ISL_2223162, EPI_ISL_2223163, EPI_ISL_2223164, EPI_ISL_2223166, EPI_ISL_2223168, EPI_ISL_2223169, EPI_ISL_2223170, EPI_ISL_2223172, EPI_ISL_2223173, EPI_ISL_2223174, EPI_ISL_2223176, EPI_ISL_2223178, EPI_ISL_2223180, EPI_ISL_2223181, EPI_ISL_2223183, EPI_ISL_2223184, EPI_ISL_2223185, EPI_ISL_2223188, EPI_ISL_2223189, EPI_ISL_2223192, EPI_ISL_2223193, EPI_ISL_2223194, EPI_ISL_2223195, EPI_ISL_2223196, EPI_ISL_2223197, EPI_ISL_2223198, EPI_ISL_2223199, EPI_ISL_2223202, EPI_ISL_2223206, EPI_ISL_2223207, EPI_ISL_2223208, EPI_ISL_2223210, EPI_ISL_2223213, EPI_ISL_2223216, EPI_ISL_2223219, EPI_ISL_2223221, EPI_ISL_2223222, EPI_ISL_2223223, EPI_ISL_2223224, EPI_ISL_2223225, EPI_ISL_2223226, EPI_ISL_2223227, EPI_ISL_2223229, EPI_ISL_2223230, EPI_ISL_2223231, EPI_ISL_2223232, EPI_ISL_2223233, EPI_ISL_2223234, EPI_ISL_2223237, EPI_ISL_2223242, EPI_ISL_2223243, EPI_ISL_2223245, EPI_ISL_2223246, EPI_ISL_2223248, EPI_ISL_2223249, EPI_ISL_2223253, EPI_ISL_2223256, EPI_ISL_2223257, EPI_ISL_2223258, EPI_ISL_2223259, EPI_ISL_2223263, EPI_ISL_2223265, EPI_ISL_2223266, EPI_ISL_2223267, EPI_ISL_2223268, EPI_ISL_2223269, EPI_ISL_2223271, EPI_ISL_2223272, EPI_ISL_2223276, EPI_ISL_2223277, EPI_ISL_2223278, EPI_ISL_2223281, EPI_ISL_2223282, EPI_ISL_2223283, EPI_ISL_2223285, EPI_ISL_2223286, EPI_ISL_2223289, EPI_ISL_2223290, EPI_ISL_2223291, EPI_ISL_2223293, EPI_ISL_2223294, EPI_ISL_2223295, EPI_ISL_2223296, EPI_ISL_2223297, EPI_ISL_2223301, EPI_ISL_2223303, EPI_ISL_2223304, EPI_ISL_2223305, EPI_ISL_2223308, EPI_ISL_2223309, EPI_ISL_2223310, EPI_ISL_2223311, EPI_ISL_2223315, EPI_ISL_2223316, EPI_ISL_2223317, EPI_ISL_2223319, EPI_ISL_2223320, EPI_ISL_2223321, EPI_ISL_2223322, EPI_ISL_2223324, EPI_ISL_2223325, EPI_ISL_2223326, EPI_ISL_2223327, EPI_ISL_2223328, EPI_ISL_2223329, EPI_ISL_2223331, EPI_ISL_2223333, EPI_ISL_2223335, EPI_ISL_2223337, EPI_ISL_2223338, EPI_ISL_2223341, EPI_ISL_2223342, EPI_ISL_2223343, EPI_ISL_2223344, EPI_ISL_2223345, EPI_ISL_2223346, EPI_ISL_2223347, EPI_ISL_2223348, EPI_ISL_2223349, EPI_ISL_2223350, EPI_ISL_2223351, EPI_ISL_2223352, EPI_ISL_2223353, EPI_ISL_2223354, EPI_ISL_2223355, EPI_ISL_2223356, EPI_ISL_2223357, EPI_ISL_2223358, EPI_ISL_2223360, EPI_ISL_2223361, EPI_ISL_2223362, EPI_ISL_2223363, EPI_ISL_2223364, EPI_ISL_2223365, EPI_ISL_2223368, EPI_ISL_2223377

| | | | |
|---|---|---|---|
| see above | Houston Methodist Hospital | Houston Methodist Hospital | Randall J. Olsen, Paul A. Christensen, S. Wesley Long, Sishir Subedi, Robert Olson, Marcus Nguyen, James J. Davis, Matthew Ojeda Saavedra, Prasanti Yerramilli, Layne Pruitt, Kristina Reppond, Madison N. Shyer, Jessica Cambric, Ryan Gadd, Ilya J. Finkelstein, Jimmy Gollihar, and James M. Musser |
| EPI_ISL_2223378 | Ohio Department of Health Laboratory | Ohio Department of Health Laboratory | Holmes, Jennifer; Eric Brandt, Keoni Omura, Glen McGillivary, Caitlin McDonnell, Jade Mowery, Stephanie Mccracken, Tyler Payne, Kirtana Ramadugu, Erica Leasure, Brent Lee, Kelsey Florek, Heather Blankenship, Allison Black, Quanta Brown, and Tammy Bannerman |

EPI_ISL_2223380, EPI_ISL_2223381, EPI_ISL_2223382, EPI_ISL_2223383, EPI_ISL_2223384, EPI_ISL_2223387, EPI_ISL_2223389, EPI_ISL_2223390, EPI_ISL_2223391, EPI_ISL_2223392, EPI_ISL_2223394, EPI_ISL_2223395, EPI_ISL_2223397, EPI_ISL_2223402, EPI_ISL_2223403, EPI_ISL_2223408, EPI_ISL_2223413, EPI_ISL_2223418, EPI_ISL_2223420, EPI_ISL_2223421, EPI_ISL_2223424, EPI_ISL_2223426, EPI_ISL_2223428, EPI_ISL_2223434, EPI_ISL_2223436, EPI_ISL_2223440, EPI_ISL_2223445, EPI_ISL_2223447, EPI_ISL_2223450, EPI_ISL_2223453, EPI_ISL_2223454, EPI_ISL_2223456, EPI_ISL_2223457, EPI_ISL_2223458, EPI_ISL_2223459, EPI_ISL_2223460, EPI_ISL_2223461, EPI_ISL_2223462, EPI_ISL_2223463, EPI_ISL_2223464, EPI_ISL_2223466, EPI_ISL_2223467, EPI_ISL_2223468, EPI_ISL_2223469, EPI_ISL_2223470, EPI_ISL_2223472, EPI_ISL_2223473, EPI_ISL_2223474, EPI_ISL_2223475, EPI_ISL_2223476, EPI_ISL_2223478, EPI_ISL_2223479, EPI_ISL_2223480, EPI_ISL_2223481, EPI_ISL_2223483, EPI_ISL_2223484, EPI_ISL_2223485, EPI_ISL_2223486, EPI_ISL_2223487, EPI_ISL_2223489, EPI_ISL_2223491, EPI_ISL_2223492, EPI_ISL_2223493, EPI_ISL_2223494, EPI_ISL_2223495, EPI_ISL_2223496, EPI_ISL_2223497, EPI_ISL_2223498, EPI_ISL_2223499, EPI_ISL_2223500, EPI_ISL_2223501, EPI_ISL_2223502, EPI_ISL_2223503, EPI_ISL_2223504, EPI_ISL_2223505, EPI_ISL_2223506, EPI_ISL_2223507, EPI_ISL_2223508, EPI_ISL_2223509, EPI_ISL_2223511, EPI_ISL_2223513, EPI_ISL_2223514, EPI_ISL_2223516, EPI_ISL_2223517, EPI_ISL_2223518, EPI_ISL_2223519, EPI_ISL_2223520, EPI_ISL_2223521, EPI_ISL_2223522, EPI_ISL_2223523, EPI_ISL_2223524, EPI_ISL_2223525, EPI_ISL_2223526, EPI_ISL_2223527, EPI_ISL_2223528, EPI_ISL_2223529, EPI_ISL_2223530, EPI_ISL_2223531, EPI_ISL_2223532, EPI_ISL_2223533, EPI_ISL_2223534, EPI_ISL_2223535, EPI_ISL_2223536, EPI_ISL_2223537, EPI_ISL_2223538, EPI_ISL_2223539,

| | | | |
|---|---|---|---|
| EPI_ISL_2223540, EPI_ISL_2223541, EPI_ISL_2223542, EPI_ISL_2223543, EPI_ISL_2223544, EPI_ISL_2223545, EPI_ISL_2223546, EPI_ISL_2223547, EPI_ISL_2223548, EPI_ISL_2223549, EPI_ISL_2223550, EPI_ISL_2223551, EPI_ISL_2223552, EPI_ISL_2223553, EPI_ISL_2223554, EPI_ISL_2223555, EPI_ISL_2223556, EPI_ISL_2223557, EPI_ISL_2223558, EPI_ISL_2223559, EPI_ISL_2223560, EPI_ISL_2223561, EPI_ISL_2223562, EPI_ISL_2223563, EPI_ISL_2223564, EPI_ISL_2223565, EPI_ISL_2223566, EPI_ISL_2223567, EPI_ISL_2223569, EPI_ISL_2223570, EPI_ISL_2223571, EPI_ISL_2223572, EPI_ISL_2223573, EPI_ISL_2223574, EPI_ISL_2223575, EPI_ISL_2223576, EPI_ISL_2223577, EPI_ISL_2223578, EPI_ISL_2223579, EPI_ISL_2223580, EPI_ISL_2223581, EPI_ISL_2223582, EPI_ISL_2223583, EPI_ISL_2223584, EPI_ISL_2223585, EPI_ISL_2223586, EPI_ISL_2223587, EPI_ISL_2223588, EPI_ISL_2223589, EPI_ISL_2223590, EPI_ISL_2223591, EPI_ISL_2223592, EPI_ISL_2223593, EPI_ISL_2223594, EPI_ISL_2223595, EPI_ISL_2223596, EPI_ISL_2223597, EPI_ISL_2223598, EPI_ISL_2223599, EPI_ISL_2223600, EPI_ISL_2223602, EPI_ISL_2223603, EPI_ISL_2223604, EPI_ISL_2223605, EPI_ISL_2223606, EPI_ISL_2223607, EPI_ISL_2223608, EPI_ISL_2223609, EPI_ISL_2223610, EPI_ISL_2223611, EPI_ISL_2223612 | | | |
| see above | Houston Methodist Hospital | Houston Methodist Hospital | Randall J. Olsen, Paul A. Christensen, S. Wesley Long, Sishir Subedi, Robert Olson, Marcus Nguyen, James J. Davis, Matthew Ojeda Saavedra, Prasanti Yerramilli, Layne Pruitt, Kristina Reppond, Madison N. Shyer, Jessica Cambric, Ryan Gadd, Ilya J. Finkelstein, Jimmy Gollihar, and James M. Musser |
| EPI_ISL_2223613 | Ohio Department of Health Laboratory | Ohio Department of Health Laboratory | Holmes, Jennifer; Eric Brandt, Keoni Omura, Glen McGillivary, Caitlin McDonnell, Jade Mowery, Stephanie Mccracken, Tyler Payne, Kirtana Ramadugu, Erica Leasure, Brent Lee, Kelsey Florek, Heather Blankenship, Allison Black, Quanta Brown, and Tammy Bannerman |
| EPI_ISL_2223614, EPI_ISL_2223615, EPI_ISL_2223616, EPI_ISL_2223617, EPI_ISL_2223618, EPI_ISL_2223619, EPI_ISL_2223620, EPI_ISL_2223622, EPI_ISL_2223623, EPI_ISL_2223624, EPI_ISL_2223625, EPI_ISL_2223626, EPI_ISL_2223627, EPI_ISL_2223628, EPI_ISL_2223629, EPI_ISL_2223630, EPI_ISL_2223631, EPI_ISL_2223632, EPI_ISL_2223633, EPI_ISL_2223634, EPI_ISL_2223635, EPI_ISL_2223636, EPI_ISL_2223637, EPI_ISL_2223638, EPI_ISL_2223639, EPI_ISL_2223640, EPI_ISL_2223641, EPI_ISL_2223642, EPI_ISL_2223643, EPI_ISL_2223644, EPI_ISL_2223645, EPI_ISL_2223646, EPI_ISL_2223647, EPI_ISL_2223648, EPI_ISL_2223649, EPI_ISL_2223650, EPI_ISL_2223651, EPI_ISL_2223652, EPI_ISL_2223653, EPI_ISL_2223654, EPI_ISL_2223655, EPI_ISL_2223656, EPI_ISL_2223657, EPI_ISL_2223658, EPI_ISL_2223659, EPI_ISL_2223660, EPI_ISL_2223661, EPI_ISL_2223662, EPI_ISL_2223663, EPI_ISL_2223664, EPI_ISL_2223665, EPI_ISL_2223666, EPI_ISL_2223667, EPI_ISL_2223668, EPI_ISL_2223669, EPI_ISL_2223670, EPI_ISL_2223671, EPI_ISL_2223672, EPI_ISL_2223673, EPI_ISL_2223674, EPI_ISL_2223675, EPI_ISL_2223676, EPI_ISL_2223677, EPI_ISL_2223678, EPI_ISL_2223679, EPI_ISL_2223680, EPI_ISL_2223681, EPI_ISL_2223682, EPI_ISL_2223683, EPI_ISL_2223684, EPI_ISL_2223685, EPI_ISL_2223686, EPI_ISL_2223687, EPI_ISL_2223688, EPI_ISL_2223689, EPI_ISL_2223690, EPI_ISL_2223691, EPI_ISL_2223692, EPI_ISL_2223693, EPI_ISL_2223694, EPI_ISL_2223695, EPI_ISL_2223696, EPI_ISL_2223697, EPI_ISL_2223698, EPI_ISL_2223699, EPI_ISL_2223700, EPI_ISL_2223701, EPI_ISL_2223702, EPI_ISL_2223703, EPI_ISL_2223704, EPI_ISL_2223705, EPI_ISL_2223706, EPI_ISL_2223707, EPI_ISL_2223708, EPI_ISL_2223709, EPI_ISL_2223710, EPI_ISL_2223711, EPI_ISL_2223712, EPI_ISL_2223713, EPI_ISL_2223714, EPI_ISL_2223715, EPI_ISL_2223716, EPI_ISL_2223718, EPI_ISL_2223719, EPI_ISL_2223720, EPI_ISL_2223721, EPI_ISL_2223722, EPI_ISL_2223723, EPI_ISL_2223724, EPI_ISL_2223725, EPI_ISL_2223726, EPI_ISL_2223727, EPI_ISL_2223728, EPI_ISL_2223729, EPI_ISL_2223730, EPI_ISL_2223731, EPI_ISL_2223732, EPI_ISL_2223733, EPI_ISL_2223734, EPI_ISL_2223735, EPI_ISL_2223736, EPI_ISL_2223737, EPI_ISL_2223738, EPI_ISL_2223739, EPI_ISL_2223740, EPI_ISL_2223741, EPI_ISL_2223742, EPI_ISL_2223743, EPI_ISL_2223744, EPI_ISL_2223745, EPI_ISL_2223746, EPI_ISL_2223747, EPI_ISL_2223748, EPI_ISL_2223749, EPI_ISL_2223750, EPI_ISL_2223751, EPI_ISL_2223752, EPI_ISL_2223753, EPI_ISL_2223754, EPI_ISL_2223755, EPI_ISL_2223756, EPI_ISL_2223757, EPI_ISL_2223758, EPI_ISL_2223759, EPI_ISL_2223760, EPI_ISL_2223761, EPI_ISL_2223762, EPI_ISL_2223763, EPI_ISL_2223764, EPI_ISL_2223765, EPI_ISL_2223766, EPI_ISL_2223767, EPI_ISL_2223768, EPI_ISL_2223769, EPI_ISL_2223770, EPI_ISL_2223771, EPI_ISL_2223772, EPI_ISL_2223773, EPI_ISL_2223774, EPI_ISL_2223775, EPI_ISL_2223776, EPI_ISL_2223777, EPI_ISL_2223778, EPI_ISL_2223779, EPI_ISL_2223780, EPI_ISL_2223781, EPI_ISL_2223782, EPI_ISL_2223783, EPI_ISL_2223784, EPI_ISL_2223785, EPI_ISL_2223786, EPI_ISL_2223787, EPI_ISL_2223788, EPI_ISL_2223789, EPI_ISL_2223790, EPI_ISL_2223791, EPI_ISL_2223792, EPI_ISL_2223793, EPI_ISL_2223794, EPI_ISL_2223795, EPI_ISL_2223796, EPI_ISL_2223797, EPI_ISL_2223798, EPI_ISL_2223799, EPI_ISL_2223800, EPI_ISL_2223801, EPI_ISL_2223802, EPI_ISL_2223803, EPI_ISL_2223804, EPI_ISL_2223805, EPI_ISL_2223806, EPI_ISL_2223807, EPI_ISL_2223808, EPI_ISL_2223809, EPI_ISL_2223810, EPI_ISL_2223811, EPI_ISL_2223812, EPI_ISL_2223813, EPI_ISL_2223814, EPI_ISL_2223815, EPI_ISL_2223816, EPI_ISL_2223817, EPI_ISL_2223818, EPI_ISL_2223819, EPI_ISL_2223820, EPI_ISL_2223821, EPI_ISL_2223822, EPI_ISL_2223823, EPI_ISL_2223824, EPI_ISL_2223825, EPI_ISL_2223826, EPI_ISL_2223827, EPI_ISL_2223828, EPI_ISL_2223829, EPI_ISL_2223830, EPI_ISL_2223831, EPI_ISL_2223832, EPI_ISL_2223833, EPI_ISL_2223834, EPI_ISL_2223835, EPI_ISL_2223836, EPI_ISL_2223837, EPI_ISL_2223838, EPI_ISL_2223839, EPI_ISL_2223840, EPI_ISL_2223841, EPI_ISL_2223842, EPI_ISL_2223843, EPI_ISL_2223844, EPI_ISL_2223845, EPI_ISL_2223846, EPI_ISL_2223847, EPI_ISL_2223848, EPI_ISL_2223849, EPI_ISL_2223850, EPI_ISL_2223851, EPI_ISL_2223852 | | | |
| see above | Houston Methodist Hospital | Houston Methodist Hospital | Randall J. Olsen, Paul A. Christensen, S. Wesley Long, Sishir Subedi, Robert Olson, Marcus Nguyen, James J. Davis, Matthew Ojeda Saavedra, Prasanti Yerramilli, Layne Pruitt, Kristina Reppond, Madison N. Shyer, Jessica Cambric, Ryan Gadd, Ilya J. Finkelstein, Jimmy Gollihar, and James M. Musser |
| EPI_ISL_2223853 | Ohio Department of Health Laboratory | Ohio Department of Health Laboratory | Holmes, Jennifer; Eric Brandt, Keoni Omura, Glen McGillivary, Caitlin McDonnell, Jade Mowery, Stephanie Mccracken, Tyler Payne, Kirtana Ramadugu, Erica Leasure, Brent Lee, Kelsey Florek, Heather Blankenship, Allison Black, Quanta Brown, and Tammy Bannerman |
| EPI_ISL_2223854, EPI_ISL_2223855, EPI_ISL_2223856, EPI_ISL_2223857, EPI_ISL_2223858, EPI_ISL_2223859, EPI_ISL_2223860, EPI_ISL_2223861, EPI_ISL_2223862, EPI_ISL_2223863, EPI_ISL_2223864, EPI_ISL_2223865, EPI_ISL_2223866, EPI_ISL_2223867, EPI_ISL_2223868, EPI_ISL_2223869, EPI_ISL_2223870, EPI_ISL_2223871, EPI_ISL_2223872, EPI_ISL_2223873, EPI_ISL_2223874, EPI_ISL_2223875, EPI_ISL_2223876, EPI_ISL_2223877, EPI_ISL_2223878, EPI_ISL_2223879, EPI_ISL_2223880, EPI_ISL_2223881, EPI_ISL_2223882, EPI_ISL_2223883, EPI_ISL_2223884, EPI_ISL_2223885, EPI_ISL_2223886, EPI_ISL_2223887, EPI_ISL_2223888, EPI_ISL_2223889, EPI_ISL_2223890, EPI_ISL_2223891, EPI_ISL_2223892, EPI_ISL_2223893, EPI_ISL_2223894, EPI_ISL_2223895, EPI_ISL_2223896, EPI_ISL_2223897, EPI_ISL_2223898, EPI_ISL_2223899, EPI_ISL_2223900, EPI_ISL_2223901, EPI_ISL_2223902, EPI_ISL_2223903, EPI_ISL_2223904, EPI_ISL_2223905, EPI_ISL_2223906, EPI_ISL_2223907, EPI_ISL_2223908, EPI_ISL_2223909, EPI_ISL_2223910, EPI_ISL_2223911, EPI_ISL_2223912, EPI_ISL_2223913, EPI_ISL_2223914, EPI_ISL_2223915, EPI_ISL_2223916, EPI_ISL_2223917, EPI_ISL_2223918, EPI_ISL_2223919, EPI_ISL_2223920, EPI_ISL_2223921, EPI_ISL_2223922, EPI_ISL_2223923, EPI_ISL_2223924, EPI_ISL_2223925, EPI_ISL_2223926, EPI_ISL_2223927, EPI_ISL_2223928, EPI_ISL_2223929, EPI_ISL_2223930, EPI_ISL_2223931, EPI_ISL_2223932, EPI_ISL_2223933, EPI_ISL_2223934, EPI_ISL_2223935, EPI_ISL_2223936, EPI_ISL_2223937, EPI_ISL_2223938, EPI_ISL_2223939, EPI_ISL_2223940, EPI_ISL_2223941, EPI_ISL_2223942, EPI_ISL_2223943, EPI_ISL_2223944, EPI_ISL_2223945, EPI_ISL_2223946, EPI_ISL_2223947, EPI_ISL_2223948, EPI_ISL_2223949, EPI_ISL_2223950, EPI_ISL_2223951, EPI_ISL_2223952, EPI_ISL_2223953, EPI_ISL_2223954, EPI_ISL_2223955, EPI_ISL_2223956, EPI_ISL_2223957, EPI_ISL_2223958, EPI_ISL_2223959, EPI_ISL_2223960, EPI_ISL_2223961, EPI_ISL_2223962, EPI_ISL_2223963, EPI_ISL_2223964, EPI_ISL_2223965, EPI_ISL_2223966, EPI_ISL_2223967, EPI_ISL_2223968, EPI_ISL_2223969, EPI_ISL_2223970, EPI_ISL_2223971, EPI_ISL_2223972, EPI_ISL_2223973, EPI_ISL_2223974, EPI_ISL_2223975, EPI_ISL_2223976, EPI_ISL_2223977, EPI_ISL_2223978, EPI_ISL_2223979, EPI_ISL_2223980, EPI_ISL_2223981, EPI_ISL_2223982, EPI_ISL_2223983, EPI_ISL_2223984, EPI_ISL_2223985, EPI_ISL_2223986, EPI_ISL_2223987, EPI_ISL_2223988, EPI_ISL_2223989, EPI_ISL_2223990, EPI_ISL_2223991, EPI_ISL_2223992, EPI_ISL_2223993, EPI_ISL_2223994, EPI_ISL_2223995, EPI_ISL_2223996, EPI_ISL_2223997, EPI_ISL_2223998, EPI_ISL_2223999, EPI_ISL_2224000, EPI_ISL_2224001, EPI_ISL_2224002, EPI_ISL_2224003, EPI_ISL_2224004, EPI_ISL_2224005, EPI_ISL_2224006, EPI_ISL_2224007, EPI_ISL_2224008, EPI_ISL_2224009, EPI_ISL_2224010, EPI_ISL_2224011, EPI_ISL_2224012, EPI_ISL_2224013, EPI_ISL_2224014, EPI_ISL_2224015, EPI_ISL_2224016, EPI_ISL_2224017, EPI_ISL_2224018, EPI_ISL_2224019, EPI_ISL_2224020, EPI_ISL_2224021, EPI_ISL_2224022, EPI_ISL_2224023, EPI_ISL_2224024, EPI_ISL_2224025, EPI_ISL_2224026, EPI_ISL_2224027, EPI_ISL_2224028, EPI_ISL_2224029, EPI_ISL_2224030, EPI_ISL_2224031, EPI_ISL_2224032, EPI_ISL_2224033, EPI_ISL_2224034, EPI_ISL_2224035, EPI_ISL_2224036, EPI_ISL_2224037, EPI_ISL_2224038, EPI_ISL_2224039, EPI_ISL_2224040, EPI_ISL_2224041, EPI_ISL_2224042, EPI_ISL_2224043, EPI_ISL_2224044, EPI_ISL_2224045, EPI_ISL_2224046, EPI_ISL_2224047, EPI_ISL_2224048, EPI_ISL_2224049, EPI_ISL_2224050, EPI_ISL_2224051, EPI_ISL_2224052, EPI_ISL_2224053, EPI_ISL_2224054, EPI_ISL_2224055, EPI_ISL_2224056, EPI_ISL_2224057, EPI_ISL_2224058, EPI_ISL_2224059, EPI_ISL_2224060, EPI_ISL_2224061, EPI_ISL_2224062, EPI_ISL_2224063, EPI_ISL_2224064, EPI_ISL_2224065, EPI_ISL_2224066, EPI_ISL_2224067, EPI_ISL_2224069, EPI_ISL_2224070, EPI_ISL_2224071, EPI_ISL_2224072, EPI_ISL_2224074, EPI_ISL_2224075, EPI_ISL_2224078, EPI_ISL_2224079, EPI_ISL_2224081, EPI_ISL_2224082, EPI_ISL_2224083, EPI_ISL_2224084, EPI_ISL_2224085, EPI_ISL_2224086, EPI_ISL_2224087, EPI_ISL_2224088, EPI_ISL_2224089, EPI_ISL_2224090, EPI_ISL_2224091, EPI_ISL_2224092, EPI_ISL_2224093, EPI_ISL_2224094, EPI_ISL_2224095, EPI_ISL_2224096, EPI_ISL_2224097, EPI_ISL_2224099, EPI_ISL_2224100, EPI_ISL_2224102, EPI_ISL_2224104, EPI_ISL_2224105, EPI_ISL_2224106, EPI_ISL_2224107, EPI_ISL_2224108, EPI_ISL_2224109, EPI_ISL_2224111, EPI_ISL_2224112, EPI_ISL_2224113, EPI_ISL_2224114, EPI_ISL_2224115, EPI_ISL_2224116, EPI_ISL_2224117, EPI_ISL_2224118, EPI_ISL_2224119, EPI_ISL_2224120, EPI_ISL_2224122, EPI_ISL_2224123, EPI_ISL_2224124, EPI_ISL_2224125, EPI_ISL_2224126, EPI_ISL_2224127, EPI_ISL_2224128, EPI_ISL_2224129, EPI_ISL_2224130, EPI_ISL_2224131, EPI_ISL_2224132, EPI_ISL_2224133, EPI_ISL_2224134, EPI_ISL_2224135, EPI_ISL_2224136, EPI_ISL_2224137, EPI_ISL_2224138, EPI_ISL_2224139, EPI_ISL_2224140, EPI_ISL_2224141, EPI_ISL_2224142, EPI_ISL_2224143, EPI_ISL_2224144, EPI_ISL_2224145, EPI_ISL_2224146, EPI_ISL_2224147, EPI_ISL_2224148, EPI_ISL_2224149, EPI_ISL_2224150, EPI_ISL_2224151, EPI_ISL_2224152, EPI_ISL_2224153, EPI_ISL_2224154, EPI_ISL_2224155, EPI_ISL_2224156, EPI_ISL_2224157, EPI_ISL_2224159, EPI_ISL_2224160, EPI_ISL_2224161, EPI_ISL_2224162, EPI_ISL_2224163, EPI_ISL_2224164, EPI_ISL_2224165, EPI_ISL_2224166, EPI_ISL_2224167, EPI_ISL_2224168, EPI_ISL_2224169, EPI_ISL_2224170, EPI_ISL_2224171, EPI_ISL_2224172, EPI_ISL_2224173, EPI_ISL_2224174, EPI_ISL_2224175, EPI_ISL_2224176, EPI_ISL_2224177, EPI_ISL_2224178, EPI_ISL_2224179, EPI_ISL_2224180, EPI_ISL_2224181, EPI_ISL_2224182, EPI_ISL_2224183, EPI_ISL_2224184, EPI_ISL_2224185, EPI_ISL_2224186, EPI_ISL_2224187, EPI_ISL_2224188, EPI_ISL_2224189, EPI_ISL_2224190, EPI_ISL_2224191, EPI_ISL_2224192, EPI_ISL_2224193, EPI_ISL_2224194, EPI_ISL_2224195, EPI_ISL_2224196, EPI_ISL_2224197, EPI_ISL_2224198, EPI_ISL_2224199, EPI_ISL_2224200, EPI_ISL_2224201, EPI_ISL_2224202, EPI_ISL_2224203, EPI_ISL_2224204, EPI_ISL_2224205, EPI_ISL_2224206, EPI_ISL_2224207, EPI_ISL_2224208, EPI_ISL_2224209, EPI_ISL_2224210, EPI_ISL_2224211, EPI_ISL_2224212, EPI_ISL_2224213, EPI_ISL_2224214, EPI_ISL_2224215, EPI_ISL_2224216, EPI_ISL_2224217, EPI_ISL_2224218, EPI_ISL_2224219, EPI_ISL_2224220, EPI_ISL_2224221, EPI_ISL_2224222, EPI_ISL_2224223, EPI_ISL_2224224, EPI_ISL_2224225, EPI_ISL_2224226, EPI_ISL_2224227, EPI_ISL_2224228, EPI_ISL_2224229, EPI_ISL_2224230, EPI_ISL_2224231, EPI_ISL_2224232, EPI_ISL_2224233, EPI_ISL_2224234, EPI_ISL_2224235, EPI_ISL_2224236, EPI_ISL_2224237, EPI_ISL_2224238, EPI_ISL_2224239, EPI_ISL_2224240, EPI_ISL_2224241, EPI_ISL_2224242, EPI_ISL_2224243, EPI_ISL_2224244, EPI_ISL_2224245, EPI_ISL_2224246, EPI_ISL_2224247, EPI_ISL_2224248, EPI_ISL_2224249, EPI_ISL_2224250, EPI_ISL_2224251, EPI_ISL_2224252, EPI_ISL_2224253, EPI_ISL_2224254, EPI_ISL_2224255, EPI_ISL_2224256, EPI_ISL_2224257, EPI_ISL_2224258, EPI_ISL_2224259, EPI_ISL_2224260, EPI_ISL_2224261, EPI_ISL_2224262, EPI_ISL_2224263, EPI_ISL_2224264, EPI_ISL_2224265, EPI_ISL_2224266, EPI_ISL_2224267, EPI_ISL_2224268, EPI_ISL_2224269, EPI_ISL_2224270, EPI_ISL_2224271, EPI_ISL_2224272, EPI_ISL_2224273, EPI_ISL_2224274, EPI_ISL_2224275, EPI_ISL_2224276, EPI_ISL_2224277, EPI_ISL_2224278, EPI_ISL_2224279, EPI_ISL_2224280, EPI_ISL_2224281, EPI_ISL_2224282, EPI_ISL_2224283, EPI_ISL_2224284, EPI_ISL_2224285, EPI_ISL_2224286, EPI_ISL_2224287, EPI_ISL_2224288, EPI_ISL_2224289, EPI_ISL_2224290, EPI_ISL_2224291, EPI_ISL_2224292, EPI_ISL_2224293, EPI_ISL_2224294, EPI_ISL_2224295, EPI_ISL_2224296, EPI_ISL_2224297, EPI_ISL_2224298, EPI_ISL_2224299, EPI_ISL_2224300, EPI_ISL_2224301, EPI_ISL_2224302, EPI_ISL_2224304 | | | |
| see above | Houston Methodist Hospital | Houston Methodist Hospital | Randall J. Olsen, Paul A. Christensen, S. Wesley Long, Sishir Subedi, Robert Olson, Marcus Nguyen, James J. Davis, Matthew Ojeda Saavedra, Prasanti Yerramilli, Layne Pruitt, Kristina Reppond, Madison N. Shyer, Jessica Cambric, Ryan Gadd, Ilya J. Finkelstein, Jimmy Gollihar, and James M. Musser |
| EPI_ISL_2224305, EPI_ISL_2224306 | University of Mississippi Medical Center, Department of Pathology | University of Mississippi Medical Center, Molecular and Genomics Core Facility | Ashley C. Johnson, Ithiel J. Frame, Krishna K. Ayyalasomayajula, Michael R. Garrett, D. Ashley Robinson |
| EPI_ISL_2224307 | Houston Methodist Hospital | Houston Methodist Hospital | Randall J. Olsen, Paul A. Christensen, S. Wesley Long, Sishir Subedi, Robert Olson, Marcus Nguyen, James J. Davis, Matthew Ojeda Saavedra, Prasanti Yerramilli, Layne Pruitt, Kristina Reppond, Madison N. Shyer, Jessica Cambric, Ryan Gadd, Ilya J. Finkelstein, Jimmy Gollihar, and James M. Musser |
| EPI_ISL_2224308, EPI_ISL_2224309 | University of Mississippi Medical Center, Department of Pathology | University of Mississippi Medical Center, Molecular and Genomics Core Facility | Ashley C. Johnson, Ithiel J. Frame, Krishna K. Ayyalasomayajula, Michael R. Garrett, D. Ashley Robinson |
| EPI_ISL_2224310 | Houston Methodist Hospital | Houston Methodist Hospital | Randall J. Olsen, Paul A. Christensen, S. Wesley Long, Sishir Subedi, Robert Olson, Marcus Nguyen, James J. Davis, Matthew Ojeda Saavedra, Prasanti Yerramilli, Layne Pruitt, Kristina Reppond, Madison N. Shyer, Jessica Cambric, Ryan Gadd, Ilya J. Finkelstein, Jimmy Gollihar, and James M. Musser |
| EPI_ISL_2224311 | University of Mississippi Medical Center, Department of Pathology | University of Mississippi Medical Center, Molecular and Genomics Core Facility | Ashley C. Johnson, Ithiel J. Frame, Krishna K. Ayyalasomayajula, Michael R. Garrett, D. Ashley Robinson |
| EPI_ISL_2224313 | Houston Methodist Hospital | Houston Methodist Hospital | Randall J. Olsen, Paul A. Christensen, S. Wesley Long, Sishir Subedi, Robert Olson, Marcus Nguyen, James J. Davis, Matthew Ojeda Saavedra, Prasanti Yerramilli, Layne Pruitt, Kristina Reppond, Madison N. Shyer, Jessica Cambric, Ryan Gadd, Ilya J. Finkelstein, Jimmy Gollihar, and James M. Musser |
| EPI_ISL_2224314 | University of Mississippi Medical Center, Department of Pathology | University of Mississippi Medical Center, Molecular and Genomics Core Facility | Ashley C. Johnson, Ithiel J. Frame, Krishna K. Ayyalasomayajula, Michael R. Garrett, D. Ashley Robinson |
| EPI_ISL_2224316 | Houston Methodist Hospital | Houston Methodist Hospital | Randall J. Olsen, Paul A. Christensen, S. Wesley Long, Sishir Subedi, Robert Olson, Marcus Nguyen, James J. Davis, Matthew Ojeda Saavedra, Prasanti Yerramilli, Layne Pruitt, Kristina Reppond, Madison N. Shyer, Jessica Cambric, Ryan Gadd, Ilya J. Finkelstein, Jimmy Gollihar, and James M. Musser |
| EPI_ISL_2224317, EPI_ISL_2224318 | University of Mississippi Medical Center, Department of Pathology | University of Mississippi Medical Center, Molecular and Genomics Core Facility | Ashley C. Johnson, Ithiel J. Frame, Krishna K. Ayyalasomayajula, Michael R. Garrett, D. Ashley Robinson |
| EPI_ISL_2224319, EPI_ISL_2224320 | Houston Methodist Hospital | Houston Methodist Hospital | Randall J. Olsen, Paul A. Christensen, S. Wesley Long, Sishir Subedi, Robert Olson, Marcus Nguyen, James J. Davis, Matthew Ojeda Saavedra, Prasanti Yerramilli, Layne Pruitt, Kristina Reppond, Madison N. Shyer, Jessica Cambric, Ryan Gadd, Ilya J. Finkelstein, Jimmy Gollihar, and James M. Musser |
| EPI_ISL_2224321 | University of Mississippi Medical Center, Department of Pathology | University of Mississippi Medical Center, Molecular and Genomics Core Facility | Ashley C. Johnson, Ithiel J. Frame, Krishna K. Ayyalasomayajula, Michael R. Garrett, D. Ashley Robinson |
| EPI_ISL_2224322 | Houston Methodist Hospital | Houston Methodist Hospital | Randall J. Olsen, Paul A. Christensen, S. Wesley Long, Sishir Subedi, Robert Olson, Marcus Nguyen, James J. Davis, Matthew Ojeda Saavedra, Prasanti Yerramilli, Layne Pruitt, Kristina Reppond, Madison N. Shyer, Jessica Cambric, Ryan Gadd, Ilya J. Finkelstein, Jimmy Gollihar, and James M. Musser |
| EPI_ISL_2224323 | University of Mississippi Medical Center, Department of Pathology | University of Mississippi Medical Center, Molecular and Genomics Core Facility | Ashley C. Johnson, Ithiel J. Frame, Krishna K. Ayyalasomayajula, Michael R. Garrett, D. Ashley Robinson |
| EPI_ISL_2224324, EPI_ISL_2224325, EPI_ISL_2224327 | Houston Methodist Hospital | Houston Methodist Hospital | Randall J. Olsen, Paul A. Christensen, S. Wesley Long, Sishir Subedi, Robert Olson, Marcus Nguyen, James J. Davis, Matthew Ojeda Saavedra, Prasanti Yerramilli, Layne Pruitt, Kristina Reppond, Madison N. Shyer, Jessica Cambric, Ryan Gadd, Ilya J. Finkelstein, Jimmy Gollihar, and James M. Musser |
| EPI_ISL_2224328 | University of Mississippi Medical Center, Department of Pathology | University of Mississippi Medical Center, Molecular and Genomics Core Facility | Ashley C. Johnson, Ithiel J. Frame, Krishna K. Ayyalasomayajula, Michael R. Garrett, D. Ashley Robinson |
| EPI_ISL_2224329, EPI_ISL_2224330 | Houston Methodist Hospital | Houston Methodist Hospital | Randall J. Olsen, Paul A. Christensen, S. Wesley Long, Sishir Subedi, Robert Olson, Marcus Nguyen, James J. Davis, Matthew Ojeda Saavedra, Prasanti Yerramilli, Layne Pruitt, Kristina Reppond, Madison N. Shyer, Jessica Cambric, Ryan Gadd, Ilya J. Finkelstein, Jimmy Gollihar, and James M. Musser |
| EPI_ISL_2224331 | University of Mississippi Medical Center, Department of Pathology | University of Mississippi Medical Center, Molecular and Genomics Core Facility | Ashley C. Johnson, Ithiel J. Frame, Krishna K. Ayyalasomayajula, Michael R. Garrett, D. Ashley Robinson |
| EPI_ISL_2224332 | Houston Methodist Hospital | Houston Methodist Hospital | Randall J. Olsen, Paul A. Christensen, S. Wesley Long, Sishir Subedi, Robert Olson, Marcus Nguyen, James J. Davis, Matthew Ojeda Saavedra, Prasanti Yerramilli, Layne Pruitt, Kristina Reppond, Madison N. Shyer, Jessica Cambric, Ryan Gadd, Ilya J. Finkelstein, Jimmy Gollihar, and James M. Musser |
| EPI_ISL_2224333 | MS Public Health Laboratory | University of Mississippi Medical Center, Molecular and Genomics Core Facility | Ashley C. Johnson, Ithiel J. Frame, Krishna K. Ayyalasomayajula, Michael R. Garrett, D. Ashley Robinson |
| EPI_ISL_2224334, EPI_ISL_2224335 | Houston Methodist Hospital | Houston Methodist Hospital | Randall J. Olsen, Paul A. Christensen, S. Wesley Long, Sishir Subedi, Robert Olson, Marcus Nguyen, James J. Davis, Matthew Ojeda Saavedra, Prasanti Yerramilli, Layne Pruitt, Kristina Reppond, Madison N. Shyer, Jessica Cambric, Ryan Gadd, Ilya J. Finkelstein, Jimmy Gollihar, and James M. Musser |
| EPI_ISL_2224336 | MS Public Health Laboratory | University of Mississippi Medical Center, Molecular and Genomics Core Facility | Ashley C. Johnson, Ithiel J. Frame, Krishna K. Ayyalasomayajula, Michael R. Garrett, D. Ashley Robinson |
| EPI_ISL_2224337 | Houston Methodist Hospital | Houston Methodist Hospital | Randall J. Olsen, Paul A. Christensen, S. Wesley Long, Sishir Subedi, Robert Olson, Marcus Nguyen, James J. Davis, Matthew Ojeda Saavedra, Prasanti Yerramilli, Layne Pruitt, Kristina Reppond, Madison N. Shyer, Jessica Cambric, Ryan Gadd, Ilya J. Finkelstein, Jimmy Gollihar, and James M. Musser |
| EPI_ISL_2224338 | MS Public Health Laboratory | University of Mississippi Medical Center, Molecular and Genomics Core Facility | Ashley C. Johnson, Ithiel J. Frame, Krishna K. Ayyalasomayajula, Michael R. Garrett, D. Ashley Robinson |
| EPI_ISL_2224339, EPI_ISL_2224340, EPI_ISL_2224342, EPI_ISL_2224343 | Houston Methodist Hospital | Houston Methodist Hospital | Randall J. Olsen, Paul A. Christensen, S. Wesley Long, Sishir Subedi, Robert Olson, Marcus Nguyen, James J. Davis, Matthew Ojeda Saavedra, Prasanti Yerramilli, Layne Pruitt, Kristina Reppond, Madison N. Shyer, Jessica Cambric, Ryan Gadd, Ilya J. Finkelstein, Jimmy Gollihar, and James M. Musser |
| EPI_ISL_2224344 | MS Public Health Laboratory | University of Mississippi Medical Center, | Ashley C. Johnson, Ithiel J. Frame, Krishna K. Ayyalasomayajula, Michael R. Garrett, D. Ashley Robinson |

| | | | |
|---|---|---|---|
| | | Molecular and Genomics Core Facility | |
| EPI_ISL_2224345, EPI_ISL_2224347, EPI_ISL_2224348, EPI_ISL_2224349, EPI_ISL_2224351, EPI_ISL_2224352, EPI_ISL_2224354 | Houston Methodist Hospital | Houston Methodist Hospital | Randall J. Olsen, Paul A. Christensen, S. Wesley Long, Sishir Subedi, Robert Olson, Marcus Nguyen, James J. Davis, Matthew Ojeda Saavedra, Prasanti Yerramilli, Layne Pruitt, Kristina Reppond, Madison N. Shyer, Jessica Cambric, Ryan Gadd, Ilya J. Finkelstein, Jimmy Gollihar, and James M. Musser |
| EPI_ISL_2224355 | MS Public Health Laboratory | University of Mississippi Medical Center, Molecular and Genomics Core Facility | Ashley C. Johnson, Ithiel J. Frame, Krishna K. Ayyalasomayajula, Michael R. Garrett, D. Ashley Robinson |
| EPI_ISL_2224356, EPI_ISL_2224357 | Houston Methodist Hospital | Houston Methodist Hospital | Randall J. Olsen, Paul A. Christensen, S. Wesley Long, Sishir Subedi, Robert Olson, Marcus Nguyen, James J. Davis, Matthew Ojeda Saavedra, Prasanti Yerramilli, Layne Pruitt, Kristina Reppond, Madison N. Shyer, Jessica Cambric, Ryan Gadd, Ilya J. Finkelstein, Jimmy Gollihar, and James M. Musser |
| EPI_ISL_2224358 | MS Public Health Laboratory | University of Mississippi Medical Center, Molecular and Genomics Core Facility | Ashley C. Johnson, Ithiel J. Frame, Krishna K. Ayyalasomayajula, Michael R. Garrett, D. Ashley Robinson |
| EPI_ISL_2224359 | Houston Methodist Hospital | Houston Methodist Hospital | Randall J. Olsen, Paul A. Christensen, S. Wesley Long, Sishir Subedi, Robert Olson, Marcus Nguyen, James J. Davis, Matthew Ojeda Saavedra, Prasanti Yerramilli, Layne Pruitt, Kristina Reppond, Madison N. Shyer, Jessica Cambric, Ryan Gadd, Ilya J. Finkelstein, Jimmy Gollihar, and James M. Musser |
| EPI_ISL_2224360 | MS Public Health Laboratory | University of Mississippi Medical Center, Molecular and Genomics Core Facility | Ashley C. Johnson, Ithiel J. Frame, Krishna K. Ayyalasomayajula, Michael R. Garrett, D. Ashley Robinson |
| EPI_ISL_2224361, EPI_ISL_2224362 | Houston Methodist Hospital | Houston Methodist Hospital | Randall J. Olsen, Paul A. Christensen, S. Wesley Long, Sishir Subedi, Robert Olson, Marcus Nguyen, James J. Davis, Matthew Ojeda Saavedra, Prasanti Yerramilli, Layne Pruitt, Kristina Reppond, Madison N. Shyer, Jessica Cambric, Ryan Gadd, Ilya J. Finkelstein, Jimmy Gollihar, and James M. Musser |
| EPI_ISL_2224363 | MS Public Health Laboratory | University of Mississippi Medical Center, Molecular and Genomics Core Facility | Ashley C. Johnson, Ithiel J. Frame, Krishna K. Ayyalasomayajula, Michael R. Garrett, D. Ashley Robinson |
| EPI_ISL_2224364, EPI_ISL_2224365, EPI_ISL_2224366, EPI_ISL_2224368, EPI_ISL_2224370, EPI_ISL_2224371 | Houston Methodist Hospital | Houston Methodist Hospital | Randall J. Olsen, Paul A. Christensen, S. Wesley Long, Sishir Subedi, Robert Olson, Marcus Nguyen, James J. Davis, Matthew Ojeda Saavedra, Prasanti Yerramilli, Layne Pruitt, Kristina Reppond, Madison N. Shyer, Jessica Cambric, Ryan Gadd, Ilya J. Finkelstein, Jimmy Gollihar, and James M. Musser |
| EPI_ISL_2224372 | MS Public Health Laboratory | University of Mississippi Medical Center, Molecular and Genomics Core Facility | Ashley C. Johnson, Ithiel J. Frame, Krishna K. Ayyalasomayajula, Michael R. Garrett, D. Ashley Robinson |
| EPI_ISL_2224373, EPI_ISL_2224374 | Houston Methodist Hospital | Houston Methodist Hospital | Randall J. Olsen, Paul A. Christensen, S. Wesley Long, Sishir Subedi, Robert Olson, Marcus Nguyen, James J. Davis, Matthew Ojeda Saavedra, Prasanti Yerramilli, Layne Pruitt, Kristina Reppond, Madison N. Shyer, Jessica Cambric, Ryan Gadd, Ilya J. Finkelstein, Jimmy Gollihar, and James M. Musser |
| EPI_ISL_2224375 | MS Public Health Laboratory | University of Mississippi Medical Center, Molecular and Genomics Core Facility | Ashley C. Johnson, Ithiel J. Frame, Krishna K. Ayyalasomayajula, Michael R. Garrett, D. Ashley Robinson |
| EPI_ISL_2224376, EPI_ISL_2224377 | Houston Methodist Hospital | Houston Methodist Hospital | Randall J. Olsen, Paul A. Christensen, S. Wesley Long, Sishir Subedi, Robert Olson, Marcus Nguyen, James J. Davis, Matthew Ojeda Saavedra, Prasanti Yerramilli, Layne Pruitt, Kristina Reppond, Madison N. Shyer, Jessica Cambric, Ryan Gadd, Ilya J. Finkelstein, Jimmy Gollihar, and James M. Musser |
| EPI_ISL_2224378 | MS Public Health Laboratory | University of Mississippi Medical Center, Molecular and Genomics Core Facility | Ashley C. Johnson, Ithiel J. Frame, Krishna K. Ayyalasomayajula, Michael R. Garrett, D. Ashley Robinson |
| EPI_ISL_2224379, EPI_ISL_2224380 | Houston Methodist Hospital | Houston Methodist Hospital | Randall J. Olsen, Paul A. Christensen, S. Wesley Long, Sishir Subedi, Robert Olson, Marcus Nguyen, James J. Davis, Matthew Ojeda Saavedra, Prasanti Yerramilli, Layne Pruitt, Kristina Reppond, Madison N. Shyer, Jessica Cambric, Ryan Gadd, Ilya J. Finkelstein, Jimmy Gollihar, and James M. Musser |
| EPI_ISL_2224381 | MS Public Health Laboratory | University of Mississippi Medical Center, Molecular and Genomics Core Facility | Ashley C. Johnson, Ithiel J. Frame, Krishna K. Ayyalasomayajula, Michael R. Garrett, D. Ashley Robinson |
| EPI_ISL_2224382 | Houston Methodist Hospital | Houston Methodist Hospital | Randall J. Olsen, Paul A. Christensen, S. Wesley Long, Sishir Subedi, Robert Olson, Marcus Nguyen, James J. Davis, Matthew Ojeda Saavedra, Prasanti Yerramilli, Layne Pruitt, Kristina Reppond, Madison N. Shyer, Jessica Cambric, Ryan Gadd, Ilya J. Finkelstein, Jimmy Gollihar, and James M. Musser |
| EPI_ISL_2224383 | MS Public Health Laboratory | University of Mississippi Medical Center, Molecular and Genomics Core Facility | Ashley C. Johnson, Ithiel J. Frame, Krishna K. Ayyalasomayajula, Michael R. Garrett, D. Ashley Robinson |
| EPI_ISL_2224384 | Houston Methodist Hospital | Houston Methodist Hospital | Randall J. Olsen, Paul A. Christensen, S. Wesley Long, Sishir Subedi, Robert Olson, Marcus Nguyen, James J. Davis, Matthew Ojeda Saavedra, Prasanti Yerramilli, Layne Pruitt, Kristina Reppond, Madison N. Shyer, Jessica Cambric, Ryan Gadd, Ilya J. Finkelstein, Jimmy Gollihar, and James M. Musser |
| EPI_ISL_2224385 | MS Public Health Laboratory | University of Mississippi Medical Center, Molecular and Genomics Core Facility | Ashley C. Johnson, Ithiel J. Frame, Krishna K. Ayyalasomayajula, Michael R. Garrett, D. Ashley Robinson |
| EPI_ISL_2224386, EPI_ISL_2224387, EPI_ISL_2224389, EPI_ISL_2224390 | Houston Methodist Hospital | Houston Methodist Hospital | Randall J. Olsen, Paul A. Christensen, S. Wesley Long, Sishir Subedi, Robert Olson, Marcus Nguyen, James J. Davis, Matthew Ojeda Saavedra, Prasanti Yerramilli, Layne Pruitt, Kristina Reppond, Madison N. Shyer, Jessica Cambric, Ryan Gadd, Ilya J. Finkelstein, Jimmy Gollihar, and James M. Musser |
| EPI_ISL_2224391 | MS Public Health Laboratory | University of Mississippi Medical Center, Molecular and Genomics Core Facility | Ashley C. Johnson, Ithiel J. Frame, Krishna K. Ayyalasomayajula, Michael R. Garrett, D. Ashley Robinson |
| EPI_ISL_2224392, EPI_ISL_2224393 | Houston Methodist Hospital | Houston Methodist Hospital | Randall J. Olsen, Paul A. Christensen, S. Wesley Long, Sishir Subedi, Robert Olson, Marcus Nguyen, James J. Davis, Matthew Ojeda Saavedra, Prasanti Yerramilli, Layne Pruitt, Kristina Reppond, Madison N. Shyer, Jessica Cambric, Ryan Gadd, Ilya J. Finkelstein, Jimmy Gollihar, and James M. Musser |
| EPI_ISL_2224394 | MS Public Health Laboratory | University of Mississippi Medical Center, Molecular and Genomics Core Facility | Ashley C. Johnson, Ithiel J. Frame, Krishna K. Ayyalasomayajula, Michael R. Garrett, D. Ashley Robinson |
| EPI_ISL_2224395, EPI_ISL_2224396 | Houston Methodist Hospital | Houston Methodist Hospital | Randall J. Olsen, Paul A. Christensen, S. Wesley Long, Sishir Subedi, Robert Olson, Marcus Nguyen, James J. Davis, Matthew Ojeda Saavedra, Prasanti Yerramilli, Layne Pruitt, Kristina Reppond, Madison N. Shyer, Jessica Cambric, Ryan Gadd, Ilya J. Finkelstein, Jimmy Gollihar, and James M. Musser |
| EPI_ISL_2224397 | MS Public Health Laboratory | University of Mississippi Medical Center, Molecular and Genomics Core Facility | Ashley C. Johnson, Ithiel J. Frame, Krishna K. Ayyalasomayajula, Michael R. Garrett, D. Ashley Robinson |
| EPI_ISL_2224398, EPI_ISL_2224399 | Houston Methodist Hospital | Houston Methodist Hospital | Randall J. Olsen, Paul A. Christensen, S. Wesley Long, Sishir Subedi, Robert Olson, Marcus Nguyen, James J. Davis, Matthew Ojeda Saavedra, Prasanti Yerramilli, Layne Pruitt, Kristina Reppond, Madison N. Shyer, Jessica Cambric, Ryan Gadd, Ilya J. Finkelstein, Jimmy Gollihar, and James M. Musser |
| EPI_ISL_2224400 | MS Public Health Laboratory | University of Mississippi Medical Center, Molecular and Genomics Core Facility | Ashley C. Johnson, Ithiel J. Frame, Krishna K. Ayyalasomayajula, Michael R. Garrett, D. Ashley Robinson |
| EPI_ISL_2224401 | Houston Methodist Hospital | Houston Methodist Hospital | Randall J. Olsen, Paul A. Christensen, S. Wesley Long, Sishir Subedi, Robert Olson, Marcus Nguyen, James J. Davis, Matthew Ojeda Saavedra, Prasanti Yerramilli, Layne Pruitt, Kristina Reppond, Madison N. Shyer, Jessica Cambric, Ryan Gadd, Ilya J. Finkelstein, Jimmy Gollihar, and James M. Musser |
| EPI_ISL_2224402 | MS Public Health Laboratory | University of Mississippi Medical Center, Molecular and Genomics Core Facility | Ashley C. Johnson, Ithiel J. Frame, Krishna K. Ayyalasomayajula, Michael R. Garrett, D. Ashley Robinson |
| EPI_ISL_2224403, EPI_ISL_2224404, EPI_ISL_2224406 | Houston Methodist Hospital | Houston Methodist Hospital | Randall J. Olsen, Paul A. Christensen, S. Wesley Long, Sishir Subedi, Robert Olson, Marcus Nguyen, James J. Davis, Matthew Ojeda Saavedra, Prasanti Yerramilli, Layne Pruitt, Kristina Reppond, Madison N. Shyer, Jessica Cambric, Ryan Gadd, Ilya J. Finkelstein, Jimmy Gollihar, and James M. Musser |
| EPI_ISL_2224407 | MS Public Health Laboratory | University of Mississippi Medical Center, Molecular and Genomics Core Facility | Ashley C. Johnson, Ithiel J. Frame, Krishna K. Ayyalasomayajula, Michael R. Garrett, D. Ashley Robinson |
| EPI_ISL_2224408, EPI_ISL_2224409, EPI_ISL_2224411, EPI_ISL_2224412 | Houston Methodist Hospital | Houston Methodist Hospital | Randall J. Olsen, Paul A. Christensen, S. Wesley Long, Sishir Subedi, Robert Olson, Marcus Nguyen, James J. Davis, Matthew Ojeda Saavedra, Prasanti Yerramilli, Layne Pruitt, Kristina Reppond, Madison N. Shyer, Jessica Cambric, Ryan Gadd, Ilya J. Finkelstein, Jimmy Gollihar, and James M. Musser |
| EPI_ISL_2224413 | MS Public Health Laboratory | University of Mississippi Medical Center, Molecular and Genomics Core Facility | Ashley C. Johnson, Ithiel J. Frame, Krishna K. Ayyalasomayajula, Michael R. Garrett, D. Ashley Robinson |
| EPI_ISL_2224414, EPI_ISL_2224415 | Houston Methodist Hospital | Houston Methodist Hospital | Randall J. Olsen, Paul A. Christensen, S. Wesley Long, Sishir Subedi, Robert Olson, Marcus Nguyen, James J. Davis, Matthew Ojeda Saavedra, Prasanti Yerramilli, Layne Pruitt, Kristina Reppond, Madison N. Shyer, Jessica Cambric, Ryan Gadd, Ilya J. Finkelstein, Jimmy Gollihar, and James M. Musser |
| EPI_ISL_2224416 | MS Public Health Laboratory | University of Mississippi Medical Center, Molecular and Genomics Core Facility | Ashley C. Johnson, Ithiel J. Frame, Krishna K. Ayyalasomayajula, Michael R. Garrett, D. Ashley Robinson |
| EPI_ISL_2224417, EPI_ISL_2224418, EPI_ISL_2224420, EPI_ISL_2224421 | Houston Methodist Hospital | Houston Methodist Hospital | Randall J. Olsen, Paul A. Christensen, S. Wesley Long, Sishir Subedi, Robert Olson, Marcus Nguyen, James J. Davis, Matthew Ojeda Saavedra, Prasanti Yerramilli, Layne Pruitt, Kristina Reppond, Madison N. Shyer, Jessica Cambric, Ryan Gadd, Ilya J. Finkelstein, Jimmy Gollihar, and James M. Musser |
| EPI_ISL_2224422 | MS Public Health Laboratory | University of Mississippi Medical Center, Molecular and Genomics Core Facility | Ashley C. Johnson, Ithiel J. Frame, Krishna K. Ayyalasomayajula, Michael R. Garrett, D. Ashley Robinson |

| | | | |
|---|---|---|---|
| EPI_ISL_2224423 | Houston Methodist Hospital | Houston Methodist Hospital | Randall J. Olsen, Paul A. Christensen, S. Wesley Long, Sishir Subedi, Robert Olson, Marcus Nguyen, James J. Davis, Matthew Ojeda Saavedra, Prasanti Yerramilli, Layne Pruitt, Kristina Reppond, Madison N. Shyer, Jessica Cambric, Ryan Gadd, Ilya J. Finkelstein, Jimmy Gollihar, and James M. Musser |
| EPI_ISL_2224424 | MS Public Health Laboratory | University of Mississippi Medical Center, Molecular and Genomics Core Facility | Ashley C. Johnson, Ithiel J. Frame, Krishna K. Ayyalasomayajula, Michael R. Garrett, D. Ashley Robinson |
| EPI_ISL_2224425, EPI_ISL_2224426, EPI_ISL_2224428, EPI_ISL_2224429 | Houston Methodist Hospital | Houston Methodist Hospital | Randall J. Olsen, Paul A. Christensen, S. Wesley Long, Sishir Subedi, Robert Olson, Marcus Nguyen, James J. Davis, Matthew Ojeda Saavedra, Prasanti Yerramilli, Layne Pruitt, Kristina Reppond, Madison N. Shyer, Jessica Cambric, Ryan Gadd, Ilya J. Finkelstein, Jimmy Gollihar, and James M. Musser |
| EPI_ISL_2224430 | MS Public Health Laboratory | University of Mississippi Medical Center, Molecular and Genomics Core Facility | Ashley C. Johnson, Ithiel J. Frame, Krishna K. Ayyalasomayajula, Michael R. Garrett, D. Ashley Robinson |
| EPI_ISL_2224431, EPI_ISL_2224432, EPI_ISL_2224434 | Houston Methodist Hospital | Houston Methodist Hospital | Randall J. Olsen, Paul A. Christensen, S. Wesley Long, Sishir Subedi, Robert Olson, Marcus Nguyen, James J. Davis, Matthew Ojeda Saavedra, Prasanti Yerramilli, Layne Pruitt, Kristina Reppond, Madison N. Shyer, Jessica Cambric, Ryan Gadd, Ilya J. Finkelstein, Jimmy Gollihar, and James M. Musser |
| EPI_ISL_2224435 | MS Public Health Laboratory | University of Mississippi Medical Center, Molecular and Genomics Core Facility | Ashley C. Johnson, Ithiel J. Frame, Krishna K. Ayyalasomayajula, Michael R. Garrett, D. Ashley Robinson |
| EPI_ISL_2224436, EPI_ISL_2224437, EPI_ISL_2224439, EPI_ISL_2224440 | Houston Methodist Hospital | Houston Methodist Hospital | Randall J. Olsen, Paul A. Christensen, S. Wesley Long, Sishir Subedi, Robert Olson, Marcus Nguyen, James J. Davis, Matthew Ojeda Saavedra, Prasanti Yerramilli, Layne Pruitt, Kristina Reppond, Madison N. Shyer, Jessica Cambric, Ryan Gadd, Ilya J. Finkelstein, Jimmy Gollihar, and James M. Musser |
| EPI_ISL_2224441 | MS Public Health Laboratory | University of Mississippi Medical Center, Molecular and Genomics Core Facility | Ashley C. Johnson, Ithiel J. Frame, Krishna K. Ayyalasomayajula, Michael R. Garrett, D. Ashley Robinson |
| EPI_ISL_2224442, EPI_ISL_2224443 | Houston Methodist Hospital | Houston Methodist Hospital | Randall J. Olsen, Paul A. Christensen, S. Wesley Long, Sishir Subedi, Robert Olson, Marcus Nguyen, James J. Davis, Matthew Ojeda Saavedra, Prasanti Yerramilli, Layne Pruitt, Kristina Reppond, Madison N. Shyer, Jessica Cambric, Ryan Gadd, Ilya J. Finkelstein, Jimmy Gollihar, and James M. Musser |
| EPI_ISL_2224444 | MS Public Health Laboratory | University of Mississippi Medical Center, Molecular and Genomics Core Facility | Ashley C. Johnson, Ithiel J. Frame, Krishna K. Ayyalasomayajula, Michael R. Garrett, D. Ashley Robinson |
| EPI_ISL_2224445, EPI_ISL_2224446, EPI_ISL_2224448, EPI_ISL_2224449 | Houston Methodist Hospital | Houston Methodist Hospital | Randall J. Olsen, Paul A. Christensen, S. Wesley Long, Sishir Subedi, Robert Olson, Marcus Nguyen, James J. Davis, Matthew Ojeda Saavedra, Prasanti Yerramilli, Layne Pruitt, Kristina Reppond, Madison N. Shyer, Jessica Cambric, Ryan Gadd, Ilya J. Finkelstein, Jimmy Gollihar, and James M. Musser |
| EPI_ISL_2224450 | MS Public Health Laboratory | University of Mississippi Medical Center, Molecular and Genomics Core Facility | Ashley C. Johnson, Ithiel J. Frame, Krishna K. Ayyalasomayajula, Michael R. Garrett, D. Ashley Robinson |
| EPI_ISL_2224451, EPI_ISL_2224452 | Houston Methodist Hospital | Houston Methodist Hospital | Randall J. Olsen, Paul A. Christensen, S. Wesley Long, Sishir Subedi, Robert Olson, Marcus Nguyen, James J. Davis, Matthew Ojeda Saavedra, Prasanti Yerramilli, Layne Pruitt, Kristina Reppond, Madison N. Shyer, Jessica Cambric, Ryan Gadd, Ilya J. Finkelstein, Jimmy Gollihar, and James M. Musser |
| EPI_ISL_2224453 | MS Public Health Laboratory | University of Mississippi Medical Center, Molecular and Genomics Core Facility | Ashley C. Johnson, Ithiel J. Frame, Krishna K. Ayyalasomayajula, Michael R. Garrett, D. Ashley Robinson |
| EPI_ISL_2224454, EPI_ISL_2224455 | Houston Methodist Hospital | Houston Methodist Hospital | Randall J. Olsen, Paul A. Christensen, S. Wesley Long, Sishir Subedi, Robert Olson, Marcus Nguyen, James J. Davis, Matthew Ojeda Saavedra, Prasanti Yerramilli, Layne Pruitt, Kristina Reppond, Madison N. Shyer, Jessica Cambric, Ryan Gadd, Ilya J. Finkelstein, Jimmy Gollihar, and James M. Musser |
| EPI_ISL_2224456 | MS Public Health Laboratory | University of Mississippi Medical Center, Molecular and Genomics Core Facility | Ashley C. Johnson, Ithiel J. Frame, Krishna K. Ayyalasomayajula, Michael R. Garrett, D. Ashley Robinson |
| EPI_ISL_2224457, EPI_ISL_2224458, EPI_ISL_2224460, EPI_ISL_2224461 | Houston Methodist Hospital | Houston Methodist Hospital | Randall J. Olsen, Paul A. Christensen, S. Wesley Long, Sishir Subedi, Robert Olson, Marcus Nguyen, James J. Davis, Matthew Ojeda Saavedra, Prasanti Yerramilli, Layne Pruitt, Kristina Reppond, Madison N. Shyer, Jessica Cambric, Ryan Gadd, Ilya J. Finkelstein, Jimmy Gollihar, and James M. Musser |
| EPI_ISL_2224462 | MS Public Health Laboratory | University of Mississippi Medical Center, Molecular and Genomics Core Facility | Ashley C. Johnson, Ithiel J. Frame, Krishna K. Ayyalasomayajula, Michael R. Garrett, D. Ashley Robinson |
| EPI_ISL_2224463, EPI_ISL_2224464 | Houston Methodist Hospital | Houston Methodist Hospital | Randall J. Olsen, Paul A. Christensen, S. Wesley Long, Sishir Subedi, Robert Olson, Marcus Nguyen, James J. Davis, Matthew Ojeda Saavedra, Prasanti Yerramilli, Layne Pruitt, Kristina Reppond, Madison N. Shyer, Jessica Cambric, Ryan Gadd, Ilya J. Finkelstein, Jimmy Gollihar, and James M. Musser |
| EPI_ISL_2224465 | MS Public Health Laboratory | University of Mississippi Medical Center, Molecular and Genomics Core Facility | Ashley C. Johnson, Ithiel J. Frame, Krishna K. Ayyalasomayajula, Michael R. Garrett, D. Ashley Robinson |
| EPI_ISL_2224466, EPI_ISL_2224467, EPI_ISL_2224469, EPI_ISL_2224470, EPI_ISL_2224472, EPI_ISL_2224474, EPI_ISL_2224475, EPI_ISL_2224476, EPI_ISL_2224477, EPI_ISL_2224479, EPI_ISL_2224480, EPI_ISL_2224481, EPI_ISL_2224482, EPI_ISL_2224483, EPI_ISL_2224484, EPI_ISL_2224485, EPI_ISL_2224486, EPI_ISL_2224487, EPI_ISL_2224488, EPI_ISL_2224489, EPI_ISL_2224490, EPI_ISL_2224491, EPI_ISL_2224492, EPI_ISL_2224493, EPI_ISL_2224494, EPI_ISL_2224495, EPI_ISL_2224496, EPI_ISL_2224497, EPI_ISL_2224498, EPI_ISL_2224499, EPI_ISL_2224500, EPI_ISL_2224501, EPI_ISL_2224502, EPI_ISL_2224503, EPI_ISL_2224504, EPI_ISL_2224505, EPI_ISL_2224506, EPI_ISL_2224507, EPI_ISL_2224508, EPI_ISL_2224509, EPI_ISL_2224510, EPI_ISL_2224511, EPI_ISL_2224512, EPI_ISL_2224513, EPI_ISL_2224514, EPI_ISL_2224515, EPI_ISL_2224516, EPI_ISL_2224517, EPI_ISL_2224518, EPI_ISL_2224519, EPI_ISL_2224520, EPI_ISL_2224521, EPI_ISL_2224522, EPI_ISL_2224523, EPI_ISL_2224524, EPI_ISL_2224525, EPI_ISL_2224526, EPI_ISL_2224527, EPI_ISL_2224528, EPI_ISL_2224529, EPI_ISL_2224530, EPI_ISL_2224531, EPI_ISL_2224532, EPI_ISL_2224533, EPI_ISL_2224534, EPI_ISL_2224535, EPI_ISL_2224536, EPI_ISL_2224537, EPI_ISL_2224538, EPI_ISL_2224539, EPI_ISL_2224540, EPI_ISL_2224541, EPI_ISL_2224542, EPI_ISL_2224543, EPI_ISL_2224544, EPI_ISL_2224545, EPI_ISL_2224546, EPI_ISL_2224547, EPI_ISL_2224548, EPI_ISL_2224549, EPI_ISL_2224550, EPI_ISL_2224551, EPI_ISL_2224552, EPI_ISL_2224553, EPI_ISL_2224554, EPI_ISL_2224555, EPI_ISL_2224556, EPI_ISL_2224557, EPI_ISL_2224558, EPI_ISL_2224559, EPI_ISL_2224560, EPI_ISL_2224561, EPI_ISL_2224562, EPI_ISL_2224563, EPI_ISL_2224564, EPI_ISL_2224565, EPI_ISL_2224566, EPI_ISL_2224567, EPI_ISL_2224568, EPI_ISL_2224569, EPI_ISL_2224570, EPI_ISL_2224571, EPI_ISL_2224572, EPI_ISL_2224573, EPI_ISL_2224574, EPI_ISL_2224575, EPI_ISL_2224576, EPI_ISL_2224577, EPI_ISL_2224578, EPI_ISL_2224579, EPI_ISL_2224580, EPI_ISL_2224581, EPI_ISL_2224582, EPI_ISL_2224583, EPI_ISL_2224584, EPI_ISL_2224585, EPI_ISL_2224586, EPI_ISL_2224587, EPI_ISL_2224588, EPI_ISL_2224589, EPI_ISL_2224590, EPI_ISL_2224591, EPI_ISL_2224592, EPI_ISL_2224593, EPI_ISL_2224594, EPI_ISL_2224595, EPI_ISL_2224596, EPI_ISL_2224597, EPI_ISL_2224598, EPI_ISL_2224599, EPI_ISL_2224600, EPI_ISL_2224601, EPI_ISL_2224602, EPI_ISL_2224603, EPI_ISL_2224604, EPI_ISL_2224605, EPI_ISL_2224606, EPI_ISL_2224607, EPI_ISL_2224608, EPI_ISL_2224609, EPI_ISL_2224610, EPI_ISL_2224611, EPI_ISL_2224612, EPI_ISL_2224613, EPI_ISL_2224614, EPI_ISL_2224615, EPI_ISL_2224616, EPI_ISL_2224617, EPI_ISL_2224618, EPI_ISL_2224619, EPI_ISL_2224620, EPI_ISL_2224621, EPI_ISL_2224622, EPI_ISL_2224623, EPI_ISL_2224624, EPI_ISL_2224625, EPI_ISL_2224626, EPI_ISL_2224627, EPI_ISL_2224628, EPI_ISL_2224629, EPI_ISL_2224630, EPI_ISL_2224631, EPI_ISL_2224632, EPI_ISL_2224633, EPI_ISL_2224634, EPI_ISL_2224635, EPI_ISL_2224636, EPI_ISL_2224637, EPI_ISL_2224638, EPI_ISL_2224639, EPI_ISL_2224640, EPI_ISL_2224641, EPI_ISL_2224642, EPI_ISL_2224643, EPI_ISL_2224644, EPI_ISL_2224645, EPI_ISL_2224647, EPI_ISL_2224648, EPI_ISL_2224649, EPI_ISL_2224650, EPI_ISL_2224651, EPI_ISL_2224652, EPI_ISL_2224653, EPI_ISL_2224655, EPI_ISL_2224656, EPI_ISL_2224657, EPI_ISL_2224658, EPI_ISL_2224659, EPI_ISL_2224660, EPI_ISL_2224661, EPI_ISL_2224662, EPI_ISL_2224663, EPI_ISL_2224664, EPI_ISL_2224665, EPI_ISL_2224666, EPI_ISL_2224667, EPI_ISL_2224668, EPI_ISL_2224669, EPI_ISL_2224670, EPI_ISL_2224671, EPI_ISL_2224672, EPI_ISL_2224673, EPI_ISL_2224674, EPI_ISL_2224675, EPI_ISL_2224676, EPI_ISL_2224677, EPI_ISL_2224678, EPI_ISL_2224679, EPI_ISL_2224680, EPI_ISL_2224681, EPI_ISL_2224682, EPI_ISL_2224683, EPI_ISL_2224684, EPI_ISL_2224685, EPI_ISL_2224686, EPI_ISL_2224687, EPI_ISL_2224688, EPI_ISL_2224689, EPI_ISL_2224690, EPI_ISL_2224691, EPI_ISL_2224692, EPI_ISL_2224693, EPI_ISL_2224694, EPI_ISL_2224695, EPI_ISL_2224696, EPI_ISL_2224697, EPI_ISL_2224698, EPI_ISL_2224699, EPI_ISL_2224700, EPI_ISL_2224701 | | | |
| see above | Houston Methodist Hospital | Houston Methodist Hospital | Randall J. Olsen, Paul A. Christensen, S. Wesley Long, Sishir Subedi, Robert Olson, Marcus Nguyen, James J. Davis, Matthew Ojeda Saavedra, Prasanti Yerramilli, Layne Pruitt, Kristina Reppond, Madison N. Shyer, Jessica Cambric, Ryan Gadd, Ilya J. Finkelstein, Jimmy Gollihar, and James M. Musser |
| EPI_ISL_2224702 | Hackensack Medical Center | New York Genome Center | Michael Zody, Andre Corvelo, Dayna M. Oschwald, Samantha Fennessey, Tom Maniatis, Liang Chen, Jose Mediavilla, Marcus Cunningham, Kaelea Composto, Kar Chow, David Perlin, Barry Kreiswirth |
| EPI_ISL_2224703 | Houston Methodist Hospital | Houston Methodist Hospital | Randall J. Olsen, Paul A. Christensen, S. Wesley Long, Sishir Subedi, Robert Olson, Marcus Nguyen, James J. Davis, Matthew Ojeda Saavedra, Prasanti Yerramilli, Layne Pruitt, Kristina Reppond, Madison N. Shyer, Jessica Cambric, Ryan Gadd, Ilya J. Finkelstein, Jimmy Gollihar, and James M. Musser |
| EPI_ISL_2224704, EPI_ISL_2224705 | Hackensack Medical Center | New York Genome Center | Michael Zody, Andre Corvelo, Dayna M. Oschwald, Samantha Fennessey, Tom Maniatis, Liang Chen, Jose Mediavilla, Marcus Cunningham, Kaelea Composto, Kar Chow, David Perlin, Barry Kreiswirth |
| EPI_ISL_2224706 | Houston Methodist Hospital | Houston Methodist Hospital | Randall J. Olsen, Paul A. Christensen, S. Wesley Long, Sishir Subedi, Robert Olson, Marcus Nguyen, James J. Davis, Matthew Ojeda Saavedra, Prasanti Yerramilli, Layne Pruitt, Kristina Reppond, Madison N. Shyer, Jessica Cambric, Ryan Gadd, Ilya J. Finkelstein, Jimmy Gollihar, and James M. Musser |
| EPI_ISL_2224707 | Hackensack Medical Center | New York Genome Center | Michael Zody, Andre Corvelo, Dayna M. Oschwald, Samantha Fennessey, Tom Maniatis, Liang Chen, Jose Mediavilla, Marcus Cunningham, Kaelea Composto, Kar Chow, David Perlin, Barry Kreiswirth |
| EPI_ISL_2224708, EPI_ISL_2224709, EPI_ISL_2224710, EPI_ISL_2224711, EPI_ISL_2224712, EPI_ISL_2224713, EPI_ISL_2224714, EPI_ISL_2224715, EPI_ISL_2224716, EPI_ISL_2224717, EPI_ISL_2224718, EPI_ISL_2224719, EPI_ISL_2224720, EPI_ISL_2224721, EPI_ISL_2224722, EPI_ISL_2224723, EPI_ISL_2224724, EPI_ISL_2224725, EPI_ISL_2224727, EPI_ISL_2224728, EPI_ISL_2224729, EPI_ISL_2224730, EPI_ISL_2224731, EPI_ISL_2224732, EPI_ISL_2224733, EPI_ISL_2224734, EPI_ISL_2224735, EPI_ISL_2224736, EPI_ISL_2224737, EPI_ISL_2224738, EPI_ISL_2224739, EPI_ISL_2224740, EPI_ISL_2224741, EPI_ISL_2224742, EPI_ISL_2224744, EPI_ISL_2224745, EPI_ISL_2224746, EPI_ISL_2224747, EPI_ISL_2224749, EPI_ISL_2224750, EPI_ISL_2224753, EPI_ISL_2224757, EPI_ISL_2224759, EPI_ISL_2224765, EPI_ISL_2224766, EPI_ISL_2224767, EPI_ISL_2224768, EPI_ISL_2224770, EPI_ISL_2224771, EPI_ISL_2224772, EPI_ISL_2224773, EPI_ISL_2224774, EPI_ISL_2224776, EPI_ISL_2224777, EPI_ISL_2224778, EPI_ISL_2224779, EPI_ISL_2224780, EPI_ISL_2224781, EPI_ISL_2224782, EPI_ISL_2224784, EPI_ISL_2224785, EPI_ISL_2224786, EPI_ISL_2224787, EPI_ISL_2224788, EPI_ISL_2224789, EPI_ISL_2224790, EPI_ISL_2224791, EPI_ISL_2224793, EPI_ISL_2224794, EPI_ISL_2224795, EPI_ISL_2224796, EPI_ISL_2224797, EPI_ISL_2224798, EPI_ISL_2224799, EPI_ISL_2224801, EPI_ISL_2224803, EPI_ISL_2224804, EPI_ISL_2224805, EPI_ISL_2224806, EPI_ISL_2224809, EPI_ISL_2224810, EPI_ISL_2224814, EPI_ISL_2224815, EPI_ISL_2224816, EPI_ISL_2224817, EPI_ISL_2224818, EPI_ISL_2224819, EPI_ISL_2224820, EPI_ISL_2224822, EPI_ISL_2224823, EPI_ISL_2224824, EPI_ISL_2224825, EPI_ISL_2224826, EPI_ISL_2224827, EPI_ISL_2224828, EPI_ISL_2224829, EPI_ISL_2224830, EPI_ISL_2224831, EPI_ISL_2224832, EPI_ISL_2224833, EPI_ISL_2224834, EPI_ISL_2224835, EPI_ISL_2224836, EPI_ISL_2224837, EPI_ISL_2224838, EPI_ISL_2224839, EPI_ISL_2224840, EPI_ISL_2224841, EPI_ISL_2224842, EPI_ISL_2224843, EPI_ISL_2224844, EPI_ISL_2224848, EPI_ISL_2224849, EPI_ISL_2224850, EPI_ISL_2224851, EPI_ISL_2224854, EPI_ISL_2224855, EPI_ISL_2224859, EPI_ISL_2224860, EPI_ISL_2224863, EPI_ISL_2224864, EPI_ISL_2224865, EPI_ISL_2224866, EPI_ISL_2224867, EPI_ISL_2224868, EPI_ISL_2224869, EPI_ISL_2224870, EPI_ISL_2224871, EPI_ISL_2224872, EPI_ISL_2224873, EPI_ISL_2224874, EPI_ISL_2224875, EPI_ISL_2224876, EPI_ISL_2224877, EPI_ISL_2224878, EPI_ISL_2224879, EPI_ISL_2224882, EPI_ISL_2224884, EPI_ISL_2224885, EPI_ISL_2224886, EPI_ISL_2224887, EPI_ISL_2224888, EPI_ISL_2224890, EPI_ISL_2224891, EPI_ISL_2224892, EPI_ISL_2224896, EPI_ISL_2224899, EPI_ISL_2224900, EPI_ISL_2224901, EPI_ISL_2224902, EPI_ISL_2224907, EPI_ISL_2224908, EPI_ISL_2224909, EPI_ISL_2224912, EPI_ISL_2224913, EPI_ISL_2224914, EPI_ISL_2224915, EPI_ISL_2224916, EPI_ISL_2224917, EPI_ISL_2224918, EPI_ISL_2224920, EPI_ISL_2224921, EPI_ISL_2224922, EPI_ISL_2224923, EPI_ISL_2224924, EPI_ISL_2224925, EPI_ISL_2224926, EPI_ISL_2224927, EPI_ISL_2224928, EPI_ISL_2224929, EPI_ISL_2224930, EPI_ISL_2224933, EPI_ISL_2224934, EPI_ISL_2224935, EPI_ISL_2224936, EPI_ISL_2224937, EPI_ISL_2224938, EPI_ISL_2224939, EPI_ISL_2224940, EPI_ISL_2224941, EPI_ISL_2224942, EPI_ISL_2224943, EPI_ISL_2224944, EPI_ISL_2224945, EPI_ISL_2224946, EPI_ISL_2224947, EPI_ISL_2224948, EPI_ISL_2224949, EPI_ISL_2224950, EPI_ISL_2224951, EPI_ISL_2224952, EPI_ISL_2224953, EPI_ISL_2224954, EPI_ISL_2224955, EPI_ISL_2224956, EPI_ISL_2224957, EPI_ISL_2224958, EPI_ISL_2224959, EPI_ISL_2224960, EPI_ISL_2224961, EPI_ISL_2224962, EPI_ISL_2224963, EPI_ISL_2224964, EPI_ISL_2224965, EPI_ISL_2224966, EPI_ISL_2224968, EPI_ISL_2224969, EPI_ISL_2224970, EPI_ISL_2224971, EPI_ISL_2224972, EPI_ISL_2224973, EPI_ISL_2224974, EPI_ISL_2224975, EPI_ISL_2224976, EPI_ISL_2224977, EPI_ISL_2224978, EPI_ISL_2224979, EPI_ISL_2224980, EPI_ISL_2224981, EPI_ISL_2224982, EPI_ISL_2224983, EPI_ISL_2224984, EPI_ISL_2224986, EPI_ISL_2224987, EPI_ISL_2224989, EPI_ISL_2224990, EPI_ISL_2224991, EPI_ISL_2224992, EPI_ISL_2224993, EPI_ISL_2224994, EPI_ISL_2224995, EPI_ISL_2224996, EPI_ISL_2224997, EPI_ISL_2224998, EPI_ISL_2224999, EPI_ISL_2225000, EPI_ISL_2225001, EPI_ISL_2225002, EPI_ISL_2225003, EPI_ISL_2225005, EPI_ISL_2225006, EPI_ISL_2225007, EPI_ISL_2225008, EPI_ISL_2225009, EPI_ISL_2225010, EPI_ISL_2225011, EPI_ISL_2225012, EPI_ISL_2225013, EPI_ISL_2225014, EPI_ISL_2225015, EPI_ISL_2225016, EPI_ISL_2225017, EPI_ISL_2225018, EPI_ISL_2225019, EPI_ISL_2225020, EPI_ISL_2225021, EPI_ISL_2225022, EPI_ISL_2225023, EPI_ISL_2225024, EPI_ISL_2225027, EPI_ISL_2225028, EPI_ISL_2225030, EPI_ISL_2225031, EPI_ISL_2225032, EPI_ISL_2225033, EPI_ISL_2225034, EPI_ISL_2225035, EPI_ISL_2225036, EPI_ISL_2225037, EPI_ISL_2225038, EPI_ISL_2225039, EPI_ISL_2225040, EPI_ISL_2225041, EPI_ISL_2225042, EPI_ISL_2225043, EPI_ISL_2225044, EPI_ISL_2225045, EPI_ISL_2225046, EPI_ISL_2225047, EPI_ISL_2225048, EPI_ISL_2225051, EPI_ISL_2225053, EPI_ISL_2225054, EPI_ISL_2225055, EPI_ISL_2225056, EPI_ISL_2225057, EPI_ISL_2225058, EPI_ISL_2225059, EPI_ISL_2225060, EPI_ISL_2225061, EPI_ISL_2225062, EPI_ISL_2225063, EPI_ISL_2225064, EPI_ISL_2225065, EPI_ISL_2225066, EPI_ISL_2225067, EPI_ISL_2225068, EPI_ISL_2225069, EPI_ISL_2225070, EPI_ISL_2225071, EPI_ISL_2225072, EPI_ISL_2225073, EPI_ISL_2225074, EPI_ISL_2225075, EPI_ISL_2225076, EPI_ISL_2225077, EPI_ISL_2225078, EPI_ISL_2225079, EPI_ISL_2225080, EPI_ISL_2225081, EPI_ISL_2225082, EPI_ISL_2225083, EPI_ISL_2225084, EPI_ISL_2225085, EPI_ISL_2225086, EPI_ISL_2225087, EPI_ISL_2225089, EPI_ISL_2225090, EPI_ISL_2225091, EPI_ISL_2225092, EPI_ISL_2225093, EPI_ISL_2225094, EPI_ISL_2225095, EPI_ISL_2225096, EPI_ISL_2225097, EPI_ISL_2225098, EPI_ISL_2225099, EPI_ISL_2225100, EPI_ISL_2225101, EPI_ISL_2225102, EPI_ISL_2225103, EPI_ISL_2225104, EPI_ISL_2225105, EPI_ISL_2225106, EPI_ISL_2225107, EPI_ISL_2225108, EPI_ISL_2225109, EPI_ISL_2225110, EPI_ISL_2225111, EPI_ISL_2225112, EPI_ISL_2225113, EPI_ISL_2225114, EPI_ISL_2225115, EPI_ISL_2225116, EPI_ISL_2225117, EPI_ISL_2225118, EPI_ISL_2225119, EPI_ISL_2225120, EPI_ISL_2225121, EPI_ISL_2225122, EPI_ISL_2225123, EPI_ISL_2225124, EPI_ISL_2225125, EPI_ISL_2225126, EPI_ISL_2225127, EPI_ISL_2225128, EPI_ISL_2225129, EPI_ISL_2225130, EPI_ISL_2225131, EPI_ISL_2225133, EPI_ISL_2225134, EPI_ISL_2225135, EPI_ISL_2225136 | | | |
| see above | Houston Methodist Hospital | Houston Methodist Hospital | Randall J. Olsen, Paul A. Christensen, S. Wesley Long, Sishir Subedi, Robert Olson, Marcus Nguyen, James J. Davis, Matthew Ojeda Saavedra, Prasanti Yerramilli, Layne Pruitt, Kristina Reppond, Madison N. Shyer, Jessica Cambric, Ryan Gadd, Ilya J. Finkelstein, Jimmy Gollihar, and James M. Musser |
| EPI_ISL_2225137 | Wyoming Public Health Laboratory | Wyoming Public Health Laboratory | Jim Mildenberger, Taylor Fearing, Channing Weber, Ashley Norberg, Chayse Rowley, Marley Goetz, Brian Dominguez, Elliot Thomasson, Brittany Oher, Sam Britz, Cari Sloma, and Rob Christensen |
| EPI_ISL_2225138 | Houston Methodist Hospital | Houston Methodist Hospital | Randall J. Olsen, Paul A. Christensen, S. Wesley Long, Sishir Subedi, Robert Olson, Marcus Nguyen, James J. Davis, Matthew Ojeda Saavedra, Prasanti Yerramilli, Layne Pruitt, Kristina Reppond, Madison N. Shyer, Jessica Cambric, Ryan Gadd, Ilya J. Finkelstein, Jimmy Gollihar, and James M. Musser |
| EPI_ISL_2225139 | Wyoming Public Health Laboratory | Wyoming Public Health Laboratory | Jim Mildenberger, Taylor Fearing, Channing Weber, Ashley Norberg, Chayse Rowley, Marley Goetz, Brian Dominguez, Elliot Thomasson, Brittany Oher, Sam Britz, Cari Sloma, and Rob Christensen |
| EPI_ISL_2225141 | Houston Methodist Hospital | Houston Methodist Hospital | Randall J. Olsen, Paul A. Christensen, S. Wesley Long, Sishir Subedi, Robert Olson, Marcus Nguyen, James J. Davis, Matthew Ojeda Saavedra, Prasanti Yerramilli, Layne Pruitt, Kristina Reppond, Madison N. Shyer, Jessica Cambric, Ryan Gadd, Ilya J. Finkelstein, Jimmy Gollihar, and James M. Musser |

| | | | |
|---|---|---|---|
| EPI_ISL_2225143, EPI_ISL_2225144 | Wyoming Public Health Laboratory | Wyoming Public Health Laboratory | Jim Mildenberger, Taylor Fearing, Channing Weber, Ashley Norberg, Chayse Rowley, Marley Goetz, Brian Dominguez, Elliot Thomasson, Brittany Oher, Sam Britz, Cari Sloma, and Rob Christensen |
| EPI_ISL_2225145 | Houston Methodist Hospital | Houston Methodist Hospital | Randall J. Olsen, Paul A. Christensen, S. Wesley Long, Sishir Subedi, Robert Olson, Marcus Nguyen, James J. Davis, Matthew Ojeda Saavedra, Prasanti Yerramilli, Layne Pruitt, Kristina Reppond, Madison N. Shyer, Jessica Cambric, Ryan Gadd, Ilya J. Finkelstein, Jimmy Gollihar, and James M. Musser |
| EPI_ISL_2225146, EPI_ISL_2225147 | Wyoming Public Health Laboratory | Wyoming Public Health Laboratory | Jim Mildenberger, Taylor Fearing, Channing Weber, Ashley Norberg, Chayse Rowley, Marley Goetz, Brian Dominguez, Elliot Thomasson, Brittany Oher, Sam Britz, Cari Sloma, and Rob Christensen |
| EPI_ISL_2225148 | Houston Methodist Hospital | Houston Methodist Hospital | Randall J. Olsen, Paul A. Christensen, S. Wesley Long, Sishir Subedi, Robert Olson, Marcus Nguyen, James J. Davis, Matthew Ojeda Saavedra, Prasanti Yerramilli, Layne Pruitt, Kristina Reppond, Madison N. Shyer, Jessica Cambric, Ryan Gadd, Ilya J. Finkelstein, Jimmy Gollihar, and James M. Musser |
| EPI_ISL_2225149 | Wyoming Public Health Laboratory | Wyoming Public Health Laboratory | Jim Mildenberger, Taylor Fearing, Channing Weber, Ashley Norberg, Chayse Rowley, Marley Goetz, Brian Dominguez, Elliot Thomasson, Brittany Oher, Sam Britz, Cari Sloma, and Rob Christensen |
| EPI_ISL_2225151 | Houston Methodist Hospital | Houston Methodist Hospital | Randall J. Olsen, Paul A. Christensen, S. Wesley Long, Sishir Subedi, Robert Olson, Marcus Nguyen, James J. Davis, Matthew Ojeda Saavedra, Prasanti Yerramilli, Layne Pruitt, Kristina Reppond, Madison N. Shyer, Jessica Cambric, Ryan Gadd, Ilya J. Finkelstein, Jimmy Gollihar, and James M. Musser |
| EPI_ISL_2225153 | Wyoming Public Health Laboratory | Wyoming Public Health Laboratory | Jim Mildenberger, Taylor Fearing, Channing Weber, Ashley Norberg, Chayse Rowley, Marley Goetz, Brian Dominguez, Elliot Thomasson, Brittany Oher, Sam Britz, Cari Sloma, and Rob Christensen |
| EPI_ISL_2225154 | Houston Methodist Hospital | Houston Methodist Hospital | Randall J. Olsen, Paul A. Christensen, S. Wesley Long, Sishir Subedi, Robert Olson, Marcus Nguyen, James J. Davis, Matthew Ojeda Saavedra, Prasanti Yerramilli, Layne Pruitt, Kristina Reppond, Madison N. Shyer, Jessica Cambric, Ryan Gadd, Ilya J. Finkelstein, Jimmy Gollihar, and James M. Musser |
| EPI_ISL_2225155, EPI_ISL_2225156 | Wyoming Public Health Laboratory | Wyoming Public Health Laboratory | Jim Mildenberger, Taylor Fearing, Channing Weber, Ashley Norberg, Chayse Rowley, Marley Goetz, Brian Dominguez, Elliot Thomasson, Brittany Oher, Sam Britz, Cari Sloma, and Rob Christensen |
| EPI_ISL_2225157 | Houston Methodist Hospital | Houston Methodist Hospital | Randall J. Olsen, Paul A. Christensen, S. Wesley Long, Sishir Subedi, Robert Olson, Marcus Nguyen, James J. Davis, Matthew Ojeda Saavedra, Prasanti Yerramilli, Layne Pruitt, Kristina Reppond, Madison N. Shyer, Jessica Cambric, Ryan Gadd, Ilya J. Finkelstein, Jimmy Gollihar, and James M. Musser |
| EPI_ISL_2225158, EPI_ISL_2225159 | Wyoming Public Health Laboratory | Wyoming Public Health Laboratory | Jim Mildenberger, Taylor Fearing, Channing Weber, Ashley Norberg, Chayse Rowley, Marley Goetz, Brian Dominguez, Elliot Thomasson, Brittany Oher, Sam Britz, Cari Sloma, and Rob Christensen |
| EPI_ISL_2225160 | Houston Methodist Hospital | Houston Methodist Hospital | Randall J. Olsen, Paul A. Christensen, S. Wesley Long, Sishir Subedi, Robert Olson, Marcus Nguyen, James J. Davis, Matthew Ojeda Saavedra, Prasanti Yerramilli, Layne Pruitt, Kristina Reppond, Madison N. Shyer, Jessica Cambric, Ryan Gadd, Ilya J. Finkelstein, Jimmy Gollihar, and James M. Musser |
| EPI_ISL_2225161, EPI_ISL_2225162 | Wyoming Public Health Laboratory | Wyoming Public Health Laboratory | Jim Mildenberger, Taylor Fearing, Channing Weber, Ashley Norberg, Chayse Rowley, Marley Goetz, Brian Dominguez, Elliot Thomasson, Brittany Oher, Sam Britz, Cari Sloma, and Rob Christensen |
| EPI_ISL_2225163 | Houston Methodist Hospital | Houston Methodist Hospital | Randall J. Olsen, Paul A. Christensen, S. Wesley Long, Sishir Subedi, Robert Olson, Marcus Nguyen, James J. Davis, Matthew Ojeda Saavedra, Prasanti Yerramilli, Layne Pruitt, Kristina Reppond, Madison N. Shyer, Jessica Cambric, Ryan Gadd, Ilya J. Finkelstein, Jimmy Gollihar, and James M. Musser |
| EPI_ISL_2225164, EPI_ISL_2225165 | Wyoming Public Health Laboratory | Wyoming Public Health Laboratory | Jim Mildenberger, Taylor Fearing, Channing Weber, Ashley Norberg, Chayse Rowley, Marley Goetz, Brian Dominguez, Elliot Thomasson, Brittany Oher, Sam Britz, Cari Sloma, and Rob Christensen |
| EPI_ISL_2225166 | Houston Methodist Hospital | Houston Methodist Hospital | Randall J. Olsen, Paul A. Christensen, S. Wesley Long, Sishir Subedi, Robert Olson, Marcus Nguyen, James J. Davis, Matthew Ojeda Saavedra, Prasanti Yerramilli, Layne Pruitt, Kristina Reppond, Madison N. Shyer, Jessica Cambric, Ryan Gadd, Ilya J. Finkelstein, Jimmy Gollihar, and James M. Musser |
| EPI_ISL_2225167, EPI_ISL_2225168 | Wyoming Public Health Laboratory | Wyoming Public Health Laboratory | Jim Mildenberger, Taylor Fearing, Channing Weber, Ashley Norberg, Chayse Rowley, Marley Goetz, Brian Dominguez, Elliot Thomasson, Brittany Oher, Sam Britz, Cari Sloma, and Rob Christensen |
| EPI_ISL_2225169 | Houston Methodist Hospital | Houston Methodist Hospital | Randall J. Olsen, Paul A. Christensen, S. Wesley Long, Sishir Subedi, Robert Olson, Marcus Nguyen, James J. Davis, Matthew Ojeda Saavedra, Prasanti Yerramilli, Layne Pruitt, Kristina Reppond, Madison N. Shyer, Jessica Cambric, Ryan Gadd, Ilya J. Finkelstein, Jimmy Gollihar, and James M. Musser |
| EPI_ISL_2225170, EPI_ISL_2225171, EPI_ISL_2225173 | Wyoming Public Health Laboratory | Wyoming Public Health Laboratory | Jim Mildenberger, Taylor Fearing, Channing Weber, Ashley Norberg, Chayse Rowley, Marley Goetz, Brian Dominguez, Elliot Thomasson, Brittany Oher, Sam Britz, Cari Sloma, and Rob Christensen |
| EPI_ISL_2225175 | Houston Methodist Hospital | Houston Methodist Hospital | Randall J. Olsen, Paul A. Christensen, S. Wesley Long, Sishir Subedi, Robert Olson, Marcus Nguyen, James J. Davis, Matthew Ojeda Saavedra, Prasanti Yerramilli, Layne Pruitt, Kristina Reppond, Madison N. Shyer, Jessica Cambric, Ryan Gadd, Ilya J. Finkelstein, Jimmy Gollihar, and James M. Musser |
| EPI_ISL_2225177 | Wyoming Public Health Laboratory | Wyoming Public Health Laboratory | Jim Mildenberger, Taylor Fearing, Channing Weber, Ashley Norberg, Chayse Rowley, Marley Goetz, Brian Dominguez, Elliot Thomasson, Brittany Oher, Sam Britz, Cari Sloma, and Rob Christensen |
| EPI_ISL_2225178 | Houston Methodist Hospital | Houston Methodist Hospital | Randall J. Olsen, Paul A. Christensen, S. Wesley Long, Sishir Subedi, Robert Olson, Marcus Nguyen, James J. Davis, Matthew Ojeda Saavedra, Prasanti Yerramilli, Layne Pruitt, Kristina Reppond, Madison N. Shyer, Jessica Cambric, Ryan Gadd, Ilya J. Finkelstein, Jimmy Gollihar, and James M. Musser |
| EPI_ISL_2225179 | Wyoming Public Health Laboratory | Wyoming Public Health Laboratory | Jim Mildenberger, Taylor Fearing, Channing Weber, Ashley Norberg, Chayse Rowley, Marley Goetz, Brian Dominguez, Elliot Thomasson, Brittany Oher, Sam Britz, Cari Sloma, and Rob Christensen |
| EPI_ISL_2225180 | Houston Methodist Hospital | Houston Methodist Hospital | Randall J. Olsen, Paul A. Christensen, S. Wesley Long, Sishir Subedi, Robert Olson, Marcus Nguyen, James J. Davis, Matthew Ojeda Saavedra, Prasanti Yerramilli, Layne Pruitt, Kristina Reppond, Madison N. Shyer, Jessica Cambric, Ryan Gadd, Ilya J. Finkelstein, Jimmy Gollihar, and James M. Musser |
| EPI_ISL_2225181 | Wyoming Public Health Laboratory | Wyoming Public Health Laboratory | Jim Mildenberger, Taylor Fearing, Channing Weber, Ashley Norberg, Chayse Rowley, Marley Goetz, Brian Dominguez, Elliot Thomasson, Brittany Oher, Sam Britz, Cari Sloma, and Rob Christensen |
| EPI_ISL_2225182, EPI_ISL_2225183 | Houston Methodist Hospital | Houston Methodist Hospital | Randall J. Olsen, Paul A. Christensen, S. Wesley Long, Sishir Subedi, Robert Olson, Marcus Nguyen, James J. Davis, Matthew Ojeda Saavedra, Prasanti Yerramilli, Layne Pruitt, Kristina Reppond, Madison N. Shyer, Jessica Cambric, Ryan Gadd, Ilya J. Finkelstein, Jimmy Gollihar, and James M. Musser |
| EPI_ISL_2225184 | Wyoming Public Health Laboratory | Wyoming Public Health Laboratory | Jim Mildenberger, Taylor Fearing, Channing Weber, Ashley Norberg, Chayse Rowley, Marley Goetz, Brian Dominguez, Elliot Thomasson, Brittany Oher, Sam Britz, Cari Sloma, and Rob Christensen |
| EPI_ISL_2225185 | Houston Methodist Hospital | Houston Methodist Hospital | Randall J. Olsen, Paul A. Christensen, S. Wesley Long, Sishir Subedi, Robert Olson, Marcus Nguyen, James J. Davis, Matthew Ojeda Saavedra, Prasanti Yerramilli, Layne Pruitt, Kristina Reppond, Madison N. Shyer, Jessica Cambric, Ryan Gadd, Ilya J. Finkelstein, Jimmy Gollihar, and James M. Musser |
| EPI_ISL_2225186 | Wyoming Public Health Laboratory | Wyoming Public Health Laboratory | Jim Mildenberger, Taylor Fearing, Channing Weber, Ashley Norberg, Chayse Rowley, Marley Goetz, Brian Dominguez, Elliot Thomasson, Brittany Oher, Sam Britz, Cari Sloma, and Rob Christensen |
| EPI_ISL_2225188 | Houston Methodist Hospital | Houston Methodist Hospital | Randall J. Olsen, Paul A. Christensen, S. Wesley Long, Sishir Subedi, Robert Olson, Marcus Nguyen, James J. Davis, Matthew Ojeda Saavedra, Prasanti Yerramilli, Layne Pruitt, Kristina Reppond, Madison N. Shyer, Jessica Cambric, Ryan Gadd, Ilya J. Finkelstein, Jimmy Gollihar, and James M. Musser |
| EPI_ISL_2225189 | Wyoming Public Health Laboratory | Wyoming Public Health Laboratory | Jim Mildenberger, Taylor Fearing, Channing Weber, Ashley Norberg, Chayse Rowley, Marley Goetz, Brian Dominguez, Elliot Thomasson, Brittany Oher, Sam Britz, Cari Sloma, and Rob Christensen |
| EPI_ISL_2225190, EPI_ISL_2225191 | Houston Methodist Hospital | Houston Methodist Hospital | Randall J. Olsen, Paul A. Christensen, S. Wesley Long, Sishir Subedi, Robert Olson, Marcus Nguyen, James J. Davis, Matthew Ojeda Saavedra, Prasanti Yerramilli, Layne Pruitt, Kristina Reppond, Madison N. Shyer, Jessica Cambric, Ryan Gadd, Ilya J. Finkelstein, Jimmy Gollihar, and James M. Musser |
| EPI_ISL_2225192 | Wyoming Public Health Laboratory | Wyoming Public Health Laboratory | Jim Mildenberger, Taylor Fearing, Channing Weber, Ashley Norberg, Chayse Rowley, Marley Goetz, Brian Dominguez, Elliot Thomasson, Brittany Oher, Sam Britz, Cari Sloma, and Rob Christensen |
| EPI_ISL_2225193, EPI_ISL_2225196, EPI_ISL_2225197, EPI_ISL_2225198, EPI_ISL_2225199, EPI_ISL_2225200, EPI_ISL_2225201, EPI_ISL_2225202, EPI_ISL_2225203, EPI_ISL_2225204, EPI_ISL_2225205, EPI_ISL_2225206, EPI_ISL_2225207, EPI_ISL_2225208, EPI_ISL_2225209, EPI_ISL_2225210, EPI_ISL_2225211, EPI_ISL_2225212, EPI_ISL_2225213, EPI_ISL_2225214, EPI_ISL_2225215, EPI_ISL_2225216, EPI_ISL_2225217, EPI_ISL_2225218, EPI_ISL_2225219, EPI_ISL_2225220, EPI_ISL_2225221, EPI_ISL_2225222, EPI_ISL_2225223, EPI_ISL_2225224, EPI_ISL_2225225, EPI_ISL_2225226, EPI_ISL_2225227, EPI_ISL_2225228, EPI_ISL_2225229, EPI_ISL_2225230, EPI_ISL_2225231, EPI_ISL_2225232, EPI_ISL_2225233, EPI_ISL_2225234, EPI_ISL_2225235, EPI_ISL_2225236, EPI_ISL_2225237, EPI_ISL_2225238, EPI_ISL_2225239, EPI_ISL_2225240, EPI_ISL_2225241, EPI_ISL_2225242, EPI_ISL_2225243, EPI_ISL_2225244, EPI_ISL_2225245, EPI_ISL_2225246, EPI_ISL_2225247, EPI_ISL_2225248, EPI_ISL_2225249, EPI_ISL_2225250, EPI_ISL_2225251, EPI_ISL_2225252, EPI_ISL_2225253, EPI_ISL_2225254, EPI_ISL_2225255, EPI_ISL_2225256, EPI_ISL_2225257, EPI_ISL_2225258, EPI_ISL_2225259, EPI_ISL_2225261, EPI_ISL_2225262, EPI_ISL_2225263, EPI_ISL_2225264, EPI_ISL_2225265, EPI_ISL_2225266, EPI_ISL_2225267, EPI_ISL_2225268, EPI_ISL_2225269, EPI_ISL_2225270, EPI_ISL_2225272, EPI_ISL_2225273, EPI_ISL_2225274, EPI_ISL_2225275, EPI_ISL_2225276, EPI_ISL_2225277, EPI_ISL_2225278, EPI_ISL_2225279, EPI_ISL_2225280, EPI_ISL_2225281, EPI_ISL_2225282, EPI_ISL_2225283, EPI_ISL_2225284, EPI_ISL_2225285, EPI_ISL_2225286, EPI_ISL_2225287, EPI_ISL_2225288, EPI_ISL_2225290, EPI_ISL_2225291, EPI_ISL_2225292, EPI_ISL_2225293, EPI_ISL_2225294, EPI_ISL_2225295, EPI_ISL_2225296, EPI_ISL_2225297, EPI_ISL_2225298, EPI_ISL_2225299, EPI_ISL_2225300, EPI_ISL_2225301, EPI_ISL_2225302, EPI_ISL_2225303, EPI_ISL_2225304, EPI_ISL_2225305, EPI_ISL_2225306, EPI_ISL_2225307, EPI_ISL_2225308, EPI_ISL_2225309, EPI_ISL_2225310, EPI_ISL_2225311, EPI_ISL_2225312, EPI_ISL_2225313, EPI_ISL_2225314, EPI_ISL_2225315, EPI_ISL_2225316, EPI_ISL_2225317, EPI_ISL_2225318, EPI_ISL_2225319, EPI_ISL_2225320, EPI_ISL_2225321, EPI_ISL_2225322, EPI_ISL_2225323, EPI_ISL_2225324, EPI_ISL_2225326, EPI_ISL_2225327, EPI_ISL_2225328, EPI_ISL_2225329, EPI_ISL_2225330, EPI_ISL_2225331, EPI_ISL_2225332, EPI_ISL_2225333, EPI_ISL_2225334, EPI_ISL_2225335, EPI_ISL_2225336, EPI_ISL_2225337, EPI_ISL_2225338, EPI_ISL_2225339, EPI_ISL_2225340, EPI_ISL_2225341, EPI_ISL_2225342, EPI_ISL_2225344, EPI_ISL_2225345, EPI_ISL_2225346, EPI_ISL_2225347, EPI_ISL_2225348, EPI_ISL_2225349, EPI_ISL_2225350, EPI_ISL_2225351, EPI_ISL_2225352, EPI_ISL_2225353, EPI_ISL_2225354, EPI_ISL_2225355, EPI_ISL_2225356, EPI_ISL_2225357, EPI_ISL_2225358, EPI_ISL_2225360 | | | |
| see above | Houston Methodist Hospital | Houston Methodist Hospital | Randall J. Olsen, Paul A. Christensen, S. Wesley Long, Sishir Subedi, Robert Olson, Marcus Nguyen, James J. Davis, Matthew Ojeda Saavedra, Prasanti Yerramilli, Layne Pruitt, Kristina Reppond, Madison N. Shyer, Jessica Cambric, Ryan Gadd, Ilya J. Finkelstein, Jimmy Gollihar, and James M. Musser |
| EPI_ISL_2225361, EPI_ISL_2225362 | Wyoming Public Health Laboratory | Wyoming Public Health Laboratory | Jim Mildenberger, Taylor Fearing, Channing Weber, Ashley Norberg, Chayse Rowley, Marley Goetz, Brian Dominguez, Elliot Thomasson, Brittany Oher, Sam Britz, Cari Sloma, and Rob Christensen |
| EPI_ISL_2225363 | Houston Methodist Hospital | Houston Methodist Hospital | Randall J. Olsen, Paul A. Christensen, S. Wesley Long, Sishir Subedi, Robert Olson, Marcus Nguyen, James J. Davis, Matthew Ojeda Saavedra, Prasanti Yerramilli, Layne Pruitt, Kristina Reppond, Madison N. Shyer, Jessica Cambric, Ryan Gadd, Ilya J. Finkelstein, Jimmy Gollihar, and James M. Musser |
| EPI_ISL_2225364, EPI_ISL_2225365 | Wyoming Public Health Laboratory | Wyoming Public Health Laboratory | Jim Mildenberger, Taylor Fearing, Channing Weber, Ashley Norberg, Chayse Rowley, Marley Goetz, Brian Dominguez, Elliot Thomasson, Brittany Oher, Sam Britz, Cari Sloma, and Rob Christensen |
| EPI_ISL_2225366, EPI_ISL_2225367 | Houston Methodist Hospital | Houston Methodist Hospital | Randall J. Olsen, Paul A. Christensen, S. Wesley Long, Sishir Subedi, Robert Olson, Marcus Nguyen, James J. Davis, Matthew Ojeda Saavedra, Prasanti Yerramilli, Layne Pruitt, Kristina Reppond, Madison N. Shyer, Jessica Cambric, Ryan Gadd, Ilya J. Finkelstein, Jimmy Gollihar, and James M. Musser |
| EPI_ISL_2225369 | Wyoming Public Health Laboratory | Wyoming Public Health Laboratory | Jim Mildenberger, Taylor Fearing, Channing Weber, Ashley Norberg, Chayse Rowley, Marley Goetz, Brian Dominguez, Elliot Thomasson, Brittany Oher, Sam Britz, Cari Sloma, and Rob Christensen |
| EPI_ISL_2225370, EPI_ISL_2225372 | Houston Methodist Hospital | Houston Methodist Hospital | Randall J. Olsen, Paul A. Christensen, S. Wesley Long, Sishir Subedi, Robert Olson, Marcus Nguyen, James J. Davis, Matthew Ojeda Saavedra, Prasanti Yerramilli, Layne Pruitt, Kristina Reppond, Madison N. Shyer, Jessica Cambric, Ryan Gadd, Ilya J. Finkelstein, Jimmy Gollihar, and James M. Musser |
| EPI_ISL_2225373 | Wyoming Public Health Laboratory | Wyoming Public Health Laboratory | Jim Mildenberger, Taylor Fearing, Channing Weber, Ashley Norberg, Chayse Rowley, Marley Goetz, Brian Dominguez, Elliot Thomasson, Brittany Oher, Sam Britz, Cari Sloma, and Rob Christensen |
| EPI_ISL_2225374, EPI_ISL_2225375, EPI_ISL_2225377, EPI_ISL_2225379, EPI_ISL_2225380, EPI_ISL_2225382 | Houston Methodist Hospital | Houston Methodist Hospital | Randall J. Olsen, Paul A. Christensen, S. Wesley Long, Sishir Subedi, Robert Olson, Marcus Nguyen, James J. Davis, Matthew Ojeda Saavedra, Prasanti Yerramilli, Layne Pruitt, Kristina Reppond, Madison N. Shyer, Jessica Cambric, Ryan Gadd, Ilya J. Finkelstein, Jimmy Gollihar, and James M. Musser |
| EPI_ISL_2225383 | Wyoming Public Health Laboratory | Wyoming Public Health Laboratory | Jim Mildenberger, Taylor Fearing, Channing Weber, Ashley Norberg, Chayse Rowley, Marley Goetz, Brian Dominguez, Elliot Thomasson, Brittany Oher, Sam Britz, Cari Sloma, and Rob Christensen |
| EPI_ISL_2225384, EPI_ISL_2225385, EPI_ISL_2225387, EPI_ISL_2225389, | Houston Methodist Hospital | Houston Methodist Hospital | Randall J. Olsen, Paul A. Christensen, S. Wesley Long, Sishir Subedi, Robert Olson, Marcus Nguyen, James J. Davis, Matthew Ojeda Saavedra, Prasanti Yerramilli, Layne Pruitt, Kristina Reppond, Madison N. Shyer, Jessica Cambric, Ryan Gadd, Ilya J. Finkelstein, Jimmy Gollihar, and James M. Musser |

| | | | |
|---|---|---|---|
| EPI_ISL_2225390 | | | |
| EPI_ISL_2225391 | Wyoming Public Health Laboratory | Wyoming Public Health Laboratory | Jim Mildenberger, Taylor Fearing, Channing Weber, Ashley Norberg, Chayse Rowley, Marley Goetz, Brian Dominguez, Elliot Thomasson, Brittany Oher, Sam Britz, Cari Sloma, and Rob Christensen |
| EPI_ISL_2225392, EPI_ISL_2225393 | Houston Methodist Hospital | Houston Methodist Hospital | Randall J. Olsen, Paul A. Christensen, S. Wesley Long, Sishir Subedi, Robert Olson, Marcus Nguyen, James J. Davis, Matthew Ojeda Saavedra, Prasanti Yerramilli, Layne Pruitt, Kristina Reppond, Madison N. Shyer, Jessica Cambric, Ryan Gadd, Ilya J. Finkelstein, Jimmy Gollihar, and James M. Musser |
| EPI_ISL_2225394 | Wyoming Public Health Laboratory | Wyoming Public Health Laboratory | Jim Mildenberger, Taylor Fearing, Channing Weber, Ashley Norberg, Chayse Rowley, Marley Goetz, Brian Dominguez, Elliot Thomasson, Brittany Oher, Sam Britz, Cari Sloma, and Rob Christensen |
| EPI_ISL_2225395 | Houston Methodist Hospital | Houston Methodist Hospital | Randall J. Olsen, Paul A. Christensen, S. Wesley Long, Sishir Subedi, Robert Olson, Marcus Nguyen, James J. Davis, Matthew Ojeda Saavedra, Prasanti Yerramilli, Layne Pruitt, Kristina Reppond, Madison N. Shyer, Jessica Cambric, Ryan Gadd, Ilya J. Finkelstein, Jimmy Gollihar, and James M. Musser |
| EPI_ISL_2225396 | Wyoming Public Health Laboratory | Wyoming Public Health Laboratory | Jim Mildenberger, Taylor Fearing, Channing Weber, Ashley Norberg, Chayse Rowley, Marley Goetz, Brian Dominguez, Elliot Thomasson, Brittany Oher, Sam Britz, Cari Sloma, and Rob Christensen |
| EPI_ISL_2225397, EPI_ISL_2225398 | Houston Methodist Hospital | Houston Methodist Hospital | Randall J. Olsen, Paul A. Christensen, S. Wesley Long, Sishir Subedi, Robert Olson, Marcus Nguyen, James J. Davis, Matthew Ojeda Saavedra, Prasanti Yerramilli, Layne Pruitt, Kristina Reppond, Madison N. Shyer, Jessica Cambric, Ryan Gadd, Ilya J. Finkelstein, Jimmy Gollihar, and James M. Musser |
| EPI_ISL_2225399 | Wyoming Public Health Laboratory | Wyoming Public Health Laboratory | Jim Mildenberger, Taylor Fearing, Channing Weber, Ashley Norberg, Chayse Rowley, Marley Goetz, Brian Dominguez, Elliot Thomasson, Brittany Oher, Sam Britz, Cari Sloma, and Rob Christensen |
| EPI_ISL_2225400, EPI_ISL_2225401, EPI_ISL_2225402, EPI_ISL_2225403, EPI_ISL_2225404, EPI_ISL_2225405, EPI_ISL_2225406, EPI_ISL_2225407, EPI_ISL_2225408, EPI_ISL_2225409, EPI_ISL_2225410, EPI_ISL_2225411, EPI_ISL_2225412, EPI_ISL_2225414, EPI_ISL_2225415, EPI_ISL_2225416, EPI_ISL_2225417, EPI_ISL_2225418, EPI_ISL_2225419, EPI_ISL_2225420, EPI_ISL_2225421, EPI_ISL_2225422, EPI_ISL_2225423, EPI_ISL_2225424, EPI_ISL_2225425, EPI_ISL_2225426, EPI_ISL_2225427, EPI_ISL_2225428, EPI_ISL_2225429, EPI_ISL_2225430, EPI_ISL_2225431, EPI_ISL_2225432, EPI_ISL_2225433, EPI_ISL_2225434, EPI_ISL_2225435, EPI_ISL_2225436, EPI_ISL_2225437, EPI_ISL_2225438, EPI_ISL_2225439, EPI_ISL_2225440, EPI_ISL_2225441, EPI_ISL_2225442, EPI_ISL_2225443, EPI_ISL_2225444, EPI_ISL_2225445, EPI_ISL_2225446, EPI_ISL_2225447, EPI_ISL_2225448, EPI_ISL_2225449, EPI_ISL_2225450, EPI_ISL_2225451, EPI_ISL_2225452, EPI_ISL_2225453, EPI_ISL_2225454, EPI_ISL_2225455, EPI_ISL_2225456, EPI_ISL_2225457, EPI_ISL_2225458, EPI_ISL_2225459, EPI_ISL_2225460, EPI_ISL_2225461, EPI_ISL_2225462, EPI_ISL_2225463, EPI_ISL_2225464, EPI_ISL_2225465, EPI_ISL_2225466, EPI_ISL_2225467, EPI_ISL_2225468, EPI_ISL_2225469, EPI_ISL_2225470, EPI_ISL_2225471, EPI_ISL_2225472, EPI_ISL_2225473, EPI_ISL_2225474, EPI_ISL_2225475, EPI_ISL_2225476, EPI_ISL_2225477, EPI_ISL_2225478, EPI_ISL_2225479, EPI_ISL_2225480, EPI_ISL_2225481, EPI_ISL_2225482, EPI_ISL_2225483, EPI_ISL_2225484, EPI_ISL_2225485, EPI_ISL_2225486, EPI_ISL_2225487, EPI_ISL_2225488, EPI_ISL_2225489, EPI_ISL_2225490, EPI_ISL_2225491, EPI_ISL_2225492, EPI_ISL_2225493, EPI_ISL_2225494, EPI_ISL_2225495, EPI_ISL_2225496, EPI_ISL_2225497, EPI_ISL_2225498, EPI_ISL_2225499, EPI_ISL_2225500, EPI_ISL_2225501, EPI_ISL_2225502, EPI_ISL_2225503, EPI_ISL_2225504, EPI_ISL_2225505, EPI_ISL_2225506, EPI_ISL_2225507, EPI_ISL_2225508, EPI_ISL_2225509, EPI_ISL_2225510, EPI_ISL_2225511, EPI_ISL_2225512, EPI_ISL_2225513 | | | |
| see above | Houston Methodist Hospital | Houston Methodist Hospital | Randall J. Olsen, Paul A. Christensen, S. Wesley Long, Sishir Subedi, Robert Olson, Marcus Nguyen, James J. Davis, Matthew Ojeda Saavedra, Prasanti Yerramilli, Layne Pruitt, Kristina Reppond, Madison N. Shyer, Jessica Cambric, Ryan Gadd, Ilya J. Finkelstein, Jimmy Gollihar, and James M. Musser |
| EPI_ISL_2225516, EPI_ISL_2225517, EPI_ISL_2225520, EPI_ISL_2225521, EPI_ISL_2225523, EPI_ISL_2225530 | Wyoming Public Health Laboratory | Wyoming Public Health Laboratory | Jim Mildenberger, Taylor Fearing, Channing Weber, Ashley Norberg, Chayse Rowley, Marley Goetz, Brian Dominguez, Elliot Thomasson, Brittany Oher, Sam Britz, Cari Sloma, and Rob Christensen |
| EPI_ISL_2225534, EPI_ISL_2225535, EPI_ISL_2225536, EPI_ISL_2225537, EPI_ISL_2225538, EPI_ISL_2225539, EPI_ISL_2225540, EPI_ISL_2225541, EPI_ISL_2225542, EPI_ISL_2225543, EPI_ISL_2225544, EPI_ISL_2225545, EPI_ISL_2225546, EPI_ISL_2225547, EPI_ISL_2225548, EPI_ISL_2225549, EPI_ISL_2225550, EPI_ISL_2225551, EPI_ISL_2225552, EPI_ISL_2225553, EPI_ISL_2225554, EPI_ISL_2225556, EPI_ISL_2225557, EPI_ISL_2225559, EPI_ISL_2225560, EPI_ISL_2225561, EPI_ISL_2225562, EPI_ISL_2225564, EPI_ISL_2225565, EPI_ISL_2225566, EPI_ISL_2225567, EPI_ISL_2225568, EPI_ISL_2225569, EPI_ISL_2225570, EPI_ISL_2225571, EPI_ISL_2225572, EPI_ISL_2225573, EPI_ISL_2225574, EPI_ISL_2225575, EPI_ISL_2225576, EPI_ISL_2225577, EPI_ISL_2225578, EPI_ISL_2225579, EPI_ISL_2225580, EPI_ISL_2225581, EPI_ISL_2225582, EPI_ISL_2225583, EPI_ISL_2225584, EPI_ISL_2225585 | | | |
| see above | University of Wisconsin-Madison AIDS Vaccine Research Laboratories | University of Wisconsin-Madison AIDS Vaccine Research Laboratories | Gage Moreno, Katarina Braun, et al. AIDS Vaccine Research Laboratories |
| EPI_ISL_2226635, EPI_ISL_2226636, EPI_ISL_2226637, EPI_ISL_2226638, EPI_ISL_2226639, EPI_ISL_2226640, EPI_ISL_2226641, EPI_ISL_2226642, EPI_ISL_2226643 | Infectious Disease Program, Broad Institute of Harvard and MIT | Infectious Disease Program, Broad Institute of Harvard and MIT | Lemieux,J.E., Siddle,K.J., Shaw,B., Adams,G., Pierce,V., Turbett,S., Anahtar,M., Branda,J., Slater,D., Harris,J., Lin,A.E., Gladden-Young,A., Lagerborg,K., Rudy,M., DeRuff,K., Carter,A., Normandin,E., Bauer,M., Reilly,S., Tomkins-Tinch,C., Loreth,C., Chaluvadi,S., Neumann,A., Cusick,C., Chapman,S.B., Gnirke,A., Flowers,K., Cerrato,F., Birren,B.W., Gallagher,G., Smole,S., Park,D.J., MacInnis,B.L., Ryan,E., LaRocque,R., Rosenberg,E., Sabeti,P.C. |
| EPI_ISL_2226644, EPI_ISL_2226645 | Laboratory Medicine, University of Washington | Laboratory Medicine, University of Washington | Roychoudhury,P., Xie,H., Shrestha,L., Bakhash,S.M., Lin,M.J., Baker,N.R., Ellis,S., Sathees,S., Huang,M.-L., Jerome,K.R., Greninger,A. |
| EPI_ISL_2226646 | Respiratory Viruses Branch, US Centers for Disease Control and Prevention | Respiratory Viruses Branch, US Centers for Disease Control and Prevention | Tao,Y., Li,Y., Zhang,J., Kelleher,A., Lynch,B., Queen,K.,Uehara,A., Cook,P., Ng,H.J.J., Marine,R., Paden,C.R., Batra,D.,Wang,H., Coalter,T., Padilla,J., Davis,M., Sheth,M., Nobles,S.,Burroughs,M., Lee,J., Retchless,A. and Tong,S. |
| EPI_ISL_2226649 | Department of General Services Division of Consolidated Laboratory Services, Virginia Division of Consolidated Laboratory Services Sequencing Submission Group | Department of General Services Division of Consolidated Laboratory Services, Virginia Division of Consolidated Laboratory Services Sequencing Submission Group | Fink,L. |
| EPI_ISL_2226650 | Department of General Services Division of Consolidated Laboratory Services, Virginia Division of Consolidated Laboratory Services Sequencing Submission Group | Department of General Services Division of Consolidated Laboratory Services, Virginia Division of Consolidated Laboratory Services Sequencing Submission Group | Fink,L. |
| EPI_ISL_2226766, EPI_ISL_2226768, EPI_ISL_2226769, EPI_ISL_2226770, EPI_ISL_2226771, EPI_ISL_2226772, EPI_ISL_2226773, EPI_ISL_2226774, EPI_ISL_2226775, EPI_ISL_2226776, EPI_ISL_2226777, EPI_ISL_2226778, EPI_ISL_2226780, EPI_ISL_2226781, EPI_ISL_2226782, EPI_ISL_2226783, EPI_ISL_2226784, EPI_ISL_2226785, EPI_ISL_2226786, EPI_ISL_2226787, EPI_ISL_2226788, EPI_ISL_2226789, EPI_ISL_2226790, EPI_ISL_2226791, EPI_ISL_2226792, EPI_ISL_2226793, EPI_ISL_2226794, EPI_ISL_2226795, EPI_ISL_2226797, EPI_ISL_2226798, EPI_ISL_2226799, EPI_ISL_2226800, EPI_ISL_2226801, EPI_ISL_2226802, EPI_ISL_2226803, EPI_ISL_2226804, EPI_ISL_2226805, EPI_ISL_2226806, EPI_ISL_2226807, EPI_ISL_2226808, EPI_ISL_2226810, EPI_ISL_2226811, EPI_ISL_2226812, EPI_ISL_2226813, EPI_ISL_2226814, EPI_ISL_2226815, EPI_ISL_2226816 | | | |
| see above | Wyoming Public Health Laboratory | Wyoming Public Health Laboratory | Jim Mildenberger, Taylor Fearing, Channing Weber, Ashley Norberg, Chayse Rowley, Marley Goetz, Brian Dominguez, Elliot Thomasson, Brittany Oher, Sam Britz, Cari Sloma, and Rob Christensen |
| EPI_ISL_2226817, EPI_ISL_2226818, EPI_ISL_2226819, EPI_ISL_2226820, EPI_ISL_2226821, EPI_ISL_2226822, EPI_ISL_2226823, EPI_ISL_2226824, EPI_ISL_2226825, EPI_ISL_2226826, EPI_ISL_2226827, EPI_ISL_2226828, EPI_ISL_2226829, EPI_ISL_2226830, EPI_ISL_2226831 | | | |
| see above | Wyoming Public Health Laboratory | Wyoming Public Health Laboratory | Jim Mildenberger, Wanda Manley, Noah Hull, Taylor Fearing, Lynette Gumbleton, Channing Weber, Ashley Norberg, Chayse Rowley, Marley Goetz, Brian Dominguez, Elliot Thomasson, Sam Britz, Cari Sloma, and Rob Christensen |
| EPI_ISL_2226832 | Department of General Services Division of Consolidated Laboratory Services, Virginia Division of Consolidated Laboratory Services Sequencing Submission Group | Department of General Services Division of Consolidated Laboratory Services, Virginia Division of Consolidated Laboratory Services Sequencing Submission Group | Fink,L. |
| EPI_ISL_2227374, EPI_ISL_2227375, EPI_ISL_2227376, EPI_ISL_2227377, EPI_ISL_2227378, EPI_ISL_2227379, EPI_ISL_2227380, EPI_ISL_2227381, EPI_ISL_2227382, EPI_ISL_2227383, EPI_ISL_2227384, EPI_ISL_2227385, EPI_ISL_2227386, EPI_ISL_2227387, EPI_ISL_2227388, EPI_ISL_2227390, EPI_ISL_2227391, EPI_ISL_2227392, EPI_ISL_2227393 | | | |
| see above | Wichita State University - Molecular Diagnostics Lab | Kansas Health and Environmental Lab | Mike Grose, Katherine Wiggins, Jonathan Barnell, Ben Olsen, and Phil Adam |
| EPI_ISL_2227394 | Kansas Health and Environmental Lab | Kansas Health and Environmental Lab | Mike Grose, Katherine Wiggins, Jonathan Barnell, Ben Olsen, and Phil Adam |
| EPI_ISL_2227396, EPI_ISL_2227397, EPI_ISL_2227399, EPI_ISL_2227400, EPI_ISL_2227401, EPI_ISL_2227402, EPI_ISL_2227403, EPI_ISL_2227404, EPI_ISL_2227405, EPI_ISL_2227406, EPI_ISL_2227407, EPI_ISL_2227408, EPI_ISL_2227409 | | | |
| see above | Wichita State University - Molecular Diagnostics Lab | Kansas Health and Environmental Lab | Mike Grose, Jonathan Barnell, Ben Olsen, and Phil Adam |
| EPI_ISL_2227410, EPI_ISL_2227411, EPI_ISL_2227412 | Kansas Health and Environmental Lab | Kansas Health and Environmental Lab | Mike Grose, Jonathan Barnell, Ben Olsen, and Phil Adam |
| EPI_ISL_2227413, EPI_ISL_2227414, EPI_ISL_2227415, EPI_ISL_2227416, EPI_ISL_2227417, EPI_ISL_2227418, EPI_ISL_2227419, EPI_ISL_2227420, EPI_ISL_2227421, EPI_ISL_2227422, EPI_ISL_2227423, EPI_ISL_2227424, EPI_ISL_2227425, EPI_ISL_2227426, EPI_ISL_2227427, EPI_ISL_2227429, EPI_ISL_2227430, EPI_ISL_2227431, EPI_ISL_2227432, EPI_ISL_2227433, EPI_ISL_2227434, EPI_ISL_2227435, EPI_ISL_2227436 | | | |
| see above | MD PHL | MD PHL | Maryland Department of Health Laboratories Administration |
| EPI_ISL_2227437, EPI_ISL_2227438, EPI_ISL_2227439, EPI_ISL_2227441 | Kansas Health and Environmental Lab | Kansas Health and Environmental Lab | Mike Grose, Katherine Wiggins, Jonathan Barnell, Ben Olsen, and Phil Adam |
| EPI_ISL_2227442, EPI_ISL_2227443, EPI_ISL_2227444, EPI_ISL_2227445, EPI_ISL_2227446, EPI_ISL_2227447, EPI_ISL_2227448, EPI_ISL_2227449, EPI_ISL_2227451, EPI_ISL_2227452, EPI_ISL_2227453, EPI_ISL_2227454, EPI_ISL_2227455, EPI_ISL_2227456 | | | |
| see above | Kansas Health and Environmental Lab | Wichita State University - Molecular Diagnostics Lab | Mike Grose, Katherine Wiggins, Jonathan Barnell, Ben Olsen, and Phil Adam |
| EPI_ISL_2227457, EPI_ISL_2227458, EPI_ISL_2227459 | Alaska State Virology Laboratory | Alaska State Virology Laboratory | Stephanie DeRonde, Elva House, Lisa Smith, Ph.D., Jack Chen, Ph.D. |
| EPI_ISL_2227462, EPI_ISL_2227463, EPI_ISL_2227464 | Alaska State Virology Laboratory | Alaska State Virology Laboratory | Stephanie DeRonde, Elva House, Jacob Zidek, Lisa Smith, Ph.D., Jack Chen, Ph.D. |
| EPI_ISL_2227466, EPI_ISL_2227467, EPI_ISL_2227469, EPI_ISL_2227470, | Oregon State University TRACE Project | OSU Center for Genome Research and Biocomputing | Oregon State University TRACE Project |

| | | | |
|---|---|---|---|
| EPI_ISL_2227471, EPI_ISL_2227472 | | | |
| EPI_ISL_2227473, EPI_ISL_2227475 | TriCore Reference Laboratories | Center for Global Health, University of New Mexico Health Sciences Center | Daryl Domman, Kurt Schwalm, Valerie Morley, Cecilia Thompson, Kendra Pesko, Karissa Culbreath, Darrell Dinwiddie |
| EPI_ISL_2227476, EPI_ISL_2227477, EPI_ISL_2227478, EPI_ISL_2227479, EPI_ISL_2227480, EPI_ISL_2227481, EPI_ISL_2227482, EPI_ISL_2227487, EPI_ISL_2227488, EPI_ISL_2227489, EPI_ISL_2227490, EPI_ISL_2227491, EPI_ISL_2227492, EPI_ISL_2227493, EPI_ISL_2227494, EPI_ISL_2227495 | | | |
| see above | Arizona State Public Health Laboratory | Arizona State Public Health Laboratory | Trung Huynh, Jessica Escobar, Katherine Fullerton, Nobuko Fukushima, Stacy White, Linda Getsinger, Victor Waddell |
| EPI_ISL_2227496, EPI_ISL_2227497, EPI_ISL_2227498, EPI_ISL_2227499, EPI_ISL_2227500, EPI_ISL_2227501, EPI_ISL_2227502, EPI_ISL_2227503, EPI_ISL_2227504, EPI_ISL_2227505, EPI_ISL_2227506, EPI_ISL_2227507, EPI_ISL_2227508, EPI_ISL_2227509, EPI_ISL_2227510, EPI_ISL_2227511, EPI_ISL_2227512, EPI_ISL_2227513, EPI_ISL_2227514, EPI_ISL_2227515, EPI_ISL_2227516, EPI_ISL_2227517, EPI_ISL_2227518, EPI_ISL_2227519, EPI_ISL_2227520, EPI_ISL_2227521, EPI_ISL_2227522, EPI_ISL_2227523, EPI_ISL_2227524, EPI_ISL_2227525, EPI_ISL_2227526, EPI_ISL_2227527, EPI_ISL_2227528, EPI_ISL_2227529, EPI_ISL_2227530, EPI_ISL_2227531, EPI_ISL_2227532, EPI_ISL_2227533, EPI_ISL_2227534, EPI_ISL_2227535, EPI_ISL_2227536, EPI_ISL_2227537, EPI_ISL_2227538, EPI_ISL_2227539, EPI_ISL_2227542, EPI_ISL_2227543, EPI_ISL_2227544, EPI_ISL_2227545, EPI_ISL_2227546, EPI_ISL_2227547, EPI_ISL_2227548, EPI_ISL_2227549 | | | |
| see above | TriCore Reference Laboratories | Center for Global Health, University of New Mexico Health Sciences Center | Daryl Domman, Kurt Schwalm, Valerie Morley, Cecilia Thompson, Kendra Pesko, Karissa Culbreath, Darrell Dinwiddie |
| EPI_ISL_2227550, EPI_ISL_2227551, EPI_ISL_2227552 | Arizona State Public Health Laboratory | Arizona State Public Health Laboratory | Trung Huynh, Jessica Escobar, Katherine Fullerton, Nobuko Fukushima, Stacy White, Linda Getsinger, Victor Waddell |
| EPI_ISL_2227556 | Arkansas Department of Health | Center for Global Health, University of New Mexico Health Sciences Center | Daryl Domman, Kurt Schwalm, Valerie Morley, Atul Kothari, Joshua L. Kennedy, Darrell Dinwiddie |
| EPI_ISL_2227566, EPI_ISL_2227567, EPI_ISL_2227568, EPI_ISL_2227574, EPI_ISL_2227575, EPI_ISL_2227576 | Arizona State Public Health Laboratory | Arizona State Public Health Laboratory | Trung Huynh, Jessica Escobar, Katherine Fullerton, Nobuko Fukushima, Stacy White, Linda Getsinger, Victor Waddell |
| EPI_ISL_2227577, EPI_ISL_2227578, EPI_ISL_2227579, EPI_ISL_2227580, EPI_ISL_2227581, EPI_ISL_2227582, EPI_ISL_2227583, EPI_ISL_2227584, EPI_ISL_2227585, EPI_ISL_2227586, EPI_ISL_2227587, EPI_ISL_2227588, EPI_ISL_2227589, EPI_ISL_2227590, EPI_ISL_2227591, EPI_ISL_2227592, EPI_ISL_2227593, EPI_ISL_2227594, EPI_ISL_2227595, EPI_ISL_2227596, EPI_ISL_2227597, EPI_ISL_2227598, EPI_ISL_2227599, EPI_ISL_2227600, EPI_ISL_2227601, EPI_ISL_2227602, EPI_ISL_2227603, EPI_ISL_2227604, EPI_ISL_2227605, EPI_ISL_2227606, EPI_ISL_2227607, EPI_ISL_2227608, EPI_ISL_2227609, EPI_ISL_2227610, EPI_ISL_2227611, EPI_ISL_2227612, EPI_ISL_2227613, EPI_ISL_2227614, EPI_ISL_2227615, EPI_ISL_2227616, EPI_ISL_2227617, EPI_ISL_2227618, EPI_ISL_2227619, EPI_ISL_2227620, EPI_ISL_2227621, EPI_ISL_2227622, EPI_ISL_2227623, EPI_ISL_2227624, EPI_ISL_2227625, EPI_ISL_2227626, EPI_ISL_2227627, EPI_ISL_2227628, EPI_ISL_2227629, EPI_ISL_2227630, EPI_ISL_2227631, EPI_ISL_2227632, EPI_ISL_2227633, EPI_ISL_2227634, EPI_ISL_2227635, EPI_ISL_2227636, EPI_ISL_2227637, EPI_ISL_2227638, EPI_ISL_2227639, EPI_ISL_2227640, EPI_ISL_2227641, EPI_ISL_2227642, EPI_ISL_2227643, EPI_ISL_2227644, EPI_ISL_2227645, EPI_ISL_2227646, EPI_ISL_2227647, EPI_ISL_2227675, EPI_ISL_2227676, EPI_ISL_2227677, EPI_ISL_2227678, EPI_ISL_2227679, EPI_ISL_2227680, EPI_ISL_2227681, EPI_ISL_2227682, EPI_ISL_2227683, EPI_ISL_2227684, EPI_ISL_2227685, EPI_ISL_2227686, EPI_ISL_2227687, EPI_ISL_2227688, EPI_ISL_2227689, EPI_ISL_2227690, EPI_ISL_2227691, EPI_ISL_2227692, EPI_ISL_2227693, EPI_ISL_2227694, EPI_ISL_2227695, EPI_ISL_2227696, EPI_ISL_2227697, EPI_ISL_2227698, EPI_ISL_2227699, EPI_ISL_2227700, EPI_ISL_2227701, EPI_ISL_2227702, EPI_ISL_2227703, EPI_ISL_2227704, EPI_ISL_2227705, EPI_ISL_2227706, EPI_ISL_2227707, EPI_ISL_2227708, EPI_ISL_2227709, EPI_ISL_2227710, EPI_ISL_2227711, EPI_ISL_2227712, EPI_ISL_2227713, EPI_ISL_2227714, EPI_ISL_2227715, EPI_ISL_2227716, EPI_ISL_2227717, EPI_ISL_2227718, EPI_ISL_2227719, EPI_ISL_2227720, EPI_ISL_2227721, EPI_ISL_2227722, EPI_ISL_2227723, EPI_ISL_2227724, EPI_ISL_2227725, EPI_ISL_2227726, EPI_ISL_2227727, EPI_ISL_2227728, EPI_ISL_2227729, EPI_ISL_2227730, EPI_ISL_2227731, EPI_ISL_2227732, EPI_ISL_2227733, EPI_ISL_2227734, EPI_ISL_2227735, EPI_ISL_2227736, EPI_ISL_2227737, EPI_ISL_2227738, EPI_ISL_2227739, EPI_ISL_2227740, EPI_ISL_2227741, EPI_ISL_2227742, EPI_ISL_2227743 | | | |
| see above | OHSU Lab Services Molecular Microbiology Lab | Oregon SARS-CoV-2 Genome Sequencing Center | Brendan L. O'Connell, Sally Grindstaff, Sonia Acharya, Cierra LaBlanc, Kayla Carter, Ruth V. Nichols, Alec J. Hirsch, Donna Hansel, Guang Fan, Xuan Qin, Daniel N. Streblow, William B. Messer, Andrew C. Adey, Benjamin N. Bimber, Brian J. O'Roak |
| EPI_ISL_2227744 | San Juan Regional Medical Center | New Mexico Department of Health Scientific Laboratory | Ellie Johnson, D'eldra Malone, Jennifer Benoit, Ratheesh Rajan, Linda Salazar, Anastacia Griego-Fisher |
| EPI_ISL_2228075 | Johns Hopkins University | Genomics and Discovery, Respiratory Viruses Branch, Division of Viral Diseases, Centers for Disease Control and Prevention | Yan Li, Jing Zhang, Anna Kelleher, Ying Tao, Brian Lynch, Krista Queen, Anna Uehara, Peter Cook, Han Jia Justin Ng, Rachel Marine, Clinton R. Paden, Dhwani Batra, Haibin Wang, Tara Coalter, Jasmine Padilla, Morgan Davis, Mili Sheth, Sarah Nobles, Mark Burroughs, Justin Lee, Adam Retchless, Suxiang Tong |
| EPI_ISL_2228088, EPI_ISL_2228089, EPI_ISL_2228090 | CT Department of Public Health | CT Department of Public Health | Claire_Pearson,Tu_N_Nguyen |
| EPI_ISL_2228091, EPI_ISL_2228092, EPI_ISL_2228093, EPI_ISL_2228094, EPI_ISL_2228095, EPI_ISL_2228097, EPI_ISL_2228098 | CDPH, Viral and Rickettsial Disease Laboratory | Pathogen Discovery, Respiratory Viruses Branch, Division of Viral Diseases, Centers for Disease Control and Prevention | Yan Li, Jing Zhang, Anna Kelleher, Ying Tao, Brian Lynch, Krista Queen, Anna Uehara, Peter Cook, Han Jia Justin Ng, Rachel Marine, Clinton R. Paden, Dhwani Batra, Haibin Wang, Tara Coalter, Jasmine Padilla, Morgan Davis, Mili Sheth, Sarah Nobles, Mark Burroughs, Justin Lee, Adam Retchless, Suxiang Tong |
| EPI_ISL_2228205, EPI_ISL_2228206, EPI_ISL_2228207, EPI_ISL_2228208, EPI_ISL_2228209, EPI_ISL_2228210, EPI_ISL_2228211, EPI_ISL_2228212, EPI_ISL_2228213, EPI_ISL_2228214, EPI_ISL_2228215, EPI_ISL_2228216, EPI_ISL_2228217, EPI_ISL_2228218, EPI_ISL_2228219, EPI_ISL_2228220, EPI_ISL_2228221, EPI_ISL_2228222, EPI_ISL_2228223, EPI_ISL_2228225, EPI_ISL_2228226, EPI_ISL_2228227, EPI_ISL_2228228, EPI_ISL_2228229, EPI_ISL_2228230, EPI_ISL_2228231, EPI_ISL_2228232, EPI_ISL_2228233, EPI_ISL_2228234, EPI_ISL_2228235, EPI_ISL_2228236, EPI_ISL_2228237, EPI_ISL_2228238, EPI_ISL_2228239, EPI_ISL_2228240, EPI_ISL_2228241, EPI_ISL_2228242, EPI_ISL_2228243, EPI_ISL_2228244, EPI_ISL_2228245 | | | |
| see above | TriCore Reference Laboratories | Center for Global Health, University of New Mexico Health Sciences Center | Daryl Domman, Kurt Schwalm, Valerie Morley, Cecilia Thompson, Kendra Pesko, Karissa Culbreath, Darrell Dinwiddie |
| EPI_ISL_2228246, EPI_ISL_2228247 | Contra Costa County Public Health Lab | Chan-Zuckerberg Biohub | CZB Cliahub Consortium |
| EPI_ISL_2228248 | CA DPH Viral and Rickettsial Disease Laboratory | Chan-Zuckerberg Biohub | CZB Cliahub Consortium |
| EPI_ISL_2228249 | Contra Costa County Public Health Lab | Chan-Zuckerberg Biohub | CZB Cliahub Consortium |
| EPI_ISL_2228250 | CA DPH Viral and Rickettsial Disease Laboratory | Chan-Zuckerberg Biohub | CZB Cliahub Consortium |
| EPI_ISL_2228251, EPI_ISL_2228252 | Contra Costa County Public Health Lab | Chan-Zuckerberg Biohub | CZB Cliahub Consortium |
| EPI_ISL_2228253, EPI_ISL_2228254 | CA DPH Viral and Rickettsial Disease Laboratory | Chan-Zuckerberg Biohub | CZB Cliahub Consortium |
| EPI_ISL_2228256, EPI_ISL_2228257 | Contra Costa County Public Health Lab | Chan-Zuckerberg Biohub | CZB Cliahub Consortium |
| EPI_ISL_2228258 | CA DPH Viral and Rickettsial Disease Laboratory | Chan-Zuckerberg Biohub | CZB Cliahub Consortium |
| EPI_ISL_2228259 | Santa Clara County Public Health Laboratory | Chan-Zuckerberg Biohub | CZB Cliahub Consortium |
| EPI_ISL_2228260, EPI_ISL_2228261 | CA DPH Viral and Rickettsial Disease Laboratory | Chan-Zuckerberg Biohub | CZB Cliahub Consortium |
| EPI_ISL_2228262 | Contra Costa County Public Health Lab | Chan-Zuckerberg Biohub | CZB Cliahub Consortium |
| EPI_ISL_2228263 | County of San Luis Obispo Public Health Laboratory | Chan-Zuckerberg Biohub | CZB Cliahub Consortium |
| EPI_ISL_2228264 | Humboldt County Public Health Laboratory | Chan-Zuckerberg Biohub | CZB Cliahub Consortium |
| EPI_ISL_2228265, EPI_ISL_2228266, EPI_ISL_2228267 | Contra Costa County Public Health Lab | Chan-Zuckerberg Biohub | CZB Cliahub Consortium |
| EPI_ISL_2228268, EPI_ISL_2228269 | CA DPH Viral and Rickettsial Disease Laboratory | Chan-Zuckerberg Biohub | CZB Cliahub Consortium |
| EPI_ISL_2228270, EPI_ISL_2228271 | Contra Costa County Public Health Lab | Chan-Zuckerberg Biohub | CZB Cliahub Consortium |
| EPI_ISL_2228272, EPI_ISL_2228273 | CA DPH Viral and Rickettsial Disease Laboratory | Chan-Zuckerberg Biohub | CZB Cliahub Consortium |
| EPI_ISL_2228274 | County of San Luis Obispo Public Health Laboratory | Chan-Zuckerberg Biohub | CZB Cliahub Consortium |
| EPI_ISL_2228275 | Contra Costa County Public Health Lab | Chan-Zuckerberg Biohub | CZB Cliahub Consortium |
| EPI_ISL_2228276 | CA DPH Viral and Rickettsial Disease Laboratory | Chan-Zuckerberg Biohub | CZB Cliahub Consortium |
| EPI_ISL_2228277 | Contra Costa County Public Health Lab | Chan-Zuckerberg Biohub | CZB Cliahub Consortium |
| EPI_ISL_2228278, EPI_ISL_2228279 | CA DPH Viral and Rickettsial Disease Laboratory | Chan-Zuckerberg Biohub | CZB Cliahub Consortium |
| EPI_ISL_2228280 | Contra Costa County Public Health Lab | Chan-Zuckerberg Biohub | CZB Cliahub Consortium |

| | | | |
|---|---|---|---|
| EPI_ISL_2228281 | CA DPH Viral and Rickettsial Disease Laboratory | Chan-Zuckerberg Biohub | CZB Cliahub Consortium |
| EPI_ISL_2228282, EPI_ISL_2228283 | Contra Costa County Public Health Lab | Chan-Zuckerberg Biohub | CZB Cliahub Consortium |
| EPI_ISL_2228284, EPI_ISL_2228285 | Humboldt County Public Health Laboratory | Chan-Zuckerberg Biohub | CZB Cliahub Consortium |
| EPI_ISL_2228286 | Contra Costa County Public Health Lab | Chan-Zuckerberg Biohub | CZB Cliahub Consortium |
| EPI_ISL_2228287 | CA DPH Viral and Rickettsial Disease Laboratory | Chan-Zuckerberg Biohub | CZB Cliahub Consortium |
| EPI_ISL_2228288, EPI_ISL_2228290 | Humboldt County Public Health Laboratory | Chan-Zuckerberg Biohub | CZB Cliahub Consortium |
| EPI_ISL_2228291 | Contra Costa County Public Health Lab | Chan-Zuckerberg Biohub | CZB Cliahub Consortium |
| EPI_ISL_2228292 | Orange County Public Health Lab | Chan-Zuckerberg Biohub | CZB Cliahub Consortium |
| EPI_ISL_2228293 | County of San Luis Obispo Public Health Laboratory | Chan-Zuckerberg Biohub | CZB Cliahub Consortium |
| EPI_ISL_2228294 | Contra Costa County Public Health Lab | Chan-Zuckerberg Biohub | CZB Cliahub Consortium |
| EPI_ISL_2228295 | Humboldt County Public Health Laboratory | Chan-Zuckerberg Biohub | CZB Cliahub Consortium |
| EPI_ISL_2228296 | Contra Costa County Public Health Lab | Chan-Zuckerberg Biohub | CZB Cliahub Consortium |
| EPI_ISL_2228297 | County of San Luis Obispo Public Health Laboratory | Chan-Zuckerberg Biohub | CZB Cliahub Consortium |
| EPI_ISL_2228298, EPI_ISL_2228299 | Contra Costa County Public Health Lab | Chan-Zuckerberg Biohub | CZB Cliahub Consortium |
| EPI_ISL_2228300 | CA DPH Viral and Rickettsial Disease Laboratory | Chan-Zuckerberg Biohub | CZB Cliahub Consortium |
| EPI_ISL_2228301 | Humboldt County Public Health Laboratory | Chan-Zuckerberg Biohub | CZB Cliahub Consortium |
| EPI_ISL_2228302 | CA DPH Viral and Rickettsial Disease Laboratory | Chan-Zuckerberg Biohub | CZB Cliahub Consortium |
| EPI_ISL_2228303, EPI_ISL_2228304 | Humboldt County Public Health Laboratory | Chan-Zuckerberg Biohub | CZB Cliahub Consortium |
| EPI_ISL_2228305 | Contra Costa County Public Health Lab | Chan-Zuckerberg Biohub | CZB Cliahub Consortium |
| EPI_ISL_2228306, EPI_ISL_2228307 | CA DPH Viral and Rickettsial Disease Laboratory | Chan-Zuckerberg Biohub | CZB Cliahub Consortium |
| EPI_ISL_2228308, EPI_ISL_2228309 | Contra Costa County Public Health Lab | Chan-Zuckerberg Biohub | CZB Cliahub Consortium |
| EPI_ISL_2228310 | Humboldt County Public Health Laboratory | Chan-Zuckerberg Biohub | CZB Cliahub Consortium |
| EPI_ISL_2228311 | County of San Luis Obispo Public Health Laboratory | Chan-Zuckerberg Biohub | CZB Cliahub Consortium |
| EPI_ISL_2228312 | Contra Costa County Public Health Lab | Chan-Zuckerberg Biohub | CZB Cliahub Consortium |
| EPI_ISL_2228313 | CA DPH Viral and Rickettsial Disease Laboratory | Chan-Zuckerberg Biohub | CZB Cliahub Consortium |
| EPI_ISL_2228314 | Humboldt County Public Health Laboratory | Chan-Zuckerberg Biohub | CZB Cliahub Consortium |
| EPI_ISL_2228315 | Contra Costa County Public Health Lab | Chan-Zuckerberg Biohub | CZB Cliahub Consortium |
| EPI_ISL_2228316 | CA DPH Viral and Rickettsial Disease Laboratory | Chan-Zuckerberg Biohub | CZB Cliahub Consortium |
| EPI_ISL_2228317 | Contra Costa County Public Health Lab | Chan-Zuckerberg Biohub | CZB Cliahub Consortium |
| EPI_ISL_2228318 | CA DPH Viral and Rickettsial Disease Laboratory | Chan-Zuckerberg Biohub | CZB Cliahub Consortium |
| EPI_ISL_2228319 | Humboldt County Public Health Laboratory | Chan-Zuckerberg Biohub | CZB Cliahub Consortium |
| EPI_ISL_2228320, EPI_ISL_2228321, EPI_ISL_2228322, EPI_ISL_2228323, EPI_ISL_2228324, EPI_ISL_2228326 | Contra Costa County Public Health Lab | Chan-Zuckerberg Biohub | CZB Cliahub Consortium |
| EPI_ISL_2228327, EPI_ISL_2228329 | CA DPH Viral and Rickettsial Disease Laboratory | Chan-Zuckerberg Biohub | CZB Cliahub Consortium |
| EPI_ISL_2228330, EPI_ISL_2228331 | Contra Costa County Public Health Lab | Chan-Zuckerberg Biohub | CZB Cliahub Consortium |
| EPI_ISL_2228332 | CA DPH Viral and Rickettsial Disease Laboratory | Chan-Zuckerberg Biohub | CZB Cliahub Consortium |
| EPI_ISL_2228333, EPI_ISL_2228334, EPI_ISL_2228335 | Contra Costa County Public Health Lab | Chan-Zuckerberg Biohub | CZB Cliahub Consortium |
| EPI_ISL_2228336 | Humboldt County Public Health Laboratory | Chan-Zuckerberg Biohub | CZB Cliahub Consortium |
| EPI_ISL_2228337 | CA DPH Viral and Rickettsial Disease Laboratory | Chan-Zuckerberg Biohub | CZB Cliahub Consortium |
| EPI_ISL_2228338 | Contra Costa County Public Health Lab | Chan-Zuckerberg Biohub | CZB Cliahub Consortium |
| EPI_ISL_2228339 | County of San Luis Obispo Public Health Laboratory | Chan-Zuckerberg Biohub | CZB Cliahub Consortium |
| EPI_ISL_2228340 | CA DPH Viral and Rickettsial Disease Laboratory | Chan-Zuckerberg Biohub | CZB Cliahub Consortium |
| EPI_ISL_2228341, EPI_ISL_2228342 | Contra Costa County Public Health Lab | Chan-Zuckerberg Biohub | CZB Cliahub Consortium |
| EPI_ISL_2228343 | Humboldt County Public Health Laboratory | Chan-Zuckerberg Biohub | CZB Cliahub Consortium |
| EPI_ISL_2228344 | Contra Costa County Public Health Lab | Chan-Zuckerberg Biohub | CZB Cliahub Consortium |
| EPI_ISL_2228345, EPI_ISL_2228346 | CA DPH Viral and Rickettsial Disease Laboratory | Chan-Zuckerberg Biohub | CZB Cliahub Consortium |
| EPI_ISL_2228347 | Contra Costa County Public Health Lab | Chan-Zuckerberg Biohub | CZB Cliahub Consortium |
| EPI_ISL_2228348, EPI_ISL_2228349, EPI_ISL_2228350 | CA DPH Viral and Rickettsial Disease Laboratory | Chan-Zuckerberg Biohub | CZB Cliahub Consortium |
| EPI_ISL_2228351 | Orange County Public Health Lab | Chan-Zuckerberg Biohub | CZB Cliahub Consortium |

| | | | |
|---|---|---|---|
| EPI_ISL_2228352 | Contra Costa County Public Health Lab | Chan-Zuckerberg Biohub | CZB Cliahub Consortium |
| EPI_ISL_2228353 | CA DPH Viral and Rickettsial Disease Laboratory | Chan-Zuckerberg Biohub | CZB Cliahub Consortium |
| EPI_ISL_2228354 | Santa Clara County Public Health Laboratory | Chan-Zuckerberg Biohub | CZB Cliahub Consortium |
| EPI_ISL_2228355, EPI_ISL_2228356 | CA DPH Viral and Rickettsial Disease Laboratory | Chan-Zuckerberg Biohub | CZB Cliahub Consortium |
| EPI_ISL_2228357 | County of San Luis Obispo Public Health Laboratory | Chan-Zuckerberg Biohub | CZB Cliahub Consortium |
| EPI_ISL_2228358 | CA DPH Viral and Rickettsial Disease Laboratory | Chan-Zuckerberg Biohub | CZB Cliahub Consortium |
| EPI_ISL_2228359 | Humboldt County Public Health Laboratory | Chan-Zuckerberg Biohub | CZB Cliahub Consortium |
| EPI_ISL_2228360, EPI_ISL_2228361 | CA DPH Viral and Rickettsial Disease Laboratory | Chan-Zuckerberg Biohub | CZB Cliahub Consortium |
| EPI_ISL_2228362 | Contra Costa County Public Health Lab | Chan-Zuckerberg Biohub | CZB Cliahub Consortium |
| EPI_ISL_2228363 | Humboldt County Public Health Laboratory | Chan-Zuckerberg Biohub | CZB Cliahub Consortium |
| EPI_ISL_2228365 | Contra Costa County Public Health Lab | Chan-Zuckerberg Biohub | CZB Cliahub Consortium |
| EPI_ISL_2228366 | Humboldt County Public Health Laboratory | Chan-Zuckerberg Biohub | CZB Cliahub Consortium |
| EPI_ISL_2228367 | County of San Luis Obispo Public Health Laboratory | Chan-Zuckerberg Biohub | CZB Cliahub Consortium |
| EPI_ISL_2228368 | Humboldt County Public Health Laboratory | Chan-Zuckerberg Biohub | CZB Cliahub Consortium |
| EPI_ISL_2228369, EPI_ISL_2228370 | CA DPH Viral and Rickettsial Disease Laboratory | Chan-Zuckerberg Biohub | CZB Cliahub Consortium |
| EPI_ISL_2228371, EPI_ISL_2228372, EPI_ISL_2228373 | Contra Costa County Public Health Lab | Chan-Zuckerberg Biohub | CZB Cliahub Consortium |
| EPI_ISL_2228374 | CA DPH Viral and Rickettsial Disease Laboratory | Chan-Zuckerberg Biohub | CZB Cliahub Consortium |
| EPI_ISL_2228375 | Contra Costa County Public Health Lab | Chan-Zuckerberg Biohub | CZB Cliahub Consortium |
| EPI_ISL_2228376 | CA DPH Viral and Rickettsial Disease Laboratory | Chan-Zuckerberg Biohub | CZB Cliahub Consortium |
| EPI_ISL_2228377 | Humboldt County Public Health Laboratory | Chan-Zuckerberg Biohub | CZB Cliahub Consortium |
| EPI_ISL_2228378, EPI_ISL_2228379 | Contra Costa County Public Health Lab | Chan-Zuckerberg Biohub | CZB Cliahub Consortium |
| EPI_ISL_2228380 | CA DPH Viral and Rickettsial Disease Laboratory | Chan-Zuckerberg Biohub | CZB Cliahub Consortium |
| EPI_ISL_2228381 | Santa Clara County Public Health Laboratory | Chan-Zuckerberg Biohub | CZB Cliahub Consortium |
| EPI_ISL_2228382 | CA DPH Viral and Rickettsial Disease Laboratory | Chan-Zuckerberg Biohub | CZB Cliahub Consortium |
| EPI_ISL_2228383 | Orange County Public Health Lab | Chan-Zuckerberg Biohub | CZB Cliahub Consortium |
| EPI_ISL_2228384 | Contra Costa County Public Health Lab | Chan-Zuckerberg Biohub | CZB Cliahub Consortium |
| EPI_ISL_2228385 | CA DPH Viral and Rickettsial Disease Laboratory | Chan-Zuckerberg Biohub | CZB Cliahub Consortium |
| EPI_ISL_2228386, EPI_ISL_2228387, EPI_ISL_2228388 | Contra Costa County Public Health Lab | Chan-Zuckerberg Biohub | CZB Cliahub Consortium |
| EPI_ISL_2228390 | County of San Luis Obispo Public Health Laboratory | Chan-Zuckerberg Biohub | CZB Cliahub Consortium |
| EPI_ISL_2228391 | CA DPH Viral and Rickettsial Disease Laboratory | Chan-Zuckerberg Biohub | CZB Cliahub Consortium |
| EPI_ISL_2228392, EPI_ISL_2228393, EPI_ISL_2228394 | Contra Costa County Public Health Lab | Chan-Zuckerberg Biohub | CZB Cliahub Consortium |
| EPI_ISL_2228395 | Humboldt County Public Health Laboratory | Chan-Zuckerberg Biohub | CZB Cliahub Consortium |
| EPI_ISL_2228396 | Santa Clara County Public Health Laboratory | Chan-Zuckerberg Biohub | CZB Cliahub Consortium |
| EPI_ISL_2228397, EPI_ISL_2228398 | Contra Costa County Public Health Lab | Chan-Zuckerberg Biohub | CZB Cliahub Consortium |
| EPI_ISL_2228399 | Humboldt County Public Health Laboratory | Chan-Zuckerberg Biohub | CZB Cliahub Consortium |
| EPI_ISL_2228400 | County of San Luis Obispo Public Health Laboratory | Chan-Zuckerberg Biohub | CZB Cliahub Consortium |
| EPI_ISL_2228401 | Contra Costa County Public Health Lab | Chan-Zuckerberg Biohub | CZB Cliahub Consortium |
| EPI_ISL_2228402 | CA DPH Viral and Rickettsial Disease Laboratory | Chan-Zuckerberg Biohub | CZB Cliahub Consortium |
| EPI_ISL_2228403 | Contra Costa County Public Health Lab | Chan-Zuckerberg Biohub | CZB Cliahub Consortium |
| EPI_ISL_2228404 | Humboldt County Public Health Laboratory | Chan-Zuckerberg Biohub | CZB Cliahub Consortium |
| EPI_ISL_2228405, EPI_ISL_2228406 | CA DPH Viral and Rickettsial Disease Laboratory | Chan-Zuckerberg Biohub | CZB Cliahub Consortium |
| EPI_ISL_2228407, EPI_ISL_2228408 | Contra Costa County Public Health Lab | Chan-Zuckerberg Biohub | CZB Cliahub Consortium |
| EPI_ISL_2228410 | CA DPH Viral and Rickettsial Disease Laboratory | Chan-Zuckerberg Biohub | CZB Cliahub Consortium |
| EPI_ISL_2228411, EPI_ISL_2228412 | Humboldt County Public Health Laboratory | Chan-Zuckerberg Biohub | CZB Cliahub Consortium |
| EPI_ISL_2228413 | Contra Costa County Public Health Lab | Chan-Zuckerberg Biohub | CZB Cliahub Consortium |
| EPI_ISL_2228414 | CA DPH Viral and Rickettsial Disease Laboratory | Chan-Zuckerberg Biohub | CZB Cliahub Consortium |
| EPI_ISL_2228415 | Santa Clara County Public Health Laboratory | Chan-Zuckerberg Biohub | CZB Cliahub Consortium |
| EPI_ISL_2228416 | Humboldt County Public Health | Chan-Zuckerberg Biohub | CZB Cliahub Consortium |

| | | | |
|---|---|---|---|
| | Laboratory | | |
| EPI_ISL_2228417 | Contra Costa County Public Health Lab | Chan-Zuckerberg Biohub | CZB Cliahub Consortium |
| EPI_ISL_2228418 | CA DPH Viral and Rickettsial Disease Laboratory | Chan-Zuckerberg Biohub | CZB Cliahub Consortium |
| EPI_ISL_2228419 | Humboldt County Public Health Laboratory | Chan-Zuckerberg Biohub | CZB Cliahub Consortium |
| EPI_ISL_2228420 | County of San Luis Obispo Public Health Laboratory | Chan-Zuckerberg Biohub | CZB Cliahub Consortium |
| EPI_ISL_2228421 | CA DPH Viral and Rickettsial Disease Laboratory | Chan-Zuckerberg Biohub | CZB Cliahub Consortium |
| EPI_ISL_2228422 | Humboldt County Public Health Laboratory | Chan-Zuckerberg Biohub | CZB Cliahub Consortium |
| EPI_ISL_2228423 | Contra Costa County Public Health Lab | Chan-Zuckerberg Biohub | CZB Cliahub Consortium |
| EPI_ISL_2228424 | Humboldt County Public Health Laboratory | Chan-Zuckerberg Biohub | CZB Cliahub Consortium |
| EPI_ISL_2228425 | Contra Costa County Public Health Lab | Chan-Zuckerberg Biohub | CZB Cliahub Consortium |
| EPI_ISL_2228426 | CA DPH Viral and Rickettsial Disease Laboratory | Chan-Zuckerberg Biohub | CZB Cliahub Consortium |
| EPI_ISL_2228427, EPI_ISL_2228428 | Contra Costa County Public Health Lab | Chan-Zuckerberg Biohub | CZB Cliahub Consortium |
| EPI_ISL_2228934, EPI_ISL_2228936, EPI_ISL_2228939, EPI_ISL_2228941, EPI_ISL_2228944, EPI_ISL_2228947, EPI_ISL_2228949, EPI_ISL_2228951, EPI_ISL_2228954, EPI_ISL_2228957 | SC Dept of Health and Env. Control-Bureau of Laboratories | Centers for Disease Control and Prevention Division of Viral Diseases, Pathogen Discovery | Mili Sheth, Sarah Nobles, Jasmine Padilla, Mark Burroughs, Shoshona Le, Katie Dillon, Peter Cook, Clinton R. Paden, Dhwani Batra, Krista Queen, Kristen Knipe, Dakota Howard, Yvette Unoarumhi, Darlene Wagner, Matthew Schmerer, Ben L. Rambo-Martin, Kristine Lacek, Sam Shepard, Alison Laufer Halpin, Dave Wentworth, Vivien Dugan, Suxiang Tong, Justin Lee |
| EPI_ISL_2228960, EPI_ISL_2228962, EPI_ISL_2228965, EPI_ISL_2228967, EPI_ISL_2228970, EPI_ISL_2228973, EPI_ISL_2228976 | DE Public Health Laboratory | Centers for Disease Control and Prevention Division of Viral Diseases, Pathogen Discovery | Mili Sheth, Sarah Nobles, Jasmine Padilla, Mark Burroughs, Shoshona Le, Katie Dillon, Peter Cook, Clinton R. Paden, Dhwani Batra, Krista Queen, Kristen Knipe, Dakota Howard, Yvette Unoarumhi, Darlene Wagner, Matthew Schmerer, Ben L. Rambo-Martin, Kristine Lacek, Sam Shepard, Alison Laufer Halpin, Dave Wentworth, Vivien Dugan, Suxiang Tong, Justin Lee |
| EPI_ISL_2228979, EPI_ISL_2228982, EPI_ISL_2228984, EPI_ISL_2228987, EPI_ISL_2228990, EPI_ISL_2228992, EPI_ISL_2228995 | SD Public Health Laboratory | Centers for Disease Control and Prevention Division of Viral Diseases, Pathogen Discovery | Mili Sheth, Sarah Nobles, Jasmine Padilla, Mark Burroughs, Shoshona Le, Katie Dillon, Peter Cook, Clinton R. Paden, Dhwani Batra, Krista Queen, Kristen Knipe, Dakota Howard, Yvette Unoarumhi, Darlene Wagner, Matthew Schmerer, Ben L. Rambo-Martin, Kristine Lacek, Sam Shepard, Alison Laufer Halpin, Dave Wentworth, Vivien Dugan, Suxiang Tong, Justin Lee |
| EPI_ISL_2228998, EPI_ISL_2229001, EPI_ISL_2229003, EPI_ISL_2229006, EPI_ISL_2229008, EPI_ISL_2229011 | ME Health and Environmental Testing Laboratory | Centers for Disease Control and Prevention Division of Viral Diseases, Pathogen Discovery | Mili Sheth, Sarah Nobles, Jasmine Padilla, Mark Burroughs, Shoshona Le, Katie Dillon, Peter Cook, Clinton R. Paden, Dhwani Batra, Krista Queen, Kristen Knipe, Dakota Howard, Yvette Unoarumhi, Darlene Wagner, Matthew Schmerer, Ben L. Rambo-Martin, Kristine Lacek, Sam Shepard, Alison Laufer Halpin, Dave Wentworth, Vivien Dugan, Suxiang Tong, Justin Lee |
| EPI_ISL_2229014, EPI_ISL_2229017, EPI_ISL_2229020, EPI_ISL_2229022, EPI_ISL_2229025, EPI_ISL_2229027, EPI_ISL_2229030, EPI_ISL_2229032, EPI_ISL_2229034, EPI_ISL_2229036, EPI_ISL_2229039, EPI_ISL_2229042, EPI_ISL_2229045, EPI_ISL_2229047, EPI_ISL_2229050, EPI_ISL_2229052, EPI_ISL_2229054 | | | |
| see above | "AK State Public Health Lab, State Health Department" | Centers for Disease Control and Prevention Division of Viral Diseases, Pathogen Discovery | Mili Sheth, Sarah Nobles, Jasmine Padilla, Mark Burroughs, Shoshona Le, Katie Dillon, Peter Cook, Clinton R. Paden, Dhwani Batra, Krista Queen, Kristen Knipe, Dakota Howard, Yvette Unoarumhi, Darlene Wagner, Matthew Schmerer, Ben L. Rambo-Martin, Kristine Lacek, Sam Shepard, Alison Laufer Halpin, Dave Wentworth, Vivien Dugan, Suxiang Tong, Justin Lee |
| EPI_ISL_2229055, EPI_ISL_2229056, EPI_ISL_2229057 | FL Bureau of Public Health Laboratories-Miami | Centers for Disease Control and Prevention Division of Viral Diseases, Pathogen Discovery | Mili Sheth, Sarah Nobles, Jasmine Padilla, Mark Burroughs, Shoshona Le, Katie Dillon, Peter Cook, Clinton R. Paden, Dhwani Batra, Krista Queen, Kristen Knipe, Dakota Howard, Yvette Unoarumhi, Darlene Wagner, Matthew Schmerer, Ben L. Rambo-Martin, Kristine Lacek, Sam Shepard, Alison Laufer Halpin, Dave Wentworth, Vivien Dugan, Suxiang Tong, Justin Lee |
| EPI_ISL_2229058, EPI_ISL_2229059, EPI_ISL_2229060, EPI_ISL_2229061, EPI_ISL_2229062, EPI_ISL_2229063 | IA State Hygienic Laboratory | Centers for Disease Control and Prevention Division of Viral Diseases, Pathogen Discovery | Mili Sheth, Sarah Nobles, Jasmine Padilla, Mark Burroughs, Shoshona Le, Katie Dillon, Peter Cook, Clinton R. Paden, Dhwani Batra, Krista Queen, Kristen Knipe, Dakota Howard, Yvette Unoarumhi, Darlene Wagner, Matthew Schmerer, Ben L. Rambo-Martin, Kristine Lacek, Sam Shepard, Alison Laufer Halpin, Dave Wentworth, Vivien Dugan, Suxiang Tong, Justin Lee |
| EPI_ISL_2229064, EPI_ISL_2229065, EPI_ISL_2229066, EPI_ISL_2229067, EPI_ISL_2229068, EPI_ISL_2229069, EPI_ISL_2229070, EPI_ISL_2229071, EPI_ISL_2229072 | FL Bureau of Public Health Laboratories-Tampa | Centers for Disease Control and Prevention Division of Viral Diseases, Pathogen Discovery | Mili Sheth, Sarah Nobles, Jasmine Padilla, Mark Burroughs, Shoshona Le, Katie Dillon, Peter Cook, Clinton R. Paden, Dhwani Batra, Krista Queen, Kristen Knipe, Dakota Howard, Yvette Unoarumhi, Darlene Wagner, Matthew Schmerer, Ben L. Rambo-Martin, Kristine Lacek, Sam Shepard, Alison Laufer Halpin, Dave Wentworth, Vivien Dugan, Suxiang Tong, Justin Lee |
| EPI_ISL_2229073 | VA-Division of Consolidated Laboratory Services | Centers for Disease Control and Prevention Division of Viral Diseases, Pathogen Discovery | Mili Sheth, Sarah Nobles, Jasmine Padilla, Mark Burroughs, Shoshona Le, Katie Dillon, Peter Cook, Clinton R. Paden, Dhwani Batra, Krista Queen, Kristen Knipe, Dakota Howard, Yvette Unoarumhi, Darlene Wagner, Matthew Schmerer, Ben L. Rambo-Martin, Kristine Lacek, Sam Shepard, Alison Laufer Halpin, Dave Wentworth, Vivien Dugan, Suxiang Tong, Justin Lee |
| EPI_ISL_2229074, EPI_ISL_2229075, EPI_ISL_2229076, EPI_ISL_2229077, EPI_ISL_2229078, EPI_ISL_2229079, EPI_ISL_2229080, EPI_ISL_2229082, EPI_ISL_2229083, EPI_ISL_2229084 | NE Public Health Laboratory | Centers for Disease Control and Prevention Division of Viral Diseases, Pathogen Discovery | Mili Sheth, Sarah Nobles, Jasmine Padilla, Mark Burroughs, Shoshona Le, Katie Dillon, Peter Cook, Clinton R. Paden, Dhwani Batra, Krista Queen, Kristen Knipe, Dakota Howard, Yvette Unoarumhi, Darlene Wagner, Matthew Schmerer, Ben L. Rambo-Martin, Kristine Lacek, Sam Shepard, Alison Laufer Halpin, Dave Wentworth, Vivien Dugan, Suxiang Tong, Justin Lee |
| EPI_ISL_2229085, EPI_ISL_2229086 | "OK Public Health Laboratory, Oklahoma State DOH" | Centers for Disease Control and Prevention Division of Viral Diseases, Pathogen Discovery | Mili Sheth, Sarah Nobles, Jasmine Padilla, Mark Burroughs, Shoshona Le, Katie Dillon, Peter Cook, Clinton R. Paden, Dhwani Batra, Krista Queen, Kristen Knipe, Dakota Howard, Yvette Unoarumhi, Darlene Wagner, Matthew Schmerer, Ben L. Rambo-Martin, Kristine Lacek, Sam Shepard, Alison Laufer Halpin, Dave Wentworth, Vivien Dugan, Suxiang Tong, Justin Lee |
| EPI_ISL_2229087, EPI_ISL_2229088, EPI_ISL_2229089, EPI_ISL_2229090, EPI_ISL_2229091, EPI_ISL_2229092, EPI_ISL_2229093, EPI_ISL_2229094, EPI_ISL_2229095, EPI_ISL_2229096, EPI_ISL_2229097, EPI_ISL_2229098, EPI_ISL_2229099, EPI_ISL_2229100, EPI_ISL_2229101, EPI_ISL_2229102, EPI_ISL_2229103, EPI_ISL_2229104, EPI_ISL_2229105 | | | |
| see above | Guam Public Health Laboratory | Centers for Disease Control and Prevention Division of Viral Diseases, Pathogen Discovery | Mili Sheth, Sarah Nobles, Jasmine Padilla, Mark Burroughs, Shoshona Le, Katie Dillon, Peter Cook, Clinton R. Paden, Dhwani Batra, Krista Queen, Kristen Knipe, Dakota Howard, Yvette Unoarumhi, Darlene Wagner, Matthew Schmerer, Ben L. Rambo-Martin, Kristine Lacek, Sam Shepard, Alison Laufer Halpin, Dave Wentworth, Vivien Dugan, Suxiang Tong, Justin Lee |
| EPI_ISL_2229106, EPI_ISL_2229107, EPI_ISL_2229108, EPI_ISL_2229109, EPI_ISL_2229110, EPI_ISL_2229111 | KS Health and Environmental Laboratories | Centers for Disease Control and Prevention Division of Viral Diseases, Pathogen Discovery | Mili Sheth, Sarah Nobles, Jasmine Padilla, Mark Burroughs, Shoshona Le, Katie Dillon, Peter Cook, Clinton R. Paden, Dhwani Batra, Krista Queen, Kristen Knipe, Dakota Howard, Yvette Unoarumhi, Darlene Wagner, Matthew Schmerer, Ben L. Rambo-Martin, Kristine Lacek, Sam Shepard, Alison Laufer Halpin, Dave Wentworth, Vivien Dugan, Suxiang Tong, Justin Lee |

| | | | |
|---|---|---|---|
| EPI_ISL_2229112, EPI_ISL_2229113, EPI_ISL_2229114, EPI_ISL_2229115, EPI_ISL_2229116 | CT-Dr. Katherine A. Kelley State Public Health Lab | Centers for Disease Control and Prevention Division of Viral Diseases, Pathogen Discovery | Mili Sheth, Sarah Nobles, Jasmine Padilla, Mark Burroughs, Shoshona Le, Katie Dillon, Peter Cook, Clinton R. Paden, Dhwani Batra, Krista Queen, Kristen Knipe, Dakota Howard, Yvette Unoarumhi, Darlene Wagner, Matthew Schmerer, Ben L. Rambo-Martin, Kristine Lacek, Sam Shepard, Alison Laufer Halpin, Dave Wentworth, Vivien Dugan, Suxiang Tong, Justin Lee |
| EPI_ISL_2229117, EPI_ISL_2229118, EPI_ISL_2229119, EPI_ISL_2229120, EPI_ISL_2229121, EPI_ISL_2229122, EPI_ISL_2229123, EPI_ISL_2229124 | "AR Dept. of Health-PHL, Molecular Diagnostics" | Centers for Disease Control and Prevention Division of Viral Diseases, Pathogen Discovery | Mili Sheth, Sarah Nobles, Jasmine Padilla, Mark Burroughs, Shoshona Le, Katie Dillon, Peter Cook, Clinton R. Paden, Dhwani Batra, Krista Queen, Kristen Knipe, Dakota Howard, Yvette Unoarumhi, Darlene Wagner, Matthew Schmerer, Ben L. Rambo-Martin, Kristine Lacek, Sam Shepard, Alison Laufer Halpin, Dave Wentworth, Vivien Dugan, Suxiang Tong, Justin Lee |
| EPI_ISL_2229125, EPI_ISL_2229126 | "TX DSHS, Lab Services Section MC 1947" | Centers for Disease Control and Prevention Division of Viral Diseases, Pathogen Discovery | Mili Sheth, Sarah Nobles, Jasmine Padilla, Mark Burroughs, Shoshona Le, Katie Dillon, Peter Cook, Clinton R. Paden, Dhwani Batra, Krista Queen, Kristen Knipe, Dakota Howard, Yvette Unoarumhi, Darlene Wagner, Matthew Schmerer, Ben L. Rambo-Martin, Kristine Lacek, Sam Shepard, Alison Laufer Halpin, Dave Wentworth, Vivien Dugan, Suxiang Tong, Justin Lee |
| EPI_ISL_2229127, EPI_ISL_2229128, EPI_ISL_2229129, EPI_ISL_2229130, EPI_ISL_2229131, EPI_ISL_2229132 | WY Public Health Laboratory | Centers for Disease Control and Prevention Division of Viral Diseases, Pathogen Discovery | Mili Sheth, Sarah Nobles, Jasmine Padilla, Mark Burroughs, Shoshona Le, Katie Dillon, Peter Cook, Clinton R. Paden, Dhwani Batra, Krista Queen, Kristen Knipe, Dakota Howard, Yvette Unoarumhi, Darlene Wagner, Matthew Schmerer, Ben L. Rambo-Martin, Kristine Lacek, Sam Shepard, Alison Laufer Halpin, Dave Wentworth, Vivien Dugan, Suxiang Tong, Justin Lee |
| EPI_ISL_2229134, EPI_ISL_2229135, EPI_ISL_2229136, EPI_ISL_2229137, EPI_ISL_2229138, EPI_ISL_2229139, EPI_ISL_2229140, EPI_ISL_2229141, EPI_ISL_2229142, EPI_ISL_2229143 | MI - Michigan Department of Health and Human Services - Bureau of Laboratories | Centers for Disease Control and Prevention Division of Viral Diseases, Pathogen Discovery | Mili Sheth, Sarah Nobles, Jasmine Padilla, Mark Burroughs, Shoshona Le, Katie Dillon, Peter Cook, Clinton R. Paden, Dhwani Batra, Krista Queen, Kristen Knipe, Dakota Howard, Yvette Unoarumhi, Darlene Wagner, Matthew Schmerer, Ben L. Rambo-Martin, Kristine Lacek, Sam Shepard, Alison Laufer Halpin, Dave Wentworth, Vivien Dugan, Suxiang Tong, Justin Lee |
| EPI_ISL_2229144, EPI_ISL_2229145, EPI_ISL_2229146, EPI_ISL_2229147, EPI_ISL_2229148, EPI_ISL_2229149, EPI_ISL_2229150, EPI_ISL_2229151, EPI_ISL_2229152, EPI_ISL_2229153, EPI_ISL_2229154, EPI_ISL_2229155, EPI_ISL_2229156, EPI_ISL_2229157 | | | |
| see above | "CDPH, Viral and Rickettsial Disease Laboratory" | Centers for Disease Control and Prevention Division of Viral Diseases, Pathogen Discovery | Mili Sheth, Sarah Nobles, Jasmine Padilla, Mark Burroughs, Shoshona Le, Katie Dillon, Peter Cook, Clinton R. Paden, Dhwani Batra, Krista Queen, Kristen Knipe, Dakota Howard, Yvette Unoarumhi, Darlene Wagner, Matthew Schmerer, Ben L. Rambo-Martin, Kristine Lacek, Sam Shepard, Alison Laufer Halpin, Dave Wentworth, Vivien Dugan, Suxiang Tong, Justin Lee |
| EPI_ISL_2229158, EPI_ISL_2229159, EPI_ISL_2229160, EPI_ISL_2229161 | PR Public Health Lab | Centers for Disease Control and Prevention Division of Viral Diseases, Pathogen Discovery | Mili Sheth, Sarah Nobles, Jasmine Padilla, Mark Burroughs, Shoshona Le, Katie Dillon, Peter Cook, Clinton R. Paden, Dhwani Batra, Krista Queen, Kristen Knipe, Dakota Howard, Yvette Unoarumhi, Darlene Wagner, Matthew Schmerer, Ben L. Rambo-Martin, Kristine Lacek, Sam Shepard, Alison Laufer Halpin, Dave Wentworth, Vivien Dugan, Suxiang Tong, Justin Lee |
| EPI_ISL_2229162, EPI_ISL_2229163 | Philadelphia Department of Public Health | Centers for Disease Control and Prevention Division of Viral Diseases, Pathogen Discovery | Mili Sheth, Sarah Nobles, Jasmine Padilla, Mark Burroughs, Shoshona Le, Katie Dillon, Peter Cook, Clinton R. Paden, Dhwani Batra, Krista Queen, Kristen Knipe, Dakota Howard, Yvette Unoarumhi, Darlene Wagner, Matthew Schmerer, Ben L. Rambo-Martin, Kristine Lacek, Sam Shepard, Alison Laufer Halpin, Dave Wentworth, Vivien Dugan, Suxiang Tong, Justin Lee |
| EPI_ISL_2229164, EPI_ISL_2229165, EPI_ISL_2229166, EPI_ISL_2229167, EPI_ISL_2229168, EPI_ISL_2229169, EPI_ISL_2229170, EPI_ISL_2229171, EPI_ISL_2229172 | RI State Health Laboratories | Centers for Disease Control and Prevention Division of Viral Diseases, Pathogen Discovery | Mili Sheth, Sarah Nobles, Jasmine Padilla, Mark Burroughs, Shoshona Le, Katie Dillon, Peter Cook, Clinton R. Paden, Dhwani Batra, Krista Queen, Kristen Knipe, Dakota Howard, Yvette Unoarumhi, Darlene Wagner, Matthew Schmerer, Ben L. Rambo-Martin, Kristine Lacek, Sam Shepard, Alison Laufer Halpin, Dave Wentworth, Vivien Dugan, Suxiang Tong, Justin Lee |
| EPI_ISL_2229173, EPI_ISL_2229174, EPI_ISL_2229175, EPI_ISL_2229176, EPI_ISL_2229177, EPI_ISL_2229178, EPI_ISL_2229179, EPI_ISL_2229180, EPI_ISL_2229181, EPI_ISL_2229182, EPI_ISL_2229183, EPI_ISL_2229184 | | | |
| see above | OH Department of Health Laboratory | Centers for Disease Control and Prevention Division of Viral Diseases, Pathogen Discovery | Mili Sheth, Sarah Nobles, Jasmine Padilla, Mark Burroughs, Shoshona Le, Katie Dillon, Peter Cook, Clinton R. Paden, Dhwani Batra, Krista Queen, Kristen Knipe, Dakota Howard, Yvette Unoarumhi, Darlene Wagner, Matthew Schmerer, Ben L. Rambo-Martin, Kristine Lacek, Sam Shepard, Alison Laufer Halpin, Dave Wentworth, Vivien Dugan, Suxiang Tong, Justin Lee |
| EPI_ISL_2229185, EPI_ISL_2229186, EPI_ISL_2229187, EPI_ISL_2229188 | WI State Laboratory of Hygiene | Centers for Disease Control and Prevention Division of Viral Diseases, Pathogen Discovery | Mili Sheth, Sarah Nobles, Jasmine Padilla, Mark Burroughs, Shoshona Le, Katie Dillon, Peter Cook, Clinton R. Paden, Dhwani Batra, Krista Queen, Kristen Knipe, Dakota Howard, Yvette Unoarumhi, Darlene Wagner, Matthew Schmerer, Ben L. Rambo-Martin, Kristine Lacek, Sam Shepard, Alison Laufer Halpin, Dave Wentworth, Vivien Dugan, Suxiang Tong, Justin Lee |
| EPI_ISL_2229189, EPI_ISL_2229190, EPI_ISL_2229191, EPI_ISL_2229192, EPI_ISL_2229193 | CA-Los Angeles County Public Health Laboratory | Centers for Disease Control and Prevention Division of Viral Diseases, Pathogen Discovery | Mili Sheth, Sarah Nobles, Jasmine Padilla, Mark Burroughs, Shoshona Le, Katie Dillon, Peter Cook, Clinton R. Paden, Dhwani Batra, Krista Queen, Kristen Knipe, Dakota Howard, Yvette Unoarumhi, Darlene Wagner, Matthew Schmerer, Ben L. Rambo-Martin, Kristine Lacek, Sam Shepard, Alison Laufer Halpin, Dave Wentworth, Vivien Dugan, Suxiang Tong, Justin Lee |
| EPI_ISL_2229194, EPI_ISL_2229195, EPI_ISL_2229196, EPI_ISL_2229197, EPI_ISL_2229198, EPI_ISL_2229199 | NH Dept. of Health and Human Services Public Health Labs | Centers for Disease Control and Prevention Division of Viral Diseases, Pathogen Discovery | Mili Sheth, Sarah Nobles, Jasmine Padilla, Mark Burroughs, Shoshona Le, Katie Dillon, Peter Cook, Clinton R. Paden, Dhwani Batra, Krista Queen, Kristen Knipe, Dakota Howard, Yvette Unoarumhi, Darlene Wagner, Matthew Schmerer, Ben L. Rambo-Martin, Kristine Lacek, Sam Shepard, Alison Laufer Halpin, Dave Wentworth, Vivien Dugan, Suxiang Tong, Justin Lee |
| EPI_ISL_2229200, EPI_ISL_2229201, EPI_ISL_2229202, EPI_ISL_2229203, EPI_ISL_2229204, EPI_ISL_2229205, EPI_ISL_2229206 | MD DOH Laboratories Administration | Centers for Disease Control and Prevention Division of Viral Diseases, Pathogen Discovery | Mili Sheth, Sarah Nobles, Jasmine Padilla, Mark Burroughs, Shoshona Le, Katie Dillon, Peter Cook, Clinton R. Paden, Dhwani Batra, Krista Queen, Kristen Knipe, Dakota Howard, Yvette Unoarumhi, Darlene Wagner, Matthew Schmerer, Ben L. Rambo-Martin, Kristine Lacek, Sam Shepard, Alison Laufer Halpin, Dave Wentworth, Vivien Dugan, Suxiang Tong, Justin Lee |
| EPI_ISL_2229207 | Idaho Bureau of Laboratories | Center for Global Health, University of New Mexico Health Sciences Center | Daryl Domman, Kurt Schwalm, Valerie Morley, Matthew Burns, Robert Voermans, Christopher Ball, Darrell Dinwiddie |
| EPI_ISL_2229208 | MD DOH Laboratories Administration | Centers for Disease Control and Prevention Division of Viral Diseases, Pathogen Discovery | Mili Sheth, Sarah Nobles, Jasmine Padilla, Mark Burroughs, Shoshona Le, Katie Dillon, Peter Cook, Clinton R. Paden, Dhwani Batra, Krista Queen, Kristen Knipe, Dakota Howard, Yvette Unoarumhi, Darlene Wagner, Matthew Schmerer, Ben L. Rambo-Martin, Kristine Lacek, Sam Shepard, Alison Laufer Halpin, Dave Wentworth, Vivien Dugan, Suxiang Tong, Justin Lee |
| EPI_ISL_2229209, EPI_ISL_2229210 | Idaho Bureau of Laboratories | Center for Global Health, University of New Mexico Health Sciences Center | Daryl Domman, Kurt Schwalm, Valerie Morley, Matthew Burns, Robert Voermans, Christopher Ball, Darrell Dinwiddie |
| EPI_ISL_2229211 | MD DOH Laboratories Administration | Centers for Disease Control and Prevention Division of Viral Diseases, Pathogen Discovery | Mili Sheth, Sarah Nobles, Jasmine Padilla, Mark Burroughs, Shoshona Le, Katie Dillon, Peter Cook, Clinton R. Paden, Dhwani Batra, Krista Queen, Kristen Knipe, Dakota Howard, Yvette Unoarumhi, Darlene Wagner, Matthew Schmerer, Ben L. Rambo-Martin, Kristine Lacek, Sam Shepard, Alison Laufer Halpin, Dave Wentworth, Vivien Dugan, Suxiang Tong, Justin Lee |
| EPI_ISL_2229212, EPI_ISL_2229213, EPI_ISL_2229214, EPI_ISL_2229215 | Idaho Bureau of Laboratories | Center for Global Health, University of New Mexico Health Sciences Center | Daryl Domman, Kurt Schwalm, Valerie Morley, Matthew Burns, Robert Voermans, Christopher Ball, Darrell Dinwiddie |
| EPI_ISL_2229216 | IL Dept. of Public Health Springfield Laboratory | Centers for Disease Control and Prevention Division of Viral Diseases, Pathogen Discovery | Mili Sheth, Sarah Nobles, Jasmine Padilla, Mark Burroughs, Shoshona Le, Katie Dillon, Peter Cook, Clinton R. Paden, Dhwani Batra, Krista Queen, Kristen Knipe, Dakota Howard, Yvette Unoarumhi, Darlene Wagner, Matthew Schmerer, Ben L. Rambo-Martin, Kristine Lacek, Sam Shepard, Alison Laufer Halpin, Dave Wentworth, Vivien Dugan, Suxiang Tong, Justin Lee |
| EPI_ISL_2229217, EPI_ISL_2229218 | Idaho Bureau of Laboratories | Center for Global Health, University of New Mexico Health Sciences Center | Daryl Domman, Kurt Schwalm, Valerie Morley, Matthew Burns, Robert Voermans, Christopher Ball, Darrell Dinwiddie |
| EPI_ISL_2229219 | IL Dept. of Public Health Springfield Laboratory | Centers for Disease Control and Prevention Division of Viral Diseases, Pathogen | Mili Sheth, Sarah Nobles, Jasmine Padilla, Mark Burroughs, Shoshona Le, Katie Dillon, Peter Cook, Clinton R. Paden, Dhwani Batra, Krista Queen, Kristen Knipe, Dakota Howard, Yvette Unoarumhi, Darlene Wagner, Matthew Schmerer, Ben L. Rambo-Martin, Kristine Lacek, Sam Shepard, Alison Laufer Halpin, Dave Wentworth, Vivien Dugan, Suxiang Tong, Justin Lee |

| | | | |
|---|---|---|---|
| | | Discovery | |
| EPI_ISL_2229220 | Idaho Bureau of Laboratories | Center for Global Health, University of New Mexico Health Sciences Center | Daryl Domman, Kurt Schwalm, Valerie Morley, Matthew Burns, Robert Voermans, Christopher Ball, Darrell Dinwiddie |
| EPI_ISL_2229221 | IL Dept. of Public Health Springfield Laboratory | Centers for Disease Control and Prevention Division of Viral Diseases, Pathogen Discovery | Mili Sheth, Sarah Nobles, Jasmine Padilla, Mark Burroughs, Shoshona Le, Katie Dillon, Peter Cook, Clinton R. Paden, Dhwani Batra, Krista Queen, Kristen Knipe, Dakota Howard, Yvette Unoarumhi, Darlene Wagner, Matthew Schmerer, Ben L. Rambo-Martin, Kristine Lacek, Sam Shepard, Alison Laufer Halpin, Dave Wentworth, Vivien Dugan, Suxiang Tong, Justin Lee |
| EPI_ISL_2229222 | Idaho Bureau of Laboratories | Center for Global Health, University of New Mexico Health Sciences Center | Daryl Domman, Kurt Schwalm, Valerie Morley, Matthew Burns, Robert Voermans, Christopher Ball, Darrell Dinwiddie |
| EPI_ISL_2229223 | IL Dept. of Public Health Springfield Laboratory | Centers for Disease Control and Prevention Division of Viral Diseases, Pathogen Discovery | Mili Sheth, Sarah Nobles, Jasmine Padilla, Mark Burroughs, Shoshona Le, Katie Dillon, Peter Cook, Clinton R. Paden, Dhwani Batra, Krista Queen, Kristen Knipe, Dakota Howard, Yvette Unoarumhi, Darlene Wagner, Matthew Schmerer, Ben L. Rambo-Martin, Kristine Lacek, Sam Shepard, Alison Laufer Halpin, Dave Wentworth, Vivien Dugan, Suxiang Tong, Justin Lee |
| EPI_ISL_2229224, EPI_ISL_2229225 | Idaho Bureau of Laboratories | Center for Global Health, University of New Mexico Health Sciences Center | Daryl Domman, Kurt Schwalm, Valerie Morley, Matthew Burns, Robert Voermans, Christopher Ball, Darrell Dinwiddie |
| EPI_ISL_2229226 | IL Dept. of Public Health Springfield Laboratory | Centers for Disease Control and Prevention Division of Viral Diseases, Pathogen Discovery | Mili Sheth, Sarah Nobles, Jasmine Padilla, Mark Burroughs, Shoshona Le, Katie Dillon, Peter Cook, Clinton R. Paden, Dhwani Batra, Krista Queen, Kristen Knipe, Dakota Howard, Yvette Unoarumhi, Darlene Wagner, Matthew Schmerer, Ben L. Rambo-Martin, Kristine Lacek, Sam Shepard, Alison Laufer Halpin, Dave Wentworth, Vivien Dugan, Suxiang Tong, Justin Lee |
| EPI_ISL_2229227, EPI_ISL_2229228 | Idaho Bureau of Laboratories | Center for Global Health, University of New Mexico Health Sciences Center | Daryl Domman, Kurt Schwalm, Valerie Morley, Matthew Burns, Robert Voermans, Christopher Ball, Darrell Dinwiddie |
| EPI_ISL_2229229 | IL Dept. of Public Health Springfield Laboratory | Centers for Disease Control and Prevention Division of Viral Diseases, Pathogen Discovery | Mili Sheth, Sarah Nobles, Jasmine Padilla, Mark Burroughs, Shoshona Le, Katie Dillon, Peter Cook, Clinton R. Paden, Dhwani Batra, Krista Queen, Kristen Knipe, Dakota Howard, Yvette Unoarumhi, Darlene Wagner, Matthew Schmerer, Ben L. Rambo-Martin, Kristine Lacek, Sam Shepard, Alison Laufer Halpin, Dave Wentworth, Vivien Dugan, Suxiang Tong, Justin Lee |
| EPI_ISL_2229230, EPI_ISL_2229231 | Idaho Bureau of Laboratories | Center for Global Health, University of New Mexico Health Sciences Center | Daryl Domman, Kurt Schwalm, Valerie Morley, Matthew Burns, Robert Voermans, Christopher Ball, Darrell Dinwiddie |
| EPI_ISL_2229232 | IL Dept. of Public Health Springfield Laboratory | Centers for Disease Control and Prevention Division of Viral Diseases, Pathogen Discovery | Mili Sheth, Sarah Nobles, Jasmine Padilla, Mark Burroughs, Shoshona Le, Katie Dillon, Peter Cook, Clinton R. Paden, Dhwani Batra, Krista Queen, Kristen Knipe, Dakota Howard, Yvette Unoarumhi, Darlene Wagner, Matthew Schmerer, Ben L. Rambo-Martin, Kristine Lacek, Sam Shepard, Alison Laufer Halpin, Dave Wentworth, Vivien Dugan, Suxiang Tong, Justin Lee |
| EPI_ISL_2229233, EPI_ISL_2229234 | Idaho Bureau of Laboratories | Center for Global Health, University of New Mexico Health Sciences Center | Daryl Domman, Kurt Schwalm, Valerie Morley, Matthew Burns, Robert Voermans, Christopher Ball, Darrell Dinwiddie |
| EPI_ISL_2229235, EPI_ISL_2229236, EPI_ISL_2229237, EPI_ISL_2229238, EPI_ISL_2229239, EPI_ISL_2229240, EPI_ISL_2229241 | IL Dept. of Public Health Springfield Laboratory | Centers for Disease Control and Prevention Division of Viral Diseases, Pathogen Discovery | Mili Sheth, Sarah Nobles, Jasmine Padilla, Mark Burroughs, Shoshona Le, Katie Dillon, Peter Cook, Clinton R. Paden, Dhwani Batra, Krista Queen, Kristen Knipe, Dakota Howard, Yvette Unoarumhi, Darlene Wagner, Matthew Schmerer, Ben L. Rambo-Martin, Kristine Lacek, Sam Shepard, Alison Laufer Halpin, Dave Wentworth, Vivien Dugan, Suxiang Tong, Justin Lee |
| EPI_ISL_2229242, EPI_ISL_2229243 | UT-Unified State Labs: Public Health Utah DOH | Centers for Disease Control and Prevention Division of Viral Diseases, Pathogen Discovery | Mili Sheth, Sarah Nobles, Jasmine Padilla, Mark Burroughs, Shoshona Le, Katie Dillon, Peter Cook, Clinton R. Paden, Dhwani Batra, Krista Queen, Kristen Knipe, Dakota Howard, Yvette Unoarumhi, Darlene Wagner, Matthew Schmerer, Ben L. Rambo-Martin, Kristine Lacek, Sam Shepard, Alison Laufer Halpin, Dave Wentworth, Vivien Dugan, Suxiang Tong, Justin Lee |
| EPI_ISL_2229244, EPI_ISL_2229245, EPI_ISL_2229246, EPI_ISL_2229247, EPI_ISL_2229248, EPI_ISL_2229249 | DC Public Health Lab/ Dept. of Forensic Sciences | Centers for Disease Control and Prevention Division of Viral Diseases, Pathogen Discovery | Mili Sheth, Sarah Nobles, Jasmine Padilla, Mark Burroughs, Shoshona Le, Katie Dillon, Peter Cook, Clinton R. Paden, Dhwani Batra, Krista Queen, Kristen Knipe, Dakota Howard, Yvette Unoarumhi, Darlene Wagner, Matthew Schmerer, Ben L. Rambo-Martin, Kristine Lacek, Sam Shepard, Alison Laufer Halpin, Dave Wentworth, Vivien Dugan, Suxiang Tong, Justin Lee |
| EPI_ISL_2229250, EPI_ISL_2229251, EPI_ISL_2229252, EPI_ISL_2229253, EPI_ISL_2229254, EPI_ISL_2229255, EPI_ISL_2229256, EPI_ISL_2229257 | NC State Laboratory of Public Health | Centers for Disease Control and Prevention Division of Viral Diseases, Pathogen Discovery | Mili Sheth, Sarah Nobles, Jasmine Padilla, Mark Burroughs, Shoshona Le, Katie Dillon, Peter Cook, Clinton R. Paden, Dhwani Batra, Krista Queen, Kristen Knipe, Dakota Howard, Yvette Unoarumhi, Darlene Wagner, Matthew Schmerer, Ben L. Rambo-Martin, Kristine Lacek, Sam Shepard, Alison Laufer Halpin, Dave Wentworth, Vivien Dugan, Suxiang Tong, Justin Lee |
| EPI_ISL_2229258, EPI_ISL_2229259, EPI_ISL_2229260 | WVDHHR - Office of Laboratory Services | Centers for Disease Control and Prevention Division of Viral Diseases, Pathogen Discovery | Mili Sheth, Sarah Nobles, Jasmine Padilla, Mark Burroughs, Shoshona Le, Katie Dillon, Peter Cook, Clinton R. Paden, Dhwani Batra, Krista Queen, Kristen Knipe, Dakota Howard, Yvette Unoarumhi, Darlene Wagner, Matthew Schmerer, Ben L. Rambo-Martin, Kristine Lacek, Sam Shepard, Alison Laufer Halpin, Dave Wentworth, Vivien Dugan, Suxiang Tong, Justin Lee |
| EPI_ISL_2229261 | MO State Public Health Laboratory | Centers for Disease Control and Prevention Division of Viral Diseases, Pathogen Discovery | Mili Sheth, Sarah Nobles, Jasmine Padilla, Mark Burroughs, Shoshona Le, Katie Dillon, Peter Cook, Clinton R. Paden, Dhwani Batra, Krista Queen, Kristen Knipe, Dakota Howard, Yvette Unoarumhi, Darlene Wagner, Matthew Schmerer, Ben L. Rambo-Martin, Kristine Lacek, Sam Shepard, Alison Laufer Halpin, Dave Wentworth, Vivien Dugan, Suxiang Tong, Justin Lee |
| EPI_ISL_2229262, EPI_ISL_2229263, EPI_ISL_2229264, EPI_ISL_2229265, EPI_ISL_2229266, EPI_ISL_2229267, EPI_ISL_2229268 | FL Bur. of Public Health Laboratories-Jacksonville | Centers for Disease Control and Prevention Division of Viral Diseases, Pathogen Discovery | Mili Sheth, Sarah Nobles, Jasmine Padilla, Mark Burroughs, Shoshona Le, Katie Dillon, Peter Cook, Clinton R. Paden, Dhwani Batra, Krista Queen, Kristen Knipe, Dakota Howard, Yvette Unoarumhi, Darlene Wagner, Matthew Schmerer, Ben L. Rambo-Martin, Kristine Lacek, Sam Shepard, Alison Laufer Halpin, Dave Wentworth, Vivien Dugan, Suxiang Tong, Justin Lee |
| EPI_ISL_2229269, EPI_ISL_2229270, EPI_ISL_2229271, EPI_ISL_2229272, EPI_ISL_2229273 | LA Office of Public Health Laboratories | Centers for Disease Control and Prevention Division of Viral Diseases, Pathogen Discovery | Mili Sheth, Sarah Nobles, Jasmine Padilla, Mark Burroughs, Shoshona Le, Katie Dillon, Peter Cook, Clinton R. Paden, Dhwani Batra, Krista Queen, Kristen Knipe, Dakota Howard, Yvette Unoarumhi, Darlene Wagner, Matthew Schmerer, Ben L. Rambo-Martin, Kristine Lacek, Sam Shepard, Alison Laufer Halpin, Dave Wentworth, Vivien Dugan, Suxiang Tong, Justin Lee |
| EPI_ISL_2229274 | "NYSDOH Wadsworth Center, Virology Lab" | Centers for Disease Control and Prevention Division of Viral Diseases, Pathogen Discovery | Mili Sheth, Sarah Nobles, Jasmine Padilla, Mark Burroughs, Shoshona Le, Katie Dillon, Peter Cook, Clinton R. Paden, Dhwani Batra, Krista Queen, Kristen Knipe, Dakota Howard, Yvette Unoarumhi, Darlene Wagner, Matthew Schmerer, Ben L. Rambo-Martin, Kristine Lacek, Sam Shepard, Alison Laufer Halpin, Dave Wentworth, Vivien Dugan, Suxiang Tong, Justin Lee |
| EPI_ISL_2229275, EPI_ISL_2229276, EPI_ISL_2229277, EPI_ISL_2229278, EPI_ISL_2229279 | "AZ SPHL, Arizona Department of Health Services" | Centers for Disease Control and Prevention Division of Viral Diseases, Pathogen Discovery | Mili Sheth, Sarah Nobles, Jasmine Padilla, Mark Burroughs, Shoshona Le, Katie Dillon, Peter Cook, Clinton R. Paden, Dhwani Batra, Krista Queen, Kristen Knipe, Dakota Howard, Yvette Unoarumhi, Darlene Wagner, Matthew Schmerer, Ben L. Rambo-Martin, Kristine Lacek, Sam Shepard, Alison Laufer Halpin, Dave Wentworth, Vivien Dugan, Suxiang Tong, Justin Lee |
| EPI_ISL_2229280, EPI_ISL_2229281, EPI_ISL_2229282 | AL Dept. of Public Health Bureau of Clinical Laboratories | Centers for Disease Control and Prevention Division of Viral Diseases, Pathogen Discovery | Mili Sheth, Sarah Nobles, Jasmine Padilla, Mark Burroughs, Shoshona Le, Katie Dillon, Peter Cook, Clinton R. Paden, Dhwani Batra, Krista Queen, Kristen Knipe, Dakota Howard, Yvette Unoarumhi, Darlene Wagner, Matthew Schmerer, Ben L. Rambo-Martin, Kristine Lacek, Sam Shepard, Alison Laufer Halpin, Dave Wentworth, Vivien Dugan, Suxiang Tong, Justin Lee |
| EPI_ISL_2229283, EPI_ISL_2229284, EPI_ISL_2229285, EPI_ISL_2229286, EPI_ISL_2229287, EPI_ISL_2229288, EPI_ISL_2229289, EPI_ISL_2229290 | "NM Dept. Health, Scientific Laboratory Division " | Centers for Disease Control and Prevention Division of Viral Diseases, Pathogen Discovery | Mili Sheth, Sarah Nobles, Jasmine Padilla, Mark Burroughs, Shoshona Le, Katie Dillon, Peter Cook, Clinton R. Paden, Dhwani Batra, Krista Queen, Kristen Knipe, Dakota Howard, Yvette Unoarumhi, Darlene Wagner, Matthew Schmerer, Ben L. Rambo-Martin, Kristine Lacek, Sam Shepard, Alison Laufer Halpin, Dave Wentworth, Vivien Dugan, Suxiang Tong, Justin Lee |
| EPI_ISL_2229291, EPI_ISL_2229292, EPI_ISL_2229293, EPI_ISL_2229294, EPI_ISL_2229295, EPI_ISL_2229296 | MT Public Health Laboratory | Centers for Disease Control and Prevention Division of Viral Diseases, Pathogen Discovery | Mili Sheth, Sarah Nobles, Jasmine Padilla, Mark Burroughs, Shoshona Le, Katie Dillon, Peter Cook, Clinton R. Paden, Dhwani Batra, Krista Queen, Kristen Knipe, Dakota Howard, Yvette Unoarumhi, Darlene Wagner, Matthew Schmerer, Ben L. Rambo-Martin, Kristine Lacek, Sam Shepard, Alison Laufer Halpin, Dave Wentworth, Vivien Dugan, Suxiang Tong, Justin Lee |

EPI_ISL_2229297, EPI_ISL_2229298, EPI_ISL_2229299, EPI_ISL_2229300, EPI_ISL_2229301, EPI_ISL_2229302, EPI_ISL_2229303, EPI_ISL_2229304, EPI_ISL_2229305, EPI_ISL_2229306, EPI_ISL_2229307, EPI_ISL_2229308, EPI_ISL_2229309, EPI_ISL_2229310, EPI_ISL_2229311, EPI_ISL_2229312, EPI_ISL_2229313, EPI_ISL_2229314, EPI_ISL_2229315, EPI_ISL_2229316, EPI_ISL_2229317, EPI_ISL_2229318, EPI_ISL_2229319, EPI_ISL_2229320, EPI_ISL_2229321, EPI_ISL_2229322, EPI_ISL_2229323, EPI_ISL_2229324

| | | | |
|---|---|---|---|
| see above | GA Department of Public Health Laboratory | Centers for Disease Control and Prevention Division of Viral Diseases, Pathogen | Mili Sheth, Sarah Nobles, Jasmine Padilla, Mark Burroughs, Shoshona Le, Katie Dillon, Peter Cook, Clinton R. Paden, Dhwani Batra, Krista Queen, Kristen Knipe, Dakota Howard, Yvette Unoarumhi, Darlene Wagner, Matthew Schmerer, Ben L. Rambo-Martin, Kristine Lacek, Sam Shepard, Alison Laufer Halpin, Dave Wentworth, Vivien Dugan, Suxiang Tong, Justin Lee |

| | | | |
|---|---|---|---|
| | | Discovery | |
| EPI_ISL_2229325, EPI_ISL_2229326, EPI_ISL_2229327, EPI_ISL_2229328, EPI_ISL_2229331 | Louisiana Office of Public Heath Laboratory | LSUHS Emerging Viral Threat Laboratory | Gregory L. Ware, Alexander Mijalis, Jeremy P. Kamil, Jennifer L. Carroll, Maarten Van Diest, Rona S. Scott, Andrew D. Yurochko, Christopher G. Kevil, John A. Vanchiere |
| EPI_ISL_2229333, EPI_ISL_2229335, EPI_ISL_2229338, EPI_ISL_2229340 | Louisiana Office of Public Heath Laboratory | LSUHS Emerging Viral Threat Laboratory | Alexander Mijalis, Jeremy P. Kamil, Jennifer L. Carroll, Maarten Van Diest, Rona S. Scott, Gregory L. Ware, Andrew D. Yurochko, Christopher G. Kevil, John A. Vanchiere |
| EPI_ISL_2229343 | LSUHS Emerging Viral Threat Laboratory | LSUHS Emerging Viral Threat Laboratory | Alexander Mijalis, Jeremy P. Kamil, Jennifer L. Carroll, Maarten Van Diest, Rona S. Scott, Gregory L. Ware, Andrew D. Yurochko, Christopher G. Kevil, John A. Vanchiere |
| EPI_ISL_2229345, EPI_ISL_2229346, EPI_ISL_2229347, EPI_ISL_2229348, EPI_ISL_2229349, EPI_ISL_2229350 | LSUHS Emerging Viral Threat Laboratory | LSUHS Emerging Viral Threat Laboratory | Jeremy P. Kamil, Jennifer L. Carroll, Maarten Van Diest, Rona S. Scott,Gregory L. Ware, Alexander Mijalis, Andrew D. Yurochko, Christopher G. Kevil, John A. Vanchiere |
| EPI_ISL_2229357, EPI_ISL_2229359, EPI_ISL_2229361, EPI_ISL_2229362, EPI_ISL_2229363, EPI_ISL_2229364 | LSUHS Emerging Viral Threat Laboratory | LSUHS Emerging Viral Threat Laboratory | Jennifer L. Carroll, Maarten Van Diest, Rona S. Scott,Gregory L. Ware, Alexander Mijalis, Jeremy P. Kamil, Andrew D. Yurochko, Christopher G. Kevil, John A. Vanchiere |
| EPI_ISL_2229366, EPI_ISL_2229367, EPI_ISL_2229368, EPI_ISL_2229370 | LSUHS Emerging Viral Threat Laboratory | LSUHS Emerging Viral Threat Laboratory | Maarten Van Diest, Rona S. Scott,Gregory L. Ware, Alexander Mijalis, Jeremy P. Kamil, Jennifer L. Carroll, Andrew D. Yurochko, Christopher G. Kevil, John A. Vanchiere |
| EPI_ISL_2229371, EPI_ISL_2229373, EPI_ISL_2229374, EPI_ISL_2229375, EPI_ISL_2229379, EPI_ISL_2229380 | Willis-Knighton Medical Center Hospital Laboratory | LSUHS Emerging Viral Threat Laboratory | Maarten Van Diest, Rona S. Scott,Gregory L. Ware, Alexander Mijalis, Jeremy P. Kamil, Jennifer L. Carroll, Andrew D. Yurochko, Christopher G. Kevil, John A. Vanchiere |
| EPI_ISL_2229381, EPI_ISL_2229382, EPI_ISL_2229383, EPI_ISL_2229384, EPI_ISL_2229385, EPI_ISL_2229386, EPI_ISL_2229387, EPI_ISL_2229388, EPI_ISL_2229389, EPI_ISL_2229390, EPI_ISL_2229392, EPI_ISL_2229393, EPI_ISL_2229394, EPI_ISL_2229396, EPI_ISL_2229398, EPI_ISL_2229399 | | | |
| see above | Willis-Knighton Medical Center Hospital Laboratory | LSUHS Emerging Viral Threat Laboratory | Rona S. Scott, Gregory L. Ware, Alexander Mijalis, Jeremy P. Kamil, Jennifer L. Carroll, Maarten Van Diest, Andrew D. Yurochko, Christopher G. Kevil, John A. Vanchiere |
| EPI_ISL_2229401, EPI_ISL_2229402, EPI_ISL_2229403, EPI_ISL_2229404, EPI_ISL_2229405, EPI_ISL_2229406, EPI_ISL_2229407, EPI_ISL_2229410, EPI_ISL_2229411, EPI_ISL_2229412, EPI_ISL_2229413, EPI_ISL_2229414, EPI_ISL_2229415, EPI_ISL_2229416, EPI_ISL_2229417, EPI_ISL_2229418, EPI_ISL_2229421, EPI_ISL_2229422, EPI_ISL_2229423, EPI_ISL_2229424, EPI_ISL_2229425, EPI_ISL_2229426, EPI_ISL_2229428, EPI_ISL_2229429, EPI_ISL_2229430, EPI_ISL_2229432, EPI_ISL_2229433, EPI_ISL_2229434, EPI_ISL_2229435, EPI_ISL_2229436, EPI_ISL_2229437, EPI_ISL_2229438, EPI_ISL_2229440, EPI_ISL_2229441, EPI_ISL_2229442, EPI_ISL_2229443, EPI_ISL_2229445, EPI_ISL_2229449, EPI_ISL_2229453, EPI_ISL_2229454, EPI_ISL_2229455, EPI_ISL_2229456, EPI_ISL_2229457, EPI_ISL_2229459, EPI_ISL_2229460, EPI_ISL_2229461, EPI_ISL_2229462, EPI_ISL_2229463, EPI_ISL_2229464, EPI_ISL_2229465, EPI_ISL_2229467, EPI_ISL_2229468, EPI_ISL_2229469, EPI_ISL_2229470, EPI_ISL_2229472, EPI_ISL_2229473, EPI_ISL_2229474, EPI_ISL_2229475, EPI_ISL_2229476, EPI_ISL_2229477, EPI_ISL_2229478, EPI_ISL_2229480 | | | |
| see above | Virginia Division of Consolidated Laboratory Services | Virginia Division of Consolidated Laboratory Services | Virginia DCLS |
| EPI_ISL_2229481, EPI_ISL_2229482, EPI_ISL_2229483, EPI_ISL_2229484, EPI_ISL_2229485, EPI_ISL_2229486, EPI_ISL_2229488, EPI_ISL_2229489, EPI_ISL_2229491, EPI_ISL_2229492, EPI_ISL_2229493, EPI_ISL_2229494, EPI_ISL_2229495, EPI_ISL_2229496, EPI_ISL_2229497, EPI_ISL_2229498, EPI_ISL_2229499, EPI_ISL_2229500, EPI_ISL_2229501, EPI_ISL_2229502, EPI_ISL_2229503, EPI_ISL_2229504, EPI_ISL_2229505, EPI_ISL_2229506, EPI_ISL_2229507, EPI_ISL_2229508, EPI_ISL_2229509, EPI_ISL_2229510, EPI_ISL_2229511, EPI_ISL_2229512, EPI_ISL_2229513, EPI_ISL_2229514, EPI_ISL_2229515, EPI_ISL_2229516, EPI_ISL_2229517, EPI_ISL_2229518, EPI_ISL_2229519, EPI_ISL_2229520, EPI_ISL_2229521, EPI_ISL_2229522, EPI_ISL_2229523, EPI_ISL_2229525, EPI_ISL_2229526, EPI_ISL_2229527, EPI_ISL_2229529, EPI_ISL_2229530, EPI_ISL_2229531, EPI_ISL_2229532, EPI_ISL_2229533, EPI_ISL_2229534, EPI_ISL_2229535, EPI_ISL_2229536, EPI_ISL_2229538 | | | |
| see above | Massachusetts State Public Health Laboratory | Massachusetts State Public Health Laboratory | Andrew Lang, Timelia Fink, Glen Gallagher, Sandra Smole |
| EPI_ISL_2229539, EPI_ISL_2229540, EPI_ISL_2229541, EPI_ISL_2229542, EPI_ISL_2229543, EPI_ISL_2229544, EPI_ISL_2229545, EPI_ISL_2229546, EPI_ISL_2229547, EPI_ISL_2229548, EPI_ISL_2229549, EPI_ISL_2229550, EPI_ISL_2229551, EPI_ISL_2229552, EPI_ISL_2229553, EPI_ISL_2229554, EPI_ISL_2229555, EPI_ISL_2229556, EPI_ISL_2229557, EPI_ISL_2229558, EPI_ISL_2229559, EPI_ISL_2229560, EPI_ISL_2229561, EPI_ISL_2229562, EPI_ISL_2229563, EPI_ISL_2229564, EPI_ISL_2229565, EPI_ISL_2229566, EPI_ISL_2229567, EPI_ISL_2229568, EPI_ISL_2229569, EPI_ISL_2229570, EPI_ISL_2229571, EPI_ISL_2229572, EPI_ISL_2229573, EPI_ISL_2229574, EPI_ISL_2229576, EPI_ISL_2229577, EPI_ISL_2229578, EPI_ISL_2229579, EPI_ISL_2229580, EPI_ISL_2229581, EPI_ISL_2229582, EPI_ISL_2229583, EPI_ISL_2229584, EPI_ISL_2229585, EPI_ISL_2229586, EPI_ISL_2229587, EPI_ISL_2229588, EPI_ISL_2229589, EPI_ISL_2229590, EPI_ISL_2229591, EPI_ISL_2229592, EPI_ISL_2229593, EPI_ISL_2229594, EPI_ISL_2229595, EPI_ISL_2229596, EPI_ISL_2229597, EPI_ISL_2229598, EPI_ISL_2229599, EPI_ISL_2229600, EPI_ISL_2229601, EPI_ISL_2229602, EPI_ISL_2229603, EPI_ISL_2229604, EPI_ISL_2229605, EPI_ISL_2229606, EPI_ISL_2229607, EPI_ISL_2229608, EPI_ISL_2229609, EPI_ISL_2229610, EPI_ISL_2229611, EPI_ISL_2229612, EPI_ISL_2229613, EPI_ISL_2229614, EPI_ISL_2229615, EPI_ISL_2229616, EPI_ISL_2229617, EPI_ISL_2229618, EPI_ISL_2229619, EPI_ISL_2229620, EPI_ISL_2229621, EPI_ISL_2229622, EPI_ISL_2229623, EPI_ISL_2229624, EPI_ISL_2229625, EPI_ISL_2229626, EPI_ISL_2229627, EPI_ISL_2229628, EPI_ISL_2229629, EPI_ISL_2229630, EPI_ISL_2229631, EPI_ISL_2229632, EPI_ISL_2229633, EPI_ISL_2229634 | | | |
| see above | Erie County Public Health (ECPHL) | University at Buffalo Genomics and Bioinformatics Core | Jonathan Bard, Natalie Lamb, Alyssa Pohlman, Brandon Marzullo, Amanda Boccolucci, Norma Nowak, Donald Yergeau, Jennifer Surtees |
| EPI_ISL_2229635, EPI_ISL_2229636, EPI_ISL_2229637, EPI_ISL_2229638, EPI_ISL_2229639, EPI_ISL_2229640, EPI_ISL_2229641, EPI_ISL_2229642, EPI_ISL_2229643, EPI_ISL_2229644, EPI_ISL_2229645, EPI_ISL_2229646, EPI_ISL_2229647, EPI_ISL_2229649, EPI_ISL_2229650, EPI_ISL_2229651, EPI_ISL_2229652, EPI_ISL_2229653, EPI_ISL_2229654, EPI_ISL_2229655, EPI_ISL_2229656, EPI_ISL_2229657, EPI_ISL_2229658, EPI_ISL_2229659, EPI_ISL_2229660, EPI_ISL_2229661, EPI_ISL_2229662, EPI_ISL_2229663, EPI_ISL_2229664, EPI_ISL_2229665, EPI_ISL_2229666, EPI_ISL_2229667, EPI_ISL_2229668, EPI_ISL_2229669, EPI_ISL_2229670, EPI_ISL_2229671, EPI_ISL_2229672, EPI_ISL_2229673, EPI_ISL_2229674, EPI_ISL_2229675, EPI_ISL_2229676, EPI_ISL_2229678, EPI_ISL_2229679, EPI_ISL_2229680, EPI_ISL_2229681, EPI_ISL_2229682, EPI_ISL_2229683, EPI_ISL_2229684, EPI_ISL_2229685, EPI_ISL_2229686, EPI_ISL_2229687, EPI_ISL_2229688, EPI_ISL_2229689, EPI_ISL_2229690, EPI_ISL_2229691, EPI_ISL_2229692, EPI_ISL_2229693, EPI_ISL_2229694, EPI_ISL_2229695, EPI_ISL_2229696, EPI_ISL_2229697, EPI_ISL_2229698, EPI_ISL_2229699, EPI_ISL_2229700, EPI_ISL_2229701, EPI_ISL_2229702, EPI_ISL_2229703, EPI_ISL_2229704, EPI_ISL_2229705, EPI_ISL_2229706, EPI_ISL_2229707, EPI_ISL_2229708, EPI_ISL_2229709, EPI_ISL_2229710, EPI_ISL_2229711, EPI_ISL_2229712, EPI_ISL_2229713, EPI_ISL_2229714, EPI_ISL_2229715, EPI_ISL_2229716, EPI_ISL_2229717, EPI_ISL_2229719, EPI_ISL_2229720, EPI_ISL_2229721, EPI_ISL_2229722, EPI_ISL_2229723, EPI_ISL_2229724, EPI_ISL_2229725, EPI_ISL_2229726, EPI_ISL_2229727, EPI_ISL_2229728, EPI_ISL_2229729, EPI_ISL_2229730, EPI_ISL_2229731, EPI_ISL_2229732, EPI_ISL_2229733, EPI_ISL_2229734, EPI_ISL_2229735, EPI_ISL_2229736, EPI_ISL_2229737, EPI_ISL_2229738, EPI_ISL_2229739, EPI_ISL_2229740, EPI_ISL_2229741, EPI_ISL_2229742, EPI_ISL_2229743, EPI_ISL_2229744, EPI_ISL_2229745, EPI_ISL_2229746, EPI_ISL_2229747, EPI_ISL_2229748, EPI_ISL_2229749, EPI_ISL_2229750, EPI_ISL_2229751, EPI_ISL_2229752, EPI_ISL_2229753, EPI_ISL_2229754, EPI_ISL_2229755, EPI_ISL_2229756, EPI_ISL_2229757, EPI_ISL_2229759, EPI_ISL_2229760, EPI_ISL_2229761, EPI_ISL_2229762, EPI_ISL_2229763, EPI_ISL_2229764 | | | |
| see above | Kaleida Health Laboratories | University at Buffalo Genomics and Bioinformatics Core | Jonathan Bard, Natalie Lamb, Alyssa Pohlman, Brandon Marzullo, Amanda Boccolucci, Norma Nowak, Donald Yergeau, Jennifer Surtees |
| EPI_ISL_2229765, EPI_ISL_2229766, EPI_ISL_2229767, EPI_ISL_2229768, EPI_ISL_2229769, EPI_ISL_2229770, EPI_ISL_2229771, EPI_ISL_2229772, EPI_ISL_2229773, EPI_ISL_2229774, EPI_ISL_2229775, EPI_ISL_2229776, EPI_ISL_2229777, EPI_ISL_2229778, EPI_ISL_2229779, EPI_ISL_2229780, EPI_ISL_2229781, EPI_ISL_2229782, EPI_ISL_2229783, EPI_ISL_2229784, EPI_ISL_2229785, EPI_ISL_2229786, EPI_ISL_2229787, EPI_ISL_2229788, EPI_ISL_2229789, EPI_ISL_2229790, EPI_ISL_2229791, EPI_ISL_2229792, EPI_ISL_2229793, EPI_ISL_2229794, EPI_ISL_2229796, EPI_ISL_2229797, EPI_ISL_2229798, EPI_ISL_2229799, EPI_ISL_2229800, EPI_ISL_2229801, EPI_ISL_2229802, EPI_ISL_2229803, EPI_ISL_2229804, EPI_ISL_2229805, EPI_ISL_2229806, EPI_ISL_2229807, EPI_ISL_2229808, EPI_ISL_2229810, EPI_ISL_2229811, EPI_ISL_2229812, EPI_ISL_2229813, EPI_ISL_2229814 | | | |
| see above | Massachusetts State Public Health Laboratory | Massachusetts State Public Health Laboratory | Andrew Lang, Timelia Fink, Glen Gallagher, Sandra Smole |
| EPI_ISL_2229815, EPI_ISL_2229816, EPI_ISL_2229817, EPI_ISL_2229818, EPI_ISL_2229819, EPI_ISL_2229820, EPI_ISL_2229821, EPI_ISL_2229822, EPI_ISL_2229823, EPI_ISL_2229824, EPI_ISL_2229825, EPI_ISL_2229826, EPI_ISL_2229827, EPI_ISL_2229828, EPI_ISL_2229829, EPI_ISL_2229830, EPI_ISL_2229831, EPI_ISL_2229832, EPI_ISL_2229833, EPI_ISL_2229834, EPI_ISL_2229835, EPI_ISL_2229836, EPI_ISL_2229837 | | | |
| see above | Nebraska Public Health Laboratory | NPHL COVID-19 Response Team | NPHL COVID-19 Response Team |
| EPI_ISL_2229842, EPI_ISL_2229843, EPI_ISL_2229844, EPI_ISL_2229845, EPI_ISL_2229846, EPI_ISL_2229847, EPI_ISL_2229848, EPI_ISL_2229849, EPI_ISL_2229850, EPI_ISL_2229851, EPI_ISL_2229852, EPI_ISL_2229853, EPI_ISL_2229854, EPI_ISL_2229855, EPI_ISL_2229856, EPI_ISL_2229857, EPI_ISL_2229858, EPI_ISL_2229859, EPI_ISL_2229860, EPI_ISL_2229861, EPI_ISL_2229862, EPI_ISL_2229863, EPI_ISL_2229864, EPI_ISL_2229865, EPI_ISL_2229866, EPI_ISL_2229867, EPI_ISL_2229868, EPI_ISL_2229869, EPI_ISL_2229870, EPI_ISL_2229871, EPI_ISL_2229872, EPI_ISL_2229873, EPI_ISL_2229874, EPI_ISL_2229875, EPI_ISL_2229876, EPI_ISL_2229877, EPI_ISL_2229878, EPI_ISL_2229879, EPI_ISL_2229880, EPI_ISL_2229881, EPI_ISL_2229882, EPI_ISL_2229883, EPI_ISL_2229884, EPI_ISL_2229885, EPI_ISL_2229886, EPI_ISL_2229887, EPI_ISL_2229888, EPI_ISL_2229889, EPI_ISL_2229890, EPI_ISL_2229891, EPI_ISL_2229892, EPI_ISL_2229893, EPI_ISL_2229894, EPI_ISL_2229895, EPI_ISL_2229896, EPI_ISL_2229897, EPI_ISL_2229898, EPI_ISL_2229899, EPI_ISL_2229900, EPI_ISL_2229901, EPI_ISL_2229902, EPI_ISL_2229903, EPI_ISL_2229904, EPI_ISL_2229905, EPI_ISL_2229906, EPI_ISL_2229907, EPI_ISL_2229908, EPI_ISL_2229909, EPI_ISL_2229910, EPI_ISL_2229911, EPI_ISL_2229912, EPI_ISL_2229913, EPI_ISL_2229914, EPI_ISL_2229915, EPI_ISL_2229916, EPI_ISL_2229917, EPI_ISL_2229918, EPI_ISL_2229919, EPI_ISL_2229920, EPI_ISL_2229921, EPI_ISL_2229922, EPI_ISL_2229923, EPI_ISL_2229924, EPI_ISL_2229925 | | | |
| see above | Hackensack Medical Center | New York Genome Center | Michael Zody, Andre Corvelo, Dayna M. Oschwald, Samantha Fennessey, Tom Maniatis, Liang Chen, Jose Mediavilla, Marcus Cunningham, Kaelea Composto, Kar Chow, David Perlin, Barry Kreiswirth |
| EPI_ISL_2229926, EPI_ISL_2229927, EPI_ISL_2229928, EPI_ISL_2229929, EPI_ISL_2229930, EPI_ISL_2229931, EPI_ISL_2229932, EPI_ISL_2229933, EPI_ISL_2229934, EPI_ISL_2229935, EPI_ISL_2229937, EPI_ISL_2229938, EPI_ISL_2229939, EPI_ISL_2229941, EPI_ISL_2229942, EPI_ISL_2229943, EPI_ISL_2229944, EPI_ISL_2229945, EPI_ISL_2229946, EPI_ISL_2229947, EPI_ISL_2229948, EPI_ISL_2229949, EPI_ISL_2229951, EPI_ISL_2229952, EPI_ISL_2229953, EPI_ISL_2229955, EPI_ISL_2229956, EPI_ISL_2229957, EPI_ISL_2229958, EPI_ISL_2229959, EPI_ISL_2229960, EPI_ISL_2229961, EPI_ISL_2229962, EPI_ISL_2229963, EPI_ISL_2229964, EPI_ISL_2229965, EPI_ISL_2229966, EPI_ISL_2229967, EPI_ISL_2229968, EPI_ISL_2229969, EPI_ISL_2229971, EPI_ISL_2229972, EPI_ISL_2229973, EPI_ISL_2229974, EPI_ISL_2229975, EPI_ISL_2229976, EPI_ISL_2229977, EPI_ISL_2229978, EPI_ISL_2229979, EPI_ISL_2229980, EPI_ISL_2229981, EPI_ISL_2229982, EPI_ISL_2229983, EPI_ISL_2229984, EPI_ISL_2229985, EPI_ISL_2229986, EPI_ISL_2229987, EPI_ISL_2229989, EPI_ISL_2229990, EPI_ISL_2229991, EPI_ISL_2229992, EPI_ISL_2229994, EPI_ISL_2229996, EPI_ISL_2229997, EPI_ISL_2229998, EPI_ISL_2229999, EPI_ISL_2230000, EPI_ISL_2230001, EPI_ISL_2230002, EPI_ISL_2230003, EPI_ISL_2230005, EPI_ISL_2230006, EPI_ISL_2230007, EPI_ISL_2230008, EPI_ISL_2230009, EPI_ISL_2230010, EPI_ISL_2230011, EPI_ISL_2230012, EPI_ISL_2230013, EPI_ISL_2230014, EPI_ISL_2230015, EPI_ISL_2230016, EPI_ISL_2230017, EPI_ISL_2230019, EPI_ISL_2230020, EPI_ISL_2230021, EPI_ISL_2230022 | | | |
| see above | Florida Bureau of Public Health Laboratories | Florida Bureau of Public Health Laboratories | Sarah Schmedes, Jason Blanton |
| EPI_ISL_2230023, EPI_ISL_2230024, EPI_ISL_2230025, EPI_ISL_2230026, EPI_ISL_2230027, EPI_ISL_2230028, EPI_ISL_2230029, EPI_ISL_2230030, EPI_ISL_2230031, EPI_ISL_2230032, EPI_ISL_2230033, EPI_ISL_2230034, EPI_ISL_2230035, EPI_ISL_2230036, EPI_ISL_2230037, EPI_ISL_2230038, EPI_ISL_2230039, EPI_ISL_2230040 | | | |
| see above | The Jackson Laboratory | The Jackson Laboratory | Long J, Bergeron D, Renzette N, Adams M, Omerza G, Kelly K, Li L |
| EPI_ISL_2230466, EPI_ISL_2230467 | Wichita State University - Molecular Diagnostics Lab | Kansas Health and Environmental Lab | Mike Grose, Katherine Wiggins, Jonathan Barnell, Ben Olsen, and Phil Adam |
| EPI_ISL_2230470, EPI_ISL_2230471, EPI_ISL_2230472, EPI_ISL_2230473, EPI_ISL_2230474, EPI_ISL_2230475, EPI_ISL_2230476, EPI_ISL_2230477 | Kansas Health and Environmental Lab | Kansas Health and Environmental Lab | Mike Grose, Katherine Wiggins, Jonathan Barnell, Ben Olsen, and Phil Adam |
| EPI_ISL_2230598, EPI_ISL_2230599, EPI_ISL_2230600, EPI_ISL_2230601, EPI_ISL_2230602, EPI_ISL_2230603, EPI_ISL_2230604, EPI_ISL_2230605, EPI_ISL_2230606, EPI_ISL_2230607, EPI_ISL_2230608, EPI_ISL_2230609, EPI_ISL_2230610, EPI_ISL_2230611, EPI_ISL_2230612, EPI_ISL_2230613, EPI_ISL_2230614, EPI_ISL_2230615, EPI_ISL_2230616, EPI_ISL_2230617, EPI_ISL_2230618, EPI_ISL_2230619, EPI_ISL_2230620 | | | |
| see above | University of Wisconsin-Madison AIDS | University of Wisconsin-Madison AIDS | Gage Moreno, Katarina Braun, et al. AIDS Vaccine Research Laboratories |

| | | | |
|---|---|---|---|
| | Vaccine Research Laboratories | Vaccine Research Laboratories | |
| EPI_ISL_2230621, EPI_ISL_2230622, EPI_ISL_2230623, EPI_ISL_2230624, EPI_ISL_2230625, EPI_ISL_2230626, EPI_ISL_2230627, EPI_ISL_2230628, EPI_ISL_2230629, EPI_ISL_2230630, EPI_ISL_2230631, EPI_ISL_2230632, EPI_ISL_2230633, EPI_ISL_2230634, EPI_ISL_2230635, EPI_ISL_2230636, EPI_ISL_2230637, EPI_ISL_2230638, EPI_ISL_2230639, EPI_ISL_2230640, EPI_ISL_2230641, EPI_ISL_2230642, EPI_ISL_2230643, EPI_ISL_2230644, EPI_ISL_2230645, EPI_ISL_2230646, EPI_ISL_2230647, EPI_ISL_2230648, EPI_ISL_2230649, EPI_ISL_2230650, EPI_ISL_2230651, EPI_ISL_2230652, EPI_ISL_2230653, EPI_ISL_2230654, EPI_ISL_2230655, EPI_ISL_2230656, EPI_ISL_2230657, EPI_ISL_2230658, EPI_ISL_2230659, EPI_ISL_2230660, EPI_ISL_2230661, EPI_ISL_2230662, EPI_ISL_2230663, EPI_ISL_2230664, EPI_ISL_2230665, EPI_ISL_2230666, EPI_ISL_2230667, EPI_ISL_2230668, EPI_ISL_2230669, EPI_ISL_2230670, EPI_ISL_2230671, EPI_ISL_2230672, EPI_ISL_2230673, EPI_ISL_2230674, EPI_ISL_2230675, EPI_ISL_2230676, EPI_ISL_2230677, EPI_ISL_2230678 | | | |
| see above | Noblis | Noblis | Lauren Brinkac, Shiela Diepold, Shane Mitchell, June Kang, Lee F. Kolakowski, William M. Nelson, Katharine Jennings |
| EPI_ISL_2230701, EPI_ISL_2230702, EPI_ISL_2230703, EPI_ISL_2230705 | Alaska State Virology Laboratory | Alaska State Virology Laboratory | Stephanie DeRonde, Elva House, Lisa Smith, Ph.D., Jack Chen, Ph.D. |
| EPI_ISL_2230706, EPI_ISL_2230708, EPI_ISL_2230709, EPI_ISL_2230710, EPI_ISL_2230711, EPI_ISL_2230712, EPI_ISL_2230713, EPI_ISL_2230714, EPI_ISL_2230715, EPI_ISL_2230716, EPI_ISL_2230717, EPI_ISL_2230718, EPI_ISL_2230719, EPI_ISL_2230720, EPI_ISL_2230721, EPI_ISL_2230722, EPI_ISL_2230723, EPI_ISL_2230724, EPI_ISL_2230725, EPI_ISL_2230726, EPI_ISL_2230727, EPI_ISL_2230728, EPI_ISL_2230729, EPI_ISL_2230730, EPI_ISL_2230732, EPI_ISL_2230733, EPI_ISL_2230734, EPI_ISL_2230735, EPI_ISL_2230736, EPI_ISL_2230737, EPI_ISL_2230738, EPI_ISL_2230739, EPI_ISL_2230740, EPI_ISL_2230741, EPI_ISL_2230742, EPI_ISL_2230743 | | | |
| see above | Southern Nevada Public Health Laboratory | Southern Nevada Public Health Laboratory | Michael Picker |
| EPI_ISL_2230744, EPI_ISL_2230745, EPI_ISL_2230746, EPI_ISL_2230747, EPI_ISL_2230748, EPI_ISL_2230749, EPI_ISL_2230750, EPI_ISL_2230751, EPI_ISL_2230752, EPI_ISL_2230753, EPI_ISL_2230754, EPI_ISL_2230755, EPI_ISL_2230756, EPI_ISL_2230757, EPI_ISL_2230758 | | | |
| see above | Kentucky State Public Health Lab | Kentucky State Public Health Lab | Anjayya Tummala, Karim George, Joshua Tobias, William Grooms, Vaneet Arora, Matthew Johnson, Rachel Zinner, Rhonda Lucas |
| EPI_ISL_2230761, EPI_ISL_2230762, EPI_ISL_2230763, EPI_ISL_2230765, EPI_ISL_2230766, EPI_ISL_2230767, EPI_ISL_2230771 | Santa Clara County public Health Laboratory | Santa Clara County public Health Laboratory | Santa Clara County Public Health Department |
| EPI_ISL_2230794, EPI_ISL_2230795, EPI_ISL_2230798, EPI_ISL_2230799, EPI_ISL_2230800 | City of Milwaukee Health Department Laboratory | City of Milwaukee Health Department Laboratory | Sanjib Bhattacharyya, Manjeet Khubbar, Amy Bauer, Jennifer Lentz, Samantha Scott |
| EPI_ISL_2230801 | InDRE | Instituto Nacional de Medicina Genomica | Hidalgo-Miranda A, Mendoza-Vargas A, Reyes-Grajeda JP, Cedro-Tanda A, Gisela Barrera-Badillo, Irma Lopez-Martinez, Jose Ernesto Ramirez González, Gonzalez-Barrera D, Rangel-DeLeon D, Munguia-Garza P, Garcia-Cardenas FJ, Gonzalez-Woge MA, Herrera-Montalvo LA. |
| EPI_ISL_2230802, EPI_ISL_2230803 | Instituto Nacional de Medicina Genomica | Instituto Nacional de Medicina Genomica | Hidalgo-Miranda A, Cedro-Tanda A, Mendoza-Vargas A, Reyes-Grajeda JP, Cisneros- Villanueva M, Gonzalez-Barrera D, Rangel-DeLeon D, Munguia-Garza P, Ramirez-Vega O, Escobar-Arrazola, M, Herrera-Montalvo LA. |
| EPI_ISL_2230804 | Salud Digna | Instituto Nacional de Medicina Genomica | Hidalgo-Miranda A, Cedro-Tanda A, Mendoza-Vargas A, Reyes-Grajeda JP, Abraham Campos-Romero, Moreno-Camacho José Luis, Rodriguez-Gallegos Jorge, Luna-Ruiz Marco, Gonzalez-Barrera D, Rangel-DeLeon D, Munguia-Garza P, Ramirez-Vega O, Escobar-Arrazola, M, Herrera-Montalvo LA. |
| EPI_ISL_2230805 | InDRE | Instituto Nacional de Medicina Genomica | Hidalgo-Miranda A, Mendoza-Vargas A, Reyes-Grajeda JP, Cedro-Tanda A, Gisela Barrera-Badillo, Irma Lopez-Martinez, Jose Ernesto Ramirez González, Gonzalez-Barrera D, Rangel-DeLeon D, Munguia-Garza P, Garcia-Cardenas FJ, Gonzalez-Woge MA, Herrera-Montalvo LA. |
| EPI_ISL_2230806 | Salud Digna | Instituto Nacional de Medicina Genomica | Hidalgo-Miranda A, Cedro-Tanda A, Mendoza-Vargas A, Reyes-Grajeda JP, Abraham Campos-Romero, Moreno-Camacho José Luis, Rodriguez-Gallegos Jorge, Luna-Ruiz Marco, Gonzalez-Barrera D, Rangel-DeLeon D, Munguia-Garza P, Ramirez-Vega O, Escobar-Arrazola, M, Herrera-Montalvo LA. |
| EPI_ISL_2230807 | Instituto Nacional de Medicina Genomica | Instituto Nacional de Medicina Genomica | Hidalgo-Miranda A, Cedro-Tanda A, Mendoza-Vargas A, Reyes-Grajeda JP, Cisneros- Villanueva M, Gonzalez-Barrera D, Rangel-DeLeon D, Munguia-Garza P, Ramirez-Vega O, Escobar-Arrazola, M, Herrera-Montalvo LA. |
| EPI_ISL_2230808 | InDRE | Instituto Nacional de Medicina Genomica | Hidalgo-Miranda A, Mendoza-Vargas A, Reyes-Grajeda JP, Cedro-Tanda A, Gisela Barrera-Badillo, Irma Lopez-Martinez, Jose Ernesto Ramirez González, Gonzalez-Barrera D, Rangel-DeLeon D, Munguia-Garza P, Garcia-Cardenas FJ, Gonzalez-Woge MA, Herrera-Montalvo LA. |
| EPI_ISL_2230809 | Instituto Nacional de Medicina Genomica | Instituto Nacional de Medicina Genomica | Hidalgo-Miranda A, Cedro-Tanda A, Mendoza-Vargas A, Reyes-Grajeda JP, Cisneros- Villanueva M, Gonzalez-Barrera D, Rangel-DeLeon D, Munguia-Garza P, Ramirez-Vega O, Escobar-Arrazola, M, Herrera-Montalvo LA. |
| EPI_ISL_2230810, EPI_ISL_2230811 | Salud Digna | Instituto Nacional de Medicina Genomica | Hidalgo-Miranda A, Cedro-Tanda A, Mendoza-Vargas A, Reyes-Grajeda JP, Abraham Campos-Romero, Moreno-Camacho José Luis, Rodriguez-Gallegos Jorge, Luna-Ruiz Marco, Gonzalez-Barrera D, Rangel-DeLeon D, Munguia-Garza P, Ramirez-Vega O, Escobar-Arrazola, M, Herrera-Montalvo LA. |
| EPI_ISL_2230812 | Instituto Nacional de Medicina Genomica | Instituto Nacional de Medicina Genomica | Hidalgo-Miranda A, Cedro-Tanda A, Mendoza-Vargas A, Reyes-Grajeda JP, Cisneros- Villanueva M, Gonzalez-Barrera D, Rangel-DeLeon D, Munguia-Garza P, Ramirez-Vega O, Escobar-Arrazola, M, Herrera-Montalvo LA. |
| EPI_ISL_2230813 | Salud Digna | Instituto Nacional de Medicina Genomica | Hidalgo-Miranda A, Cedro-Tanda A, Mendoza-Vargas A, Reyes-Grajeda JP, Abraham Campos-Romero, Moreno-Camacho José Luis, Rodriguez-Gallegos Jorge, Luna-Ruiz Marco, Gonzalez-Barrera D, Rangel-DeLeon D, Munguia-Garza P, Ramirez-Vega O, Escobar-Arrazola, M, Herrera-Montalvo LA. |
| EPI_ISL_2230814 | InDRE | Instituto Nacional de Medicina Genomica | Hidalgo-Miranda A, Mendoza-Vargas A, Reyes-Grajeda JP, Cedro-Tanda A, Gisela Barrera-Badillo, Irma Lopez-Martinez, Jose Ernesto Ramirez González, Gonzalez-Barrera D, Rangel-DeLeon D, Munguia-Garza P, Garcia-Cardenas FJ, Gonzalez-Woge MA, Herrera-Montalvo LA. |
| EPI_ISL_2230815 | Instituto Nacional de Medicina Genomica | Instituto Nacional de Medicina Genomica | Hidalgo-Miranda A, Cedro-Tanda A, Mendoza-Vargas A, Reyes-Grajeda JP, Cisneros- Villanueva M, Gonzalez-Barrera D, Rangel-DeLeon D, Munguia-Garza P, Ramirez-Vega O, Escobar-Arrazola, M, Herrera-Montalvo LA. |
| EPI_ISL_2230816, EPI_ISL_2230817 | Salud Digna | Instituto Nacional de Medicina Genomica | Hidalgo-Miranda A, Cedro-Tanda A, Mendoza-Vargas A, Reyes-Grajeda JP, Abraham Campos-Romero, Moreno-Camacho José Luis, Rodriguez-Gallegos Jorge, Luna-Ruiz Marco, Gonzalez-Barrera D, Rangel-DeLeon D, Munguia-Garza P, Ramirez-Vega O, Escobar-Arrazola, M, Herrera-Montalvo LA. |
| EPI_ISL_2230818, EPI_ISL_2230819, EPI_ISL_2230820 | Instituto Nacional de Medicina Genomica | Instituto Nacional de Medicina Genomica | Hidalgo-Miranda A, Cedro-Tanda A, Mendoza-Vargas A, Reyes-Grajeda JP, Cisneros- Villanueva M, Gonzalez-Barrera D, Rangel-DeLeon D, Munguia-Garza P, Ramirez-Vega O, Escobar-Arrazola, M, Herrera-Montalvo LA. |
| EPI_ISL_2230821, EPI_ISL_2230822, EPI_ISL_2230823, EPI_ISL_2230824 | InDRE | Instituto Nacional de Medicina Genomica | Hidalgo-Miranda A, Mendoza-Vargas A, Reyes-Grajeda JP, Cedro-Tanda A, Gisela Barrera-Badillo, Irma Lopez-Martinez, Jose Ernesto Ramirez González, Gonzalez-Barrera D, Rangel-DeLeon D, Munguia-Garza P, Garcia-Cardenas FJ, Gonzalez-Woge MA, Herrera-Montalvo LA. |
| EPI_ISL_2230825 | Salud Digna | Instituto Nacional de Medicina Genomica | Hidalgo-Miranda A, Cedro-Tanda A, Mendoza-Vargas A, Reyes-Grajeda JP, Abraham Campos-Romero, Moreno-Camacho José Luis, Rodriguez-Gallegos Jorge, Luna-Ruiz Marco, Gonzalez-Barrera D, Rangel-DeLeon D, Munguia-Garza P, Ramirez-Vega O, Escobar-Arrazola, M, Herrera-Montalvo LA. |
| EPI_ISL_2230826 | Instituto Nacional de Medicina Genomica | Instituto Nacional de Medicina Genomica | Hidalgo-Miranda A, Cedro-Tanda A, Mendoza-Vargas A, Reyes-Grajeda JP, Cisneros- Villanueva M, Gonzalez-Barrera D, Rangel-DeLeon D, Munguia-Garza P, Ramirez-Vega O, Escobar-Arrazola, M, Herrera-Montalvo LA. |
| EPI_ISL_2230827 | InDRE | Instituto Nacional de Medicina Genomica | Hidalgo-Miranda A, Mendoza-Vargas A, Reyes-Grajeda JP, Cedro-Tanda A, Gisela Barrera-Badillo, Irma Lopez-Martinez, Jose Ernesto Ramirez González, Gonzalez-Barrera D, Rangel-DeLeon D, Munguia-Garza P, Garcia-Cardenas FJ, Gonzalez-Woge MA, Herrera-Montalvo LA. |
| EPI_ISL_2230828 | Salud Digna | Instituto Nacional de Medicina Genomica | Hidalgo-Miranda A, Cedro-Tanda A, Mendoza-Vargas A, Reyes-Grajeda JP, Abraham Campos-Romero, Moreno-Camacho José Luis, Rodriguez-Gallegos Jorge, Luna-Ruiz Marco, Gonzalez-Barrera D, Rangel-DeLeon D, Munguia-Garza P, Ramirez-Vega O, Escobar-Arrazola, M, Herrera-Montalvo LA. |
| EPI_ISL_2230829 | Instituto Nacional de Medicina Genomica | Instituto Nacional de Medicina Genomica | Hidalgo-Miranda A, Cedro-Tanda A, Mendoza-Vargas A, Reyes-Grajeda JP, Cisneros- Villanueva M, Gonzalez-Barrera D, Rangel-DeLeon D, Munguia-Garza P, Ramirez-Vega O, Escobar-Arrazola, M, Herrera-Montalvo LA. |
| EPI_ISL_2230830, EPI_ISL_2230831, EPI_ISL_2230832 | Salud Digna | Instituto Nacional de Medicina Genomica | Hidalgo-Miranda A, Cedro-Tanda A, Mendoza-Vargas A, Reyes-Grajeda JP, Abraham Campos-Romero, Moreno-Camacho José Luis, Rodriguez-Gallegos Jorge, Luna-Ruiz Marco, Gonzalez-Barrera D, Rangel-DeLeon D, Munguia-Garza P, Ramirez-Vega O, Escobar-Arrazola, M, Herrera-Montalvo LA. |
| EPI_ISL_2230833 | InDRE | Instituto Nacional de Medicina Genomica | Hidalgo-Miranda A, Mendoza-Vargas A, Reyes-Grajeda JP, Cedro-Tanda A, Gisela Barrera-Badillo, Irma Lopez-Martinez, Jose Ernesto Ramirez González, Gonzalez-Barrera D, Rangel-DeLeon D, Munguia-Garza P, Garcia-Cardenas FJ, Gonzalez-Woge MA, Herrera-Montalvo LA. |
| EPI_ISL_2230835 | Salud Digna | Instituto Nacional de Medicina Genomica | Hidalgo-Miranda A, Cedro-Tanda A, Mendoza-Vargas A, Reyes-Grajeda JP, Abraham Campos-Romero, Moreno-Camacho José Luis, Rodriguez-Gallegos Jorge, Luna-Ruiz Marco, Gonzalez-Barrera D, Rangel-DeLeon D, Munguia-Garza P, Ramirez-Vega O, Escobar-Arrazola, M, Herrera-Montalvo LA. |
| EPI_ISL_2230836 | Instituto Nacional de Medicina Genomica | Instituto Nacional de Medicina Genomica | Hidalgo-Miranda A, Cedro-Tanda A, Mendoza-Vargas A, Reyes-Grajeda JP, Cisneros- Villanueva M, Gonzalez-Barrera D, Rangel-DeLeon D, Munguia-Garza P, Ramirez-Vega O, Escobar-Arrazola, M, Herrera-Montalvo LA. |
| EPI_ISL_2230837 | InDRE | Instituto Nacional de Medicina Genomica | Hidalgo-Miranda A, Mendoza-Vargas A, Reyes-Grajeda JP, Cedro-Tanda A, Gisela Barrera-Badillo, Irma Lopez-Martinez, Jose Ernesto Ramirez González, Gonzalez-Barrera D, Rangel-DeLeon D, Munguia-Garza P, Garcia-Cardenas FJ, Gonzalez-Woge MA, Herrera-Montalvo LA. |
| EPI_ISL_2230838 | Salud Digna | Instituto Nacional de Medicina Genomica | Hidalgo-Miranda A, Cedro-Tanda A, Mendoza-Vargas A, Reyes-Grajeda JP, Abraham Campos-Romero, Moreno-Camacho José Luis, Rodriguez-Gallegos Jorge, Luna-Ruiz Marco, Gonzalez-Barrera D, Rangel-DeLeon D, Munguia-Garza P, Ramirez-Vega O, Escobar-Arrazola, M, Herrera-Montalvo LA. |
| EPI_ISL_2230839 | Instituto Nacional de Medicina Genomica | Instituto Nacional de Medicina Genomica | Hidalgo-Miranda A, Cedro-Tanda A, Mendoza-Vargas A, Reyes-Grajeda JP, Cisneros- Villanueva M, Gonzalez-Barrera D, Rangel-DeLeon D, Munguia-Garza P, Ramirez-Vega O, Escobar-Arrazola, M, Herrera-Montalvo LA. |
| EPI_ISL_2230840 | InDRE | Instituto Nacional de Medicina Genomica | Hidalgo-Miranda A, Mendoza-Vargas A, Reyes-Grajeda JP, Cedro-Tanda A, Gisela Barrera-Badillo, Irma Lopez-Martinez, Jose Ernesto Ramirez González, Gonzalez-Barrera D, Rangel-DeLeon D, Munguia-Garza P, Garcia-Cardenas FJ, Gonzalez-Woge MA, Herrera-Montalvo LA. |
| EPI_ISL_2230841, EPI_ISL_2230842, EPI_ISL_2230843, EPI_ISL_2230844 | Salud Digna | Instituto Nacional de Medicina Genomica | Hidalgo-Miranda A, Cedro-Tanda A, Mendoza-Vargas A, Reyes-Grajeda JP, Abraham Campos-Romero, Moreno-Camacho José Luis, Rodriguez-Gallegos Jorge, Luna-Ruiz Marco, Gonzalez-Barrera D, Rangel-DeLeon D, Munguia-Garza P, Ramirez-Vega O, Escobar-Arrazola, M, Herrera-Montalvo LA. |
| EPI_ISL_2230845, EPI_ISL_2230846 | InDRE | Instituto Nacional de Medicina Genomica | Hidalgo-Miranda A, Mendoza-Vargas A, Reyes-Grajeda JP, Cedro-Tanda A, Gisela Barrera-Badillo, Irma Lopez-Martinez, Jose Ernesto Ramirez González, Gonzalez-Barrera D, Rangel-DeLeon D, Munguia-Garza P, Garcia-Cardenas FJ, Gonzalez-Woge MA, Herrera-Montalvo LA. |
| EPI_ISL_2230847, EPI_ISL_2230848 | Instituto Nacional de Medicina Genomica | Instituto Nacional de Medicina Genomica | Hidalgo-Miranda A, Cedro-Tanda A, Mendoza-Vargas A, Reyes-Grajeda JP, Cisneros- Villanueva M, Gonzalez-Barrera D, Rangel-DeLeon D, Munguia-Garza P, Ramirez-Vega O, Escobar-Arrazola, M, Herrera-Montalvo LA. |
| EPI_ISL_2230849, EPI_ISL_2230850, | Salud Digna | Instituto Nacional de Medicina Genomica | Hidalgo-Miranda A, Cedro-Tanda A, Mendoza-Vargas A, Reyes-Grajeda JP, Abraham Campos-Romero, Moreno-Camacho José Luis, Rodriguez-Gallegos Jorge, Luna-Ruiz Marco, Gonzalez-Barrera D, Rangel-DeLeon D, Munguia-Garza P, Ramirez-Vega O, Escobar-Arrazola, M, Herrera-Montalvo LA. |

| | | | |
|---|---|---|---|
| EPI_ISL_2230851 | | | |
| EPI_ISL_2230852 | InDRE | Instituto Nacional de Medicina Genomica | Hidalgo-Miranda A, Mendoza-Vargas A, Reyes-Grajeda JP, Cedro-Tanda A, Gisela Barrera-Badillo, Irma Lopez-Martinez, Jose Ernesto Ramirez González, Gonzalez-Barrera D, Rangel-DeLeon D, Munguia-Garza P, Garcia-Cardenas FJ, Gonzalez-Woge MA, Herrera-Montalvo LA. |
| EPI_ISL_2230853, EPI_ISL_2230854 | Salud Digna | Instituto Nacional de Medicina Genomica | Hidalgo-Miranda A, Cedro-Tanda A, Mendoza-Vargas A, Reyes-Grajeda JP, Abraham Campos-Romero, Moreno-Camacho José Luis, Rodriguez-Gallegos Jorge, Luna-Ruiz Marco, Gonzalez-Barrera D, Rangel-DeLeon D, Munguia-Garza P, Ramirez-Vega O, Escobar-Arrazola, M, Herrera-Montalvo LA. |
| EPI_ISL_2230855 | InDRE | Instituto Nacional de Medicina Genomica | Hidalgo-Miranda A, Mendoza-Vargas A, Reyes-Grajeda JP, Cedro-Tanda A, Gisela Barrera-Badillo, Irma Lopez-Martinez, Jose Ernesto Ramirez González, Gonzalez-Barrera D, Rangel-DeLeon D, Munguia-Garza P, Garcia-Cardenas FJ, Gonzalez-Woge MA, Herrera-Montalvo LA. |
| EPI_ISL_2230856, EPI_ISL_2230857 | Salud Digna | Instituto Nacional de Medicina Genomica | Hidalgo-Miranda A, Cedro-Tanda A, Mendoza-Vargas A, Reyes-Grajeda JP, Abraham Campos-Romero, Moreno-Camacho José Luis, Rodriguez-Gallegos Jorge, Luna-Ruiz Marco, Gonzalez-Barrera D, Rangel-DeLeon D, Munguia-Garza P, Ramirez-Vega O, Escobar-Arrazola, M, Herrera-Montalvo LA. |
| EPI_ISL_2230858 | Instituto Nacional de Medicina Genomica | Instituto Nacional de Medicina Genomica | Hidalgo-Miranda A, Cedro-Tanda A, Mendoza-Vargas A, Reyes-Grajeda JP, Cisneros- Villanueva M, Gonzalez-Barrera D, Rangel-DeLeon D, Munguia-Garza P, Ramirez-Vega O, Escobar-Arrazola, M, Herrera-Montalvo LA. |
| EPI_ISL_2230859 | Salud Digna | Instituto Nacional de Medicina Genomica | Hidalgo-Miranda A, Cedro-Tanda A, Mendoza-Vargas A, Reyes-Grajeda JP, Abraham Campos-Romero, Moreno-Camacho José Luis, Rodriguez-Gallegos Jorge, Luna-Ruiz Marco, Gonzalez-Barrera D, Rangel-DeLeon D, Munguia-Garza P, Ramirez-Vega O, Escobar-Arrazola, M, Herrera-Montalvo LA. |
| EPI_ISL_2230860 | InDRE | Instituto Nacional de Medicina Genomica | Hidalgo-Miranda A, Mendoza-Vargas A, Reyes-Grajeda JP, Cedro-Tanda A, Gisela Barrera-Badillo, Irma Lopez-Martinez, Jose Ernesto Ramirez González, Gonzalez-Barrera D, Rangel-DeLeon D, Munguia-Garza P, Garcia-Cardenas FJ, Gonzalez-Woge MA, Herrera-Montalvo LA. |
| EPI_ISL_2230861 | Instituto Nacional de Medicina Genomica | Instituto Nacional de Medicina Genomica | Hidalgo-Miranda A, Cedro-Tanda A, Mendoza-Vargas A, Reyes-Grajeda JP, Cisneros- Villanueva M, Gonzalez-Barrera D, Rangel-DeLeon D, Munguia-Garza P, Ramirez-Vega O, Escobar-Arrazola, M, Herrera-Montalvo LA. |
| EPI_ISL_2230862 | Salud Digna | Instituto Nacional de Medicina Genomica | Hidalgo-Miranda A, Cedro-Tanda A, Mendoza-Vargas A, Reyes-Grajeda JP, Abraham Campos-Romero, Moreno-Camacho José Luis, Rodriguez-Gallegos Jorge, Luna-Ruiz Marco, Gonzalez-Barrera D, Rangel-DeLeon D, Munguia-Garza P, Ramirez-Vega O, Escobar-Arrazola, M, Herrera-Montalvo LA. |
| EPI_ISL_2230863, EPI_ISL_2230864 | InDRE | Instituto Nacional de Medicina Genomica | Hidalgo-Miranda A, Mendoza-Vargas A, Reyes-Grajeda JP, Cedro-Tanda A, Gisela Barrera-Badillo, Irma Lopez-Martinez, Jose Ernesto Ramirez González, Gonzalez-Barrera D, Rangel-DeLeon D, Munguia-Garza P, Garcia-Cardenas FJ, Gonzalez-Woge MA, Herrera-Montalvo LA. |
| EPI_ISL_2230865, EPI_ISL_2230866, EPI_ISL_2230867, EPI_ISL_2230868, EPI_ISL_2230869, EPI_ISL_2230870, EPI_ISL_2230871, EPI_ISL_2230872, EPI_ISL_2230873, EPI_ISL_2230874, EPI_ISL_2230875, EPI_ISL_2230876, EPI_ISL_2230877, EPI_ISL_2230878, EPI_ISL_2230879 | | | |
| see above | Salud Digna | Instituto Nacional de Medicina Genomica | Hidalgo-Miranda A, Cedro-Tanda A, Mendoza-Vargas A, Reyes-Grajeda JP, Abraham Campos-Romero, Moreno-Camacho José Luis, Rodriguez-Gallegos Jorge, Luna-Ruiz Marco, Gonzalez-Barrera D, Rangel-DeLeon D, Munguia-Garza P, Ramirez-Vega O, Escobar-Arrazola, M, Herrera-Montalvo LA. |
| EPI_ISL_2230880 | InDRE | Instituto Nacional de Medicina Genomica | Hidalgo-Miranda A, Mendoza-Vargas A, Reyes-Grajeda JP, Cedro-Tanda A, Gisela Barrera-Badillo, Irma Lopez-Martinez, Jose Ernesto Ramirez González, Gonzalez-Barrera D, Rangel-DeLeon D, Munguia-Garza P, Garcia-Cardenas FJ, Gonzalez-Woge MA, Herrera-Montalvo LA. |
| EPI_ISL_2230881, EPI_ISL_2230882, EPI_ISL_2230883, EPI_ISL_2230884, EPI_ISL_2230885, EPI_ISL_2230886 | Salud Digna | Instituto Nacional de Medicina Genomica | Hidalgo-Miranda A, Cedro-Tanda A, Mendoza-Vargas A, Reyes-Grajeda JP, Abraham Campos-Romero, Moreno-Camacho José Luis, Rodriguez-Gallegos Jorge, Luna-Ruiz Marco, Gonzalez-Barrera D, Rangel-DeLeon D, Munguia-Garza P, Ramirez-Vega O, Escobar-Arrazola, M, Herrera-Montalvo LA. |
| EPI_ISL_2230897 | InDRE | Instituto Nacional de Medicina Genomica | Hidalgo-Miranda A, Mendoza-Vargas A, Reyes-Grajeda JP, Cedro-Tanda A, Gisela Barrera-Badillo, Irma Lopez-Martinez, Jose Ernesto Ramirez González, Gonzalez-Barrera D, Rangel-DeLeon D, Munguia-Garza P, Garcia-Cardenas FJ, Gonzalez-Woge MA, Herrera-Montalvo LA. |
| EPI_ISL_2230898, EPI_ISL_2230899, EPI_ISL_2230900, EPI_ISL_2230901, EPI_ISL_2230902, EPI_ISL_2230903 | Salud Digna | Instituto Nacional de Medicina Genomica | Hidalgo-Miranda A, Cedro-Tanda A, Mendoza-Vargas A, Reyes-Grajeda JP, Abraham Campos-Romero, Moreno-Camacho José Luis, Rodriguez-Gallegos Jorge, Luna-Ruiz Marco, Gonzalez-Barrera D, Rangel-DeLeon D, Munguia-Garza P, Ramirez-Vega O, Escobar-Arrazola, M, Herrera-Montalvo LA. |
| EPI_ISL_2230905 | InDRE | Instituto Nacional de Medicina Genomica | Hidalgo-Miranda A, Mendoza-Vargas A, Reyes-Grajeda JP, Cedro-Tanda A, Gisela Barrera-Badillo, Irma Lopez-Martinez, Jose Ernesto Ramirez González, Gonzalez-Barrera D, Rangel-DeLeon D, Munguia-Garza P, Garcia-Cardenas FJ, Gonzalez-Woge MA, Herrera-Montalvo LA. |
| EPI_ISL_2230906 | Salud Digna | Instituto Nacional de Medicina Genomica | Hidalgo-Miranda A, Cedro-Tanda A, Mendoza-Vargas A, Reyes-Grajeda JP, Abraham Campos-Romero, Moreno-Camacho José Luis, Rodriguez-Gallegos Jorge, Luna-Ruiz Marco, Gonzalez-Barrera D, Rangel-DeLeon D, Munguia-Garza P, Ramirez-Vega O, Escobar-Arrazola, M, Herrera-Montalvo LA. |
| EPI_ISL_2230907, EPI_ISL_2230908, EPI_ISL_2230909, EPI_ISL_2230910 | InDRE | Instituto Nacional de Medicina Genomica | Hidalgo-Miranda A, Mendoza-Vargas A, Reyes-Grajeda JP, Cedro-Tanda A, Gisela Barrera-Badillo, Irma Lopez-Martinez, Jose Ernesto Ramirez González, Gonzalez-Barrera D, Rangel-DeLeon D, Munguia-Garza P, Garcia-Cardenas FJ, Gonzalez-Woge MA, Herrera-Montalvo LA. |
| EPI_ISL_2230911, EPI_ISL_2230912 | Salud Digna | Instituto Nacional de Medicina Genomica | Hidalgo-Miranda A, Cedro-Tanda A, Mendoza-Vargas A, Reyes-Grajeda JP, Abraham Campos-Romero, Moreno-Camacho José Luis, Rodriguez-Gallegos Jorge, Luna-Ruiz Marco, Gonzalez-Barrera D, Rangel-DeLeon D, Munguia-Garza P, Ramirez-Vega O, Escobar-Arrazola, M, Herrera-Montalvo LA. |
| EPI_ISL_2230913 | Instituto Nacional de Medicina Genomica | Instituto Nacional de Medicina Genomica | Hidalgo-Miranda A, Cedro-Tanda A, Mendoza-Vargas A, Reyes-Grajeda JP, Cisneros- Villanueva M, Gonzalez-Barrera D, Rangel-DeLeon D, Munguia-Garza P, Ramirez-Vega O, Escobar-Arrazola, M, Herrera-Montalvo LA. |
| EPI_ISL_2230914, EPI_ISL_2230915 | Salud Digna | Instituto Nacional de Medicina Genomica | Hidalgo-Miranda A, Cedro-Tanda A, Mendoza-Vargas A, Reyes-Grajeda JP, Abraham Campos-Romero, Moreno-Camacho José Luis, Rodriguez-Gallegos Jorge, Luna-Ruiz Marco, Gonzalez-Barrera D, Rangel-DeLeon D, Munguia-Garza P, Ramirez-Vega O, Escobar-Arrazola, M, Herrera-Montalvo LA. |
| EPI_ISL_2230916 | InDRE | Instituto Nacional de Medicina Genomica | Hidalgo-Miranda A, Mendoza-Vargas A, Reyes-Grajeda JP, Cedro-Tanda A, Gisela Barrera-Badillo, Irma Lopez-Martinez, Jose Ernesto Ramirez González, Gonzalez-Barrera D, Rangel-DeLeon D, Munguia-Garza P, Garcia-Cardenas FJ, Gonzalez-Woge MA, Herrera-Montalvo LA. |
| EPI_ISL_2230917, EPI_ISL_2230918, EPI_ISL_2230919 | Salud Digna | Instituto Nacional de Medicina Genomica | Hidalgo-Miranda A, Cedro-Tanda A, Mendoza-Vargas A, Reyes-Grajeda JP, Abraham Campos-Romero, Moreno-Camacho José Luis, Rodriguez-Gallegos Jorge, Luna-Ruiz Marco, Gonzalez-Barrera D, Rangel-DeLeon D, Munguia-Garza P, Ramirez-Vega O, Escobar-Arrazola, M, Herrera-Montalvo LA. |
| EPI_ISL_2230920, EPI_ISL_2230921 | Instituto Nacional de Medicina Genomica | Instituto Nacional de Medicina Genomica | Hidalgo-Miranda A, Cedro-Tanda A, Mendoza-Vargas A, Reyes-Grajeda JP, Cisneros- Villanueva M, Gonzalez-Barrera D, Rangel-DeLeon D, Munguia-Garza P, Ramirez-Vega O, Escobar-Arrazola, M, Herrera-Montalvo LA. |
| EPI_ISL_2230922, EPI_ISL_2230923, EPI_ISL_2230924, EPI_ISL_2230925 | Salud Digna | Instituto Nacional de Medicina Genomica | Hidalgo-Miranda A, Cedro-Tanda A, Mendoza-Vargas A, Reyes-Grajeda JP, Abraham Campos-Romero, Moreno-Camacho José Luis, Rodriguez-Gallegos Jorge, Luna-Ruiz Marco, Gonzalez-Barrera D, Rangel-DeLeon D, Munguia-Garza P, Ramirez-Vega O, Escobar-Arrazola, M, Herrera-Montalvo LA. |
| EPI_ISL_2230926, EPI_ISL_2230927, EPI_ISL_2230928, EPI_ISL_2230929, EPI_ISL_2230930, EPI_ISL_2230931, EPI_ISL_2230932, EPI_ISL_2230933, EPI_ISL_2230934, EPI_ISL_2230935, EPI_ISL_2230936, EPI_ISL_2230937, EPI_ISL_2230938, EPI_ISL_2230939, EPI_ISL_2230940, EPI_ISL_2230941, EPI_ISL_2230942 | | | |
| see above | Instituto Nacional de Medicina Genomica | Instituto Nacional de Medicina Genomica | Hidalgo-Miranda A, Cedro-Tanda A, Mendoza-Vargas A, Reyes-Grajeda JP, Cisneros- Villanueva M, Gonzalez-Barrera D, Rangel-DeLeon D, Munguia-Garza P, Ramirez-Vega O, Escobar-Arrazola, M, Herrera-Montalvo LA. |
| EPI_ISL_2230943, EPI_ISL_2230944, EPI_ISL_2230945, EPI_ISL_2230946, EPI_ISL_2230947, EPI_ISL_2230948, EPI_ISL_2230949, EPI_ISL_2230950, EPI_ISL_2230951, EPI_ISL_2230952, EPI_ISL_2230953, EPI_ISL_2230954, EPI_ISL_2230955, EPI_ISL_2230956, EPI_ISL_2230957, EPI_ISL_2230958, EPI_ISL_2230959, EPI_ISL_2230960, EPI_ISL_2230961, EPI_ISL_2230962, EPI_ISL_2230963, EPI_ISL_2230964, EPI_ISL_2230965, EPI_ISL_2230966, EPI_ISL_2230967, EPI_ISL_2230968, EPI_ISL_2230969, EPI_ISL_2230970, EPI_ISL_2230971, EPI_ISL_2230972, EPI_ISL_2230973, EPI_ISL_2230974 | | | |
| see above | InDRE | Instituto Nacional de Medicina Genomica | Hidalgo-Miranda A, Mendoza-Vargas A, Reyes-Grajeda JP, Cedro-Tanda A, Gisela Barrera-Badillo, Irma Lopez-Martinez, Jose Ernesto Ramirez González, Gonzalez-Barrera D, Rangel-DeLeon D, Munguia-Garza P, Garcia-Cardenas FJ, Gonzalez-Woge MA, Herrera-Montalvo LA. |
| EPI_ISL_2230975, EPI_ISL_2230976, EPI_ISL_2230977, EPI_ISL_2230978, EPI_ISL_2230979, EPI_ISL_2230980, EPI_ISL_2230981, EPI_ISL_2230982, EPI_ISL_2230983, EPI_ISL_2230984, EPI_ISL_2230985, EPI_ISL_2230986, EPI_ISL_2230987, EPI_ISL_2230988, EPI_ISL_2230989, EPI_ISL_2230990, EPI_ISL_2230991 | | | |
| see above | Salud Digna | Instituto Nacional de Medicina Genomica | Hidalgo-Miranda A, Cedro-Tanda A, Mendoza-Vargas A, Reyes-Grajeda JP, Abraham Campos-Romero, Moreno-Camacho José Luis, Rodriguez-Gallegos Jorge, Luna-Ruiz Marco, Gonzalez-Barrera D, Rangel-DeLeon D, Munguia-Garza P, Ramirez-Vega O, Escobar-Arrazola, M, Herrera-Montalvo LA. |
| EPI_ISL_2230992, EPI_ISL_2230993, EPI_ISL_2230994 | Instituto Nacional de Medicina Genomica | Instituto Nacional de Medicina Genomica | Hidalgo-Miranda A, Cedro-Tanda A, Mendoza-Vargas A, Reyes-Grajeda JP, Cisneros- Villanueva M, Gonzalez-Barrera D, Rangel-DeLeon D, Munguia-Garza P, Ramirez-Vega O, Escobar-Arrazola, M, Herrera-Montalvo LA. |
| EPI_ISL_2230995, EPI_ISL_2230996, EPI_ISL_2230997, EPI_ISL_2230998, EPI_ISL_2230999, EPI_ISL_2231000, EPI_ISL_2231001, EPI_ISL_2231002, EPI_ISL_2231003, EPI_ISL_2231004, EPI_ISL_2231005, EPI_ISL_2231006, EPI_ISL_2231007, EPI_ISL_2231008, EPI_ISL_2231009, EPI_ISL_2231010, EPI_ISL_2231011, EPI_ISL_2231012, EPI_ISL_2231013, EPI_ISL_2231014, EPI_ISL_2231015, EPI_ISL_2231016, EPI_ISL_2231017, EPI_ISL_2231018, EPI_ISL_2231019, EPI_ISL_2231020, EPI_ISL_2231021, EPI_ISL_2231022, EPI_ISL_2231023, EPI_ISL_2231024, EPI_ISL_2231025, EPI_ISL_2231026, EPI_ISL_2231027, EPI_ISL_2231028, EPI_ISL_2231029, EPI_ISL_2231030, EPI_ISL_2231031, EPI_ISL_2231032, EPI_ISL_2231033, EPI_ISL_2231034, EPI_ISL_2231035, EPI_ISL_2231036, EPI_ISL_2231037 | | | |
| see above | Salud Digna | Instituto Nacional de Medicina Genomica | Hidalgo-Miranda A, Cedro-Tanda A, Mendoza-Vargas A, Reyes-Grajeda JP, Abraham Campos-Romero, Moreno-Camacho José Luis, Rodriguez-Gallegos Jorge, Luna-Ruiz Marco, Gonzalez-Barrera D, Rangel-DeLeon D, Munguia-Garza P, Ramirez-Vega O, Escobar-Arrazola, M, Herrera-Montalvo LA. |
| EPI_ISL_2231038, EPI_ISL_2231039, EPI_ISL_2231040 | San Diego County Public Health Laboratory | Andersen lab at Scripps Research | SEARCH Alliance San Diego with Ashleigh Murphy, Jovan Shephard, Brett Austin |
| EPI_ISL_2231041 | Sharp HealthCare Laboratory | Andersen lab at Scripps Research | SEARCH Alliance San Diego with Aaron Harding, Jacquelyn Berumen, Cathy Woerle, Liam McGinnis, Art Mendoza, Omid Bakhtar |
| EPI_ISL_2231042, EPI_ISL_2231043 | San Diego County Public Health Laboratory | Andersen lab at Scripps Research | SEARCH Alliance San Diego with Ashleigh Murphy, Jovan Shephard, Brett Austin |
| EPI_ISL_2231044 | Sharp HealthCare Laboratory | Andersen lab at Scripps Research | SEARCH Alliance San Diego with Aaron Harding, Jacquelyn Berumen, Cathy Woerle, Liam McGinnis, Art Mendoza, Omid Bakhtar |
| EPI_ISL_2231045 | San Diego County Public Health Laboratory | Andersen lab at Scripps Research | SEARCH Alliance San Diego with Ashleigh Murphy, Jovan Shephard, Brett Austin |
| EPI_ISL_2231046, EPI_ISL_2231047 | Sharp HealthCare Laboratory | Andersen lab at Scripps Research | SEARCH Alliance San Diego with Aaron Harding, Jacquelyn Berumen, Cathy Woerle, Liam McGinnis, Art Mendoza, Omid Bakhtar |
| EPI_ISL_2231048 | San Diego County Public Health Laboratory | Andersen lab at Scripps Research | SEARCH Alliance San Diego with Ashleigh Murphy, Jovan Shephard, Brett Austin |
| EPI_ISL_2231049 | Sharp HealthCare Laboratory | Andersen lab at Scripps Research | SEARCH Alliance San Diego with Aaron Harding, Jacquelyn Berumen, Cathy Woerle, Liam McGinnis, Art Mendoza, Omid Bakhtar |
| EPI_ISL_2231050 | San Diego County Public Health Laboratory | Andersen lab at Scripps Research | SEARCH Alliance San Diego with Ashleigh Murphy, Jovan Shephard, Brett Austin |

| | | | |
|---|---|---|---|
| EPI_ISL_2231051 | Sharp HealthCare Laboratory | Andersen lab at Scripps Research | SEARCH Alliance San Diego with Aaron Harding, Jacquelyn Berumen, Cathy Woerle, Liam McGinnis, Art Mendoza, Omid Bakhtar |
| EPI_ISL_2231052, EPI_ISL_2231053 | San Diego County Public Health Laboratory | Andersen lab at Scripps Research | SEARCH Alliance San Diego with Ashleigh Murphy, Jovan Shephard, Brett Austin |
| EPI_ISL_2231054 | Sharp HealthCare Laboratory | Andersen lab at Scripps Research | SEARCH Alliance San Diego with Aaron Harding, Jacquelyn Berumen, Cathy Woerle, Liam McGinnis, Art Mendoza, Omid Bakhtar |
| EPI_ISL_2231055 | San Diego County Public Health Laboratory | Andersen lab at Scripps Research | SEARCH Alliance San Diego with Ashleigh Murphy, Jovan Shephard, Brett Austin |
| EPI_ISL_2231056, EPI_ISL_2231057, EPI_ISL_2231058, EPI_ISL_2231059 | Sharp HealthCare Laboratory | Andersen lab at Scripps Research | SEARCH Alliance San Diego with Aaron Harding, Jacquelyn Berumen, Cathy Woerle, Liam McGinnis, Art Mendoza, Omid Bakhtar |
| EPI_ISL_2231060, EPI_ISL_2231061 | San Diego County Public Health Laboratory | Andersen lab at Scripps Research | SEARCH Alliance San Diego with Ashleigh Murphy, Jovan Shephard, Brett Austin |
| EPI_ISL_2231062 | Sharp HealthCare Laboratory | Andersen lab at Scripps Research | SEARCH Alliance San Diego with Aaron Harding, Jacquelyn Berumen, Cathy Woerle, Liam McGinnis, Art Mendoza, Omid Bakhtar |
| EPI_ISL_2231063 | San Diego County Public Health Laboratory | Andersen lab at Scripps Research | SEARCH Alliance San Diego with Ashleigh Murphy, Jovan Shephard, Brett Austin |
| EPI_ISL_2231064 | Sharp HealthCare Laboratory | Andersen lab at Scripps Research | SEARCH Alliance San Diego with Aaron Harding, Jacquelyn Berumen, Cathy Woerle, Liam McGinnis, Art Mendoza, Omid Bakhtar |
| EPI_ISL_2231065 | San Diego County Public Health Laboratory | Andersen lab at Scripps Research | SEARCH Alliance San Diego with Ashleigh Murphy, Jovan Shephard, Brett Austin |
| EPI_ISL_2231066 | Sharp HealthCare Laboratory | Andersen lab at Scripps Research | SEARCH Alliance San Diego with Aaron Harding, Jacquelyn Berumen, Cathy Woerle, Liam McGinnis, Art Mendoza, Omid Bakhtar |
| EPI_ISL_2231067 | San Diego County Public Health Laboratory | Andersen lab at Scripps Research | SEARCH Alliance San Diego with Ashleigh Murphy, Jovan Shephard, Brett Austin |
| EPI_ISL_2231068 | Sharp HealthCare Laboratory | Andersen lab at Scripps Research | SEARCH Alliance San Diego with Aaron Harding, Jacquelyn Berumen, Cathy Woerle, Liam McGinnis, Art Mendoza, Omid Bakhtar |
| EPI_ISL_2231069, EPI_ISL_2231070, EPI_ISL_2231071, EPI_ISL_2231072 | San Diego County Public Health Laboratory | Andersen lab at Scripps Research | SEARCH Alliance San Diego with Ashleigh Murphy, Jovan Shephard, Brett Austin |
| EPI_ISL_2231073, EPI_ISL_2231074 | Sharp HealthCare Laboratory | Andersen lab at Scripps Research | SEARCH Alliance San Diego with Aaron Harding, Jacquelyn Berumen, Cathy Woerle, Liam McGinnis, Art Mendoza, Omid Bakhtar |
| EPI_ISL_2231075 | San Diego County Public Health Laboratory | Andersen lab at Scripps Research | SEARCH Alliance San Diego with Ashleigh Murphy, Jovan Shephard, Brett Austin |
| EPI_ISL_2231076 | Sharp HealthCare Laboratory | Andersen lab at Scripps Research | SEARCH Alliance San Diego with Aaron Harding, Jacquelyn Berumen, Cathy Woerle, Liam McGinnis, Art Mendoza, Omid Bakhtar |
| EPI_ISL_2231077, EPI_ISL_2231078 | San Diego County Public Health Laboratory | Andersen lab at Scripps Research | SEARCH Alliance San Diego with Ashleigh Murphy, Jovan Shephard, Brett Austin |
| EPI_ISL_2231079, EPI_ISL_2231080 | Sharp HealthCare Laboratory | Andersen lab at Scripps Research | SEARCH Alliance San Diego with Aaron Harding, Jacquelyn Berumen, Cathy Woerle, Liam McGinnis, Art Mendoza, Omid Bakhtar |
| EPI_ISL_2231081 | San Diego County Public Health Laboratory | Andersen lab at Scripps Research | SEARCH Alliance San Diego with Ashleigh Murphy, Jovan Shephard, Brett Austin |
| EPI_ISL_2231082 | Centro de Diagnostico COVID-19 UABC Tijuana | Andersen lab at Scripps Research | SEARCH Alliance San Diego with Idanya Rubi Serafin Higuera, Manuel Sanchez Alavez, Jorge Luis Jimenez Niebla, German Ibarra, Jonathan Vincent Baena, Oscar Efren Zazueta Fierro |
| EPI_ISL_2231083, EPI_ISL_2231084 | San Diego County Public Health Laboratory | Andersen lab at Scripps Research | SEARCH Alliance San Diego with Ashleigh Murphy, Jovan Shephard, Brett Austin |
| EPI_ISL_2231085 | Sharp HealthCare Laboratory | Andersen lab at Scripps Research | SEARCH Alliance San Diego with Aaron Harding, Jacquelyn Berumen, Cathy Woerle, Liam McGinnis, Art Mendoza, Omid Bakhtar |
| EPI_ISL_2231086, EPI_ISL_2231087 | San Diego County Public Health Laboratory | Andersen lab at Scripps Research | SEARCH Alliance San Diego with Ashleigh Murphy, Jovan Shephard, Brett Austin |
| EPI_ISL_2231088 | Sharp HealthCare Laboratory | Andersen lab at Scripps Research | SEARCH Alliance San Diego with Aaron Harding, Jacquelyn Berumen, Cathy Woerle, Liam McGinnis, Art Mendoza, Omid Bakhtar |
| EPI_ISL_2231089 | San Diego County Public Health Laboratory | Andersen lab at Scripps Research | SEARCH Alliance San Diego with Ashleigh Murphy, Jovan Shephard, Brett Austin |
| EPI_ISL_2231090, EPI_ISL_2231091, EPI_ISL_2231092, EPI_ISL_2231093, EPI_ISL_2231094, EPI_ISL_2231095, EPI_ISL_2231096, EPI_ISL_2231097, EPI_ISL_2231098, EPI_ISL_2231099, EPI_ISL_2231100, EPI_ISL_2231101, EPI_ISL_2231102, EPI_ISL_2231103 | | | |
| see above | Sharp HealthCare Laboratory | Andersen lab at Scripps Research | SEARCH Alliance San Diego with Aaron Harding, Jacquelyn Berumen, Cathy Woerle, Liam McGinnis, Art Mendoza, Omid Bakhtar |
| EPI_ISL_2231104, EPI_ISL_2231105, EPI_ISL_2231106, EPI_ISL_2231107, EPI_ISL_2231108, EPI_ISL_2231109, EPI_ISL_2231110, EPI_ISL_2231111, EPI_ISL_2231112, EPI_ISL_2231113, EPI_ISL_2231114, EPI_ISL_2231115, EPI_ISL_2231116, EPI_ISL_2231117, EPI_ISL_2231118, EPI_ISL_2231119, EPI_ISL_2231120, EPI_ISL_2231121, EPI_ISL_2231122, EPI_ISL_2231123, EPI_ISL_2231124, EPI_ISL_2231125, EPI_ISL_2231126, EPI_ISL_2231127, EPI_ISL_2231128 | | | |
| see above | San Diego County Public Health Laboratory | Andersen lab at Scripps Research | SEARCH Alliance San Diego with Ashleigh Murphy, Jovan Shephard, Brett Austin |
| EPI_ISL_2231129, EPI_ISL_2231130, EPI_ISL_2231131, EPI_ISL_2231132, EPI_ISL_2231133, EPI_ISL_2231134, EPI_ISL_2231135, EPI_ISL_2231136, EPI_ISL_2231137, EPI_ISL_2231138, EPI_ISL_2231139, EPI_ISL_2231140, EPI_ISL_2231141, EPI_ISL_2231142, EPI_ISL_2231143, EPI_ISL_2231144, EPI_ISL_2231145, EPI_ISL_2231146, EPI_ISL_2231147, EPI_ISL_2231148, EPI_ISL_2231149, EPI_ISL_2231150, EPI_ISL_2231151 | | | |
| see above | Sharp HealthCare Laboratory | Andersen lab at Scripps Research | SEARCH Alliance San Diego with Aaron Harding, Jacquelyn Berumen, Cathy Woerle, Liam McGinnis, Art Mendoza, Omid Bakhtar |
| EPI_ISL_2231152, EPI_ISL_2231153 | Centro de Diagnostico COVID-19 UABC Tijuana | Andersen lab at Scripps Research | SEARCH Alliance San Diego with Idanya Rubi Serafin Higuera, Manuel Sanchez Alavez, Jorge Luis Jimenez Niebla, German Ibarra, Jonathan Vincent Baena, Oscar Efren Zazueta Fierro |
| EPI_ISL_2231154, EPI_ISL_2231155, EPI_ISL_2231156, EPI_ISL_2231157, EPI_ISL_2231158, EPI_ISL_2231159, EPI_ISL_2231160 | Sharp HealthCare Laboratory | Andersen lab at Scripps Research | SEARCH Alliance San Diego with Aaron Harding, Jacquelyn Berumen, Cathy Woerle, Liam McGinnis, Art Mendoza, Omid Bakhtar |
| EPI_ISL_2231161 | San Diego County Public Health Laboratory | Andersen lab at Scripps Research | SEARCH Alliance San Diego with Ashleigh Murphy, Jovan Shephard, Brett Austin |
| EPI_ISL_2231162, EPI_ISL_2231163, EPI_ISL_2231164, EPI_ISL_2231165, EPI_ISL_2231166, EPI_ISL_2231167, EPI_ISL_2231168, EPI_ISL_2231169, EPI_ISL_2231170 | Sharp HealthCare Laboratory | Andersen lab at Scripps Research | SEARCH Alliance San Diego with Aaron Harding, Jacquelyn Berumen, Cathy Woerle, Liam McGinnis, Art Mendoza, Omid Bakhtar |
| EPI_ISL_2231171, EPI_ISL_2231172 | Centro de Diagnostico COVID-19 UABC Tijuana | Andersen lab at Scripps Research | SEARCH Alliance San Diego with Idanya Rubi Serafin Higuera, Manuel Sanchez Alavez, Jorge Luis Jimenez Niebla, German Ibarra, Jonathan Vincent Baena, Oscar Efren Zazueta Fierro |
| EPI_ISL_2231173 | San Diego County Public Health Laboratory | Andersen lab at Scripps Research | SEARCH Alliance San Diego with Ashleigh Murphy, Jovan Shephard, Brett Austin |
| EPI_ISL_2231174 | Centro de Diagnostico COVID-19 UABC Tijuana | Andersen lab at Scripps Research | SEARCH Alliance San Diego with Idanya Rubi Serafin Higuera, Manuel Sanchez Alavez, Jorge Luis Jimenez Niebla, German Ibarra, Jonathan Vincent Baena, Oscar Efren Zazueta Fierro |
| EPI_ISL_2231175, EPI_ISL_2231176, EPI_ISL_2231177 | Sharp HealthCare Laboratory | Andersen lab at Scripps Research | SEARCH Alliance San Diego with Aaron Harding, Jacquelyn Berumen, Cathy Woerle, Liam McGinnis, Art Mendoza, Omid Bakhtar |
| EPI_ISL_2231178 | San Diego County Public Health Laboratory | Andersen lab at Scripps Research | SEARCH Alliance San Diego with Ashleigh Murphy, Jovan Shephard, Brett Austin |
| EPI_ISL_2231179, EPI_ISL_2231180, EPI_ISL_2231181, EPI_ISL_2231182, EPI_ISL_2231183, EPI_ISL_2231184, EPI_ISL_2231185, EPI_ISL_2231186, EPI_ISL_2231187, EPI_ISL_2231188, EPI_ISL_2231189 | | | |
| see above | Sharp HealthCare Laboratory | Andersen lab at Scripps Research | SEARCH Alliance San Diego with Aaron Harding, Jacquelyn Berumen, Cathy Woerle, Liam McGinnis, Art Mendoza, Omid Bakhtar |
| EPI_ISL_2231190, EPI_ISL_2231191, | Centro de Diagnostico COVID-19 UABC Tijuana | Andersen lab at Scripps Research | SEARCH Alliance San Diego with Idanya Rubi Serafin Higuera, Manuel Sanchez Alavez, Jorge Luis Jimenez Niebla, German Ibarra, Jonathan Vincent Baena, Oscar Efren Zazueta Fierro |

| | | | |
|---|---|---|---|
| EPI_ISL_2231192 | | | |
| EPI_ISL_2231193 | San Diego County Public Health Laboratory | Andersen lab at Scripps Research | SEARCH Alliance San Diego with Ashleigh Murphy, Jovan Shephard, Brett Austin |
| EPI_ISL_2231194, EPI_ISL_2231195, EPI_ISL_2231196, EPI_ISL_2231197, EPI_ISL_2231198 | Sharp HealthCare Laboratory | Andersen lab at Scripps Research | SEARCH Alliance San Diego with Aaron Harding, Jacquelyn Berumen, Cathy Woerle, Liam McGinnis, Art Mendoza, Omid Bakhtar |
| EPI_ISL_2231199, EPI_ISL_2231200 | Centro de Diagnostico COVID-19 UABC Tijuana | Andersen lab at Scripps Research | SEARCH Alliance San Diego with Idanya Rubi Serafin Higuera, Manuel Sanchez Alavez, Jorge Luis Jimenez Niebla, German Ibarra, Jonathan Vincent Baena, Oscar Efren Zazueta Fierro |
| EPI_ISL_2231201, EPI_ISL_2231202, EPI_ISL_2231203, EPI_ISL_2231204 | San Diego County Public Health Laboratory | Andersen lab at Scripps Research | SEARCH Alliance San Diego with Ashleigh Murphy, Jovan Shephard, Brett Austin |
| EPI_ISL_2231205, EPI_ISL_2231206 | Sharp HealthCare Laboratory | Andersen lab at Scripps Research | SEARCH Alliance San Diego with Aaron Harding, Jacquelyn Berumen, Cathy Woerle, Liam McGinnis, Art Mendoza, Omid Bakhtar |
| EPI_ISL_2231207, EPI_ISL_2231208, EPI_ISL_2231209, EPI_ISL_2231210, EPI_ISL_2231211, EPI_ISL_2231212, EPI_ISL_2231213, EPI_ISL_2231214, EPI_ISL_2231215, EPI_ISL_2231216, EPI_ISL_2231217, EPI_ISL_2231218, EPI_ISL_2231219, EPI_ISL_2231220, EPI_ISL_2231221, EPI_ISL_2231222, EPI_ISL_2231223, EPI_ISL_2231224, EPI_ISL_2231225, EPI_ISL_2231226, EPI_ISL_2231227, EPI_ISL_2231228, EPI_ISL_2231229, EPI_ISL_2231230, EPI_ISL_2231231, EPI_ISL_2231232, EPI_ISL_2231233, EPI_ISL_2231234, EPI_ISL_2231235, EPI_ISL_2231236, EPI_ISL_2231237, EPI_ISL_2231238, EPI_ISL_2231239, EPI_ISL_2231240, EPI_ISL_2231241, EPI_ISL_2231242, EPI_ISL_2231243, EPI_ISL_2231244, EPI_ISL_2231245, EPI_ISL_2231246, EPI_ISL_2231247, EPI_ISL_2231248, EPI_ISL_2231249, EPI_ISL_2231250, EPI_ISL_2231251, EPI_ISL_2231252, EPI_ISL_2231253, EPI_ISL_2231254, EPI_ISL_2231255, EPI_ISL_2231256, EPI_ISL_2231257, EPI_ISL_2231258, EPI_ISL_2231259, EPI_ISL_2231260, EPI_ISL_2231261, EPI_ISL_2231262, EPI_ISL_2231263, EPI_ISL_2231264, EPI_ISL_2231265, EPI_ISL_2231266, EPI_ISL_2231267, EPI_ISL_2231268, EPI_ISL_2231269 | | | |
| see above | San Diego County Public Health Laboratory | Andersen lab at Scripps Research | SEARCH Alliance San Diego with Ashleigh Murphy, Jovan Shephard, Brett Austin |
| EPI_ISL_2231270, EPI_ISL_2231271, EPI_ISL_2231272, EPI_ISL_2231273, EPI_ISL_2231274, EPI_ISL_2231275, EPI_ISL_2231276, EPI_ISL_2231277, EPI_ISL_2231278, EPI_ISL_2231279, EPI_ISL_2231280, EPI_ISL_2231281, EPI_ISL_2231282, EPI_ISL_2231283, EPI_ISL_2231284, EPI_ISL_2231285, EPI_ISL_2231286, EPI_ISL_2231287, EPI_ISL_2231288, EPI_ISL_2231289, EPI_ISL_2231290, EPI_ISL_2231291, EPI_ISL_2231292, EPI_ISL_2231293, EPI_ISL_2231294, EPI_ISL_2231295, EPI_ISL_2231296, EPI_ISL_2231297, EPI_ISL_2231298, EPI_ISL_2231299, EPI_ISL_2231300, EPI_ISL_2231301, EPI_ISL_2231302, EPI_ISL_2231303, EPI_ISL_2231304, EPI_ISL_2231305, EPI_ISL_2231306, EPI_ISL_2231307, EPI_ISL_2231308, EPI_ISL_2231309, EPI_ISL_2231310, EPI_ISL_2231311, EPI_ISL_2231312, EPI_ISL_2231313, EPI_ISL_2231314, EPI_ISL_2231315, EPI_ISL_2231316, EPI_ISL_2231317, EPI_ISL_2231318, EPI_ISL_2231319, EPI_ISL_2231320 | | | |
| see above | Sharp HealthCare Laboratory | Andersen lab at Scripps Research | SEARCH Alliance San Diego with Aaron Harding, Jacquelyn Berumen, Cathy Woerle, Liam McGinnis, Art Mendoza, Omid Bakhtar |
| EPI_ISL_2231321, EPI_ISL_2231322, EPI_ISL_2231323, EPI_ISL_2231324, EPI_ISL_2231325, EPI_ISL_2231326, EPI_ISL_2231327, EPI_ISL_2231328, EPI_ISL_2231329, EPI_ISL_2231330, EPI_ISL_2231331, EPI_ISL_2231332, EPI_ISL_2231333, EPI_ISL_2231334, EPI_ISL_2231335, EPI_ISL_2231336, EPI_ISL_2231337, EPI_ISL_2231338, EPI_ISL_2231339, EPI_ISL_2231340, EPI_ISL_2231341, EPI_ISL_2231342, EPI_ISL_2231343, EPI_ISL_2231344, EPI_ISL_2231345, EPI_ISL_2231346, EPI_ISL_2231347, EPI_ISL_2231348, EPI_ISL_2231349, EPI_ISL_2231350, EPI_ISL_2231351, EPI_ISL_2231352, EPI_ISL_2231353, EPI_ISL_2231354, EPI_ISL_2231355, EPI_ISL_2231356, EPI_ISL_2231357, EPI_ISL_2231358, EPI_ISL_2231359, EPI_ISL_2231360, EPI_ISL_2231361, EPI_ISL_2231362, EPI_ISL_2231363, EPI_ISL_2231364, EPI_ISL_2231365, EPI_ISL_2231366, EPI_ISL_2231367, EPI_ISL_2231368, EPI_ISL_2231369, EPI_ISL_2231370, EPI_ISL_2231371, EPI_ISL_2231372, EPI_ISL_2231373, EPI_ISL_2231374, EPI_ISL_2231375, EPI_ISL_2231376, EPI_ISL_2231377, EPI_ISL_2231378, EPI_ISL_2231379, EPI_ISL_2231380, EPI_ISL_2231381, EPI_ISL_2231382, EPI_ISL_2231383, EPI_ISL_2231384, EPI_ISL_2231385, EPI_ISL_2231386, EPI_ISL_2231387, EPI_ISL_2231388, EPI_ISL_2231389, EPI_ISL_2231390, EPI_ISL_2231391, EPI_ISL_2231392, EPI_ISL_2231393, EPI_ISL_2231394, EPI_ISL_2231395, EPI_ISL_2231396, EPI_ISL_2231397, EPI_ISL_2231398, EPI_ISL_2231399, EPI_ISL_2231400, EPI_ISL_2231401, EPI_ISL_2231402, EPI_ISL_2231403, EPI_ISL_2231404, EPI_ISL_2231405, EPI_ISL_2231406, EPI_ISL_2231407, EPI_ISL_2231408, EPI_ISL_2231409, EPI_ISL_2231410, EPI_ISL_2231411, EPI_ISL_2231412, EPI_ISL_2231413, EPI_ISL_2231414, EPI_ISL_2231415, EPI_ISL_2231416, EPI_ISL_2231417, EPI_ISL_2231418, EPI_ISL_2231419, EPI_ISL_2231420, EPI_ISL_2231421, EPI_ISL_2231422, EPI_ISL_2231423, EPI_ISL_2231424, EPI_ISL_2231425, EPI_ISL_2231426, EPI_ISL_2231427, EPI_ISL_2231428, EPI_ISL_2231429, EPI_ISL_2231430, EPI_ISL_2231431, EPI_ISL_2231432, EPI_ISL_2231433, EPI_ISL_2231434 | | | |
| see above | Centro de Diagnostico COVID-19 UABC Tijuana | Andersen lab at Scripps Research | SEARCH Alliance San Diego with Idanya Rubi Serafin Higuera, Manuel Sanchez Alavez, Jorge Luis Jimenez Niebla, German Ibarra, Jonathan Vincent Baena, Oscar Efren Zazueta Fierro |
| EPI_ISL_2231435, EPI_ISL_2231436, EPI_ISL_2231437, EPI_ISL_2231438, EPI_ISL_2231439, EPI_ISL_2231440, EPI_ISL_2231441, EPI_ISL_2231442, EPI_ISL_2231443, EPI_ISL_2231444, EPI_ISL_2231445, EPI_ISL_2231446, EPI_ISL_2231447, EPI_ISL_2231448, EPI_ISL_2231449, EPI_ISL_2231450, EPI_ISL_2231451, EPI_ISL_2231452, EPI_ISL_2231453, EPI_ISL_2231454, EPI_ISL_2231455, EPI_ISL_2231456, EPI_ISL_2231457, EPI_ISL_2231458, EPI_ISL_2231459, EPI_ISL_2231460, EPI_ISL_2231461, EPI_ISL_2231462, EPI_ISL_2231463, EPI_ISL_2231464, EPI_ISL_2231465 | | | |
| see above | San Diego County Public Health Laboratory | Andersen lab at Scripps Research | SEARCH Alliance San Diego with Ashleigh Murphy, Jovan Shephard, Brett Austin |
| EPI_ISL_2231466, EPI_ISL_2231467 | Centro de Diagnostico COVID-19 UABC Tijuana | Andersen lab at Scripps Research | SEARCH Alliance San Diego with Idanya Rubi Serafin Higuera, Manuel Sanchez Alavez, Jorge Luis Jimenez Niebla, German Ibarra, Jonathan Vincent Baena, Oscar Efren Zazueta Fierro |
| EPI_ISL_2231468, EPI_ISL_2231469, EPI_ISL_2231470, EPI_ISL_2231471 | San Diego County Public Health Laboratory | Andersen lab at Scripps Research | SEARCH Alliance San Diego with Ashleigh Murphy, Jovan Shephard, Brett Austin |
| EPI_ISL_2231472 | Centro de Diagnostico COVID-19 UABC Tijuana | Andersen lab at Scripps Research | SEARCH Alliance San Diego with Idanya Rubi Serafin Higuera, Manuel Sanchez Alavez, Jorge Luis Jimenez Niebla, German Ibarra, Jonathan Vincent Baena, Oscar Efren Zazueta Fierro |
| EPI_ISL_2231473, EPI_ISL_2231474, EPI_ISL_2231475 | San Diego County Public Health Laboratory | Andersen lab at Scripps Research | SEARCH Alliance San Diego with Ashleigh Murphy, Jovan Shephard, Brett Austin |
| EPI_ISL_2231476 | Centro de Diagnostico COVID-19 UABC Tijuana | Andersen lab at Scripps Research | SEARCH Alliance San Diego with Idanya Rubi Serafin Higuera, Manuel Sanchez Alavez, Jorge Luis Jimenez Niebla, German Ibarra, Jonathan Vincent Baena, Oscar Efren Zazueta Fierro |
| EPI_ISL_2231477, EPI_ISL_2231478 | San Diego County Public Health Laboratory | Andersen lab at Scripps Research | SEARCH Alliance San Diego with Ashleigh Murphy, Jovan Shephard, Brett Austin |
| EPI_ISL_2231479 | Centro de Diagnostico COVID-19 UABC Tijuana | Andersen lab at Scripps Research | SEARCH Alliance San Diego with Idanya Rubi Serafin Higuera, Manuel Sanchez Alavez, Jorge Luis Jimenez Niebla, German Ibarra, Jonathan Vincent Baena, Oscar Efren Zazueta Fierro |
| EPI_ISL_2231480, EPI_ISL_2231481, EPI_ISL_2231482, EPI_ISL_2231483, EPI_ISL_2231484, EPI_ISL_2231485 | San Diego County Public Health Laboratory | Andersen lab at Scripps Research | SEARCH Alliance San Diego with Ashleigh Murphy, Jovan Shephard, Brett Austin |
| EPI_ISL_2231486 | Centro de Diagnostico COVID-19 UABC Tijuana | Andersen lab at Scripps Research | SEARCH Alliance San Diego with Idanya Rubi Serafin Higuera, Manuel Sanchez Alavez, Jorge Luis Jimenez Niebla, German Ibarra, Jonathan Vincent Baena, Oscar Efren Zazueta Fierro |
| EPI_ISL_2231487, EPI_ISL_2231488, EPI_ISL_2231489, EPI_ISL_2231490, EPI_ISL_2231491, EPI_ISL_2231492, EPI_ISL_2231493 | San Diego County Public Health Laboratory | Andersen lab at Scripps Research | SEARCH Alliance San Diego with Ashleigh Murphy, Jovan Shephard, Brett Austin |
| EPI_ISL_2231494 | Centro de Diagnostico COVID-19 UABC Tijuana | Andersen lab at Scripps Research | SEARCH Alliance San Diego with Idanya Rubi Serafin Higuera, Manuel Sanchez Alavez, Jorge Luis Jimenez Niebla, German Ibarra, Jonathan Vincent Baena, Oscar Efren Zazueta Fierro |
| EPI_ISL_2231495, EPI_ISL_2231496, EPI_ISL_2231497 | San Diego County Public Health Laboratory | Andersen lab at Scripps Research | SEARCH Alliance San Diego with Ashleigh Murphy, Jovan Shephard, Brett Austin |
| EPI_ISL_2231498, EPI_ISL_2231499 | Centro de Diagnostico COVID-19 UABC Tijuana | Andersen lab at Scripps Research | SEARCH Alliance San Diego with Idanya Rubi Serafin Higuera, Manuel Sanchez Alavez, Jorge Luis Jimenez Niebla, German Ibarra, Jonathan Vincent Baena, Oscar Efren Zazueta Fierro |
| EPI_ISL_2231500, EPI_ISL_2231501 | San Diego County Public Health Laboratory | Andersen lab at Scripps Research | SEARCH Alliance San Diego with Ashleigh Murphy, Jovan Shephard, Brett Austin |
| EPI_ISL_2231502, EPI_ISL_2231503 | Centro de Diagnostico COVID-19 UABC Tijuana | Andersen lab at Scripps Research | SEARCH Alliance San Diego with Idanya Rubi Serafin Higuera, Manuel Sanchez Alavez, Jorge Luis Jimenez Niebla, German Ibarra, Jonathan Vincent Baena, Oscar Efren Zazueta Fierro |
| EPI_ISL_2231504, EPI_ISL_2231505, EPI_ISL_2231506, EPI_ISL_2231507 | San Diego County Public Health Laboratory | Andersen lab at Scripps Research | SEARCH Alliance San Diego with Ashleigh Murphy, Jovan Shephard, Brett Austin |
| EPI_ISL_2231508, EPI_ISL_2231509 | Centro de Diagnostico COVID-19 UABC Tijuana | Andersen lab at Scripps Research | SEARCH Alliance San Diego with Idanya Rubi Serafin Higuera, Manuel Sanchez Alavez, Jorge Luis Jimenez Niebla, German Ibarra, Jonathan Vincent Baena, Oscar Efren Zazueta Fierro |
| EPI_ISL_2231510, EPI_ISL_2231511, EPI_ISL_2231512, EPI_ISL_2231513, EPI_ISL_2231514 | San Diego County Public Health Laboratory | Andersen lab at Scripps Research | SEARCH Alliance San Diego with Ashleigh Murphy, Jovan Shephard, Brett Austin |
| EPI_ISL_2231515, EPI_ISL_2231516, EPI_ISL_2231517 | Centro de Diagnostico COVID-19 UABC Tijuana | Andersen lab at Scripps Research | SEARCH Alliance San Diego with Idanya Rubi Serafin Higuera, Manuel Sanchez Alavez, Jorge Luis Jimenez Niebla, German Ibarra, Jonathan Vincent Baena, Oscar Efren Zazueta Fierro |
| EPI_ISL_2231518, EPI_ISL_2231519, | San Diego County Public Health Laboratory | Andersen lab at Scripps Research | SEARCH Alliance San Diego with Ashleigh Murphy, Jovan Shephard, Brett Austin |

| | | | |
|---|---|---|---|
| EPI_ISL_2231520, EPI_ISL_2231521, EPI_ISL_2231522 | | | |
| EPI_ISL_2231523, EPI_ISL_2231524, EPI_ISL_2231525 | Centro de Diagnostico COVID-19 UABC Tijuana | Andersen lab at Scripps Research | SEARCH Alliance San Diego with Idanya Rubi Serafin Higuera, Manuel Sanchez Alavez, Jorge Luis Jimenez Niebla, German Ibarra, Jonathan Vincent Baena, Oscar Efren Zazueta Fierro |
| EPI_ISL_2231526, EPI_ISL_2231527, EPI_ISL_2231528, EPI_ISL_2231529, EPI_ISL_2231530, EPI_ISL_2231531, EPI_ISL_2231532, EPI_ISL_2231533, EPI_ISL_2231534, EPI_ISL_2231535, EPI_ISL_2231536, EPI_ISL_2231537, EPI_ISL_2231538, EPI_ISL_2231539, EPI_ISL_2231540, EPI_ISL_2231541, EPI_ISL_2231542, EPI_ISL_2231543, EPI_ISL_2231544, EPI_ISL_2231545, EPI_ISL_2231546, EPI_ISL_2231547, EPI_ISL_2231548, EPI_ISL_2231549, EPI_ISL_2231550, EPI_ISL_2231551, EPI_ISL_2231552, EPI_ISL_2231553, EPI_ISL_2231554, EPI_ISL_2231555, EPI_ISL_2231556, EPI_ISL_2231557, EPI_ISL_2231558, EPI_ISL_2231559, EPI_ISL_2231560, EPI_ISL_2231561, EPI_ISL_2231562, EPI_ISL_2231563, EPI_ISL_2231564 | | | |
| see above | Sharp HealthCare Laboratory | Andersen lab at Scripps Research | SEARCH Alliance San Diego with Aaron Harding, Jacquelyn Berumen, Cathy Woerle, Liam McGinnis, Art Mendoza, Omid Bakhtar |
| EPI_ISL_2232209, EPI_ISL_2232210, EPI_ISL_2232211, EPI_ISL_2232213, EPI_ISL_2232218, EPI_ISL_2232219, EPI_ISL_2232220, EPI_ISL_2232222, EPI_ISL_2232223, EPI_ISL_2232224, EPI_ISL_2232225, EPI_ISL_2232226, EPI_ISL_2232227, EPI_ISL_2232228, EPI_ISL_2232229, EPI_ISL_2232230, EPI_ISL_2232231, EPI_ISL_2232232, EPI_ISL_2232233, EPI_ISL_2232234, EPI_ISL_2232235, EPI_ISL_2232236, EPI_ISL_2232237, EPI_ISL_2232238, EPI_ISL_2232241, EPI_ISL_2232245, EPI_ISL_2232246, EPI_ISL_2232247, EPI_ISL_2232249, EPI_ISL_2232251 | | | |
| see above | City of Milwaukee Health Department Laboratory | City of Milwaukee Health Department Laboratory | Sanjib Bhattacharyya, Manjeet Khubbar, Amy Bauer, Jennifer Lentz, Samantha Scott |
| EPI_ISL_2232408, EPI_ISL_2232409, EPI_ISL_2232410, EPI_ISL_2232411, EPI_ISL_2232412, EPI_ISL_2232413, EPI_ISL_2232414, EPI_ISL_2232415, EPI_ISL_2232416, EPI_ISL_2232417, EPI_ISL_2232418, EPI_ISL_2232419, EPI_ISL_2232420, EPI_ISL_2232421, EPI_ISL_2232422, EPI_ISL_2232423, EPI_ISL_2232424, EPI_ISL_2232425, EPI_ISL_2232426, EPI_ISL_2232427, EPI_ISL_2232428, EPI_ISL_2232429, EPI_ISL_2232430, EPI_ISL_2232431, EPI_ISL_2232432, EPI_ISL_2232433, EPI_ISL_2232434, EPI_ISL_2232435, EPI_ISL_2232436, EPI_ISL_2232437, EPI_ISL_2232439, EPI_ISL_2232440, EPI_ISL_2232441, EPI_ISL_2232442, EPI_ISL_2232443, EPI_ISL_2232444, EPI_ISL_2232445, EPI_ISL_2232446, EPI_ISL_2232447, EPI_ISL_2232448, EPI_ISL_2232449, EPI_ISL_2232451, EPI_ISL_2232452, EPI_ISL_2232453, EPI_ISL_2232454, EPI_ISL_2232455, EPI_ISL_2232456, EPI_ISL_2232457, EPI_ISL_2232458, EPI_ISL_2232459, EPI_ISL_2232460, EPI_ISL_2232461, EPI_ISL_2232462, EPI_ISL_2232463, EPI_ISL_2232464, EPI_ISL_2232465, EPI_ISL_2232466, EPI_ISL_2232467, EPI_ISL_2232468, EPI_ISL_2232469, EPI_ISL_2232470, EPI_ISL_2232471, EPI_ISL_2232472, EPI_ISL_2232473, EPI_ISL_2232474, EPI_ISL_2232475, EPI_ISL_2232476, EPI_ISL_2232477, EPI_ISL_2232478, EPI_ISL_2232479, EPI_ISL_2232480, EPI_ISL_2232481, EPI_ISL_2232482, EPI_ISL_2232483, EPI_ISL_2232484, EPI_ISL_2232485, EPI_ISL_2232486, EPI_ISL_2232487, EPI_ISL_2232488, EPI_ISL_2232489, EPI_ISL_2232490, EPI_ISL_2232491, EPI_ISL_2232492, EPI_ISL_2232493, EPI_ISL_2232494, EPI_ISL_2232495, EPI_ISL_2232496, EPI_ISL_2232497, EPI_ISL_2232498, EPI_ISL_2232499, EPI_ISL_2232500, EPI_ISL_2232501, EPI_ISL_2232502, EPI_ISL_2232503, EPI_ISL_2232504, EPI_ISL_2232505, EPI_ISL_2232506, EPI_ISL_2232507, EPI_ISL_2232508, EPI_ISL_2232509, EPI_ISL_2232510, EPI_ISL_2232511, EPI_ISL_2232512, EPI_ISL_2232513, EPI_ISL_2232514, EPI_ISL_2232515, EPI_ISL_2232516, EPI_ISL_2232517, EPI_ISL_2232518, EPI_ISL_2232519, EPI_ISL_2232520, EPI_ISL_2232521, EPI_ISL_2232522, EPI_ISL_2232523, EPI_ISL_2232524, EPI_ISL_2232525, EPI_ISL_2232526, EPI_ISL_2232527, EPI_ISL_2232528, EPI_ISL_2232529, EPI_ISL_2232530, EPI_ISL_2232531, EPI_ISL_2232532, EPI_ISL_2232533, EPI_ISL_2232534, EPI_ISL_2232535, EPI_ISL_2232536, EPI_ISL_2232537, EPI_ISL_2232538, EPI_ISL_2232539, EPI_ISL_2232540, EPI_ISL_2232541 | | | |
| see above | Rhode Island Department of Health | Infectious Disease Program, Broad Institute of Harvard and MIT | Siddle,K.J., Azevedo,K., Miller,A., Adams,G., Pearlman,L., Gladden-Young,A., Lagerborg,K., Rudy,M., DeRuff,K., Carter,A., Normandin,E., Bauer,M., Reilly,S., Tomkins-Tinch,C., Loreth,C., Chaluvadi,S., Lemieux,J.E., Birren,B.W., Sabeti,P.C., Huard,R., King,E., Park,D.J., and MacInnis,B.L. |
| EPI_ISL_2232542, EPI_ISL_2232543, EPI_ISL_2232544, EPI_ISL_2232545, EPI_ISL_2232546, EPI_ISL_2232547, EPI_ISL_2232548, EPI_ISL_2232549, EPI_ISL_2232550, EPI_ISL_2232551, EPI_ISL_2232552, EPI_ISL_2232553, EPI_ISL_2232554, EPI_ISL_2232555, EPI_ISL_2232556, EPI_ISL_2232558, EPI_ISL_2232559, EPI_ISL_2232560, EPI_ISL_2232561, EPI_ISL_2232562, EPI_ISL_2232563, EPI_ISL_2232564, EPI_ISL_2232565, EPI_ISL_2232566, EPI_ISL_2232567, EPI_ISL_2232568, EPI_ISL_2232569, EPI_ISL_2232570, EPI_ISL_2232571, EPI_ISL_2232572, EPI_ISL_2232573, EPI_ISL_2232574 | | | |
| see above | Minnesota Department of Health, Public Health Laboratory | Minnesota Department of Health, Public Health Laboratory | Alexandra Lorentz, Jacob Garfin, Matt Plumb, and Xiong Wang |
| EPI_ISL_2232575, EPI_ISL_2232576, EPI_ISL_2232577, EPI_ISL_2232578 | Centracare Laboratory Services | Minnesota Department of Health, Public Health Laboratory | Alexandra Lorentz, Jacob Garfin, Matt Plumb, and Xiong Wang |
| EPI_ISL_2232579, EPI_ISL_2232580, EPI_ISL_2232581, EPI_ISL_2232582, EPI_ISL_2232583, EPI_ISL_2232584, EPI_ISL_2232585, EPI_ISL_2232586, EPI_ISL_2232588, EPI_ISL_2232589 | Minnesota Department of Health, Public Health Laboratory | Minnesota Department of Health, Public Health Laboratory | Alexandra Lorentz, Jacob Garfin, Matt Plumb, and Xiong Wang |
| EPI_ISL_2232590, EPI_ISL_2232591, EPI_ISL_2232593, EPI_ISL_2232594, EPI_ISL_2232595, EPI_ISL_2232596, EPI_ISL_2232598, EPI_ISL_2232599, EPI_ISL_2232600, EPI_ISL_2232601, EPI_ISL_2232602, EPI_ISL_2232603, EPI_ISL_2232604, EPI_ISL_2232605, EPI_ISL_2232606 | | | |
| see above | Premier Medical Lab | Minnesota Department of Health, Public Health Laboratory | Alexandra Lorentz, Jacob Garfin, Matt Plumb, and Xiong Wang |
| EPI_ISL_2232607, EPI_ISL_2232608, EPI_ISL_2232610, EPI_ISL_2232611 | M Health Fairview | Minnesota Department of Health, Public Health Laboratory | Alexandra Lorentz, Jacob Garfin, Matt Plumb, and Xiong Wang |
| EPI_ISL_2232989, EPI_ISL_2232990, EPI_ISL_2232991, EPI_ISL_2232992, EPI_ISL_2232993, EPI_ISL_2232994, EPI_ISL_2232995, EPI_ISL_2232996, EPI_ISL_2232997, EPI_ISL_2232998, EPI_ISL_2232999, EPI_ISL_2233000, EPI_ISL_2233001, EPI_ISL_2233003, EPI_ISL_2233004, EPI_ISL_2233005, EPI_ISL_2233006, EPI_ISL_2233007, EPI_ISL_2233008, EPI_ISL_2233009, EPI_ISL_2233010, EPI_ISL_2233011, EPI_ISL_2233012, EPI_ISL_2233013, EPI_ISL_2233014, EPI_ISL_2233015, EPI_ISL_2233017, EPI_ISL_2233018, EPI_ISL_2233019, EPI_ISL_2233021, EPI_ISL_2233023, EPI_ISL_2233024 | | | |
| see above | Curative | New Mexico Department of Health Scientific Laboratory | Ellie Johnson, D'eldra Malone, Jennifer Benoit, Ratheesh Rajan, Linda Salazar, Keila Gutierrez, Anastacia Griego-Fisher |
| EPI_ISL_2233025, EPI_ISL_2233026, EPI_ISL_2233027, EPI_ISL_2233028, EPI_ISL_2233029, EPI_ISL_2233030, EPI_ISL_2233031, EPI_ISL_2233032, EPI_ISL_2233033 | TriCore Reference Laboratory Inc. | New Mexico Department of Health Scientific Laboratory | Ellie Johnson, D'eldra Malone, Jennifer Benoit, Ratheesh Rajan, Linda Salazar, Keila Gutierrez, Anastacia Griego-Fisher |
| EPI_ISL_2233036, EPI_ISL_2233037, EPI_ISL_2233038, EPI_ISL_2233040, EPI_ISL_2233041, EPI_ISL_2233042, EPI_ISL_2233043, EPI_ISL_2233044, EPI_ISL_2233046, EPI_ISL_2233049, EPI_ISL_2233050, EPI_ISL_2233051, EPI_ISL_2233052, EPI_ISL_2233053, EPI_ISL_2233054, EPI_ISL_2233055, EPI_ISL_2233056, EPI_ISL_2233057, EPI_ISL_2233058, EPI_ISL_2233059 | | | |
| see above | UTGSAF | UTGSAF | Jessica Podnar, Sylvie Beaudenon, Audrey Kelly, Zachary Carver, Anna Battenhouse, Andreas Matouschek |
| EPI_ISL_2233060, EPI_ISL_2233061, EPI_ISL_2233062, EPI_ISL_2233063, EPI_ISL_2233064, EPI_ISL_2233065, EPI_ISL_2233066, EPI_ISL_2233067, EPI_ISL_2233068, EPI_ISL_2233069, EPI_ISL_2233070, EPI_ISL_2233071, EPI_ISL_2233072, EPI_ISL_2233073, EPI_ISL_2233074, EPI_ISL_2233075, EPI_ISL_2233076, EPI_ISL_2233077, EPI_ISL_2233078, EPI_ISL_2233079, EPI_ISL_2233080, EPI_ISL_2233081, EPI_ISL_2233082, EPI_ISL_2233083 | | | |
| see above | Nebraska Public Health Laboratory | NPHL COVID-19 Response Team | NPHL COVID-19 Response Team |
| EPI_ISL_2233110, EPI_ISL_2233111, EPI_ISL_2233112, EPI_ISL_2233113, EPI_ISL_2233114, EPI_ISL_2233115, EPI_ISL_2233116, EPI_ISL_2233117, EPI_ISL_2233118, EPI_ISL_2233119, EPI_ISL_2233120, EPI_ISL_2233121, EPI_ISL_2233122, EPI_ISL_2233123, EPI_ISL_2233124, EPI_ISL_2233125, EPI_ISL_2233126, EPI_ISL_2233127, EPI_ISL_2233128, EPI_ISL_2233129, EPI_ISL_2233130, EPI_ISL_2233131, EPI_ISL_2233132, EPI_ISL_2233133, EPI_ISL_2233134, EPI_ISL_2233135, EPI_ISL_2233136, EPI_ISL_2233137, EPI_ISL_2233138, EPI_ISL_2233139, EPI_ISL_2233140, EPI_ISL_2233141, EPI_ISL_2233142, EPI_ISL_2233143, EPI_ISL_2233144, EPI_ISL_2233149, EPI_ISL_2233150, EPI_ISL_2233151, EPI_ISL_2233152, EPI_ISL_2233153, EPI_ISL_2233155, EPI_ISL_2233156, EPI_ISL_2233157, EPI_ISL_2233158, EPI_ISL_2233159, EPI_ISL_2233160, EPI_ISL_2233161, EPI_ISL_2233162, EPI_ISL_2233163, EPI_ISL_2233164, EPI_ISL_2233165, EPI_ISL_2233166, EPI_ISL_2233167, EPI_ISL_2233168, EPI_ISL_2233169, EPI_ISL_2233170, EPI_ISL_2233171, EPI_ISL_2233172, EPI_ISL_2233173, EPI_ISL_2233174, EPI_ISL_2233175, EPI_ISL_2233176, EPI_ISL_2233177, EPI_ISL_2233178, EPI_ISL_2233179, EPI_ISL_2233180, EPI_ISL_2233181, EPI_ISL_2233182, EPI_ISL_2233183, EPI_ISL_2233184, EPI_ISL_2233185, EPI_ISL_2233186, EPI_ISL_2233187, EPI_ISL_2233188, EPI_ISL_2233189, EPI_ISL_2233190, EPI_ISL_2233191, EPI_ISL_2233193, EPI_ISL_2233194, EPI_ISL_2233195, EPI_ISL_2233196, EPI_ISL_2233197, EPI_ISL_2233198, EPI_ISL_2233199, EPI_ISL_2233201, EPI_ISL_2233202, EPI_ISL_2233203, EPI_ISL_2233204, EPI_ISL_2233205, EPI_ISL_2233206, EPI_ISL_2233207, EPI_ISL_2233209, EPI_ISL_2233210, EPI_ISL_2233211, EPI_ISL_2233212, EPI_ISL_2233213, EPI_ISL_2233214, EPI_ISL_2233215, EPI_ISL_2233216, EPI_ISL_2233217, EPI_ISL_2233218, EPI_ISL_2233219, EPI_ISL_2233220, EPI_ISL_2233221, EPI_ISL_2233222, EPI_ISL_2233223, EPI_ISL_2233224, EPI_ISL_2233225, EPI_ISL_2233226, EPI_ISL_2233227, EPI_ISL_2233228, EPI_ISL_2233231, EPI_ISL_2233232, EPI_ISL_2233245, EPI_ISL_2233246, EPI_ISL_2233247, EPI_ISL_2233249, EPI_ISL_2233250, EPI_ISL_2233253, EPI_ISL_2233254, EPI_ISL_2233255, EPI_ISL_2233256, EPI_ISL_2233257, EPI_ISL_2233258, EPI_ISL_2233259, EPI_ISL_2233260, EPI_ISL_2233261, EPI_ISL_2233262, EPI_ISL_2233263, EPI_ISL_2233264, EPI_ISL_2233267, EPI_ISL_2233268, EPI_ISL_2233269, EPI_ISL_2233270, EPI_ISL_2233273, EPI_ISL_2233274, EPI_ISL_2233275, EPI_ISL_2233276, EPI_ISL_2233277, EPI_ISL_2233278, EPI_ISL_2233279, EPI_ISL_2233280, EPI_ISL_2233281, EPI_ISL_2233282, EPI_ISL_2233283, EPI_ISL_2233284, EPI_ISL_2233285, EPI_ISL_2233286, EPI_ISL_2233288, EPI_ISL_2233290, EPI_ISL_2233291, EPI_ISL_2233292, EPI_ISL_2233293, EPI_ISL_2233294, EPI_ISL_2233295, EPI_ISL_2233296, EPI_ISL_2233297, EPI_ISL_2233298, EPI_ISL_2233299, EPI_ISL_2233301, EPI_ISL_2233302, EPI_ISL_2233303, EPI_ISL_2233304, EPI_ISL_2233305, EPI_ISL_2233306, EPI_ISL_2233307, EPI_ISL_2233308, EPI_ISL_2233309, EPI_ISL_2233310, EPI_ISL_2233311, EPI_ISL_2233312, EPI_ISL_2233313, EPI_ISL_2233314, EPI_ISL_2233315, EPI_ISL_2233318, EPI_ISL_2233319, EPI_ISL_2233320, EPI_ISL_2233322, EPI_ISL_2233323, EPI_ISL_2233324, EPI_ISL_2233325, EPI_ISL_2233327, EPI_ISL_2233328, EPI_ISL_2233329, EPI_ISL_2233330, EPI_ISL_2233331, EPI_ISL_2233332, EPI_ISL_2233333, EPI_ISL_2233335, EPI_ISL_2233336, EPI_ISL_2233337 | | | |
| see above | UW Virology Lab | UW Virology Lab | Pavitra Roychoudhury, Hong Xie, Lasata Shrestha, Tien V. Nguyen, Shah Mohamed Bakhash, Michelle Lin, Noah R. Baker, Sean Ellis, Meei-Li Huang, Keith R Jerome, Alexander Greninger |
| EPI_ISL_2233669 | City of Milwaukee Health Department Laboratory | City of Milwaukee Health Department Laboratory | Sanjib Bhattacharyya, Manjeet Khubbar, Amy Bauer, Jennifer Lentz, Samantha Scott |
| EPI_ISL_2233910, EPI_ISL_2233911, EPI_ISL_2233912, EPI_ISL_2233913, EPI_ISL_2233914, EPI_ISL_2233915, EPI_ISL_2233916, EPI_ISL_2233917, EPI_ISL_2233918, EPI_ISL_2233919, EPI_ISL_2233920, EPI_ISL_2233921, EPI_ISL_2233922, EPI_ISL_2233923, EPI_ISL_2233924, EPI_ISL_2233925, EPI_ISL_2233926, EPI_ISL_2233927, EPI_ISL_2233928, EPI_ISL_2233929, EPI_ISL_2233930, EPI_ISL_2233931, EPI_ISL_2233932, EPI_ISL_2233933, EPI_ISL_2233934, EPI_ISL_2233935, EPI_ISL_2233936, EPI_ISL_2233937, EPI_ISL_2233938, EPI_ISL_2233939, EPI_ISL_2233940, EPI_ISL_2233941, EPI_ISL_2233942, EPI_ISL_2233943, EPI_ISL_2233944, EPI_ISL_2233945, EPI_ISL_2233946, EPI_ISL_2233947, EPI_ISL_2233948, EPI_ISL_2233949, EPI_ISL_2233950, EPI_ISL_2233951, EPI_ISL_2233952, EPI_ISL_2233953, EPI_ISL_2233954, EPI_ISL_2233955, EPI_ISL_2233956, EPI_ISL_2233957, EPI_ISL_2233958, EPI_ISL_2233959, EPI_ISL_2233960, EPI_ISL_2233961, EPI_ISL_2233962, EPI_ISL_2233963, EPI_ISL_2233964, EPI_ISL_2233965, EPI_ISL_2233966, EPI_ISL_2233967, EPI_ISL_2233968, EPI_ISL_2233969, EPI_ISL_2233970, EPI_ISL_2233971, EPI_ISL_2233972, EPI_ISL_2233973, EPI_ISL_2233974, EPI_ISL_2233975, EPI_ISL_2233976, EPI_ISL_2233977, EPI_ISL_2233978, EPI_ISL_2233979, EPI_ISL_2233980, EPI_ISL_2233981, EPI_ISL_2233982, EPI_ISL_2233983, EPI_ISL_2233984, EPI_ISL_2233985, EPI_ISL_2233986, EPI_ISL_2233988, EPI_ISL_2233989, EPI_ISL_2233990, EPI_ISL_2233991, EPI_ISL_2233992, EPI_ISL_2233994, EPI_ISL_2233996, EPI_ISL_2233997, EPI_ISL_2233998, EPI_ISL_2233999, EPI_ISL_2234000, EPI_ISL_2234001, EPI_ISL_2234002, EPI_ISL_2234003, EPI_ISL_2234004, EPI_ISL_2234005, EPI_ISL_2234006, EPI_ISL_2234007, EPI_ISL_2234008, EPI_ISL_2234009, EPI_ISL_2234010, EPI_ISL_2234011, EPI_ISL_2234012, EPI_ISL_2234013, EPI_ISL_2234014, EPI_ISL_2234015, EPI_ISL_2234016, EPI_ISL_2234017, EPI_ISL_2234018, EPI_ISL_2234019, EPI_ISL_2234020, EPI_ISL_2234021, EPI_ISL_2234022, EPI_ISL_2234023, EPI_ISL_2234024, EPI_ISL_2234025, EPI_ISL_2234026, EPI_ISL_2234027, EPI_ISL_2234028, EPI_ISL_2234029, EPI_ISL_2234030, EPI_ISL_2234031, EPI_ISL_2234032, EPI_ISL_2234033, EPI_ISL_2234034, EPI_ISL_2234035, EPI_ISL_2234036, EPI_ISL_2234037, EPI_ISL_2234038, EPI_ISL_2234039, EPI_ISL_2234040, EPI_ISL_2234041, EPI_ISL_2234042, EPI_ISL_2234043, EPI_ISL_2234044, EPI_ISL_2234045, EPI_ISL_2234046, EPI_ISL_2234047, EPI_ISL_2234048, EPI_ISL_2234049, EPI_ISL_2234050, EPI_ISL_2234051, EPI_ISL_2234052, EPI_ISL_2234053, EPI_ISL_2234054, EPI_ISL_2234055, EPI_ISL_2234056, EPI_ISL_2234057, EPI_ISL_2234058, EPI_ISL_2234059, EPI_ISL_2234060, EPI_ISL_2234061, EPI_ISL_2234062, EPI_ISL_2234063, EPI_ISL_2234064, EPI_ISL_2234065, EPI_ISL_2234066, EPI_ISL_2234067, EPI_ISL_2234068, EPI_ISL_2234069, EPI_ISL_2234070, EPI_ISL_2234071, EPI_ISL_2234072, EPI_ISL_2234073, EPI_ISL_2234075, EPI_ISL_2234076, EPI_ISL_2234077, EPI_ISL_2234078, EPI_ISL_2234079, EPI_ISL_2234080, EPI_ISL_2234081, EPI_ISL_2234082, EPI_ISL_2234083, EPI_ISL_2234084, EPI_ISL_2234085, EPI_ISL_2234086, EPI_ISL_2234087, EPI_ISL_2234088, EPI_ISL_2234090, EPI_ISL_2234091, EPI_ISL_2234092, EPI_ISL_2234093, EPI_ISL_2234094, EPI_ISL_2234095, EPI_ISL_2234096, EPI_ISL_2234097, EPI_ISL_2234098, EPI_ISL_2234099, EPI_ISL_2234100, EPI_ISL_2234101, EPI_ISL_2234102, EPI_ISL_2234103, EPI_ISL_2234104, EPI_ISL_2234105, EPI_ISL_2234106, EPI_ISL_2234107, EPI_ISL_2234109, EPI_ISL_2234110, EPI_ISL_2234111, EPI_ISL_2234112, EPI_ISL_2234113, EPI_ISL_2234114, EPI_ISL_2234115, EPI_ISL_2234116, EPI_ISL_2234117, EPI_ISL_2234118, EPI_ISL_2234119, EPI_ISL_2234120, EPI_ISL_2234121, EPI_ISL_2234122, EPI_ISL_2234123, EPI_ISL_2234124, EPI_ISL_2234125, EPI_ISL_2234126, EPI_ISL_2234127, EPI_ISL_2234128, EPI_ISL_2234129, EPI_ISL_2234130, EPI_ISL_2234131, EPI_ISL_2234132, EPI_ISL_2234133, EPI_ISL_2234134, EPI_ISL_2234135, EPI_ISL_2234136, EPI_ISL_2234137, EPI_ISL_2234138, EPI_ISL_2234139, EPI_ISL_2234140, EPI_ISL_2234141, EPI_ISL_2234142, EPI_ISL_2234143, EPI_ISL_2234144, EPI_ISL_2234145, EPI_ISL_2234146, EPI_ISL_2234147, EPI_ISL_2234148, EPI_ISL_2234149, EPI_ISL_2234150, EPI_ISL_2234151, EPI_ISL_2234152, EPI_ISL_2234153, EPI_ISL_2234154, EPI_ISL_2234155, EPI_ISL_2234156, EPI_ISL_2234157, EPI_ISL_2234158, EPI_ISL_2234159, EPI_ISL_2234160, EPI_ISL_2234161, EPI_ISL_2234162, EPI_ISL_2234163, EPI_ISL_2234164, EPI_ISL_2234165, EPI_ISL_2234166, EPI_ISL_2234167, EPI_ISL_2234168, EPI_ISL_2234169, EPI_ISL_2234170, EPI_ISL_2234171, EPI_ISL_2234172, EPI_ISL_2234173, EPI_ISL_2234174, EPI_ISL_2234175, EPI_ISL_2234176, EPI_ISL_2234177, EPI_ISL_2234178, EPI_ISL_2234179, EPI_ISL_2234180, EPI_ISL_2234181, EPI_ISL_2234182, EPI_ISL_2234183, EPI_ISL_2234184, EPI_ISL_2234185, EPI_ISL_2234186, EPI_ISL_2234187, EPI_ISL_2234188, EPI_ISL_2234189, EPI_ISL_2234190, EPI_ISL_2234191, EPI_ISL_2234192, EPI_ISL_2234193, EPI_ISL_2234194, EPI_ISL_2234195, EPI_ISL_2234196, EPI_ISL_2234197, EPI_ISL_2234198, EPI_ISL_2234199, EPI_ISL_2234200, EPI_ISL_2234201, EPI_ISL_2234202, EPI_ISL_2234203, EPI_ISL_2234204, EPI_ISL_2234205, EPI_ISL_2234206, EPI_ISL_2234207, EPI_ISL_2234208, EPI_ISL_2234209, EPI_ISL_2234210, EPI_ISL_2234211, EPI_ISL_2234212, EPI_ISL_2234213, EPI_ISL_2234214, EPI_ISL_2234215, EPI_ISL_2234216, EPI_ISL_2234217, EPI_ISL_2234218, EPI_ISL_2234219, EPI_ISL_2234220, EPI_ISL_2234221, EPI_ISL_2234222, EPI_ISL_2234223, EPI_ISL_2234224, EPI_ISL_2234225, EPI_ISL_2234226, EPI_ISL_2234227, EPI_ISL_2234228, EPI_ISL_2234229, EPI_ISL_2234230, EPI_ISL_2234231, EPI_ISL_2234232, EPI_ISL_2234233, EPI_ISL_2234234, EPI_ISL_2234235, EPI_ISL_2234236, EPI_ISL_2234237, EPI_ISL_2234238, EPI_ISL_2234239, EPI_ISL_2234240, EPI_ISL_2234241, EPI_ISL_2234242, EPI_ISL_2234243, EPI_ISL_2234244, EPI_ISL_2234245, EPI_ISL_2234246, EPI_ISL_2234247, EPI_ISL_2234248, EPI_ISL_2234249, EPI_ISL_2234250, EPI_ISL_2234251, EPI_ISL_2234252, EPI_ISL_2234253, EPI_ISL_2234254, EPI_ISL_2234255, EPI_ISL_2234256, EPI_ISL_2234257, EPI_ISL_2234258, EPI_ISL_2234259, EPI_ISL_2234260, EPI_ISL_2234261, EPI_ISL_2234262, EPI_ISL_2234263, EPI_ISL_2234264, EPI_ISL_2234265, EPI_ISL_2234266, EPI_ISL_2234267, EPI_ISL_2234268, EPI_ISL_2234269, EPI_ISL_2234270, EPI_ISL_2234271, EPI_ISL_2234272, EPI_ISL_2234273, EPI_ISL_2234274, EPI_ISL_2234275, EPI_ISL_2234276, EPI_ISL_2234277, EPI_ISL_2234278, EPI_ISL_2234279, EPI_ISL_2234280, EPI_ISL_2234281, EPI_ISL_2234282, EPI_ISL_2234283, EPI_ISL_2234284, EPI_ISL_2234285, EPI_ISL_2234286, EPI_ISL_2234287, EPI_ISL_2234288, EPI_ISL_2234289, EPI_ISL_2234290, EPI_ISL_2234291, EPI_ISL_2234292, EPI_ISL_2234293, EPI_ISL_2234294, EPI_ISL_2234295, EPI_ISL_2234296, EPI_ISL_2234297, EPI_ISL_2234298, EPI_ISL_2234299, EPI_ISL_2234300, EPI_ISL_2234301, EPI_ISL_2234302, EPI_ISL_2234304, EPI_ISL_2234305, EPI_ISL_2234306, EPI_ISL_2234307, EPI_ISL_2234308, EPI_ISL_2234309, EPI_ISL_2234310, EPI_ISL_2234311, EPI_ISL_2234313, EPI_ISL_2234314, EPI_ISL_2234315, EPI_ISL_2234316, EPI_ISL_2234317, EPI_ISL_2234318, EPI_ISL_2234319, EPI_ISL_2234320, EPI_ISL_2234321, EPI_ISL_2234323, EPI_ISL_2234325, EPI_ISL_2234326, EPI_ISL_2234327, EPI_ISL_2234328, EPI_ISL_2234329, EPI_ISL_2234330, EPI_ISL_2234331, EPI_ISL_2234332, EPI_ISL_2234333, EPI_ISL_2234334, EPI_ISL_2234335, EPI_ISL_2234336, EPI_ISL_2234337, EPI_ISL_2234338, EPI_ISL_2234339, EPI_ISL_2234340, EPI_ISL_2234341, EPI_ISL_2234342, EPI_ISL_2234343, EPI_ISL_2234344, EPI_ISL_2234345, EPI_ISL_2234346, EPI_ISL_2234347, EPI_ISL_2234348, EPI_ISL_2234349, EPI_ISL_2234350, EPI_ISL_2234351, EPI_ISL_2234352, EPI_ISL_2234353, EPI_ISL_2234354, EPI_ISL_2234355, EPI_ISL_2234356 | | | |
| see above | California Department of Public Health | California Department of Public Health | CDPH IDLB COVIDNet et al |
| EPI_ISL_2234832, EPI_ISL_2234833, EPI_ISL_2234834, EPI_ISL_2234835, EPI_ISL_2234836, EPI_ISL_2234837, EPI_ISL_2234838, EPI_ISL_2234839, EPI_ISL_2234840, EPI_ISL_2234841, EPI_ISL_2234842, EPI_ISL_2234843, EPI_ISL_2234844, EPI_ISL_2234845, EPI_ISL_2234846, EPI_ISL_2234847, EPI_ISL_2234848, EPI_ISL_2234849, EPI_ISL_2234851, EPI_ISL_2234852, EPI_ISL_2234853, EPI_ISL_2234858, | | | |

EPI_ISL_2234859, EPI_ISL_2234860, EPI_ISL_2234861, EPI_ISL_2234864, EPI_ISL_2234865, EPI_ISL_2234866, EPI_ISL_2234868, EPI_ISL_2234869

| | | | |
|---|---|---|---|
| see above | City of Milwaukee Health Department Laboratory | City of Milwaukee Health Department Laboratory | Sanjib Bhattacharyya, Manjeet Khubbar, Amy Bauer, Jennifer Lentz, Samantha Scott |

EPI_ISL_2234909, EPI_ISL_2234911, EPI_ISL_2234912, EPI_ISL_2234913, EPI_ISL_2234914, EPI_ISL_2234916, EPI_ISL_2234917, EPI_ISL_2234918, EPI_ISL_2234920, EPI_ISL_2234921, EPI_ISL_2234923, EPI_ISL_2234924, EPI_ISL_2234925, EPI_ISL_2234926, EPI_ISL_2234927, EPI_ISL_2234928, EPI_ISL_2234929, EPI_ISL_2234930, EPI_ISL_2234931, EPI_ISL_2234932, EPI_ISL_2234933, EPI_ISL_2234935, EPI_ISL_2234936, EPI_ISL_2234937, EPI_ISL_2234938, EPI_ISL_2234939, EPI_ISL_2234940, EPI_ISL_2234941, EPI_ISL_2234942, EPI_ISL_2234943, EPI_ISL_2234944, EPI_ISL_2234945, EPI_ISL_2234946, EPI_ISL_2234947, EPI_ISL_2234948, EPI_ISL_2234949, EPI_ISL_2234950, EPI_ISL_2234951, EPI_ISL_2234952, EPI_ISL_2234953, EPI_ISL_2234954, EPI_ISL_2234955, EPI_ISL_2234956, EPI_ISL_2234957, EPI_ISL_2234958, EPI_ISL_2234959, EPI_ISL_2234960, EPI_ISL_2234961, EPI_ISL_2234962, EPI_ISL_2234963, EPI_ISL_2234964, EPI_ISL_2234965, EPI_ISL_2234966, EPI_ISL_2234967, EPI_ISL_2234968, EPI_ISL_2234969, EPI_ISL_2234970, EPI_ISL_2234971, EPI_ISL_2234972, EPI_ISL_2234973, EPI_ISL_2234975, EPI_ISL_2234976, EPI_ISL_2234978, EPI_ISL_2234979, EPI_ISL_2234980, EPI_ISL_2234981, EPI_ISL_2234982, EPI_ISL_2234983, EPI_ISL_2234984, EPI_ISL_2234986, EPI_ISL_2234987, EPI_ISL_2234988, EPI_ISL_2234989, EPI_ISL_2234991, EPI_ISL_2234992, EPI_ISL_2234993, EPI_ISL_2234995, EPI_ISL_2234997, EPI_ISL_2234999, EPI_ISL_2235000, EPI_ISL_2235001, EPI_ISL_2235003, EPI_ISL_2235004, EPI_ISL_2235005, EPI_ISL_2235006, EPI_ISL_2235007, EPI_ISL_2235008, EPI_ISL_2235009, EPI_ISL_2235010, EPI_ISL_2235011, EPI_ISL_2235012, EPI_ISL_2235013, EPI_ISL_2235014, EPI_ISL_2235015, EPI_ISL_2235016, EPI_ISL_2235017, EPI_ISL_2235019, EPI_ISL_2235020, EPI_ISL_2235021, EPI_ISL_2235022, EPI_ISL_2235023, EPI_ISL_2235024, EPI_ISL_2235025, EPI_ISL_2235026, EPI_ISL_2235027, EPI_ISL_2235028, EPI_ISL_2235029, EPI_ISL_2235030, EPI_ISL_2235031, EPI_ISL_2235032, EPI_ISL_2235033, EPI_ISL_2235034, EPI_ISL_2235035, EPI_ISL_2235036, EPI_ISL_2235037, EPI_ISL_2235040, EPI_ISL_2235041, EPI_ISL_2235042, EPI_ISL_2235043, EPI_ISL_2235044, EPI_ISL_2235045, EPI_ISL_2235046, EPI_ISL_2235047, EPI_ISL_2235048, EPI_ISL_2235049, EPI_ISL_2235050, EPI_ISL_2235051, EPI_ISL_2235052, EPI_ISL_2235053, EPI_ISL_2235055, EPI_ISL_2235057, EPI_ISL_2235058, EPI_ISL_2235059, EPI_ISL_2235060, EPI_ISL_2235061, EPI_ISL_2235062, EPI_ISL_2235063, EPI_ISL_2235064, EPI_ISL_2235065, EPI_ISL_2235066, EPI_ISL_2235068, EPI_ISL_2235070, EPI_ISL_2235071, EPI_ISL_2235072, EPI_ISL_2235073, EPI_ISL_2235074, EPI_ISL_2235075, EPI_ISL_2235076, EPI_ISL_2235077, EPI_ISL_2235078, EPI_ISL_2235079, EPI_ISL_2235080, EPI_ISL_2235082, EPI_ISL_2235083, EPI_ISL_2235084, EPI_ISL_2235085, EPI_ISL_2235086, EPI_ISL_2235087, EPI_ISL_2235088, EPI_ISL_2235089, EPI_ISL_2235090, EPI_ISL_2235091, EPI_ISL_2235092, EPI_ISL_2235093, EPI_ISL_2235095, EPI_ISL_2235096, EPI_ISL_2235097, EPI_ISL_2235098, EPI_ISL_2235099, EPI_ISL_2235100, EPI_ISL_2235101, EPI_ISL_2235102, EPI_ISL_2235103, EPI_ISL_2235104, EPI_ISL_2235105, EPI_ISL_2235106, EPI_ISL_2235107, EPI_ISL_2235108, EPI_ISL_2235109, EPI_ISL_2235110, EPI_ISL_2235111, EPI_ISL_2235112, EPI_ISL_2235113, EPI_ISL_2235114, EPI_ISL_2235115, EPI_ISL_2235116, EPI_ISL_2235117, EPI_ISL_2235118, EPI_ISL_2235119, EPI_ISL_2235120, EPI_ISL_2235121, EPI_ISL_2235123, EPI_ISL_2235124, EPI_ISL_2235125, EPI_ISL_2235126, EPI_ISL_2235127, EPI_ISL_2235129, EPI_ISL_2235130, EPI_ISL_2235131, EPI_ISL_2235132, EPI_ISL_2235133, EPI_ISL_2235134, EPI_ISL_2235135, EPI_ISL_2235136, EPI_ISL_2235137, EPI_ISL_2235138, EPI_ISL_2235140, EPI_ISL_2235141, EPI_ISL_2235142, EPI_ISL_2235143, EPI_ISL_2235144, EPI_ISL_2235146, EPI_ISL_2235147, EPI_ISL_2235148, EPI_ISL_2235149, EPI_ISL_2235150, EPI_ISL_2235151, EPI_ISL_2235153, EPI_ISL_2235154, EPI_ISL_2235155, EPI_ISL_2235156, EPI_ISL_2235157, EPI_ISL_2235158, EPI_ISL_2235159, EPI_ISL_2235160, EPI_ISL_2235161, EPI_ISL_2235162, EPI_ISL_2235163, EPI_ISL_2235164, EPI_ISL_2235166, EPI_ISL_2235168, EPI_ISL_2235169, EPI_ISL_2235170, EPI_ISL_2235171, EPI_ISL_2235172, EPI_ISL_2235173, EPI_ISL_2235174, EPI_ISL_2235175, EPI_ISL_2235176, EPI_ISL_2235177, EPI_ISL_2235178, EPI_ISL_2235180, EPI_ISL_2235181, EPI_ISL_2235182, EPI_ISL_2235183, EPI_ISL_2235184, EPI_ISL_2235185, EPI_ISL_2235186, EPI_ISL_2235187, EPI_ISL_2235188, EPI_ISL_2235189, EPI_ISL_2235190, EPI_ISL_2235191, EPI_ISL_2235192, EPI_ISL_2235193, EPI_ISL_2235194, EPI_ISL_2235195, EPI_ISL_2235196, EPI_ISL_2235197, EPI_ISL_2235198, EPI_ISL_2235199, EPI_ISL_2235202, EPI_ISL_2235203, EPI_ISL_2235204, EPI_ISL_2235205, EPI_ISL_2235206, EPI_ISL_2235207, EPI_ISL_2235208, EPI_ISL_2235209, EPI_ISL_2235210, EPI_ISL_2235211, EPI_ISL_2235212, EPI_ISL_2235213, EPI_ISL_2235215, EPI_ISL_2235216, EPI_ISL_2235217, EPI_ISL_2235218

| | | | |
|---|---|---|---|
| see above | Pandemic Response Lab - NYC | Pandemic Response Lab, R&D | Henry Lee, Michael Hammerling, Melissa Hopkins, Cybill del Castillo, Shinyoung Clair Kang, William Ward, Pradeep Bugga, Sol Rey, Dylan Law, Katharine Nelson, Haiping Hao, Jon Laurent |
| EPI_ISL_2240781 | Ohio Department of Health Laboratory | Ohio Department of Health Laboratory | Holmes, Jennifer; Eric Brandt, Keoni Omura, Glen McGillivary, Caitlin McDonnell, Jade Mowery, Stephanie Mccracken, Tyler Payne, Kirtana Ramadugu, Erica Leasure, Brent Lee, Kelsey Florek, Heather Blankenship, Allison Black, Quanta Brown, and Tammy Bannerman |
| EPI_ISL_2241626 | Fulgent Genetics | Centers for Disease Control and Prevention Division of Viral Diseases, Pathogen Discovery | Dakota Howard, Dhwani Batra, Peter W. Cook, Kara Moser, Adrian Paskey, Jason Caravas, Benjamin Rambo-Martin, Shatavia Morrison, Christopher Gulvick, Scott Sammons, Yvette Unoarumhi, Darlene Wagner, Matthew Schmerer, Harry Gao, Mickey Li, John Gao, Joseph Fierro, Benafsh Sapra, Becky Tsai, Yan Meng, Doreen Ng, James Xie, Clinton R. Paden, Duncan MacCannell |

EPI_ISL_2241627, EPI_ISL_2241628, EPI_ISL_2241629, EPI_ISL_2241630, EPI_ISL_2241631, EPI_ISL_2241632, EPI_ISL_2241633, EPI_ISL_2241634, EPI_ISL_2241635, EPI_ISL_2241636, EPI_ISL_2241637, EPI_ISL_2241638, EPI_ISL_2241639, EPI_ISL_2241640, EPI_ISL_2241641, EPI_ISL_2241642

| | | | |
|---|---|---|---|
| see above | Quest Diagnostics Incorporated | Centers for Disease Control and Prevention Division of Viral Diseases, Pathogen Discovery | Dakota Howard, Dhwani Batra, Peter W. Cook, Kara Moser, Adrian Paskey, Jason Caravas, Benjamin Rambo-Martin, Shatavia Morrison, Christopher Gulvick, Scott Sammons, Yvette Unoarumhi, Darlene Wagner, Matthew Schmerer, S. H. Rosenthal, A. Gerasimova, R. M. Kagan, B. Anderson, M. Hua, Y. Liu, L.E. Bernstein, K.E. Livingston, A. Perez, I. A. Shlyakhter, R. V. Rolando, R. Owen, P. Tanpaiboon, F. Lacbawan, Clinton R. Paden, Duncan MacCannell |

EPI_ISL_2241643, EPI_ISL_2241644, EPI_ISL_2241645, EPI_ISL_2241646, EPI_ISL_2241647, EPI_ISL_2241648, EPI_ISL_2241649, EPI_ISL_2241650, EPI_ISL_2241651, EPI_ISL_2241652, EPI_ISL_2241653, EPI_ISL_2241654, EPI_ISL_2241655, EPI_ISL_2241656, EPI_ISL_2241657, EPI_ISL_2241658, EPI_ISL_2241659, EPI_ISL_2241660, EPI_ISL_2241661, EPI_ISL_2241662, EPI_ISL_2241663, EPI_ISL_2241664, EPI_ISL_2241666, EPI_ISL_2241667, EPI_ISL_2241668, EPI_ISL_2241669, EPI_ISL_2241670, EPI_ISL_2241671, EPI_ISL_2241672, EPI_ISL_2241673, EPI_ISL_2241674, EPI_ISL_2241675, EPI_ISL_2241676, EPI_ISL_2241677, EPI_ISL_2241678, EPI_ISL_2241680, EPI_ISL_2241681, EPI_ISL_2241682, EPI_ISL_2241683, EPI_ISL_2241684, EPI_ISL_2241685, EPI_ISL_2241686, EPI_ISL_2241687, EPI_ISL_2241688, EPI_ISL_2241689

| | | | |
|---|---|---|---|
| see above | Aegis Sciences Corporation | Centers for Disease Control and Prevention Division of Viral Diseases, Pathogen Discovery | Dakota Howard, Dhwani Batra, Peter W. Cook, Kara Moser, Adrian Paskey, Jason Caravas, Benjamin Rambo-Martin, Shatavia Morrison, Christopher Gulvick, Scott Sammons, Yvette Unoarumhi, Darlene Wagner, Matthew Schmerer, Cyndi Clark, Patrick Campbell, Rob Case, Vikramsinha Ghorpade, Holly Houdeshell, Ola Kvalvaag, Dillon Nall, Ethan Sanders, Alec Vest, Shaun Westlund, Matthew Hardison, Clinton R. Paden, Duncan MacCannell |
| EPI_ISL_2241690, EPI_ISL_2241691, EPI_ISL_2241692, EPI_ISL_2241693 | Laboratory Corporation of America | Centers for Disease Control and Prevention Division of Viral Diseases, Pathogen Discovery | Dakota Howard, Dhwani Batra, Peter W. Cook, Kara Moser, Adrian Paskey, Jason Caravas, Benjamin Rambo-Martin, Shatavia Morrison, Christopher Gulvick, Scott Sammons, Yvette Unoarumhi, Darlene Wagner, Matthew Schmerer, Minoo Agarwal, Eyad Almasri, Debbie Boles, Ayla Burns, Nuthawin Charoensri, Oren Cohen, Susan Countryman, Mary Ann Cristobal, Bobbi Croy, Suzanne Dale, Hrushikesh Deshmukh, Amanda Douglas, Vincent Drouillon, Marcia Eisenberg, Howard Engler, Rama Ghatti, Prashant Gupta, Susan Hicks, Jake Humphrey, Lax Iyer, Manoj Jain, Mohan Kolli, Brian Krueger, Tim Kuphal, Stanley Letovsky, Michael Levandoski, Craig Lukasik, Jonathan Meltzer, Brian Norvell, Mindy Nye, Scott Parker, Christos Petropoulos, John Pruitt, Steven Ragan, Scott Ryan, Mike Sapeta, Jana Schroth, Suresh Babu Selvaraju, Goran Stevovic, Amanda Suchanek, Andrea Throop, Lyndon Tilson, Thomas Urban, Joe Voshell, Kimberly Wagner, Jonathan Williams, Mary Williamson, Qian Zeng, Tricia Zwiefelhofer, Clinton R. Paden, Duncan MacCannell |

EPI_ISL_2241694, EPI_ISL_2241695, EPI_ISL_2241696, EPI_ISL_2241697, EPI_ISL_2241698, EPI_ISL_2241699, EPI_ISL_2241700, EPI_ISL_2241701, EPI_ISL_2241702, EPI_ISL_2241703, EPI_ISL_2241704, EPI_ISL_2241705, EPI_ISL_2241706, EPI_ISL_2241707, EPI_ISL_2241708, EPI_ISL_2241709, EPI_ISL_2241710, EPI_ISL_2241711, EPI_ISL_2241712, EPI_ISL_2241713, EPI_ISL_2241714, EPI_ISL_2241715, EPI_ISL_2241716, EPI_ISL_2241717, EPI_ISL_2241718, EPI_ISL_2241719, EPI_ISL_2241720, EPI_ISL_2241721, EPI_ISL_2241722, EPI_ISL_2241723, EPI_ISL_2241724, EPI_ISL_2241725, EPI_ISL_2241726, EPI_ISL_2241727, EPI_ISL_2241728, EPI_ISL_2241729, EPI_ISL_2241730, EPI_ISL_2241731, EPI_ISL_2241732, EPI_ISL_2241733, EPI_ISL_2241734, EPI_ISL_2241735, EPI_ISL_2241736, EPI_ISL_2241737, EPI_ISL_2241738, EPI_ISL_2241739, EPI_ISL_2241740, EPI_ISL_2241741, EPI_ISL_2241742, EPI_ISL_2241743, EPI_ISL_2241744, EPI_ISL_2241745, EPI_ISL_2241746, EPI_ISL_2241747, EPI_ISL_2241748, EPI_ISL_2241749, EPI_ISL_2241750, EPI_ISL_2241751, EPI_ISL_2241752, EPI_ISL_2241753, EPI_ISL_2241754, EPI_ISL_2241755, EPI_ISL_2241757, EPI_ISL_2241758, EPI_ISL_2241759, EPI_ISL_2241760, EPI_ISL_2241761, EPI_ISL_2241762, EPI_ISL_2241763, EPI_ISL_2241764, EPI_ISL_2241765, EPI_ISL_2241766, EPI_ISL_2241767, EPI_ISL_2241768, EPI_ISL_2241769, EPI_ISL_2241770, EPI_ISL_2241771, EPI_ISL_2241772, EPI_ISL_2241773, EPI_ISL_2241774, EPI_ISL_2241775, EPI_ISL_2241776, EPI_ISL_2241777, EPI_ISL_2241779, EPI_ISL_2241780, EPI_ISL_2241781, EPI_ISL_2241782, EPI_ISL_2241783, EPI_ISL_2241784, EPI_ISL_2241785, EPI_ISL_2241786, EPI_ISL_2241788, EPI_ISL_2241789, EPI_ISL_2241790, EPI_ISL_2241791, EPI_ISL_2241792, EPI_ISL_2241793, EPI_ISL_2241794, EPI_ISL_2241795, EPI_ISL_2241796, EPI_ISL_2241797, EPI_ISL_2241798, EPI_ISL_2241799, EPI_ISL_2241800, EPI_ISL_2241801, EPI_ISL_2241802, EPI_ISL_2241803, EPI_ISL_2241804, EPI_ISL_2241805, EPI_ISL_2241806, EPI_ISL_2241807, EPI_ISL_2241808, EPI_ISL_2241810, EPI_ISL_2241811, EPI_ISL_2241812, EPI_ISL_2241813, EPI_ISL_2241814, EPI_ISL_2241815, EPI_ISL_2241816, EPI_ISL_2241817, EPI_ISL_2241818, EPI_ISL_2241819, EPI_ISL_2241820, EPI_ISL_2241821, EPI_ISL_2241822, EPI_ISL_2241823, EPI_ISL_2241824, EPI_ISL_2241825, EPI_ISL_2241826, EPI_ISL_2241827, EPI_ISL_2241828, EPI_ISL_2241829, EPI_ISL_2241830, EPI_ISL_2241831, EPI_ISL_2241832, EPI_ISL_2241833, EPI_ISL_2241834, EPI_ISL_2241835, EPI_ISL_2241836, EPI_ISL_2241837, EPI_ISL_2241838, EPI_ISL_2241839, EPI_ISL_2241840, EPI_ISL_2241841, EPI_ISL_2241842, EPI_ISL_2241843, EPI_ISL_2241844, EPI_ISL_2241846, EPI_ISL_2241847, EPI_ISL_2241848, EPI_ISL_2241849, EPI_ISL_2241850, EPI_ISL_2241851, EPI_ISL_2241852, EPI_ISL_2241853, EPI_ISL_2241854, EPI_ISL_2241855, EPI_ISL_2241856, EPI_ISL_2241857, EPI_ISL_2241858, EPI_ISL_2241859, EPI_ISL_2241860, EPI_ISL_2241861, EPI_ISL_2241862, EPI_ISL_2241863, EPI_ISL_2241864, EPI_ISL_2241865, EPI_ISL_2241866, EPI_ISL_2241867, EPI_ISL_2241868, EPI_ISL_2241869, EPI_ISL_2241870, EPI_ISL_2241871, EPI_ISL_2241872, EPI_ISL_2241873, EPI_ISL_2241874, EPI_ISL_2241875, EPI_ISL_2241876, EPI_ISL_2241877, EPI_ISL_2241878, EPI_ISL_2241879, EPI_ISL_2241881, EPI_ISL_2241882, EPI_ISL_2241883, EPI_ISL_2241884, EPI_ISL_2241885, EPI_ISL_2241886, EPI_ISL_2241887, EPI_ISL_2241888, EPI_ISL_2241889, EPI_ISL_2241890, EPI_ISL_2241891, EPI_ISL_2241892, EPI_ISL_2241893, EPI_ISL_2241894, EPI_ISL_2241895, EPI_ISL_2241896, EPI_ISL_2241897, EPI_ISL_2241898, EPI_ISL_2241899, EPI_ISL_2241900, EPI_ISL_2241901, EPI_ISL_2241902, EPI_ISL_2241903, EPI_ISL_2241904, EPI_ISL_2241905, EPI_ISL_2241906, EPI_ISL_2241907, EPI_ISL_2241908, EPI_ISL_2241909, EPI_ISL_2241910, EPI_ISL_2241911, EPI_ISL_2241912, EPI_ISL_2241913, EPI_ISL_2241914, EPI_ISL_2241915, EPI_ISL_2241916, EPI_ISL_2241917, EPI_ISL_2241918, EPI_ISL_2241919, EPI_ISL_2241920, EPI_ISL_2241921, EPI_ISL_2241922, EPI_ISL_2241923, EPI_ISL_2241924, EPI_ISL_2241925, EPI_ISL_2241926, EPI_ISL_2241927, EPI_ISL_2241928, EPI_ISL_2241929, EPI_ISL_2241930, EPI_ISL_2241931, EPI_ISL_2241932, EPI_ISL_2241933, EPI_ISL_2241934, EPI_ISL_2241935, EPI_ISL_2241936, EPI_ISL_2241937, EPI_ISL_2241938, EPI_ISL_2241939, EPI_ISL_2241940, EPI_ISL_2241941, EPI_ISL_2241942, EPI_ISL_2241943, EPI_ISL_2241944, EPI_ISL_2241945, EPI_ISL_2241946, EPI_ISL_2241947, EPI_ISL_2241948, EPI_ISL_2241949, EPI_ISL_2241950, EPI_ISL_2241951, EPI_ISL_2241952, EPI_ISL_2241953, EPI_ISL_2241954, EPI_ISL_2241955, EPI_ISL_2241956, EPI_ISL_2241957, EPI_ISL_2241958, EPI_ISL_2241959, EPI_ISL_2241960, EPI_ISL_2241961, EPI_ISL_2241962, EPI_ISL_2241963, EPI_ISL_2241964, EPI_ISL_2241965, EPI_ISL_2241966, EPI_ISL_2241968, EPI_ISL_2241969, EPI_ISL_2241970, EPI_ISL_2241971, EPI_ISL_2241972, EPI_ISL_2241973, EPI_ISL_2241974, EPI_ISL_2241975, EPI_ISL_2241976, EPI_ISL_2241977, EPI_ISL_2241978, EPI_ISL_2241979, EPI_ISL_2241980, EPI_ISL_2241982, EPI_ISL_2241983, EPI_ISL_2241984, EPI_ISL_2241985, EPI_ISL_2241986, EPI_ISL_2241987, EPI_ISL_2241988, EPI_ISL_2241989, EPI_ISL_2241990, EPI_ISL_2241991, EPI_ISL_2241992, EPI_ISL_2241993, EPI_ISL_2241994, EPI_ISL_2241995, EPI_ISL_2241996, EPI_ISL_2241997, EPI_ISL_2241998, EPI_ISL_2241999, EPI_ISL_2242000, EPI_ISL_2242001, EPI_ISL_2242002, EPI_ISL_2242003, EPI_ISL_2242004, EPI_ISL_2242005, EPI_ISL_2242006, EPI_ISL_2242007, EPI_ISL_2242008, EPI_ISL_2242009, EPI_ISL_2242010, EPI_ISL_2242011, EPI_ISL_2242012, EPI_ISL_2242013, EPI_ISL_2242014, EPI_ISL_2242015, EPI_ISL_2242016, EPI_ISL_2242017, EPI_ISL_2242018, EPI_ISL_2242019, EPI_ISL_2242020, EPI_ISL_2242021, EPI_ISL_2242022, EPI_ISL_2242023, EPI_ISL_2242024, EPI_ISL_2242025, EPI_ISL_2242026, EPI_ISL_2242027, EPI_ISL_2242028, EPI_ISL_2242029, EPI_ISL_2242030, EPI_ISL_2242031, EPI_ISL_2242032, EPI_ISL_2242033, EPI_ISL_2242034, EPI_ISL_2242035, EPI_ISL_2242036, EPI_ISL_2242037, EPI_ISL_2242038, EPI_ISL_2242039, EPI_ISL_2242040, EPI_ISL_2242041, EPI_ISL_2242042, EPI_ISL_2242043, EPI_ISL_2242044, EPI_ISL_2242045, EPI_ISL_2242046, EPI_ISL_2242047, EPI_ISL_2242048, EPI_ISL_2242049, EPI_ISL_2242050, EPI_ISL_2242051, EPI_ISL_2242052, EPI_ISL_2242053, EPI_ISL_2242054, EPI_ISL_2242055, EPI_ISL_2242056, EPI_ISL_2242057, EPI_ISL_2242058, EPI_ISL_2242059, EPI_ISL_2242060, EPI_ISL_2242061, EPI_ISL_2242062, EPI_ISL_2242063, EPI_ISL_2242064, EPI_ISL_2242065, EPI_ISL_2242066, EPI_ISL_2242067, EPI_ISL_2242068, EPI_ISL_2242069, EPI_ISL_2242070, EPI_ISL_2242071, EPI_ISL_2242072, EPI_ISL_2242073, EPI_ISL_2242074, EPI_ISL_2242075, EPI_ISL_2242076, EPI_ISL_2242077, EPI_ISL_2242078, EPI_ISL_2242079, EPI_ISL_2242080, EPI_ISL_2242081, EPI_ISL_2242082, EPI_ISL_2242083, EPI_ISL_2242084, EPI_ISL_2242085, EPI_ISL_2242086, EPI_ISL_2242087, EPI_ISL_2242088, EPI_ISL_2242089, EPI_ISL_2242090, EPI_ISL_2242091, EPI_ISL_2242092, EPI_ISL_2242093, EPI_ISL_2242094, EPI_ISL_2242095, EPI_ISL_2242096, EPI_ISL_2242097, EPI_ISL_2242098, EPI_ISL_2242099, EPI_ISL_2242100, EPI_ISL_2242101, EPI_ISL_2242102, EPI_ISL_2242103, EPI_ISL_2242104, EPI_ISL_2242105, EPI_ISL_2242106, EPI_ISL_2242107, EPI_ISL_2242108, EPI_ISL_2242109, EPI_ISL_2242110, EPI_ISL_2242111, EPI_ISL_2242112, EPI_ISL_2242113, EPI_ISL_2242114, EPI_ISL_2242115, EPI_ISL_2242116, EPI_ISL_2242117, EPI_ISL_2242118, EPI_ISL_2242119, EPI_ISL_2242120, EPI_ISL_2242121, EPI_ISL_2242122, EPI_ISL_2242123, EPI_ISL_2242124, EPI_ISL_2242125, EPI_ISL_2242126, EPI_ISL_2242127, EPI_ISL_2242128, EPI_ISL_2242129, EPI_ISL_2242130, EPI_ISL_2242131, EPI_ISL_2242132, EPI_ISL_2242133, EPI_ISL_2242134, EPI_ISL_2242135, EPI_ISL_2242136, EPI_ISL_2242137, EPI_ISL_2242138, EPI_ISL_2242139, EPI_ISL_2242140, EPI_ISL_2242141, EPI_ISL_2242142, EPI_ISL_2242143, EPI_ISL_2242144, EPI_ISL_2242145, EPI_ISL_2242146, EPI_ISL_2242147, EPI_ISL_2242148, EPI_ISL_2242149, EPI_ISL_2242150, EPI_ISL_2242151, EPI_ISL_2242152, EPI_ISL_2242153, EPI_ISL_2242154, EPI_ISL_2242155, EPI_ISL_2242156, EPI_ISL_2242157, EPI_ISL_2242158, EPI_ISL_2242159, EPI_ISL_2242160, EPI_ISL_2242161, EPI_ISL_2242162, EPI_ISL_2242163, EPI_ISL_2242164, EPI_ISL_2242165, EPI_ISL_2242166, EPI_ISL_2242167, EPI_ISL_2242168, EPI_ISL_2242169, EPI_ISL_2242170, EPI_ISL_2242171, EPI_ISL_2242172, EPI_ISL_2242173, EPI_ISL_2242174, EPI_ISL_2242175, EPI_ISL_2242176, EPI_ISL_2242177, EPI_ISL_2242178, EPI_ISL_2242179, EPI_ISL_2242180, EPI_ISL_2242181, EPI_ISL_2242182, EPI_ISL_2242184, EPI_ISL_2242185, EPI_ISL_2242186, EPI_ISL_2242187, EPI_ISL_2242188, EPI_ISL_2242189, EPI_ISL_2242190, EPI_ISL_2242191, EPI_ISL_2242192, EPI_ISL_2242193, EPI_ISL_2242194, EPI_ISL_2242195, EPI_ISL_2242196, EPI_ISL_2242197, EPI_ISL_2242198, EPI_ISL_2242199, EPI_ISL_2242200, EPI_ISL_2242201, EPI_ISL_2242202, EPI_ISL_2242203, EPI_ISL_2242204, EPI_ISL_2242205, EPI_ISL_2242206, EPI_ISL_2242207, EPI_ISL_2242208, EPI_ISL_2242209, EPI_ISL_2242210, EPI_ISL_2242211, EPI_ISL_2242212, EPI_ISL_2242213, EPI_ISL_2242214, EPI_ISL_2242215, EPI_ISL_2242216, EPI_ISL_2242217, EPI_ISL_2242218, EPI_ISL_2242219, EPI_ISL_2242220, EPI_ISL_2242221, EPI_ISL_2242222, EPI_ISL_2242223, EPI_ISL_2242224, EPI_ISL_2242225, EPI_ISL_2242226, EPI_ISL_2242227, EPI_ISL_2242228, EPI_ISL_2242229, EPI_ISL_2242230, EPI_ISL_2242231, EPI_ISL_2242232, EPI_ISL_2242233, EPI_ISL_2242234, EPI_ISL_2242235, EPI_ISL_2242236, EPI_ISL_2242237, EPI_ISL_2242238, EPI_ISL_2242239, EPI_ISL_2242240, EPI_ISL_2242241, EPI_ISL_2242242, EPI_ISL_2242243, EPI_ISL_2242244, EPI_ISL_2242245, EPI_ISL_2242246, EPI_ISL_2242247, EPI_ISL_2242248, EPI_ISL_2242249, EPI_ISL_2242250, EPI_ISL_2242251, EPI_ISL_2242252, EPI_ISL_2242253, EPI_ISL_2242254, EPI_ISL_2242255, EPI_ISL_2242256, EPI_ISL_2242257, EPI_ISL_2242258, EPI_ISL_2242259, EPI_ISL_2242260, EPI_ISL_2242261, EPI_ISL_2242262, EPI_ISL_2242263, EPI_ISL_2242264, EPI_ISL_2242265, EPI_ISL_2242266, EPI_ISL_2242267, EPI_ISL_2242268, EPI_ISL_2242269, EPI_ISL_2242270, EPI_ISL_2242271, EPI_ISL_2242272, EPI_ISL_2242273, EPI_ISL_2242274, EPI_ISL_2242275, EPI_ISL_2242276, EPI_ISL_2242277, EPI_ISL_2242278, EPI_ISL_2242279, EPI_ISL_2242280, EPI_ISL_2242281, EPI_ISL_2242282, EPI_ISL_2242283, EPI_ISL_2242284, EPI_ISL_2242285, EPI_ISL_2242286, EPI_ISL_2242287, EPI_ISL_2242288, EPI_ISL_2242289, EPI_ISL_2242290, EPI_ISL_2242291, EPI_ISL_2242292, EPI_ISL_2242293, EPI_ISL_2242294, EPI_ISL_2242295, EPI_ISL_2242296, EPI_ISL_2242297, EPI_ISL_2242298, EPI_ISL_2242299, EPI_ISL_2242300, EPI_ISL_2242301, EPI_ISL_2242302, EPI_ISL_2242303, EPI_ISL_2242304, EPI_ISL_2242305, EPI_ISL_2242306, EPI_ISL_2242307, EPI_ISL_2242308, EPI_ISL_2242309, EPI_ISL_2242310, EPI_ISL_2242311, EPI_ISL_2242312, EPI_ISL_2242313, EPI_ISL_2242314, EPI_ISL_2242315, EPI_ISL_2242316, EPI_ISL_2242317, EPI_ISL_2242318, EPI_ISL_2242319, EPI_ISL_2242320, EPI_ISL_2242321, EPI_ISL_2242322, EPI_ISL_2242323, EPI_ISL_2242324, EPI_ISL_2242325, EPI_ISL_2242326, EPI_ISL_2242327, EPI_ISL_2242328, EPI_ISL_2242329, EPI_ISL_2242330, EPI_ISL_2242331, EPI_ISL_2242332, EPI_ISL_2242333, EPI_ISL_2242334, EPI_ISL_2242335, EPI_ISL_2242336, EPI_ISL_2242337, EPI_ISL_2242338, EPI_ISL_2242339, EPI_ISL_2242340, EPI_ISL_2242341, EPI_ISL_2242342, EPI_ISL_2242343, EPI_ISL_2242344, EPI_ISL_2242345, EPI_ISL_2242346, EPI_ISL_2242347, EPI_ISL_2242348, EPI_ISL_2242349, EPI_ISL_2242350, EPI_ISL_2242351, EPI_ISL_2242352, EPI_ISL_2242353, EPI_ISL_2242354, EPI_ISL_2242355, EPI_ISL_2242356, EPI_ISL_2242357, EPI_ISL_2242358, EPI_ISL_2242359, EPI_ISL_2242360, EPI_ISL_2242361, EPI_ISL_2242362, EPI_ISL_2242363, EPI_ISL_2242364, EPI_ISL_2242365, EPI_ISL_2242366, EPI_ISL_2242367, EPI_ISL_2242368, EPI_ISL_2242369, EPI_ISL_2242370, EPI_ISL_2242371, EPI_ISL_2242372, EPI_ISL_2242373, EPI_ISL_2242374, EPI_ISL_2242375, EPI_ISL_2242376, EPI_ISL_2242377, EPI_ISL_2242378, EPI_ISL_2242379, EPI_ISL_2242380, EPI_ISL_2242381, EPI_ISL_2242382, EPI_ISL_2242383, EPI_ISL_2242384, EPI_ISL_2242385, EPI_ISL_2242386, EPI_ISL_2242387, EPI_ISL_2242388, EPI_ISL_2242389, EPI_ISL_2242390, EPI_ISL_2242391, EPI_ISL_2242392, EPI_ISL_2242393, EPI_ISL_2242394, EPI_ISL_2242395, EPI_ISL_2242396, EPI_ISL_2242397, EPI_ISL_2242398, EPI_ISL_2242399, EPI_ISL_2242400, EPI_ISL_2242401, EPI_ISL_2242402, EPI_ISL_2242403, EPI_ISL_2242404, EPI_ISL_2242405, EPI_ISL_2242406, EPI_ISL_2242407, EPI_ISL_2242408, EPI_ISL_2242409, EPI_ISL_2242410, EPI_ISL_2242411, EPI_ISL_2242412, EPI_ISL_2242413, EPI_ISL_2242414, EPI_ISL_2242415, EPI_ISL_2242416, EPI_ISL_2242417, EPI_ISL_2242418, EPI_ISL_2242419, EPI_ISL_2242420, EPI_ISL_2242421, EPI_ISL_2242422, EPI_ISL_2242423, EPI_ISL_2242424, EPI_ISL_2242425, EPI_ISL_2242426, EPI_ISL_2242427, EPI_ISL_2242428, EPI_ISL_2242429, EPI_ISL_2242430, EPI_ISL_2242431, EPI_ISL_2242432, EPI_ISL_2242433, EPI_ISL_2242434, EPI_ISL_2242435, EPI_ISL_2242436, EPI_ISL_2242437, EPI_ISL_2242438, EPI_ISL_2242439, EPI_ISL_2242440, EPI_ISL_2242441, EPI_ISL_2242442, EPI_ISL_2242443, EPI_ISL_2242444, EPI_ISL_2242445, EPI_ISL_2242446, EPI_ISL_2242447, EPI_ISL_2242448, EPI_ISL_2242449, EPI_ISL_2242450, EPI_ISL_2242451, EPI_ISL_2242452, EPI_ISL_2242453, EPI_ISL_2242454, EPI_ISL_2242455, EPI_ISL_2242456, EPI_ISL_2242457, EPI_ISL_2242458, EPI_ISL_2242459, EPI_ISL_2242460, EPI_ISL_2242461, EPI_ISL_2242462, EPI_ISL_2242463, EPI_ISL_2242464, EPI_ISL_2242465, EPI_ISL_2242466, EPI_ISL_2242467, EPI_ISL_2242468, EPI_ISL_2242469, EPI_ISL_2242470, EPI_ISL_2242471, EPI_ISL_2242472, EPI_ISL_2242473, EPI_ISL_2242474, EPI_ISL_2242475, EPI_ISL_2242476, EPI_ISL_2242477, EPI_ISL_2242478, EPI_ISL_2242479, EPI_ISL_2242480, EPI_ISL_2242481, EPI_ISL_2242482, EPI_ISL_2242483, EPI_ISL_2242484, EPI_ISL_2242485, EPI_ISL_2242486, EPI_ISL_2242487, EPI_ISL_2242488, EPI_ISL_2242489, EPI_ISL_2242490, EPI_ISL_2242491, EPI_ISL_2242492, EPI_ISL_2242493, EPI_ISL_2242494, EPI_ISL_2242495, EPI_ISL_2242496, EPI_ISL_2242497, EPI_ISL_2242498, EPI_ISL_2242499, EPI_ISL_2242500, EPI_ISL_2242501, EPI_ISL_2242502, EPI_ISL_2242503, EPI_ISL_2242504, EPI_ISL_2242505, EPI_ISL_2242507, EPI_ISL_2242508

| | | | |
|---|---|---|---|
| see above | Aegis Sciences Corporation | Centers for Disease Control and Prevention Division of Viral Diseases, Pathogen Discovery | Dakota Howard, Dhwani Batra, Peter W. Cook, Kara Moser, Adrian Paskey, Jason Caravas, Benjamin Rambo-Martin, Shatavia Morrison, Christopher Gulvick, Scott Sammons, Yvette Unoarumhi, Darlene Wagner, Matthew Schmerer, Cyndi Clark, Patrick Campbell, Rob Case, Vikramsinha Ghorpade, Holly Houdeshell, Ola Kvalvaag, Dillon Nall, Ethan Sanders, Alec Vest, Shaun Westlund, Matthew Hardison, Clinton R. Paden, Duncan MacCannell |
| EPI_ISL_2242509 | IN State Department of Health Laboratory Services | IN State Department of Health Laboratory Services | Cassandra Campion, Jamie Yeadon, Brian Pope, Lixia Liu, Kyle Brownlee, Melissa Hindenlang, Mark Glazier |
| EPI_ISL_2242510 | Aegis Sciences Corporation | Centers for Disease Control and Prevention Division of Viral Diseases, Pathogen Discovery | Dakota Howard, Dhwani Batra, Peter W. Cook, Kara Moser, Adrian Paskey, Jason Caravas, Benjamin Rambo-Martin, Shatavia Morrison, Christopher Gulvick, Scott Sammons, Yvette Unoarumhi, Darlene Wagner, Matthew Schmerer, Cyndi Clark, Patrick Campbell, Rob Case, Vikramsinha Ghorpade, Holly Houdeshell, Ola Kvalvaag, Dillon Nall, Ethan Sanders, Alec Vest, Shaun Westlund, Matthew Hardison, Clinton R. Paden, Duncan MacCannell |
| EPI_ISL_2242511 | IN State Department of Health Laboratory Services | IN State Department of Health Laboratory Services | Cassandra Campion, Jamie Yeadon, Brian Pope, Lixia Liu, Kyle Brownlee, Melissa Hindenlang, Mark Glazier |
| EPI_ISL_2242512 | Aegis Sciences Corporation | Centers for Disease Control and Prevention Division of Viral Diseases, Pathogen Discovery | Dakota Howard, Dhwani Batra, Peter W. Cook, Kara Moser, Adrian Paskey, Jason Caravas, Benjamin Rambo-Martin, Shatavia Morrison, Christopher Gulvick, Scott Sammons, Yvette Unoarumhi, Darlene Wagner, Matthew Schmerer, Cyndi Clark, Patrick Campbell, Rob Case, Vikramsinha Ghorpade, Holly Houdeshell, Ola Kvalvaag, Dillon Nall, Ethan Sanders, Alec Vest, Shaun Westlund, Matthew Hardison, Clinton R. Paden, Duncan MacCannell |
| EPI_ISL_2242513 | IN State Department of Health Laboratory Services | IN State Department of Health Laboratory Services | Cassandra Campion, Jamie Yeadon, Brian Pope, Lixia Liu, Kyle Brownlee, Melissa Hindenlang, Mark Glazier |
| EPI_ISL_2242514 | Aegis Sciences Corporation | Centers for Disease Control and Prevention Division of Viral Diseases, Pathogen Discovery | Dakota Howard, Dhwani Batra, Peter W. Cook, Kara Moser, Adrian Paskey, Jason Caravas, Benjamin Rambo-Martin, Shatavia Morrison, Christopher Gulvick, Scott Sammons, Yvette Unoarumhi, Darlene Wagner, Matthew Schmerer, Cyndi Clark, Patrick Campbell, Rob Case, Vikramsinha Ghorpade, Holly Houdeshell, Ola Kvalvaag, Dillon Nall, Ethan Sanders, Alec Vest, Shaun Westlund, Matthew Hardison, Clinton R. Paden, Duncan MacCannell |
| EPI_ISL_2242515 | IN State Department of Health Laboratory Services | IN State Department of Health Laboratory Services | Cassandra Campion, Jamie Yeadon, Brian Pope, Lixia Liu, Kyle Brownlee, Melissa Hindenlang, Mark Glazier |
| EPI_ISL_2242516 | Aegis Sciences Corporation | Centers for Disease Control and Prevention Division of Viral Diseases, Pathogen Discovery | Dakota Howard, Dhwani Batra, Peter W. Cook, Kara Moser, Adrian Paskey, Jason Caravas, Benjamin Rambo-Martin, Shatavia Morrison, Christopher Gulvick, Scott Sammons, Yvette Unoarumhi, Darlene Wagner, Matthew Schmerer, Cyndi Clark, Patrick Campbell, Rob Case, Vikramsinha Ghorpade, Holly Houdeshell, Ola Kvalvaag, Dillon Nall, Ethan Sanders, Alec Vest, Shaun Westlund, Matthew Hardison, Clinton R. Paden, Duncan MacCannell |
| EPI_ISL_2242517 | IN State Department of Health Laboratory Services | IN State Department of Health Laboratory Services | Cassandra Campion, Jamie Yeadon, Brian Pope, Lixia Liu, Kyle Brownlee, Melissa Hindenlang, Mark Glazier |
| EPI_ISL_2242518 | Aegis Sciences Corporation | Centers for Disease Control and Prevention Division of Viral Diseases, Pathogen Discovery | Dakota Howard, Dhwani Batra, Peter W. Cook, Kara Moser, Adrian Paskey, Jason Caravas, Benjamin Rambo-Martin, Shatavia Morrison, Christopher Gulvick, Scott Sammons, Yvette Unoarumhi, Darlene Wagner, Matthew Schmerer, Cyndi Clark, Patrick Campbell, Rob Case, Vikramsinha Ghorpade, Holly Houdeshell, Ola Kvalvaag, Dillon Nall, Ethan Sanders, Alec Vest, Shaun Westlund, Matthew Hardison, Clinton R. Paden, Duncan MacCannell |

| | | | |
|---|---|---|---|
| EPI_ISL_2242519 | IN State Department of Health Laboratory Services | IN State Department of Health Laboratory Services | Cassandra Campion, Jamie Yeadon, Brian Pope, Lixia Liu, Kyle Brownlee, Melissa Hindenlang, Mark Glazier |
| EPI_ISL_2242520, EPI_ISL_2242522, EPI_ISL_2242524 | Aegis Sciences Corporation | Centers for Disease Control and Prevention Division of Viral Diseases, Pathogen Discovery | Dakota Howard, Dhwani Batra, Peter W. Cook, Kara Moser, Adrian Paskey, Jason Caravas, Benjamin Rambo-Martin, Shatavia Morrison, Christopher Gulvick, Scott Sammons, Yvette Unoarumhi, Darlene Wagner, Matthew Schmerer, Cyndi Clark, Patrick Campbell, Rob Case, Vikramsinha Ghorpade, Holly Houdeshell, Ola Kvalvaag, Dillon Nall, Ethan Sanders, Alec Vest, Shaun Westlund, Matthew Hardison, Clinton R. Paden, Duncan MacCannell |
| EPI_ISL_2242525 | IN State Department of Health Laboratory Services | IN State Department of Health Laboratory Services | Cassandra Campion, Jamie Yeadon, Brian Pope, Lixia Liu, Kyle Brownlee, Melissa Hindenlang, Mark Glazier |
| EPI_ISL_2242526, EPI_ISL_2242528, EPI_ISL_2242530 | Aegis Sciences Corporation | Centers for Disease Control and Prevention Division of Viral Diseases, Pathogen Discovery | Dakota Howard, Dhwani Batra, Peter W. Cook, Kara Moser, Adrian Paskey, Jason Caravas, Benjamin Rambo-Martin, Shatavia Morrison, Christopher Gulvick, Scott Sammons, Yvette Unoarumhi, Darlene Wagner, Matthew Schmerer, Cyndi Clark, Patrick Campbell, Rob Case, Vikramsinha Ghorpade, Holly Houdeshell, Ola Kvalvaag, Dillon Nall, Ethan Sanders, Alec Vest, Shaun Westlund, Matthew Hardison, Clinton R. Paden, Duncan MacCannell |
| EPI_ISL_2242531 | IN State Department of Health Laboratory Services | IN State Department of Health Laboratory Services | Cassandra Campion, Jamie Yeadon, Brian Pope, Lixia Liu, Kyle Brownlee, Melissa Hindenlang, Mark Glazier |
| EPI_ISL_2242532 | Aegis Sciences Corporation | Centers for Disease Control and Prevention Division of Viral Diseases, Pathogen Discovery | Dakota Howard, Dhwani Batra, Peter W. Cook, Kara Moser, Adrian Paskey, Jason Caravas, Benjamin Rambo-Martin, Shatavia Morrison, Christopher Gulvick, Scott Sammons, Yvette Unoarumhi, Darlene Wagner, Matthew Schmerer, Cyndi Clark, Patrick Campbell, Rob Case, Vikramsinha Ghorpade, Holly Houdeshell, Ola Kvalvaag, Dillon Nall, Ethan Sanders, Alec Vest, Shaun Westlund, Matthew Hardison, Clinton R. Paden, Duncan MacCannell |
| EPI_ISL_2242533 | IN State Department of Health Laboratory Services | IN State Department of Health Laboratory Services | Cassandra Campion, Jamie Yeadon, Brian Pope, Lixia Liu, Kyle Brownlee, Melissa Hindenlang, Mark Glazier |
| EPI_ISL_2242534, EPI_ISL_2242535, EPI_ISL_2242537 | Aegis Sciences Corporation | Centers for Disease Control and Prevention Division of Viral Diseases, Pathogen Discovery | Dakota Howard, Dhwani Batra, Peter W. Cook, Kara Moser, Adrian Paskey, Jason Caravas, Benjamin Rambo-Martin, Shatavia Morrison, Christopher Gulvick, Scott Sammons, Yvette Unoarumhi, Darlene Wagner, Matthew Schmerer, Cyndi Clark, Patrick Campbell, Rob Case, Vikramsinha Ghorpade, Holly Houdeshell, Ola Kvalvaag, Dillon Nall, Ethan Sanders, Alec Vest, Shaun Westlund, Matthew Hardison, Clinton R. Paden, Duncan MacCannell |
| EPI_ISL_2242538 | IN State Department of Health Laboratory Services | IN State Department of Health Laboratory Services | Cassandra Campion, Jamie Yeadon, Brian Pope, Lixia Liu, Kyle Brownlee, Melissa Hindenlang, Mark Glazier |
| EPI_ISL_2242539 | Aegis Sciences Corporation | Centers for Disease Control and Prevention Division of Viral Diseases, Pathogen Discovery | Dakota Howard, Dhwani Batra, Peter W. Cook, Kara Moser, Adrian Paskey, Jason Caravas, Benjamin Rambo-Martin, Shatavia Morrison, Christopher Gulvick, Scott Sammons, Yvette Unoarumhi, Darlene Wagner, Matthew Schmerer, Cyndi Clark, Patrick Campbell, Rob Case, Vikramsinha Ghorpade, Holly Houdeshell, Ola Kvalvaag, Dillon Nall, Ethan Sanders, Alec Vest, Shaun Westlund, Matthew Hardison, Clinton R. Paden, Duncan MacCannell |
| EPI_ISL_2242540 | IN State Department of Health Laboratory Services | IN State Department of Health Laboratory Services | Cassandra Campion, Jamie Yeadon, Brian Pope, Lixia Liu, Kyle Brownlee, Melissa Hindenlang, Mark Glazier |
| EPI_ISL_2242541 | Aegis Sciences Corporation | Centers for Disease Control and Prevention Division of Viral Diseases, Pathogen Discovery | Dakota Howard, Dhwani Batra, Peter W. Cook, Kara Moser, Adrian Paskey, Jason Caravas, Benjamin Rambo-Martin, Shatavia Morrison, Christopher Gulvick, Scott Sammons, Yvette Unoarumhi, Darlene Wagner, Matthew Schmerer, Cyndi Clark, Patrick Campbell, Rob Case, Vikramsinha Ghorpade, Holly Houdeshell, Ola Kvalvaag, Dillon Nall, Ethan Sanders, Alec Vest, Shaun Westlund, Matthew Hardison, Clinton R. Paden, Duncan MacCannell |
| EPI_ISL_2242542 | IN State Department of Health Laboratory Services | IN State Department of Health Laboratory Services | Cassandra Campion, Jamie Yeadon, Brian Pope, Lixia Liu, Kyle Brownlee, Melissa Hindenlang, Mark Glazier |
| EPI_ISL_2242543 | Aegis Sciences Corporation | Centers for Disease Control and Prevention Division of Viral Diseases, Pathogen Discovery | Dakota Howard, Dhwani Batra, Peter W. Cook, Kara Moser, Adrian Paskey, Jason Caravas, Benjamin Rambo-Martin, Shatavia Morrison, Christopher Gulvick, Scott Sammons, Yvette Unoarumhi, Darlene Wagner, Matthew Schmerer, Cyndi Clark, Patrick Campbell, Rob Case, Vikramsinha Ghorpade, Holly Houdeshell, Ola Kvalvaag, Dillon Nall, Ethan Sanders, Alec Vest, Shaun Westlund, Matthew Hardison, Clinton R. Paden, Duncan MacCannell |
| EPI_ISL_2242544 | IN State Department of Health Laboratory Services | IN State Department of Health Laboratory Services | Cassandra Campion, Jamie Yeadon, Brian Pope, Lixia Liu, Kyle Brownlee, Melissa Hindenlang, Mark Glazier |
| EPI_ISL_2242545 | Aegis Sciences Corporation | Centers for Disease Control and Prevention Division of Viral Diseases, Pathogen Discovery | Dakota Howard, Dhwani Batra, Peter W. Cook, Kara Moser, Adrian Paskey, Jason Caravas, Benjamin Rambo-Martin, Shatavia Morrison, Christopher Gulvick, Scott Sammons, Yvette Unoarumhi, Darlene Wagner, Matthew Schmerer, Cyndi Clark, Patrick Campbell, Rob Case, Vikramsinha Ghorpade, Holly Houdeshell, Ola Kvalvaag, Dillon Nall, Ethan Sanders, Alec Vest, Shaun Westlund, Matthew Hardison, Clinton R. Paden, Duncan MacCannell |
| EPI_ISL_2242546, EPI_ISL_2242547 | IN State Department of Health Laboratory Services | IN State Department of Health Laboratory Services | Cassandra Campion, Jamie Yeadon, Brian Pope, Lixia Liu, Kyle Brownlee, Melissa Hindenlang, Mark Glazier |
| EPI_ISL_2242548, EPI_ISL_2242549, EPI_ISL_2242550 | Aegis Sciences Corporation | Centers for Disease Control and Prevention Division of Viral Diseases, Pathogen Discovery | Dakota Howard, Dhwani Batra, Peter W. Cook, Kara Moser, Adrian Paskey, Jason Caravas, Benjamin Rambo-Martin, Shatavia Morrison, Christopher Gulvick, Scott Sammons, Yvette Unoarumhi, Darlene Wagner, Matthew Schmerer, Cyndi Clark, Patrick Campbell, Rob Case, Vikramsinha Ghorpade, Holly Houdeshell, Ola Kvalvaag, Dillon Nall, Ethan Sanders, Alec Vest, Shaun Westlund, Matthew Hardison, Clinton R. Paden, Duncan MacCannell |
| EPI_ISL_2242551 | IN State Department of Health Laboratory Services | IN State Department of Health Laboratory Services | Cassandra Campion, Jamie Yeadon, Brian Pope, Lixia Liu, Kyle Brownlee, Melissa Hindenlang, Mark Glazier |
| EPI_ISL_2242552, EPI_ISL_2242553 | Aegis Sciences Corporation | Centers for Disease Control and Prevention Division of Viral Diseases, Pathogen Discovery | Dakota Howard, Dhwani Batra, Peter W. Cook, Kara Moser, Adrian Paskey, Jason Caravas, Benjamin Rambo-Martin, Shatavia Morrison, Christopher Gulvick, Scott Sammons, Yvette Unoarumhi, Darlene Wagner, Matthew Schmerer, Cyndi Clark, Patrick Campbell, Rob Case, Vikramsinha Ghorpade, Holly Houdeshell, Ola Kvalvaag, Dillon Nall, Ethan Sanders, Alec Vest, Shaun Westlund, Matthew Hardison, Clinton R. Paden, Duncan MacCannell |
| EPI_ISL_2242554 | IN State Department of Health Laboratory Services | IN State Department of Health Laboratory Services | Cassandra Campion, Jamie Yeadon, Brian Pope, Lixia Liu, Kyle Brownlee, Melissa Hindenlang, Mark Glazier |
| EPI_ISL_2242555 | Aegis Sciences Corporation | Centers for Disease Control and Prevention Division of Viral Diseases, Pathogen Discovery | Dakota Howard, Dhwani Batra, Peter W. Cook, Kara Moser, Adrian Paskey, Jason Caravas, Benjamin Rambo-Martin, Shatavia Morrison, Christopher Gulvick, Scott Sammons, Yvette Unoarumhi, Darlene Wagner, Matthew Schmerer, Cyndi Clark, Patrick Campbell, Rob Case, Vikramsinha Ghorpade, Holly Houdeshell, Ola Kvalvaag, Dillon Nall, Ethan Sanders, Alec Vest, Shaun Westlund, Matthew Hardison, Clinton R. Paden, Duncan MacCannell |
| EPI_ISL_2242556 | IN State Department of Health Laboratory Services | IN State Department of Health Laboratory Services | Cassandra Campion, Jamie Yeadon, Brian Pope, Lixia Liu, Kyle Brownlee, Melissa Hindenlang, Mark Glazier |
| EPI_ISL_2242557 | Aegis Sciences Corporation | Centers for Disease Control and Prevention Division of Viral Diseases, Pathogen Discovery | Dakota Howard, Dhwani Batra, Peter W. Cook, Kara Moser, Adrian Paskey, Jason Caravas, Benjamin Rambo-Martin, Shatavia Morrison, Christopher Gulvick, Scott Sammons, Yvette Unoarumhi, Darlene Wagner, Matthew Schmerer, Cyndi Clark, Patrick Campbell, Rob Case, Vikramsinha Ghorpade, Holly Houdeshell, Ola Kvalvaag, Dillon Nall, Ethan Sanders, Alec Vest, Shaun Westlund, Matthew Hardison, Clinton R. Paden, Duncan MacCannell |
| EPI_ISL_2242558, EPI_ISL_2242559 | IN State Department of Health Laboratory Services | IN State Department of Health Laboratory Services | Cassandra Campion, Jamie Yeadon, Brian Pope, Lixia Liu, Kyle Brownlee, Melissa Hindenlang, Mark Glazier |
| EPI_ISL_2242560 | Aegis Sciences Corporation | Centers for Disease Control and Prevention Division of Viral Diseases, Pathogen Discovery | Dakota Howard, Dhwani Batra, Peter W. Cook, Kara Moser, Adrian Paskey, Jason Caravas, Benjamin Rambo-Martin, Shatavia Morrison, Christopher Gulvick, Scott Sammons, Yvette Unoarumhi, Darlene Wagner, Matthew Schmerer, Cyndi Clark, Patrick Campbell, Rob Case, Vikramsinha Ghorpade, Holly Houdeshell, Ola Kvalvaag, Dillon Nall, Ethan Sanders, Alec Vest, Shaun Westlund, Matthew Hardison, Clinton R. Paden, Duncan MacCannell |
| EPI_ISL_2242561 | IN State Department of Health Laboratory Services | IN State Department of Health Laboratory Services | Cassandra Campion, Jamie Yeadon, Brian Pope, Lixia Liu, Kyle Brownlee, Melissa Hindenlang, Mark Glazier |
| EPI_ISL_2242562 | Aegis Sciences Corporation | Centers for Disease Control and Prevention Division of Viral Diseases, Pathogen Discovery | Dakota Howard, Dhwani Batra, Peter W. Cook, Kara Moser, Adrian Paskey, Jason Caravas, Benjamin Rambo-Martin, Shatavia Morrison, Christopher Gulvick, Scott Sammons, Yvette Unoarumhi, Darlene Wagner, Matthew Schmerer, Cyndi Clark, Patrick Campbell, Rob Case, Vikramsinha Ghorpade, Holly Houdeshell, Ola Kvalvaag, Dillon Nall, Ethan Sanders, Alec Vest, Shaun Westlund, Matthew Hardison, Clinton R. Paden, Duncan MacCannell |
| EPI_ISL_2242563 | IN State Department of Health Laboratory Services | IN State Department of Health Laboratory Services | Cassandra Campion, Jamie Yeadon, Brian Pope, Lixia Liu, Kyle Brownlee, Melissa Hindenlang, Mark Glazier |
| EPI_ISL_2242564 | Aegis Sciences Corporation | Centers for Disease Control and Prevention Division of Viral Diseases, Pathogen Discovery | Dakota Howard, Dhwani Batra, Peter W. Cook, Kara Moser, Adrian Paskey, Jason Caravas, Benjamin Rambo-Martin, Shatavia Morrison, Christopher Gulvick, Scott Sammons, Yvette Unoarumhi, Darlene Wagner, Matthew Schmerer, Cyndi Clark, Patrick Campbell, Rob Case, Vikramsinha Ghorpade, Holly Houdeshell, Ola Kvalvaag, Dillon Nall, Ethan Sanders, Alec Vest, Shaun Westlund, Matthew Hardison, Clinton R. Paden, Duncan MacCannell |
| EPI_ISL_2242565 | IN State Department of Health Laboratory Services | IN State Department of Health Laboratory Services | Cassandra Campion, Jamie Yeadon, Brian Pope, Lixia Liu, Kyle Brownlee, Melissa Hindenlang, Mark Glazier |
| EPI_ISL_2242566, EPI_ISL_2242568, EPI_ISL_2242570, EPI_ISL_2242571 | Aegis Sciences Corporation | Centers for Disease Control and Prevention Division of Viral Diseases, Pathogen Discovery | Dakota Howard, Dhwani Batra, Peter W. Cook, Kara Moser, Adrian Paskey, Jason Caravas, Benjamin Rambo-Martin, Shatavia Morrison, Christopher Gulvick, Scott Sammons, Yvette Unoarumhi, Darlene Wagner, Matthew Schmerer, Cyndi Clark, Patrick Campbell, Rob Case, Vikramsinha Ghorpade, Holly Houdeshell, Ola Kvalvaag, Dillon Nall, Ethan Sanders, Alec Vest, Shaun Westlund, Matthew Hardison, Clinton R. Paden, Duncan MacCannell |
| EPI_ISL_2242572 | IN State Department of Health Laboratory Services | IN State Department of Health Laboratory Services | Cassandra Campion, Jamie Yeadon, Brian Pope, Lixia Liu, Kyle Brownlee, Melissa Hindenlang, Mark Glazier |
| EPI_ISL_2242573 | Aegis Sciences Corporation | Centers for Disease Control and Prevention Division of Viral Diseases, Pathogen Discovery | Dakota Howard, Dhwani Batra, Peter W. Cook, Kara Moser, Adrian Paskey, Jason Caravas, Benjamin Rambo-Martin, Shatavia Morrison, Christopher Gulvick, Scott Sammons, Yvette Unoarumhi, Darlene Wagner, Matthew Schmerer, Cyndi Clark, Patrick Campbell, Rob Case, Vikramsinha Ghorpade, Holly Houdeshell, Ola Kvalvaag, Dillon Nall, Ethan Sanders, Alec Vest, Shaun Westlund, Matthew Hardison, Clinton R. Paden, Duncan MacCannell |
| EPI_ISL_2242574 | IN State Department of Health Laboratory Services | IN State Department of Health Laboratory Services | Cassandra Campion, Jamie Yeadon, Brian Pope, Lixia Liu, Kyle Brownlee, Melissa Hindenlang, Mark Glazier |
| EPI_ISL_2242575 | Aegis Sciences Corporation | Centers for Disease Control and Prevention Division of Viral Diseases, Pathogen Discovery | Dakota Howard, Dhwani Batra, Peter W. Cook, Kara Moser, Adrian Paskey, Jason Caravas, Benjamin Rambo-Martin, Shatavia Morrison, Christopher Gulvick, Scott Sammons, Yvette Unoarumhi, Darlene Wagner, Matthew Schmerer, Cyndi Clark, Patrick Campbell, Rob Case, Vikramsinha Ghorpade, Holly Houdeshell, Ola Kvalvaag, Dillon Nall, Ethan Sanders, Alec Vest, Shaun Westlund, Matthew Hardison, Clinton R. Paden, Duncan MacCannell |
| EPI_ISL_2242576, EPI_ISL_2242577, | IN State Department of Health Laboratory Services | IN State Department of Health Laboratory Services | Cassandra Campion, Jamie Yeadon, Brian Pope, Lixia Liu, Kyle Brownlee, Melissa Hindenlang, Mark Glazier |

| | | | |
|---|---|---|---|
| EPI_ISL_2242579 | | | |
| EPI_ISL_2242580, EPI_ISL_2242582 | Aegis Sciences Corporation | Centers for Disease Control and Prevention Division of Viral Diseases, Pathogen Discovery | Dakota Howard, Dhwani Batra, Peter W. Cook, Kara Moser, Adrian Paskey, Jason Caravas, Benjamin Rambo-Martin, Shatavia Morrison, Christopher Gulvick, Scott Sammons, Yvette Unoarumhi, Darlene Wagner, Matthew Schmerer, Cyndi Clark, Patrick Campbell, Rob Case, Vikramsinha Ghorpade, Holly Houdeshell, Ola Kvalvaag, Dillon Nall, Ethan Sanders, Alec Vest, Shaun Westlund, Matthew Hardison, Clinton R. Paden, Duncan MacCannell |
| EPI_ISL_2242583 | IN State Department of Health Laboratory Services | IN State Department of Health Laboratory Services | Cassandra Campion, Jamie Yeadon, Brian Pope, Lixia Liu, Kyle Brownlee, Melissa Hindenlang, Mark Glazier |
| EPI_ISL_2242584, EPI_ISL_2242586 | Aegis Sciences Corporation | Centers for Disease Control and Prevention Division of Viral Diseases, Pathogen Discovery | Dakota Howard, Dhwani Batra, Peter W. Cook, Kara Moser, Adrian Paskey, Jason Caravas, Benjamin Rambo-Martin, Shatavia Morrison, Christopher Gulvick, Scott Sammons, Yvette Unoarumhi, Darlene Wagner, Matthew Schmerer, Cyndi Clark, Patrick Campbell, Rob Case, Vikramsinha Ghorpade, Holly Houdeshell, Ola Kvalvaag, Dillon Nall, Ethan Sanders, Alec Vest, Shaun Westlund, Matthew Hardison, Clinton R. Paden, Duncan MacCannell |
| EPI_ISL_2242587 | IN State Department of Health Laboratory Services | IN State Department of Health Laboratory Services | Cassandra Campion, Jamie Yeadon, Brian Pope, Lixia Liu, Kyle Brownlee, Melissa Hindenlang, Mark Glazier |
| EPI_ISL_2242588 | Aegis Sciences Corporation | Centers for Disease Control and Prevention Division of Viral Diseases, Pathogen Discovery | Dakota Howard, Dhwani Batra, Peter W. Cook, Kara Moser, Adrian Paskey, Jason Caravas, Benjamin Rambo-Martin, Shatavia Morrison, Christopher Gulvick, Scott Sammons, Yvette Unoarumhi, Darlene Wagner, Matthew Schmerer, Cyndi Clark, Patrick Campbell, Rob Case, Vikramsinha Ghorpade, Holly Houdeshell, Ola Kvalvaag, Dillon Nall, Ethan Sanders, Alec Vest, Shaun Westlund, Matthew Hardison, Clinton R. Paden, Duncan MacCannell |
| EPI_ISL_2242589 | IN State Department of Health Laboratory Services | IN State Department of Health Laboratory Services | Cassandra Campion, Jamie Yeadon, Brian Pope, Lixia Liu, Kyle Brownlee, Melissa Hindenlang, Mark Glazier |
| EPI_ISL_2242590 | Aegis Sciences Corporation | Centers for Disease Control and Prevention Division of Viral Diseases, Pathogen Discovery | Dakota Howard, Dhwani Batra, Peter W. Cook, Kara Moser, Adrian Paskey, Jason Caravas, Benjamin Rambo-Martin, Shatavia Morrison, Christopher Gulvick, Scott Sammons, Yvette Unoarumhi, Darlene Wagner, Matthew Schmerer, Cyndi Clark, Patrick Campbell, Rob Case, Vikramsinha Ghorpade, Holly Houdeshell, Ola Kvalvaag, Dillon Nall, Ethan Sanders, Alec Vest, Shaun Westlund, Matthew Hardison, Clinton R. Paden, Duncan MacCannell |
| EPI_ISL_2242591 | IN State Department of Health Laboratory Services | IN State Department of Health Laboratory Services | Cassandra Campion, Jamie Yeadon, Brian Pope, Lixia Liu, Kyle Brownlee, Melissa Hindenlang, Mark Glazier |
| EPI_ISL_2242592, EPI_ISL_2242593, EPI_ISL_2242595 | Aegis Sciences Corporation | Centers for Disease Control and Prevention Division of Viral Diseases, Pathogen Discovery | Dakota Howard, Dhwani Batra, Peter W. Cook, Kara Moser, Adrian Paskey, Jason Caravas, Benjamin Rambo-Martin, Shatavia Morrison, Christopher Gulvick, Scott Sammons, Yvette Unoarumhi, Darlene Wagner, Matthew Schmerer, Cyndi Clark, Patrick Campbell, Rob Case, Vikramsinha Ghorpade, Holly Houdeshell, Ola Kvalvaag, Dillon Nall, Ethan Sanders, Alec Vest, Shaun Westlund, Matthew Hardison, Clinton R. Paden, Duncan MacCannell |
| EPI_ISL_2242596 | IN State Department of Health Laboratory Services | IN State Department of Health Laboratory Services | Cassandra Campion, Jamie Yeadon, Brian Pope, Lixia Liu, Kyle Brownlee, Melissa Hindenlang, Mark Glazier |
| EPI_ISL_2242597 | Aegis Sciences Corporation | Centers for Disease Control and Prevention Division of Viral Diseases, Pathogen Discovery | Dakota Howard, Dhwani Batra, Peter W. Cook, Kara Moser, Adrian Paskey, Jason Caravas, Benjamin Rambo-Martin, Shatavia Morrison, Christopher Gulvick, Scott Sammons, Yvette Unoarumhi, Darlene Wagner, Matthew Schmerer, Cyndi Clark, Patrick Campbell, Rob Case, Vikramsinha Ghorpade, Holly Houdeshell, Ola Kvalvaag, Dillon Nall, Ethan Sanders, Alec Vest, Shaun Westlund, Matthew Hardison, Clinton R. Paden, Duncan MacCannell |
| EPI_ISL_2242598 | IN State Department of Health Laboratory Services | IN State Department of Health Laboratory Services | Cassandra Campion, Jamie Yeadon, Brian Pope, Lixia Liu, Kyle Brownlee, Melissa Hindenlang, Mark Glazier |
| EPI_ISL_2242599 | Aegis Sciences Corporation | Centers for Disease Control and Prevention Division of Viral Diseases, Pathogen Discovery | Dakota Howard, Dhwani Batra, Peter W. Cook, Kara Moser, Adrian Paskey, Jason Caravas, Benjamin Rambo-Martin, Shatavia Morrison, Christopher Gulvick, Scott Sammons, Yvette Unoarumhi, Darlene Wagner, Matthew Schmerer, Cyndi Clark, Patrick Campbell, Rob Case, Vikramsinha Ghorpade, Holly Houdeshell, Ola Kvalvaag, Dillon Nall, Ethan Sanders, Alec Vest, Shaun Westlund, Matthew Hardison, Clinton R. Paden, Duncan MacCannell |
| EPI_ISL_2242600 | IN State Department of Health Laboratory Services | IN State Department of Health Laboratory Services | Cassandra Campion, Jamie Yeadon, Brian Pope, Lixia Liu, Kyle Brownlee, Melissa Hindenlang, Mark Glazier |

EPI_ISL_2242601, EPI_ISL_2242603, EPI_ISL_2242604, EPI_ISL_2242605, EPI_ISL_2242607, EPI_ISL_2242608, EPI_ISL_2242609, EPI_ISL_2242610, EPI_ISL_2242611, EPI_ISL_2242612, EPI_ISL_2242613, EPI_ISL_2242614, EPI_ISL_2242615, EPI_ISL_2242616, EPI_ISL_2242617, EPI_ISL_2242618, EPI_ISL_2242619, EPI_ISL_2242620, EPI_ISL_2242621, EPI_ISL_2242622, EPI_ISL_2242623, EPI_ISL_2242624,
EPI_ISL_2242625, EPI_ISL_2242626, EPI_ISL_2242627, EPI_ISL_2242628, EPI_ISL_2242629, EPI_ISL_2242630, EPI_ISL_2242631, EPI_ISL_2242632, EPI_ISL_2242633, EPI_ISL_2242634, EPI_ISL_2242635, EPI_ISL_2242637, EPI_ISL_2242638, EPI_ISL_2242639, EPI_ISL_2242640, EPI_ISL_2242641, EPI_ISL_2242642, EPI_ISL_2242643, EPI_ISL_2242644, EPI_ISL_2242645, EPI_ISL_2242646, EPI_ISL_2242647,
EPI_ISL_2242648, EPI_ISL_2242649, EPI_ISL_2242650, EPI_ISL_2242651, EPI_ISL_2242652, EPI_ISL_2242653, EPI_ISL_2242654, EPI_ISL_2242655, EPI_ISL_2242656, EPI_ISL_2242657, EPI_ISL_2242658, EPI_ISL_2242660, EPI_ISL_2242661, EPI_ISL_2242662, EPI_ISL_2242663, EPI_ISL_2242664, EPI_ISL_2242665, EPI_ISL_2242666, EPI_ISL_2242667, EPI_ISL_2242668, EPI_ISL_2242669, EPI_ISL_2242670,
EPI_ISL_2242671, EPI_ISL_2242672, EPI_ISL_2242673, EPI_ISL_2242674, EPI_ISL_2242675, EPI_ISL_2242676, EPI_ISL_2242677, EPI_ISL_2242678, EPI_ISL_2242679, EPI_ISL_2242680, EPI_ISL_2242681, EPI_ISL_2242682, EPI_ISL_2242683, EPI_ISL_2242684, EPI_ISL_2242685, EPI_ISL_2242686, EPI_ISL_2242687, EPI_ISL_2242688, EPI_ISL_2242689, EPI_ISL_2242690, EPI_ISL_2242691, EPI_ISL_2242692,
EPI_ISL_2242694, EPI_ISL_2242695, EPI_ISL_2242696, EPI_ISL_2242697, EPI_ISL_2242698, EPI_ISL_2242699, EPI_ISL_2242700, EPI_ISL_2242701, EPI_ISL_2242703, EPI_ISL_2242704, EPI_ISL_2242705, EPI_ISL_2242706, EPI_ISL_2242707, EPI_ISL_2242708, EPI_ISL_2242709, EPI_ISL_2242710, EPI_ISL_2242711, EPI_ISL_2242712, EPI_ISL_2242713, EPI_ISL_2242714, EPI_ISL_2242716, EPI_ISL_2242717,
EPI_ISL_2242718, EPI_ISL_2242719, EPI_ISL_2242720, EPI_ISL_2242721, EPI_ISL_2242722, EPI_ISL_2242724, EPI_ISL_2242725, EPI_ISL_2242726, EPI_ISL_2242727, EPI_ISL_2242728, EPI_ISL_2242729, EPI_ISL_2242730, EPI_ISL_2242732, EPI_ISL_2242733, EPI_ISL_2242734, EPI_ISL_2242735, EPI_ISL_2242736, EPI_ISL_2242737, EPI_ISL_2242738, EPI_ISL_2242739, EPI_ISL_2242740, EPI_ISL_2242743,
EPI_ISL_2242744, EPI_ISL_2242745, EPI_ISL_2242746, EPI_ISL_2242747, EPI_ISL_2242748, EPI_ISL_2242749, EPI_ISL_2242750, EPI_ISL_2242751, EPI_ISL_2242752, EPI_ISL_2242753, EPI_ISL_2242754, EPI_ISL_2242755, EPI_ISL_2242756, EPI_ISL_2242757, EPI_ISL_2242758, EPI_ISL_2242759, EPI_ISL_2242760, EPI_ISL_2242761, EPI_ISL_2242762, EPI_ISL_2242763, EPI_ISL_2242764, EPI_ISL_2242765,
EPI_ISL_2242766, EPI_ISL_2242767, EPI_ISL_2242769, EPI_ISL_2242771, EPI_ISL_2242772, EPI_ISL_2242773, EPI_ISL_2242774, EPI_ISL_2242775, EPI_ISL_2242776, EPI_ISL_2242777, EPI_ISL_2242778, EPI_ISL_2242779, EPI_ISL_2242780, EPI_ISL_2242781, EPI_ISL_2242782, EPI_ISL_2242783, EPI_ISL_2242784, EPI_ISL_2242785, EPI_ISL_2242786, EPI_ISL_2242788, EPI_ISL_2242789, EPI_ISL_2242790,
EPI_ISL_2242791, EPI_ISL_2242792, EPI_ISL_2242793, EPI_ISL_2242794, EPI_ISL_2242795, EPI_ISL_2242797, EPI_ISL_2242798, EPI_ISL_2242799, EPI_ISL_2242800, EPI_ISL_2242801, EPI_ISL_2242802, EPI_ISL_2242803, EPI_ISL_2242804, EPI_ISL_2242805, EPI_ISL_2242806, EPI_ISL_2242807, EPI_ISL_2242808, EPI_ISL_2242810, EPI_ISL_2242811, EPI_ISL_2242812, EPI_ISL_2242813, EPI_ISL_2242814,
EPI_ISL_2242815, EPI_ISL_2242816, EPI_ISL_2242817, EPI_ISL_2242818, EPI_ISL_2242819, EPI_ISL_2242820, EPI_ISL_2242821, EPI_ISL_2242822, EPI_ISL_2242823, EPI_ISL_2242824, EPI_ISL_2242825, EPI_ISL_2242826, EPI_ISL_2242827, EPI_ISL_2242828, EPI_ISL_2242829, EPI_ISL_2242830, EPI_ISL_2242831, EPI_ISL_2242832, EPI_ISL_2242833, EPI_ISL_2242834, EPI_ISL_2242835, EPI_ISL_2242836,
EPI_ISL_2242837, EPI_ISL_2242838, EPI_ISL_2242839, EPI_ISL_2242840, EPI_ISL_2242841, EPI_ISL_2242842, EPI_ISL_2242843, EPI_ISL_2242844, EPI_ISL_2242845, EPI_ISL_2242846, EPI_ISL_2242847, EPI_ISL_2242848, EPI_ISL_2242849, EPI_ISL_2242850, EPI_ISL_2242851, EPI_ISL_2242852, EPI_ISL_2242853, EPI_ISL_2242854, EPI_ISL_2242855, EPI_ISL_2242856, EPI_ISL_2242857, EPI_ISL_2242858,
EPI_ISL_2242859, EPI_ISL_2242860, EPI_ISL_2242862, EPI_ISL_2242863, EPI_ISL_2242864, EPI_ISL_2242865, EPI_ISL_2242866, EPI_ISL_2242867, EPI_ISL_2242868, EPI_ISL_2242869, EPI_ISL_2242870, EPI_ISL_2242871, EPI_ISL_2242872, EPI_ISL_2242873, EPI_ISL_2242874, EPI_ISL_2242875, EPI_ISL_2242876, EPI_ISL_2242877, EPI_ISL_2242878, EPI_ISL_2242879, EPI_ISL_2242880, EPI_ISL_2242881,
EPI_ISL_2242882, EPI_ISL_2242883, EPI_ISL_2242884, EPI_ISL_2242885, EPI_ISL_2242886, EPI_ISL_2242887, EPI_ISL_2242888, EPI_ISL_2242889, EPI_ISL_2242890, EPI_ISL_2242891, EPI_ISL_2242892, EPI_ISL_2242893, EPI_ISL_2242894, EPI_ISL_2242895, EPI_ISL_2242896, EPI_ISL_2242897, EPI_ISL_2242898, EPI_ISL_2242899, EPI_ISL_2242900, EPI_ISL_2242901, EPI_ISL_2242902, EPI_ISL_2242903,
EPI_ISL_2242904, EPI_ISL_2242905, EPI_ISL_2242906, EPI_ISL_2242907, EPI_ISL_2242908, EPI_ISL_2242909, EPI_ISL_2242910, EPI_ISL_2242911, EPI_ISL_2242912, EPI_ISL_2242913, EPI_ISL_2242914, EPI_ISL_2242915, EPI_ISL_2242916, EPI_ISL_2242917, EPI_ISL_2242918, EPI_ISL_2242919, EPI_ISL_2242920, EPI_ISL_2242921, EPI_ISL_2242922, EPI_ISL_2242923, EPI_ISL_2242924, EPI_ISL_2242925,
EPI_ISL_2242926, EPI_ISL_2242927, EPI_ISL_2242928, EPI_ISL_2242929, EPI_ISL_2242930, EPI_ISL_2242931, EPI_ISL_2242932, EPI_ISL_2242933, EPI_ISL_2242934, EPI_ISL_2242935, EPI_ISL_2242936, EPI_ISL_2242937, EPI_ISL_2242938, EPI_ISL_2242939, EPI_ISL_2242940, EPI_ISL_2242941, EPI_ISL_2242942, EPI_ISL_2242943, EPI_ISL_2242944, EPI_ISL_2242945, EPI_ISL_2242946, EPI_ISL_2242947,
EPI_ISL_2242948, EPI_ISL_2242949, EPI_ISL_2242950, EPI_ISL_2242951, EPI_ISL_2242952, EPI_ISL_2242953, EPI_ISL_2242954, EPI_ISL_2242955, EPI_ISL_2242956, EPI_ISL_2242957, EPI_ISL_2242958, EPI_ISL_2242959, EPI_ISL_2242960, EPI_ISL_2242961, EPI_ISL_2242962, EPI_ISL_2242963, EPI_ISL_2242964, EPI_ISL_2242965, EPI_ISL_2242966, EPI_ISL_2242968, EPI_ISL_2242969, EPI_ISL_2242970,
EPI_ISL_2242971, EPI_ISL_2242972, EPI_ISL_2242973, EPI_ISL_2242974, EPI_ISL_2242975, EPI_ISL_2242976, EPI_ISL_2242977, EPI_ISL_2242978, EPI_ISL_2242979, EPI_ISL_2242980, EPI_ISL_2242981, EPI_ISL_2242982, EPI_ISL_2242983, EPI_ISL_2242984, EPI_ISL_2242985, EPI_ISL_2242986, EPI_ISL_2242987, EPI_ISL_2242988, EPI_ISL_2242989, EPI_ISL_2242990, EPI_ISL_2242991, EPI_ISL_2242992,
EPI_ISL_2242993, EPI_ISL_2242994, EPI_ISL_2242995, EPI_ISL_2242996, EPI_ISL_2242997, EPI_ISL_2242998, EPI_ISL_2242999, EPI_ISL_2243000, EPI_ISL_2243001, EPI_ISL_2243002, EPI_ISL_2243003, EPI_ISL_2243004, EPI_ISL_2243005, EPI_ISL_2243006, EPI_ISL_2243007, EPI_ISL_2243008, EPI_ISL_2243009, EPI_ISL_2243010, EPI_ISL_2243011, EPI_ISL_2243012, EPI_ISL_2243013, EPI_ISL_2243014,
EPI_ISL_2243015, EPI_ISL_2243016, EPI_ISL_2243017, EPI_ISL_2243018, EPI_ISL_2243019, EPI_ISL_2243021, EPI_ISL_2243022, EPI_ISL_2243023, EPI_ISL_2243024, EPI_ISL_2243025, EPI_ISL_2243026, EPI_ISL_2243027, EPI_ISL_2243028, EPI_ISL_2243029, EPI_ISL_2243030, EPI_ISL_2243031, EPI_ISL_2243032, EPI_ISL_2243033, EPI_ISL_2243034, EPI_ISL_2243035, EPI_ISL_2243036, EPI_ISL_2243037,
EPI_ISL_2243038, EPI_ISL_2243039, EPI_ISL_2243040, EPI_ISL_2243041, EPI_ISL_2243042, EPI_ISL_2243043, EPI_ISL_2243044, EPI_ISL_2243045, EPI_ISL_2243046, EPI_ISL_2243047, EPI_ISL_2243048, EPI_ISL_2243049, EPI_ISL_2243050, EPI_ISL_2243051, EPI_ISL_2243052, EPI_ISL_2243053, EPI_ISL_2243054, EPI_ISL_2243055, EPI_ISL_2243056, EPI_ISL_2243057, EPI_ISL_2243058, EPI_ISL_2243059,
EPI_ISL_2243060, EPI_ISL_2243061, EPI_ISL_2243062, EPI_ISL_2243063, EPI_ISL_2243064, EPI_ISL_2243065, EPI_ISL_2243066, EPI_ISL_2243067, EPI_ISL_2243068, EPI_ISL_2243069, EPI_ISL_2243070, EPI_ISL_2243071, EPI_ISL_2243072, EPI_ISL_2243073, EPI_ISL_2243074, EPI_ISL_2243075, EPI_ISL_2243076, EPI_ISL_2243077, EPI_ISL_2243078, EPI_ISL_2243079, EPI_ISL_2243080, EPI_ISL_2243081,
EPI_ISL_2243082, EPI_ISL_2243083, EPI_ISL_2243084, EPI_ISL_2243085, EPI_ISL_2243086, EPI_ISL_2243087, EPI_ISL_2243088, EPI_ISL_2243089, EPI_ISL_2243090, EPI_ISL_2243091, EPI_ISL_2243092, EPI_ISL_2243093, EPI_ISL_2243094, EPI_ISL_2243095, EPI_ISL_2243096, EPI_ISL_2243097, EPI_ISL_2243098, EPI_ISL_2243099, EPI_ISL_2243100, EPI_ISL_2243101, EPI_ISL_2243102, EPI_ISL_2243103,
EPI_ISL_2243104, EPI_ISL_2243105, EPI_ISL_2243106, EPI_ISL_2243107, EPI_ISL_2243108, EPI_ISL_2243109, EPI_ISL_2243110, EPI_ISL_2243111, EPI_ISL_2243112, EPI_ISL_2243113, EPI_ISL_2243114, EPI_ISL_2243115, EPI_ISL_2243116, EPI_ISL_2243117, EPI_ISL_2243118, EPI_ISL_2243119, EPI_ISL_2243120, EPI_ISL_2243121, EPI_ISL_2243122, EPI_ISL_2243123, EPI_ISL_2243124, EPI_ISL_2243125,
EPI_ISL_2243126, EPI_ISL_2243127, EPI_ISL_2243128, EPI_ISL_2243129, EPI_ISL_2243130, EPI_ISL_2243131, EPI_ISL_2243132, EPI_ISL_2243133, EPI_ISL_2243134, EPI_ISL_2243135, EPI_ISL_2243136, EPI_ISL_2243137, EPI_ISL_2243138, EPI_ISL_2243139, EPI_ISL_2243140, EPI_ISL_2243141, EPI_ISL_2243142, EPI_ISL_2243143, EPI_ISL_2243144, EPI_ISL_2243145, EPI_ISL_2243146, EPI_ISL_2243147,
EPI_ISL_2243148, EPI_ISL_2243149, EPI_ISL_2243150, EPI_ISL_2243151, EPI_ISL_2243152, EPI_ISL_2243153, EPI_ISL_2243154, EPI_ISL_2243155, EPI_ISL_2243156, EPI_ISL_2243157, EPI_ISL_2243158, EPI_ISL_2243159, EPI_ISL_2243160, EPI_ISL_2243161, EPI_ISL_2243162, EPI_ISL_2243163, EPI_ISL_2243164, EPI_ISL_2243165, EPI_ISL_2243166, EPI_ISL_2243167, EPI_ISL_2243168, EPI_ISL_2243169,
EPI_ISL_2243170, EPI_ISL_2243171, EPI_ISL_2243172, EPI_ISL_2243173, EPI_ISL_2243174, EPI_ISL_2243175, EPI_ISL_2243176, EPI_ISL_2243177, EPI_ISL_2243178, EPI_ISL_2243179, EPI_ISL_2243180, EPI_ISL_2243181, EPI_ISL_2243182, EPI_ISL_2243183, EPI_ISL_2243184, EPI_ISL_2243185, EPI_ISL_2243186, EPI_ISL_2243187, EPI_ISL_2243188, EPI_ISL_2243189, EPI_ISL_2243190, EPI_ISL_2243191,
EPI_ISL_2243192, EPI_ISL_2243193, EPI_ISL_2243194, EPI_ISL_2243195, EPI_ISL_2243196, EPI_ISL_2243197, EPI_ISL_2243198, EPI_ISL_2243199, EPI_ISL_2243200, EPI_ISL_2243201, EPI_ISL_2243202, EPI_ISL_2243203, EPI_ISL_2243204, EPI_ISL_2243205, EPI_ISL_2243206, EPI_ISL_2243207, EPI_ISL_2243208, EPI_ISL_2243209, EPI_ISL_2243210, EPI_ISL_2243211, EPI_ISL_2243212, EPI_ISL_2243213,
EPI_ISL_2243214, EPI_ISL_2243215, EPI_ISL_2243216, EPI_ISL_2243217, EPI_ISL_2243218, EPI_ISL_2243219, EPI_ISL_2243220, EPI_ISL_2243221, EPI_ISL_2243222, EPI_ISL_2243224, EPI_ISL_2243225, EPI_ISL_2243226, EPI_ISL_2243227, EPI_ISL_2243228, EPI_ISL_2243229, EPI_ISL_2243230, EPI_ISL_2243231, EPI_ISL_2243232, EPI_ISL_2243233, EPI_ISL_2243234, EPI_ISL_2243235, EPI_ISL_2243236,
EPI_ISL_2243237, EPI_ISL_2243238, EPI_ISL_2243239, EPI_ISL_2243240, EPI_ISL_2243241, EPI_ISL_2243242, EPI_ISL_2243243, EPI_ISL_2243244, EPI_ISL_2243245, EPI_ISL_2243246, EPI_ISL_2243247, EPI_ISL_2243248, EPI_ISL_2243249, EPI_ISL_2243250, EPI_ISL_2243251, EPI_ISL_2243252, EPI_ISL_2243253, EPI_ISL_2243254, EPI_ISL_2243255, EPI_ISL_2243256, EPI_ISL_2243257, EPI_ISL_2243258,
EPI_ISL_2243259, EPI_ISL_2243260, EPI_ISL_2243261, EPI_ISL_2243262, EPI_ISL_2243263, EPI_ISL_2243264, EPI_ISL_2243265, EPI_ISL_2243266, EPI_ISL_2243267, EPI_ISL_2243268, EPI_ISL_2243269, EPI_ISL_2243270, EPI_ISL_2243271, EPI_ISL_2243272, EPI_ISL_2243273, EPI_ISL_2243274, EPI_ISL_2243275, EPI_ISL_2243276, EPI_ISL_2243277, EPI_ISL_2243278, EPI_ISL_2243279, EPI_ISL_2243280,
EPI_ISL_2243281, EPI_ISL_2243282, EPI_ISL_2243283, EPI_ISL_2243284, EPI_ISL_2243285, EPI_ISL_2243286, EPI_ISL_2243287, EPI_ISL_2243288, EPI_ISL_2243289, EPI_ISL_2243290, EPI_ISL_2243291, EPI_ISL_2243292, EPI_ISL_2243293, EPI_ISL_2243294, EPI_ISL_2243295, EPI_ISL_2243296, EPI_ISL_2243297, EPI_ISL_2243298, EPI_ISL_2243299, EPI_ISL_2243300, EPI_ISL_2243301, EPI_ISL_2243302,
EPI_ISL_2243303, EPI_ISL_2243304, EPI_ISL_2243305, EPI_ISL_2243306, EPI_ISL_2243307, EPI_ISL_2243308, EPI_ISL_2243309, EPI_ISL_2243310, EPI_ISL_2243311, EPI_ISL_2243312, EPI_ISL_2243313, EPI_ISL_2243314, EPI_ISL_2243315, EPI_ISL_2243316, EPI_ISL_2243317, EPI_ISL_2243319, EPI_ISL_2243320, EPI_ISL_2243321, EPI_ISL_2243322, EPI_ISL_2243323, EPI_ISL_2243324, EPI_ISL_2243325,
EPI_ISL_2243326, EPI_ISL_2243327, EPI_ISL_2243328, EPI_ISL_2243329, EPI_ISL_2243330, EPI_ISL_2243332, EPI_ISL_2243333, EPI_ISL_2243334, EPI_ISL_2243335, EPI_ISL_2243336, EPI_ISL_2243337, EPI_ISL_2243338, EPI_ISL_2243339, EPI_ISL_2243340, EPI_ISL_2243341, EPI_ISL_2243342, EPI_ISL_2243343, EPI_ISL_2243344, EPI_ISL_2243345, EPI_ISL_2243346, EPI_ISL_2243347, EPI_ISL_2243348,
EPI_ISL_2243350, EPI_ISL_2243351, EPI_ISL_2243352, EPI_ISL_2243353, EPI_ISL_2243354, EPI_ISL_2243355, EPI_ISL_2243356, EPI_ISL_2243357, EPI_ISL_2243358, EPI_ISL_2243359, EPI_ISL_2243360, EPI_ISL_2243362, EPI_ISL_2243363, EPI_ISL_2243364, EPI_ISL_2243365, EPI_ISL_2243367, EPI_ISL_2243368, EPI_ISL_2243369, EPI_ISL_2243370, EPI_ISL_2243371, EPI_ISL_2243372, EPI_ISL_2243373,
EPI_ISL_2243374, EPI_ISL_2243375, EPI_ISL_2243376, EPI_ISL_2243377, EPI_ISL_2243378, EPI_ISL_2243379, EPI_ISL_2243380, EPI_ISL_2243381, EPI_ISL_2243382, EPI_ISL_2243383, EPI_ISL_2243384, EPI_ISL_2243385, EPI_ISL_2243386, EPI_ISL_2243387, EPI_ISL_2243388, EPI_ISL_2243389, EPI_ISL_2243390, EPI_ISL_2243391, EPI_ISL_2243392, EPI_ISL_2243393, EPI_ISL_2243394, EPI_ISL_2243395,
EPI_ISL_2243396, EPI_ISL_2243397, EPI_ISL_2243398, EPI_ISL_2243399, EPI_ISL_2243400, EPI_ISL_2243401, EPI_ISL_2243402, EPI_ISL_2243403, EPI_ISL_2243404, EPI_ISL_2243405, EPI_ISL_2243406, EPI_ISL_2243407, EPI_ISL_2243409, EPI_ISL_2243410, EPI_ISL_2243411, EPI_ISL_2243412, EPI_ISL_2243413, EPI_ISL_2243414, EPI_ISL_2243415, EPI_ISL_2243416, EPI_ISL_2243417, EPI_ISL_2243418,
EPI_ISL_2243419, EPI_ISL_2243420, EPI_ISL_2243421, EPI_ISL_2243422, EPI_ISL_2243423, EPI_ISL_2243424, EPI_ISL_2243425, EPI_ISL_2243426, EPI_ISL_2243427, EPI_ISL_2243428, EPI_ISL_2243429, EPI_ISL_2243430, EPI_ISL_2243431, EPI_ISL_2243432, EPI_ISL_2243433, EPI_ISL_2243434, EPI_ISL_2243435, EPI_ISL_2243437, EPI_ISL_2243438, EPI_ISL_2243439, EPI_ISL_2243440, EPI_ISL_2243441,
EPI_ISL_2243442, EPI_ISL_2243443, EPI_ISL_2243444, EPI_ISL_2243445, EPI_ISL_2243446, EPI_ISL_2243447, EPI_ISL_2243448, EPI_ISL_2243449, EPI_ISL_2243450, EPI_ISL_2243451, EPI_ISL_2243452, EPI_ISL_2243454, EPI_ISL_2243455, EPI_ISL_2243456, EPI_ISL_2243457, EPI_ISL_2243458, EPI_ISL_2243459, EPI_ISL_2243460, EPI_ISL_2243461, EPI_ISL_2243462, EPI_ISL_2243463, EPI_ISL_2243464,
EPI_ISL_2243465, EPI_ISL_2243466, EPI_ISL_2243467, EPI_ISL_2243468, EPI_ISL_2243469, EPI_ISL_2243470, EPI_ISL_2243471, EPI_ISL_2243472, EPI_ISL_2243473, EPI_ISL_2243474, EPI_ISL_2243475, EPI_ISL_2243476, EPI_ISL_2243477, EPI_ISL_2243478, EPI_ISL_2243479, EPI_ISL_2243480, EPI_ISL_2243481, EPI_ISL_2243482, EPI_ISL_2243483, EPI_ISL_2243484, EPI_ISL_2243485, EPI_ISL_2243486,
EPI_ISL_2243487, EPI_ISL_2243488, EPI_ISL_2243489, EPI_ISL_2243490, EPI_ISL_2243491, EPI_ISL_2243492, EPI_ISL_2243493, EPI_ISL_2243494, EPI_ISL_2243495, EPI_ISL_2243496, EPI_ISL_2243497, EPI_ISL_2243498, EPI_ISL_2243499, EPI_ISL_2243500, EPI_ISL_2243501, EPI_ISL_2243502, EPI_ISL_2243503, EPI_ISL_2243504, EPI_ISL_2243505, EPI_ISL_2243506, EPI_ISL_2243507, EPI_ISL_2243508,
EPI_ISL_2243510, EPI_ISL_2243512, EPI_ISL_2243513, EPI_ISL_2243514, EPI_ISL_2243515, EPI_ISL_2243516, EPI_ISL_2243517, EPI_ISL_2243518, EPI_ISL_2243519, EPI_ISL_2243520, EPI_ISL_2243521, EPI_ISL_2243522, EPI_ISL_2243523, EPI_ISL_2243524, EPI_ISL_2243525, EPI_ISL_2243526, EPI_ISL_2243527, EPI_ISL_2243528, EPI_ISL_2243529, EPI_ISL_2243530, EPI_ISL_2243531, EPI_ISL_2243532,
EPI_ISL_2243533, EPI_ISL_2243534, EPI_ISL_2243535, EPI_ISL_2243536, EPI_ISL_2243537, EPI_ISL_2243538, EPI_ISL_2243539, EPI_ISL_2243540, EPI_ISL_2243541, EPI_ISL_2243542, EPI_ISL_2243543, EPI_ISL_2243544, EPI_ISL_2243545, EPI_ISL_2243546, EPI_ISL_2243548, EPI_ISL_2243549, EPI_ISL_2243551, EPI_ISL_2243552, EPI_ISL_2243553, EPI_ISL_2243554, EPI_ISL_2243555, EPI_ISL_2243556,
EPI_ISL_2243557, EPI_ISL_2243558, EPI_ISL_2243559, EPI_ISL_2243560, EPI_ISL_2243561, EPI_ISL_2243562, EPI_ISL_2243564, EPI_ISL_2243565, EPI_ISL_2243566, EPI_ISL_2243567, EPI_ISL_2243568, EPI_ISL_2243569, EPI_ISL_2243570, EPI_ISL_2243571, EPI_ISL_2243572, EPI_ISL_2243573, EPI_ISL_2243574, EPI_ISL_2243575, EPI_ISL_2243576, EPI_ISL_2243577, EPI_ISL_2243578, EPI_ISL_2243579,
EPI_ISL_2243580, EPI_ISL_2243581, EPI_ISL_2243582, EPI_ISL_2243583, EPI_ISL_2243584, EPI_ISL_2243585, EPI_ISL_2243586, EPI_ISL_2243587, EPI_ISL_2243588, EPI_ISL_2243589, EPI_ISL_2243590, EPI_ISL_2243591, EPI_ISL_2243592, EPI_ISL_2243593, EPI_ISL_2243594, EPI_ISL_2243595, EPI_ISL_2243596, EPI_ISL_2243598, EPI_ISL_2243599, EPI_ISL_2243600, EPI_ISL_2243601, EPI_ISL_2243602,
EPI_ISL_2243603, EPI_ISL_2243604, EPI_ISL_2243605, EPI_ISL_2243606, EPI_ISL_2243607, EPI_ISL_2243608, EPI_ISL_2243609, EPI_ISL_2243610, EPI_ISL_2243611, EPI_ISL_2243612, EPI_ISL_2243613, EPI_ISL_2243614, EPI_ISL_2243615, EPI_ISL_2243616, EPI_ISL_2243617, EPI_ISL_2243618, EPI_ISL_2243619, EPI_ISL_2243620, EPI_ISL_2243621, EPI_ISL_2243623, EPI_ISL_2243624, EPI_ISL_2243625,
EPI_ISL_2243626, EPI_ISL_2243627, EPI_ISL_2243628, EPI_ISL_2243629, EPI_ISL_2243630, EPI_ISL_2243631, EPI_ISL_2243632, EPI_ISL_2243633, EPI_ISL_2243634, EPI_ISL_2243635, EPI_ISL_2243636, EPI_ISL_2243637, EPI_ISL_2243638, EPI_ISL_2243639, EPI_ISL_2243640, EPI_ISL_2243641, EPI_ISL_2243642, EPI_ISL_2243644, EPI_ISL_2243645, EPI_ISL_2243646, EPI_ISL_2243647, EPI_ISL_2243648,
EPI_ISL_2243649, EPI_ISL_2243650, EPI_ISL_2243651, EPI_ISL_2243652, EPI_ISL_2243653, EPI_ISL_2243654, EPI_ISL_2243655, EPI_ISL_2243656, EPI_ISL_2243657, EPI_ISL_2243658, EPI_ISL_2243659, EPI_ISL_2243660, EPI_ISL_2243661, EPI_ISL_2243662, EPI_ISL_2243663, EPI_ISL_2243664, EPI_ISL_2243665, EPI_ISL_2243666, EPI_ISL_2243667, EPI_ISL_2243668, EPI_ISL_2243669, EPI_ISL_2243670,
EPI_ISL_2243671, EPI_ISL_2243672, EPI_ISL_2243673, EPI_ISL_2243674, EPI_ISL_2243675, EPI_ISL_2243676, EPI_ISL_2243677, EPI_ISL_2243678, EPI_ISL_2243679, EPI_ISL_2243680, EPI_ISL_2243681, EPI_ISL_2243682, EPI_ISL_2243683, EPI_ISL_2243684, EPI_ISL_2243685, EPI_ISL_2243686, EPI_ISL_2243687, EPI_ISL_2243688, EPI_ISL_2243689, EPI_ISL_2243690, EPI_ISL_2243691, EPI_ISL_2243692,
EPI_ISL_2243693, EPI_ISL_2243694, EPI_ISL_2243695, EPI_ISL_2243696, EPI_ISL_2243697, EPI_ISL_2243698, EPI_ISL_2243699, EPI_ISL_2243701, EPI_ISL_2243702, EPI_ISL_2243703, EPI_ISL_2243704, EPI_ISL_2243705, EPI_ISL_2243706, EPI_ISL_2243707, EPI_ISL_2243708, EPI_ISL_2243709, EPI_ISL_2243710, EPI_ISL_2243711, EPI_ISL_2243712, EPI_ISL_2243713, EPI_ISL_2243714, EPI_ISL_2243715,
EPI_ISL_2243716, EPI_ISL_2243717, EPI_ISL_2243718, EPI_ISL_2243719, EPI_ISL_2243720, EPI_ISL_2243721, EPI_ISL_2243722, EPI_ISL_2243723, EPI_ISL_2243724, EPI_ISL_2243725, EPI_ISL_2243726, EPI_ISL_2243727, EPI_ISL_2243728, EPI_ISL_2243729, EPI_ISL_2243730, EPI_ISL_2243731, EPI_ISL_2243732, EPI_ISL_2243733, EPI_ISL_2243734, EPI_ISL_2243735, EPI_ISL_2243736, EPI_ISL_2243737,
EPI_ISL_2243738, EPI_ISL_2243739, EPI_ISL_2243740, EPI_ISL_2243741, EPI_ISL_2243742, EPI_ISL_2243743, EPI_ISL_2243744, EPI_ISL_2243745, EPI_ISL_2243746, EPI_ISL_2243747, EPI_ISL_2243748, EPI_ISL_2243749, EPI_ISL_2243750, EPI_ISL_2243751, EPI_ISL_2243752, EPI_ISL_2243753, EPI_ISL_2243754, EPI_ISL_2243755, EPI_ISL_2243756, EPI_ISL_2243757, EPI_ISL_2243758, EPI_ISL_2243759,
EPI_ISL_2243760, EPI_ISL_2243761, EPI_ISL_2243762, EPI_ISL_2243763, EPI_ISL_2243764, EPI_ISL_2243765, EPI_ISL_2243767, EPI_ISL_2243768, EPI_ISL_2243769, EPI_ISL_2243770, EPI_ISL_2243771, EPI_ISL_2243773, EPI_ISL_2243774, EPI_ISL_2243775, EPI_ISL_2243776, EPI_ISL_2243777, EPI_ISL_2243778, EPI_ISL_2243779, EPI_ISL_2243780, EPI_ISL_2243781, EPI_ISL_2243783, EPI_ISL_2243784,
EPI_ISL_2243785, EPI_ISL_2243786, EPI_ISL_2243787, EPI_ISL_2243788, EPI_ISL_2243789, EPI_ISL_2243790, EPI_ISL_2243791, EPI_ISL_2243792, EPI_ISL_2243793, EPI_ISL_2243794, EPI_ISL_2243795, EPI_ISL_2243796, EPI_ISL_2243797, EPI_ISL_2243798, EPI_ISL_2243799, EPI_ISL_2243800, EPI_ISL_2243801, EPI_ISL_2243802, EPI_ISL_2243803, EPI_ISL_2243804, EPI_ISL_2243805, EPI_ISL_2243806,
EPI_ISL_2243807, EPI_ISL_2243808, EPI_ISL_2243809, EPI_ISL_2243810, EPI_ISL_2243811, EPI_ISL_2243812, EPI_ISL_2243813, EPI_ISL_2243815, EPI_ISL_2243816, EPI_ISL_2243817, EPI_ISL_2243818, EPI_ISL_2243819, EPI_ISL_2243820, EPI_ISL_2243821, EPI_ISL_2243822, EPI_ISL_2243823, EPI_ISL_2243824, EPI_ISL_2243825, EPI_ISL_2243826, EPI_ISL_2243827, EPI_ISL_2243828, EPI_ISL_2243829,
EPI_ISL_2243830, EPI_ISL_2243831, EPI_ISL_2243832, EPI_ISL_2243834, EPI_ISL_2243835, EPI_ISL_2243836, EPI_ISL_2243837, EPI_ISL_2243838, EPI_ISL_2243839, EPI_ISL_2243840, EPI_ISL_2243841, EPI_ISL_2243842, EPI_ISL_2243843, EPI_ISL_2243844, EPI_ISL_2243845, EPI_ISL_2243846, EPI_ISL_2243847, EPI_ISL_2243848, EPI_ISL_2243849, EPI_ISL_2243850, EPI_ISL_2243851, EPI_ISL_2243852,
EPI_ISL_2243853, EPI_ISL_2243854, EPI_ISL_2243855, EPI_ISL_2243856, EPI_ISL_2243857, EPI_ISL_2243858, EPI_ISL_2243859, EPI_ISL_2243860, EPI_ISL_2243861, EPI_ISL_2243862, EPI_ISL_2243863, EPI_ISL_2243864, EPI_ISL_2243865, EPI_ISL_2243866, EPI_ISL_2243867, EPI_ISL_2243868, EPI_ISL_2243869, EPI_ISL_2243870, EPI_ISL_2243871, EPI_ISL_2243872, EPI_ISL_2243873, EPI_ISL_2243874,
EPI_ISL_2243875, EPI_ISL_2243876, EPI_ISL_2243877, EPI_ISL_2243878, EPI_ISL_2243879, EPI_ISL_2243880, EPI_ISL_2243881, EPI_ISL_2243882, EPI_ISL_2243883, EPI_ISL_2243884, EPI_ISL_2243885, EPI_ISL_2243886, EPI_ISL_2243887, EPI_ISL_2243888, EPI_ISL_2243889, EPI_ISL_2243890, EPI_ISL_2243891, EPI_ISL_2243892, EPI_ISL_2243893, EPI_ISL_2243894, EPI_ISL_2243895, EPI_ISL_2243896,
EPI_ISL_2243897, EPI_ISL_2243898, EPI_ISL_2243899, EPI_ISL_2243900, EPI_ISL_2243901, EPI_ISL_2243902, EPI_ISL_2243903, EPI_ISL_2243904, EPI_ISL_2243905, EPI_ISL_2243906, EPI_ISL_2243907, EPI_ISL_2243908, EPI_ISL_2243909, EPI_ISL_2243910, EPI_ISL_2243911, EPI_ISL_2243912, EPI_ISL_2243913, EPI_ISL_2243914, EPI_ISL_2243915, EPI_ISL_2243916, EPI_ISL_2243917, EPI_ISL_2243918,
EPI_ISL_2243919, EPI_ISL_2243920, EPI_ISL_2243921, EPI_ISL_2243922, EPI_ISL_2243923, EPI_ISL_2243924, EPI_ISL_2243925, EPI_ISL_2243926, EPI_ISL_2243927, EPI_ISL_2243928, EPI_ISL_2243929, EPI_ISL_2243930, EPI_ISL_2243931, EPI_ISL_2243933, EPI_ISL_2243934, EPI_ISL_2243935, EPI_ISL_2243936, EPI_ISL_2243937, EPI_ISL_2243938, EPI_ISL_2243939, EPI_ISL_2243940, EPI_ISL_2243941,
EPI_ISL_2243942, EPI_ISL_2243943, EPI_ISL_2243944, EPI_ISL_2243945, EPI_ISL_2243946, EPI_ISL_2243947, EPI_ISL_2243948, EPI_ISL_2243949, EPI_ISL_2243950, EPI_ISL_2243951, EPI_ISL_2243952, EPI_ISL_2243953, EPI_ISL_2243954, EPI_ISL_2243955, EPI_ISL_2243956, EPI_ISL_2243957, EPI_ISL_2243958, EPI_ISL_2243959, EPI_ISL_2243961, EPI_ISL_2243962, EPI_ISL_2243963, EPI_ISL_2243964,
EPI_ISL_2243965, EPI_ISL_2243966, EPI_ISL_2243967, EPI_ISL_2243968, EPI_ISL_2243969, EPI_ISL_2243970, EPI_ISL_2243971, EPI_ISL_2243972, EPI_ISL_2243973, EPI_ISL_2243974, EPI_ISL_2243975, EPI_ISL_2243976, EPI_ISL_2243977, EPI_ISL_2243978, EPI_ISL_2243980, EPI_ISL_2243981, EPI_ISL_2243982, EPI_ISL_2243983, EPI_ISL_2243984, EPI_ISL_2243985, EPI_ISL_2243986, EPI_ISL_2243987,
EPI_ISL_2243988, EPI_ISL_2243989, EPI_ISL_2243990, EPI_ISL_2243991, EPI_ISL_2243992, EPI_ISL_2243993, EPI_ISL_2243994, EPI_ISL_2243995, EPI_ISL_2243996, EPI_ISL_2243997, EPI_ISL_2243998, EPI_ISL_2243999, EPI_ISL_2244000, EPI_ISL_2244001, EPI_ISL_2244002, EPI_ISL_2244003, EPI_ISL_2244004, EPI_ISL_2244005, EPI_ISL_2244006, EPI_ISL_2244007, EPI_ISL_2244008, EPI_ISL_2244009,
EPI_ISL_2244010, EPI_ISL_2244011, EPI_ISL_2244012, EPI_ISL_2244013, EPI_ISL_2244014, EPI_ISL_2244015, EPI_ISL_2244016, EPI_ISL_2244017, EPI_ISL_2244018, EPI_ISL_2244019, EPI_ISL_2244020, EPI_ISL_2244021, EPI_ISL_2244022, EPI_ISL_2244023, EPI_ISL_2244024, EPI_ISL_2244025, EPI_ISL_2244026, EPI_ISL_2244028, EPI_ISL_2244029, EPI_ISL_2244030, EPI_ISL_2244031, EPI_ISL_2244032,
EPI_ISL_2244033, EPI_ISL_2244034, EPI_ISL_2244035, EPI_ISL_2244037, EPI_ISL_2244038, EPI_ISL_2244039, EPI_ISL_2244040, EPI_ISL_2244041, EPI_ISL_2244042, EPI_ISL_2244043, EPI_ISL_2244044, EPI_ISL_2244045, EPI_ISL_2244046, EPI_ISL_2244047, EPI_ISL_2244048, EPI_ISL_2244049, EPI_ISL_2244050, EPI_ISL_2244051, EPI_ISL_2244052, EPI_ISL_2244053, EPI_ISL_2244054, EPI_ISL_2244055,
EPI_ISL_2244056, EPI_ISL_2244057, EPI_ISL_2244059, EPI_ISL_2244060, EPI_ISL_2244061, EPI_ISL_2244062, EPI_ISL_2244063, EPI_ISL_2244064, EPI_ISL_2244065, EPI_ISL_2244066, EPI_ISL_2244067, EPI_ISL_2244068, EPI_ISL_2244069, EPI_ISL_2244070, EPI_ISL_2244071, EPI_ISL_2244073, EPI_ISL_2244074, EPI_ISL_2244075, EPI_ISL_2244076, EPI_ISL_2244077, EPI_ISL_2244078, EPI_ISL_2244079,
EPI_ISL_2244080, EPI_ISL_2244081, EPI_ISL_2244082, EPI_ISL_2244083, EPI_ISL_2244084, EPI_ISL_2244085, EPI_ISL_2244086, EPI_ISL_2244087, EPI_ISL_2244088, EPI_ISL_2244089, EPI_ISL_2244090, EPI_ISL_2244091, EPI_ISL_2244092, EPI_ISL_2244093, EPI_ISL_2244095, EPI_ISL_2244096, EPI_ISL_2244097, EPI_ISL_2244098, EPI_ISL_2244099, EPI_ISL_2244100, EPI_ISL_2244101, EPI_ISL_2244102,
EPI_ISL_2244103, EPI_ISL_2244104, EPI_ISL_2244106, EPI_ISL_2244107, EPI_ISL_2244108, EPI_ISL_2244109, EPI_ISL_2244110, EPI_ISL_2244111, EPI_ISL_2244112, EPI_ISL_2244113, EPI_ISL_2244114, EPI_ISL_2244115, EPI_ISL_2244117, EPI_ISL_2244118, EPI_ISL_2244119, EPI_ISL_2244120, EPI_ISL_2244121, EPI_ISL_2244123, EPI_ISL_2244124, EPI_ISL_2244125, EPI_ISL_2244126, EPI_ISL_2244127,

| | | | |
|---|---|---|---|
| EPI_ISL_2244128, EPI_ISL_2244129, EPI_ISL_2244130, EPI_ISL_2244131, EPI_ISL_2244132, EPI_ISL_2244133, EPI_ISL_2244134, EPI_ISL_2244135, EPI_ISL_2244136, EPI_ISL_2244137, EPI_ISL_2244138, EPI_ISL_2244139, EPI_ISL_2244140, EPI_ISL_2244141, EPI_ISL_2244142, EPI_ISL_2244143, EPI_ISL_2244144, EPI_ISL_2244145, EPI_ISL_2244146, EPI_ISL_2244147, EPI_ISL_2244148, EPI_ISL_2244149, EPI_ISL_2244151, EPI_ISL_2244152, EPI_ISL_2244153, EPI_ISL_2244154, EPI_ISL_2244155, EPI_ISL_2244156, EPI_ISL_2244157, EPI_ISL_2244158, EPI_ISL_2244159, EPI_ISL_2244160, EPI_ISL_2244161, EPI_ISL_2244162, EPI_ISL_2244163, EPI_ISL_2244164, EPI_ISL_2244165, EPI_ISL_2244167, EPI_ISL_2244168, EPI_ISL_2244169, EPI_ISL_2244170, EPI_ISL_2244171, EPI_ISL_2244172, EPI_ISL_2244173, EPI_ISL_2244174, EPI_ISL_2244175, EPI_ISL_2244176, EPI_ISL_2244177, EPI_ISL_2244178, EPI_ISL_2244179, EPI_ISL_2244180, EPI_ISL_2244181, EPI_ISL_2244182, EPI_ISL_2244183, EPI_ISL_2244184, EPI_ISL_2244185, EPI_ISL_2244186, EPI_ISL_2244188, EPI_ISL_2244189, EPI_ISL_2244190, EPI_ISL_2244191, EPI_ISL_2244192, EPI_ISL_2244193, EPI_ISL_2244194, EPI_ISL_2244195, EPI_ISL_2244196, EPI_ISL_2244197, EPI_ISL_2244198, EPI_ISL_2244199, EPI_ISL_2244200, EPI_ISL_2244201, EPI_ISL_2244202, EPI_ISL_2244203, EPI_ISL_2244204, EPI_ISL_2244205, EPI_ISL_2244206, EPI_ISL_2244207, EPI_ISL_2244208, EPI_ISL_2244209, EPI_ISL_2244210, EPI_ISL_2244211, EPI_ISL_2244212, EPI_ISL_2244213, EPI_ISL_2244214, EPI_ISL_2244215, EPI_ISL_2244216, EPI_ISL_2244217, EPI_ISL_2244218, EPI_ISL_2244219, EPI_ISL_2244220, EPI_ISL_2244221, EPI_ISL_2244222, EPI_ISL_2244223, EPI_ISL_2244224, EPI_ISL_2244225, EPI_ISL_2244226, EPI_ISL_2244227, EPI_ISL_2244228, EPI_ISL_2244229, EPI_ISL_2244230, EPI_ISL_2244231, EPI_ISL_2244232, EPI_ISL_2244233, EPI_ISL_2244234, EPI_ISL_2244235, EPI_ISL_2244236, EPI_ISL_2244237, EPI_ISL_2244238, EPI_ISL_2244239, EPI_ISL_2244240, EPI_ISL_2244241, EPI_ISL_2244242, EPI_ISL_2244243, EPI_ISL_2244244, EPI_ISL_2244245, EPI_ISL_2244246, EPI_ISL_2244247, EPI_ISL_2244248, EPI_ISL_2244249, EPI_ISL_2244250, EPI_ISL_2244251, EPI_ISL_2244252, EPI_ISL_2244253, EPI_ISL_2244254, EPI_ISL_2244255, EPI_ISL_2244256, EPI_ISL_2244257, EPI_ISL_2244258, EPI_ISL_2244259, EPI_ISL_2244260, EPI_ISL_2244261, EPI_ISL_2244262, EPI_ISL_2244263, EPI_ISL_2244264, EPI_ISL_2244265, EPI_ISL_2244266, EPI_ISL_2244267, EPI_ISL_2244268, EPI_ISL_2244269, EPI_ISL_2244270, EPI_ISL_2244271, EPI_ISL_2244272, EPI_ISL_2244273, EPI_ISL_2244274, EPI_ISL_2244275, EPI_ISL_2244276, EPI_ISL_2244277, EPI_ISL_2244278, EPI_ISL_2244279, EPI_ISL_2244280, EPI_ISL_2244281, EPI_ISL_2244282, EPI_ISL_2244283, EPI_ISL_2244284, EPI_ISL_2244285, EPI_ISL_2244286, EPI_ISL_2244287, EPI_ISL_2244288, EPI_ISL_2244289, EPI_ISL_2244290, EPI_ISL_2244291, EPI_ISL_2244292, EPI_ISL_2244293, EPI_ISL_2244294, EPI_ISL_2244295, EPI_ISL_2244296, EPI_ISL_2244297, EPI_ISL_2244298, EPI_ISL_2244299, EPI_ISL_2244300, EPI_ISL_2244301, EPI_ISL_2244302, EPI_ISL_2244303, EPI_ISL_2244305, EPI_ISL_2244306, EPI_ISL_2244307, EPI_ISL_2244308, EPI_ISL_2244309, EPI_ISL_2244310, EPI_ISL_2244311, EPI_ISL_2244312, EPI_ISL_2244314, EPI_ISL_2244315, EPI_ISL_2244316, EPI_ISL_2244317, EPI_ISL_2244318, EPI_ISL_2244319, EPI_ISL_2244320, EPI_ISL_2244321, EPI_ISL_2244322, EPI_ISL_2244323, EPI_ISL_2244324, EPI_ISL_2244325, EPI_ISL_2244326, EPI_ISL_2244327, EPI_ISL_2244328, EPI_ISL_2244329, EPI_ISL_2244330, EPI_ISL_2244331, EPI_ISL_2244332, EPI_ISL_2244333, EPI_ISL_2244334, EPI_ISL_2244335, EPI_ISL_2244336, EPI_ISL_2244337, EPI_ISL_2244338, EPI_ISL_2244339, EPI_ISL_2244340, EPI_ISL_2244341, EPI_ISL_2244342, EPI_ISL_2244343, EPI_ISL_2244344, EPI_ISL_2244345, EPI_ISL_2244346, EPI_ISL_2244347, EPI_ISL_2244348, EPI_ISL_2244349, EPI_ISL_2244350, EPI_ISL_2244351, EPI_ISL_2244352, EPI_ISL_2244353, EPI_ISL_2244354, EPI_ISL_2244355, EPI_ISL_2244356, EPI_ISL_2244357, EPI_ISL_2244358, EPI_ISL_2244359, EPI_ISL_2244360, EPI_ISL_2244361, EPI_ISL_2244362, EPI_ISL_2244363, EPI_ISL_2244364, EPI_ISL_2244365, EPI_ISL_2244366, EPI_ISL_2244367, EPI_ISL_2244368, EPI_ISL_2244369, EPI_ISL_2244370, EPI_ISL_2244371, EPI_ISL_2244372, EPI_ISL_2244373, EPI_ISL_2244374, EPI_ISL_2244375, EPI_ISL_2244376, EPI_ISL_2244377, EPI_ISL_2244378, EPI_ISL_2244379, EPI_ISL_2244380, EPI_ISL_2244381, EPI_ISL_2244382, EPI_ISL_2244383, EPI_ISL_2244384, EPI_ISL_2244385, EPI_ISL_2244386, EPI_ISL_2244387, EPI_ISL_2244388, EPI_ISL_2244389, EPI_ISL_2244390, EPI_ISL_2244391, EPI_ISL_2244392, EPI_ISL_2244393, EPI_ISL_2244394, EPI_ISL_2244395, EPI_ISL_2244396, EPI_ISL_2244397, EPI_ISL_2244398, EPI_ISL_2244399, EPI_ISL_2244400, EPI_ISL_2244401, EPI_ISL_2244402, EPI_ISL_2244403, EPI_ISL_2244404, EPI_ISL_2244405, EPI_ISL_2244406, EPI_ISL_2244407, EPI_ISL_2244408, EPI_ISL_2244409, EPI_ISL_2244410, EPI_ISL_2244411, EPI_ISL_2244412, EPI_ISL_2244413, EPI_ISL_2244414, EPI_ISL_2244415, EPI_ISL_2244416, EPI_ISL_2244417, EPI_ISL_2244418, EPI_ISL_2244419, EPI_ISL_2244420, EPI_ISL_2244421, EPI_ISL_2244422, EPI_ISL_2244423, EPI_ISL_2244424, EPI_ISL_2244425, EPI_ISL_2244426, EPI_ISL_2244427, EPI_ISL_2244428, EPI_ISL_2244429, EPI_ISL_2244430, EPI_ISL_2244431, EPI_ISL_2244432, EPI_ISL_2244433, EPI_ISL_2244434, EPI_ISL_2244435, EPI_ISL_2244436, EPI_ISL_2244437, EPI_ISL_2244438, EPI_ISL_2244439, EPI_ISL_2244440, EPI_ISL_2244441, EPI_ISL_2244442, EPI_ISL_2244443, EPI_ISL_2244444, EPI_ISL_2244445, EPI_ISL_2244446, EPI_ISL_2244447, EPI_ISL_2244448, EPI_ISL_2244449, EPI_ISL_2244450, EPI_ISL_2244451, EPI_ISL_2244452, EPI_ISL_2244453, EPI_ISL_2244454, EPI_ISL_2244455, EPI_ISL_2244456, EPI_ISL_2244457, EPI_ISL_2244458, EPI_ISL_2244459, EPI_ISL_2244460, EPI_ISL_2244461, EPI_ISL_2244462, EPI_ISL_2244463, EPI_ISL_2244464, EPI_ISL_2244465, EPI_ISL_2244466, EPI_ISL_2244467, EPI_ISL_2244468, EPI_ISL_2244469, EPI_ISL_2244470, EPI_ISL_2244471, EPI_ISL_2244472, EPI_ISL_2244473, EPI_ISL_2244474, EPI_ISL_2244475, EPI_ISL_2244476, EPI_ISL_2244477, EPI_ISL_2244478, EPI_ISL_2244479, EPI_ISL_2244480, EPI_ISL_2244481, EPI_ISL_2244482, EPI_ISL_2244483, EPI_ISL_2244484, EPI_ISL_2244485, EPI_ISL_2244486, EPI_ISL_2244487, EPI_ISL_2244488, EPI_ISL_2244489, EPI_ISL_2244490, EPI_ISL_2244491, EPI_ISL_2244492, EPI_ISL_2244493, EPI_ISL_2244494, EPI_ISL_2244495, EPI_ISL_2244496, EPI_ISL_2244497, EPI_ISL_2244498, EPI_ISL_2244499, EPI_ISL_2244500, EPI_ISL_2244501, EPI_ISL_2244502, EPI_ISL_2244503, EPI_ISL_2244504, EPI_ISL_2244505, EPI_ISL_2244506, EPI_ISL_2244507, EPI_ISL_2244508, EPI_ISL_2244509, EPI_ISL_2244510, EPI_ISL_2244511, EPI_ISL_2244512, EPI_ISL_2244513, EPI_ISL_2244514, EPI_ISL_2244515, EPI_ISL_2244516, EPI_ISL_2244517, EPI_ISL_2244518, EPI_ISL_2244519, EPI_ISL_2244520, EPI_ISL_2244521, EPI_ISL_2244522, EPI_ISL_2244523, EPI_ISL_2244524, EPI_ISL_2244525, EPI_ISL_2244526, EPI_ISL_2244527, EPI_ISL_2244528, EPI_ISL_2244529, EPI_ISL_2244530, EPI_ISL_2244531, EPI_ISL_2244532, EPI_ISL_2244533, EPI_ISL_2244534, EPI_ISL_2244535, EPI_ISL_2244536, EPI_ISL_2244537, EPI_ISL_2244538, EPI_ISL_2244539, EPI_ISL_2244540, EPI_ISL_2244541, EPI_ISL_2244542, EPI_ISL_2244543, EPI_ISL_2244544, EPI_ISL_2244545, EPI_ISL_2244546, EPI_ISL_2244547, EPI_ISL_2244548, EPI_ISL_2244549, EPI_ISL_2244550, EPI_ISL_2244551, EPI_ISL_2244552, EPI_ISL_2244553, EPI_ISL_2244554, EPI_ISL_2244555, EPI_ISL_2244556, EPI_ISL_2244557, EPI_ISL_2244558, EPI_ISL_2244559, EPI_ISL_2244560, EPI_ISL_2244561, EPI_ISL_2244562, EPI_ISL_2244563, EPI_ISL_2244564, EPI_ISL_2244565, EPI_ISL_2244566, EPI_ISL_2244567, EPI_ISL_2244568, EPI_ISL_2244569, EPI_ISL_2244570, EPI_ISL_2244571, EPI_ISL_2244572, EPI_ISL_2244573, EPI_ISL_2244574, EPI_ISL_2244575, EPI_ISL_2244576, EPI_ISL_2244577, EPI_ISL_2244578, EPI_ISL_2244579, EPI_ISL_2244580, EPI_ISL_2244581, EPI_ISL_2244582, EPI_ISL_2244583, EPI_ISL_2244584, EPI_ISL_2244585, EPI_ISL_2244586, EPI_ISL_2244587, EPI_ISL_2244588, EPI_ISL_2244589, EPI_ISL_2244590, EPI_ISL_2244591, EPI_ISL_2244592, EPI_ISL_2244593, EPI_ISL_2244594, EPI_ISL_2244595, EPI_ISL_2244596, EPI_ISL_2244597, EPI_ISL_2244598, EPI_ISL_2244599, EPI_ISL_2244600, EPI_ISL_2244601, EPI_ISL_2244602, EPI_ISL_2244603, EPI_ISL_2244604, EPI_ISL_2244605, EPI_ISL_2244606, EPI_ISL_2244607, EPI_ISL_2244608, EPI_ISL_2244609, EPI_ISL_2244610, EPI_ISL_2244611, EPI_ISL_2244612, EPI_ISL_2244613, EPI_ISL_2244614, EPI_ISL_2244615, EPI_ISL_2244616, EPI_ISL_2244617, EPI_ISL_2244618, EPI_ISL_2244619, EPI_ISL_2244620, EPI_ISL_2244621, EPI_ISL_2244622, EPI_ISL_2244623, EPI_ISL_2244624, EPI_ISL_2244625, EPI_ISL_2244627, EPI_ISL_2244629, EPI_ISL_2244630, EPI_ISL_2244631, EPI_ISL_2244632, EPI_ISL_2244633, EPI_ISL_2244634, EPI_ISL_2244635, EPI_ISL_2244636, EPI_ISL_2244637, EPI_ISL_2244638, EPI_ISL_2244639, EPI_ISL_2244640, EPI_ISL_2244641, EPI_ISL_2244642, EPI_ISL_2244643, EPI_ISL_2244644, EPI_ISL_2244645, EPI_ISL_2244646, EPI_ISL_2244647, EPI_ISL_2244648, EPI_ISL_2244649, EPI_ISL_2244650, EPI_ISL_2244651, EPI_ISL_2244652, EPI_ISL_2244653, EPI_ISL_2244654, EPI_ISL_2244655, EPI_ISL_2244656, EPI_ISL_2244657, EPI_ISL_2244658, EPI_ISL_2244659, EPI_ISL_2244660, EPI_ISL_2244661, EPI_ISL_2244662, EPI_ISL_2244663, EPI_ISL_2244664, EPI_ISL_2244665, EPI_ISL_2244666, EPI_ISL_2244667, EPI_ISL_2244668, EPI_ISL_2244669, EPI_ISL_2244670, EPI_ISL_2244671, EPI_ISL_2244673, EPI_ISL_2244674, EPI_ISL_2244675, EPI_ISL_2244676, EPI_ISL_2244677, EPI_ISL_2244678, EPI_ISL_2244679, EPI_ISL_2244680, EPI_ISL_2244681, EPI_ISL_2244682, EPI_ISL_2244683, EPI_ISL_2244684, EPI_ISL_2244685, EPI_ISL_2244686, EPI_ISL_2244687, EPI_ISL_2244688, EPI_ISL_2244689, EPI_ISL_2244690, EPI_ISL_2244691, EPI_ISL_2244692, EPI_ISL_2244693, EPI_ISL_2244694, EPI_ISL_2244695, EPI_ISL_2244696, EPI_ISL_2244697, EPI_ISL_2244698, EPI_ISL_2244699, EPI_ISL_2244700, EPI_ISL_2244701, EPI_ISL_2244702, EPI_ISL_2244703, EPI_ISL_2244704, EPI_ISL_2244705, EPI_ISL_2244706, EPI_ISL_2244707, EPI_ISL_2244708, EPI_ISL_2244709, EPI_ISL_2244710, EPI_ISL_2244711, EPI_ISL_2244712, EPI_ISL_2244713, EPI_ISL_2244714, EPI_ISL_2244715, EPI_ISL_2244716, EPI_ISL_2244717, EPI_ISL_2244718, EPI_ISL_2244719, EPI_ISL_2244720, EPI_ISL_2244721, EPI_ISL_2244722, EPI_ISL_2244723, EPI_ISL_2244724, EPI_ISL_2244725, EPI_ISL_2244726, EPI_ISL_2244727, EPI_ISL_2244728, EPI_ISL_2244729, EPI_ISL_2244730, EPI_ISL_2244731, EPI_ISL_2244732, EPI_ISL_2244733, EPI_ISL_2244734, EPI_ISL_2244735, EPI_ISL_2244736, EPI_ISL_2244737, EPI_ISL_2244738, EPI_ISL_2244739, EPI_ISL_2244740, EPI_ISL_2244741, EPI_ISL_2244742, EPI_ISL_2244743, EPI_ISL_2244744, EPI_ISL_2244745, EPI_ISL_2244746, EPI_ISL_2244747, EPI_ISL_2244748, EPI_ISL_2244749, EPI_ISL_2244750, EPI_ISL_2244751, EPI_ISL_2244752, EPI_ISL_2244753, EPI_ISL_2244754, EPI_ISL_2244755, EPI_ISL_2244756, EPI_ISL_2244757, EPI_ISL_2244758, EPI_ISL_2244759, EPI_ISL_2244760, EPI_ISL_2244761, EPI_ISL_2244762, EPI_ISL_2244763, EPI_ISL_2244764, EPI_ISL_2244765, EPI_ISL_2244766, EPI_ISL_2244767, EPI_ISL_2244768, EPI_ISL_2244769, EPI_ISL_2244770, EPI_ISL_2244771, EPI_ISL_2244772, EPI_ISL_2244773, EPI_ISL_2244774, EPI_ISL_2244776, EPI_ISL_2244777, EPI_ISL_2244778, EPI_ISL_2244779, EPI_ISL_2244780, EPI_ISL_2244781, EPI_ISL_2244782, EPI_ISL_2244784, EPI_ISL_2244785, EPI_ISL_2244786, EPI_ISL_2244787, EPI_ISL_2244788, EPI_ISL_2244789, EPI_ISL_2244790, EPI_ISL_2244791, EPI_ISL_2244792, EPI_ISL_2244793, EPI_ISL_2244794, EPI_ISL_2244795, EPI_ISL_2244796, EPI_ISL_2244797, EPI_ISL_2244798, EPI_ISL_2244799, EPI_ISL_2244800, EPI_ISL_2244801, EPI_ISL_2244802, EPI_ISL_2244803, EPI_ISL_2244804, EPI_ISL_2244805, EPI_ISL_2244806, EPI_ISL_2244807, EPI_ISL_2244808, EPI_ISL_2244809, EPI_ISL_2244810, EPI_ISL_2244811, EPI_ISL_2244812, EPI_ISL_2244813, EPI_ISL_2244814, EPI_ISL_2244815, EPI_ISL_2244816, EPI_ISL_2244817, EPI_ISL_2244818, EPI_ISL_2244819, EPI_ISL_2244820, EPI_ISL_2244821, EPI_ISL_2244822, EPI_ISL_2244823, EPI_ISL_2244824, EPI_ISL_2244825, EPI_ISL_2244826, EPI_ISL_2244827, EPI_ISL_2244828, EPI_ISL_2244829, EPI_ISL_2244830, EPI_ISL_2244831, EPI_ISL_2244832, EPI_ISL_2244833, EPI_ISL_2244834, EPI_ISL_2244835, EPI_ISL_2244836, EPI_ISL_2244837, EPI_ISL_2244838, EPI_ISL_2244839, EPI_ISL_2244840, EPI_ISL_2244841, EPI_ISL_2244842, EPI_ISL_2244843, EPI_ISL_2244844, EPI_ISL_2244845, EPI_ISL_2244846, EPI_ISL_2244847, EPI_ISL_2244848, EPI_ISL_2244849, EPI_ISL_2244850, EPI_ISL_2244851, EPI_ISL_2244852, EPI_ISL_2244853, EPI_ISL_2244854, EPI_ISL_2244855, EPI_ISL_2244856, EPI_ISL_2244857, EPI_ISL_2244858, EPI_ISL_2244859, EPI_ISL_2244860, EPI_ISL_2244861, EPI_ISL_2244862, EPI_ISL_2244863, EPI_ISL_2244864, EPI_ISL_2244865, EPI_ISL_2244866, EPI_ISL_2244867, EPI_ISL_2244868, EPI_ISL_2244869, EPI_ISL_2244870, EPI_ISL_2244871, EPI_ISL_2244872, EPI_ISL_2244873, EPI_ISL_2244874, EPI_ISL_2244875, EPI_ISL_2244876, EPI_ISL_2244877, EPI_ISL_2244878, EPI_ISL_2244879, EPI_ISL_2244880, EPI_ISL_2244881, EPI_ISL_2244882, EPI_ISL_2244883, EPI_ISL_2244884, EPI_ISL_2244885, EPI_ISL_2244886, EPI_ISL_2244887, EPI_ISL_2244888, EPI_ISL_2244889, EPI_ISL_2244890, EPI_ISL_2244891, EPI_ISL_2244892, EPI_ISL_2244893, EPI_ISL_2244894, EPI_ISL_2244895, EPI_ISL_2244896 | | | |
| see above | Aegis Sciences Corporation | Centers for Disease Control and Prevention Division of Viral Diseases, Pathogen Discovery | Dakota Howard, Dhwani Batra, Peter W. Cook, Kara Moser, Adrian Paskey, Jason Caravas, Benjamin Rambo-Martin, Shatavia Morrison, Christopher Gulvick, Scott Sammons, Yvette Unoarumhi, Darlene Wagner, Matthew Schmerer, Cyndi Clark, Patrick Campbell, Rob Case, Vikramsinha Ghorpade, Holly Houdeshell, Ola Kvalvaag, Dillon Nall, Ethan Sanders, Alec Vest, Shaun Westlund, Matthew Hardison, Clinton R. Paden, Duncan MacCannell |
| EPI_ISL_2244900, EPI_ISL_2244901, EPI_ISL_2244902, EPI_ISL_2244903, EPI_ISL_2244905, EPI_ISL_2244906, EPI_ISL_2244907, EPI_ISL_2244908 | Infinity Biologix | Centers for Disease Control and Prevention Division of Viral Diseases, Pathogen Discovery | Dakota Howard, Dhwani Batra, Peter W. Cook, Kara Moser, Adrian Paskey, Jason Caravas, Benjamin Rambo-Martin, Shatavia Morrison, Christopher Gulvick, Scott Sammons, Yvette Unoarumhi, Darlene Wagner, Matthew Schmerer, Christian Bixby, Yihe Wang, Jonathan Schultz, Chirayu Goswami, Russ Hager, Robin Grimwood, Clinton R. Paden, Duncan MacCannell |
| EPI_ISL_2244913, EPI_ISL_2244914, EPI_ISL_2244915 | Minnesota Department of Health, Public Health Laboratory | Minnesota Department of Health, Public Health Laboratory | Alexandra Lorentz, Jacob Garfin, Matt Plumb, and Xiong Wang |
| EPI_ISL_2244916, EPI_ISL_2244917, EPI_ISL_2244918 | Mayo Clinic & Mayo Clinic Laboratories | Minnesota Department of Health, Public Health Laboratory | Alexandra Lorentz, Jacob Garfin, Matt Plumb, and Xiong Wang |
| EPI_ISL_2244919, EPI_ISL_2244920, EPI_ISL_2244921 | Minnesota Department of Health, Public Health Laboratory | Minnesota Department of Health, Public Health Laboratory | Alexandra Lorentz, Jacob Garfin, Matt Plumb, and Xiong Wang |
| EPI_ISL_2244922, EPI_ISL_2244923, EPI_ISL_2244924, EPI_ISL_2244925, EPI_ISL_2244926, EPI_ISL_2244927, EPI_ISL_2244928, EPI_ISL_2244929, EPI_ISL_2244930, EPI_ISL_2244931, EPI_ISL_2244932, EPI_ISL_2244933, EPI_ISL_2244934, EPI_ISL_2244935, EPI_ISL_2244936, EPI_ISL_2244937, EPI_ISL_2244938, EPI_ISL_2244939, EPI_ISL_2244940, EPI_ISL_2244941, EPI_ISL_2244943, EPI_ISL_2244944, EPI_ISL_2244945, EPI_ISL_2244946, EPI_ISL_2244947, EPI_ISL_2244948, EPI_ISL_2244949, EPI_ISL_2244950, EPI_ISL_2244951, EPI_ISL_2244952, EPI_ISL_2244953, EPI_ISL_2244954, EPI_ISL_2244955, EPI_ISL_2244956, EPI_ISL_2244957, EPI_ISL_2244958, EPI_ISL_2244959, EPI_ISL_2244960, EPI_ISL_2244961, EPI_ISL_2244962, EPI_ISL_2244963, EPI_ISL_2244964, EPI_ISL_2244965, EPI_ISL_2244966, EPI_ISL_2244968, EPI_ISL_2244969, EPI_ISL_2244970, EPI_ISL_2244971, EPI_ISL_2244972, EPI_ISL_2244973, EPI_ISL_2244974, EPI_ISL_2244975, EPI_ISL_2244976, EPI_ISL_2244977, EPI_ISL_2244978, EPI_ISL_2244979, EPI_ISL_2244980, EPI_ISL_2244981, EPI_ISL_2244982, EPI_ISL_2244983, EPI_ISL_2244984, EPI_ISL_2244985, EPI_ISL_2244986, EPI_ISL_2244987, EPI_ISL_2244988, EPI_ISL_2244989, EPI_ISL_2244990, EPI_ISL_2244991, EPI_ISL_2244992, EPI_ISL_2244993, EPI_ISL_2244994, EPI_ISL_2244995, EPI_ISL_2244996, EPI_ISL_2244997, EPI_ISL_2244998, EPI_ISL_2244999, EPI_ISL_2245000, EPI_ISL_2245001 | | | |
| see above | M Health Fairview | Minnesota Department of Health, Public Health Laboratory | Alexandra Lorentz, Jacob Garfin, Matt Plumb, and Xiong Wang |
| EPI_ISL_2245002, EPI_ISL_2245003, EPI_ISL_2245004, EPI_ISL_2245007, EPI_ISL_2245008, EPI_ISL_2245009, EPI_ISL_2245010, EPI_ISL_2245011, EPI_ISL_2245012, EPI_ISL_2245013 | Florida Bureau of Public Health Laboratories | Florida Bureau of Public Health Laboratories | Sarah Schmedes, Jason Blanton |
| EPI_ISL_2245468, EPI_ISL_2245469, EPI_ISL_2245470, EPI_ISL_2245471 | University of Wisconsin-Madison AIDS Vaccine Research Laboratories | University of Wisconsin-Madison AIDS Vaccine Research Laboratories | Gage Moreno, Katarina Braun, et al. AIDS Vaccine Research Laboratories |
| EPI_ISL_2245472, EPI_ISL_2245473, EPI_ISL_2245474, EPI_ISL_2245475, EPI_ISL_2245476, EPI_ISL_2245477, EPI_ISL_2245478, EPI_ISL_2245479, EPI_ISL_2245480, EPI_ISL_2245481, EPI_ISL_2245482, EPI_ISL_2245483, EPI_ISL_2245484, EPI_ISL_2245486, EPI_ISL_2245487, EPI_ISL_2245488, EPI_ISL_2245489, EPI_ISL_2245490, EPI_ISL_2245491, EPI_ISL_2245492, EPI_ISL_2245493, EPI_ISL_2245494, EPI_ISL_2245495, EPI_ISL_2245497, EPI_ISL_2245498, EPI_ISL_2245500, EPI_ISL_2245502, EPI_ISL_2245503, EPI_ISL_2245504, EPI_ISL_2245505, EPI_ISL_2245506, EPI_ISL_2245507, EPI_ISL_2245508, EPI_ISL_2245509, EPI_ISL_2245510, EPI_ISL_2245511, EPI_ISL_2245512, EPI_ISL_2245513, EPI_ISL_2245514, EPI_ISL_2245515, EPI_ISL_2245517, EPI_ISL_2245518, EPI_ISL_2245519, EPI_ISL_2245520, EPI_ISL_2245521, EPI_ISL_2245523, EPI_ISL_2245524, EPI_ISL_2245525, EPI_ISL_2245526, EPI_ISL_2245527, EPI_ISL_2245528, EPI_ISL_2245529, EPI_ISL_2245530, EPI_ISL_2245531, EPI_ISL_2245532, EPI_ISL_2245533, EPI_ISL_2245534, EPI_ISL_2245535, EPI_ISL_2245536, EPI_ISL_2245537, EPI_ISL_2245538, EPI_ISL_2245539, EPI_ISL_2245540, EPI_ISL_2245541, EPI_ISL_2245542, EPI_ISL_2245544, EPI_ISL_2245545, EPI_ISL_2245546, EPI_ISL_2245547, EPI_ISL_2245548, EPI_ISL_2245549, EPI_ISL_2245550, EPI_ISL_2245551, EPI_ISL_2245552, EPI_ISL_2245553, EPI_ISL_2245554, EPI_ISL_2245555, EPI_ISL_2245556, EPI_ISL_2245557, EPI_ISL_2245558, EPI_ISL_2245559, EPI_ISL_2245560, EPI_ISL_2245561, EPI_ISL_2245563, EPI_ISL_2245564, EPI_ISL_2245565, EPI_ISL_2245566, EPI_ISL_2245567, EPI_ISL_2245568, EPI_ISL_2245569, EPI_ISL_2245570, EPI_ISL_2245571, EPI_ISL_2245572, EPI_ISL_2245573, EPI_ISL_2245575, EPI_ISL_2245576, EPI_ISL_2245578, EPI_ISL_2245579, EPI_ISL_2245580, EPI_ISL_2245581, EPI_ISL_2245582, EPI_ISL_2245583, EPI_ISL_2245584, EPI_ISL_2245585, EPI_ISL_2245586, EPI_ISL_2245587, EPI_ISL_2245588, EPI_ISL_2245589, EPI_ISL_2245590, EPI_ISL_2245591, EPI_ISL_2245592, EPI_ISL_2245593, EPI_ISL_2245594, EPI_ISL_2245595, EPI_ISL_2245596, EPI_ISL_2245597, EPI_ISL_2245598, EPI_ISL_2245599, EPI_ISL_2245600, EPI_ISL_2245601, EPI_ISL_2245603, EPI_ISL_2245604, EPI_ISL_2245605, EPI_ISL_2245606, EPI_ISL_2245607, EPI_ISL_2245609, EPI_ISL_2245610, EPI_ISL_2245611, EPI_ISL_2245612, EPI_ISL_2245613, EPI_ISL_2245614, EPI_ISL_2245615, EPI_ISL_2245616, EPI_ISL_2245617, EPI_ISL_2245618, EPI_ISL_2245619, EPI_ISL_2245620, EPI_ISL_2245621, EPI_ISL_2245622, EPI_ISL_2245623, EPI_ISL_2245624, EPI_ISL_2245625, EPI_ISL_2245626, EPI_ISL_2245627, EPI_ISL_2245628, EPI_ISL_2245629, EPI_ISL_2245630, EPI_ISL_2245631, EPI_ISL_2245632, EPI_ISL_2245633 | | | |
| see above | Maine Health and Environmental Testing Laboratory | Tewhey Lab, The Jackson Laboratory | Matluk,N., Dewey,H., Iosue,F., Barter,M., Lynch,R., Munger,H. and Tewhey,R. |
| EPI_ISL_2246200, EPI_ISL_2246201, EPI_ISL_2246202, EPI_ISL_2246203, EPI_ISL_2246204, EPI_ISL_2246205, EPI_ISL_2246207, EPI_ISL_2246208, EPI_ISL_2246209, EPI_ISL_2246210, EPI_ISL_2246211, EPI_ISL_2246212, EPI_ISL_2246213, EPI_ISL_2246214, EPI_ISL_2246215, EPI_ISL_2246216, EPI_ISL_2246218, EPI_ISL_2246219, EPI_ISL_2246220, EPI_ISL_2246221, EPI_ISL_2246222 | | | |
| see above | DC Public Health Lab/ Dept. of Forensic Sciences | DC Public Health Lab/ Dept. of Forensic Sciences | Janis Doss, Scott Nguyen, Elizabeth Zelaya, Sarah Scott, Connie Maza, Monica Mann, Brittany Hamilton, David Payne, Jocelyn Hauser |
| EPI_ISL_2246331, EPI_ISL_2246340, EPI_ISL_2246382, EPI_ISL_2246398, EPI_ISL_2246400, EPI_ISL_2246401, EPI_ISL_2246402, EPI_ISL_2246403, EPI_ISL_2246404, EPI_ISL_2246405, EPI_ISL_2246406, EPI_ISL_2246407, EPI_ISL_2246408, EPI_ISL_2246409, EPI_ISL_2246410, EPI_ISL_2246411, EPI_ISL_2246412, EPI_ISL_2246413, EPI_ISL_2246414, EPI_ISL_2246415, EPI_ISL_2246417, EPI_ISL_2246418, EPI_ISL_2246419, EPI_ISL_2246420, EPI_ISL_2246421, EPI_ISL_2246422, EPI_ISL_2246423, EPI_ISL_2246424, EPI_ISL_2246425, EPI_ISL_2246426, EPI_ISL_2246427, EPI_ISL_2246428, EPI_ISL_2246429, EPI_ISL_2246430, EPI_ISL_2246431, EPI_ISL_2246432, EPI_ISL_2246433, EPI_ISL_2246434, EPI_ISL_2246435, EPI_ISL_2246436, EPI_ISL_2246437, EPI_ISL_2246438, EPI_ISL_2246439, EPI_ISL_2246440, EPI_ISL_2246441, EPI_ISL_2246442, EPI_ISL_2246443, EPI_ISL_2246444, EPI_ISL_2246445, EPI_ISL_2246447, EPI_ISL_2246450, EPI_ISL_2246451, EPI_ISL_2246452, EPI_ISL_2246453, EPI_ISL_2246454, EPI_ISL_2246455, EPI_ISL_2246456, EPI_ISL_2246457, EPI_ISL_2246458, EPI_ISL_2246459, EPI_ISL_2246460, EPI_ISL_2246461, EPI_ISL_2246462, EPI_ISL_2246463, EPI_ISL_2246464, EPI_ISL_2246465, EPI_ISL_2246467, EPI_ISL_2246468, EPI_ISL_2246469, EPI_ISL_2246470, EPI_ISL_2246471, EPI_ISL_2246473, EPI_ISL_2246474, EPI_ISL_2246475, EPI_ISL_2246476, EPI_ISL_2246477, EPI_ISL_2246478, EPI_ISL_2246479, EPI_ISL_2246480, EPI_ISL_2246481, EPI_ISL_2246482, EPI_ISL_2246483, EPI_ISL_2246484, EPI_ISL_2246485, EPI_ISL_2246486, EPI_ISL_2246487, EPI_ISL_2246488, EPI_ISL_2246491, EPI_ISL_2246492, EPI_ISL_2246495, EPI_ISL_2246496, EPI_ISL_2246500, EPI_ISL_2246501, EPI_ISL_2246502, EPI_ISL_2246503, EPI_ISL_2246504, EPI_ISL_2246505, EPI_ISL_2246507, EPI_ISL_2246508, EPI_ISL_2246512, EPI_ISL_2246514, EPI_ISL_2246515, EPI_ISL_2246517, EPI_ISL_2246525, EPI_ISL_2246526, EPI_ISL_2246527, EPI_ISL_2246530, EPI_ISL_2246531, EPI_ISL_2246538, EPI_ISL_2246539, EPI_ISL_2246545, EPI_ISL_2246547, EPI_ISL_2246548, EPI_ISL_2246551, EPI_ISL_2246553, EPI_ISL_2246556, EPI_ISL_2246559, EPI_ISL_2246560, EPI_ISL_2246562, EPI_ISL_2246566, EPI_ISL_2246572, EPI_ISL_2246573, EPI_ISL_2246576 | | | |
| see above | Michigan Department of Health and Human Services, Bureau of Laboratories | Michigan Department of Health and Human Services, Bureau of Laboratories | Blankenship HM, Riner D, Soehnlen MK |
| EPI_ISL_2246579, EPI_ISL_2246580, EPI_ISL_2246581, EPI_ISL_2246582, EPI_ISL_2246583, EPI_ISL_2246584, EPI_ISL_2246585, EPI_ISL_2246586, EPI_ISL_2246587, EPI_ISL_2246588, EPI_ISL_2246589, EPI_ISL_2246590, EPI_ISL_2246591, EPI_ISL_2246592, EPI_ISL_2246593, EPI_ISL_2246594, EPI_ISL_2246595, EPI_ISL_2246596, EPI_ISL_2246597, EPI_ISL_2246598, EPI_ISL_2246599, EPI_ISL_2246600, EPI_ISL_2246601, EPI_ISL_2246602, EPI_ISL_2246603, EPI_ISL_2246604, EPI_ISL_2246605, EPI_ISL_2246606, EPI_ISL_2246607, EPI_ISL_2246608, EPI_ISL_2246609, EPI_ISL_2246610, EPI_ISL_2246611, EPI_ISL_2246613, EPI_ISL_2246614, EPI_ISL_2246615, EPI_ISL_2246616, EPI_ISL_2246617, EPI_ISL_2246618, EPI_ISL_2246619, EPI_ISL_2246620, EPI_ISL_2246621, EPI_ISL_2246622, EPI_ISL_2246623, EPI_ISL_2246624, EPI_ISL_2246626, EPI_ISL_2246627, EPI_ISL_2246628, EPI_ISL_2246629, EPI_ISL_2246630, EPI_ISL_2246631, EPI_ISL_2246632, EPI_ISL_2246633, EPI_ISL_2246634, EPI_ISL_2246635, EPI_ISL_2246636, EPI_ISL_2246637, EPI_ISL_2246638, EPI_ISL_2246639, EPI_ISL_2246640, EPI_ISL_2246641, EPI_ISL_2246642, EPI_ISL_2246643, EPI_ISL_2246644, EPI_ISL_2246645, EPI_ISL_2246646, EPI_ISL_2246647, EPI_ISL_2246649, EPI_ISL_2246650, EPI_ISL_2246652, EPI_ISL_2246653, EPI_ISL_2246654, EPI_ISL_2246655, EPI_ISL_2246657, EPI_ISL_2246658, EPI_ISL_2246659, EPI_ISL_2246660, EPI_ISL_2246661, EPI_ISL_2246662, EPI_ISL_2246663, EPI_ISL_2246664, EPI_ISL_2246665, EPI_ISL_2246666, EPI_ISL_2246667, EPI_ISL_2246668, EPI_ISL_2246669, EPI_ISL_2246670, EPI_ISL_2246671, EPI_ISL_2246672, EPI_ISL_2246673, EPI_ISL_2246674, EPI_ISL_2246675, EPI_ISL_2246676, EPI_ISL_2246677, EPI_ISL_2246678, EPI_ISL_2246679, EPI_ISL_2246680, EPI_ISL_2246681, EPI_ISL_2246682, EPI_ISL_2246683, EPI_ISL_2246684, EPI_ISL_2246685, EPI_ISL_2246686, EPI_ISL_2246687, EPI_ISL_2246688, EPI_ISL_2246689, EPI_ISL_2246690, EPI_ISL_2246691, EPI_ISL_2246692, EPI_ISL_2246693, EPI_ISL_2246694, EPI_ISL_2246695, EPI_ISL_2246696, EPI_ISL_2246697, EPI_ISL_2246698 | | | |
| see above | University of Chicago Medicine | RIPHL at Rush University Medical Center | Stefan Green, Kevin Kunstman, Max Kolton, Marieta Hyde, Laura Furtado, Felix Araujo Perez, Cindy Bethel, Lauren Megger |

| | | | |
|---|---|---|---|
| EPI_ISL_2246699, EPI_ISL_2246700, EPI_ISL_2246701, EPI_ISL_2246702, EPI_ISL_2246703, EPI_ISL_2246704, EPI_ISL_2246705, EPI_ISL_2246706, EPI_ISL_2246707, EPI_ISL_2246708, EPI_ISL_2246709, EPI_ISL_2246710, EPI_ISL_2246711, EPI_ISL_2246712, EPI_ISL_2246713 | | | |
| see above | Northwestern Memorial Hospital | RIPHL at Rush University Medical Center | Stefan Green, Kevin Kunstman, Max Kolton, Marieta Hyde, Laura Furtado, Felix Araujo Perez, Chao Qi, Amber Kimble |
| EPI_ISL_2246714, EPI_ISL_2246715, EPI_ISL_2246716, EPI_ISL_2246717, EPI_ISL_2246718, EPI_ISL_2246719, EPI_ISL_2246720, EPI_ISL_2246721, EPI_ISL_2246722, EPI_ISL_2246723, EPI_ISL_2246724, EPI_ISL_2246725, EPI_ISL_2246726, EPI_ISL_2246727, EPI_ISL_2246728, EPI_ISL_2246729, EPI_ISL_2246730, EPI_ISL_2246731, EPI_ISL_2246732, EPI_ISL_2246733, EPI_ISL_2246734, EPI_ISL_2246735, EPI_ISL_2246736, EPI_ISL_2246737 | | | |
| see above | John H Stroger Jr Hospital of Cook County | RIPHL at Rush University Medical Center | Stefan Green, Kevin Kunstman, Max Kolton, Marieta Hyde, Laura Furtado, Felix Araujo Perez, Sharon Welbel, Ah-Young Sung |
| EPI_ISL_2246738, EPI_ISL_2246739, EPI_ISL_2246740, EPI_ISL_2246742, EPI_ISL_2246744, EPI_ISL_2246745, EPI_ISL_2246747, EPI_ISL_2246749, EPI_ISL_2246751, EPI_ISL_2246752, EPI_ISL_2246753, EPI_ISL_2246754, EPI_ISL_2246755, EPI_ISL_2246756, EPI_ISL_2246757, EPI_ISL_2246760, EPI_ISL_2246761, EPI_ISL_2246762, EPI_ISL_2246763, EPI_ISL_2246764, EPI_ISL_2246768, EPI_ISL_2246769, EPI_ISL_2246770, EPI_ISL_2246771, EPI_ISL_2246773, EPI_ISL_2246774, EPI_ISL_2246775, EPI_ISL_2246779, EPI_ISL_2246781, EPI_ISL_2246782 | | | |
| see above | Wyoming Public Health Laboratory | Wyoming Public Health Laboratory | Jim Mildenberger, Taylor Fearing, Channing Weber, Ashley Norberg, Chayse Rowley, Marley Goetz, Brian Dominguez, Elliot Thomasson, Brittany Oher, Sam Britz, Cari Sloma, and Rob Christensen |
| EPI_ISL_2246783, EPI_ISL_2246784, EPI_ISL_2246786 | WLA VAMC | Pathogen Discovery, Respiratory Viruses Branch, Division of Viral Diseases, Centers for Disease Control and Prevention | Yan Li, Jing Zhang, Anna Kelleher, Ying Tao, Brian Lynch, Krista Queen, Anna Uehara, Peter Cook, Han Jia Justin Ng, Rachel Marine, Clinton R. Paden, Dhwani Batra, Haibin Wang, Tara Coalter, Jasmine Padilla, Morgan Davis, Mili Sheth, Sarah Nobles, Mark Burroughs, Justin Lee, Adam Retchless, Suxiang Tong |
| EPI_ISL_2246787, EPI_ISL_2246788, EPI_ISL_2246790, EPI_ISL_2246791, EPI_ISL_2246792, EPI_ISL_2246794, EPI_ISL_2246795, EPI_ISL_2246796, EPI_ISL_2246797, EPI_ISL_2246799, EPI_ISL_2246800, EPI_ISL_2246801, EPI_ISL_2246802, EPI_ISL_2246803, EPI_ISL_2246805, EPI_ISL_2246806, EPI_ISL_2246809, EPI_ISL_2246811, EPI_ISL_2246812 | | | |
| see above | Wyoming Public Health Laboratory | Wyoming Public Health Laboratory | Jim Mildenberger, Taylor Fearing, Channing Weber, Ashley Norberg, Chayse Rowley, Marley Goetz, Brian Dominguez, Elliot Thomasson, Brittany Oher, Sam Britz, Cari Sloma, and Rob Christensen |
| EPI_ISL_2246827, EPI_ISL_2246828 | LESP Ciudad de Mexico /LANS | Instituto de Diagnostico y Referencia Epidemiologicos (INDRE) | Claudia Wong-Arambula, Abril Rodriguez-Maldonado, Vanessa Rivero-Arredondo, Ariadna Medina-Benitez, Joaquin Quiroz-Mercado, Sergio Rangel-Guerrero, Natividad Cruz-Ortiz, Tatiana Nunez-Garcia, Gisela Barrera-Badillo, Lucia Hernandez-Rivas, Irma Lopez-Martinez, Ernesto Ramirez-Gonzalez. |
| EPI_ISL_2246829, EPI_ISL_2246830, EPI_ISL_2246831, EPI_ISL_2246832, EPI_ISL_2246833 | LESP Michoacan | Instituto de Diagnostico y Referencia Epidemiologicos (INDRE) | Claudia Wong-Arambula, Abril Rodriguez-Maldonado, Vanessa Rivero-Arredondo, Ariadna Medina-Benitez, Joaquin Quiroz-Mercado, Sergio Rangel-Guerrero, Natividad Cruz-Ortiz, Tatiana Nunez-Garcia, Gisela Barrera-Badillo, Lucia Hernandez-Rivas, Irma Lopez-Martinez, Ernesto Ramirez-Gonzalez. |
| EPI_ISL_2246834, EPI_ISL_2246835, EPI_ISL_2246836, EPI_ISL_2246837 | LESP Sinaloa | Instituto de Diagnostico y Referencia Epidemiologicos (INDRE) | Claudia Wong-Arambula, Abril Rodriguez-Maldonado, Vanessa Rivero-Arredondo, Ariadna Medina-Benitez, Joaquin Quiroz-Mercado, Sergio Rangel-Guerrero, Natividad Cruz-Ortiz, Tatiana Nunez-Garcia, Gisela Barrera-Badillo, Lucia Hernandez-Rivas, Irma Lopez-Martinez, Ernesto Ramirez-Gonzalez. |
| EPI_ISL_2246838 | LESP Ciudad de Mexico | Instituto de Diagnostico y Referencia Epidemiologicos (INDRE) | Claudia Wong-Arambula, Abril Rodriguez-Maldonado, Vanessa Rivero-Arredondo, Ariadna Medina-Benitez, Joaquin Quiroz-Mercado, Sergio Rangel-Guerrero, Natividad Cruz-Ortiz, Tatiana Nunez-Garcia, Gisela Barrera-Badillo, Lucia Hernandez-Rivas, Irma Lopez-Martinez, Ernesto Ramirez-Gonzalez. |
| EPI_ISL_2246839 | LESP Nuevo Leon | Instituto de Diagnostico y Referencia Epidemiologicos (INDRE) | Claudia Wong-Arambula, Abril Rodriguez-Maldonado, Vanessa Rivero-Arredondo, Ariadna Medina-Benitez, Joaquin Quiroz-Mercado, Sergio Rangel-Guerrero, Natividad Cruz-Ortiz, Tatiana Nunez-Garcia, Gisela Barrera-Badillo, Lucia Hernandez-Rivas, Irma Lopez-Martinez, Ernesto Ramirez-Gonzalez. |
| EPI_ISL_2246840 | LESP Tamaulipas | Instituto de Diagnostico y Referencia Epidemiologicos (INDRE) | Claudia Wong-Arambula, Abril Rodriguez-Maldonado, Vanessa Rivero-Arredondo, Ariadna Medina-Benitez, Joaquin Quiroz-Mercado, Sergio Rangel-Guerrero, Natividad Cruz-Ortiz, Tatiana Nunez-Garcia, Gisela Barrera-Badillo, Lucia Hernandez-Rivas, Irma Lopez-Martinez, Ernesto Ramirez-Gonzalez. |
| EPI_ISL_2246841, EPI_ISL_2246842, EPI_ISL_2246843, EPI_ISL_2246844, EPI_ISL_2246845, EPI_ISL_2246846, EPI_ISL_2246847, EPI_ISL_2246848, EPI_ISL_2246850, EPI_ISL_2246851, EPI_ISL_2246852, EPI_ISL_2246853, EPI_ISL_2246854, EPI_ISL_2246855, EPI_ISL_2246856, EPI_ISL_2246857, EPI_ISL_2246858, EPI_ISL_2246859, EPI_ISL_2246860, EPI_ISL_2246861, EPI_ISL_2246862, EPI_ISL_2246863, EPI_ISL_2246865, EPI_ISL_2246866, EPI_ISL_2246867, EPI_ISL_2246868, EPI_ISL_2246869, EPI_ISL_2246870, EPI_ISL_2246871, EPI_ISL_2246874, EPI_ISL_2246876, EPI_ISL_2246877, EPI_ISL_2246878, EPI_ISL_2246879, EPI_ISL_2246880, EPI_ISL_2246881, EPI_ISL_2246882, EPI_ISL_2246883, EPI_ISL_2246884, EPI_ISL_2246886 | | | |
| see above | Wyoming Public Health Laboratory | Wyoming Public Health Laboratory | Jim Mildenberger, Taylor Fearing, Channing Weber, Ashley Norberg, Chayse Rowley, Marley Goetz, Brian Dominguez, Elliot Thomasson, Brittany Oher, Sam Britz, Cari Sloma, and Rob Christensen |
| EPI_ISL_2246887, EPI_ISL_2246888, EPI_ISL_2246889 | Florida Bureau of Public Health Laboratories | Florida Bureau of Public Health Laboratories | Sarah Schmedes, Jason Blanton |
| EPI_ISL_2247060, EPI_ISL_2247061, EPI_ISL_2247062, EPI_ISL_2247063, EPI_ISL_2247064, EPI_ISL_2247065, EPI_ISL_2247066, EPI_ISL_2247067, EPI_ISL_2247068, EPI_ISL_2247069, EPI_ISL_2247070, EPI_ISL_2247071, EPI_ISL_2247072, EPI_ISL_2247073, EPI_ISL_2247074, EPI_ISL_2247075, EPI_ISL_2247076, EPI_ISL_2247077, EPI_ISL_2247078, EPI_ISL_2247079, EPI_ISL_2247080, EPI_ISL_2247081, EPI_ISL_2247082, EPI_ISL_2247083, EPI_ISL_2247084, EPI_ISL_2247085, EPI_ISL_2247086, EPI_ISL_2247087, EPI_ISL_2247088, EPI_ISL_2247089, EPI_ISL_2247090, EPI_ISL_2247091, EPI_ISL_2247092, EPI_ISL_2247093, EPI_ISL_2247094, EPI_ISL_2247095, EPI_ISL_2247096, EPI_ISL_2247097, EPI_ISL_2247098, EPI_ISL_2247099, EPI_ISL_2247100, EPI_ISL_2247101, EPI_ISL_2247102, EPI_ISL_2247103, EPI_ISL_2247104, EPI_ISL_2247105, EPI_ISL_2247106, EPI_ISL_2247107, EPI_ISL_2247108, EPI_ISL_2247109, EPI_ISL_2247110, EPI_ISL_2247111, EPI_ISL_2247112, EPI_ISL_2247113, EPI_ISL_2247114, EPI_ISL_2247115, EPI_ISL_2247116, EPI_ISL_2247117, EPI_ISL_2247118, EPI_ISL_2247119, EPI_ISL_2247120, EPI_ISL_2247121, EPI_ISL_2247122, EPI_ISL_2247123, EPI_ISL_2247124, EPI_ISL_2247125, EPI_ISL_2247126, EPI_ISL_2247127, EPI_ISL_2247128, EPI_ISL_2247129, EPI_ISL_2247130, EPI_ISL_2247131, EPI_ISL_2247132, EPI_ISL_2247133, EPI_ISL_2247134, EPI_ISL_2247135, EPI_ISL_2247136, EPI_ISL_2247137, EPI_ISL_2247138, EPI_ISL_2247139, EPI_ISL_2247140, EPI_ISL_2247141, EPI_ISL_2247142, EPI_ISL_2247143, EPI_ISL_2247144, EPI_ISL_2247145, EPI_ISL_2247146, EPI_ISL_2247147, EPI_ISL_2247148, EPI_ISL_2247149, EPI_ISL_2247150, EPI_ISL_2247151, EPI_ISL_2247152, EPI_ISL_2247153, EPI_ISL_2247154, EPI_ISL_2247155, EPI_ISL_2247156, EPI_ISL_2247157, EPI_ISL_2247158, EPI_ISL_2247159, EPI_ISL_2247160, EPI_ISL_2247161, EPI_ISL_2247162, EPI_ISL_2247163, EPI_ISL_2247164, EPI_ISL_2247165, EPI_ISL_2247166, EPI_ISL_2247167, EPI_ISL_2247168, EPI_ISL_2247169, EPI_ISL_2247170, EPI_ISL_2247171, EPI_ISL_2247172, EPI_ISL_2247173, EPI_ISL_2247174, EPI_ISL_2247175, EPI_ISL_2247176, EPI_ISL_2247177, EPI_ISL_2247178, EPI_ISL_2247179, EPI_ISL_2247180, EPI_ISL_2247181, EPI_ISL_2247182, EPI_ISL_2247183, EPI_ISL_2247184, EPI_ISL_2247185, EPI_ISL_2247186, EPI_ISL_2247187, EPI_ISL_2247188, EPI_ISL_2247189, EPI_ISL_2247190, EPI_ISL_2247191, EPI_ISL_2247192, EPI_ISL_2247193, EPI_ISL_2247194, EPI_ISL_2247195, EPI_ISL_2247196, EPI_ISL_2247197, EPI_ISL_2247198, EPI_ISL_2247199, EPI_ISL_2247200, EPI_ISL_2247201, EPI_ISL_2247202, EPI_ISL_2247203, EPI_ISL_2247204, EPI_ISL_2247205, EPI_ISL_2247206, EPI_ISL_2247207, EPI_ISL_2247208, EPI_ISL_2247209, EPI_ISL_2247210, EPI_ISL_2247211, EPI_ISL_2247212, EPI_ISL_2247213, EPI_ISL_2247214, EPI_ISL_2247215, EPI_ISL_2247216, EPI_ISL_2247217, EPI_ISL_2247218, EPI_ISL_2247219, EPI_ISL_2247220, EPI_ISL_2247221, EPI_ISL_2247222, EPI_ISL_2247223, EPI_ISL_2247224, EPI_ISL_2247225, EPI_ISL_2247226, EPI_ISL_2247227, EPI_ISL_2247228, EPI_ISL_2247229, EPI_ISL_2247230, EPI_ISL_2247231, EPI_ISL_2247232, EPI_ISL_2247233, EPI_ISL_2247234, EPI_ISL_2247235, EPI_ISL_2247236, EPI_ISL_2247237, EPI_ISL_2247238, EPI_ISL_2247239, EPI_ISL_2247240, EPI_ISL_2247241, EPI_ISL_2247242, EPI_ISL_2247243, EPI_ISL_2247244, EPI_ISL_2247245, EPI_ISL_2247246, EPI_ISL_2247247, EPI_ISL_2247248, EPI_ISL_2247249, EPI_ISL_2247250, EPI_ISL_2247251, EPI_ISL_2247252, EPI_ISL_2247253, EPI_ISL_2247254, EPI_ISL_2247255, EPI_ISL_2247256, EPI_ISL_2247257, EPI_ISL_2247258, EPI_ISL_2247259, EPI_ISL_2247260, EPI_ISL_2247261, EPI_ISL_2247262, EPI_ISL_2247263, EPI_ISL_2247264, EPI_ISL_2247265, EPI_ISL_2247266, EPI_ISL_2247267, EPI_ISL_2247268, EPI_ISL_2247269, EPI_ISL_2247270, EPI_ISL_2247271, EPI_ISL_2247272, EPI_ISL_2247273, EPI_ISL_2247274, EPI_ISL_2247275, EPI_ISL_2247276, EPI_ISL_2247277, EPI_ISL_2247278, EPI_ISL_2247279, EPI_ISL_2247280, EPI_ISL_2247281, EPI_ISL_2247282, EPI_ISL_2247283, EPI_ISL_2247284, EPI_ISL_2247285, EPI_ISL_2247286, EPI_ISL_2247287, EPI_ISL_2247288, EPI_ISL_2247289, EPI_ISL_2247290, EPI_ISL_2247291, EPI_ISL_2247292, EPI_ISL_2247293, EPI_ISL_2247294, EPI_ISL_2247295, EPI_ISL_2247296, EPI_ISL_2247297, EPI_ISL_2247298, EPI_ISL_2247299, EPI_ISL_2247300, EPI_ISL_2247301, EPI_ISL_2247302, EPI_ISL_2247303, EPI_ISL_2247304, EPI_ISL_2247305, EPI_ISL_2247306, EPI_ISL_2247307, EPI_ISL_2247308, EPI_ISL_2247309, EPI_ISL_2247310, EPI_ISL_2247311, EPI_ISL_2247312, EPI_ISL_2247313, EPI_ISL_2247314, EPI_ISL_2247315, EPI_ISL_2247316, EPI_ISL_2247317, EPI_ISL_2247318, EPI_ISL_2247319, EPI_ISL_2247320, EPI_ISL_2247321, EPI_ISL_2247322, EPI_ISL_2247323, EPI_ISL_2247324, EPI_ISL_2247325, EPI_ISL_2247326, EPI_ISL_2247327, EPI_ISL_2247328, EPI_ISL_2247329, EPI_ISL_2247330, EPI_ISL_2247331, EPI_ISL_2247332, EPI_ISL_2247333, EPI_ISL_2247334, EPI_ISL_2247335, EPI_ISL_2247336, EPI_ISL_2247337, EPI_ISL_2247338, EPI_ISL_2247339, EPI_ISL_2247340 | | | |
| see above | Quest Diagnostics Incorporated | Centers for Disease Control and Prevention Division of Viral Diseases, Pathogen Discovery | Dakota Howard, Dhwani Batra, Peter W. Cook, Kara Moser, Adrian Paskey, Jason Caravas, Benjamin Rambo-Martin, Shatavia Morrison, Christopher Gulvick, Scott Sammons, Yvette Unoarumhi, Darlene Wagner, Matthew Schmerer, S. H. Rosenthal, A. Gerasimova, R. M. Kagan, B. Anderson, M. Hua, Y. Liu, L.E. Bernstein, K.E. Livingston, A. Perez, I. A. Shlyakhter, R. V. Rolando, R. Owen, P. Tanpaiboon, F. Lacbawan, Clinton R. Paden, Duncan MacCannell |
| EPI_ISL_2247341, EPI_ISL_2247342, EPI_ISL_2247343, EPI_ISL_2247344, EPI_ISL_2247345, EPI_ISL_2247346, EPI_ISL_2247347, EPI_ISL_2247348, EPI_ISL_2247349, EPI_ISL_2247350, EPI_ISL_2247351, EPI_ISL_2247352, EPI_ISL_2247353, EPI_ISL_2247354, EPI_ISL_2247355, EPI_ISL_2247356, EPI_ISL_2247357, EPI_ISL_2247358, EPI_ISL_2247359, EPI_ISL_2247360, EPI_ISL_2247361, EPI_ISL_2247362, EPI_ISL_2247363, EPI_ISL_2247364, EPI_ISL_2247365, EPI_ISL_2247366, EPI_ISL_2247367, EPI_ISL_2247368, EPI_ISL_2247369, EPI_ISL_2247370, EPI_ISL_2247371, EPI_ISL_2247372, EPI_ISL_2247373, EPI_ISL_2247374, EPI_ISL_2247375, EPI_ISL_2247376, EPI_ISL_2247377, EPI_ISL_2247378, EPI_ISL_2247379, EPI_ISL_2247380, EPI_ISL_2247381, EPI_ISL_2247382, EPI_ISL_2247383, EPI_ISL_2247384, EPI_ISL_2247385, EPI_ISL_2247386, EPI_ISL_2247387, EPI_ISL_2247388, EPI_ISL_2247389, EPI_ISL_2247390, EPI_ISL_2247391, EPI_ISL_2247392, EPI_ISL_2247393, EPI_ISL_2247394, EPI_ISL_2247395, EPI_ISL_2247396, EPI_ISL_2247397, EPI_ISL_2247398, EPI_ISL_2247399, EPI_ISL_2247400, EPI_ISL_2247401, EPI_ISL_2247402, EPI_ISL_2247403, EPI_ISL_2247404, EPI_ISL_2247405, EPI_ISL_2247406, EPI_ISL_2247407, EPI_ISL_2247408, EPI_ISL_2247409, EPI_ISL_2247410, EPI_ISL_2247411, EPI_ISL_2247412, EPI_ISL_2247413, EPI_ISL_2247414, EPI_ISL_2247415, EPI_ISL_2247416, EPI_ISL_2247417, EPI_ISL_2247418, EPI_ISL_2247419, EPI_ISL_2247420, EPI_ISL_2247421, EPI_ISL_2247422, EPI_ISL_2247423, EPI_ISL_2247424, EPI_ISL_2247425, EPI_ISL_2247426, EPI_ISL_2247427, EPI_ISL_2247428, EPI_ISL_2247429, EPI_ISL_2247430, EPI_ISL_2247431, EPI_ISL_2247432, EPI_ISL_2247433, EPI_ISL_2247434, EPI_ISL_2247435, EPI_ISL_2247436, EPI_ISL_2247437, EPI_ISL_2247438, EPI_ISL_2247439, EPI_ISL_2247440, EPI_ISL_2247441, EPI_ISL_2247442, EPI_ISL_2247443, EPI_ISL_2247444, EPI_ISL_2247445, EPI_ISL_2247446, EPI_ISL_2247447, EPI_ISL_2247448, EPI_ISL_2247449, EPI_ISL_2247450, EPI_ISL_2247451, EPI_ISL_2247452, EPI_ISL_2247453, EPI_ISL_2247454, EPI_ISL_2247455, EPI_ISL_2247456, EPI_ISL_2247457, EPI_ISL_2247458, EPI_ISL_2247459, EPI_ISL_2247460, EPI_ISL_2247461, EPI_ISL_2247462, EPI_ISL_2247463, EPI_ISL_2247464, EPI_ISL_2247465, EPI_ISL_2247466, EPI_ISL_2247467, EPI_ISL_2247468, EPI_ISL_2247469, EPI_ISL_2247470, EPI_ISL_2247471, EPI_ISL_2247472, EPI_ISL_2247473, EPI_ISL_2247474, EPI_ISL_2247475, EPI_ISL_2247476, EPI_ISL_2247477, EPI_ISL_2247478, EPI_ISL_2247479, EPI_ISL_2247480, EPI_ISL_2247481, EPI_ISL_2247482, EPI_ISL_2247483, EPI_ISL_2247484, EPI_ISL_2247485, EPI_ISL_2247486, EPI_ISL_2247487, EPI_ISL_2247488, EPI_ISL_2247489, EPI_ISL_2247490, EPI_ISL_2247491, EPI_ISL_2247492, EPI_ISL_2247493, EPI_ISL_2247494, EPI_ISL_2247495, EPI_ISL_2247496, EPI_ISL_2247497, EPI_ISL_2247498, EPI_ISL_2247499, EPI_ISL_2247500, EPI_ISL_2247501, EPI_ISL_2247502, EPI_ISL_2247503, EPI_ISL_2247504, EPI_ISL_2247505, EPI_ISL_2247506, EPI_ISL_2247507, EPI_ISL_2247508, EPI_ISL_2247509, EPI_ISL_2247510, EPI_ISL_2247511, EPI_ISL_2247512, EPI_ISL_2247513, EPI_ISL_2247514, EPI_ISL_2247515, EPI_ISL_2247516, EPI_ISL_2247517, EPI_ISL_2247518, EPI_ISL_2247519, EPI_ISL_2247520, EPI_ISL_2247521, EPI_ISL_2247522, EPI_ISL_2247523, EPI_ISL_2247524, EPI_ISL_2247525, EPI_ISL_2247526, EPI_ISL_2247527, EPI_ISL_2247528, EPI_ISL_2247529, EPI_ISL_2247530, EPI_ISL_2247531, EPI_ISL_2247532, EPI_ISL_2247533, EPI_ISL_2247534, EPI_ISL_2247535, EPI_ISL_2247536, EPI_ISL_2247537, EPI_ISL_2247538, EPI_ISL_2247539, EPI_ISL_2247540, EPI_ISL_2247541, EPI_ISL_2247542, EPI_ISL_2247543, EPI_ISL_2247544, EPI_ISL_2247545, EPI_ISL_2247546, EPI_ISL_2247547, EPI_ISL_2247548, EPI_ISL_2247549, EPI_ISL_2247550, EPI_ISL_2247551, EPI_ISL_2247552, EPI_ISL_2247553, EPI_ISL_2247554, EPI_ISL_2247555, EPI_ISL_2247556, EPI_ISL_2247557, EPI_ISL_2247558, EPI_ISL_2247559, EPI_ISL_2247560, EPI_ISL_2247561, EPI_ISL_2247562, EPI_ISL_2247563, EPI_ISL_2247564, EPI_ISL_2247565, EPI_ISL_2247566, EPI_ISL_2247567, EPI_ISL_2247568, EPI_ISL_2247569, EPI_ISL_2247570, EPI_ISL_2247571, EPI_ISL_2247572, EPI_ISL_2247573, EPI_ISL_2247574, EPI_ISL_2247575, EPI_ISL_2247576, EPI_ISL_2247577, EPI_ISL_2247578, EPI_ISL_2247579, EPI_ISL_2247580, EPI_ISL_2247581, EPI_ISL_2247582, EPI_ISL_2247583, EPI_ISL_2247584, EPI_ISL_2247585, EPI_ISL_2247586, EPI_ISL_2247587, EPI_ISL_2247588, EPI_ISL_2247589, EPI_ISL_2247590, EPI_ISL_2247591, EPI_ISL_2247592, EPI_ISL_2247593, EPI_ISL_2247594, EPI_ISL_2247595, EPI_ISL_2247596, EPI_ISL_2247598, EPI_ISL_2247599, EPI_ISL_2247600, EPI_ISL_2247601, EPI_ISL_2247603, EPI_ISL_2247604, EPI_ISL_2247605, EPI_ISL_2247606, EPI_ISL_2247607, EPI_ISL_2247608, EPI_ISL_2247609, EPI_ISL_2247610, EPI_ISL_2247611, EPI_ISL_2247612, EPI_ISL_2247613, EPI_ISL_2247614, EPI_ISL_2247615, EPI_ISL_2247616, EPI_ISL_2247617, EPI_ISL_2247618, EPI_ISL_2247619, EPI_ISL_2247620, EPI_ISL_2247621, EPI_ISL_2247622, EPI_ISL_2247623, EPI_ISL_2247624, EPI_ISL_2247625, EPI_ISL_2247626, EPI_ISL_2247627, EPI_ISL_2247628, EPI_ISL_2247629, EPI_ISL_2247630, EPI_ISL_2247631, EPI_ISL_2247632, EPI_ISL_2247633, EPI_ISL_2247634, EPI_ISL_2247635, EPI_ISL_2247636, EPI_ISL_2247637, EPI_ISL_2247638, EPI_ISL_2247639, EPI_ISL_2247640, EPI_ISL_2247641, EPI_ISL_2247642, EPI_ISL_2247643, EPI_ISL_2247644, EPI_ISL_2247645, EPI_ISL_2247646, EPI_ISL_2247647, EPI_ISL_2247648, EPI_ISL_2247649, EPI_ISL_2247650, EPI_ISL_2247651, EPI_ISL_2247652, EPI_ISL_2247653, EPI_ISL_2247655, EPI_ISL_2247656, EPI_ISL_2247657, EPI_ISL_2247658, EPI_ISL_2247659, EPI_ISL_2247660, EPI_ISL_2247661, EPI_ISL_2247662, EPI_ISL_2247663, EPI_ISL_2247664, EPI_ISL_2247665, EPI_ISL_2247666, EPI_ISL_2247667, EPI_ISL_2247668, EPI_ISL_2247669, EPI_ISL_2247670, EPI_ISL_2247671, EPI_ISL_2247672, EPI_ISL_2247673, EPI_ISL_2247674, EPI_ISL_2247675, EPI_ISL_2247676, EPI_ISL_2247677, EPI_ISL_2247678, EPI_ISL_2247679, EPI_ISL_2247680, EPI_ISL_2247681, EPI_ISL_2247682, EPI_ISL_2247683, EPI_ISL_2247684, EPI_ISL_2247685, EPI_ISL_2247686, EPI_ISL_2247687, EPI_ISL_2247688, EPI_ISL_2247689, EPI_ISL_2247690, EPI_ISL_2247691, EPI_ISL_2247692, EPI_ISL_2247693, EPI_ISL_2247694, EPI_ISL_2247695, EPI_ISL_2247696, EPI_ISL_2247697, EPI_ISL_2247698, EPI_ISL_2247699, EPI_ISL_2247700, EPI_ISL_2247701, EPI_ISL_2247702, EPI_ISL_2247703, EPI_ISL_2247704, EPI_ISL_2247705, EPI_ISL_2247706, EPI_ISL_2247707, EPI_ISL_2247708, EPI_ISL_2247709, EPI_ISL_2247710, EPI_ISL_2247711, EPI_ISL_2247712, EPI_ISL_2247713, EPI_ISL_2247714, EPI_ISL_2247715, EPI_ISL_2247716, EPI_ISL_2247717, EPI_ISL_2247718, EPI_ISL_2247719, EPI_ISL_2247720, EPI_ISL_2247721, EPI_ISL_2247722, EPI_ISL_2247723, EPI_ISL_2247724, EPI_ISL_2247725, EPI_ISL_2247726, EPI_ISL_2247727, EPI_ISL_2247728, EPI_ISL_2247729, EPI_ISL_2247730, EPI_ISL_2247731, EPI_ISL_2247732, EPI_ISL_2247733, EPI_ISL_2247734, EPI_ISL_2247735, EPI_ISL_2247736, EPI_ISL_2247737, EPI_ISL_2247738, EPI_ISL_2247739, EPI_ISL_2247740, EPI_ISL_2247742, EPI_ISL_2247743, EPI_ISL_2247744, EPI_ISL_2247745, EPI_ISL_2247746, EPI_ISL_2247747, EPI_ISL_2247748, EPI_ISL_2247749, EPI_ISL_2247750, EPI_ISL_2247751, EPI_ISL_2247752, EPI_ISL_2247753, EPI_ISL_2247754, EPI_ISL_2247755, EPI_ISL_2247756, EPI_ISL_2247757, EPI_ISL_2247758, EPI_ISL_2247759, EPI_ISL_2247761, EPI_ISL_2247762, EPI_ISL_2247763, EPI_ISL_2247764, EPI_ISL_2247765, EPI_ISL_2247766, EPI_ISL_2247767, EPI_ISL_2247768, EPI_ISL_2247769, EPI_ISL_2247770, EPI_ISL_2247771, EPI_ISL_2247772, EPI_ISL_2247773, EPI_ISL_2247774, EPI_ISL_2247775, EPI_ISL_2247776, EPI_ISL_2247777, EPI_ISL_2247778, EPI_ISL_2247779, EPI_ISL_2247780, EPI_ISL_2247781, EPI_ISL_2247782, EPI_ISL_2247783, EPI_ISL_2247784, EPI_ISL_2247785, EPI_ISL_2247786, EPI_ISL_2247787, EPI_ISL_2247788, EPI_ISL_2247789, EPI_ISL_2247790, EPI_ISL_2247791, EPI_ISL_2247792, EPI_ISL_2247793, EPI_ISL_2247794, EPI_ISL_2247795, EPI_ISL_2247796, EPI_ISL_2247797, EPI_ISL_2247798, EPI_ISL_2247799, EPI_ISL_2247800, EPI_ISL_2247801, EPI_ISL_2247802, EPI_ISL_2247803, EPI_ISL_2247804, EPI_ISL_2247805, EPI_ISL_2247806, EPI_ISL_2247807, EPI_ISL_2247809, EPI_ISL_2247810, EPI_ISL_2247811, EPI_ISL_2247812, EPI_ISL_2247813, EPI_ISL_2247814, EPI_ISL_2247815, EPI_ISL_2247816, EPI_ISL_2247817, EPI_ISL_2247818, EPI_ISL_2247819, EPI_ISL_2247820, EPI_ISL_2247821, EPI_ISL_2247822, EPI_ISL_2247823, EPI_ISL_2247824, EPI_ISL_2247825, EPI_ISL_2247826, EPI_ISL_2247827, EPI_ISL_2247828, EPI_ISL_2247829, EPI_ISL_2247830, EPI_ISL_2247831, EPI_ISL_2247832, EPI_ISL_2247833, EPI_ISL_2247834, EPI_ISL_2247835, EPI_ISL_2247836, EPI_ISL_2247837, EPI_ISL_2247838, EPI_ISL_2247839, EPI_ISL_2247840, EPI_ISL_2247841, EPI_ISL_2247842, EPI_ISL_2247843, EPI_ISL_2247844, EPI_ISL_2247845, EPI_ISL_2247846, EPI_ISL_2247847, EPI_ISL_2247848, EPI_ISL_2247849, EPI_ISL_2247850, EPI_ISL_2247851, EPI_ISL_2247852, EPI_ISL_2247853, EPI_ISL_2247854, EPI_ISL_2247855, EPI_ISL_2247856, EPI_ISL_2247857, EPI_ISL_2247858, EPI_ISL_2247859, EPI_ISL_2247860, EPI_ISL_2247861, EPI_ISL_2247862, EPI_ISL_2247863, EPI_ISL_2247864, EPI_ISL_2247865, EPI_ISL_2247866, EPI_ISL_2247867, EPI_ISL_2247868, EPI_ISL_2247869, EPI_ISL_2247870, EPI_ISL_2247871, EPI_ISL_2247872, EPI_ISL_2247873, EPI_ISL_2247874, EPI_ISL_2247875, EPI_ISL_2247876, EPI_ISL_2247877, EPI_ISL_2247878, EPI_ISL_2247879, EPI_ISL_2247880, EPI_ISL_2247881, EPI_ISL_2247882, EPI_ISL_2247883, EPI_ISL_2247884, EPI_ISL_2247885, EPI_ISL_2247886, EPI_ISL_2247887, EPI_ISL_2247888, EPI_ISL_2247889, EPI_ISL_2247890, EPI_ISL_2247891, EPI_ISL_2247892, EPI_ISL_2247893, EPI_ISL_2247894, EPI_ISL_2247895, EPI_ISL_2247896, EPI_ISL_2247897, EPI_ISL_2247898, EPI_ISL_2247899, EPI_ISL_2247900, EPI_ISL_2247901, EPI_ISL_2247902, EPI_ISL_2247903, EPI_ISL_2247904, EPI_ISL_2247905, EPI_ISL_2247906, EPI_ISL_2247907, EPI_ISL_2247908, EPI_ISL_2247909, EPI_ISL_2247910, EPI_ISL_2247911, EPI_ISL_2247912, EPI_ISL_2247913, EPI_ISL_2247914, EPI_ISL_2247915, EPI_ISL_2247916, EPI_ISL_2247917, EPI_ISL_2247918, EPI_ISL_2247919, EPI_ISL_2247920, EPI_ISL_2247921, EPI_ISL_2247922, EPI_ISL_2247923, EPI_ISL_2247924, EPI_ISL_2247925, EPI_ISL_2247926, EPI_ISL_2247927, EPI_ISL_2247928, EPI_ISL_2247929, EPI_ISL_2247930, EPI_ISL_2247931, EPI_ISL_2247932, EPI_ISL_2247933, EPI_ISL_2247934, EPI_ISL_2247935, EPI_ISL_2247936, EPI_ISL_2247937, EPI_ISL_2247938, EPI_ISL_2247939, EPI_ISL_2247940, EPI_ISL_2247941, EPI_ISL_2247942, EPI_ISL_2247943, EPI_ISL_2247944, EPI_ISL_2247945, EPI_ISL_2247946, EPI_ISL_2247947, EPI_ISL_2247948, EPI_ISL_2247949, EPI_ISL_2247950, EPI_ISL_2247951, EPI_ISL_2247952, EPI_ISL_2247953, EPI_ISL_2247954, EPI_ISL_2247955, EPI_ISL_2247956, EPI_ISL_2247957, EPI_ISL_2247958 | | | |
| see above | Helix/Illumina | Centers for Disease Control and Prevention Division of Viral Diseases, Pathogen Discovery | Dakota Howard, Dhwani Batra, Peter W. Cook, Kara Moser, Adrian Paskey, Jason Caravas, Benjamin Rambo-Martin, Shatavia Morrison, Christopher Gulvick, Scott Sammons, Yvette Unoarumhi, Darlene Wagner, Matthew Schmerer, Eileen de Feo, Jan Antico, Christine Tran, Matthew Tolentino, Shannon Wickline, Kim Gietzen, Brad Sickler, Jingtao Liu, Eric Allen, Phil Febbo, Nicole L. Washington, Simon White, Geraint Levan, Kelly Schiabor Barrett, Elizabeth Cirulli, Alexandre Bolze, Ary Ascencio, Charlotte Rivera-Garcia, Ryan Cho, Jason Nguyen, Sherry Wang, Jimmy Ramirez, Tyler Cassens, Efren Sandoval, Magnus Isaksson, William Lee, David Becker, Marc Laurent, James Lu, Clinton R. Paden, Duncan MacCannell |
| EPI_ISL_2247959 | Wyoming Public Health Laboratory | Wyoming Public Health Laboratory | Jim Mildenberger, Taylor Fearing, Channing Weber, Ashley Norberg, Chayse Rowley, Marley Goetz, Brian Dominguez, Elliot Thomasson, Brittany Oher, Sam Britz, Cari Sloma, and Rob Christensen |
| EPI_ISL_2247960 | Helix/Illumina | Centers for Disease Control and Prevention Division of Viral Diseases, Pathogen Discovery | Dakota Howard, Dhwani Batra, Peter W. Cook, Kara Moser, Adrian Paskey, Jason Caravas, Benjamin Rambo-Martin, Shatavia Morrison, Christopher Gulvick, Scott Sammons, Yvette Unoarumhi, Darlene Wagner, Matthew Schmerer, Eileen de Feo, Jan Antico, Christine Tran, Matthew Tolentino, Shannon Wickline, Kim Gietzen, Brad Sickler, Jingtao Liu, Eric Allen, Phil Febbo, Nicole L. Washington, Simon White, Geraint Levan, Kelly Schiabor Barrett, Elizabeth Cirulli, Alexandre Bolze, Ary Ascencio, Charlotte Rivera-Garcia, Ryan Cho, Jason Nguyen, Sherry Wang, Jimmy Ramirez, Tyler Cassens, Efren Sandoval, Magnus Isaksson, William Lee, David Becker, Marc Laurent, James Lu, Clinton R. Paden, Duncan MacCannell |
| EPI_ISL_2247961, EPI_ISL_2247962 | Wyoming Public Health Laboratory | Wyoming Public Health Laboratory | Jim Mildenberger, Taylor Fearing, Channing Weber, Ashley Norberg, Chayse Rowley, Marley Goetz, Brian Dominguez, Elliot Thomasson, Brittany Oher, Sam Britz, Cari Sloma, and Rob Christensen |
| EPI_ISL_2247963 | Helix/Illumina | Centers for Disease Control and Prevention Division of Viral Diseases, Pathogen Discovery | Dakota Howard, Dhwani Batra, Peter W. Cook, Kara Moser, Adrian Paskey, Jason Caravas, Benjamin Rambo-Martin, Shatavia Morrison, Christopher Gulvick, Scott Sammons, Yvette Unoarumhi, Darlene Wagner, Matthew Schmerer, Eileen de Feo, Jan Antico, Christine Tran, Matthew Tolentino, Shannon Wickline, Kim Gietzen, Brad Sickler, Jingtao Liu, Eric Allen, Phil Febbo, Nicole L. Washington, Simon White, Geraint Levan, Kelly Schiabor Barrett, Elizabeth Cirulli, Alexandre Bolze, Ary Ascencio, Charlotte Rivera-Garcia, Ryan Cho, Jason Nguyen, Sherry Wang, Jimmy Ramirez, Tyler Cassens, Efren Sandoval, Magnus Isaksson, William Lee, David Becker, Marc Laurent, James Lu, Clinton R. Paden, Duncan MacCannell |
| EPI_ISL_2247964 | Wyoming Public Health Laboratory | Wyoming Public Health Laboratory | Jim Mildenberger, Taylor Fearing, Channing Weber, Ashley Norberg, Chayse Rowley, Marley Goetz, Brian Dominguez, Elliot Thomasson, Brittany Oher, Sam Britz, Cari Sloma, and Rob Christensen |
| EPI_ISL_2247965 | Helix/Illumina | Centers for Disease Control and Prevention Division of Viral Diseases, Pathogen Discovery | Dakota Howard, Dhwani Batra, Peter W. Cook, Kara Moser, Adrian Paskey, Jason Caravas, Benjamin Rambo-Martin, Shatavia Morrison, Christopher Gulvick, Scott Sammons, Yvette Unoarumhi, Darlene Wagner, Matthew Schmerer, Eileen de Feo, Jan Antico, Christine Tran, Matthew Tolentino, Shannon Wickline, Kim Gietzen, Brad Sickler, Jingtao Liu, Eric Allen, Phil Febbo, Nicole L. Washington, Simon White, Geraint Levan, Kelly Schiabor Barrett, Elizabeth Cirulli, Alexandre Bolze, Ary Ascencio, Charlotte Rivera-Garcia, Ryan Cho, Jason Nguyen, Sherry Wang, Jimmy Ramirez, Tyler Cassens, Efren Sandoval, Magnus Isaksson, William Lee, David Becker, Marc Laurent, James Lu, Clinton R. Paden, Duncan MacCannell |
| EPI_ISL_2247966, | Wyoming Public Health Laboratory | Wyoming Public Health Laboratory | Jim Mildenberger, Taylor Fearing, Channing Weber, Ashley Norberg, Chayse Rowley, Marley Goetz, Brian Dominguez, Elliot Thomasson, Brittany Oher, Sam Britz, Cari Sloma, and Rob Christensen |

| | | | |
|---|---|---|---|
| EPI_ISL_2247967 | | | |
| EPI_ISL_2247968, EPI_ISL_2247969 | Helix/Illumina | Centers for Disease Control and Prevention Division of Viral Diseases, Pathogen Discovery | Dakota Howard, Dhwani Batra, Peter W. Cook, Kara Moser, Adrian Paskey, Jason Caravas, Benjamin Rambo-Martin, Shatavia Morrison, Christopher Gulvick, Scott Sammons, Yvette Unoarumhi, Darlene Wagner, Matthew Schmerer, Eileen de Feo, Jan Antico, Christine Tran, Matthew Tolentino, Shannon Wickline, Kim Gietzen, Brad Sickler, Jingtao Liu, Eric Allen, Phil Febbo, Nicole L. Washington, Simon White, Geraint Levan, Kelly Schiabor Barrett, Elizabeth Cirulli, Alexandre Bolze, Ary Ascencio, Charlotte Rivera-Garcia, Ryan Cho, Jason Nguyen, Sherry Wang, Jimmy Ramirez, Tyler Cassens, Efren Sandoval, Magnus Isaksson, William Lee, David Becker, Marc Laurent, James Lu, Clinton R. Paden, Duncan MacCannell |
| EPI_ISL_2247970, EPI_ISL_2247971, EPI_ISL_2247972 | Wyoming Public Health Laboratory | Wyoming Public Health Laboratory | Jim Mildenberger, Taylor Fearing, Channing Weber, Ashley Norberg, Chayse Rowley, Marley Goetz, Brian Dominguez, Elliot Thomasson, Brittany Oher, Sam Britz, Cari Sloma, and Rob Christensen |
| EPI_ISL_2247973 | Helix/Illumina | Centers for Disease Control and Prevention Division of Viral Diseases, Pathogen Discovery | Dakota Howard, Dhwani Batra, Peter W. Cook, Kara Moser, Adrian Paskey, Jason Caravas, Benjamin Rambo-Martin, Shatavia Morrison, Christopher Gulvick, Scott Sammons, Yvette Unoarumhi, Darlene Wagner, Matthew Schmerer, Eileen de Feo, Jan Antico, Christine Tran, Matthew Tolentino, Shannon Wickline, Kim Gietzen, Brad Sickler, Jingtao Liu, Eric Allen, Phil Febbo, Nicole L. Washington, Simon White, Geraint Levan, Kelly Schiabor Barrett, Elizabeth Cirulli, Alexandre Bolze, Ary Ascencio, Charlotte Rivera-Garcia, Ryan Cho, Jason Nguyen, Sherry Wang, Jimmy Ramirez, Tyler Cassens, Efren Sandoval, Magnus Isaksson, William Lee, David Becker, Marc Laurent, James Lu, Clinton R. Paden, Duncan MacCannell |
| EPI_ISL_2247974, EPI_ISL_2247975 | Wyoming Public Health Laboratory | Wyoming Public Health Laboratory | Jim Mildenberger, Taylor Fearing, Channing Weber, Ashley Norberg, Chayse Rowley, Marley Goetz, Brian Dominguez, Elliot Thomasson, Brittany Oher, Sam Britz, Cari Sloma, and Rob Christensen |
| EPI_ISL_2247976 | Helix/Illumina | Centers for Disease Control and Prevention Division of Viral Diseases, Pathogen Discovery | Dakota Howard, Dhwani Batra, Peter W. Cook, Kara Moser, Adrian Paskey, Jason Caravas, Benjamin Rambo-Martin, Shatavia Morrison, Christopher Gulvick, Scott Sammons, Yvette Unoarumhi, Darlene Wagner, Matthew Schmerer, Eileen de Feo, Jan Antico, Christine Tran, Matthew Tolentino, Shannon Wickline, Kim Gietzen, Brad Sickler, Jingtao Liu, Eric Allen, Phil Febbo, Nicole L. Washington, Simon White, Geraint Levan, Kelly Schiabor Barrett, Elizabeth Cirulli, Alexandre Bolze, Ary Ascencio, Charlotte Rivera-Garcia, Ryan Cho, Jason Nguyen, Sherry Wang, Jimmy Ramirez, Tyler Cassens, Efren Sandoval, Magnus Isaksson, William Lee, David Becker, Marc Laurent, James Lu, Clinton R. Paden, Duncan MacCannell |
| EPI_ISL_2247977, EPI_ISL_2247978 | Wyoming Public Health Laboratory | Wyoming Public Health Laboratory | Jim Mildenberger, Taylor Fearing, Channing Weber, Ashley Norberg, Chayse Rowley, Marley Goetz, Brian Dominguez, Elliot Thomasson, Brittany Oher, Sam Britz, Cari Sloma, and Rob Christensen |
| EPI_ISL_2247979 | Helix/Illumina | Centers for Disease Control and Prevention Division of Viral Diseases, Pathogen Discovery | Dakota Howard, Dhwani Batra, Peter W. Cook, Kara Moser, Adrian Paskey, Jason Caravas, Benjamin Rambo-Martin, Shatavia Morrison, Christopher Gulvick, Scott Sammons, Yvette Unoarumhi, Darlene Wagner, Matthew Schmerer, Eileen de Feo, Jan Antico, Christine Tran, Matthew Tolentino, Shannon Wickline, Kim Gietzen, Brad Sickler, Jingtao Liu, Eric Allen, Phil Febbo, Nicole L. Washington, Simon White, Geraint Levan, Kelly Schiabor Barrett, Elizabeth Cirulli, Alexandre Bolze, Ary Ascencio, Charlotte Rivera-Garcia, Ryan Cho, Jason Nguyen, Sherry Wang, Jimmy Ramirez, Tyler Cassens, Efren Sandoval, Magnus Isaksson, William Lee, David Becker, Marc Laurent, James Lu, Clinton R. Paden, Duncan MacCannell |
| EPI_ISL_2247980 | Wyoming Public Health Laboratory | Wyoming Public Health Laboratory | Jim Mildenberger, Taylor Fearing, Channing Weber, Ashley Norberg, Chayse Rowley, Marley Goetz, Brian Dominguez, Elliot Thomasson, Brittany Oher, Sam Britz, Cari Sloma, and Rob Christensen |
| EPI_ISL_2247981 | Helix/Illumina | Centers for Disease Control and Prevention Division of Viral Diseases, Pathogen Discovery | Dakota Howard, Dhwani Batra, Peter W. Cook, Kara Moser, Adrian Paskey, Jason Caravas, Benjamin Rambo-Martin, Shatavia Morrison, Christopher Gulvick, Scott Sammons, Yvette Unoarumhi, Darlene Wagner, Matthew Schmerer, Eileen de Feo, Jan Antico, Christine Tran, Matthew Tolentino, Shannon Wickline, Kim Gietzen, Brad Sickler, Jingtao Liu, Eric Allen, Phil Febbo, Nicole L. Washington, Simon White, Geraint Levan, Kelly Schiabor Barrett, Elizabeth Cirulli, Alexandre Bolze, Ary Ascencio, Charlotte Rivera-Garcia, Ryan Cho, Jason Nguyen, Sherry Wang, Jimmy Ramirez, Tyler Cassens, Efren Sandoval, Magnus Isaksson, William Lee, David Becker, Marc Laurent, James Lu, Clinton R. Paden, Duncan MacCannell |
| EPI_ISL_2247982, EPI_ISL_2247983 | Wyoming Public Health Laboratory | Wyoming Public Health Laboratory | Jim Mildenberger, Taylor Fearing, Channing Weber, Ashley Norberg, Chayse Rowley, Marley Goetz, Brian Dominguez, Elliot Thomasson, Brittany Oher, Sam Britz, Cari Sloma, and Rob Christensen |
| EPI_ISL_2247984 | Helix/Illumina | Centers for Disease Control and Prevention Division of Viral Diseases, Pathogen Discovery | Dakota Howard, Dhwani Batra, Peter W. Cook, Kara Moser, Adrian Paskey, Jason Caravas, Benjamin Rambo-Martin, Shatavia Morrison, Christopher Gulvick, Scott Sammons, Yvette Unoarumhi, Darlene Wagner, Matthew Schmerer, Eileen de Feo, Jan Antico, Christine Tran, Matthew Tolentino, Shannon Wickline, Kim Gietzen, Brad Sickler, Jingtao Liu, Eric Allen, Phil Febbo, Nicole L. Washington, Simon White, Geraint Levan, Kelly Schiabor Barrett, Elizabeth Cirulli, Alexandre Bolze, Ary Ascencio, Charlotte Rivera-Garcia, Ryan Cho, Jason Nguyen, Sherry Wang, Jimmy Ramirez, Tyler Cassens, Efren Sandoval, Magnus Isaksson, William Lee, David Becker, Marc Laurent, James Lu, Clinton R. Paden, Duncan MacCannell |
| EPI_ISL_2247985, EPI_ISL_2247986, EPI_ISL_2247987, EPI_ISL_2247988 | Wyoming Public Health Laboratory | Wyoming Public Health Laboratory | Jim Mildenberger, Taylor Fearing, Channing Weber, Ashley Norberg, Chayse Rowley, Marley Goetz, Brian Dominguez, Elliot Thomasson, Brittany Oher, Sam Britz, Cari Sloma, and Rob Christensen |
| EPI_ISL_2247989 | Helix/Illumina | Centers for Disease Control and Prevention Division of Viral Diseases, Pathogen Discovery | Dakota Howard, Dhwani Batra, Peter W. Cook, Kara Moser, Adrian Paskey, Jason Caravas, Benjamin Rambo-Martin, Shatavia Morrison, Christopher Gulvick, Scott Sammons, Yvette Unoarumhi, Darlene Wagner, Matthew Schmerer, Eileen de Feo, Jan Antico, Christine Tran, Matthew Tolentino, Shannon Wickline, Kim Gietzen, Brad Sickler, Jingtao Liu, Eric Allen, Phil Febbo, Nicole L. Washington, Simon White, Geraint Levan, Kelly Schiabor Barrett, Elizabeth Cirulli, Alexandre Bolze, Ary Ascencio, Charlotte Rivera-Garcia, Ryan Cho, Jason Nguyen, Sherry Wang, Jimmy Ramirez, Tyler Cassens, Efren Sandoval, Magnus Isaksson, William Lee, David Becker, Marc Laurent, James Lu, Clinton R. Paden, Duncan MacCannell |
| EPI_ISL_2247990, EPI_ISL_2247991 | Wyoming Public Health Laboratory | Wyoming Public Health Laboratory | Jim Mildenberger, Taylor Fearing, Channing Weber, Ashley Norberg, Chayse Rowley, Marley Goetz, Brian Dominguez, Elliot Thomasson, Brittany Oher, Sam Britz, Cari Sloma, and Rob Christensen |
| EPI_ISL_2247992 | Helix/Illumina | Centers for Disease Control and Prevention Division of Viral Diseases, Pathogen Discovery | Dakota Howard, Dhwani Batra, Peter W. Cook, Kara Moser, Adrian Paskey, Jason Caravas, Benjamin Rambo-Martin, Shatavia Morrison, Christopher Gulvick, Scott Sammons, Yvette Unoarumhi, Darlene Wagner, Matthew Schmerer, Eileen de Feo, Jan Antico, Christine Tran, Matthew Tolentino, Shannon Wickline, Kim Gietzen, Brad Sickler, Jingtao Liu, Eric Allen, Phil Febbo, Nicole L. Washington, Simon White, Geraint Levan, Kelly Schiabor Barrett, Elizabeth Cirulli, Alexandre Bolze, Ary Ascencio, Charlotte Rivera-Garcia, Ryan Cho, Jason Nguyen, Sherry Wang, Jimmy Ramirez, Tyler Cassens, Efren Sandoval, Magnus Isaksson, William Lee, David Becker, Marc Laurent, James Lu, Clinton R. Paden, Duncan MacCannell |
| EPI_ISL_2247993 | Wyoming Public Health Laboratory | Wyoming Public Health Laboratory | Jim Mildenberger, Taylor Fearing, Channing Weber, Ashley Norberg, Chayse Rowley, Marley Goetz, Brian Dominguez, Elliot Thomasson, Brittany Oher, Sam Britz, Cari Sloma, and Rob Christensen |
| EPI_ISL_2247994 | Helix/Illumina | Centers for Disease Control and Prevention Division of Viral Diseases, Pathogen Discovery | Dakota Howard, Dhwani Batra, Peter W. Cook, Kara Moser, Adrian Paskey, Jason Caravas, Benjamin Rambo-Martin, Shatavia Morrison, Christopher Gulvick, Scott Sammons, Yvette Unoarumhi, Darlene Wagner, Matthew Schmerer, Eileen de Feo, Jan Antico, Christine Tran, Matthew Tolentino, Shannon Wickline, Kim Gietzen, Brad Sickler, Jingtao Liu, Eric Allen, Phil Febbo, Nicole L. Washington, Simon White, Geraint Levan, Kelly Schiabor Barrett, Elizabeth Cirulli, Alexandre Bolze, Ary Ascencio, Charlotte Rivera-Garcia, Ryan Cho, Jason Nguyen, Sherry Wang, Jimmy Ramirez, Tyler Cassens, Efren Sandoval, Magnus Isaksson, William Lee, David Becker, Marc Laurent, James Lu, Clinton R. Paden, Duncan MacCannell |
| EPI_ISL_2247995 | Wyoming Public Health Laboratory | Wyoming Public Health Laboratory | Jim Mildenberger, Taylor Fearing, Channing Weber, Ashley Norberg, Chayse Rowley, Marley Goetz, Brian Dominguez, Elliot Thomasson, Brittany Oher, Sam Britz, Cari Sloma, and Rob Christensen |
| EPI_ISL_2247997 | Helix/Illumina | Centers for Disease Control and Prevention Division of Viral Diseases, Pathogen Discovery | Dakota Howard, Dhwani Batra, Peter W. Cook, Kara Moser, Adrian Paskey, Jason Caravas, Benjamin Rambo-Martin, Shatavia Morrison, Christopher Gulvick, Scott Sammons, Yvette Unoarumhi, Darlene Wagner, Matthew Schmerer, Eileen de Feo, Jan Antico, Christine Tran, Matthew Tolentino, Shannon Wickline, Kim Gietzen, Brad Sickler, Jingtao Liu, Eric Allen, Phil Febbo, Nicole L. Washington, Simon White, Geraint Levan, Kelly Schiabor Barrett, Elizabeth Cirulli, Alexandre Bolze, Ary Ascencio, Charlotte Rivera-Garcia, Ryan Cho, Jason Nguyen, Sherry Wang, Jimmy Ramirez, Tyler Cassens, Efren Sandoval, Magnus Isaksson, William Lee, David Becker, Marc Laurent, James Lu, Clinton R. Paden, Duncan MacCannell |
| EPI_ISL_2247999 | Wyoming Public Health Laboratory | Wyoming Public Health Laboratory | Jim Mildenberger, Taylor Fearing, Channing Weber, Ashley Norberg, Chayse Rowley, Marley Goetz, Brian Dominguez, Elliot Thomasson, Brittany Oher, Sam Britz, Cari Sloma, and Rob Christensen |
| EPI_ISL_2248000 | Helix/Illumina | Centers for Disease Control and Prevention Division of Viral Diseases, Pathogen Discovery | Dakota Howard, Dhwani Batra, Peter W. Cook, Kara Moser, Adrian Paskey, Jason Caravas, Benjamin Rambo-Martin, Shatavia Morrison, Christopher Gulvick, Scott Sammons, Yvette Unoarumhi, Darlene Wagner, Matthew Schmerer, Eileen de Feo, Jan Antico, Christine Tran, Matthew Tolentino, Shannon Wickline, Kim Gietzen, Brad Sickler, Jingtao Liu, Eric Allen, Phil Febbo, Nicole L. Washington, Simon White, Geraint Levan, Kelly Schiabor Barrett, Elizabeth Cirulli, Alexandre Bolze, Ary Ascencio, Charlotte Rivera-Garcia, Ryan Cho, Jason Nguyen, Sherry Wang, Jimmy Ramirez, Tyler Cassens, Efren Sandoval, Magnus Isaksson, William Lee, David Becker, Marc Laurent, James Lu, Clinton R. Paden, Duncan MacCannell |
| EPI_ISL_2248001 | Wyoming Public Health Laboratory | Wyoming Public Health Laboratory | Jim Mildenberger, Taylor Fearing, Channing Weber, Ashley Norberg, Chayse Rowley, Marley Goetz, Brian Dominguez, Elliot Thomasson, Brittany Oher, Sam Britz, Cari Sloma, and Rob Christensen |
| EPI_ISL_2248002 | Helix/Illumina | Centers for Disease Control and Prevention Division of Viral Diseases, Pathogen Discovery | Dakota Howard, Dhwani Batra, Peter W. Cook, Kara Moser, Adrian Paskey, Jason Caravas, Benjamin Rambo-Martin, Shatavia Morrison, Christopher Gulvick, Scott Sammons, Yvette Unoarumhi, Darlene Wagner, Matthew Schmerer, Eileen de Feo, Jan Antico, Christine Tran, Matthew Tolentino, Shannon Wickline, Kim Gietzen, Brad Sickler, Jingtao Liu, Eric Allen, Phil Febbo, Nicole L. Washington, Simon White, Geraint Levan, Kelly Schiabor Barrett, Elizabeth Cirulli, Alexandre Bolze, Ary Ascencio, Charlotte Rivera-Garcia, Ryan Cho, Jason Nguyen, Sherry Wang, Jimmy Ramirez, Tyler Cassens, Efren Sandoval, Magnus Isaksson, William Lee, David Becker, Marc Laurent, James Lu, Clinton R. Paden, Duncan MacCannell |
| EPI_ISL_2248003, EPI_ISL_2248004 | Wyoming Public Health Laboratory | Wyoming Public Health Laboratory | Jim Mildenberger, Taylor Fearing, Channing Weber, Ashley Norberg, Chayse Rowley, Marley Goetz, Brian Dominguez, Elliot Thomasson, Brittany Oher, Sam Britz, Cari Sloma, and Rob Christensen |
| EPI_ISL_2248005, EPI_ISL_2248006, EPI_ISL_2248007, EPI_ISL_2248008, EPI_ISL_2248009, EPI_ISL_2248010, EPI_ISL_2248012, EPI_ISL_2248013, EPI_ISL_2248014, EPI_ISL_2248015, EPI_ISL_2248016, EPI_ISL_2248017, EPI_ISL_2248018, EPI_ISL_2248019, EPI_ISL_2248020, EPI_ISL_2248021, EPI_ISL_2248022, EPI_ISL_2248023, EPI_ISL_2248024, EPI_ISL_2248025, EPI_ISL_2248026, EPI_ISL_2248027, EPI_ISL_2248028, EPI_ISL_2248029, EPI_ISL_2248030, EPI_ISL_2248031, EPI_ISL_2248032, EPI_ISL_2248033, EPI_ISL_2248034, EPI_ISL_2248035, EPI_ISL_2248036, EPI_ISL_2248037, EPI_ISL_2248038, EPI_ISL_2248039, EPI_ISL_2248040, EPI_ISL_2248041, EPI_ISL_2248042, EPI_ISL_2248043, EPI_ISL_2248044, EPI_ISL_2248045, EPI_ISL_2248046, EPI_ISL_2248047, EPI_ISL_2248048, EPI_ISL_2248049, EPI_ISL_2248050, EPI_ISL_2248051, EPI_ISL_2248052, EPI_ISL_2248053, EPI_ISL_2248054, EPI_ISL_2248055, EPI_ISL_2248056, EPI_ISL_2248057, EPI_ISL_2248058, EPI_ISL_2248059, EPI_ISL_2248060, EPI_ISL_2248061, EPI_ISL_2248062, EPI_ISL_2248063, EPI_ISL_2248064, EPI_ISL_2248065, EPI_ISL_2248066, EPI_ISL_2248068, EPI_ISL_2248069, EPI_ISL_2248070, EPI_ISL_2248071, EPI_ISL_2248072, EPI_ISL_2248073, EPI_ISL_2248074, EPI_ISL_2248075, EPI_ISL_2248076, EPI_ISL_2248077, EPI_ISL_2248078, EPI_ISL_2248079, EPI_ISL_2248080, EPI_ISL_2248081, EPI_ISL_2248082, EPI_ISL_2248083, EPI_ISL_2248084, EPI_ISL_2248085, EPI_ISL_2248086, EPI_ISL_2248087, EPI_ISL_2248088, EPI_ISL_2248089, EPI_ISL_2248090, EPI_ISL_2248091, EPI_ISL_2248092, EPI_ISL_2248093, EPI_ISL_2248094, EPI_ISL_2248096, EPI_ISL_2248097, EPI_ISL_2248098, EPI_ISL_2248099, EPI_ISL_2248100, EPI_ISL_2248101, EPI_ISL_2248102, EPI_ISL_2248103, EPI_ISL_2248104, EPI_ISL_2248105, EPI_ISL_2248106, EPI_ISL_2248107, EPI_ISL_2248108, EPI_ISL_2248109, EPI_ISL_2248110, EPI_ISL_2248111, EPI_ISL_2248112, EPI_ISL_2248113, EPI_ISL_2248114, EPI_ISL_2248115, EPI_ISL_2248116, EPI_ISL_2248117, EPI_ISL_2248118, EPI_ISL_2248119, EPI_ISL_2248120, EPI_ISL_2248121, EPI_ISL_2248122, EPI_ISL_2248123, EPI_ISL_2248124, EPI_ISL_2248125, EPI_ISL_2248126, EPI_ISL_2248127, EPI_ISL_2248128, EPI_ISL_2248129, EPI_ISL_2248130, EPI_ISL_2248131, EPI_ISL_2248132, EPI_ISL_2248133, EPI_ISL_2248134, EPI_ISL_2248135, EPI_ISL_2248136, EPI_ISL_2248137, EPI_ISL_2248138, EPI_ISL_2248139, EPI_ISL_2248140, EPI_ISL_2248142, EPI_ISL_2248143, EPI_ISL_2248144, EPI_ISL_2248145, EPI_ISL_2248146, EPI_ISL_2248147, EPI_ISL_2248148, EPI_ISL_2248149, EPI_ISL_2248150, EPI_ISL_2248151, EPI_ISL_2248152, EPI_ISL_2248153, EPI_ISL_2248154, EPI_ISL_2248155, EPI_ISL_2248156, EPI_ISL_2248157, EPI_ISL_2248158, EPI_ISL_2248159, EPI_ISL_2248160, EPI_ISL_2248161, EPI_ISL_2248162, EPI_ISL_2248163, EPI_ISL_2248164, EPI_ISL_2248165, EPI_ISL_2248219, EPI_ISL_2248220, EPI_ISL_2248221, EPI_ISL_2248222, EPI_ISL_2248223, EPI_ISL_2248224, EPI_ISL_2248225, EPI_ISL_2248226, EPI_ISL_2248227, EPI_ISL_2248228, EPI_ISL_2248229, EPI_ISL_2248230, EPI_ISL_2248231, EPI_ISL_2248232, EPI_ISL_2248233, EPI_ISL_2248234, EPI_ISL_2248235, EPI_ISL_2248236, EPI_ISL_2248237, EPI_ISL_2248238, EPI_ISL_2248239, EPI_ISL_2248240, EPI_ISL_2248241, EPI_ISL_2248242, EPI_ISL_2248243, EPI_ISL_2248244, EPI_ISL_2248245, EPI_ISL_2248246, EPI_ISL_2248247, EPI_ISL_2248248, EPI_ISL_2248249, EPI_ISL_2248250, EPI_ISL_2248251, EPI_ISL_2248252, EPI_ISL_2248253, EPI_ISL_2248254, EPI_ISL_2248255, EPI_ISL_2248256, EPI_ISL_2248257, EPI_ISL_2248258, EPI_ISL_2248259, EPI_ISL_2248260, EPI_ISL_2248261, EPI_ISL_2248262, EPI_ISL_2248263, EPI_ISL_2248264, EPI_ISL_2248265, EPI_ISL_2248266, EPI_ISL_2248267, EPI_ISL_2248268, EPI_ISL_2248269, EPI_ISL_2248270, EPI_ISL_2248271, EPI_ISL_2248272, EPI_ISL_2248273, EPI_ISL_2248274, EPI_ISL_2248275, EPI_ISL_2248276, EPI_ISL_2248277, EPI_ISL_2248278, EPI_ISL_2248279, EPI_ISL_2248280, EPI_ISL_2248281, EPI_ISL_2248282, EPI_ISL_2248283, EPI_ISL_2248284, EPI_ISL_2248285, EPI_ISL_2248286, EPI_ISL_2248287, EPI_ISL_2248288, EPI_ISL_2248289, EPI_ISL_2248290, EPI_ISL_2248291, EPI_ISL_2248292, EPI_ISL_2248293, EPI_ISL_2248294, EPI_ISL_2248295, EPI_ISL_2248296, EPI_ISL_2248297, EPI_ISL_2248298, EPI_ISL_2248299, EPI_ISL_2248300, EPI_ISL_2248301, EPI_ISL_2248302, EPI_ISL_2248303, EPI_ISL_2248304, EPI_ISL_2248305, EPI_ISL_2248306, EPI_ISL_2248307, EPI_ISL_2248308, EPI_ISL_2248309, EPI_ISL_2248310, EPI_ISL_2248311, EPI_ISL_2248312, EPI_ISL_2248313, EPI_ISL_2248314, EPI_ISL_2248315, EPI_ISL_2248316, EPI_ISL_2248317, EPI_ISL_2248318, EPI_ISL_2248319, EPI_ISL_2248320, EPI_ISL_2248321, EPI_ISL_2248322, EPI_ISL_2248323, EPI_ISL_2248324, EPI_ISL_2248325, EPI_ISL_2248326, EPI_ISL_2248327, EPI_ISL_2248328, EPI_ISL_2248329, EPI_ISL_2248330, EPI_ISL_2248331, EPI_ISL_2248332, EPI_ISL_2248333, EPI_ISL_2248334, EPI_ISL_2248335, EPI_ISL_2248336, EPI_ISL_2248337, EPI_ISL_2248338, EPI_ISL_2248339, EPI_ISL_2248340, EPI_ISL_2248341, EPI_ISL_2248342, EPI_ISL_2248343, EPI_ISL_2248344, EPI_ISL_2248345, EPI_ISL_2248346, EPI_ISL_2248347, EPI_ISL_2248348, EPI_ISL_2248349, EPI_ISL_2248350, EPI_ISL_2248351, EPI_ISL_2248352, EPI_ISL_2248353, EPI_ISL_2248354, EPI_ISL_2248355, EPI_ISL_2248356, EPI_ISL_2248357, EPI_ISL_2248358, EPI_ISL_2248359, EPI_ISL_2248362, EPI_ISL_2248363, EPI_ISL_2248364, EPI_ISL_2248365, EPI_ISL_2248366, EPI_ISL_2248367, EPI_ISL_2248368, EPI_ISL_2248369, EPI_ISL_2248370, EPI_ISL_2248371, EPI_ISL_2248372, EPI_ISL_2248373, EPI_ISL_2248374, EPI_ISL_2248375, EPI_ISL_2248376, EPI_ISL_2248377, EPI_ISL_2248378, EPI_ISL_2248379, EPI_ISL_2248380, EPI_ISL_2248381, EPI_ISL_2248382, EPI_ISL_2248383, EPI_ISL_2248384, EPI_ISL_2248385, EPI_ISL_2248386, EPI_ISL_2248387, EPI_ISL_2248388, EPI_ISL_2248389, EPI_ISL_2248390, EPI_ISL_2248391, EPI_ISL_2248392, EPI_ISL_2248393, EPI_ISL_2248394, EPI_ISL_2248395, EPI_ISL_2248396, EPI_ISL_2248397, EPI_ISL_2248398, EPI_ISL_2248399, EPI_ISL_2248400, EPI_ISL_2248401, EPI_ISL_2248402, EPI_ISL_2248403, EPI_ISL_2248404, EPI_ISL_2248405, EPI_ISL_2248406, EPI_ISL_2248407, EPI_ISL_2248408, EPI_ISL_2248409, EPI_ISL_2248410, EPI_ISL_2248411, EPI_ISL_2248412, EPI_ISL_2248413, EPI_ISL_2248414, EPI_ISL_2248415, EPI_ISL_2248416, EPI_ISL_2248417, EPI_ISL_2248418, EPI_ISL_2248419, EPI_ISL_2248420, EPI_ISL_2248421, EPI_ISL_2248422, EPI_ISL_2248423, EPI_ISL_2248424, EPI_ISL_2248425, EPI_ISL_2248426, EPI_ISL_2248427, EPI_ISL_2248428, EPI_ISL_2248429, EPI_ISL_2248430, EPI_ISL_2248431, EPI_ISL_2248432, EPI_ISL_2248433, EPI_ISL_2248434, EPI_ISL_2248435, EPI_ISL_2248436, EPI_ISL_2248437, EPI_ISL_2248438, EPI_ISL_2248439, EPI_ISL_2248440, EPI_ISL_2248441, EPI_ISL_2248442, EPI_ISL_2248443, EPI_ISL_2248444, EPI_ISL_2248445, EPI_ISL_2248446, EPI_ISL_2248447, EPI_ISL_2248448, EPI_ISL_2248449, EPI_ISL_2248450, EPI_ISL_2248451, EPI_ISL_2248452, EPI_ISL_2248453, EPI_ISL_2248454, EPI_ISL_2248455, EPI_ISL_2248456, EPI_ISL_2248457, EPI_ISL_2248458, EPI_ISL_2248459 | | | |
| see above | Helix/Illumina | Centers for Disease Control and Prevention Division of Viral Diseases, Pathogen Discovery | Dakota Howard, Dhwani Batra, Peter W. Cook, Kara Moser, Adrian Paskey, Jason Caravas, Benjamin Rambo-Martin, Shatavia Morrison, Christopher Gulvick, Scott Sammons, Yvette Unoarumhi, Darlene Wagner, Matthew Schmerer, Eileen de Feo, Jan Antico, Christine Tran, Matthew Tolentino, Shannon Wickline, Kim Gietzen, Brad Sickler, Jingtao Liu, Eric Allen, Phil Febbo, Nicole L. Washington, Simon White, Geraint Levan, Kelly Schiabor Barrett, Elizabeth Cirulli, Alexandre Bolze, Ary Ascencio, Charlotte Rivera-Garcia, Ryan Cho, Jason Nguyen, Sherry Wang, Jimmy Ramirez, Tyler Cassens, Efren Sandoval, Magnus Isaksson, William Lee, David Becker, Marc Laurent, James Lu, Clinton R. Paden, Duncan MacCannell |
| EPI_ISL_2248460, EPI_ISL_2248461, EPI_ISL_2248462, EPI_ISL_2248463, EPI_ISL_2248464, EPI_ISL_2248465, EPI_ISL_2248466, EPI_ISL_2248467, EPI_ISL_2248468, EPI_ISL_2248469, EPI_ISL_2248470, EPI_ISL_2248471, EPI_ISL_2248472, EPI_ISL_2248473, EPI_ISL_2248474, EPI_ISL_2248475, EPI_ISL_2248476, EPI_ISL_2248477, EPI_ISL_2248478, EPI_ISL_2248479, EPI_ISL_2248480, EPI_ISL_2248481, EPI_ISL_2248482, EPI_ISL_2248483, EPI_ISL_2248484, EPI_ISL_2248485, EPI_ISL_2248486, EPI_ISL_2248487, EPI_ISL_2248488, EPI_ISL_2248489, EPI_ISL_2248490, EPI_ISL_2248491, EPI_ISL_2248492, EPI_ISL_2248493, EPI_ISL_2248494, EPI_ISL_2248495, EPI_ISL_2248496, EPI_ISL_2248497, EPI_ISL_2248498, EPI_ISL_2248499, EPI_ISL_2248500, EPI_ISL_2248501, EPI_ISL_2248502, EPI_ISL_2248503, EPI_ISL_2248504, EPI_ISL_2248505, EPI_ISL_2248506, EPI_ISL_2248507, EPI_ISL_2248508, EPI_ISL_2248509, EPI_ISL_2248510, EPI_ISL_2248511, EPI_ISL_2248512, EPI_ISL_2248513, EPI_ISL_2248514, EPI_ISL_2248515, EPI_ISL_2248516, EPI_ISL_2248517, EPI_ISL_2248518, EPI_ISL_2248519, EPI_ISL_2248520, EPI_ISL_2248521, EPI_ISL_2248522, EPI_ISL_2248523, EPI_ISL_2248524, EPI_ISL_2248525, EPI_ISL_2248526, EPI_ISL_2248527, EPI_ISL_2248528, EPI_ISL_2248529, EPI_ISL_2248530, EPI_ISL_2248531, EPI_ISL_2248532, EPI_ISL_2248533, EPI_ISL_2248534, EPI_ISL_2248535, EPI_ISL_2248536, EPI_ISL_2248537, EPI_ISL_2248538, EPI_ISL_2248539, EPI_ISL_2248540, EPI_ISL_2248541, EPI_ISL_2248542, EPI_ISL_2248543, EPI_ISL_2248544, EPI_ISL_2248545, EPI_ISL_2248546, EPI_ISL_2248547, EPI_ISL_2248548, EPI_ISL_2248549, EPI_ISL_2248550, EPI_ISL_2248551, EPI_ISL_2248552, EPI_ISL_2248553, EPI_ISL_2248554, EPI_ISL_2248555, EPI_ISL_2248556, EPI_ISL_2248557, EPI_ISL_2248558, EPI_ISL_2248559, EPI_ISL_2248560, EPI_ISL_2248561, EPI_ISL_2248562, EPI_ISL_2248563, EPI_ISL_2248564, EPI_ISL_2248565, EPI_ISL_2248566, EPI_ISL_2248567, EPI_ISL_2248568, EPI_ISL_2248569, EPI_ISL_2248570, EPI_ISL_2248571, EPI_ISL_2248572, EPI_ISL_2248573, EPI_ISL_2248574, EPI_ISL_2248575, EPI_ISL_2248576, EPI_ISL_2248577, EPI_ISL_2248578, EPI_ISL_2248579, EPI_ISL_2248580, EPI_ISL_2248581, EPI_ISL_2248582, EPI_ISL_2248583, EPI_ISL_2248584, EPI_ISL_2248585, EPI_ISL_2248586, EPI_ISL_2248587, EPI_ISL_2248588, EPI_ISL_2248589, EPI_ISL_2248590, EPI_ISL_2248591, EPI_ISL_2248592, EPI_ISL_2248593, EPI_ISL_2248594, EPI_ISL_2248595, EPI_ISL_2248596, EPI_ISL_2248597, EPI_ISL_2248598, EPI_ISL_2248599, EPI_ISL_2248600, EPI_ISL_2248601, EPI_ISL_2248602, EPI_ISL_2248603, EPI_ISL_2248604, EPI_ISL_2248605, EPI_ISL_2248606, EPI_ISL_2248607, EPI_ISL_2248608, EPI_ISL_2248609, EPI_ISL_2248610, EPI_ISL_2248611, EPI_ISL_2248612, EPI_ISL_2248613, EPI_ISL_2248614, EPI_ISL_2248615, EPI_ISL_2248616, EPI_ISL_2248617, EPI_ISL_2248618, EPI_ISL_2248619, EPI_ISL_2248620, EPI_ISL_2248621, EPI_ISL_2248622, EPI_ISL_2248623, EPI_ISL_2248624, EPI_ISL_2248625, EPI_ISL_2248626, EPI_ISL_2248627, EPI_ISL_2248628, EPI_ISL_2248629, EPI_ISL_2248630, EPI_ISL_2248631, EPI_ISL_2248632, EPI_ISL_2248633, EPI_ISL_2248634, EPI_ISL_2248635, EPI_ISL_2248636, EPI_ISL_2248637, EPI_ISL_2248638, EPI_ISL_2248639, EPI_ISL_2248640, EPI_ISL_2248641, EPI_ISL_2248642, EPI_ISL_2248643, EPI_ISL_2248644, EPI_ISL_2248645, EPI_ISL_2248646, EPI_ISL_2248647, EPI_ISL_2248648, EPI_ISL_2248649, EPI_ISL_2248650, EPI_ISL_2248651, EPI_ISL_2248652, EPI_ISL_2248653, EPI_ISL_2248654, EPI_ISL_2248655, EPI_ISL_2248656, EPI_ISL_2248657, EPI_ISL_2248658, EPI_ISL_2248659, EPI_ISL_2248660, EPI_ISL_2248661, EPI_ISL_2248662, EPI_ISL_2248663, EPI_ISL_2248664, EPI_ISL_2248665, EPI_ISL_2248666, EPI_ISL_2248667, EPI_ISL_2248668, EPI_ISL_2248669, EPI_ISL_2248670, EPI_ISL_2248671, EPI_ISL_2248672, EPI_ISL_2248673, EPI_ISL_2248674, EPI_ISL_2248675, EPI_ISL_2248676, EPI_ISL_2248677, EPI_ISL_2248678, EPI_ISL_2248679, EPI_ISL_2248680, EPI_ISL_2248681, EPI_ISL_2248682, EPI_ISL_2248683, EPI_ISL_2248684, EPI_ISL_2248685, EPI_ISL_2248686, EPI_ISL_2248687, EPI_ISL_2248688, EPI_ISL_2248689, EPI_ISL_2248690, EPI_ISL_2248691, EPI_ISL_2248692, EPI_ISL_2248693, EPI_ISL_2248694, EPI_ISL_2248695, EPI_ISL_2248696, EPI_ISL_2248697, EPI_ISL_2248698, EPI_ISL_2248699, EPI_ISL_2248700, EPI_ISL_2248701, EPI_ISL_2248702, EPI_ISL_2248703 | | | |
| see above | Quest Diagnostics Incorporated | Centers for Disease Control and Prevention Division of Viral Diseases, Pathogen Discovery | Dakota Howard, Dhwani Batra, Peter W. Cook, Kara Moser, Adrian Paskey, Jason Caravas, Benjamin Rambo-Martin, Shatavia Morrison, Christopher Gulvick, Scott Sammons, Yvette Unoarumhi, Darlene Wagner, Matthew Schmerer, S. H. Rosenthal, A. Gerasimova, R. M. Kagan, B. Anderson, M. Hua, Y. Liu, L.E. Bernstein, K.E. Livingston, A. Perez, I. A. Shlyakhter, R. V. Rolando, R. Owen, P. Tanpaiboon, F. Lacbawan, Clinton R. Paden, Duncan MacCannell |
| EPI_ISL_2248704, EPI_ISL_2248705, EPI_ISL_2248706, EPI_ISL_2248707, EPI_ISL_2248708, EPI_ISL_2248709, EPI_ISL_2248710, EPI_ISL_2248711, EPI_ISL_2248712, EPI_ISL_2248713, EPI_ISL_2248714, EPI_ISL_2248715, EPI_ISL_2248716, EPI_ISL_2248717, EPI_ISL_2248718 | | | |

| | | | |
|---|---|---|---|
| see above | Univeristy of New Mexico Hospital | Center for Global Health, University of New Mexico Health Sciences Center | Daryl Domman, Kurt Schwalm, Valerie Morley, Justin Bacca, Jon Femling, Darrell Dinwiddie |
| EPI_ISL_2248719, EPI_ISL_2248720, EPI_ISL_2248721, EPI_ISL_2248722, EPI_ISL_2248723, EPI_ISL_2248724, EPI_ISL_2248725 | Wyoming Public Health Laboratory | Wyoming Public Health Laboratory | Jim Mildenberger, Wanda Manley, Noah Hull, Taylor Fearing, Lynette Gumbleton, Channing Weber, Ashley Norberg, Chayse Rowley, Marley Goetz, Brian Dominguez, Elliot Thomasson, Sam Britz, Cari Sloma, and Rob Christensen |
| EPI_ISL_2248726 | ME Health and Environmental Testing Laboratory | Centers for Disease Control and Prevention Division of Viral Diseases, Pathogen Discovery | Mili Sheth, Sarah Nobles, Jasmine Padilla, Mark Burroughs, Shoshona Le, Katie Dillon, Peter Cook, Clinton R. Paden, Dhwani Batra, Krista Queen, Kristen Knipe, Dakota Howard, Yvette Unoarumhi, Darlene Wagner, Matthew Schmerer, Ben L. Rambo-Martin, Kristine Lacek, Sam Shepard, Alison Laufer Halpin, Dave Wentworth, Vivien Dugan, Suxiang Tong, Justin Lee |
| EPI_ISL_2248727 | "CDPH, Viral and Rickettsial Disease Laboratory" | Centers for Disease Control and Prevention Division of Viral Diseases, Pathogen Discovery | Mili Sheth, Sarah Nobles, Jasmine Padilla, Mark Burroughs, Shoshona Le, Katie Dillon, Peter Cook, Clinton R. Paden, Dhwani Batra, Krista Queen, Kristen Knipe, Dakota Howard, Yvette Unoarumhi, Darlene Wagner, Matthew Schmerer, Ben L. Rambo-Martin, Kristine Lacek, Sam Shepard, Alison Laufer Halpin, Dave Wentworth, Vivien Dugan, Suxiang Tong, Justin Lee |
| EPI_ISL_2248728 | RI State Health Laboratories | Centers for Disease Control and Prevention Division of Viral Diseases, Pathogen Discovery | Mili Sheth, Sarah Nobles, Jasmine Padilla, Mark Burroughs, Shoshona Le, Katie Dillon, Peter Cook, Clinton R. Paden, Dhwani Batra, Krista Queen, Kristen Knipe, Dakota Howard, Yvette Unoarumhi, Darlene Wagner, Matthew Schmerer, Ben L. Rambo-Martin, Kristine Lacek, Sam Shepard, Alison Laufer Halpin, Dave Wentworth, Vivien Dugan, Suxiang Tong, Justin Lee |
| EPI_ISL_2248729 | WI State Laboratory of Hygiene | Centers for Disease Control and Prevention Division of Viral Diseases, Pathogen Discovery | Mili Sheth, Sarah Nobles, Jasmine Padilla, Mark Burroughs, Shoshona Le, Katie Dillon, Peter Cook, Clinton R. Paden, Dhwani Batra, Krista Queen, Kristen Knipe, Dakota Howard, Yvette Unoarumhi, Darlene Wagner, Matthew Schmerer, Ben L. Rambo-Martin, Kristine Lacek, Sam Shepard, Alison Laufer Halpin, Dave Wentworth, Vivien Dugan, Suxiang Tong, Justin Lee |
| EPI_ISL_2248730 | CA-Los Angeles County Public Health Laboratory | Centers for Disease Control and Prevention Division of Viral Diseases, Pathogen Discovery | Mili Sheth, Sarah Nobles, Jasmine Padilla, Mark Burroughs, Shoshona Le, Katie Dillon, Peter Cook, Clinton R. Paden, Dhwani Batra, Krista Queen, Kristen Knipe, Dakota Howard, Yvette Unoarumhi, Darlene Wagner, Matthew Schmerer, Ben L. Rambo-Martin, Kristine Lacek, Sam Shepard, Alison Laufer Halpin, Dave Wentworth, Vivien Dugan, Suxiang Tong, Justin Lee |
| EPI_ISL_2248731 | MD DOH Laboratories Administration | Centers for Disease Control and Prevention Division of Viral Diseases, Pathogen Discovery | Mili Sheth, Sarah Nobles, Jasmine Padilla, Mark Burroughs, Shoshona Le, Katie Dillon, Peter Cook, Clinton R. Paden, Dhwani Batra, Krista Queen, Kristen Knipe, Dakota Howard, Yvette Unoarumhi, Darlene Wagner, Matthew Schmerer, Ben L. Rambo-Martin, Kristine Lacek, Sam Shepard, Alison Laufer Halpin, Dave Wentworth, Vivien Dugan, Suxiang Tong, Justin Lee |
| EPI_ISL_2248732 | IL Dept. of Public Health Springfield Laboratory | Centers for Disease Control and Prevention Division of Viral Diseases, Pathogen Discovery | Mili Sheth, Sarah Nobles, Jasmine Padilla, Mark Burroughs, Shoshona Le, Katie Dillon, Peter Cook, Clinton R. Paden, Dhwani Batra, Krista Queen, Kristen Knipe, Dakota Howard, Yvette Unoarumhi, Darlene Wagner, Matthew Schmerer, Ben L. Rambo-Martin, Kristine Lacek, Sam Shepard, Alison Laufer Halpin, Dave Wentworth, Vivien Dugan, Suxiang Tong, Justin Lee |
| EPI_ISL_2248733 | "NM Dept. Health, Scientific Laboratory Division " | Centers for Disease Control and Prevention Division of Viral Diseases, Pathogen Discovery | Mili Sheth, Sarah Nobles, Jasmine Padilla, Mark Burroughs, Shoshona Le, Katie Dillon, Peter Cook, Clinton R. Paden, Dhwani Batra, Krista Queen, Kristen Knipe, Dakota Howard, Yvette Unoarumhi, Darlene Wagner, Matthew Schmerer, Ben L. Rambo-Martin, Kristine Lacek, Sam Shepard, Alison Laufer Halpin, Dave Wentworth, Vivien Dugan, Suxiang Tong, Justin Lee |
| EPI_ISL_2248734, EPI_ISL_2248735, EPI_ISL_2248736, EPI_ISL_2248737, EPI_ISL_2248738, EPI_ISL_2248739, EPI_ISL_2248740, EPI_ISL_2248741, EPI_ISL_2248742 | GA Department of Public Health | GA Department of Public Health | Stacy Reeves, Jonathan Edwards, Cynthia Dixey, Tonia Parrott, Aliyah Fields, Taylor Smith |
| EPI_ISL_2248743 | Kansas Health and Environmental Lab | Kansas Health and Environmental Lab | Mike Grose, Katherine Wiggins, Jonathan Barnell, Ben Olsen, and Phil Adam |
| EPI_ISL_2248744, EPI_ISL_2248745, EPI_ISL_2248746, EPI_ISL_2248747, EPI_ISL_2248749, EPI_ISL_2248750, EPI_ISL_2248751, EPI_ISL_2248752, EPI_ISL_2248753 | Kansas Health and Environmental Lab | Kansas Health and Environmental Lab | Mike Grose, Jonathan Barnell, Ben Olsen, and Phil Adam |
| EPI_ISL_2248754, EPI_ISL_2248755, EPI_ISL_2248756, EPI_ISL_2248757 | Wichita State University - Molecular Diagnostics Lab | Kansas Health and Environmental Lab | Mike Grose, Jonathan Barnell, Ben Olsen, and Phil Adam |
| EPI_ISL_2249206, EPI_ISL_2249207, EPI_ISL_2249208, EPI_ISL_2249209, EPI_ISL_2249210, EPI_ISL_2249211, EPI_ISL_2249212, EPI_ISL_2249213, EPI_ISL_2249214, EPI_ISL_2249215, EPI_ISL_2249216, EPI_ISL_2249217, EPI_ISL_2249218, EPI_ISL_2249219, EPI_ISL_2249220, EPI_ISL_2249221, EPI_ISL_2249222, EPI_ISL_2249223, EPI_ISL_2249224, EPI_ISL_2249225, EPI_ISL_2249226 | | | |
| see above | Gundersen Clinical Microbiology Laboratory | Kabara Cancer Research Institute | Craig S. Richmond, Paraic A. Kenny |
| EPI_ISL_2249227 | Hospital Sharp | Microbial Genomics Laboratory | Bruno Gomez-Gil, Julissa Enciso-Ibarra, Alejandra Garcia-Gasca, Daniel Fregoso-Rueda |
| EPI_ISL_2249228, EPI_ISL_2249229 | Hospital Margarita Maza de Juárez | Microbial Genomics Laboratory | Bruno Gomez-Gil, Julissa Enciso-Ibarra, Alejandra Garcia-Gasca, Daniel Fregoso-Rueda |
| EPI_ISL_2249230, EPI_ISL_2249231 | Hospital Sharp | Microbial Genomics Laboratory | Bruno Gomez-Gil, Julissa Enciso-Ibarra, Alejandra Garcia-Gasca, Daniel Fregoso-Rueda |
| EPI_ISL_2249233 | Hospital Margarita Maza de Juárez | Microbial Genomics Laboratory | Bruno Gomez-Gil, Julissa Enciso-Ibarra, Alejandra Garcia-Gasca, Daniel Fregoso-Rueda |
| EPI_ISL_2249234 | Imágenes Diagnósticas de Mazatlán | Microbial Genomics Laboratory | Bruno Gomez-Gil, Julissa Enciso-Ibarra, Alejandra Garcia-Gasca, Daniel Fregoso-Rueda |
| EPI_ISL_2249235 | Hospital Margarita Maza de Juárez | Microbial Genomics Laboratory | Bruno Gomez-Gil, Julissa Enciso-Ibarra, Alejandra Garcia-Gasca, Daniel Fregoso-Rueda |
| EPI_ISL_2249236, EPI_ISL_2249237, EPI_ISL_2249238 | Hospital Sharp | Microbial Genomics Laboratory | Bruno Gomez-Gil, Julissa Enciso-Ibarra, Alejandra Garcia-Gasca, Daniel Fregoso-Rueda |
| EPI_ISL_2249239 | Hospital Margarita Maza de Juárez | Microbial Genomics Laboratory | Bruno Gomez-Gil, Julissa Enciso-Ibarra, Alejandra Garcia-Gasca, Daniel Fregoso-Rueda |
| EPI_ISL_2249240 | Hospital Sharp | Microbial Genomics Laboratory | Bruno Gomez-Gil, Julissa Enciso-Ibarra, Alejandra Garcia-Gasca, Daniel Fregoso-Rueda |
| EPI_ISL_2249241, EPI_ISL_2249242, EPI_ISL_2249243, EPI_ISL_2249244, EPI_ISL_2249245 | Hospital Margarita Maza de Juárez | Microbial Genomics Laboratory | Bruno Gomez-Gil, Julissa Enciso-Ibarra, Alejandra Garcia-Gasca, Daniel Fregoso-Rueda |
| EPI_ISL_2249246, EPI_ISL_2249247, EPI_ISL_2249248, EPI_ISL_2249249 | Hospital Sharp | Microbial Genomics Laboratory | Bruno Gomez-Gil, Julissa Enciso-Ibarra, Alejandra Garcia-Gasca, Daniel Fregoso-Rueda |
| EPI_ISL_2249250 | Hospital Margarita Maza de Juárez | Microbial Genomics Laboratory | Bruno Gomez-Gil, Julissa Enciso-Ibarra, Alejandra Garcia-Gasca, Daniel Fregoso-Rueda |
| EPI_ISL_2249251, EPI_ISL_2249252, EPI_ISL_2249253 | Hospital Sharp | Microbial Genomics Laboratory | Bruno Gomez-Gil, Julissa Enciso-Ibarra, Alejandra Garcia-Gasca, Daniel Fregoso-Rueda |
| EPI_ISL_2249254 | Hospital Margarita Maza de Juárez | Microbial Genomics Laboratory | Bruno Gomez-Gil, Julissa Enciso-Ibarra, Alejandra Garcia-Gasca, Daniel Fregoso-Rueda |
| EPI_ISL_2249255 | Hospital Sharp | Microbial Genomics Laboratory | Bruno Gomez-Gil, Julissa Enciso-Ibarra, Alejandra Garcia-Gasca, Daniel Fregoso-Rueda |
| EPI_ISL_2249259, EPI_ISL_2249260, EPI_ISL_2249261, EPI_ISL_2249262, EPI_ISL_2249263, EPI_ISL_2249264, EPI_ISL_2249265, EPI_ISL_2249266, EPI_ISL_2249267, EPI_ISL_2249268, EPI_ISL_2249269, EPI_ISL_2249270, EPI_ISL_2249271, EPI_ISL_2249272, EPI_ISL_2249273, EPI_ISL_2249274, EPI_ISL_2249275, EPI_ISL_2249276, EPI_ISL_2249278, EPI_ISL_2249279, EPI_ISL_2249280, EPI_ISL_2249281, EPI_ISL_2249282, EPI_ISL_2249283, EPI_ISL_2249284, EPI_ISL_2249285, EPI_ISL_2249286, EPI_ISL_2249287, EPI_ISL_2249288, EPI_ISL_2249289, EPI_ISL_2249290, EPI_ISL_2249291, EPI_ISL_2249292, EPI_ISL_2249293, EPI_ISL_2249294, EPI_ISL_2249295, EPI_ISL_2249296, EPI_ISL_2249297, EPI_ISL_2249298 | | | |
| see above | New Mexico Department of Health Scientific Laboratory | Center for Global Health, University of New Mexico Health Sciences Center | Daryl Domman, Kurt Schwalm, Twila Kunde, Joseph Hicks, Anastacia Griego, Michael Edwards, Valerie Morley, Darrell Dinwiddie |
| EPI_ISL_2249299, EPI_ISL_2249300, EPI_ISL_2249301, EPI_ISL_2249302, EPI_ISL_2249303, EPI_ISL_2249304, EPI_ISL_2249305, EPI_ISL_2249306, EPI_ISL_2249307, EPI_ISL_2249308, EPI_ISL_2249309, EPI_ISL_2249310, EPI_ISL_2249311, EPI_ISL_2249312, EPI_ISL_2249313, EPI_ISL_2249314, EPI_ISL_2249315, EPI_ISL_2249316, EPI_ISL_2249317, EPI_ISL_2249318, EPI_ISL_2249319, EPI_ISL_2249320, EPI_ISL_2249321, EPI_ISL_2249322, EPI_ISL_2249323, EPI_ISL_2249324, EPI_ISL_2249325, EPI_ISL_2249326, EPI_ISL_2249327, EPI_ISL_2249328, EPI_ISL_2249329, EPI_ISL_2249330, EPI_ISL_2249331, EPI_ISL_2249332 | | | |
| see above | TriCore Reference Laboratories | Center for Global Health, University of New Mexico Health Sciences Center | Daryl Domman, Kurt Schwalm, Valerie Morley, Cecilia Thompson, Kendra Pesko, Karissa Culbreath, Darrell Dinwiddie |

| | | | |
|---|---|---|---|
| EPI_ISL_2249466, EPI_ISL_2249467, EPI_ISL_2249469 | Wyoming Public Health Laboratory | Wyoming Public Health Laboratory | Jim Mildenberger, Taylor Fearing, Channing Weber, Ashley Norberg, Chayse Rowley, Marley Goetz, Brian Dominguez, Elliot Thomasson, Brittany Oher, Sam Britz, Cari Sloma, and Rob Christensen |
| EPI_ISL_2249474, EPI_ISL_2249475, EPI_ISL_2249476, EPI_ISL_2249477, EPI_ISL_2249478, EPI_ISL_2249479, EPI_ISL_2249480, EPI_ISL_2249481, EPI_ISL_2249482, EPI_ISL_2249483, EPI_ISL_2249484, EPI_ISL_2249485, EPI_ISL_2249486, EPI_ISL_2249487, EPI_ISL_2249488, EPI_ISL_2249489, EPI_ISL_2249490, EPI_ISL_2249491, EPI_ISL_2249492, EPI_ISL_2249493, EPI_ISL_2249494, EPI_ISL_2249495, EPI_ISL_2249496, EPI_ISL_2249497, EPI_ISL_2249498 | | | |
| see above | Arizona State Public Health Laboratory | Arizona State Public Health Laboratory | Trung Huynh, Jessica Escobar, Katherine Fullerton, Nobuko Fukushima, Stacy White, Linda Getsinger, Victor Waddell |
| EPI_ISL_2249501, EPI_ISL_2249502, EPI_ISL_2249503, EPI_ISL_2249504, EPI_ISL_2249505, EPI_ISL_2249506, EPI_ISL_2249507, EPI_ISL_2249508, EPI_ISL_2249509, EPI_ISL_2249510, EPI_ISL_2249511, EPI_ISL_2249512, EPI_ISL_2249513 | | | |
| see above | Wyoming Public Health Laboratory | Wyoming Public Health Laboratory | Jim Mildenberger, Taylor Fearing, Channing Weber, Ashley Norberg, Chayse Rowley, Marley Goetz, Brian Dominguez, Elliot Thomasson, Brittany Oher, Sam Britz, Cari Sloma, and Rob Christensen |
| EPI_ISL_2249518, EPI_ISL_2249519, EPI_ISL_2249520, EPI_ISL_2249521, EPI_ISL_2249522, EPI_ISL_2249523, EPI_ISL_2249524, EPI_ISL_2249525, EPI_ISL_2249526, EPI_ISL_2249527, EPI_ISL_2249528 | | | |
| see above | Minnesota Department of Health, Public Health Laboratory | Minnesota Department of Health, Public Health Laboratory | Alexandra Lorentz, Jacob Garfin, Matt Plumb, and Xiong Wang |
| EPI_ISL_2249529, EPI_ISL_2249530 | Mayo Clinic & Mayo Clinic Laboratories | Minnesota Department of Health, Public Health Laboratory | Alexandra Lorentz, Jacob Garfin, Matt Plumb, and Xiong Wang |
| EPI_ISL_2249531, EPI_ISL_2249532, EPI_ISL_2249533, EPI_ISL_2249534, EPI_ISL_2249535, EPI_ISL_2249536, EPI_ISL_2249537, EPI_ISL_2249538, EPI_ISL_2249539, EPI_ISL_2249540, EPI_ISL_2249541, EPI_ISL_2249542, EPI_ISL_2249543, EPI_ISL_2249544, EPI_ISL_2249545, EPI_ISL_2249546, EPI_ISL_2249547, EPI_ISL_2249548, EPI_ISL_2249549, EPI_ISL_2249550, EPI_ISL_2249551 | | | |
| see above | Minnesota Department of Health, Public Health Laboratory | Minnesota Department of Health, Public Health Laboratory | Alexandra Lorentz, Jacob Garfin, Matt Plumb, and Xiong Wang |
| EPI_ISL_2249552, EPI_ISL_2249553, EPI_ISL_2249554, EPI_ISL_2249555, EPI_ISL_2249556, EPI_ISL_2249557, EPI_ISL_2249558, EPI_ISL_2249559, EPI_ISL_2249560, EPI_ISL_2249561, EPI_ISL_2249562, EPI_ISL_2249563, EPI_ISL_2249564, EPI_ISL_2249565, EPI_ISL_2249566, EPI_ISL_2249567, EPI_ISL_2249568, EPI_ISL_2249569, EPI_ISL_2249570, EPI_ISL_2249571, EPI_ISL_2249572, EPI_ISL_2249573, EPI_ISL_2249574, EPI_ISL_2249575, EPI_ISL_2249576, EPI_ISL_2249577, EPI_ISL_2249578, EPI_ISL_2249579, EPI_ISL_2249580, EPI_ISL_2249581, EPI_ISL_2249582, EPI_ISL_2249583, EPI_ISL_2249584, EPI_ISL_2249585, EPI_ISL_2249586, EPI_ISL_2249587, EPI_ISL_2249588, EPI_ISL_2249589, EPI_ISL_2249590, EPI_ISL_2249591, EPI_ISL_2249592, EPI_ISL_2249593, EPI_ISL_2249594, EPI_ISL_2249595, EPI_ISL_2249596 | | | |
| see above | M Health Fairview | Minnesota Department of Health, Public Health Laboratory | Alexandra Lorentz, Jacob Garfin, Matt Plumb, and Xiong Wang |
| EPI_ISL_2249598, EPI_ISL_2249599, EPI_ISL_2249601, EPI_ISL_2249602, EPI_ISL_2249604, EPI_ISL_2249605, EPI_ISL_2249606, EPI_ISL_2249607, EPI_ISL_2249608, EPI_ISL_2249609, EPI_ISL_2249610, EPI_ISL_2249611, EPI_ISL_2249612, EPI_ISL_2249614, EPI_ISL_2249616, EPI_ISL_2249618, EPI_ISL_2249619, EPI_ISL_2249621, EPI_ISL_2249622, EPI_ISL_2249623, EPI_ISL_2249625, EPI_ISL_2249626 | | | |
| see above | Minnesota Department of Health, Public Health Laboratory | Minnesota Department of Health, Public Health Laboratory | Alexandra Lorentz, Jacob Garfin, Matt Plumb, and Xiong Wang |
| EPI_ISL_2249627, EPI_ISL_2249628, EPI_ISL_2249629, EPI_ISL_2249630, EPI_ISL_2249631, EPI_ISL_2249632, EPI_ISL_2249633, EPI_ISL_2249634, EPI_ISL_2249635, EPI_ISL_2249639, EPI_ISL_2249640, EPI_ISL_2249641, EPI_ISL_2249642, EPI_ISL_2249643, EPI_ISL_2249644, EPI_ISL_2249645, EPI_ISL_2249646, EPI_ISL_2249648 | | | |
| see above | M Health Fairview | Minnesota Department of Health, Public Health Laboratory | Alexandra Lorentz, Jacob Garfin, Matt Plumb, and Xiong Wang |
| EPI_ISL_2249654, EPI_ISL_2249657, EPI_ISL_2249658, EPI_ISL_2249659, EPI_ISL_2249660, EPI_ISL_2249661, EPI_ISL_2249662, EPI_ISL_2249663, EPI_ISL_2249664, EPI_ISL_2249665, EPI_ISL_2249666, EPI_ISL_2249672, EPI_ISL_2249674, EPI_ISL_2249675, EPI_ISL_2249676, EPI_ISL_2249678, EPI_ISL_2249679, EPI_ISL_2249682, EPI_ISL_2249683, EPI_ISL_2249684, EPI_ISL_2249685, EPI_ISL_2249686, EPI_ISL_2249689, EPI_ISL_2249690, EPI_ISL_2249691, EPI_ISL_2249692, EPI_ISL_2249693, EPI_ISL_2249694, EPI_ISL_2249695, EPI_ISL_2249696, EPI_ISL_2249697, EPI_ISL_2249698, EPI_ISL_2249699, EPI_ISL_2249701, EPI_ISL_2249702, EPI_ISL_2249703, EPI_ISL_2249704, EPI_ISL_2249705, EPI_ISL_2249706, EPI_ISL_2249707, EPI_ISL_2249708, EPI_ISL_2249709, EPI_ISL_2249710, EPI_ISL_2249711, EPI_ISL_2249713, EPI_ISL_2249714, EPI_ISL_2249715, EPI_ISL_2249716, EPI_ISL_2249717, EPI_ISL_2249718, EPI_ISL_2249719, EPI_ISL_2249720, EPI_ISL_2249721, EPI_ISL_2249722, EPI_ISL_2249723, EPI_ISL_2249724, EPI_ISL_2249725, EPI_ISL_2249726, EPI_ISL_2249727, EPI_ISL_2249728, EPI_ISL_2249729, EPI_ISL_2249731, EPI_ISL_2249732, EPI_ISL_2249733, EPI_ISL_2249734, EPI_ISL_2249735, EPI_ISL_2249736, EPI_ISL_2249737, EPI_ISL_2249738, EPI_ISL_2249739, EPI_ISL_2249740, EPI_ISL_2249742, EPI_ISL_2249743, EPI_ISL_2249744, EPI_ISL_2249745, EPI_ISL_2249746, EPI_ISL_2249747, EPI_ISL_2249748, EPI_ISL_2249749, EPI_ISL_2249750, EPI_ISL_2249751, EPI_ISL_2249752, EPI_ISL_2249753, EPI_ISL_2249754, EPI_ISL_2249755, EPI_ISL_2249756, EPI_ISL_2249757, EPI_ISL_2249758, EPI_ISL_2249759, EPI_ISL_2249760, EPI_ISL_2249761, EPI_ISL_2249764, EPI_ISL_2249765, EPI_ISL_2249766, EPI_ISL_2249768, EPI_ISL_2249769, EPI_ISL_2249770, EPI_ISL_2249771, EPI_ISL_2249772, EPI_ISL_2249773, EPI_ISL_2249774, EPI_ISL_2249775, EPI_ISL_2249776, EPI_ISL_2249777, EPI_ISL_2249778, EPI_ISL_2249779, EPI_ISL_2249780, EPI_ISL_2249782, EPI_ISL_2249783, EPI_ISL_2249784, EPI_ISL_2249785, EPI_ISL_2249786, EPI_ISL_2249787, EPI_ISL_2249788, EPI_ISL_2249789, EPI_ISL_2249790, EPI_ISL_2249791, EPI_ISL_2249792, EPI_ISL_2249793, EPI_ISL_2249794, EPI_ISL_2249795, EPI_ISL_2249796, EPI_ISL_2249797, EPI_ISL_2249798, EPI_ISL_2249799, EPI_ISL_2249800, EPI_ISL_2249801, EPI_ISL_2249802, EPI_ISL_2249803, EPI_ISL_2249804, EPI_ISL_2249805, EPI_ISL_2249806, EPI_ISL_2249807, EPI_ISL_2249808, EPI_ISL_2249809, EPI_ISL_2249810, EPI_ISL_2249811, EPI_ISL_2249812, EPI_ISL_2249813, EPI_ISL_2249814, EPI_ISL_2249815, EPI_ISL_2249816, EPI_ISL_2249817, EPI_ISL_2249818, EPI_ISL_2249819, EPI_ISL_2249820, EPI_ISL_2249821, EPI_ISL_2249822, EPI_ISL_2249823, EPI_ISL_2249824, EPI_ISL_2249825, EPI_ISL_2249826, EPI_ISL_2249827, EPI_ISL_2249828, EPI_ISL_2249829, EPI_ISL_2249830, EPI_ISL_2249831, EPI_ISL_2249832, EPI_ISL_2249833, EPI_ISL_2249834, EPI_ISL_2249837, EPI_ISL_2249838, EPI_ISL_2249839, EPI_ISL_2249840, EPI_ISL_2249841, EPI_ISL_2249842, EPI_ISL_2249843, EPI_ISL_2249844, EPI_ISL_2249845, EPI_ISL_2249846, EPI_ISL_2249847, EPI_ISL_2249848, EPI_ISL_2249849, EPI_ISL_2249850, EPI_ISL_2249851, EPI_ISL_2249852, EPI_ISL_2249853, EPI_ISL_2249854, EPI_ISL_2249855, EPI_ISL_2249856, EPI_ISL_2249857, EPI_ISL_2249858, EPI_ISL_2249859, EPI_ISL_2249860, EPI_ISL_2249861, EPI_ISL_2249862, EPI_ISL_2249863, EPI_ISL_2249864, EPI_ISL_2249865, EPI_ISL_2249866, EPI_ISL_2249867, EPI_ISL_2249868, EPI_ISL_2249869, EPI_ISL_2249870, EPI_ISL_2249871, EPI_ISL_2249872, EPI_ISL_2249873, EPI_ISL_2249874, EPI_ISL_2249875, EPI_ISL_2249876, EPI_ISL_2249877, EPI_ISL_2249878, EPI_ISL_2249879, EPI_ISL_2249880, EPI_ISL_2249881, EPI_ISL_2249882, EPI_ISL_2249887, EPI_ISL_2249888, EPI_ISL_2249889, EPI_ISL_2249890, EPI_ISL_2249891, EPI_ISL_2249892, EPI_ISL_2249893, EPI_ISL_2249894, EPI_ISL_2249895, EPI_ISL_2249897, EPI_ISL_2249898, EPI_ISL_2249899, EPI_ISL_2249900, EPI_ISL_2249902, EPI_ISL_2249903, EPI_ISL_2249904, EPI_ISL_2249905, EPI_ISL_2249906, EPI_ISL_2249907, EPI_ISL_2249908, EPI_ISL_2249909, EPI_ISL_2249910, EPI_ISL_2249911, EPI_ISL_2249912, EPI_ISL_2249913, EPI_ISL_2249914, EPI_ISL_2249915, EPI_ISL_2249916, EPI_ISL_2249917, EPI_ISL_2249918, EPI_ISL_2249919, EPI_ISL_2249921, EPI_ISL_2249922, EPI_ISL_2249923, EPI_ISL_2249924, EPI_ISL_2249925, EPI_ISL_2249926, EPI_ISL_2249927, EPI_ISL_2249928, EPI_ISL_2249929, EPI_ISL_2249930, EPI_ISL_2249931, EPI_ISL_2249932, EPI_ISL_2249933, EPI_ISL_2249934, EPI_ISL_2249935, EPI_ISL_2249936, EPI_ISL_2249937, EPI_ISL_2249938, EPI_ISL_2249939, EPI_ISL_2249940, EPI_ISL_2249941, EPI_ISL_2249942, EPI_ISL_2249943, EPI_ISL_2249944, EPI_ISL_2249945, EPI_ISL_2249946, EPI_ISL_2249947, EPI_ISL_2249948, EPI_ISL_2249949, EPI_ISL_2249950, EPI_ISL_2249951, EPI_ISL_2249952, EPI_ISL_2249953, EPI_ISL_2249954, EPI_ISL_2249955, EPI_ISL_2249956, EPI_ISL_2249957, EPI_ISL_2249958, EPI_ISL_2249959, EPI_ISL_2249960, EPI_ISL_2249961, EPI_ISL_2249962, EPI_ISL_2249963, EPI_ISL_2249964, EPI_ISL_2249965, EPI_ISL_2249966, EPI_ISL_2249967, EPI_ISL_2249968, EPI_ISL_2249969, EPI_ISL_2249971, EPI_ISL_2249972, EPI_ISL_2249973, EPI_ISL_2249975, EPI_ISL_2249976, EPI_ISL_2249977, EPI_ISL_2249978, EPI_ISL_2249979, EPI_ISL_2249980, EPI_ISL_2249981, EPI_ISL_2249982, EPI_ISL_2249983, EPI_ISL_2249984, EPI_ISL_2249985, EPI_ISL_2249986, EPI_ISL_2249987, EPI_ISL_2249988, EPI_ISL_2249989, EPI_ISL_2249990, EPI_ISL_2249991, EPI_ISL_2249992, EPI_ISL_2249993, EPI_ISL_2249994, EPI_ISL_2249995, EPI_ISL_2249996, EPI_ISL_2249998, EPI_ISL_2249999, EPI_ISL_2250000, EPI_ISL_2250001, EPI_ISL_2250002, EPI_ISL_2250003, EPI_ISL_2250004, EPI_ISL_2250005, EPI_ISL_2250006, EPI_ISL_2250007, EPI_ISL_2250008, EPI_ISL_2250009, EPI_ISL_2250011, EPI_ISL_2250012, EPI_ISL_2250013, EPI_ISL_2250014, EPI_ISL_2250015 | | | |
| see above | Reditus Laboratories | Reditus Laboratories | Joshua J. Geltz, Ph.D., Robert M. Sgambelluri, Ph.D., Cassy Philips, M.S., Alexa Eichelberger, M.S. |
| EPI_ISL_2250017, EPI_ISL_2250018, EPI_ISL_2250020, EPI_ISL_2250021, EPI_ISL_2250022, EPI_ISL_2250023, EPI_ISL_2250024, EPI_ISL_2250026, EPI_ISL_2250027, EPI_ISL_2250030, EPI_ISL_2250032, EPI_ISL_2250036, EPI_ISL_2250038, EPI_ISL_2250039, EPI_ISL_2250040, EPI_ISL_2250042, EPI_ISL_2250044, EPI_ISL_2250045, EPI_ISL_2250047, EPI_ISL_2250049, EPI_ISL_2250050, EPI_ISL_2250052, EPI_ISL_2250054, EPI_ISL_2250055, EPI_ISL_2250056, EPI_ISL_2250057, EPI_ISL_2250059, EPI_ISL_2250060, EPI_ISL_2250061, EPI_ISL_2250062, EPI_ISL_2250063, EPI_ISL_2250064, EPI_ISL_2250065, EPI_ISL_2250066, EPI_ISL_2250067, EPI_ISL_2250069, EPI_ISL_2250070, EPI_ISL_2250072, EPI_ISL_2250073, EPI_ISL_2250078, EPI_ISL_2250079, EPI_ISL_2250082, EPI_ISL_2250084, EPI_ISL_2250085, EPI_ISL_2250086, EPI_ISL_2250088, EPI_ISL_2250089, EPI_ISL_2250090, EPI_ISL_2250091, EPI_ISL_2250092, EPI_ISL_2250093, EPI_ISL_2250098, EPI_ISL_2250100, EPI_ISL_2250101, EPI_ISL_2250102, EPI_ISL_2250103, EPI_ISL_2250104, EPI_ISL_2250105, EPI_ISL_2250106, EPI_ISL_2250107, EPI_ISL_2250108, EPI_ISL_2250109, EPI_ISL_2250110, EPI_ISL_2250112, EPI_ISL_2250113, EPI_ISL_2250114, EPI_ISL_2250115, EPI_ISL_2250117, EPI_ISL_2250118, EPI_ISL_2250119, EPI_ISL_2250120, EPI_ISL_2250121, EPI_ISL_2250122, EPI_ISL_2250123, EPI_ISL_2250124, EPI_ISL_2250126, EPI_ISL_2250127, EPI_ISL_2250128, EPI_ISL_2250129, EPI_ISL_2250130, EPI_ISL_2250132, EPI_ISL_2250133, EPI_ISL_2250134, EPI_ISL_2250135, EPI_ISL_2250136, EPI_ISL_2250137, EPI_ISL_2250138, EPI_ISL_2250139, EPI_ISL_2250141, EPI_ISL_2250142, EPI_ISL_2250143, EPI_ISL_2250144, EPI_ISL_2250145, EPI_ISL_2250146, EPI_ISL_2250147, EPI_ISL_2250148, EPI_ISL_2250149 | | | |
| see above | Arizona State University | Arizona State University | Peter T. Skidmore, LaRinda A. Holland, Matthew F. Smith, Rabia Maqsood, Nicholas J. Mellor, Joy M. Blain, Valerie Harris, Joshua LaBaer, Vel Murugan, Efrem S. Lim |
| EPI_ISL_2250153, EPI_ISL_2250154, EPI_ISL_2250155, EPI_ISL_2250156, EPI_ISL_2250157, EPI_ISL_2250158, EPI_ISL_2250159, EPI_ISL_2250160, EPI_ISL_2250161, EPI_ISL_2250162, EPI_ISL_2250163, EPI_ISL_2250164, EPI_ISL_2250165, EPI_ISL_2250166, EPI_ISL_2250167, EPI_ISL_2250168, EPI_ISL_2250169, EPI_ISL_2250170, EPI_ISL_2250171, EPI_ISL_2250172, EPI_ISL_2250173, EPI_ISL_2250174, EPI_ISL_2250175, EPI_ISL_2250176, EPI_ISL_2250177, EPI_ISL_2250178, EPI_ISL_2250179, EPI_ISL_2250180, EPI_ISL_2250181, EPI_ISL_2250182, EPI_ISL_2250183 | | | |
| see above | Oregon State Public Health Laboratory | Oregon State Public Health Laboratory | Rafia Razzaque, Eugene Yeboah, Vanda Makris, Laura Tsaknaridis, John Fontana and Shane Sevey |
| EPI_ISL_2253127, EPI_ISL_2253128, EPI_ISL_2253129, EPI_ISL_2253130, EPI_ISL_2253131, EPI_ISL_2253132, EPI_ISL_2253133, EPI_ISL_2253134, EPI_ISL_2253135, EPI_ISL_2253136, EPI_ISL_2253137, EPI_ISL_2253138, EPI_ISL_2253139, EPI_ISL_2253140, EPI_ISL_2253141, EPI_ISL_2253142, EPI_ISL_2253143, EPI_ISL_2253144, EPI_ISL_2253145, EPI_ISL_2253146, EPI_ISL_2253147, EPI_ISL_2253148, EPI_ISL_2253149, EPI_ISL_2253150, EPI_ISL_2253151, EPI_ISL_2253152, EPI_ISL_2253153, EPI_ISL_2253154, EPI_ISL_2253155, EPI_ISL_2253156, EPI_ISL_2253157, EPI_ISL_2253158, EPI_ISL_2253159, EPI_ISL_2253160, EPI_ISL_2253161, EPI_ISL_2253162, EPI_ISL_2253163, EPI_ISL_2253164, EPI_ISL_2253165 | | | |
| see above | Respiratory Viruses Branch, Centers for Disease Control and Prevention | Respiratory Viruses Branch, Centers for Disease Control and Prevention | Howard,D., Batra,D., Cook,P.W., Moser,K., Paskey,A., Caravas,J., Rambo-Martin,B., Morrison,S., Gulvick,C., Sammons,S., Unoarumhi,Y., Wagner,D., Schmerer,M., Rosenthal,S.H., Gerasimova,A., Kagan,R.M., Anderson,B., Hua,M., Liu,Y., Bernstein,L., Livingston,K., Perez,A., Shlyakhter,I.A., Rolando,R.V., Owen,R., Tanpaiboon,P., Lacbawan,F., Paden,C.R., MacCannell,D. |
| EPI_ISL_2253166, EPI_ISL_2253167, EPI_ISL_2253168, EPI_ISL_2253169, EPI_ISL_2253170, EPI_ISL_2253171, EPI_ISL_2253172, EPI_ISL_2253173, EPI_ISL_2253174, EPI_ISL_2253175 | Infectious Diseases, Chan-Zuckerberg Biohub Covidtracker | Infectious Diseases, Chan-Zuckerberg Biohub Covidtracker | CZB Cliahub Consortium |
| EPI_ISL_2253185, EPI_ISL_2253186, EPI_ISL_2253187, EPI_ISL_2253188, EPI_ISL_2253189, EPI_ISL_2253195, EPI_ISL_2253196 | Laboratory Medicine, University of Washington | Laboratory Medicine, University of Washington | Roychoudhury,P., Xie,H., Shrestha,L., Bakhash,S.M., Lin,M.J., Baker,N.R., Ellis,S., Sathees,S., Huang,M.-L., Jerome,K.R., Greninger,A. |
| EPI_ISL_2253199 | Respiratory Viruses Branch, Centers for Disease Control and Prevention | Respiratory Viruses Branch, Centers for Disease Control and Prevention | Howard,D., Batra,D., Cook,P.W., Moser,K., Paskey,A., Caravas,J.,Rambo-Martin,B., Morrison,S., Gulvick,C., Sammons,S., Unoarumhi,Y.,Schmerer,M., Roychoudhury,P., Xie,H., Shrestha,L., Nguyen,T.V.,Bakhash,S.M., Lin,M., Baker,N.R., Perez,R., Ellis,S., Breit,N.,Livingston,R.J., Huang,M.-L., Jerome,K.R., Mathias,P.,Greninger,A., Paden,C.R. and MacCannell,D. |
| EPI_ISL_2253201, EPI_ISL_2253202, EPI_ISL_2253203 | Respiratory Viruses Branch, Centers for Disease Control and Prevention | Respiratory Viruses Branch, Centers for Disease Control and Prevention | Howard,D., Batra,D., Cook,P.W., Moser,K., Paskey,A., Caravas,J., Rambo-Martin,B., Morrison,S., Gulvick,C., Sammons,S., Unoarumhi,Y., Wagner,D., Schmerer,M., Gao,H., Li,M., Gao,J., Fierro,J., Sapra,B., Tsai,B., Meng,Y., Ng,D., Xie,J., Paden,C.R., MacCannell,D. |
| EPI_ISL_2253206, EPI_ISL_2253207, EPI_ISL_2253208, EPI_ISL_2253209, EPI_ISL_2253210 | Respiratory Viruses Branch, Centers for Disease Control and Prevention | Respiratory Viruses Branch, Centers for Disease Control and Prevention | Howard,D., Batra,D., Cook,P.W., Moser,K., Paskey,A., Caravas,J., Rambo-Martin,B., Morrison,S., Gulvick,C., Sammons,S., Unoarumhi,Y., Wagner,D., Schmerer,M., Agarwal,M., Almasri,E., Boles,D., Burns,A., Charoensri,N., Cohen,O., Countryman,S., Cristobal,M.A., Croy,B., Dale,S., Deshmukh,H., Douglas,A., Drouillon,V., Eisenberg,M., Engler,H., Ghatti,R., Gupta,P., Hicks,S., Humphrey,J., Iyer,L., Jain,M., Kolli,M., Krueger,B., Kuphal,T., Letovsky,S., Levandoski,M., Lukasik,C., Meltzer,J., Norvell,B., Nye,M., Parker,S., Petropoulos,C., Pruitt,J., Ragan,S., Ryan,S., Sapeta,M., Schroth,J., Selvaraju,S.B., Stevovic,G., Suchanek,A., Throop,A., Tilson,L., Urban,T., Voshell,J., Wagner,K., Williams,J., Williamson,M., Zeng,Q., Zwiefelhofer,T., Paden,C.R., MacCannell,D. |
| EPI_ISL_2254263, EPI_ISL_2254264, EPI_ISL_2254265, EPI_ISL_2254266, EPI_ISL_2254267, EPI_ISL_2254268, EPI_ISL_2254269, EPI_ISL_2254271, EPI_ISL_2254272, EPI_ISL_2254273, EPI_ISL_2254274, EPI_ISL_2254275, EPI_ISL_2254276, EPI_ISL_2254277, EPI_ISL_2254278, EPI_ISL_2254279, EPI_ISL_2254280, EPI_ISL_2254281, EPI_ISL_2254282, EPI_ISL_2254283, EPI_ISL_2254284, EPI_ISL_2254285, EPI_ISL_2254286, EPI_ISL_2254287, EPI_ISL_2254288, EPI_ISL_2254290, EPI_ISL_2254291, EPI_ISL_2254292, EPI_ISL_2254293, EPI_ISL_2254295, EPI_ISL_2254296, EPI_ISL_2254297, EPI_ISL_2254298, EPI_ISL_2254299, EPI_ISL_2254300, EPI_ISL_2254301, EPI_ISL_2254302, EPI_ISL_2254303, EPI_ISL_2254304, EPI_ISL_2254305, EPI_ISL_2254306, EPI_ISL_2254307, EPI_ISL_2254309, EPI_ISL_2254310, EPI_ISL_2254311, EPI_ISL_2254312, EPI_ISL_2254313, EPI_ISL_2254314, EPI_ISL_2254315, EPI_ISL_2254316, EPI_ISL_2254317, EPI_ISL_2254318, EPI_ISL_2254319, EPI_ISL_2254320, EPI_ISL_2254321, EPI_ISL_2254322, EPI_ISL_2254323, EPI_ISL_2254324, EPI_ISL_2254325, EPI_ISL_2254326, EPI_ISL_2254327, EPI_ISL_2254328, EPI_ISL_2254329, EPI_ISL_2254330, EPI_ISL_2254331, EPI_ISL_2254332, EPI_ISL_2254333, EPI_ISL_2254334, EPI_ISL_2254335, EPI_ISL_2254337, EPI_ISL_2254338, EPI_ISL_2254339, EPI_ISL_2254340, EPI_ISL_2254341, EPI_ISL_2254342, EPI_ISL_2254343, EPI_ISL_2254344, EPI_ISL_2254345, EPI_ISL_2254346, EPI_ISL_2254347, EPI_ISL_2254348, EPI_ISL_2254351, EPI_ISL_2254352, EPI_ISL_2254353, EPI_ISL_2254354, EPI_ISL_2254355, EPI_ISL_2254356, EPI_ISL_2254357, EPI_ISL_2254358, EPI_ISL_2254359, EPI_ISL_2254360, EPI_ISL_2254361, EPI_ISL_2254362, EPI_ISL_2254363, EPI_ISL_2254364, EPI_ISL_2254365, EPI_ISL_2254366, EPI_ISL_2254367, EPI_ISL_2254368, EPI_ISL_2254369, EPI_ISL_2254370, EPI_ISL_2254371, EPI_ISL_2254372, EPI_ISL_2254373, EPI_ISL_2254374, EPI_ISL_2254375, EPI_ISL_2254376, EPI_ISL_2254377, EPI_ISL_2254378, EPI_ISL_2254379, EPI_ISL_2254380, EPI_ISL_2254381, EPI_ISL_2254382, EPI_ISL_2254383, EPI_ISL_2254384, EPI_ISL_2254385, EPI_ISL_2254386, EPI_ISL_2254387, EPI_ISL_2254388, EPI_ISL_2254389, EPI_ISL_2254390, EPI_ISL_2254391, EPI_ISL_2254392, EPI_ISL_2254393, EPI_ISL_2254394, EPI_ISL_2254395, EPI_ISL_2254396, EPI_ISL_2254397, EPI_ISL_2254398, EPI_ISL_2254400, EPI_ISL_2254401, EPI_ISL_2254402, EPI_ISL_2254403, EPI_ISL_2254404, EPI_ISL_2254405, EPI_ISL_2254406, EPI_ISL_2254407, EPI_ISL_2254408, EPI_ISL_2254409, EPI_ISL_2254410, EPI_ISL_2254411, EPI_ISL_2254412, EPI_ISL_2254413, EPI_ISL_2254414, EPI_ISL_2254415, EPI_ISL_2254416, EPI_ISL_2254417, EPI_ISL_2254418, EPI_ISL_2254419, EPI_ISL_2254420, EPI_ISL_2254421, EPI_ISL_2254422, EPI_ISL_2254423, EPI_ISL_2254424, EPI_ISL_2254425, EPI_ISL_2254426, EPI_ISL_2254427, EPI_ISL_2254428, EPI_ISL_2254429, EPI_ISL_2254430, EPI_ISL_2254431, EPI_ISL_2254432, EPI_ISL_2254433, EPI_ISL_2254434, EPI_ISL_2254435, EPI_ISL_2254436, EPI_ISL_2254438, EPI_ISL_2254439, EPI_ISL_2254440, EPI_ISL_2254441, EPI_ISL_2254442, EPI_ISL_2254443, EPI_ISL_2254444, EPI_ISL_2254445, EPI_ISL_2254446, EPI_ISL_2254447, EPI_ISL_2254448, EPI_ISL_2254449, EPI_ISL_2254450, EPI_ISL_2254451, EPI_ISL_2254453, EPI_ISL_2254454, EPI_ISL_2254455, EPI_ISL_2254456, EPI_ISL_2254457, EPI_ISL_2254458, EPI_ISL_2254459, EPI_ISL_2254460, EPI_ISL_2254461, EPI_ISL_2254462, EPI_ISL_2254463, EPI_ISL_2254464, EPI_ISL_2254465, EPI_ISL_2254466, EPI_ISL_2254467, EPI_ISL_2254468, EPI_ISL_2254469, EPI_ISL_2254470, EPI_ISL_2254472, EPI_ISL_2254473, EPI_ISL_2254474, EPI_ISL_2254475, EPI_ISL_2254476, EPI_ISL_2254477, EPI_ISL_2254478, EPI_ISL_2254479, EPI_ISL_2254480, EPI_ISL_2254482, EPI_ISL_2254483, EPI_ISL_2254484, EPI_ISL_2254485, EPI_ISL_2254486, EPI_ISL_2254487, EPI_ISL_2254488, EPI_ISL_2254489, EPI_ISL_2254490, EPI_ISL_2254491, EPI_ISL_2254492, EPI_ISL_2254493, EPI_ISL_2254494 | | | |
| see above | Respiratory Viruses Branch, Centers for Disease Control and Prevention | Respiratory Viruses Branch, Centers for Disease Control and Prevention | Howard,D., Batra,D., Cook,P.W., Moser,K., Paskey,A., Caravas,J., Rambo-Martin,B., Morrison,S., Gulvick,C., Sammons,S., Unoarumhi,Y., Wagner,D., Schmerer,M., Clark,C., Campbell,P., Case,R., Ghorpade,V., Houdeshell,H., Kvalvaag,O., Nall,D., Sanders,E., Vest,A., Westlund,S., Hardison,M., Paden,C.R., MacCannell,D. |
| EPI_ISL_2254502, EPI_ISL_2254507, EPI_ISL_2254509, EPI_ISL_2254514, EPI_ISL_2254517, EPI_ISL_2254518, EPI_ISL_2254522, EPI_ISL_2254523, EPI_ISL_2254524, EPI_ISL_2254526, EPI_ISL_2254527, EPI_ISL_2254528, EPI_ISL_2254529, EPI_ISL_2254530, EPI_ISL_2254531, EPI_ISL_2254534, EPI_ISL_2254535, EPI_ISL_2254537, EPI_ISL_2254538, EPI_ISL_2254539, EPI_ISL_2254540, EPI_ISL_2254543, | | | |

| | | | |
|---|---|---|---|
| EPI_ISL_2254544, EPI_ISL_2254545, EPI_ISL_2254546, EPI_ISL_2254547, EPI_ISL_2254548, EPI_ISL_2254550, EPI_ISL_2254551, EPI_ISL_2254552, EPI_ISL_2254553, EPI_ISL_2254557, EPI_ISL_2254558, EPI_ISL_2254560, EPI_ISL_2254561, EPI_ISL_2254562, EPI_ISL_2254563, EPI_ISL_2254565, EPI_ISL_2254566, EPI_ISL_2254568, EPI_ISL_2254569, EPI_ISL_2254570, EPI_ISL_2254571, EPI_ISL_2254572, EPI_ISL_2254573, EPI_ISL_2254574, EPI_ISL_2254575, EPI_ISL_2254576, EPI_ISL_2254582, EPI_ISL_2254588, EPI_ISL_2254589, EPI_ISL_2254590, EPI_ISL_2254592, EPI_ISL_2254593, EPI_ISL_2254594, EPI_ISL_2254595, EPI_ISL_2254596, EPI_ISL_2254597, EPI_ISL_2254598, EPI_ISL_2254599, EPI_ISL_2254600, EPI_ISL_2254601, EPI_ISL_2254602, EPI_ISL_2254604, EPI_ISL_2254605, EPI_ISL_2254606, EPI_ISL_2254607, EPI_ISL_2254608, EPI_ISL_2254611, EPI_ISL_2254613, EPI_ISL_2254614, EPI_ISL_2254617, EPI_ISL_2254618, EPI_ISL_2254619, EPI_ISL_2254620, EPI_ISL_2254621, EPI_ISL_2254622, EPI_ISL_2254623, EPI_ISL_2254624, EPI_ISL_2254625, EPI_ISL_2254628, EPI_ISL_2254629, EPI_ISL_2254630, EPI_ISL_2254631, EPI_ISL_2254632, EPI_ISL_2254633, EPI_ISL_2254635, EPI_ISL_2254636, EPI_ISL_2254637, EPI_ISL_2254638, EPI_ISL_2254639, EPI_ISL_2254640, EPI_ISL_2254641, EPI_ISL_2254642, EPI_ISL_2254643, EPI_ISL_2254645, EPI_ISL_2254646, EPI_ISL_2254647, EPI_ISL_2254648, EPI_ISL_2254649, EPI_ISL_2254650, EPI_ISL_2254651, EPI_ISL_2254652, EPI_ISL_2254653, EPI_ISL_2254655, EPI_ISL_2254656, EPI_ISL_2254660, EPI_ISL_2254661, EPI_ISL_2254663, EPI_ISL_2254664, EPI_ISL_2254665, EPI_ISL_2254666, EPI_ISL_2254667, EPI_ISL_2254668, EPI_ISL_2254669, EPI_ISL_2254673, EPI_ISL_2254674, EPI_ISL_2254675, EPI_ISL_2254676, EPI_ISL_2254677, EPI_ISL_2254678, EPI_ISL_2254679, EPI_ISL_2254680, EPI_ISL_2254681, EPI_ISL_2254682, EPI_ISL_2254683, EPI_ISL_2254684, EPI_ISL_2254685, EPI_ISL_2254686, EPI_ISL_2254688, EPI_ISL_2254689, EPI_ISL_2254690, EPI_ISL_2254691, EPI_ISL_2254692, EPI_ISL_2254695, EPI_ISL_2254696, EPI_ISL_2254697, EPI_ISL_2254698, EPI_ISL_2254699, EPI_ISL_2254700, EPI_ISL_2254701, EPI_ISL_2254702, EPI_ISL_2254704, EPI_ISL_2254705, EPI_ISL_2254706, EPI_ISL_2254707, EPI_ISL_2254708, EPI_ISL_2254709, EPI_ISL_2254710, EPI_ISL_2254711, EPI_ISL_2254712, EPI_ISL_2254714, EPI_ISL_2254716 | | | |
| see above | Duke Center for Genomic and Computational Biology (GCB), Duke University | Duke Center for Genomic and Computational Biology (GCB), Duke University | Berrio,A., Devos,N., Wray,G.A., Haase,S.B., Denny,T.N. |
| EPI_ISL_2258223, EPI_ISL_2258224, EPI_ISL_2258228, EPI_ISL_2258229, EPI_ISL_2258230, EPI_ISL_2258231, EPI_ISL_2258233, EPI_ISL_2258234, EPI_ISL_2258235, EPI_ISL_2258236, EPI_ISL_2258237, EPI_ISL_2258239, EPI_ISL_2258240, EPI_ISL_2258241, EPI_ISL_2258242, EPI_ISL_2258243, EPI_ISL_2258244, EPI_ISL_2258245, EPI_ISL_2258247, EPI_ISL_2258249, EPI_ISL_2258250, EPI_ISL_2258251, EPI_ISL_2258252, EPI_ISL_2258254, EPI_ISL_2258255, EPI_ISL_2258256, EPI_ISL_2258257, EPI_ISL_2258258, EPI_ISL_2258259, EPI_ISL_2258260, EPI_ISL_2258263, EPI_ISL_2258264, EPI_ISL_2258265, EPI_ISL_2258268, EPI_ISL_2258269, EPI_ISL_2258270 | | | |
| see above | City of Milwaukee Health Department Laboratory | City of Milwaukee Health Department Laboratory | Sanjib Bhattacharyya, Manjeet Khubbar, Amy Bauer, Jennifer Lentz, Samantha Scott |
| EPI_ISL_2259228 | UCLA Clinical Micro Lab | Los Angeles County Public Health Laboratories | P. Hemarajata et al. |
| EPI_ISL_2259229, EPI_ISL_2259230, EPI_ISL_2259231, EPI_ISL_2259232, EPI_ISL_2259233, EPI_ISL_2259234, EPI_ISL_2259235 | Pomona Valley Hospital Medical Center | Los Angeles County Public Health Laboratories | P. Hemarajata et al. |
| EPI_ISL_2259237, EPI_ISL_2259238, EPI_ISL_2259240, EPI_ISL_2259241 | Huntington Memorial Hospital | Los Angeles County Public Health Laboratories | P. Hemarajata et al. |
| EPI_ISL_2259242 | Martin Luther King Jr. Community Hospital | Los Angeles County Public Health Laboratories | P. Hemarajata et al. |
| EPI_ISL_2259243, EPI_ISL_2259244, EPI_ISL_2259245, EPI_ISL_2259246, EPI_ISL_2259248 | Los Angeles County Public Health Laboratories | Los Angeles County Public Health Laboratories | P. Hemarajata et al. |
| EPI_ISL_2259249 | USC Clinical Lab | Los Angeles County Public Health Laboratories | P. Hemarajata et al. |
| EPI_ISL_2259250 | Flow Diagnostics Lab | Los Angeles County Public Health Laboratories | P. Hemarajata et al. |
| EPI_ISL_2259251 | Quest Diagnostics | Los Angeles County Public Health Laboratories | P. Hemarajata et al. |
| EPI_ISL_2259412 | DC Public Health Lab/ Dept. of Forensic Sciences | DC Public Health Lab/ Dept. of Forensic Sciences | Janis Doss, Scott Nguyen, Elizabeth Zelaya, Sarah Scott, Connie Maza, Monica Mann, Brittany Hamilton, David Payne, Jocelyn Hauser |
| EPI_ISL_2259427, EPI_ISL_2259428, EPI_ISL_2259429, EPI_ISL_2259430, EPI_ISL_2259431 | Pomona Valley Hospital Medical Center | Los Angeles County Public Health Laboratories | P. Hemarajata et al. |
| EPI_ISL_2259432, EPI_ISL_2259433, EPI_ISL_2259434, EPI_ISL_2259435 | USC Clinical Lab | Los Angeles County Public Health Laboratories | P. Hemarajata et al. |
| EPI_ISL_2259436 | UCLA Clinical Micro Lab | Los Angeles County Public Health Laboratories | P. Hemarajata et al. |
| EPI_ISL_2259437 | Pomona Valley Hospital Medical Center | Los Angeles County Public Health Laboratories | P. Hemarajata et al. |
| EPI_ISL_2259438 | Flow Diagnostics Lab | Los Angeles County Public Health Laboratories | P. Hemarajata et al. |
| EPI_ISL_2259439 | Providence Little Company of Mary Medical Center | Los Angeles County Public Health Laboratories | P. Hemarajata et al. |
| EPI_ISL_2260370, EPI_ISL_2260371, EPI_ISL_2260373, EPI_ISL_2260375, EPI_ISL_2260377, EPI_ISL_2260379, EPI_ISL_2260381, EPI_ISL_2260385, EPI_ISL_2260386, EPI_ISL_2260388, EPI_ISL_2260390, EPI_ISL_2260392 | | | |
| see above | USC Clinical Lab | Los Angeles County Public Health Laboratories | P. Hemarajata et al. |
| EPI_ISL_2260394, EPI_ISL_2260396 | Flow Diagnostics Lab | Los Angeles County Public Health Laboratories | P. Hemarajata et al. |
| EPI_ISL_2261789, EPI_ISL_2261791, EPI_ISL_2261793, EPI_ISL_2261795, EPI_ISL_2261797, EPI_ISL_2261799, EPI_ISL_2261801, EPI_ISL_2261805, EPI_ISL_2261807, EPI_ISL_2261809, EPI_ISL_2261811, EPI_ISL_2261813, EPI_ISL_2261825, EPI_ISL_2261828, EPI_ISL_2261829 | | | |
| see above | Wichita State University - Molecular Diagnostics Lab | Kansas Health and Environmental Lab | Mike Grose, Jonathan Barnell, Ben Olsen, and Phil Adam |
| EPI_ISL_2262493, EPI_ISL_2262495 | DOHMH Riverside | New York City Public Health Laboratory | Jade Wang, et al. |
| EPI_ISL_2262497 | DOHMH Chelsea | New York City Public Health Laboratory | Jade Wang, et al. |
| EPI_ISL_2262499 | DOHMH Morrisania | New York City Public Health Laboratory | Jade Wang, et al. |
| EPI_ISL_2262500 | DOHMH Jamaica | New York City Public Health Laboratory | Jade Wang, et al. |
| EPI_ISL_2262502 | DOHMH Corona | New York City Public Health Laboratory | Jade Wang, et al. |
| EPI_ISL_2262504 | DOHMH Riverside | New York City Public Health Laboratory | Jade Wang, et al. |
| EPI_ISL_2262506, EPI_ISL_2262508 | DOHMH Crown Heights | New York City Public Health Laboratory | Jade Wang, et al. |
| EPI_ISL_2262510 | DOHMH Jamaica | New York City Public Health Laboratory | Jade Wang, et al. |
| EPI_ISL_2262511 | DOHMH Morrisania | New York City Public Health Laboratory | Jade Wang, et al. |
| EPI_ISL_2262513 | DOHMH Fort Greene | New York City Public Health Laboratory | Jade Wang, et al. |
| EPI_ISL_2262515, EPI_ISL_2262517 | DOHMH PHL | New York City Public Health Laboratory | Jade Wang, et al. |
| EPI_ISL_2262519, EPI_ISL_2262521 | DOHMH Jamaica | New York City Public Health Laboratory | Jade Wang, et al. |
| EPI_ISL_2262522 | DOHMH Corona | New York City Public Health Laboratory | Jade Wang, et al. |
| EPI_ISL_2262524 | DOHMH Central Harlem | New York City Public Health Laboratory | Jade Wang, et al. |
| EPI_ISL_2262526, EPI_ISL_2262528 | DOHMH Riverside | New York City Public Health Laboratory | Jade Wang, et al. |
| EPI_ISL_2262529 | DOHMH Central Harlem | New York City Public Health Laboratory | Jade Wang, et al. |
| EPI_ISL_2262531 | DOHMH Crown Heights | New York City Public Health Laboratory | Jade Wang, et al. |
| EPI_ISL_2262533, EPI_ISL_2262535 | DOHMH Chelsea | New York City Public Health Laboratory | Jade Wang, et al. |
| EPI_ISL_2262537, EPI_ISL_2262539 | DOHMH Jamaica | New York City Public Health Laboratory | Jade Wang, et al. |

| | | | |
|---|---|---|---|
| EPI_ISL_2262540 | DOHMH PHL | New York City Public Health Laboratory | Jade Wang, et al. |
| EPI_ISL_2262542 | DOHMH Morrisania | New York City Public Health Laboratory | Jade Wang, et al. |
| EPI_ISL_2262544 | DOHMH Central Harlem | New York City Public Health Laboratory | Jade Wang, et al. |
| EPI_ISL_2262546, EPI_ISL_2262548, EPI_ISL_2262550, EPI_ISL_2262552, EPI_ISL_2262554 | OCME Office Of Chief Medical Examiner | New York City Public Health Laboratory | Jade Wang, et al. |
| EPI_ISL_2262555 | DOHMH Jamaica | New York City Public Health Laboratory | Jade Wang, et al. |
| EPI_ISL_2262557, EPI_ISL_2262559 | DOHMH Central Harlem | New York City Public Health Laboratory | Jade Wang, et al. |
| EPI_ISL_2262561 | DOHMH Chelsea | New York City Public Health Laboratory | Jade Wang, et al. |
| EPI_ISL_2262563 | DOHMH Jamaica | New York City Public Health Laboratory | Jade Wang, et al. |
| EPI_ISL_2262564, EPI_ISL_2262566 | OCME Office Of Chief Medical Examiner | New York City Public Health Laboratory | Jade Wang, et al. |
| EPI_ISL_2262568, EPI_ISL_2262570, EPI_ISL_2262572, EPI_ISL_2262573 | Poplar Healthcare | New York City Public Health Laboratory | Jade Wang, et al. |
| EPI_ISL_2262575 | OCME Office Of Chief Medical Examiner | New York City Public Health Laboratory | Jade Wang, et al. |
| EPI_ISL_2262577, EPI_ISL_2262579, EPI_ISL_2262581 | DOHMH Chelsea | New York City Public Health Laboratory | Jade Wang, et al. |
| EPI_ISL_2262582, EPI_ISL_2262584 | DOHMH Central Harlem | New York City Public Health Laboratory | Jade Wang, et al. |
| EPI_ISL_2262586, EPI_ISL_2262588 | DOHMH Jamaica | New York City Public Health Laboratory | Jade Wang, et al. |
| EPI_ISL_2262590 | DOHMH Fort Greene | New York City Public Health Laboratory | Jade Wang, et al. |
| EPI_ISL_2262592 | DOHMH Central Harlem | New York City Public Health Laboratory | Jade Wang, et al. |
| EPI_ISL_2262594, EPI_ISL_2262595, EPI_ISL_2262597 | DOHMH Morrisania | New York City Public Health Laboratory | Jade Wang, et al. |
| EPI_ISL_2262599 | DOHMH Jamaica | New York City Public Health Laboratory | Jade Wang, et al. |
| EPI_ISL_2262601, EPI_ISL_2262602 | DOHMH Corona | New York City Public Health Laboratory | Jade Wang, et al. |
| EPI_ISL_2262604, EPI_ISL_2262606, EPI_ISL_2262607 | DOHMH PHL | New York City Public Health Laboratory | Jade Wang, et al. |
| EPI_ISL_2262609 | DOHMH Riverside | New York City Public Health Laboratory | Jade Wang, et al. |
| EPI_ISL_2262612 | DOHMH Fort Greene | New York City Public Health Laboratory | Jade Wang, et al. |
| EPI_ISL_2262613 | DOHMH Crown Heights | New York City Public Health Laboratory | Jade Wang, et al. |
| EPI_ISL_2262615 | OCME Office Of Chief Medical Examiner | New York City Public Health Laboratory | Jade Wang, et al. |
| EPI_ISL_2262617 | DOHMH Jamaica | New York City Public Health Laboratory | Jade Wang, et al. |
| EPI_ISL_2262618 | DOHMH Morrisania | New York City Public Health Laboratory | Jade Wang, et al. |
| EPI_ISL_2262620, EPI_ISL_2262622 | DOHMH PHL | New York City Public Health Laboratory | Jade Wang, et al. |
| EPI_ISL_2262624 | DOHMH Jamaica | New York City Public Health Laboratory | Jade Wang, et al. |
| EPI_ISL_2262626 | DOHMH Crown Heights | New York City Public Health Laboratory | Jade Wang, et al. |
| EPI_ISL_2262628, EPI_ISL_2262629 | DOHMH Morrisania | New York City Public Health Laboratory | Jade Wang, et al. |
| EPI_ISL_2262631, EPI_ISL_2262633 | DOHMH Central Harlem | New York City Public Health Laboratory | Jade Wang, et al. |
| EPI_ISL_2262635, EPI_ISL_2262636, EPI_ISL_2262638, EPI_ISL_2262642, EPI_ISL_2262644, EPI_ISL_2262645, EPI_ISL_2262647 | DOHMH Jamaica | New York City Public Health Laboratory | Jade Wang, et al. |
| EPI_ISL_2262649 | DOHMH Central Harlem | New York City Public Health Laboratory | Jade Wang, et al. |
| EPI_ISL_2262651, EPI_ISL_2262652 | DOHMH Fort Greene | New York City Public Health Laboratory | Jade Wang, et al. |
| EPI_ISL_2262654 | DOHMH PHL | New York City Public Health Laboratory | Jade Wang, et al. |
| EPI_ISL_2262655, EPI_ISL_2262657, EPI_ISL_2262659 | DOHMH Morrisania | New York City Public Health Laboratory | Jade Wang, et al. |
| EPI_ISL_2262661 | DOHMH Jamaica | New York City Public Health Laboratory | Jade Wang, et al. |
| EPI_ISL_2262663 | Department of Homeless Services | New York City Public Health Laboratory | Jade Wang, et al. |
| EPI_ISL_2262665 | OCME Office Of Chief Medical Examiner | New York City Public Health Laboratory | Jade Wang, et al. |
| EPI_ISL_2262666 | DOHMH PHL | New York City Public Health Laboratory | Jade Wang, et al. |
| EPI_ISL_2262668 | DOHMH Corona | New York City Public Health Laboratory | Jade Wang, et al. |
| EPI_ISL_2262670, EPI_ISL_2262672 | DOHMH Chelsea | New York City Public Health Laboratory | Jade Wang, et al. |
| EPI_ISL_2262674 | DOHMH Riverside | New York City Public Health Laboratory | Jade Wang, et al. |
| EPI_ISL_2262675, EPI_ISL_2262676, EPI_ISL_2262677 | DOHMH Morrisania | New York City Public Health Laboratory | Jade Wang, et al. |
| EPI_ISL_2262679 | DOHMH Central Harlem | New York City Public Health Laboratory | Jade Wang, et al. |
| EPI_ISL_2262680 | DOHMH Riverside | New York City Public Health Laboratory | Jade Wang, et al. |
| EPI_ISL_2262682 | DOHMH Central Harlem | New York City Public Health Laboratory | Jade Wang, et al. |
| EPI_ISL_2262684 | Department of Homeless Services | New York City Public Health Laboratory | Jade Wang, et al. |
| EPI_ISL_2262686 | OCME Office Of Chief Medical Examiner | New York City Public Health Laboratory | Jade Wang, et al. |
| EPI_ISL_2263450, EPI_ISL_2263452, EPI_ISL_2263453, EPI_ISL_2263457, EPI_ISL_2263459, EPI_ISL_2263462, EPI_ISL_2263464, EPI_ISL_2263469, EPI_ISL_2263477, EPI_ISL_2263479, EPI_ISL_2263482, EPI_ISL_2263487, EPI_ISL_2263490 | | | |
| see above | Arizona State Public Health Laboratory | Arizona State Public Health Laboratory | Trung Huynh, Jessica Escobar, Katherine Fullerton, Nobuko Fukushima, Stacy White, Linda Getsinger, Victor Waddell |
| EPI_ISL_2264408, EPI_ISL_2264410, EPI_ISL_2264412, EPI_ISL_2264417, EPI_ISL_2264419, EPI_ISL_2264421, EPI_ISL_2264423, EPI_ISL_2264424, EPI_ISL_2264428, EPI_ISL_2264431, EPI_ISL_2264432, EPI_ISL_2264433, EPI_ISL_2264434, EPI_ISL_2264435, EPI_ISL_2264437, EPI_ISL_2264439 | | | |
| see above | World Reference Center of Emerging | ClickSeq Lab, University of Texas Medical | Jaworski E., Langsjoen R.M., Judy B., Newman P., Plante J.A., Plante K.S., Miller A.L., Zhou Y., Swetnam D., Dong J., Ren P., Pyles R.B., Ksiazek T., Menachery V.D., Weaver S.C, Routh A. |

| | | |
|---|---|---|
| | Viruses and Arboviruses (WRCEVA), University of Texas Medical Branch (UTMB), Galveston | Branch, UTMB, Galveston |

EPI_ISL_2264443, EPI_ISL_2264446, EPI_ISL_2264449, EPI_ISL_2264452, EPI_ISL_2264454, EPI_ISL_2264456, EPI_ISL_2264458, EPI_ISL_2264461, EPI_ISL_2264464, EPI_ISL_2264467, EPI_ISL_2264470, EPI_ISL_2264474, EPI_ISL_2264476, EPI_ISL_2264479, EPI_ISL_2264482, EPI_ISL_2264485, EPI_ISL_2264488, EPI_ISL_2264491, EPI_ISL_2264494, EPI_ISL_2264496, EPI_ISL_2264499, EPI_ISL_2264502, EPI_ISL_2264505

| | | | |
|---|---|---|---|
| see above | Clinical Microbiology, University of Texas Medical Branch (UTMB), Galveston | ClickSeq Lab, University of Texas Medical Branch, UTMB, Galveston | Jaworski E., Langsjoen R.M., Judy B., Newman P., Plante J.A., Plante K.S., Miller A.L., Zhou Y., Swetnam D., Dong J., Ren P., Pyles R.B., Ksiazek T., Menachery V.D., Weaver S.C, Routh A. |

EPI_ISL_2267988, EPI_ISL_2267991, EPI_ISL_2267993, EPI_ISL_2267996, EPI_ISL_2267998, EPI_ISL_2268001, EPI_ISL_2268003, EPI_ISL_2268005, EPI_ISL_2268007, EPI_ISL_2268009, EPI_ISL_2268011, EPI_ISL_2268014, EPI_ISL_2268016, EPI_ISL_2268019, EPI_ISL_2268021, EPI_ISL_2268024, EPI_ISL_2268026, EPI_ISL_2268030, EPI_ISL_2268032, EPI_ISL_2268035, EPI_ISL_2268037, EPI_ISL_2268040, EPI_ISL_2268042, EPI_ISL_2268045, EPI_ISL_2268048, EPI_ISL_2268050, EPI_ISL_2268053, EPI_ISL_2268055, EPI_ISL_2268057, EPI_ISL_2268060, EPI_ISL_2268063, EPI_ISL_2268065, EPI_ISL_2268068, EPI_ISL_2268070, EPI_ISL_2268073, EPI_ISL_2268076, EPI_ISL_2268078, EPI_ISL_2268080, EPI_ISL_2268085, EPI_ISL_2268087, EPI_ISL_2268089, EPI_ISL_2268092, EPI_ISL_2268094, EPI_ISL_2268097, EPI_ISL_2268099, EPI_ISL_2268101, EPI_ISL_2268104, EPI_ISL_2268106, EPI_ISL_2268109, EPI_ISL_2268111, EPI_ISL_2268113, EPI_ISL_2268115, EPI_ISL_2268118, EPI_ISL_2268120, EPI_ISL_2268123, EPI_ISL_2268125, EPI_ISL_2268128, EPI_ISL_2268130, EPI_ISL_2268133

| | | | |
|---|---|---|---|
| see above | Aegis Sciences Corporation | Centers for Disease Control and Prevention Division of Viral Diseases, Pathogen Discovery | Dakota Howard, Dhwani Batra, Peter W. Cook, Kara Moser, Adrian Paskey, Jason Caravas, Benjamin Rambo-Martin, Shatavia Morrison, Christopher Gulvick, Scott Sammons, Yvette Unoarumhi, Darlene Wagner, Matthew Schmerer, Cyndi Clark, Patrick Campbell, Rob Case, Vikramsinha Ghorpade, Holly Houdeshell, Ola Kvalvaag, Dillon Nall, Ethan Sanders, Alec Vest, Shaun Westlund, Matthew Hardison, Clinton R. Paden, Duncan MacCannell |

EPI_ISL_2268144, EPI_ISL_2268146, EPI_ISL_2268148, EPI_ISL_2268152, EPI_ISL_2268154, EPI_ISL_2268156, EPI_ISL_2268159, EPI_ISL_2268161, EPI_ISL_2268164, EPI_ISL_2268166, EPI_ISL_2268169, EPI_ISL_2268171, EPI_ISL_2268174, EPI_ISL_2268176, EPI_ISL_2268178, EPI_ISL_2268181, EPI_ISL_2268184, EPI_ISL_2268186, EPI_ISL_2268189

| | | | |
|---|---|---|---|
| see above | Quest Diagnostics Incorporated | Centers for Disease Control and Prevention Division of Viral Diseases, Pathogen Discovery | Dakota Howard, Dhwani Batra, Peter W. Cook, Kara Moser, Adrian Paskey, Jason Caravas, Benjamin Rambo-Martin, Shatavia Morrison, Christopher Gulvick, Scott Sammons, Yvette Unoarumhi, Darlene Wagner, Matthew Schmerer, S. H. Rosenthal, A. Gerasimova, R. M. Kagan, B. Anderson, M. Hua, Y. Liu, L.E. Bernstein, K.E. Livingston, A. Perez, I. A. Shlyakhter, R. V. Rolando, R. Owen, P. Tanpaiboon, F. Lacbawan, Clinton R. Paden, Duncan MacCannell |

EPI_ISL_2268201, EPI_ISL_2268203, EPI_ISL_2268206, EPI_ISL_2268208, EPI_ISL_2268211, EPI_ISL_2268213, EPI_ISL_2268216, EPI_ISL_2268218, EPI_ISL_2268220, EPI_ISL_2268223, EPI_ISL_2268225, EPI_ISL_2268228, EPI_ISL_2268230, EPI_ISL_2268233, EPI_ISL_2268235, EPI_ISL_2268238, EPI_ISL_2268242, EPI_ISL_2268245, EPI_ISL_2268247, EPI_ISL_2268252, EPI_ISL_2268256, EPI_ISL_2268259

| | | | |
|---|---|---|---|
| see above | Aegis Sciences Corporation | Centers for Disease Control and Prevention Division of Viral Diseases, Pathogen Discovery | Dakota Howard, Dhwani Batra, Peter W. Cook, Kara Moser, Adrian Paskey, Jason Caravas, Benjamin Rambo-Martin, Shatavia Morrison, Christopher Gulvick, Scott Sammons, Yvette Unoarumhi, Darlene Wagner, Matthew Schmerer, Cyndi Clark, Patrick Campbell, Rob Case, Vikramsinha Ghorpade, Holly Houdeshell, Ola Kvalvaag, Dillon Nall, Ethan Sanders, Alec Vest, Shaun Westlund, Matthew Hardison, Clinton R. Paden, Duncan MacCannell |

EPI_ISL_2268271, EPI_ISL_2268273, EPI_ISL_2268275, EPI_ISL_2268278, EPI_ISL_2268280, EPI_ISL_2268282, EPI_ISL_2268287, EPI_ISL_2268291, EPI_ISL_2268293, EPI_ISL_2268295, EPI_ISL_2268297, EPI_ISL_2268299, EPI_ISL_2268302, EPI_ISL_2268304, EPI_ISL_2268307, EPI_ISL_2268309, EPI_ISL_2268311, EPI_ISL_2268314, EPI_ISL_2268316, EPI_ISL_2268319, EPI_ISL_2268321, EPI_ISL_2268323, EPI_ISL_2268325, EPI_ISL_2268328, EPI_ISL_2268331, EPI_ISL_2268333, EPI_ISL_2268335, EPI_ISL_2268336

| | | | |
|---|---|---|---|
| see above | Quest Diagnostics Incorporated | Centers for Disease Control and Prevention Division of Viral Diseases, Pathogen Discovery | Dakota Howard, Dhwani Batra, Peter W. Cook, Kara Moser, Adrian Paskey, Jason Caravas, Benjamin Rambo-Martin, Shatavia Morrison, Christopher Gulvick, Scott Sammons, Yvette Unoarumhi, Darlene Wagner, Matthew Schmerer, S. H. Rosenthal, A. Gerasimova, R. M. Kagan, B. Anderson, M. Hua, Y. Liu, L.E. Bernstein, K.E. Livingston, A. Perez, I. A. Shlyakhter, R. V. Rolando, R. Owen, P. Tanpaiboon, F. Lacbawan, Clinton R. Paden, Duncan MacCannell |

EPI_ISL_2268337, EPI_ISL_2268338, EPI_ISL_2268339, EPI_ISL_2268340, EPI_ISL_2268341, EPI_ISL_2268342, EPI_ISL_2268343, EPI_ISL_2268344, EPI_ISL_2268345, EPI_ISL_2268346, EPI_ISL_2268347, EPI_ISL_2268348, EPI_ISL_2268349, EPI_ISL_2268350, EPI_ISL_2268351, EPI_ISL_2268352, EPI_ISL_2268353, EPI_ISL_2268354, EPI_ISL_2268355, EPI_ISL_2268356, EPI_ISL_2268357, EPI_ISL_2268361, EPI_ISL_2268362, EPI_ISL_2268363, EPI_ISL_2268364, EPI_ISL_2268365

| | | | |
|---|---|---|---|
| see above | Helix/Illumina | Centers for Disease Control and Prevention Division of Viral Diseases, Pathogen Discovery | Dakota Howard, Dhwani Batra, Peter W. Cook, Kara Moser, Adrian Paskey, Jason Caravas, Benjamin Rambo-Martin, Shatavia Morrison, Christopher Gulvick, Scott Sammons, Yvette Unoarumhi, Darlene Wagner, Matthew Schmerer, Eileen de Feo, Jan Antico, Christine Tran, Matthew Tolentino, Shannon Wickline, Kim Gietzen, Brad Sickler, Jingtao Liu, Eric Allen, Phil Febbo, Nicole L. Washington, Simon White, Geraint Levan, Kelly Schiabor Barrett, Elizabeth Cirulli, Alexandre Bolze, Ary Ascencio, Charlotte Rivera-Garcia, Ryan Cho, Jason Nguyen, Sherry Wang, Jimmy Ramirez, Tyler Cassens, Efren Sandoval, Magnus Isaksson, William Lee, David Becker, Marc Laurent, James Lu, Clinton R. Paden, Duncan MacCannell |

EPI_ISL_2268366, EPI_ISL_2268367, EPI_ISL_2268368, EPI_ISL_2268369, EPI_ISL_2268370, EPI_ISL_2268371, EPI_ISL_2268372, EPI_ISL_2268373, EPI_ISL_2268374, EPI_ISL_2268375, EPI_ISL_2268376, EPI_ISL_2268377, EPI_ISL_2268378, EPI_ISL_2268379, EPI_ISL_2268380, EPI_ISL_2268381, EPI_ISL_2268382, EPI_ISL_2268383, EPI_ISL_2268384, EPI_ISL_2268385, EPI_ISL_2268386, EPI_ISL_2268387, EPI_ISL_2268388, EPI_ISL_2268389, EPI_ISL_2268390, EPI_ISL_2268391, EPI_ISL_2268392, EPI_ISL_2268393, EPI_ISL_2268394, EPI_ISL_2268395, EPI_ISL_2268396, EPI_ISL_2268397, EPI_ISL_2268398, EPI_ISL_2268399, EPI_ISL_2268400, EPI_ISL_2268401, EPI_ISL_2268402, EPI_ISL_2268403, EPI_ISL_2268404, EPI_ISL_2268405, EPI_ISL_2268406, EPI_ISL_2268407, EPI_ISL_2268408, EPI_ISL_2268409, EPI_ISL_2268410, EPI_ISL_2268411, EPI_ISL_2268412, EPI_ISL_2268413, EPI_ISL_2268414, EPI_ISL_2268415, EPI_ISL_2268416, EPI_ISL_2268417, EPI_ISL_2268418, EPI_ISL_2268419, EPI_ISL_2268420, EPI_ISL_2268421, EPI_ISL_2268422, EPI_ISL_2268423, EPI_ISL_2268424, EPI_ISL_2268425, EPI_ISL_2268426, EPI_ISL_2268427, EPI_ISL_2268428, EPI_ISL_2268429, EPI_ISL_2268430, EPI_ISL_2268431, EPI_ISL_2268432, EPI_ISL_2268433, EPI_ISL_2268434, EPI_ISL_2268435, EPI_ISL_2268436, EPI_ISL_2268437, EPI_ISL_2268438, EPI_ISL_2268439, EPI_ISL_2268440, EPI_ISL_2268441, EPI_ISL_2268442, EPI_ISL_2268443, EPI_ISL_2268444, EPI_ISL_2268445, EPI_ISL_2268446, EPI_ISL_2268447, EPI_ISL_2268448, EPI_ISL_2268449, EPI_ISL_2268450, EPI_ISL_2268451, EPI_ISL_2268452, EPI_ISL_2268453, EPI_ISL_2268454, EPI_ISL_2268455, EPI_ISL_2268456, EPI_ISL_2268457, EPI_ISL_2268458, EPI_ISL_2268459, EPI_ISL_2268460, EPI_ISL_2268461, EPI_ISL_2268462, EPI_ISL_2268463, EPI_ISL_2268464, EPI_ISL_2268465, EPI_ISL_2268466, EPI_ISL_2268467, EPI_ISL_2268468, EPI_ISL_2268469, EPI_ISL_2268470, EPI_ISL_2268471, EPI_ISL_2268472, EPI_ISL_2268473, EPI_ISL_2268474, EPI_ISL_2268475, EPI_ISL_2268476, EPI_ISL_2268477, EPI_ISL_2268478, EPI_ISL_2268479, EPI_ISL_2268480, EPI_ISL_2268481, EPI_ISL_2268482, EPI_ISL_2268483, EPI_ISL_2268484, EPI_ISL_2268485, EPI_ISL_2268486, EPI_ISL_2268487, EPI_ISL_2268488, EPI_ISL_2268489, EPI_ISL_2268490, EPI_ISL_2268491, EPI_ISL_2268492, EPI_ISL_2268493, EPI_ISL_2268494, EPI_ISL_2268495, EPI_ISL_2268496, EPI_ISL_2268497, EPI_ISL_2268498, EPI_ISL_2268499, EPI_ISL_2268500, EPI_ISL_2268501, EPI_ISL_2268502, EPI_ISL_2268503, EPI_ISL_2268504, EPI_ISL_2268505, EPI_ISL_2268506, EPI_ISL_2268507, EPI_ISL_2268508, EPI_ISL_2268509, EPI_ISL_2268510, EPI_ISL_2268511, EPI_ISL_2268512, EPI_ISL_2268513, EPI_ISL_2268514, EPI_ISL_2268515, EPI_ISL_2268516, EPI_ISL_2268517, EPI_ISL_2268518, EPI_ISL_2268519, EPI_ISL_2268520, EPI_ISL_2268521, EPI_ISL_2268522, EPI_ISL_2268523, EPI_ISL_2268524, EPI_ISL_2268525, EPI_ISL_2268526, EPI_ISL_2268527, EPI_ISL_2268528, EPI_ISL_2268529, EPI_ISL_2268530, EPI_ISL_2268531, EPI_ISL_2268532, EPI_ISL_2268533, EPI_ISL_2268534, EPI_ISL_2268535, EPI_ISL_2268536, EPI_ISL_2268537, EPI_ISL_2268538, EPI_ISL_2268539, EPI_ISL_2268540, EPI_ISL_2268541, EPI_ISL_2268542, EPI_ISL_2268543, EPI_ISL_2268544, EPI_ISL_2268545, EPI_ISL_2268546, EPI_ISL_2268547, EPI_ISL_2268548, EPI_ISL_2268549, EPI_ISL_2268550, EPI_ISL_2268551, EPI_ISL_2268552, EPI_ISL_2268553, EPI_ISL_2268554, EPI_ISL_2268555, EPI_ISL_2268556, EPI_ISL_2268557, EPI_ISL_2268558, EPI_ISL_2268559, EPI_ISL_2268560, EPI_ISL_2268561, EPI_ISL_2268562, EPI_ISL_2268563, EPI_ISL_2268564, EPI_ISL_2268565, EPI_ISL_2268566, EPI_ISL_2268567, EPI_ISL_2268568, EPI_ISL_2268569, EPI_ISL_2268570, EPI_ISL_2268571, EPI_ISL_2268572, EPI_ISL_2268573, EPI_ISL_2268574, EPI_ISL_2268575, EPI_ISL_2268576, EPI_ISL_2268577, EPI_ISL_2268578, EPI_ISL_2268579, EPI_ISL_2268580, EPI_ISL_2268581, EPI_ISL_2268582, EPI_ISL_2268583, EPI_ISL_2268584, EPI_ISL_2268585, EPI_ISL_2268586, EPI_ISL_2268587, EPI_ISL_2268588, EPI_ISL_2268589, EPI_ISL_2268590, EPI_ISL_2268591, EPI_ISL_2268592, EPI_ISL_2268593, EPI_ISL_2268594, EPI_ISL_2268595, EPI_ISL_2268596, EPI_ISL_2268597, EPI_ISL_2268598, EPI_ISL_2268599, EPI_ISL_2268600, EPI_ISL_2268601, EPI_ISL_2268602, EPI_ISL_2268603, EPI_ISL_2268604, EPI_ISL_2268605, EPI_ISL_2268606, EPI_ISL_2268607, EPI_ISL_2268608, EPI_ISL_2268609, EPI_ISL_2268610, EPI_ISL_2268611, EPI_ISL_2268612, EPI_ISL_2268613, EPI_ISL_2268614, EPI_ISL_2268615, EPI_ISL_2268616, EPI_ISL_2268617, EPI_ISL_2268618, EPI_ISL_2268619, EPI_ISL_2268620, EPI_ISL_2268621, EPI_ISL_2268622, EPI_ISL_2268623, EPI_ISL_2268624, EPI_ISL_2268625, EPI_ISL_2268626, EPI_ISL_2268627, EPI_ISL_2268628, EPI_ISL_2268629, EPI_ISL_2268630, EPI_ISL_2268631, EPI_ISL_2268632, EPI_ISL_2268633, EPI_ISL_2268634, EPI_ISL_2268635, EPI_ISL_2268636, EPI_ISL_2268637, EPI_ISL_2268638, EPI_ISL_2268639, EPI_ISL_2268640, EPI_ISL_2268641, EPI_ISL_2268642, EPI_ISL_2268643, EPI_ISL_2268644, EPI_ISL_2268645, EPI_ISL_2268646, EPI_ISL_2268647, EPI_ISL_2268648, EPI_ISL_2268649, EPI_ISL_2268650, EPI_ISL_2268651, EPI_ISL_2268652, EPI_ISL_2268653, EPI_ISL_2268654, EPI_ISL_2268655, EPI_ISL_2268656, EPI_ISL_2268657, EPI_ISL_2268658, EPI_ISL_2268659, EPI_ISL_2268660, EPI_ISL_2268661, EPI_ISL_2268662, EPI_ISL_2268663, EPI_ISL_2268664, EPI_ISL_2268665, EPI_ISL_2268666, EPI_ISL_2268667, EPI_ISL_2268668, EPI_ISL_2268669, EPI_ISL_2268670, EPI_ISL_2268671, EPI_ISL_2268672, EPI_ISL_2268673, EPI_ISL_2268674, EPI_ISL_2268675, EPI_ISL_2268676, EPI_ISL_2268677, EPI_ISL_2268678, EPI_ISL_2268679, EPI_ISL_2268680, EPI_ISL_2268681, EPI_ISL_2268682, EPI_ISL_2268683, EPI_ISL_2268684, EPI_ISL_2268685, EPI_ISL_2268686, EPI_ISL_2268687, EPI_ISL_2268688, EPI_ISL_2268689, EPI_ISL_2268690, EPI_ISL_2268691, EPI_ISL_2268692, EPI_ISL_2268693, EPI_ISL_2268694, EPI_ISL_2268695, EPI_ISL_2268696, EPI_ISL_2268697, EPI_ISL_2268698, EPI_ISL_2268699, EPI_ISL_2268700, EPI_ISL_2268701, EPI_ISL_2268702, EPI_ISL_2268703, EPI_ISL_2268704, EPI_ISL_2268705, EPI_ISL_2268706, EPI_ISL_2268707, EPI_ISL_2268708, EPI_ISL_2268709, EPI_ISL_2268710, EPI_ISL_2268711, EPI_ISL_2268712, EPI_ISL_2268713, EPI_ISL_2268714, EPI_ISL_2268715, EPI_ISL_2268716, EPI_ISL_2268717, EPI_ISL_2268718, EPI_ISL_2268719, EPI_ISL_2268720, EPI_ISL_2268721, EPI_ISL_2268722, EPI_ISL_2268723, EPI_ISL_2268724, EPI_ISL_2268725, EPI_ISL_2268726, EPI_ISL_2268727, EPI_ISL_2268728, EPI_ISL_2268729, EPI_ISL_2268730, EPI_ISL_2268731, EPI_ISL_2268732, EPI_ISL_2268733, EPI_ISL_2268734, EPI_ISL_2268735, EPI_ISL_2268736, EPI_ISL_2268737, EPI_ISL_2268738, EPI_ISL_2268739, EPI_ISL_2268740, EPI_ISL_2268741, EPI_ISL_2268742, EPI_ISL_2268743, EPI_ISL_2268744, EPI_ISL_2268745, EPI_ISL_2268746, EPI_ISL_2268747, EPI_ISL_2268748, EPI_ISL_2268749, EPI_ISL_2268750, EPI_ISL_2268751, EPI_ISL_2268752, EPI_ISL_2268753, EPI_ISL_2268754, EPI_ISL_2268755, EPI_ISL_2268756, EPI_ISL_2268757, EPI_ISL_2268758, EPI_ISL_2268759, EPI_ISL_2268760, EPI_ISL_2268761, EPI_ISL_2268762, EPI_ISL_2268763, EPI_ISL_2268764, EPI_ISL_2268765, EPI_ISL_2268766, EPI_ISL_2268767, EPI_ISL_2268768, EPI_ISL_2268769, EPI_ISL_2268770, EPI_ISL_2268771, EPI_ISL_2268772, EPI_ISL_2268773, EPI_ISL_2268774, EPI_ISL_2268775, EPI_ISL_2268776, EPI_ISL_2268777, EPI_ISL_2268778, EPI_ISL_2268779, EPI_ISL_2268780, EPI_ISL_2268781, EPI_ISL_2268782, EPI_ISL_2268783, EPI_ISL_2268784, EPI_ISL_2268785, EPI_ISL_2268786, EPI_ISL_2268787, EPI_ISL_2268788, EPI_ISL_2268789, EPI_ISL_2268790, EPI_ISL_2268791, EPI_ISL_2268792, EPI_ISL_2268793, EPI_ISL_2268794, EPI_ISL_2268795, EPI_ISL_2268796, EPI_ISL_2268797, EPI_ISL_2268798, EPI_ISL_2268799, EPI_ISL_2268800, EPI_ISL_2268801, EPI_ISL_2268802, EPI_ISL_2268803, EPI_ISL_2268804, EPI_ISL_2268805, EPI_ISL_2268806, EPI_ISL_2268807, EPI_ISL_2268808, EPI_ISL_2268809, EPI_ISL_2268810, EPI_ISL_2268811, EPI_ISL_2268812, EPI_ISL_2268813, EPI_ISL_2268814, EPI_ISL_2268815, EPI_ISL_2268816, EPI_ISL_2268817, EPI_ISL_2268818, EPI_ISL_2268819, EPI_ISL_2268820, EPI_ISL_2268821, EPI_ISL_2268822, EPI_ISL_2268823, EPI_ISL_2268824, EPI_ISL_2268825, EPI_ISL_2268826, EPI_ISL_2268827, EPI_ISL_2268828, EPI_ISL_2268829, EPI_ISL_2268830, EPI_ISL_2268831, EPI_ISL_2268832, EPI_ISL_2268833, EPI_ISL_2268834, EPI_ISL_2268835, EPI_ISL_2268836, EPI_ISL_2268837, EPI_ISL_2268838, EPI_ISL_2268839, EPI_ISL_2268840, EPI_ISL_2268841, EPI_ISL_2268842, EPI_ISL_2268843, EPI_ISL_2268844, EPI_ISL_2268845, EPI_ISL_2268846, EPI_ISL_2268847, EPI_ISL_2268848, EPI_ISL_2268849, EPI_ISL_2268850, EPI_ISL_2268851, EPI_ISL_2268852, EPI_ISL_2268853, EPI_ISL_2268854, EPI_ISL_2268855, EPI_ISL_2268856, EPI_ISL_2268857, EPI_ISL_2268858, EPI_ISL_2268859, EPI_ISL_2268860, EPI_ISL_2268861, EPI_ISL_2268862, EPI_ISL_2268863, EPI_ISL_2268864, EPI_ISL_2268865, EPI_ISL_2268866, EPI_ISL_2268867, EPI_ISL_2268868, EPI_ISL_2268869, EPI_ISL_2268870, EPI_ISL_2268871, EPI_ISL_2268872, EPI_ISL_2268873, EPI_ISL_2268874, EPI_ISL_2268875, EPI_ISL_2268876, EPI_ISL_2268877, EPI_ISL_2268878, EPI_ISL_2268879, EPI_ISL_2268880, EPI_ISL_2268881, EPI_ISL_2268882, EPI_ISL_2268883, EPI_ISL_2268884, EPI_ISL_2268885, EPI_ISL_2268886, EPI_ISL_2268887, EPI_ISL_2268888, EPI_ISL_2268889, EPI_ISL_2268890, EPI_ISL_2268891, EPI_ISL_2268892, EPI_ISL_2268893, EPI_ISL_2268894, EPI_ISL_2268895, EPI_ISL_2268896, EPI_ISL_2268897, EPI_ISL_2268898, EPI_ISL_2268899, EPI_ISL_2268900, EPI_ISL_2268901, EPI_ISL_2268902, EPI_ISL_2268903, EPI_ISL_2268904, EPI_ISL_2268905, EPI_ISL_2268906, EPI_ISL_2268907, EPI_ISL_2268908, EPI_ISL_2268909, EPI_ISL_2268910, EPI_ISL_2268911, EPI_ISL_2268912, EPI_ISL_2268913, EPI_ISL_2268914, EPI_ISL_2268915, EPI_ISL_2268916, EPI_ISL_2268917, EPI_ISL_2268918, EPI_ISL_2268919, EPI_ISL_2268920, EPI_ISL_2268921, EPI_ISL_2268922, EPI_ISL_2268923, EPI_ISL_2268924, EPI_ISL_2268925, EPI_ISL_2268926, EPI_ISL_2268927, EPI_ISL_2268928, EPI_ISL_2268929, EPI_ISL_2268930, EPI_ISL_2268931, EPI_ISL_2268932, EPI_ISL_2268933, EPI_ISL_2268934, EPI_ISL_2268935, EPI_ISL_2268936, EPI_ISL_2268937, EPI_ISL_2268938, EPI_ISL_2268939, EPI_ISL_2268940, EPI_ISL_2268941, EPI_ISL_2268942, EPI_ISL_2268943, EPI_ISL_2268944, EPI_ISL_2268945, EPI_ISL_2268946, EPI_ISL_2268947, EPI_ISL_2268948, EPI_ISL_2268949, EPI_ISL_2268950, EPI_ISL_2268951, EPI_ISL_2268952, EPI_ISL_2268953, EPI_ISL_2268954, EPI_ISL_2268955, EPI_ISL_2268956, EPI_ISL_2268957, EPI_ISL_2268958, EPI_ISL_2268959, EPI_ISL_2268960, EPI_ISL_2268961, EPI_ISL_2268962, EPI_ISL_2268963, EPI_ISL_2268964, EPI_ISL_2268965, EPI_ISL_2268966, EPI_ISL_2268967, EPI_ISL_2268968, EPI_ISL_2268969, EPI_ISL_2268970, EPI_ISL_2268971, EPI_ISL_2268972, EPI_ISL_2268973, EPI_ISL_2268974, EPI_ISL_2268975, EPI_ISL_2268976, EPI_ISL_2268977, EPI_ISL_2268978, EPI_ISL_2268979, EPI_ISL_2268980, EPI_ISL_2268981, EPI_ISL_2268982, EPI_ISL_2268983, EPI_ISL_2268984, EPI_ISL_2268985, EPI_ISL_2268986, EPI_ISL_2268987, EPI_ISL_2268988, EPI_ISL_2268989, EPI_ISL_2268990, EPI_ISL_2268991, EPI_ISL_2268992, EPI_ISL_2268993, EPI_ISL_2268994, EPI_ISL_2268995, EPI_ISL_2268996, EPI_ISL_2268997, EPI_ISL_2268998, EPI_ISL_2268999, EPI_ISL_2269000, EPI_ISL_2269001, EPI_ISL_2269002, EPI_ISL_2269003, EPI_ISL_2269004, EPI_ISL_2269005, EPI_ISL_2269006, EPI_ISL_2269007, EPI_ISL_2269008, EPI_ISL_2269009, EPI_ISL_2269010, EPI_ISL_2269011, EPI_ISL_2269012, EPI_ISL_2269013, EPI_ISL_2269014, EPI_ISL_2269015, EPI_ISL_2269016, EPI_ISL_2269017, EPI_ISL_2269018, EPI_ISL_2269019, EPI_ISL_2269020, EPI_ISL_2269021, EPI_ISL_2269022, EPI_ISL_2269023, EPI_ISL_2269024, EPI_ISL_2269025, EPI_ISL_2269026, EPI_ISL_2269027, EPI_ISL_2269028, EPI_ISL_2269029, EPI_ISL_2269030, EPI_ISL_2269031, EPI_ISL_2269032, EPI_ISL_2269033, EPI_ISL_2269034, EPI_ISL_2269035, EPI_ISL_2269036, EPI_ISL_2269037, EPI_ISL_2269038, EPI_ISL_2269039, EPI_ISL_2269040, EPI_ISL_2269041, EPI_ISL_2269042, EPI_ISL_2269043, EPI_ISL_2269044, EPI_ISL_2269045, EPI_ISL_2269046, EPI_ISL_2269047, EPI_ISL_2269048, EPI_ISL_2269049, EPI_ISL_2269050, EPI_ISL_2269051, EPI_ISL_2269052, EPI_ISL_2269053, EPI_ISL_2269054, EPI_ISL_2269055, EPI_ISL_2269056, EPI_ISL_2269057, EPI_ISL_2269058, EPI_ISL_2269059, EPI_ISL_2269060, EPI_ISL_2269061, EPI_ISL_2269062, EPI_ISL_2269063, EPI_ISL_2269064, EPI_ISL_2269065, EPI_ISL_2269066, EPI_ISL_2269067, EPI_ISL_2269068, EPI_ISL_2269069, EPI_ISL_2269070, EPI_ISL_2269071, EPI_ISL_2269072, EPI_ISL_2269073, EPI_ISL_2269074, EPI_ISL_2269075, EPI_ISL_2269076, EPI_ISL_2269077, EPI_ISL_2269078, EPI_ISL_2269079, EPI_ISL_2269080, EPI_ISL_2269081, EPI_ISL_2269082, EPI_ISL_2269083, EPI_ISL_2269084, EPI_ISL_2269085, EPI_ISL_2269086, EPI_ISL_2269087, EPI_ISL_2269088, EPI_ISL_2269089, EPI_ISL_2269090, EPI_ISL_2269091, EPI_ISL_2269092, EPI_ISL_2269093, EPI_ISL_2269094, EPI_ISL_2269095, EPI_ISL_2269096, EPI_ISL_2269097, EPI_ISL_2269098, EPI_ISL_2269099, EPI_ISL_2269100, EPI_ISL_2269101, EPI_ISL_2269102, EPI_ISL_2269103, EPI_ISL_2269104, EPI_ISL_2269105, EPI_ISL_2269106, EPI_ISL_2269107, EPI_ISL_2269108, EPI_ISL_2269109, EPI_ISL_2269110, EPI_ISL_2269111, EPI_ISL_2269112, EPI_ISL_2269113, EPI_ISL_2269114, EPI_ISL_2269115, EPI_ISL_2269116, EPI_ISL_2269117, EPI_ISL_2269118, EPI_ISL_2269119, EPI_ISL_2269120, EPI_ISL_2269121, EPI_ISL_2269122, EPI_ISL_2269123, EPI_ISL_2269124, EPI_ISL_2269125, EPI_ISL_2269126, EPI_ISL_2269127, EPI_ISL_2269128, EPI_ISL_2269129, EPI_ISL_2269130, EPI_ISL_2269131, EPI_ISL_2269132, EPI_ISL_2269133, EPI_ISL_2269134, EPI_ISL_2269135, EPI_ISL_2269136, EPI_ISL_2269137, EPI_ISL_2269138, EPI_ISL_2269139, EPI_ISL_2269140, EPI_ISL_2269141, EPI_ISL_2269142, EPI_ISL_2269143, EPI_ISL_2269144, EPI_ISL_2269145, EPI_ISL_2269146, EPI_ISL_2269147, EPI_ISL_2269148, EPI_ISL_2269149, EPI_ISL_2269150, EPI_ISL_2269151, EPI_ISL_2269152, EPI_ISL_2269153, EPI_ISL_2269154, EPI_ISL_2269155, EPI_ISL_2269156, EPI_ISL_2269157, EPI_ISL_2269158, EPI_ISL_2269159, EPI_ISL_2269160, EPI_ISL_2269161, EPI_ISL_2269162, EPI_ISL_2269163, EPI_ISL_2269164, EPI_ISL_2269165, EPI_ISL_2269166, EPI_ISL_2269167, EPI_ISL_2269168, EPI_ISL_2269169, EPI_ISL_2269170, EPI_ISL_2269171, EPI_ISL_2269172, EPI_ISL_2269173, EPI_ISL_2269174, EPI_ISL_2269175, EPI_ISL_2269176, EPI_ISL_2269177, EPI_ISL_2269178, EPI_ISL_2269179, EPI_ISL_2269180, EPI_ISL_2269181, EPI_ISL_2269182, EPI_ISL_2269183, EPI_ISL_2269184, EPI_ISL_2269185, EPI_ISL_2269186, EPI_ISL_2269187, EPI_ISL_2269188, EPI_ISL_2269189, EPI_ISL_2269190, EPI_ISL_2269191, EPI_ISL_2269192, EPI_ISL_2269193, EPI_ISL_2269194, EPI_ISL_2269195, EPI_ISL_2269196, EPI_ISL_2269197, EPI_ISL_2269198, EPI_ISL_2269199, EPI_ISL_2269200, EPI_ISL_2269201, EPI_ISL_2269202, EPI_ISL_2269203, EPI_ISL_2269204, EPI_ISL_2269205, EPI_ISL_2269206, EPI_ISL_2269207, EPI_ISL_2269208, EPI_ISL_2269209, EPI_ISL_2269210, EPI_ISL_2269211, EPI_ISL_2269212, EPI_ISL_2269213, EPI_ISL_2269214, EPI_ISL_2269215

| | | | |
|---|---|---|---|
| see above | Quest Diagnostics Incorporated | Centers for Disease Control and Prevention Division of Viral Diseases, Pathogen Discovery | Dakota Howard, Dhwani Batra, Peter W. Cook, Kara Moser, Adrian Paskey, Jason Caravas, Benjamin Rambo-Martin, Shatavia Morrison, Christopher Gulvick, Scott Sammons, Yvette Unoarumhi, Darlene Wagner, Matthew Schmerer, S. H. Rosenthal, A. Gerasimova, R. M. Kagan, B. Anderson, M. Hua, Y. Liu, L.E. Bernstein, K.E. Livingston, A. Perez, I. A. Shlyakhter, R. V. Rolando, R. Owen, P. Tanpaiboon, F. Lacbawan, Clinton R. Paden, Duncan MacCannell |

EPI_ISL_2269216, EPI_ISL_2269217, EPI_ISL_2269218, EPI_ISL_2269220, EPI_ISL_2269221, EPI_ISL_2269222, EPI_ISL_2269223, EPI_ISL_2269224, EPI_ISL_2269225, EPI_ISL_2269226, EPI_ISL_2269227, EPI_ISL_2269228, EPI_ISL_2269229, EPI_ISL_2269230, EPI_ISL_2269231, EPI_ISL_2269232, EPI_ISL_2269233, EPI_ISL_2269234, EPI_ISL_2269235, EPI_ISL_2269236, EPI_ISL_2269237, EPI_ISL_2269239, EPI_ISL_2269240, EPI_ISL_2269241, EPI_ISL_2269242, EPI_ISL_2269243, EPI_ISL_2269244, EPI_ISL_2269245, EPI_ISL_2269246, EPI_ISL_2269247, EPI_ISL_2269248, EPI_ISL_2269249, EPI_ISL_2269250, EPI_ISL_2269251, EPI_ISL_2269252, EPI_ISL_2269253, EPI_ISL_2269254, EPI_ISL_2269255, EPI_ISL_2269256, EPI_ISL_2269257, EPI_ISL_2269258, EPI_ISL_2269259, EPI_ISL_2269260, EPI_ISL_2269261, EPI_ISL_2269262, EPI_ISL_2269264, EPI_ISL_2269265, EPI_ISL_2269266, EPI_ISL_2269267, EPI_ISL_2269268, EPI_ISL_2269269, EPI_ISL_2269270, EPI_ISL_2269271, EPI_ISL_2269272, EPI_ISL_2269273, EPI_ISL_2269274, EPI_ISL_2269275, EPI_ISL_2269276, EPI_ISL_2269277, EPI_ISL_2269278, EPI_ISL_2269279, EPI_ISL_2269280, EPI_ISL_2269281, EPI_ISL_2269282, EPI_ISL_2269283, EPI_ISL_2269284, EPI_ISL_2269285, EPI_ISL_2269286, EPI_ISL_2269287, EPI_ISL_2269288, EPI_ISL_2269289, EPI_ISL_2269290, EPI_ISL_2269291, EPI_ISL_2269292, EPI_ISL_2269293, EPI_ISL_2269294, EPI_ISL_2269295, EPI_ISL_2269296, EPI_ISL_2269297, EPI_ISL_2269298, EPI_ISL_2269299, EPI_ISL_2269300, EPI_ISL_2269301, EPI_ISL_2269302, EPI_ISL_2269303, EPI_ISL_2269304, EPI_ISL_2269305, EPI_ISL_2269307, EPI_ISL_2269308, EPI_ISL_2269309, EPI_ISL_2269310, EPI_ISL_2269311, EPI_ISL_2269312, EPI_ISL_2269313, EPI_ISL_2269314, EPI_ISL_2269315, EPI_ISL_2269316, EPI_ISL_2269317, EPI_ISL_2269318, EPI_ISL_2269319, EPI_ISL_2269320, EPI_ISL_2269321, EPI_ISL_2269322, EPI_ISL_2269323, EPI_ISL_2269324, EPI_ISL_2269325, EPI_ISL_2269326, EPI_ISL_2269327, EPI_ISL_2269328, EPI_ISL_2269329, EPI_ISL_2269330, EPI_ISL_2269331, EPI_ISL_2269332, EPI_ISL_2269333, EPI_ISL_2269334, EPI_ISL_2269335, EPI_ISL_2269336, EPI_ISL_2269337, EPI_ISL_2269338, EPI_ISL_2269339, EPI_ISL_2269340, EPI_ISL_2269341, EPI_ISL_2269342, EPI_ISL_2269343, EPI_ISL_2269344, EPI_ISL_2269346, EPI_ISL_2269347, EPI_ISL_2269348, EPI_ISL_2269349, EPI_ISL_2269350, EPI_ISL_2269351, EPI_ISL_2269352, EPI_ISL_2269353, EPI_ISL_2269354, EPI_ISL_2269356, EPI_ISL_2269357, EPI_ISL_2269358, EPI_ISL_2269359, EPI_ISL_2269360, EPI_ISL_2269361, EPI_ISL_2269362, EPI_ISL_2269363, EPI_ISL_2269364, EPI_ISL_2269366, EPI_ISL_2269368, EPI_ISL_2269370, EPI_ISL_2269371, EPI_ISL_2269372, EPI_ISL_2269373, EPI_ISL_2269374, EPI_ISL_2269376, EPI_ISL_2269377, EPI_ISL_2269378, EPI_ISL_2269379, EPI_ISL_2269380, EPI_ISL_2269381, EPI_ISL_2269382, EPI_ISL_2269383, EPI_ISL_2269384, EPI_ISL_2269385, EPI_ISL_2269386, EPI_ISL_2269387, EPI_ISL_2269388, EPI_ISL_2269389, EPI_ISL_2269390, EPI_ISL_2269391, EPI_ISL_2269392, EPI_ISL_2269393, EPI_ISL_2269394, EPI_ISL_2269395, EPI_ISL_2269396, EPI_ISL_2269399, EPI_ISL_2269400, EPI_ISL_2269401, EPI_ISL_2269402, EPI_ISL_2269403, EPI_ISL_2269404, EPI_ISL_2269405, EPI_ISL_2269406, EPI_ISL_2269407, EPI_ISL_2269408, EPI_ISL_2269409, EPI_ISL_2269410, EPI_ISL_2269411, EPI_ISL_2269412, EPI_ISL_2269413, EPI_ISL_2269414, EPI_ISL_2269415, EPI_ISL_2269416, EPI_ISL_2269417, EPI_ISL_2269418, EPI_ISL_2269419, EPI_ISL_2269420, EPI_ISL_2269421, EPI_ISL_2269422, EPI_ISL_2269423, EPI_ISL_2269424, EPI_ISL_2269426, EPI_ISL_2269427, EPI_ISL_2269429, EPI_ISL_2269430, EPI_ISL_2269431, EPI_ISL_2269433, EPI_ISL_2269434, EPI_ISL_2269435, EPI_ISL_2269436, EPI_ISL_2269437, EPI_ISL_2269438, EPI_ISL_2269439, EPI_ISL_2269440, EPI_ISL_2269441, EPI_ISL_2269442, EPI_ISL_2269443, EPI_ISL_2269445, EPI_ISL_2269446, EPI_ISL_2269447, EPI_ISL_2269448, EPI_ISL_2269449, EPI_ISL_2269450, EPI_ISL_2269451, EPI_ISL_2269452, EPI_ISL_2269453, EPI_ISL_2269454, EPI_ISL_2269455, EPI_ISL_2269456, EPI_ISL_2269457, EPI_ISL_2269458, EPI_ISL_2269459, EPI_ISL_2269460, EPI_ISL_2269461, EPI_ISL_2269462, EPI_ISL_2269463, EPI_ISL_2269464, EPI_ISL_2269465, EPI_ISL_2269466, EPI_ISL_2269467, EPI_ISL_2269468, EPI_ISL_2269469, EPI_ISL_2269470, EPI_ISL_2269472, EPI_ISL_2269473, EPI_ISL_2269474, EPI_ISL_2269475, EPI_ISL_2269476, EPI_ISL_2269477, EPI_ISL_2269480, EPI_ISL_2269481, EPI_ISL_2269482, EPI_ISL_2269483, EPI_ISL_2269484, EPI_ISL_2269485, EPI_ISL_2269486, EPI_ISL_2269487, EPI_ISL_2269488, EPI_ISL_2269489, EPI_ISL_2269490, EPI_ISL_2269492, EPI_ISL_2269493, EPI_ISL_2269494, EPI_ISL_2269495, EPI_ISL_2269496, EPI_ISL_2269497, EPI_ISL_2269498, EPI_ISL_2269499, EPI_ISL_2269500, EPI_ISL_2269501, EPI_ISL_2269502, EPI_ISL_2269503, EPI_ISL_2269504, EPI_ISL_2269505, EPI_ISL_2269506, EPI_ISL_2269507, EPI_ISL_2269508, EPI_ISL_2269509, EPI_ISL_2269510, EPI_ISL_2269511, EPI_ISL_2269513, EPI_ISL_2269515, EPI_ISL_2269516, EPI_ISL_2269517, EPI_ISL_2269518, EPI_ISL_2269519, EPI_ISL_2269520, EPI_ISL_2269521, EPI_ISL_2269523, EPI_ISL_2269524, EPI_ISL_2269525, EPI_ISL_2269526, EPI_ISL_2269527, EPI_ISL_2269528, EPI_ISL_2269529, EPI_ISL_2269530, EPI_ISL_2269531, EPI_ISL_2269532, EPI_ISL_2269533, EPI_ISL_2269534, EPI_ISL_2269535, EPI_ISL_2269536, EPI_ISL_2269537, EPI_ISL_2269538, EPI_ISL_2269539, EPI_ISL_2269540, EPI_ISL_2269541, EPI_ISL_2269542, EPI_ISL_2269544, EPI_ISL_2269545, EPI_ISL_2269546, EPI_ISL_2269547, EPI_ISL_2269548, EPI_ISL_2269549, EPI_ISL_2269550, EPI_ISL_2269551, EPI_ISL_2269552, EPI_ISL_2269553, EPI_ISL_2269554, EPI_ISL_2269555, EPI_ISL_2269556, EPI_ISL_2269557, EPI_ISL_2269558, EPI_ISL_2269559, EPI_ISL_2269560, EPI_ISL_2269561, EPI_ISL_2269562, EPI_ISL_2269563, EPI_ISL_2269564, EPI_ISL_2269565, EPI_ISL_2269566, EPI_ISL_2269567, EPI_ISL_2269568, EPI_ISL_2269569, EPI_ISL_2269571, EPI_ISL_2269572, EPI_ISL_2269573, EPI_ISL_2269574, EPI_ISL_2269575, EPI_ISL_2269576, EPI_ISL_2269577, EPI_ISL_2269578, EPI_ISL_2269579, EPI_ISL_2269580, EPI_ISL_2269581, EPI_ISL_2269582, EPI_ISL_2269583, EPI_ISL_2269584, EPI_ISL_2269586, EPI_ISL_2269587, EPI_ISL_2269588, EPI_ISL_2269589, EPI_ISL_2269590, EPI_ISL_2269591, EPI_ISL_2269592, EPI_ISL_2269593, EPI_ISL_2269594, EPI_ISL_2269595, EPI_ISL_2269596, EPI_ISL_2269597, EPI_ISL_2269598, EPI_ISL_2269599, EPI_ISL_2269600, EPI_ISL_2269601, EPI_ISL_2269602, EPI_ISL_2269603, EPI_ISL_2269605, EPI_ISL_2269607, EPI_ISL_2269608, EPI_ISL_2269609, EPI_ISL_2269610, EPI_ISL_2269611

| | | | |
|---|---|---|---|
| see above | Fulgent Genetics | Centers for Disease Control and Prevention Division of Viral Diseases, Pathogen Discovery | Dakota Howard, Dhwani Batra, Peter W. Cook, Kara Moser, Adrian Paskey, Jason Caravas, Benjamin Rambo-Martin, Shatavia Morrison, Christopher Gulvick, Scott Sammons, Yvette Unoarumhi, Darlene Wagner, Matthew Schmerer, Harry Gao, Mickey Li, John Gao, Joseph Fierro, Benafsh Sapra, Becky Tsai, Yan Meng, Doreen Ng, James Xie, Clinton R. Paden, Duncan MacCannell |

EPI_ISL_2269612, EPI_ISL_2269613, EPI_ISL_2269614, EPI_ISL_2269615, EPI_ISL_2269616, EPI_ISL_2269617, EPI_ISL_2269618, EPI_ISL_2269619, EPI_ISL_2269620, EPI_ISL_2269621, EPI_ISL_2269622, EPI_ISL_2269623, EPI_ISL_2269624, EPI_ISL_2269625, EPI_ISL_2269626, EPI_ISL_2269627, EPI_ISL_2269628, EPI_ISL_2269629, EPI_ISL_2269630, EPI_ISL_2269631, EPI_ISL_2269632, EPI_ISL_2269633, EPI_ISL_2269634, EPI_ISL_2269635, EPI_ISL_2269636, EPI_ISL_2269637, EPI_ISL_2269638, EPI_ISL_2269640, EPI_ISL_2269641, EPI_ISL_2269643, EPI_ISL_2269644, EPI_ISL_2269645, EPI_ISL_2269646, EPI_ISL_2269647, EPI_ISL_2269648, EPI_ISL_2269649, EPI_ISL_2269650, EPI_ISL_2269651, EPI_ISL_2269652, EPI_ISL_2269653, EPI_ISL_2269654, EPI_ISL_2269655, EPI_ISL_2269656, EPI_ISL_2269657, EPI_ISL_2269658, EPI_ISL_2269659, EPI_ISL_2269660, EPI_ISL_2269661, EPI_ISL_2269662, EPI_ISL_2269663, EPI_ISL_2269664, EPI_ISL_2269665, EPI_ISL_2269667, EPI_ISL_2269668, EPI_ISL_2269669, EPI_ISL_2269670, EPI_ISL_2269671, EPI_ISL_2269672, EPI_ISL_2269673, EPI_ISL_2269674, EPI_ISL_2269675, EPI_ISL_2269676, EPI_ISL_2269677, EPI_ISL_2269678, EPI_ISL_2269679, EPI_ISL_2269680, EPI_ISL_2269681, EPI_ISL_2269682, EPI_ISL_2269683, EPI_ISL_2269684, EPI_ISL_2269685, EPI_ISL_2269686, EPI_ISL_2269687, EPI_ISL_2269688, EPI_ISL_2269689, EPI_ISL_2269690, EPI_ISL_2269691, EPI_ISL_2269692, EPI_ISL_2269693, EPI_ISL_2269694, EPI_ISL_2269695, EPI_ISL_2269696, EPI_ISL_2269697, EPI_ISL_2269698, EPI_ISL_2269699, EPI_ISL_2269700, EPI_ISL_2269701, EPI_ISL_2269702, EPI_ISL_2269703, EPI_ISL_2269704, EPI_ISL_2269705, EPI_ISL_2269706, EPI_ISL_2269707, EPI_ISL_2269708, EPI_ISL_2269709, EPI_ISL_2269710, EPI_ISL_2269711, EPI_ISL_2269712, EPI_ISL_2269713, EPI_ISL_2269714, EPI_ISL_2269715, EPI_ISL_2269716, EPI_ISL_2269717, EPI_ISL_2269718, EPI_ISL_2269719, EPI_ISL_2269720, EPI_ISL_2269721, EPI_ISL_2269722, EPI_ISL_2269723, EPI_ISL_2269724, EPI_ISL_2269725, EPI_ISL_2269726, EPI_ISL_2269727, EPI_ISL_2269728, EPI_ISL_2269729, EPI_ISL_2269730, EPI_ISL_2269731, EPI_ISL_2269732, EPI_ISL_2269733, EPI_ISL_2269734, EPI_ISL_2269735, EPI_ISL_2269736, EPI_ISL_2269737, EPI_ISL_2269738, EPI_ISL_2269739, EPI_ISL_2269740, EPI_ISL_2269741, EPI_ISL_2269742, EPI_ISL_2269743, EPI_ISL_2269744, EPI_ISL_2269745, EPI_ISL_2269746, EPI_ISL_2269747, EPI_ISL_2269748, EPI_ISL_2269749, EPI_ISL_2269750, EPI_ISL_2269751, EPI_ISL_2269752, EPI_ISL_2269753, EPI_ISL_2269754, EPI_ISL_2269755, EPI_ISL_2269756, EPI_ISL_2269757, EPI_ISL_2269758, EPI_ISL_2269759, EPI_ISL_2269760, EPI_ISL_2269761, EPI_ISL_2269762, EPI_ISL_2269763, EPI_ISL_2269764, EPI_ISL_2269765, EPI_ISL_2269766, EPI_ISL_2269767, EPI_ISL_2269768, EPI_ISL_2269769, EPI_ISL_2269770, EPI_ISL_2269771, EPI_ISL_2269772, EPI_ISL_2269773, EPI_ISL_2269774, EPI_ISL_2269775, EPI_ISL_2269776, EPI_ISL_2269777, EPI_ISL_2269778, EPI_ISL_2269779, EPI_ISL_2269780, EPI_ISL_2269781, EPI_ISL_2269782, EPI_ISL_2269783, EPI_ISL_2269784, EPI_ISL_2269785, EPI_ISL_2269786, EPI_ISL_2269787, EPI_ISL_2269788, EPI_ISL_2269789, EPI_ISL_2269790, EPI_ISL_2269791, EPI_ISL_2269792, EPI_ISL_2269793, EPI_ISL_2269794, EPI_ISL_2269795, EPI_ISL_2269796, EPI_ISL_2269797, EPI_ISL_2269798, EPI_ISL_2269799, EPI_ISL_2269800, EPI_ISL_2269801, EPI_ISL_2269802, EPI_ISL_2269803, EPI_ISL_2269804, EPI_ISL_2269805, EPI_ISL_2269806, EPI_ISL_2269807, EPI_ISL_2269808, EPI_ISL_2269809, EPI_ISL_2269810, EPI_ISL_2269811, EPI_ISL_2269812,

| | | | |
|---|---|---|---|
| EPI_ISL_2269813, EPI_ISL_2269814, EPI_ISL_2269815, EPI_ISL_2269816, EPI_ISL_2269817, EPI_ISL_2269818, EPI_ISL_2269819, EPI_ISL_2269820, EPI_ISL_2269821, EPI_ISL_2269822, EPI_ISL_2269823, EPI_ISL_2269824, EPI_ISL_2269825, EPI_ISL_2269826, EPI_ISL_2269827, EPI_ISL_2269828, EPI_ISL_2269829, EPI_ISL_2269830, EPI_ISL_2269831, EPI_ISL_2269832, EPI_ISL_2269833, EPI_ISL_2269834, EPI_ISL_2269835, EPI_ISL_2269836, EPI_ISL_2269837, EPI_ISL_2269838, EPI_ISL_2269839, EPI_ISL_2269840, EPI_ISL_2269841, EPI_ISL_2269842, EPI_ISL_2269843, EPI_ISL_2269844, EPI_ISL_2269845, EPI_ISL_2269846, EPI_ISL_2269847, EPI_ISL_2269848, EPI_ISL_2269849, EPI_ISL_2269850, EPI_ISL_2269851, EPI_ISL_2269852, EPI_ISL_2269853, EPI_ISL_2269854, EPI_ISL_2269855, EPI_ISL_2269856, EPI_ISL_2269857, EPI_ISL_2269858, EPI_ISL_2269859, EPI_ISL_2269860, EPI_ISL_2269861, EPI_ISL_2269862, EPI_ISL_2269863, EPI_ISL_2269864, EPI_ISL_2269865, EPI_ISL_2269866, EPI_ISL_2269867, EPI_ISL_2269868, EPI_ISL_2269869, EPI_ISL_2269870, EPI_ISL_2269871, EPI_ISL_2269872, EPI_ISL_2269873, EPI_ISL_2269874, EPI_ISL_2269875, EPI_ISL_2269876, EPI_ISL_2269877, EPI_ISL_2269878, EPI_ISL_2269879, EPI_ISL_2269880, EPI_ISL_2269881, EPI_ISL_2269882, EPI_ISL_2269883, EPI_ISL_2269884, EPI_ISL_2269885, EPI_ISL_2269886, EPI_ISL_2269887, EPI_ISL_2269888, EPI_ISL_2269889, EPI_ISL_2269890, EPI_ISL_2269891, EPI_ISL_2269892, EPI_ISL_2269893, EPI_ISL_2269894, EPI_ISL_2269895, EPI_ISL_2269896, EPI_ISL_2269897, EPI_ISL_2269898, EPI_ISL_2269899, EPI_ISL_2269900, EPI_ISL_2269901, EPI_ISL_2269902, EPI_ISL_2269903, EPI_ISL_2269904, EPI_ISL_2269905, EPI_ISL_2269906, EPI_ISL_2269907, EPI_ISL_2269908, EPI_ISL_2269909, EPI_ISL_2269910, EPI_ISL_2269911, EPI_ISL_2269912, EPI_ISL_2269913, EPI_ISL_2269914, EPI_ISL_2269915, EPI_ISL_2269916, EPI_ISL_2269917, EPI_ISL_2269918, EPI_ISL_2269919, EPI_ISL_2269920, EPI_ISL_2269921, EPI_ISL_2269922, EPI_ISL_2269923, EPI_ISL_2269924, EPI_ISL_2269925, EPI_ISL_2269926, EPI_ISL_2269927, EPI_ISL_2269928, EPI_ISL_2269929, EPI_ISL_2269930, EPI_ISL_2269931, EPI_ISL_2269932, EPI_ISL_2269933, EPI_ISL_2269934, EPI_ISL_2269935, EPI_ISL_2269936, EPI_ISL_2269937, EPI_ISL_2269938, EPI_ISL_2269939, EPI_ISL_2269940, EPI_ISL_2269941, EPI_ISL_2269942, EPI_ISL_2269943, EPI_ISL_2269944, EPI_ISL_2269945, EPI_ISL_2269946, EPI_ISL_2269947, EPI_ISL_2269948, EPI_ISL_2269949, EPI_ISL_2269950, EPI_ISL_2269951, EPI_ISL_2269952, EPI_ISL_2269953, EPI_ISL_2269954, EPI_ISL_2269955, EPI_ISL_2269956, EPI_ISL_2269957, EPI_ISL_2269958, EPI_ISL_2269959, EPI_ISL_2269960, EPI_ISL_2269961, EPI_ISL_2269962, EPI_ISL_2269963, EPI_ISL_2269964, EPI_ISL_2269965, EPI_ISL_2269966, EPI_ISL_2269967, EPI_ISL_2269968, EPI_ISL_2269969, EPI_ISL_2269970, EPI_ISL_2269971, EPI_ISL_2269972, EPI_ISL_2269973, EPI_ISL_2269974, EPI_ISL_2269975, EPI_ISL_2269976, EPI_ISL_2269977, EPI_ISL_2269978, EPI_ISL_2269979, EPI_ISL_2269980, EPI_ISL_2269981, EPI_ISL_2269982, EPI_ISL_2269983, EPI_ISL_2269984, EPI_ISL_2269985, EPI_ISL_2269986, EPI_ISL_2269987, EPI_ISL_2269988, EPI_ISL_2269989, EPI_ISL_2269990, EPI_ISL_2269991, EPI_ISL_2269992, EPI_ISL_2269993, EPI_ISL_2269994, EPI_ISL_2269995, EPI_ISL_2269996, EPI_ISL_2269997, EPI_ISL_2269998, EPI_ISL_2269999, EPI_ISL_2270000, EPI_ISL_2270001, EPI_ISL_2270002, EPI_ISL_2270003, EPI_ISL_2270004, EPI_ISL_2270005, EPI_ISL_2270006, EPI_ISL_2270007, EPI_ISL_2270008, EPI_ISL_2270009, EPI_ISL_2270010, EPI_ISL_2270011, EPI_ISL_2270012, EPI_ISL_2270013, EPI_ISL_2270014, EPI_ISL_2270015, EPI_ISL_2270016, EPI_ISL_2270017, EPI_ISL_2270018, EPI_ISL_2270019, EPI_ISL_2270020, EPI_ISL_2270021, EPI_ISL_2270022, EPI_ISL_2270023, EPI_ISL_2270024, EPI_ISL_2270025, EPI_ISL_2270026, EPI_ISL_2270027, EPI_ISL_2270028, EPI_ISL_2270029, EPI_ISL_2270030, EPI_ISL_2270031, EPI_ISL_2270032, EPI_ISL_2270033, EPI_ISL_2270034, EPI_ISL_2270035, EPI_ISL_2270036, EPI_ISL_2270037, EPI_ISL_2270039, EPI_ISL_2270042, EPI_ISL_2270044, EPI_ISL_2270047, EPI_ISL_2270050, EPI_ISL_2270055, EPI_ISL_2270058, EPI_ISL_2270061, EPI_ISL_2270063, EPI_ISL_2270066, EPI_ISL_2270069, EPI_ISL_2270071, EPI_ISL_2270073, EPI_ISL_2270075, EPI_ISL_2270078, EPI_ISL_2270080, EPI_ISL_2270083, EPI_ISL_2270087, EPI_ISL_2270089, EPI_ISL_2270092, EPI_ISL_2270094, EPI_ISL_2270097, EPI_ISL_2270100, EPI_ISL_2270103, EPI_ISL_2270105, EPI_ISL_2270109, EPI_ISL_2270111, EPI_ISL_2270114, EPI_ISL_2270116, EPI_ISL_2270119, EPI_ISL_2270122, EPI_ISL_2270124, EPI_ISL_2270127, EPI_ISL_2270130, EPI_ISL_2270132, EPI_ISL_2270135, EPI_ISL_2270138, EPI_ISL_2270140, EPI_ISL_2270143, EPI_ISL_2270146, EPI_ISL_2270148, EPI_ISL_2270151, EPI_ISL_2270154, EPI_ISL_2270156, EPI_ISL_2270159, EPI_ISL_2270162, EPI_ISL_2270164, EPI_ISL_2270166, EPI_ISL_2270167, EPI_ISL_2270168, EPI_ISL_2270169, EPI_ISL_2270170, EPI_ISL_2270171, EPI_ISL_2270172, EPI_ISL_2270173, EPI_ISL_2270174, EPI_ISL_2270175, EPI_ISL_2270176, EPI_ISL_2270177, EPI_ISL_2270178, EPI_ISL_2270179, EPI_ISL_2270180, EPI_ISL_2270181, EPI_ISL_2270182, EPI_ISL_2270183, EPI_ISL_2270184, EPI_ISL_2270185, EPI_ISL_2270186, EPI_ISL_2270187, EPI_ISL_2270188, EPI_ISL_2270189, EPI_ISL_2270190, EPI_ISL_2270191, EPI_ISL_2270192, EPI_ISL_2270193, EPI_ISL_2270194, EPI_ISL_2270195, EPI_ISL_2270196, EPI_ISL_2270197, EPI_ISL_2270198, EPI_ISL_2270199, EPI_ISL_2270200, EPI_ISL_2270201, EPI_ISL_2270202, EPI_ISL_2270203, EPI_ISL_2270204, EPI_ISL_2270205, EPI_ISL_2270206, EPI_ISL_2270207, EPI_ISL_2270208, EPI_ISL_2270209, EPI_ISL_2270210, EPI_ISL_2270211, EPI_ISL_2270212, EPI_ISL_2270213, EPI_ISL_2270214, EPI_ISL_2270215, EPI_ISL_2270216, EPI_ISL_2270217, EPI_ISL_2270218, EPI_ISL_2270219, EPI_ISL_2270220, EPI_ISL_2270221, EPI_ISL_2270222, EPI_ISL_2270223, EPI_ISL_2270224, EPI_ISL_2270225, EPI_ISL_2270226, EPI_ISL_2270227, EPI_ISL_2270228, EPI_ISL_2270229, EPI_ISL_2270230, EPI_ISL_2270231, EPI_ISL_2270232, EPI_ISL_2270233, EPI_ISL_2270234, EPI_ISL_2270235, EPI_ISL_2270236, EPI_ISL_2270237, EPI_ISL_2270238, EPI_ISL_2270239, EPI_ISL_2270241, EPI_ISL_2270242, EPI_ISL_2270243, EPI_ISL_2270244, EPI_ISL_2270245, EPI_ISL_2270246, EPI_ISL_2270247, EPI_ISL_2270248, EPI_ISL_2270249, EPI_ISL_2270250, EPI_ISL_2270251, EPI_ISL_2270252, EPI_ISL_2270253, EPI_ISL_2270254, EPI_ISL_2270255, EPI_ISL_2270256, EPI_ISL_2270257, EPI_ISL_2270258, EPI_ISL_2270259, EPI_ISL_2270260, EPI_ISL_2270261, EPI_ISL_2270262, EPI_ISL_2270263, EPI_ISL_2270264, EPI_ISL_2270265, EPI_ISL_2270266, EPI_ISL_2270267, EPI_ISL_2270268, EPI_ISL_2270269, EPI_ISL_2270270, EPI_ISL_2270271, EPI_ISL_2270272, EPI_ISL_2270273, EPI_ISL_2270274, EPI_ISL_2270275, EPI_ISL_2270276, EPI_ISL_2270277, EPI_ISL_2270278, EPI_ISL_2270279, EPI_ISL_2270280, EPI_ISL_2270281, EPI_ISL_2270282, EPI_ISL_2270283, EPI_ISL_2270284, EPI_ISL_2270285, EPI_ISL_2270286, EPI_ISL_2270287, EPI_ISL_2270288, EPI_ISL_2270289, EPI_ISL_2270290, EPI_ISL_2270291, EPI_ISL_2270292, EPI_ISL_2270293, EPI_ISL_2270294, EPI_ISL_2270295, EPI_ISL_2270296, EPI_ISL_2270297, EPI_ISL_2270298, EPI_ISL_2270299, EPI_ISL_2270300, EPI_ISL_2270301, EPI_ISL_2270302, EPI_ISL_2270303, EPI_ISL_2270304, EPI_ISL_2270305, EPI_ISL_2270306, EPI_ISL_2270307, EPI_ISL_2270309, EPI_ISL_2270310, EPI_ISL_2270311, EPI_ISL_2270312, EPI_ISL_2270313, EPI_ISL_2270314, EPI_ISL_2270315, EPI_ISL_2270316, EPI_ISL_2270317, EPI_ISL_2270318, EPI_ISL_2270319, EPI_ISL_2270320, EPI_ISL_2270321, EPI_ISL_2270322, EPI_ISL_2270323, EPI_ISL_2270324, EPI_ISL_2270325, EPI_ISL_2270326, EPI_ISL_2270327, EPI_ISL_2270328, EPI_ISL_2270329, EPI_ISL_2270330, EPI_ISL_2270331, EPI_ISL_2270332, EPI_ISL_2270333, EPI_ISL_2270334, EPI_ISL_2270335, EPI_ISL_2270336, EPI_ISL_2270337, EPI_ISL_2270338, EPI_ISL_2270339, EPI_ISL_2270340, EPI_ISL_2270341, EPI_ISL_2270342, EPI_ISL_2270343, EPI_ISL_2270344, EPI_ISL_2270345, EPI_ISL_2270346, EPI_ISL_2270347, EPI_ISL_2270348, EPI_ISL_2270350, EPI_ISL_2270351, EPI_ISL_2270352, EPI_ISL_2270353, EPI_ISL_2270354 | | | |
| see above | Helix/Illumina | Centers for Disease Control and Prevention Division of Viral Diseases, Pathogen Discovery | Dakota Howard, Dhwani Batra, Peter W. Cook, Kara Moser, Adrian Paskey, Jason Caravas, Benjamin Rambo-Martin, Shatavia Morrison, Christopher Gulvick, Scott Sammons, Yvette Unoarumhi, Darlene Wagner, Matthew Schmerer, Eileen de Feo, Jan Antico, Christine Tran, Matthew Tolentino, Shannon Wickline, Kim Gietzen, Brad Sickler, Jingtao Liu, Eric Allen, Phil Febbo, Nicole L. Washington, Simon White, Geraint Levan, Kelly Schiabor Barrett, Elizabeth Cirulli, Alexandre Bolze, Ary Ascencio, Charlotte Rivera-Garcia, Ryan Cho, Jason Nguyen, Sherry Wang, Jimmy Ramirez, Tyler Cassens, Efren Sandoval, Magnus Isaksson, William Lee, David Becker, Marc Laurent, James Lu, Clinton R. Paden, Duncan MacCannell |
| EPI_ISL_2270355 | Mobile County Health Department | Synergy Laboratories | Megan Cornwell |
| EPI_ISL_2270356, EPI_ISL_2270357, EPI_ISL_2270358, EPI_ISL_2270359, EPI_ISL_2270360, EPI_ISL_2270361, EPI_ISL_2270362, EPI_ISL_2270363, EPI_ISL_2270364, EPI_ISL_2270365, EPI_ISL_2270366, EPI_ISL_2270367, EPI_ISL_2270368, EPI_ISL_2270369, EPI_ISL_2270370, EPI_ISL_2270371, EPI_ISL_2270372, EPI_ISL_2270373, EPI_ISL_2270374, EPI_ISL_2270375, EPI_ISL_2270377, EPI_ISL_2270378, EPI_ISL_2270379, EPI_ISL_2270380, EPI_ISL_2270381, EPI_ISL_2270382, EPI_ISL_2270383, EPI_ISL_2270384, EPI_ISL_2270385, EPI_ISL_2270386, EPI_ISL_2270387, EPI_ISL_2270388, EPI_ISL_2270389, EPI_ISL_2270390, EPI_ISL_2270391, EPI_ISL_2270392, EPI_ISL_2270393, EPI_ISL_2270394, EPI_ISL_2270395, EPI_ISL_2270396, EPI_ISL_2270397, EPI_ISL_2270398, EPI_ISL_2270399, EPI_ISL_2270400, EPI_ISL_2270401, EPI_ISL_2270402, EPI_ISL_2270403, EPI_ISL_2270405, EPI_ISL_2270406, EPI_ISL_2270408, EPI_ISL_2270409, EPI_ISL_2270410, EPI_ISL_2270411, EPI_ISL_2270412, EPI_ISL_2270413, EPI_ISL_2270414, EPI_ISL_2270415, EPI_ISL_2270416, EPI_ISL_2270417, EPI_ISL_2270418, EPI_ISL_2270419, EPI_ISL_2270420, EPI_ISL_2270421, EPI_ISL_2270422, EPI_ISL_2270423, EPI_ISL_2270424, EPI_ISL_2270425, EPI_ISL_2270426, EPI_ISL_2270427, EPI_ISL_2270428, EPI_ISL_2270429, EPI_ISL_2270430, EPI_ISL_2270431, EPI_ISL_2270432, EPI_ISL_2270433, EPI_ISL_2270434, EPI_ISL_2270435, EPI_ISL_2270436, EPI_ISL_2270437, EPI_ISL_2270438, EPI_ISL_2270439, EPI_ISL_2270440, EPI_ISL_2270441, EPI_ISL_2270442, EPI_ISL_2270443, EPI_ISL_2270444, EPI_ISL_2270445, EPI_ISL_2270446, EPI_ISL_2270448, EPI_ISL_2270449, EPI_ISL_2270450, EPI_ISL_2270451, EPI_ISL_2270452, EPI_ISL_2270453, EPI_ISL_2270454, EPI_ISL_2270455, EPI_ISL_2270456, EPI_ISL_2270457, EPI_ISL_2270458, EPI_ISL_2270459, EPI_ISL_2270460, EPI_ISL_2270461, EPI_ISL_2270462, EPI_ISL_2270463, EPI_ISL_2270464, EPI_ISL_2270465, EPI_ISL_2270466, EPI_ISL_2270467, EPI_ISL_2270468, EPI_ISL_2270469, EPI_ISL_2270470, EPI_ISL_2270471, EPI_ISL_2270472, EPI_ISL_2270473, EPI_ISL_2270474, EPI_ISL_2270475, EPI_ISL_2270476, EPI_ISL_2270477, EPI_ISL_2270478, EPI_ISL_2270479, EPI_ISL_2270480, EPI_ISL_2270481, EPI_ISL_2270482, EPI_ISL_2270483, EPI_ISL_2270484, EPI_ISL_2270485, EPI_ISL_2270486, EPI_ISL_2270487, EPI_ISL_2270488, EPI_ISL_2270489, EPI_ISL_2270490, EPI_ISL_2270491, EPI_ISL_2270492, EPI_ISL_2270493, EPI_ISL_2270494, EPI_ISL_2270495, EPI_ISL_2270496, EPI_ISL_2270497, EPI_ISL_2270498, EPI_ISL_2270499, EPI_ISL_2270500, EPI_ISL_2270501, EPI_ISL_2270502, EPI_ISL_2270503, EPI_ISL_2270504, EPI_ISL_2270505, EPI_ISL_2270506, EPI_ISL_2270507, EPI_ISL_2270508, EPI_ISL_2270509, EPI_ISL_2270510, EPI_ISL_2270511, EPI_ISL_2270512, EPI_ISL_2270513, EPI_ISL_2270514, EPI_ISL_2270515, EPI_ISL_2270516, EPI_ISL_2270517, EPI_ISL_2270518, EPI_ISL_2270519, EPI_ISL_2270520, EPI_ISL_2270521, EPI_ISL_2270522, EPI_ISL_2270524, EPI_ISL_2270525, EPI_ISL_2270526, EPI_ISL_2270527, EPI_ISL_2270528, EPI_ISL_2270529, EPI_ISL_2270530, EPI_ISL_2270531, EPI_ISL_2270532, EPI_ISL_2270533, EPI_ISL_2270534, EPI_ISL_2270535, EPI_ISL_2270536, EPI_ISL_2270537, EPI_ISL_2270538, EPI_ISL_2270539, EPI_ISL_2270540, EPI_ISL_2270541, EPI_ISL_2270542, EPI_ISL_2270543, EPI_ISL_2270544, EPI_ISL_2270545, EPI_ISL_2270546, EPI_ISL_2270547, EPI_ISL_2270548, EPI_ISL_2270549, EPI_ISL_2270550, EPI_ISL_2270551, EPI_ISL_2270552, EPI_ISL_2270553, EPI_ISL_2270554, EPI_ISL_2270555, EPI_ISL_2270556, EPI_ISL_2270557, EPI_ISL_2270558, EPI_ISL_2270559, EPI_ISL_2270560, EPI_ISL_2270561, EPI_ISL_2270562, EPI_ISL_2270563, EPI_ISL_2270564, EPI_ISL_2270565, EPI_ISL_2270566, EPI_ISL_2270567, EPI_ISL_2270568, EPI_ISL_2270569, EPI_ISL_2270570, EPI_ISL_2270571, EPI_ISL_2270572, EPI_ISL_2270573, EPI_ISL_2270574, EPI_ISL_2270575, EPI_ISL_2270576, EPI_ISL_2270577, EPI_ISL_2270578, EPI_ISL_2270579, EPI_ISL_2270580, EPI_ISL_2270581, EPI_ISL_2270582, EPI_ISL_2270583, EPI_ISL_2270584, EPI_ISL_2270585, EPI_ISL_2270586, EPI_ISL_2270587, EPI_ISL_2270588, EPI_ISL_2270589, EPI_ISL_2270590, EPI_ISL_2270591, EPI_ISL_2270592, EPI_ISL_2270593, EPI_ISL_2270594, EPI_ISL_2270595, EPI_ISL_2270596, EPI_ISL_2270597, EPI_ISL_2270598, EPI_ISL_2270599, EPI_ISL_2270600, EPI_ISL_2270601, EPI_ISL_2270602, EPI_ISL_2270603, EPI_ISL_2270604, EPI_ISL_2270605, EPI_ISL_2270606, EPI_ISL_2270607, EPI_ISL_2270608, EPI_ISL_2270609, EPI_ISL_2270610, EPI_ISL_2270611, EPI_ISL_2270613, EPI_ISL_2270614, EPI_ISL_2270615, EPI_ISL_2270616, EPI_ISL_2270617, EPI_ISL_2270618, EPI_ISL_2270619, EPI_ISL_2270620, EPI_ISL_2270621, EPI_ISL_2270622, EPI_ISL_2270623, EPI_ISL_2270624, EPI_ISL_2270625, EPI_ISL_2270626, EPI_ISL_2270627, EPI_ISL_2270628, EPI_ISL_2270629, EPI_ISL_2270630, EPI_ISL_2270631, EPI_ISL_2270632, EPI_ISL_2270633, EPI_ISL_2270634, EPI_ISL_2270635, EPI_ISL_2270636, EPI_ISL_2270637, EPI_ISL_2270638, EPI_ISL_2270639, EPI_ISL_2270640, EPI_ISL_2270641, EPI_ISL_2270642, EPI_ISL_2270643, EPI_ISL_2270645, EPI_ISL_2270646, EPI_ISL_2270647, EPI_ISL_2270648, EPI_ISL_2270649, EPI_ISL_2270650, EPI_ISL_2270651, EPI_ISL_2270652, EPI_ISL_2270653, EPI_ISL_2270654, EPI_ISL_2270655, EPI_ISL_2270656, EPI_ISL_2270657, EPI_ISL_2270658, EPI_ISL_2270659, EPI_ISL_2270660, EPI_ISL_2270661, EPI_ISL_2270662, EPI_ISL_2270663, EPI_ISL_2270664, EPI_ISL_2270665, EPI_ISL_2270666, EPI_ISL_2270667, EPI_ISL_2270668, EPI_ISL_2270669, EPI_ISL_2270670, EPI_ISL_2270671, EPI_ISL_2270672, EPI_ISL_2270673, EPI_ISL_2270674, EPI_ISL_2270675, EPI_ISL_2270676, EPI_ISL_2270677, EPI_ISL_2270678, EPI_ISL_2270679, EPI_ISL_2270680, EPI_ISL_2270681, EPI_ISL_2270682, EPI_ISL_2270683, EPI_ISL_2270684, EPI_ISL_2270685, EPI_ISL_2270686, EPI_ISL_2270687, EPI_ISL_2270688, EPI_ISL_2270689, EPI_ISL_2270690, EPI_ISL_2270691, EPI_ISL_2270692, EPI_ISL_2270693, EPI_ISL_2270694, EPI_ISL_2270695, EPI_ISL_2270696, EPI_ISL_2270697, EPI_ISL_2270698, EPI_ISL_2270699, EPI_ISL_2270700, EPI_ISL_2270701, EPI_ISL_2270702, EPI_ISL_2270703, EPI_ISL_2270704, EPI_ISL_2270705, EPI_ISL_2270706, EPI_ISL_2270707, EPI_ISL_2270708, EPI_ISL_2270709, EPI_ISL_2270710, EPI_ISL_2270711, EPI_ISL_2270712, EPI_ISL_2270713, EPI_ISL_2270714, EPI_ISL_2270715, EPI_ISL_2270716, EPI_ISL_2270717, EPI_ISL_2270718, EPI_ISL_2270719, EPI_ISL_2270720, EPI_ISL_2270721, EPI_ISL_2270722, EPI_ISL_2270723, EPI_ISL_2270724, EPI_ISL_2270725, EPI_ISL_2270726, EPI_ISL_2270727, EPI_ISL_2270728, EPI_ISL_2270729, EPI_ISL_2270730, EPI_ISL_2270731, EPI_ISL_2270732, EPI_ISL_2270733, EPI_ISL_2270734, EPI_ISL_2270735, EPI_ISL_2270736, EPI_ISL_2270737, EPI_ISL_2270738, EPI_ISL_2270739, EPI_ISL_2270740, EPI_ISL_2270741, EPI_ISL_2270742, EPI_ISL_2270743, EPI_ISL_2270744, EPI_ISL_2270745, EPI_ISL_2270746, EPI_ISL_2270747, EPI_ISL_2270748, EPI_ISL_2270749, EPI_ISL_2270750, EPI_ISL_2270751, EPI_ISL_2270752, EPI_ISL_2270753, EPI_ISL_2270754, EPI_ISL_2270755, EPI_ISL_2270756, EPI_ISL_2270757, EPI_ISL_2270758, EPI_ISL_2270759, EPI_ISL_2270760, EPI_ISL_2270761, EPI_ISL_2270763, EPI_ISL_2270764, EPI_ISL_2270765, EPI_ISL_2270766, EPI_ISL_2270767, EPI_ISL_2270768, EPI_ISL_2270769, EPI_ISL_2270770, EPI_ISL_2270771, EPI_ISL_2270772, EPI_ISL_2270773, EPI_ISL_2270774, EPI_ISL_2270775, EPI_ISL_2270776, EPI_ISL_2270777, EPI_ISL_2270778, EPI_ISL_2270779, EPI_ISL_2270781, EPI_ISL_2270782, EPI_ISL_2270783, EPI_ISL_2270784, EPI_ISL_2270785, EPI_ISL_2270786, EPI_ISL_2270787, EPI_ISL_2270788, EPI_ISL_2270789, EPI_ISL_2270790, EPI_ISL_2270791, EPI_ISL_2270792, EPI_ISL_2270793, EPI_ISL_2270794, EPI_ISL_2270795, EPI_ISL_2270796, EPI_ISL_2270797, EPI_ISL_2270798, EPI_ISL_2270799, EPI_ISL_2270800, EPI_ISL_2270801, EPI_ISL_2270802, EPI_ISL_2270803, EPI_ISL_2270804, EPI_ISL_2270805, EPI_ISL_2270806, EPI_ISL_2270807, EPI_ISL_2270808, EPI_ISL_2270809, EPI_ISL_2270810, EPI_ISL_2270811, EPI_ISL_2270812, EPI_ISL_2270813, EPI_ISL_2270815, EPI_ISL_2270816, EPI_ISL_2270817, EPI_ISL_2270818, EPI_ISL_2270819, EPI_ISL_2270820, EPI_ISL_2270821, EPI_ISL_2270822, EPI_ISL_2270823, EPI_ISL_2270824, EPI_ISL_2270825, EPI_ISL_2270826, EPI_ISL_2270827, EPI_ISL_2270828, EPI_ISL_2270829, EPI_ISL_2270830, EPI_ISL_2270831, EPI_ISL_2270832, EPI_ISL_2270833, EPI_ISL_2270834 | | | |
| see above | Helix/Illumina | Centers for Disease Control and Prevention Division of Viral Diseases, Pathogen Discovery | Dakota Howard, Dhwani Batra, Peter W. Cook, Kara Moser, Adrian Paskey, Jason Caravas, Benjamin Rambo-Martin, Shatavia Morrison, Christopher Gulvick, Scott Sammons, Yvette Unoarumhi, Darlene Wagner, Matthew Schmerer, Eileen de Feo, Jan Antico, Christine Tran, Matthew Tolentino, Shannon Wickline, Kim Gietzen, Brad Sickler, Jingtao Liu, Eric Allen, Phil Febbo, Nicole L. Washington, Simon White, Geraint Levan, Kelly Schiabor Barrett, Elizabeth Cirulli, Alexandre Bolze, Ary Ascencio, Charlotte Rivera-Garcia, Ryan Cho, Jason Nguyen, Sherry Wang, Jimmy Ramirez, Tyler Cassens, Efren Sandoval, Magnus Isaksson, William Lee, David Becker, Marc Laurent, James Lu, Clinton R. Paden, Duncan MacCannell |
| EPI_ISL_2270930, EPI_ISL_2270931, EPI_ISL_2270932, EPI_ISL_2270933, EPI_ISL_2270934, EPI_ISL_2270935, EPI_ISL_2270936, EPI_ISL_2270937, EPI_ISL_2270938, EPI_ISL_2270939, EPI_ISL_2270940, EPI_ISL_2270941, EPI_ISL_2270942, EPI_ISL_2270943, EPI_ISL_2270944, EPI_ISL_2270945, EPI_ISL_2270946, EPI_ISL_2270947, EPI_ISL_2270948, EPI_ISL_2270949, EPI_ISL_2270951, EPI_ISL_2270952, EPI_ISL_2270953, EPI_ISL_2270954, EPI_ISL_2270955, EPI_ISL_2270957, EPI_ISL_2270958, EPI_ISL_2270959, EPI_ISL_2270960, EPI_ISL_2270961, EPI_ISL_2270962, EPI_ISL_2270963, EPI_ISL_2270964, EPI_ISL_2270965, EPI_ISL_2270966, EPI_ISL_2270967, EPI_ISL_2270968, EPI_ISL_2270969, EPI_ISL_2270970, EPI_ISL_2270971, EPI_ISL_2270972, EPI_ISL_2270973, EPI_ISL_2270974, EPI_ISL_2270975, EPI_ISL_2270976, EPI_ISL_2270977, EPI_ISL_2270978, EPI_ISL_2270979, EPI_ISL_2270980, EPI_ISL_2270981, EPI_ISL_2270982, EPI_ISL_2270983, EPI_ISL_2270984, EPI_ISL_2270985, EPI_ISL_2270986, EPI_ISL_2270992, EPI_ISL_2271002, EPI_ISL_2271010, EPI_ISL_2271012, EPI_ISL_2271015, EPI_ISL_2271017, EPI_ISL_2271020, EPI_ISL_2271021, EPI_ISL_2271022, EPI_ISL_2271023, EPI_ISL_2271025, EPI_ISL_2271027, EPI_ISL_2271028, EPI_ISL_2271029 | | | |
| see above | Wyoming Public Health Laboratory | Wyoming Public Health Laboratory | Jim Mildenberger, Taylor Fearing, Channing Weber, Ashley Norberg, Chayse Rowley, Marley Goetz, Brian Dominguez, Elliot Thomasson, Brittany Oher, Sam Britz, Cari Sloma, and Rob Christensen |
| EPI_ISL_2271710, EPI_ISL_2271711, EPI_ISL_2271712, EPI_ISL_2271713, EPI_ISL_2271714, EPI_ISL_2271715, EPI_ISL_2271716, EPI_ISL_2271717, EPI_ISL_2271718, EPI_ISL_2271719, EPI_ISL_2271721, EPI_ISL_2271722 | | | |
| see above | Kansas Health and Environmental Lab | Kansas Health and Environmental Lab | Mike Grose, Katherine Wiggins, Jonathan Barnell, Ben Olsen, and Phil Adam |
| EPI_ISL_2271723, EPI_ISL_2271724, EPI_ISL_2271725, EPI_ISL_2271726, EPI_ISL_2271727, EPI_ISL_2271728, EPI_ISL_2271729, EPI_ISL_2271730, EPI_ISL_2271731, EPI_ISL_2271732, EPI_ISL_2271734, EPI_ISL_2271735, EPI_ISL_2271736, EPI_ISL_2271737, EPI_ISL_2271738, EPI_ISL_2271739, EPI_ISL_2271740, EPI_ISL_2271741, EPI_ISL_2271742, EPI_ISL_2271743, EPI_ISL_2271745, EPI_ISL_2271746, EPI_ISL_2271747, EPI_ISL_2271748, EPI_ISL_2271749, EPI_ISL_2271750, EPI_ISL_2271751, EPI_ISL_2271752, EPI_ISL_2271753, EPI_ISL_2271754, EPI_ISL_2271755, EPI_ISL_2271756, EPI_ISL_2271757, EPI_ISL_2271758, EPI_ISL_2271759, EPI_ISL_2271760, EPI_ISL_2271761, EPI_ISL_2271762, EPI_ISL_2271763, EPI_ISL_2271764, EPI_ISL_2271765, EPI_ISL_2271766, EPI_ISL_2271767, EPI_ISL_2271768, EPI_ISL_2271769, EPI_ISL_2271770, EPI_ISL_2271771, EPI_ISL_2271772, EPI_ISL_2271773, EPI_ISL_2271774, EPI_ISL_2271775, EPI_ISL_2271776, EPI_ISL_2271777, EPI_ISL_2271778, EPI_ISL_2271779, EPI_ISL_2271780, EPI_ISL_2271781, EPI_ISL_2271782, EPI_ISL_2271784, EPI_ISL_2271785, EPI_ISL_2271786, EPI_ISL_2271787, EPI_ISL_2271788, EPI_ISL_2271789, EPI_ISL_2271790, EPI_ISL_2271791, EPI_ISL_2271792, EPI_ISL_2271793, EPI_ISL_2271794, EPI_ISL_2271795, EPI_ISL_2271796, EPI_ISL_2271797, EPI_ISL_2271798, EPI_ISL_2271799, EPI_ISL_2271800, EPI_ISL_2271801, EPI_ISL_2271802, EPI_ISL_2271803, EPI_ISL_2271804, EPI_ISL_2271805, EPI_ISL_2271806, EPI_ISL_2271807, EPI_ISL_2271808, EPI_ISL_2271809, EPI_ISL_2271810, EPI_ISL_2271811, EPI_ISL_2271812, EPI_ISL_2271813, EPI_ISL_2271814, EPI_ISL_2271815, EPI_ISL_2271816, EPI_ISL_2271817, EPI_ISL_2271818, EPI_ISL_2271820, EPI_ISL_2271821, EPI_ISL_2271822, EPI_ISL_2271823, EPI_ISL_2271824, EPI_ISL_2271825, EPI_ISL_2271827, EPI_ISL_2271830, EPI_ISL_2271831, EPI_ISL_2271832, EPI_ISL_2271833, EPI_ISL_2271834, EPI_ISL_2271835, EPI_ISL_2271836, EPI_ISL_2271837, EPI_ISL_2271838, EPI_ISL_2271839, EPI_ISL_2271840, EPI_ISL_2271842, EPI_ISL_2271843, EPI_ISL_2271844, EPI_ISL_2271845, EPI_ISL_2271846, EPI_ISL_2271848, EPI_ISL_2271850, EPI_ISL_2271851, EPI_ISL_2271852, EPI_ISL_2271854, EPI_ISL_2271855, EPI_ISL_2271856, EPI_ISL_2271857, EPI_ISL_2271859, EPI_ISL_2271860, EPI_ISL_2271861, EPI_ISL_2271862, EPI_ISL_2271864, EPI_ISL_2271865, EPI_ISL_2271866, EPI_ISL_2271867, EPI_ISL_2271868, EPI_ISL_2271870, EPI_ISL_2271871, EPI_ISL_2271872, EPI_ISL_2271873, EPI_ISL_2271874 | | | |
| see above | Michigan Department of Health and Human Services, Bureau of Laboratories | Michigan Department of Health and Human Services, Bureau of Laboratories | Blankenship HM, Riner D, Soehnlen MK |
| EPI_ISL_2271875, EPI_ISL_2271876, EPI_ISL_2271877, EPI_ISL_2271878, EPI_ISL_2271879, EPI_ISL_2271880, EPI_ISL_2271881, EPI_ISL_2271882, EPI_ISL_2271884, EPI_ISL_2271885, EPI_ISL_2271886, EPI_ISL_2271887, EPI_ISL_2271888, EPI_ISL_2271889, EPI_ISL_2271890, EPI_ISL_2271891, EPI_ISL_2271892, EPI_ISL_2271893, EPI_ISL_2271894, EPI_ISL_2271895, EPI_ISL_2271896, EPI_ISL_2271897, EPI_ISL_2271898, EPI_ISL_2271899, EPI_ISL_2271900, EPI_ISL_2271901 | | | |
| see above | MD PHL | MD PHL | Maryland Department of Health Laboratories Administration |
| EPI_ISL_2272498, EPI_ISL_2272499, EPI_ISL_2272500 | LANIIA | LANIIA | Cristobal Chaidez, Bruno Gomez-Gil, Jean Pierre González-Gómez, Juan Daniel Lira-Morales, Irvin González-López, Julissa Enciso-Ibarra, Célida Isabel Martínez-Rodríguez |
| EPI_ISL_2272501, EPI_ISL_2272502, EPI_ISL_2272503, EPI_ISL_2272505, EPI_ISL_2272506, EPI_ISL_2272507, EPI_ISL_2272508, EPI_ISL_2272509, EPI_ISL_2272510, EPI_ISL_2272511, EPI_ISL_2272512, EPI_ISL_2272513, EPI_ISL_2272514, EPI_ISL_2272515, EPI_ISL_2272516, EPI_ISL_2272517, EPI_ISL_2272518, EPI_ISL_2272519, EPI_ISL_2272521 | | | |
| see above | Servicio Diagnóstico Clínico | LANIIA | Cristobal Chaidez, Bruno Gomez-Gil, Jean Pierre González-Gómez, Juan Daniel Lira-Morales, Irvin González-López, Julissa Enciso-Ibarra, Célida Isabel Martínez-Rodríguez |
| EPI_ISL_2272522, EPI_ISL_2272523, EPI_ISL_2272524, EPI_ISL_2272525, EPI_ISL_2272526, EPI_ISL_2272527, EPI_ISL_2272528, EPI_ISL_2272529, EPI_ISL_2272530, EPI_ISL_2272531, EPI_ISL_2272532, EPI_ISL_2272533, EPI_ISL_2272534, EPI_ISL_2272535, EPI_ISL_2272536, EPI_ISL_2272537, EPI_ISL_2272538, EPI_ISL_2272539, EPI_ISL_2272540, EPI_ISL_2272541, EPI_ISL_2272542, EPI_ISL_2272543, EPI_ISL_2272544, EPI_ISL_2272545, EPI_ISL_2272546, EPI_ISL_2272547, EPI_ISL_2272548, EPI_ISL_2272549, EPI_ISL_2272550, EPI_ISL_2272551, EPI_ISL_2272552, EPI_ISL_2272553, EPI_ISL_2272554, EPI_ISL_2272555, EPI_ISL_2272556, EPI_ISL_2272557 | | | |
| see above | Alaska State Virology Laboratory | Alaska State Virology Laboratory | Stephanie DeRonde, Elva House, Jacob Zidek, Lisa Smith, Ph.D., Jack Chen, Ph.D. |
| EPI_ISL_2272558, EPI_ISL_2272559, EPI_ISL_2272560, EPI_ISL_2272561, EPI_ISL_2272562, EPI_ISL_2272563, EPI_ISL_2272564, EPI_ISL_2272565, EPI_ISL_2272566, EPI_ISL_2272567, EPI_ISL_2272568, EPI_ISL_2272569, EPI_ISL_2272570, EPI_ISL_2272571, EPI_ISL_2272572, EPI_ISL_2272573, EPI_ISL_2272574, EPI_ISL_2272575, EPI_ISL_2272576, EPI_ISL_2272577, EPI_ISL_2272578, EPI_ISL_2272579, EPI_ISL_2272580, EPI_ISL_2272581, EPI_ISL_2272582, EPI_ISL_2272583, EPI_ISL_2272584, EPI_ISL_2272585, EPI_ISL_2272586, EPI_ISL_2272587, EPI_ISL_2272588, EPI_ISL_2272589, EPI_ISL_2272590, EPI_ISL_2272591, EPI_ISL_2272592, EPI_ISL_2272593, EPI_ISL_2272594, EPI_ISL_2272595, EPI_ISL_2272596 | | | |
| see above | GA Department of Public Health | GA Department of Public Health | Stacy Reeves, Jonathan Edwards, Cynthia Dixey, Tonia Parrott, Aliyah Fields, Taylor Smith |
| EPI_ISL_2272603 | City of Milwaukee Health Department Laboratory | City of Milwaukee Health Department Laboratory | Sanjib Bhattacharyya, Manjeet Khubbar, Amy Bauer, Jennifer Lentz, Samantha Scott |
| EPI_ISL_2272604, EPI_ISL_2272605, EPI_ISL_2272606, EPI_ISL_2272607, EPI_ISL_2272608, EPI_ISL_2272609, EPI_ISL_2272610, EPI_ISL_2272611, EPI_ISL_2272612, EPI_ISL_2272613, EPI_ISL_2272614, EPI_ISL_2272615, EPI_ISL_2272616, EPI_ISL_2272617, EPI_ISL_2272618, EPI_ISL_2272619, EPI_ISL_2272620, EPI_ISL_2272621, EPI_ISL_2272622, EPI_ISL_2272623, EPI_ISL_2272624, EPI_ISL_2272625, EPI_ISL_2272626, EPI_ISL_2272627, EPI_ISL_2272628, EPI_ISL_2272629, EPI_ISL_2272630, EPI_ISL_2272631, EPI_ISL_2272632, EPI_ISL_2272633, EPI_ISL_2272634, EPI_ISL_2272635, EPI_ISL_2272636, EPI_ISL_2272637, EPI_ISL_2272638, EPI_ISL_2272639, EPI_ISL_2272640, EPI_ISL_2272641, EPI_ISL_2272642, EPI_ISL_2272643, EPI_ISL_2272644, EPI_ISL_2272645, EPI_ISL_2272646, EPI_ISL_2272647, EPI_ISL_2272648, EPI_ISL_2272649, EPI_ISL_2272650, EPI_ISL_2272651, EPI_ISL_2272652, EPI_ISL_2272653, EPI_ISL_2272654, EPI_ISL_2272655, EPI_ISL_2272656, EPI_ISL_2272657, EPI_ISL_2272658, EPI_ISL_2272659, EPI_ISL_2272660, EPI_ISL_2272661, EPI_ISL_2272662, EPI_ISL_2272663, EPI_ISL_2272664, EPI_ISL_2272665, EPI_ISL_2272666, EPI_ISL_2272667, EPI_ISL_2272668, EPI_ISL_2272669 | | | |
| see above | UW Virology Lab | UW Virology Lab | Pavitra Roychoudhury, Hong Xie, Lasata Shrestha, Tien V. Nguyen, Shah Mohamed Bakhash, Michelle Lin, Noah R. Baker, Ricardo Perez, Sean Ellis, Nathan Breit, Robert J. Livingston, Meei-Li Huang, Keith R Jerome, Patrick Mathias, Alexander Greninger |
| EPI_ISL_2272938, EPI_ISL_2272939, EPI_ISL_2272941, EPI_ISL_2272942, EPI_ISL_2272943, EPI_ISL_2272944, EPI_ISL_2272945, EPI_ISL_2272946, EPI_ISL_2272947, EPI_ISL_2272948, EPI_ISL_2272949, EPI_ISL_2272950, EPI_ISL_2272951, EPI_ISL_2272952, EPI_ISL_2272953, EPI_ISL_2272954, EPI_ISL_2272955, EPI_ISL_2272956, EPI_ISL_2272957 | | | |
| see above | Wichita State University - Molecular Diagnostics Lab | Kansas Health and Environmental Lab | Mike Grose, Katherine Wiggins, Jonathan Barnell, Ben Olsen, and Phil Adam |
| EPI_ISL_2273092, EPI_ISL_2273093, EPI_ISL_2273094, EPI_ISL_2273095, EPI_ISL_2273096, EPI_ISL_2273097, EPI_ISL_2273098, EPI_ISL_2273099, EPI_ISL_2273100, EPI_ISL_2273101, EPI_ISL_2273102, EPI_ISL_2273103, EPI_ISL_2273104, EPI_ISL_2273106, EPI_ISL_2273107, EPI_ISL_2273108, EPI_ISL_2273109, EPI_ISL_2273110, EPI_ISL_2273111, EPI_ISL_2273112, EPI_ISL_2273113, EPI_ISL_2273114, EPI_ISL_2273115, EPI_ISL_2273116, EPI_ISL_2273118, EPI_ISL_2273120, EPI_ISL_2273121, EPI_ISL_2273123, EPI_ISL_2273126, EPI_ISL_2273130, EPI_ISL_2273131, EPI_ISL_2273132, EPI_ISL_2273133, EPI_ISL_2273134, EPI_ISL_2273136, EPI_ISL_2273137 | | | |
| see above | Blue Water Diagnostic Laboratory, LLC | University of Louisville Laboratory of Melissa Smith | Elizabeth Hudson, Matt Seetin, Erik Korte, Alonzo Shepherd, Corey T. Watson, Eric C. Rouchka, Melissa L. Smith |
| EPI_ISL_2273727, EPI_ISL_2273729, EPI_ISL_2273730, EPI_ISL_2273731, EPI_ISL_2273732, EPI_ISL_2273733, EPI_ISL_2273734, EPI_ISL_2273735, EPI_ISL_2273736, EPI_ISL_2273737, EPI_ISL_2273738, EPI_ISL_2273739, EPI_ISL_2273740, EPI_ISL_2273742, EPI_ISL_2273743, EPI_ISL_2273744, EPI_ISL_2273745, EPI_ISL_2273746, EPI_ISL_2273747, EPI_ISL_2273748, EPI_ISL_2273749, EPI_ISL_2273750, EPI_ISL_2273751, EPI_ISL_2273752, EPI_ISL_2273753, EPI_ISL_2273754, EPI_ISL_2273755, EPI_ISL_2273756, EPI_ISL_2273757, EPI_ISL_2273758, EPI_ISL_2273759, EPI_ISL_2273760, EPI_ISL_2273761, EPI_ISL_2273762, EPI_ISL_2273763, EPI_ISL_2273764, EPI_ISL_2273765, EPI_ISL_2273766, EPI_ISL_2273767, EPI_ISL_2273768, EPI_ISL_2273769, EPI_ISL_2273770, EPI_ISL_2273771, EPI_ISL_2273772, EPI_ISL_2273773, EPI_ISL_2273774, EPI_ISL_2273775, EPI_ISL_2273776, EPI_ISL_2273777, EPI_ISL_2273778, EPI_ISL_2273779, EPI_ISL_2273780, EPI_ISL_2273781, EPI_ISL_2273782, EPI_ISL_2273783, EPI_ISL_2273784, EPI_ISL_2273785, EPI_ISL_2273786, EPI_ISL_2273787, EPI_ISL_2273788, EPI_ISL_2273789, EPI_ISL_2273790, EPI_ISL_2273791, EPI_ISL_2273792, EPI_ISL_2273793, EPI_ISL_2273794, EPI_ISL_2273795, EPI_ISL_2273796, EPI_ISL_2273797, EPI_ISL_2273798, EPI_ISL_2273799, EPI_ISL_2273800, EPI_ISL_2273801, EPI_ISL_2273802, EPI_ISL_2273803, EPI_ISL_2273804, EPI_ISL_2273805, EPI_ISL_2273806, EPI_ISL_2273807, EPI_ISL_2273808, EPI_ISL_2273809, EPI_ISL_2273811, EPI_ISL_2273812, EPI_ISL_2273813, EPI_ISL_2273814, EPI_ISL_2273815, EPI_ISL_2273816, EPI_ISL_2273817, | | | |

| | | | |
|---|---|---|---|
| EPI_ISL_2273818, EPI_ISL_2273819, EPI_ISL_2273820, EPI_ISL_2273821, EPI_ISL_2273822, EPI_ISL_2273823, EPI_ISL_2273824, EPI_ISL_2273825, EPI_ISL_2273826, EPI_ISL_2273827, EPI_ISL_2273828, EPI_ISL_2273829, EPI_ISL_2273830, EPI_ISL_2273831, EPI_ISL_2273832, EPI_ISL_2273833, EPI_ISL_2273834, EPI_ISL_2273835, EPI_ISL_2273836, EPI_ISL_2273837 | | | |
| see above | UW Virology Lab | UW Virology Lab | Pavitra Roychoudhury, Hong Xie, Lasata Shrestha, Tien V. Nguyen, Shah Mohamed Bakhash, Michelle Lin, Noah R. Baker, Ricardo Perez, Sean Ellis, Nathan Breit, Robert J. Livingston, Meei-Li Huang, Keith R Jerome, Patrick Mathias, Alexander Greninger |
| EPI_ISL_2273838, EPI_ISL_2273839, EPI_ISL_2273840, EPI_ISL_2273841, EPI_ISL_2273843, EPI_ISL_2273849, EPI_ISL_2273850, EPI_ISL_2273851, EPI_ISL_2273853, EPI_ISL_2273858, EPI_ISL_2273859, EPI_ISL_2273860, EPI_ISL_2273863, EPI_ISL_2273864, EPI_ISL_2273865, EPI_ISL_2273866, EPI_ISL_2273868 | | | |
| see above | City of Milwaukee Health Department Laboratory | City of Milwaukee Health Department Laboratory | Sanjib Bhattacharyya, Manjeet Khubbar, Amy Bauer, Jennifer Lentz, Samantha Scott |
| EPI_ISL_2273871, EPI_ISL_2273872, EPI_ISL_2273873, EPI_ISL_2273874, EPI_ISL_2273875, EPI_ISL_2273876, EPI_ISL_2273877, EPI_ISL_2273878, EPI_ISL_2273879, EPI_ISL_2273880, EPI_ISL_2273881, EPI_ISL_2273882, EPI_ISL_2273883, EPI_ISL_2273884, EPI_ISL_2273885, EPI_ISL_2273886, EPI_ISL_2273887, EPI_ISL_2273888, EPI_ISL_2273889, EPI_ISL_2273890, EPI_ISL_2273891, EPI_ISL_2273892, EPI_ISL_2273893, EPI_ISL_2273894, EPI_ISL_2273895, EPI_ISL_2273896, EPI_ISL_2273897, EPI_ISL_2273898, EPI_ISL_2273899, EPI_ISL_2273900, EPI_ISL_2273901, EPI_ISL_2273902, EPI_ISL_2273903, EPI_ISL_2273904, EPI_ISL_2273905, EPI_ISL_2273906, EPI_ISL_2273907, EPI_ISL_2273908, EPI_ISL_2273909, EPI_ISL_2273910, EPI_ISL_2273911, EPI_ISL_2273912, EPI_ISL_2273913, EPI_ISL_2273914, EPI_ISL_2273915, EPI_ISL_2273916, EPI_ISL_2273917, EPI_ISL_2273918, EPI_ISL_2273919, EPI_ISL_2273920, EPI_ISL_2273921, EPI_ISL_2273922, EPI_ISL_2273923, EPI_ISL_2273924, EPI_ISL_2273925, EPI_ISL_2273926, EPI_ISL_2273927, EPI_ISL_2273928, EPI_ISL_2273929, EPI_ISL_2273930, EPI_ISL_2273931, EPI_ISL_2273932, EPI_ISL_2273933, EPI_ISL_2273934, EPI_ISL_2273935, EPI_ISL_2273936, EPI_ISL_2273937, EPI_ISL_2273938, EPI_ISL_2273939, EPI_ISL_2273940, EPI_ISL_2273941, EPI_ISL_2273942, EPI_ISL_2273943, EPI_ISL_2273944, EPI_ISL_2273945, EPI_ISL_2273946, EPI_ISL_2273947, EPI_ISL_2273948, EPI_ISL_2273949, EPI_ISL_2273950, EPI_ISL_2273951, EPI_ISL_2273952, EPI_ISL_2273953, EPI_ISL_2273954, EPI_ISL_2273955, EPI_ISL_2273956, EPI_ISL_2273957, EPI_ISL_2273958, EPI_ISL_2273959, EPI_ISL_2273960, EPI_ISL_2273961, EPI_ISL_2273962, EPI_ISL_2273963, EPI_ISL_2273964, EPI_ISL_2273965, EPI_ISL_2273966, EPI_ISL_2273967, EPI_ISL_2273968, EPI_ISL_2273969, EPI_ISL_2273970, EPI_ISL_2273971, EPI_ISL_2273972, EPI_ISL_2273973, EPI_ISL_2273974, EPI_ISL_2273975, EPI_ISL_2273976, EPI_ISL_2273977, EPI_ISL_2273978, EPI_ISL_2273979, EPI_ISL_2273980, EPI_ISL_2273981, EPI_ISL_2273982, EPI_ISL_2273983, EPI_ISL_2273984, EPI_ISL_2273985, EPI_ISL_2273986, EPI_ISL_2273987, EPI_ISL_2273988, EPI_ISL_2273989, EPI_ISL_2273990, EPI_ISL_2273991, EPI_ISL_2273992, EPI_ISL_2273993, EPI_ISL_2273994, EPI_ISL_2273995, EPI_ISL_2273996, EPI_ISL_2273997, EPI_ISL_2273998 | | | |
| see above | Weill Cornell Medicine | New York Genome Center | Michael Zody, Andre Corvelo, Dayna M. Oschwald, Samantha Fennessey, Tom Maniatis, Melissa Cushing, Olivier Elemento, Margaret Elizabeth Ross, Chris Mason, Priya Velu, Hanna Rennert, Arryn Craney, Lars F Westblade |
| EPI_ISL_2274141, EPI_ISL_2274144, EPI_ISL_2274145, EPI_ISL_2274146, EPI_ISL_2274147, EPI_ISL_2274148, EPI_ISL_2274149, EPI_ISL_2274150, EPI_ISL_2274151, EPI_ISL_2274152, EPI_ISL_2274153, EPI_ISL_2274154, EPI_ISL_2274155, EPI_ISL_2274156, EPI_ISL_2274157, EPI_ISL_2274158, EPI_ISL_2274159, EPI_ISL_2274160, EPI_ISL_2274161, EPI_ISL_2274164, EPI_ISL_2274165, EPI_ISL_2274166, EPI_ISL_2274167, EPI_ISL_2274169, EPI_ISL_2274171, EPI_ISL_2274172, EPI_ISL_2274173, EPI_ISL_2274174, EPI_ISL_2274175, EPI_ISL_2274176, EPI_ISL_2274178, EPI_ISL_2274181, EPI_ISL_2274182, EPI_ISL_2274185, EPI_ISL_2274186, EPI_ISL_2274187, EPI_ISL_2274188, EPI_ISL_2274189, EPI_ISL_2274190, EPI_ISL_2274191, EPI_ISL_2274192, EPI_ISL_2274193, EPI_ISL_2274194, EPI_ISL_2274195, EPI_ISL_2274196, EPI_ISL_2274197, EPI_ISL_2274198, EPI_ISL_2274199, EPI_ISL_2274200, EPI_ISL_2274201, EPI_ISL_2274202, EPI_ISL_2274203, EPI_ISL_2274204, EPI_ISL_2274205, EPI_ISL_2274206, EPI_ISL_2274207, EPI_ISL_2274208, EPI_ISL_2274209, EPI_ISL_2274210, EPI_ISL_2274211, EPI_ISL_2274212, EPI_ISL_2274213, EPI_ISL_2274214, EPI_ISL_2274215, EPI_ISL_2274216, EPI_ISL_2274217, EPI_ISL_2274218, EPI_ISL_2274220, EPI_ISL_2274221, EPI_ISL_2274222, EPI_ISL_2274223, EPI_ISL_2274225, EPI_ISL_2274226, EPI_ISL_2274227, EPI_ISL_2274228, EPI_ISL_2274229, EPI_ISL_2274230, EPI_ISL_2274231, EPI_ISL_2274232, EPI_ISL_2274233, EPI_ISL_2274234, EPI_ISL_2274235, EPI_ISL_2274236, EPI_ISL_2274237, EPI_ISL_2274238, EPI_ISL_2274239, EPI_ISL_2274240, EPI_ISL_2274241, EPI_ISL_2274242 | | | |
| see above | Johns Hopkins Hospital Department of Pathology | Johns Hopkins Hospital Department of Pathology | C. Paul Morris, Chun Huai Luo, Adannaya Amadi, Matthew Schwartz, Nicholas Gallagher, Heba H. Mostafa |
| EPI_ISL_2274492 | NORTHWELL HEALTH LABORATORIES | Wadsworth Center, New York State Department of Health | Kirsten St. George, Daryl M. Lamson, Alexis Russell, Matthew Shudt, Melissa A Leisner, Jonathan Plitnick, Catharine Prussing, Navjot Singh, John Kelly, Erasmus Schneider, Erica Lasek-Nesselquist |
| EPI_ISL_2274493, EPI_ISL_2274494 | Columbia University Irving Medical Center | Wadsworth Center, New York State Department of Health | Kirsten St. George, Daryl M. Lamson, Alexis Russell, Matthew Shudt, Melissa A Leisner, Jonathan Plitnick, Catharine Prussing, Navjot Singh, John Kelly, Erasmus Schneider, Erica Lasek-Nesselquist |
| EPI_ISL_2274495 | SUNY UPSTATE MEDICAL UNIVERSITY | Wadsworth Center, New York State Department of Health | Kirsten St. George, Daryl M. Lamson, Alexis Russell, Matthew Shudt, Melissa A Leisner, Jonathan Plitnick, Catharine Prussing, Navjot Singh, John Kelly, Erasmus Schneider, Erica Lasek-Nesselquist |
| EPI_ISL_2274496, EPI_ISL_2274497, EPI_ISL_2274498 | URMC LABS | Wadsworth Center, New York State Department of Health | Kirsten St. George, Daryl M. Lamson, Alexis Russell, Matthew Shudt, Melissa A Leisner, Jonathan Plitnick, Catharine Prussing, Navjot Singh, John Kelly, Erasmus Schneider, Erica Lasek-Nesselquist |
| EPI_ISL_2274499 | WESTCHESTER MEDICAL CENTER | Wadsworth Center, New York State Department of Health | Kirsten St. George, Daryl M. Lamson, Alexis Russell, Matthew Shudt, Melissa A Leisner, Jonathan Plitnick, Catharine Prussing, Navjot Singh, John Kelly, Erasmus Schneider, Erica Lasek-Nesselquist |
| EPI_ISL_2274500 | URMC LABS | Wadsworth Center, New York State Department of Health | Kirsten St. George, Daryl M. Lamson, Alexis Russell, Matthew Shudt, Melissa A Leisner, Jonathan Plitnick, Catharine Prussing, Navjot Singh, John Kelly, Erasmus Schneider, Erica Lasek-Nesselquist |
| EPI_ISL_2274501, EPI_ISL_2274502 | MEMORIAL SLOAN KETTERING CANCER CENTER | Wadsworth Center, New York State Department of Health | Kirsten St. George, Daryl M. Lamson, Alexis Russell, Matthew Shudt, Melissa A Leisner, Jonathan Plitnick, Catharine Prussing, Navjot Singh, John Kelly, Erasmus Schneider, Erica Lasek-Nesselquist |
| EPI_ISL_2274503, EPI_ISL_2274504 | MONTEFIORE MEDICAL CENTER LABORATORIES | Wadsworth Center, New York State Department of Health | Kirsten St. George, Daryl M. Lamson, Alexis Russell, Matthew Shudt, Melissa A Leisner, Jonathan Plitnick, Catharine Prussing, Navjot Singh, John Kelly, Erasmus Schneider, Erica Lasek-Nesselquist |
| EPI_ISL_2274505 | URMC LABS | Wadsworth Center, New York State Department of Health | Kirsten St. George, Daryl M. Lamson, Alexis Russell, Matthew Shudt, Melissa A Leisner, Jonathan Plitnick, Catharine Prussing, Navjot Singh, John Kelly, Erasmus Schneider, Erica Lasek-Nesselquist |
| EPI_ISL_2274506, EPI_ISL_2274507, EPI_ISL_2274508, EPI_ISL_2274509 | Columbia University Irving Medical Center | Wadsworth Center, New York State Department of Health | Kirsten St. George, Daryl M. Lamson, Alexis Russell, Matthew Shudt, Melissa A Leisner, Jonathan Plitnick, Catharine Prussing, Navjot Singh, John Kelly, Erasmus Schneider, Erica Lasek-Nesselquist |
| EPI_ISL_2274510 | New York Presbyterian Hospital | Wadsworth Center, New York State Department of Health | Kirsten St. George, Daryl M. Lamson, Alexis Russell, Matthew Shudt, Melissa A Leisner, Jonathan Plitnick, Catharine Prussing, Navjot Singh, John Kelly, Erasmus Schneider, Erica Lasek-Nesselquist |
| EPI_ISL_2274511 | NORTHWELL HEALTH LABORATORIES | Wadsworth Center, New York State Department of Health | Kirsten St. George, Daryl M. Lamson, Alexis Russell, Matthew Shudt, Melissa A Leisner, Jonathan Plitnick, Catharine Prussing, Navjot Singh, John Kelly, Erasmus Schneider, Erica Lasek-Nesselquist |
| EPI_ISL_2274512, EPI_ISL_2274513, EPI_ISL_2274514 | WESTCHESTER MEDICAL CENTER | Wadsworth Center, New York State Department of Health | Kirsten St. George, Daryl M. Lamson, Alexis Russell, Matthew Shudt, Melissa A Leisner, Jonathan Plitnick, Catharine Prussing, Navjot Singh, John Kelly, Erasmus Schneider, Erica Lasek-Nesselquist |
| EPI_ISL_2274515 | NORTHWELL HEALTH LABORATORIES | Wadsworth Center, New York State Department of Health | Kirsten St. George, Daryl M. Lamson, Alexis Russell, Matthew Shudt, Melissa A Leisner, Jonathan Plitnick, Catharine Prussing, Navjot Singh, John Kelly, Erasmus Schneider, Erica Lasek-Nesselquist |
| EPI_ISL_2274516, EPI_ISL_2274517 | Columbia University Irving Medical Center | Wadsworth Center, New York State Department of Health | Kirsten St. George, Daryl M. Lamson, Alexis Russell, Matthew Shudt, Melissa A Leisner, Jonathan Plitnick, Catharine Prussing, Navjot Singh, John Kelly, Erasmus Schneider, Erica Lasek-Nesselquist |
| EPI_ISL_2274518, EPI_ISL_2274519, EPI_ISL_2274520, EPI_ISL_2274521 | URMC LABS | Wadsworth Center, New York State Department of Health | Kirsten St. George, Daryl M. Lamson, Alexis Russell, Matthew Shudt, Melissa A Leisner, Jonathan Plitnick, Catharine Prussing, Navjot Singh, John Kelly, Erasmus Schneider, Erica Lasek-Nesselquist |
| EPI_ISL_2274522 | SUNY UPSTATE MEDICAL UNIVERSITY | Wadsworth Center, New York State Department of Health | Kirsten St. George, Daryl M. Lamson, Alexis Russell, Matthew Shudt, Melissa A Leisner, Jonathan Plitnick, Catharine Prussing, Navjot Singh, John Kelly, Erasmus Schneider, Erica Lasek-Nesselquist |
| EPI_ISL_2274523, EPI_ISL_2274524, EPI_ISL_2274525, EPI_ISL_2274526, EPI_ISL_2274527, EPI_ISL_2274528, EPI_ISL_2274529, EPI_ISL_2274530, EPI_ISL_2274531, EPI_ISL_2274532, EPI_ISL_2274533 | | | |
| see above | WESTCHESTER MEDICAL CENTER | Wadsworth Center, New York State Department of Health | Kirsten St. George, Daryl M. Lamson, Alexis Russell, Matthew Shudt, Melissa A Leisner, Jonathan Plitnick, Catharine Prussing, Navjot Singh, John Kelly, Erasmus Schneider, Erica Lasek-Nesselquist |
| EPI_ISL_2274534, EPI_ISL_2274535, EPI_ISL_2274536 | NORTHWELL HEALTH LABORATORIES | Wadsworth Center, New York State Department of Health | Kirsten St. George, Daryl M. Lamson, Alexis Russell, Matthew Shudt, Melissa A Leisner, Jonathan Plitnick, Catharine Prussing, Navjot Singh, John Kelly, Erasmus Schneider, Erica Lasek-Nesselquist |
| EPI_ISL_2274537 | Columbia University Irving Medical Center | Wadsworth Center, New York State Department of Health | Kirsten St. George, Daryl M. Lamson, Alexis Russell, Matthew Shudt, Melissa A Leisner, Jonathan Plitnick, Catharine Prussing, Navjot Singh, John Kelly, Erasmus Schneider, Erica Lasek-Nesselquist |
| EPI_ISL_2274538 | WHITE PLAINS HOSPITAL CENTER LABORATORY | Wadsworth Center, New York State Department of Health | Kirsten St. George, Daryl M. Lamson, Alexis Russell, Matthew Shudt, Melissa A Leisner, Jonathan Plitnick, Catharine Prussing, Navjot Singh, John Kelly, Erasmus Schneider, Erica Lasek-Nesselquist |
| EPI_ISL_2274539, EPI_ISL_2274540, EPI_ISL_2274541, EPI_ISL_2274542, EPI_ISL_2274543 | WESTCHESTER MEDICAL CENTER | Wadsworth Center, New York State Department of Health | Kirsten St. George, Daryl M. Lamson, Alexis Russell, Matthew Shudt, Melissa A Leisner, Jonathan Plitnick, Catharine Prussing, Navjot Singh, John Kelly, Erasmus Schneider, Erica Lasek-Nesselquist |
| EPI_ISL_2274544 | URMC LABS | Wadsworth Center, New York State Department of Health | Kirsten St. George, Daryl M. Lamson, Alexis Russell, Matthew Shudt, Melissa A Leisner, Jonathan Plitnick, Catharine Prussing, Navjot Singh, John Kelly, Erasmus Schneider, Erica Lasek-Nesselquist |
| EPI_ISL_2274545 | WHITE PLAINS HOSPITAL CENTER LABORATORY | Wadsworth Center, New York State Department of Health | Kirsten St. George, Daryl M. Lamson, Alexis Russell, Matthew Shudt, Melissa A Leisner, Jonathan Plitnick, Catharine Prussing, Navjot Singh, John Kelly, Erasmus Schneider, Erica Lasek-Nesselquist |
| EPI_ISL_2274546 | URMC LABS | Wadsworth Center, New York State Department of Health | Kirsten St. George, Daryl M. Lamson, Alexis Russell, Matthew Shudt, Melissa A Leisner, Jonathan Plitnick, Catharine Prussing, Navjot Singh, John Kelly, Erasmus Schneider, Erica Lasek-Nesselquist |
| EPI_ISL_2274547, EPI_ISL_2274548, EPI_ISL_2274549, EPI_ISL_2274550, EPI_ISL_2274551, EPI_ISL_2274552, EPI_ISL_2274553, EPI_ISL_2274554, EPI_ISL_2274555 | WESTCHESTER MEDICAL CENTER | Wadsworth Center, New York State Department of Health | Kirsten St. George, Daryl M. Lamson, Alexis Russell, Matthew Shudt, Melissa A Leisner, Jonathan Plitnick, Catharine Prussing, Navjot Singh, John Kelly, Erasmus Schneider, Erica Lasek-Nesselquist |
| EPI_ISL_2274556, EPI_ISL_2274557 | Columbia University Irving Medical Center | Wadsworth Center, New York State Department of Health | Kirsten St. George, Daryl M. Lamson, Alexis Russell, Matthew Shudt, Melissa A Leisner, Jonathan Plitnick, Catharine Prussing, Navjot Singh, John Kelly, Erasmus Schneider, Erica Lasek-Nesselquist |
| EPI_ISL_2274558, EPI_ISL_2274559 | WESTCHESTER MEDICAL CENTER | Wadsworth Center, New York State Department of Health | Kirsten St. George, Daryl M. Lamson, Alexis Russell, Matthew Shudt, Melissa A Leisner, Jonathan Plitnick, Catharine Prussing, Navjot Singh, John Kelly, Erasmus Schneider, Erica Lasek-Nesselquist |

| | | | |
|---|---|---|---|
| EPI_ISL_2274560 | URMC LABS | Wadsworth Center, New York State Department of Health | Kirsten St. George, Daryl M. Lamson, Alexis Russell, Matthew Shudt, Melissa A Leisner, Jonathan Plitnick, Catharine Prussing, Navjot Singh, John Kelly, Erasmus Schneider, Erica Lasek-Nesselquist |
| EPI_ISL_2274561 | SARATOGA HOSPITAL LABORATORY | Wadsworth Center, New York State Department of Health | Kirsten St. George, Daryl M. Lamson, Alexis Russell, Matthew Shudt, Melissa A Leisner, Jonathan Plitnick, Catharine Prussing, Navjot Singh, John Kelly, Erasmus Schneider, Erica Lasek-Nesselquist |
| EPI_ISL_2274562, EPI_ISL_2274563 | URMC LABS | Wadsworth Center, New York State Department of Health | Kirsten St. George, Daryl M. Lamson, Alexis Russell, Matthew Shudt, Melissa A Leisner, Jonathan Plitnick, Catharine Prussing, Navjot Singh, John Kelly, Erasmus Schneider, Erica Lasek-Nesselquist |
| EPI_ISL_2274564, EPI_ISL_2274565 | WESTCHESTER MEDICAL CENTER | Wadsworth Center, New York State Department of Health | Kirsten St. George, Daryl M. Lamson, Alexis Russell, Matthew Shudt, Melissa A Leisner, Jonathan Plitnick, Catharine Prussing, Navjot Singh, John Kelly, Erasmus Schneider, Erica Lasek-Nesselquist |
| EPI_ISL_2274566, EPI_ISL_2274567 | Columbia University Irving Medical Center | Wadsworth Center, New York State Department of Health | Kirsten St. George, Daryl M. Lamson, Alexis Russell, Matthew Shudt, Melissa A Leisner, Jonathan Plitnick, Catharine Prussing, Navjot Singh, John Kelly, Erasmus Schneider, Erica Lasek-Nesselquist |
| EPI_ISL_2274568, EPI_ISL_2274569 | URMC LABS | Wadsworth Center, New York State Department of Health | Kirsten St. George, Daryl M. Lamson, Alexis Russell, Matthew Shudt, Melissa A Leisner, Jonathan Plitnick, Catharine Prussing, Navjot Singh, John Kelly, Erasmus Schneider, Erica Lasek-Nesselquist |
| EPI_ISL_2274570 | SUNY UPSTATE MEDICAL UNIVERSITY | Wadsworth Center, New York State Department of Health | Kirsten St. George, Daryl M. Lamson, Alexis Russell, Matthew Shudt, Melissa A Leisner, Jonathan Plitnick, Catharine Prussing, Navjot Singh, John Kelly, Erasmus Schneider, Erica Lasek-Nesselquist |
| EPI_ISL_2274571, EPI_ISL_2274572 | NORTHWELL HEALTH LABORATORIES | Wadsworth Center, New York State Department of Health | Kirsten St. George, Daryl M. Lamson, Alexis Russell, Matthew Shudt, Melissa A Leisner, Jonathan Plitnick, Catharine Prussing, Navjot Singh, John Kelly, Erasmus Schneider, Erica Lasek-Nesselquist |
| EPI_ISL_2274574 | URMC LABS | Wadsworth Center, New York State Department of Health | Kirsten St. George, Daryl M. Lamson, Alexis Russell, Matthew Shudt, Melissa A Leisner, Jonathan Plitnick, Catharine Prussing, Navjot Singh, John Kelly, Erasmus Schneider, Erica Lasek-Nesselquist |
| EPI_ISL_2274575 | SUNY UPSTATE MEDICAL UNIVERSITY | Wadsworth Center, New York State Department of Health | Kirsten St. George, Daryl M. Lamson, Alexis Russell, Matthew Shudt, Melissa A Leisner, Jonathan Plitnick, Catharine Prussing, Navjot Singh, John Kelly, Erasmus Schneider, Erica Lasek-Nesselquist |
| EPI_ISL_2274576, EPI_ISL_2274577, EPI_ISL_2274578, EPI_ISL_2274579, EPI_ISL_2274580, EPI_ISL_2274581, EPI_ISL_2274582, EPI_ISL_2274583, EPI_ISL_2274584, EPI_ISL_2274585 | STONY BROOK UNIVERSITY HOSPITAL | Wadsworth Center, New York State Department of Health | Kirsten St. George, Daryl M. Lamson, Alexis Russell, Matthew Shudt, Melissa A Leisner, Jonathan Plitnick, Catharine Prussing, Navjot Singh, John Kelly, Erasmus Schneider, Erica Lasek-Nesselquist |
| EPI_ISL_2274586 | OSWEGO HOSPITAL LABORATORY | Wadsworth Center, New York State Department of Health | Kirsten St. George, Daryl M. Lamson, Alexis Russell, Matthew Shudt, Melissa A Leisner, Jonathan Plitnick, Catharine Prussing, Navjot Singh, John Kelly, Erasmus Schneider, Erica Lasek-Nesselquist |
| EPI_ISL_2274587, EPI_ISL_2274588, EPI_ISL_2274589, EPI_ISL_2274590, EPI_ISL_2274591, EPI_ISL_2274592, EPI_ISL_2274593, EPI_ISL_2274594, EPI_ISL_2274595, EPI_ISL_2274596, EPI_ISL_2274597, EPI_ISL_2274598, EPI_ISL_2274599, EPI_ISL_2274600, EPI_ISL_2274601, EPI_ISL_2274602, EPI_ISL_2274603, EPI_ISL_2274604, EPI_ISL_2274605, EPI_ISL_2274606 | | | |
| see above | NYC Pandemic Response Lab | Wadsworth Center, New York State Department of Health | Kirsten St. George, Daryl M. Lamson, Alexis Russell, Matthew Shudt, Melissa A Leisner, Jonathan Plitnick, Catharine Prussing, Navjot Singh, John Kelly, Erasmus Schneider, Erica Lasek-Nesselquist |
| EPI_ISL_2274607, EPI_ISL_2274608, EPI_ISL_2274609, EPI_ISL_2274610, EPI_ISL_2274611, EPI_ISL_2274612, EPI_ISL_2274613, EPI_ISL_2274614, EPI_ISL_2274615, EPI_ISL_2274616, EPI_ISL_2274617 | | | |
| see above | MONTEFIORE MEDICAL CENTER LABORATORIES | Wadsworth Center, New York State Department of Health | Kirsten St. George, Daryl M. Lamson, Alexis Russell, Matthew Shudt, Melissa A Leisner, Jonathan Plitnick, Catharine Prussing, Navjot Singh, John Kelly, Erasmus Schneider, Erica Lasek-Nesselquist |
| EPI_ISL_2274618, EPI_ISL_2274619, EPI_ISL_2274620, EPI_ISL_2274621, EPI_ISL_2274622, EPI_ISL_2274623, EPI_ISL_2274624, EPI_ISL_2274625, EPI_ISL_2274626, EPI_ISL_2274627, EPI_ISL_2274628, EPI_ISL_2274629, EPI_ISL_2274630 | | | |
| see above | ALBANY MEDICAL CENTER | Wadsworth Center, New York State Department of Health | Kirsten St. George, Daryl M. Lamson, Alexis Russell, Matthew Shudt, Melissa A Leisner, Jonathan Plitnick, Catharine Prussing, Navjot Singh, John Kelly, Erasmus Schneider, Erica Lasek-Nesselquist |
| EPI_ISL_2274631, EPI_ISL_2274632, EPI_ISL_2274633, EPI_ISL_2274634, EPI_ISL_2274635, EPI_ISL_2274636, EPI_ISL_2274637, EPI_ISL_2274638, EPI_ISL_2274639, EPI_ISL_2274640, EPI_ISL_2274641, EPI_ISL_2274642, EPI_ISL_2274643, EPI_ISL_2274644, EPI_ISL_2274645, EPI_ISL_2274646 | | | |
| see above | URMC LABS | Wadsworth Center, New York State Department of Health | Kirsten St. George, Daryl M. Lamson, Alexis Russell, Matthew Shudt, Melissa A Leisner, Jonathan Plitnick, Catharine Prussing, Navjot Singh, John Kelly, Erasmus Schneider, Erica Lasek-Nesselquist |
| EPI_ISL_2274647, EPI_ISL_2274648, EPI_ISL_2274649, EPI_ISL_2274650, EPI_ISL_2274651, EPI_ISL_2274652, EPI_ISL_2274653, EPI_ISL_2274654 | WESTCHESTER MEDICAL CENTER | Wadsworth Center, New York State Department of Health | Kirsten St. George, Daryl M. Lamson, Alexis Russell, Matthew Shudt, Melissa A Leisner, Jonathan Plitnick, Catharine Prussing, Navjot Singh, John Kelly, Erasmus Schneider, Erica Lasek-Nesselquist |
| EPI_ISL_2274657, EPI_ISL_2274658, EPI_ISL_2274659, EPI_ISL_2274660, EPI_ISL_2274661, EPI_ISL_2274662, EPI_ISL_2274663 | SUNY UPSTATE MEDICAL UNIVERSITY | Wadsworth Center, New York State Department of Health | Kirsten St. George, Daryl M. Lamson, Alexis Russell, Matthew Shudt, Melissa A Leisner, Jonathan Plitnick, Catharine Prussing, Navjot Singh, John Kelly, Erasmus Schneider, Erica Lasek-Nesselquist |
| EPI_ISL_2274664 | ALBANY MEDICAL CENTER | Wadsworth Center, New York State Department of Health | Kirsten St. George, Daryl M. Lamson, Alexis Russell, Matthew Shudt, Melissa A Leisner, Jonathan Plitnick, Catharine Prussing, Navjot Singh, John Kelly, Erasmus Schneider, Erica Lasek-Nesselquist |
| EPI_ISL_2274665, EPI_ISL_2274666, EPI_ISL_2274667, EPI_ISL_2274668, EPI_ISL_2274669, EPI_ISL_2274670, EPI_ISL_2274671, EPI_ISL_2274672, EPI_ISL_2274673, EPI_ISL_2274674, EPI_ISL_2274675 | | | |
| see above | URMC LABS | Wadsworth Center, New York State Department of Health | Kirsten St. George, Daryl M. Lamson, Alexis Russell, Matthew Shudt, Melissa A Leisner, Jonathan Plitnick, Catharine Prussing, Navjot Singh, John Kelly, Erasmus Schneider, Erica Lasek-Nesselquist |
| EPI_ISL_2274676, EPI_ISL_2274677, EPI_ISL_2274678, EPI_ISL_2274679, EPI_ISL_2274680, EPI_ISL_2274681, EPI_ISL_2274682, EPI_ISL_2274683, EPI_ISL_2274684, EPI_ISL_2274685, EPI_ISL_2274686, EPI_ISL_2274687, EPI_ISL_2274688, EPI_ISL_2274689, EPI_ISL_2274690, EPI_ISL_2274691, EPI_ISL_2274692, EPI_ISL_2274693, EPI_ISL_2274694, EPI_ISL_2274695, EPI_ISL_2274696, EPI_ISL_2274697, EPI_ISL_2274698, EPI_ISL_2274699, EPI_ISL_2274700, EPI_ISL_2274701, EPI_ISL_2274702, EPI_ISL_2274703, EPI_ISL_2274704, EPI_ISL_2274705, EPI_ISL_2274706, EPI_ISL_2274707, EPI_ISL_2274708, EPI_ISL_2274709, EPI_ISL_2274710, EPI_ISL_2274711, EPI_ISL_2274712, EPI_ISL_2274713, EPI_ISL_2274714 | | | |
| see above | NYC Pandemic Response Lab | Wadsworth Center, New York State Department of Health | Kirsten St. George, Daryl M. Lamson, Alexis Russell, Matthew Shudt, Melissa A Leisner, Jonathan Plitnick, Catharine Prussing, Navjot Singh, John Kelly, Erasmus Schneider, Erica Lasek-Nesselquist |
| EPI_ISL_2276081 | Southern Nevada Public Health Laboratory | Southern Nevada Public Health Laboratory | Michael Picker |
| EPI_ISL_2276083, EPI_ISL_2276084, EPI_ISL_2276085, EPI_ISL_2276086, EPI_ISL_2276087, EPI_ISL_2276088, EPI_ISL_2276089, EPI_ISL_2276091, EPI_ISL_2276092, EPI_ISL_2276093 | Wichita State University - Molecular Diagnostics Lab | Kansas Health and Environmental Lab | Mike Grose, Katherine Wiggins, Jonathan Barnell, Ben Olsen, and Phil Adam |
| EPI_ISL_2276094, EPI_ISL_2276095, EPI_ISL_2276096, EPI_ISL_2276097, EPI_ISL_2276098, EPI_ISL_2276099, EPI_ISL_2276100 | Kansas Health and Environmental Lab | Kansas Health and Environmental Lab | Mike Grose, Katherine Wiggins, Jonathan Barnell, Ben Olsen, and Phil Adam |
| EPI_ISL_2276102 | City of Milwaukee Health Department Laboratory | City of Milwaukee Health Department Laboratory | Sanjib Bhattacharyya, Manjeet Khubbar, Amy Bauer, Jennifer Lentz, Samantha Scott |
| EPI_ISL_2276104, EPI_ISL_2276105 | Instituto Nacional de Medicina Genomica | Instituto Nacional de Medicina Genomica | Hidalgo-Miranda A, Cedro-Tanda A, Mendoza-Vargas A, Reyes-Grajeda JP, Cisneros- Villanueva M, Orjuela-Rodríguez M, Gonzalez-Barrera D, Rangel-DeLeon D, Munguia-Garza P, Ramirez-Vega O, Escobar-Arrazola, M, Herrera-Montalvo LA. |
| EPI_ISL_2276106 | InDRE | Instituto Nacional de Medicina Genomica | Hidalgo-Miranda A, Mendoza-Vargas A, Reyes-Grajeda JP, Cedro-Tanda A, Cisneros-Villanueva M, Orjuela-Rodríguez M, Gisela Barrera-Badillo, Irma Lopez-Martinez, Jose Ernesto Ramirez González, Gonzalez-Barrera D, Rangel-DeLeon D, Munguia-Garza P, Garcia-Cardenas FJ, Gonzalez-Woge MA, Herrera-Montalvo LA. |
| EPI_ISL_2276107 | Salud Digna | Instituto Nacional de Medicina Genomica | Hidalgo-Miranda A, Cedro-Tanda A, Mendoza-Vargas A, Reyes-Grajeda JP, Cisneros-Villanueva M, Orjuela-Rodríguez M, Abraham Campos-Romero, Moreno-Camacho José Luis, Rodriguez-Gallegos Jorge, Luna-Ruiz Marco, Gonzalez-Barrera D, Rangel-DeLeon D, Munguia-Garza P, Ramirez-Vega O, Escobar-Arrazola, M, Herrera-Montalvo LA. |

| | | | |
|---|---|---|---|
| EPI_ISL_2276108 | Instituto Nacional de Medicina Genomica | Instituto Nacional de Medicina Genomica | Hidalgo-Miranda A, Cedro-Tanda A, Mendoza-Vargas A, Reyes-Grajeda JP, Cisneros- Villanueva M, Orjuela-Rodríguez M, Gonzalez-Barrera D, Rangel-DeLeon D, Munguia-Garza P, Ramirez-Vega O, Escobar-Arrazola, M, Herrera-Montalvo LA. |
| EPI_ISL_2276109, EPI_ISL_2276110 | Salud Digna | Instituto Nacional de Medicina Genomica | Hidalgo-Miranda A, Cedro-Tanda A, Mendoza-Vargas A, Reyes-Grajeda JP, Cisneros-Villanueva M, Orjuela-Rodríguez M, Abraham Campos-Romero, Moreno-Camacho José Luis, Rodriguez-Gallegos Jorge, Luna-Ruiz Marco, Gonzalez-Barrera D, Rangel-DeLeon D, Munguia-Garza P, Ramirez-Vega O, Escobar-Arrazola, M, Herrera-Montalvo LA. |
| EPI_ISL_2276120, EPI_ISL_2276121, EPI_ISL_2276122, EPI_ISL_2276123, EPI_ISL_2276130 | City of Milwaukee Health Department Laboratory | City of Milwaukee Health Department Laboratory | Sanjib Bhattacharyya, Manjeet Khubbar, Amy Bauer, Jennifer Lentz, Samantha Scott |
| EPI_ISL_2276140, EPI_ISL_2276141, EPI_ISL_2276142, EPI_ISL_2276143, EPI_ISL_2276162, EPI_ISL_2276191, EPI_ISL_2276192, EPI_ISL_2276194, EPI_ISL_2276195, EPI_ISL_2276196, EPI_ISL_2276197, EPI_ISL_2276198, EPI_ISL_2276199, EPI_ISL_2276201, EPI_ISL_2276202, EPI_ISL_2276203, EPI_ISL_2276205, EPI_ISL_2276207, EPI_ISL_2276208, EPI_ISL_2276211, EPI_ISL_2276214, EPI_ISL_2276215, EPI_ISL_2276216, EPI_ISL_2276219, EPI_ISL_2276222, EPI_ISL_2276223, EPI_ISL_2276225, EPI_ISL_2276230, EPI_ISL_2276231, EPI_ISL_2276232, EPI_ISL_2276234, EPI_ISL_2276236, EPI_ISL_2276237, EPI_ISL_2276238, EPI_ISL_2276239, EPI_ISL_2276241, EPI_ISL_2276242, EPI_ISL_2276244, EPI_ISL_2276245, EPI_ISL_2276246, EPI_ISL_2276248, EPI_ISL_2276249, EPI_ISL_2276251, EPI_ISL_2276252, EPI_ISL_2276253, EPI_ISL_2276255, EPI_ISL_2276260, EPI_ISL_2276261, EPI_ISL_2276262, EPI_ISL_2276263, EPI_ISL_2276264, EPI_ISL_2276265, EPI_ISL_2276266, EPI_ISL_2276267, EPI_ISL_2276268, EPI_ISL_2276269, EPI_ISL_2276270, EPI_ISL_2276271, EPI_ISL_2276272, EPI_ISL_2276274, EPI_ISL_2276275, EPI_ISL_2276276, EPI_ISL_2276277, EPI_ISL_2276278, EPI_ISL_2276279, EPI_ISL_2276280, EPI_ISL_2276281, EPI_ISL_2276283, EPI_ISL_2276285, EPI_ISL_2276286, EPI_ISL_2276287, EPI_ISL_2276295, EPI_ISL_2276299, EPI_ISL_2276300, EPI_ISL_2276301, EPI_ISL_2276302, EPI_ISL_2276303, EPI_ISL_2276305, EPI_ISL_2276306, EPI_ISL_2276307, EPI_ISL_2276308, EPI_ISL_2276309, EPI_ISL_2276311, EPI_ISL_2276312, EPI_ISL_2276314, EPI_ISL_2276317, EPI_ISL_2276320, EPI_ISL_2276321, EPI_ISL_2276322, EPI_ISL_2276323, EPI_ISL_2276325, EPI_ISL_2276326, EPI_ISL_2276327, EPI_ISL_2276328, EPI_ISL_2276329, EPI_ISL_2276331, EPI_ISL_2276333, EPI_ISL_2276334, EPI_ISL_2276341, EPI_ISL_2276342, EPI_ISL_2276386, EPI_ISL_2276388, EPI_ISL_2276389, EPI_ISL_2276390, EPI_ISL_2276391, EPI_ISL_2276392, EPI_ISL_2276393, EPI_ISL_2276394, EPI_ISL_2276395, EPI_ISL_2276396, EPI_ISL_2276397, EPI_ISL_2276398, EPI_ISL_2276399, EPI_ISL_2276401, EPI_ISL_2276402, EPI_ISL_2276414, EPI_ISL_2276422, EPI_ISL_2276431, EPI_ISL_2276434, EPI_ISL_2276443, EPI_ISL_2276444, EPI_ISL_2276447, EPI_ISL_2276472, EPI_ISL_2276473, EPI_ISL_2276475, EPI_ISL_2276476, EPI_ISL_2276478, EPI_ISL_2276479, EPI_ISL_2276482, EPI_ISL_2276484, EPI_ISL_2276486, EPI_ISL_2276487, EPI_ISL_2276490, EPI_ISL_2276492, EPI_ISL_2276493, EPI_ISL_2276494, EPI_ISL_2276496, EPI_ISL_2276498, EPI_ISL_2276499, EPI_ISL_2276502, EPI_ISL_2276506, EPI_ISL_2276507, EPI_ISL_2276515, EPI_ISL_2276517, EPI_ISL_2276519, EPI_ISL_2276524, EPI_ISL_2276529, EPI_ISL_2276532, EPI_ISL_2276536, EPI_ISL_2276538, EPI_ISL_2276539, EPI_ISL_2276540, EPI_ISL_2276546, EPI_ISL_2276547, EPI_ISL_2276548, EPI_ISL_2276549, EPI_ISL_2276551, EPI_ISL_2276554, EPI_ISL_2276556, EPI_ISL_2276561, EPI_ISL_2276563, EPI_ISL_2276564, EPI_ISL_2276565, EPI_ISL_2276568, EPI_ISL_2276569, EPI_ISL_2276571, EPI_ISL_2276572, EPI_ISL_2276573, EPI_ISL_2276574, EPI_ISL_2276577, EPI_ISL_2276579, EPI_ISL_2276581, EPI_ISL_2276582, EPI_ISL_2276586, EPI_ISL_2276588, EPI_ISL_2276589, EPI_ISL_2276590, EPI_ISL_2276593, EPI_ISL_2276594, EPI_ISL_2276597, EPI_ISL_2276598, EPI_ISL_2276599, EPI_ISL_2276602, EPI_ISL_2276603, EPI_ISL_2276604, EPI_ISL_2276607, EPI_ISL_2276608, EPI_ISL_2276609, EPI_ISL_2276610, EPI_ISL_2276611, EPI_ISL_2276612, EPI_ISL_2276613, EPI_ISL_2276614, EPI_ISL_2276615, EPI_ISL_2276617, EPI_ISL_2276618, EPI_ISL_2276620, EPI_ISL_2276622, EPI_ISL_2276623, EPI_ISL_2276624, EPI_ISL_2276625, EPI_ISL_2276626, EPI_ISL_2276628, EPI_ISL_2276629, EPI_ISL_2276630, EPI_ISL_2276631, EPI_ISL_2276632, EPI_ISL_2276633, EPI_ISL_2276634, EPI_ISL_2276635, EPI_ISL_2276636, EPI_ISL_2276640, EPI_ISL_2276641, EPI_ISL_2276643, EPI_ISL_2276655, EPI_ISL_2276656, EPI_ISL_2276657, EPI_ISL_2276659, EPI_ISL_2276660, EPI_ISL_2276661, EPI_ISL_2276663, EPI_ISL_2276664, EPI_ISL_2276665, EPI_ISL_2276667, EPI_ISL_2276668, EPI_ISL_2276669, EPI_ISL_2276670, EPI_ISL_2276672, EPI_ISL_2276674, EPI_ISL_2276675, EPI_ISL_2276676, EPI_ISL_2276677, EPI_ISL_2276678, EPI_ISL_2276683, EPI_ISL_2276684, EPI_ISL_2276685, EPI_ISL_2276686, EPI_ISL_2276687, EPI_ISL_2276688, EPI_ISL_2276689, EPI_ISL_2276691, EPI_ISL_2276693, EPI_ISL_2276694, EPI_ISL_2276695, EPI_ISL_2276696, EPI_ISL_2276698, EPI_ISL_2276699, EPI_ISL_2276700, EPI_ISL_2276701, EPI_ISL_2276702, EPI_ISL_2276704, EPI_ISL_2276709, EPI_ISL_2276711, EPI_ISL_2276712, EPI_ISL_2276713, EPI_ISL_2276715, EPI_ISL_2276716, EPI_ISL_2276717, EPI_ISL_2276718, EPI_ISL_2276719, EPI_ISL_2276720, EPI_ISL_2276721, EPI_ISL_2276722, EPI_ISL_2276723, EPI_ISL_2276724, EPI_ISL_2276726, EPI_ISL_2276728, EPI_ISL_2276732, EPI_ISL_2276733, EPI_ISL_2276734, EPI_ISL_2276735, EPI_ISL_2276736, EPI_ISL_2276737, EPI_ISL_2276739, EPI_ISL_2276740, EPI_ISL_2276741, EPI_ISL_2276744, EPI_ISL_2276746, EPI_ISL_2276747, EPI_ISL_2276748, EPI_ISL_2276749, EPI_ISL_2276751, EPI_ISL_2276752, EPI_ISL_2276753, EPI_ISL_2276755, EPI_ISL_2276756, EPI_ISL_2276757, EPI_ISL_2276758, EPI_ISL_2276759, EPI_ISL_2276764, EPI_ISL_2276765, EPI_ISL_2276767, EPI_ISL_2276773, EPI_ISL_2276774, EPI_ISL_2276776, EPI_ISL_2276780, EPI_ISL_2276782, EPI_ISL_2276783, EPI_ISL_2276787, EPI_ISL_2276788, EPI_ISL_2276789, EPI_ISL_2276791, EPI_ISL_2276792, EPI_ISL_2276793, EPI_ISL_2276794, EPI_ISL_2276795, EPI_ISL_2276796, EPI_ISL_2276797, EPI_ISL_2276798, EPI_ISL_2276801, EPI_ISL_2276804, EPI_ISL_2276805, EPI_ISL_2276806, EPI_ISL_2276811, EPI_ISL_2276812, EPI_ISL_2276815, EPI_ISL_2276817, EPI_ISL_2276819, EPI_ISL_2276820, EPI_ISL_2276822, EPI_ISL_2276823, EPI_ISL_2276826, EPI_ISL_2276832, EPI_ISL_2276834, EPI_ISL_2276835, EPI_ISL_2276836, EPI_ISL_2276839, EPI_ISL_2276841, EPI_ISL_2276842, EPI_ISL_2276843, EPI_ISL_2276844, EPI_ISL_2276845, EPI_ISL_2276847, EPI_ISL_2276848, EPI_ISL_2276849, EPI_ISL_2276850, EPI_ISL_2276852, EPI_ISL_2276854, EPI_ISL_2276855, EPI_ISL_2276856, EPI_ISL_2276857, EPI_ISL_2276858, EPI_ISL_2276859, EPI_ISL_2276860, EPI_ISL_2276861, EPI_ISL_2276862, EPI_ISL_2276863, EPI_ISL_2276864, EPI_ISL_2276867, EPI_ISL_2276868, EPI_ISL_2276870, EPI_ISL_2276873, EPI_ISL_2276874, EPI_ISL_2276875, EPI_ISL_2276876, EPI_ISL_2276877, EPI_ISL_2276882, EPI_ISL_2276884, EPI_ISL_2276887, EPI_ISL_2276888, EPI_ISL_2276889, EPI_ISL_2276890, EPI_ISL_2276892, EPI_ISL_2276896, EPI_ISL_2276897, EPI_ISL_2276899, EPI_ISL_2276901, EPI_ISL_2276903, EPI_ISL_2276904, EPI_ISL_2276905, EPI_ISL_2276906, EPI_ISL_2276907, EPI_ISL_2276908, EPI_ISL_2276909, EPI_ISL_2276911, EPI_ISL_2276914, EPI_ISL_2276929, EPI_ISL_2276932, EPI_ISL_2276934, EPI_ISL_2276938, EPI_ISL_2276939, EPI_ISL_2276944, EPI_ISL_2276946, EPI_ISL_2276948, EPI_ISL_2276950, EPI_ISL_2276953, EPI_ISL_2276954, EPI_ISL_2276956, EPI_ISL_2276959, EPI_ISL_2276962, EPI_ISL_2276964, EPI_ISL_2276965, EPI_ISL_2276967, EPI_ISL_2276968, EPI_ISL_2276970, EPI_ISL_2276971, EPI_ISL_2276973, EPI_ISL_2276974, EPI_ISL_2276976, EPI_ISL_2276978, EPI_ISL_2276980, EPI_ISL_2276981, EPI_ISL_2276983, EPI_ISL_2276984, EPI_ISL_2276986, EPI_ISL_2276987, EPI_ISL_2276988, EPI_ISL_2276990, EPI_ISL_2276991, EPI_ISL_2276993, EPI_ISL_2276994, EPI_ISL_2276995, EPI_ISL_2276996, EPI_ISL_2276999, EPI_ISL_2277000, EPI_ISL_2277001, EPI_ISL_2277002, EPI_ISL_2277005, EPI_ISL_2277006, EPI_ISL_2277008, EPI_ISL_2277009, EPI_ISL_2277012, EPI_ISL_2277013, EPI_ISL_2277015, EPI_ISL_2277016, EPI_ISL_2277018, EPI_ISL_2277023, EPI_ISL_2277024, EPI_ISL_2277025, EPI_ISL_2277026, EPI_ISL_2277028, EPI_ISL_2277029, EPI_ISL_2277030, EPI_ISL_2277031, EPI_ISL_2277033, EPI_ISL_2277034, EPI_ISL_2277035, EPI_ISL_2277036, EPI_ISL_2277040, EPI_ISL_2277043, EPI_ISL_2277045, EPI_ISL_2277046, EPI_ISL_2277047, EPI_ISL_2277048, EPI_ISL_2277049, EPI_ISL_2277050, EPI_ISL_2277051, EPI_ISL_2277052, EPI_ISL_2277055, EPI_ISL_2277057, EPI_ISL_2277058, EPI_ISL_2277060, EPI_ISL_2277063, EPI_ISL_2277064, EPI_ISL_2277067, EPI_ISL_2277069, EPI_ISL_2277073, EPI_ISL_2277081, EPI_ISL_2277084, EPI_ISL_2277087, EPI_ISL_2277089, EPI_ISL_2277090, EPI_ISL_2277093, EPI_ISL_2277094, EPI_ISL_2277096, EPI_ISL_2277097, EPI_ISL_2277098, EPI_ISL_2277099, EPI_ISL_2277100, EPI_ISL_2277101, EPI_ISL_2277102, EPI_ISL_2277103, EPI_ISL_2277104, EPI_ISL_2277105, EPI_ISL_2277106, EPI_ISL_2277116, EPI_ISL_2277126, EPI_ISL_2277129, EPI_ISL_2277135, EPI_ISL_2277136, EPI_ISL_2277139, EPI_ISL_2277142, EPI_ISL_2277145, EPI_ISL_2277146, EPI_ISL_2277148, EPI_ISL_2277151, EPI_ISL_2277153, EPI_ISL_2277154, EPI_ISL_2277156, EPI_ISL_2277157, EPI_ISL_2277171, EPI_ISL_2277172, EPI_ISL_2277173, EPI_ISL_2277174, EPI_ISL_2277175, EPI_ISL_2277176, EPI_ISL_2277177, EPI_ISL_2277181, EPI_ISL_2277185, EPI_ISL_2277189, EPI_ISL_2277190, EPI_ISL_2277191, EPI_ISL_2277194, EPI_ISL_2277197, EPI_ISL_2277199, EPI_ISL_2277200, EPI_ISL_2277201, EPI_ISL_2277202, EPI_ISL_2277203, EPI_ISL_2277205, EPI_ISL_2277209, EPI_ISL_2277210, EPI_ISL_2277211, EPI_ISL_2277212, EPI_ISL_2277215, EPI_ISL_2277217, EPI_ISL_2277219, EPI_ISL_2277220, EPI_ISL_2277221, EPI_ISL_2277226, EPI_ISL_2277230, EPI_ISL_2277233, EPI_ISL_2277234, EPI_ISL_2277236, EPI_ISL_2277237, EPI_ISL_2277239, EPI_ISL_2277240, EPI_ISL_2277241, EPI_ISL_2277242, EPI_ISL_2277244, EPI_ISL_2277245, EPI_ISL_2277249, EPI_ISL_2277251, EPI_ISL_2277252, EPI_ISL_2277253, EPI_ISL_2277255, EPI_ISL_2277257, EPI_ISL_2277258, EPI_ISL_2277259, EPI_ISL_2277260, EPI_ISL_2277264, EPI_ISL_2277265, EPI_ISL_2277267, EPI_ISL_2277268, EPI_ISL_2277269, EPI_ISL_2277271, EPI_ISL_2277274, EPI_ISL_2277277, EPI_ISL_2277278, EPI_ISL_2277279, EPI_ISL_2277280, EPI_ISL_2277281, EPI_ISL_2277283, EPI_ISL_2277284, EPI_ISL_2277285, EPI_ISL_2277286, EPI_ISL_2277289, EPI_ISL_2277290, EPI_ISL_2277291, EPI_ISL_2277293, EPI_ISL_2277294, EPI_ISL_2277295, EPI_ISL_2277296, EPI_ISL_2277299, EPI_ISL_2277300, EPI_ISL_2277301, EPI_ISL_2277303, EPI_ISL_2277304, EPI_ISL_2277305, EPI_ISL_2277307, EPI_ISL_2277308, EPI_ISL_2277309, EPI_ISL_2277313, EPI_ISL_2277316, EPI_ISL_2277317, EPI_ISL_2277318, EPI_ISL_2277321, EPI_ISL_2277322, EPI_ISL_2277323, EPI_ISL_2277324, EPI_ISL_2277325, EPI_ISL_2277326, EPI_ISL_2277327, EPI_ISL_2277328, EPI_ISL_2277329, EPI_ISL_2277336, EPI_ISL_2277337, EPI_ISL_2277338, EPI_ISL_2277339, EPI_ISL_2277340, EPI_ISL_2277341, EPI_ISL_2277342, EPI_ISL_2277343, EPI_ISL_2277344, EPI_ISL_2277345, EPI_ISL_2277350, EPI_ISL_2277353, EPI_ISL_2277355, EPI_ISL_2277356, EPI_ISL_2277357, EPI_ISL_2277358, EPI_ISL_2277359, EPI_ISL_2277360, EPI_ISL_2277361, EPI_ISL_2277362, EPI_ISL_2277363, EPI_ISL_2277364, EPI_ISL_2277365, EPI_ISL_2277366, EPI_ISL_2277367, EPI_ISL_2277368, EPI_ISL_2277369, EPI_ISL_2277371, EPI_ISL_2277373, EPI_ISL_2277374, EPI_ISL_2277375, EPI_ISL_2277376, EPI_ISL_2277377, EPI_ISL_2277379, EPI_ISL_2277381, EPI_ISL_2277383, EPI_ISL_2277384, EPI_ISL_2277385, EPI_ISL_2277386, EPI_ISL_2277398, EPI_ISL_2277400, EPI_ISL_2277401, EPI_ISL_2277404, EPI_ISL_2277407, EPI_ISL_2277408, EPI_ISL_2277410, EPI_ISL_2277411, EPI_ISL_2277416, EPI_ISL_2277417, EPI_ISL_2277418, EPI_ISL_2277419, EPI_ISL_2277420, EPI_ISL_2277421, EPI_ISL_2277422, EPI_ISL_2277423, EPI_ISL_2277424, EPI_ISL_2277425, EPI_ISL_2277426, EPI_ISL_2277427, EPI_ISL_2277428, EPI_ISL_2277430, EPI_ISL_2277432, EPI_ISL_2277435, EPI_ISL_2277437, EPI_ISL_2277438, EPI_ISL_2277441, EPI_ISL_2277450, EPI_ISL_2277471, EPI_ISL_2277475, EPI_ISL_2277477, EPI_ISL_2277480, EPI_ISL_2277493, EPI_ISL_2277494, EPI_ISL_2277496, EPI_ISL_2277497, EPI_ISL_2277500, EPI_ISL_2277505, EPI_ISL_2277506, EPI_ISL_2277507, EPI_ISL_2277509, EPI_ISL_2277511, EPI_ISL_2277514, EPI_ISL_2277518, EPI_ISL_2277519, EPI_ISL_2277520, EPI_ISL_2277521, EPI_ISL_2277522, EPI_ISL_2277526, EPI_ISL_2277528, EPI_ISL_2277531, EPI_ISL_2277532, EPI_ISL_2277533, EPI_ISL_2277534, EPI_ISL_2277543, EPI_ISL_2277544, EPI_ISL_2277545, EPI_ISL_2277546, EPI_ISL_2277547, EPI_ISL_2277552, EPI_ISL_2277553, EPI_ISL_2277556, EPI_ISL_2277557, EPI_ISL_2277558, EPI_ISL_2277559, EPI_ISL_2277560, EPI_ISL_2277561, EPI_ISL_2277562, EPI_ISL_2277564, EPI_ISL_2277565, EPI_ISL_2277567, EPI_ISL_2277568, EPI_ISL_2277570, EPI_ISL_2277571, EPI_ISL_2277572, EPI_ISL_2277574, EPI_ISL_2277575, EPI_ISL_2277576, EPI_ISL_2277577, EPI_ISL_2277578, EPI_ISL_2277579, EPI_ISL_2277580, EPI_ISL_2277581, EPI_ISL_2277582, EPI_ISL_2277584, EPI_ISL_2277585, EPI_ISL_2277587, EPI_ISL_2277588, EPI_ISL_2277591, EPI_ISL_2277592, EPI_ISL_2277593, EPI_ISL_2277594, EPI_ISL_2277595, EPI_ISL_2277597, EPI_ISL_2277600, EPI_ISL_2277601, EPI_ISL_2277602, EPI_ISL_2277603, EPI_ISL_2277604, EPI_ISL_2277607, EPI_ISL_2277608, EPI_ISL_2277614, EPI_ISL_2277619 | | | |
| see above | Colorado Department of Public Health and Environment | Colorado Department of Public Health and Environment | Laura Bankers, Molly C. Hetherington-Rauth, Diana Ir, Alexandria Rossheim, Shannon R. Matzinger, Sarah Elizabeth Totten, Emily A. Travanty |
| EPI_ISL_2277634, EPI_ISL_2277636, EPI_ISL_2277638 | Nevada State Public Health Laboratory | Nevada State Public Health Laboratory | Andrew Gorzalski, Mark Pandori |
| EPI_ISL_2277639, EPI_ISL_2277640, EPI_ISL_2277641 | Stony Brook Medicine | New York Genome Center | Michael Zody, Andre Corvelo, Dayna M. Oschwald, Samantha Fennessey, Tom Maniatis , Sarah A. Halbohn, Alaa B. Salaheldin, Sarah A. Malik, Josue M. Flores Acuna, Dimitri Gnatenko, Silvia G. Spitzer, Margaret M. McGovern, Eric D. Spitzer, Richard R. Kew, Kenneth R. Shroyer, Jiang Chen, Susan V. Donelan |
| EPI_ISL_2277642 | Hospital Sharp | Microbial Genomics Laboratory | Bruno Gomez-Gil, Julissa Enciso-Ibarra, Alejandra Garcia-Gasca, Daniel Fregoso-Rueda |
| EPI_ISL_2277645, EPI_ISL_2277646, EPI_ISL_2277647, EPI_ISL_2277648, EPI_ISL_2277649, EPI_ISL_2277650, EPI_ISL_2277651, EPI_ISL_2277652, EPI_ISL_2277653, EPI_ISL_2277654, EPI_ISL_2277656, EPI_ISL_2277658, EPI_ISL_2277660, EPI_ISL_2277662, EPI_ISL_2277664, EPI_ISL_2277666, EPI_ISL_2277667, EPI_ISL_2277669, EPI_ISL_2277671, EPI_ISL_2277673, EPI_ISL_2277675, EPI_ISL_2277677, EPI_ISL_2277679, EPI_ISL_2277681, EPI_ISL_2277683, EPI_ISL_2277685, EPI_ISL_2277687, EPI_ISL_2277689, EPI_ISL_2277691, EPI_ISL_2277692, EPI_ISL_2277694, EPI_ISL_2277696, EPI_ISL_2277698, EPI_ISL_2277700, EPI_ISL_2277702, EPI_ISL_2277704, EPI_ISL_2277706, EPI_ISL_2277708, EPI_ISL_2277710, EPI_ISL_2277713, EPI_ISL_2277714, EPI_ISL_2277716, EPI_ISL_2277718, EPI_ISL_2277720, EPI_ISL_2277722, EPI_ISL_2277724, EPI_ISL_2277726, EPI_ISL_2277728, EPI_ISL_2277730, EPI_ISL_2277732, EPI_ISL_2277733, EPI_ISL_2277734, EPI_ISL_2277735, EPI_ISL_2277736, EPI_ISL_2277737, EPI_ISL_2277738, EPI_ISL_2277739, EPI_ISL_2277740, EPI_ISL_2277741, EPI_ISL_2277742, EPI_ISL_2277743, EPI_ISL_2277744, EPI_ISL_2277745, EPI_ISL_2277746, EPI_ISL_2277747, EPI_ISL_2277748, EPI_ISL_2277749, EPI_ISL_2277750, EPI_ISL_2277751, EPI_ISL_2277752, EPI_ISL_2277753, EPI_ISL_2277754, EPI_ISL_2277755, EPI_ISL_2277756, EPI_ISL_2277757, EPI_ISL_2277758, EPI_ISL_2277759, EPI_ISL_2277760, EPI_ISL_2277761, EPI_ISL_2277762, EPI_ISL_2277763, EPI_ISL_2277764, EPI_ISL_2277765, EPI_ISL_2277766, EPI_ISL_2277767, EPI_ISL_2277768, EPI_ISL_2277769, EPI_ISL_2277770, EPI_ISL_2277771, EPI_ISL_2277772, EPI_ISL_2277773, EPI_ISL_2277774, EPI_ISL_2277775, EPI_ISL_2277776, EPI_ISL_2277777, EPI_ISL_2277778, EPI_ISL_2277779, EPI_ISL_2277780, EPI_ISL_2277781, EPI_ISL_2277782, EPI_ISL_2277783, EPI_ISL_2277784, EPI_ISL_2277785, EPI_ISL_2277786, EPI_ISL_2277787, EPI_ISL_2277788, EPI_ISL_2277789, EPI_ISL_2277790, EPI_ISL_2277791, EPI_ISL_2277792, EPI_ISL_2277793, EPI_ISL_2277794, EPI_ISL_2277795, EPI_ISL_2277796, EPI_ISL_2277797, EPI_ISL_2277798, EPI_ISL_2277799, EPI_ISL_2277800, EPI_ISL_2277801, EPI_ISL_2277802, EPI_ISL_2277803, EPI_ISL_2277804, EPI_ISL_2277805, EPI_ISL_2277806, EPI_ISL_2277807, EPI_ISL_2277808, EPI_ISL_2277809, EPI_ISL_2277810, EPI_ISL_2277811, EPI_ISL_2277812, EPI_ISL_2277813, EPI_ISL_2277814, EPI_ISL_2277815, EPI_ISL_2277816, EPI_ISL_2277817, EPI_ISL_2277818, EPI_ISL_2277819, EPI_ISL_2277820, EPI_ISL_2277821, EPI_ISL_2277822, EPI_ISL_2277823, EPI_ISL_2277824, EPI_ISL_2277825, EPI_ISL_2277826, EPI_ISL_2277827, EPI_ISL_2277828, EPI_ISL_2277829, EPI_ISL_2277830, EPI_ISL_2277831, EPI_ISL_2277832, EPI_ISL_2277833, EPI_ISL_2277834, EPI_ISL_2277835, EPI_ISL_2277836, EPI_ISL_2277837, EPI_ISL_2277838, EPI_ISL_2277839, EPI_ISL_2277840, EPI_ISL_2277841, EPI_ISL_2277842, EPI_ISL_2277843, EPI_ISL_2277844, EPI_ISL_2277845, EPI_ISL_2277846, EPI_ISL_2277847, EPI_ISL_2277848, EPI_ISL_2277849, EPI_ISL_2277850, EPI_ISL_2277851, EPI_ISL_2277852, EPI_ISL_2277853, EPI_ISL_2277854, EPI_ISL_2277855, EPI_ISL_2277856, EPI_ISL_2277857, EPI_ISL_2277858, EPI_ISL_2277859, EPI_ISL_2277860, EPI_ISL_2277861, EPI_ISL_2277862, EPI_ISL_2277863, EPI_ISL_2277864, EPI_ISL_2277865, EPI_ISL_2277866, EPI_ISL_2277867, EPI_ISL_2277868, EPI_ISL_2277869, EPI_ISL_2277870, EPI_ISL_2277871, EPI_ISL_2277872, EPI_ISL_2277873, EPI_ISL_2277874, EPI_ISL_2277875, EPI_ISL_2277876, EPI_ISL_2277877, EPI_ISL_2277878, EPI_ISL_2277879, EPI_ISL_2277880, EPI_ISL_2277881, EPI_ISL_2277882, EPI_ISL_2277883, EPI_ISL_2277884, EPI_ISL_2277885, EPI_ISL_2277886, EPI_ISL_2277887, EPI_ISL_2277888, EPI_ISL_2277889, EPI_ISL_2277890, EPI_ISL_2277891, EPI_ISL_2277892, EPI_ISL_2277893, EPI_ISL_2277894, EPI_ISL_2277895, EPI_ISL_2277896, EPI_ISL_2277897, EPI_ISL_2277898, EPI_ISL_2277899, EPI_ISL_2277900, EPI_ISL_2277901, EPI_ISL_2277902, EPI_ISL_2277903, EPI_ISL_2277904, EPI_ISL_2277905, EPI_ISL_2277906, EPI_ISL_2277907, EPI_ISL_2277955, EPI_ISL_2277956, EPI_ISL_2277957, EPI_ISL_2277958, EPI_ISL_2277959, EPI_ISL_2277960, EPI_ISL_2277961, EPI_ISL_2277962, EPI_ISL_2277963, EPI_ISL_2277964, EPI_ISL_2277965, EPI_ISL_2277966, EPI_ISL_2277967, EPI_ISL_2277968, EPI_ISL_2277969, EPI_ISL_2277970, EPI_ISL_2277971, EPI_ISL_2277972, EPI_ISL_2277973, EPI_ISL_2277974, EPI_ISL_2277975, EPI_ISL_2277976, EPI_ISL_2277977, EPI_ISL_2277978, EPI_ISL_2277979, EPI_ISL_2277980, EPI_ISL_2277981, EPI_ISL_2277982, EPI_ISL_2277983, EPI_ISL_2277984, EPI_ISL_2277985, EPI_ISL_2277986, EPI_ISL_2277987, EPI_ISL_2277988, EPI_ISL_2277989, EPI_ISL_2277990, EPI_ISL_2277991, EPI_ISL_2277992, EPI_ISL_2277993, EPI_ISL_2277994, EPI_ISL_2277995, EPI_ISL_2277996, EPI_ISL_2277997, EPI_ISL_2277998, EPI_ISL_2277999, EPI_ISL_2278000, EPI_ISL_2278001, EPI_ISL_2278002, EPI_ISL_2278003, EPI_ISL_2278004, EPI_ISL_2278005, EPI_ISL_2278006, EPI_ISL_2278007, EPI_ISL_2278008, EPI_ISL_2278009 | | | |
| see above | Florida Bureau of Public Health Laboratories | Florida Bureau of Public Health Laboratories | Sarah Schmedes, Jason Blanton |
| EPI_ISL_2278731 | MONTEFIORE MEDICAL CENTER LABORATORIES | Wadsworth Center, New York State Department of Health | Kirsten St. George, Daryl M. Lamson, Alexis Russell, Matthew Shudt, Melissa A Leisner, Jonathan Plitnick, Catharine Prussing, Navjot Singh, John Kelly, Erasmus Schneider, Erica Lasek-Nesselquist |
| EPI_ISL_2278732, EPI_ISL_2278733, EPI_ISL_2278734 | SUNY UPSTATE MEDICAL UNIVERSITY | Wadsworth Center, New York State Department of Health | Kirsten St. George, Daryl M. Lamson, Alexis Russell, Matthew Shudt, Melissa A Leisner, Jonathan Plitnick, Catharine Prussing, Navjot Singh, John Kelly, Erasmus Schneider, Erica Lasek-Nesselquist |
| EPI_ISL_2278735 | URMC LABS | Wadsworth Center, New York State Department of Health | Kirsten St. George, Daryl M. Lamson, Alexis Russell, Matthew Shudt, Melissa A Leisner, Jonathan Plitnick, Catharine Prussing, Navjot Singh, John Kelly, Erasmus Schneider, Erica Lasek-Nesselquist |
| EPI_ISL_2278736 | MONTEFIORE MEDICAL CENTER LABORATORIES | Wadsworth Center, New York State Department of Health | Kirsten St. George, Daryl M. Lamson, Alexis Russell, Matthew Shudt, Melissa A Leisner, Jonathan Plitnick, Catharine Prussing, Navjot Singh, John Kelly, Erasmus Schneider, Erica Lasek-Nesselquist |
| EPI_ISL_2278737, EPI_ISL_2278738 | ALBANY MEDICAL CENTER | Wadsworth Center, New York State Department of Health | Kirsten St. George, Daryl M. Lamson, Alexis Russell, Matthew Shudt, Melissa A Leisner, Jonathan Plitnick, Catharine Prussing, Navjot Singh, John Kelly, Erasmus Schneider, Erica Lasek-Nesselquist |
| EPI_ISL_2278739 | URMC LABS | Wadsworth Center, New York State Department of Health | Kirsten St. George, Daryl M. Lamson, Alexis Russell, Matthew Shudt, Melissa A Leisner, Jonathan Plitnick, Catharine Prussing, Navjot Singh, John Kelly, Erasmus Schneider, Erica Lasek-Nesselquist |
| EPI_ISL_2278740, EPI_ISL_2278741 | SUNY UPSTATE MEDICAL UNIVERSITY | Wadsworth Center, New York State Department of Health | Kirsten St. George, Daryl M. Lamson, Alexis Russell, Matthew Shudt, Melissa A Leisner, Jonathan Plitnick, Catharine Prussing, Navjot Singh, John Kelly, Erasmus Schneider, Erica Lasek-Nesselquist |
| EPI_ISL_2278742 | ALBANY MEDICAL CENTER | Wadsworth Center, New York State Department of Health | Kirsten St. George, Daryl M. Lamson, Alexis Russell, Matthew Shudt, Melissa A Leisner, Jonathan Plitnick, Catharine Prussing, Navjot Singh, John Kelly, Erasmus Schneider, Erica Lasek-Nesselquist |
| EPI_ISL_2278743, EPI_ISL_2278744, EPI_ISL_2278745, EPI_ISL_2278746, EPI_ISL_2278747, EPI_ISL_2278748, EPI_ISL_2278749, EPI_ISL_2278750, EPI_ISL_2278751, EPI_ISL_2278752, EPI_ISL_2278753, EPI_ISL_2278754, EPI_ISL_2278755, EPI_ISL_2278756, EPI_ISL_2278757, EPI_ISL_2278758, EPI_ISL_2278759, EPI_ISL_2278760, EPI_ISL_2278761, EPI_ISL_2278762, EPI_ISL_2278763, EPI_ISL_2278764, EPI_ISL_2278765, EPI_ISL_2278766, EPI_ISL_2278767, EPI_ISL_2278768, EPI_ISL_2278769, EPI_ISL_2278770, EPI_ISL_2278771, EPI_ISL_2278772, EPI_ISL_2278773, EPI_ISL_2278774, EPI_ISL_2278775, EPI_ISL_2278776, EPI_ISL_2278777, EPI_ISL_2278778, EPI_ISL_2278779, EPI_ISL_2278780, EPI_ISL_2278781, EPI_ISL_2278782, EPI_ISL_2278783, EPI_ISL_2278784, EPI_ISL_2278785, EPI_ISL_2278786, EPI_ISL_2278787, EPI_ISL_2278788, EPI_ISL_2278789, EPI_ISL_2278790, EPI_ISL_2278791, EPI_ISL_2278792, EPI_ISL_2278793, EPI_ISL_2278794, EPI_ISL_2278795, EPI_ISL_2278796 | | | |
| see above | URMC LABS | Wadsworth Center, New York State Department of Health | Kirsten St. George, Daryl M. Lamson, Alexis Russell, Matthew Shudt, Melissa A Leisner, Jonathan Plitnick, Catharine Prussing, Navjot Singh, John Kelly, Erasmus Schneider, Erica Lasek-Nesselquist |
| EPI_ISL_2278797 | Maimonides Medical Center | Wadsworth Center, New York State Department of Health | Kirsten St. George, Daryl M. Lamson, Alexis Russell, Matthew Shudt, Melissa A Leisner, Jonathan Plitnick, Catharine Prussing, Navjot Singh, John Kelly, Erasmus Schneider, Erica Lasek-Nesselquist |
| EPI_ISL_2278798, EPI_ISL_2278799 | URMC LABS | Wadsworth Center, New York State Department of Health | Kirsten St. George, Daryl M. Lamson, Alexis Russell, Matthew Shudt, Melissa A Leisner, Jonathan Plitnick, Catharine Prussing, Navjot Singh, John Kelly, Erasmus Schneider, Erica Lasek-Nesselquist |
| EPI_ISL_2278975, EPI_ISL_2278976, EPI_ISL_2278977, EPI_ISL_2278978, EPI_ISL_2278979, EPI_ISL_2278980, EPI_ISL_2278981, EPI_ISL_2278982, EPI_ISL_2278983, EPI_ISL_2278984, EPI_ISL_2278985, EPI_ISL_2278986, EPI_ISL_2278987, EPI_ISL_2278988, EPI_ISL_2278989, EPI_ISL_2278990, EPI_ISL_2278991, EPI_ISL_2278992, EPI_ISL_2278993, EPI_ISL_2278994, EPI_ISL_2278995, EPI_ISL_2278996, EPI_ISL_2278997, EPI_ISL_2278998, EPI_ISL_2278999, EPI_ISL_2279000, EPI_ISL_2279001, EPI_ISL_2279002, EPI_ISL_2279003, EPI_ISL_2279004, EPI_ISL_2279005, EPI_ISL_2279006, EPI_ISL_2279007, EPI_ISL_2279008, EPI_ISL_2279010, EPI_ISL_2279011, EPI_ISL_2279012, EPI_ISL_2279013, EPI_ISL_2279014, EPI_ISL_2279016, EPI_ISL_2279017, EPI_ISL_2279018, EPI_ISL_2279019, EPI_ISL_2279020, EPI_ISL_2279021, EPI_ISL_2279022, EPI_ISL_2279024, EPI_ISL_2279025, EPI_ISL_2279026, EPI_ISL_2279027, EPI_ISL_2279028, EPI_ISL_2279029, EPI_ISL_2279030, EPI_ISL_2279031, EPI_ISL_2279032, EPI_ISL_2279033, EPI_ISL_2279034, EPI_ISL_2279035, EPI_ISL_2279036, EPI_ISL_2279037, EPI_ISL_2279038, EPI_ISL_2279039, EPI_ISL_2279040, EPI_ISL_2279041, EPI_ISL_2279042, EPI_ISL_2279043, EPI_ISL_2279044, EPI_ISL_2279045, EPI_ISL_2279046, EPI_ISL_2279047, EPI_ISL_2279048, EPI_ISL_2279050, EPI_ISL_2279051, EPI_ISL_2279052, EPI_ISL_2279053, EPI_ISL_2279054, EPI_ISL_2279055, EPI_ISL_2279056, EPI_ISL_2279057, EPI_ISL_2279058, EPI_ISL_2279059, EPI_ISL_2279060, EPI_ISL_2279061, EPI_ISL_2279065, EPI_ISL_2279067, EPI_ISL_2279069, EPI_ISL_2279070, EPI_ISL_2279071, EPI_ISL_2279072, EPI_ISL_2279073, EPI_ISL_2279074, EPI_ISL_2279075, EPI_ISL_2279076, EPI_ISL_2279077, EPI_ISL_2279078, EPI_ISL_2279079, EPI_ISL_2279080, EPI_ISL_2279081, EPI_ISL_2279085, EPI_ISL_2279087, EPI_ISL_2279089, EPI_ISL_2279092, EPI_ISL_2279093, EPI_ISL_2279094, EPI_ISL_2279095, EPI_ISL_2279096, EPI_ISL_2279097, EPI_ISL_2279099, EPI_ISL_2279101, EPI_ISL_2279102, EPI_ISL_2279103, EPI_ISL_2279104, EPI_ISL_2279105, EPI_ISL_2279106, EPI_ISL_2279107, EPI_ISL_2279109, EPI_ISL_2279111, EPI_ISL_2279112, EPI_ISL_2279113, EPI_ISL_2279114, EPI_ISL_2279115, EPI_ISL_2279117, EPI_ISL_2279118, EPI_ISL_2279119, EPI_ISL_2279120, EPI_ISL_2279122, EPI_ISL_2279123, EPI_ISL_2279124, EPI_ISL_2279125, EPI_ISL_2279126, EPI_ISL_2279127, EPI_ISL_2279128, EPI_ISL_2279129, EPI_ISL_2279130, EPI_ISL_2279131, EPI_ISL_2279132, EPI_ISL_2279133, EPI_ISL_2279134, EPI_ISL_2279135, EPI_ISL_2279136, EPI_ISL_2279137, EPI_ISL_2279138, EPI_ISL_2279139, EPI_ISL_2279140, EPI_ISL_2279141, EPI_ISL_2279142, EPI_ISL_2279143, EPI_ISL_2279144, EPI_ISL_2279145, EPI_ISL_2279146, EPI_ISL_2279147, EPI_ISL_2279148, EPI_ISL_2279149, EPI_ISL_2279150, | | | |

| | | | |
|---|---|---|---|
| EPI_ISL_2279151, EPI_ISL_2279152, EPI_ISL_2279153, EPI_ISL_2279154, EPI_ISL_2279156, EPI_ISL_2279157, EPI_ISL_2279158, EPI_ISL_2279159, EPI_ISL_2279160, EPI_ISL_2279161, EPI_ISL_2279162, EPI_ISL_2279163, EPI_ISL_2279164, EPI_ISL_2279165, EPI_ISL_2279166, EPI_ISL_2279167, EPI_ISL_2279168, EPI_ISL_2279169, EPI_ISL_2279170, EPI_ISL_2279171, EPI_ISL_2279172, EPI_ISL_2279173, EPI_ISL_2279174, EPI_ISL_2279176, EPI_ISL_2279177, EPI_ISL_2279178, EPI_ISL_2279179, EPI_ISL_2279180, EPI_ISL_2279181, EPI_ISL_2279182, EPI_ISL_2279183, EPI_ISL_2279184, EPI_ISL_2279185, EPI_ISL_2279186, EPI_ISL_2279187, EPI_ISL_2279188, EPI_ISL_2279189, EPI_ISL_2279190, EPI_ISL_2279191, EPI_ISL_2279192, EPI_ISL_2279193, EPI_ISL_2279194, EPI_ISL_2279195, EPI_ISL_2279196, EPI_ISL_2279197, EPI_ISL_2279198, EPI_ISL_2279199, EPI_ISL_2279200, EPI_ISL_2279201, EPI_ISL_2279202, EPI_ISL_2279203, EPI_ISL_2279204, EPI_ISL_2279205, EPI_ISL_2279206, EPI_ISL_2279207, EPI_ISL_2279208, EPI_ISL_2279209, EPI_ISL_2279210, EPI_ISL_2279211, EPI_ISL_2279212, EPI_ISL_2279213, EPI_ISL_2279214, EPI_ISL_2279215, EPI_ISL_2279216, EPI_ISL_2279217, EPI_ISL_2279218, EPI_ISL_2279219, EPI_ISL_2279220, EPI_ISL_2279221, EPI_ISL_2279222, EPI_ISL_2279223, EPI_ISL_2279224, EPI_ISL_2279227, EPI_ISL_2279228, EPI_ISL_2279229, EPI_ISL_2279230, EPI_ISL_2279231, EPI_ISL_2279233, EPI_ISL_2279234, EPI_ISL_2279235, EPI_ISL_2279236, EPI_ISL_2279237, EPI_ISL_2279238, EPI_ISL_2279239, EPI_ISL_2279240, EPI_ISL_2279241, EPI_ISL_2279242, EPI_ISL_2279243, EPI_ISL_2279244, EPI_ISL_2279245, EPI_ISL_2279246, EPI_ISL_2279247, EPI_ISL_2279248, EPI_ISL_2279249, EPI_ISL_2279250, EPI_ISL_2279251, EPI_ISL_2279252, EPI_ISL_2279253, EPI_ISL_2279254, EPI_ISL_2279255, EPI_ISL_2279256, EPI_ISL_2279257, EPI_ISL_2279258, EPI_ISL_2279259, EPI_ISL_2279260, EPI_ISL_2279261, EPI_ISL_2279262, EPI_ISL_2279263, EPI_ISL_2279264, EPI_ISL_2279265, EPI_ISL_2279266, EPI_ISL_2279267, EPI_ISL_2279268, EPI_ISL_2279269, EPI_ISL_2279270, EPI_ISL_2279271, EPI_ISL_2279272, EPI_ISL_2279273, EPI_ISL_2279274, EPI_ISL_2279275, EPI_ISL_2279276, EPI_ISL_2279277, EPI_ISL_2279278, EPI_ISL_2279279, EPI_ISL_2279280, EPI_ISL_2279281, EPI_ISL_2279282, EPI_ISL_2279283, EPI_ISL_2279284, EPI_ISL_2279285, EPI_ISL_2279286, EPI_ISL_2279287, EPI_ISL_2279288, EPI_ISL_2279289, EPI_ISL_2279290, EPI_ISL_2279291, EPI_ISL_2279292, EPI_ISL_2279293, EPI_ISL_2279294, EPI_ISL_2279296, EPI_ISL_2279297, EPI_ISL_2279298, EPI_ISL_2279299, EPI_ISL_2279300, EPI_ISL_2279301, EPI_ISL_2279302, EPI_ISL_2279303, EPI_ISL_2279304, EPI_ISL_2279305, EPI_ISL_2279306, EPI_ISL_2279307, EPI_ISL_2279308, EPI_ISL_2279309, EPI_ISL_2279310, EPI_ISL_2279311, EPI_ISL_2279312, EPI_ISL_2279313, EPI_ISL_2279314, EPI_ISL_2279315, EPI_ISL_2279316, EPI_ISL_2279317, EPI_ISL_2279318, EPI_ISL_2279319, EPI_ISL_2279320, EPI_ISL_2279322, EPI_ISL_2279323, EPI_ISL_2279324, EPI_ISL_2279325, EPI_ISL_2279326, EPI_ISL_2279327, EPI_ISL_2279328, EPI_ISL_2279329, EPI_ISL_2279330, EPI_ISL_2279331, EPI_ISL_2279332, EPI_ISL_2279333, EPI_ISL_2279334, EPI_ISL_2279335, EPI_ISL_2279336, EPI_ISL_2279337, EPI_ISL_2279338, EPI_ISL_2279339, EPI_ISL_2279340, EPI_ISL_2279341, EPI_ISL_2279342, EPI_ISL_2279343, EPI_ISL_2279344, EPI_ISL_2279345, EPI_ISL_2279346, EPI_ISL_2279347, EPI_ISL_2279348, EPI_ISL_2279349, EPI_ISL_2279350, EPI_ISL_2279351, EPI_ISL_2279352, EPI_ISL_2279353, EPI_ISL_2279354, EPI_ISL_2279355, EPI_ISL_2279356, EPI_ISL_2279357, EPI_ISL_2279358, EPI_ISL_2279359, EPI_ISL_2279360, EPI_ISL_2279361, EPI_ISL_2279364, EPI_ISL_2279366, EPI_ISL_2279367, EPI_ISL_2279368, EPI_ISL_2279369, EPI_ISL_2279370, EPI_ISL_2279371, EPI_ISL_2279373, EPI_ISL_2279375, EPI_ISL_2279376, EPI_ISL_2279377, EPI_ISL_2279379, EPI_ISL_2279381, EPI_ISL_2279382, EPI_ISL_2279384, EPI_ISL_2279386, EPI_ISL_2279387, EPI_ISL_2279388, EPI_ISL_2279389, EPI_ISL_2279390, EPI_ISL_2279391, EPI_ISL_2279392, EPI_ISL_2279393, EPI_ISL_2279394, EPI_ISL_2279399, EPI_ISL_2279401, EPI_ISL_2279402, EPI_ISL_2279403, EPI_ISL_2279404, EPI_ISL_2279405, EPI_ISL_2279407, EPI_ISL_2279408, EPI_ISL_2279409, EPI_ISL_2279411, EPI_ISL_2279414, EPI_ISL_2279415, EPI_ISL_2279416, EPI_ISL_2279417, EPI_ISL_2279419, EPI_ISL_2279420, EPI_ISL_2279421, EPI_ISL_2279422, EPI_ISL_2279423, EPI_ISL_2279424, EPI_ISL_2279425, EPI_ISL_2279426, EPI_ISL_2279427, EPI_ISL_2279428, EPI_ISL_2279429, EPI_ISL_2279430, EPI_ISL_2279431, EPI_ISL_2279432, EPI_ISL_2279433, EPI_ISL_2279434, EPI_ISL_2279435, EPI_ISL_2279436, EPI_ISL_2279437, EPI_ISL_2279438, EPI_ISL_2279439, EPI_ISL_2279440, EPI_ISL_2279441, EPI_ISL_2279442, EPI_ISL_2279443, EPI_ISL_2279444, EPI_ISL_2279445, EPI_ISL_2279446, EPI_ISL_2279447, EPI_ISL_2279448, EPI_ISL_2279449, EPI_ISL_2279450, EPI_ISL_2279451, EPI_ISL_2279453, EPI_ISL_2279454, EPI_ISL_2279455, EPI_ISL_2279456, EPI_ISL_2279457, EPI_ISL_2279458, EPI_ISL_2279459, EPI_ISL_2279460, EPI_ISL_2279461, EPI_ISL_2279462, EPI_ISL_2279463, EPI_ISL_2279464, EPI_ISL_2279465, EPI_ISL_2279466, EPI_ISL_2279467, EPI_ISL_2279468, EPI_ISL_2279469, EPI_ISL_2279470, EPI_ISL_2279471, EPI_ISL_2279472, EPI_ISL_2279473, EPI_ISL_2279474, EPI_ISL_2279475, EPI_ISL_2279476, EPI_ISL_2279477, EPI_ISL_2279478, EPI_ISL_2279480, EPI_ISL_2279481, EPI_ISL_2279482, EPI_ISL_2279483, EPI_ISL_2279484, EPI_ISL_2279485, EPI_ISL_2279486, EPI_ISL_2279487, EPI_ISL_2279488, EPI_ISL_2279489, EPI_ISL_2279490, EPI_ISL_2279491, EPI_ISL_2279492, EPI_ISL_2279493, EPI_ISL_2279494, EPI_ISL_2279495, EPI_ISL_2279496, EPI_ISL_2279497 | | | |
| see above | Wisconsin State Laboratory of Hygiene Communicable Disease Division | Wisconsin State Laboratory of Hygiene Communicable Disease Division | Abigail C. Shockey, Alicia J. Mooney, Erika M. Hanson, Tonya Danz, Richard Griesser, Sara Wagner, Kelsey R. Florek |
| EPI_ISL_2279499, EPI_ISL_2279500, EPI_ISL_2279501, EPI_ISL_2279502 | SUNY UPSTATE MEDICAL UNIVERSITY | Wadsworth Center, New York State Department of Health | Kirsten St. George, Daryl M. Lamson, Alexis Russell, Matthew Shudt, Melissa A Leisner, Jonathan Plitnick, Catharine Prussing, Navjot Singh, John Kelly, Erasmus Schneider, Erica Lasek-Nesselquist |
| EPI_ISL_2279503, EPI_ISL_2279504, EPI_ISL_2279505 | ALBANY MEDICAL CENTER | Wadsworth Center, New York State Department of Health | Kirsten St. George, Daryl M. Lamson, Alexis Russell, Matthew Shudt, Melissa A Leisner, Jonathan Plitnick, Catharine Prussing, Navjot Singh, John Kelly, Erasmus Schneider, Erica Lasek-Nesselquist |
| EPI_ISL_2279506, EPI_ISL_2279507, EPI_ISL_2279508, EPI_ISL_2279509, EPI_ISL_2279510, EPI_ISL_2279511, EPI_ISL_2279512, EPI_ISL_2279513, EPI_ISL_2279514, EPI_ISL_2279515, EPI_ISL_2279516, EPI_ISL_2279517, EPI_ISL_2279518, EPI_ISL_2279519, EPI_ISL_2279520, EPI_ISL_2279521, EPI_ISL_2279522, EPI_ISL_2279523, EPI_ISL_2279524, EPI_ISL_2279525, EPI_ISL_2279526, EPI_ISL_2279527, EPI_ISL_2279528, EPI_ISL_2279529, EPI_ISL_2279530, EPI_ISL_2279531, EPI_ISL_2279532, EPI_ISL_2279533, EPI_ISL_2279534, EPI_ISL_2279535, EPI_ISL_2279536, EPI_ISL_2279537, EPI_ISL_2279538, EPI_ISL_2279539, EPI_ISL_2279540, EPI_ISL_2279541, EPI_ISL_2279542, EPI_ISL_2279543, EPI_ISL_2279544, EPI_ISL_2279545, EPI_ISL_2279546, EPI_ISL_2279547, EPI_ISL_2279548, EPI_ISL_2279549, EPI_ISL_2279550, EPI_ISL_2279551, EPI_ISL_2279552, EPI_ISL_2279553, EPI_ISL_2279554, EPI_ISL_2279555, EPI_ISL_2279556, EPI_ISL_2279557, EPI_ISL_2279558, EPI_ISL_2279559, EPI_ISL_2279560, EPI_ISL_2279561, EPI_ISL_2279562, EPI_ISL_2279563, EPI_ISL_2279564, EPI_ISL_2279565, EPI_ISL_2279566, EPI_ISL_2279567, EPI_ISL_2279568, EPI_ISL_2279569, EPI_ISL_2279570, EPI_ISL_2279571, EPI_ISL_2279572, EPI_ISL_2279573, EPI_ISL_2279574, EPI_ISL_2279575, EPI_ISL_2279576, EPI_ISL_2279577, EPI_ISL_2279578, EPI_ISL_2279579, EPI_ISL_2279580, EPI_ISL_2279581, EPI_ISL_2279582, EPI_ISL_2279583, EPI_ISL_2279584 | | | |
| see above | URMC LABS | Wadsworth Center, New York State Department of Health | Kirsten St. George, Daryl M. Lamson, Alexis Russell, Matthew Shudt, Melissa A Leisner, Jonathan Plitnick, Catharine Prussing, Navjot Singh, John Kelly, Erasmus Schneider, Erica Lasek-Nesselquist |
| EPI_ISL_2279585 | BIO-REFERENCE LABORATORIES | Wadsworth Center, New York State Department of Health | Kirsten St. George, Daryl M. Lamson, Alexis Russell, Matthew Shudt, Melissa A Leisner, Jonathan Plitnick, Catharine Prussing, Navjot Singh, John Kelly, Erasmus Schneider, Erica Lasek-Nesselquist |
| EPI_ISL_2279699, EPI_ISL_2279700, EPI_ISL_2279701, EPI_ISL_2279703, EPI_ISL_2279704, EPI_ISL_2279705, EPI_ISL_2279706, EPI_ISL_2279707, EPI_ISL_2279708, EPI_ISL_2279709, EPI_ISL_2279710, EPI_ISL_2279711, EPI_ISL_2279712, EPI_ISL_2279713, EPI_ISL_2279714, EPI_ISL_2279715, EPI_ISL_2279716, EPI_ISL_2279717, EPI_ISL_2279718, EPI_ISL_2279720, EPI_ISL_2279722, EPI_ISL_2279723, EPI_ISL_2279726, EPI_ISL_2279727, EPI_ISL_2279729, EPI_ISL_2279730, EPI_ISL_2279731, EPI_ISL_2279732, EPI_ISL_2279733, EPI_ISL_2279734, EPI_ISL_2279735, EPI_ISL_2279736, EPI_ISL_2279737, EPI_ISL_2279738, EPI_ISL_2279739, EPI_ISL_2279740, EPI_ISL_2279741, EPI_ISL_2279742, EPI_ISL_2279743, EPI_ISL_2279744, EPI_ISL_2279745, EPI_ISL_2279746, EPI_ISL_2279747, EPI_ISL_2279748, EPI_ISL_2279749, EPI_ISL_2279750, EPI_ISL_2279751, EPI_ISL_2279752, EPI_ISL_2279753, EPI_ISL_2279754, EPI_ISL_2279756, EPI_ISL_2279757, EPI_ISL_2279758, EPI_ISL_2279759, EPI_ISL_2279761, EPI_ISL_2279762, EPI_ISL_2279763, EPI_ISL_2279764, EPI_ISL_2279765, EPI_ISL_2279766, EPI_ISL_2279767, EPI_ISL_2279768, EPI_ISL_2279769, EPI_ISL_2279770, EPI_ISL_2279771, EPI_ISL_2279772, EPI_ISL_2279773, EPI_ISL_2279774, EPI_ISL_2279775, EPI_ISL_2279776, EPI_ISL_2279777, EPI_ISL_2279778, EPI_ISL_2279779, EPI_ISL_2279780, EPI_ISL_2279781, EPI_ISL_2279782, EPI_ISL_2279783, EPI_ISL_2279784, EPI_ISL_2279785, EPI_ISL_2279786, EPI_ISL_2279787, EPI_ISL_2279788, EPI_ISL_2279789, EPI_ISL_2279790, EPI_ISL_2279791, EPI_ISL_2279792, EPI_ISL_2279793, EPI_ISL_2279794, EPI_ISL_2279795, EPI_ISL_2279796, EPI_ISL_2279797, EPI_ISL_2279798, EPI_ISL_2279799, EPI_ISL_2279800, EPI_ISL_2279801, EPI_ISL_2279802, EPI_ISL_2279803, EPI_ISL_2279804, EPI_ISL_2279805, EPI_ISL_2279806, EPI_ISL_2279807, EPI_ISL_2279808, EPI_ISL_2279809, EPI_ISL_2279810, EPI_ISL_2279811, EPI_ISL_2279812, EPI_ISL_2279813, EPI_ISL_2279814, EPI_ISL_2279815, EPI_ISL_2279816, EPI_ISL_2279817, EPI_ISL_2279818, EPI_ISL_2279819, EPI_ISL_2279820, EPI_ISL_2279821, EPI_ISL_2279822, EPI_ISL_2279823, EPI_ISL_2279824, EPI_ISL_2279825, EPI_ISL_2279826, EPI_ISL_2279827, EPI_ISL_2279828, EPI_ISL_2279829, EPI_ISL_2279830, EPI_ISL_2279831, EPI_ISL_2279832, EPI_ISL_2279833, EPI_ISL_2279834, EPI_ISL_2279835, EPI_ISL_2279836, EPI_ISL_2279837, EPI_ISL_2279838, EPI_ISL_2279839, EPI_ISL_2279840, EPI_ISL_2279841, EPI_ISL_2279842, EPI_ISL_2279843, EPI_ISL_2279844, EPI_ISL_2279846, EPI_ISL_2279847, EPI_ISL_2279848, EPI_ISL_2279849, EPI_ISL_2279850, EPI_ISL_2279851, EPI_ISL_2279852, EPI_ISL_2279853, EPI_ISL_2279854, EPI_ISL_2279855, EPI_ISL_2279856, EPI_ISL_2279857, EPI_ISL_2279858, EPI_ISL_2279859, EPI_ISL_2279860, EPI_ISL_2279861, EPI_ISL_2279862, EPI_ISL_2279863, EPI_ISL_2279864, EPI_ISL_2279865, EPI_ISL_2279866, EPI_ISL_2279867, EPI_ISL_2279868, EPI_ISL_2279869, EPI_ISL_2279870, EPI_ISL_2279871, EPI_ISL_2279872, EPI_ISL_2279873, EPI_ISL_2279874, EPI_ISL_2279875, EPI_ISL_2279876, EPI_ISL_2279877, EPI_ISL_2279878, EPI_ISL_2279879, EPI_ISL_2279880, EPI_ISL_2279881, EPI_ISL_2279882, EPI_ISL_2279883, EPI_ISL_2279884, EPI_ISL_2279885, EPI_ISL_2279886, EPI_ISL_2279887, EPI_ISL_2279888, EPI_ISL_2279889, EPI_ISL_2279890, EPI_ISL_2279891, EPI_ISL_2279892, EPI_ISL_2279893, EPI_ISL_2279894, EPI_ISL_2279895, EPI_ISL_2279897, EPI_ISL_2279898, EPI_ISL_2279899, EPI_ISL_2279900, EPI_ISL_2279901, EPI_ISL_2279902, EPI_ISL_2279903, EPI_ISL_2279904, EPI_ISL_2279905, EPI_ISL_2279906, EPI_ISL_2279907, EPI_ISL_2279908, EPI_ISL_2279909, EPI_ISL_2279911, EPI_ISL_2279912, EPI_ISL_2279913, EPI_ISL_2279914, EPI_ISL_2279915, EPI_ISL_2279916, EPI_ISL_2279917, EPI_ISL_2279918, EPI_ISL_2279919, EPI_ISL_2279921, EPI_ISL_2279922, EPI_ISL_2279923, EPI_ISL_2279924, EPI_ISL_2279925, EPI_ISL_2279926, EPI_ISL_2279927, EPI_ISL_2279928, EPI_ISL_2279930, EPI_ISL_2279931, EPI_ISL_2279932, EPI_ISL_2279933, EPI_ISL_2279934, EPI_ISL_2279935, EPI_ISL_2279936, EPI_ISL_2279937, EPI_ISL_2279938, EPI_ISL_2279939, EPI_ISL_2279941, EPI_ISL_2279942, EPI_ISL_2279943, EPI_ISL_2279944, EPI_ISL_2279945, EPI_ISL_2279946, EPI_ISL_2279947, EPI_ISL_2279948, EPI_ISL_2279949, EPI_ISL_2279950, EPI_ISL_2279951, EPI_ISL_2279952, EPI_ISL_2279954, EPI_ISL_2279956, EPI_ISL_2279957, EPI_ISL_2279958, EPI_ISL_2279959, EPI_ISL_2279960, EPI_ISL_2279961, EPI_ISL_2279962, EPI_ISL_2279963, EPI_ISL_2279964, EPI_ISL_2279965, EPI_ISL_2279966, EPI_ISL_2279967, EPI_ISL_2279968, EPI_ISL_2279969, EPI_ISL_2279970, EPI_ISL_2279971, EPI_ISL_2279972, EPI_ISL_2279973, EPI_ISL_2279974, EPI_ISL_2279975, EPI_ISL_2279976, EPI_ISL_2279977, EPI_ISL_2279978, EPI_ISL_2279979, EPI_ISL_2279980, EPI_ISL_2279981, EPI_ISL_2279982, EPI_ISL_2279983, EPI_ISL_2279984, EPI_ISL_2279985, EPI_ISL_2279987, EPI_ISL_2279988, EPI_ISL_2279989, EPI_ISL_2279990, EPI_ISL_2279991, EPI_ISL_2279992, EPI_ISL_2279993, EPI_ISL_2279994, EPI_ISL_2279995, EPI_ISL_2279997, EPI_ISL_2279998, EPI_ISL_2279999, EPI_ISL_2280000, EPI_ISL_2280001 | | | |
| see above | Utah Public Health Laboratory | Utah Public Health Laboratory | Erin L. Young, Kelly F. Oakeson, Tara Gallagher |
| EPI_ISL_2280002, EPI_ISL_2280003, EPI_ISL_2280004, EPI_ISL_2280005, EPI_ISL_2280006, EPI_ISL_2280007, EPI_ISL_2280008, EPI_ISL_2280009, EPI_ISL_2280010, EPI_ISL_2280011, EPI_ISL_2280012, EPI_ISL_2280013, EPI_ISL_2280014, EPI_ISL_2280015, EPI_ISL_2280016, EPI_ISL_2280017, EPI_ISL_2280018, EPI_ISL_2280019, EPI_ISL_2280021, EPI_ISL_2280022, EPI_ISL_2280023, EPI_ISL_2280024, EPI_ISL_2280026, EPI_ISL_2280027, EPI_ISL_2280028, EPI_ISL_2280029, EPI_ISL_2280030, EPI_ISL_2280032, EPI_ISL_2280033, EPI_ISL_2280034, EPI_ISL_2280036, EPI_ISL_2280037, EPI_ISL_2280038, EPI_ISL_2280039, EPI_ISL_2280040, EPI_ISL_2280041, EPI_ISL_2280042, EPI_ISL_2280043, EPI_ISL_2280044, EPI_ISL_2280045, EPI_ISL_2280046, EPI_ISL_2280047, EPI_ISL_2280048, EPI_ISL_2280049, EPI_ISL_2280050, EPI_ISL_2280051, EPI_ISL_2280052, EPI_ISL_2280053, EPI_ISL_2280054, EPI_ISL_2280055, EPI_ISL_2280058, EPI_ISL_2280059, EPI_ISL_2280060, EPI_ISL_2280061, EPI_ISL_2280062, EPI_ISL_2280063, EPI_ISL_2280064, EPI_ISL_2280065, EPI_ISL_2280067, EPI_ISL_2280068, EPI_ISL_2280069, EPI_ISL_2280070, EPI_ISL_2280071, EPI_ISL_2280072, EPI_ISL_2280075, EPI_ISL_2280076 | | | |
| see above | TXDSHS | TXDSHS | Rashmi Tuladhar, Bonnie Oh, Jenny Zhang, Maliha Rahman, Mayela Pedrueza, Anita Pokharel, Karen Bobier, Lorraine Rodriguez, Myong Koag, Chun Wang, Rachel Lee, Grace Kubin |
| EPI_ISL_2280080, EPI_ISL_2280081, EPI_ISL_2280082, EPI_ISL_2280083, EPI_ISL_2280084, EPI_ISL_2280085, EPI_ISL_2280086, EPI_ISL_2280087, EPI_ISL_2280088, EPI_ISL_2280089, EPI_ISL_2280090, EPI_ISL_2280091, EPI_ISL_2280092, EPI_ISL_2280093, EPI_ISL_2280094, EPI_ISL_2280095, EPI_ISL_2280096, EPI_ISL_2280097, EPI_ISL_2280098, EPI_ISL_2280099, EPI_ISL_2280100, EPI_ISL_2280101, EPI_ISL_2280102, EPI_ISL_2280103, EPI_ISL_2280104, EPI_ISL_2280105, EPI_ISL_2280106, EPI_ISL_2280107, EPI_ISL_2280108, EPI_ISL_2280109, EPI_ISL_2280110, EPI_ISL_2280111, EPI_ISL_2280112, EPI_ISL_2280113, EPI_ISL_2280114, EPI_ISL_2280115, EPI_ISL_2280116, EPI_ISL_2280117, EPI_ISL_2280118, EPI_ISL_2280119, EPI_ISL_2280120, EPI_ISL_2280121, EPI_ISL_2280122, EPI_ISL_2280123, EPI_ISL_2280124, EPI_ISL_2280125, EPI_ISL_2280126, EPI_ISL_2280127, EPI_ISL_2280128, EPI_ISL_2280129, EPI_ISL_2280130, EPI_ISL_2280131, EPI_ISL_2280132, EPI_ISL_2280133, EPI_ISL_2280134, EPI_ISL_2280135, EPI_ISL_2280136, EPI_ISL_2280137, EPI_ISL_2280138, EPI_ISL_2280139, EPI_ISL_2280140, EPI_ISL_2280141, EPI_ISL_2280142, EPI_ISL_2280143, EPI_ISL_2280144, EPI_ISL_2280145, EPI_ISL_2280146, EPI_ISL_2280147, EPI_ISL_2280148, EPI_ISL_2280149, EPI_ISL_2280150, EPI_ISL_2280151, EPI_ISL_2280152, EPI_ISL_2280153, EPI_ISL_2280154, EPI_ISL_2280155, EPI_ISL_2280156, EPI_ISL_2280157, EPI_ISL_2280158, EPI_ISL_2280159, EPI_ISL_2280160, EPI_ISL_2280161, EPI_ISL_2280162, EPI_ISL_2280163, EPI_ISL_2280164, EPI_ISL_2280165, EPI_ISL_2280166, EPI_ISL_2280167, EPI_ISL_2280168, EPI_ISL_2280169, EPI_ISL_2280170, EPI_ISL_2280171, EPI_ISL_2280172, EPI_ISL_2280173, EPI_ISL_2280174, EPI_ISL_2280175 | | | |
| see above | Quest Diagnostics Incorporated | Centers for Disease Control and Prevention Division of Viral Diseases, Pathogen Discovery | Dakota Howard, Dhwani Batra, Peter W. Cook, Kara Moser, Adrian Paskey, Jason Caravas, Benjamin Rambo-Martin, Shatavia Morrison, Christopher Gulvick, Scott Sammons, Yvette Unoarumhi, Darlene Wagner, Matthew Schmerer, S. H. Rosenthal, A. Gerasimova, R. M. Kagan, B. Anderson, M. Hua, Y. Liu, L.E. Bernstein, K.E. Livingston, A. Perez, I. A. Shlyakhter, R. V. Rolando, R. Owen, P. Tanpaiboon, F. Lacbawan, Clinton R. Paden, Duncan MacCannell |
| EPI_ISL_2280176, EPI_ISL_2280177, EPI_ISL_2280178, EPI_ISL_2280179, EPI_ISL_2280180, EPI_ISL_2280181, EPI_ISL_2280182, EPI_ISL_2280183, EPI_ISL_2280184, EPI_ISL_2280185, EPI_ISL_2280186, EPI_ISL_2280187, EPI_ISL_2280188, EPI_ISL_2280189, EPI_ISL_2280191, EPI_ISL_2280192, EPI_ISL_2280193, EPI_ISL_2280194, EPI_ISL_2280195, EPI_ISL_2280196, EPI_ISL_2280197, EPI_ISL_2280198, EPI_ISL_2280199, EPI_ISL_2280200, EPI_ISL_2280201, EPI_ISL_2280202, EPI_ISL_2280203, EPI_ISL_2280204, EPI_ISL_2280205, EPI_ISL_2280206, EPI_ISL_2280207, EPI_ISL_2280208, EPI_ISL_2280209, EPI_ISL_2280210, EPI_ISL_2280211, EPI_ISL_2280212, EPI_ISL_2280213, EPI_ISL_2280214, EPI_ISL_2280215, EPI_ISL_2280216, EPI_ISL_2280217, EPI_ISL_2280218, EPI_ISL_2280219, EPI_ISL_2280220, EPI_ISL_2280221, EPI_ISL_2280222, EPI_ISL_2280223, EPI_ISL_2280224, EPI_ISL_2280225, EPI_ISL_2280226 | | | |
| see above | Helix/Illumina | Centers for Disease Control and Prevention Division of Viral Diseases, Pathogen Discovery | Dakota Howard, Dhwani Batra, Peter W. Cook, Kara Moser, Adrian Paskey, Jason Caravas, Benjamin Rambo-Martin, Shatavia Morrison, Christopher Gulvick, Scott Sammons, Yvette Unoarumhi, Darlene Wagner, Matthew Schmerer, Eileen de Feo, Jan Antico, Christine Tran, Matthew Tolentino, Shannon Wickline, Kim Gietzen, Brad Sickler, Jingtao Liu, Eric Allen, Phil Febbo, Nicole L. Washington, Simon White, Geraint Levan, Kelly Schiabor Barrett, Elizabeth Cirulli, Alexandre Bolze, Ary Ascencio, Charlotte Rivera-Garcia, Ryan Cho, Jason Nguyen, Sherry Wang, Jimmy Ramirez, Tyler Cassens, Efren Sandoval, Magnus Isaksson, William Lee, David Becker, Marc Laurent, James Lu, Clinton R. Paden, Duncan MacCannell |
| EPI_ISL_2280227, EPI_ISL_2280228, EPI_ISL_2280229, EPI_ISL_2280230, EPI_ISL_2280232, EPI_ISL_2280234, EPI_ISL_2280235, EPI_ISL_2280236, EPI_ISL_2280237, EPI_ISL_2280238, EPI_ISL_2280239, EPI_ISL_2280240, EPI_ISL_2280241, EPI_ISL_2280242, EPI_ISL_2280243, EPI_ISL_2280244, EPI_ISL_2280245, EPI_ISL_2280246, EPI_ISL_2280248, EPI_ISL_2280249, EPI_ISL_2280250, EPI_ISL_2280251, EPI_ISL_2280252, EPI_ISL_2280253, EPI_ISL_2280254, EPI_ISL_2280255, EPI_ISL_2280256, EPI_ISL_2280257, EPI_ISL_2280258, EPI_ISL_2280259, EPI_ISL_2280260, EPI_ISL_2280261, EPI_ISL_2280262, EPI_ISL_2280263, EPI_ISL_2280264, EPI_ISL_2280265, EPI_ISL_2280266, EPI_ISL_2280267, EPI_ISL_2280268, EPI_ISL_2280269, EPI_ISL_2280270, EPI_ISL_2280271, EPI_ISL_2280272, EPI_ISL_2280273, EPI_ISL_2280275, EPI_ISL_2280276, EPI_ISL_2280277, EPI_ISL_2280278, EPI_ISL_2280279, EPI_ISL_2280280, EPI_ISL_2280281, EPI_ISL_2280282 | | | |
| see above | Fulgent Genetics | Centers for Disease Control and Prevention Division of Viral Diseases, Pathogen Discovery | Dakota Howard, Dhwani Batra, Peter W. Cook, Kara Moser, Adrian Paskey, Jason Caravas, Benjamin Rambo-Martin, Shatavia Morrison, Christopher Gulvick, Scott Sammons, Yvette Unoarumhi, Darlene Wagner, Matthew Schmerer, Harry Gao, Mickey Li, John Gao, Joseph Fierro, Benafsh Sapra, Becky Tsai, Yan Meng, Doreen Ng, James Xie, Clinton R. Paden, Duncan MacCannell |
| EPI_ISL_2280285, EPI_ISL_2280286, EPI_ISL_2280287, EPI_ISL_2280288, EPI_ISL_2280289, EPI_ISL_2280290, EPI_ISL_2280292, EPI_ISL_2280293, EPI_ISL_2280294, EPI_ISL_2280295, EPI_ISL_2280296, EPI_ISL_2280298, EPI_ISL_2280300, EPI_ISL_2280301, EPI_ISL_2280302, EPI_ISL_2280303, EPI_ISL_2280305, EPI_ISL_2280306, EPI_ISL_2280307, EPI_ISL_2280308, EPI_ISL_2280309, EPI_ISL_2280310, EPI_ISL_2280311, EPI_ISL_2280312, EPI_ISL_2280313, EPI_ISL_2280314, EPI_ISL_2280315, EPI_ISL_2280316, EPI_ISL_2280317, EPI_ISL_2280318, EPI_ISL_2280319, EPI_ISL_2280320, EPI_ISL_2280321, EPI_ISL_2280322, EPI_ISL_2280323, EPI_ISL_2280324, EPI_ISL_2280325, EPI_ISL_2280326, EPI_ISL_2280327, EPI_ISL_2280328, EPI_ISL_2280329, EPI_ISL_2280330, EPI_ISL_2280331, EPI_ISL_2280332, EPI_ISL_2280333, EPI_ISL_2280334, EPI_ISL_2280335, EPI_ISL_2280336, EPI_ISL_2280337, EPI_ISL_2280338, EPI_ISL_2280339, EPI_ISL_2280340, EPI_ISL_2280341, EPI_ISL_2280342, EPI_ISL_2280343, EPI_ISL_2280344, EPI_ISL_2280345, EPI_ISL_2280346, EPI_ISL_2280347, EPI_ISL_2280348, EPI_ISL_2280349, EPI_ISL_2280350, EPI_ISL_2280351, EPI_ISL_2280352, EPI_ISL_2280353, EPI_ISL_2280354, EPI_ISL_2280355, EPI_ISL_2280356, EPI_ISL_2280357, EPI_ISL_2280358, EPI_ISL_2280359, EPI_ISL_2280360, EPI_ISL_2280361, EPI_ISL_2280362, EPI_ISL_2280363, EPI_ISL_2280364, EPI_ISL_2280365, EPI_ISL_2280366, EPI_ISL_2280367, EPI_ISL_2280368, EPI_ISL_2280369, EPI_ISL_2280370, EPI_ISL_2280371, EPI_ISL_2280372, EPI_ISL_2280373, EPI_ISL_2280374, EPI_ISL_2280375, EPI_ISL_2280376, EPI_ISL_2280377, EPI_ISL_2280378, EPI_ISL_2280379, EPI_ISL_2280380, EPI_ISL_2280381, EPI_ISL_2280382, EPI_ISL_2280383, EPI_ISL_2280384, EPI_ISL_2280385, EPI_ISL_2280386, EPI_ISL_2280387, EPI_ISL_2280388, EPI_ISL_2280389, EPI_ISL_2280390, EPI_ISL_2280391, EPI_ISL_2280392, EPI_ISL_2280393, EPI_ISL_2280394, EPI_ISL_2280395, EPI_ISL_2280396, EPI_ISL_2280397, EPI_ISL_2280398, EPI_ISL_2280399, EPI_ISL_2280400, EPI_ISL_2280401, EPI_ISL_2280402, EPI_ISL_2280403, EPI_ISL_2280404, EPI_ISL_2280405, EPI_ISL_2280406, EPI_ISL_2280407, EPI_ISL_2280408, EPI_ISL_2280409, EPI_ISL_2280410, EPI_ISL_2280411, EPI_ISL_2280412, EPI_ISL_2280413, EPI_ISL_2280414, EPI_ISL_2280415, EPI_ISL_2280416, EPI_ISL_2280417, EPI_ISL_2280418, EPI_ISL_2280419, EPI_ISL_2280420, EPI_ISL_2280421, EPI_ISL_2280422, EPI_ISL_2280423, EPI_ISL_2280424, EPI_ISL_2280425, EPI_ISL_2280426, EPI_ISL_2280427, EPI_ISL_2280428, EPI_ISL_2280429, EPI_ISL_2280430, EPI_ISL_2280431, EPI_ISL_2280432, EPI_ISL_2280433, EPI_ISL_2280434, EPI_ISL_2280435, EPI_ISL_2280436, EPI_ISL_2280437, EPI_ISL_2280438, EPI_ISL_2280440, EPI_ISL_2280441, EPI_ISL_2280442, EPI_ISL_2280443, EPI_ISL_2280444, EPI_ISL_2280445, EPI_ISL_2280446, EPI_ISL_2280447, EPI_ISL_2280448, EPI_ISL_2280449, EPI_ISL_2280450, EPI_ISL_2280451, EPI_ISL_2280452, EPI_ISL_2280453, EPI_ISL_2280454, EPI_ISL_2280455, EPI_ISL_2280456, EPI_ISL_2280457, EPI_ISL_2280458, EPI_ISL_2280459, EPI_ISL_2280460, EPI_ISL_2280461, EPI_ISL_2280462, EPI_ISL_2280463, EPI_ISL_2280464, EPI_ISL_2280465, EPI_ISL_2280466, EPI_ISL_2280467, EPI_ISL_2280468, EPI_ISL_2280469, EPI_ISL_2280470, EPI_ISL_2280471, EPI_ISL_2280472, EPI_ISL_2280473, EPI_ISL_2280474, EPI_ISL_2280475, EPI_ISL_2280476, EPI_ISL_2280477, EPI_ISL_2280478, EPI_ISL_2280479, EPI_ISL_2280480, EPI_ISL_2280481, EPI_ISL_2280482, EPI_ISL_2280483, EPI_ISL_2280484, EPI_ISL_2280486, EPI_ISL_2280487, EPI_ISL_2280488, EPI_ISL_2280489, EPI_ISL_2280490, EPI_ISL_2280491, EPI_ISL_2280492, EPI_ISL_2280493, EPI_ISL_2280494, EPI_ISL_2280495, EPI_ISL_2280496, EPI_ISL_2280497, EPI_ISL_2280498, EPI_ISL_2280499, EPI_ISL_2280500, EPI_ISL_2280501, EPI_ISL_2280502, EPI_ISL_2280503, EPI_ISL_2280504, EPI_ISL_2280505, EPI_ISL_2280506, EPI_ISL_2280507, EPI_ISL_2280508, EPI_ISL_2280509, EPI_ISL_2280510, EPI_ISL_2280511, EPI_ISL_2280512, EPI_ISL_2280513, EPI_ISL_2280514, EPI_ISL_2280515, EPI_ISL_2280516, EPI_ISL_2280517, EPI_ISL_2280518, EPI_ISL_2280519, EPI_ISL_2280520, EPI_ISL_2280521, EPI_ISL_2280522, EPI_ISL_2280523, EPI_ISL_2280524, EPI_ISL_2280526, EPI_ISL_2280527, EPI_ISL_2280528, EPI_ISL_2280529, EPI_ISL_2280530, EPI_ISL_2280531, EPI_ISL_2280532, EPI_ISL_2280533, EPI_ISL_2280534, EPI_ISL_2280535, EPI_ISL_2280536, EPI_ISL_2280537, EPI_ISL_2280538, EPI_ISL_2280539, EPI_ISL_2280540, EPI_ISL_2280541, EPI_ISL_2280542, EPI_ISL_2280543, EPI_ISL_2280544, EPI_ISL_2280545, EPI_ISL_2280546, EPI_ISL_2280547, EPI_ISL_2280548, EPI_ISL_2280549, EPI_ISL_2280550, EPI_ISL_2280551, EPI_ISL_2280552, EPI_ISL_2280553, EPI_ISL_2280554, EPI_ISL_2280555, EPI_ISL_2280556, EPI_ISL_2280557, EPI_ISL_2280558, EPI_ISL_2280559, EPI_ISL_2280560, EPI_ISL_2280561, EPI_ISL_2280562, EPI_ISL_2280563, EPI_ISL_2280564, EPI_ISL_2280565, EPI_ISL_2280566, EPI_ISL_2280567, EPI_ISL_2280568, EPI_ISL_2280569, EPI_ISL_2280570, EPI_ISL_2280571, EPI_ISL_2280572, EPI_ISL_2280573, EPI_ISL_2280574, EPI_ISL_2280575, EPI_ISL_2280576, EPI_ISL_2280577, EPI_ISL_2280578, EPI_ISL_2280579, EPI_ISL_2280580, EPI_ISL_2280581, EPI_ISL_2280582, EPI_ISL_2280583, EPI_ISL_2280584, EPI_ISL_2280585, EPI_ISL_2280586, EPI_ISL_2280587, EPI_ISL_2280588, EPI_ISL_2280589, EPI_ISL_2280590, EPI_ISL_2280591, EPI_ISL_2280592, EPI_ISL_2280593, EPI_ISL_2280594, EPI_ISL_2280595, EPI_ISL_2280596, EPI_ISL_2280597, EPI_ISL_2280598, EPI_ISL_2280599, EPI_ISL_2280600, EPI_ISL_2280601, EPI_ISL_2280602, EPI_ISL_2280603, EPI_ISL_2280604, EPI_ISL_2280605, EPI_ISL_2280607, EPI_ISL_2280608, EPI_ISL_2280609, EPI_ISL_2280610, EPI_ISL_2280611, EPI_ISL_2280612, EPI_ISL_2280613, EPI_ISL_2280614, EPI_ISL_2280615, EPI_ISL_2280616, EPI_ISL_2280618, EPI_ISL_2280619, EPI_ISL_2280620, EPI_ISL_2280621, EPI_ISL_2280622, EPI_ISL_2280623, EPI_ISL_2280624, EPI_ISL_2280625, EPI_ISL_2280626, EPI_ISL_2280627, EPI_ISL_2280628, EPI_ISL_2280629, EPI_ISL_2280630, EPI_ISL_2280631, EPI_ISL_2280632, EPI_ISL_2280633, EPI_ISL_2280634, EPI_ISL_2280635, EPI_ISL_2280636, EPI_ISL_2280637, EPI_ISL_2280638, EPI_ISL_2280639, EPI_ISL_2280640, EPI_ISL_2280641, EPI_ISL_2280642, EPI_ISL_2280643, EPI_ISL_2280644, EPI_ISL_2280645, EPI_ISL_2280646, EPI_ISL_2280647, EPI_ISL_2280648, EPI_ISL_2280649, EPI_ISL_2280650, EPI_ISL_2280651, EPI_ISL_2280652, EPI_ISL_2280653, EPI_ISL_2280654, EPI_ISL_2280655, EPI_ISL_2280656, EPI_ISL_2280657, EPI_ISL_2280658, EPI_ISL_2280659, EPI_ISL_2280660, EPI_ISL_2280661, EPI_ISL_2280662, EPI_ISL_2280663, EPI_ISL_2280664, EPI_ISL_2280665, EPI_ISL_2280666, EPI_ISL_2280667, EPI_ISL_2280668, EPI_ISL_2280669, EPI_ISL_2280671, EPI_ISL_2280672, EPI_ISL_2280673, EPI_ISL_2280674, EPI_ISL_2280675, EPI_ISL_2280676, EPI_ISL_2280677, EPI_ISL_2280678, EPI_ISL_2280679, EPI_ISL_2280680, EPI_ISL_2280681, EPI_ISL_2280682, EPI_ISL_2280683, EPI_ISL_2280684, EPI_ISL_2280685, EPI_ISL_2280686, EPI_ISL_2280687, EPI_ISL_2280688, EPI_ISL_2280689, EPI_ISL_2280690, EPI_ISL_2280691, EPI_ISL_2280692, EPI_ISL_2280693, EPI_ISL_2280694, EPI_ISL_2280695, EPI_ISL_2280696, EPI_ISL_2280697, EPI_ISL_2280698, EPI_ISL_2280699, EPI_ISL_2280700, EPI_ISL_2280701, EPI_ISL_2280702, EPI_ISL_2280703, EPI_ISL_2280704, EPI_ISL_2280705, EPI_ISL_2280706, EPI_ISL_2280707, EPI_ISL_2280708, EPI_ISL_2280709, EPI_ISL_2280710, EPI_ISL_2280711, EPI_ISL_2280712, EPI_ISL_2280713, EPI_ISL_2280714, EPI_ISL_2280715, EPI_ISL_2280716, EPI_ISL_2280717, EPI_ISL_2280718, EPI_ISL_2280719, EPI_ISL_2280720, EPI_ISL_2280721, EPI_ISL_2280722, EPI_ISL_2280723, EPI_ISL_2280724, EPI_ISL_2280725, EPI_ISL_2280726, EPI_ISL_2280727, EPI_ISL_2280728, EPI_ISL_2280729, EPI_ISL_2280730, EPI_ISL_2280731, EPI_ISL_2280732, EPI_ISL_2280733, EPI_ISL_2280734, EPI_ISL_2280736, EPI_ISL_2280737, EPI_ISL_2280738, EPI_ISL_2280739, EPI_ISL_2280740, EPI_ISL_2280741, EPI_ISL_2280742, EPI_ISL_2280743, EPI_ISL_2280744, EPI_ISL_2280745, EPI_ISL_2280746, EPI_ISL_2280747, EPI_ISL_2280748, EPI_ISL_2280749, EPI_ISL_2280750, EPI_ISL_2280751, EPI_ISL_2280752, EPI_ISL_2280753, EPI_ISL_2280754, EPI_ISL_2280755, EPI_ISL_2280756, EPI_ISL_2280757, EPI_ISL_2280758, EPI_ISL_2280759, EPI_ISL_2280760, EPI_ISL_2280761, EPI_ISL_2280762, EPI_ISL_2280763, EPI_ISL_2280764, EPI_ISL_2280765, EPI_ISL_2280766, EPI_ISL_2280767, EPI_ISL_2280768, EPI_ISL_2280769, EPI_ISL_2280770, EPI_ISL_2280771, EPI_ISL_2280772, EPI_ISL_2280773, EPI_ISL_2280774, EPI_ISL_2280775, EPI_ISL_2280776, EPI_ISL_2280777, EPI_ISL_2280778, EPI_ISL_2280779, EPI_ISL_2280780, EPI_ISL_2280781, EPI_ISL_2280782, EPI_ISL_2280783, EPI_ISL_2280784, EPI_ISL_2280785, EPI_ISL_2280786, EPI_ISL_2280787, EPI_ISL_2280788, EPI_ISL_2280789, EPI_ISL_2280790, EPI_ISL_2280791, EPI_ISL_2280792, EPI_ISL_2280793, EPI_ISL_2280794, EPI_ISL_2280795, EPI_ISL_2280796, EPI_ISL_2280797, EPI_ISL_2280798, EPI_ISL_2280799, EPI_ISL_2280800, EPI_ISL_2280801, EPI_ISL_2280802, EPI_ISL_2280803, EPI_ISL_2280804, EPI_ISL_2280805, EPI_ISL_2280806, EPI_ISL_2280807, EPI_ISL_2280808, EPI_ISL_2280809, EPI_ISL_2280811, EPI_ISL_2280812, EPI_ISL_2280813, EPI_ISL_2280814, EPI_ISL_2280815, EPI_ISL_2280816, EPI_ISL_2280817, EPI_ISL_2280818, EPI_ISL_2280819, EPI_ISL_2280820, EPI_ISL_2280821, EPI_ISL_2280822, EPI_ISL_2280823, EPI_ISL_2280824, EPI_ISL_2280825, EPI_ISL_2280826, EPI_ISL_2280827, EPI_ISL_2280828, EPI_ISL_2280829, EPI_ISL_2280830, EPI_ISL_2280831, EPI_ISL_2280832, EPI_ISL_2280833, EPI_ISL_2280834, EPI_ISL_2280835, EPI_ISL_2280836, EPI_ISL_2280837, EPI_ISL_2280838, EPI_ISL_2280839, EPI_ISL_2280840, EPI_ISL_2280841, EPI_ISL_2280842, EPI_ISL_2280843, EPI_ISL_2280844, EPI_ISL_2280845, EPI_ISL_2280846, EPI_ISL_2280847, EPI_ISL_2280848, EPI_ISL_2280849, EPI_ISL_2280850, EPI_ISL_2280852, EPI_ISL_2280853, EPI_ISL_2280854, EPI_ISL_2280855, EPI_ISL_2280856, EPI_ISL_2280857, EPI_ISL_2280858, EPI_ISL_2280859, EPI_ISL_2280860, EPI_ISL_2280861, EPI_ISL_2280862, EPI_ISL_2280863, EPI_ISL_2280864, EPI_ISL_2280865, EPI_ISL_2280866, EPI_ISL_2280867, EPI_ISL_2280868, EPI_ISL_2280869, EPI_ISL_2280870, EPI_ISL_2280871, EPI_ISL_2280872, EPI_ISL_2280873, EPI_ISL_2280874, EPI_ISL_2280875, EPI_ISL_2280876, EPI_ISL_2280877, EPI_ISL_2280878, EPI_ISL_2280879, EPI_ISL_2280880, EPI_ISL_2280881, EPI_ISL_2280882, EPI_ISL_2280883, EPI_ISL_2280884, EPI_ISL_2280886, EPI_ISL_2280887, EPI_ISL_2280888, EPI_ISL_2280889, EPI_ISL_2280890, EPI_ISL_2280891, EPI_ISL_2280892, EPI_ISL_2280893, EPI_ISL_2280894, EPI_ISL_2280895, EPI_ISL_2280896, EPI_ISL_2280897, EPI_ISL_2280898, EPI_ISL_2280899, EPI_ISL_2280900, EPI_ISL_2280902, EPI_ISL_2280903, EPI_ISL_2280904, EPI_ISL_2280905, EPI_ISL_2280906, EPI_ISL_2280907, EPI_ISL_2280908, EPI_ISL_2280910, EPI_ISL_2280911, EPI_ISL_2280912, EPI_ISL_2280913, EPI_ISL_2280914, EPI_ISL_2280915, EPI_ISL_2280916, EPI_ISL_2280917, EPI_ISL_2280918, EPI_ISL_2280919, EPI_ISL_2280921, EPI_ISL_2280922, EPI_ISL_2280923, EPI_ISL_2280924, EPI_ISL_2280925, EPI_ISL_2280926, EPI_ISL_2280927, EPI_ISL_2280928, EPI_ISL_2280929, EPI_ISL_2280930, EPI_ISL_2280931, EPI_ISL_2280932, EPI_ISL_2280933, EPI_ISL_2280934, EPI_ISL_2280935, EPI_ISL_2280936, EPI_ISL_2280937, EPI_ISL_2280938, EPI_ISL_2280939, EPI_ISL_2280940, EPI_ISL_2280941, EPI_ISL_2280942, EPI_ISL_2280943, EPI_ISL_2280944, EPI_ISL_2280945, EPI_ISL_2280946, EPI_ISL_2280947, EPI_ISL_2280948, EPI_ISL_2280950, EPI_ISL_2280951, EPI_ISL_2280953, EPI_ISL_2280954, EPI_ISL_2280955, EPI_ISL_2280956, EPI_ISL_2280957, EPI_ISL_2280958, EPI_ISL_2280959, EPI_ISL_2280960, EPI_ISL_2280961, EPI_ISL_2280962, EPI_ISL_2280963, EPI_ISL_2280964, EPI_ISL_2280965, EPI_ISL_2280966, EPI_ISL_2280967, EPI_ISL_2280968, EPI_ISL_2280969, EPI_ISL_2280970, EPI_ISL_2280971, EPI_ISL_2280972, EPI_ISL_2280973, EPI_ISL_2280974, EPI_ISL_2280975, EPI_ISL_2280976, EPI_ISL_2280977, EPI_ISL_2280978, EPI_ISL_2280979, EPI_ISL_2280980, EPI_ISL_2280981, EPI_ISL_2280982, EPI_ISL_2280983, EPI_ISL_2280984, EPI_ISL_2280985, EPI_ISL_2280986, EPI_ISL_2280987, EPI_ISL_2280988, EPI_ISL_2280989, EPI_ISL_2280990, EPI_ISL_2280991, EPI_ISL_2280992, EPI_ISL_2280993, EPI_ISL_2280994, EPI_ISL_2280995, EPI_ISL_2280996, EPI_ISL_2280997, EPI_ISL_2280998, EPI_ISL_2280999, EPI_ISL_2281000, EPI_ISL_2281001, EPI_ISL_2281002, EPI_ISL_2281003, EPI_ISL_2281004, EPI_ISL_2281005, EPI_ISL_2281007, EPI_ISL_2281008, | | | |

| | | | |
|---|---|---|---|
| EPI_ISL_2281009, EPI_ISL_2281010, EPI_ISL_2281011, EPI_ISL_2281013, EPI_ISL_2281014, EPI_ISL_2281015, EPI_ISL_2281016, EPI_ISL_2281017, EPI_ISL_2281018, EPI_ISL_2281019, EPI_ISL_2281020, EPI_ISL_2281022, EPI_ISL_2281023, EPI_ISL_2281024, EPI_ISL_2281025, EPI_ISL_2281026, EPI_ISL_2281027, EPI_ISL_2281028, EPI_ISL_2281029, EPI_ISL_2281030, EPI_ISL_2281031, EPI_ISL_2281032, EPI_ISL_2281033, EPI_ISL_2281034, EPI_ISL_2281035, EPI_ISL_2281036, EPI_ISL_2281037, EPI_ISL_2281038, EPI_ISL_2281039, EPI_ISL_2281040, EPI_ISL_2281041, EPI_ISL_2281042, EPI_ISL_2281044, EPI_ISL_2281045, EPI_ISL_2281046, EPI_ISL_2281047, EPI_ISL_2281048, EPI_ISL_2281049, EPI_ISL_2281050, EPI_ISL_2281051, EPI_ISL_2281052, EPI_ISL_2281053, EPI_ISL_2281054, EPI_ISL_2281055, EPI_ISL_2281056, EPI_ISL_2281057, EPI_ISL_2281058, EPI_ISL_2281059, EPI_ISL_2281060, EPI_ISL_2281061, EPI_ISL_2281062, EPI_ISL_2281063, EPI_ISL_2281064, EPI_ISL_2281065, EPI_ISL_2281066, EPI_ISL_2281067, EPI_ISL_2281068, EPI_ISL_2281069, EPI_ISL_2281070, EPI_ISL_2281071, EPI_ISL_2281072, EPI_ISL_2281073, EPI_ISL_2281074, EPI_ISL_2281075, EPI_ISL_2281076, EPI_ISL_2281077, EPI_ISL_2281078, EPI_ISL_2281079, EPI_ISL_2281080, EPI_ISL_2281081, EPI_ISL_2281082, EPI_ISL_2281083, EPI_ISL_2281084, EPI_ISL_2281085, EPI_ISL_2281086, EPI_ISL_2281087, EPI_ISL_2281088, EPI_ISL_2281089, EPI_ISL_2281090, EPI_ISL_2281091, EPI_ISL_2281092, EPI_ISL_2281093, EPI_ISL_2281094, EPI_ISL_2281095, EPI_ISL_2281096, EPI_ISL_2281097, EPI_ISL_2281098, EPI_ISL_2281099, EPI_ISL_2281100, EPI_ISL_2281101, EPI_ISL_2281102, EPI_ISL_2281103, EPI_ISL_2281105, EPI_ISL_2281106, EPI_ISL_2281107, EPI_ISL_2281108, EPI_ISL_2281109, EPI_ISL_2281110, EPI_ISL_2281111, EPI_ISL_2281112, EPI_ISL_2281113, EPI_ISL_2281114, EPI_ISL_2281115, EPI_ISL_2281116, EPI_ISL_2281117, EPI_ISL_2281118, EPI_ISL_2281119, EPI_ISL_2281120, EPI_ISL_2281121, EPI_ISL_2281122, EPI_ISL_2281123, EPI_ISL_2281124, EPI_ISL_2281125, EPI_ISL_2281126, EPI_ISL_2281127, EPI_ISL_2281129, EPI_ISL_2281130, EPI_ISL_2281131, EPI_ISL_2281132, EPI_ISL_2281133, EPI_ISL_2281134, EPI_ISL_2281135, EPI_ISL_2281136, EPI_ISL_2281137, EPI_ISL_2281138, EPI_ISL_2281139, EPI_ISL_2281140, EPI_ISL_2281142, EPI_ISL_2281143, EPI_ISL_2281144, EPI_ISL_2281145, EPI_ISL_2281146, EPI_ISL_2281147, EPI_ISL_2281148, EPI_ISL_2281149, EPI_ISL_2281150, EPI_ISL_2281152, EPI_ISL_2281153, EPI_ISL_2281154, EPI_ISL_2281155, EPI_ISL_2281156, EPI_ISL_2281157, EPI_ISL_2281158, EPI_ISL_2281159, EPI_ISL_2281160, EPI_ISL_2281161, EPI_ISL_2281162, EPI_ISL_2281163, EPI_ISL_2281164, EPI_ISL_2281165, EPI_ISL_2281166, EPI_ISL_2281167, EPI_ISL_2281168, EPI_ISL_2281169, EPI_ISL_2281170, EPI_ISL_2281171, EPI_ISL_2281172, EPI_ISL_2281174, EPI_ISL_2281175, EPI_ISL_2281176, EPI_ISL_2281177, EPI_ISL_2281178, EPI_ISL_2281179, EPI_ISL_2281180, EPI_ISL_2281181, EPI_ISL_2281182, EPI_ISL_2281183, EPI_ISL_2281184, EPI_ISL_2281185, EPI_ISL_2281186, EPI_ISL_2281187, EPI_ISL_2281188, EPI_ISL_2281189, EPI_ISL_2281190, EPI_ISL_2281191, EPI_ISL_2281192, EPI_ISL_2281193, EPI_ISL_2281194, EPI_ISL_2281195, EPI_ISL_2281196, EPI_ISL_2281197, EPI_ISL_2281198, EPI_ISL_2281199, EPI_ISL_2281200, EPI_ISL_2281201, EPI_ISL_2281202, EPI_ISL_2281203, EPI_ISL_2281204, EPI_ISL_2281205, EPI_ISL_2281206, EPI_ISL_2281207, EPI_ISL_2281208, EPI_ISL_2281209, EPI_ISL_2281210, EPI_ISL_2281211, EPI_ISL_2281212, EPI_ISL_2281214, EPI_ISL_2281215, EPI_ISL_2281216, EPI_ISL_2281217, EPI_ISL_2281218, EPI_ISL_2281219, EPI_ISL_2281220, EPI_ISL_2281221, EPI_ISL_2281222, EPI_ISL_2281223, EPI_ISL_2281224, EPI_ISL_2281226 | | | |
| see above | Aegis Sciences Corporation | Centers for Disease Control and Prevention Division of Viral Diseases, Pathogen Discovery | Dakota Howard, Dhwani Batra, Peter W. Cook, Kara Moser, Adrian Paskey, Jason Caravas, Benjamin Rambo-Martin, Shatavia Morrison, Christopher Gulvick, Scott Sammons, Yvette Unoarumhi, Darlene Wagner, Matthew Schmerer, Cyndi Clark, Patrick Campbell, Rob Case, Vikramsinha Ghorpade, Holly Houdeshell, Ola Kvalvaag, Dillon Nall, Ethan Sanders, Alec Vest, Shaun Westlund, Matthew Hardison, Clinton R. Paden, Duncan MacCannell |
| EPI_ISL_2281306, EPI_ISL_2281339, EPI_ISL_2281344, EPI_ISL_2281347, EPI_ISL_2281348, EPI_ISL_2281352, EPI_ISL_2281358, EPI_ISL_2281361, EPI_ISL_2281367, EPI_ISL_2281370, EPI_ISL_2281383, EPI_ISL_2281392, EPI_ISL_2281413, EPI_ISL_2281415, EPI_ISL_2281417, EPI_ISL_2281418, EPI_ISL_2281420, EPI_ISL_2281421, EPI_ISL_2281422, EPI_ISL_2281424, EPI_ISL_2281427, EPI_ISL_2281430, EPI_ISL_2281431, EPI_ISL_2281432, EPI_ISL_2281437, EPI_ISL_2281441, EPI_ISL_2281444, EPI_ISL_2281449 | | | |
| see above | TGen North | TGen North | "Jolene Bowers, Heather Centner, Chris French, Hayley Yaglom, Ashlyn Pfeiffer, Darrin Lemmer, Dave Engelthaler, The Arizona COVID Genomics Union (ACGU)" |
| EPI_ISL_2281451 | TGen North | TGen North | Jolene Bowers, Heather Centner, Chris French, Hayley Yaglom, Ashlyn Pfeiffer, Darrin Lemmer, Dave Engelthaler, The Arizona COVID Genomics Union (ACGU) |
| EPI_ISL_2281456, EPI_ISL_2281457, EPI_ISL_2281458, EPI_ISL_2281459, EPI_ISL_2281463, EPI_ISL_2281464, EPI_ISL_2281466, EPI_ISL_2281470, EPI_ISL_2281474, EPI_ISL_2281478, EPI_ISL_2281479, EPI_ISL_2281481, EPI_ISL_2281483 | | | |
| see above | TGen North | TGen North | "Jolene Bowers, Heather Centner, Chris French, Hayley Yaglom, Ashlyn Pfeiffer, Darrin Lemmer, Dave Engelthaler, The Arizona COVID Genomics Union (ACGU)" |
| EPI_ISL_2281485 | TGen North | TGen North | Jolene Bowers, Heather Centner, Chris French, Hayley Yaglom, Ashlyn Pfeiffer, Darrin Lemmer, Dave Engelthaler, The Arizona COVID Genomics Union (ACGU) |
| EPI_ISL_2281488, EPI_ISL_2281490, EPI_ISL_2281491, EPI_ISL_2281492, EPI_ISL_2281494, EPI_ISL_2281496, EPI_ISL_2281498, EPI_ISL_2281500, EPI_ISL_2281501, EPI_ISL_2281503, EPI_ISL_2281509, EPI_ISL_2281510, EPI_ISL_2281512, EPI_ISL_2281513, EPI_ISL_2281515, EPI_ISL_2281516, EPI_ISL_2281517, EPI_ISL_2281519, EPI_ISL_2281520, EPI_ISL_2281521, EPI_ISL_2281526, EPI_ISL_2281528, EPI_ISL_2281532, EPI_ISL_2281533, EPI_ISL_2281535, EPI_ISL_2281536, EPI_ISL_2281537, EPI_ISL_2281539, EPI_ISL_2281541, EPI_ISL_2281542, EPI_ISL_2281543, EPI_ISL_2281546, EPI_ISL_2281549, EPI_ISL_2281552, EPI_ISL_2281553, EPI_ISL_2281554, EPI_ISL_2281556, EPI_ISL_2281561, EPI_ISL_2281567, EPI_ISL_2281569, EPI_ISL_2281570, EPI_ISL_2281571, EPI_ISL_2281579, EPI_ISL_2281580, EPI_ISL_2281582, EPI_ISL_2281585, EPI_ISL_2281590, EPI_ISL_2281594, EPI_ISL_2281597, EPI_ISL_2281598, EPI_ISL_2281599, EPI_ISL_2281600, EPI_ISL_2281601, EPI_ISL_2281604, EPI_ISL_2281607, EPI_ISL_2281608, EPI_ISL_2281609, EPI_ISL_2281613, EPI_ISL_2281621, EPI_ISL_2281622, EPI_ISL_2281623, EPI_ISL_2281624, EPI_ISL_2281629, EPI_ISL_2281633, EPI_ISL_2281636, EPI_ISL_2281638, EPI_ISL_2281639, EPI_ISL_2281641, EPI_ISL_2281642, EPI_ISL_2281643, EPI_ISL_2281646, EPI_ISL_2281652, EPI_ISL_2281653, EPI_ISL_2281654, EPI_ISL_2281661, EPI_ISL_2281662, EPI_ISL_2281663, EPI_ISL_2281665, EPI_ISL_2281668, EPI_ISL_2281670, EPI_ISL_2281672, EPI_ISL_2281676, EPI_ISL_2281679, EPI_ISL_2281681, EPI_ISL_2281682, EPI_ISL_2281689, EPI_ISL_2281690, EPI_ISL_2281691, EPI_ISL_2281698, EPI_ISL_2281700, EPI_ISL_2281705, EPI_ISL_2281706, EPI_ISL_2281708, EPI_ISL_2281709, EPI_ISL_2281710, EPI_ISL_2281714, EPI_ISL_2281716, EPI_ISL_2281717, EPI_ISL_2281719, EPI_ISL_2281720, EPI_ISL_2281721, EPI_ISL_2281731, EPI_ISL_2281732, EPI_ISL_2281733, EPI_ISL_2281734, EPI_ISL_2281736, EPI_ISL_2281737, EPI_ISL_2281739, EPI_ISL_2281740, EPI_ISL_2281742, EPI_ISL_2281748, EPI_ISL_2281749, EPI_ISL_2281751, EPI_ISL_2281753, EPI_ISL_2281754 | | | |
| see above | TGen North | TGen North | "Jolene Bowers, Heather Centner, Chris French, Hayley Yaglom, Ashlyn Pfeiffer, Darrin Lemmer, Dave Engelthaler, The Arizona COVID Genomics Union (ACGU)" |
| EPI_ISL_2281846, EPI_ISL_2281847, EPI_ISL_2281848, EPI_ISL_2281849, EPI_ISL_2281850, EPI_ISL_2281851, EPI_ISL_2281852, EPI_ISL_2281853, EPI_ISL_2281854, EPI_ISL_2281855, EPI_ISL_2281856, EPI_ISL_2281857, EPI_ISL_2281859, EPI_ISL_2281860, EPI_ISL_2281861, EPI_ISL_2281862, EPI_ISL_2281863, EPI_ISL_2281864, EPI_ISL_2281866, EPI_ISL_2281867, EPI_ISL_2281868, EPI_ISL_2281869, EPI_ISL_2281870, EPI_ISL_2281871, EPI_ISL_2281873, EPI_ISL_2281875, EPI_ISL_2281876, EPI_ISL_2281877, EPI_ISL_2281878, EPI_ISL_2281879, EPI_ISL_2281880, EPI_ISL_2281881, EPI_ISL_2281882, EPI_ISL_2281883, EPI_ISL_2281884, EPI_ISL_2281887, EPI_ISL_2281888, EPI_ISL_2281889 | | | |
| see above | NJDOH, Public Health and Environmental Laboratories | NJ_PHEL | Lindsey Bodnar, Shiv K. Verma, Jacquelyn Deverell, Ryan Pachucki, Dana Woell, Byeong Jeong |
| EPI_ISL_2282015 | NJDOH, Public Health and Environmental Laboratories | NJ_PHEL | Lindsey Bodnar, Shiv K. Verma, Jacquelyn Deverell, Ryan Pachuchi, Dana Woell, Byeong Jeong |
| EPI_ISL_2282016, EPI_ISL_2282018, EPI_ISL_2282020, EPI_ISL_2282022, EPI_ISL_2282024, EPI_ISL_2282026, EPI_ISL_2282028, EPI_ISL_2282029, EPI_ISL_2282031, EPI_ISL_2282035, EPI_ISL_2282037, EPI_ISL_2282038, EPI_ISL_2282040, EPI_ISL_2282042, EPI_ISL_2282044, EPI_ISL_2282046, EPI_ISL_2282047 | | | |
| see above | NJDOH, Public Health and Environmental Laboratories | NJ_PHEL | Lindsey Bodnar, Shiv K. Verma, Jacquelyn Deverell, Matthew Scarnati, Dana Woell, Byeong Jeong |
| EPI_ISL_2282235, EPI_ISL_2282236, EPI_ISL_2282237, EPI_ISL_2282238, EPI_ISL_2282239, EPI_ISL_2282240, EPI_ISL_2282241, EPI_ISL_2282242, EPI_ISL_2282243, EPI_ISL_2282244, EPI_ISL_2282245, EPI_ISL_2282246 | | | |
| see above | Weill Cornell Medicine | New York Genome Center | Michael Zody, Andre Corvelo, Dayna M. Oschwald, Samantha Fennessey, Tom Maniatis, Melissa Cushing, Olivier Elemento, Margaret Elizabeth Ross, Chris Mason, Priya Velu, Hanna Rennert, Arryn Craney, Lars F Westblade |
| EPI_ISL_2282249, EPI_ISL_2282250, EPI_ISL_2282251, EPI_ISL_2282252, EPI_ISL_2282253, EPI_ISL_2282254, EPI_ISL_2282255, EPI_ISL_2282256, EPI_ISL_2282257, EPI_ISL_2282258, EPI_ISL_2282259, EPI_ISL_2282260, EPI_ISL_2282261, EPI_ISL_2282262, EPI_ISL_2282263, EPI_ISL_2282265, EPI_ISL_2282266, EPI_ISL_2282267, EPI_ISL_2282269, EPI_ISL_2282270, EPI_ISL_2282271, EPI_ISL_2282272, EPI_ISL_2282273, EPI_ISL_2282274, EPI_ISL_2282275, EPI_ISL_2282277, EPI_ISL_2282278, EPI_ISL_2282279, EPI_ISL_2282280, EPI_ISL_2282281, EPI_ISL_2282282, EPI_ISL_2282283, EPI_ISL_2282285, EPI_ISL_2282286, EPI_ISL_2282287, EPI_ISL_2282288, EPI_ISL_2282289, EPI_ISL_2282290, EPI_ISL_2282291, EPI_ISL_2282292, EPI_ISL_2282293, EPI_ISL_2282294, EPI_ISL_2282295, EPI_ISL_2282296, EPI_ISL_2282297, EPI_ISL_2282299, EPI_ISL_2282300, EPI_ISL_2282301, EPI_ISL_2282303, EPI_ISL_2282304, EPI_ISL_2282306, EPI_ISL_2282307, EPI_ISL_2282308, EPI_ISL_2282309, EPI_ISL_2282310, EPI_ISL_2282311, EPI_ISL_2282314, EPI_ISL_2282315, EPI_ISL_2282316, EPI_ISL_2282317, EPI_ISL_2282319, EPI_ISL_2282320, EPI_ISL_2282321, EPI_ISL_2282322, EPI_ISL_2282323, EPI_ISL_2282324, EPI_ISL_2282325, EPI_ISL_2282326, EPI_ISL_2282327, EPI_ISL_2282328, EPI_ISL_2282329, EPI_ISL_2282330, EPI_ISL_2282331, EPI_ISL_2282332, EPI_ISL_2282333, EPI_ISL_2282334, EPI_ISL_2282335, EPI_ISL_2282336, EPI_ISL_2282337, EPI_ISL_2282338, EPI_ISL_2282339, EPI_ISL_2282340, EPI_ISL_2282341, EPI_ISL_2282342, EPI_ISL_2282343, EPI_ISL_2282344, EPI_ISL_2282345, EPI_ISL_2282348, EPI_ISL_2282349, EPI_ISL_2282350, EPI_ISL_2282351, EPI_ISL_2282352, EPI_ISL_2282353, EPI_ISL_2282355, EPI_ISL_2282356, EPI_ISL_2282357, EPI_ISL_2282358, EPI_ISL_2282359, EPI_ISL_2282360, EPI_ISL_2282361, EPI_ISL_2282362, EPI_ISL_2282363, EPI_ISL_2282365, EPI_ISL_2282366, EPI_ISL_2282367, EPI_ISL_2282368, EPI_ISL_2282370, EPI_ISL_2282371, EPI_ISL_2282372, EPI_ISL_2282373, EPI_ISL_2282374, EPI_ISL_2282375, EPI_ISL_2282376, EPI_ISL_2282377, EPI_ISL_2282379, EPI_ISL_2282381, EPI_ISL_2282383, EPI_ISL_2282384, EPI_ISL_2282385, EPI_ISL_2282386, EPI_ISL_2282387, EPI_ISL_2282388, EPI_ISL_2282389, EPI_ISL_2282390, EPI_ISL_2282391, EPI_ISL_2282392, EPI_ISL_2282393, EPI_ISL_2282394, EPI_ISL_2282395, EPI_ISL_2282396, EPI_ISL_2282398, EPI_ISL_2282399, EPI_ISL_2282400, EPI_ISL_2282401, EPI_ISL_2282402, EPI_ISL_2282403, EPI_ISL_2282404, EPI_ISL_2282405, EPI_ISL_2282406, EPI_ISL_2282407, EPI_ISL_2282408, EPI_ISL_2282409, EPI_ISL_2282410, EPI_ISL_2282411, EPI_ISL_2282412, EPI_ISL_2282413, EPI_ISL_2282414, EPI_ISL_2282415, EPI_ISL_2282416, EPI_ISL_2282417, EPI_ISL_2282418, EPI_ISL_2282419, EPI_ISL_2282420, EPI_ISL_2282421, EPI_ISL_2282422, EPI_ISL_2282423, EPI_ISL_2282424, EPI_ISL_2282426, EPI_ISL_2282427, EPI_ISL_2282428, EPI_ISL_2282429, EPI_ISL_2282431, EPI_ISL_2282432, EPI_ISL_2282433, EPI_ISL_2282434, EPI_ISL_2282436, EPI_ISL_2282437, EPI_ISL_2282438, EPI_ISL_2282439, EPI_ISL_2282440, EPI_ISL_2282441, EPI_ISL_2282442, EPI_ISL_2282443, EPI_ISL_2282444, EPI_ISL_2282445, EPI_ISL_2282446, EPI_ISL_2282447, EPI_ISL_2282448, EPI_ISL_2282449, EPI_ISL_2282450, EPI_ISL_2282451, EPI_ISL_2282452, EPI_ISL_2282453, EPI_ISL_2282454, EPI_ISL_2282455, EPI_ISL_2282456, EPI_ISL_2282457, EPI_ISL_2282458, EPI_ISL_2282459, EPI_ISL_2282460, EPI_ISL_2282461, EPI_ISL_2282462, EPI_ISL_2282463, EPI_ISL_2282464, EPI_ISL_2282465, EPI_ISL_2282467, EPI_ISL_2282468, EPI_ISL_2282469, EPI_ISL_2282470, EPI_ISL_2282471, EPI_ISL_2282472, EPI_ISL_2282473, EPI_ISL_2282474, EPI_ISL_2282475, EPI_ISL_2282476, EPI_ISL_2282477, EPI_ISL_2282478, EPI_ISL_2282479, EPI_ISL_2282480, EPI_ISL_2282481, EPI_ISL_2282482, EPI_ISL_2282483, EPI_ISL_2282484, EPI_ISL_2282485, EPI_ISL_2282486, EPI_ISL_2282487, EPI_ISL_2282489, EPI_ISL_2282490, EPI_ISL_2282491, EPI_ISL_2282492, EPI_ISL_2282493, EPI_ISL_2282495, EPI_ISL_2282496, EPI_ISL_2282497, EPI_ISL_2282498, EPI_ISL_2282499, EPI_ISL_2282500, EPI_ISL_2282501, EPI_ISL_2282502, EPI_ISL_2282503, EPI_ISL_2282504, EPI_ISL_2282506, EPI_ISL_2282507, EPI_ISL_2282508, EPI_ISL_2282509, EPI_ISL_2282510, EPI_ISL_2282511, EPI_ISL_2282512, EPI_ISL_2282513, EPI_ISL_2282514, EPI_ISL_2282515, EPI_ISL_2282516, EPI_ISL_2282517, EPI_ISL_2282518, EPI_ISL_2282519, EPI_ISL_2282520, EPI_ISL_2282521, EPI_ISL_2282522, EPI_ISL_2282523, EPI_ISL_2282524, EPI_ISL_2282525, EPI_ISL_2282527, EPI_ISL_2282528, EPI_ISL_2282529, EPI_ISL_2282531, EPI_ISL_2282532, EPI_ISL_2282533, EPI_ISL_2282534, EPI_ISL_2282535, EPI_ISL_2282536, EPI_ISL_2282537, EPI_ISL_2282538, EPI_ISL_2282539, EPI_ISL_2282540, EPI_ISL_2282541, EPI_ISL_2282542, EPI_ISL_2282543, EPI_ISL_2282544, EPI_ISL_2282545, EPI_ISL_2282546, EPI_ISL_2282547, EPI_ISL_2282549, EPI_ISL_2282550, EPI_ISL_2282551, EPI_ISL_2282552, EPI_ISL_2282553, EPI_ISL_2282554, EPI_ISL_2282555, EPI_ISL_2282557, EPI_ISL_2282558, EPI_ISL_2282559, EPI_ISL_2282561, EPI_ISL_2282562, EPI_ISL_2282563, EPI_ISL_2282564, EPI_ISL_2282565, EPI_ISL_2282566, EPI_ISL_2282567, EPI_ISL_2282568, EPI_ISL_2282569, EPI_ISL_2282570, EPI_ISL_2282571, EPI_ISL_2282572, EPI_ISL_2282573, EPI_ISL_2282574, EPI_ISL_2282575, EPI_ISL_2282576, EPI_ISL_2282577, EPI_ISL_2282578, EPI_ISL_2282579, EPI_ISL_2282580, EPI_ISL_2282581, EPI_ISL_2282582, EPI_ISL_2282583, EPI_ISL_2282584, EPI_ISL_2282585, EPI_ISL_2282586, EPI_ISL_2282587, EPI_ISL_2282588, EPI_ISL_2282589, EPI_ISL_2282590, EPI_ISL_2282591, EPI_ISL_2282592, EPI_ISL_2282593, EPI_ISL_2282594, EPI_ISL_2282595, EPI_ISL_2282596, EPI_ISL_2282598, EPI_ISL_2282599, EPI_ISL_2282600, EPI_ISL_2282601, EPI_ISL_2282602, EPI_ISL_2282603, EPI_ISL_2282604, EPI_ISL_2282606, EPI_ISL_2282607, EPI_ISL_2282608, EPI_ISL_2282609, EPI_ISL_2282610, EPI_ISL_2282611, EPI_ISL_2282612, EPI_ISL_2282613, EPI_ISL_2282614, EPI_ISL_2282615, EPI_ISL_2282616, EPI_ISL_2282617, EPI_ISL_2282618, EPI_ISL_2282619, EPI_ISL_2282620, EPI_ISL_2282621, EPI_ISL_2282622, EPI_ISL_2282623, EPI_ISL_2282624, EPI_ISL_2282625, EPI_ISL_2282626, EPI_ISL_2282627, EPI_ISL_2282628, EPI_ISL_2282629, EPI_ISL_2282630, EPI_ISL_2282631, EPI_ISL_2282632, EPI_ISL_2282634, EPI_ISL_2282635, EPI_ISL_2282638, EPI_ISL_2282639, EPI_ISL_2282640, EPI_ISL_2282641, EPI_ISL_2282642, EPI_ISL_2282643, EPI_ISL_2282644, EPI_ISL_2282645, EPI_ISL_2282646, EPI_ISL_2282648, EPI_ISL_2282649, EPI_ISL_2282650, EPI_ISL_2282651, EPI_ISL_2282652, EPI_ISL_2282653, EPI_ISL_2282654, EPI_ISL_2282655, EPI_ISL_2282656, EPI_ISL_2282657, EPI_ISL_2282658, EPI_ISL_2282659, EPI_ISL_2282660, EPI_ISL_2282661, EPI_ISL_2282662, EPI_ISL_2282663, EPI_ISL_2282664, EPI_ISL_2282665, EPI_ISL_2282666, EPI_ISL_2282667, EPI_ISL_2282668, EPI_ISL_2282669, EPI_ISL_2282670, EPI_ISL_2282671, EPI_ISL_2282672, EPI_ISL_2282673, EPI_ISL_2282674, EPI_ISL_2282675, EPI_ISL_2282676, EPI_ISL_2282677, EPI_ISL_2282679, EPI_ISL_2282680, EPI_ISL_2282681, EPI_ISL_2282682, EPI_ISL_2282683, EPI_ISL_2282684, EPI_ISL_2282685, EPI_ISL_2282686, EPI_ISL_2282687, EPI_ISL_2282688, EPI_ISL_2282689, EPI_ISL_2282690, EPI_ISL_2282691, EPI_ISL_2282692, EPI_ISL_2282693, EPI_ISL_2282694, EPI_ISL_2282695, EPI_ISL_2282696, EPI_ISL_2282697, EPI_ISL_2282698, EPI_ISL_2282699, EPI_ISL_2282700, EPI_ISL_2282702, EPI_ISL_2282704, EPI_ISL_2282705, EPI_ISL_2282706, EPI_ISL_2282707, EPI_ISL_2282708, EPI_ISL_2282709, EPI_ISL_2282710, EPI_ISL_2282711, EPI_ISL_2282712, EPI_ISL_2282713, EPI_ISL_2282714, EPI_ISL_2282715, EPI_ISL_2282716, EPI_ISL_2282717, EPI_ISL_2282718, EPI_ISL_2282719, EPI_ISL_2282720, EPI_ISL_2282721, EPI_ISL_2282722, EPI_ISL_2282723, EPI_ISL_2282724, EPI_ISL_2282725, EPI_ISL_2282726, EPI_ISL_2282727, EPI_ISL_2282728, EPI_ISL_2282729, EPI_ISL_2282730, EPI_ISL_2282731, EPI_ISL_2282732, EPI_ISL_2282733, EPI_ISL_2282734, EPI_ISL_2282735, EPI_ISL_2282736, EPI_ISL_2282737, EPI_ISL_2282738, EPI_ISL_2282739, EPI_ISL_2282740, EPI_ISL_2282741, EPI_ISL_2282742, EPI_ISL_2282743, EPI_ISL_2282744, EPI_ISL_2282745, EPI_ISL_2282746, EPI_ISL_2282747, EPI_ISL_2282748, EPI_ISL_2282749, EPI_ISL_2282750, EPI_ISL_2282751, EPI_ISL_2282752, EPI_ISL_2282753, EPI_ISL_2282754, EPI_ISL_2282756, EPI_ISL_2282757, EPI_ISL_2282758, EPI_ISL_2282759, EPI_ISL_2282760, EPI_ISL_2282761, EPI_ISL_2282762, EPI_ISL_2282763, EPI_ISL_2282764, EPI_ISL_2282765, EPI_ISL_2282766, EPI_ISL_2282767, EPI_ISL_2282768, EPI_ISL_2282769, EPI_ISL_2282770, EPI_ISL_2282771, EPI_ISL_2282772, EPI_ISL_2282773, EPI_ISL_2282774, EPI_ISL_2282775, EPI_ISL_2282776, EPI_ISL_2282777, EPI_ISL_2282778, EPI_ISL_2282779, EPI_ISL_2282780, EPI_ISL_2282781, EPI_ISL_2282783, EPI_ISL_2282785, EPI_ISL_2282786, EPI_ISL_2282787, EPI_ISL_2282788, EPI_ISL_2282789, EPI_ISL_2282790, EPI_ISL_2282791, EPI_ISL_2282792, EPI_ISL_2282793, EPI_ISL_2282794, EPI_ISL_2282795, EPI_ISL_2282796, EPI_ISL_2282797, EPI_ISL_2282798, EPI_ISL_2282799, EPI_ISL_2282800, EPI_ISL_2282801, EPI_ISL_2282803, EPI_ISL_2282804, EPI_ISL_2282805, EPI_ISL_2282806, EPI_ISL_2282807, EPI_ISL_2282808, EPI_ISL_2282809, EPI_ISL_2282810, EPI_ISL_2282811, EPI_ISL_2282812, EPI_ISL_2282813, EPI_ISL_2282814, EPI_ISL_2282815, EPI_ISL_2282817, EPI_ISL_2282818, EPI_ISL_2282819, EPI_ISL_2282820, EPI_ISL_2282821, EPI_ISL_2282822, EPI_ISL_2282823, EPI_ISL_2282824, EPI_ISL_2282825, EPI_ISL_2282826, EPI_ISL_2282827, EPI_ISL_2282828, EPI_ISL_2282829, EPI_ISL_2282830, EPI_ISL_2282831, EPI_ISL_2282832, EPI_ISL_2282833, EPI_ISL_2282834, EPI_ISL_2282835, EPI_ISL_2282836, EPI_ISL_2282837, EPI_ISL_2282838, EPI_ISL_2282839, EPI_ISL_2282840, EPI_ISL_2282841, EPI_ISL_2282842, EPI_ISL_2282843, EPI_ISL_2282844, EPI_ISL_2282846, EPI_ISL_2282847, EPI_ISL_2282848, EPI_ISL_2282849, EPI_ISL_2282850, EPI_ISL_2282851, EPI_ISL_2282852, EPI_ISL_2282853, EPI_ISL_2282854, EPI_ISL_2282855, EPI_ISL_2282856, EPI_ISL_2282857, EPI_ISL_2282858, EPI_ISL_2282859, EPI_ISL_2282860, EPI_ISL_2282861, EPI_ISL_2282862, EPI_ISL_2282863, EPI_ISL_2282864, EPI_ISL_2282865, EPI_ISL_2282866, EPI_ISL_2282867, EPI_ISL_2282868, EPI_ISL_2282869, EPI_ISL_2282870, EPI_ISL_2282872, EPI_ISL_2282873, EPI_ISL_2282874, EPI_ISL_2282875, EPI_ISL_2282876, EPI_ISL_2282877, EPI_ISL_2282878, EPI_ISL_2282879, EPI_ISL_2282880, EPI_ISL_2282882, EPI_ISL_2282883, EPI_ISL_2282884, EPI_ISL_2282885, EPI_ISL_2282886, EPI_ISL_2282888, EPI_ISL_2282889, EPI_ISL_2282892, EPI_ISL_2282894, EPI_ISL_2282895, EPI_ISL_2282896, EPI_ISL_2282897, EPI_ISL_2282899, EPI_ISL_2282901, EPI_ISL_2282902, EPI_ISL_2282903, EPI_ISL_2282904, EPI_ISL_2282905, EPI_ISL_2282906, EPI_ISL_2282908, EPI_ISL_2282909, EPI_ISL_2282910, EPI_ISL_2282912, EPI_ISL_2282913, EPI_ISL_2282914, EPI_ISL_2282915, EPI_ISL_2282916, EPI_ISL_2282917, EPI_ISL_2282918, EPI_ISL_2282919, EPI_ISL_2282920, EPI_ISL_2282921, EPI_ISL_2282922, EPI_ISL_2282923, EPI_ISL_2282924, EPI_ISL_2282925, EPI_ISL_2282926, EPI_ISL_2282927, EPI_ISL_2282928, EPI_ISL_2282929, EPI_ISL_2282930, EPI_ISL_2282931, EPI_ISL_2282932, EPI_ISL_2282933, EPI_ISL_2282934, EPI_ISL_2282935, EPI_ISL_2282936, EPI_ISL_2282937, EPI_ISL_2282938, EPI_ISL_2282939, EPI_ISL_2282940, EPI_ISL_2282942, EPI_ISL_2282943, EPI_ISL_2282944, EPI_ISL_2282945, EPI_ISL_2282946, EPI_ISL_2282947, EPI_ISL_2282949, EPI_ISL_2282950, EPI_ISL_2282951, EPI_ISL_2282952, EPI_ISL_2282953, EPI_ISL_2282954, EPI_ISL_2282955, EPI_ISL_2282956, EPI_ISL_2282957, EPI_ISL_2282958, EPI_ISL_2282959, EPI_ISL_2282960, EPI_ISL_2282961, EPI_ISL_2282963, EPI_ISL_2282964, EPI_ISL_2282965, EPI_ISL_2282966, EPI_ISL_2282967, EPI_ISL_2282968, EPI_ISL_2282969, EPI_ISL_2282970, EPI_ISL_2282971, EPI_ISL_2282972, EPI_ISL_2282973, EPI_ISL_2282974, EPI_ISL_2282975, EPI_ISL_2282976, EPI_ISL_2282977, EPI_ISL_2282978, EPI_ISL_2282979, EPI_ISL_2282980, EPI_ISL_2282981, EPI_ISL_2282982, EPI_ISL_2282983, EPI_ISL_2282984, EPI_ISL_2282985, EPI_ISL_2282986, EPI_ISL_2282987, EPI_ISL_2282988, EPI_ISL_2282990, EPI_ISL_2282991, EPI_ISL_2282992, EPI_ISL_2282993, EPI_ISL_2282994, EPI_ISL_2282995, EPI_ISL_2282996, EPI_ISL_2282997, EPI_ISL_2282998, EPI_ISL_2282999, EPI_ISL_2283000, EPI_ISL_2283001, EPI_ISL_2283002, EPI_ISL_2283003, EPI_ISL_2283004, EPI_ISL_2283005, EPI_ISL_2283006, EPI_ISL_2283007, EPI_ISL_2283008, EPI_ISL_2283009, EPI_ISL_2283010, EPI_ISL_2283011, EPI_ISL_2283012, EPI_ISL_2283013, EPI_ISL_2283014, EPI_ISL_2283016, EPI_ISL_2283017, EPI_ISL_2283018, EPI_ISL_2283019, EPI_ISL_2283020, EPI_ISL_2283021, EPI_ISL_2283022, EPI_ISL_2283023, EPI_ISL_2283024, EPI_ISL_2283025, EPI_ISL_2283026, EPI_ISL_2283027, EPI_ISL_2283028, EPI_ISL_2283029, EPI_ISL_2283030, EPI_ISL_2283031, EPI_ISL_2283032, EPI_ISL_2283033, EPI_ISL_2283034, EPI_ISL_2283035, EPI_ISL_2283036, EPI_ISL_2283037, EPI_ISL_2283038, EPI_ISL_2283039, EPI_ISL_2283040, EPI_ISL_2283041, EPI_ISL_2283042, EPI_ISL_2283043, EPI_ISL_2283044, EPI_ISL_2283045, EPI_ISL_2283046, EPI_ISL_2283047, EPI_ISL_2283048, EPI_ISL_2283049, EPI_ISL_2283050, EPI_ISL_2283051, EPI_ISL_2283052, EPI_ISL_2283053, EPI_ISL_2283054, EPI_ISL_2283055, EPI_ISL_2283056, EPI_ISL_2283057, EPI_ISL_2283059, EPI_ISL_2283060, EPI_ISL_2283061, EPI_ISL_2283062, EPI_ISL_2283063, EPI_ISL_2283064, EPI_ISL_2283065, EPI_ISL_2283066, EPI_ISL_2283067, EPI_ISL_2283068, EPI_ISL_2283069, EPI_ISL_2283070, EPI_ISL_2283071, EPI_ISL_2283072, EPI_ISL_2283073, EPI_ISL_2283074, EPI_ISL_2283075, EPI_ISL_2283076, EPI_ISL_2283077, EPI_ISL_2283079, EPI_ISL_2283080, EPI_ISL_2283081, EPI_ISL_2283082, EPI_ISL_2283083, EPI_ISL_2283084, EPI_ISL_2283085, EPI_ISL_2283086, EPI_ISL_2283087, EPI_ISL_2283088, EPI_ISL_2283089, EPI_ISL_2283090, EPI_ISL_2283091, EPI_ISL_2283093, EPI_ISL_2283094, EPI_ISL_2283095, EPI_ISL_2283096, EPI_ISL_2283097, EPI_ISL_2283098, EPI_ISL_2283099, EPI_ISL_2283100, EPI_ISL_2283101, EPI_ISL_2283102, EPI_ISL_2283103, EPI_ISL_2283104, EPI_ISL_2283105, EPI_ISL_2283106, EPI_ISL_2283107, EPI_ISL_2283108, EPI_ISL_2283109, EPI_ISL_2283110, EPI_ISL_2283111, EPI_ISL_2283112, EPI_ISL_2283113 | | | |
| see above | Infinity Biologix | Centers for Disease Control and Prevention Division of Viral Diseases, Pathogen Discovery | Dakota Howard, Dhwani Batra, Peter W. Cook, Kara Moser, Adrian Paskey, Jason Caravas, Benjamin Rambo-Martin, Shatavia Morrison, Christopher Gulvick, Scott Sammons, Yvette Unoarumhi, Darlene Wagner, Matthew Schmerer, Christian Bixby, Yihe Wang, Jonathan Schultz, Chirayu Goswami, Russ Hager, Robin Grimwood, Clinton R. Paden, Duncan MacCannell |
| EPI_ISL_2283114 | Weill Cornell Medicine | New York Genome Center | Michael Zody, Andre Corvelo, Dayna M. Oschwald, Samantha Fennessey, Tom Maniatis, Melissa Cushing, Olivier Elemento, Margaret Elizabeth Ross, Chris Mason, Priya Velu, Hanna Rennert, Arryn Craney, Lars F Westblade |
| EPI_ISL_2283115 | Infinity Biologix | Centers for Disease Control and Prevention Division of Viral Diseases, Pathogen Discovery | Dakota Howard, Dhwani Batra, Peter W. Cook, Kara Moser, Adrian Paskey, Jason Caravas, Benjamin Rambo-Martin, Shatavia Morrison, Christopher Gulvick, Scott Sammons, Yvette Unoarumhi, Darlene Wagner, Matthew Schmerer, Christian Bixby, Yihe Wang, Jonathan Schultz, Chirayu Goswami, Russ Hager, Robin Grimwood, Clinton R. Paden, Duncan MacCannell |
| EPI_ISL_2283116, EPI_ISL_2283117 | Weill Cornell Medicine | New York Genome Center | Michael Zody, Andre Corvelo, Dayna M. Oschwald, Samantha Fennessey, Tom Maniatis, Melissa Cushing, Olivier Elemento, Margaret Elizabeth Ross, Chris Mason, Priya Velu, Hanna Rennert, Arryn Craney, Lars F Westblade |
| EPI_ISL_2283118 | Infinity Biologix | Centers for Disease Control and Prevention Division of Viral Diseases, Pathogen Discovery | Dakota Howard, Dhwani Batra, Peter W. Cook, Kara Moser, Adrian Paskey, Jason Caravas, Benjamin Rambo-Martin, Shatavia Morrison, Christopher Gulvick, Scott Sammons, Yvette Unoarumhi, Darlene Wagner, Matthew Schmerer, Christian Bixby, Yihe Wang, Jonathan Schultz, Chirayu Goswami, Russ Hager, Robin Grimwood, Clinton R. Paden, Duncan MacCannell |
| EPI_ISL_2283119 | Weill Cornell Medicine | New York Genome Center | Michael Zody, Andre Corvelo, Dayna M. Oschwald, Samantha Fennessey, Tom Maniatis, Melissa Cushing, Olivier Elemento, Margaret Elizabeth Ross, Chris Mason, Priya Velu, Hanna Rennert, Arryn Craney, Lars F Westblade |
| EPI_ISL_2283120 | Infinity Biologix | Centers for Disease Control and Prevention Division of Viral Diseases, Pathogen Discovery | Dakota Howard, Dhwani Batra, Peter W. Cook, Kara Moser, Adrian Paskey, Jason Caravas, Benjamin Rambo-Martin, Shatavia Morrison, Christopher Gulvick, Scott Sammons, Yvette Unoarumhi, Darlene Wagner, Matthew Schmerer, Christian Bixby, Yihe Wang, Jonathan Schultz, Chirayu Goswami, Russ Hager, Robin Grimwood, Clinton R. Paden, Duncan MacCannell |
| EPI_ISL_2283121, EPI_ISL_2283122 | Weill Cornell Medicine | New York Genome Center | Michael Zody, Andre Corvelo, Dayna M. Oschwald, Samantha Fennessey, Tom Maniatis, Melissa Cushing, Olivier Elemento, Margaret Elizabeth Ross, Chris Mason, Priya Velu, Hanna Rennert, Arryn Craney, Lars F Westblade |
| EPI_ISL_2283123 | Infinity Biologix | Centers for Disease Control and Prevention Division of Viral Diseases, Pathogen Discovery | Dakota Howard, Dhwani Batra, Peter W. Cook, Kara Moser, Adrian Paskey, Jason Caravas, Benjamin Rambo-Martin, Shatavia Morrison, Christopher Gulvick, Scott Sammons, Yvette Unoarumhi, Darlene Wagner, Matthew Schmerer, Christian Bixby, Yihe Wang, Jonathan Schultz, Chirayu Goswami, Russ Hager, Robin Grimwood, Clinton R. Paden, Duncan MacCannell |
| EPI_ISL_2283124 | Weill Cornell Medicine | New York Genome Center | Michael Zody, Andre Corvelo, Dayna M. Oschwald, Samantha Fennessey, Tom Maniatis, Melissa Cushing, Olivier Elemento, Margaret Elizabeth Ross, Chris Mason, Priya Velu, Hanna Rennert, Arryn Craney, Lars F Westblade |
| EPI_ISL_2283125 | Infinity Biologix | Centers for Disease Control and Prevention | Dakota Howard, Dhwani Batra, Peter W. Cook, Kara Moser, Adrian Paskey, Jason Caravas, Benjamin Rambo-Martin, Shatavia Morrison, Christopher Gulvick, Scott Sammons, Yvette Unoarumhi, Darlene Wagner, Matthew Schmerer, Christian Bixby, Yihe Wang, Jonathan Schultz, Chirayu |

| | | | |
|---|---|---|---|
| | | Division of Viral Diseases, Pathogen Discovery | Goswami, Russ Hager, Robin Grimwood, Clinton R. Paden, Duncan MacCannell |
| EPI_ISL_2283126, EPI_ISL_2283127 | Weill Cornell Medicine | New York Genome Center | Michael Zody, Andre Corvelo, Dayna M. Oschwald, Samantha Fennessey, Tom Maniatis, Melissa Cushing, Olivier Elemento, Margaret Elizabeth Ross, Chris Mason, Priya Velu, Hanna Rennert, Arryn Craney, Lars F Westblade |
| EPI_ISL_2283128 | Infinity Biologix | Centers for Disease Control and Prevention Division of Viral Diseases, Pathogen Discovery | Dakota Howard, Dhwani Batra, Peter W. Cook, Kara Moser, Adrian Paskey, Jason Caravas, Benjamin Rambo-Martin, Shatavia Morrison, Christopher Gulvick, Scott Sammons, Yvette Unoarumhi, Darlene Wagner, Matthew Schmerer, Christian Bixby, Yihe Wang, Jonathan Schultz, Chirayu Goswami, Russ Hager, Robin Grimwood, Clinton R. Paden, Duncan MacCannell |
| EPI_ISL_2283129, EPI_ISL_2283130 | Weill Cornell Medicine | New York Genome Center | Michael Zody, Andre Corvelo, Dayna M. Oschwald, Samantha Fennessey, Tom Maniatis, Melissa Cushing, Olivier Elemento, Margaret Elizabeth Ross, Chris Mason, Priya Velu, Hanna Rennert, Arryn Craney, Lars F Westblade |
| EPI_ISL_2283131 | Infinity Biologix | Centers for Disease Control and Prevention Division of Viral Diseases, Pathogen Discovery | Dakota Howard, Dhwani Batra, Peter W. Cook, Kara Moser, Adrian Paskey, Jason Caravas, Benjamin Rambo-Martin, Shatavia Morrison, Christopher Gulvick, Scott Sammons, Yvette Unoarumhi, Darlene Wagner, Matthew Schmerer, Christian Bixby, Yihe Wang, Jonathan Schultz, Chirayu Goswami, Russ Hager, Robin Grimwood, Clinton R. Paden, Duncan MacCannell |
| EPI_ISL_2283132 | Weill Cornell Medicine | New York Genome Center | Michael Zody, Andre Corvelo, Dayna M. Oschwald, Samantha Fennessey, Tom Maniatis, Melissa Cushing, Olivier Elemento, Margaret Elizabeth Ross, Chris Mason, Priya Velu, Hanna Rennert, Arryn Craney, Lars F Westblade |
| EPI_ISL_2283133 | Infinity Biologix | Centers for Disease Control and Prevention Division of Viral Diseases, Pathogen Discovery | Dakota Howard, Dhwani Batra, Peter W. Cook, Kara Moser, Adrian Paskey, Jason Caravas, Benjamin Rambo-Martin, Shatavia Morrison, Christopher Gulvick, Scott Sammons, Yvette Unoarumhi, Darlene Wagner, Matthew Schmerer, Christian Bixby, Yihe Wang, Jonathan Schultz, Chirayu Goswami, Russ Hager, Robin Grimwood, Clinton R. Paden, Duncan MacCannell |
| EPI_ISL_2283134 | Weill Cornell Medicine | New York Genome Center | Michael Zody, Andre Corvelo, Dayna M. Oschwald, Samantha Fennessey, Tom Maniatis, Melissa Cushing, Olivier Elemento, Margaret Elizabeth Ross, Chris Mason, Priya Velu, Hanna Rennert, Arryn Craney, Lars F Westblade |
| EPI_ISL_2283135 | Infinity Biologix | Centers for Disease Control and Prevention Division of Viral Diseases, Pathogen Discovery | Dakota Howard, Dhwani Batra, Peter W. Cook, Kara Moser, Adrian Paskey, Jason Caravas, Benjamin Rambo-Martin, Shatavia Morrison, Christopher Gulvick, Scott Sammons, Yvette Unoarumhi, Darlene Wagner, Matthew Schmerer, Christian Bixby, Yihe Wang, Jonathan Schultz, Chirayu Goswami, Russ Hager, Robin Grimwood, Clinton R. Paden, Duncan MacCannell |
| EPI_ISL_2283136 | Weill Cornell Medicine | New York Genome Center | Michael Zody, Andre Corvelo, Dayna M. Oschwald, Samantha Fennessey, Tom Maniatis, Melissa Cushing, Olivier Elemento, Margaret Elizabeth Ross, Chris Mason, Priya Velu, Hanna Rennert, Arryn Craney, Lars F Westblade |
| EPI_ISL_2283137 | Infinity Biologix | Centers for Disease Control and Prevention Division of Viral Diseases, Pathogen Discovery | Dakota Howard, Dhwani Batra, Peter W. Cook, Kara Moser, Adrian Paskey, Jason Caravas, Benjamin Rambo-Martin, Shatavia Morrison, Christopher Gulvick, Scott Sammons, Yvette Unoarumhi, Darlene Wagner, Matthew Schmerer, Christian Bixby, Yihe Wang, Jonathan Schultz, Chirayu Goswami, Russ Hager, Robin Grimwood, Clinton R. Paden, Duncan MacCannell |
| EPI_ISL_2283138, EPI_ISL_2283139 | Weill Cornell Medicine | New York Genome Center | Michael Zody, Andre Corvelo, Dayna M. Oschwald, Samantha Fennessey, Tom Maniatis, Melissa Cushing, Olivier Elemento, Margaret Elizabeth Ross, Chris Mason, Priya Velu, Hanna Rennert, Arryn Craney, Lars F Westblade |
| EPI_ISL_2283140, EPI_ISL_2283141 | Infinity Biologix | Centers for Disease Control and Prevention Division of Viral Diseases, Pathogen Discovery | Dakota Howard, Dhwani Batra, Peter W. Cook, Kara Moser, Adrian Paskey, Jason Caravas, Benjamin Rambo-Martin, Shatavia Morrison, Christopher Gulvick, Scott Sammons, Yvette Unoarumhi, Darlene Wagner, Matthew Schmerer, Christian Bixby, Yihe Wang, Jonathan Schultz, Chirayu Goswami, Russ Hager, Robin Grimwood, Clinton R. Paden, Duncan MacCannell |
| EPI_ISL_2283142 | Weill Cornell Medicine | New York Genome Center | Michael Zody, Andre Corvelo, Dayna M. Oschwald, Samantha Fennessey, Tom Maniatis, Melissa Cushing, Olivier Elemento, Margaret Elizabeth Ross, Chris Mason, Priya Velu, Hanna Rennert, Arryn Craney, Lars F Westblade |
| EPI_ISL_2283143, EPI_ISL_2283144 | Infinity Biologix | Centers for Disease Control and Prevention Division of Viral Diseases, Pathogen Discovery | Dakota Howard, Dhwani Batra, Peter W. Cook, Kara Moser, Adrian Paskey, Jason Caravas, Benjamin Rambo-Martin, Shatavia Morrison, Christopher Gulvick, Scott Sammons, Yvette Unoarumhi, Darlene Wagner, Matthew Schmerer, Christian Bixby, Yihe Wang, Jonathan Schultz, Chirayu Goswami, Russ Hager, Robin Grimwood, Clinton R. Paden, Duncan MacCannell |
| EPI_ISL_2283145, EPI_ISL_2283146 | Weill Cornell Medicine | New York Genome Center | Michael Zody, Andre Corvelo, Dayna M. Oschwald, Samantha Fennessey, Tom Maniatis, Melissa Cushing, Olivier Elemento, Margaret Elizabeth Ross, Chris Mason, Priya Velu, Hanna Rennert, Arryn Craney, Lars F Westblade |
| EPI_ISL_2283147 | Infinity Biologix | Centers for Disease Control and Prevention Division of Viral Diseases, Pathogen Discovery | Dakota Howard, Dhwani Batra, Peter W. Cook, Kara Moser, Adrian Paskey, Jason Caravas, Benjamin Rambo-Martin, Shatavia Morrison, Christopher Gulvick, Scott Sammons, Yvette Unoarumhi, Darlene Wagner, Matthew Schmerer, Christian Bixby, Yihe Wang, Jonathan Schultz, Chirayu Goswami, Russ Hager, Robin Grimwood, Clinton R. Paden, Duncan MacCannell |
| EPI_ISL_2283148 | Weill Cornell Medicine | New York Genome Center | Michael Zody, Andre Corvelo, Dayna M. Oschwald, Samantha Fennessey, Tom Maniatis, Melissa Cushing, Olivier Elemento, Margaret Elizabeth Ross, Chris Mason, Priya Velu, Hanna Rennert, Arryn Craney, Lars F Westblade |
| EPI_ISL_2283149 | Infinity Biologix | Centers for Disease Control and Prevention Division of Viral Diseases, Pathogen Discovery | Dakota Howard, Dhwani Batra, Peter W. Cook, Kara Moser, Adrian Paskey, Jason Caravas, Benjamin Rambo-Martin, Shatavia Morrison, Christopher Gulvick, Scott Sammons, Yvette Unoarumhi, Darlene Wagner, Matthew Schmerer, Christian Bixby, Yihe Wang, Jonathan Schultz, Chirayu Goswami, Russ Hager, Robin Grimwood, Clinton R. Paden, Duncan MacCannell |
| EPI_ISL_2283150, EPI_ISL_2283151 | Weill Cornell Medicine | New York Genome Center | Michael Zody, Andre Corvelo, Dayna M. Oschwald, Samantha Fennessey, Tom Maniatis, Melissa Cushing, Olivier Elemento, Margaret Elizabeth Ross, Chris Mason, Priya Velu, Hanna Rennert, Arryn Craney, Lars F Westblade |
| EPI_ISL_2283152 | Infinity Biologix | Centers for Disease Control and Prevention Division of Viral Diseases, Pathogen Discovery | Dakota Howard, Dhwani Batra, Peter W. Cook, Kara Moser, Adrian Paskey, Jason Caravas, Benjamin Rambo-Martin, Shatavia Morrison, Christopher Gulvick, Scott Sammons, Yvette Unoarumhi, Darlene Wagner, Matthew Schmerer, Christian Bixby, Yihe Wang, Jonathan Schultz, Chirayu Goswami, Russ Hager, Robin Grimwood, Clinton R. Paden, Duncan MacCannell |
| EPI_ISL_2283153, EPI_ISL_2283154 | Weill Cornell Medicine | New York Genome Center | Michael Zody, Andre Corvelo, Dayna M. Oschwald, Samantha Fennessey, Tom Maniatis, Melissa Cushing, Olivier Elemento, Margaret Elizabeth Ross, Chris Mason, Priya Velu, Hanna Rennert, Arryn Craney, Lars F Westblade |
| EPI_ISL_2283155 | Infinity Biologix | Centers for Disease Control and Prevention Division of Viral Diseases, Pathogen Discovery | Dakota Howard, Dhwani Batra, Peter W. Cook, Kara Moser, Adrian Paskey, Jason Caravas, Benjamin Rambo-Martin, Shatavia Morrison, Christopher Gulvick, Scott Sammons, Yvette Unoarumhi, Darlene Wagner, Matthew Schmerer, Christian Bixby, Yihe Wang, Jonathan Schultz, Chirayu Goswami, Russ Hager, Robin Grimwood, Clinton R. Paden, Duncan MacCannell |
| EPI_ISL_2283156 | Weill Cornell Medicine | New York Genome Center | Michael Zody, Andre Corvelo, Dayna M. Oschwald, Samantha Fennessey, Tom Maniatis, Melissa Cushing, Olivier Elemento, Margaret Elizabeth Ross, Chris Mason, Priya Velu, Hanna Rennert, Arryn Craney, Lars F Westblade |
| EPI_ISL_2283157 | Infinity Biologix | Centers for Disease Control and Prevention Division of Viral Diseases, Pathogen Discovery | Dakota Howard, Dhwani Batra, Peter W. Cook, Kara Moser, Adrian Paskey, Jason Caravas, Benjamin Rambo-Martin, Shatavia Morrison, Christopher Gulvick, Scott Sammons, Yvette Unoarumhi, Darlene Wagner, Matthew Schmerer, Christian Bixby, Yihe Wang, Jonathan Schultz, Chirayu Goswami, Russ Hager, Robin Grimwood, Clinton R. Paden, Duncan MacCannell |
| EPI_ISL_2283158, EPI_ISL_2283159 | Weill Cornell Medicine | New York Genome Center | Michael Zody, Andre Corvelo, Dayna M. Oschwald, Samantha Fennessey, Tom Maniatis, Melissa Cushing, Olivier Elemento, Margaret Elizabeth Ross, Chris Mason, Priya Velu, Hanna Rennert, Arryn Craney, Lars F Westblade |
| EPI_ISL_2283160 | Infinity Biologix | Centers for Disease Control and Prevention Division of Viral Diseases, Pathogen Discovery | Dakota Howard, Dhwani Batra, Peter W. Cook, Kara Moser, Adrian Paskey, Jason Caravas, Benjamin Rambo-Martin, Shatavia Morrison, Christopher Gulvick, Scott Sammons, Yvette Unoarumhi, Darlene Wagner, Matthew Schmerer, Christian Bixby, Yihe Wang, Jonathan Schultz, Chirayu Goswami, Russ Hager, Robin Grimwood, Clinton R. Paden, Duncan MacCannell |
| EPI_ISL_2283161, EPI_ISL_2283162 | Weill Cornell Medicine | New York Genome Center | Michael Zody, Andre Corvelo, Dayna M. Oschwald, Samantha Fennessey, Tom Maniatis, Melissa Cushing, Olivier Elemento, Margaret Elizabeth Ross, Chris Mason, Priya Velu, Hanna Rennert, Arryn Craney, Lars F Westblade |
| EPI_ISL_2283163 | Infinity Biologix | Centers for Disease Control and Prevention Division of Viral Diseases, Pathogen Discovery | Dakota Howard, Dhwani Batra, Peter W. Cook, Kara Moser, Adrian Paskey, Jason Caravas, Benjamin Rambo-Martin, Shatavia Morrison, Christopher Gulvick, Scott Sammons, Yvette Unoarumhi, Darlene Wagner, Matthew Schmerer, Christian Bixby, Yihe Wang, Jonathan Schultz, Chirayu Goswami, Russ Hager, Robin Grimwood, Clinton R. Paden, Duncan MacCannell |
| EPI_ISL_2283164 | Weill Cornell Medicine | New York Genome Center | Michael Zody, Andre Corvelo, Dayna M. Oschwald, Samantha Fennessey, Tom Maniatis, Melissa Cushing, Olivier Elemento, Margaret Elizabeth Ross, Chris Mason, Priya Velu, Hanna Rennert, Arryn Craney, Lars F Westblade |
| EPI_ISL_2283165 | Infinity Biologix | Centers for Disease Control and Prevention Division of Viral Diseases, Pathogen Discovery | Dakota Howard, Dhwani Batra, Peter W. Cook, Kara Moser, Adrian Paskey, Jason Caravas, Benjamin Rambo-Martin, Shatavia Morrison, Christopher Gulvick, Scott Sammons, Yvette Unoarumhi, Darlene Wagner, Matthew Schmerer, Christian Bixby, Yihe Wang, Jonathan Schultz, Chirayu Goswami, Russ Hager, Robin Grimwood, Clinton R. Paden, Duncan MacCannell |
| EPI_ISL_2283166, EPI_ISL_2283167 | Weill Cornell Medicine | New York Genome Center | Michael Zody, Andre Corvelo, Dayna M. Oschwald, Samantha Fennessey, Tom Maniatis, Melissa Cushing, Olivier Elemento, Margaret Elizabeth Ross, Chris Mason, Priya Velu, Hanna Rennert, Arryn Craney, Lars F Westblade |
| EPI_ISL_2283168 | Infinity Biologix | Centers for Disease Control and Prevention Division of Viral Diseases, Pathogen Discovery | Dakota Howard, Dhwani Batra, Peter W. Cook, Kara Moser, Adrian Paskey, Jason Caravas, Benjamin Rambo-Martin, Shatavia Morrison, Christopher Gulvick, Scott Sammons, Yvette Unoarumhi, Darlene Wagner, Matthew Schmerer, Christian Bixby, Yihe Wang, Jonathan Schultz, Chirayu Goswami, Russ Hager, Robin Grimwood, Clinton R. Paden, Duncan MacCannell |
| EPI_ISL_2283169, EPI_ISL_2283170 | Weill Cornell Medicine | New York Genome Center | Michael Zody, Andre Corvelo, Dayna M. Oschwald, Samantha Fennessey, Tom Maniatis, Melissa Cushing, Olivier Elemento, Margaret Elizabeth Ross, Chris Mason, Priya Velu, Hanna Rennert, Arryn Craney, Lars F Westblade |
| EPI_ISL_2283171 | Infinity Biologix | Centers for Disease Control and Prevention Division of Viral Diseases, Pathogen Discovery | Dakota Howard, Dhwani Batra, Peter W. Cook, Kara Moser, Adrian Paskey, Jason Caravas, Benjamin Rambo-Martin, Shatavia Morrison, Christopher Gulvick, Scott Sammons, Yvette Unoarumhi, Darlene Wagner, Matthew Schmerer, Christian Bixby, Yihe Wang, Jonathan Schultz, Chirayu Goswami, Russ Hager, Robin Grimwood, Clinton R. Paden, Duncan MacCannell |
| EPI_ISL_2283172, EPI_ISL_2283174, EPI_ISL_2283175 | Weill Cornell Medicine | New York Genome Center | Michael Zody, Andre Corvelo, Dayna M. Oschwald, Samantha Fennessey, Tom Maniatis, Melissa Cushing, Olivier Elemento, Margaret Elizabeth Ross, Chris Mason, Priya Velu, Hanna Rennert, Arryn Craney, Lars F Westblade |
| EPI_ISL_2283176 | Infinity Biologix | Centers for Disease Control and Prevention Division of Viral Diseases, Pathogen Discovery | Dakota Howard, Dhwani Batra, Peter W. Cook, Kara Moser, Adrian Paskey, Jason Caravas, Benjamin Rambo-Martin, Shatavia Morrison, Christopher Gulvick, Scott Sammons, Yvette Unoarumhi, Darlene Wagner, Matthew Schmerer, Christian Bixby, Yihe Wang, Jonathan Schultz, Chirayu Goswami, Russ Hager, Robin Grimwood, Clinton R. Paden, Duncan MacCannell |
| EPI_ISL_2283177, EPI_ISL_2283178 | Weill Cornell Medicine | New York Genome Center | Michael Zody, Andre Corvelo, Dayna M. Oschwald, Samantha Fennessey, Tom Maniatis, Melissa Cushing, Olivier Elemento, Margaret Elizabeth Ross, Chris Mason, Priya Velu, Hanna Rennert, Arryn Craney, Lars F Westblade |
| EPI_ISL_2283179 | Infinity Biologix | Centers for Disease Control and Prevention Division of Viral Diseases, Pathogen Discovery | Dakota Howard, Dhwani Batra, Peter W. Cook, Kara Moser, Adrian Paskey, Jason Caravas, Benjamin Rambo-Martin, Shatavia Morrison, Christopher Gulvick, Scott Sammons, Yvette Unoarumhi, Darlene Wagner, Matthew Schmerer, Christian Bixby, Yihe Wang, Jonathan Schultz, Chirayu Goswami, Russ Hager, Robin Grimwood, Clinton R. Paden, Duncan MacCannell |
| EPI_ISL_2283180 | Weill Cornell Medicine | New York Genome Center | Michael Zody, Andre Corvelo, Dayna M. Oschwald, Samantha Fennessey, Tom Maniatis, Melissa Cushing, Olivier Elemento, Margaret Elizabeth Ross, Chris Mason, Priya Velu, Hanna Rennert, Arryn Craney, Lars F Westblade |
| EPI_ISL_2283181 | Infinity Biologix | Centers for Disease Control and Prevention | Dakota Howard, Dhwani Batra, Peter W. Cook, Kara Moser, Adrian Paskey, Jason Caravas, Benjamin Rambo-Martin, Shatavia Morrison, Christopher Gulvick, Scott Sammons, Yvette Unoarumhi, Darlene Wagner, Matthew Schmerer, Christian Bixby, Yihe Wang, Jonathan Schultz, Chirayu |

| | | | |
|---|---|---|---|
| | | Division of Viral Diseases, Pathogen Discovery | Goswami, Russ Hager, Robin Grimwood, Clinton R. Paden, Duncan MacCannell |
| EPI_ISL_2283182 | Weill Cornell Medicine | New York Genome Center | Michael Zody, Andre Corvelo, Dayna M. Oschwald, Samantha Fennessey, Tom Maniatis, Melissa Cushing, Olivier Elemento, Margaret Elizabeth Ross, Chris Mason, Priya Velu, Hanna Rennert, Arryn Craney, Lars F Westblade |
| EPI_ISL_2283183 | Infinity Biologix | Centers for Disease Control and Prevention Division of Viral Diseases, Pathogen Discovery | Dakota Howard, Dhwani Batra, Peter W. Cook, Kara Moser, Adrian Paskey, Jason Caravas, Benjamin Rambo-Martin, Shatavia Morrison, Christopher Gulvick, Scott Sammons, Yvette Unoarumhi, Darlene Wagner, Matthew Schmerer, Christian Bixby, Yihe Wang, Jonathan Schultz, Chirayu Goswami, Russ Hager, Robin Grimwood, Clinton R. Paden, Duncan MacCannell |
| EPI_ISL_2283184 | Weill Cornell Medicine | New York Genome Center | Michael Zody, Andre Corvelo, Dayna M. Oschwald, Samantha Fennessey, Tom Maniatis, Melissa Cushing, Olivier Elemento, Margaret Elizabeth Ross, Chris Mason, Priya Velu, Hanna Rennert, Arryn Craney, Lars F Westblade |
| EPI_ISL_2283185 | Infinity Biologix | Centers for Disease Control and Prevention Division of Viral Diseases, Pathogen Discovery | Dakota Howard, Dhwani Batra, Peter W. Cook, Kara Moser, Adrian Paskey, Jason Caravas, Benjamin Rambo-Martin, Shatavia Morrison, Christopher Gulvick, Scott Sammons, Yvette Unoarumhi, Darlene Wagner, Matthew Schmerer, Christian Bixby, Yihe Wang, Jonathan Schultz, Chirayu Goswami, Russ Hager, Robin Grimwood, Clinton R. Paden, Duncan MacCannell |
| EPI_ISL_2283186, EPI_ISL_2283187 | Weill Cornell Medicine | New York Genome Center | Michael Zody, Andre Corvelo, Dayna M. Oschwald, Samantha Fennessey, Tom Maniatis, Melissa Cushing, Olivier Elemento, Margaret Elizabeth Ross, Chris Mason, Priya Velu, Hanna Rennert, Arryn Craney, Lars F Westblade |
| EPI_ISL_2283188 | Infinity Biologix | Centers for Disease Control and Prevention Division of Viral Diseases, Pathogen Discovery | Dakota Howard, Dhwani Batra, Peter W. Cook, Kara Moser, Adrian Paskey, Jason Caravas, Benjamin Rambo-Martin, Shatavia Morrison, Christopher Gulvick, Scott Sammons, Yvette Unoarumhi, Darlene Wagner, Matthew Schmerer, Christian Bixby, Yihe Wang, Jonathan Schultz, Chirayu Goswami, Russ Hager, Robin Grimwood, Clinton R. Paden, Duncan MacCannell |
| EPI_ISL_2283189, EPI_ISL_2283190 | Weill Cornell Medicine | New York Genome Center | Michael Zody, Andre Corvelo, Dayna M. Oschwald, Samantha Fennessey, Tom Maniatis, Melissa Cushing, Olivier Elemento, Margaret Elizabeth Ross, Chris Mason, Priya Velu, Hanna Rennert, Arryn Craney, Lars F Westblade |
| EPI_ISL_2283191 | Infinity Biologix | Centers for Disease Control and Prevention Division of Viral Diseases, Pathogen Discovery | Dakota Howard, Dhwani Batra, Peter W. Cook, Kara Moser, Adrian Paskey, Jason Caravas, Benjamin Rambo-Martin, Shatavia Morrison, Christopher Gulvick, Scott Sammons, Yvette Unoarumhi, Darlene Wagner, Matthew Schmerer, Christian Bixby, Yihe Wang, Jonathan Schultz, Chirayu Goswami, Russ Hager, Robin Grimwood, Clinton R. Paden, Duncan MacCannell |
| EPI_ISL_2283192 | Weill Cornell Medicine | New York Genome Center | Michael Zody, Andre Corvelo, Dayna M. Oschwald, Samantha Fennessey, Tom Maniatis, Melissa Cushing, Olivier Elemento, Margaret Elizabeth Ross, Chris Mason, Priya Velu, Hanna Rennert, Arryn Craney, Lars F Westblade |
| EPI_ISL_2283193 | Infinity Biologix | Centers for Disease Control and Prevention Division of Viral Diseases, Pathogen Discovery | Dakota Howard, Dhwani Batra, Peter W. Cook, Kara Moser, Adrian Paskey, Jason Caravas, Benjamin Rambo-Martin, Shatavia Morrison, Christopher Gulvick, Scott Sammons, Yvette Unoarumhi, Darlene Wagner, Matthew Schmerer, Christian Bixby, Yihe Wang, Jonathan Schultz, Chirayu Goswami, Russ Hager, Robin Grimwood, Clinton R. Paden, Duncan MacCannell |
| EPI_ISL_2283194, EPI_ISL_2283195 | Weill Cornell Medicine | New York Genome Center | Michael Zody, Andre Corvelo, Dayna M. Oschwald, Samantha Fennessey, Tom Maniatis, Melissa Cushing, Olivier Elemento, Margaret Elizabeth Ross, Chris Mason, Priya Velu, Hanna Rennert, Arryn Craney, Lars F Westblade |
| EPI_ISL_2283196 | Infinity Biologix | Centers for Disease Control and Prevention Division of Viral Diseases, Pathogen Discovery | Dakota Howard, Dhwani Batra, Peter W. Cook, Kara Moser, Adrian Paskey, Jason Caravas, Benjamin Rambo-Martin, Shatavia Morrison, Christopher Gulvick, Scott Sammons, Yvette Unoarumhi, Darlene Wagner, Matthew Schmerer, Christian Bixby, Yihe Wang, Jonathan Schultz, Chirayu Goswami, Russ Hager, Robin Grimwood, Clinton R. Paden, Duncan MacCannell |
| EPI_ISL_2283197 | Weill Cornell Medicine | New York Genome Center | Michael Zody, Andre Corvelo, Dayna M. Oschwald, Samantha Fennessey, Tom Maniatis, Melissa Cushing, Olivier Elemento, Margaret Elizabeth Ross, Chris Mason, Priya Velu, Hanna Rennert, Arryn Craney, Lars F Westblade |
| EPI_ISL_2283198 | Infinity Biologix | Centers for Disease Control and Prevention Division of Viral Diseases, Pathogen Discovery | Dakota Howard, Dhwani Batra, Peter W. Cook, Kara Moser, Adrian Paskey, Jason Caravas, Benjamin Rambo-Martin, Shatavia Morrison, Christopher Gulvick, Scott Sammons, Yvette Unoarumhi, Darlene Wagner, Matthew Schmerer, Christian Bixby, Yihe Wang, Jonathan Schultz, Chirayu Goswami, Russ Hager, Robin Grimwood, Clinton R. Paden, Duncan MacCannell |
| EPI_ISL_2283199, EPI_ISL_2283200 | Weill Cornell Medicine | New York Genome Center | Michael Zody, Andre Corvelo, Dayna M. Oschwald, Samantha Fennessey, Tom Maniatis, Melissa Cushing, Olivier Elemento, Margaret Elizabeth Ross, Chris Mason, Priya Velu, Hanna Rennert, Arryn Craney, Lars F Westblade |
| EPI_ISL_2283201 | Infinity Biologix | Centers for Disease Control and Prevention Division of Viral Diseases, Pathogen Discovery | Dakota Howard, Dhwani Batra, Peter W. Cook, Kara Moser, Adrian Paskey, Jason Caravas, Benjamin Rambo-Martin, Shatavia Morrison, Christopher Gulvick, Scott Sammons, Yvette Unoarumhi, Darlene Wagner, Matthew Schmerer, Christian Bixby, Yihe Wang, Jonathan Schultz, Chirayu Goswami, Russ Hager, Robin Grimwood, Clinton R. Paden, Duncan MacCannell |
| EPI_ISL_2283202 | Weill Cornell Medicine | New York Genome Center | Michael Zody, Andre Corvelo, Dayna M. Oschwald, Samantha Fennessey, Tom Maniatis, Melissa Cushing, Olivier Elemento, Margaret Elizabeth Ross, Chris Mason, Priya Velu, Hanna Rennert, Arryn Craney, Lars F Westblade |
| EPI_ISL_2283203 | Infinity Biologix | Centers for Disease Control and Prevention Division of Viral Diseases, Pathogen Discovery | Dakota Howard, Dhwani Batra, Peter W. Cook, Kara Moser, Adrian Paskey, Jason Caravas, Benjamin Rambo-Martin, Shatavia Morrison, Christopher Gulvick, Scott Sammons, Yvette Unoarumhi, Darlene Wagner, Matthew Schmerer, Christian Bixby, Yihe Wang, Jonathan Schultz, Chirayu Goswami, Russ Hager, Robin Grimwood, Clinton R. Paden, Duncan MacCannell |
| EPI_ISL_2283204, EPI_ISL_2283205 | Weill Cornell Medicine | New York Genome Center | Michael Zody, Andre Corvelo, Dayna M. Oschwald, Samantha Fennessey, Tom Maniatis, Melissa Cushing, Olivier Elemento, Margaret Elizabeth Ross, Chris Mason, Priya Velu, Hanna Rennert, Arryn Craney, Lars F Westblade |
| EPI_ISL_2283206 | Infinity Biologix | Centers for Disease Control and Prevention Division of Viral Diseases, Pathogen Discovery | Dakota Howard, Dhwani Batra, Peter W. Cook, Kara Moser, Adrian Paskey, Jason Caravas, Benjamin Rambo-Martin, Shatavia Morrison, Christopher Gulvick, Scott Sammons, Yvette Unoarumhi, Darlene Wagner, Matthew Schmerer, Christian Bixby, Yihe Wang, Jonathan Schultz, Chirayu Goswami, Russ Hager, Robin Grimwood, Clinton R. Paden, Duncan MacCannell |
| EPI_ISL_2283207, EPI_ISL_2283208 | Weill Cornell Medicine | New York Genome Center | Michael Zody, Andre Corvelo, Dayna M. Oschwald, Samantha Fennessey, Tom Maniatis, Melissa Cushing, Olivier Elemento, Margaret Elizabeth Ross, Chris Mason, Priya Velu, Hanna Rennert, Arryn Craney, Lars F Westblade |
| EPI_ISL_2283209 | Infinity Biologix | Centers for Disease Control and Prevention Division of Viral Diseases, Pathogen Discovery | Dakota Howard, Dhwani Batra, Peter W. Cook, Kara Moser, Adrian Paskey, Jason Caravas, Benjamin Rambo-Martin, Shatavia Morrison, Christopher Gulvick, Scott Sammons, Yvette Unoarumhi, Darlene Wagner, Matthew Schmerer, Christian Bixby, Yihe Wang, Jonathan Schultz, Chirayu Goswami, Russ Hager, Robin Grimwood, Clinton R. Paden, Duncan MacCannell |
| EPI_ISL_2283210 | Weill Cornell Medicine | New York Genome Center | Michael Zody, Andre Corvelo, Dayna M. Oschwald, Samantha Fennessey, Tom Maniatis, Melissa Cushing, Olivier Elemento, Margaret Elizabeth Ross, Chris Mason, Priya Velu, Hanna Rennert, Arryn Craney, Lars F Westblade |
| EPI_ISL_2283211 | Infinity Biologix | Centers for Disease Control and Prevention Division of Viral Diseases, Pathogen Discovery | Dakota Howard, Dhwani Batra, Peter W. Cook, Kara Moser, Adrian Paskey, Jason Caravas, Benjamin Rambo-Martin, Shatavia Morrison, Christopher Gulvick, Scott Sammons, Yvette Unoarumhi, Darlene Wagner, Matthew Schmerer, Christian Bixby, Yihe Wang, Jonathan Schultz, Chirayu Goswami, Russ Hager, Robin Grimwood, Clinton R. Paden, Duncan MacCannell |
| EPI_ISL_2283212, EPI_ISL_2283213 | Weill Cornell Medicine | New York Genome Center | Michael Zody, Andre Corvelo, Dayna M. Oschwald, Samantha Fennessey, Tom Maniatis, Melissa Cushing, Olivier Elemento, Margaret Elizabeth Ross, Chris Mason, Priya Velu, Hanna Rennert, Arryn Craney, Lars F Westblade |
| EPI_ISL_2283214 | Infinity Biologix | Centers for Disease Control and Prevention Division of Viral Diseases, Pathogen Discovery | Dakota Howard, Dhwani Batra, Peter W. Cook, Kara Moser, Adrian Paskey, Jason Caravas, Benjamin Rambo-Martin, Shatavia Morrison, Christopher Gulvick, Scott Sammons, Yvette Unoarumhi, Darlene Wagner, Matthew Schmerer, Christian Bixby, Yihe Wang, Jonathan Schultz, Chirayu Goswami, Russ Hager, Robin Grimwood, Clinton R. Paden, Duncan MacCannell |
| EPI_ISL_2283215 | Weill Cornell Medicine | New York Genome Center | Michael Zody, Andre Corvelo, Dayna M. Oschwald, Samantha Fennessey, Tom Maniatis, Melissa Cushing, Olivier Elemento, Margaret Elizabeth Ross, Chris Mason, Priya Velu, Hanna Rennert, Arryn Craney, Lars F Westblade |
| EPI_ISL_2283216 | Infinity Biologix | Centers for Disease Control and Prevention Division of Viral Diseases, Pathogen Discovery | Dakota Howard, Dhwani Batra, Peter W. Cook, Kara Moser, Adrian Paskey, Jason Caravas, Benjamin Rambo-Martin, Shatavia Morrison, Christopher Gulvick, Scott Sammons, Yvette Unoarumhi, Darlene Wagner, Matthew Schmerer, Christian Bixby, Yihe Wang, Jonathan Schultz, Chirayu Goswami, Russ Hager, Robin Grimwood, Clinton R. Paden, Duncan MacCannell |
| EPI_ISL_2283217, EPI_ISL_2283218 | Weill Cornell Medicine | New York Genome Center | Michael Zody, Andre Corvelo, Dayna M. Oschwald, Samantha Fennessey, Tom Maniatis, Melissa Cushing, Olivier Elemento, Margaret Elizabeth Ross, Chris Mason, Priya Velu, Hanna Rennert, Arryn Craney, Lars F Westblade |
| EPI_ISL_2283219 | Infinity Biologix | Centers for Disease Control and Prevention Division of Viral Diseases, Pathogen Discovery | Dakota Howard, Dhwani Batra, Peter W. Cook, Kara Moser, Adrian Paskey, Jason Caravas, Benjamin Rambo-Martin, Shatavia Morrison, Christopher Gulvick, Scott Sammons, Yvette Unoarumhi, Darlene Wagner, Matthew Schmerer, Christian Bixby, Yihe Wang, Jonathan Schultz, Chirayu Goswami, Russ Hager, Robin Grimwood, Clinton R. Paden, Duncan MacCannell |
| EPI_ISL_2283220, EPI_ISL_2283221 | Weill Cornell Medicine | New York Genome Center | Michael Zody, Andre Corvelo, Dayna M. Oschwald, Samantha Fennessey, Tom Maniatis, Melissa Cushing, Olivier Elemento, Margaret Elizabeth Ross, Chris Mason, Priya Velu, Hanna Rennert, Arryn Craney, Lars F Westblade |
| EPI_ISL_2283222 | Infinity Biologix | Centers for Disease Control and Prevention Division of Viral Diseases, Pathogen Discovery | Dakota Howard, Dhwani Batra, Peter W. Cook, Kara Moser, Adrian Paskey, Jason Caravas, Benjamin Rambo-Martin, Shatavia Morrison, Christopher Gulvick, Scott Sammons, Yvette Unoarumhi, Darlene Wagner, Matthew Schmerer, Christian Bixby, Yihe Wang, Jonathan Schultz, Chirayu Goswami, Russ Hager, Robin Grimwood, Clinton R. Paden, Duncan MacCannell |
| EPI_ISL_2283223 | Weill Cornell Medicine | New York Genome Center | Michael Zody, Andre Corvelo, Dayna M. Oschwald, Samantha Fennessey, Tom Maniatis, Melissa Cushing, Olivier Elemento, Margaret Elizabeth Ross, Chris Mason, Priya Velu, Hanna Rennert, Arryn Craney, Lars F Westblade |
| EPI_ISL_2283224 | Infinity Biologix | Centers for Disease Control and Prevention Division of Viral Diseases, Pathogen Discovery | Dakota Howard, Dhwani Batra, Peter W. Cook, Kara Moser, Adrian Paskey, Jason Caravas, Benjamin Rambo-Martin, Shatavia Morrison, Christopher Gulvick, Scott Sammons, Yvette Unoarumhi, Darlene Wagner, Matthew Schmerer, Christian Bixby, Yihe Wang, Jonathan Schultz, Chirayu Goswami, Russ Hager, Robin Grimwood, Clinton R. Paden, Duncan MacCannell |
| EPI_ISL_2283225, EPI_ISL_2283226 | Weill Cornell Medicine | New York Genome Center | Michael Zody, Andre Corvelo, Dayna M. Oschwald, Samantha Fennessey, Tom Maniatis, Melissa Cushing, Olivier Elemento, Margaret Elizabeth Ross, Chris Mason, Priya Velu, Hanna Rennert, Arryn Craney, Lars F Westblade |
| EPI_ISL_2283227 | Infinity Biologix | Centers for Disease Control and Prevention Division of Viral Diseases, Pathogen Discovery | Dakota Howard, Dhwani Batra, Peter W. Cook, Kara Moser, Adrian Paskey, Jason Caravas, Benjamin Rambo-Martin, Shatavia Morrison, Christopher Gulvick, Scott Sammons, Yvette Unoarumhi, Darlene Wagner, Matthew Schmerer, Christian Bixby, Yihe Wang, Jonathan Schultz, Chirayu Goswami, Russ Hager, Robin Grimwood, Clinton R. Paden, Duncan MacCannell |
| EPI_ISL_2283228, EPI_ISL_2283229 | Weill Cornell Medicine | New York Genome Center | Michael Zody, Andre Corvelo, Dayna M. Oschwald, Samantha Fennessey, Tom Maniatis, Melissa Cushing, Olivier Elemento, Margaret Elizabeth Ross, Chris Mason, Priya Velu, Hanna Rennert, Arryn Craney, Lars F Westblade |
| EPI_ISL_2283230 | Infinity Biologix | Centers for Disease Control and Prevention Division of Viral Diseases, Pathogen Discovery | Dakota Howard, Dhwani Batra, Peter W. Cook, Kara Moser, Adrian Paskey, Jason Caravas, Benjamin Rambo-Martin, Shatavia Morrison, Christopher Gulvick, Scott Sammons, Yvette Unoarumhi, Darlene Wagner, Matthew Schmerer, Christian Bixby, Yihe Wang, Jonathan Schultz, Chirayu Goswami, Russ Hager, Robin Grimwood, Clinton R. Paden, Duncan MacCannell |
| EPI_ISL_2283231 | Weill Cornell Medicine | New York Genome Center | Michael Zody, Andre Corvelo, Dayna M. Oschwald, Samantha Fennessey, Tom Maniatis, Melissa Cushing, Olivier Elemento, Margaret Elizabeth Ross, Chris Mason, Priya Velu, Hanna Rennert, Arryn Craney, Lars F Westblade |
| EPI_ISL_2283232 | Infinity Biologix | Centers for Disease Control and Prevention | Dakota Howard, Dhwani Batra, Peter W. Cook, Kara Moser, Adrian Paskey, Jason Caravas, Benjamin Rambo-Martin, Shatavia Morrison, Christopher Gulvick, Scott Sammons, Yvette Unoarumhi, Darlene Wagner, Matthew Schmerer, Christian Bixby, Yihe Wang, Jonathan Schultz, Chirayu |

| | | | |
|---|---|---|---|
| | | Division of Viral Diseases, Pathogen Discovery | Goswami, Russ Hager, Robin Grimwood, Clinton R. Paden, Duncan MacCannell |
| EPI_ISL_2283233, EPI_ISL_2283234 | Weill Cornell Medicine | New York Genome Center | Michael Zody, Andre Corvelo, Dayna M. Oschwald, Samantha Fennessey, Tom Maniatis, Melissa Cushing, Olivier Elemento, Margaret Elizabeth Ross, Chris Mason, Priya Velu, Hanna Rennert, Arryn Craney, Lars F Westblade |
| EPI_ISL_2283235 | Infinity Biologix | Centers for Disease Control and Prevention Division of Viral Diseases, Pathogen Discovery | Dakota Howard, Dhwani Batra, Peter W. Cook, Kara Moser, Adrian Paskey, Jason Caravas, Benjamin Rambo-Martin, Shatavia Morrison, Christopher Gulvick, Scott Sammons, Yvette Unoarumhi, Darlene Wagner, Matthew Schmerer, Christian Bixby, Yihe Wang, Jonathan Schultz, Chirayu Goswami, Russ Hager, Robin Grimwood, Clinton R. Paden, Duncan MacCannell |
| EPI_ISL_2283236, EPI_ISL_2283237 | Weill Cornell Medicine | New York Genome Center | Michael Zody, Andre Corvelo, Dayna M. Oschwald, Samantha Fennessey, Tom Maniatis, Melissa Cushing, Olivier Elemento, Margaret Elizabeth Ross, Chris Mason, Priya Velu, Hanna Rennert, Arryn Craney, Lars F Westblade |
| EPI_ISL_2283238 | Infinity Biologix | Centers for Disease Control and Prevention Division of Viral Diseases, Pathogen Discovery | Dakota Howard, Dhwani Batra, Peter W. Cook, Kara Moser, Adrian Paskey, Jason Caravas, Benjamin Rambo-Martin, Shatavia Morrison, Christopher Gulvick, Scott Sammons, Yvette Unoarumhi, Darlene Wagner, Matthew Schmerer, Christian Bixby, Yihe Wang, Jonathan Schultz, Chirayu Goswami, Russ Hager, Robin Grimwood, Clinton R. Paden, Duncan MacCannell |
| EPI_ISL_2283239, EPI_ISL_2283240 | Weill Cornell Medicine | New York Genome Center | Michael Zody, Andre Corvelo, Dayna M. Oschwald, Samantha Fennessey, Tom Maniatis, Melissa Cushing, Olivier Elemento, Margaret Elizabeth Ross, Chris Mason, Priya Velu, Hanna Rennert, Arryn Craney, Lars F Westblade |
| EPI_ISL_2283241 | Infinity Biologix | Centers for Disease Control and Prevention Division of Viral Diseases, Pathogen Discovery | Dakota Howard, Dhwani Batra, Peter W. Cook, Kara Moser, Adrian Paskey, Jason Caravas, Benjamin Rambo-Martin, Shatavia Morrison, Christopher Gulvick, Scott Sammons, Yvette Unoarumhi, Darlene Wagner, Matthew Schmerer, Christian Bixby, Yihe Wang, Jonathan Schultz, Chirayu Goswami, Russ Hager, Robin Grimwood, Clinton R. Paden, Duncan MacCannell |
| EPI_ISL_2283242 | Weill Cornell Medicine | New York Genome Center | Michael Zody, Andre Corvelo, Dayna M. Oschwald, Samantha Fennessey, Tom Maniatis, Melissa Cushing, Olivier Elemento, Margaret Elizabeth Ross, Chris Mason, Priya Velu, Hanna Rennert, Arryn Craney, Lars F Westblade |
| EPI_ISL_2283243 | Infinity Biologix | Centers for Disease Control and Prevention Division of Viral Diseases, Pathogen Discovery | Dakota Howard, Dhwani Batra, Peter W. Cook, Kara Moser, Adrian Paskey, Jason Caravas, Benjamin Rambo-Martin, Shatavia Morrison, Christopher Gulvick, Scott Sammons, Yvette Unoarumhi, Darlene Wagner, Matthew Schmerer, Christian Bixby, Yihe Wang, Jonathan Schultz, Chirayu Goswami, Russ Hager, Robin Grimwood, Clinton R. Paden, Duncan MacCannell |
| EPI_ISL_2283244, EPI_ISL_2283245 | Weill Cornell Medicine | New York Genome Center | Michael Zody, Andre Corvelo, Dayna M. Oschwald, Samantha Fennessey, Tom Maniatis, Melissa Cushing, Olivier Elemento, Margaret Elizabeth Ross, Chris Mason, Priya Velu, Hanna Rennert, Arryn Craney, Lars F Westblade |
| EPI_ISL_2283246 | Infinity Biologix | Centers for Disease Control and Prevention Division of Viral Diseases, Pathogen Discovery | Dakota Howard, Dhwani Batra, Peter W. Cook, Kara Moser, Adrian Paskey, Jason Caravas, Benjamin Rambo-Martin, Shatavia Morrison, Christopher Gulvick, Scott Sammons, Yvette Unoarumhi, Darlene Wagner, Matthew Schmerer, Christian Bixby, Yihe Wang, Jonathan Schultz, Chirayu Goswami, Russ Hager, Robin Grimwood, Clinton R. Paden, Duncan MacCannell |
| EPI_ISL_2283247, EPI_ISL_2283248 | Weill Cornell Medicine | New York Genome Center | Michael Zody, Andre Corvelo, Dayna M. Oschwald, Samantha Fennessey, Tom Maniatis, Melissa Cushing, Olivier Elemento, Margaret Elizabeth Ross, Chris Mason, Priya Velu, Hanna Rennert, Arryn Craney, Lars F Westblade |
| EPI_ISL_2283249 | Infinity Biologix | Centers for Disease Control and Prevention Division of Viral Diseases, Pathogen Discovery | Dakota Howard, Dhwani Batra, Peter W. Cook, Kara Moser, Adrian Paskey, Jason Caravas, Benjamin Rambo-Martin, Shatavia Morrison, Christopher Gulvick, Scott Sammons, Yvette Unoarumhi, Darlene Wagner, Matthew Schmerer, Christian Bixby, Yihe Wang, Jonathan Schultz, Chirayu Goswami, Russ Hager, Robin Grimwood, Clinton R. Paden, Duncan MacCannell |
| EPI_ISL_2283250, EPI_ISL_2283251 | Weill Cornell Medicine | New York Genome Center | Michael Zody, Andre Corvelo, Dayna M. Oschwald, Samantha Fennessey, Tom Maniatis, Melissa Cushing, Olivier Elemento, Margaret Elizabeth Ross, Chris Mason, Priya Velu, Hanna Rennert, Arryn Craney, Lars F Westblade |
| EPI_ISL_2283252 | Infinity Biologix | Centers for Disease Control and Prevention Division of Viral Diseases, Pathogen Discovery | Dakota Howard, Dhwani Batra, Peter W. Cook, Kara Moser, Adrian Paskey, Jason Caravas, Benjamin Rambo-Martin, Shatavia Morrison, Christopher Gulvick, Scott Sammons, Yvette Unoarumhi, Darlene Wagner, Matthew Schmerer, Christian Bixby, Yihe Wang, Jonathan Schultz, Chirayu Goswami, Russ Hager, Robin Grimwood, Clinton R. Paden, Duncan MacCannell |
| EPI_ISL_2283253, EPI_ISL_2283254 | Weill Cornell Medicine | New York Genome Center | Michael Zody, Andre Corvelo, Dayna M. Oschwald, Samantha Fennessey, Tom Maniatis, Melissa Cushing, Olivier Elemento, Margaret Elizabeth Ross, Chris Mason, Priya Velu, Hanna Rennert, Arryn Craney, Lars F Westblade |
| EPI_ISL_2283255 | Infinity Biologix | Centers for Disease Control and Prevention Division of Viral Diseases, Pathogen Discovery | Dakota Howard, Dhwani Batra, Peter W. Cook, Kara Moser, Adrian Paskey, Jason Caravas, Benjamin Rambo-Martin, Shatavia Morrison, Christopher Gulvick, Scott Sammons, Yvette Unoarumhi, Darlene Wagner, Matthew Schmerer, Christian Bixby, Yihe Wang, Jonathan Schultz, Chirayu Goswami, Russ Hager, Robin Grimwood, Clinton R. Paden, Duncan MacCannell |
| EPI_ISL_2283256, EPI_ISL_2283257 | Weill Cornell Medicine | New York Genome Center | Michael Zody, Andre Corvelo, Dayna M. Oschwald, Samantha Fennessey, Tom Maniatis, Melissa Cushing, Olivier Elemento, Margaret Elizabeth Ross, Chris Mason, Priya Velu, Hanna Rennert, Arryn Craney, Lars F Westblade |
| EPI_ISL_2283258 | Infinity Biologix | Centers for Disease Control and Prevention Division of Viral Diseases, Pathogen Discovery | Dakota Howard, Dhwani Batra, Peter W. Cook, Kara Moser, Adrian Paskey, Jason Caravas, Benjamin Rambo-Martin, Shatavia Morrison, Christopher Gulvick, Scott Sammons, Yvette Unoarumhi, Darlene Wagner, Matthew Schmerer, Christian Bixby, Yihe Wang, Jonathan Schultz, Chirayu Goswami, Russ Hager, Robin Grimwood, Clinton R. Paden, Duncan MacCannell |
| EPI_ISL_2283259, EPI_ISL_2283260 | Weill Cornell Medicine | New York Genome Center | Michael Zody, Andre Corvelo, Dayna M. Oschwald, Samantha Fennessey, Tom Maniatis, Melissa Cushing, Olivier Elemento, Margaret Elizabeth Ross, Chris Mason, Priya Velu, Hanna Rennert, Arryn Craney, Lars F Westblade |
| EPI_ISL_2283261 | Infinity Biologix | Centers for Disease Control and Prevention Division of Viral Diseases, Pathogen Discovery | Dakota Howard, Dhwani Batra, Peter W. Cook, Kara Moser, Adrian Paskey, Jason Caravas, Benjamin Rambo-Martin, Shatavia Morrison, Christopher Gulvick, Scott Sammons, Yvette Unoarumhi, Darlene Wagner, Matthew Schmerer, Christian Bixby, Yihe Wang, Jonathan Schultz, Chirayu Goswami, Russ Hager, Robin Grimwood, Clinton R. Paden, Duncan MacCannell |
| EPI_ISL_2283262, EPI_ISL_2283263 | Weill Cornell Medicine | New York Genome Center | Michael Zody, Andre Corvelo, Dayna M. Oschwald, Samantha Fennessey, Tom Maniatis, Melissa Cushing, Olivier Elemento, Margaret Elizabeth Ross, Chris Mason, Priya Velu, Hanna Rennert, Arryn Craney, Lars F Westblade |
| EPI_ISL_2283264 | Infinity Biologix | Centers for Disease Control and Prevention Division of Viral Diseases, Pathogen Discovery | Dakota Howard, Dhwani Batra, Peter W. Cook, Kara Moser, Adrian Paskey, Jason Caravas, Benjamin Rambo-Martin, Shatavia Morrison, Christopher Gulvick, Scott Sammons, Yvette Unoarumhi, Darlene Wagner, Matthew Schmerer, Christian Bixby, Yihe Wang, Jonathan Schultz, Chirayu Goswami, Russ Hager, Robin Grimwood, Clinton R. Paden, Duncan MacCannell |
| EPI_ISL_2283265 | Weill Cornell Medicine | New York Genome Center | Michael Zody, Andre Corvelo, Dayna M. Oschwald, Samantha Fennessey, Tom Maniatis, Melissa Cushing, Olivier Elemento, Margaret Elizabeth Ross, Chris Mason, Priya Velu, Hanna Rennert, Arryn Craney, Lars F Westblade |
| EPI_ISL_2283266 | Infinity Biologix | Centers for Disease Control and Prevention Division of Viral Diseases, Pathogen Discovery | Dakota Howard, Dhwani Batra, Peter W. Cook, Kara Moser, Adrian Paskey, Jason Caravas, Benjamin Rambo-Martin, Shatavia Morrison, Christopher Gulvick, Scott Sammons, Yvette Unoarumhi, Darlene Wagner, Matthew Schmerer, Christian Bixby, Yihe Wang, Jonathan Schultz, Chirayu Goswami, Russ Hager, Robin Grimwood, Clinton R. Paden, Duncan MacCannell |
| EPI_ISL_2283267 | Weill Cornell Medicine | New York Genome Center | Michael Zody, Andre Corvelo, Dayna M. Oschwald, Samantha Fennessey, Tom Maniatis, Melissa Cushing, Olivier Elemento, Margaret Elizabeth Ross, Chris Mason, Priya Velu, Hanna Rennert, Arryn Craney, Lars F Westblade |
| EPI_ISL_2283268 | Infinity Biologix | Centers for Disease Control and Prevention Division of Viral Diseases, Pathogen Discovery | Dakota Howard, Dhwani Batra, Peter W. Cook, Kara Moser, Adrian Paskey, Jason Caravas, Benjamin Rambo-Martin, Shatavia Morrison, Christopher Gulvick, Scott Sammons, Yvette Unoarumhi, Darlene Wagner, Matthew Schmerer, Christian Bixby, Yihe Wang, Jonathan Schultz, Chirayu Goswami, Russ Hager, Robin Grimwood, Clinton R. Paden, Duncan MacCannell |
| EPI_ISL_2283269, EPI_ISL_2283270 | Weill Cornell Medicine | New York Genome Center | Michael Zody, Andre Corvelo, Dayna M. Oschwald, Samantha Fennessey, Tom Maniatis, Melissa Cushing, Olivier Elemento, Margaret Elizabeth Ross, Chris Mason, Priya Velu, Hanna Rennert, Arryn Craney, Lars F Westblade |
| EPI_ISL_2283271 | Infinity Biologix | Centers for Disease Control and Prevention Division of Viral Diseases, Pathogen Discovery | Dakota Howard, Dhwani Batra, Peter W. Cook, Kara Moser, Adrian Paskey, Jason Caravas, Benjamin Rambo-Martin, Shatavia Morrison, Christopher Gulvick, Scott Sammons, Yvette Unoarumhi, Darlene Wagner, Matthew Schmerer, Christian Bixby, Yihe Wang, Jonathan Schultz, Chirayu Goswami, Russ Hager, Robin Grimwood, Clinton R. Paden, Duncan MacCannell |
| EPI_ISL_2283272, EPI_ISL_2283273 | Weill Cornell Medicine | New York Genome Center | Michael Zody, Andre Corvelo, Dayna M. Oschwald, Samantha Fennessey, Tom Maniatis, Melissa Cushing, Olivier Elemento, Margaret Elizabeth Ross, Chris Mason, Priya Velu, Hanna Rennert, Arryn Craney, Lars F Westblade |
| EPI_ISL_2283274 | Infinity Biologix | Centers for Disease Control and Prevention Division of Viral Diseases, Pathogen Discovery | Dakota Howard, Dhwani Batra, Peter W. Cook, Kara Moser, Adrian Paskey, Jason Caravas, Benjamin Rambo-Martin, Shatavia Morrison, Christopher Gulvick, Scott Sammons, Yvette Unoarumhi, Darlene Wagner, Matthew Schmerer, Christian Bixby, Yihe Wang, Jonathan Schultz, Chirayu Goswami, Russ Hager, Robin Grimwood, Clinton R. Paden, Duncan MacCannell |
| EPI_ISL_2283275, EPI_ISL_2283276 | Weill Cornell Medicine | New York Genome Center | Michael Zody, Andre Corvelo, Dayna M. Oschwald, Samantha Fennessey, Tom Maniatis, Melissa Cushing, Olivier Elemento, Margaret Elizabeth Ross, Chris Mason, Priya Velu, Hanna Rennert, Arryn Craney, Lars F Westblade |
| EPI_ISL_2283277 | Infinity Biologix | Centers for Disease Control and Prevention Division of Viral Diseases, Pathogen Discovery | Dakota Howard, Dhwani Batra, Peter W. Cook, Kara Moser, Adrian Paskey, Jason Caravas, Benjamin Rambo-Martin, Shatavia Morrison, Christopher Gulvick, Scott Sammons, Yvette Unoarumhi, Darlene Wagner, Matthew Schmerer, Christian Bixby, Yihe Wang, Jonathan Schultz, Chirayu Goswami, Russ Hager, Robin Grimwood, Clinton R. Paden, Duncan MacCannell |
| EPI_ISL_2283278, EPI_ISL_2283279 | Weill Cornell Medicine | New York Genome Center | Michael Zody, Andre Corvelo, Dayna M. Oschwald, Samantha Fennessey, Tom Maniatis, Melissa Cushing, Olivier Elemento, Margaret Elizabeth Ross, Chris Mason, Priya Velu, Hanna Rennert, Arryn Craney, Lars F Westblade |
| EPI_ISL_2283280 | Infinity Biologix | Centers for Disease Control and Prevention Division of Viral Diseases, Pathogen Discovery | Dakota Howard, Dhwani Batra, Peter W. Cook, Kara Moser, Adrian Paskey, Jason Caravas, Benjamin Rambo-Martin, Shatavia Morrison, Christopher Gulvick, Scott Sammons, Yvette Unoarumhi, Darlene Wagner, Matthew Schmerer, Christian Bixby, Yihe Wang, Jonathan Schultz, Chirayu Goswami, Russ Hager, Robin Grimwood, Clinton R. Paden, Duncan MacCannell |
| EPI_ISL_2283281 | Weill Cornell Medicine | New York Genome Center | Michael Zody, Andre Corvelo, Dayna M. Oschwald, Samantha Fennessey, Tom Maniatis, Melissa Cushing, Olivier Elemento, Margaret Elizabeth Ross, Chris Mason, Priya Velu, Hanna Rennert, Arryn Craney, Lars F Westblade |
| EPI_ISL_2283282 | Infinity Biologix | Centers for Disease Control and Prevention Division of Viral Diseases, Pathogen Discovery | Dakota Howard, Dhwani Batra, Peter W. Cook, Kara Moser, Adrian Paskey, Jason Caravas, Benjamin Rambo-Martin, Shatavia Morrison, Christopher Gulvick, Scott Sammons, Yvette Unoarumhi, Darlene Wagner, Matthew Schmerer, Christian Bixby, Yihe Wang, Jonathan Schultz, Chirayu Goswami, Russ Hager, Robin Grimwood, Clinton R. Paden, Duncan MacCannell |
| EPI_ISL_2283283 | Weill Cornell Medicine | New York Genome Center | Michael Zody, Andre Corvelo, Dayna M. Oschwald, Samantha Fennessey, Tom Maniatis, Melissa Cushing, Olivier Elemento, Margaret Elizabeth Ross, Chris Mason, Priya Velu, Hanna Rennert, Arryn Craney, Lars F Westblade |

EPI_ISL_2283284, EPI_ISL_2283285, EPI_ISL_2283286, EPI_ISL_2283287, EPI_ISL_2283289, EPI_ISL_2283291, EPI_ISL_2283292, EPI_ISL_2283293, EPI_ISL_2283294, EPI_ISL_2283295, EPI_ISL_2283296, EPI_ISL_2283297, EPI_ISL_2283298, EPI_ISL_2283299, EPI_ISL_2283300, EPI_ISL_2283301, EPI_ISL_2283303, EPI_ISL_2283304, EPI_ISL_2283305, EPI_ISL_2283306, EPI_ISL_2283307, EPI_ISL_2283308, EPI_ISL_2283309, EPI_ISL_2283310, EPI_ISL_2283311, EPI_ISL_2283312, EPI_ISL_2283313, EPI_ISL_2283314, EPI_ISL_2283315, EPI_ISL_2283316, EPI_ISL_2283317, EPI_ISL_2283318, EPI_ISL_2283319, EPI_ISL_2283320, EPI_ISL_2283321, EPI_ISL_2283322, EPI_ISL_2283323, EPI_ISL_2283324, EPI_ISL_2283325, EPI_ISL_2283326, EPI_ISL_2283327, EPI_ISL_2283328, EPI_ISL_2283329, EPI_ISL_2283330, EPI_ISL_2283331, EPI_ISL_2283332, EPI_ISL_2283333, EPI_ISL_2283334, EPI_ISL_2283335, EPI_ISL_2283336, EPI_ISL_2283337, EPI_ISL_2283338, EPI_ISL_2283339, EPI_ISL_2283340, EPI_ISL_2283341, EPI_ISL_2283342, EPI_ISL_2283343, EPI_ISL_2283344, EPI_ISL_2283345, EPI_ISL_2283346, EPI_ISL_2283347, EPI_ISL_2283348, EPI_ISL_2283349, EPI_ISL_2283350, EPI_ISL_2283352, EPI_ISL_2283353, EPI_ISL_2283354, EPI_ISL_2283355, EPI_ISL_2283356, EPI_ISL_2283357, EPI_ISL_2283358, EPI_ISL_2283359, EPI_ISL_2283360, EPI_ISL_2283361, EPI_ISL_2283362, EPI_ISL_2283363, EPI_ISL_2283364, EPI_ISL_2283365, EPI_ISL_2283366, EPI_ISL_2283367, EPI_ISL_2283368, EPI_ISL_2283369, EPI_ISL_2283370, EPI_ISL_2283371, EPI_ISL_2283372, EPI_ISL_2283373, EPI_ISL_2283374, EPI_ISL_2283375,

| | | | |
|---|---|---|---|
| EPI_ISL_2283376, EPI_ISL_2283377, EPI_ISL_2283378, EPI_ISL_2283379, EPI_ISL_2283380, EPI_ISL_2283381, EPI_ISL_2283382, EPI_ISL_2283383, EPI_ISL_2283384, EPI_ISL_2283385, EPI_ISL_2283386, EPI_ISL_2283387, EPI_ISL_2283388, EPI_ISL_2283389, EPI_ISL_2283390, EPI_ISL_2283391, EPI_ISL_2283392, EPI_ISL_2283393, EPI_ISL_2283394, EPI_ISL_2283395, EPI_ISL_2283396, EPI_ISL_2283397, EPI_ISL_2283398, EPI_ISL_2283399, EPI_ISL_2283400, EPI_ISL_2283401, EPI_ISL_2283402, EPI_ISL_2283403, EPI_ISL_2283404, EPI_ISL_2283405, EPI_ISL_2283406, EPI_ISL_2283407, EPI_ISL_2283408, EPI_ISL_2283409, EPI_ISL_2283410, EPI_ISL_2283411, EPI_ISL_2283412, EPI_ISL_2283413, EPI_ISL_2283414, EPI_ISL_2283415, EPI_ISL_2283416, EPI_ISL_2283417, EPI_ISL_2283418, EPI_ISL_2283419, EPI_ISL_2283420, EPI_ISL_2283421, EPI_ISL_2283422, EPI_ISL_2283423, EPI_ISL_2283424, EPI_ISL_2283425, EPI_ISL_2283426, EPI_ISL_2283427, EPI_ISL_2283428, EPI_ISL_2283429, EPI_ISL_2283430, EPI_ISL_2283431, EPI_ISL_2283432, EPI_ISL_2283433, EPI_ISL_2283434, EPI_ISL_2283435, EPI_ISL_2283436, EPI_ISL_2283437, EPI_ISL_2283438, EPI_ISL_2283439, EPI_ISL_2283440, EPI_ISL_2283441, EPI_ISL_2283442, EPI_ISL_2283443, EPI_ISL_2283444, EPI_ISL_2283445, EPI_ISL_2283446, EPI_ISL_2283447, EPI_ISL_2283448, EPI_ISL_2283449, EPI_ISL_2283450, EPI_ISL_2283451, EPI_ISL_2283452, EPI_ISL_2283453, EPI_ISL_2283454, EPI_ISL_2283455, EPI_ISL_2283456, EPI_ISL_2283457, EPI_ISL_2283458, EPI_ISL_2283459, EPI_ISL_2283460, EPI_ISL_2283461, EPI_ISL_2283462, EPI_ISL_2283463, EPI_ISL_2283464, EPI_ISL_2283465, EPI_ISL_2283466, EPI_ISL_2283467, EPI_ISL_2283468, EPI_ISL_2283469, EPI_ISL_2283470, EPI_ISL_2283471, EPI_ISL_2283472, EPI_ISL_2283473, EPI_ISL_2283474, EPI_ISL_2283475, EPI_ISL_2283476, EPI_ISL_2283477, EPI_ISL_2283478, EPI_ISL_2283479, EPI_ISL_2283480, EPI_ISL_2283481, EPI_ISL_2283482, EPI_ISL_2283483, EPI_ISL_2283484, EPI_ISL_2283485, EPI_ISL_2283486, EPI_ISL_2283487, EPI_ISL_2283488, EPI_ISL_2283489, EPI_ISL_2283490, EPI_ISL_2283491, EPI_ISL_2283492, EPI_ISL_2283493, EPI_ISL_2283494, EPI_ISL_2283495, EPI_ISL_2283496, EPI_ISL_2283497, EPI_ISL_2283498, EPI_ISL_2283499, EPI_ISL_2283500, EPI_ISL_2283501, EPI_ISL_2283502, EPI_ISL_2283503, EPI_ISL_2283504, EPI_ISL_2283505, EPI_ISL_2283506, EPI_ISL_2283507, EPI_ISL_2283508, EPI_ISL_2283509, EPI_ISL_2283510, EPI_ISL_2283511, EPI_ISL_2283512, EPI_ISL_2283513, EPI_ISL_2283514, EPI_ISL_2283515, EPI_ISL_2283516, EPI_ISL_2283517, EPI_ISL_2283518, EPI_ISL_2283519, EPI_ISL_2283520, EPI_ISL_2283521, EPI_ISL_2283522, EPI_ISL_2283523, EPI_ISL_2283524, EPI_ISL_2283525, EPI_ISL_2283526, EPI_ISL_2283527, EPI_ISL_2283528, EPI_ISL_2283529, EPI_ISL_2283530, EPI_ISL_2283531, EPI_ISL_2283532, EPI_ISL_2283533, EPI_ISL_2283534, EPI_ISL_2283535, EPI_ISL_2283536, EPI_ISL_2283537, EPI_ISL_2283538, EPI_ISL_2283539, EPI_ISL_2283540, EPI_ISL_2283541, EPI_ISL_2283542, EPI_ISL_2283543, EPI_ISL_2283544, EPI_ISL_2283547, EPI_ISL_2283548, EPI_ISL_2283549, EPI_ISL_2283550, EPI_ISL_2283551, EPI_ISL_2283552, EPI_ISL_2283553, EPI_ISL_2283554, EPI_ISL_2283555, EPI_ISL_2283556, EPI_ISL_2283557, EPI_ISL_2283558, EPI_ISL_2283559, EPI_ISL_2283560, EPI_ISL_2283561, EPI_ISL_2283562, EPI_ISL_2283563, EPI_ISL_2283564, EPI_ISL_2283565, EPI_ISL_2283566, EPI_ISL_2283567, EPI_ISL_2283568, EPI_ISL_2283569, EPI_ISL_2283570, EPI_ISL_2283571, EPI_ISL_2283572, EPI_ISL_2283573, EPI_ISL_2283574, EPI_ISL_2283575, EPI_ISL_2283576, EPI_ISL_2283577, EPI_ISL_2283578, EPI_ISL_2283579, EPI_ISL_2283580, EPI_ISL_2283581, EPI_ISL_2283582, EPI_ISL_2283583, EPI_ISL_2283584, EPI_ISL_2283585, EPI_ISL_2283586, EPI_ISL_2283587, EPI_ISL_2283588, EPI_ISL_2283589, EPI_ISL_2283590, EPI_ISL_2283591, EPI_ISL_2283592, EPI_ISL_2283593, EPI_ISL_2283594, EPI_ISL_2283595, EPI_ISL_2283596, EPI_ISL_2283597, EPI_ISL_2283598, EPI_ISL_2283599, EPI_ISL_2283600, EPI_ISL_2283601, EPI_ISL_2283602, EPI_ISL_2283603, EPI_ISL_2283604, EPI_ISL_2283605, EPI_ISL_2283606, EPI_ISL_2283607, EPI_ISL_2283608, EPI_ISL_2283609, EPI_ISL_2283610, EPI_ISL_2283611, EPI_ISL_2283612, EPI_ISL_2283613, EPI_ISL_2283614, EPI_ISL_2283615 | | | |
| see above | Infinity Biologix | Centers for Disease Control and Prevention Division of Viral Diseases, Pathogen Discovery | Dakota Howard, Dhwani Batra, Peter W. Cook, Kara Moser, Adrian Paskey, Jason Caravas, Benjamin Rambo-Martin, Shatavia Morrison, Christopher Gulvick, Scott Sammons, Yvette Unoarumhi, Darlene Wagner, Matthew Schmerer, Christian Bixby, Yihe Wang, Jonathan Schultz, Chirayu Goswami, Russ Hager, Robin Grimwood, Clinton R. Paden, Duncan MacCannell |
| EPI_ISL_2283616, EPI_ISL_2283617, EPI_ISL_2283618, EPI_ISL_2283619, EPI_ISL_2283620, EPI_ISL_2283621, EPI_ISL_2283622, EPI_ISL_2283623, EPI_ISL_2283624, EPI_ISL_2283625, EPI_ISL_2283626, EPI_ISL_2283627, EPI_ISL_2283628, EPI_ISL_2283629, EPI_ISL_2283630, EPI_ISL_2283631, EPI_ISL_2283632, EPI_ISL_2283633, EPI_ISL_2283634, EPI_ISL_2283635, EPI_ISL_2283636 | | | |
| see above | Wyoming Public Health Laboratory | Wyoming Public Health Laboratory | Jim Mildenberger, Taylor Fearing, Channing Weber, Ashley Norberg, Chayse Rowley, Marley Goetz, Brian Dominguez, Elliot Thomasson, Brittany Oher, Sam Britz, Cari Sloma, and Rob Christensen |
| EPI_ISL_2283681, EPI_ISL_2283682, EPI_ISL_2283683, EPI_ISL_2283684, EPI_ISL_2283685 | LESP Oaxaca | Instituto de Diagnostico y Referencia Epidemiologicos (INDRE) | Claudia Wong-Arambula, Abril Rodriguez-Maldonado, Vanessa Rivero-Arredondo, Ariadna Medina-Benitez, Joaquin Quiroz-Mercado, Sergio Rangel-Guerrero, Natividad Cruz-Ortiz, Tatiana Nunez-Garcia, Gisela Barrera-Badillo, Lucia Hernandez-Rivas, Irma Lopez-Martinez, Ernesto Ramirez-Gonzalez. |
| EPI_ISL_2283686 | Lesp Jalisco /Unidad de Patologia | Instituto de Diagnostico y Referencia Epidemiologicos (INDRE) | Claudia Wong-Arambula, Abril Rodriguez-Maldonado, Vanessa Rivero-Arredondo, Ariadna Medina-Benitez, Joaquin Quiroz-Mercado, Sergio Rangel-Guerrero, Natividad Cruz-Ortiz, Tatiana Nunez-Garcia, Gisela Barrera-Badillo, Lucia Hernandez-Rivas, Irma Lopez-Martinez, Ernesto Ramirez-Gonzalez. |
| EPI_ISL_2283687 | LESP Tamaulipas | Instituto de Diagnostico y Referencia Epidemiologicos (INDRE) | Claudia Wong-Arambula, Abril Rodriguez-Maldonado, Vanessa Rivero-Arredondo, Ariadna Medina-Benitez, Joaquin Quiroz-Mercado, Sergio Rangel-Guerrero, Natividad Cruz-Ortiz, Tatiana Nunez-Garcia, Gisela Barrera-Badillo, Lucia Hernandez-Rivas, Irma Lopez-Martinez, Ernesto Ramirez-Gonzalez. |
| EPI_ISL_2283688 | LESP Hidalgo | Instituto de Diagnostico y Referencia Epidemiologicos (INDRE) | Claudia Wong-Arambula, Abril Rodriguez-Maldonado, Vanessa Rivero-Arredondo, Ariadna Medina-Benitez, Joaquin Quiroz-Mercado, Sergio Rangel-Guerrero, Natividad Cruz-Ortiz, Tatiana Nunez-Garcia, Gisela Barrera-Badillo, Lucia Hernandez-Rivas, Irma Lopez-Martinez, Ernesto Ramirez-Gonzalez. |
| EPI_ISL_2283689 | LESP Campeche | Instituto de Diagnostico y Referencia Epidemiologicos (INDRE) | Claudia Wong-Arambula, Abril Rodriguez-Maldonado, Vanessa Rivero-Arredondo, Ariadna Medina-Benitez, Joaquin Quiroz-Mercado, Sergio Rangel-Guerrero, Natividad Cruz-Ortiz, Tatiana Nunez-Garcia, Gisela Barrera-Badillo, Lucia Hernandez-Rivas, Irma Lopez-Martinez, Ernesto Ramirez-Gonzalez. |
| EPI_ISL_2283690 | LESP Hidalgo | Instituto de Diagnostico y Referencia Epidemiologicos (INDRE) | Claudia Wong-Arambula, Abril Rodriguez-Maldonado, Vanessa Rivero-Arredondo, Ariadna Medina-Benitez, Joaquin Quiroz-Mercado, Sergio Rangel-Guerrero, Natividad Cruz-Ortiz, Tatiana Nunez-Garcia, Gisela Barrera-Badillo, Lucia Hernandez-Rivas, Irma Lopez-Martinez, Ernesto Ramirez-Gonzalez. |
| EPI_ISL_2283691, EPI_ISL_2283692 | LESP Chihuahua | Instituto de Diagnostico y Referencia Epidemiologicos (INDRE) | Claudia Wong-Arambula, Abril Rodriguez-Maldonado, Vanessa Rivero-Arredondo, Ariadna Medina-Benitez, Joaquin Quiroz-Mercado, Sergio Rangel-Guerrero, Natividad Cruz-Ortiz, Tatiana Nunez-Garcia, Gisela Barrera-Badillo, Lucia Hernandez-Rivas, Irma Lopez-Martinez, Ernesto Ramirez-Gonzalez. |
| EPI_ISL_2283693 | LESP Tamaulipas | Instituto de Diagnostico y Referencia Epidemiologicos (INDRE) | Claudia Wong-Arambula, Abril Rodriguez-Maldonado, Vanessa Rivero-Arredondo, Ariadna Medina-Benitez, Joaquin Quiroz-Mercado, Sergio Rangel-Guerrero, Natividad Cruz-Ortiz, Tatiana Nunez-Garcia, Gisela Barrera-Badillo, Lucia Hernandez-Rivas, Irma Lopez-Martinez, Ernesto Ramirez-Gonzalez. |
| EPI_ISL_2283694 | LESP Oaxaca | Instituto de Diagnostico y Referencia Epidemiologicos (INDRE) | Claudia Wong-Arambula, Abril Rodriguez-Maldonado, Vanessa Rivero-Arredondo, Ariadna Medina-Benitez, Joaquin Quiroz-Mercado, Sergio Rangel-Guerrero, Natividad Cruz-Ortiz, Tatiana Nunez-Garcia, Gisela Barrera-Badillo, Lucia Hernandez-Rivas, Irma Lopez-Martinez, Ernesto Ramirez-Gonzalez. |
| EPI_ISL_2283695, EPI_ISL_2283696 | LESP Michoacan | Instituto de Diagnostico y Referencia Epidemiologicos (INDRE) | Claudia Wong-Arambula, Abril Rodriguez-Maldonado, Vanessa Rivero-Arredondo, Ariadna Medina-Benitez, Joaquin Quiroz-Mercado, Sergio Rangel-Guerrero, Natividad Cruz-Ortiz, Tatiana Nunez-Garcia, Gisela Barrera-Badillo, Lucia Hernandez-Rivas, Irma Lopez-Martinez, Ernesto Ramirez-Gonzalez. |
| EPI_ISL_2283697 | LESP Colima | Instituto de Diagnostico y Referencia Epidemiologicos (INDRE) | Claudia Wong-Arambula, Abril Rodriguez-Maldonado, Vanessa Rivero-Arredondo, Ariadna Medina-Benitez, Joaquin Quiroz-Mercado, Sergio Rangel-Guerrero, Natividad Cruz-Ortiz, Tatiana Nunez-Garcia, Gisela Barrera-Badillo, Lucia Hernandez-Rivas, Irma Lopez-Martinez, Ernesto Ramirez-Gonzalez. |
| EPI_ISL_2283698, EPI_ISL_2283699 | LESP Yucatan | Instituto de Diagnostico y Referencia Epidemiologicos (INDRE) | Claudia Wong-Arambula, Abril Rodriguez-Maldonado, Vanessa Rivero-Arredondo, Ariadna Medina-Benitez, Joaquin Quiroz-Mercado, Sergio Rangel-Guerrero, Natividad Cruz-Ortiz, Tatiana Nunez-Garcia, Gisela Barrera-Badillo, Lucia Hernandez-Rivas, Irma Lopez-Martinez, Ernesto Ramirez-Gonzalez. |
| EPI_ISL_2283700, EPI_ISL_2283701, EPI_ISL_2283702, EPI_ISL_2283703 | LESP Sinaloa | Instituto de Diagnostico y Referencia Epidemiologicos (INDRE) | Claudia Wong-Arambula, Abril Rodriguez-Maldonado, Vanessa Rivero-Arredondo, Ariadna Medina-Benitez, Joaquin Quiroz-Mercado, Sergio Rangel-Guerrero, Natividad Cruz-Ortiz, Tatiana Nunez-Garcia, Gisela Barrera-Badillo, Lucia Hernandez-Rivas, Irma Lopez-Martinez, Ernesto Ramirez-Gonzalez. |
| EPI_ISL_2283704, EPI_ISL_2283705, EPI_ISL_2283706, EPI_ISL_2283707, EPI_ISL_2283708 | LESP Ciudad de Mexico | Instituto de Diagnostico y Referencia Epidemiologicos (INDRE) | Claudia Wong-Arambula, Abril Rodriguez-Maldonado, Vanessa Rivero-Arredondo, Ariadna Medina-Benitez, Joaquin Quiroz-Mercado, Sergio Rangel-Guerrero, Natividad Cruz-Ortiz, Tatiana Nunez-Garcia, Gisela Barrera-Badillo, Lucia Hernandez-Rivas, Irma Lopez-Martinez, Ernesto Ramirez-Gonzalez. |
| EPI_ISL_2283709, EPI_ISL_2283710 | LESP Sinaloa | Instituto de Diagnostico y Referencia Epidemiologicos (INDRE) | Claudia Wong-Arambula, Abril Rodriguez-Maldonado, Vanessa Rivero-Arredondo, Ariadna Medina-Benitez, Joaquin Quiroz-Mercado, Sergio Rangel-Guerrero, Natividad Cruz-Ortiz, Tatiana Nunez-Garcia, Gisela Barrera-Badillo, Lucia Hernandez-Rivas, Irma Lopez-Martinez, Ernesto Ramirez-Gonzalez. |
| EPI_ISL_2283711, EPI_ISL_2283712, EPI_ISL_2283713, EPI_ISL_2283714, EPI_ISL_2283715, EPI_ISL_2283716 | LESP Tamaulipas | Instituto de Diagnostico y Referencia Epidemiologicos (INDRE) | Claudia Wong-Arambula, Abril Rodriguez-Maldonado, Vanessa Rivero-Arredondo, Ariadna Medina-Benitez, Joaquin Quiroz-Mercado, Sergio Rangel-Guerrero, Natividad Cruz-Ortiz, Tatiana Nunez-Garcia, Gisela Barrera-Badillo, Lucia Hernandez-Rivas, Irma Lopez-Martinez, Ernesto Ramirez-Gonzalez. |
| EPI_ISL_2283717, EPI_ISL_2283718 | LESP Guanajuato | Instituto de Diagnostico y Referencia Epidemiologicos (INDRE) | Claudia Wong-Arambula, Abril Rodriguez-Maldonado, Vanessa Rivero-Arredondo, Ariadna Medina-Benitez, Joaquin Quiroz-Mercado, Sergio Rangel-Guerrero, Natividad Cruz-Ortiz, Tatiana Nunez-Garcia, Gisela Barrera-Badillo, Lucia Hernandez-Rivas, Irma Lopez-Martinez, Ernesto Ramirez-Gonzalez. |
| EPI_ISL_2283719 | LESP Yucatan | Instituto de Diagnostico y Referencia Epidemiologicos (INDRE) | Claudia Wong-Arambula, Abril Rodriguez-Maldonado, Vanessa Rivero-Arredondo, Ariadna Medina-Benitez, Joaquin Quiroz-Mercado, Sergio Rangel-Guerrero, Natividad Cruz-Ortiz, Tatiana Nunez-Garcia, Gisela Barrera-Badillo, Lucia Hernandez-Rivas, Irma Lopez-Martinez, Ernesto Ramirez-Gonzalez. |
| EPI_ISL_2283720 | LESP Queretaro | Instituto de Diagnostico y Referencia Epidemiologicos (INDRE) | Claudia Wong-Arambula, Abril Rodriguez-Maldonado, Vanessa Rivero-Arredondo, Ariadna Medina-Benitez, Joaquin Quiroz-Mercado, Sergio Rangel-Guerrero, Natividad Cruz-Ortiz, Tatiana Nunez-Garcia, Gisela Barrera-Badillo, Lucia Hernandez-Rivas, Irma Lopez-Martinez, Ernesto Ramirez-Gonzalez. |
| EPI_ISL_2283721, EPI_ISL_2283722 | LESP Guanajuato | Instituto de Diagnostico y Referencia Epidemiologicos (INDRE) | Claudia Wong-Arambula, Abril Rodriguez-Maldonado, Vanessa Rivero-Arredondo, Ariadna Medina-Benitez, Joaquin Quiroz-Mercado, Sergio Rangel-Guerrero, Natividad Cruz-Ortiz, Tatiana Nunez-Garcia, Gisela Barrera-Badillo, Lucia Hernandez-Rivas, Irma Lopez-Martinez, Ernesto Ramirez-Gonzalez. |
| EPI_ISL_2283723, EPI_ISL_2283724 | LESP Nuevo Leon | Instituto de Diagnostico y Referencia Epidemiologicos (INDRE) | Claudia Wong-Arambula, Abril Rodriguez-Maldonado, Vanessa Rivero-Arredondo, Ariadna Medina-Benitez, Joaquin Quiroz-Mercado, Sergio Rangel-Guerrero, Natividad Cruz-Ortiz, Tatiana Nunez-Garcia, Gisela Barrera-Badillo, Lucia Hernandez-Rivas, Irma Lopez-Martinez, Ernesto Ramirez-Gonzalez. |
| EPI_ISL_2283725 | LESP Chihuahua | Instituto de Diagnostico y Referencia Epidemiologicos (INDRE) | Claudia Wong-Arambula, Abril Rodriguez-Maldonado, Vanessa Rivero-Arredondo, Ariadna Medina-Benitez, Joaquin Quiroz-Mercado, Sergio Rangel-Guerrero, Natividad Cruz-Ortiz, Tatiana Nunez-Garcia, Gisela Barrera-Badillo, Lucia Hernandez-Rivas, Irma Lopez-Martinez, Ernesto Ramirez-Gonzalez. |
| EPI_ISL_2283812, EPI_ISL_2283813, EPI_ISL_2283814, EPI_ISL_2283815, EPI_ISL_2283816, EPI_ISL_2283817, EPI_ISL_2283818, EPI_ISL_2283819, EPI_ISL_2283820, EPI_ISL_2283821, EPI_ISL_2283822, EPI_ISL_2283823, EPI_ISL_2283824, EPI_ISL_2283825, EPI_ISL_2283826, EPI_ISL_2283828, EPI_ISL_2283829, EPI_ISL_2283830, EPI_ISL_2283831, EPI_ISL_2283832, EPI_ISL_2283833, EPI_ISL_2283834, EPI_ISL_2283835, EPI_ISL_2283836, EPI_ISL_2283837, EPI_ISL_2283838, EPI_ISL_2283839, EPI_ISL_2283840, EPI_ISL_2283842, EPI_ISL_2283843, EPI_ISL_2283844, EPI_ISL_2283845, EPI_ISL_2283846, EPI_ISL_2283847, EPI_ISL_2283848, EPI_ISL_2283849, EPI_ISL_2283850, EPI_ISL_2283851, EPI_ISL_2283852, EPI_ISL_2283853, EPI_ISL_2283854, EPI_ISL_2283855, EPI_ISL_2283856, EPI_ISL_2283857, EPI_ISL_2283858, EPI_ISL_2283859, EPI_ISL_2283860, EPI_ISL_2283861, EPI_ISL_2283863, EPI_ISL_2283864, EPI_ISL_2283865, EPI_ISL_2283866, EPI_ISL_2283867, EPI_ISL_2283868, EPI_ISL_2283869, EPI_ISL_2283870, EPI_ISL_2283871, EPI_ISL_2283872, EPI_ISL_2283873, EPI_ISL_2283874, EPI_ISL_2283875, EPI_ISL_2283876, EPI_ISL_2283877, EPI_ISL_2283878, EPI_ISL_2283881, EPI_ISL_2283882, EPI_ISL_2283884, EPI_ISL_2283885, EPI_ISL_2283886, EPI_ISL_2283887, EPI_ISL_2283888, EPI_ISL_2283889 | | | |
| see above | Aegis Sciences Corporation | Centers for Disease Control and Prevention Division of Viral Diseases, Pathogen Discovery | Dakota Howard, Dhwani Batra, Peter W. Cook, Kara Moser, Adrian Paskey, Jason Caravas, Benjamin Rambo-Martin, Shatavia Morrison, Christopher Gulvick, Scott Sammons, Yvette Unoarumhi, Darlene Wagner, Matthew Schmerer, Cyndi Clark, Patrick Campbell, Rob Case, Vikramsinha Ghorpade, Holly Houdeshell, Ola Kvalvaag, Dillon Nall, Ethan Sanders, Alec Vest, Shaun Westlund, Matthew Hardison, Clinton R. Paden, Duncan MacCannell |
| EPI_ISL_2283890, EPI_ISL_2283892, EPI_ISL_2283893, EPI_ISL_2283894, EPI_ISL_2283896, EPI_ISL_2283897, EPI_ISL_2283899, EPI_ISL_2283900, EPI_ISL_2283901, EPI_ISL_2283902, EPI_ISL_2283904, EPI_ISL_2283907, EPI_ISL_2283908, EPI_ISL_2283909, EPI_ISL_2283910, EPI_ISL_2283911, EPI_ISL_2283912, EPI_ISL_2283913, EPI_ISL_2283914, EPI_ISL_2283915, EPI_ISL_2283916, EPI_ISL_2283917, EPI_ISL_2283918, EPI_ISL_2283920, EPI_ISL_2283921, EPI_ISL_2283922, EPI_ISL_2283923, EPI_ISL_2283924, EPI_ISL_2283925, EPI_ISL_2283926, EPI_ISL_2283927, EPI_ISL_2283928, EPI_ISL_2283929, EPI_ISL_2283930, EPI_ISL_2283931, EPI_ISL_2283932, EPI_ISL_2283933, EPI_ISL_2283935, EPI_ISL_2283936, EPI_ISL_2283937, EPI_ISL_2283939, EPI_ISL_2283940, EPI_ISL_2283941, EPI_ISL_2283942, EPI_ISL_2283943, EPI_ISL_2283944, EPI_ISL_2283945, EPI_ISL_2283946, EPI_ISL_2283947, EPI_ISL_2283948, EPI_ISL_2283949, EPI_ISL_2283950, EPI_ISL_2283951, EPI_ISL_2283952, EPI_ISL_2283953, EPI_ISL_2283954, EPI_ISL_2283955, EPI_ISL_2283956, EPI_ISL_2283957, EPI_ISL_2283958, EPI_ISL_2283959, EPI_ISL_2283960, EPI_ISL_2283961, EPI_ISL_2283962, EPI_ISL_2283963, EPI_ISL_2283964, EPI_ISL_2283965, EPI_ISL_2283966, EPI_ISL_2283967, EPI_ISL_2283969, EPI_ISL_2283970, EPI_ISL_2283972, EPI_ISL_2283973, EPI_ISL_2283974, EPI_ISL_2283975, EPI_ISL_2283976, EPI_ISL_2283979, EPI_ISL_2283980, EPI_ISL_2283981, EPI_ISL_2283982, EPI_ISL_2283984, EPI_ISL_2283985, EPI_ISL_2283986, EPI_ISL_2283987, EPI_ISL_2283988, EPI_ISL_2283989, EPI_ISL_2283990, EPI_ISL_2283991, EPI_ISL_2283992, EPI_ISL_2283994, EPI_ISL_2283995, EPI_ISL_2283996, EPI_ISL_2283998, EPI_ISL_2283999, EPI_ISL_2284000, EPI_ISL_2284002, EPI_ISL_2284003, EPI_ISL_2284004, EPI_ISL_2284005, EPI_ISL_2284006, EPI_ISL_2284007, EPI_ISL_2284008, EPI_ISL_2284009, EPI_ISL_2284010, EPI_ISL_2284011, EPI_ISL_2284012, EPI_ISL_2284013, EPI_ISL_2284014, EPI_ISL_2284015, EPI_ISL_2284016, EPI_ISL_2284017, EPI_ISL_2284018, EPI_ISL_2284019, EPI_ISL_2284020, EPI_ISL_2284021, EPI_ISL_2284022, EPI_ISL_2284023, EPI_ISL_2284025, EPI_ISL_2284026, EPI_ISL_2284027, EPI_ISL_2284028, EPI_ISL_2284029, EPI_ISL_2284030, EPI_ISL_2284031, EPI_ISL_2284033, EPI_ISL_2284034, EPI_ISL_2284035, EPI_ISL_2284036, EPI_ISL_2284037, EPI_ISL_2284038, EPI_ISL_2284039, EPI_ISL_2284040, EPI_ISL_2284041, EPI_ISL_2284042, EPI_ISL_2284043, EPI_ISL_2284044, EPI_ISL_2284046, EPI_ISL_2284047, EPI_ISL_2284048, EPI_ISL_2284049, EPI_ISL_2284050, EPI_ISL_2284051, EPI_ISL_2284052, EPI_ISL_2284053, EPI_ISL_2284054, EPI_ISL_2284055, EPI_ISL_2284056, EPI_ISL_2284057, EPI_ISL_2284058, EPI_ISL_2284059, EPI_ISL_2284060, EPI_ISL_2284061, EPI_ISL_2284062, EPI_ISL_2284063, EPI_ISL_2284064, EPI_ISL_2284065, EPI_ISL_2284066, EPI_ISL_2284067, EPI_ISL_2284068, EPI_ISL_2284069, EPI_ISL_2284070, EPI_ISL_2284071, EPI_ISL_2284073, EPI_ISL_2284074, EPI_ISL_2284076, EPI_ISL_2284077, EPI_ISL_2284078, EPI_ISL_2284079, EPI_ISL_2284080, EPI_ISL_2284081, EPI_ISL_2284082, EPI_ISL_2284083, EPI_ISL_2284084, EPI_ISL_2284085, EPI_ISL_2284086, EPI_ISL_2284087, EPI_ISL_2284089, EPI_ISL_2284090, EPI_ISL_2284091, EPI_ISL_2284092, EPI_ISL_2284093, EPI_ISL_2284094, EPI_ISL_2284095, EPI_ISL_2284096, EPI_ISL_2284097, EPI_ISL_2284098, EPI_ISL_2284099, EPI_ISL_2284100, EPI_ISL_2284101, EPI_ISL_2284102, EPI_ISL_2284103, EPI_ISL_2284104, EPI_ISL_2284105, EPI_ISL_2284106, EPI_ISL_2284107, EPI_ISL_2284108, EPI_ISL_2284109, EPI_ISL_2284110, EPI_ISL_2284111, EPI_ISL_2284112, EPI_ISL_2284113, EPI_ISL_2284114, EPI_ISL_2284115, EPI_ISL_2284116, EPI_ISL_2284117, EPI_ISL_2284118, EPI_ISL_2284119, EPI_ISL_2284120, EPI_ISL_2284121, EPI_ISL_2284122, EPI_ISL_2284123, EPI_ISL_2284124, EPI_ISL_2284125, EPI_ISL_2284126, EPI_ISL_2284127, EPI_ISL_2284129, EPI_ISL_2284130, EPI_ISL_2284131, EPI_ISL_2284132, EPI_ISL_2284133, EPI_ISL_2284134, EPI_ISL_2284135, EPI_ISL_2284137, EPI_ISL_2284138, EPI_ISL_2284139, EPI_ISL_2284140, EPI_ISL_2284141, EPI_ISL_2284142, EPI_ISL_2284143, EPI_ISL_2284144, EPI_ISL_2284145, EPI_ISL_2284146, EPI_ISL_2284147, EPI_ISL_2284148, EPI_ISL_2284149, EPI_ISL_2284150, EPI_ISL_2284151, EPI_ISL_2284152, EPI_ISL_2284153, EPI_ISL_2284154, EPI_ISL_2284155, EPI_ISL_2284156, EPI_ISL_2284157, EPI_ISL_2284158, EPI_ISL_2284159, EPI_ISL_2284160, EPI_ISL_2284161, EPI_ISL_2284162, EPI_ISL_2284163, EPI_ISL_2284164, EPI_ISL_2284165, EPI_ISL_2284166, EPI_ISL_2284167, EPI_ISL_2284168, EPI_ISL_2284170, EPI_ISL_2284171, EPI_ISL_2284174, EPI_ISL_2284175, EPI_ISL_2284176, EPI_ISL_2284177, EPI_ISL_2284178, EPI_ISL_2284179, EPI_ISL_2284180, EPI_ISL_2284181, EPI_ISL_2284182, EPI_ISL_2284183, EPI_ISL_2284184, EPI_ISL_2284185, EPI_ISL_2284186, EPI_ISL_2284187, EPI_ISL_2284188, EPI_ISL_2284189, EPI_ISL_2284190, EPI_ISL_2284191, EPI_ISL_2284192, EPI_ISL_2284193, EPI_ISL_2284194, EPI_ISL_2284195, EPI_ISL_2284196, EPI_ISL_2284197, EPI_ISL_2284198 | | | |
| see above | VA Connecticut Healthcare System | Yale Center for Genomic Analysis | Shrikant Mane, Kaya Bilguvar, Curt Scharfe, Irina Tikhonova, Brooke Sullivan |
| EPI_ISL_2284199, EPI_ISL_2284200, EPI_ISL_2284201, EPI_ISL_2284202, EPI_ISL_2284204, EPI_ISL_2284205, EPI_ISL_2284206, EPI_ISL_2284207, EPI_ISL_2284208, EPI_ISL_2284209, EPI_ISL_2284210, EPI_ISL_2284211, EPI_ISL_2284212, EPI_ISL_2284213, EPI_ISL_2284214, EPI_ISL_2284215, EPI_ISL_2284216, EPI_ISL_2284217, EPI_ISL_2284218, EPI_ISL_2284219, EPI_ISL_2284220, EPI_ISL_2284221, EPI_ISL_2284222 | | | |
| see above | Infinity Biologix | Centers for Disease Control and Prevention Division of Viral Diseases, Pathogen | Dakota Howard, Dhwani Batra, Peter W. Cook, Kara Moser, Adrian Paskey, Jason Caravas, Benjamin Rambo-Martin, Shatavia Morrison, Christopher Gulvick, Scott Sammons, Yvette Unoarumhi, Darlene Wagner, Matthew Schmerer, Christian Bixby, Yihe Wang, Jonathan Schultz, Chirayu Goswami, Russ Hager, Robin Grimwood, Clinton R. Paden, Duncan MacCannell |

| | | | |
|---|---|---|---|
| | | Discovery | |
| EPI_ISL_2284693, EPI_ISL_2284695, EPI_ISL_2284700, EPI_ISL_2284702, EPI_ISL_2284704, EPI_ISL_2284705, EPI_ISL_2284706, EPI_ISL_2284707, EPI_ISL_2284710, EPI_ISL_2284711, EPI_ISL_2284713, EPI_ISL_2284714, EPI_ISL_2284715, EPI_ISL_2284716, EPI_ISL_2284717, EPI_ISL_2284718, EPI_ISL_2284719, EPI_ISL_2284720, EPI_ISL_2284722, EPI_ISL_2284723, EPI_ISL_2284725, EPI_ISL_2284726, EPI_ISL_2284727, EPI_ISL_2284728, EPI_ISL_2284729, EPI_ISL_2284731, EPI_ISL_2284732, EPI_ISL_2284733, EPI_ISL_2284734, EPI_ISL_2284735, EPI_ISL_2284738, EPI_ISL_2284739, EPI_ISL_2284740, EPI_ISL_2284741, EPI_ISL_2284743, EPI_ISL_2284745, EPI_ISL_2284746, EPI_ISL_2284747, EPI_ISL_2284750, EPI_ISL_2284751, EPI_ISL_2284753, EPI_ISL_2284754, EPI_ISL_2284755, EPI_ISL_2284756, EPI_ISL_2284757, EPI_ISL_2284758, EPI_ISL_2284759, EPI_ISL_2284760, EPI_ISL_2284761, EPI_ISL_2284762, EPI_ISL_2284763, EPI_ISL_2284765, EPI_ISL_2284766, EPI_ISL_2284767, EPI_ISL_2284768, EPI_ISL_2284769, EPI_ISL_2284771, EPI_ISL_2284772, EPI_ISL_2284773, EPI_ISL_2284775, EPI_ISL_2284776, EPI_ISL_2284778, EPI_ISL_2284779, EPI_ISL_2284780, EPI_ISL_2284781, EPI_ISL_2284782, EPI_ISL_2284783, EPI_ISL_2284784, EPI_ISL_2284786, EPI_ISL_2284787, EPI_ISL_2284788, EPI_ISL_2284789, EPI_ISL_2284791, EPI_ISL_2284792, EPI_ISL_2284793, EPI_ISL_2284795, EPI_ISL_2284797, EPI_ISL_2284798, EPI_ISL_2284799, EPI_ISL_2284800, EPI_ISL_2284801, EPI_ISL_2284802, EPI_ISL_2284803, EPI_ISL_2284804, EPI_ISL_2284805, EPI_ISL_2284806, EPI_ISL_2284807, EPI_ISL_2284808, EPI_ISL_2284809, EPI_ISL_2284811, EPI_ISL_2284812, EPI_ISL_2284813, EPI_ISL_2284814, EPI_ISL_2284815, EPI_ISL_2284816, EPI_ISL_2284817, EPI_ISL_2284818, EPI_ISL_2284819, EPI_ISL_2284820, EPI_ISL_2284822, EPI_ISL_2284823, EPI_ISL_2284824, EPI_ISL_2284825, EPI_ISL_2284826, EPI_ISL_2284827, EPI_ISL_2284828, EPI_ISL_2284830, EPI_ISL_2284831, EPI_ISL_2284832, EPI_ISL_2284833, EPI_ISL_2284837, EPI_ISL_2284839, EPI_ISL_2284842, EPI_ISL_2284843, EPI_ISL_2284844, EPI_ISL_2284846, EPI_ISL_2284847, EPI_ISL_2284848, EPI_ISL_2284851, EPI_ISL_2284852, EPI_ISL_2284853 | | | |
| see above | Utah Public Health Laboratory | Utah Public Health Laboratory | Erin L. Young, Kelly F. Oakeson, Tara Gallagher |
| EPI_ISL_2284854, EPI_ISL_2284855, EPI_ISL_2284856, EPI_ISL_2284857, EPI_ISL_2284858, EPI_ISL_2284859, EPI_ISL_2284860, EPI_ISL_2284861, EPI_ISL_2284863, EPI_ISL_2284864, EPI_ISL_2284865, EPI_ISL_2284866, EPI_ISL_2284867 | | | |
| see above | Wichita State University - Molecular Diagnostics Lab | Kansas Health and Environmental Lab | Mike Grose, Katherine Wiggins, Jonathan Barnell, Ben Olsen, and Phil Adam |
| EPI_ISL_2284979, EPI_ISL_2284980, EPI_ISL_2284981, EPI_ISL_2284982, EPI_ISL_2284983, EPI_ISL_2284984, EPI_ISL_2284985, EPI_ISL_2284986, EPI_ISL_2284987, EPI_ISL_2284988, EPI_ISL_2284989, EPI_ISL_2284990, EPI_ISL_2284991, EPI_ISL_2284992, EPI_ISL_2284993, EPI_ISL_2284994, EPI_ISL_2284995, EPI_ISL_2284997, EPI_ISL_2284998, EPI_ISL_2284999, EPI_ISL_2285000, EPI_ISL_2285002, EPI_ISL_2285003, EPI_ISL_2285004, EPI_ISL_2285005, EPI_ISL_2285006, EPI_ISL_2285007, EPI_ISL_2285008, EPI_ISL_2285009, EPI_ISL_2285010, EPI_ISL_2285011, EPI_ISL_2285012, EPI_ISL_2285013, EPI_ISL_2285014, EPI_ISL_2285015, EPI_ISL_2285016, EPI_ISL_2285018, EPI_ISL_2285019, EPI_ISL_2285020, EPI_ISL_2285021, EPI_ISL_2285022, EPI_ISL_2285023, EPI_ISL_2285025, EPI_ISL_2285026, EPI_ISL_2285027, EPI_ISL_2285029, EPI_ISL_2285030, EPI_ISL_2285031, EPI_ISL_2285032, EPI_ISL_2285033, EPI_ISL_2285034, EPI_ISL_2285035, EPI_ISL_2285036, EPI_ISL_2285037, EPI_ISL_2285038, EPI_ISL_2285040, EPI_ISL_2285042, EPI_ISL_2285043, EPI_ISL_2285044, EPI_ISL_2285045, EPI_ISL_2285046, EPI_ISL_2285047, EPI_ISL_2285048, EPI_ISL_2285050, EPI_ISL_2285053, EPI_ISL_2285054, EPI_ISL_2285055, EPI_ISL_2285056, EPI_ISL_2285057, EPI_ISL_2285058, EPI_ISL_2285059, EPI_ISL_2285060, EPI_ISL_2285061, EPI_ISL_2285062, EPI_ISL_2285063, EPI_ISL_2285064, EPI_ISL_2285065, EPI_ISL_2285066, EPI_ISL_2285068, EPI_ISL_2285069, EPI_ISL_2285070, EPI_ISL_2285071, EPI_ISL_2285072, EPI_ISL_2285073, EPI_ISL_2285074, EPI_ISL_2285075, EPI_ISL_2285076, EPI_ISL_2285077, EPI_ISL_2285078, EPI_ISL_2285079, EPI_ISL_2285080, EPI_ISL_2285081, EPI_ISL_2285082, EPI_ISL_2285083, EPI_ISL_2285084, EPI_ISL_2285085, EPI_ISL_2285086, EPI_ISL_2285087, EPI_ISL_2285088, EPI_ISL_2285089, EPI_ISL_2285090, EPI_ISL_2285091, EPI_ISL_2285092, EPI_ISL_2285093, EPI_ISL_2285094, EPI_ISL_2285095, EPI_ISL_2285096, EPI_ISL_2285097, EPI_ISL_2285099, EPI_ISL_2285100, EPI_ISL_2285101, EPI_ISL_2285102, EPI_ISL_2285103 | | | |
| see above | Mako Medical | Centers for Disease Control and Prevention Division of Viral Diseases, Pathogen Discovery | Dakota Howard, Dhwani Batra, Peter W. Cook, Kara Moser, Adrian Paskey, Jason Caravas, Benjamin Rambo-Martin, Shatavia Morrison, Christopher Gulvick, Scott Sammons, Yvette Unoarumhi, Darlene Wagner, Matthew Schmerer,Matthew Tugwell, Lauren Moon, Clinton R. Paden, Duncan MacCannell |
| EPI_ISL_2285734, EPI_ISL_2285735, EPI_ISL_2285737, EPI_ISL_2285738, EPI_ISL_2285739, EPI_ISL_2285740, EPI_ISL_2285742, EPI_ISL_2285743, EPI_ISL_2285744, EPI_ISL_2285745, EPI_ISL_2285748, EPI_ISL_2285749, EPI_ISL_2285750, EPI_ISL_2285751, EPI_ISL_2285753, EPI_ISL_2285754, EPI_ISL_2285755, EPI_ISL_2285762, EPI_ISL_2285763, EPI_ISL_2285764, EPI_ISL_2285765, EPI_ISL_2285766, EPI_ISL_2285767, EPI_ISL_2285768, EPI_ISL_2285770, EPI_ISL_2285771, EPI_ISL_2285773, EPI_ISL_2285774, EPI_ISL_2285775, EPI_ISL_2285777, EPI_ISL_2285779, EPI_ISL_2285781, EPI_ISL_2285782, EPI_ISL_2285783, EPI_ISL_2285787, EPI_ISL_2285788, EPI_ISL_2285789, EPI_ISL_2285791, EPI_ISL_2285792, EPI_ISL_2285793, EPI_ISL_2285794, EPI_ISL_2285795, EPI_ISL_2285796, EPI_ISL_2285797, EPI_ISL_2285798 | | | |
| see above | Wyoming Public Health Laboratory | Wyoming Public Health Laboratory | Jim Mildenberger, Taylor Fearing, Channing Weber, Ashley Norberg, Chayse Rowley, Marley Goetz, Brian Dominguez, Elliot Thomasson, Brittany Oher, Sam Britz, Cari Sloma, and Rob Christensen |
| EPI_ISL_2285826, EPI_ISL_2285827, EPI_ISL_2285828, EPI_ISL_2285829, EPI_ISL_2285830, EPI_ISL_2285831, EPI_ISL_2285832, EPI_ISL_2285833, EPI_ISL_2285834, EPI_ISL_2285835, EPI_ISL_2285836, EPI_ISL_2285837, EPI_ISL_2285838, EPI_ISL_2285839 | | | |
| see above | Kansas Health and Environmental Lab | Kansas Health and Environmental Lab | Mike Grose, Katherine Wiggins, Jonathan Barnell, Ben Olsen, and Phil Adam |
| EPI_ISL_2285982, EPI_ISL_2285983, EPI_ISL_2285984, EPI_ISL_2285985, EPI_ISL_2285986, EPI_ISL_2285987, EPI_ISL_2285989, EPI_ISL_2285990, EPI_ISL_2285991, EPI_ISL_2285992, EPI_ISL_2285993, EPI_ISL_2285994, EPI_ISL_2285995, EPI_ISL_2285996, EPI_ISL_2285997, EPI_ISL_2285998, EPI_ISL_2285999, EPI_ISL_2286000, EPI_ISL_2286001 | | | |
| see above | Arizona State Public Health Laboratory | Arizona State Public Health Laboratory | Trung Huynh, Jessica Escobar, Katherine Fullerton, Nobuko Fukushima, Stacy White, Linda Getsinger, Victor Waddell |
| EPI_ISL_2287804, EPI_ISL_2287807, EPI_ISL_2287810, EPI_ISL_2287813 | San Juan Regional Medical Center | New Mexico Department of Health Scientific Laboratory | Ellie Johnson, Anastacia Griego-Fisher, D'eldra Malone, Jennifer Benoit |
| EPI_ISL_2287816, EPI_ISL_2287820, EPI_ISL_2287822, EPI_ISL_2287824, EPI_ISL_2287827, EPI_ISL_2287830 | New Mexico Department of Health Scientific Laboratory | New Mexico Department of Health Scientific Laboratory | Ellie Johnson, Anastacia Griego-Fisher, D'eldra Malone, Jennifer Benoit |
| EPI_ISL_2287834, EPI_ISL_2287837, EPI_ISL_2287840, EPI_ISL_2287843, EPI_ISL_2287846, EPI_ISL_2287849 | San Juan Regional Medical Center | New Mexico Department of Health Scientific Laboratory | Ellie Johnson, Anastacia Griego-Fisher, D'eldra Malone, Jennifer Benoit |
| EPI_ISL_2287853, EPI_ISL_2287856, EPI_ISL_2287859, EPI_ISL_2287865, EPI_ISL_2287869, EPI_ISL_2287872, EPI_ISL_2287874, EPI_ISL_2287875 | New Mexico Department of Health Scientific Laboratory | New Mexico Department of Health Scientific Laboratory | Ellie Johnson, Anastacia Griego-Fisher, D'eldra Malone, Jennifer Benoit |
| EPI_ISL_2287876, EPI_ISL_2287880, EPI_ISL_2287883 | San Juan Regional Medical Center | New Mexico Department of Health Scientific Laboratory | Ellie Johnson, Anastacia Griego-Fisher, D'eldra Malone, Jennifer Benoit |
| EPI_ISL_2287886 | New Mexico Department of Health Scientific Laboratory | New Mexico Department of Health Scientific Laboratory | Ellie Johnson, Anastacia Griego-Fisher, D'eldra Malone, Jennifer Benoit |
| EPI_ISL_2287889, EPI_ISL_2287893, EPI_ISL_2287896 | San Juan Regional Medical Center | New Mexico Department of Health Scientific Laboratory | Ellie Johnson, Anastacia Griego-Fisher, D'eldra Malone, Jennifer Benoit |
| EPI_ISL_2287899 | New Mexico Department of Health Scientific Laboratory | New Mexico Department of Health Scientific Laboratory | Ellie Johnson, Anastacia Griego-Fisher, D'eldra Malone, Jennifer Benoit |
| EPI_ISL_2287902 | Sierra Vista Hospital | New Mexico Department of Health Scientific Laboratory | Ellie Johnson, Anastacia Griego-Fisher, D'eldra Malone, Jennifer Benoit |
| EPI_ISL_2287906, EPI_ISL_2287908, EPI_ISL_2287915, EPI_ISL_2287918, EPI_ISL_2287921 | New Mexico Department of Health Scientific Laboratory | New Mexico Department of Health Scientific Laboratory | Ellie Johnson, Anastacia Griego-Fisher, D'eldra Malone, Jennifer Benoit |
| EPI_ISL_2287924, EPI_ISL_2287928, EPI_ISL_2287931 | San Juan Regional Medical Center | New Mexico Department of Health Scientific Laboratory | Ellie Johnson, Anastacia Griego-Fisher, D'eldra Malone, Jennifer Benoit |
| EPI_ISL_2287933 | New Mexico Department of Health Scientific Laboratory | New Mexico Department of Health Scientific Laboratory | Ellie Johnson, Anastacia Griego-Fisher, D'eldra Malone, Jennifer Benoit |
| EPI_ISL_2287937, EPI_ISL_2287940 | TriCore Reference Laboratory Inc. | New Mexico Department of Health Scientific Laboratory | Ellie Johnson, Anastacia Griego-Fisher, D'eldra Malone, Jennifer Benoit |
| EPI_ISL_2287943 | New Mexico Department of Health Scientific Laboratory | New Mexico Department of Health Scientific Laboratory | Ellie Johnson, Anastacia Griego-Fisher, D'eldra Malone, Jennifer Benoit |
| EPI_ISL_2289071, EPI_ISL_2289072, EPI_ISL_2289073, EPI_ISL_2289074 | Kansas Health and Environmental Lab | Kansas Health and Environmental Lab | Mike Grose, Katherine Wiggins, Jonathan Barnell, Ben Olsen, and Phil Adam |
| EPI_ISL_2289075, EPI_ISL_2289076, EPI_ISL_2289077, EPI_ISL_2289078, EPI_ISL_2289079, EPI_ISL_2289080, EPI_ISL_2289081, EPI_ISL_2289085 | Wichita State University - Molecular Diagnostics Lab | Kansas Health and Environmental Lab | Mike Grose, Katherine Wiggins, Jonathan Barnell, Ben Olsen, and Phil Adam |
| EPI_ISL_2289145, EPI_ISL_2289146, EPI_ISL_2289147, EPI_ISL_2289148, EPI_ISL_2289149, EPI_ISL_2289150, EPI_ISL_2289151, EPI_ISL_2289152, EPI_ISL_2289153, EPI_ISL_2289154, EPI_ISL_2289155, EPI_ISL_2289156, EPI_ISL_2289157, EPI_ISL_2289158, EPI_ISL_2289159, EPI_ISL_2289160, EPI_ISL_2289161, EPI_ISL_2289162, EPI_ISL_2289163, EPI_ISL_2289164, EPI_ISL_2289165, EPI_ISL_2289166, EPI_ISL_2289167, EPI_ISL_2289169, EPI_ISL_2289170, EPI_ISL_2289171, EPI_ISL_2289172, EPI_ISL_2289173, EPI_ISL_2289175, EPI_ISL_2289176, EPI_ISL_2289177 | | | |
| see above | Microbiology Division, SC DHEC | Microbiology Division, SC DHEC | Flores,H., Freeman,J. |
| EPI_ISL_2289768, EPI_ISL_2289769, EPI_ISL_2289770, EPI_ISL_2289771, EPI_ISL_2289773, EPI_ISL_2289775, EPI_ISL_2289776, EPI_ISL_2289779, EPI_ISL_2289780, EPI_ISL_2289781, EPI_ISL_2289782, EPI_ISL_2289783, EPI_ISL_2289784, EPI_ISL_2289785, EPI_ISL_2289786, EPI_ISL_2289787, EPI_ISL_2289788, EPI_ISL_2289789, EPI_ISL_2289790, EPI_ISL_2289791, EPI_ISL_2289792, EPI_ISL_2289797, EPI_ISL_2289798, EPI_ISL_2289799, EPI_ISL_2289800, EPI_ISL_2289801, EPI_ISL_2289802, EPI_ISL_2289804, EPI_ISL_2289805, EPI_ISL_2289806, EPI_ISL_2289809, EPI_ISL_2289810, EPI_ISL_2289812, EPI_ISL_2289813, EPI_ISL_2289814, EPI_ISL_2289816, EPI_ISL_2289818, EPI_ISL_2289819, EPI_ISL_2289820, EPI_ISL_2289821, EPI_ISL_2289822, EPI_ISL_2289823, EPI_ISL_2289824, EPI_ISL_2289826, EPI_ISL_2289827, EPI_ISL_2289828, EPI_ISL_2289829, EPI_ISL_2289830, EPI_ISL_2289831, EPI_ISL_2289832, EPI_ISL_2289833, EPI_ISL_2289834, EPI_ISL_2289835, EPI_ISL_2289837, EPI_ISL_2289838, EPI_ISL_2289839, EPI_ISL_2289840, EPI_ISL_2289844, EPI_ISL_2289845, EPI_ISL_2289846, EPI_ISL_2289847, EPI_ISL_2289850, EPI_ISL_2289851, EPI_ISL_2289852, EPI_ISL_2289853, EPI_ISL_2289854, EPI_ISL_2289855, EPI_ISL_2289857, EPI_ISL_2289858, EPI_ISL_2289859, EPI_ISL_2289860, EPI_ISL_2289861, EPI_ISL_2289863, EPI_ISL_2289865, EPI_ISL_2289866, EPI_ISL_2289867, EPI_ISL_2289869, EPI_ISL_2289870, EPI_ISL_2289874, EPI_ISL_2289882, EPI_ISL_2289883, EPI_ISL_2289885, EPI_ISL_2289892, EPI_ISL_2289893, EPI_ISL_2289895, EPI_ISL_2289896, EPI_ISL_2289902, EPI_ISL_2289903, | | | |

EPI_ISL_2289904, EPI_ISL_2289906, EPI_ISL_2289908, EPI_ISL_2289909, EPI_ISL_2289910, EPI_ISL_2289912, EPI_ISL_2289913, EPI_ISL_2289914, EPI_ISL_2289915, EPI_ISL_2289916, EPI_ISL_2289917, EPI_ISL_2289919, EPI_ISL_2289920, EPI_ISL_2289929, EPI_ISL_2289930, EPI_ISL_2289931, EPI_ISL_2289932, EPI_ISL_2289933, EPI_ISL_2289936, EPI_ISL_2289937, EPI_ISL_2289938, EPI_ISL_2289939, EPI_ISL_2289940, EPI_ISL_2289941, EPI_ISL_2289943, EPI_ISL_2289944, EPI_ISL_2289945, EPI_ISL_2289949, EPI_ISL_2289952, EPI_ISL_2289953, EPI_ISL_2289954, EPI_ISL_2289956, EPI_ISL_2289957, EPI_ISL_2289958, EPI_ISL_2289959, EPI_ISL_2289960, EPI_ISL_2289961, EPI_ISL_2289962, EPI_ISL_2289963, EPI_ISL_2289964, EPI_ISL_2289965, EPI_ISL_2289966, EPI_ISL_2289967, EPI_ISL_2289968, EPI_ISL_2289969, EPI_ISL_2289970, EPI_ISL_2289971, EPI_ISL_2289972, EPI_ISL_2289973, EPI_ISL_2289974, EPI_ISL_2289975, EPI_ISL_2289976, EPI_ISL_2289977, EPI_ISL_2289979, EPI_ISL_2289980, EPI_ISL_2289981, EPI_ISL_2289983, EPI_ISL_2289984, EPI_ISL_2289985, EPI_ISL_2289986, EPI_ISL_2289987, EPI_ISL_2289989, EPI_ISL_2289990, EPI_ISL_2289991, EPI_ISL_2289992, EPI_ISL_2289993, EPI_ISL_2289994, EPI_ISL_2289995, EPI_ISL_2289996, EPI_ISL_2289997, EPI_ISL_2289998, EPI_ISL_2289999, EPI_ISL_2290000, EPI_ISL_2290001, EPI_ISL_2290002, EPI_ISL_2290003, EPI_ISL_2290004, EPI_ISL_2290005, EPI_ISL_2290006, EPI_ISL_2290007, EPI_ISL_2290009, EPI_ISL_2290010, EPI_ISL_2290011, EPI_ISL_2290013, EPI_ISL_2290014, EPI_ISL_2290015, EPI_ISL_2290016, EPI_ISL_2290017, EPI_ISL_2290018, EPI_ISL_2290019, EPI_ISL_2290020, EPI_ISL_2290021, EPI_ISL_2290022, EPI_ISL_2290023, EPI_ISL_2290024, EPI_ISL_2290025, EPI_ISL_2290026, EPI_ISL_2290028, EPI_ISL_2290029, EPI_ISL_2290030, EPI_ISL_2290032, EPI_ISL_2290033, EPI_ISL_2290036, EPI_ISL_2290038, EPI_ISL_2290039, EPI_ISL_2290040, EPI_ISL_2290041, EPI_ISL_2290044, EPI_ISL_2290045, EPI_ISL_2290046, EPI_ISL_2290047, EPI_ISL_2290048, EPI_ISL_2290049, EPI_ISL_2290050, EPI_ISL_2290052, EPI_ISL_2290053, EPI_ISL_2290054, EPI_ISL_2290055, EPI_ISL_2290056, EPI_ISL_2290057, EPI_ISL_2290058, EPI_ISL_2290059, EPI_ISL_2290064, EPI_ISL_2290065, EPI_ISL_2290066, EPI_ISL_2290068, EPI_ISL_2290069, EPI_ISL_2290070, EPI_ISL_2290071, EPI_ISL_2290072, EPI_ISL_2290073, EPI_ISL_2290074, EPI_ISL_2290075, EPI_ISL_2290077, EPI_ISL_2290078, EPI_ISL_2290079, EPI_ISL_2290081, EPI_ISL_2290083, EPI_ISL_2290086, EPI_ISL_2290088, EPI_ISL_2290089, EPI_ISL_2290090, EPI_ISL_2290092, EPI_ISL_2290093, EPI_ISL_2290094, EPI_ISL_2290095, EPI_ISL_2290096, EPI_ISL_2290097, EPI_ISL_2290098, EPI_ISL_2290100, EPI_ISL_2290101, EPI_ISL_2290103, EPI_ISL_2290104, EPI_ISL_2290105, EPI_ISL_2290107, EPI_ISL_2290110, EPI_ISL_2290111, EPI_ISL_2290115, EPI_ISL_2290116, EPI_ISL_2290118, EPI_ISL_2290119, EPI_ISL_2290120, EPI_ISL_2290122, EPI_ISL_2290123, EPI_ISL_2290124, EPI_ISL_2290125, EPI_ISL_2290129, EPI_ISL_2290130, EPI_ISL_2290131, EPI_ISL_2290133, EPI_ISL_2290134, EPI_ISL_2290135, EPI_ISL_2290136, EPI_ISL_2290138, EPI_ISL_2290140, EPI_ISL_2290143, EPI_ISL_2290162, EPI_ISL_2290163, EPI_ISL_2290164, EPI_ISL_2290165, EPI_ISL_2290166, EPI_ISL_2290167, EPI_ISL_2290168, EPI_ISL_2290169, EPI_ISL_2290170, EPI_ISL_2290171, EPI_ISL_2290172, EPI_ISL_2290173, EPI_ISL_2290174, EPI_ISL_2290175, EPI_ISL_2290176, EPI_ISL_2290177, EPI_ISL_2290178, EPI_ISL_2290179, EPI_ISL_2290180, EPI_ISL_2290181, EPI_ISL_2290182, EPI_ISL_2290183, EPI_ISL_2290184, EPI_ISL_2290185, EPI_ISL_2290186, EPI_ISL_2290187, EPI_ISL_2290188, EPI_ISL_2290189, EPI_ISL_2290190, EPI_ISL_2290191, EPI_ISL_2290192, EPI_ISL_2290193, EPI_ISL_2290194, EPI_ISL_2290195, EPI_ISL_2290196, EPI_ISL_2290197, EPI_ISL_2290198, EPI_ISL_2290199, EPI_ISL_2290200, EPI_ISL_2290201, EPI_ISL_2290202, EPI_ISL_2290203, EPI_ISL_2290204, EPI_ISL_2290205, EPI_ISL_2290206, EPI_ISL_2290207, EPI_ISL_2290208, EPI_ISL_2290209, EPI_ISL_2290210, EPI_ISL_2290211, EPI_ISL_2290212, EPI_ISL_2290213, EPI_ISL_2290214, EPI_ISL_2290215, EPI_ISL_2290216, EPI_ISL_2290217, EPI_ISL_2290218, EPI_ISL_2290219, EPI_ISL_2290220, EPI_ISL_2290221, EPI_ISL_2290222, EPI_ISL_2290223, EPI_ISL_2290224, EPI_ISL_2290225, EPI_ISL_2290226, EPI_ISL_2290227, EPI_ISL_2290228, EPI_ISL_2290229, EPI_ISL_2290230, EPI_ISL_2290231, EPI_ISL_2290232, EPI_ISL_2290233, EPI_ISL_2290234, EPI_ISL_2290235, EPI_ISL_2290236, EPI_ISL_2290237, EPI_ISL_2290238, EPI_ISL_2290239, EPI_ISL_2290240, EPI_ISL_2290241, EPI_ISL_2290242, EPI_ISL_2290243, EPI_ISL_2290244, EPI_ISL_2290245, EPI_ISL_2290246, EPI_ISL_2290247, EPI_ISL_2290248, EPI_ISL_2290249, EPI_ISL_2290250, EPI_ISL_2290251, EPI_ISL_2290252, EPI_ISL_2290253, EPI_ISL_2290254, EPI_ISL_2290255, EPI_ISL_2290256, EPI_ISL_2290257, EPI_ISL_2290258, EPI_ISL_2290259, EPI_ISL_2290260, EPI_ISL_2290261, EPI_ISL_2290262, EPI_ISL_2290263, EPI_ISL_2290264, EPI_ISL_2290265, EPI_ISL_2290266, EPI_ISL_2290267, EPI_ISL_2290268, EPI_ISL_2290269, EPI_ISL_2290270, EPI_ISL_2290271, EPI_ISL_2290272, EPI_ISL_2290273, EPI_ISL_2290274, EPI_ISL_2290275, EPI_ISL_2290276, EPI_ISL_2290277, EPI_ISL_2290278, EPI_ISL_2290279, EPI_ISL_2290280, EPI_ISL_2290281, EPI_ISL_2290282, EPI_ISL_2290283, EPI_ISL_2290284, EPI_ISL_2290285, EPI_ISL_2290286, EPI_ISL_2290287, EPI_ISL_2290288, EPI_ISL_2290289, EPI_ISL_2290290, EPI_ISL_2290291, EPI_ISL_2290292, EPI_ISL_2290293, EPI_ISL_2290294, EPI_ISL_2290295, EPI_ISL_2290296, EPI_ISL_2290297, EPI_ISL_2290298, EPI_ISL_2290299, EPI_ISL_2290300, EPI_ISL_2290301, EPI_ISL_2290302, EPI_ISL_2290303, EPI_ISL_2290304, EPI_ISL_2290305, EPI_ISL_2290306, EPI_ISL_2290307, EPI_ISL_2290308, EPI_ISL_2290309, EPI_ISL_2290310, EPI_ISL_2290311, EPI_ISL_2290312, EPI_ISL_2290313, EPI_ISL_2290314, EPI_ISL_2290315, EPI_ISL_2290316, EPI_ISL_2290317, EPI_ISL_2290318, EPI_ISL_2290319, EPI_ISL_2290320, EPI_ISL_2290321, EPI_ISL_2290322, EPI_ISL_2290323, EPI_ISL_2290324, EPI_ISL_2290325, EPI_ISL_2290326, EPI_ISL_2290327, EPI_ISL_2290328, EPI_ISL_2290329, EPI_ISL_2290330, EPI_ISL_2290331, EPI_ISL_2290332, EPI_ISL_2290333, EPI_ISL_2290334, EPI_ISL_2290335, EPI_ISL_2290336, EPI_ISL_2290337, EPI_ISL_2290338, EPI_ISL_2290339, EPI_ISL_2290340, EPI_ISL_2290341, EPI_ISL_2290342, EPI_ISL_2290343, EPI_ISL_2290344, EPI_ISL_2290345, EPI_ISL_2290346, EPI_ISL_2290347, EPI_ISL_2290348, EPI_ISL_2290349, EPI_ISL_2290350, EPI_ISL_2290351, EPI_ISL_2290352, EPI_ISL_2290353, EPI_ISL_2290354, EPI_ISL_2290355, EPI_ISL_2290356, EPI_ISL_2290357, EPI_ISL_2290358, EPI_ISL_2290359, EPI_ISL_2290360, EPI_ISL_2290361, EPI_ISL_2290362, EPI_ISL_2290363, EPI_ISL_2290364, EPI_ISL_2290365, EPI_ISL_2290366, EPI_ISL_2290367, EPI_ISL_2290368, EPI_ISL_2290369, EPI_ISL_2290370, EPI_ISL_2290371, EPI_ISL_2290372, EPI_ISL_2290373, EPI_ISL_2290374, EPI_ISL_2290375, EPI_ISL_2290376, EPI_ISL_2290377, EPI_ISL_2290378, EPI_ISL_2290379, EPI_ISL_2290380, EPI_ISL_2290381, EPI_ISL_2290382, EPI_ISL_2290383, EPI_ISL_2290384, EPI_ISL_2290385, EPI_ISL_2290386, EPI_ISL_2290387, EPI_ISL_2290388, EPI_ISL_2290389, EPI_ISL_2290390, EPI_ISL_2290391, EPI_ISL_2290392, EPI_ISL_2290393, EPI_ISL_2290394, EPI_ISL_2290395, EPI_ISL_2290396, EPI_ISL_2290397, EPI_ISL_2290398, EPI_ISL_2290399, EPI_ISL_2290400, EPI_ISL_2290401, EPI_ISL_2290402, EPI_ISL_2290403, EPI_ISL_2290404, EPI_ISL_2290405, EPI_ISL_2290406, EPI_ISL_2290407, EPI_ISL_2290408, EPI_ISL_2290409, EPI_ISL_2290410, EPI_ISL_2290411, EPI_ISL_2290412, EPI_ISL_2290413, EPI_ISL_2290414, EPI_ISL_2290415, EPI_ISL_2290416, EPI_ISL_2290417, EPI_ISL_2290418, EPI_ISL_2290419, EPI_ISL_2290420, EPI_ISL_2290421, EPI_ISL_2290422, EPI_ISL_2290423, EPI_ISL_2290424, EPI_ISL_2290425, EPI_ISL_2290426, EPI_ISL_2290427, EPI_ISL_2290428, EPI_ISL_2290429, EPI_ISL_2290430, EPI_ISL_2290431, EPI_ISL_2290432, EPI_ISL_2290433, EPI_ISL_2290434, EPI_ISL_2290435, EPI_ISL_2290436, EPI_ISL_2290437, EPI_ISL_2290438, EPI_ISL_2290439, EPI_ISL_2290440, EPI_ISL_2290441, EPI_ISL_2290442, EPI_ISL_2290443, EPI_ISL_2290444, EPI_ISL_2290445, EPI_ISL_2290446, EPI_ISL_2290447, EPI_ISL_2290448, EPI_ISL_2290449, EPI_ISL_2290450, EPI_ISL_2290451, EPI_ISL_2290452, EPI_ISL_2290453, EPI_ISL_2290454, EPI_ISL_2290455, EPI_ISL_2290456, EPI_ISL_2290457, EPI_ISL_2290458, EPI_ISL_2290459, EPI_ISL_2290460, EPI_ISL_2290461, EPI_ISL_2290462, EPI_ISL_2290463, EPI_ISL_2290464, EPI_ISL_2290465, EPI_ISL_2290466, EPI_ISL_2290467, EPI_ISL_2290468, EPI_ISL_2290469, EPI_ISL_2290470, EPI_ISL_2290471, EPI_ISL_2290472, EPI_ISL_2290473, EPI_ISL_2290474, EPI_ISL_2290475, EPI_ISL_2290476, EPI_ISL_2290478, EPI_ISL_2290479, EPI_ISL_2290480, EPI_ISL_2290481, EPI_ISL_2290482, EPI_ISL_2290483, EPI_ISL_2290484, EPI_ISL_2290485, EPI_ISL_2290486, EPI_ISL_2290487, EPI_ISL_2290488, EPI_ISL_2290489, EPI_ISL_2290490, EPI_ISL_2290491, EPI_ISL_2290492, EPI_ISL_2290493, EPI_ISL_2290494, EPI_ISL_2290495, EPI_ISL_2290496, EPI_ISL_2290497, EPI_ISL_2290498, EPI_ISL_2290499, EPI_ISL_2290500, EPI_ISL_2290501, EPI_ISL_2290502, EPI_ISL_2290503, EPI_ISL_2290504, EPI_ISL_2290505, EPI_ISL_2290506, EPI_ISL_2290507, EPI_ISL_2290508, EPI_ISL_2290509, EPI_ISL_2290510, EPI_ISL_2290511, EPI_ISL_2290512, EPI_ISL_2290513, EPI_ISL_2290514, EPI_ISL_2290515, EPI_ISL_2290516, EPI_ISL_2290517, EPI_ISL_2290518, EPI_ISL_2290519, EPI_ISL_2290520, EPI_ISL_2290521, EPI_ISL_2290522, EPI_ISL_2290523, EPI_ISL_2290524, EPI_ISL_2290525, EPI_ISL_2290526, EPI_ISL_2290527, EPI_ISL_2290528, EPI_ISL_2290529, EPI_ISL_2290530, EPI_ISL_2290531, EPI_ISL_2290532, EPI_ISL_2290533, EPI_ISL_2290534, EPI_ISL_2290535, EPI_ISL_2290536, EPI_ISL_2290537, EPI_ISL_2290538, EPI_ISL_2290539, EPI_ISL_2290540, EPI_ISL_2290541, EPI_ISL_2290542, EPI_ISL_2290543, EPI_ISL_2290544, EPI_ISL_2290545, EPI_ISL_2290546, EPI_ISL_2290547, EPI_ISL_2290548, EPI_ISL_2290549, EPI_ISL_2290550, EPI_ISL_2290551, EPI_ISL_2290552, EPI_ISL_2290553, EPI_ISL_2290554, EPI_ISL_2290555, EPI_ISL_2290556, EPI_ISL_2290557, EPI_ISL_2290558, EPI_ISL_2290559, EPI_ISL_2290560, EPI_ISL_2290561, EPI_ISL_2290562, EPI_ISL_2290563, EPI_ISL_2290564, EPI_ISL_2290565, EPI_ISL_2290566, EPI_ISL_2290567, EPI_ISL_2290568, EPI_ISL_2290569, EPI_ISL_2290570, EPI_ISL_2290571, EPI_ISL_2290572, EPI_ISL_2290573, EPI_ISL_2290574, EPI_ISL_2290575, EPI_ISL_2290576, EPI_ISL_2290577, EPI_ISL_2290578

| | | | |
|---|---|---|---|
| see above | Utah Public Health Laboratory | Utah Public Health Laboratory | Erin L. Young, Kelly F. Oakeson, Tara Gallagher |
| EPI_ISL_2290768, EPI_ISL_2290769 | MSHS Clinical Microbiology Laboratories | MSHS Pathogen Surveillance Program | Ana S. Gonzalez-Reiche, Hala Alshammary, Mitchell J. Sullivan, Brianne Ciferri, Ajay Obla, Angela Amoako, Mahmoud Awawda, Daniel Floda, Julia Matthews, Ashley Salimbangon, Levy Sominsky, Katherine Beach, Kayla Russo, Charles Gleason, Shelcie Fabre, Giulio Kleiner, Zenab Khan, Bremy Alburquerque, Adriana van de Guchte, Komal Srivastava, Matthew M. Hernandez, Jayeeta Dutta, Denise Jurczyszak, Nancy Francoeur, Betsaida Salom Melo, Irina Oussenko, Gintaras Deikus, Juan Soto, Shwetha Hara Sridhar, Ying-Chih Wang, Kathryn Twyman, Deena R. Altman, Robert Sebra, Adolfo García-Sastre, Marta Luksza, Gopi Patel, Sarah Schaefer, Melissa Gitman, Michael D. Nowak, Alberto Paniz-Mondolfi, Emilia Mia Sordillo, Viviana Simon, Harm van Bakel |

EPI_ISL_2290770, EPI_ISL_2290771, EPI_ISL_2290772, EPI_ISL_2290773, EPI_ISL_2290774, EPI_ISL_2290775, EPI_ISL_2290776, EPI_ISL_2290777, EPI_ISL_2290778, EPI_ISL_2290779, EPI_ISL_2290780, EPI_ISL_2290781, EPI_ISL_2290782, EPI_ISL_2290783, EPI_ISL_2290784, EPI_ISL_2290785, EPI_ISL_2290786, EPI_ISL_2290787, EPI_ISL_2290788, EPI_ISL_2290789, EPI_ISL_2290790, EPI_ISL_2290791, EPI_ISL_2290792, EPI_ISL_2290793, EPI_ISL_2290794, EPI_ISL_2290795, EPI_ISL_2290796, EPI_ISL_2290797, EPI_ISL_2290798, EPI_ISL_2290799, EPI_ISL_2290800, EPI_ISL_2290801, EPI_ISL_2290802, EPI_ISL_2290803, EPI_ISL_2290804, EPI_ISL_2290805, EPI_ISL_2290806, EPI_ISL_2290807, EPI_ISL_2290808, EPI_ISL_2290809, EPI_ISL_2290810, EPI_ISL_2290811, EPI_ISL_2290812, EPI_ISL_2290813, EPI_ISL_2290814, EPI_ISL_2290815, EPI_ISL_2290816, EPI_ISL_2290817, EPI_ISL_2290818, EPI_ISL_2290819, EPI_ISL_2290820, EPI_ISL_2290821, EPI_ISL_2290822, EPI_ISL_2290823, EPI_ISL_2290824, EPI_ISL_2290825, EPI_ISL_2290826, EPI_ISL_2290827, EPI_ISL_2290828, EPI_ISL_2290829, EPI_ISL_2290830, EPI_ISL_2290831, EPI_ISL_2290832, EPI_ISL_2290833, EPI_ISL_2290834, EPI_ISL_2290835, EPI_ISL_2290836, EPI_ISL_2290837, EPI_ISL_2290838, EPI_ISL_2290839, EPI_ISL_2290840, EPI_ISL_2290841, EPI_ISL_2290842, EPI_ISL_2290843, EPI_ISL_2290844, EPI_ISL_2290845, EPI_ISL_2290846, EPI_ISL_2290847, EPI_ISL_2290848, EPI_ISL_2290849, EPI_ISL_2290850, EPI_ISL_2290851, EPI_ISL_2290852, EPI_ISL_2290853, EPI_ISL_2290854, EPI_ISL_2290855, EPI_ISL_2290856, EPI_ISL_2290857, EPI_ISL_2290858, EPI_ISL_2290859, EPI_ISL_2290860, EPI_ISL_2290861, EPI_ISL_2290862, EPI_ISL_2290863, EPI_ISL_2290864, EPI_ISL_2290865, EPI_ISL_2290866, EPI_ISL_2290867, EPI_ISL_2290868, EPI_ISL_2290869, EPI_ISL_2290870, EPI_ISL_2290871, EPI_ISL_2290872, EPI_ISL_2290873, EPI_ISL_2290874, EPI_ISL_2290875, EPI_ISL_2290876, EPI_ISL_2290877, EPI_ISL_2290878, EPI_ISL_2290879, EPI_ISL_2290880, EPI_ISL_2290881, EPI_ISL_2290882, EPI_ISL_2290883, EPI_ISL_2290884, EPI_ISL_2290885, EPI_ISL_2290886, EPI_ISL_2290887, EPI_ISL_2290888, EPI_ISL_2290889, EPI_ISL_2290890, EPI_ISL_2290891, EPI_ISL_2290892, EPI_ISL_2290893, EPI_ISL_2290894, EPI_ISL_2290895, EPI_ISL_2290896, EPI_ISL_2290897, EPI_ISL_2290898, EPI_ISL_2290899, EPI_ISL_2290900, EPI_ISL_2290901, EPI_ISL_2290902, EPI_ISL_2290903, EPI_ISL_2290904, EPI_ISL_2290905, EPI_ISL_2290906, EPI_ISL_2290907, EPI_ISL_2290908, EPI_ISL_2290909, EPI_ISL_2290910, EPI_ISL_2290911, EPI_ISL_2290912, EPI_ISL_2290913, EPI_ISL_2290914, EPI_ISL_2290915, EPI_ISL_2290916, EPI_ISL_2290917, EPI_ISL_2290918, EPI_ISL_2290919, EPI_ISL_2290920, EPI_ISL_2290921, EPI_ISL_2290922, EPI_ISL_2290973, EPI_ISL_2290974, EPI_ISL_2290975, EPI_ISL_2290976, EPI_ISL_2290977, EPI_ISL_2290978, EPI_ISL_2290979, EPI_ISL_2290980, EPI_ISL_2290981, EPI_ISL_2290982, EPI_ISL_2290983, EPI_ISL_2290984, EPI_ISL_2290985, EPI_ISL_2290986, EPI_ISL_2290987, EPI_ISL_2290988, EPI_ISL_2290989, EPI_ISL_2290990, EPI_ISL_2290991, EPI_ISL_2290992, EPI_ISL_2290993, EPI_ISL_2290994, EPI_ISL_2290995, EPI_ISL_2290996, EPI_ISL_2290997, EPI_ISL_2290998, EPI_ISL_2290999, EPI_ISL_2291000, EPI_ISL_2291001, EPI_ISL_2291002, EPI_ISL_2291003, EPI_ISL_2291004, EPI_ISL_2291005, EPI_ISL_2291006, EPI_ISL_2291007, EPI_ISL_2291008, EPI_ISL_2291009, EPI_ISL_2291010, EPI_ISL_2291011, EPI_ISL_2291012, EPI_ISL_2291013, EPI_ISL_2291014, EPI_ISL_2291015, EPI_ISL_2291016, EPI_ISL_2291017, EPI_ISL_2291018, EPI_ISL_2291019, EPI_ISL_2291020, EPI_ISL_2291021, EPI_ISL_2291022, EPI_ISL_2291023, EPI_ISL_2291024, EPI_ISL_2291025, EPI_ISL_2291026, EPI_ISL_2291027, EPI_ISL_2291028, EPI_ISL_2291029, EPI_ISL_2291030, EPI_ISL_2291031, EPI_ISL_2291032, EPI_ISL_2291033, EPI_ISL_2291034, EPI_ISL_2291035, EPI_ISL_2291036, EPI_ISL_2291037, EPI_ISL_2291038, EPI_ISL_2291039, EPI_ISL_2291040, EPI_ISL_2291041, EPI_ISL_2291042, EPI_ISL_2291043, EPI_ISL_2291044, EPI_ISL_2291045, EPI_ISL_2291046, EPI_ISL_2291047, EPI_ISL_2291048, EPI_ISL_2291049, EPI_ISL_2291050, EPI_ISL_2291051, EPI_ISL_2291052, EPI_ISL_2291053, EPI_ISL_2291054, EPI_ISL_2291055, EPI_ISL_2291056, EPI_ISL_2291057, EPI_ISL_2291058, EPI_ISL_2291059, EPI_ISL_2291060, EPI_ISL_2291061, EPI_ISL_2291062, EPI_ISL_2291063, EPI_ISL_2291064, EPI_ISL_2291065, EPI_ISL_2291066, EPI_ISL_2291067, EPI_ISL_2291068, EPI_ISL_2291069, EPI_ISL_2291070, EPI_ISL_2291071, EPI_ISL_2291072, EPI_ISL_2291073, EPI_ISL_2291074, EPI_ISL_2291075, EPI_ISL_2291076, EPI_ISL_2291077, EPI_ISL_2291078, EPI_ISL_2291079, EPI_ISL_2291080, EPI_ISL_2291081, EPI_ISL_2291082, EPI_ISL_2291083, EPI_ISL_2291084, EPI_ISL_2291085, EPI_ISL_2291086, EPI_ISL_2291087, EPI_ISL_2291088, EPI_ISL_2291089, EPI_ISL_2291090, EPI_ISL_2291091, EPI_ISL_2291092, EPI_ISL_2291093, EPI_ISL_2291094, EPI_ISL_2291095, EPI_ISL_2291096, EPI_ISL_2291097, EPI_ISL_2291098, EPI_ISL_2291099, EPI_ISL_2291100, EPI_ISL_2291101, EPI_ISL_2291102, EPI_ISL_2291103, EPI_ISL_2291104, EPI_ISL_2291105, EPI_ISL_2291106, EPI_ISL_2291107, EPI_ISL_2291108, EPI_ISL_2291109, EPI_ISL_2291110, EPI_ISL_2291111, EPI_ISL_2291112, EPI_ISL_2291113, EPI_ISL_2291114, EPI_ISL_2291115, EPI_ISL_2291116, EPI_ISL_2291117, EPI_ISL_2291118, EPI_ISL_2291119, EPI_ISL_2291120, EPI_ISL_2291121, EPI_ISL_2291122, EPI_ISL_2291123, EPI_ISL_2291124, EPI_ISL_2291125, EPI_ISL_2291126, EPI_ISL_2291127, EPI_ISL_2291128, EPI_ISL_2291129, EPI_ISL_2291130, EPI_ISL_2291131, EPI_ISL_2291132, EPI_ISL_2291133, EPI_ISL_2291134, EPI_ISL_2291135, EPI_ISL_2291136, EPI_ISL_2291137, EPI_ISL_2291138, EPI_ISL_2291139, EPI_ISL_2291140, EPI_ISL_2291141, EPI_ISL_2291142, EPI_ISL_2291143, EPI_ISL_2291144, EPI_ISL_2291145, EPI_ISL_2291146, EPI_ISL_2291147, EPI_ISL_2291148, EPI_ISL_2291149, EPI_ISL_2291150, EPI_ISL_2291151, EPI_ISL_2291152, EPI_ISL_2291153, EPI_ISL_2291154, EPI_ISL_2291155, EPI_ISL_2291156, EPI_ISL_2291157, EPI_ISL_2291158, EPI_ISL_2291159, EPI_ISL_2291160, EPI_ISL_2291161, EPI_ISL_2291162, EPI_ISL_2291163, EPI_ISL_2291164, EPI_ISL_2291165, EPI_ISL_2291166, EPI_ISL_2291167, EPI_ISL_2291168, EPI_ISL_2291169, EPI_ISL_2291170, EPI_ISL_2291171, EPI_ISL_2291172, EPI_ISL_2291173, EPI_ISL_2291174, EPI_ISL_2291175, EPI_ISL_2291176, EPI_ISL_2291209, EPI_ISL_2291210, EPI_ISL_2291211, EPI_ISL_2291212, EPI_ISL_2291213, EPI_ISL_2291214, EPI_ISL_2291215, EPI_ISL_2291216, EPI_ISL_2291217, EPI_ISL_2291218, EPI_ISL_2291219, EPI_ISL_2291220, EPI_ISL_2291221, EPI_ISL_2291222, EPI_ISL_2291223, EPI_ISL_2291224, EPI_ISL_2291225, EPI_ISL_2291226, EPI_ISL_2291227, EPI_ISL_2291228, EPI_ISL_2291229, EPI_ISL_2291230, EPI_ISL_2291231, EPI_ISL_2291232, EPI_ISL_2291233, EPI_ISL_2291234, EPI_ISL_2291235, EPI_ISL_2291236, EPI_ISL_2291237, EPI_ISL_2291238, EPI_ISL_2291239, EPI_ISL_2291240, EPI_ISL_2291241, EPI_ISL_2291242, EPI_ISL_2291243, EPI_ISL_2291244, EPI_ISL_2291245, EPI_ISL_2291246, EPI_ISL_2291247, EPI_ISL_2291248, EPI_ISL_2291249, EPI_ISL_2291250, EPI_ISL_2291251, EPI_ISL_2291252, EPI_ISL_2291253, EPI_ISL_2291254, EPI_ISL_2291255, EPI_ISL_2291256, EPI_ISL_2291257, EPI_ISL_2291258, EPI_ISL_2291259, EPI_ISL_2291260, EPI_ISL_2291261, EPI_ISL_2291262, EPI_ISL_2291263, EPI_ISL_2291264, EPI_ISL_2291265, EPI_ISL_2291266, EPI_ISL_2291267, EPI_ISL_2291268, EPI_ISL_2291269, EPI_ISL_2291270, EPI_ISL_2291271, EPI_ISL_2291272, EPI_ISL_2291273, EPI_ISL_2291274, EPI_ISL_2291275, EPI_ISL_2291276, EPI_ISL_2291277, EPI_ISL_2291278, EPI_ISL_2291279, EPI_ISL_2291280, EPI_ISL_2291281, EPI_ISL_2291282, EPI_ISL_2291283, EPI_ISL_2291284, EPI_ISL_2291285, EPI_ISL_2291286, EPI_ISL_2291287, EPI_ISL_2291288, EPI_ISL_2291289, EPI_ISL_2291290, EPI_ISL_2291291, EPI_ISL_2291292, EPI_ISL_2291293, EPI_ISL_2291294, EPI_ISL_2291295, EPI_ISL_2291296, EPI_ISL_2291297, EPI_ISL_2291298, EPI_ISL_2291299, EPI_ISL_2291300, EPI_ISL_2291301, EPI_ISL_2291302, EPI_ISL_2291303, EPI_ISL_2291304, EPI_ISL_2291305, EPI_ISL_2291306, EPI_ISL_2291307, EPI_ISL_2291308, EPI_ISL_2291309, EPI_ISL_2291310, EPI_ISL_2291311, EPI_ISL_2291312, EPI_ISL_2291313, EPI_ISL_2291314, EPI_ISL_2291315, EPI_ISL_2291316, EPI_ISL_2291317, EPI_ISL_2291318, EPI_ISL_2291319, EPI_ISL_2291320, EPI_ISL_2291321, EPI_ISL_2291322, EPI_ISL_2291323, EPI_ISL_2291324, EPI_ISL_2291325, EPI_ISL_2291326, EPI_ISL_2291327, EPI_ISL_2291328, EPI_ISL_2291329, EPI_ISL_2291330, EPI_ISL_2291331, EPI_ISL_2291332, EPI_ISL_2291333, EPI_ISL_2291334, EPI_ISL_2291335, EPI_ISL_2291336, EPI_ISL_2291337, EPI_ISL_2291338, EPI_ISL_2291407, EPI_ISL_2291408, EPI_ISL_2291409, EPI_ISL_2291410, EPI_ISL_2291411, EPI_ISL_2291412, EPI_ISL_2291413, EPI_ISL_2291414, EPI_ISL_2291415, EPI_ISL_2291416, EPI_ISL_2291417, EPI_ISL_2291418, EPI_ISL_2291419, EPI_ISL_2291420, EPI_ISL_2291421, EPI_ISL_2291422, EPI_ISL_2291423, EPI_ISL_2291424, EPI_ISL_2291425, EPI_ISL_2291426, EPI_ISL_2291427, EPI_ISL_2291428, EPI_ISL_2291429, EPI_ISL_2291430, EPI_ISL_2291431, EPI_ISL_2291432, EPI_ISL_2291433, EPI_ISL_2291434, EPI_ISL_2291435, EPI_ISL_2291436, EPI_ISL_2291437, EPI_ISL_2291438, EPI_ISL_2291439, EPI_ISL_2291440, EPI_ISL_2291441, EPI_ISL_2291442, EPI_ISL_2291443, EPI_ISL_2291444, EPI_ISL_2291445, EPI_ISL_2291446, EPI_ISL_2291447, EPI_ISL_2291448, EPI_ISL_2291449, EPI_ISL_2291450, EPI_ISL_2291451, EPI_ISL_2291452, EPI_ISL_2291453, EPI_ISL_2291454, EPI_ISL_2291455, EPI_ISL_2291456, EPI_ISL_2291457, EPI_ISL_2291458, EPI_ISL_2291459, EPI_ISL_2291460, EPI_ISL_2291461, EPI_ISL_2291462, EPI_ISL_2291463, EPI_ISL_2291464, EPI_ISL_2291465, EPI_ISL_2291466, EPI_ISL_2291467, EPI_ISL_2291468, EPI_ISL_2291469, EPI_ISL_2291470, EPI_ISL_2291471, EPI_ISL_2291472, EPI_ISL_2291473, EPI_ISL_2291474, EPI_ISL_2291475, EPI_ISL_2291476, EPI_ISL_2291477, EPI_ISL_2291478, EPI_ISL_2291479, EPI_ISL_2291480, EPI_ISL_2291481, EPI_ISL_2291482, EPI_ISL_2291483, EPI_ISL_2291484, EPI_ISL_2291485, EPI_ISL_2291486, EPI_ISL_2291487, EPI_ISL_2291488, EPI_ISL_2291489, EPI_ISL_2291490, EPI_ISL_2291491, EPI_ISL_2291492, EPI_ISL_2291493, EPI_ISL_2291494, EPI_ISL_2291495, EPI_ISL_2291496, EPI_ISL_2291497, EPI_ISL_2291498, EPI_ISL_2291499, EPI_ISL_2291500, EPI_ISL_2291501, EPI_ISL_2291502, EPI_ISL_2291503, EPI_ISL_2291504, EPI_ISL_2291505, EPI_ISL_2291506, EPI_ISL_2291507, EPI_ISL_2291508, EPI_ISL_2291509, EPI_ISL_2291510, EPI_ISL_2291511, EPI_ISL_2291512, EPI_ISL_2291513, EPI_ISL_2291514, EPI_ISL_2291515, EPI_ISL_2291516, EPI_ISL_2291517, EPI_ISL_2291518, EPI_ISL_2291519, EPI_ISL_2291520, EPI_ISL_2291521, EPI_ISL_2291522, EPI_ISL_2291523, EPI_ISL_2291524, EPI_ISL_2291525, EPI_ISL_2291526, EPI_ISL_2291527, EPI_ISL_2291528, EPI_ISL_2291529, EPI_ISL_2291530, EPI_ISL_2291531, EPI_ISL_2291532, EPI_ISL_2291533, EPI_ISL_2291534, EPI_ISL_2291535, EPI_ISL_2291536, EPI_ISL_2291537, EPI_ISL_2291538, EPI_ISL_2291540, EPI_ISL_2291541, EPI_ISL_2291542, EPI_ISL_2291543, EPI_ISL_2291544, EPI_ISL_2291545, EPI_ISL_2291546, EPI_ISL_2291547, EPI_ISL_2291548, EPI_ISL_2291549, EPI_ISL_2291550, EPI_ISL_2291551, EPI_ISL_2291552, EPI_ISL_2291553, EPI_ISL_2291554, EPI_ISL_2291555, EPI_ISL_2291556, EPI_ISL_2291557, EPI_ISL_2291558, EPI_ISL_2291559, EPI_ISL_2291560, EPI_ISL_2291561, EPI_ISL_2291562, EPI_ISL_2291563, EPI_ISL_2291564, EPI_ISL_2291565, EPI_ISL_2291566, EPI_ISL_2291567, EPI_ISL_2291568, EPI_ISL_2291569, EPI_ISL_2291570, EPI_ISL_2291571, EPI_ISL_2291572, EPI_ISL_2291573, EPI_ISL_2291574, EPI_ISL_2291575, EPI_ISL_2291576, EPI_ISL_2291577, EPI_ISL_2291578, EPI_ISL_2291579, EPI_ISL_2291580, EPI_ISL_2291581, EPI_ISL_2291582, EPI_ISL_2291583, EPI_ISL_2291584, EPI_ISL_2291585, EPI_ISL_2291586, EPI_ISL_2291587, EPI_ISL_2291588, EPI_ISL_2291589, EPI_ISL_2291590, EPI_ISL_2291591, EPI_ISL_2291592, EPI_ISL_2291593, EPI_ISL_2291594, EPI_ISL_2291595, EPI_ISL_2291596, EPI_ISL_2291597, EPI_ISL_2291598, EPI_ISL_2291600, EPI_ISL_2291634, EPI_ISL_2291635, EPI_ISL_2291636, EPI_ISL_2291637, EPI_ISL_2291638, EPI_ISL_2291639, EPI_ISL_2291640, EPI_ISL_2291641, EPI_ISL_2291642, EPI_ISL_2291643, EPI_ISL_2291644, EPI_ISL_2291645, EPI_ISL_2291646, EPI_ISL_2291647, EPI_ISL_2291648, EPI_ISL_2291649, EPI_ISL_2291650, EPI_ISL_2291651, EPI_ISL_2291652, EPI_ISL_2291653, EPI_ISL_2291654, EPI_ISL_2291655, EPI_ISL_2291656, EPI_ISL_2291657, EPI_ISL_2291658, EPI_ISL_2291659, EPI_ISL_2291660, EPI_ISL_2291661, EPI_ISL_2291662, EPI_ISL_2291663, EPI_ISL_2291664, EPI_ISL_2291665, EPI_ISL_2291666, EPI_ISL_2291667, EPI_ISL_2291668, EPI_ISL_2291669, EPI_ISL_2291670, EPI_ISL_2291671, EPI_ISL_2291672, EPI_ISL_2291673, EPI_ISL_2291674, EPI_ISL_2291675, EPI_ISL_2291676, EPI_ISL_2291677, EPI_ISL_2291678, EPI_ISL_2291679, EPI_ISL_2291680, EPI_ISL_2291681, EPI_ISL_2291682, EPI_ISL_2291683, EPI_ISL_2291684, EPI_ISL_2291685, EPI_ISL_2291686, EPI_ISL_2291687, EPI_ISL_2291688, EPI_ISL_2291689, EPI_ISL_2291690, EPI_ISL_2291691, EPI_ISL_2291692, EPI_ISL_2291693, EPI_ISL_2291694, EPI_ISL_2291695, EPI_ISL_2291696, EPI_ISL_2291697, EPI_ISL_2291698, EPI_ISL_2291699, EPI_ISL_2291700, EPI_ISL_2291701, EPI_ISL_2291702, EPI_ISL_2291703, EPI_ISL_2291704, EPI_ISL_2291705, EPI_ISL_2291706, EPI_ISL_2291707, EPI_ISL_2291708, EPI_ISL_2291709, EPI_ISL_2291710, EPI_ISL_2291711, EPI_ISL_2291712, EPI_ISL_2291713, EPI_ISL_2291714, EPI_ISL_2291715, EPI_ISL_2291716, EPI_ISL_2291717, EPI_ISL_2291718, EPI_ISL_2291719, EPI_ISL_2291720, EPI_ISL_2291721, EPI_ISL_2291722, EPI_ISL_2291723, EPI_ISL_2291724, EPI_ISL_2291725, EPI_ISL_2291726, EPI_ISL_2291727, EPI_ISL_2291728, EPI_ISL_2291729, EPI_ISL_2291730, EPI_ISL_2291731, EPI_ISL_2291732, EPI_ISL_2291733, EPI_ISL_2291734, EPI_ISL_2291735, EPI_ISL_2291736, EPI_ISL_2291737, EPI_ISL_2291738, EPI_ISL_2291739, EPI_ISL_2291740, EPI_ISL_2291741, EPI_ISL_2291742, EPI_ISL_2291743, EPI_ISL_2291744, EPI_ISL_2291745, EPI_ISL_2291746, EPI_ISL_2291747, EPI_ISL_2291748, EPI_ISL_2291749, EPI_ISL_2291750, EPI_ISL_2291751, EPI_ISL_2291752, EPI_ISL_2291753, EPI_ISL_2291754, EPI_ISL_2291755, EPI_ISL_2291756, EPI_ISL_2291757, EPI_ISL_2291758, EPI_ISL_2291759, EPI_ISL_2291760, EPI_ISL_2291761, EPI_ISL_2291762, EPI_ISL_2291763, EPI_ISL_2291764, EPI_ISL_2291765, EPI_ISL_2291766, EPI_ISL_2291767, EPI_ISL_2291768, EPI_ISL_2291769, EPI_ISL_2291770, EPI_ISL_2291771, EPI_ISL_2291772, EPI_ISL_2291773, EPI_ISL_2291774, EPI_ISL_2291775, EPI_ISL_2291776, EPI_ISL_2291777, EPI_ISL_2291778, EPI_ISL_2291779, EPI_ISL_2291780, EPI_ISL_2291781, EPI_ISL_2291782, EPI_ISL_2291783, EPI_ISL_2291784, EPI_ISL_2291785, EPI_ISL_2291786, EPI_ISL_2291787, EPI_ISL_2291788, EPI_ISL_2291789, EPI_ISL_2291790, EPI_ISL_2291791, EPI_ISL_2291792, EPI_ISL_2291793, EPI_ISL_2291794, EPI_ISL_2291795, EPI_ISL_2291796, EPI_ISL_2291797, EPI_ISL_2291798, EPI_ISL_2291799

| | | | |
|---|---|---|---|
| see above | Utah Public Health Laboratory | Utah Public Health Laboratory | Erin L. Young, Kelly F. Oakeson, Tara Gallagher |
| EPI_ISL_2291826, EPI_ISL_2291827, EPI_ISL_2291828, EPI_ISL_2291829, EPI_ISL_2291830, EPI_ISL_2291831 | MD PHL | MD PHL | Maryland Department of Health Laboratories Administration |

EPI_ISL_2291832, EPI_ISL_2291833, EPI_ISL_2291834, EPI_ISL_2291835, EPI_ISL_2291836, EPI_ISL_2291837, EPI_ISL_2291838, EPI_ISL_2291839, EPI_ISL_2291840, EPI_ISL_2291841, EPI_ISL_2291842, EPI_ISL_2291843, EPI_ISL_2291844, EPI_ISL_2291845, EPI_ISL_2291846, EPI_ISL_2291847, EPI_ISL_2291848, EPI_ISL_2291849, EPI_ISL_2291850, EPI_ISL_2291851, EPI_ISL_2291852, EPI_ISL_2291853, EPI_ISL_2291854, EPI_ISL_2291855, EPI_ISL_2291856, EPI_ISL_2291857, EPI_ISL_2291858, EPI_ISL_2291859, EPI_ISL_2291860, EPI_ISL_2291861, EPI_ISL_2291862, EPI_ISL_2291863, EPI_ISL_2291864, EPI_ISL_2291865, EPI_ISL_2291866, EPI_ISL_2291867, EPI_ISL_2291868, EPI_ISL_2291869, EPI_ISL_2291870, EPI_ISL_2291871, EPI_ISL_2291872, EPI_ISL_2291873, EPI_ISL_2291874, EPI_ISL_2291875, EPI_ISL_2291876, EPI_ISL_2291877, EPI_ISL_2291878, EPI_ISL_2291879, EPI_ISL_2291880, EPI_ISL_2291881, EPI_ISL_2291882, EPI_ISL_2291883, EPI_ISL_2291884, EPI_ISL_2291885, EPI_ISL_2291886, EPI_ISL_2291887, EPI_ISL_2291888, EPI_ISL_2291889, EPI_ISL_2291890, EPI_ISL_2291891, EPI_ISL_2291892, EPI_ISL_2291893, EPI_ISL_2291894, EPI_ISL_2291895, EPI_ISL_2291896, EPI_ISL_2291897, EPI_ISL_2291898, EPI_ISL_2291899, EPI_ISL_2291900, EPI_ISL_2291901, EPI_ISL_2291902, EPI_ISL_2291903, EPI_ISL_2291904, EPI_ISL_2291905, EPI_ISL_2291906, EPI_ISL_2291907, EPI_ISL_2291908, EPI_ISL_2291909, EPI_ISL_2291910, EPI_ISL_2291911, EPI_ISL_2291912, EPI_ISL_2291913, EPI_ISL_2291914, EPI_ISL_2291915, EPI_ISL_2291916, EPI_ISL_2291917, EPI_ISL_2291918, EPI_ISL_2291919, EPI_ISL_2291920, EPI_ISL_2291921, EPI_ISL_2291922, EPI_ISL_2291923, EPI_ISL_2291924, EPI_ISL_2291925, EPI_ISL_2291926, EPI_ISL_2291927, EPI_ISL_2291928, EPI_ISL_2291929, EPI_ISL_2291930, EPI_ISL_2291931, EPI_ISL_2291932, EPI_ISL_2291933, EPI_ISL_2291934, EPI_ISL_2291935, EPI_ISL_2291936, EPI_ISL_2291937, EPI_ISL_2291938, EPI_ISL_2291939, EPI_ISL_2291940, EPI_ISL_2291941, EPI_ISL_2291942, EPI_ISL_2291943, EPI_ISL_2291944, EPI_ISL_2291945, EPI_ISL_2291946, EPI_ISL_2291947, EPI_ISL_2291948, EPI_ISL_2291949, EPI_ISL_2291950, EPI_ISL_2291951, EPI_ISL_2291952, EPI_ISL_2291953, EPI_ISL_2291954, EPI_ISL_2291955, EPI_ISL_2291956, EPI_ISL_2291957, EPI_ISL_2291958, EPI_ISL_2291959, EPI_ISL_2291960, EPI_ISL_2291961, EPI_ISL_2291962, EPI_ISL_2291963, EPI_ISL_2291964, EPI_ISL_2291965, EPI_ISL_2291966, EPI_ISL_2291967, EPI_ISL_2291968, EPI_ISL_2291969, EPI_ISL_2291970, EPI_ISL_2291971, EPI_ISL_2291972, EPI_ISL_2291973, EPI_ISL_2291974, EPI_ISL_2291975, EPI_ISL_2291976, EPI_ISL_2291977, EPI_ISL_2291978, EPI_ISL_2291979, EPI_ISL_2291980, EPI_ISL_2291981, EPI_ISL_2291982, EPI_ISL_2291983, EPI_ISL_2291984, EPI_ISL_2291985, EPI_ISL_2291986, EPI_ISL_2291987, EPI_ISL_2291988, EPI_ISL_2291989, EPI_ISL_2291990, EPI_ISL_2291991, EPI_ISL_2291992, EPI_ISL_2291993, EPI_ISL_2292096, EPI_ISL_2292097, EPI_ISL_2292098, EPI_ISL_2292099, EPI_ISL_2292100, EPI_ISL_2292101, EPI_ISL_2292102, EPI_ISL_2292103, EPI_ISL_2292104, EPI_ISL_2292105, EPI_ISL_2292106, EPI_ISL_2292107, EPI_ISL_2292108, EPI_ISL_2292109, EPI_ISL_2292110, EPI_ISL_2292111, EPI_ISL_2292112, EPI_ISL_2292113, EPI_ISL_2292114, EPI_ISL_2292115, EPI_ISL_2292116, EPI_ISL_2292117, EPI_ISL_2292118, EPI_ISL_2292119, EPI_ISL_2292120, EPI_ISL_2292121, EPI_ISL_2292122, EPI_ISL_2292123, EPI_ISL_2292124, EPI_ISL_2292125, EPI_ISL_2292126, EPI_ISL_2292127, EPI_ISL_2292128, EPI_ISL_2292129, EPI_ISL_2292130, EPI_ISL_2292131, EPI_ISL_2292132, EPI_ISL_2292133, EPI_ISL_2292134, EPI_ISL_2292135, EPI_ISL_2292136, EPI_ISL_2292137, EPI_ISL_2292138, EPI_ISL_2292139, EPI_ISL_2292140, EPI_ISL_2292141, EPI_ISL_2292142, EPI_ISL_2292143, EPI_ISL_2292144, EPI_ISL_2292145, EPI_ISL_2292146, EPI_ISL_2292147, EPI_ISL_2292148, EPI_ISL_2292149, EPI_ISL_2292150, EPI_ISL_2292151, EPI_ISL_2292152, EPI_ISL_2292153, EPI_ISL_2292154, EPI_ISL_2292155, EPI_ISL_2292156, EPI_ISL_2292157, EPI_ISL_2292158, EPI_ISL_2292159, EPI_ISL_2292160, EPI_ISL_2292161, EPI_ISL_2292162, EPI_ISL_2292163, EPI_ISL_2292164, EPI_ISL_2292165, EPI_ISL_2292166, EPI_ISL_2292167, EPI_ISL_2292168, EPI_ISL_2292169, EPI_ISL_2292170, EPI_ISL_2292171, EPI_ISL_2292172, EPI_ISL_2292173, EPI_ISL_2292174, EPI_ISL_2292175, EPI_ISL_2292176, EPI_ISL_2292177, EPI_ISL_2292178, EPI_ISL_2292179, EPI_ISL_2292180, EPI_ISL_2292181, EPI_ISL_2292182, EPI_ISL_2292183, EPI_ISL_2292184, EPI_ISL_2292185, EPI_ISL_2292186, EPI_ISL_2292187, EPI_ISL_2292188, EPI_ISL_2292189, EPI_ISL_2292190, EPI_ISL_2292191, EPI_ISL_2292192, EPI_ISL_2292193, EPI_ISL_2292194, EPI_ISL_2292195, EPI_ISL_2292196, EPI_ISL_2292197, EPI_ISL_2292198, EPI_ISL_2292199, EPI_ISL_2292200, EPI_ISL_2292201, EPI_ISL_2292202, EPI_ISL_2292203, EPI_ISL_2292204, EPI_ISL_2292205, EPI_ISL_2292206, EPI_ISL_2292207, EPI_ISL_2292208, EPI_ISL_2292209, EPI_ISL_2292210, EPI_ISL_2292211, EPI_ISL_2292212, EPI_ISL_2292213, EPI_ISL_2292214, EPI_ISL_2292215, EPI_ISL_2292216, EPI_ISL_2292217, EPI_ISL_2292218, EPI_ISL_2292219, EPI_ISL_2292220, EPI_ISL_2292221, EPI_ISL_2292222, EPI_ISL_2292223, EPI_ISL_2292224, EPI_ISL_2292225, EPI_ISL_2292226, EPI_ISL_2292227, EPI_ISL_2292228, EPI_ISL_2292229, EPI_ISL_2292230, EPI_ISL_2292231, EPI_ISL_2292232, EPI_ISL_2292233, EPI_ISL_2292234, EPI_ISL_2292235, EPI_ISL_2292236, EPI_ISL_2292237, EPI_ISL_2292238, EPI_ISL_2292239, EPI_ISL_2292240, EPI_ISL_2292241, EPI_ISL_2292242, EPI_ISL_2292243, EPI_ISL_2292244, EPI_ISL_2292245, EPI_ISL_2292246, EPI_ISL_2292247, EPI_ISL_2292248, EPI_ISL_2292249, EPI_ISL_2292250, EPI_ISL_2292251, EPI_ISL_2292252, EPI_ISL_2292253, EPI_ISL_2292254, EPI_ISL_2292255, EPI_ISL_2292256, EPI_ISL_2292257, EPI_ISL_2292258, EPI_ISL_2292259, EPI_ISL_2292260, EPI_ISL_2292261, EPI_ISL_2292262, EPI_ISL_2292263, EPI_ISL_2292264, EPI_ISL_2292265, EPI_ISL_2292266, EPI_ISL_2292267, EPI_ISL_2292268, EPI_ISL_2292269, EPI_ISL_2292270, EPI_ISL_2292271, EPI_ISL_2292272, EPI_ISL_2292273, EPI_ISL_2292274, EPI_ISL_2292275, EPI_ISL_2292276, EPI_ISL_2292277, EPI_ISL_2292278, EPI_ISL_2292279, EPI_ISL_2292280, EPI_ISL_2292281, EPI_ISL_2292282, EPI_ISL_2292283, EPI_ISL_2292284, EPI_ISL_2292285, EPI_ISL_2292286, EPI_ISL_2292287, EPI_ISL_2292288, EPI_ISL_2292289, EPI_ISL_2292290, EPI_ISL_2292291, EPI_ISL_2292292, EPI_ISL_2292293, EPI_ISL_2292294, EPI_ISL_2292295, EPI_ISL_2292296, EPI_ISL_2292297, EPI_ISL_2292298, EPI_ISL_2292299, EPI_ISL_2292300, EPI_ISL_2292301, EPI_ISL_2292302, EPI_ISL_2292303, EPI_ISL_2292304, EPI_ISL_2292305, EPI_ISL_2292306, EPI_ISL_2292307, EPI_ISL_2292308, EPI_ISL_2292309, EPI_ISL_2292310, EPI_ISL_2292311, EPI_ISL_2292312, EPI_ISL_2292313, EPI_ISL_2292314, EPI_ISL_2292315, EPI_ISL_2292316, EPI_ISL_2292317, EPI_ISL_2292318, EPI_ISL_2292319, EPI_ISL_2292320, EPI_ISL_2292321, EPI_ISL_2292322, EPI_ISL_2292323, EPI_ISL_2292324, EPI_ISL_2292325, EPI_ISL_2292326, EPI_ISL_2292327, EPI_ISL_2292328, EPI_ISL_2292329, EPI_ISL_2292330, EPI_ISL_2292331, EPI_ISL_2292332, EPI_ISL_2292333, EPI_ISL_2292334, EPI_ISL_2292335, EPI_ISL_2292336, EPI_ISL_2292337, EPI_ISL_2292338, EPI_ISL_2292339, EPI_ISL_2292340, EPI_ISL_2292341, EPI_ISL_2292342, EPI_ISL_2292343, EPI_ISL_2292344, EPI_ISL_2292345, EPI_ISL_2292346, EPI_ISL_2292347, EPI_ISL_2292348, EPI_ISL_2292349, EPI_ISL_2292350, EPI_ISL_2292351, EPI_ISL_2292352, EPI_ISL_2292353, EPI_ISL_2292354, EPI_ISL_2292355, EPI_ISL_2292356, EPI_ISL_2292357, EPI_ISL_2292358, EPI_ISL_2292359, EPI_ISL_2292360, EPI_ISL_2292361, EPI_ISL_2292362, EPI_ISL_2292363, EPI_ISL_2292364, EPI_ISL_2292365, EPI_ISL_2292366, EPI_ISL_2292367, EPI_ISL_2292368, EPI_ISL_2292369, EPI_ISL_2292370, EPI_ISL_2292371, EPI_ISL_2292372, EPI_ISL_2292373, EPI_ISL_2292374, EPI_ISL_2292375, EPI_ISL_2292376, EPI_ISL_2292377, EPI_ISL_2292378, EPI_ISL_2292379, EPI_ISL_2292380, EPI_ISL_2292381, EPI_ISL_2292382, EPI_ISL_2292383, EPI_ISL_2292384, EPI_ISL_2292385, EPI_ISL_2292386, EPI_ISL_2292387, EPI_ISL_2292388, EPI_ISL_2292389, EPI_ISL_2292390, EPI_ISL_2292391, EPI_ISL_2292392, EPI_ISL_2292393, EPI_ISL_2292394, EPI_ISL_2292395, EPI_ISL_2292396, EPI_ISL_2292397, EPI_ISL_2292398, EPI_ISL_2292399, EPI_ISL_2292400, EPI_ISL_2292401, EPI_ISL_2292402, EPI_ISL_2292403, EPI_ISL_2292404, EPI_ISL_2292405, EPI_ISL_2292406, EPI_ISL_2292407, EPI_ISL_2292408, EPI_ISL_2292409, EPI_ISL_2292410, EPI_ISL_2292411, EPI_ISL_2292412, EPI_ISL_2292413, EPI_ISL_2292414, EPI_ISL_2292415, EPI_ISL_2292416, EPI_ISL_2292417, EPI_ISL_2292418, EPI_ISL_2292419, EPI_ISL_2292420, EPI_ISL_2292421, EPI_ISL_2292422, EPI_ISL_2292423, EPI_ISL_2292424, EPI_ISL_2292425, EPI_ISL_2292426, EPI_ISL_2292427, EPI_ISL_2292428, EPI_ISL_2292429, EPI_ISL_2292430, EPI_ISL_2292431, EPI_ISL_2292432, EPI_ISL_2292433, EPI_ISL_2292434, EPI_ISL_2292435, EPI_ISL_2292436, EPI_ISL_2292437, EPI_ISL_2292438, EPI_ISL_2292439, EPI_ISL_2292440, EPI_ISL_2292441, EPI_ISL_2292442, EPI_ISL_2292443, EPI_ISL_2292444, EPI_ISL_2292445, EPI_ISL_2292446, EPI_ISL_2292447, EPI_ISL_2292448, EPI_ISL_2292449, EPI_ISL_2292450, EPI_ISL_2292451, EPI_ISL_2292452, EPI_ISL_2292453, EPI_ISL_2292454, EPI_ISL_2292455, EPI_ISL_2292456, EPI_ISL_2292457, EPI_ISL_2292458, EPI_ISL_2292459, EPI_ISL_2292460, EPI_ISL_2292461, EPI_ISL_2292462, EPI_ISL_2292463, EPI_ISL_2292464, EPI_ISL_2292465, EPI_ISL_2292466, EPI_ISL_2292467, EPI_ISL_2292468, EPI_ISL_2292469, EPI_ISL_2292470, EPI_ISL_2292471, EPI_ISL_2292472, EPI_ISL_2292473, EPI_ISL_2292474, EPI_ISL_2292475, EPI_ISL_2292476, EPI_ISL_2292477, EPI_ISL_2292478, EPI_ISL_2292479, EPI_ISL_2292480, EPI_ISL_2292481, EPI_ISL_2292482, EPI_ISL_2292483, EPI_ISL_2292484, EPI_ISL_2292485, EPI_ISL_2292486, EPI_ISL_2292487, EPI_ISL_2292488, EPI_ISL_2292489, EPI_ISL_2292490, EPI_ISL_2292491, EPI_ISL_2292492, EPI_ISL_2292493, EPI_ISL_2292494, EPI_ISL_2292495, EPI_ISL_2292496, EPI_ISL_2292497, EPI_ISL_2292498, EPI_ISL_2292499, EPI_ISL_2292500, EPI_ISL_2292501, EPI_ISL_2292502, EPI_ISL_2292503, EPI_ISL_2292504, EPI_ISL_2292505, EPI_ISL_2292506, EPI_ISL_2292507, EPI_ISL_2292508, EPI_ISL_2292509, EPI_ISL_2292510, EPI_ISL_2292511, EPI_ISL_2292512, EPI_ISL_2292513, EPI_ISL_2292514, EPI_ISL_2292515, EPI_ISL_2292516, EPI_ISL_2292517, EPI_ISL_2292518, EPI_ISL_2292519, EPI_ISL_2292520, EPI_ISL_2292521, EPI_ISL_2292522, EPI_ISL_2292523, EPI_ISL_2292524, EPI_ISL_2292525, EPI_ISL_2292526, EPI_ISL_2292527, EPI_ISL_2292528, EPI_ISL_2292529, EPI_ISL_2292530, EPI_ISL_2292531, EPI_ISL_2292532, EPI_ISL_2292533, EPI_ISL_2292534, EPI_ISL_2292535, EPI_ISL_2292536, EPI_ISL_2292537, EPI_ISL_2292538, EPI_ISL_2292539, EPI_ISL_2292540, EPI_ISL_2292541, EPI_ISL_2292542, EPI_ISL_2292543, EPI_ISL_2292544, EPI_ISL_2292545, EPI_ISL_2292546, EPI_ISL_2292547, EPI_ISL_2292548, EPI_ISL_2292549, EPI_ISL_2292550, EPI_ISL_2292551, EPI_ISL_2292552, EPI_ISL_2292553, EPI_ISL_2292554, EPI_ISL_2292555, EPI_ISL_2292556, EPI_ISL_2292557, EPI_ISL_2292558, EPI_ISL_2292559, EPI_ISL_2292560, EPI_ISL_2292561, EPI_ISL_2292562, EPI_ISL_2292563, EPI_ISL_2292564, EPI_ISL_2292565, EPI_ISL_2292566, EPI_ISL_2292567, EPI_ISL_2292568, EPI_ISL_2292569, EPI_ISL_2292570, EPI_ISL_2292571, EPI_ISL_2292572, EPI_ISL_2292573, EPI_ISL_2292574, EPI_ISL_2292575, EPI_ISL_2292576, EPI_ISL_2292577, EPI_ISL_2292578, EPI_ISL_2292579, EPI_ISL_2292580, EPI_ISL_2292581, EPI_ISL_2292582, EPI_ISL_2292583, EPI_ISL_2292584, EPI_ISL_2292585, EPI_ISL_2292586, EPI_ISL_2292587, EPI_ISL_2292588, EPI_ISL_2292589, EPI_ISL_2292590, EPI_ISL_2292591, EPI_ISL_2292592, EPI_ISL_2292593, EPI_ISL_2292594, EPI_ISL_2292595, EPI_ISL_2292596, EPI_ISL_2292597, EPI_ISL_2292598, EPI_ISL_2292599, EPI_ISL_2292600, EPI_ISL_2292601, EPI_ISL_2292602, EPI_ISL_2292603, EPI_ISL_2292604, EPI_ISL_2292605, EPI_ISL_2292606, EPI_ISL_2292729, EPI_ISL_2292730, EPI_ISL_2292731, EPI_ISL_2292732, EPI_ISL_2292733, EPI_ISL_2292734, EPI_ISL_2292735, EPI_ISL_2292736, EPI_ISL_2292737,

EPI_ISL_2292738, EPI_ISL_2292739, EPI_ISL_2292740, EPI_ISL_2292741, EPI_ISL_2292742, EPI_ISL_2292743, EPI_ISL_2292744, EPI_ISL_2292745, EPI_ISL_2292746, EPI_ISL_2292747, EPI_ISL_2292748, EPI_ISL_2292749, EPI_ISL_2292750, EPI_ISL_2292751, EPI_ISL_2292752, EPI_ISL_2292753, EPI_ISL_2292754, EPI_ISL_2292755, EPI_ISL_2292756, EPI_ISL_2292757, EPI_ISL_2292758, EPI_ISL_2292759, EPI_ISL_2292760, EPI_ISL_2292761, EPI_ISL_2292762, EPI_ISL_2292763, EPI_ISL_2292764, EPI_ISL_2292765, EPI_ISL_2292766, EPI_ISL_2292767, EPI_ISL_2292768, EPI_ISL_2292769, EPI_ISL_2292770, EPI_ISL_2292771, EPI_ISL_2292772, EPI_ISL_2292773, EPI_ISL_2292774, EPI_ISL_2292775, EPI_ISL_2292776, EPI_ISL_2292777, EPI_ISL_2292778, EPI_ISL_2292779, EPI_ISL_2292780, EPI_ISL_2292781, EPI_ISL_2292782, EPI_ISL_2292783, EPI_ISL_2292784, EPI_ISL_2292785, EPI_ISL_2292786, EPI_ISL_2292787, EPI_ISL_2292788, EPI_ISL_2292789, EPI_ISL_2292790, EPI_ISL_2292791, EPI_ISL_2292792, EPI_ISL_2292793, EPI_ISL_2292794, EPI_ISL_2292795, EPI_ISL_2292796, EPI_ISL_2292797, EPI_ISL_2292798, EPI_ISL_2292799, EPI_ISL_2292800, EPI_ISL_2292801, EPI_ISL_2292802, EPI_ISL_2292803, EPI_ISL_2292804, EPI_ISL_2292805, EPI_ISL_2292806, EPI_ISL_2292807, EPI_ISL_2292808, EPI_ISL_2292809, EPI_ISL_2292810, EPI_ISL_2292811, EPI_ISL_2292812, EPI_ISL_2292813, EPI_ISL_2292814, EPI_ISL_2292815, EPI_ISL_2292816, EPI_ISL_2292817, EPI_ISL_2292818, EPI_ISL_2292819, EPI_ISL_2292820, EPI_ISL_2292821, EPI_ISL_2292822, EPI_ISL_2292823, EPI_ISL_2292824, EPI_ISL_2292825, EPI_ISL_2292826, EPI_ISL_2292827, EPI_ISL_2292828, EPI_ISL_2292829, EPI_ISL_2292830, EPI_ISL_2292831, EPI_ISL_2292832, EPI_ISL_2292833, EPI_ISL_2292834, EPI_ISL_2292835, EPI_ISL_2292836, EPI_ISL_2292837, EPI_ISL_2292838, EPI_ISL_2292839, EPI_ISL_2292840, EPI_ISL_2292841, EPI_ISL_2292842, EPI_ISL_2292843, EPI_ISL_2292844, EPI_ISL_2292845, EPI_ISL_2292846, EPI_ISL_2292847, EPI_ISL_2292848, EPI_ISL_2292849, EPI_ISL_2292850, EPI_ISL_2292851, EPI_ISL_2292852, EPI_ISL_2292853, EPI_ISL_2292854, EPI_ISL_2292855, EPI_ISL_2292856, EPI_ISL_2292857, EPI_ISL_2292858, EPI_ISL_2292859, EPI_ISL_2292860, EPI_ISL_2292861, EPI_ISL_2292862, EPI_ISL_2292863, EPI_ISL_2292864, EPI_ISL_2292865, EPI_ISL_2292866, EPI_ISL_2292867, EPI_ISL_2292868, EPI_ISL_2292869, EPI_ISL_2292870, EPI_ISL_2292871, EPI_ISL_2292872, EPI_ISL_2292873, EPI_ISL_2292874, EPI_ISL_2292875, EPI_ISL_2292876, EPI_ISL_2292877, EPI_ISL_2292878, EPI_ISL_2292879, EPI_ISL_2292880, EPI_ISL_2292881, EPI_ISL_2292882, EPI_ISL_2292883, EPI_ISL_2292884, EPI_ISL_2292885, EPI_ISL_2292886, EPI_ISL_2292887, EPI_ISL_2292888, EPI_ISL_2292889, EPI_ISL_2292890, EPI_ISL_2292891, EPI_ISL_2292892, EPI_ISL_2292893, EPI_ISL_2292894, EPI_ISL_2292895, EPI_ISL_2292896, EPI_ISL_2292897, EPI_ISL_2292898, EPI_ISL_2292899, EPI_ISL_2292900, EPI_ISL_2292901, EPI_ISL_2292902, EPI_ISL_2292903, EPI_ISL_2292904, EPI_ISL_2292905, EPI_ISL_2292906, EPI_ISL_2292907, EPI_ISL_2292908, EPI_ISL_2292909, EPI_ISL_2292910, EPI_ISL_2292911, EPI_ISL_2292912, EPI_ISL_2292913, EPI_ISL_2292914, EPI_ISL_2292915, EPI_ISL_2292916, EPI_ISL_2292917, EPI_ISL_2292918, EPI_ISL_2292919, EPI_ISL_2292920, EPI_ISL_2292921, EPI_ISL_2292922, EPI_ISL_2292923, EPI_ISL_2292924, EPI_ISL_2292925, EPI_ISL_2292926, EPI_ISL_2292927, EPI_ISL_2292928, EPI_ISL_2292929, EPI_ISL_2292930, EPI_ISL_2292931, EPI_ISL_2292932, EPI_ISL_2292933, EPI_ISL_2292934, EPI_ISL_2292935, EPI_ISL_2292936, EPI_ISL_2292937, EPI_ISL_2292938, EPI_ISL_2292939, EPI_ISL_2292940, EPI_ISL_2292941, EPI_ISL_2292942, EPI_ISL_2292943, EPI_ISL_2292944, EPI_ISL_2292945, EPI_ISL_2292946, EPI_ISL_2292947, EPI_ISL_2292948, EPI_ISL_2292949, EPI_ISL_2292950, EPI_ISL_2292951, EPI_ISL_2292952, EPI_ISL_2292953, EPI_ISL_2292954, EPI_ISL_2292955, EPI_ISL_2292956, EPI_ISL_2292957, EPI_ISL_2292958, EPI_ISL_2292959, EPI_ISL_2292960, EPI_ISL_2292961, EPI_ISL_2292962, EPI_ISL_2292963, EPI_ISL_2292964, EPI_ISL_2292965, EPI_ISL_2292966, EPI_ISL_2292967, EPI_ISL_2292968, EPI_ISL_2292969, EPI_ISL_2292970, EPI_ISL_2292971

| | | | |
|---|---|---|---|
| see above | Utah Public Health Laboratory | Utah Public Health Laboratory | Erin L. Young, Kelly F. Oakeson, Tara Gallagher |
| EPI_ISL_2292991, EPI_ISL_2292992 | Wyoming Public Health Laboratory | Wyoming Public Health Laboratory | Jim Mildenberger, Wanda Manley, Noah Hull, Taylor Fearing, Lynette Gumbleton, Channing Weber, Ashley Norberg, Chayse Rowley, Marley Goetz, Brian Dominguez, Elliot Thomasson, Sam Britz, Cari Sloma, and Rob Christensen |
| EPI_ISL_2293023, EPI_ISL_2293024 | LANIIA | LANIIA | Cristobal Chaidez, Bruno Gomez-Gil, Jean Pierre González-Gómez, Juan Daniel Lira-Morales, Irvin González-López, Julissa Enciso-Ibarra, Célida Isabel Martínez-Rodríguez |

EPI_ISL_2293043, EPI_ISL_2293044, EPI_ISL_2293045, EPI_ISL_2293046, EPI_ISL_2293047, EPI_ISL_2293048, EPI_ISL_2293049, EPI_ISL_2293050, EPI_ISL_2293051, EPI_ISL_2293052, EPI_ISL_2293053, EPI_ISL_2293054, EPI_ISL_2293055, EPI_ISL_2293056

| | | | |
|---|---|---|---|
| see above | Public Health Laboratory, Minnesota Department of Health | University of Minnesota Genomics Center | Daryl M. Gohl, Benjamin Auch, John Garbe, Jaquelyn Kuriger-Laber, Corbin Dirkx, Andy Nelson, and Sean Wang |

EPI_ISL_2293057, EPI_ISL_2293058, EPI_ISL_2293059, EPI_ISL_2293060, EPI_ISL_2293061, EPI_ISL_2293062, EPI_ISL_2293063, EPI_ISL_2293064, EPI_ISL_2293065, EPI_ISL_2293066, EPI_ISL_2293067, EPI_ISL_2293068, EPI_ISL_2293069, EPI_ISL_2293070, EPI_ISL_2293071, EPI_ISL_2293072, EPI_ISL_2293073, EPI_ISL_2293074, EPI_ISL_2293075, EPI_ISL_2293076, EPI_ISL_2293077, EPI_ISL_2293078, EPI_ISL_2293079, EPI_ISL_2293080, EPI_ISL_2293081, EPI_ISL_2293082, EPI_ISL_2293083, EPI_ISL_2293084, EPI_ISL_2293085, EPI_ISL_2293086, EPI_ISL_2293087, EPI_ISL_2293088, EPI_ISL_2293089, EPI_ISL_2293090, EPI_ISL_2293091, EPI_ISL_2293092, EPI_ISL_2293093, EPI_ISL_2293094, EPI_ISL_2293095, EPI_ISL_2293096, EPI_ISL_2293097, EPI_ISL_2293098, EPI_ISL_2293099, EPI_ISL_2293100, EPI_ISL_2293101, EPI_ISL_2293102, EPI_ISL_2293103, EPI_ISL_2293104, EPI_ISL_2293105, EPI_ISL_2293106, EPI_ISL_2293107, EPI_ISL_2293108, EPI_ISL_2293109, EPI_ISL_2293110, EPI_ISL_2293111, EPI_ISL_2293112, EPI_ISL_2293113, EPI_ISL_2293114, EPI_ISL_2293115, EPI_ISL_2293116, EPI_ISL_2293117, EPI_ISL_2293118, EPI_ISL_2293119, EPI_ISL_2293120, EPI_ISL_2293121, EPI_ISL_2293122, EPI_ISL_2293123, EPI_ISL_2293124, EPI_ISL_2293125, EPI_ISL_2293126, EPI_ISL_2293127, EPI_ISL_2293128, EPI_ISL_2293129, EPI_ISL_2293130, EPI_ISL_2293131, EPI_ISL_2293132, EPI_ISL_2293133, EPI_ISL_2293134, EPI_ISL_2293135, EPI_ISL_2293136, EPI_ISL_2293137, EPI_ISL_2293138, EPI_ISL_2293139, EPI_ISL_2293140, EPI_ISL_2293141, EPI_ISL_2293142, EPI_ISL_2293143, EPI_ISL_2293144, EPI_ISL_2293145, EPI_ISL_2293146, EPI_ISL_2293147, EPI_ISL_2293148, EPI_ISL_2293149, EPI_ISL_2293150, EPI_ISL_2293151, EPI_ISL_2293152, EPI_ISL_2293153, EPI_ISL_2293154, EPI_ISL_2293155, EPI_ISL_2293156, EPI_ISL_2293157, EPI_ISL_2293158, EPI_ISL_2293159, EPI_ISL_2293160, EPI_ISL_2293161, EPI_ISL_2293162, EPI_ISL_2293163, EPI_ISL_2293164, EPI_ISL_2293165, EPI_ISL_2293166, EPI_ISL_2293167, EPI_ISL_2293168, EPI_ISL_2293169, EPI_ISL_2293170, EPI_ISL_2293171, EPI_ISL_2293172, EPI_ISL_2293173, EPI_ISL_2293174, EPI_ISL_2293175, EPI_ISL_2293176, EPI_ISL_2293177, EPI_ISL_2293178, EPI_ISL_2293179, EPI_ISL_2293180, EPI_ISL_2293181, EPI_ISL_2293182, EPI_ISL_2293183, EPI_ISL_2293184, EPI_ISL_2293185, EPI_ISL_2293186, EPI_ISL_2293187, EPI_ISL_2293188, EPI_ISL_2293189, EPI_ISL_2293190, EPI_ISL_2293191, EPI_ISL_2293192, EPI_ISL_2293193, EPI_ISL_2293194, EPI_ISL_2293195, EPI_ISL_2293196, EPI_ISL_2293197, EPI_ISL_2293198, EPI_ISL_2293199, EPI_ISL_2293200, EPI_ISL_2293201, EPI_ISL_2293202, EPI_ISL_2293203, EPI_ISL_2293204, EPI_ISL_2293205, EPI_ISL_2293206, EPI_ISL_2293207, EPI_ISL_2293208, EPI_ISL_2293209, EPI_ISL_2293210, EPI_ISL_2293211, EPI_ISL_2293212, EPI_ISL_2293213, EPI_ISL_2293214, EPI_ISL_2293215, EPI_ISL_2293216, EPI_ISL_2293217, EPI_ISL_2293218, EPI_ISL_2293219, EPI_ISL_2293220, EPI_ISL_2293221, EPI_ISL_2293222, EPI_ISL_2293223, EPI_ISL_2293224, EPI_ISL_2293225, EPI_ISL_2293226, EPI_ISL_2293227, EPI_ISL_2293228, EPI_ISL_2293229, EPI_ISL_2293230, EPI_ISL_2293231, EPI_ISL_2293232, EPI_ISL_2293233, EPI_ISL_2293234, EPI_ISL_2293235, EPI_ISL_2293236, EPI_ISL_2293237, EPI_ISL_2293238, EPI_ISL_2293239, EPI_ISL_2293240, EPI_ISL_2293241, EPI_ISL_2293242, EPI_ISL_2293243, EPI_ISL_2293244

| | | | |
|---|---|---|---|
| see above | Public Health Laboratory, Minnesota Department of Health | University of Minnesota Genomics Center | Daryl M. Gohl, Benjamin Auch, John Garbe, Jaquelyn Kuriger-Laber, Corbin Dirkx, and Chris Carlson |

EPI_ISL_2293387, EPI_ISL_2293388, EPI_ISL_2293389, EPI_ISL_2293391, EPI_ISL_2293392, EPI_ISL_2293393, EPI_ISL_2293394, EPI_ISL_2293395, EPI_ISL_2293396, EPI_ISL_2293397, EPI_ISL_2293398, EPI_ISL_2293399, EPI_ISL_2293401, EPI_ISL_2293402, EPI_ISL_2293403, EPI_ISL_2293404, EPI_ISL_2293405, EPI_ISL_2293406, EPI_ISL_2293407, EPI_ISL_2293408, EPI_ISL_2293409, EPI_ISL_2293410, EPI_ISL_2293411, EPI_ISL_2293412, EPI_ISL_2293413, EPI_ISL_2293414, EPI_ISL_2293649, EPI_ISL_2293650, EPI_ISL_2293651, EPI_ISL_2293652, EPI_ISL_2293653, EPI_ISL_2293654, EPI_ISL_2293655, EPI_ISL_2293656, EPI_ISL_2293658, EPI_ISL_2293661, EPI_ISL_2293662, EPI_ISL_2293663, EPI_ISL_2293664, EPI_ISL_2293666, EPI_ISL_2293668, EPI_ISL_2293669, EPI_ISL_2293670, EPI_ISL_2293671, EPI_ISL_2293672, EPI_ISL_2293673, EPI_ISL_2293674, EPI_ISL_2293675, EPI_ISL_2293676, EPI_ISL_2293677, EPI_ISL_2293678, EPI_ISL_2293679, EPI_ISL_2293680, EPI_ISL_2293681, EPI_ISL_2293682, EPI_ISL_2293683, EPI_ISL_2293684, EPI_ISL_2293685, EPI_ISL_2293686, EPI_ISL_2293687, EPI_ISL_2293688, EPI_ISL_2293689

| | | | |
|---|---|---|---|
| see above | Microbiology Division, SC DHEC | Microbiology Division, SC DHEC | Freeman,J., Flores,H. |

EPI_ISL_2294378, EPI_ISL_2294379, EPI_ISL_2294380, EPI_ISL_2294381, EPI_ISL_2294382, EPI_ISL_2294383, EPI_ISL_2294384, EPI_ISL_2294385, EPI_ISL_2294386, EPI_ISL_2294387, EPI_ISL_2294388, EPI_ISL_2294389, EPI_ISL_2294390, EPI_ISL_2294391, EPI_ISL_2294392, EPI_ISL_2294393, EPI_ISL_2294394, EPI_ISL_2294395, EPI_ISL_2294396, EPI_ISL_2294397, EPI_ISL_2294398, EPI_ISL_2294399, EPI_ISL_2294400, EPI_ISL_2294401, EPI_ISL_2294402, EPI_ISL_2294403, EPI_ISL_2294404, EPI_ISL_2294405, EPI_ISL_2294406, EPI_ISL_2294407, EPI_ISL_2294408, EPI_ISL_2294409, EPI_ISL_2294410, EPI_ISL_2294411, EPI_ISL_2294412, EPI_ISL_2294413, EPI_ISL_2294414, EPI_ISL_2294415, EPI_ISL_2294416, EPI_ISL_2294417, EPI_ISL_2294418, EPI_ISL_2294419, EPI_ISL_2294420, EPI_ISL_2294421, EPI_ISL_2294422, EPI_ISL_2294423, EPI_ISL_2294424, EPI_ISL_2294425, EPI_ISL_2294426, EPI_ISL_2294427, EPI_ISL_2294428, EPI_ISL_2294429, EPI_ISL_2294430, EPI_ISL_2294431, EPI_ISL_2294432, EPI_ISL_2294433, EPI_ISL_2294434, EPI_ISL_2294435, EPI_ISL_2294436, EPI_ISL_2294437, EPI_ISL_2294438, EPI_ISL_2294439, EPI_ISL_2294440, EPI_ISL_2294441, EPI_ISL_2294442, EPI_ISL_2294443, EPI_ISL_2294444, EPI_ISL_2294445, EPI_ISL_2294446, EPI_ISL_2294447, EPI_ISL_2294448, EPI_ISL_2294449, EPI_ISL_2294450, EPI_ISL_2294451, EPI_ISL_2294452, EPI_ISL_2294453, EPI_ISL_2294454, EPI_ISL_2294455, EPI_ISL_2294456, EPI_ISL_2294457, EPI_ISL_2294458, EPI_ISL_2294459, EPI_ISL_2294460, EPI_ISL_2294461, EPI_ISL_2294462, EPI_ISL_2294463, EPI_ISL_2294464, EPI_ISL_2294465, EPI_ISL_2294466, EPI_ISL_2294467, EPI_ISL_2294468, EPI_ISL_2294469, EPI_ISL_2294470, EPI_ISL_2294471, EPI_ISL_2294472, EPI_ISL_2294473, EPI_ISL_2294474, EPI_ISL_2294475, EPI_ISL_2294476, EPI_ISL_2294477, EPI_ISL_2294478, EPI_ISL_2294479, EPI_ISL_2294480, EPI_ISL_2294481, EPI_ISL_2294482, EPI_ISL_2294483, EPI_ISL_2294484, EPI_ISL_2294485, EPI_ISL_2294486, EPI_ISL_2294487, EPI_ISL_2294488, EPI_ISL_2294489, EPI_ISL_2294490, EPI_ISL_2294491, EPI_ISL_2294492, EPI_ISL_2294493, EPI_ISL_2294494, EPI_ISL_2294495, EPI_ISL_2294496, EPI_ISL_2294497, EPI_ISL_2294498, EPI_ISL_2294499, EPI_ISL_2294500, EPI_ISL_2294501, EPI_ISL_2294502, EPI_ISL_2294503, EPI_ISL_2294504, EPI_ISL_2294505, EPI_ISL_2294506, EPI_ISL_2294507, EPI_ISL_2294508, EPI_ISL_2294509, EPI_ISL_2294510, EPI_ISL_2294511, EPI_ISL_2294512, EPI_ISL_2294513, EPI_ISL_2294514, EPI_ISL_2294515, EPI_ISL_2294516, EPI_ISL_2294517, EPI_ISL_2294518, EPI_ISL_2294519, EPI_ISL_2294520, EPI_ISL_2294521, EPI_ISL_2294522, EPI_ISL_2294523, EPI_ISL_2294524, EPI_ISL_2294525, EPI_ISL_2294526, EPI_ISL_2294527, EPI_ISL_2294528, EPI_ISL_2294529, EPI_ISL_2294530, EPI_ISL_2294531, EPI_ISL_2294532, EPI_ISL_2294533, EPI_ISL_2294535, EPI_ISL_2294536, EPI_ISL_2294537, EPI_ISL_2294538, EPI_ISL_2294539, EPI_ISL_2294540, EPI_ISL_2294541, EPI_ISL_2294542, EPI_ISL_2294543, EPI_ISL_2294544, EPI_ISL_2294545, EPI_ISL_2294546, EPI_ISL_2294547, EPI_ISL_2294548, EPI_ISL_2294549, EPI_ISL_2294550, EPI_ISL_2294551, EPI_ISL_2294552, EPI_ISL_2294553, EPI_ISL_2294554, EPI_ISL_2294555, EPI_ISL_2294556, EPI_ISL_2294557, EPI_ISL_2294558, EPI_ISL_2294559, EPI_ISL_2294560, EPI_ISL_2294561, EPI_ISL_2294562, EPI_ISL_2294563, EPI_ISL_2294564, EPI_ISL_2294565, EPI_ISL_2294566, EPI_ISL_2294567, EPI_ISL_2294568, EPI_ISL_2294569, EPI_ISL_2294570, EPI_ISL_2294571, EPI_ISL_2294572, EPI_ISL_2294573

| | | | |
|---|---|---|---|
| see above | Public Health Laboratory, Minnesota Department of Health | University of Minnesota Genomics Center | Daryl M. Gohl, John Garbe, Jaquelyn Kuriger-Laber, Corbin Dirkx, and Sean Wang |

EPI_ISL_2294595, EPI_ISL_2294596, EPI_ISL_2294597, EPI_ISL_2294598, EPI_ISL_2294599, EPI_ISL_2294600, EPI_ISL_2294601, EPI_ISL_2294602, EPI_ISL_2294603, EPI_ISL_2294604, EPI_ISL_2294605, EPI_ISL_2294606, EPI_ISL_2294607, EPI_ISL_2294609, EPI_ISL_2294610, EPI_ISL_2294611, EPI_ISL_2294613, EPI_ISL_2294614, EPI_ISL_2294616, EPI_ISL_2294617, EPI_ISL_2294618, EPI_ISL_2294619, EPI_ISL_2294620, EPI_ISL_2294621, EPI_ISL_2294622, EPI_ISL_2294623, EPI_ISL_2294624, EPI_ISL_2294625, EPI_ISL_2294626, EPI_ISL_2294627, EPI_ISL_2294628, EPI_ISL_2294629, EPI_ISL_2294630, EPI_ISL_2294631, EPI_ISL_2294632, EPI_ISL_2294633, EPI_ISL_2294634, EPI_ISL_2294635, EPI_ISL_2294636, EPI_ISL_2294637, EPI_ISL_2294638, EPI_ISL_2294639, EPI_ISL_2294640, EPI_ISL_2294641, EPI_ISL_2294642, EPI_ISL_2294643, EPI_ISL_2294644, EPI_ISL_2294645, EPI_ISL_2294646, EPI_ISL_2294647, EPI_ISL_2294648, EPI_ISL_2294649, EPI_ISL_2294650, EPI_ISL_2294651, EPI_ISL_2294652, EPI_ISL_2294653, EPI_ISL_2294654, EPI_ISL_2294655, EPI_ISL_2294656, EPI_ISL_2294657, EPI_ISL_2294658, EPI_ISL_2294659, EPI_ISL_2294660, EPI_ISL_2294661, EPI_ISL_2294662, EPI_ISL_2294663, EPI_ISL_2294664, EPI_ISL_2294665, EPI_ISL_2294666, EPI_ISL_2294667, EPI_ISL_2294668, EPI_ISL_2294669, EPI_ISL_2294670, EPI_ISL_2294671, EPI_ISL_2294672, EPI_ISL_2294673, EPI_ISL_2294674, EPI_ISL_2294675, EPI_ISL_2294676, EPI_ISL_2294677, EPI_ISL_2294678, EPI_ISL_2294679, EPI_ISL_2294680, EPI_ISL_2294681, EPI_ISL_2294682, EPI_ISL_2294683, EPI_ISL_2294684, EPI_ISL_2294685, EPI_ISL_2294686, EPI_ISL_2294687, EPI_ISL_2294688, EPI_ISL_2294689, EPI_ISL_2294690, EPI_ISL_2294691, EPI_ISL_2294692, EPI_ISL_2294693, EPI_ISL_2294694, EPI_ISL_2294695, EPI_ISL_2294696, EPI_ISL_2294697, EPI_ISL_2294698, EPI_ISL_2294699, EPI_ISL_2294700, EPI_ISL_2294701, EPI_ISL_2294702, EPI_ISL_2294703, EPI_ISL_2294704, EPI_ISL_2294705, EPI_ISL_2294706, EPI_ISL_2294707, EPI_ISL_2294708, EPI_ISL_2294709, EPI_ISL_2294710, EPI_ISL_2294711, EPI_ISL_2294712, EPI_ISL_2294713, EPI_ISL_2294714, EPI_ISL_2294715, EPI_ISL_2294716, EPI_ISL_2294717, EPI_ISL_2294718, EPI_ISL_2294719, EPI_ISL_2294720, EPI_ISL_2294721, EPI_ISL_2294722, EPI_ISL_2294723, EPI_ISL_2294724, EPI_ISL_2294725, EPI_ISL_2294726, EPI_ISL_2294727, EPI_ISL_2294729, EPI_ISL_2294730, EPI_ISL_2294731, EPI_ISL_2294732, EPI_ISL_2294733, EPI_ISL_2294734, EPI_ISL_2294735, EPI_ISL_2294736, EPI_ISL_2294737, EPI_ISL_2294738, EPI_ISL_2294740, EPI_ISL_2294742, EPI_ISL_2294743, EPI_ISL_2294744, EPI_ISL_2294745, EPI_ISL_2294747, EPI_ISL_2294748, EPI_ISL_2294749, EPI_ISL_2294750, EPI_ISL_2294751, EPI_ISL_2294752, EPI_ISL_2294753, EPI_ISL_2294754, EPI_ISL_2294755, EPI_ISL_2294756, EPI_ISL_2294757, EPI_ISL_2294758, EPI_ISL_2294759, EPI_ISL_2294760, EPI_ISL_2294761, EPI_ISL_2294762, EPI_ISL_2294763, EPI_ISL_2294764, EPI_ISL_2294765, EPI_ISL_2294766, EPI_ISL_2294767, EPI_ISL_2294768, EPI_ISL_2294769, EPI_ISL_2294770, EPI_ISL_2294771, EPI_ISL_2294772, EPI_ISL_2294774, EPI_ISL_2294775, EPI_ISL_2294776, EPI_ISL_2294777, EPI_ISL_2294778, EPI_ISL_2294779, EPI_ISL_2294780, EPI_ISL_2294781, EPI_ISL_2294782, EPI_ISL_2294783, EPI_ISL_2294784, EPI_ISL_2294785, EPI_ISL_2294786, EPI_ISL_2294787, EPI_ISL_2294788, EPI_ISL_2294789, EPI_ISL_2294790, EPI_ISL_2294791, EPI_ISL_2294792, EPI_ISL_2294793, EPI_ISL_2294794, EPI_ISL_2294795, EPI_ISL_2294796, EPI_ISL_2294797, EPI_ISL_2294798, EPI_ISL_2294799, EPI_ISL_2294800, EPI_ISL_2294801, EPI_ISL_2294802, EPI_ISL_2294803, EPI_ISL_2294804, EPI_ISL_2294805, EPI_ISL_2294806, EPI_ISL_2294807, EPI_ISL_2294808, EPI_ISL_2294809, EPI_ISL_2294810, EPI_ISL_2294811, EPI_ISL_2294812, EPI_ISL_2294813, EPI_ISL_2294814, EPI_ISL_2294815, EPI_ISL_2294816, EPI_ISL_2294817, EPI_ISL_2294818, EPI_ISL_2294819, EPI_ISL_2294820, EPI_ISL_2294821, EPI_ISL_2294822, EPI_ISL_2294823, EPI_ISL_2294824, EPI_ISL_2294825, EPI_ISL_2294826, EPI_ISL_2294827, EPI_ISL_2294828, EPI_ISL_2294829, EPI_ISL_2294830, EPI_ISL_2294831, EPI_ISL_2294832, EPI_ISL_2294833, EPI_ISL_2294834, EPI_ISL_2294835, EPI_ISL_2294836, EPI_ISL_2294837, EPI_ISL_2294838, EPI_ISL_2294839, EPI_ISL_2294840, EPI_ISL_2294841, EPI_ISL_2294842, EPI_ISL_2294843, EPI_ISL_2294844, EPI_ISL_2294845, EPI_ISL_2294846, EPI_ISL_2294847, EPI_ISL_2294848, EPI_ISL_2294849, EPI_ISL_2294850, EPI_ISL_2294851, EPI_ISL_2294852, EPI_ISL_2294853, EPI_ISL_2294854, EPI_ISL_2294856, EPI_ISL_2294857, EPI_ISL_2294858, EPI_ISL_2294859, EPI_ISL_2294860, EPI_ISL_2294861, EPI_ISL_2294862, EPI_ISL_2294863, EPI_ISL_2294864, EPI_ISL_2294865, EPI_ISL_2294866, EPI_ISL_2294867, EPI_ISL_2294868, EPI_ISL_2294869, EPI_ISL_2294870, EPI_ISL_2294871, EPI_ISL_2294872, EPI_ISL_2294873, EPI_ISL_2294874, EPI_ISL_2294875, EPI_ISL_2294876, EPI_ISL_2294877, EPI_ISL_2294878, EPI_ISL_2294879, EPI_ISL_2294880, EPI_ISL_2294881, EPI_ISL_2294882, EPI_ISL_2294883, EPI_ISL_2294884, EPI_ISL_2294885, EPI_ISL_2294886, EPI_ISL_2294887, EPI_ISL_2294888, EPI_ISL_2294889, EPI_ISL_2294890, EPI_ISL_2294891, EPI_ISL_2294892, EPI_ISL_2294893, EPI_ISL_2294894, EPI_ISL_2294895, EPI_ISL_2294896, EPI_ISL_2294897, EPI_ISL_2294898, EPI_ISL_2294899, EPI_ISL_2294900, EPI_ISL_2294901, EPI_ISL_2294902, EPI_ISL_2294903, EPI_ISL_2294904, EPI_ISL_2294905, EPI_ISL_2294906, EPI_ISL_2294907, EPI_ISL_2294908, EPI_ISL_2294909, EPI_ISL_2294910, EPI_ISL_2294911, EPI_ISL_2294912, EPI_ISL_2294913, EPI_ISL_2294914, EPI_ISL_2294915, EPI_ISL_2294916, EPI_ISL_2294917, EPI_ISL_2294918, EPI_ISL_2294919, EPI_ISL_2294920, EPI_ISL_2294921, EPI_ISL_2294922, EPI_ISL_2294923, EPI_ISL_2294924, EPI_ISL_2294925, EPI_ISL_2294926, EPI_ISL_2294927, EPI_ISL_2294928, EPI_ISL_2294929, EPI_ISL_2294930, EPI_ISL_2294931, EPI_ISL_2294932, EPI_ISL_2294933, EPI_ISL_2294934, EPI_ISL_2294935, EPI_ISL_2294936, EPI_ISL_2294937, EPI_ISL_2294938, EPI_ISL_2294939, EPI_ISL_2294940, EPI_ISL_2294941, EPI_ISL_2294942, EPI_ISL_2294943, EPI_ISL_2294944, EPI_ISL_2294945, EPI_ISL_2294946, EPI_ISL_2294947, EPI_ISL_2294948, EPI_ISL_2294949, EPI_ISL_2294950, EPI_ISL_2294951, EPI_ISL_2294952, EPI_ISL_2294953, EPI_ISL_2294954, EPI_ISL_2294955, EPI_ISL_2294956, EPI_ISL_2294957, EPI_ISL_2294958, EPI_ISL_2294959, EPI_ISL_2294960, EPI_ISL_2294961, EPI_ISL_2294962, EPI_ISL_2294963, EPI_ISL_2294964, EPI_ISL_2294965, EPI_ISL_2294966, EPI_ISL_2294967, EPI_ISL_2294968, EPI_ISL_2294969, EPI_ISL_2294970, EPI_ISL_2294971, EPI_ISL_2294972, EPI_ISL_2294973, EPI_ISL_2294974, EPI_ISL_2294975, EPI_ISL_2294976, EPI_ISL_2294977, EPI_ISL_2294978, EPI_ISL_2294979, EPI_ISL_2294980, EPI_ISL_2294981, EPI_ISL_2294982, EPI_ISL_2294983, EPI_ISL_2294984, EPI_ISL_2294985, EPI_ISL_2294986, EPI_ISL_2294987, EPI_ISL_2294988, EPI_ISL_2294989, EPI_ISL_2294990, EPI_ISL_2294991, EPI_ISL_2294992, EPI_ISL_2294993, EPI_ISL_2294994, EPI_ISL_2294995, EPI_ISL_2294996, EPI_ISL_2294997, EPI_ISL_2294998, EPI_ISL_2294999, EPI_ISL_2295000, EPI_ISL_2295001, EPI_ISL_2295002, EPI_ISL_2295003, EPI_ISL_2295004, EPI_ISL_2295005, EPI_ISL_2295006, EPI_ISL_2295007, EPI_ISL_2295008, EPI_ISL_2295009, EPI_ISL_2295010, EPI_ISL_2295011, EPI_ISL_2295012, EPI_ISL_2295013, EPI_ISL_2295014, EPI_ISL_2295015, EPI_ISL_2295016, EPI_ISL_2295017, EPI_ISL_2295018, EPI_ISL_2295019, EPI_ISL_2295020, EPI_ISL_2295021, EPI_ISL_2295022, EPI_ISL_2295023, EPI_ISL_2295024, EPI_ISL_2295025, EPI_ISL_2295026, EPI_ISL_2295027, EPI_ISL_2295028, EPI_ISL_2295029, EPI_ISL_2295030, EPI_ISL_2295031, EPI_ISL_2295032, EPI_ISL_2295033, EPI_ISL_2295034, EPI_ISL_2295035, EPI_ISL_2295036, EPI_ISL_2295037, EPI_ISL_2295038, EPI_ISL_2295039, EPI_ISL_2295040, EPI_ISL_2295041, EPI_ISL_2295042, EPI_ISL_2295043, EPI_ISL_2295044, EPI_ISL_2295045, EPI_ISL_2295046, EPI_ISL_2295047, EPI_ISL_2295048, EPI_ISL_2295049, EPI_ISL_2295050, EPI_ISL_2295051, EPI_ISL_2295052, EPI_ISL_2295053, EPI_ISL_2295054, EPI_ISL_2295055, EPI_ISL_2295056, EPI_ISL_2295057, EPI_ISL_2295058, EPI_ISL_2295059, EPI_ISL_2295060, EPI_ISL_2295061, EPI_ISL_2295062, EPI_ISL_2295063, EPI_ISL_2295064, EPI_ISL_2295065, EPI_ISL_2295066, EPI_ISL_2295067, EPI_ISL_2295068, EPI_ISL_2295069, EPI_ISL_2295070, EPI_ISL_2295071, EPI_ISL_2295072, EPI_ISL_2295073, EPI_ISL_2295074, EPI_ISL_2295075, EPI_ISL_2295077, EPI_ISL_2295078, EPI_ISL_2295079, EPI_ISL_2295080, EPI_ISL_2295081, EPI_ISL_2295082, EPI_ISL_2295083, EPI_ISL_2295084, EPI_ISL_2295085, EPI_ISL_2295086, EPI_ISL_2295087, EPI_ISL_2295088, EPI_ISL_2295089, EPI_ISL_2295090, EPI_ISL_2295091, EPI_ISL_2295092, EPI_ISL_2295093, EPI_ISL_2295094, EPI_ISL_2295095, EPI_ISL_2295096, EPI_ISL_2295097, EPI_ISL_2295098, EPI_ISL_2295099, EPI_ISL_2295100, EPI_ISL_2295101, EPI_ISL_2295102, EPI_ISL_2295103, EPI_ISL_2295104, EPI_ISL_2295105, EPI_ISL_2295106, EPI_ISL_2295107, EPI_ISL_2295108, EPI_ISL_2295109, EPI_ISL_2295110, EPI_ISL_2295111, EPI_ISL_2295112, EPI_ISL_2295113, EPI_ISL_2295114, EPI_ISL_2295115, EPI_ISL_2295116, EPI_ISL_2295117, EPI_ISL_2295118, EPI_ISL_2295119, EPI_ISL_2295120, EPI_ISL_2295121, EPI_ISL_2295122, EPI_ISL_2295123, EPI_ISL_2295124, EPI_ISL_2295125, EPI_ISL_2295126, EPI_ISL_2295127, EPI_ISL_2295128, EPI_ISL_2295129, EPI_ISL_2295130, EPI_ISL_2295131, EPI_ISL_2295132, EPI_ISL_2295133, EPI_ISL_2295134, EPI_ISL_2295135, EPI_ISL_2295136, EPI_ISL_2295137, EPI_ISL_2295138, EPI_ISL_2295139, EPI_ISL_2295140, EPI_ISL_2295141, EPI_ISL_2295142, EPI_ISL_2295143, EPI_ISL_2295144, EPI_ISL_2295145, EPI_ISL_2295146, EPI_ISL_2295147, EPI_ISL_2295148, EPI_ISL_2295149, EPI_ISL_2295150, EPI_ISL_2295151, EPI_ISL_2295152, EPI_ISL_2295153, EPI_ISL_2295154, EPI_ISL_2295155, EPI_ISL_2295156, EPI_ISL_2295157, EPI_ISL_2295158, EPI_ISL_2295159, EPI_ISL_2295160, EPI_ISL_2295161, EPI_ISL_2295162, EPI_ISL_2295163, EPI_ISL_2295164, EPI_ISL_2295165, EPI_ISL_2295166, EPI_ISL_2295167, EPI_ISL_2295168, EPI_ISL_2295169, EPI_ISL_2295170, EPI_ISL_2295171, EPI_ISL_2295172, EPI_ISL_2295173, EPI_ISL_2295174, EPI_ISL_2295175, EPI_ISL_2295176, EPI_ISL_2295177, EPI_ISL_2295178, EPI_ISL_2295179, EPI_ISL_2295180, EPI_ISL_2295181, EPI_ISL_2295182, EPI_ISL_2295183, EPI_ISL_2295184, EPI_ISL_2295185, EPI_ISL_2295186, EPI_ISL_2295187, EPI_ISL_2295188, EPI_ISL_2295189, EPI_ISL_2295190, EPI_ISL_2295191, EPI_ISL_2295192, EPI_ISL_2295193, EPI_ISL_2295194, EPI_ISL_2295195, EPI_ISL_2295196, EPI_ISL_2295197, EPI_ISL_2295198, EPI_ISL_2295199, EPI_ISL_2295200, EPI_ISL_2295201, EPI_ISL_2295202, EPI_ISL_2295203, EPI_ISL_2295204, EPI_ISL_2295205, EPI_ISL_2295206, EPI_ISL_2295207, EPI_ISL_2295208, EPI_ISL_2295209, EPI_ISL_2295210, EPI_ISL_2295211, EPI_ISL_2295212, EPI_ISL_2295213, EPI_ISL_2295214, EPI_ISL_2295215, EPI_ISL_2295216, EPI_ISL_2295217, EPI_ISL_2295218, EPI_ISL_2295219, EPI_ISL_2295220, EPI_ISL_2295221, EPI_ISL_2295222, EPI_ISL_2295223, EPI_ISL_2295224, EPI_ISL_2295225, EPI_ISL_2295226, EPI_ISL_2295227, EPI_ISL_2295228, EPI_ISL_2295229, EPI_ISL_2295230, EPI_ISL_2295231, EPI_ISL_2295232, EPI_ISL_2295233, EPI_ISL_2295234, EPI_ISL_2295235, EPI_ISL_2295236, EPI_ISL_2295237, EPI_ISL_2295238, EPI_ISL_2295239, EPI_ISL_2295240, EPI_ISL_2295241, EPI_ISL_2295242, EPI_ISL_2295243, EPI_ISL_2295244, EPI_ISL_2295245, EPI_ISL_2295246, EPI_ISL_2295247, EPI_ISL_2295248, EPI_ISL_2295249, EPI_ISL_2295250, EPI_ISL_2295251, EPI_ISL_2295252, EPI_ISL_2295253, EPI_ISL_2295254, EPI_ISL_2295255, EPI_ISL_2295256, EPI_ISL_2295257, EPI_ISL_2295258, EPI_ISL_2295259, EPI_ISL_2295260, EPI_ISL_2295261, EPI_ISL_2295262, EPI_ISL_2295263, EPI_ISL_2295264, EPI_ISL_2295265, EPI_ISL_2295266, EPI_ISL_2295267, EPI_ISL_2295268, EPI_ISL_2295269, EPI_ISL_2295270, EPI_ISL_2295271, EPI_ISL_2295272, EPI_ISL_2295273, EPI_ISL_2295274, EPI_ISL_2295275, EPI_ISL_2295276, EPI_ISL_2295277, EPI_ISL_2295278, EPI_ISL_2295279, EPI_ISL_2295280, EPI_ISL_2295281, EPI_ISL_2295282, EPI_ISL_2295283

| | | | |
|---|---|---|---|
| see above | University of Wisconsin-Madison AIDS Vaccine Research Laboratories | University of Wisconsin-Madison AIDS Vaccine Research Laboratories | Gage Moreno, Katarina Braun, et al. AIDS Vaccine Research Laboratories |
| EPI_ISL_2295284 | AR Dept. of Health-PHL, Molecular Diagnostics | Genomics and Discovery, Respiratory Viruses Branch, Division of Viral Diseases, Centers for Disease Control and Prevention | Jing Zhang, Anna Kelleher, Ying Tao, Yan Li, Brian Lynch, Krista Queen, Anna Uehara, Peter Cook, Han Jia Justin Ng, Rachel Marine, Clinton R. Paden, Dhwani Batra, Haibin Wang, Tara Coalter, Jasmine Padilla, Morgan Davis, Mili Sheth, Sarah Nobles, Mark Burroughs, Justin Lee, Adam Retchless, Suxiang Tong |
| EPI_ISL_2295285 | OK Public Health Laboratory, Oklahoma State DOH | Genomics and Discovery, Respiratory Viruses Branch, Division of Viral Diseases, Centers for Disease Control and Prevention | Jing Zhang, Anna Kelleher, Ying Tao, Yan Li, Brian Lynch, Krista Queen, Anna Uehara, Peter Cook, Han Jia Justin Ng, Rachel Marine, Clinton R. Paden, Dhwani Batra, Haibin Wang, Tara Coalter, Jasmine Padilla, Morgan Davis, Mili Sheth, Sarah Nobles, Mark Burroughs, Justin Lee, Adam Retchless, Suxiang Tong |
| EPI_ISL_2295287 | NC State Laboratory of Public Health | Genomics and Discovery, Respiratory Viruses Branch, Division of Viral Diseases, Centers for Disease Control and Prevention | Jing Zhang, Anna Kelleher, Ying Tao, Yan Li, Brian Lynch, Krista Queen, Anna Uehara, Peter Cook, Han Jia Justin Ng, Rachel Marine, Clinton R. Paden, Dhwani Batra, Haibin Wang, Tara Coalter, Jasmine Padilla, Morgan Davis, Mili Sheth, Sarah Nobles, Mark Burroughs, Justin Lee, Adam Retchless, Suxiang Tong |
| EPI_ISL_2295288, EPI_ISL_2295289 | TN Division of Laboratory Services | Genomics and Discovery, Respiratory Viruses Branch, Division of Viral Diseases, Centers for Disease Control and Prevention | Jing Zhang, Anna Kelleher, Ying Tao, Yan Li, Brian Lynch, Krista Queen, Anna Uehara, Peter Cook, Han Jia Justin Ng, Rachel Marine, Clinton R. Paden, Dhwani Batra, Haibin Wang, Tara Coalter, Jasmine Padilla, Morgan Davis, Mili Sheth, Sarah Nobles, Mark Burroughs, Justin Lee, Adam Retchless, Suxiang Tong |

EPI_ISL_2295290, EPI_ISL_2295291, EPI_ISL_2295293, EPI_ISL_2295294, EPI_ISL_2295295, EPI_ISL_2295296, EPI_ISL_2295297, EPI_ISL_2295298, EPI_ISL_2295299, EPI_ISL_2295300, EPI_ISL_2295301, EPI_ISL_2295302, EPI_ISL_2295303, EPI_ISL_2295304, EPI_ISL_2295305, EPI_ISL_2295306, EPI_ISL_2295307, EPI_ISL_2295308, EPI_ISL_2295309, EPI_ISL_2295310, EPI_ISL_2295311, EPI_ISL_2295312, EPI_ISL_2295313, EPI_ISL_2295314, EPI_ISL_2295315, EPI_ISL_2295316, EPI_ISL_2295317, EPI_ISL_2295318, EPI_ISL_2295319, EPI_ISL_2295320, EPI_ISL_2295321, EPI_ISL_2295322, EPI_ISL_2295323, EPI_ISL_2295324, EPI_ISL_2295325, EPI_ISL_2295326, EPI_ISL_2295327, EPI_ISL_2295328, EPI_ISL_2295329, EPI_ISL_2295331, EPI_ISL_2295332, EPI_ISL_2295333, EPI_ISL_2295334, EPI_ISL_2295335, EPI_ISL_2295337, EPI_ISL_2295338, EPI_ISL_2295339, EPI_ISL_2295340, EPI_ISL_2295341, EPI_ISL_2295342, EPI_ISL_2295343, EPI_ISL_2295344, EPI_ISL_2295345, EPI_ISL_2295346, EPI_ISL_2295347, EPI_ISL_2295348, EPI_ISL_2295349, EPI_ISL_2295350, EPI_ISL_2295351, EPI_ISL_2295352, EPI_ISL_2295353, EPI_ISL_2295354, EPI_ISL_2295355, EPI_ISL_2295356, EPI_ISL_2295357, EPI_ISL_2295359, EPI_ISL_2295360, EPI_ISL_2295361, EPI_ISL_2295362, EPI_ISL_2295363, EPI_ISL_2295364, EPI_ISL_2295365, EPI_ISL_2295366, EPI_ISL_2295367, EPI_ISL_2295368, EPI_ISL_2295369, EPI_ISL_2295370, EPI_ISL_2295371, EPI_ISL_2295372, EPI_ISL_2295373, EPI_ISL_2295374, EPI_ISL_2295376, EPI_ISL_2295378, EPI_ISL_2295379, EPI_ISL_2295382, EPI_ISL_2295383, EPI_ISL_2295385

| | | | |
|---|---|---|---|
| see above | WLA VAMC | Genomics and Discovery, Respiratory Viruses Branch, Division of Viral Diseases, Centers for Disease Control and Prevention | Jing Zhang, Anna Kelleher, Ying Tao, Yan Li, Brian Lynch, Krista Queen, Anna Uehara, Peter Cook, Han Jia Justin Ng, Rachel Marine, Clinton R. Paden, Dhwani Batra, Haibin Wang, Tara Coalter, Jasmine Padilla, Morgan Davis, Mili Sheth, Sarah Nobles, Mark Burroughs, Justin Lee, Adam Retchless, Suxiang Tong |

EPI_ISL_2295453, EPI_ISL_2295454, EPI_ISL_2295455, EPI_ISL_2295456, EPI_ISL_2295457, EPI_ISL_2295458, EPI_ISL_2295459, EPI_ISL_2295460, EPI_ISL_2295461, EPI_ISL_2295462, EPI_ISL_2295463, EPI_ISL_2295464, EPI_ISL_2295465, EPI_ISL_2295466, EPI_ISL_2295467, EPI_ISL_2295468, EPI_ISL_2295469, EPI_ISL_2295470, EPI_ISL_2295471, EPI_ISL_2295472, EPI_ISL_2295473, EPI_ISL_2295474, EPI_ISL_2295475, EPI_ISL_2295476, EPI_ISL_2295477, EPI_ISL_2295478, EPI_ISL_2295479, EPI_ISL_2295480, EPI_ISL_2295481, EPI_ISL_2295482, EPI_ISL_2295483, EPI_ISL_2295484, EPI_ISL_2295485, EPI_ISL_2295486, EPI_ISL_2295487, EPI_ISL_2295488, EPI_ISL_2295489, EPI_ISL_2295490, EPI_ISL_2295491, EPI_ISL_2295492, EPI_ISL_2295493, EPI_ISL_2295494, EPI_ISL_2295495, EPI_ISL_2295496, EPI_ISL_2295497, EPI_ISL_2295498, EPI_ISL_2295499, EPI_ISL_2295500, EPI_ISL_2295501, EPI_ISL_2295502, EPI_ISL_2295503, EPI_ISL_2295504, EPI_ISL_2295505, EPI_ISL_2295506, EPI_ISL_2295507, EPI_ISL_2295508, EPI_ISL_2295509, EPI_ISL_2295510, EPI_ISL_2295511, EPI_ISL_2295512, EPI_ISL_2295513, EPI_ISL_2295514, EPI_ISL_2295515, EPI_ISL_2295516, EPI_ISL_2295517, EPI_ISL_2295518, EPI_ISL_2295519, EPI_ISL_2295520, EPI_ISL_2295521, EPI_ISL_2295522, EPI_ISL_2295523, EPI_ISL_2295524, EPI_ISL_2295525, EPI_ISL_2295526, EPI_ISL_2295527, EPI_ISL_2295528, EPI_ISL_2295529, EPI_ISL_2295530, EPI_ISL_2295531, EPI_ISL_2295532, EPI_ISL_2295533

| | | | |
|---|---|---|---|
| see above | UCSC Genomics Institute | UCSC Genomics Institute | Mark Akeson, Ikenna Anigbogu, Eric Beraut, Isabel Bjork, Molly Cassatt-Johnstone, Terren Chang, Russell Corbett-Detig, Namrita Dhillon, Maximilian Haeussler, David Haussler, Angie Hinrichs, Miten Jain, Joshua Kapp, A. Marm Kilpatrick, Jakob McBroome, Hugh Olsen, Jeremy Sanford, Beth Shapiro, Michael Stone, Bryan Thornlow, Yatish Turakhia, Ciara Wanket |
| EPI_ISL_2295534, EPI_ISL_2295535, EPI_ISL_2295536, EPI_ISL_2295537, EPI_ISL_2295538, EPI_ISL_2295539, EPI_ISL_2295541, EPI_ISL_2295542, EPI_ISL_2295543, EPI_ISL_2295544, EPI_ISL_2295545, EPI_ISL_2295546, EPI_ISL_2295547, EPI_ISL_2295548, EPI_ISL_2295549, EPI_ISL_2295550, EPI_ISL_2295551, EPI_ISL_2295552, EPI_ISL_2295553, EPI_ISL_2295554, EPI_ISL_2295555, EPI_ISL_2295556, EPI_ISL_2295557, EPI_ISL_2295558, EPI_ISL_2295560, EPI_ISL_2295561, EPI_ISL_2295562, EPI_ISL_2295563, EPI_ISL_2295565, EPI_ISL_2295566, EPI_ISL_2295567, EPI_ISL_2295568, EPI_ISL_2295569, EPI_ISL_2295570, EPI_ISL_2295572, EPI_ISL_2295573, EPI_ISL_2295574, EPI_ISL_2295575, EPI_ISL_2295576, EPI_ISL_2295577, EPI_ISL_2295578, EPI_ISL_2295579, EPI_ISL_2295580, EPI_ISL_2295581, EPI_ISL_2295582, EPI_ISL_2295583, EPI_ISL_2295584, EPI_ISL_2295587 | | | |
| see above | UTMG Pathology LLC, University Clinical Health | UTMG Pathology LLC, University Clinical Health | Kumaravelu,P., Hodge,T., Matande,K., Hymon,T., Baselski,V., Azouz,A. |
| EPI_ISL_2295588, EPI_ISL_2295589 | LESP Ciudad de Mexico | Instituto de Diagnostico y Referencia Epidemiologicos (INDRE) | Claudia Wong-Arambula, Abril Rodriguez-Maldonado, Vanessa Rivero-Arredondo, Ariadna Medina-Benitez, Joaquin Quiroz-Mercado, Sergio Rangel-Guerrero, Natividad Cruz-Ortiz, Tatiana Nunez-Garcia, Gisela Barrera-Badillo, Lucia Hernandez-Rivas, Irma Lopez-Martinez, Ernesto Ramirez-Gonzalez. |
| EPI_ISL_2295590 | LESP Guanajuato | Instituto de Diagnostico y Referencia Epidemiologicos (INDRE) | Claudia Wong-Arambula, Abril Rodriguez-Maldonado, Vanessa Rivero-Arredondo, Ariadna Medina-Benitez, Joaquin Quiroz-Mercado, Sergio Rangel-Guerrero, Natividad Cruz-Ortiz, Tatiana Nunez-Garcia, Gisela Barrera-Badillo, Lucia Hernandez-Rivas, Irma Lopez-Martinez, Ernesto Ramirez-Gonzalez. |
| EPI_ISL_2295592, EPI_ISL_2295595, EPI_ISL_2295596 | Florida Bureau of Public Health Laboratories | Florida Bureau of Public Health Laboratories | Sarah Schmedes, Jason Blanton |
| EPI_ISL_2295599, EPI_ISL_2295600, EPI_ISL_2295601, EPI_ISL_2295602, EPI_ISL_2295603, EPI_ISL_2295604, EPI_ISL_2295605, EPI_ISL_2295606, EPI_ISL_2295607, EPI_ISL_2295608, EPI_ISL_2295609, EPI_ISL_2295610, EPI_ISL_2295611, EPI_ISL_2295612, EPI_ISL_2295613, EPI_ISL_2295614, EPI_ISL_2295615, EPI_ISL_2295616, EPI_ISL_2295617, EPI_ISL_2295618, EPI_ISL_2295619, EPI_ISL_2295620, EPI_ISL_2295621, EPI_ISL_2295622, EPI_ISL_2295623, EPI_ISL_2295624, EPI_ISL_2295625, EPI_ISL_2295626, EPI_ISL_2295627, EPI_ISL_2295628, EPI_ISL_2295629, EPI_ISL_2295630, EPI_ISL_2295631, EPI_ISL_2295632, EPI_ISL_2295633, EPI_ISL_2295634, EPI_ISL_2295635, EPI_ISL_2295636, EPI_ISL_2295637, EPI_ISL_2295638, EPI_ISL_2295639, EPI_ISL_2295640, EPI_ISL_2295641, EPI_ISL_2295642, EPI_ISL_2295643, EPI_ISL_2295644, EPI_ISL_2295645, EPI_ISL_2295646, EPI_ISL_2295647, EPI_ISL_2295648, EPI_ISL_2295649, EPI_ISL_2295650, EPI_ISL_2295651, EPI_ISL_2295652, EPI_ISL_2295653, EPI_ISL_2295654, EPI_ISL_2295655, EPI_ISL_2295656, EPI_ISL_2295657, EPI_ISL_2295658, EPI_ISL_2295659, EPI_ISL_2295660, EPI_ISL_2295661, EPI_ISL_2295662, EPI_ISL_2295663, EPI_ISL_2295664, EPI_ISL_2295665, EPI_ISL_2295666, EPI_ISL_2295667, EPI_ISL_2295668, EPI_ISL_2295669, EPI_ISL_2295670, EPI_ISL_2295671, EPI_ISL_2295672, EPI_ISL_2295673, EPI_ISL_2295674, EPI_ISL_2295675, EPI_ISL_2295676, EPI_ISL_2295677, EPI_ISL_2295678, EPI_ISL_2295679, EPI_ISL_2295680, EPI_ISL_2295681, EPI_ISL_2295682, EPI_ISL_2295683, EPI_ISL_2295684, EPI_ISL_2295685, EPI_ISL_2295686, EPI_ISL_2295687, EPI_ISL_2295688, EPI_ISL_2295689, EPI_ISL_2295690, EPI_ISL_2295691, EPI_ISL_2295692, EPI_ISL_2295693, EPI_ISL_2295694, EPI_ISL_2295695, EPI_ISL_2295696, EPI_ISL_2295697, EPI_ISL_2295698, EPI_ISL_2295699, EPI_ISL_2295700, EPI_ISL_2295701, EPI_ISL_2295702, EPI_ISL_2295703, EPI_ISL_2295704, EPI_ISL_2295705, EPI_ISL_2295706, EPI_ISL_2295707, EPI_ISL_2295708, EPI_ISL_2295709, EPI_ISL_2295710, EPI_ISL_2295711, EPI_ISL_2295712, EPI_ISL_2295713, EPI_ISL_2295714, EPI_ISL_2295715, EPI_ISL_2295716, EPI_ISL_2295717, EPI_ISL_2295718, EPI_ISL_2295719, EPI_ISL_2295720, EPI_ISL_2295721, EPI_ISL_2295722, EPI_ISL_2295723, EPI_ISL_2295724, EPI_ISL_2295725, EPI_ISL_2295726, EPI_ISL_2295727, EPI_ISL_2295728, EPI_ISL_2295729, EPI_ISL_2295730, EPI_ISL_2295731, EPI_ISL_2295732, EPI_ISL_2295733, EPI_ISL_2295734, EPI_ISL_2295735, EPI_ISL_2295736, EPI_ISL_2295737, EPI_ISL_2295738, EPI_ISL_2295739, EPI_ISL_2295740, EPI_ISL_2295741, EPI_ISL_2295742, EPI_ISL_2295743, EPI_ISL_2295744, EPI_ISL_2295745, EPI_ISL_2295746, EPI_ISL_2295747, EPI_ISL_2295748, EPI_ISL_2295749, EPI_ISL_2295750, EPI_ISL_2295751, EPI_ISL_2295752, EPI_ISL_2295753, EPI_ISL_2295754, EPI_ISL_2295755, EPI_ISL_2295756, EPI_ISL_2295757, EPI_ISL_2295758, EPI_ISL_2295759, EPI_ISL_2295760, EPI_ISL_2295761, EPI_ISL_2295762, EPI_ISL_2295763, EPI_ISL_2295764, EPI_ISL_2295765, EPI_ISL_2295766, EPI_ISL_2295767, EPI_ISL_2295768, EPI_ISL_2295769, EPI_ISL_2295770, EPI_ISL_2295771, EPI_ISL_2295772, EPI_ISL_2295773, EPI_ISL_2295774, EPI_ISL_2295775, EPI_ISL_2295776, EPI_ISL_2295777, EPI_ISL_2295778, EPI_ISL_2295779, EPI_ISL_2295780, EPI_ISL_2295781, EPI_ISL_2295782, EPI_ISL_2295783 | | | |
| see above | Public Health Laboratory, Minnesota Department of Health | University of Minnesota Genomics Center | Daryl M. Gohl, John Garbe, Jaquelyn Kuriger-Laber, Corbin Dirkx, and Sean Wang |
| EPI_ISL_2295786, EPI_ISL_2295787, EPI_ISL_2295789, EPI_ISL_2295790, EPI_ISL_2295791, EPI_ISL_2295792, EPI_ISL_2295793, EPI_ISL_2295794, EPI_ISL_2295795, EPI_ISL_2295796, EPI_ISL_2295797, EPI_ISL_2295799, EPI_ISL_2295801, EPI_ISL_2295802, EPI_ISL_2295803, EPI_ISL_2295804, EPI_ISL_2295805, EPI_ISL_2295806, EPI_ISL_2295808, EPI_ISL_2295809, EPI_ISL_2295810, EPI_ISL_2295811, EPI_ISL_2295812, EPI_ISL_2295813, EPI_ISL_2295815 | | | |
| see above | Oregon State University TRACE Project | OSU Center for Genome Research and Biocomputing | Oregon State University TRACE Project |
| EPI_ISL_2295818, EPI_ISL_2295819, EPI_ISL_2295820, EPI_ISL_2295821, EPI_ISL_2295822, EPI_ISL_2295823, EPI_ISL_2295824, EPI_ISL_2295825, EPI_ISL_2295826, EPI_ISL_2295827, EPI_ISL_2295828, EPI_ISL_2295829, EPI_ISL_2295830, EPI_ISL_2295832, EPI_ISL_2295833, EPI_ISL_2295834, EPI_ISL_2295835, EPI_ISL_2295836, EPI_ISL_2295837, EPI_ISL_2295838, EPI_ISL_2295839, EPI_ISL_2295840, EPI_ISL_2295842, EPI_ISL_2295843, EPI_ISL_2295844, EPI_ISL_2295845, EPI_ISL_2295846, EPI_ISL_2295847, EPI_ISL_2295848, EPI_ISL_2295849, EPI_ISL_2295850, EPI_ISL_2295851, EPI_ISL_2295852, EPI_ISL_2295853, EPI_ISL_2295854, EPI_ISL_2295855, EPI_ISL_2295856, EPI_ISL_2295858, EPI_ISL_2295859, EPI_ISL_2295860, EPI_ISL_2295861, EPI_ISL_2295862, EPI_ISL_2295863, EPI_ISL_2295864, EPI_ISL_2295866, EPI_ISL_2295867, EPI_ISL_2295869, EPI_ISL_2295870, EPI_ISL_2295871, EPI_ISL_2295872, EPI_ISL_2295873, EPI_ISL_2295874, EPI_ISL_2295875, EPI_ISL_2295876, EPI_ISL_2295877, EPI_ISL_2295878, EPI_ISL_2295879, EPI_ISL_2295880, EPI_ISL_2295881, EPI_ISL_2295882, EPI_ISL_2295883, EPI_ISL_2295884, EPI_ISL_2295886, EPI_ISL_2295887, EPI_ISL_2295888, EPI_ISL_2295889, EPI_ISL_2295890, EPI_ISL_2295891, EPI_ISL_2295892, EPI_ISL_2295893, EPI_ISL_2295894, EPI_ISL_2295895, EPI_ISL_2295896, EPI_ISL_2295897, EPI_ISL_2295898, EPI_ISL_2295899, EPI_ISL_2295900, EPI_ISL_2295901, EPI_ISL_2295902, EPI_ISL_2295903, EPI_ISL_2295904, EPI_ISL_2295905, EPI_ISL_2295906, EPI_ISL_2295907, EPI_ISL_2295908, EPI_ISL_2295909, EPI_ISL_2295910, EPI_ISL_2295911, EPI_ISL_2295912, EPI_ISL_2295913, EPI_ISL_2295914, EPI_ISL_2295915, EPI_ISL_2295916, EPI_ISL_2295917, EPI_ISL_2295918, EPI_ISL_2295919, EPI_ISL_2295920, EPI_ISL_2295921, EPI_ISL_2295922, EPI_ISL_2295923, EPI_ISL_2295924, EPI_ISL_2295925, EPI_ISL_2295926 | | | |
| see above | Illinois Department of Public Health | Gagnon Lab, Southern Illinois University | Keith Gagnon |
| EPI_ISL_2295927, EPI_ISL_2295928, EPI_ISL_2295929, EPI_ISL_2295930, EPI_ISL_2295931, EPI_ISL_2295932, EPI_ISL_2295933, EPI_ISL_2295934, EPI_ISL_2295935, EPI_ISL_2295936, EPI_ISL_2295937, EPI_ISL_2295938, EPI_ISL_2295939, EPI_ISL_2295940, EPI_ISL_2295942, EPI_ISL_2295943, EPI_ISL_2295944, EPI_ISL_2295945, EPI_ISL_2295946, EPI_ISL_2295947, EPI_ISL_2295948, EPI_ISL_2295949, EPI_ISL_2295950, EPI_ISL_2295951, EPI_ISL_2295952, EPI_ISL_2295953, EPI_ISL_2295954, EPI_ISL_2295955, EPI_ISL_2295956, EPI_ISL_2295957, EPI_ISL_2295958, EPI_ISL_2295960, EPI_ISL_2295962, EPI_ISL_2295963, EPI_ISL_2295964, EPI_ISL_2295965, EPI_ISL_2295966, EPI_ISL_2295967, EPI_ISL_2295968, EPI_ISL_2295969, EPI_ISL_2295970, EPI_ISL_2295971, EPI_ISL_2295972, EPI_ISL_2295973, EPI_ISL_2295975, EPI_ISL_2295976, EPI_ISL_2295978, EPI_ISL_2295979, EPI_ISL_2295981, EPI_ISL_2295982, EPI_ISL_2295983, EPI_ISL_2295984, EPI_ISL_2295985, EPI_ISL_2295986, EPI_ISL_2295988, EPI_ISL_2295989, EPI_ISL_2295991, EPI_ISL_2295992, EPI_ISL_2295993, EPI_ISL_2295994, EPI_ISL_2295995, EPI_ISL_2295997, EPI_ISL_2295998, EPI_ISL_2295999, EPI_ISL_2296000, EPI_ISL_2296003, EPI_ISL_2296004, EPI_ISL_2296005, EPI_ISL_2296006, EPI_ISL_2296007, EPI_ISL_2296008, EPI_ISL_2296009, EPI_ISL_2296011, EPI_ISL_2296012, EPI_ISL_2296013, EPI_ISL_2296014, EPI_ISL_2296017, EPI_ISL_2296018, EPI_ISL_2296019, EPI_ISL_2296020, EPI_ISL_2296021, EPI_ISL_2296022, EPI_ISL_2296023, EPI_ISL_2296024, EPI_ISL_2296025, EPI_ISL_2296027, EPI_ISL_2296028, EPI_ISL_2296029, EPI_ISL_2296030, EPI_ISL_2296031, EPI_ISL_2296033, EPI_ISL_2296034, EPI_ISL_2296035, EPI_ISL_2296038, EPI_ISL_2296039, EPI_ISL_2296040, EPI_ISL_2296041, EPI_ISL_2296043, EPI_ISL_2296044, EPI_ISL_2296045, EPI_ISL_2296046 | | | |
| see above | Simple Laboratories | Gagnon Lab, Southern Illinois University | Keith Gagnon |
| EPI_ISL_2296051, EPI_ISL_2296054 | Arizona State Public Health Laboratory | Arizona State Public Health Laboratory | Trung Huynh, Jessica Escobar, Katherine Fullerton, Nobuko Fukushima, Stacy White, Linda Getsinger, Victor Waddell |
| EPI_ISL_2296059 | LESP Oaxaca | Instituto de Diagnostico y Referencia Epidemiologicos (INDRE) | Claudia Wong-Arambula, Abril Rodriguez-Maldonado, Vanessa Rivero-Arredondo, Ariadna Medina-Benitez, Joaquin Quiroz-Mercado, Sergio Rangel-Guerrero, Natividad Cruz-Ortiz, Tatiana Nunez-Garcia, Gisela Barrera-Badillo, Lucia Hernandez-Rivas, Irma Lopez-Martinez, Ernesto Ramirez-Gonzalez. |
| EPI_ISL_2296060 | LESP Ciudad de Mexico | Instituto de Diagnostico y Referencia Epidemiologicos (INDRE) | Claudia Wong-Arambula, Abril Rodriguez-Maldonado, Vanessa Rivero-Arredondo, Ariadna Medina-Benitez, Joaquin Quiroz-Mercado, Sergio Rangel-Guerrero, Natividad Cruz-Ortiz, Tatiana Nunez-Garcia, Gisela Barrera-Badillo, Lucia Hernandez-Rivas, Irma Lopez-Martinez, Ernesto Ramirez-Gonzalez. |
| EPI_ISL_2296061, EPI_ISL_2296062 | LESP Quintana Roo | Instituto de Diagnostico y Referencia Epidemiologicos (INDRE) | Claudia Wong-Arambula, Abril Rodriguez-Maldonado, Vanessa Rivero-Arredondo, Ariadna Medina-Benitez, Joaquin Quiroz-Mercado, Sergio Rangel-Guerrero, Natividad Cruz-Ortiz, Tatiana Nunez-Garcia, Gisela Barrera-Badillo, Lucia Hernandez-Rivas, Irma Lopez-Martinez, Ernesto Ramirez-Gonzalez. |
| EPI_ISL_2296063 | LESP Hidalgo | Instituto de Diagnostico y Referencia Epidemiologicos (INDRE) | Claudia Wong-Arambula, Abril Rodriguez-Maldonado, Vanessa Rivero-Arredondo, Ariadna Medina-Benitez, Joaquin Quiroz-Mercado, Sergio Rangel-Guerrero, Natividad Cruz-Ortiz, Tatiana Nunez-Garcia, Gisela Barrera-Badillo, Lucia Hernandez-Rivas, Irma Lopez-Martinez, Ernesto Ramirez-Gonzalez. |
| EPI_ISL_2296064 | LESP Baja California | Instituto de Diagnostico y Referencia Epidemiologicos (INDRE) | Claudia Wong-Arambula, Abril Rodriguez-Maldonado, Vanessa Rivero-Arredondo, Ariadna Medina-Benitez, Joaquin Quiroz-Mercado, Sergio Rangel-Guerrero, Natividad Cruz-Ortiz, Tatiana Nunez-Garcia, Gisela Barrera-Badillo, Lucia Hernandez-Rivas, Irma Lopez-Martinez, Ernesto Ramirez-Gonzalez. |
| EPI_ISL_2296065 | LESP Tamaulipas | Instituto de Diagnostico y Referencia Epidemiologicos (INDRE) | Claudia Wong-Arambula, Abril Rodriguez-Maldonado, Vanessa Rivero-Arredondo, Ariadna Medina-Benitez, Joaquin Quiroz-Mercado, Sergio Rangel-Guerrero, Natividad Cruz-Ortiz, Tatiana Nunez-Garcia, Gisela Barrera-Badillo, Lucia Hernandez-Rivas, Irma Lopez-Martinez, Ernesto Ramirez-Gonzalez. |
| EPI_ISL_2296066 | LESP Michoacan | Instituto de Diagnostico y Referencia Epidemiologicos (INDRE) | Claudia Wong-Arambula, Abril Rodriguez-Maldonado, Vanessa Rivero-Arredondo, Ariadna Medina-Benitez, Joaquin Quiroz-Mercado, Sergio Rangel-Guerrero, Natividad Cruz-Ortiz, Tatiana Nunez-Garcia, Gisela Barrera-Badillo, Lucia Hernandez-Rivas, Irma Lopez-Martinez, Ernesto Ramirez-Gonzalez. |
| EPI_ISL_2296067 | LESP Colima | Instituto de Diagnostico y Referencia Epidemiologicos (INDRE) | Claudia Wong-Arambula, Abril Rodriguez-Maldonado, Vanessa Rivero-Arredondo, Ariadna Medina-Benitez, Joaquin Quiroz-Mercado, Sergio Rangel-Guerrero, Natividad Cruz-Ortiz, Tatiana Nunez-Garcia, Gisela Barrera-Badillo, Lucia Hernandez-Rivas, Irma Lopez-Martinez, Ernesto Ramirez-Gonzalez. |
| EPI_ISL_2296068 | LESP Nuevo Leon | Instituto de Diagnostico y Referencia Epidemiologicos (INDRE) | Claudia Wong-Arambula, Abril Rodriguez-Maldonado, Vanessa Rivero-Arredondo, Ariadna Medina-Benitez, Joaquin Quiroz-Mercado, Sergio Rangel-Guerrero, Natividad Cruz-Ortiz, Tatiana Nunez-Garcia, Gisela Barrera-Badillo, Lucia Hernandez-Rivas, Irma Lopez-Martinez, Ernesto Ramirez-Gonzalez. |
| EPI_ISL_2296069 | LESP Guanajuato | Instituto de Diagnostico y Referencia Epidemiologicos (INDRE) | Claudia Wong-Arambula, Abril Rodriguez-Maldonado, Vanessa Rivero-Arredondo, Ariadna Medina-Benitez, Joaquin Quiroz-Mercado, Sergio Rangel-Guerrero, Natividad Cruz-Ortiz, Tatiana Nunez-Garcia, Gisela Barrera-Badillo, Lucia Hernandez-Rivas, Irma Lopez-Martinez, Ernesto Ramirez-Gonzalez. |
| EPI_ISL_2296070 | LESP Jalisco | Instituto de Diagnostico y Referencia Epidemiologicos (INDRE) | Claudia Wong-Arambula, Abril Rodriguez-Maldonado, Vanessa Rivero-Arredondo, Ariadna Medina-Benitez, Joaquin Quiroz-Mercado, Sergio Rangel-Guerrero, Natividad Cruz-Ortiz, Tatiana Nunez-Garcia, Gisela Barrera-Badillo, Lucia Hernandez-Rivas, Irma Lopez-Martinez, Ernesto Ramirez-Gonzalez. |
| EPI_ISL_2296071 | LESP Sinaloa | Instituto de Diagnostico y Referencia Epidemiologicos (INDRE) | Claudia Wong-Arambula, Abril Rodriguez-Maldonado, Vanessa Rivero-Arredondo, Ariadna Medina-Benitez, Joaquin Quiroz-Mercado, Sergio Rangel-Guerrero, Natividad Cruz-Ortiz, Tatiana Nunez-Garcia, Gisela Barrera-Badillo, Lucia Hernandez-Rivas, Irma Lopez-Martinez, Ernesto Ramirez-Gonzalez. |
| EPI_ISL_2296072 | LESP Ciudad de Mexico | Instituto de Diagnostico y Referencia Epidemiologicos (INDRE) | Claudia Wong-Arambula, Abril Rodriguez-Maldonado, Vanessa Rivero-Arredondo, Ariadna Medina-Benitez, Joaquin Quiroz-Mercado, Sergio Rangel-Guerrero, Natividad Cruz-Ortiz, Tatiana Nunez-Garcia, Gisela Barrera-Badillo, Lucia Hernandez-Rivas, Irma Lopez-Martinez, Ernesto Ramirez-Gonzalez. |
| EPI_ISL_2296073 | LESP Estado de Mexico | Instituto de Diagnostico y Referencia Epidemiologicos (INDRE) | Claudia Wong-Arambula, Abril Rodriguez-Maldonado, Vanessa Rivero-Arredondo, Ariadna Medina-Benitez, Joaquin Quiroz-Mercado, Sergio Rangel-Guerrero, Natividad Cruz-Ortiz, Tatiana Nunez-Garcia, Gisela Barrera-Badillo, Lucia Hernandez-Rivas, Irma Lopez-Martinez, Ernesto Ramirez-Gonzalez. |
| EPI_ISL_2296074, EPI_ISL_2296075, EPI_ISL_2296076, EPI_ISL_2296077, EPI_ISL_2296078, EPI_ISL_2296079, EPI_ISL_2296080, EPI_ISL_2296081 | LESP Ciudad de Mexico | Instituto de Diagnostico y Referencia Epidemiologicos (INDRE) | Claudia Wong-Arambula, Abril Rodriguez-Maldonado, Vanessa Rivero-Arredondo, Ariadna Medina-Benitez, Joaquin Quiroz-Mercado, Sergio Rangel-Guerrero, Natividad Cruz-Ortiz, Tatiana Nunez-Garcia, Gisela Barrera-Badillo, Lucia Hernandez-Rivas, Irma Lopez-Martinez, Ernesto Ramirez-Gonzalez. |
| EPI_ISL_2296082 | LESP Estado de Mexico | Instituto de Diagnostico y Referencia Epidemiologicos (INDRE) | Claudia Wong-Arambula, Abril Rodriguez-Maldonado, Vanessa Rivero-Arredondo, Ariadna Medina-Benitez, Joaquin Quiroz-Mercado, Sergio Rangel-Guerrero, Natividad Cruz-Ortiz, Tatiana Nunez-Garcia, Gisela Barrera-Badillo, Lucia Hernandez-Rivas, Irma Lopez-Martinez, Ernesto Ramirez-Gonzalez. |
| EPI_ISL_2296083 | LESP Ciudad de Mexico | Instituto de Diagnostico y Referencia Epidemiologicos (INDRE) | Claudia Wong-Arambula, Abril Rodriguez-Maldonado, Vanessa Rivero-Arredondo, Ariadna Medina-Benitez, Joaquin Quiroz-Mercado, Sergio Rangel-Guerrero, Natividad Cruz-Ortiz, Tatiana Nunez-Garcia, Gisela Barrera-Badillo, Lucia Hernandez-Rivas, Irma Lopez-Martinez, Ernesto Ramirez-Gonzalez. |
| EPI_ISL_2296084 | LESP Tamaulipas | Instituto de Diagnostico y Referencia Epidemiologicos (INDRE) | Claudia Wong-Arambula, Abril Rodriguez-Maldonado, Vanessa Rivero-Arredondo, Ariadna Medina-Benitez, Joaquin Quiroz-Mercado, Sergio Rangel-Guerrero, Natividad Cruz-Ortiz, Tatiana Nunez-Garcia, Gisela Barrera-Badillo, Lucia Hernandez-Rivas, Irma Lopez-Martinez, Ernesto Ramirez-Gonzalez. |
| EPI_ISL_2296085 | LESP Chiapas | Instituto de Diagnostico y Referencia Epidemiologicos (INDRE) | Claudia Wong-Arambula, Abril Rodriguez-Maldonado, Vanessa Rivero-Arredondo, Ariadna Medina-Benitez, Joaquin Quiroz-Mercado, Sergio Rangel-Guerrero, Natividad Cruz-Ortiz, Tatiana Nunez-Garcia, Gisela Barrera-Badillo, Lucia Hernandez-Rivas, Irma Lopez-Martinez, Ernesto Ramirez-Gonzalez. |
| EPI_ISL_2296086 | LESP Baja California | Instituto de Diagnostico y Referencia Epidemiologicos (INDRE) | Claudia Wong-Arambula, Abril Rodriguez-Maldonado, Vanessa Rivero-Arredondo, Ariadna Medina-Benitez, Joaquin Quiroz-Mercado, Sergio Rangel-Guerrero, Natividad Cruz-Ortiz, Tatiana Nunez-Garcia, Gisela Barrera-Badillo, Lucia Hernandez-Rivas, Irma Lopez-Martinez, Ernesto Ramirez-Gonzalez. |
| EPI_ISL_2296087 | LESP Quintana Roo | Instituto de Diagnostico y Referencia Epidemiologicos (INDRE) | Claudia Wong-Arambula, Abril Rodriguez-Maldonado, Vanessa Rivero-Arredondo, Ariadna Medina-Benitez, Joaquin Quiroz-Mercado, Sergio Rangel-Guerrero, Natividad Cruz-Ortiz, Tatiana Nunez-Garcia, Gisela Barrera-Badillo, Lucia Hernandez-Rivas, Irma Lopez-Martinez, Ernesto Ramirez-Gonzalez. |
| EPI_ISL_2296088, EPI_ISL_2296089, EPI_ISL_2296090, | LESP Yucatan | Instituto de Diagnostico y Referencia Epidemiologicos (INDRE) | Claudia Wong-Arambula, Abril Rodriguez-Maldonado, Vanessa Rivero-Arredondo, Ariadna Medina-Benitez, Joaquin Quiroz-Mercado, Sergio Rangel-Guerrero, Natividad Cruz-Ortiz, Tatiana Nunez-Garcia, Gisela Barrera-Badillo, Lucia Hernandez-Rivas, Irma Lopez-Martinez, Ernesto Ramirez-Gonzalez. |

| | | | |
|---|---|---|---|
| EPI_ISL_2296091 | | | |
| EPI_ISL_2296092 | LESP Chihuahua | Instituto de Diagnostico y Referencia Epidemiologicos (INDRE) | Claudia Wong-Arambula, Abril Rodriguez-Maldonado, Vanessa Rivero-Arredondo, Ariadna Medina-Benitez, Joaquin Quiroz-Mercado, Sergio Rangel-Guerrero, Natividad Cruz-Ortiz, Tatiana Nunez-Garcia, Gisela Barrera-Badillo, Lucia Hernandez-Rivas, Irma Lopez-Martinez, Ernesto Ramirez-Gonzalez. |
| EPI_ISL_2296093 | LESP Guanajuato | Instituto de Diagnostico y Referencia Epidemiologicos (INDRE) | Claudia Wong-Arambula, Abril Rodriguez-Maldonado, Vanessa Rivero-Arredondo, Ariadna Medina-Benitez, Joaquin Quiroz-Mercado, Sergio Rangel-Guerrero, Natividad Cruz-Ortiz, Tatiana Nunez-Garcia, Gisela Barrera-Badillo, Lucia Hernandez-Rivas, Irma Lopez-Martinez, Ernesto Ramirez-Gonzalez. |
| EPI_ISL_2296094 | LESP Morelos | Instituto de Diagnostico y Referencia Epidemiologicos (INDRE) | Claudia Wong-Arambula, Abril Rodriguez-Maldonado, Vanessa Rivero-Arredondo, Ariadna Medina-Benitez, Joaquin Quiroz-Mercado, Sergio Rangel-Guerrero, Natividad Cruz-Ortiz, Tatiana Nunez-Garcia, Gisela Barrera-Badillo, Lucia Hernandez-Rivas, Irma Lopez-Martinez, Ernesto Ramirez-Gonzalez. |
| EPI_ISL_2296095, EPI_ISL_2296096 | LESP Guanajuato | Instituto de Diagnostico y Referencia Epidemiologicos (INDRE) | Claudia Wong-Arambula, Abril Rodriguez-Maldonado, Vanessa Rivero-Arredondo, Ariadna Medina-Benitez, Joaquin Quiroz-Mercado, Sergio Rangel-Guerrero, Natividad Cruz-Ortiz, Tatiana Nunez-Garcia, Gisela Barrera-Badillo, Lucia Hernandez-Rivas, Irma Lopez-Martinez, Ernesto Ramirez-Gonzalez. |
| EPI_ISL_2296097 | LESP Hidalgo | Instituto de Diagnostico y Referencia Epidemiologicos (INDRE) | Claudia Wong-Arambula, Abril Rodriguez-Maldonado, Vanessa Rivero-Arredondo, Ariadna Medina-Benitez, Joaquin Quiroz-Mercado, Sergio Rangel-Guerrero, Natividad Cruz-Ortiz, Tatiana Nunez-Garcia, Gisela Barrera-Badillo, Lucia Hernandez-Rivas, Irma Lopez-Martinez, Ernesto Ramirez-Gonzalez. |
| EPI_ISL_2296098, EPI_ISL_2296099 | LESP Michoacan | Instituto de Diagnostico y Referencia Epidemiologicos (INDRE) | Claudia Wong-Arambula, Abril Rodriguez-Maldonado, Vanessa Rivero-Arredondo, Ariadna Medina-Benitez, Joaquin Quiroz-Mercado, Sergio Rangel-Guerrero, Natividad Cruz-Ortiz, Tatiana Nunez-Garcia, Gisela Barrera-Badillo, Lucia Hernandez-Rivas, Irma Lopez-Martinez, Ernesto Ramirez-Gonzalez. |
| EPI_ISL_2296100 | LESP Colima | Instituto de Diagnostico y Referencia Epidemiologicos (INDRE) | Claudia Wong-Arambula, Abril Rodriguez-Maldonado, Vanessa Rivero-Arredondo, Ariadna Medina-Benitez, Joaquin Quiroz-Mercado, Sergio Rangel-Guerrero, Natividad Cruz-Ortiz, Tatiana Nunez-Garcia, Gisela Barrera-Badillo, Lucia Hernandez-Rivas, Irma Lopez-Martinez, Ernesto Ramirez-Gonzalez. |
| EPI_ISL_2296101 | LESP Guanajuato | Instituto de Diagnostico y Referencia Epidemiologicos (INDRE) | Claudia Wong-Arambula, Abril Rodriguez-Maldonado, Vanessa Rivero-Arredondo, Ariadna Medina-Benitez, Joaquin Quiroz-Mercado, Sergio Rangel-Guerrero, Natividad Cruz-Ortiz, Tatiana Nunez-Garcia, Gisela Barrera-Badillo, Lucia Hernandez-Rivas, Irma Lopez-Martinez, Ernesto Ramirez-Gonzalez. |
| EPI_ISL_2296102, EPI_ISL_2296103 | LESP Sinaloa | Instituto de Diagnostico y Referencia Epidemiologicos (INDRE) | Claudia Wong-Arambula, Abril Rodriguez-Maldonado, Vanessa Rivero-Arredondo, Ariadna Medina-Benitez, Joaquin Quiroz-Mercado, Sergio Rangel-Guerrero, Natividad Cruz-Ortiz, Tatiana Nunez-Garcia, Gisela Barrera-Badillo, Lucia Hernandez-Rivas, Irma Lopez-Martinez, Ernesto Ramirez-Gonzalez. |
| EPI_ISL_2296104 | LESP Baja California | Instituto de Diagnostico y Referencia Epidemiologicos (INDRE) | Claudia Wong-Arambula, Abril Rodriguez-Maldonado, Vanessa Rivero-Arredondo, Ariadna Medina-Benitez, Joaquin Quiroz-Mercado, Sergio Rangel-Guerrero, Natividad Cruz-Ortiz, Tatiana Nunez-Garcia, Gisela Barrera-Badillo, Lucia Hernandez-Rivas, Irma Lopez-Martinez, Ernesto Ramirez-Gonzalez. |
| EPI_ISL_2296105 | LESP Sinaloa | Instituto de Diagnostico y Referencia Epidemiologicos (INDRE) | Claudia Wong-Arambula, Abril Rodriguez-Maldonado, Vanessa Rivero-Arredondo, Ariadna Medina-Benitez, Joaquin Quiroz-Mercado, Sergio Rangel-Guerrero, Natividad Cruz-Ortiz, Tatiana Nunez-Garcia, Gisela Barrera-Badillo, Lucia Hernandez-Rivas, Irma Lopez-Martinez, Ernesto Ramirez-Gonzalez. |
| EPI_ISL_2296106, EPI_ISL_2296107 | LESP Guanajuato | Instituto de Diagnostico y Referencia Epidemiologicos (INDRE) | Claudia Wong-Arambula, Abril Rodriguez-Maldonado, Vanessa Rivero-Arredondo, Ariadna Medina-Benitez, Joaquin Quiroz-Mercado, Sergio Rangel-Guerrero, Natividad Cruz-Ortiz, Tatiana Nunez-Garcia, Gisela Barrera-Badillo, Lucia Hernandez-Rivas, Irma Lopez-Martinez, Ernesto Ramirez-Gonzalez. |
| EPI_ISL_2296108 | LESP Yucatan | Instituto de Diagnostico y Referencia Epidemiologicos (INDRE) | Claudia Wong-Arambula, Abril Rodriguez-Maldonado, Vanessa Rivero-Arredondo, Ariadna Medina-Benitez, Joaquin Quiroz-Mercado, Sergio Rangel-Guerrero, Natividad Cruz-Ortiz, Tatiana Nunez-Garcia, Gisela Barrera-Badillo, Lucia Hernandez-Rivas, Irma Lopez-Martinez, Ernesto Ramirez-Gonzalez. |
| EPI_ISL_2296109 | LESP Jalisco | Instituto de Diagnostico y Referencia Epidemiologicos (INDRE) | Claudia Wong-Arambula, Abril Rodriguez-Maldonado, Vanessa Rivero-Arredondo, Ariadna Medina-Benitez, Joaquin Quiroz-Mercado, Sergio Rangel-Guerrero, Natividad Cruz-Ortiz, Tatiana Nunez-Garcia, Gisela Barrera-Badillo, Lucia Hernandez-Rivas, Irma Lopez-Martinez, Ernesto Ramirez-Gonzalez. |
| EPI_ISL_2296110 | LESP Ciudad de Mexico | Instituto de Diagnostico y Referencia Epidemiologicos (INDRE) | Claudia Wong-Arambula, Abril Rodriguez-Maldonado, Vanessa Rivero-Arredondo, Ariadna Medina-Benitez, Joaquin Quiroz-Mercado, Sergio Rangel-Guerrero, Natividad Cruz-Ortiz, Tatiana Nunez-Garcia, Gisela Barrera-Badillo, Lucia Hernandez-Rivas, Irma Lopez-Martinez, Ernesto Ramirez-Gonzalez. |
| EPI_ISL_2296381, EPI_ISL_2296382, EPI_ISL_2296383, EPI_ISL_2296384, EPI_ISL_2296385, EPI_ISL_2296386, EPI_ISL_2296387, EPI_ISL_2296388, EPI_ISL_2296389, EPI_ISL_2296390, EPI_ISL_2296391, EPI_ISL_2296392, EPI_ISL_2296393, EPI_ISL_2296394, EPI_ISL_2296395, EPI_ISL_2296396, EPI_ISL_2296397, EPI_ISL_2296398, EPI_ISL_2296399, EPI_ISL_2296400, EPI_ISL_2296401, EPI_ISL_2296402, EPI_ISL_2296403, EPI_ISL_2296404, EPI_ISL_2296405, EPI_ISL_2296406, EPI_ISL_2296407, EPI_ISL_2296408, EPI_ISL_2296409, EPI_ISL_2296410, EPI_ISL_2296411, EPI_ISL_2296412, EPI_ISL_2296413, EPI_ISL_2296414, EPI_ISL_2296415, EPI_ISL_2296416, EPI_ISL_2296417, EPI_ISL_2296418, EPI_ISL_2296419, EPI_ISL_2296420, EPI_ISL_2296421, EPI_ISL_2296422, EPI_ISL_2296423, EPI_ISL_2296424, EPI_ISL_2296425, EPI_ISL_2296426, EPI_ISL_2296427, EPI_ISL_2296428, EPI_ISL_2296429, EPI_ISL_2296430, EPI_ISL_2296431, EPI_ISL_2296432, EPI_ISL_2296433, EPI_ISL_2296434, EPI_ISL_2296435, EPI_ISL_2296436, EPI_ISL_2296437, EPI_ISL_2296438, EPI_ISL_2296439, EPI_ISL_2296440, EPI_ISL_2296442, EPI_ISL_2296443, EPI_ISL_2296444, EPI_ISL_2296445, EPI_ISL_2296446, EPI_ISL_2296447, EPI_ISL_2296448, EPI_ISL_2296449, EPI_ISL_2296450, EPI_ISL_2296451, EPI_ISL_2296452, EPI_ISL_2296453, EPI_ISL_2296454, EPI_ISL_2296455, EPI_ISL_2296456, EPI_ISL_2296457, EPI_ISL_2296458, EPI_ISL_2296459, EPI_ISL_2296460, EPI_ISL_2296461, EPI_ISL_2296462, EPI_ISL_2296463, EPI_ISL_2296464, EPI_ISL_2296465, EPI_ISL_2296466 | | | |
| see above | Yale Clinical Virology Lab | Grubaugh Lab - Yale School of Public Health | Joseph Fauver, Mallery Breban, Isabel Ott, Tara Alpert, Mary Petrone, Anderson Brito, Chantal Vogels, Annie Watkins, Chaney Kalinich, Jessica Rothman, Marie L. Landry, Nathan Grubaugh |
| EPI_ISL_2296467, EPI_ISL_2296468, EPI_ISL_2296469, EPI_ISL_2296470 | Yale Pathology Lab | Grubaugh Lab - Yale School of Public Health | Joseph Fauver, Mallery Breban, Isabel Ott, Tara Alpert, Mary Petrone, Anderson Brito, Chantal Vogels, Annie Watkins, Chaney Kalinich, Jessica Rothman, Jianhui Wang, Chen Liu, Pei Hui, Nathan Grubaugh |
| EPI_ISL_2296473, EPI_ISL_2296474, EPI_ISL_2296475, EPI_ISL_2296476, EPI_ISL_2296477 | Minnesota Department of Health, Public Health Laboratory | Minnesota Department of Health, Public Health Laboratory | Alexandra Lorentz, Jacob Garfin, Matt Plumb, and Xiong Wang |
| EPI_ISL_2296478 | Centracare Laboratory Services | Minnesota Department of Health, Public Health Laboratory | Alexandra Lorentz, Jacob Garfin, Matt Plumb, and Xiong Wang |
| EPI_ISL_2296479, EPI_ISL_2296480, EPI_ISL_2296481, EPI_ISL_2296482, EPI_ISL_2296483, EPI_ISL_2296484, EPI_ISL_2296485, EPI_ISL_2296486, EPI_ISL_2296487, EPI_ISL_2296488, EPI_ISL_2296489, EPI_ISL_2296490, EPI_ISL_2296491, EPI_ISL_2296492, EPI_ISL_2296493, EPI_ISL_2296494, EPI_ISL_2296496, EPI_ISL_2296497, EPI_ISL_2296498 | | | |
| see above | Minnesota Department of Health, Public Health Laboratory | Minnesota Department of Health, Public Health Laboratory | Alexandra Lorentz, Jacob Garfin, Matt Plumb, and Xiong Wang |
| EPI_ISL_2296499, EPI_ISL_2296500, EPI_ISL_2296501 | Centracare Laboratory Services | Minnesota Department of Health, Public Health Laboratory | Alexandra Lorentz, Jacob Garfin, Matt Plumb, and Xiong Wang |
| EPI_ISL_2296503, EPI_ISL_2296504, EPI_ISL_2296505 | Minnesota Department of Health, Public Health Laboratory | Minnesota Department of Health, Public Health Laboratory | Alexandra Lorentz, Jacob Garfin, Matt Plumb, and Xiong Wang |
| EPI_ISL_2296506, EPI_ISL_2296507, EPI_ISL_2296508, EPI_ISL_2296509, EPI_ISL_2296510, EPI_ISL_2296511, EPI_ISL_2296512, EPI_ISL_2296513, EPI_ISL_2296514, EPI_ISL_2296515, EPI_ISL_2296516, EPI_ISL_2296517, EPI_ISL_2296518, EPI_ISL_2296519, EPI_ISL_2296520 | | | |
| see above | Centracare Laboratory Services | Minnesota Department of Health, Public Health Laboratory | Alexandra Lorentz, Jacob Garfin, Matt Plumb, and Xiong Wang |
| EPI_ISL_2296521, EPI_ISL_2296522, EPI_ISL_2296523, EPI_ISL_2296524 | Premier Medical Lab | Minnesota Department of Health, Public Health Laboratory | Alexandra Lorentz, Jacob Garfin, Matt Plumb, and Xiong Wang |
| EPI_ISL_2298008, EPI_ISL_2298620 | Ohio Department of Health Laboratory | Ohio Department of Health Laboratory | Holmes, Jennifer; Eric Brandt, Keoni Omura, Glen McGillivary, Caitlin McDonnell, Jade Mowery, Stephanie Mccracken, Tyler Payne, Kirtana Ramadugu, Erica Leasure, Brent Lee, Kelsey Florek, Heather Blankenship, Allison Black, Quanta Brown, and Tammy Bannerman |
| EPI_ISL_2299296, EPI_ISL_2299297, EPI_ISL_2299299, EPI_ISL_2299301, EPI_ISL_2299303, EPI_ISL_2299304, EPI_ISL_2299306, EPI_ISL_2299307, EPI_ISL_2299309, EPI_ISL_2299311, EPI_ISL_2299313, EPI_ISL_2299315, EPI_ISL_2299317, EPI_ISL_2299318, EPI_ISL_2299320, EPI_ISL_2299322, EPI_ISL_2299323, EPI_ISL_2299326, EPI_ISL_2299328, EPI_ISL_2299330, EPI_ISL_2299331, EPI_ISL_2299332, EPI_ISL_2299334, EPI_ISL_2299336, EPI_ISL_2299338, EPI_ISL_2299339, EPI_ISL_2299341, EPI_ISL_2299343, EPI_ISL_2299345, EPI_ISL_2299347, EPI_ISL_2299349, EPI_ISL_2299350, EPI_ISL_2299351, EPI_ISL_2299353, EPI_ISL_2299355, EPI_ISL_2299356, EPI_ISL_2299358, EPI_ISL_2299359, EPI_ISL_2299361, EPI_ISL_2299362, EPI_ISL_2299364, EPI_ISL_2299365, EPI_ISL_2299366, EPI_ISL_2299368, EPI_ISL_2299370, EPI_ISL_2299371, EPI_ISL_2299373, EPI_ISL_2299375, EPI_ISL_2299377, EPI_ISL_2299379, EPI_ISL_2299380, EPI_ISL_2299382, EPI_ISL_2299384 | | | |
| see above | UW Virology Lab | UW Virology Lab | Pavitra Roychoudhury, Hong Xie, Lasata Shrestha, Tien V. Nguyen, Shah Mohamed Bakhash, Michelle Lin, Noah R. Baker, Sean Ellis, Meei-Li Huang, Keith R Jerome, Alexander Greninger |
| EPI_ISL_2299386 | Ohio Department of Health Laboratory | Ohio Department of Health Laboratory | Holmes, Jennifer; Eric Brandt, Keoni Omura, Glen McGillivary, Caitlin McDonnell, Jade Mowery, Stephanie Mccracken, Tyler Payne, Kirtana Ramadugu, Erica Leasure, Brent Lee, Kelsey Florek, Heather Blankenship, Allison Black, Quanta Brown, and Tammy Bannerman |
| EPI_ISL_2299387, EPI_ISL_2299388, EPI_ISL_2299390, EPI_ISL_2299392, EPI_ISL_2299394, EPI_ISL_2299395, EPI_ISL_2299396, EPI_ISL_2299398, EPI_ISL_2299400, EPI_ISL_2299402, EPI_ISL_2299404, EPI_ISL_2299405, EPI_ISL_2299410, EPI_ISL_2299412, EPI_ISL_2299414, EPI_ISL_2299416, EPI_ISL_2299417, EPI_ISL_2299419, EPI_ISL_2299421, EPI_ISL_2299423, EPI_ISL_2299425, EPI_ISL_2299427, EPI_ISL_2299428, EPI_ISL_2299429, EPI_ISL_2299431, EPI_ISL_2299434, EPI_ISL_2299435, EPI_ISL_2299437, EPI_ISL_2299439, EPI_ISL_2299441, EPI_ISL_2299443, EPI_ISL_2299445, EPI_ISL_2299446, EPI_ISL_2299448, EPI_ISL_2299449, EPI_ISL_2299451, EPI_ISL_2299452, EPI_ISL_2299454, EPI_ISL_2299456, EPI_ISL_2299458, EPI_ISL_2299459, EPI_ISL_2299461, EPI_ISL_2299463, EPI_ISL_2299466, EPI_ISL_2299467, EPI_ISL_2299469, EPI_ISL_2299470, EPI_ISL_2299471, EPI_ISL_2299472, EPI_ISL_2299474, EPI_ISL_2299476, EPI_ISL_2299480, EPI_ISL_2299482, EPI_ISL_2299483, EPI_ISL_2299485, EPI_ISL_2299487, EPI_ISL_2299488, EPI_ISL_2299490, EPI_ISL_2299492, EPI_ISL_2299494, EPI_ISL_2299495, EPI_ISL_2299497, EPI_ISL_2299499, EPI_ISL_2299500, EPI_ISL_2299502, EPI_ISL_2299506, EPI_ISL_2299510, EPI_ISL_2299511, EPI_ISL_2299513, EPI_ISL_2299514, EPI_ISL_2299516, EPI_ISL_2299518, EPI_ISL_2299519, EPI_ISL_2299521, EPI_ISL_2299522 | | | |
| see above | UW Virology Lab | UW Virology Lab | Pavitra Roychoudhury, Hong Xie, Lasata Shrestha, Tien V. Nguyen, Shah Mohamed Bakhash, Michelle Lin, Noah R. Baker, Sean Ellis, Meei-Li Huang, Keith R Jerome, Alexander Greninger |
| EPI_ISL_2300099 | Ohio Department of Health Laboratory | Ohio Department of Health Laboratory | Holmes, Jennifer; Eric Brandt, Keoni Omura, Glen McGillivary, Caitlin McDonnell, Jade Mowery, Stephanie Mccracken, Tyler Payne, Kirtana Ramadugu, Erica Leasure, Brent Lee, Kelsey Florek, Heather Blankenship, Allison Black, Quanta Brown, and Tammy Bannerman |
| EPI_ISL_2300283, EPI_ISL_2300286, EPI_ISL_2300289, EPI_ISL_2300291, EPI_ISL_2300295, EPI_ISL_2300297, EPI_ISL_2300298, EPI_ISL_2300300, EPI_ISL_2300302, EPI_ISL_2300304, EPI_ISL_2300305, EPI_ISL_2300308, EPI_ISL_2300314, EPI_ISL_2300317, EPI_ISL_2300323, EPI_ISL_2300328, EPI_ISL_2300331, EPI_ISL_2300334, EPI_ISL_2300337, EPI_ISL_2300339, EPI_ISL_2300342, EPI_ISL_2300345, EPI_ISL_2300351, EPI_ISL_2300353, EPI_ISL_2300355, EPI_ISL_2300357, EPI_ISL_2300360, EPI_ISL_2300361, EPI_ISL_2300364, EPI_ISL_2300367, EPI_ISL_2300370, EPI_ISL_2300374, EPI_ISL_2300379, EPI_ISL_2300381, EPI_ISL_2300384, EPI_ISL_2300387, EPI_ISL_2300389, EPI_ISL_2300392, EPI_ISL_2300394, EPI_ISL_2300396, EPI_ISL_2300398, EPI_ISL_2300401, EPI_ISL_2300404, EPI_ISL_2300407, EPI_ISL_2300414, EPI_ISL_2300419, EPI_ISL_2300422, EPI_ISL_2300429, EPI_ISL_2300431, EPI_ISL_2300432, EPI_ISL_2300433, EPI_ISL_2300436, EPI_ISL_2300439, EPI_ISL_2300441, EPI_ISL_2300442, EPI_ISL_2300446, EPI_ISL_2300448, EPI_ISL_2300451, EPI_ISL_2300453, EPI_ISL_2300455, EPI_ISL_2300459, EPI_ISL_2300461, EPI_ISL_2300462, EPI_ISL_2300465, EPI_ISL_2300467, EPI_ISL_2300472, EPI_ISL_2300476, EPI_ISL_2300478, EPI_ISL_2300482, EPI_ISL_2300483, EPI_ISL_2300486, EPI_ISL_2300491, EPI_ISL_2300496, EPI_ISL_2300498, EPI_ISL_2300501, EPI_ISL_2300504, EPI_ISL_2300508, EPI_ISL_2300510, EPI_ISL_2300513, EPI_ISL_2300516, EPI_ISL_2300519, EPI_ISL_2300523, EPI_ISL_2300525, EPI_ISL_2300527, EPI_ISL_2300529, EPI_ISL_2300531, EPI_ISL_2300533, EPI_ISL_2300536, EPI_ISL_2300540, EPI_ISL_2300541, EPI_ISL_2300543, EPI_ISL_2300545, EPI_ISL_2300547, EPI_ISL_2300551, EPI_ISL_2300554, EPI_ISL_2300556, EPI_ISL_2300559, EPI_ISL_2300561, EPI_ISL_2300562, EPI_ISL_2300565, EPI_ISL_2300567, EPI_ISL_2300570, EPI_ISL_2300573, EPI_ISL_2300575, EPI_ISL_2300577, EPI_ISL_2300580, EPI_ISL_2300582, EPI_ISL_2300585, EPI_ISL_2300587, EPI_ISL_2300590, EPI_ISL_2300593, EPI_ISL_2300597, EPI_ISL_2300604, EPI_ISL_2300606, EPI_ISL_2300609, EPI_ISL_2300611, EPI_ISL_2300614, EPI_ISL_2300615, EPI_ISL_2300617, EPI_ISL_2300620, EPI_ISL_2300623, EPI_ISL_2300626, EPI_ISL_2300631 | | | |
| see above | Illinois Department of Public Health | Illinois Department of Public Health - Chicago Lab | Vineet K. Dhiman, Ira Heimler, Joel Price |
| EPI_ISL_2300671, EPI_ISL_2300673, EPI_ISL_2300677, EPI_ISL_2300679, EPI_ISL_2300681, EPI_ISL_2300685, EPI_ISL_2300688, EPI_ISL_2300690, EPI_ISL_2300692, EPI_ISL_2300693, EPI_ISL_2300694, EPI_ISL_2300696, EPI_ISL_2300699, EPI_ISL_2300701, EPI_ISL_2300703, EPI_ISL_2300706, EPI_ISL_2300709, EPI_ISL_2300712 | | | |
| see above | UW Virology Lab | UW Virology Lab | Pavitra Roychoudhury, Hong Xie, Lasata Shrestha, Tien V. Nguyen, Shah Mohamed Bakhash, Michelle Lin, Noah R. Baker, Sean Ellis, Meei-Li Huang, Keith R Jerome, Alexander Greninger |
| EPI_ISL_2301431, EPI_ISL_2301432, EPI_ISL_2301436, EPI_ISL_2301437, EPI_ISL_2301438, EPI_ISL_2301439, EPI_ISL_2301440, EPI_ISL_2301441, EPI_ISL_2301442, EPI_ISL_2301443, EPI_ISL_2301444, EPI_ISL_2301445, EPI_ISL_2301446, EPI_ISL_2301447, EPI_ISL_2301448, EPI_ISL_2301449, EPI_ISL_2301450, EPI_ISL_2301451, EPI_ISL_2301452, EPI_ISL_2301453, EPI_ISL_2301454, EPI_ISL_2301455, EPI_ISL_2301456, EPI_ISL_2301457, EPI_ISL_2301458, EPI_ISL_2301459, EPI_ISL_2301460, EPI_ISL_2301461, EPI_ISL_2301462, EPI_ISL_2301463, EPI_ISL_2301464, EPI_ISL_2301465, EPI_ISL_2301466, EPI_ISL_2301467, EPI_ISL_2301468, EPI_ISL_2301469, EPI_ISL_2301470 | | | |
| see above | WVT Laboratory, Corvallis, Oregon | OSU Center for Genome Research and Biocomputing | Oregon State University TRACE Project |
| EPI_ISL_2303998, EPI_ISL_2303999, EPI_ISL_2304000, EPI_ISL_2304001, EPI_ISL_2304002, EPI_ISL_2304003, EPI_ISL_2304004, EPI_ISL_2304005, EPI_ISL_2304006, EPI_ISL_2304007, EPI_ISL_2304008, EPI_ISL_2304009, EPI_ISL_2304010, EPI_ISL_2304011, EPI_ISL_2304012, EPI_ISL_2304013, EPI_ISL_2304014, EPI_ISL_2304015, EPI_ISL_2304016, EPI_ISL_2304017, EPI_ISL_2304022, EPI_ISL_2304023, EPI_ISL_2304024, EPI_ISL_2304025, EPI_ISL_2304026, EPI_ISL_2304027, EPI_ISL_2304028 | | | |

| | | | |
|---|---|---|---|
| see above | University of Oregon COVID-19 MAP Laboratory | University of Oregon Genomics and Cell Charactericzation Core Facility (GC3F) | Douglas Turnbull, Ariana White, Jeff Bishop, Jason Carriere, Jason Sydes, Peter Batzel |
| EPI_ISL_2304192, EPI_ISL_2304193, EPI_ISL_2304194, EPI_ISL_2304195, EPI_ISL_2304196, EPI_ISL_2304197, EPI_ISL_2304198, EPI_ISL_2304199, EPI_ISL_2304200, EPI_ISL_2304202, EPI_ISL_2304203, EPI_ISL_2304205, EPI_ISL_2304206, EPI_ISL_2304207, EPI_ISL_2304208, EPI_ISL_2304210, EPI_ISL_2304211, EPI_ISL_2304213, EPI_ISL_2304215, EPI_ISL_2304216, EPI_ISL_2304217, EPI_ISL_2304219, EPI_ISL_2304220, EPI_ISL_2304221, EPI_ISL_2304222, EPI_ISL_2304223, EPI_ISL_2304224, EPI_ISL_2304225, EPI_ISL_2304226, EPI_ISL_2304227, EPI_ISL_2304228, EPI_ISL_2304229, EPI_ISL_2304230, EPI_ISL_2304231, EPI_ISL_2304232, EPI_ISL_2304233, EPI_ISL_2304234, EPI_ISL_2304235, EPI_ISL_2304236, EPI_ISL_2304237, EPI_ISL_2304238, EPI_ISL_2304240, EPI_ISL_2304241, EPI_ISL_2304242, EPI_ISL_2304243, EPI_ISL_2304245, EPI_ISL_2304246, EPI_ISL_2304247, EPI_ISL_2304248, EPI_ISL_2304249, EPI_ISL_2304250, EPI_ISL_2304251, EPI_ISL_2304252, EPI_ISL_2304253, EPI_ISL_2304254, EPI_ISL_2304255, EPI_ISL_2304256, EPI_ISL_2304257, EPI_ISL_2304258, EPI_ISL_2304259, EPI_ISL_2304260, EPI_ISL_2304261, EPI_ISL_2304262, EPI_ISL_2304263, EPI_ISL_2304264, EPI_ISL_2304265, EPI_ISL_2304266, EPI_ISL_2304267, EPI_ISL_2304268, EPI_ISL_2304269, EPI_ISL_2304270, EPI_ISL_2304271 | | | |
| see above | Missouri State Public Health Laboratory | Missouri State Public Health Laboratory | Matthew Sinn, Joshua Barry, Ashley New |
| EPI_ISL_2304307, EPI_ISL_2304308 | Alameda County Public Health Department | Alameda County Public Health Department | Kristina Hsieh, Maria Elma, Kevin Libuit |
| EPI_ISL_2304309 | Alameda County Public Health Department | Alameda County Public Health Department | Kristina Hsieh, Maria Elma, Kevin Libuit |
| EPI_ISL_2304310, EPI_ISL_2304311, EPI_ISL_2304312, EPI_ISL_2304313, EPI_ISL_2304314, EPI_ISL_2304315, EPI_ISL_2304316 | Alameda County Public Health Department | Alameda County Public Health Department | Kristina Hsieh, Maria Elma, Kevin Libuit |
| EPI_ISL_2304317, EPI_ISL_2304318, EPI_ISL_2304319, EPI_ISL_2304320, EPI_ISL_2304321, EPI_ISL_2304322, EPI_ISL_2304323, EPI_ISL_2304324, EPI_ISL_2304325, EPI_ISL_2304326, EPI_ISL_2304327 | | | |
| see above | Alameda County Public Health Department | Alameda County Public Health Department | Kristina Hsieh, Maria Elma, Kevin Libuit |
| EPI_ISL_2304328, EPI_ISL_2304329, EPI_ISL_2304331, EPI_ISL_2304332, EPI_ISL_2304333, EPI_ISL_2304334, EPI_ISL_2304335, EPI_ISL_2304336 | Alameda County Public Health Department | Alameda County Public Health Department | Kristna Hsieh, Maria Elma, Kevin Libuit |
| EPI_ISL_2304337 | Alameda County Public Health Department | Alameda County Public Health Department | Kristina Hsieh, Maria Elma, Kevin Libuit |
| EPI_ISL_2304338 | Alameda County Public Health Department | Alameda County Public Health Department | Kristna Hsieh, Maria Elma, Kevin Libuit |
| EPI_ISL_2304339 | Alameda County Public Health Department | Alameda County Public Health Department | Kristina Hsieh, Maria Elma, Kevin Libuit |
| EPI_ISL_2304340 | Alameda County Public Health Department | Alameda County Public Health Department | Kristina Hsieh, Maria Elma, Kevin Libuit |
| EPI_ISL_2304341, EPI_ISL_2304342, EPI_ISL_2304343, EPI_ISL_2304344 | Alameda County Public Health Department | Alameda County Public Health Department | Kristna Hsieh, Maria Elma, Kevin Libuit |
| EPI_ISL_2304345, EPI_ISL_2304346, EPI_ISL_2304347, EPI_ISL_2304348, EPI_ISL_2304349, EPI_ISL_2304350, EPI_ISL_2304351, EPI_ISL_2304352, EPI_ISL_2304353, EPI_ISL_2304354, EPI_ISL_2304355, EPI_ISL_2304356, EPI_ISL_2304357, EPI_ISL_2304358, EPI_ISL_2304359, EPI_ISL_2304360, EPI_ISL_2304361, EPI_ISL_2304362, EPI_ISL_2304363, EPI_ISL_2304364, EPI_ISL_2304366, EPI_ISL_2304367, EPI_ISL_2304368, EPI_ISL_2304369, EPI_ISL_2304370, EPI_ISL_2304371, EPI_ISL_2304372, EPI_ISL_2304373, EPI_ISL_2304374, EPI_ISL_2304375, EPI_ISL_2304376, EPI_ISL_2304377, EPI_ISL_2304378, EPI_ISL_2304379, EPI_ISL_2304380, EPI_ISL_2304381, EPI_ISL_2304382, EPI_ISL_2304383, EPI_ISL_2304384, EPI_ISL_2304385, EPI_ISL_2304386, EPI_ISL_2304387, EPI_ISL_2304388, EPI_ISL_2304389, EPI_ISL_2304390, EPI_ISL_2304391, EPI_ISL_2304392, EPI_ISL_2304393, EPI_ISL_2304394, EPI_ISL_2304395, EPI_ISL_2304396, EPI_ISL_2304397, EPI_ISL_2304398, EPI_ISL_2304399, EPI_ISL_2304400, EPI_ISL_2304401, EPI_ISL_2304402, EPI_ISL_2304403, EPI_ISL_2304404, EPI_ISL_2304405, EPI_ISL_2304406, EPI_ISL_2304407, EPI_ISL_2304408, EPI_ISL_2304409, EPI_ISL_2304410, EPI_ISL_2304411, EPI_ISL_2304412, EPI_ISL_2304413, EPI_ISL_2304414, EPI_ISL_2304416, EPI_ISL_2304417, EPI_ISL_2304418, EPI_ISL_2304419, EPI_ISL_2304420, EPI_ISL_2304421, EPI_ISL_2304422, EPI_ISL_2304423, EPI_ISL_2304424, EPI_ISL_2304425, EPI_ISL_2304426, EPI_ISL_2304427, EPI_ISL_2304428, EPI_ISL_2304429, EPI_ISL_2304430, EPI_ISL_2304431, EPI_ISL_2304432, EPI_ISL_2304433, EPI_ISL_2304434, EPI_ISL_2304435, EPI_ISL_2304436, EPI_ISL_2304437, EPI_ISL_2304438, EPI_ISL_2304439, EPI_ISL_2304440, EPI_ISL_2304441, EPI_ISL_2304442, EPI_ISL_2304443, EPI_ISL_2304444, EPI_ISL_2304445, EPI_ISL_2304446, EPI_ISL_2304447, EPI_ISL_2304448, EPI_ISL_2304449, EPI_ISL_2304450, EPI_ISL_2304451, EPI_ISL_2304452, EPI_ISL_2304453, EPI_ISL_2304454, EPI_ISL_2304455, EPI_ISL_2304456, EPI_ISL_2304457, EPI_ISL_2304458, EPI_ISL_2304459, EPI_ISL_2304460 | | | |
| see above | Indiana Animal Disease Diagnostic Laboratory | Carpi Laboratory - Purdue University | Jack Dorman, Ilinca I Ciubotariu, Nicole M Perry, Jobin J Kattoor, Lev Gorenstein, Abebe A Fola, G Kenitra Hendrix, Rebecca P Wilkes, Giovanna Carpi |
| EPI_ISL_2304472, EPI_ISL_2304473, EPI_ISL_2304474, EPI_ISL_2304475, EPI_ISL_2304477, EPI_ISL_2304478, EPI_ISL_2304479, EPI_ISL_2304480, EPI_ISL_2304481, EPI_ISL_2304482, EPI_ISL_2304483, EPI_ISL_2304484, EPI_ISL_2304485, EPI_ISL_2304487, EPI_ISL_2304489, EPI_ISL_2304491, EPI_ISL_2304493, EPI_ISL_2304494, EPI_ISL_2304496, EPI_ISL_2304497, EPI_ISL_2304498, EPI_ISL_2304499 | | | |
| see above | IN State Department of Health Laboratory Services | IN State Department of Health Laboratory Services | Cassandra Campion, Jamie Yeadon, Brian Pope, Lixia Liu, Kyle Brownlee, Melissa Hindenlang, Mark Glazier |
| EPI_ISL_2304500, EPI_ISL_2304502, EPI_ISL_2304503, EPI_ISL_2304504, EPI_ISL_2304505, EPI_ISL_2304506, EPI_ISL_2304507, EPI_ISL_2304508, EPI_ISL_2304509, EPI_ISL_2304510, EPI_ISL_2304511, EPI_ISL_2304512, EPI_ISL_2304513, EPI_ISL_2304514, EPI_ISL_2304515, EPI_ISL_2304516, EPI_ISL_2304517, EPI_ISL_2304518, EPI_ISL_2304519, EPI_ISL_2304520, EPI_ISL_2304521, EPI_ISL_2304522, EPI_ISL_2304523, EPI_ISL_2304524, EPI_ISL_2304525, EPI_ISL_2304526, EPI_ISL_2304527, EPI_ISL_2304528, EPI_ISL_2304529, EPI_ISL_2304530, EPI_ISL_2304531, EPI_ISL_2304532, EPI_ISL_2304533, EPI_ISL_2304534, EPI_ISL_2304535, EPI_ISL_2304536, EPI_ISL_2304537, EPI_ISL_2304538, EPI_ISL_2304539, EPI_ISL_2304540, EPI_ISL_2304541, EPI_ISL_2304542, EPI_ISL_2304543, EPI_ISL_2304544, EPI_ISL_2304545, EPI_ISL_2304546, EPI_ISL_2304547, EPI_ISL_2304548, EPI_ISL_2304549, EPI_ISL_2304550, EPI_ISL_2304551, EPI_ISL_2304552, EPI_ISL_2304553, EPI_ISL_2304554, EPI_ISL_2304555, EPI_ISL_2304556, EPI_ISL_2304557, EPI_ISL_2304558, EPI_ISL_2304559, EPI_ISL_2304560, EPI_ISL_2304561, EPI_ISL_2304562, EPI_ISL_2304563, EPI_ISL_2304564, EPI_ISL_2304565, EPI_ISL_2304566, EPI_ISL_2304567, EPI_ISL_2304568, EPI_ISL_2304569, EPI_ISL_2304570, EPI_ISL_2304571, EPI_ISL_2304572, EPI_ISL_2304573, EPI_ISL_2304574, EPI_ISL_2304575, EPI_ISL_2304576, EPI_ISL_2304577, EPI_ISL_2304578, EPI_ISL_2304579, EPI_ISL_2304580, EPI_ISL_2304581, EPI_ISL_2304582, EPI_ISL_2304583, EPI_ISL_2304584, EPI_ISL_2304585, EPI_ISL_2304586, EPI_ISL_2304587, EPI_ISL_2304588, EPI_ISL_2304589, EPI_ISL_2304590, EPI_ISL_2304591, EPI_ISL_2304592, EPI_ISL_2304593, EPI_ISL_2304594, EPI_ISL_2304595, EPI_ISL_2304596, EPI_ISL_2304597, EPI_ISL_2304598, EPI_ISL_2304599, EPI_ISL_2304600, EPI_ISL_2304601, EPI_ISL_2304602, EPI_ISL_2304603, EPI_ISL_2304604, EPI_ISL_2304605, EPI_ISL_2304606, EPI_ISL_2304607, EPI_ISL_2304608, EPI_ISL_2304609, EPI_ISL_2304610, EPI_ISL_2304611, EPI_ISL_2304612, EPI_ISL_2304613, EPI_ISL_2304614, EPI_ISL_2304615, EPI_ISL_2304616, EPI_ISL_2304617, EPI_ISL_2304618, EPI_ISL_2304619, EPI_ISL_2304620, EPI_ISL_2304621, EPI_ISL_2304622, EPI_ISL_2304623, EPI_ISL_2304624, EPI_ISL_2304625, EPI_ISL_2304626, EPI_ISL_2304627, EPI_ISL_2304628, EPI_ISL_2304629, EPI_ISL_2304630, EPI_ISL_2304631, EPI_ISL_2304632, EPI_ISL_2304633, EPI_ISL_2304634, EPI_ISL_2304635, EPI_ISL_2304636, EPI_ISL_2304637, EPI_ISL_2304638, EPI_ISL_2304639, EPI_ISL_2304640, EPI_ISL_2304641, EPI_ISL_2304642, EPI_ISL_2304643, EPI_ISL_2304644, EPI_ISL_2304645, EPI_ISL_2304646, EPI_ISL_2304647, EPI_ISL_2304648, EPI_ISL_2304649, EPI_ISL_2304650, EPI_ISL_2304651, EPI_ISL_2304652, EPI_ISL_2304653, EPI_ISL_2304654, EPI_ISL_2304655, EPI_ISL_2304656, EPI_ISL_2304657, EPI_ISL_2304658, EPI_ISL_2304659, EPI_ISL_2304660, EPI_ISL_2304661, EPI_ISL_2304662, EPI_ISL_2304663, EPI_ISL_2304664, EPI_ISL_2304665, EPI_ISL_2304666, EPI_ISL_2304667, EPI_ISL_2304668, EPI_ISL_2304669, EPI_ISL_2304670, EPI_ISL_2304671, EPI_ISL_2304672, EPI_ISL_2304673, EPI_ISL_2304674, EPI_ISL_2304675, EPI_ISL_2304676, EPI_ISL_2304677, EPI_ISL_2304678, EPI_ISL_2304679, EPI_ISL_2304680, EPI_ISL_2304681, EPI_ISL_2304682, EPI_ISL_2304683, EPI_ISL_2304684, EPI_ISL_2304685, EPI_ISL_2304686, EPI_ISL_2304687, EPI_ISL_2304688, EPI_ISL_2304689, EPI_ISL_2304690, EPI_ISL_2304691, EPI_ISL_2304692, EPI_ISL_2304693, EPI_ISL_2304694, EPI_ISL_2304695, EPI_ISL_2304696, EPI_ISL_2304697, EPI_ISL_2304698, EPI_ISL_2304699, EPI_ISL_2304700, EPI_ISL_2304701, EPI_ISL_2304702, EPI_ISL_2304703, EPI_ISL_2304704, EPI_ISL_2304705, EPI_ISL_2304706, EPI_ISL_2304707, EPI_ISL_2304708, EPI_ISL_2304709, EPI_ISL_2304710, EPI_ISL_2304711, EPI_ISL_2304712, EPI_ISL_2304713, EPI_ISL_2304714, EPI_ISL_2304715, EPI_ISL_2304716, EPI_ISL_2304717, EPI_ISL_2304718, EPI_ISL_2304719, EPI_ISL_2304720, EPI_ISL_2304721, EPI_ISL_2304722, EPI_ISL_2304723, EPI_ISL_2304724, EPI_ISL_2304725, EPI_ISL_2304726, EPI_ISL_2304727, EPI_ISL_2304728, EPI_ISL_2304729, EPI_ISL_2304730, EPI_ISL_2304731, EPI_ISL_2304732, EPI_ISL_2304733, EPI_ISL_2304734, EPI_ISL_2304735, EPI_ISL_2304736, EPI_ISL_2304737, EPI_ISL_2304738, EPI_ISL_2304739, EPI_ISL_2304740, EPI_ISL_2304741, EPI_ISL_2304742, EPI_ISL_2304743, EPI_ISL_2304744, EPI_ISL_2304745, EPI_ISL_2304746, EPI_ISL_2304747, EPI_ISL_2304748, EPI_ISL_2304749, EPI_ISL_2304750, EPI_ISL_2304751, EPI_ISL_2304752, EPI_ISL_2304753, EPI_ISL_2304754, EPI_ISL_2304755, EPI_ISL_2304756, EPI_ISL_2304757, EPI_ISL_2304758, EPI_ISL_2304759, EPI_ISL_2304760, EPI_ISL_2304761, EPI_ISL_2304762, EPI_ISL_2304763, EPI_ISL_2304764, EPI_ISL_2304765, EPI_ISL_2304766, EPI_ISL_2304767, EPI_ISL_2304768, EPI_ISL_2304769, EPI_ISL_2304770, EPI_ISL_2304771, EPI_ISL_2304772, EPI_ISL_2304773, EPI_ISL_2304774, EPI_ISL_2304775, EPI_ISL_2304776, EPI_ISL_2304777, EPI_ISL_2304778, EPI_ISL_2304779, EPI_ISL_2304780, EPI_ISL_2304781, EPI_ISL_2304782, EPI_ISL_2304783, EPI_ISL_2304784, EPI_ISL_2304785, EPI_ISL_2304786, EPI_ISL_2304787, EPI_ISL_2304788, EPI_ISL_2304789, EPI_ISL_2304790, EPI_ISL_2304791, EPI_ISL_2304792, EPI_ISL_2304793, EPI_ISL_2304794, EPI_ISL_2304795, EPI_ISL_2304796, EPI_ISL_2304797, EPI_ISL_2304798, EPI_ISL_2304799 | | | |
| see above | Quest Diagnostics Incorporated | Centers for Disease Control and Prevention Division of Viral Diseases, Pathogen Discovery | Dakota Howard, Dhwani Batra, Peter W. Cook, Kara Moser, Adrian Paskey, Jason Caravas, Benjamin Rambo-Martin, Shatavia Morrison, Christopher Gulvick, Scott Sammons, Yvette Unoarumhi, Darlene Wagner, Matthew Schmerer, S. H. Rosenthal, A. Gerasimova, R. M. Kagan, B. Anderson, M. Hua, Y. Liu, L.E. Bernstein, K.E. Livingston, A. Perez, I. A. Shlyakhter, R. V. Rolando, R. Owen, P. Tanpaiboon, F. Lacbawan, Clinton R. Paden, Duncan MacCannell |
| EPI_ISL_2304800, EPI_ISL_2304801, EPI_ISL_2304802, EPI_ISL_2304803, EPI_ISL_2304804, EPI_ISL_2304806, EPI_ISL_2304807, EPI_ISL_2304808, EPI_ISL_2304811, EPI_ISL_2304812, EPI_ISL_2304813, EPI_ISL_2304814, EPI_ISL_2304815, EPI_ISL_2304816, EPI_ISL_2304817, EPI_ISL_2304818, EPI_ISL_2304819, EPI_ISL_2304820, EPI_ISL_2304821, EPI_ISL_2304823, EPI_ISL_2304824, EPI_ISL_2304825, EPI_ISL_2304827, EPI_ISL_2304828, EPI_ISL_2304829, EPI_ISL_2304830, EPI_ISL_2304831, EPI_ISL_2304832, EPI_ISL_2304833, EPI_ISL_2304834, EPI_ISL_2304835, EPI_ISL_2304836, EPI_ISL_2304837, EPI_ISL_2304839, EPI_ISL_2304840, EPI_ISL_2304841, EPI_ISL_2304842, EPI_ISL_2304843, EPI_ISL_2304844, EPI_ISL_2304845, EPI_ISL_2304846, EPI_ISL_2304848, EPI_ISL_2304849, EPI_ISL_2304851, EPI_ISL_2304852, EPI_ISL_2304853, EPI_ISL_2304854, EPI_ISL_2304856, EPI_ISL_2304857, EPI_ISL_2304858, EPI_ISL_2304859, EPI_ISL_2304861, EPI_ISL_2304862, EPI_ISL_2304863, EPI_ISL_2304864, EPI_ISL_2304865, EPI_ISL_2304866, EPI_ISL_2304867, EPI_ISL_2304868, EPI_ISL_2304870, EPI_ISL_2304871, EPI_ISL_2304872, EPI_ISL_2304873, EPI_ISL_2304875, EPI_ISL_2304876, EPI_ISL_2304879, EPI_ISL_2304880, EPI_ISL_2304881, EPI_ISL_2304882, EPI_ISL_2304883, EPI_ISL_2304884, EPI_ISL_2304885, EPI_ISL_2304886, EPI_ISL_2304887, EPI_ISL_2304888, EPI_ISL_2304889, EPI_ISL_2304890, EPI_ISL_2304891, EPI_ISL_2304892, EPI_ISL_2304893, EPI_ISL_2304894, EPI_ISL_2304895, EPI_ISL_2304896, EPI_ISL_2304897, EPI_ISL_2304898, EPI_ISL_2304899, EPI_ISL_2304900, EPI_ISL_2304901, EPI_ISL_2304902, EPI_ISL_2304903, EPI_ISL_2304904, EPI_ISL_2304905, EPI_ISL_2304906, EPI_ISL_2304907, EPI_ISL_2304908, EPI_ISL_2304909, EPI_ISL_2304910, EPI_ISL_2304911, EPI_ISL_2304912, EPI_ISL_2304914, EPI_ISL_2304915, EPI_ISL_2304916, EPI_ISL_2304917, EPI_ISL_2304918, EPI_ISL_2304919, EPI_ISL_2304920, EPI_ISL_2304922, EPI_ISL_2304923, EPI_ISL_2304924, EPI_ISL_2304925, EPI_ISL_2304926, EPI_ISL_2304927, EPI_ISL_2304928, EPI_ISL_2304929, EPI_ISL_2304930, EPI_ISL_2304931, EPI_ISL_2304932, EPI_ISL_2304933, EPI_ISL_2304934, EPI_ISL_2304935, EPI_ISL_2304936, EPI_ISL_2304937, EPI_ISL_2304938, EPI_ISL_2304940, EPI_ISL_2304942, EPI_ISL_2304943, EPI_ISL_2304947, EPI_ISL_2304948, EPI_ISL_2304949, EPI_ISL_2304950, EPI_ISL_2304951, EPI_ISL_2304952, EPI_ISL_2304953, EPI_ISL_2304954, EPI_ISL_2304955, EPI_ISL_2304956, EPI_ISL_2304957, EPI_ISL_2304958, EPI_ISL_2304959, EPI_ISL_2304960, EPI_ISL_2304961, EPI_ISL_2304962, EPI_ISL_2304963, EPI_ISL_2304964, EPI_ISL_2304965, EPI_ISL_2304966, EPI_ISL_2304967, EPI_ISL_2304970, EPI_ISL_2304971, EPI_ISL_2304974, EPI_ISL_2304976, EPI_ISL_2304977, EPI_ISL_2304978, EPI_ISL_2304979, EPI_ISL_2304980, EPI_ISL_2304981, EPI_ISL_2304982, EPI_ISL_2304983, EPI_ISL_2304984, EPI_ISL_2304986, EPI_ISL_2304987, EPI_ISL_2304988, EPI_ISL_2304989, EPI_ISL_2304990, EPI_ISL_2304991, EPI_ISL_2304992, EPI_ISL_2304993, EPI_ISL_2304994, EPI_ISL_2304995, EPI_ISL_2304996, EPI_ISL_2304997, EPI_ISL_2304998, EPI_ISL_2304999, EPI_ISL_2305000, EPI_ISL_2305001, EPI_ISL_2305002, EPI_ISL_2305004, EPI_ISL_2305005, EPI_ISL_2305007, EPI_ISL_2305008, EPI_ISL_2305010, EPI_ISL_2305011, EPI_ISL_2305012, EPI_ISL_2305013, EPI_ISL_2305014, EPI_ISL_2305015, EPI_ISL_2305016, EPI_ISL_2305018, EPI_ISL_2305019, EPI_ISL_2305021, EPI_ISL_2305022, EPI_ISL_2305023, EPI_ISL_2305024, EPI_ISL_2305026, EPI_ISL_2305027, EPI_ISL_2305028, EPI_ISL_2305029, EPI_ISL_2305030, EPI_ISL_2305034, EPI_ISL_2305036, EPI_ISL_2305037, EPI_ISL_2305038, EPI_ISL_2305039, EPI_ISL_2305040, EPI_ISL_2305041, EPI_ISL_2305042, EPI_ISL_2305044, EPI_ISL_2305045, EPI_ISL_2305046, EPI_ISL_2305047, EPI_ISL_2305048, EPI_ISL_2305049, EPI_ISL_2305050, EPI_ISL_2305051, EPI_ISL_2305052, EPI_ISL_2305054, EPI_ISL_2305055, EPI_ISL_2305056, EPI_ISL_2305057, EPI_ISL_2305058, EPI_ISL_2305059, EPI_ISL_2305060, EPI_ISL_2305061, EPI_ISL_2305062, EPI_ISL_2305063, EPI_ISL_2305065, EPI_ISL_2305066, EPI_ISL_2305067, EPI_ISL_2305068, EPI_ISL_2305069, EPI_ISL_2305070, EPI_ISL_2305071, EPI_ISL_2305072, EPI_ISL_2305073, EPI_ISL_2305074, EPI_ISL_2305075, EPI_ISL_2305076, EPI_ISL_2305077, EPI_ISL_2305079, EPI_ISL_2305080, EPI_ISL_2305081, EPI_ISL_2305082, EPI_ISL_2305083, EPI_ISL_2305084, EPI_ISL_2305086, EPI_ISL_2305087, EPI_ISL_2305088, EPI_ISL_2305089, EPI_ISL_2305092, EPI_ISL_2305093, EPI_ISL_2305095, EPI_ISL_2305096, EPI_ISL_2305097, EPI_ISL_2305098, EPI_ISL_2305099, EPI_ISL_2305100, EPI_ISL_2305101, EPI_ISL_2305102, EPI_ISL_2305103, EPI_ISL_2305106, EPI_ISL_2305107, EPI_ISL_2305108, EPI_ISL_2305110, EPI_ISL_2305111, EPI_ISL_2305112, EPI_ISL_2305113, EPI_ISL_2305115, EPI_ISL_2305117, EPI_ISL_2305118, EPI_ISL_2305119, EPI_ISL_2305120, EPI_ISL_2305121, EPI_ISL_2305122, EPI_ISL_2305123, EPI_ISL_2305124, EPI_ISL_2305125, EPI_ISL_2305126, EPI_ISL_2305128, EPI_ISL_2305129, EPI_ISL_2305131, EPI_ISL_2305132, EPI_ISL_2305133, EPI_ISL_2305134, EPI_ISL_2305135, EPI_ISL_2305137, EPI_ISL_2305138, EPI_ISL_2305139, EPI_ISL_2305140, EPI_ISL_2305141, EPI_ISL_2305142, EPI_ISL_2305143, EPI_ISL_2305144, EPI_ISL_2305145, EPI_ISL_2305146, EPI_ISL_2305147, EPI_ISL_2305149, EPI_ISL_2305150, EPI_ISL_2305151, EPI_ISL_2305152, EPI_ISL_2305153, EPI_ISL_2305154, EPI_ISL_2305156, EPI_ISL_2305157, EPI_ISL_2305158, EPI_ISL_2305161, EPI_ISL_2305162, EPI_ISL_2305163, EPI_ISL_2305164, EPI_ISL_2305165, EPI_ISL_2305166, EPI_ISL_2305167, EPI_ISL_2305169, EPI_ISL_2305170, EPI_ISL_2305171, EPI_ISL_2305172, EPI_ISL_2305173, EPI_ISL_2305174, EPI_ISL_2305175, EPI_ISL_2305176, EPI_ISL_2305177, EPI_ISL_2305178, EPI_ISL_2305179, EPI_ISL_2305180, EPI_ISL_2305182, EPI_ISL_2305183, EPI_ISL_2305184, EPI_ISL_2305185, EPI_ISL_2305186, EPI_ISL_2305188, EPI_ISL_2305190, EPI_ISL_2305191, EPI_ISL_2305192, EPI_ISL_2305193, EPI_ISL_2305194, EPI_ISL_2305195, EPI_ISL_2305196, EPI_ISL_2305198, EPI_ISL_2305199, EPI_ISL_2305200, EPI_ISL_2305201, EPI_ISL_2305202, EPI_ISL_2305204, EPI_ISL_2305205, EPI_ISL_2305206, EPI_ISL_2305207, EPI_ISL_2305208, EPI_ISL_2305209, EPI_ISL_2305210, EPI_ISL_2305212, EPI_ISL_2305213, EPI_ISL_2305215, EPI_ISL_2305216, EPI_ISL_2305218, EPI_ISL_2305219, EPI_ISL_2305220, EPI_ISL_2305221, EPI_ISL_2305222, EPI_ISL_2305223, EPI_ISL_2305224, EPI_ISL_2305225, EPI_ISL_2305226, EPI_ISL_2305227, EPI_ISL_2305230, EPI_ISL_2305231, EPI_ISL_2305232, EPI_ISL_2305233, EPI_ISL_2305234, EPI_ISL_2305235, EPI_ISL_2305236, EPI_ISL_2305238, EPI_ISL_2305240, EPI_ISL_2305241, EPI_ISL_2305242, EPI_ISL_2305243, EPI_ISL_2305244, EPI_ISL_2305246, EPI_ISL_2305247, EPI_ISL_2305249, EPI_ISL_2305250, EPI_ISL_2305251, EPI_ISL_2305252, EPI_ISL_2305253, EPI_ISL_2305255, EPI_ISL_2305256, EPI_ISL_2305257, EPI_ISL_2305258, EPI_ISL_2305261, EPI_ISL_2305262, EPI_ISL_2305263, EPI_ISL_2305264, EPI_ISL_2305267, EPI_ISL_2305268, EPI_ISL_2305269, EPI_ISL_2305270, EPI_ISL_2305271, EPI_ISL_2305272, EPI_ISL_2305273, EPI_ISL_2305274, EPI_ISL_2305275, EPI_ISL_2305276, EPI_ISL_2305278, EPI_ISL_2305279, EPI_ISL_2305280, EPI_ISL_2305281, EPI_ISL_2305282, EPI_ISL_2305283, EPI_ISL_2305284, EPI_ISL_2305285, EPI_ISL_2305287, EPI_ISL_2305290, EPI_ISL_2305291, EPI_ISL_2305292, EPI_ISL_2305293, EPI_ISL_2305294, EPI_ISL_2305295, EPI_ISL_2305296, EPI_ISL_2305297, EPI_ISL_2305298, EPI_ISL_2305299, EPI_ISL_2305301, EPI_ISL_2305303, EPI_ISL_2305304, EPI_ISL_2305305, EPI_ISL_2305307, EPI_ISL_2305309, EPI_ISL_2305310, EPI_ISL_2305311, EPI_ISL_2305312, EPI_ISL_2305313, EPI_ISL_2305314, EPI_ISL_2305316, EPI_ISL_2305317, EPI_ISL_2305318, EPI_ISL_2305319, EPI_ISL_2305320, EPI_ISL_2305321, EPI_ISL_2305322, EPI_ISL_2305323, EPI_ISL_2305324, EPI_ISL_2305325, EPI_ISL_2305326, EPI_ISL_2305327, EPI_ISL_2305328, EPI_ISL_2305329, EPI_ISL_2305330, EPI_ISL_2305331, EPI_ISL_2305332, EPI_ISL_2305333, EPI_ISL_2305335, EPI_ISL_2305336, EPI_ISL_2305337, EPI_ISL_2305338, EPI_ISL_2305339, EPI_ISL_2305340, EPI_ISL_2305341, EPI_ISL_2305342, EPI_ISL_2305343, EPI_ISL_2305344, EPI_ISL_2305345, EPI_ISL_2305346, EPI_ISL_2305347, EPI_ISL_2305348, EPI_ISL_2305349, EPI_ISL_2305350, EPI_ISL_2305351, EPI_ISL_2305352, EPI_ISL_2305353, EPI_ISL_2305354, EPI_ISL_2305355, EPI_ISL_2305357, EPI_ISL_2305358, EPI_ISL_2305359, EPI_ISL_2305360, EPI_ISL_2305361, EPI_ISL_2305362, EPI_ISL_2305363, EPI_ISL_2305364, EPI_ISL_2305365, EPI_ISL_2305366, EPI_ISL_2305367, EPI_ISL_2305368, EPI_ISL_2305369, EPI_ISL_2305372, EPI_ISL_2305373, EPI_ISL_2305375, EPI_ISL_2305376, EPI_ISL_2305377, EPI_ISL_2305380, EPI_ISL_2305381, EPI_ISL_2305384, EPI_ISL_2305387, EPI_ISL_2305388, EPI_ISL_2305389, EPI_ISL_2305390, EPI_ISL_2305391, EPI_ISL_2305392, EPI_ISL_2305394, EPI_ISL_2305395, EPI_ISL_2305396, EPI_ISL_2305397, EPI_ISL_2305398, EPI_ISL_2305399, EPI_ISL_2305401, EPI_ISL_2305402, EPI_ISL_2305403, EPI_ISL_2305405, EPI_ISL_2305408, EPI_ISL_2305409, EPI_ISL_2305411, EPI_ISL_2305413, EPI_ISL_2305415, EPI_ISL_2305416, EPI_ISL_2305417, EPI_ISL_2305418, EPI_ISL_2305420, EPI_ISL_2305422, EPI_ISL_2305423, EPI_ISL_2305424, EPI_ISL_2305425, EPI_ISL_2305426, EPI_ISL_2305435, EPI_ISL_2305436, EPI_ISL_2305438, EPI_ISL_2305439, EPI_ISL_2305442, EPI_ISL_2305443, EPI_ISL_2305445, EPI_ISL_2305446, EPI_ISL_2305449, EPI_ISL_2305450, EPI_ISL_2305452, EPI_ISL_2305453, EPI_ISL_2305456, EPI_ISL_2305457, EPI_ISL_2305460, EPI_ISL_2305462, EPI_ISL_2305463, EPI_ISL_2305464, EPI_ISL_2305465, EPI_ISL_2305467, EPI_ISL_2305471, EPI_ISL_2305472, EPI_ISL_2305474, EPI_ISL_2305478, EPI_ISL_2305483, EPI_ISL_2305484, EPI_ISL_2305485, EPI_ISL_2305486, EPI_ISL_2305488, EPI_ISL_2305491, EPI_ISL_2305492, EPI_ISL_2305493, EPI_ISL_2305495, EPI_ISL_2305497, EPI_ISL_2305498, EPI_ISL_2305499, EPI_ISL_2305500, EPI_ISL_2305501, EPI_ISL_2305502, EPI_ISL_2305505, EPI_ISL_2305506, EPI_ISL_2305508, EPI_ISL_2305509, EPI_ISL_2305511, EPI_ISL_2305512, EPI_ISL_2305513, EPI_ISL_2305514, EPI_ISL_2305515, EPI_ISL_2305516, EPI_ISL_2305517, EPI_ISL_2305519, EPI_ISL_2305521, EPI_ISL_2305522, EPI_ISL_2305523, EPI_ISL_2305524, EPI_ISL_2305526, EPI_ISL_2305527, EPI_ISL_2305528, EPI_ISL_2305530, EPI_ISL_2305533, EPI_ISL_2305534, EPI_ISL_2305537, EPI_ISL_2305540, EPI_ISL_2305541, EPI_ISL_2305542, EPI_ISL_2305543, EPI_ISL_2305544, EPI_ISL_2305547, EPI_ISL_2305550, EPI_ISL_2305552, EPI_ISL_2305553, EPI_ISL_2305554, EPI_ISL_2305556, EPI_ISL_2305558, EPI_ISL_2305559, EPI_ISL_2305560, EPI_ISL_2305562, EPI_ISL_2305564, EPI_ISL_2305565, EPI_ISL_2305566, EPI_ISL_2305567, EPI_ISL_2305568, EPI_ISL_2305572, EPI_ISL_2305573, EPI_ISL_2305574, EPI_ISL_2305575, EPI_ISL_2305577, EPI_ISL_2305578, EPI_ISL_2305580, EPI_ISL_2305581, EPI_ISL_2305587, EPI_ISL_2305588, EPI_ISL_2305589, EPI_ISL_2305592, EPI_ISL_2305595, EPI_ISL_2305596, EPI_ISL_2305601, EPI_ISL_2305602, EPI_ISL_2305603, EPI_ISL_2305605, EPI_ISL_2305607, EPI_ISL_2305608, EPI_ISL_2305610, EPI_ISL_2305613, EPI_ISL_2305615, EPI_ISL_2305616, EPI_ISL_2305617, EPI_ISL_2305619, EPI_ISL_2305623, EPI_ISL_2305625, EPI_ISL_2305626, EPI_ISL_2305629, EPI_ISL_2305630, EPI_ISL_2305631, EPI_ISL_2305633, EPI_ISL_2305634, EPI_ISL_2305635, EPI_ISL_2305636, EPI_ISL_2305637, EPI_ISL_2305640, EPI_ISL_2305642, EPI_ISL_2305643, EPI_ISL_2305646, EPI_ISL_2305647, EPI_ISL_2305649, EPI_ISL_2305653, EPI_ISL_2305655, EPI_ISL_2305658, EPI_ISL_2305660, EPI_ISL_2305662, EPI_ISL_2305664, EPI_ISL_2305666, EPI_ISL_2305667, EPI_ISL_2305670, EPI_ISL_2305671, EPI_ISL_2305672, EPI_ISL_2305673, EPI_ISL_2305674, EPI_ISL_2305675, EPI_ISL_2305676, EPI_ISL_2305678, EPI_ISL_2305679, EPI_ISL_2305680, EPI_ISL_2305681, EPI_ISL_2305682, EPI_ISL_2305683, EPI_ISL_2305684, EPI_ISL_2305686, EPI_ISL_2305687, EPI_ISL_2305688, EPI_ISL_2305689, EPI_ISL_2305690, EPI_ISL_2305691, EPI_ISL_2305693, EPI_ISL_2305694, EPI_ISL_2305695, EPI_ISL_2305697, EPI_ISL_2305698, EPI_ISL_2305699, EPI_ISL_2305702, EPI_ISL_2305703, EPI_ISL_2305704, EPI_ISL_2305705, EPI_ISL_2305707, EPI_ISL_2305708, EPI_ISL_2305709, EPI_ISL_2305710, EPI_ISL_2305713, EPI_ISL_2305714, EPI_ISL_2305715, EPI_ISL_2305716, EPI_ISL_2305717, EPI_ISL_2305718, EPI_ISL_2305721, EPI_ISL_2305722, EPI_ISL_2305724, EPI_ISL_2305725, EPI_ISL_2305727, EPI_ISL_2305728, EPI_ISL_2305729, EPI_ISL_2305730, EPI_ISL_2305731, EPI_ISL_2305733, | | | |

| | | | |
|---|---|---|---|
| EPI_ISL_2305734, EPI_ISL_2305735, EPI_ISL_2305736, EPI_ISL_2305737, EPI_ISL_2305738, EPI_ISL_2305740, EPI_ISL_2305741, EPI_ISL_2305742, EPI_ISL_2305743, EPI_ISL_2305744, EPI_ISL_2305745, EPI_ISL_2305748, EPI_ISL_2305750, EPI_ISL_2305751, EPI_ISL_2305754, EPI_ISL_2305755, EPI_ISL_2305759, EPI_ISL_2305761, EPI_ISL_2305762, EPI_ISL_2305763, EPI_ISL_2305764, EPI_ISL_2305765, EPI_ISL_2305766, EPI_ISL_2305767, EPI_ISL_2305769, EPI_ISL_2305770, EPI_ISL_2305771, EPI_ISL_2305772, EPI_ISL_2305773, EPI_ISL_2305775, EPI_ISL_2305776, EPI_ISL_2305777, EPI_ISL_2305778, EPI_ISL_2305779, EPI_ISL_2305782, EPI_ISL_2305784, EPI_ISL_2305785, EPI_ISL_2305786, EPI_ISL_2305788, EPI_ISL_2305790, EPI_ISL_2305791, EPI_ISL_2305792, EPI_ISL_2305793, EPI_ISL_2305795, EPI_ISL_2305798, EPI_ISL_2305799, EPI_ISL_2305800, EPI_ISL_2305803, EPI_ISL_2305804, EPI_ISL_2305805, EPI_ISL_2305806, EPI_ISL_2305807, EPI_ISL_2305808, EPI_ISL_2305810, EPI_ISL_2305812, EPI_ISL_2305813, EPI_ISL_2305814, EPI_ISL_2305815, EPI_ISL_2305817, EPI_ISL_2305818, EPI_ISL_2305819, EPI_ISL_2305820, EPI_ISL_2305821, EPI_ISL_2305822, EPI_ISL_2305824, EPI_ISL_2305825, EPI_ISL_2305826, EPI_ISL_2305827, EPI_ISL_2305828, EPI_ISL_2305831, EPI_ISL_2305832, EPI_ISL_2305834, EPI_ISL_2305835, EPI_ISL_2305836, EPI_ISL_2305838, EPI_ISL_2305839, EPI_ISL_2305840, EPI_ISL_2305842, EPI_ISL_2305843, EPI_ISL_2305844, EPI_ISL_2305845, EPI_ISL_2305846, EPI_ISL_2305849, EPI_ISL_2305850, EPI_ISL_2305852, EPI_ISL_2305853, EPI_ISL_2305854, EPI_ISL_2305855, EPI_ISL_2305856, EPI_ISL_2305857, EPI_ISL_2305858, EPI_ISL_2305860, EPI_ISL_2305861, EPI_ISL_2305862, EPI_ISL_2305863, EPI_ISL_2305865, EPI_ISL_2305866, EPI_ISL_2305867, EPI_ISL_2305868, EPI_ISL_2305869, EPI_ISL_2305871, EPI_ISL_2305872, EPI_ISL_2305873, EPI_ISL_2305875, EPI_ISL_2305876, EPI_ISL_2305878, EPI_ISL_2305879, EPI_ISL_2305882, EPI_ISL_2305883, EPI_ISL_2305884, EPI_ISL_2305885, EPI_ISL_2305886, EPI_ISL_2305888, EPI_ISL_2305889, EPI_ISL_2305890, EPI_ISL_2305892, EPI_ISL_2305894, EPI_ISL_2305895, EPI_ISL_2305896, EPI_ISL_2305897, EPI_ISL_2305898, EPI_ISL_2305899, EPI_ISL_2305901, EPI_ISL_2305902, EPI_ISL_2305904, EPI_ISL_2305905, EPI_ISL_2305906, EPI_ISL_2305907, EPI_ISL_2305908, EPI_ISL_2305909, EPI_ISL_2305910, EPI_ISL_2305911, EPI_ISL_2305912, EPI_ISL_2305913, EPI_ISL_2305914, EPI_ISL_2305915, EPI_ISL_2305916, EPI_ISL_2305918, EPI_ISL_2305919, EPI_ISL_2305920, EPI_ISL_2305921, EPI_ISL_2305922, EPI_ISL_2305923, EPI_ISL_2305924, EPI_ISL_2305925, EPI_ISL_2305926, EPI_ISL_2305927, EPI_ISL_2305929, EPI_ISL_2305930, EPI_ISL_2305932, EPI_ISL_2305933, EPI_ISL_2305935, EPI_ISL_2305937, EPI_ISL_2305938, EPI_ISL_2305939, EPI_ISL_2305940, EPI_ISL_2305941, EPI_ISL_2305943, EPI_ISL_2305944, EPI_ISL_2305945, EPI_ISL_2305946, EPI_ISL_2305947, EPI_ISL_2305948, EPI_ISL_2305949, EPI_ISL_2305950, EPI_ISL_2305951, EPI_ISL_2305952, EPI_ISL_2305953, EPI_ISL_2305954, EPI_ISL_2305955, EPI_ISL_2305956, EPI_ISL_2305957, EPI_ISL_2305958, EPI_ISL_2305959, EPI_ISL_2305960, EPI_ISL_2305961, EPI_ISL_2305962, EPI_ISL_2305963, EPI_ISL_2305964, EPI_ISL_2305965, EPI_ISL_2305966, EPI_ISL_2305967, EPI_ISL_2305968, EPI_ISL_2305969, EPI_ISL_2305970, EPI_ISL_2305971, EPI_ISL_2305972, EPI_ISL_2305973, EPI_ISL_2305974, EPI_ISL_2305975, EPI_ISL_2305976, EPI_ISL_2305977, EPI_ISL_2305978, EPI_ISL_2305979, EPI_ISL_2305980, EPI_ISL_2305981, EPI_ISL_2305982, EPI_ISL_2305983, EPI_ISL_2305984, EPI_ISL_2305985, EPI_ISL_2305986, EPI_ISL_2305987, EPI_ISL_2305988, EPI_ISL_2305989, EPI_ISL_2305991, EPI_ISL_2305992, EPI_ISL_2305993, EPI_ISL_2305994, EPI_ISL_2305995, EPI_ISL_2305996, EPI_ISL_2305997, EPI_ISL_2305998, EPI_ISL_2305999, EPI_ISL_2306000, EPI_ISL_2306001, EPI_ISL_2306002, EPI_ISL_2306003, EPI_ISL_2306004, EPI_ISL_2306005, EPI_ISL_2306006, EPI_ISL_2306007, EPI_ISL_2306008, EPI_ISL_2306009, EPI_ISL_2306010, EPI_ISL_2306011, EPI_ISL_2306012, EPI_ISL_2306013, EPI_ISL_2306014, EPI_ISL_2306015, EPI_ISL_2306016, EPI_ISL_2306017, EPI_ISL_2306018, EPI_ISL_2306019, EPI_ISL_2306020, EPI_ISL_2306021, EPI_ISL_2306022, EPI_ISL_2306023, EPI_ISL_2306024, EPI_ISL_2306025, EPI_ISL_2306026, EPI_ISL_2306027, EPI_ISL_2306028, EPI_ISL_2306029, EPI_ISL_2306030, EPI_ISL_2306031, EPI_ISL_2306032, EPI_ISL_2306033, EPI_ISL_2306034, EPI_ISL_2306035, EPI_ISL_2306036, EPI_ISL_2306037, EPI_ISL_2306038, EPI_ISL_2306039, EPI_ISL_2306040, EPI_ISL_2306041, EPI_ISL_2306042, EPI_ISL_2306043, EPI_ISL_2306044, EPI_ISL_2306045, EPI_ISL_2306046, EPI_ISL_2306047, EPI_ISL_2306048, EPI_ISL_2306049, EPI_ISL_2306050, EPI_ISL_2306051, EPI_ISL_2306052, EPI_ISL_2306053, EPI_ISL_2306054, EPI_ISL_2306055, EPI_ISL_2306056, EPI_ISL_2306057, EPI_ISL_2306058, EPI_ISL_2306059, EPI_ISL_2306060, EPI_ISL_2306061, EPI_ISL_2306062, EPI_ISL_2306063, EPI_ISL_2306064, EPI_ISL_2306065, EPI_ISL_2306066, EPI_ISL_2306067, EPI_ISL_2306068, EPI_ISL_2306069, EPI_ISL_2306070, EPI_ISL_2306071, EPI_ISL_2306073, EPI_ISL_2306074, EPI_ISL_2306075, EPI_ISL_2306076, EPI_ISL_2306077, EPI_ISL_2306078, EPI_ISL_2306079, EPI_ISL_2306080, EPI_ISL_2306081, EPI_ISL_2306082, EPI_ISL_2306083, EPI_ISL_2306084, EPI_ISL_2306085, EPI_ISL_2306086, EPI_ISL_2306087, EPI_ISL_2306088, EPI_ISL_2306089, EPI_ISL_2306090, EPI_ISL_2306091, EPI_ISL_2306092, EPI_ISL_2306093, EPI_ISL_2306094, EPI_ISL_2306095, EPI_ISL_2306096, EPI_ISL_2306097, EPI_ISL_2306098, EPI_ISL_2306099, EPI_ISL_2306101, EPI_ISL_2306102, EPI_ISL_2306103, EPI_ISL_2306104, EPI_ISL_2306105, EPI_ISL_2306107, EPI_ISL_2306108, EPI_ISL_2306109, EPI_ISL_2306110, EPI_ISL_2306111, EPI_ISL_2306113, EPI_ISL_2306114, EPI_ISL_2306115, EPI_ISL_2306116, EPI_ISL_2306117, EPI_ISL_2306118, EPI_ISL_2306119, EPI_ISL_2306120, EPI_ISL_2306121, EPI_ISL_2306122, EPI_ISL_2306123, EPI_ISL_2306124, EPI_ISL_2306125, EPI_ISL_2306126, EPI_ISL_2306128, EPI_ISL_2306129, EPI_ISL_2306130, EPI_ISL_2306131, EPI_ISL_2306132, EPI_ISL_2306133, EPI_ISL_2306134, EPI_ISL_2306135, EPI_ISL_2306136, EPI_ISL_2306137, EPI_ISL_2306138, EPI_ISL_2306139, EPI_ISL_2306140, EPI_ISL_2306141, EPI_ISL_2306142, EPI_ISL_2306143, EPI_ISL_2306144, EPI_ISL_2306145, EPI_ISL_2306146, EPI_ISL_2306147, EPI_ISL_2306148, EPI_ISL_2306149, EPI_ISL_2306150, EPI_ISL_2306151, EPI_ISL_2306152, EPI_ISL_2306153, EPI_ISL_2306154, EPI_ISL_2306155, EPI_ISL_2306156, EPI_ISL_2306157, EPI_ISL_2306158, EPI_ISL_2306159, EPI_ISL_2306160, EPI_ISL_2306161, EPI_ISL_2306162, EPI_ISL_2306163, EPI_ISL_2306164, EPI_ISL_2306165, EPI_ISL_2306166, EPI_ISL_2306168, EPI_ISL_2306169, EPI_ISL_2306170, EPI_ISL_2306171, EPI_ISL_2306172, EPI_ISL_2306174, EPI_ISL_2306175, EPI_ISL_2306176, EPI_ISL_2306177, EPI_ISL_2306178, EPI_ISL_2306179, EPI_ISL_2306180, EPI_ISL_2306181, EPI_ISL_2306182, EPI_ISL_2306183, EPI_ISL_2306184, EPI_ISL_2306185, EPI_ISL_2306186, EPI_ISL_2306187, EPI_ISL_2306188, EPI_ISL_2306189, EPI_ISL_2306190, EPI_ISL_2306192, EPI_ISL_2306193, EPI_ISL_2306194, EPI_ISL_2306195, EPI_ISL_2306196, EPI_ISL_2306197, EPI_ISL_2306198, EPI_ISL_2306199, EPI_ISL_2306200, EPI_ISL_2306201, EPI_ISL_2306202, EPI_ISL_2306203, EPI_ISL_2306204, EPI_ISL_2306205, EPI_ISL_2306206, EPI_ISL_2306207, EPI_ISL_2306208, EPI_ISL_2306209, EPI_ISL_2306210, EPI_ISL_2306211, EPI_ISL_2306212, EPI_ISL_2306213, EPI_ISL_2306214, EPI_ISL_2306215, EPI_ISL_2306216, EPI_ISL_2306217, EPI_ISL_2306218, EPI_ISL_2306219, EPI_ISL_2306220, EPI_ISL_2306221, EPI_ISL_2306222, EPI_ISL_2306223, EPI_ISL_2306224, EPI_ISL_2306225, EPI_ISL_2306226, EPI_ISL_2306227, EPI_ISL_2306228, EPI_ISL_2306229, EPI_ISL_2306230, EPI_ISL_2306231, EPI_ISL_2306232, EPI_ISL_2306233, EPI_ISL_2306234, EPI_ISL_2306235, EPI_ISL_2306236, EPI_ISL_2306237, EPI_ISL_2306238, EPI_ISL_2306239, EPI_ISL_2306240, EPI_ISL_2306241, EPI_ISL_2306242, EPI_ISL_2306243, EPI_ISL_2306244, EPI_ISL_2306245, EPI_ISL_2306246, EPI_ISL_2306247, EPI_ISL_2306248, EPI_ISL_2306249, EPI_ISL_2306250, EPI_ISL_2306251, EPI_ISL_2306252, EPI_ISL_2306253, EPI_ISL_2306254, EPI_ISL_2306255, EPI_ISL_2306256, EPI_ISL_2306257, EPI_ISL_2306258, EPI_ISL_2306259, EPI_ISL_2306260, EPI_ISL_2306261, EPI_ISL_2306262, EPI_ISL_2306263, EPI_ISL_2306264, EPI_ISL_2306265, EPI_ISL_2306266, EPI_ISL_2306267, EPI_ISL_2306268, EPI_ISL_2306269, EPI_ISL_2306270, EPI_ISL_2306271, EPI_ISL_2306272, EPI_ISL_2306273, EPI_ISL_2306274, EPI_ISL_2306275, EPI_ISL_2306276, EPI_ISL_2306277, EPI_ISL_2306278, EPI_ISL_2306279, EPI_ISL_2306280, EPI_ISL_2306281, EPI_ISL_2306282, EPI_ISL_2306283, EPI_ISL_2306284, EPI_ISL_2306285, EPI_ISL_2306286, EPI_ISL_2306288, EPI_ISL_2306289, EPI_ISL_2306290, EPI_ISL_2306291, EPI_ISL_2306292, EPI_ISL_2306293, EPI_ISL_2306294, EPI_ISL_2306295, EPI_ISL_2306296, EPI_ISL_2306297, EPI_ISL_2306298, EPI_ISL_2306299, EPI_ISL_2306300, EPI_ISL_2306301, EPI_ISL_2306302, EPI_ISL_2306303, EPI_ISL_2306304, EPI_ISL_2306305, EPI_ISL_2306306, EPI_ISL_2306307, EPI_ISL_2306308, EPI_ISL_2306309, EPI_ISL_2306310, EPI_ISL_2306311, EPI_ISL_2306312, EPI_ISL_2306313, EPI_ISL_2306314, EPI_ISL_2306315, EPI_ISL_2306316, EPI_ISL_2306317, EPI_ISL_2306318, EPI_ISL_2306319, EPI_ISL_2306321, EPI_ISL_2306322, EPI_ISL_2306323, EPI_ISL_2306324, EPI_ISL_2306325, EPI_ISL_2306326, EPI_ISL_2306327, EPI_ISL_2306328, EPI_ISL_2306329, EPI_ISL_2306330, EPI_ISL_2306331, EPI_ISL_2306332, EPI_ISL_2306333, EPI_ISL_2306334, EPI_ISL_2306335, EPI_ISL_2306336, EPI_ISL_2306337, EPI_ISL_2306338, EPI_ISL_2306339, EPI_ISL_2306340, EPI_ISL_2306341, EPI_ISL_2306342, EPI_ISL_2306343, EPI_ISL_2306344, EPI_ISL_2306345, EPI_ISL_2306346, EPI_ISL_2306347, EPI_ISL_2306348, EPI_ISL_2306349, EPI_ISL_2306350, EPI_ISL_2306351, EPI_ISL_2306352, EPI_ISL_2306353, EPI_ISL_2306354, EPI_ISL_2306355, EPI_ISL_2306356, EPI_ISL_2306357, EPI_ISL_2306358, EPI_ISL_2306359, EPI_ISL_2306360, EPI_ISL_2306361, EPI_ISL_2306362, EPI_ISL_2306364, EPI_ISL_2306365, EPI_ISL_2306366, EPI_ISL_2306367, EPI_ISL_2306368, EPI_ISL_2306369, EPI_ISL_2306370, EPI_ISL_2306371, EPI_ISL_2306372, EPI_ISL_2306373, EPI_ISL_2306374, EPI_ISL_2306375, EPI_ISL_2306376, EPI_ISL_2306377, EPI_ISL_2306378, EPI_ISL_2306379, EPI_ISL_2306380, EPI_ISL_2306382, EPI_ISL_2306383, EPI_ISL_2306384, EPI_ISL_2306385, EPI_ISL_2306386, EPI_ISL_2306387, EPI_ISL_2306388, EPI_ISL_2306389, EPI_ISL_2306390, EPI_ISL_2306391, EPI_ISL_2306392, EPI_ISL_2306393, EPI_ISL_2306394, EPI_ISL_2306395, EPI_ISL_2306396, EPI_ISL_2306397, EPI_ISL_2306398, EPI_ISL_2306399, EPI_ISL_2306400, EPI_ISL_2306402, EPI_ISL_2306403, EPI_ISL_2306404, EPI_ISL_2306405, EPI_ISL_2306406, EPI_ISL_2306407, EPI_ISL_2306408, EPI_ISL_2306409, EPI_ISL_2306410, EPI_ISL_2306411, EPI_ISL_2306412, EPI_ISL_2306413, EPI_ISL_2306414, EPI_ISL_2306415, EPI_ISL_2306416, EPI_ISL_2306417, EPI_ISL_2306418, EPI_ISL_2306419, EPI_ISL_2306420, EPI_ISL_2306421, EPI_ISL_2306422, EPI_ISL_2306423, EPI_ISL_2306424, EPI_ISL_2306425, EPI_ISL_2306426, EPI_ISL_2306427, EPI_ISL_2306428, EPI_ISL_2306429, EPI_ISL_2306430, EPI_ISL_2306431, EPI_ISL_2306432, EPI_ISL_2306433, EPI_ISL_2306434, EPI_ISL_2306435, EPI_ISL_2306436, EPI_ISL_2306437, EPI_ISL_2306438, EPI_ISL_2306439, EPI_ISL_2306440, EPI_ISL_2306441, EPI_ISL_2306442, EPI_ISL_2306443, EPI_ISL_2306444, EPI_ISL_2306445, EPI_ISL_2306446, EPI_ISL_2306447, EPI_ISL_2306448, EPI_ISL_2306449, EPI_ISL_2306451, EPI_ISL_2306452, EPI_ISL_2306453, EPI_ISL_2306454, EPI_ISL_2306455, EPI_ISL_2306456, EPI_ISL_2306457, EPI_ISL_2306458, EPI_ISL_2306460, EPI_ISL_2306461, EPI_ISL_2306462, EPI_ISL_2306463, EPI_ISL_2306464, EPI_ISL_2306465, EPI_ISL_2306466, EPI_ISL_2306467, EPI_ISL_2306468, EPI_ISL_2306469, EPI_ISL_2306470, EPI_ISL_2306471, EPI_ISL_2306472, EPI_ISL_2306473, EPI_ISL_2306474, EPI_ISL_2306475, EPI_ISL_2306476, EPI_ISL_2306477, EPI_ISL_2306478, EPI_ISL_2306479, EPI_ISL_2306480, EPI_ISL_2306481, EPI_ISL_2306482, EPI_ISL_2306483, EPI_ISL_2306484, EPI_ISL_2306485, EPI_ISL_2306486, EPI_ISL_2306488, EPI_ISL_2306489, EPI_ISL_2306491, EPI_ISL_2306492, EPI_ISL_2306493, EPI_ISL_2306494, EPI_ISL_2306495, EPI_ISL_2306496, EPI_ISL_2306497, EPI_ISL_2306498, EPI_ISL_2306499, EPI_ISL_2306500, EPI_ISL_2306501, EPI_ISL_2306502, EPI_ISL_2306503, EPI_ISL_2306504, EPI_ISL_2306505, EPI_ISL_2306506, EPI_ISL_2306507, EPI_ISL_2306508, EPI_ISL_2306509, EPI_ISL_2306510, EPI_ISL_2306511, EPI_ISL_2306512, EPI_ISL_2306513, EPI_ISL_2306514, EPI_ISL_2306515, EPI_ISL_2306516, EPI_ISL_2306517, EPI_ISL_2306518, EPI_ISL_2306519, EPI_ISL_2306520, EPI_ISL_2306521, EPI_ISL_2306522, EPI_ISL_2306523, EPI_ISL_2306524, EPI_ISL_2306525, EPI_ISL_2306526, EPI_ISL_2306527, EPI_ISL_2306528, EPI_ISL_2306529, EPI_ISL_2306530, EPI_ISL_2306531, EPI_ISL_2306532, EPI_ISL_2306533, EPI_ISL_2306534, EPI_ISL_2306535, EPI_ISL_2306536, EPI_ISL_2306537, EPI_ISL_2306538, EPI_ISL_2306539, EPI_ISL_2306540, EPI_ISL_2306541, EPI_ISL_2306542, EPI_ISL_2306544, EPI_ISL_2306545, EPI_ISL_2306546, EPI_ISL_2306547, EPI_ISL_2306548, EPI_ISL_2306549, EPI_ISL_2306550, EPI_ISL_2306551, EPI_ISL_2306552, EPI_ISL_2306553, EPI_ISL_2306554, EPI_ISL_2306555, EPI_ISL_2306556, EPI_ISL_2306557, EPI_ISL_2306558, EPI_ISL_2306559, EPI_ISL_2306560, EPI_ISL_2306563, EPI_ISL_2306564, EPI_ISL_2306565, EPI_ISL_2306566, EPI_ISL_2306568, EPI_ISL_2306569, EPI_ISL_2306570, EPI_ISL_2306571, EPI_ISL_2306572, EPI_ISL_2306573, EPI_ISL_2306574, EPI_ISL_2306575, EPI_ISL_2306576, EPI_ISL_2306577, EPI_ISL_2306578, EPI_ISL_2306579, EPI_ISL_2306581, EPI_ISL_2306582, EPI_ISL_2306583, EPI_ISL_2306584, EPI_ISL_2306585, EPI_ISL_2306586, EPI_ISL_2306587, EPI_ISL_2306588, EPI_ISL_2306589, EPI_ISL_2306590, EPI_ISL_2306591, EPI_ISL_2306592, EPI_ISL_2306593, EPI_ISL_2306594, EPI_ISL_2306595, EPI_ISL_2306596, EPI_ISL_2306597, EPI_ISL_2306598, EPI_ISL_2306599, EPI_ISL_2306600, EPI_ISL_2306601, EPI_ISL_2306602, EPI_ISL_2306603, EPI_ISL_2306604, EPI_ISL_2306606, EPI_ISL_2306607, EPI_ISL_2306608, EPI_ISL_2306609, EPI_ISL_2306610, EPI_ISL_2306611, EPI_ISL_2306612, EPI_ISL_2306613, EPI_ISL_2306614, EPI_ISL_2306615, EPI_ISL_2306616, EPI_ISL_2306617, EPI_ISL_2306618, EPI_ISL_2306619, EPI_ISL_2306620, EPI_ISL_2306621, EPI_ISL_2306622, EPI_ISL_2306623, EPI_ISL_2306624, EPI_ISL_2306625, EPI_ISL_2306626, EPI_ISL_2306627, EPI_ISL_2306628, EPI_ISL_2306629, EPI_ISL_2306630, EPI_ISL_2306631, EPI_ISL_2306632, EPI_ISL_2306633, EPI_ISL_2306634, EPI_ISL_2306635, EPI_ISL_2306636, EPI_ISL_2306637, EPI_ISL_2306638, EPI_ISL_2306640, EPI_ISL_2306641, EPI_ISL_2306642, EPI_ISL_2306643, EPI_ISL_2306644, EPI_ISL_2306645, EPI_ISL_2306646, EPI_ISL_2306647, EPI_ISL_2306648, EPI_ISL_2306649, EPI_ISL_2306650, EPI_ISL_2306651, EPI_ISL_2306652, EPI_ISL_2306653, EPI_ISL_2306654, EPI_ISL_2306655, EPI_ISL_2306656, EPI_ISL_2306657, EPI_ISL_2306658, EPI_ISL_2306659, EPI_ISL_2306660, EPI_ISL_2306661, EPI_ISL_2306662, EPI_ISL_2306663, EPI_ISL_2306664, EPI_ISL_2306665, EPI_ISL_2306666, EPI_ISL_2306667, EPI_ISL_2306668, EPI_ISL_2306669, EPI_ISL_2306670, EPI_ISL_2306671, EPI_ISL_2306672, EPI_ISL_2306673, EPI_ISL_2306674, EPI_ISL_2306675, EPI_ISL_2306676, EPI_ISL_2306677, EPI_ISL_2306678, EPI_ISL_2306679, EPI_ISL_2306680, EPI_ISL_2306681, EPI_ISL_2306682, EPI_ISL_2306683, EPI_ISL_2306684, EPI_ISL_2306685, EPI_ISL_2306686, EPI_ISL_2306687, EPI_ISL_2306688, EPI_ISL_2306689, EPI_ISL_2306690, EPI_ISL_2306691, EPI_ISL_2306692, EPI_ISL_2306693, EPI_ISL_2306695, EPI_ISL_2306696, EPI_ISL_2306697, EPI_ISL_2306698, EPI_ISL_2306699, EPI_ISL_2306700, EPI_ISL_2306701, EPI_ISL_2306702, EPI_ISL_2306703, EPI_ISL_2306704, EPI_ISL_2306705, EPI_ISL_2306706, EPI_ISL_2306707, EPI_ISL_2306708, EPI_ISL_2306709, EPI_ISL_2306710, EPI_ISL_2306711, EPI_ISL_2306712, EPI_ISL_2306713, EPI_ISL_2306714, EPI_ISL_2306716, EPI_ISL_2306717, EPI_ISL_2306718, EPI_ISL_2306719, EPI_ISL_2306720, EPI_ISL_2306721, EPI_ISL_2306724, EPI_ISL_2306725, EPI_ISL_2306726, EPI_ISL_2306727, EPI_ISL_2306728, EPI_ISL_2306729, EPI_ISL_2306730, EPI_ISL_2306731, EPI_ISL_2306732, EPI_ISL_2306733, EPI_ISL_2306734, EPI_ISL_2306735, EPI_ISL_2306736, EPI_ISL_2306737, EPI_ISL_2306738, EPI_ISL_2306739, EPI_ISL_2306740, EPI_ISL_2306741, EPI_ISL_2306742, EPI_ISL_2306743, EPI_ISL_2306744, EPI_ISL_2306745, EPI_ISL_2306746, EPI_ISL_2306747, EPI_ISL_2306748, EPI_ISL_2306749, EPI_ISL_2306750, EPI_ISL_2306751, EPI_ISL_2306752, EPI_ISL_2306753, EPI_ISL_2306754, EPI_ISL_2306755, EPI_ISL_2306756, EPI_ISL_2306757, EPI_ISL_2306758, EPI_ISL_2306759, EPI_ISL_2306761, EPI_ISL_2306762, EPI_ISL_2306763, EPI_ISL_2306764, EPI_ISL_2306765, EPI_ISL_2306766, EPI_ISL_2306767, EPI_ISL_2306768, EPI_ISL_2306769, EPI_ISL_2306770, EPI_ISL_2306771, EPI_ISL_2306772, EPI_ISL_2306773, EPI_ISL_2306774, EPI_ISL_2306775, EPI_ISL_2306776, EPI_ISL_2306777, EPI_ISL_2306778, EPI_ISL_2306779, EPI_ISL_2306780, EPI_ISL_2306781, EPI_ISL_2306782, EPI_ISL_2306783, EPI_ISL_2306784, EPI_ISL_2306785, EPI_ISL_2306786, EPI_ISL_2306787, EPI_ISL_2306788, EPI_ISL_2306789, EPI_ISL_2306790, EPI_ISL_2306791, EPI_ISL_2306792, EPI_ISL_2306793, EPI_ISL_2306794, EPI_ISL_2306795, EPI_ISL_2306796, EPI_ISL_2306797, EPI_ISL_2306798, EPI_ISL_2306799, EPI_ISL_2306800, EPI_ISL_2306801, EPI_ISL_2306802, EPI_ISL_2306803, EPI_ISL_2306804, EPI_ISL_2306805, EPI_ISL_2306806, EPI_ISL_2306807, EPI_ISL_2306808, EPI_ISL_2306809, EPI_ISL_2306810, EPI_ISL_2306811, EPI_ISL_2306812, EPI_ISL_2306813, EPI_ISL_2306814, EPI_ISL_2306815, EPI_ISL_2306816, EPI_ISL_2306817, EPI_ISL_2306818, EPI_ISL_2306819, EPI_ISL_2306820, EPI_ISL_2306821, EPI_ISL_2306822, EPI_ISL_2306823, EPI_ISL_2306824, EPI_ISL_2306825, EPI_ISL_2306826, EPI_ISL_2306827, EPI_ISL_2306828, EPI_ISL_2306829, EPI_ISL_2306830, EPI_ISL_2306831, EPI_ISL_2306832, EPI_ISL_2306833, EPI_ISL_2306834, EPI_ISL_2306835, EPI_ISL_2306836, EPI_ISL_2306837, EPI_ISL_2306838, EPI_ISL_2306839, EPI_ISL_2306840, EPI_ISL_2306841, EPI_ISL_2306842, EPI_ISL_2306843, EPI_ISL_2306844, EPI_ISL_2306845, EPI_ISL_2306846, EPI_ISL_2306847, EPI_ISL_2306848, EPI_ISL_2306849, EPI_ISL_2306850, EPI_ISL_2306851, EPI_ISL_2306852, EPI_ISL_2306853, EPI_ISL_2306854, EPI_ISL_2306855, EPI_ISL_2306856, EPI_ISL_2306857, EPI_ISL_2306858, EPI_ISL_2306859, EPI_ISL_2306860, EPI_ISL_2306861, EPI_ISL_2306862, EPI_ISL_2306863, EPI_ISL_2306864, EPI_ISL_2306865, EPI_ISL_2306866, EPI_ISL_2306867, EPI_ISL_2306868, EPI_ISL_2306869, EPI_ISL_2306870, EPI_ISL_2306871, EPI_ISL_2306872, EPI_ISL_2306873, EPI_ISL_2306874, EPI_ISL_2306875, EPI_ISL_2306876, EPI_ISL_2306877, EPI_ISL_2306878, EPI_ISL_2306879, EPI_ISL_2306880, EPI_ISL_2306881, EPI_ISL_2306882, EPI_ISL_2306883, EPI_ISL_2306884, EPI_ISL_2306885, EPI_ISL_2306886, EPI_ISL_2306887, EPI_ISL_2306888, EPI_ISL_2306889, EPI_ISL_2306890, EPI_ISL_2306891, EPI_ISL_2306892, EPI_ISL_2306893, EPI_ISL_2306894, EPI_ISL_2306895, EPI_ISL_2306897, EPI_ISL_2306898, EPI_ISL_2306899, EPI_ISL_2306900, EPI_ISL_2306901, EPI_ISL_2306902, EPI_ISL_2306903, EPI_ISL_2306905, EPI_ISL_2306906, EPI_ISL_2306907, EPI_ISL_2306908, EPI_ISL_2306909, EPI_ISL_2306910, EPI_ISL_2306911, EPI_ISL_2306912, EPI_ISL_2306913, EPI_ISL_2306914, EPI_ISL_2306915, EPI_ISL_2306916, EPI_ISL_2306917, EPI_ISL_2306918, EPI_ISL_2306919, EPI_ISL_2306920, EPI_ISL_2306921, EPI_ISL_2306922, EPI_ISL_2306923, EPI_ISL_2306924, EPI_ISL_2306925, EPI_ISL_2306926, EPI_ISL_2306927, EPI_ISL_2306928, EPI_ISL_2306929, EPI_ISL_2306930, EPI_ISL_2306931, EPI_ISL_2306932, EPI_ISL_2306933, EPI_ISL_2306934, EPI_ISL_2306935, EPI_ISL_2306936, EPI_ISL_2306937, EPI_ISL_2306938, EPI_ISL_2306939, EPI_ISL_2306940, EPI_ISL_2306941, EPI_ISL_2306942, EPI_ISL_2306943, EPI_ISL_2306944, EPI_ISL_2306945, EPI_ISL_2306946, EPI_ISL_2306947, EPI_ISL_2306948, EPI_ISL_2306950, EPI_ISL_2306951, EPI_ISL_2306952, EPI_ISL_2306953, EPI_ISL_2306954, EPI_ISL_2306955, EPI_ISL_2306956, EPI_ISL_2306957, EPI_ISL_2306958, EPI_ISL_2306959, EPI_ISL_2306960, EPI_ISL_2306962, EPI_ISL_2306963, EPI_ISL_2306964, EPI_ISL_2306965, EPI_ISL_2306966, EPI_ISL_2306967, EPI_ISL_2306968, EPI_ISL_2306969, EPI_ISL_2306970, EPI_ISL_2306971, EPI_ISL_2306972, EPI_ISL_2306973, EPI_ISL_2306974, EPI_ISL_2306975, EPI_ISL_2306976, EPI_ISL_2306977, EPI_ISL_2306978, EPI_ISL_2306979, EPI_ISL_2306980, EPI_ISL_2306981, EPI_ISL_2306982, EPI_ISL_2306983, EPI_ISL_2306984, EPI_ISL_2306985, EPI_ISL_2306986, EPI_ISL_2306987, EPI_ISL_2306988, EPI_ISL_2306989, EPI_ISL_2306990, EPI_ISL_2306991, EPI_ISL_2306992, EPI_ISL_2306993, EPI_ISL_2306994, EPI_ISL_2306995, EPI_ISL_2306996, EPI_ISL_2306997, EPI_ISL_2306998, EPI_ISL_2306999, EPI_ISL_2307000, EPI_ISL_2307001, EPI_ISL_2307002, EPI_ISL_2307003, EPI_ISL_2307004, EPI_ISL_2307005, EPI_ISL_2307006, EPI_ISL_2307007, EPI_ISL_2307008, EPI_ISL_2307010, EPI_ISL_2307011, EPI_ISL_2307012, EPI_ISL_2307013, EPI_ISL_2307014, EPI_ISL_2307015, EPI_ISL_2307016, EPI_ISL_2307017, EPI_ISL_2307018, EPI_ISL_2307019, EPI_ISL_2307020, EPI_ISL_2307021, EPI_ISL_2307022, EPI_ISL_2307023, EPI_ISL_2307024, EPI_ISL_2307025, EPI_ISL_2307027, EPI_ISL_2307028, EPI_ISL_2307029, EPI_ISL_2307030, EPI_ISL_2307031, EPI_ISL_2307032, EPI_ISL_2307033, EPI_ISL_2307034, EPI_ISL_2307035, EPI_ISL_2307036, EPI_ISL_2307037, EPI_ISL_2307038, EPI_ISL_2307039, EPI_ISL_2307040, EPI_ISL_2307041, EPI_ISL_2307042, EPI_ISL_2307043, EPI_ISL_2307044, EPI_ISL_2307045, EPI_ISL_2307046, EPI_ISL_2307047, EPI_ISL_2307048, EPI_ISL_2307049, EPI_ISL_2307050, EPI_ISL_2307051, EPI_ISL_2307052, EPI_ISL_2307053, EPI_ISL_2307054, EPI_ISL_2307055, EPI_ISL_2307056, EPI_ISL_2307057, EPI_ISL_2307058, EPI_ISL_2307059, EPI_ISL_2307060, EPI_ISL_2307061, EPI_ISL_2307062, EPI_ISL_2307063, EPI_ISL_2307064, EPI_ISL_2307065, EPI_ISL_2307066, EPI_ISL_2307067, EPI_ISL_2307068, EPI_ISL_2307069, EPI_ISL_2307070, EPI_ISL_2307072, EPI_ISL_2307073, EPI_ISL_2307074, EPI_ISL_2307075, EPI_ISL_2307076, EPI_ISL_2307077, EPI_ISL_2307078, EPI_ISL_2307079 | | | |
| see above | Laboratory Corporation of America | Centers for Disease Control and Prevention Division of Viral Diseases, Pathogen Discovery | Dakota Howard, Dhwani Batra, Peter W. Cook, Kara Moser, Adrian Paskey, Jason Caravas, Benjamin Rambo-Martin, Shatavia Morrison, Christopher Gulvick, Scott Sammons, Yvette Unoarumhi, Darlene Wagner, Matthew Schmerer, Minoo Agarwal, Eyad Almasri, Debbie Boles, Ayla Burns, Nuthawin Charoensri, Oren Cohen, Susan Countryman, Mary Ann Cristobal, Bobbi Croy, Suzanne Dale, Hrushikesh Deshmukh, Amanda Douglas, Vincent Drouillon, Marcia Eisenberg, Howard Engler, Rama Ghatti, Prashant Gupta, Susan Hicks, Jake Humphrey, Lax Iyer, Manoj Jain, Mohan Kolli, Brian Krueger, Tim Kuphal, Stanley Letovsky, Michael Levandoski, Craig Lukasik, Jonathan Meltzer, Brian Norvell, Mindy Nye, Scott Parker, Christos Petropoulos, John Pruitt, Steven Ragan, Scott Ryan, Mike Sapeta, Jana Schroth, Suresh Babu Selvaraju, Goran Stevovic, Amanda Suchanek, Andrea Throop, Lyndon Tilson, Thomas Urban, Joe Voshell, Kimberly Wagner, Jonathan Williams, Mary Williamson, Qian Zeng, Tricia Zwiefelhofer, Clinton R. Paden, Duncan MacCannell |
| EPI_ISL_2307081, EPI_ISL_2307082, EPI_ISL_2307083, EPI_ISL_2307084 | Ohio Department of Health Laboratory | Ohio Department of Health Laboratory | Holmes, Jennifer; Eric Brandt, Keoni Omura, Glen McGillivary, Caitlin McDonnell, Jade Mowery, Stephanie Mccracken, Tyler Payne, Kirtana Ramadugu, Erica Leasure, Brent Lee, Kelsey Florek, Heather Blankenship, Allison Black, Quanta Brown, and Tammy Bannerman |
| EPI_ISL_2307137, EPI_ISL_2307139 | TGen North | TGen North | "Jolene Bowers, Heather Centner, Chris French, Hayley Yaglom, Ashlyn Pfeiffer, Darrin Lemmer, Dave Engelthaler, The Arizona COVID Genomics Union (ACGU)" |
| EPI_ISL_2307140, EPI_ISL_2307141, EPI_ISL_2307142, EPI_ISL_2307143, EPI_ISL_2307144, EPI_ISL_2307145, EPI_ISL_2307146, EPI_ISL_2307147, EPI_ISL_2307148, EPI_ISL_2307149, EPI_ISL_2307150, EPI_ISL_2307151, EPI_ISL_2307152, EPI_ISL_2307153, EPI_ISL_2307154, EPI_ISL_2307155, EPI_ISL_2307156, EPI_ISL_2307157, EPI_ISL_2307158, EPI_ISL_2307159, EPI_ISL_2307160, EPI_ISL_2307161, EPI_ISL_2307162, EPI_ISL_2307163, EPI_ISL_2307164 | | | |
| see above | Quest Diagnostics Incorporated | Centers for Disease Control and Prevention Division of Viral Diseases, Pathogen Discovery | Dakota Howard, Dhwani Batra, Peter W. Cook, Kara Moser, Adrian Paskey, Jason Caravas, Benjamin Rambo-Martin, Shatavia Morrison, Christopher Gulvick, Scott Sammons, Yvette Unoarumhi, Darlene Wagner, Matthew Schmerer, S. H. Rosenthal, A. Gerasimova, R. M. Kagan, B. Anderson, M. Hua, Y. Liu, L.E. Bernstein, K.E. Livingston, A. Perez, I. A. Shlyakhter, R. V. Rolando, R. Owen, P. Tanpaiboon, F. Lacbawan, Clinton R. Paden, Duncan MacCannell |
| EPI_ISL_2307166, EPI_ISL_2307167 | Laboratory Corporation of America | Centers for Disease Control and Prevention Division of Viral Diseases, Pathogen Discovery | Dakota Howard, Dhwani Batra, Peter W. Cook, Kara Moser, Adrian Paskey, Jason Caravas, Benjamin Rambo-Martin, Shatavia Morrison, Christopher Gulvick, Scott Sammons, Yvette Unoarumhi, Darlene Wagner, Matthew Schmerer, Minoo Agarwal, Eyad Almasri, Debbie Boles, Ayla Burns, Nuthawin Charoensri, Oren Cohen, Susan Countryman, Mary Ann Cristobal, Bobbi Croy, Suzanne Dale, Hrushikesh Deshmukh, Amanda Douglas, Vincent Drouillon, Marcia Eisenberg, Howard Engler, Rama Ghatti, Prashant Gupta, Susan Hicks, Jake Humphrey, Lax Iyer, Manoj Jain, Mohan Kolli, Brian Krueger, Tim Kuphal, Stanley Letovsky, Michael Levandoski, Craig Lukasik, Jonathan Meltzer, Brian Norvell, Mindy Nye, Scott Parker, Christos Petropoulos, John Pruitt, Steven Ragan, Scott Ryan, Mike Sapeta, Jana Schroth, Suresh Babu Selvaraju, Goran Stevovic, Amanda Suchanek, Andrea Throop, Lyndon Tilson, Thomas Urban, Joe Voshell, Kimberly Wagner, Jonathan Williams, Mary Williamson, Qian Zeng, Tricia Zwiefelhofer, Clinton R. Paden, Duncan MacCannell |
| EPI_ISL_2307168, EPI_ISL_2307169, EPI_ISL_2307170, EPI_ISL_2307171, EPI_ISL_2307172, EPI_ISL_2307173, EPI_ISL_2307174, EPI_ISL_2307175, EPI_ISL_2307176, EPI_ISL_2307177, EPI_ISL_2307178, EPI_ISL_2307179, EPI_ISL_2307180, EPI_ISL_2307181, EPI_ISL_2307182, EPI_ISL_2307183, EPI_ISL_2307184, EPI_ISL_2307185, EPI_ISL_2307186, EPI_ISL_2307187, EPI_ISL_2307188, EPI_ISL_2307189, EPI_ISL_2307190, EPI_ISL_2307191, EPI_ISL_2307192, EPI_ISL_2307193, EPI_ISL_2307194, EPI_ISL_2307195, EPI_ISL_2307196, EPI_ISL_2307197, EPI_ISL_2307198, EPI_ISL_2307199, EPI_ISL_2307200, EPI_ISL_2307201, EPI_ISL_2307202, EPI_ISL_2307203, EPI_ISL_2307204, EPI_ISL_2307205, EPI_ISL_2307206, EPI_ISL_2307207, EPI_ISL_2307208, EPI_ISL_2307210, EPI_ISL_2307211, EPI_ISL_2307212, EPI_ISL_2307213, EPI_ISL_2307214, EPI_ISL_2307215, EPI_ISL_2307216, EPI_ISL_2307217, EPI_ISL_2307218, EPI_ISL_2307219, EPI_ISL_2307220, EPI_ISL_2307221, EPI_ISL_2307222, EPI_ISL_2307223, EPI_ISL_2307224, EPI_ISL_2307225, EPI_ISL_2307226, EPI_ISL_2307227, EPI_ISL_2307228, EPI_ISL_2307229, EPI_ISL_2307230, EPI_ISL_2307231, EPI_ISL_2307232, EPI_ISL_2307233, EPI_ISL_2307234, EPI_ISL_2307235, EPI_ISL_2307236, EPI_ISL_2307237, EPI_ISL_2307238, EPI_ISL_2307239, EPI_ISL_2307240, EPI_ISL_2307241, EPI_ISL_2307242, EPI_ISL_2307243, EPI_ISL_2307244, EPI_ISL_2307245, EPI_ISL_2307246, EPI_ISL_2307247, EPI_ISL_2307248, EPI_ISL_2307249, EPI_ISL_2307250, EPI_ISL_2307251, EPI_ISL_2307252, EPI_ISL_2307253, EPI_ISL_2307254, EPI_ISL_2307255, EPI_ISL_2307256, EPI_ISL_2307257, EPI_ISL_2307258, EPI_ISL_2307259, EPI_ISL_2307260, EPI_ISL_2307261, EPI_ISL_2307262, EPI_ISL_2307263, EPI_ISL_2307264, EPI_ISL_2307265, EPI_ISL_2307267, EPI_ISL_2307268, EPI_ISL_2307269, EPI_ISL_2307272, EPI_ISL_2307273, EPI_ISL_2307274, EPI_ISL_2307275, EPI_ISL_2307276, EPI_ISL_2307277, EPI_ISL_2307278, EPI_ISL_2307279, EPI_ISL_2307281, EPI_ISL_2307282, EPI_ISL_2307283, EPI_ISL_2307284, EPI_ISL_2307285, EPI_ISL_2307286, EPI_ISL_2307287, EPI_ISL_2307288, EPI_ISL_2307289, EPI_ISL_2307290, EPI_ISL_2307291, EPI_ISL_2307292, EPI_ISL_2307293, EPI_ISL_2307294, EPI_ISL_2307296, EPI_ISL_2307297, EPI_ISL_2307298, EPI_ISL_2307299, EPI_ISL_2307300, EPI_ISL_2307301, EPI_ISL_2307302, EPI_ISL_2307303, EPI_ISL_2307304, EPI_ISL_2307305, EPI_ISL_2307306, EPI_ISL_2307307, EPI_ISL_2307310, EPI_ISL_2307311, EPI_ISL_2307312, EPI_ISL_2307313, EPI_ISL_2307314, EPI_ISL_2307315, EPI_ISL_2307317, EPI_ISL_2307322, EPI_ISL_2307323, EPI_ISL_2307324 | | | |
| see above | Mako Medical | Centers for Disease Control and Prevention Division of Viral Diseases, Pathogen Discovery | Dakota Howard, Dhwani Batra, Peter W. Cook, Kara Moser, Adrian Paskey, Jason Caravas, Benjamin Rambo-Martin, Shatavia Morrison, Christopher Gulvick, Scott Sammons, Yvette Unoarumhi, Darlene Wagner, Matthew Schmerer,Matthew Tugwell, Lauren Moon, Clinton R. Paden, Duncan MacCannell |
| EPI_ISL_2308217, EPI_ISL_2308218, EPI_ISL_2308219, EPI_ISL_2308221, EPI_ISL_2308222, EPI_ISL_2308223, EPI_ISL_2308224, EPI_ISL_2308225, EPI_ISL_2308227, EPI_ISL_2308228, EPI_ISL_2308230, EPI_ISL_2308231, EPI_ISL_2308232, EPI_ISL_2308233 | | | |
| see above | Wyoming Public Health Laboratory | Wyoming Public Health Laboratory | Jim Mildenberger, Wanda Manley, Noah Hull, Taylor Fearing, Lynette Gumbleton, Channing Weber, Ashley Norberg, Chayse Rowley, Marley Goetz, Brian Dominguez, Elliot Thomasson, Sam Britz, Cari Sloma, and Rob Christensen |
| EPI_ISL_2308282, EPI_ISL_2308283, EPI_ISL_2308284, EPI_ISL_2308285, EPI_ISL_2308286, EPI_ISL_2308287, EPI_ISL_2308288, EPI_ISL_2308289, EPI_ISL_2308290, EPI_ISL_2308291, EPI_ISL_2308292, EPI_ISL_2308293, EPI_ISL_2308294, EPI_ISL_2308295, EPI_ISL_2308296, EPI_ISL_2308297, EPI_ISL_2308298, EPI_ISL_2308299, EPI_ISL_2308300, EPI_ISL_2308301 | | | |
| see above | Monterey County Public Health Laboratory | Monterey County Public Health Laboratory | Monterey County Public Health Laboratory |
| EPI_ISL_2308302, EPI_ISL_2308303, EPI_ISL_2308304, EPI_ISL_2308305, EPI_ISL_2308306, EPI_ISL_2308307, EPI_ISL_2308308, EPI_ISL_2308309, EPI_ISL_2308310, EPI_ISL_2308311, EPI_ISL_2308312, EPI_ISL_2308313, EPI_ISL_2308314, EPI_ISL_2308315, EPI_ISL_2308316, EPI_ISL_2308317, EPI_ISL_2308318, EPI_ISL_2308319, EPI_ISL_2308320, EPI_ISL_2308321, EPI_ISL_2308322, EPI_ISL_2308323, EPI_ISL_2308324, EPI_ISL_2308325, EPI_ISL_2308326, EPI_ISL_2308327, EPI_ISL_2308328, EPI_ISL_2308329, EPI_ISL_2308330 | | | |
| see above | unknown | Noblis | Lauren Brinkac, Shiela Diepold, Shane Mitchell, June Kang, Lee F. Kolakowski, William M. Nelson, Katharine Jennings |
| EPI_ISL_2308331, EPI_ISL_2308332, EPI_ISL_2308333, EPI_ISL_2308334, EPI_ISL_2308335, EPI_ISL_2308336, EPI_ISL_2308337, EPI_ISL_2308338, EPI_ISL_2308339, EPI_ISL_2308340, EPI_ISL_2308341, EPI_ISL_2308342, EPI_ISL_2308343, EPI_ISL_2308344, EPI_ISL_2308345, EPI_ISL_2308347, EPI_ISL_2308348, EPI_ISL_2308349, EPI_ISL_2308350 | | | |
| see above | Montana Public Health Laboratory | Montana Public Health Laboratory | Joy Ritter, Michelle Mozer, Carrie Biskupiak, Deborah Gibson, Michael Dills |
| EPI_ISL_2308352 | Nevada State Public Health Laboratory | Nevada State Public Health Laboratory | Andrew Gorzalski, Mark Pandori |
| EPI_ISL_2308353, EPI_ISL_2308354, EPI_ISL_2308355, EPI_ISL_2308356, EPI_ISL_2308357, EPI_ISL_2308358, EPI_ISL_2308359, EPI_ISL_2308360, EPI_ISL_2308361, EPI_ISL_2308362, EPI_ISL_2308364, EPI_ISL_2308365, EPI_ISL_2308366, EPI_ISL_2308367, EPI_ISL_2308368, EPI_ISL_2308369 | | | |
| see above | Monterey County Public Health Laboratory | Monterey County Public Health Laboratory | Monterey County Public Health Laboratory |

| | | | |
|---|---|---|---|
| EPI_ISL_2308371, EPI_ISL_2308372, EPI_ISL_2308373, EPI_ISL_2308374, EPI_ISL_2308375, EPI_ISL_2308376, EPI_ISL_2308377, EPI_ISL_2308378, EPI_ISL_2308379, EPI_ISL_2308380, EPI_ISL_2308381, EPI_ISL_2308382, EPI_ISL_2308383, EPI_ISL_2308384, EPI_ISL_2308385, EPI_ISL_2308386, EPI_ISL_2308387, EPI_ISL_2308388, EPI_ISL_2308389, EPI_ISL_2308390, EPI_ISL_2308391, EPI_ISL_2308392, EPI_ISL_2308393, EPI_ISL_2308394, EPI_ISL_2308395, EPI_ISL_2308396, EPI_ISL_2308397, EPI_ISL_2308398 | | | |
| see above | Nebraska Public Health Laboratory | NPHL COVID-19 Response Team | NPHL COVID-19 Response Team |
| EPI_ISL_2308496, EPI_ISL_2308497, EPI_ISL_2308498, EPI_ISL_2308499, EPI_ISL_2308500, EPI_ISL_2308501, EPI_ISL_2308502, EPI_ISL_2308503, EPI_ISL_2308504, EPI_ISL_2308505, EPI_ISL_2308506, EPI_ISL_2308507 | | | |
| see above | Kansas Health and Environmental Lab | Kansas Health and Environmental Lab | Mike Grose, Katherine Wiggins, Jonathan Barnell, Ben Olsen, and Phil Adam |
| EPI_ISL_2308509, EPI_ISL_2308510, EPI_ISL_2308511, EPI_ISL_2308512, EPI_ISL_2308513, EPI_ISL_2308514, EPI_ISL_2308515, EPI_ISL_2308516, EPI_ISL_2308517, EPI_ISL_2308518, EPI_ISL_2308519, EPI_ISL_2308520 | | | |
| see above | Wyoming Public Health Laboratory | Wyoming Public Health Laboratory | Jim Mildenberger, Taylor Fearing, Channing Weber, Ashley Norberg, Chayse Rowley, Marley Goetz, Brian Dominguez, Elliot Thomasson, Brittany Oher, Sam Britz, Cari Sloma, and Rob Christensen |
| EPI_ISL_2308523, EPI_ISL_2308524, EPI_ISL_2308525, EPI_ISL_2308526, EPI_ISL_2308527, EPI_ISL_2308528, EPI_ISL_2308529, EPI_ISL_2308530, EPI_ISL_2308531, EPI_ISL_2308532, EPI_ISL_2308533, EPI_ISL_2308534, EPI_ISL_2308535, EPI_ISL_2308536, EPI_ISL_2308537, EPI_ISL_2308538, EPI_ISL_2308539, EPI_ISL_2308540, EPI_ISL_2308541, EPI_ISL_2308542, EPI_ISL_2308543, EPI_ISL_2308544, EPI_ISL_2308545, EPI_ISL_2308546, EPI_ISL_2308547 | | | |
| see above | Nebraska Public Health Laboratory | NPHL COVID-19 Response Team | NPHL COVID-19 Response Team |
| EPI_ISL_2308645, EPI_ISL_2308646, EPI_ISL_2308647, EPI_ISL_2308648, EPI_ISL_2308649, EPI_ISL_2308650, EPI_ISL_2308651, EPI_ISL_2308652, EPI_ISL_2308653, EPI_ISL_2308654, EPI_ISL_2308655, EPI_ISL_2308656, EPI_ISL_2308657, EPI_ISL_2308658, EPI_ISL_2308659, EPI_ISL_2308660, EPI_ISL_2308661, EPI_ISL_2308662, EPI_ISL_2308663, EPI_ISL_2308664, EPI_ISL_2308665, EPI_ISL_2308666, EPI_ISL_2308667, EPI_ISL_2308668, EPI_ISL_2308669, EPI_ISL_2308670 | | | |
| see above | Alaska State Virology Laboratory | Alaska State Virology Laboratory | Stephanie DeRonde, Elva House, Jacob Zidek, Lisa Smith, Ph.D., Jack Chen, Ph.D. |
| EPI_ISL_2308671, EPI_ISL_2308672, EPI_ISL_2308673, EPI_ISL_2308674, EPI_ISL_2308675, EPI_ISL_2308676, EPI_ISL_2308677, EPI_ISL_2308679, EPI_ISL_2308680, EPI_ISL_2308681 | Kansas Health and Environmental Lab | Kansas Health and Environmental Lab | Mike Grose, Katherine Wiggins, Jonathan Barnell, Ben Olsen, and Phil Adam |
| EPI_ISL_2308682 | Clinical Reference Laboratory | Kansas Health and Environmental Lab | Mike Grose, Katherine Wiggins, Jonathan Barnell, Ben Olsen, and Phil Adam |
| EPI_ISL_2308721 | Wyoming Public Health Laboratory | Wyoming Public Health Laboratory | Jim Mildenberger, Wanda Manley, Noah Hull, Taylor Fearing, Lynette Gumbleton, Channing Weber, Ashley Norberg, Chayse Rowley, Marley Goetz, Brian Dominguez, Elliot Thomasson, Sam Britz, Cari Sloma, and Rob Christensen |
| EPI_ISL_2308755, EPI_ISL_2308758, EPI_ISL_2308762, EPI_ISL_2308763, EPI_ISL_2308766, EPI_ISL_2308768, EPI_ISL_2308769 | Wyoming Public Health Laboratory | Wyoming Public Health Laboratory | Jim Mildenberger, Taylor Fearing, Channing Weber, Ashley Norberg, Chayse Rowley, Marley Goetz, Brian Dominguez, Elliot Thomasson, Brittany Oher, Sam Britz, Cari Sloma, and Rob Christensen |
| EPI_ISL_2308781 | New York Presbyterian Hospital | Wadsworth Center, New York State Department of Health | Kirsten St. George, Daryl M. Lamson, Alexis Russell, Matthew Shudt, Melissa A Leisner, Jonathan Plitnick, Catharine Prussing, Navjot Singh, John Kelly, Erasmus Schneider, Erica Lasek-Nesselquist |
| EPI_ISL_2308782, EPI_ISL_2308783 | WESTCHESTER MEDICAL CENTER | Wadsworth Center, New York State Department of Health | Kirsten St. George, Daryl M. Lamson, Alexis Russell, Matthew Shudt, Melissa A Leisner, Jonathan Plitnick, Catharine Prussing, Navjot Singh, John Kelly, Erasmus Schneider, Erica Lasek-Nesselquist |
| EPI_ISL_2308784, EPI_ISL_2308785 | MEMORIAL SLOAN KETTERING CANCER CENTER | Wadsworth Center, New York State Department of Health | Kirsten St. George, Daryl M. Lamson, Alexis Russell, Matthew Shudt, Melissa A Leisner, Jonathan Plitnick, Catharine Prussing, Navjot Singh, John Kelly, Erasmus Schneider, Erica Lasek-Nesselquist |
| EPI_ISL_2308786 | Columbia University Irving Medical Center | Wadsworth Center, New York State Department of Health | Kirsten St. George, Daryl M. Lamson, Alexis Russell, Matthew Shudt, Melissa A Leisner, Jonathan Plitnick, Catharine Prussing, Navjot Singh, John Kelly, Erasmus Schneider, Erica Lasek-Nesselquist |
| EPI_ISL_2308787 | NYC Pandemic Response Lab | Wadsworth Center, New York State Department of Health | Kirsten St. George, Daryl M. Lamson, Alexis Russell, Matthew Shudt, Melissa A Leisner, Jonathan Plitnick, Catharine Prussing, Navjot Singh, John Kelly, Erasmus Schneider, Erica Lasek-Nesselquist |
| EPI_ISL_2308788, EPI_ISL_2308789 | MONTEFIORE MEDICAL CENTER LABORATORIES | Wadsworth Center, New York State Department of Health | Kirsten St. George, Daryl M. Lamson, Alexis Russell, Matthew Shudt, Melissa A Leisner, Jonathan Plitnick, Catharine Prussing, Navjot Singh, John Kelly, Erasmus Schneider, Erica Lasek-Nesselquist |
| EPI_ISL_2308790, EPI_ISL_2308791 | URMC LABS | Wadsworth Center, New York State Department of Health | Kirsten St. George, Daryl M. Lamson, Alexis Russell, Matthew Shudt, Melissa A Leisner, Jonathan Plitnick, Catharine Prussing, Navjot Singh, John Kelly, Erasmus Schneider, Erica Lasek-Nesselquist |
| EPI_ISL_2308792 | WESTCHESTER MEDICAL CENTER | Wadsworth Center, New York State Department of Health | Kirsten St. George, Daryl M. Lamson, Alexis Russell, Matthew Shudt, Melissa A Leisner, Jonathan Plitnick, Catharine Prussing, Navjot Singh, John Kelly, Erasmus Schneider, Erica Lasek-Nesselquist |
| EPI_ISL_2308793, EPI_ISL_2308794 | URMC LABS | Wadsworth Center, New York State Department of Health | Kirsten St. George, Daryl M. Lamson, Alexis Russell, Matthew Shudt, Melissa A Leisner, Jonathan Plitnick, Catharine Prussing, Navjot Singh, John Kelly, Erasmus Schneider, Erica Lasek-Nesselquist |
| EPI_ISL_2308795, EPI_ISL_2308796, EPI_ISL_2308797, EPI_ISL_2308798 | NYC Pandemic Response Lab | Wadsworth Center, New York State Department of Health | Kirsten St. George, Daryl M. Lamson, Alexis Russell, Matthew Shudt, Melissa A Leisner, Jonathan Plitnick, Catharine Prussing, Navjot Singh, John Kelly, Erasmus Schneider, Erica Lasek-Nesselquist |
| EPI_ISL_2308799, EPI_ISL_2308800, EPI_ISL_2308801 | URMC LABS | Wadsworth Center, New York State Department of Health | Kirsten St. George, Daryl M. Lamson, Alexis Russell, Matthew Shudt, Melissa A Leisner, Jonathan Plitnick, Catharine Prussing, Navjot Singh, John Kelly, Erasmus Schneider, Erica Lasek-Nesselquist |
| EPI_ISL_2308802 | ALBANY MEDICAL CENTER | Wadsworth Center, New York State Department of Health | Kirsten St. George, Daryl M. Lamson, Alexis Russell, Matthew Shudt, Melissa A Leisner, Jonathan Plitnick, Catharine Prussing, Navjot Singh, John Kelly, Erasmus Schneider, Erica Lasek-Nesselquist |
| EPI_ISL_2308803, EPI_ISL_2308804, EPI_ISL_2308805, EPI_ISL_2308806 | URMC LABS | Wadsworth Center, New York State Department of Health | Kirsten St. George, Daryl M. Lamson, Alexis Russell, Matthew Shudt, Melissa A Leisner, Jonathan Plitnick, Catharine Prussing, Navjot Singh, John Kelly, Erasmus Schneider, Erica Lasek-Nesselquist |
| EPI_ISL_2308954, EPI_ISL_2308956, EPI_ISL_2308958, EPI_ISL_2308960, EPI_ISL_2308963, EPI_ISL_2308964, EPI_ISL_2308965, EPI_ISL_2308966, EPI_ISL_2308967, EPI_ISL_2308969, EPI_ISL_2308970, EPI_ISL_2308971, EPI_ISL_2308973, EPI_ISL_2308976, EPI_ISL_2308977, EPI_ISL_2308978, EPI_ISL_2308979, EPI_ISL_2308981, EPI_ISL_2308982, EPI_ISL_2308983, EPI_ISL_2308984, EPI_ISL_2308985, EPI_ISL_2308988, EPI_ISL_2308989, EPI_ISL_2308990, EPI_ISL_2308992, EPI_ISL_2308993, EPI_ISL_2308995, EPI_ISL_2308996, EPI_ISL_2308997, EPI_ISL_2308998, EPI_ISL_2309002, EPI_ISL_2309004, EPI_ISL_2309006, EPI_ISL_2309007, EPI_ISL_2309011, EPI_ISL_2309013, EPI_ISL_2309014, EPI_ISL_2309015, EPI_ISL_2309016, EPI_ISL_2309017, EPI_ISL_2309018, EPI_ISL_2309019, EPI_ISL_2309020, EPI_ISL_2309021, EPI_ISL_2309024, EPI_ISL_2309027, EPI_ISL_2309029, EPI_ISL_2309031, EPI_ISL_2309033, EPI_ISL_2309035, EPI_ISL_2309036, EPI_ISL_2309040, EPI_ISL_2309041, EPI_ISL_2309042, EPI_ISL_2309045, EPI_ISL_2309046, EPI_ISL_2309053, EPI_ISL_2309054, EPI_ISL_2309055, EPI_ISL_2309056, EPI_ISL_2309059, EPI_ISL_2309060, EPI_ISL_2309062, EPI_ISL_2309064, EPI_ISL_2309065, EPI_ISL_2309066, EPI_ISL_2309068, EPI_ISL_2309069, EPI_ISL_2309070, EPI_ISL_2309071, EPI_ISL_2309073, EPI_ISL_2309074, EPI_ISL_2309077, EPI_ISL_2309080, EPI_ISL_2309082, EPI_ISL_2309083, EPI_ISL_2309085, EPI_ISL_2309087, EPI_ISL_2309089, EPI_ISL_2309093, EPI_ISL_2309094, EPI_ISL_2309096, EPI_ISL_2309103, EPI_ISL_2309104, EPI_ISL_2309105, EPI_ISL_2309106, EPI_ISL_2309108, EPI_ISL_2309109, EPI_ISL_2309111, EPI_ISL_2309112, EPI_ISL_2309115, EPI_ISL_2309116, EPI_ISL_2309117, EPI_ISL_2309121, EPI_ISL_2309124, EPI_ISL_2309125, EPI_ISL_2309129, EPI_ISL_2309132, EPI_ISL_2309137, EPI_ISL_2309138, EPI_ISL_2309139, EPI_ISL_2309140, EPI_ISL_2309141, EPI_ISL_2309145, EPI_ISL_2309146, EPI_ISL_2309148, EPI_ISL_2309149, EPI_ISL_2309153, EPI_ISL_2309154, EPI_ISL_2309158, EPI_ISL_2309159, EPI_ISL_2309160, EPI_ISL_2309162, EPI_ISL_2309163, EPI_ISL_2309165, EPI_ISL_2309166, EPI_ISL_2309170, EPI_ISL_2309172, EPI_ISL_2309173, EPI_ISL_2309178, EPI_ISL_2309181, EPI_ISL_2309189, EPI_ISL_2309190, EPI_ISL_2309191, EPI_ISL_2309193, EPI_ISL_2309194, EPI_ISL_2309195, EPI_ISL_2309198, EPI_ISL_2309201, EPI_ISL_2309202, EPI_ISL_2309206, EPI_ISL_2309207, EPI_ISL_2309215, EPI_ISL_2309217, EPI_ISL_2309218, EPI_ISL_2309219, EPI_ISL_2309221, EPI_ISL_2309227, EPI_ISL_2309228, EPI_ISL_2309229, EPI_ISL_2309231, EPI_ISL_2309232, EPI_ISL_2309233, EPI_ISL_2309234, EPI_ISL_2309235, EPI_ISL_2309236, EPI_ISL_2309238, EPI_ISL_2309240, EPI_ISL_2309242, EPI_ISL_2309243, EPI_ISL_2309246, EPI_ISL_2309249, EPI_ISL_2309252, EPI_ISL_2309253, EPI_ISL_2309255, EPI_ISL_2309256, EPI_ISL_2309257, EPI_ISL_2309258, EPI_ISL_2309261, EPI_ISL_2309262, EPI_ISL_2309263, EPI_ISL_2309264, EPI_ISL_2309266, EPI_ISL_2309268, EPI_ISL_2309270, EPI_ISL_2309272, EPI_ISL_2309273, EPI_ISL_2309275, EPI_ISL_2309276, EPI_ISL_2309277, EPI_ISL_2309278, EPI_ISL_2309279, EPI_ISL_2309282, EPI_ISL_2309283, EPI_ISL_2309286, EPI_ISL_2309289, EPI_ISL_2309290, EPI_ISL_2309292, EPI_ISL_2309293, EPI_ISL_2309294, EPI_ISL_2309295, EPI_ISL_2309299, EPI_ISL_2309300, EPI_ISL_2309302, EPI_ISL_2309303, EPI_ISL_2309304, EPI_ISL_2309305, EPI_ISL_2309310, EPI_ISL_2309311, EPI_ISL_2309312, EPI_ISL_2309314, EPI_ISL_2309315, EPI_ISL_2309316, EPI_ISL_2309317, EPI_ISL_2309318, EPI_ISL_2309319, EPI_ISL_2309321, EPI_ISL_2309324, EPI_ISL_2309327, EPI_ISL_2309328, EPI_ISL_2309331, EPI_ISL_2309332, EPI_ISL_2309335, EPI_ISL_2309336, EPI_ISL_2309337, EPI_ISL_2309338, EPI_ISL_2309339, EPI_ISL_2309341, EPI_ISL_2309344, EPI_ISL_2309345, EPI_ISL_2309346, EPI_ISL_2309347, EPI_ISL_2309348, EPI_ISL_2309352, EPI_ISL_2309357, EPI_ISL_2309358, EPI_ISL_2309359, EPI_ISL_2309361, EPI_ISL_2309364, EPI_ISL_2309365, EPI_ISL_2309366, EPI_ISL_2309367, EPI_ISL_2309368, EPI_ISL_2309369, EPI_ISL_2309374, EPI_ISL_2309376, EPI_ISL_2309377, EPI_ISL_2309378, EPI_ISL_2309379, EPI_ISL_2309380, EPI_ISL_2309381, EPI_ISL_2309385, EPI_ISL_2309386, EPI_ISL_2309387, EPI_ISL_2309388, EPI_ISL_2309391, EPI_ISL_2309393, EPI_ISL_2309394, EPI_ISL_2309396, EPI_ISL_2309398, EPI_ISL_2309399, EPI_ISL_2309400, EPI_ISL_2309401, EPI_ISL_2309402, EPI_ISL_2309404, EPI_ISL_2309405, EPI_ISL_2309406, EPI_ISL_2309407, EPI_ISL_2309408, EPI_ISL_2309410, EPI_ISL_2309412, EPI_ISL_2309413, EPI_ISL_2309414, EPI_ISL_2309415, EPI_ISL_2309416, EPI_ISL_2309417, EPI_ISL_2309418, EPI_ISL_2309420, EPI_ISL_2309421, EPI_ISL_2309422, EPI_ISL_2309423, EPI_ISL_2309424, EPI_ISL_2309425, EPI_ISL_2309426, EPI_ISL_2309427, EPI_ISL_2309429, EPI_ISL_2309430, EPI_ISL_2309433, EPI_ISL_2309434, EPI_ISL_2309435, EPI_ISL_2309438, EPI_ISL_2309440, EPI_ISL_2309441, EPI_ISL_2309442, EPI_ISL_2309443, EPI_ISL_2309445, EPI_ISL_2309446, EPI_ISL_2309450, EPI_ISL_2309453, EPI_ISL_2309454, EPI_ISL_2309456, EPI_ISL_2309458, EPI_ISL_2309459, EPI_ISL_2309461, EPI_ISL_2309462, EPI_ISL_2309465, EPI_ISL_2309466, EPI_ISL_2309467, EPI_ISL_2309468, EPI_ISL_2309469, EPI_ISL_2309472, EPI_ISL_2309473, EPI_ISL_2309474, EPI_ISL_2309475, EPI_ISL_2309476, EPI_ISL_2309477, EPI_ISL_2309478, EPI_ISL_2309482, EPI_ISL_2309483, EPI_ISL_2309485, EPI_ISL_2309486, EPI_ISL_2309488, EPI_ISL_2309489, EPI_ISL_2309490, EPI_ISL_2309491, EPI_ISL_2309492, EPI_ISL_2309493, EPI_ISL_2309496, EPI_ISL_2309497, EPI_ISL_2309498, EPI_ISL_2309499, EPI_ISL_2309501, EPI_ISL_2309505, EPI_ISL_2309506, EPI_ISL_2309507, EPI_ISL_2309508, EPI_ISL_2309509, EPI_ISL_2309510, EPI_ISL_2309511, EPI_ISL_2309513, EPI_ISL_2309514, EPI_ISL_2309515, EPI_ISL_2309516, EPI_ISL_2309517, EPI_ISL_2309518, EPI_ISL_2309519, EPI_ISL_2309521, EPI_ISL_2309522, EPI_ISL_2309523, EPI_ISL_2309524, EPI_ISL_2309525, EPI_ISL_2309530, EPI_ISL_2309531, EPI_ISL_2309532, EPI_ISL_2309533, EPI_ISL_2309535, EPI_ISL_2309536, EPI_ISL_2309537, EPI_ISL_2309539, EPI_ISL_2309541, EPI_ISL_2309542, EPI_ISL_2309543, EPI_ISL_2309544, EPI_ISL_2309545, EPI_ISL_2309547, EPI_ISL_2309548, EPI_ISL_2309551, EPI_ISL_2309554, EPI_ISL_2309555, EPI_ISL_2309556, EPI_ISL_2309557, EPI_ISL_2309559, EPI_ISL_2309560, EPI_ISL_2309561, EPI_ISL_2309564, EPI_ISL_2309566, EPI_ISL_2309567, EPI_ISL_2309569, EPI_ISL_2309570, EPI_ISL_2309571, EPI_ISL_2309572, EPI_ISL_2309575, EPI_ISL_2309576, EPI_ISL_2309578, EPI_ISL_2309579, EPI_ISL_2309580, EPI_ISL_2309583, EPI_ISL_2309584, EPI_ISL_2309588, EPI_ISL_2309589, EPI_ISL_2309591, EPI_ISL_2309592, EPI_ISL_2309594, EPI_ISL_2309595, EPI_ISL_2309597, EPI_ISL_2309599, EPI_ISL_2309601, EPI_ISL_2309602, EPI_ISL_2309603, EPI_ISL_2309604, EPI_ISL_2309606, EPI_ISL_2309608, EPI_ISL_2309610, EPI_ISL_2309611, EPI_ISL_2309612, EPI_ISL_2309613, EPI_ISL_2309614, EPI_ISL_2309615, EPI_ISL_2309616, EPI_ISL_2309618, EPI_ISL_2309619, EPI_ISL_2309620, EPI_ISL_2309621, EPI_ISL_2309627, EPI_ISL_2309629, EPI_ISL_2309632, EPI_ISL_2309634, EPI_ISL_2309635, EPI_ISL_2309638, EPI_ISL_2309639, EPI_ISL_2309643, EPI_ISL_2309644, EPI_ISL_2309647, EPI_ISL_2309658, EPI_ISL_2309659, EPI_ISL_2309662, EPI_ISL_2309664, EPI_ISL_2309667, EPI_ISL_2309669, EPI_ISL_2309671, EPI_ISL_2309673, EPI_ISL_2309675, EPI_ISL_2309679, EPI_ISL_2309680, EPI_ISL_2309681, EPI_ISL_2309682, EPI_ISL_2309683, EPI_ISL_2309685, EPI_ISL_2309693, EPI_ISL_2309694, EPI_ISL_2309697, EPI_ISL_2309698, EPI_ISL_2309701, EPI_ISL_2309703, EPI_ISL_2309704, EPI_ISL_2309712, EPI_ISL_2309716, EPI_ISL_2309720, EPI_ISL_2309723, EPI_ISL_2309727, EPI_ISL_2309728, EPI_ISL_2309729, EPI_ISL_2309735, EPI_ISL_2309738, EPI_ISL_2309742, EPI_ISL_2309745, EPI_ISL_2309746, EPI_ISL_2309747, EPI_ISL_2309748, EPI_ISL_2309749, EPI_ISL_2309750, EPI_ISL_2309752, EPI_ISL_2309755, EPI_ISL_2309756, EPI_ISL_2309763, EPI_ISL_2309766, EPI_ISL_2309767, EPI_ISL_2309770, EPI_ISL_2309771, EPI_ISL_2309777, EPI_ISL_2309781, EPI_ISL_2309788, EPI_ISL_2309790, EPI_ISL_2309792, EPI_ISL_2309793, EPI_ISL_2309794, EPI_ISL_2309795, EPI_ISL_2309796, EPI_ISL_2309797, EPI_ISL_2309798, EPI_ISL_2309801, EPI_ISL_2309802, EPI_ISL_2309803, EPI_ISL_2309805, EPI_ISL_2309806, EPI_ISL_2309808, EPI_ISL_2309810, EPI_ISL_2309815, EPI_ISL_2309816, EPI_ISL_2309819, EPI_ISL_2309820, EPI_ISL_2309821, EPI_ISL_2309825, EPI_ISL_2309827, EPI_ISL_2309828, EPI_ISL_2309830, EPI_ISL_2309833, EPI_ISL_2309834, EPI_ISL_2309837, EPI_ISL_2309840, EPI_ISL_2309844, EPI_ISL_2309846, EPI_ISL_2309847, EPI_ISL_2309849, EPI_ISL_2309850, EPI_ISL_2309853, EPI_ISL_2309854, EPI_ISL_2309855, EPI_ISL_2309857, EPI_ISL_2309858, EPI_ISL_2309860, EPI_ISL_2309861, EPI_ISL_2309862, EPI_ISL_2309863, EPI_ISL_2309866, EPI_ISL_2309867, EPI_ISL_2309870, EPI_ISL_2309873, EPI_ISL_2309874, EPI_ISL_2309876, EPI_ISL_2309878, EPI_ISL_2309879, EPI_ISL_2309880, EPI_ISL_2309881, EPI_ISL_2309882, EPI_ISL_2309884, EPI_ISL_2309887, EPI_ISL_2309888, EPI_ISL_2309889, EPI_ISL_2309892, EPI_ISL_2309893, EPI_ISL_2309894, EPI_ISL_2309895, EPI_ISL_2309896, EPI_ISL_2309897, EPI_ISL_2309898, EPI_ISL_2309899, EPI_ISL_2309901, EPI_ISL_2309902, EPI_ISL_2309903, EPI_ISL_2309906, EPI_ISL_2309908, EPI_ISL_2309909, EPI_ISL_2309910, EPI_ISL_2309911, EPI_ISL_2309912, EPI_ISL_2309914, EPI_ISL_2309915, EPI_ISL_2309917, EPI_ISL_2309918, EPI_ISL_2309919, EPI_ISL_2309920, EPI_ISL_2309922, EPI_ISL_2309923, EPI_ISL_2309925, EPI_ISL_2309926, EPI_ISL_2309927, EPI_ISL_2309929, EPI_ISL_2309931, EPI_ISL_2309934, EPI_ISL_2309935, EPI_ISL_2309936, EPI_ISL_2309937, EPI_ISL_2309939, EPI_ISL_2309940, EPI_ISL_2309941, EPI_ISL_2309942, EPI_ISL_2309943, EPI_ISL_2309946, EPI_ISL_2309947, EPI_ISL_2309948, EPI_ISL_2309949, EPI_ISL_2309950, EPI_ISL_2309951, EPI_ISL_2309952, EPI_ISL_2309953, EPI_ISL_2309954, EPI_ISL_2309956, EPI_ISL_2309957, EPI_ISL_2309958, EPI_ISL_2309960, EPI_ISL_2309961, EPI_ISL_2309962, EPI_ISL_2309963, EPI_ISL_2309965, EPI_ISL_2309966, EPI_ISL_2309967, EPI_ISL_2309969, EPI_ISL_2309970, EPI_ISL_2309972, EPI_ISL_2309973, EPI_ISL_2309974, EPI_ISL_2309975, EPI_ISL_2309978, EPI_ISL_2309979, EPI_ISL_2309981, EPI_ISL_2309983, EPI_ISL_2309984, EPI_ISL_2309985, EPI_ISL_2309986, EPI_ISL_2309987, EPI_ISL_2309988, EPI_ISL_2309989, EPI_ISL_2309990, EPI_ISL_2309991, EPI_ISL_2309993, EPI_ISL_2309996, EPI_ISL_2309999, EPI_ISL_2310000, EPI_ISL_2310001, EPI_ISL_2310002, EPI_ISL_2310003, EPI_ISL_2310005, EPI_ISL_2310007, EPI_ISL_2310008, EPI_ISL_2310009, EPI_ISL_2310010, EPI_ISL_2310012, EPI_ISL_2310015, EPI_ISL_2310020, EPI_ISL_2310021, EPI_ISL_2310022, EPI_ISL_2310023, EPI_ISL_2310025, EPI_ISL_2310027, EPI_ISL_2310028, EPI_ISL_2310029, EPI_ISL_2310030, EPI_ISL_2310031, EPI_ISL_2310032, EPI_ISL_2310033, EPI_ISL_2310034, EPI_ISL_2310036, EPI_ISL_2310037, EPI_ISL_2310038, EPI_ISL_2310039, EPI_ISL_2310041, EPI_ISL_2310043, EPI_ISL_2310044, EPI_ISL_2310045, EPI_ISL_2310047, EPI_ISL_2310048, EPI_ISL_2310050, EPI_ISL_2310051, EPI_ISL_2310052, EPI_ISL_2310053, EPI_ISL_2310055, EPI_ISL_2310056, EPI_ISL_2310057, EPI_ISL_2310058, EPI_ISL_2310059, EPI_ISL_2310061, EPI_ISL_2310062, EPI_ISL_2310063, EPI_ISL_2310066, EPI_ISL_2310067, EPI_ISL_2310069, EPI_ISL_2310071, EPI_ISL_2310072, EPI_ISL_2310073, EPI_ISL_2310074, EPI_ISL_2310075, EPI_ISL_2310076, EPI_ISL_2310077, EPI_ISL_2310079, EPI_ISL_2310080, EPI_ISL_2310081, EPI_ISL_2310083, EPI_ISL_2310085, EPI_ISL_2310087, EPI_ISL_2310088, EPI_ISL_2310089, EPI_ISL_2310090, EPI_ISL_2310094, EPI_ISL_2310096, EPI_ISL_2310103, EPI_ISL_2310104, EPI_ISL_2310106, EPI_ISL_2310110, EPI_ISL_2310112, EPI_ISL_2310115, EPI_ISL_2310116, EPI_ISL_2310118, EPI_ISL_2310119, EPI_ISL_2310121, EPI_ISL_2310122, EPI_ISL_2310123, EPI_ISL_2310124, EPI_ISL_2310126, EPI_ISL_2310127, EPI_ISL_2310129, EPI_ISL_2310130, EPI_ISL_2310131, EPI_ISL_2310134, EPI_ISL_2310136, EPI_ISL_2310138, EPI_ISL_2310139, EPI_ISL_2310140, EPI_ISL_2310141, EPI_ISL_2310142, EPI_ISL_2310146, EPI_ISL_2310149, EPI_ISL_2310151, EPI_ISL_2310154, EPI_ISL_2310155, EPI_ISL_2310156, EPI_ISL_2310157, EPI_ISL_2310158, EPI_ISL_2310159, EPI_ISL_2310160, EPI_ISL_2310161, EPI_ISL_2310165, EPI_ISL_2310166, EPI_ISL_2310167, EPI_ISL_2310168, EPI_ISL_2310169, EPI_ISL_2310171, EPI_ISL_2310173, EPI_ISL_2310174, EPI_ISL_2310175, EPI_ISL_2310176, EPI_ISL_2310177, EPI_ISL_2310178, EPI_ISL_2310181, EPI_ISL_2310183, EPI_ISL_2310186, EPI_ISL_2310187, EPI_ISL_2310191, EPI_ISL_2310192, EPI_ISL_2310193, EPI_ISL_2310194, EPI_ISL_2310195, EPI_ISL_2310196, EPI_ISL_2310199, EPI_ISL_2310200, EPI_ISL_2310202, EPI_ISL_2310203, EPI_ISL_2310204, EPI_ISL_2310205, EPI_ISL_2310206, EPI_ISL_2310208, EPI_ISL_2310211, EPI_ISL_2310213, EPI_ISL_2310215, EPI_ISL_2310216, EPI_ISL_2310217, EPI_ISL_2310218, EPI_ISL_2310220, EPI_ISL_2310228, EPI_ISL_2310229, EPI_ISL_2310230, EPI_ISL_2310232, EPI_ISL_2310233, EPI_ISL_2310234, EPI_ISL_2310237, EPI_ISL_2310238, EPI_ISL_2310239, EPI_ISL_2310240, EPI_ISL_2310241, EPI_ISL_2310242, EPI_ISL_2310243, EPI_ISL_2310244, EPI_ISL_2310245, EPI_ISL_2310253, EPI_ISL_2310255, EPI_ISL_2310256, EPI_ISL_2310257, EPI_ISL_2310263, EPI_ISL_2310265, EPI_ISL_2310276, EPI_ISL_2310278, EPI_ISL_2310279, EPI_ISL_2310282, EPI_ISL_2310284, EPI_ISL_2310286, EPI_ISL_2310289, EPI_ISL_2310291, EPI_ISL_2310295, EPI_ISL_2310296, EPI_ISL_2310317, EPI_ISL_2310329, EPI_ISL_2310334, EPI_ISL_2310370, EPI_ISL_2310371, EPI_ISL_2310372, EPI_ISL_2310373, EPI_ISL_2310374, EPI_ISL_2310375, EPI_ISL_2310376, | | | |

EPI_ISL_2310377, EPI_ISL_2310378, EPI_ISL_2310382, EPI_ISL_2310383, EPI_ISL_2310387, EPI_ISL_2310388, EPI_ISL_2310389, EPI_ISL_2310390, EPI_ISL_2310392, EPI_ISL_2310393, EPI_ISL_2310395, EPI_ISL_2310398, EPI_ISL_2310400, EPI_ISL_2310401, EPI_ISL_2310402, EPI_ISL_2310403, EPI_ISL_2310404, EPI_ISL_2310407, EPI_ISL_2310410, EPI_ISL_2310411, EPI_ISL_2310412, EPI_ISL_2310414, EPI_ISL_2310417, EPI_ISL_2310419, EPI_ISL_2310420, EPI_ISL_2310422, EPI_ISL_2310423, EPI_ISL_2310426, EPI_ISL_2310427, EPI_ISL_2310433, EPI_ISL_2310436, EPI_ISL_2310437, EPI_ISL_2310438, EPI_ISL_2310439, EPI_ISL_2310440, EPI_ISL_2310441, EPI_ISL_2310442, EPI_ISL_2310443, EPI_ISL_2310444, EPI_ISL_2310446, EPI_ISL_2310447, EPI_ISL_2310448, EPI_ISL_2310449, EPI_ISL_2310450, EPI_ISL_2310451, EPI_ISL_2310452, EPI_ISL_2310453, EPI_ISL_2310455, EPI_ISL_2310456, EPI_ISL_2310457, EPI_ISL_2310458, EPI_ISL_2310459, EPI_ISL_2310460, EPI_ISL_2310461, EPI_ISL_2310463, EPI_ISL_2310464, EPI_ISL_2310465, EPI_ISL_2310466, EPI_ISL_2310468, EPI_ISL_2310470, EPI_ISL_2310475, EPI_ISL_2310476, EPI_ISL_2310477, EPI_ISL_2310480, EPI_ISL_2310482, EPI_ISL_2310483, EPI_ISL_2310485, EPI_ISL_2310486, EPI_ISL_2310488, EPI_ISL_2310489, EPI_ISL_2310490, EPI_ISL_2310491, EPI_ISL_2310492, EPI_ISL_2310493, EPI_ISL_2310494, EPI_ISL_2310496, EPI_ISL_2310500, EPI_ISL_2310501, EPI_ISL_2310502, EPI_ISL_2310503, EPI_ISL_2310505, EPI_ISL_2310506, EPI_ISL_2310507, EPI_ISL_2310509, EPI_ISL_2310510, EPI_ISL_2310512, EPI_ISL_2310514, EPI_ISL_2310515, EPI_ISL_2310516, EPI_ISL_2310519, EPI_ISL_2310520, EPI_ISL_2310523, EPI_ISL_2310524, EPI_ISL_2310525, EPI_ISL_2310526, EPI_ISL_2310527, EPI_ISL_2310530, EPI_ISL_2310531, EPI_ISL_2310533, EPI_ISL_2310536, EPI_ISL_2310537, EPI_ISL_2310539, EPI_ISL_2310540, EPI_ISL_2310543, EPI_ISL_2310544, EPI_ISL_2310545, EPI_ISL_2310549, EPI_ISL_2310550, EPI_ISL_2310551, EPI_ISL_2310552, EPI_ISL_2310553, EPI_ISL_2310557, EPI_ISL_2310559, EPI_ISL_2310560, EPI_ISL_2310561, EPI_ISL_2310563, EPI_ISL_2310564, EPI_ISL_2310565, EPI_ISL_2310566, EPI_ISL_2310568, EPI_ISL_2310569, EPI_ISL_2310570, EPI_ISL_2310571, EPI_ISL_2310572, EPI_ISL_2310575, EPI_ISL_2310577, EPI_ISL_2310579, EPI_ISL_2310581, EPI_ISL_2310582, EPI_ISL_2310584, EPI_ISL_2310586, EPI_ISL_2310587, EPI_ISL_2310588, EPI_ISL_2310590, EPI_ISL_2310591, EPI_ISL_2310592, EPI_ISL_2310593, EPI_ISL_2310594, EPI_ISL_2310595, EPI_ISL_2310596, EPI_ISL_2310599, EPI_ISL_2310600, EPI_ISL_2310601, EPI_ISL_2310602, EPI_ISL_2310603, EPI_ISL_2310604, EPI_ISL_2310606, EPI_ISL_2310607, EPI_ISL_2310609, EPI_ISL_2310612, EPI_ISL_2310613, EPI_ISL_2310615, EPI_ISL_2310616, EPI_ISL_2310618, EPI_ISL_2310620, EPI_ISL_2310621, EPI_ISL_2310622, EPI_ISL_2310623, EPI_ISL_2310624, EPI_ISL_2310625, EPI_ISL_2310626, EPI_ISL_2310627, EPI_ISL_2310628, EPI_ISL_2310629, EPI_ISL_2310630, EPI_ISL_2310631, EPI_ISL_2310632, EPI_ISL_2310633, EPI_ISL_2310634, EPI_ISL_2310635, EPI_ISL_2310639, EPI_ISL_2310641, EPI_ISL_2310642, EPI_ISL_2310644, EPI_ISL_2310646, EPI_ISL_2310647, EPI_ISL_2310648, EPI_ISL_2310652, EPI_ISL_2310653, EPI_ISL_2310654, EPI_ISL_2310656, EPI_ISL_2310658, EPI_ISL_2310659, EPI_ISL_2310660, EPI_ISL_2310661, EPI_ISL_2310662, EPI_ISL_2310663, EPI_ISL_2310664, EPI_ISL_2310665, EPI_ISL_2310666, EPI_ISL_2310667, EPI_ISL_2310669, EPI_ISL_2310670, EPI_ISL_2310671, EPI_ISL_2310672, EPI_ISL_2310673, EPI_ISL_2310676, EPI_ISL_2310677, EPI_ISL_2310678, EPI_ISL_2310679, EPI_ISL_2310680, EPI_ISL_2310681, EPI_ISL_2310682, EPI_ISL_2310683, EPI_ISL_2310684, EPI_ISL_2310685, EPI_ISL_2310690, EPI_ISL_2310691, EPI_ISL_2310692, EPI_ISL_2310693, EPI_ISL_2310694, EPI_ISL_2310695, EPI_ISL_2310696, EPI_ISL_2310698, EPI_ISL_2310699, EPI_ISL_2310701, EPI_ISL_2310702, EPI_ISL_2310703, EPI_ISL_2310704, EPI_ISL_2310705, EPI_ISL_2310706, EPI_ISL_2310707, EPI_ISL_2310708, EPI_ISL_2310709, EPI_ISL_2310710, EPI_ISL_2310711, EPI_ISL_2310712, EPI_ISL_2310713, EPI_ISL_2310714, EPI_ISL_2310716, EPI_ISL_2310717, EPI_ISL_2310718, EPI_ISL_2310719, EPI_ISL_2310720, EPI_ISL_2310721, EPI_ISL_2310722, EPI_ISL_2310723, EPI_ISL_2310725, EPI_ISL_2310727, EPI_ISL_2310729, EPI_ISL_2310732, EPI_ISL_2310736, EPI_ISL_2310738, EPI_ISL_2310745, EPI_ISL_2310747, EPI_ISL_2310748, EPI_ISL_2310749, EPI_ISL_2310750, EPI_ISL_2310751, EPI_ISL_2310753, EPI_ISL_2310754, EPI_ISL_2310755, EPI_ISL_2310756, EPI_ISL_2310757, EPI_ISL_2310759, EPI_ISL_2310762, EPI_ISL_2310765, EPI_ISL_2310766, EPI_ISL_2310767, EPI_ISL_2310768, EPI_ISL_2310769, EPI_ISL_2310770, EPI_ISL_2310771, EPI_ISL_2310774, EPI_ISL_2310776, EPI_ISL_2310780, EPI_ISL_2310781, EPI_ISL_2310782, EPI_ISL_2310783, EPI_ISL_2310784, EPI_ISL_2310785, EPI_ISL_2310787, EPI_ISL_2310788, EPI_ISL_2310789, EPI_ISL_2310790, EPI_ISL_2310792, EPI_ISL_2310793, EPI_ISL_2310795, EPI_ISL_2310796, EPI_ISL_2310797, EPI_ISL_2310798, EPI_ISL_2310799, EPI_ISL_2310800, EPI_ISL_2310801, EPI_ISL_2310802, EPI_ISL_2310803, EPI_ISL_2310805, EPI_ISL_2310807, EPI_ISL_2310808, EPI_ISL_2310809, EPI_ISL_2310811, EPI_ISL_2310813, EPI_ISL_2310814, EPI_ISL_2310815, EPI_ISL_2310816, EPI_ISL_2310817, EPI_ISL_2310818, EPI_ISL_2310819, EPI_ISL_2310821, EPI_ISL_2310822, EPI_ISL_2310823, EPI_ISL_2310824, EPI_ISL_2310827, EPI_ISL_2310830, EPI_ISL_2310831, EPI_ISL_2310832, EPI_ISL_2310834, EPI_ISL_2310835, EPI_ISL_2310836, EPI_ISL_2310837, EPI_ISL_2310838, EPI_ISL_2310839, EPI_ISL_2310840, EPI_ISL_2310841, EPI_ISL_2310842, EPI_ISL_2310843, EPI_ISL_2310844, EPI_ISL_2310848, EPI_ISL_2310849, EPI_ISL_2310850, EPI_ISL_2310852, EPI_ISL_2310853, EPI_ISL_2310854, EPI_ISL_2310855, EPI_ISL_2310857, EPI_ISL_2310859, EPI_ISL_2310860, EPI_ISL_2310861, EPI_ISL_2310862, EPI_ISL_2310864, EPI_ISL_2310866, EPI_ISL_2310867, EPI_ISL_2310869, EPI_ISL_2310870, EPI_ISL_2310873, EPI_ISL_2310875, EPI_ISL_2310877, EPI_ISL_2310878, EPI_ISL_2310879, EPI_ISL_2310881, EPI_ISL_2310882, EPI_ISL_2310887, EPI_ISL_2310889, EPI_ISL_2310890, EPI_ISL_2310893, EPI_ISL_2310894, EPI_ISL_2310897, EPI_ISL_2310900, EPI_ISL_2310902, EPI_ISL_2310903, EPI_ISL_2310906, EPI_ISL_2310909, EPI_ISL_2310911, EPI_ISL_2310912, EPI_ISL_2310914, EPI_ISL_2310915, EPI_ISL_2310916, EPI_ISL_2310917, EPI_ISL_2310920, EPI_ISL_2310922, EPI_ISL_2310923, EPI_ISL_2310927, EPI_ISL_2310930, EPI_ISL_2310931, EPI_ISL_2310935, EPI_ISL_2310936, EPI_ISL_2310938, EPI_ISL_2310940, EPI_ISL_2310944, EPI_ISL_2310947, EPI_ISL_2310948, EPI_ISL_2310949, EPI_ISL_2310953, EPI_ISL_2310954, EPI_ISL_2310955, EPI_ISL_2310956, EPI_ISL_2310957, EPI_ISL_2310958, EPI_ISL_2310961, EPI_ISL_2310963, EPI_ISL_2310964, EPI_ISL_2310965, EPI_ISL_2310967, EPI_ISL_2310969, EPI_ISL_2310970, EPI_ISL_2310971, EPI_ISL_2310972, EPI_ISL_2310975, EPI_ISL_2310977, EPI_ISL_2310981, EPI_ISL_2310982, EPI_ISL_2310983, EPI_ISL_2310984, EPI_ISL_2310985, EPI_ISL_2310986, EPI_ISL_2310987, EPI_ISL_2310990, EPI_ISL_2310991, EPI_ISL_2310994, EPI_ISL_2310995, EPI_ISL_2310996, EPI_ISL_2310997, EPI_ISL_2310999, EPI_ISL_2311000, EPI_ISL_2311001, EPI_ISL_2311002, EPI_ISL_2311005, EPI_ISL_2311006, EPI_ISL_2311008, EPI_ISL_2311009, EPI_ISL_2311010, EPI_ISL_2311011, EPI_ISL_2311012, EPI_ISL_2311013, EPI_ISL_2311014, EPI_ISL_2311015, EPI_ISL_2311016, EPI_ISL_2311018, EPI_ISL_2311021, EPI_ISL_2311022, EPI_ISL_2311023, EPI_ISL_2311025, EPI_ISL_2311026, EPI_ISL_2311028, EPI_ISL_2311029, EPI_ISL_2311031, EPI_ISL_2311033, EPI_ISL_2311034, EPI_ISL_2311035, EPI_ISL_2311037, EPI_ISL_2311038, EPI_ISL_2311039, EPI_ISL_2311040, EPI_ISL_2311041, EPI_ISL_2311042, EPI_ISL_2311043, EPI_ISL_2311044, EPI_ISL_2311045, EPI_ISL_2311046, EPI_ISL_2311047, EPI_ISL_2311048, EPI_ISL_2311050, EPI_ISL_2311051, EPI_ISL_2311052, EPI_ISL_2311053, EPI_ISL_2311055, EPI_ISL_2311057, EPI_ISL_2311058, EPI_ISL_2311059, EPI_ISL_2311060, EPI_ISL_2311061, EPI_ISL_2311062, EPI_ISL_2311064, EPI_ISL_2311065, EPI_ISL_2311067, EPI_ISL_2311068, EPI_ISL_2311069, EPI_ISL_2311071, EPI_ISL_2311072, EPI_ISL_2311073, EPI_ISL_2311075, EPI_ISL_2311076, EPI_ISL_2311077, EPI_ISL_2311080, EPI_ISL_2311081, EPI_ISL_2311082, EPI_ISL_2311083, EPI_ISL_2311084, EPI_ISL_2311085, EPI_ISL_2311086, EPI_ISL_2311087, EPI_ISL_2311088, EPI_ISL_2311089, EPI_ISL_2311090, EPI_ISL_2311091, EPI_ISL_2311092, EPI_ISL_2311093, EPI_ISL_2311095, EPI_ISL_2311096, EPI_ISL_2311098, EPI_ISL_2311099, EPI_ISL_2311100, EPI_ISL_2311101, EPI_ISL_2311103, EPI_ISL_2311105, EPI_ISL_2311107, EPI_ISL_2311108, EPI_ISL_2311109, EPI_ISL_2311112, EPI_ISL_2311114, EPI_ISL_2311115, EPI_ISL_2311116, EPI_ISL_2311117, EPI_ISL_2311118, EPI_ISL_2311120, EPI_ISL_2311121, EPI_ISL_2311122, EPI_ISL_2311123, EPI_ISL_2311125, EPI_ISL_2311127, EPI_ISL_2311128, EPI_ISL_2311130, EPI_ISL_2311131, EPI_ISL_2311132, EPI_ISL_2311133, EPI_ISL_2311134, EPI_ISL_2311135, EPI_ISL_2311136, EPI_ISL_2311137, EPI_ISL_2311138, EPI_ISL_2311139, EPI_ISL_2311140, EPI_ISL_2311141, EPI_ISL_2311142, EPI_ISL_2311143

| | | | |
|---|---|---|---|
| see above | Colorado Department of Public Health and Environment | Colorado Department of Public Health and Environment | Laura Bankers, Molly C. Hetherington-Rauth, Diana Ir, Alexandria Rossheim, Shannon R. Matzinger, Sarah Elizabeth Totten, Emily A. Travanty |
| EPI_ISL_2311744, EPI_ISL_2311745, EPI_ISL_2311746 | WVDHHR - Office of Laboratory Services | Genomics and Discovery, Respiratory Viruses Branch, Division of Viral Diseases, Centers for Disease Control and Prevention | Jing Zhang, Anna Kelleher, Ying Tao, Yan Li, Brian Lynch, Krista Queen, Anna Uehara, Peter Cook, Han Jia Justin Ng, Rachel Marine, Clinton R. Paden, Dhwani Batra, Haibin Wang, Tara Coalter, Jasmine Padilla, Morgan Davis, Mili Sheth, Sarah Nobles, Mark Burroughs, Justin Lee, Adam Retchless, Suxiang Tong |
| EPI_ISL_2311747, EPI_ISL_2311748, EPI_ISL_2311749, EPI_ISL_2311750, EPI_ISL_2311751 | GA Department of Public Health Laboratory | Genomics and Discovery, Respiratory Viruses Branch, Division of Viral Diseases, Centers for Disease Control and Prevention | Jing Zhang, Anna Kelleher, Ying Tao, Yan Li, Brian Lynch, Krista Queen, Anna Uehara, Peter Cook, Han Jia Justin Ng, Rachel Marine, Clinton R. Paden, Dhwani Batra, Haibin Wang, Tara Coalter, Jasmine Padilla, Morgan Davis, Mili Sheth, Sarah Nobles, Mark Burroughs, Justin Lee, Adam Retchless, Suxiang Tong |
| EPI_ISL_2311752 | AL Dept. of Public Health Bureau of Clinical Laboratories | Genomics and Discovery, Respiratory Viruses Branch, Division of Viral Diseases, Centers for Disease Control and Prevention | Jing Zhang, Anna Kelleher, Ying Tao, Yan Li, Brian Lynch, Krista Queen, Anna Uehara, Peter Cook, Han Jia Justin Ng, Rachel Marine, Clinton R. Paden, Dhwani Batra, Haibin Wang, Tara Coalter, Jasmine Padilla, Morgan Davis, Mili Sheth, Sarah Nobles, Mark Burroughs, Justin Lee, Adam Retchless, Suxiang Tong |
| EPI_ISL_2311753, EPI_ISL_2311754 | NC State Laboratory of Public Health | Genomics and Discovery, Respiratory Viruses Branch, Division of Viral Diseases, Centers for Disease Control and Prevention | Jing Zhang, Anna Kelleher, Ying Tao, Yan Li, Brian Lynch, Krista Queen, Anna Uehara, Peter Cook, Han Jia Justin Ng, Rachel Marine, Clinton R. Paden, Dhwani Batra, Haibin Wang, Tara Coalter, Jasmine Padilla, Morgan Davis, Mili Sheth, Sarah Nobles, Mark Burroughs, Justin Lee, Adam Retchless, Suxiang Tong |
| EPI_ISL_2311755, EPI_ISL_2311756, EPI_ISL_2311757 | AR Dept. of Health-PHL, Molecular Diagnostics | Genomics and Discovery, Respiratory Viruses Branch, Division of Viral Diseases, Centers for Disease Control and Prevention | Jing Zhang, Anna Kelleher, Ying Tao, Yan Li, Brian Lynch, Krista Queen, Anna Uehara, Peter Cook, Han Jia Justin Ng, Rachel Marine, Clinton R. Paden, Dhwani Batra, Haibin Wang, Tara Coalter, Jasmine Padilla, Morgan Davis, Mili Sheth, Sarah Nobles, Mark Burroughs, Justin Lee, Adam Retchless, Suxiang Tong |
| EPI_ISL_2311758 | LA Office of Public Health Laboratories | Genomics and Discovery, Respiratory Viruses Branch, Division of Viral Diseases, Centers for Disease Control and Prevention | Jing Zhang, Anna Kelleher, Ying Tao, Yan Li, Brian Lynch, Krista Queen, Anna Uehara, Peter Cook, Han Jia Justin Ng, Rachel Marine, Clinton R. Paden, Dhwani Batra, Haibin Wang, Tara Coalter, Jasmine Padilla, Morgan Davis, Mili Sheth, Sarah Nobles, Mark Burroughs, Justin Lee, Adam Retchless, Suxiang Tong |
| EPI_ISL_2311759, EPI_ISL_2311760, EPI_ISL_2311761, EPI_ISL_2311762, EPI_ISL_2311763, EPI_ISL_2311764, EPI_ISL_2311765 | GA Department of Public Health Laboratory | Genomics and Discovery, Respiratory Viruses Branch, Division of Viral Diseases, Centers for Disease Control and Prevention | Jing Zhang, Anna Kelleher, Ying Tao, Yan Li, Brian Lynch, Krista Queen, Anna Uehara, Peter Cook, Han Jia Justin Ng, Rachel Marine, Clinton R. Paden, Dhwani Batra, Haibin Wang, Tara Coalter, Jasmine Padilla, Morgan Davis, Mili Sheth, Sarah Nobles, Mark Burroughs, Justin Lee, Adam Retchless, Suxiang Tong |
| EPI_ISL_2311766, EPI_ISL_2311767 | TN Division of Laboratory Services | Genomics and Discovery, Respiratory Viruses Branch, Division of Viral Diseases, Centers for Disease Control and Prevention | Jing Zhang, Anna Kelleher, Ying Tao, Yan Li, Brian Lynch, Krista Queen, Anna Uehara, Peter Cook, Han Jia Justin Ng, Rachel Marine, Clinton R. Paden, Dhwani Batra, Haibin Wang, Tara Coalter, Jasmine Padilla, Morgan Davis, Mili Sheth, Sarah Nobles, Mark Burroughs, Justin Lee, Adam Retchless, Suxiang Tong |
| EPI_ISL_2311768, EPI_ISL_2311769, EPI_ISL_2311770, EPI_ISL_2311771, EPI_ISL_2311772, EPI_ISL_2311773, EPI_ISL_2311774 | OK Public Health Laboratory, Oklahoma State DOH | Genomics and Discovery, Respiratory Viruses Branch, Division of Viral Diseases, Centers for Disease Control and Prevention | Jing Zhang, Anna Kelleher, Ying Tao, Yan Li, Brian Lynch, Krista Queen, Anna Uehara, Peter Cook, Han Jia Justin Ng, Rachel Marine, Clinton R. Paden, Dhwani Batra, Haibin Wang, Tara Coalter, Jasmine Padilla, Morgan Davis, Mili Sheth, Sarah Nobles, Mark Burroughs, Justin Lee, Adam Retchless, Suxiang Tong |
| EPI_ISL_2311862 | Monterey County Public Health Laboratory | Monterey County Public Health Laboratory | Monterey County Public Health Laboratory |

EPI_ISL_2311863, EPI_ISL_2311864, EPI_ISL_2311865, EPI_ISL_2311866, EPI_ISL_2311867, EPI_ISL_2311869, EPI_ISL_2311870, EPI_ISL_2311871, EPI_ISL_2311872, EPI_ISL_2311873, EPI_ISL_2311874, EPI_ISL_2311875, EPI_ISL_2311876, EPI_ISL_2311877, EPI_ISL_2311878, EPI_ISL_2311879, EPI_ISL_2311880, EPI_ISL_2311881, EPI_ISL_2311882, EPI_ISL_2311883, EPI_ISL_2311884, EPI_ISL_2311885, EPI_ISL_2311886, EPI_ISL_2311887, EPI_ISL_2311888, EPI_ISL_2311889, EPI_ISL_2311890, EPI_ISL_2311891, EPI_ISL_2311892, EPI_ISL_2311893, EPI_ISL_2311894, EPI_ISL_2311895, EPI_ISL_2311897, EPI_ISL_2311898, EPI_ISL_2311899, EPI_ISL_2311900, EPI_ISL_2311901, EPI_ISL_2311902, EPI_ISL_2311903, EPI_ISL_2311904, EPI_ISL_2311905, EPI_ISL_2311906, EPI_ISL_2311907, EPI_ISL_2311908, EPI_ISL_2311909, EPI_ISL_2311911, EPI_ISL_2311912, EPI_ISL_2311913, EPI_ISL_2311914, EPI_ISL_2311915, EPI_ISL_2311916, EPI_ISL_2311917, EPI_ISL_2311918, EPI_ISL_2311919, EPI_ISL_2311920, EPI_ISL_2311921, EPI_ISL_2311922, EPI_ISL_2311923, EPI_ISL_2311925, EPI_ISL_2311926, EPI_ISL_2311927, EPI_ISL_2311928, EPI_ISL_2311929, EPI_ISL_2311930, EPI_ISL_2311931, EPI_ISL_2311932, EPI_ISL_2311933, EPI_ISL_2311934, EPI_ISL_2311935, EPI_ISL_2311936, EPI_ISL_2311937, EPI_ISL_2311938, EPI_ISL_2311939, EPI_ISL_2311940, EPI_ISL_2311941, EPI_ISL_2311942, EPI_ISL_2311943, EPI_ISL_2311944, EPI_ISL_2311945, EPI_ISL_2311946, EPI_ISL_2311947, EPI_ISL_2311948, EPI_ISL_2311949, EPI_ISL_2311950, EPI_ISL_2311951, EPI_ISL_2311952, EPI_ISL_2311953, EPI_ISL_2311954, EPI_ISL_2311955, EPI_ISL_2311956, EPI_ISL_2311957, EPI_ISL_2311958, EPI_ISL_2311959, EPI_ISL_2311960, EPI_ISL_2311961, EPI_ISL_2311962, EPI_ISL_2311963, EPI_ISL_2311964, EPI_ISL_2311965, EPI_ISL_2311966, EPI_ISL_2311968, EPI_ISL_2311969, EPI_ISL_2311970, EPI_ISL_2311971, EPI_ISL_2311972, EPI_ISL_2311973, EPI_ISL_2311974, EPI_ISL_2311975, EPI_ISL_2311976, EPI_ISL_2311977, EPI_ISL_2311978, EPI_ISL_2311979, EPI_ISL_2311980, EPI_ISL_2311981, EPI_ISL_2311982, EPI_ISL_2311983, EPI_ISL_2311985, EPI_ISL_2311986, EPI_ISL_2311987, EPI_ISL_2311988, EPI_ISL_2311989, EPI_ISL_2311990, EPI_ISL_2311991, EPI_ISL_2311992, EPI_ISL_2311993, EPI_ISL_2311994, EPI_ISL_2311995, EPI_ISL_2311996, EPI_ISL_2311997, EPI_ISL_2311998, EPI_ISL_2311999, EPI_ISL_2312000, EPI_ISL_2312001, EPI_ISL_2312002, EPI_ISL_2312003, EPI_ISL_2312004, EPI_ISL_2312005, EPI_ISL_2312006, EPI_ISL_2312007, EPI_ISL_2312008, EPI_ISL_2312009, EPI_ISL_2312010, EPI_ISL_2312011, EPI_ISL_2312012, EPI_ISL_2312013, EPI_ISL_2312014, EPI_ISL_2312015, EPI_ISL_2312016, EPI_ISL_2312017, EPI_ISL_2312018, EPI_ISL_2312019, EPI_ISL_2312020, EPI_ISL_2312021, EPI_ISL_2312022, EPI_ISL_2312023, EPI_ISL_2312024, EPI_ISL_2312025, EPI_ISL_2312026, EPI_ISL_2312027, EPI_ISL_2312028, EPI_ISL_2312029, EPI_ISL_2312030, EPI_ISL_2312031, EPI_ISL_2312032, EPI_ISL_2312033, EPI_ISL_2312034, EPI_ISL_2312035, EPI_ISL_2312036, EPI_ISL_2312037, EPI_ISL_2312038, EPI_ISL_2312039, EPI_ISL_2312040, EPI_ISL_2312041, EPI_ISL_2312042, EPI_ISL_2312043, EPI_ISL_2312044, EPI_ISL_2312045, EPI_ISL_2312046, EPI_ISL_2312047, EPI_ISL_2312048, EPI_ISL_2312049, EPI_ISL_2312050, EPI_ISL_2312051, EPI_ISL_2312052, EPI_ISL_2312053, EPI_ISL_2312054, EPI_ISL_2312055, EPI_ISL_2312056, EPI_ISL_2312057, EPI_ISL_2312058, EPI_ISL_2312059, EPI_ISL_2312060, EPI_ISL_2312062, EPI_ISL_2312063, EPI_ISL_2312064, EPI_ISL_2312065, EPI_ISL_2312066, EPI_ISL_2312067, EPI_ISL_2312068, EPI_ISL_2312069, EPI_ISL_2312070, EPI_ISL_2312071, EPI_ISL_2312072, EPI_ISL_2312073, EPI_ISL_2312074, EPI_ISL_2312075, EPI_ISL_2312076, EPI_ISL_2312077, EPI_ISL_2312078, EPI_ISL_2312079, EPI_ISL_2312080, EPI_ISL_2312081, EPI_ISL_2312082, EPI_ISL_2312083, EPI_ISL_2312084, EPI_ISL_2312085, EPI_ISL_2312086, EPI_ISL_2312087, EPI_ISL_2312088, EPI_ISL_2312089, EPI_ISL_2312090, EPI_ISL_2312091, EPI_ISL_2312093, EPI_ISL_2312094, EPI_ISL_2312095, EPI_ISL_2312096, EPI_ISL_2312097, EPI_ISL_2312098, EPI_ISL_2312099, EPI_ISL_2312100, EPI_ISL_2312101, EPI_ISL_2312102, EPI_ISL_2312103, EPI_ISL_2312104, EPI_ISL_2312105, EPI_ISL_2312107, EPI_ISL_2312108, EPI_ISL_2312109, EPI_ISL_2312110, EPI_ISL_2312111, EPI_ISL_2312112, EPI_ISL_2312113, EPI_ISL_2312114, EPI_ISL_2312115, EPI_ISL_2312116, EPI_ISL_2312117, EPI_ISL_2312118, EPI_ISL_2312119, EPI_ISL_2312120, EPI_ISL_2312121, EPI_ISL_2312122, EPI_ISL_2312123, EPI_ISL_2312124, EPI_ISL_2312125, EPI_ISL_2312126, EPI_ISL_2312127, EPI_ISL_2312128, EPI_ISL_2312129, EPI_ISL_2312130, EPI_ISL_2312131, EPI_ISL_2312132, EPI_ISL_2312133, EPI_ISL_2312134, EPI_ISL_2312135, EPI_ISL_2312136, EPI_ISL_2312137, EPI_ISL_2312139, EPI_ISL_2312140, EPI_ISL_2312141, EPI_ISL_2312142, EPI_ISL_2312143, EPI_ISL_2312144, EPI_ISL_2312145, EPI_ISL_2312146, EPI_ISL_2312147, EPI_ISL_2312148, EPI_ISL_2312149, EPI_ISL_2312150, EPI_ISL_2312151, EPI_ISL_2312152, EPI_ISL_2312153, EPI_ISL_2312154, EPI_ISL_2312155, EPI_ISL_2312156, EPI_ISL_2312157, EPI_ISL_2312158, EPI_ISL_2312159, EPI_ISL_2312160, EPI_ISL_2312161, EPI_ISL_2312163, EPI_ISL_2312164, EPI_ISL_2312165, EPI_ISL_2312166, EPI_ISL_2312167, EPI_ISL_2312168, EPI_ISL_2312169, EPI_ISL_2312170, EPI_ISL_2312171, EPI_ISL_2312172, EPI_ISL_2312173, EPI_ISL_2312174, EPI_ISL_2312175, EPI_ISL_2312176, EPI_ISL_2312177, EPI_ISL_2312178, EPI_ISL_2312179, EPI_ISL_2312180, EPI_ISL_2312181, EPI_ISL_2312182, EPI_ISL_2312183, EPI_ISL_2312184, EPI_ISL_2312185, EPI_ISL_2312186, EPI_ISL_2312187, EPI_ISL_2312188, EPI_ISL_2312189, EPI_ISL_2312190, EPI_ISL_2312191, EPI_ISL_2312192, EPI_ISL_2312193, EPI_ISL_2312195, EPI_ISL_2312196, EPI_ISL_2312197, EPI_ISL_2312198, EPI_ISL_2312199, EPI_ISL_2312200, EPI_ISL_2312201, EPI_ISL_2312202, EPI_ISL_2312203, EPI_ISL_2312204, EPI_ISL_2312205, EPI_ISL_2312206, EPI_ISL_2312207, EPI_ISL_2312209, EPI_ISL_2312210, EPI_ISL_2312211, EPI_ISL_2312212, EPI_ISL_2312213, EPI_ISL_2312214, EPI_ISL_2312215, EPI_ISL_2312216, EPI_ISL_2312217, EPI_ISL_2312218, EPI_ISL_2312219, EPI_ISL_2312220, EPI_ISL_2312221, EPI_ISL_2312222, EPI_ISL_2312223, EPI_ISL_2312225, EPI_ISL_2312226, EPI_ISL_2312227, EPI_ISL_2312228, EPI_ISL_2312229, EPI_ISL_2312230, EPI_ISL_2312231, EPI_ISL_2312232, EPI_ISL_2312233, EPI_ISL_2312234, EPI_ISL_2312235, EPI_ISL_2312236, EPI_ISL_2312237, EPI_ISL_2312238, EPI_ISL_2312239, EPI_ISL_2312240, EPI_ISL_2312241, EPI_ISL_2312242, EPI_ISL_2312243, EPI_ISL_2312244, EPI_ISL_2312245, EPI_ISL_2312247, EPI_ISL_2312248, EPI_ISL_2312249, EPI_ISL_2312250, EPI_ISL_2312251, EPI_ISL_2312252, EPI_ISL_2312253, EPI_ISL_2312254, EPI_ISL_2312255, EPI_ISL_2312256, EPI_ISL_2312257, EPI_ISL_2312258, EPI_ISL_2312259, EPI_ISL_2312260, EPI_ISL_2312261, EPI_ISL_2312262, EPI_ISL_2312263, EPI_ISL_2312264, EPI_ISL_2312265, EPI_ISL_2312266, EPI_ISL_2312267, EPI_ISL_2312268, EPI_ISL_2312269, EPI_ISL_2312270, EPI_ISL_2312271, EPI_ISL_2312272, EPI_ISL_2312273, EPI_ISL_2312274, EPI_ISL_2312275, EPI_ISL_2312276, EPI_ISL_2312278, EPI_ISL_2312279, EPI_ISL_2312280, EPI_ISL_2312281, EPI_ISL_2312283, EPI_ISL_2312284, EPI_ISL_2312285, EPI_ISL_2312286, EPI_ISL_2312287, EPI_ISL_2312288, EPI_ISL_2312289, EPI_ISL_2312290, EPI_ISL_2312291, EPI_ISL_2312292, EPI_ISL_2312293, EPI_ISL_2312294, EPI_ISL_2312295, EPI_ISL_2312296, EPI_ISL_2312297, EPI_ISL_2312298, EPI_ISL_2312299, EPI_ISL_2312300, EPI_ISL_2312301, EPI_ISL_2312302, EPI_ISL_2312303, EPI_ISL_2312304, EPI_ISL_2312305, EPI_ISL_2312306, EPI_ISL_2312307, EPI_ISL_2312308, EPI_ISL_2312309, EPI_ISL_2312310, EPI_ISL_2312311, EPI_ISL_2312312, EPI_ISL_2312313, EPI_ISL_2312314, EPI_ISL_2312315, EPI_ISL_2312316, EPI_ISL_2312317, EPI_ISL_2312318, EPI_ISL_2312319, EPI_ISL_2312320, EPI_ISL_2312321, EPI_ISL_2312322, EPI_ISL_2312323, EPI_ISL_2312324, EPI_ISL_2312325, EPI_ISL_2312326, EPI_ISL_2312327, EPI_ISL_2312328, EPI_ISL_2312329, EPI_ISL_2312330, EPI_ISL_2312331, EPI_ISL_2312332, EPI_ISL_2312333, EPI_ISL_2312334, EPI_ISL_2312335, EPI_ISL_2312336, EPI_ISL_2312337, EPI_ISL_2312338, EPI_ISL_2312339, EPI_ISL_2312340, EPI_ISL_2312341, EPI_ISL_2312342, EPI_ISL_2312343, EPI_ISL_2312344, EPI_ISL_2312345, EPI_ISL_2312347, EPI_ISL_2312348, EPI_ISL_2312349, EPI_ISL_2312350, EPI_ISL_2312351, EPI_ISL_2312352, EPI_ISL_2312353, EPI_ISL_2312354, EPI_ISL_2312355, EPI_ISL_2312356, EPI_ISL_2312357, EPI_ISL_2312358, EPI_ISL_2312359, EPI_ISL_2312360, EPI_ISL_2312361, EPI_ISL_2312362, EPI_ISL_2312363, EPI_ISL_2312364, EPI_ISL_2312365, EPI_ISL_2312366, EPI_ISL_2312367, EPI_ISL_2312368, EPI_ISL_2312369, EPI_ISL_2312370, EPI_ISL_2312372, EPI_ISL_2312373, EPI_ISL_2312374, EPI_ISL_2312375, EPI_ISL_2312376, EPI_ISL_2312377, EPI_ISL_2312379, EPI_ISL_2312380, EPI_ISL_2312381, EPI_ISL_2312382, EPI_ISL_2312383, EPI_ISL_2312384, EPI_ISL_2312385, EPI_ISL_2312386, EPI_ISL_2312387, EPI_ISL_2312388, EPI_ISL_2312389, EPI_ISL_2312390, EPI_ISL_2312391, EPI_ISL_2312392, EPI_ISL_2312393, EPI_ISL_2312394, EPI_ISL_2312395, EPI_ISL_2312396, EPI_ISL_2312397, EPI_ISL_2312398, EPI_ISL_2312399, EPI_ISL_2312400, EPI_ISL_2312401, EPI_ISL_2312402, EPI_ISL_2312403, EPI_ISL_2312404, EPI_ISL_2312405, EPI_ISL_2312406, EPI_ISL_2312407, EPI_ISL_2312408, EPI_ISL_2312409, EPI_ISL_2312410, EPI_ISL_2312411, EPI_ISL_2312412, EPI_ISL_2312413, EPI_ISL_2312414, EPI_ISL_2312415, EPI_ISL_2312416, EPI_ISL_2312417, EPI_ISL_2312418, EPI_ISL_2312419, EPI_ISL_2312420, EPI_ISL_2312421, EPI_ISL_2312422, EPI_ISL_2312423, EPI_ISL_2312424, EPI_ISL_2312425, EPI_ISL_2312426, EPI_ISL_2312427, EPI_ISL_2312428, EPI_ISL_2312429, EPI_ISL_2312430, EPI_ISL_2312431, EPI_ISL_2312432, EPI_ISL_2312433, EPI_ISL_2312434, EPI_ISL_2312435, EPI_ISL_2312436, EPI_ISL_2312437, EPI_ISL_2312438, EPI_ISL_2312439, EPI_ISL_2312440, EPI_ISL_2312441, EPI_ISL_2312442, EPI_ISL_2312443, EPI_ISL_2312444, EPI_ISL_2312445, EPI_ISL_2312446, EPI_ISL_2312447, EPI_ISL_2312448, EPI_ISL_2312449, EPI_ISL_2312450, EPI_ISL_2312451, EPI_ISL_2312452, EPI_ISL_2312453, EPI_ISL_2312454, EPI_ISL_2312455, EPI_ISL_2312456, EPI_ISL_2312457, EPI_ISL_2312458, EPI_ISL_2312459, EPI_ISL_2312460, EPI_ISL_2312461, EPI_ISL_2312462, EPI_ISL_2312463, EPI_ISL_2312464, EPI_ISL_2312465, EPI_ISL_2312466, EPI_ISL_2312467, EPI_ISL_2312468, EPI_ISL_2312469, EPI_ISL_2312470, EPI_ISL_2312471, EPI_ISL_2312472, EPI_ISL_2312473, EPI_ISL_2312474, EPI_ISL_2312475, EPI_ISL_2312476, EPI_ISL_2312477, EPI_ISL_2312478, EPI_ISL_2312479, EPI_ISL_2312480, EPI_ISL_2312481, EPI_ISL_2312483, EPI_ISL_2312484, EPI_ISL_2312485, EPI_ISL_2312486, EPI_ISL_2312488, EPI_ISL_2312489, EPI_ISL_2312490, EPI_ISL_2312491

| | | | |
|---|---|---|---|
| see above | Broad Institute Clinical Research Sequencing Platform | Infectious Disease Program, Broad Institute of Harvard and MIT | Siddle,K.J., Adams,G., Pearlman,L., Gladden-Young,A., Vicente,G., Blumenstiel,B., DeFelice,M., Lee,M., McGovern,S., Lagerborg,K., Rudy,M., DeRuff,K., Carter,A., Normandin,E., Bauer,M., Reilly,S., Tomkins-Tinch,C., Loreth,C., Chaluvadi,S., Meldrim,J., Granger,B., Lemieux,J.E., Birren,B.W., Sabeti,P.C., Larkin,K., Dodge,S., Lennon,N., Madoff,L., Brown,C., Gallagher,G., Smole,S., Park,D.J., Gabriel,S., and MacInnis,B.L. |

EPI_ISL_2312728, EPI_ISL_2312729, EPI_ISL_2312730, EPI_ISL_2312731, EPI_ISL_2312732, EPI_ISL_2312733, EPI_ISL_2312734, EPI_ISL_2312735, EPI_ISL_2312736, EPI_ISL_2312737, EPI_ISL_2312738, EPI_ISL_2312739, EPI_ISL_2312740, EPI_ISL_2312741, EPI_ISL_2312742, EPI_ISL_2312743, EPI_ISL_2312744, EPI_ISL_2312745, EPI_ISL_2312746, EPI_ISL_2312747, EPI_ISL_2312748, EPI_ISL_2312749, EPI_ISL_2312750, EPI_ISL_2312751, EPI_ISL_2312753, EPI_ISL_2312754, EPI_ISL_2312755, EPI_ISL_2312756, EPI_ISL_2312757, EPI_ISL_2312758, EPI_ISL_2312759, EPI_ISL_2312760, EPI_ISL_2312761, EPI_ISL_2312763, EPI_ISL_2312764, EPI_ISL_2312765, EPI_ISL_2312766, EPI_ISL_2312767, EPI_ISL_2312768, EPI_ISL_2312769, EPI_ISL_2312770, EPI_ISL_2312771, EPI_ISL_2312772, EPI_ISL_2312773, EPI_ISL_2312774, EPI_ISL_2312775, EPI_ISL_2312776, EPI_ISL_2312777, EPI_ISL_2312778, EPI_ISL_2312780, EPI_ISL_2312781, EPI_ISL_2312782, EPI_ISL_2312783, EPI_ISL_2312784, EPI_ISL_2312785, EPI_ISL_2312786, EPI_ISL_2312787, EPI_ISL_2312788, EPI_ISL_2312789, EPI_ISL_2312790, EPI_ISL_2312791, EPI_ISL_2312792, EPI_ISL_2312793, EPI_ISL_2312794, EPI_ISL_2312795, EPI_ISL_2312796, EPI_ISL_2312797, EPI_ISL_2312798, EPI_ISL_2312799, EPI_ISL_2312800, EPI_ISL_2312801, EPI_ISL_2312802, EPI_ISL_2312803, EPI_ISL_2312804, EPI_ISL_2312805, EPI_ISL_2312806, EPI_ISL_2312807, EPI_ISL_2312809, EPI_ISL_2312810, EPI_ISL_2312811, EPI_ISL_2312812, EPI_ISL_2312813, EPI_ISL_2312815, EPI_ISL_2312816, EPI_ISL_2312817, EPI_ISL_2312818, EPI_ISL_2312819, EPI_ISL_2312820, EPI_ISL_2312821, EPI_ISL_2312822, EPI_ISL_2312823, EPI_ISL_2312824, EPI_ISL_2312825, EPI_ISL_2312826, EPI_ISL_2312827, EPI_ISL_2312828, EPI_ISL_2312829, EPI_ISL_2312830, EPI_ISL_2312831, EPI_ISL_2312832, EPI_ISL_2312833, EPI_ISL_2312834, EPI_ISL_2312835, EPI_ISL_2312836, EPI_ISL_2312837, EPI_ISL_2312838, EPI_ISL_2312839, EPI_ISL_2312840, EPI_ISL_2312841, EPI_ISL_2312842, EPI_ISL_2312843, EPI_ISL_2312844, EPI_ISL_2312845, EPI_ISL_2312846, EPI_ISL_2312847, EPI_ISL_2312848, EPI_ISL_2312849, EPI_ISL_2312850, EPI_ISL_2312851, EPI_ISL_2312852, EPI_ISL_2312853, EPI_ISL_2312854, EPI_ISL_2312855, EPI_ISL_2312856, EPI_ISL_2312857, EPI_ISL_2312858, EPI_ISL_2312859, EPI_ISL_2312860, EPI_ISL_2312861, EPI_ISL_2312862, EPI_ISL_2312863, EPI_ISL_2312864, EPI_ISL_2312865, EPI_ISL_2312866, EPI_ISL_2312867, EPI_ISL_2312868, EPI_ISL_2312869, EPI_ISL_2312870, EPI_ISL_2312871, EPI_ISL_2312872, EPI_ISL_2312873, EPI_ISL_2312874, EPI_ISL_2312875, EPI_ISL_2312876, EPI_ISL_2312877, EPI_ISL_2312878, EPI_ISL_2312879, EPI_ISL_2312880, EPI_ISL_2312881, EPI_ISL_2312882, EPI_ISL_2312883, EPI_ISL_2312884, EPI_ISL_2312885, EPI_ISL_2312886, EPI_ISL_2312887, EPI_ISL_2312888, EPI_ISL_2312889, EPI_ISL_2312890, EPI_ISL_2312891, EPI_ISL_2312892, EPI_ISL_2312893, EPI_ISL_2312894, EPI_ISL_2312895, EPI_ISL_2312896, EPI_ISL_2312897, EPI_ISL_2312898, EPI_ISL_2312899, EPI_ISL_2312900, EPI_ISL_2312901, EPI_ISL_2312902, EPI_ISL_2312903, EPI_ISL_2312904, EPI_ISL_2312905, EPI_ISL_2312906, EPI_ISL_2312907, EPI_ISL_2312908, EPI_ISL_2312909, EPI_ISL_2312910, EPI_ISL_2312911, EPI_ISL_2312912, EPI_ISL_2312913, EPI_ISL_2312914, EPI_ISL_2312915, EPI_ISL_2312916, EPI_ISL_2312917, EPI_ISL_2312918, EPI_ISL_2312919, EPI_ISL_2312920, EPI_ISL_2312921, EPI_ISL_2312922, EPI_ISL_2312923, EPI_ISL_2312924, EPI_ISL_2312925, EPI_ISL_2312926,

| | | | |
|---|---|---|---|
| EPI_ISL_2312927, EPI_ISL_2312928, EPI_ISL_2312929, EPI_ISL_2312930, EPI_ISL_2312931, EPI_ISL_2312932, EPI_ISL_2312933, EPI_ISL_2312934, EPI_ISL_2312935, EPI_ISL_2312936, EPI_ISL_2312937, EPI_ISL_2312938, EPI_ISL_2312939, EPI_ISL_2312940, EPI_ISL_2312941, EPI_ISL_2312942, EPI_ISL_2312943, EPI_ISL_2312944, EPI_ISL_2312945, EPI_ISL_2312946, EPI_ISL_2312947, EPI_ISL_2312948, EPI_ISL_2312949, EPI_ISL_2312950, EPI_ISL_2312951, EPI_ISL_2312952, EPI_ISL_2312954, EPI_ISL_2312955, EPI_ISL_2312956, EPI_ISL_2312957, EPI_ISL_2312958, EPI_ISL_2312959, EPI_ISL_2312960 | | | |
| see above | The Ohio State University Applied Microbiology Services Laboratory | The Ohio State University Applied Microbiology Services Laboratory | Seth A. Faith PhD |
| EPI_ISL_2312961, EPI_ISL_2312962, EPI_ISL_2312963, EPI_ISL_2312964, EPI_ISL_2312966, EPI_ISL_2312967, EPI_ISL_2312968, EPI_ISL_2312969, EPI_ISL_2312970, EPI_ISL_2312971, EPI_ISL_2312972, EPI_ISL_2312973, EPI_ISL_2312974, EPI_ISL_2312975, EPI_ISL_2312976, EPI_ISL_2312977, EPI_ISL_2312978, EPI_ISL_2312979, EPI_ISL_2312980, EPI_ISL_2312981 | | | |
| see above | Microbiology Division, SC DHEC | Microbiology Division, SC DHEC | Freeman,J., Flores,H. |
| EPI_ISL_2312982, EPI_ISL_2312983, EPI_ISL_2312984, EPI_ISL_2312985, EPI_ISL_2312986, EPI_ISL_2312987, EPI_ISL_2312988, EPI_ISL_2312989, EPI_ISL_2312990, EPI_ISL_2312991 | Microbiology Division, SC DHEC | Microbiology Division, SC DHEC | Freeman,J. |
| EPI_ISL_2312992, EPI_ISL_2312993, EPI_ISL_2312994, EPI_ISL_2312995, EPI_ISL_2312997, EPI_ISL_2312998, EPI_ISL_2312999, EPI_ISL_2313000, EPI_ISL_2313001, EPI_ISL_2313002, EPI_ISL_2313003, EPI_ISL_2313004, EPI_ISL_2313005, EPI_ISL_2313006, EPI_ISL_2313007, EPI_ISL_2313008, EPI_ISL_2313009, EPI_ISL_2313010, EPI_ISL_2313011, EPI_ISL_2313012, EPI_ISL_2313013, EPI_ISL_2313014, EPI_ISL_2313016, EPI_ISL_2313017, EPI_ISL_2313018, EPI_ISL_2313019, EPI_ISL_2313020, EPI_ISL_2313021, EPI_ISL_2313022, EPI_ISL_2313023, EPI_ISL_2313024, EPI_ISL_2313026, EPI_ISL_2313027, EPI_ISL_2313028, EPI_ISL_2313029, EPI_ISL_2313030, EPI_ISL_2313032, EPI_ISL_2313033, EPI_ISL_2313034, EPI_ISL_2313035, EPI_ISL_2313036, EPI_ISL_2313037, EPI_ISL_2313038, EPI_ISL_2313039, EPI_ISL_2313040, EPI_ISL_2313041, EPI_ISL_2313042, EPI_ISL_2313043, EPI_ISL_2313044 | | | |
| see above | Microbiology Division, SC DHEC | Microbiology Division, SC DHEC | Freeman,J., Flores,H. |
| EPI_ISL_2313087, EPI_ISL_2313088, EPI_ISL_2313089, EPI_ISL_2313090, EPI_ISL_2313091, EPI_ISL_2313092, EPI_ISL_2313093, EPI_ISL_2313094, EPI_ISL_2313095, EPI_ISL_2313096, EPI_ISL_2313097 | | | |
| see above | Houston Health Department, Disease Prevention and Control | Houston Health Department, Disease Prevention and Control | Penn,R., Brown,P., Lara,A., Lai,Y. |
| EPI_ISL_2313101, EPI_ISL_2313102, EPI_ISL_2313103, EPI_ISL_2313104, EPI_ISL_2313105, EPI_ISL_2313106, EPI_ISL_2313107, EPI_ISL_2313108, EPI_ISL_2313109, EPI_ISL_2313110, EPI_ISL_2313111 | | | |
| see above | Laboratory Medicine, University of Washington | Laboratory Medicine, University of Washington | Roychoudhury,P., Xie,H., Shrestha,L., Bakhash,S.M., Lin,M.J., Baker,N.R., Ellis,S., Sathees,S., Huang,M.-L., Jerome,K.R., Greninger,A. |
| EPI_ISL_2313477, EPI_ISL_2313479, EPI_ISL_2313481, EPI_ISL_2313483, EPI_ISL_2313485, EPI_ISL_2313486, EPI_ISL_2313488, EPI_ISL_2313490, EPI_ISL_2313492, EPI_ISL_2313494, EPI_ISL_2313495, EPI_ISL_2313497, EPI_ISL_2313499, EPI_ISL_2313501, EPI_ISL_2313502, EPI_ISL_2313504, EPI_ISL_2313506, EPI_ISL_2313508, EPI_ISL_2313510, EPI_ISL_2313511, EPI_ISL_2313513, EPI_ISL_2313515, EPI_ISL_2313517, EPI_ISL_2313518, EPI_ISL_2313520, EPI_ISL_2313521, EPI_ISL_2313523, EPI_ISL_2313525, EPI_ISL_2313527, EPI_ISL_2313528, EPI_ISL_2313532, EPI_ISL_2313534, EPI_ISL_2313535, EPI_ISL_2313539, EPI_ISL_2313541, EPI_ISL_2313542, EPI_ISL_2313544, EPI_ISL_2313546, EPI_ISL_2313548, EPI_ISL_2313550, EPI_ISL_2313551, EPI_ISL_2313553, EPI_ISL_2313555, EPI_ISL_2313557, EPI_ISL_2313558, EPI_ISL_2313560, EPI_ISL_2313562, EPI_ISL_2313564, EPI_ISL_2313565, EPI_ISL_2313567, EPI_ISL_2313568, EPI_ISL_2313570, EPI_ISL_2313572, EPI_ISL_2313573, EPI_ISL_2313575, EPI_ISL_2313576, EPI_ISL_2313578, EPI_ISL_2313580, EPI_ISL_2313582, EPI_ISL_2313584, EPI_ISL_2313586, EPI_ISL_2313587, EPI_ISL_2313589, EPI_ISL_2313591, EPI_ISL_2313592, EPI_ISL_2313594, EPI_ISL_2313596, EPI_ISL_2313597, EPI_ISL_2313599, EPI_ISL_2313601, EPI_ISL_2313602, EPI_ISL_2313604, EPI_ISL_2313606, EPI_ISL_2313608, EPI_ISL_2313610, EPI_ISL_2313612, EPI_ISL_2313613, EPI_ISL_2313617, EPI_ISL_2313619, EPI_ISL_2313621, EPI_ISL_2313623, EPI_ISL_2313624, EPI_ISL_2313626, EPI_ISL_2313628, EPI_ISL_2313630, EPI_ISL_2313632, EPI_ISL_2313633, EPI_ISL_2313635, EPI_ISL_2313637, EPI_ISL_2313640, EPI_ISL_2313642, EPI_ISL_2313644, EPI_ISL_2313646, EPI_ISL_2313647, EPI_ISL_2313649, EPI_ISL_2313651, EPI_ISL_2313653, EPI_ISL_2313655, EPI_ISL_2313657, EPI_ISL_2313658, EPI_ISL_2313660, EPI_ISL_2313662, EPI_ISL_2313663, EPI_ISL_2313665, EPI_ISL_2313667, EPI_ISL_2313668, EPI_ISL_2313670, EPI_ISL_2313672, EPI_ISL_2313674, EPI_ISL_2313675, EPI_ISL_2313677, EPI_ISL_2313679, EPI_ISL_2313680, EPI_ISL_2313681, EPI_ISL_2313682, EPI_ISL_2313684, EPI_ISL_2313686, EPI_ISL_2313687, EPI_ISL_2313689, EPI_ISL_2313691, EPI_ISL_2313695, EPI_ISL_2313696, EPI_ISL_2313698, EPI_ISL_2313699, EPI_ISL_2313701, EPI_ISL_2313703, EPI_ISL_2313705, EPI_ISL_2313707, EPI_ISL_2313709, EPI_ISL_2313711, EPI_ISL_2313713, EPI_ISL_2313715, EPI_ISL_2313716, EPI_ISL_2313718, EPI_ISL_2313720, EPI_ISL_2313722, EPI_ISL_2313723, EPI_ISL_2313725, EPI_ISL_2313727, EPI_ISL_2313728, EPI_ISL_2313730, EPI_ISL_2313732, EPI_ISL_2313734, EPI_ISL_2313736, EPI_ISL_2313737, EPI_ISL_2313739, EPI_ISL_2313741, EPI_ISL_2313743, EPI_ISL_2313744, EPI_ISL_2313746, EPI_ISL_2313748, EPI_ISL_2313749, EPI_ISL_2313751, EPI_ISL_2313753, EPI_ISL_2313755, EPI_ISL_2313756, EPI_ISL_2313758, EPI_ISL_2313760, EPI_ISL_2313762, EPI_ISL_2313764, EPI_ISL_2313765, EPI_ISL_2313767, EPI_ISL_2313771, EPI_ISL_2313773, EPI_ISL_2313775, EPI_ISL_2313777, EPI_ISL_2313778, EPI_ISL_2313780, EPI_ISL_2313783, EPI_ISL_2313787, EPI_ISL_2313789, EPI_ISL_2313791, EPI_ISL_2313792, EPI_ISL_2313793, EPI_ISL_2313794, EPI_ISL_2313795, EPI_ISL_2313796, EPI_ISL_2313797, EPI_ISL_2313799, EPI_ISL_2313800, EPI_ISL_2313802, EPI_ISL_2313803, EPI_ISL_2313805, EPI_ISL_2313806, EPI_ISL_2313808, EPI_ISL_2313810, EPI_ISL_2313812, EPI_ISL_2313814, EPI_ISL_2313816, EPI_ISL_2313818, EPI_ISL_2313819, EPI_ISL_2313821, EPI_ISL_2313823, EPI_ISL_2313825, EPI_ISL_2313827, EPI_ISL_2313828, EPI_ISL_2313830, EPI_ISL_2313832, EPI_ISL_2313834, EPI_ISL_2313836, EPI_ISL_2313838, EPI_ISL_2313840, EPI_ISL_2313841, EPI_ISL_2313843, EPI_ISL_2313844, EPI_ISL_2313846, EPI_ISL_2313848, EPI_ISL_2313850, EPI_ISL_2313852, EPI_ISL_2313854, EPI_ISL_2313855, EPI_ISL_2313857, EPI_ISL_2313859, EPI_ISL_2313861, EPI_ISL_2313862, EPI_ISL_2313864, EPI_ISL_2313865, EPI_ISL_2313867, EPI_ISL_2313869, EPI_ISL_2313870, EPI_ISL_2313871, EPI_ISL_2313873, EPI_ISL_2313874, EPI_ISL_2313876, EPI_ISL_2313878 | | | |
| see above | Respiratory Viruses Branch, Centers for Disease Control and Prevention | Respiratory Viruses Branch, Centers for Disease Control and Prevention | Howard,D., Batra,D., Cook,P.W., Moser,K., Paskey,A., Caravas,J., Rambo-Martin,B., Morrison,S., Gulvick,C., Sammons,S., Unoarumhi,Y., Wagner,D., Schmerer,M., Bixby,C., Wang,Y., Schultz,J., Goswami,C., Hager,R., Grimwood,R., Paden,C.R., MacCannell,D. |
| EPI_ISL_2319171, EPI_ISL_2319172, EPI_ISL_2319173, EPI_ISL_2319174, EPI_ISL_2319175, EPI_ISL_2319176, EPI_ISL_2319177, EPI_ISL_2319178, EPI_ISL_2319179, EPI_ISL_2319180, EPI_ISL_2319181, EPI_ISL_2319182, EPI_ISL_2319183, EPI_ISL_2319184, EPI_ISL_2319185, EPI_ISL_2319186, EPI_ISL_2319187, EPI_ISL_2319188, EPI_ISL_2319189, EPI_ISL_2319190, EPI_ISL_2319191, EPI_ISL_2319192, EPI_ISL_2319194, EPI_ISL_2319195, EPI_ISL_2319196, EPI_ISL_2319197, EPI_ISL_2319198, EPI_ISL_2319199, EPI_ISL_2319200, EPI_ISL_2319201, EPI_ISL_2319202, EPI_ISL_2319203, EPI_ISL_2319204, EPI_ISL_2319205, EPI_ISL_2319206, EPI_ISL_2319207, EPI_ISL_2319208, EPI_ISL_2319209, EPI_ISL_2319210, EPI_ISL_2319211, EPI_ISL_2319212, EPI_ISL_2319213, EPI_ISL_2319214, EPI_ISL_2319215, EPI_ISL_2319216, EPI_ISL_2319217, EPI_ISL_2319218, EPI_ISL_2319219, EPI_ISL_2319220, EPI_ISL_2319221, EPI_ISL_2319222, EPI_ISL_2319224, EPI_ISL_2319225, EPI_ISL_2319226, EPI_ISL_2319227, EPI_ISL_2319228, EPI_ISL_2319229, EPI_ISL_2319230, EPI_ISL_2319231, EPI_ISL_2319232, EPI_ISL_2319233, EPI_ISL_2319234, EPI_ISL_2319235, EPI_ISL_2319236, EPI_ISL_2319238, EPI_ISL_2319239, EPI_ISL_2319240, EPI_ISL_2319241, EPI_ISL_2319242, EPI_ISL_2319243, EPI_ISL_2319244, EPI_ISL_2319245, EPI_ISL_2319246, EPI_ISL_2319247, EPI_ISL_2319248, EPI_ISL_2319249, EPI_ISL_2319250, EPI_ISL_2319251, EPI_ISL_2319252, EPI_ISL_2319253, EPI_ISL_2319254, EPI_ISL_2319255, EPI_ISL_2319256, EPI_ISL_2319257, EPI_ISL_2319259, EPI_ISL_2319260, EPI_ISL_2319261, EPI_ISL_2319262, EPI_ISL_2319263, EPI_ISL_2319264, EPI_ISL_2319266, EPI_ISL_2319267, EPI_ISL_2319268, EPI_ISL_2319269, EPI_ISL_2319270, EPI_ISL_2319271, EPI_ISL_2319272, EPI_ISL_2319273, EPI_ISL_2319274, EPI_ISL_2319275, EPI_ISL_2319276, EPI_ISL_2319277, EPI_ISL_2319278, EPI_ISL_2319279, EPI_ISL_2319280, EPI_ISL_2319281, EPI_ISL_2319282, EPI_ISL_2319284, EPI_ISL_2319285, EPI_ISL_2319286, EPI_ISL_2319287, EPI_ISL_2319288, EPI_ISL_2319289, EPI_ISL_2319291, EPI_ISL_2319293, EPI_ISL_2319294, EPI_ISL_2319295, EPI_ISL_2319296, EPI_ISL_2319297, EPI_ISL_2319298, EPI_ISL_2319299, EPI_ISL_2319300, EPI_ISL_2319301, EPI_ISL_2319302, EPI_ISL_2319303, EPI_ISL_2319304, EPI_ISL_2319306, EPI_ISL_2319307, EPI_ISL_2319308, EPI_ISL_2319309, EPI_ISL_2319310, EPI_ISL_2319311, EPI_ISL_2319312 | | | |
| see above | Salud Digna | Instituto Nacional de Medicina Genomica | Hidalgo-Miranda A, Cedro-Tanda A, Mendoza-Vargas A, Reyes-Grajeda JP, Abraham Campos-Romero, Moreno-Camacho José Luis, Rodriguez-Gallegos Jorge, Luna-Ruiz Marco, Gonzalez-Barrera D, Rangel-DeLeon D, Munguia-Garza P, Ramirez-Vega O, Escobar-Arrazola, M, Herrera-Montalvo LA. |
| EPI_ISL_2319313, EPI_ISL_2319314, EPI_ISL_2319315, EPI_ISL_2319316, EPI_ISL_2319317, EPI_ISL_2319319, EPI_ISL_2319320, EPI_ISL_2319321, EPI_ISL_2319322, EPI_ISL_2319323, EPI_ISL_2319324, EPI_ISL_2319326, EPI_ISL_2319327, EPI_ISL_2319328, EPI_ISL_2319329, EPI_ISL_2319331, EPI_ISL_2319332, EPI_ISL_2319333, EPI_ISL_2319334, EPI_ISL_2319335, EPI_ISL_2319336, EPI_ISL_2319337, EPI_ISL_2319338, EPI_ISL_2319339, EPI_ISL_2319341, EPI_ISL_2319342, EPI_ISL_2319343, EPI_ISL_2319344, EPI_ISL_2319345, EPI_ISL_2319346, EPI_ISL_2319347, EPI_ISL_2319348, EPI_ISL_2319349, EPI_ISL_2319350, EPI_ISL_2319351, EPI_ISL_2319352, EPI_ISL_2319353, EPI_ISL_2319354, EPI_ISL_2319355, EPI_ISL_2319356, EPI_ISL_2319357, EPI_ISL_2319358, EPI_ISL_2319359, EPI_ISL_2319360, EPI_ISL_2319361, EPI_ISL_2319362, EPI_ISL_2319363, EPI_ISL_2319364, EPI_ISL_2319365, EPI_ISL_2319366, EPI_ISL_2319367, EPI_ISL_2319368, EPI_ISL_2319369, EPI_ISL_2319370, EPI_ISL_2319371, EPI_ISL_2319372, EPI_ISL_2319373, EPI_ISL_2319376, EPI_ISL_2319377, EPI_ISL_2319378, EPI_ISL_2319379, EPI_ISL_2319380, EPI_ISL_2319381 | | | |
| see above | INDRE | Instituto Nacional de Medicina Genomica | Hidalgo-Miranda A, Mendoza-Vargas A, Reyes-Grajeda JP, Cedro-Tanda A, Gisela Barrera-Badillo, Irma Lopez-Martinez, Jose Ernesto Ramirez González, Gonzalez-Barrera D, Rangel-DeLeon D, Munguia-Garza P, Garcia-Cardenas FJ, Gonzalez-Woge MA, Herrera-Montalvo LA. |
| EPI_ISL_2319382, EPI_ISL_2319383, EPI_ISL_2319384, EPI_ISL_2319386 | Salud Digna | Instituto Nacional de Medicina Genomica | Hidalgo-Miranda A, Cedro-Tanda A, Mendoza-Vargas A, Reyes-Grajeda JP, Abraham Campos-Romero, Moreno-Camacho José Luis, Rodriguez-Gallegos Jorge, Luna-Ruiz Marco, Gonzalez-Barrera D, Rangel-DeLeon D, Munguia-Garza P, Ramirez-Vega O, Escobar-Arrazola, M, Herrera-Montalvo LA. |
| EPI_ISL_2319390 | Instituto Nacional de Medicina Genomica | Instituto Nacional de Medicina Genomica | Hidalgo-Miranda A, Cedro-Tanda A, Mendoza-Vargas A, Reyes-Grajeda JP, Cisneros- Villanueva M, Gonzalez-Barrera D, Rangel-DeLeon D, Munguia-Garza P, Ramirez-Vega O, Escobar-Arrazola, M, Herrera-Montalvo LA. |
| EPI_ISL_2319391, EPI_ISL_2319392, EPI_ISL_2319393, EPI_ISL_2319394, EPI_ISL_2319396, EPI_ISL_2319398, EPI_ISL_2319399, EPI_ISL_2319400, EPI_ISL_2319401, EPI_ISL_2319403, EPI_ISL_2319407, EPI_ISL_2319409, EPI_ISL_2319410, EPI_ISL_2319411, EPI_ISL_2319412 | | | |
| see above | Salud Digna | Instituto Nacional de Medicina Genomica | Hidalgo-Miranda A, Cedro-Tanda A, Mendoza-Vargas A, Reyes-Grajeda JP, Abraham Campos-Romero, Moreno-Camacho José Luis, Rodriguez-Gallegos Jorge, Luna-Ruiz Marco, Gonzalez-Barrera D, Rangel-DeLeon D, Munguia-Garza P, Ramirez-Vega O, Escobar-Arrazola, M, Herrera-Montalvo LA. |
| EPI_ISL_2319413, EPI_ISL_2319414 | Ohio Department of Health Laboratory | Ohio Department of Health Laboratory | Holmes, Jennifer; Eric Brandt, Keoni Omura, Glen McGillivary, Caitlin McDonnell, Jade Mowery, Stephanie Mccracken, Tyler Payne, Kirtana Ramadugu, Erica Leasure, Brent Lee, Kelsey Florek, Heather Blankenship, Allison Black, Quanta Brown, and Tammy Bannerman |
| EPI_ISL_2319415, EPI_ISL_2319416, EPI_ISL_2319417, EPI_ISL_2319418, EPI_ISL_2319419, EPI_ISL_2319420, EPI_ISL_2319421, EPI_ISL_2319422, EPI_ISL_2319423, EPI_ISL_2319424, EPI_ISL_2319425, EPI_ISL_2319426, EPI_ISL_2319427, EPI_ISL_2319428, EPI_ISL_2319429, EPI_ISL_2319430, EPI_ISL_2319431, EPI_ISL_2319432, EPI_ISL_2319433, EPI_ISL_2319434, EPI_ISL_2319435, EPI_ISL_2319436, EPI_ISL_2319437, EPI_ISL_2319438, EPI_ISL_2319439, EPI_ISL_2319440, EPI_ISL_2319441, EPI_ISL_2319442, EPI_ISL_2319443, EPI_ISL_2319444, EPI_ISL_2319445, EPI_ISL_2319446, EPI_ISL_2319447, EPI_ISL_2319448, EPI_ISL_2319449, EPI_ISL_2319450, EPI_ISL_2319451, EPI_ISL_2319452, EPI_ISL_2319453, EPI_ISL_2319454, EPI_ISL_2319455, EPI_ISL_2319456, EPI_ISL_2319457, EPI_ISL_2319458, EPI_ISL_2319459, EPI_ISL_2319460, EPI_ISL_2319461, EPI_ISL_2319462, EPI_ISL_2319463, EPI_ISL_2319464, EPI_ISL_2319465, EPI_ISL_2319466, EPI_ISL_2319467, EPI_ISL_2319468, EPI_ISL_2319469, EPI_ISL_2319470, EPI_ISL_2319471, EPI_ISL_2319472, EPI_ISL_2319473, EPI_ISL_2319474, EPI_ISL_2319475, EPI_ISL_2319476, EPI_ISL_2319477, EPI_ISL_2319478, EPI_ISL_2319479, EPI_ISL_2319480, EPI_ISL_2319481, EPI_ISL_2319482, EPI_ISL_2319483, EPI_ISL_2319484, EPI_ISL_2319485, EPI_ISL_2319486, EPI_ISL_2319487, EPI_ISL_2319488, EPI_ISL_2319489, EPI_ISL_2319490, EPI_ISL_2319491, EPI_ISL_2319492, EPI_ISL_2319493, EPI_ISL_2319494, EPI_ISL_2319495, EPI_ISL_2319496, EPI_ISL_2319497, EPI_ISL_2319498, EPI_ISL_2319499, EPI_ISL_2319500, EPI_ISL_2319501, EPI_ISL_2319502, EPI_ISL_2319503, EPI_ISL_2319504, EPI_ISL_2319505, EPI_ISL_2319506, EPI_ISL_2319507, EPI_ISL_2319508, EPI_ISL_2319509, EPI_ISL_2319510, EPI_ISL_2319511, EPI_ISL_2319512, EPI_ISL_2319513, EPI_ISL_2319514, EPI_ISL_2319515, EPI_ISL_2319516, EPI_ISL_2319517, EPI_ISL_2319518, EPI_ISL_2319519, EPI_ISL_2319520, EPI_ISL_2319521, EPI_ISL_2319522, EPI_ISL_2319523, EPI_ISL_2319524, EPI_ISL_2319525, EPI_ISL_2319526, EPI_ISL_2319527, EPI_ISL_2319528, EPI_ISL_2319529, EPI_ISL_2319530, EPI_ISL_2319531, EPI_ISL_2319532, EPI_ISL_2319533, EPI_ISL_2319534, EPI_ISL_2319536, EPI_ISL_2319537, EPI_ISL_2319538, EPI_ISL_2319539, EPI_ISL_2319540, EPI_ISL_2319541, EPI_ISL_2319542, EPI_ISL_2319543, EPI_ISL_2319544, EPI_ISL_2319545, EPI_ISL_2319546, EPI_ISL_2319547, EPI_ISL_2319548, EPI_ISL_2319550, EPI_ISL_2319551, EPI_ISL_2319552, EPI_ISL_2319553, EPI_ISL_2319554, EPI_ISL_2319555, EPI_ISL_2319556, EPI_ISL_2319557, EPI_ISL_2319558, EPI_ISL_2319559, EPI_ISL_2319560, EPI_ISL_2319561, EPI_ISL_2319562, EPI_ISL_2319563, EPI_ISL_2319564, EPI_ISL_2319565, EPI_ISL_2319566, EPI_ISL_2319567, EPI_ISL_2319568, EPI_ISL_2319569, EPI_ISL_2319570, EPI_ISL_2319571, EPI_ISL_2319572, EPI_ISL_2319573, EPI_ISL_2319574, EPI_ISL_2319575, EPI_ISL_2319576, EPI_ISL_2319577, EPI_ISL_2319578, EPI_ISL_2319579, EPI_ISL_2319580, EPI_ISL_2319581, EPI_ISL_2319582, EPI_ISL_2319583, EPI_ISL_2319584, EPI_ISL_2319585, EPI_ISL_2319586, EPI_ISL_2319587, EPI_ISL_2319588, EPI_ISL_2319589, EPI_ISL_2319590, EPI_ISL_2319591, EPI_ISL_2319592, EPI_ISL_2319593, EPI_ISL_2319594, EPI_ISL_2319595, EPI_ISL_2319596, EPI_ISL_2319597, EPI_ISL_2319598, EPI_ISL_2319599 | | | |
| see above | Maryland Genomics, Institute for Genome Sciences, University of Maryland School of Medicine | Maryland Genomics, Institute for Genome Sciences, University of Maryland School of Medicine | Tallon, Luke J; Sadzewicz, Lisa D; Humphrys, Mike; Ott, Sandra; Roussey, Holly; Mehta, Aditya; Vavikolanu, Kranthi; Fraser, Claire M; Ravel, Jacques |
| EPI_ISL_2319600, EPI_ISL_2319601, EPI_ISL_2319602, EPI_ISL_2319628 | Ohio Department of Health Laboratory | Ohio Department of Health Laboratory | Holmes, Jennifer; Eric Brandt, Keoni Omura, Glen McGillivary, Caitlin McDonnell, Jade Mowery, Stephanie Mccracken, Tyler Payne, Kirtana Ramadugu, Erica Leasure, Brent Lee, Kelsey Florek, Heather Blankenship, Allison Black, Quanta Brown, and Tammy Bannerman |
| EPI_ISL_2319629, EPI_ISL_2319630, EPI_ISL_2319631, EPI_ISL_2319632, EPI_ISL_2319633, EPI_ISL_2319634, EPI_ISL_2319635, EPI_ISL_2319636, EPI_ISL_2319637, EPI_ISL_2319638, EPI_ISL_2319639, EPI_ISL_2319640, EPI_ISL_2319641, EPI_ISL_2319642, EPI_ISL_2319643, EPI_ISL_2319644, EPI_ISL_2319645, EPI_ISL_2319646, EPI_ISL_2319648, EPI_ISL_2319649, EPI_ISL_2319651, EPI_ISL_2319652, EPI_ISL_2319653, EPI_ISL_2319654, EPI_ISL_2319655, EPI_ISL_2319656, EPI_ISL_2319657, EPI_ISL_2319658, EPI_ISL_2319659, EPI_ISL_2319660, EPI_ISL_2319661, EPI_ISL_2319662, EPI_ISL_2319663, EPI_ISL_2319664, EPI_ISL_2319665, EPI_ISL_2319666, EPI_ISL_2319667, EPI_ISL_2319668, EPI_ISL_2319669, EPI_ISL_2319670, EPI_ISL_2319671, EPI_ISL_2319672, EPI_ISL_2319673, EPI_ISL_2319674, EPI_ISL_2319675, EPI_ISL_2319676, EPI_ISL_2319677, EPI_ISL_2319678, EPI_ISL_2319679, EPI_ISL_2319680, EPI_ISL_2319681, EPI_ISL_2319682, EPI_ISL_2319683, EPI_ISL_2319685, EPI_ISL_2319686, EPI_ISL_2319687, EPI_ISL_2319688, EPI_ISL_2319689, EPI_ISL_2319690, EPI_ISL_2319691, EPI_ISL_2319693, EPI_ISL_2319694, EPI_ISL_2319695, EPI_ISL_2319696, EPI_ISL_2319697, EPI_ISL_2319698, EPI_ISL_2319699, EPI_ISL_2319700, EPI_ISL_2319701, EPI_ISL_2319702, EPI_ISL_2319703, EPI_ISL_2319704, EPI_ISL_2319705, EPI_ISL_2319706, EPI_ISL_2319707, EPI_ISL_2319708, EPI_ISL_2319709, EPI_ISL_2319710, EPI_ISL_2319711, EPI_ISL_2319713, EPI_ISL_2319714, EPI_ISL_2319715, EPI_ISL_2319716, EPI_ISL_2319717, EPI_ISL_2319719, EPI_ISL_2319720, EPI_ISL_2319721, EPI_ISL_2319722, EPI_ISL_2319723, EPI_ISL_2319724, EPI_ISL_2319725, EPI_ISL_2319726, EPI_ISL_2319727, EPI_ISL_2319728, EPI_ISL_2319729, EPI_ISL_2319731, EPI_ISL_2319732, EPI_ISL_2319733, EPI_ISL_2319734, EPI_ISL_2319735, EPI_ISL_2319736, EPI_ISL_2319737, EPI_ISL_2319738, EPI_ISL_2319739, EPI_ISL_2319740, EPI_ISL_2319741, EPI_ISL_2319742, EPI_ISL_2319743, EPI_ISL_2319744, EPI_ISL_2319746, EPI_ISL_2319747, EPI_ISL_2319748, EPI_ISL_2319749, EPI_ISL_2319750, EPI_ISL_2319752, EPI_ISL_2319753, EPI_ISL_2319754, EPI_ISL_2319755, EPI_ISL_2319756, EPI_ISL_2319757, EPI_ISL_2319758, EPI_ISL_2319759, EPI_ISL_2319760, EPI_ISL_2319761, EPI_ISL_2319763, EPI_ISL_2319764, EPI_ISL_2319765, EPI_ISL_2319766, EPI_ISL_2319767, EPI_ISL_2319768, EPI_ISL_2319769, EPI_ISL_2319771, EPI_ISL_2319772, EPI_ISL_2319774, EPI_ISL_2319775, EPI_ISL_2319776, EPI_ISL_2319777, EPI_ISL_2319778, EPI_ISL_2319779, EPI_ISL_2319781, EPI_ISL_2319783, EPI_ISL_2319784, EPI_ISL_2319785, EPI_ISL_2319786, EPI_ISL_2319787, EPI_ISL_2319789, EPI_ISL_2319791, EPI_ISL_2319792, EPI_ISL_2319794, EPI_ISL_2319795, EPI_ISL_2319796, EPI_ISL_2319797, EPI_ISL_2319798, EPI_ISL_2319799, EPI_ISL_2319800, EPI_ISL_2319801, EPI_ISL_2319802, EPI_ISL_2319803, EPI_ISL_2319804, EPI_ISL_2319805, EPI_ISL_2319806, EPI_ISL_2319808, EPI_ISL_2319809, EPI_ISL_2319810, EPI_ISL_2319811, EPI_ISL_2319812, EPI_ISL_2319813, EPI_ISL_2319814, EPI_ISL_2319817, EPI_ISL_2319818, EPI_ISL_2319819, EPI_ISL_2319820, EPI_ISL_2319821, EPI_ISL_2319822, EPI_ISL_2319823, EPI_ISL_2319824, EPI_ISL_2319825, EPI_ISL_2319826, EPI_ISL_2319830, EPI_ISL_2319833, EPI_ISL_2319834, EPI_ISL_2319835, EPI_ISL_2319837, EPI_ISL_2319839, EPI_ISL_2319840, EPI_ISL_2319841, EPI_ISL_2319842, EPI_ISL_2319843, EPI_ISL_2319844, EPI_ISL_2319845, EPI_ISL_2319846, EPI_ISL_2319847, EPI_ISL_2319848, EPI_ISL_2319849, EPI_ISL_2319850, EPI_ISL_2319851, EPI_ISL_2319852, EPI_ISL_2319853, EPI_ISL_2319854, EPI_ISL_2319856, EPI_ISL_2319857, EPI_ISL_2319858, EPI_ISL_2319859, EPI_ISL_2319860, EPI_ISL_2319861, EPI_ISL_2319862, EPI_ISL_2319863, EPI_ISL_2319864, EPI_ISL_2319865, EPI_ISL_2319866, EPI_ISL_2319867, EPI_ISL_2319868, EPI_ISL_2319869, EPI_ISL_2319870, EPI_ISL_2319871, EPI_ISL_2319872, EPI_ISL_2319873, EPI_ISL_2319874, EPI_ISL_2319875, EPI_ISL_2319876, EPI_ISL_2319877, EPI_ISL_2319878, EPI_ISL_2319879, EPI_ISL_2319880, EPI_ISL_2319881, EPI_ISL_2319882, EPI_ISL_2319883, EPI_ISL_2319884, EPI_ISL_2319885, EPI_ISL_2319886, EPI_ISL_2319887, EPI_ISL_2319888, EPI_ISL_2319889, EPI_ISL_2319890, EPI_ISL_2319891, EPI_ISL_2319892, EPI_ISL_2319893, EPI_ISL_2319894, EPI_ISL_2319895, EPI_ISL_2319896, EPI_ISL_2319897, EPI_ISL_2319898, EPI_ISL_2319899, EPI_ISL_2319900, EPI_ISL_2319901, EPI_ISL_2319902, EPI_ISL_2319903, EPI_ISL_2319904, EPI_ISL_2319905, EPI_ISL_2319906, EPI_ISL_2319907, EPI_ISL_2319908, EPI_ISL_2319909, EPI_ISL_2319910, EPI_ISL_2319911, EPI_ISL_2319912, EPI_ISL_2319913, EPI_ISL_2319914, EPI_ISL_2319915, EPI_ISL_2319916, EPI_ISL_2319917, EPI_ISL_2319918, EPI_ISL_2319919, EPI_ISL_2319920, EPI_ISL_2319921, EPI_ISL_2319922, EPI_ISL_2319923, EPI_ISL_2319924, EPI_ISL_2319925, EPI_ISL_2319926, EPI_ISL_2319927, EPI_ISL_2319928, EPI_ISL_2319929, EPI_ISL_2319930, EPI_ISL_2319932, EPI_ISL_2319933, EPI_ISL_2319934, EPI_ISL_2319935, EPI_ISL_2319936, EPI_ISL_2319937, EPI_ISL_2319938, EPI_ISL_2319941, EPI_ISL_2319942, EPI_ISL_2319943, EPI_ISL_2319944, EPI_ISL_2319945, EPI_ISL_2319946, EPI_ISL_2319947, EPI_ISL_2319948, EPI_ISL_2319949, EPI_ISL_2319950, EPI_ISL_2319951, EPI_ISL_2319952, EPI_ISL_2319953, EPI_ISL_2319954, EPI_ISL_2319955, EPI_ISL_2319956, EPI_ISL_2319957, EPI_ISL_2319958, EPI_ISL_2319959, EPI_ISL_2319960, EPI_ISL_2319961, EPI_ISL_2319962, EPI_ISL_2319963, EPI_ISL_2319964, EPI_ISL_2319965, EPI_ISL_2319966, EPI_ISL_2319967, EPI_ISL_2319968, EPI_ISL_2319969, EPI_ISL_2319970, EPI_ISL_2319971, EPI_ISL_2319972, EPI_ISL_2319973, EPI_ISL_2319974, EPI_ISL_2319975, EPI_ISL_2319976, EPI_ISL_2319977, EPI_ISL_2319978, EPI_ISL_2319979, EPI_ISL_2319980, EPI_ISL_2319981, EPI_ISL_2319982 | | | |
| see above | Reditus Laboratories | Reditus Laboratories | Joshua J. Geltz, Ph.D., Robert M. Sgambelluri, Ph.D., Rex Dyer, Ph.D., Cassy Philips, M.S., Alexa Eichelberger, M.S. |
| EPI_ISL_2319983, EPI_ISL_2319985 | Ohio Department of Health Laboratory | Ohio Department of Health Laboratory | Holmes, Jennifer; Eric Brandt, Keoni Omura, Glen McGillivary, Caitlin McDonnell, Jade Mowery, Stephanie Mccracken, Tyler Payne, Kirtana Ramadugu, Erica Leasure, Brent Lee, Kelsey Florek, Heather Blankenship, Allison Black, Quanta Brown, and Tammy Bannerman |

| | | | |
|---|---|---|---|
| EPI_ISL_2319986, EPI_ISL_2319987, EPI_ISL_2319988, EPI_ISL_2319989, EPI_ISL_2319990, EPI_ISL_2319991, EPI_ISL_2319992, EPI_ISL_2319993, EPI_ISL_2319994, EPI_ISL_2319995, EPI_ISL_2319996, EPI_ISL_2319997, EPI_ISL_2319998, EPI_ISL_2319999, EPI_ISL_2320000, EPI_ISL_2320001, EPI_ISL_2320002, EPI_ISL_2320003, EPI_ISL_2320004, EPI_ISL_2320005, EPI_ISL_2320006, EPI_ISL_2320008, EPI_ISL_2320009, EPI_ISL_2320010, EPI_ISL_2320011, EPI_ISL_2320012, EPI_ISL_2320013, EPI_ISL_2320014, EPI_ISL_2320015, EPI_ISL_2320016, EPI_ISL_2320017, EPI_ISL_2320018, EPI_ISL_2320019, EPI_ISL_2320020, EPI_ISL_2320021, EPI_ISL_2320022, EPI_ISL_2320023, EPI_ISL_2320024, EPI_ISL_2320025, EPI_ISL_2320026, EPI_ISL_2320027, EPI_ISL_2320028, EPI_ISL_2320029, EPI_ISL_2320030, EPI_ISL_2320031, EPI_ISL_2320032, EPI_ISL_2320033, EPI_ISL_2320034, EPI_ISL_2320035, EPI_ISL_2320036, EPI_ISL_2320037, EPI_ISL_2320038, EPI_ISL_2320039, EPI_ISL_2320040, EPI_ISL_2320041, EPI_ISL_2320042, EPI_ISL_2320043, EPI_ISL_2320044, EPI_ISL_2320045, EPI_ISL_2320046, EPI_ISL_2320047, EPI_ISL_2320048, EPI_ISL_2320049, EPI_ISL_2320050, EPI_ISL_2320051, EPI_ISL_2320052, EPI_ISL_2320053, EPI_ISL_2320054, EPI_ISL_2320055, EPI_ISL_2320056, EPI_ISL_2320057, EPI_ISL_2320058, EPI_ISL_2320059, EPI_ISL_2320060, EPI_ISL_2320061, EPI_ISL_2320062, EPI_ISL_2320063, EPI_ISL_2320064, EPI_ISL_2320065, EPI_ISL_2320066, EPI_ISL_2320067, EPI_ISL_2320068, EPI_ISL_2320069, EPI_ISL_2320070, EPI_ISL_2320071, EPI_ISL_2320072, EPI_ISL_2320073, EPI_ISL_2320074, EPI_ISL_2320075, EPI_ISL_2320076, EPI_ISL_2320077, EPI_ISL_2320078, EPI_ISL_2320079, EPI_ISL_2320080, EPI_ISL_2320081, EPI_ISL_2320082, EPI_ISL_2320083, EPI_ISL_2320084, EPI_ISL_2320085, EPI_ISL_2320086, EPI_ISL_2320087, EPI_ISL_2320088, EPI_ISL_2320089, EPI_ISL_2320090, EPI_ISL_2320091, EPI_ISL_2320092 | | | |
| see above | Helix/Illumina | Centers for Disease Control and Prevention Division of Viral Diseases, Pathogen Discovery | Dakota Howard, Dhwani Batra, Peter W. Cook, Kara Moser, Adrian Paskey, Jason Caravas, Benjamin Rambo-Martin, Shatavia Morrison, Christopher Gulvick, Scott Sammons, Yvette Unoarumhi, Darlene Wagner, Matthew Schmerer, Eileen de Feo, Jan Antico, Christine Tran, Matthew Tolentino, Shannon Wickline, Kim Gietzen, Brad Sickler, Jingtao Liu, Eric Allen, Phil Febbo, Nicole L. Washington, Simon White, Geraint Levan, Kelly Schiabor Barrett, Elizabeth Cirulli, Alexandre Bolze, Ary Ascencio, Charlotte Rivera-Garcia, Ryan Cho, Jason Nguyen, Sherry Wang, Jimmy Ramirez, Tyler Cassens, Efren Sandoval, Magnus Isaksson, William Lee, David Becker, Marc Laurent, James Lu, Clinton R. Paden, Duncan MacCannell |
| EPI_ISL_2320093 | Wexner Medical Center | Wexner Medical Center | Koenig,S., Seminetta,J. |
| EPI_ISL_2320094, EPI_ISL_2320095, EPI_ISL_2320096, EPI_ISL_2320097, EPI_ISL_2320098, EPI_ISL_2320099, EPI_ISL_2320100, EPI_ISL_2320101, EPI_ISL_2320102, EPI_ISL_2320103, EPI_ISL_2320104 | | | |
| see above | Helix/Illumina | Centers for Disease Control and Prevention Division of Viral Diseases, Pathogen Discovery | Dakota Howard, Dhwani Batra, Peter W. Cook, Kara Moser, Adrian Paskey, Jason Caravas, Benjamin Rambo-Martin, Shatavia Morrison, Christopher Gulvick, Scott Sammons, Yvette Unoarumhi, Darlene Wagner, Matthew Schmerer, Eileen de Feo, Jan Antico, Christine Tran, Matthew Tolentino, Shannon Wickline, Kim Gietzen, Brad Sickler, Jingtao Liu, Eric Allen, Phil Febbo, Nicole L. Washington, Simon White, Geraint Levan, Kelly Schiabor Barrett, Elizabeth Cirulli, Alexandre Bolze, Ary Ascencio, Charlotte Rivera-Garcia, Ryan Cho, Jason Nguyen, Sherry Wang, Jimmy Ramirez, Tyler Cassens, Efren Sandoval, Magnus Isaksson, William Lee, David Becker, Marc Laurent, James Lu, Clinton R. Paden, Duncan MacCannell |
| EPI_ISL_2320105 | Ohio Department of Health Laboratory | Ohio Department of Health Laboratory | Holmes, Jennifer; Eric Brandt, Keoni Omura, Glen McGillivary, Caitlin McDonnell, Jade Mowery, Stephanie Mccracken, Tyler Payne, Kirtana Ramadugu, Erica Leasure, Brent Lee, Kelsey Florek, Heather Blankenship, Allison Black, Quanta Brown, and Tammy Bannerman |
| EPI_ISL_2320106, EPI_ISL_2320107, EPI_ISL_2320108, EPI_ISL_2320109, EPI_ISL_2320110, EPI_ISL_2320111, EPI_ISL_2320112, EPI_ISL_2320114, EPI_ISL_2320115, EPI_ISL_2320116, EPI_ISL_2320117, EPI_ISL_2320118, EPI_ISL_2320119, EPI_ISL_2320120, EPI_ISL_2320121, EPI_ISL_2320122, EPI_ISL_2320123, EPI_ISL_2320124, EPI_ISL_2320125, EPI_ISL_2320126, EPI_ISL_2320127, EPI_ISL_2320128, EPI_ISL_2320129, EPI_ISL_2320130, EPI_ISL_2320132, EPI_ISL_2320133, EPI_ISL_2320134, EPI_ISL_2320135, EPI_ISL_2320136, EPI_ISL_2320137, EPI_ISL_2320138, EPI_ISL_2320139, EPI_ISL_2320140, EPI_ISL_2320141, EPI_ISL_2320142, EPI_ISL_2320143, EPI_ISL_2320144, EPI_ISL_2320145, EPI_ISL_2320146, EPI_ISL_2320147, EPI_ISL_2320148, EPI_ISL_2320149, EPI_ISL_2320150, EPI_ISL_2320151, EPI_ISL_2320152, EPI_ISL_2320153, EPI_ISL_2320154, EPI_ISL_2320155, EPI_ISL_2320156, EPI_ISL_2320157, EPI_ISL_2320158, EPI_ISL_2320159, EPI_ISL_2320160, EPI_ISL_2320161, EPI_ISL_2320162, EPI_ISL_2320163, EPI_ISL_2320164, EPI_ISL_2320165, EPI_ISL_2320166, EPI_ISL_2320167, EPI_ISL_2320168, EPI_ISL_2320169, EPI_ISL_2320170, EPI_ISL_2320171, EPI_ISL_2320172, EPI_ISL_2320173, EPI_ISL_2320174, EPI_ISL_2320175, EPI_ISL_2320176, EPI_ISL_2320177, EPI_ISL_2320178, EPI_ISL_2320179, EPI_ISL_2320180, EPI_ISL_2320181, EPI_ISL_2320182, EPI_ISL_2320183, EPI_ISL_2320185, EPI_ISL_2320186, EPI_ISL_2320187, EPI_ISL_2320188, EPI_ISL_2320189, EPI_ISL_2320190, EPI_ISL_2320191, EPI_ISL_2320192, EPI_ISL_2320193, EPI_ISL_2320194, EPI_ISL_2320195, EPI_ISL_2320196, EPI_ISL_2320197, EPI_ISL_2320198, EPI_ISL_2320199, EPI_ISL_2320200, EPI_ISL_2320201, EPI_ISL_2320202, EPI_ISL_2320203, EPI_ISL_2320204, EPI_ISL_2320205, EPI_ISL_2320206, EPI_ISL_2320207, EPI_ISL_2320208, EPI_ISL_2320209, EPI_ISL_2320210, EPI_ISL_2320211, EPI_ISL_2320212, EPI_ISL_2320213, EPI_ISL_2320214, EPI_ISL_2320215, EPI_ISL_2320216, EPI_ISL_2320217, EPI_ISL_2320218, EPI_ISL_2320219, EPI_ISL_2320220, EPI_ISL_2320221, EPI_ISL_2320222, EPI_ISL_2320223, EPI_ISL_2320224, EPI_ISL_2320225, EPI_ISL_2320226, EPI_ISL_2320227, EPI_ISL_2320228, EPI_ISL_2320229, EPI_ISL_2320230, EPI_ISL_2320231, EPI_ISL_2320232, EPI_ISL_2320233, EPI_ISL_2320234, EPI_ISL_2320235, EPI_ISL_2320236, EPI_ISL_2320237, EPI_ISL_2320238, EPI_ISL_2320239, EPI_ISL_2320240, EPI_ISL_2320241, EPI_ISL_2320242, EPI_ISL_2320243, EPI_ISL_2320244, EPI_ISL_2320245, EPI_ISL_2320246, EPI_ISL_2320247, EPI_ISL_2320248, EPI_ISL_2320249, EPI_ISL_2320250, EPI_ISL_2320251, EPI_ISL_2320252, EPI_ISL_2320253, EPI_ISL_2320254, EPI_ISL_2320255, EPI_ISL_2320256, EPI_ISL_2320257, EPI_ISL_2320258, EPI_ISL_2320259, EPI_ISL_2320260, EPI_ISL_2320261, EPI_ISL_2320262, EPI_ISL_2320263, EPI_ISL_2320264, EPI_ISL_2320265, EPI_ISL_2320266, EPI_ISL_2320267, EPI_ISL_2320268, EPI_ISL_2320269, EPI_ISL_2320270, EPI_ISL_2320271, EPI_ISL_2320272, EPI_ISL_2320273, EPI_ISL_2320274, EPI_ISL_2320275, EPI_ISL_2320276, EPI_ISL_2320277, EPI_ISL_2320278, EPI_ISL_2320279, EPI_ISL_2320280, EPI_ISL_2320281, EPI_ISL_2320282, EPI_ISL_2320283, EPI_ISL_2320284, EPI_ISL_2320285, EPI_ISL_2320286, EPI_ISL_2320287, EPI_ISL_2320288, EPI_ISL_2320289, EPI_ISL_2320290, EPI_ISL_2320291, EPI_ISL_2320292, EPI_ISL_2320293, EPI_ISL_2320294, EPI_ISL_2320295, EPI_ISL_2320296, EPI_ISL_2320297, EPI_ISL_2320298, EPI_ISL_2320299, EPI_ISL_2320300, EPI_ISL_2320301, EPI_ISL_2320302, EPI_ISL_2320303, EPI_ISL_2320304, EPI_ISL_2320305, EPI_ISL_2320306, EPI_ISL_2320307, EPI_ISL_2320308, EPI_ISL_2320309, EPI_ISL_2320310, EPI_ISL_2320311, EPI_ISL_2320312, EPI_ISL_2320313, EPI_ISL_2320314, EPI_ISL_2320315, EPI_ISL_2320316, EPI_ISL_2320317, EPI_ISL_2320318, EPI_ISL_2320319, EPI_ISL_2320320, EPI_ISL_2320321, EPI_ISL_2320322, EPI_ISL_2320323, EPI_ISL_2320324, EPI_ISL_2320325, EPI_ISL_2320326, EPI_ISL_2320327, EPI_ISL_2320328, EPI_ISL_2320329, EPI_ISL_2320330, EPI_ISL_2320331, EPI_ISL_2320332, EPI_ISL_2320333, EPI_ISL_2320334, EPI_ISL_2320335, EPI_ISL_2320336, EPI_ISL_2320337, EPI_ISL_2320338, EPI_ISL_2320339, EPI_ISL_2320341, EPI_ISL_2320342, EPI_ISL_2320343, EPI_ISL_2320344, EPI_ISL_2320345, EPI_ISL_2320346, EPI_ISL_2320347, EPI_ISL_2320348, EPI_ISL_2320349, EPI_ISL_2320350, EPI_ISL_2320351, EPI_ISL_2320352, EPI_ISL_2320353, EPI_ISL_2320354, EPI_ISL_2320355, EPI_ISL_2320356, EPI_ISL_2320357, EPI_ISL_2320358, EPI_ISL_2320359, EPI_ISL_2320360, EPI_ISL_2320361, EPI_ISL_2320362, EPI_ISL_2320363, EPI_ISL_2320364, EPI_ISL_2320365, EPI_ISL_2320366, EPI_ISL_2320367, EPI_ISL_2320368, EPI_ISL_2320369, EPI_ISL_2320370, EPI_ISL_2320371, EPI_ISL_2320372, EPI_ISL_2320373, EPI_ISL_2320374, EPI_ISL_2320375, EPI_ISL_2320376, EPI_ISL_2320377, EPI_ISL_2320378, EPI_ISL_2320379, EPI_ISL_2320380, EPI_ISL_2320381, EPI_ISL_2320382, EPI_ISL_2320383, EPI_ISL_2320384, EPI_ISL_2320385, EPI_ISL_2320386, EPI_ISL_2320387, EPI_ISL_2320388, EPI_ISL_2320389, EPI_ISL_2320390, EPI_ISL_2320391, EPI_ISL_2320392, EPI_ISL_2320393, EPI_ISL_2320394, EPI_ISL_2320395, EPI_ISL_2320396, EPI_ISL_2320397, EPI_ISL_2320398, EPI_ISL_2320399, EPI_ISL_2320400, EPI_ISL_2320401, EPI_ISL_2320402, EPI_ISL_2320403, EPI_ISL_2320404, EPI_ISL_2320405, EPI_ISL_2320406, EPI_ISL_2320407, EPI_ISL_2320408, EPI_ISL_2320409, EPI_ISL_2320410, EPI_ISL_2320411 | | | |
| see above | Helix/Illumina | Centers for Disease Control and Prevention Division of Viral Diseases, Pathogen Discovery | Dakota Howard, Dhwani Batra, Peter W. Cook, Kara Moser, Adrian Paskey, Jason Caravas, Benjamin Rambo-Martin, Shatavia Morrison, Christopher Gulvick, Scott Sammons, Yvette Unoarumhi, Darlene Wagner, Matthew Schmerer, Eileen de Feo, Jan Antico, Christine Tran, Matthew Tolentino, Shannon Wickline, Kim Gietzen, Brad Sickler, Jingtao Liu, Eric Allen, Phil Febbo, Nicole L. Washington, Simon White, Geraint Levan, Kelly Schiabor Barrett, Elizabeth Cirulli, Alexandre Bolze, Ary Ascencio, Charlotte Rivera-Garcia, Ryan Cho, Jason Nguyen, Sherry Wang, Jimmy Ramirez, Tyler Cassens, Efren Sandoval, Magnus Isaksson, William Lee, David Becker, Marc Laurent, James Lu, Clinton R. Paden, Duncan MacCannell |
| EPI_ISL_2320412 | Ohio Department of Health Laboratory | Ohio Department of Health Laboratory | Holmes, Jennifer; Eric Brandt, Keoni Omura, Glen McGillivary, Caitlin McDonnell, Jade Mowery, Stephanie Mccracken, Tyler Payne, Kirtana Ramadugu, Erica Leasure, Brent Lee, Kelsey Florek, Heather Blankenship, Allison Black, Quanta Brown, and Tammy Bannerman |
| EPI_ISL_2320413, EPI_ISL_2320414, EPI_ISL_2320415, EPI_ISL_2320416, EPI_ISL_2320417, EPI_ISL_2320418, EPI_ISL_2320419, EPI_ISL_2320420, EPI_ISL_2320421, EPI_ISL_2320422, EPI_ISL_2320423, EPI_ISL_2320424, EPI_ISL_2320425, EPI_ISL_2320426, EPI_ISL_2320427, EPI_ISL_2320428, EPI_ISL_2320429, EPI_ISL_2320430, EPI_ISL_2320431, EPI_ISL_2320432, EPI_ISL_2320433, EPI_ISL_2320434, EPI_ISL_2320435, EPI_ISL_2320436, EPI_ISL_2320437, EPI_ISL_2320438, EPI_ISL_2320439, EPI_ISL_2320440, EPI_ISL_2320441, EPI_ISL_2320442, EPI_ISL_2320443, EPI_ISL_2320444, EPI_ISL_2320445, EPI_ISL_2320446, EPI_ISL_2320447, EPI_ISL_2320448, EPI_ISL_2320449, EPI_ISL_2320450, EPI_ISL_2320451, EPI_ISL_2320452, EPI_ISL_2320453, EPI_ISL_2320454, EPI_ISL_2320455, EPI_ISL_2320456, EPI_ISL_2320457, EPI_ISL_2320458, EPI_ISL_2320459, EPI_ISL_2320460, EPI_ISL_2320461, EPI_ISL_2320462, EPI_ISL_2320463, EPI_ISL_2320464, EPI_ISL_2320465, EPI_ISL_2320466, EPI_ISL_2320467, EPI_ISL_2320468, EPI_ISL_2320470, EPI_ISL_2320471, EPI_ISL_2320472, EPI_ISL_2320473, EPI_ISL_2320474, EPI_ISL_2320475, EPI_ISL_2320476, EPI_ISL_2320477, EPI_ISL_2320478, EPI_ISL_2320479, EPI_ISL_2320480, EPI_ISL_2320481, EPI_ISL_2320482, EPI_ISL_2320483, EPI_ISL_2320484, EPI_ISL_2320485, EPI_ISL_2320486, EPI_ISL_2320488, EPI_ISL_2320489, EPI_ISL_2320490, EPI_ISL_2320491, EPI_ISL_2320492, EPI_ISL_2320493, EPI_ISL_2320494, EPI_ISL_2320495, EPI_ISL_2320496, EPI_ISL_2320497, EPI_ISL_2320498, EPI_ISL_2320499, EPI_ISL_2320500, EPI_ISL_2320501, EPI_ISL_2320502, EPI_ISL_2320503, EPI_ISL_2320504, EPI_ISL_2320505, EPI_ISL_2320506, EPI_ISL_2320507, EPI_ISL_2320508, EPI_ISL_2320509, EPI_ISL_2320510, EPI_ISL_2320511, EPI_ISL_2320512, EPI_ISL_2320513, EPI_ISL_2320514, EPI_ISL_2320515, EPI_ISL_2320516, EPI_ISL_2320517, EPI_ISL_2320518, EPI_ISL_2320520, EPI_ISL_2320521, EPI_ISL_2320522, EPI_ISL_2320523, EPI_ISL_2320524, EPI_ISL_2320525, EPI_ISL_2320526, EPI_ISL_2320527, EPI_ISL_2320528, EPI_ISL_2320529, EPI_ISL_2320530, EPI_ISL_2320531, EPI_ISL_2320532, EPI_ISL_2320533, EPI_ISL_2320534, EPI_ISL_2320535, EPI_ISL_2320536, EPI_ISL_2320537, EPI_ISL_2320538, EPI_ISL_2320539, EPI_ISL_2320540, EPI_ISL_2320541, EPI_ISL_2320542, EPI_ISL_2320543, EPI_ISL_2320544, EPI_ISL_2320545, EPI_ISL_2320546, EPI_ISL_2320547, EPI_ISL_2320548, EPI_ISL_2320549, EPI_ISL_2320550, EPI_ISL_2320551, EPI_ISL_2320552, EPI_ISL_2320553, EPI_ISL_2320554, EPI_ISL_2320555, EPI_ISL_2320556, EPI_ISL_2320557, EPI_ISL_2320558, EPI_ISL_2320559, EPI_ISL_2320560, EPI_ISL_2320561, EPI_ISL_2320562, EPI_ISL_2320563, EPI_ISL_2320564, EPI_ISL_2320565, EPI_ISL_2320566, EPI_ISL_2320567, EPI_ISL_2320568, EPI_ISL_2320569, EPI_ISL_2320570, EPI_ISL_2320571, EPI_ISL_2320572, EPI_ISL_2320573, EPI_ISL_2320574, EPI_ISL_2320575, EPI_ISL_2320576, EPI_ISL_2320577, EPI_ISL_2320578, EPI_ISL_2320579, EPI_ISL_2320580, EPI_ISL_2320581, EPI_ISL_2320582, EPI_ISL_2320583, EPI_ISL_2320584, EPI_ISL_2320585, EPI_ISL_2320586, EPI_ISL_2320587, EPI_ISL_2320588, EPI_ISL_2320589, EPI_ISL_2320590, EPI_ISL_2320591, EPI_ISL_2320592, EPI_ISL_2320593, EPI_ISL_2320594, EPI_ISL_2320595, EPI_ISL_2320596, EPI_ISL_2320597, EPI_ISL_2320598, EPI_ISL_2320599, EPI_ISL_2320600, EPI_ISL_2320601, EPI_ISL_2320602, EPI_ISL_2320603, EPI_ISL_2320604, EPI_ISL_2320605, EPI_ISL_2320606, EPI_ISL_2320607, EPI_ISL_2320608, EPI_ISL_2320609, EPI_ISL_2320610, EPI_ISL_2320611, EPI_ISL_2320612, EPI_ISL_2320613, EPI_ISL_2320614, EPI_ISL_2320615, EPI_ISL_2320616, EPI_ISL_2320617, EPI_ISL_2320618, EPI_ISL_2320619, EPI_ISL_2320620, EPI_ISL_2320621, EPI_ISL_2320622, EPI_ISL_2320623, EPI_ISL_2320624, EPI_ISL_2320625, EPI_ISL_2320626, EPI_ISL_2320627, EPI_ISL_2320628, EPI_ISL_2320629, EPI_ISL_2320630 | | | |
| see above | Helix/Illumina | Centers for Disease Control and Prevention Division of Viral Diseases, Pathogen Discovery | Dakota Howard, Dhwani Batra, Peter W. Cook, Kara Moser, Adrian Paskey, Jason Caravas, Benjamin Rambo-Martin, Shatavia Morrison, Christopher Gulvick, Scott Sammons, Yvette Unoarumhi, Darlene Wagner, Matthew Schmerer, Eileen de Feo, Jan Antico, Christine Tran, Matthew Tolentino, Shannon Wickline, Kim Gietzen, Brad Sickler, Jingtao Liu, Eric Allen, Phil Febbo, Nicole L. Washington, Simon White, Geraint Levan, Kelly Schiabor Barrett, Elizabeth Cirulli, Alexandre Bolze, Ary Ascencio, Charlotte Rivera-Garcia, Ryan Cho, Jason Nguyen, Sherry Wang, Jimmy Ramirez, Tyler Cassens, Efren Sandoval, Magnus Isaksson, William Lee, David Becker, Marc Laurent, James Lu, Clinton R. Paden, Duncan MacCannell |
| EPI_ISL_2320631 | Ohio Department of Health Laboratory | Ohio Department of Health Laboratory | Holmes, Jennifer; Eric Brandt, Keoni Omura, Glen McGillivary, Caitlin McDonnell, Jade Mowery, Stephanie Mccracken, Tyler Payne, Kirtana Ramadugu, Erica Leasure, Brent Lee, Kelsey Florek, Heather Blankenship, Allison Black, Quanta Brown, and Tammy Bannerman |
| EPI_ISL_2320632, EPI_ISL_2320633, EPI_ISL_2320634, EPI_ISL_2320635, EPI_ISL_2320636, EPI_ISL_2320637, EPI_ISL_2320638, EPI_ISL_2320639, EPI_ISL_2320640, EPI_ISL_2320641, EPI_ISL_2320642, EPI_ISL_2320643, EPI_ISL_2320644, EPI_ISL_2320645, EPI_ISL_2320646, EPI_ISL_2320647, EPI_ISL_2320648, EPI_ISL_2320649, EPI_ISL_2320650, EPI_ISL_2320651, EPI_ISL_2320652, EPI_ISL_2320653, EPI_ISL_2320654, EPI_ISL_2320655, EPI_ISL_2320656, EPI_ISL_2320657, EPI_ISL_2320658, EPI_ISL_2320659, EPI_ISL_2320660, EPI_ISL_2320661, EPI_ISL_2320662, EPI_ISL_2320663, EPI_ISL_2320664, EPI_ISL_2320665, EPI_ISL_2320666, EPI_ISL_2320667, EPI_ISL_2320668, EPI_ISL_2320669, EPI_ISL_2320670, EPI_ISL_2320671, EPI_ISL_2320672, EPI_ISL_2320673, EPI_ISL_2320674, EPI_ISL_2320675, EPI_ISL_2320676, EPI_ISL_2320677, EPI_ISL_2320678, EPI_ISL_2320679, EPI_ISL_2320680, EPI_ISL_2320681, EPI_ISL_2320682, EPI_ISL_2320683, EPI_ISL_2320684, EPI_ISL_2320685, EPI_ISL_2320686, EPI_ISL_2320687, EPI_ISL_2320688, EPI_ISL_2320689, EPI_ISL_2320690, EPI_ISL_2320691, EPI_ISL_2320692, EPI_ISL_2320693, EPI_ISL_2320694, EPI_ISL_2320695, EPI_ISL_2320696, EPI_ISL_2320697, EPI_ISL_2320698, EPI_ISL_2320699, EPI_ISL_2320700, EPI_ISL_2320701, EPI_ISL_2320702, EPI_ISL_2320703, EPI_ISL_2320704, EPI_ISL_2320705, EPI_ISL_2320706, EPI_ISL_2320707, EPI_ISL_2320708, EPI_ISL_2320709, EPI_ISL_2320710, EPI_ISL_2320711, EPI_ISL_2320712, EPI_ISL_2320713, EPI_ISL_2320714, EPI_ISL_2320715, EPI_ISL_2320716, EPI_ISL_2320717, EPI_ISL_2320718, EPI_ISL_2320719, EPI_ISL_2320720, EPI_ISL_2320721, EPI_ISL_2320722, EPI_ISL_2320723, EPI_ISL_2320724, EPI_ISL_2320725, EPI_ISL_2320726, EPI_ISL_2320727, EPI_ISL_2320728, EPI_ISL_2320729, EPI_ISL_2320730, EPI_ISL_2320731, EPI_ISL_2320732, EPI_ISL_2320733, EPI_ISL_2320734, EPI_ISL_2320735, EPI_ISL_2320736, EPI_ISL_2320737, EPI_ISL_2320738, EPI_ISL_2320739, EPI_ISL_2320740, EPI_ISL_2320741, EPI_ISL_2320742, EPI_ISL_2320743, EPI_ISL_2320744, EPI_ISL_2320745, EPI_ISL_2320746, EPI_ISL_2320747, EPI_ISL_2320748, EPI_ISL_2320749, EPI_ISL_2320750, EPI_ISL_2320751, EPI_ISL_2320752, EPI_ISL_2320753, EPI_ISL_2320754, EPI_ISL_2320755, EPI_ISL_2320756, EPI_ISL_2320757, EPI_ISL_2320758, EPI_ISL_2320759, EPI_ISL_2320760, EPI_ISL_2320761, EPI_ISL_2320762, EPI_ISL_2320763, EPI_ISL_2320764, EPI_ISL_2320765, EPI_ISL_2320766, EPI_ISL_2320767, EPI_ISL_2320768, EPI_ISL_2320769, EPI_ISL_2320770, EPI_ISL_2320771, EPI_ISL_2320772, EPI_ISL_2320773, EPI_ISL_2320774, EPI_ISL_2320775, EPI_ISL_2320776, EPI_ISL_2320777, EPI_ISL_2320778, EPI_ISL_2320779, EPI_ISL_2320780, EPI_ISL_2320781, EPI_ISL_2320782, EPI_ISL_2320783, EPI_ISL_2320784, EPI_ISL_2320785, EPI_ISL_2320786, EPI_ISL_2320787, EPI_ISL_2320788, EPI_ISL_2320789, EPI_ISL_2320790, EPI_ISL_2320791, EPI_ISL_2320792, EPI_ISL_2320793, EPI_ISL_2320794, EPI_ISL_2320795, EPI_ISL_2320796, EPI_ISL_2320797, EPI_ISL_2320798, EPI_ISL_2320799, EPI_ISL_2320800, EPI_ISL_2320801, EPI_ISL_2320802, EPI_ISL_2320803, EPI_ISL_2320804, EPI_ISL_2320805, EPI_ISL_2320806, EPI_ISL_2320807, EPI_ISL_2320808, EPI_ISL_2320809, EPI_ISL_2320810, EPI_ISL_2320811, EPI_ISL_2320812, EPI_ISL_2320813, EPI_ISL_2320814, EPI_ISL_2320815, EPI_ISL_2320816, EPI_ISL_2320817, EPI_ISL_2320818, EPI_ISL_2320819, EPI_ISL_2320820, EPI_ISL_2320821, EPI_ISL_2320822, EPI_ISL_2320823, EPI_ISL_2320824, EPI_ISL_2320825, EPI_ISL_2320826, EPI_ISL_2320827, EPI_ISL_2320828, EPI_ISL_2320829, EPI_ISL_2320830, EPI_ISL_2320831, EPI_ISL_2320832, EPI_ISL_2320833, EPI_ISL_2320834, EPI_ISL_2320835, EPI_ISL_2320836, EPI_ISL_2320837, EPI_ISL_2320838, EPI_ISL_2320839, EPI_ISL_2320840, EPI_ISL_2320841, EPI_ISL_2320842, EPI_ISL_2320843, EPI_ISL_2320844, EPI_ISL_2320845, EPI_ISL_2320846, EPI_ISL_2320847, EPI_ISL_2320848, EPI_ISL_2320849, EPI_ISL_2320850, EPI_ISL_2320851, EPI_ISL_2320852, EPI_ISL_2320853, EPI_ISL_2320854, EPI_ISL_2320855, EPI_ISL_2320856, EPI_ISL_2320857, EPI_ISL_2320858, EPI_ISL_2320859, EPI_ISL_2320860, EPI_ISL_2320861, EPI_ISL_2320862, EPI_ISL_2320863, EPI_ISL_2320864, EPI_ISL_2320865, EPI_ISL_2320866, EPI_ISL_2320867, EPI_ISL_2320868, EPI_ISL_2320869, EPI_ISL_2320870, EPI_ISL_2320871, EPI_ISL_2320872, EPI_ISL_2320873, EPI_ISL_2320874, EPI_ISL_2320875, EPI_ISL_2320876, EPI_ISL_2320877, EPI_ISL_2320878, EPI_ISL_2320879, EPI_ISL_2320880, EPI_ISL_2320881, EPI_ISL_2320882, EPI_ISL_2320883, EPI_ISL_2320884, EPI_ISL_2320885, EPI_ISL_2320886, EPI_ISL_2320887, EPI_ISL_2320888, EPI_ISL_2320889, EPI_ISL_2320890, EPI_ISL_2320891, EPI_ISL_2320892, EPI_ISL_2320893, EPI_ISL_2320894, EPI_ISL_2320895, EPI_ISL_2320896, EPI_ISL_2320897, EPI_ISL_2320898, EPI_ISL_2320899, EPI_ISL_2320900, EPI_ISL_2320901, EPI_ISL_2320902, EPI_ISL_2320903, EPI_ISL_2320904, EPI_ISL_2320905, EPI_ISL_2320906, EPI_ISL_2320907, EPI_ISL_2320908, EPI_ISL_2320909, EPI_ISL_2320910, EPI_ISL_2320911, EPI_ISL_2320912, EPI_ISL_2320913, EPI_ISL_2320914, EPI_ISL_2320915, EPI_ISL_2320916, EPI_ISL_2320917, EPI_ISL_2320918, EPI_ISL_2320919, EPI_ISL_2320920, EPI_ISL_2320921, EPI_ISL_2320922, EPI_ISL_2320923, EPI_ISL_2320924, EPI_ISL_2320925, EPI_ISL_2320926, EPI_ISL_2320927, EPI_ISL_2320928, EPI_ISL_2320929, EPI_ISL_2320930, EPI_ISL_2320931, EPI_ISL_2320932, EPI_ISL_2320933, EPI_ISL_2320934, EPI_ISL_2320935, EPI_ISL_2320936, EPI_ISL_2320937, EPI_ISL_2320938, EPI_ISL_2320939, EPI_ISL_2320940, EPI_ISL_2320941, EPI_ISL_2320942, EPI_ISL_2320943, EPI_ISL_2320944, EPI_ISL_2320945, EPI_ISL_2320946, EPI_ISL_2320947, EPI_ISL_2320948, EPI_ISL_2320949, EPI_ISL_2320950, EPI_ISL_2320951, EPI_ISL_2320952, EPI_ISL_2320953, EPI_ISL_2320954, EPI_ISL_2320955, EPI_ISL_2320956, EPI_ISL_2320957, EPI_ISL_2320958, EPI_ISL_2320959, EPI_ISL_2320960, EPI_ISL_2320961, EPI_ISL_2320962, EPI_ISL_2320963, EPI_ISL_2320964, EPI_ISL_2320965, EPI_ISL_2320966, EPI_ISL_2320967, EPI_ISL_2320968, EPI_ISL_2320969, EPI_ISL_2320970, EPI_ISL_2320971, EPI_ISL_2320972, EPI_ISL_2320973, EPI_ISL_2320974, EPI_ISL_2320975, EPI_ISL_2320976, EPI_ISL_2320977, EPI_ISL_2320978, EPI_ISL_2320979, EPI_ISL_2320980, EPI_ISL_2320981, EPI_ISL_2320982, EPI_ISL_2320983, EPI_ISL_2320984, EPI_ISL_2320985, EPI_ISL_2320986, EPI_ISL_2320987, EPI_ISL_2320988, EPI_ISL_2320989, EPI_ISL_2320990, EPI_ISL_2320991, EPI_ISL_2320992, EPI_ISL_2320993, EPI_ISL_2320994, EPI_ISL_2320995, EPI_ISL_2320996, EPI_ISL_2320997, EPI_ISL_2320998, EPI_ISL_2320999, EPI_ISL_2321000, EPI_ISL_2321001, EPI_ISL_2321002, EPI_ISL_2321003, EPI_ISL_2321004, EPI_ISL_2321005, EPI_ISL_2321006, EPI_ISL_2321007, EPI_ISL_2321008, EPI_ISL_2321009, EPI_ISL_2321010, EPI_ISL_2321011, EPI_ISL_2321012, EPI_ISL_2321013, EPI_ISL_2321014, EPI_ISL_2321015, EPI_ISL_2321016, EPI_ISL_2321017, EPI_ISL_2321018, EPI_ISL_2321019, EPI_ISL_2321020, EPI_ISL_2321021, EPI_ISL_2321022, EPI_ISL_2321023, EPI_ISL_2321024, EPI_ISL_2321025, EPI_ISL_2321026, EPI_ISL_2321027, EPI_ISL_2321028, EPI_ISL_2321029, EPI_ISL_2321030, EPI_ISL_2321031, EPI_ISL_2321032, EPI_ISL_2321033, EPI_ISL_2321034, EPI_ISL_2321035, EPI_ISL_2321036, EPI_ISL_2321037, EPI_ISL_2321038, EPI_ISL_2321039, EPI_ISL_2321040, EPI_ISL_2321041, EPI_ISL_2321042, EPI_ISL_2321043, EPI_ISL_2321044, EPI_ISL_2321045, EPI_ISL_2321046, EPI_ISL_2321047, EPI_ISL_2321048, EPI_ISL_2321049, EPI_ISL_2321050, EPI_ISL_2321051, EPI_ISL_2321052, EPI_ISL_2321053, EPI_ISL_2321054, EPI_ISL_2321055, EPI_ISL_2321056, EPI_ISL_2321057, EPI_ISL_2321058, EPI_ISL_2321059, EPI_ISL_2321060, EPI_ISL_2321061, EPI_ISL_2321062, EPI_ISL_2321063, EPI_ISL_2321064, EPI_ISL_2321065, EPI_ISL_2321066, EPI_ISL_2321067, EPI_ISL_2321068, EPI_ISL_2321069, EPI_ISL_2321070, EPI_ISL_2321072, EPI_ISL_2321073, EPI_ISL_2321074, EPI_ISL_2321075, EPI_ISL_2321076, EPI_ISL_2321077, EPI_ISL_2321078, EPI_ISL_2321079, EPI_ISL_2321080, EPI_ISL_2321081, EPI_ISL_2321082, EPI_ISL_2321083, EPI_ISL_2321084, EPI_ISL_2321085, EPI_ISL_2321086, EPI_ISL_2321087, EPI_ISL_2321088, EPI_ISL_2321089, EPI_ISL_2321090, EPI_ISL_2321091, EPI_ISL_2321092, EPI_ISL_2321093, EPI_ISL_2321094, EPI_ISL_2321095, EPI_ISL_2321096, EPI_ISL_2321097, EPI_ISL_2321098, EPI_ISL_2321099, EPI_ISL_2321100, EPI_ISL_2321101, EPI_ISL_2321102, EPI_ISL_2321103, EPI_ISL_2321104, EPI_ISL_2321105, EPI_ISL_2321106, EPI_ISL_2321107, EPI_ISL_2321108, EPI_ISL_2321109, EPI_ISL_2321110, EPI_ISL_2321111, EPI_ISL_2321112, EPI_ISL_2321113, EPI_ISL_2321114, EPI_ISL_2321115, EPI_ISL_2321116, EPI_ISL_2321117, EPI_ISL_2321118, EPI_ISL_2321119, EPI_ISL_2321120, EPI_ISL_2321121, EPI_ISL_2321122, EPI_ISL_2321123, EPI_ISL_2321124, EPI_ISL_2321125, EPI_ISL_2321126, EPI_ISL_2321127, EPI_ISL_2321128, EPI_ISL_2321129, EPI_ISL_2321130, EPI_ISL_2321131, EPI_ISL_2321132, EPI_ISL_2321133, EPI_ISL_2321134, EPI_ISL_2321135, EPI_ISL_2321136, EPI_ISL_2321137, EPI_ISL_2321138, EPI_ISL_2321139, EPI_ISL_2321140, EPI_ISL_2321141, EPI_ISL_2321142, EPI_ISL_2321143, EPI_ISL_2321144, EPI_ISL_2321145, EPI_ISL_2321146, EPI_ISL_2321147, EPI_ISL_2321148, EPI_ISL_2321149, EPI_ISL_2321150, EPI_ISL_2321151, EPI_ISL_2321152, EPI_ISL_2321153 | | | |
| see above | Helix/Illumina | Centers for Disease Control and Prevention Division of Viral Diseases, Pathogen Discovery | Dakota Howard, Dhwani Batra, Peter W. Cook, Kara Moser, Adrian Paskey, Jason Caravas, Benjamin Rambo-Martin, Shatavia Morrison, Christopher Gulvick, Scott Sammons, Yvette Unoarumhi, Darlene Wagner, Matthew Schmerer, Eileen de Feo, Jan Antico, Christine Tran, Matthew Tolentino, Shannon Wickline, Kim Gietzen, Brad Sickler, Jingtao Liu, Eric Allen, Phil Febbo, Nicole L. Washington, Simon White, Geraint Levan, Kelly Schiabor Barrett, Elizabeth Cirulli, Alexandre Bolze, Ary Ascencio, Charlotte Rivera-Garcia, Ryan Cho, Jason Nguyen, Sherry Wang, Jimmy Ramirez, Tyler Cassens, Efren Sandoval, Magnus Isaksson, William Lee, David Becker, Marc Laurent, James Lu, Clinton R. Paden, Duncan MacCannell |
| EPI_ISL_2321155, EPI_ISL_2321156, EPI_ISL_2321231, EPI_ISL_2321232, EPI_ISL_2321233, EPI_ISL_2321234, EPI_ISL_2321235, EPI_ISL_2321236, EPI_ISL_2321237, EPI_ISL_2321238, EPI_ISL_2321239, EPI_ISL_2321240, EPI_ISL_2321241, EPI_ISL_2321242, EPI_ISL_2321244, EPI_ISL_2321245, EPI_ISL_2321246, EPI_ISL_2321247, EPI_ISL_2321248, EPI_ISL_2321249, EPI_ISL_2321250, EPI_ISL_2321251, EPI_ISL_2321252, EPI_ISL_2321253, EPI_ISL_2321254, EPI_ISL_2321255, EPI_ISL_2321256, EPI_ISL_2321257, EPI_ISL_2321258, EPI_ISL_2321259, EPI_ISL_2321260, EPI_ISL_2321262, EPI_ISL_2321263, EPI_ISL_2321264, EPI_ISL_2321265, EPI_ISL_2321266, EPI_ISL_2321267, EPI_ISL_2321268, EPI_ISL_2321269, EPI_ISL_2321270, EPI_ISL_2321271, EPI_ISL_2321272, EPI_ISL_2321273, EPI_ISL_2321274, EPI_ISL_2321275, EPI_ISL_2321276, EPI_ISL_2321277, EPI_ISL_2321278, EPI_ISL_2321279, EPI_ISL_2321280, EPI_ISL_2321281, EPI_ISL_2321282, EPI_ISL_2321283, EPI_ISL_2321284, EPI_ISL_2321285, EPI_ISL_2321286, EPI_ISL_2321287, EPI_ISL_2321288, EPI_ISL_2321289, EPI_ISL_2321290, EPI_ISL_2321293, EPI_ISL_2321294, EPI_ISL_2321295, EPI_ISL_2321296, EPI_ISL_2321297, EPI_ISL_2321299, EPI_ISL_2321300, EPI_ISL_2321301, EPI_ISL_2321302, EPI_ISL_2321303, EPI_ISL_2321304, EPI_ISL_2321305, EPI_ISL_2321306, EPI_ISL_2321307, EPI_ISL_2321308, EPI_ISL_2321309, EPI_ISL_2321311, EPI_ISL_2321312, EPI_ISL_2321313, EPI_ISL_2321314, EPI_ISL_2321315, EPI_ISL_2321316, EPI_ISL_2321317, EPI_ISL_2321318, EPI_ISL_2321319, EPI_ISL_2321320, EPI_ISL_2321321, EPI_ISL_2321322, EPI_ISL_2321323, EPI_ISL_2321324, EPI_ISL_2321325, EPI_ISL_2321326, EPI_ISL_2321327, EPI_ISL_2321328, EPI_ISL_2321329, EPI_ISL_2321330, EPI_ISL_2321332, EPI_ISL_2321333, EPI_ISL_2321334, EPI_ISL_2321335, EPI_ISL_2321336, EPI_ISL_2321337, EPI_ISL_2321338, EPI_ISL_2321339, EPI_ISL_2321340, EPI_ISL_2321343, EPI_ISL_2321344, EPI_ISL_2321345, EPI_ISL_2321346, EPI_ISL_2321347, EPI_ISL_2321348, EPI_ISL_2321349, EPI_ISL_2321350, EPI_ISL_2321351, EPI_ISL_2321352, EPI_ISL_2321353, EPI_ISL_2321354, EPI_ISL_2321355, EPI_ISL_2321357, EPI_ISL_2321358, EPI_ISL_2321360, EPI_ISL_2321361, EPI_ISL_2321362, EPI_ISL_2321363, EPI_ISL_2321364, EPI_ISL_2321365, EPI_ISL_2321366, EPI_ISL_2321367, EPI_ISL_2321368, EPI_ISL_2321369, EPI_ISL_2321370, EPI_ISL_2321371, EPI_ISL_2321372, EPI_ISL_2321373, EPI_ISL_2321374, EPI_ISL_2321375, EPI_ISL_2321376, EPI_ISL_2321377, EPI_ISL_2321378, EPI_ISL_2321379, EPI_ISL_2321380, EPI_ISL_2321381, EPI_ISL_2321382, EPI_ISL_2321383, EPI_ISL_2321384, EPI_ISL_2321385, EPI_ISL_2321386, EPI_ISL_2321387 | | | |
| see above | Aegis Sciences Corporation | Centers for Disease Control and Prevention Division of Viral Diseases, Pathogen Discovery | Dakota Howard, Dhwani Batra, Peter W. Cook, Kara Moser, Adrian Paskey, Jason Caravas, Benjamin Rambo-Martin, Shatavia Morrison, Christopher Gulvick, Scott Sammons, Yvette Unoarumhi, Darlene Wagner, Matthew Schmerer, Cyndi Clark, Patrick Campbell, Rob Case, Vikramsinha Ghorpade, Holly Houdeshell, Ola Kvalvaag, Dillon Nall, Ethan Sanders, Alec Vest, Shaun Westlund, Matthew Hardison, Clinton R. Paden, Duncan MacCannell |
| EPI_ISL_2321388 | Tripler Army Medical Center | Tripler Army Medical Center | Jonathan A. D'Ambrozio, Rachael Downham, Timothy S. Horseman, Lindsey L. O'Neal, Zakariae Bram, and Catherine F. Uyehara |
| EPI_ISL_2321389, EPI_ISL_2321390, EPI_ISL_2321391, EPI_ISL_2321392, EPI_ISL_2321393, EPI_ISL_2321394, EPI_ISL_2321395, EPI_ISL_2321396, EPI_ISL_2321397, EPI_ISL_2321398, EPI_ISL_2321399, EPI_ISL_2321400, EPI_ISL_2321401, EPI_ISL_2321402, EPI_ISL_2321403, EPI_ISL_2321404, EPI_ISL_2321405, EPI_ISL_2321406, EPI_ISL_2321407, EPI_ISL_2321408, EPI_ISL_2321409, EPI_ISL_2321410, EPI_ISL_2321411, EPI_ISL_2321412, EPI_ISL_2321413, EPI_ISL_2321414, EPI_ISL_2321415, EPI_ISL_2321416, EPI_ISL_2321417, EPI_ISL_2321418, EPI_ISL_2321419, EPI_ISL_2321420, EPI_ISL_2321421, EPI_ISL_2321422, EPI_ISL_2321423, EPI_ISL_2321424, EPI_ISL_2321425, EPI_ISL_2321426, EPI_ISL_2321427, EPI_ISL_2321428, EPI_ISL_2321429, EPI_ISL_2321430, EPI_ISL_2321431, EPI_ISL_2321432, EPI_ISL_2321433, EPI_ISL_2321434, EPI_ISL_2321435, EPI_ISL_2321436, EPI_ISL_2321437, EPI_ISL_2321438, EPI_ISL_2321439, EPI_ISL_2321440, EPI_ISL_2321441, EPI_ISL_2321442, EPI_ISL_2321443, EPI_ISL_2321444, EPI_ISL_2321445, EPI_ISL_2321446, EPI_ISL_2321447, EPI_ISL_2321448, EPI_ISL_2321449, EPI_ISL_2321450, EPI_ISL_2321451, EPI_ISL_2321452, EPI_ISL_2321453, EPI_ISL_2321454, EPI_ISL_2321455, EPI_ISL_2321456, EPI_ISL_2321457, EPI_ISL_2321458, EPI_ISL_2321459, EPI_ISL_2321460, EPI_ISL_2321461, EPI_ISL_2321462, EPI_ISL_2321463, EPI_ISL_2321464, EPI_ISL_2321465, EPI_ISL_2321466, EPI_ISL_2321467, EPI_ISL_2321468, EPI_ISL_2321469, EPI_ISL_2321470, EPI_ISL_2321471, EPI_ISL_2321472, EPI_ISL_2321473, EPI_ISL_2321474, EPI_ISL_2321475, EPI_ISL_2321476, EPI_ISL_2321477, EPI_ISL_2321478, EPI_ISL_2321479, EPI_ISL_2321480, EPI_ISL_2321481, EPI_ISL_2321482, EPI_ISL_2321483, EPI_ISL_2321484, EPI_ISL_2321485, EPI_ISL_2321486, EPI_ISL_2321487, EPI_ISL_2321488, EPI_ISL_2321489, EPI_ISL_2321490, EPI_ISL_2321491, EPI_ISL_2321492, EPI_ISL_2321493, EPI_ISL_2321494, EPI_ISL_2321495, EPI_ISL_2321496, EPI_ISL_2321497, EPI_ISL_2321498, EPI_ISL_2321499, EPI_ISL_2321500, EPI_ISL_2321501, EPI_ISL_2321502, EPI_ISL_2321503, EPI_ISL_2321504, EPI_ISL_2321505, EPI_ISL_2321506, EPI_ISL_2321507, EPI_ISL_2321508, EPI_ISL_2321509, EPI_ISL_2321510, EPI_ISL_2321511, EPI_ISL_2321512, EPI_ISL_2321513, EPI_ISL_2321514, EPI_ISL_2321515, EPI_ISL_2321516, EPI_ISL_2321517, EPI_ISL_2321518, EPI_ISL_2321519, EPI_ISL_2321520, EPI_ISL_2321521, EPI_ISL_2321522, EPI_ISL_2321523, EPI_ISL_2321524, EPI_ISL_2321525, EPI_ISL_2321526, EPI_ISL_2321527, EPI_ISL_2321528, EPI_ISL_2321529, EPI_ISL_2321530, EPI_ISL_2321531, EPI_ISL_2321532, EPI_ISL_2321533, EPI_ISL_2321534, EPI_ISL_2321535, EPI_ISL_2321536, EPI_ISL_2321537, EPI_ISL_2321538, EPI_ISL_2321539, EPI_ISL_2321540, EPI_ISL_2321541, EPI_ISL_2321542, EPI_ISL_2321543, EPI_ISL_2321544, EPI_ISL_2321545, EPI_ISL_2321546, EPI_ISL_2321547, EPI_ISL_2321548, EPI_ISL_2321549, EPI_ISL_2321550, EPI_ISL_2321551, EPI_ISL_2321552, EPI_ISL_2321553, EPI_ISL_2321554, EPI_ISL_2321555, EPI_ISL_2321556, EPI_ISL_2321557, EPI_ISL_2321558, EPI_ISL_2321559, EPI_ISL_2321560, EPI_ISL_2321561, EPI_ISL_2321562, EPI_ISL_2321563, EPI_ISL_2321564, EPI_ISL_2321565, EPI_ISL_2321566, EPI_ISL_2321567, EPI_ISL_2321568, EPI_ISL_2321569, EPI_ISL_2321570, EPI_ISL_2321571 | | | |
| see above | Emory University | Centers for Disease Control and Prevention Division of Viral Diseases, Pathogen Discovery | Mili Sheth, Sarah Nobles, Jasmine Padilla, Mark Burroughs, Shoshona Le, Katie Dillon, Peter Cook, Clinton R. Paden, Dhwani Batra, Krista Queen, Kristen Knipe, Dakota Howard, Yvette Unoarumhi, Darlene Wagner, Matthew Schmerer, Ben L. Rambo-Martin, Kristine Lacek, Sam Shepard, Alison Laufer Halpin, Dave Wentworth, Vivien Dugan, Suxiang Tong, Justin Lee |
| EPI_ISL_2322716 | Ohio Department of Health Laboratory | Ohio Department of Health Laboratory | Holmes, Jennifer; Eric Brandt, Keoni Omura, Glen McGillivary, Caitlin McDonnell, Jade Mowery, Stephanie Mccracken, Tyler Payne, Kirtana Ramadugu, Erica Leasure, Brent Lee, Kelsey Florek, Heather Blankenship, Allison Black, Quanta Brown, and Tammy Bannerman |
| EPI_ISL_2322717, EPI_ISL_2322719, EPI_ISL_2322720, EPI_ISL_2322721, EPI_ISL_2322722, EPI_ISL_2322723, EPI_ISL_2322724, EPI_ISL_2322725, EPI_ISL_2322726, EPI_ISL_2322727, EPI_ISL_2322728, EPI_ISL_2322729, EPI_ISL_2322730, EPI_ISL_2322731, EPI_ISL_2322732, EPI_ISL_2322733, EPI_ISL_2322734, EPI_ISL_2322735, EPI_ISL_2322736, EPI_ISL_2322737, EPI_ISL_2322738, EPI_ISL_2322739, EPI_ISL_2322740, EPI_ISL_2322741 | | | |
| see above | MD PHL | MD PHL | Maryland Department of Health Laboratories Administration |
| EPI_ISL_2322743 | Ohio Department of Health Laboratory | Ohio Department of Health Laboratory | Holmes, Jennifer; Eric Brandt, Keoni Omura, Glen McGillivary, Caitlin McDonnell, Jade Mowery, Stephanie Mccracken, Tyler Payne, Kirtana Ramadugu, Erica Leasure, Brent Lee, Kelsey Florek, Heather Blankenship, Allison Black, Quanta Brown, and Tammy Bannerman |

| | | | |
|---|---|---|---|
| EPI_ISL_2322752 | Washington State Department of Health Public Health Laboratories | Washington State Department of Health Public Health Laboratories | Drew MacKellar, Philip Dykema, Denny Russell, Joenice Gonzalez, Hannah Gray, Geoff Melly, Vanessa De Los Santos, Darren Lucas, JohnAric Peterson, Avi Singh, Rebecca Cao |
| EPI_ISL_2322753 | AR Dept. of Health-PHL, Molecular Diagnostics | Genomics and Discovery, Respiratory Viruses Branch, Division of Viral Diseases, Centers for Disease Control and Prevention | Jing Zhang, Anna Kelleher, Ying Tao, Yan Li, Brian Lynch, Krista Queen, Anna Uehara, Peter Cook, Han Jia Justin Ng, Rachel Marine, Clinton R. Paden, Dhwani Batra, Haibin Wang, Tara Coalter, Jasmine Padilla, Morgan Davis, Mili Sheth, Sarah Nobles, Mark Burroughs, Justin Lee, Adam Retchless, Suxiang Tong |
| EPI_ISL_2322754 | TN Division of Laboratory Services | Genomics and Discovery, Respiratory Viruses Branch, Division of Viral Diseases, Centers for Disease Control and Prevention | Jing Zhang, Anna Kelleher, Ying Tao, Yan Li, Brian Lynch, Krista Queen, Anna Uehara, Peter Cook, Han Jia Justin Ng, Rachel Marine, Clinton R. Paden, Dhwani Batra, Haibin Wang, Tara Coalter, Jasmine Padilla, Morgan Davis, Mili Sheth, Sarah Nobles, Mark Burroughs, Justin Lee, Adam Retchless, Suxiang Tong |
| EPI_ISL_2322755, EPI_ISL_2322756 | Wexner Medical Center | The Ohio State University College of Medicine | Koenig,S., Seminetta,J. |
| EPI_ISL_2322764, EPI_ISL_2322765, EPI_ISL_2322766, EPI_ISL_2322767, EPI_ISL_2322768, EPI_ISL_2322769, EPI_ISL_2322770, EPI_ISL_2322771, EPI_ISL_2322772, EPI_ISL_2322773, EPI_ISL_2322774, EPI_ISL_2322775, EPI_ISL_2322776, EPI_ISL_2322777, EPI_ISL_2322778, EPI_ISL_2322779, EPI_ISL_2322780, EPI_ISL_2322781, EPI_ISL_2322782, EPI_ISL_2322783, EPI_ISL_2322784, EPI_ISL_2322785, EPI_ISL_2322786, EPI_ISL_2322787, EPI_ISL_2322788, EPI_ISL_2322789, EPI_ISL_2322790, EPI_ISL_2322791, EPI_ISL_2322792, EPI_ISL_2322793, EPI_ISL_2322794, EPI_ISL_2322796, EPI_ISL_2322797, EPI_ISL_2322798, EPI_ISL_2322799, EPI_ISL_2322800, EPI_ISL_2322801, EPI_ISL_2322803, EPI_ISL_2322804, EPI_ISL_2322805, EPI_ISL_2322806, EPI_ISL_2322807, EPI_ISL_2322808, EPI_ISL_2322809, EPI_ISL_2322810, EPI_ISL_2322811, EPI_ISL_2322812, EPI_ISL_2322813, EPI_ISL_2322814, EPI_ISL_2322815, EPI_ISL_2322816, EPI_ISL_2322817, EPI_ISL_2322818, EPI_ISL_2322819, EPI_ISL_2322821, EPI_ISL_2322822, EPI_ISL_2322823, EPI_ISL_2322824, EPI_ISL_2322825, EPI_ISL_2322826, EPI_ISL_2322827, EPI_ISL_2322828, EPI_ISL_2322829, EPI_ISL_2322830, EPI_ISL_2322831, EPI_ISL_2322832, EPI_ISL_2322834, EPI_ISL_2322835, EPI_ISL_2322836, EPI_ISL_2322837, EPI_ISL_2322838, EPI_ISL_2322839, EPI_ISL_2322840 | | | |
| see above | New Mexico Department of Health Scientific Laboratory | New Mexico Department of Health Scientific Laboratory | Ellie Johnson, Anastacia Griego-Fisher, D'eldra Malone, Jennifer Benoit |
| EPI_ISL_2322841, EPI_ISL_2322842 | WLA VAMC | Genomics and Discovery, Respiratory Viruses Branch, Division of Viral Diseases, Centers for Disease Control and Prevention | Jing Zhang, Anna Kelleher, Ying Tao, Yan Li, Brian Lynch, Krista Queen, Anna Uehara, Peter Cook, Han Jia Justin Ng, Rachel Marine, Clinton R. Paden, Dhwani Batra, Haibin Wang, Tara Coalter, Jasmine Padilla, Morgan Davis, Mili Sheth, Sarah Nobles, Mark Burroughs, Justin Lee, Adam Retchless, Suxiang Tong |
| EPI_ISL_2322843 | New Mexico Department of Health Scientific Laboratory | New Mexico Department of Health Scientific Laboratory | Ellie Johnson, Anastacia Griego-Fisher, D'eldra Malone, Jennifer Benoit |
| EPI_ISL_2322844 | WLA VAMC | Genomics and Discovery, Respiratory Viruses Branch, Division of Viral Diseases, Centers for Disease Control and Prevention | Jing Zhang, Anna Kelleher, Ying Tao, Yan Li, Brian Lynch, Krista Queen, Anna Uehara, Peter Cook, Han Jia Justin Ng, Rachel Marine, Clinton R. Paden, Dhwani Batra, Haibin Wang, Tara Coalter, Jasmine Padilla, Morgan Davis, Mili Sheth, Sarah Nobles, Mark Burroughs, Justin Lee, Adam Retchless, Suxiang Tong |
| EPI_ISL_2322845, EPI_ISL_2322846, EPI_ISL_2322847, EPI_ISL_2322848, EPI_ISL_2322849, EPI_ISL_2322851, EPI_ISL_2322853, EPI_ISL_2322854, EPI_ISL_2322855, EPI_ISL_2322856, EPI_ISL_2322857, EPI_ISL_2322858 | | | |
| see above | New Mexico Department of Health Scientific Laboratory | New Mexico Department of Health Scientific Laboratory | Ellie Johnson, Anastacia Griego-Fisher, D'eldra Malone, Jennifer Benoit |
| EPI_ISL_2322859 | GA Department of Public Health Laboratory | Genomics and Discovery, Respiratory Viruses Branch, Division of Viral Diseases, Centers for Disease Control and Prevention | Jing Zhang, Anna Kelleher, Ying Tao, Yan Li, Brian Lynch, Krista Queen, Anna Uehara, Peter Cook, Han Jia Justin Ng, Rachel Marine, Clinton R. Paden, Dhwani Batra, Haibin Wang, Tara Coalter, Jasmine Padilla, Morgan Davis, Mili Sheth, Sarah Nobles, Mark Burroughs, Justin Lee, Adam Retchless, Suxiang Tong |
| EPI_ISL_2322860, EPI_ISL_2322861, EPI_ISL_2322862 | AR Dept. of Health-PHL, Molecular Diagnostics | Genomics and Discovery, Respiratory Viruses Branch, Division of Viral Diseases, Centers for Disease Control and Prevention | Jing Zhang, Anna Kelleher, Ying Tao, Yan Li, Brian Lynch, Krista Queen, Anna Uehara, Peter Cook, Han Jia Justin Ng, Rachel Marine, Clinton R. Paden, Dhwani Batra, Haibin Wang, Tara Coalter, Jasmine Padilla, Morgan Davis, Mili Sheth, Sarah Nobles, Mark Burroughs, Justin Lee, Adam Retchless, Suxiang Tong |
| EPI_ISL_2322863, EPI_ISL_2322864, EPI_ISL_2322866, EPI_ISL_2322867, EPI_ISL_2322869, EPI_ISL_2322870, EPI_ISL_2322871, EPI_ISL_2322874, EPI_ISL_2322875, EPI_ISL_2322876, EPI_ISL_2322877, EPI_ISL_2322879, EPI_ISL_2322882, EPI_ISL_2322883, EPI_ISL_2322885, EPI_ISL_2322886, EPI_ISL_2322887, EPI_ISL_2322888, EPI_ISL_2322889, EPI_ISL_2322890, EPI_ISL_2322891, EPI_ISL_2322892, EPI_ISL_2322893, EPI_ISL_2322894, EPI_ISL_2322895, EPI_ISL_2322897 | | | |
| see above | Wyoming Public Health Laboratory | Wyoming Public Health Laboratory | Jim Mildenberger, Taylor Fearing, Channing Weber, Ashley Norberg, Chayse Rowley, Marley Goetz, Brian Dominguez, Elliot Thomasson, Brittany Oher, Sam Britz, Cari Sloma, and Rob Christensen |
| EPI_ISL_2322900, EPI_ISL_2322901 | DOHMH Morrisania | New York City Public Health Laboratory | Jade Wang, et al. |
| EPI_ISL_2322902 | DOHMH Fort Greene | New York City Public Health Laboratory | Jade Wang, et al. |
| EPI_ISL_2322903, EPI_ISL_2322904, EPI_ISL_2322905 | DOHMH Corona | New York City Public Health Laboratory | Jade Wang, et al. |
| EPI_ISL_2322906, EPI_ISL_2322907 | DOHMH Crown Heights | New York City Public Health Laboratory | Jade Wang, et al. |
| EPI_ISL_2322908, EPI_ISL_2322909, EPI_ISL_2322910 | DOHMH PHL | New York City Public Health Laboratory | Jade Wang, et al. |
| EPI_ISL_2322911 | DOHMH Fort Greene | New York City Public Health Laboratory | Jade Wang, et al. |
| EPI_ISL_2322912, EPI_ISL_2322913 | DOHMH Central Harlem | New York City Public Health Laboratory | Jade Wang, et al. |
| EPI_ISL_2322914, EPI_ISL_2322915, EPI_ISL_2322916, EPI_ISL_2322917, EPI_ISL_2322918 | DOHMH Jamaica | New York City Public Health Laboratory | Jade Wang, et al. |
| EPI_ISL_2322919 | DOHMH Riverside | New York City Public Health Laboratory | Jade Wang, et al. |
| EPI_ISL_2322920 | OCME Office Of Chief Medical Examiner | New York City Public Health Laboratory | Jade Wang, et al. |
| EPI_ISL_2323151, EPI_ISL_2323152, EPI_ISL_2323153, EPI_ISL_2323154, EPI_ISL_2323155, EPI_ISL_2323156, EPI_ISL_2323157, EPI_ISL_2323158, EPI_ISL_2323159, EPI_ISL_2323160, EPI_ISL_2323161, EPI_ISL_2323162, EPI_ISL_2323163, EPI_ISL_2323164, EPI_ISL_2323165, EPI_ISL_2323166, EPI_ISL_2323167, EPI_ISL_2323168, EPI_ISL_2323169, EPI_ISL_2323170, EPI_ISL_2323171, EPI_ISL_2323172, EPI_ISL_2323173 | | | |
| see above | Nebraska Public Health Laboratory | NPHL COVID-19 Response Team | NPHL COVID-19 Response Team |
| EPI_ISL_2323452 | GA Department of Public Health Laboratory | Genomics and Discovery, Respiratory Viruses Branch, Division of Viral Diseases, Centers for Disease Control and Prevention | Jing Zhang, Anna Kelleher, Ying Tao, Yan Li, Brian Lynch, Krista Queen, Anna Uehara, Peter Cook, Han Jia Justin Ng, Rachel Marine, Clinton R. Paden, Dhwani Batra, Haibin Wang, Tara Coalter, Jasmine Padilla, Morgan Davis, Mili Sheth, Sarah Nobles, Mark Burroughs, Justin Lee, Adam Retchless, Suxiang Tong |
| EPI_ISL_2323453, EPI_ISL_2323454, EPI_ISL_2323455 | AR Dept. of Health-PHL, Molecular Diagnostics | Genomics and Discovery, Respiratory Viruses Branch, Division of Viral Diseases, Centers for Disease Control and Prevention | Jing Zhang, Anna Kelleher, Ying Tao, Yan Li, Brian Lynch, Krista Queen, Anna Uehara, Peter Cook, Han Jia Justin Ng, Rachel Marine, Clinton R. Paden, Dhwani Batra, Haibin Wang, Tara Coalter, Jasmine Padilla, Morgan Davis, Mili Sheth, Sarah Nobles, Mark Burroughs, Justin Lee, Adam Retchless, Suxiang Tong |
| EPI_ISL_2323456 | OK Public Health Laboratory, Oklahoma State DOH | Genomics and Discovery, Respiratory Viruses Branch, Division of Viral Diseases, Centers for Disease Control and Prevention | Jing Zhang, Anna Kelleher, Ying Tao, Yan Li, Brian Lynch, Krista Queen, Anna Uehara, Peter Cook, Han Jia Justin Ng, Rachel Marine, Clinton R. Paden, Dhwani Batra, Haibin Wang, Tara Coalter, Jasmine Padilla, Morgan Davis, Mili Sheth, Sarah Nobles, Mark Burroughs, Justin Lee, Adam Retchless, Suxiang Tong |
| EPI_ISL_2323457, EPI_ISL_2323458, EPI_ISL_2323460, EPI_ISL_2323461, EPI_ISL_2323462, EPI_ISL_2323466, EPI_ISL_2323468, EPI_ISL_2323471, EPI_ISL_2323473, EPI_ISL_2323475, EPI_ISL_2323476, EPI_ISL_2323483, EPI_ISL_2323484, EPI_ISL_2323485, EPI_ISL_2323486, EPI_ISL_2323487, EPI_ISL_2323488, EPI_ISL_2323489, EPI_ISL_2323491, EPI_ISL_2323492, EPI_ISL_2323494, EPI_ISL_2323496, EPI_ISL_2323497, EPI_ISL_2323498, EPI_ISL_2323499, EPI_ISL_2323500, EPI_ISL_2323501, EPI_ISL_2323502, EPI_ISL_2323503, EPI_ISL_2323504, EPI_ISL_2323505, EPI_ISL_2323506, EPI_ISL_2323507, EPI_ISL_2323508, EPI_ISL_2323510, EPI_ISL_2323511, EPI_ISL_2323512, EPI_ISL_2323513, EPI_ISL_2323514, EPI_ISL_2323515, EPI_ISL_2323516, EPI_ISL_2323517, EPI_ISL_2323518, EPI_ISL_2323519, EPI_ISL_2323520, EPI_ISL_2323521, EPI_ISL_2323522, EPI_ISL_2323523, EPI_ISL_2323525, EPI_ISL_2323526, EPI_ISL_2323527, EPI_ISL_2323528, EPI_ISL_2323529, EPI_ISL_2323530, EPI_ISL_2323531, EPI_ISL_2323532, EPI_ISL_2323533, EPI_ISL_2323535, EPI_ISL_2323536, EPI_ISL_2323537, EPI_ISL_2323538, EPI_ISL_2323539, EPI_ISL_2323540, EPI_ISL_2323541, EPI_ISL_2323542, EPI_ISL_2323543, EPI_ISL_2323544, EPI_ISL_2323545, EPI_ISL_2323546, EPI_ISL_2323547, EPI_ISL_2323548, EPI_ISL_2323549, EPI_ISL_2323551, EPI_ISL_2323552, EPI_ISL_2323553, EPI_ISL_2323554, EPI_ISL_2323555, EPI_ISL_2323557, EPI_ISL_2323558, EPI_ISL_2323559, EPI_ISL_2323560, EPI_ISL_2323561, EPI_ISL_2323562, EPI_ISL_2323563, EPI_ISL_2323564, EPI_ISL_2323566, EPI_ISL_2323567, EPI_ISL_2323569, EPI_ISL_2323570, EPI_ISL_2323571, EPI_ISL_2323572, EPI_ISL_2323573, EPI_ISL_2323574, EPI_ISL_2323575, EPI_ISL_2323576, EPI_ISL_2323577, EPI_ISL_2323578, EPI_ISL_2323579, EPI_ISL_2323580, EPI_ISL_2323581, EPI_ISL_2323582, EPI_ISL_2323584, EPI_ISL_2323585, EPI_ISL_2323586, EPI_ISL_2323587, EPI_ISL_2323588, EPI_ISL_2323589, EPI_ISL_2323590, EPI_ISL_2323591, EPI_ISL_2323592, EPI_ISL_2323593, EPI_ISL_2323594, EPI_ISL_2323595, EPI_ISL_2323596, EPI_ISL_2323597, EPI_ISL_2323598, EPI_ISL_2323599, EPI_ISL_2323600, EPI_ISL_2323601, EPI_ISL_2323602, EPI_ISL_2323603, EPI_ISL_2323605, EPI_ISL_2323606, EPI_ISL_2323607, EPI_ISL_2323608, EPI_ISL_2323609, EPI_ISL_2323610, EPI_ISL_2323611, EPI_ISL_2323612, EPI_ISL_2323613, EPI_ISL_2323614, EPI_ISL_2323615, EPI_ISL_2323616, EPI_ISL_2323617, EPI_ISL_2323618, EPI_ISL_2323619, EPI_ISL_2323620, EPI_ISL_2323621, EPI_ISL_2323622, EPI_ISL_2323624, EPI_ISL_2323627, EPI_ISL_2323628, EPI_ISL_2323629, EPI_ISL_2323630, EPI_ISL_2323631, EPI_ISL_2323632, EPI_ISL_2323633, EPI_ISL_2323634, EPI_ISL_2323635, EPI_ISL_2323637, EPI_ISL_2323638, EPI_ISL_2323639, EPI_ISL_2323640, EPI_ISL_2323641, EPI_ISL_2323642, EPI_ISL_2323643, EPI_ISL_2323644, EPI_ISL_2323645, EPI_ISL_2323646, EPI_ISL_2323647, EPI_ISL_2323648, EPI_ISL_2323649, EPI_ISL_2323650, EPI_ISL_2323651, EPI_ISL_2323652, EPI_ISL_2323653, EPI_ISL_2323654, EPI_ISL_2323655, EPI_ISL_2323656, EPI_ISL_2323657, EPI_ISL_2323658, EPI_ISL_2323659, EPI_ISL_2323660, EPI_ISL_2323661, EPI_ISL_2323663, EPI_ISL_2323664, EPI_ISL_2323665, EPI_ISL_2323666, EPI_ISL_2323667, EPI_ISL_2323668, EPI_ISL_2323669, EPI_ISL_2323670, EPI_ISL_2323671, EPI_ISL_2323672, EPI_ISL_2323673, EPI_ISL_2323674, EPI_ISL_2323675, EPI_ISL_2323676, EPI_ISL_2323677, EPI_ISL_2323678, EPI_ISL_2323679, EPI_ISL_2323680, EPI_ISL_2323681, EPI_ISL_2323682, EPI_ISL_2323683, EPI_ISL_2323684, EPI_ISL_2323685, EPI_ISL_2323686, EPI_ISL_2323687, EPI_ISL_2323688, EPI_ISL_2323689, EPI_ISL_2323690, EPI_ISL_2323691, EPI_ISL_2323692, EPI_ISL_2323693, EPI_ISL_2323694, EPI_ISL_2323695, EPI_ISL_2323696, EPI_ISL_2323698, EPI_ISL_2323699, EPI_ISL_2323701, EPI_ISL_2323702, EPI_ISL_2323703, EPI_ISL_2323704, EPI_ISL_2323705, EPI_ISL_2323706, EPI_ISL_2323709, EPI_ISL_2323710, EPI_ISL_2323711, EPI_ISL_2323712, EPI_ISL_2323713, EPI_ISL_2323714, EPI_ISL_2323715, EPI_ISL_2323716, EPI_ISL_2323717, EPI_ISL_2323718, EPI_ISL_2323719, EPI_ISL_2323720, EPI_ISL_2323721, EPI_ISL_2323723, EPI_ISL_2323724, EPI_ISL_2323725, EPI_ISL_2323726, EPI_ISL_2323727, EPI_ISL_2323728, EPI_ISL_2323729, EPI_ISL_2323730, EPI_ISL_2323731, EPI_ISL_2323732, EPI_ISL_2323733, EPI_ISL_2323734, EPI_ISL_2323735, EPI_ISL_2323736, EPI_ISL_2323737, EPI_ISL_2323738, EPI_ISL_2323739, EPI_ISL_2323741, EPI_ISL_2323742, EPI_ISL_2323743, EPI_ISL_2323744, EPI_ISL_2323745, EPI_ISL_2323746, EPI_ISL_2323747, EPI_ISL_2323748, EPI_ISL_2323749, EPI_ISL_2323750, EPI_ISL_2323751, EPI_ISL_2323752, EPI_ISL_2323753, EPI_ISL_2323754, EPI_ISL_2323755, EPI_ISL_2323756, EPI_ISL_2323757, EPI_ISL_2323758, EPI_ISL_2323759, EPI_ISL_2323760, EPI_ISL_2323761, EPI_ISL_2323762, EPI_ISL_2323763, EPI_ISL_2323764, EPI_ISL_2323765, EPI_ISL_2323766, EPI_ISL_2323767, EPI_ISL_2323768, EPI_ISL_2323769, EPI_ISL_2323770, EPI_ISL_2323771, EPI_ISL_2323772, EPI_ISL_2323773, EPI_ISL_2323774, EPI_ISL_2323775, EPI_ISL_2323776, EPI_ISL_2323777, EPI_ISL_2323778, EPI_ISL_2323779, EPI_ISL_2323780, EPI_ISL_2323781, EPI_ISL_2323782, EPI_ISL_2323783, EPI_ISL_2323784, EPI_ISL_2323785, EPI_ISL_2323786, EPI_ISL_2323787, EPI_ISL_2323788, EPI_ISL_2323789, EPI_ISL_2323790, EPI_ISL_2323791, EPI_ISL_2323792, EPI_ISL_2323793, EPI_ISL_2323794, EPI_ISL_2323795, EPI_ISL_2323796, EPI_ISL_2323797, EPI_ISL_2323798, EPI_ISL_2323799, EPI_ISL_2323800, EPI_ISL_2323801, EPI_ISL_2323802, EPI_ISL_2323803, EPI_ISL_2323804, EPI_ISL_2323805, EPI_ISL_2323806, EPI_ISL_2323807, EPI_ISL_2323808, EPI_ISL_2323809, EPI_ISL_2323810, EPI_ISL_2323811, EPI_ISL_2323812, EPI_ISL_2323813, EPI_ISL_2323814, EPI_ISL_2323815, EPI_ISL_2323816, EPI_ISL_2323817, EPI_ISL_2323818, EPI_ISL_2323819, EPI_ISL_2323820, EPI_ISL_2323822, EPI_ISL_2323824, EPI_ISL_2323825, EPI_ISL_2323827, EPI_ISL_2323828, EPI_ISL_2323829, EPI_ISL_2323830, EPI_ISL_2323831, EPI_ISL_2323832, EPI_ISL_2323833, EPI_ISL_2323834, EPI_ISL_2323835, EPI_ISL_2323837, EPI_ISL_2323838, EPI_ISL_2323839, EPI_ISL_2323841, EPI_ISL_2323843, EPI_ISL_2323844, EPI_ISL_2323845, EPI_ISL_2323846, EPI_ISL_2323847, EPI_ISL_2323848, EPI_ISL_2323849, EPI_ISL_2323850, EPI_ISL_2323851, EPI_ISL_2323852, EPI_ISL_2323853, EPI_ISL_2323854, EPI_ISL_2323855, EPI_ISL_2323856, EPI_ISL_2323857, EPI_ISL_2323858, EPI_ISL_2323859, EPI_ISL_2323860, EPI_ISL_2323861, EPI_ISL_2323862, EPI_ISL_2323863, EPI_ISL_2323864, EPI_ISL_2323865, EPI_ISL_2323866, EPI_ISL_2323867, EPI_ISL_2323868, EPI_ISL_2323869, EPI_ISL_2323870, EPI_ISL_2323871, EPI_ISL_2323872, EPI_ISL_2323873, EPI_ISL_2323874, EPI_ISL_2323875, EPI_ISL_2323876, EPI_ISL_2323877, EPI_ISL_2323878, EPI_ISL_2323879, EPI_ISL_2323880, EPI_ISL_2323881, EPI_ISL_2323882, EPI_ISL_2323883, EPI_ISL_2323884, EPI_ISL_2323885, EPI_ISL_2323886, EPI_ISL_2323887, EPI_ISL_2323888, EPI_ISL_2323889, EPI_ISL_2323890, EPI_ISL_2323892, EPI_ISL_2323893, EPI_ISL_2323894, EPI_ISL_2323895, EPI_ISL_2323896, EPI_ISL_2323897, EPI_ISL_2323898, EPI_ISL_2323899, EPI_ISL_2323900, EPI_ISL_2323901, EPI_ISL_2323902, EPI_ISL_2323903, EPI_ISL_2323904, EPI_ISL_2323905, EPI_ISL_2323906, EPI_ISL_2323907, EPI_ISL_2323908, EPI_ISL_2323909, EPI_ISL_2323910, EPI_ISL_2323911, EPI_ISL_2323912, EPI_ISL_2323913, EPI_ISL_2323914, EPI_ISL_2323915, EPI_ISL_2323916, EPI_ISL_2323917, EPI_ISL_2323918, EPI_ISL_2323919, EPI_ISL_2323920, EPI_ISL_2323921, EPI_ISL_2323922, EPI_ISL_2323923, EPI_ISL_2323924, EPI_ISL_2323925, EPI_ISL_2323926, EPI_ISL_2323927, EPI_ISL_2323929, EPI_ISL_2323930, EPI_ISL_2323931, EPI_ISL_2323932, EPI_ISL_2323933, EPI_ISL_2323934, EPI_ISL_2323935, EPI_ISL_2323936, EPI_ISL_2323937, EPI_ISL_2323938, EPI_ISL_2323939, EPI_ISL_2323941, EPI_ISL_2323942, EPI_ISL_2323943, EPI_ISL_2323944, EPI_ISL_2323945, EPI_ISL_2323946, EPI_ISL_2323947, EPI_ISL_2323948, EPI_ISL_2323949, EPI_ISL_2323950, EPI_ISL_2323951, EPI_ISL_2323952, EPI_ISL_2323953, EPI_ISL_2323954, EPI_ISL_2323955, EPI_ISL_2323956, EPI_ISL_2323957, EPI_ISL_2323958, EPI_ISL_2323959, EPI_ISL_2323960, EPI_ISL_2323961, EPI_ISL_2323962, EPI_ISL_2323963, EPI_ISL_2323964, EPI_ISL_2323965, EPI_ISL_2323966, EPI_ISL_2323967, EPI_ISL_2323968, EPI_ISL_2323969, EPI_ISL_2323970, EPI_ISL_2323971, EPI_ISL_2323972, EPI_ISL_2323973, EPI_ISL_2323974, EPI_ISL_2323975, EPI_ISL_2323976, EPI_ISL_2323978, EPI_ISL_2323980, EPI_ISL_2323981, EPI_ISL_2323982, EPI_ISL_2323983, EPI_ISL_2323984, EPI_ISL_2323985, EPI_ISL_2323986, EPI_ISL_2323987, EPI_ISL_2323988, EPI_ISL_2323989, EPI_ISL_2323990, EPI_ISL_2323991, EPI_ISL_2323992, EPI_ISL_2323994, EPI_ISL_2323995, EPI_ISL_2323996, EPI_ISL_2323997, EPI_ISL_2323998, EPI_ISL_2323999, EPI_ISL_2324000, EPI_ISL_2324001, EPI_ISL_2324002, EPI_ISL_2324003, EPI_ISL_2324005, EPI_ISL_2324006, EPI_ISL_2324007, EPI_ISL_2324008, EPI_ISL_2324010, EPI_ISL_2324011, EPI_ISL_2324012, EPI_ISL_2324013, EPI_ISL_2324014, EPI_ISL_2324015, EPI_ISL_2324016, EPI_ISL_2324017, EPI_ISL_2324018, EPI_ISL_2324019, EPI_ISL_2324020, EPI_ISL_2324021, EPI_ISL_2324022, EPI_ISL_2324023, EPI_ISL_2324025, EPI_ISL_2324026, EPI_ISL_2324027, EPI_ISL_2324028, EPI_ISL_2324029, EPI_ISL_2324030, EPI_ISL_2324031, EPI_ISL_2324032, EPI_ISL_2324033, EPI_ISL_2324034, EPI_ISL_2324035, EPI_ISL_2324036, EPI_ISL_2324037, EPI_ISL_2324038, EPI_ISL_2324039, EPI_ISL_2324040, EPI_ISL_2324041, EPI_ISL_2324042, EPI_ISL_2324043, EPI_ISL_2324044, EPI_ISL_2324045, EPI_ISL_2324046, EPI_ISL_2324047, EPI_ISL_2324048, EPI_ISL_2324049, EPI_ISL_2324050, EPI_ISL_2324051, EPI_ISL_2324053, EPI_ISL_2324054, EPI_ISL_2324055, EPI_ISL_2324056, EPI_ISL_2324057, EPI_ISL_2324059, EPI_ISL_2324060, EPI_ISL_2324061, EPI_ISL_2324062, EPI_ISL_2324063, EPI_ISL_2324064, EPI_ISL_2324065, EPI_ISL_2324066, EPI_ISL_2324067, EPI_ISL_2324068, EPI_ISL_2324069, EPI_ISL_2324070, EPI_ISL_2324071, EPI_ISL_2324072, EPI_ISL_2324073, EPI_ISL_2324074, EPI_ISL_2324075, EPI_ISL_2324077, EPI_ISL_2324078, EPI_ISL_2324079, EPI_ISL_2324080, EPI_ISL_2324081, EPI_ISL_2324082, EPI_ISL_2324083, EPI_ISL_2324084, EPI_ISL_2324085, EPI_ISL_2324086, EPI_ISL_2324087, EPI_ISL_2324088, EPI_ISL_2324089, EPI_ISL_2324090, EPI_ISL_2324091, EPI_ISL_2324092, EPI_ISL_2324093, EPI_ISL_2324094, EPI_ISL_2324096, EPI_ISL_2324097, EPI_ISL_2324099, EPI_ISL_2324100, EPI_ISL_2324101, EPI_ISL_2324102, EPI_ISL_2324103, EPI_ISL_2324104, EPI_ISL_2324105, EPI_ISL_2324106, EPI_ISL_2324107, EPI_ISL_2324108, | | | |

| | | | |
|---|---|---|---|
| EPI_ISL_2324109, EPI_ISL_2324110, EPI_ISL_2324111, EPI_ISL_2324112, EPI_ISL_2324114, EPI_ISL_2324115, EPI_ISL_2324116, EPI_ISL_2324117, EPI_ISL_2324118, EPI_ISL_2324120, EPI_ISL_2324121, EPI_ISL_2324122, EPI_ISL_2324123, EPI_ISL_2324124, EPI_ISL_2324125, EPI_ISL_2324127, EPI_ISL_2324128, EPI_ISL_2324129, EPI_ISL_2324130, EPI_ISL_2324131, EPI_ISL_2324132, EPI_ISL_2324133, EPI_ISL_2324134, EPI_ISL_2324135, EPI_ISL_2324136, EPI_ISL_2324137, EPI_ISL_2324138, EPI_ISL_2324139, EPI_ISL_2324140, EPI_ISL_2324141, EPI_ISL_2324142, EPI_ISL_2324143, EPI_ISL_2324144, EPI_ISL_2324145, EPI_ISL_2324146, EPI_ISL_2324147, EPI_ISL_2324148, EPI_ISL_2324149, EPI_ISL_2324150, EPI_ISL_2324151, EPI_ISL_2324152, EPI_ISL_2324153, EPI_ISL_2324154, EPI_ISL_2324155, EPI_ISL_2324156, EPI_ISL_2324157, EPI_ISL_2324158, EPI_ISL_2324159, EPI_ISL_2324160, EPI_ISL_2324161, EPI_ISL_2324162, EPI_ISL_2324163, EPI_ISL_2324165, EPI_ISL_2324166, EPI_ISL_2324167, EPI_ISL_2324168 | | | |
| see above | Infinity Biologix | Centers for Disease Control and Prevention Division of Viral Diseases, Pathogen Discovery | Dakota Howard, Dhwani Batra, Peter W. Cook, Kara Moser, Adrian Paskey, Jason Caravas, Benjamin Rambo-Martin, Shatavia Morrison, Christopher Gulvick, Scott Sammons, Yvette Unoarumhi, Darlene Wagner, Matthew Schmerer, Christian Bixby, Yihe Wang, Jonathan Schultz, Chirayu Goswami, Russ Hager, Robin Grimwood, Clinton R. Paden, Duncan MacCannell |
| EPI_ISL_2324440, EPI_ISL_2324441, EPI_ISL_2324442, EPI_ISL_2324443, EPI_ISL_2324444 | Tulare County Public Health Laboratory | Tulare County Public Health Laboratory | Denise L Lopez, Joshua Kootstra |
| EPI_ISL_2324445, EPI_ISL_2324446, EPI_ISL_2324447, EPI_ISL_2324448, EPI_ISL_2324449, EPI_ISL_2324450, EPI_ISL_2324451, EPI_ISL_2324452, EPI_ISL_2324453, EPI_ISL_2324455, EPI_ISL_2324456, EPI_ISL_2324457, EPI_ISL_2324458, EPI_ISL_2324459, EPI_ISL_2324460, EPI_ISL_2324461, EPI_ISL_2324462, EPI_ISL_2324463, EPI_ISL_2324464, EPI_ISL_2324466, EPI_ISL_2324467, EPI_ISL_2324468, EPI_ISL_2324469, EPI_ISL_2324470, EPI_ISL_2324471, EPI_ISL_2324472, EPI_ISL_2324473, EPI_ISL_2324474, EPI_ISL_2324475, EPI_ISL_2324476, EPI_ISL_2324477, EPI_ISL_2324478, EPI_ISL_2324479, EPI_ISL_2324480, EPI_ISL_2324481, EPI_ISL_2324482, EPI_ISL_2324483, EPI_ISL_2324484, EPI_ISL_2324485, EPI_ISL_2324486, EPI_ISL_2324487, EPI_ISL_2324488, EPI_ISL_2324489, EPI_ISL_2324490, EPI_ISL_2324492, EPI_ISL_2324493, EPI_ISL_2324494, EPI_ISL_2324495, EPI_ISL_2324496, EPI_ISL_2324497, EPI_ISL_2324498, EPI_ISL_2324499, EPI_ISL_2324500, EPI_ISL_2324501, EPI_ISL_2324502, EPI_ISL_2324503, EPI_ISL_2324504, EPI_ISL_2324505, EPI_ISL_2324506, EPI_ISL_2324507, EPI_ISL_2324508, EPI_ISL_2324509, EPI_ISL_2324510, EPI_ISL_2324511, EPI_ISL_2324512, EPI_ISL_2324513, EPI_ISL_2324514, EPI_ISL_2324515, EPI_ISL_2324516, EPI_ISL_2324517, EPI_ISL_2324518, EPI_ISL_2324519, EPI_ISL_2324520, EPI_ISL_2324521, EPI_ISL_2324522, EPI_ISL_2324523, EPI_ISL_2324524, EPI_ISL_2324525, EPI_ISL_2324527, EPI_ISL_2324528, EPI_ISL_2324529, EPI_ISL_2324530, EPI_ISL_2324531, EPI_ISL_2324532, EPI_ISL_2324533, EPI_ISL_2324534, EPI_ISL_2324535, EPI_ISL_2324536, EPI_ISL_2324537, EPI_ISL_2324538, EPI_ISL_2324539, EPI_ISL_2324540, EPI_ISL_2324542, EPI_ISL_2324543, EPI_ISL_2324544, EPI_ISL_2324545, EPI_ISL_2324546, EPI_ISL_2324547, EPI_ISL_2324548, EPI_ISL_2324549, EPI_ISL_2324550, EPI_ISL_2324551, EPI_ISL_2324552, EPI_ISL_2324553, EPI_ISL_2324554, EPI_ISL_2324555, EPI_ISL_2324556, EPI_ISL_2324557, EPI_ISL_2324558, EPI_ISL_2324559, EPI_ISL_2324560, EPI_ISL_2324561, EPI_ISL_2324562, EPI_ISL_2324564, EPI_ISL_2324565, EPI_ISL_2324566, EPI_ISL_2324567, EPI_ISL_2324568, EPI_ISL_2324569, EPI_ISL_2324570, EPI_ISL_2324571, EPI_ISL_2324572, EPI_ISL_2324573, EPI_ISL_2324574, EPI_ISL_2324575, EPI_ISL_2324576, EPI_ISL_2324577, EPI_ISL_2324578, EPI_ISL_2324579, EPI_ISL_2324580, EPI_ISL_2324581, EPI_ISL_2324582, EPI_ISL_2324583, EPI_ISL_2324584, EPI_ISL_2324585, EPI_ISL_2324586, EPI_ISL_2324587, EPI_ISL_2324588, EPI_ISL_2324589, EPI_ISL_2324590, EPI_ISL_2324591, EPI_ISL_2324592 | | | |
| see above | Laboratory Corporation of America | Centers for Disease Control and Prevention Division of Viral Diseases, Pathogen Discovery | Dakota Howard, Dhwani Batra, Peter W. Cook, Kara Moser, Adrian Paskey, Jason Caravas, Benjamin Rambo-Martin, Shatavia Morrison, Christopher Gulvick, Scott Sammons, Yvette Unoarumhi, Darlene Wagner, Matthew Schmerer, Minoo Agarwal, Eyad Almasri, Debbie Boles, Ayla Burns, Nuthawin Charoensri, Oren Cohen, Susan Countryman, Mary Ann Cristobal, Bobbi Croy, Suzanne Dale, Hrushikesh Deshmukh, Amanda Douglas, Vincent Drouillon, Marcia Eisenberg, Howard Engler, Rama Ghatti, Prashant Gupta, Susan Hicks, Jake Humphrey, Lax Iyer, Lisa Pfefferle, Manoj Jain, Matthew Robinson, Mohan Kolli, Brian Krueger, Tim Kuphal, Stanley Letovsky, Michael Levandoski, Craig Lukasik, Jonathan Meltzer, Brian Norvell, Mindy Nye, Scott Parker, Christos Petropoulos, John Pruitt, Steven Ragan, Scott Ryan, Mike Sapeta, Jana Schroth, Suresh Babu Selvaraju, Goran Stevovic, Amanda Suchanek, Andrea Throop, Lyndon Tilson, Thomas Urban, Joe Voshell, Kimberly Wagner, Jonathan Williams, Mary Williamson, Qian Zeng, Tricia Zwiefelhofer, Clinton R. Paden, Duncan MacCannell |
| EPI_ISL_2325062, EPI_ISL_2325063, EPI_ISL_2325064, EPI_ISL_2325065, EPI_ISL_2325066, EPI_ISL_2325067, EPI_ISL_2325068, EPI_ISL_2325069, EPI_ISL_2325070, EPI_ISL_2325071, EPI_ISL_2325072, EPI_ISL_2325073, EPI_ISL_2325074, EPI_ISL_2325075, EPI_ISL_2325076, EPI_ISL_2325077, EPI_ISL_2325078 | | | |
| see above | Minnesota Department of Health, Public Health Laboratory | Minnesota Department of Health, Public Health Laboratory | Alexandra Lorentz, Jacob Garfin, Matt Plumb, and Xiong Wang |
| EPI_ISL_2325079, EPI_ISL_2325080 | Centracare Laboratory Services | Minnesota Department of Health, Public Health Laboratory | Alexandra Lorentz, Jacob Garfin, Matt Plumb, and Xiong Wang |
| EPI_ISL_2325081, EPI_ISL_2325082, EPI_ISL_2325083, EPI_ISL_2325084, EPI_ISL_2325085, EPI_ISL_2325086, EPI_ISL_2325087, EPI_ISL_2325088, EPI_ISL_2325089, EPI_ISL_2325090 | Minnesota Department of Health, Public Health Laboratory | Minnesota Department of Health, Public Health Laboratory | Alexandra Lorentz, Jacob Garfin, Matt Plumb, and Xiong Wang |
| EPI_ISL_2325091, EPI_ISL_2325092, EPI_ISL_2325093, EPI_ISL_2325094 | Mayo Clinic & Mayo Clinic Laboratories | Minnesota Department of Health, Public Health Laboratory | Alexandra Lorentz, Jacob Garfin, Matt Plumb, and Xiong Wang |
| EPI_ISL_2325095, EPI_ISL_2325097, EPI_ISL_2325098, EPI_ISL_2325099, EPI_ISL_2325100, EPI_ISL_2325101, EPI_ISL_2325102, EPI_ISL_2325103, EPI_ISL_2325104, EPI_ISL_2325106, EPI_ISL_2325107, EPI_ISL_2325111, EPI_ISL_2325112, EPI_ISL_2325113, EPI_ISL_2325114, EPI_ISL_2325115, EPI_ISL_2325116, EPI_ISL_2325117, EPI_ISL_2325118 | | | |
| see above | Minnesota Department of Health, Public Health Laboratory | Minnesota Department of Health, Public Health Laboratory | Alexandra Lorentz, Jacob Garfin, Matt Plumb, and Xiong Wang |
| EPI_ISL_2325130, EPI_ISL_2325131, EPI_ISL_2325132, EPI_ISL_2325133, EPI_ISL_2325134, EPI_ISL_2325135, EPI_ISL_2325136, EPI_ISL_2325137, EPI_ISL_2325138, EPI_ISL_2325139, EPI_ISL_2325140, EPI_ISL_2325141, EPI_ISL_2325142, EPI_ISL_2325143, EPI_ISL_2325144, EPI_ISL_2325145, EPI_ISL_2325146, EPI_ISL_2325147, EPI_ISL_2325148, EPI_ISL_2325149, EPI_ISL_2325150, EPI_ISL_2325151, EPI_ISL_2325152, EPI_ISL_2325153, EPI_ISL_2325154, EPI_ISL_2325155, EPI_ISL_2325156, EPI_ISL_2325157, EPI_ISL_2325158, EPI_ISL_2325159, EPI_ISL_2325160, EPI_ISL_2325161, EPI_ISL_2325162, EPI_ISL_2325163, EPI_ISL_2325164, EPI_ISL_2325165, EPI_ISL_2325166, EPI_ISL_2325167, EPI_ISL_2325168, EPI_ISL_2325169, EPI_ISL_2325170, EPI_ISL_2325171, EPI_ISL_2325172, EPI_ISL_2325173, EPI_ISL_2325174, EPI_ISL_2325175, EPI_ISL_2325176 | | | |
| see above | Arizona State Public Health Laboratory | Arizona State Public Health Laboratory | Trung Huynh, Jessica Escobar, Katherine Fullerton, Nobuko Fukushima, Stacy White, Linda Getsinger, Victor Waddell |
| EPI_ISL_2329056, EPI_ISL_2329060, EPI_ISL_2329061, EPI_ISL_2329064, EPI_ISL_2329067, EPI_ISL_2329071, EPI_ISL_2329074, EPI_ISL_2329078, EPI_ISL_2329082, EPI_ISL_2329086, EPI_ISL_2329089, EPI_ISL_2329093, EPI_ISL_2329096, EPI_ISL_2329103, EPI_ISL_2329107, EPI_ISL_2329110, EPI_ISL_2329113, EPI_ISL_2329116, EPI_ISL_2329121, EPI_ISL_2329125, EPI_ISL_2329128, EPI_ISL_2329132, EPI_ISL_2329135, EPI_ISL_2329138, EPI_ISL_2329141, EPI_ISL_2329145, EPI_ISL_2329148, EPI_ISL_2329151, EPI_ISL_2329155, EPI_ISL_2329158, EPI_ISL_2329162, EPI_ISL_2329164, EPI_ISL_2329170, EPI_ISL_2329172, EPI_ISL_2329176, EPI_ISL_2329180, EPI_ISL_2329183, EPI_ISL_2329187, EPI_ISL_2329191, EPI_ISL_2329194, EPI_ISL_2329197, EPI_ISL_2329201, EPI_ISL_2329204, EPI_ISL_2329207, EPI_ISL_2329210, EPI_ISL_2329214, EPI_ISL_2329217, EPI_ISL_2329221, EPI_ISL_2329224, EPI_ISL_2329227, EPI_ISL_2329230, EPI_ISL_2329233, EPI_ISL_2329236, EPI_ISL_2329239, EPI_ISL_2329240, EPI_ISL_2329241, EPI_ISL_2329242, EPI_ISL_2329243, EPI_ISL_2329244, EPI_ISL_2329245, EPI_ISL_2329247, EPI_ISL_2329248, EPI_ISL_2329249, EPI_ISL_2329250, EPI_ISL_2329251, EPI_ISL_2329252, EPI_ISL_2329253, EPI_ISL_2329254, EPI_ISL_2329255, EPI_ISL_2329256, EPI_ISL_2329257, EPI_ISL_2329258, EPI_ISL_2329259, EPI_ISL_2329260, EPI_ISL_2329261, EPI_ISL_2329262, EPI_ISL_2329263, EPI_ISL_2329264, EPI_ISL_2329265, EPI_ISL_2329266, EPI_ISL_2329267, EPI_ISL_2329268, EPI_ISL_2329269, EPI_ISL_2329270, EPI_ISL_2329271, EPI_ISL_2329272, EPI_ISL_2329273, EPI_ISL_2329274, EPI_ISL_2329275, EPI_ISL_2329276, EPI_ISL_2329277, EPI_ISL_2329278, EPI_ISL_2329279, EPI_ISL_2329280, EPI_ISL_2329281, EPI_ISL_2329282, EPI_ISL_2329284 | | | |
| see above | Centro de Diagnostico COVID-19 UABC Tijuana | Andersen lab at Scripps Research | SEARCH Alliance San Diego with Idanya Rubi Serafin Higuera, Manuel Sanchez Alavez, Jorge Luis Jimenez Niebla, German Ibarra, Jonathan Vincent Baena, Oscar Efren Zazueta Fierro |
| EPI_ISL_2329286, EPI_ISL_2329287, EPI_ISL_2329288, EPI_ISL_2329289, EPI_ISL_2329290, EPI_ISL_2329291, EPI_ISL_2329292, EPI_ISL_2329293, EPI_ISL_2329294, EPI_ISL_2329295, EPI_ISL_2329296, EPI_ISL_2329297, EPI_ISL_2329298, EPI_ISL_2329299, EPI_ISL_2329300, EPI_ISL_2329301, EPI_ISL_2329302, EPI_ISL_2329303, EPI_ISL_2329304, EPI_ISL_2329305, EPI_ISL_2329306, EPI_ISL_2329307, EPI_ISL_2329308, EPI_ISL_2329309, EPI_ISL_2329310, EPI_ISL_2329311, EPI_ISL_2329312, EPI_ISL_2329313, EPI_ISL_2329314, EPI_ISL_2329315, EPI_ISL_2329316, EPI_ISL_2329317, EPI_ISL_2329318, EPI_ISL_2329319, EPI_ISL_2329320, EPI_ISL_2329321, EPI_ISL_2329322, EPI_ISL_2329323, EPI_ISL_2329324, EPI_ISL_2329325, EPI_ISL_2329326, EPI_ISL_2329327, EPI_ISL_2329328, EPI_ISL_2329329, EPI_ISL_2329330, EPI_ISL_2329331, EPI_ISL_2329332, EPI_ISL_2329333, EPI_ISL_2329334 | | | |
| see above | Sharp HealthCare Laboratory | Andersen lab at Scripps Research | SEARCH Alliance San Diego with Aaron Harding, Jacquelyn Berumen, Cathy Woerle, Liam McGinnis, Art Mendoza, Omid Bakhtar |
| EPI_ISL_2329335, EPI_ISL_2329336, EPI_ISL_2329337, EPI_ISL_2329338, EPI_ISL_2329339, EPI_ISL_2329340, EPI_ISL_2329341, EPI_ISL_2329342, EPI_ISL_2329343, EPI_ISL_2329344, EPI_ISL_2329345, EPI_ISL_2329346, EPI_ISL_2329347, EPI_ISL_2329348, EPI_ISL_2329349, EPI_ISL_2329350, EPI_ISL_2329351, EPI_ISL_2329352, EPI_ISL_2329353, EPI_ISL_2329354, EPI_ISL_2329355, EPI_ISL_2329356, EPI_ISL_2329358, EPI_ISL_2329359, EPI_ISL_2329360, EPI_ISL_2329361, EPI_ISL_2329362, EPI_ISL_2329363, EPI_ISL_2329365, EPI_ISL_2329366, EPI_ISL_2329367, EPI_ISL_2329368, EPI_ISL_2329369, EPI_ISL_2329370, EPI_ISL_2329371, EPI_ISL_2329372, EPI_ISL_2329373, EPI_ISL_2329374, EPI_ISL_2329375, EPI_ISL_2329377, EPI_ISL_2329378, EPI_ISL_2329379, EPI_ISL_2329380, EPI_ISL_2329381, EPI_ISL_2329382, EPI_ISL_2329383, EPI_ISL_2329384, EPI_ISL_2329385, EPI_ISL_2329386, EPI_ISL_2329387, EPI_ISL_2329388, EPI_ISL_2329389, EPI_ISL_2329390, EPI_ISL_2329391, EPI_ISL_2329392, EPI_ISL_2329393, EPI_ISL_2329394, EPI_ISL_2329396, EPI_ISL_2329397, EPI_ISL_2329398, EPI_ISL_2329399, EPI_ISL_2329400, EPI_ISL_2329401, EPI_ISL_2329402, EPI_ISL_2329403, EPI_ISL_2329404, EPI_ISL_2329405, EPI_ISL_2329406, EPI_ISL_2329407, EPI_ISL_2329408 | | | |
| see above | San Diego County Public Health Laboratory | Andersen lab at Scripps Research | SEARCH Alliance San Diego with Ashleigh Murphy, Jovan Shephard, Brett Austin |
| EPI_ISL_2329409, EPI_ISL_2329410, EPI_ISL_2329411, EPI_ISL_2329412, EPI_ISL_2329413, EPI_ISL_2329414, EPI_ISL_2329415, EPI_ISL_2329416, EPI_ISL_2329417, EPI_ISL_2329418, EPI_ISL_2329419, EPI_ISL_2329420, EPI_ISL_2329421, EPI_ISL_2329422, EPI_ISL_2329423, EPI_ISL_2329424, EPI_ISL_2329425, EPI_ISL_2329426, EPI_ISL_2329427, EPI_ISL_2329428, EPI_ISL_2329429, EPI_ISL_2329431, EPI_ISL_2329432, EPI_ISL_2329434, EPI_ISL_2329435, EPI_ISL_2329436, EPI_ISL_2329437, EPI_ISL_2329438, EPI_ISL_2329439, EPI_ISL_2329440, EPI_ISL_2329441, EPI_ISL_2329442, EPI_ISL_2329443, EPI_ISL_2329444, EPI_ISL_2329445, EPI_ISL_2329446, EPI_ISL_2329447, EPI_ISL_2329448, EPI_ISL_2329449, EPI_ISL_2329450, EPI_ISL_2329451, EPI_ISL_2329452, EPI_ISL_2329453, EPI_ISL_2329454, EPI_ISL_2329455, EPI_ISL_2329456, EPI_ISL_2329457, EPI_ISL_2329458, EPI_ISL_2329459, EPI_ISL_2329460, EPI_ISL_2329461, EPI_ISL_2329462, EPI_ISL_2329463, EPI_ISL_2329464, EPI_ISL_2329465 | | | |
| see above | Sharp HealthCare Laboratory | Andersen lab at Scripps Research | SEARCH Alliance San Diego with Aaron Harding, Jacquelyn Berumen, Cathy Woerle, Liam McGinnis, Art Mendoza, Omid Bakhtar |
| EPI_ISL_2329466, EPI_ISL_2329467, EPI_ISL_2329468, EPI_ISL_2329469, EPI_ISL_2329471, EPI_ISL_2329472, EPI_ISL_2329473, EPI_ISL_2329474, EPI_ISL_2329475, EPI_ISL_2329477, EPI_ISL_2329478, EPI_ISL_2329479, EPI_ISL_2329480, EPI_ISL_2329481, EPI_ISL_2329482, EPI_ISL_2329483, EPI_ISL_2329484, EPI_ISL_2329485, EPI_ISL_2329486, EPI_ISL_2329487, EPI_ISL_2329489, EPI_ISL_2329490, EPI_ISL_2329491, EPI_ISL_2329492, EPI_ISL_2329493, EPI_ISL_2329494, EPI_ISL_2329495, EPI_ISL_2329496, EPI_ISL_2329497, EPI_ISL_2329498, EPI_ISL_2329499, EPI_ISL_2329500, EPI_ISL_2329501, EPI_ISL_2329502, EPI_ISL_2329503, EPI_ISL_2329504, EPI_ISL_2329505, EPI_ISL_2329506, EPI_ISL_2329507, EPI_ISL_2329508, EPI_ISL_2329509, EPI_ISL_2329510, EPI_ISL_2329511, EPI_ISL_2329512, EPI_ISL_2329513, EPI_ISL_2329514, EPI_ISL_2329515, EPI_ISL_2329516, EPI_ISL_2329517, EPI_ISL_2329518, EPI_ISL_2329519, EPI_ISL_2329520, EPI_ISL_2329521, EPI_ISL_2329522, EPI_ISL_2329523, EPI_ISL_2329524, EPI_ISL_2329525, EPI_ISL_2329526, EPI_ISL_2329527, EPI_ISL_2329528, EPI_ISL_2329529, EPI_ISL_2329530, EPI_ISL_2329531, EPI_ISL_2329532, EPI_ISL_2329534, EPI_ISL_2329535, EPI_ISL_2329536, EPI_ISL_2329537, EPI_ISL_2329538, EPI_ISL_2329539, EPI_ISL_2329540, EPI_ISL_2329541, EPI_ISL_2329542, EPI_ISL_2329543, EPI_ISL_2329544, EPI_ISL_2329545, EPI_ISL_2329546, EPI_ISL_2329547, EPI_ISL_2329548, EPI_ISL_2329549, EPI_ISL_2329550, EPI_ISL_2329551, EPI_ISL_2329552, EPI_ISL_2329553, EPI_ISL_2329554, EPI_ISL_2329555, EPI_ISL_2329556, EPI_ISL_2329557, EPI_ISL_2329558, EPI_ISL_2329559, EPI_ISL_2329560, EPI_ISL_2329561, EPI_ISL_2329562, EPI_ISL_2329563, EPI_ISL_2329564, EPI_ISL_2329565, EPI_ISL_2329566, EPI_ISL_2329567, EPI_ISL_2329568, EPI_ISL_2329569, EPI_ISL_2329570, EPI_ISL_2329571, EPI_ISL_2329572, EPI_ISL_2329573, EPI_ISL_2329574, EPI_ISL_2329575, EPI_ISL_2329576, EPI_ISL_2329577, EPI_ISL_2329578, EPI_ISL_2329579, EPI_ISL_2329580, EPI_ISL_2329581, EPI_ISL_2329582, EPI_ISL_2329583, EPI_ISL_2329584, EPI_ISL_2329585, EPI_ISL_2329586, EPI_ISL_2329587, EPI_ISL_2329588, EPI_ISL_2329589, EPI_ISL_2329590, EPI_ISL_2329591, EPI_ISL_2329592, EPI_ISL_2329593, EPI_ISL_2329594, EPI_ISL_2329595, EPI_ISL_2329596, EPI_ISL_2329597, EPI_ISL_2329598, EPI_ISL_2329599, EPI_ISL_2329600, EPI_ISL_2329601, EPI_ISL_2329602, EPI_ISL_2329603, EPI_ISL_2329604, EPI_ISL_2329605, EPI_ISL_2329606, EPI_ISL_2329608 | | | |
| see above | San Diego County Public Health Laboratory | Andersen lab at Scripps Research | SEARCH Alliance San Diego with Ashleigh Murphy, Jovan Shephard, Brett Austin |
| EPI_ISL_2331465, EPI_ISL_2331466, EPI_ISL_2331467, EPI_ISL_2331468, EPI_ISL_2331469, EPI_ISL_2331470, EPI_ISL_2331471, EPI_ISL_2331472, EPI_ISL_2331473, EPI_ISL_2331474, EPI_ISL_2331475, EPI_ISL_2331477, EPI_ISL_2331478, EPI_ISL_2331479, EPI_ISL_2331480, EPI_ISL_2331481, EPI_ISL_2331482, EPI_ISL_2331483, EPI_ISL_2331484, EPI_ISL_2331485, EPI_ISL_2331486, EPI_ISL_2331487, EPI_ISL_2331488, EPI_ISL_2331489, EPI_ISL_2331490, EPI_ISL_2331491, EPI_ISL_2331492, EPI_ISL_2331493, EPI_ISL_2331494, EPI_ISL_2331495, EPI_ISL_2331496, EPI_ISL_2331497, EPI_ISL_2331498, EPI_ISL_2331499, EPI_ISL_2331500, EPI_ISL_2331501, EPI_ISL_2331502, EPI_ISL_2331503, EPI_ISL_2331504, EPI_ISL_2331505, EPI_ISL_2331506, EPI_ISL_2331507, EPI_ISL_2331508, EPI_ISL_2331509, EPI_ISL_2331510, EPI_ISL_2331511, EPI_ISL_2331513, EPI_ISL_2331514, EPI_ISL_2331515, EPI_ISL_2331516, EPI_ISL_2331518, EPI_ISL_2331519, EPI_ISL_2331520, EPI_ISL_2331521, EPI_ISL_2331522, EPI_ISL_2331523, EPI_ISL_2331524, EPI_ISL_2331525, EPI_ISL_2331526, EPI_ISL_2331528, EPI_ISL_2331529, EPI_ISL_2331531, EPI_ISL_2331532, EPI_ISL_2331533, EPI_ISL_2331534, EPI_ISL_2331535, EPI_ISL_2331536, EPI_ISL_2331537, EPI_ISL_2331538, EPI_ISL_2331539, EPI_ISL_2331541, EPI_ISL_2331542, EPI_ISL_2331543, EPI_ISL_2331544, EPI_ISL_2331545, EPI_ISL_2331546, EPI_ISL_2331547, EPI_ISL_2331548, EPI_ISL_2331549, EPI_ISL_2331550, EPI_ISL_2331551, EPI_ISL_2331552, EPI_ISL_2331553, EPI_ISL_2331554, EPI_ISL_2331555, EPI_ISL_2331556, EPI_ISL_2331557, EPI_ISL_2331558, EPI_ISL_2331559, EPI_ISL_2331560, EPI_ISL_2331561, EPI_ISL_2331562, EPI_ISL_2331563, EPI_ISL_2331564, EPI_ISL_2331565, EPI_ISL_2331566, EPI_ISL_2331567, EPI_ISL_2331568, EPI_ISL_2331569, EPI_ISL_2331570, EPI_ISL_2331571, EPI_ISL_2331572, EPI_ISL_2331573, EPI_ISL_2331574, EPI_ISL_2331575, EPI_ISL_2331576, EPI_ISL_2331577, EPI_ISL_2331578, EPI_ISL_2331579, EPI_ISL_2331580, EPI_ISL_2331581, EPI_ISL_2331582, EPI_ISL_2331583, EPI_ISL_2331584, EPI_ISL_2331585, EPI_ISL_2331586, EPI_ISL_2331587, EPI_ISL_2331588, EPI_ISL_2331589, EPI_ISL_2331590, EPI_ISL_2331591, EPI_ISL_2331592, EPI_ISL_2331593, EPI_ISL_2331595, EPI_ISL_2331596, EPI_ISL_2331597, EPI_ISL_2331598, EPI_ISL_2331599, EPI_ISL_2331600, EPI_ISL_2331601, EPI_ISL_2331602, EPI_ISL_2331603, EPI_ISL_2331604, EPI_ISL_2331605, EPI_ISL_2331606, EPI_ISL_2331607, EPI_ISL_2331608, EPI_ISL_2331609, EPI_ISL_2331610, EPI_ISL_2331611, EPI_ISL_2331612, EPI_ISL_2331613, EPI_ISL_2331615, EPI_ISL_2331616, EPI_ISL_2331617, EPI_ISL_2331618, EPI_ISL_2331619, EPI_ISL_2331620, EPI_ISL_2331621, EPI_ISL_2331622, EPI_ISL_2331623, EPI_ISL_2331624, EPI_ISL_2331625, EPI_ISL_2331626, EPI_ISL_2331627, EPI_ISL_2331628, EPI_ISL_2331629, EPI_ISL_2331630, EPI_ISL_2331632, EPI_ISL_2331633, EPI_ISL_2331634, EPI_ISL_2331635, EPI_ISL_2331636, EPI_ISL_2331637, EPI_ISL_2331638, EPI_ISL_2331639, EPI_ISL_2331640, EPI_ISL_2331641, EPI_ISL_2331642, EPI_ISL_2331643, EPI_ISL_2331644, EPI_ISL_2331645, EPI_ISL_2331646, EPI_ISL_2331647, EPI_ISL_2331648, EPI_ISL_2331649, EPI_ISL_2331650, EPI_ISL_2331651, EPI_ISL_2331652, EPI_ISL_2331653, EPI_ISL_2331654, EPI_ISL_2331655, EPI_ISL_2331656, EPI_ISL_2331657, EPI_ISL_2331658, EPI_ISL_2331659, EPI_ISL_2331660, EPI_ISL_2331661, EPI_ISL_2331662, EPI_ISL_2331663, EPI_ISL_2331664, EPI_ISL_2331665, EPI_ISL_2331666, EPI_ISL_2331667, EPI_ISL_2331668, EPI_ISL_2331669, EPI_ISL_2331670, EPI_ISL_2331671, EPI_ISL_2331672, EPI_ISL_2331673, EPI_ISL_2331674 | | | |
| see above | Johns Hopkins Hospital Department of Pathology | Johns Hopkins Hospital Department of Pathology | C. Paul Morris, Chun Huai Luo, Adannaya Amadi, Matthew Schwartz, Nicholas Gallagher, Heba H. Mostafa |
| EPI_ISL_2332647, EPI_ISL_2332648, EPI_ISL_2332649 | Minnesota Department of Health, Public Health Laboratory | Minnesota Department of Health, Public Health Laboratory | Alexandra Lorentz, Jacob Garfin, Matt Plumb, and Xiong Wang |
| EPI_ISL_2332650 | Centracare Laboratory Services | Minnesota Department of Health, Public Health Laboratory | Alexandra Lorentz, Jacob Garfin, Matt Plumb, and Xiong Wang |
| EPI_ISL_2332651 | Premier Medical Lab | Minnesota Department of Health, Public Health Laboratory | Alexandra Lorentz, Jacob Garfin, Matt Plumb, and Xiong Wang |

| | | | |
|---|---|---|---|
| EPI_ISL_2332652, EPI_ISL_2332653 | Centracare Laboratory Services | Minnesota Department of Health, Public Health Laboratory | Alexandra Lorentz, Jacob Garfin, Matt Plumb, and Xiong Wang |
| EPI_ISL_2332654 | M Health Fairview | Minnesota Department of Health, Public Health Laboratory | Alexandra Lorentz, Jacob Garfin, Matt Plumb, and Xiong Wang |
| EPI_ISL_2332655 | Maple Grove Hospital | Minnesota Department of Health, Public Health Laboratory | Alexandra Lorentz, Jacob Garfin, Matt Plumb, and Xiong Wang |
| EPI_ISL_2332656 | Minnesota Department of Health, Public Health Laboratory | Minnesota Department of Health, Public Health Laboratory | Alexandra Lorentz, Jacob Garfin, Matt Plumb, and Xiong Wang |
| EPI_ISL_2332657, EPI_ISL_2332658, EPI_ISL_2332659 | Centracare Laboratory Services | Minnesota Department of Health, Public Health Laboratory | Alexandra Lorentz, Jacob Garfin, Matt Plumb, and Xiong Wang |
| EPI_ISL_2332660 | Maple Grove Hospital | Minnesota Department of Health, Public Health Laboratory | Alexandra Lorentz, Jacob Garfin, Matt Plumb, and Xiong Wang |
| EPI_ISL_2332661 | Minnesota Department of Health, Public Health Laboratory | Minnesota Department of Health, Public Health Laboratory | Alexandra Lorentz, Jacob Garfin, Matt Plumb, and Xiong Wang |
| EPI_ISL_2332662 | M Health Fairview | Minnesota Department of Health, Public Health Laboratory | Alexandra Lorentz, Jacob Garfin, Matt Plumb, and Xiong Wang |
| EPI_ISL_2332663, EPI_ISL_2332664 | Centracare Laboratory Services | Minnesota Department of Health, Public Health Laboratory | Alexandra Lorentz, Jacob Garfin, Matt Plumb, and Xiong Wang |
| EPI_ISL_2332665, EPI_ISL_2332666 | Premier Medical Lab | Minnesota Department of Health, Public Health Laboratory | Alexandra Lorentz, Jacob Garfin, Matt Plumb, and Xiong Wang |
| EPI_ISL_2332667, EPI_ISL_2332668 | Maple Grove Hospital | Minnesota Department of Health, Public Health Laboratory | Alexandra Lorentz, Jacob Garfin, Matt Plumb, and Xiong Wang |
| EPI_ISL_2332669 | Minnesota Department of Health, Public Health Laboratory | Minnesota Department of Health, Public Health Laboratory | Alexandra Lorentz, Jacob Garfin, Matt Plumb, and Xiong Wang |
| EPI_ISL_2332670, EPI_ISL_2332671 | Centracare Laboratory Services | Minnesota Department of Health, Public Health Laboratory | Alexandra Lorentz, Jacob Garfin, Matt Plumb, and Xiong Wang |
| EPI_ISL_2332672 | M Health Fairview | Minnesota Department of Health, Public Health Laboratory | Alexandra Lorentz, Jacob Garfin, Matt Plumb, and Xiong Wang |
| EPI_ISL_2332673, EPI_ISL_2332674, EPI_ISL_2332675, EPI_ISL_2332676, EPI_ISL_2332677, EPI_ISL_2332678 | Centracare Laboratory Services | Minnesota Department of Health, Public Health Laboratory | Alexandra Lorentz, Jacob Garfin, Matt Plumb, and Xiong Wang |
| EPI_ISL_2332679, EPI_ISL_2332680, EPI_ISL_2332681, EPI_ISL_2332682, EPI_ISL_2332683 | Maple Grove Hospital | Minnesota Department of Health, Public Health Laboratory | Alexandra Lorentz, Jacob Garfin, Matt Plumb, and Xiong Wang |
| EPI_ISL_2332684, EPI_ISL_2332685, EPI_ISL_2332686 | Minnesota Department of Health, Public Health Laboratory | Minnesota Department of Health, Public Health Laboratory | Alexandra Lorentz, Jacob Garfin, Matt Plumb, and Xiong Wang |
| EPI_ISL_2332687, EPI_ISL_2332688 | Centracare Laboratory Services | Minnesota Department of Health, Public Health Laboratory | Alexandra Lorentz, Jacob Garfin, Matt Plumb, and Xiong Wang |
| EPI_ISL_2332689 | Premier Medical Lab | Minnesota Department of Health, Public Health Laboratory | Alexandra Lorentz, Jacob Garfin, Matt Plumb, and Xiong Wang |
| EPI_ISL_2332690, EPI_ISL_2332691, EPI_ISL_2332692 | Minnesota Department of Health, Public Health Laboratory | Minnesota Department of Health, Public Health Laboratory | Alexandra Lorentz, Jacob Garfin, Matt Plumb, and Xiong Wang |
| EPI_ISL_2332693, EPI_ISL_2332694, EPI_ISL_2332695 | Centracare Laboratory Services | Minnesota Department of Health, Public Health Laboratory | Alexandra Lorentz, Jacob Garfin, Matt Plumb, and Xiong Wang |
| EPI_ISL_2332698 | Minnesota Department of Health, Public Health Laboratory | Minnesota Department of Health, Public Health Laboratory | Alexandra Lorentz, Jacob Garfin, Matt Plumb, and Xiong Wang |
| EPI_ISL_2332699 | Maple Grove Hospital | Minnesota Department of Health, Public Health Laboratory | Alexandra Lorentz, Jacob Garfin, Matt Plumb, and Xiong Wang |
| EPI_ISL_2332700 | Premier Medical Lab | Minnesota Department of Health, Public Health Laboratory | Alexandra Lorentz, Jacob Garfin, Matt Plumb, and Xiong Wang |
| EPI_ISL_2332701 | Minnesota Department of Health, Public Health Laboratory | Minnesota Department of Health, Public Health Laboratory | Alexandra Lorentz, Jacob Garfin, Matt Plumb, and Xiong Wang |
| EPI_ISL_2332702 | M Health Fairview | Minnesota Department of Health, Public Health Laboratory | Alexandra Lorentz, Jacob Garfin, Matt Plumb, and Xiong Wang |
| EPI_ISL_2332703 | Centracare Laboratory Services | Minnesota Department of Health, Public Health Laboratory | Alexandra Lorentz, Jacob Garfin, Matt Plumb, and Xiong Wang |
| EPI_ISL_2332812, EPI_ISL_2332813, EPI_ISL_2332814, EPI_ISL_2332815, EPI_ISL_2332816, EPI_ISL_2332817, EPI_ISL_2332818, EPI_ISL_2332819, EPI_ISL_2332820, EPI_ISL_2332821, EPI_ISL_2332822, EPI_ISL_2332823, EPI_ISL_2332824, EPI_ISL_2332825, EPI_ISL_2332826, EPI_ISL_2332827, EPI_ISL_2332828, EPI_ISL_2332829, EPI_ISL_2332830, EPI_ISL_2332831 | | | |
| see above | DC Public Health Lab/ Dept. of Forensic Sciences | DC Public Health Lab/ Dept. of Forensic Sciences | Janis Doss, Scott Nguyen, Elizabeth Zelaya, Sarah Scott, Connie Maza, Monica Mann, Brittany Hamilton, David Payne, Jocelyn Hauser |
| EPI_ISL_2332832, EPI_ISL_2332833, EPI_ISL_2332834, EPI_ISL_2332835, EPI_ISL_2332836, EPI_ISL_2332837, EPI_ISL_2332838, EPI_ISL_2332839, EPI_ISL_2332840, EPI_ISL_2332841, EPI_ISL_2332842, EPI_ISL_2332843, EPI_ISL_2332844, EPI_ISL_2332845, EPI_ISL_2332846, EPI_ISL_2332847, EPI_ISL_2332848, EPI_ISL_2332849, EPI_ISL_2332850, EPI_ISL_2332851, EPI_ISL_2332852, EPI_ISL_2332853, EPI_ISL_2332854, EPI_ISL_2332855, EPI_ISL_2332856, EPI_ISL_2332857, EPI_ISL_2332858, EPI_ISL_2332859, EPI_ISL_2332860, EPI_ISL_2332861, EPI_ISL_2332862, EPI_ISL_2332863, EPI_ISL_2332864, EPI_ISL_2332865, EPI_ISL_2332866, EPI_ISL_2332867, EPI_ISL_2332868, EPI_ISL_2332869, EPI_ISL_2332870, EPI_ISL_2332871, EPI_ISL_2332872, EPI_ISL_2332873, EPI_ISL_2332874, EPI_ISL_2332875, EPI_ISL_2332876, EPI_ISL_2332878, EPI_ISL_2332879, EPI_ISL_2332880, EPI_ISL_2332881, EPI_ISL_2332882, EPI_ISL_2332883, EPI_ISL_2332884, EPI_ISL_2332885, EPI_ISL_2332886, EPI_ISL_2332887, EPI_ISL_2332888, EPI_ISL_2332889, EPI_ISL_2332890, EPI_ISL_2332891, EPI_ISL_2332892, EPI_ISL_2332893, EPI_ISL_2332894, EPI_ISL_2332895, EPI_ISL_2332896, EPI_ISL_2332897, EPI_ISL_2332898, EPI_ISL_2332899, EPI_ISL_2332900, EPI_ISL_2332901, EPI_ISL_2332902, EPI_ISL_2332903, EPI_ISL_2332904, EPI_ISL_2332905, EPI_ISL_2332906, EPI_ISL_2332907, EPI_ISL_2332908, EPI_ISL_2332909, EPI_ISL_2332910, EPI_ISL_2332911, EPI_ISL_2332912, EPI_ISL_2332913, EPI_ISL_2332914, EPI_ISL_2332915, EPI_ISL_2332916, EPI_ISL_2332917, EPI_ISL_2332918, EPI_ISL_2332919, EPI_ISL_2332920, EPI_ISL_2332921, EPI_ISL_2332922, EPI_ISL_2332923, EPI_ISL_2332924, EPI_ISL_2332925, EPI_ISL_2332926, EPI_ISL_2332927, EPI_ISL_2332929, EPI_ISL_2332930, EPI_ISL_2332931, EPI_ISL_2332933, EPI_ISL_2332934, EPI_ISL_2332935, EPI_ISL_2332936, EPI_ISL_2332937, EPI_ISL_2332938, EPI_ISL_2332939, EPI_ISL_2332940, EPI_ISL_2332941, EPI_ISL_2332942, EPI_ISL_2332943, EPI_ISL_2332944, EPI_ISL_2332945, EPI_ISL_2332946, EPI_ISL_2332947, EPI_ISL_2332948, EPI_ISL_2332949, EPI_ISL_2332950, EPI_ISL_2332951, EPI_ISL_2332952, EPI_ISL_2332953, EPI_ISL_2332954, EPI_ISL_2332955, EPI_ISL_2332956, EPI_ISL_2332957, EPI_ISL_2332958, EPI_ISL_2332959, EPI_ISL_2332960, EPI_ISL_2332961, EPI_ISL_2332962, EPI_ISL_2332963, EPI_ISL_2332964, EPI_ISL_2332965, EPI_ISL_2332966, EPI_ISL_2332967, EPI_ISL_2332968, EPI_ISL_2332969, EPI_ISL_2332970, EPI_ISL_2332971, EPI_ISL_2332973, EPI_ISL_2332974, EPI_ISL_2332975, EPI_ISL_2332976, EPI_ISL_2332978, EPI_ISL_2332979, EPI_ISL_2332980, EPI_ISL_2332981, EPI_ISL_2332982, EPI_ISL_2332983, EPI_ISL_2332984, EPI_ISL_2332985, EPI_ISL_2332986, EPI_ISL_2332987, EPI_ISL_2332988, EPI_ISL_2332989, EPI_ISL_2332990, EPI_ISL_2332991, EPI_ISL_2332992, EPI_ISL_2332993, EPI_ISL_2332994, EPI_ISL_2332995, EPI_ISL_2332996, EPI_ISL_2332997, EPI_ISL_2332998, EPI_ISL_2332999, EPI_ISL_2333000, EPI_ISL_2333001, EPI_ISL_2333002, EPI_ISL_2333003, EPI_ISL_2333004, EPI_ISL_2333005, EPI_ISL_2333006, EPI_ISL_2333007, EPI_ISL_2333008, EPI_ISL_2333009, EPI_ISL_2333010, EPI_ISL_2333011, EPI_ISL_2333012, EPI_ISL_2333013, EPI_ISL_2333014, EPI_ISL_2333015, EPI_ISL_2333016, EPI_ISL_2333017, EPI_ISL_2333018, EPI_ISL_2333019, EPI_ISL_2333020, EPI_ISL_2333021, EPI_ISL_2333022, EPI_ISL_2333023, EPI_ISL_2333024, EPI_ISL_2333025, EPI_ISL_2333026, EPI_ISL_2333027, EPI_ISL_2333028, EPI_ISL_2333030, EPI_ISL_2333031, EPI_ISL_2333032, EPI_ISL_2333033, EPI_ISL_2333034, EPI_ISL_2333035, EPI_ISL_2333036, EPI_ISL_2333037, EPI_ISL_2333039, EPI_ISL_2333040, EPI_ISL_2333041, EPI_ISL_2333042, EPI_ISL_2333043, EPI_ISL_2333044, EPI_ISL_2333045, EPI_ISL_2333046, EPI_ISL_2333047, EPI_ISL_2333048, EPI_ISL_2333049, EPI_ISL_2333050, EPI_ISL_2333051, EPI_ISL_2333052, EPI_ISL_2333053, EPI_ISL_2333054, EPI_ISL_2333055, EPI_ISL_2333056, EPI_ISL_2333057, EPI_ISL_2333058, EPI_ISL_2333059, EPI_ISL_2333060, EPI_ISL_2333061, EPI_ISL_2333063, EPI_ISL_2333064, EPI_ISL_2333065, EPI_ISL_2333066, EPI_ISL_2333067, EPI_ISL_2333068, EPI_ISL_2333069, EPI_ISL_2333070 | | | |
| see above | UW Virology Lab | UW Virology Lab | Pavitra Roychoudhury, Hong Xie, Lasata Shrestha, Tien V. Nguyen, Shah Mohamed Bakhash, Michelle Lin, Noah R. Baker, Ricardo Perez, Sean Ellis, Nathan Breit, Robert J. Livingston, Meei-Li Huang, Keith R Jerome, Patrick Mathias, Alexander Greninger |
| EPI_ISL_2333901, EPI_ISL_2333902 | MS Public Health Laboratory | University of Mississippi Medical Center, Molecular and Genomics Core Facility | Ashley C. Johnson, Ithiel J. Frame, Krishna K. Ayyalasomayajula, Michael R. Garrett, D. Ashley Robinson |
| EPI_ISL_2333903, EPI_ISL_2333904 | University of Mississippi Medical Center, Department of Pathology | University of Mississippi Medical Center, Molecular and Genomics Core Facility | Ashley C. Johnson, Ithiel J. Frame, Krishna K. Ayyalasomayajula, Michael R. Garrett, D. Ashley Robinson |
| EPI_ISL_2333905 | MS Public Health Laboratory | University of Mississippi Medical Center, Molecular and Genomics Core Facility | Ashley C. Johnson, Ithiel J. Frame, Krishna K. Ayyalasomayajula, Michael R. Garrett, D. Ashley Robinson |
| EPI_ISL_2333906, EPI_ISL_2333907 | University of Mississippi Medical Center, Department of Pathology | University of Mississippi Medical Center, Molecular and Genomics Core Facility | Ashley C. Johnson, Ithiel J. Frame, Krishna K. Ayyalasomayajula, Michael R. Garrett, D. Ashley Robinson |
| EPI_ISL_2333908 | MS Public Health Laboratory | University of Mississippi Medical Center, Molecular and Genomics Core Facility | Ashley C. Johnson, Ithiel J. Frame, Krishna K. Ayyalasomayajula, Michael R. Garrett, D. Ashley Robinson |
| EPI_ISL_2333909, EPI_ISL_2333910, EPI_ISL_2333911, EPI_ISL_2333912, | University of Mississippi Medical Center, Department of Pathology | University of Mississippi Medical Center, Molecular and Genomics Core Facility | Ashley C. Johnson, Ithiel J. Frame, Krishna K. Ayyalasomayajula, Michael R. Garrett, D. Ashley Robinson |

| | | | |
|---|---|---|---|
| EPI_ISL_2333913, EPI_ISL_2333914 | | | |
| EPI_ISL_2333915 | MS Public Health Laboratory | University of Mississippi Medical Center, Molecular and Genomics Core Facility | Ashley C. Johnson, Ithiel J. Frame, Krishna K. Ayyalasomayajula, Michael R. Garrett, D. Ashley Robinson |
| EPI_ISL_2333916, EPI_ISL_2333917, EPI_ISL_2333918, EPI_ISL_2333926 | University of Mississippi Medical Center, Department of Pathology | University of Mississippi Medical Center, Molecular and Genomics Core Facility | Ashley C. Johnson, Ithiel J. Frame, Krishna K. Ayyalasomayajula, Michael R. Garrett, D. Ashley Robinson |
| EPI_ISL_2334059, EPI_ISL_2334060, EPI_ISL_2334061, EPI_ISL_2334062, EPI_ISL_2334063, EPI_ISL_2334064, EPI_ISL_2334065, EPI_ISL_2334066, EPI_ISL_2334067, EPI_ISL_2334069, EPI_ISL_2334070, EPI_ISL_2334071, EPI_ISL_2334072, EPI_ISL_2334073, EPI_ISL_2334074, EPI_ISL_2334075, EPI_ISL_2334076, EPI_ISL_2334077, EPI_ISL_2334078, EPI_ISL_2334079, EPI_ISL_2334080, EPI_ISL_2334081, EPI_ISL_2334082, EPI_ISL_2334083, EPI_ISL_2334084, EPI_ISL_2334085, EPI_ISL_2334086, EPI_ISL_2334087, EPI_ISL_2334088, EPI_ISL_2334089, EPI_ISL_2334090, EPI_ISL_2334091, EPI_ISL_2334092, EPI_ISL_2334093 | | | |
| see above | TXDSHS | TXDSHS | Rashmi Tuladhar, Bonnie Oh, Jenny Zhang, Maliha Rahman, Mayela Pedrueza, Anita Pokharel, Karen Bobier, Lorraine Rodriguez, Myong Koag, Chun Wang, Rachel Lee, Grace Kubin |
| EPI_ISL_2334518, EPI_ISL_2334519, EPI_ISL_2334520, EPI_ISL_2334521, EPI_ISL_2334522, EPI_ISL_2334523, EPI_ISL_2334524, EPI_ISL_2334525, EPI_ISL_2334526, EPI_ISL_2334527, EPI_ISL_2334528, EPI_ISL_2334529, EPI_ISL_2334530, EPI_ISL_2334533, EPI_ISL_2334534, EPI_ISL_2334535, EPI_ISL_2334536, EPI_ISL_2334537, EPI_ISL_2334538, EPI_ISL_2334539, EPI_ISL_2334540, EPI_ISL_2334541, EPI_ISL_2334542, EPI_ISL_2334543, EPI_ISL_2334544, EPI_ISL_2334545, EPI_ISL_2334546, EPI_ISL_2334547, EPI_ISL_2334548, EPI_ISL_2334549, EPI_ISL_2334550, EPI_ISL_2334551, EPI_ISL_2334552, EPI_ISL_2334553, EPI_ISL_2334554, EPI_ISL_2334556, EPI_ISL_2334557, EPI_ISL_2334558, EPI_ISL_2334559, EPI_ISL_2334560, EPI_ISL_2334561, EPI_ISL_2334562, EPI_ISL_2334563, EPI_ISL_2334564, EPI_ISL_2334565, EPI_ISL_2334566, EPI_ISL_2334567, EPI_ISL_2334568, EPI_ISL_2334569, EPI_ISL_2334570, EPI_ISL_2334572, EPI_ISL_2334573, EPI_ISL_2334574, EPI_ISL_2334576, EPI_ISL_2334577, EPI_ISL_2334579, EPI_ISL_2334580, EPI_ISL_2334581, EPI_ISL_2334582, EPI_ISL_2334583, EPI_ISL_2334584, EPI_ISL_2334585, EPI_ISL_2334586, EPI_ISL_2335022, EPI_ISL_2335023, EPI_ISL_2335025, EPI_ISL_2335026, EPI_ISL_2335027, EPI_ISL_2335028, EPI_ISL_2335029, EPI_ISL_2335030, EPI_ISL_2335031, EPI_ISL_2335032, EPI_ISL_2335033, EPI_ISL_2335034, EPI_ISL_2335035, EPI_ISL_2335036, EPI_ISL_2335038, EPI_ISL_2335039, EPI_ISL_2335040, EPI_ISL_2335041, EPI_ISL_2335043, EPI_ISL_2335044, EPI_ISL_2335045, EPI_ISL_2335047, EPI_ISL_2335048, EPI_ISL_2335049, EPI_ISL_2335050, EPI_ISL_2335051, EPI_ISL_2335052, EPI_ISL_2335053, EPI_ISL_2335054, EPI_ISL_2335055, EPI_ISL_2335056, EPI_ISL_2335057, EPI_ISL_2335058, EPI_ISL_2335059, EPI_ISL_2335060, EPI_ISL_2335061, EPI_ISL_2335062, EPI_ISL_2335064, EPI_ISL_2335065, EPI_ISL_2335067, EPI_ISL_2335068, EPI_ISL_2335070, EPI_ISL_2335071, EPI_ISL_2335072, EPI_ISL_2335073, EPI_ISL_2335074, EPI_ISL_2335075, EPI_ISL_2335076, EPI_ISL_2335078, EPI_ISL_2335079, EPI_ISL_2335081, EPI_ISL_2335082, EPI_ISL_2335083, EPI_ISL_2335084, EPI_ISL_2335085, EPI_ISL_2335086, EPI_ISL_2335087, EPI_ISL_2335089 | | | |
| see above | North Dakota Department of Health, Public Health Laboratory | North Dakota Department of Health, Public Health Laboratory | Lisa Wingerter |
| EPI_ISL_2335091, EPI_ISL_2335092, EPI_ISL_2335093, EPI_ISL_2335094 | University of Michigan Clinical Microbiology Laboratory | Lauring Lab, University of Michigan, Department of Microbiology and Immunology | Gilbert |
| EPI_ISL_2335095, EPI_ISL_2335096, EPI_ISL_2335097, EPI_ISL_2335098, EPI_ISL_2335099, EPI_ISL_2335101, EPI_ISL_2335102, EPI_ISL_2335103, EPI_ISL_2335105, EPI_ISL_2335106, EPI_ISL_2335107, EPI_ISL_2335108, EPI_ISL_2335109, EPI_ISL_2335110, EPI_ISL_2335111, EPI_ISL_2335113, EPI_ISL_2335114, EPI_ISL_2335115, EPI_ISL_2335116, EPI_ISL_2335117, EPI_ISL_2335118, EPI_ISL_2335119, EPI_ISL_2335120, EPI_ISL_2335121, EPI_ISL_2335123, EPI_ISL_2335124, EPI_ISL_2335125, EPI_ISL_2335126, EPI_ISL_2335127, EPI_ISL_2335128, EPI_ISL_2335130, EPI_ISL_2335131, EPI_ISL_2335132, EPI_ISL_2335133, EPI_ISL_2335136, EPI_ISL_2335137, EPI_ISL_2335138, EPI_ISL_2335139, EPI_ISL_2335140, EPI_ISL_2335141, EPI_ISL_2335143, EPI_ISL_2335144 | | | |
| see above | IVY3 Central Lab, Vanderbilt University Medical Center | Lauring Lab, University of Michigan, Department of Microbiology and Immunology | Gilbert |
| EPI_ISL_2336152, EPI_ISL_2336154, EPI_ISL_2336155, EPI_ISL_2336156, EPI_ISL_2336157, EPI_ISL_2336158, EPI_ISL_2336159, EPI_ISL_2336160, EPI_ISL_2336161, EPI_ISL_2336162, EPI_ISL_2336163, EPI_ISL_2336164, EPI_ISL_2336165, EPI_ISL_2336166, EPI_ISL_2336167, EPI_ISL_2336168, EPI_ISL_2336169, EPI_ISL_2336170, EPI_ISL_2336171, EPI_ISL_2336172, EPI_ISL_2336173, EPI_ISL_2336174, EPI_ISL_2336175, EPI_ISL_2336176, EPI_ISL_2336177, EPI_ISL_2336178, EPI_ISL_2336179, EPI_ISL_2336180, EPI_ISL_2336181, EPI_ISL_2336182, EPI_ISL_2336183, EPI_ISL_2336184, EPI_ISL_2336185, EPI_ISL_2336186, EPI_ISL_2336187, EPI_ISL_2336188, EPI_ISL_2336189, EPI_ISL_2336190, EPI_ISL_2336191, EPI_ISL_2336192, EPI_ISL_2336193, EPI_ISL_2336194, EPI_ISL_2336195, EPI_ISL_2336196, EPI_ISL_2336197, EPI_ISL_2336198, EPI_ISL_2336199, EPI_ISL_2336200, EPI_ISL_2336201, EPI_ISL_2336202, EPI_ISL_2336203, EPI_ISL_2336204, EPI_ISL_2336205, EPI_ISL_2336206, EPI_ISL_2336207, EPI_ISL_2336208, EPI_ISL_2336209, EPI_ISL_2336210, EPI_ISL_2336211, EPI_ISL_2336212, EPI_ISL_2336213, EPI_ISL_2336214, EPI_ISL_2336215, EPI_ISL_2336216, EPI_ISL_2336217, EPI_ISL_2336218, EPI_ISL_2336219, EPI_ISL_2336220, EPI_ISL_2336221, EPI_ISL_2336222, EPI_ISL_2336223, EPI_ISL_2336224, EPI_ISL_2336225, EPI_ISL_2336226, EPI_ISL_2336227, EPI_ISL_2336228, EPI_ISL_2336229, EPI_ISL_2336230, EPI_ISL_2336231, EPI_ISL_2336232, EPI_ISL_2336233, EPI_ISL_2336234, EPI_ISL_2336235, EPI_ISL_2336236, EPI_ISL_2336237, EPI_ISL_2336238, EPI_ISL_2336239, EPI_ISL_2336240, EPI_ISL_2336241, EPI_ISL_2336242, EPI_ISL_2336243, EPI_ISL_2336244, EPI_ISL_2336245, EPI_ISL_2336246, EPI_ISL_2336247, EPI_ISL_2336248, EPI_ISL_2336249, EPI_ISL_2336250, EPI_ISL_2336251, EPI_ISL_2336252, EPI_ISL_2336253 | | | |
| see above | Washington State Department of Health Public Health Laboratories | Washington State Department of Health Public Health Laboratories | Drew MacKellar, Philip Dykema, Denny Russell, Joenice Gonzalez, Hannah Gray, Geoff Melly, Vanessa De Los Santos, Darren Lucas, JohnAric Peterson, Avi Singh, Rebecca Cao |
| EPI_ISL_2339275, EPI_ISL_2339276, EPI_ISL_2339277, EPI_ISL_2339278, EPI_ISL_2339279, EPI_ISL_2339280, EPI_ISL_2339281, EPI_ISL_2339282, EPI_ISL_2339283, EPI_ISL_2339284, EPI_ISL_2339285, EPI_ISL_2339286, EPI_ISL_2339287, EPI_ISL_2339288, EPI_ISL_2339289, EPI_ISL_2339290, EPI_ISL_2339291, EPI_ISL_2339292, EPI_ISL_2339293, EPI_ISL_2339294, EPI_ISL_2339295, EPI_ISL_2339296, EPI_ISL_2339301, EPI_ISL_2339302, EPI_ISL_2339303, EPI_ISL_2339304 | | | |
| see above | Arizona State University | Arizona State University | Peter T. Skidmore, LaRinda A. Holland, Matthew F. Smith, Rabia Maqsood, Nicholas J. Mellor, Joy M. Blain, Valerie Harris, Joshua LaBaer, Vel Murugan, Efrem S. Lim |
| EPI_ISL_2339306, EPI_ISL_2339307, EPI_ISL_2339308, EPI_ISL_2339309, EPI_ISL_2339310, EPI_ISL_2339311, EPI_ISL_2339312, EPI_ISL_2339313, EPI_ISL_2339314, EPI_ISL_2339315, EPI_ISL_2339316, EPI_ISL_2339317, EPI_ISL_2339318, EPI_ISL_2339319, EPI_ISL_2339320, EPI_ISL_2339321, EPI_ISL_2339322, EPI_ISL_2339323, EPI_ISL_2339324, EPI_ISL_2339325, EPI_ISL_2339326, EPI_ISL_2339327, EPI_ISL_2339328, EPI_ISL_2339329, EPI_ISL_2339330, EPI_ISL_2339331, EPI_ISL_2339332, EPI_ISL_2339333, EPI_ISL_2339334, EPI_ISL_2339335, EPI_ISL_2339336, EPI_ISL_2339337, EPI_ISL_2339338, EPI_ISL_2339339, EPI_ISL_2339340, EPI_ISL_2339341, EPI_ISL_2339342 | | | |
| see above | Oregon State Public Health Laboratory | Oregon State Public Health Laboratory | Rafia Razzaque, Eugene Yeboah, Vanda Makris, Laura Tsaknaridis, John Fontana and Shane Sevey |
| EPI_ISL_2339343, EPI_ISL_2339344, EPI_ISL_2339345, EPI_ISL_2339346, EPI_ISL_2339347, EPI_ISL_2339348, EPI_ISL_2339349, EPI_ISL_2339350, EPI_ISL_2339351, EPI_ISL_2339352, EPI_ISL_2339353, EPI_ISL_2339354, EPI_ISL_2339355, EPI_ISL_2339356, EPI_ISL_2339357, EPI_ISL_2339358, EPI_ISL_2339359, EPI_ISL_2339360, EPI_ISL_2339361, EPI_ISL_2339362, EPI_ISL_2339363, EPI_ISL_2339364, EPI_ISL_2339366, EPI_ISL_2339367, EPI_ISL_2339368, EPI_ISL_2339369, EPI_ISL_2339370, EPI_ISL_2339371, EPI_ISL_2339372, EPI_ISL_2339373, EPI_ISL_2339374, EPI_ISL_2339375, EPI_ISL_2339377, EPI_ISL_2339378, EPI_ISL_2339379, EPI_ISL_2339380, EPI_ISL_2339381, EPI_ISL_2339382, EPI_ISL_2339383, EPI_ISL_2339384, EPI_ISL_2339385, EPI_ISL_2339386, EPI_ISL_2339387, EPI_ISL_2339388, EPI_ISL_2339389, EPI_ISL_2339390, EPI_ISL_2339391, EPI_ISL_2339392, EPI_ISL_2339393, EPI_ISL_2339394, EPI_ISL_2339395, EPI_ISL_2339396, EPI_ISL_2339397, EPI_ISL_2339398, EPI_ISL_2339399, EPI_ISL_2339400, EPI_ISL_2339401, EPI_ISL_2339402, EPI_ISL_2339404, EPI_ISL_2339405, EPI_ISL_2339406, EPI_ISL_2339407, EPI_ISL_2339408, EPI_ISL_2339409, EPI_ISL_2339410, EPI_ISL_2339411, EPI_ISL_2339412, EPI_ISL_2339413, EPI_ISL_2339414, EPI_ISL_2339415, EPI_ISL_2339416, EPI_ISL_2339417, EPI_ISL_2339418, EPI_ISL_2339419, EPI_ISL_2339420, EPI_ISL_2339421, EPI_ISL_2339422, EPI_ISL_2339423, EPI_ISL_2339425, EPI_ISL_2339426, EPI_ISL_2339427, EPI_ISL_2339428, EPI_ISL_2339430, EPI_ISL_2339431, EPI_ISL_2339432, EPI_ISL_2339433, EPI_ISL_2339434, EPI_ISL_2339435 | | | |
| see above | Illinois Department of Public Health - Springfield Lab | Illinois Department of Public Health - Springfield Lab | Bryan Sim, Gordon McCall |
| EPI_ISL_2339910, EPI_ISL_2339911, EPI_ISL_2339913, EPI_ISL_2339915, EPI_ISL_2339916, EPI_ISL_2339918, EPI_ISL_2339919, EPI_ISL_2339920, EPI_ISL_2339921, EPI_ISL_2339923, EPI_ISL_2339924, EPI_ISL_2339925, EPI_ISL_2339926, EPI_ISL_2339928, EPI_ISL_2339930, EPI_ISL_2339931, EPI_ISL_2339932, EPI_ISL_2339933, EPI_ISL_2339934, EPI_ISL_2339935, EPI_ISL_2339936, EPI_ISL_2339937, EPI_ISL_2339938, EPI_ISL_2339939, EPI_ISL_2339940, EPI_ISL_2339942, EPI_ISL_2339943, EPI_ISL_2339944, EPI_ISL_2339946, EPI_ISL_2339947, EPI_ISL_2339948, EPI_ISL_2339949, EPI_ISL_2339950, EPI_ISL_2339951, EPI_ISL_2339952, EPI_ISL_2339953, EPI_ISL_2339954, EPI_ISL_2339955, EPI_ISL_2339956, EPI_ISL_2339957, EPI_ISL_2339959, EPI_ISL_2339960, EPI_ISL_2339962, EPI_ISL_2339963, EPI_ISL_2339964, EPI_ISL_2339966, EPI_ISL_2339967, EPI_ISL_2339969, EPI_ISL_2339970, EPI_ISL_2339971, EPI_ISL_2339972, EPI_ISL_2339974, EPI_ISL_2339976, EPI_ISL_2339977, EPI_ISL_2339978, EPI_ISL_2339979, EPI_ISL_2339980, EPI_ISL_2339981, EPI_ISL_2339982, EPI_ISL_2339983, EPI_ISL_2339985, EPI_ISL_2339986, EPI_ISL_2339987, EPI_ISL_2339989, EPI_ISL_2339991, EPI_ISL_2339992, EPI_ISL_2339993, EPI_ISL_2339994, EPI_ISL_2339995, EPI_ISL_2339996, EPI_ISL_2339997, EPI_ISL_2339998, EPI_ISL_2339999, EPI_ISL_2340001, EPI_ISL_2340002, EPI_ISL_2340003, EPI_ISL_2340007, EPI_ISL_2340008, EPI_ISL_2340009, EPI_ISL_2340010, EPI_ISL_2340011, EPI_ISL_2340012, EPI_ISL_2340013, EPI_ISL_2340015, EPI_ISL_2340016, EPI_ISL_2340017, EPI_ISL_2340018, EPI_ISL_2340019, EPI_ISL_2340020, EPI_ISL_2340021, EPI_ISL_2340022, EPI_ISL_2340023, EPI_ISL_2340024, EPI_ISL_2340025, EPI_ISL_2340026, EPI_ISL_2340027, EPI_ISL_2340028, EPI_ISL_2340029, EPI_ISL_2340030, EPI_ISL_2340032, EPI_ISL_2340033, EPI_ISL_2340034, EPI_ISL_2340035, EPI_ISL_2340036, EPI_ISL_2340037, EPI_ISL_2340038, EPI_ISL_2340039, EPI_ISL_2340040, EPI_ISL_2340041, EPI_ISL_2340042, EPI_ISL_2340043, EPI_ISL_2340044, EPI_ISL_2340045 | | | |
| see above | North Dakota Department of Health, Public Health Laboratory | North Dakota Department of Health, Public Health Laboratory | Lisa Wingerter |
| EPI_ISL_2340546, EPI_ISL_2340547, EPI_ISL_2340548, EPI_ISL_2340549, EPI_ISL_2340550, EPI_ISL_2340551, EPI_ISL_2340552, EPI_ISL_2340553, EPI_ISL_2340554, EPI_ISL_2340555, EPI_ISL_2340556, EPI_ISL_2340557, EPI_ISL_2340558, EPI_ISL_2340559, EPI_ISL_2340560, EPI_ISL_2340561, EPI_ISL_2340562, EPI_ISL_2340563, EPI_ISL_2340564, EPI_ISL_2340565, EPI_ISL_2340566, EPI_ISL_2340567, EPI_ISL_2340568, EPI_ISL_2340569, EPI_ISL_2340570, EPI_ISL_2340571, EPI_ISL_2340572, EPI_ISL_2340573, EPI_ISL_2340574, EPI_ISL_2340575, EPI_ISL_2340576, EPI_ISL_2340577, EPI_ISL_2340578 | | | |
| see above | GHRC | TIGSS | Benjamin W. Neuman, Melissa M. Kahl-Mcdonagh, Kurt A. Zuelke, Charlie D. Johnson, Allison Ficht, Sankar P. Chaki, Sierra Guidry, Joshua Hill, Richard Metz, Marcel Brun |
| EPI_ISL_2340579, EPI_ISL_2340580, EPI_ISL_2340581, EPI_ISL_2340582, EPI_ISL_2340583, EPI_ISL_2340584, EPI_ISL_2340585, EPI_ISL_2340586, EPI_ISL_2340588, EPI_ISL_2340589, EPI_ISL_2340590, EPI_ISL_2340591, EPI_ISL_2340592, EPI_ISL_2340593, EPI_ISL_2340594, EPI_ISL_2340595, EPI_ISL_2340596, EPI_ISL_2340597, EPI_ISL_2340599, EPI_ISL_2340600, EPI_ISL_2340601, EPI_ISL_2340602, EPI_ISL_2340603, EPI_ISL_2340604, EPI_ISL_2340605, EPI_ISL_2340606, EPI_ISL_2340608, EPI_ISL_2340609, EPI_ISL_2340610, EPI_ISL_2340611, EPI_ISL_2340612, EPI_ISL_2340613, EPI_ISL_2340614, EPI_ISL_2340615, EPI_ISL_2340616, EPI_ISL_2340617, EPI_ISL_2340618, EPI_ISL_2340619, EPI_ISL_2340620, EPI_ISL_2340621, EPI_ISL_2340622, EPI_ISL_2340623, EPI_ISL_2340624, EPI_ISL_2340625, EPI_ISL_2340626, EPI_ISL_2340627, EPI_ISL_2340628, EPI_ISL_2340629, EPI_ISL_2340630, EPI_ISL_2340631, EPI_ISL_2340632, EPI_ISL_2340633, EPI_ISL_2340634, EPI_ISL_2340635, EPI_ISL_2340636, EPI_ISL_2340637, EPI_ISL_2340638, EPI_ISL_2340639, EPI_ISL_2340640, EPI_ISL_2340641, EPI_ISL_2340642, EPI_ISL_2340643, EPI_ISL_2340644, EPI_ISL_2340645, EPI_ISL_2340646, EPI_ISL_2340647, EPI_ISL_2340648, EPI_ISL_2340649, EPI_ISL_2340650, EPI_ISL_2340651, EPI_ISL_2340652, EPI_ISL_2340653, EPI_ISL_2340654, EPI_ISL_2340657, EPI_ISL_2340658, EPI_ISL_2340659, EPI_ISL_2340660, EPI_ISL_2340661, EPI_ISL_2340662, EPI_ISL_2340663, EPI_ISL_2340664, EPI_ISL_2340665 | | | |
| see above | GHRC | TxGen | Benjamin W. Neuman, Melissa M. Kahl-Mcdonagh, Kurt A. Zuelke, Charlie D. Johnson, Allison Ficht, Sankar P. Chaki, Sierra Guidry, Joshua Hill, Richard Metz, Marcel Brun |
| EPI_ISL_2340666, EPI_ISL_2340667, EPI_ISL_2340668, EPI_ISL_2340669, EPI_ISL_2340670, EPI_ISL_2340671, EPI_ISL_2340672, EPI_ISL_2340673, EPI_ISL_2340674, EPI_ISL_2340676, EPI_ISL_2340677, EPI_ISL_2340678, EPI_ISL_2340679, EPI_ISL_2340680, EPI_ISL_2340681, EPI_ISL_2340682, EPI_ISL_2340683, EPI_ISL_2340684 | | | |
| see above | University of Wisconsin-Madison AIDS Vaccine Research Laboratories | University of Wisconsin-Madison AIDS Vaccine Research Laboratories | Gage Moreno, Katarina Braun, et al. AIDS Vaccine Research Laboratories |
| EPI_ISL_2340685, EPI_ISL_2340686, EPI_ISL_2340688, EPI_ISL_2340690, EPI_ISL_2340691, EPI_ISL_2340692, EPI_ISL_2340695, EPI_ISL_2340696, EPI_ISL_2340697, EPI_ISL_2340698, EPI_ISL_2340699, EPI_ISL_2340700, EPI_ISL_2340701, EPI_ISL_2340702, EPI_ISL_2340703, EPI_ISL_2340704, EPI_ISL_2340705, EPI_ISL_2340706, EPI_ISL_2340707, EPI_ISL_2340708, EPI_ISL_2340709, EPI_ISL_2340710, EPI_ISL_2340711, EPI_ISL_2340712, EPI_ISL_2340713, EPI_ISL_2340714, EPI_ISL_2340715, EPI_ISL_2340716, EPI_ISL_2340717, EPI_ISL_2340718, EPI_ISL_2340719, EPI_ISL_2340720, EPI_ISL_2340721, EPI_ISL_2340722, EPI_ISL_2340723, EPI_ISL_2340724, EPI_ISL_2340725, EPI_ISL_2340727, EPI_ISL_2340728, EPI_ISL_2340729, EPI_ISL_2340730, EPI_ISL_2340731, EPI_ISL_2340732, EPI_ISL_2340733, EPI_ISL_2340734, EPI_ISL_2340735, EPI_ISL_2340736, EPI_ISL_2340737, EPI_ISL_2340738, EPI_ISL_2340740, EPI_ISL_2340741, EPI_ISL_2340742, EPI_ISL_2340743, EPI_ISL_2340744, EPI_ISL_2340745, EPI_ISL_2340746, EPI_ISL_2340747, EPI_ISL_2340748, EPI_ISL_2340749, EPI_ISL_2340752, EPI_ISL_2340753, EPI_ISL_2340754, EPI_ISL_2340755, EPI_ISL_2340756, EPI_ISL_2340757, EPI_ISL_2340758, EPI_ISL_2340759, EPI_ISL_2340760, EPI_ISL_2340761, EPI_ISL_2340762, EPI_ISL_2340763, EPI_ISL_2340764, EPI_ISL_2340765, EPI_ISL_2340766, EPI_ISL_2340767, EPI_ISL_2340768, EPI_ISL_2340769, EPI_ISL_2340770, EPI_ISL_2340771, EPI_ISL_2340772, EPI_ISL_2340773, EPI_ISL_2340774, EPI_ISL_2340775, EPI_ISL_2340776, EPI_ISL_2340777, EPI_ISL_2340778, EPI_ISL_2340779, EPI_ISL_2340780, EPI_ISL_2340781, EPI_ISL_2340782, EPI_ISL_2340783, EPI_ISL_2340784, EPI_ISL_2340785, EPI_ISL_2340786, EPI_ISL_2340787, EPI_ISL_2340788, EPI_ISL_2340789, EPI_ISL_2340790, EPI_ISL_2340791, EPI_ISL_2340792, EPI_ISL_2340793, EPI_ISL_2340794, EPI_ISL_2340795, EPI_ISL_2340796, EPI_ISL_2340797, EPI_ISL_2340798, EPI_ISL_2340799, EPI_ISL_2340800, EPI_ISL_2340801, EPI_ISL_2340802, EPI_ISL_2340803, EPI_ISL_2340804, EPI_ISL_2340805, EPI_ISL_2340806, EPI_ISL_2340807, EPI_ISL_2340808, EPI_ISL_2340809, EPI_ISL_2340810, EPI_ISL_2340811, EPI_ISL_2340812, EPI_ISL_2340813, EPI_ISL_2340814, EPI_ISL_2340815, EPI_ISL_2340816, EPI_ISL_2340817, EPI_ISL_2340818, EPI_ISL_2340819, EPI_ISL_2340820, EPI_ISL_2340821, EPI_ISL_2340822, EPI_ISL_2340823, EPI_ISL_2340824, EPI_ISL_2340825, EPI_ISL_2340826 | | | |
| see above | UW Virology Lab | UW Virology Lab | Pavitra Roychoudhury, Hong Xie, Lasata Shrestha, Tien V. Nguyen, Shah Mohamed Bakhash, Michelle Lin, Noah R. Baker, Sean Ellis, Meei-Li Huang, Keith R Jerome, Alexander Greninger |
| EPI_ISL_2340827, EPI_ISL_2340828, EPI_ISL_2340829, EPI_ISL_2340830, EPI_ISL_2340831, EPI_ISL_2340832, EPI_ISL_2340833, EPI_ISL_2340834, EPI_ISL_2340835, EPI_ISL_2340836, EPI_ISL_2340837, EPI_ISL_2340838, EPI_ISL_2340839, EPI_ISL_2340840 | | | |
| see above | University of Wisconsin-Madison AIDS Vaccine Research Laboratories | University of Wisconsin-Madison AIDS Vaccine Research Laboratories | Gage Moreno, Katarina Braun, et al. AIDS Vaccine Research Laboratories |
| EPI_ISL_2340843, EPI_ISL_2340844, EPI_ISL_2340845, EPI_ISL_2340846, EPI_ISL_2340847, EPI_ISL_2340848, EPI_ISL_2340849, EPI_ISL_2340850, EPI_ISL_2340851, EPI_ISL_2340852, EPI_ISL_2340853, EPI_ISL_2340854, EPI_ISL_2340855, EPI_ISL_2340856, EPI_ISL_2340857, EPI_ISL_2340858, EPI_ISL_2340859 | | | |
| see above | GA Department of Public Health | GA Department of Public Health | Stacy Reeves, Jonathan Edwards, Cynthia Dixey, Tonia Parrott, Aliyah Fields, Taylor Smith |
| EPI_ISL_2340860 | LESP Guanajuato | Instituto de Diagnostico y Referencia Epidemiologicos (INDRE) | Claudia Wong-Arambula, Abril Rodriguez-Maldonado, Vanessa Rivero-Arredondo, Ariadna Medina-Benitez, Joaquin Quiroz-Mercado, Sergio Rangel-Guerrero, Natividad Cruz-Ortiz, Tatiana Nunez-Garcia, Gisela Barrera-Badillo, Lucia Hernandez-Rivas, Irma Lopez-Martinez, Ernesto Ramirez-Gonzalez. |
| EPI_ISL_2340861 | LESP Jalisco | Instituto de Diagnostico y Referencia Epidemiologicos (INDRE) | Claudia Wong-Arambula, Abril Rodriguez-Maldonado, Vanessa Rivero-Arredondo, Ariadna Medina-Benitez, Joaquin Quiroz-Mercado, Sergio Rangel-Guerrero, Natividad Cruz-Ortiz, Tatiana Nunez-Garcia, Gisela Barrera-Badillo, Lucia Hernandez-Rivas, Irma Lopez-Martinez, Ernesto Ramirez-Gonzalez. |
| EPI_ISL_2340862 | LESP Michoacan | Instituto de Diagnostico y Referencia Epidemiologicos (INDRE) | Claudia Wong-Arambula, Abril Rodriguez-Maldonado, Vanessa Rivero-Arredondo, Ariadna Medina-Benitez, Joaquin Quiroz-Mercado, Sergio Rangel-Guerrero, Natividad Cruz-Ortiz, Tatiana Nunez-Garcia, Gisela Barrera-Badillo, Lucia Hernandez-Rivas, Irma Lopez-Martinez, Ernesto Ramirez-Gonzalez. |
| EPI_ISL_2340863, EPI_ISL_2340864 | LESP Guanajuato | Instituto de Diagnostico y Referencia Epidemiologicos (INDRE) | Claudia Wong-Arambula, Abril Rodriguez-Maldonado, Vanessa Rivero-Arredondo, Ariadna Medina-Benitez, Joaquin Quiroz-Mercado, Sergio Rangel-Guerrero, Natividad Cruz-Ortiz, Tatiana Nunez-Garcia, Gisela Barrera-Badillo, Lucia Hernandez-Rivas, Irma Lopez-Martinez, Ernesto Ramirez-Gonzalez. |
| EPI_ISL_2340865 | LESP Michoacan | Instituto de Diagnostico y Referencia Epidemiologicos (INDRE) | Claudia Wong-Arambula, Abril Rodriguez-Maldonado, Vanessa Rivero-Arredondo, Ariadna Medina-Benitez, Joaquin Quiroz-Mercado, Sergio Rangel-Guerrero, Natividad Cruz-Ortiz, Tatiana Nunez-Garcia, Gisela Barrera-Badillo, Lucia Hernandez-Rivas, Irma Lopez-Martinez, Ernesto Ramirez-Gonzalez. |
| EPI_ISL_2340866 | LESP Guanajuato | Instituto de Diagnostico y Referencia Epidemiologicos (INDRE) | Claudia Wong-Arambula, Abril Rodriguez-Maldonado, Vanessa Rivero-Arredondo, Ariadna Medina-Benitez, Joaquin Quiroz-Mercado, Sergio Rangel-Guerrero, Natividad Cruz-Ortiz, Tatiana Nunez-Garcia, Gisela Barrera-Badillo, Lucia Hernandez-Rivas, Irma Lopez-Martinez, Ernesto Ramirez-Gonzalez. |
| EPI_ISL_2340867 | LESP Nuevo Leon | Instituto de Diagnostico y Referencia Epidemiologicos (INDRE) | Claudia Wong-Arambula, Abril Rodriguez-Maldonado, Vanessa Rivero-Arredondo, Ariadna Medina-Benitez, Joaquin Quiroz-Mercado, Sergio Rangel-Guerrero, Natividad Cruz-Ortiz, Tatiana Nunez-Garcia, Gisela Barrera-Badillo, Lucia Hernandez-Rivas, Irma Lopez-Martinez, Ernesto Ramirez-Gonzalez. |
| EPI_ISL_2340868, | LESP Michoacan | Instituto de Diagnostico y Referencia | Claudia Wong-Arambula, Abril Rodriguez-Maldonado, Vanessa Rivero-Arredondo, Ariadna Medina-Benitez, Joaquin Quiroz-Mercado, Sergio Rangel-Guerrero, Natividad Cruz-Ortiz, Tatiana Nunez-Garcia, Gisela Barrera-Badillo, Lucia Hernandez-Rivas, Irma Lopez-Martinez, Ernesto |

| | | | |
|---|---|---|---|
| EPI_ISL_2340869, EPI_ISL_2340870 | | Epidemiologicos (INDRE) | Ramirez-Gonzalez. |
| EPI_ISL_2340871 | LESP Oaxaca | Instituto de Diagnostico y Referencia Epidemiologicos (INDRE) | Claudia Wong-Arambula, Abril Rodriguez-Maldonado, Vanessa Rivero-Arredondo, Ariadna Medina-Benitez, Joaquin Quiroz-Mercado, Natividad Cruz-Ortiz, Tatiana Nunez-Garcia, Gisela Barrera-Badillo, Lucia Hernandez-Rivas, Irma Lopez-Martinez, Ernesto Ramirez-Gonzalez. |
| EPI_ISL_2340872, EPI_ISL_2340873 | LESP Michoacan | Instituto de Diagnostico y Referencia Epidemiologicos (INDRE) | Claudia Wong-Arambula, Abril Rodriguez-Maldonado, Vanessa Rivero-Arredondo, Ariadna Medina-Benitez, Joaquin Quiroz-Mercado, Sergio Rangel-Guerrero, Natividad Cruz-Ortiz, Tatiana Nunez-Garcia, Gisela Barrera-Badillo, Lucia Hernandez-Rivas, Irma Lopez-Martinez, Ernesto Ramirez-Gonzalez. |
| EPI_ISL_2340874 | LESP Yucatan | Instituto de Diagnostico y Referencia Epidemiologicos (INDRE) | Claudia Wong-Arambula, Abril Rodriguez-Maldonado, Vanessa Rivero-Arredondo, Ariadna Medina-Benitez, Joaquin Quiroz-Mercado, Sergio Rangel-Guerrero, Natividad Cruz-Ortiz, Tatiana Nunez-Garcia, Gisela Barrera-Badillo, Lucia Hernandez-Rivas, Irma Lopez-Martinez, Ernesto Ramirez-Gonzalez. |
| EPI_ISL_2340875 | LESP Sinaloa | Instituto de Diagnostico y Referencia Epidemiologicos (INDRE) | Claudia Wong-Arambula, Abril Rodriguez-Maldonado, Vanessa Rivero-Arredondo, Ariadna Medina-Benitez, Joaquin Quiroz-Mercado, Sergio Rangel-Guerrero, Natividad Cruz-Ortiz, Tatiana Nunez-Garcia, Gisela Barrera-Badillo, Lucia Hernandez-Rivas, Irma Lopez-Martinez, Ernesto Ramirez-Gonzalez. |
| EPI_ISL_2340876 | LESP Queretaro | Instituto de Diagnostico y Referencia Epidemiologicos (INDRE) | Claudia Wong-Arambula, Abril Rodriguez-Maldonado, Vanessa Rivero-Arredondo, Ariadna Medina-Benitez, Joaquin Quiroz-Mercado, Sergio Rangel-Guerrero, Natividad Cruz-Ortiz, Tatiana Nunez-Garcia, Gisela Barrera-Badillo, Lucia Hernandez-Rivas, Irma Lopez-Martinez, Ernesto Ramirez-Gonzalez. |
| EPI_ISL_2340877, EPI_ISL_2340878 | LESP Tamaulipas | Instituto de Diagnostico y Referencia Epidemiologicos (INDRE) | Claudia Wong-Arambula, Abril Rodriguez-Maldonado, Vanessa Rivero-Arredondo, Ariadna Medina-Benitez, Joaquin Quiroz-Mercado, Sergio Rangel-Guerrero, Natividad Cruz-Ortiz, Tatiana Nunez-Garcia, Gisela Barrera-Badillo, Lucia Hernandez-Rivas, Irma Lopez-Martinez, Ernesto Ramirez-Gonzalez. |
| EPI_ISL_2340879, EPI_ISL_2340880 | LESP Guanajuato | Instituto de Diagnostico y Referencia Epidemiologicos (INDRE) | Claudia Wong-Arambula, Abril Rodriguez-Maldonado, Vanessa Rivero-Arredondo, Ariadna Medina-Benitez, Joaquin Quiroz-Mercado, Sergio Rangel-Guerrero, Natividad Cruz-Ortiz, Tatiana Nunez-Garcia, Gisela Barrera-Badillo, Lucia Hernandez-Rivas, Irma Lopez-Martinez, Ernesto Ramirez-Gonzalez. |
| EPI_ISL_2340881 | LESP Queretaro | Instituto de Diagnostico y Referencia Epidemiologicos (INDRE) | Claudia Wong-Arambula, Abril Rodriguez-Maldonado, Vanessa Rivero-Arredondo, Ariadna Medina-Benitez, Joaquin Quiroz-Mercado, Sergio Rangel-Guerrero, Natividad Cruz-Ortiz, Tatiana Nunez-Garcia, Gisela Barrera-Badillo, Lucia Hernandez-Rivas, Irma Lopez-Martinez, Ernesto Ramirez-Gonzalez. |
| EPI_ISL_2340882 | LESP Ciudad de Mexico | Instituto de Diagnostico y Referencia Epidemiologicos (INDRE) | Claudia Wong-Arambula, Abril Rodriguez-Maldonado, Vanessa Rivero-Arredondo, Ariadna Medina-Benitez, Joaquin Quiroz-Mercado, Sergio Rangel-Guerrero, Natividad Cruz-Ortiz, Tatiana Nunez-Garcia, Gisela Barrera-Badillo, Lucia Hernandez-Rivas, Irma Lopez-Martinez, Ernesto Ramirez-Gonzalez. |
| EPI_ISL_2340883 | LESP Queretaro | Instituto de Diagnostico y Referencia Epidemiologicos (INDRE) | Claudia Wong-Arambula, Abril Rodriguez-Maldonado, Vanessa Rivero-Arredondo, Ariadna Medina-Benitez, Joaquin Quiroz-Mercado, Sergio Rangel-Guerrero, Natividad Cruz-Ortiz, Tatiana Nunez-Garcia, Gisela Barrera-Badillo, Lucia Hernandez-Rivas, Irma Lopez-Martinez, Ernesto Ramirez-Gonzalez. |
| EPI_ISL_2340884, EPI_ISL_2340885 | LESP Sinaloa | Instituto de Diagnostico y Referencia Epidemiologicos (INDRE) | Claudia Wong-Arambula, Abril Rodriguez-Maldonado, Vanessa Rivero-Arredondo, Ariadna Medina-Benitez, Joaquin Quiroz-Mercado, Sergio Rangel-Guerrero, Natividad Cruz-Ortiz, Tatiana Nunez-Garcia, Gisela Barrera-Badillo, Lucia Hernandez-Rivas, Irma Lopez-Martinez, Ernesto Ramirez-Gonzalez. |
| EPI_ISL_2340886 | LESP Tamaulipas | Instituto de Diagnostico y Referencia Epidemiologicos (INDRE) | Claudia Wong-Arambula, Abril Rodriguez-Maldonado, Vanessa Rivero-Arredondo, Ariadna Medina-Benitez, Joaquin Quiroz-Mercado, Sergio Rangel-Guerrero, Natividad Cruz-Ortiz, Tatiana Nunez-Garcia, Gisela Barrera-Badillo, Lucia Hernandez-Rivas, Irma Lopez-Martinez, Ernesto Ramirez-Gonzalez. |
| EPI_ISL_2340887 | LESP Campeche | Instituto de Diagnostico y Referencia Epidemiologicos (INDRE) | Claudia Wong-Arambula, Abril Rodriguez-Maldonado, Vanessa Rivero-Arredondo, Ariadna Medina-Benitez, Joaquin Quiroz-Mercado, Sergio Rangel-Guerrero, Natividad Cruz-Ortiz, Tatiana Nunez-Garcia, Gisela Barrera-Badillo, Lucia Hernandez-Rivas, Irma Lopez-Martinez, Ernesto Ramirez-Gonzalez. |
| EPI_ISL_2340888, EPI_ISL_2340889 | LESP Chihuahua | Instituto de Diagnostico y Referencia Epidemiologicos (INDRE) | Claudia Wong-Arambula, Abril Rodriguez-Maldonado, Vanessa Rivero-Arredondo, Ariadna Medina-Benitez, Joaquin Quiroz-Mercado, Sergio Rangel-Guerrero, Natividad Cruz-Ortiz, Tatiana Nunez-Garcia, Gisela Barrera-Badillo, Lucia Hernandez-Rivas, Irma Lopez-Martinez, Ernesto Ramirez-Gonzalez. |
| EPI_ISL_2340890, EPI_ISL_2340891, EPI_ISL_2340892, EPI_ISL_2340893, EPI_ISL_2340894, EPI_ISL_2340895, EPI_ISL_2340896, EPI_ISL_2340897, EPI_ISL_2340898, EPI_ISL_2340899, EPI_ISL_2340900, EPI_ISL_2340901, EPI_ISL_2340902 | | | |
| see above | LESP Michoacan | Instituto de Diagnostico y Referencia Epidemiologicos (INDRE) | Claudia Wong-Arambula, Abril Rodriguez-Maldonado, Vanessa Rivero-Arredondo, Ariadna Medina-Benitez, Joaquin Quiroz-Mercado, Sergio Rangel-Guerrero, Natividad Cruz-Ortiz, Tatiana Nunez-Garcia, Gisela Barrera-Badillo, Lucia Hernandez-Rivas, Irma Lopez-Martinez, Ernesto Ramirez-Gonzalez. |
| EPI_ISL_2340903, EPI_ISL_2340904, EPI_ISL_2340905, EPI_ISL_2340906, EPI_ISL_2340907, EPI_ISL_2340908, EPI_ISL_2340909, EPI_ISL_2340910, EPI_ISL_2340911, EPI_ISL_2340912, EPI_ISL_2340913, EPI_ISL_2340914, EPI_ISL_2340915, EPI_ISL_2340916, EPI_ISL_2340917, EPI_ISL_2340918, EPI_ISL_2340919, EPI_ISL_2340920, EPI_ISL_2340921, EPI_ISL_2340922, EPI_ISL_2340923, EPI_ISL_2340924, EPI_ISL_2340925, EPI_ISL_2340926, EPI_ISL_2340927, EPI_ISL_2340928, EPI_ISL_2340929, EPI_ISL_2340930, EPI_ISL_2340931, EPI_ISL_2340932, EPI_ISL_2340933, EPI_ISL_2340934, EPI_ISL_2340935, EPI_ISL_2340936, EPI_ISL_2340937, EPI_ISL_2340938, EPI_ISL_2340939, EPI_ISL_2340940, EPI_ISL_2340941 | | | |
| see above | LESP Yucatan | Instituto de Diagnostico y Referencia Epidemiologicos (INDRE) | Claudia Wong-Arambula, Abril Rodriguez-Maldonado, Vanessa Rivero-Arredondo, Ariadna Medina-Benitez, Joaquin Quiroz-Mercado, Sergio Rangel-Guerrero, Natividad Cruz-Ortiz, Tatiana Nunez-Garcia, Gisela Barrera-Badillo, Lucia Hernandez-Rivas, Irma Lopez-Martinez, Ernesto Ramirez-Gonzalez. |
| EPI_ISL_2340942, EPI_ISL_2340943, EPI_ISL_2340944, EPI_ISL_2340945, EPI_ISL_2340946, EPI_ISL_2340947, EPI_ISL_2340948, EPI_ISL_2340949 | LESP Guanajuato | Instituto de Diagnostico y Referencia Epidemiologicos (INDRE) | Claudia Wong-Arambula, Abril Rodriguez-Maldonado, Vanessa Rivero-Arredondo, Ariadna Medina-Benitez, Joaquin Quiroz-Mercado, Sergio Rangel-Guerrero, Natividad Cruz-Ortiz, Tatiana Nunez-Garcia, Gisela Barrera-Badillo, Lucia Hernandez-Rivas, Irma Lopez-Martinez, Ernesto Ramirez-Gonzalez. |
| EPI_ISL_2340950, EPI_ISL_2340951, EPI_ISL_2340952, EPI_ISL_2340953, EPI_ISL_2340954 | LESP Puebla | Instituto de Diagnostico y Referencia Epidemiologicos (INDRE) | Claudia Wong-Arambula, Abril Rodriguez-Maldonado, Vanessa Rivero-Arredondo, Ariadna Medina-Benitez, Joaquin Quiroz-Mercado, Sergio Rangel-Guerrero, Natividad Cruz-Ortiz, Tatiana Nunez-Garcia, Gisela Barrera-Badillo, Lucia Hernandez-Rivas, Irma Lopez-Martinez, Ernesto Ramirez-Gonzalez. |
| EPI_ISL_2340955, EPI_ISL_2340956, EPI_ISL_2340957, EPI_ISL_2340958 | LESP Hidalgo | Instituto de Diagnostico y Referencia Epidemiologicos (INDRE) | Claudia Wong-Arambula, Abril Rodriguez-Maldonado, Vanessa Rivero-Arredondo, Ariadna Medina-Benitez, Joaquin Quiroz-Mercado, Sergio Rangel-Guerrero, Natividad Cruz-Ortiz, Tatiana Nunez-Garcia, Gisela Barrera-Badillo, Lucia Hernandez-Rivas, Irma Lopez-Martinez, Ernesto Ramirez-Gonzalez. |
| EPI_ISL_2340959 | LESP Queretaro | Instituto de Diagnostico y Referencia Epidemiologicos (INDRE) | Claudia Wong-Arambula, Abril Rodriguez-Maldonado, Vanessa Rivero-Arredondo, Ariadna Medina-Benitez, Joaquin Quiroz-Mercado, Sergio Rangel-Guerrero, Natividad Cruz-Ortiz, Tatiana Nunez-Garcia, Gisela Barrera-Badillo, Lucia Hernandez-Rivas, Irma Lopez-Martinez, Ernesto Ramirez-Gonzalez. |
| EPI_ISL_2340960, EPI_ISL_2340961 | LESP Sinaloa | Instituto de Diagnostico y Referencia Epidemiologicos (INDRE) | Claudia Wong-Arambula, Abril Rodriguez-Maldonado, Vanessa Rivero-Arredondo, Ariadna Medina-Benitez, Joaquin Quiroz-Mercado, Sergio Rangel-Guerrero, Natividad Cruz-Ortiz, Tatiana Nunez-Garcia, Gisela Barrera-Badillo, Lucia Hernandez-Rivas, Irma Lopez-Martinez, Ernesto Ramirez-Gonzalez. |
| EPI_ISL_2340962, EPI_ISL_2340963, EPI_ISL_2340964, EPI_ISL_2340965, EPI_ISL_2340966, EPI_ISL_2340967, EPI_ISL_2340968 | LESP Queretaro | Instituto de Diagnostico y Referencia Epidemiologicos (INDRE) | Claudia Wong-Arambula, Abril Rodriguez-Maldonado, Vanessa Rivero-Arredondo, Ariadna Medina-Benitez, Joaquin Quiroz-Mercado, Sergio Rangel-Guerrero, Natividad Cruz-Ortiz, Tatiana Nunez-Garcia, Gisela Barrera-Badillo, Lucia Hernandez-Rivas, Irma Lopez-Martinez, Ernesto Ramirez-Gonzalez. |
| EPI_ISL_2340969, EPI_ISL_2340970, EPI_ISL_2340971 | LESP Veracruz | Instituto de Diagnostico y Referencia Epidemiologicos (INDRE) | Claudia Wong-Arambula, Abril Rodriguez-Maldonado, Vanessa Rivero-Arredondo, Ariadna Medina-Benitez, Joaquin Quiroz-Mercado, Sergio Rangel-Guerrero, Natividad Cruz-Ortiz, Tatiana Nunez-Garcia, Gisela Barrera-Badillo, Lucia Hernandez-Rivas, Irma Lopez-Martinez, Ernesto Ramirez-Gonzalez. |
| EPI_ISL_2340972, EPI_ISL_2340973 | LESP Sinaloa | Instituto de Diagnostico y Referencia Epidemiologicos (INDRE) | Claudia Wong-Arambula, Abril Rodriguez-Maldonado, Vanessa Rivero-Arredondo, Ariadna Medina-Benitez, Joaquin Quiroz-Mercado, Sergio Rangel-Guerrero, Natividad Cruz-Ortiz, Tatiana Nunez-Garcia, Gisela Barrera-Badillo, Lucia Hernandez-Rivas, Irma Lopez-Martinez, Ernesto Ramirez-Gonzalez. |
| EPI_ISL_2340974, EPI_ISL_2340975 | LESP Tamaulipas | Instituto de Diagnostico y Referencia Epidemiologicos (INDRE) | Claudia Wong-Arambula, Abril Rodriguez-Maldonado, Vanessa Rivero-Arredondo, Ariadna Medina-Benitez, Joaquin Quiroz-Mercado, Sergio Rangel-Guerrero, Natividad Cruz-Ortiz, Tatiana Nunez-Garcia, Gisela Barrera-Badillo, Lucia Hernandez-Rivas, Irma Lopez-Martinez, Ernesto Ramirez-Gonzalez. |
| EPI_ISL_2340976, EPI_ISL_2340977 | LESP Ciudad de Mexico | Instituto de Diagnostico y Referencia Epidemiologicos (INDRE) | Claudia Wong-Arambula, Abril Rodriguez-Maldonado, Vanessa Rivero-Arredondo, Ariadna Medina-Benitez, Joaquin Quiroz-Mercado, Sergio Rangel-Guerrero, Natividad Cruz-Ortiz, Tatiana Nunez-Garcia, Gisela Barrera-Badillo, Lucia Hernandez-Rivas, Irma Lopez-Martinez, Ernesto Ramirez-Gonzalez. |
| EPI_ISL_2340978 | LESP Sonora | Instituto de Diagnostico y Referencia Epidemiologicos (INDRE) | Claudia Wong-Arambula, Abril Rodriguez-Maldonado, Vanessa Rivero-Arredondo, Ariadna Medina-Benitez, Joaquin Quiroz-Mercado, Sergio Rangel-Guerrero, Natividad Cruz-Ortiz, Tatiana Nunez-Garcia, Gisela Barrera-Badillo, Lucia Hernandez-Rivas, Irma Lopez-Martinez, Ernesto Ramirez-Gonzalez. |
| EPI_ISL_2340979 | LESP Baja California | Instituto de Diagnostico y Referencia Epidemiologicos (INDRE) | Claudia Wong-Arambula, Abril Rodriguez-Maldonado, Vanessa Rivero-Arredondo, Ariadna Medina-Benitez, Joaquin Quiroz-Mercado, Sergio Rangel-Guerrero, Natividad Cruz-Ortiz, Tatiana Nunez-Garcia, Gisela Barrera-Badillo, Lucia Hernandez-Rivas, Irma Lopez-Martinez, Ernesto Ramirez-Gonzalez. |
| EPI_ISL_2340980, EPI_ISL_2340981, EPI_ISL_2340982, EPI_ISL_2340983, EPI_ISL_2340984, EPI_ISL_2340985, EPI_ISL_2340986, EPI_ISL_2340987, EPI_ISL_2340988, EPI_ISL_2340989, EPI_ISL_2340990, EPI_ISL_2340991, EPI_ISL_2340992, EPI_ISL_2340993, EPI_ISL_2340994, EPI_ISL_2340995, EPI_ISL_2340996 | | | |
| see above | LESP Guanajuato | Instituto de Diagnostico y Referencia Epidemiologicos (INDRE) | Claudia Wong-Arambula, Abril Rodriguez-Maldonado, Vanessa Rivero-Arredondo, Ariadna Medina-Benitez, Joaquin Quiroz-Mercado, Sergio Rangel-Guerrero, Natividad Cruz-Ortiz, Tatiana Nunez-Garcia, Gisela Barrera-Badillo, Lucia Hernandez-Rivas, Irma Lopez-Martinez, Ernesto Ramirez-Gonzalez. |
| EPI_ISL_2340997 | LESP Tamaulipas | Instituto de Diagnostico y Referencia Epidemiologicos (INDRE) | Claudia Wong-Arambula, Abril Rodriguez-Maldonado, Vanessa Rivero-Arredondo, Ariadna Medina-Benitez, Joaquin Quiroz-Mercado, Sergio Rangel-Guerrero, Natividad Cruz-Ortiz, Tatiana Nunez-Garcia, Gisela Barrera-Badillo, Lucia Hernandez-Rivas, Irma Lopez-Martinez, Ernesto Ramirez-Gonzalez. |
| EPI_ISL_2340998, EPI_ISL_2340999, EPI_ISL_2341000, EPI_ISL_2341001, EPI_ISL_2341002, EPI_ISL_2341003, EPI_ISL_2341004, EPI_ISL_2341005 | LESP Yucatan | Instituto de Diagnostico y Referencia Epidemiologicos (INDRE) | Claudia Wong-Arambula, Abril Rodriguez-Maldonado, Vanessa Rivero-Arredondo, Ariadna Medina-Benitez, Joaquin Quiroz-Mercado, Sergio Rangel-Guerrero, Natividad Cruz-Ortiz, Tatiana Nunez-Garcia, Gisela Barrera-Badillo, Lucia Hernandez-Rivas, Irma Lopez-Martinez, Ernesto Ramirez-Gonzalez. |
| EPI_ISL_2341006 | LESP Ciudad de Mexico | Instituto de Diagnostico y Referencia Epidemiologicos (INDRE) | Claudia Wong-Arambula, Abril Rodriguez-Maldonado, Vanessa Rivero-Arredondo, Ariadna Medina-Benitez, Joaquin Quiroz-Mercado, Sergio Rangel-Guerrero, Natividad Cruz-Ortiz, Tatiana Nunez-Garcia, Gisela Barrera-Badillo, Lucia Hernandez-Rivas, Irma Lopez-Martinez, Ernesto Ramirez-Gonzalez. |
| EPI_ISL_2341007 | LESP Jalisco | Instituto de Diagnostico y Referencia | Claudia Wong-Arambula, Abril Rodriguez-Maldonado, Vanessa Rivero-Arredondo, Ariadna Medina-Benitez, Joaquin Quiroz-Mercado, Sergio Rangel-Guerrero, Natividad Cruz-Ortiz, Tatiana Nunez-Garcia, Gisela Barrera-Badillo, Lucia Hernandez-Rivas, Irma Lopez-Martinez, Ernesto |

| | | | |
|---|---|---|---|
| | | Epidemiologicos (INDRE) | Ramirez-Gonzalez. |
| EPI_ISL_2341008 | LESP Queretaro | Instituto de Diagnostico y Referencia Epidemiologicos (INDRE) | Claudia Wong-Arambula, Abril Rodriguez-Maldonado, Vanessa Rivero-Arredondo, Ariadna Medina-Benitez, Joaquin Quiroz-Mercado, Sergio Rangel-Guerrero, Natividad Cruz-Ortiz, Tatiana Nunez-Garcia, Gisela Barrera-Badillo, Lucia Hernandez-Rivas, Irma Lopez-Martinez, Ernesto Ramirez-Gonzalez. |
| EPI_ISL_2341009, EPI_ISL_2341010, EPI_ISL_2341011, EPI_ISL_2341012 | LESP Guanajuato | Instituto de Diagnostico y Referencia Epidemiologicos (INDRE) | Claudia Wong-Arambula, Abril Rodriguez-Maldonado, Vanessa Rivero-Arredondo, Ariadna Medina-Benitez, Joaquin Quiroz-Mercado, Sergio Rangel-Guerrero, Natividad Cruz-Ortiz, Tatiana Nunez-Garcia, Gisela Barrera-Badillo, Lucia Hernandez-Rivas, Irma Lopez-Martinez, Ernesto Ramirez-Gonzalez. |
| EPI_ISL_2341013 | LESP Baja California Sur | Instituto de Diagnostico y Referencia Epidemiologicos (INDRE) | Claudia Wong-Arambula, Abril Rodriguez-Maldonado, Vanessa Rivero-Arredondo, Ariadna Medina-Benitez, Joaquin Quiroz-Mercado, Sergio Rangel-Guerrero, Natividad Cruz-Ortiz, Tatiana Nunez-Garcia, Gisela Barrera-Badillo, Lucia Hernandez-Rivas, Irma Lopez-Martinez, Ernesto Ramirez-Gonzalez. |
| EPI_ISL_2341014, EPI_ISL_2341015 | LESP Morelos | Instituto de Diagnostico y Referencia Epidemiologicos (INDRE) | Claudia Wong-Arambula, Abril Rodriguez-Maldonado, Vanessa Rivero-Arredondo, Ariadna Medina-Benitez, Joaquin Quiroz-Mercado, Sergio Rangel-Guerrero, Natividad Cruz-Ortiz, Tatiana Nunez-Garcia, Gisela Barrera-Badillo, Lucia Hernandez-Rivas, Irma Lopez-Martinez, Ernesto Ramirez-Gonzalez. |
| EPI_ISL_2341016, EPI_ISL_2341017, EPI_ISL_2341018, EPI_ISL_2341019, EPI_ISL_2341020 | LESP Guanajuato | Instituto de Diagnostico y Referencia Epidemiologicos (INDRE) | Claudia Wong-Arambula, Abril Rodriguez-Maldonado, Vanessa Rivero-Arredondo, Ariadna Medina-Benitez, Joaquin Quiroz-Mercado, Sergio Rangel-Guerrero, Natividad Cruz-Ortiz, Tatiana Nunez-Garcia, Gisela Barrera-Badillo, Lucia Hernandez-Rivas, Irma Lopez-Martinez, Ernesto Ramirez-Gonzalez. |
| EPI_ISL_2341021, EPI_ISL_2341022 | LESP Nuevo Leon | Instituto de Diagnostico y Referencia Epidemiologicos (INDRE) | Claudia Wong-Arambula, Abril Rodriguez-Maldonado, Vanessa Rivero-Arredondo, Ariadna Medina-Benitez, Joaquin Quiroz-Mercado, Sergio Rangel-Guerrero, Natividad Cruz-Ortiz, Tatiana Nunez-Garcia, Gisela Barrera-Badillo, Lucia Hernandez-Rivas, Irma Lopez-Martinez, Ernesto Ramirez-Gonzalez. |
| EPI_ISL_2341023 | LESP Campeche | Instituto de Diagnostico y Referencia Epidemiologicos (INDRE) | Claudia Wong-Arambula, Abril Rodriguez-Maldonado, Vanessa Rivero-Arredondo, Ariadna Medina-Benitez, Joaquin Quiroz-Mercado, Sergio Rangel-Guerrero, Natividad Cruz-Ortiz, Tatiana Nunez-Garcia, Gisela Barrera-Badillo, Lucia Hernandez-Rivas, Irma Lopez-Martinez, Ernesto Ramirez-Gonzalez. |
| EPI_ISL_2341024, EPI_ISL_2341025, EPI_ISL_2341026, EPI_ISL_2341027 | LESP Yucatan | Instituto de Diagnostico y Referencia Epidemiologicos (INDRE) | Claudia Wong-Arambula, Abril Rodriguez-Maldonado, Vanessa Rivero-Arredondo, Ariadna Medina-Benitez, Joaquin Quiroz-Mercado, Sergio Rangel-Guerrero, Natividad Cruz-Ortiz, Tatiana Nunez-Garcia, Gisela Barrera-Badillo, Lucia Hernandez-Rivas, Irma Lopez-Martinez, Ernesto Ramirez-Gonzalez. |
| EPI_ISL_2341028 | Salud Digna | Instituto Nacional de Medicina Genomica | Hidalgo-Miranda A, Cedro-Tanda A, Cisneros-Villanueva M, Orjuela-Rodríguez M, Mendoza-Vargas A, Reyes-Grajeda JP, Abraham Campos-Romero, Moreno-Camacho José Luis, Rodriguez-Gallegos Jorge, Luna-Ruiz Marco, Gonzalez-Barrera D, Rangel-DeLeon D, Munguia-Garza P, Ramirez-Vega O, Escobar-Arrazola, M, Herrera-Montalvo LA. |
| EPI_ISL_2341029 | INDRE | Instituto Nacional de Medicina Genomica | Hidalgo-Miranda A, Mendoza-Vargas A, Reyes-Grajeda JP, Cedro-Tanda A, Cisneros-Villanueva M, Orjuela-Rodríguez M, Gisela Barrera-Badillo, Irma Lopez-Martinez, Jose Ernesto Ramirez González, Gonzalez-Barrera D, Rangel-DeLeon D, Munguia-Garza P, Garcia-Cardenas FJ, Gonzalez-Woge MA, Herrera-Montalvo LA. |
| EPI_ISL_2341030 | Salud Digna | Instituto Nacional de Medicina Genomica | Hidalgo-Miranda A, Cedro-Tanda A, Cisneros-Villanueva M, Orjuela-Rodríguez M, Mendoza-Vargas A, Reyes-Grajeda JP, Abraham Campos-Romero, Moreno-Camacho José Luis, Rodriguez-Gallegos Jorge, Luna-Ruiz Marco, Gonzalez-Barrera D, Rangel-DeLeon D, Munguia-Garza P, Ramirez-Vega O, Escobar-Arrazola, M, Herrera-Montalvo LA. |
| EPI_ISL_2341031 | INDRE | Instituto Nacional de Medicina Genomica | Hidalgo-Miranda A, Mendoza-Vargas A, Reyes-Grajeda JP, Cedro-Tanda A, Cisneros-Villanueva M, Orjuela-Rodríguez M, Gisela Barrera-Badillo, Irma Lopez-Martinez, Jose Ernesto Ramirez González, Gonzalez-Barrera D, Rangel-DeLeon D, Munguia-Garza P, Garcia-Cardenas FJ, Gonzalez-Woge MA, Herrera-Montalvo LA. |
| EPI_ISL_2341032, EPI_ISL_2341035, EPI_ISL_2341036, EPI_ISL_2341037, EPI_ISL_2341038, EPI_ISL_2341039 | Salud Digna | Instituto Nacional de Medicina Genomica | Hidalgo-Miranda A, Cedro-Tanda A, Cisneros-Villanueva M, Orjuela-Rodríguez M, Mendoza-Vargas A, Reyes-Grajeda JP, Abraham Campos-Romero, Moreno-Camacho José Luis, Rodriguez-Gallegos Jorge, Luna-Ruiz Marco, Gonzalez-Barrera D, Rangel-DeLeon D, Munguia-Garza P, Ramirez-Vega O, Escobar-Arrazola, M, Herrera-Montalvo LA. |
| EPI_ISL_2341040, EPI_ISL_2341041 | INDRE | Instituto Nacional de Medicina Genomica | Hidalgo-Miranda A, Mendoza-Vargas A, Reyes-Grajeda JP, Cedro-Tanda A, Cisneros-Villanueva M, Orjuela-Rodríguez M, Gisela Barrera-Badillo, Irma Lopez-Martinez, Jose Ernesto Ramirez González, Gonzalez-Barrera D, Rangel-DeLeon D, Munguia-Garza P, Garcia-Cardenas FJ, Gonzalez-Woge MA, Herrera-Montalvo LA. |
| EPI_ISL_2341042, EPI_ISL_2341043, EPI_ISL_2341044, EPI_ISL_2341045 | Salud Digna | Instituto Nacional de Medicina Genomica | Hidalgo-Miranda A, Cedro-Tanda A, Cisneros-Villanueva M, Orjuela-Rodríguez M, Mendoza-Vargas A, Reyes-Grajeda JP, Abraham Campos-Romero, Moreno-Camacho José Luis, Rodriguez-Gallegos Jorge, Luna-Ruiz Marco, Gonzalez-Barrera D, Rangel-DeLeon D, Munguia-Garza P, Ramirez-Vega O, Escobar-Arrazola, M, Herrera-Montalvo LA. |
| EPI_ISL_2341046, EPI_ISL_2341047, EPI_ISL_2341048, EPI_ISL_2341049, EPI_ISL_2341050, EPI_ISL_2341051, EPI_ISL_2341052, EPI_ISL_2341053, EPI_ISL_2341054, EPI_ISL_2341055, EPI_ISL_2341056, EPI_ISL_2341057, EPI_ISL_2341058, EPI_ISL_2341059, EPI_ISL_2341060, EPI_ISL_2341061, EPI_ISL_2341062, EPI_ISL_2341063, EPI_ISL_2341064, EPI_ISL_2341065, EPI_ISL_2341066, EPI_ISL_2341067, EPI_ISL_2341068, EPI_ISL_2341069, EPI_ISL_2341070, EPI_ISL_2341071, EPI_ISL_2341072, EPI_ISL_2341073, EPI_ISL_2341074, EPI_ISL_2341075, EPI_ISL_2341076, EPI_ISL_2341077, EPI_ISL_2341078, EPI_ISL_2341079, EPI_ISL_2341080, EPI_ISL_2341081, EPI_ISL_2341082, EPI_ISL_2341083, EPI_ISL_2341084, EPI_ISL_2341085, EPI_ISL_2341086, EPI_ISL_2341087, EPI_ISL_2341088, EPI_ISL_2341089, EPI_ISL_2341090, EPI_ISL_2341091, EPI_ISL_2341092, EPI_ISL_2341093, EPI_ISL_2341094, EPI_ISL_2341095, EPI_ISL_2341096, EPI_ISL_2341097, EPI_ISL_2341098, EPI_ISL_2341099, EPI_ISL_2341100, EPI_ISL_2341101, EPI_ISL_2341102, EPI_ISL_2341103, EPI_ISL_2341104, EPI_ISL_2341105, EPI_ISL_2341106, EPI_ISL_2341107, EPI_ISL_2341108, EPI_ISL_2341109, EPI_ISL_2341110, EPI_ISL_2341111, EPI_ISL_2341112, EPI_ISL_2341113, EPI_ISL_2341114, EPI_ISL_2341115, EPI_ISL_2341116, EPI_ISL_2341117, EPI_ISL_2341118, EPI_ISL_2341119, EPI_ISL_2341120, EPI_ISL_2341121, EPI_ISL_2341122, EPI_ISL_2341123, EPI_ISL_2341124, EPI_ISL_2341125, EPI_ISL_2341126, EPI_ISL_2341127, EPI_ISL_2341128, EPI_ISL_2341129, EPI_ISL_2341130, EPI_ISL_2341131, EPI_ISL_2341132, EPI_ISL_2341133, EPI_ISL_2341134, EPI_ISL_2341135, EPI_ISL_2341136, EPI_ISL_2341137, EPI_ISL_2341138, EPI_ISL_2341139, EPI_ISL_2341140, EPI_ISL_2341141, EPI_ISL_2341142, EPI_ISL_2341143, EPI_ISL_2341144, EPI_ISL_2341145, EPI_ISL_2341146, EPI_ISL_2341147, EPI_ISL_2341148, EPI_ISL_2341149, EPI_ISL_2341150, EPI_ISL_2341151, EPI_ISL_2341152, EPI_ISL_2341153, EPI_ISL_2341154, EPI_ISL_2341155, EPI_ISL_2341156, EPI_ISL_2341157, EPI_ISL_2341158, EPI_ISL_2341159, EPI_ISL_2341160, EPI_ISL_2341161, EPI_ISL_2341162, EPI_ISL_2341163, EPI_ISL_2341164, EPI_ISL_2341165, EPI_ISL_2341166, EPI_ISL_2341167, EPI_ISL_2341168, EPI_ISL_2341169, EPI_ISL_2341170, EPI_ISL_2341171, EPI_ISL_2341172, EPI_ISL_2341173, EPI_ISL_2341174, EPI_ISL_2341175, EPI_ISL_2341176, EPI_ISL_2341177, EPI_ISL_2341178, EPI_ISL_2341179, EPI_ISL_2341180, EPI_ISL_2341181, EPI_ISL_2341182, EPI_ISL_2341183, EPI_ISL_2341184, EPI_ISL_2341185, EPI_ISL_2341186, EPI_ISL_2341187, EPI_ISL_2341188, EPI_ISL_2341189, EPI_ISL_2341190, EPI_ISL_2341191, EPI_ISL_2341192, EPI_ISL_2341193, EPI_ISL_2341194, EPI_ISL_2341195, EPI_ISL_2341196, EPI_ISL_2341197, EPI_ISL_2341198, EPI_ISL_2341199, EPI_ISL_2341200, EPI_ISL_2341201, EPI_ISL_2341202, EPI_ISL_2341203, EPI_ISL_2341204, EPI_ISL_2341205, EPI_ISL_2341206, EPI_ISL_2341207, EPI_ISL_2341208, EPI_ISL_2341209, EPI_ISL_2341210, EPI_ISL_2341211, EPI_ISL_2341212, EPI_ISL_2341213, EPI_ISL_2341214, EPI_ISL_2341215, EPI_ISL_2341216, EPI_ISL_2341217, EPI_ISL_2341218, EPI_ISL_2341219, EPI_ISL_2341220, EPI_ISL_2341221, EPI_ISL_2341222, EPI_ISL_2341223, EPI_ISL_2341224, EPI_ISL_2341225, EPI_ISL_2341226, EPI_ISL_2341227, EPI_ISL_2341228, EPI_ISL_2341229, EPI_ISL_2341230, EPI_ISL_2341231, EPI_ISL_2341232, EPI_ISL_2341233, EPI_ISL_2341234, EPI_ISL_2341235, EPI_ISL_2341236, EPI_ISL_2341237, EPI_ISL_2341238, EPI_ISL_2341239, EPI_ISL_2341240, EPI_ISL_2341241, EPI_ISL_2341242, EPI_ISL_2341243, EPI_ISL_2341244, EPI_ISL_2341245, EPI_ISL_2341246, EPI_ISL_2341247, EPI_ISL_2341248, EPI_ISL_2341249, EPI_ISL_2341250, EPI_ISL_2341251, EPI_ISL_2341252, EPI_ISL_2341253, EPI_ISL_2341254, EPI_ISL_2341255, EPI_ISL_2341256, EPI_ISL_2341257, EPI_ISL_2341258, EPI_ISL_2341259, EPI_ISL_2341260, EPI_ISL_2341261, EPI_ISL_2341262, EPI_ISL_2341263, EPI_ISL_2341264, EPI_ISL_2341265, EPI_ISL_2341266, EPI_ISL_2341267, EPI_ISL_2341268, EPI_ISL_2341269, EPI_ISL_2341270, EPI_ISL_2341271, EPI_ISL_2341272, EPI_ISL_2341273, EPI_ISL_2341274, EPI_ISL_2341275, EPI_ISL_2341276, EPI_ISL_2341277, EPI_ISL_2341278, EPI_ISL_2341279, EPI_ISL_2341280, EPI_ISL_2341281, EPI_ISL_2341282, EPI_ISL_2341283, EPI_ISL_2341284, EPI_ISL_2341285, EPI_ISL_2341286, EPI_ISL_2341287, EPI_ISL_2341288, EPI_ISL_2341289, EPI_ISL_2341290, EPI_ISL_2341291, EPI_ISL_2341292, EPI_ISL_2341293, EPI_ISL_2341294, EPI_ISL_2341295, EPI_ISL_2341296, EPI_ISL_2341297, EPI_ISL_2341298, EPI_ISL_2341299, EPI_ISL_2341300, EPI_ISL_2341301, EPI_ISL_2341302, EPI_ISL_2341303, EPI_ISL_2341304, EPI_ISL_2341305, EPI_ISL_2341306, EPI_ISL_2341307, EPI_ISL_2341308, EPI_ISL_2341309, EPI_ISL_2341310, EPI_ISL_2341311, EPI_ISL_2341312, EPI_ISL_2341313, EPI_ISL_2341314 | | | |
| see above | Tulare County Public Health Laboratory | Tulare County Public Health Laboratory | Denise L Lopez, Joshua Kootstra |
| EPI_ISL_2341315, EPI_ISL_2341316, EPI_ISL_2341317, EPI_ISL_2341318, EPI_ISL_2341319, EPI_ISL_2341320, EPI_ISL_2341321, EPI_ISL_2341322, EPI_ISL_2341323, EPI_ISL_2341324, EPI_ISL_2341325, EPI_ISL_2341326, EPI_ISL_2341327, EPI_ISL_2341328, EPI_ISL_2341329, EPI_ISL_2341330, EPI_ISL_2341331, EPI_ISL_2341332, EPI_ISL_2341333, EPI_ISL_2341334, EPI_ISL_2341335, EPI_ISL_2341336, EPI_ISL_2341337, EPI_ISL_2341338, EPI_ISL_2341339, EPI_ISL_2341340, EPI_ISL_2341341, EPI_ISL_2341342, EPI_ISL_2341343, EPI_ISL_2341344, EPI_ISL_2341345, EPI_ISL_2341346 | | | |
| see above | Instituto Nacional de Medicina Genomica | Centro de Investigación en Enfermedades Infecciosas (CIENI), Instituto Nacional de Enfermedades Respiratorias (INER) | Matias-Florentino M, Perez-Garcia M, Hidalgo-Miranda A, Mendoza-Vargas A, Reyes-Grajeda JP, Cedro-Tanda A, Herrera-Montalvo LA, Reyes-Teran G, Avila-Rios S, Boukadida C |
| EPI_ISL_2341347, EPI_ISL_2341348, EPI_ISL_2341349, EPI_ISL_2341350, EPI_ISL_2341351, EPI_ISL_2341352, EPI_ISL_2341353, EPI_ISL_2341354, EPI_ISL_2341355, EPI_ISL_2341356, EPI_ISL_2341357, EPI_ISL_2341358, EPI_ISL_2341359, EPI_ISL_2341360, EPI_ISL_2341361, EPI_ISL_2341362, EPI_ISL_2341363, EPI_ISL_2341364, EPI_ISL_2341365, EPI_ISL_2341366, EPI_ISL_2341367, EPI_ISL_2341368, EPI_ISL_2341369, EPI_ISL_2341370, EPI_ISL_2341371, EPI_ISL_2341372, EPI_ISL_2341373, EPI_ISL_2341374, EPI_ISL_2341375, EPI_ISL_2341376, EPI_ISL_2341377, EPI_ISL_2341378, EPI_ISL_2341379, EPI_ISL_2341380, EPI_ISL_2341381, EPI_ISL_2341382, EPI_ISL_2341383, EPI_ISL_2341384, EPI_ISL_2341385, EPI_ISL_2341386, EPI_ISL_2341387, EPI_ISL_2341388, EPI_ISL_2341389, EPI_ISL_2341390, EPI_ISL_2341391, EPI_ISL_2341392, EPI_ISL_2341393, EPI_ISL_2341394, EPI_ISL_2341395, EPI_ISL_2341396, EPI_ISL_2341397, EPI_ISL_2341398, EPI_ISL_2341399, EPI_ISL_2341400, EPI_ISL_2341401, EPI_ISL_2341402, EPI_ISL_2341403, EPI_ISL_2341404, EPI_ISL_2341405, EPI_ISL_2341406, EPI_ISL_2341407, EPI_ISL_2341408, EPI_ISL_2341409, EPI_ISL_2341410, EPI_ISL_2341411 | | | |
| see above | The Ohio State University Applied Microbiology Services Laboratory | The Ohio State University Applied Microbiology Services Laboratory | Seth A. Faith PhD |
| EPI_ISL_2341412, EPI_ISL_2341413 | UCSC Genomics Institute | UCSC Genomics Institute | Mark Akeson, Ikenna Anigbogu, Eric Beraut, Isabel Bjork, Molly Cassatt-Johnstone, Terren Chang, Russell Corbett-Detig, Namrita Dhillon, Maximilian Haeussler, David Haussler, Angie Hinrichs, Miten Jain, Joshua Kapp, A. Marm Kilpatrick, Jakob McBroome, Hugh Olsen, Jeremy Sanford, Beth Shapiro, Michael Stone, Bryan Thornlow, Yatish Turakhia, Ciara Wanket |
| EPI_ISL_2341414 | Contra Costa County Public Health Lab | Chan-Zuckerberg Biohub | CZB Cliahub Consortium |
| EPI_ISL_2341415 | Humboldt County Public Health Laboratory | Chan-Zuckerberg Biohub | CZB Cliahub Consortium |
| EPI_ISL_2341416, EPI_ISL_2341417, EPI_ISL_2341418, EPI_ISL_2341419, EPI_ISL_2341420 | CA DPH Viral and Rickettsial Disease Laboratory | Chan-Zuckerberg Biohub | CZB Cliahub Consortium |
| EPI_ISL_2341421 | Humboldt County Public Health Laboratory | Chan-Zuckerberg Biohub | CZB Cliahub Consortium |
| EPI_ISL_2341422 | CA DPH Viral and Rickettsial Disease Laboratory | Chan-Zuckerberg Biohub | CZB Cliahub Consortium |
| EPI_ISL_2341423, EPI_ISL_2341424 | Contra Costa County Public Health Lab | Chan-Zuckerberg Biohub | CZB Cliahub Consortium |
| EPI_ISL_2341425, EPI_ISL_2341426 | CA DPH Viral and Rickettsial Disease Laboratory | Chan-Zuckerberg Biohub | CZB Cliahub Consortium |
| EPI_ISL_2341427 | Contra Costa County Public Health Lab | Chan-Zuckerberg Biohub | CZB Cliahub Consortium |
| EPI_ISL_2341428 | CA DPH Viral and Rickettsial Disease Laboratory | Chan-Zuckerberg Biohub | CZB Cliahub Consortium |
| EPI_ISL_2341429 | Contra Costa County Public Health Lab | Chan-Zuckerberg Biohub | CZB Cliahub Consortium |
| EPI_ISL_2341430 | CA DPH Viral and Rickettsial Disease Laboratory | Chan-Zuckerberg Biohub | CZB Cliahub Consortium |
| EPI_ISL_2341431, EPI_ISL_2341432 | Contra Costa County Public Health Lab | Chan-Zuckerberg Biohub | CZB Cliahub Consortium |
| EPI_ISL_2341433, | CA DPH Viral and Rickettsial Disease | Chan-Zuckerberg Biohub | CZB Cliahub Consortium |

| | | | |
|---|---|---|---|
| EPI_ISL_2341434, EPI_ISL_2341435 | Laboratory | | |
| EPI_ISL_2341436 | Humboldt County Public Health Laboratory | Chan-Zuckerberg Biohub | CZB Cliahub Consortium |
| EPI_ISL_2341437 | Contra Costa County Public Health Lab | Chan-Zuckerberg Biohub | CZB Cliahub Consortium |
| EPI_ISL_2341438 | Santa Clara County Public Health Laboratory | Chan-Zuckerberg Biohub | CZB Cliahub Consortium |
| EPI_ISL_2341439 | Contra Costa County Public Health Lab | Chan-Zuckerberg Biohub | CZB Cliahub Consortium |
| EPI_ISL_2341440, EPI_ISL_2341441 | CA DPH Viral and Rickettsial Disease Laboratory | Chan-Zuckerberg Biohub | CZB Cliahub Consortium |
| EPI_ISL_2341442 | Humboldt County Public Health Laboratory | Chan-Zuckerberg Biohub | CZB Cliahub Consortium |
| EPI_ISL_2341443 | CA DPH Viral and Rickettsial Disease Laboratory | Chan-Zuckerberg Biohub | CZB Cliahub Consortium |
| EPI_ISL_2341444 | Contra Costa County Public Health Lab | Chan-Zuckerberg Biohub | CZB Cliahub Consortium |
| EPI_ISL_2341445, EPI_ISL_2341446 | CA DPH Viral and Rickettsial Disease Laboratory | Chan-Zuckerberg Biohub | CZB Cliahub Consortium |
| EPI_ISL_2341447 | Contra Costa County Public Health Lab | Chan-Zuckerberg Biohub | CZB Cliahub Consortium |
| EPI_ISL_2341449 | Humboldt County Public Health Laboratory | Chan-Zuckerberg Biohub | CZB Cliahub Consortium |
| EPI_ISL_2341450, EPI_ISL_2341451, EPI_ISL_2341452 | Contra Costa County Public Health Lab | Chan-Zuckerberg Biohub | CZB Cliahub Consortium |
| EPI_ISL_2341453 | CA DPH Viral and Rickettsial Disease Laboratory | Chan-Zuckerberg Biohub | CZB Cliahub Consortium |
| EPI_ISL_2341454, EPI_ISL_2341455, EPI_ISL_2341456, EPI_ISL_2341457 | Contra Costa County Public Health Lab | Chan-Zuckerberg Biohub | CZB Cliahub Consortium |
| EPI_ISL_2341458 | Humboldt County Public Health Laboratory | Chan-Zuckerberg Biohub | CZB Cliahub Consortium |
| EPI_ISL_2341459 | Contra Costa County Public Health Lab | Chan-Zuckerberg Biohub | CZB Cliahub Consortium |
| EPI_ISL_2341460 | Santa Clara County Public Health Laboratory | Chan-Zuckerberg Biohub | CZB Cliahub Consortium |
| EPI_ISL_2341462 | CA DPH Viral and Rickettsial Disease Laboratory | Chan-Zuckerberg Biohub | CZB Cliahub Consortium |
| EPI_ISL_2341463 | Humboldt County Public Health Laboratory | Chan-Zuckerberg Biohub | CZB Cliahub Consortium |
| EPI_ISL_2341464, EPI_ISL_2341465, EPI_ISL_2341466, EPI_ISL_2341467 | CA DPH Viral and Rickettsial Disease Laboratory | Chan-Zuckerberg Biohub | CZB Cliahub Consortium |
| EPI_ISL_2341468 | Contra Costa County Public Health Lab | Chan-Zuckerberg Biohub | CZB Cliahub Consortium |
| EPI_ISL_2341469 | Santa Clara County Public Health Laboratory | Chan-Zuckerberg Biohub | CZB Cliahub Consortium |
| EPI_ISL_2341470 | CA DPH Viral and Rickettsial Disease Laboratory | Chan-Zuckerberg Biohub | CZB Cliahub Consortium |
| EPI_ISL_2341471, EPI_ISL_2341472 | Humboldt County Public Health Laboratory | Chan-Zuckerberg Biohub | CZB Cliahub Consortium |
| EPI_ISL_2341473 | Santa Clara County Public Health Laboratory | Chan-Zuckerberg Biohub | CZB Cliahub Consortium |
| EPI_ISL_2341474, EPI_ISL_2341475, EPI_ISL_2341476, EPI_ISL_2341477 | CA DPH Viral and Rickettsial Disease Laboratory | Chan-Zuckerberg Biohub | CZB Cliahub Consortium |
| EPI_ISL_2341478 | Humboldt County Public Health Laboratory | Chan-Zuckerberg Biohub | CZB Cliahub Consortium |
| EPI_ISL_2341479 | CA DPH Viral and Rickettsial Disease Laboratory | Chan-Zuckerberg Biohub | CZB Cliahub Consortium |
| EPI_ISL_2341480 | Humboldt County Public Health Laboratory | Chan-Zuckerberg Biohub | CZB Cliahub Consortium |
| EPI_ISL_2341481 | Contra Costa County Public Health Lab | Chan-Zuckerberg Biohub | CZB Cliahub Consortium |
| EPI_ISL_2341482 | Santa Clara County Public Health Laboratory | Chan-Zuckerberg Biohub | CZB Cliahub Consortium |
| EPI_ISL_2341483, EPI_ISL_2341484, EPI_ISL_2341485, EPI_ISL_2341486 | CA DPH Viral and Rickettsial Disease Laboratory | Chan-Zuckerberg Biohub | CZB Cliahub Consortium |
| EPI_ISL_2341487, EPI_ISL_2341488 | Humboldt County Public Health Laboratory | Chan-Zuckerberg Biohub | CZB Cliahub Consortium |
| EPI_ISL_2341489 | Santa Clara County Public Health Laboratory | Chan-Zuckerberg Biohub | CZB Cliahub Consortium |
| EPI_ISL_2341490 | Contra Costa County Public Health Lab | Chan-Zuckerberg Biohub | CZB Cliahub Consortium |
| EPI_ISL_2341492 | CA DPH Viral and Rickettsial Disease Laboratory | Chan-Zuckerberg Biohub | CZB Cliahub Consortium |
| EPI_ISL_2341493, EPI_ISL_2341494 | Contra Costa County Public Health Lab | Chan-Zuckerberg Biohub | CZB Cliahub Consortium |
| EPI_ISL_2341495, EPI_ISL_2341496 | Santa Clara County Public Health Laboratory | Chan-Zuckerberg Biohub | CZB Cliahub Consortium |
| EPI_ISL_2341497 | Contra Costa County Public Health Lab | Chan-Zuckerberg Biohub | CZB Cliahub Consortium |
| EPI_ISL_2341498 | Humboldt County Public Health Laboratory | Chan-Zuckerberg Biohub | CZB Cliahub Consortium |
| EPI_ISL_2341499, EPI_ISL_2341500 | Contra Costa County Public Health Lab | Chan-Zuckerberg Biohub | CZB Cliahub Consortium |
| EPI_ISL_2341501 | Humboldt County Public Health Laboratory | Chan-Zuckerberg Biohub | CZB Cliahub Consortium |
| EPI_ISL_2341502, EPI_ISL_2341503, EPI_ISL_2341504 | CA DPH Viral and Rickettsial Disease Laboratory | Chan-Zuckerberg Biohub | CZB Cliahub Consortium |
| EPI_ISL_2341505 | Contra Costa County Public Health Lab | Chan-Zuckerberg Biohub | CZB Cliahub Consortium |

| | | | |
|---|---|---|---|
| EPI_ISL_2341506, EPI_ISL_2341507 | CA DPH Viral and Rickettsial Disease Laboratory | Chan-Zuckerberg Biohub | CZB Cliahub Consortium |
| EPI_ISL_2341508 | Contra Costa County Public Health Lab | Chan-Zuckerberg Biohub | CZB Cliahub Consortium |
| EPI_ISL_2341509 | Humboldt County Public Health Laboratory | Chan-Zuckerberg Biohub | CZB Cliahub Consortium |
| EPI_ISL_2341510 | CA DPH Viral and Rickettsial Disease Laboratory | Chan-Zuckerberg Biohub | CZB Cliahub Consortium |
| EPI_ISL_2341511 | Santa Clara County Public Health Laboratory | Chan-Zuckerberg Biohub | CZB Cliahub Consortium |
| EPI_ISL_2341512 | Contra Costa County Public Health Lab | Chan-Zuckerberg Biohub | CZB Cliahub Consortium |
| EPI_ISL_2341513, EPI_ISL_2341514 | CA DPH Viral and Rickettsial Disease Laboratory | Chan-Zuckerberg Biohub | CZB Cliahub Consortium |
| EPI_ISL_2341515, EPI_ISL_2341516 | Contra Costa County Public Health Lab | Chan-Zuckerberg Biohub | CZB Cliahub Consortium |
| EPI_ISL_2341517 | Santa Clara County Public Health Laboratory | Chan-Zuckerberg Biohub | CZB Cliahub Consortium |
| EPI_ISL_2341518 | Contra Costa County Public Health Lab | Chan-Zuckerberg Biohub | CZB Cliahub Consortium |
| EPI_ISL_2341519 | CA DPH Viral and Rickettsial Disease Laboratory | Chan-Zuckerberg Biohub | CZB Cliahub Consortium |
| EPI_ISL_2341520 | Contra Costa County Public Health Lab | Chan-Zuckerberg Biohub | CZB Cliahub Consortium |
| EPI_ISL_2341521, EPI_ISL_2341522, EPI_ISL_2341523 | CA DPH Viral and Rickettsial Disease Laboratory | Chan-Zuckerberg Biohub | CZB Cliahub Consortium |
| EPI_ISL_2341524, EPI_ISL_2341525, EPI_ISL_2341526 | Contra Costa County Public Health Lab | Chan-Zuckerberg Biohub | CZB Cliahub Consortium |
| EPI_ISL_2341527, EPI_ISL_2341528, EPI_ISL_2341529, EPI_ISL_2341530 | CA DPH Viral and Rickettsial Disease Laboratory | Chan-Zuckerberg Biohub | CZB Cliahub Consortium |
| EPI_ISL_2341531 | Santa Clara County Public Health Laboratory | Chan-Zuckerberg Biohub | CZB Cliahub Consortium |
| EPI_ISL_2341532 | CA DPH Viral and Rickettsial Disease Laboratory | Chan-Zuckerberg Biohub | CZB Cliahub Consortium |
| EPI_ISL_2341533 | Santa Clara County Public Health Laboratory | Chan-Zuckerberg Biohub | CZB Cliahub Consortium |
| EPI_ISL_2341534, EPI_ISL_2341535 | Contra Costa County Public Health Lab | Chan-Zuckerberg Biohub | CZB Cliahub Consortium |
| EPI_ISL_2341536 | Santa Clara County Public Health Laboratory | Chan-Zuckerberg Biohub | CZB Cliahub Consortium |
| EPI_ISL_2341537, EPI_ISL_2341538, EPI_ISL_2341539 | Contra Costa County Public Health Lab | Chan-Zuckerberg Biohub | CZB Cliahub Consortium |
| EPI_ISL_2341540 | CA DPH Viral and Rickettsial Disease Laboratory | Chan-Zuckerberg Biohub | CZB Cliahub Consortium |
| EPI_ISL_2341541 | Contra Costa County Public Health Lab | Chan-Zuckerberg Biohub | CZB Cliahub Consortium |
| EPI_ISL_2341542, EPI_ISL_2341543 | Humboldt County Public Health Laboratory | Chan-Zuckerberg Biohub | CZB Cliahub Consortium |
| EPI_ISL_2341544 | Contra Costa County Public Health Lab | Chan-Zuckerberg Biohub | CZB Cliahub Consortium |
| EPI_ISL_2341545, EPI_ISL_2341546 | CA DPH Viral and Rickettsial Disease Laboratory | Chan-Zuckerberg Biohub | CZB Cliahub Consortium |
| EPI_ISL_2341547 | Contra Costa County Public Health Lab | Chan-Zuckerberg Biohub | CZB Cliahub Consortium |
| EPI_ISL_2341548, EPI_ISL_2341549, EPI_ISL_2341550 | CA DPH Viral and Rickettsial Disease Laboratory | Chan-Zuckerberg Biohub | CZB Cliahub Consortium |
| EPI_ISL_2341551 | Contra Costa County Public Health Lab | Chan-Zuckerberg Biohub | CZB Cliahub Consortium |
| EPI_ISL_2341552 | CA DPH Viral and Rickettsial Disease Laboratory | Chan-Zuckerberg Biohub | CZB Cliahub Consortium |
| EPI_ISL_2341553, EPI_ISL_2341554, EPI_ISL_2341555 | Contra Costa County Public Health Lab | Chan-Zuckerberg Biohub | CZB Cliahub Consortium |
| EPI_ISL_2341556 | County of San Luis Obispo Public Health Laboratory | Chan-Zuckerberg Biohub | CZB Cliahub Consortium |
| EPI_ISL_2341557 | Santa Clara County Public Health Laboratory | Chan-Zuckerberg Biohub | CZB Cliahub Consortium |
| EPI_ISL_2341558 | Contra Costa County Public Health Lab | Chan-Zuckerberg Biohub | CZB Cliahub Consortium |
| EPI_ISL_2341559 | CA DPH Viral and Rickettsial Disease Laboratory | Chan-Zuckerberg Biohub | CZB Cliahub Consortium |
| EPI_ISL_2341560 | Santa Clara County Public Health Laboratory | Chan-Zuckerberg Biohub | CZB Cliahub Consortium |
| EPI_ISL_2341561 | Contra Costa County Public Health Lab | Chan-Zuckerberg Biohub | CZB Cliahub Consortium |
| EPI_ISL_2341562 | CA DPH Viral and Rickettsial Disease Laboratory | Chan-Zuckerberg Biohub | CZB Cliahub Consortium |
| EPI_ISL_2341563, EPI_ISL_2341565 | Humboldt County Public Health Laboratory | Chan-Zuckerberg Biohub | CZB Cliahub Consortium |
| EPI_ISL_2341566 | CA DPH Viral and Rickettsial Disease Laboratory | Chan-Zuckerberg Biohub | CZB Cliahub Consortium |
| EPI_ISL_2341567 | Humboldt County Public Health Laboratory | Chan-Zuckerberg Biohub | CZB Cliahub Consortium |
| EPI_ISL_2341568, EPI_ISL_2341569 | CA DPH Viral and Rickettsial Disease Laboratory | Chan-Zuckerberg Biohub | CZB Cliahub Consortium |
| EPI_ISL_2341570 | Contra Costa County Public Health Lab | Chan-Zuckerberg Biohub | CZB Cliahub Consortium |
| EPI_ISL_2341571 | CA DPH Viral and Rickettsial Disease Laboratory | Chan-Zuckerberg Biohub | CZB Cliahub Consortium |
| EPI_ISL_2341572 | Humboldt County Public Health Laboratory | Chan-Zuckerberg Biohub | CZB Cliahub Consortium |
| EPI_ISL_2341573, EPI_ISL_2341574, EPI_ISL_2341575, EPI_ISL_2341576 | CA DPH Viral and Rickettsial Disease Laboratory | Chan-Zuckerberg Biohub | CZB Cliahub Consortium |

| | | | |
|---|---|---|---|
| EPI_ISL_2341577 | Contra Costa County Public Health Lab | Chan-Zuckerberg Biohub | CZB Cliahub Consortium |
| EPI_ISL_2341578, EPI_ISL_2341579, EPI_ISL_2341580, EPI_ISL_2341581, EPI_ISL_2341582, EPI_ISL_2341583 | CA DPH Viral and Rickettsial Disease Laboratory | Chan-Zuckerberg Biohub | CZB Cliahub Consortium |
| EPI_ISL_2341584 | Santa Clara County Public Health Laboratory | Chan-Zuckerberg Biohub | CZB Cliahub Consortium |
| EPI_ISL_2341585 | CA DPH Viral and Rickettsial Disease Laboratory | Chan-Zuckerberg Biohub | CZB Cliahub Consortium |
| EPI_ISL_2341586 | Contra Costa County Public Health Lab | Chan-Zuckerberg Biohub | CZB Cliahub Consortium |
| EPI_ISL_2341587, EPI_ISL_2341588 | CA DPH Viral and Rickettsial Disease Laboratory | Chan-Zuckerberg Biohub | CZB Cliahub Consortium |
| EPI_ISL_2341589, EPI_ISL_2341590 | Contra Costa County Public Health Lab | Chan-Zuckerberg Biohub | CZB Cliahub Consortium |
| EPI_ISL_2341591 | CA DPH Viral and Rickettsial Disease Laboratory | Chan-Zuckerberg Biohub | CZB Cliahub Consortium |
| EPI_ISL_2341592 | Santa Clara County Public Health Laboratory | Chan-Zuckerberg Biohub | CZB Cliahub Consortium |
| EPI_ISL_2341593 | Contra Costa County Public Health Lab | Chan-Zuckerberg Biohub | CZB Cliahub Consortium |
| EPI_ISL_2341594 | CA DPH Viral and Rickettsial Disease Laboratory | Chan-Zuckerberg Biohub | CZB Cliahub Consortium |
| EPI_ISL_2341595 | Humboldt County Public Health Laboratory | Chan-Zuckerberg Biohub | CZB Cliahub Consortium |
| EPI_ISL_2341596 | CA DPH Viral and Rickettsial Disease Laboratory | Chan-Zuckerberg Biohub | CZB Cliahub Consortium |
| EPI_ISL_2341597 | Contra Costa County Public Health Lab | Chan-Zuckerberg Biohub | CZB Cliahub Consortium |
| EPI_ISL_2341598 | CA DPH Viral and Rickettsial Disease Laboratory | Chan-Zuckerberg Biohub | CZB Cliahub Consortium |
| EPI_ISL_2341599, EPI_ISL_2341600 | Contra Costa County Public Health Lab | Chan-Zuckerberg Biohub | CZB Cliahub Consortium |
| EPI_ISL_2341601 | CA DPH Viral and Rickettsial Disease Laboratory | Chan-Zuckerberg Biohub | CZB Cliahub Consortium |
| EPI_ISL_2341602 | Contra Costa County Public Health Lab | Chan-Zuckerberg Biohub | CZB Cliahub Consortium |
| EPI_ISL_2341604 | CA DPH Viral and Rickettsial Disease Laboratory | Chan-Zuckerberg Biohub | CZB Cliahub Consortium |
| EPI_ISL_2341605, EPI_ISL_2341606 | Contra Costa County Public Health Lab | Chan-Zuckerberg Biohub | CZB Cliahub Consortium |
| EPI_ISL_2341607 | CA DPH Viral and Rickettsial Disease Laboratory | Chan-Zuckerberg Biohub | CZB Cliahub Consortium |
| EPI_ISL_2341608 | Contra Costa County Public Health Lab | Chan-Zuckerberg Biohub | CZB Cliahub Consortium |
| EPI_ISL_2341609 | CA DPH Viral and Rickettsial Disease Laboratory | Chan-Zuckerberg Biohub | CZB Cliahub Consortium |
| EPI_ISL_2341610, EPI_ISL_2341611, EPI_ISL_2341612 | Contra Costa County Public Health Lab | Chan-Zuckerberg Biohub | CZB Cliahub Consortium |
| EPI_ISL_2341613 | Humboldt County Public Health Laboratory | Chan-Zuckerberg Biohub | CZB Cliahub Consortium |
| EPI_ISL_2341614, EPI_ISL_2341615 | Contra Costa County Public Health Lab | Chan-Zuckerberg Biohub | CZB Cliahub Consortium |
| EPI_ISL_2341616 | Humboldt County Public Health Laboratory | Chan-Zuckerberg Biohub | CZB Cliahub Consortium |
| EPI_ISL_2341617, EPI_ISL_2341618, EPI_ISL_2341619 | CA DPH Viral and Rickettsial Disease Laboratory | Chan-Zuckerberg Biohub | CZB Cliahub Consortium |
| EPI_ISL_2341621 | Humboldt County Public Health Laboratory | Chan-Zuckerberg Biohub | CZB Cliahub Consortium |
| EPI_ISL_2341622 | Contra Costa County Public Health Lab | Chan-Zuckerberg Biohub | CZB Cliahub Consortium |
| EPI_ISL_2341623 | Santa Clara County Public Health Laboratory | Chan-Zuckerberg Biohub | CZB Cliahub Consortium |
| EPI_ISL_2341624 | Contra Costa County Public Health Lab | Chan-Zuckerberg Biohub | CZB Cliahub Consortium |
| EPI_ISL_2341625 | Humboldt County Public Health Laboratory | Chan-Zuckerberg Biohub | CZB Cliahub Consortium |
| EPI_ISL_2341626 | CA DPH Viral and Rickettsial Disease Laboratory | Chan-Zuckerberg Biohub | CZB Cliahub Consortium |
| EPI_ISL_2341627 | Humboldt County Public Health Laboratory | Chan-Zuckerberg Biohub | CZB Cliahub Consortium |
| EPI_ISL_2341628 | Contra Costa County Public Health Lab | Chan-Zuckerberg Biohub | CZB Cliahub Consortium |
| EPI_ISL_2341629 | Humboldt County Public Health Laboratory | Chan-Zuckerberg Biohub | CZB Cliahub Consortium |
| EPI_ISL_2341630 | Contra Costa County Public Health Lab | Chan-Zuckerberg Biohub | CZB Cliahub Consortium |
| EPI_ISL_2341631 | CA DPH Viral and Rickettsial Disease Laboratory | Chan-Zuckerberg Biohub | CZB Cliahub Consortium |
| EPI_ISL_2341632, EPI_ISL_2341633, EPI_ISL_2341634 | Contra Costa County Public Health Lab | Chan-Zuckerberg Biohub | CZB Cliahub Consortium |
| EPI_ISL_2341635 | CA DPH Viral and Rickettsial Disease Laboratory | Chan-Zuckerberg Biohub | CZB Cliahub Consortium |
| EPI_ISL_2341636 | Santa Clara County Public Health Laboratory | Chan-Zuckerberg Biohub | CZB Cliahub Consortium |
| EPI_ISL_2341638 | Contra Costa County Public Health Lab | Chan-Zuckerberg Biohub | CZB Cliahub Consortium |
| EPI_ISL_2341639 | CA DPH Viral and Rickettsial Disease Laboratory | Chan-Zuckerberg Biohub | CZB Cliahub Consortium |
| EPI_ISL_2341640 | Contra Costa County Public Health Lab | Chan-Zuckerberg Biohub | CZB Cliahub Consortium |
| EPI_ISL_2341641 | Santa Clara County Public Health Laboratory | Chan-Zuckerberg Biohub | CZB Cliahub Consortium |
| EPI_ISL_2341642, EPI_ISL_2341643 | Contra Costa County Public Health Lab | Chan-Zuckerberg Biohub | CZB Cliahub Consortium |

| | | | |
|---|---|---|---|
| EPI_ISL_2341644 | CA DPH Viral and Rickettsial Disease Laboratory | Chan-Zuckerberg Biohub | CZB Cliahub Consortium |
| EPI_ISL_2341645 | Contra Costa County Public Health Lab | Chan-Zuckerberg Biohub | CZB Cliahub Consortium |
| EPI_ISL_2341646, EPI_ISL_2341647, EPI_ISL_2341648 | CA DPH Viral and Rickettsial Disease Laboratory | Chan-Zuckerberg Biohub | CZB Cliahub Consortium |
| EPI_ISL_2341649 | Santa Clara County Public Health Laboratory | Chan-Zuckerberg Biohub | CZB Cliahub Consortium |
| EPI_ISL_2341650 | Humboldt County Public Health Laboratory | Chan-Zuckerberg Biohub | CZB Cliahub Consortium |
| EPI_ISL_2341651, EPI_ISL_2341652 | CA DPH Viral and Rickettsial Disease Laboratory | Chan-Zuckerberg Biohub | CZB Cliahub Consortium |
| EPI_ISL_2341654 | Contra Costa County Public Health Lab | Chan-Zuckerberg Biohub | CZB Cliahub Consortium |
| EPI_ISL_2341655 | Humboldt County Public Health Laboratory | Chan-Zuckerberg Biohub | CZB Cliahub Consortium |
| EPI_ISL_2341656, EPI_ISL_2341657 | Contra Costa County Public Health Lab | Chan-Zuckerberg Biohub | CZB Cliahub Consortium |
| EPI_ISL_2341658 | CA DPH Viral and Rickettsial Disease Laboratory | Chan-Zuckerberg Biohub | CZB Cliahub Consortium |
| EPI_ISL_2341659 | Contra Costa County Public Health Lab | Chan-Zuckerberg Biohub | CZB Cliahub Consortium |
| EPI_ISL_2341660 | CA DPH Viral and Rickettsial Disease Laboratory | Chan-Zuckerberg Biohub | CZB Cliahub Consortium |
| EPI_ISL_2341661, EPI_ISL_2341662, EPI_ISL_2341663, EPI_ISL_2341664 | Contra Costa County Public Health Lab | Chan-Zuckerberg Biohub | CZB Cliahub Consortium |
| EPI_ISL_2342365, EPI_ISL_2342366, EPI_ISL_2342367, EPI_ISL_2342368, EPI_ISL_2342369, EPI_ISL_2342370, EPI_ISL_2342371, EPI_ISL_2342372, EPI_ISL_2342373, EPI_ISL_2342374, EPI_ISL_2342375, EPI_ISL_2342376, EPI_ISL_2342377, EPI_ISL_2342378, EPI_ISL_2342379, EPI_ISL_2342380, EPI_ISL_2342381, EPI_ISL_2342382, EPI_ISL_2342383, EPI_ISL_2342384, EPI_ISL_2342385, EPI_ISL_2342386, EPI_ISL_2342387, EPI_ISL_2342388, EPI_ISL_2342389, EPI_ISL_2342390, EPI_ISL_2342391, EPI_ISL_2342392, EPI_ISL_2342393, EPI_ISL_2342394, EPI_ISL_2342395, EPI_ISL_2342396, EPI_ISL_2342397, EPI_ISL_2342398, EPI_ISL_2342399, EPI_ISL_2342400, EPI_ISL_2342401, EPI_ISL_2342402, EPI_ISL_2342403, EPI_ISL_2342404, EPI_ISL_2342405, EPI_ISL_2342406, EPI_ISL_2342407, EPI_ISL_2342408, EPI_ISL_2342409, EPI_ISL_2342410, EPI_ISL_2342411, EPI_ISL_2342412, EPI_ISL_2342413, EPI_ISL_2342414, EPI_ISL_2342415, EPI_ISL_2342416, EPI_ISL_2342417, EPI_ISL_2342418, EPI_ISL_2342419, EPI_ISL_2342420, EPI_ISL_2342421, EPI_ISL_2342422, EPI_ISL_2342423, EPI_ISL_2342424, EPI_ISL_2342425, EPI_ISL_2342426, EPI_ISL_2342427, EPI_ISL_2342428, EPI_ISL_2342429, EPI_ISL_2342430, EPI_ISL_2342431, EPI_ISL_2342432, EPI_ISL_2342433, EPI_ISL_2342434, EPI_ISL_2342435, EPI_ISL_2342436, EPI_ISL_2342437, EPI_ISL_2342438, EPI_ISL_2342439, EPI_ISL_2342440, EPI_ISL_2342441, EPI_ISL_2342442, EPI_ISL_2342443, EPI_ISL_2342444, EPI_ISL_2342445, EPI_ISL_2342446, EPI_ISL_2342447, EPI_ISL_2342448, EPI_ISL_2342449, EPI_ISL_2342450, EPI_ISL_2342451, EPI_ISL_2342452, EPI_ISL_2342453, EPI_ISL_2342454, EPI_ISL_2342455, EPI_ISL_2342456, EPI_ISL_2342457, EPI_ISL_2342458, EPI_ISL_2342459, EPI_ISL_2342460, EPI_ISL_2342461, EPI_ISL_2342462, EPI_ISL_2342463, EPI_ISL_2342464 | | | |
| see above | Utah Public Health Laboratory | Utah Public Health Laboratory | Erin L. Young, Kelly F. Oakeson, Tara Gallagher |
| EPI_ISL_2342981 | LESP Queretaro | Instituto de Diagnostico y Referencia Epidemiologicos (INDRE) | Claudia Wong-Arambula, Abril Rodriguez-Maldonado, Vanessa Rivero-Arredondo, Ariadna Medina-Benitez, Joaquin Quiroz-Mercado, Sergio Rangel-Guerrero, Natividad Cruz-Ortiz, Tatiana Nunez-Garcia, Gisela Barrera-Badillo, Lucia Hernandez-Rivas, Irma Lopez-Martinez, Ernesto Ramirez-Gonzalez. |
| EPI_ISL_2342982 | LESP Yucatan | Instituto de Diagnostico y Referencia Epidemiologicos (INDRE) | Claudia Wong-Arambula, Abril Rodriguez-Maldonado, Vanessa Rivero-Arredondo, Ariadna Medina-Benitez, Joaquin Quiroz-Mercado, Sergio Rangel-Guerrero, Natividad Cruz-Ortiz, Tatiana Nunez-Garcia, Gisela Barrera-Badillo, Lucia Hernandez-Rivas, Irma Lopez-Martinez, Ernesto Ramirez-Gonzalez. |
| EPI_ISL_2342983 | LESP Guanajuato | Instituto de Diagnostico y Referencia Epidemiologicos (INDRE) | Claudia Wong-Arambula, Abril Rodriguez-Maldonado, Vanessa Rivero-Arredondo, Ariadna Medina-Benitez, Joaquin Quiroz-Mercado, Sergio Rangel-Guerrero, Natividad Cruz-Ortiz, Tatiana Nunez-Garcia, Gisela Barrera-Badillo, Lucia Hernandez-Rivas, Irma Lopez-Martinez, Ernesto Ramirez-Gonzalez. |
| EPI_ISL_2342984 | LESP Tamaulipas | Instituto de Diagnostico y Referencia Epidemiologicos (INDRE) | Claudia Wong-Arambula, Abril Rodriguez-Maldonado, Vanessa Rivero-Arredondo, Ariadna Medina-Benitez, Joaquin Quiroz-Mercado, Sergio Rangel-Guerrero, Natividad Cruz-Ortiz, Tatiana Nunez-Garcia, Gisela Barrera-Badillo, Lucia Hernandez-Rivas, Irma Lopez-Martinez, Ernesto Ramirez-Gonzalez. |
| EPI_ISL_2342985 | LESP Sinaloa | Instituto de Diagnostico y Referencia Epidemiologicos (INDRE) | Claudia Wong-Arambula, Abril Rodriguez-Maldonado, Vanessa Rivero-Arredondo, Ariadna Medina-Benitez, Joaquin Quiroz-Mercado, Sergio Rangel-Guerrero, Natividad Cruz-Ortiz, Tatiana Nunez-Garcia, Gisela Barrera-Badillo, Lucia Hernandez-Rivas, Irma Lopez-Martinez, Ernesto Ramirez-Gonzalez. |
| EPI_ISL_2342986, EPI_ISL_2342987 | LESP Yucatan | Instituto de Diagnostico y Referencia Epidemiologicos (INDRE) | Claudia Wong-Arambula, Abril Rodriguez-Maldonado, Vanessa Rivero-Arredondo, Ariadna Medina-Benitez, Joaquin Quiroz-Mercado, Sergio Rangel-Guerrero, Natividad Cruz-Ortiz, Tatiana Nunez-Garcia, Gisela Barrera-Badillo, Lucia Hernandez-Rivas, Irma Lopez-Martinez, Ernesto Ramirez-Gonzalez. |
| EPI_ISL_2342988 | LESP Guanajuato | Instituto de Diagnostico y Referencia Epidemiologicos (INDRE) | Claudia Wong-Arambula, Abril Rodriguez-Maldonado, Vanessa Rivero-Arredondo, Ariadna Medina-Benitez, Joaquin Quiroz-Mercado, Sergio Rangel-Guerrero, Natividad Cruz-Ortiz, Tatiana Nunez-Garcia, Gisela Barrera-Badillo, Lucia Hernandez-Rivas, Irma Lopez-Martinez, Ernesto Ramirez-Gonzalez. |
| EPI_ISL_2342989, EPI_ISL_2342990 | LESP Ciudad de Mexico | Instituto de Diagnostico y Referencia Epidemiologicos (INDRE) | Claudia Wong-Arambula, Abril Rodriguez-Maldonado, Vanessa Rivero-Arredondo, Ariadna Medina-Benitez, Joaquin Quiroz-Mercado, Sergio Rangel-Guerrero, Natividad Cruz-Ortiz, Tatiana Nunez-Garcia, Gisela Barrera-Badillo, Lucia Hernandez-Rivas, Irma Lopez-Martinez, Ernesto Ramirez-Gonzalez. |
| EPI_ISL_2342991, EPI_ISL_2342992 | LESP Michoacan | Instituto de Diagnostico y Referencia Epidemiologicos (INDRE) | Claudia Wong-Arambula, Abril Rodriguez-Maldonado, Vanessa Rivero-Arredondo, Ariadna Medina-Benitez, Joaquin Quiroz-Mercado, Sergio Rangel-Guerrero, Natividad Cruz-Ortiz, Tatiana Nunez-Garcia, Gisela Barrera-Badillo, Lucia Hernandez-Rivas, Irma Lopez-Martinez, Ernesto Ramirez-Gonzalez. |
| EPI_ISL_2342993 | LESP Sinaloa | Instituto de Diagnostico y Referencia Epidemiologicos (INDRE) | Claudia Wong-Arambula, Abril Rodriguez-Maldonado, Vanessa Rivero-Arredondo, Ariadna Medina-Benitez, Joaquin Quiroz-Mercado, Sergio Rangel-Guerrero, Natividad Cruz-Ortiz, Tatiana Nunez-Garcia, Gisela Barrera-Badillo, Lucia Hernandez-Rivas, Irma Lopez-Martinez, Ernesto Ramirez-Gonzalez. |
| EPI_ISL_2342994 | LESP Queretaro | Instituto de Diagnostico y Referencia Epidemiologicos (INDRE) | Claudia Wong-Arambula, Abril Rodriguez-Maldonado, Vanessa Rivero-Arredondo, Ariadna Medina-Benitez, Joaquin Quiroz-Mercado, Sergio Rangel-Guerrero, Natividad Cruz-Ortiz, Tatiana Nunez-Garcia, Gisela Barrera-Badillo, Lucia Hernandez-Rivas, Irma Lopez-Martinez, Ernesto Ramirez-Gonzalez. |
| EPI_ISL_2342995 | LESP Yucatan | Instituto de Diagnostico y Referencia Epidemiologicos (INDRE) | Claudia Wong-Arambula, Abril Rodriguez-Maldonado, Vanessa Rivero-Arredondo, Ariadna Medina-Benitez, Joaquin Quiroz-Mercado, Sergio Rangel-Guerrero, Natividad Cruz-Ortiz, Tatiana Nunez-Garcia, Gisela Barrera-Badillo, Lucia Hernandez-Rivas, Irma Lopez-Martinez, Ernesto Ramirez-Gonzalez. |
| EPI_ISL_2342996, EPI_ISL_2342997 | LESP Ciudad de Mexico | Instituto de Diagnostico y Referencia Epidemiologicos (INDRE) | Claudia Wong-Arambula, Abril Rodriguez-Maldonado, Vanessa Rivero-Arredondo, Ariadna Medina-Benitez, Joaquin Quiroz-Mercado, Sergio Rangel-Guerrero, Natividad Cruz-Ortiz, Tatiana Nunez-Garcia, Gisela Barrera-Badillo, Lucia Hernandez-Rivas, Irma Lopez-Martinez, Ernesto Ramirez-Gonzalez. |
| EPI_ISL_2342998 | LESP Queretaro | Instituto de Diagnostico y Referencia Epidemiologicos (INDRE) | Claudia Wong-Arambula, Abril Rodriguez-Maldonado, Vanessa Rivero-Arredondo, Ariadna Medina-Benitez, Joaquin Quiroz-Mercado, Sergio Rangel-Guerrero, Natividad Cruz-Ortiz, Tatiana Nunez-Garcia, Gisela Barrera-Badillo, Lucia Hernandez-Rivas, Irma Lopez-Martinez, Ernesto Ramirez-Gonzalez. |
| EPI_ISL_2342999 | LESP Michoacan/LANS | Instituto de Diagnostico y Referencia Epidemiologicos (INDRE) | Claudia Wong-Arambula, Abril Rodriguez-Maldonado, Vanessa Rivero-Arredondo, Ariadna Medina-Benitez, Joaquin Quiroz-Mercado, Sergio Rangel-Guerrero, Natividad Cruz-Ortiz, Tatiana Nunez-Garcia, Gisela Barrera-Badillo, Lucia Hernandez-Rivas, Irma Lopez-Martinez, Ernesto Ramirez-Gonzalez. |
| EPI_ISL_2343000, EPI_ISL_2343001, EPI_ISL_2343002, EPI_ISL_2343003, EPI_ISL_2343004 | LESP Yucatan | Instituto de Diagnostico y Referencia Epidemiologicos (INDRE) | Claudia Wong-Arambula, Abril Rodriguez-Maldonado, Vanessa Rivero-Arredondo, Ariadna Medina-Benitez, Joaquin Quiroz-Mercado, Sergio Rangel-Guerrero, Natividad Cruz-Ortiz, Tatiana Nunez-Garcia, Gisela Barrera-Badillo, Lucia Hernandez-Rivas, Irma Lopez-Martinez, Ernesto Ramirez-Gonzalez. |
| EPI_ISL_2343005 | LESP Sinaloa | Instituto de Diagnostico y Referencia Epidemiologicos (INDRE) | Claudia Wong-Arambula, Abril Rodriguez-Maldonado, Vanessa Rivero-Arredondo, Ariadna Medina-Benitez, Joaquin Quiroz-Mercado, Sergio Rangel-Guerrero, Natividad Cruz-Ortiz, Tatiana Nunez-Garcia, Gisela Barrera-Badillo, Lucia Hernandez-Rivas, Irma Lopez-Martinez, Ernesto Ramirez-Gonzalez. |
| EPI_ISL_2343006, EPI_ISL_2343007 | LESP Ciudad de Mexico | Instituto de Diagnostico y Referencia Epidemiologicos (INDRE) | Claudia Wong-Arambula, Abril Rodriguez-Maldonado, Vanessa Rivero-Arredondo, Ariadna Medina-Benitez, Joaquin Quiroz-Mercado, Sergio Rangel-Guerrero, Natividad Cruz-Ortiz, Tatiana Nunez-Garcia, Gisela Barrera-Badillo, Lucia Hernandez-Rivas, Irma Lopez-Martinez, Ernesto Ramirez-Gonzalez. |
| EPI_ISL_2343008 | LESP Tamaulipas | Instituto de Diagnostico y Referencia Epidemiologicos (INDRE) | Claudia Wong-Arambula, Abril Rodriguez-Maldonado, Vanessa Rivero-Arredondo, Ariadna Medina-Benitez, Joaquin Quiroz-Mercado, Sergio Rangel-Guerrero, Natividad Cruz-Ortiz, Tatiana Nunez-Garcia, Gisela Barrera-Badillo, Lucia Hernandez-Rivas, Irma Lopez-Martinez, Ernesto Ramirez-Gonzalez. |
| EPI_ISL_2343009 | LESP Ciudad de Mexico | Instituto de Diagnostico y Referencia Epidemiologicos (INDRE) | Claudia Wong-Arambula, Abril Rodriguez-Maldonado, Vanessa Rivero-Arredondo, Ariadna Medina-Benitez, Joaquin Quiroz-Mercado, Sergio Rangel-Guerrero, Natividad Cruz-Ortiz, Tatiana Nunez-Garcia, Gisela Barrera-Badillo, Lucia Hernandez-Rivas, Irma Lopez-Martinez, Ernesto Ramirez-Gonzalez. |
| EPI_ISL_2343010 | LESP Hidalgo | Instituto de Diagnostico y Referencia Epidemiologicos (INDRE) | Claudia Wong-Arambula, Abril Rodriguez-Maldonado, Vanessa Rivero-Arredondo, Ariadna Medina-Benitez, Joaquin Quiroz-Mercado, Sergio Rangel-Guerrero, Natividad Cruz-Ortiz, Tatiana Nunez-Garcia, Gisela Barrera-Badillo, Lucia Hernandez-Rivas, Irma Lopez-Martinez, Ernesto Ramirez-Gonzalez. |
| EPI_ISL_2343011, EPI_ISL_2343012, EPI_ISL_2343013, EPI_ISL_2343014, EPI_ISL_2343015, EPI_ISL_2343016 | LESP Michoacan | Instituto de Diagnostico y Referencia Epidemiologicos (INDRE) | Claudia Wong-Arambula, Abril Rodriguez-Maldonado, Vanessa Rivero-Arredondo, Ariadna Medina-Benitez, Joaquin Quiroz-Mercado, Sergio Rangel-Guerrero, Natividad Cruz-Ortiz, Tatiana Nunez-Garcia, Gisela Barrera-Badillo, Lucia Hernandez-Rivas, Irma Lopez-Martinez, Ernesto Ramirez-Gonzalez. |
| EPI_ISL_2343017 | LESP Colima | Instituto de Diagnostico y Referencia Epidemiologicos (INDRE) | Claudia Wong-Arambula, Abril Rodriguez-Maldonado, Vanessa Rivero-Arredondo, Ariadna Medina-Benitez, Joaquin Quiroz-Mercado, Sergio Rangel-Guerrero, Natividad Cruz-Ortiz, Tatiana Nunez-Garcia, Gisela Barrera-Badillo, Lucia Hernandez-Rivas, Irma Lopez-Martinez, Ernesto Ramirez-Gonzalez. |
| EPI_ISL_2343018 | LESP Yucatan | Instituto de Diagnostico y Referencia Epidemiologicos (INDRE) | Claudia Wong-Arambula, Abril Rodriguez-Maldonado, Vanessa Rivero-Arredondo, Ariadna Medina-Benitez, Joaquin Quiroz-Mercado, Sergio Rangel-Guerrero, Natividad Cruz-Ortiz, Tatiana Nunez-Garcia, Gisela Barrera-Badillo, Lucia Hernandez-Rivas, Irma Lopez-Martinez, Ernesto Ramirez-Gonzalez. |
| EPI_ISL_2343019 | LESP Queretaro | Instituto de Diagnostico y Referencia Epidemiologicos (INDRE) | Claudia Wong-Arambula, Abril Rodriguez-Maldonado, Vanessa Rivero-Arredondo, Ariadna Medina-Benitez, Joaquin Quiroz-Mercado, Sergio Rangel-Guerrero, Natividad Cruz-Ortiz, Tatiana Nunez-Garcia, Gisela Barrera-Badillo, Lucia Hernandez-Rivas, Irma Lopez-Martinez, Ernesto Ramirez-Gonzalez. |
| EPI_ISL_2343020 | LESP Ciudad de Mexico /Hospital Angeles | Instituto de Diagnostico y Referencia Epidemiologicos (INDRE) | Claudia Wong-Arambula, Abril Rodriguez-Maldonado, Vanessa Rivero-Arredondo, Ariadna Medina-Benitez, Joaquin Quiroz-Mercado, Sergio Rangel-Guerrero, Natividad Cruz-Ortiz, Tatiana Nunez-Garcia, Gisela Barrera-Badillo, Lucia Hernandez-Rivas, Irma Lopez-Martinez, Ernesto Ramirez-Gonzalez. |

| | | | |
|---|---|---|---|
| EPI_ISL_2343021 | LESP Veracruz | Instituto de Diagnostico y Referencia Epidemiologicos (INDRE) | Claudia Wong-Arambula, Abril Rodriguez-Maldonado, Vanessa Rivero-Arredondo, Ariadna Medina-Benitez, Joaquin Quiroz-Mercado, Sergio Rangel-Guerrero, Natividad Cruz-Ortiz, Tatiana Nunez-Garcia, Gisela Barrera-Badillo, Lucia Hernandez-Rivas, Irma Lopez-Martinez, Ernesto Ramirez-Gonzalez. |
| EPI_ISL_2343022 | LESP Sinaloa | Instituto de Diagnostico y Referencia Epidemiologicos (INDRE) | Claudia Wong-Arambula, Abril Rodriguez-Maldonado, Vanessa Rivero-Arredondo, Ariadna Medina-Benitez, Joaquin Quiroz-Mercado, Sergio Rangel-Guerrero, Natividad Cruz-Ortiz, Tatiana Nunez-Garcia, Gisela Barrera-Badillo, Lucia Hernandez-Rivas, Irma Lopez-Martinez, Ernesto Ramirez-Gonzalez. |
| EPI_ISL_2343023 | LESP Tamaulipas | Instituto de Diagnostico y Referencia Epidemiologicos (INDRE) | Claudia Wong-Arambula, Abril Rodriguez-Maldonado, Vanessa Rivero-Arredondo, Ariadna Medina-Benitez, Joaquin Quiroz-Mercado, Sergio Rangel-Guerrero, Natividad Cruz-Ortiz, Tatiana Nunez-Garcia, Gisela Barrera-Badillo, Lucia Hernandez-Rivas, Irma Lopez-Martinez, Ernesto Ramirez-Gonzalez. |
| EPI_ISL_2343024 | LESP Chiapas | Instituto de Diagnostico y Referencia Epidemiologicos (INDRE) | Claudia Wong-Arambula, Abril Rodriguez-Maldonado, Vanessa Rivero-Arredondo, Ariadna Medina-Benitez, Joaquin Quiroz-Mercado, Sergio Rangel-Guerrero, Natividad Cruz-Ortiz, Tatiana Nunez-Garcia, Gisela Barrera-Badillo, Lucia Hernandez-Rivas, Irma Lopez-Martinez, Ernesto Ramirez-Gonzalez. |
| EPI_ISL_2343025 | LESP Sonora | Instituto de Diagnostico y Referencia Epidemiologicos (INDRE) | Claudia Wong-Arambula, Abril Rodriguez-Maldonado, Vanessa Rivero-Arredondo, Ariadna Medina-Benitez, Joaquin Quiroz-Mercado, Sergio Rangel-Guerrero, Natividad Cruz-Ortiz, Tatiana Nunez-Garcia, Gisela Barrera-Badillo, Lucia Hernandez-Rivas, Irma Lopez-Martinez, Ernesto Ramirez-Gonzalez. |
| EPI_ISL_2343026, EPI_ISL_2343027, EPI_ISL_2343028 | LESP Guanajuato | Instituto de Diagnostico y Referencia Epidemiologicos (INDRE) | Claudia Wong-Arambula, Abril Rodriguez-Maldonado, Vanessa Rivero-Arredondo, Ariadna Medina-Benitez, Joaquin Quiroz-Mercado, Sergio Rangel-Guerrero, Natividad Cruz-Ortiz, Tatiana Nunez-Garcia, Gisela Barrera-Badillo, Lucia Hernandez-Rivas, Irma Lopez-Martinez, Ernesto Ramirez-Gonzalez. |
| EPI_ISL_2343029 | LESP Yucatan | Instituto de Diagnostico y Referencia Epidemiologicos (INDRE) | Claudia Wong-Arambula, Abril Rodriguez-Maldonado, Vanessa Rivero-Arredondo, Ariadna Medina-Benitez, Joaquin Quiroz-Mercado, Sergio Rangel-Guerrero, Natividad Cruz-Ortiz, Tatiana Nunez-Garcia, Gisela Barrera-Badillo, Lucia Hernandez-Rivas, Irma Lopez-Martinez, Ernesto Ramirez-Gonzalez. |
| EPI_ISL_2343253, EPI_ISL_2343255, EPI_ISL_2343256, EPI_ISL_2343257, EPI_ISL_2343258, EPI_ISL_2343259, EPI_ISL_2343260, EPI_ISL_2343262, EPI_ISL_2343264, EPI_ISL_2343265, EPI_ISL_2343266, EPI_ISL_2343267, EPI_ISL_2343269 | | | |
| see above | Willis-Knighton Medical Center Hospital Laboratory | LSUHS Emerging Viral Threat Laboratory | April N. Johnson, Lorie M. Atkins, Gregory L. Ware, Alexander Mijalis, Jeremy P. Kamil, Jennifer L. Carroll, Maarten Van Diest, Rona S. Scott, Andrew D. Yurochko, Christopher G. Kevil, John A. Vanchiere, Joseph A. Bocchini |
| EPI_ISL_2343270, EPI_ISL_2343271, EPI_ISL_2343273, EPI_ISL_2343274, EPI_ISL_2343275, EPI_ISL_2343276, EPI_ISL_2343277, EPI_ISL_2343278, EPI_ISL_2343279, EPI_ISL_2343280, EPI_ISL_2343282, EPI_ISL_2343283, EPI_ISL_2343284, EPI_ISL_2343285, EPI_ISL_2343286, EPI_ISL_2343287, EPI_ISL_2343288, EPI_ISL_2343289, EPI_ISL_2343290 | | | |
| see above | Willis-Knighton Medical Center Hospital Laboratory | LSUHS Emerging Viral Threat Laboratory | Lorie M. Atkins, April N. Johnson, Gregory L. Ware, Alexander Mijalis, Jeremy P. Kamil, Jennifer L. Carroll, Maarten Van Diest, Rona S. Scott, Andrew D. Yurochko, Christopher G. Kevil, John A. Vanchiere, Joseph A. Bocchini |
| EPI_ISL_2343297, EPI_ISL_2343298, EPI_ISL_2343299, EPI_ISL_2343300, EPI_ISL_2343301, EPI_ISL_2343302, EPI_ISL_2343303, EPI_ISL_2343304, EPI_ISL_2343306, EPI_ISL_2343307, EPI_ISL_2343308, EPI_ISL_2343309, EPI_ISL_2343310, EPI_ISL_2343311, EPI_ISL_2343312, EPI_ISL_2343315, EPI_ISL_2343316 | | | |
| see above | Louisiana Office of Public Heath Laboratory | LSUHS Emerging Viral Threat Laboratory | Gregory L. Ware, Alexander Mijalis, Jeremy P. Kamil, Jennifer L. Carroll, Maarten Van Diest, Rona S. Scott, Andrew D. Yurochko, Christopher G. Kevil, John A. Vanchiere |
| EPI_ISL_2344462, EPI_ISL_2344463, EPI_ISL_2344464, EPI_ISL_2344465, EPI_ISL_2344466, EPI_ISL_2344467, EPI_ISL_2344468, EPI_ISL_2344469, EPI_ISL_2344470, EPI_ISL_2344471, EPI_ISL_2344472, EPI_ISL_2344473, EPI_ISL_2344474, EPI_ISL_2344475, EPI_ISL_2344476, EPI_ISL_2344477, EPI_ISL_2344478, EPI_ISL_2344479, EPI_ISL_2344480, EPI_ISL_2344481, EPI_ISL_2344482, EPI_ISL_2344483, EPI_ISL_2344484, EPI_ISL_2344485, EPI_ISL_2344486, EPI_ISL_2344487, EPI_ISL_2344488, EPI_ISL_2344489, EPI_ISL_2344490, EPI_ISL_2344491, EPI_ISL_2344492, EPI_ISL_2344493, EPI_ISL_2344494, EPI_ISL_2344495, EPI_ISL_2344496, EPI_ISL_2344497, EPI_ISL_2344498, EPI_ISL_2344499, EPI_ISL_2344500, EPI_ISL_2344501, EPI_ISL_2344502, EPI_ISL_2344503, EPI_ISL_2344504, EPI_ISL_2344505, EPI_ISL_2344506, EPI_ISL_2344507, EPI_ISL_2344508, EPI_ISL_2344509, EPI_ISL_2344510, EPI_ISL_2344511, EPI_ISL_2344512, EPI_ISL_2344513, EPI_ISL_2344514, EPI_ISL_2344515, EPI_ISL_2344516, EPI_ISL_2344517, EPI_ISL_2344518, EPI_ISL_2344519, EPI_ISL_2344520, EPI_ISL_2344521, EPI_ISL_2344522, EPI_ISL_2344523, EPI_ISL_2344524, EPI_ISL_2344525, EPI_ISL_2344526, EPI_ISL_2344527, EPI_ISL_2344528, EPI_ISL_2344529, EPI_ISL_2344530, EPI_ISL_2344531, EPI_ISL_2344532, EPI_ISL_2344533, EPI_ISL_2344534, EPI_ISL_2344535, EPI_ISL_2344536, EPI_ISL_2344537, EPI_ISL_2344538, EPI_ISL_2344539 | | | |
| see above | Virginia Division of Consolidated Laboratory Services | Virginia Division of Consolidated Laboratory Services | Virginia DCLS |
| EPI_ISL_2346100, EPI_ISL_2346101, EPI_ISL_2346102, EPI_ISL_2346103, EPI_ISL_2346104, EPI_ISL_2346105, EPI_ISL_2346106, EPI_ISL_2346107, EPI_ISL_2346108, EPI_ISL_2346109, EPI_ISL_2346110, EPI_ISL_2346111, EPI_ISL_2346112, EPI_ISL_2346113, EPI_ISL_2346114, EPI_ISL_2346115, EPI_ISL_2346116, EPI_ISL_2346117, EPI_ISL_2346118, EPI_ISL_2346119, EPI_ISL_2346120, EPI_ISL_2346121, EPI_ISL_2346122, EPI_ISL_2346123 | | | |
| see above | Oregon State Public Health Laboratory | Oregon State Public Health Laboratory | Rafia Razzaque, Eugene Yeboah, Vanda Makris, Laura Tsaknaridis, John Fontana and Shane Sevey |
| EPI_ISL_2346124, EPI_ISL_2346125 | Emory Molecular Diagnostics Laboratory, Emory Healthcare | Piantadosi Lab, Emory Department of Pathology | Ahmed Babiker, Anne Piantadosi |
| EPI_ISL_2346126 | Emory Molecular Diagnostics Laboratory, Emory Healthcare | Piantadosi Lab, Emory Department of Pathology | Jesse Waggoner, Anne Piantadosi |
| EPI_ISL_2346127, EPI_ISL_2346128, EPI_ISL_2346129 | Emory Molecular Diagnostics Laboratory, Emory Healthcare | Piantadosi Lab, Emory Department of Pathology | Ahmed Babiker, Anne Piantadosi |
| EPI_ISL_2346130, EPI_ISL_2346131, EPI_ISL_2346132, EPI_ISL_2346133 | Grady Memorial Hospital, Clinical Microbiology Laboratory | Piantadosi Lab, Emory Department of Pathology | Yun F (Wayne) Wang, Anne Piantadosi |
| EPI_ISL_2346134, EPI_ISL_2346135, EPI_ISL_2346136, EPI_ISL_2346137, EPI_ISL_2346138, EPI_ISL_2346139, EPI_ISL_2346140, EPI_ISL_2346141, EPI_ISL_2346142, EPI_ISL_2346143 | Emory Molecular Diagnostics Laboratory, Emory Healthcare | Piantadosi Lab, Emory Department of Pathology | Ahmed Babiker, Anne Piantadosi |
| EPI_ISL_2346144 | Grady Memorial Hospital, Clinical Microbiology Laboratory | Piantadosi Lab, Emory Department of Pathology | Yun F (Wayne) Wang, Anne Piantadosi |
| EPI_ISL_2346145, EPI_ISL_2346146, EPI_ISL_2346147, EPI_ISL_2346148, EPI_ISL_2346149, EPI_ISL_2346150, EPI_ISL_2346154, EPI_ISL_2346155 | Emory Molecular Diagnostics Laboratory, Emory Healthcare | Piantadosi Lab, Emory Department of Pathology | Ahmed Babiker, Anne Piantadosi |
| EPI_ISL_2346156 | Grady Memorial Hospital, Clinical Microbiology Laboratory | Piantadosi Lab, Emory Department of Pathology | Yun F (Wayne) Wang, Anne Piantadosi |
| EPI_ISL_2346158 | Emory Molecular Diagnostics Laboratory, Emory Healthcare | Piantadosi Lab, Emory Department of Pathology | Ahmed Babiker, Anne Piantadosi |
| EPI_ISL_2346159 | Grady Memorial Hospital, Clinical Microbiology Laboratory | Piantadosi Lab, Emory Department of Pathology | Yun F (Wayne) Wang, Anne Piantadosi |
| EPI_ISL_2346160, EPI_ISL_2346161 | Emory Molecular Diagnostics Laboratory, Emory Healthcare | Piantadosi Lab, Emory Department of Pathology | Ahmed Babiker, Anne Piantadosi |
| EPI_ISL_2346173, EPI_ISL_2346178, EPI_ISL_2346185, EPI_ISL_2346187, EPI_ISL_2346207, EPI_ISL_2346219, EPI_ISL_2346221, EPI_ISL_2346222, EPI_ISL_2346229, EPI_ISL_2346230, EPI_ISL_2346234, EPI_ISL_2346253, EPI_ISL_2346259, EPI_ISL_2346280 | | | |
| see above | Notre Dame COVID Response and Testing Laboratory | Notre Dame Genomics & Bioinformatics Core Facility | Michael Pfrender, Joseph Sarro, Melissa Stephens |
| EPI_ISL_2346323, EPI_ISL_2346324, EPI_ISL_2346325, EPI_ISL_2346326, EPI_ISL_2346327, EPI_ISL_2346328, EPI_ISL_2346329, EPI_ISL_2346330, EPI_ISL_2346331, EPI_ISL_2346332, EPI_ISL_2346333, EPI_ISL_2346334, EPI_ISL_2346335, EPI_ISL_2346336, EPI_ISL_2346337, EPI_ISL_2346338 | | | |
| see above | The Jackson Laboratory | The Jackson Laboratory | Long J, Bergeron D, Renzette N, Adams M, Omerza G, Kelly K, Li L |
| EPI_ISL_2346436, EPI_ISL_2346437, EPI_ISL_2346438, EPI_ISL_2346439, EPI_ISL_2346440, EPI_ISL_2346441, EPI_ISL_2346442, EPI_ISL_2346443, EPI_ISL_2346444, EPI_ISL_2346445, EPI_ISL_2346446, EPI_ISL_2346447, EPI_ISL_2346448, EPI_ISL_2346449, EPI_ISL_2346450, EPI_ISL_2346451, EPI_ISL_2346452, EPI_ISL_2346453, EPI_ISL_2346454, EPI_ISL_2346455, EPI_ISL_2346456, EPI_ISL_2346457, EPI_ISL_2346458, EPI_ISL_2346459, EPI_ISL_2346460, EPI_ISL_2346461, EPI_ISL_2346462, EPI_ISL_2346463, EPI_ISL_2346464, EPI_ISL_2346465, EPI_ISL_2346466, EPI_ISL_2346467, EPI_ISL_2346468, EPI_ISL_2346469, EPI_ISL_2346470 | | | |
| see above | Instituto Nacional de Enfermedades Respiratorias (INER) | Centro de Investigación en Enfermedades Infecciosas (CIENI), Instituto Nacional de Enfermedades Respiratorias (INER) | Consorcio Mexicano de Vigilancia Genómica (CoViGen-Mex). Authors (in alphabetical order): Julio Elias Alvarado-Yaah, Carlos F. Arias, Santiago Ávila-Ríos, Eduardo Becerril-Vargas, Víctor Hugo Borja-Aburto, Celia Boukadida, Juan Bautista Chale-Dzul, Célida Duque Molina, José Antonio Enciso-Moreno, Gloria Elena Espinosa-Ayala, Fernando Fontove-Herrera, Victor Eduardo Garcia-Arias, Concepción Grajales-Muñiz, Ricardo Grande, Alejandra Hernández-Terán, Alfredo Herrera-Estrella, Carla Ivón Herrera-Najera, Pavel Isa, Brenda Irasema Maldonado-Meza, Bernardo Martínez-Miguel, José Arturo Martínez-Orozco, Margarita Matías-Florentino, Fidencio Mejía-Nepomuceno, María Guadalupe de Jesús Mireles-Rivera, Gloria María Molina-Salinas, Hector Montoya-Fuentes, Mario Mújica-Sánchez, José Esteban Muñoz-Medina, José de Jesús Nuñez-Contreras, Alicia Ocaña-Mondragón, Luis Alberto Ochoa-Carrera, Hector Esteban Paz-Juárez, Francisco Pulido, Helen Haydee Fernanda Ramírez-Plascencia, Angel Gustavo Salas-Lais, Alejandro Sanchez-Flores, Clara Esperanza Santacruz-Tinoco, María Guadalupe Santiago-Mauricio, Jorge Salas-Hernández, Nelly Sélem-Mojica, Blanca Taboada, Gloria Vazquez, Joel Armando Vázquez-Pérez |
| EPI_ISL_2348498 | Nationwide Children's Hospital Clinical Microbiology Lab | James Molecular Laboratory | Huanyu Wang, Dan Jones, Sophonie Jean, Richard Eltringham, John Madison, Pamela Snyder, Huolin Tu, Amy L. Leber |
| EPI_ISL_2348837 | Salud Digna | Instituto Nacional de Medicina Genomica | Hidalgo-Miranda A, Cedro-Tanda A, Mendoza-Vargas A, Reyes-Grajeda JP, Abraham Campos-Romero, Moreno-Camacho José Luis, Rodriguez-Gallegos Jorge, Luna-Ruiz Marco, Gonzalez-Barrera D, Rangel-DeLeon D, Munguia-Garza P, Ramirez-Vega O, Escobar-Arrazola, M, Herrera-Montalvo LA. |
| EPI_ISL_2348839 | Utah Public Health Laboratory, Utah Public Health Laboratory Infectious Disease submission group | Utah Public Health Laboratory, Utah Public Health Laboratory Infectious Disease submission group | Gallagher,T., Young,E.L. and Oakeson,K.F. |
| EPI_ISL_2348926 | Respiratory Viruses Branch, Centers for Disease Control and Prevention | Respiratory Viruses Branch, Centers for Disease Control and Prevention | Howard,D., Batra,D., Cook,P.W., Moser,K., Paskey,A., Caravas,J.,Rambo-Martin,B., Morrison,S., Gulvick,C., Sammons,S., Unoarumhi,Y.,Schmerer,M., Roychoudhury,P., Xie,H., Shrestha,L., Nguyen,T.V.,Bakhash,S.M., Lin,M., Baker,N.R., Perez,R., Ellis,S., Breit,N.,Livingston,R.J., Huang,M.-L., Jerome,K.R., Mathias,P.,Greninger,A., Paden,C.R. and MacCannell,D. |

| | | | |
|---|---|---|---|
| EPI_ISL_2348927, EPI_ISL_2348928, EPI_ISL_2348929, EPI_ISL_2348930, EPI_ISL_2348931, EPI_ISL_2348932, EPI_ISL_2348933, EPI_ISL_2348934, EPI_ISL_2348935, EPI_ISL_2348936, EPI_ISL_2348937, EPI_ISL_2348938, EPI_ISL_2348939, EPI_ISL_2348940, EPI_ISL_2348941, EPI_ISL_2348942, EPI_ISL_2348943, EPI_ISL_2348944, EPI_ISL_2348945, EPI_ISL_2348946, EPI_ISL_2348947, EPI_ISL_2348948, EPI_ISL_2348949, EPI_ISL_2348950, EPI_ISL_2348951, EPI_ISL_2348952, EPI_ISL_2348953 | | | |
| see above | Oregon State Public Health Laboratory | Oregon State Public Health Laboratory | Rafia Razzaque, Eugene Yeboah, Vanda Makris, Laura Tsaknaridis, John Fontana and Shane Sevey |
| EPI_ISL_2348954, EPI_ISL_2348955, EPI_ISL_2348956, EPI_ISL_2348957, EPI_ISL_2348958, EPI_ISL_2348959 | Alaska State Virology Lab, Alaska State Public Health Labs | Alaska State Virology Lab, Alaska State Public Health Labs | DeRonde,S., House,E., Zidek,J., Redlinger,M., Smith,L., Chen,J. |
| EPI_ISL_2348960, EPI_ISL_2348961, EPI_ISL_2348962 | Laboratory Medicine, University of Washington | Laboratory Medicine, University of Washington | Roychoudhury,P., Xie,H., Shrestha,L., Bakhash,S.M., Rachleff,V.M., Lin,M.J., Baker,N.R., Ellis,S., Sathees,S., Huang,M.-L., Jerome,K.R., Greninger,A. |
| EPI_ISL_2348963, EPI_ISL_2348964, EPI_ISL_2348965, EPI_ISL_2348966 | Laboratory Medicine, University of Washington | Laboratory Medicine, University of Washington | Roychoudhury,P., Xie,H., Shrestha,L., Bakhash,S.M., Lin,M.J., Baker,N.R., Ellis,S., Sathees,S., Huang,M.-L., Jerome,K.R., Greninger,A. |
| EPI_ISL_2348967 | Laboratory Medicine, University of Washington | Laboratory Medicine, University of Washington | Roychoudhury,P., Xie,H., Shrestha,L., Bakhash,S.M., Lin,M.J., Baker,N.R., Ellis,S., Sathees,S., Rachleff,V.M., Huang,M.-L., Jerome,K.R., Greninger,A. |
| EPI_ISL_2348968, EPI_ISL_2348969, EPI_ISL_2348970, EPI_ISL_2348971 | Laboratory Medicine, University of Washington | Laboratory Medicine, University of Washington | Roychoudhury,P., Xie,H., Shrestha,L., Bakhash,S.M., Lin,M.J., Baker,N.R., Ellis,S., Huang,M.-L., Jerome,K.R., Greninger,A. |
| EPI_ISL_2348972, EPI_ISL_2348973, EPI_ISL_2348974 | Laboratory Medicine, University of Washington | Laboratory Medicine, University of Washington | Roychoudhury,P., Xie,H., Shrestha,L., Bakhash,S.M., Rachleff,V.M., Lin,M.J., Baker,N.R., Ellis,S., Sathees,S., Huang,M.-L., Jerome,K.R., Greninger,A. |
| EPI_ISL_2348975, EPI_ISL_2348976 | Laboratory Medicine, University of Washington | Laboratory Medicine, University of Washington | Roychoudhury,P., Xie,H., Shrestha,L., Bakhash,S.M., Lin,M.J., Baker,N.R., Ellis,S., Huang,M.-L., Jerome,K.R., Greninger,A. |
| EPI_ISL_2348977 | Laboratory Medicine, University of Washington | Laboratory Medicine, University of Washington | Roychoudhury,P., Xie,H., Shrestha,L., Bakhash,S.M., Lin,M.J., Baker,N.R., Ellis,S., Sathees,S., Huang,M.-L., Jerome,K.R., Greninger,A. |
| EPI_ISL_2349269 | Respiratory Viruses Branch, Centers for Disease Control and Prevention | Respiratory Viruses Branch, Centers for Disease Control and Prevention | Howard,D., Batra,D., Cook,P.W., Moser,K., Paskey,A., Caravas,J.,Rambo-Martin,B., Morrison,S., Gulvick,C., Sammons,S., Unoarumhi,Y.,Schmerer,M., Roychoudhury,P., Xie,H., Shrestha,L., Nguyen,T.V.,Bakhash,S.M., Lin,M., Baker,N.R., Perez,R., Ellis,S., Breit,N.,Livingston,R.J., Huang,M.-L., Jerome,K.R., Mathias,P.,Greninger,A., Paden,C.R. and MacCannell,D. |
| EPI_ISL_2349270, EPI_ISL_2349271 | Utah Public Health Laboratory, Utah Public Health Laboratory Infectious Disease submission group | Utah Public Health Laboratory, Utah Public Health Laboratory Infectious Disease submission group | Gallagher,T., Young,E.L., Oakeson,K.F. |
| EPI_ISL_2349615, EPI_ISL_2349616, EPI_ISL_2349617, EPI_ISL_2349618, EPI_ISL_2349619, EPI_ISL_2349620, EPI_ISL_2349623, EPI_ISL_2349624, EPI_ISL_2349625, EPI_ISL_2349626, EPI_ISL_2349627, EPI_ISL_2349628, EPI_ISL_2349629, EPI_ISL_2349631, EPI_ISL_2349633, EPI_ISL_2349634, EPI_ISL_2349635, EPI_ISL_2349636, EPI_ISL_2349637, EPI_ISL_2349638, EPI_ISL_2349639, EPI_ISL_2349640, EPI_ISL_2349641, EPI_ISL_2349642, EPI_ISL_2349643, EPI_ISL_2349644, EPI_ISL_2349645, EPI_ISL_2349646, EPI_ISL_2349648, EPI_ISL_2349650, EPI_ISL_2349651, EPI_ISL_2349652, EPI_ISL_2349653, EPI_ISL_2349654, EPI_ISL_2349655, EPI_ISL_2349656, EPI_ISL_2349657, EPI_ISL_2349658, EPI_ISL_2349659, EPI_ISL_2349660, EPI_ISL_2349661, EPI_ISL_2349662, EPI_ISL_2349663, EPI_ISL_2349664, EPI_ISL_2349665, EPI_ISL_2349668, EPI_ISL_2349669, EPI_ISL_2349670, EPI_ISL_2349671, EPI_ISL_2349672, EPI_ISL_2349673, EPI_ISL_2349674, EPI_ISL_2349675, EPI_ISL_2349676, EPI_ISL_2349677, EPI_ISL_2349678, EPI_ISL_2349679, EPI_ISL_2349680, EPI_ISL_2349681, EPI_ISL_2349682, EPI_ISL_2349683, EPI_ISL_2349684, EPI_ISL_2349685, EPI_ISL_2349686, EPI_ISL_2349688, EPI_ISL_2349690, EPI_ISL_2349691 | | | |
| see above | Pandemic Response Lab - NYC | Pandemic Response Lab, R&D | Henry Lee, Michael Hammerling, Melissa Hopkins, Cybill del Castillo, Shinyoung Clair Kang, William Ward, Pradeep Bugga, Sol Rey, Dylan Law, Katharine Nelson, Haiping Hao, Jon Laurent |
| EPI_ISL_2349881, EPI_ISL_2349882, EPI_ISL_2349883, EPI_ISL_2349884, EPI_ISL_2349885, EPI_ISL_2349886, EPI_ISL_2349888, EPI_ISL_2349889, EPI_ISL_2349890, EPI_ISL_2349891, EPI_ISL_2349892, EPI_ISL_2349893, EPI_ISL_2349894, EPI_ISL_2349895, EPI_ISL_2349896, EPI_ISL_2349897, EPI_ISL_2349898, EPI_ISL_2349899, EPI_ISL_2349900, EPI_ISL_2349901, EPI_ISL_2349902, EPI_ISL_2349903, EPI_ISL_2349904, EPI_ISL_2349906, EPI_ISL_2349907, EPI_ISL_2349908, EPI_ISL_2349909, EPI_ISL_2349910, EPI_ISL_2349911, EPI_ISL_2349912, EPI_ISL_2349913, EPI_ISL_2349914, EPI_ISL_2349915, EPI_ISL_2349916, EPI_ISL_2349917, EPI_ISL_2349918, EPI_ISL_2349919, EPI_ISL_2349921, EPI_ISL_2349922, EPI_ISL_2349923, EPI_ISL_2349924, EPI_ISL_2349925, EPI_ISL_2349926, EPI_ISL_2349928, EPI_ISL_2349929, EPI_ISL_2349931, EPI_ISL_2349932, EPI_ISL_2349933, EPI_ISL_2349934, EPI_ISL_2349935, EPI_ISL_2349938, EPI_ISL_2349940, EPI_ISL_2349941, EPI_ISL_2349942, EPI_ISL_2349943, EPI_ISL_2349944, EPI_ISL_2349945, EPI_ISL_2349946, EPI_ISL_2349947, EPI_ISL_2349948, EPI_ISL_2349949, EPI_ISL_2349950, EPI_ISL_2349951, EPI_ISL_2349952, EPI_ISL_2349953, EPI_ISL_2349954, EPI_ISL_2349955, EPI_ISL_2349956, EPI_ISL_2349957, EPI_ISL_2349958, EPI_ISL_2349959, EPI_ISL_2349961, EPI_ISL_2349962, EPI_ISL_2349964, EPI_ISL_2349965, EPI_ISL_2349966, EPI_ISL_2349967, EPI_ISL_2349968, EPI_ISL_2349970, EPI_ISL_2349971, EPI_ISL_2349972, EPI_ISL_2349973, EPI_ISL_2349974, EPI_ISL_2349975, EPI_ISL_2349976, EPI_ISL_2349978, EPI_ISL_2349979, EPI_ISL_2349980, EPI_ISL_2349981, EPI_ISL_2349983, EPI_ISL_2349984, EPI_ISL_2349985, EPI_ISL_2349986, EPI_ISL_2349987, EPI_ISL_2349988, EPI_ISL_2349989, EPI_ISL_2349990, EPI_ISL_2349993, EPI_ISL_2349994, EPI_ISL_2349995, EPI_ISL_2349996, EPI_ISL_2349997, EPI_ISL_2349998, EPI_ISL_2349999, EPI_ISL_2350000, EPI_ISL_2350002, EPI_ISL_2350004, EPI_ISL_2350005, EPI_ISL_2350006, EPI_ISL_2350008, EPI_ISL_2350009, EPI_ISL_2350010, EPI_ISL_2350011, EPI_ISL_2350012, EPI_ISL_2350014, EPI_ISL_2350015, EPI_ISL_2350016, EPI_ISL_2350018, EPI_ISL_2350019, EPI_ISL_2350021, EPI_ISL_2350022, EPI_ISL_2350023, EPI_ISL_2350024, EPI_ISL_2350025, EPI_ISL_2350026, EPI_ISL_2350027, EPI_ISL_2350028, EPI_ISL_2350029, EPI_ISL_2350030, EPI_ISL_2350031, EPI_ISL_2350032, EPI_ISL_2350033, EPI_ISL_2350035, EPI_ISL_2350036, EPI_ISL_2350037, EPI_ISL_2350038, EPI_ISL_2350039, EPI_ISL_2350040, EPI_ISL_2350041, EPI_ISL_2350042, EPI_ISL_2350043, EPI_ISL_2350044, EPI_ISL_2350048, EPI_ISL_2350051, EPI_ISL_2350052, EPI_ISL_2350054, EPI_ISL_2350055, EPI_ISL_2350056, EPI_ISL_2350057, EPI_ISL_2350058, EPI_ISL_2350060, EPI_ISL_2350061, EPI_ISL_2350062, EPI_ISL_2350063, EPI_ISL_2350064, EPI_ISL_2350065, EPI_ISL_2350066, EPI_ISL_2350067, EPI_ISL_2350068, EPI_ISL_2350069, EPI_ISL_2350072, EPI_ISL_2350075, EPI_ISL_2350076, EPI_ISL_2350077, EPI_ISL_2350078, EPI_ISL_2350079, EPI_ISL_2350081, EPI_ISL_2350082, EPI_ISL_2350084, EPI_ISL_2350085, EPI_ISL_2350086, EPI_ISL_2350087, EPI_ISL_2350088, EPI_ISL_2350089, EPI_ISL_2350093 | | | |
| see above | Salud Digna | Instituto Nacional de Medicina Genomica | Hidalgo-Miranda A, Cedro-Tanda A, Mendoza-Vargas A, Reyes-Grajeda JP, Abraham Campos-Romero, Moreno-Camacho José Luis, Rodriguez-Gallegos Jorge, Luna-Ruiz Marco, Gonzalez-Barrera D, Rangel-DeLeon D, Munguia-Garza P, Ramirez-Vega O, Escobar-Arrazola, M, Herrera-Montalvo LA. |
| EPI_ISL_2350095, EPI_ISL_2350096, EPI_ISL_2350098, EPI_ISL_2350099, EPI_ISL_2350100, EPI_ISL_2350101, EPI_ISL_2350102, EPI_ISL_2350104, EPI_ISL_2350106, EPI_ISL_2350107, EPI_ISL_2350108, EPI_ISL_2350109, EPI_ISL_2350110, EPI_ISL_2350111, EPI_ISL_2350112, EPI_ISL_2350113 | | | |
| see above | Instituto Nacional de Medicina Genomica | Instituto Nacional de Medicina Genomica | Hidalgo-Miranda A, Cedro-Tanda A, Mendoza-Vargas A, Reyes-Grajeda JP, Cisneros- Villanueva M, Gonzalez-Barrera D, Rangel-DeLeon D, Munguia-Garza P, Ramirez-Vega O, Escobar-Arrazola, M, Herrera-Montalvo LA. |
| EPI_ISL_2350114, EPI_ISL_2350115, EPI_ISL_2350117, EPI_ISL_2350119, EPI_ISL_2350120, EPI_ISL_2350121, EPI_ISL_2350122, EPI_ISL_2350123, EPI_ISL_2350124, EPI_ISL_2350125, EPI_ISL_2350126, EPI_ISL_2350127, EPI_ISL_2350128, EPI_ISL_2350129, EPI_ISL_2350130, EPI_ISL_2350131, EPI_ISL_2350132, EPI_ISL_2350133, EPI_ISL_2350134, EPI_ISL_2350135, EPI_ISL_2350136, EPI_ISL_2350137, EPI_ISL_2350139, EPI_ISL_2350140, EPI_ISL_2350141 | | | |
| see above | Salud Digna | Instituto Nacional de Medicina Genomica | Hidalgo-Miranda A, Cedro-Tanda A, Mendoza-Vargas A, Reyes-Grajeda JP, Abraham Campos-Romero, Moreno-Camacho José Luis, Rodriguez-Gallegos Jorge, Luna-Ruiz Marco, Gonzalez-Barrera D, Rangel-DeLeon D, Munguia-Garza P, Ramirez-Vega O, Escobar-Arrazola, M, Herrera-Montalvo LA. |
| EPI_ISL_2350143, EPI_ISL_2350144, EPI_ISL_2350145, EPI_ISL_2350146, EPI_ISL_2350147, EPI_ISL_2350149, EPI_ISL_2350150, EPI_ISL_2350151, EPI_ISL_2350152, EPI_ISL_2350153, EPI_ISL_2350154, EPI_ISL_2350155, EPI_ISL_2350156, EPI_ISL_2350157, EPI_ISL_2350158, EPI_ISL_2350160, EPI_ISL_2350162, EPI_ISL_2350163, EPI_ISL_2350164, EPI_ISL_2350165, EPI_ISL_2350169, EPI_ISL_2350170, EPI_ISL_2350171, EPI_ISL_2350172, EPI_ISL_2350173, EPI_ISL_2350175, EPI_ISL_2350176, EPI_ISL_2350177, EPI_ISL_2350178, EPI_ISL_2350179, EPI_ISL_2350181, EPI_ISL_2350182, EPI_ISL_2350183, EPI_ISL_2350184 | | | |
| see above | Molecular Biology, New Mexico Department of Health Scientific Laboratory | Molecular Biology, New Mexico Department of Health Scientific Laboratory | Johnson,E.J., Malone,D., Benoit,J., Rajan,R., Salazar,L., Gutierrez,K., Griego-Fisher,A.M. |
| EPI_ISL_2350365, EPI_ISL_2350366, EPI_ISL_2350367, EPI_ISL_2350368, EPI_ISL_2350369, EPI_ISL_2350370, EPI_ISL_2350371, EPI_ISL_2350372, EPI_ISL_2350373, EPI_ISL_2350374, EPI_ISL_2350375, EPI_ISL_2350376, EPI_ISL_2350377, EPI_ISL_2350378, EPI_ISL_2350379, EPI_ISL_2350380, EPI_ISL_2350381, EPI_ISL_2350382, EPI_ISL_2350383, EPI_ISL_2350384, EPI_ISL_2350385, EPI_ISL_2350386, EPI_ISL_2350387, EPI_ISL_2350388, EPI_ISL_2350389, EPI_ISL_2350390, EPI_ISL_2350391, EPI_ISL_2350392, EPI_ISL_2350393, EPI_ISL_2350394, EPI_ISL_2350395, EPI_ISL_2350396, EPI_ISL_2350397, EPI_ISL_2350398, EPI_ISL_2350399, EPI_ISL_2350400, EPI_ISL_2350401, EPI_ISL_2350402 | | | |
| see above | CSL, Maryland Department of Health | CSL, Maryland Department of Health | Keller,E., Washington,Y. |
| EPI_ISL_2354483, EPI_ISL_2354484, EPI_ISL_2354485, EPI_ISL_2354486, EPI_ISL_2354487, EPI_ISL_2354488, EPI_ISL_2354490, EPI_ISL_2354491, EPI_ISL_2354493, EPI_ISL_2354494, EPI_ISL_2354495, EPI_ISL_2354496, EPI_ISL_2354497 | | | |
| see above | Microbiology Division, SC DHEC | Microbiology Division, SC DHEC | Flores,H., Freeman,J. |
| EPI_ISL_2361362 | LESP Guanajuato | Instituto de diagnóstico y Referencia Epidemiologicos (INDRE) Departamento de Virología | Claudia Wong-Arambula, Abril Rodriguez-Maldonado, Vanessa Rivero-Arredondo, Ariadna Medina-Benitez, Joaquin Quiroz-Mercado, Sergio Rangel-Guerrero, Natividad Cruz-Ortiz, Tatiana Nunez-Garcia, Gisela Barrera-Badillo, Lucia Hernandez-Rivas, Irma Lopez-Martinez, Ernesto Ramirez-Gonzalez. |
| EPI_ISL_2361366, EPI_ISL_2361367, EPI_ISL_2361369, EPI_ISL_2361370, EPI_ISL_2361371, EPI_ISL_2361372, EPI_ISL_2361373, EPI_ISL_2361374, EPI_ISL_2361375, EPI_ISL_2361376, EPI_ISL_2361377, EPI_ISL_2361378, EPI_ISL_2361379, EPI_ISL_2361380, EPI_ISL_2361381, EPI_ISL_2361382, EPI_ISL_2361383, EPI_ISL_2361384, EPI_ISL_2361385, EPI_ISL_2361386, EPI_ISL_2361387, EPI_ISL_2361388, EPI_ISL_2361389, EPI_ISL_2361390, EPI_ISL_2361391, EPI_ISL_2361392, EPI_ISL_2361393, EPI_ISL_2361394, EPI_ISL_2361395, EPI_ISL_2361396, EPI_ISL_2361397, EPI_ISL_2361398, EPI_ISL_2361399, EPI_ISL_2361400, EPI_ISL_2361401, EPI_ISL_2361402, EPI_ISL_2361403, EPI_ISL_2361404, EPI_ISL_2361405, EPI_ISL_2361406, EPI_ISL_2361407, EPI_ISL_2361408, EPI_ISL_2361409, EPI_ISL_2361410, EPI_ISL_2361411, EPI_ISL_2361412, EPI_ISL_2361413, EPI_ISL_2361415, EPI_ISL_2361416, EPI_ISL_2361417, EPI_ISL_2361418, EPI_ISL_2361419, EPI_ISL_2361420, EPI_ISL_2361421, EPI_ISL_2361422, EPI_ISL_2361423, EPI_ISL_2361424, EPI_ISL_2361425, EPI_ISL_2361426, EPI_ISL_2361427, EPI_ISL_2361428, EPI_ISL_2361429, EPI_ISL_2361430, EPI_ISL_2361431, EPI_ISL_2361432, EPI_ISL_2361433, EPI_ISL_2361434, EPI_ISL_2361436, EPI_ISL_2361437, EPI_ISL_2361438, EPI_ISL_2361439, EPI_ISL_2361440, EPI_ISL_2361442, EPI_ISL_2361445, EPI_ISL_2361446, EPI_ISL_2361447, EPI_ISL_2361449, EPI_ISL_2361451, EPI_ISL_2361452, EPI_ISL_2361453, EPI_ISL_2361454 | | | |
| see above | IN State Department of Health Laboratory Services | IN State Department of Health Laboratory Services | Cassandra Campion, Jamie Yeadon, Brian Pope, Lixia Liu, Kyle Brownlee, Melissa Hindenlang, Mark Glazier |
| EPI_ISL_2361879 | Instituto Nacional de Enfermedades Respiratorias (INER) | Centro de Investigación en Enfermedades Infecciosas (CIENI), Instituto Nacional de Enfermedades Respiratorias (INER) | Consorcio Mexicano de Vigilancia Genómica (CoViGen-Mex). Authors (in alphabetical order): Julio Elias Alvarado-Yaah, Carlos F. Arias, Santiago Ávila-Ríos, Eduardo Becerril-Vargas, Víctor Hugo Borja-Aburto, Celia Boukadida, Juan Bautista Chale-Dzul, Célida Duque Molina, José Antonio Enciso-Moreno, Gloria Elena Espinosa-Ayala, Fernando Fontove-Herrera, Victor Eduardo Garcia-Arias, Concepción Grajales-Muñiz, Ricardo Grande, Alejandra Hernández-Terán, Alfredo Herrera-Estrella, Carla Ivón Herrera-Najera, Pavel Isa, Brenda Irasema Maldonado-Meza, Bernardo Martínez-Miguel, José Arturo Martínez-Orozco, Margarita Matías-Florentino, Fidencio Mejía-Nepomuceno, María Guadalupe de Jesús Mireles-Rivera, Gloria María Molina-Salinas, Hector Montoya-Fuentes, Mario Mújica-Sánchez, José Esteban Muñoz-Medina, José de Jesús Nuñez-Contreras, Alicia Ocaña-Mondragón, Luis Alberto Ochoa-Carrera, Hector Esteban Paz-Juárez, Francisco Pulido, Helen Haydee Fernanda Ramírez-Plascencia, Angel Gustavo Salas-Lais, Alejandro Sanchez-Flores, Clara Esperanza Santacruz-Tinoco, María Guadalupe Santiago-Mauricio, Jorge Salas-Hernández, Nelly Sélem-Mojica, Blanca Taboada, Gloria Vazquez, Joel Armando Vázquez-Pérez |
| EPI_ISL_2362006, EPI_ISL_2362007, EPI_ISL_2362008, EPI_ISL_2362009, EPI_ISL_2362010, EPI_ISL_2362011, EPI_ISL_2362012, EPI_ISL_2362013, EPI_ISL_2362014, EPI_ISL_2362015, EPI_ISL_2362016, EPI_ISL_2362017, EPI_ISL_2362018, EPI_ISL_2362019, EPI_ISL_2362020, EPI_ISL_2362021, EPI_ISL_2362022, EPI_ISL_2362023, EPI_ISL_2362024, EPI_ISL_2362025, EPI_ISL_2362026, EPI_ISL_2362027, EPI_ISL_2362028, EPI_ISL_2362029, EPI_ISL_2362030, EPI_ISL_2362031, EPI_ISL_2362032, EPI_ISL_2362033, EPI_ISL_2362034, EPI_ISL_2362035, EPI_ISL_2362036, EPI_ISL_2362037, EPI_ISL_2362038, EPI_ISL_2362039, EPI_ISL_2362040 | | | |
| see above | Virginia Division of Consolidated Laboratory Services | Virginia Division of Consolidated Laboratory Services | Virginia DCLS |
| EPI_ISL_2362354 | Flow Health | Infectious Disease Program, Broad Institute of Harvard and MIT | Lemieux,J.E., Siddle,K.J., Adams,G., Gladden-Young,A., Lagerborg,K., Rudy,M., DeRuff,K., Carter,A., Normandin,E., Bauer,M., Reilly,S., Tomkins-Tinch,C., Loreth,C., Chaluvadi,S., Birren,B.W., Gallagher,G., Smole,S., Park,D.J., MacInnis,B.L., and Sabeti,P.C. |
| EPI_ISL_2362355, EPI_ISL_2362356, EPI_ISL_2362357, EPI_ISL_2362358, EPI_ISL_2362359, EPI_ISL_2362360, EPI_ISL_2362361, EPI_ISL_2362362, EPI_ISL_2362365, EPI_ISL_2362366, EPI_ISL_2362367, EPI_ISL_2362368, EPI_ISL_2362369, EPI_ISL_2362370, EPI_ISL_2362371, EPI_ISL_2362372, EPI_ISL_2362373, EPI_ISL_2362374, EPI_ISL_2362375 | | | |
| see above | Massachusetts General Hospital | Infectious Disease Program, Broad Institute of Harvard and MIT | Lemieux,J.E., Siddle,K.J., Shaw,B., Adams,G., Pierce,V., Turbett,S., Anahtar,M., Branda,J., Slater,D., Harris,J., Lin,A.E., Gladden-Young,A., Lagerborg,K., Rudy,M., DeRuff,K., Carter,A., Normandin,E., Bauer,M., Reilly,S., Tomkins-Tinch,C., Loreth,C., Chaluvadi,S., Neumann,A., Cusick,C., Chapman,S.B., Gnirke,A., Flowers,K., Cerrato,F., Birren,B.W., Gallagher,G., Smole,S., Park,D.J., MacInnis,B.L., Ryan,E., LaRocque,R., Rosenberg,E. and Sabeti,P.C. |
| EPI_ISL_2362376, EPI_ISL_2362377, EPI_ISL_2362378, EPI_ISL_2362379, EPI_ISL_2362380, EPI_ISL_2362381, EPI_ISL_2362382, EPI_ISL_2362383, EPI_ISL_2362384, EPI_ISL_2362385, EPI_ISL_2362386, EPI_ISL_2362387, EPI_ISL_2362388, EPI_ISL_2362389, EPI_ISL_2362390, EPI_ISL_2362391, EPI_ISL_2362393, EPI_ISL_2362394, EPI_ISL_2362395, EPI_ISL_2362396, EPI_ISL_2362397, EPI_ISL_2362398, EPI_ISL_2362399, EPI_ISL_2362400, EPI_ISL_2362401, EPI_ISL_2362402, EPI_ISL_2362403, EPI_ISL_2362404, EPI_ISL_2362405, EPI_ISL_2362406, EPI_ISL_2362408, EPI_ISL_2362409, EPI_ISL_2362410, EPI_ISL_2362411, EPI_ISL_2362412, EPI_ISL_2362413, EPI_ISL_2362414, EPI_ISL_2362415, EPI_ISL_2362416, EPI_ISL_2362417, EPI_ISL_2362418, EPI_ISL_2362419, EPI_ISL_2362420, EPI_ISL_2362421, EPI_ISL_2362422, EPI_ISL_2362424, EPI_ISL_2362425, EPI_ISL_2362426, EPI_ISL_2362427, EPI_ISL_2362428, EPI_ISL_2362429, EPI_ISL_2362430, EPI_ISL_2362431, EPI_ISL_2362432, EPI_ISL_2362433, EPI_ISL_2362435, EPI_ISL_2362436, EPI_ISL_2362437, EPI_ISL_2362438, EPI_ISL_2362439, EPI_ISL_2362440, EPI_ISL_2362442, EPI_ISL_2362443, EPI_ISL_2362444, EPI_ISL_2362445, EPI_ISL_2362446, EPI_ISL_2362448, EPI_ISL_2362451, EPI_ISL_2362453, EPI_ISL_2362454, EPI_ISL_2362456, EPI_ISL_2362460, EPI_ISL_2362461, EPI_ISL_2362462, EPI_ISL_2362466, EPI_ISL_2362467, EPI_ISL_2362468, EPI_ISL_2362469, EPI_ISL_2362470, EPI_ISL_2362471, EPI_ISL_2362474, EPI_ISL_2362475, EPI_ISL_2362476, EPI_ISL_2362477, EPI_ISL_2362478, EPI_ISL_2362479, EPI_ISL_2362480, EPI_ISL_2362481, EPI_ISL_2362482 | | | |

| | | | |
|---|---|---|---|
| see above | Rhode Island Department of Health | Infectious Disease Program, Broad Institute of Harvard and MIT | Siddle,K.J., Azevedo,K., Miller,A., Adams,G., Pearlman,L., Gladden-Young,A., Lagerborg,K., Rudy,M., DeRuff,K., Carter,A., Normandin,E., Bauer,M., Reilly,S., Tomkins-Tinch,C., Loreth,C., Chaluvadi,S., Lemieux,J.E., Birren,B.W., Sabeti,P.C., Huard,R., King,E., Park,D.J., and MacInnis,B.L. |
| EPI_ISL_2362485, EPI_ISL_2362487 | Flow Health | Infectious Disease Program, Broad Institute of Harvard and MIT | Lemieux,J.E., Siddle,K.J., Adams,G., Gladden-Young,A., Lagerborg,K., Rudy,M., DeRuff,K., Carter,A., Normandin,E., Bauer,M., Reilly,S., Tomkins-Tinch,C., Loreth,C., Chaluvadi,S., Birren,B.W., Gallagher,G., Smole,S., Park,D.J., MacInnis,B.L., and Sabeti,P.C. |
| EPI_ISL_2362687 | LESP Yucatan | Instituto de Diagnostico y Referencia Epidemiologicos (INDRE) | Claudia Wong-Arambula, Abril Rodriguez-Maldonado, Vanessa Rivero-Arredondo, Ariadna Medina-Benitez, Joaquin Quiroz-Mercado, Sergio Rangel-Guerrero, Natividad Cruz-Ortiz, Tatiana Nunez-Garcia, Gisela Barrera-Badillo, Lucia Hernandez-Rivas, Irma Lopez-Martinez, Ernesto Ramirez-Gonzalez. |
| EPI_ISL_2362688, EPI_ISL_2362689, EPI_ISL_2362690, EPI_ISL_2362691, EPI_ISL_2362692 | LESP Guanajuato | Instituto de Diagnostico y Referencia Epidemiologicos (INDRE) | Claudia Wong-Arambula, Abril Rodriguez-Maldonado, Vanessa Rivero-Arredondo, Ariadna Medina-Benitez, Joaquin Quiroz-Mercado, Sergio Rangel-Guerrero, Natividad Cruz-Ortiz, Tatiana Nunez-Garcia, Gisela Barrera-Badillo, Lucia Hernandez-Rivas, Irma Lopez-Martinez, Ernesto Ramirez-Gonzalez. |
| EPI_ISL_2362693 | LESP Jalisco | Instituto de Diagnostico y Referencia Epidemiologicos (INDRE) | Claudia Wong-Arambula, Abril Rodriguez-Maldonado, Vanessa Rivero-Arredondo, Ariadna Medina-Benitez, Joaquin Quiroz-Mercado, Sergio Rangel-Guerrero, Natividad Cruz-Ortiz, Tatiana Nunez-Garcia, Gisela Barrera-Badillo, Lucia Hernandez-Rivas, Irma Lopez-Martinez, Ernesto Ramirez-Gonzalez. |
| EPI_ISL_2362694 | LESP Ciudad de Mexico | Instituto de Diagnostico y Referencia Epidemiologicos (INDRE) | Claudia Wong-Arambula, Abril Rodriguez-Maldonado, Vanessa Rivero-Arredondo, Ariadna Medina-Benitez, Joaquin Quiroz-Mercado, Sergio Rangel-Guerrero, Natividad Cruz-Ortiz, Tatiana Nunez-Garcia, Gisela Barrera-Badillo, Lucia Hernandez-Rivas, Irma Lopez-Martinez, Ernesto Ramirez-Gonzalez. |
| EPI_ISL_2362695, EPI_ISL_2362696 | LESP Tamaulipas | Instituto de Diagnostico y Referencia Epidemiologicos (INDRE) | Claudia Wong-Arambula, Abril Rodriguez-Maldonado, Vanessa Rivero-Arredondo, Ariadna Medina-Benitez, Joaquin Quiroz-Mercado, Sergio Rangel-Guerrero, Natividad Cruz-Ortiz, Tatiana Nunez-Garcia, Gisela Barrera-Badillo, Lucia Hernandez-Rivas, Irma Lopez-Martinez, Ernesto Ramirez-Gonzalez. |
| EPI_ISL_2362697 | LESP Sonora | Instituto de Diagnostico y Referencia Epidemiologicos (INDRE) | Claudia Wong-Arambula, Abril Rodriguez-Maldonado, Vanessa Rivero-Arredondo, Ariadna Medina-Benitez, Joaquin Quiroz-Mercado, Sergio Rangel-Guerrero, Natividad Cruz-Ortiz, Tatiana Nunez-Garcia, Gisela Barrera-Badillo, Lucia Hernandez-Rivas, Irma Lopez-Martinez, Ernesto Ramirez-Gonzalez. |
| EPI_ISL_2362698 | LESP Tamaulipas | Instituto de Diagnostico y Referencia Epidemiologicos (INDRE) | Claudia Wong-Arambula, Abril Rodriguez-Maldonado, Vanessa Rivero-Arredondo, Ariadna Medina-Benitez, Joaquin Quiroz-Mercado, Sergio Rangel-Guerrero, Natividad Cruz-Ortiz, Tatiana Nunez-Garcia, Gisela Barrera-Badillo, Lucia Hernandez-Rivas, Irma Lopez-Martinez, Ernesto Ramirez-Gonzalez. |
| EPI_ISL_2362700 | Laboratory Medicine, University of Washington | Laboratory Medicine, University of Washington | Roychoudhury,P., Xie,H., Shrestha,L., Bakhash,S.M., Lin,M.J.,Baker,N.R., Ellis,S., Sathees,S., Huang,M.-L., Jerome,K.R. and Greninger,A. |
| EPI_ISL_2364620, EPI_ISL_2364622, EPI_ISL_2364623, EPI_ISL_2364625, EPI_ISL_2364627, EPI_ISL_2364628, EPI_ISL_2364630, EPI_ISL_2364631, EPI_ISL_2364633, EPI_ISL_2364634, EPI_ISL_2364636, EPI_ISL_2364637, EPI_ISL_2364639, EPI_ISL_2364640, EPI_ISL_2364642, EPI_ISL_2364643, EPI_ISL_2364645, EPI_ISL_2364647, EPI_ISL_2364648, EPI_ISL_2364649, EPI_ISL_2364651, EPI_ISL_2364652, EPI_ISL_2364654, EPI_ISL_2364655, EPI_ISL_2364657, EPI_ISL_2364658, EPI_ISL_2364660, EPI_ISL_2364661, EPI_ISL_2364662, EPI_ISL_2364664, EPI_ISL_2364665, EPI_ISL_2364667, EPI_ISL_2364668, EPI_ISL_2364670, EPI_ISL_2364671, EPI_ISL_2364673, EPI_ISL_2364674, EPI_ISL_2364676, EPI_ISL_2364677, EPI_ISL_2364678, EPI_ISL_2364679, EPI_ISL_2364681, EPI_ISL_2364682, EPI_ISL_2364683, EPI_ISL_2364684, EPI_ISL_2364685, EPI_ISL_2364686, EPI_ISL_2364687, EPI_ISL_2364688, EPI_ISL_2364689, EPI_ISL_2364690, EPI_ISL_2364691, EPI_ISL_2364692, EPI_ISL_2364693, EPI_ISL_2364694, EPI_ISL_2364695, EPI_ISL_2364696, EPI_ISL_2364697, EPI_ISL_2364698, EPI_ISL_2364699, EPI_ISL_2364700, EPI_ISL_2364701, EPI_ISL_2364702, EPI_ISL_2364703, EPI_ISL_2364704, EPI_ISL_2364705, EPI_ISL_2364706, EPI_ISL_2364707, EPI_ISL_2364708, EPI_ISL_2364709, EPI_ISL_2364710, EPI_ISL_2364711, EPI_ISL_2364712, EPI_ISL_2364713, EPI_ISL_2364714, EPI_ISL_2364715, EPI_ISL_2364716, EPI_ISL_2364717, EPI_ISL_2364718, EPI_ISL_2364719, EPI_ISL_2364720, EPI_ISL_2364721, EPI_ISL_2364722, EPI_ISL_2364723, EPI_ISL_2364724, EPI_ISL_2364725, EPI_ISL_2364726, EPI_ISL_2364727, EPI_ISL_2364728, EPI_ISL_2364729, EPI_ISL_2364730, EPI_ISL_2364731, EPI_ISL_2364732, EPI_ISL_2364733, EPI_ISL_2364734, EPI_ISL_2364735, EPI_ISL_2364736, EPI_ISL_2364737, EPI_ISL_2364738, EPI_ISL_2364739, EPI_ISL_2364740, EPI_ISL_2364741, EPI_ISL_2364742, EPI_ISL_2364744, EPI_ISL_2364745, EPI_ISL_2364746, EPI_ISL_2364747, EPI_ISL_2364748, EPI_ISL_2364749, EPI_ISL_2364750, EPI_ISL_2364751, EPI_ISL_2364752, EPI_ISL_2364754, EPI_ISL_2364755, EPI_ISL_2364756, EPI_ISL_2364757, EPI_ISL_2364758, EPI_ISL_2364759, EPI_ISL_2364760, EPI_ISL_2364761, EPI_ISL_2364762, EPI_ISL_2364763, EPI_ISL_2364764, EPI_ISL_2364765, EPI_ISL_2364766, EPI_ISL_2364767, EPI_ISL_2364768, EPI_ISL_2364769, EPI_ISL_2364770, EPI_ISL_2364771, EPI_ISL_2364772, EPI_ISL_2364773, EPI_ISL_2364774, EPI_ISL_2364775, EPI_ISL_2364776, EPI_ISL_2364777, EPI_ISL_2364778, EPI_ISL_2364779, EPI_ISL_2364780, EPI_ISL_2364781, EPI_ISL_2364782, EPI_ISL_2364783, EPI_ISL_2364784, EPI_ISL_2364785, EPI_ISL_2364786, EPI_ISL_2364787 | | | |
| see above | Pathology and Laboratory Medicine Institute, Cleveland Clinic, Ohio, USA | Pathology and Laboratory Medicine Institute, Cleveland Clinic, Ohio, USA | Jennifer Starbuck, Concetta Peck, Joy Nakitandwe, Kristen McDonnell, David Plunkett, Zheng Jin Tu, Jay Brock, Yu-Wei Cheng, Gary Procop, Daniel Rhoads, Daniel H. Farkas, David Bosler |
| EPI_ISL_2367564, EPI_ISL_2367606 | Ohio Department of Health Laboratory | Ohio Department of Health Laboratory | Holmes, Jennifer; Eric Brandt, Keoni Omura, Glen McGillivary, Caitlin McDonnell, Jade Mowery, Stephanie Mccracken, Tyler Payne, Kirtana Ramadugu, Erica Leasure, Brent Lee, Kelsey Florek, Heather Blankenship, Allison Black, Quanta Brown, and Tammy Bannerman |
| EPI_ISL_2367607, EPI_ISL_2367608, EPI_ISL_2367609, EPI_ISL_2367610, EPI_ISL_2367611, EPI_ISL_2367612, EPI_ISL_2367613, EPI_ISL_2367614, EPI_ISL_2367615, EPI_ISL_2367616, EPI_ISL_2367617, EPI_ISL_2367618, EPI_ISL_2367619, EPI_ISL_2367620, EPI_ISL_2367621, EPI_ISL_2367622, EPI_ISL_2367623, EPI_ISL_2367624, EPI_ISL_2367625, EPI_ISL_2367626, EPI_ISL_2367627, EPI_ISL_2367628, EPI_ISL_2367629, EPI_ISL_2367630, EPI_ISL_2367631, EPI_ISL_2367632, EPI_ISL_2367633, EPI_ISL_2367634, EPI_ISL_2367635, EPI_ISL_2367636, EPI_ISL_2367637, EPI_ISL_2367638, EPI_ISL_2367639, EPI_ISL_2367640, EPI_ISL_2367641, EPI_ISL_2367642, EPI_ISL_2367643, EPI_ISL_2367644, EPI_ISL_2367645, EPI_ISL_2367646, EPI_ISL_2367647, EPI_ISL_2367648, EPI_ISL_2367649, EPI_ISL_2367650, EPI_ISL_2367651, EPI_ISL_2367652 | | | |
| see above | Helix/Illumina | Centers for Disease Control and Prevention Division of Viral Diseases, Pathogen Discovery | Dakota Howard, Dhwani Batra, Peter W. Cook, Kara Moser, Adrian Paskey, Jason Caravas, Benjamin Rambo-Martin, Shatavia Morrison, Christopher Gulvick, Scott Sammons, Yvette Unoarumhi, Darlene Wagner, Matthew Schmerer, Eileen de Feo, Jan Antico, Christine Tran, Matthew Tolentino, Shannon Wickline, Kim Gietzen, Brad Sickler, Jingtao Liu, Eric Allen, Phil Febbo, Nicole L. Washington, Simon White, Geraint Levan, Kelly Schiabor Barrett, Elizabeth Cirulli, Alexandre Bolze, Ary Ascencio, Charlotte Rivera-Garcia, Ryan Cho, Jason Nguyen, Sherry Wang, Jimmy Ramirez, Tyler Cassens, Efren Sandoval, Magnus Isaksson, William Lee, David Becker, Marc Laurent, James Lu, Clinton R. Paden, Duncan MacCannell |
| EPI_ISL_2367653, EPI_ISL_2367654, EPI_ISL_2367655, EPI_ISL_2367656, EPI_ISL_2367657, EPI_ISL_2367658, EPI_ISL_2367659, EPI_ISL_2367660, EPI_ISL_2367661, EPI_ISL_2367662, EPI_ISL_2367663, EPI_ISL_2367664, EPI_ISL_2367665, EPI_ISL_2367666, EPI_ISL_2367667, EPI_ISL_2367668, EPI_ISL_2367669, EPI_ISL_2367671, EPI_ISL_2367672, EPI_ISL_2367673, EPI_ISL_2367674, EPI_ISL_2367675, EPI_ISL_2367676, EPI_ISL_2367677 | | | |
| see above | Aegis Sciences Corporation | Centers for Disease Control and Prevention Division of Viral Diseases, Pathogen Discovery | Dakota Howard, Dhwani Batra, Peter W. Cook, Kara Moser, Adrian Paskey, Jason Caravas, Benjamin Rambo-Martin, Shatavia Morrison, Christopher Gulvick, Scott Sammons, Yvette Unoarumhi, Darlene Wagner, Matthew Schmerer, Cyndi Clark, Patrick Campbell, Rob Case, Vikramsinha Ghorpade, Holly Houdeshell, Ola Kvalvaag, Dillon Nall, Ethan Sanders, Alec Vest, Shaun Westlund, Matthew Hardison, Clinton R. Paden, Duncan MacCannell |
| EPI_ISL_2367678, EPI_ISL_2367679, EPI_ISL_2367680, EPI_ISL_2367681, EPI_ISL_2367682, EPI_ISL_2367683, EPI_ISL_2367684, EPI_ISL_2367685, EPI_ISL_2367686, EPI_ISL_2367687, EPI_ISL_2367688, EPI_ISL_2367689, EPI_ISL_2367690, EPI_ISL_2367691, EPI_ISL_2367692, EPI_ISL_2367693, EPI_ISL_2367694, EPI_ISL_2367695, EPI_ISL_2367696, EPI_ISL_2367697, EPI_ISL_2367698, EPI_ISL_2367699, EPI_ISL_2367700, EPI_ISL_2367701, EPI_ISL_2367702, EPI_ISL_2367703, EPI_ISL_2367704, EPI_ISL_2367705, EPI_ISL_2367706, EPI_ISL_2367707, EPI_ISL_2367708, EPI_ISL_2367709, EPI_ISL_2367710, EPI_ISL_2367711, EPI_ISL_2367712, EPI_ISL_2367713, EPI_ISL_2367714, EPI_ISL_2367715, EPI_ISL_2367716, EPI_ISL_2367717, EPI_ISL_2367718, EPI_ISL_2367719, EPI_ISL_2367720, EPI_ISL_2367721, EPI_ISL_2367722, EPI_ISL_2367723, EPI_ISL_2367724, EPI_ISL_2367725, EPI_ISL_2367726, EPI_ISL_2367727, EPI_ISL_2367728, EPI_ISL_2367729, EPI_ISL_2367730, EPI_ISL_2367731, EPI_ISL_2367732, EPI_ISL_2367733, EPI_ISL_2367734, EPI_ISL_2367735, EPI_ISL_2367736 | | | |
| see above | Quest Diagnostics Incorporated | Centers for Disease Control and Prevention Division of Viral Diseases, Pathogen Discovery | Dakota Howard, Dhwani Batra, Peter W. Cook, Kara Moser, Adrian Paskey, Jason Caravas, Benjamin Rambo-Martin, Shatavia Morrison, Christopher Gulvick, Scott Sammons, Yvette Unoarumhi, Darlene Wagner, Matthew Schmerer, S. H. Rosenthal, A. Gerasimova, R. M. Kagan, B. Anderson, M. Hua, Y. Liu, L.E. Bernstein, K.E. Livingston, A. Perez, I. A. Shlyakhter, R. V. Rolando, R. Owen, P. Tanpaiboon, F. Lacbawan, Clinton R. Paden, Duncan MacCannell |
| EPI_ISL_2367737 | Ohio Department of Health Laboratory | Ohio Department of Health Laboratory | Holmes, Jennifer; Eric Brandt, Keoni Omura, Glen McGillivary, Caitlin McDonnell, Jade Mowery, Stephanie Mccracken, Tyler Payne, Kirtana Ramadugu, Erica Leasure, Brent Lee, Kelsey Florek, Heather Blankenship, Allison Black, Quanta Brown, and Tammy Bannerman |
| EPI_ISL_2367738, EPI_ISL_2367739, EPI_ISL_2367740, EPI_ISL_2367741, EPI_ISL_2367742, EPI_ISL_2367743, EPI_ISL_2367744, EPI_ISL_2367745, EPI_ISL_2367746, EPI_ISL_2367747, EPI_ISL_2367748, EPI_ISL_2367749, EPI_ISL_2367750, EPI_ISL_2367751, EPI_ISL_2367752, EPI_ISL_2367753, EPI_ISL_2367754, EPI_ISL_2367755, EPI_ISL_2367756, EPI_ISL_2367757, EPI_ISL_2367758, EPI_ISL_2367759, EPI_ISL_2367760, EPI_ISL_2367761, EPI_ISL_2367764, EPI_ISL_2367766, EPI_ISL_2367768, EPI_ISL_2367769, EPI_ISL_2367772, EPI_ISL_2367773, EPI_ISL_2367776, EPI_ISL_2367778, EPI_ISL_2367780, EPI_ISL_2367782, EPI_ISL_2367784, EPI_ISL_2367786, EPI_ISL_2367788, EPI_ISL_2367789, EPI_ISL_2367792, EPI_ISL_2367793, EPI_ISL_2367794, EPI_ISL_2367795, EPI_ISL_2367796, EPI_ISL_2367797, EPI_ISL_2367798, EPI_ISL_2367799, EPI_ISL_2367800, EPI_ISL_2367801, EPI_ISL_2367802, EPI_ISL_2367803, EPI_ISL_2367804, EPI_ISL_2367805, EPI_ISL_2367806, EPI_ISL_2367807, EPI_ISL_2367808, EPI_ISL_2367809, EPI_ISL_2367810, EPI_ISL_2367811, EPI_ISL_2367812, EPI_ISL_2367813, EPI_ISL_2367814, EPI_ISL_2367815, EPI_ISL_2367816, EPI_ISL_2367817, EPI_ISL_2367818, EPI_ISL_2367819, EPI_ISL_2367820, EPI_ISL_2367821, EPI_ISL_2367822, EPI_ISL_2367823, EPI_ISL_2367824, EPI_ISL_2367825, EPI_ISL_2367826, EPI_ISL_2367827, EPI_ISL_2367828, EPI_ISL_2367829, EPI_ISL_2367830, EPI_ISL_2367831, EPI_ISL_2367832, EPI_ISL_2367833, EPI_ISL_2367834, EPI_ISL_2367835, EPI_ISL_2367836, EPI_ISL_2367837, EPI_ISL_2367838, EPI_ISL_2367839, EPI_ISL_2367840, EPI_ISL_2367841, EPI_ISL_2367842, EPI_ISL_2367843, EPI_ISL_2367844, EPI_ISL_2367845, EPI_ISL_2367846, EPI_ISL_2367847, EPI_ISL_2367848, EPI_ISL_2367849, EPI_ISL_2367850, EPI_ISL_2367851, EPI_ISL_2367852, EPI_ISL_2367853, EPI_ISL_2367854, EPI_ISL_2367855, EPI_ISL_2367856, EPI_ISL_2367857, EPI_ISL_2367858, EPI_ISL_2367859, EPI_ISL_2367860, EPI_ISL_2367861, EPI_ISL_2367862, EPI_ISL_2367863, EPI_ISL_2367864, EPI_ISL_2367865, EPI_ISL_2367866, EPI_ISL_2367867, EPI_ISL_2367868, EPI_ISL_2367869, EPI_ISL_2367870, EPI_ISL_2367871, EPI_ISL_2367872, EPI_ISL_2367873, EPI_ISL_2367874, EPI_ISL_2367875, EPI_ISL_2367876, EPI_ISL_2367877, EPI_ISL_2367878, EPI_ISL_2367879, EPI_ISL_2367880, EPI_ISL_2367881, EPI_ISL_2367882, EPI_ISL_2367883, EPI_ISL_2367884, EPI_ISL_2367885, EPI_ISL_2367886, EPI_ISL_2367887, EPI_ISL_2367888, EPI_ISL_2367889, EPI_ISL_2367890, EPI_ISL_2367891, EPI_ISL_2367892, EPI_ISL_2367893, EPI_ISL_2367894, EPI_ISL_2367895, EPI_ISL_2367896, EPI_ISL_2367897, EPI_ISL_2367898, EPI_ISL_2367899, EPI_ISL_2367900, EPI_ISL_2367901, EPI_ISL_2367902, EPI_ISL_2367903, EPI_ISL_2367904, EPI_ISL_2367905, EPI_ISL_2367906, EPI_ISL_2367907, EPI_ISL_2367908, EPI_ISL_2367909, EPI_ISL_2367910, EPI_ISL_2367911, EPI_ISL_2367912, EPI_ISL_2367913, EPI_ISL_2367914, EPI_ISL_2367915, EPI_ISL_2367916, EPI_ISL_2367917, EPI_ISL_2367918, EPI_ISL_2367919, EPI_ISL_2367920, EPI_ISL_2367921, EPI_ISL_2367922, EPI_ISL_2367923, EPI_ISL_2367924, EPI_ISL_2367925, EPI_ISL_2367926, EPI_ISL_2367927, EPI_ISL_2367928, EPI_ISL_2367929, EPI_ISL_2367930, EPI_ISL_2367931, EPI_ISL_2367932, EPI_ISL_2367933, EPI_ISL_2367934, EPI_ISL_2367935, EPI_ISL_2367936, EPI_ISL_2367937, EPI_ISL_2367938, EPI_ISL_2367939, EPI_ISL_2367940, EPI_ISL_2367941, EPI_ISL_2367942, EPI_ISL_2367943, EPI_ISL_2367944, EPI_ISL_2367945, EPI_ISL_2367946, EPI_ISL_2367947, EPI_ISL_2367948, EPI_ISL_2367949, EPI_ISL_2367950, EPI_ISL_2367951, EPI_ISL_2367952, EPI_ISL_2367953, EPI_ISL_2367954, EPI_ISL_2367955, EPI_ISL_2367956, EPI_ISL_2367957, EPI_ISL_2367958, EPI_ISL_2367959, EPI_ISL_2367960, EPI_ISL_2367961, EPI_ISL_2367962, EPI_ISL_2367963, EPI_ISL_2367964, EPI_ISL_2367965, EPI_ISL_2367966, EPI_ISL_2367967, EPI_ISL_2367968, EPI_ISL_2367969, EPI_ISL_2367970, EPI_ISL_2367971, EPI_ISL_2367972, EPI_ISL_2367973, EPI_ISL_2367974, EPI_ISL_2367975, EPI_ISL_2367976, EPI_ISL_2367977, EPI_ISL_2367978, EPI_ISL_2367979, EPI_ISL_2367980, EPI_ISL_2367981, EPI_ISL_2367982, EPI_ISL_2367983, EPI_ISL_2367984, EPI_ISL_2367985, EPI_ISL_2367986, EPI_ISL_2367987, EPI_ISL_2367988, EPI_ISL_2367989, EPI_ISL_2367990, EPI_ISL_2367991, EPI_ISL_2367992, EPI_ISL_2367993, EPI_ISL_2367994, EPI_ISL_2367995, EPI_ISL_2367996, EPI_ISL_2367997, EPI_ISL_2367998, EPI_ISL_2367999, EPI_ISL_2368000, EPI_ISL_2368001, EPI_ISL_2368002, EPI_ISL_2368003, EPI_ISL_2368004, EPI_ISL_2368005, EPI_ISL_2368006, EPI_ISL_2368007, EPI_ISL_2368008, EPI_ISL_2368009, EPI_ISL_2368010, EPI_ISL_2368011, EPI_ISL_2368012, EPI_ISL_2368013, EPI_ISL_2368014, EPI_ISL_2368015, EPI_ISL_2368016, EPI_ISL_2368017, EPI_ISL_2368018, EPI_ISL_2368019, EPI_ISL_2368020, EPI_ISL_2368021, EPI_ISL_2368022, EPI_ISL_2368023, EPI_ISL_2368024, EPI_ISL_2368025, EPI_ISL_2368026, EPI_ISL_2368027, EPI_ISL_2368028, EPI_ISL_2368029, EPI_ISL_2368030, EPI_ISL_2368031, EPI_ISL_2368032, EPI_ISL_2368033, EPI_ISL_2368034, EPI_ISL_2368035, EPI_ISL_2368036, EPI_ISL_2368037, EPI_ISL_2368038, EPI_ISL_2368039, EPI_ISL_2368040, EPI_ISL_2368041, EPI_ISL_2368042, EPI_ISL_2368043, EPI_ISL_2368044, EPI_ISL_2368045, EPI_ISL_2368046, EPI_ISL_2368047, EPI_ISL_2368048, EPI_ISL_2368049, EPI_ISL_2368050, EPI_ISL_2368051, EPI_ISL_2368052, EPI_ISL_2368053, EPI_ISL_2368054, EPI_ISL_2368055, EPI_ISL_2368056, EPI_ISL_2368057, EPI_ISL_2368058, EPI_ISL_2368059, EPI_ISL_2368060, EPI_ISL_2368061, EPI_ISL_2368062, EPI_ISL_2368063, EPI_ISL_2368064, EPI_ISL_2368065, EPI_ISL_2368066 | | | |
| see above | Quest Diagnostics Incorporated | Centers for Disease Control and Prevention Division of Viral Diseases, Pathogen Discovery | Dakota Howard, Dhwani Batra, Peter W. Cook, Kara Moser, Adrian Paskey, Jason Caravas, Benjamin Rambo-Martin, Shatavia Morrison, Christopher Gulvick, Scott Sammons, Yvette Unoarumhi, Darlene Wagner, Matthew Schmerer, S. H. Rosenthal, A. Gerasimova, R. M. Kagan, B. Anderson, M. Hua, Y. Liu, L.E. Bernstein, K.E. Livingston, A. Perez, I. A. Shlyakhter, R. V. Rolando, R. Owen, P. Tanpaiboon, F. Lacbawan, Clinton R. Paden, Duncan MacCannell |
| EPI_ISL_2368067 | Ohio Department of Health Laboratory | Ohio Department of Health Laboratory | Holmes, Jennifer; Eric Brandt, Keoni Omura, Glen McGillivary, Caitlin McDonnell, Jade Mowery, Stephanie Mccracken, Tyler Payne, Kirtana Ramadugu, Erica Leasure, Brent Lee, Kelsey Florek, Heather Blankenship, Allison Black, Quanta Brown, and Tammy Bannerman |
| EPI_ISL_2368068, EPI_ISL_2368069, EPI_ISL_2368070, EPI_ISL_2368071, EPI_ISL_2368072, EPI_ISL_2368073, EPI_ISL_2368074, EPI_ISL_2368075, EPI_ISL_2368076, EPI_ISL_2368077, EPI_ISL_2368078, EPI_ISL_2368080, EPI_ISL_2368081, EPI_ISL_2368083, EPI_ISL_2368084, EPI_ISL_2368085, EPI_ISL_2368086, EPI_ISL_2368087, EPI_ISL_2368088, EPI_ISL_2368089, EPI_ISL_2368090, EPI_ISL_2368091, EPI_ISL_2368092, EPI_ISL_2368094, EPI_ISL_2368095, EPI_ISL_2368096, EPI_ISL_2368097, EPI_ISL_2368098, EPI_ISL_2368099, EPI_ISL_2368100, EPI_ISL_2368101, EPI_ISL_2368102, EPI_ISL_2368103, EPI_ISL_2368104, EPI_ISL_2368105, EPI_ISL_2368106, EPI_ISL_2368107, EPI_ISL_2368108, EPI_ISL_2368109, EPI_ISL_2368110, EPI_ISL_2368111, EPI_ISL_2368112, EPI_ISL_2368113, EPI_ISL_2368114, EPI_ISL_2368115, EPI_ISL_2368116, EPI_ISL_2368118, EPI_ISL_2368119, EPI_ISL_2368121, EPI_ISL_2368122, EPI_ISL_2368123, EPI_ISL_2368124, EPI_ISL_2368125, EPI_ISL_2368126, EPI_ISL_2368127, EPI_ISL_2368128, EPI_ISL_2368129, EPI_ISL_2368131, EPI_ISL_2368132, EPI_ISL_2368133, EPI_ISL_2368134, EPI_ISL_2368135, EPI_ISL_2368136, EPI_ISL_2368137, EPI_ISL_2368138, EPI_ISL_2368139, EPI_ISL_2368140, EPI_ISL_2368141, EPI_ISL_2368142, EPI_ISL_2368143, EPI_ISL_2368144, EPI_ISL_2368145, EPI_ISL_2368146, EPI_ISL_2368147, EPI_ISL_2368148, EPI_ISL_2368150, EPI_ISL_2368152, EPI_ISL_2368153, EPI_ISL_2368154, EPI_ISL_2368156, EPI_ISL_2368157, EPI_ISL_2368158, EPI_ISL_2368159, EPI_ISL_2368160 | | | |
| see above | Fulgent Genetics | Centers for Disease Control and Prevention Division of Viral Diseases, Pathogen Discovery | Dakota Howard, Dhwani Batra, Peter W. Cook, Kara Moser, Adrian Paskey, Jason Caravas, Benjamin Rambo-Martin, Shatavia Morrison, Christopher Gulvick, Scott Sammons, Yvette Unoarumhi, Darlene Wagner, Matthew Schmerer, Harry Gao, Mickey Li, John Gao, Joseph Fierro, Benafsh Sapra, Becky Tsai, Yan Meng, Doreen Ng, James Xie, Clinton R. Paden, Duncan MacCannell |
| EPI_ISL_2368161 | Ohio Department of Health Laboratory | Ohio Department of Health Laboratory | Holmes, Jennifer; Eric Brandt, Keoni Omura, Glen McGillivary, Caitlin McDonnell, Jade Mowery, Stephanie Mccracken, Tyler Payne, Kirtana Ramadugu, Erica Leasure, Brent Lee, Kelsey Florek, Heather Blankenship, Allison Black, Quanta Brown, and Tammy Bannerman |
| EPI_ISL_2368162, EPI_ISL_2368163, EPI_ISL_2368165, EPI_ISL_2368166, EPI_ISL_2368167, EPI_ISL_2368168, EPI_ISL_2368170, EPI_ISL_2368171, EPI_ISL_2368172, EPI_ISL_2368173, EPI_ISL_2368174, EPI_ISL_2368176, EPI_ISL_2368177, EPI_ISL_2368178, EPI_ISL_2368179, EPI_ISL_2368180, EPI_ISL_2368181, EPI_ISL_2368182, EPI_ISL_2368183, EPI_ISL_2368184, EPI_ISL_2368186, EPI_ISL_2368187, EPI_ISL_2368188, EPI_ISL_2368191, EPI_ISL_2368192, EPI_ISL_2368193, EPI_ISL_2368194, EPI_ISL_2368195, EPI_ISL_2368196, EPI_ISL_2368197, EPI_ISL_2368199, EPI_ISL_2368200, EPI_ISL_2368201, EPI_ISL_2368202, EPI_ISL_2368203, EPI_ISL_2368204 | | | |
| see above | Fulgent Genetics | Centers for Disease Control and Prevention Division of Viral Diseases, Pathogen Discovery | Dakota Howard, Dhwani Batra, Peter W. Cook, Kara Moser, Adrian Paskey, Jason Caravas, Benjamin Rambo-Martin, Shatavia Morrison, Christopher Gulvick, Scott Sammons, Yvette Unoarumhi, Darlene Wagner, Matthew Schmerer, Harry Gao, Mickey Li, John Gao, Joseph Fierro, Benafsh Sapra, Becky Tsai, Yan Meng, Doreen Ng, James Xie, Clinton R. Paden, Duncan MacCannell |
| EPI_ISL_2368205, EPI_ISL_2368206, EPI_ISL_2368207, EPI_ISL_2368208, EPI_ISL_2368209, EPI_ISL_2368210, EPI_ISL_2368211, EPI_ISL_2368212, EPI_ISL_2368213, EPI_ISL_2368214, EPI_ISL_2368215, EPI_ISL_2368216, EPI_ISL_2368217, EPI_ISL_2368218, EPI_ISL_2368219, EPI_ISL_2368220, EPI_ISL_2368221, EPI_ISL_2368222, EPI_ISL_2368223, EPI_ISL_2368224, EPI_ISL_2368225, EPI_ISL_2368226, EPI_ISL_2368227, EPI_ISL_2368228, EPI_ISL_2368229, EPI_ISL_2368230, EPI_ISL_2368231, EPI_ISL_2368232, EPI_ISL_2368233, EPI_ISL_2368234, EPI_ISL_2368235, EPI_ISL_2368236, EPI_ISL_2368237, EPI_ISL_2368238, EPI_ISL_2368239, EPI_ISL_2368240, EPI_ISL_2368241, EPI_ISL_2368242, EPI_ISL_2368243, EPI_ISL_2368244, EPI_ISL_2368245, EPI_ISL_2368246, EPI_ISL_2368247, EPI_ISL_2368248, EPI_ISL_2368249, EPI_ISL_2368250, EPI_ISL_2368251, EPI_ISL_2368252, EPI_ISL_2368253, EPI_ISL_2368254, EPI_ISL_2368255, EPI_ISL_2368256, EPI_ISL_2368257, EPI_ISL_2368258, EPI_ISL_2368259, EPI_ISL_2368260, EPI_ISL_2368261, EPI_ISL_2368262, EPI_ISL_2368263, EPI_ISL_2368265, EPI_ISL_2368266, EPI_ISL_2368267, EPI_ISL_2368268, EPI_ISL_2368269, EPI_ISL_2368270, EPI_ISL_2368271, EPI_ISL_2368272, EPI_ISL_2368273, EPI_ISL_2368274, EPI_ISL_2368275, EPI_ISL_2368276, EPI_ISL_2368277, EPI_ISL_2368278, EPI_ISL_2368279, EPI_ISL_2368280, EPI_ISL_2368282, EPI_ISL_2368283, EPI_ISL_2368284, EPI_ISL_2368285, EPI_ISL_2368286, EPI_ISL_2368287, EPI_ISL_2368288, EPI_ISL_2368289, EPI_ISL_2368290, EPI_ISL_2368291, EPI_ISL_2368292, EPI_ISL_2368293, EPI_ISL_2368294, EPI_ISL_2368295, EPI_ISL_2368296, EPI_ISL_2368297, EPI_ISL_2368298, EPI_ISL_2368299, EPI_ISL_2368300, EPI_ISL_2368301, EPI_ISL_2368302, EPI_ISL_2368303, EPI_ISL_2368304, EPI_ISL_2368305, EPI_ISL_2368306, EPI_ISL_2368307, EPI_ISL_2368308, EPI_ISL_2368310, EPI_ISL_2368311, EPI_ISL_2368312, EPI_ISL_2368313, EPI_ISL_2368314, EPI_ISL_2368315, EPI_ISL_2368316, EPI_ISL_2368317, EPI_ISL_2368318, EPI_ISL_2368319, EPI_ISL_2368320, EPI_ISL_2368321, EPI_ISL_2368322, EPI_ISL_2368324, EPI_ISL_2368325, EPI_ISL_2368326, EPI_ISL_2368327, EPI_ISL_2368328, EPI_ISL_2368329, EPI_ISL_2368330, EPI_ISL_2368332, EPI_ISL_2368333, EPI_ISL_2368334, EPI_ISL_2368335, EPI_ISL_2368336, EPI_ISL_2368337, EPI_ISL_2368338, EPI_ISL_2368339, EPI_ISL_2368340, EPI_ISL_2368341, EPI_ISL_2368342, EPI_ISL_2368343, EPI_ISL_2368344, EPI_ISL_2368345, EPI_ISL_2368346, EPI_ISL_2368347, EPI_ISL_2368348, EPI_ISL_2368349, EPI_ISL_2368350, EPI_ISL_2368351, EPI_ISL_2368352, EPI_ISL_2368353, EPI_ISL_2368354, EPI_ISL_2368355, EPI_ISL_2368356, EPI_ISL_2368357, EPI_ISL_2368358, EPI_ISL_2368359, EPI_ISL_2368360, EPI_ISL_2368361, EPI_ISL_2368362, EPI_ISL_2368363, EPI_ISL_2368364, EPI_ISL_2368365, EPI_ISL_2368366, EPI_ISL_2368367, EPI_ISL_2368368, EPI_ISL_2368369, EPI_ISL_2368370, EPI_ISL_2368371, EPI_ISL_2368372, EPI_ISL_2368373, EPI_ISL_2368374, EPI_ISL_2368375, EPI_ISL_2368376, EPI_ISL_2368377, EPI_ISL_2368378, EPI_ISL_2368379, EPI_ISL_2368380, EPI_ISL_2368381, EPI_ISL_2368382, EPI_ISL_2368383, EPI_ISL_2368384, EPI_ISL_2368385, EPI_ISL_2368386, EPI_ISL_2368387, EPI_ISL_2368388, EPI_ISL_2368389, EPI_ISL_2368390, EPI_ISL_2368391, EPI_ISL_2368392, EPI_ISL_2368393, EPI_ISL_2368394, EPI_ISL_2368395, EPI_ISL_2368396, EPI_ISL_2368397, EPI_ISL_2368398, EPI_ISL_2368399, EPI_ISL_2368400, EPI_ISL_2368402, EPI_ISL_2368403, EPI_ISL_2368404, EPI_ISL_2368405, EPI_ISL_2368406, EPI_ISL_2368407, EPI_ISL_2368408, EPI_ISL_2368409, EPI_ISL_2368410, EPI_ISL_2368411, EPI_ISL_2368412, EPI_ISL_2368413, EPI_ISL_2368414, EPI_ISL_2368415, EPI_ISL_2368416, EPI_ISL_2368417, EPI_ISL_2368418, EPI_ISL_2368419, EPI_ISL_2368420, EPI_ISL_2368424, EPI_ISL_2368425, EPI_ISL_2368426, EPI_ISL_2368427, EPI_ISL_2368428, EPI_ISL_2368429, EPI_ISL_2368430, EPI_ISL_2368431, EPI_ISL_2368432, EPI_ISL_2368433, EPI_ISL_2368434, EPI_ISL_2368436, EPI_ISL_2368437, EPI_ISL_2368438, EPI_ISL_2368439, EPI_ISL_2368440, EPI_ISL_2368441, EPI_ISL_2368442, EPI_ISL_2368443, EPI_ISL_2368444, EPI_ISL_2368445, EPI_ISL_2368446, EPI_ISL_2368447, EPI_ISL_2368448, EPI_ISL_2368449, EPI_ISL_2368450, EPI_ISL_2368451, EPI_ISL_2368452, EPI_ISL_2368453, EPI_ISL_2368454, EPI_ISL_2368455, EPI_ISL_2368456, EPI_ISL_2368457, EPI_ISL_2368458, EPI_ISL_2368459, EPI_ISL_2368460, EPI_ISL_2368461, EPI_ISL_2368462, EPI_ISL_2368463, EPI_ISL_2368464, EPI_ISL_2368465, EPI_ISL_2368466, EPI_ISL_2368467, EPI_ISL_2368468, EPI_ISL_2368469, EPI_ISL_2368470, EPI_ISL_2368471, EPI_ISL_2368472, EPI_ISL_2368473, EPI_ISL_2368474, EPI_ISL_2368475, EPI_ISL_2368476, EPI_ISL_2368477, EPI_ISL_2368478, EPI_ISL_2368479, EPI_ISL_2368480, EPI_ISL_2368481, EPI_ISL_2368482, EPI_ISL_2368483, EPI_ISL_2368484, EPI_ISL_2368485, EPI_ISL_2368486, EPI_ISL_2368487, EPI_ISL_2368488, EPI_ISL_2368489, EPI_ISL_2368490, EPI_ISL_2368491, EPI_ISL_2368492, EPI_ISL_2368493, EPI_ISL_2368495, EPI_ISL_2368498, EPI_ISL_2368501, EPI_ISL_2368507, EPI_ISL_2368509, EPI_ISL_2368512, EPI_ISL_2368516, | | | |

EPI_ISL_2368519, EPI_ISL_2368522, EPI_ISL_2368524, EPI_ISL_2368527, EPI_ISL_2368531, EPI_ISL_2368533, EPI_ISL_2368536, EPI_ISL_2368539, EPI_ISL_2368542, EPI_ISL_2368545, EPI_ISL_2368548, EPI_ISL_2368551, EPI_ISL_2368557, EPI_ISL_2368559, EPI_ISL_2368563, EPI_ISL_2368565, EPI_ISL_2368569, EPI_ISL_2368572, EPI_ISL_2368574, EPI_ISL_2368576, EPI_ISL_2368579, EPI_ISL_2368582,
EPI_ISL_2368591, EPI_ISL_2368597, EPI_ISL_2368600, EPI_ISL_2368603, EPI_ISL_2368606, EPI_ISL_2368608, EPI_ISL_2368611, EPI_ISL_2368613, EPI_ISL_2368616, EPI_ISL_2368619, EPI_ISL_2368622, EPI_ISL_2368625, EPI_ISL_2368628, EPI_ISL_2368630, EPI_ISL_2368636, EPI_ISL_2368638, EPI_ISL_2368641, EPI_ISL_2368644, EPI_ISL_2368646, EPI_ISL_2368649, EPI_ISL_2368652, EPI_ISL_2368655,
EPI_ISL_2368657, EPI_ISL_2368661, EPI_ISL_2368664, EPI_ISL_2368666, EPI_ISL_2368670, EPI_ISL_2368673, EPI_ISL_2368675, EPI_ISL_2368678, EPI_ISL_2368681, EPI_ISL_2368684, EPI_ISL_2368687, EPI_ISL_2368690, EPI_ISL_2368693, EPI_ISL_2368698, EPI_ISL_2368699, EPI_ISL_2368707, EPI_ISL_2368709, EPI_ISL_2368712, EPI_ISL_2368715, EPI_ISL_2368717, EPI_ISL_2368718, EPI_ISL_2368723,
EPI_ISL_2368726, EPI_ISL_2368727, EPI_ISL_2368730, EPI_ISL_2368733, EPI_ISL_2368739, EPI_ISL_2368742, EPI_ISL_2368744, EPI_ISL_2368747, EPI_ISL_2368750, EPI_ISL_2368753, EPI_ISL_2368756, EPI_ISL_2368758, EPI_ISL_2368761, EPI_ISL_2368764, EPI_ISL_2368767, EPI_ISL_2368769, EPI_ISL_2368772, EPI_ISL_2368775, EPI_ISL_2368778, EPI_ISL_2368781, EPI_ISL_2368784, EPI_ISL_2368786,
EPI_ISL_2368789, EPI_ISL_2368792, EPI_ISL_2368794, EPI_ISL_2368796, EPI_ISL_2368797, EPI_ISL_2368800, EPI_ISL_2368803, EPI_ISL_2368806, EPI_ISL_2368809, EPI_ISL_2368812, EPI_ISL_2368814, EPI_ISL_2368817, EPI_ISL_2368820, EPI_ISL_2368823, EPI_ISL_2368826, EPI_ISL_2368827, EPI_ISL_2368829, EPI_ISL_2368836, EPI_ISL_2368838, EPI_ISL_2368841, EPI_ISL_2368843, EPI_ISL_2368845,
EPI_ISL_2368848, EPI_ISL_2368851, EPI_ISL_2368854, EPI_ISL_2368857, EPI_ISL_2368859, EPI_ISL_2368862, EPI_ISL_2368865, EPI_ISL_2368868, EPI_ISL_2368870, EPI_ISL_2368873, EPI_ISL_2368876, EPI_ISL_2368879, EPI_ISL_2368882, EPI_ISL_2368884, EPI_ISL_2368887, EPI_ISL_2368889, EPI_ISL_2368892, EPI_ISL_2368895, EPI_ISL_2368898, EPI_ISL_2368900, EPI_ISL_2368903, EPI_ISL_2368906,
EPI_ISL_2368909, EPI_ISL_2368912, EPI_ISL_2368915, EPI_ISL_2368922, EPI_ISL_2368925, EPI_ISL_2368927, EPI_ISL_2368928, EPI_ISL_2368931, EPI_ISL_2368934, EPI_ISL_2368936, EPI_ISL_2368939, EPI_ISL_2368945, EPI_ISL_2368948, EPI_ISL_2368954, EPI_ISL_2368957, EPI_ISL_2368963, EPI_ISL_2368966, EPI_ISL_2368969, EPI_ISL_2368971, EPI_ISL_2368973, EPI_ISL_2368979, EPI_ISL_2368981

see above | Aegis Sciences Corporation | Centers for Disease Control and Prevention Division of Viral Diseases, Pathogen Discovery | Dakota Howard, Dhwani Batra, Peter W. Cook, Kara Moser, Adrian Paskey, Jason Caravas, Benjamin Rambo-Martin, Shatavia Morrison, Christopher Gulvick, Scott Sammons, Yvette Unoarumhi, Darlene Wagner, Matthew Schmerer, Cyndi Clark, Patrick Campbell, Rob Case, Vikramsinha Ghorpade, Holly Houdeshell, Ola Kvalvaag, Dillon Nall, Ethan Sanders, Alec Vest, Shaun Westlund, Matthew Hardison, Clinton R. Paden, Duncan MacCannell

EPI_ISL_2369005, EPI_ISL_2369008, EPI_ISL_2369011, EPI_ISL_2369014, EPI_ISL_2369017, EPI_ISL_2369020, EPI_ISL_2369022, EPI_ISL_2369025, EPI_ISL_2369028, EPI_ISL_2369031, EPI_ISL_2369034, EPI_ISL_2369035, EPI_ISL_2369037, EPI_ISL_2369040, EPI_ISL_2369043, EPI_ISL_2369046, EPI_ISL_2369049, EPI_ISL_2369051, EPI_ISL_2369054, EPI_ISL_2369057, EPI_ISL_2369059, EPI_ISL_2369062,
EPI_ISL_2369065, EPI_ISL_2369067, EPI_ISL_2369070, EPI_ISL_2369073, EPI_ISL_2369076, EPI_ISL_2369078, EPI_ISL_2369081, EPI_ISL_2369084, EPI_ISL_2369087, EPI_ISL_2369090, EPI_ISL_2369093, EPI_ISL_2369095, EPI_ISL_2369098, EPI_ISL_2369101, EPI_ISL_2369103, EPI_ISL_2369106, EPI_ISL_2369109, EPI_ISL_2369111, EPI_ISL_2369114, EPI_ISL_2369116, EPI_ISL_2369119, EPI_ISL_2369122,
EPI_ISL_2369124, EPI_ISL_2369127, EPI_ISL_2369130, EPI_ISL_2369132, EPI_ISL_2369135, EPI_ISL_2369139, EPI_ISL_2369141, EPI_ISL_2369144, EPI_ISL_2369147, EPI_ISL_2369149, EPI_ISL_2369152, EPI_ISL_2369155, EPI_ISL_2369158, EPI_ISL_2369161, EPI_ISL_2369164, EPI_ISL_2369167, EPI_ISL_2369170, EPI_ISL_2369172, EPI_ISL_2369175, EPI_ISL_2369178, EPI_ISL_2369181, EPI_ISL_2369184,
EPI_ISL_2369186, EPI_ISL_2369189, EPI_ISL_2369192, EPI_ISL_2369195, EPI_ISL_2369197, EPI_ISL_2369200, EPI_ISL_2369203, EPI_ISL_2369206, EPI_ISL_2369209, EPI_ISL_2369212, EPI_ISL_2369215, EPI_ISL_2369216, EPI_ISL_2369218, EPI_ISL_2369220, EPI_ISL_2369223, EPI_ISL_2369226, EPI_ISL_2369228, EPI_ISL_2369231, EPI_ISL_2369234, EPI_ISL_2369237, EPI_ISL_2369239, EPI_ISL_2369242,
EPI_ISL_2369244, EPI_ISL_2369247, EPI_ISL_2369250, EPI_ISL_2369253, EPI_ISL_2369255, EPI_ISL_2369258, EPI_ISL_2369261, EPI_ISL_2369264, EPI_ISL_2369267, EPI_ISL_2369270, EPI_ISL_2369273, EPI_ISL_2369276, EPI_ISL_2369279, EPI_ISL_2369282, EPI_ISL_2369285, EPI_ISL_2369287, EPI_ISL_2369290, EPI_ISL_2369293, EPI_ISL_2369296, EPI_ISL_2369299, EPI_ISL_2369302, EPI_ISL_2369304,
EPI_ISL_2369307, EPI_ISL_2369309, EPI_ISL_2369312, EPI_ISL_2369315, EPI_ISL_2369317, EPI_ISL_2369320, EPI_ISL_2369322, EPI_ISL_2369325, EPI_ISL_2369328, EPI_ISL_2369330, EPI_ISL_2369332, EPI_ISL_2369335, EPI_ISL_2369338, EPI_ISL_2369341, EPI_ISL_2369344, EPI_ISL_2369347, EPI_ISL_2369351, EPI_ISL_2369354, EPI_ISL_2369357, EPI_ISL_2369360, EPI_ISL_2369363, EPI_ISL_2369365,
EPI_ISL_2369368, EPI_ISL_2369372, EPI_ISL_2369375, EPI_ISL_2369378, EPI_ISL_2369381, EPI_ISL_2369384, EPI_ISL_2369386, EPI_ISL_2369388, EPI_ISL_2369390, EPI_ISL_2369393, EPI_ISL_2369396, EPI_ISL_2369399, EPI_ISL_2369402, EPI_ISL_2369405, EPI_ISL_2369407, EPI_ISL_2369410, EPI_ISL_2369413, EPI_ISL_2369416, EPI_ISL_2369418, EPI_ISL_2369421, EPI_ISL_2369424

see above | Quest Diagnostics Incorporated | Centers for Disease Control and Prevention Division of Viral Diseases, Pathogen Discovery | Dakota Howard, Dhwani Batra, Peter W. Cook, Kara Moser, Adrian Paskey, Jason Caravas, Benjamin Rambo-Martin, Shatavia Morrison, Christopher Gulvick, Scott Sammons, Yvette Unoarumhi, Darlene Wagner, Matthew Schmerer, S. H. Rosenthal, A. Gerasimova, R. M. Kagan, B. Anderson, M. Hua, Y. Liu, L.E. Bernstein, K.E. Livingston, A. Perez, I. A. Shlyakhter, R. V. Rolando, R. Owen, P. Tanpaiboon, F. Lacbawan, Clinton R. Paden, Duncan MacCannell

EPI_ISL_2369942, EPI_ISL_2369945, EPI_ISL_2369947, EPI_ISL_2369952, EPI_ISL_2369953, EPI_ISL_2369955, EPI_ISL_2369957, EPI_ISL_2369960, EPI_ISL_2369963, EPI_ISL_2369964, EPI_ISL_2369966, EPI_ISL_2369968, EPI_ISL_2369970, EPI_ISL_2369972, EPI_ISL_2369973, EPI_ISL_2369975, EPI_ISL_2369977, EPI_ISL_2369979, EPI_ISL_2369981, EPI_ISL_2369983, EPI_ISL_2369986, EPI_ISL_2369988,
EPI_ISL_2369989, EPI_ISL_2369991, EPI_ISL_2369993, EPI_ISL_2369996, EPI_ISL_2369998, EPI_ISL_2370000, EPI_ISL_2370002, EPI_ISL_2370004, EPI_ISL_2370006, EPI_ISL_2370008, EPI_ISL_2370010, EPI_ISL_2370012, EPI_ISL_2370014, EPI_ISL_2370016, EPI_ISL_2370018, EPI_ISL_2370022, EPI_ISL_2370024, EPI_ISL_2370026, EPI_ISL_2370027, EPI_ISL_2370030, EPI_ISL_2370031, EPI_ISL_2370034,
EPI_ISL_2370036, EPI_ISL_2370038, EPI_ISL_2370040, EPI_ISL_2370042, EPI_ISL_2370044, EPI_ISL_2370046, EPI_ISL_2370048, EPI_ISL_2370050, EPI_ISL_2370052, EPI_ISL_2370054, EPI_ISL_2370056, EPI_ISL_2370059, EPI_ISL_2370062, EPI_ISL_2370065, EPI_ISL_2370067, EPI_ISL_2370070, EPI_ISL_2370072, EPI_ISL_2370074, EPI_ISL_2370076, EPI_ISL_2370079, EPI_ISL_2370081, EPI_ISL_2370083,
EPI_ISL_2370086, EPI_ISL_2370088, EPI_ISL_2370090, EPI_ISL_2370092, EPI_ISL_2370094, EPI_ISL_2370096, EPI_ISL_2370098, EPI_ISL_2370099, EPI_ISL_2370102, EPI_ISL_2370105, EPI_ISL_2370107, EPI_ISL_2370113, EPI_ISL_2370115, EPI_ISL_2370117, EPI_ISL_2370120, EPI_ISL_2370123, EPI_ISL_2370125, EPI_ISL_2370128, EPI_ISL_2370130, EPI_ISL_2370133, EPI_ISL_2370135, EPI_ISL_2370138,
EPI_ISL_2370139, EPI_ISL_2370142, EPI_ISL_2370144, EPI_ISL_2370146, EPI_ISL_2370148, EPI_ISL_2370150, EPI_ISL_2370152, EPI_ISL_2370154, EPI_ISL_2370156, EPI_ISL_2370158, EPI_ISL_2370160, EPI_ISL_2370162, EPI_ISL_2370164, EPI_ISL_2370166, EPI_ISL_2370168, EPI_ISL_2370170, EPI_ISL_2370174, EPI_ISL_2370176, EPI_ISL_2370178, EPI_ISL_2370180, EPI_ISL_2370181, EPI_ISL_2370183,
EPI_ISL_2370185, EPI_ISL_2370187, EPI_ISL_2370188, EPI_ISL_2370190, EPI_ISL_2370192, EPI_ISL_2370194, EPI_ISL_2370196, EPI_ISL_2370198, EPI_ISL_2370200, EPI_ISL_2370203, EPI_ISL_2370205, EPI_ISL_2370207, EPI_ISL_2370209, EPI_ISL_2370211, EPI_ISL_2370213, EPI_ISL_2370215, EPI_ISL_2370217, EPI_ISL_2370219, EPI_ISL_2370221, EPI_ISL_2370223, EPI_ISL_2370225, EPI_ISL_2370227,
EPI_ISL_2370229, EPI_ISL_2370231, EPI_ISL_2370233, EPI_ISL_2370235, EPI_ISL_2370237, EPI_ISL_2370239, EPI_ISL_2370241, EPI_ISL_2370243, EPI_ISL_2370245, EPI_ISL_2370247, EPI_ISL_2370249, EPI_ISL_2370251, EPI_ISL_2370253, EPI_ISL_2370255, EPI_ISL_2370257, EPI_ISL_2370259, EPI_ISL_2370261, EPI_ISL_2370263, EPI_ISL_2370264, EPI_ISL_2370267, EPI_ISL_2370268, EPI_ISL_2370270,
EPI_ISL_2370272, EPI_ISL_2370274, EPI_ISL_2370276, EPI_ISL_2370278, EPI_ISL_2370280, EPI_ISL_2370282, EPI_ISL_2370284, EPI_ISL_2370289, EPI_ISL_2370290, EPI_ISL_2370293, EPI_ISL_2370295, EPI_ISL_2370297, EPI_ISL_2370301, EPI_ISL_2370303, EPI_ISL_2370305, EPI_ISL_2370307, EPI_ISL_2370309, EPI_ISL_2370310, EPI_ISL_2370312, EPI_ISL_2370314, EPI_ISL_2370316, EPI_ISL_2370318,
EPI_ISL_2370322, EPI_ISL_2370324, EPI_ISL_2370326, EPI_ISL_2370328, EPI_ISL_2370330, EPI_ISL_2370332, EPI_ISL_2370334, EPI_ISL_2370336, EPI_ISL_2370338, EPI_ISL_2370340, EPI_ISL_2370343, EPI_ISL_2370345, EPI_ISL_2370348, EPI_ISL_2370351, EPI_ISL_2370353, EPI_ISL_2370355, EPI_ISL_2370356, EPI_ISL_2370359, EPI_ISL_2370360, EPI_ISL_2370362, EPI_ISL_2370364, EPI_ISL_2370366,
EPI_ISL_2370368, EPI_ISL_2370370, EPI_ISL_2370372, EPI_ISL_2370374, EPI_ISL_2370376, EPI_ISL_2370378, EPI_ISL_2370379, EPI_ISL_2370381, EPI_ISL_2370382, EPI_ISL_2370384, EPI_ISL_2370386, EPI_ISL_2370387, EPI_ISL_2370389, EPI_ISL_2370391, EPI_ISL_2370392, EPI_ISL_2370394, EPI_ISL_2370396, EPI_ISL_2370398, EPI_ISL_2370399, EPI_ISL_2370401, EPI_ISL_2370402, EPI_ISL_2370406,
EPI_ISL_2370408, EPI_ISL_2370409, EPI_ISL_2370411, EPI_ISL_2370413, EPI_ISL_2370417, EPI_ISL_2370419, EPI_ISL_2370420, EPI_ISL_2370422, EPI_ISL_2370424, EPI_ISL_2370426, EPI_ISL_2370429, EPI_ISL_2370431, EPI_ISL_2370434, EPI_ISL_2370437, EPI_ISL_2370439, EPI_ISL_2370442, EPI_ISL_2370445, EPI_ISL_2370448, EPI_ISL_2370450, EPI_ISL_2370453, EPI_ISL_2370455, EPI_ISL_2370458,
EPI_ISL_2370460, EPI_ISL_2370463, EPI_ISL_2370465, EPI_ISL_2370470, EPI_ISL_2370473, EPI_ISL_2370475, EPI_ISL_2370477, EPI_ISL_2370480, EPI_ISL_2370485, EPI_ISL_2370488, EPI_ISL_2370489, EPI_ISL_2370494, EPI_ISL_2370496, EPI_ISL_2370498, EPI_ISL_2370500, EPI_ISL_2370502, EPI_ISL_2370504, EPI_ISL_2370507, EPI_ISL_2370509, EPI_ISL_2370511, EPI_ISL_2370513, EPI_ISL_2370514,
EPI_ISL_2370516, EPI_ISL_2370518, EPI_ISL_2370521, EPI_ISL_2370523, EPI_ISL_2370525, EPI_ISL_2370528, EPI_ISL_2370530, EPI_ISL_2370533, EPI_ISL_2370536, EPI_ISL_2370538, EPI_ISL_2370541, EPI_ISL_2370543, EPI_ISL_2370546, EPI_ISL_2370548, EPI_ISL_2370550, EPI_ISL_2370555, EPI_ISL_2370558, EPI_ISL_2370560, EPI_ISL_2370562, EPI_ISL_2370564, EPI_ISL_2370566, EPI_ISL_2370568,
EPI_ISL_2370571, EPI_ISL_2370574, EPI_ISL_2370576, EPI_ISL_2370579, EPI_ISL_2370582, EPI_ISL_2370584, EPI_ISL_2370585, EPI_ISL_2370587, EPI_ISL_2370589, EPI_ISL_2370592, EPI_ISL_2370596, EPI_ISL_2370598, EPI_ISL_2370601, EPI_ISL_2370602, EPI_ISL_2370604, EPI_ISL_2370606, EPI_ISL_2370608, EPI_ISL_2370609, EPI_ISL_2370611, EPI_ISL_2370613, EPI_ISL_2370616, EPI_ISL_2370619,
EPI_ISL_2370622, EPI_ISL_2370625, EPI_ISL_2370627, EPI_ISL_2370630, EPI_ISL_2370632, EPI_ISL_2370635, EPI_ISL_2370638, EPI_ISL_2370640, EPI_ISL_2370643, EPI_ISL_2370645, EPI_ISL_2370648, EPI_ISL_2370650, EPI_ISL_2370653, EPI_ISL_2370656, EPI_ISL_2370659, EPI_ISL_2370661, EPI_ISL_2370664, EPI_ISL_2370667, EPI_ISL_2370669, EPI_ISL_2370672, EPI_ISL_2370675, EPI_ISL_2370678,
EPI_ISL_2370680, EPI_ISL_2370683, EPI_ISL_2370686, EPI_ISL_2370689, EPI_ISL_2370692, EPI_ISL_2370695, EPI_ISL_2370698, EPI_ISL_2370701, EPI_ISL_2370704, EPI_ISL_2370706, EPI_ISL_2370709, EPI_ISL_2370713, EPI_ISL_2370718

see above | Aegis Sciences Corporation | Centers for Disease Control and Prevention Division of Viral Diseases, Pathogen Discovery | Dakota Howard, Dhwani Batra, Peter W. Cook, Kara Moser, Adrian Paskey, Jason Caravas, Benjamin Rambo-Martin, Shatavia Morrison, Christopher Gulvick, Scott Sammons, Yvette Unoarumhi, Darlene Wagner, Matthew Schmerer, Cyndi Clark, Patrick Campbell, Rob Case, Vikramsinha Ghorpade, Holly Houdeshell, Ola Kvalvaag, Dillon Nall, Ethan Sanders, Alec Vest, Shaun Westlund, Matthew Hardison, Clinton R. Paden, Duncan MacCannell

EPI_ISL_2371407, EPI_ISL_2371408, EPI_ISL_2371409, EPI_ISL_2371410, EPI_ISL_2371411, EPI_ISL_2371412, EPI_ISL_2371413, EPI_ISL_2371414, EPI_ISL_2371415, EPI_ISL_2371416, EPI_ISL_2371417, EPI_ISL_2371418, EPI_ISL_2371419, EPI_ISL_2371420, EPI_ISL_2371421, EPI_ISL_2371422, EPI_ISL_2371423, EPI_ISL_2371424, EPI_ISL_2371425, EPI_ISL_2371426, EPI_ISL_2371427, EPI_ISL_2371428,
EPI_ISL_2371429, EPI_ISL_2371430, EPI_ISL_2371431, EPI_ISL_2371432, EPI_ISL_2371433, EPI_ISL_2371434, EPI_ISL_2371435, EPI_ISL_2371436, EPI_ISL_2371437, EPI_ISL_2371438, EPI_ISL_2371439, EPI_ISL_2371440, EPI_ISL_2371441, EPI_ISL_2371442, EPI_ISL_2371443, EPI_ISL_2371444, EPI_ISL_2371445, EPI_ISL_2371446, EPI_ISL_2371447, EPI_ISL_2371448, EPI_ISL_2371449, EPI_ISL_2371450,
EPI_ISL_2371451, EPI_ISL_2371452, EPI_ISL_2371453, EPI_ISL_2371454, EPI_ISL_2371455, EPI_ISL_2371456, EPI_ISL_2371457, EPI_ISL_2371458, EPI_ISL_2371459, EPI_ISL_2371460, EPI_ISL_2371461, EPI_ISL_2371462, EPI_ISL_2371463, EPI_ISL_2371464, EPI_ISL_2371465, EPI_ISL_2371466, EPI_ISL_2371467, EPI_ISL_2371468, EPI_ISL_2371469, EPI_ISL_2371470, EPI_ISL_2371471, EPI_ISL_2371472,
EPI_ISL_2371473, EPI_ISL_2371474, EPI_ISL_2371475, EPI_ISL_2371476, EPI_ISL_2371477, EPI_ISL_2371478, EPI_ISL_2371479, EPI_ISL_2371480, EPI_ISL_2371481, EPI_ISL_2371482, EPI_ISL_2371483, EPI_ISL_2371484, EPI_ISL_2371485, EPI_ISL_2371486, EPI_ISL_2371487, EPI_ISL_2371488, EPI_ISL_2371489, EPI_ISL_2371490, EPI_ISL_2371491, EPI_ISL_2371492, EPI_ISL_2371493, EPI_ISL_2371494,
EPI_ISL_2371495, EPI_ISL_2371496, EPI_ISL_2371497, EPI_ISL_2371498, EPI_ISL_2371499, EPI_ISL_2371500, EPI_ISL_2371501, EPI_ISL_2371502, EPI_ISL_2371503, EPI_ISL_2371504, EPI_ISL_2371505, EPI_ISL_2371506, EPI_ISL_2371507, EPI_ISL_2371508, EPI_ISL_2371509, EPI_ISL_2371510, EPI_ISL_2371511, EPI_ISL_2371512, EPI_ISL_2371513

see above | Mako Medical | Centers for Disease Control and Prevention Division of Viral Diseases, Pathogen Discovery | Dakota Howard, Dhwani Batra, Peter W. Cook, Kara Moser, Adrian Paskey, Jason Caravas, Benjamin Rambo-Martin, Shatavia Morrison, Christopher Gulvick, Scott Sammons, Yvette Unoarumhi, Darlene Wagner, Matthew Schmerer,Matthew Tugwell, Lauren Moon, Clinton R. Paden, Duncan MacCannell

EPI_ISL_2371514, EPI_ISL_2371515, EPI_ISL_2371516, EPI_ISL_2371517, EPI_ISL_2371518, EPI_ISL_2371519, EPI_ISL_2371520, EPI_ISL_2371522, EPI_ISL_2371523, EPI_ISL_2371524, EPI_ISL_2371525, EPI_ISL_2371526, EPI_ISL_2371527, EPI_ISL_2371528, EPI_ISL_2371529, EPI_ISL_2371530, EPI_ISL_2371531, EPI_ISL_2371532, EPI_ISL_2371533, EPI_ISL_2371534, EPI_ISL_2371535, EPI_ISL_2371536,
EPI_ISL_2371537, EPI_ISL_2371538, EPI_ISL_2371539, EPI_ISL_2371540, EPI_ISL_2371541, EPI_ISL_2371542, EPI_ISL_2371543, EPI_ISL_2371544, EPI_ISL_2371546, EPI_ISL_2371547, EPI_ISL_2371548, EPI_ISL_2371549, EPI_ISL_2371550, EPI_ISL_2371551, EPI_ISL_2371552, EPI_ISL_2371553, EPI_ISL_2371554, EPI_ISL_2371555, EPI_ISL_2371556, EPI_ISL_2371557, EPI_ISL_2371558, EPI_ISL_2371559,
EPI_ISL_2371560, EPI_ISL_2371561, EPI_ISL_2371563, EPI_ISL_2371564, EPI_ISL_2371565, EPI_ISL_2371566, EPI_ISL_2371567, EPI_ISL_2371568, EPI_ISL_2371569, EPI_ISL_2371570, EPI_ISL_2371571, EPI_ISL_2371572, EPI_ISL_2371573, EPI_ISL_2371574, EPI_ISL_2371575, EPI_ISL_2371576, EPI_ISL_2371578, EPI_ISL_2371579, EPI_ISL_2371580, EPI_ISL_2371581, EPI_ISL_2371582, EPI_ISL_2371583,
EPI_ISL_2371584, EPI_ISL_2371585, EPI_ISL_2371586, EPI_ISL_2371587, EPI_ISL_2371588, EPI_ISL_2371589, EPI_ISL_2371590, EPI_ISL_2371591, EPI_ISL_2371592, EPI_ISL_2371593, EPI_ISL_2371594, EPI_ISL_2371595, EPI_ISL_2371598, EPI_ISL_2371599, EPI_ISL_2371600, EPI_ISL_2371601, EPI_ISL_2371602, EPI_ISL_2371603, EPI_ISL_2371604, EPI_ISL_2371605, EPI_ISL_2371606, EPI_ISL_2371607,
EPI_ISL_2371608, EPI_ISL_2371610, EPI_ISL_2371611, EPI_ISL_2371613, EPI_ISL_2371614, EPI_ISL_2371615, EPI_ISL_2371616, EPI_ISL_2371617, EPI_ISL_2371618, EPI_ISL_2371619, EPI_ISL_2371620, EPI_ISL_2371621, EPI_ISL_2371622, EPI_ISL_2371623, EPI_ISL_2371624, EPI_ISL_2371625, EPI_ISL_2371626, EPI_ISL_2371627, EPI_ISL_2371628, EPI_ISL_2371629, EPI_ISL_2371631, EPI_ISL_2371632,
EPI_ISL_2371633, EPI_ISL_2371634, EPI_ISL_2371635, EPI_ISL_2371636, EPI_ISL_2371637, EPI_ISL_2371638, EPI_ISL_2371639, EPI_ISL_2371640, EPI_ISL_2371641, EPI_ISL_2371642, EPI_ISL_2371643, EPI_ISL_2371644, EPI_ISL_2371645, EPI_ISL_2371646, EPI_ISL_2371647, EPI_ISL_2371648, EPI_ISL_2371649, EPI_ISL_2371650, EPI_ISL_2371651, EPI_ISL_2371652, EPI_ISL_2371653, EPI_ISL_2371654,
EPI_ISL_2371655, EPI_ISL_2371656, EPI_ISL_2371657, EPI_ISL_2371658, EPI_ISL_2371659, EPI_ISL_2371660, EPI_ISL_2371661, EPI_ISL_2371663, EPI_ISL_2371664, EPI_ISL_2371665, EPI_ISL_2371666, EPI_ISL_2371667, EPI_ISL_2371668, EPI_ISL_2371669, EPI_ISL_2371671, EPI_ISL_2371672, EPI_ISL_2371673, EPI_ISL_2371674, EPI_ISL_2371675, EPI_ISL_2371676, EPI_ISL_2371677, EPI_ISL_2371678,
EPI_ISL_2371679, EPI_ISL_2371680, EPI_ISL_2371681, EPI_ISL_2371682, EPI_ISL_2371683, EPI_ISL_2371684, EPI_ISL_2371685, EPI_ISL_2371686, EPI_ISL_2371687, EPI_ISL_2371688, EPI_ISL_2371689, EPI_ISL_2371690

see above | Fulgent Genetics | Centers for Disease Control and Prevention Division of Viral Diseases, Pathogen Discovery | Dakota Howard, Dhwani Batra, Peter W. Cook, Kara Moser, Adrian Paskey, Jason Caravas, Benjamin Rambo-Martin, Shatavia Morrison, Christopher Gulvick, Scott Sammons, Yvette Unoarumhi, Darlene Wagner, Matthew Schmerer, Harry Gao, Mickey Li, John Gao, Joseph Fierro, Benafsh Sapra, Becky Tsai, Yan Meng, Doreen Ng, James Xie, Clinton R. Paden, Duncan MacCannell

EPI_ISL_2371691, EPI_ISL_2371692, EPI_ISL_2371693, EPI_ISL_2371694, EPI_ISL_2371695, EPI_ISL_2371696, EPI_ISL_2371697, EPI_ISL_2371698, EPI_ISL_2371699, EPI_ISL_2371700, EPI_ISL_2371701, EPI_ISL_2371702, EPI_ISL_2371703, EPI_ISL_2371704, EPI_ISL_2371705, EPI_ISL_2371706, EPI_ISL_2371707, EPI_ISL_2371708, EPI_ISL_2371709, EPI_ISL_2371710, EPI_ISL_2371711, EPI_ISL_2371712,
EPI_ISL_2371713, EPI_ISL_2371714, EPI_ISL_2371715, EPI_ISL_2371716, EPI_ISL_2371717, EPI_ISL_2371718, EPI_ISL_2371719, EPI_ISL_2371720, EPI_ISL_2371721, EPI_ISL_2371722, EPI_ISL_2371723, EPI_ISL_2371724, EPI_ISL_2371725, EPI_ISL_2371726, EPI_ISL_2371727, EPI_ISL_2371728, EPI_ISL_2371729, EPI_ISL_2371730, EPI_ISL_2371731, EPI_ISL_2371732, EPI_ISL_2371733, EPI_ISL_2371734,
EPI_ISL_2371735, EPI_ISL_2371736, EPI_ISL_2371737, EPI_ISL_2371738, EPI_ISL_2371739, EPI_ISL_2371740, EPI_ISL_2371741, EPI_ISL_2371742, EPI_ISL_2371743, EPI_ISL_2371744, EPI_ISL_2371745, EPI_ISL_2371746, EPI_ISL_2371747, EPI_ISL_2371748, EPI_ISL_2371749, EPI_ISL_2371750, EPI_ISL_2371751, EPI_ISL_2371752, EPI_ISL_2371753, EPI_ISL_2371754, EPI_ISL_2371755, EPI_ISL_2371756,
EPI_ISL_2371757, EPI_ISL_2371758, EPI_ISL_2371759, EPI_ISL_2371760, EPI_ISL_2371761, EPI_ISL_2371762, EPI_ISL_2371763, EPI_ISL_2371764, EPI_ISL_2371765, EPI_ISL_2371766, EPI_ISL_2371767, EPI_ISL_2371768, EPI_ISL_2371769, EPI_ISL_2371770, EPI_ISL_2371771, EPI_ISL_2371772, EPI_ISL_2371773, EPI_ISL_2371774, EPI_ISL_2371775, EPI_ISL_2371776, EPI_ISL_2371777, EPI_ISL_2371778,
EPI_ISL_2371779, EPI_ISL_2371780, EPI_ISL_2371781, EPI_ISL_2371782, EPI_ISL_2371783, EPI_ISL_2371784, EPI_ISL_2371785, EPI_ISL_2371786, EPI_ISL_2371787, EPI_ISL_2371788, EPI_ISL_2371789, EPI_ISL_2371790, EPI_ISL_2371791, EPI_ISL_2371792, EPI_ISL_2371793, EPI_ISL_2371794, EPI_ISL_2371795, EPI_ISL_2371796, EPI_ISL_2371797, EPI_ISL_2371798, EPI_ISL_2371799, EPI_ISL_2371800,
EPI_ISL_2371801, EPI_ISL_2371802, EPI_ISL_2371803, EPI_ISL_2371804, EPI_ISL_2371805, EPI_ISL_2371806, EPI_ISL_2371807, EPI_ISL_2371808, EPI_ISL_2371809, EPI_ISL_2371810, EPI_ISL_2371811, EPI_ISL_2371812, EPI_ISL_2371813, EPI_ISL_2371814, EPI_ISL_2371815, EPI_ISL_2371816, EPI_ISL_2371817, EPI_ISL_2371818, EPI_ISL_2371819, EPI_ISL_2371820, EPI_ISL_2371821, EPI_ISL_2371822,
EPI_ISL_2371823, EPI_ISL_2371824, EPI_ISL_2371825, EPI_ISL_2371826, EPI_ISL_2371827, EPI_ISL_2371828, EPI_ISL_2371829, EPI_ISL_2371830, EPI_ISL_2371831, EPI_ISL_2371832, EPI_ISL_2371833, EPI_ISL_2371834, EPI_ISL_2371835, EPI_ISL_2371836, EPI_ISL_2371837, EPI_ISL_2371838, EPI_ISL_2371839, EPI_ISL_2371840, EPI_ISL_2371841, EPI_ISL_2371842, EPI_ISL_2371843, EPI_ISL_2371844,
EPI_ISL_2371845, EPI_ISL_2371846, EPI_ISL_2371847, EPI_ISL_2371849, EPI_ISL_2371852, EPI_ISL_2371854, EPI_ISL_2371857, EPI_ISL_2371860, EPI_ISL_2371863, EPI_ISL_2371867, EPI_ISL_2371868, EPI_ISL_2371869, EPI_ISL_2371870, EPI_ISL_2371871, EPI_ISL_2371872, EPI_ISL_2371873, EPI_ISL_2371874, EPI_ISL_2371875, EPI_ISL_2371876, EPI_ISL_2371877, EPI_ISL_2371878, EPI_ISL_2371879,
EPI_ISL_2371880, EPI_ISL_2371881, EPI_ISL_2371882, EPI_ISL_2371883, EPI_ISL_2371884, EPI_ISL_2371885, EPI_ISL_2371886, EPI_ISL_2371887, EPI_ISL_2371888, EPI_ISL_2371889, EPI_ISL_2371890, EPI_ISL_2371891, EPI_ISL_2371892, EPI_ISL_2371893, EPI_ISL_2371894, EPI_ISL_2371895, EPI_ISL_2371896, EPI_ISL_2371897, EPI_ISL_2371898, EPI_ISL_2371899, EPI_ISL_2371900, EPI_ISL_2371901,
EPI_ISL_2371902, EPI_ISL_2371903, EPI_ISL_2371904, EPI_ISL_2371905, EPI_ISL_2371906, EPI_ISL_2371907, EPI_ISL_2371908, EPI_ISL_2371909, EPI_ISL_2371910, EPI_ISL_2371911, EPI_ISL_2371912, EPI_ISL_2371913, EPI_ISL_2371914, EPI_ISL_2371915, EPI_ISL_2371916, EPI_ISL_2371917, EPI_ISL_2371918, EPI_ISL_2371919, EPI_ISL_2371920, EPI_ISL_2371921, EPI_ISL_2371922, EPI_ISL_2371923,
EPI_ISL_2371924, EPI_ISL_2371925, EPI_ISL_2371926, EPI_ISL_2371927, EPI_ISL_2371928, EPI_ISL_2371929, EPI_ISL_2371930, EPI_ISL_2371931, EPI_ISL_2371932, EPI_ISL_2371933, EPI_ISL_2371934, EPI_ISL_2371935, EPI_ISL_2371936, EPI_ISL_2371937, EPI_ISL_2371938, EPI_ISL_2371939, EPI_ISL_2371940, EPI_ISL_2371941, EPI_ISL_2371942, EPI_ISL_2371943, EPI_ISL_2371944, EPI_ISL_2371945,
EPI_ISL_2371946, EPI_ISL_2371947, EPI_ISL_2371948, EPI_ISL_2371949, EPI_ISL_2371950, EPI_ISL_2371951, EPI_ISL_2371952, EPI_ISL_2371953, EPI_ISL_2371954, EPI_ISL_2371955, EPI_ISL_2371956, EPI_ISL_2371957, EPI_ISL_2371958, EPI_ISL_2371959, EPI_ISL_2371960, EPI_ISL_2371961, EPI_ISL_2371962, EPI_ISL_2371963, EPI_ISL_2371964, EPI_ISL_2371965, EPI_ISL_2371966, EPI_ISL_2371967,
EPI_ISL_2371968, EPI_ISL_2371969, EPI_ISL_2371970, EPI_ISL_2371971, EPI_ISL_2371972, EPI_ISL_2371973, EPI_ISL_2371974, EPI_ISL_2371975, EPI_ISL_2371976, EPI_ISL_2371977, EPI_ISL_2371978, EPI_ISL_2371979, EPI_ISL_2371980, EPI_ISL_2371981, EPI_ISL_2371982, EPI_ISL_2371983, EPI_ISL_2371984, EPI_ISL_2371985, EPI_ISL_2371986, EPI_ISL_2371987, EPI_ISL_2371988, EPI_ISL_2371989,
EPI_ISL_2371990, EPI_ISL_2371991, EPI_ISL_2371992, EPI_ISL_2371993, EPI_ISL_2371994, EPI_ISL_2371995, EPI_ISL_2371996, EPI_ISL_2371997, EPI_ISL_2371998, EPI_ISL_2371999, EPI_ISL_2372000, EPI_ISL_2372001, EPI_ISL_2372002, EPI_ISL_2372003, EPI_ISL_2372004, EPI_ISL_2372005, EPI_ISL_2372006, EPI_ISL_2372007, EPI_ISL_2372008, EPI_ISL_2372009, EPI_ISL_2372010, EPI_ISL_2372011,
EPI_ISL_2372012, EPI_ISL_2372013, EPI_ISL_2372014, EPI_ISL_2372015, EPI_ISL_2372016, EPI_ISL_2372017, EPI_ISL_2372018, EPI_ISL_2372019, EPI_ISL_2372020, EPI_ISL_2372021, EPI_ISL_2372022, EPI_ISL_2372023, EPI_ISL_2372024, EPI_ISL_2372025, EPI_ISL_2372026, EPI_ISL_2372027, EPI_ISL_2372028, EPI_ISL_2372029, EPI_ISL_2372030, EPI_ISL_2372031, EPI_ISL_2372032, EPI_ISL_2372033,
EPI_ISL_2372034, EPI_ISL_2372035, EPI_ISL_2372036, EPI_ISL_2372037, EPI_ISL_2372038, EPI_ISL_2372039, EPI_ISL_2372040, EPI_ISL_2372041, EPI_ISL_2372042, EPI_ISL_2372043, EPI_ISL_2372044, EPI_ISL_2372045, EPI_ISL_2372046, EPI_ISL_2372047, EPI_ISL_2372048, EPI_ISL_2372049, EPI_ISL_2372050, EPI_ISL_2372051, EPI_ISL_2372052, EPI_ISL_2372053, EPI_ISL_2372054, EPI_ISL_2372055,
EPI_ISL_2372056, EPI_ISL_2372057, EPI_ISL_2372058, EPI_ISL_2372059, EPI_ISL_2372060, EPI_ISL_2372061, EPI_ISL_2372062, EPI_ISL_2372063, EPI_ISL_2372064, EPI_ISL_2372065, EPI_ISL_2372066, EPI_ISL_2372067, EPI_ISL_2372068, EPI_ISL_2372069, EPI_ISL_2372070, EPI_ISL_2372071, EPI_ISL_2372072, EPI_ISL_2372073, EPI_ISL_2372074, EPI_ISL_2372075, EPI_ISL_2372076, EPI_ISL_2372077,
EPI_ISL_2372078, EPI_ISL_2372079, EPI_ISL_2372080, EPI_ISL_2372081, EPI_ISL_2372082, EPI_ISL_2372083, EPI_ISL_2372084, EPI_ISL_2372085, EPI_ISL_2372086, EPI_ISL_2372087, EPI_ISL_2372088, EPI_ISL_2372089, EPI_ISL_2372090, EPI_ISL_2372091, EPI_ISL_2372092, EPI_ISL_2372093, EPI_ISL_2372094, EPI_ISL_2372095, EPI_ISL_2372096, EPI_ISL_2372097, EPI_ISL_2372098, EPI_ISL_2372099,
EPI_ISL_2372100, EPI_ISL_2372101, EPI_ISL_2372102, EPI_ISL_2372103, EPI_ISL_2372104, EPI_ISL_2372105, EPI_ISL_2372106, EPI_ISL_2372107, EPI_ISL_2372108, EPI_ISL_2372109, EPI_ISL_2372110, EPI_ISL_2372111, EPI_ISL_2372112, EPI_ISL_2372113, EPI_ISL_2372114, EPI_ISL_2372115, EPI_ISL_2372116, EPI_ISL_2372117, EPI_ISL_2372118, EPI_ISL_2372119, EPI_ISL_2372120, EPI_ISL_2372121,
EPI_ISL_2372122, EPI_ISL_2372123, EPI_ISL_2372124, EPI_ISL_2372125, EPI_ISL_2372126, EPI_ISL_2372127, EPI_ISL_2372128, EPI_ISL_2372129, EPI_ISL_2372130, EPI_ISL_2372131, EPI_ISL_2372132, EPI_ISL_2372133, EPI_ISL_2372134, EPI_ISL_2372135, EPI_ISL_2372136, EPI_ISL_2372137, EPI_ISL_2372138, EPI_ISL_2372139, EPI_ISL_2372140, EPI_ISL_2372141, EPI_ISL_2372142, EPI_ISL_2372143,
EPI_ISL_2372144, EPI_ISL_2372145, EPI_ISL_2372146, EPI_ISL_2372147, EPI_ISL_2372148, EPI_ISL_2372149, EPI_ISL_2372150, EPI_ISL_2372151, EPI_ISL_2372152, EPI_ISL_2372153, EPI_ISL_2372154, EPI_ISL_2372155, EPI_ISL_2372156, EPI_ISL_2372157, EPI_ISL_2372158, EPI_ISL_2372159, EPI_ISL_2372160, EPI_ISL_2372161, EPI_ISL_2372162, EPI_ISL_2372163, EPI_ISL_2372164, EPI_ISL_2372165,
EPI_ISL_2372166, EPI_ISL_2372167, EPI_ISL_2372168, EPI_ISL_2372169, EPI_ISL_2372170, EPI_ISL_2372171, EPI_ISL_2372172, EPI_ISL_2372173, EPI_ISL_2372174, EPI_ISL_2372175, EPI_ISL_2372176, EPI_ISL_2372177, EPI_ISL_2372178, EPI_ISL_2372179, EPI_ISL_2372180, EPI_ISL_2372181, EPI_ISL_2372182, EPI_ISL_2372183, EPI_ISL_2372184, EPI_ISL_2372185, EPI_ISL_2372186, EPI_ISL_2372187,
EPI_ISL_2372188, EPI_ISL_2372189, EPI_ISL_2372190, EPI_ISL_2372191, EPI_ISL_2372192, EPI_ISL_2372193, EPI_ISL_2372194, EPI_ISL_2372195, EPI_ISL_2372196, EPI_ISL_2372197, EPI_ISL_2372198, EPI_ISL_2372199, EPI_ISL_2372200, EPI_ISL_2372201, EPI_ISL_2372202, EPI_ISL_2372203, EPI_ISL_2372204, EPI_ISL_2372205, EPI_ISL_2372206, EPI_ISL_2372207, EPI_ISL_2372208, EPI_ISL_2372209,
EPI_ISL_2372210, EPI_ISL_2372211, EPI_ISL_2372212, EPI_ISL_2372213, EPI_ISL_2372214, EPI_ISL_2372215, EPI_ISL_2372216, EPI_ISL_2372217, EPI_ISL_2372218, EPI_ISL_2372219, EPI_ISL_2372220, EPI_ISL_2372221, EPI_ISL_2372222, EPI_ISL_2372223, EPI_ISL_2372224, EPI_ISL_2372225, EPI_ISL_2372226, EPI_ISL_2372227, EPI_ISL_2372228, EPI_ISL_2372229, EPI_ISL_2372230, EPI_ISL_2372231,
EPI_ISL_2372232, EPI_ISL_2372233, EPI_ISL_2372234, EPI_ISL_2372235, EPI_ISL_2372236, EPI_ISL_2372237, EPI_ISL_2372238, EPI_ISL_2372239, EPI_ISL_2372240, EPI_ISL_2372241, EPI_ISL_2372242, EPI_ISL_2372243, EPI_ISL_2372244, EPI_ISL_2372245, EPI_ISL_2372246, EPI_ISL_2372247, EPI_ISL_2372248, EPI_ISL_2372249, EPI_ISL_2372250, EPI_ISL_2372251, EPI_ISL_2372252, EPI_ISL_2372253,
EPI_ISL_2372254, EPI_ISL_2372255, EPI_ISL_2372256, EPI_ISL_2372257, EPI_ISL_2372258, EPI_ISL_2372259, EPI_ISL_2372260, EPI_ISL_2372261, EPI_ISL_2372262, EPI_ISL_2372263, EPI_ISL_2372264, EPI_ISL_2372265

see above | Quest Diagnostics Incorporated | Centers for Disease Control and Prevention Division of Viral Diseases, Pathogen Discovery | Dakota Howard, Dhwani Batra, Peter W. Cook, Kara Moser, Adrian Paskey, Jason Caravas, Benjamin Rambo-Martin, Shatavia Morrison, Christopher Gulvick, Scott Sammons, Yvette Unoarumhi, Darlene Wagner, Matthew Schmerer, S. H. Rosenthal, A. Gerasimova, R. M. Kagan, B. Anderson, M. Hua, Y. Liu, L.E. Bernstein, K.E. Livingston, A. Perez, I. A. Shlyakhter, R. V. Rolando, R. Owen, P. Tanpaiboon, F. Lacbawan, Clinton R. Paden, Duncan MacCannell

EPI_ISL_2372298, EPI_ISL_2372300, EPI_ISL_2372302, EPI_ISL_2372304, EPI_ISL_2372306, EPI_ISL_2372308, EPI_ISL_2372310, EPI_ISL_2372312, EPI_ISL_2372314, EPI_ISL_2372316, EPI_ISL_2372318, EPI_ISL_2372320, EPI_ISL_2372322, EPI_ISL_2372324, EPI_ISL_2372326, EPI_ISL_2372328, EPI_ISL_2372330, EPI_ISL_2372332, EPI_ISL_2372334, EPI_ISL_2372335, EPI_ISL_2372337, EPI_ISL_2372339,
EPI_ISL_2372341, EPI_ISL_2372343, EPI_ISL_2372345, EPI_ISL_2372347, EPI_ISL_2372349, EPI_ISL_2372351, EPI_ISL_2372353, EPI_ISL_2372355, EPI_ISL_2372357, EPI_ISL_2372359, EPI_ISL_2372360, EPI_ISL_2372362, EPI_ISL_2372364, EPI_ISL_2372366, EPI_ISL_2372368, EPI_ISL_2372370, EPI_ISL_2372372, EPI_ISL_2372374, EPI_ISL_2372376, EPI_ISL_2372378, EPI_ISL_2372380, EPI_ISL_2372382,
EPI_ISL_2372384, EPI_ISL_2372386, EPI_ISL_2372388, EPI_ISL_2372391, EPI_ISL_2372393, EPI_ISL_2372394, EPI_ISL_2372396, EPI_ISL_2372397, EPI_ISL_2372399, EPI_ISL_2372401

see above | Aegis Sciences Corporation | Centers for Disease Control and Prevention Division of Viral Diseases, Pathogen Discovery | Dakota Howard, Dhwani Batra, Peter W. Cook, Kara Moser, Adrian Paskey, Jason Caravas, Benjamin Rambo-Martin, Shatavia Morrison, Christopher Gulvick, Scott Sammons, Yvette Unoarumhi, Darlene Wagner, Matthew Schmerer, Cyndi Clark, Patrick Campbell, Rob Case, Vikramsinha Ghorpade, Holly Houdeshell, Ola Kvalvaag, Dillon Nall, Ethan Sanders, Alec Vest, Shaun Westlund, Matthew Hardison, Clinton R. Paden, Duncan MacCannell

EPI_ISL_2372410, EPI_ISL_2372411, EPI_ISL_2372412, EPI_ISL_2372413, EPI_ISL_2372414, EPI_ISL_2372415, EPI_ISL_2372416, EPI_ISL_2372417, EPI_ISL_2372418, EPI_ISL_2372419, EPI_ISL_2372420, EPI_ISL_2372421, EPI_ISL_2372422, EPI_ISL_2372423

see above | Quest Diagnostics Incorporated | Centers for Disease Control and Prevention Division of Viral Diseases, Pathogen Discovery | Dakota Howard, Dhwani Batra, Peter W. Cook, Kara Moser, Adrian Paskey, Jason Caravas, Benjamin Rambo-Martin, Shatavia Morrison, Christopher Gulvick, Scott Sammons, Yvette Unoarumhi, Darlene Wagner, Matthew Schmerer, S. H. Rosenthal, A. Gerasimova, R. M. Kagan, B. Anderson, M. Hua, Y. Liu, L.E. Bernstein, K.E. Livingston, A. Perez, I. A. Shlyakhter, R. V. Rolando, R. Owen, P. Tanpaiboon, F. Lacbawan, Clinton R. Paden, Duncan MacCannell

EPI_ISL_2372424, EPI_ISL_2372426, EPI_ISL_2372427, EPI_ISL_2372428, EPI_ISL_2372429, EPI_ISL_2372430, EPI_ISL_2372431, EPI_ISL_2372432, EPI_ISL_2372433, EPI_ISL_2372434, EPI_ISL_2372435, EPI_ISL_2372436, EPI_ISL_2372437, EPI_ISL_2372438, EPI_ISL_2372439, EPI_ISL_2372440, EPI_ISL_2372442, EPI_ISL_2372443, EPI_ISL_2372444, EPI_ISL_2372445, EPI_ISL_2372446, EPI_ISL_2372447,
EPI_ISL_2372448, EPI_ISL_2372449, EPI_ISL_2372450, EPI_ISL_2372451, EPI_ISL_2372452, EPI_ISL_2372453, EPI_ISL_2372454, EPI_ISL_2372455, EPI_ISL_2372456, EPI_ISL_2372457, EPI_ISL_2372458, EPI_ISL_2372459, EPI_ISL_2372461, EPI_ISL_2372462, EPI_ISL_2372463, EPI_ISL_2372464, EPI_ISL_2372465, EPI_ISL_2372466

see above | Aegis Sciences Corporation | Centers for Disease Control and Prevention Division of Viral Diseases, Pathogen Discovery | Dakota Howard, Dhwani Batra, Peter W. Cook, Kara Moser, Adrian Paskey, Jason Caravas, Benjamin Rambo-Martin, Shatavia Morrison, Christopher Gulvick, Scott Sammons, Yvette Unoarumhi, Darlene Wagner, Matthew Schmerer, Cyndi Clark, Patrick Campbell, Rob Case, Vikramsinha Ghorpade, Holly Houdeshell, Ola Kvalvaag, Dillon Nall, Ethan Sanders, Alec Vest, Shaun Westlund, Matthew Hardison, Clinton R. Paden, Duncan MacCannell

| | | | |
|---|---|---|---|
| EPI_ISL_2372467, EPI_ISL_2372468, EPI_ISL_2372469, EPI_ISL_2372470, EPI_ISL_2372471, EPI_ISL_2372472, EPI_ISL_2372473, EPI_ISL_2372474, EPI_ISL_2372475, EPI_ISL_2372476, EPI_ISL_2372477, EPI_ISL_2372478, EPI_ISL_2372479, EPI_ISL_2372480, EPI_ISL_2372481, EPI_ISL_2372482, EPI_ISL_2372483, EPI_ISL_2372484, EPI_ISL_2372485, EPI_ISL_2372486, EPI_ISL_2372487, EPI_ISL_2372488, EPI_ISL_2372489, EPI_ISL_2372490, EPI_ISL_2372491, EPI_ISL_2372492, EPI_ISL_2372493, EPI_ISL_2372494, EPI_ISL_2372495, EPI_ISL_2372496, EPI_ISL_2372497, EPI_ISL_2372498, EPI_ISL_2372499, EPI_ISL_2372500, EPI_ISL_2372501, EPI_ISL_2372502, EPI_ISL_2372503, EPI_ISL_2372504, EPI_ISL_2372505, EPI_ISL_2372506, EPI_ISL_2372507, EPI_ISL_2372508, EPI_ISL_2372509, EPI_ISL_2372510, EPI_ISL_2372511, EPI_ISL_2372512, EPI_ISL_2372513, EPI_ISL_2372514, EPI_ISL_2372515, EPI_ISL_2372516, EPI_ISL_2372517, EPI_ISL_2372518, EPI_ISL_2372519, EPI_ISL_2372520, EPI_ISL_2372521, EPI_ISL_2372522, EPI_ISL_2372523, EPI_ISL_2372524, EPI_ISL_2372525, EPI_ISL_2372526, EPI_ISL_2372527, EPI_ISL_2372528, EPI_ISL_2372529, EPI_ISL_2372530, EPI_ISL_2372531, EPI_ISL_2372532, EPI_ISL_2372533, EPI_ISL_2372534, EPI_ISL_2372535, EPI_ISL_2372536, EPI_ISL_2372537, EPI_ISL_2372538, EPI_ISL_2372539, EPI_ISL_2372540, EPI_ISL_2372541, EPI_ISL_2372542, EPI_ISL_2372544, EPI_ISL_2372545, EPI_ISL_2372547, EPI_ISL_2372548, EPI_ISL_2372549, EPI_ISL_2372550, EPI_ISL_2372551, EPI_ISL_2372552, EPI_ISL_2372554, EPI_ISL_2372555, EPI_ISL_2372556, EPI_ISL_2372557, EPI_ISL_2372559, EPI_ISL_2372560, EPI_ISL_2372561, EPI_ISL_2372562, EPI_ISL_2372563, EPI_ISL_2372564, EPI_ISL_2372565, EPI_ISL_2372566, EPI_ISL_2372567, EPI_ISL_2372568, EPI_ISL_2372569, EPI_ISL_2372570, EPI_ISL_2372571, EPI_ISL_2372572, EPI_ISL_2372573, EPI_ISL_2372574, EPI_ISL_2372575, EPI_ISL_2372576, EPI_ISL_2372577, EPI_ISL_2372578, EPI_ISL_2372579, EPI_ISL_2372580, EPI_ISL_2372581, EPI_ISL_2372583, EPI_ISL_2372584, EPI_ISL_2372585, EPI_ISL_2372586, EPI_ISL_2372587, EPI_ISL_2372588, EPI_ISL_2372589, EPI_ISL_2372590, EPI_ISL_2372591, EPI_ISL_2372592, EPI_ISL_2372593, EPI_ISL_2372594, EPI_ISL_2372595, EPI_ISL_2372596, EPI_ISL_2372597, EPI_ISL_2372598, EPI_ISL_2372599, EPI_ISL_2372600, EPI_ISL_2372601, EPI_ISL_2372602, EPI_ISL_2372603, EPI_ISL_2372604, EPI_ISL_2372605, EPI_ISL_2372606, EPI_ISL_2372607, EPI_ISL_2372608, EPI_ISL_2372609, EPI_ISL_2372610, EPI_ISL_2372611, EPI_ISL_2372612, EPI_ISL_2372613, EPI_ISL_2372614, EPI_ISL_2372615, EPI_ISL_2372616, EPI_ISL_2372617, EPI_ISL_2372618, EPI_ISL_2372619, EPI_ISL_2372621, EPI_ISL_2372622, EPI_ISL_2372623, EPI_ISL_2372624, EPI_ISL_2372625, EPI_ISL_2372626, EPI_ISL_2372627, EPI_ISL_2372628, EPI_ISL_2372629, EPI_ISL_2372630, EPI_ISL_2372631, EPI_ISL_2372632, EPI_ISL_2372633, EPI_ISL_2372634, EPI_ISL_2372635, EPI_ISL_2372636, EPI_ISL_2372637, EPI_ISL_2372638, EPI_ISL_2372639, EPI_ISL_2372640, EPI_ISL_2372641, EPI_ISL_2372642, EPI_ISL_2372643, EPI_ISL_2372644, EPI_ISL_2372645, EPI_ISL_2372646, EPI_ISL_2372647, EPI_ISL_2372648, EPI_ISL_2372649, EPI_ISL_2372650, EPI_ISL_2372651, EPI_ISL_2372652, EPI_ISL_2372653, EPI_ISL_2372654, EPI_ISL_2372655, EPI_ISL_2372656, EPI_ISL_2372657, EPI_ISL_2372658, EPI_ISL_2372659, EPI_ISL_2372660, EPI_ISL_2372661, EPI_ISL_2372662, EPI_ISL_2372663, EPI_ISL_2372664, EPI_ISL_2372665, EPI_ISL_2372666, EPI_ISL_2372667, EPI_ISL_2372668, EPI_ISL_2372669, EPI_ISL_2372670, EPI_ISL_2372671, EPI_ISL_2372672, EPI_ISL_2372673, EPI_ISL_2372674, EPI_ISL_2372675, EPI_ISL_2372676, EPI_ISL_2372677, EPI_ISL_2372678, EPI_ISL_2372679, EPI_ISL_2372680, EPI_ISL_2372681, EPI_ISL_2372683, EPI_ISL_2372684, EPI_ISL_2372685, EPI_ISL_2372686 | | | |
| see above | Mako Medical | Centers for Disease Control and Prevention Division of Viral Diseases, Pathogen Discovery | Dakota Howard, Dhwani Batra, Peter W. Cook, Kara Moser, Adrian Paskey, Jason Caravas, Benjamin Rambo-Martin, Shatavia Morrison, Christopher Gulvick, Scott Sammons, Yvette Unoarumhi, Darlene Wagner, Matthew Schmerer,Matthew Tugwell, Lauren Moon, Clinton R. Paden, Duncan MacCannell |
| EPI_ISL_2372783, EPI_ISL_2372784, EPI_ISL_2372786, EPI_ISL_2372787, EPI_ISL_2372788, EPI_ISL_2372789, EPI_ISL_2372790, EPI_ISL_2372791, EPI_ISL_2372792, EPI_ISL_2372793, EPI_ISL_2372794, EPI_ISL_2372795, EPI_ISL_2372796, EPI_ISL_2372797, EPI_ISL_2372798, EPI_ISL_2372799, EPI_ISL_2372800, EPI_ISL_2372801, EPI_ISL_2372802, EPI_ISL_2372803 | | | |
| see above | Infinity Biologix | Centers for Disease Control and Prevention Division of Viral Diseases, Pathogen Discovery | Dakota Howard, Dhwani Batra, Peter W. Cook, Kara Moser, Adrian Paskey, Jason Caravas, Benjamin Rambo-Martin, Shatavia Morrison, Christopher Gulvick, Scott Sammons, Yvette Unoarumhi, Darlene Wagner, Matthew Schmerer, Christian Bixby, Yihe Wang, Jonathan Schultz, Chirayu Goswami, Russ Hager, Robin Grimwood, Clinton R. Paden, Duncan MacCannell |
| EPI_ISL_2372899, EPI_ISL_2372900, EPI_ISL_2372901, EPI_ISL_2372902, EPI_ISL_2372903, EPI_ISL_2372904, EPI_ISL_2372905, EPI_ISL_2372906, EPI_ISL_2372907, EPI_ISL_2372908, EPI_ISL_2372909, EPI_ISL_2372910, EPI_ISL_2372911, EPI_ISL_2372912, EPI_ISL_2372913, EPI_ISL_2372914, EPI_ISL_2372915, EPI_ISL_2372916, EPI_ISL_2372917, EPI_ISL_2372918, EPI_ISL_2372919, EPI_ISL_2372920, EPI_ISL_2372921, EPI_ISL_2372922, EPI_ISL_2372923, EPI_ISL_2372924, EPI_ISL_2372925, EPI_ISL_2372926, EPI_ISL_2372927, EPI_ISL_2372928, EPI_ISL_2372929, EPI_ISL_2372930, EPI_ISL_2372931, EPI_ISL_2372932, EPI_ISL_2372933, EPI_ISL_2372934, EPI_ISL_2372935, EPI_ISL_2372936, EPI_ISL_2372937, EPI_ISL_2372938, EPI_ISL_2372939, EPI_ISL_2372940, EPI_ISL_2372941, EPI_ISL_2372942, EPI_ISL_2372943, EPI_ISL_2372944, EPI_ISL_2372945, EPI_ISL_2372946, EPI_ISL_2372947, EPI_ISL_2372948, EPI_ISL_2372949, EPI_ISL_2372950, EPI_ISL_2372951, EPI_ISL_2372952 | | | |
| see above | Quest Diagnostics Incorporated | Centers for Disease Control and Prevention Division of Viral Diseases, Pathogen Discovery | Dakota Howard, Dhwani Batra, Peter W. Cook, Kara Moser, Adrian Paskey, Jason Caravas, Benjamin Rambo-Martin, Shatavia Morrison, Christopher Gulvick, Scott Sammons, Yvette Unoarumhi, Darlene Wagner, Matthew Schmerer, S. H. Rosenthal, A. Gerasimova, R. M. Kagan, B. Anderson, M. Hua, Y. Liu, L.E. Bernstein, K.E. Livingston, A. Perez, I. A. Shlyakhter, R. V. Rolando, R. Owen, P. Tanpaiboon, F. Lacbawan, Clinton R. Paden, Duncan MacCannell |
| EPI_ISL_2372953, EPI_ISL_2372954, EPI_ISL_2372955, EPI_ISL_2372956, EPI_ISL_2372957, EPI_ISL_2372958, EPI_ISL_2372959, EPI_ISL_2372960, EPI_ISL_2372961, EPI_ISL_2372962, EPI_ISL_2372963, EPI_ISL_2372964, EPI_ISL_2372965, EPI_ISL_2372966, EPI_ISL_2372967, EPI_ISL_2372968, EPI_ISL_2372969, EPI_ISL_2372970, EPI_ISL_2372971, EPI_ISL_2372972, EPI_ISL_2372973, EPI_ISL_2372974, EPI_ISL_2372975, EPI_ISL_2372976, EPI_ISL_2372977, EPI_ISL_2372978, EPI_ISL_2372979, EPI_ISL_2372980, EPI_ISL_2372981, EPI_ISL_2372982, EPI_ISL_2372983, EPI_ISL_2372984, EPI_ISL_2372985, EPI_ISL_2372986, EPI_ISL_2372987, EPI_ISL_2372988, EPI_ISL_2372989, EPI_ISL_2372990, EPI_ISL_2372991, EPI_ISL_2372992, EPI_ISL_2372993, EPI_ISL_2372994, EPI_ISL_2372995, EPI_ISL_2372996, EPI_ISL_2372997, EPI_ISL_2372998, EPI_ISL_2372999, EPI_ISL_2373000, EPI_ISL_2373001, EPI_ISL_2373002, EPI_ISL_2373003, EPI_ISL_2373004, EPI_ISL_2373005, EPI_ISL_2373006, EPI_ISL_2373007, EPI_ISL_2373008, EPI_ISL_2373009, EPI_ISL_2373010, EPI_ISL_2373011, EPI_ISL_2373012, EPI_ISL_2373013, EPI_ISL_2373014, EPI_ISL_2373015, EPI_ISL_2373016, EPI_ISL_2373017, EPI_ISL_2373018, EPI_ISL_2373019, EPI_ISL_2373020, EPI_ISL_2373021, EPI_ISL_2373022, EPI_ISL_2373023, EPI_ISL_2373024, EPI_ISL_2373025, EPI_ISL_2373026, EPI_ISL_2373027, EPI_ISL_2373028, EPI_ISL_2373029, EPI_ISL_2373030, EPI_ISL_2373031, EPI_ISL_2373032, EPI_ISL_2373033, EPI_ISL_2373034, EPI_ISL_2373035, EPI_ISL_2373036, EPI_ISL_2373037, EPI_ISL_2373038, EPI_ISL_2373040, EPI_ISL_2373041, EPI_ISL_2373042, EPI_ISL_2373043, EPI_ISL_2373044, EPI_ISL_2373045, EPI_ISL_2373046, EPI_ISL_2373047, EPI_ISL_2373048, EPI_ISL_2373049, EPI_ISL_2373050 | | | |
| see above | Northwestern Memorial Hospital | Northwestern University - Ozer Lab | Ramon Lorenzo-Redondo, Lacy M. Simons, Taylor J. Dean, Chad J. Achenbach, Lawrence J. Jennings, Chao Qi, Michael G. Ison, Judd F. Hultquist, Egon A. Ozer |
| EPI_ISL_2373051, EPI_ISL_2373052, EPI_ISL_2373053, EPI_ISL_2373054, EPI_ISL_2373055, EPI_ISL_2373056, EPI_ISL_2373057, EPI_ISL_2373058, EPI_ISL_2373059, EPI_ISL_2373060, EPI_ISL_2373061, EPI_ISL_2373062, EPI_ISL_2373063, EPI_ISL_2373064, EPI_ISL_2373065, EPI_ISL_2373066, EPI_ISL_2373067, EPI_ISL_2373068, EPI_ISL_2373069, EPI_ISL_2373070, EPI_ISL_2373071, EPI_ISL_2373072, EPI_ISL_2373073, EPI_ISL_2373074, EPI_ISL_2373075, EPI_ISL_2373076, EPI_ISL_2373077, EPI_ISL_2373078, EPI_ISL_2373079, EPI_ISL_2373080, EPI_ISL_2373081, EPI_ISL_2373082, EPI_ISL_2373083, EPI_ISL_2373084, EPI_ISL_2373085, EPI_ISL_2373086, EPI_ISL_2373087, EPI_ISL_2373088, EPI_ISL_2373089, EPI_ISL_2373090, EPI_ISL_2373091, EPI_ISL_2373092, EPI_ISL_2373093, EPI_ISL_2373094, EPI_ISL_2373095, EPI_ISL_2373096, EPI_ISL_2373097, EPI_ISL_2373098, EPI_ISL_2373099, EPI_ISL_2373100, EPI_ISL_2373101, EPI_ISL_2373102, EPI_ISL_2373103, EPI_ISL_2373104, EPI_ISL_2373105, EPI_ISL_2373106, EPI_ISL_2373107 | | | |
| see above | Lurie Children's Hospital of Chicago | Northwestern University - Ozer Lab | Ramon Lorenzo-Redondo, Lacy M. Simons, Taylor J. Dean, Michael G. Ison, Xiaotian, Zheng, William J. Muller, Larry K. Kociolek, Judd F. Hultquist, Egon A. Ozer |
| EPI_ISL_2374152, EPI_ISL_2374153, EPI_ISL_2374308, EPI_ISL_2374560 | Ohio Department of Health Laboratory | Ohio Department of Health Laboratory | Holmes, Jennifer; Eric Brandt, Keoni Omura, Glen McGillivary, Caitlin McDonnell, Jade Mowery, Stephanie Mccracken, Tyler Payne, Kirtana Ramadugu, Erica Leasure, Brent Lee, Kelsey Florek, Heather Blankenship, Allison Black, Quanta Brown, and Tammy Bannerman |
| EPI_ISL_2374561, EPI_ISL_2374562, EPI_ISL_2374563, EPI_ISL_2374564, EPI_ISL_2374565, EPI_ISL_2374566, EPI_ISL_2374567, EPI_ISL_2374568, EPI_ISL_2374569, EPI_ISL_2374570, EPI_ISL_2374571, EPI_ISL_2374573, EPI_ISL_2374574, EPI_ISL_2374575, EPI_ISL_2374576, EPI_ISL_2374577, EPI_ISL_2374578, EPI_ISL_2374579, EPI_ISL_2374580, EPI_ISL_2374581, EPI_ISL_2374582, EPI_ISL_2374583, EPI_ISL_2374584, EPI_ISL_2374585, EPI_ISL_2374586, EPI_ISL_2374587, EPI_ISL_2374588, EPI_ISL_2374589, EPI_ISL_2374590, EPI_ISL_2374591, EPI_ISL_2374592, EPI_ISL_2374593, EPI_ISL_2374594, EPI_ISL_2374595, EPI_ISL_2374596, EPI_ISL_2374598, EPI_ISL_2374599, EPI_ISL_2374600, EPI_ISL_2374601, EPI_ISL_2374602, EPI_ISL_2374603, EPI_ISL_2374604, EPI_ISL_2374605, EPI_ISL_2374606, EPI_ISL_2374607, EPI_ISL_2374608, EPI_ISL_2374609, EPI_ISL_2374610, EPI_ISL_2374611, EPI_ISL_2374612, EPI_ISL_2374614, EPI_ISL_2374615, EPI_ISL_2374616, EPI_ISL_2374617, EPI_ISL_2374618, EPI_ISL_2374619, EPI_ISL_2374620, EPI_ISL_2374621, EPI_ISL_2374622, EPI_ISL_2374623, EPI_ISL_2374624, EPI_ISL_2374625, EPI_ISL_2374626, EPI_ISL_2374627, EPI_ISL_2374628, EPI_ISL_2374629, EPI_ISL_2374630, EPI_ISL_2374631, EPI_ISL_2374632, EPI_ISL_2374633, EPI_ISL_2374634, EPI_ISL_2374635, EPI_ISL_2374636, EPI_ISL_2374637, EPI_ISL_2374638, EPI_ISL_2374639, EPI_ISL_2374640, EPI_ISL_2374641, EPI_ISL_2374642, EPI_ISL_2374643, EPI_ISL_2374644, EPI_ISL_2374645, EPI_ISL_2374646, EPI_ISL_2374647, EPI_ISL_2374648, EPI_ISL_2374649, EPI_ISL_2374650, EPI_ISL_2374651, EPI_ISL_2374652, EPI_ISL_2374653 | | | |
| see above | Michigan Department of Health and Human Services, Bureau of Laboratories | Michigan Department of Health and Human Services, Bureau of Laboratories | Blankenship HM, Riner D, Soehnlen MK |
| EPI_ISL_2374654 | Ohio Department of Health Laboratory | Ohio Department of Health Laboratory | Holmes, Jennifer; Eric Brandt, Keoni Omura, Glen McGillivary, Caitlin McDonnell, Jade Mowery, Stephanie Mccracken, Tyler Payne, Kirtana Ramadugu, Erica Leasure, Brent Lee, Kelsey Florek, Heather Blankenship, Allison Black, Quanta Brown, and Tammy Bannerman |
| EPI_ISL_2374655, EPI_ISL_2374656, EPI_ISL_2374657, EPI_ISL_2374658, EPI_ISL_2374659, EPI_ISL_2374660, EPI_ISL_2374661, EPI_ISL_2374662, EPI_ISL_2374663, EPI_ISL_2374664, EPI_ISL_2374665, EPI_ISL_2374666, EPI_ISL_2374667, EPI_ISL_2374668, EPI_ISL_2374669, EPI_ISL_2374671, EPI_ISL_2374672, EPI_ISL_2374673, EPI_ISL_2374674, EPI_ISL_2374675, EPI_ISL_2374676, EPI_ISL_2374677, EPI_ISL_2374678, EPI_ISL_2374679, EPI_ISL_2374680, EPI_ISL_2374681, EPI_ISL_2374682, EPI_ISL_2374683, EPI_ISL_2374684, EPI_ISL_2374685, EPI_ISL_2374686, EPI_ISL_2374687, EPI_ISL_2374689, EPI_ISL_2374690, EPI_ISL_2374692, EPI_ISL_2374693, EPI_ISL_2374694, EPI_ISL_2374695, EPI_ISL_2374696, EPI_ISL_2374697, EPI_ISL_2374698, EPI_ISL_2374699, EPI_ISL_2374700, EPI_ISL_2374701, EPI_ISL_2374702, EPI_ISL_2374703, EPI_ISL_2374704, EPI_ISL_2374705, EPI_ISL_2374706, EPI_ISL_2374707, EPI_ISL_2374708, EPI_ISL_2374709, EPI_ISL_2374710, EPI_ISL_2374711, EPI_ISL_2374712, EPI_ISL_2374713, EPI_ISL_2374714, EPI_ISL_2374715, EPI_ISL_2374716, EPI_ISL_2374717, EPI_ISL_2374718, EPI_ISL_2374719, EPI_ISL_2374720, EPI_ISL_2374721, EPI_ISL_2374722, EPI_ISL_2374723, EPI_ISL_2374724, EPI_ISL_2374725, EPI_ISL_2374726, EPI_ISL_2374732, EPI_ISL_2374738, EPI_ISL_2374739, EPI_ISL_2374743, EPI_ISL_2374744, EPI_ISL_2374745, EPI_ISL_2374748, EPI_ISL_2374749, EPI_ISL_2374750, EPI_ISL_2374752, EPI_ISL_2374754, EPI_ISL_2374759, EPI_ISL_2374763, EPI_ISL_2374766, EPI_ISL_2374768, EPI_ISL_2374773, EPI_ISL_2374775, EPI_ISL_2374777, EPI_ISL_2374780, EPI_ISL_2374781, EPI_ISL_2374782, EPI_ISL_2374784, EPI_ISL_2374785, EPI_ISL_2374787, EPI_ISL_2374788, EPI_ISL_2374789, EPI_ISL_2374790, EPI_ISL_2374791, EPI_ISL_2374792, EPI_ISL_2374794, EPI_ISL_2374796, EPI_ISL_2374797, EPI_ISL_2374798, EPI_ISL_2374800, EPI_ISL_2374802, EPI_ISL_2374803, EPI_ISL_2374804, EPI_ISL_2374806, EPI_ISL_2374807, EPI_ISL_2374808, EPI_ISL_2374810, EPI_ISL_2374812, EPI_ISL_2374813, EPI_ISL_2374814, EPI_ISL_2374819, EPI_ISL_2374820, EPI_ISL_2374824, EPI_ISL_2374825, EPI_ISL_2374829, EPI_ISL_2374836, EPI_ISL_2374837, EPI_ISL_2374842, EPI_ISL_2374843, EPI_ISL_2374844, EPI_ISL_2374846, EPI_ISL_2374850, EPI_ISL_2374851, EPI_ISL_2374853, EPI_ISL_2374854, EPI_ISL_2374855, EPI_ISL_2374856, EPI_ISL_2374857, EPI_ISL_2374858, EPI_ISL_2374859, EPI_ISL_2374860, EPI_ISL_2374861, EPI_ISL_2374862, EPI_ISL_2374863, EPI_ISL_2374864, EPI_ISL_2374865, EPI_ISL_2374867, EPI_ISL_2374868, EPI_ISL_2374869, EPI_ISL_2374870, EPI_ISL_2374872, EPI_ISL_2374873, EPI_ISL_2374874, EPI_ISL_2374875, EPI_ISL_2374876, EPI_ISL_2374877, EPI_ISL_2374878, EPI_ISL_2374879, EPI_ISL_2374880, EPI_ISL_2374881, EPI_ISL_2374883, EPI_ISL_2374884, EPI_ISL_2374885, EPI_ISL_2374886, EPI_ISL_2374887, EPI_ISL_2374889, EPI_ISL_2374890, EPI_ISL_2374891, EPI_ISL_2374893, EPI_ISL_2374894, EPI_ISL_2374895, EPI_ISL_2374896, EPI_ISL_2374897, EPI_ISL_2374898, EPI_ISL_2374899, EPI_ISL_2374900, EPI_ISL_2374901, EPI_ISL_2374902, EPI_ISL_2374903, EPI_ISL_2374904, EPI_ISL_2374905, EPI_ISL_2374906, EPI_ISL_2374907, EPI_ISL_2374908, EPI_ISL_2374909, EPI_ISL_2374910, EPI_ISL_2374911, EPI_ISL_2374912, EPI_ISL_2374913, EPI_ISL_2374914, EPI_ISL_2374915, EPI_ISL_2374916, EPI_ISL_2374917, EPI_ISL_2374918, EPI_ISL_2374919, EPI_ISL_2374921, EPI_ISL_2374922, EPI_ISL_2374923, EPI_ISL_2374924, EPI_ISL_2374925, EPI_ISL_2374926 | | | |
| see above | Michigan Department of Health and Human Services, Bureau of Laboratories | Michigan Department of Health and Human Services, Bureau of Laboratories | Blankenship HM, Riner D, Soehnlen MK |
| EPI_ISL_2374929, EPI_ISL_2374930, EPI_ISL_2375304 | Ohio Department of Health Laboratory | Ohio Department of Health Laboratory | Holmes, Jennifer; Eric Brandt, Keoni Omura, Glen McGillivary, Caitlin McDonnell, Jade Mowery, Stephanie Mccracken, Tyler Payne, Kirtana Ramadugu, Erica Leasure, Brent Lee, Kelsey Florek, Heather Blankenship, Allison Black, Quanta Brown, and Tammy Bannerman |
| EPI_ISL_2375450, EPI_ISL_2375451, EPI_ISL_2375452, EPI_ISL_2375453, EPI_ISL_2375454, EPI_ISL_2375455, EPI_ISL_2375456, EPI_ISL_2375457, EPI_ISL_2375458, EPI_ISL_2375459, EPI_ISL_2375460, EPI_ISL_2375461 | | | |
| see above | NCSLPH | NCSLPH | Chase K, Miller MC, Greene S, Glover W |
| EPI_ISL_2375495, EPI_ISL_2375496, EPI_ISL_2375497 | Ohio Department of Health Laboratory | Ohio Department of Health Laboratory | Holmes, Jennifer; Eric Brandt, Keoni Omura, Glen McGillivary, Caitlin McDonnell, Jade Mowery, Stephanie Mccracken, Tyler Payne, Kirtana Ramadugu, Erica Leasure, Brent Lee, Kelsey Florek, Heather Blankenship, Allison Black, Quanta Brown, and Tammy Bannerman |
| EPI_ISL_2375498, EPI_ISL_2375499 | DC Public Health Lab/ Dept. of Forensic Sciences | DC Public Health Lab/ Dept. of Forensic Sciences | Janis Doss, Scott Nguyen, Elizabeth Zelaya, Sarah Scott, Connie Maza, Monica Mann, Brittany Hamilton, David Payne, Jocelyn Hauser |
| EPI_ISL_2375500, EPI_ISL_2375501, EPI_ISL_2375502, EPI_ISL_2375512 | Ohio Department of Health Laboratory | Ohio Department of Health Laboratory | Holmes, Jennifer; Eric Brandt, Keoni Omura, Glen McGillivary, Caitlin McDonnell, Jade Mowery, Stephanie Mccracken, Tyler Payne, Kirtana Ramadugu, Erica Leasure, Brent Lee, Kelsey Florek, Heather Blankenship, Allison Black, Quanta Brown, and Tammy Bannerman |
| EPI_ISL_2375513, EPI_ISL_2375514, EPI_ISL_2375515, EPI_ISL_2375516, EPI_ISL_2375517, EPI_ISL_2375518, EPI_ISL_2375519, EPI_ISL_2375520, EPI_ISL_2375521, EPI_ISL_2375522, EPI_ISL_2375523, EPI_ISL_2375524, EPI_ISL_2375525, EPI_ISL_2375526, EPI_ISL_2375527, EPI_ISL_2375528, EPI_ISL_2375529, EPI_ISL_2375530, EPI_ISL_2375531, EPI_ISL_2375532, EPI_ISL_2375533, EPI_ISL_2375534, EPI_ISL_2375535, EPI_ISL_2375536 | | | |
| see above | Nebraska Public Health Laboratory | NPHL COVID-19 Response Team | NPHL COVID-19 Response Team |
| EPI_ISL_2375537, EPI_ISL_2375538, EPI_ISL_2375539, EPI_ISL_2375540, EPI_ISL_2375541, EPI_ISL_2375542, EPI_ISL_2375543, EPI_ISL_2375544, EPI_ISL_2375545, EPI_ISL_2375546, EPI_ISL_2375547, EPI_ISL_2375548, EPI_ISL_2375549, EPI_ISL_2375550, EPI_ISL_2375551, EPI_ISL_2375552, EPI_ISL_2375553, EPI_ISL_2375554, EPI_ISL_2375555, EPI_ISL_2375556, EPI_ISL_2375557, EPI_ISL_2375558, EPI_ISL_2375559, EPI_ISL_2375560, EPI_ISL_2375561, EPI_ISL_2375562, EPI_ISL_2375563, EPI_ISL_2375564, EPI_ISL_2375565, EPI_ISL_2375566, EPI_ISL_2375567, EPI_ISL_2375568, EPI_ISL_2375569, EPI_ISL_2375570, EPI_ISL_2375571, EPI_ISL_2375572, EPI_ISL_2375573, EPI_ISL_2375574, EPI_ISL_2375575, EPI_ISL_2375576, EPI_ISL_2375578, EPI_ISL_2375579, EPI_ISL_2375580, EPI_ISL_2375581, EPI_ISL_2375582, EPI_ISL_2375583, EPI_ISL_2375584, EPI_ISL_2375585, EPI_ISL_2375586, EPI_ISL_2375587, EPI_ISL_2375588, EPI_ISL_2375589, EPI_ISL_2375590, EPI_ISL_2375591, EPI_ISL_2375592, EPI_ISL_2375593, EPI_ISL_2375594, EPI_ISL_2375595, EPI_ISL_2375596, EPI_ISL_2375597, EPI_ISL_2375598, EPI_ISL_2375599, EPI_ISL_2375600, EPI_ISL_2375601, EPI_ISL_2375602, EPI_ISL_2375603, EPI_ISL_2375604, EPI_ISL_2375605, EPI_ISL_2375606, EPI_ISL_2375607, EPI_ISL_2375609, EPI_ISL_2375610, EPI_ISL_2375611, EPI_ISL_2375612, EPI_ISL_2375613, EPI_ISL_2375614, EPI_ISL_2375615, EPI_ISL_2375616, EPI_ISL_2375617, EPI_ISL_2375618, EPI_ISL_2375619, EPI_ISL_2375620, EPI_ISL_2375621, EPI_ISL_2375622, EPI_ISL_2375623, EPI_ISL_2375625, EPI_ISL_2375626, EPI_ISL_2375627, EPI_ISL_2375628, EPI_ISL_2375629, EPI_ISL_2375630, EPI_ISL_2375631, EPI_ISL_2375632, EPI_ISL_2375633, EPI_ISL_2375634, EPI_ISL_2375635, EPI_ISL_2375636, EPI_ISL_2375637, EPI_ISL_2375638, EPI_ISL_2375639, EPI_ISL_2375640, EPI_ISL_2375641, EPI_ISL_2375642, EPI_ISL_2375643, EPI_ISL_2375644, EPI_ISL_2375645, EPI_ISL_2375646, EPI_ISL_2375647, EPI_ISL_2375648, EPI_ISL_2375649, EPI_ISL_2375650, EPI_ISL_2375651, EPI_ISL_2375652, EPI_ISL_2375653, EPI_ISL_2375654, EPI_ISL_2375655, EPI_ISL_2375656, EPI_ISL_2375657, EPI_ISL_2375658, EPI_ISL_2375659, EPI_ISL_2375660, EPI_ISL_2375661, EPI_ISL_2375662, EPI_ISL_2375663, EPI_ISL_2375664, EPI_ISL_2375665, EPI_ISL_2375666, EPI_ISL_2375667, EPI_ISL_2375668, EPI_ISL_2375669, EPI_ISL_2375670, EPI_ISL_2375671, EPI_ISL_2375672, EPI_ISL_2375673, EPI_ISL_2375674, EPI_ISL_2375675, EPI_ISL_2375676, EPI_ISL_2375677, EPI_ISL_2375678, EPI_ISL_2375679, EPI_ISL_2375680, EPI_ISL_2375681, EPI_ISL_2375682, EPI_ISL_2375683, EPI_ISL_2375684, EPI_ISL_2375685, EPI_ISL_2375686, EPI_ISL_2375687, EPI_ISL_2375688, EPI_ISL_2375689, EPI_ISL_2375690, EPI_ISL_2375691, EPI_ISL_2375692, EPI_ISL_2375693, EPI_ISL_2375694, EPI_ISL_2375695, EPI_ISL_2375696, EPI_ISL_2375697, EPI_ISL_2375698, EPI_ISL_2375699, EPI_ISL_2375700, EPI_ISL_2375701, EPI_ISL_2375702, EPI_ISL_2375703, EPI_ISL_2375704, EPI_ISL_2375705, EPI_ISL_2375706, EPI_ISL_2375707, EPI_ISL_2375708, EPI_ISL_2375709, EPI_ISL_2375710, EPI_ISL_2375711, EPI_ISL_2375712, EPI_ISL_2375713, EPI_ISL_2375714, EPI_ISL_2375715, EPI_ISL_2375716, EPI_ISL_2375717, EPI_ISL_2375718, EPI_ISL_2375719, EPI_ISL_2375720, EPI_ISL_2375721, EPI_ISL_2375722, EPI_ISL_2375723, EPI_ISL_2375724, EPI_ISL_2375725, EPI_ISL_2375726, EPI_ISL_2375727, EPI_ISL_2375728, EPI_ISL_2375729, EPI_ISL_2375730, EPI_ISL_2375731, EPI_ISL_2375732, EPI_ISL_2375733, EPI_ISL_2375734, EPI_ISL_2375735, EPI_ISL_2375736, EPI_ISL_2375737, EPI_ISL_2375738, EPI_ISL_2375739, EPI_ISL_2375740, EPI_ISL_2375741, EPI_ISL_2375742, EPI_ISL_2375743, EPI_ISL_2375744, EPI_ISL_2375745, EPI_ISL_2375746, EPI_ISL_2375747, EPI_ISL_2375748, EPI_ISL_2375749, EPI_ISL_2375751, EPI_ISL_2375754, EPI_ISL_2375755, EPI_ISL_2375756, EPI_ISL_2375757, EPI_ISL_2375758, EPI_ISL_2375759, EPI_ISL_2375760, EPI_ISL_2375761, EPI_ISL_2375762, EPI_ISL_2375763, EPI_ISL_2375764, EPI_ISL_2375765, EPI_ISL_2375766, EPI_ISL_2375767, EPI_ISL_2375768, EPI_ISL_2375769, EPI_ISL_2375770, EPI_ISL_2375771, EPI_ISL_2375772, EPI_ISL_2375773, EPI_ISL_2375774, EPI_ISL_2375775, EPI_ISL_2375776, EPI_ISL_2375777, EPI_ISL_2375778, EPI_ISL_2375779, EPI_ISL_2375780, EPI_ISL_2375782, EPI_ISL_2375783, EPI_ISL_2375784, EPI_ISL_2375785, EPI_ISL_2375786, EPI_ISL_2375787, EPI_ISL_2375788, EPI_ISL_2375789, EPI_ISL_2375790, EPI_ISL_2375791, EPI_ISL_2375792, EPI_ISL_2375793, EPI_ISL_2375794, EPI_ISL_2375795 | | | |
| see above | Infinity Biologix | Centers for Disease Control and Prevention Division of Viral Diseases, Pathogen Discovery | Dakota Howard, Dhwani Batra, Peter W. Cook, Kara Moser, Adrian Paskey, Jason Caravas, Benjamin Rambo-Martin, Shatavia Morrison, Christopher Gulvick, Scott Sammons, Yvette Unoarumhi, Darlene Wagner, Matthew Schmerer, Christian Bixby, Yihe Wang, Jonathan Schultz, Chirayu Goswami, Russ Hager, Robin Grimwood, Clinton R. Paden, Duncan MacCannell |
| EPI_ISL_2375802, EPI_ISL_2375803, EPI_ISL_2375804, EPI_ISL_2375805, EPI_ISL_2375806, EPI_ISL_2375807, EPI_ISL_2375808, EPI_ISL_2375809, EPI_ISL_2375810, EPI_ISL_2375811, EPI_ISL_2375812, EPI_ISL_2375813, EPI_ISL_2375814, EPI_ISL_2375815, EPI_ISL_2375816, EPI_ISL_2375817, EPI_ISL_2375818, EPI_ISL_2375819, EPI_ISL_2375820, EPI_ISL_2375821, EPI_ISL_2375822, EPI_ISL_2375823, EPI_ISL_2375824, EPI_ISL_2375825, EPI_ISL_2375826, EPI_ISL_2375827, EPI_ISL_2375828, EPI_ISL_2375829, EPI_ISL_2375830, EPI_ISL_2375831, EPI_ISL_2375832, EPI_ISL_2375833, EPI_ISL_2375834, EPI_ISL_2375835, EPI_ISL_2375836, EPI_ISL_2375837, EPI_ISL_2375996, EPI_ISL_2375997, EPI_ISL_2375998, EPI_ISL_2375999, EPI_ISL_2376000, EPI_ISL_2376001, EPI_ISL_2376002, EPI_ISL_2376003, EPI_ISL_2376004, EPI_ISL_2376005, EPI_ISL_2376006, EPI_ISL_2376007, EPI_ISL_2376008, EPI_ISL_2376009, EPI_ISL_2376010, EPI_ISL_2376011, EPI_ISL_2376012, EPI_ISL_2376013, EPI_ISL_2376014, EPI_ISL_2376015, EPI_ISL_2376016, EPI_ISL_2376017, EPI_ISL_2376018, EPI_ISL_2376019, EPI_ISL_2376020, EPI_ISL_2376021, EPI_ISL_2376022, EPI_ISL_2376023, EPI_ISL_2376024, EPI_ISL_2376025, EPI_ISL_2376026 | | | |
| see above | Alaska State Virology Laboratory | Alaska State Virology Laboratory | Stephanie DeRonde, Elva House, Jacob Zidek, Lisa Smith, Ph.D., Jack Chen, Ph.D. |
| EPI_ISL_2376035, EPI_ISL_2376036, EPI_ISL_2376037, EPI_ISL_2376038, EPI_ISL_2376039, EPI_ISL_2376040, EPI_ISL_2376041, EPI_ISL_2376042, EPI_ISL_2376043, EPI_ISL_2376044, EPI_ISL_2376045, EPI_ISL_2376046, EPI_ISL_2376047, EPI_ISL_2376048, EPI_ISL_2376049, EPI_ISL_2376050, EPI_ISL_2376051, EPI_ISL_2376052, EPI_ISL_2376053, EPI_ISL_2376054, EPI_ISL_2376055, EPI_ISL_2376056, EPI_ISL_2376057, EPI_ISL_2376058, EPI_ISL_2376059, EPI_ISL_2376060, EPI_ISL_2376061, EPI_ISL_2376062, EPI_ISL_2376063, EPI_ISL_2376064, EPI_ISL_2376065, EPI_ISL_2376066, EPI_ISL_2376067 | | | |
| see above | University of Michigan Clinical Microbiology Laboratory | Lauring Lab, University of Michigan, Department of Microbiology and Immunology | Gilbert |

| | | | |
|---|---|---|---|
| EPI_ISL_2376068, EPI_ISL_2376069, EPI_ISL_2376070, EPI_ISL_2376071, EPI_ISL_2376072, EPI_ISL_2376073, EPI_ISL_2376074, EPI_ISL_2376075, EPI_ISL_2376076, EPI_ISL_2376077, EPI_ISL_2376078, EPI_ISL_2376079, EPI_ISL_2376080, EPI_ISL_2376081, EPI_ISL_2376082, EPI_ISL_2376083, EPI_ISL_2376084, EPI_ISL_2376085, EPI_ISL_2376086, EPI_ISL_2376087, EPI_ISL_2376088, EPI_ISL_2376089, EPI_ISL_2376090, EPI_ISL_2376091, EPI_ISL_2376092, EPI_ISL_2376093, EPI_ISL_2376094, EPI_ISL_2376095, EPI_ISL_2376096, EPI_ISL_2376097, EPI_ISL_2376098, EPI_ISL_2376099 | | | |
| see above | Alaska State Virology Laboratory | Alaska State Virology Laboratory | Stephanie DeRonde, Elva House, Jacob Zidek, Lisa Smith, Ph.D., Jack Chen, Ph.D. |
| EPI_ISL_2376100, EPI_ISL_2376101, EPI_ISL_2376102, EPI_ISL_2376103, EPI_ISL_2376104, EPI_ISL_2376105, EPI_ISL_2376106, EPI_ISL_2376107, EPI_ISL_2376108, EPI_ISL_2376109, EPI_ISL_2376110, EPI_ISL_2376111, EPI_ISL_2376112, EPI_ISL_2376113, EPI_ISL_2376114, EPI_ISL_2376115, EPI_ISL_2376117, EPI_ISL_2376118, EPI_ISL_2376119, EPI_ISL_2376120, EPI_ISL_2376121, EPI_ISL_2376122, EPI_ISL_2376123, EPI_ISL_2376124, EPI_ISL_2376125, EPI_ISL_2376126, EPI_ISL_2376127, EPI_ISL_2376128, EPI_ISL_2376129, EPI_ISL_2376130, EPI_ISL_2376131, EPI_ISL_2376132, EPI_ISL_2376133, EPI_ISL_2376134, EPI_ISL_2376135, EPI_ISL_2376136, EPI_ISL_2376137 | | | |
| see above | NJDOH, Public Health and Environmental Laboratories | NJ_PHEL | Lindsey Bodnar, Shiv K. Verma, Jacquelyn Deverell, Dana Woell, Allison Roder, Byeong Jeong |
| EPI_ISL_2376140, EPI_ISL_2376141, EPI_ISL_2376142, EPI_ISL_2376143, EPI_ISL_2376144, EPI_ISL_2376145, EPI_ISL_2376146, EPI_ISL_2376147, EPI_ISL_2376148, EPI_ISL_2376149, EPI_ISL_2376150, EPI_ISL_2376151, EPI_ISL_2376152, EPI_ISL_2376153, EPI_ISL_2376154, EPI_ISL_2376155, EPI_ISL_2376156, EPI_ISL_2376157, EPI_ISL_2376158, EPI_ISL_2376159, EPI_ISL_2376160, EPI_ISL_2376161, EPI_ISL_2376162, EPI_ISL_2376163, EPI_ISL_2376164, EPI_ISL_2376165, EPI_ISL_2376166, EPI_ISL_2376167, EPI_ISL_2376168, EPI_ISL_2376169, EPI_ISL_2376170, EPI_ISL_2376171, EPI_ISL_2376172, EPI_ISL_2376173, EPI_ISL_2376174, EPI_ISL_2376175, EPI_ISL_2376176, EPI_ISL_2376177, EPI_ISL_2376178, EPI_ISL_2376179, EPI_ISL_2376180, EPI_ISL_2376181, EPI_ISL_2376182, EPI_ISL_2376183, EPI_ISL_2376184, EPI_ISL_2376185, EPI_ISL_2376186, EPI_ISL_2376187, EPI_ISL_2376188, EPI_ISL_2376189 | | | |
| see above | Montana Public Health Laboratory | Montana Public Health Laboratory | Joy Ritter, Michelle Mozer, Carrie Biskupiak, Deborah Gibson, Michael Dills |
| EPI_ISL_2376192 | Notre Dame COVID Response and Testing Laboratory | Notre Dame Genomics & Bioinformatics Core Facility | Michael Pfrender, Joseph Sarro, Melissa Stephens |
| EPI_ISL_2376213, EPI_ISL_2376214 | NCSLPH | NCSLPH | Chase K, Miller MC, Greene S, Glover W |
| EPI_ISL_2376234, EPI_ISL_2376235, EPI_ISL_2376236, EPI_ISL_2376237, EPI_ISL_2376238, EPI_ISL_2376239, EPI_ISL_2376240, EPI_ISL_2376241, EPI_ISL_2376242, EPI_ISL_2376243, EPI_ISL_2376244, EPI_ISL_2376245, EPI_ISL_2376246, EPI_ISL_2376247, EPI_ISL_2376248, EPI_ISL_2376249, EPI_ISL_2376250, EPI_ISL_2376251, EPI_ISL_2376252, EPI_ISL_2376253, EPI_ISL_2376254, EPI_ISL_2376255, EPI_ISL_2376256, EPI_ISL_2376257, EPI_ISL_2376258, EPI_ISL_2376259 | | | |
| see above | Gundersen Clinical Microbiology Laboratory | Kabara Cancer Research Institute | Craig S. Richmond, Paraic A. Kenny |
| EPI_ISL_2376260, EPI_ISL_2376261, EPI_ISL_2376262, EPI_ISL_2376263, EPI_ISL_2376264, EPI_ISL_2376265, EPI_ISL_2376266, EPI_ISL_2376267, EPI_ISL_2376268, EPI_ISL_2376269, EPI_ISL_2376270, EPI_ISL_2376271, EPI_ISL_2376272, EPI_ISL_2376273, EPI_ISL_2376274, EPI_ISL_2376275, EPI_ISL_2376276, EPI_ISL_2376277, EPI_ISL_2376278, EPI_ISL_2376279, EPI_ISL_2376280, EPI_ISL_2376281, EPI_ISL_2376282, EPI_ISL_2376283, EPI_ISL_2376284 | | | |
| see above | Wexner Medical Center | The Ohio State University College of Medicine | Koenig,S., Seminetta,J. |
| EPI_ISL_2376422, EPI_ISL_2376423 | Omega Diagnostics at Mounes | Omega Diagnostics at Mounes | Ashleigh McGrail,Cynthia Corley,Latira Haynes-Jacob, Vivek Khare MD |
| EPI_ISL_2376424, EPI_ISL_2376425 | GA Department of Public Health Laboratory | Genomics and Discovery, Respiratory Viruses Branch, Division of Viral Diseases, Centers for Disease Control and Prevention | Anna Kelleher, Ying Tao, Yan Li, Jing Zhang, Brian Lynch, Krista Queen, Anna Uehara, Peter Cook, Han Jia Justin Ng, Rachel Marine, Clinton R. Paden, Dhwani Batra, Haibin Wang, Tara Coalter, Jasmine Padilla, Morgan Davis, Mili Sheth, Sarah Nobles, Mark Burroughs, Justin Lee, Adam Retchless, Suxiang Tong |
| EPI_ISL_2376426 | LA Office of Public Health Laboratories | Genomics and Discovery, Respiratory Viruses Branch, Division of Viral Diseases, Centers for Disease Control and Prevention | Anna Kelleher, Ying Tao, Yan Li, Jing Zhang, Brian Lynch, Krista Queen, Anna Uehara, Peter Cook, Han Jia Justin Ng, Rachel Marine, Clinton R. Paden, Dhwani Batra, Haibin Wang, Tara Coalter, Jasmine Padilla, Morgan Davis, Mili Sheth, Sarah Nobles, Mark Burroughs, Justin Lee, Adam Retchless, Suxiang Tong |
| EPI_ISL_2376427 | GA Department of Public Health Laboratory | Genomics and Discovery, Respiratory Viruses Branch, Division of Viral Diseases, Centers for Disease Control and Prevention | Anna Kelleher, Ying Tao, Yan Li, Jing Zhang, Brian Lynch, Krista Queen, Anna Uehara, Peter Cook, Han Jia Justin Ng, Rachel Marine, Clinton R. Paden, Dhwani Batra, Haibin Wang, Tara Coalter, Jasmine Padilla, Morgan Davis, Mili Sheth, Sarah Nobles, Mark Burroughs, Justin Lee, Adam Retchless, Suxiang Tong |
| EPI_ISL_2376428 | Omega Diagnostics at Mounes | Omega Diagnostics at Mounes | Ashleigh McGrail,Cynthia Corley,Latira Haynes-Jacob, Vivek Khare MD |
| EPI_ISL_2376429, EPI_ISL_2376430, EPI_ISL_2376431, EPI_ISL_2376432, EPI_ISL_2376433, EPI_ISL_2376434, EPI_ISL_2376435 | Wyoming Public Health Laboratory | Wyoming Public Health Laboratory | Jim Mildenberger, Wanda Manley, Noah Hull, Taylor Fearing, Lynette Gumbleton, Channing Weber, Ashley Norberg, Chayse Rowley, Marley Goetz, Brian Dominguez, Elliot Thomasson, Sam Britz, Cari Sloma, and Rob Christensen |
| EPI_ISL_2376437, EPI_ISL_2376438 | Omega Diagnostics at Mounes | Omega Diagnostics at Mounes | Ashleigh McGrail,Cynthia Corley,Latira Haynes-Jacob, Vivek Khare MD |
| EPI_ISL_2376439, EPI_ISL_2376440 | Omega Diagnostics at Mounes | Omega Diagnotics at Mounes | Ashleigh McGrail,Cynthia Corley,Latira Haynes-Jacob, Vivek Khare MD |
| EPI_ISL_2376441, EPI_ISL_2376442, EPI_ISL_2376443, EPI_ISL_2376444, EPI_ISL_2376445, EPI_ISL_2376446, EPI_ISL_2376447, EPI_ISL_2376448, EPI_ISL_2376449, EPI_ISL_2376450, EPI_ISL_2376451, EPI_ISL_2376452, EPI_ISL_2376453, EPI_ISL_2376454, EPI_ISL_2376455, EPI_ISL_2376456, EPI_ISL_2376457, EPI_ISL_2376458, EPI_ISL_2376459, EPI_ISL_2376460, EPI_ISL_2376461, EPI_ISL_2376462, EPI_ISL_2376463, EPI_ISL_2376464, EPI_ISL_2376465, EPI_ISL_2376466, EPI_ISL_2376467, EPI_ISL_2376468, EPI_ISL_2376469, EPI_ISL_2376470, EPI_ISL_2376471, EPI_ISL_2376472, EPI_ISL_2376473, EPI_ISL_2376474, EPI_ISL_2376475, EPI_ISL_2376476, EPI_ISL_2376477, EPI_ISL_2376478, EPI_ISL_2376479, EPI_ISL_2376480, EPI_ISL_2376481, EPI_ISL_2376482, EPI_ISL_2376483, EPI_ISL_2376484, EPI_ISL_2376485, EPI_ISL_2376486, EPI_ISL_2376487, EPI_ISL_2376488, EPI_ISL_2376489, EPI_ISL_2376490, EPI_ISL_2376491, EPI_ISL_2376492, EPI_ISL_2376493, EPI_ISL_2376494, EPI_ISL_2376495, EPI_ISL_2376496, EPI_ISL_2376497, EPI_ISL_2376498, EPI_ISL_2376499, EPI_ISL_2376500, EPI_ISL_2376501, EPI_ISL_2376502, EPI_ISL_2376503 | | | |
| see above | State Testing Facility | Altius Institute for Biomedical Research | Daniel Bates, Rebecca Bruders, Michael Buckley, Mark Frerker, Amanda Gale, Clem Green, Muhammad Halimun, Kneshay Harper, Matt Hartman, Alex Isner, Audra Johnson, Jessica Kunder, Lauren Mitchell, Jemma Nelson, Alex Nguyen, Sofia Olsson, Sadie Patraw, Tobias Ragoczy, Joshua Richards, Jean Robinston, Jacob Rodriguez, John Stamatoyannopoulos, Eric Thorland, Julia Wald |
| EPI_ISL_2376504 | Omega Diagnotics at Mounes | Omega Diagnotics at Mounes | Ashleigh McGrail,Cynthia Corley,Latira Haynes-Jacob, Vivek Khare MD |
| EPI_ISL_2376505, EPI_ISL_2376506, EPI_ISL_2376507, EPI_ISL_2376508, EPI_ISL_2376509, EPI_ISL_2376510, EPI_ISL_2376511, EPI_ISL_2376512, EPI_ISL_2376513, EPI_ISL_2376514, EPI_ISL_2376515, EPI_ISL_2376516, EPI_ISL_2376517, EPI_ISL_2376518, EPI_ISL_2376519, EPI_ISL_2376520, EPI_ISL_2376521, EPI_ISL_2376522, EPI_ISL_2376523, EPI_ISL_2376524, EPI_ISL_2376525, EPI_ISL_2376526, EPI_ISL_2376527, EPI_ISL_2376528 | | | |
| see above | State Testing Facility | Altius Institute for Biomedical Research | Daniel Bates, Rebecca Bruders, Michael Buckley, Mark Frerker, Amanda Gale, Clem Green, Muhammad Halimun, Kneshay Harper, Matt Hartman, Alex Isner, Audra Johnson, Jessica Kunder, Lauren Mitchell, Jemma Nelson, Alex Nguyen, Sofia Olsson, Sadie Patraw, Tobias Ragoczy, Joshua Richards, Jean Robinston, Jacob Rodriguez, John Stamatoyannopoulos, Eric Thorland, Julia Wald |
| EPI_ISL_2376530 | Omega Diagnostics at Mounes | Omega Diagnostics at Mounes | Ashleigh McGrail,Cynthia Corley,Latira Haynes-Jacob, Vivek Khare MD |
| EPI_ISL_2376531 | Omega Diagnostics at Mounes | Omega Diagnotics at Mounes | Ashleigh McGrail,Cynthia Corley,Latira Haynes-Jacob, Vivek Khare MD |
| EPI_ISL_2376532, EPI_ISL_2376533 | Omega Diagnostics at Mounes | Omega Diagnostics at Mounes | Ashleigh McGrail,Cynthia Corley,Latira Haynes-Jacob, Vivek Khare MD |
| EPI_ISL_2376534, EPI_ISL_2376535, EPI_ISL_2376536, EPI_ISL_2376537, EPI_ISL_2376538, EPI_ISL_2376539, EPI_ISL_2376540, EPI_ISL_2376541, EPI_ISL_2376542, EPI_ISL_2376543, EPI_ISL_2376544, EPI_ISL_2376545, EPI_ISL_2376546, EPI_ISL_2376547, EPI_ISL_2376548 | | | |
| see above | Brien Lab, St. Louis University | Brien Lab, St. Louis University | James D. Brien, Amelia K. Pinto, Mariah Hassert, Tara Steffen, Alexandria M. Dickson, Colleen R. Bocke, Elizabeth Geerling, E. Taylor Stone, Robin R. Chamberland, Alexis Guenette DO |
| EPI_ISL_2376549, EPI_ISL_2376550, EPI_ISL_2376551, EPI_ISL_2376552, EPI_ISL_2376553, EPI_ISL_2376554, EPI_ISL_2376555, EPI_ISL_2376556, EPI_ISL_2376557, EPI_ISL_2376558, EPI_ISL_2376559, EPI_ISL_2376560, EPI_ISL_2376561, EPI_ISL_2376562, EPI_ISL_2376563, EPI_ISL_2376564, EPI_ISL_2376565, EPI_ISL_2376566, EPI_ISL_2376567, EPI_ISL_2376568, EPI_ISL_2376569, EPI_ISL_2376570, EPI_ISL_2376571 | | | |
| see above | Minnesota Department of Health, Public Health Laboratory | Minnesota Department of Health, Public Health Laboratory | Alexandra Lorentz, Jacob Garfin, Matt Plumb, and Xiong Wang |
| EPI_ISL_2376572 | M Health Fairview | Minnesota Department of Health, Public Health Laboratory | Alexandra Lorentz, Jacob Garfin, Matt Plumb, and Xiong Wang |
| EPI_ISL_2376573, EPI_ISL_2376574, EPI_ISL_2376575 | Mayo Clinic & Mayo Clinic Laboratories | Minnesota Department of Health, Public Health Laboratory | Alexandra Lorentz, Jacob Garfin, Matt Plumb, and Xiong Wang |
| EPI_ISL_2376576, EPI_ISL_2376577, EPI_ISL_2376578, EPI_ISL_2376579, EPI_ISL_2376580, EPI_ISL_2376581, EPI_ISL_2376583, EPI_ISL_2376584, EPI_ISL_2376585, EPI_ISL_2376586, EPI_ISL_2376587, EPI_ISL_2376588, EPI_ISL_2376589, EPI_ISL_2376590, EPI_ISL_2376591, EPI_ISL_2376592, EPI_ISL_2376593, EPI_ISL_2376594, EPI_ISL_2376595, EPI_ISL_2376596, EPI_ISL_2376597, EPI_ISL_2376598, EPI_ISL_2376599, EPI_ISL_2376600, EPI_ISL_2376601, EPI_ISL_2376602, EPI_ISL_2376603, EPI_ISL_2376604, EPI_ISL_2376605, EPI_ISL_2376606, EPI_ISL_2376607, EPI_ISL_2376608, EPI_ISL_2376609, EPI_ISL_2376610, EPI_ISL_2376611, EPI_ISL_2376612, EPI_ISL_2376613, EPI_ISL_2376614, EPI_ISL_2376615, EPI_ISL_2376616 | | | |
| see above | M Health Fairview | Minnesota Department of Health, Public Health Laboratory | Alexandra Lorentz, Jacob Garfin, Matt Plumb, and Xiong Wang |
| EPI_ISL_2376618 | GA Department of Public Health Laboratory | Genomics and Discovery, Respiratory Viruses Branch, Division of Viral Diseases, Centers for Disease Control and Prevention | Anna Kelleher, Ying Tao, Yan Li, Jing Zhang, Brian Lynch, Krista Queen, Anna Uehara, Peter Cook, Han Jia Justin Ng, Rachel Marine, Clinton R. Paden, Dhwani Batra, Haibin Wang, Tara Coalter, Jasmine Padilla, Morgan Davis, Mili Sheth, Sarah Nobles, Mark Burroughs, Justin Lee, Adam Retchless, Suxiang Tong |
| EPI_ISL_2377585, EPI_ISL_2377586, EPI_ISL_2377587, EPI_ISL_2377588, EPI_ISL_2377589, EPI_ISL_2377590, EPI_ISL_2377591, EPI_ISL_2377592, EPI_ISL_2377593, EPI_ISL_2377594, EPI_ISL_2377595, EPI_ISL_2377596, EPI_ISL_2377597, EPI_ISL_2377598, EPI_ISL_2377599, EPI_ISL_2377600, EPI_ISL_2377601, EPI_ISL_2377602, EPI_ISL_2377603, EPI_ISL_2377604, EPI_ISL_2377605, EPI_ISL_2377606, EPI_ISL_2377607, EPI_ISL_2377609, EPI_ISL_2377611, EPI_ISL_2377612, EPI_ISL_2377613, EPI_ISL_2377614, EPI_ISL_2377615, EPI_ISL_2377616, EPI_ISL_2377617, EPI_ISL_2377619, EPI_ISL_2377620, EPI_ISL_2377622, EPI_ISL_2377623, EPI_ISL_2377624, EPI_ISL_2377625, EPI_ISL_2377626, EPI_ISL_2377627, EPI_ISL_2377629, EPI_ISL_2377630, EPI_ISL_2377631, EPI_ISL_2377632, EPI_ISL_2377635, EPI_ISL_2377636, EPI_ISL_2377637, EPI_ISL_2377638, EPI_ISL_2377640, EPI_ISL_2377641, EPI_ISL_2377642, EPI_ISL_2377643, EPI_ISL_2377644, EPI_ISL_2377645, EPI_ISL_2377646, EPI_ISL_2377647, EPI_ISL_2377648, EPI_ISL_2377649, EPI_ISL_2377650, EPI_ISL_2377651, EPI_ISL_2377652, EPI_ISL_2377653, EPI_ISL_2377654, EPI_ISL_2377655, EPI_ISL_2377656, EPI_ISL_2377657, EPI_ISL_2377658, EPI_ISL_2377659, EPI_ISL_2377660, EPI_ISL_2377661, EPI_ISL_2377662, EPI_ISL_2377663, EPI_ISL_2377664, EPI_ISL_2377666, EPI_ISL_2377668, EPI_ISL_2377669, EPI_ISL_2377670, EPI_ISL_2377671, EPI_ISL_2377672, EPI_ISL_2377673, EPI_ISL_2377674, EPI_ISL_2377675, EPI_ISL_2377677, EPI_ISL_2377678, EPI_ISL_2377679, EPI_ISL_2377681, EPI_ISL_2377682, EPI_ISL_2377683 | | | |
| see above | Maryland Genomics, Institute for Genome Sciences, University of Maryland School of Medicine | Maryland Genomics, Institute for Genome Sciences, University of Maryland School of Medicine | Tallon, Luke J; Sadzewicz, Lisa D; Humphrys, Mike; Ott, Sandra; Roussey, Holly; Mehta, Aditya; Vavikolanu, Kranthi; Fraser, Claire M; Ravel, Jacques |
| EPI_ISL_2377698, EPI_ISL_2377699, EPI_ISL_2377700, EPI_ISL_2377701, EPI_ISL_2377702, EPI_ISL_2377703, EPI_ISL_2377704, EPI_ISL_2377705, EPI_ISL_2377706, EPI_ISL_2377707, EPI_ISL_2377708, EPI_ISL_2377709, EPI_ISL_2377710, EPI_ISL_2377711, EPI_ISL_2377712, EPI_ISL_2377713, EPI_ISL_2377714, EPI_ISL_2377715, EPI_ISL_2377716, EPI_ISL_2377717, EPI_ISL_2377718, EPI_ISL_2377719, EPI_ISL_2377720, EPI_ISL_2377721, EPI_ISL_2377722, EPI_ISL_2377723, EPI_ISL_2377724, EPI_ISL_2377725, EPI_ISL_2377726, EPI_ISL_2377727, EPI_ISL_2377728, EPI_ISL_2377729, EPI_ISL_2377730, EPI_ISL_2377731, EPI_ISL_2377732, EPI_ISL_2377733, EPI_ISL_2377734, EPI_ISL_2377735, EPI_ISL_2377736, EPI_ISL_2377737 | | | |
| see above | Texas Department of State Health Services (TXDSHS) | Texas Department of State Health Services (TXDSHS) | Rashmi Tuladhar, Bonnie Oh, Jenny Zhang, Maliha Rahman, Mayela Pedrueza, Anita Pokharel, Karen Bobier, Lorraine Rodriguez, Myong Koag, Chun Wang, Rachel Lee, Grace Kubin |
| EPI_ISL_2377896, EPI_ISL_2377898, EPI_ISL_2377899, | Wexner Medical Center | The Ohio State University College of Medicine | Koenig,S., Seminetta,J. |

| | | | |
|---|---|---|---|
| EPI_ISL_2377900, EPI_ISL_2377901, EPI_ISL_2377902, EPI_ISL_2377903, EPI_ISL_2377904, EPI_ISL_2377905 | | | |
| EPI_ISL_2377906, EPI_ISL_2377907, EPI_ISL_2377908, EPI_ISL_2377909, EPI_ISL_2377910, EPI_ISL_2377911, EPI_ISL_2377912, EPI_ISL_2377913, EPI_ISL_2377914, EPI_ISL_2377915, EPI_ISL_2377916, EPI_ISL_2377917, EPI_ISL_2377918, EPI_ISL_2377919, EPI_ISL_2377920, EPI_ISL_2377921, EPI_ISL_2377922, EPI_ISL_2377923, EPI_ISL_2377924, EPI_ISL_2377925, EPI_ISL_2377926, EPI_ISL_2377927, EPI_ISL_2377928, EPI_ISL_2377929, EPI_ISL_2377930, EPI_ISL_2377931, EPI_ISL_2377932, EPI_ISL_2377933, EPI_ISL_2377934, EPI_ISL_2377935, EPI_ISL_2377936, EPI_ISL_2377937, EPI_ISL_2377938, EPI_ISL_2377939, EPI_ISL_2377940, EPI_ISL_2377941, EPI_ISL_2377942, EPI_ISL_2377943, EPI_ISL_2377944, EPI_ISL_2377945, EPI_ISL_2377946, EPI_ISL_2377947, EPI_ISL_2377948, EPI_ISL_2377949, EPI_ISL_2377950, EPI_ISL_2377951, EPI_ISL_2377952, EPI_ISL_2377953, EPI_ISL_2377954, EPI_ISL_2377955, EPI_ISL_2377956, EPI_ISL_2377957, EPI_ISL_2377958, EPI_ISL_2377959, EPI_ISL_2377960, EPI_ISL_2377961, EPI_ISL_2377962, EPI_ISL_2377963, EPI_ISL_2377964, EPI_ISL_2377965, EPI_ISL_2377966, EPI_ISL_2377967, EPI_ISL_2377968, EPI_ISL_2377969, EPI_ISL_2377970, EPI_ISL_2377971, EPI_ISL_2377972, EPI_ISL_2377973, EPI_ISL_2377974, EPI_ISL_2377975, EPI_ISL_2377976, EPI_ISL_2377977, EPI_ISL_2377978, EPI_ISL_2377979, EPI_ISL_2377980, EPI_ISL_2377981, EPI_ISL_2377982, EPI_ISL_2377983, EPI_ISL_2377984, EPI_ISL_2377985, EPI_ISL_2377986, EPI_ISL_2377987, EPI_ISL_2377988, EPI_ISL_2377989, EPI_ISL_2377990, EPI_ISL_2377991, EPI_ISL_2377992, EPI_ISL_2377993, EPI_ISL_2377994, EPI_ISL_2377995, EPI_ISL_2377997, EPI_ISL_2377998, EPI_ISL_2377999, EPI_ISL_2378000, EPI_ISL_2378001, EPI_ISL_2378002, EPI_ISL_2378003, EPI_ISL_2378004, EPI_ISL_2378005, EPI_ISL_2378006, EPI_ISL_2378007, EPI_ISL_2378008, EPI_ISL_2378009, EPI_ISL_2378010, EPI_ISL_2378011, EPI_ISL_2378012, EPI_ISL_2378013, EPI_ISL_2378014, EPI_ISL_2378015, EPI_ISL_2378016, EPI_ISL_2378017, EPI_ISL_2378019, EPI_ISL_2378020, EPI_ISL_2378021, EPI_ISL_2378022, EPI_ISL_2378023, EPI_ISL_2378024, EPI_ISL_2378025, EPI_ISL_2378026, EPI_ISL_2378027, EPI_ISL_2378028, EPI_ISL_2378029, EPI_ISL_2378030, EPI_ISL_2378031, EPI_ISL_2378032, EPI_ISL_2378033 | | | |
| see above | State Testing Facility | Altius Institute for Biomedical Research | Daniel Bates, Rebecca Bruders, Michael Buckley, Mark Frerker, Amanda Gale, Clem Green, Muhammad Halimun, Kneshay Harper, Matt Hartman, Alex Isner, Audra Johnson, Jessica Kunder, Lauren Mitchell, Jemma Nelson, Alex Nguyen, Sofia Olsson, Sadie Patraw, Tobias Ragoczy, Joshua Richards, Jean Robinston, Jacob Rodriguez, John Stamatoyannopoulos, Eric Thorland, Julia Wald |
| EPI_ISL_2378034 | Seattle Academy | Altius Institute for Biomedical Research | Daniel Bates, Rebecca Bruders, Michael Buckley, Mark Frerker, Amanda Gale, Clem Green, Muhammad Halimun, Kneshay Harper, Matt Hartman, Alex Isner, Audra Johnson, Jessica Kunder, Lauren Mitchell, Jemma Nelson, Alex Nguyen, Sofia Olsson, Sadie Patraw, Tobias Ragoczy, Joshua Richards, Jean Robinston, Jacob Rodriguez, John Stamatoyannopoulos, Eric Thorland, Julia Wald |
| EPI_ISL_2378035, EPI_ISL_2378036, EPI_ISL_2378037, EPI_ISL_2378038, EPI_ISL_2378039, EPI_ISL_2378040, EPI_ISL_2378041, EPI_ISL_2378042, EPI_ISL_2378043, EPI_ISL_2378044, EPI_ISL_2378045, EPI_ISL_2378046, EPI_ISL_2378047, EPI_ISL_2378048, EPI_ISL_2378049, EPI_ISL_2378050, EPI_ISL_2378051, EPI_ISL_2378052, EPI_ISL_2378053, EPI_ISL_2378054, EPI_ISL_2378055, EPI_ISL_2378056, EPI_ISL_2378057 | | | |
| see above | State Testing Facility | Altius Institute for Biomedical Research | Daniel Bates, Rebecca Bruders, Michael Buckley, Mark Frerker, Amanda Gale, Clem Green, Muhammad Halimun, Kneshay Harper, Matt Hartman, Alex Isner, Audra Johnson, Jessica Kunder, Lauren Mitchell, Jemma Nelson, Alex Nguyen, Sofia Olsson, Sadie Patraw, Tobias Ragoczy, Joshua Richards, Jean Robinston, Jacob Rodriguez, John Stamatoyannopoulos, Eric Thorland, Julia Wald |
| EPI_ISL_2378058 | Seattle Academy | Altius Institute for Biomedical Research | Daniel Bates, Rebecca Bruders, Michael Buckley, Mark Frerker, Amanda Gale, Clem Green, Muhammad Halimun, Kneshay Harper, Matt Hartman, Alex Isner, Audra Johnson, Jessica Kunder, Lauren Mitchell, Jemma Nelson, Alex Nguyen, Sofia Olsson, Sadie Patraw, Tobias Ragoczy, Joshua Richards, Jean Robinston, Jacob Rodriguez, John Stamatoyannopoulos, Eric Thorland, Julia Wald |
| EPI_ISL_2378059, EPI_ISL_2378060, EPI_ISL_2378061, EPI_ISL_2378062, EPI_ISL_2378063, EPI_ISL_2378064, EPI_ISL_2378065, EPI_ISL_2378066, EPI_ISL_2378067, EPI_ISL_2378068, EPI_ISL_2378069, EPI_ISL_2378070, EPI_ISL_2378071, EPI_ISL_2378072, EPI_ISL_2378073, EPI_ISL_2378074, EPI_ISL_2378075, EPI_ISL_2378076, EPI_ISL_2378077, EPI_ISL_2378078, EPI_ISL_2378079, EPI_ISL_2378080, EPI_ISL_2378081 | | | |
| see above | State Testing Facility | Altius Institute for Biomedical Research | Daniel Bates, Rebecca Bruders, Michael Buckley, Mark Frerker, Amanda Gale, Clem Green, Muhammad Halimun, Kneshay Harper, Matt Hartman, Alex Isner, Audra Johnson, Jessica Kunder, Lauren Mitchell, Jemma Nelson, Alex Nguyen, Sofia Olsson, Sadie Patraw, Tobias Ragoczy, Joshua Richards, Jean Robinston, Jacob Rodriguez, John Stamatoyannopoulos, Eric Thorland, Julia Wald |
| EPI_ISL_2378082 | Bertschi School | Altius Institute for Biomedical Research | Daniel Bates, Rebecca Bruders, Michael Buckley, Mark Frerker, Amanda Gale, Clem Green, Muhammad Halimun, Kneshay Harper, Matt Hartman, Alex Isner, Audra Johnson, Jessica Kunder, Lauren Mitchell, Jemma Nelson, Alex Nguyen, Sofia Olsson, Sadie Patraw, Tobias Ragoczy, Joshua Richards, Jean Robinston, Jacob Rodriguez, John Stamatoyannopoulos, Eric Thorland, Julia Wald |
| EPI_ISL_2378083, EPI_ISL_2378084, EPI_ISL_2378085, EPI_ISL_2378086, EPI_ISL_2378087, EPI_ISL_2378088, EPI_ISL_2378089, EPI_ISL_2378090, EPI_ISL_2378091, EPI_ISL_2378092, EPI_ISL_2378093, EPI_ISL_2378094, EPI_ISL_2378095, EPI_ISL_2378096, EPI_ISL_2378097, EPI_ISL_2378098, EPI_ISL_2378099, EPI_ISL_2378100, EPI_ISL_2378101, EPI_ISL_2378102, EPI_ISL_2378103, EPI_ISL_2378104, EPI_ISL_2378105, EPI_ISL_2378106, EPI_ISL_2378107, EPI_ISL_2378108, EPI_ISL_2378109, EPI_ISL_2378110, EPI_ISL_2378111, EPI_ISL_2378112, EPI_ISL_2378113, EPI_ISL_2378114, EPI_ISL_2378115, EPI_ISL_2378116, EPI_ISL_2378117, EPI_ISL_2378118, EPI_ISL_2378119, EPI_ISL_2378120, EPI_ISL_2378121, EPI_ISL_2378122, EPI_ISL_2378123, EPI_ISL_2378124, EPI_ISL_2378125, EPI_ISL_2378126, EPI_ISL_2378127, EPI_ISL_2378128, EPI_ISL_2378129, EPI_ISL_2378130, EPI_ISL_2378131, EPI_ISL_2378132, EPI_ISL_2378133, EPI_ISL_2378134, EPI_ISL_2378135, EPI_ISL_2378136, EPI_ISL_2378137, EPI_ISL_2378138, EPI_ISL_2378139, EPI_ISL_2378140, EPI_ISL_2378141, EPI_ISL_2378142, EPI_ISL_2378143, EPI_ISL_2378144, EPI_ISL_2378145, EPI_ISL_2378146, EPI_ISL_2378147, EPI_ISL_2378148, EPI_ISL_2378149, EPI_ISL_2378150, EPI_ISL_2378151, EPI_ISL_2378152, EPI_ISL_2378153, EPI_ISL_2378154, EPI_ISL_2378155, EPI_ISL_2378156, EPI_ISL_2378157, EPI_ISL_2378158, EPI_ISL_2378159, EPI_ISL_2378160, EPI_ISL_2378161, EPI_ISL_2378162, EPI_ISL_2378163, EPI_ISL_2378164, EPI_ISL_2378165, EPI_ISL_2378166, EPI_ISL_2378167, EPI_ISL_2378168, EPI_ISL_2378169, EPI_ISL_2378170, EPI_ISL_2378171, EPI_ISL_2378172, EPI_ISL_2378173, EPI_ISL_2378174, EPI_ISL_2378175, EPI_ISL_2378176, EPI_ISL_2378177, EPI_ISL_2378178, EPI_ISL_2378179, EPI_ISL_2378180, EPI_ISL_2378181, EPI_ISL_2378182, EPI_ISL_2378183, EPI_ISL_2378185, EPI_ISL_2378186, EPI_ISL_2378187, EPI_ISL_2378188, EPI_ISL_2378189, EPI_ISL_2378190, EPI_ISL_2378191, EPI_ISL_2378192, EPI_ISL_2378193, EPI_ISL_2378194, EPI_ISL_2378195, EPI_ISL_2378196, EPI_ISL_2378197, EPI_ISL_2378198, EPI_ISL_2378199, EPI_ISL_2378200, EPI_ISL_2378201, EPI_ISL_2378202, EPI_ISL_2378203, EPI_ISL_2378204, EPI_ISL_2378205, EPI_ISL_2378206, EPI_ISL_2378207, EPI_ISL_2378208, EPI_ISL_2378209, EPI_ISL_2378210, EPI_ISL_2378211, EPI_ISL_2378212, EPI_ISL_2378213, EPI_ISL_2378214, EPI_ISL_2378215, EPI_ISL_2378216, EPI_ISL_2378217, EPI_ISL_2378218, EPI_ISL_2378219, EPI_ISL_2378220, EPI_ISL_2378221, EPI_ISL_2378222, EPI_ISL_2378223, EPI_ISL_2378224, EPI_ISL_2378225, EPI_ISL_2378226, EPI_ISL_2378227, EPI_ISL_2378228, EPI_ISL_2378229, EPI_ISL_2378230, EPI_ISL_2378231, EPI_ISL_2378232, EPI_ISL_2378233, EPI_ISL_2378234, EPI_ISL_2378235, EPI_ISL_2378236, EPI_ISL_2378237, EPI_ISL_2378238, EPI_ISL_2378239, EPI_ISL_2378240, EPI_ISL_2378241, EPI_ISL_2378242, EPI_ISL_2378243, EPI_ISL_2378244, EPI_ISL_2378245, EPI_ISL_2378246, EPI_ISL_2378247, EPI_ISL_2378248, EPI_ISL_2378249, EPI_ISL_2378250, EPI_ISL_2378251, EPI_ISL_2378252, EPI_ISL_2378253, EPI_ISL_2378254, EPI_ISL_2378255, EPI_ISL_2378256, EPI_ISL_2378257, EPI_ISL_2378258 | | | |
| see above | State Testing Facility | Altius Institute for Biomedical Research | Daniel Bates, Rebecca Bruders, Michael Buckley, Mark Frerker, Amanda Gale, Clem Green, Muhammad Halimun, Kneshay Harper, Matt Hartman, Alex Isner, Audra Johnson, Jessica Kunder, Lauren Mitchell, Jemma Nelson, Alex Nguyen, Sofia Olsson, Sadie Patraw, Tobias Ragoczy, Joshua Richards, Jean Robinston, Jacob Rodriguez, John Stamatoyannopoulos, Eric Thorland, Julia Wald |
| EPI_ISL_2378259 | Seattle University Athletics | Altius Institute for Biomedical Research | Daniel Bates, Rebecca Bruders, Michael Buckley, Mark Frerker, Amanda Gale, Clem Green, Muhammad Halimun, Kneshay Harper, Matt Hartman, Alex Isner, Audra Johnson, Jessica Kunder, Lauren Mitchell, Jemma Nelson, Alex Nguyen, Sofia Olsson, Sadie Patraw, Tobias Ragoczy, Joshua Richards, Jean Robinston, Jacob Rodriguez, John Stamatoyannopoulos, Eric Thorland, Julia Wald |
| EPI_ISL_2378260, EPI_ISL_2378261, EPI_ISL_2378262, EPI_ISL_2378263, EPI_ISL_2378264, EPI_ISL_2378265, EPI_ISL_2378266, EPI_ISL_2378267, EPI_ISL_2378268, EPI_ISL_2378269, EPI_ISL_2378270, EPI_ISL_2378271, EPI_ISL_2378272, EPI_ISL_2378273, EPI_ISL_2378274, EPI_ISL_2378275, EPI_ISL_2378276, EPI_ISL_2378277, EPI_ISL_2378278, EPI_ISL_2378279, EPI_ISL_2378280, EPI_ISL_2378281, EPI_ISL_2378282, EPI_ISL_2378283, EPI_ISL_2378284, EPI_ISL_2378285, EPI_ISL_2378286, EPI_ISL_2378287, EPI_ISL_2378288, EPI_ISL_2378289, EPI_ISL_2378290, EPI_ISL_2378291, EPI_ISL_2378292, EPI_ISL_2378293, EPI_ISL_2378294, EPI_ISL_2378295, EPI_ISL_2378296, EPI_ISL_2378297, EPI_ISL_2378298, EPI_ISL_2378299, EPI_ISL_2378300, EPI_ISL_2378301, EPI_ISL_2378302, EPI_ISL_2378303, EPI_ISL_2378304, EPI_ISL_2378305, EPI_ISL_2378306, EPI_ISL_2378307, EPI_ISL_2378308, EPI_ISL_2378309, EPI_ISL_2378310, EPI_ISL_2378311, EPI_ISL_2378312, EPI_ISL_2378313, EPI_ISL_2378314, EPI_ISL_2378315, EPI_ISL_2378316, EPI_ISL_2378317, EPI_ISL_2378318, EPI_ISL_2378319, EPI_ISL_2378320, EPI_ISL_2378321, EPI_ISL_2378322, EPI_ISL_2378323, EPI_ISL_2378324, EPI_ISL_2378325, EPI_ISL_2378326, EPI_ISL_2378327, EPI_ISL_2378328, EPI_ISL_2378329, EPI_ISL_2378330, EPI_ISL_2378331, EPI_ISL_2378332, EPI_ISL_2378333, EPI_ISL_2378334, EPI_ISL_2378335, EPI_ISL_2378336, EPI_ISL_2378337, EPI_ISL_2378338, EPI_ISL_2378339, EPI_ISL_2378340, EPI_ISL_2378341, EPI_ISL_2378342, EPI_ISL_2378343, EPI_ISL_2378344, EPI_ISL_2378345, EPI_ISL_2378346, EPI_ISL_2378347, EPI_ISL_2378348, EPI_ISL_2378349, EPI_ISL_2378350, EPI_ISL_2378351, EPI_ISL_2378352, EPI_ISL_2378353, EPI_ISL_2378354, EPI_ISL_2378355, EPI_ISL_2378356, EPI_ISL_2378357, EPI_ISL_2378358, EPI_ISL_2378359, EPI_ISL_2378360, EPI_ISL_2378361, EPI_ISL_2378362, EPI_ISL_2378363, EPI_ISL_2378364, EPI_ISL_2378365, EPI_ISL_2378366, EPI_ISL_2378367, EPI_ISL_2378368, EPI_ISL_2378369, EPI_ISL_2378370, EPI_ISL_2378371, EPI_ISL_2378372, EPI_ISL_2378373, EPI_ISL_2378374, EPI_ISL_2378375, EPI_ISL_2378376, EPI_ISL_2378377, EPI_ISL_2378378, EPI_ISL_2378379, EPI_ISL_2378380, EPI_ISL_2378381, EPI_ISL_2378382, EPI_ISL_2378383, EPI_ISL_2378384, EPI_ISL_2378385, EPI_ISL_2378386, EPI_ISL_2378387, EPI_ISL_2378388, EPI_ISL_2378389, EPI_ISL_2378390, EPI_ISL_2378391, EPI_ISL_2378392, EPI_ISL_2378393, EPI_ISL_2378394, EPI_ISL_2378395, EPI_ISL_2378396, EPI_ISL_2378397, EPI_ISL_2378398, EPI_ISL_2378399, EPI_ISL_2378400, EPI_ISL_2378401, EPI_ISL_2378402, EPI_ISL_2378403, EPI_ISL_2378404, EPI_ISL_2378405, EPI_ISL_2378406, EPI_ISL_2378407, EPI_ISL_2378408, EPI_ISL_2378409, EPI_ISL_2378410, EPI_ISL_2378411, EPI_ISL_2378412, EPI_ISL_2378413, EPI_ISL_2378414, EPI_ISL_2378415, EPI_ISL_2378416, EPI_ISL_2378417, EPI_ISL_2378418, EPI_ISL_2378419, EPI_ISL_2378420, EPI_ISL_2378421, EPI_ISL_2378422, EPI_ISL_2378423, EPI_ISL_2378424, EPI_ISL_2378425, EPI_ISL_2378426, EPI_ISL_2378427, EPI_ISL_2378428, EPI_ISL_2378429, EPI_ISL_2378430, EPI_ISL_2378431, EPI_ISL_2378432, EPI_ISL_2378433, EPI_ISL_2378434, EPI_ISL_2378435, EPI_ISL_2378436, EPI_ISL_2378437, EPI_ISL_2378438, EPI_ISL_2378439, EPI_ISL_2378440, EPI_ISL_2378441, EPI_ISL_2378442, EPI_ISL_2378443, EPI_ISL_2378444, EPI_ISL_2378445, EPI_ISL_2378446, EPI_ISL_2378447, EPI_ISL_2378448 | | | |
| see above | State Testing Facility | Altius Institute for Biomedical Research | Daniel Bates, Rebecca Bruders, Michael Buckley, Mark Frerker, Amanda Gale, Clem Green, Muhammad Halimun, Kneshay Harper, Matt Hartman, Alex Isner, Audra Johnson, Jessica Kunder, Lauren Mitchell, Jemma Nelson, Alex Nguyen, Sofia Olsson, Sadie Patraw, Tobias Ragoczy, Joshua Richards, Jean Robinston, Jacob Rodriguez, John Stamatoyannopoulos, Eric Thorland, Julia Wald |
| EPI_ISL_2378449 | Seattle University Athletics | Altius Institute for Biomedical Research | Daniel Bates, Rebecca Bruders, Michael Buckley, Mark Frerker, Amanda Gale, Clem Green, Muhammad Halimun, Kneshay Harper, Matt Hartman, Alex Isner, Audra Johnson, Jessica Kunder, Lauren Mitchell, Jemma Nelson, Alex Nguyen, Sofia Olsson, Sadie Patraw, Tobias Ragoczy, Joshua Richards, Jean Robinston, Jacob Rodriguez, John Stamatoyannopoulos, Eric Thorland, Julia Wald |
| EPI_ISL_2378450, EPI_ISL_2378451, EPI_ISL_2378452, EPI_ISL_2378453, EPI_ISL_2378454, EPI_ISL_2378455, EPI_ISL_2378456, EPI_ISL_2378457, EPI_ISL_2378458, EPI_ISL_2378459, EPI_ISL_2378460 | | | |
| see above | State Testing Facility | Altius Institute for Biomedical Research | Daniel Bates, Rebecca Bruders, Michael Buckley, Mark Frerker, Amanda Gale, Clem Green, Muhammad Halimun, Kneshay Harper, Matt Hartman, Alex Isner, Audra Johnson, Jessica Kunder, Lauren Mitchell, Jemma Nelson, Alex Nguyen, Sofia Olsson, Sadie Patraw, Tobias Ragoczy, Joshua Richards, Jean Robinston, Jacob Rodriguez, John Stamatoyannopoulos, Eric Thorland, Julia Wald |
| EPI_ISL_2378461 | Seattle University Athletics | Altius Institute for Biomedical Research | Daniel Bates, Rebecca Bruders, Michael Buckley, Mark Frerker, Amanda Gale, Clem Green, Muhammad Halimun, Kneshay Harper, Matt Hartman, Alex Isner, Audra Johnson, Jessica Kunder, Lauren Mitchell, Jemma Nelson, Alex Nguyen, Sofia Olsson, Sadie Patraw, Tobias Ragoczy, Joshua Richards, Jean Robinston, Jacob Rodriguez, John Stamatoyannopoulos, Eric Thorland, Julia Wald |
| EPI_ISL_2378462, EPI_ISL_2378463, EPI_ISL_2378464, EPI_ISL_2378465, EPI_ISL_2378466, EPI_ISL_2378467, EPI_ISL_2378468, EPI_ISL_2378469, EPI_ISL_2378470, EPI_ISL_2378471, EPI_ISL_2378472, EPI_ISL_2378473, EPI_ISL_2378474, EPI_ISL_2378475, EPI_ISL_2378476, EPI_ISL_2378477, EPI_ISL_2378478, EPI_ISL_2378479, EPI_ISL_2378480, EPI_ISL_2378481, EPI_ISL_2378482, EPI_ISL_2378483, EPI_ISL_2378484, EPI_ISL_2378485, EPI_ISL_2378486, EPI_ISL_2378487, EPI_ISL_2378488, EPI_ISL_2378489, EPI_ISL_2378490, EPI_ISL_2378491, EPI_ISL_2378492, EPI_ISL_2378493, EPI_ISL_2378494, EPI_ISL_2378495, EPI_ISL_2378496, EPI_ISL_2378497, EPI_ISL_2378498, EPI_ISL_2378499, EPI_ISL_2378500, EPI_ISL_2378501 | | | |
| see above | State Testing Facility | Altius Institute for Biomedical Research | Daniel Bates, Rebecca Bruders, Michael Buckley, Mark Frerker, Amanda Gale, Clem Green, Muhammad Halimun, Kneshay Harper, Matt Hartman, Alex Isner, Audra Johnson, Jessica Kunder, Lauren Mitchell, Jemma Nelson, Alex Nguyen, Sofia Olsson, Sadie Patraw, Tobias Ragoczy, Joshua Richards, Jean Robinston, Jacob Rodriguez, John Stamatoyannopoulos, Eric Thorland, Julia Wald |
| EPI_ISL_2378502 | Seattle University | Altius Institute for Biomedical Research | Daniel Bates, Rebecca Bruders, Michael Buckley, Mark Frerker, Amanda Gale, Clem Green, Muhammad Halimun, Kneshay Harper, Matt Hartman, Alex Isner, Audra Johnson, Jessica Kunder, Lauren Mitchell, Jemma Nelson, Alex Nguyen, Sofia Olsson, Sadie Patraw, Tobias Ragoczy, Joshua Richards, Jean Robinston, Jacob Rodriguez, John Stamatoyannopoulos, Eric Thorland, Julia Wald |
| EPI_ISL_2378503, EPI_ISL_2378504, EPI_ISL_2378505, EPI_ISL_2378506, EPI_ISL_2378507, EPI_ISL_2378508, EPI_ISL_2378509, EPI_ISL_2378510, EPI_ISL_2378511, EPI_ISL_2378512, EPI_ISL_2378513, EPI_ISL_2378514, EPI_ISL_2378515, EPI_ISL_2378516, EPI_ISL_2378517, EPI_ISL_2378518, EPI_ISL_2378519, EPI_ISL_2378520, EPI_ISL_2378521, EPI_ISL_2378522, EPI_ISL_2378523, EPI_ISL_2378524, EPI_ISL_2378525, EPI_ISL_2378526, EPI_ISL_2378527, EPI_ISL_2378528, EPI_ISL_2378529, EPI_ISL_2378530, EPI_ISL_2378531, EPI_ISL_2378532, EPI_ISL_2378533, EPI_ISL_2378534, EPI_ISL_2378535, EPI_ISL_2378536, EPI_ISL_2378537, EPI_ISL_2378538, EPI_ISL_2378539, EPI_ISL_2378540, EPI_ISL_2378541, EPI_ISL_2378542, EPI_ISL_2378543, EPI_ISL_2378544, EPI_ISL_2378545, EPI_ISL_2378546, EPI_ISL_2378547, EPI_ISL_2378548, EPI_ISL_2378549, EPI_ISL_2378550, EPI_ISL_2378551, EPI_ISL_2378552, EPI_ISL_2378553, EPI_ISL_2378554, EPI_ISL_2378555, EPI_ISL_2378556, EPI_ISL_2378557, EPI_ISL_2378558, EPI_ISL_2378559, EPI_ISL_2378560, EPI_ISL_2378561, EPI_ISL_2378562, EPI_ISL_2378563, EPI_ISL_2378564, EPI_ISL_2378565, EPI_ISL_2378566, EPI_ISL_2378567, EPI_ISL_2378568, EPI_ISL_2378569, EPI_ISL_2378570, EPI_ISL_2378571, EPI_ISL_2378572, EPI_ISL_2378573, EPI_ISL_2378574, EPI_ISL_2378575, EPI_ISL_2378576, EPI_ISL_2378577, EPI_ISL_2378578, EPI_ISL_2378579, EPI_ISL_2378580, EPI_ISL_2378581, EPI_ISL_2378582, EPI_ISL_2378583, EPI_ISL_2378584, EPI_ISL_2378585, EPI_ISL_2378586, EPI_ISL_2378587, EPI_ISL_2378588, EPI_ISL_2378589, EPI_ISL_2378590, EPI_ISL_2378591, EPI_ISL_2378592, EPI_ISL_2378593, EPI_ISL_2378594, EPI_ISL_2378595, EPI_ISL_2378596, EPI_ISL_2378597, EPI_ISL_2378598, EPI_ISL_2378599, EPI_ISL_2378600, EPI_ISL_2378601, EPI_ISL_2378602, EPI_ISL_2378603, EPI_ISL_2378604, EPI_ISL_2378605, EPI_ISL_2378606, EPI_ISL_2378607, EPI_ISL_2378609, EPI_ISL_2378610, EPI_ISL_2378611, EPI_ISL_2378612, EPI_ISL_2378613, EPI_ISL_2378614, EPI_ISL_2378615, EPI_ISL_2378616, EPI_ISL_2378617, EPI_ISL_2378618, EPI_ISL_2378619, EPI_ISL_2378620, EPI_ISL_2378621, EPI_ISL_2378622, EPI_ISL_2378623, EPI_ISL_2378624, EPI_ISL_2378625, EPI_ISL_2378626, EPI_ISL_2378627, EPI_ISL_2378628, EPI_ISL_2378629, EPI_ISL_2378630, EPI_ISL_2378631 | | | |
| see above | State Testing Facility | Altius Institute for Biomedical Research | Daniel Bates, Rebecca Bruders, Michael Buckley, Mark Frerker, Amanda Gale, Clem Green, Muhammad Halimun, Kneshay Harper, Matt Hartman, Alex Isner, Audra Johnson, Jessica Kunder, Lauren Mitchell, Jemma Nelson, Alex Nguyen, Sofia Olsson, Sadie Patraw, Tobias Ragoczy, Joshua Richards, Jean Robinston, Jacob Rodriguez, John Stamatoyannopoulos, Eric Thorland, Julia Wald |
| EPI_ISL_2378632, EPI_ISL_2378634, EPI_ISL_2378635, EPI_ISL_2378636, EPI_ISL_2378637, EPI_ISL_2378638, EPI_ISL_2378639 | The Ohio State University Applied Microbiology Services Laboratory | The Ohio State University Applied Microbiology Services Laboratory | Seth A. Faith PhD |
| EPI_ISL_2378711, EPI_ISL_2378712, EPI_ISL_2378713, EPI_ISL_2378714, EPI_ISL_2378715, EPI_ISL_2378716, EPI_ISL_2378717, EPI_ISL_2378718, EPI_ISL_2378719, EPI_ISL_2378720, EPI_ISL_2378721, EPI_ISL_2378722, EPI_ISL_2378723 | | | |
| see above | Maryland Genomics, Institute for Genome Sciences, University of Maryland School of Medicine | Maryland Genomics, Institute for Genome Sciences, University of Maryland School of Medicine | Tallon, Luke J; Sadzewicz, Lisa D; Humphrys, Mike; Ott, Sandra; Roussey, Holly; Mehta, Aditya; Vavikolanu, Kranthi; Fraser, Claire M; Ravel, Jacques |
| EPI_ISL_2378755, EPI_ISL_2378756, EPI_ISL_2378757, EPI_ISL_2378758, EPI_ISL_2378759, EPI_ISL_2378760, EPI_ISL_2378761, EPI_ISL_2378763, EPI_ISL_2378768, EPI_ISL_2378769, EPI_ISL_2378770, EPI_ISL_2378771, EPI_ISL_2378772, EPI_ISL_2378774, EPI_ISL_2378775, EPI_ISL_2378776, EPI_ISL_2378777, EPI_ISL_2378780, EPI_ISL_2378781, EPI_ISL_2378783, EPI_ISL_2378785, EPI_ISL_2378789, EPI_ISL_2378790, EPI_ISL_2378791, EPI_ISL_2378792, EPI_ISL_2378795, EPI_ISL_2378796, EPI_ISL_2378797, EPI_ISL_2378798, EPI_ISL_2378800, EPI_ISL_2378801, EPI_ISL_2378802, EPI_ISL_2378804, EPI_ISL_2378806, EPI_ISL_2378807, EPI_ISL_2378809, EPI_ISL_2378811, EPI_ISL_2378813, EPI_ISL_2378815, EPI_ISL_2378816, EPI_ISL_2378817, EPI_ISL_2378819, EPI_ISL_2378820, EPI_ISL_2378822 | | | |
| see above | UCSC Genomics Institute | UCSC Genomics Institute | Mark Akeson, Ikenna Anigbogu, Eric Beraut, Isabel Bjork, Molly Cassatt-Johnstone, Terren Chang, Russell Corbett-Detig, Namrita Dhillon, Maximilian Haeussler, David Haussler, Angie Hinrichs, Miten Jain, Joshua Kapp, A. Marm Kilpatrick, Jakob McBroome, Hugh Olsen, Jeremy Sanford, Beth Shapiro, Michael Stone, Bryan Thornlow, Yatish Turakhia, Ciara Wanket |
| EPI_ISL_2378824, EPI_ISL_2378825, EPI_ISL_2378826, EPI_ISL_2378827, EPI_ISL_2378828, EPI_ISL_2378829, EPI_ISL_2378830, EPI_ISL_2378831, EPI_ISL_2378832, EPI_ISL_2378833, EPI_ISL_2378834, EPI_ISL_2378835, EPI_ISL_2378836, EPI_ISL_2378837, EPI_ISL_2378838, EPI_ISL_2378839, EPI_ISL_2378840, EPI_ISL_2378841, EPI_ISL_2378842, EPI_ISL_2378843, EPI_ISL_2378844, EPI_ISL_2378845, EPI_ISL_2378846, EPI_ISL_2378847, EPI_ISL_2378848, EPI_ISL_2378849, EPI_ISL_2378850, EPI_ISL_2378851, EPI_ISL_2378852, EPI_ISL_2378853, EPI_ISL_2378854, EPI_ISL_2378855, EPI_ISL_2378856, EPI_ISL_2378857, EPI_ISL_2378858, EPI_ISL_2378859, EPI_ISL_2378860, EPI_ISL_2378861, EPI_ISL_2378862, EPI_ISL_2378863, EPI_ISL_2378864, EPI_ISL_2378865, EPI_ISL_2378866, EPI_ISL_2378867, EPI_ISL_2378868, EPI_ISL_2378869, EPI_ISL_2378870, EPI_ISL_2378871, EPI_ISL_2378872, EPI_ISL_2378873, EPI_ISL_2378874, EPI_ISL_2378875, EPI_ISL_2378876, EPI_ISL_2378877, EPI_ISL_2378878, EPI_ISL_2378879, EPI_ISL_2378880, EPI_ISL_2378881, EPI_ISL_2378882, EPI_ISL_2378883, EPI_ISL_2378884, EPI_ISL_2378885, EPI_ISL_2378886, EPI_ISL_2378887, EPI_ISL_2378888, EPI_ISL_2378889, EPI_ISL_2378890, EPI_ISL_2378891 | | | |
| see above | Washington State Department of Health Public Health Laboratories | Washington State Department of Health Public Health Laboratories | Drew MacKellar, Philip Dykema, Denny Russell, Joenice Gonzalez, Hannah Gray, Geoff Melly, Vanessa De Los Santos, Darren Lucas, JohnAric Peterson, Avi Singh, Rebecca Cao |
| EPI_ISL_2378897, EPI_ISL_2378898, EPI_ISL_2378899, EPI_ISL_2378902, EPI_ISL_2378903, EPI_ISL_2378904, EPI_ISL_2378906, EPI_ISL_2378907, EPI_ISL_2378908, EPI_ISL_2378909, EPI_ISL_2378910, EPI_ISL_2378911, EPI_ISL_2378912, EPI_ISL_2378913, EPI_ISL_2378914, EPI_ISL_2378915, EPI_ISL_2378916, EPI_ISL_2378917, EPI_ISL_2378918, EPI_ISL_2378920, EPI_ISL_2378921, EPI_ISL_2378923, EPI_ISL_2378924, EPI_ISL_2378925, EPI_ISL_2378926, EPI_ISL_2378927, EPI_ISL_2378928, EPI_ISL_2378929, EPI_ISL_2378930, EPI_ISL_2378931, EPI_ISL_2378932, EPI_ISL_2378933, EPI_ISL_2378934, EPI_ISL_2378935, EPI_ISL_2378936, EPI_ISL_2378937, EPI_ISL_2378938, EPI_ISL_2378939, EPI_ISL_2378940, EPI_ISL_2378941, EPI_ISL_2378942, EPI_ISL_2378943, EPI_ISL_2378944, EPI_ISL_2378945, EPI_ISL_2378946, EPI_ISL_2378948, EPI_ISL_2378949, EPI_ISL_2378950, EPI_ISL_2378951, EPI_ISL_2378952, EPI_ISL_2378954, EPI_ISL_2378955, EPI_ISL_2378956, EPI_ISL_2378957, EPI_ISL_2378958, EPI_ISL_2378959, EPI_ISL_2378960, EPI_ISL_2378961, EPI_ISL_2378962, EPI_ISL_2378964, EPI_ISL_2378966, EPI_ISL_2378967, EPI_ISL_2378968, EPI_ISL_2378969, EPI_ISL_2378970, EPI_ISL_2378971, | | | |

| | | | |
|---|---|---|---|
| EPI_ISL_2378973, EPI_ISL_2378974, EPI_ISL_2378975, EPI_ISL_2378976, EPI_ISL_2378977, EPI_ISL_2378978, EPI_ISL_2378979, EPI_ISL_2378980, EPI_ISL_2378981, EPI_ISL_2378982, EPI_ISL_2378983 | | | |
| see above | OHSU Lab Services Molecular Microbiology Lab | Oregon SARS-CoV-2 Genome Sequencing Center | Brendan L. O'Connell, Sally Grindstaff, Sonia Acharya, Cierra LaBlanc, Kayla Carter, Ruth V. Nichols, Alec J. Hirsch, Donna Hansel, Guang Fan, Xuan Qin, Daniel N. Streblow, William B. Messer, Andrew C. Adey, Benjamin N. Bimber, Brian J. O'Roak |
| EPI_ISL_2379247, EPI_ISL_2379248, EPI_ISL_2379249, EPI_ISL_2379250, EPI_ISL_2379251, EPI_ISL_2379252, EPI_ISL_2379253, EPI_ISL_2379254, EPI_ISL_2379255, EPI_ISL_2379256, EPI_ISL_2379257, EPI_ISL_2379258, EPI_ISL_2379259, EPI_ISL_2379260, EPI_ISL_2379261, EPI_ISL_2379262, EPI_ISL_2379263 | | | |
| see above | University of Pittsburgh | Genomics and Discovery, Respiratory Viruses Branch, Division of Viral Diseases, Centers for Disease Control and Prevention | Anna Kelleher, Ying Tao, Yan Li, Jing Zhang, Brian Lynch, Krista Queen, Anna Uehara, Peter Cook, Han Jia Justin Ng, Rachel Marine, Clinton R. Paden, Dhwani Batra, Haibin Wang, Tara Coalter, Jasmine Padilla, Morgan Davis, Mili Sheth, Sarah Nobles, Mark Burroughs, Justin Lee, Adam Retchless, Suxiang Tong |
| EPI_ISL_2380769, EPI_ISL_2380770, EPI_ISL_2380771, EPI_ISL_2380772, EPI_ISL_2380773, EPI_ISL_2380774, EPI_ISL_2380775, EPI_ISL_2380776, EPI_ISL_2380777, EPI_ISL_2380778, EPI_ISL_2380779, EPI_ISL_2380780, EPI_ISL_2380781, EPI_ISL_2380782, EPI_ISL_2380783, EPI_ISL_2380784, EPI_ISL_2380785, EPI_ISL_2380786, EPI_ISL_2380787, EPI_ISL_2380788, EPI_ISL_2380789, EPI_ISL_2380790, EPI_ISL_2380791, EPI_ISL_2380792, EPI_ISL_2380793, EPI_ISL_2380794, EPI_ISL_2380795, EPI_ISL_2380796, EPI_ISL_2380797, EPI_ISL_2380798, EPI_ISL_2380799, EPI_ISL_2380800, EPI_ISL_2380801, EPI_ISL_2380802, EPI_ISL_2380803, EPI_ISL_2380804, EPI_ISL_2380805, EPI_ISL_2380806, EPI_ISL_2380807, EPI_ISL_2380808, EPI_ISL_2380810, EPI_ISL_2380811, EPI_ISL_2380813, EPI_ISL_2380814, EPI_ISL_2380815, EPI_ISL_2380816, EPI_ISL_2380817, EPI_ISL_2380818, EPI_ISL_2380819, EPI_ISL_2380821, EPI_ISL_2380822, EPI_ISL_2380824, EPI_ISL_2380825, EPI_ISL_2380826, EPI_ISL_2380827, EPI_ISL_2380828, EPI_ISL_2380829, EPI_ISL_2380833, EPI_ISL_2380834, EPI_ISL_2380837, EPI_ISL_2380838, EPI_ISL_2380839, EPI_ISL_2380840, EPI_ISL_2380841, EPI_ISL_2380842, EPI_ISL_2380845, EPI_ISL_2380847, EPI_ISL_2380848, EPI_ISL_2380849, EPI_ISL_2380850, EPI_ISL_2380852, EPI_ISL_2380853, EPI_ISL_2380854, EPI_ISL_2380855, EPI_ISL_2380856, EPI_ISL_2380857 | | | |
| see above | Clinical Molecular Microbiology Laboratory, UNC Hospitals | Jeremy Wang | Jeremy Wang, Alexander Rubinsteyn, Colleen Rice, Jason Smedberg, Shawn Hawken, Melissa Miller, Corbin Jones, Robert Hagan |
| EPI_ISL_2381022, EPI_ISL_2381025, EPI_ISL_2381028 | Infinity Biologix | Centers for Disease Control and Prevention Division of Viral Diseases, Pathogen Discovery | Dakota Howard, Dhwani Batra, Peter W. Cook, Kara Moser, Adrian Paskey, Jason Caravas, Benjamin Rambo-Martin, Shatavia Morrison, Christopher Gulvick, Scott Sammons, Yvette Unoarumhi, Darlene Wagner, Matthew Schmerer, Christian Bixby, Yihe Wang, Jonathan Schultz, Chirayu Goswami, Russ Hager, Robin Grimwood, Clinton R. Paden, Duncan MacCannell |
| EPI_ISL_2381900 | Curative | New Mexico Department of Health Scientific Laboratory | Ellie Johnson,D'eldra Malone, Jennifer Benoit, Ratheesh Rajan, Linda Salazar, Keila Gutierrez, Anastacia Griego-Fisher |
| EPI_ISL_2381901, EPI_ISL_2381902, EPI_ISL_2381903, EPI_ISL_2381904, EPI_ISL_2381905, EPI_ISL_2381906, EPI_ISL_2381907, EPI_ISL_2381908, EPI_ISL_2381909, EPI_ISL_2381910, EPI_ISL_2381911, EPI_ISL_2381912, EPI_ISL_2381913, EPI_ISL_2381914, EPI_ISL_2381915, EPI_ISL_2381916, EPI_ISL_2381917, EPI_ISL_2381918, EPI_ISL_2381919, EPI_ISL_2381920, EPI_ISL_2381921, EPI_ISL_2381922, EPI_ISL_2381923, EPI_ISL_2381924, EPI_ISL_2381925, EPI_ISL_2381926, EPI_ISL_2381927, EPI_ISL_2381928, EPI_ISL_2381929, EPI_ISL_2381930, EPI_ISL_2381931, EPI_ISL_2381932, EPI_ISL_2381933, EPI_ISL_2381934, EPI_ISL_2381935, EPI_ISL_2381936, EPI_ISL_2381937, EPI_ISL_2381939, EPI_ISL_2381940, EPI_ISL_2381941, EPI_ISL_2381942, EPI_ISL_2381943, EPI_ISL_2381944, EPI_ISL_2381945, EPI_ISL_2381946, EPI_ISL_2381947, EPI_ISL_2381948, EPI_ISL_2381949, EPI_ISL_2381950, EPI_ISL_2381951, EPI_ISL_2381952, EPI_ISL_2381953, EPI_ISL_2381954, EPI_ISL_2381955, EPI_ISL_2381956, EPI_ISL_2381957, EPI_ISL_2381958, EPI_ISL_2381959, EPI_ISL_2381960, EPI_ISL_2381961, EPI_ISL_2381962, EPI_ISL_2381963, EPI_ISL_2381964, EPI_ISL_2381965, EPI_ISL_2381966, EPI_ISL_2381967, EPI_ISL_2381968, EPI_ISL_2381969, EPI_ISL_2381970, EPI_ISL_2381971, EPI_ISL_2381972, EPI_ISL_2381973, EPI_ISL_2381974, EPI_ISL_2381975, EPI_ISL_2381976, EPI_ISL_2381977, EPI_ISL_2381978, EPI_ISL_2381979, EPI_ISL_2381981, EPI_ISL_2381982, EPI_ISL_2381983, EPI_ISL_2381984, EPI_ISL_2381985, EPI_ISL_2381986, EPI_ISL_2381987, EPI_ISL_2381988, EPI_ISL_2381989, EPI_ISL_2381990, EPI_ISL_2381991 | | | |
| see above | New Mexico Department of Health Scientific Laboratory | New Mexico Department of Health Scientific Laboratory | Ellie Johnson,D'eldra Malone, Jennifer Benoit, Ratheesh Rajan, Linda Salazar, Keila Gutierrez, Anastacia Griego-Fisher |
| EPI_ISL_2382009, EPI_ISL_2382072, EPI_ISL_2382080 | Ohio Department of Health Laboratory | Ohio Department of Health Laboratory | Holmes, Jennifer; Eric Brandt, Keoni Omura, Glen McGillivary, Caitlin McDonnell, Jade Mowery, Stephanie Mccracken, Tyler Payne, Kirtana Ramadugu, Erica Leasure, Brent Lee, Kelsey Florek, Heather Blankenship, Allison Black, Quanta Brown, and Tammy Bannerman |
| EPI_ISL_2382081, EPI_ISL_2382082, EPI_ISL_2382083, EPI_ISL_2382084, EPI_ISL_2382085, EPI_ISL_2382086, EPI_ISL_2382087, EPI_ISL_2382088, EPI_ISL_2382089, EPI_ISL_2382090, EPI_ISL_2382091, EPI_ISL_2382093, EPI_ISL_2382094, EPI_ISL_2382095, EPI_ISL_2382096, EPI_ISL_2382097, EPI_ISL_2382098, EPI_ISL_2382099, EPI_ISL_2382100, EPI_ISL_2382101, EPI_ISL_2382102, EPI_ISL_2382103, EPI_ISL_2382104, EPI_ISL_2382105, EPI_ISL_2382106, EPI_ISL_2382107, EPI_ISL_2382108, EPI_ISL_2382109, EPI_ISL_2382110, EPI_ISL_2382111, EPI_ISL_2382112, EPI_ISL_2382113, EPI_ISL_2382114, EPI_ISL_2382115, EPI_ISL_2382116, EPI_ISL_2382117, EPI_ISL_2382118, EPI_ISL_2382119, EPI_ISL_2382120, EPI_ISL_2382121, EPI_ISL_2382122, EPI_ISL_2382123, EPI_ISL_2382124, EPI_ISL_2382125, EPI_ISL_2382126, EPI_ISL_2382127, EPI_ISL_2382128, EPI_ISL_2382129, EPI_ISL_2382132, EPI_ISL_2382133, EPI_ISL_2382135, EPI_ISL_2382136, EPI_ISL_2382137, EPI_ISL_2382138, EPI_ISL_2382139, EPI_ISL_2382140, EPI_ISL_2382141, EPI_ISL_2382142, EPI_ISL_2382143, EPI_ISL_2382144, EPI_ISL_2382145, EPI_ISL_2382146, EPI_ISL_2382147, EPI_ISL_2382148, EPI_ISL_2382149, EPI_ISL_2382151, EPI_ISL_2382153, EPI_ISL_2382154, EPI_ISL_2382155, EPI_ISL_2382158, EPI_ISL_2382159, EPI_ISL_2382160, EPI_ISL_2382161, EPI_ISL_2382162, EPI_ISL_2382163, EPI_ISL_2382164, EPI_ISL_2382165, EPI_ISL_2382166, EPI_ISL_2382167, EPI_ISL_2382168, EPI_ISL_2382169, EPI_ISL_2382170, EPI_ISL_2382171, EPI_ISL_2382172, EPI_ISL_2382173, EPI_ISL_2382174, EPI_ISL_2382175, EPI_ISL_2382176, EPI_ISL_2382177, EPI_ISL_2382178, EPI_ISL_2382179, EPI_ISL_2382180, EPI_ISL_2382182, EPI_ISL_2382183, EPI_ISL_2382184, EPI_ISL_2382185, EPI_ISL_2382186, EPI_ISL_2382187, EPI_ISL_2382188, EPI_ISL_2382189 | | | |
| see above | Mako Medical | Centers for Disease Control and Prevention Division of Viral Diseases, Pathogen Discovery | Dakota Howard, Dhwani Batra, Peter W. Cook, Kara Moser, Adrian Paskey, Jason Caravas, Benjamin Rambo-Martin, Shatavia Morrison, Christopher Gulvick, Scott Sammons, Yvette Unoarumhi, Darlene Wagner, Matthew Schmerer,Matthew Tugwell, Lauren Moon, Clinton R. Paden, Duncan MacCannell |
| EPI_ISL_2382267 | Ohio Department of Health Laboratory | Ohio Department of Health Laboratory | Holmes, Jennifer; Eric Brandt, Keoni Omura, Glen McGillivary, Caitlin McDonnell, Jade Mowery, Stephanie Mccracken, Tyler Payne, Kirtana Ramadugu, Erica Leasure, Brent Lee, Kelsey Florek, Heather Blankenship, Allison Black, Quanta Brown, and Tammy Bannerman |
| EPI_ISL_2382295, EPI_ISL_2382296, EPI_ISL_2382297, EPI_ISL_2382298, EPI_ISL_2382299, EPI_ISL_2382300, EPI_ISL_2382301, EPI_ISL_2382302, EPI_ISL_2382303, EPI_ISL_2382304, EPI_ISL_2382305 | | | |
| see above | Uconn Health Clinical Microbiology | Brenton Graveley Lab, UConn Health | Lori Avery, Christina Caron, Stacey Ferraro, Kimberly Hayes, Michael Senh, Sara Olson, Xintao Wei, Brenton Graveley |
| EPI_ISL_2382306, EPI_ISL_2382307, EPI_ISL_2382308, EPI_ISL_2382309, EPI_ISL_2382310 | Wexner Medical Center | The Ohio State University College of Medicine | Koenig,S., Seminetta,J. |
| EPI_ISL_2382311, EPI_ISL_2382312 | Blue Water Diagnostic Laboratory, LLC | University of Louisville Laboratory of Melissa Smith | Elizabeth Hudson, Matt Seetin, Erik Korte, Alonzo Shepherd, Corey T. Watson, Eric C. Rouchka, Melissa L. Smith |
| EPI_ISL_2382313 | University of Pittsburgh | Genomics and Discovery, Respiratory Viruses Branch, Division of Viral Diseases, Centers for Disease Control and Prevention | Anna Kelleher, Ying Tao, Yan Li, Jing Zhang, Brian Lynch, Krista Queen, Anna Uehara, Peter Cook, Han Jia Justin Ng, Rachel Marine, Clinton R. Paden, Dhwani Batra, Haibin Wang, Tara Coalter, Jasmine Padilla, Morgan Davis, Mili Sheth, Sarah Nobles, Mark Burroughs, Justin Lee, Adam Retchless, Suxiang Tong |
| EPI_ISL_2382314, EPI_ISL_2382315, EPI_ISL_2382316, EPI_ISL_2382317, EPI_ISL_2382318, EPI_ISL_2382319, EPI_ISL_2382320, EPI_ISL_2382321, EPI_ISL_2382322, EPI_ISL_2382323, EPI_ISL_2382324, EPI_ISL_2382325, EPI_ISL_2382326, EPI_ISL_2382327, EPI_ISL_2382328, EPI_ISL_2382329, EPI_ISL_2382330, EPI_ISL_2382331, EPI_ISL_2382332, EPI_ISL_2382333, EPI_ISL_2382334, EPI_ISL_2382335, EPI_ISL_2382336, EPI_ISL_2382337, EPI_ISL_2382338, EPI_ISL_2382339, EPI_ISL_2382340, EPI_ISL_2382341, EPI_ISL_2382342, EPI_ISL_2382343, EPI_ISL_2382344 | | | |
| see above | Wexner Medical Center | The Ohio State University College of Medicine | Koenig,S., Seminetta,J. |
| EPI_ISL_2382366, EPI_ISL_2382367, EPI_ISL_2382368, EPI_ISL_2382370, EPI_ISL_2382371, EPI_ISL_2382372, EPI_ISL_2382373, EPI_ISL_2382374, EPI_ISL_2382375, EPI_ISL_2382376, EPI_ISL_2382377, EPI_ISL_2382378, EPI_ISL_2382379, EPI_ISL_2382380, EPI_ISL_2382381, EPI_ISL_2382382, EPI_ISL_2382383, EPI_ISL_2382384, EPI_ISL_2382385, EPI_ISL_2382387, EPI_ISL_2382389, EPI_ISL_2382390, EPI_ISL_2382391, EPI_ISL_2382392, EPI_ISL_2382393, EPI_ISL_2382395, EPI_ISL_2382396, EPI_ISL_2382397, EPI_ISL_2382398, EPI_ISL_2382399, EPI_ISL_2382400, EPI_ISL_2382401 | | | |
| see above | Arizona State Public Health Laboratory | Arizona State Public Health Laboratory | Trung Huynh, Jessica Escobar, Katherine Fullerton, Nobuko Fukushima, Stacy White, Linda Getsinger, Victor Waddell |
| EPI_ISL_2382424, EPI_ISL_2382425, EPI_ISL_2382426, EPI_ISL_2382427, EPI_ISL_2382428, EPI_ISL_2382429, EPI_ISL_2382430, EPI_ISL_2382431, EPI_ISL_2382432, EPI_ISL_2382433, EPI_ISL_2382434, EPI_ISL_2382435, EPI_ISL_2382436, EPI_ISL_2382437, EPI_ISL_2382438, EPI_ISL_2382439, EPI_ISL_2382440, EPI_ISL_2382441, EPI_ISL_2382442, EPI_ISL_2382443, EPI_ISL_2382444, EPI_ISL_2382445, EPI_ISL_2382446, EPI_ISL_2382447, EPI_ISL_2382448, EPI_ISL_2382449, EPI_ISL_2382450, EPI_ISL_2382451 | | | |
| see above | Nebraska Public Health Laboratory | NPHL COVID-19 Response Team | NPHL COVID-19 Response Team |
| EPI_ISL_2382452, EPI_ISL_2382453, EPI_ISL_2382454, EPI_ISL_2382455, EPI_ISL_2382456, EPI_ISL_2382457, EPI_ISL_2382458, EPI_ISL_2382459, EPI_ISL_2382460, EPI_ISL_2382461, EPI_ISL_2382462, EPI_ISL_2382463, EPI_ISL_2382464, EPI_ISL_2382465, EPI_ISL_2382466, EPI_ISL_2382467, EPI_ISL_2382468, EPI_ISL_2382469, EPI_ISL_2382470, EPI_ISL_2382471, EPI_ISL_2382472, EPI_ISL_2382473, EPI_ISL_2382474, EPI_ISL_2382475, EPI_ISL_2382476, EPI_ISL_2382477, EPI_ISL_2382478, EPI_ISL_2382479, EPI_ISL_2382480, EPI_ISL_2382481, EPI_ISL_2382482, EPI_ISL_2382483, EPI_ISL_2382484, EPI_ISL_2382485, EPI_ISL_2382486, EPI_ISL_2382487, EPI_ISL_2382488, EPI_ISL_2382489, EPI_ISL_2382490, EPI_ISL_2382491, EPI_ISL_2382492, EPI_ISL_2382493, EPI_ISL_2382494, EPI_ISL_2382495, EPI_ISL_2382496, EPI_ISL_2382497, EPI_ISL_2382498, EPI_ISL_2382499, EPI_ISL_2382500, EPI_ISL_2382501, EPI_ISL_2382502, EPI_ISL_2382503, EPI_ISL_2382505, EPI_ISL_2382506, EPI_ISL_2382507, EPI_ISL_2382508, EPI_ISL_2382509, EPI_ISL_2382510, EPI_ISL_2382512, EPI_ISL_2382513, EPI_ISL_2382514, EPI_ISL_2382515, EPI_ISL_2382516, EPI_ISL_2382518, EPI_ISL_2382519, EPI_ISL_2382520, EPI_ISL_2382521 | | | |
| see above | The Ohio State University Applied Microbiology Services Laboratory | The Ohio State University Applied Microbiology Services Laboratory | Seth A. Faith PhD |
| EPI_ISL_2382522 | State Testing Facility | Altius Institute for Biomedical Research | Daniel Bates, Rebecca Bruders, Michael Buckley, Mark Frerker, Amanda Gale, Clem Green, Muhammad Halimun, Kneshay Harper, Matt Hartman, Alex Isner, Audra Johnson, Jessica Kunder, Lauren Mitchell, Jemma Nelson, Alex Nguyen, Sofia Olsson, Sadie Patraw, Tobias Ragoczy, Joshua Richards, Jean Robinston, Jacob Rodriguez, John Stamatoyannopoulos, Eric Thorland, Julia Wald |
| EPI_ISL_2382523, EPI_ISL_2382524, EPI_ISL_2382525, EPI_ISL_2382526, EPI_ISL_2382527, EPI_ISL_2382528, EPI_ISL_2382529, EPI_ISL_2382530, EPI_ISL_2382531, EPI_ISL_2382532, EPI_ISL_2382533 | | | |
| see above | Oregon State Public Health Laboratory | Oregon State Public Health Laboratory | Rafia Razzaque, Eugene Yeboah, Vanda Makris, Laura Tsaknaridis, John Fontana and Shane Sevey |
| EPI_ISL_2382542 | DC Public Health Lab/ Dept. of Forensic Sciences | DC Public Health Lab/ Dept. of Forensic Sciences | Janis Doss, Scott Nguyen, Elizabeth Zelaya, Sarah Scott, Connie Maza, Monica Mann, Brittany Hamilton, David Payne, Jocelyn Hauser |
| EPI_ISL_2382544, EPI_ISL_2382545, EPI_ISL_2382546, EPI_ISL_2382547, EPI_ISL_2382548, EPI_ISL_2382549, EPI_ISL_2382550 | Wichita State University- Molecular Diagonstic lab | Kansas Health and Environmental Lab | Katherine Wiggins, Mike Grose, Jonathan Barnell, Ben Olsen, and Phil Adam |
| EPI_ISL_2382551, EPI_ISL_2382552, EPI_ISL_2382553, EPI_ISL_2382554, EPI_ISL_2382555, EPI_ISL_2382556, EPI_ISL_2382557 | Kansas Health and Environmental Lab | Kansas Health and Environmental Lab | Katherine Wiggins, Mike Grose, Jonathan Barnell, Ben Olsen, and Phil Adam |
| EPI_ISL_2382558, EPI_ISL_2382560, EPI_ISL_2382561, EPI_ISL_2382562, EPI_ISL_2382563, EPI_ISL_2382564, EPI_ISL_2382565, EPI_ISL_2382566, EPI_ISL_2382567, EPI_ISL_2382569 | Wichita State University- Molecular Diagonstic lab | Kansas Health and Environmental Lab | Katherine Wiggins, Mike Grose, Jonathan Barnell, Ben Olsen, and Phil Adam |

| | | | |
|---|---|---|---|
| EPI_ISL_2382728, EPI_ISL_2382729, EPI_ISL_2382730, EPI_ISL_2382731, EPI_ISL_2382732 | University of Wisconsin-Madison AIDS Vaccine Research Laboratories | University of Wisconsin-Madison AIDS Vaccine Research Laboratories | Gage Moreno, Katarina Braun, et al. AIDS Vaccine Research Laboratories |
| EPI_ISL_2382734, EPI_ISL_2382735, EPI_ISL_2382736, EPI_ISL_2382737, EPI_ISL_2382738, EPI_ISL_2382739, EPI_ISL_2382740, EPI_ISL_2382741, EPI_ISL_2382742, EPI_ISL_2382743, EPI_ISL_2382744, EPI_ISL_2382745, EPI_ISL_2382746, EPI_ISL_2382747, EPI_ISL_2382748 | | | |
| see above | GA Department of Public Health | GA Department of Public Health | Stacy Reeves, Jonathan Edwards, Cynthia Dixey, Tonia Parrott, Aliyah Fields, Taylor Smith |
| EPI_ISL_2382749, EPI_ISL_2382750, EPI_ISL_2382751, EPI_ISL_2382752, EPI_ISL_2382753, EPI_ISL_2382754, EPI_ISL_2382755, EPI_ISL_2382756, EPI_ISL_2382757, EPI_ISL_2382758, EPI_ISL_2382759, EPI_ISL_2382760, EPI_ISL_2382761, EPI_ISL_2382762, EPI_ISL_2382763, EPI_ISL_2382764, EPI_ISL_2382765, EPI_ISL_2382766, EPI_ISL_2382767, EPI_ISL_2382768, EPI_ISL_2382769, EPI_ISL_2382770, EPI_ISL_2382771, EPI_ISL_2382772, EPI_ISL_2382773, EPI_ISL_2382774, EPI_ISL_2382775, EPI_ISL_2382776, EPI_ISL_2382777, EPI_ISL_2382778, EPI_ISL_2382779, EPI_ISL_2382780, EPI_ISL_2382781, EPI_ISL_2382782, EPI_ISL_2382783, EPI_ISL_2382784, EPI_ISL_2382785, EPI_ISL_2382786, EPI_ISL_2382787, EPI_ISL_2382788, EPI_ISL_2382789, EPI_ISL_2382790, EPI_ISL_2382791, EPI_ISL_2382792, EPI_ISL_2382793, EPI_ISL_2382794, EPI_ISL_2382795, EPI_ISL_2382796, EPI_ISL_2382797, EPI_ISL_2382798, EPI_ISL_2382799, EPI_ISL_2382800, EPI_ISL_2382801, EPI_ISL_2382802, EPI_ISL_2382803, EPI_ISL_2382804 | | | |
| see above | The Ohio State University Applied Microbiology Services Laboratory | The Ohio State University Applied Microbiology Services Laboratory | Seth A. Faith PhD |
| EPI_ISL_2383008, EPI_ISL_2383009, EPI_ISL_2383010, EPI_ISL_2383011, EPI_ISL_2383012, EPI_ISL_2383013, EPI_ISL_2383014, EPI_ISL_2383016, EPI_ISL_2383017, EPI_ISL_2383018 | Wichita State University- Molecular Diagonstic lab | Kansas Health and Environmental Lab | Katherine Wiggins, Mike Grose, Jonathan Barnell, Ben Olsen, and Phil Adam |
| EPI_ISL_2383019, EPI_ISL_2383020, EPI_ISL_2383021 | Kansas Health and Environmental Lab | Kansas Health and Environmental Lab | Katherine Wiggins, Mike Grose, Jonathan Barnell, Ben Olsen, and Phil Adam |
| EPI_ISL_2383022, EPI_ISL_2383023, EPI_ISL_2383024, EPI_ISL_2383025, EPI_ISL_2383027, EPI_ISL_2383029, EPI_ISL_2383030, EPI_ISL_2383031 | Wichita State University- Molecular Diagonstic lab | Kansas Health and Environmental Lab | Katherine Wiggins, Mike Grose, Jonathan Barnell, Ben Olsen, and Phil Adam |
| EPI_ISL_2383036, EPI_ISL_2383037, EPI_ISL_2383038, EPI_ISL_2383039, EPI_ISL_2383040, EPI_ISL_2383041, EPI_ISL_2383042, EPI_ISL_2383043, EPI_ISL_2383044 | unknown | Noblis | Lauren Brinkac, Shiela Diepold, Shane Mitchell, June Kang, Lee F. Kolakowski, William M. Nelson, Katharine Jennings |
| EPI_ISL_2383045, EPI_ISL_2383046, EPI_ISL_2383047, EPI_ISL_2383048, EPI_ISL_2383049, EPI_ISL_2383050, EPI_ISL_2383051, EPI_ISL_2383052, EPI_ISL_2383053, EPI_ISL_2383054, EPI_ISL_2383055 | | | |
| see above | GA Department of Public Health | GA Department of Public Health | Stacy Reeves, Jonathan Edwards, Cynthia Dixey, Tonia Parrott, Aliyah Fields, Taylor Smith |
| EPI_ISL_2383059, EPI_ISL_2383062, EPI_ISL_2383066, EPI_ISL_2383068, EPI_ISL_2383071, EPI_ISL_2383072, EPI_ISL_2383074, EPI_ISL_2383076 | Arizona State Public Health Laboratory | Arizona State Public Health Laboratory | Trung Huynh, Jessica Escobar, Katherine Fullerton, Nobuko Fukushima, Stacy White, Linda Getsinger, Victor Waddell |
| EPI_ISL_2383077, EPI_ISL_2383078, EPI_ISL_2383079, EPI_ISL_2383080, EPI_ISL_2383081, EPI_ISL_2383082, EPI_ISL_2383083, EPI_ISL_2383084, EPI_ISL_2383085, EPI_ISL_2383086, EPI_ISL_2383087, EPI_ISL_2383088, EPI_ISL_2383089, EPI_ISL_2383090, EPI_ISL_2383091, EPI_ISL_2383092, EPI_ISL_2383093, EPI_ISL_2383094, EPI_ISL_2383095, EPI_ISL_2383096, EPI_ISL_2383097, EPI_ISL_2383098, EPI_ISL_2383099, EPI_ISL_2383100, EPI_ISL_2383101, EPI_ISL_2383102, EPI_ISL_2383103, EPI_ISL_2383104, EPI_ISL_2383105, EPI_ISL_2383106, EPI_ISL_2383107, EPI_ISL_2383108, EPI_ISL_2383109, EPI_ISL_2383110, EPI_ISL_2383111, EPI_ISL_2383112, EPI_ISL_2383113, EPI_ISL_2383114, EPI_ISL_2383115, EPI_ISL_2383116, EPI_ISL_2383117, EPI_ISL_2383118, EPI_ISL_2383119, EPI_ISL_2383120, EPI_ISL_2383121, EPI_ISL_2383122, EPI_ISL_2383123, EPI_ISL_2383124, EPI_ISL_2383125 | | | |
| see above | Tampa General Hospital Esoteric Lab | Tampa General Hospital Esoteric Research & Development Lab | Grant Vestal, Deanna Becker, Dominic Uy, Vicki Healer, Amorce Lima, Suzane Silbert |
| EPI_ISL_2383126, EPI_ISL_2383127, EPI_ISL_2383128, EPI_ISL_2383129, EPI_ISL_2383130, EPI_ISL_2383131, EPI_ISL_2383132, EPI_ISL_2383133, EPI_ISL_2383134, EPI_ISL_2383136, EPI_ISL_2383137, EPI_ISL_2383138, EPI_ISL_2383139, EPI_ISL_2383140, EPI_ISL_2383141, EPI_ISL_2383142, EPI_ISL_2383143, EPI_ISL_2383144, EPI_ISL_2383145, EPI_ISL_2383146, EPI_ISL_2383148, EPI_ISL_2383149, EPI_ISL_2383150, EPI_ISL_2383151, EPI_ISL_2383152 | | | |
| see above | Wyoming Public Health Laboratory | Wyoming Public Health Laboratory | Jim Mildenberger, Wanda Manley, Noah Hull, Taylor Fearing, Lynette Gumbleton, Channing Weber, Ashley Norberg, Chayse Rowley, Marley Goetz, Brian Dominguez, Elliot Thomasson, Sam Britz, Cari Sloma, and Rob Christensen |
| EPI_ISL_2383154, EPI_ISL_2383155, EPI_ISL_2383159, EPI_ISL_2383160, EPI_ISL_2383161, EPI_ISL_2383162, EPI_ISL_2383164, EPI_ISL_2383166, EPI_ISL_2383167, EPI_ISL_2383169, EPI_ISL_2383171, EPI_ISL_2383174, EPI_ISL_2383176, EPI_ISL_2383177, EPI_ISL_2383179, EPI_ISL_2383180 | | | |
| see above | Willis-Knighton Medical Center Hospital Laboratory | LSUHS Emerging Viral Threat Laboratory | Lorie M. Atkins, April N. Johnson, Gregory L. Ware, Alexander Mijalis, Jeremy P. Kamil, Jennifer L. Carroll, Maarten Van Diest, Rona S. Scott, Andrew D. Yurochko, Christopher G. Kevil, John A. Vanchiere, Joseph A. Bocchini |
| EPI_ISL_2383181, EPI_ISL_2383183, EPI_ISL_2383184, EPI_ISL_2383188, EPI_ISL_2383189, EPI_ISL_2383190 | LSUHS Emerging Viral Threat Laboratory | LSUHS Emerging Viral Threat Laboratory | Gregory L. Ware, Alexander Mijalis, Jeremy P. Kamil, Jennifer L. Carroll, Maarten Van Diest, Rona S. Scott, Andrew D. Yurochko, Christopher G. Kevil, John A. Vanchiere |
| EPI_ISL_2383191, EPI_ISL_2383192, EPI_ISL_2383193, EPI_ISL_2383196, EPI_ISL_2383197, EPI_ISL_2383198, EPI_ISL_2383199, EPI_ISL_2383200, EPI_ISL_2383201, EPI_ISL_2383203, EPI_ISL_2383204 | | | |
| see above | Louisiana Office of Public Heath Laboratory | LSUHS Emerging Viral Threat Laboratory | Gregory L. Ware, Alexander Mijalis, Jeremy P. Kamil, Jennifer L. Carroll, Maarten Van Diest, Rona S. Scott, Andrew D. Yurochko, Christopher G. Kevil, John A. Vanchiere |
| EPI_ISL_2383207, EPI_ISL_2383208, EPI_ISL_2383209, EPI_ISL_2383210, EPI_ISL_2383211, EPI_ISL_2383212, EPI_ISL_2383213, EPI_ISL_2383215, EPI_ISL_2383216, EPI_ISL_2383217, EPI_ISL_2383218, EPI_ISL_2383219, EPI_ISL_2383220, EPI_ISL_2383221, EPI_ISL_2383222, EPI_ISL_2383223, EPI_ISL_2383226, EPI_ISL_2383227, EPI_ISL_2383228, EPI_ISL_2383229, EPI_ISL_2383230, EPI_ISL_2383231, EPI_ISL_2383232, EPI_ISL_2383233, EPI_ISL_2383234, EPI_ISL_2383235 | | | |
| see above | State Testing Facility | Altius Institute for Biomedical Research | Daniel Bates, Rebecca Bruders, Michael Buckley, Mark Frerker, Amanda Gale, Clem Green, Muhammad Halimun, Kneshay Harper, Matt Hartman, Alex Isner, Audra Johnson, Jessica Kunder, Lauren Mitchell, Jemma Nelson, Alex Nguyen, Sofia Olsson, Sadie Patraw, Tobias Ragoczy, Joshua Richards, Jean Robinston, Jacob Rodriguez, John Stamatoyannopoulos, Eric Thorland, Julia Wald |
| EPI_ISL_2383320, EPI_ISL_2383321, EPI_ISL_2383324, EPI_ISL_2383328, EPI_ISL_2383329, EPI_ISL_2383330, EPI_ISL_2383331, EPI_ISL_2383332, EPI_ISL_2383333, EPI_ISL_2383334, EPI_ISL_2383335, EPI_ISL_2383336, EPI_ISL_2383337, EPI_ISL_2383338, EPI_ISL_2383339, EPI_ISL_2383340, EPI_ISL_2383341, EPI_ISL_2383342, EPI_ISL_2383343, EPI_ISL_2383344, EPI_ISL_2383345, EPI_ISL_2383346, EPI_ISL_2383347, EPI_ISL_2383348, EPI_ISL_2383351, EPI_ISL_2383352, EPI_ISL_2383353, EPI_ISL_2383354, EPI_ISL_2383355, EPI_ISL_2383356, EPI_ISL_2383357, EPI_ISL_2383359, EPI_ISL_2383360, EPI_ISL_2383361, EPI_ISL_2383362, EPI_ISL_2383363, EPI_ISL_2383364, EPI_ISL_2383365, EPI_ISL_2383366, EPI_ISL_2383367, EPI_ISL_2383368, EPI_ISL_2383370, EPI_ISL_2383371, EPI_ISL_2383372, EPI_ISL_2383373, EPI_ISL_2383374, EPI_ISL_2383375, EPI_ISL_2383376, EPI_ISL_2383378, EPI_ISL_2383380, EPI_ISL_2383383, EPI_ISL_2383384, EPI_ISL_2383385, EPI_ISL_2383386, EPI_ISL_2383387, EPI_ISL_2383388, EPI_ISL_2383389, EPI_ISL_2383390, EPI_ISL_2383391, EPI_ISL_2383393, EPI_ISL_2383396, EPI_ISL_2383397, EPI_ISL_2383398, EPI_ISL_2383399, EPI_ISL_2383400, EPI_ISL_2383401, EPI_ISL_2383402, EPI_ISL_2383404, EPI_ISL_2383405, EPI_ISL_2383407, EPI_ISL_2383408, EPI_ISL_2383409, EPI_ISL_2383411, EPI_ISL_2383468, EPI_ISL_2383469, EPI_ISL_2383470, EPI_ISL_2383474, EPI_ISL_2383475, EPI_ISL_2383476, EPI_ISL_2383477, EPI_ISL_2383478, EPI_ISL_2383479, EPI_ISL_2383480, EPI_ISL_2383481, EPI_ISL_2383483, EPI_ISL_2383484, EPI_ISL_2383485, EPI_ISL_2383486, EPI_ISL_2383487, EPI_ISL_2383488, EPI_ISL_2383489, EPI_ISL_2383491, EPI_ISL_2383492, EPI_ISL_2383493, EPI_ISL_2383497, EPI_ISL_2383498, EPI_ISL_2383499, EPI_ISL_2383500, EPI_ISL_2383501, EPI_ISL_2383503, EPI_ISL_2383505, EPI_ISL_2383507, EPI_ISL_2383508, EPI_ISL_2383509, EPI_ISL_2383510, EPI_ISL_2383511, EPI_ISL_2383513, EPI_ISL_2383514, EPI_ISL_2383515, EPI_ISL_2383516, EPI_ISL_2383517, EPI_ISL_2383518, EPI_ISL_2383519, EPI_ISL_2383520, EPI_ISL_2383521, EPI_ISL_2383523, EPI_ISL_2383524, EPI_ISL_2383525, EPI_ISL_2383526, EPI_ISL_2383527, EPI_ISL_2383529, EPI_ISL_2383531, EPI_ISL_2383532, EPI_ISL_2383533, EPI_ISL_2383534, EPI_ISL_2383535, EPI_ISL_2383536, EPI_ISL_2383537, EPI_ISL_2383538, EPI_ISL_2383539, EPI_ISL_2383540, EPI_ISL_2383541 | | | |
| see above | US Air Force School of Aerospace Medicine | US Air Force School of Aerospace Medicine | Anthony Fries, Jennifer Meyer, William Gruner, Amanda Javorina, Carol Garrett, Sarah Purves, Clarise Starr, Elizabeth Macias |
| EPI_ISL_2383572, EPI_ISL_2383573, EPI_ISL_2383575, EPI_ISL_2383576, EPI_ISL_2383577, EPI_ISL_2383578, EPI_ISL_2383579, EPI_ISL_2383580, EPI_ISL_2383581, EPI_ISL_2383582, EPI_ISL_2383583, EPI_ISL_2383584, EPI_ISL_2383585, EPI_ISL_2383586, EPI_ISL_2383587, EPI_ISL_2383588, EPI_ISL_2383589, EPI_ISL_2383590, EPI_ISL_2383591, EPI_ISL_2383592, EPI_ISL_2383593, EPI_ISL_2383594, EPI_ISL_2383595, EPI_ISL_2383596, EPI_ISL_2383597, EPI_ISL_2383598, EPI_ISL_2383600, EPI_ISL_2383601, EPI_ISL_2383602, EPI_ISL_2383603, EPI_ISL_2383605, EPI_ISL_2383606, EPI_ISL_2383607, EPI_ISL_2383608, EPI_ISL_2383612, EPI_ISL_2383613, EPI_ISL_2383614, EPI_ISL_2383615, EPI_ISL_2383616, EPI_ISL_2383617, EPI_ISL_2383619, EPI_ISL_2383620, EPI_ISL_2383622, EPI_ISL_2383623, EPI_ISL_2383624, EPI_ISL_2383625, EPI_ISL_2383626, EPI_ISL_2383627, EPI_ISL_2383628, EPI_ISL_2383629, EPI_ISL_2383630, EPI_ISL_2383631, EPI_ISL_2383632, EPI_ISL_2383633, EPI_ISL_2383634, EPI_ISL_2383636, EPI_ISL_2383637, EPI_ISL_2383638, EPI_ISL_2383639, EPI_ISL_2383640, EPI_ISL_2383641, EPI_ISL_2383642, EPI_ISL_2383643, EPI_ISL_2383644, EPI_ISL_2383645, EPI_ISL_2383646, EPI_ISL_2383647, EPI_ISL_2383648, EPI_ISL_2383649, EPI_ISL_2383650, EPI_ISL_2383651, EPI_ISL_2383652, EPI_ISL_2383653, EPI_ISL_2383654, EPI_ISL_2383655, EPI_ISL_2383656, EPI_ISL_2383657, EPI_ISL_2383658, EPI_ISL_2383659, EPI_ISL_2383661, EPI_ISL_2383663, EPI_ISL_2383664, EPI_ISL_2383666, EPI_ISL_2383668, EPI_ISL_2383669, EPI_ISL_2383670, EPI_ISL_2383671, EPI_ISL_2383672, EPI_ISL_2383675, EPI_ISL_2383676, EPI_ISL_2383677, EPI_ISL_2383679, EPI_ISL_2383680, EPI_ISL_2383681, EPI_ISL_2383682, EPI_ISL_2383684, EPI_ISL_2383686, EPI_ISL_2383687, EPI_ISL_2383688, EPI_ISL_2383689, EPI_ISL_2383690, EPI_ISL_2383691, EPI_ISL_2383692, EPI_ISL_2383693, EPI_ISL_2383694, EPI_ISL_2383696, EPI_ISL_2383697, EPI_ISL_2383698, EPI_ISL_2383699, EPI_ISL_2383701, EPI_ISL_2383702, EPI_ISL_2383703, EPI_ISL_2383704, EPI_ISL_2383705, EPI_ISL_2383706, EPI_ISL_2383707, EPI_ISL_2383708, EPI_ISL_2383709, EPI_ISL_2383710, EPI_ISL_2383711, EPI_ISL_2383712, EPI_ISL_2383713, EPI_ISL_2383714, EPI_ISL_2383715, EPI_ISL_2383716, EPI_ISL_2383717, EPI_ISL_2383718, EPI_ISL_2383719, EPI_ISL_2383720, EPI_ISL_2383721, EPI_ISL_2383723, EPI_ISL_2383724, EPI_ISL_2383726, EPI_ISL_2383727, EPI_ISL_2383728, EPI_ISL_2383729, EPI_ISL_2383730, EPI_ISL_2383731, EPI_ISL_2383732, EPI_ISL_2383733, EPI_ISL_2383734, EPI_ISL_2383735, EPI_ISL_2383737, EPI_ISL_2383738, EPI_ISL_2383739, EPI_ISL_2383740, EPI_ISL_2383741, EPI_ISL_2383742, EPI_ISL_2383744, EPI_ISL_2383745, EPI_ISL_2383746, EPI_ISL_2383747, EPI_ISL_2383748, EPI_ISL_2383749, EPI_ISL_2383750, EPI_ISL_2383751, EPI_ISL_2383752, EPI_ISL_2383753, EPI_ISL_2383754, EPI_ISL_2383757, EPI_ISL_2383759, EPI_ISL_2383760, EPI_ISL_2383762, EPI_ISL_2383763, EPI_ISL_2383764, EPI_ISL_2383766, EPI_ISL_2383767, EPI_ISL_2383768, EPI_ISL_2383769, EPI_ISL_2383770, EPI_ISL_2383771, EPI_ISL_2383772, EPI_ISL_2383773, EPI_ISL_2383774, EPI_ISL_2383775, EPI_ISL_2383777, EPI_ISL_2383778, EPI_ISL_2383779, EPI_ISL_2383780, EPI_ISL_2383781, EPI_ISL_2383782, EPI_ISL_2383783, EPI_ISL_2383784, EPI_ISL_2383785, EPI_ISL_2383786, EPI_ISL_2383787, EPI_ISL_2383788, EPI_ISL_2383789, EPI_ISL_2383790, EPI_ISL_2383791, EPI_ISL_2383792, EPI_ISL_2383793, EPI_ISL_2383794, EPI_ISL_2383795, EPI_ISL_2383796, EPI_ISL_2383797, EPI_ISL_2383798, EPI_ISL_2383799, EPI_ISL_2383800, EPI_ISL_2383801, EPI_ISL_2383802, EPI_ISL_2383803, EPI_ISL_2383804, EPI_ISL_2383805, EPI_ISL_2383806, EPI_ISL_2383807, EPI_ISL_2383809, EPI_ISL_2383810, EPI_ISL_2383811, EPI_ISL_2383812, EPI_ISL_2383813, EPI_ISL_2383814, EPI_ISL_2383815, EPI_ISL_2383816, EPI_ISL_2383817, EPI_ISL_2383818, EPI_ISL_2383819, EPI_ISL_2383820, EPI_ISL_2383821, EPI_ISL_2383822, EPI_ISL_2383823, EPI_ISL_2383824, EPI_ISL_2383825, EPI_ISL_2383826, EPI_ISL_2383827, EPI_ISL_2383828, EPI_ISL_2383829, EPI_ISL_2383830, EPI_ISL_2383832, EPI_ISL_2383833, EPI_ISL_2383834, EPI_ISL_2383836, EPI_ISL_2383837, EPI_ISL_2383838, EPI_ISL_2383839, EPI_ISL_2383840, EPI_ISL_2383841, EPI_ISL_2383843, EPI_ISL_2383844, EPI_ISL_2383847, EPI_ISL_2383848, EPI_ISL_2383849, EPI_ISL_2383850, EPI_ISL_2383851, EPI_ISL_2383852, EPI_ISL_2383853, EPI_ISL_2383854, EPI_ISL_2383855, EPI_ISL_2383856, EPI_ISL_2383858, EPI_ISL_2383859, EPI_ISL_2383860, EPI_ISL_2383861, EPI_ISL_2383862, EPI_ISL_2383863, EPI_ISL_2383864, EPI_ISL_2383865, EPI_ISL_2383866, EPI_ISL_2383867, EPI_ISL_2383868, EPI_ISL_2383869, EPI_ISL_2383870, EPI_ISL_2383871, EPI_ISL_2383872, EPI_ISL_2383873, EPI_ISL_2383874, EPI_ISL_2383875, EPI_ISL_2383876, EPI_ISL_2383877, EPI_ISL_2383878, EPI_ISL_2383879, EPI_ISL_2383881, EPI_ISL_2383882, EPI_ISL_2383883, EPI_ISL_2383884, EPI_ISL_2383885, EPI_ISL_2383887, EPI_ISL_2383888, EPI_ISL_2383889, EPI_ISL_2383890, EPI_ISL_2383891, EPI_ISL_2383892, EPI_ISL_2383893 | | | |
| see above | DPHL | Delaware Public Health Lab | Rebecca Savage |

| | | | |
|---|---|---|---|
| EPI_ISL_2383894, EPI_ISL_2383895 | CT-Dr. Katherine A. Kelley State Public Health Lab | Centers for Disease Control and Prevention Division of Viral Diseases, Pathogen Discovery | Mili Sheth, Sarah Nobles, Jasmine Padilla, Mark Burroughs, Shoshona Le, Katie Dillon, Peter Cook, Clinton R. Paden, Dhwani Batra, Krista Queen, Kristen Knipe, Dakota Howard, Yvette Unoarumhi, Darlene Wagner, Matthew Schmerer, Ben L. Rambo-Martin, Kristine Lacek, Sam Shepard, Alison Laufer Halpin, Dave Wentworth, Vivien Dugan, Suxiang Tong, Justin Lee |
| EPI_ISL_2383896 | Commonwealth Healthcare Center | Centers for Disease Control and Prevention Division of Viral Diseases, Pathogen Discovery | Mili Sheth, Sarah Nobles, Jasmine Padilla, Mark Burroughs, Shoshona Le, Katie Dillon, Peter Cook, Clinton R. Paden, Dhwani Batra, Krista Queen, Kristen Knipe, Dakota Howard, Yvette Unoarumhi, Darlene Wagner, Matthew Schmerer, Ben L. Rambo-Martin, Kristine Lacek, Sam Shepard, Alison Laufer Halpin, Dave Wentworth, Vivien Dugan, Suxiang Tong, Justin Lee |
| EPI_ISL_2383897, EPI_ISL_2383898 | WVDHHR - Office of Laboratory Services | Centers for Disease Control and Prevention Division of Viral Diseases, Pathogen Discovery | Mili Sheth, Sarah Nobles, Jasmine Padilla, Mark Burroughs, Shoshona Le, Katie Dillon, Peter Cook, Clinton R. Paden, Dhwani Batra, Krista Queen, Kristen Knipe, Dakota Howard, Yvette Unoarumhi, Darlene Wagner, Matthew Schmerer, Ben L. Rambo-Martin, Kristine Lacek, Sam Shepard, Alison Laufer Halpin, Dave Wentworth, Vivien Dugan, Suxiang Tong, Justin Lee |
| EPI_ISL_2383899, EPI_ISL_2383900, EPI_ISL_2383901, EPI_ISL_2383902, EPI_ISL_2383903, EPI_ISL_2383904, EPI_ISL_2383905, EPI_ISL_2383906, EPI_ISL_2383907, EPI_ISL_2383908, EPI_ISL_2383909, EPI_ISL_2383910, EPI_ISL_2383911, EPI_ISL_2383912, EPI_ISL_2383913, EPI_ISL_2383914, EPI_ISL_2383915 | | | |
| see above | CO Dept. of Public Health and Environment, Lab Services Division | Centers for Disease Control and Prevention Division of Viral Diseases, Pathogen Discovery | Mili Sheth, Sarah Nobles, Jasmine Padilla, Mark Burroughs, Shoshona Le, Katie Dillon, Peter Cook, Clinton R. Paden, Dhwani Batra, Krista Queen, Kristen Knipe, Dakota Howard, Yvette Unoarumhi, Darlene Wagner, Matthew Schmerer, Ben L. Rambo-Martin, Kristine Lacek, Sam Shepard, Alison Laufer Halpin, Dave Wentworth, Vivien Dugan, Suxiang Tong, Justin Lee |
| EPI_ISL_2383916, EPI_ISL_2383917, EPI_ISL_2383918, EPI_ISL_2383919, EPI_ISL_2383920, EPI_ISL_2383921, EPI_ISL_2383922, EPI_ISL_2383923, EPI_ISL_2383924, EPI_ISL_2383925, EPI_ISL_2383926, EPI_ISL_2383927, EPI_ISL_2383928, EPI_ISL_2383929, EPI_ISL_2383930, EPI_ISL_2383931, EPI_ISL_2383932, EPI_ISL_2383933, EPI_ISL_2383934, EPI_ISL_2383935, EPI_ISL_2383936, EPI_ISL_2383937, EPI_ISL_2383938, EPI_ISL_2383939, EPI_ISL_2383940, EPI_ISL_2383941, EPI_ISL_2383942, EPI_ISL_2383943, EPI_ISL_2383944 | | | |
| see above | ME Health and Environmental Testing Laboratory | Centers for Disease Control and Prevention Division of Viral Diseases, Pathogen Discovery | Mili Sheth, Sarah Nobles, Jasmine Padilla, Mark Burroughs, Shoshona Le, Katie Dillon, Peter Cook, Clinton R. Paden, Dhwani Batra, Krista Queen, Kristen Knipe, Dakota Howard, Yvette Unoarumhi, Darlene Wagner, Matthew Schmerer, Ben L. Rambo-Martin, Kristine Lacek, Sam Shepard, Alison Laufer Halpin, Dave Wentworth, Vivien Dugan, Suxiang Tong, Justin Lee |
| EPI_ISL_2383945, EPI_ISL_2383946, EPI_ISL_2383947 | LA Office of Public Health Laboratories | Centers for Disease Control and Prevention Division of Viral Diseases, Pathogen Discovery | Mili Sheth, Sarah Nobles, Jasmine Padilla, Mark Burroughs, Shoshona Le, Katie Dillon, Peter Cook, Clinton R. Paden, Dhwani Batra, Krista Queen, Kristen Knipe, Dakota Howard, Yvette Unoarumhi, Darlene Wagner, Matthew Schmerer, Ben L. Rambo-Martin, Kristine Lacek, Sam Shepard, Alison Laufer Halpin, Dave Wentworth, Vivien Dugan, Suxiang Tong, Justin Lee |
| EPI_ISL_2383948, EPI_ISL_2383949, EPI_ISL_2383950, EPI_ISL_2383951, EPI_ISL_2383952, EPI_ISL_2383953, EPI_ISL_2383954, EPI_ISL_2383955 | SC Dept of Health and Env. Control-Bureau of Laboratories | Centers for Disease Control and Prevention Division of Viral Diseases, Pathogen Discovery | Mili Sheth, Sarah Nobles, Jasmine Padilla, Mark Burroughs, Shoshona Le, Katie Dillon, Peter Cook, Clinton R. Paden, Dhwani Batra, Krista Queen, Kristen Knipe, Dakota Howard, Yvette Unoarumhi, Darlene Wagner, Matthew Schmerer, Ben L. Rambo-Martin, Kristine Lacek, Sam Shepard, Alison Laufer Halpin, Dave Wentworth, Vivien Dugan, Suxiang Tong, Justin Lee |
| EPI_ISL_2383956, EPI_ISL_2383957, EPI_ISL_2383958, EPI_ISL_2383959, EPI_ISL_2383960 | VA-Division of Consolidated Laboratory Services | Centers for Disease Control and Prevention Division of Viral Diseases, Pathogen Discovery | Mili Sheth, Sarah Nobles, Jasmine Padilla, Mark Burroughs, Shoshona Le, Katie Dillon, Peter Cook, Clinton R. Paden, Dhwani Batra, Krista Queen, Kristen Knipe, Dakota Howard, Yvette Unoarumhi, Darlene Wagner, Matthew Schmerer, Ben L. Rambo-Martin, Kristine Lacek, Sam Shepard, Alison Laufer Halpin, Dave Wentworth, Vivien Dugan, Suxiang Tong, Justin Lee |
| EPI_ISL_2383961, EPI_ISL_2383962, EPI_ISL_2383963, EPI_ISL_2383964, EPI_ISL_2383965, EPI_ISL_2383966, EPI_ISL_2383967, EPI_ISL_2383968, EPI_ISL_2383969, EPI_ISL_2383970, EPI_ISL_2383971 | | | |
| see above | ID Bureau of Laboratories | Centers for Disease Control and Prevention Division of Viral Diseases, Pathogen Discovery | Mili Sheth, Sarah Nobles, Jasmine Padilla, Mark Burroughs, Shoshona Le, Katie Dillon, Peter Cook, Clinton R. Paden, Dhwani Batra, Krista Queen, Kristen Knipe, Dakota Howard, Yvette Unoarumhi, Darlene Wagner, Matthew Schmerer, Ben L. Rambo-Martin, Kristine Lacek, Sam Shepard, Alison Laufer Halpin, Dave Wentworth, Vivien Dugan, Suxiang Tong, Justin Lee |
| EPI_ISL_2383972, EPI_ISL_2383973, EPI_ISL_2383974, EPI_ISL_2383975, EPI_ISL_2383976, EPI_ISL_2383977, EPI_ISL_2383978, EPI_ISL_2383979, EPI_ISL_2383980, EPI_ISL_2383981 | KY State Public Health Lab | Centers for Disease Control and Prevention Division of Viral Diseases, Pathogen Discovery | Mili Sheth, Sarah Nobles, Jasmine Padilla, Mark Burroughs, Shoshona Le, Katie Dillon, Peter Cook, Clinton R. Paden, Dhwani Batra, Krista Queen, Kristen Knipe, Dakota Howard, Yvette Unoarumhi, Darlene Wagner, Matthew Schmerer, Ben L. Rambo-Martin, Kristine Lacek, Sam Shepard, Alison Laufer Halpin, Dave Wentworth, Vivien Dugan, Suxiang Tong, Justin Lee |
| EPI_ISL_2383982, EPI_ISL_2383983, EPI_ISL_2383984, EPI_ISL_2383985 | AL Dept. of Public Health Bureau of Clinical Laboratories | Centers for Disease Control and Prevention Division of Viral Diseases, Pathogen Discovery | Mili Sheth, Sarah Nobles, Jasmine Padilla, Mark Burroughs, Shoshona Le, Katie Dillon, Peter Cook, Clinton R. Paden, Dhwani Batra, Krista Queen, Kristen Knipe, Dakota Howard, Yvette Unoarumhi, Darlene Wagner, Matthew Schmerer, Ben L. Rambo-Martin, Kristine Lacek, Sam Shepard, Alison Laufer Halpin, Dave Wentworth, Vivien Dugan, Suxiang Tong, Justin Lee |
| EPI_ISL_2383986, EPI_ISL_2383987 | CA-Los Angeles County Public Health Laboratory | Centers for Disease Control and Prevention Division of Viral Diseases, Pathogen Discovery | Mili Sheth, Sarah Nobles, Jasmine Padilla, Mark Burroughs, Shoshona Le, Katie Dillon, Peter Cook, Clinton R. Paden, Dhwani Batra, Krista Queen, Kristen Knipe, Dakota Howard, Yvette Unoarumhi, Darlene Wagner, Matthew Schmerer, Ben L. Rambo-Martin, Kristine Lacek, Sam Shepard, Alison Laufer Halpin, Dave Wentworth, Vivien Dugan, Suxiang Tong, Justin Lee |
| EPI_ISL_2383988, EPI_ISL_2383989, EPI_ISL_2383990, EPI_ISL_2383991, EPI_ISL_2383992 | UT-Unified State Labs: Public Health Utah DOH | Centers for Disease Control and Prevention Division of Viral Diseases, Pathogen Discovery | Mili Sheth, Sarah Nobles, Jasmine Padilla, Mark Burroughs, Shoshona Le, Katie Dillon, Peter Cook, Clinton R. Paden, Dhwani Batra, Krista Queen, Kristen Knipe, Dakota Howard, Yvette Unoarumhi, Darlene Wagner, Matthew Schmerer, Ben L. Rambo-Martin, Kristine Lacek, Sam Shepard, Alison Laufer Halpin, Dave Wentworth, Vivien Dugan, Suxiang Tong, Justin Lee |
| EPI_ISL_2383993, EPI_ISL_2383994, EPI_ISL_2383995, EPI_ISL_2383996, EPI_ISL_2383997, EPI_ISL_2383998, EPI_ISL_2383999, EPI_ISL_2384000, EPI_ISL_2384001 | DC Public Health Lab/ Dept. of Forensic Sciences | Centers for Disease Control and Prevention Division of Viral Diseases, Pathogen Discovery | Mili Sheth, Sarah Nobles, Jasmine Padilla, Mark Burroughs, Shoshona Le, Katie Dillon, Peter Cook, Clinton R. Paden, Dhwani Batra, Krista Queen, Kristen Knipe, Dakota Howard, Yvette Unoarumhi, Darlene Wagner, Matthew Schmerer, Ben L. Rambo-Martin, Kristine Lacek, Sam Shepard, Alison Laufer Halpin, Dave Wentworth, Vivien Dugan, Suxiang Tong, Justin Lee |
| EPI_ISL_2384002, EPI_ISL_2384003, EPI_ISL_2384004, EPI_ISL_2384005, EPI_ISL_2384006, EPI_ISL_2384007, EPI_ISL_2384008 | NH Dept. of Health and Human Services Public Health Labs | Centers for Disease Control and Prevention Division of Viral Diseases, Pathogen Discovery | Mili Sheth, Sarah Nobles, Jasmine Padilla, Mark Burroughs, Shoshona Le, Katie Dillon, Peter Cook, Clinton R. Paden, Dhwani Batra, Krista Queen, Kristen Knipe, Dakota Howard, Yvette Unoarumhi, Darlene Wagner, Matthew Schmerer, Ben L. Rambo-Martin, Kristine Lacek, Sam Shepard, Alison Laufer Halpin, Dave Wentworth, Vivien Dugan, Suxiang Tong, Justin Lee |
| EPI_ISL_2384009, EPI_ISL_2384010, EPI_ISL_2384011, EPI_ISL_2384012, EPI_ISL_2384013, EPI_ISL_2384014, EPI_ISL_2384015 | MT Public Health Laboratory | Centers for Disease Control and Prevention Division of Viral Diseases, Pathogen Discovery | Mili Sheth, Sarah Nobles, Jasmine Padilla, Mark Burroughs, Shoshona Le, Katie Dillon, Peter Cook, Clinton R. Paden, Dhwani Batra, Krista Queen, Kristen Knipe, Dakota Howard, Yvette Unoarumhi, Darlene Wagner, Matthew Schmerer, Ben L. Rambo-Martin, Kristine Lacek, Sam Shepard, Alison Laufer Halpin, Dave Wentworth, Vivien Dugan, Suxiang Tong, Justin Lee |
| EPI_ISL_2384016, EPI_ISL_2384017, EPI_ISL_2384018, EPI_ISL_2384019, EPI_ISL_2384020, EPI_ISL_2384021, EPI_ISL_2384022, EPI_ISL_2384023, EPI_ISL_2384024, EPI_ISL_2384025 | PR Public Health Lab | Centers for Disease Control and Prevention Division of Viral Diseases, Pathogen Discovery | Mili Sheth, Sarah Nobles, Jasmine Padilla, Mark Burroughs, Shoshona Le, Katie Dillon, Peter Cook, Clinton R. Paden, Dhwani Batra, Krista Queen, Kristen Knipe, Dakota Howard, Yvette Unoarumhi, Darlene Wagner, Matthew Schmerer, Ben L. Rambo-Martin, Kristine Lacek, Sam Shepard, Alison Laufer Halpin, Dave Wentworth, Vivien Dugan, Suxiang Tong, Justin Lee |
| EPI_ISL_2384026, EPI_ISL_2384027, EPI_ISL_2384028 | OK Public Health Laboratory, Oklahoma State DOH | Centers for Disease Control and Prevention Division of Viral Diseases, Pathogen Discovery | Mili Sheth, Sarah Nobles, Jasmine Padilla, Mark Burroughs, Shoshona Le, Katie Dillon, Peter Cook, Clinton R. Paden, Dhwani Batra, Krista Queen, Kristen Knipe, Dakota Howard, Yvette Unoarumhi, Darlene Wagner, Matthew Schmerer, Ben L. Rambo-Martin, Kristine Lacek, Sam Shepard, Alison Laufer Halpin, Dave Wentworth, Vivien Dugan, Suxiang Tong, Justin Lee |
| EPI_ISL_2384029, EPI_ISL_2384030, EPI_ISL_2384031, EPI_ISL_2384032, | ND Dept. of Health Laboratory Services-Microbiology | Centers for Disease Control and Prevention Division of Viral Diseases, Pathogen Discovery | Mili Sheth, Sarah Nobles, Jasmine Padilla, Mark Burroughs, Shoshona Le, Katie Dillon, Peter Cook, Clinton R. Paden, Dhwani Batra, Krista Queen, Kristen Knipe, Dakota Howard, Yvette Unoarumhi, Darlene Wagner, Matthew Schmerer, Ben L. Rambo-Martin, Kristine Lacek, Sam Shepard, Alison Laufer Halpin, Dave Wentworth, Vivien Dugan, Suxiang Tong, Justin Lee |

| | | | |
|---|---|---|---|
| EPI_ISL_2384033, EPI_ISL_2384034, EPI_ISL_2384035, EPI_ISL_2384036 | | | |
| EPI_ISL_2384037, EPI_ISL_2384038, EPI_ISL_2384039, EPI_ISL_2384040, EPI_ISL_2384041, EPI_ISL_2384042, EPI_ISL_2384043, EPI_ISL_2384044, EPI_ISL_2384045, EPI_ISL_2384046 | NM Dept. Health, Scientific Laboratory Division | Centers for Disease Control and Prevention Division of Viral Diseases, Pathogen Discovery | Mili Sheth, Sarah Nobles, Jasmine Padilla, Mark Burroughs, Shoshona Le, Katie Dillon, Peter Cook, Clinton R. Paden, Dhwani Batra, Krista Queen, Kristen Knipe, Dakota Howard, Yvette Unoarumhi, Darlene Wagner, Matthew Schmerer, Ben L. Rambo-Martin, Kristine Lacek, Sam Shepard, Alison Laufer Halpin, Dave Wentworth, Vivien Dugan, Suxiang Tong, Justin Lee |
| EPI_ISL_2384047, EPI_ISL_2384048, EPI_ISL_2384049, EPI_ISL_2384050, EPI_ISL_2384051, EPI_ISL_2384052, EPI_ISL_2384053, EPI_ISL_2384054, EPI_ISL_2384055 | IA State Hygienic Laboratory | Centers for Disease Control and Prevention Division of Viral Diseases, Pathogen Discovery | Mili Sheth, Sarah Nobles, Jasmine Padilla, Mark Burroughs, Shoshona Le, Katie Dillon, Peter Cook, Clinton R. Paden, Dhwani Batra, Krista Queen, Kristen Knipe, Dakota Howard, Yvette Unoarumhi, Darlene Wagner, Matthew Schmerer, Ben L. Rambo-Martin, Kristine Lacek, Sam Shepard, Alison Laufer Halpin, Dave Wentworth, Vivien Dugan, Suxiang Tong, Justin Lee |
| EPI_ISL_2384056, EPI_ISL_2384057, EPI_ISL_2384058, EPI_ISL_2384059, EPI_ISL_2384060, EPI_ISL_2384061, EPI_ISL_2384062, EPI_ISL_2384063 | WY Public Health Laboratory | Centers for Disease Control and Prevention Division of Viral Diseases, Pathogen Discovery | Mili Sheth, Sarah Nobles, Jasmine Padilla, Mark Burroughs, Shoshona Le, Katie Dillon, Peter Cook, Clinton R. Paden, Dhwani Batra, Krista Queen, Kristen Knipe, Dakota Howard, Yvette Unoarumhi, Darlene Wagner, Matthew Schmerer, Ben L. Rambo-Martin, Kristine Lacek, Sam Shepard, Alison Laufer Halpin, Dave Wentworth, Vivien Dugan, Suxiang Tong, Justin Lee |
| EPI_ISL_2384064, EPI_ISL_2384065, EPI_ISL_2384066, EPI_ISL_2384067, EPI_ISL_2384068, EPI_ISL_2384069 | CT-Dr. Katherine A. Kelley State Public Health Lab | Centers for Disease Control and Prevention Division of Viral Diseases, Pathogen Discovery | Mili Sheth, Sarah Nobles, Jasmine Padilla, Mark Burroughs, Shoshona Le, Katie Dillon, Peter Cook, Clinton R. Paden, Dhwani Batra, Krista Queen, Kristen Knipe, Dakota Howard, Yvette Unoarumhi, Darlene Wagner, Matthew Schmerer, Ben L. Rambo-Martin, Kristine Lacek, Sam Shepard, Alison Laufer Halpin, Dave Wentworth, Vivien Dugan, Suxiang Tong, Justin Lee |
| EPI_ISL_2384070, EPI_ISL_2384071, EPI_ISL_2384072, EPI_ISL_2384073, EPI_ISL_2384074 | SD Public Health Laboratory | Centers for Disease Control and Prevention Division of Viral Diseases, Pathogen Discovery | Mili Sheth, Sarah Nobles, Jasmine Padilla, Mark Burroughs, Shoshona Le, Katie Dillon, Peter Cook, Clinton R. Paden, Dhwani Batra, Krista Queen, Kristen Knipe, Dakota Howard, Yvette Unoarumhi, Darlene Wagner, Matthew Schmerer, Ben L. Rambo-Martin, Kristine Lacek, Sam Shepard, Alison Laufer Halpin, Dave Wentworth, Vivien Dugan, Suxiang Tong, Justin Lee |
| EPI_ISL_2384075, EPI_ISL_2384076, EPI_ISL_2384077, EPI_ISL_2384078, EPI_ISL_2384079, EPI_ISL_2384080, EPI_ISL_2384081, EPI_ISL_2384082, EPI_ISL_2384083, EPI_ISL_2384084, EPI_ISL_2384085, EPI_ISL_2384086, EPI_ISL_2384087 | | | |
| see above | OH Department of Health Laboratory | Centers for Disease Control and Prevention Division of Viral Diseases, Pathogen Discovery | Mili Sheth, Sarah Nobles, Jasmine Padilla, Mark Burroughs, Shoshona Le, Katie Dillon, Peter Cook, Clinton R. Paden, Dhwani Batra, Krista Queen, Kristen Knipe, Dakota Howard, Yvette Unoarumhi, Darlene Wagner, Matthew Schmerer, Ben L. Rambo-Martin, Kristine Lacek, Sam Shepard, Alison Laufer Halpin, Dave Wentworth, Vivien Dugan, Suxiang Tong, Justin Lee |
| EPI_ISL_2384088, EPI_ISL_2384091, EPI_ISL_2384093, EPI_ISL_2384096, EPI_ISL_2384099, EPI_ISL_2384102, EPI_ISL_2384105, EPI_ISL_2384107, EPI_ISL_2384110 | HI Dept. of Health, State Laboratories Division | Centers for Disease Control and Prevention Division of Viral Diseases, Pathogen Discovery | Mili Sheth, Sarah Nobles, Jasmine Padilla, Mark Burroughs, Shoshona Le, Katie Dillon, Peter Cook, Clinton R. Paden, Dhwani Batra, Krista Queen, Kristen Knipe, Dakota Howard, Yvette Unoarumhi, Darlene Wagner, Matthew Schmerer, Ben L. Rambo-Martin, Kristine Lacek, Sam Shepard, Alison Laufer Halpin, Dave Wentworth, Vivien Dugan, Suxiang Tong, Justin Lee |
| EPI_ISL_2384113, EPI_ISL_2384115, EPI_ISL_2384118, EPI_ISL_2384121 | PA Department of Health, Bureau of Laboratories | Centers for Disease Control and Prevention Division of Viral Diseases, Pathogen Discovery | Mili Sheth, Sarah Nobles, Jasmine Padilla, Mark Burroughs, Shoshona Le, Katie Dillon, Peter Cook, Clinton R. Paden, Dhwani Batra, Krista Queen, Kristen Knipe, Dakota Howard, Yvette Unoarumhi, Darlene Wagner, Matthew Schmerer, Ben L. Rambo-Martin, Kristine Lacek, Sam Shepard, Alison Laufer Halpin, Dave Wentworth, Vivien Dugan, Suxiang Tong, Justin Lee |
| EPI_ISL_2384123, EPI_ISL_2384126, EPI_ISL_2384128, EPI_ISL_2384131, EPI_ISL_2384134, EPI_ISL_2384137 | RI State Health Laboratories | Centers for Disease Control and Prevention Division of Viral Diseases, Pathogen Discovery | Mili Sheth, Sarah Nobles, Jasmine Padilla, Mark Burroughs, Shoshona Le, Katie Dillon, Peter Cook, Clinton R. Paden, Dhwani Batra, Krista Queen, Kristen Knipe, Dakota Howard, Yvette Unoarumhi, Darlene Wagner, Matthew Schmerer, Ben L. Rambo-Martin, Kristine Lacek, Sam Shepard, Alison Laufer Halpin, Dave Wentworth, Vivien Dugan, Suxiang Tong, Justin Lee |
| EPI_ISL_2384139 | Alaska State Virology Lab, Alaska State Public Health Labs | Alaska State Virology Lab, Alaska State Public Health Labs | DeRonde,S., House,E., Zidek,J., Redlinger,M., Smith,L., Chen,J. |
| EPI_ISL_2384140, EPI_ISL_2384143 | RI State Health Laboratories | Centers for Disease Control and Prevention Division of Viral Diseases, Pathogen Discovery | Mili Sheth, Sarah Nobles, Jasmine Padilla, Mark Burroughs, Shoshona Le, Katie Dillon, Peter Cook, Clinton R. Paden, Dhwani Batra, Krista Queen, Kristen Knipe, Dakota Howard, Yvette Unoarumhi, Darlene Wagner, Matthew Schmerer, Ben L. Rambo-Martin, Kristine Lacek, Sam Shepard, Alison Laufer Halpin, Dave Wentworth, Vivien Dugan, Suxiang Tong, Justin Lee |
| EPI_ISL_2384145 | Alaska State Virology Lab, Alaska State Public Health Labs | Alaska State Virology Lab, Alaska State Public Health Labs | DeRonde,S., House,E., Zidek,J., Redlinger,M., Smith,L., Chen,J. |
| EPI_ISL_2384146, EPI_ISL_2384147, EPI_ISL_2384148, EPI_ISL_2384149, EPI_ISL_2384150, EPI_ISL_2384151, EPI_ISL_2384152, EPI_ISL_2384153, EPI_ISL_2384154, EPI_ISL_2384155, EPI_ISL_2384156, EPI_ISL_2384157, EPI_ISL_2384158 | | | |
| see above | RI State Health Laboratories | Centers for Disease Control and Prevention Division of Viral Diseases, Pathogen Discovery | Mili Sheth, Sarah Nobles, Jasmine Padilla, Mark Burroughs, Shoshona Le, Katie Dillon, Peter Cook, Clinton R. Paden, Dhwani Batra, Krista Queen, Kristen Knipe, Dakota Howard, Yvette Unoarumhi, Darlene Wagner, Matthew Schmerer, Ben L. Rambo-Martin, Kristine Lacek, Sam Shepard, Alison Laufer Halpin, Dave Wentworth, Vivien Dugan, Suxiang Tong, Justin Lee |
| EPI_ISL_2384159, EPI_ISL_2384160, EPI_ISL_2384161, EPI_ISL_2384162, EPI_ISL_2384163, EPI_ISL_2384164, EPI_ISL_2384165, EPI_ISL_2384166, EPI_ISL_2384167, EPI_ISL_2384168, EPI_ISL_2384170, EPI_ISL_2384171, EPI_ISL_2384172, EPI_ISL_2384173, EPI_ISL_2384174, EPI_ISL_2384175, EPI_ISL_2384176, EPI_ISL_2384177, EPI_ISL_2384178, EPI_ISL_2384179, EPI_ISL_2384180, EPI_ISL_2384181, EPI_ISL_2384182, EPI_ISL_2384183, EPI_ISL_2384184 | | | |
| see above | AR Dept. of Health-PHL, Molecular Diagnostics | Centers for Disease Control and Prevention Division of Viral Diseases, Pathogen Discovery | Mili Sheth, Sarah Nobles, Jasmine Padilla, Mark Burroughs, Shoshona Le, Katie Dillon, Peter Cook, Clinton R. Paden, Dhwani Batra, Krista Queen, Kristen Knipe, Dakota Howard, Yvette Unoarumhi, Darlene Wagner, Matthew Schmerer, Ben L. Rambo-Martin, Kristine Lacek, Sam Shepard, Alison Laufer Halpin, Dave Wentworth, Vivien Dugan, Suxiang Tong, Justin Lee |
| EPI_ISL_2384185, EPI_ISL_2384186, EPI_ISL_2384187, EPI_ISL_2384188, EPI_ISL_2384189, EPI_ISL_2384190, EPI_ISL_2384191, EPI_ISL_2384192, EPI_ISL_2384193, EPI_ISL_2384194, EPI_ISL_2384195, EPI_ISL_2384196, EPI_ISL_2384197, EPI_ISL_2384198, EPI_ISL_2384199, EPI_ISL_2384200 | | | |
| see above | MI - Michigan Department of Health and Human Services - Bureau of Laboratories | Centers for Disease Control and Prevention Division of Viral Diseases, Pathogen Discovery | Mili Sheth, Sarah Nobles, Jasmine Padilla, Mark Burroughs, Shoshona Le, Katie Dillon, Peter Cook, Clinton R. Paden, Dhwani Batra, Krista Queen, Kristen Knipe, Dakota Howard, Yvette Unoarumhi, Darlene Wagner, Matthew Schmerer, Ben L. Rambo-Martin, Kristine Lacek, Sam Shepard, Alison Laufer Halpin, Dave Wentworth, Vivien Dugan, Suxiang Tong, Justin Lee |
| EPI_ISL_2384201, EPI_ISL_2384202, EPI_ISL_2384203, EPI_ISL_2384204, EPI_ISL_2384205, EPI_ISL_2384206, EPI_ISL_2384207, EPI_ISL_2384208, EPI_ISL_2384209 | KS Health and Environmental Laboratories | Centers for Disease Control and Prevention Division of Viral Diseases, Pathogen Discovery | Mili Sheth, Sarah Nobles, Jasmine Padilla, Mark Burroughs, Shoshona Le, Katie Dillon, Peter Cook, Clinton R. Paden, Dhwani Batra, Krista Queen, Kristen Knipe, Dakota Howard, Yvette Unoarumhi, Darlene Wagner, Matthew Schmerer, Ben L. Rambo-Martin, Kristine Lacek, Sam Shepard, Alison Laufer Halpin, Dave Wentworth, Vivien Dugan, Suxiang Tong, Justin Lee |
| EPI_ISL_2384210, EPI_ISL_2384211, EPI_ISL_2384212, EPI_ISL_2384213, EPI_ISL_2384215, EPI_ISL_2384216, EPI_ISL_2384217, | OR State PHL-Virology/Immunology Section | Centers for Disease Control and Prevention Division of Viral Diseases, Pathogen Discovery | Mili Sheth, Sarah Nobles, Jasmine Padilla, Mark Burroughs, Shoshona Le, Katie Dillon, Peter Cook, Clinton R. Paden, Dhwani Batra, Krista Queen, Kristen Knipe, Dakota Howard, Yvette Unoarumhi, Darlene Wagner, Matthew Schmerer, Ben L. Rambo-Martin, Kristine Lacek, Sam Shepard, Alison Laufer Halpin, Dave Wentworth, Vivien Dugan, Suxiang Tong, Justin Lee |

| | | | |
|---|---|---|---|
| EPI_ISL_2384218, EPI_ISL_2384219, EPI_ISL_2384220 | | | |
| EPI_ISL_2384221, EPI_ISL_2384222, EPI_ISL_2384223 | NC State Laboratory of Public Health | Centers for Disease Control and Prevention Division of Viral Diseases, Pathogen Discovery | Mili Sheth, Sarah Nobles, Jasmine Padilla, Mark Burroughs, Shoshona Le, Katie Dillon, Peter Cook, Clinton R. Paden, Dhwani Batra, Krista Queen, Kristen Knipe, Dakota Howard, Yvette Unoarumhi, Darlene Wagner, Matthew Schmerer, Ben L. Rambo-Martin, Kristine Lacek, Sam Shepard, Alison Laufer Halpin, Dave Wentworth, Vivien Dugan, Suxiang Tong, Justin Lee |
| EPI_ISL_2384224, EPI_ISL_2384225, EPI_ISL_2384226, EPI_ISL_2384227, EPI_ISL_2384228, EPI_ISL_2384229, EPI_ISL_2384230, EPI_ISL_2384231, EPI_ISL_2384232 | VI-US Virgin Islands Department of Health | Centers for Disease Control and Prevention Division of Viral Diseases, Pathogen Discovery | Mili Sheth, Sarah Nobles, Jasmine Padilla, Mark Burroughs, Shoshona Le, Katie Dillon, Peter Cook, Clinton R. Paden, Dhwani Batra, Krista Queen, Kristen Knipe, Dakota Howard, Yvette Unoarumhi, Darlene Wagner, Matthew Schmerer, Ben L. Rambo-Martin, Kristine Lacek, Sam Shepard, Alison Laufer Halpin, Dave Wentworth, Vivien Dugan, Suxiang Tong, Justin Lee |
| EPI_ISL_2384233, EPI_ISL_2384235, EPI_ISL_2384238, EPI_ISL_2384239, EPI_ISL_2384240, EPI_ISL_2384242, EPI_ISL_2384243, EPI_ISL_2384244, EPI_ISL_2384245, EPI_ISL_2384246, EPI_ISL_2384250, EPI_ISL_2384251, EPI_ISL_2384253, EPI_ISL_2384254, EPI_ISL_2384255, EPI_ISL_2384256, EPI_ISL_2384257, EPI_ISL_2384258, EPI_ISL_2384259, EPI_ISL_2384260, EPI_ISL_2384261, EPI_ISL_2384262, EPI_ISL_2384263, EPI_ISL_2384264, EPI_ISL_2384265, EPI_ISL_2384266, EPI_ISL_2384267, EPI_ISL_2384270, EPI_ISL_2384271, EPI_ISL_2384273, EPI_ISL_2384275, EPI_ISL_2384277, EPI_ISL_2384278, EPI_ISL_2384279, EPI_ISL_2384280, EPI_ISL_2384282, EPI_ISL_2384283, EPI_ISL_2384284, EPI_ISL_2384290, EPI_ISL_2384291, EPI_ISL_2384293, EPI_ISL_2384294, EPI_ISL_2384295, EPI_ISL_2384299, EPI_ISL_2384302, EPI_ISL_2384304, EPI_ISL_2384306, EPI_ISL_2384307, EPI_ISL_2384311, EPI_ISL_2384312, EPI_ISL_2384314, EPI_ISL_2384317, EPI_ISL_2384318, EPI_ISL_2384320, EPI_ISL_2384321, EPI_ISL_2384322, EPI_ISL_2384324, EPI_ISL_2384325, EPI_ISL_2384328, EPI_ISL_2384329, EPI_ISL_2384330, EPI_ISL_2384333, EPI_ISL_2384334, EPI_ISL_2384335, EPI_ISL_2384336, EPI_ISL_2384338, EPI_ISL_2384339, EPI_ISL_2384341, EPI_ISL_2384342, EPI_ISL_2384343, EPI_ISL_2384344, EPI_ISL_2384345, EPI_ISL_2384349, EPI_ISL_2384351, EPI_ISL_2384352, EPI_ISL_2384356, EPI_ISL_2384357, EPI_ISL_2384359, EPI_ISL_2384360, EPI_ISL_2384361, EPI_ISL_2384362, EPI_ISL_2384363, EPI_ISL_2384364, EPI_ISL_2384365, EPI_ISL_2384366, EPI_ISL_2384368 | | | |
| see above | North Dakota Department of Health, Public Health Laboratory | North Dakota Department of Health, Public Health Laboratory | Lisa Wingerter |
| EPI_ISL_2388777 | URMC LABS | URMC LABS | Dwight Hardy, Andrew Cameron, Stepan Voljehn, Nicole Pecora |
| EPI_ISL_2391507 | Centro de Investigación Biomédica del Noreste (CIBIN) | Instituto de Biotecnología de la UNAM | Consorcio Mexicano de Vigilancia Genómica (CoViGen-Mex). Authors (in alphabetical order): Julio Elias Alvarado-Yaah, Carlos F. Arias, Santiago Ávila-Ríos, Eduardo Becerril-Vargas, Víctor Hugo Borja-Aburto, Celia Boukadida, Cristóbal Cháidez-Quiróz, Juan Bautista Chale-Dzul, Ricardo Ciria Merce, Andreu Comas-García, Célida Duque Molina, Julissa Enciso-Ibarra, José Antonio Enciso-Moreno, Gloria Elena Espinosa-Ayala, Fernando Fontove-Herrera, Daniel Fregoso-Rueda, Victor Eduardo Garcia-Arias, Alejandra Garcia-Gasca, Bruno Gomez-Gil, Jean Pierre González, Irvin González-López, Concepción Grajales-Muñiz, Ricardo Grande, Rosa María Gutierrez Rios, Alejandra Hernández-Terán, Alfredo Herrera-Estrella, Carla Ivón Herrera-Najera, Pavel Isa, Daniel Lira Morales, Susana Lopez, Antonio Loza Román, Brenda Irasema Maldonado-Meza, Bernardo Martínez-Miguel, José Arturo Martínez-Orozco, Célida Martínez- Rodríguez, Margarita Matías-Florentino, Fidencio Mejía-Nepomuceno, María Guadalupe de Jesús Mireles-Rivera, Gloria María Molina-Salinas, Hector Montoya-Fuentes, , Mario Mújica-Sánchez, José Esteban Muñoz-Medina, José de Jesús Nuñez-Contreras, Alicia Ocaña-Mondragón, Luis Alberto Ochoa-Carrera, Hector Esteban Paz-Juárez, Francisco Pulido, Helen Haydee Fernanda Ramírez-Plascencia, Jorge Salas-Hernández, Angel Gustavo Salas-Lais, Alejandro Sanchez-Flores, Clara Esperanza Santacruz-Tinoco, María Guadalupe Santiago-Mauricio, Selene Zárate, Nelly Sélem-Mojica, Blanca Taboada, Gloria Vazquez, Joel Armando Vázquez-Pérez. |
| EPI_ISL_2391508, EPI_ISL_2391509, EPI_ISL_2391510, EPI_ISL_2391511 | Laboratorio Central de Epidemiología (LCE) | Instituto de Biotecnología de la UNAM | Consorcio Mexicano de Vigilancia Genómica (CoViGen-Mex). Authors (in alphabetical order): Julio Elias Alvarado-Yaah, Carlos F. Arias, Santiago Ávila-Ríos, Eduardo Becerril-Vargas, Víctor Hugo Borja-Aburto, Celia Boukadida, Cristóbal Cháidez-Quiróz, Juan Bautista Chale-Dzul, Ricardo Ciria Merce, Andreu Comas-García, Célida Duque Molina, Julissa Enciso-Ibarra, José Antonio Enciso-Moreno, Gloria Elena Espinosa-Ayala, Fernando Fontove-Herrera, Daniel Fregoso-Rueda, Victor Eduardo Garcia-Arias, Alejandra Garcia-Gasca, Bruno Gomez-Gil, Jean Pierre González, Irvin González-López, Concepción Grajales-Muñiz, Ricardo Grande, Rosa María Gutierrez Rios, Alejandra Hernández-Terán, Alfredo Herrera-Estrella, Carla Ivón Herrera-Najera, Pavel Isa, Daniel Lira Morales, Susana Lopez, Antonio Loza Román, Brenda Irasema Maldonado-Meza, Bernardo Martínez-Miguel, José Arturo Martínez-Orozco, Célida Martínez- Rodríguez, Margarita Matías-Florentino, Fidencio Mejía-Nepomuceno, María Guadalupe de Jesús Mireles-Rivera, Gloria María Molina-Salinas, Hector Montoya-Fuentes, , Mario Mújica-Sánchez, José Esteban Muñoz-Medina, José de Jesús Nuñez-Contreras, Alicia Ocaña-Mondragón, Luis Alberto Ochoa-Carrera, Hector Esteban Paz-Juárez, Francisco Pulido, Helen Haydee Fernanda Ramírez-Plascencia, Jorge Salas-Hernández, Angel Gustavo Salas-Lais, Alejandro Sanchez-Flores, Clara Esperanza Santacruz-Tinoco, María Guadalupe Santiago-Mauricio, Selene Zárate, Nelly Sélem-Mojica, Blanca Taboada, Gloria Vazquez, Joel Armando Vázquez-Pérez. |
| EPI_ISL_2391512 | Unidad de Investigación Médica de Yucatán (UIMY) | Instituto de Biotecnología de la UNAM | Consorcio Mexicano de Vigilancia Genómica (CoViGen-Mex). Authors (in alphabetical order): Julio Elias Alvarado-Yaah, Carlos F. Arias, Santiago Ávila-Ríos, Eduardo Becerril-Vargas, Víctor Hugo Borja-Aburto, Celia Boukadida, Cristóbal Cháidez-Quiróz, Juan Bautista Chale-Dzul, Ricardo Ciria Merce, Andreu Comas-García, Célida Duque Molina, Julissa Enciso-Ibarra, José Antonio Enciso-Moreno, Gloria Elena Espinosa-Ayala, Fernando Fontove-Herrera, Daniel Fregoso-Rueda, Victor Eduardo Garcia-Arias, Alejandra Garcia-Gasca, Bruno Gomez-Gil, Jean Pierre González, Irvin González-López, Concepción Grajales-Muñiz, Ricardo Grande, Rosa María Gutierrez Rios, Alejandra Hernández-Terán, Alfredo Herrera-Estrella, Carla Ivón Herrera-Najera, Pavel Isa, Daniel Lira Morales, Susana Lopez, Antonio Loza Román, Brenda Irasema Maldonado-Meza, Bernardo Martínez-Miguel, José Arturo Martínez-Orozco, Célida Martínez- Rodríguez, Margarita Matías-Florentino, Fidencio Mejía-Nepomuceno, María Guadalupe de Jesús Mireles-Rivera, Gloria María Molina-Salinas, Hector Montoya-Fuentes, , Mario Mújica-Sánchez, José Esteban Muñoz-Medina, José de Jesús Nuñez-Contreras, Alicia Ocaña-Mondragón, Luis Alberto Ochoa-Carrera, Hector Esteban Paz-Juárez, Francisco Pulido, Helen Haydee Fernanda Ramírez-Plascencia, Jorge Salas-Hernández, Angel Gustavo Salas-Lais, Alejandro Sanchez-Flores, Clara Esperanza Santacruz-Tinoco, María Guadalupe Santiago-Mauricio, Selene Zárate, Nelly Sélem-Mojica, Blanca Taboada, Gloria Vazquez, Joel Armando Vázquez-Pérez. |
| EPI_ISL_2391513, EPI_ISL_2391514, EPI_ISL_2391515, EPI_ISL_2391516, EPI_ISL_2391517, EPI_ISL_2391518, EPI_ISL_2391519, EPI_ISL_2391520, EPI_ISL_2391521, EPI_ISL_2391522 | Laboratorio Central de Epidemiología (LCE) | Instituto de Biotecnología de la UNAM | Consorcio Mexicano de Vigilancia Genómica (CoViGen-Mex). Authors (in alphabetical order): Julio Elias Alvarado-Yaah, Carlos F. Arias, Santiago Ávila-Ríos, Eduardo Becerril-Vargas, Víctor Hugo Borja-Aburto, Celia Boukadida, Cristóbal Cháidez-Quiróz, Juan Bautista Chale-Dzul, Ricardo Ciria Merce, Andreu Comas-García, Célida Duque Molina, Julissa Enciso-Ibarra, José Antonio Enciso-Moreno, Gloria Elena Espinosa-Ayala, Fernando Fontove-Herrera, Daniel Fregoso-Rueda, Victor Eduardo Garcia-Arias, Alejandra Garcia-Gasca, Bruno Gomez-Gil, Jean Pierre González, Irvin González-López, Concepción Grajales-Muñiz, Ricardo Grande, Rosa María Gutierrez Rios, Alejandra Hernández-Terán, Alfredo Herrera-Estrella, Carla Ivón Herrera-Najera, Pavel Isa, Daniel Lira Morales, Susana Lopez, Antonio Loza Román, Brenda Irasema Maldonado-Meza, Bernardo Martínez-Miguel, José Arturo Martínez-Orozco, Célida Martínez- Rodríguez, Margarita Matías-Florentino, Fidencio Mejía-Nepomuceno, María Guadalupe de Jesús Mireles-Rivera, Gloria María Molina-Salinas, Hector Montoya-Fuentes, , Mario Mújica-Sánchez, José Esteban Muñoz-Medina, José de Jesús Nuñez-Contreras, Alicia Ocaña-Mondragón, Luis Alberto Ochoa-Carrera, Hector Esteban Paz-Juárez, Francisco Pulido, Helen Haydee Fernanda Ramírez-Plascencia, Jorge Salas-Hernández, Angel Gustavo Salas-Lais, Alejandro Sanchez-Flores, Clara Esperanza Santacruz-Tinoco, María Guadalupe Santiago-Mauricio, Selene Zárate, Nelly Sélem-Mojica, Blanca Taboada, Gloria Vazquez, Joel Armando Vázquez-Pérez. |
| EPI_ISL_2391523 | Centro de Investigación Biomédica del Noreste (CIBIN) | Instituto de Biotecnología de la UNAM | Consorcio Mexicano de Vigilancia Genómica (CoViGen-Mex). Authors (in alphabetical order): Julio Elias Alvarado-Yaah, Carlos F. Arias, Santiago Ávila-Ríos, Eduardo Becerril-Vargas, Víctor Hugo Borja-Aburto, Celia Boukadida, Cristóbal Cháidez-Quiróz, Juan Bautista Chale-Dzul, Ricardo Ciria Merce, Andreu Comas-García, Célida Duque Molina, Julissa Enciso-Ibarra, José Antonio Enciso-Moreno, Gloria Elena Espinosa-Ayala, Fernando Fontove-Herrera, Daniel Fregoso-Rueda, Victor Eduardo Garcia-Arias, Alejandra Garcia-Gasca, Bruno Gomez-Gil, Jean Pierre González, Irvin González-López, Concepción Grajales-Muñiz, Ricardo Grande, Rosa María Gutierrez Rios, Alejandra Hernández-Terán, Alfredo Herrera-Estrella, Carla Ivón Herrera-Najera, Pavel Isa, Daniel Lira Morales, Susana Lopez, Antonio Loza Román, Brenda Irasema Maldonado-Meza, Bernardo Martínez-Miguel, José Arturo Martínez-Orozco, Célida Martínez- Rodríguez, Margarita Matías-Florentino, Fidencio Mejía-Nepomuceno, María Guadalupe de Jesús Mireles-Rivera, Gloria María Molina-Salinas, Hector Montoya-Fuentes, , Mario Mújica-Sánchez, José Esteban Muñoz-Medina, José de Jesús Nuñez-Contreras, Alicia Ocaña-Mondragón, Luis Alberto Ochoa-Carrera, Hector Esteban Paz-Juárez, Francisco Pulido, Helen Haydee Fernanda Ramírez-Plascencia, Jorge Salas-Hernández, Angel Gustavo Salas-Lais, Alejandro Sanchez-Flores, Clara Esperanza Santacruz-Tinoco, María Guadalupe Santiago-Mauricio, Selene Zárate, Nelly Sélem-Mojica, Blanca Taboada, Gloria Vazquez, Joel Armando Vázquez-Pérez. |
| EPI_ISL_2391524, EPI_ISL_2391525, EPI_ISL_2391526, EPI_ISL_2391527, EPI_ISL_2391528, EPI_ISL_2391529, EPI_ISL_2391530, EPI_ISL_2391531, EPI_ISL_2391532 | Laboratorio Central de Epidemiología (LCE) | Instituto de Biotecnología de la UNAM | Consorcio Mexicano de Vigilancia Genómica (CoViGen-Mex). Authors (in alphabetical order): Julio Elias Alvarado-Yaah, Carlos F. Arias, Santiago Ávila-Ríos, Eduardo Becerril-Vargas, Víctor Hugo Borja-Aburto, Celia Boukadida, Cristóbal Cháidez-Quiróz, Juan Bautista Chale-Dzul, Ricardo Ciria Merce, Andreu Comas-García, Célida Duque Molina, Julissa Enciso-Ibarra, José Antonio Enciso-Moreno, Gloria Elena Espinosa-Ayala, Fernando Fontove-Herrera, Daniel Fregoso-Rueda, Victor Eduardo Garcia-Arias, Alejandra Garcia-Gasca, Bruno Gomez-Gil, Jean Pierre González, Irvin González-López, Concepción Grajales-Muñiz, Ricardo Grande, Rosa María Gutierrez Rios, Alejandra Hernández-Terán, Alfredo Herrera-Estrella, Carla Ivón Herrera-Najera, Pavel Isa, Daniel Lira Morales, Susana Lopez, Antonio Loza Román, Brenda Irasema Maldonado-Meza, Bernardo Martínez-Miguel, José Arturo Martínez-Orozco, Célida Martínez- Rodríguez, Margarita Matías-Florentino, Fidencio Mejía-Nepomuceno, María Guadalupe de Jesús Mireles-Rivera, Gloria María Molina-Salinas, Hector Montoya-Fuentes, , Mario Mújica-Sánchez, José Esteban Muñoz-Medina, José de Jesús Nuñez-Contreras, Alicia Ocaña-Mondragón, Luis Alberto Ochoa-Carrera, Hector Esteban Paz-Juárez, Francisco Pulido, Helen Haydee Fernanda Ramírez-Plascencia, Jorge Salas-Hernández, Angel Gustavo Salas-Lais, Alejandro Sanchez-Flores, Clara Esperanza Santacruz-Tinoco, María Guadalupe Santiago-Mauricio, Selene Zárate, Nelly Sélem-Mojica, Blanca Taboada, Gloria Vazquez, Joel Armando Vázquez-Pérez. |
| EPI_ISL_2391533 | Centro de Investigación Biomédica del Noreste (CIBIN) | Instituto de Biotecnología de la UNAM | Consorcio Mexicano de Vigilancia Genómica (CoViGen-Mex). Authors (in alphabetical order): Julio Elias Alvarado-Yaah, Carlos F. Arias, Santiago Ávila-Ríos, Eduardo Becerril-Vargas, Víctor Hugo Borja-Aburto, Celia Boukadida, Cristóbal Cháidez-Quiróz, Juan Bautista Chale-Dzul, Ricardo Ciria Merce, Andreu Comas-García, Célida Duque Molina, Julissa Enciso-Ibarra, José Antonio Enciso-Moreno, Gloria Elena Espinosa-Ayala, Fernando Fontove-Herrera, Daniel Fregoso-Rueda, Victor Eduardo Garcia-Arias, Alejandra Garcia-Gasca, Bruno Gomez-Gil, Jean Pierre González, Irvin González-López, Concepción Grajales-Muñiz, Ricardo Grande, Rosa María Gutierrez Rios, Alejandra Hernández-Terán, Alfredo Herrera-Estrella, Carla Ivón Herrera-Najera, Pavel Isa, Daniel Lira Morales, Susana Lopez, Antonio Loza Román, Brenda Irasema Maldonado-Meza, Bernardo Martínez-Miguel, José Arturo Martínez-Orozco, Célida Martínez- Rodríguez, Margarita Matías-Florentino, Fidencio Mejía-Nepomuceno, María Guadalupe de Jesús Mireles-Rivera, Gloria María Molina-Salinas, Hector Montoya-Fuentes, , Mario Mújica-Sánchez, José Esteban Muñoz-Medina, José de Jesús Nuñez-Contreras, Alicia Ocaña-Mondragón, Luis Alberto Ochoa-Carrera, Hector Esteban Paz-Juárez, Francisco Pulido, Helen Haydee Fernanda Ramírez-Plascencia, Jorge Salas-Hernández, Angel Gustavo Salas-Lais, Alejandro Sanchez-Flores, Clara Esperanza Santacruz-Tinoco, María Guadalupe Santiago-Mauricio, Selene Zárate, Nelly Sélem-Mojica, Blanca Taboada, Gloria Vazquez, Joel Armando Vázquez-Pérez. |
| EPI_ISL_2391534, EPI_ISL_2391535 | Laboratorio Central de Epidemiología (LCE) | Instituto de Biotecnología de la UNAM | Consorcio Mexicano de Vigilancia Genómica (CoViGen-Mex). Authors (in alphabetical order): Julio Elias Alvarado-Yaah, Carlos F. Arias, Santiago Ávila-Ríos, Eduardo Becerril-Vargas, Víctor Hugo Borja-Aburto, Celia Boukadida, Cristóbal Cháidez-Quiróz, Juan Bautista Chale-Dzul, Ricardo Ciria Merce, Andreu Comas-García, Célida Duque Molina, Julissa Enciso-Ibarra, José Antonio Enciso-Moreno, Gloria Elena Espinosa-Ayala, Fernando Fontove-Herrera, Daniel Fregoso-Rueda, Victor Eduardo Garcia-Arias, Alejandra Garcia-Gasca, Bruno Gomez-Gil, Jean Pierre González, Irvin González-López, Concepción Grajales-Muñiz, Ricardo Grande, Rosa María Gutierrez Rios, Alejandra Hernández-Terán, Alfredo Herrera-Estrella, Carla Ivón Herrera-Najera, Pavel Isa, Daniel Lira Morales, Susana Lopez, Antonio Loza Román, Brenda Irasema Maldonado-Meza, Bernardo Martínez-Miguel, José Arturo Martínez-Orozco, Célida Martínez- Rodríguez, Margarita Matías-Florentino, Fidencio Mejía-Nepomuceno, María Guadalupe de Jesús Mireles-Rivera, Gloria María Molina-Salinas, Hector Montoya-Fuentes, , Mario Mújica-Sánchez, José Esteban Muñoz-Medina, José de Jesús Nuñez-Contreras, Alicia Ocaña-Mondragón, Luis Alberto Ochoa-Carrera, Hector Esteban Paz-Juárez, Francisco Pulido, Helen Haydee Fernanda Ramírez-Plascencia, Jorge Salas-Hernández, Angel Gustavo Salas-Lais, Alejandro Sanchez-Flores, Clara Esperanza Santacruz-Tinoco, María Guadalupe Santiago-Mauricio, Selene Zárate, Nelly Sélem-Mojica, Blanca Taboada, Gloria Vazquez, Joel Armando Vázquez-Pérez. |
| EPI_ISL_2391536 | Centro de Investigación Biomédica de Occidente (CIBO) | Instituto de Biotecnología de la UNAM | Consorcio Mexicano de Vigilancia Genómica (CoViGen-Mex). Authors (in alphabetical order): Julio Elias Alvarado-Yaah, Carlos F. Arias, Santiago Ávila-Ríos, Eduardo Becerril-Vargas, Víctor Hugo Borja-Aburto, Celia Boukadida, Cristóbal Cháidez-Quiróz, Juan Bautista Chale-Dzul, Ricardo Ciria Merce, Andreu Comas-García, Célida Duque Molina, Julissa Enciso-Ibarra, José Antonio Enciso-Moreno, Gloria Elena Espinosa-Ayala, Fernando Fontove-Herrera, Daniel Fregoso-Rueda, Victor Eduardo Garcia-Arias, Alejandra Garcia-Gasca, Bruno Gomez-Gil, Jean Pierre González, Irvin González-López, Concepción Grajales-Muñiz, Ricardo Grande, Rosa María Gutierrez Rios, Alejandra Hernández-Terán, Alfredo Herrera-Estrella, Carla Ivón Herrera-Najera, Pavel Isa, Daniel Lira Morales, Susana Lopez, Antonio Loza Román, Brenda Irasema Maldonado-Meza, Bernardo Martínez-Miguel, José Arturo Martínez-Orozco, Célida Martínez- Rodríguez, Margarita Matías-Florentino, Fidencio Mejía-Nepomuceno, María Guadalupe de Jesús Mireles-Rivera, Gloria María Molina-Salinas, Hector Montoya-Fuentes, , Mario Mújica-Sánchez, José Esteban Muñoz-Medina, José de Jesús Nuñez-Contreras, Alicia Ocaña-Mondragón, Luis Alberto Ochoa-Carrera, Hector Esteban Paz-Juárez, Francisco Pulido, Helen Haydee Fernanda Ramírez-Plascencia, Jorge Salas-Hernández, Angel Gustavo Salas-Lais, Alejandro Sanchez-Flores, Clara Esperanza Santacruz-Tinoco, María Guadalupe Santiago-Mauricio, Selene Zárate, Nelly Sélem-Mojica, Blanca Taboada, Gloria Vazquez, Joel Armando Vázquez-Pérez. |
| EPI_ISL_2391537 | Unidad de Investigación Médica de Yucatán (UIMY) | Instituto de Biotecnología de la UNAM | Consorcio Mexicano de Vigilancia Genómica (CoViGen-Mex). Authors (in alphabetical order): Julio Elias Alvarado-Yaah, Carlos F. Arias, Santiago Ávila-Ríos, Eduardo Becerril-Vargas, Víctor Hugo Borja-Aburto, Celia Boukadida, Cristóbal Cháidez-Quiróz, Juan Bautista Chale-Dzul, Ricardo Ciria Merce, Andreu Comas-García, Célida Duque Molina, Julissa Enciso-Ibarra, José Antonio Enciso-Moreno, Gloria Elena Espinosa-Ayala, Fernando Fontove-Herrera, Daniel Fregoso-Rueda, Victor Eduardo Garcia-Arias, Alejandra Garcia-Gasca, Bruno Gomez-Gil, Jean Pierre González, Irvin González-López, Concepción Grajales-Muñiz, Ricardo Grande, Rosa María Gutierrez Rios, Alejandra Hernández-Terán, Alfredo Herrera-Estrella, Carla Ivón Herrera-Najera, Pavel Isa, Daniel Lira Morales, Susana Lopez, Antonio Loza Román, Brenda Irasema Maldonado-Meza, Bernardo Martínez-Miguel, José Arturo Martínez-Orozco, Célida Martínez- Rodríguez, Margarita Matías-Florentino, Fidencio Mejía-Nepomuceno, María Guadalupe de Jesús Mireles-Rivera, Gloria María Molina-Salinas, Hector Montoya-Fuentes, , Mario Mújica-Sánchez, José Esteban Muñoz-Medina, José de Jesús Nuñez-Contreras, Alicia Ocaña-Mondragón, Luis Alberto Ochoa-Carrera, Hector Esteban Paz-Juárez, Francisco Pulido, Helen Haydee Fernanda Ramírez-Plascencia, Jorge Salas-Hernández, Angel Gustavo Salas-Lais, Alejandro Sanchez-Flores, Clara Esperanza Santacruz-Tinoco, María Guadalupe Santiago-Mauricio, Selene Zárate, Nelly Sélem-Mojica, Blanca Taboada, Gloria Vazquez, Joel Armando Vázquez-Pérez. |
| EPI_ISL_2391538, EPI_ISL_2391539 | Laboratorio Central de Epidemiología (LCE) | Instituto de Biotecnología de la UNAM | Consorcio Mexicano de Vigilancia Genómica (CoViGen-Mex). Authors (in alphabetical order): Julio Elias Alvarado-Yaah, Carlos F. Arias, Santiago Ávila-Ríos, Eduardo Becerril-Vargas, Víctor Hugo Borja-Aburto, Celia Boukadida, Cristóbal Cháidez-Quiróz, Juan Bautista Chale-Dzul, Ricardo Ciria Merce, Andreu Comas-García, Célida Duque Molina, Julissa Enciso-Ibarra, José Antonio Enciso-Moreno, Gloria Elena Espinosa-Ayala, Fernando Fontove-Herrera, Daniel Fregoso-Rueda, Victor Eduardo Garcia-Arias, Alejandra Garcia-Gasca, Bruno Gomez-Gil, Jean Pierre González, Irvin González-López, Concepción Grajales-Muñiz, Ricardo Grande, Rosa María Gutierrez Rios, Alejandra Hernández-Terán, Alfredo Herrera-Estrella, Carla Ivón Herrera-Najera, Pavel Isa, Daniel Lira Morales, Susana Lopez, Antonio Loza Román, Brenda Irasema Maldonado-Meza, Bernardo Martínez-Miguel, José Arturo Martínez-Orozco, Célida Martínez- Rodríguez, Margarita Matías-Florentino, Fidencio Mejía-Nepomuceno, María Guadalupe de Jesús Mireles-Rivera, Gloria María Molina-Salinas, Hector Montoya-Fuentes, , Mario Mújica-Sánchez, José Esteban Muñoz-Medina, José de Jesús Nuñez-Contreras, Alicia Ocaña-Mondragón, Luis Alberto Ochoa-Carrera, Hector Esteban Paz-Juárez, Francisco Pulido, Helen Haydee Fernanda Ramírez-Plascencia, Jorge Salas-Hernández, Angel Gustavo Salas-Lais, Alejandro Sanchez-Flores, Clara Esperanza Santacruz-Tinoco, María Guadalupe Santiago-Mauricio, Selene Zárate, Nelly Sélem-Mojica, Blanca Taboada, Gloria Vazquez, Joel Armando Vázquez-Pérez. |
| EPI_ISL_2391540 | Unidad de Investigación Biomédica de Zacatecas (UIBZ) | Instituto de Biotecnología de la UNAM | Consorcio Mexicano de Vigilancia Genómica (CoViGen-Mex). Authors (in alphabetical order): Julio Elias Alvarado-Yaah, Carlos F. Arias, Santiago Ávila-Ríos, Eduardo Becerril-Vargas, Víctor Hugo Borja-Aburto, Celia Boukadida, Cristóbal Cháidez-Quiróz, Juan Bautista Chale-Dzul, Ricardo Ciria Merce, Andreu Comas-García, Célida Duque Molina, Julissa Enciso-Ibarra, José Antonio Enciso-Moreno, Gloria Elena Espinosa-Ayala, Fernando Fontove-Herrera, Daniel Fregoso-Rueda, Victor Eduardo Garcia-Arias, Alejandra Garcia-Gasca, Bruno Gomez-Gil, Jean Pierre González, Irvin González-López, Concepción Grajales-Muñiz, Ricardo Grande, Rosa María Gutierrez Rios, Alejandra Hernández-Terán, Alfredo Herrera-Estrella, Carla Ivón Herrera-Najera, Pavel Isa, Daniel Lira Morales, Susana Lopez, Antonio Loza Román, Brenda Irasema Maldonado-Meza, Bernardo Martínez-Miguel, José Arturo Martínez-Orozco, Célida Martínez- Rodríguez, Margarita Matías-Florentino, Fidencio Mejía-Nepomuceno, María Guadalupe de Jesús Mireles-Rivera, Gloria María Molina-Salinas, Hector Montoya-Fuentes, , Mario Mújica-Sánchez, José Esteban Muñoz-Medina, José de Jesús Nuñez-Contreras, Alicia Ocaña-Mondragón, Luis Alberto Ochoa-Carrera, Hector Esteban Paz-Juárez, Francisco Pulido, Helen Haydee Fernanda Ramírez-Plascencia, Jorge Salas-Hernández, Angel Gustavo Salas-Lais, Alejandro Sanchez-Flores, Clara Esperanza Santacruz-Tinoco, María Guadalupe Santiago-Mauricio, Selene Zárate, Nelly Sélem-Mojica, Blanca Taboada, Gloria Vazquez, Joel Armando Vázquez-Pérez. |
| EPI_ISL_2391541, | Laboratorio Central de Epidemiología | Instituto de Biotecnología de la UNAM | Consorcio Mexicano de Vigilancia Genómica (CoViGen-Mex). Authors (in alphabetical order): Julio Elias Alvarado-Yaah, Carlos F. Arias, Santiago Ávila-Ríos, Eduardo Becerril-Vargas, Víctor Hugo Borja-Aburto, Celia Boukadida, Cristóbal Cháidez-Quiróz, Juan Bautista Chale-Dzul, Ricardo |

| | | | |
|---|---|---|---|
| EPI_ISL_2391542, EPI_ISL_2391543 | (LCE) | | Ciria Merce, Andreu Comas-García, Célida Duque Molina, Julissa Enciso-Ibarra, José Antonio Enciso-Moreno, Gloria Elena Espinosa-Ayala, Fernando Fontove-Herrera, Daniel Fregoso-Rueda, Victor Eduardo Garcia-Arias, Alejandra Garcia-Gasca, Bruno Gomez-Gil, Jean Pierre González, Irvin González-López, Concepción Grajales-Muñiz, Ricardo Grande, Rosa María Gutierrez Rios, Alejandra Hernández-Terán, Alfredo Herrera-Estrella, Carla Ivón Herrera-Najera, Pavel Isa, Daniel Lira Morales, Susana Lopez, Antonio Loza Román, Brenda Irasema Maldonado-Meza, Bernardo Martínez-Miguel, José Arturo Martínez-Orozco, Célida Martínez- Rodríguez, Margarita Matías-Florentino, Fidencio Mejía-Nepomuceno, María Guadalupe de Jesús Mireles-Rivera, Gloria María Molina-Salinas, Hector Montoya-Fuentes, , Mario Mújica-Sánchez, José Esteban Muñoz-Medina, José de Jesús Nuñez-Contreras, Alicia Ocaña-Mondragón, Luis Alberto Ochoa-Carrera, Hector Esteban Paz-Juárez, Francisco Pulido, Helen Haydee Fernanda Ramírez-Plascencia, Jorge Salas-Hernández, Angel Gustavo Salas-Lais, Alejandro Sanchez-Flores, Clara Esperanza Santacruz-Tinoco, María Guadalupe Santiago-Mauricio, Selene Zárate, Nelly Sélem-Mojica, Blanca Taboada, Gloria Vazquez, Joel Armando Vázquez-Pérez. |
| EPI_ISL_2391544 | Centro de Investigación Biomédica de Occidente (CIBO) | Instituto de Biotecnología de la UNAM | Consorcio Mexicano de Vigilancia Genómica (CoViGen-Mex). Authors (in alphabetical order): Julio Elias Alvarado-Yaah, Carlos F. Arias, Santiago Ávila-Ríos, Eduardo Becerril-Vargas, Víctor Hugo Borja-Aburto, Celia Boukadida, Cristóbal Cháidez-Quiróz, Juan Bautista Chale-Dzul, Ricardo Ciria Merce, Andreu Comas-García, Célida Duque Molina, Julissa Enciso-Ibarra, José Antonio Enciso-Moreno, Gloria Elena Espinosa-Ayala, Fernando Fontove-Herrera, Daniel Fregoso-Rueda, Victor Eduardo Garcia-Arias, Alejandra Garcia-Gasca, Bruno Gomez-Gil, Jean Pierre González, Irvin González-López, Concepción Grajales-Muñiz, Ricardo Grande, Rosa María Gutierrez Rios, Alejandra Hernández-Terán, Alfredo Herrera-Estrella, Carla Ivón Herrera-Najera, Pavel Isa, Daniel Lira Morales, Susana Lopez, Antonio Loza Román, Brenda Irasema Maldonado-Meza, Bernardo Martínez-Miguel, José Arturo Martínez-Orozco, Célida Martínez- Rodríguez, Margarita Matías-Florentino, Fidencio Mejía-Nepomuceno, María Guadalupe de Jesús Mireles-Rivera, Gloria María Molina-Salinas, Hector Montoya-Fuentes, , Mario Mújica-Sánchez, José Esteban Muñoz-Medina, José de Jesús Nuñez-Contreras, Alicia Ocaña-Mondragón, Luis Alberto Ochoa-Carrera, Hector Esteban Paz-Juárez, Francisco Pulido, Helen Haydee Fernanda Ramírez-Plascencia, Jorge Salas-Hernández, Angel Gustavo Salas-Lais, Alejandro Sanchez-Flores, Clara Esperanza Santacruz-Tinoco, María Guadalupe Santiago-Mauricio, Selene Zárate, Nelly Sélem-Mojica, Blanca Taboada, Gloria Vazquez, Joel Armando Vázquez-Pérez. |
| EPI_ISL_2391545, EPI_ISL_2391546, EPI_ISL_2391547, EPI_ISL_2391548, EPI_ISL_2391549, EPI_ISL_2391550, EPI_ISL_2391551, EPI_ISL_2391552, EPI_ISL_2391553, EPI_ISL_2391554, EPI_ISL_2391555, EPI_ISL_2391556, EPI_ISL_2391557, EPI_ISL_2391558, EPI_ISL_2391559, EPI_ISL_2391560, EPI_ISL_2391561, EPI_ISL_2391562, EPI_ISL_2391563, EPI_ISL_2391564 | | | |
| see above | Laboratorio Central de Epidemiología (LCE) | Instituto de Biotecnología de la UNAM | Consorcio Mexicano de Vigilancia Genómica (CoViGen-Mex). Authors (in alphabetical order): Julio Elias Alvarado-Yaah, Carlos F. Arias, Santiago Ávila-Ríos, Eduardo Becerril-Vargas, Víctor Hugo Borja-Aburto, Celia Boukadida, Cristóbal Cháidez-Quiróz, Juan Bautista Chale-Dzul, Ricardo Ciria Merce, Andreu Comas-García, Célida Duque Molina, Julissa Enciso-Ibarra, José Antonio Enciso-Moreno, Gloria Elena Espinosa-Ayala, Fernando Fontove-Herrera, Daniel Fregoso-Rueda, Victor Eduardo Garcia-Arias, Alejandra Garcia-Gasca, Bruno Gomez-Gil, Jean Pierre González, Irvin González-López, Concepción Grajales-Muñiz, Ricardo Grande, Rosa María Gutierrez Rios, Alejandra Hernández-Terán, Alfredo Herrera-Estrella, Carla Ivón Herrera-Najera, Pavel Isa, Daniel Lira Morales, Susana Lopez, Antonio Loza Román, Brenda Irasema Maldonado-Meza, Bernardo Martínez-Miguel, José Arturo Martínez-Orozco, Célida Martínez- Rodríguez, Margarita Matías-Florentino, Fidencio Mejía-Nepomuceno, María Guadalupe de Jesús Mireles-Rivera, Gloria María Molina-Salinas, Hector Montoya-Fuentes, , Mario Mújica-Sánchez, José Esteban Muñoz-Medina, José de Jesús Nuñez-Contreras, Alicia Ocaña-Mondragón, Luis Alberto Ochoa-Carrera, Hector Esteban Paz-Juárez, Francisco Pulido, Helen Haydee Fernanda Ramírez-Plascencia, Jorge Salas-Hernández, Angel Gustavo Salas-Lais, Alejandro Sanchez-Flores, Clara Esperanza Santacruz-Tinoco, María Guadalupe Santiago-Mauricio, Selene Zárate, Nelly Sélem-Mojica, Blanca Taboada, Gloria Vazquez, Joel Armando Vázquez-Pérez. |
| EPI_ISL_2391565 | Centro de Investigación Biomédica de Occidente (CIBO) | Instituto de Biotecnología de la UNAM | Consorcio Mexicano de Vigilancia Genómica (CoViGen-Mex). Authors (in alphabetical order): Julio Elias Alvarado-Yaah, Carlos F. Arias, Santiago Ávila-Ríos, Eduardo Becerril-Vargas, Víctor Hugo Borja-Aburto, Celia Boukadida, Cristóbal Cháidez-Quiróz, Juan Bautista Chale-Dzul, Ricardo Ciria Merce, Andreu Comas-García, Célida Duque Molina, Julissa Enciso-Ibarra, José Antonio Enciso-Moreno, Gloria Elena Espinosa-Ayala, Fernando Fontove-Herrera, Daniel Fregoso-Rueda, Victor Eduardo Garcia-Arias, Alejandra Garcia-Gasca, Bruno Gomez-Gil, Jean Pierre González, Irvin González-López, Concepción Grajales-Muñiz, Ricardo Grande, Rosa María Gutierrez Rios, Alejandra Hernández-Terán, Alfredo Herrera-Estrella, Carla Ivón Herrera-Najera, Pavel Isa, Daniel Lira Morales, Susana Lopez, Antonio Loza Román, Brenda Irasema Maldonado-Meza, Bernardo Martínez-Miguel, José Arturo Martínez-Orozco, Célida Martínez- Rodríguez, Margarita Matías-Florentino, Fidencio Mejía-Nepomuceno, María Guadalupe de Jesús Mireles-Rivera, Gloria María Molina-Salinas, Hector Montoya-Fuentes, , Mario Mújica-Sánchez, José Esteban Muñoz-Medina, José de Jesús Nuñez-Contreras, Alicia Ocaña-Mondragón, Luis Alberto Ochoa-Carrera, Hector Esteban Paz-Juárez, Francisco Pulido, Helen Haydee Fernanda Ramírez-Plascencia, Jorge Salas-Hernández, Angel Gustavo Salas-Lais, Alejandro Sanchez-Flores, Clara Esperanza Santacruz-Tinoco, María Guadalupe Santiago-Mauricio, Selene Zárate, Nelly Sélem-Mojica, Blanca Taboada, Gloria Vazquez, Joel Armando Vázquez-Pérez. |
| EPI_ISL_2391566, EPI_ISL_2391567 | Laboratorio Central de Epidemiología (LCE) | Instituto de Biotecnología de la UNAM | Consorcio Mexicano de Vigilancia Genómica (CoViGen-Mex). Authors (in alphabetical order): Julio Elias Alvarado-Yaah, Carlos F. Arias, Santiago Ávila-Ríos, Eduardo Becerril-Vargas, Víctor Hugo Borja-Aburto, Celia Boukadida, Cristóbal Cháidez-Quiróz, Juan Bautista Chale-Dzul, Ricardo Ciria Merce, Andreu Comas-García, Célida Duque Molina, Julissa Enciso-Ibarra, José Antonio Enciso-Moreno, Gloria Elena Espinosa-Ayala, Fernando Fontove-Herrera, Daniel Fregoso-Rueda, Victor Eduardo Garcia-Arias, Alejandra Garcia-Gasca, Bruno Gomez-Gil, Jean Pierre González, Irvin González-López, Concepción Grajales-Muñiz, Ricardo Grande, Rosa María Gutierrez Rios, Alejandra Hernández-Terán, Alfredo Herrera-Estrella, Carla Ivón Herrera-Najera, Pavel Isa, Daniel Lira Morales, Susana Lopez, Antonio Loza Román, Brenda Irasema Maldonado-Meza, Bernardo Martínez-Miguel, José Arturo Martínez-Orozco, Célida Martínez- Rodríguez, Margarita Matías-Florentino, Fidencio Mejía-Nepomuceno, María Guadalupe de Jesús Mireles-Rivera, Gloria María Molina-Salinas, Hector Montoya-Fuentes, , Mario Mújica-Sánchez, José Esteban Muñoz-Medina, José de Jesús Nuñez-Contreras, Alicia Ocaña-Mondragón, Luis Alberto Ochoa-Carrera, Hector Esteban Paz-Juárez, Francisco Pulido, Helen Haydee Fernanda Ramírez-Plascencia, Jorge Salas-Hernández, Angel Gustavo Salas-Lais, Alejandro Sanchez-Flores, Clara Esperanza Santacruz-Tinoco, María Guadalupe Santiago-Mauricio, Selene Zárate, Nelly Sélem-Mojica, Blanca Taboada, Gloria Vazquez, Joel Armando Vázquez-Pérez. |
| EPI_ISL_2391568 | Centro de Investigación Biomédica de Occidente (CIBO) | Instituto de Biotecnología de la UNAM | Consorcio Mexicano de Vigilancia Genómica (CoViGen-Mex). Authors (in alphabetical order): Julio Elias Alvarado-Yaah, Carlos F. Arias, Santiago Ávila-Ríos, Eduardo Becerril-Vargas, Víctor Hugo Borja-Aburto, Celia Boukadida, Cristóbal Cháidez-Quiróz, Juan Bautista Chale-Dzul, Ricardo Ciria Merce, Andreu Comas-García, Célida Duque Molina, Julissa Enciso-Ibarra, José Antonio Enciso-Moreno, Gloria Elena Espinosa-Ayala, Fernando Fontove-Herrera, Daniel Fregoso-Rueda, Victor Eduardo Garcia-Arias, Alejandra Garcia-Gasca, Bruno Gomez-Gil, Jean Pierre González, Irvin González-López, Concepción Grajales-Muñiz, Ricardo Grande, Rosa María Gutierrez Rios, Alejandra Hernández-Terán, Alfredo Herrera-Estrella, Carla Ivón Herrera-Najera, Pavel Isa, Daniel Lira Morales, Susana Lopez, Antonio Loza Román, Brenda Irasema Maldonado-Meza, Bernardo Martínez-Miguel, José Arturo Martínez-Orozco, Célida Martínez- Rodríguez, Margarita Matías-Florentino, Fidencio Mejía-Nepomuceno, María Guadalupe de Jesús Mireles-Rivera, Gloria María Molina-Salinas, Hector Montoya-Fuentes, , Mario Mújica-Sánchez, José Esteban Muñoz-Medina, José de Jesús Nuñez-Contreras, Alicia Ocaña-Mondragón, Luis Alberto Ochoa-Carrera, Hector Esteban Paz-Juárez, Francisco Pulido, Helen Haydee Fernanda Ramírez-Plascencia, Jorge Salas-Hernández, Angel Gustavo Salas-Lais, Alejandro Sanchez-Flores, Clara Esperanza Santacruz-Tinoco, María Guadalupe Santiago-Mauricio, Selene Zárate, Nelly Sélem-Mojica, Blanca Taboada, Gloria Vazquez, Joel Armando Vázquez-Pérez. |
| EPI_ISL_2391569, EPI_ISL_2391570, EPI_ISL_2391571, EPI_ISL_2391572, EPI_ISL_2391573, EPI_ISL_2391574 | Laboratorio Central de Epidemiología (LCE) | Instituto de Biotecnología de la UNAM | Consorcio Mexicano de Vigilancia Genómica (CoViGen-Mex). Authors (in alphabetical order): Julio Elias Alvarado-Yaah, Carlos F. Arias, Santiago Ávila-Ríos, Eduardo Becerril-Vargas, Víctor Hugo Borja-Aburto, Celia Boukadida, Cristóbal Cháidez-Quiróz, Juan Bautista Chale-Dzul, Ricardo Ciria Merce, Andreu Comas-García, Célida Duque Molina, Julissa Enciso-Ibarra, José Antonio Enciso-Moreno, Gloria Elena Espinosa-Ayala, Fernando Fontove-Herrera, Daniel Fregoso-Rueda, Victor Eduardo Garcia-Arias, Alejandra Garcia-Gasca, Bruno Gomez-Gil, Jean Pierre González, Irvin González-López, Concepción Grajales-Muñiz, Ricardo Grande, Rosa María Gutierrez Rios, Alejandra Hernández-Terán, Alfredo Herrera-Estrella, Carla Ivón Herrera-Najera, Pavel Isa, Daniel Lira Morales, Susana Lopez, Antonio Loza Román, Brenda Irasema Maldonado-Meza, Bernardo Martínez-Miguel, José Arturo Martínez-Orozco, Célida Martínez- Rodríguez, Margarita Matías-Florentino, Fidencio Mejía-Nepomuceno, María Guadalupe de Jesús Mireles-Rivera, Gloria María Molina-Salinas, Hector Montoya-Fuentes, , Mario Mújica-Sánchez, José Esteban Muñoz-Medina, José de Jesús Nuñez-Contreras, Alicia Ocaña-Mondragón, Luis Alberto Ochoa-Carrera, Hector Esteban Paz-Juárez, Francisco Pulido, Helen Haydee Fernanda Ramírez-Plascencia, Jorge Salas-Hernández, Angel Gustavo Salas-Lais, Alejandro Sanchez-Flores, Clara Esperanza Santacruz-Tinoco, María Guadalupe Santiago-Mauricio, Selene Zárate, Nelly Sélem-Mojica, Blanca Taboada, Gloria Vazquez, Joel Armando Vázquez-Pérez. |
| EPI_ISL_2391575 | Centro de Investigación Biomédica del Noreste (CIBIN) | Instituto de Biotecnología de la UNAM | Consorcio Mexicano de Vigilancia Genómica (CoViGen-Mex). Authors (in alphabetical order): Julio Elias Alvarado-Yaah, Carlos F. Arias, Santiago Ávila-Ríos, Eduardo Becerril-Vargas, Víctor Hugo Borja-Aburto, Celia Boukadida, Cristóbal Cháidez-Quiróz, Juan Bautista Chale-Dzul, Ricardo Ciria Merce, Andreu Comas-García, Célida Duque Molina, Julissa Enciso-Ibarra, José Antonio Enciso-Moreno, Gloria Elena Espinosa-Ayala, Fernando Fontove-Herrera, Daniel Fregoso-Rueda, Victor Eduardo Garcia-Arias, Alejandra Garcia-Gasca, Bruno Gomez-Gil, Jean Pierre González, Irvin González-López, Concepción Grajales-Muñiz, Ricardo Grande, Rosa María Gutierrez Rios, Alejandra Hernández-Terán, Alfredo Herrera-Estrella, Carla Ivón Herrera-Najera, Pavel Isa, Daniel Lira Morales, Susana Lopez, Antonio Loza Román, Brenda Irasema Maldonado-Meza, Bernardo Martínez-Miguel, José Arturo Martínez-Orozco, Célida Martínez- Rodríguez, Margarita Matías-Florentino, Fidencio Mejía-Nepomuceno, María Guadalupe de Jesús Mireles-Rivera, Gloria María Molina-Salinas, Hector Montoya-Fuentes, , Mario Mújica-Sánchez, José Esteban Muñoz-Medina, José de Jesús Nuñez-Contreras, Alicia Ocaña-Mondragón, Luis Alberto Ochoa-Carrera, Hector Esteban Paz-Juárez, Francisco Pulido, Helen Haydee Fernanda Ramírez-Plascencia, Jorge Salas-Hernández, Angel Gustavo Salas-Lais, Alejandro Sanchez-Flores, Clara Esperanza Santacruz-Tinoco, María Guadalupe Santiago-Mauricio, Selene Zárate, Nelly Sélem-Mojica, Blanca Taboada, Gloria Vazquez, Joel Armando Vázquez-Pérez. |
| EPI_ISL_2391576, EPI_ISL_2391577 | Laboratorio Central de Epidemiología (LCE) | Instituto de Biotecnología de la UNAM | Consorcio Mexicano de Vigilancia Genómica (CoViGen-Mex). Authors (in alphabetical order): Julio Elias Alvarado-Yaah, Carlos F. Arias, Santiago Ávila-Ríos, Eduardo Becerril-Vargas, Víctor Hugo Borja-Aburto, Celia Boukadida, Cristóbal Cháidez-Quiróz, Juan Bautista Chale-Dzul, Ricardo Ciria Merce, Andreu Comas-García, Célida Duque Molina, Julissa Enciso-Ibarra, José Antonio Enciso-Moreno, Gloria Elena Espinosa-Ayala, Fernando Fontove-Herrera, Daniel Fregoso-Rueda, Victor Eduardo Garcia-Arias, Alejandra Garcia-Gasca, Bruno Gomez-Gil, Jean Pierre González, Irvin González-López, Concepción Grajales-Muñiz, Ricardo Grande, Rosa María Gutierrez Rios, Alejandra Hernández-Terán, Alfredo Herrera-Estrella, Carla Ivón Herrera-Najera, Pavel Isa, Daniel Lira Morales, Susana Lopez, Antonio Loza Román, Brenda Irasema Maldonado-Meza, Bernardo Martínez-Miguel, José Arturo Martínez-Orozco, Célida Martínez- Rodríguez, Margarita Matías-Florentino, Fidencio Mejía-Nepomuceno, María Guadalupe de Jesús Mireles-Rivera, Gloria María Molina-Salinas, Hector Montoya-Fuentes, , Mario Mújica-Sánchez, José Esteban Muñoz-Medina, José de Jesús Nuñez-Contreras, Alicia Ocaña-Mondragón, Luis Alberto Ochoa-Carrera, Hector Esteban Paz-Juárez, Francisco Pulido, Helen Haydee Fernanda Ramírez-Plascencia, Jorge Salas-Hernández, Angel Gustavo Salas-Lais, Alejandro Sanchez-Flores, Clara Esperanza Santacruz-Tinoco, María Guadalupe Santiago-Mauricio, Selene Zárate, Nelly Sélem-Mojica, Blanca Taboada, Gloria Vazquez, Joel Armando Vázquez-Pérez. |
| EPI_ISL_2391578 | Unidad de Investigación Médica de Yucatán (UIMY) | Instituto de Biotecnología de la UNAM | Consorcio Mexicano de Vigilancia Genómica (CoViGen-Mex). Authors (in alphabetical order): Julio Elias Alvarado-Yaah, Carlos F. Arias, Santiago Ávila-Ríos, Eduardo Becerril-Vargas, Víctor Hugo Borja-Aburto, Celia Boukadida, Cristóbal Cháidez-Quiróz, Juan Bautista Chale-Dzul, Ricardo Ciria Merce, Andreu Comas-García, Célida Duque Molina, Julissa Enciso-Ibarra, José Antonio Enciso-Moreno, Gloria Elena Espinosa-Ayala, Fernando Fontove-Herrera, Daniel Fregoso-Rueda, Victor Eduardo Garcia-Arias, Alejandra Garcia-Gasca, Bruno Gomez-Gil, Jean Pierre González, Irvin González-López, Concepción Grajales-Muñiz, Ricardo Grande, Rosa María Gutierrez Rios, Alejandra Hernández-Terán, Alfredo Herrera-Estrella, Carla Ivón Herrera-Najera, Pavel Isa, Daniel Lira Morales, Susana Lopez, Antonio Loza Román, Brenda Irasema Maldonado-Meza, Bernardo Martínez-Miguel, José Arturo Martínez-Orozco, Célida Martínez- Rodríguez, Margarita Matías-Florentino, Fidencio Mejía-Nepomuceno, María Guadalupe de Jesús Mireles-Rivera, Gloria María Molina-Salinas, Hector Montoya-Fuentes, , Mario Mújica-Sánchez, José Esteban Muñoz-Medina, José de Jesús Nuñez-Contreras, Alicia Ocaña-Mondragón, Luis Alberto Ochoa-Carrera, Hector Esteban Paz-Juárez, Francisco Pulido, Helen Haydee Fernanda Ramírez-Plascencia, Jorge Salas-Hernández, Angel Gustavo Salas-Lais, Alejandro Sanchez-Flores, Clara Esperanza Santacruz-Tinoco, María Guadalupe Santiago-Mauricio, Selene Zárate, Nelly Sélem-Mojica, Blanca Taboada, Gloria Vazquez, Joel Armando Vázquez-Pérez. |
| EPI_ISL_2391579, EPI_ISL_2391580 | Laboratorio Central de Epidemiología (LCE) | Instituto de Biotecnología de la UNAM | Consorcio Mexicano de Vigilancia Genómica (CoViGen-Mex). Authors (in alphabetical order): Julio Elias Alvarado-Yaah, Carlos F. Arias, Santiago Ávila-Ríos, Eduardo Becerril-Vargas, Víctor Hugo Borja-Aburto, Celia Boukadida, Cristóbal Cháidez-Quiróz, Juan Bautista Chale-Dzul, Ricardo Ciria Merce, Andreu Comas-García, Célida Duque Molina, Julissa Enciso-Ibarra, José Antonio Enciso-Moreno, Gloria Elena Espinosa-Ayala, Fernando Fontove-Herrera, Daniel Fregoso-Rueda, Victor Eduardo Garcia-Arias, Alejandra Garcia-Gasca, Bruno Gomez-Gil, Jean Pierre González, Irvin González-López, Concepción Grajales-Muñiz, Ricardo Grande, Rosa María Gutierrez Rios, Alejandra Hernández-Terán, Alfredo Herrera-Estrella, Carla Ivón Herrera-Najera, Pavel Isa, Daniel Lira Morales, Susana Lopez, Antonio Loza Román, Brenda Irasema Maldonado-Meza, Bernardo Martínez-Miguel, José Arturo Martínez-Orozco, Célida Martínez- Rodríguez, Margarita Matías-Florentino, Fidencio Mejía-Nepomuceno, María Guadalupe de Jesús Mireles-Rivera, Gloria María Molina-Salinas, Hector Montoya-Fuentes, , Mario Mújica-Sánchez, José Esteban Muñoz-Medina, José de Jesús Nuñez-Contreras, Alicia Ocaña-Mondragón, Luis Alberto Ochoa-Carrera, Hector Esteban Paz-Juárez, Francisco Pulido, Helen Haydee Fernanda Ramírez-Plascencia, Jorge Salas-Hernández, Angel Gustavo Salas-Lais, Alejandro Sanchez-Flores, Clara Esperanza Santacruz-Tinoco, María Guadalupe Santiago-Mauricio, Selene Zárate, Nelly Sélem-Mojica, Blanca Taboada, Gloria Vazquez, Joel Armando Vázquez-Pérez. |
| EPI_ISL_2391581 | Unidad de Investigación Médica de Yucatán (UIMY) | Instituto de Biotecnología de la UNAM | Consorcio Mexicano de Vigilancia Genómica (CoViGen-Mex). Authors (in alphabetical order): Julio Elias Alvarado-Yaah, Carlos F. Arias, Santiago Ávila-Ríos, Eduardo Becerril-Vargas, Víctor Hugo Borja-Aburto, Celia Boukadida, Cristóbal Cháidez-Quiróz, Juan Bautista Chale-Dzul, Ricardo Ciria Merce, Andreu Comas-García, Célida Duque Molina, Julissa Enciso-Ibarra, José Antonio Enciso-Moreno, Gloria Elena Espinosa-Ayala, Fernando Fontove-Herrera, Daniel Fregoso-Rueda, Victor Eduardo Garcia-Arias, Alejandra Garcia-Gasca, Bruno Gomez-Gil, Jean Pierre González, Irvin González-López, Concepción Grajales-Muñiz, Ricardo Grande, Rosa María Gutierrez Rios, Alejandra Hernández-Terán, Alfredo Herrera-Estrella, Carla Ivón Herrera-Najera, Pavel Isa, Daniel Lira Morales, Susana Lopez, Antonio Loza Román, Brenda Irasema Maldonado-Meza, Bernardo Martínez-Miguel, José Arturo Martínez-Orozco, Célida Martínez- Rodríguez, Margarita Matías-Florentino, Fidencio Mejía-Nepomuceno, María Guadalupe de Jesús Mireles-Rivera, Gloria María Molina-Salinas, Hector Montoya-Fuentes, , Mario Mújica-Sánchez, José Esteban Muñoz-Medina, José de Jesús Nuñez-Contreras, Alicia Ocaña-Mondragón, Luis Alberto Ochoa-Carrera, Hector Esteban Paz-Juárez, Francisco Pulido, Helen Haydee Fernanda Ramírez-Plascencia, Jorge Salas-Hernández, Angel Gustavo Salas-Lais, Alejandro Sanchez-Flores, Clara Esperanza Santacruz-Tinoco, María Guadalupe Santiago-Mauricio, Selene Zárate, Nelly Sélem-Mojica, Blanca Taboada, Gloria Vazquez, Joel Armando Vázquez-Pérez. |
| EPI_ISL_2391582, EPI_ISL_2391583, EPI_ISL_2391584, EPI_ISL_2391585, EPI_ISL_2391586, EPI_ISL_2391587 | Laboratorio Central de Epidemiología (LCE) | Instituto de Biotecnología de la UNAM | Consorcio Mexicano de Vigilancia Genómica (CoViGen-Mex). Authors (in alphabetical order): Julio Elias Alvarado-Yaah, Carlos F. Arias, Santiago Ávila-Ríos, Eduardo Becerril-Vargas, Víctor Hugo Borja-Aburto, Celia Boukadida, Cristóbal Cháidez-Quiróz, Juan Bautista Chale-Dzul, Ricardo Ciria Merce, Andreu Comas-García, Célida Duque Molina, Julissa Enciso-Ibarra, José Antonio Enciso-Moreno, Gloria Elena Espinosa-Ayala, Fernando Fontove-Herrera, Daniel Fregoso-Rueda, Victor Eduardo Garcia-Arias, Alejandra Garcia-Gasca, Bruno Gomez-Gil, Jean Pierre González, Irvin González-López, Concepción Grajales-Muñiz, Ricardo Grande, Rosa María Gutierrez Rios, Alejandra Hernández-Terán, Alfredo Herrera-Estrella, Carla Ivón Herrera-Najera, Pavel Isa, Daniel Lira Morales, Susana Lopez, Antonio Loza Román, Brenda Irasema Maldonado-Meza, Bernardo Martínez-Miguel, José Arturo Martínez-Orozco, Célida Martínez- Rodríguez, Margarita Matías-Florentino, Fidencio Mejía-Nepomuceno, María Guadalupe de Jesús Mireles-Rivera, Gloria María Molina-Salinas, Hector Montoya-Fuentes, , Mario Mújica-Sánchez, José Esteban Muñoz-Medina, José de Jesús Nuñez-Contreras, Alicia Ocaña-Mondragón, Luis Alberto Ochoa-Carrera, Hector Esteban Paz-Juárez, Francisco Pulido, Helen Haydee Fernanda Ramírez-Plascencia, Jorge Salas-Hernández, Angel Gustavo Salas-Lais, Alejandro Sanchez-Flores, Clara Esperanza Santacruz-Tinoco, María Guadalupe Santiago-Mauricio, Selene Zárate, Nelly Sélem-Mojica, Blanca Taboada, Gloria Vazquez, Joel Armando Vázquez-Pérez. |
| EPI_ISL_2391588 | Centro de Investigación Biomédica del Noreste (CIBIN) | Instituto de Biotecnología de la UNAM | Consorcio Mexicano de Vigilancia Genómica (CoViGen-Mex). Authors (in alphabetical order): Julio Elias Alvarado-Yaah, Carlos F. Arias, Santiago Ávila-Ríos, Eduardo Becerril-Vargas, Víctor Hugo Borja-Aburto, Celia Boukadida, Cristóbal Cháidez-Quiróz, Juan Bautista Chale-Dzul, Ricardo Ciria Merce, Andreu Comas-García, Célida Duque Molina, Julissa Enciso-Ibarra, José Antonio Enciso-Moreno, Gloria Elena Espinosa-Ayala, Fernando Fontove-Herrera, Daniel Fregoso-Rueda, Victor Eduardo Garcia-Arias, Alejandra Garcia-Gasca, Bruno Gomez-Gil, Jean Pierre González, Irvin González-López, Concepción Grajales-Muñiz, Ricardo Grande, Rosa María Gutierrez Rios, Alejandra Hernández-Terán, Alfredo Herrera-Estrella, Carla Ivón Herrera-Najera, Pavel Isa, Daniel Lira Morales, Susana Lopez, Antonio Loza Román, Brenda Irasema Maldonado-Meza, Bernardo Martínez-Miguel, José Arturo Martínez-Orozco, Célida Martínez- Rodríguez, Margarita Matías-Florentino, Fidencio Mejía-Nepomuceno, María Guadalupe de Jesús Mireles-Rivera, Gloria María Molina-Salinas, Hector Montoya-Fuentes, , Mario Mújica-Sánchez, José Esteban Muñoz-Medina, José de Jesús Nuñez-Contreras, Alicia Ocaña-Mondragón, Luis Alberto Ochoa-Carrera, Hector Esteban Paz-Juárez, Francisco Pulido, Helen Haydee Fernanda Ramírez-Plascencia, Jorge Salas-Hernández, Angel Gustavo Salas-Lais, Alejandro Sanchez-Flores, Clara Esperanza Santacruz-Tinoco, María Guadalupe Santiago-Mauricio, Selene Zárate, Nelly Sélem-Mojica, Blanca Taboada, Gloria Vazquez, Joel Armando Vázquez-Pérez. |
| EPI_ISL_2391589, EPI_ISL_2391590, EPI_ISL_2391591, EPI_ISL_2391592, EPI_ISL_2391593, EPI_ISL_2391594, EPI_ISL_2391595, EPI_ISL_2391596, EPI_ISL_2391597 | Laboratorio Central de Epidemiología (LCE) | Instituto de Biotecnología de la UNAM | Consorcio Mexicano de Vigilancia Genómica (CoViGen-Mex). Authors (in alphabetical order): Julio Elias Alvarado-Yaah, Carlos F. Arias, Santiago Ávila-Ríos, Eduardo Becerril-Vargas, Víctor Hugo Borja-Aburto, Celia Boukadida, Cristóbal Cháidez-Quiróz, Juan Bautista Chale-Dzul, Ricardo Ciria Merce, Andreu Comas-García, Célida Duque Molina, Julissa Enciso-Ibarra, José Antonio Enciso-Moreno, Gloria Elena Espinosa-Ayala, Fernando Fontove-Herrera, Daniel Fregoso-Rueda, Victor Eduardo Garcia-Arias, Alejandra Garcia-Gasca, Bruno Gomez-Gil, Jean Pierre González, Irvin González-López, Concepción Grajales-Muñiz, Ricardo Grande, Rosa María Gutierrez Rios, Alejandra Hernández-Terán, Alfredo Herrera-Estrella, Carla Ivón Herrera-Najera, Pavel Isa, Daniel Lira Morales, Susana Lopez, Antonio Loza Román, Brenda Irasema Maldonado-Meza, Bernardo Martínez-Miguel, José Arturo Martínez-Orozco, Célida Martínez- Rodríguez, Margarita Matías-Florentino, Fidencio Mejía-Nepomuceno, María Guadalupe de Jesús Mireles-Rivera, Gloria María Molina-Salinas, Hector Montoya-Fuentes, , Mario Mújica-Sánchez, José Esteban Muñoz-Medina, José de Jesús Nuñez-Contreras, Alicia Ocaña-Mondragón, Luis Alberto Ochoa-Carrera, Hector Esteban Paz-Juárez, Francisco Pulido, Helen Haydee Fernanda Ramírez-Plascencia, Jorge Salas-Hernández, Angel Gustavo Salas-Lais, Alejandro Sanchez-Flores, Clara Esperanza Santacruz-Tinoco, María Guadalupe Santiago-Mauricio, Selene Zárate, Nelly Sélem-Mojica, Blanca Taboada, Gloria Vazquez, Joel Armando Vázquez-Pérez. |
| EPI_ISL_2391598 | Centro de Investigación Biomédica del Noreste (CIBIN) | Instituto de Biotecnología de la UNAM | Consorcio Mexicano de Vigilancia Genómica (CoViGen-Mex). Authors (in alphabetical order): Julio Elias Alvarado-Yaah, Carlos F. Arias, Santiago Ávila-Ríos, Eduardo Becerril-Vargas, Víctor Hugo Borja-Aburto, Celia Boukadida, Cristóbal Cháidez-Quiróz, Juan Bautista Chale-Dzul, Ricardo Ciria Merce, Andreu Comas-García, Célida Duque Molina, Julissa Enciso-Ibarra, José Antonio Enciso-Moreno, Gloria Elena Espinosa-Ayala, Fernando Fontove-Herrera, Daniel Fregoso-Rueda, Victor Eduardo Garcia-Arias, Alejandra Garcia-Gasca, Bruno Gomez-Gil, Jean Pierre González, Irvin González-López, Concepción Grajales-Muñiz, Ricardo Grande, Rosa María Gutierrez Rios, Alejandra Hernández-Terán, Alfredo Herrera-Estrella, Carla Ivón Herrera-Najera, Pavel Isa, Daniel Lira Morales, Susana Lopez, Antonio Loza Román, Brenda Irasema Maldonado-Meza, Bernardo Martínez-Miguel, José Arturo Martínez-Orozco, Célida Martínez- Rodríguez, Margarita Matías-Florentino, Fidencio Mejía-Nepomuceno, María Guadalupe de Jesús Mireles-Rivera, Gloria María Molina-Salinas, Hector Montoya-Fuentes, , Mario Mújica-Sánchez, José Esteban Muñoz-Medina, José de Jesús Nuñez-Contreras, Alicia Ocaña-Mondragón, Luis Alberto Ochoa-Carrera, Hector Esteban Paz-Juárez, Francisco Pulido, Helen Haydee Fernanda Ramírez-Plascencia, Jorge Salas-Hernández, Angel Gustavo Salas-Lais, Alejandro Sanchez-Flores, Clara Esperanza Santacruz-Tinoco, María Guadalupe Santiago-Mauricio, Selene Zárate, Nelly Sélem-Mojica, Blanca Taboada, Gloria Vazquez, Joel Armando Vázquez-Pérez. |
| EPI_ISL_2391599, EPI_ISL_2391600, EPI_ISL_2391601, EPI_ISL_2391602, EPI_ISL_2391603, EPI_ISL_2391604, EPI_ISL_2391605, EPI_ISL_2391606, EPI_ISL_2391607, EPI_ISL_2391608, EPI_ISL_2391609, EPI_ISL_2391610, EPI_ISL_2391611, EPI_ISL_2391612, EPI_ISL_2391613, EPI_ISL_2391614, EPI_ISL_2391615, EPI_ISL_2391616, EPI_ISL_2391617, EPI_ISL_2391618, EPI_ISL_2391619, EPI_ISL_2391620, EPI_ISL_2391621, EPI_ISL_2391622, EPI_ISL_2391623, EPI_ISL_2391624, EPI_ISL_2391625, EPI_ISL_2391626, EPI_ISL_2391627, EPI_ISL_2391628, EPI_ISL_2391629, EPI_ISL_2391630, EPI_ISL_2391631, EPI_ISL_2391632, EPI_ISL_2391633, EPI_ISL_2391634, EPI_ISL_2391635, EPI_ISL_2391636, EPI_ISL_2391637, EPI_ISL_2391638 | | | |

| | | | |
|---|---|---|---|
| see above | Laboratorio Central de Epidemiología (LCE) | Instituto de Biotecnología de la UNAM | Consorcio Mexicano de Vigilancia Genómica (CoViGen-Mex). Authors (in alphabetical order): Julio Elias Alvarado-Yaah, Carlos F. Arias, Santiago Ávila-Ríos, Eduardo Becerril-Vargas, Víctor Hugo Borja-Aburto, Celia Boukadida, Cristóbal Cháidez-Quiróz, Juan Bautista Chale-Dzul, Ricardo Ciria Merce, Andreu Comas-García, Célida Duque Molina, Julissa Enciso-Ibarra, José Antonio Enciso-Moreno, Gloria Elena Espinosa-Ayala, Fernando Fontove-Herrera, Daniel Fregoso-Rueda, Victor Eduardo Garcia-Arias, Alejandra Garcia-Gasca, Bruno Gomez-Gil, Jean Pierre González, Irvin González-López, Concepción Grajales-Muñiz, Ricardo Grande, Rosa María Gutierrez Rios, Alejandra Hernández-Terán, Alfredo Herrera-Estrella, Carla Ivón Herrera-Najera, Pavel Isa, Daniel Lira Morales, Susana Lopez, Antonio Loza Román, Brenda Irasema Maldonado-Meza, Bernardo Martínez-Miguel, José Arturo Martínez-Orozco, Célida Martínez- Rodríguez, Margarita Matías-Florentino, Fidencio Mejía-Nepomuceno, María Guadalupe de Jesús Mireles-Rivera, Gloria María Molina-Salinas, Hector Montoya-Fuentes, , Mario Mújica-Sánchez, José Esteban Muñoz-Medina, José de Jesús Nuñez-Contreras, Alicia Ocaña-Mondragón, Luis Alberto Ochoa-Carrera, Hector Esteban Paz-Juárez, Francisco Pulido, Helen Haydee Fernanda Ramírez-Plascencia, Jorge Salas-Hernández, Angel Gustavo Salas-Lais, Alejandro Sanchez-Flores, Clara Esperanza Santacruz-Tinoco, María Guadalupe Santiago-Mauricio, Selene Zárate, Nelly Sélem-Mojica, Blanca Taboada, Gloria Vazquez, Joel Armando Vázquez-Pérez. |
| EPI_ISL_2391639 | Centro de Investigación Biomédica del Noreste (CIBIN) | Instituto de Biotecnología de la UNAM | Consorcio Mexicano de Vigilancia Genómica (CoViGen-Mex). Authors (in alphabetical order): Julio Elias Alvarado-Yaah, Carlos F. Arias, Santiago Ávila-Ríos, Eduardo Becerril-Vargas, Víctor Hugo Borja-Aburto, Celia Boukadida, Cristóbal Cháidez-Quiróz, Juan Bautista Chale-Dzul, Ricardo Ciria Merce, Andreu Comas-García, Célida Duque Molina, Julissa Enciso-Ibarra, José Antonio Enciso-Moreno, Gloria Elena Espinosa-Ayala, Fernando Fontove-Herrera, Daniel Fregoso-Rueda, Victor Eduardo Garcia-Arias, Alejandra Garcia-Gasca, Bruno Gomez-Gil, Jean Pierre González, Irvin González-López, Concepción Grajales-Muñiz, Ricardo Grande, Rosa María Gutierrez Rios, Alejandra Hernández-Terán, Alfredo Herrera-Estrella, Carla Ivón Herrera-Najera, Pavel Isa, Daniel Lira Morales, Susana Lopez, Antonio Loza Román, Brenda Irasema Maldonado-Meza, Bernardo Martínez-Miguel, José Arturo Martínez-Orozco, Célida Martínez- Rodríguez, Margarita Matías-Florentino, Fidencio Mejía-Nepomuceno, María Guadalupe de Jesús Mireles-Rivera, Gloria María Molina-Salinas, Hector Montoya-Fuentes, , Mario Mújica-Sánchez, José Esteban Muñoz-Medina, José de Jesús Nuñez-Contreras, Alicia Ocaña-Mondragón, Luis Alberto Ochoa-Carrera, Hector Esteban Paz-Juárez, Francisco Pulido, Helen Haydee Fernanda Ramírez-Plascencia, Jorge Salas-Hernández, Angel Gustavo Salas-Lais, Alejandro Sanchez-Flores, Clara Esperanza Santacruz-Tinoco, María Guadalupe Santiago-Mauricio, Selene Zárate, Nelly Sélem-Mojica, Blanca Taboada, Gloria Vazquez, Joel Armando Vázquez-Pérez. |
| EPI_ISL_2391640, EPI_ISL_2391641, EPI_ISL_2391642, EPI_ISL_2391643, EPI_ISL_2391644, EPI_ISL_2391645, EPI_ISL_2391646, EPI_ISL_2391647, EPI_ISL_2391648, EPI_ISL_2391649 | Laboratorio Central de Epidemiología (LCE) | Instituto de Biotecnología de la UNAM | Consorcio Mexicano de Vigilancia Genómica (CoViGen-Mex). Authors (in alphabetical order): Julio Elias Alvarado-Yaah, Carlos F. Arias, Santiago Ávila-Ríos, Eduardo Becerril-Vargas, Víctor Hugo Borja-Aburto, Celia Boukadida, Cristóbal Cháidez-Quiróz, Juan Bautista Chale-Dzul, Ricardo Ciria Merce, Andreu Comas-García, Célida Duque Molina, Julissa Enciso-Ibarra, José Antonio Enciso-Moreno, Gloria Elena Espinosa-Ayala, Fernando Fontove-Herrera, Daniel Fregoso-Rueda, Victor Eduardo Garcia-Arias, Alejandra Garcia-Gasca, Bruno Gomez-Gil, Jean Pierre González, Irvin González-López, Concepción Grajales-Muñiz, Ricardo Grande, Rosa María Gutierrez Rios, Alejandra Hernández-Terán, Alfredo Herrera-Estrella, Carla Ivón Herrera-Najera, Pavel Isa, Daniel Lira Morales, Susana Lopez, Antonio Loza Román, Brenda Irasema Maldonado-Meza, Bernardo Martínez-Miguel, José Arturo Martínez-Orozco, Célida Martínez- Rodríguez, Margarita Matías-Florentino, Fidencio Mejía-Nepomuceno, María Guadalupe de Jesús Mireles-Rivera, Gloria María Molina-Salinas, Hector Montoya-Fuentes, , Mario Mújica-Sánchez, José Esteban Muñoz-Medina, José de Jesús Nuñez-Contreras, Alicia Ocaña-Mondragón, Luis Alberto Ochoa-Carrera, Hector Esteban Paz-Juárez, Francisco Pulido, Helen Haydee Fernanda Ramírez-Plascencia, Jorge Salas-Hernández, Angel Gustavo Salas-Lais, Alejandro Sanchez-Flores, Clara Esperanza Santacruz-Tinoco, María Guadalupe Santiago-Mauricio, Selene Zárate, Nelly Sélem-Mojica, Blanca Taboada, Gloria Vazquez, Joel Armando Vázquez-Pérez. |
| EPI_ISL_2391650 | Unidad de Investigación Médica de Yucatán (UIMY) | Instituto de Biotecnología de la UNAM | Consorcio Mexicano de Vigilancia Genómica (CoViGen-Mex). Authors (in alphabetical order): Julio Elias Alvarado-Yaah, Carlos F. Arias, Santiago Ávila-Ríos, Eduardo Becerril-Vargas, Víctor Hugo Borja-Aburto, Celia Boukadida, Cristóbal Cháidez-Quiróz, Juan Bautista Chale-Dzul, Ricardo Ciria Merce, Andreu Comas-García, Célida Duque Molina, Julissa Enciso-Ibarra, José Antonio Enciso-Moreno, Gloria Elena Espinosa-Ayala, Fernando Fontove-Herrera, Daniel Fregoso-Rueda, Victor Eduardo Garcia-Arias, Alejandra Garcia-Gasca, Bruno Gomez-Gil, Jean Pierre González, Irvin González-López, Concepción Grajales-Muñiz, Ricardo Grande, Rosa María Gutierrez Rios, Alejandra Hernández-Terán, Alfredo Herrera-Estrella, Carla Ivón Herrera-Najera, Pavel Isa, Daniel Lira Morales, Susana Lopez, Antonio Loza Román, Brenda Irasema Maldonado-Meza, Bernardo Martínez-Miguel, José Arturo Martínez-Orozco, Célida Martínez- Rodríguez, Margarita Matías-Florentino, Fidencio Mejía-Nepomuceno, María Guadalupe de Jesús Mireles-Rivera, Gloria María Molina-Salinas, Hector Montoya-Fuentes, , Mario Mújica-Sánchez, José Esteban Muñoz-Medina, José de Jesús Nuñez-Contreras, Alicia Ocaña-Mondragón, Luis Alberto Ochoa-Carrera, Hector Esteban Paz-Juárez, Francisco Pulido, Helen Haydee Fernanda Ramírez-Plascencia, Jorge Salas-Hernández, Angel Gustavo Salas-Lais, Alejandro Sanchez-Flores, Clara Esperanza Santacruz-Tinoco, María Guadalupe Santiago-Mauricio, Selene Zárate, Nelly Sélem-Mojica, Blanca Taboada, Gloria Vazquez, Joel Armando Vázquez-Pérez. |
| EPI_ISL_2391651, EPI_ISL_2391652, EPI_ISL_2391653, EPI_ISL_2391654 | Laboratorio Central de Epidemiología (LCE) | Instituto de Biotecnología de la UNAM | Consorcio Mexicano de Vigilancia Genómica (CoViGen-Mex). Authors (in alphabetical order): Julio Elias Alvarado-Yaah, Carlos F. Arias, Santiago Ávila-Ríos, Eduardo Becerril-Vargas, Víctor Hugo Borja-Aburto, Celia Boukadida, Cristóbal Cháidez-Quiróz, Juan Bautista Chale-Dzul, Ricardo Ciria Merce, Andreu Comas-García, Célida Duque Molina, Julissa Enciso-Ibarra, José Antonio Enciso-Moreno, Gloria Elena Espinosa-Ayala, Fernando Fontove-Herrera, Daniel Fregoso-Rueda, Victor Eduardo Garcia-Arias, Alejandra Garcia-Gasca, Bruno Gomez-Gil, Jean Pierre González, Irvin González-López, Concepción Grajales-Muñiz, Ricardo Grande, Rosa María Gutierrez Rios, Alejandra Hernández-Terán, Alfredo Herrera-Estrella, Carla Ivón Herrera-Najera, Pavel Isa, Daniel Lira Morales, Susana Lopez, Antonio Loza Román, Brenda Irasema Maldonado-Meza, Bernardo Martínez-Miguel, José Arturo Martínez-Orozco, Célida Martínez- Rodríguez, Margarita Matías-Florentino, Fidencio Mejía-Nepomuceno, María Guadalupe de Jesús Mireles-Rivera, Gloria María Molina-Salinas, Hector Montoya-Fuentes, , Mario Mújica-Sánchez, José Esteban Muñoz-Medina, José de Jesús Nuñez-Contreras, Alicia Ocaña-Mondragón, Luis Alberto Ochoa-Carrera, Hector Esteban Paz-Juárez, Francisco Pulido, Helen Haydee Fernanda Ramírez-Plascencia, Jorge Salas-Hernández, Angel Gustavo Salas-Lais, Alejandro Sanchez-Flores, Clara Esperanza Santacruz-Tinoco, María Guadalupe Santiago-Mauricio, Selene Zárate, Nelly Sélem-Mojica, Blanca Taboada, Gloria Vazquez, Joel Armando Vázquez-Pérez. |
| EPI_ISL_2391655 | Centro de Investigación Biomédica del Noreste (CIBIN) | Instituto de Biotecnología de la UNAM | Consorcio Mexicano de Vigilancia Genómica (CoViGen-Mex). Authors (in alphabetical order): Julio Elias Alvarado-Yaah, Carlos F. Arias, Santiago Ávila-Ríos, Eduardo Becerril-Vargas, Víctor Hugo Borja-Aburto, Celia Boukadida, Cristóbal Cháidez-Quiróz, Juan Bautista Chale-Dzul, Ricardo Ciria Merce, Andreu Comas-García, Célida Duque Molina, Julissa Enciso-Ibarra, José Antonio Enciso-Moreno, Gloria Elena Espinosa-Ayala, Fernando Fontove-Herrera, Daniel Fregoso-Rueda, Victor Eduardo Garcia-Arias, Alejandra Garcia-Gasca, Bruno Gomez-Gil, Jean Pierre González, Irvin González-López, Concepción Grajales-Muñiz, Ricardo Grande, Rosa María Gutierrez Rios, Alejandra Hernández-Terán, Alfredo Herrera-Estrella, Carla Ivón Herrera-Najera, Pavel Isa, Daniel Lira Morales, Susana Lopez, Antonio Loza Román, Brenda Irasema Maldonado-Meza, Bernardo Martínez-Miguel, José Arturo Martínez-Orozco, Célida Martínez- Rodríguez, Margarita Matías-Florentino, Fidencio Mejía-Nepomuceno, María Guadalupe de Jesús Mireles-Rivera, Gloria María Molina-Salinas, Hector Montoya-Fuentes, , Mario Mújica-Sánchez, José Esteban Muñoz-Medina, José de Jesús Nuñez-Contreras, Alicia Ocaña-Mondragón, Luis Alberto Ochoa-Carrera, Hector Esteban Paz-Juárez, Francisco Pulido, Helen Haydee Fernanda Ramírez-Plascencia, Jorge Salas-Hernández, Angel Gustavo Salas-Lais, Alejandro Sanchez-Flores, Clara Esperanza Santacruz-Tinoco, María Guadalupe Santiago-Mauricio, Selene Zárate, Nelly Sélem-Mojica, Blanca Taboada, Gloria Vazquez, Joel Armando Vázquez-Pérez. |
| EPI_ISL_2391656, EPI_ISL_2391657, EPI_ISL_2391658, EPI_ISL_2391659, EPI_ISL_2391660 | Laboratorio Central de Epidemiología (LCE) | Instituto de Biotecnología de la UNAM | Consorcio Mexicano de Vigilancia Genómica (CoViGen-Mex). Authors (in alphabetical order): Julio Elias Alvarado-Yaah, Carlos F. Arias, Santiago Ávila-Ríos, Eduardo Becerril-Vargas, Víctor Hugo Borja-Aburto, Celia Boukadida, Cristóbal Cháidez-Quiróz, Juan Bautista Chale-Dzul, Ricardo Ciria Merce, Andreu Comas-García, Célida Duque Molina, Julissa Enciso-Ibarra, José Antonio Enciso-Moreno, Gloria Elena Espinosa-Ayala, Fernando Fontove-Herrera, Daniel Fregoso-Rueda, Victor Eduardo Garcia-Arias, Alejandra Garcia-Gasca, Bruno Gomez-Gil, Jean Pierre González, Irvin González-López, Concepción Grajales-Muñiz, Ricardo Grande, Rosa María Gutierrez Rios, Alejandra Hernández-Terán, Alfredo Herrera-Estrella, Carla Ivón Herrera-Najera, Pavel Isa, Daniel Lira Morales, Susana Lopez, Antonio Loza Román, Brenda Irasema Maldonado-Meza, Bernardo Martínez-Miguel, José Arturo Martínez-Orozco, Célida Martínez- Rodríguez, Margarita Matías-Florentino, Fidencio Mejía-Nepomuceno, María Guadalupe de Jesús Mireles-Rivera, Gloria María Molina-Salinas, Hector Montoya-Fuentes, , Mario Mújica-Sánchez, José Esteban Muñoz-Medina, José de Jesús Nuñez-Contreras, Alicia Ocaña-Mondragón, Luis Alberto Ochoa-Carrera, Hector Esteban Paz-Juárez, Francisco Pulido, Helen Haydee Fernanda Ramírez-Plascencia, Jorge Salas-Hernández, Angel Gustavo Salas-Lais, Alejandro Sanchez-Flores, Clara Esperanza Santacruz-Tinoco, María Guadalupe Santiago-Mauricio, Selene Zárate, Nelly Sélem-Mojica, Blanca Taboada, Gloria Vazquez, Joel Armando Vázquez-Pérez. |
| EPI_ISL_2391661 | Centro de Investigación Biomédica de Occidente (CIBO) | Instituto de Biotecnología de la UNAM | Consorcio Mexicano de Vigilancia Genómica (CoViGen-Mex). Authors (in alphabetical order): Julio Elias Alvarado-Yaah, Carlos F. Arias, Santiago Ávila-Ríos, Eduardo Becerril-Vargas, Víctor Hugo Borja-Aburto, Celia Boukadida, Cristóbal Cháidez-Quiróz, Juan Bautista Chale-Dzul, Ricardo Ciria Merce, Andreu Comas-García, Célida Duque Molina, Julissa Enciso-Ibarra, José Antonio Enciso-Moreno, Gloria Elena Espinosa-Ayala, Fernando Fontove-Herrera, Daniel Fregoso-Rueda, Victor Eduardo Garcia-Arias, Alejandra Garcia-Gasca, Bruno Gomez-Gil, Jean Pierre González, Irvin González-López, Concepción Grajales-Muñiz, Ricardo Grande, Rosa María Gutierrez Rios, Alejandra Hernández-Terán, Alfredo Herrera-Estrella, Carla Ivón Herrera-Najera, Pavel Isa, Daniel Lira Morales, Susana Lopez, Antonio Loza Román, Brenda Irasema Maldonado-Meza, Bernardo Martínez-Miguel, José Arturo Martínez-Orozco, Célida Martínez- Rodríguez, Margarita Matías-Florentino, Fidencio Mejía-Nepomuceno, María Guadalupe de Jesús Mireles-Rivera, Gloria María Molina-Salinas, Hector Montoya-Fuentes, , Mario Mújica-Sánchez, José Esteban Muñoz-Medina, José de Jesús Nuñez-Contreras, Alicia Ocaña-Mondragón, Luis Alberto Ochoa-Carrera, Hector Esteban Paz-Juárez, Francisco Pulido, Helen Haydee Fernanda Ramírez-Plascencia, Jorge Salas-Hernández, Angel Gustavo Salas-Lais, Alejandro Sanchez-Flores, Clara Esperanza Santacruz-Tinoco, María Guadalupe Santiago-Mauricio, Selene Zárate, Nelly Sélem-Mojica, Blanca Taboada, Gloria Vazquez, Joel Armando Vázquez-Pérez. |
| EPI_ISL_2391662, EPI_ISL_2391663, EPI_ISL_2391664, EPI_ISL_2391665, EPI_ISL_2391666, EPI_ISL_2391667, EPI_ISL_2391668, EPI_ISL_2391669, EPI_ISL_2391670, EPI_ISL_2391671, EPI_ISL_2391672, EPI_ISL_2391673 | | | |
| see above | Laboratorio Central de Epidemiología (LCE) | Instituto de Biotecnología de la UNAM | Consorcio Mexicano de Vigilancia Genómica (CoViGen-Mex). Authors (in alphabetical order): Julio Elias Alvarado-Yaah, Carlos F. Arias, Santiago Ávila-Ríos, Eduardo Becerril-Vargas, Víctor Hugo Borja-Aburto, Celia Boukadida, Cristóbal Cháidez-Quiróz, Juan Bautista Chale-Dzul, Ricardo Ciria Merce, Andreu Comas-García, Célida Duque Molina, Julissa Enciso-Ibarra, José Antonio Enciso-Moreno, Gloria Elena Espinosa-Ayala, Fernando Fontove-Herrera, Daniel Fregoso-Rueda, Victor Eduardo Garcia-Arias, Alejandra Garcia-Gasca, Bruno Gomez-Gil, Jean Pierre González, Irvin González-López, Concepción Grajales-Muñiz, Ricardo Grande, Rosa María Gutierrez Rios, Alejandra Hernández-Terán, Alfredo Herrera-Estrella, Carla Ivón Herrera-Najera, Pavel Isa, Daniel Lira Morales, Susana Lopez, Antonio Loza Román, Brenda Irasema Maldonado-Meza, Bernardo Martínez-Miguel, José Arturo Martínez-Orozco, Célida Martínez- Rodríguez, Margarita Matías-Florentino, Fidencio Mejía-Nepomuceno, María Guadalupe de Jesús Mireles-Rivera, Gloria María Molina-Salinas, Hector Montoya-Fuentes, , Mario Mújica-Sánchez, José Esteban Muñoz-Medina, José de Jesús Nuñez-Contreras, Alicia Ocaña-Mondragón, Luis Alberto Ochoa-Carrera, Hector Esteban Paz-Juárez, Francisco Pulido, Helen Haydee Fernanda Ramírez-Plascencia, Jorge Salas-Hernández, Angel Gustavo Salas-Lais, Alejandro Sanchez-Flores, Clara Esperanza Santacruz-Tinoco, María Guadalupe Santiago-Mauricio, Selene Zárate, Nelly Sélem-Mojica, Blanca Taboada, Gloria Vazquez, Joel Armando Vázquez-Pérez. |
| EPI_ISL_2391674 | Centro de Investigación Biomédica del Noreste (CIBIN) | Instituto de Biotecnología de la UNAM | Consorcio Mexicano de Vigilancia Genómica (CoViGen-Mex). Authors (in alphabetical order): Julio Elias Alvarado-Yaah, Carlos F. Arias, Santiago Ávila-Ríos, Eduardo Becerril-Vargas, Víctor Hugo Borja-Aburto, Celia Boukadida, Cristóbal Cháidez-Quiróz, Juan Bautista Chale-Dzul, Ricardo Ciria Merce, Andreu Comas-García, Célida Duque Molina, Julissa Enciso-Ibarra, José Antonio Enciso-Moreno, Gloria Elena Espinosa-Ayala, Fernando Fontove-Herrera, Daniel Fregoso-Rueda, Victor Eduardo Garcia-Arias, Alejandra Garcia-Gasca, Bruno Gomez-Gil, Jean Pierre González, Irvin González-López, Concepción Grajales-Muñiz, Ricardo Grande, Rosa María Gutierrez Rios, Alejandra Hernández-Terán, Alfredo Herrera-Estrella, Carla Ivón Herrera-Najera, Pavel Isa, Daniel Lira Morales, Susana Lopez, Antonio Loza Román, Brenda Irasema Maldonado-Meza, Bernardo Martínez-Miguel, José Arturo Martínez-Orozco, Célida Martínez- Rodríguez, Margarita Matías-Florentino, Fidencio Mejía-Nepomuceno, María Guadalupe de Jesús Mireles-Rivera, Gloria María Molina-Salinas, Hector Montoya-Fuentes, , Mario Mújica-Sánchez, José Esteban Muñoz-Medina, José de Jesús Nuñez-Contreras, Alicia Ocaña-Mondragón, Luis Alberto Ochoa-Carrera, Hector Esteban Paz-Juárez, Francisco Pulido, Helen Haydee Fernanda Ramírez-Plascencia, Jorge Salas-Hernández, Angel Gustavo Salas-Lais, Alejandro Sanchez-Flores, Clara Esperanza Santacruz-Tinoco, María Guadalupe Santiago-Mauricio, Selene Zárate, Nelly Sélem-Mojica, Blanca Taboada, Gloria Vazquez, Joel Armando Vázquez-Pérez. |
| EPI_ISL_2391675, EPI_ISL_2391676, EPI_ISL_2391677, EPI_ISL_2391678, EPI_ISL_2391679, EPI_ISL_2391680, EPI_ISL_2391681, EPI_ISL_2391682, EPI_ISL_2391683, EPI_ISL_2391684, EPI_ISL_2391685, EPI_ISL_2391686, EPI_ISL_2391687 | | | |
| see above | Laboratorio Central de Epidemiología (LCE) | Instituto de Biotecnología de la UNAM | Consorcio Mexicano de Vigilancia Genómica (CoViGen-Mex). Authors (in alphabetical order): Julio Elias Alvarado-Yaah, Carlos F. Arias, Santiago Ávila-Ríos, Eduardo Becerril-Vargas, Víctor Hugo Borja-Aburto, Celia Boukadida, Cristóbal Cháidez-Quiróz, Juan Bautista Chale-Dzul, Ricardo Ciria Merce, Andreu Comas-García, Célida Duque Molina, Julissa Enciso-Ibarra, José Antonio Enciso-Moreno, Gloria Elena Espinosa-Ayala, Fernando Fontove-Herrera, Daniel Fregoso-Rueda, Victor Eduardo Garcia-Arias, Alejandra Garcia-Gasca, Bruno Gomez-Gil, Jean Pierre González, Irvin González-López, Concepción Grajales-Muñiz, Ricardo Grande, Rosa María Gutierrez Rios, Alejandra Hernández-Terán, Alfredo Herrera-Estrella, Carla Ivón Herrera-Najera, Pavel Isa, Daniel Lira Morales, Susana Lopez, Antonio Loza Román, Brenda Irasema Maldonado-Meza, Bernardo Martínez-Miguel, José Arturo Martínez-Orozco, Célida Martínez- Rodríguez, Margarita Matías-Florentino, Fidencio Mejía-Nepomuceno, María Guadalupe de Jesús Mireles-Rivera, Gloria María Molina-Salinas, Hector Montoya-Fuentes, , Mario Mújica-Sánchez, José Esteban Muñoz-Medina, José de Jesús Nuñez-Contreras, Alicia Ocaña-Mondragón, Luis Alberto Ochoa-Carrera, Hector Esteban Paz-Juárez, Francisco Pulido, Helen Haydee Fernanda Ramírez-Plascencia, Jorge Salas-Hernández, Angel Gustavo Salas-Lais, Alejandro Sanchez-Flores, Clara Esperanza Santacruz-Tinoco, María Guadalupe Santiago-Mauricio, Selene Zárate, Nelly Sélem-Mojica, Blanca Taboada, Gloria Vazquez, Joel Armando Vázquez-Pérez. |
| EPI_ISL_2391688 | Unidad de Investigación Biomédica de Zacatecas (UIBZ) | Instituto de Biotecnología de la UNAM | Consorcio Mexicano de Vigilancia Genómica (CoViGen-Mex). Authors (in alphabetical order): Julio Elias Alvarado-Yaah, Carlos F. Arias, Santiago Ávila-Ríos, Eduardo Becerril-Vargas, Víctor Hugo Borja-Aburto, Celia Boukadida, Cristóbal Cháidez-Quiróz, Juan Bautista Chale-Dzul, Ricardo Ciria Merce, Andreu Comas-García, Célida Duque Molina, Julissa Enciso-Ibarra, José Antonio Enciso-Moreno, Gloria Elena Espinosa-Ayala, Fernando Fontove-Herrera, Daniel Fregoso-Rueda, Victor Eduardo Garcia-Arias, Alejandra Garcia-Gasca, Bruno Gomez-Gil, Jean Pierre González, Irvin González-López, Concepción Grajales-Muñiz, Ricardo Grande, Rosa María Gutierrez Rios, Alejandra Hernández-Terán, Alfredo Herrera-Estrella, Carla Ivón Herrera-Najera, Pavel Isa, Daniel Lira Morales, Susana Lopez, Antonio Loza Román, Brenda Irasema Maldonado-Meza, Bernardo Martínez-Miguel, José Arturo Martínez-Orozco, Célida Martínez- Rodríguez, Margarita Matías-Florentino, Fidencio Mejía-Nepomuceno, María Guadalupe de Jesús Mireles-Rivera, Gloria María Molina-Salinas, Hector Montoya-Fuentes, , Mario Mújica-Sánchez, José Esteban Muñoz-Medina, José de Jesús Nuñez-Contreras, Alicia Ocaña-Mondragón, Luis Alberto Ochoa-Carrera, Hector Esteban Paz-Juárez, Francisco Pulido, Helen Haydee Fernanda Ramírez-Plascencia, Jorge Salas-Hernández, Angel Gustavo Salas-Lais, Alejandro Sanchez-Flores, Clara Esperanza Santacruz-Tinoco, María Guadalupe Santiago-Mauricio, Selene Zárate, Nelly Sélem-Mojica, Blanca Taboada, Gloria Vazquez, Joel Armando Vázquez-Pérez. |
| EPI_ISL_2391689, EPI_ISL_2391690 | Laboratorio Central de Epidemiología (LCE) | Instituto de Biotecnología de la UNAM | Consorcio Mexicano de Vigilancia Genómica (CoViGen-Mex). Authors (in alphabetical order): Julio Elias Alvarado-Yaah, Carlos F. Arias, Santiago Ávila-Ríos, Eduardo Becerril-Vargas, Víctor Hugo Borja-Aburto, Celia Boukadida, Cristóbal Cháidez-Quiróz, Juan Bautista Chale-Dzul, Ricardo Ciria Merce, Andreu Comas-García, Célida Duque Molina, Julissa Enciso-Ibarra, José Antonio Enciso-Moreno, Gloria Elena Espinosa-Ayala, Fernando Fontove-Herrera, Daniel Fregoso-Rueda, Victor Eduardo Garcia-Arias, Alejandra Garcia-Gasca, Bruno Gomez-Gil, Jean Pierre González, Irvin González-López, Concepción Grajales-Muñiz, Ricardo Grande, Rosa María Gutierrez Rios, Alejandra Hernández-Terán, Alfredo Herrera-Estrella, Carla Ivón Herrera-Najera, Pavel Isa, Daniel Lira Morales, Susana Lopez, Antonio Loza Román, Brenda Irasema Maldonado-Meza, Bernardo Martínez-Miguel, José Arturo Martínez-Orozco, Célida Martínez- Rodríguez, Margarita Matías-Florentino, Fidencio Mejía-Nepomuceno, María Guadalupe de Jesús Mireles-Rivera, Gloria María Molina-Salinas, Hector Montoya-Fuentes, , Mario Mújica-Sánchez, José Esteban Muñoz-Medina, José de Jesús Nuñez-Contreras, Alicia Ocaña-Mondragón, Luis Alberto Ochoa-Carrera, Hector Esteban Paz-Juárez, Francisco Pulido, Helen Haydee Fernanda Ramírez-Plascencia, Jorge Salas-Hernández, Angel Gustavo Salas-Lais, Alejandro Sanchez-Flores, Clara Esperanza Santacruz-Tinoco, María Guadalupe Santiago-Mauricio, Selene Zárate, Nelly Sélem-Mojica, Blanca Taboada, Gloria Vazquez, Joel Armando Vázquez-Pérez. |
| EPI_ISL_2391691 | Centro de Investigación Biomédica del Noreste (CIBIN) | Instituto de Biotecnología de la UNAM | Consorcio Mexicano de Vigilancia Genómica (CoViGen-Mex). Authors (in alphabetical order): Julio Elias Alvarado-Yaah, Carlos F. Arias, Santiago Ávila-Ríos, Eduardo Becerril-Vargas, Víctor Hugo Borja-Aburto, Celia Boukadida, Cristóbal Cháidez-Quiróz, Juan Bautista Chale-Dzul, Ricardo Ciria Merce, Andreu Comas-García, Célida Duque Molina, Julissa Enciso-Ibarra, José Antonio Enciso-Moreno, Gloria Elena Espinosa-Ayala, Fernando Fontove-Herrera, Daniel Fregoso-Rueda, Victor Eduardo Garcia-Arias, Alejandra Garcia-Gasca, Bruno Gomez-Gil, Jean Pierre González, Irvin González-López, Concepción Grajales-Muñiz, Ricardo Grande, Rosa María Gutierrez Rios, Alejandra Hernández-Terán, Alfredo Herrera-Estrella, Carla Ivón Herrera-Najera, Pavel Isa, Daniel Lira Morales, Susana Lopez, Antonio Loza Román, Brenda Irasema Maldonado-Meza, Bernardo Martínez-Miguel, José Arturo Martínez-Orozco, Célida Martínez- Rodríguez, Margarita Matías-Florentino, Fidencio Mejía-Nepomuceno, María Guadalupe de Jesús Mireles-Rivera, Gloria María Molina-Salinas, Hector Montoya-Fuentes, , Mario Mújica-Sánchez, José Esteban Muñoz-Medina, José de Jesús Nuñez-Contreras, Alicia Ocaña-Mondragón, Luis Alberto Ochoa-Carrera, Hector Esteban Paz-Juárez, Francisco Pulido, Helen Haydee Fernanda Ramírez-Plascencia, Jorge Salas-Hernández, Angel Gustavo Salas-Lais, Alejandro Sanchez-Flores, Clara Esperanza Santacruz-Tinoco, María Guadalupe Santiago-Mauricio, Selene Zárate, Nelly Sélem-Mojica, Blanca Taboada, Gloria Vazquez, Joel Armando Vázquez-Pérez. |
| EPI_ISL_2391692, EPI_ISL_2391693, EPI_ISL_2391694, EPI_ISL_2391695, EPI_ISL_2391696, EPI_ISL_2391697, EPI_ISL_2391698, EPI_ISL_2391699, EPI_ISL_2391700, EPI_ISL_2391701, EPI_ISL_2391702, EPI_ISL_2391703, EPI_ISL_2391704, EPI_ISL_2391705, EPI_ISL_2391706, EPI_ISL_2391707, EPI_ISL_2391708 | | | |
| see above | Laboratorio Central de Epidemiología (LCE) | Instituto de Biotecnología de la UNAM | Consorcio Mexicano de Vigilancia Genómica (CoViGen-Mex). Authors (in alphabetical order): Julio Elias Alvarado-Yaah, Carlos F. Arias, Santiago Ávila-Ríos, Eduardo Becerril-Vargas, Víctor Hugo Borja-Aburto, Celia Boukadida, Cristóbal Cháidez-Quiróz, Juan Bautista Chale-Dzul, Ricardo Ciria Merce, Andreu Comas-García, Célida Duque Molina, Julissa Enciso-Ibarra, José Antonio Enciso-Moreno, Gloria Elena Espinosa-Ayala, Fernando Fontove-Herrera, Daniel Fregoso-Rueda, Victor Eduardo Garcia-Arias, Alejandra Garcia-Gasca, Bruno Gomez-Gil, Jean Pierre González, Irvin González-López, Concepción Grajales-Muñiz, Ricardo Grande, Rosa María Gutierrez Rios, Alejandra Hernández-Terán, Alfredo Herrera-Estrella, Carla Ivón Herrera-Najera, Pavel Isa, Daniel Lira Morales, Susana Lopez, Antonio Loza Román, Brenda Irasema Maldonado-Meza, Bernardo Martínez-Miguel, José Arturo Martínez-Orozco, Célida Martínez- Rodríguez, Margarita Matías-Florentino, Fidencio Mejía-Nepomuceno, María Guadalupe de Jesús Mireles-Rivera, Gloria María Molina-Salinas, Hector Montoya-Fuentes, , Mario Mújica-Sánchez, José Esteban Muñoz-Medina, José de Jesús Nuñez-Contreras, Alicia Ocaña-Mondragón, Luis Alberto Ochoa-Carrera, Hector Esteban Paz-Juárez, Francisco Pulido, Helen Haydee Fernanda Ramírez-Plascencia, Jorge Salas-Hernández, Angel Gustavo Salas-Lais, Alejandro Sanchez-Flores, Clara Esperanza Santacruz-Tinoco, María Guadalupe Santiago-Mauricio, Selene Zárate, Nelly Sélem-Mojica, Blanca Taboada, Gloria Vazquez, Joel Armando Vázquez-Pérez. |
| EPI_ISL_2391709 | Centro de Investigación Biomédica del Noreste (CIBIN) | Instituto de Biotecnología de la UNAM | Consorcio Mexicano de Vigilancia Genómica (CoViGen-Mex). Authors (in alphabetical order): Julio Elias Alvarado-Yaah, Carlos F. Arias, Santiago Ávila-Ríos, Eduardo Becerril-Vargas, Víctor Hugo Borja-Aburto, Celia Boukadida, Cristóbal Cháidez-Quiróz, Juan Bautista Chale-Dzul, Ricardo Ciria Merce, Andreu Comas-García, Célida Duque Molina, Julissa Enciso-Ibarra, José Antonio Enciso-Moreno, Gloria Elena Espinosa-Ayala, Fernando Fontove-Herrera, Daniel Fregoso-Rueda, Victor Eduardo Garcia-Arias, Alejandra Garcia-Gasca, Bruno Gomez-Gil, Jean Pierre González, Irvin González-López, Concepción Grajales-Muñiz, Ricardo Grande, Rosa María Gutierrez Rios, Alejandra Hernández-Terán, Alfredo Herrera-Estrella, Carla Ivón Herrera-Najera, Pavel Isa, Daniel Lira Morales, Susana Lopez, Antonio Loza Román, Brenda Irasema Maldonado-Meza, Bernardo Martínez-Miguel, José Arturo Martínez-Orozco, Célida Martínez- Rodríguez, Margarita Matías-Florentino, Fidencio Mejía-Nepomuceno, María Guadalupe de Jesús Mireles-Rivera, Gloria María Molina-Salinas, Hector Montoya-Fuentes, , Mario Mújica-Sánchez, José Esteban Muñoz-Medina, José de Jesús Nuñez-Contreras, Alicia Ocaña-Mondragón, Luis Alberto Ochoa-Carrera, Hector Esteban Paz-Juárez, Francisco Pulido, Helen Haydee Fernanda Ramírez-Plascencia, Jorge Salas-Hernández, Angel Gustavo Salas-Lais, Alejandro Sanchez-Flores, Clara Esperanza Santacruz-Tinoco, |

| | | | |
|---|---|---|---|
| | | | María Guadalupe Santiago-Mauricio, Selene Zárate, Nelly Sélem-Mojica, Blanca Taboada, Gloria Vazquez, Joel Armando Vázquez-Pérez. |
| EPI_ISL_2391710, EPI_ISL_2391711 | Laboratorio Central de Epidemiología (LCE) | Instituto de Biotecnología de la UNAM | Consorcio Mexicano de Vigilancia Genómica (CoViGen-Mex). Authors (in alphabetical order): Julio Elias Alvarado-Yaah, Carlos F. Arias, Santiago Ávila-Ríos, Eduardo Becerril-Vargas, Víctor Hugo Borja-Aburto, Celia Boukadida, Cristóbal Cháidez-Quiróz, Juan Bautista Chale-Dzul, Ricardo Ciria Merce, Andreu Comas-García, Célida Duque Molina, Julissa Enciso-Ibarra, José Antonio Enciso-Moreno, Gloria Elena Espinosa-Ayala, Fernando Fontove-Herrera, Daniel Fregoso-Rueda, Victor Eduardo Garcia-Arias, Alejandra Garcia-Gasca, Bruno Gomez-Gil, Jean Pierre González, Irvin González-López, Concepción Grajales-Muñiz, Ricardo Grande, Rosa María Gutierrez Rios, Alejandra Hernández-Terán, Alfredo Herrera-Estrella, Carla Ivón Herrera-Najera, Pavel Isa, Daniel Lira Morales, Susana Lopez, Antonio Loza Román, Brenda Irasema Maldonado-Meza, Bernardo Martínez-Miguel, José Arturo Martínez-Orozco, Célida Martínez- Rodríguez, Margarita Matías-Florentino, Fidencio Mejía-Nepomuceno, María Guadalupe de Jesús Mireles-Rivera, Gloria María Molina-Salinas, Hector Montoya-Fuentes, , Mario Mújica-Sánchez, José Esteban Muñoz-Medina, José de Jesús Nuñez-Contreras, Alicia Ocaña-Mondragón, Luis Alberto Ochoa-Carrera, Hector Esteban Paz-Juárez, Francisco Pulido, Helen Haydee Fernanda Ramírez-Plascencia, Jorge Salas-Hernández, Angel Gustavo Salas-Lais, Alejandro Sanchez-Flores, Clara Esperanza Santacruz-Tinoco, María Guadalupe Santiago-Mauricio, Selene Zárate, Nelly Sélem-Mojica, Blanca Taboada, Gloria Vazquez, Joel Armando Vázquez-Pérez. |
| EPI_ISL_2391712 | Centro de Investigación Biomédica de Occidente (CIBO) | Instituto de Biotecnología de la UNAM | Consorcio Mexicano de Vigilancia Genómica (CoViGen-Mex). Authors (in alphabetical order): Julio Elias Alvarado-Yaah, Carlos F. Arias, Santiago Ávila-Ríos, Eduardo Becerril-Vargas, Víctor Hugo Borja-Aburto, Celia Boukadida, Cristóbal Cháidez-Quiróz, Juan Bautista Chale-Dzul, Ricardo Ciria Merce, Andreu Comas-García, Célida Duque Molina, Julissa Enciso-Ibarra, José Antonio Enciso-Moreno, Gloria Elena Espinosa-Ayala, Fernando Fontove-Herrera, Daniel Fregoso-Rueda, Victor Eduardo Garcia-Arias, Alejandra Garcia-Gasca, Bruno Gomez-Gil, Jean Pierre González, Irvin González-López, Concepción Grajales-Muñiz, Ricardo Grande, Rosa María Gutierrez Rios, Alejandra Hernández-Terán, Alfredo Herrera-Estrella, Carla Ivón Herrera-Najera, Pavel Isa, Daniel Lira Morales, Susana Lopez, Antonio Loza Román, Brenda Irasema Maldonado-Meza, Bernardo Martínez-Miguel, José Arturo Martínez-Orozco, Célida Martínez- Rodríguez, Margarita Matías-Florentino, Fidencio Mejía-Nepomuceno, María Guadalupe de Jesús Mireles-Rivera, Gloria María Molina-Salinas, Hector Montoya-Fuentes, , Mario Mújica-Sánchez, José Esteban Muñoz-Medina, José de Jesús Nuñez-Contreras, Alicia Ocaña-Mondragón, Luis Alberto Ochoa-Carrera, Hector Esteban Paz-Juárez, Francisco Pulido, Helen Haydee Fernanda Ramírez-Plascencia, Jorge Salas-Hernández, Angel Gustavo Salas-Lais, Alejandro Sanchez-Flores, Clara Esperanza Santacruz-Tinoco, María Guadalupe Santiago-Mauricio, Selene Zárate, Nelly Sélem-Mojica, Blanca Taboada, Gloria Vazquez, Joel Armando Vázquez-Pérez. |
| EPI_ISL_2391713, EPI_ISL_2391714, EPI_ISL_2391715, EPI_ISL_2391716, EPI_ISL_2391717, EPI_ISL_2391718 | Laboratorio Central de Epidemiología (LCE) | Instituto de Biotecnología de la UNAM | Consorcio Mexicano de Vigilancia Genómica (CoViGen-Mex). Authors (in alphabetical order): Julio Elias Alvarado-Yaah, Carlos F. Arias, Santiago Ávila-Ríos, Eduardo Becerril-Vargas, Víctor Hugo Borja-Aburto, Celia Boukadida, Cristóbal Cháidez-Quiróz, Juan Bautista Chale-Dzul, Ricardo Ciria Merce, Andreu Comas-García, Célida Duque Molina, Julissa Enciso-Ibarra, José Antonio Enciso-Moreno, Gloria Elena Espinosa-Ayala, Fernando Fontove-Herrera, Daniel Fregoso-Rueda, Victor Eduardo Garcia-Arias, Alejandra Garcia-Gasca, Bruno Gomez-Gil, Jean Pierre González, Irvin González-López, Concepción Grajales-Muñiz, Ricardo Grande, Rosa María Gutierrez Rios, Alejandra Hernández-Terán, Alfredo Herrera-Estrella, Carla Ivón Herrera-Najera, Pavel Isa, Daniel Lira Morales, Susana Lopez, Antonio Loza Román, Brenda Irasema Maldonado-Meza, Bernardo Martínez-Miguel, José Arturo Martínez-Orozco, Célida Martínez- Rodríguez, Margarita Matías-Florentino, Fidencio Mejía-Nepomuceno, María Guadalupe de Jesús Mireles-Rivera, Gloria María Molina-Salinas, Hector Montoya-Fuentes, , Mario Mújica-Sánchez, José Esteban Muñoz-Medina, José de Jesús Nuñez-Contreras, Alicia Ocaña-Mondragón, Luis Alberto Ochoa-Carrera, Hector Esteban Paz-Juárez, Francisco Pulido, Helen Haydee Fernanda Ramírez-Plascencia, Jorge Salas-Hernández, Angel Gustavo Salas-Lais, Alejandro Sanchez-Flores, Clara Esperanza Santacruz-Tinoco, María Guadalupe Santiago-Mauricio, Selene Zárate, Nelly Sélem-Mojica, Blanca Taboada, Gloria Vazquez, Joel Armando Vázquez-Pérez. |
| EPI_ISL_2391719 | Centro de Investigación Biomédica del Noreste (CIBIN) | Instituto de Biotecnología de la UNAM | Consorcio Mexicano de Vigilancia Genómica (CoViGen-Mex). Authors (in alphabetical order): Julio Elias Alvarado-Yaah, Carlos F. Arias, Santiago Ávila-Ríos, Eduardo Becerril-Vargas, Víctor Hugo Borja-Aburto, Celia Boukadida, Cristóbal Cháidez-Quiróz, Juan Bautista Chale-Dzul, Ricardo Ciria Merce, Andreu Comas-García, Célida Duque Molina, Julissa Enciso-Ibarra, José Antonio Enciso-Moreno, Gloria Elena Espinosa-Ayala, Fernando Fontove-Herrera, Daniel Fregoso-Rueda, Victor Eduardo Garcia-Arias, Alejandra Garcia-Gasca, Bruno Gomez-Gil, Jean Pierre González, Irvin González-López, Concepción Grajales-Muñiz, Ricardo Grande, Rosa María Gutierrez Rios, Alejandra Hernández-Terán, Alfredo Herrera-Estrella, Carla Ivón Herrera-Najera, Pavel Isa, Daniel Lira Morales, Susana Lopez, Antonio Loza Román, Brenda Irasema Maldonado-Meza, Bernardo Martínez-Miguel, José Arturo Martínez-Orozco, Célida Martínez- Rodríguez, Margarita Matías-Florentino, Fidencio Mejía-Nepomuceno, María Guadalupe de Jesús Mireles-Rivera, Gloria María Molina-Salinas, Hector Montoya-Fuentes, , Mario Mújica-Sánchez, José Esteban Muñoz-Medina, José de Jesús Nuñez-Contreras, Alicia Ocaña-Mondragón, Luis Alberto Ochoa-Carrera, Hector Esteban Paz-Juárez, Francisco Pulido, Helen Haydee Fernanda Ramírez-Plascencia, Jorge Salas-Hernández, Angel Gustavo Salas-Lais, Alejandro Sanchez-Flores, Clara Esperanza Santacruz-Tinoco, María Guadalupe Santiago-Mauricio, Selene Zárate, Nelly Sélem-Mojica, Blanca Taboada, Gloria Vazquez, Joel Armando Vázquez-Pérez. |
| EPI_ISL_2391720 | Unidad de Investigación Médica de Yucatán (UIMY) | Instituto de Biotecnología de la UNAM | Consorcio Mexicano de Vigilancia Genómica (CoViGen-Mex). Authors (in alphabetical order): Julio Elias Alvarado-Yaah, Carlos F. Arias, Santiago Ávila-Ríos, Eduardo Becerril-Vargas, Víctor Hugo Borja-Aburto, Celia Boukadida, Cristóbal Cháidez-Quiróz, Juan Bautista Chale-Dzul, Ricardo Ciria Merce, Andreu Comas-García, Célida Duque Molina, Julissa Enciso-Ibarra, José Antonio Enciso-Moreno, Gloria Elena Espinosa-Ayala, Fernando Fontove-Herrera, Daniel Fregoso-Rueda, Victor Eduardo Garcia-Arias, Alejandra Garcia-Gasca, Bruno Gomez-Gil, Jean Pierre González, Irvin González-López, Concepción Grajales-Muñiz, Ricardo Grande, Rosa María Gutierrez Rios, Alejandra Hernández-Terán, Alfredo Herrera-Estrella, Carla Ivón Herrera-Najera, Pavel Isa, Daniel Lira Morales, Susana Lopez, Antonio Loza Román, Brenda Irasema Maldonado-Meza, Bernardo Martínez-Miguel, José Arturo Martínez-Orozco, Célida Martínez- Rodríguez, Margarita Matías-Florentino, Fidencio Mejía-Nepomuceno, María Guadalupe de Jesús Mireles-Rivera, Gloria María Molina-Salinas, Hector Montoya-Fuentes, , Mario Mújica-Sánchez, José Esteban Muñoz-Medina, José de Jesús Nuñez-Contreras, Alicia Ocaña-Mondragón, Luis Alberto Ochoa-Carrera, Hector Esteban Paz-Juárez, Francisco Pulido, Helen Haydee Fernanda Ramírez-Plascencia, Jorge Salas-Hernández, Angel Gustavo Salas-Lais, Alejandro Sanchez-Flores, Clara Esperanza Santacruz-Tinoco, María Guadalupe Santiago-Mauricio, Selene Zárate, Nelly Sélem-Mojica, Blanca Taboada, Gloria Vazquez, Joel Armando Vázquez-Pérez. |
| EPI_ISL_2391721, EPI_ISL_2391722, EPI_ISL_2391723, EPI_ISL_2391724, EPI_ISL_2391725, EPI_ISL_2391726 | Laboratorio Central de Epidemiología (LCE) | Instituto de Biotecnología de la UNAM | Consorcio Mexicano de Vigilancia Genómica (CoViGen-Mex). Authors (in alphabetical order): Julio Elias Alvarado-Yaah, Carlos F. Arias, Santiago Ávila-Ríos, Eduardo Becerril-Vargas, Víctor Hugo Borja-Aburto, Celia Boukadida, Cristóbal Cháidez-Quiróz, Juan Bautista Chale-Dzul, Ricardo Ciria Merce, Andreu Comas-García, Célida Duque Molina, Julissa Enciso-Ibarra, José Antonio Enciso-Moreno, Gloria Elena Espinosa-Ayala, Fernando Fontove-Herrera, Daniel Fregoso-Rueda, Victor Eduardo Garcia-Arias, Alejandra Garcia-Gasca, Bruno Gomez-Gil, Jean Pierre González, Irvin González-López, Concepción Grajales-Muñiz, Ricardo Grande, Rosa María Gutierrez Rios, Alejandra Hernández-Terán, Alfredo Herrera-Estrella, Carla Ivón Herrera-Najera, Pavel Isa, Daniel Lira Morales, Susana Lopez, Antonio Loza Román, Brenda Irasema Maldonado-Meza, Bernardo Martínez-Miguel, José Arturo Martínez-Orozco, Célida Martínez- Rodríguez, Margarita Matías-Florentino, Fidencio Mejía-Nepomuceno, María Guadalupe de Jesús Mireles-Rivera, Gloria María Molina-Salinas, Hector Montoya-Fuentes, , Mario Mújica-Sánchez, José Esteban Muñoz-Medina, José de Jesús Nuñez-Contreras, Alicia Ocaña-Mondragón, Luis Alberto Ochoa-Carrera, Hector Esteban Paz-Juárez, Francisco Pulido, Helen Haydee Fernanda Ramírez-Plascencia, Jorge Salas-Hernández, Angel Gustavo Salas-Lais, Alejandro Sanchez-Flores, Clara Esperanza Santacruz-Tinoco, María Guadalupe Santiago-Mauricio, Selene Zárate, Nelly Sélem-Mojica, Blanca Taboada, Gloria Vazquez, Joel Armando Vázquez-Pérez. |
| EPI_ISL_2391727 | Centro de Investigación Biomédica del Noreste (CIBIN) | Instituto de Biotecnología de la UNAM | Consorcio Mexicano de Vigilancia Genómica (CoViGen-Mex). Authors (in alphabetical order): Julio Elias Alvarado-Yaah, Carlos F. Arias, Santiago Ávila-Ríos, Eduardo Becerril-Vargas, Víctor Hugo Borja-Aburto, Celia Boukadida, Cristóbal Cháidez-Quiróz, Juan Bautista Chale-Dzul, Ricardo Ciria Merce, Andreu Comas-García, Célida Duque Molina, Julissa Enciso-Ibarra, José Antonio Enciso-Moreno, Gloria Elena Espinosa-Ayala, Fernando Fontove-Herrera, Daniel Fregoso-Rueda, Victor Eduardo Garcia-Arias, Alejandra Garcia-Gasca, Bruno Gomez-Gil, Jean Pierre González, Irvin González-López, Concepción Grajales-Muñiz, Ricardo Grande, Rosa María Gutierrez Rios, Alejandra Hernández-Terán, Alfredo Herrera-Estrella, Carla Ivón Herrera-Najera, Pavel Isa, Daniel Lira Morales, Susana Lopez, Antonio Loza Román, Brenda Irasema Maldonado-Meza, Bernardo Martínez-Miguel, José Arturo Martínez-Orozco, Célida Martínez- Rodríguez, Margarita Matías-Florentino, Fidencio Mejía-Nepomuceno, María Guadalupe de Jesús Mireles-Rivera, Gloria María Molina-Salinas, Hector Montoya-Fuentes, , Mario Mújica-Sánchez, José Esteban Muñoz-Medina, José de Jesús Nuñez-Contreras, Alicia Ocaña-Mondragón, Luis Alberto Ochoa-Carrera, Hector Esteban Paz-Juárez, Francisco Pulido, Helen Haydee Fernanda Ramírez-Plascencia, Jorge Salas-Hernández, Angel Gustavo Salas-Lais, Alejandro Sanchez-Flores, Clara Esperanza Santacruz-Tinoco, María Guadalupe Santiago-Mauricio, Selene Zárate, Nelly Sélem-Mojica, Blanca Taboada, Gloria Vazquez, Joel Armando Vázquez-Pérez. |
| EPI_ISL_2391728 | Unidad de Investigación Médica de Yucatán (UIMY) | Instituto de Biotecnología de la UNAM | Consorcio Mexicano de Vigilancia Genómica (CoViGen-Mex). Authors (in alphabetical order): Julio Elias Alvarado-Yaah, Carlos F. Arias, Santiago Ávila-Ríos, Eduardo Becerril-Vargas, Víctor Hugo Borja-Aburto, Celia Boukadida, Cristóbal Cháidez-Quiróz, Juan Bautista Chale-Dzul, Ricardo Ciria Merce, Andreu Comas-García, Célida Duque Molina, Julissa Enciso-Ibarra, José Antonio Enciso-Moreno, Gloria Elena Espinosa-Ayala, Fernando Fontove-Herrera, Daniel Fregoso-Rueda, Victor Eduardo Garcia-Arias, Alejandra Garcia-Gasca, Bruno Gomez-Gil, Jean Pierre González, Irvin González-López, Concepción Grajales-Muñiz, Ricardo Grande, Rosa María Gutierrez Rios, Alejandra Hernández-Terán, Alfredo Herrera-Estrella, Carla Ivón Herrera-Najera, Pavel Isa, Daniel Lira Morales, Susana Lopez, Antonio Loza Román, Brenda Irasema Maldonado-Meza, Bernardo Martínez-Miguel, José Arturo Martínez-Orozco, Célida Martínez- Rodríguez, Margarita Matías-Florentino, Fidencio Mejía-Nepomuceno, María Guadalupe de Jesús Mireles-Rivera, Gloria María Molina-Salinas, Hector Montoya-Fuentes, , Mario Mújica-Sánchez, José Esteban Muñoz-Medina, José de Jesús Nuñez-Contreras, Alicia Ocaña-Mondragón, Luis Alberto Ochoa-Carrera, Hector Esteban Paz-Juárez, Francisco Pulido, Helen Haydee Fernanda Ramírez-Plascencia, Jorge Salas-Hernández, Angel Gustavo Salas-Lais, Alejandro Sanchez-Flores, Clara Esperanza Santacruz-Tinoco, María Guadalupe Santiago-Mauricio, Selene Zárate, Nelly Sélem-Mojica, Blanca Taboada, Gloria Vazquez, Joel Armando Vázquez-Pérez. |
| EPI_ISL_2391729, EPI_ISL_2391730, EPI_ISL_2391731 | Laboratorio Central de Epidemiología (LCE) | Instituto de Biotecnología de la UNAM | Consorcio Mexicano de Vigilancia Genómica (CoViGen-Mex). Authors (in alphabetical order): Julio Elias Alvarado-Yaah, Carlos F. Arias, Santiago Ávila-Ríos, Eduardo Becerril-Vargas, Víctor Hugo Borja-Aburto, Celia Boukadida, Cristóbal Cháidez-Quiróz, Juan Bautista Chale-Dzul, Ricardo Ciria Merce, Andreu Comas-García, Célida Duque Molina, Julissa Enciso-Ibarra, José Antonio Enciso-Moreno, Gloria Elena Espinosa-Ayala, Fernando Fontove-Herrera, Daniel Fregoso-Rueda, Victor Eduardo Garcia-Arias, Alejandra Garcia-Gasca, Bruno Gomez-Gil, Jean Pierre González, Irvin González-López, Concepción Grajales-Muñiz, Ricardo Grande, Rosa María Gutierrez Rios, Alejandra Hernández-Terán, Alfredo Herrera-Estrella, Carla Ivón Herrera-Najera, Pavel Isa, Daniel Lira Morales, Susana Lopez, Antonio Loza Román, Brenda Irasema Maldonado-Meza, Bernardo Martínez-Miguel, José Arturo Martínez-Orozco, Célida Martínez- Rodríguez, Margarita Matías-Florentino, Fidencio Mejía-Nepomuceno, María Guadalupe de Jesús Mireles-Rivera, Gloria María Molina-Salinas, Hector Montoya-Fuentes, , Mario Mújica-Sánchez, José Esteban Muñoz-Medina, José de Jesús Nuñez-Contreras, Alicia Ocaña-Mondragón, Luis Alberto Ochoa-Carrera, Hector Esteban Paz-Juárez, Francisco Pulido, Helen Haydee Fernanda Ramírez-Plascencia, Jorge Salas-Hernández, Angel Gustavo Salas-Lais, Alejandro Sanchez-Flores, Clara Esperanza Santacruz-Tinoco, María Guadalupe Santiago-Mauricio, Selene Zárate, Nelly Sélem-Mojica, Blanca Taboada, Gloria Vazquez, Joel Armando Vázquez-Pérez. |
| EPI_ISL_2391732 | Centro de Investigación Biomédica del Noreste (CIBIN) | Instituto de Biotecnología de la UNAM | Consorcio Mexicano de Vigilancia Genómica (CoViGen-Mex). Authors (in alphabetical order): Julio Elias Alvarado-Yaah, Carlos F. Arias, Santiago Ávila-Ríos, Eduardo Becerril-Vargas, Víctor Hugo Borja-Aburto, Celia Boukadida, Cristóbal Cháidez-Quiróz, Juan Bautista Chale-Dzul, Ricardo Ciria Merce, Andreu Comas-García, Célida Duque Molina, Julissa Enciso-Ibarra, José Antonio Enciso-Moreno, Gloria Elena Espinosa-Ayala, Fernando Fontove-Herrera, Daniel Fregoso-Rueda, Victor Eduardo Garcia-Arias, Alejandra Garcia-Gasca, Bruno Gomez-Gil, Jean Pierre González, Irvin González-López, Concepción Grajales-Muñiz, Ricardo Grande, Rosa María Gutierrez Rios, Alejandra Hernández-Terán, Alfredo Herrera-Estrella, Carla Ivón Herrera-Najera, Pavel Isa, Daniel Lira Morales, Susana Lopez, Antonio Loza Román, Brenda Irasema Maldonado-Meza, Bernardo Martínez-Miguel, José Arturo Martínez-Orozco, Célida Martínez- Rodríguez, Margarita Matías-Florentino, Fidencio Mejía-Nepomuceno, María Guadalupe de Jesús Mireles-Rivera, Gloria María Molina-Salinas, Hector Montoya-Fuentes, , Mario Mújica-Sánchez, José Esteban Muñoz-Medina, José de Jesús Nuñez-Contreras, Alicia Ocaña-Mondragón, Luis Alberto Ochoa-Carrera, Hector Esteban Paz-Juárez, Francisco Pulido, Helen Haydee Fernanda Ramírez-Plascencia, Jorge Salas-Hernández, Angel Gustavo Salas-Lais, Alejandro Sanchez-Flores, Clara Esperanza Santacruz-Tinoco, María Guadalupe Santiago-Mauricio, Selene Zárate, Nelly Sélem-Mojica, Blanca Taboada, Gloria Vazquez, Joel Armando Vázquez-Pérez. |
| EPI_ISL_2391733 | Unidad de Investigación Médica de Yucatán (UIMY) | Instituto de Biotecnología de la UNAM | Consorcio Mexicano de Vigilancia Genómica (CoViGen-Mex). Authors (in alphabetical order): Julio Elias Alvarado-Yaah, Carlos F. Arias, Santiago Ávila-Ríos, Eduardo Becerril-Vargas, Víctor Hugo Borja-Aburto, Celia Boukadida, Cristóbal Cháidez-Quiróz, Juan Bautista Chale-Dzul, Ricardo Ciria Merce, Andreu Comas-García, Célida Duque Molina, Julissa Enciso-Ibarra, José Antonio Enciso-Moreno, Gloria Elena Espinosa-Ayala, Fernando Fontove-Herrera, Daniel Fregoso-Rueda, Victor Eduardo Garcia-Arias, Alejandra Garcia-Gasca, Bruno Gomez-Gil, Jean Pierre González, Irvin González-López, Concepción Grajales-Muñiz, Ricardo Grande, Rosa María Gutierrez Rios, Alejandra Hernández-Terán, Alfredo Herrera-Estrella, Carla Ivón Herrera-Najera, Pavel Isa, Daniel Lira Morales, Susana Lopez, Antonio Loza Román, Brenda Irasema Maldonado-Meza, Bernardo Martínez-Miguel, José Arturo Martínez-Orozco, Célida Martínez- Rodríguez, Margarita Matías-Florentino, Fidencio Mejía-Nepomuceno, María Guadalupe de Jesús Mireles-Rivera, Gloria María Molina-Salinas, Hector Montoya-Fuentes, , Mario Mújica-Sánchez, José Esteban Muñoz-Medina, José de Jesús Nuñez-Contreras, Alicia Ocaña-Mondragón, Luis Alberto Ochoa-Carrera, Hector Esteban Paz-Juárez, Francisco Pulido, Helen Haydee Fernanda Ramírez-Plascencia, Jorge Salas-Hernández, Angel Gustavo Salas-Lais, Alejandro Sanchez-Flores, Clara Esperanza Santacruz-Tinoco, María Guadalupe Santiago-Mauricio, Selene Zárate, Nelly Sélem-Mojica, Blanca Taboada, Gloria Vazquez, Joel Armando Vázquez-Pérez. |
| EPI_ISL_2391734 | Laboratorio Central de Epidemiología (LCE) | Instituto de Biotecnología de la UNAM | Consorcio Mexicano de Vigilancia Genómica (CoViGen-Mex). Authors (in alphabetical order): Julio Elias Alvarado-Yaah, Carlos F. Arias, Santiago Ávila-Ríos, Eduardo Becerril-Vargas, Víctor Hugo Borja-Aburto, Celia Boukadida, Cristóbal Cháidez-Quiróz, Juan Bautista Chale-Dzul, Ricardo Ciria Merce, Andreu Comas-García, Célida Duque Molina, Julissa Enciso-Ibarra, José Antonio Enciso-Moreno, Gloria Elena Espinosa-Ayala, Fernando Fontove-Herrera, Daniel Fregoso-Rueda, Victor Eduardo Garcia-Arias, Alejandra Garcia-Gasca, Bruno Gomez-Gil, Jean Pierre González, Irvin González-López, Concepción Grajales-Muñiz, Ricardo Grande, Rosa María Gutierrez Rios, Alejandra Hernández-Terán, Alfredo Herrera-Estrella, Carla Ivón Herrera-Najera, Pavel Isa, Daniel Lira Morales, Susana Lopez, Antonio Loza Román, Brenda Irasema Maldonado-Meza, Bernardo Martínez-Miguel, José Arturo Martínez-Orozco, Célida Martínez- Rodríguez, Margarita Matías-Florentino, Fidencio Mejía-Nepomuceno, María Guadalupe de Jesús Mireles-Rivera, Gloria María Molina-Salinas, Hector Montoya-Fuentes, , Mario Mújica-Sánchez, José Esteban Muñoz-Medina, José de Jesús Nuñez-Contreras, Alicia Ocaña-Mondragón, Luis Alberto Ochoa-Carrera, Hector Esteban Paz-Juárez, Francisco Pulido, Helen Haydee Fernanda Ramírez-Plascencia, Jorge Salas-Hernández, Angel Gustavo Salas-Lais, Alejandro Sanchez-Flores, Clara Esperanza Santacruz-Tinoco, María Guadalupe Santiago-Mauricio, Selene Zárate, Nelly Sélem-Mojica, Blanca Taboada, Gloria Vazquez, Joel Armando Vázquez-Pérez. |
| EPI_ISL_2391735 | Centro de Investigación Biomédica de Occidente (CIBO) | Instituto de Biotecnología de la UNAM | Consorcio Mexicano de Vigilancia Genómica (CoViGen-Mex). Authors (in alphabetical order): Julio Elias Alvarado-Yaah, Carlos F. Arias, Santiago Ávila-Ríos, Eduardo Becerril-Vargas, Víctor Hugo Borja-Aburto, Celia Boukadida, Cristóbal Cháidez-Quiróz, Juan Bautista Chale-Dzul, Ricardo Ciria Merce, Andreu Comas-García, Célida Duque Molina, Julissa Enciso-Ibarra, José Antonio Enciso-Moreno, Gloria Elena Espinosa-Ayala, Fernando Fontove-Herrera, Daniel Fregoso-Rueda, Victor Eduardo Garcia-Arias, Alejandra Garcia-Gasca, Bruno Gomez-Gil, Jean Pierre González, Irvin González-López, Concepción Grajales-Muñiz, Ricardo Grande, Rosa María Gutierrez Rios, Alejandra Hernández-Terán, Alfredo Herrera-Estrella, Carla Ivón Herrera-Najera, Pavel Isa, Daniel Lira Morales, Susana Lopez, Antonio Loza Román, Brenda Irasema Maldonado-Meza, Bernardo Martínez-Miguel, José Arturo Martínez-Orozco, Célida Martínez- Rodríguez, Margarita Matías-Florentino, Fidencio Mejía-Nepomuceno, María Guadalupe de Jesús Mireles-Rivera, Gloria María Molina-Salinas, Hector Montoya-Fuentes, , Mario Mújica-Sánchez, José Esteban Muñoz-Medina, José de Jesús Nuñez-Contreras, Alicia Ocaña-Mondragón, Luis Alberto Ochoa-Carrera, Hector Esteban Paz-Juárez, Francisco Pulido, Helen Haydee Fernanda Ramírez-Plascencia, Jorge Salas-Hernández, Angel Gustavo Salas-Lais, Alejandro Sanchez-Flores, Clara Esperanza Santacruz-Tinoco, María Guadalupe Santiago-Mauricio, Selene Zárate, Nelly Sélem-Mojica, Blanca Taboada, Gloria Vazquez, Joel Armando Vázquez-Pérez. |
| EPI_ISL_2391736 | Unidad de Investigación Biomédica de Zacatecas (UIBZ) | Instituto de Biotecnología de la UNAM | Consorcio Mexicano de Vigilancia Genómica (CoViGen-Mex). Authors (in alphabetical order): Julio Elias Alvarado-Yaah, Carlos F. Arias, Santiago Ávila-Ríos, Eduardo Becerril-Vargas, Víctor Hugo Borja-Aburto, Celia Boukadida, Cristóbal Cháidez-Quiróz, Juan Bautista Chale-Dzul, Ricardo Ciria Merce, Andreu Comas-García, Célida Duque Molina, Julissa Enciso-Ibarra, José Antonio Enciso-Moreno, Gloria Elena Espinosa-Ayala, Fernando Fontove-Herrera, Daniel Fregoso-Rueda, Victor Eduardo Garcia-Arias, Alejandra Garcia-Gasca, Bruno Gomez-Gil, Jean Pierre González, Irvin González-López, Concepción Grajales-Muñiz, Ricardo Grande, Rosa María Gutierrez Rios, Alejandra Hernández-Terán, Alfredo Herrera-Estrella, Carla Ivón Herrera-Najera, Pavel Isa, Daniel Lira Morales, Susana Lopez, Antonio Loza Román, Brenda Irasema Maldonado-Meza, Bernardo Martínez-Miguel, José Arturo Martínez-Orozco, Célida Martínez- Rodríguez, Margarita Matías-Florentino, Fidencio Mejía-Nepomuceno, María Guadalupe de Jesús Mireles-Rivera, Gloria María Molina-Salinas, Hector Montoya-Fuentes, , Mario Mújica-Sánchez, José Esteban Muñoz-Medina, José de Jesús Nuñez-Contreras, Alicia Ocaña-Mondragón, Luis Alberto Ochoa-Carrera, Hector Esteban Paz-Juárez, Francisco Pulido, Helen Haydee Fernanda Ramírez-Plascencia, Jorge Salas-Hernández, Angel Gustavo Salas-Lais, Alejandro Sanchez-Flores, Clara Esperanza Santacruz-Tinoco, María Guadalupe Santiago-Mauricio, Selene Zárate, Nelly Sélem-Mojica, Blanca Taboada, Gloria Vazquez, Joel Armando Vázquez-Pérez. |
| EPI_ISL_2391737 | Laboratorio Central de Epidemiología (LCE) | Instituto de Biotecnología de la UNAM | Consorcio Mexicano de Vigilancia Genómica (CoViGen-Mex). Authors (in alphabetical order): Julio Elias Alvarado-Yaah, Carlos F. Arias, Santiago Ávila-Ríos, Eduardo Becerril-Vargas, Víctor Hugo Borja-Aburto, Celia Boukadida, Cristóbal Cháidez-Quiróz, Juan Bautista Chale-Dzul, Ricardo Ciria Merce, Andreu Comas-García, Célida Duque Molina, Julissa Enciso-Ibarra, José Antonio Enciso-Moreno, Gloria Elena Espinosa-Ayala, Fernando Fontove-Herrera, Daniel Fregoso-Rueda, Victor Eduardo Garcia-Arias, Alejandra Garcia-Gasca, Bruno Gomez-Gil, Jean Pierre González, Irvin González-López, Concepción Grajales-Muñiz, Ricardo Grande, Rosa María Gutierrez Rios, Alejandra Hernández-Terán, Alfredo Herrera-Estrella, Carla Ivón Herrera-Najera, Pavel Isa, Daniel Lira Morales, Susana Lopez, Antonio Loza Román, Brenda Irasema Maldonado-Meza, Bernardo Martínez-Miguel, José Arturo Martínez-Orozco, Célida Martínez- Rodríguez, Margarita Matías-Florentino, Fidencio Mejía-Nepomuceno, María Guadalupe de Jesús Mireles-Rivera, Gloria María Molina-Salinas, Hector Montoya-Fuentes, , Mario Mújica-Sánchez, José Esteban Muñoz-Medina, José de Jesús Nuñez-Contreras, Alicia Ocaña-Mondragón, Luis Alberto Ochoa-Carrera, Hector Esteban Paz-Juárez, Francisco Pulido, Helen Haydee Fernanda Ramírez-Plascencia, Jorge Salas-Hernández, Angel Gustavo Salas-Lais, Alejandro Sanchez-Flores, Clara Esperanza Santacruz-Tinoco, María Guadalupe Santiago-Mauricio, Selene Zárate, Nelly Sélem-Mojica, Blanca Taboada, Gloria Vazquez, Joel Armando Vázquez-Pérez. |
| EPI_ISL_2391738 | Centro de Investigación Biomédica de Occidente (CIBO) | Instituto de Biotecnología de la UNAM | Consorcio Mexicano de Vigilancia Genómica (CoViGen-Mex). Authors (in alphabetical order): Julio Elias Alvarado-Yaah, Carlos F. Arias, Santiago Ávila-Ríos, Eduardo Becerril-Vargas, Víctor Hugo Borja-Aburto, Celia Boukadida, Cristóbal Cháidez-Quiróz, Juan Bautista Chale-Dzul, Ricardo Ciria Merce, Andreu Comas-García, Célida Duque Molina, Julissa Enciso-Ibarra, José Antonio Enciso-Moreno, Gloria Elena Espinosa-Ayala, Fernando Fontove-Herrera, Daniel Fregoso-Rueda, Victor Eduardo Garcia-Arias, Alejandra Garcia-Gasca, Bruno Gomez-Gil, Jean Pierre González, Irvin González-López, Concepción Grajales-Muñiz, Ricardo Grande, Rosa María Gutierrez Rios, Alejandra Hernández-Terán, Alfredo Herrera-Estrella, Carla Ivón Herrera-Najera, Pavel Isa, Daniel Lira Morales, Susana Lopez, Antonio Loza Román, Brenda Irasema Maldonado-Meza, Bernardo Martínez-Miguel, José Arturo Martínez-Orozco, Célida Martínez- Rodríguez, Margarita Matías-Florentino, Fidencio Mejía-Nepomuceno, María Guadalupe de Jesús Mireles-Rivera, Gloria María Molina-Salinas, Hector Montoya-Fuentes, , Mario Mújica-Sánchez, José Esteban Muñoz-Medina, José de Jesús Nuñez-Contreras, Alicia Ocaña-Mondragón, Luis Alberto Ochoa-Carrera, Hector Esteban Paz-Juárez, Francisco Pulido, Helen Haydee Fernanda Ramírez-Plascencia, Jorge Salas-Hernández, Angel Gustavo Salas-Lais, Alejandro Sanchez-Flores, Clara Esperanza Santacruz-Tinoco, María Guadalupe Santiago-Mauricio, Selene Zárate, Nelly Sélem-Mojica, Blanca Taboada, Gloria Vazquez, Joel Armando Vázquez-Pérez. |
| EPI_ISL_2391739 | Centro de Investigación Biomédica del Noreste (CIBIN) | Instituto de Biotecnología de la UNAM | Consorcio Mexicano de Vigilancia Genómica (CoViGen-Mex). Authors (in alphabetical order): Julio Elias Alvarado-Yaah, Carlos F. Arias, Santiago Ávila-Ríos, Eduardo Becerril-Vargas, Víctor Hugo Borja-Aburto, Celia Boukadida, Cristóbal Cháidez-Quiróz, Juan Bautista Chale-Dzul, Ricardo Ciria Merce, Andreu Comas-García, Célida Duque Molina, Julissa Enciso-Ibarra, José Antonio Enciso-Moreno, Gloria Elena Espinosa-Ayala, Fernando Fontove-Herrera, Daniel Fregoso-Rueda, Victor Eduardo Garcia-Arias, Alejandra Garcia-Gasca, Bruno Gomez-Gil, Jean Pierre González, Irvin González-López, Concepción Grajales-Muñiz, Ricardo Grande, Rosa María Gutierrez Rios, Alejandra Hernández-Terán, Alfredo Herrera-Estrella, Carla Ivón Herrera-Najera, Pavel Isa, Daniel Lira Morales, Susana Lopez, Antonio Loza Román, Brenda Irasema Maldonado-Meza, Bernardo Martínez-Miguel, José Arturo Martínez-Orozco, Célida Martínez- Rodríguez, Margarita Matías-Florentino, Fidencio Mejía-Nepomuceno, María Guadalupe de Jesús Mireles-Rivera, Gloria María Molina-Salinas, Hector Montoya-Fuentes, , Mario Mújica-Sánchez, José Esteban Muñoz-Medina, José de Jesús Nuñez-Contreras, Alicia Ocaña-Mondragón, Luis Alberto Ochoa-Carrera, Hector Esteban Paz-Juárez, Francisco Pulido, Helen Haydee Fernanda Ramírez-Plascencia, Jorge Salas-Hernández, Angel Gustavo Salas-Lais, Alejandro Sanchez-Flores, Clara Esperanza Santacruz-Tinoco, |

| | | | |
|---|---|---|---|
| | | | María Guadalupe Santiago-Mauricio, Selene Zárate, Nelly Sélem-Mojica, Blanca Taboada, Gloria Vazquez, Joel Armando Vázquez-Pérez. |
| EPI_ISL_2391740, EPI_ISL_2391741, EPI_ISL_2391742, EPI_ISL_2391743, EPI_ISL_2391744 | Laboratorio Central de Epidemiología (LCE) | Instituto de Biotecnología de la UNAM | Consorcio Mexicano de Vigilancia Genómica (CoViGen-Mex). Authors (in alphabetical order): Julio Elias Alvarado-Yaah, Carlos F. Arias, Santiago Ávila-Ríos, Eduardo Becerril-Vargas, Víctor Hugo Borja-Aburto, Celia Boukadida, Cristóbal Cháidez-Quiróz, Juan Bautista Chale-Dzul, Ricardo Ciria Merce, Andreu Comas-García, Célida Duque Molina, Julissa Enciso-Ibarra, José Antonio Enciso-Moreno, Gloria Elena Espinosa-Ayala, Fernando Fontove-Herrera, Daniel Fregoso-Rueda, Victor Eduardo Garcia-Arias, Alejandra Garcia-Gasca, Bruno Gomez-Gil, Jean Pierre González, Irvin González-López, Concepción Grajales-Muñiz, Ricardo Grande, Rosa María Gutierrez Rios, Alejandra Hernández-Terán, Alfredo Herrera-Estrella, Carla Ivón Herrera-Najera, Pavel Isa, Daniel Lira Morales, Susana Lopez, Antonio Loza Román, Brenda Irasema Maldonado-Meza, Bernardo Martínez-Miguel, José Arturo Martínez-Orozco, Célida Martínez- Rodríguez, Margarita Matías-Florentino, Fidencio Mejía-Nepomuceno, María Guadalupe de Jesús Mireles-Rivera, Gloria María Molina-Salinas, Hector Montoya-Fuentes, , Mario Mújica-Sánchez, José Esteban Muñoz-Medina, José de Jesús Nuñez-Contreras, Alicia Ocaña-Mondragón, Luis Alberto Ochoa-Carrera, Hector Esteban Paz-Juárez, Francisco Pulido, Helen Haydee Fernanda Ramírez-Plascencia, Jorge Salas-Hernández, Angel Gustavo Salas-Lais, Alejandro Sanchez-Flores, Clara Esperanza Santacruz-Tinoco, María Guadalupe Santiago-Mauricio, Selene Zárate, Nelly Sélem-Mojica, Blanca Taboada, Gloria Vazquez, Joel Armando Vázquez-Pérez. |
| EPI_ISL_2391745 | Unidad de Investigación Biomédica de Zacatecas (UIBZ) | Instituto de Biotecnología de la UNAM | Consorcio Mexicano de Vigilancia Genómica (CoViGen-Mex). Authors (in alphabetical order): Julio Elias Alvarado-Yaah, Carlos F. Arias, Santiago Ávila-Ríos, Eduardo Becerril-Vargas, Víctor Hugo Borja-Aburto, Celia Boukadida, Cristóbal Cháidez-Quiróz, Juan Bautista Chale-Dzul, Ricardo Ciria Merce, Andreu Comas-García, Célida Duque Molina, Julissa Enciso-Ibarra, José Antonio Enciso-Moreno, Gloria Elena Espinosa-Ayala, Fernando Fontove-Herrera, Daniel Fregoso-Rueda, Victor Eduardo Garcia-Arias, Alejandra Garcia-Gasca, Bruno Gomez-Gil, Jean Pierre González, Irvin González-López, Concepción Grajales-Muñiz, Ricardo Grande, Rosa María Gutierrez Rios, Alejandra Hernández-Terán, Alfredo Herrera-Estrella, Carla Ivón Herrera-Najera, Pavel Isa, Daniel Lira Morales, Susana Lopez, Antonio Loza Román, Brenda Irasema Maldonado-Meza, Bernardo Martínez-Miguel, José Arturo Martínez-Orozco, Célida Martínez- Rodríguez, Margarita Matías-Florentino, Fidencio Mejía-Nepomuceno, María Guadalupe de Jesús Mireles-Rivera, Gloria María Molina-Salinas, Hector Montoya-Fuentes, , Mario Mújica-Sánchez, José Esteban Muñoz-Medina, José de Jesús Nuñez-Contreras, Alicia Ocaña-Mondragón, Luis Alberto Ochoa-Carrera, Hector Esteban Paz-Juárez, Francisco Pulido, Helen Haydee Fernanda Ramírez-Plascencia, Jorge Salas-Hernández, Angel Gustavo Salas-Lais, Alejandro Sanchez-Flores, Clara Esperanza Santacruz-Tinoco, María Guadalupe Santiago-Mauricio, Selene Zárate, Nelly Sélem-Mojica, Blanca Taboada, Gloria Vazquez, Joel Armando Vázquez-Pérez. |
| EPI_ISL_2391746 | Centro de Investigación Biomédica del Noreste (CIBIN) | Instituto de Biotecnología de la UNAM | Consorcio Mexicano de Vigilancia Genómica (CoViGen-Mex). Authors (in alphabetical order): Julio Elias Alvarado-Yaah, Carlos F. Arias, Santiago Ávila-Ríos, Eduardo Becerril-Vargas, Víctor Hugo Borja-Aburto, Celia Boukadida, Cristóbal Cháidez-Quiróz, Juan Bautista Chale-Dzul, Ricardo Ciria Merce, Andreu Comas-García, Célida Duque Molina, Julissa Enciso-Ibarra, José Antonio Enciso-Moreno, Gloria Elena Espinosa-Ayala, Fernando Fontove-Herrera, Daniel Fregoso-Rueda, Victor Eduardo Garcia-Arias, Alejandra Garcia-Gasca, Bruno Gomez-Gil, Jean Pierre González, Irvin González-López, Concepción Grajales-Muñiz, Ricardo Grande, Rosa María Gutierrez Rios, Alejandra Hernández-Terán, Alfredo Herrera-Estrella, Carla Ivón Herrera-Najera, Pavel Isa, Daniel Lira Morales, Susana Lopez, Antonio Loza Román, Brenda Irasema Maldonado-Meza, Bernardo Martínez-Miguel, José Arturo Martínez-Orozco, Célida Martínez- Rodríguez, Margarita Matías-Florentino, Fidencio Mejía-Nepomuceno, María Guadalupe de Jesús Mireles-Rivera, Gloria María Molina-Salinas, Hector Montoya-Fuentes, , Mario Mújica-Sánchez, José Esteban Muñoz-Medina, José de Jesús Nuñez-Contreras, Alicia Ocaña-Mondragón, Luis Alberto Ochoa-Carrera, Hector Esteban Paz-Juárez, Francisco Pulido, Helen Haydee Fernanda Ramírez-Plascencia, Jorge Salas-Hernández, Angel Gustavo Salas-Lais, Alejandro Sanchez-Flores, Clara Esperanza Santacruz-Tinoco, María Guadalupe Santiago-Mauricio, Selene Zárate, Nelly Sélem-Mojica, Blanca Taboada, Gloria Vazquez, Joel Armando Vázquez-Pérez. |
| EPI_ISL_2391747, EPI_ISL_2391748 | Laboratorio Central de Epidemiología (LCE) | Instituto de Biotecnología de la UNAM | Consorcio Mexicano de Vigilancia Genómica (CoViGen-Mex). Authors (in alphabetical order): Julio Elias Alvarado-Yaah, Carlos F. Arias, Santiago Ávila-Ríos, Eduardo Becerril-Vargas, Víctor Hugo Borja-Aburto, Celia Boukadida, Cristóbal Cháidez-Quiróz, Juan Bautista Chale-Dzul, Ricardo Ciria Merce, Andreu Comas-García, Célida Duque Molina, Julissa Enciso-Ibarra, José Antonio Enciso-Moreno, Gloria Elena Espinosa-Ayala, Fernando Fontove-Herrera, Daniel Fregoso-Rueda, Victor Eduardo Garcia-Arias, Alejandra Garcia-Gasca, Bruno Gomez-Gil, Jean Pierre González, Irvin González-López, Concepción Grajales-Muñiz, Ricardo Grande, Rosa María Gutierrez Rios, Alejandra Hernández-Terán, Alfredo Herrera-Estrella, Carla Ivón Herrera-Najera, Pavel Isa, Daniel Lira Morales, Susana Lopez, Antonio Loza Román, Brenda Irasema Maldonado-Meza, Bernardo Martínez-Miguel, José Arturo Martínez-Orozco, Célida Martínez- Rodríguez, Margarita Matías-Florentino, Fidencio Mejía-Nepomuceno, María Guadalupe de Jesús Mireles-Rivera, Gloria María Molina-Salinas, Hector Montoya-Fuentes, , Mario Mújica-Sánchez, José Esteban Muñoz-Medina, José de Jesús Nuñez-Contreras, Alicia Ocaña-Mondragón, Luis Alberto Ochoa-Carrera, Hector Esteban Paz-Juárez, Francisco Pulido, Helen Haydee Fernanda Ramírez-Plascencia, Jorge Salas-Hernández, Angel Gustavo Salas-Lais, Alejandro Sanchez-Flores, Clara Esperanza Santacruz-Tinoco, María Guadalupe Santiago-Mauricio, Selene Zárate, Nelly Sélem-Mojica, Blanca Taboada, Gloria Vazquez, Joel Armando Vázquez-Pérez. |
| EPI_ISL_2391749, EPI_ISL_2391750, EPI_ISL_2391751, EPI_ISL_2391752 | Pomona Valley Hospital Medical Center | Los Angeles County Public Health Laboratories | P. Hemarajata et al. |
| EPI_ISL_2391753 | California Hospital Medical Center | Los Angeles County Public Health Laboratories | P. Hemarajata et al. |
| EPI_ISL_2391755, EPI_ISL_2391756, EPI_ISL_2391757, EPI_ISL_2391758, EPI_ISL_2391759 | Flow Diagnostics Lab | Los Angeles County Public Health Laboratories | P. Hemarajata et al. |
| EPI_ISL_2391760, EPI_ISL_2391761, EPI_ISL_2391762, EPI_ISL_2391763, EPI_ISL_2391764 | UCLA Clinical Micro Lab | Los Angeles County Public Health Laboratories | P. Hemarajata et al. |
| EPI_ISL_2391780 | Pomona Valley Hospital Medical Center | Los Angeles County Public Health Laboratories | P. Hemarajata et al. |
| EPI_ISL_2391781, EPI_ISL_2391782, EPI_ISL_2391783, EPI_ISL_2391784, EPI_ISL_2391785, EPI_ISL_2391786, EPI_ISL_2391787 | USC Clinical Lab | Los Angeles County Public Health Laboratories | P. Hemarajata et al. |
| EPI_ISL_2391788 | Huntington Memorial Hospital | Los Angeles County Public Health Laboratories | P. Hemarajata et al. |
| EPI_ISL_2391789 | Flow Diagnostics Lab | Los Angeles County Public Health Laboratories | P. Hemarajata et al. |
| EPI_ISL_2391791 | Sun Clinical Laboratory | Los Angeles County Public Health Laboratories | P. Hemarajata et al. |
| EPI_ISL_2391792 | Providence Saint Joseph Medical Center | Los Angeles County Public Health Laboratories | P. Hemarajata et al. |
| EPI_ISL_2391793 | City of Hope Comprehensive Cancer Center | Los Angeles County Public Health Laboratories | P. Hemarajata et al. |
| EPI_ISL_2391795, EPI_ISL_2391796, EPI_ISL_2391797, EPI_ISL_2391798, EPI_ISL_2391799, EPI_ISL_2391800, EPI_ISL_2391801, EPI_ISL_2391802, EPI_ISL_2391803, EPI_ISL_2391804, EPI_ISL_2391805 | | | |
| see above | Flow Diagnostics Lab | Los Angeles County Public Health Laboratories | P. Hemarajata et al. |
| EPI_ISL_2391807 | Huntington Memorial Hospital | Los Angeles County Public Health Laboratories | P. Hemarajata et al. |
| EPI_ISL_2391808, EPI_ISL_2391809, EPI_ISL_2391810, EPI_ISL_2391811, EPI_ISL_2391812, EPI_ISL_2391813 | UCLA Clinical Micro Lab | Los Angeles County Public Health Laboratories | P. Hemarajata et al. |
| EPI_ISL_2392167, EPI_ISL_2392168 | Pomona Valley Hospital Medical Center | Los Angeles County Public Health Laboratories | P. Hemarajata et al. |
| EPI_ISL_2392169, EPI_ISL_2392170 | Flow Diagnostics Lab | Los Angeles County Public Health Laboratories | P. Hemarajata et al. |
| EPI_ISL_2392171, EPI_ISL_2392172, EPI_ISL_2392173, EPI_ISL_2392174, EPI_ISL_2392175, EPI_ISL_2392176, EPI_ISL_2392177, EPI_ISL_2392178, EPI_ISL_2392179, EPI_ISL_2392180, EPI_ISL_2392191, EPI_ISL_2392192, EPI_ISL_2392193, EPI_ISL_2392194 | | | |
| see above | UCLA Clinical Micro Lab | Los Angeles County Public Health Laboratories | P. Hemarajata et al. |
| EPI_ISL_2397325 | Rhode Island State Health Laboratory | Rhode Island State Health Laboratory | Ewa King, Richard C. Huard, Kristin Carpenter-Azevedo |
| EPI_ISL_2397329, EPI_ISL_2397330, EPI_ISL_2397331, EPI_ISL_2397332, EPI_ISL_2397333, EPI_ISL_2397334, EPI_ISL_2397335, EPI_ISL_2397336, EPI_ISL_2397337, EPI_ISL_2397338, EPI_ISL_2397339, EPI_ISL_2397340, EPI_ISL_2397341, EPI_ISL_2397342, EPI_ISL_2397343, EPI_ISL_2397344, EPI_ISL_2397345, EPI_ISL_2397346, EPI_ISL_2397347, EPI_ISL_2397348, EPI_ISL_2397349, EPI_ISL_2397350, EPI_ISL_2397353, EPI_ISL_2397356, EPI_ISL_2397359, EPI_ISL_2397361, EPI_ISL_2397363, EPI_ISL_2397364, EPI_ISL_2397365, EPI_ISL_2397366, EPI_ISL_2397367, EPI_ISL_2397368, EPI_ISL_2397369, EPI_ISL_2397370, EPI_ISL_2397371, EPI_ISL_2397372, EPI_ISL_2397373, EPI_ISL_2397374, EPI_ISL_2397375, EPI_ISL_2397376, EPI_ISL_2397377, EPI_ISL_2397378, EPI_ISL_2397379, EPI_ISL_2397380, EPI_ISL_2397381, EPI_ISL_2397382, EPI_ISL_2397383, EPI_ISL_2397384, EPI_ISL_2397385, EPI_ISL_2397386, EPI_ISL_2397387, EPI_ISL_2397388, EPI_ISL_2397389, EPI_ISL_2397390, EPI_ISL_2397391, EPI_ISL_2397392, EPI_ISL_2397393, EPI_ISL_2397394, EPI_ISL_2397395, EPI_ISL_2397396, EPI_ISL_2397397, EPI_ISL_2397398, EPI_ISL_2397399, EPI_ISL_2397400, EPI_ISL_2397401, EPI_ISL_2397402, EPI_ISL_2397403, EPI_ISL_2397404, EPI_ISL_2397405, EPI_ISL_2397406, EPI_ISL_2397407, EPI_ISL_2397408, EPI_ISL_2397409, EPI_ISL_2397410, EPI_ISL_2397411, EPI_ISL_2397412, EPI_ISL_2397413, EPI_ISL_2397414, EPI_ISL_2397415, EPI_ISL_2397416, EPI_ISL_2397417, EPI_ISL_2397418, EPI_ISL_2397419, EPI_ISL_2397420, EPI_ISL_2397421, EPI_ISL_2397422, EPI_ISL_2397423, EPI_ISL_2397424, EPI_ISL_2397425, EPI_ISL_2397426, EPI_ISL_2397427, EPI_ISL_2397428, EPI_ISL_2397429, EPI_ISL_2397430, EPI_ISL_2397431, EPI_ISL_2397432, EPI_ISL_2397433, EPI_ISL_2397434, EPI_ISL_2397435, EPI_ISL_2397436, EPI_ISL_2397437, EPI_ISL_2397438, EPI_ISL_2397439, EPI_ISL_2397440, EPI_ISL_2397441, EPI_ISL_2397442, EPI_ISL_2397443, EPI_ISL_2397444, EPI_ISL_2397445, EPI_ISL_2397446, EPI_ISL_2397447, EPI_ISL_2397448, EPI_ISL_2397449, EPI_ISL_2397450, EPI_ISL_2397451, EPI_ISL_2397452, EPI_ISL_2397453, EPI_ISL_2397454, EPI_ISL_2397455, EPI_ISL_2397456, EPI_ISL_2397457, EPI_ISL_2397458, EPI_ISL_2397459, EPI_ISL_2397460, EPI_ISL_2397461, EPI_ISL_2397462, EPI_ISL_2397463, EPI_ISL_2397464, EPI_ISL_2397465, EPI_ISL_2397466, EPI_ISL_2397467, EPI_ISL_2397468, EPI_ISL_2397469, EPI_ISL_2397470, EPI_ISL_2397471, EPI_ISL_2397472, EPI_ISL_2397473, EPI_ISL_2397474, EPI_ISL_2397475, EPI_ISL_2397476, EPI_ISL_2397477, EPI_ISL_2397478, EPI_ISL_2397479, EPI_ISL_2397480, EPI_ISL_2397481, EPI_ISL_2397482, EPI_ISL_2397483, EPI_ISL_2397484, EPI_ISL_2397485, EPI_ISL_2397486, EPI_ISL_2397487, EPI_ISL_2397488, EPI_ISL_2397489, EPI_ISL_2397490, EPI_ISL_2397491, EPI_ISL_2397492, EPI_ISL_2397493, EPI_ISL_2397494, EPI_ISL_2397495, EPI_ISL_2397496, EPI_ISL_2397497, EPI_ISL_2397498, EPI_ISL_2397499, EPI_ISL_2397500, EPI_ISL_2397501, EPI_ISL_2397502, EPI_ISL_2397503, EPI_ISL_2397504, EPI_ISL_2397505, EPI_ISL_2397506, EPI_ISL_2397507, EPI_ISL_2397508, EPI_ISL_2397509, EPI_ISL_2397510, EPI_ISL_2397511, EPI_ISL_2397512, EPI_ISL_2397513, EPI_ISL_2397514, EPI_ISL_2397516, EPI_ISL_2397517, EPI_ISL_2397518, EPI_ISL_2397519, EPI_ISL_2397520, EPI_ISL_2397521, EPI_ISL_2397522, EPI_ISL_2397523, EPI_ISL_2397524, EPI_ISL_2397525, EPI_ISL_2397526, EPI_ISL_2397527, EPI_ISL_2397528, EPI_ISL_2397529, EPI_ISL_2397530, EPI_ISL_2397531, EPI_ISL_2397532, EPI_ISL_2397533, EPI_ISL_2397534, EPI_ISL_2397535, EPI_ISL_2397536, EPI_ISL_2397537, EPI_ISL_2397538, EPI_ISL_2397539, EPI_ISL_2397540, EPI_ISL_2397541, EPI_ISL_2397542, EPI_ISL_2397543, EPI_ISL_2397544, EPI_ISL_2397545, EPI_ISL_2397546, EPI_ISL_2397547, EPI_ISL_2397548, EPI_ISL_2397549, EPI_ISL_2397550, EPI_ISL_2397551, EPI_ISL_2397552, EPI_ISL_2397553, EPI_ISL_2397554, EPI_ISL_2397555, EPI_ISL_2397556, EPI_ISL_2397557, EPI_ISL_2397558, EPI_ISL_2397559, EPI_ISL_2397560, EPI_ISL_2397561, EPI_ISL_2397562, EPI_ISL_2397563, EPI_ISL_2397564, EPI_ISL_2397565, EPI_ISL_2397566, EPI_ISL_2397567, EPI_ISL_2397568, EPI_ISL_2397569, EPI_ISL_2397570, EPI_ISL_2397571, EPI_ISL_2397572, EPI_ISL_2397573, EPI_ISL_2397574, EPI_ISL_2397575, EPI_ISL_2397576, EPI_ISL_2397577, EPI_ISL_2397578, EPI_ISL_2397579, EPI_ISL_2397580, EPI_ISL_2397581, EPI_ISL_2397582, EPI_ISL_2397583, EPI_ISL_2397584, EPI_ISL_2397585, EPI_ISL_2397586, EPI_ISL_2397587, EPI_ISL_2397588, EPI_ISL_2397589, EPI_ISL_2397590, EPI_ISL_2397591, EPI_ISL_2397592, EPI_ISL_2397593, EPI_ISL_2397594, EPI_ISL_2397595, EPI_ISL_2397596, EPI_ISL_2397597, EPI_ISL_2397598, EPI_ISL_2397599, EPI_ISL_2397600, EPI_ISL_2397601, EPI_ISL_2397602, EPI_ISL_2397603, EPI_ISL_2397604, EPI_ISL_2397605, EPI_ISL_2397606, EPI_ISL_2397607, EPI_ISL_2397608, EPI_ISL_2397609, EPI_ISL_2397610, EPI_ISL_2397611, EPI_ISL_2397612, EPI_ISL_2397613, EPI_ISL_2397614, EPI_ISL_2397615, EPI_ISL_2397616, EPI_ISL_2397617, EPI_ISL_2397618, EPI_ISL_2397619, EPI_ISL_2397620, EPI_ISL_2397621, EPI_ISL_2397622, EPI_ISL_2397623, EPI_ISL_2397624, EPI_ISL_2397625, EPI_ISL_2397626, EPI_ISL_2397627, EPI_ISL_2397628, EPI_ISL_2397629, EPI_ISL_2397630, EPI_ISL_2397631, EPI_ISL_2397632, EPI_ISL_2397633, EPI_ISL_2397634, EPI_ISL_2397635, EPI_ISL_2397636, EPI_ISL_2397637, EPI_ISL_2397638, EPI_ISL_2397639, EPI_ISL_2397640, EPI_ISL_2397641, EPI_ISL_2397642, EPI_ISL_2397643, EPI_ISL_2397644, EPI_ISL_2397645, EPI_ISL_2397646, EPI_ISL_2397647, EPI_ISL_2397648, EPI_ISL_2397649, EPI_ISL_2397650, EPI_ISL_2397651, EPI_ISL_2397652, EPI_ISL_2397653, EPI_ISL_2397654, EPI_ISL_2397655, EPI_ISL_2397656, EPI_ISL_2397657, EPI_ISL_2397658, EPI_ISL_2397659, EPI_ISL_2397660, EPI_ISL_2397661, EPI_ISL_2397662, EPI_ISL_2397663, EPI_ISL_2397664, EPI_ISL_2397665, EPI_ISL_2397666, EPI_ISL_2397667, EPI_ISL_2397668, EPI_ISL_2397669, EPI_ISL_2397670, EPI_ISL_2397671, EPI_ISL_2397672, EPI_ISL_2397673, EPI_ISL_2397674, EPI_ISL_2397675, EPI_ISL_2397676, EPI_ISL_2397677, EPI_ISL_2397678, EPI_ISL_2397679, EPI_ISL_2397680, EPI_ISL_2397681, EPI_ISL_2397682, EPI_ISL_2397683, EPI_ISL_2397684, EPI_ISL_2397685, EPI_ISL_2397686, EPI_ISL_2397687, EPI_ISL_2397688, EPI_ISL_2397689, EPI_ISL_2397690, EPI_ISL_2397691, EPI_ISL_2397692, EPI_ISL_2397693, EPI_ISL_2397694, EPI_ISL_2397695, EPI_ISL_2397696, EPI_ISL_2397697, EPI_ISL_2397698, EPI_ISL_2397699, EPI_ISL_2397700, EPI_ISL_2397701, EPI_ISL_2397702, EPI_ISL_2397703, EPI_ISL_2397704, EPI_ISL_2397705, EPI_ISL_2397706, EPI_ISL_2397707, EPI_ISL_2397708, EPI_ISL_2397709, EPI_ISL_2397710, EPI_ISL_2397711, EPI_ISL_2397712, EPI_ISL_2397713, EPI_ISL_2397714, EPI_ISL_2397715, EPI_ISL_2397716, EPI_ISL_2397717, EPI_ISL_2397718, EPI_ISL_2397719, EPI_ISL_2397720, EPI_ISL_2397721, EPI_ISL_2397722, EPI_ISL_2397723, EPI_ISL_2397724, EPI_ISL_2397725, EPI_ISL_2397726, EPI_ISL_2397727, EPI_ISL_2397728, EPI_ISL_2397729, EPI_ISL_2397730, EPI_ISL_2397731, EPI_ISL_2397732, EPI_ISL_2397733, EPI_ISL_2397734, EPI_ISL_2397735, EPI_ISL_2397736, EPI_ISL_2397737, EPI_ISL_2397738, EPI_ISL_2397739, EPI_ISL_2397740, EPI_ISL_2397741, EPI_ISL_2397742, EPI_ISL_2397743, EPI_ISL_2397744, EPI_ISL_2397745, EPI_ISL_2397746, EPI_ISL_2397747, EPI_ISL_2397748, EPI_ISL_2397749, EPI_ISL_2397750, EPI_ISL_2397751, EPI_ISL_2397752, EPI_ISL_2397753, EPI_ISL_2397754, EPI_ISL_2397755, EPI_ISL_2397756, EPI_ISL_2397757, EPI_ISL_2397758, EPI_ISL_2397759, EPI_ISL_2397760, EPI_ISL_2397761, EPI_ISL_2397762, EPI_ISL_2397763, EPI_ISL_2397764, EPI_ISL_2397765, EPI_ISL_2397766, EPI_ISL_2397767, EPI_ISL_2397768, EPI_ISL_2397769, EPI_ISL_2397770, EPI_ISL_2397771, EPI_ISL_2397772, EPI_ISL_2397773, EPI_ISL_2397774, EPI_ISL_2397775, EPI_ISL_2397776, EPI_ISL_2397777, | | | |

| | | | |
|---|---|---|---|
| EPI_ISL_2397778, EPI_ISL_2397779, EPI_ISL_2397780, EPI_ISL_2397781, EPI_ISL_2397782, EPI_ISL_2397783, EPI_ISL_2397784, EPI_ISL_2397785, EPI_ISL_2397786, EPI_ISL_2397787, EPI_ISL_2397788, EPI_ISL_2397789, EPI_ISL_2397790, EPI_ISL_2397791, EPI_ISL_2397792, EPI_ISL_2397793, EPI_ISL_2397794, EPI_ISL_2397795, EPI_ISL_2397796, EPI_ISL_2397797 | | | |
| see above | Quest Diagnostics Incorporated | Centers for Disease Control and Prevention Division of Viral Diseases, Pathogen Discovery | Dakota Howard, Dhwani Batra, Peter W. Cook, Kara Moser, Adrian Paskey, Jason Caravas, Benjamin Rambo-Martin, Shatavia Morrison, Christopher Gulvick, Scott Sammons, Yvette Unoarumhi, Darlene Wagner, Matthew Schmerer, S. H. Rosenthal, A. Gerasimova, R. M. Kagan, B. Anderson, M. Hua, Y. Liu, L.E. Bernstein, K.E. Livingston, A. Perez, I. A. Shlyakhter, R. V. Rolando, R. Owen, P. Tanpaiboon, F. Lacbawan, Clinton R. Paden, Duncan MacCannell |
| EPI_ISL_2397798, EPI_ISL_2397799, EPI_ISL_2397800, EPI_ISL_2397801, EPI_ISL_2397802, EPI_ISL_2397803, EPI_ISL_2397804, EPI_ISL_2397805, EPI_ISL_2397806, EPI_ISL_2397807, EPI_ISL_2397811, EPI_ISL_2397813, EPI_ISL_2397814, EPI_ISL_2397815, EPI_ISL_2397816, EPI_ISL_2397817, EPI_ISL_2397818, EPI_ISL_2397819, EPI_ISL_2397820, EPI_ISL_2397821, EPI_ISL_2397822, EPI_ISL_2397823, EPI_ISL_2397824, EPI_ISL_2397825, EPI_ISL_2397826, EPI_ISL_2397827, EPI_ISL_2397828, EPI_ISL_2397830, EPI_ISL_2397831, EPI_ISL_2397832, EPI_ISL_2397833, EPI_ISL_2397834, EPI_ISL_2397835, EPI_ISL_2397837, EPI_ISL_2397840, EPI_ISL_2397841, EPI_ISL_2397844, EPI_ISL_2397845, EPI_ISL_2397846, EPI_ISL_2397847, EPI_ISL_2397848, EPI_ISL_2397849, EPI_ISL_2397850, EPI_ISL_2397852, EPI_ISL_2397853, EPI_ISL_2397854, EPI_ISL_2397855, EPI_ISL_2397857, EPI_ISL_2397858, EPI_ISL_2397859, EPI_ISL_2397860, EPI_ISL_2397861, EPI_ISL_2397862, EPI_ISL_2397864, EPI_ISL_2397866, EPI_ISL_2397868, EPI_ISL_2397869, EPI_ISL_2397870, EPI_ISL_2397871, EPI_ISL_2397872, EPI_ISL_2397873, EPI_ISL_2397874, EPI_ISL_2397875, EPI_ISL_2397876, EPI_ISL_2397877, EPI_ISL_2397878, EPI_ISL_2397879, EPI_ISL_2397882, EPI_ISL_2397884, EPI_ISL_2397885, EPI_ISL_2397886, EPI_ISL_2397888, EPI_ISL_2397890, EPI_ISL_2397891, EPI_ISL_2397892, EPI_ISL_2397895, EPI_ISL_2397896, EPI_ISL_2397897, EPI_ISL_2397900, EPI_ISL_2397901, EPI_ISL_2397902, EPI_ISL_2397903, EPI_ISL_2397904, EPI_ISL_2397905, EPI_ISL_2397906, EPI_ISL_2397907, EPI_ISL_2397908, EPI_ISL_2397909, EPI_ISL_2397910, EPI_ISL_2397911, EPI_ISL_2397912, EPI_ISL_2397914, EPI_ISL_2397915, EPI_ISL_2397916, EPI_ISL_2397917, EPI_ISL_2397919, EPI_ISL_2397920, EPI_ISL_2397921, EPI_ISL_2397922, EPI_ISL_2397924, EPI_ISL_2397925, EPI_ISL_2397926, EPI_ISL_2397927, EPI_ISL_2397928, EPI_ISL_2397929, EPI_ISL_2397930, EPI_ISL_2397931, EPI_ISL_2397932, EPI_ISL_2397934, EPI_ISL_2397936, EPI_ISL_2397937, EPI_ISL_2397938, EPI_ISL_2397939, EPI_ISL_2397940, EPI_ISL_2397941, EPI_ISL_2397942, EPI_ISL_2397943, EPI_ISL_2397944, EPI_ISL_2397946, EPI_ISL_2397948, EPI_ISL_2397949, EPI_ISL_2397950, EPI_ISL_2397951, EPI_ISL_2397952, EPI_ISL_2397953, EPI_ISL_2397955, EPI_ISL_2397956, EPI_ISL_2397957, EPI_ISL_2397958, EPI_ISL_2397959, EPI_ISL_2397960, EPI_ISL_2397961, EPI_ISL_2397964, EPI_ISL_2397966, EPI_ISL_2397967, EPI_ISL_2397968, EPI_ISL_2397969, EPI_ISL_2397970, EPI_ISL_2397971, EPI_ISL_2397972, EPI_ISL_2397973, EPI_ISL_2397974, EPI_ISL_2397975, EPI_ISL_2397976, EPI_ISL_2397977, EPI_ISL_2397980, EPI_ISL_2397982, EPI_ISL_2397983, EPI_ISL_2397984, EPI_ISL_2397985, EPI_ISL_2397986, EPI_ISL_2397988, EPI_ISL_2397989, EPI_ISL_2397990, EPI_ISL_2397991, EPI_ISL_2397992, EPI_ISL_2397994, EPI_ISL_2397995, EPI_ISL_2397996, EPI_ISL_2397997, EPI_ISL_2397998, EPI_ISL_2397999, EPI_ISL_2398000, EPI_ISL_2398001, EPI_ISL_2398002, EPI_ISL_2398003, EPI_ISL_2398004, EPI_ISL_2398006, EPI_ISL_2398007, EPI_ISL_2398008, EPI_ISL_2398009, EPI_ISL_2398012, EPI_ISL_2398013, EPI_ISL_2398014, EPI_ISL_2398015, EPI_ISL_2398016, EPI_ISL_2398018, EPI_ISL_2398020, EPI_ISL_2398021, EPI_ISL_2398022, EPI_ISL_2398023, EPI_ISL_2398025, EPI_ISL_2398026, EPI_ISL_2398027, EPI_ISL_2398028, EPI_ISL_2398029, EPI_ISL_2398030, EPI_ISL_2398031, EPI_ISL_2398032, EPI_ISL_2398033, EPI_ISL_2398034, EPI_ISL_2398035, EPI_ISL_2398036, EPI_ISL_2398037, EPI_ISL_2398038, EPI_ISL_2398039, EPI_ISL_2398040, EPI_ISL_2398041, EPI_ISL_2398042, EPI_ISL_2398043, EPI_ISL_2398044, EPI_ISL_2398045, EPI_ISL_2398046, EPI_ISL_2398047, EPI_ISL_2398049, EPI_ISL_2398050, EPI_ISL_2398051, EPI_ISL_2398052, EPI_ISL_2398053, EPI_ISL_2398054, EPI_ISL_2398055, EPI_ISL_2398057, EPI_ISL_2398058, EPI_ISL_2398059, EPI_ISL_2398060, EPI_ISL_2398061, EPI_ISL_2398062, EPI_ISL_2398063, EPI_ISL_2398064, EPI_ISL_2398065, EPI_ISL_2398067, EPI_ISL_2398068, EPI_ISL_2398069, EPI_ISL_2398070, EPI_ISL_2398072, EPI_ISL_2398073, EPI_ISL_2398074, EPI_ISL_2398075, EPI_ISL_2398076, EPI_ISL_2398077, EPI_ISL_2398078, EPI_ISL_2398079, EPI_ISL_2398080, EPI_ISL_2398081, EPI_ISL_2398082, EPI_ISL_2398083, EPI_ISL_2398084, EPI_ISL_2398085, EPI_ISL_2398086, EPI_ISL_2398087, EPI_ISL_2398088, EPI_ISL_2398089, EPI_ISL_2398090, EPI_ISL_2398092, EPI_ISL_2398093, EPI_ISL_2398094, EPI_ISL_2398095, EPI_ISL_2398096, EPI_ISL_2398097, EPI_ISL_2398098, EPI_ISL_2398099, EPI_ISL_2398101, EPI_ISL_2398102, EPI_ISL_2398103, EPI_ISL_2398104, EPI_ISL_2398105, EPI_ISL_2398106, EPI_ISL_2398107, EPI_ISL_2398108, EPI_ISL_2398109, EPI_ISL_2398110, EPI_ISL_2398111, EPI_ISL_2398112, EPI_ISL_2398113, EPI_ISL_2398114, EPI_ISL_2398115, EPI_ISL_2398117, EPI_ISL_2398118, EPI_ISL_2398119, EPI_ISL_2398120, EPI_ISL_2398121, EPI_ISL_2398122, EPI_ISL_2398123, EPI_ISL_2398126, EPI_ISL_2398127, EPI_ISL_2398128, EPI_ISL_2398129, EPI_ISL_2398130, EPI_ISL_2398131, EPI_ISL_2398132, EPI_ISL_2398133, EPI_ISL_2398134, EPI_ISL_2398135, EPI_ISL_2398136, EPI_ISL_2398137, EPI_ISL_2398138, EPI_ISL_2398139, EPI_ISL_2398140, EPI_ISL_2398141, EPI_ISL_2398142, EPI_ISL_2398143, EPI_ISL_2398144, EPI_ISL_2398145, EPI_ISL_2398146, EPI_ISL_2398147, EPI_ISL_2398148, EPI_ISL_2398149, EPI_ISL_2398150, EPI_ISL_2398152, EPI_ISL_2398153, EPI_ISL_2398154, EPI_ISL_2398155, EPI_ISL_2398156, EPI_ISL_2398157, EPI_ISL_2398158, EPI_ISL_2398159, EPI_ISL_2398160, EPI_ISL_2398161, EPI_ISL_2398162, EPI_ISL_2398163, EPI_ISL_2398164, EPI_ISL_2398165, EPI_ISL_2398166, EPI_ISL_2398167, EPI_ISL_2398168, EPI_ISL_2398169, EPI_ISL_2398170, EPI_ISL_2398171, EPI_ISL_2398172, EPI_ISL_2398173, EPI_ISL_2398174, EPI_ISL_2398175, EPI_ISL_2398176, EPI_ISL_2398177, EPI_ISL_2398178, EPI_ISL_2398179, EPI_ISL_2398180, EPI_ISL_2398181, EPI_ISL_2398182, EPI_ISL_2398183, EPI_ISL_2398184, EPI_ISL_2398185, EPI_ISL_2398186, EPI_ISL_2398187, EPI_ISL_2398188, EPI_ISL_2398189, EPI_ISL_2398190, EPI_ISL_2398191, EPI_ISL_2398192, EPI_ISL_2398193, EPI_ISL_2398194, EPI_ISL_2398195, EPI_ISL_2398196, EPI_ISL_2398197, EPI_ISL_2398198, EPI_ISL_2398199, EPI_ISL_2398200, EPI_ISL_2398202, EPI_ISL_2398203, EPI_ISL_2398204, EPI_ISL_2398205, EPI_ISL_2398206, EPI_ISL_2398207, EPI_ISL_2398208, EPI_ISL_2398209, EPI_ISL_2398210, EPI_ISL_2398211, EPI_ISL_2398212, EPI_ISL_2398213, EPI_ISL_2398214, EPI_ISL_2398215, EPI_ISL_2398216, EPI_ISL_2398217, EPI_ISL_2398218, EPI_ISL_2398219, EPI_ISL_2398220, EPI_ISL_2398222, EPI_ISL_2398223, EPI_ISL_2398225, EPI_ISL_2398226, EPI_ISL_2398227, EPI_ISL_2398228, EPI_ISL_2398230, EPI_ISL_2398231, EPI_ISL_2398232, EPI_ISL_2398233, EPI_ISL_2398234, EPI_ISL_2398235, EPI_ISL_2398236, EPI_ISL_2398237, EPI_ISL_2398239, EPI_ISL_2398240, EPI_ISL_2398241, EPI_ISL_2398242, EPI_ISL_2398243, EPI_ISL_2398244, EPI_ISL_2398245, EPI_ISL_2398246, EPI_ISL_2398247, EPI_ISL_2398248, EPI_ISL_2398249, EPI_ISL_2398250, EPI_ISL_2398251, EPI_ISL_2398252, EPI_ISL_2398253, EPI_ISL_2398254, EPI_ISL_2398255, EPI_ISL_2398256, EPI_ISL_2398257, EPI_ISL_2398258, EPI_ISL_2398259, EPI_ISL_2398260, EPI_ISL_2398261, EPI_ISL_2398262, EPI_ISL_2398264, EPI_ISL_2398265, EPI_ISL_2398266, EPI_ISL_2398267, EPI_ISL_2398268, EPI_ISL_2398269, EPI_ISL_2398270, EPI_ISL_2398271, EPI_ISL_2398272, EPI_ISL_2398274, EPI_ISL_2398275, EPI_ISL_2398276, EPI_ISL_2398277, EPI_ISL_2398278, EPI_ISL_2398280, EPI_ISL_2398281, EPI_ISL_2398283, EPI_ISL_2398284, EPI_ISL_2398285, EPI_ISL_2398286, EPI_ISL_2398287, EPI_ISL_2398289, EPI_ISL_2398292, EPI_ISL_2398295, EPI_ISL_2398297, EPI_ISL_2398300, EPI_ISL_2398302, EPI_ISL_2398303, EPI_ISL_2398304, EPI_ISL_2398305, EPI_ISL_2398306, EPI_ISL_2398307, EPI_ISL_2398308, EPI_ISL_2398309, EPI_ISL_2398310, EPI_ISL_2398311, EPI_ISL_2398312, EPI_ISL_2398313, EPI_ISL_2398314, EPI_ISL_2398315, EPI_ISL_2398316, EPI_ISL_2398317, EPI_ISL_2398318, EPI_ISL_2398319, EPI_ISL_2398320, EPI_ISL_2398321, EPI_ISL_2398322, EPI_ISL_2398324, EPI_ISL_2398325, EPI_ISL_2398326, EPI_ISL_2398327, EPI_ISL_2398328, EPI_ISL_2398329, EPI_ISL_2398330, EPI_ISL_2398332, EPI_ISL_2398333, EPI_ISL_2398334, EPI_ISL_2398335, EPI_ISL_2398336, EPI_ISL_2398337, EPI_ISL_2398338, EPI_ISL_2398339, EPI_ISL_2398340, EPI_ISL_2398342, EPI_ISL_2398343, EPI_ISL_2398344, EPI_ISL_2398346, EPI_ISL_2398347, EPI_ISL_2398348, EPI_ISL_2398349, EPI_ISL_2398350, EPI_ISL_2398351, EPI_ISL_2398352, EPI_ISL_2398353, EPI_ISL_2398354, EPI_ISL_2398355, EPI_ISL_2398356, EPI_ISL_2398357, EPI_ISL_2398358, EPI_ISL_2398359, EPI_ISL_2398360, EPI_ISL_2398361, EPI_ISL_2398362, EPI_ISL_2398363, EPI_ISL_2398364, EPI_ISL_2398365, EPI_ISL_2398366, EPI_ISL_2398367, EPI_ISL_2398368, EPI_ISL_2398369, EPI_ISL_2398370, EPI_ISL_2398371, EPI_ISL_2398372, EPI_ISL_2398373, EPI_ISL_2398374, EPI_ISL_2398375, EPI_ISL_2398376, EPI_ISL_2398377, EPI_ISL_2398378, EPI_ISL_2398379, EPI_ISL_2398380, EPI_ISL_2398381, EPI_ISL_2398382, EPI_ISL_2398383, EPI_ISL_2398384, EPI_ISL_2398385, EPI_ISL_2398386, EPI_ISL_2398387, EPI_ISL_2398388, EPI_ISL_2398389, EPI_ISL_2398390, EPI_ISL_2398391, EPI_ISL_2398392, EPI_ISL_2398393, EPI_ISL_2398394, EPI_ISL_2398395, EPI_ISL_2398396, EPI_ISL_2398397, EPI_ISL_2398398, EPI_ISL_2398399, EPI_ISL_2398400, EPI_ISL_2398401, EPI_ISL_2398402, EPI_ISL_2398404, EPI_ISL_2398405, EPI_ISL_2398406, EPI_ISL_2398407, EPI_ISL_2398408, EPI_ISL_2398409, EPI_ISL_2398410, EPI_ISL_2398411, EPI_ISL_2398412, EPI_ISL_2398413, EPI_ISL_2398415, EPI_ISL_2398416, EPI_ISL_2398417, EPI_ISL_2398418, EPI_ISL_2398419, EPI_ISL_2398420, EPI_ISL_2398422, EPI_ISL_2398423, EPI_ISL_2398424, EPI_ISL_2398425, EPI_ISL_2398427, EPI_ISL_2398428, EPI_ISL_2398429, EPI_ISL_2398430, EPI_ISL_2398431, EPI_ISL_2398432, EPI_ISL_2398433, EPI_ISL_2398434, EPI_ISL_2398435, EPI_ISL_2398437, EPI_ISL_2398438, EPI_ISL_2398439, EPI_ISL_2398440, EPI_ISL_2398441, EPI_ISL_2398442, EPI_ISL_2398443, EPI_ISL_2398444, EPI_ISL_2398445, EPI_ISL_2398446, EPI_ISL_2398447, EPI_ISL_2398448, EPI_ISL_2398449, EPI_ISL_2398450, EPI_ISL_2398451, EPI_ISL_2398452, EPI_ISL_2398453, EPI_ISL_2398454, EPI_ISL_2398455, EPI_ISL_2398456, EPI_ISL_2398457, EPI_ISL_2398458, EPI_ISL_2398459, EPI_ISL_2398460, EPI_ISL_2398461, EPI_ISL_2398463, EPI_ISL_2398464, EPI_ISL_2398465, EPI_ISL_2398466, EPI_ISL_2398467, EPI_ISL_2398468, EPI_ISL_2398469, EPI_ISL_2398470, EPI_ISL_2398471, EPI_ISL_2398472, EPI_ISL_2398473, EPI_ISL_2398474, EPI_ISL_2398475, EPI_ISL_2398476, EPI_ISL_2398477, EPI_ISL_2398478, EPI_ISL_2398479, EPI_ISL_2398480, EPI_ISL_2398481, EPI_ISL_2398482, EPI_ISL_2398483, EPI_ISL_2398484, EPI_ISL_2398485, EPI_ISL_2398486, EPI_ISL_2398487, EPI_ISL_2398488, EPI_ISL_2398489, EPI_ISL_2398490, EPI_ISL_2398491, EPI_ISL_2398492, EPI_ISL_2398494, EPI_ISL_2398495, EPI_ISL_2398496, EPI_ISL_2398497, EPI_ISL_2398498, EPI_ISL_2398499, EPI_ISL_2398500, EPI_ISL_2398501, EPI_ISL_2398502, EPI_ISL_2398503, EPI_ISL_2398504, EPI_ISL_2398505, EPI_ISL_2398506, EPI_ISL_2398507, EPI_ISL_2398508, EPI_ISL_2398509, EPI_ISL_2398510, EPI_ISL_2398511, EPI_ISL_2398512, EPI_ISL_2398513, EPI_ISL_2398514, EPI_ISL_2398515, EPI_ISL_2398516, EPI_ISL_2398517, EPI_ISL_2398518, EPI_ISL_2398519, EPI_ISL_2398520, EPI_ISL_2398521, EPI_ISL_2398522, EPI_ISL_2398524, EPI_ISL_2398525, EPI_ISL_2398526, EPI_ISL_2398527, EPI_ISL_2398528, EPI_ISL_2398529, EPI_ISL_2398530, EPI_ISL_2398531, EPI_ISL_2398532, EPI_ISL_2398533, EPI_ISL_2398535, EPI_ISL_2398536, EPI_ISL_2398537, EPI_ISL_2398538, EPI_ISL_2398539, EPI_ISL_2398540, EPI_ISL_2398541, EPI_ISL_2398542, EPI_ISL_2398543, EPI_ISL_2398544, EPI_ISL_2398545, EPI_ISL_2398546, EPI_ISL_2398547, EPI_ISL_2398548, EPI_ISL_2398549, EPI_ISL_2398550, EPI_ISL_2398551, EPI_ISL_2398552, EPI_ISL_2398553, EPI_ISL_2398554, EPI_ISL_2398555, EPI_ISL_2398556, EPI_ISL_2398557, EPI_ISL_2398558, EPI_ISL_2398559, EPI_ISL_2398560, EPI_ISL_2398561, EPI_ISL_2398562, EPI_ISL_2398563, EPI_ISL_2398564, EPI_ISL_2398565, EPI_ISL_2398566, EPI_ISL_2398567, EPI_ISL_2398568, EPI_ISL_2398569, EPI_ISL_2398570, EPI_ISL_2398571, EPI_ISL_2398572, EPI_ISL_2398573, EPI_ISL_2398574, EPI_ISL_2398575, EPI_ISL_2398576, EPI_ISL_2398577, EPI_ISL_2398578, EPI_ISL_2398579, EPI_ISL_2398580, EPI_ISL_2398581, EPI_ISL_2398582, EPI_ISL_2398583, EPI_ISL_2398584, EPI_ISL_2398585, EPI_ISL_2398586, EPI_ISL_2398587, EPI_ISL_2398588, EPI_ISL_2398589, EPI_ISL_2398590, EPI_ISL_2398591, EPI_ISL_2398592, EPI_ISL_2398593, EPI_ISL_2398594, EPI_ISL_2398595, EPI_ISL_2398596, EPI_ISL_2398597, EPI_ISL_2398598, EPI_ISL_2398599, EPI_ISL_2398600, EPI_ISL_2398602, EPI_ISL_2398603, EPI_ISL_2398604, EPI_ISL_2398605, EPI_ISL_2398606, EPI_ISL_2398607, EPI_ISL_2398608, EPI_ISL_2398609, EPI_ISL_2398610, EPI_ISL_2398611, EPI_ISL_2398612, EPI_ISL_2398613, EPI_ISL_2398614, EPI_ISL_2398615, EPI_ISL_2398616, EPI_ISL_2398617, EPI_ISL_2398618, EPI_ISL_2398619, EPI_ISL_2398620, EPI_ISL_2398621, EPI_ISL_2398622, EPI_ISL_2398623, EPI_ISL_2398625, EPI_ISL_2398626, EPI_ISL_2398627, EPI_ISL_2398631, EPI_ISL_2398633, EPI_ISL_2398634, EPI_ISL_2398635, EPI_ISL_2398636, EPI_ISL_2398637, EPI_ISL_2398638, EPI_ISL_2398639, EPI_ISL_2398640, EPI_ISL_2398641, EPI_ISL_2398642, EPI_ISL_2398643, EPI_ISL_2398644, EPI_ISL_2398645, EPI_ISL_2398646, EPI_ISL_2398647, EPI_ISL_2398648, EPI_ISL_2398649, EPI_ISL_2398650, EPI_ISL_2398651, EPI_ISL_2398652, EPI_ISL_2398653, EPI_ISL_2398654, EPI_ISL_2398655, EPI_ISL_2398656, EPI_ISL_2398658, EPI_ISL_2398659, EPI_ISL_2398660, EPI_ISL_2398661, EPI_ISL_2398662, EPI_ISL_2398663, EPI_ISL_2398664, EPI_ISL_2398665, EPI_ISL_2398666, EPI_ISL_2398667, EPI_ISL_2398668, EPI_ISL_2398669, EPI_ISL_2398670, EPI_ISL_2398671, EPI_ISL_2398672, EPI_ISL_2398673, EPI_ISL_2398674, EPI_ISL_2398675, EPI_ISL_2398676, EPI_ISL_2398677, EPI_ISL_2398678, EPI_ISL_2398679, EPI_ISL_2398680, EPI_ISL_2398681, EPI_ISL_2398682, EPI_ISL_2398683, EPI_ISL_2398684, EPI_ISL_2398686, EPI_ISL_2398687, EPI_ISL_2398688, EPI_ISL_2398689, EPI_ISL_2398690, EPI_ISL_2398691, EPI_ISL_2398692, EPI_ISL_2398694, EPI_ISL_2398695, EPI_ISL_2398696, EPI_ISL_2398698, EPI_ISL_2398699, EPI_ISL_2398700, EPI_ISL_2398701, EPI_ISL_2398702, EPI_ISL_2398703, EPI_ISL_2398704, EPI_ISL_2398705, EPI_ISL_2398706, EPI_ISL_2398707, EPI_ISL_2398708, EPI_ISL_2398709, EPI_ISL_2398710, EPI_ISL_2398711, EPI_ISL_2398712, EPI_ISL_2398713, EPI_ISL_2398716, EPI_ISL_2398717, EPI_ISL_2398718, EPI_ISL_2398719, EPI_ISL_2398720, EPI_ISL_2398721, EPI_ISL_2398722, EPI_ISL_2398723, EPI_ISL_2398724, EPI_ISL_2398725, EPI_ISL_2398726, EPI_ISL_2398728, EPI_ISL_2398729, EPI_ISL_2398730, EPI_ISL_2398731, EPI_ISL_2398732, EPI_ISL_2398733, EPI_ISL_2398734, EPI_ISL_2398735, EPI_ISL_2398736, EPI_ISL_2398738, EPI_ISL_2398739, EPI_ISL_2398740, EPI_ISL_2398741, EPI_ISL_2398742, EPI_ISL_2398743, EPI_ISL_2398744, EPI_ISL_2398745, EPI_ISL_2398746, EPI_ISL_2398747, EPI_ISL_2398748, EPI_ISL_2398749, EPI_ISL_2398750, EPI_ISL_2398751, EPI_ISL_2398752, EPI_ISL_2398753, EPI_ISL_2398754, EPI_ISL_2398755, EPI_ISL_2398756, EPI_ISL_2398757, EPI_ISL_2398758, EPI_ISL_2398759, EPI_ISL_2398760, EPI_ISL_2398761, EPI_ISL_2398762, EPI_ISL_2398763, EPI_ISL_2398764, EPI_ISL_2398765, EPI_ISL_2398766, EPI_ISL_2398767, EPI_ISL_2398768, EPI_ISL_2398769, EPI_ISL_2398770, EPI_ISL_2398771, EPI_ISL_2398772, EPI_ISL_2398773, EPI_ISL_2398774, EPI_ISL_2398775, EPI_ISL_2398776, EPI_ISL_2398777, EPI_ISL_2398778, EPI_ISL_2398779, EPI_ISL_2398780, EPI_ISL_2398781, EPI_ISL_2398782, EPI_ISL_2398783, EPI_ISL_2398784, EPI_ISL_2398785, EPI_ISL_2398786, EPI_ISL_2398787, EPI_ISL_2398788, EPI_ISL_2398789, EPI_ISL_2398790, EPI_ISL_2398791, EPI_ISL_2398792, EPI_ISL_2398793, EPI_ISL_2398794, EPI_ISL_2398795, EPI_ISL_2398796, EPI_ISL_2398797, EPI_ISL_2398798, EPI_ISL_2398799, EPI_ISL_2398801, EPI_ISL_2398802, EPI_ISL_2398803, EPI_ISL_2398804, EPI_ISL_2398805, EPI_ISL_2398806, EPI_ISL_2398807, EPI_ISL_2398808, EPI_ISL_2398809, EPI_ISL_2398810, EPI_ISL_2398811, EPI_ISL_2398812, EPI_ISL_2398813, EPI_ISL_2398815, EPI_ISL_2398816, EPI_ISL_2398817, EPI_ISL_2398818, EPI_ISL_2398819, EPI_ISL_2398820, EPI_ISL_2398821, EPI_ISL_2398822, EPI_ISL_2398823, EPI_ISL_2398824, EPI_ISL_2398825, EPI_ISL_2398826, EPI_ISL_2398827, EPI_ISL_2398828, EPI_ISL_2398829, EPI_ISL_2398830, EPI_ISL_2398831, EPI_ISL_2398832, EPI_ISL_2398833, EPI_ISL_2398834, EPI_ISL_2398835, EPI_ISL_2398836, EPI_ISL_2398837, EPI_ISL_2398838, EPI_ISL_2398839, EPI_ISL_2398840, EPI_ISL_2398841, EPI_ISL_2398842, EPI_ISL_2398843, EPI_ISL_2398845, EPI_ISL_2398846, EPI_ISL_2398847, EPI_ISL_2398848, EPI_ISL_2398849, EPI_ISL_2398850, EPI_ISL_2398851, EPI_ISL_2398852, EPI_ISL_2398853, EPI_ISL_2398855, EPI_ISL_2398856, EPI_ISL_2398857, EPI_ISL_2398858, EPI_ISL_2398859, EPI_ISL_2398860, EPI_ISL_2398861, EPI_ISL_2398862, EPI_ISL_2398863, EPI_ISL_2398864, EPI_ISL_2398865, EPI_ISL_2398866, EPI_ISL_2398867, EPI_ISL_2398868, EPI_ISL_2398869, EPI_ISL_2398870, EPI_ISL_2398871, EPI_ISL_2398872, EPI_ISL_2398873, EPI_ISL_2398874, EPI_ISL_2398875, EPI_ISL_2398876, EPI_ISL_2398877, EPI_ISL_2398878, EPI_ISL_2398879, EPI_ISL_2398880, EPI_ISL_2398881, EPI_ISL_2398883, EPI_ISL_2398884, EPI_ISL_2398885, EPI_ISL_2398886, EPI_ISL_2398887, EPI_ISL_2398888, EPI_ISL_2398889, EPI_ISL_2398890, EPI_ISL_2398891, EPI_ISL_2398892, EPI_ISL_2398893, EPI_ISL_2398894, EPI_ISL_2398895, EPI_ISL_2398896, EPI_ISL_2398897, EPI_ISL_2398898, EPI_ISL_2398899, EPI_ISL_2398900, EPI_ISL_2398901, EPI_ISL_2398902, EPI_ISL_2398903, EPI_ISL_2398904, EPI_ISL_2398905, EPI_ISL_2398906, EPI_ISL_2398907, EPI_ISL_2398909, EPI_ISL_2398911, EPI_ISL_2398912, EPI_ISL_2398913, EPI_ISL_2398914, EPI_ISL_2398915, EPI_ISL_2398916, EPI_ISL_2398917, EPI_ISL_2398918, EPI_ISL_2398919, EPI_ISL_2398920, EPI_ISL_2398921, EPI_ISL_2398922, EPI_ISL_2398923, EPI_ISL_2398924, EPI_ISL_2398925, EPI_ISL_2398926, EPI_ISL_2398927, EPI_ISL_2398928, EPI_ISL_2398931, EPI_ISL_2398932, EPI_ISL_2398933, EPI_ISL_2398934, EPI_ISL_2398935, EPI_ISL_2398936, EPI_ISL_2398937, EPI_ISL_2398938, EPI_ISL_2398939, EPI_ISL_2398940, EPI_ISL_2398941, EPI_ISL_2398942, EPI_ISL_2398943, EPI_ISL_2398944, EPI_ISL_2398945, EPI_ISL_2398946, EPI_ISL_2398947, EPI_ISL_2398948, EPI_ISL_2398949, EPI_ISL_2398950, EPI_ISL_2398951, EPI_ISL_2398952, EPI_ISL_2398953, EPI_ISL_2398954, EPI_ISL_2398955, EPI_ISL_2398956, EPI_ISL_2398957, EPI_ISL_2398958, EPI_ISL_2398959, EPI_ISL_2398960, EPI_ISL_2398961, EPI_ISL_2398962, EPI_ISL_2398963, EPI_ISL_2398965, EPI_ISL_2398966, EPI_ISL_2398967, EPI_ISL_2398968, EPI_ISL_2398969, EPI_ISL_2398972, EPI_ISL_2398973, EPI_ISL_2398974, EPI_ISL_2398976, EPI_ISL_2398977, EPI_ISL_2398978, EPI_ISL_2398979, EPI_ISL_2398980, EPI_ISL_2398981, EPI_ISL_2398982, EPI_ISL_2398983, EPI_ISL_2398984, EPI_ISL_2398985, EPI_ISL_2398986, EPI_ISL_2398987, EPI_ISL_2398988, EPI_ISL_2398989, EPI_ISL_2398990, EPI_ISL_2398991, EPI_ISL_2398992, EPI_ISL_2398993, EPI_ISL_2398994, EPI_ISL_2398995, EPI_ISL_2398996, EPI_ISL_2398997, EPI_ISL_2398998, EPI_ISL_2399000, EPI_ISL_2399001, EPI_ISL_2399002, EPI_ISL_2399003, EPI_ISL_2399004, EPI_ISL_2399005, EPI_ISL_2399007, EPI_ISL_2399008, EPI_ISL_2399009, EPI_ISL_2399010, EPI_ISL_2399012, EPI_ISL_2399015, EPI_ISL_2399018, EPI_ISL_2399020, EPI_ISL_2399022, EPI_ISL_2399024, EPI_ISL_2399026, EPI_ISL_2399028, EPI_ISL_2399030, EPI_ISL_2399033, EPI_ISL_2399035, EPI_ISL_2399036, EPI_ISL_2399038, EPI_ISL_2399041, EPI_ISL_2399042, EPI_ISL_2399043, EPI_ISL_2399044, EPI_ISL_2399045, EPI_ISL_2399046, EPI_ISL_2399047, EPI_ISL_2399048, EPI_ISL_2399050, EPI_ISL_2399051, EPI_ISL_2399052, EPI_ISL_2399053, EPI_ISL_2399054, EPI_ISL_2399055, EPI_ISL_2399056, EPI_ISL_2399057, EPI_ISL_2399058, EPI_ISL_2399059, EPI_ISL_2399060, EPI_ISL_2399061, EPI_ISL_2399062, EPI_ISL_2399063, EPI_ISL_2399064, EPI_ISL_2399065, EPI_ISL_2399066, EPI_ISL_2399067, EPI_ISL_2399068, EPI_ISL_2399069, EPI_ISL_2399070, EPI_ISL_2399071, EPI_ISL_2399072, EPI_ISL_2399073, EPI_ISL_2399074, EPI_ISL_2399075, EPI_ISL_2399076, EPI_ISL_2399077, EPI_ISL_2399078, EPI_ISL_2399079, EPI_ISL_2399080, EPI_ISL_2399081, EPI_ISL_2399082, EPI_ISL_2399083, EPI_ISL_2399084, EPI_ISL_2399085, EPI_ISL_2399086, EPI_ISL_2399087, EPI_ISL_2399088, EPI_ISL_2399089, EPI_ISL_2399090, EPI_ISL_2399091, EPI_ISL_2399092, EPI_ISL_2399093, EPI_ISL_2399094, EPI_ISL_2399095, EPI_ISL_2399096, EPI_ISL_2399097, EPI_ISL_2399099, EPI_ISL_2399100, EPI_ISL_2399101, EPI_ISL_2399102, EPI_ISL_2399103, EPI_ISL_2399104, EPI_ISL_2399105, EPI_ISL_2399106, EPI_ISL_2399107, EPI_ISL_2399108, EPI_ISL_2399109, EPI_ISL_2399111, EPI_ISL_2399112, EPI_ISL_2399113, EPI_ISL_2399114, EPI_ISL_2399116, EPI_ISL_2399117, EPI_ISL_2399119, EPI_ISL_2399120, EPI_ISL_2399121, EPI_ISL_2399122, EPI_ISL_2399123, EPI_ISL_2399124, EPI_ISL_2399125, EPI_ISL_2399126, EPI_ISL_2399127, EPI_ISL_2399128, EPI_ISL_2399129, EPI_ISL_2399130, EPI_ISL_2399131, EPI_ISL_2399132, EPI_ISL_2399133, EPI_ISL_2399135, EPI_ISL_2399136, EPI_ISL_2399137, EPI_ISL_2399138, EPI_ISL_2399140, EPI_ISL_2399141, EPI_ISL_2399142, EPI_ISL_2399143, EPI_ISL_2399144, EPI_ISL_2399145, EPI_ISL_2399146, EPI_ISL_2399147, EPI_ISL_2399148, EPI_ISL_2399149, EPI_ISL_2399150, EPI_ISL_2399151, EPI_ISL_2399152, EPI_ISL_2399154, EPI_ISL_2399155, EPI_ISL_2399156, EPI_ISL_2399157, EPI_ISL_2399158, EPI_ISL_2399159, EPI_ISL_2399160, EPI_ISL_2399161, EPI_ISL_2399162, EPI_ISL_2399163, EPI_ISL_2399164, EPI_ISL_2399165, EPI_ISL_2399166, EPI_ISL_2399167, EPI_ISL_2399168, EPI_ISL_2399169, EPI_ISL_2399170, EPI_ISL_2399171, EPI_ISL_2399172, EPI_ISL_2399173, EPI_ISL_2399174, EPI_ISL_2399175, EPI_ISL_2399176, EPI_ISL_2399177, EPI_ISL_2399178, EPI_ISL_2399179, EPI_ISL_2399180, EPI_ISL_2399181, EPI_ISL_2399182, EPI_ISL_2399183, EPI_ISL_2399184, EPI_ISL_2399185, EPI_ISL_2399186, EPI_ISL_2399187, EPI_ISL_2399188, EPI_ISL_2399189, EPI_ISL_2399190, EPI_ISL_2399191, EPI_ISL_2399192, EPI_ISL_2399193, EPI_ISL_2399194, EPI_ISL_2399195, EPI_ISL_2399196, EPI_ISL_2399197, EPI_ISL_2399198, EPI_ISL_2399199, EPI_ISL_2399200, EPI_ISL_2399201, EPI_ISL_2399202, EPI_ISL_2399203, EPI_ISL_2399204, EPI_ISL_2399205, EPI_ISL_2399206, EPI_ISL_2399207, EPI_ISL_2399208, EPI_ISL_2399209, EPI_ISL_2399210, EPI_ISL_2399211, EPI_ISL_2399212, EPI_ISL_2399213, EPI_ISL_2399214, EPI_ISL_2399215, EPI_ISL_2399216, EPI_ISL_2399217, EPI_ISL_2399218, EPI_ISL_2399219, EPI_ISL_2399220, EPI_ISL_2399221, EPI_ISL_2399222, EPI_ISL_2399223, EPI_ISL_2399224, EPI_ISL_2399225, EPI_ISL_2399226, EPI_ISL_2399227, EPI_ISL_2399228, EPI_ISL_2399229, EPI_ISL_2399230, EPI_ISL_2399231, EPI_ISL_2399232, EPI_ISL_2399233, EPI_ISL_2399234, EPI_ISL_2399235, EPI_ISL_2399236, EPI_ISL_2399237, EPI_ISL_2399238, EPI_ISL_2399239, EPI_ISL_2399240, EPI_ISL_2399241, EPI_ISL_2399242, EPI_ISL_2399243, EPI_ISL_2399244, EPI_ISL_2399245, EPI_ISL_2399246, EPI_ISL_2399247, EPI_ISL_2399248, EPI_ISL_2399249, EPI_ISL_2399250, EPI_ISL_2399251, EPI_ISL_2399252, EPI_ISL_2399253, EPI_ISL_2399254, EPI_ISL_2399255, EPI_ISL_2399256, EPI_ISL_2399257, EPI_ISL_2399258, EPI_ISL_2399259, EPI_ISL_2399260, EPI_ISL_2399261, EPI_ISL_2399262, EPI_ISL_2399263, EPI_ISL_2399264, EPI_ISL_2399265, EPI_ISL_2399266, EPI_ISL_2399267, EPI_ISL_2399268, EPI_ISL_2399269, EPI_ISL_2399270, EPI_ISL_2399271, EPI_ISL_2399272, EPI_ISL_2399273, EPI_ISL_2399274, EPI_ISL_2399275, EPI_ISL_2399276, EPI_ISL_2399277, EPI_ISL_2399278, EPI_ISL_2399279, EPI_ISL_2399280, EPI_ISL_2399281, EPI_ISL_2399282, EPI_ISL_2399283, EPI_ISL_2399284, EPI_ISL_2399285, EPI_ISL_2399286, EPI_ISL_2399287, EPI_ISL_2399288, EPI_ISL_2399289, EPI_ISL_2399290, EPI_ISL_2399291, EPI_ISL_2399292, EPI_ISL_2399293, EPI_ISL_2399294, EPI_ISL_2399295, EPI_ISL_2399296, EPI_ISL_2399297, EPI_ISL_2399298, EPI_ISL_2399299, EPI_ISL_2399300, EPI_ISL_2399301, EPI_ISL_2399302, EPI_ISL_2399303, EPI_ISL_2399304, EPI_ISL_2399305, EPI_ISL_2399306, EPI_ISL_2399307, EPI_ISL_2399308, EPI_ISL_2399309, EPI_ISL_2399310, EPI_ISL_2399311, EPI_ISL_2399312, EPI_ISL_2399314, EPI_ISL_2399315, EPI_ISL_2399316, EPI_ISL_2399317, EPI_ISL_2399319, EPI_ISL_2399320, EPI_ISL_2399322, EPI_ISL_2399323, EPI_ISL_2399324, EPI_ISL_2399325, EPI_ISL_2399326, EPI_ISL_2399327, EPI_ISL_2399328, EPI_ISL_2399329, EPI_ISL_2399330, EPI_ISL_2399331, EPI_ISL_2399332, EPI_ISL_2399333, EPI_ISL_2399334, EPI_ISL_2399335, EPI_ISL_2399336, EPI_ISL_2399337, EPI_ISL_2399338, EPI_ISL_2399339, EPI_ISL_2399340, EPI_ISL_2399341, EPI_ISL_2399342, EPI_ISL_2399343, EPI_ISL_2399344, EPI_ISL_2399345, EPI_ISL_2399346, EPI_ISL_2399347, EPI_ISL_2399348, EPI_ISL_2399349, EPI_ISL_2399350, EPI_ISL_2399351, EPI_ISL_2399352, EPI_ISL_2399353, EPI_ISL_2399354, EPI_ISL_2399355, EPI_ISL_2399356, EPI_ISL_2399357, EPI_ISL_2399358, EPI_ISL_2399359, EPI_ISL_2399360, EPI_ISL_2399361, EPI_ISL_2399362, EPI_ISL_2399363, EPI_ISL_2399364, EPI_ISL_2399365, EPI_ISL_2399366, EPI_ISL_2399367, EPI_ISL_2399368, EPI_ISL_2399369, EPI_ISL_2399370, EPI_ISL_2399371, EPI_ISL_2399372, EPI_ISL_2399373, EPI_ISL_2399374, EPI_ISL_2399375, EPI_ISL_2399376, EPI_ISL_2399377, EPI_ISL_2399378, EPI_ISL_2399379, EPI_ISL_2399380, EPI_ISL_2399381, EPI_ISL_2399382, EPI_ISL_2399383, EPI_ISL_2399384, EPI_ISL_2399385, EPI_ISL_2399386, EPI_ISL_2399387, EPI_ISL_2399388, EPI_ISL_2399389, EPI_ISL_2399390, EPI_ISL_2399391, EPI_ISL_2399392, EPI_ISL_2399393, EPI_ISL_2399394, EPI_ISL_2399395, EPI_ISL_2399396, EPI_ISL_2399397, EPI_ISL_2399398, EPI_ISL_2399399, EPI_ISL_2399400, EPI_ISL_2399401, EPI_ISL_2399402, EPI_ISL_2399403, EPI_ISL_2399404, EPI_ISL_2399405, EPI_ISL_2399406, EPI_ISL_2399407, EPI_ISL_2399408, EPI_ISL_2399409, EPI_ISL_2399410, EPI_ISL_2399411, EPI_ISL_2399412, EPI_ISL_2399413, EPI_ISL_2399414, EPI_ISL_2399415, EPI_ISL_2399416, EPI_ISL_2399417 | | | |
| see above | Laboratory Corporation of America | Centers for Disease Control and Prevention Division of Viral Diseases, Pathogen Discovery | Dakota Howard, Dhwani Batra, Peter W. Cook, Kara Moser, Adrian Paskey, Jason Caravas, Benjamin Rambo-Martin, Shatavia Morrison, Christopher Gulvick, Scott Sammons, Yvette Unoarumhi, Darlene Wagner, Matthew Schmerer, Minoo Agarwal, Eyad Almasri, Debbie Boles, Ayla Burns, Nuthawin Charoensri, Oren Cohen, Susan Countryman, Mary Ann Cristobal, Bobbi Croy, Suzanne Dale, Hrushikesh Deshmukh, Amanda Douglas, Vincent Drouillon, Marcia Eisenberg, Howard Engler, Rama Ghatti, Prashant Gupta, Susan Hicks, Jake Humphrey, Lax Iyer, Lisa Pfefferle, Manoj Jain, Matthew Robinson, Mohan Kolli, Brian Krueger, Tim Kuphal, Stanley Letovsky, Michael Levandoski, Craig Lukasik, Jonathan Meltzer, Brian Norvell, Mindy Nye, Scott Parker, Christos Petropoulos, John Pruitt, Steven Ragan, Scott Ryan, Mike Sapeta, Jana Schroth, Suresh Babu Selvaraju, Goran Stevovic, Amanda Suchanek, Andrea Throop, Lyndon Tilson, Thomas Urban, Joe Voshell, Kimberly Wagner, Jonathan Williams, Mary Williamson, Qian Zeng, Tricia Zwiefelhofer, Clinton R. Paden, Duncan MacCannell |
| EPI_ISL_2399427 | UT-Unified State Labs: Public Health Utah DOH | Centers for Disease Control and Prevention Division of Viral Diseases, Pathogen Discovery | Mili Sheth, Sarah Nobles, Jasmine Padilla, Mark Burroughs, Shoshona Le, Katie Dillon, Peter Cook, Clinton R. Paden, Dhwani Batra, Krista Queen, Kristen Knipe, Dakota Howard, Yvette Unoarumhi, Darlene Wagner, Matthew Schmerer, Ben L. Rambo-Martin, Kristine Lacek, Sam Shepard, Alison Laufer Halpin, Dave Wentworth, Vivien Dugan, Suxiang Tong, Justin Lee |
| EPI_ISL_2399428 | RI State Health Laboratories | Centers for Disease Control and Prevention Division of Viral Diseases, Pathogen Discovery | Mili Sheth, Sarah Nobles, Jasmine Padilla, Mark Burroughs, Shoshona Le, Katie Dillon, Peter Cook, Clinton R. Paden, Dhwani Batra, Krista Queen, Kristen Knipe, Dakota Howard, Yvette Unoarumhi, Darlene Wagner, Matthew Schmerer, Ben L. Rambo-Martin, Kristine Lacek, Sam Shepard, Alison Laufer Halpin, Dave Wentworth, Vivien Dugan, Suxiang Tong, Justin Lee |
| EPI_ISL_2399429 | OR State PHL-Virology/Immunology Section | Centers for Disease Control and Prevention Division of Viral Diseases, Pathogen Discovery | Mili Sheth, Sarah Nobles, Jasmine Padilla, Mark Burroughs, Shoshona Le, Katie Dillon, Peter Cook, Clinton R. Paden, Dhwani Batra, Krista Queen, Kristen Knipe, Dakota Howard, Yvette Unoarumhi, Darlene Wagner, Matthew Schmerer, Ben L. Rambo-Martin, Kristine Lacek, Sam Shepard, Alison Laufer Halpin, Dave Wentworth, Vivien Dugan, Suxiang Tong, Justin Lee |
| EPI_ISL_2399430 | VI-US Virgin Islands Department of Health | Centers for Disease Control and Prevention Division of Viral Diseases, Pathogen Discovery | Mili Sheth, Sarah Nobles, Jasmine Padilla, Mark Burroughs, Shoshona Le, Katie Dillon, Peter Cook, Clinton R. Paden, Dhwani Batra, Krista Queen, Kristen Knipe, Dakota Howard, Yvette Unoarumhi, Darlene Wagner, Matthew Schmerer, Ben L. Rambo-Martin, Kristine Lacek, Sam Shepard, Alison Laufer Halpin, Dave Wentworth, Vivien Dugan, Suxiang Tong, Justin Lee |
| EPI_ISL_2399463, EPI_ISL_2399464, EPI_ISL_2399465, EPI_ISL_2399466, EPI_ISL_2399467, EPI_ISL_2399468, EPI_ISL_2399469, EPI_ISL_2399470, EPI_ISL_2399471, EPI_ISL_2399472, EPI_ISL_2399473, EPI_ISL_2399474, EPI_ISL_2399475, EPI_ISL_2399476, EPI_ISL_2399477, EPI_ISL_2399478, EPI_ISL_2399479, EPI_ISL_2399480, EPI_ISL_2399481, EPI_ISL_2399482, EPI_ISL_2399483, EPI_ISL_2399484, EPI_ISL_2399485, EPI_ISL_2399486, EPI_ISL_2399487, EPI_ISL_2399488, EPI_ISL_2399489, EPI_ISL_2399490, EPI_ISL_2399491, EPI_ISL_2399492, EPI_ISL_2399493, EPI_ISL_2399494, EPI_ISL_2399495, EPI_ISL_2399496, EPI_ISL_2399497, EPI_ISL_2399498, EPI_ISL_2399499, EPI_ISL_2399500, EPI_ISL_2399501, EPI_ISL_2399502, EPI_ISL_2399503, EPI_ISL_2399504, EPI_ISL_2399505, EPI_ISL_2399506, EPI_ISL_2399507, EPI_ISL_2399508, EPI_ISL_2399509, EPI_ISL_2399510, EPI_ISL_2399511, EPI_ISL_2399512, EPI_ISL_2399513, EPI_ISL_2399514, EPI_ISL_2399515 | | | |
| see above | Quest Diagnostics Incorporated | Centers for Disease Control and Prevention Division of Viral Diseases, Pathogen Discovery | Dakota Howard, Dhwani Batra, Peter W. Cook, Kara Moser, Adrian Paskey, Jason Caravas, Benjamin Rambo-Martin, Shatavia Morrison, Christopher Gulvick, Scott Sammons, Yvette Unoarumhi, Darlene Wagner, Matthew Schmerer, S. H. Rosenthal, A. Gerasimova, R. M. Kagan, B. Anderson, M. Hua, Y. Liu, L.E. Bernstein, K.E. Livingston, A. Perez, I. A. Shlyakhter, R. V. Rolando, R. Owen, P. Tanpaiboon, F. Lacbawan, Clinton R. Paden, Duncan MacCannell |
| EPI_ISL_2399520, EPI_ISL_2399521, EPI_ISL_2399522, EPI_ISL_2399523, EPI_ISL_2399524, EPI_ISL_2399525, EPI_ISL_2399526, EPI_ISL_2399527, EPI_ISL_2399528, EPI_ISL_2399529, EPI_ISL_2399530, EPI_ISL_2399531, EPI_ISL_2399532, EPI_ISL_2399533, EPI_ISL_2399534, EPI_ISL_2399535, EPI_ISL_2399536, EPI_ISL_2399537, EPI_ISL_2399538, EPI_ISL_2399539, EPI_ISL_2399540, EPI_ISL_2399541, EPI_ISL_2399542, EPI_ISL_2399543 | | | |
| see above | Wexner Medical Center | The Ohio State University College of Medicine | Koenig,S., Seminetta,J. |
| EPI_ISL_2399547, EPI_ISL_2399548, EPI_ISL_2399549, EPI_ISL_2399551, EPI_ISL_2399552, EPI_ISL_2399553, EPI_ISL_2399554, EPI_ISL_2399555, EPI_ISL_2399556, EPI_ISL_2399557, EPI_ISL_2399558, EPI_ISL_2399559, EPI_ISL_2399560, EPI_ISL_2399561, EPI_ISL_2399562, EPI_ISL_2399563, EPI_ISL_2399564, EPI_ISL_2399565, EPI_ISL_2399566, EPI_ISL_2399567, EPI_ISL_2399568, EPI_ISL_2399569, EPI_ISL_2399570, EPI_ISL_2399571, EPI_ISL_2399572, EPI_ISL_2399573, EPI_ISL_2399574, EPI_ISL_2399575, EPI_ISL_2399576, EPI_ISL_2399577, EPI_ISL_2399578, EPI_ISL_2399579, EPI_ISL_2399580, EPI_ISL_2399581, EPI_ISL_2399582, EPI_ISL_2399583, EPI_ISL_2399584, EPI_ISL_2399585, EPI_ISL_2399586, EPI_ISL_2399587, EPI_ISL_2399588, EPI_ISL_2399589, EPI_ISL_2399590, EPI_ISL_2399591, EPI_ISL_2399592, EPI_ISL_2399593, EPI_ISL_2399594, EPI_ISL_2399595, EPI_ISL_2399596, EPI_ISL_2399597, EPI_ISL_2399598, EPI_ISL_2399599, EPI_ISL_2399600, EPI_ISL_2399601, EPI_ISL_2399602, EPI_ISL_2399603, EPI_ISL_2399604, EPI_ISL_2399605, EPI_ISL_2399606, EPI_ISL_2399607, EPI_ISL_2399608, EPI_ISL_2399609, EPI_ISL_2399610 | | | |
| see above | The Ohio State University Applied Microbiology Services Laboratory | The Ohio State University Applied Microbiology Services Laboratory | Seth A. Faith PhD |
| EPI_ISL_2399612, | UCLA Clinical Micro Lab | Los Angeles County Public Health | P. Hemarajata et al. |

| | | | |
|---|---|---|---|
| EPI_ISL_2399613, EPI_ISL_2399614, EPI_ISL_2399615, EPI_ISL_2399616 | | Laboratories | |
| EPI_ISL_2399618 | Flow Diagnostics Lab | Los Angeles County Public Health Laboratories | P. Hemarajata et al. |
| EPI_ISL_2399619, EPI_ISL_2399620 | UCLA Clinical Micro Lab | Los Angeles County Public Health Laboratories | P. Hemarajata et al. |
| EPI_ISL_2399621, EPI_ISL_2399622, EPI_ISL_2399623, EPI_ISL_2399624, EPI_ISL_2399625, EPI_ISL_2399626, EPI_ISL_2399627, EPI_ISL_2399628, EPI_ISL_2399629, EPI_ISL_2399630, EPI_ISL_2399631, EPI_ISL_2399632, EPI_ISL_2399633, EPI_ISL_2399634, EPI_ISL_2399635, EPI_ISL_2399636, EPI_ISL_2399637, EPI_ISL_2399638, EPI_ISL_2399639, EPI_ISL_2399640, EPI_ISL_2399641, EPI_ISL_2399642, EPI_ISL_2399643, EPI_ISL_2399644, EPI_ISL_2399645, EPI_ISL_2399646, EPI_ISL_2399647, EPI_ISL_2399648, EPI_ISL_2399649, EPI_ISL_2399650, EPI_ISL_2399651, EPI_ISL_2399652, EPI_ISL_2399653, EPI_ISL_2399654, EPI_ISL_2399655, EPI_ISL_2399656, EPI_ISL_2399657, EPI_ISL_2399658, EPI_ISL_2399659, EPI_ISL_2399660, EPI_ISL_2399661, EPI_ISL_2399662, EPI_ISL_2399663, EPI_ISL_2399664, EPI_ISL_2399665, EPI_ISL_2399666, EPI_ISL_2399667, EPI_ISL_2399668, EPI_ISL_2399669, EPI_ISL_2399670, EPI_ISL_2399671, EPI_ISL_2399672, EPI_ISL_2399673, EPI_ISL_2399674, EPI_ISL_2399675, EPI_ISL_2399676, EPI_ISL_2399677, EPI_ISL_2399678, EPI_ISL_2399679, EPI_ISL_2399680, EPI_ISL_2399681, EPI_ISL_2399682, EPI_ISL_2399683, EPI_ISL_2399684, EPI_ISL_2399685, EPI_ISL_2399686, EPI_ISL_2399687, EPI_ISL_2399688, EPI_ISL_2399689, EPI_ISL_2399690, EPI_ISL_2399691, EPI_ISL_2399692, EPI_ISL_2399693, EPI_ISL_2399694, EPI_ISL_2399695, EPI_ISL_2399696, EPI_ISL_2399697, EPI_ISL_2399698, EPI_ISL_2399699, EPI_ISL_2399700, EPI_ISL_2399701, EPI_ISL_2399702, EPI_ISL_2399703, EPI_ISL_2399704, EPI_ISL_2399705, EPI_ISL_2399706, EPI_ISL_2399707, EPI_ISL_2399708, EPI_ISL_2399709, EPI_ISL_2399710, EPI_ISL_2399711, EPI_ISL_2399712, EPI_ISL_2399713, EPI_ISL_2399714, EPI_ISL_2399715, EPI_ISL_2399716, EPI_ISL_2399717, EPI_ISL_2399718, EPI_ISL_2399719, EPI_ISL_2399720, EPI_ISL_2399721, EPI_ISL_2399722, EPI_ISL_2399723, EPI_ISL_2399724, EPI_ISL_2399725, EPI_ISL_2399726, EPI_ISL_2399727, EPI_ISL_2399728, EPI_ISL_2399729, EPI_ISL_2399730, EPI_ISL_2399731, EPI_ISL_2399732, EPI_ISL_2399733, EPI_ISL_2399734, EPI_ISL_2399735, EPI_ISL_2399736, EPI_ISL_2399737, EPI_ISL_2399738, EPI_ISL_2399739, EPI_ISL_2399740, EPI_ISL_2399741, EPI_ISL_2399742, EPI_ISL_2399743, EPI_ISL_2399744, EPI_ISL_2399745, EPI_ISL_2399746, EPI_ISL_2399747, EPI_ISL_2399749, EPI_ISL_2399750, EPI_ISL_2399751, EPI_ISL_2399752, EPI_ISL_2399753, EPI_ISL_2399754, EPI_ISL_2399756, EPI_ISL_2399757 | | | |
| see above | The Ohio State University Applied Microbiology Services Laboratory | The Ohio State University Applied Microbiology Services Laboratory | Seth A. Faith PhD |
| EPI_ISL_2399758, EPI_ISL_2399759, EPI_ISL_2399760, EPI_ISL_2399761, EPI_ISL_2399762, EPI_ISL_2399763, EPI_ISL_2399764, EPI_ISL_2399765, EPI_ISL_2399766, EPI_ISL_2399767, EPI_ISL_2399768, EPI_ISL_2399769, EPI_ISL_2399770, EPI_ISL_2399771, EPI_ISL_2399772, EPI_ISL_2399773, EPI_ISL_2399774, EPI_ISL_2399775, EPI_ISL_2399776, EPI_ISL_2399777, EPI_ISL_2399778, EPI_ISL_2399779, EPI_ISL_2399780, EPI_ISL_2399781, EPI_ISL_2399782, EPI_ISL_2399783, EPI_ISL_2399784, EPI_ISL_2399785 | | | |
| see above | Wexner Medical Center | The Ohio State University College of Medicine | Koenig,S., Seminetta,J. |
| EPI_ISL_2400091, EPI_ISL_2400092, EPI_ISL_2400093, EPI_ISL_2400094, EPI_ISL_2400095, EPI_ISL_2400096, EPI_ISL_2400097, EPI_ISL_2400098, EPI_ISL_2400099, EPI_ISL_2400100, EPI_ISL_2400101, EPI_ISL_2400102, EPI_ISL_2400103, EPI_ISL_2400104, EPI_ISL_2400105, EPI_ISL_2400106, EPI_ISL_2400107, EPI_ISL_2400108, EPI_ISL_2400109, EPI_ISL_2400110, EPI_ISL_2400111, EPI_ISL_2400112, EPI_ISL_2400113, EPI_ISL_2400114, EPI_ISL_2400115, EPI_ISL_2400116, EPI_ISL_2400117, EPI_ISL_2400118, EPI_ISL_2400119, EPI_ISL_2400120 | | | |
| see above | Wyoming Public Health Laboratory | Wyoming Public Health Laboratory | Jim Mildenberger, Taylor Fearing, Channing Weber, Ashley Norberg, Chayse Rowley, Marley Goetz, Brian Dominguez, Elliot Thomasson, Brittany Oher, Sam Britz, Cari Sloma, and Rob Christensen |
| EPI_ISL_2400121, EPI_ISL_2400122, EPI_ISL_2400123, EPI_ISL_2400124, EPI_ISL_2400125, EPI_ISL_2400126, EPI_ISL_2400127, EPI_ISL_2400128, EPI_ISL_2400129, EPI_ISL_2400130, EPI_ISL_2400131, EPI_ISL_2400132, EPI_ISL_2400133, EPI_ISL_2400134 | | | |
| see above | Kansas Health and Environmental Lab | Kansas Health and Environmental Lab | Mike Grose, Katherine Wiggins, Jonathan Barnell, Ben Olsen, and Phil Adam |
| EPI_ISL_2400226, EPI_ISL_2400227, EPI_ISL_2400228, EPI_ISL_2400229, EPI_ISL_2400230, EPI_ISL_2400231, EPI_ISL_2400232 | Wexner Medical Center | The Ohio State University College of Medicine | Koenig,S., Seminetta,J. |
| EPI_ISL_2400233, EPI_ISL_2400234, EPI_ISL_2400235, EPI_ISL_2400236, EPI_ISL_2400237, EPI_ISL_2400238, EPI_ISL_2400239, EPI_ISL_2400240, EPI_ISL_2400241, EPI_ISL_2400242, EPI_ISL_2400243 | | | |
| see above | Wichita State University - Molecular Diagnostics Lab | Kansas Health and Environmental Lab | Mike Grose, Katherine Wiggins, Jonathan Barnell, Ben Olsen, and Phil Adam |
| EPI_ISL_2400245, EPI_ISL_2400246, EPI_ISL_2400247, EPI_ISL_2400248, EPI_ISL_2400249, EPI_ISL_2400250, EPI_ISL_2400251, EPI_ISL_2400252, EPI_ISL_2400253, EPI_ISL_2400254, EPI_ISL_2400255, EPI_ISL_2400256, EPI_ISL_2400257, EPI_ISL_2400258, EPI_ISL_2400259, EPI_ISL_2400261, EPI_ISL_2400262, EPI_ISL_2400263 | | | |
| see above | Massachusetts State Public Health Laboratory | Massachusetts State Public Health Laboratory | Andrew Lang, Timelia Fink, Glen Gallagher, Sandra Smole |
| EPI_ISL_2400264, EPI_ISL_2400265, EPI_ISL_2400266, EPI_ISL_2400267, EPI_ISL_2400268, EPI_ISL_2400269, EPI_ISL_2400270, EPI_ISL_2400271, EPI_ISL_2400272, EPI_ISL_2400273, EPI_ISL_2400274, EPI_ISL_2400275, EPI_ISL_2400276, EPI_ISL_2400277, EPI_ISL_2400278, EPI_ISL_2400279, EPI_ISL_2400280, EPI_ISL_2400281, EPI_ISL_2400282, EPI_ISL_2400283, EPI_ISL_2400284, EPI_ISL_2400285, EPI_ISL_2400286 | | | |
| see above | Nebraska Public Health Laboratory | NPHL COVID-19 Response Team | NPHL COVID-19 Response Team |
| EPI_ISL_2400287 | NYU Langone Health | Departments of Pathology and Medicine, New York University School of Medicine | Adriana Heguy, Dacia Dimartino, Emily Guzman, Christian Marier, Peter Meyn, Sitharam Ramaswami, Gael Westby, Paul Zappile, Yutong Zhang, Paolo Cotzia, Guiqing Wang |
| EPI_ISL_2400288, EPI_ISL_2400289, EPI_ISL_2400290, EPI_ISL_2400291, EPI_ISL_2400292, EPI_ISL_2400293, EPI_ISL_2400295, EPI_ISL_2400296, EPI_ISL_2400297, EPI_ISL_2400298, EPI_ISL_2400299, EPI_ISL_2400333, EPI_ISL_2400335 | | | |
| see above | Kansas Health and Environmental Lab | Kansas Health and Environmental Lab | Mike Grose, Katherine Wiggins, Jonathan Barnell, Ben Olsen, and Phil Adam |
| EPI_ISL_2400336 | NYU Langone Health | Departments of Pathology and Medicine, New York University School of Medicine | Adriana Heguy, Dacia Dimartino, Emily Guzman, Christian Marier, Peter Meyn, Sitharam Ramaswami, Gael Westby, Paul Zappile, Yutong Zhang, Paolo Cotzia, Guiqing Wang |
| EPI_ISL_2400337, EPI_ISL_2400338, EPI_ISL_2400339, EPI_ISL_2400340, EPI_ISL_2400341, EPI_ISL_2400343, EPI_ISL_2400344, EPI_ISL_2400345 | Kansas Health and Environmental Lab | Kansas Health and Environmental Lab | Mike Grose, Katherine Wiggins, Jonathan Barnell, Ben Olsen, and Phil Adam |
| EPI_ISL_2400357, EPI_ISL_2400358, EPI_ISL_2400360, EPI_ISL_2400361, EPI_ISL_2400362, EPI_ISL_2400363, EPI_ISL_2400364, EPI_ISL_2400366, EPI_ISL_2400367, EPI_ISL_2400368, EPI_ISL_2400369, EPI_ISL_2400370, EPI_ISL_2400371, EPI_ISL_2400372, EPI_ISL_2400373, EPI_ISL_2400374, EPI_ISL_2400384 | | | |
| see above | University of Michigan Clinical Microbiology Laboratory | Lauring Lab, University of Michigan, Department of Microbiology and Immunology | Gilbert |
| EPI_ISL_2400385, EPI_ISL_2400386 | IVY3 Central Lab, Vanderbilt University Medical Center | Lauring Lab, University of Michigan, Department of Microbiology and Immunology | Gilbert |
| EPI_ISL_2400387, EPI_ISL_2400388, EPI_ISL_2400389, EPI_ISL_2400390, EPI_ISL_2400391, EPI_ISL_2400392 | DC Public Health Lab/ Dept. of Forensic Sciences | DC Public Health Lab/ Dept. of Forensic Sciences | Janis Doss, Scott Nguyen, Elizabeth Zelaya, Sarah Scott, Connie Maza, Monica Mann, Brittany Hamilton, David Payne, Jocelyn Hauser |
| EPI_ISL_2400416 | Wyoming Public Health Laboratory | Wyoming Public Health Laboratory | Jim Mildenberger, Wanda Manley, Noah Hull, Taylor Fearing, Lynette Gumbleton, Channing Weber, Ashley Norberg, Chayse Rowley, Marley Goetz, Brian Dominguez, Elliot Thomasson, Sam Britz, Cari Sloma, and Rob Christensen |
| EPI_ISL_2400425 | DOHMH Morrisania | New York City Public Health Laboratory | Jade Wang, et al. |
| EPI_ISL_2400426 | DOHMH Chelsea | New York City Public Health Laboratory | Jade Wang, et al. |
| EPI_ISL_2400427 | Department of Homeless Services | New York City Public Health Laboratory | Jade Wang, et al. |
| EPI_ISL_2400428, EPI_ISL_2400429 | OCME Office Of Chief Medical Examiner | New York City Public Health Laboratory | Jade Wang, et al. |
| EPI_ISL_2400430 | DOHMH Crown Heights | New York City Public Health Laboratory | Jade Wang, et al. |
| EPI_ISL_2400431 | DOHMH PHL | New York City Public Health Laboratory | Jade Wang, et al. |
| EPI_ISL_2400432 | DOHMH Central Harlem | New York City Public Health Laboratory | Jade Wang, et al. |
| EPI_ISL_2400433 | DOHMH Morrisania | New York City Public Health Laboratory | Jade Wang, et al. |
| EPI_ISL_2400434, EPI_ISL_2400435 | DOHMH Corona | New York City Public Health Laboratory | Jade Wang, et al. |
| EPI_ISL_2400436 | DOHMH Chelsea | New York City Public Health Laboratory | Jade Wang, et al. |
| EPI_ISL_2400437, EPI_ISL_2400438, EPI_ISL_2400439, EPI_ISL_2400440, EPI_ISL_2400441 | Quest Diagnostics | New York City Public Health Laboratory | Jade Wang, et al. |
| EPI_ISL_2400442 | OCME Office Of Chief Medical Examiner | New York City Public Health Laboratory | Jade Wang, et al. |
| EPI_ISL_2400443, EPI_ISL_2400444, EPI_ISL_2400445, EPI_ISL_2400446, EPI_ISL_2400447, EPI_ISL_2400448, EPI_ISL_2400449, EPI_ISL_2400450, EPI_ISL_2400451, EPI_ISL_2400452, EPI_ISL_2400453, EPI_ISL_2400454, EPI_ISL_2400455, EPI_ISL_2400456, EPI_ISL_2400457, EPI_ISL_2400458, EPI_ISL_2400459 | | | |
| see above | Molecular Biology Laboratory, Westchester Medical Center | Genomics Core Laboratory, New York Medical College | Weihua Huang, Utsav Pandey, Jian Zhuge, Changhong Yin |
| EPI_ISL_2400468, EPI_ISL_2400469, EPI_ISL_2400470, EPI_ISL_2400471, EPI_ISL_2400472, EPI_ISL_2400473, EPI_ISL_2400474, EPI_ISL_2400475, EPI_ISL_2400476, EPI_ISL_2400477, EPI_ISL_2400478, EPI_ISL_2400479, EPI_ISL_2400480, EPI_ISL_2400481, EPI_ISL_2400482, EPI_ISL_2400483, EPI_ISL_2400484, EPI_ISL_2400485, EPI_ISL_2400486, EPI_ISL_2400487, EPI_ISL_2400488, EPI_ISL_2400489, EPI_ISL_2400490, EPI_ISL_2400491, EPI_ISL_2400492 | | | |

| | | | |
|---|---|---|---|
| see above | Maryland Genomics, Institute for Genome Sciences, University of Maryland School of Medicine | Maryland Genomics, Institute for Genome Sciences, University of Maryland School of Medicine | Tallon, Luke J; Sadzewicz, Lisa D; Humphrys, Mike; Ott, Sandra; Roussey, Holly; Mehta, Aditya; Vavikolanu, Kranthi; Fraser, Claire M; Ravel, Jacques |
| EPI_ISL_2400493, EPI_ISL_2400494, EPI_ISL_2400495, EPI_ISL_2400496, EPI_ISL_2400497, EPI_ISL_2400498 | Azarian Lab at Burnett School of Biomedical Sciences | Azarian Lab at Burnett School of Biomedical Sciences | Catherine Johnston, Eleonora Cella, Taj Azarian |
| EPI_ISL_2400511, EPI_ISL_2400513 | NCSLPH | NCSLPH | Chase K, Miller MC, Greene S, Glover W |
| EPI_ISL_2400514, EPI_ISL_2400516 | Notre Dame COVID Response and Testing Laboratory | Notre Dame Genomics & Bioinformatics Core Facility | Michael Pfrender, Joseph Sarro, Melissa Stephens |
| EPI_ISL_2401933, EPI_ISL_2401934, EPI_ISL_2401935, EPI_ISL_2401936, EPI_ISL_2401937, EPI_ISL_2401938, EPI_ISL_2401939, EPI_ISL_2401940, EPI_ISL_2401941, EPI_ISL_2401942, EPI_ISL_2401943, EPI_ISL_2401944, EPI_ISL_2401945, EPI_ISL_2401946, EPI_ISL_2401947, EPI_ISL_2401948, EPI_ISL_2401949, EPI_ISL_2401950, EPI_ISL_2401951, EPI_ISL_2401952, EPI_ISL_2401953, EPI_ISL_2401954, EPI_ISL_2401955, EPI_ISL_2401956, EPI_ISL_2401957, EPI_ISL_2401958, EPI_ISL_2401959, EPI_ISL_2401960, EPI_ISL_2401961, EPI_ISL_2401962, EPI_ISL_2401963, EPI_ISL_2401964, EPI_ISL_2401965, EPI_ISL_2401966, EPI_ISL_2401967, EPI_ISL_2401968, EPI_ISL_2401969, EPI_ISL_2401970, EPI_ISL_2401971, EPI_ISL_2401973, EPI_ISL_2401974, EPI_ISL_2401975, EPI_ISL_2401976, EPI_ISL_2401977, EPI_ISL_2401978, EPI_ISL_2401979, EPI_ISL_2401980, EPI_ISL_2401981, EPI_ISL_2401982, EPI_ISL_2401983, EPI_ISL_2401984, EPI_ISL_2401985, EPI_ISL_2401986, EPI_ISL_2401987, EPI_ISL_2401988, EPI_ISL_2401989, EPI_ISL_2401990, EPI_ISL_2401991, EPI_ISL_2401992, EPI_ISL_2401993, EPI_ISL_2401994, EPI_ISL_2401995, EPI_ISL_2401996, EPI_ISL_2401997, EPI_ISL_2401998, EPI_ISL_2401999, EPI_ISL_2402000, EPI_ISL_2402001, EPI_ISL_2402002, EPI_ISL_2402003, EPI_ISL_2402004, EPI_ISL_2402005, EPI_ISL_2402006, EPI_ISL_2402007, EPI_ISL_2402008, EPI_ISL_2402009, EPI_ISL_2402010, EPI_ISL_2402011, EPI_ISL_2402012, EPI_ISL_2402013, EPI_ISL_2402014 | | | |
| see above | Centro de Investigación Biomédica del Noreste (CIBIN) | Unidad de Genomica Avanzada | Consorcio Mexicano de Vigilancia Genomica (CoViGen-Mex). Authors (in alphabetical order): Julio Elias Alvarado-Yaah, Carlos F. Arias, Santiago Avila-Rios, Victor Hugo Borja-Aburto, Celia Boukadida, Juan Bautista Chale-Dzul , Jose Antonio Enciso-Moreno, Gloria Elena Espinoza-Ayala, Fernando Fontove-Herrera, Concepcion Grajales-Muniz, Ricardo Grande, Alfredo Herrera-Estrella, Carla Ivon Herrera-Najera, Pavel Isa, Brenda Irasema Maldonado-Meza, Bernardo Martinez-Miguel, Margarita Matias-Florentino, Maria Guadalupe de Jesus Mireles-Rivera, Gloria Maria Molina-Salinas, Hector Montoya-Fuentes, Jose Esteban Munoz-Medina, Jose de Jesus Nunez-Contreras, Alicia Ocana-Mondragon, Luis Alberto Ochoa-Carrera, Hector Esteban Paz-Juarez, Francisco Pulido, Helen Haydee Fernanda Ramirez-Plascencia, Angel Gustavo Salas-Lais, Jorge Ivan Salinal-Nevarez, Alejandro Sanchez-Flores, Clara Esperanza Santacruz-Tinoco, Maria Guadalupe Santiago-Mauricio, Nelly Selem-Mojica, Blanca Taboada, Gloria Vazquez |
| EPI_ISL_2402015, EPI_ISL_2402016, EPI_ISL_2402017, EPI_ISL_2402018, EPI_ISL_2402019, EPI_ISL_2402020, EPI_ISL_2402021, EPI_ISL_2402022, EPI_ISL_2402023, EPI_ISL_2402024, EPI_ISL_2402025, EPI_ISL_2402026, EPI_ISL_2402027, EPI_ISL_2402028, EPI_ISL_2402029, EPI_ISL_2402030, EPI_ISL_2402031, EPI_ISL_2402032, EPI_ISL_2402033, EPI_ISL_2402034, EPI_ISL_2402035, EPI_ISL_2402036, EPI_ISL_2402037, EPI_ISL_2402038, EPI_ISL_2402039, EPI_ISL_2402040, EPI_ISL_2402041, EPI_ISL_2402042, EPI_ISL_2402043, EPI_ISL_2402044, EPI_ISL_2402045, EPI_ISL_2402046, EPI_ISL_2402047, EPI_ISL_2402048, EPI_ISL_2402049, EPI_ISL_2402050, EPI_ISL_2402051, EPI_ISL_2402052, EPI_ISL_2402053, EPI_ISL_2402054, EPI_ISL_2402055, EPI_ISL_2402056, EPI_ISL_2402057, EPI_ISL_2402058, EPI_ISL_2402059, EPI_ISL_2402060, EPI_ISL_2402061, EPI_ISL_2402062, EPI_ISL_2402063, EPI_ISL_2402064, EPI_ISL_2402065 | | | |
| see above | Unidad de Investigación Médica de Yucatán (UIMY) | Unidad de Genomica Avanzada | Consorcio Mexicano de Vigilancia Genomica (CoViGen-Mex). Authors (in alphabetical order): Julio Elias Alvarado-Yaah, Carlos F. Arias, Santiago Avila-Rios, Victor Hugo Borja-Aburto, Celia Boukadida, Juan Bautista Chale-Dzul , Jose Antonio Enciso-Moreno, Gloria Elena Espinoza-Ayala, Fernando Fontove-Herrera, Concepcion Grajales-Muniz, Ricardo Grande, Alfredo Herrera-Estrella, Carla Ivon Herrera-Najera, Pavel Isa, Brenda Irasema Maldonado-Meza, Bernardo Martinez-Miguel, Margarita Matias-Florentino, Maria Guadalupe de Jesus Mireles-Rivera, Gloria Maria Molina-Salinas, Hector Montoya-Fuentes, Jose Esteban Munoz-Medina, Jose de Jesus Nunez-Contreras, Alicia Ocana-Mondragon, Luis Alberto Ochoa-Carrera, Hector Esteban Paz-Juarez, Francisco Pulido, Helen Haydee Fernanda Ramirez-Plascencia, Angel Gustavo Salas-Lais, Jorge Ivan Salinal-Nevarez, Alejandro Sanchez-Flores, Clara Esperanza Santacruz-Tinoco, Maria Guadalupe Santiago-Mauricio, Nelly Selem-Mojica, Blanca Taboada, Gloria Vazquez |
| EPI_ISL_2402066, EPI_ISL_2402067, EPI_ISL_2402068, EPI_ISL_2402069, EPI_ISL_2402070, EPI_ISL_2402071, EPI_ISL_2402072, EPI_ISL_2402073, EPI_ISL_2402074, EPI_ISL_2402075, EPI_ISL_2402076, EPI_ISL_2402077, EPI_ISL_2402078, EPI_ISL_2402079, EPI_ISL_2402080, EPI_ISL_2402081, EPI_ISL_2402082, EPI_ISL_2402083, EPI_ISL_2402084, EPI_ISL_2402085, EPI_ISL_2402086, EPI_ISL_2402087, EPI_ISL_2402088, EPI_ISL_2402089, EPI_ISL_2402090, EPI_ISL_2402091, EPI_ISL_2402092, EPI_ISL_2402093, EPI_ISL_2402094, EPI_ISL_2402095, EPI_ISL_2402096, EPI_ISL_2402097, EPI_ISL_2402098, EPI_ISL_2402099, EPI_ISL_2402100, EPI_ISL_2402101, EPI_ISL_2402102, EPI_ISL_2402103, EPI_ISL_2402104, EPI_ISL_2402105, EPI_ISL_2402106, EPI_ISL_2402107, EPI_ISL_2402108, EPI_ISL_2402109, EPI_ISL_2402110, EPI_ISL_2402111, EPI_ISL_2402112, EPI_ISL_2402113, EPI_ISL_2402114, EPI_ISL_2402115, EPI_ISL_2402116, EPI_ISL_2402117, EPI_ISL_2402118, EPI_ISL_2402119, EPI_ISL_2402120, EPI_ISL_2402121, EPI_ISL_2402122, EPI_ISL_2402123, EPI_ISL_2402124, EPI_ISL_2402125, EPI_ISL_2402126, EPI_ISL_2402127, EPI_ISL_2402128, EPI_ISL_2402129, EPI_ISL_2402130, EPI_ISL_2402131 | | | |
| see above | Unidad de Investigación Biomédica de Zacatecas (UIBZ) | Unidad de Genomica Avanzada | Consorcio Mexicano de Vigilancia Genomica (CoViGen-Mex). Authors (in alphabetical order): Julio Elias Alvarado-Yaah, Carlos F. Arias, Santiago Avila-Rios, Victor Hugo Borja-Aburto, Celia Boukadida, Juan Bautista Chale-Dzul , Jose Antonio Enciso-Moreno, Gloria Elena Espinoza-Ayala, Fernando Fontove-Herrera, Concepcion Grajales-Muniz, Ricardo Grande, Alfredo Herrera-Estrella, Carla Ivon Herrera-Najera, Pavel Isa, Brenda Irasema Maldonado-Meza, Bernardo Martinez-Miguel, Margarita Matias-Florentino, Maria Guadalupe de Jesus Mireles-Rivera, Gloria Maria Molina-Salinas, Hector Montoya-Fuentes, Jose Esteban Munoz-Medina, Jose de Jesus Nunez-Contreras, Alicia Ocana-Mondragon, Luis Alberto Ochoa-Carrera, Hector Esteban Paz-Juarez, Francisco Pulido, Helen Haydee Fernanda Ramirez-Plascencia, Angel Gustavo Salas-Lais, Jorge Ivan Salinal-Nevarez, Alejandro Sanchez-Flores, Clara Esperanza Santacruz-Tinoco, Maria Guadalupe Santiago-Mauricio, Nelly Selem-Mojica, Blanca Taboada, Gloria Vazquez |
| EPI_ISL_2402132, EPI_ISL_2402133, EPI_ISL_2402134, EPI_ISL_2402135, EPI_ISL_2402136, EPI_ISL_2402137, EPI_ISL_2402138, EPI_ISL_2402139, EPI_ISL_2402140, EPI_ISL_2402141, EPI_ISL_2402142, EPI_ISL_2402143, EPI_ISL_2402144, EPI_ISL_2402145, EPI_ISL_2402146, EPI_ISL_2402147, EPI_ISL_2402148, EPI_ISL_2402149, EPI_ISL_2402150, EPI_ISL_2402151, EPI_ISL_2402152, EPI_ISL_2402153, EPI_ISL_2402154, EPI_ISL_2402155, EPI_ISL_2402156, EPI_ISL_2402157, EPI_ISL_2402158, EPI_ISL_2402159, EPI_ISL_2402160, EPI_ISL_2402161, EPI_ISL_2402162, EPI_ISL_2402163, EPI_ISL_2402164, EPI_ISL_2402165, EPI_ISL_2402166, EPI_ISL_2402167, EPI_ISL_2402168, EPI_ISL_2402169, EPI_ISL_2402170, EPI_ISL_2402171, EPI_ISL_2402172, EPI_ISL_2402173, EPI_ISL_2402174, EPI_ISL_2402175, EPI_ISL_2402176, EPI_ISL_2402177, EPI_ISL_2402178 | | | |
| see above | Centro de Investigación Biomédica de Occidente (CIBO) | Unidad de Genomica Avanzada | Consorcio Mexicano de Vigilancia Genomica (CoViGen-Mex). Authors (in alphabetical order): Julio Elias Alvarado-Yaah, Carlos F. Arias, Santiago Avila-Rios, Victor Hugo Borja-Aburto, Celia Boukadida, Juan Bautista Chale-Dzul , Jose Antonio Enciso-Moreno, Gloria Elena Espinoza-Ayala, Fernando Fontove-Herrera, Concepcion Grajales-Muniz, Ricardo Grande, Alfredo Herrera-Estrella, Carla Ivon Herrera-Najera, Pavel Isa, Brenda Irasema Maldonado-Meza, Bernardo Martinez-Miguel, Margarita Matias-Florentino, Maria Guadalupe de Jesus Mireles-Rivera, Gloria Maria Molina-Salinas, Hector Montoya-Fuentes, Jose Esteban Munoz-Medina, Jose de Jesus Nunez-Contreras, Alicia Ocana-Mondragon, Luis Alberto Ochoa-Carrera, Hector Esteban Paz-Juarez, Francisco Pulido, Helen Haydee Fernanda Ramirez-Plascencia, Angel Gustavo Salas-Lais, Jorge Ivan Salinal-Nevarez, Alejandro Sanchez-Flores, Clara Esperanza Santacruz-Tinoco, Maria Guadalupe Santiago-Mauricio, Nelly Selem-Mojica, Blanca Taboada, Gloria Vazquez |
| EPI_ISL_2402179, EPI_ISL_2402180, EPI_ISL_2402181, EPI_ISL_2402183, EPI_ISL_2402184, EPI_ISL_2402185, EPI_ISL_2402186, EPI_ISL_2402187, EPI_ISL_2402188, EPI_ISL_2402189, EPI_ISL_2402190, EPI_ISL_2402191, EPI_ISL_2402192, EPI_ISL_2402193, EPI_ISL_2402194, EPI_ISL_2402195, EPI_ISL_2402196, EPI_ISL_2402197, EPI_ISL_2402198, EPI_ISL_2402199, EPI_ISL_2402200, EPI_ISL_2402201, EPI_ISL_2402202, EPI_ISL_2402203, EPI_ISL_2402204, EPI_ISL_2402205, EPI_ISL_2402206, EPI_ISL_2402207, EPI_ISL_2402208, EPI_ISL_2402209, EPI_ISL_2402210, EPI_ISL_2402211, EPI_ISL_2402212, EPI_ISL_2402213, EPI_ISL_2402214, EPI_ISL_2402215, EPI_ISL_2402216, EPI_ISL_2402217, EPI_ISL_2402218, EPI_ISL_2402219, EPI_ISL_2402220, EPI_ISL_2402221, EPI_ISL_2402222, EPI_ISL_2402223, EPI_ISL_2402224, EPI_ISL_2402225, EPI_ISL_2402226, EPI_ISL_2402227, EPI_ISL_2402228 | | | |
| see above | Laboratorio Central de Epidemiología (LCE) | Unidad de Genomica Avanzada | Consorcio Mexicano de Vigilancia Genomica (CoViGen-Mex). Authors (in alphabetical order): Julio Elias Alvarado-Yaah, Carlos F. Arias, Santiago Avila-Rios, Victor Hugo Borja-Aburto, Celia Boukadida, Juan Bautista Chale-Dzul , Jose Antonio Enciso-Moreno, Gloria Elena Espinoza-Ayala, Fernando Fontove-Herrera, Concepcion Grajales-Muniz, Ricardo Grande, Alfredo Herrera-Estrella, Carla Ivon Herrera-Najera, Pavel Isa, Brenda Irasema Maldonado-Meza, Bernardo Martinez-Miguel, Margarita Matias-Florentino, Maria Guadalupe de Jesus Mireles-Rivera, Gloria Maria Molina-Salinas, Hector Montoya-Fuentes, Jose Esteban Munoz-Medina, Jose de Jesus Nunez-Contreras, Alicia Ocana-Mondragon, Luis Alberto Ochoa-Carrera, Hector Esteban Paz-Juarez, Francisco Pulido, Helen Haydee Fernanda Ramirez-Plascencia, Angel Gustavo Salas-Lais, Jorge Ivan Salinal-Nevarez, Alejandro Sanchez-Flores, Clara Esperanza Santacruz-Tinoco, Maria Guadalupe Santiago-Mauricio, Nelly Selem-Mojica, Blanca Taboada, Gloria Vazquez |
| EPI_ISL_2402229, EPI_ISL_2402230, EPI_ISL_2402231, EPI_ISL_2402232, EPI_ISL_2402233, EPI_ISL_2402234, EPI_ISL_2402235, EPI_ISL_2402236, EPI_ISL_2402237, EPI_ISL_2402238, EPI_ISL_2402239, EPI_ISL_2402240, EPI_ISL_2402241, EPI_ISL_2402242, EPI_ISL_2402243, EPI_ISL_2402244, EPI_ISL_2402245, EPI_ISL_2402246, EPI_ISL_2402247, EPI_ISL_2402249, EPI_ISL_2402250, EPI_ISL_2402251, EPI_ISL_2402252, EPI_ISL_2402253, EPI_ISL_2402254, EPI_ISL_2402255, EPI_ISL_2402256, EPI_ISL_2402257, EPI_ISL_2402259, EPI_ISL_2402260, EPI_ISL_2402261, EPI_ISL_2402262, EPI_ISL_2402263, EPI_ISL_2402265, EPI_ISL_2402266, EPI_ISL_2402267, EPI_ISL_2402268, EPI_ISL_2402269, EPI_ISL_2402270, EPI_ISL_2402271, EPI_ISL_2402272, EPI_ISL_2402273, EPI_ISL_2402274, EPI_ISL_2402275, EPI_ISL_2402276, EPI_ISL_2402277, EPI_ISL_2402278, EPI_ISL_2402279, EPI_ISL_2402280, EPI_ISL_2402281, EPI_ISL_2402282, EPI_ISL_2402283, EPI_ISL_2402284, EPI_ISL_2402285, EPI_ISL_2402286, EPI_ISL_2402287, EPI_ISL_2402288, EPI_ISL_2402290, EPI_ISL_2402292, EPI_ISL_2402293, EPI_ISL_2402294, EPI_ISL_2402295, EPI_ISL_2402297, EPI_ISL_2402298, EPI_ISL_2402299, EPI_ISL_2402300, EPI_ISL_2402301, EPI_ISL_2402302, EPI_ISL_2402303, EPI_ISL_2402304 | | | |
| see above | US Air Force School of Aerospace Medicine | US Air Force School of Aerospace Medicine | Anthony Fries, Jennifer Meyer, William Gruner, Amanda Javorina, Carol Garrett, Sarah Purves, Clarise Starr, Elizabeth Macias |
| EPI_ISL_2402331, EPI_ISL_2402333, EPI_ISL_2402334, EPI_ISL_2402335, EPI_ISL_2402336, EPI_ISL_2402337, EPI_ISL_2402338, EPI_ISL_2402339, EPI_ISL_2402340, EPI_ISL_2402341, EPI_ISL_2402342, EPI_ISL_2402346, EPI_ISL_2402351, EPI_ISL_2402352, EPI_ISL_2402353, EPI_ISL_2402354, EPI_ISL_2402355, EPI_ISL_2402356, EPI_ISL_2402357, EPI_ISL_2402358, EPI_ISL_2402359, EPI_ISL_2402360, EPI_ISL_2402362, EPI_ISL_2402363, EPI_ISL_2402364, EPI_ISL_2402366, EPI_ISL_2402367, EPI_ISL_2402368, EPI_ISL_2402369, EPI_ISL_2402370 | | | |
| see above | Wyoming Public Health Laboratory | Wyoming Public Health Laboratory | Jim Mildenberger, Taylor Fearing, Channing Weber, Ashley Norberg, Chayse Rowley, Marley Goetz, Brian Dominguez, Elliot Thomasson, Brittany Oher, Sam Britz, Cari Sloma, and Rob Christensen |
| EPI_ISL_2402572, EPI_ISL_2402574, EPI_ISL_2402575, EPI_ISL_2402577, EPI_ISL_2402578, EPI_ISL_2402579, EPI_ISL_2402580, EPI_ISL_2402581, EPI_ISL_2402582, EPI_ISL_2402583, EPI_ISL_2402584, EPI_ISL_2402586, EPI_ISL_2402588, EPI_ISL_2402589, EPI_ISL_2402590, EPI_ISL_2402591, EPI_ISL_2402592, EPI_ISL_2402593, EPI_ISL_2402594, EPI_ISL_2402596, EPI_ISL_2402597, EPI_ISL_2402598, EPI_ISL_2402599, EPI_ISL_2402600, EPI_ISL_2402601, EPI_ISL_2402602 | | | |
| see above | Minnesota Department of Health, Public Health Laboratory | Minnesota Department of Health, Public Health Laboratory | Alexandra Lorentz, Jacob Garfin, Matt Plumb, and Xiong Wang |
| EPI_ISL_2402603, EPI_ISL_2402604, EPI_ISL_2402605, EPI_ISL_2402606, EPI_ISL_2402607, EPI_ISL_2402608, EPI_ISL_2402609, EPI_ISL_2402610, EPI_ISL_2402611, EPI_ISL_2402612, EPI_ISL_2402613, EPI_ISL_2402614, EPI_ISL_2402615, EPI_ISL_2402616, EPI_ISL_2402617, EPI_ISL_2402618, EPI_ISL_2402619, EPI_ISL_2402620, EPI_ISL_2402621, EPI_ISL_2402623, EPI_ISL_2402624, EPI_ISL_2402625, EPI_ISL_2402626, EPI_ISL_2402627, EPI_ISL_2402628, EPI_ISL_2402629, EPI_ISL_2402630, EPI_ISL_2402631, EPI_ISL_2402632, EPI_ISL_2402811, EPI_ISL_2402812 | | | |
| see above | Centracare Laboratory Services | Minnesota Department of Health, Public Health Laboratory | Alexandra Lorentz, Jacob Garfin, Matt Plumb, and Xiong Wang |
| EPI_ISL_2402813, EPI_ISL_2402814, EPI_ISL_2402815, EPI_ISL_2402816, EPI_ISL_2402817, EPI_ISL_2402818, EPI_ISL_2402819, EPI_ISL_2402820, EPI_ISL_2402821, EPI_ISL_2402822, EPI_ISL_2402823, EPI_ISL_2402824, EPI_ISL_2402825, EPI_ISL_2402826, EPI_ISL_2402827, EPI_ISL_2402828, EPI_ISL_2402829, EPI_ISL_2402830, EPI_ISL_2402831, EPI_ISL_2402832, EPI_ISL_2402833, EPI_ISL_2402834, EPI_ISL_2402835, EPI_ISL_2402836, EPI_ISL_2402837, EPI_ISL_2402838, EPI_ISL_2402839, EPI_ISL_2402840, EPI_ISL_2402841, EPI_ISL_2402842, EPI_ISL_2402843, EPI_ISL_2402844, EPI_ISL_2402845, EPI_ISL_2402846, EPI_ISL_2402847, EPI_ISL_2402848, EPI_ISL_2402849, EPI_ISL_2402850, EPI_ISL_2402851, EPI_ISL_2402852, EPI_ISL_2402853, EPI_ISL_2402854, EPI_ISL_2402855, EPI_ISL_2402856, EPI_ISL_2402857, EPI_ISL_2402858, EPI_ISL_2402859, EPI_ISL_2402860, EPI_ISL_2402861, EPI_ISL_2402862, EPI_ISL_2402863, EPI_ISL_2402864, EPI_ISL_2402865, EPI_ISL_2402866, EPI_ISL_2402867, EPI_ISL_2402868, EPI_ISL_2402869, EPI_ISL_2402870, EPI_ISL_2402871, EPI_ISL_2402872, EPI_ISL_2402873, EPI_ISL_2402874, EPI_ISL_2402875, EPI_ISL_2402876, EPI_ISL_2402877, EPI_ISL_2402878, EPI_ISL_2402879, EPI_ISL_2402880, EPI_ISL_2402881, EPI_ISL_2402882, EPI_ISL_2402883, EPI_ISL_2402884, EPI_ISL_2402885, EPI_ISL_2402886, EPI_ISL_2402887, EPI_ISL_2402888, EPI_ISL_2402889, EPI_ISL_2402890, EPI_ISL_2402891, EPI_ISL_2402892, EPI_ISL_2402893, EPI_ISL_2402894, EPI_ISL_2402895, EPI_ISL_2402896, EPI_ISL_2402897, EPI_ISL_2402898, EPI_ISL_2402899, EPI_ISL_2402900, EPI_ISL_2402901, EPI_ISL_2402902, EPI_ISL_2402903, EPI_ISL_2402904, EPI_ISL_2402905, EPI_ISL_2402906, EPI_ISL_2402907, EPI_ISL_2402908, EPI_ISL_2402909, EPI_ISL_2402910, EPI_ISL_2402911, EPI_ISL_2402912, EPI_ISL_2402913, EPI_ISL_2402914, EPI_ISL_2402915, EPI_ISL_2402916, EPI_ISL_2402917, EPI_ISL_2402918, EPI_ISL_2402919, EPI_ISL_2402920, EPI_ISL_2402921, EPI_ISL_2402922, EPI_ISL_2402923, EPI_ISL_2402924, EPI_ISL_2402925, EPI_ISL_2402926, EPI_ISL_2402927, EPI_ISL_2402928, EPI_ISL_2402929, EPI_ISL_2402930, EPI_ISL_2402931, EPI_ISL_2402932, EPI_ISL_2402933, EPI_ISL_2402934, EPI_ISL_2402935 | | | |
| see above | Weill Cornell Medicine | New York Genome Center | Michael Zody, Andre Corvelo, Dayna M. Oschwald, Samantha Fennessey, Tom Maniatis, Melissa Cushing, Olivier Elemento, Margaret Elizabeth Ross, Chris Mason, Priya Velu, Hanna Rennert, Arryn Craney, Lars F Westblade |
| EPI_ISL_2402939, EPI_ISL_2402940, EPI_ISL_2402941, EPI_ISL_2402943, EPI_ISL_2402944, EPI_ISL_2402945, EPI_ISL_2402947, EPI_ISL_2402948, EPI_ISL_2402949, EPI_ISL_2402950, EPI_ISL_2402951, EPI_ISL_2402952, EPI_ISL_2402954, EPI_ISL_2402956, EPI_ISL_2402957, EPI_ISL_2402960, EPI_ISL_2402962, EPI_ISL_2402963, EPI_ISL_2402965, EPI_ISL_2402966, EPI_ISL_2402967, EPI_ISL_2402968, EPI_ISL_2402969, EPI_ISL_2402970, EPI_ISL_2402971, EPI_ISL_2402972, EPI_ISL_2402973, EPI_ISL_2402974, EPI_ISL_2402975, EPI_ISL_2402977, EPI_ISL_2402978, EPI_ISL_2402979, EPI_ISL_2402981, EPI_ISL_2402982, EPI_ISL_2402984, EPI_ISL_2402985, EPI_ISL_2402986, EPI_ISL_2402987, EPI_ISL_2402988, EPI_ISL_2402989, EPI_ISL_2402991, EPI_ISL_2402992, EPI_ISL_2402993, EPI_ISL_2402994, EPI_ISL_2402995, EPI_ISL_2402997, EPI_ISL_2402998, EPI_ISL_2402999, EPI_ISL_2403000, EPI_ISL_2403001, EPI_ISL_2403003, EPI_ISL_2403004, EPI_ISL_2403007 | | | |
| see above | North Dakota Department of Health, Public Health Laboratory | North Dakota Department of Health, Public Health Laboratory | Lisa Wingerter |
| EPI_ISL_2403009, EPI_ISL_2403010, EPI_ISL_2403011, EPI_ISL_2403012, EPI_ISL_2403013, EPI_ISL_2403014, EPI_ISL_2403015, EPI_ISL_2403016, EPI_ISL_2403017, EPI_ISL_2403018, EPI_ISL_2403019, EPI_ISL_2403020, EPI_ISL_2403021, EPI_ISL_2403022, EPI_ISL_2403023, EPI_ISL_2403024, EPI_ISL_2403025, EPI_ISL_2403026, EPI_ISL_2403027, EPI_ISL_2403028, EPI_ISL_2403029, EPI_ISL_2403030, EPI_ISL_2403031, EPI_ISL_2403032, EPI_ISL_2403033, EPI_ISL_2403034, EPI_ISL_2403035, EPI_ISL_2403036, EPI_ISL_2403037, EPI_ISL_2403038, EPI_ISL_2403039, EPI_ISL_2403040, EPI_ISL_2403041, EPI_ISL_2403042, EPI_ISL_2403043, EPI_ISL_2403044, EPI_ISL_2403046, EPI_ISL_2403047, EPI_ISL_2403048, EPI_ISL_2403049, EPI_ISL_2403050, EPI_ISL_2403051, EPI_ISL_2403052, EPI_ISL_2403053, EPI_ISL_2403054, EPI_ISL_2403055, EPI_ISL_2403056, EPI_ISL_2403057, EPI_ISL_2403058, EPI_ISL_2403059, EPI_ISL_2403060, EPI_ISL_2403061, EPI_ISL_2403062, EPI_ISL_2403063, EPI_ISL_2403064, EPI_ISL_2403065, EPI_ISL_2403066, EPI_ISL_2403067, EPI_ISL_2403068, EPI_ISL_2403069, EPI_ISL_2403070, EPI_ISL_2403071, EPI_ISL_2403072, EPI_ISL_2403073, EPI_ISL_2403074, EPI_ISL_2403075, EPI_ISL_2403076, EPI_ISL_2403077, EPI_ISL_2403078, EPI_ISL_2403079, EPI_ISL_2403080, EPI_ISL_2403081, EPI_ISL_2403082, EPI_ISL_2403083, EPI_ISL_2403084, EPI_ISL_2403085, EPI_ISL_2403086, EPI_ISL_2403087, EPI_ISL_2403088, EPI_ISL_2403089, EPI_ISL_2403090, EPI_ISL_2403091, EPI_ISL_2403092, EPI_ISL_2403093, EPI_ISL_2403094, EPI_ISL_2403095, EPI_ISL_2403096, EPI_ISL_2403097, EPI_ISL_2403098, EPI_ISL_2403101, EPI_ISL_2403102, EPI_ISL_2403103, EPI_ISL_2403104, EPI_ISL_2403105, EPI_ISL_2403106, EPI_ISL_2403107, EPI_ISL_2403108, EPI_ISL_2403109, EPI_ISL_2403110, EPI_ISL_2403111, EPI_ISL_2403112, EPI_ISL_2403113, EPI_ISL_2403114, EPI_ISL_2403115, EPI_ISL_2403116, EPI_ISL_2403117, EPI_ISL_2403118, EPI_ISL_2403119, EPI_ISL_2403120, EPI_ISL_2403121, EPI_ISL_2403122, EPI_ISL_2403123, EPI_ISL_2403124, EPI_ISL_2403125, EPI_ISL_2403127, EPI_ISL_2403129, EPI_ISL_2403130, EPI_ISL_2403131, EPI_ISL_2403132, EPI_ISL_2403133, EPI_ISL_2403134, EPI_ISL_2403135, EPI_ISL_2403136, EPI_ISL_2403137, EPI_ISL_2403138, EPI_ISL_2403139, EPI_ISL_2403140, EPI_ISL_2403141, EPI_ISL_2403142, EPI_ISL_2403144, EPI_ISL_2403145, EPI_ISL_2403146, EPI_ISL_2403147, EPI_ISL_2403148, EPI_ISL_2403149, EPI_ISL_2403150, EPI_ISL_2403151, EPI_ISL_2403152, EPI_ISL_2403153, EPI_ISL_2403154, EPI_ISL_2403155, EPI_ISL_2403156, EPI_ISL_2403157, EPI_ISL_2403158, EPI_ISL_2403159, EPI_ISL_2403160, EPI_ISL_2403161, EPI_ISL_2403162, EPI_ISL_2403163, EPI_ISL_2403164, EPI_ISL_2403165, EPI_ISL_2403166, EPI_ISL_2403167, EPI_ISL_2403168, EPI_ISL_2403169, EPI_ISL_2403170, EPI_ISL_2403171, EPI_ISL_2403172, EPI_ISL_2403173, EPI_ISL_2403174, EPI_ISL_2403175, EPI_ISL_2403176, EPI_ISL_2403177, EPI_ISL_2403178, EPI_ISL_2403179, EPI_ISL_2403180, EPI_ISL_2403181, EPI_ISL_2403182, EPI_ISL_2403183, EPI_ISL_2403184, EPI_ISL_2403185, EPI_ISL_2403186 | | | |
| see above | Illinois Department of Public Health - Springfield Lab | Illinois Department of Public Health - Springfield Lab | Bryan Sim, Gordon McCall |
| EPI_ISL_2403257 | Wyoming Public Health Laboratory | Wyoming Public Health Laboratory | Jim Mildenberger, Taylor Fearing, Channing Weber, Ashley Norberg, Chayse Rowley, Marley Goetz, Brian Dominguez, Elliot Thomasson,Sam Britz, Cari Sloma, Robert Petit, and Rob Christensen |
| EPI_ISL_2403263, EPI_ISL_2403264, EPI_ISL_2403265, EPI_ISL_2403268, EPI_ISL_2403269 | Kansas Health and Environmental Lab | Kansas Health and Environmental Lab | Katherine Wiggins, Mike Grose, Jonathan Barnell, Ben Olsen, and Phil Adam |
| EPI_ISL_2403657, EPI_ISL_2403658 | Minnesota Department of Health, Public Health Laboratory | Minnesota Department of Health, Public Health Laboratory | Alexandra Lorentz, Jacob Garfin, Matt Plumb, and Xiong Wang |

| | | | |
|---|---|---|---|
| EPI_ISL_2403659, EPI_ISL_2403660, EPI_ISL_2403661, EPI_ISL_2403662, EPI_ISL_2403663, EPI_ISL_2403664, EPI_ISL_2403665, EPI_ISL_2403666, EPI_ISL_2403667, EPI_ISL_2403668, EPI_ISL_2403669, EPI_ISL_2403670, EPI_ISL_2403671, EPI_ISL_2403672, EPI_ISL_2403673, EPI_ISL_2403674, EPI_ISL_2403675, EPI_ISL_2403676, EPI_ISL_2403677, EPI_ISL_2403678, EPI_ISL_2403679, EPI_ISL_2403680, EPI_ISL_2403681, EPI_ISL_2403682, EPI_ISL_2403684, EPI_ISL_2403685, EPI_ISL_2403686, EPI_ISL_2403687, EPI_ISL_2403689, EPI_ISL_2403690 | | | |
| see above | Hennepin County Medical Center | Minnesota Department of Health, Public Health Laboratory | Alexandra Lorentz, Jacob Garfin, Matt Plumb, and Xiong Wang |
| EPI_ISL_2403691 | Minnesota Department of Health, Public Health Laboratory | Minnesota Department of Health, Public Health Laboratory | Alexandra Lorentz, Jacob Garfin, Matt Plumb, and Xiong Wang |
| EPI_ISL_2403692, EPI_ISL_2403693, EPI_ISL_2403694, EPI_ISL_2403695, EPI_ISL_2403696, EPI_ISL_2403697, EPI_ISL_2403698, EPI_ISL_2403699, EPI_ISL_2403700, EPI_ISL_2403701, EPI_ISL_2403702 | | | |
| see above | Hennepin County Medical Center | Minnesota Department of Health, Public Health Laboratory | Alexandra Lorentz, Jacob Garfin, Matt Plumb, and Xiong Wang |
| EPI_ISL_2403703, EPI_ISL_2403704, EPI_ISL_2403705, EPI_ISL_2403706, EPI_ISL_2403707, EPI_ISL_2403708, EPI_ISL_2403709, EPI_ISL_2403710, EPI_ISL_2403711, EPI_ISL_2403712, EPI_ISL_2403713, EPI_ISL_2403714, EPI_ISL_2403715, EPI_ISL_2403716, EPI_ISL_2403717, EPI_ISL_2403718, EPI_ISL_2403719, EPI_ISL_2403720, EPI_ISL_2403721, EPI_ISL_2403722, EPI_ISL_2403723, EPI_ISL_2403724 | | | |
| see above | Minnesota Department of Health, Public Health Laboratory | Minnesota Department of Health, Public Health Laboratory | Alexandra Lorentz, Jacob Garfin, Matt Plumb, and Xiong Wang |
| EPI_ISL_2403725, EPI_ISL_2403726, EPI_ISL_2403727, EPI_ISL_2403728, EPI_ISL_2403731, EPI_ISL_2403732, EPI_ISL_2403733, EPI_ISL_2403734, EPI_ISL_2403735 | Kansas Health and Environmental Lab | Kansas Health and Environmental Lab | Katheirne Wiggins, Mike Grose, Jonathan Barnell, Ben Olsen, and Phil Adam |
| EPI_ISL_2404690, EPI_ISL_2404691, EPI_ISL_2404692, EPI_ISL_2404693, EPI_ISL_2404694, EPI_ISL_2404695, EPI_ISL_2404696, EPI_ISL_2404697, EPI_ISL_2404698, EPI_ISL_2404699, EPI_ISL_2404700, EPI_ISL_2404702, EPI_ISL_2404704, EPI_ISL_2404705, EPI_ISL_2404706, EPI_ISL_2404707, EPI_ISL_2404708, EPI_ISL_2404709, EPI_ISL_2404710, EPI_ISL_2404711, EPI_ISL_2404713, EPI_ISL_2404714, EPI_ISL_2404715, EPI_ISL_2404716, EPI_ISL_2404717, EPI_ISL_2404718, EPI_ISL_2404720, EPI_ISL_2404721, EPI_ISL_2404722, EPI_ISL_2404723, EPI_ISL_2404724, EPI_ISL_2404725, EPI_ISL_2404727 | | | |
| see above | Wyoming Public Health Laboratory | Wyoming Public Health Laboratory | Jim Mildenberger, Taylor Fearing, Channing Weber, Ashley Norberg, Chayse Rowley, Marley Goetz, Brian Dominguez, Elliot Thomasson, Brittany Oher, Sam Britz, Cari Sloma, and Rob Christensen |
| EPI_ISL_2404733, EPI_ISL_2404734, EPI_ISL_2404735, EPI_ISL_2404736, EPI_ISL_2404737, EPI_ISL_2404738, EPI_ISL_2404739, EPI_ISL_2404740, EPI_ISL_2404741, EPI_ISL_2404742, EPI_ISL_2404743, EPI_ISL_2404744, EPI_ISL_2404745, EPI_ISL_2404746, EPI_ISL_2404747, EPI_ISL_2404748, EPI_ISL_2404749, EPI_ISL_2404750, EPI_ISL_2404751, EPI_ISL_2404752, EPI_ISL_2404753, EPI_ISL_2404754, EPI_ISL_2404755, EPI_ISL_2404756, EPI_ISL_2404757, EPI_ISL_2404758 | | | |
| see above | Oregon State Public Health Laboratory | Oregon State Public Health Laboratory | Rafia Razzaque, Eugene Yeboah, Vanda Makris, Laura Tsaknaridis, John Fontana and Shane Sevey |
| EPI_ISL_2404759, EPI_ISL_2404760, EPI_ISL_2404761, EPI_ISL_2404762, EPI_ISL_2404763 | Oregon State University TRACE Project | OSU Center for Genome Research and Biocomputing | Oregon State University TRACE Project |
| EPI_ISL_2404764, EPI_ISL_2404765, EPI_ISL_2404766, EPI_ISL_2404767, EPI_ISL_2404768, EPI_ISL_2404769, EPI_ISL_2404770, EPI_ISL_2404771, EPI_ISL_2404772, EPI_ISL_2404773, EPI_ISL_2404774, EPI_ISL_2404775, EPI_ISL_2404776, EPI_ISL_2404777, EPI_ISL_2404778, EPI_ISL_2404779, EPI_ISL_2404780, EPI_ISL_2404781, EPI_ISL_2404782, EPI_ISL_2404783, EPI_ISL_2404784, EPI_ISL_2404785, EPI_ISL_2404786, EPI_ISL_2404787, EPI_ISL_2404790, EPI_ISL_2404791, EPI_ISL_2404793, EPI_ISL_2404794 | | | |
| see above | Wyoming Public Health Laboratory | Wyoming Public Health Laboratory | Jim Mildenberger, Taylor Fearing, Channing Weber, Ashley Norberg, Chayse Rowley, Marley Goetz, Brian Dominguez, Elliot Thomasson, Brittany Oher, Sam Britz, Cari Sloma, and Rob Christensen |
| EPI_ISL_2404795, EPI_ISL_2404796, EPI_ISL_2404797, EPI_ISL_2404798, EPI_ISL_2404799 | Weill Cornell Medicine | New York Genome Center | Michael Zody, Andre Corvelo, Dayna M. Oschwald, Samantha Fennessey, Tom Maniatis, Melissa Cushing, Olivier Elemento, Margaret Elizabeth Ross, Chris Mason, Priya Velu, Hanna Rennert, Arryn Craney, Lars F Westblade |
| EPI_ISL_2404801 | GOOD SAMARITAN HOSPTIAL | Santa Clara County public Health Laboratory | Santa Clara County Public Health Department |
| EPI_ISL_2404809, EPI_ISL_2404812, EPI_ISL_2404813, EPI_ISL_2404814, EPI_ISL_2404815, EPI_ISL_2404816, EPI_ISL_2404820, EPI_ISL_2404821, EPI_ISL_2404823, EPI_ISL_2404824, EPI_ISL_2404825 | | | |
| see above | VMC | Santa Clara County public Health Laboratory | Santa Clara County Public Health Department |
| EPI_ISL_2404827 | PAMF PALO ALTO | Santa Clara County public Health Laboratory | Santa Clara County Public Health Department |
| EPI_ISL_2404828, EPI_ISL_2404830, EPI_ISL_2404831, EPI_ISL_2404832, EPI_ISL_2404833, EPI_ISL_2404838, EPI_ISL_2404839 | VMC | Santa Clara County public Health Laboratory | Santa Clara County Public Health Department |
| EPI_ISL_2404840, EPI_ISL_2404841, EPI_ISL_2404842, EPI_ISL_2404843, EPI_ISL_2404844, EPI_ISL_2404845, EPI_ISL_2404846 | University of Oregon COVID-19 MAP Laboratory | University of Oregon Genomics and Cell Charactericzation Core Facility (GC3F) | Douglas Turnbull, Ariana White, Jeff Bishop, Jason Carriere, Jason Sydes, Peter Batzel |
| EPI_ISL_2404847 | Pomona Valley Hospital Medical Center | Los Angeles County Public Health Laboratories | P. Hemarajata et al. |
| EPI_ISL_2404848, EPI_ISL_2404849, EPI_ISL_2404850, EPI_ISL_2404851, EPI_ISL_2404852, EPI_ISL_2404853, EPI_ISL_2404854, EPI_ISL_2404855, EPI_ISL_2404856, EPI_ISL_2404857, EPI_ISL_2404858, EPI_ISL_2404859, EPI_ISL_2404860, EPI_ISL_2404861, EPI_ISL_2404862, EPI_ISL_2404863, EPI_ISL_2404864, EPI_ISL_2404865, EPI_ISL_2404866, EPI_ISL_2404867, EPI_ISL_2404868, EPI_ISL_2404869 | | | |
| see above | UCLA Clinical Micro Lab | Los Angeles County Public Health Laboratories | P. Hemarajata et al. |
| EPI_ISL_2404993, EPI_ISL_2404994, EPI_ISL_2404995, EPI_ISL_2404996, EPI_ISL_2404997, EPI_ISL_2404998, EPI_ISL_2404999, EPI_ISL_2405000, EPI_ISL_2405001, EPI_ISL_2405002, EPI_ISL_2405003, EPI_ISL_2405004, EPI_ISL_2405005, EPI_ISL_2405006, EPI_ISL_2405007, EPI_ISL_2405008, EPI_ISL_2405009, EPI_ISL_2405010, EPI_ISL_2405011, EPI_ISL_2405012, EPI_ISL_2405013, EPI_ISL_2405014, EPI_ISL_2405015, EPI_ISL_2405016, EPI_ISL_2405017, EPI_ISL_2405018, EPI_ISL_2405019, EPI_ISL_2405020, EPI_ISL_2405021, EPI_ISL_2405022, EPI_ISL_2405023, EPI_ISL_2405024, EPI_ISL_2405025, EPI_ISL_2405026, EPI_ISL_2405027, EPI_ISL_2405028, EPI_ISL_2405029, EPI_ISL_2405030, EPI_ISL_2405031, EPI_ISL_2405032, EPI_ISL_2405033, EPI_ISL_2405034, EPI_ISL_2405035, EPI_ISL_2405036, EPI_ISL_2405037, EPI_ISL_2405038, EPI_ISL_2405039, EPI_ISL_2405040, EPI_ISL_2405041, EPI_ISL_2405042, EPI_ISL_2405043, EPI_ISL_2405044, EPI_ISL_2405045, EPI_ISL_2405046, EPI_ISL_2405047, EPI_ISL_2405048, EPI_ISL_2405049, EPI_ISL_2405050, EPI_ISL_2405051, EPI_ISL_2405052, EPI_ISL_2405053, EPI_ISL_2405054, EPI_ISL_2405055, EPI_ISL_2405056, EPI_ISL_2405057, EPI_ISL_2405058, EPI_ISL_2405059, EPI_ISL_2405060, EPI_ISL_2405061, EPI_ISL_2405062, EPI_ISL_2405063, EPI_ISL_2405064, EPI_ISL_2405065, EPI_ISL_2405066, EPI_ISL_2405067, EPI_ISL_2405068, EPI_ISL_2405069, EPI_ISL_2405070, EPI_ISL_2405071, EPI_ISL_2405072, EPI_ISL_2405073, EPI_ISL_2405074, EPI_ISL_2405075, EPI_ISL_2405076, EPI_ISL_2405077, EPI_ISL_2405078, EPI_ISL_2405079, EPI_ISL_2405080, EPI_ISL_2405081, EPI_ISL_2405082, EPI_ISL_2405083, EPI_ISL_2405084, EPI_ISL_2405085, EPI_ISL_2405086, EPI_ISL_2405087, EPI_ISL_2405088, EPI_ISL_2405089, EPI_ISL_2405090, EPI_ISL_2405091, EPI_ISL_2405092, EPI_ISL_2405093 | | | |
| see above | UW Virology Lab | UW Virology Lab | Pavitra Roychoudhury, Hong Xie, Lasata Shrestha, Shah Mohamed Bakhash, Noah R. Baker, Sean Ellis, Saraswathi Sathees, Meei-Li Huang, Keith R Jerome, Alexander Greninger |
| EPI_ISL_2405094, EPI_ISL_2405095, EPI_ISL_2405096, EPI_ISL_2405097, EPI_ISL_2405098, EPI_ISL_2405099, EPI_ISL_2405100, EPI_ISL_2405101, EPI_ISL_2405102, EPI_ISL_2405103, EPI_ISL_2405104, EPI_ISL_2405105, EPI_ISL_2405106, EPI_ISL_2405107, EPI_ISL_2405108 | | | |
| see above | Molecular Biology Laboratory, Westchester Medical Center | Genomics Core Laboratory, New York Medical College | Weihua Huang, Utsav Pandey, Jian Zhuge, Changhong Yin |
| EPI_ISL_2405167 | UCLA Clinical Micro Lab | Los Angeles County Public Health Laboratories | P. Hemarajata et al. |
| EPI_ISL_2405178 | Pomona Valley Hospital Medical Center | Los Angeles County Public Health Laboratories | P. Hemarajata et al. |
| EPI_ISL_2405179 | California Hospital Medical Center | Los Angeles County Public Health Laboratories | P. Hemarajata et al. |
| EPI_ISL_2405180, EPI_ISL_2405181 | USC Clinical Lab | Los Angeles County Public Health Laboratories | P. Hemarajata et al. |
| EPI_ISL_2405182 | Huntington Memorial Hospital | Los Angeles County Public Health Laboratories | P. Hemarajata et al. |
| EPI_ISL_2405183 | UCLA Clinical Micro Lab | Los Angeles County Public Health Laboratories | P. Hemarajata et al. |
| EPI_ISL_2405184, EPI_ISL_2405185 | Flow Diagnostics Lab | Los Angeles County Public Health Laboratories | P. Hemarajata et al. |
| EPI_ISL_2405186, EPI_ISL_2405187, EPI_ISL_2405188, EPI_ISL_2405189, EPI_ISL_2405190, EPI_ISL_2405191, EPI_ISL_2405192, EPI_ISL_2405193, EPI_ISL_2405194, EPI_ISL_2405195, EPI_ISL_2405196, EPI_ISL_2405197, EPI_ISL_2405198, EPI_ISL_2405199, EPI_ISL_2405200 | | | |
| see above | USC Clinical Lab | Los Angeles County Public Health Laboratories | P. Hemarajata et al. |
| EPI_ISL_2405202, EPI_ISL_2405203, EPI_ISL_2405204 | Pomona Valley Hospital Medical Center | Los Angeles County Public Health Laboratories | P. Hemarajata et al. |
| EPI_ISL_2405205 | California Hospital Medical Center | Los Angeles County Public Health | P. Hemarajata et al. |

| | | | |
|---|---|---|---|
| | | Laboratories | |
| EPI_ISL_2405206 | Providence Saint Joseph Medical Center | Los Angeles County Public Health Laboratories | P. Hemarajata et al. |
| EPI_ISL_2405207, EPI_ISL_2405208, EPI_ISL_2405210, EPI_ISL_2405212, EPI_ISL_2405213, EPI_ISL_2405214, EPI_ISL_2405215 | Flow Diagnostics Lab | Los Angeles County Public Health Laboratories | P. Hemarajata et al. |
| EPI_ISL_2420130, EPI_ISL_2420133, EPI_ISL_2420136, EPI_ISL_2420139, EPI_ISL_2420142, EPI_ISL_2420144, EPI_ISL_2420149, EPI_ISL_2420152, EPI_ISL_2420155, EPI_ISL_2420158, EPI_ISL_2420160, EPI_ISL_2420163, EPI_ISL_2420165, EPI_ISL_2420168, EPI_ISL_2420170, EPI_ISL_2420173, EPI_ISL_2420182, EPI_ISL_2420185, EPI_ISL_2420188, EPI_ISL_2420194, EPI_ISL_2420197, EPI_ISL_2420200, EPI_ISL_2420208, EPI_ISL_2420211, EPI_ISL_2420213, EPI_ISL_2420216, EPI_ISL_2420219, EPI_ISL_2420222, EPI_ISL_2420225, EPI_ISL_2420228, EPI_ISL_2420233, EPI_ISL_2420236, EPI_ISL_2420239, EPI_ISL_2420242, EPI_ISL_2420244, EPI_ISL_2420245 | | | |
| see above | IN State Department of Health Laboratory Services | IN State Department of Health Laboratory Services | Cassandra Campion, Jamie Yeadon, Brian Pope, Lixia Liu, Kyle Brownlee, Melissa Hindenlang, Mark Glazier |
| EPI_ISL_2420985, EPI_ISL_2420986, EPI_ISL_2420987, EPI_ISL_2420988, EPI_ISL_2420989, EPI_ISL_2420990, EPI_ISL_2420991, EPI_ISL_2420992, EPI_ISL_2420993, EPI_ISL_2420994, EPI_ISL_2420995, EPI_ISL_2420996, EPI_ISL_2420997, EPI_ISL_2420998, EPI_ISL_2420999, EPI_ISL_2421000, EPI_ISL_2421001, EPI_ISL_2421002, EPI_ISL_2421003, EPI_ISL_2421004, EPI_ISL_2421005, EPI_ISL_2421006, EPI_ISL_2421007, EPI_ISL_2421008, EPI_ISL_2421009, EPI_ISL_2421010, EPI_ISL_2421011, EPI_ISL_2421012, EPI_ISL_2421013, EPI_ISL_2421014, EPI_ISL_2421015, EPI_ISL_2421016, EPI_ISL_2421017, EPI_ISL_2421018 | | | |
| see above | Montana Public Health Laboratory | Montana Public Health Laboratory | Joy Ritter, Michelle Mozer, Carrie Biskupiak, Deborah Gibson, Michael Dills |
| EPI_ISL_2421019 | Mako Medical | Centers for Disease Control and Prevention Division of Viral Diseases, Pathogen Discovery | Dakota Howard, Dhwani Batra, Peter W. Cook, Kara Moser, Adrian Paskey, Jason Caravas, Benjamin Rambo-Martin, Shatavia Morrison, Christopher Gulvick, Scott Sammons, Yvette Unoarumhi, Darlene Wagner, Matthew Schmerer,Matthew Tugwell, Lauren Moon, Clinton R. Paden, Duncan MacCannell |
| EPI_ISL_2421021, EPI_ISL_2421022, EPI_ISL_2421023, EPI_ISL_2421024, EPI_ISL_2421025, EPI_ISL_2421026, EPI_ISL_2421027, EPI_ISL_2421028, EPI_ISL_2421029, EPI_ISL_2421030, EPI_ISL_2421031, EPI_ISL_2421032, EPI_ISL_2421033, EPI_ISL_2421034, EPI_ISL_2421035, EPI_ISL_2421036, EPI_ISL_2421037, EPI_ISL_2421038, EPI_ISL_2421039, EPI_ISL_2421040, EPI_ISL_2421041, EPI_ISL_2421042 | | | |
| see above | Montana Public Health Laboratory | Montana Public Health Laboratory | Joy Ritter, Michelle Mozer, Carrie Biskupiak, Deborah Gibson, Michael Dills |
| EPI_ISL_2421044 | NORTHWELL HEALTH LABORATORIES | Wadsworth Center, New York State Department of Health | Kirsten St. George, Daryl M. Lamson, Alexis Russell, Matthew Shudt, Melissa A Leisner, Jonathan Plitnick, Catharine Prussing, Navjot Singh, John Kelly, Erasmus Schneider, Erica Lasek-Nesselquist |
| EPI_ISL_2421045, EPI_ISL_2421046, EPI_ISL_2421047, EPI_ISL_2421048, EPI_ISL_2421049, EPI_ISL_2421050, EPI_ISL_2421051, EPI_ISL_2421052, EPI_ISL_2421053 | STONY BROOK UNIVERSITY HOSPITAL | Wadsworth Center, New York State Department of Health | Kirsten St. George, Daryl M. Lamson, Alexis Russell, Matthew Shudt, Melissa A Leisner, Jonathan Plitnick, Catharine Prussing, Navjot Singh, John Kelly, Erasmus Schneider, Erica Lasek-Nesselquist |
| EPI_ISL_2421054, EPI_ISL_2421055, EPI_ISL_2421056, EPI_ISL_2421057, EPI_ISL_2421058, EPI_ISL_2421059, EPI_ISL_2421060, EPI_ISL_2421061, EPI_ISL_2421062, EPI_ISL_2421063, EPI_ISL_2421064, EPI_ISL_2421065, EPI_ISL_2421066, EPI_ISL_2421067, EPI_ISL_2421068, EPI_ISL_2421069, EPI_ISL_2421070, EPI_ISL_2421071, EPI_ISL_2421072, EPI_ISL_2421073, EPI_ISL_2421074, EPI_ISL_2421075, EPI_ISL_2421076, EPI_ISL_2421077, EPI_ISL_2421078, EPI_ISL_2421079, EPI_ISL_2421080, EPI_ISL_2421081 | | | |
| see above | SUNY UPSTATE MEDICAL UNIVERSITY | Wadsworth Center, New York State Department of Health | Kirsten St. George, Daryl M. Lamson, Alexis Russell, Matthew Shudt, Melissa A Leisner, Jonathan Plitnick, Catharine Prussing, Navjot Singh, John Kelly, Erasmus Schneider, Erica Lasek-Nesselquist |
| EPI_ISL_2421082, EPI_ISL_2421083, EPI_ISL_2421084, EPI_ISL_2421085, EPI_ISL_2421086, EPI_ISL_2421087, EPI_ISL_2421088, EPI_ISL_2421089, EPI_ISL_2421090, EPI_ISL_2421091, EPI_ISL_2421092, EPI_ISL_2421093, EPI_ISL_2421094, EPI_ISL_2421095 | | | |
| see above | BOSTON HEART DIAGNOSTICS CORP | Wadsworth Center, New York State Department of Health | Kirsten St. George, Daryl M. Lamson, Alexis Russell, Matthew Shudt, Melissa A Leisner, Jonathan Plitnick, Catharine Prussing, Navjot Singh, John Kelly, Erasmus Schneider, Erica Lasek-Nesselquist |
| EPI_ISL_2421096, EPI_ISL_2421097, EPI_ISL_2421098, EPI_ISL_2421099, EPI_ISL_2421100, EPI_ISL_2421101, EPI_ISL_2421102, EPI_ISL_2421103, EPI_ISL_2421104 | MONTEFIORE MEDICAL CENTER LABORATORIES | Wadsworth Center, New York State Department of Health | Kirsten St. George, Daryl M. Lamson, Alexis Russell, Matthew Shudt, Melissa A Leisner, Jonathan Plitnick, Catharine Prussing, Navjot Singh, John Kelly, Erasmus Schneider, Erica Lasek-Nesselquist |
| EPI_ISL_2421105, EPI_ISL_2421106, EPI_ISL_2421107 | WESTCHESTER MEDICAL CENTER | Wadsworth Center, New York State Department of Health | Kirsten St. George, Daryl M. Lamson, Alexis Russell, Matthew Shudt, Melissa A Leisner, Jonathan Plitnick, Catharine Prussing, Navjot Singh, John Kelly, Erasmus Schneider, Erica Lasek-Nesselquist |
| EPI_ISL_2421140, EPI_ISL_2421141, EPI_ISL_2421142, EPI_ISL_2421143 | ALBANY MEDICAL CENTER | Wadsworth Center, New York State Department of Health | Kirsten St. George, Daryl M. Lamson, Alexis Russell, Matthew Shudt, Melissa A Leisner, Jonathan Plitnick, Catharine Prussing, Navjot Singh, John Kelly, Erasmus Schneider, Erica Lasek-Nesselquist |
| EPI_ISL_2421144, EPI_ISL_2421145, EPI_ISL_2421146, EPI_ISL_2421147, EPI_ISL_2421148, EPI_ISL_2421149, EPI_ISL_2421150, EPI_ISL_2421151, EPI_ISL_2421152, EPI_ISL_2421153, EPI_ISL_2421154, EPI_ISL_2421155, EPI_ISL_2421156 | | | |
| see above | URMC LABS | Wadsworth Center, New York State Department of Health | Kirsten St. George, Daryl M. Lamson, Alexis Russell, Matthew Shudt, Melissa A Leisner, Jonathan Plitnick, Catharine Prussing, Navjot Singh, John Kelly, Erasmus Schneider, Erica Lasek-Nesselquist |
| EPI_ISL_2421157, EPI_ISL_2421158, EPI_ISL_2421159, EPI_ISL_2421160 | GLENS FALLS HOSPITAL LABORATORY | Wadsworth Center, New York State Department of Health | Kirsten St. George, Daryl M. Lamson, Alexis Russell, Matthew Shudt, Melissa A Leisner, Jonathan Plitnick, Catharine Prussing, Navjot Singh, John Kelly, Erasmus Schneider, Erica Lasek-Nesselquist |
| EPI_ISL_2421161, EPI_ISL_2421162, EPI_ISL_2421163, EPI_ISL_2421164, EPI_ISL_2421165, EPI_ISL_2421166, EPI_ISL_2421167, EPI_ISL_2421168, EPI_ISL_2421169, EPI_ISL_2421170, EPI_ISL_2421171, EPI_ISL_2421172, EPI_ISL_2421173, EPI_ISL_2421174, EPI_ISL_2421175, EPI_ISL_2421176, EPI_ISL_2421177, EPI_ISL_2421178, EPI_ISL_2421179, EPI_ISL_2421180, EPI_ISL_2421181, EPI_ISL_2421182, EPI_ISL_2421183, EPI_ISL_2421184, EPI_ISL_2421185, EPI_ISL_2421186, EPI_ISL_2421187, EPI_ISL_2421188 | | | |
| see above | TEMPUS LABS INC | Wadsworth Center, New York State Department of Health | Kirsten St. George, Daryl M. Lamson, Alexis Russell, Matthew Shudt, Melissa A Leisner, Jonathan Plitnick, Catharine Prussing, Navjot Singh, John Kelly, Erasmus Schneider, Erica Lasek-Nesselquist |
| EPI_ISL_2421189, EPI_ISL_2421190, EPI_ISL_2421191, EPI_ISL_2421192, EPI_ISL_2421193, EPI_ISL_2421194, EPI_ISL_2421195, EPI_ISL_2421196, EPI_ISL_2421197, EPI_ISL_2421198, EPI_ISL_2421199, EPI_ISL_2421200, EPI_ISL_2421201, EPI_ISL_2421202, EPI_ISL_2421203, EPI_ISL_2421204, EPI_ISL_2421205, EPI_ISL_2421206, EPI_ISL_2421207, EPI_ISL_2421208, EPI_ISL_2421209, EPI_ISL_2421210, EPI_ISL_2421211 | | | |
| see above | BOSTON HEART DIAGNOSTICS CORP | Wadsworth Center, New York State Department of Health | Kirsten St. George, Daryl M. Lamson, Alexis Russell, Matthew Shudt, Melissa A Leisner, Jonathan Plitnick, Catharine Prussing, Navjot Singh, John Kelly, Erasmus Schneider, Erica Lasek-Nesselquist |
| EPI_ISL_2421214 | WESTCHESTER MEDICAL CENTER | Wadsworth Center, New York State Department of Health | Kirsten St. George, Daryl M. Lamson, Alexis Russell, Matthew Shudt, Melissa A Leisner, Jonathan Plitnick, Catharine Prussing, Navjot Singh, John Kelly, Erasmus Schneider, Erica Lasek-Nesselquist |
| EPI_ISL_2421215 | NYC Pandemic Response Lab | Wadsworth Center, New York State Department of Health | Kirsten St. George, Daryl M. Lamson, Alexis Russell, Matthew Shudt, Melissa A Leisner, Jonathan Plitnick, Catharine Prussing, Navjot Singh, John Kelly, Erasmus Schneider, Erica Lasek-Nesselquist |
| EPI_ISL_2421216 | Columbia University Irving Medical Center | Wadsworth Center, New York State Department of Health | Kirsten St. George, Daryl M. Lamson, Alexis Russell, Matthew Shudt, Melissa A Leisner, Jonathan Plitnick, Catharine Prussing, Navjot Singh, John Kelly, Erasmus Schneider, Erica Lasek-Nesselquist |
| EPI_ISL_2421217 | MONTEFIORE MEDICAL CENTER LABORATORIES | Wadsworth Center, New York State Department of Health | Kirsten St. George, Daryl M. Lamson, Alexis Russell, Matthew Shudt, Melissa A Leisner, Jonathan Plitnick, Catharine Prussing, Navjot Singh, John Kelly, Erasmus Schneider, Erica Lasek-Nesselquist |
| EPI_ISL_2421218, EPI_ISL_2421219 | Columbia University Irving Medical Center | Wadsworth Center, New York State Department of Health | Kirsten St. George, Daryl M. Lamson, Alexis Russell, Matthew Shudt, Melissa A Leisner, Jonathan Plitnick, Catharine Prussing, Navjot Singh, John Kelly, Erasmus Schneider, Erica Lasek-Nesselquist |
| EPI_ISL_2421220 | SUNY UPSTATE MEDICAL UNIVERSITY | Wadsworth Center, New York State Department of Health | Kirsten St. George, Daryl M. Lamson, Alexis Russell, Matthew Shudt, Melissa A Leisner, Jonathan Plitnick, Catharine Prussing, Navjot Singh, John Kelly, Erasmus Schneider, Erica Lasek-Nesselquist |
| EPI_ISL_2421221, EPI_ISL_2421222 | NORTHWELL HEALTH LABORATORIES | Wadsworth Center, New York State Department of Health | Kirsten St. George, Daryl M. Lamson, Alexis Russell, Matthew Shudt, Melissa A Leisner, Jonathan Plitnick, Catharine Prussing, Navjot Singh, John Kelly, Erasmus Schneider, Erica Lasek-Nesselquist |
| EPI_ISL_2421223, EPI_ISL_2421224, EPI_ISL_2421225, EPI_ISL_2421226, EPI_ISL_2421227 | URMC LABS | Wadsworth Center, New York State Department of Health | Kirsten St. George, Daryl M. Lamson, Alexis Russell, Matthew Shudt, Melissa A Leisner, Jonathan Plitnick, Catharine Prussing, Navjot Singh, John Kelly, Erasmus Schneider, Erica Lasek-Nesselquist |
| EPI_ISL_2421228 | Garnet Health Medical Center Laboratory | Wadsworth Center, New York State Department of Health | Kirsten St. George, Daryl M. Lamson, Alexis Russell, Matthew Shudt, Melissa A Leisner, Jonathan Plitnick, Catharine Prussing, Navjot Singh, John Kelly, Erasmus Schneider, Erica Lasek-Nesselquist |
| EPI_ISL_2421229 | GLENS FALLS HOSPITAL LABORATORY | Wadsworth Center, New York State Department of Health | Kirsten St. George, Daryl M. Lamson, Alexis Russell, Matthew Shudt, Melissa A Leisner, Jonathan Plitnick, Catharine Prussing, Navjot Singh, John Kelly, Erasmus Schneider, Erica Lasek-Nesselquist |

| | | | |
|---|---|---|---|
| EPI_ISL_2421230, EPI_ISL_2421231, EPI_ISL_2421232, EPI_ISL_2421233, EPI_ISL_2421234, EPI_ISL_2421235, EPI_ISL_2421236, EPI_ISL_2421237, EPI_ISL_2421238, EPI_ISL_2421239, EPI_ISL_2421240, EPI_ISL_2421241, EPI_ISL_2421242, EPI_ISL_2421243, EPI_ISL_2421244, EPI_ISL_2421245, EPI_ISL_2421246, EPI_ISL_2421247, EPI_ISL_2421248 | | | |
| see above | BOSTON HEART DIAGNOSTICS CORP | Wadsworth Center, New York State Department of Health | Kirsten St. George, Daryl M. Lamson, Alexis Russell, Matthew Shudt, Melissa A Leisner, Jonathan Plitnick, Catharine Prussing, Navjot Singh, John Kelly, Erasmus Schneider, Erica Lasek-Nesselquist |
| EPI_ISL_2421249, EPI_ISL_2421250, EPI_ISL_2421251, EPI_ISL_2421252 | Saratoga Hospital Laboratory | Wadsworth Center, New York State Department of Health | Kirsten St. George, Daryl M. Lamson, Alexis Russell, Matthew Shudt, Melissa A Leisner, Jonathan Plitnick, Catharine Prussing, Navjot Singh, John Kelly, Erasmus Schneider, Erica Lasek-Nesselquist |
| EPI_ISL_2421253 | Vermont DoH | Wadsworth Center, New York State Department of Health | Kirsten St. George, Daryl M. Lamson, Alexis Russell, Matthew Shudt, Melissa A Leisner, Jonathan Plitnick, Catharine Prussing, Navjot Singh, John Kelly, Erasmus Schneider, Erica Lasek-Nesselquist |
| EPI_ISL_2421254, EPI_ISL_2421255, EPI_ISL_2421256, EPI_ISL_2421257, EPI_ISL_2421258, EPI_ISL_2421259, EPI_ISL_2421260, EPI_ISL_2421261, EPI_ISL_2421262, EPI_ISL_2421263, EPI_ISL_2421264, EPI_ISL_2421265, EPI_ISL_2421266, EPI_ISL_2421267, EPI_ISL_2421268, EPI_ISL_2421269, EPI_ISL_2421270, EPI_ISL_2421271, EPI_ISL_2421272, EPI_ISL_2421273, EPI_ISL_2421274, EPI_ISL_2421275, EPI_ISL_2421276, EPI_ISL_2421277 | | | |
| see above | Montana Public Health Laboratory | Montana Public Health Laboratory | Joy Ritter, Michelle Mozer, Carrie Biskupiak, Deborah Gibson, Michael Dills |
| EPI_ISL_2421289, EPI_ISL_2421290, EPI_ISL_2421291, EPI_ISL_2421292, EPI_ISL_2421293, EPI_ISL_2421294, EPI_ISL_2421295, EPI_ISL_2421296, EPI_ISL_2421297, EPI_ISL_2421298, EPI_ISL_2421299, EPI_ISL_2421300, EPI_ISL_2421301, EPI_ISL_2421302, EPI_ISL_2421303, EPI_ISL_2421304, EPI_ISL_2421305, EPI_ISL_2421306, EPI_ISL_2421307, EPI_ISL_2421308, EPI_ISL_2421309, EPI_ISL_2421310, EPI_ISL_2421311, EPI_ISL_2421312, EPI_ISL_2421313, EPI_ISL_2421314, EPI_ISL_2421315, EPI_ISL_2421316, EPI_ISL_2421317, EPI_ISL_2421318, EPI_ISL_2421319, EPI_ISL_2421320, EPI_ISL_2421321, EPI_ISL_2421322, EPI_ISL_2421323, EPI_ISL_2421324, EPI_ISL_2421325, EPI_ISL_2421326, EPI_ISL_2421327, EPI_ISL_2421328, EPI_ISL_2421329, EPI_ISL_2421330, EPI_ISL_2421331, EPI_ISL_2421332, EPI_ISL_2421333, EPI_ISL_2421334, EPI_ISL_2421335, EPI_ISL_2421336, EPI_ISL_2421337, EPI_ISL_2421338, EPI_ISL_2421339, EPI_ISL_2421340, EPI_ISL_2421341, EPI_ISL_2421342, EPI_ISL_2421343, EPI_ISL_2421344, EPI_ISL_2421345, EPI_ISL_2421346, EPI_ISL_2421347, EPI_ISL_2421348, EPI_ISL_2421349, EPI_ISL_2421350, EPI_ISL_2421351, EPI_ISL_2421352, EPI_ISL_2421353, EPI_ISL_2421354, EPI_ISL_2421355, EPI_ISL_2421356, EPI_ISL_2421357, EPI_ISL_2421358, EPI_ISL_2421359, EPI_ISL_2421360, EPI_ISL_2421361, EPI_ISL_2421362, EPI_ISL_2421363, EPI_ISL_2421364, EPI_ISL_2421365, EPI_ISL_2421366, EPI_ISL_2421367, EPI_ISL_2421368, EPI_ISL_2421369, EPI_ISL_2421370, EPI_ISL_2421371, EPI_ISL_2421372, EPI_ISL_2421373, EPI_ISL_2421374, EPI_ISL_2421375, EPI_ISL_2421376, EPI_ISL_2421377, EPI_ISL_2421378, EPI_ISL_2421379, EPI_ISL_2421380, EPI_ISL_2421381, EPI_ISL_2421382, EPI_ISL_2421384, EPI_ISL_2421385, EPI_ISL_2421386, EPI_ISL_2421387, EPI_ISL_2421388, EPI_ISL_2421389, EPI_ISL_2421390, EPI_ISL_2421391, EPI_ISL_2421392, EPI_ISL_2421393, EPI_ISL_2421394, EPI_ISL_2421395, EPI_ISL_2421396, EPI_ISL_2421397, EPI_ISL_2421398, EPI_ISL_2421399, EPI_ISL_2421400, EPI_ISL_2421403, EPI_ISL_2421404, EPI_ISL_2421405, EPI_ISL_2421406, EPI_ISL_2421407, EPI_ISL_2421408, EPI_ISL_2421409, EPI_ISL_2421410, EPI_ISL_2421411, EPI_ISL_2421412, EPI_ISL_2421413, EPI_ISL_2421414, EPI_ISL_2421415, EPI_ISL_2421416, EPI_ISL_2421418, EPI_ISL_2421419, EPI_ISL_2421420, EPI_ISL_2421421, EPI_ISL_2421422, EPI_ISL_2421423, EPI_ISL_2421424, EPI_ISL_2421425, EPI_ISL_2421426, EPI_ISL_2421427, EPI_ISL_2421428, EPI_ISL_2421429, EPI_ISL_2421430, EPI_ISL_2421431, EPI_ISL_2421432, EPI_ISL_2421433, EPI_ISL_2421434, EPI_ISL_2421435, EPI_ISL_2421436, EPI_ISL_2421437, EPI_ISL_2421438, EPI_ISL_2421439, EPI_ISL_2421440, EPI_ISL_2421441, EPI_ISL_2421442, EPI_ISL_2421443, EPI_ISL_2421444, EPI_ISL_2421445, EPI_ISL_2421446, EPI_ISL_2421447, EPI_ISL_2421448, EPI_ISL_2421449, EPI_ISL_2421450, EPI_ISL_2421451, EPI_ISL_2421452, EPI_ISL_2421453, EPI_ISL_2421454, EPI_ISL_2421455, EPI_ISL_2421456, EPI_ISL_2421457, EPI_ISL_2421458, EPI_ISL_2421459, EPI_ISL_2421460, EPI_ISL_2421461, EPI_ISL_2421462, EPI_ISL_2421463, EPI_ISL_2421464, EPI_ISL_2421465, EPI_ISL_2421466, EPI_ISL_2421467, EPI_ISL_2421468, EPI_ISL_2421469, EPI_ISL_2421470, EPI_ISL_2421471, EPI_ISL_2421472, EPI_ISL_2421473, EPI_ISL_2421474, EPI_ISL_2421475, EPI_ISL_2421476, EPI_ISL_2421477, EPI_ISL_2421478, EPI_ISL_2421479, EPI_ISL_2421480, EPI_ISL_2421481, EPI_ISL_2421482, EPI_ISL_2421483, EPI_ISL_2421484, EPI_ISL_2421485, EPI_ISL_2421486, EPI_ISL_2421487, EPI_ISL_2421488, EPI_ISL_2421489, EPI_ISL_2421490, EPI_ISL_2421491, EPI_ISL_2421492, EPI_ISL_2421493, EPI_ISL_2421494, EPI_ISL_2421495, EPI_ISL_2421496, EPI_ISL_2421497, EPI_ISL_2421498, EPI_ISL_2421499, EPI_ISL_2421500, EPI_ISL_2421501, EPI_ISL_2421502, EPI_ISL_2421503, EPI_ISL_2421504, EPI_ISL_2421505, EPI_ISL_2421506, EPI_ISL_2421507, EPI_ISL_2421508, EPI_ISL_2421509, EPI_ISL_2421510, EPI_ISL_2421511, EPI_ISL_2421512, EPI_ISL_2421513, EPI_ISL_2421514, EPI_ISL_2421515, EPI_ISL_2421516, EPI_ISL_2421517, EPI_ISL_2421518, EPI_ISL_2421519, EPI_ISL_2421520, EPI_ISL_2421521, EPI_ISL_2421522, EPI_ISL_2421523, EPI_ISL_2421524, EPI_ISL_2421525, EPI_ISL_2421526, EPI_ISL_2421527, EPI_ISL_2421528, EPI_ISL_2421529, EPI_ISL_2421530, EPI_ISL_2421531, EPI_ISL_2421532, EPI_ISL_2421533, EPI_ISL_2421534, EPI_ISL_2421535, EPI_ISL_2421536, EPI_ISL_2421537, EPI_ISL_2421538, EPI_ISL_2421539, EPI_ISL_2421540, EPI_ISL_2421542, EPI_ISL_2421543, EPI_ISL_2421544, EPI_ISL_2421545, EPI_ISL_2421546, EPI_ISL_2421547, EPI_ISL_2421548, EPI_ISL_2421549, EPI_ISL_2421550, EPI_ISL_2421551, EPI_ISL_2421552, EPI_ISL_2421553, EPI_ISL_2421554, EPI_ISL_2421555, EPI_ISL_2421556, EPI_ISL_2421557, EPI_ISL_2421559, EPI_ISL_2421560, EPI_ISL_2421561, EPI_ISL_2421562, EPI_ISL_2421563, EPI_ISL_2421564, EPI_ISL_2421565, EPI_ISL_2421566, EPI_ISL_2421567, EPI_ISL_2421568, EPI_ISL_2421569, EPI_ISL_2421570, EPI_ISL_2421571, EPI_ISL_2421572, EPI_ISL_2421573, EPI_ISL_2421574, EPI_ISL_2421575, EPI_ISL_2421576, EPI_ISL_2421577, EPI_ISL_2421578, EPI_ISL_2421579, EPI_ISL_2421580, EPI_ISL_2421581, EPI_ISL_2421582, EPI_ISL_2421583, EPI_ISL_2421584, EPI_ISL_2421585, EPI_ISL_2421586, EPI_ISL_2421587, EPI_ISL_2421588, EPI_ISL_2421589, EPI_ISL_2421590, EPI_ISL_2421591, EPI_ISL_2421592, EPI_ISL_2421593, EPI_ISL_2421594, EPI_ISL_2421595, EPI_ISL_2421596, EPI_ISL_2421597, EPI_ISL_2421598, EPI_ISL_2421599, EPI_ISL_2421600, EPI_ISL_2421601, EPI_ISL_2421602, EPI_ISL_2421603, EPI_ISL_2421604, EPI_ISL_2421605, EPI_ISL_2421606, EPI_ISL_2421607, EPI_ISL_2421608, EPI_ISL_2421610, EPI_ISL_2421611, EPI_ISL_2421612, EPI_ISL_2421613, EPI_ISL_2421614, EPI_ISL_2421615, EPI_ISL_2421616, EPI_ISL_2421617, EPI_ISL_2421618, EPI_ISL_2421619, EPI_ISL_2421620, EPI_ISL_2421621, EPI_ISL_2421622, EPI_ISL_2421623, EPI_ISL_2421624, EPI_ISL_2421625, EPI_ISL_2421626, EPI_ISL_2421628, EPI_ISL_2421629, EPI_ISL_2421630, EPI_ISL_2421631, EPI_ISL_2421632, EPI_ISL_2421633, EPI_ISL_2421634, EPI_ISL_2421635, EPI_ISL_2421636, EPI_ISL_2421637, EPI_ISL_2421638, EPI_ISL_2421639, EPI_ISL_2421640, EPI_ISL_2421641, EPI_ISL_2421642, EPI_ISL_2421643, EPI_ISL_2421644, EPI_ISL_2421645, EPI_ISL_2421646, EPI_ISL_2421647, EPI_ISL_2421648, EPI_ISL_2421649, EPI_ISL_2421650, EPI_ISL_2421651, EPI_ISL_2421652, EPI_ISL_2421653, EPI_ISL_2421654, EPI_ISL_2421655, EPI_ISL_2421656, EPI_ISL_2421657, EPI_ISL_2421658, EPI_ISL_2421659, EPI_ISL_2421660, EPI_ISL_2421661, EPI_ISL_2421662, EPI_ISL_2421663, EPI_ISL_2421664, EPI_ISL_2421665, EPI_ISL_2421666, EPI_ISL_2421667, EPI_ISL_2421668, EPI_ISL_2421669, EPI_ISL_2421670, EPI_ISL_2421671, EPI_ISL_2421672, EPI_ISL_2421673, EPI_ISL_2421674, EPI_ISL_2421675, EPI_ISL_2421676, EPI_ISL_2421677, EPI_ISL_2421678, EPI_ISL_2421679, EPI_ISL_2421680, EPI_ISL_2421681, EPI_ISL_2421682, EPI_ISL_2421683 | | | |
| see above | Aegis Sciences Corporation | Centers for Disease Control and Prevention Division of Viral Diseases, Pathogen Discovery | Dakota Howard, Dhwani Batra, Peter W. Cook, Kara Moser, Adrian Paskey, Jason Caravas, Benjamin Rambo-Martin, Shatavia Morrison, Christopher Gulvick, Scott Sammons, Yvette Unoarumhi, Darlene Wagner, Matthew Schmerer, Cyndi Clark, Patrick Campbell, Rob Case, Vikramsinha Ghorpade, Holly Houdeshell, Ola Kvalvaag, Dillon Nall, Ethan Sanders, Alec Vest, Shaun Westlund, Matthew Hardison, Clinton R. Paden, Duncan MacCannell |
| EPI_ISL_2421684, EPI_ISL_2421685 | Alaska State Virology Laboratory | Alaska State Virology Laboratory | Stephanie DeRonde, Elva House, Jacob Zidek, Lisa Smith, Ph.D., Jack Chen, Ph.D. |
| EPI_ISL_2421686 | Aegis Sciences Corporation | Centers for Disease Control and Prevention Division of Viral Diseases, Pathogen Discovery | Dakota Howard, Dhwani Batra, Peter W. Cook, Kara Moser, Adrian Paskey, Jason Caravas, Benjamin Rambo-Martin, Shatavia Morrison, Christopher Gulvick, Scott Sammons, Yvette Unoarumhi, Darlene Wagner, Matthew Schmerer, Cyndi Clark, Patrick Campbell, Rob Case, Vikramsinha Ghorpade, Holly Houdeshell, Ola Kvalvaag, Dillon Nall, Ethan Sanders, Alec Vest, Shaun Westlund, Matthew Hardison, Clinton R. Paden, Duncan MacCannell |
| EPI_ISL_2421687, EPI_ISL_2421688 | Alaska State Virology Laboratory | Alaska State Virology Laboratory | Stephanie DeRonde, Elva House, Jacob Zidek, Lisa Smith, Ph.D., Jack Chen, Ph.D. |
| EPI_ISL_2421689 | Aegis Sciences Corporation | Centers for Disease Control and Prevention Division of Viral Diseases, Pathogen Discovery | Dakota Howard, Dhwani Batra, Peter W. Cook, Kara Moser, Adrian Paskey, Jason Caravas, Benjamin Rambo-Martin, Shatavia Morrison, Christopher Gulvick, Scott Sammons, Yvette Unoarumhi, Darlene Wagner, Matthew Schmerer, Cyndi Clark, Patrick Campbell, Rob Case, Vikramsinha Ghorpade, Holly Houdeshell, Ola Kvalvaag, Dillon Nall, Ethan Sanders, Alec Vest, Shaun Westlund, Matthew Hardison, Clinton R. Paden, Duncan MacCannell |
| EPI_ISL_2421690, EPI_ISL_2421691 | Alaska State Virology Laboratory | Alaska State Virology Laboratory | Stephanie DeRonde, Elva House, Jacob Zidek, Lisa Smith, Ph.D., Jack Chen, Ph.D. |
| EPI_ISL_2421692 | Aegis Sciences Corporation | Centers for Disease Control and Prevention Division of Viral Diseases, Pathogen Discovery | Dakota Howard, Dhwani Batra, Peter W. Cook, Kara Moser, Adrian Paskey, Jason Caravas, Benjamin Rambo-Martin, Shatavia Morrison, Christopher Gulvick, Scott Sammons, Yvette Unoarumhi, Darlene Wagner, Matthew Schmerer, Cyndi Clark, Patrick Campbell, Rob Case, Vikramsinha Ghorpade, Holly Houdeshell, Ola Kvalvaag, Dillon Nall, Ethan Sanders, Alec Vest, Shaun Westlund, Matthew Hardison, Clinton R. Paden, Duncan MacCannell |
| EPI_ISL_2421693, EPI_ISL_2421694 | Alaska State Virology Laboratory | Alaska State Virology Laboratory | Stephanie DeRonde, Elva House, Jacob Zidek, Lisa Smith, Ph.D., Jack Chen, Ph.D. |
| EPI_ISL_2421695 | Aegis Sciences Corporation | Centers for Disease Control and Prevention Division of Viral Diseases, Pathogen Discovery | Dakota Howard, Dhwani Batra, Peter W. Cook, Kara Moser, Adrian Paskey, Jason Caravas, Benjamin Rambo-Martin, Shatavia Morrison, Christopher Gulvick, Scott Sammons, Yvette Unoarumhi, Darlene Wagner, Matthew Schmerer, Cyndi Clark, Patrick Campbell, Rob Case, Vikramsinha Ghorpade, Holly Houdeshell, Ola Kvalvaag, Dillon Nall, Ethan Sanders, Alec Vest, Shaun Westlund, Matthew Hardison, Clinton R. Paden, Duncan MacCannell |
| EPI_ISL_2421696, EPI_ISL_2421697 | Alaska State Virology Laboratory | Alaska State Virology Laboratory | Stephanie DeRonde, Elva House, Jacob Zidek, Lisa Smith, Ph.D., Jack Chen, Ph.D. |
| EPI_ISL_2421698 | Aegis Sciences Corporation | Centers for Disease Control and Prevention Division of Viral Diseases, Pathogen Discovery | Dakota Howard, Dhwani Batra, Peter W. Cook, Kara Moser, Adrian Paskey, Jason Caravas, Benjamin Rambo-Martin, Shatavia Morrison, Christopher Gulvick, Scott Sammons, Yvette Unoarumhi, Darlene Wagner, Matthew Schmerer, Cyndi Clark, Patrick Campbell, Rob Case, Vikramsinha Ghorpade, Holly Houdeshell, Ola Kvalvaag, Dillon Nall, Ethan Sanders, Alec Vest, Shaun Westlund, Matthew Hardison, Clinton R. Paden, Duncan MacCannell |
| EPI_ISL_2421699, EPI_ISL_2421700, EPI_ISL_2421701 | Alaska State Virology Laboratory | Alaska State Virology Laboratory | Stephanie DeRonde, Elva House, Jacob Zidek, Lisa Smith, Ph.D., Jack Chen, Ph.D. |
| EPI_ISL_2421702 | Aegis Sciences Corporation | Centers for Disease Control and Prevention Division of Viral Diseases, Pathogen Discovery | Dakota Howard, Dhwani Batra, Peter W. Cook, Kara Moser, Adrian Paskey, Jason Caravas, Benjamin Rambo-Martin, Shatavia Morrison, Christopher Gulvick, Scott Sammons, Yvette Unoarumhi, Darlene Wagner, Matthew Schmerer, Cyndi Clark, Patrick Campbell, Rob Case, Vikramsinha Ghorpade, Holly Houdeshell, Ola Kvalvaag, Dillon Nall, Ethan Sanders, Alec Vest, Shaun Westlund, Matthew Hardison, Clinton R. Paden, Duncan MacCannell |
| EPI_ISL_2421703 | Alaska State Virology Laboratory | Alaska State Virology Laboratory | Stephanie DeRonde, Elva House, Jacob Zidek, Lisa Smith, Ph.D., Jack Chen, Ph.D. |
| EPI_ISL_2421704 | Aegis Sciences Corporation | Centers for Disease Control and Prevention Division of Viral Diseases, Pathogen Discovery | Dakota Howard, Dhwani Batra, Peter W. Cook, Kara Moser, Adrian Paskey, Jason Caravas, Benjamin Rambo-Martin, Shatavia Morrison, Christopher Gulvick, Scott Sammons, Yvette Unoarumhi, Darlene Wagner, Matthew Schmerer, Cyndi Clark, Patrick Campbell, Rob Case, Vikramsinha Ghorpade, Holly Houdeshell, Ola Kvalvaag, Dillon Nall, Ethan Sanders, Alec Vest, Shaun Westlund, Matthew Hardison, Clinton R. Paden, Duncan MacCannell |
| EPI_ISL_2421705, EPI_ISL_2421706, EPI_ISL_2421707 | Alaska State Virology Laboratory | Alaska State Virology Laboratory | Stephanie DeRonde, Elva House, Jacob Zidek, Lisa Smith, Ph.D., Jack Chen, Ph.D. |
| EPI_ISL_2421708 | Aegis Sciences Corporation | Centers for Disease Control and Prevention Division of Viral Diseases, Pathogen Discovery | Dakota Howard, Dhwani Batra, Peter W. Cook, Kara Moser, Adrian Paskey, Jason Caravas, Benjamin Rambo-Martin, Shatavia Morrison, Christopher Gulvick, Scott Sammons, Yvette Unoarumhi, Darlene Wagner, Matthew Schmerer, Cyndi Clark, Patrick Campbell, Rob Case, Vikramsinha Ghorpade, Holly Houdeshell, Ola Kvalvaag, Dillon Nall, Ethan Sanders, Alec Vest, Shaun Westlund, Matthew Hardison, Clinton R. Paden, Duncan MacCannell |
| EPI_ISL_2421709, EPI_ISL_2421710 | Alaska State Virology Laboratory | Alaska State Virology Laboratory | Stephanie DeRonde, Elva House, Jacob Zidek, Lisa Smith, Ph.D., Jack Chen, Ph.D. |
| EPI_ISL_2421711 | Aegis Sciences Corporation | Centers for Disease Control and Prevention Division of Viral Diseases, Pathogen Discovery | Dakota Howard, Dhwani Batra, Peter W. Cook, Kara Moser, Adrian Paskey, Jason Caravas, Benjamin Rambo-Martin, Shatavia Morrison, Christopher Gulvick, Scott Sammons, Yvette Unoarumhi, Darlene Wagner, Matthew Schmerer, Cyndi Clark, Patrick Campbell, Rob Case, Vikramsinha Ghorpade, Holly Houdeshell, Ola Kvalvaag, Dillon Nall, Ethan Sanders, Alec Vest, Shaun Westlund, Matthew Hardison, Clinton R. Paden, Duncan MacCannell |
| EPI_ISL_2421712 | Alaska State Virology Laboratory | Alaska State Virology Laboratory | Stephanie DeRonde, Elva House, Jacob Zidek, Lisa Smith, Ph.D., Jack Chen, Ph.D. |
| EPI_ISL_2421713 | Aegis Sciences Corporation | Centers for Disease Control and Prevention Division of Viral Diseases, Pathogen Discovery | Dakota Howard, Dhwani Batra, Peter W. Cook, Kara Moser, Adrian Paskey, Jason Caravas, Benjamin Rambo-Martin, Shatavia Morrison, Christopher Gulvick, Scott Sammons, Yvette Unoarumhi, Darlene Wagner, Matthew Schmerer, Cyndi Clark, Patrick Campbell, Rob Case, Vikramsinha Ghorpade, Holly Houdeshell, Ola Kvalvaag, Dillon Nall, Ethan Sanders, Alec Vest, Shaun Westlund, Matthew Hardison, Clinton R. Paden, Duncan MacCannell |
| EPI_ISL_2421714, EPI_ISL_2421715 | Alaska State Virology Laboratory | Alaska State Virology Laboratory | Stephanie DeRonde, Elva House, Jacob Zidek, Lisa Smith, Ph.D., Jack Chen, Ph.D. |
| EPI_ISL_2421716 | Aegis Sciences Corporation | Centers for Disease Control and Prevention Division of Viral Diseases, Pathogen Discovery | Dakota Howard, Dhwani Batra, Peter W. Cook, Kara Moser, Adrian Paskey, Jason Caravas, Benjamin Rambo-Martin, Shatavia Morrison, Christopher Gulvick, Scott Sammons, Yvette Unoarumhi, Darlene Wagner, Matthew Schmerer, Cyndi Clark, Patrick Campbell, Rob Case, Vikramsinha Ghorpade, Holly Houdeshell, Ola Kvalvaag, Dillon Nall, Ethan Sanders, Alec Vest, Shaun Westlund, Matthew Hardison, Clinton R. Paden, Duncan MacCannell |
| EPI_ISL_2421717, EPI_ISL_2421718 | Alaska State Virology Laboratory | Alaska State Virology Laboratory | Stephanie DeRonde, Elva House, Jacob Zidek, Lisa Smith, Ph.D., Jack Chen, Ph.D. |
| EPI_ISL_2421719 | Aegis Sciences Corporation | Centers for Disease Control and Prevention Division of Viral Diseases, Pathogen Discovery | Dakota Howard, Dhwani Batra, Peter W. Cook, Kara Moser, Adrian Paskey, Jason Caravas, Benjamin Rambo-Martin, Shatavia Morrison, Christopher Gulvick, Scott Sammons, Yvette Unoarumhi, Darlene Wagner, Matthew Schmerer, Cyndi Clark, Patrick Campbell, Rob Case, Vikramsinha Ghorpade, Holly Houdeshell, Ola Kvalvaag, Dillon Nall, Ethan Sanders, Alec Vest, Shaun Westlund, Matthew Hardison, Clinton R. Paden, Duncan MacCannell |
| EPI_ISL_2421720, EPI_ISL_2421721 | Alaska State Virology Laboratory | Alaska State Virology Laboratory | Stephanie DeRonde, Elva House, Jacob Zidek, Lisa Smith, Ph.D., Jack Chen, Ph.D. |

| | | | |
|---|---|---|---|
| EPI_ISL_2421722 | Aegis Sciences Corporation | Centers for Disease Control and Prevention Division of Viral Diseases, Pathogen Discovery | Dakota Howard, Dhwani Batra, Peter W. Cook, Kara Moser, Adrian Paskey, Jason Caravas, Benjamin Rambo-Martin, Shatavia Morrison, Christopher Gulvick, Scott Sammons, Yvette Unoarumhi, Darlene Wagner, Matthew Schmerer, Cyndi Clark, Patrick Campbell, Rob Case, Vikramsinha Ghorpade, Holly Houdeshell, Ola Kvalvaag, Dillon Nall, Ethan Sanders, Alec Vest, Shaun Westlund, Matthew Hardison, Clinton R. Paden, Duncan MacCannell |
| EPI_ISL_2421723, EPI_ISL_2421724 | Alaska State Virology Laboratory | Alaska State Virology Laboratory | Stephanie DeRonde, Elva House, Jacob Zidek, Lisa Smith, Ph.D., Jack Chen, Ph.D. |
| EPI_ISL_2421725 | Aegis Sciences Corporation | Centers for Disease Control and Prevention Division of Viral Diseases, Pathogen Discovery | Dakota Howard, Dhwani Batra, Peter W. Cook, Kara Moser, Adrian Paskey, Jason Caravas, Benjamin Rambo-Martin, Shatavia Morrison, Christopher Gulvick, Scott Sammons, Yvette Unoarumhi, Darlene Wagner, Matthew Schmerer, Cyndi Clark, Patrick Campbell, Rob Case, Vikramsinha Ghorpade, Holly Houdeshell, Ola Kvalvaag, Dillon Nall, Ethan Sanders, Alec Vest, Shaun Westlund, Matthew Hardison, Clinton R. Paden, Duncan MacCannell |
| EPI_ISL_2421726, EPI_ISL_2421727 | Alaska State Virology Laboratory | Alaska State Virology Laboratory | Stephanie DeRonde, Elva House, Jacob Zidek, Lisa Smith, Ph.D., Jack Chen, Ph.D. |
| EPI_ISL_2421728, EPI_ISL_2421729, EPI_ISL_2421730, EPI_ISL_2421731, EPI_ISL_2421732, EPI_ISL_2421733, EPI_ISL_2421734, EPI_ISL_2421735, EPI_ISL_2421736, EPI_ISL_2421737, EPI_ISL_2421738, EPI_ISL_2421739, EPI_ISL_2421740, EPI_ISL_2421741, EPI_ISL_2421742, EPI_ISL_2421743, EPI_ISL_2421744, EPI_ISL_2421745, EPI_ISL_2421746, EPI_ISL_2421747, EPI_ISL_2421748, EPI_ISL_2421749, EPI_ISL_2421750, EPI_ISL_2421751, EPI_ISL_2421752, EPI_ISL_2421753, EPI_ISL_2421754, EPI_ISL_2421755, EPI_ISL_2421756, EPI_ISL_2421757, EPI_ISL_2421758, EPI_ISL_2421759, EPI_ISL_2421761, EPI_ISL_2421762, EPI_ISL_2421763, EPI_ISL_2421764, EPI_ISL_2421766, EPI_ISL_2421767, EPI_ISL_2421768, EPI_ISL_2421769, EPI_ISL_2421770, EPI_ISL_2421771, EPI_ISL_2421772, EPI_ISL_2421773, EPI_ISL_2421774, EPI_ISL_2421775, EPI_ISL_2421776, EPI_ISL_2421777, EPI_ISL_2421778, EPI_ISL_2421779, EPI_ISL_2421780, EPI_ISL_2421781, EPI_ISL_2421782, EPI_ISL_2421783, EPI_ISL_2421784, EPI_ISL_2421785, EPI_ISL_2421786, EPI_ISL_2421787, EPI_ISL_2421788, EPI_ISL_2421789, EPI_ISL_2421790, EPI_ISL_2421791, EPI_ISL_2421792, EPI_ISL_2421793, EPI_ISL_2421794, EPI_ISL_2421795, EPI_ISL_2421796, EPI_ISL_2421797, EPI_ISL_2421798, EPI_ISL_2421799, EPI_ISL_2421800, EPI_ISL_2421801, EPI_ISL_2421802, EPI_ISL_2421803, EPI_ISL_2421804, EPI_ISL_2421805, EPI_ISL_2421806, EPI_ISL_2421807, EPI_ISL_2421808, EPI_ISL_2421809, EPI_ISL_2421810, EPI_ISL_2421811, EPI_ISL_2421812, EPI_ISL_2421813, EPI_ISL_2421814, EPI_ISL_2421815, EPI_ISL_2421816, EPI_ISL_2421817, EPI_ISL_2421818, EPI_ISL_2421819, EPI_ISL_2421820, EPI_ISL_2421821, EPI_ISL_2421822, EPI_ISL_2421823, EPI_ISL_2421824, EPI_ISL_2421825, EPI_ISL_2421826, EPI_ISL_2421827, EPI_ISL_2421828, EPI_ISL_2421829, EPI_ISL_2421830, EPI_ISL_2421831, EPI_ISL_2421832, EPI_ISL_2421833, EPI_ISL_2421834, EPI_ISL_2421835, EPI_ISL_2421836, EPI_ISL_2421837, EPI_ISL_2421838, EPI_ISL_2421839, EPI_ISL_2421840, EPI_ISL_2421841, EPI_ISL_2421842, EPI_ISL_2421843, EPI_ISL_2421844, EPI_ISL_2421845, EPI_ISL_2421846, EPI_ISL_2421847, EPI_ISL_2421848, EPI_ISL_2421849, EPI_ISL_2421850, EPI_ISL_2421851, EPI_ISL_2421852, EPI_ISL_2421853, EPI_ISL_2421854, EPI_ISL_2421855, EPI_ISL_2421856, EPI_ISL_2421857, EPI_ISL_2421858, EPI_ISL_2421859, EPI_ISL_2421860, EPI_ISL_2421861, EPI_ISL_2421862, EPI_ISL_2421863, EPI_ISL_2421864, EPI_ISL_2421865, EPI_ISL_2421866, EPI_ISL_2421868, EPI_ISL_2421869, EPI_ISL_2421870, EPI_ISL_2421871, EPI_ISL_2421872, EPI_ISL_2421873, EPI_ISL_2421874, EPI_ISL_2421875, EPI_ISL_2421876, EPI_ISL_2421877, EPI_ISL_2421878, EPI_ISL_2421879, EPI_ISL_2421880, EPI_ISL_2421881, EPI_ISL_2421882, EPI_ISL_2421883, EPI_ISL_2421884, EPI_ISL_2421885, EPI_ISL_2421886, EPI_ISL_2421887, EPI_ISL_2421888, EPI_ISL_2421889, EPI_ISL_2421890, EPI_ISL_2421891, EPI_ISL_2421892, EPI_ISL_2421893, EPI_ISL_2421894, EPI_ISL_2421895, EPI_ISL_2421896, EPI_ISL_2421897, EPI_ISL_2421898, EPI_ISL_2421899, EPI_ISL_2421900, EPI_ISL_2421901, EPI_ISL_2421902, EPI_ISL_2421903, EPI_ISL_2421904, EPI_ISL_2421905, EPI_ISL_2421906, EPI_ISL_2421907, EPI_ISL_2421908, EPI_ISL_2421909, EPI_ISL_2421910, EPI_ISL_2421911, EPI_ISL_2421912, EPI_ISL_2421913, EPI_ISL_2421914, EPI_ISL_2421915, EPI_ISL_2421916, EPI_ISL_2421917, EPI_ISL_2421918, EPI_ISL_2421919, EPI_ISL_2421920, EPI_ISL_2421921, EPI_ISL_2421922, EPI_ISL_2421923, EPI_ISL_2421924, EPI_ISL_2421925, EPI_ISL_2421926, EPI_ISL_2421927, EPI_ISL_2421929, EPI_ISL_2421930, EPI_ISL_2421931, EPI_ISL_2421932, EPI_ISL_2421933, EPI_ISL_2421934, EPI_ISL_2421935, EPI_ISL_2421936, EPI_ISL_2421937, EPI_ISL_2421938, EPI_ISL_2421939, EPI_ISL_2421940, EPI_ISL_2421941, EPI_ISL_2421942, EPI_ISL_2421943, EPI_ISL_2421944, EPI_ISL_2421945, EPI_ISL_2421946, EPI_ISL_2421947, EPI_ISL_2421948, EPI_ISL_2421949, EPI_ISL_2421950, EPI_ISL_2421951, EPI_ISL_2421952, EPI_ISL_2421953, EPI_ISL_2421954, EPI_ISL_2421955, EPI_ISL_2421956, EPI_ISL_2421957, EPI_ISL_2421958, EPI_ISL_2421959, EPI_ISL_2421961, EPI_ISL_2421962, EPI_ISL_2421963, EPI_ISL_2421964, EPI_ISL_2421965, EPI_ISL_2421966, EPI_ISL_2421967, EPI_ISL_2421968, EPI_ISL_2421969, EPI_ISL_2421970, EPI_ISL_2421971, EPI_ISL_2421972, EPI_ISL_2421973, EPI_ISL_2421974, EPI_ISL_2421975, EPI_ISL_2421976, EPI_ISL_2421977, EPI_ISL_2421978, EPI_ISL_2421979, EPI_ISL_2421981, EPI_ISL_2421982, EPI_ISL_2421983, EPI_ISL_2421984, EPI_ISL_2421985, EPI_ISL_2421986, EPI_ISL_2421987, EPI_ISL_2421988, EPI_ISL_2421989, EPI_ISL_2421990, EPI_ISL_2421991, EPI_ISL_2421992, EPI_ISL_2421993, EPI_ISL_2421994, EPI_ISL_2421995, EPI_ISL_2421996, EPI_ISL_2421997, EPI_ISL_2421998, EPI_ISL_2421999, EPI_ISL_2422000, EPI_ISL_2422001, EPI_ISL_2422002, EPI_ISL_2422003, EPI_ISL_2422004, EPI_ISL_2422005, EPI_ISL_2422006, EPI_ISL_2422007, EPI_ISL_2422008, EPI_ISL_2422010, EPI_ISL_2422011, EPI_ISL_2422012, EPI_ISL_2422013, EPI_ISL_2422014, EPI_ISL_2422015, EPI_ISL_2422016, EPI_ISL_2422017, EPI_ISL_2422018, EPI_ISL_2422019, EPI_ISL_2422020, EPI_ISL_2422021, EPI_ISL_2422022, EPI_ISL_2422023, EPI_ISL_2422024, EPI_ISL_2422025, EPI_ISL_2422026, EPI_ISL_2422027, EPI_ISL_2422028, EPI_ISL_2422029, EPI_ISL_2422030, EPI_ISL_2422031, EPI_ISL_2422032, EPI_ISL_2422033, EPI_ISL_2422034, EPI_ISL_2422035, EPI_ISL_2422036, EPI_ISL_2422037, EPI_ISL_2422038, EPI_ISL_2422039, EPI_ISL_2422040, EPI_ISL_2422041, EPI_ISL_2422042, EPI_ISL_2422043, EPI_ISL_2422044, EPI_ISL_2422045, EPI_ISL_2422046 | | | |
| see above | Aegis Sciences Corporation | Centers for Disease Control and Prevention Division of Viral Diseases, Pathogen Discovery | Dakota Howard, Dhwani Batra, Peter W. Cook, Kara Moser, Adrian Paskey, Jason Caravas, Benjamin Rambo-Martin, Shatavia Morrison, Christopher Gulvick, Scott Sammons, Yvette Unoarumhi, Darlene Wagner, Matthew Schmerer, Cyndi Clark, Patrick Campbell, Rob Case, Vikramsinha Ghorpade, Holly Houdeshell, Ola Kvalvaag, Dillon Nall, Ethan Sanders, Alec Vest, Shaun Westlund, Matthew Hardison, Clinton R. Paden, Duncan MacCannell |
| EPI_ISL_2422047, EPI_ISL_2422048, EPI_ISL_2422050, EPI_ISL_2422051 | Alaska State Virology Laboratory | Alaska State Virology Laboratory | Stephanie DeRonde, Elva House, Jacob Zidek, Lisa Smith, Ph.D., Jack Chen, Ph.D. |
| EPI_ISL_2422052 | Aegis Sciences Corporation | Centers for Disease Control and Prevention Division of Viral Diseases, Pathogen Discovery | Dakota Howard, Dhwani Batra, Peter W. Cook, Kara Moser, Adrian Paskey, Jason Caravas, Benjamin Rambo-Martin, Shatavia Morrison, Christopher Gulvick, Scott Sammons, Yvette Unoarumhi, Darlene Wagner, Matthew Schmerer, Cyndi Clark, Patrick Campbell, Rob Case, Vikramsinha Ghorpade, Holly Houdeshell, Ola Kvalvaag, Dillon Nall, Ethan Sanders, Alec Vest, Shaun Westlund, Matthew Hardison, Clinton R. Paden, Duncan MacCannell |
| EPI_ISL_2422053, EPI_ISL_2422054 | Alaska State Virology Laboratory | Alaska State Virology Laboratory | Stephanie DeRonde, Elva House, Jacob Zidek, Lisa Smith, Ph.D., Jack Chen, Ph.D. |
| EPI_ISL_2422055 | Aegis Sciences Corporation | Centers for Disease Control and Prevention Division of Viral Diseases, Pathogen Discovery | Dakota Howard, Dhwani Batra, Peter W. Cook, Kara Moser, Adrian Paskey, Jason Caravas, Benjamin Rambo-Martin, Shatavia Morrison, Christopher Gulvick, Scott Sammons, Yvette Unoarumhi, Darlene Wagner, Matthew Schmerer, Cyndi Clark, Patrick Campbell, Rob Case, Vikramsinha Ghorpade, Holly Houdeshell, Ola Kvalvaag, Dillon Nall, Ethan Sanders, Alec Vest, Shaun Westlund, Matthew Hardison, Clinton R. Paden, Duncan MacCannell |
| EPI_ISL_2422056, EPI_ISL_2422057 | Alaska State Virology Laboratory | Alaska State Virology Laboratory | Stephanie DeRonde, Elva House, Jacob Zidek, Lisa Smith, Ph.D., Jack Chen, Ph.D. |
| EPI_ISL_2422058 | Aegis Sciences Corporation | Centers for Disease Control and Prevention Division of Viral Diseases, Pathogen Discovery | Dakota Howard, Dhwani Batra, Peter W. Cook, Kara Moser, Adrian Paskey, Jason Caravas, Benjamin Rambo-Martin, Shatavia Morrison, Christopher Gulvick, Scott Sammons, Yvette Unoarumhi, Darlene Wagner, Matthew Schmerer, Cyndi Clark, Patrick Campbell, Rob Case, Vikramsinha Ghorpade, Holly Houdeshell, Ola Kvalvaag, Dillon Nall, Ethan Sanders, Alec Vest, Shaun Westlund, Matthew Hardison, Clinton R. Paden, Duncan MacCannell |
| EPI_ISL_2422059, EPI_ISL_2422060 | Alaska State Virology Laboratory | Alaska State Virology Laboratory | Stephanie DeRonde, Elva House, Jacob Zidek, Lisa Smith, Ph.D., Jack Chen, Ph.D. |
| EPI_ISL_2422061 | Aegis Sciences Corporation | Centers for Disease Control and Prevention Division of Viral Diseases, Pathogen Discovery | Dakota Howard, Dhwani Batra, Peter W. Cook, Kara Moser, Adrian Paskey, Jason Caravas, Benjamin Rambo-Martin, Shatavia Morrison, Christopher Gulvick, Scott Sammons, Yvette Unoarumhi, Darlene Wagner, Matthew Schmerer, Cyndi Clark, Patrick Campbell, Rob Case, Vikramsinha Ghorpade, Holly Houdeshell, Ola Kvalvaag, Dillon Nall, Ethan Sanders, Alec Vest, Shaun Westlund, Matthew Hardison, Clinton R. Paden, Duncan MacCannell |
| EPI_ISL_2422062, EPI_ISL_2422063 | Alaska State Virology Laboratory | Alaska State Virology Laboratory | Stephanie DeRonde, Elva House, Jacob Zidek, Lisa Smith, Ph.D., Jack Chen, Ph.D. |
| EPI_ISL_2422064 | Aegis Sciences Corporation | Centers for Disease Control and Prevention Division of Viral Diseases, Pathogen Discovery | Dakota Howard, Dhwani Batra, Peter W. Cook, Kara Moser, Adrian Paskey, Jason Caravas, Benjamin Rambo-Martin, Shatavia Morrison, Christopher Gulvick, Scott Sammons, Yvette Unoarumhi, Darlene Wagner, Matthew Schmerer, Cyndi Clark, Patrick Campbell, Rob Case, Vikramsinha Ghorpade, Holly Houdeshell, Ola Kvalvaag, Dillon Nall, Ethan Sanders, Alec Vest, Shaun Westlund, Matthew Hardison, Clinton R. Paden, Duncan MacCannell |
| EPI_ISL_2422065, EPI_ISL_2422066 | Alaska State Virology Laboratory | Alaska State Virology Laboratory | Stephanie DeRonde, Elva House, Jacob Zidek, Lisa Smith, Ph.D., Jack Chen, Ph.D. |
| EPI_ISL_2422067 | Aegis Sciences Corporation | Centers for Disease Control and Prevention Division of Viral Diseases, Pathogen Discovery | Dakota Howard, Dhwani Batra, Peter W. Cook, Kara Moser, Adrian Paskey, Jason Caravas, Benjamin Rambo-Martin, Shatavia Morrison, Christopher Gulvick, Scott Sammons, Yvette Unoarumhi, Darlene Wagner, Matthew Schmerer, Cyndi Clark, Patrick Campbell, Rob Case, Vikramsinha Ghorpade, Holly Houdeshell, Ola Kvalvaag, Dillon Nall, Ethan Sanders, Alec Vest, Shaun Westlund, Matthew Hardison, Clinton R. Paden, Duncan MacCannell |
| EPI_ISL_2422068, EPI_ISL_2422069 | Alaska State Virology Laboratory | Alaska State Virology Laboratory | Stephanie DeRonde, Elva House, Jacob Zidek, Lisa Smith, Ph.D., Jack Chen, Ph.D. |
| EPI_ISL_2422070 | Aegis Sciences Corporation | Centers for Disease Control and Prevention Division of Viral Diseases, Pathogen Discovery | Dakota Howard, Dhwani Batra, Peter W. Cook, Kara Moser, Adrian Paskey, Jason Caravas, Benjamin Rambo-Martin, Shatavia Morrison, Christopher Gulvick, Scott Sammons, Yvette Unoarumhi, Darlene Wagner, Matthew Schmerer, Cyndi Clark, Patrick Campbell, Rob Case, Vikramsinha Ghorpade, Holly Houdeshell, Ola Kvalvaag, Dillon Nall, Ethan Sanders, Alec Vest, Shaun Westlund, Matthew Hardison, Clinton R. Paden, Duncan MacCannell |
| EPI_ISL_2422071, EPI_ISL_2422072 | Alaska State Virology Laboratory | Alaska State Virology Laboratory | Stephanie DeRonde, Elva House, Jacob Zidek, Lisa Smith, Ph.D., Jack Chen, Ph.D. |
| EPI_ISL_2422073 | Aegis Sciences Corporation | Centers for Disease Control and Prevention Division of Viral Diseases, Pathogen Discovery | Dakota Howard, Dhwani Batra, Peter W. Cook, Kara Moser, Adrian Paskey, Jason Caravas, Benjamin Rambo-Martin, Shatavia Morrison, Christopher Gulvick, Scott Sammons, Yvette Unoarumhi, Darlene Wagner, Matthew Schmerer, Cyndi Clark, Patrick Campbell, Rob Case, Vikramsinha Ghorpade, Holly Houdeshell, Ola Kvalvaag, Dillon Nall, Ethan Sanders, Alec Vest, Shaun Westlund, Matthew Hardison, Clinton R. Paden, Duncan MacCannell |
| EPI_ISL_2422074, EPI_ISL_2422075 | Alaska State Virology Laboratory | Alaska State Virology Laboratory | Stephanie DeRonde, Elva House, Jacob Zidek, Lisa Smith, Ph.D., Jack Chen, Ph.D. |
| EPI_ISL_2422076 | Aegis Sciences Corporation | Centers for Disease Control and Prevention Division of Viral Diseases, Pathogen Discovery | Dakota Howard, Dhwani Batra, Peter W. Cook, Kara Moser, Adrian Paskey, Jason Caravas, Benjamin Rambo-Martin, Shatavia Morrison, Christopher Gulvick, Scott Sammons, Yvette Unoarumhi, Darlene Wagner, Matthew Schmerer, Cyndi Clark, Patrick Campbell, Rob Case, Vikramsinha Ghorpade, Holly Houdeshell, Ola Kvalvaag, Dillon Nall, Ethan Sanders, Alec Vest, Shaun Westlund, Matthew Hardison, Clinton R. Paden, Duncan MacCannell |
| EPI_ISL_2422077, EPI_ISL_2422078 | Alaska State Virology Laboratory | Alaska State Virology Laboratory | Stephanie DeRonde, Elva House, Jacob Zidek, Lisa Smith, Ph.D., Jack Chen, Ph.D. |
| EPI_ISL_2422079 | Aegis Sciences Corporation | Centers for Disease Control and Prevention Division of Viral Diseases, Pathogen Discovery | Dakota Howard, Dhwani Batra, Peter W. Cook, Kara Moser, Adrian Paskey, Jason Caravas, Benjamin Rambo-Martin, Shatavia Morrison, Christopher Gulvick, Scott Sammons, Yvette Unoarumhi, Darlene Wagner, Matthew Schmerer, Cyndi Clark, Patrick Campbell, Rob Case, Vikramsinha Ghorpade, Holly Houdeshell, Ola Kvalvaag, Dillon Nall, Ethan Sanders, Alec Vest, Shaun Westlund, Matthew Hardison, Clinton R. Paden, Duncan MacCannell |
| EPI_ISL_2422080 | Alaska State Virology Laboratory | Alaska State Virology Laboratory | Stephanie DeRonde, Elva House, Jacob Zidek, Lisa Smith, Ph.D., Jack Chen, Ph.D. |
| EPI_ISL_2422081 | Aegis Sciences Corporation | Centers for Disease Control and Prevention Division of Viral Diseases, Pathogen Discovery | Dakota Howard, Dhwani Batra, Peter W. Cook, Kara Moser, Adrian Paskey, Jason Caravas, Benjamin Rambo-Martin, Shatavia Morrison, Christopher Gulvick, Scott Sammons, Yvette Unoarumhi, Darlene Wagner, Matthew Schmerer, Cyndi Clark, Patrick Campbell, Rob Case, Vikramsinha Ghorpade, Holly Houdeshell, Ola Kvalvaag, Dillon Nall, Ethan Sanders, Alec Vest, Shaun Westlund, Matthew Hardison, Clinton R. Paden, Duncan MacCannell |
| EPI_ISL_2422082, EPI_ISL_2422083 | Alaska State Virology Laboratory | Alaska State Virology Laboratory | Stephanie DeRonde, Elva House, Jacob Zidek, Lisa Smith, Ph.D., Jack Chen, Ph.D. |
| EPI_ISL_2422084 | Aegis Sciences Corporation | Centers for Disease Control and Prevention Division of Viral Diseases, Pathogen Discovery | Dakota Howard, Dhwani Batra, Peter W. Cook, Kara Moser, Adrian Paskey, Jason Caravas, Benjamin Rambo-Martin, Shatavia Morrison, Christopher Gulvick, Scott Sammons, Yvette Unoarumhi, Darlene Wagner, Matthew Schmerer, Cyndi Clark, Patrick Campbell, Rob Case, Vikramsinha Ghorpade, Holly Houdeshell, Ola Kvalvaag, Dillon Nall, Ethan Sanders, Alec Vest, Shaun Westlund, Matthew Hardison, Clinton R. Paden, Duncan MacCannell |
| EPI_ISL_2422085, EPI_ISL_2422086 | Alaska State Virology Laboratory | Alaska State Virology Laboratory | Stephanie DeRonde, Elva House, Jacob Zidek, Lisa Smith, Ph.D., Jack Chen, Ph.D. |
| EPI_ISL_2422087 | Aegis Sciences Corporation | Centers for Disease Control and Prevention Division of Viral Diseases, Pathogen Discovery | Dakota Howard, Dhwani Batra, Peter W. Cook, Kara Moser, Adrian Paskey, Jason Caravas, Benjamin Rambo-Martin, Shatavia Morrison, Christopher Gulvick, Scott Sammons, Yvette Unoarumhi, Darlene Wagner, Matthew Schmerer, Cyndi Clark, Patrick Campbell, Rob Case, Vikramsinha Ghorpade, Holly Houdeshell, Ola Kvalvaag, Dillon Nall, Ethan Sanders, Alec Vest, Shaun Westlund, Matthew Hardison, Clinton R. Paden, Duncan MacCannell |

| | | | |
|---|---|---|---|
| EPI_ISL_2422088, EPI_ISL_2422089 | Alaska State Virology Laboratory | Alaska State Virology Laboratory | Stephanie DeRonde, Elva House, Jacob Zidek, Lisa Smith, Ph.D., Jack Chen, Ph.D. |
| EPI_ISL_2422090 | Aegis Sciences Corporation | Centers for Disease Control and Prevention Division of Viral Diseases, Pathogen Discovery | Dakota Howard, Dhwani Batra, Peter W. Cook, Kara Moser, Adrian Paskey, Jason Caravas, Benjamin Rambo-Martin, Shatavia Morrison, Christopher Gulvick, Scott Sammons, Yvette Unoarumhi, Darlene Wagner, Matthew Schmerer, Cyndi Clark, Patrick Campbell, Rob Case, Vikramsinha Ghorpade, Holly Houdeshell, Ola Kvalvaag, Dillon Nall, Ethan Sanders, Alec Vest, Shaun Westlund, Matthew Hardison, Clinton R. Paden, Duncan MacCannell |
| EPI_ISL_2422091, EPI_ISL_2422092 | Alaska State Virology Laboratory | Alaska State Virology Laboratory | Stephanie DeRonde, Elva House, Jacob Zidek, Lisa Smith, Ph.D., Jack Chen, Ph.D. |
| EPI_ISL_2422093 | Aegis Sciences Corporation | Centers for Disease Control and Prevention Division of Viral Diseases, Pathogen Discovery | Dakota Howard, Dhwani Batra, Peter W. Cook, Kara Moser, Adrian Paskey, Jason Caravas, Benjamin Rambo-Martin, Shatavia Morrison, Christopher Gulvick, Scott Sammons, Yvette Unoarumhi, Darlene Wagner, Matthew Schmerer, Cyndi Clark, Patrick Campbell, Rob Case, Vikramsinha Ghorpade, Holly Houdeshell, Ola Kvalvaag, Dillon Nall, Ethan Sanders, Alec Vest, Shaun Westlund, Matthew Hardison, Clinton R. Paden, Duncan MacCannell |
| EPI_ISL_2422094, EPI_ISL_2422095 | Alaska State Virology Laboratory | Alaska State Virology Laboratory | Stephanie DeRonde, Elva House, Jacob Zidek, Lisa Smith, Ph.D., Jack Chen, Ph.D. |
| EPI_ISL_2422096 | Aegis Sciences Corporation | Centers for Disease Control and Prevention Division of Viral Diseases, Pathogen Discovery | Dakota Howard, Dhwani Batra, Peter W. Cook, Kara Moser, Adrian Paskey, Jason Caravas, Benjamin Rambo-Martin, Shatavia Morrison, Christopher Gulvick, Scott Sammons, Yvette Unoarumhi, Darlene Wagner, Matthew Schmerer, Cyndi Clark, Patrick Campbell, Rob Case, Vikramsinha Ghorpade, Holly Houdeshell, Ola Kvalvaag, Dillon Nall, Ethan Sanders, Alec Vest, Shaun Westlund, Matthew Hardison, Clinton R. Paden, Duncan MacCannell |
| EPI_ISL_2422097, EPI_ISL_2422098 | Alaska State Virology Laboratory | Alaska State Virology Laboratory | Stephanie DeRonde, Elva House, Jacob Zidek, Lisa Smith, Ph.D., Jack Chen, Ph.D. |
| EPI_ISL_2422099 | Aegis Sciences Corporation | Centers for Disease Control and Prevention Division of Viral Diseases, Pathogen Discovery | Dakota Howard, Dhwani Batra, Peter W. Cook, Kara Moser, Adrian Paskey, Jason Caravas, Benjamin Rambo-Martin, Shatavia Morrison, Christopher Gulvick, Scott Sammons, Yvette Unoarumhi, Darlene Wagner, Matthew Schmerer, Cyndi Clark, Patrick Campbell, Rob Case, Vikramsinha Ghorpade, Holly Houdeshell, Ola Kvalvaag, Dillon Nall, Ethan Sanders, Alec Vest, Shaun Westlund, Matthew Hardison, Clinton R. Paden, Duncan MacCannell |
| EPI_ISL_2422100, EPI_ISL_2422101, EPI_ISL_2422103, EPI_ISL_2422104 | Alaska State Virology Laboratory | Alaska State Virology Laboratory | Stephanie DeRonde, Elva House, Jacob Zidek, Lisa Smith, Ph.D., Jack Chen, Ph.D. |
| EPI_ISL_2422105 | Aegis Sciences Corporation | Centers for Disease Control and Prevention Division of Viral Diseases, Pathogen Discovery | Dakota Howard, Dhwani Batra, Peter W. Cook, Kara Moser, Adrian Paskey, Jason Caravas, Benjamin Rambo-Martin, Shatavia Morrison, Christopher Gulvick, Scott Sammons, Yvette Unoarumhi, Darlene Wagner, Matthew Schmerer, Cyndi Clark, Patrick Campbell, Rob Case, Vikramsinha Ghorpade, Holly Houdeshell, Ola Kvalvaag, Dillon Nall, Ethan Sanders, Alec Vest, Shaun Westlund, Matthew Hardison, Clinton R. Paden, Duncan MacCannell |
| EPI_ISL_2422106 | Alaska State Virology Laboratory | Alaska State Virology Laboratory | Stephanie DeRonde, Elva House, Jacob Zidek, Lisa Smith, Ph.D., Jack Chen, Ph.D. |
| EPI_ISL_2422107 | Aegis Sciences Corporation | Centers for Disease Control and Prevention Division of Viral Diseases, Pathogen Discovery | Dakota Howard, Dhwani Batra, Peter W. Cook, Kara Moser, Adrian Paskey, Jason Caravas, Benjamin Rambo-Martin, Shatavia Morrison, Christopher Gulvick, Scott Sammons, Yvette Unoarumhi, Darlene Wagner, Matthew Schmerer, Cyndi Clark, Patrick Campbell, Rob Case, Vikramsinha Ghorpade, Holly Houdeshell, Ola Kvalvaag, Dillon Nall, Ethan Sanders, Alec Vest, Shaun Westlund, Matthew Hardison, Clinton R. Paden, Duncan MacCannell |
| EPI_ISL_2422108 | Alaska State Virology Laboratory | Alaska State Virology Laboratory | Stephanie DeRonde, Elva House, Jacob Zidek, Lisa Smith, Ph.D., Jack Chen, Ph.D. |
| EPI_ISL_2422109, EPI_ISL_2422110, EPI_ISL_2422111, EPI_ISL_2422112, EPI_ISL_2422113, EPI_ISL_2422114, EPI_ISL_2422115, EPI_ISL_2422116, EPI_ISL_2422117, EPI_ISL_2422118, EPI_ISL_2422119 | | | |
| see above | Aegis Sciences Corporation | Centers for Disease Control and Prevention Division of Viral Diseases, Pathogen Discovery | Dakota Howard, Dhwani Batra, Peter W. Cook, Kara Moser, Adrian Paskey, Jason Caravas, Benjamin Rambo-Martin, Shatavia Morrison, Christopher Gulvick, Scott Sammons, Yvette Unoarumhi, Darlene Wagner, Matthew Schmerer, Cyndi Clark, Patrick Campbell, Rob Case, Vikramsinha Ghorpade, Holly Houdeshell, Ola Kvalvaag, Dillon Nall, Ethan Sanders, Alec Vest, Shaun Westlund, Matthew Hardison, Clinton R. Paden, Duncan MacCannell |
| EPI_ISL_2422227, EPI_ISL_2422228, EPI_ISL_2422229, EPI_ISL_2422230, EPI_ISL_2422231, EPI_ISL_2422232, EPI_ISL_2422233, EPI_ISL_2422234, EPI_ISL_2422235, EPI_ISL_2422236, EPI_ISL_2422237, EPI_ISL_2422238, EPI_ISL_2422239, EPI_ISL_2422240, EPI_ISL_2422241, EPI_ISL_2422242, EPI_ISL_2422243, EPI_ISL_2422244, EPI_ISL_2422245 | | | |
| see above | Kansas Health and Environmental Lab | Kansas Health and Environmental Lab | Katherine Wiggins, Mike Grose, Jonathan Barnell, Ben Olsen, and Phil Adam |
| EPI_ISL_2422246 | Washington University | Genomics and Discovery, Respiratory Viruses Branch, Division of Viral Diseases, Centers for Disease Control and Prevention | Anna Kelleher, Ying Tao, Yan Li, Jing Zhang, Brian Lynch, Krista Queen, Anna Uehara, Peter Cook, Han Jia Justin Ng, Rachel Marine, Clinton R. Paden, Dhwani Batra, Haibin Wang, Tara Coalter, Jasmine Padilla, Morgan Davis, Mili Sheth, Sarah Nobles, Mark Burroughs, Justin Lee, Adam Retchless, Suxiang Tong |
| EPI_ISL_2422247, EPI_ISL_2422248, EPI_ISL_2422249, EPI_ISL_2422250, EPI_ISL_2422251, EPI_ISL_2422252, EPI_ISL_2422254 | Springfield Greene County Health Dept. | Genomics and Discovery, Respiratory Viruses Branch, Division of Viral Diseases, Centers for Disease Control and Prevention | Anna Kelleher, Ying Tao, Yan Li, Jing Zhang, Brian Lynch, Krista Queen, Anna Uehara, Peter Cook, Han Jia Justin Ng, Rachel Marine, Clinton R. Paden, Dhwani Batra, Haibin Wang, Tara Coalter, Jasmine Padilla, Morgan Davis, Mili Sheth, Sarah Nobles, Mark Burroughs, Justin Lee, Adam Retchless, Suxiang Tong |
| EPI_ISL_2422255, EPI_ISL_2422256 | Washington University | Genomics and Discovery, Respiratory Viruses Branch, Division of Viral Diseases, Centers for Disease Control and Prevention | Anna Kelleher, Ying Tao, Yan Li, Jing Zhang, Brian Lynch, Krista Queen, Anna Uehara, Peter Cook, Han Jia Justin Ng, Rachel Marine, Clinton R. Paden, Dhwani Batra, Haibin Wang, Tara Coalter, Jasmine Padilla, Morgan Davis, Mili Sheth, Sarah Nobles, Mark Burroughs, Justin Lee, Adam Retchless, Suxiang Tong |
| EPI_ISL_2422257 | MO State Public Health Laboratory | Genomics and Discovery, Respiratory Viruses Branch, Division of Viral Diseases, Centers for Disease Control and Prevention | Anna Kelleher, Ying Tao, Yan Li, Jing Zhang, Brian Lynch, Krista Queen, Anna Uehara, Peter Cook, Han Jia Justin Ng, Rachel Marine, Clinton R. Paden, Dhwani Batra, Haibin Wang, Tara Coalter, Jasmine Padilla, Morgan Davis, Mili Sheth, Sarah Nobles, Mark Burroughs, Justin Lee, Adam Retchless, Suxiang Tong |
| EPI_ISL_2422283, EPI_ISL_2422284, EPI_ISL_2422285, EPI_ISL_2422286, EPI_ISL_2422287, EPI_ISL_2422288, EPI_ISL_2422289, EPI_ISL_2422290, EPI_ISL_2422291, EPI_ISL_2422292, EPI_ISL_2422293, EPI_ISL_2422294, EPI_ISL_2422295, EPI_ISL_2422296, EPI_ISL_2422297, EPI_ISL_2422298, EPI_ISL_2422299, EPI_ISL_2422300, EPI_ISL_2422301, EPI_ISL_2422302, EPI_ISL_2422303, EPI_ISL_2422304, EPI_ISL_2422305, EPI_ISL_2422306, EPI_ISL_2422307, EPI_ISL_2422308, EPI_ISL_2422309, EPI_ISL_2422310, EPI_ISL_2422311, EPI_ISL_2422312 | | | |
| see above | SUNY UPSTATE MEDICAL UNIVERSITY | Wadsworth Center, New York State Department of Health | Kirsten St. George, Daryl M. Lamson, Alexis Russell, Matthew Shudt, Melissa A Leisner, Jonathan Plitnick, Catharine Prussing, Navjot Singh, John Kelly, Erasmus Schneider, Erica Lasek-Nesselquist |
| EPI_ISL_2422313 | ALBANY MEDICAL CENTER | Wadsworth Center, New York State Department of Health | Kirsten St. George, Daryl M. Lamson, Alexis Russell, Matthew Shudt, Melissa A Leisner, Jonathan Plitnick, Catharine Prussing, Navjot Singh, John Kelly, Erasmus Schneider, Erica Lasek-Nesselquist |
| EPI_ISL_2422314, EPI_ISL_2422315, EPI_ISL_2422316, EPI_ISL_2422317, EPI_ISL_2422318, EPI_ISL_2422319, EPI_ISL_2422320, EPI_ISL_2422321, EPI_ISL_2422322 | STONY BROOK UNIVERSITY HOSPITAL | Wadsworth Center, New York State Department of Health | Kirsten St. George, Daryl M. Lamson, Alexis Russell, Matthew Shudt, Melissa A Leisner, Jonathan Plitnick, Catharine Prussing, Navjot Singh, John Kelly, Erasmus Schneider, Erica Lasek-Nesselquist |
| EPI_ISL_2422323, EPI_ISL_2422324, EPI_ISL_2422325, EPI_ISL_2422326 | MONTEFIORE MEDICAL CENTER LABORATORIES | Wadsworth Center, New York State Department of Health | Kirsten St. George, Daryl M. Lamson, Alexis Russell, Matthew Shudt, Melissa A Leisner, Jonathan Plitnick, Catharine Prussing, Navjot Singh, John Kelly, Erasmus Schneider, Erica Lasek-Nesselquist |
| EPI_ISL_2422327, EPI_ISL_2422328, EPI_ISL_2422329, EPI_ISL_2422330, EPI_ISL_2422331, EPI_ISL_2422332, EPI_ISL_2422333, EPI_ISL_2422334, EPI_ISL_2422335, EPI_ISL_2422336, EPI_ISL_2422337, EPI_ISL_2422338, EPI_ISL_2422339, EPI_ISL_2422340, EPI_ISL_2422341, EPI_ISL_2422342, EPI_ISL_2422343, EPI_ISL_2422344, EPI_ISL_2422345, EPI_ISL_2422346, EPI_ISL_2422347, EPI_ISL_2422348, EPI_ISL_2422349, EPI_ISL_2422350, EPI_ISL_2422351, EPI_ISL_2422352, EPI_ISL_2422353, EPI_ISL_2422354, EPI_ISL_2422355, EPI_ISL_2422356, EPI_ISL_2422357, EPI_ISL_2422358, EPI_ISL_2422359, EPI_ISL_2422360 | | | |
| see above | URMC LABS | Wadsworth Center, New York State Department of Health | Kirsten St. George, Daryl M. Lamson, Alexis Russell, Matthew Shudt, Melissa A Leisner, Jonathan Plitnick, Catharine Prussing, Navjot Singh, John Kelly, Erasmus Schneider, Erica Lasek-Nesselquist |
| EPI_ISL_2422433 | LA Office of Public Health Laboratories | Genomics and Discovery, Respiratory Viruses Branch, Division of Viral Diseases, Centers for Disease Control and Prevention | Anna Kelleher, Ying Tao, Yan Li, Jing Zhang, Brian Lynch, Krista Queen, Anna Uehara, Peter Cook, Han Jia Justin Ng, Rachel Marine, Clinton R. Paden, Dhwani Batra, Haibin Wang, Tara Coalter, Jasmine Padilla, Morgan Davis, Mili Sheth, Sarah Nobles, Mark Burroughs, Justin Lee, Adam Retchless, Suxiang Tong |
| EPI_ISL_2422434 | NC State Laboratory of Public Health | Genomics and Discovery, Respiratory Viruses Branch, Division of Viral Diseases, Centers for Disease Control and Prevention | Anna Kelleher, Ying Tao, Yan Li, Jing Zhang, Brian Lynch, Krista Queen, Anna Uehara, Peter Cook, Han Jia Justin Ng, Rachel Marine, Clinton R. Paden, Dhwani Batra, Haibin Wang, Tara Coalter, Jasmine Padilla, Morgan Davis, Mili Sheth, Sarah Nobles, Mark Burroughs, Justin Lee, Adam Retchless, Suxiang Tong |
| EPI_ISL_2422435, EPI_ISL_2422436 | SD Public Health Laboratory | Genomics and Discovery, Respiratory Viruses Branch, Division of Viral Diseases, Centers for Disease Control and Prevention | Anna Kelleher, Ying Tao, Yan Li, Jing Zhang, Brian Lynch, Krista Queen, Anna Uehara, Peter Cook, Han Jia Justin Ng, Rachel Marine, Clinton R. Paden, Dhwani Batra, Haibin Wang, Tara Coalter, Jasmine Padilla, Morgan Davis, Mili Sheth, Sarah Nobles, Mark Burroughs, Justin Lee, Adam Retchless, Suxiang Tong |
| EPI_ISL_2422437, EPI_ISL_2422438, EPI_ISL_2422439, EPI_ISL_2422440, EPI_ISL_2422441, EPI_ISL_2422442, EPI_ISL_2422443 | AR Dept. of Health-PHL, Molecular Diagnostics | Genomics and Discovery, Respiratory Viruses Branch, Division of Viral Diseases, Centers for Disease Control and Prevention | Anna Kelleher, Ying Tao, Yan Li, Jing Zhang, Brian Lynch, Krista Queen, Anna Uehara, Peter Cook, Han Jia Justin Ng, Rachel Marine, Clinton R. Paden, Dhwani Batra, Haibin Wang, Tara Coalter, Jasmine Padilla, Morgan Davis, Mili Sheth, Sarah Nobles, Mark Burroughs, Justin Lee, Adam Retchless, Suxiang Tong |
| EPI_ISL_2422444, EPI_ISL_2422445, EPI_ISL_2422446, | OK Public Health Laboratory, Oklahoma State DOH | Genomics and Discovery, Respiratory Viruses Branch, Division of Viral Diseases, Centers for Disease Control and Prevention | Anna Kelleher, Ying Tao, Yan Li, Jing Zhang, Brian Lynch, Krista Queen, Anna Uehara, Peter Cook, Han Jia Justin Ng, Rachel Marine, Clinton R. Paden, Dhwani Batra, Haibin Wang, Tara Coalter, Jasmine Padilla, Morgan Davis, Mili Sheth, Sarah Nobles, Mark Burroughs, Justin Lee, Adam Retchless, Suxiang Tong |

| | | | |
|---|---|---|---|
| EPI_ISL_2422447, EPI_ISL_2422448, EPI_ISL_2422449, EPI_ISL_2422450 | | | |
| EPI_ISL_2422451, EPI_ISL_2422452, EPI_ISL_2422453, EPI_ISL_2422454, EPI_ISL_2422455, EPI_ISL_2422456 | WVDHHR - Office of Laboratory Services | Genomics and Discovery, Respiratory Viruses Branch, Division of Viral Diseases, Centers for Disease Control and Prevention | Anna Kelleher, Ying Tao, Yan Li, Jing Zhang, Brian Lynch, Krista Queen, Anna Uehara, Peter Cook, Han Jia Justin Ng, Rachel Marine, Clinton R. Paden, Dhwani Batra, Haibin Wang, Tara Coalter, Jasmine Padilla, Morgan Davis, Mili Sheth, Sarah Nobles, Mark Burroughs, Justin Lee, Adam Retchless, Suxiang Tong |
| EPI_ISL_2422457, EPI_ISL_2422458, EPI_ISL_2422459, EPI_ISL_2422460, EPI_ISL_2422461 | GA Department of Public Health Laboratory | Genomics and Discovery, Respiratory Viruses Branch, Division of Viral Diseases, Centers for Disease Control and Prevention | Anna Kelleher, Ying Tao, Yan Li, Jing Zhang, Brian Lynch, Krista Queen, Anna Uehara, Peter Cook, Han Jia Justin Ng, Rachel Marine, Clinton R. Paden, Dhwani Batra, Haibin Wang, Tara Coalter, Jasmine Padilla, Morgan Davis, Mili Sheth, Sarah Nobles, Mark Burroughs, Justin Lee, Adam Retchless, Suxiang Tong |
| EPI_ISL_2422462, EPI_ISL_2422463 | TN Division of Laboratory Services | Genomics and Discovery, Respiratory Viruses Branch, Division of Viral Diseases, Centers for Disease Control and Prevention | Anna Kelleher, Ying Tao, Yan Li, Jing Zhang, Brian Lynch, Krista Queen, Anna Uehara, Peter Cook, Han Jia Justin Ng, Rachel Marine, Clinton R. Paden, Dhwani Batra, Haibin Wang, Tara Coalter, Jasmine Padilla, Morgan Davis, Mili Sheth, Sarah Nobles, Mark Burroughs, Justin Lee, Adam Retchless, Suxiang Tong |
| EPI_ISL_2422464, EPI_ISL_2422465, EPI_ISL_2422466, EPI_ISL_2422467, EPI_ISL_2422468, EPI_ISL_2422469, EPI_ISL_2422470, EPI_ISL_2422471, EPI_ISL_2422472, EPI_ISL_2422473, EPI_ISL_2422474, EPI_ISL_2422475, EPI_ISL_2422476, EPI_ISL_2422477, EPI_ISL_2422478, EPI_ISL_2422479, EPI_ISL_2422480, EPI_ISL_2422481, EPI_ISL_2422482, EPI_ISL_2422483, EPI_ISL_2422484, EPI_ISL_2422485, EPI_ISL_2422486, EPI_ISL_2422487, EPI_ISL_2422488, EPI_ISL_2422489, EPI_ISL_2422490, EPI_ISL_2422491, EPI_ISL_2422492, EPI_ISL_2422493, EPI_ISL_2422494 | | | |
| see above | Wyoming Public Health Laboratory | Wyoming Public Health Laboratory | Jim Mildenberger, Wanda Manley, Noah Hull, Taylor Fearing, Lynette Gumbleton, Channing Weber, Ashley Norberg, Chayse Rowley, Marley Goetz, Brian Dominguez, Elliot Thomasson, Sam Britz, Cari Sloma, and Rob Christensen |
| EPI_ISL_2422573, EPI_ISL_2422576, EPI_ISL_2422580, EPI_ISL_2422584, EPI_ISL_2422587, EPI_ISL_2422590, EPI_ISL_2422592, EPI_ISL_2422596, EPI_ISL_2422599, EPI_ISL_2422602, EPI_ISL_2422606, EPI_ISL_2422609, EPI_ISL_2422613, EPI_ISL_2422616, EPI_ISL_2422620, EPI_ISL_2422623, EPI_ISL_2422626, EPI_ISL_2422629, EPI_ISL_2422631, EPI_ISL_2422632, EPI_ISL_2422633, EPI_ISL_2422634, EPI_ISL_2422635, EPI_ISL_2422636, EPI_ISL_2422637, EPI_ISL_2422638, EPI_ISL_2422639, EPI_ISL_2422640, EPI_ISL_2422641, EPI_ISL_2422642, EPI_ISL_2422643, EPI_ISL_2422644, EPI_ISL_2422645, EPI_ISL_2422646, EPI_ISL_2422647, EPI_ISL_2422648, EPI_ISL_2422649 | | | |
| see above | San Diego County Public Health Laboratory | Andersen lab at Scripps Research | SEARCH Alliance San Diego with Ashleigh Murphy, Jovan Shephard, Brett Austin |
| EPI_ISL_2422650 | Wyoming Public Health Laboratory | Wyoming Public Health Laboratory | Jim Mildenberger, Wanda Manley, Noah Hull, Taylor Fearing, Lynette Gumbleton, Channing Weber, Ashley Norberg, Chayse Rowley, Marley Goetz, Brian Dominguez, Elliot Thomasson, Sam Britz, Cari Sloma, and Rob Christensen |
| EPI_ISL_2422651, EPI_ISL_2422652, EPI_ISL_2422653, EPI_ISL_2422654, EPI_ISL_2422655, EPI_ISL_2422656, EPI_ISL_2422657, EPI_ISL_2422658, EPI_ISL_2422659, EPI_ISL_2422660, EPI_ISL_2422661, EPI_ISL_2422662, EPI_ISL_2422663, EPI_ISL_2422664, EPI_ISL_2422665, EPI_ISL_2422666, EPI_ISL_2422667, EPI_ISL_2422668 | | | |
| see above | San Diego County Public Health Laboratory | Andersen lab at Scripps Research | SEARCH Alliance San Diego with Ashleigh Murphy, Jovan Shephard, Brett Austin |
| EPI_ISL_2422669, EPI_ISL_2422670, EPI_ISL_2422671, EPI_ISL_2422672, EPI_ISL_2422673, EPI_ISL_2422674, EPI_ISL_2422675, EPI_ISL_2422676, EPI_ISL_2422677, EPI_ISL_2422678, EPI_ISL_2422679, EPI_ISL_2422680, EPI_ISL_2422681, EPI_ISL_2422682, EPI_ISL_2422683, EPI_ISL_2422684, EPI_ISL_2422685, EPI_ISL_2422686, EPI_ISL_2422687, EPI_ISL_2422688, EPI_ISL_2422689, EPI_ISL_2422690, EPI_ISL_2422691, EPI_ISL_2422692, EPI_ISL_2422693, EPI_ISL_2422694, EPI_ISL_2422695, EPI_ISL_2422696, EPI_ISL_2422697, EPI_ISL_2422698, EPI_ISL_2422699, EPI_ISL_2422700, EPI_ISL_2422701, EPI_ISL_2422702, EPI_ISL_2422703, EPI_ISL_2422704, EPI_ISL_2422705, EPI_ISL_2422706, EPI_ISL_2422707, EPI_ISL_2422708, EPI_ISL_2422709, EPI_ISL_2422710, EPI_ISL_2422711, EPI_ISL_2422712, EPI_ISL_2422713, EPI_ISL_2422714, EPI_ISL_2422715, EPI_ISL_2422716, EPI_ISL_2422717, EPI_ISL_2422718, EPI_ISL_2422719 | | | |
| see above | Sharp HealthCare Laboratory | Andersen lab at Scripps Research | SEARCH Alliance San Diego with Aaron Harding, Jacquelyn Berumen, Cathy Woerle, Liam McGinnis, Art Mendoza, Omid Bakhtar |
| EPI_ISL_2422720, EPI_ISL_2422722, EPI_ISL_2422723, EPI_ISL_2422724, EPI_ISL_2422725, EPI_ISL_2422726, EPI_ISL_2422727, EPI_ISL_2422728, EPI_ISL_2422729, EPI_ISL_2422730, EPI_ISL_2422731, EPI_ISL_2422732, EPI_ISL_2422733, EPI_ISL_2422734, EPI_ISL_2422735, EPI_ISL_2422737, EPI_ISL_2422738, EPI_ISL_2422739, EPI_ISL_2422740, EPI_ISL_2422741, EPI_ISL_2422742, EPI_ISL_2422743, EPI_ISL_2422744, EPI_ISL_2422745, EPI_ISL_2422746, EPI_ISL_2422747, EPI_ISL_2422748, EPI_ISL_2422749, EPI_ISL_2422750, EPI_ISL_2422751, EPI_ISL_2422752, EPI_ISL_2422753, EPI_ISL_2422754, EPI_ISL_2422755, EPI_ISL_2422756, EPI_ISL_2422757, EPI_ISL_2422758, EPI_ISL_2422759, EPI_ISL_2422760, EPI_ISL_2422761, EPI_ISL_2422762, EPI_ISL_2422763, EPI_ISL_2422764, EPI_ISL_2422765, EPI_ISL_2422766, EPI_ISL_2422767, EPI_ISL_2422768, EPI_ISL_2422769, EPI_ISL_2422770, EPI_ISL_2422771, EPI_ISL_2422772, EPI_ISL_2422773, EPI_ISL_2422774, EPI_ISL_2422775, EPI_ISL_2422776, EPI_ISL_2422777, EPI_ISL_2422778, EPI_ISL_2422779, EPI_ISL_2422780, EPI_ISL_2422781, EPI_ISL_2422782 | | | |
| see above | San Diego County Public Health Laboratory | Andersen lab at Scripps Research | SEARCH Alliance San Diego with Ashleigh Murphy, Jovan Shephard, Brett Austin |
| EPI_ISL_2422783, EPI_ISL_2422784, EPI_ISL_2422785, EPI_ISL_2422786, EPI_ISL_2422787, EPI_ISL_2422788, EPI_ISL_2422789, EPI_ISL_2422790, EPI_ISL_2422791, EPI_ISL_2422792, EPI_ISL_2422793, EPI_ISL_2422794, EPI_ISL_2422795, EPI_ISL_2422796, EPI_ISL_2422797, EPI_ISL_2422798, EPI_ISL_2422799, EPI_ISL_2422800, EPI_ISL_2422801, EPI_ISL_2422802, EPI_ISL_2422803, EPI_ISL_2422804, EPI_ISL_2422805, EPI_ISL_2422806, EPI_ISL_2422807, EPI_ISL_2422808, EPI_ISL_2422809, EPI_ISL_2422810, EPI_ISL_2422811, EPI_ISL_2422812, EPI_ISL_2422813, EPI_ISL_2422814, EPI_ISL_2422815, EPI_ISL_2422816, EPI_ISL_2422817, EPI_ISL_2422818, EPI_ISL_2422819, EPI_ISL_2422820, EPI_ISL_2422821, EPI_ISL_2422822, EPI_ISL_2422823, EPI_ISL_2422824, EPI_ISL_2422825, EPI_ISL_2422826, EPI_ISL_2422827, EPI_ISL_2422828, EPI_ISL_2422829, EPI_ISL_2422830, EPI_ISL_2422831, EPI_ISL_2422832, EPI_ISL_2422833, EPI_ISL_2422834, EPI_ISL_2422835, EPI_ISL_2422836, EPI_ISL_2422837, EPI_ISL_2422838, EPI_ISL_2422839, EPI_ISL_2422840, EPI_ISL_2422841, EPI_ISL_2422842, EPI_ISL_2422843, EPI_ISL_2422844, EPI_ISL_2422845, EPI_ISL_2422846, EPI_ISL_2422847, EPI_ISL_2422848, EPI_ISL_2422849, EPI_ISL_2422850, EPI_ISL_2422851, EPI_ISL_2422852, EPI_ISL_2422853, EPI_ISL_2422854, EPI_ISL_2422855, EPI_ISL_2422856, EPI_ISL_2422857, EPI_ISL_2422858, EPI_ISL_2422859, EPI_ISL_2422860, EPI_ISL_2422861, EPI_ISL_2422862, EPI_ISL_2422863, EPI_ISL_2422864, EPI_ISL_2422865, EPI_ISL_2422866, EPI_ISL_2422867, EPI_ISL_2422868, EPI_ISL_2422869, EPI_ISL_2422870, EPI_ISL_2422871, EPI_ISL_2422872, EPI_ISL_2422873, EPI_ISL_2422874, EPI_ISL_2422875, EPI_ISL_2422876, EPI_ISL_2422877, EPI_ISL_2422878, EPI_ISL_2422879, EPI_ISL_2422880, EPI_ISL_2422881, EPI_ISL_2422882, EPI_ISL_2422883, EPI_ISL_2422884, EPI_ISL_2422885, EPI_ISL_2422886, EPI_ISL_2422887, EPI_ISL_2422888, EPI_ISL_2422889, EPI_ISL_2422890, EPI_ISL_2422891, EPI_ISL_2422892, EPI_ISL_2422893, EPI_ISL_2422894, EPI_ISL_2422895, EPI_ISL_2422896 | | | |
| see above | Sharp HealthCare Laboratory | Andersen lab at Scripps Research | SEARCH Alliance San Diego with Aaron Harding, Jacquelyn Berumen, Cathy Woerle, Liam McGinnis, Art Mendoza, Omid Bakhtar |
| EPI_ISL_2422897, EPI_ISL_2422898, EPI_ISL_2422899, EPI_ISL_2422900, EPI_ISL_2422901, EPI_ISL_2422902, EPI_ISL_2422903, EPI_ISL_2422905, EPI_ISL_2422906, EPI_ISL_2422907, EPI_ISL_2422908, EPI_ISL_2422909, EPI_ISL_2422910, EPI_ISL_2422911, EPI_ISL_2422912, EPI_ISL_2422913, EPI_ISL_2422914, EPI_ISL_2422915, EPI_ISL_2422916, EPI_ISL_2422917, EPI_ISL_2422918, EPI_ISL_2422919, EPI_ISL_2422920, EPI_ISL_2422921, EPI_ISL_2422922, EPI_ISL_2422923, EPI_ISL_2422924, EPI_ISL_2422925, EPI_ISL_2422926, EPI_ISL_2422927, EPI_ISL_2422928, EPI_ISL_2422929, EPI_ISL_2422930, EPI_ISL_2422931, EPI_ISL_2422932, EPI_ISL_2422933, EPI_ISL_2422934, EPI_ISL_2422935, EPI_ISL_2422936, EPI_ISL_2422937, EPI_ISL_2422938, EPI_ISL_2422939, EPI_ISL_2422940, EPI_ISL_2422941, EPI_ISL_2422942, EPI_ISL_2422943, EPI_ISL_2422944, EPI_ISL_2422945, EPI_ISL_2422946, EPI_ISL_2422947, EPI_ISL_2422948, EPI_ISL_2422949, EPI_ISL_2422950, EPI_ISL_2422951, EPI_ISL_2422952, EPI_ISL_2422953, EPI_ISL_2422954, EPI_ISL_2422955, EPI_ISL_2422956, EPI_ISL_2422957, EPI_ISL_2422958, EPI_ISL_2422959, EPI_ISL_2422960, EPI_ISL_2422961, EPI_ISL_2422962, EPI_ISL_2422963, EPI_ISL_2422964, EPI_ISL_2422965, EPI_ISL_2422966, EPI_ISL_2422967, EPI_ISL_2422969, EPI_ISL_2422970, EPI_ISL_2422971, EPI_ISL_2422972, EPI_ISL_2422973, EPI_ISL_2422974, EPI_ISL_2422975, EPI_ISL_2422976, EPI_ISL_2422977, EPI_ISL_2422978, EPI_ISL_2422979, EPI_ISL_2422980, EPI_ISL_2422981, EPI_ISL_2422982, EPI_ISL_2422983, EPI_ISL_2422984, EPI_ISL_2422985, EPI_ISL_2422986, EPI_ISL_2422987, EPI_ISL_2422988, EPI_ISL_2422989, EPI_ISL_2422990, EPI_ISL_2422991, EPI_ISL_2422992, EPI_ISL_2422993, EPI_ISL_2422995, EPI_ISL_2422997, EPI_ISL_2422998, EPI_ISL_2423003, EPI_ISL_2423004, EPI_ISL_2423005, EPI_ISL_2423006, EPI_ISL_2423007, EPI_ISL_2423010, EPI_ISL_2423011, EPI_ISL_2423013, EPI_ISL_2423014, EPI_ISL_2423015, EPI_ISL_2423016, EPI_ISL_2423017, EPI_ISL_2423018, EPI_ISL_2423019, EPI_ISL_2423020, EPI_ISL_2423021, EPI_ISL_2423022, EPI_ISL_2423023, EPI_ISL_2423025, EPI_ISL_2423026, EPI_ISL_2423027, EPI_ISL_2423029, EPI_ISL_2423031, EPI_ISL_2423032 | | | |
| see above | San Diego County Public Health Laboratory | Andersen lab at Scripps Research | SEARCH Alliance San Diego with Ashleigh Murphy, Jovan Shephard, Brett Austin |
| EPI_ISL_2423035, EPI_ISL_2423037, EPI_ISL_2423038, EPI_ISL_2423039, EPI_ISL_2423040, EPI_ISL_2423041, EPI_ISL_2423042, EPI_ISL_2423043, EPI_ISL_2423044, EPI_ISL_2423045, EPI_ISL_2423046, EPI_ISL_2423047, EPI_ISL_2423048, EPI_ISL_2423050, EPI_ISL_2423051, EPI_ISL_2423052, EPI_ISL_2423053, EPI_ISL_2423054, EPI_ISL_2423055, EPI_ISL_2423056, EPI_ISL_2423057, EPI_ISL_2423058, EPI_ISL_2423059, EPI_ISL_2423060, EPI_ISL_2423061, EPI_ISL_2423062, EPI_ISL_2423063, EPI_ISL_2423064, EPI_ISL_2423065, EPI_ISL_2423066, EPI_ISL_2423067, EPI_ISL_2423068, EPI_ISL_2423069, EPI_ISL_2423070, EPI_ISL_2423071, EPI_ISL_2423072, EPI_ISL_2423074, EPI_ISL_2423075, EPI_ISL_2423076, EPI_ISL_2423077, EPI_ISL_2423078, EPI_ISL_2423079 | | | |
| see above | Wyoming Public Health Laboratory | Wyoming Public Health Laboratory | Jim Mildenberger, Taylor Fearing, Channing Weber, Ashley Norberg, Chayse Rowley, Marley Goetz, Brian Dominguez, Elliot Thomasson, Brittany Oher, Sam Britz, Cari Sloma, and Rob Christensen |
| EPI_ISL_2423156, EPI_ISL_2423157, EPI_ISL_2423158, EPI_ISL_2423159, EPI_ISL_2423160, EPI_ISL_2423161, EPI_ISL_2423162, EPI_ISL_2423163, EPI_ISL_2423164, EPI_ISL_2423165, EPI_ISL_2423166, EPI_ISL_2423167, EPI_ISL_2423168, EPI_ISL_2423169, EPI_ISL_2423170, EPI_ISL_2423171, EPI_ISL_2423172 | | | |
| see above | Wyoming Public Health Laboratory | Wyoming Public Health Laboratory | Jim Mildenberger, Taylor Fearing, Channing Weber, Ashley Norberg, Chayse Rowley, Marley Goetz, Brian Dominguez, Elliot Thomasson,Sam Britz, Cari Sloma, Robert Petit, and Rob Christensen |
| EPI_ISL_2423318, EPI_ISL_2423319, EPI_ISL_2423321, EPI_ISL_2423322, EPI_ISL_2423323, EPI_ISL_2423324, EPI_ISL_2423325, EPI_ISL_2423326, EPI_ISL_2423327, EPI_ISL_2423328, EPI_ISL_2423330, EPI_ISL_2423331, EPI_ISL_2423333, EPI_ISL_2423335, EPI_ISL_2423336 | | | |
| see above | Wyoming Public Health Laboratory | Wyoming Public Health Laboratory | Jim Mildenberger, Taylor Fearing, Channing Weber, Ashley Norberg, Chayse Rowley, Marley Goetz, Brian Dominguez, Elliot Thomasson, Brittany Oher, Sam Britz, Cari Sloma, and Rob Christensen |
| EPI_ISL_2423409 | Wyoming Public Health Laboratory | Wyoming Public Health Laboratory | Jim Mildenberger, Wanda Manley, Noah Hull, Taylor Fearing, Lynette Gumbleton, Channing Weber, Ashley Norberg, Chayse Rowley, Marley Goetz, Brian Dominguez, Elliot Thomasson, Sam Britz, Cari Sloma, and Rob Christensen |
| EPI_ISL_2423412, EPI_ISL_2423413, EPI_ISL_2423414, EPI_ISL_2423415, EPI_ISL_2423416, EPI_ISL_2423417 | Alaska State Virology Laboratory | Alaska State Virology Laboratory | Stephanie DeRonde, Elva House, Jacob Zidek, Lisa Smith, Ph.D., Jack Chen, Ph.D. |
| EPI_ISL_2423508, EPI_ISL_2423509, EPI_ISL_2423510, EPI_ISL_2423511, EPI_ISL_2423512, EPI_ISL_2423513, EPI_ISL_2423514, EPI_ISL_2423515, EPI_ISL_2423516, EPI_ISL_2423517 | Oregon State Public Health Laboratory | Oregon State Public Health Laboratory | Rafia Razzaque, Eugene Yeboah, Vanda Makris, Laura Tsaknaridis, John Fontana and Shane Sevey |
| EPI_ISL_2423518, EPI_ISL_2423519, EPI_ISL_2423520, EPI_ISL_2423521, EPI_ISL_2423522, EPI_ISL_2423523, EPI_ISL_2423524, EPI_ISL_2423525, EPI_ISL_2423526, EPI_ISL_2423527, EPI_ISL_2423528, EPI_ISL_2423529, EPI_ISL_2423530, EPI_ISL_2423531, EPI_ISL_2423532, EPI_ISL_2423533, EPI_ISL_2423534 | | | |
| see above | Atlanta VA Medical Center | Genomics and Discovery, Respiratory Viruses Branch, Division of Viral Diseases, Centers for Disease Control and Prevention | Anna Kelleher, Ying Tao, Yan Li, Jing Zhang, Brian Lynch, Krista Queen, Anna Uehara, Peter Cook, Han Jia Justin Ng, Rachel Marine, Clinton R. Paden, Dhwani Batra, Haibin Wang, Tara Coalter, Jasmine Padilla, Morgan Davis, Mili Sheth, Sarah Nobles, Mark Burroughs, Justin Lee, Adam Retchless, Suxiang Tong |
| EPI_ISL_2423536, EPI_ISL_2423537, EPI_ISL_2423538, EPI_ISL_2423539, EPI_ISL_2423540, EPI_ISL_2423541, EPI_ISL_2423542, EPI_ISL_2423543, EPI_ISL_2423544, EPI_ISL_2423545, EPI_ISL_2423546, EPI_ISL_2423547, EPI_ISL_2423548, EPI_ISL_2423549, EPI_ISL_2423550, EPI_ISL_2423551, EPI_ISL_2423552, EPI_ISL_2423553, EPI_ISL_2423554 | | | |
| see above | TXDSHS | TXDSHS | Rashmi Tuladhar, Bonnie Oh, Jenny Zhang, Maliha Rahman, Mayela Pedrueza, Anita Pokharel, Karen Bobier, Lorraine Rodriguez, Myong Koag, Chun Wang, Rachel Lee, Grace Kubin |
| EPI_ISL_2423559, EPI_ISL_2423560, EPI_ISL_2423561, EPI_ISL_2423562, EPI_ISL_2423563, EPI_ISL_2423567, EPI_ISL_2423568, EPI_ISL_2423570, EPI_ISL_2423571, EPI_ISL_2423575, EPI_ISL_2423578, EPI_ISL_2423579, EPI_ISL_2423580, EPI_ISL_2423581, EPI_ISL_2423582, EPI_ISL_2423583, EPI_ISL_2423585, EPI_ISL_2423586, EPI_ISL_2423587, EPI_ISL_2423589, EPI_ISL_2423590, EPI_ISL_2423592, EPI_ISL_2423594, EPI_ISL_2423597, EPI_ISL_2423598, EPI_ISL_2423599, EPI_ISL_2423600, EPI_ISL_2423601, EPI_ISL_2423605, EPI_ISL_2423606, EPI_ISL_2423607, EPI_ISL_2423608, EPI_ISL_2423610, EPI_ISL_2423614, EPI_ISL_2423619, EPI_ISL_2423620, EPI_ISL_2423621, EPI_ISL_2423622, EPI_ISL_2423627, EPI_ISL_2423635, EPI_ISL_2423636, EPI_ISL_2423638, EPI_ISL_2423639, EPI_ISL_2423640, EPI_ISL_2423641, EPI_ISL_2423642, EPI_ISL_2423643, EPI_ISL_2423646, EPI_ISL_2423651, EPI_ISL_2423652, EPI_ISL_2423653, EPI_ISL_2423655, EPI_ISL_2423656, EPI_ISL_2423662, EPI_ISL_2423666, EPI_ISL_2423668, EPI_ISL_2423670, EPI_ISL_2423671, EPI_ISL_2423673, EPI_ISL_2423675, EPI_ISL_2423678, EPI_ISL_2423679, EPI_ISL_2423681, EPI_ISL_2423682, EPI_ISL_2423683, EPI_ISL_2423686, EPI_ISL_2423690, EPI_ISL_2423692, EPI_ISL_2423693, EPI_ISL_2423694, EPI_ISL_2423698, EPI_ISL_2423699, EPI_ISL_2423700, EPI_ISL_2423701, EPI_ISL_2423702, EPI_ISL_2423703, EPI_ISL_2423704, EPI_ISL_2423705, EPI_ISL_2423706, EPI_ISL_2423707, EPI_ISL_2423709, EPI_ISL_2423710, EPI_ISL_2423711, EPI_ISL_2423713, EPI_ISL_2423715, EPI_ISL_2423716, EPI_ISL_2423717, EPI_ISL_2423719, EPI_ISL_2423722, EPI_ISL_2423723, EPI_ISL_2423727, EPI_ISL_2423728, EPI_ISL_2423729, EPI_ISL_2423730, EPI_ISL_2423731, EPI_ISL_2423736, EPI_ISL_2423737, EPI_ISL_2423741, EPI_ISL_2423742, EPI_ISL_2423744, EPI_ISL_2423747, EPI_ISL_2423748, EPI_ISL_2423749, EPI_ISL_2423750, EPI_ISL_2423751, EPI_ISL_2423752, EPI_ISL_2423754, EPI_ISL_2423756, EPI_ISL_2423758, EPI_ISL_2423761, EPI_ISL_2423762, EPI_ISL_2423764, EPI_ISL_2423767, EPI_ISL_2423769, EPI_ISL_2423775, EPI_ISL_2423776, EPI_ISL_2423777, EPI_ISL_2423778, EPI_ISL_2423779, EPI_ISL_2423782, EPI_ISL_2423783, EPI_ISL_2423786, EPI_ISL_2423788, EPI_ISL_2423789, EPI_ISL_2423790, EPI_ISL_2423791, EPI_ISL_2423793, EPI_ISL_2423794, EPI_ISL_2423795, EPI_ISL_2423796, EPI_ISL_2423797, EPI_ISL_2423799, EPI_ISL_2423800, EPI_ISL_2423801, EPI_ISL_2423803, EPI_ISL_2423804, EPI_ISL_2423805, EPI_ISL_2423806, EPI_ISL_2423807, EPI_ISL_2423808, EPI_ISL_2423810, EPI_ISL_2423814, EPI_ISL_2423815, EPI_ISL_2423816, EPI_ISL_2423818, EPI_ISL_2423819, EPI_ISL_2423821, EPI_ISL_2423824, EPI_ISL_2423826, EPI_ISL_2423828, EPI_ISL_2423832, EPI_ISL_2423834, EPI_ISL_2423835, EPI_ISL_2423844, EPI_ISL_2423847, EPI_ISL_2423849, EPI_ISL_2423853, EPI_ISL_2423854, EPI_ISL_2423856, EPI_ISL_2423857, EPI_ISL_2423858, EPI_ISL_2423859, EPI_ISL_2423861, EPI_ISL_2423864, EPI_ISL_2423866, EPI_ISL_2423867, EPI_ISL_2423876, EPI_ISL_2423878, EPI_ISL_2423879, EPI_ISL_2423882, EPI_ISL_2423883, EPI_ISL_2423885, EPI_ISL_2423886, EPI_ISL_2423887, EPI_ISL_2423889, EPI_ISL_2423891, EPI_ISL_2423892, EPI_ISL_2423893, EPI_ISL_2423901, EPI_ISL_2423902, EPI_ISL_2423905, EPI_ISL_2423906, EPI_ISL_2423910, EPI_ISL_2423911, EPI_ISL_2423913, EPI_ISL_2423918, EPI_ISL_2423919, EPI_ISL_2423921, EPI_ISL_2423922, EPI_ISL_2423923, EPI_ISL_2423925, EPI_ISL_2423926, EPI_ISL_2423927, EPI_ISL_2423928, EPI_ISL_2423929, EPI_ISL_2423932, EPI_ISL_2423933, EPI_ISL_2423934, EPI_ISL_2423935, EPI_ISL_2423936, EPI_ISL_2423937, EPI_ISL_2423938, EPI_ISL_2423941, EPI_ISL_2423945, EPI_ISL_2423947, EPI_ISL_2423950, EPI_ISL_2423951, EPI_ISL_2423954, EPI_ISL_2423955, EPI_ISL_2423956, EPI_ISL_2423957, EPI_ISL_2423958, EPI_ISL_2423960, EPI_ISL_2423968, EPI_ISL_2423971, EPI_ISL_2423972, EPI_ISL_2423973, EPI_ISL_2423978, EPI_ISL_2423979, EPI_ISL_2423980, EPI_ISL_2423981, EPI_ISL_2423982, EPI_ISL_2423983, EPI_ISL_2423985, EPI_ISL_2423986, EPI_ISL_2423987, EPI_ISL_2423989, EPI_ISL_2423990, EPI_ISL_2423996, EPI_ISL_2423997, EPI_ISL_2423998, EPI_ISL_2423999, EPI_ISL_2424002, EPI_ISL_2424003, EPI_ISL_2424004, EPI_ISL_2424005, EPI_ISL_2424006, EPI_ISL_2424008, EPI_ISL_2424010, EPI_ISL_2424011, EPI_ISL_2424012, EPI_ISL_2424013, EPI_ISL_2424014, EPI_ISL_2424018, EPI_ISL_2424019, EPI_ISL_2424020, EPI_ISL_2424021, EPI_ISL_2424023, EPI_ISL_2424027, EPI_ISL_2424028, EPI_ISL_2424029, EPI_ISL_2424032, EPI_ISL_2424033, EPI_ISL_2424035, EPI_ISL_2424038, EPI_ISL_2424039, EPI_ISL_2424041, EPI_ISL_2424045, EPI_ISL_2424047 | | | |
| see above | TGen North | TGen North | "Jolene Bowers, Heather Centner, Chris French, Hayley Yaglom, Ashlyn Pfeiffer, Darrin Lemmer, Dave Engelthaler, The Arizona COVID Genomics Union (ACGU)" |

| | | | |
|---|---|---|---|
| EPI_ISL_2424048, EPI_ISL_2424050, EPI_ISL_2424051, EPI_ISL_2424053, EPI_ISL_2424054, EPI_ISL_2424058, EPI_ISL_2424064, EPI_ISL_2424065, EPI_ISL_2424066, EPI_ISL_2424067, EPI_ISL_2424074 | | | |
| see above | TGen North | TGen North | Jolene Bowers, Heather Centner, Chris French, Hayley Yaglom, Ashlyn Pfeiffer, Darrin Lemmer, Dave Engelthaler, The Arizona COVID Genomics Union (ACGU) |
| EPI_ISL_2424078 | TGen North | TGen North | "Jolene Bowers, Heather Centner, Chris French, Hayley Yaglom, Ashlyn Pfeiffer, Darrin Lemmer, Dave Engelthaler, The Arizona COVID Genomics Union (ACGU)" |
| EPI_ISL_2424081, EPI_ISL_2424083, EPI_ISL_2424084, EPI_ISL_2424086, EPI_ISL_2424087, EPI_ISL_2424088, EPI_ISL_2424092, EPI_ISL_2424093, EPI_ISL_2424098, EPI_ISL_2424099, EPI_ISL_2424100, EPI_ISL_2424103, EPI_ISL_2424104, EPI_ISL_2424105, EPI_ISL_2424106, EPI_ISL_2424108, EPI_ISL_2424109, EPI_ISL_2424111, EPI_ISL_2424112, EPI_ISL_2424113, EPI_ISL_2424114, EPI_ISL_2424117, EPI_ISL_2424119, EPI_ISL_2424120, EPI_ISL_2424121, EPI_ISL_2424122, EPI_ISL_2424124, EPI_ISL_2424125, EPI_ISL_2424127, EPI_ISL_2424128, EPI_ISL_2424129, EPI_ISL_2424134 | | | |
| see above | TGen North | TGen North | Jolene Bowers, Heather Centner, Chris French, Hayley Yaglom, Ashlyn Pfeiffer, Darrin Lemmer, Dave Engelthaler, The Arizona COVID Genomics Union (ACGU) |
| EPI_ISL_2424166 | UCLA Clinical Micro Lab | Los Angeles County Public Health Laboratories | P. Hemarajata et al. |
| EPI_ISL_2424167 | USC Clinical Lab | Los Angeles County Public Health Laboratories | P. Hemarajata et al. |
| EPI_ISL_2424168, EPI_ISL_2424169, EPI_ISL_2424170, EPI_ISL_2424171, EPI_ISL_2424172, EPI_ISL_2424173 | UCLA Clinical Micro Lab | Los Angeles County Public Health Laboratories | P. Hemarajata et al. |
| EPI_ISL_2424174 | USC Clinical Lab | Los Angeles County Public Health Laboratories | P. Hemarajata et al. |
| EPI_ISL_2424175, EPI_ISL_2424177, EPI_ISL_2424178, EPI_ISL_2424179, EPI_ISL_2424180, EPI_ISL_2424181, EPI_ISL_2424182, EPI_ISL_2424183 | UCLA Clinical Micro Lab | Los Angeles County Public Health Laboratories | P. Hemarajata et al. |
| EPI_ISL_2424184, EPI_ISL_2424185, EPI_ISL_2424186, EPI_ISL_2424187 | USC Clinical Lab | Los Angeles County Public Health Laboratories | P. Hemarajata et al. |
| EPI_ISL_2424188, EPI_ISL_2424189, EPI_ISL_2424190 | UCLA Clinical Micro Lab | Los Angeles County Public Health Laboratories | P. Hemarajata et al. |
| EPI_ISL_2424191 | Flow Diagnostics Lab | Los Angeles County Public Health Laboratories | P. Hemarajata et al. |
| EPI_ISL_2424192, EPI_ISL_2424193, EPI_ISL_2424194, EPI_ISL_2424195, EPI_ISL_2424196, EPI_ISL_2424197, EPI_ISL_2424198, EPI_ISL_2424199, EPI_ISL_2424200, EPI_ISL_2424201, EPI_ISL_2424202, EPI_ISL_2424204, EPI_ISL_2424205, EPI_ISL_2424206, EPI_ISL_2424207, EPI_ISL_2424208, EPI_ISL_2424209, EPI_ISL_2424210, EPI_ISL_2424211 | | | |
| see above | UCLA Clinical Micro Lab | Los Angeles County Public Health Laboratories | P. Hemarajata et al. |
| EPI_ISL_2424212 | Martin Luther King Jr. Community Hospital | Los Angeles County Public Health Laboratories | P. Hemarajata et al. |
| EPI_ISL_2424213, EPI_ISL_2424214, EPI_ISL_2424215, EPI_ISL_2424216, EPI_ISL_2424217, EPI_ISL_2424218, EPI_ISL_2424219, EPI_ISL_2424220, EPI_ISL_2424221, EPI_ISL_2424222, EPI_ISL_2424223, EPI_ISL_2424224, EPI_ISL_2424225, EPI_ISL_2424226, EPI_ISL_2424227, EPI_ISL_2424228, EPI_ISL_2424229, EPI_ISL_2424230, EPI_ISL_2424231, EPI_ISL_2424232, EPI_ISL_2424233 | | | |
| see above | USC Clinical Lab | Los Angeles County Public Health Laboratories | P. Hemarajata et al. |
| EPI_ISL_2424234 | UCLA Clinical Micro Lab | Los Angeles County Public Health Laboratories | P. Hemarajata et al. |
| EPI_ISL_2424235, EPI_ISL_2424236, EPI_ISL_2424237, EPI_ISL_2424238, EPI_ISL_2424239, EPI_ISL_2424240, EPI_ISL_2424241, EPI_ISL_2424242, EPI_ISL_2424243, EPI_ISL_2424244, EPI_ISL_2424245, EPI_ISL_2424246, EPI_ISL_2424247, EPI_ISL_2424248, EPI_ISL_2424249, EPI_ISL_2424250, EPI_ISL_2424251, EPI_ISL_2424252, EPI_ISL_2424253, EPI_ISL_2424254, EPI_ISL_2424255, EPI_ISL_2424256, EPI_ISL_2424257, EPI_ISL_2424258, EPI_ISL_2424259, EPI_ISL_2424260, EPI_ISL_2424261, EPI_ISL_2424262, EPI_ISL_2424263, EPI_ISL_2424264 | | | |
| see above | Oregon State Public Health Laboratory | Oregon State Public Health Laboratory | Rafia Razzaque, Eugene Yeboah, Vanda Makris, Laura Tsaknaridis, John Fontana and Shane Sevey |
| EPI_ISL_2425344 | Minnesota Department of Health, Public Health Laboratory | Minnesota Department of Health, Public Health Laboratory | Alexandra Lorentz, Jacob Garfin, Matt Plumb, and Xiong Wang |
| EPI_ISL_2425345 | M Health Fairview | Minnesota Department of Health, Public Health Laboratory | Alexandra Lorentz, Jacob Garfin, Matt Plumb, and Xiong Wang |
| EPI_ISL_2425346, EPI_ISL_2425347 | Minnesota Department of Health, Public Health Laboratory | Minnesota Department of Health, Public Health Laboratory | Alexandra Lorentz, Jacob Garfin, Matt Plumb, and Xiong Wang |
| EPI_ISL_2425348 | M Health Fairview | Minnesota Department of Health, Public Health Laboratory | Alexandra Lorentz, Jacob Garfin, Matt Plumb, and Xiong Wang |
| EPI_ISL_2425349 | Minnesota Department of Health, Public Health Laboratory | Minnesota Department of Health, Public Health Laboratory | Alexandra Lorentz, Jacob Garfin, Matt Plumb, and Xiong Wang |
| EPI_ISL_2425350 | M Health Fairview | Minnesota Department of Health, Public Health Laboratory | Alexandra Lorentz, Jacob Garfin, Matt Plumb, and Xiong Wang |
| EPI_ISL_2425351 | Minnesota Department of Health, Public Health Laboratory | Minnesota Department of Health, Public Health Laboratory | Alexandra Lorentz, Jacob Garfin, Matt Plumb, and Xiong Wang |
| EPI_ISL_2425352, EPI_ISL_2425353, EPI_ISL_2425354, EPI_ISL_2425355, EPI_ISL_2425356, EPI_ISL_2425357 | M Health Fairview | Minnesota Department of Health, Public Health Laboratory | Alexandra Lorentz, Jacob Garfin, Matt Plumb, and Xiong Wang |
| EPI_ISL_2425358 | Minnesota Department of Health, Public Health Laboratory | Minnesota Department of Health, Public Health Laboratory | Alexandra Lorentz, Jacob Garfin, Matt Plumb, and Xiong Wang |
| EPI_ISL_2425359, EPI_ISL_2425360 | Centracare Laboratory Services | Minnesota Department of Health, Public Health Laboratory | Alexandra Lorentz, Jacob Garfin, Matt Plumb, and Xiong Wang |
| EPI_ISL_2425361, EPI_ISL_2425362, EPI_ISL_2425363 | Minnesota Department of Health, Public Health Laboratory | Minnesota Department of Health, Public Health Laboratory | Alexandra Lorentz, Jacob Garfin, Matt Plumb, and Xiong Wang |
| EPI_ISL_2425364, EPI_ISL_2425365, EPI_ISL_2425366, EPI_ISL_2425367, EPI_ISL_2425368, EPI_ISL_2425369, EPI_ISL_2425370, EPI_ISL_2425371, EPI_ISL_2425372, EPI_ISL_2425373, EPI_ISL_2425374 | | | |
| see above | M Health Fairview | Minnesota Department of Health, Public Health Laboratory | Alexandra Lorentz, Jacob Garfin, Matt Plumb, and Xiong Wang |
| EPI_ISL_2425375, EPI_ISL_2425376, EPI_ISL_2425377 | Minnesota Department of Health, Public Health Laboratory | Minnesota Department of Health, Public Health Laboratory | Alexandra Lorentz, Jacob Garfin, Matt Plumb, and Xiong Wang |
| EPI_ISL_2425378, EPI_ISL_2425379, EPI_ISL_2425380, EPI_ISL_2425381, EPI_ISL_2425382 | M Health Fairview | Minnesota Department of Health, Public Health Laboratory | Alexandra Lorentz, Jacob Garfin, Matt Plumb, and Xiong Wang |
| EPI_ISL_2425383, EPI_ISL_2425394 | Minnesota Department of Health, Public Health Laboratory | Minnesota Department of Health, Public Health Laboratory | Alexandra Lorentz, Jacob Garfin, Matt Plumb, and Xiong Wang |
| EPI_ISL_2425395, EPI_ISL_2425396, EPI_ISL_2425397 | M Health Fairview | Minnesota Department of Health, Public Health Laboratory | Alexandra Lorentz, Jacob Garfin, Matt Plumb, and Xiong Wang |
| EPI_ISL_2425398, EPI_ISL_2425399 | Minnesota Department of Health, Public Health Laboratory | Minnesota Department of Health, Public Health Laboratory | Alexandra Lorentz, Jacob Garfin, Matt Plumb, and Xiong Wang |

| | | | |
|---|---|---|---|
| EPI_ISL_2425400 | M Health Fairview | Minnesota Department of Health, Public Health Laboratory | Alexandra Lorentz, Jacob Garfin, Matt Plumb, and Xiong Wang |
| EPI_ISL_2425401 | Minnesota Department of Health, Public Health Laboratory | Minnesota Department of Health, Public Health Laboratory | Alexandra Lorentz, Jacob Garfin, Matt Plumb, and Xiong Wang |
| EPI_ISL_2425402, EPI_ISL_2425403, EPI_ISL_2425404, EPI_ISL_2425405 | M Health Fairview | Minnesota Department of Health, Public Health Laboratory | Alexandra Lorentz, Jacob Garfin, Matt Plumb, and Xiong Wang |
| EPI_ISL_2425406, EPI_ISL_2425407, EPI_ISL_2425409, EPI_ISL_2425410, EPI_ISL_2425411, EPI_ISL_2425412, EPI_ISL_2425413, EPI_ISL_2425414, EPI_ISL_2425415, EPI_ISL_2425417, EPI_ISL_2425418, EPI_ISL_2425419, EPI_ISL_2425421, EPI_ISL_2425422, EPI_ISL_2425424, EPI_ISL_2425425, EPI_ISL_2425426, EPI_ISL_2425427, EPI_ISL_2425428, EPI_ISL_2425429, EPI_ISL_2425430, EPI_ISL_2425431, EPI_ISL_2425432 | | | |
| see above | MD PHL | MD PHL | Maryland Department of Health Laboratories Administration |
| EPI_ISL_2425444, EPI_ISL_2425445, EPI_ISL_2425446, EPI_ISL_2425447, EPI_ISL_2425448, EPI_ISL_2425449, EPI_ISL_2425450, EPI_ISL_2425451, EPI_ISL_2425452, EPI_ISL_2425453 | DC Public Health Lab/ Dept. of Forensic Sciences | DC Public Health Lab/ Dept. of Forensic Sciences | Janis Doss, Scott Nguyen, Elizabeth Zelaya, Sarah Scott, Connie Maza, Monica Mann, Brittany Hamilton, David Payne, Jocelyn Hauser |
| EPI_ISL_2425454, EPI_ISL_2425455, EPI_ISL_2425456, EPI_ISL_2425457, EPI_ISL_2425458, EPI_ISL_2425459, EPI_ISL_2425460, EPI_ISL_2425461, EPI_ISL_2425462, EPI_ISL_2425463, EPI_ISL_2425464, EPI_ISL_2425465, EPI_ISL_2425466, EPI_ISL_2425467, EPI_ISL_2425468, EPI_ISL_2425469, EPI_ISL_2425470, EPI_ISL_2425471, EPI_ISL_2425472, EPI_ISL_2425473, EPI_ISL_2425474, EPI_ISL_2425475, EPI_ISL_2425476, EPI_ISL_2425477, EPI_ISL_2425478, EPI_ISL_2425479, EPI_ISL_2425480, EPI_ISL_2425481, EPI_ISL_2425482, EPI_ISL_2425483, EPI_ISL_2425484, EPI_ISL_2425485, EPI_ISL_2425486, EPI_ISL_2425487, EPI_ISL_2425488 | | | |
| see above | Virginia Division of Consolidated Laboratory Services | Virginia Division of Consolidated Laboratory Services | Virginia DCLS |
| EPI_ISL_2425490, EPI_ISL_2425491, EPI_ISL_2425492, EPI_ISL_2425493, EPI_ISL_2425494, EPI_ISL_2425495, EPI_ISL_2425497, EPI_ISL_2425498, EPI_ISL_2425499, EPI_ISL_2425500, EPI_ISL_2425501, EPI_ISL_2425502, EPI_ISL_2425503, EPI_ISL_2425504, EPI_ISL_2425505, EPI_ISL_2425506, EPI_ISL_2425507, EPI_ISL_2425508, EPI_ISL_2425509, EPI_ISL_2425510, EPI_ISL_2425511, EPI_ISL_2425512, EPI_ISL_2425513, EPI_ISL_2425514, EPI_ISL_2425515, EPI_ISL_2425516, EPI_ISL_2425517, EPI_ISL_2425518, EPI_ISL_2425519, EPI_ISL_2425520, EPI_ISL_2425521, EPI_ISL_2425522, EPI_ISL_2425523, EPI_ISL_2425524, EPI_ISL_2425525, EPI_ISL_2425526, EPI_ISL_2425527 | | | |
| see above | Rhode Island State Health Laboratory | Rhode Island State Health Laboratory | Ewa King, Richard C. Huard, Kristin Carpenter-Azevedo |
| EPI_ISL_2426099, EPI_ISL_2426100, EPI_ISL_2426101, EPI_ISL_2426102, EPI_ISL_2426103, EPI_ISL_2426104, EPI_ISL_2426105, EPI_ISL_2426106, EPI_ISL_2426107, EPI_ISL_2426108, EPI_ISL_2426109, EPI_ISL_2426110, EPI_ISL_2426111, EPI_ISL_2426112, EPI_ISL_2426113, EPI_ISL_2426114, EPI_ISL_2426115, EPI_ISL_2426116, EPI_ISL_2426117, EPI_ISL_2426118, EPI_ISL_2426119, EPI_ISL_2426120, EPI_ISL_2426121, EPI_ISL_2426122, EPI_ISL_2426123, EPI_ISL_2426124, EPI_ISL_2426125, EPI_ISL_2426126, EPI_ISL_2426127, EPI_ISL_2426128, EPI_ISL_2426129, EPI_ISL_2426130, EPI_ISL_2426131, EPI_ISL_2426132, EPI_ISL_2426133, EPI_ISL_2426134, EPI_ISL_2426135, EPI_ISL_2426136, EPI_ISL_2426137, EPI_ISL_2426138, EPI_ISL_2426139, EPI_ISL_2426140, EPI_ISL_2426141 | | | |
| see above | Yale Clinical Virology Lab | Grubaugh Lab - Yale School of Public Health | Joseph Fauver, Mallery Breban, Isabel Ott, Tara Alpert, Mary Petrone, Anderson Brito, Chantal Vogels, Annie Watkins, Chaney Kalinich, Jessica Rothman, Marie L. Landry, Nathan Grubaugh |
| EPI_ISL_2426142 | Yale Pathology Lab | Grubaugh Lab - Yale School of Public Health | Joseph Fauver, Mallery Breban, Isabel Ott, Tara Alpert, Mary Petrone, Anderson Brito, Chantal Vogels, Annie Watkins, Chaney Kalinich, Jessica Rothman, Jianhui Wang, Chen Liu, Pei Hui, Nathan Grubaugh |
| EPI_ISL_2426260, EPI_ISL_2426261, EPI_ISL_2426262, EPI_ISL_2426263, EPI_ISL_2426264, EPI_ISL_2426265, EPI_ISL_2426266, EPI_ISL_2426267, EPI_ISL_2426268, EPI_ISL_2426269, EPI_ISL_2426270, EPI_ISL_2426271, EPI_ISL_2426272, EPI_ISL_2426273, EPI_ISL_2426274, EPI_ISL_2426275, EPI_ISL_2426276, EPI_ISL_2426277, EPI_ISL_2426278, EPI_ISL_2426279, EPI_ISL_2426280, EPI_ISL_2426281, EPI_ISL_2426282, EPI_ISL_2426283, EPI_ISL_2426284, EPI_ISL_2426285, EPI_ISL_2426287, EPI_ISL_2426288, EPI_ISL_2426289, EPI_ISL_2426290, EPI_ISL_2426291, EPI_ISL_2426292, EPI_ISL_2426293, EPI_ISL_2426294, EPI_ISL_2426295, EPI_ISL_2426296, EPI_ISL_2426298, EPI_ISL_2426299, EPI_ISL_2426300, EPI_ISL_2426301, EPI_ISL_2426302, EPI_ISL_2426303, EPI_ISL_2426304, EPI_ISL_2426305, EPI_ISL_2426306, EPI_ISL_2426307, EPI_ISL_2426308, EPI_ISL_2426309, EPI_ISL_2426310, EPI_ISL_2426311, EPI_ISL_2426312, EPI_ISL_2426313, EPI_ISL_2426314, EPI_ISL_2426315, EPI_ISL_2426316, EPI_ISL_2426317, EPI_ISL_2426318, EPI_ISL_2426319, EPI_ISL_2426320, EPI_ISL_2426321, EPI_ISL_2426322, EPI_ISL_2426323, EPI_ISL_2426324, EPI_ISL_2426325, EPI_ISL_2426326, EPI_ISL_2426327, EPI_ISL_2426328, EPI_ISL_2426329, EPI_ISL_2426330, EPI_ISL_2426331, EPI_ISL_2426332, EPI_ISL_2426333, EPI_ISL_2426334, EPI_ISL_2426335, EPI_ISL_2426336, EPI_ISL_2426337, EPI_ISL_2426338, EPI_ISL_2426339, EPI_ISL_2426340, EPI_ISL_2426341, EPI_ISL_2426342, EPI_ISL_2426343, EPI_ISL_2426344, EPI_ISL_2426346, EPI_ISL_2426347, EPI_ISL_2426348, EPI_ISL_2426349, EPI_ISL_2426350, EPI_ISL_2426351, EPI_ISL_2426352, EPI_ISL_2426353, EPI_ISL_2426354, EPI_ISL_2426355, EPI_ISL_2426356, EPI_ISL_2426357, EPI_ISL_2426358, EPI_ISL_2426359, EPI_ISL_2426360, EPI_ISL_2426361, EPI_ISL_2426362, EPI_ISL_2426364, EPI_ISL_2426365, EPI_ISL_2426366, EPI_ISL_2426367, EPI_ISL_2426368, EPI_ISL_2426369, EPI_ISL_2426370, EPI_ISL_2426371, EPI_ISL_2426372, EPI_ISL_2426373, EPI_ISL_2426374, EPI_ISL_2426375, EPI_ISL_2426376, EPI_ISL_2426377, EPI_ISL_2426378, EPI_ISL_2426379, EPI_ISL_2426380, EPI_ISL_2426381, EPI_ISL_2426382, EPI_ISL_2426383, EPI_ISL_2426384, EPI_ISL_2426385, EPI_ISL_2426386, EPI_ISL_2426387, EPI_ISL_2426388, EPI_ISL_2426389, EPI_ISL_2426390, EPI_ISL_2426391, EPI_ISL_2426392, EPI_ISL_2426393, EPI_ISL_2426394, EPI_ISL_2426396, EPI_ISL_2426397, EPI_ISL_2426398 | | | |
| see above | UW Virology Lab | UW Virology Lab | Pavitra Roychoudhury, Hong Xie, Lasata Shrestha, Tien V. Nguyen, Shah Mohamed Bakhash, Michelle Lin, Noah R. Baker, Ricardo Perez, Sean Ellis, Nathan Breit, Robert J. Livingston, Meei-Li Huang, Keith R Jerome, Patrick Mathias, Alexander Greninger |
| EPI_ISL_2426399, EPI_ISL_2426400, EPI_ISL_2426401, EPI_ISL_2426402, EPI_ISL_2426403, EPI_ISL_2426404, EPI_ISL_2426405, EPI_ISL_2426406, EPI_ISL_2426407, EPI_ISL_2426408, EPI_ISL_2426409, EPI_ISL_2426410, EPI_ISL_2426411, EPI_ISL_2426412, EPI_ISL_2426414, EPI_ISL_2426415, EPI_ISL_2426416, EPI_ISL_2426417, EPI_ISL_2426418, EPI_ISL_2426419, EPI_ISL_2426420, EPI_ISL_2426421, EPI_ISL_2426422, EPI_ISL_2426423, EPI_ISL_2426424, EPI_ISL_2426425, EPI_ISL_2426426, EPI_ISL_2426427, EPI_ISL_2426428, EPI_ISL_2426429, EPI_ISL_2426430, EPI_ISL_2426431, EPI_ISL_2426432, EPI_ISL_2426433, EPI_ISL_2426434, EPI_ISL_2426435, EPI_ISL_2426436, EPI_ISL_2426437, EPI_ISL_2426439, EPI_ISL_2426440, EPI_ISL_2426441, EPI_ISL_2426442, EPI_ISL_2426443, EPI_ISL_2426444, EPI_ISL_2426445, EPI_ISL_2426446, EPI_ISL_2426447, EPI_ISL_2426448, EPI_ISL_2426449, EPI_ISL_2426450, EPI_ISL_2426451, EPI_ISL_2426452, EPI_ISL_2426453, EPI_ISL_2426454, EPI_ISL_2426455, EPI_ISL_2426456, EPI_ISL_2426457, EPI_ISL_2426458, EPI_ISL_2426459, EPI_ISL_2426461, EPI_ISL_2426462, EPI_ISL_2426463, EPI_ISL_2426464, EPI_ISL_2426465, EPI_ISL_2426467, EPI_ISL_2426468, EPI_ISL_2426469, EPI_ISL_2426470, EPI_ISL_2426471, EPI_ISL_2426473, EPI_ISL_2426474, EPI_ISL_2426475, EPI_ISL_2426476, EPI_ISL_2426477, EPI_ISL_2426478, EPI_ISL_2426479, EPI_ISL_2426480, EPI_ISL_2426481, EPI_ISL_2426483, EPI_ISL_2426484, EPI_ISL_2426485, EPI_ISL_2426486, EPI_ISL_2426487, EPI_ISL_2426488, EPI_ISL_2426489, EPI_ISL_2426490, EPI_ISL_2426491, EPI_ISL_2426492, EPI_ISL_2426494, EPI_ISL_2426495, EPI_ISL_2426496, EPI_ISL_2426497, EPI_ISL_2426498, EPI_ISL_2426499, EPI_ISL_2426500, EPI_ISL_2426501, EPI_ISL_2426502, EPI_ISL_2426503, EPI_ISL_2426504, EPI_ISL_2426505, EPI_ISL_2426506, EPI_ISL_2426507, EPI_ISL_2426508, EPI_ISL_2426509, EPI_ISL_2426510, EPI_ISL_2426511, EPI_ISL_2426512, EPI_ISL_2426513, EPI_ISL_2426514, EPI_ISL_2426515, EPI_ISL_2426516, EPI_ISL_2426517, EPI_ISL_2426518, EPI_ISL_2426519, EPI_ISL_2426520, EPI_ISL_2426521, EPI_ISL_2426522, EPI_ISL_2426523, EPI_ISL_2426524, EPI_ISL_2426525, EPI_ISL_2426526, EPI_ISL_2426528, EPI_ISL_2426529, EPI_ISL_2426530, EPI_ISL_2426531, EPI_ISL_2426533, EPI_ISL_2426534, EPI_ISL_2426535, EPI_ISL_2426536, EPI_ISL_2426537, EPI_ISL_2426538, EPI_ISL_2426539, EPI_ISL_2426540, EPI_ISL_2426541, EPI_ISL_2426542, EPI_ISL_2426543, EPI_ISL_2426545, EPI_ISL_2426546, EPI_ISL_2426547, EPI_ISL_2426548, EPI_ISL_2426549, EPI_ISL_2426550, EPI_ISL_2426551, EPI_ISL_2426552, EPI_ISL_2426553, EPI_ISL_2426554, EPI_ISL_2426555, EPI_ISL_2426556, EPI_ISL_2426557, EPI_ISL_2426558, EPI_ISL_2426559, EPI_ISL_2426560, EPI_ISL_2426561, EPI_ISL_2426563, EPI_ISL_2426564, EPI_ISL_2426566, EPI_ISL_2426567, EPI_ISL_2426568, EPI_ISL_2426569, EPI_ISL_2426570, EPI_ISL_2426571, EPI_ISL_2426572, EPI_ISL_2426573, EPI_ISL_2426574, EPI_ISL_2426575, EPI_ISL_2426576, EPI_ISL_2426577 | | | |
| see above | UW Virology Lab | UW Virology Lab | Pavitra Roychoudhury, Hong Xie, Lasata Shrestha, Tien V. Nguyen, Shah Mohamed Bakhash, Michelle Lin, Noah R. Baker, Sean Ellis, Meei-Li Huang, Keith R Jerome, Alexander Greninger |
| EPI_ISL_2426659, EPI_ISL_2426660, EPI_ISL_2426661, EPI_ISL_2426662, EPI_ISL_2426663, EPI_ISL_2426664, EPI_ISL_2426665, EPI_ISL_2426666, EPI_ISL_2426667, EPI_ISL_2426668, EPI_ISL_2426669, EPI_ISL_2426670, EPI_ISL_2426671, EPI_ISL_2426672, EPI_ISL_2426673, EPI_ISL_2426674, EPI_ISL_2426675, EPI_ISL_2426676, EPI_ISL_2426677, EPI_ISL_2426678, EPI_ISL_2426679, EPI_ISL_2426680, EPI_ISL_2426681, EPI_ISL_2426682, EPI_ISL_2426683, EPI_ISL_2426684, EPI_ISL_2426685, EPI_ISL_2426687, EPI_ISL_2426688, EPI_ISL_2426689, EPI_ISL_2426690, EPI_ISL_2426691, EPI_ISL_2426692, EPI_ISL_2426693 | | | |
| see above | Kansas Health and Environmental Lab | Kansas Health and Environmental Lab | Katherine Wiggins, Mike Grose, Jonathan Barnell, Ben Olsen, and Phil Adam |
| EPI_ISL_2426695, EPI_ISL_2426696, EPI_ISL_2426699, EPI_ISL_2426700, EPI_ISL_2426701, EPI_ISL_2426702, EPI_ISL_2426703, EPI_ISL_2426705, EPI_ISL_2426707, EPI_ISL_2426709, EPI_ISL_2426710, EPI_ISL_2426711, EPI_ISL_2426712, EPI_ISL_2426713, EPI_ISL_2426714, EPI_ISL_2426718, EPI_ISL_2426720, EPI_ISL_2426721, EPI_ISL_2426722, EPI_ISL_2426723, EPI_ISL_2426726, EPI_ISL_2426727, EPI_ISL_2426731, EPI_ISL_2426732, EPI_ISL_2426733, EPI_ISL_2426734, EPI_ISL_2426735, EPI_ISL_2426736, EPI_ISL_2426737, EPI_ISL_2426738, EPI_ISL_2426739, EPI_ISL_2426741, EPI_ISL_2426742, EPI_ISL_2426743, EPI_ISL_2426744, EPI_ISL_2426745, EPI_ISL_2426746, EPI_ISL_2426747, EPI_ISL_2426748, EPI_ISL_2426749, EPI_ISL_2426752, EPI_ISL_2426753, EPI_ISL_2426754, EPI_ISL_2426755, EPI_ISL_2426756, EPI_ISL_2426757, EPI_ISL_2426758, EPI_ISL_2426772, EPI_ISL_2426773, EPI_ISL_2426774, EPI_ISL_2426775, EPI_ISL_2426778, EPI_ISL_2426781, EPI_ISL_2426782, EPI_ISL_2426783 | | | |
| see above | US Air Force School of Aerospace Medicine | US Air Force School of Aerospace Medicine | Anthony Fries, Jennifer Meyer, William Gruner, Amanda Javorina, Carol Garrett, Sarah Purves, Clarise Starr, Elizabeth Macias |
| EPI_ISL_2426784, EPI_ISL_2426785 | MS Public Health Laboratory | University of Mississippi Medical Center, Molecular and Genomics Core Facility | Ashley C. Johnson, Ithiel J. Frame, Krishna K. Ayyalasomayajula, Michael R. Garrett, D. Ashley Robinson |
| EPI_ISL_2426786, EPI_ISL_2426787 | University of Mississippi Medical Center, Department of Pathology | University of Mississippi Medical Center, Molecular and Genomics Core Facility | Ashley C. Johnson, Ithiel J. Frame, Krishna K. Ayyalasomayajula, Michael R. Garrett, D. Ashley Robinson |
| EPI_ISL_2426788, EPI_ISL_2426789 | MS Public Health Laboratory | University of Mississippi Medical Center, Molecular and Genomics Core Facility | Ashley C. Johnson, Ithiel J. Frame, Krishna K. Ayyalasomayajula, Michael R. Garrett, D. Ashley Robinson |
| EPI_ISL_2426790 | University of Mississippi Medical Center, Department of Pathology | University of Mississippi Medical Center, Molecular and Genomics Core Facility | Ashley C. Johnson, Ithiel J. Frame, Krishna K. Ayyalasomayajula, Michael R. Garrett, D. Ashley Robinson |
| EPI_ISL_2426791, EPI_ISL_2426792 | MS Public Health Laboratory | University of Mississippi Medical Center, Molecular and Genomics Core Facility | Ashley C. Johnson, Ithiel J. Frame, Krishna K. Ayyalasomayajula, Michael R. Garrett, D. Ashley Robinson |
| EPI_ISL_2426793, EPI_ISL_2426794 | University of Mississippi Medical Center, Department of Pathology | University of Mississippi Medical Center, Molecular and Genomics Core Facility | Ashley C. Johnson, Ithiel J. Frame, Krishna K. Ayyalasomayajula, Michael R. Garrett, D. Ashley Robinson |
| EPI_ISL_2426795, EPI_ISL_2426796 | MS Public Health Laboratory | University of Mississippi Medical Center, Molecular and Genomics Core Facility | Ashley C. Johnson, Ithiel J. Frame, Krishna K. Ayyalasomayajula, Michael R. Garrett, D. Ashley Robinson |
| EPI_ISL_2426797 | University of Mississippi Medical Center, Department of Pathology | University of Mississippi Medical Center, Molecular and Genomics Core Facility | Ashley C. Johnson, Ithiel J. Frame, Krishna K. Ayyalasomayajula, Michael R. Garrett, D. Ashley Robinson |
| EPI_ISL_2426798 | MS Public Health Laboratory | University of Mississippi Medical Center, Molecular and Genomics Core Facility | Ashley C. Johnson, Ithiel J. Frame, Krishna K. Ayyalasomayajula, Michael R. Garrett, D. Ashley Robinson |
| EPI_ISL_2426799, EPI_ISL_2426800, EPI_ISL_2426801, EPI_ISL_2426802, EPI_ISL_2426803, EPI_ISL_2426804, EPI_ISL_2426805, EPI_ISL_2426806, EPI_ISL_2426807, EPI_ISL_2426808, EPI_ISL_2426809, EPI_ISL_2426810 | | | |
| see above | University of Mississippi Medical Center, Department of Pathology | University of Mississippi Medical Center, Molecular and Genomics Core Facility | Ashley C. Johnson, Ithiel J. Frame, Krishna K. Ayyalasomayajula, Michael R. Garrett, D. Ashley Robinson |
| EPI_ISL_2426814, EPI_ISL_2426815, EPI_ISL_2426816, EPI_ISL_2426817, EPI_ISL_2426818, EPI_ISL_2426819, EPI_ISL_2426820, EPI_ISL_2426821, EPI_ISL_2426822, EPI_ISL_2426823, EPI_ISL_2426824, EPI_ISL_2426825, EPI_ISL_2426826, EPI_ISL_2426827, EPI_ISL_2426828, EPI_ISL_2426829, EPI_ISL_2426830, EPI_ISL_2426831, EPI_ISL_2426832, EPI_ISL_2426833, EPI_ISL_2426834, EPI_ISL_2426835, EPI_ISL_2426836, EPI_ISL_2426837, EPI_ISL_2426838, EPI_ISL_2426839, EPI_ISL_2426840, EPI_ISL_2426841, EPI_ISL_2426842, EPI_ISL_2426843, EPI_ISL_2426844, EPI_ISL_2426845, EPI_ISL_2426847, EPI_ISL_2426848, EPI_ISL_2426849, EPI_ISL_2426850, EPI_ISL_2426851, EPI_ISL_2426852 | | | |
| see above | Rhode Island State Health Laboratory | Rhode Island State Health Laboratory | Ewa King, Richard C. Huard, Kristin Carpenter-Azevedo |
| EPI_ISL_2426853, EPI_ISL_2426854, EPI_ISL_2426855, EPI_ISL_2426856, EPI_ISL_2426857, EPI_ISL_2426858, EPI_ISL_2426859, EPI_ISL_2426860 | Centers for Disease Control and Prevention, Dengue Branch | Centers for Disease Control and Prevention, Dengue Branch | Gilberto A. Santiago, Glenda Gonzalez, Betzabel Flores, Keyla Charriez, Gabriela Paz-Bailey, Jorge L. Munoz-Jordan |
| EPI_ISL_2426861, EPI_ISL_2426862, EPI_ISL_2426863, EPI_ISL_2426864, EPI_ISL_2426865 | Puerto Rico Department of Health | Centers for Disease Control and Prevention, Dengue Branch | Gilberto A. Santiago, Glenda Gonzalez, Betzabel Flores, Keyla Charriez, Gabriela Paz-Bailey, Jorge L. Munoz-Jordan |

| | | | |
|---|---|---|---|
| EPI_ISL_2426866, EPI_ISL_2426867, EPI_ISL_2426868, EPI_ISL_2426869, EPI_ISL_2426870, EPI_ISL_2426871, EPI_ISL_2426872, EPI_ISL_2426873, EPI_ISL_2426874, EPI_ISL_2426875, EPI_ISL_2426876, EPI_ISL_2426877, EPI_ISL_2426878, EPI_ISL_2426879 | | | |
| see above | Centers for Disease Control and Prevention, Dengue Branch | Centers for Disease Control and Prevention, Dengue Branch | Gilberto A. Santiago, Glenda Gonzalez, Betzabel Flores, Keyla Charriez, Gabriela Paz-Bailey, Jorge L. Munoz-Jordan |
| EPI_ISL_2426880, EPI_ISL_2426881 | Puerto Rico Department of Health | Centers for Disease Control and Prevention, Dengue Branch | Gilberto A. Santiago, Glenda Gonzalez, Betzabel Flores, Keyla Charriez, Gabriela Paz-Bailey, Jorge L. Munoz-Jordan |
| EPI_ISL_2426882, EPI_ISL_2426883, EPI_ISL_2426884, EPI_ISL_2426885, EPI_ISL_2426886, EPI_ISL_2426887, EPI_ISL_2426888, EPI_ISL_2426889, EPI_ISL_2426890, EPI_ISL_2426891 | Centers for Disease Control and Prevention, Dengue Branch | Centers for Disease Control and Prevention, Dengue Branch | Gilberto A. Santiago, Glenda Gonzalez, Betzabel Flores, Keyla Charriez, Gabriela Paz-Bailey, Jorge L. Munoz-Jordan |
| EPI_ISL_2426892 | Puerto Rico Department of Health | Centers for Disease Control and Prevention, Dengue Branch | Gilberto A. Santiago, Glenda Gonzalez, Betzabel Flores, Keyla Charriez, Gabriela Paz-Bailey, Jorge L. Munoz-Jordan |
| EPI_ISL_2426893, EPI_ISL_2426894, EPI_ISL_2426895 | Centers for Disease Control and Prevention, Dengue Branch | Centers for Disease Control and Prevention, Dengue Branch | Gilberto A. Santiago, Glenda Gonzalez, Betzabel Flores, Keyla Charriez, Gabriela Paz-Bailey, Jorge L. Munoz-Jordan |
| EPI_ISL_2426896, EPI_ISL_2426897 | Puerto Rico Department of Health | Centers for Disease Control and Prevention, Dengue Branch | Gilberto A. Santiago, Glenda Gonzalez, Betzabel Flores, Keyla Charriez, Gabriela Paz-Bailey, Jorge L. Munoz-Jordan |
| EPI_ISL_2426898, EPI_ISL_2426899, EPI_ISL_2426900, EPI_ISL_2426901, EPI_ISL_2426902, EPI_ISL_2426903, EPI_ISL_2426904, EPI_ISL_2426905, EPI_ISL_2426906, EPI_ISL_2426907, EPI_ISL_2426908, EPI_ISL_2426909, EPI_ISL_2426910, EPI_ISL_2426911, EPI_ISL_2426912, EPI_ISL_2426913, EPI_ISL_2426914, EPI_ISL_2426915, EPI_ISL_2426916, EPI_ISL_2426917, EPI_ISL_2426918, EPI_ISL_2426919, EPI_ISL_2426920, EPI_ISL_2426921, EPI_ISL_2426922, EPI_ISL_2426923, EPI_ISL_2426924, EPI_ISL_2426925, EPI_ISL_2426926, EPI_ISL_2426927, EPI_ISL_2426928, EPI_ISL_2426929, EPI_ISL_2426930, EPI_ISL_2426931, EPI_ISL_2426932, EPI_ISL_2426933, EPI_ISL_2426934, EPI_ISL_2426935, EPI_ISL_2426936, EPI_ISL_2426937, EPI_ISL_2426938, EPI_ISL_2426939, EPI_ISL_2426940, EPI_ISL_2426941, EPI_ISL_2426942, EPI_ISL_2426943 | | | |
| see above | Centers for Disease Control and Prevention, Dengue Branch | Centers for Disease Control and Prevention, Dengue Branch | Gilberto A. Santiago, Glenda Gonzalez, Betzabel Flores, Keyla Charriez, Gabriela Paz-Bailey, Jorge L. Munoz-Jordan |
| EPI_ISL_2426985, EPI_ISL_2426986, EPI_ISL_2426987, EPI_ISL_2426988, EPI_ISL_2426989, EPI_ISL_2426990, EPI_ISL_2426993, EPI_ISL_2426994 | UC Davis Genome Center and Healthy Davis Together | UC Davis Genome Center | UC Davis |
| EPI_ISL_2427230, EPI_ISL_2427231, EPI_ISL_2427232, EPI_ISL_2427233, EPI_ISL_2427234, EPI_ISL_2427235, EPI_ISL_2427236, EPI_ISL_2427238, EPI_ISL_2427239, EPI_ISL_2427240, EPI_ISL_2427241, EPI_ISL_2427242, EPI_ISL_2427243, EPI_ISL_2427244, EPI_ISL_2427246, EPI_ISL_2427247, EPI_ISL_2427248, EPI_ISL_2427249, EPI_ISL_2427250, EPI_ISL_2427251, EPI_ISL_2427252, EPI_ISL_2427253, EPI_ISL_2427254, EPI_ISL_2427255, EPI_ISL_2427256, EPI_ISL_2427257, EPI_ISL_2427258 | | | |
| see above | South Dakota Public Health Laboratory | South Dakota Public Health Laboratory | Jacob Garfin and Chris Carlson |
| EPI_ISL_2427259, EPI_ISL_2427260, EPI_ISL_2427261, EPI_ISL_2427262, EPI_ISL_2427263, EPI_ISL_2427264, EPI_ISL_2427265, EPI_ISL_2427266, EPI_ISL_2427267, EPI_ISL_2427268, EPI_ISL_2427269, EPI_ISL_2427270, EPI_ISL_2427271, EPI_ISL_2427272, EPI_ISL_2427273, EPI_ISL_2427274, EPI_ISL_2427275, EPI_ISL_2427276, EPI_ISL_2427277, EPI_ISL_2427278, EPI_ISL_2427279, EPI_ISL_2427280, EPI_ISL_2427281, EPI_ISL_2427282, EPI_ISL_2427283, EPI_ISL_2427284, EPI_ISL_2427285, EPI_ISL_2427286, EPI_ISL_2427287, EPI_ISL_2427288, EPI_ISL_2427289, EPI_ISL_2427290, EPI_ISL_2427291, EPI_ISL_2427292, EPI_ISL_2427293, EPI_ISL_2427294, EPI_ISL_2427295, EPI_ISL_2427296, EPI_ISL_2427297, EPI_ISL_2427298, EPI_ISL_2427299, EPI_ISL_2427300, EPI_ISL_2427301, EPI_ISL_2427302, EPI_ISL_2427309, EPI_ISL_2427310, EPI_ISL_2427311, EPI_ISL_2427312, EPI_ISL_2427313, EPI_ISL_2427314, EPI_ISL_2427315, EPI_ISL_2427316, EPI_ISL_2427317, EPI_ISL_2427318, EPI_ISL_2427319, EPI_ISL_2427320, EPI_ISL_2427321 | | | |
| see above | IVY3 Central Lab, Vanderbilt University Medical Center | Lauring Lab, University of Michigan, Department of Microbiology and Immunology | Gilbert |
| EPI_ISL_2427403, EPI_ISL_2427404, EPI_ISL_2427405, EPI_ISL_2427406, EPI_ISL_2427408, EPI_ISL_2427409, EPI_ISL_2427410, EPI_ISL_2427411, EPI_ISL_2427412, EPI_ISL_2427413, EPI_ISL_2427414, EPI_ISL_2427415, EPI_ISL_2427416, EPI_ISL_2427417, EPI_ISL_2427418, EPI_ISL_2427419, EPI_ISL_2427420, EPI_ISL_2427421, EPI_ISL_2427422, EPI_ISL_2427423, EPI_ISL_2427424, EPI_ISL_2427425, EPI_ISL_2427426, EPI_ISL_2427427, EPI_ISL_2427428, EPI_ISL_2427429, EPI_ISL_2427430, EPI_ISL_2427431, EPI_ISL_2427432, EPI_ISL_2427433, EPI_ISL_2427434, EPI_ISL_2427435, EPI_ISL_2427436, EPI_ISL_2427437, EPI_ISL_2427438, EPI_ISL_2427439, EPI_ISL_2427440, EPI_ISL_2427441, EPI_ISL_2427442, EPI_ISL_2427443, EPI_ISL_2427444, EPI_ISL_2427445, EPI_ISL_2427446, EPI_ISL_2427447, EPI_ISL_2427448, EPI_ISL_2427449, EPI_ISL_2427450, EPI_ISL_2427451, EPI_ISL_2427452, EPI_ISL_2427453, EPI_ISL_2427454, EPI_ISL_2427455, EPI_ISL_2427456, EPI_ISL_2427457, EPI_ISL_2427458, EPI_ISL_2427459, EPI_ISL_2427460, EPI_ISL_2427461, EPI_ISL_2427462, EPI_ISL_2427463, EPI_ISL_2427464, EPI_ISL_2427465, EPI_ISL_2427466, EPI_ISL_2427467, EPI_ISL_2427468, EPI_ISL_2427469, EPI_ISL_2427470, EPI_ISL_2427471, EPI_ISL_2427472, EPI_ISL_2427473, EPI_ISL_2427474, EPI_ISL_2427475, EPI_ISL_2427476, EPI_ISL_2427477, EPI_ISL_2427478, EPI_ISL_2427479, EPI_ISL_2427480, EPI_ISL_2427481, EPI_ISL_2427482, EPI_ISL_2427483, EPI_ISL_2427484, EPI_ISL_2427485, EPI_ISL_2427486, EPI_ISL_2427488, EPI_ISL_2427489, EPI_ISL_2427490, EPI_ISL_2427491, EPI_ISL_2427493, EPI_ISL_2427495, EPI_ISL_2427496, EPI_ISL_2427499, EPI_ISL_2427500 | | | |
| see above | NYU Langone Health | Departments of Pathology and Medicine, New York University School of Medicine | Adriana Heguy, Dacia Dimartino, Emily Guzman, Christian Marier, Peter Meyn, Sitharam Ramaswami, Gael Westby, Paul Zappile, Yutong Zhang, Paolo Cotzia, Guiqing Wang |
| EPI_ISL_2429084 | Azarian Lab at Burnett School of Biomedical Sciences | Azarian Lab at Burnett School of Biomedical Sciences | Catherine Johnston, Eleonora Cella, Taj Azarian |
| EPI_ISL_2429085, EPI_ISL_2429086, EPI_ISL_2429087, EPI_ISL_2429088, EPI_ISL_2429089, EPI_ISL_2429090, EPI_ISL_2429091, EPI_ISL_2429092, EPI_ISL_2429094, EPI_ISL_2429095, EPI_ISL_2429096, EPI_ISL_2429097, EPI_ISL_2429098, EPI_ISL_2429099, EPI_ISL_2429100, EPI_ISL_2429101, EPI_ISL_2429102, EPI_ISL_2429103, EPI_ISL_2429104, EPI_ISL_2429105, EPI_ISL_2429106, EPI_ISL_2429107, EPI_ISL_2429108, EPI_ISL_2429109, EPI_ISL_2429110, EPI_ISL_2429111, EPI_ISL_2429112, EPI_ISL_2429113, EPI_ISL_2429114, EPI_ISL_2429115, EPI_ISL_2429116, EPI_ISL_2429117, EPI_ISL_2429118, EPI_ISL_2429119, EPI_ISL_2429120, EPI_ISL_2429121, EPI_ISL_2429122 | | | |
| see above | Alaska State Virology Laboratory | Alaska State Virology Laboratory | Stephanie DeRonde, Elva House, Jacob Zidek, Lisa Smith, Ph.D., Jack Chen, Ph.D. |
| EPI_ISL_2432594, EPI_ISL_2432595, EPI_ISL_2432598, EPI_ISL_2432599, EPI_ISL_2432600, EPI_ISL_2432601, EPI_ISL_2432602, EPI_ISL_2432603, EPI_ISL_2432604, EPI_ISL_2432605, EPI_ISL_2432606, EPI_ISL_2432607, EPI_ISL_2432608, EPI_ISL_2432609, EPI_ISL_2432610, EPI_ISL_2432611, EPI_ISL_2432612, EPI_ISL_2432613, EPI_ISL_2432615, EPI_ISL_2432616, EPI_ISL_2432617, EPI_ISL_2432618, EPI_ISL_2432619, EPI_ISL_2432620, EPI_ISL_2432621, EPI_ISL_2432622, EPI_ISL_2432623, EPI_ISL_2432624, EPI_ISL_2432625, EPI_ISL_2432626, EPI_ISL_2432627, EPI_ISL_2432628, EPI_ISL_2432629, EPI_ISL_2432630, EPI_ISL_2432631, EPI_ISL_2432632, EPI_ISL_2432633, EPI_ISL_2432634, EPI_ISL_2432635, EPI_ISL_2432636, EPI_ISL_2432637, EPI_ISL_2432638, EPI_ISL_2432639, EPI_ISL_2432640, EPI_ISL_2432641, EPI_ISL_2432642, EPI_ISL_2432643, EPI_ISL_2432644, EPI_ISL_2432645, EPI_ISL_2432646, EPI_ISL_2432647, EPI_ISL_2432648, EPI_ISL_2432649, EPI_ISL_2432650, EPI_ISL_2432651, EPI_ISL_2432652, EPI_ISL_2432653, EPI_ISL_2432654, EPI_ISL_2432655, EPI_ISL_2432656, EPI_ISL_2432657, EPI_ISL_2432658, EPI_ISL_2432659, EPI_ISL_2432660, EPI_ISL_2432661, EPI_ISL_2432662, EPI_ISL_2432663, EPI_ISL_2432664, EPI_ISL_2432665, EPI_ISL_2432666, EPI_ISL_2432667, EPI_ISL_2432668, EPI_ISL_2432669, EPI_ISL_2432670, EPI_ISL_2432671, EPI_ISL_2432672, EPI_ISL_2432673, EPI_ISL_2432674, EPI_ISL_2432675, EPI_ISL_2432676, EPI_ISL_2432677, EPI_ISL_2432678, EPI_ISL_2432679, EPI_ISL_2432680, EPI_ISL_2432681, EPI_ISL_2432682, EPI_ISL_2432683, EPI_ISL_2432684, EPI_ISL_2432685, EPI_ISL_2432686, EPI_ISL_2432687, EPI_ISL_2432688, EPI_ISL_2432689, EPI_ISL_2432690, EPI_ISL_2432691, EPI_ISL_2432692, EPI_ISL_2432693, EPI_ISL_2432694, EPI_ISL_2432695, EPI_ISL_2432696, EPI_ISL_2432697, EPI_ISL_2432698, EPI_ISL_2432699, EPI_ISL_2432700, EPI_ISL_2432701, EPI_ISL_2432702, EPI_ISL_2432703, EPI_ISL_2432704, EPI_ISL_2432705, EPI_ISL_2432706, EPI_ISL_2432707, EPI_ISL_2432708, EPI_ISL_2432709, EPI_ISL_2432710, EPI_ISL_2432711, EPI_ISL_2432712, EPI_ISL_2432714, EPI_ISL_2432715, EPI_ISL_2432716, EPI_ISL_2432717, EPI_ISL_2432718, EPI_ISL_2432719, EPI_ISL_2432720, EPI_ISL_2432721, EPI_ISL_2432722, EPI_ISL_2432723, EPI_ISL_2432724, EPI_ISL_2432725, EPI_ISL_2432726, EPI_ISL_2432727, EPI_ISL_2432728, EPI_ISL_2432729, EPI_ISL_2432730, EPI_ISL_2432731, EPI_ISL_2432732, EPI_ISL_2432733, EPI_ISL_2432734, EPI_ISL_2432735, EPI_ISL_2432736, EPI_ISL_2432737, EPI_ISL_2432738, EPI_ISL_2432739, EPI_ISL_2432740, EPI_ISL_2432741, EPI_ISL_2432742, EPI_ISL_2432743, EPI_ISL_2432744, EPI_ISL_2432745, EPI_ISL_2432746, EPI_ISL_2432747, EPI_ISL_2432748, EPI_ISL_2432749, EPI_ISL_2432750, EPI_ISL_2432751, EPI_ISL_2432752, EPI_ISL_2432753, EPI_ISL_2432754, EPI_ISL_2432755, EPI_ISL_2432756, EPI_ISL_2432757, EPI_ISL_2432758, EPI_ISL_2432759, EPI_ISL_2432760, EPI_ISL_2432761, EPI_ISL_2432762, EPI_ISL_2432763, EPI_ISL_2432764, EPI_ISL_2432765, EPI_ISL_2432766, EPI_ISL_2432767, EPI_ISL_2432768, EPI_ISL_2432769, EPI_ISL_2432770, EPI_ISL_2432771, EPI_ISL_2432772, EPI_ISL_2432773, EPI_ISL_2432774, EPI_ISL_2432775, EPI_ISL_2432776, EPI_ISL_2432777, EPI_ISL_2432778, EPI_ISL_2432779, EPI_ISL_2432780, EPI_ISL_2432781, EPI_ISL_2432782, EPI_ISL_2432783, EPI_ISL_2432784, EPI_ISL_2432785, EPI_ISL_2432786, EPI_ISL_2432787, EPI_ISL_2432788, EPI_ISL_2432789, EPI_ISL_2432790, EPI_ISL_2432791, EPI_ISL_2432792, EPI_ISL_2432793, EPI_ISL_2432794, EPI_ISL_2432795, EPI_ISL_2432796, EPI_ISL_2432797, EPI_ISL_2432798, EPI_ISL_2432799, EPI_ISL_2432800, EPI_ISL_2432801, EPI_ISL_2432802, EPI_ISL_2432803, EPI_ISL_2432804, EPI_ISL_2432805, EPI_ISL_2432806, EPI_ISL_2432807, EPI_ISL_2432808, EPI_ISL_2432809, EPI_ISL_2432810, EPI_ISL_2432811, EPI_ISL_2432812, EPI_ISL_2432813, EPI_ISL_2432814, EPI_ISL_2432815, EPI_ISL_2432816, EPI_ISL_2432817, EPI_ISL_2432818, EPI_ISL_2432819, EPI_ISL_2432820, EPI_ISL_2432821, EPI_ISL_2432822, EPI_ISL_2432823, EPI_ISL_2432824, EPI_ISL_2432825, EPI_ISL_2432826, EPI_ISL_2432827, EPI_ISL_2432828, EPI_ISL_2432829, EPI_ISL_2432830, EPI_ISL_2432831, EPI_ISL_2432832, EPI_ISL_2432833, EPI_ISL_2432834, EPI_ISL_2432835, EPI_ISL_2432836, EPI_ISL_2432837 | | | |
| see above | UW Virology Lab | UW Virology Lab | Pavitra Roychoudhury, Hong Xie, Lasata Shrestha, Tien V. Nguyen, Shah Mohamed Bakhash, Michelle Lin, Noah R. Baker, Ricardo Perez, Sean Ellis, Nathan Breit, Robert J. Livingston, Meei-Li Huang, Keith R Jerome, Patrick Mathias, Alexander Greninger |
| EPI_ISL_2432839, EPI_ISL_2432840, EPI_ISL_2432841, EPI_ISL_2432842, EPI_ISL_2432843, EPI_ISL_2432844, EPI_ISL_2432845, EPI_ISL_2432847, EPI_ISL_2432848, EPI_ISL_2432849, EPI_ISL_2432850, EPI_ISL_2432851, EPI_ISL_2432852, EPI_ISL_2432853, EPI_ISL_2432854, EPI_ISL_2432855, EPI_ISL_2432856, EPI_ISL_2432857, EPI_ISL_2432858, EPI_ISL_2432859, EPI_ISL_2432860, EPI_ISL_2432861, EPI_ISL_2432863, EPI_ISL_2432864, EPI_ISL_2432865, EPI_ISL_2432866, EPI_ISL_2432867, EPI_ISL_2432868, EPI_ISL_2432869, EPI_ISL_2432870, EPI_ISL_2432871, EPI_ISL_2432872, EPI_ISL_2432873, EPI_ISL_2432874, EPI_ISL_2432875, EPI_ISL_2432876, EPI_ISL_2432877, EPI_ISL_2432878, EPI_ISL_2432880, EPI_ISL_2432881, EPI_ISL_2432883, EPI_ISL_2432884, EPI_ISL_2432885, EPI_ISL_2432886, EPI_ISL_2432887, EPI_ISL_2432888, EPI_ISL_2432889, EPI_ISL_2432890, EPI_ISL_2432891, EPI_ISL_2432892, EPI_ISL_2432893, EPI_ISL_2432894, EPI_ISL_2432895, EPI_ISL_2432896, EPI_ISL_2432897, EPI_ISL_2432898, EPI_ISL_2432899, EPI_ISL_2432900, EPI_ISL_2432901, EPI_ISL_2432902, EPI_ISL_2432903, EPI_ISL_2432904, EPI_ISL_2432905, EPI_ISL_2432907, EPI_ISL_2432908, EPI_ISL_2432909, EPI_ISL_2432910, EPI_ISL_2432911, EPI_ISL_2432912, EPI_ISL_2432913, EPI_ISL_2432917, EPI_ISL_2432918, EPI_ISL_2432919, EPI_ISL_2432921 | | | |
| see above | UW Virology Lab | UW Virology Lab | Pavitra Roychoudhury, Hong Xie, Lasata Shrestha, Tien V. Nguyen, Shah Mohamed Bakhash, Michelle Lin, Noah R. Baker, Sean Ellis, Meei-Li Huang, Keith R Jerome, Alexander Greninger |
| EPI_ISL_2432958, EPI_ISL_2432959, EPI_ISL_2432960, EPI_ISL_2432961, EPI_ISL_2432962, EPI_ISL_2432963, EPI_ISL_2432964, EPI_ISL_2432965, EPI_ISL_2432966, EPI_ISL_2432967, EPI_ISL_2432968, EPI_ISL_2432969, EPI_ISL_2432970, EPI_ISL_2432971, EPI_ISL_2432972, EPI_ISL_2432973, EPI_ISL_2432974, EPI_ISL_2432975, EPI_ISL_2432976, EPI_ISL_2432977, EPI_ISL_2432978, EPI_ISL_2432979, EPI_ISL_2432980, EPI_ISL_2432981, EPI_ISL_2432982, EPI_ISL_2432983, EPI_ISL_2432984, EPI_ISL_2432985, EPI_ISL_2432986, EPI_ISL_2432987, EPI_ISL_2432988, EPI_ISL_2432989, EPI_ISL_2432990, EPI_ISL_2432991, EPI_ISL_2432992, EPI_ISL_2432993, EPI_ISL_2432994, EPI_ISL_2432995, EPI_ISL_2432996, EPI_ISL_2432997, EPI_ISL_2432998, EPI_ISL_2432999, EPI_ISL_2433000, EPI_ISL_2433001, EPI_ISL_2433002, EPI_ISL_2433003, EPI_ISL_2433004, EPI_ISL_2433005, EPI_ISL_2433006, EPI_ISL_2433007, EPI_ISL_2433008, EPI_ISL_2433009, EPI_ISL_2433010, EPI_ISL_2433011, EPI_ISL_2433012, EPI_ISL_2433013, EPI_ISL_2433014, EPI_ISL_2433015, EPI_ISL_2433016, EPI_ISL_2433017, EPI_ISL_2433018, EPI_ISL_2433019, EPI_ISL_2433020, EPI_ISL_2433021, EPI_ISL_2433022, EPI_ISL_2433023, EPI_ISL_2433024, EPI_ISL_2433025, EPI_ISL_2433026, EPI_ISL_2433027, EPI_ISL_2433028, EPI_ISL_2433029, EPI_ISL_2433030, EPI_ISL_2433031, EPI_ISL_2433032, EPI_ISL_2433033, EPI_ISL_2433034, EPI_ISL_2433035, EPI_ISL_2433036, EPI_ISL_2433037, EPI_ISL_2433038, EPI_ISL_2433039, EPI_ISL_2433040, EPI_ISL_2433041, EPI_ISL_2433042, EPI_ISL_2433043, EPI_ISL_2433044, EPI_ISL_2433045, EPI_ISL_2433046, EPI_ISL_2433047, EPI_ISL_2433048, EPI_ISL_2433049, EPI_ISL_2433050, EPI_ISL_2433051, EPI_ISL_2433052, EPI_ISL_2433053, EPI_ISL_2433054, EPI_ISL_2433055, EPI_ISL_2433056, EPI_ISL_2433057, EPI_ISL_2433058, EPI_ISL_2433059, EPI_ISL_2433060, EPI_ISL_2433061, EPI_ISL_2433062, EPI_ISL_2433063, EPI_ISL_2433064, EPI_ISL_2433065, EPI_ISL_2433066, EPI_ISL_2433067, EPI_ISL_2433068, EPI_ISL_2433069, EPI_ISL_2433070, EPI_ISL_2433071, EPI_ISL_2433072, EPI_ISL_2433079, EPI_ISL_2433080, EPI_ISL_2433081, EPI_ISL_2433082, EPI_ISL_2433083, EPI_ISL_2433084, EPI_ISL_2433085, EPI_ISL_2433086, EPI_ISL_2433087, EPI_ISL_2433088, EPI_ISL_2433089, EPI_ISL_2433090, EPI_ISL_2433091, EPI_ISL_2433092, EPI_ISL_2433093, EPI_ISL_2433094, EPI_ISL_2433095, EPI_ISL_2433096, EPI_ISL_2433097, EPI_ISL_2433098, EPI_ISL_2433099, EPI_ISL_2433100, EPI_ISL_2433101, EPI_ISL_2433102, EPI_ISL_2433103, EPI_ISL_2433104, EPI_ISL_2433105, EPI_ISL_2433106, EPI_ISL_2433107, EPI_ISL_2433108, EPI_ISL_2433109, EPI_ISL_2433110, EPI_ISL_2433111, EPI_ISL_2433112, EPI_ISL_2433113, EPI_ISL_2433114, EPI_ISL_2433115, EPI_ISL_2433116, EPI_ISL_2433117, EPI_ISL_2433118, EPI_ISL_2433119, EPI_ISL_2433120, EPI_ISL_2433121, EPI_ISL_2433122, EPI_ISL_2433123, EPI_ISL_2433124, EPI_ISL_2433125, EPI_ISL_2433126, EPI_ISL_2433127, EPI_ISL_2433128, EPI_ISL_2433129, EPI_ISL_2433130, EPI_ISL_2433131, EPI_ISL_2433132, EPI_ISL_2433133, EPI_ISL_2433134, EPI_ISL_2433135, EPI_ISL_2433136, EPI_ISL_2433137, EPI_ISL_2433138, EPI_ISL_2433139, EPI_ISL_2433140, EPI_ISL_2433141, EPI_ISL_2433142, EPI_ISL_2433143, EPI_ISL_2433144, EPI_ISL_2433145, EPI_ISL_2433146, EPI_ISL_2433147, EPI_ISL_2433148, EPI_ISL_2433149, EPI_ISL_2433150, EPI_ISL_2433151, EPI_ISL_2433152, EPI_ISL_2433153, EPI_ISL_2433154, EPI_ISL_2433155, EPI_ISL_2433156, EPI_ISL_2433157, EPI_ISL_2433158, EPI_ISL_2433159, EPI_ISL_2433160, EPI_ISL_2433161, EPI_ISL_2433162, EPI_ISL_2433163, EPI_ISL_2433164, EPI_ISL_2433165, EPI_ISL_2433166, EPI_ISL_2433167, EPI_ISL_2433168, EPI_ISL_2433169, EPI_ISL_2433170, EPI_ISL_2433171, EPI_ISL_2433172, EPI_ISL_2433173, EPI_ISL_2433174, EPI_ISL_2433175, EPI_ISL_2433176, EPI_ISL_2433177, EPI_ISL_2433178, EPI_ISL_2433179, EPI_ISL_2433180, EPI_ISL_2433181, EPI_ISL_2433182, EPI_ISL_2433183, EPI_ISL_2433184, EPI_ISL_2433185, EPI_ISL_2433186, EPI_ISL_2433187, EPI_ISL_2433188, EPI_ISL_2433189, EPI_ISL_2433190, EPI_ISL_2433191, EPI_ISL_2433192, EPI_ISL_2433193, EPI_ISL_2433194, EPI_ISL_2433195, EPI_ISL_2433196, EPI_ISL_2433197, EPI_ISL_2433198, EPI_ISL_2433199, EPI_ISL_2433200, EPI_ISL_2433201, EPI_ISL_2433202, EPI_ISL_2433203, EPI_ISL_2433204, EPI_ISL_2433205, EPI_ISL_2433206, EPI_ISL_2433207, EPI_ISL_2433208, EPI_ISL_2433209, EPI_ISL_2433210, EPI_ISL_2433211, EPI_ISL_2433212, EPI_ISL_2433213, EPI_ISL_2433214, EPI_ISL_2433215, EPI_ISL_2433216, EPI_ISL_2433217, EPI_ISL_2433218, EPI_ISL_2433219, EPI_ISL_2433220, EPI_ISL_2433221, EPI_ISL_2433222, EPI_ISL_2433223, EPI_ISL_2433224, EPI_ISL_2433225, EPI_ISL_2433226, EPI_ISL_2433227, EPI_ISL_2433228, EPI_ISL_2433229, EPI_ISL_2433230, EPI_ISL_2433231, EPI_ISL_2433232, EPI_ISL_2433238, EPI_ISL_2433239, EPI_ISL_2433240, EPI_ISL_2433241, EPI_ISL_2433242, EPI_ISL_2433243, EPI_ISL_2433244, EPI_ISL_2433245, EPI_ISL_2433246, EPI_ISL_2433247, EPI_ISL_2433248, EPI_ISL_2433249, EPI_ISL_2433250, EPI_ISL_2433251, EPI_ISL_2433252, EPI_ISL_2433253, EPI_ISL_2433254, EPI_ISL_2433255, EPI_ISL_2433256, EPI_ISL_2433257, EPI_ISL_2434124, EPI_ISL_2434125, EPI_ISL_2434126, EPI_ISL_2434127, EPI_ISL_2434128, EPI_ISL_2434129 | | | |
| see above | Pathology and Laboratory Medicine Institute, Cleveland Clinic, Ohio, USA | Pathology and Laboratory Medicine Institute, Cleveland Clinic, Ohio, USA | Concetta Peck, Jennifer Starbuck, Joy Nakitandwe, Kristen McDonnell, David Plunkett, Zheng Jin Tu, Jay Brock, Yu-Wei Cheng, Gary Procop, Daniel Rhoads, Daniel H. Farkas, David Bosler |
| EPI_ISL_2434989, EPI_ISL_2436469, EPI_ISL_2437300 | The Ohio State University Applied Microbiology Services Laboratory | The Ohio State University Applied Microbiology Services Laboratory | Seth A. Faith PhD |
| EPI_ISL_2439137, EPI_ISL_2439139, EPI_ISL_2439140, EPI_ISL_2439141, EPI_ISL_2439143, EPI_ISL_2439144, EPI_ISL_2439145, EPI_ISL_2439146, EPI_ISL_2439147, EPI_ISL_2439148, EPI_ISL_2439149, EPI_ISL_2439150, EPI_ISL_2439152, EPI_ISL_2439153, EPI_ISL_2439154, EPI_ISL_2439155, EPI_ISL_2439156, EPI_ISL_2439157, EPI_ISL_2439158, EPI_ISL_2439161, EPI_ISL_2439162, EPI_ISL_2439165, EPI_ISL_2439166, EPI_ISL_2439167, EPI_ISL_2439168, EPI_ISL_2439169, EPI_ISL_2439170, EPI_ISL_2439171, EPI_ISL_2439172, EPI_ISL_2439173, EPI_ISL_2439175, EPI_ISL_2439177, EPI_ISL_2439178, EPI_ISL_2439179, EPI_ISL_2439180, EPI_ISL_2439181 | | | |
| see above | IN State Department of Health Laboratory Services | IN State Department of Health Laboratory Services | Cassandra Campion, Jamie Yeadon, Brian Pope, Lixia Liu, Kyle Brownlee, Melissa Hindenlang, Mark Glazier |
| EPI_ISL_2439296, EPI_ISL_2439297, EPI_ISL_2439298, EPI_ISL_2439299, EPI_ISL_2439300, EPI_ISL_2439301, EPI_ISL_2439302, EPI_ISL_2439303, EPI_ISL_2439304, EPI_ISL_2439305, EPI_ISL_2439306, EPI_ISL_2439307, EPI_ISL_2439308, EPI_ISL_2439309, EPI_ISL_2439310, EPI_ISL_2439311, EPI_ISL_2439312, EPI_ISL_2439313, EPI_ISL_2439314, EPI_ISL_2439315, EPI_ISL_2439316, EPI_ISL_2439317, EPI_ISL_2439318, EPI_ISL_2439319, EPI_ISL_2439320, EPI_ISL_2439321, EPI_ISL_2439322, EPI_ISL_2439323, EPI_ISL_2439324, EPI_ISL_2439325, EPI_ISL_2439326, EPI_ISL_2439327, EPI_ISL_2439328, EPI_ISL_2439329, EPI_ISL_2439330, EPI_ISL_2439331, EPI_ISL_2439332, EPI_ISL_2439333, EPI_ISL_2439334, EPI_ISL_2439335, EPI_ISL_2439336, EPI_ISL_2439337, EPI_ISL_2439338, EPI_ISL_2439339, EPI_ISL_2439340, EPI_ISL_2439341, EPI_ISL_2439342, EPI_ISL_2439343, EPI_ISL_2439344, EPI_ISL_2439345, EPI_ISL_2439346, EPI_ISL_2439347, EPI_ISL_2439348, EPI_ISL_2439349, EPI_ISL_2439350, EPI_ISL_2439351, EPI_ISL_2439352, EPI_ISL_2439353, EPI_ISL_2439354, EPI_ISL_2439355, EPI_ISL_2439356, EPI_ISL_2439357, EPI_ISL_2439358, EPI_ISL_2439359, EPI_ISL_2439360, EPI_ISL_2439361, | | | |

| | | | |
|---|---|---|---|
| EPI_ISL_2439362, EPI_ISL_2439363, EPI_ISL_2439364, EPI_ISL_2439365, EPI_ISL_2439366, EPI_ISL_2439367, EPI_ISL_2439368, EPI_ISL_2439369, EPI_ISL_2439370, EPI_ISL_2439371, EPI_ISL_2439372, EPI_ISL_2439373, EPI_ISL_2439374, EPI_ISL_2439375, EPI_ISL_2439376, EPI_ISL_2439377, EPI_ISL_2439378, EPI_ISL_2439379, EPI_ISL_2439380, EPI_ISL_2439381, EPI_ISL_2439382, EPI_ISL_2439383, EPI_ISL_2439384, EPI_ISL_2439385, EPI_ISL_2439386, EPI_ISL_2439387, EPI_ISL_2439388, EPI_ISL_2439389, EPI_ISL_2439390, EPI_ISL_2439391, EPI_ISL_2439392, EPI_ISL_2439393, EPI_ISL_2439394, EPI_ISL_2439395, EPI_ISL_2439396, EPI_ISL_2439397, EPI_ISL_2439398, EPI_ISL_2439399, EPI_ISL_2439400, EPI_ISL_2439401, EPI_ISL_2439402, EPI_ISL_2439403, EPI_ISL_2439404, EPI_ISL_2439405, EPI_ISL_2439406, EPI_ISL_2439407, EPI_ISL_2439408, EPI_ISL_2439409, EPI_ISL_2439410, EPI_ISL_2439411, EPI_ISL_2439412, EPI_ISL_2439413, EPI_ISL_2439414, EPI_ISL_2439415, EPI_ISL_2439416, EPI_ISL_2439417, EPI_ISL_2439418, EPI_ISL_2439419, EPI_ISL_2439420, EPI_ISL_2439421, EPI_ISL_2439422, EPI_ISL_2439423, EPI_ISL_2439424, EPI_ISL_2439425, EPI_ISL_2439426, EPI_ISL_2439427, EPI_ISL_2439428, EPI_ISL_2439429, EPI_ISL_2439430, EPI_ISL_2439431, EPI_ISL_2439432, EPI_ISL_2439433, EPI_ISL_2439434, EPI_ISL_2439435, EPI_ISL_2439436, EPI_ISL_2439437, EPI_ISL_2439438, EPI_ISL_2439439, EPI_ISL_2439440, EPI_ISL_2439441, EPI_ISL_2439442, EPI_ISL_2439443, EPI_ISL_2439444, EPI_ISL_2439445, EPI_ISL_2439446, EPI_ISL_2439447, EPI_ISL_2439448, EPI_ISL_2439449, EPI_ISL_2439450, EPI_ISL_2439451, EPI_ISL_2439452, EPI_ISL_2439453, EPI_ISL_2439454, EPI_ISL_2439455, EPI_ISL_2439456, EPI_ISL_2439457, EPI_ISL_2439458, EPI_ISL_2439459, EPI_ISL_2439460, EPI_ISL_2439461, EPI_ISL_2439462, EPI_ISL_2439463, EPI_ISL_2439464, EPI_ISL_2439465, EPI_ISL_2439466, EPI_ISL_2439467, EPI_ISL_2439468, EPI_ISL_2439469, EPI_ISL_2439470, EPI_ISL_2439471, EPI_ISL_2439472, EPI_ISL_2439473, EPI_ISL_2439474, EPI_ISL_2439475, EPI_ISL_2439476, EPI_ISL_2439477, EPI_ISL_2439478, EPI_ISL_2439479, EPI_ISL_2439480, EPI_ISL_2439481, EPI_ISL_2439482, EPI_ISL_2439483, EPI_ISL_2439484, EPI_ISL_2439485, EPI_ISL_2439486, EPI_ISL_2439487, EPI_ISL_2439488, EPI_ISL_2439489, EPI_ISL_2439490, EPI_ISL_2439491, EPI_ISL_2439492, EPI_ISL_2439493, EPI_ISL_2439494, EPI_ISL_2439495, EPI_ISL_2439496, EPI_ISL_2439497, EPI_ISL_2439498, EPI_ISL_2439499, EPI_ISL_2439500, EPI_ISL_2439501, EPI_ISL_2439502, EPI_ISL_2439503, EPI_ISL_2439504, EPI_ISL_2439505, EPI_ISL_2439506, EPI_ISL_2439507, EPI_ISL_2439508, EPI_ISL_2439509, EPI_ISL_2439510, EPI_ISL_2439511, EPI_ISL_2439512, EPI_ISL_2439513, EPI_ISL_2439514, EPI_ISL_2439515, EPI_ISL_2439516, EPI_ISL_2439517, EPI_ISL_2439518, EPI_ISL_2439519, EPI_ISL_2439520, EPI_ISL_2439521, EPI_ISL_2439522, EPI_ISL_2439523, EPI_ISL_2439524, EPI_ISL_2439525, EPI_ISL_2439526, EPI_ISL_2439527, EPI_ISL_2439528, EPI_ISL_2439529, EPI_ISL_2439530, EPI_ISL_2439531, EPI_ISL_2439532, EPI_ISL_2439533, EPI_ISL_2439534, EPI_ISL_2439535, EPI_ISL_2439536, EPI_ISL_2439537, EPI_ISL_2439538, EPI_ISL_2439539, EPI_ISL_2439540, EPI_ISL_2439541, EPI_ISL_2439542, EPI_ISL_2439543, EPI_ISL_2439544, EPI_ISL_2439545, EPI_ISL_2439546, EPI_ISL_2439547, EPI_ISL_2439548, EPI_ISL_2439549, EPI_ISL_2439550, EPI_ISL_2439551, EPI_ISL_2439552, EPI_ISL_2439553, EPI_ISL_2439554, EPI_ISL_2439555, EPI_ISL_2439556, EPI_ISL_2439557, EPI_ISL_2439558, EPI_ISL_2439559, EPI_ISL_2439560, EPI_ISL_2439561, EPI_ISL_2439562, EPI_ISL_2439563, EPI_ISL_2439564, EPI_ISL_2439565, EPI_ISL_2439566, EPI_ISL_2439567, EPI_ISL_2439568, EPI_ISL_2439569, EPI_ISL_2439570, EPI_ISL_2439571, EPI_ISL_2439572, EPI_ISL_2439573, EPI_ISL_2439574, EPI_ISL_2439575, EPI_ISL_2439576, EPI_ISL_2439577, EPI_ISL_2439578, EPI_ISL_2439579, EPI_ISL_2439580, EPI_ISL_2439581, EPI_ISL_2439582, EPI_ISL_2439583, EPI_ISL_2439584, EPI_ISL_2439585, EPI_ISL_2439586, EPI_ISL_2439587, EPI_ISL_2439588, EPI_ISL_2439589, EPI_ISL_2439590, EPI_ISL_2439591, EPI_ISL_2439592, EPI_ISL_2439593, EPI_ISL_2439594, EPI_ISL_2439595, EPI_ISL_2439596, EPI_ISL_2439597, EPI_ISL_2439598, EPI_ISL_2439599, EPI_ISL_2439600, EPI_ISL_2439601, EPI_ISL_2439602, EPI_ISL_2439603, EPI_ISL_2439604, EPI_ISL_2439605, EPI_ISL_2439606, EPI_ISL_2439607, EPI_ISL_2439608, EPI_ISL_2439609, EPI_ISL_2439610, EPI_ISL_2439611, EPI_ISL_2439612, EPI_ISL_2439613, EPI_ISL_2439614, EPI_ISL_2439615, EPI_ISL_2439616, EPI_ISL_2439617, EPI_ISL_2439618, EPI_ISL_2439619, EPI_ISL_2439620, EPI_ISL_2439621, EPI_ISL_2439622, EPI_ISL_2439623, EPI_ISL_2439624, EPI_ISL_2439625, EPI_ISL_2439626, EPI_ISL_2439627, EPI_ISL_2439628, EPI_ISL_2439629, EPI_ISL_2439630, EPI_ISL_2439631, EPI_ISL_2439632, EPI_ISL_2439633, EPI_ISL_2439634, EPI_ISL_2439635, EPI_ISL_2439636, EPI_ISL_2439637, EPI_ISL_2439638, EPI_ISL_2439639, EPI_ISL_2439640, EPI_ISL_2439641, EPI_ISL_2439642, EPI_ISL_2439643, EPI_ISL_2439644, EPI_ISL_2439645, EPI_ISL_2439646, EPI_ISL_2439647, EPI_ISL_2439648, EPI_ISL_2439649, EPI_ISL_2439650, EPI_ISL_2439651, EPI_ISL_2439652, EPI_ISL_2439653, EPI_ISL_2439654, EPI_ISL_2439655, EPI_ISL_2439656, EPI_ISL_2439657, EPI_ISL_2439658, EPI_ISL_2439659, EPI_ISL_2439660, EPI_ISL_2439661, EPI_ISL_2439662, EPI_ISL_2439663, EPI_ISL_2439664, EPI_ISL_2439665, EPI_ISL_2439666, EPI_ISL_2439667, EPI_ISL_2439668, EPI_ISL_2439669, EPI_ISL_2439670, EPI_ISL_2439671, EPI_ISL_2439672, EPI_ISL_2439673, EPI_ISL_2439674, EPI_ISL_2439675, EPI_ISL_2439676, EPI_ISL_2439677, EPI_ISL_2439678, EPI_ISL_2439679, EPI_ISL_2439680, EPI_ISL_2439681, EPI_ISL_2439682, EPI_ISL_2439683, EPI_ISL_2439684, EPI_ISL_2439685, EPI_ISL_2439686, EPI_ISL_2439687, EPI_ISL_2439688, EPI_ISL_2439689, EPI_ISL_2439690, EPI_ISL_2439691, EPI_ISL_2439692, EPI_ISL_2439693, EPI_ISL_2439694, EPI_ISL_2439695, EPI_ISL_2439696, EPI_ISL_2439697, EPI_ISL_2439698, EPI_ISL_2439699, EPI_ISL_2439700, EPI_ISL_2439701, EPI_ISL_2439702, EPI_ISL_2439703, EPI_ISL_2439704, EPI_ISL_2439705, EPI_ISL_2439706, EPI_ISL_2439707, EPI_ISL_2439708, EPI_ISL_2439709, EPI_ISL_2439710, EPI_ISL_2439711, EPI_ISL_2439712, EPI_ISL_2439713, EPI_ISL_2439714, EPI_ISL_2439715, EPI_ISL_2439716, EPI_ISL_2439717, EPI_ISL_2439718, EPI_ISL_2439719, EPI_ISL_2439720, EPI_ISL_2439721, EPI_ISL_2439722, EPI_ISL_2439723, EPI_ISL_2439724, EPI_ISL_2439725, EPI_ISL_2439726, EPI_ISL_2439727, EPI_ISL_2439728, EPI_ISL_2439729, EPI_ISL_2439730, EPI_ISL_2439731, EPI_ISL_2439732, EPI_ISL_2439733, EPI_ISL_2439734, EPI_ISL_2439735, EPI_ISL_2439736, EPI_ISL_2439737, EPI_ISL_2439738, EPI_ISL_2439739, EPI_ISL_2439740, EPI_ISL_2439741, EPI_ISL_2439742, EPI_ISL_2439743, EPI_ISL_2439744, EPI_ISL_2439745, EPI_ISL_2439746, EPI_ISL_2439747, EPI_ISL_2439748, EPI_ISL_2439749, EPI_ISL_2439750, EPI_ISL_2439751, EPI_ISL_2439752, EPI_ISL_2439753, EPI_ISL_2439754, EPI_ISL_2439755, EPI_ISL_2439756, EPI_ISL_2439757, EPI_ISL_2439758, EPI_ISL_2439759, EPI_ISL_2439760, EPI_ISL_2439761, EPI_ISL_2439762, EPI_ISL_2439763, EPI_ISL_2439764, EPI_ISL_2439765, EPI_ISL_2439766, EPI_ISL_2439767, EPI_ISL_2439768, EPI_ISL_2439769, EPI_ISL_2439770, EPI_ISL_2439771, EPI_ISL_2439772, EPI_ISL_2439773, EPI_ISL_2439774, EPI_ISL_2439775, EPI_ISL_2439776, EPI_ISL_2439777, EPI_ISL_2439778, EPI_ISL_2439779, EPI_ISL_2439780, EPI_ISL_2439781, EPI_ISL_2439782, EPI_ISL_2439783, EPI_ISL_2439784, EPI_ISL_2439785, EPI_ISL_2439786, EPI_ISL_2439787, EPI_ISL_2439788, EPI_ISL_2439789, EPI_ISL_2439790, EPI_ISL_2439791, EPI_ISL_2439792, EPI_ISL_2439793, EPI_ISL_2439794, EPI_ISL_2439795, EPI_ISL_2439796, EPI_ISL_2439797, EPI_ISL_2439798, EPI_ISL_2439799, EPI_ISL_2439800, EPI_ISL_2439801, EPI_ISL_2439802, EPI_ISL_2439803, EPI_ISL_2439804, EPI_ISL_2439805, EPI_ISL_2439806, EPI_ISL_2439807, EPI_ISL_2439808, EPI_ISL_2439809, EPI_ISL_2439810, EPI_ISL_2439811, EPI_ISL_2439812, EPI_ISL_2439813, EPI_ISL_2439814, EPI_ISL_2439815, EPI_ISL_2439816, EPI_ISL_2439817, EPI_ISL_2439818, EPI_ISL_2439819, EPI_ISL_2439820, EPI_ISL_2439821, EPI_ISL_2439822, EPI_ISL_2439823, EPI_ISL_2439824, EPI_ISL_2439825, EPI_ISL_2439826, EPI_ISL_2439827, EPI_ISL_2439828, EPI_ISL_2439829, EPI_ISL_2439830, EPI_ISL_2439831, EPI_ISL_2439832, EPI_ISL_2439833, EPI_ISL_2439834, EPI_ISL_2439835, EPI_ISL_2439836, EPI_ISL_2439837, EPI_ISL_2439838, EPI_ISL_2439839, EPI_ISL_2439840, EPI_ISL_2439841, EPI_ISL_2439842, EPI_ISL_2439843, EPI_ISL_2439844, EPI_ISL_2439845, EPI_ISL_2439846, EPI_ISL_2439847, EPI_ISL_2439848, EPI_ISL_2439849, EPI_ISL_2439850, EPI_ISL_2439851, EPI_ISL_2439852, EPI_ISL_2439853, EPI_ISL_2439854, EPI_ISL_2439855, EPI_ISL_2439856, EPI_ISL_2439857, EPI_ISL_2439858, EPI_ISL_2439859, EPI_ISL_2439860, EPI_ISL_2439861, EPI_ISL_2439862, EPI_ISL_2439863, EPI_ISL_2439864, EPI_ISL_2439865, EPI_ISL_2439866, EPI_ISL_2439867, EPI_ISL_2439868, EPI_ISL_2439869, EPI_ISL_2439870, EPI_ISL_2439871, EPI_ISL_2439872, EPI_ISL_2439873, EPI_ISL_2439874, EPI_ISL_2439875, EPI_ISL_2439876, EPI_ISL_2439877, EPI_ISL_2439878, EPI_ISL_2439879, EPI_ISL_2439880, EPI_ISL_2439881, EPI_ISL_2439882, EPI_ISL_2439883, EPI_ISL_2439884, EPI_ISL_2439885, EPI_ISL_2439886, EPI_ISL_2439887, EPI_ISL_2439888, EPI_ISL_2439889, EPI_ISL_2439890, EPI_ISL_2439891, EPI_ISL_2439892, EPI_ISL_2439893, EPI_ISL_2439894, EPI_ISL_2439895, EPI_ISL_2439896, EPI_ISL_2439897, EPI_ISL_2439898, EPI_ISL_2439899, EPI_ISL_2439900, EPI_ISL_2439901, EPI_ISL_2439902, EPI_ISL_2439903, EPI_ISL_2439904, EPI_ISL_2439905, EPI_ISL_2439906, EPI_ISL_2439907, EPI_ISL_2439908, EPI_ISL_2439909, EPI_ISL_2439910, EPI_ISL_2439911, EPI_ISL_2439912, EPI_ISL_2439913, EPI_ISL_2439914, EPI_ISL_2439915, EPI_ISL_2439916, EPI_ISL_2439917, EPI_ISL_2439918, EPI_ISL_2439919, EPI_ISL_2439920, EPI_ISL_2439921, EPI_ISL_2439922, EPI_ISL_2439923, EPI_ISL_2439924, EPI_ISL_2439925, EPI_ISL_2439926, EPI_ISL_2439927, EPI_ISL_2439928, EPI_ISL_2439929, EPI_ISL_2439930, EPI_ISL_2439931, EPI_ISL_2439932, EPI_ISL_2439933, EPI_ISL_2439934, EPI_ISL_2439935, EPI_ISL_2439936, EPI_ISL_2439937, EPI_ISL_2439938, EPI_ISL_2439939, EPI_ISL_2439940, EPI_ISL_2439941, EPI_ISL_2439942, EPI_ISL_2439943, EPI_ISL_2439944, EPI_ISL_2439945, EPI_ISL_2439946, EPI_ISL_2439947, EPI_ISL_2439948, EPI_ISL_2439949, EPI_ISL_2439950, EPI_ISL_2439951, EPI_ISL_2439952, EPI_ISL_2439953, EPI_ISL_2439954, EPI_ISL_2439955, EPI_ISL_2439956, EPI_ISL_2439957, EPI_ISL_2439958, EPI_ISL_2439959, EPI_ISL_2439960, EPI_ISL_2439961, EPI_ISL_2439962, EPI_ISL_2439963, EPI_ISL_2439964, EPI_ISL_2439965, EPI_ISL_2439966, EPI_ISL_2439967, EPI_ISL_2439968, EPI_ISL_2439969, EPI_ISL_2439970, EPI_ISL_2439971, EPI_ISL_2439972, EPI_ISL_2439973, EPI_ISL_2439974, EPI_ISL_2439975, EPI_ISL_2439976, EPI_ISL_2439977, EPI_ISL_2439978, EPI_ISL_2439979, EPI_ISL_2439980, EPI_ISL_2439981, EPI_ISL_2439982, EPI_ISL_2439983 | | | |
| see above | Quest Diagnostics Incorporated | Centers for Disease Control and Prevention Division of Viral Diseases, Pathogen Discovery | Dakota Howard, Dhwani Batra, Peter W. Cook, Kara Moser, Adrian Paskey, Jason Caravas, Benjamin Rambo-Martin, Shatavia Morrison, Christopher Gulvick, Scott Sammons, Yvette Unoarumhi, Darlene Wagner, Matthew Schmerer, S. H. Rosenthal, A. Gerasimova, R. M. Kagan, B. Anderson, M. Hua, Y. Liu, L.E. Bernstein, K.E. Livingston, A. Perez, I. A. Shlyakhter, R. V. Rolando, R. Owen, P. Tanpaiboon, F. Lacbawan, Clinton R. Paden, Duncan MacCannell |
| EPI_ISL_2440054, EPI_ISL_2440055, EPI_ISL_2440056, EPI_ISL_2440057, EPI_ISL_2440058, EPI_ISL_2440059, EPI_ISL_2440060, EPI_ISL_2440061, EPI_ISL_2440062, EPI_ISL_2440063, EPI_ISL_2440064, EPI_ISL_2440066, EPI_ISL_2440067, EPI_ISL_2440068, EPI_ISL_2440069, EPI_ISL_2440070, EPI_ISL_2440071, EPI_ISL_2440072, EPI_ISL_2440073, EPI_ISL_2440074, EPI_ISL_2440075, EPI_ISL_2440076, EPI_ISL_2440077, EPI_ISL_2440078, EPI_ISL_2440079, EPI_ISL_2440080, EPI_ISL_2440081, EPI_ISL_2440082, EPI_ISL_2440083, EPI_ISL_2440084, EPI_ISL_2440085, EPI_ISL_2440086, EPI_ISL_2440087, EPI_ISL_2440088, EPI_ISL_2440089, EPI_ISL_2440090, EPI_ISL_2440091, EPI_ISL_2440092, EPI_ISL_2440093, EPI_ISL_2440094, EPI_ISL_2440095, EPI_ISL_2440096, EPI_ISL_2440097, EPI_ISL_2440098, EPI_ISL_2440099, EPI_ISL_2440100, EPI_ISL_2440101, EPI_ISL_2440102, EPI_ISL_2440103, EPI_ISL_2440104, EPI_ISL_2440105, EPI_ISL_2440106, EPI_ISL_2440107, EPI_ISL_2440108, EPI_ISL_2440109, EPI_ISL_2440110, EPI_ISL_2440111, EPI_ISL_2440112, EPI_ISL_2440113, EPI_ISL_2440114, EPI_ISL_2440115, EPI_ISL_2440116, EPI_ISL_2440117, EPI_ISL_2440118, EPI_ISL_2440119, EPI_ISL_2440120, EPI_ISL_2440121, EPI_ISL_2440122, EPI_ISL_2440123, EPI_ISL_2440124, EPI_ISL_2440125, EPI_ISL_2440126, EPI_ISL_2440127, EPI_ISL_2440128, EPI_ISL_2440129, EPI_ISL_2440130, EPI_ISL_2440131, EPI_ISL_2440132, EPI_ISL_2440133, EPI_ISL_2440134, EPI_ISL_2440135, EPI_ISL_2440136, EPI_ISL_2440137, EPI_ISL_2440138, EPI_ISL_2440139, EPI_ISL_2440140, EPI_ISL_2440141, EPI_ISL_2440142, EPI_ISL_2440143, EPI_ISL_2440144, EPI_ISL_2440145, EPI_ISL_2440146, EPI_ISL_2440147, EPI_ISL_2440148, EPI_ISL_2440149, EPI_ISL_2440150, EPI_ISL_2440151, EPI_ISL_2440152, EPI_ISL_2440153, EPI_ISL_2440154, EPI_ISL_2440155, EPI_ISL_2440156, EPI_ISL_2440157, EPI_ISL_2440158, EPI_ISL_2440159, EPI_ISL_2440160, EPI_ISL_2440161, EPI_ISL_2440162, EPI_ISL_2440163, EPI_ISL_2440164, EPI_ISL_2440165, EPI_ISL_2440166, EPI_ISL_2440167, EPI_ISL_2440168, EPI_ISL_2440169, EPI_ISL_2440170, EPI_ISL_2440171, EPI_ISL_2440172, EPI_ISL_2440173, EPI_ISL_2440174, EPI_ISL_2440175, EPI_ISL_2440176, EPI_ISL_2440177, EPI_ISL_2440178, EPI_ISL_2440179, EPI_ISL_2440180, EPI_ISL_2440181, EPI_ISL_2440182, EPI_ISL_2440183, EPI_ISL_2440184, EPI_ISL_2440185, EPI_ISL_2440186, EPI_ISL_2440187, EPI_ISL_2440188, EPI_ISL_2440189, EPI_ISL_2440190, EPI_ISL_2440191, EPI_ISL_2440192, EPI_ISL_2440193, EPI_ISL_2440194, EPI_ISL_2440195, EPI_ISL_2440196, EPI_ISL_2440197, EPI_ISL_2440198, EPI_ISL_2440199, EPI_ISL_2440200, EPI_ISL_2440201, EPI_ISL_2440202, EPI_ISL_2440203, EPI_ISL_2440205, EPI_ISL_2440206, EPI_ISL_2440207, EPI_ISL_2440208, EPI_ISL_2440209, EPI_ISL_2440210, EPI_ISL_2440211, EPI_ISL_2440212, EPI_ISL_2440213, EPI_ISL_2440214, EPI_ISL_2440215, EPI_ISL_2440216, EPI_ISL_2440217, EPI_ISL_2440218, EPI_ISL_2440219, EPI_ISL_2440220, EPI_ISL_2440221, EPI_ISL_2440222, EPI_ISL_2440223, EPI_ISL_2440224, EPI_ISL_2440225, EPI_ISL_2440226, EPI_ISL_2440227, EPI_ISL_2440228, EPI_ISL_2440229, EPI_ISL_2440230, EPI_ISL_2440231, EPI_ISL_2440232, EPI_ISL_2440233, EPI_ISL_2440234, EPI_ISL_2440235, EPI_ISL_2440236, EPI_ISL_2440237, EPI_ISL_2440238, EPI_ISL_2440239, EPI_ISL_2440240, EPI_ISL_2440241, EPI_ISL_2440242, EPI_ISL_2440243, EPI_ISL_2440244, EPI_ISL_2440245, EPI_ISL_2440246, EPI_ISL_2440247, EPI_ISL_2440248, EPI_ISL_2440249, EPI_ISL_2440250, EPI_ISL_2440251, EPI_ISL_2440252, EPI_ISL_2440253, EPI_ISL_2440254, EPI_ISL_2440255, EPI_ISL_2440256, EPI_ISL_2440257, EPI_ISL_2440258, EPI_ISL_2440259, EPI_ISL_2440260, EPI_ISL_2440261, EPI_ISL_2440262, EPI_ISL_2440263, EPI_ISL_2440264, EPI_ISL_2440265, EPI_ISL_2440266, EPI_ISL_2440267, EPI_ISL_2440268, EPI_ISL_2440269, EPI_ISL_2440270, EPI_ISL_2440271, EPI_ISL_2440272, EPI_ISL_2440273, EPI_ISL_2440274, EPI_ISL_2440275, EPI_ISL_2440276, EPI_ISL_2440277, EPI_ISL_2440278, EPI_ISL_2440279, EPI_ISL_2440280, EPI_ISL_2440281, EPI_ISL_2440282, EPI_ISL_2440283, EPI_ISL_2440284, EPI_ISL_2440285, EPI_ISL_2440286, EPI_ISL_2440287, EPI_ISL_2440288, EPI_ISL_2440289, EPI_ISL_2440290, EPI_ISL_2440291, EPI_ISL_2440292, EPI_ISL_2440293, EPI_ISL_2440294, EPI_ISL_2440295, EPI_ISL_2440296, EPI_ISL_2440297, EPI_ISL_2440298, EPI_ISL_2440299, EPI_ISL_2440300, EPI_ISL_2440301, EPI_ISL_2440302, EPI_ISL_2440303, EPI_ISL_2440305, EPI_ISL_2440306, EPI_ISL_2440307, EPI_ISL_2440308, EPI_ISL_2440309, EPI_ISL_2440310, EPI_ISL_2440311, EPI_ISL_2440312, EPI_ISL_2440313, EPI_ISL_2440314, EPI_ISL_2440315, EPI_ISL_2440316, EPI_ISL_2440317, EPI_ISL_2440318, EPI_ISL_2440319, EPI_ISL_2440320, EPI_ISL_2440321, EPI_ISL_2440322, EPI_ISL_2440323, EPI_ISL_2440324, EPI_ISL_2440325, EPI_ISL_2440326, EPI_ISL_2440327, EPI_ISL_2440328, EPI_ISL_2440329, EPI_ISL_2440330, EPI_ISL_2440331, EPI_ISL_2440332, EPI_ISL_2440333, EPI_ISL_2440334, EPI_ISL_2440335, EPI_ISL_2440336, EPI_ISL_2440337, EPI_ISL_2440338, EPI_ISL_2440339, EPI_ISL_2440340, EPI_ISL_2440341, EPI_ISL_2440342, EPI_ISL_2440343, EPI_ISL_2440344, EPI_ISL_2440345, EPI_ISL_2440346, EPI_ISL_2440347, EPI_ISL_2440348, EPI_ISL_2440349, EPI_ISL_2440350, EPI_ISL_2440351, EPI_ISL_2440352, EPI_ISL_2440353, EPI_ISL_2440354, EPI_ISL_2440355, EPI_ISL_2440356, EPI_ISL_2440357, EPI_ISL_2440358, EPI_ISL_2440359, EPI_ISL_2440360, EPI_ISL_2440361, EPI_ISL_2440362, EPI_ISL_2440363, EPI_ISL_2440364, EPI_ISL_2440365, EPI_ISL_2440366, EPI_ISL_2440367, EPI_ISL_2440368, EPI_ISL_2440369, EPI_ISL_2440370, EPI_ISL_2440371, EPI_ISL_2440372, EPI_ISL_2440373, EPI_ISL_2440374, EPI_ISL_2440375, EPI_ISL_2440376, EPI_ISL_2440377, EPI_ISL_2440378, EPI_ISL_2440379, EPI_ISL_2440380, EPI_ISL_2440381, EPI_ISL_2440382, EPI_ISL_2440383, EPI_ISL_2440384, EPI_ISL_2440385, EPI_ISL_2440386, EPI_ISL_2440387, EPI_ISL_2440388, EPI_ISL_2440389, EPI_ISL_2440390, EPI_ISL_2440391, EPI_ISL_2440392, EPI_ISL_2440393, EPI_ISL_2440394, EPI_ISL_2440395, EPI_ISL_2440396, EPI_ISL_2440397, EPI_ISL_2440398, EPI_ISL_2440399, EPI_ISL_2440400, EPI_ISL_2440401, EPI_ISL_2440402, EPI_ISL_2440403, EPI_ISL_2440404, EPI_ISL_2440405, EPI_ISL_2440406, EPI_ISL_2440407, EPI_ISL_2440408, EPI_ISL_2440409, EPI_ISL_2440410, EPI_ISL_2440411, EPI_ISL_2440412, EPI_ISL_2440413, EPI_ISL_2440414, EPI_ISL_2440415, EPI_ISL_2440416, EPI_ISL_2440417, EPI_ISL_2440418, EPI_ISL_2440419, EPI_ISL_2440420, EPI_ISL_2440421, EPI_ISL_2440422, EPI_ISL_2440423, EPI_ISL_2440424, EPI_ISL_2440425, EPI_ISL_2440426, EPI_ISL_2440427, EPI_ISL_2440428, EPI_ISL_2440429, EPI_ISL_2440430, EPI_ISL_2440431, EPI_ISL_2440432, EPI_ISL_2440433, EPI_ISL_2440434, EPI_ISL_2440435, EPI_ISL_2440436, EPI_ISL_2440437, EPI_ISL_2440438, EPI_ISL_2440439, EPI_ISL_2440440, EPI_ISL_2440441, EPI_ISL_2440442, EPI_ISL_2440443, EPI_ISL_2440444, EPI_ISL_2440445, EPI_ISL_2440446, EPI_ISL_2440447, EPI_ISL_2440448, EPI_ISL_2440449, EPI_ISL_2440450, EPI_ISL_2440451, EPI_ISL_2440452, EPI_ISL_2440453, EPI_ISL_2440454, EPI_ISL_2440455, EPI_ISL_2440456, EPI_ISL_2440457, EPI_ISL_2440458, EPI_ISL_2440459, EPI_ISL_2440460, EPI_ISL_2440461, EPI_ISL_2440462, EPI_ISL_2440463, EPI_ISL_2440464, EPI_ISL_2440465, EPI_ISL_2440466, EPI_ISL_2440467, EPI_ISL_2440468, EPI_ISL_2440469, EPI_ISL_2440470, EPI_ISL_2440471, EPI_ISL_2440472, EPI_ISL_2440473, EPI_ISL_2440474, EPI_ISL_2440475, EPI_ISL_2440476, EPI_ISL_2440477, EPI_ISL_2440478, EPI_ISL_2440479, EPI_ISL_2440480, EPI_ISL_2440481, EPI_ISL_2440482, EPI_ISL_2440483, EPI_ISL_2440484, EPI_ISL_2440485, EPI_ISL_2440486, EPI_ISL_2440487, EPI_ISL_2440488, EPI_ISL_2440489, EPI_ISL_2440490, EPI_ISL_2440491, EPI_ISL_2440492, EPI_ISL_2440493, EPI_ISL_2440494, EPI_ISL_2440495, EPI_ISL_2440496, EPI_ISL_2440497, EPI_ISL_2440498, EPI_ISL_2440499, EPI_ISL_2440500, EPI_ISL_2440501, EPI_ISL_2440502, EPI_ISL_2440503, EPI_ISL_2440504, EPI_ISL_2440505, EPI_ISL_2440506, EPI_ISL_2440507, EPI_ISL_2440508, EPI_ISL_2440509, EPI_ISL_2440510, EPI_ISL_2440511, EPI_ISL_2440512, EPI_ISL_2440513, EPI_ISL_2440514, EPI_ISL_2440515, EPI_ISL_2440516, EPI_ISL_2440517, EPI_ISL_2440518, EPI_ISL_2440519, EPI_ISL_2440520, EPI_ISL_2440521, EPI_ISL_2440522, EPI_ISL_2440523, EPI_ISL_2440524, EPI_ISL_2440525, EPI_ISL_2440526, EPI_ISL_2440527, EPI_ISL_2440528 | | | |
| see above | Helix/Illumina | Centers for Disease Control and Prevention Division of Viral Diseases, Pathogen Discovery | Dakota Howard, Dhwani Batra, Peter W. Cook, Kara Moser, Adrian Paskey, Jason Caravas, Benjamin Rambo-Martin, Shatavia Morrison, Christopher Gulvick, Scott Sammons, Yvette Unoarumhi, Darlene Wagner, Matthew Schmerer, Eileen de Feo, Jan Antico, Christine Tran, Matthew Tolentino, Shannon Wickline, Kim Gietzen, Brad Sickler, Jingtao Liu, Eric Allen, Phil Febbo, Nicole L. Washington, Simon White, Geraint Levan, Kelly Schiabor Barrett, Elizabeth Cirulli, Alexandre Bolze, Ary Ascencio, Charlotte Rivera-Garcia, Ryan Cho, Jason Nguyen, Sherry Wang, Jimmy Ramirez, Tyler Cassens, Efren Sandoval, Magnus Isaksson, William Lee, David Becker, Marc Laurent, James Lu, Clinton R. Paden, Duncan MacCannell |
| EPI_ISL_2440529, EPI_ISL_2440530, EPI_ISL_2440531, EPI_ISL_2440532, EPI_ISL_2440533, EPI_ISL_2440534, EPI_ISL_2440535, EPI_ISL_2440536, EPI_ISL_2440537, EPI_ISL_2440538, EPI_ISL_2440539, EPI_ISL_2440540, EPI_ISL_2440541, EPI_ISL_2440542, EPI_ISL_2440543, EPI_ISL_2440544, EPI_ISL_2440545, EPI_ISL_2440546, EPI_ISL_2440547, EPI_ISL_2440548, EPI_ISL_2440549, EPI_ISL_2440550, EPI_ISL_2440551, EPI_ISL_2440552, EPI_ISL_2440553, EPI_ISL_2440554, EPI_ISL_2440555, EPI_ISL_2440556, EPI_ISL_2440557, EPI_ISL_2440558, EPI_ISL_2440559, EPI_ISL_2440560, EPI_ISL_2440561, EPI_ISL_2440562, EPI_ISL_2440563, EPI_ISL_2440564, EPI_ISL_2440565, EPI_ISL_2440566, EPI_ISL_2440567, EPI_ISL_2440568, EPI_ISL_2440569, EPI_ISL_2440570, EPI_ISL_2440571, EPI_ISL_2440572, EPI_ISL_2440573, EPI_ISL_2440574, EPI_ISL_2440575, EPI_ISL_2440576, EPI_ISL_2440577, EPI_ISL_2440578, EPI_ISL_2440579, EPI_ISL_2440580, EPI_ISL_2440581, EPI_ISL_2440582, EPI_ISL_2440583, EPI_ISL_2440584, EPI_ISL_2440585, EPI_ISL_2440586, EPI_ISL_2440587, EPI_ISL_2440588, EPI_ISL_2440589, EPI_ISL_2440590, EPI_ISL_2440591, EPI_ISL_2440592, EPI_ISL_2440593, EPI_ISL_2440594, EPI_ISL_2440595, EPI_ISL_2440596, EPI_ISL_2440597, EPI_ISL_2440598, EPI_ISL_2440599, EPI_ISL_2440600, EPI_ISL_2440601, EPI_ISL_2440602, EPI_ISL_2440603, EPI_ISL_2440604, EPI_ISL_2440605, EPI_ISL_2440606, EPI_ISL_2440607, EPI_ISL_2440608, EPI_ISL_2440609, EPI_ISL_2440610, EPI_ISL_2440611, EPI_ISL_2440612, EPI_ISL_2440613, EPI_ISL_2440614, EPI_ISL_2440615, EPI_ISL_2440616, EPI_ISL_2440617, EPI_ISL_2440618, EPI_ISL_2440619, EPI_ISL_2440620, EPI_ISL_2440621, EPI_ISL_2440622, EPI_ISL_2440623, EPI_ISL_2440624, EPI_ISL_2440625, EPI_ISL_2440626, EPI_ISL_2440627, EPI_ISL_2440628, EPI_ISL_2440629, EPI_ISL_2440630, EPI_ISL_2440631, EPI_ISL_2440632, EPI_ISL_2440633, EPI_ISL_2440634, EPI_ISL_2440635, EPI_ISL_2440636, EPI_ISL_2440637, EPI_ISL_2440638, EPI_ISL_2440639, EPI_ISL_2440640, EPI_ISL_2440641, EPI_ISL_2440642, EPI_ISL_2440643, EPI_ISL_2440644, EPI_ISL_2440645, EPI_ISL_2440646, EPI_ISL_2440647, EPI_ISL_2440648, EPI_ISL_2440649, EPI_ISL_2440650, EPI_ISL_2440651, EPI_ISL_2440652, EPI_ISL_2440653, EPI_ISL_2440654, EPI_ISL_2440655, EPI_ISL_2440656, EPI_ISL_2440657, EPI_ISL_2440658, EPI_ISL_2440659, EPI_ISL_2440660, EPI_ISL_2440661, EPI_ISL_2440662, EPI_ISL_2440663, EPI_ISL_2440664, EPI_ISL_2440665, EPI_ISL_2440666, EPI_ISL_2440667, EPI_ISL_2440668, EPI_ISL_2440669, EPI_ISL_2440670, EPI_ISL_2440671, EPI_ISL_2440672, EPI_ISL_2440673, EPI_ISL_2440674, EPI_ISL_2440675, EPI_ISL_2440676, EPI_ISL_2440677, EPI_ISL_2440678, EPI_ISL_2440679, EPI_ISL_2440680, EPI_ISL_2440681, EPI_ISL_2440682, EPI_ISL_2440683, EPI_ISL_2440684, EPI_ISL_2440685, EPI_ISL_2440686, EPI_ISL_2440687, EPI_ISL_2440688, EPI_ISL_2440689, EPI_ISL_2440690, EPI_ISL_2440691, EPI_ISL_2440692, EPI_ISL_2440693, EPI_ISL_2440694, EPI_ISL_2440695, EPI_ISL_2440696, EPI_ISL_2440697, EPI_ISL_2440698, EPI_ISL_2440699, EPI_ISL_2440700, EPI_ISL_2440701, EPI_ISL_2440702, EPI_ISL_2440703, EPI_ISL_2440704, EPI_ISL_2440705, EPI_ISL_2440706, EPI_ISL_2440707, EPI_ISL_2440708, EPI_ISL_2440709, EPI_ISL_2440710, EPI_ISL_2440711, EPI_ISL_2440712, EPI_ISL_2440713, EPI_ISL_2440714, EPI_ISL_2440715, EPI_ISL_2440716, EPI_ISL_2440717, EPI_ISL_2440718, EPI_ISL_2440719, EPI_ISL_2440720, EPI_ISL_2440721, EPI_ISL_2440722, EPI_ISL_2440723, EPI_ISL_2440724, EPI_ISL_2440725, EPI_ISL_2440726, EPI_ISL_2440727, EPI_ISL_2440728, EPI_ISL_2440729, EPI_ISL_2440730, EPI_ISL_2440731, EPI_ISL_2440732, EPI_ISL_2440733, EPI_ISL_2440734, EPI_ISL_2440735, EPI_ISL_2440736, EPI_ISL_2440737, EPI_ISL_2440738, EPI_ISL_2440739, EPI_ISL_2440740, EPI_ISL_2440741, EPI_ISL_2440742, EPI_ISL_2440743, EPI_ISL_2440744, EPI_ISL_2440745, EPI_ISL_2440746, EPI_ISL_2440747, EPI_ISL_2440748, EPI_ISL_2440749, EPI_ISL_2440750, EPI_ISL_2440751, EPI_ISL_2440752, EPI_ISL_2440753, EPI_ISL_2440754, EPI_ISL_2440755, EPI_ISL_2440756, EPI_ISL_2440757, EPI_ISL_2440758, EPI_ISL_2440759, EPI_ISL_2440760, EPI_ISL_2440761, EPI_ISL_2440762, EPI_ISL_2440763, EPI_ISL_2440764, EPI_ISL_2440765, EPI_ISL_2440766, EPI_ISL_2440767, EPI_ISL_2440768, EPI_ISL_2440769, EPI_ISL_2440770, EPI_ISL_2440771, EPI_ISL_2440772, EPI_ISL_2440773, EPI_ISL_2440774, EPI_ISL_2440775, EPI_ISL_2440776, EPI_ISL_2440777, EPI_ISL_2440778, EPI_ISL_2440779, EPI_ISL_2440780, EPI_ISL_2440781, EPI_ISL_2440782, EPI_ISL_2440783, EPI_ISL_2440784, EPI_ISL_2440785, EPI_ISL_2440786, EPI_ISL_2440787, EPI_ISL_2440788, EPI_ISL_2440789, EPI_ISL_2440790, EPI_ISL_2440791, EPI_ISL_2440792, EPI_ISL_2440793, EPI_ISL_2440794, EPI_ISL_2440795, EPI_ISL_2440796, EPI_ISL_2440797, EPI_ISL_2440798, EPI_ISL_2440799, EPI_ISL_2440800, EPI_ISL_2440801, EPI_ISL_2440802, EPI_ISL_2440803, EPI_ISL_2440804, EPI_ISL_2440805, EPI_ISL_2440806, EPI_ISL_2440807, EPI_ISL_2440808, EPI_ISL_2440809, EPI_ISL_2440810, EPI_ISL_2440811, EPI_ISL_2440812, EPI_ISL_2440813, EPI_ISL_2440814, EPI_ISL_2440815, EPI_ISL_2440816, EPI_ISL_2440817, EPI_ISL_2440818, EPI_ISL_2440819, EPI_ISL_2440820, EPI_ISL_2440821, EPI_ISL_2440822, EPI_ISL_2440823, EPI_ISL_2440824, EPI_ISL_2440825, EPI_ISL_2440826, EPI_ISL_2440827, EPI_ISL_2440828, EPI_ISL_2440829, EPI_ISL_2440830, EPI_ISL_2440831, EPI_ISL_2440832, EPI_ISL_2440833, EPI_ISL_2440834, EPI_ISL_2440835, EPI_ISL_2440836, EPI_ISL_2440837, EPI_ISL_2440838, EPI_ISL_2440839, EPI_ISL_2440840, EPI_ISL_2440841, EPI_ISL_2440842, EPI_ISL_2440843, EPI_ISL_2440844, EPI_ISL_2440845, EPI_ISL_2440846, EPI_ISL_2440847, EPI_ISL_2440848, EPI_ISL_2440849, EPI_ISL_2440850, EPI_ISL_2440851, EPI_ISL_2440852, EPI_ISL_2440853, EPI_ISL_2440854, EPI_ISL_2440855, EPI_ISL_2440856, EPI_ISL_2440857, EPI_ISL_2440858, EPI_ISL_2440859, EPI_ISL_2440860, EPI_ISL_2440861, EPI_ISL_2440862, EPI_ISL_2440863, EPI_ISL_2440864, EPI_ISL_2440865, EPI_ISL_2440866, EPI_ISL_2440867, EPI_ISL_2440868, EPI_ISL_2440869, EPI_ISL_2440870, EPI_ISL_2440871, EPI_ISL_2440872, EPI_ISL_2440873, EPI_ISL_2440874, EPI_ISL_2440875, EPI_ISL_2440876, EPI_ISL_2440877, EPI_ISL_2440878, EPI_ISL_2440879, EPI_ISL_2440880, EPI_ISL_2440881, EPI_ISL_2440882, EPI_ISL_2440883, EPI_ISL_2440884, EPI_ISL_2440885, EPI_ISL_2440886, EPI_ISL_2440887, EPI_ISL_2440888, EPI_ISL_2440889, EPI_ISL_2440890, EPI_ISL_2440891, EPI_ISL_2440892, EPI_ISL_2440893, EPI_ISL_2440894 | | | |
| see above | Quest Diagnostics Incorporated | Centers for Disease Control and Prevention Division of Viral Diseases, Pathogen Discovery | Dakota Howard, Dhwani Batra, Peter W. Cook, Kara Moser, Adrian Paskey, Jason Caravas, Benjamin Rambo-Martin, Shatavia Morrison, Christopher Gulvick, Scott Sammons, Yvette Unoarumhi, Darlene Wagner, Matthew Schmerer, S. H. Rosenthal, A. Gerasimova, R. M. Kagan, B. Anderson, M. Hua, Y. Liu, L.E. Bernstein, K.E. Livingston, A. Perez, I. A. Shlyakhter, R. V. Rolando, R. Owen, P. Tanpaiboon, F. Lacbawan, Clinton R. Paden, Duncan MacCannell |
| EPI_ISL_2440895, EPI_ISL_2440896, EPI_ISL_2440897, EPI_ISL_2440898, EPI_ISL_2440899, EPI_ISL_2440900, EPI_ISL_2440901, EPI_ISL_2440902, EPI_ISL_2440903, EPI_ISL_2440904, EPI_ISL_2440905, EPI_ISL_2440906, EPI_ISL_2440907, EPI_ISL_2440908, EPI_ISL_2440909, EPI_ISL_2440910, EPI_ISL_2440911, EPI_ISL_2440912, EPI_ISL_2440913, EPI_ISL_2440914, EPI_ISL_2440915, EPI_ISL_2440916, EPI_ISL_2440917, EPI_ISL_2440918, EPI_ISL_2440919, EPI_ISL_2440920, EPI_ISL_2440921, EPI_ISL_2440922, EPI_ISL_2440923, EPI_ISL_2440924, EPI_ISL_2440925, EPI_ISL_2440926, EPI_ISL_2440927, EPI_ISL_2440928, EPI_ISL_2440929, EPI_ISL_2440930, EPI_ISL_2440931, EPI_ISL_2440932, EPI_ISL_2440933, EPI_ISL_2440934, EPI_ISL_2440935, EPI_ISL_2440936, EPI_ISL_2440937, EPI_ISL_2440938, EPI_ISL_2440939, EPI_ISL_2440940, EPI_ISL_2440941, EPI_ISL_2440942, EPI_ISL_2440943, EPI_ISL_2440944, EPI_ISL_2440945, EPI_ISL_2440946, EPI_ISL_2440947, EPI_ISL_2440948, EPI_ISL_2440949, EPI_ISL_2440950, EPI_ISL_2440951, EPI_ISL_2440952, EPI_ISL_2440953, EPI_ISL_2440954, EPI_ISL_2440955, EPI_ISL_2440956, EPI_ISL_2440957, EPI_ISL_2440958, EPI_ISL_2440959, EPI_ISL_2440960, EPI_ISL_2440961, EPI_ISL_2440962, EPI_ISL_2440963, EPI_ISL_2440964, EPI_ISL_2440965, EPI_ISL_2440966, EPI_ISL_2440967, EPI_ISL_2440968, EPI_ISL_2440969, EPI_ISL_2440970, EPI_ISL_2440971, EPI_ISL_2440972, EPI_ISL_2440973, EPI_ISL_2440974, EPI_ISL_2440975, EPI_ISL_2440976, EPI_ISL_2440977, EPI_ISL_2440978, EPI_ISL_2440979, EPI_ISL_2440980, EPI_ISL_2440981, EPI_ISL_2440982, EPI_ISL_2440983, EPI_ISL_2440984, EPI_ISL_2440985, EPI_ISL_2440986, EPI_ISL_2440987, EPI_ISL_2440988, EPI_ISL_2440989, EPI_ISL_2440990, EPI_ISL_2440991, EPI_ISL_2440992, EPI_ISL_2440993, EPI_ISL_2440994, EPI_ISL_2440995, EPI_ISL_2440996, EPI_ISL_2440997, EPI_ISL_2440998, EPI_ISL_2440999, EPI_ISL_2441000, EPI_ISL_2441001, EPI_ISL_2441002, EPI_ISL_2441003, EPI_ISL_2441004, EPI_ISL_2441005, EPI_ISL_2441006, EPI_ISL_2441007, EPI_ISL_2441008, EPI_ISL_2441009, EPI_ISL_2441010, EPI_ISL_2441011, EPI_ISL_2441012, EPI_ISL_2441013, EPI_ISL_2441014, EPI_ISL_2441015, EPI_ISL_2441016, EPI_ISL_2441017, EPI_ISL_2441018, EPI_ISL_2441019, EPI_ISL_2441020, EPI_ISL_2441021, EPI_ISL_2441022, EPI_ISL_2441023, EPI_ISL_2441024, EPI_ISL_2441025, EPI_ISL_2441026, EPI_ISL_2441027, EPI_ISL_2441028, EPI_ISL_2441029, EPI_ISL_2441030, EPI_ISL_2441031, EPI_ISL_2441032, EPI_ISL_2441033, EPI_ISL_2441034, EPI_ISL_2441035, EPI_ISL_2441036, EPI_ISL_2441037, EPI_ISL_2441038, EPI_ISL_2441039, EPI_ISL_2441040, EPI_ISL_2441041, EPI_ISL_2441042, EPI_ISL_2441043, EPI_ISL_2441044, EPI_ISL_2441045, EPI_ISL_2441046, EPI_ISL_2441047, EPI_ISL_2441048, EPI_ISL_2441049, EPI_ISL_2441050, EPI_ISL_2441051, EPI_ISL_2441052, EPI_ISL_2441053, EPI_ISL_2441054, EPI_ISL_2441056, EPI_ISL_2441057, EPI_ISL_2441058, EPI_ISL_2441059, EPI_ISL_2441061, EPI_ISL_2441063, EPI_ISL_2441064, EPI_ISL_2441065, EPI_ISL_2441066, EPI_ISL_2441067, EPI_ISL_2441068, EPI_ISL_2441069, EPI_ISL_2441070, EPI_ISL_2441071, EPI_ISL_2441073, EPI_ISL_2441075, EPI_ISL_2441076 | | | |
| see above | Mako Medical | Centers for Disease Control and Prevention Division of Viral Diseases, Pathogen Discovery | Dakota Howard, Dhwani Batra, Peter W. Cook, Kara Moser, Adrian Paskey, Jason Caravas, Benjamin Rambo-Martin, Shatavia Morrison, Christopher Gulvick, Scott Sammons, Yvette Unoarumhi, Darlene Wagner, Matthew Schmerer,Matthew Tugwell, Lauren Moon, Clinton R. Paden, Duncan MacCannell |
| EPI_ISL_2441773 | Springfield Greene County Health Dept. | Genomics and Discovery, Respiratory Viruses Branch, Division of Viral Diseases, Centers for Disease Control and Prevention | Anna Kelleher, Ying Tao, Yan Li, Jing Zhang, Brian Lynch, Krista Queen, Anna Uehara, Peter Cook, Han Jia Justin Ng, Rachel Marine, Clinton R. Paden, Dhwani Batra, Haibin Wang, Tara Coalter, Jasmine Padilla, Morgan Davis, Mili Sheth, Sarah Nobles, Mark Burroughs, Justin Lee, Adam Retchless, Suxiang Tong |
| EPI_ISL_2441774 | Atlanta VA Medical Center | Genomics and Discovery, Respiratory Viruses Branch, Division of Viral Diseases, Centers for Disease Control and Prevention | Anna Kelleher, Ying Tao, Yan Li, Jing Zhang, Brian Lynch, Krista Queen, Anna Uehara, Peter Cook, Han Jia Justin Ng, Rachel Marine, Clinton R. Paden, Dhwani Batra, Haibin Wang, Tara Coalter, Jasmine Padilla, Morgan Davis, Mili Sheth, Sarah Nobles, Mark Burroughs, Justin Lee, Adam Retchless, Suxiang Tong |
| EPI_ISL_2441775 | AR Dept. of Health-PHL, Molecular Diagnostics | Genomics and Discovery, Respiratory Viruses Branch, Division of Viral Diseases, Centers for Disease Control and Prevention | Anna Kelleher, Ying Tao, Yan Li, Jing Zhang, Brian Lynch, Krista Queen, Anna Uehara, Peter Cook, Han Jia Justin Ng, Rachel Marine, Clinton R. Paden, Dhwani Batra, Haibin Wang, Tara Coalter, Jasmine Padilla, Morgan Davis, Mili Sheth, Sarah Nobles, Mark Burroughs, Justin Lee, Adam Retchless, Suxiang Tong |
| EPI_ISL_2441776 | WVDHHR - Office of Laboratory Services | Genomics and Discovery, Respiratory Viruses Branch, Division of Viral Diseases, Centers for Disease Control and Prevention | Anna Kelleher, Ying Tao, Yan Li, Jing Zhang, Brian Lynch, Krista Queen, Anna Uehara, Peter Cook, Han Jia Justin Ng, Rachel Marine, Clinton R. Paden, Dhwani Batra, Haibin Wang, Tara Coalter, Jasmine Padilla, Morgan Davis, Mili Sheth, Sarah Nobles, Mark Burroughs, Justin Lee, Adam Retchless, Suxiang Tong |
| EPI_ISL_2441777, EPI_ISL_2441778, EPI_ISL_2441779 | Atlanta VA Medical Center | Genomics and Discovery, Respiratory Viruses Branch, Division of Viral Diseases, Centers for Disease Control and Prevention | Ying Tao, Yan Li, Jing Zhang, Anna Kelleher, Brian Lynch, Krista Queen, Anna Uehara, Peter Cook, Han Jia Justin Ng, Rachel Marine, Clinton R. Paden, Dhwani Batra, Haibin Wang, Tara Coalter, Jasmine Padilla, Morgan Davis, Mili Sheth, Sarah Nobles, Mark Burroughs, Justin Lee, Adam Retchless, Suxiang Tong |

| | | | |
| --- | --- | --- | --- |
| EPI_ISL_2441819, EPI_ISL_2441820, EPI_ISL_2441821, EPI_ISL_2441822, EPI_ISL_2441824, EPI_ISL_2441825, EPI_ISL_2441826, EPI_ISL_2441827, EPI_ISL_2441828, EPI_ISL_2441829, EPI_ISL_2441830, EPI_ISL_2441831, EPI_ISL_2441834, EPI_ISL_2441835, EPI_ISL_2441836, EPI_ISL_2441838, EPI_ISL_2441839, EPI_ISL_2441840, EPI_ISL_2441841, EPI_ISL_2441842, EPI_ISL_2441844, EPI_ISL_2441845, EPI_ISL_2441846, EPI_ISL_2441847, EPI_ISL_2441848, EPI_ISL_2441850, EPI_ISL_2441851, EPI_ISL_2441852, EPI_ISL_2441853, EPI_ISL_2441855, EPI_ISL_2441856, EPI_ISL_2441857, EPI_ISL_2441858, EPI_ISL_2441860, EPI_ISL_2441861, EPI_ISL_2441862, EPI_ISL_2441863, EPI_ISL_2441864, EPI_ISL_2441865, EPI_ISL_2441866, EPI_ISL_2441867, EPI_ISL_2441868, EPI_ISL_2441869, EPI_ISL_2441870, EPI_ISL_2441871, EPI_ISL_2441872, EPI_ISL_2441873, EPI_ISL_2441874, EPI_ISL_2441875, EPI_ISL_2441876, EPI_ISL_2441877, EPI_ISL_2441878, EPI_ISL_2441879, EPI_ISL_2441880, EPI_ISL_2441881, EPI_ISL_2441882, EPI_ISL_2441883, EPI_ISL_2441884, EPI_ISL_2441885, EPI_ISL_2441886, EPI_ISL_2441887, EPI_ISL_2441888, EPI_ISL_2441889, EPI_ISL_2441890, EPI_ISL_2441891, EPI_ISL_2441892, EPI_ISL_2441893, EPI_ISL_2441894, EPI_ISL_2441895, EPI_ISL_2441896, EPI_ISL_2441897, EPI_ISL_2441898, EPI_ISL_2441899, EPI_ISL_2441901, EPI_ISL_2441902, EPI_ISL_2441903, EPI_ISL_2441904, EPI_ISL_2441905, EPI_ISL_2441906, EPI_ISL_2441907, EPI_ISL_2441908, EPI_ISL_2441909, EPI_ISL_2441910, EPI_ISL_2441911, EPI_ISL_2441912, EPI_ISL_2441913, EPI_ISL_2441917, EPI_ISL_2441918, EPI_ISL_2441919, EPI_ISL_2441920, EPI_ISL_2441921, EPI_ISL_2441922, EPI_ISL_2441924, EPI_ISL_2441925, EPI_ISL_2441926, EPI_ISL_2441927, EPI_ISL_2441928, EPI_ISL_2441929, EPI_ISL_2441930, EPI_ISL_2441931, EPI_ISL_2441932, EPI_ISL_2441933, EPI_ISL_2441934, EPI_ISL_2441935, EPI_ISL_2441936, EPI_ISL_2441937, EPI_ISL_2441938, EPI_ISL_2441939, EPI_ISL_2441940, EPI_ISL_2441941, EPI_ISL_2441942, EPI_ISL_2441943, EPI_ISL_2441944, EPI_ISL_2441945, EPI_ISL_2441947, EPI_ISL_2441948, EPI_ISL_2441949, EPI_ISL_2441950, EPI_ISL_2441951, EPI_ISL_2441952, EPI_ISL_2441953, EPI_ISL_2441954, EPI_ISL_2441955, EPI_ISL_2441956, EPI_ISL_2441958, EPI_ISL_2441959, EPI_ISL_2441960, EPI_ISL_2441961, EPI_ISL_2441962 | | | |
| see above | Fulgent Genetics | Centers for Disease Control and Prevention Division of Viral Diseases, Pathogen Discovery | Dakota Howard, Dhwani Batra, Peter W. Cook, Kara Moser, Adrian Paskey, Jason Caravas, Benjamin Rambo-Martin, Shatavia Morrison, Christopher Gulvick, Scott Sammons, Yvette Unoarumhi, Darlene Wagner, Matthew Schmerer, Harry Gao, Mickey Li, John Gao, Joseph Fierro, Benafsh Sapra, Becky Tsai, Yan Meng, Doreen Ng, James Xie, Clinton R. Paden, Duncan MacCannell |
| EPI_ISL_2442280, EPI_ISL_2442281, EPI_ISL_2442282, EPI_ISL_2442283, EPI_ISL_2442284, EPI_ISL_2442285, EPI_ISL_2442286, EPI_ISL_2442287, EPI_ISL_2442288, EPI_ISL_2442289, EPI_ISL_2442290, EPI_ISL_2442291, EPI_ISL_2442292, EPI_ISL_2442293, EPI_ISL_2442294, EPI_ISL_2442295, EPI_ISL_2442296, EPI_ISL_2442297, EPI_ISL_2442298 | | | |
| see above | Wexner Medical Center | The Ohio State University College of Medicine | Koenig,S., Seminetta,J. |
| EPI_ISL_2442299, EPI_ISL_2442300, EPI_ISL_2442301, EPI_ISL_2442302, EPI_ISL_2442303, EPI_ISL_2442304, EPI_ISL_2442305, EPI_ISL_2442306, EPI_ISL_2442307, EPI_ISL_2442308, EPI_ISL_2442309, EPI_ISL_2442310, EPI_ISL_2442311, EPI_ISL_2442312, EPI_ISL_2442313, EPI_ISL_2442314, EPI_ISL_2442315, EPI_ISL_2442316, EPI_ISL_2442317, EPI_ISL_2442318, EPI_ISL_2442319, EPI_ISL_2442320, EPI_ISL_2442321, EPI_ISL_2442322, EPI_ISL_2442323, EPI_ISL_2442324, EPI_ISL_2442325, EPI_ISL_2442326, EPI_ISL_2442327, EPI_ISL_2442328, EPI_ISL_2442329, EPI_ISL_2442331, EPI_ISL_2442332, EPI_ISL_2442333, EPI_ISL_2442334, EPI_ISL_2442335, EPI_ISL_2442336, EPI_ISL_2442337 | | | |
| see above | Aegis Sciences Corporation | Centers for Disease Control and Prevention Division of Viral Diseases, Pathogen Discovery | Dakota Howard, Dhwani Batra, Peter W. Cook, Kara Moser, Adrian Paskey, Jason Caravas, Benjamin Rambo-Martin, Shatavia Morrison, Christopher Gulvick, Scott Sammons, Yvette Unoarumhi, Darlene Wagner, Matthew Schmerer, Cyndi Clark, Patrick Campbell, Rob Case, Vikramsinha Ghorpade, Holly Houdeshell, Ola Kvalvaag, Dillon Nall, Ethan Sanders, Alec Vest, Shaun Westlund, Matthew Hardison, Clinton R. Paden, Duncan MacCannell |
| EPI_ISL_2442339, EPI_ISL_2442340, EPI_ISL_2442341, EPI_ISL_2442342, EPI_ISL_2442343, EPI_ISL_2442344, EPI_ISL_2442345, EPI_ISL_2442346, EPI_ISL_2442348 | Wyoming Public Health Laboratory | Wyoming Public Health Laboratory | Jim Mildenberger, Taylor Fearing, Channing Weber, Ashley Norberg, Chayse Rowley, Marley Goetz, Brian Dominguez, Elliot Thomasson, Brittany Oher, Sam Britz, Cari Sloma, and Rob Christensen |
| EPI_ISL_2442349, EPI_ISL_2442350, EPI_ISL_2442351, EPI_ISL_2442352, EPI_ISL_2442353, EPI_ISL_2442354, EPI_ISL_2442355, EPI_ISL_2442356, EPI_ISL_2442357, EPI_ISL_2442358, EPI_ISL_2442359, EPI_ISL_2442360, EPI_ISL_2442361 | | | |
| see above | Nebraska Public Health Laboratory | NPHL COVID-19 Response Team | NPHL COVID-19 Response Team |
| EPI_ISL_2442362 | Wexner Medical Center | The Ohio State University College of Medicine | Koenig,S., Seminetta,J. |
| EPI_ISL_2442385 | Kansas Health and Environmental Lab | Kansas Health and Environmental Lab | Katherine Wiggins, Mike Grose, Jonathan Barnell, Ben Olsen, and Phil Adam |
| EPI_ISL_2442386, EPI_ISL_2442387, EPI_ISL_2442388, EPI_ISL_2442389 | Wexner Medical Center | The Ohio State University College of Medicine | Koenig,S., Seminetta,J. |
| EPI_ISL_2442390, EPI_ISL_2442391, EPI_ISL_2442392, EPI_ISL_2442393, EPI_ISL_2442394, EPI_ISL_2442395, EPI_ISL_2442396, EPI_ISL_2442397, EPI_ISL_2442398, EPI_ISL_2442399 | Nebraska Public Health Laboratory | NPHL COVID-19 Response Team | NPHL COVID-19 Response Team |
| EPI_ISL_2442400, EPI_ISL_2442401, EPI_ISL_2442402, EPI_ISL_2442403, EPI_ISL_2442404, EPI_ISL_2442405, EPI_ISL_2442406, EPI_ISL_2442407, EPI_ISL_2442408, EPI_ISL_2442409, EPI_ISL_2442410, EPI_ISL_2442411, EPI_ISL_2442412, EPI_ISL_2442413, EPI_ISL_2442414, EPI_ISL_2442415, EPI_ISL_2442416, EPI_ISL_2442417, EPI_ISL_2442418, EPI_ISL_2442419, EPI_ISL_2442420, EPI_ISL_2442421, EPI_ISL_2442422, EPI_ISL_2442423, EPI_ISL_2442424, EPI_ISL_2442425, EPI_ISL_2442426, EPI_ISL_2442427, EPI_ISL_2442428, EPI_ISL_2442429, EPI_ISL_2442430, EPI_ISL_2442431, EPI_ISL_2442432, EPI_ISL_2442433, EPI_ISL_2442434, EPI_ISL_2442435, EPI_ISL_2442436, EPI_ISL_2442437, EPI_ISL_2442438, EPI_ISL_2442439, EPI_ISL_2442442, EPI_ISL_2442443, EPI_ISL_2442444, EPI_ISL_2442445, EPI_ISL_2442446, EPI_ISL_2442447, EPI_ISL_2442448, EPI_ISL_2442449, EPI_ISL_2442451, EPI_ISL_2442453, EPI_ISL_2442454, EPI_ISL_2442456, EPI_ISL_2442457, EPI_ISL_2442458, EPI_ISL_2442459, EPI_ISL_2442460, EPI_ISL_2442461, EPI_ISL_2442462, EPI_ISL_2442463, EPI_ISL_2442464, EPI_ISL_2442465, EPI_ISL_2442466, EPI_ISL_2442467, EPI_ISL_2442468, EPI_ISL_2442469, EPI_ISL_2442470, EPI_ISL_2442471, EPI_ISL_2442472, EPI_ISL_2442473, EPI_ISL_2442474, EPI_ISL_2442475, EPI_ISL_2442476, EPI_ISL_2442477, EPI_ISL_2442478, EPI_ISL_2442479, EPI_ISL_2442480, EPI_ISL_2442481, EPI_ISL_2442482, EPI_ISL_2442483, EPI_ISL_2442484, EPI_ISL_2442486, EPI_ISL_2442487, EPI_ISL_2442488, EPI_ISL_2442489, EPI_ISL_2442490, EPI_ISL_2442491, EPI_ISL_2442492, EPI_ISL_2442493, EPI_ISL_2442494, EPI_ISL_2442495, EPI_ISL_2442496, EPI_ISL_2442497, EPI_ISL_2442498, EPI_ISL_2442499, EPI_ISL_2442500, EPI_ISL_2442501, EPI_ISL_2442503, EPI_ISL_2442504, EPI_ISL_2442505, EPI_ISL_2442506, EPI_ISL_2442507, EPI_ISL_2442508, EPI_ISL_2442509, EPI_ISL_2442510, EPI_ISL_2442511, EPI_ISL_2442512, EPI_ISL_2442513, EPI_ISL_2442514, EPI_ISL_2442515, EPI_ISL_2442516, EPI_ISL_2442517, EPI_ISL_2442518, EPI_ISL_2442519, EPI_ISL_2442520, EPI_ISL_2442521, EPI_ISL_2442522, EPI_ISL_2442523, EPI_ISL_2442524, EPI_ISL_2442525, EPI_ISL_2442526, EPI_ISL_2442527, EPI_ISL_2442528, EPI_ISL_2442529, EPI_ISL_2442530, EPI_ISL_2442531, EPI_ISL_2442532, EPI_ISL_2442533, EPI_ISL_2442535, EPI_ISL_2442536, EPI_ISL_2442537, EPI_ISL_2442538, EPI_ISL_2442539, EPI_ISL_2442540, EPI_ISL_2442541, EPI_ISL_2442542, EPI_ISL_2442543, EPI_ISL_2442545, EPI_ISL_2442546, EPI_ISL_2442547, EPI_ISL_2442548, EPI_ISL_2442549, EPI_ISL_2442550, EPI_ISL_2442551, EPI_ISL_2442552, EPI_ISL_2442553, EPI_ISL_2442554, EPI_ISL_2442555, EPI_ISL_2442556, EPI_ISL_2442557, EPI_ISL_2442558, EPI_ISL_2442559, EPI_ISL_2442560, EPI_ISL_2442561, EPI_ISL_2442562, EPI_ISL_2442563, EPI_ISL_2442564, EPI_ISL_2442565, EPI_ISL_2442566, EPI_ISL_2442567, EPI_ISL_2442568, EPI_ISL_2442569, EPI_ISL_2442570, EPI_ISL_2442571, EPI_ISL_2442572, EPI_ISL_2442573, EPI_ISL_2442574, EPI_ISL_2442575, EPI_ISL_2442576, EPI_ISL_2442577, EPI_ISL_2442578, EPI_ISL_2442579, EPI_ISL_2442580, EPI_ISL_2442581, EPI_ISL_2442582, EPI_ISL_2442583, EPI_ISL_2442584, EPI_ISL_2442586, EPI_ISL_2442587, EPI_ISL_2442588, EPI_ISL_2442589, EPI_ISL_2442590, EPI_ISL_2442591, EPI_ISL_2442592, EPI_ISL_2442593, EPI_ISL_2442594, EPI_ISL_2442596, EPI_ISL_2442597, EPI_ISL_2442600, EPI_ISL_2442601, EPI_ISL_2442602, EPI_ISL_2442604, EPI_ISL_2442605, EPI_ISL_2442606, EPI_ISL_2442607, EPI_ISL_2442608, EPI_ISL_2442609, EPI_ISL_2442610, EPI_ISL_2442611, EPI_ISL_2442613, EPI_ISL_2442614, EPI_ISL_2442615, EPI_ISL_2442616, EPI_ISL_2442617, EPI_ISL_2442618, EPI_ISL_2442619, EPI_ISL_2442620, EPI_ISL_2442621, EPI_ISL_2442622, EPI_ISL_2442623, EPI_ISL_2442624, EPI_ISL_2442625, EPI_ISL_2442626, EPI_ISL_2442627, EPI_ISL_2442628, EPI_ISL_2442629, EPI_ISL_2442631, EPI_ISL_2442632, EPI_ISL_2442633, EPI_ISL_2442634, EPI_ISL_2442635, EPI_ISL_2442636, EPI_ISL_2442637, EPI_ISL_2442638, EPI_ISL_2442639, EPI_ISL_2442640, EPI_ISL_2442641, EPI_ISL_2442642, EPI_ISL_2442643, EPI_ISL_2442644, EPI_ISL_2442645, EPI_ISL_2442646, EPI_ISL_2442647, EPI_ISL_2442648, EPI_ISL_2442649, EPI_ISL_2442650, EPI_ISL_2442651, EPI_ISL_2442652, EPI_ISL_2442653, EPI_ISL_2442654, EPI_ISL_2442655, EPI_ISL_2442656, EPI_ISL_2442657, EPI_ISL_2442658, EPI_ISL_2442659, EPI_ISL_2442661, EPI_ISL_2442662, EPI_ISL_2442663, EPI_ISL_2442664, EPI_ISL_2442665, EPI_ISL_2442666, EPI_ISL_2442667, EPI_ISL_2442668, EPI_ISL_2442669, EPI_ISL_2442670, EPI_ISL_2442671, EPI_ISL_2442672, EPI_ISL_2442673, EPI_ISL_2442674, EPI_ISL_2442675, EPI_ISL_2442676, EPI_ISL_2442677, EPI_ISL_2442678, EPI_ISL_2442679, EPI_ISL_2442680, EPI_ISL_2442681, EPI_ISL_2442682, EPI_ISL_2442683, EPI_ISL_2442684, EPI_ISL_2442685, EPI_ISL_2442686, EPI_ISL_2442687, EPI_ISL_2442689, EPI_ISL_2442690, EPI_ISL_2442691, EPI_ISL_2442692, EPI_ISL_2442693, EPI_ISL_2442694, EPI_ISL_2442695, EPI_ISL_2442696, EPI_ISL_2442697, EPI_ISL_2442698, EPI_ISL_2442699, EPI_ISL_2442700, EPI_ISL_2442701, EPI_ISL_2442702, EPI_ISL_2442703, EPI_ISL_2442704, EPI_ISL_2442706, EPI_ISL_2442707, EPI_ISL_2442709, EPI_ISL_2442710, EPI_ISL_2442711, EPI_ISL_2442712, EPI_ISL_2442713, EPI_ISL_2442714, EPI_ISL_2442715, EPI_ISL_2442716, EPI_ISL_2442717, EPI_ISL_2442718, EPI_ISL_2442719, EPI_ISL_2442720, EPI_ISL_2442721, EPI_ISL_2442722, EPI_ISL_2442723, EPI_ISL_2442725, EPI_ISL_2442727, EPI_ISL_2442728, EPI_ISL_2442729, EPI_ISL_2442730, EPI_ISL_2442731, EPI_ISL_2442732, EPI_ISL_2442733, EPI_ISL_2442734, EPI_ISL_2442735, EPI_ISL_2442736, EPI_ISL_2442737, EPI_ISL_2442738, EPI_ISL_2442739, EPI_ISL_2442740, EPI_ISL_2442741, EPI_ISL_2442742, EPI_ISL_2442743, EPI_ISL_2442744, EPI_ISL_2442745, EPI_ISL_2442747, EPI_ISL_2442748, EPI_ISL_2442749, EPI_ISL_2442751, EPI_ISL_2442752, EPI_ISL_2442753, EPI_ISL_2442754, EPI_ISL_2442756, EPI_ISL_2442757, EPI_ISL_2442758, EPI_ISL_2442759, EPI_ISL_2442760, EPI_ISL_2442761, EPI_ISL_2442763, EPI_ISL_2442765, EPI_ISL_2442768, EPI_ISL_2442769, EPI_ISL_2442770, EPI_ISL_2442771, EPI_ISL_2442772, EPI_ISL_2442773, EPI_ISL_2442774, EPI_ISL_2442775, EPI_ISL_2442776, EPI_ISL_2442777, EPI_ISL_2442778, EPI_ISL_2442779, EPI_ISL_2442780, EPI_ISL_2442781, EPI_ISL_2442782, EPI_ISL_2442783, EPI_ISL_2442784, EPI_ISL_2442785, EPI_ISL_2442786, EPI_ISL_2442787, EPI_ISL_2442789, EPI_ISL_2442790, EPI_ISL_2442791, EPI_ISL_2442792, EPI_ISL_2442793, EPI_ISL_2442794, EPI_ISL_2442795, EPI_ISL_2442796, EPI_ISL_2442797, EPI_ISL_2442798, EPI_ISL_2442799, EPI_ISL_2442800, EPI_ISL_2442801, EPI_ISL_2442802, EPI_ISL_2442803, EPI_ISL_2442804, EPI_ISL_2442805, EPI_ISL_2442806, EPI_ISL_2442807, EPI_ISL_2442808, EPI_ISL_2442809, EPI_ISL_2442810, EPI_ISL_2442811, EPI_ISL_2442812, EPI_ISL_2442813, EPI_ISL_2442814, EPI_ISL_2442815, EPI_ISL_2442816, EPI_ISL_2442817, EPI_ISL_2442818, EPI_ISL_2442819, EPI_ISL_2442820, EPI_ISL_2442821, EPI_ISL_2442822, EPI_ISL_2442823, EPI_ISL_2442824, EPI_ISL_2442825, EPI_ISL_2442826, EPI_ISL_2442827, EPI_ISL_2442828, EPI_ISL_2442829, EPI_ISL_2442830, EPI_ISL_2442831, EPI_ISL_2442832, EPI_ISL_2442833, EPI_ISL_2442834, EPI_ISL_2442836, EPI_ISL_2442839, EPI_ISL_2442840, EPI_ISL_2442841, EPI_ISL_2442842, EPI_ISL_2442844, EPI_ISL_2442846, EPI_ISL_2442847, EPI_ISL_2442848, EPI_ISL_2442851, EPI_ISL_2442852, EPI_ISL_2442853, EPI_ISL_2442854, EPI_ISL_2442855, EPI_ISL_2442856, EPI_ISL_2442857, EPI_ISL_2442858, EPI_ISL_2442859, EPI_ISL_2442860, EPI_ISL_2442861, EPI_ISL_2442862, EPI_ISL_2442863, EPI_ISL_2442864, EPI_ISL_2442865, EPI_ISL_2442866, EPI_ISL_2442867, EPI_ISL_2442868, EPI_ISL_2442869, EPI_ISL_2442870, EPI_ISL_2442871, EPI_ISL_2442872, EPI_ISL_2442873, EPI_ISL_2442875, EPI_ISL_2442876, EPI_ISL_2442877, EPI_ISL_2442878, EPI_ISL_2442879, EPI_ISL_2442881, EPI_ISL_2442882, EPI_ISL_2442883, EPI_ISL_2442884, EPI_ISL_2442885, EPI_ISL_2442886, EPI_ISL_2442887, EPI_ISL_2442888, EPI_ISL_2442889, EPI_ISL_2442890, EPI_ISL_2442891, EPI_ISL_2442892, EPI_ISL_2442893, EPI_ISL_2442894, EPI_ISL_2442895, EPI_ISL_2442896, EPI_ISL_2442897, EPI_ISL_2442899, EPI_ISL_2442900, EPI_ISL_2442901, EPI_ISL_2442903, EPI_ISL_2442904, EPI_ISL_2442905, EPI_ISL_2442906, EPI_ISL_2442907, EPI_ISL_2442908, EPI_ISL_2442909, EPI_ISL_2442910, EPI_ISL_2442911, EPI_ISL_2442912, EPI_ISL_2442915, EPI_ISL_2442916, EPI_ISL_2442917, EPI_ISL_2442918, EPI_ISL_2442919, EPI_ISL_2442920, EPI_ISL_2442921, EPI_ISL_2442922, EPI_ISL_2442923, EPI_ISL_2442924, EPI_ISL_2442925, EPI_ISL_2442927, EPI_ISL_2442928, EPI_ISL_2442929, EPI_ISL_2442930, EPI_ISL_2442931, EPI_ISL_2442932, EPI_ISL_2442933, EPI_ISL_2442934, EPI_ISL_2442935, EPI_ISL_2442936, EPI_ISL_2442937, EPI_ISL_2442939, EPI_ISL_2442940, EPI_ISL_2442941, EPI_ISL_2442942, EPI_ISL_2442943, EPI_ISL_2442944, EPI_ISL_2442945, EPI_ISL_2442946, EPI_ISL_2442947, EPI_ISL_2442948, EPI_ISL_2442949, EPI_ISL_2442950, EPI_ISL_2442951, EPI_ISL_2442952, EPI_ISL_2442953, EPI_ISL_2442954, EPI_ISL_2442955, EPI_ISL_2442956, EPI_ISL_2442957, EPI_ISL_2442958, EPI_ISL_2442959, EPI_ISL_2442961, EPI_ISL_2442962, EPI_ISL_2442963, EPI_ISL_2442964, EPI_ISL_2442965, EPI_ISL_2442966, EPI_ISL_2442967, EPI_ISL_2442968, EPI_ISL_2442969, EPI_ISL_2442970, EPI_ISL_2442971, EPI_ISL_2442972, EPI_ISL_2442974, EPI_ISL_2442975, EPI_ISL_2442976, EPI_ISL_2442977, EPI_ISL_2442978, EPI_ISL_2442979, EPI_ISL_2442980, EPI_ISL_2442981, EPI_ISL_2442982, EPI_ISL_2442983, EPI_ISL_2442984, EPI_ISL_2442986, EPI_ISL_2442988, EPI_ISL_2442989, EPI_ISL_2442990, EPI_ISL_2442991, EPI_ISL_2442992, EPI_ISL_2442993, EPI_ISL_2442994, EPI_ISL_2442995, EPI_ISL_2442996, EPI_ISL_2442997, EPI_ISL_2442998, EPI_ISL_2442999, EPI_ISL_2443000, EPI_ISL_2443001, EPI_ISL_2443003, EPI_ISL_2443004, EPI_ISL_2443005, EPI_ISL_2443006, EPI_ISL_2443007, EPI_ISL_2443008, EPI_ISL_2443009, EPI_ISL_2443010, EPI_ISL_2443011, EPI_ISL_2443012, EPI_ISL_2443015, EPI_ISL_2443016, EPI_ISL_2443017, EPI_ISL_2443018, EPI_ISL_2443020, EPI_ISL_2443021, EPI_ISL_2443022, EPI_ISL_2443023, EPI_ISL_2443024, EPI_ISL_2443025, EPI_ISL_2443026, EPI_ISL_2443027, EPI_ISL_2443028, EPI_ISL_2443029, EPI_ISL_2443030, EPI_ISL_2443031, EPI_ISL_2443032, EPI_ISL_2443033, EPI_ISL_2443034, EPI_ISL_2443035, EPI_ISL_2443036, EPI_ISL_2443037, EPI_ISL_2443038, EPI_ISL_2443039, EPI_ISL_2443041, EPI_ISL_2443042, EPI_ISL_2443043, EPI_ISL_2443044, EPI_ISL_2443045 | | | |
| see above | Florida Bureau of Public Health Laboratories | Florida Bureau of Public Health Laboratories | Sarah Schmedes, Jason Blanton |
| EPI_ISL_2443047 | LESP Baja California | Instituto de Diagnostico y Referencia Epidemiologicos (INDRE) | Claudia Wong-Arambula, Abril Rodriguez-Maldonado, Vanessa Rivero-Arredondo, Ariadna Medina-Benitez, Joaquin Quiroz-Mercado, Sergio Rangel-Guerrero, Natividad Cruz-Ortiz, Tatiana Nunez-Garcia, Gisela Barrera-Badillo, Lucia Hernandez-Rivas, Irma Lopez-Martinez, Ernesto Ramirez-Gonzalez. |
| EPI_ISL_2443048 | LESP Ciudad de Mexico | Instituto de Diagnostico y Referencia Epidemiologicos (INDRE) | Claudia Wong-Arambula, Abril Rodriguez-Maldonado, Vanessa Rivero-Arredondo, Ariadna Medina-Benitez, Joaquin Quiroz-Mercado, Sergio Rangel-Guerrero, Natividad Cruz-Ortiz, Tatiana Nunez-Garcia, Gisela Barrera-Badillo, Lucia Hernandez-Rivas, Irma Lopez-Martinez, Ernesto Ramirez-Gonzalez. |
| EPI_ISL_2443049 | LESP Quintana Roo | Instituto de Diagnostico y Referencia Epidemiologicos (INDRE) | Claudia Wong-Arambula, Abril Rodriguez-Maldonado, Vanessa Rivero-Arredondo, Ariadna Medina-Benitez, Joaquin Quiroz-Mercado, Sergio Rangel-Guerrero, Natividad Cruz-Ortiz, Tatiana Nunez-Garcia, Gisela Barrera-Badillo, Lucia Hernandez-Rivas, Irma Lopez-Martinez, Ernesto Ramirez-Gonzalez. |
| EPI_ISL_2443050 | LESP Michoacan | Instituto de Diagnostico y Referencia Epidemiologicos (INDRE) | Claudia Wong-Arambula, Abril Rodriguez-Maldonado, Vanessa Rivero-Arredondo, Ariadna Medina-Benitez, Joaquin Quiroz-Mercado, Sergio Rangel-Guerrero, Natividad Cruz-Ortiz, Tatiana Nunez-Garcia, Gisela Barrera-Badillo, Lucia Hernandez-Rivas, Irma Lopez-Martinez, Ernesto Ramirez-Gonzalez. |
| EPI_ISL_2443052 | University of Pittsburgh | Genomics and Discovery, Respiratory Viruses Branch, Division of Viral Diseases, Centers for Disease Control and Prevention | Ying Tao, Yan Li, Jing Zhang, Anna Kelleher, Brian Lynch, Krista Queen, Anna Uehara, Peter Cook, Han Jia Justin Ng, Rachel Marine, Clinton R. Paden, Dhwani Batra, Haibin Wang, Tara Coalter, Jasmine Padilla, Morgan Davis, Mili Sheth, Sarah Nobles, Mark Burroughs, Justin Lee, Adam Retchless, Suxiang Tong |
| EPI_ISL_2443054 | GA Department of Public Health Laboratory | Genomics and Discovery, Respiratory Viruses Branch, Division of Viral Diseases, Centers for Disease Control and Prevention | Ying Tao, Yan Li, Jing Zhang, Anna Kelleher, Brian Lynch, Krista Queen, Anna Uehara, Peter Cook, Han Jia Justin Ng, Rachel Marine, Clinton R. Paden, Dhwani Batra, Haibin Wang, Tara Coalter, Jasmine Padilla, Morgan Davis, Mili Sheth, Sarah Nobles, Mark Burroughs, Justin Lee, Adam Retchless, Suxiang Tong |
| EPI_ISL_2443056 | TN Division of Laboratory Services | Genomics and Discovery, Respiratory Viruses Branch, Division of Viral Diseases, Centers for Disease Control and Prevention | Ying Tao, Yan Li, Jing Zhang, Anna Kelleher, Brian Lynch, Krista Queen, Anna Uehara, Peter Cook, Han Jia Justin Ng, Rachel Marine, Clinton R. Paden, Dhwani Batra, Haibin Wang, Tara Coalter, Jasmine Padilla, Morgan Davis, Mili Sheth, Sarah Nobles, Mark Burroughs, Justin Lee, Adam Retchless, Suxiang Tong |
| EPI_ISL_2443066, EPI_ISL_2443067, EPI_ISL_2443068 | NYU Langone Health | Departments of Pathology and Medicine, New York University School of Medicine | Adriana Heguy, Dacia Dimartino, Emily Guzman, Christian Marier, Peter Meyn, Sitharam Ramaswami, Gael Westby, Paul Zappile, Yutong Zhang, Paolo Cotzia, Guiqing Wang |
| EPI_ISL_2443071, EPI_ISL_2443072, EPI_ISL_2443073, EPI_ISL_2443074, EPI_ISL_2443075, EPI_ISL_2443076, | Salud Digna | Instituto Nacional de Medicina Genomica | Hidalgo-Miranda A, Cedro-Tanda A, Cisneros-Villanueva M, Orjuela-Rodríguez M, Mendoza-Vargas A, Reyes-Grajeda JP, Abraham Campos-Romero, Moreno-Camacho José Luis, Rodriguez-Gallegos Jorge, Luna-Ruiz Marco, Gonzalez-Barrera D, Rangel-DeLeon D, Munguia-Garza P, Ramirez-Vega O, Escobar-Arrazola, M, Herrera-Montalvo LA. |

| | | | |
|---|---|---|---|
| EPI_ISL_2443077, EPI_ISL_2443078, EPI_ISL_2443079 | | | |
| EPI_ISL_2443080 | Salud Digna | Instituto Nacional de Medicina Genomica | Hidalgo-Miranda A, Cedro-Tanda A, Cisneros-Villanueva M, Orjuela-Rodríguez M, Mendoza-Vargas A, Reyes-Grajeda JP, Abraham Campos-Romero, Moreno-Camacho José Luis, Rodriguez-Gallegos Jorge, Luna-Ruiz Marco, Gonzalez-Barrera D, Rangel-DeLeon D, Munguia-Garza P, Ramirez-Vega O, Escobar-Arrazola, M, Herrera-Montalvo LA. |
| EPI_ISL_2443081 | Salud Digna | Instituto Nacional de Medicina Genomica | Hidalgo-Miranda A, Cedro-Tanda A, Cisneros-Villanueva M, Orjuela-Rodríguez M, Mendoza-Vargas A, Reyes-Grajeda JP, Abraham Campos-Romero, Moreno-Camacho José Luis, Rodriguez-Gallegos Jorge, Luna-Ruiz Marco, Gonzalez-Barrera D, Rangel-DeLeon D, Munguia-Garza P, Ramirez-Vega O, Escobar-Arrazola, M, Herrera-Montalvo LA. |
| EPI_ISL_2443082, EPI_ISL_2443083 | Instituto Nacional de Medicina Genomica | Instituto Nacional de Medicina Genomica | Hidalgo-Miranda A, Cedro-Tanda A, Cisneros-Villanueva M, Orjuela-Rodríguez M, Mendoza-Vargas A, Reyes-Grajeda JP, Gonzalez-Barrera D, Rangel-DeLeon D, Munguia-Garza P, Ramirez-Vega O, Escobar-Arrazola, M, Herrera-Montalvo LA. |
| EPI_ISL_2443084, EPI_ISL_2443085 | Salud Digna | Instituto Nacional de Medicina Genomica | Hidalgo-Miranda A, Cedro-Tanda A, Cisneros-Villanueva M, Orjuela-Rodríguez M, Mendoza-Vargas A, Reyes-Grajeda JP, Abraham Campos-Romero, Moreno-Camacho José Luis, Rodriguez-Gallegos Jorge, Luna-Ruiz Marco, Gonzalez-Barrera D, Rangel-DeLeon D, Munguia-Garza P, Ramirez-Vega O, Escobar-Arrazola, M, Herrera-Montalvo LA. |
| EPI_ISL_2443086, EPI_ISL_2443088, EPI_ISL_2443089, EPI_ISL_2443090, EPI_ISL_2443091, EPI_ISL_2443092, EPI_ISL_2443093, EPI_ISL_2443094, EPI_ISL_2443095, EPI_ISL_2443096, EPI_ISL_2443097, EPI_ISL_2443098, EPI_ISL_2443099, EPI_ISL_2443100, EPI_ISL_2443101, EPI_ISL_2443102, EPI_ISL_2443103, EPI_ISL_2443104, EPI_ISL_2443105, EPI_ISL_2443106, EPI_ISL_2443107, EPI_ISL_2443108, EPI_ISL_2443109, EPI_ISL_2443110, EPI_ISL_2443111, EPI_ISL_2443112, EPI_ISL_2443113, EPI_ISL_2443114, EPI_ISL_2443115, EPI_ISL_2443116, EPI_ISL_2443117, EPI_ISL_2443118, EPI_ISL_2443119, EPI_ISL_2443120, EPI_ISL_2443121, EPI_ISL_2443122, EPI_ISL_2443123, EPI_ISL_2443124, EPI_ISL_2443125, EPI_ISL_2443126, EPI_ISL_2443128, EPI_ISL_2443129, EPI_ISL_2443130 | | | |
| see above | NJDOH, Public Health and Environmental Laboratories | NJ_PHEL | Lindsey Bodnar, Shiv K. Verma, Ryan Pachucki, Chelsea San Filippo, Dana Woell, Allison Roder, Byeong Jeong |
| EPI_ISL_2443131, EPI_ISL_2443132, EPI_ISL_2443133, EPI_ISL_2443134, EPI_ISL_2443135, EPI_ISL_2443136, EPI_ISL_2443137, EPI_ISL_2443138, EPI_ISL_2443139, EPI_ISL_2443142, EPI_ISL_2443143, EPI_ISL_2443144, EPI_ISL_2443145, EPI_ISL_2443146, EPI_ISL_2443147, EPI_ISL_2443148, EPI_ISL_2443149, EPI_ISL_2443150, EPI_ISL_2443151, EPI_ISL_2443152, EPI_ISL_2443153, EPI_ISL_2443154, EPI_ISL_2443155, EPI_ISL_2443156, EPI_ISL_2443157, EPI_ISL_2443158, EPI_ISL_2443159, EPI_ISL_2443160 | | | |
| see above | Kansas Health and Environmental Lab | Kansas Health and Environmental Lab | Katherine Wiggins, Mike Grose, Jonathan Barnell, Ben Olsen, and Phil Adam |
| EPI_ISL_2443164, EPI_ISL_2443165, EPI_ISL_2443166, EPI_ISL_2443167, EPI_ISL_2443168, EPI_ISL_2443169, EPI_ISL_2443170, EPI_ISL_2443171, EPI_ISL_2443174, EPI_ISL_2443175 | IN State Department of Health Laboratory Services | IN State Department of Health Laboratory Services | Cassandra Campion, Jamie Yeadon, Brian Pope, Lixia Liu, Kyle Brownlee, Melissa Hindenlang, Mark Glazier |
| EPI_ISL_2443192, EPI_ISL_2443193, EPI_ISL_2443194, EPI_ISL_2443195, EPI_ISL_2443196, EPI_ISL_2443198, EPI_ISL_2443199, EPI_ISL_2443200, EPI_ISL_2443201, EPI_ISL_2443202, EPI_ISL_2443206, EPI_ISL_2443208 | | | |
| see above | Kansas Health and Environmental Lab | Kansas Health and Environmental Lab | Katherine Wiggins, Mike Grose, Jonathan Barnell, Ben Olsen, and Phil Adam |
| EPI_ISL_2443288, EPI_ISL_2443289, EPI_ISL_2443290, EPI_ISL_2443291, EPI_ISL_2443292, EPI_ISL_2443293, EPI_ISL_2443294, EPI_ISL_2443295, EPI_ISL_2443296, EPI_ISL_2443298, EPI_ISL_2443300, EPI_ISL_2443301, EPI_ISL_2443302, EPI_ISL_2443303, EPI_ISL_2443304, EPI_ISL_2443306, EPI_ISL_2443307, EPI_ISL_2443308, EPI_ISL_2443309, EPI_ISL_2443310, EPI_ISL_2443311, EPI_ISL_2443312, EPI_ISL_2443313, EPI_ISL_2443314, EPI_ISL_2443315, EPI_ISL_2443316, EPI_ISL_2443317, EPI_ISL_2443318, EPI_ISL_2443319, EPI_ISL_2443320, EPI_ISL_2443321, EPI_ISL_2443322, EPI_ISL_2443323, EPI_ISL_2443324 | | | |
| see above | TXDSHS | TXDSHS | Rashmi Tuladhar, Bonnie Oh, Jenny Zhang, Maliha Rahman, Mayela Pedrueza, Anita Pokharel, Karen Bobier, Lorraine Rodriguez, Myong Koag, Chun Wang, Rachel Lee, Grace Kubin |
| EPI_ISL_2443340, EPI_ISL_2443349, EPI_ISL_2443351, EPI_ISL_2443357 | Wyoming Public Health Laboratory | Wyoming Public Health Laboratory | Jim Mildenberger, Taylor Fearing, Channing Weber, Ashley Norberg, Chayse Rowley, Marley Goetz, Brian Dominguez, Elliot Thomasson, Sam Britz, Cari Sloma, Robert Petit, and Rob Christensen |
| EPI_ISL_2443465 | Washington State Department of Health Public Health Laboratories | Washington State Department of Health Public Health Laboratories | Drew MacKellar, Philip Dykema, Denny Russell, Joenice Gonzalez, Hannah Gray, Geoff Melly, Vanessa De Los Santos, Darren Lucas, JohnAric Peterson, Avi Singh, Rebecca Cao |
| EPI_ISL_2443466, EPI_ISL_2443467, EPI_ISL_2443468, EPI_ISL_2443469, EPI_ISL_2443470, EPI_ISL_2443471, EPI_ISL_2443472, EPI_ISL_2443474, EPI_ISL_2443475, EPI_ISL_2443476, EPI_ISL_2443477, EPI_ISL_2443478, EPI_ISL_2443479, EPI_ISL_2443480, EPI_ISL_2443481, EPI_ISL_2443482 | | | |
| see above | Baylor Scott & White-Temple | Baylor Scott & White-Temple | Ari Rao, Linden Morales, Kimberly Walker, Marcus Volz, Shelby Johnson |
| EPI_ISL_2443483 | Baylor Scott & White-Grapevine | Baylor Scott & White-Temple | Ari Rao, Linden Morales, Kimberly Walker, Marcus Volz, Shelby Johnson |
| EPI_ISL_2443484, EPI_ISL_2443485, EPI_ISL_2443486, EPI_ISL_2443487, EPI_ISL_2443488, EPI_ISL_2443489, EPI_ISL_2443490, EPI_ISL_2443491, EPI_ISL_2443492, EPI_ISL_2443493, EPI_ISL_2443494, EPI_ISL_2443495, EPI_ISL_2443496, EPI_ISL_2443497, EPI_ISL_2443498, EPI_ISL_2443499, EPI_ISL_2443500, EPI_ISL_2443501, EPI_ISL_2443502, EPI_ISL_2443504, EPI_ISL_2443505, EPI_ISL_2443506, EPI_ISL_2443507, EPI_ISL_2443508, EPI_ISL_2443509, EPI_ISL_2443510, EPI_ISL_2443511, EPI_ISL_2443512, EPI_ISL_2443513, EPI_ISL_2443514, EPI_ISL_2443516, EPI_ISL_2443517, EPI_ISL_2443518, EPI_ISL_2443519, EPI_ISL_2443520, EPI_ISL_2443521, EPI_ISL_2443522, EPI_ISL_2443523, EPI_ISL_2443524, EPI_ISL_2443525, EPI_ISL_2443526, EPI_ISL_2443527 | | | |
| see above | Baylor Scott & White-Temple | Baylor Scott & White-Temple | Ari Rao, Linden Morales, Kimberly Walker, Marcus Volz, Shelby Johnson |
| EPI_ISL_2443713, EPI_ISL_2443714, EPI_ISL_2443715, EPI_ISL_2443717, EPI_ISL_2443718, EPI_ISL_2443719, EPI_ISL_2443720, EPI_ISL_2443721, EPI_ISL_2443722, EPI_ISL_2443723, EPI_ISL_2443724, EPI_ISL_2443725, EPI_ISL_2443726, EPI_ISL_2443728, EPI_ISL_2443729, EPI_ISL_2443730, EPI_ISL_2443732, EPI_ISL_2443735, EPI_ISL_2443736, EPI_ISL_2443737, EPI_ISL_2443738, EPI_ISL_2443742, EPI_ISL_2443743, EPI_ISL_2443744, EPI_ISL_2443745, EPI_ISL_2443746, EPI_ISL_2443747, EPI_ISL_2443748, EPI_ISL_2443749, EPI_ISL_2443750, EPI_ISL_2443751, EPI_ISL_2443752, EPI_ISL_2443753, EPI_ISL_2443754, EPI_ISL_2443755, EPI_ISL_2443756, EPI_ISL_2443757, EPI_ISL_2443758, EPI_ISL_2443759, EPI_ISL_2443761, EPI_ISL_2443762, EPI_ISL_2443763, EPI_ISL_2443764, EPI_ISL_2443765, EPI_ISL_2443766, EPI_ISL_2443767, EPI_ISL_2443768, EPI_ISL_2443769, EPI_ISL_2443771, EPI_ISL_2443772, EPI_ISL_2443773, EPI_ISL_2443774, EPI_ISL_2443776, EPI_ISL_2443777 | | | |
| see above | Orange County Public Health Laboratory | University of California, Irvine | Amanda N. Pinski, Ilhem Messaoudi |
| EPI_ISL_2443778, EPI_ISL_2443779, EPI_ISL_2443780, EPI_ISL_2443781 | The Ohio State University Applied Microbiology Services Laboratory | The Ohio State University Applied Microbiology Services Laboratory | Seth A. Faith PhD |
| EPI_ISL_2443782 | Stony Brook Medicine | New York Genome Center | Michael Zody, Andre Corvelo, Dayna M. Oschwald, Samantha Fennessey, Tom Maniatis , Sarah A. Halbohn, Alaa B. Salaheldin, Sarah A. Malik, Josue M. Flores Acuna, Dimitri Gnatenko, Silvia G. Spitzer, Margaret M. McGovern, Eric D. Spitzer, Richard R. Kew, Kenneth R. Shroyer, Jiang Chen, Susan V. Donelan |
| EPI_ISL_2443783, EPI_ISL_2443784, EPI_ISL_2443785, EPI_ISL_2443786, EPI_ISL_2443788, EPI_ISL_2443789, EPI_ISL_2443790, EPI_ISL_2443791, EPI_ISL_2443792 | Weill Cornell Medicine | New York Genome Center | Michael Zody, Andre Corvelo, Dayna M. Oschwald, Samantha Fennessey, Tom Maniatis, Melissa Cushing, Olivier Elemento, Margaret Elizabeth Ross, Chris Mason, Priya Velu, Hanna Rennert, Arryn Craney, Lars F Westblade |
| EPI_ISL_2443793, EPI_ISL_2443794 | Hackensack Medical Center | New York Genome Center | Michael Zody, Andre Corvelo, Dayna M. Oschwald, Samantha Fennessey, Tom Maniatis, Liang Chen, Jose Mediavilla, Marcus Cunningham, Kaelea Composto, Kar Chow, David Perlin, Barry Kreiswirth |
| EPI_ISL_2443795 | Weill Cornell Medicine | New York Genome Center | Michael Zody, Andre Corvelo, Dayna M. Oschwald, Samantha Fennessey, Tom Maniatis, Melissa Cushing, Olivier Elemento, Margaret Elizabeth Ross, Chris Mason, Priya Velu, Hanna Rennert, Arryn Craney, Lars F Westblade |
| EPI_ISL_2443796 | Maryland Genomics, Institute for Genome Sciences, University of Maryland School of Medicine | Maryland Genomics, Institute for Genome Sciences, University of Maryland School of Medicine | Tallon, Luke J; Sadzewicz, Lisa D; Humphrys, Mike; Ott, Sandra; Roussey, Holly; Mehta, Aditya; Vavikolanu, Kranthi; Fraser, Claire M; Ravel, Jacques |
| EPI_ISL_2443797 | Minnesota Department of Health, Public Health Laboratory | Minnesota Department of Health, Public Health Laboratory | Alexandra Lorentz, Jacob Garfin, Matt Plumb, and Xiong Wang |
| EPI_ISL_2443798 | M Health Fairview | Minnesota Department of Health, Public Health Laboratory | Alexandra Lorentz, Jacob Garfin, Matt Plumb, and Xiong Wang |
| EPI_ISL_2443799 | Minnesota Department of Health, Public Health Laboratory | Minnesota Department of Health, Public Health Laboratory | Alexandra Lorentz, Jacob Garfin, Matt Plumb, and Xiong Wang |
| EPI_ISL_2443800 | M Health Fairview | Minnesota Department of Health, Public Health Laboratory | Alexandra Lorentz, Jacob Garfin, Matt Plumb, and Xiong Wang |
| EPI_ISL_2443801 | Centracare Laboratory Services | Minnesota Department of Health, Public Health Laboratory | Alexandra Lorentz, Jacob Garfin, Matt Plumb, and Xiong Wang |
| EPI_ISL_2443802, EPI_ISL_2443803 | Minnesota Department of Health, Public Health Laboratory | Minnesota Department of Health, Public Health Laboratory | Alexandra Lorentz, Jacob Garfin, Matt Plumb, and Xiong Wang |
| EPI_ISL_2443804, EPI_ISL_2443805 | M Health Fairview | Minnesota Department of Health, Public Health Laboratory | Alexandra Lorentz, Jacob Garfin, Matt Plumb, and Xiong Wang |
| EPI_ISL_2443806, EPI_ISL_2443807, EPI_ISL_2443808, EPI_ISL_2443809, EPI_ISL_2443810 | Minnesota Department of Health, Public Health Laboratory | Minnesota Department of Health, Public Health Laboratory | Alexandra Lorentz, Jacob Garfin, Matt Plumb, and Xiong Wang |
| EPI_ISL_2443811, EPI_ISL_2443812, EPI_ISL_2443813, EPI_ISL_2443814, EPI_ISL_2443815, EPI_ISL_2443816, EPI_ISL_2443817, EPI_ISL_2443818, EPI_ISL_2443819, EPI_ISL_2443820, EPI_ISL_2443821, EPI_ISL_2443822, EPI_ISL_2443823, EPI_ISL_2443824 | | | |
| see above | M Health Fairview | Minnesota Department of Health, Public Health Laboratory | Alexandra Lorentz, Jacob Garfin, Matt Plumb, and Xiong Wang |
| EPI_ISL_2443825, | Minnesota Department of Health, Public | Minnesota Department of Health, Public | Alexandra Lorentz, Jacob Garfin, Matt Plumb, and Xiong Wang |

| | | | |
|---|---|---|---|
| EPI_ISL_2443826, EPI_ISL_2443827, EPI_ISL_2443828 | Health Laboratory | Health Laboratory | |
| EPI_ISL_2443829 | Centracare Laboratory Services | Minnesota Department of Health, Public Health Laboratory | Alexandra Lorentz, Jacob Garfin, Matt Plumb, and Xiong Wang |
| EPI_ISL_2443830 | Minnesota Department of Health, Public Health Laboratory | Minnesota Department of Health, Public Health Laboratory | Alexandra Lorentz, Jacob Garfin, Matt Plumb, and Xiong Wang |
| EPI_ISL_2443831, EPI_ISL_2443832, EPI_ISL_2443833, EPI_ISL_2443834, EPI_ISL_2443835 | M Health Fairview | Minnesota Department of Health, Public Health Laboratory | Alexandra Lorentz, Jacob Garfin, Matt Plumb, and Xiong Wang |
| EPI_ISL_2443836, EPI_ISL_2443840 | Minnesota Department of Health, Public Health Laboratory | Minnesota Department of Health, Public Health Laboratory | Alexandra Lorentz, Jacob Garfin, Matt Plumb, and Xiong Wang |
| EPI_ISL_2443841 | M Health Fairview | Minnesota Department of Health, Public Health Laboratory | Alexandra Lorentz, Jacob Garfin, Matt Plumb, and Xiong Wang |
| EPI_ISL_2443842 | Minnesota Department of Health, Public Health Laboratory | Minnesota Department of Health, Public Health Laboratory | Alexandra Lorentz, Jacob Garfin, Matt Plumb, and Xiong Wang |
| EPI_ISL_2443843, EPI_ISL_2443844, EPI_ISL_2443845, EPI_ISL_2443846, EPI_ISL_2443847, EPI_ISL_2443848, EPI_ISL_2443849, EPI_ISL_2443850 | M Health Fairview | Minnesota Department of Health, Public Health Laboratory | Alexandra Lorentz, Jacob Garfin, Matt Plumb, and Xiong Wang |
| EPI_ISL_2443851, EPI_ISL_2443852 | Minnesota Department of Health, Public Health Laboratory | Minnesota Department of Health, Public Health Laboratory | Alexandra Lorentz, Jacob Garfin, Matt Plumb, and Xiong Wang |
| EPI_ISL_2443853 | M Health Fairview | Minnesota Department of Health, Public Health Laboratory | Alexandra Lorentz, Jacob Garfin, Matt Plumb, and Xiong Wang |
| EPI_ISL_2443854 | Minnesota Department of Health, Public Health Laboratory | Minnesota Department of Health, Public Health Laboratory | Alexandra Lorentz, Jacob Garfin, Matt Plumb, and Xiong Wang |
| EPI_ISL_2443855, EPI_ISL_2443856, EPI_ISL_2443857, EPI_ISL_2443858, EPI_ISL_2443859, EPI_ISL_2443860, EPI_ISL_2443861 | Hackensack Medical Center | New York Genome Center | Michael Zody, Andre Corvelo, Dayna M. Oschwald, Samantha Fennessey, Tom Maniatis, Liang Chen, Jose Mediavilla, Marcus Cunningham, Kaelea Composto, Kar Chow, David Perlin, Barry Kreiswirth |
| EPI_ISL_2443874, EPI_ISL_2443875, EPI_ISL_2443876, EPI_ISL_2443877, EPI_ISL_2443879, EPI_ISL_2443880, EPI_ISL_2443885, EPI_ISL_2443892, EPI_ISL_2443902, EPI_ISL_2443904, EPI_ISL_2443908, EPI_ISL_2443909, EPI_ISL_2443917, EPI_ISL_2443922, EPI_ISL_2443929, EPI_ISL_2443930, EPI_ISL_2443932, EPI_ISL_2443934, EPI_ISL_2443936, EPI_ISL_2443942, EPI_ISL_2443948, EPI_ISL_2443954, EPI_ISL_2443961, EPI_ISL_2443963, EPI_ISL_2443965, EPI_ISL_2443972, EPI_ISL_2443973, EPI_ISL_2443974, EPI_ISL_2443983, EPI_ISL_2443984, EPI_ISL_2443988, EPI_ISL_2443993 | | | |
| see above | Notre Dame COVID Response and Testing Laboratory | Notre Dame Genomics & Bioinformatics Core Facility | Michael Pfrender, Joseph Sarro, Melissa Stephens |
| EPI_ISL_2444223, EPI_ISL_2444224, EPI_ISL_2444225, EPI_ISL_2444226, EPI_ISL_2444227, EPI_ISL_2444228, EPI_ISL_2444229, EPI_ISL_2444230, EPI_ISL_2444231, EPI_ISL_2444232, EPI_ISL_2444233, EPI_ISL_2444234, EPI_ISL_2444235, EPI_ISL_2444236, EPI_ISL_2444237, EPI_ISL_2444238, EPI_ISL_2444239, EPI_ISL_2444240, EPI_ISL_2444241, EPI_ISL_2444242, EPI_ISL_2444243, EPI_ISL_2444244 | | | |
| see above | Indiana State Department of Health | Quantigen Biosciences | Paul J. Chidress |
| EPI_ISL_2444398, EPI_ISL_2444399, EPI_ISL_2444400, EPI_ISL_2444401, EPI_ISL_2444402, EPI_ISL_2444403, EPI_ISL_2444404, EPI_ISL_2444405, EPI_ISL_2444406, EPI_ISL_2444407, EPI_ISL_2444409, EPI_ISL_2444410, EPI_ISL_2444411, EPI_ISL_2444412, EPI_ISL_2444413, EPI_ISL_2444414, EPI_ISL_2444415, EPI_ISL_2444416, EPI_ISL_2444417, EPI_ISL_2444418, EPI_ISL_2444419, EPI_ISL_2444420, EPI_ISL_2444421, EPI_ISL_2444422, EPI_ISL_2444424, EPI_ISL_2444425, EPI_ISL_2444426, EPI_ISL_2444427, EPI_ISL_2444428, EPI_ISL_2444429, EPI_ISL_2444430, EPI_ISL_2444431, EPI_ISL_2444432, EPI_ISL_2444433, EPI_ISL_2444435, EPI_ISL_2444436, EPI_ISL_2444437, EPI_ISL_2444438, EPI_ISL_2444439, EPI_ISL_2444440, EPI_ISL_2444441, EPI_ISL_2444442, EPI_ISL_2444443, EPI_ISL_2444444, EPI_ISL_2444445, EPI_ISL_2444446, EPI_ISL_2444447, EPI_ISL_2444448, EPI_ISL_2444449, EPI_ISL_2444452, EPI_ISL_2444453, EPI_ISL_2444454, EPI_ISL_2444455, EPI_ISL_2444456, EPI_ISL_2444457, EPI_ISL_2444459, EPI_ISL_2444460, EPI_ISL_2444461, EPI_ISL_2444462, EPI_ISL_2444463, EPI_ISL_2444466, EPI_ISL_2444467, EPI_ISL_2444468, EPI_ISL_2444469, EPI_ISL_2444470, EPI_ISL_2444472, EPI_ISL_2444474, EPI_ISL_2444475, EPI_ISL_2444476, EPI_ISL_2444477, EPI_ISL_2444478, EPI_ISL_2444479, EPI_ISL_2444480, EPI_ISL_2444481, EPI_ISL_2444482, EPI_ISL_2444483, EPI_ISL_2444484, EPI_ISL_2444485, EPI_ISL_2444486, EPI_ISL_2444487, EPI_ISL_2444489, EPI_ISL_2444490, EPI_ISL_2444491, EPI_ISL_2444492, EPI_ISL_2444493, EPI_ISL_2444494, EPI_ISL_2444495, EPI_ISL_2444496, EPI_ISL_2444498, EPI_ISL_2444499, EPI_ISL_2444500, EPI_ISL_2444501, EPI_ISL_2444502, EPI_ISL_2444504, EPI_ISL_2444505, EPI_ISL_2444506, EPI_ISL_2444507, EPI_ISL_2444508, EPI_ISL_2444509, EPI_ISL_2444510, EPI_ISL_2444511, EPI_ISL_2444512, EPI_ISL_2444513, EPI_ISL_2444514, EPI_ISL_2444515, EPI_ISL_2444516, EPI_ISL_2444518, EPI_ISL_2444519, EPI_ISL_2444520, EPI_ISL_2444522, EPI_ISL_2444523, EPI_ISL_2444524, EPI_ISL_2444525, EPI_ISL_2444526, EPI_ISL_2444527, EPI_ISL_2444528, EPI_ISL_2444530, EPI_ISL_2444531, EPI_ISL_2444532, EPI_ISL_2444533, EPI_ISL_2444534, EPI_ISL_2444535, EPI_ISL_2444536, EPI_ISL_2444538, EPI_ISL_2444539, EPI_ISL_2444540, EPI_ISL_2444542, EPI_ISL_2444543, EPI_ISL_2444544, EPI_ISL_2444545, EPI_ISL_2444546, EPI_ISL_2444547, EPI_ISL_2444549, EPI_ISL_2444550, EPI_ISL_2444551, EPI_ISL_2444552, EPI_ISL_2444554, EPI_ISL_2444556, EPI_ISL_2444558, EPI_ISL_2444559 | | | |
| see above | Salud Digna | Instituto Nacional de Medicina Genomica | Hidalgo-Miranda A, Cedro-Tanda A, Mendoza-Vargas A, Reyes-Grajeda JP, Abraham Campos-Romero, Moreno-Camacho José Luis, Rodriguez-Gallegos Jorge, Luna-Ruiz Marco, Gonzalez-Barrera D, Rangel-DeLeon D, Munguia-Garza P, Ramirez-Vega O, Escobar-Arrazola, M, Herrera-Montalvo LA. |
| EPI_ISL_2444563, EPI_ISL_2444564, EPI_ISL_2444566, EPI_ISL_2444570, EPI_ISL_2444571, EPI_ISL_2444572, EPI_ISL_2444573, EPI_ISL_2444574, EPI_ISL_2444577, EPI_ISL_2444578, EPI_ISL_2444579, EPI_ISL_2444580, EPI_ISL_2444581, EPI_ISL_2444582, EPI_ISL_2444583, EPI_ISL_2444584, EPI_ISL_2444585, EPI_ISL_2444586, EPI_ISL_2444587, EPI_ISL_2444588, EPI_ISL_2444589, EPI_ISL_2444590, EPI_ISL_2444591, EPI_ISL_2444592, EPI_ISL_2444593, EPI_ISL_2444595, EPI_ISL_2444596, EPI_ISL_2444598, EPI_ISL_2444599, EPI_ISL_2444600, EPI_ISL_2444601, EPI_ISL_2444602, EPI_ISL_2444603, EPI_ISL_2444604, EPI_ISL_2444605, EPI_ISL_2444606, EPI_ISL_2444607, EPI_ISL_2444608, EPI_ISL_2444609, EPI_ISL_2444610, EPI_ISL_2444611, EPI_ISL_2444612, EPI_ISL_2444613, EPI_ISL_2444614, EPI_ISL_2444615, EPI_ISL_2444616, EPI_ISL_2444617, EPI_ISL_2444618, EPI_ISL_2444619, EPI_ISL_2444622, EPI_ISL_2444623, EPI_ISL_2444624, EPI_ISL_2444625, EPI_ISL_2444626, EPI_ISL_2444627, EPI_ISL_2444628, EPI_ISL_2444629, EPI_ISL_2444630, EPI_ISL_2444631, EPI_ISL_2444632, EPI_ISL_2444634, EPI_ISL_2444636, EPI_ISL_2444638, EPI_ISL_2444639, EPI_ISL_2444641, EPI_ISL_2444642, EPI_ISL_2444643, EPI_ISL_2444644, EPI_ISL_2444645, EPI_ISL_2444646, EPI_ISL_2444648, EPI_ISL_2444649, EPI_ISL_2444650, EPI_ISL_2444651, EPI_ISL_2444652, EPI_ISL_2444653, EPI_ISL_2444655, EPI_ISL_2444656, EPI_ISL_2444657, EPI_ISL_2444658, EPI_ISL_2444659, EPI_ISL_2444660, EPI_ISL_2444661, EPI_ISL_2444662, EPI_ISL_2444663, EPI_ISL_2444664, EPI_ISL_2444665, EPI_ISL_2444666, EPI_ISL_2444667, EPI_ISL_2444668, EPI_ISL_2444669, EPI_ISL_2444670, EPI_ISL_2444671, EPI_ISL_2444673, EPI_ISL_2444674, EPI_ISL_2444675, EPI_ISL_2444676, EPI_ISL_2444677, EPI_ISL_2444678, EPI_ISL_2444679, EPI_ISL_2444681, EPI_ISL_2444682, EPI_ISL_2444683, EPI_ISL_2444684, EPI_ISL_2444688, EPI_ISL_2444690, EPI_ISL_2444691, EPI_ISL_2444692, EPI_ISL_2444693, EPI_ISL_2444694, EPI_ISL_2444695, EPI_ISL_2444696, EPI_ISL_2444697, EPI_ISL_2444698, EPI_ISL_2444699, EPI_ISL_2444703, EPI_ISL_2444705, EPI_ISL_2444706, EPI_ISL_2444707, EPI_ISL_2444708, EPI_ISL_2444709, EPI_ISL_2444711, EPI_ISL_2444712, EPI_ISL_2444713, EPI_ISL_2444715, EPI_ISL_2444717, EPI_ISL_2444719, EPI_ISL_2444720, EPI_ISL_2444721, EPI_ISL_2444722, EPI_ISL_2444723, EPI_ISL_2444724, EPI_ISL_2444726, EPI_ISL_2444727, EPI_ISL_2444728, EPI_ISL_2444729, EPI_ISL_2444730, EPI_ISL_2444731, EPI_ISL_2444732, EPI_ISL_2444733, EPI_ISL_2444735, EPI_ISL_2444738, EPI_ISL_2444739, EPI_ISL_2444740, EPI_ISL_2444742, EPI_ISL_2444744, EPI_ISL_2444746, EPI_ISL_2444747, EPI_ISL_2444752, EPI_ISL_2444753, EPI_ISL_2444755, EPI_ISL_2444756, EPI_ISL_2444757, EPI_ISL_2444758, EPI_ISL_2444759, EPI_ISL_2444760, EPI_ISL_2444761, EPI_ISL_2444763, EPI_ISL_2444764, EPI_ISL_2444766, EPI_ISL_2444767, EPI_ISL_2444768, EPI_ISL_2444769, EPI_ISL_2444771, EPI_ISL_2444772, EPI_ISL_2444773, EPI_ISL_2444774 | | | |
| see above | Wyoming Public Health Laboratory | Wyoming Public Health Laboratory | Jim Mildenberger, Taylor Fearing, Channing Weber, Ashley Norberg, Chayse Rowley, Marley Goetz, Brian Dominguez, Elliot Thomasson, Brittany Oher, Sam Britz, Cari Sloma, and Rob Christensen |
| EPI_ISL_2445013, EPI_ISL_2445015, EPI_ISL_2445016, EPI_ISL_2445025, EPI_ISL_2445026, EPI_ISL_2445028, EPI_ISL_2445036, EPI_ISL_2445040, EPI_ISL_2445041, EPI_ISL_2445045 | Arizona State Public Health Laboratory | Arizona State Public Health Laboratory | Trung Huynh, Jessica Escobar, Katherine Fullerton, Nobuko Fukushima, Stacy White, Linda Getsinger, Victor Waddell |
| EPI_ISL_2445617, EPI_ISL_2445618, EPI_ISL_2445619, EPI_ISL_2445620, EPI_ISL_2445621, EPI_ISL_2445622, EPI_ISL_2445623, EPI_ISL_2445624, EPI_ISL_2445625, EPI_ISL_2445626, EPI_ISL_2445627, EPI_ISL_2445628, EPI_ISL_2445629, EPI_ISL_2445630, EPI_ISL_2445631, EPI_ISL_2445632, EPI_ISL_2445633 | | | |
| see above | Humboldt County Public Health Laboratory | Humboldt County Public Health Laboratory | Jeremy Corrigan |
| EPI_ISL_2445967, EPI_ISL_2445968, EPI_ISL_2445970, EPI_ISL_2445971 | TriCore Reference Laboratories | Center for Global Health, University of New Mexico Health Sciences Center | Daryl Domman, Kurt Schwalm, Valerie Morley, Cecilia Thompson, Kendra Pesko, Karissa Culbreath, Darrell Dinwiddie |
| EPI_ISL_2445972, EPI_ISL_2445973, EPI_ISL_2445974 | Univeristy of New Mexico Hospital | Center for Global Health, University of New Mexico Health Sciences Center | Daryl Domman, Kurt Schwalm, Valerie Morley, Justin Bacca, Jon Femling, Darrell Dinwiddie |
| EPI_ISL_2446035, EPI_ISL_2446036, EPI_ISL_2446037, EPI_ISL_2446038, EPI_ISL_2446039, EPI_ISL_2446040, EPI_ISL_2446041, EPI_ISL_2446042, EPI_ISL_2446043, EPI_ISL_2446044, EPI_ISL_2446045, EPI_ISL_2446046, EPI_ISL_2446047, EPI_ISL_2446048, EPI_ISL_2446049, EPI_ISL_2446050, EPI_ISL_2446051, EPI_ISL_2446052, EPI_ISL_2446053, EPI_ISL_2446054, EPI_ISL_2446055, EPI_ISL_2446056, EPI_ISL_2446057, EPI_ISL_2446058, EPI_ISL_2446059, EPI_ISL_2446060, EPI_ISL_2446061, EPI_ISL_2446062, EPI_ISL_2446063, EPI_ISL_2446064, EPI_ISL_2446065, EPI_ISL_2446066, EPI_ISL_2446067, EPI_ISL_2446068, EPI_ISL_2446069, EPI_ISL_2446070, EPI_ISL_2446071, EPI_ISL_2446072, EPI_ISL_2446073, EPI_ISL_2446074, EPI_ISL_2446075, EPI_ISL_2446076, EPI_ISL_2446077, EPI_ISL_2446078, EPI_ISL_2446079, EPI_ISL_2446080, EPI_ISL_2446081, EPI_ISL_2446082, EPI_ISL_2446084, EPI_ISL_2446085, EPI_ISL_2446086, EPI_ISL_2446087, EPI_ISL_2446088, EPI_ISL_2446089, EPI_ISL_2446090, EPI_ISL_2446091, EPI_ISL_2446092, EPI_ISL_2446093, EPI_ISL_2446094, EPI_ISL_2446096, EPI_ISL_2446099, EPI_ISL_2446102, EPI_ISL_2446105, EPI_ISL_2446107, EPI_ISL_2446111, EPI_ISL_2446114, EPI_ISL_2446117, EPI_ISL_2446120 | | | |
| see above | Pandemic Response Lab - NYC | Pandemic Response Lab, R&D | Henry Lee, Michael Hammerling, Melissa Hopkins, Cybill del Castillo, Shinyoung Clair Kang, William Ward, Pradeep Bugga, Sol Rey, Dylan Law, Katharine Nelson, Haiping Hao, Jon Laurent |
| EPI_ISL_2448298 | MONTEFIORE MEDICAL CENTER LABORATORIES | Wadsworth Center, New York State Department of Health | Kirsten St. George, Daryl M. Lamson, Alexis Russell, Matthew Shudt, Melissa A Leisner, Jonathan Plitnick, Catharine Prussing, Navjot Singh, John Kelly, Erasmus Schneider, Erica Lasek-Nesselquist |
| EPI_ISL_2448299 | SUNY UPSTATE MEDICAL UNIVERSITY | Wadsworth Center, New York State Department of Health | Kirsten St. George, Daryl M. Lamson, Alexis Russell, Matthew Shudt, Melissa A Leisner, Jonathan Plitnick, Catharine Prussing, Navjot Singh, John Kelly, Erasmus Schneider, Erica Lasek-Nesselquist |
| EPI_ISL_2448300 | TEMPUS LABS INC | Wadsworth Center, New York State Department of Health | Kirsten St. George, Daryl M. Lamson, Alexis Russell, Matthew Shudt, Melissa A Leisner, Jonathan Plitnick, Catharine Prussing, Navjot Singh, John Kelly, Erasmus Schneider, Erica Lasek-Nesselquist |
| EPI_ISL_2448301 | WESTCHESTER MEDICAL CENTER | Wadsworth Center, New York State Department of Health | Kirsten St. George, Daryl M. Lamson, Alexis Russell, Matthew Shudt, Melissa A Leisner, Jonathan Plitnick, Catharine Prussing, Navjot Singh, John Kelly, Erasmus Schneider, Erica Lasek-Nesselquist |

| | | | |
|---|---|---|---|
| EPI_ISL_2448302 | MONTEFIORE MEDICAL CENTER LABORATORIES | Wadsworth Center, New York State Department of Health | Kirsten St. George, Daryl M. Lamson, Alexis Russell, Matthew Shudt, Melissa A Leisner, Jonathan Plitnick, Catharine Prussing, Navjot Singh, John Kelly, Erasmus Schneider, Erica Lasek-Nesselquist |
| EPI_ISL_2448303 | URMC LABS | Wadsworth Center, New York State Department of Health | Kirsten St. George, Daryl M. Lamson, Alexis Russell, Matthew Shudt, Melissa A Leisner, Jonathan Plitnick, Catharine Prussing, Navjot Singh, John Kelly, Erasmus Schneider, Erica Lasek-Nesselquist |
| EPI_ISL_2448304 | BOSTON HEART DIAGNOSTICS CORP | Wadsworth Center, New York State Department of Health | Kirsten St. George, Daryl M. Lamson, Alexis Russell, Matthew Shudt, Melissa A Leisner, Jonathan Plitnick, Catharine Prussing, Navjot Singh, John Kelly, Erasmus Schneider, Erica Lasek-Nesselquist |
| EPI_ISL_2448305 | TEMPUS LABS INC | Wadsworth Center, New York State Department of Health | Kirsten St. George, Daryl M. Lamson, Alexis Russell, Matthew Shudt, Melissa A Leisner, Jonathan Plitnick, Catharine Prussing, Navjot Singh, John Kelly, Erasmus Schneider, Erica Lasek-Nesselquist |
| EPI_ISL_2448306, EPI_ISL_2448307 | URMC LABS | Wadsworth Center, New York State Department of Health | Kirsten St. George, Daryl M. Lamson, Alexis Russell, Matthew Shudt, Melissa A Leisner, Jonathan Plitnick, Catharine Prussing, Navjot Singh, John Kelly, Erasmus Schneider, Erica Lasek-Nesselquist |
| EPI_ISL_2448308, EPI_ISL_2448309 | BOSTON HEART DIAGNOSTICS CORP | Wadsworth Center, New York State Department of Health | Kirsten St. George, Daryl M. Lamson, Alexis Russell, Matthew Shudt, Melissa A Leisner, Jonathan Plitnick, Catharine Prussing, Navjot Singh, John Kelly, Erasmus Schneider, Erica Lasek-Nesselquist |
| EPI_ISL_2448310 | STONY BROOK UNIVERSITY HOSPITAL | Wadsworth Center, New York State Department of Health | Kirsten St. George, Daryl M. Lamson, Alexis Russell, Matthew Shudt, Melissa A Leisner, Jonathan Plitnick, Catharine Prussing, Navjot Singh, John Kelly, Erasmus Schneider, Erica Lasek-Nesselquist |
| EPI_ISL_2448311 | BOSTON HEART DIAGNOSTICS CORP | Wadsworth Center, New York State Department of Health | Kirsten St. George, Daryl M. Lamson, Alexis Russell, Matthew Shudt, Melissa A Leisner, Jonathan Plitnick, Catharine Prussing, Navjot Singh, John Kelly, Erasmus Schneider, Erica Lasek-Nesselquist |
| EPI_ISL_2448312 | ALBANY MEDICAL CENTER | Wadsworth Center, New York State Department of Health | Kirsten St. George, Daryl M. Lamson, Alexis Russell, Matthew Shudt, Melissa A Leisner, Jonathan Plitnick, Catharine Prussing, Navjot Singh, John Kelly, Erasmus Schneider, Erica Lasek-Nesselquist |
| EPI_ISL_2448313, EPI_ISL_2448314, EPI_ISL_2448315, EPI_ISL_2448316, EPI_ISL_2448317 | BOSTON HEART DIAGNOSTICS CORP | Wadsworth Center, New York State Department of Health | Kirsten St. George, Daryl M. Lamson, Alexis Russell, Matthew Shudt, Melissa A Leisner, Jonathan Plitnick, Catharine Prussing, Navjot Singh, John Kelly, Erasmus Schneider, Erica Lasek-Nesselquist |
| EPI_ISL_2448318 | URMC LABS | Wadsworth Center, New York State Department of Health | Kirsten St. George, Daryl M. Lamson, Alexis Russell, Matthew Shudt, Melissa A Leisner, Jonathan Plitnick, Catharine Prussing, Navjot Singh, John Kelly, Erasmus Schneider, Erica Lasek-Nesselquist |
| EPI_ISL_2448319, EPI_ISL_2448320 | BOSTON HEART DIAGNOSTICS CORP | Wadsworth Center, New York State Department of Health | Kirsten St. George, Daryl M. Lamson, Alexis Russell, Matthew Shudt, Melissa A Leisner, Jonathan Plitnick, Catharine Prussing, Navjot Singh, John Kelly, Erasmus Schneider, Erica Lasek-Nesselquist |
| EPI_ISL_2448321 | URMC LABS | Wadsworth Center, New York State Department of Health | Kirsten St. George, Daryl M. Lamson, Alexis Russell, Matthew Shudt, Melissa A Leisner, Jonathan Plitnick, Catharine Prussing, Navjot Singh, John Kelly, Erasmus Schneider, Erica Lasek-Nesselquist |
| EPI_ISL_2450366 | TN Division of Laboratory Services | Genomics and Discovery, Respiratory Viruses Branch, Division of Viral Diseases, Centers for Disease Control and Prevention | Ying Tao, Yan Li, Jing Zhang, Anna Kelleher, Brian Lynch, Krista Queen, Anna Uehara, Peter Cook, Han Jia Justin Ng, Rachel Marine, Clinton R. Paden, Dhwani Batra, Haibin Wang, Tara Coalter, Jasmine Padilla, Morgan Davis, Mili Sheth, Sarah Nobles, Mark Burroughs, Justin Lee, Adam Retchless, Suxiang Tong |
| EPI_ISL_2450379, EPI_ISL_2450380, EPI_ISL_2450381, EPI_ISL_2450386, EPI_ISL_2450387, EPI_ISL_2450388, EPI_ISL_2450389, EPI_ISL_2450390, EPI_ISL_2450391, EPI_ISL_2450392, EPI_ISL_2450393, EPI_ISL_2450394, EPI_ISL_2450395, EPI_ISL_2450396, EPI_ISL_2450397, EPI_ISL_2450398, EPI_ISL_2450399, EPI_ISL_2450400, EPI_ISL_2450401, EPI_ISL_2450402, EPI_ISL_2450403, EPI_ISL_2450404, EPI_ISL_2450405, EPI_ISL_2450406, EPI_ISL_2450408, EPI_ISL_2450409, EPI_ISL_2450410, EPI_ISL_2450411, EPI_ISL_2450412, EPI_ISL_2450413, EPI_ISL_2450414, EPI_ISL_2450415 | | | |
| see above | US Air Force School of Aerospace Medicine | US Air Force School of Aerospace Medicine | Anthony Fries, Jennifer Meyer, William Gruner, Amanda Javorina, Carol Garrett, Sarah Purves, Clarise Starr, Elizabeth Macias |
| EPI_ISL_2450617, EPI_ISL_2450618, EPI_ISL_2450619, EPI_ISL_2450620, EPI_ISL_2450621, EPI_ISL_2450622, EPI_ISL_2450623, EPI_ISL_2450624, EPI_ISL_2450625, EPI_ISL_2450626, EPI_ISL_2450627, EPI_ISL_2450628, EPI_ISL_2450629, EPI_ISL_2450630, EPI_ISL_2450631, EPI_ISL_2450632, EPI_ISL_2450633, EPI_ISL_2450634, EPI_ISL_2450635, EPI_ISL_2450636, EPI_ISL_2450637, EPI_ISL_2450638, EPI_ISL_2450639, EPI_ISL_2450640, EPI_ISL_2450641, EPI_ISL_2450642, EPI_ISL_2450643, EPI_ISL_2450644, EPI_ISL_2450645, EPI_ISL_2450646, EPI_ISL_2450647, EPI_ISL_2450648, EPI_ISL_2450649, EPI_ISL_2450650, EPI_ISL_2450651, EPI_ISL_2450652, EPI_ISL_2450653, EPI_ISL_2450654, EPI_ISL_2450655, EPI_ISL_2450656, EPI_ISL_2450657, EPI_ISL_2450658, EPI_ISL_2450659, EPI_ISL_2450660, EPI_ISL_2450661, EPI_ISL_2450662, EPI_ISL_2450663, EPI_ISL_2450664, EPI_ISL_2450665, EPI_ISL_2450666, EPI_ISL_2450667, EPI_ISL_2450668, EPI_ISL_2450669, EPI_ISL_2450670, EPI_ISL_2450671, EPI_ISL_2450672, EPI_ISL_2450673, EPI_ISL_2450674, EPI_ISL_2450675, EPI_ISL_2450676, EPI_ISL_2450677, EPI_ISL_2450678, EPI_ISL_2450679, EPI_ISL_2450680, EPI_ISL_2450681, EPI_ISL_2450682, EPI_ISL_2450683, EPI_ISL_2450684, EPI_ISL_2450685, EPI_ISL_2450686, EPI_ISL_2450687, EPI_ISL_2450688, EPI_ISL_2450689, EPI_ISL_2450690 | | | |
| see above | Weill Cornell Medicine | New York Genome Center | Michael Zody, Andre Corvelo, Dayna M. Oschwald, Samantha Fennessey, Tom Maniatis, Melissa Cushing, Olivier Elemento, Margaret Elizabeth Ross, Chris Mason, Priya Velu, Hanna Rennert, Arryn Craney, Lars F Westblade |
| EPI_ISL_2450853, EPI_ISL_2450854, EPI_ISL_2450855, EPI_ISL_2450856, EPI_ISL_2450857, EPI_ISL_2450858, EPI_ISL_2450859, EPI_ISL_2450860, EPI_ISL_2450861, EPI_ISL_2450862, EPI_ISL_2450863, EPI_ISL_2450864, EPI_ISL_2450865, EPI_ISL_2450866, EPI_ISL_2450867, EPI_ISL_2450868, EPI_ISL_2450869, EPI_ISL_2450870, EPI_ISL_2450871, EPI_ISL_2450872, EPI_ISL_2450873, EPI_ISL_2450874, EPI_ISL_2450875, EPI_ISL_2450876, EPI_ISL_2450877, EPI_ISL_2450878, EPI_ISL_2450879, EPI_ISL_2450880, EPI_ISL_2450881, EPI_ISL_2450882, EPI_ISL_2450883, EPI_ISL_2450884, EPI_ISL_2450885, EPI_ISL_2450886, EPI_ISL_2450887, EPI_ISL_2450888, EPI_ISL_2450889, EPI_ISL_2450890, EPI_ISL_2450891, EPI_ISL_2450892, EPI_ISL_2450893, EPI_ISL_2450894, EPI_ISL_2450895, EPI_ISL_2450896, EPI_ISL_2450897, EPI_ISL_2450898, EPI_ISL_2450899, EPI_ISL_2450900, EPI_ISL_2450901, EPI_ISL_2450902, EPI_ISL_2450903, EPI_ISL_2450904, EPI_ISL_2450905, EPI_ISL_2450906, EPI_ISL_2450907, EPI_ISL_2450908, EPI_ISL_2450909, EPI_ISL_2450910, EPI_ISL_2450911, EPI_ISL_2450912, EPI_ISL_2450913, EPI_ISL_2450914, EPI_ISL_2450915, EPI_ISL_2450916, EPI_ISL_2450917, EPI_ISL_2450918, EPI_ISL_2450919, EPI_ISL_2450920, EPI_ISL_2450921, EPI_ISL_2450922, EPI_ISL_2450923, EPI_ISL_2450924, EPI_ISL_2450925, EPI_ISL_2450926, EPI_ISL_2450927, EPI_ISL_2450928, EPI_ISL_2450929, EPI_ISL_2450930, EPI_ISL_2450931, EPI_ISL_2450932, EPI_ISL_2450934, EPI_ISL_2450936, EPI_ISL_2450937, EPI_ISL_2450938, EPI_ISL_2450939, EPI_ISL_2450941, EPI_ISL_2450942, EPI_ISL_2450943, EPI_ISL_2450944, EPI_ISL_2450945, EPI_ISL_2450946, EPI_ISL_2450947, EPI_ISL_2450948, EPI_ISL_2450949, EPI_ISL_2450950, EPI_ISL_2450951, EPI_ISL_2450952, EPI_ISL_2450953, EPI_ISL_2450954, EPI_ISL_2450955, EPI_ISL_2450956, EPI_ISL_2450957, EPI_ISL_2450958, EPI_ISL_2450959, EPI_ISL_2450960, EPI_ISL_2450961, EPI_ISL_2450962, EPI_ISL_2450963, EPI_ISL_2450964, EPI_ISL_2450965, EPI_ISL_2450966, EPI_ISL_2450967, EPI_ISL_2450968, EPI_ISL_2450969, EPI_ISL_2450970, EPI_ISL_2450971, EPI_ISL_2450972, EPI_ISL_2450973, EPI_ISL_2450974, EPI_ISL_2450975, EPI_ISL_2450976, EPI_ISL_2450977, EPI_ISL_2450978, EPI_ISL_2450979, EPI_ISL_2450980, EPI_ISL_2450981, EPI_ISL_2450982, EPI_ISL_2450983, EPI_ISL_2450984, EPI_ISL_2450985, EPI_ISL_2450986, EPI_ISL_2450987, EPI_ISL_2450988, EPI_ISL_2450989, EPI_ISL_2450990, EPI_ISL_2450991, EPI_ISL_2450992, EPI_ISL_2450993, EPI_ISL_2450994, EPI_ISL_2450995, EPI_ISL_2450996, EPI_ISL_2450997, EPI_ISL_2450998, EPI_ISL_2450999, EPI_ISL_2451000, EPI_ISL_2451001, EPI_ISL_2451002, EPI_ISL_2451003, EPI_ISL_2451004, EPI_ISL_2451005, EPI_ISL_2451006, EPI_ISL_2451007, EPI_ISL_2451008, EPI_ISL_2451009, EPI_ISL_2451010, EPI_ISL_2451011, EPI_ISL_2451012, EPI_ISL_2451013, EPI_ISL_2451014, EPI_ISL_2451015, EPI_ISL_2451016, EPI_ISL_2451017, EPI_ISL_2451018, EPI_ISL_2451019, EPI_ISL_2451020, EPI_ISL_2451021, EPI_ISL_2451022, EPI_ISL_2451023, EPI_ISL_2451024, EPI_ISL_2451025, EPI_ISL_2451026, EPI_ISL_2451027, EPI_ISL_2451028, EPI_ISL_2451029, EPI_ISL_2451030, EPI_ISL_2451031, EPI_ISL_2451032, EPI_ISL_2451033, EPI_ISL_2451034, EPI_ISL_2451035, EPI_ISL_2451036, EPI_ISL_2451037, EPI_ISL_2451038, EPI_ISL_2451039, EPI_ISL_2451040, EPI_ISL_2451041, EPI_ISL_2451042, EPI_ISL_2451043, EPI_ISL_2451044, EPI_ISL_2451045, EPI_ISL_2451046, EPI_ISL_2451047, EPI_ISL_2451048, EPI_ISL_2451049, EPI_ISL_2451050, EPI_ISL_2451051, EPI_ISL_2451052, EPI_ISL_2451053, EPI_ISL_2451054, EPI_ISL_2451055, EPI_ISL_2451056, EPI_ISL_2451057, EPI_ISL_2451058, EPI_ISL_2451059, EPI_ISL_2451060, EPI_ISL_2451061, EPI_ISL_2451062, EPI_ISL_2451063, EPI_ISL_2451064, EPI_ISL_2451065, EPI_ISL_2451066, EPI_ISL_2451067, EPI_ISL_2451068, EPI_ISL_2451069, EPI_ISL_2451070, EPI_ISL_2451071, EPI_ISL_2451072, EPI_ISL_2451073, EPI_ISL_2451074, EPI_ISL_2451075, EPI_ISL_2451076, EPI_ISL_2451077, EPI_ISL_2451078, EPI_ISL_2451079, EPI_ISL_2451080, EPI_ISL_2451081, EPI_ISL_2451082, EPI_ISL_2451083, EPI_ISL_2451084, EPI_ISL_2451085, EPI_ISL_2451086, EPI_ISL_2451087, EPI_ISL_2451088, EPI_ISL_2451089, EPI_ISL_2451090, EPI_ISL_2451091, EPI_ISL_2451093, EPI_ISL_2451094, EPI_ISL_2451095, EPI_ISL_2451096, EPI_ISL_2451097, EPI_ISL_2451098, EPI_ISL_2451099, EPI_ISL_2451100, EPI_ISL_2451102, EPI_ISL_2451103, EPI_ISL_2451104, EPI_ISL_2451107, EPI_ISL_2451108, EPI_ISL_2451109, EPI_ISL_2451111, EPI_ISL_2451112, EPI_ISL_2451113, EPI_ISL_2451115, EPI_ISL_2451118, EPI_ISL_2451119, EPI_ISL_2451121, EPI_ISL_2451122, EPI_ISL_2451123, EPI_ISL_2451124, EPI_ISL_2451125, EPI_ISL_2451126, EPI_ISL_2451127, EPI_ISL_2451128, EPI_ISL_2451129, EPI_ISL_2451130, EPI_ISL_2451131, EPI_ISL_2451132, EPI_ISL_2451133, EPI_ISL_2451134, EPI_ISL_2451135, EPI_ISL_2451136, EPI_ISL_2451137, EPI_ISL_2451138, EPI_ISL_2451139, EPI_ISL_2451140, EPI_ISL_2451141, EPI_ISL_2451142, EPI_ISL_2451143, EPI_ISL_2451144, EPI_ISL_2451145, EPI_ISL_2451146, EPI_ISL_2451147, EPI_ISL_2451148, EPI_ISL_2451149, EPI_ISL_2451150, EPI_ISL_2451151, EPI_ISL_2451152, EPI_ISL_2451153, EPI_ISL_2451154, EPI_ISL_2451155, EPI_ISL_2451156, EPI_ISL_2451157, EPI_ISL_2451158, EPI_ISL_2451159, EPI_ISL_2451160, EPI_ISL_2451161, EPI_ISL_2451162, EPI_ISL_2451163, EPI_ISL_2451165, EPI_ISL_2451166, EPI_ISL_2451168, EPI_ISL_2451169, EPI_ISL_2451171, EPI_ISL_2451172, EPI_ISL_2451173, EPI_ISL_2451174, EPI_ISL_2451175, EPI_ISL_2451176, EPI_ISL_2451177, EPI_ISL_2451178, EPI_ISL_2451179, EPI_ISL_2451180, EPI_ISL_2451181, EPI_ISL_2451182, EPI_ISL_2451183, EPI_ISL_2451184, EPI_ISL_2451185, EPI_ISL_2451186, EPI_ISL_2451187, EPI_ISL_2451188, EPI_ISL_2451189, EPI_ISL_2451190, EPI_ISL_2451191, EPI_ISL_2451192, EPI_ISL_2451193, EPI_ISL_2451194, EPI_ISL_2451195, EPI_ISL_2451196, EPI_ISL_2451197, EPI_ISL_2451198, EPI_ISL_2451199, EPI_ISL_2451200, EPI_ISL_2451201, EPI_ISL_2451202, EPI_ISL_2451203, EPI_ISL_2451204, EPI_ISL_2451205, EPI_ISL_2451206, EPI_ISL_2451207, EPI_ISL_2451208, EPI_ISL_2451209, EPI_ISL_2451211, EPI_ISL_2451212, EPI_ISL_2451214, EPI_ISL_2451215, EPI_ISL_2451216, EPI_ISL_2451217, EPI_ISL_2451218, EPI_ISL_2451219, EPI_ISL_2451220, EPI_ISL_2451221, EPI_ISL_2451222, EPI_ISL_2451223, EPI_ISL_2451224, EPI_ISL_2451225, EPI_ISL_2451226, EPI_ISL_2451227, EPI_ISL_2451228, EPI_ISL_2451229, EPI_ISL_2451230, EPI_ISL_2451231, EPI_ISL_2451232, EPI_ISL_2451233, EPI_ISL_2451234, EPI_ISL_2451235, EPI_ISL_2451236, EPI_ISL_2451237, EPI_ISL_2451238, EPI_ISL_2451239, EPI_ISL_2451240, EPI_ISL_2451241, EPI_ISL_2451242, EPI_ISL_2451243, EPI_ISL_2451244, EPI_ISL_2451245, EPI_ISL_2451246, EPI_ISL_2451247, EPI_ISL_2451248, EPI_ISL_2451249, EPI_ISL_2451250, EPI_ISL_2451251, EPI_ISL_2451252, EPI_ISL_2451253, EPI_ISL_2451254, EPI_ISL_2451255, EPI_ISL_2451256, EPI_ISL_2451257, EPI_ISL_2451258, EPI_ISL_2451259, EPI_ISL_2451260, EPI_ISL_2451261, EPI_ISL_2451262, EPI_ISL_2451263, EPI_ISL_2451264 | | | |
| see above | Mako Medical | Centers for Disease Control and Prevention Division of Viral Diseases, Pathogen Discovery | Dakota Howard, Dhwani Batra, Peter W. Cook, Kara Moser, Adrian Paskey, Jason Caravas, Benjamin Rambo-Martin, Shatavia Morrison, Christopher Gulvick, Scott Sammons, Yvette Unoarumhi, Darlene Wagner, Matthew Schmerer,Matthew Tugwell, Lauren Moon, Clinton R. Paden, Duncan MacCannell |
| EPI_ISL_2451265, EPI_ISL_2451266, EPI_ISL_2451267, EPI_ISL_2451268, EPI_ISL_2451269, EPI_ISL_2451270, EPI_ISL_2451271, EPI_ISL_2451272, EPI_ISL_2451273, EPI_ISL_2451274, EPI_ISL_2451275, EPI_ISL_2451276, EPI_ISL_2451277, EPI_ISL_2451278, EPI_ISL_2451279, EPI_ISL_2451280, EPI_ISL_2451281, EPI_ISL_2451282, EPI_ISL_2451283, EPI_ISL_2451284, EPI_ISL_2451285, EPI_ISL_2451286, EPI_ISL_2451287, EPI_ISL_2451288, EPI_ISL_2451289, EPI_ISL_2451290, EPI_ISL_2451291, EPI_ISL_2451292, EPI_ISL_2451293, EPI_ISL_2451294, EPI_ISL_2451295, EPI_ISL_2451296, EPI_ISL_2451297, EPI_ISL_2451298, EPI_ISL_2451299, EPI_ISL_2451300, EPI_ISL_2451301, EPI_ISL_2451302, EPI_ISL_2451303, EPI_ISL_2451304, EPI_ISL_2451305, EPI_ISL_2451306, EPI_ISL_2451307, EPI_ISL_2451308, EPI_ISL_2451309, EPI_ISL_2451310, EPI_ISL_2451311, EPI_ISL_2451312, EPI_ISL_2451313, EPI_ISL_2451314, EPI_ISL_2451315, EPI_ISL_2451316, EPI_ISL_2451317, EPI_ISL_2451318, EPI_ISL_2451319, EPI_ISL_2451320, EPI_ISL_2451321, EPI_ISL_2451322, EPI_ISL_2451323, EPI_ISL_2451324, EPI_ISL_2451325, EPI_ISL_2451326, EPI_ISL_2451327, EPI_ISL_2451328, EPI_ISL_2451329, EPI_ISL_2451330, EPI_ISL_2451331, EPI_ISL_2451332, EPI_ISL_2451333, EPI_ISL_2451334, EPI_ISL_2451335, EPI_ISL_2451336, EPI_ISL_2451337, EPI_ISL_2451338, EPI_ISL_2451339, EPI_ISL_2451340, EPI_ISL_2451341, EPI_ISL_2451342, EPI_ISL_2451343, EPI_ISL_2451344, EPI_ISL_2451345, EPI_ISL_2451346, EPI_ISL_2451347, EPI_ISL_2451348, EPI_ISL_2451349, EPI_ISL_2451350, EPI_ISL_2451351, EPI_ISL_2451352, EPI_ISL_2451353, EPI_ISL_2451354, EPI_ISL_2451355, EPI_ISL_2451356, EPI_ISL_2451357, EPI_ISL_2451358, EPI_ISL_2451359, EPI_ISL_2451360, EPI_ISL_2451361, EPI_ISL_2451362, EPI_ISL_2451363, EPI_ISL_2451364, EPI_ISL_2451365, EPI_ISL_2451366, EPI_ISL_2451367, EPI_ISL_2451368, EPI_ISL_2451369, EPI_ISL_2451370, EPI_ISL_2451371, EPI_ISL_2451372, EPI_ISL_2451373, EPI_ISL_2451374, EPI_ISL_2451375, EPI_ISL_2451376, EPI_ISL_2451377, EPI_ISL_2451378, EPI_ISL_2451379, EPI_ISL_2451380, EPI_ISL_2451381, EPI_ISL_2451382, EPI_ISL_2451383, EPI_ISL_2451384, EPI_ISL_2451385, EPI_ISL_2451386, EPI_ISL_2451387, EPI_ISL_2451388, EPI_ISL_2451389, EPI_ISL_2451390, EPI_ISL_2451391, EPI_ISL_2451392, EPI_ISL_2451393, EPI_ISL_2451394, EPI_ISL_2451395, EPI_ISL_2451396, EPI_ISL_2451397, EPI_ISL_2451398, EPI_ISL_2451399, EPI_ISL_2451400, EPI_ISL_2451401, EPI_ISL_2451402, EPI_ISL_2451403, EPI_ISL_2451404, EPI_ISL_2451405, EPI_ISL_2451406, EPI_ISL_2451407, EPI_ISL_2451408, EPI_ISL_2451409, EPI_ISL_2451410, EPI_ISL_2451411, EPI_ISL_2451412, EPI_ISL_2451413, EPI_ISL_2451414, EPI_ISL_2451415, EPI_ISL_2451416, EPI_ISL_2451417, EPI_ISL_2451418, EPI_ISL_2451419, EPI_ISL_2451420, EPI_ISL_2451421, EPI_ISL_2451422, EPI_ISL_2451423, EPI_ISL_2451424, EPI_ISL_2451425, EPI_ISL_2451426, EPI_ISL_2451427, EPI_ISL_2451428, EPI_ISL_2451429, EPI_ISL_2451430, EPI_ISL_2451431, EPI_ISL_2451432, EPI_ISL_2451433, EPI_ISL_2451434, EPI_ISL_2451435, EPI_ISL_2451436, EPI_ISL_2451437, EPI_ISL_2451438, EPI_ISL_2451439, EPI_ISL_2451440, EPI_ISL_2451441, EPI_ISL_2451442, EPI_ISL_2451443, EPI_ISL_2451444, EPI_ISL_2451445, EPI_ISL_2451446, EPI_ISL_2451447, EPI_ISL_2451448, EPI_ISL_2451449, EPI_ISL_2451450, EPI_ISL_2451451, EPI_ISL_2451452, EPI_ISL_2451453, EPI_ISL_2451454, EPI_ISL_2451455, EPI_ISL_2451456, EPI_ISL_2451457, EPI_ISL_2451458, EPI_ISL_2451459, EPI_ISL_2451460, EPI_ISL_2451461, EPI_ISL_2451462, EPI_ISL_2451463, EPI_ISL_2451464, EPI_ISL_2451465, EPI_ISL_2451466, EPI_ISL_2451467, EPI_ISL_2451468, EPI_ISL_2451469, EPI_ISL_2451470, EPI_ISL_2451471, EPI_ISL_2451472, EPI_ISL_2451473, EPI_ISL_2451474, EPI_ISL_2451475, EPI_ISL_2451476, EPI_ISL_2451477, EPI_ISL_2451478, EPI_ISL_2451479, EPI_ISL_2451480, EPI_ISL_2451481, EPI_ISL_2451482, EPI_ISL_2451483, EPI_ISL_2451484, EPI_ISL_2451485, EPI_ISL_2451486, EPI_ISL_2451487, EPI_ISL_2451488, EPI_ISL_2451489, EPI_ISL_2451490 | | | |
| see above | Quest Diagnostics Incorporated | Centers for Disease Control and Prevention Division of Viral Diseases, Pathogen Discovery | Dakota Howard, Dhwani Batra, Peter W. Cook, Kara Moser, Adrian Paskey, Jason Caravas, Benjamin Rambo-Martin, Shatavia Morrison, Christopher Gulvick, Scott Sammons, Yvette Unoarumhi, Darlene Wagner, Matthew Schmerer, S. H. Rosenthal, A. Gerasimova, R. M. Kagan, B. Anderson, M. Hua, Y. Liu, L.E. Bernstein, K.E. Livingston, A. Perez, I. A. Shlyakhter, R. V. Rolando, R. Owen, P. Tanpaiboon, F. Lacbawan, Clinton R. Paden, Duncan MacCannell |
| EPI_ISL_2451491, EPI_ISL_2451492, EPI_ISL_2451493, EPI_ISL_2451494, EPI_ISL_2451495, EPI_ISL_2451496, EPI_ISL_2451497, EPI_ISL_2451498, EPI_ISL_2451499, EPI_ISL_2451500, EPI_ISL_2451501, EPI_ISL_2451502, EPI_ISL_2451503, EPI_ISL_2451504, EPI_ISL_2451505, EPI_ISL_2451506 | | | |
| see above | Helix/Illumina | Centers for Disease Control and Prevention Division of Viral Diseases, Pathogen Discovery | Dakota Howard, Dhwani Batra, Peter W. Cook, Kara Moser, Adrian Paskey, Jason Caravas, Benjamin Rambo-Martin, Shatavia Morrison, Christopher Gulvick, Scott Sammons, Yvette Unoarumhi, Darlene Wagner, Matthew Schmerer, Eileen de Feo, Jan Antico, Christine Tran, Matthew Tolentino, Shannon Wickline, Kim Gietzen, Brad Sickler, Jingtao Liu, Eric Allen, Phil Febbo, Nicole L. Washington, Simon White, Geraint Levan, Kelly Schiabor Barrett, Elizabeth Cirulli, Alexandre Bolze, Ary Ascencio, Charlotte Rivera-Garcia, Ryan Cho, Jason Nguyen, Sherry Wang, Jimmy Ramirez, Tyler Cassens, Efren Sandoval, Magnus Isaksson, William Lee, David Becker, Marc Laurent, James Lu, Clinton R. Paden, Duncan MacCannell |
| EPI_ISL_2451507, EPI_ISL_2451508, EPI_ISL_2451509, EPI_ISL_2451510, EPI_ISL_2451511, EPI_ISL_2451512, EPI_ISL_2451513, EPI_ISL_2451514, EPI_ISL_2451515, EPI_ISL_2451516, EPI_ISL_2451517, EPI_ISL_2451518, EPI_ISL_2451519, EPI_ISL_2451520, EPI_ISL_2451521, EPI_ISL_2451522, EPI_ISL_2451523, EPI_ISL_2451524, EPI_ISL_2451525, EPI_ISL_2451526, EPI_ISL_2451527, EPI_ISL_2451528, EPI_ISL_2451529, EPI_ISL_2451530, EPI_ISL_2451531, EPI_ISL_2451532, EPI_ISL_2451533, EPI_ISL_2451534, EPI_ISL_2451535, EPI_ISL_2451536, EPI_ISL_2451537, EPI_ISL_2451538, EPI_ISL_2451539, EPI_ISL_2451540, EPI_ISL_2451541, EPI_ISL_2451542, EPI_ISL_2451543, EPI_ISL_2451544, EPI_ISL_2451545, EPI_ISL_2451546, EPI_ISL_2451547, EPI_ISL_2451548, EPI_ISL_2451549, EPI_ISL_2451550, EPI_ISL_2451551, EPI_ISL_2451552, EPI_ISL_2451553, EPI_ISL_2451554, EPI_ISL_2451555, EPI_ISL_2451556, EPI_ISL_2451557, EPI_ISL_2451558, EPI_ISL_2451559, EPI_ISL_2451560, EPI_ISL_2451561, EPI_ISL_2451562, EPI_ISL_2451563, EPI_ISL_2451564, EPI_ISL_2451565, EPI_ISL_2451566 | | | |
| see above | Quest Diagnostics Incorporated | Centers for Disease Control and Prevention Division of Viral Diseases, Pathogen Discovery | Dakota Howard, Dhwani Batra, Peter W. Cook, Kara Moser, Adrian Paskey, Jason Caravas, Benjamin Rambo-Martin, Shatavia Morrison, Christopher Gulvick, Scott Sammons, Yvette Unoarumhi, Darlene Wagner, Matthew Schmerer, S. H. Rosenthal, A. Gerasimova, R. M. Kagan, B. Anderson, M. Hua, Y. Liu, L.E. Bernstein, K.E. Livingston, A. Perez, I. A. Shlyakhter, R. V. Rolando, R. Owen, P. Tanpaiboon, F. Lacbawan, Clinton R. Paden, Duncan MacCannell |
| EPI_ISL_2451567, EPI_ISL_2451568, EPI_ISL_2451569, EPI_ISL_2451570, EPI_ISL_2451571, EPI_ISL_2451572, EPI_ISL_2451573 | MN PHL Division, Minnesota Department of Health | Centers for Disease Control and Prevention Division of Viral Diseases, Pathogen Discovery | Mili Sheth, Sarah Nobles, Jasmine Padilla, Mark Burroughs, Shoshona Le, Katie Dillon, Peter Cook, Clinton R. Paden, Dhwani Batra, Krista Queen, Kristen Knipe, Dakota Howard, Yvette Unoarumhi, Darlene Wagner, Matthew Schmerer, Ben L. Rambo-Martin, Kristine Lacek, Sam Shepard, Alison Laufer Halpin, Dave Wentworth, Vivien Dugan, Suxiang Tong, Justin Lee |
| EPI_ISL_2451574, EPI_ISL_2451575 | OK Public Health Laboratory, Oklahoma State DOH | Centers for Disease Control and Prevention Division of Viral Diseases, Pathogen Discovery | Mili Sheth, Sarah Nobles, Jasmine Padilla, Mark Burroughs, Shoshona Le, Katie Dillon, Peter Cook, Clinton R. Paden, Dhwani Batra, Krista Queen, Kristen Knipe, Dakota Howard, Yvette Unoarumhi, Darlene Wagner, Matthew Schmerer, Ben L. Rambo-Martin, Kristine Lacek, Sam Shepard, Alison Laufer Halpin, Dave Wentworth, Vivien Dugan, Suxiang Tong, Justin Lee |
| EPI_ISL_2451576, EPI_ISL_2451577, | KS Health and Environmental Laboratories | Centers for Disease Control and Prevention Division of Viral Diseases, Pathogen | Mili Sheth, Sarah Nobles, Jasmine Padilla, Mark Burroughs, Shoshona Le, Katie Dillon, Peter Cook, Clinton R. Paden, Dhwani Batra, Krista Queen, Kristen Knipe, Dakota Howard, Yvette Unoarumhi, Darlene Wagner, Matthew Schmerer, Ben L. Rambo-Martin, Kristine Lacek, Sam Shepard, Alison Laufer Halpin, Dave Wentworth, Vivien Dugan, Suxiang Tong, Justin Lee |

| | | | |
|---|---|---|---|
| EPI_ISL_2451578, EPI_ISL_2451579, EPI_ISL_2451580, EPI_ISL_2451581, EPI_ISL_2451582, EPI_ISL_2451583 | | Discovery | |
| EPI_ISL_2451584 | AZ SPHL, Arizona Department of Health Services | Centers for Disease Control and Prevention Division of Viral Diseases, Pathogen Discovery | Mili Sheth, Sarah Nobles, Jasmine Padilla, Mark Burroughs, Shoshona Le, Katie Dillon, Peter Cook, Clinton R. Paden, Dhwani Batra, Krista Queen, Kristen Knipe, Dakota Howard, Yvette Unoarumhi, Darlene Wagner, Matthew Schmerer, Ben L. Rambo-Martin, Kristine Lacek, Sam Shepard, Alison Laufer Halpin, Dave Wentworth, Vivien Dugan, Suxiang Tong, Justin Lee |
| EPI_ISL_2451585, EPI_ISL_2451586, EPI_ISL_2451587, EPI_ISL_2451588, EPI_ISL_2451589, EPI_ISL_2451590, EPI_ISL_2451591, EPI_ISL_2451592, EPI_ISL_2451593, EPI_ISL_2451594, EPI_ISL_2451595, EPI_ISL_2451596, EPI_ISL_2451597, EPI_ISL_2451598, EPI_ISL_2451599, EPI_ISL_2451600, EPI_ISL_2451601, EPI_ISL_2451602, EPI_ISL_2451603 | | | |
| see above | NV-Southern Nevada Public Health Laboratory | Centers for Disease Control and Prevention Division of Viral Diseases, Pathogen Discovery | Mili Sheth, Sarah Nobles, Jasmine Padilla, Mark Burroughs, Shoshona Le, Katie Dillon, Peter Cook, Clinton R. Paden, Dhwani Batra, Krista Queen, Kristen Knipe, Dakota Howard, Yvette Unoarumhi, Darlene Wagner, Matthew Schmerer, Ben L. Rambo-Martin, Kristine Lacek, Sam Shepard, Alison Laufer Halpin, Dave Wentworth, Vivien Dugan, Suxiang Tong, Justin Lee |
| EPI_ISL_2451604, EPI_ISL_2451605, EPI_ISL_2451606 | Philadelphia Department of Public Health | Centers for Disease Control and Prevention Division of Viral Diseases, Pathogen Discovery | Mili Sheth, Sarah Nobles, Jasmine Padilla, Mark Burroughs, Shoshona Le, Katie Dillon, Peter Cook, Clinton R. Paden, Dhwani Batra, Krista Queen, Kristen Knipe, Dakota Howard, Yvette Unoarumhi, Darlene Wagner, Matthew Schmerer, Ben L. Rambo-Martin, Kristine Lacek, Sam Shepard, Alison Laufer Halpin, Dave Wentworth, Vivien Dugan, Suxiang Tong, Justin Lee |
| EPI_ISL_2451607, EPI_ISL_2451608, EPI_ISL_2451609, EPI_ISL_2451610, EPI_ISL_2451611, EPI_ISL_2451612, EPI_ISL_2451613, EPI_ISL_2451614, EPI_ISL_2451615, EPI_ISL_2451616, EPI_ISL_2451617, EPI_ISL_2451618, EPI_ISL_2451619, EPI_ISL_2451621, EPI_ISL_2451624 | | | |
| see above | MI - Michigan Department of Health and Human Services - Bureau of Laboratories | Centers for Disease Control and Prevention Division of Viral Diseases, Pathogen Discovery | Mili Sheth, Sarah Nobles, Jasmine Padilla, Mark Burroughs, Shoshona Le, Katie Dillon, Peter Cook, Clinton R. Paden, Dhwani Batra, Krista Queen, Kristen Knipe, Dakota Howard, Yvette Unoarumhi, Darlene Wagner, Matthew Schmerer, Ben L. Rambo-Martin, Kristine Lacek, Sam Shepard, Alison Laufer Halpin, Dave Wentworth, Vivien Dugan, Suxiang Tong, Justin Lee |
| EPI_ISL_2451627, EPI_ISL_2451631, EPI_ISL_2451634, EPI_ISL_2451637, EPI_ISL_2451640, EPI_ISL_2451643, EPI_ISL_2451646 | MT Public Health Laboratory | Centers for Disease Control and Prevention Division of Viral Diseases, Pathogen Discovery | Mili Sheth, Sarah Nobles, Jasmine Padilla, Mark Burroughs, Shoshona Le, Katie Dillon, Peter Cook, Clinton R. Paden, Dhwani Batra, Krista Queen, Kristen Knipe, Dakota Howard, Yvette Unoarumhi, Darlene Wagner, Matthew Schmerer, Ben L. Rambo-Martin, Kristine Lacek, Sam Shepard, Alison Laufer Halpin, Dave Wentworth, Vivien Dugan, Suxiang Tong, Justin Lee |
| EPI_ISL_2451649, EPI_ISL_2451652, EPI_ISL_2451655, EPI_ISL_2451659, EPI_ISL_2451662, EPI_ISL_2451664, EPI_ISL_2451667, EPI_ISL_2451671, EPI_ISL_2451674, EPI_ISL_2451676, EPI_ISL_2451679, EPI_ISL_2451682, EPI_ISL_2451686, EPI_ISL_2451688, EPI_ISL_2451690, EPI_ISL_2451693, EPI_ISL_2451697 | | | |
| see above | Guam Public Health Laboratory | Centers for Disease Control and Prevention Division of Viral Diseases, Pathogen Discovery | Mili Sheth, Sarah Nobles, Jasmine Padilla, Mark Burroughs, Shoshona Le, Katie Dillon, Peter Cook, Clinton R. Paden, Dhwani Batra, Krista Queen, Kristen Knipe, Dakota Howard, Yvette Unoarumhi, Darlene Wagner, Matthew Schmerer, Ben L. Rambo-Martin, Kristine Lacek, Sam Shepard, Alison Laufer Halpin, Dave Wentworth, Vivien Dugan, Suxiang Tong, Justin Lee |
| EPI_ISL_2451700, EPI_ISL_2451703, EPI_ISL_2451706, EPI_ISL_2451710, EPI_ISL_2451713, EPI_ISL_2451716, EPI_ISL_2451720 | ME Health and Environmental Testing Laboratory | Centers for Disease Control and Prevention Division of Viral Diseases, Pathogen Discovery | Mili Sheth, Sarah Nobles, Jasmine Padilla, Mark Burroughs, Shoshona Le, Katie Dillon, Peter Cook, Clinton R. Paden, Dhwani Batra, Krista Queen, Kristen Knipe, Dakota Howard, Yvette Unoarumhi, Darlene Wagner, Matthew Schmerer, Ben L. Rambo-Martin, Kristine Lacek, Sam Shepard, Alison Laufer Halpin, Dave Wentworth, Vivien Dugan, Suxiang Tong, Justin Lee |
| EPI_ISL_2451723, EPI_ISL_2451726, EPI_ISL_2451729 | TX-Houston Health Department Laboratory | Centers for Disease Control and Prevention Division of Viral Diseases, Pathogen Discovery | Mili Sheth, Sarah Nobles, Jasmine Padilla, Mark Burroughs, Shoshona Le, Katie Dillon, Peter Cook, Clinton R. Paden, Dhwani Batra, Krista Queen, Kristen Knipe, Dakota Howard, Yvette Unoarumhi, Darlene Wagner, Matthew Schmerer, Ben L. Rambo-Martin, Kristine Lacek, Sam Shepard, Alison Laufer Halpin, Dave Wentworth, Vivien Dugan, Suxiang Tong, Justin Lee |
| EPI_ISL_2451732, EPI_ISL_2451736 | SD Public Health Laboratory | Centers for Disease Control and Prevention Division of Viral Diseases, Pathogen Discovery | Mili Sheth, Sarah Nobles, Jasmine Padilla, Mark Burroughs, Shoshona Le, Katie Dillon, Peter Cook, Clinton R. Paden, Dhwani Batra, Krista Queen, Kristen Knipe, Dakota Howard, Yvette Unoarumhi, Darlene Wagner, Matthew Schmerer, Ben L. Rambo-Martin, Kristine Lacek, Sam Shepard, Alison Laufer Halpin, Dave Wentworth, Vivien Dugan, Suxiang Tong, Justin Lee |
| EPI_ISL_2451739, EPI_ISL_2451741, EPI_ISL_2451743, EPI_ISL_2451744, EPI_ISL_2451746, EPI_ISL_2451748, EPI_ISL_2451750 | CT-Dr. Katherine A. Kelley State Public Health Lab | Centers for Disease Control and Prevention Division of Viral Diseases, Pathogen Discovery | Mili Sheth, Sarah Nobles, Jasmine Padilla, Mark Burroughs, Shoshona Le, Katie Dillon, Peter Cook, Clinton R. Paden, Dhwani Batra, Krista Queen, Kristen Knipe, Dakota Howard, Yvette Unoarumhi, Darlene Wagner, Matthew Schmerer, Ben L. Rambo-Martin, Kristine Lacek, Sam Shepard, Alison Laufer Halpin, Dave Wentworth, Vivien Dugan, Suxiang Tong, Justin Lee |
| EPI_ISL_2451752, EPI_ISL_2451754, EPI_ISL_2451755, EPI_ISL_2451757 | FL Bur. of Public Health Laboratories-Jacksonville | Centers for Disease Control and Prevention Division of Viral Diseases, Pathogen Discovery | Mili Sheth, Sarah Nobles, Jasmine Padilla, Mark Burroughs, Shoshona Le, Katie Dillon, Peter Cook, Clinton R. Paden, Dhwani Batra, Krista Queen, Kristen Knipe, Dakota Howard, Yvette Unoarumhi, Darlene Wagner, Matthew Schmerer, Ben L. Rambo-Martin, Kristine Lacek, Sam Shepard, Alison Laufer Halpin, Dave Wentworth, Vivien Dugan, Suxiang Tong, Justin Lee |
| EPI_ISL_2451759, EPI_ISL_2451761, EPI_ISL_2451763, EPI_ISL_2451765, EPI_ISL_2451766, EPI_ISL_2451768, EPI_ISL_2451770, EPI_ISL_2451772, EPI_ISL_2451775 | DC Public Health Lab/ Dept. of Forensic Sciences | Centers for Disease Control and Prevention Division of Viral Diseases, Pathogen Discovery | Mili Sheth, Sarah Nobles, Jasmine Padilla, Mark Burroughs, Shoshona Le, Katie Dillon, Peter Cook, Clinton R. Paden, Dhwani Batra, Krista Queen, Kristen Knipe, Dakota Howard, Yvette Unoarumhi, Darlene Wagner, Matthew Schmerer, Ben L. Rambo-Martin, Kristine Lacek, Sam Shepard, Alison Laufer Halpin, Dave Wentworth, Vivien Dugan, Suxiang Tong, Justin Lee |
| EPI_ISL_2451777, EPI_ISL_2451779, EPI_ISL_2451780, EPI_ISL_2451782, EPI_ISL_2451784, EPI_ISL_2451785 | AR Dept. of Health-PHL, Molecular Diagnostics | Centers for Disease Control and Prevention Division of Viral Diseases, Pathogen Discovery | Mili Sheth, Sarah Nobles, Jasmine Padilla, Mark Burroughs, Shoshona Le, Katie Dillon, Peter Cook, Clinton R. Paden, Dhwani Batra, Krista Queen, Kristen Knipe, Dakota Howard, Yvette Unoarumhi, Darlene Wagner, Matthew Schmerer, Ben L. Rambo-Martin, Kristine Lacek, Sam Shepard, Alison Laufer Halpin, Dave Wentworth, Vivien Dugan, Suxiang Tong, Justin Lee |
| EPI_ISL_2451787, EPI_ISL_2451789, EPI_ISL_2451791, EPI_ISL_2451792, EPI_ISL_2451794, EPI_ISL_2451796, EPI_ISL_2451798, EPI_ISL_2451799, EPI_ISL_2451801, EPI_ISL_2451803, EPI_ISL_2451804, EPI_ISL_2451806, EPI_ISL_2451808, EPI_ISL_2451809, EPI_ISL_2451811 | | | |
| see above | IL Dept. of Public Health Springfield Laboratory | Centers for Disease Control and Prevention Division of Viral Diseases, Pathogen Discovery | Mili Sheth, Sarah Nobles, Jasmine Padilla, Mark Burroughs, Shoshona Le, Katie Dillon, Peter Cook, Clinton R. Paden, Dhwani Batra, Krista Queen, Kristen Knipe, Dakota Howard, Yvette Unoarumhi, Darlene Wagner, Matthew Schmerer, Ben L. Rambo-Martin, Kristine Lacek, Sam Shepard, Alison Laufer Halpin, Dave Wentworth, Vivien Dugan, Suxiang Tong, Justin Lee |
| EPI_ISL_2451813, EPI_ISL_2451815, EPI_ISL_2451818, EPI_ISL_2451820, EPI_ISL_2451821, EPI_ISL_2451823, EPI_ISL_2451825, EPI_ISL_2451827, EPI_ISL_2451829 | MA State Public Health Laboratory | Centers for Disease Control and Prevention Division of Viral Diseases, Pathogen Discovery | Mili Sheth, Sarah Nobles, Jasmine Padilla, Mark Burroughs, Shoshona Le, Katie Dillon, Peter Cook, Clinton R. Paden, Dhwani Batra, Krista Queen, Kristen Knipe, Dakota Howard, Yvette Unoarumhi, Darlene Wagner, Matthew Schmerer, Ben L. Rambo-Martin, Kristine Lacek, Sam Shepard, Alison Laufer Halpin, Dave Wentworth, Vivien Dugan, Suxiang Tong, Justin Lee |
| EPI_ISL_2451831, EPI_ISL_2451833, EPI_ISL_2451834, EPI_ISL_2451836, EPI_ISL_2451838, EPI_ISL_2451839, EPI_ISL_2451841, EPI_ISL_2451843, EPI_ISL_2451845, EPI_ISL_2451846, EPI_ISL_2451848, EPI_ISL_2451850, EPI_ISL_2451852, EPI_ISL_2451854, EPI_ISL_2451856, EPI_ISL_2451858, EPI_ISL_2451859, EPI_ISL_2451861, EPI_ISL_2451862, EPI_ISL_2451864, EPI_ISL_2451866, EPI_ISL_2451868, EPI_ISL_2451869, EPI_ISL_2451871, EPI_ISL_2451873 | | | |
| see above | VA-Division of Consolidated Laboratory Services | Centers for Disease Control and Prevention Division of Viral Diseases, Pathogen Discovery | Mili Sheth, Sarah Nobles, Jasmine Padilla, Mark Burroughs, Shoshona Le, Katie Dillon, Peter Cook, Clinton R. Paden, Dhwani Batra, Krista Queen, Kristen Knipe, Dakota Howard, Yvette Unoarumhi, Darlene Wagner, Matthew Schmerer, Ben L. Rambo-Martin, Kristine Lacek, Sam Shepard, Alison Laufer Halpin, Dave Wentworth, Vivien Dugan, Suxiang Tong, Justin Lee |
| EPI_ISL_2451874, EPI_ISL_2451876, EPI_ISL_2451878 | NJ Public Health and Environmental Laboratories | Centers for Disease Control and Prevention Division of Viral Diseases, Pathogen Discovery | Mili Sheth, Sarah Nobles, Jasmine Padilla, Mark Burroughs, Shoshona Le, Katie Dillon, Peter Cook, Clinton R. Paden, Dhwani Batra, Krista Queen, Kristen Knipe, Dakota Howard, Yvette Unoarumhi, Darlene Wagner, Matthew Schmerer, Ben L. Rambo-Martin, Kristine Lacek, Sam Shepard, Alison Laufer Halpin, Dave Wentworth, Vivien Dugan, Suxiang Tong, Justin Lee |
| EPI_ISL_2451879 | WVDHHR - Office of Laboratory Services | Centers for Disease Control and Prevention Division of Viral Diseases, Pathogen Discovery | Mili Sheth, Sarah Nobles, Jasmine Padilla, Mark Burroughs, Shoshona Le, Katie Dillon, Peter Cook, Clinton R. Paden, Dhwani Batra, Krista Queen, Kristen Knipe, Dakota Howard, Yvette Unoarumhi, Darlene Wagner, Matthew Schmerer, Ben L. Rambo-Martin, Kristine Lacek, Sam Shepard, Alison Laufer Halpin, Dave Wentworth, Vivien Dugan, Suxiang Tong, Justin Lee |
| EPI_ISL_2451881, EPI_ISL_2451883, EPI_ISL_2451884, EPI_ISL_2451886, EPI_ISL_2451888, EPI_ISL_2451890, | CA-Los Angeles County Public Health Laboratory | Centers for Disease Control and Prevention Division of Viral Diseases, Pathogen Discovery | Mili Sheth, Sarah Nobles, Jasmine Padilla, Mark Burroughs, Shoshona Le, Katie Dillon, Peter Cook, Clinton R. Paden, Dhwani Batra, Krista Queen, Kristen Knipe, Dakota Howard, Yvette Unoarumhi, Darlene Wagner, Matthew Schmerer, Ben L. Rambo-Martin, Kristine Lacek, Sam Shepard, Alison Laufer Halpin, Dave Wentworth, Vivien Dugan, Suxiang Tong, Justin Lee |

| | | | |
|---|---|---|---|
| EPI_ISL_2451891, EPI_ISL_2451893, EPI_ISL_2451895 | | | |
| EPI_ISL_2451896, EPI_ISL_2451898, EPI_ISL_2451900, EPI_ISL_2451902, EPI_ISL_2451903, EPI_ISL_2451905, EPI_ISL_2451907, EPI_ISL_2451908, EPI_ISL_2451910 | ND Dept. of Health Laboratory Services-Microbiology | Centers for Disease Control and Prevention Division of Viral Diseases, Pathogen Discovery | Mili Sheth, Sarah Nobles, Jasmine Padilla, Mark Burroughs, Shoshona Le, Katie Dillon, Peter Cook, Clinton R. Paden, Dhwani Batra, Krista Queen, Kristen Knipe, Dakota Howard, Yvette Unoarumhi, Darlene Wagner, Matthew Schmerer, Ben L. Rambo-Martin, Kristine Lacek, Sam Shepard, Alison Laufer Halpin, Dave Wentworth, Vivien Dugan, Suxiang Tong, Justin Lee |
| EPI_ISL_2451912 | TX DSHS, Lab Services Section MC 1947 | Centers for Disease Control and Prevention Division of Viral Diseases, Pathogen Discovery | Mili Sheth, Sarah Nobles, Jasmine Padilla, Mark Burroughs, Shoshona Le, Katie Dillon, Peter Cook, Clinton R. Paden, Dhwani Batra, Krista Queen, Kristen Knipe, Dakota Howard, Yvette Unoarumhi, Darlene Wagner, Matthew Schmerer, Ben L. Rambo-Martin, Kristine Lacek, Sam Shepard, Alison Laufer Halpin, Dave Wentworth, Vivien Dugan, Suxiang Tong, Justin Lee |
| EPI_ISL_2451913, EPI_ISL_2451915, EPI_ISL_2451917, EPI_ISL_2451918, EPI_ISL_2451920, EPI_ISL_2451922, EPI_ISL_2451923, EPI_ISL_2451925, EPI_ISL_2451926, EPI_ISL_2451928, EPI_ISL_2451929, EPI_ISL_2451930, EPI_ISL_2451932, EPI_ISL_2451934, EPI_ISL_2451936 | | | |
| see above | NE Public Health Laboratory | Centers for Disease Control and Prevention Division of Viral Diseases, Pathogen Discovery | Mili Sheth, Sarah Nobles, Jasmine Padilla, Mark Burroughs, Shoshona Le, Katie Dillon, Peter Cook, Clinton R. Paden, Dhwani Batra, Krista Queen, Kristen Knipe, Dakota Howard, Yvette Unoarumhi, Darlene Wagner, Matthew Schmerer, Ben L. Rambo-Martin, Kristine Lacek, Sam Shepard, Alison Laufer Halpin, Dave Wentworth, Vivien Dugan, Suxiang Tong, Justin Lee |
| EPI_ISL_2451937, EPI_ISL_2451938, EPI_ISL_2451940, EPI_ISL_2451942, EPI_ISL_2451943, EPI_ISL_2451945, EPI_ISL_2451947, EPI_ISL_2451948 | AK State Public Health Lab, State Health Department | Centers for Disease Control and Prevention Division of Viral Diseases, Pathogen Discovery | Mili Sheth, Sarah Nobles, Jasmine Padilla, Mark Burroughs, Shoshona Le, Katie Dillon, Peter Cook, Clinton R. Paden, Dhwani Batra, Krista Queen, Kristen Knipe, Dakota Howard, Yvette Unoarumhi, Darlene Wagner, Matthew Schmerer, Ben L. Rambo-Martin, Kristine Lacek, Sam Shepard, Alison Laufer Halpin, Dave Wentworth, Vivien Dugan, Suxiang Tong, Justin Lee |
| EPI_ISL_2451950, EPI_ISL_2451952, EPI_ISL_2451953, EPI_ISL_2451955, EPI_ISL_2451957 | WA State Department of Health | Centers for Disease Control and Prevention Division of Viral Diseases, Pathogen Discovery | Mili Sheth, Sarah Nobles, Jasmine Padilla, Mark Burroughs, Shoshona Le, Katie Dillon, Peter Cook, Clinton R. Paden, Dhwani Batra, Krista Queen, Kristen Knipe, Dakota Howard, Yvette Unoarumhi, Darlene Wagner, Matthew Schmerer, Ben L. Rambo-Martin, Kristine Lacek, Sam Shepard, Alison Laufer Halpin, Dave Wentworth, Vivien Dugan, Suxiang Tong, Justin Lee |
| EPI_ISL_2451959, EPI_ISL_2451960, EPI_ISL_2451962, EPI_ISL_2451964, EPI_ISL_2451965, EPI_ISL_2451967 | FL Bureau of Public Health Laboratories-Tampa | Centers for Disease Control and Prevention Division of Viral Diseases, Pathogen Discovery | Mili Sheth, Sarah Nobles, Jasmine Padilla, Mark Burroughs, Shoshona Le, Katie Dillon, Peter Cook, Clinton R. Paden, Dhwani Batra, Krista Queen, Kristen Knipe, Dakota Howard, Yvette Unoarumhi, Darlene Wagner, Matthew Schmerer, Ben L. Rambo-Martin, Kristine Lacek, Sam Shepard, Alison Laufer Halpin, Dave Wentworth, Vivien Dugan, Suxiang Tong, Justin Lee |
| EPI_ISL_2451969, EPI_ISL_2451970, EPI_ISL_2451972, EPI_ISL_2451974, EPI_ISL_2451975, EPI_ISL_2451977, EPI_ISL_2451979 | SC Dept of Health and Env. Control-Bureau of Laboratories | Centers for Disease Control and Prevention Division of Viral Diseases, Pathogen Discovery | Mili Sheth, Sarah Nobles, Jasmine Padilla, Mark Burroughs, Shoshona Le, Katie Dillon, Peter Cook, Clinton R. Paden, Dhwani Batra, Krista Queen, Kristen Knipe, Dakota Howard, Yvette Unoarumhi, Darlene Wagner, Matthew Schmerer, Ben L. Rambo-Martin, Kristine Lacek, Sam Shepard, Alison Laufer Halpin, Dave Wentworth, Vivien Dugan, Suxiang Tong, Justin Lee |
| EPI_ISL_2451980, EPI_ISL_2451982, EPI_ISL_2451984, EPI_ISL_2451985, EPI_ISL_2451987, EPI_ISL_2451989, EPI_ISL_2451990, EPI_ISL_2451992, EPI_ISL_2451994, EPI_ISL_2451996 | OR State PHL-Virology/Immunology Section | Centers for Disease Control and Prevention Division of Viral Diseases, Pathogen Discovery | Mili Sheth, Sarah Nobles, Jasmine Padilla, Mark Burroughs, Shoshona Le, Katie Dillon, Peter Cook, Clinton R. Paden, Dhwani Batra, Krista Queen, Kristen Knipe, Dakota Howard, Yvette Unoarumhi, Darlene Wagner, Matthew Schmerer, Ben L. Rambo-Martin, Kristine Lacek, Sam Shepard, Alison Laufer Halpin, Dave Wentworth, Vivien Dugan, Suxiang Tong, Justin Lee |
| EPI_ISL_2451998, EPI_ISL_2451999, EPI_ISL_2452001 | LA Office of Public Health Laboratories | Centers for Disease Control and Prevention Division of Viral Diseases, Pathogen Discovery | Mili Sheth, Sarah Nobles, Jasmine Padilla, Mark Burroughs, Shoshona Le, Katie Dillon, Peter Cook, Clinton R. Paden, Dhwani Batra, Krista Queen, Kristen Knipe, Dakota Howard, Yvette Unoarumhi, Darlene Wagner, Matthew Schmerer, Ben L. Rambo-Martin, Kristine Lacek, Sam Shepard, Alison Laufer Halpin, Dave Wentworth, Vivien Dugan, Suxiang Tong, Justin Lee |
| EPI_ISL_2452003, EPI_ISL_2452004, EPI_ISL_2452006, EPI_ISL_2452008, EPI_ISL_2452009, EPI_ISL_2452011 | DE Public Health Laboratory | Centers for Disease Control and Prevention Division of Viral Diseases, Pathogen Discovery | Mili Sheth, Sarah Nobles, Jasmine Padilla, Mark Burroughs, Shoshona Le, Katie Dillon, Peter Cook, Clinton R. Paden, Dhwani Batra, Krista Queen, Kristen Knipe, Dakota Howard, Yvette Unoarumhi, Darlene Wagner, Matthew Schmerer, Ben L. Rambo-Martin, Kristine Lacek, Sam Shepard, Alison Laufer Halpin, Dave Wentworth, Vivien Dugan, Suxiang Tong, Justin Lee |
| EPI_ISL_2452013, EPI_ISL_2452015, EPI_ISL_2452016, EPI_ISL_2452018 | WI State Laboratory of Hygiene | Centers for Disease Control and Prevention Division of Viral Diseases, Pathogen Discovery | Mili Sheth, Sarah Nobles, Jasmine Padilla, Mark Burroughs, Shoshona Le, Katie Dillon, Peter Cook, Clinton R. Paden, Dhwani Batra, Krista Queen, Kristen Knipe, Dakota Howard, Yvette Unoarumhi, Darlene Wagner, Matthew Schmerer, Ben L. Rambo-Martin, Kristine Lacek, Sam Shepard, Alison Laufer Halpin, Dave Wentworth, Vivien Dugan, Suxiang Tong, Justin Lee |
| EPI_ISL_2452020 | NC State Laboratory of Public Health | Centers for Disease Control and Prevention Division of Viral Diseases, Pathogen Discovery | Mili Sheth, Sarah Nobles, Jasmine Padilla, Mark Burroughs, Shoshona Le, Katie Dillon, Peter Cook, Clinton R. Paden, Dhwani Batra, Krista Queen, Kristen Knipe, Dakota Howard, Yvette Unoarumhi, Darlene Wagner, Matthew Schmerer, Ben L. Rambo-Martin, Kristine Lacek, Sam Shepard, Alison Laufer Halpin, Dave Wentworth, Vivien Dugan, Suxiang Tong, Justin Lee |
| EPI_ISL_2452021, EPI_ISL_2452023, EPI_ISL_2452025, EPI_ISL_2452026 | VI-US Virgin Islands Department of Health | Centers for Disease Control and Prevention Division of Viral Diseases, Pathogen Discovery | Mili Sheth, Sarah Nobles, Jasmine Padilla, Mark Burroughs, Shoshona Le, Katie Dillon, Peter Cook, Clinton R. Paden, Dhwani Batra, Krista Queen, Kristen Knipe, Dakota Howard, Yvette Unoarumhi, Darlene Wagner, Matthew Schmerer, Ben L. Rambo-Martin, Kristine Lacek, Sam Shepard, Alison Laufer Halpin, Dave Wentworth, Vivien Dugan, Suxiang Tong, Justin Lee |
| EPI_ISL_2452028, EPI_ISL_2452030, EPI_ISL_2452031, EPI_ISL_2452033, EPI_ISL_2452035, EPI_ISL_2452037, EPI_ISL_2452038, EPI_ISL_2452040, EPI_ISL_2452042, EPI_ISL_2452043, EPI_ISL_2452045 | | | |
| see above | CDPH, Viral and Rickettsial Disease Laboratory | Centers for Disease Control and Prevention Division of Viral Diseases, Pathogen Discovery | Mili Sheth, Sarah Nobles, Jasmine Padilla, Mark Burroughs, Shoshona Le, Katie Dillon, Peter Cook, Clinton R. Paden, Dhwani Batra, Krista Queen, Kristen Knipe, Dakota Howard, Yvette Unoarumhi, Darlene Wagner, Matthew Schmerer, Ben L. Rambo-Martin, Kristine Lacek, Sam Shepard, Alison Laufer Halpin, Dave Wentworth, Vivien Dugan, Suxiang Tong, Justin Lee |
| EPI_ISL_2452047, EPI_ISL_2452048, EPI_ISL_2452050, EPI_ISL_2452052, EPI_ISL_2452054, EPI_ISL_2452055, EPI_ISL_2452057, EPI_ISL_2452060, EPI_ISL_2452063, EPI_ISL_2452064, EPI_ISL_2452066, EPI_ISL_2452067, EPI_ISL_2452069, EPI_ISL_2452071, EPI_ISL_2452073 | | | |
| see above | NYC Department of Health and Mental Hygiene | Centers for Disease Control and Prevention Division of Viral Diseases, Pathogen Discovery | Mili Sheth, Sarah Nobles, Jasmine Padilla, Mark Burroughs, Shoshona Le, Katie Dillon, Peter Cook, Clinton R. Paden, Dhwani Batra, Krista Queen, Kristen Knipe, Dakota Howard, Yvette Unoarumhi, Darlene Wagner, Matthew Schmerer, Ben L. Rambo-Martin, Kristine Lacek, Sam Shepard, Alison Laufer Halpin, Dave Wentworth, Vivien Dugan, Suxiang Tong, Justin Lee |
| EPI_ISL_2452074, EPI_ISL_2452076, EPI_ISL_2452078, EPI_ISL_2452079, EPI_ISL_2452081, EPI_ISL_2452083 | ID Bureau of Laboratories | Centers for Disease Control and Prevention Division of Viral Diseases, Pathogen Discovery | Mili Sheth, Sarah Nobles, Jasmine Padilla, Mark Burroughs, Shoshona Le, Katie Dillon, Peter Cook, Clinton R. Paden, Dhwani Batra, Krista Queen, Kristen Knipe, Dakota Howard, Yvette Unoarumhi, Darlene Wagner, Matthew Schmerer, Ben L. Rambo-Martin, Kristine Lacek, Sam Shepard, Alison Laufer Halpin, Dave Wentworth, Vivien Dugan, Suxiang Tong, Justin Lee |
| EPI_ISL_2452084, EPI_ISL_2452085, EPI_ISL_2452087, EPI_ISL_2452088, EPI_ISL_2452090, EPI_ISL_2452092, EPI_ISL_2452093, EPI_ISL_2452095, EPI_ISL_2452097 | WY Public Health Laboratory | Centers for Disease Control and Prevention Division of Viral Diseases, Pathogen Discovery | Mili Sheth, Sarah Nobles, Jasmine Padilla, Mark Burroughs, Shoshona Le, Katie Dillon, Peter Cook, Clinton R. Paden, Dhwani Batra, Krista Queen, Kristen Knipe, Dakota Howard, Yvette Unoarumhi, Darlene Wagner, Matthew Schmerer, Ben L. Rambo-Martin, Kristine Lacek, Sam Shepard, Alison Laufer Halpin, Dave Wentworth, Vivien Dugan, Suxiang Tong, Justin Lee |
| EPI_ISL_2452099, EPI_ISL_2452100, | NM Dept. Health, Scientific Laboratory Division | Centers for Disease Control and Prevention Division of Viral Diseases, Pathogen | Mili Sheth, Sarah Nobles, Jasmine Padilla, Mark Burroughs, Shoshona Le, Katie Dillon, Peter Cook, Clinton R. Paden, Dhwani Batra, Krista Queen, Kristen Knipe, Dakota Howard, Yvette Unoarumhi, Darlene Wagner, Matthew Schmerer, Ben L. Rambo-Martin, Kristine Lacek, Sam Shepard, Alison Laufer Halpin, Dave Wentworth, Vivien Dugan, Suxiang Tong, Justin Lee |

| | | | |
|---|---|---|---|
| EPI_ISL_2452102, EPI_ISL_2452103, EPI_ISL_2452105, EPI_ISL_2452107, EPI_ISL_2452108 | | Discovery | |
| EPI_ISL_2452110, EPI_ISL_2452112, EPI_ISL_2452113, EPI_ISL_2452115, EPI_ISL_2452117, EPI_ISL_2452119, EPI_ISL_2452120, EPI_ISL_2452122, EPI_ISL_2452124, EPI_ISL_2452125, EPI_ISL_2452127, EPI_ISL_2452129, EPI_ISL_2452130, EPI_ISL_2452132, EPI_ISL_2452134, EPI_ISL_2452135, EPI_ISL_2452137, EPI_ISL_2452139, EPI_ISL_2452140, EPI_ISL_2452142, EPI_ISL_2452144 | | | |
| see above | HI Dept. of Health, State Laboratories Division | Centers for Disease Control and Prevention Division of Viral Diseases, Pathogen Discovery | Mili Sheth, Sarah Nobles, Jasmine Padilla, Mark Burroughs, Shoshona Le, Katie Dillon, Peter Cook, Clinton R. Paden, Dhwani Batra, Krista Queen, Kristen Knipe, Dakota Howard, Yvette Unoarumhi, Darlene Wagner, Matthew Schmerer, Ben L. Rambo-Martin, Kristine Lacek, Sam Shepard, Alison Laufer Halpin, Dave Wentworth, Vivien Dugan, Suxiang Tong, Justin Lee |
| EPI_ISL_2452145, EPI_ISL_2452147, EPI_ISL_2452149, EPI_ISL_2452150, EPI_ISL_2452152, EPI_ISL_2452154, EPI_ISL_2452155, EPI_ISL_2452157, EPI_ISL_2452159, EPI_ISL_2452160, EPI_ISL_2452162, EPI_ISL_2452164 | | | |
| see above | IN State Department of Health Laboratory Services | Centers for Disease Control and Prevention Division of Viral Diseases, Pathogen Discovery | Mili Sheth, Sarah Nobles, Jasmine Padilla, Mark Burroughs, Shoshona Le, Katie Dillon, Peter Cook, Clinton R. Paden, Dhwani Batra, Krista Queen, Kristen Knipe, Dakota Howard, Yvette Unoarumhi, Darlene Wagner, Matthew Schmerer, Ben L. Rambo-Martin, Kristine Lacek, Sam Shepard, Alison Laufer Halpin, Dave Wentworth, Vivien Dugan, Suxiang Tong, Justin Lee |
| EPI_ISL_2452165, EPI_ISL_2452167, EPI_ISL_2452169, EPI_ISL_2452171 | AL Dept. of Public Health Bureau of Clinical Laboratories | Centers for Disease Control and Prevention Division of Viral Diseases, Pathogen Discovery | Mili Sheth, Sarah Nobles, Jasmine Padilla, Mark Burroughs, Shoshona Le, Katie Dillon, Peter Cook, Clinton R. Paden, Dhwani Batra, Krista Queen, Kristen Knipe, Dakota Howard, Yvette Unoarumhi, Darlene Wagner, Matthew Schmerer, Ben L. Rambo-Martin, Kristine Lacek, Sam Shepard, Alison Laufer Halpin, Dave Wentworth, Vivien Dugan, Suxiang Tong, Justin Lee |
| EPI_ISL_2452172, EPI_ISL_2452174, EPI_ISL_2452176, EPI_ISL_2452177, EPI_ISL_2452179, EPI_ISL_2452180, EPI_ISL_2452182 | IA State Hygienic Laboratory | Centers for Disease Control and Prevention Division of Viral Diseases, Pathogen Discovery | Mili Sheth, Sarah Nobles, Jasmine Padilla, Mark Burroughs, Shoshona Le, Katie Dillon, Peter Cook, Clinton R. Paden, Dhwani Batra, Krista Queen, Kristen Knipe, Dakota Howard, Yvette Unoarumhi, Darlene Wagner, Matthew Schmerer, Ben L. Rambo-Martin, Kristine Lacek, Sam Shepard, Alison Laufer Halpin, Dave Wentworth, Vivien Dugan, Suxiang Tong, Justin Lee |
| EPI_ISL_2452184, EPI_ISL_2452186, EPI_ISL_2452187, EPI_ISL_2452189, EPI_ISL_2452191, EPI_ISL_2452192, EPI_ISL_2452194, EPI_ISL_2452196, EPI_ISL_2452197 | NH Dept. of Health and Human Services Public Health Labs | Centers for Disease Control and Prevention Division of Viral Diseases, Pathogen Discovery | Mili Sheth, Sarah Nobles, Jasmine Padilla, Mark Burroughs, Shoshona Le, Katie Dillon, Peter Cook, Clinton R. Paden, Dhwani Batra, Krista Queen, Kristen Knipe, Dakota Howard, Yvette Unoarumhi, Darlene Wagner, Matthew Schmerer, Ben L. Rambo-Martin, Kristine Lacek, Sam Shepard, Alison Laufer Halpin, Dave Wentworth, Vivien Dugan, Suxiang Tong, Justin Lee |
| EPI_ISL_2452199, EPI_ISL_2452201, EPI_ISL_2452203, EPI_ISL_2452204, EPI_ISL_2452206, EPI_ISL_2452208, EPI_ISL_2452209, EPI_ISL_2452211, EPI_ISL_2452213 | OH Department of Health Laboratory | Centers for Disease Control and Prevention Division of Viral Diseases, Pathogen Discovery | Mili Sheth, Sarah Nobles, Jasmine Padilla, Mark Burroughs, Shoshona Le, Katie Dillon, Peter Cook, Clinton R. Paden, Dhwani Batra, Krista Queen, Kristen Knipe, Dakota Howard, Yvette Unoarumhi, Darlene Wagner, Matthew Schmerer, Ben L. Rambo-Martin, Kristine Lacek, Sam Shepard, Alison Laufer Halpin, Dave Wentworth, Vivien Dugan, Suxiang Tong, Justin Lee |
| EPI_ISL_2452214, EPI_ISL_2452216, EPI_ISL_2452218, EPI_ISL_2452219, EPI_ISL_2452221, EPI_ISL_2452222, EPI_ISL_2452224, EPI_ISL_2452226, EPI_ISL_2452227, EPI_ISL_2452229 | GA Department of Public Health Laboratory | Centers for Disease Control and Prevention Division of Viral Diseases, Pathogen Discovery | Mili Sheth, Sarah Nobles, Jasmine Padilla, Mark Burroughs, Shoshona Le, Katie Dillon, Peter Cook, Clinton R. Paden, Dhwani Batra, Krista Queen, Kristen Knipe, Dakota Howard, Yvette Unoarumhi, Darlene Wagner, Matthew Schmerer, Ben L. Rambo-Martin, Kristine Lacek, Sam Shepard, Alison Laufer Halpin, Dave Wentworth, Vivien Dugan, Suxiang Tong, Justin Lee |
| EPI_ISL_2452230, EPI_ISL_2452233, EPI_ISL_2452236, EPI_ISL_2452237, EPI_ISL_2452239, EPI_ISL_2452241, EPI_ISL_2452242, EPI_ISL_2452244, EPI_ISL_2452246, EPI_ISL_2452248, EPI_ISL_2452249, EPI_ISL_2452251, EPI_ISL_2452253, EPI_ISL_2452254, EPI_ISL_2452256, EPI_ISL_2452258, EPI_ISL_2452260, EPI_ISL_2452262, EPI_ISL_2452263, EPI_ISL_2452265, EPI_ISL_2452267, EPI_ISL_2452268, EPI_ISL_2452270, EPI_ISL_2452271, EPI_ISL_2452273, EPI_ISL_2452275 | | | |
| see above | RI State Health Laboratories | Centers for Disease Control and Prevention Division of Viral Diseases, Pathogen Discovery | Mili Sheth, Sarah Nobles, Jasmine Padilla, Mark Burroughs, Shoshona Le, Katie Dillon, Peter Cook, Clinton R. Paden, Dhwani Batra, Krista Queen, Kristen Knipe, Dakota Howard, Yvette Unoarumhi, Darlene Wagner, Matthew Schmerer, Ben L. Rambo-Martin, Kristine Lacek, Sam Shepard, Alison Laufer Halpin, Dave Wentworth, Vivien Dugan, Suxiang Tong, Justin Lee |
| EPI_ISL_2452277, EPI_ISL_2452279, EPI_ISL_2452280, EPI_ISL_2452282, EPI_ISL_2452284, EPI_ISL_2452285, EPI_ISL_2452287, EPI_ISL_2452289, EPI_ISL_2452291, EPI_ISL_2452292, EPI_ISL_2452294, EPI_ISL_2452296 | | | |
| see above | MD DOH Laboratories Administration | Centers for Disease Control and Prevention Division of Viral Diseases, Pathogen Discovery | Mili Sheth, Sarah Nobles, Jasmine Padilla, Mark Burroughs, Shoshona Le, Katie Dillon, Peter Cook, Clinton R. Paden, Dhwani Batra, Krista Queen, Kristen Knipe, Dakota Howard, Yvette Unoarumhi, Darlene Wagner, Matthew Schmerer, Ben L. Rambo-Martin, Kristine Lacek, Sam Shepard, Alison Laufer Halpin, Dave Wentworth, Vivien Dugan, Suxiang Tong, Justin Lee |
| EPI_ISL_2453318, EPI_ISL_2453319, EPI_ISL_2453320, EPI_ISL_2453321, EPI_ISL_2453322, EPI_ISL_2453323, EPI_ISL_2453324, EPI_ISL_2453325, EPI_ISL_2453326, EPI_ISL_2453327, EPI_ISL_2453328, EPI_ISL_2453329, EPI_ISL_2453330, EPI_ISL_2453331, EPI_ISL_2453332, EPI_ISL_2453333, EPI_ISL_2453334, EPI_ISL_2453335, EPI_ISL_2453336, EPI_ISL_2453337, EPI_ISL_2453338, EPI_ISL_2453339, EPI_ISL_2453340, EPI_ISL_2453341, EPI_ISL_2454294, EPI_ISL_2454295, EPI_ISL_2454296, EPI_ISL_2454297, EPI_ISL_2454298 | | | |
| see above | Wexner Medical Center | The Ohio State University College of Medicine | Koenig,S., Seminetta,J. |
| EPI_ISL_2454303, EPI_ISL_2454304, EPI_ISL_2454305, EPI_ISL_2454306, EPI_ISL_2454307, EPI_ISL_2454308, EPI_ISL_2454309, EPI_ISL_2454310, EPI_ISL_2454311, EPI_ISL_2454312, EPI_ISL_2454313, EPI_ISL_2454314, EPI_ISL_2454315, EPI_ISL_2454316, EPI_ISL_2454317, EPI_ISL_2454318, EPI_ISL_2454319, EPI_ISL_2454320, EPI_ISL_2454321, EPI_ISL_2454322, EPI_ISL_2454323, EPI_ISL_2454324, EPI_ISL_2454325, EPI_ISL_2454326, EPI_ISL_2454327, EPI_ISL_2454328, EPI_ISL_2454329, EPI_ISL_2454330, EPI_ISL_2454331, EPI_ISL_2454332, EPI_ISL_2454333, EPI_ISL_2454334, EPI_ISL_2454335, EPI_ISL_2454337, EPI_ISL_2454338, EPI_ISL_2454339, EPI_ISL_2454340, EPI_ISL_2454341, EPI_ISL_2454343, EPI_ISL_2454344, EPI_ISL_2454345, EPI_ISL_2454346, EPI_ISL_2454347, EPI_ISL_2454348, EPI_ISL_2454349, EPI_ISL_2454350, EPI_ISL_2454351, EPI_ISL_2454352, EPI_ISL_2454353, EPI_ISL_2454354, EPI_ISL_2454356, EPI_ISL_2454357, EPI_ISL_2454358, EPI_ISL_2454359, EPI_ISL_2454360, EPI_ISL_2454361, EPI_ISL_2454362 | | | |
| see above | TriCore Reference Laboratories | Center for Global Health, University of New Mexico Health Sciences Center | Daryl Domman, Kurt Schwalm, Valerie Morley, Cecilia Thompson, Kendra Pesko, Karissa Culbreath, Darrell Dinwiddie |
| EPI_ISL_2454363, EPI_ISL_2454365, EPI_ISL_2454366, EPI_ISL_2454367, EPI_ISL_2454368, EPI_ISL_2454369, EPI_ISL_2454370, EPI_ISL_2454371 | New Mexico Department of Health Scientific Laboratory | Center for Global Health, University of New Mexico Health Sciences Center | Daryl Domman, Kurt Schwalm, Twila Kunde, Joseph Hicks, Anastacia Griego, Michael Edwards, Valerie Morley, Darrell Dinwiddie |
| EPI_ISL_2454511, EPI_ISL_2454513, EPI_ISL_2454514, EPI_ISL_2454515, EPI_ISL_2454516, EPI_ISL_2454517, EPI_ISL_2454522, EPI_ISL_2454525, EPI_ISL_2454527, EPI_ISL_2454528, EPI_ISL_2454529 | | | |
| see above | School of Pharmacy, Shenandoah University | School of Pharmacy, Shenandoah University | Adams,S.M., Harralson,A.F., Kidd,R.S., Sawyer,G.W. |
| EPI_ISL_2454530, EPI_ISL_2454531, EPI_ISL_2454532, EPI_ISL_2454533, EPI_ISL_2454534, EPI_ISL_2454535, EPI_ISL_2454536, EPI_ISL_2454537, EPI_ISL_2454538, EPI_ISL_2454539, EPI_ISL_2454540, EPI_ISL_2454541, EPI_ISL_2454542, EPI_ISL_2454543, EPI_ISL_2454544, EPI_ISL_2454545, EPI_ISL_2454546, EPI_ISL_2454547, EPI_ISL_2454548, EPI_ISL_2454549, EPI_ISL_2454550 | | | |
| see above | Wexner Medical Center | The Ohio State University College of Medicine | Koenig,S., Seminetta,J. |
| EPI_ISL_2454551, EPI_ISL_2454552, EPI_ISL_2454553, EPI_ISL_2454554, EPI_ISL_2454555, EPI_ISL_2454556, EPI_ISL_2454557, EPI_ISL_2454558, EPI_ISL_2454559, EPI_ISL_2454560, EPI_ISL_2454561, EPI_ISL_2454562, EPI_ISL_2454563, EPI_ISL_2454564, EPI_ISL_2454565, EPI_ISL_2454566, EPI_ISL_2454567, EPI_ISL_2454568, EPI_ISL_2454569, EPI_ISL_2454570, EPI_ISL_2454571, EPI_ISL_2454572, EPI_ISL_2454573, EPI_ISL_2454574, EPI_ISL_2454575, EPI_ISL_2454576, EPI_ISL_2454577, EPI_ISL_2454578, EPI_ISL_2454579, EPI_ISL_2454580, EPI_ISL_2454581 | | | |
| see above | Alaska State Virology Laboratory | Alaska State Virology Laboratory | Stephanie DeRonde, Elva House, Jacob Zidek, Lisa Smith, Ph.D., Jack Chen, Ph.D. |
| EPI_ISL_2454585, EPI_ISL_2454590, EPI_ISL_2454596, EPI_ISL_2454601, EPI_ISL_2454603, EPI_ISL_2454606, EPI_ISL_2454607, EPI_ISL_2454609, EPI_ISL_2454615, EPI_ISL_2454619, EPI_ISL_2454620, EPI_ISL_2454622 | | | |
| see above | TGen North | TGen North | Jolene Bowers, Heather Centner, Chris French, Hayley Yaglom, Ashlyn Pfeiffer, Darrin Lemmer, Dave Engelthaler, The Arizona COVID Genomics Union (ACGU) |
| EPI_ISL_2455049, EPI_ISL_2455050, EPI_ISL_2455051, EPI_ISL_2455052, EPI_ISL_2455053, EPI_ISL_2455054, EPI_ISL_2455055, EPI_ISL_2455056, EPI_ISL_2455057, EPI_ISL_2455058, EPI_ISL_2455059, EPI_ISL_2455060, EPI_ISL_2455061, EPI_ISL_2455062, EPI_ISL_2455063, EPI_ISL_2455064, EPI_ISL_2455065, EPI_ISL_2455066, EPI_ISL_2455067, EPI_ISL_2455068, EPI_ISL_2455069, EPI_ISL_2455070 | | | |
| see above | Montana Public Health Laboratory | Montana Public Health Laboratory | Joy Ritter, Michelle Mozer, Carrie Biskupiak, Deborah Gibson, Michael Dills |
| EPI_ISL_2455071, EPI_ISL_2455072, EPI_ISL_2455073, EPI_ISL_2455074 | Wexner Medical Center | The Ohio State University College of Medicine | Koenig,S., Seminetta,J. |
| EPI_ISL_2455080, EPI_ISL_2455081, | Houston Health Dept. | Houston Health Dept. | Ryker Penn, Pamela Brown, Adolpho Lara, Yanlai Lai |

| | | | |
|---|---|---|---|
| EPI_ISL_2455082, EPI_ISL_2455083, EPI_ISL_2455084 | | | |
| EPI_ISL_2455237 | CICESE-BSL3 | CICESE-BSL3 | Cervantes Karla, Gonzalez-Sanchez Ricardo, Martinez Miguel, Galindo Clara and Licea Alexei |
| EPI_ISL_2455241 | CICESE-BSL3 | CICESE-BSL3 | Cervantes Karla, Gonzalez Ricardo, Martinez Miguel, Galindo Clara and Licea Alexei |
| EPI_ISL_2455242 | CICESE-BSL3 | CICESE-BSL3 | Karla Cervantes, Ricardo Gonzalez, Miguel Martinez, Clara Galindo and Alexei Licea |
| EPI_ISL_2455243 | CICESE-BSL3 | CICESE-BSL3 | Karla Cervantes, Ricardo Gonzalez, Miguel Martinez, Clara Galindo and Alexei Licea |
| EPI_ISL_2455265, EPI_ISL_2455266, EPI_ISL_2455267, EPI_ISL_2455268, EPI_ISL_2455269, EPI_ISL_2455270, EPI_ISL_2455271, EPI_ISL_2455272, EPI_ISL_2455273, EPI_ISL_2455274, EPI_ISL_2455275, EPI_ISL_2455276, EPI_ISL_2455277, EPI_ISL_2455278, EPI_ISL_2455279, EPI_ISL_2455280, EPI_ISL_2455281, EPI_ISL_2455282, EPI_ISL_2455283, EPI_ISL_2455284, EPI_ISL_2455285, EPI_ISL_2455286, EPI_ISL_2455287, EPI_ISL_2455288, EPI_ISL_2455289, EPI_ISL_2455290, EPI_ISL_2455291, EPI_ISL_2455292, EPI_ISL_2455293, EPI_ISL_2455294, EPI_ISL_2455295, EPI_ISL_2455296, EPI_ISL_2455297, EPI_ISL_2455298, EPI_ISL_2455299, EPI_ISL_2455300, EPI_ISL_2455301, EPI_ISL_2455303, EPI_ISL_2455304, EPI_ISL_2455305, EPI_ISL_2455306, EPI_ISL_2455308, EPI_ISL_2455309, EPI_ISL_2455310, EPI_ISL_2455312, EPI_ISL_2455313, EPI_ISL_2455315, EPI_ISL_2455317, EPI_ISL_2455318, EPI_ISL_2455319, EPI_ISL_2455320, EPI_ISL_2455321, EPI_ISL_2455322, EPI_ISL_2455324, EPI_ISL_2455325, EPI_ISL_2455326 | | | |
| see above | Johns Hopkins Hospital Department of Pathology | Johns Hopkins Hospital Department of Pathology | C. Paul Morris, Chun Huai Luo, Adannaya Amadi, Matthew Schwartz, Nicholas Gallagher, Heba H. Mostafa |
| EPI_ISL_2455341, EPI_ISL_2455342, EPI_ISL_2455343, EPI_ISL_2455344, EPI_ISL_2455345, EPI_ISL_2455346, EPI_ISL_2455347, EPI_ISL_2455348 | Minnesota Department of Health, Public Health Laboratory | Minnesota Department of Health, Public Health Laboratory | Alexandra Lorentz, Jacob Garfin, Matt Plumb, and Xiong Wang |
| EPI_ISL_2455349 | Centracare Laboratory Services | Minnesota Department of Health, Public Health Laboratory | Alexandra Lorentz, Jacob Garfin, Matt Plumb, and Xiong Wang |
| EPI_ISL_2455350, EPI_ISL_2455351, EPI_ISL_2455352, EPI_ISL_2455353, EPI_ISL_2455354, EPI_ISL_2455355, EPI_ISL_2455356, EPI_ISL_2455358, EPI_ISL_2455359, EPI_ISL_2455361, EPI_ISL_2455362, EPI_ISL_2455363, EPI_ISL_2455364, EPI_ISL_2455365, EPI_ISL_2455366, EPI_ISL_2455367, EPI_ISL_2455368 | | | |
| see above | Minnesota Department of Health, Public Health Laboratory | Minnesota Department of Health, Public Health Laboratory | Alexandra Lorentz, Jacob Garfin, Matt Plumb, and Xiong Wang |
| EPI_ISL_2455370, EPI_ISL_2455371, EPI_ISL_2455372, EPI_ISL_2455374, EPI_ISL_2455375, EPI_ISL_2455376, EPI_ISL_2455377, EPI_ISL_2455378, EPI_ISL_2455379, EPI_ISL_2455380, EPI_ISL_2455381 | | | |
| see above | HealthPartners Central Lab | Minnesota Department of Health, Public Health Laboratory | Alexandra Lorentz, Jacob Garfin, Matt Plumb, and Xiong Wang |
| EPI_ISL_2455383, EPI_ISL_2455384, EPI_ISL_2455385, EPI_ISL_2455387, EPI_ISL_2455388, EPI_ISL_2455389, EPI_ISL_2455390 | Premier Medical Lab | Minnesota Department of Health, Public Health Laboratory | Alexandra Lorentz, Jacob Garfin, Matt Plumb, and Xiong Wang |
| EPI_ISL_2455391, EPI_ISL_2455392, EPI_ISL_2455393, EPI_ISL_2455394, EPI_ISL_2455395 | Weill Cornell Medicine | New York Genome Center | Michael Zody, Andre Corvelo, Dayna M. Oschwald, Samantha Fennessey, Tom Maniatis, Melissa Cushing, Olivier Elemento, Margaret Elizabeth Ross, Chris Mason, Priya Velu, Hanna Rennert, Arryn Craney, Lars F Westblade |
| EPI_ISL_2455417, EPI_ISL_2455418, EPI_ISL_2455419, EPI_ISL_2455420, EPI_ISL_2455421, EPI_ISL_2455422, EPI_ISL_2455423, EPI_ISL_2455424, EPI_ISL_2455425, EPI_ISL_2455427, EPI_ISL_2455428, EPI_ISL_2455429, EPI_ISL_2455430, EPI_ISL_2455431, EPI_ISL_2455432, EPI_ISL_2455433, EPI_ISL_2455434, EPI_ISL_2455435, EPI_ISL_2455436, EPI_ISL_2455437, EPI_ISL_2455438, EPI_ISL_2455439, EPI_ISL_2455442, EPI_ISL_2455443, EPI_ISL_2455444, EPI_ISL_2455445, EPI_ISL_2455446, EPI_ISL_2455447, EPI_ISL_2455448, EPI_ISL_2455449, EPI_ISL_2455450 | | | |
| see above | Alaska State Virology Laboratory | Alaska State Virology Laboratory | Stephanie DeRonde, Elva House, Jacob Zidek, Lisa Smith, Ph.D., Jack Chen, Ph.D. |
| EPI_ISL_2455452, EPI_ISL_2455453, EPI_ISL_2455454, EPI_ISL_2455455, EPI_ISL_2455457, EPI_ISL_2455458, EPI_ISL_2455459, EPI_ISL_2455460, EPI_ISL_2455461, EPI_ISL_2455462, EPI_ISL_2455463, EPI_ISL_2455464, EPI_ISL_2455465, EPI_ISL_2455466, EPI_ISL_2455467, EPI_ISL_2455468, EPI_ISL_2455469, EPI_ISL_2455470, EPI_ISL_2455471, EPI_ISL_2455472, EPI_ISL_2455473, EPI_ISL_2455474, EPI_ISL_2455475, EPI_ISL_2455476, EPI_ISL_2455477, EPI_ISL_2455478 | | | |
| see above | Kansas Health and Environmental Lab | Kansas Health and Environmental Lab | Katherine Wiggins, Mike Grose, Jonathan Barnell, Ben Olsen, and Phil Adam |
| EPI_ISL_2455510, EPI_ISL_2455511, EPI_ISL_2455512, EPI_ISL_2455515, EPI_ISL_2455516, EPI_ISL_2455518, EPI_ISL_2455519, EPI_ISL_2455520, EPI_ISL_2455521, EPI_ISL_2455523, EPI_ISL_2455524, EPI_ISL_2455525, EPI_ISL_2455527, EPI_ISL_2455531, EPI_ISL_2455540, EPI_ISL_2455541, EPI_ISL_2455544, EPI_ISL_2455548, EPI_ISL_2455550, EPI_ISL_2455551, EPI_ISL_2455552 | | | |
| see above | LSUHS Emerging Viral Threat Laboratory | LSUHS Emerging Viral Threat Laboratory | Gregory L. Ware, Alexander Mijalis, Jeremy P. Kamil, Jennifer L. Carroll, Maarten Van Diest, Rona S. Scott, Andrew D. Yurochko, Christopher G. Kevil, John A. Vanchiere |
| EPI_ISL_2455569, EPI_ISL_2455571, EPI_ISL_2455572, EPI_ISL_2455573, EPI_ISL_2455575, EPI_ISL_2455581 | Arizona State Public Health Laboratory | Arizona State Public Health Laboratory | Trung Huynh, Jessica Escobar, Katherine Fullerton, Nobuko Fukushima, Stacy White, Linda Getsinger, Victor Waddell |
| EPI_ISL_2455896, EPI_ISL_2455897, EPI_ISL_2455898, EPI_ISL_2455899, EPI_ISL_2455900, EPI_ISL_2455901, EPI_ISL_2455902, EPI_ISL_2455903, EPI_ISL_2455904, EPI_ISL_2455905, EPI_ISL_2455906, EPI_ISL_2455907, EPI_ISL_2455908, EPI_ISL_2455909, EPI_ISL_2455910, EPI_ISL_2455911, EPI_ISL_2455912 | | | |
| see above | LESP Queretaro | Instituto de Diagnostico y Referencia Epidemiologicos (INDRE) | Claudia Wong-Arambula, Abril Rodriguez-Maldonado, Vanessa Rivero-Arredondo, Ariadna Medina-Benitez, Joaquin Quiroz-Mercado, Sergio Rangel-Guerrero, Natividad Cruz-Ortiz, Tatiana Nunez-Garcia, Gisela Barrera-Badillo, Lucia Hernandez-Rivas, Irma Lopez-Martinez, Ernesto Ramirez-Gonzalez. |
| EPI_ISL_2455913 | LESP Campeche | Instituto de Diagnostico y Referencia Epidemiologicos (INDRE) | Claudia Wong-Arambula, Abril Rodriguez-Maldonado, Vanessa Rivero-Arredondo, Ariadna Medina-Benitez, Joaquin Quiroz-Mercado, Sergio Rangel-Guerrero, Natividad Cruz-Ortiz, Tatiana Nunez-Garcia, Gisela Barrera-Badillo, Lucia Hernandez-Rivas, Irma Lopez-Martinez, Ernesto Ramirez-Gonzalez. |
| EPI_ISL_2455914, EPI_ISL_2455915 | LESP Chihuahua | Instituto de Diagnostico y Referencia Epidemiologicos (INDRE) | Claudia Wong-Arambula, Abril Rodriguez-Maldonado, Vanessa Rivero-Arredondo, Ariadna Medina-Benitez, Joaquin Quiroz-Mercado, Sergio Rangel-Guerrero, Natividad Cruz-Ortiz, Tatiana Nunez-Garcia, Gisela Barrera-Badillo, Lucia Hernandez-Rivas, Irma Lopez-Martinez, Ernesto Ramirez-Gonzalez. |
| EPI_ISL_2455916 | LESP Tamaulipas | Instituto de Diagnostico y Referencia Epidemiologicos (INDRE) | Claudia Wong-Arambula, Abril Rodriguez-Maldonado, Vanessa Rivero-Arredondo, Ariadna Medina-Benitez, Joaquin Quiroz-Mercado, Sergio Rangel-Guerrero, Natividad Cruz-Ortiz, Tatiana Nunez-Garcia, Gisela Barrera-Badillo, Lucia Hernandez-Rivas, Irma Lopez-Martinez, Ernesto Ramirez-Gonzalez. |
| EPI_ISL_2455917 | LESP Chihuahua | Instituto de Diagnostico y Referencia Epidemiologicos (INDRE) | Claudia Wong-Arambula, Abril Rodriguez-Maldonado, Vanessa Rivero-Arredondo, Ariadna Medina-Benitez, Joaquin Quiroz-Mercado, Sergio Rangel-Guerrero, Natividad Cruz-Ortiz, Tatiana Nunez-Garcia, Gisela Barrera-Badillo, Lucia Hernandez-Rivas, Irma Lopez-Martinez, Ernesto Ramirez-Gonzalez. |
| EPI_ISL_2455918 | LESP Tamaulipas | Instituto de Diagnostico y Referencia Epidemiologicos (INDRE) | Claudia Wong-Arambula, Abril Rodriguez-Maldonado, Vanessa Rivero-Arredondo, Ariadna Medina-Benitez, Joaquin Quiroz-Mercado, Sergio Rangel-Guerrero, Natividad Cruz-Ortiz, Tatiana Nunez-Garcia, Gisela Barrera-Badillo, Lucia Hernandez-Rivas, Irma Lopez-Martinez, Ernesto Ramirez-Gonzalez. |
| EPI_ISL_2455919, EPI_ISL_2455920 | LESP Chihuahua | Instituto de Diagnostico y Referencia Epidemiologicos (INDRE) | Claudia Wong-Arambula, Abril Rodriguez-Maldonado, Vanessa Rivero-Arredondo, Ariadna Medina-Benitez, Joaquin Quiroz-Mercado, Sergio Rangel-Guerrero, Natividad Cruz-Ortiz, Tatiana Nunez-Garcia, Gisela Barrera-Badillo, Lucia Hernandez-Rivas, Irma Lopez-Martinez, Ernesto Ramirez-Gonzalez. |
| EPI_ISL_2455921, EPI_ISL_2455922 | LESP Oaxaca | Instituto de Diagnostico y Referencia Epidemiologicos (INDRE) | Claudia Wong-Arambula, Abril Rodriguez-Maldonado, Vanessa Rivero-Arredondo, Ariadna Medina-Benitez, Joaquin Quiroz-Mercado, Sergio Rangel-Guerrero, Natividad Cruz-Ortiz, Tatiana Nunez-Garcia, Gisela Barrera-Badillo, Lucia Hernandez-Rivas, Irma Lopez-Martinez, Ernesto Ramirez-Gonzalez. |
| EPI_ISL_2455923, EPI_ISL_2455924, EPI_ISL_2455925 | LESP Tamaulipas | Instituto de Diagnostico y Referencia Epidemiologicos (INDRE) | Claudia Wong-Arambula, Abril Rodriguez-Maldonado, Vanessa Rivero-Arredondo, Ariadna Medina-Benitez, Joaquin Quiroz-Mercado, Sergio Rangel-Guerrero, Natividad Cruz-Ortiz, Tatiana Nunez-Garcia, Gisela Barrera-Badillo, Lucia Hernandez-Rivas, Irma Lopez-Martinez, Ernesto Ramirez-Gonzalez. |
| EPI_ISL_2455926, EPI_ISL_2455927, EPI_ISL_2455928, EPI_ISL_2455929 | LESP Chihuahua | Instituto de Diagnostico y Referencia Epidemiologicos (INDRE) | Claudia Wong-Arambula, Abril Rodriguez-Maldonado, Vanessa Rivero-Arredondo, Ariadna Medina-Benitez, Joaquin Quiroz-Mercado, Sergio Rangel-Guerrero, Natividad Cruz-Ortiz, Tatiana Nunez-Garcia, Gisela Barrera-Badillo, Lucia Hernandez-Rivas, Irma Lopez-Martinez, Ernesto Ramirez-Gonzalez. |
| EPI_ISL_2455930, EPI_ISL_2455931, EPI_ISL_2455932, EPI_ISL_2455933, EPI_ISL_2455935, EPI_ISL_2455936, EPI_ISL_2455937, EPI_ISL_2455938, EPI_ISL_2455939, EPI_ISL_2455940, EPI_ISL_2455941, EPI_ISL_2455942, EPI_ISL_2455943, EPI_ISL_2455944, EPI_ISL_2455945, EPI_ISL_2455946, EPI_ISL_2455947, EPI_ISL_2455948, EPI_ISL_2455949, EPI_ISL_2455950, EPI_ISL_2455951, EPI_ISL_2455952 | | | |
| see above | LESP Tamaulipas | Instituto de Diagnostico y Referencia Epidemiologicos (INDRE) | Claudia Wong-Arambula, Abril Rodriguez-Maldonado, Vanessa Rivero-Arredondo, Ariadna Medina-Benitez, Joaquin Quiroz-Mercado, Sergio Rangel-Guerrero, Natividad Cruz-Ortiz, Tatiana Nunez-Garcia, Gisela Barrera-Badillo, Lucia Hernandez-Rivas, Irma Lopez-Martinez, Ernesto Ramirez-Gonzalez. |
| EPI_ISL_2455953 | LESP San Luis Potosi | Instituto de Diagnostico y Referencia Epidemiologicos (INDRE) | Claudia Wong-Arambula, Abril Rodriguez-Maldonado, Vanessa Rivero-Arredondo, Ariadna Medina-Benitez, Joaquin Quiroz-Mercado, Sergio Rangel-Guerrero, Natividad Cruz-Ortiz, Tatiana Nunez-Garcia, Gisela Barrera-Badillo, Lucia Hernandez-Rivas, Irma Lopez-Martinez, Ernesto Ramirez-Gonzalez. |
| EPI_ISL_2455954, EPI_ISL_2455955, EPI_ISL_2455956, EPI_ISL_2455957 | LESP Tamaulipas | Instituto de Diagnostico y Referencia Epidemiologicos (INDRE) | Claudia Wong-Arambula, Abril Rodriguez-Maldonado, Vanessa Rivero-Arredondo, Ariadna Medina-Benitez, Joaquin Quiroz-Mercado, Sergio Rangel-Guerrero, Natividad Cruz-Ortiz, Tatiana Nunez-Garcia, Gisela Barrera-Badillo, Lucia Hernandez-Rivas, Irma Lopez-Martinez, Ernesto Ramirez-Gonzalez. |
| EPI_ISL_2455958 | LESP Jalisco | Instituto de Diagnostico y Referencia Epidemiologicos (INDRE) | Claudia Wong-Arambula, Abril Rodriguez-Maldonado, Vanessa Rivero-Arredondo, Ariadna Medina-Benitez, Joaquin Quiroz-Mercado, Sergio Rangel-Guerrero, Natividad Cruz-Ortiz, Tatiana Nunez-Garcia, Gisela Barrera-Badillo, Lucia Hernandez-Rivas, Irma Lopez-Martinez, Ernesto Ramirez-Gonzalez. |
| EPI_ISL_2455959 | LESP Ciudad de Mexico | Instituto de Diagnostico y Referencia Epidemiologicos (INDRE) | Claudia Wong-Arambula, Abril Rodriguez-Maldonado, Vanessa Rivero-Arredondo, Ariadna Medina-Benitez, Joaquin Quiroz-Mercado, Sergio Rangel-Guerrero, Natividad Cruz-Ortiz, Tatiana Nunez-Garcia, Gisela Barrera-Badillo, Lucia Hernandez-Rivas, Irma Lopez-Martinez, Ernesto Ramirez-Gonzalez. |
| EPI_ISL_2455960, | LESP Queretaro | Instituto de Diagnostico y Referencia | Claudia Wong-Arambula, Abril Rodriguez-Maldonado, Vanessa Rivero-Arredondo, Ariadna Medina-Benitez, Joaquin Quiroz-Mercado, Sergio Rangel-Guerrero, Natividad Cruz-Ortiz, Tatiana Nunez-Garcia, Gisela Barrera-Badillo, Lucia Hernandez-Rivas, Irma Lopez-Martinez, Ernesto |

| | | | |
|---|---|---|---|
| EPI_ISL_2455961, EPI_ISL_2455962, EPI_ISL_2455963 | | Epidemiologicos (INDRE) | Ramirez-Gonzalez. |
| EPI_ISL_2455964, EPI_ISL_2455965, EPI_ISL_2455966, EPI_ISL_2455967, EPI_ISL_2455968, EPI_ISL_2455969, EPI_ISL_2455970, EPI_ISL_2455971, EPI_ISL_2455972, EPI_ISL_2455973, EPI_ISL_2455974, EPI_ISL_2455975 | | | |
| see above | LESP Hidalgo | Instituto de Diagnostico y Referencia Epidemiologicos (INDRE) | Claudia Wong-Arambula, Abril Rodriguez-Maldonado, Vanessa Rivero-Arredondo, Ariadna Medina-Benitez, Joaquin Quiroz-Mercado, Sergio Rangel-Guerrero, Natividad Cruz-Ortiz, Tatiana Nunez-Garcia, Gisela Barrera-Badillo, Lucia Hernandez-Rivas, Irma Lopez-Martinez, Ernesto Ramirez-Gonzalez. |
| EPI_ISL_2455977, EPI_ISL_2455978 | LESP Sinaloa | Instituto de Diagnostico y Referencia Epidemiologicos (INDRE) | Claudia Wong-Arambula, Abril Rodriguez-Maldonado, Vanessa Rivero-Arredondo, Ariadna Medina-Benitez, Joaquin Quiroz-Mercado, Sergio Rangel-Guerrero, Natividad Cruz-Ortiz, Tatiana Nunez-Garcia, Gisela Barrera-Badillo, Lucia Hernandez-Rivas, Irma Lopez-Martinez, Ernesto Ramirez-Gonzalez. |
| EPI_ISL_2455979 | LESP Morelos | Instituto de Diagnostico y Referencia Epidemiologicos (INDRE) | Claudia Wong-Arambula, Abril Rodriguez-Maldonado, Vanessa Rivero-Arredondo, Ariadna Medina-Benitez, Joaquin Quiroz-Mercado, Sergio Rangel-Guerrero, Natividad Cruz-Ortiz, Tatiana Nunez-Garcia, Gisela Barrera-Badillo, Lucia Hernandez-Rivas, Irma Lopez-Martinez, Ernesto Ramirez-Gonzalez. |
| EPI_ISL_2455980 | LESP Colima | Instituto de Diagnostico y Referencia Epidemiologicos (INDRE) | Claudia Wong-Arambula, Abril Rodriguez-Maldonado, Vanessa Rivero-Arredondo, Ariadna Medina-Benitez, Joaquin Quiroz-Mercado, Sergio Rangel-Guerrero, Natividad Cruz-Ortiz, Tatiana Nunez-Garcia, Gisela Barrera-Badillo, Lucia Hernandez-Rivas, Irma Lopez-Martinez, Ernesto Ramirez-Gonzalez. |
| EPI_ISL_2455981 | LESP Queretaro | Instituto de Diagnostico y Referencia Epidemiologicos (INDRE) | Claudia Wong-Arambula, Abril Rodriguez-Maldonado, Vanessa Rivero-Arredondo, Ariadna Medina-Benitez, Joaquin Quiroz-Mercado, Sergio Rangel-Guerrero, Natividad Cruz-Ortiz, Tatiana Nunez-Garcia, Gisela Barrera-Badillo, Lucia Hernandez-Rivas, Irma Lopez-Martinez, Ernesto Ramirez-Gonzalez. |
| EPI_ISL_2455982, EPI_ISL_2455983, EPI_ISL_2455984, EPI_ISL_2455985 | LESP Nuevo Leon | Instituto de Diagnostico y Referencia Epidemiologicos (INDRE) | Claudia Wong-Arambula, Abril Rodriguez-Maldonado, Vanessa Rivero-Arredondo, Ariadna Medina-Benitez, Joaquin Quiroz-Mercado, Sergio Rangel-Guerrero, Natividad Cruz-Ortiz, Tatiana Nunez-Garcia, Gisela Barrera-Badillo, Lucia Hernandez-Rivas, Irma Lopez-Martinez, Ernesto Ramirez-Gonzalez. |
| EPI_ISL_2455986, EPI_ISL_2455987, EPI_ISL_2455988 | LESP Guanajuato | Instituto de Diagnostico y Referencia Epidemiologicos (INDRE) | Claudia Wong-Arambula, Abril Rodriguez-Maldonado, Vanessa Rivero-Arredondo, Ariadna Medina-Benitez, Joaquin Quiroz-Mercado, Sergio Rangel-Guerrero, Natividad Cruz-Ortiz, Tatiana Nunez-Garcia, Gisela Barrera-Badillo, Lucia Hernandez-Rivas, Irma Lopez-Martinez, Ernesto Ramirez-Gonzalez. |
| EPI_ISL_2455989 | LESP Queretaro | Instituto de Diagnostico y Referencia Epidemiologicos (INDRE) | Claudia Wong-Arambula, Abril Rodriguez-Maldonado, Vanessa Rivero-Arredondo, Ariadna Medina-Benitez, Joaquin Quiroz-Mercado, Sergio Rangel-Guerrero, Natividad Cruz-Ortiz, Tatiana Nunez-Garcia, Gisela Barrera-Badillo, Lucia Hernandez-Rivas, Irma Lopez-Martinez, Ernesto Ramirez-Gonzalez. |
| EPI_ISL_2455990, EPI_ISL_2455991 | LESP Guanajuato | Instituto de Diagnostico y Referencia Epidemiologicos (INDRE) | Claudia Wong-Arambula, Abril Rodriguez-Maldonado, Vanessa Rivero-Arredondo, Ariadna Medina-Benitez, Joaquin Quiroz-Mercado, Sergio Rangel-Guerrero, Natividad Cruz-Ortiz, Tatiana Nunez-Garcia, Gisela Barrera-Badillo, Lucia Hernandez-Rivas, Irma Lopez-Martinez, Ernesto Ramirez-Gonzalez. |
| EPI_ISL_2455992 | LESP Estado de Mexico | Instituto de Diagnostico y Referencia Epidemiologicos (INDRE) | Claudia Wong-Arambula, Abril Rodriguez-Maldonado, Vanessa Rivero-Arredondo, Ariadna Medina-Benitez, Joaquin Quiroz-Mercado, Sergio Rangel-Guerrero, Natividad Cruz-Ortiz, Tatiana Nunez-Garcia, Gisela Barrera-Badillo, Lucia Hernandez-Rivas, Irma Lopez-Martinez, Ernesto Ramirez-Gonzalez. |
| EPI_ISL_2455993, EPI_ISL_2455994, EPI_ISL_2455995 | LESP Puebla | Instituto de Diagnostico y Referencia Epidemiologicos (INDRE) | Claudia Wong-Arambula, Abril Rodriguez-Maldonado, Vanessa Rivero-Arredondo, Ariadna Medina-Benitez, Joaquin Quiroz-Mercado, Sergio Rangel-Guerrero, Natividad Cruz-Ortiz, Tatiana Nunez-Garcia, Gisela Barrera-Badillo, Lucia Hernandez-Rivas, Irma Lopez-Martinez, Ernesto Ramirez-Gonzalez. |
| EPI_ISL_2455996 | LESP Hidalgo | Instituto de Diagnostico y Referencia Epidemiologicos (INDRE) | Claudia Wong-Arambula, Abril Rodriguez-Maldonado, Vanessa Rivero-Arredondo, Ariadna Medina-Benitez, Joaquin Quiroz-Mercado, Sergio Rangel-Guerrero, Natividad Cruz-Ortiz, Tatiana Nunez-Garcia, Gisela Barrera-Badillo, Lucia Hernandez-Rivas, Irma Lopez-Martinez, Ernesto Ramirez-Gonzalez. |
| EPI_ISL_2455997 | LESP Quintana Roo | Instituto de Diagnostico y Referencia Epidemiologicos (INDRE) | Claudia Wong-Arambula, Abril Rodriguez-Maldonado, Vanessa Rivero-Arredondo, Ariadna Medina-Benitez, Joaquin Quiroz-Mercado, Sergio Rangel-Guerrero, Natividad Cruz-Ortiz, Tatiana Nunez-Garcia, Gisela Barrera-Badillo, Lucia Hernandez-Rivas, Irma Lopez-Martinez, Ernesto Ramirez-Gonzalez. |
| EPI_ISL_2455998, EPI_ISL_2455999, EPI_ISL_2456000 | LESP Baja California | Instituto de Diagnostico y Referencia Epidemiologicos (INDRE) | Claudia Wong-Arambula, Abril Rodriguez-Maldonado, Vanessa Rivero-Arredondo, Ariadna Medina-Benitez, Joaquin Quiroz-Mercado, Sergio Rangel-Guerrero, Natividad Cruz-Ortiz, Tatiana Nunez-Garcia, Gisela Barrera-Badillo, Lucia Hernandez-Rivas, Irma Lopez-Martinez, Ernesto Ramirez-Gonzalez. |
| EPI_ISL_2456001 | LESP Colima | Instituto de Diagnostico y Referencia Epidemiologicos (INDRE) | Claudia Wong-Arambula, Abril Rodriguez-Maldonado, Vanessa Rivero-Arredondo, Ariadna Medina-Benitez, Joaquin Quiroz-Mercado, Sergio Rangel-Guerrero, Natividad Cruz-Ortiz, Tatiana Nunez-Garcia, Gisela Barrera-Badillo, Lucia Hernandez-Rivas, Irma Lopez-Martinez, Ernesto Ramirez-Gonzalez. |
| EPI_ISL_2456002 | LESP Queretaro | Instituto de Diagnostico y Referencia Epidemiologicos (INDRE) | Claudia Wong-Arambula, Abril Rodriguez-Maldonado, Vanessa Rivero-Arredondo, Ariadna Medina-Benitez, Joaquin Quiroz-Mercado, Sergio Rangel-Guerrero, Natividad Cruz-Ortiz, Tatiana Nunez-Garcia, Gisela Barrera-Badillo, Lucia Hernandez-Rivas, Irma Lopez-Martinez, Ernesto Ramirez-Gonzalez. |
| EPI_ISL_2456111, EPI_ISL_2456114, EPI_ISL_2456117, EPI_ISL_2456120, EPI_ISL_2456123, EPI_ISL_2456126, EPI_ISL_2456129, EPI_ISL_2456132, EPI_ISL_2456135, EPI_ISL_2456138, EPI_ISL_2456141, EPI_ISL_2456143, EPI_ISL_2456146, EPI_ISL_2456149, EPI_ISL_2456153, EPI_ISL_2456156, EPI_ISL_2456160, EPI_ISL_2456163, EPI_ISL_2456166, EPI_ISL_2456169, EPI_ISL_2456172, EPI_ISL_2456175, EPI_ISL_2456179, EPI_ISL_2456182, EPI_ISL_2456185, EPI_ISL_2456188, EPI_ISL_2456191, EPI_ISL_2456193, EPI_ISL_2456196, EPI_ISL_2456200, EPI_ISL_2456203, EPI_ISL_2456206, EPI_ISL_2456209, EPI_ISL_2456212, EPI_ISL_2456215, EPI_ISL_2456218, EPI_ISL_2456222, EPI_ISL_2456225, EPI_ISL_2456228, EPI_ISL_2456231, EPI_ISL_2456234, EPI_ISL_2456237, EPI_ISL_2456240, EPI_ISL_2456244, EPI_ISL_2456247, EPI_ISL_2456250, EPI_ISL_2456253, EPI_ISL_2456255, EPI_ISL_2456258, EPI_ISL_2456261, EPI_ISL_2456264, EPI_ISL_2456267, EPI_ISL_2456273, EPI_ISL_2456276, EPI_ISL_2456279, EPI_ISL_2456282, EPI_ISL_2456285, EPI_ISL_2456288, EPI_ISL_2456291, EPI_ISL_2456294, EPI_ISL_2456297, EPI_ISL_2456300, EPI_ISL_2456303, EPI_ISL_2456307, EPI_ISL_2456310, EPI_ISL_2456313, EPI_ISL_2456316, EPI_ISL_2456318, EPI_ISL_2456321, EPI_ISL_2456324, EPI_ISL_2456327, EPI_ISL_2456331, EPI_ISL_2456334, EPI_ISL_2456337, EPI_ISL_2456340, EPI_ISL_2456343, EPI_ISL_2456346, EPI_ISL_2456350, EPI_ISL_2456353, EPI_ISL_2456356, EPI_ISL_2456359, EPI_ISL_2456362, EPI_ISL_2456365, EPI_ISL_2456367, EPI_ISL_2456370, EPI_ISL_2456374, EPI_ISL_2456377, EPI_ISL_2456380, EPI_ISL_2456383, EPI_ISL_2456386, EPI_ISL_2456389, EPI_ISL_2456392, EPI_ISL_2456395, EPI_ISL_2456398, EPI_ISL_2456401, EPI_ISL_2456404, EPI_ISL_2456407, EPI_ISL_2456410, EPI_ISL_2456413, EPI_ISL_2456415, EPI_ISL_2456418, EPI_ISL_2456422, EPI_ISL_2456425, EPI_ISL_2456428, EPI_ISL_2456430, EPI_ISL_2456434, EPI_ISL_2456438, EPI_ISL_2456441, EPI_ISL_2456444, EPI_ISL_2456447, EPI_ISL_2456449, EPI_ISL_2456453, EPI_ISL_2456455, EPI_ISL_2456458, EPI_ISL_2456462, EPI_ISL_2456465, EPI_ISL_2456468, EPI_ISL_2456470, EPI_ISL_2456474, EPI_ISL_2456477, EPI_ISL_2456480, EPI_ISL_2456483, EPI_ISL_2456486, EPI_ISL_2456489, EPI_ISL_2456492, EPI_ISL_2456496, EPI_ISL_2456499, EPI_ISL_2456502, EPI_ISL_2456505, EPI_ISL_2456508, EPI_ISL_2456510, EPI_ISL_2456513, EPI_ISL_2456516, EPI_ISL_2456519, EPI_ISL_2456522, EPI_ISL_2456525, EPI_ISL_2456528, EPI_ISL_2456531, EPI_ISL_2456534, EPI_ISL_2456537, EPI_ISL_2456541, EPI_ISL_2456544, EPI_ISL_2456547, EPI_ISL_2456550, EPI_ISL_2456553, EPI_ISL_2456556, EPI_ISL_2456558, EPI_ISL_2456561, EPI_ISL_2456565, EPI_ISL_2456568, EPI_ISL_2456571, EPI_ISL_2456574, EPI_ISL_2456577, EPI_ISL_2456580, EPI_ISL_2456583, EPI_ISL_2456586, EPI_ISL_2456589, EPI_ISL_2456592, EPI_ISL_2456595, EPI_ISL_2456598, EPI_ISL_2456601, EPI_ISL_2456604, EPI_ISL_2456607, EPI_ISL_2456609, EPI_ISL_2456612, EPI_ISL_2456616, EPI_ISL_2456618, EPI_ISL_2456622, EPI_ISL_2456625, EPI_ISL_2456629, EPI_ISL_2456632, EPI_ISL_2456635, EPI_ISL_2456638, EPI_ISL_2456641, EPI_ISL_2456645, EPI_ISL_2456649, EPI_ISL_2456652, EPI_ISL_2456654, EPI_ISL_2456657, EPI_ISL_2456660, EPI_ISL_2456663, EPI_ISL_2456666, EPI_ISL_2456670, EPI_ISL_2456673, EPI_ISL_2456676, EPI_ISL_2456678, EPI_ISL_2456681, EPI_ISL_2456685, EPI_ISL_2456688, EPI_ISL_2456691, EPI_ISL_2456694, EPI_ISL_2456697, EPI_ISL_2456698, EPI_ISL_2456699, EPI_ISL_2456700, EPI_ISL_2456701, EPI_ISL_2456702, EPI_ISL_2456703, EPI_ISL_2456704, EPI_ISL_2456705, EPI_ISL_2456706, EPI_ISL_2456707, EPI_ISL_2456708, EPI_ISL_2456709, EPI_ISL_2456710, EPI_ISL_2456711, EPI_ISL_2456712, EPI_ISL_2456713, EPI_ISL_2456714, EPI_ISL_2456715, EPI_ISL_2456716, EPI_ISL_2456717, EPI_ISL_2456718, EPI_ISL_2456719, EPI_ISL_2456720, EPI_ISL_2456721, EPI_ISL_2456722, EPI_ISL_2456723, EPI_ISL_2456724, EPI_ISL_2456725, EPI_ISL_2456726, EPI_ISL_2456727, EPI_ISL_2456728, EPI_ISL_2456729, EPI_ISL_2456730, EPI_ISL_2456731, EPI_ISL_2456732, EPI_ISL_2456733, EPI_ISL_2456734, EPI_ISL_2456735, EPI_ISL_2456736 | | | |
| see above | Utah Public Health Laboratory | Utah Public Health Laboratory | Erin L. Young, Kelly F. Oakeson, Tara Gallagher |
| EPI_ISL_2457058, EPI_ISL_2457061 | Stanford Health Care | Stanford University School of Medicine, Clinical Virology Laboratory | Daniel Solis, Mamdouh Sibai, Bernadette Troung, Becky Jiang, Michelle Verghese, ChunHong Huang, James Zehnder, Malaya K. Sahoo, and Benjamin A. Pinsky |
| EPI_ISL_2458161, EPI_ISL_2458162, EPI_ISL_2458163, EPI_ISL_2458164, EPI_ISL_2458165, EPI_ISL_2458166, EPI_ISL_2458167, EPI_ISL_2458168, EPI_ISL_2458169, EPI_ISL_2458170, EPI_ISL_2458171, EPI_ISL_2458172, EPI_ISL_2458173, EPI_ISL_2458174, EPI_ISL_2458175, EPI_ISL_2458176, EPI_ISL_2458177, EPI_ISL_2458178, EPI_ISL_2458179, EPI_ISL_2458180, EPI_ISL_2458198, EPI_ISL_2458199, EPI_ISL_2458200, EPI_ISL_2458201, EPI_ISL_2458202, EPI_ISL_2458203, EPI_ISL_2458204, EPI_ISL_2458205, EPI_ISL_2458206, EPI_ISL_2458207, EPI_ISL_2458208, EPI_ISL_2458209, EPI_ISL_2458210, EPI_ISL_2458211, EPI_ISL_2458212, EPI_ISL_2458213, EPI_ISL_2458214, EPI_ISL_2458215, EPI_ISL_2458216, EPI_ISL_2458217, EPI_ISL_2458218, EPI_ISL_2458219, EPI_ISL_2458220, EPI_ISL_2458221, EPI_ISL_2458222, EPI_ISL_2458223, EPI_ISL_2458224 | | | |
| see above | NYU Langone Health | Departments of Pathology and Medicine, New York University School of Medicine | Adriana Heguy, Dacia Dimartino, Emily Guzman, Christian Marier, Peter Meyn, Sitharam Ramaswami, Gael Westby, Paul Zappile, Yutong Zhang, Paolo Cotzia, Guiqing Wang |
| EPI_ISL_2458225, EPI_ISL_2458226, EPI_ISL_2458227 | Alaska State Virology Laboratory | Alaska State Virology Laboratory | Stephanie DeRonde, Elva House, Jacob Zidek, Lisa Smith, Ph.D., Jack Chen, Ph.D. |
| EPI_ISL_2458228, EPI_ISL_2458229, EPI_ISL_2458230, EPI_ISL_2458231, EPI_ISL_2458232, EPI_ISL_2458233, EPI_ISL_2458234, EPI_ISL_2458235, EPI_ISL_2458236, EPI_ISL_2458237, EPI_ISL_2458238, EPI_ISL_2458239, EPI_ISL_2458240, EPI_ISL_2458241, EPI_ISL_2458242, EPI_ISL_2458243, EPI_ISL_2458244, EPI_ISL_2458245, EPI_ISL_2458246, EPI_ISL_2458247, EPI_ISL_2458248, EPI_ISL_2458249, EPI_ISL_2458250, EPI_ISL_2458251, EPI_ISL_2458252, EPI_ISL_2458253, EPI_ISL_2458254, EPI_ISL_2458255, EPI_ISL_2458256, EPI_ISL_2458257, EPI_ISL_2458258, EPI_ISL_2458259, EPI_ISL_2458260, EPI_ISL_2458261, EPI_ISL_2458262, EPI_ISL_2458263, EPI_ISL_2458264, EPI_ISL_2458265, EPI_ISL_2458266, EPI_ISL_2458267, EPI_ISL_2458268, EPI_ISL_2458269, EPI_ISL_2458270, EPI_ISL_2458271, EPI_ISL_2458272, EPI_ISL_2458273, EPI_ISL_2458274, EPI_ISL_2458275, EPI_ISL_2458276, EPI_ISL_2458277, EPI_ISL_2458278, EPI_ISL_2458279, EPI_ISL_2458280, EPI_ISL_2458281, EPI_ISL_2458282, EPI_ISL_2458283, EPI_ISL_2458284, EPI_ISL_2458285, EPI_ISL_2458286, EPI_ISL_2458287, EPI_ISL_2458288, EPI_ISL_2458289, EPI_ISL_2458290, EPI_ISL_2458291, EPI_ISL_2458292, EPI_ISL_2458293, EPI_ISL_2458294, EPI_ISL_2458295, EPI_ISL_2458296, EPI_ISL_2458297, EPI_ISL_2458298, EPI_ISL_2458299, EPI_ISL_2458300, EPI_ISL_2458301, EPI_ISL_2458302, EPI_ISL_2458303, EPI_ISL_2458304, EPI_ISL_2458305, EPI_ISL_2458306, EPI_ISL_2458307, EPI_ISL_2458308, EPI_ISL_2458309, EPI_ISL_2458310, EPI_ISL_2458311, EPI_ISL_2458312, EPI_ISL_2458313, EPI_ISL_2458314, EPI_ISL_2458315, EPI_ISL_2458316, EPI_ISL_2458317, EPI_ISL_2458318, EPI_ISL_2458319, EPI_ISL_2458320, EPI_ISL_2458321, EPI_ISL_2458322, EPI_ISL_2458323, EPI_ISL_2458324, EPI_ISL_2458325, EPI_ISL_2458326, EPI_ISL_2458327, EPI_ISL_2458328, EPI_ISL_2458329, EPI_ISL_2458330, EPI_ISL_2458331, EPI_ISL_2458332, EPI_ISL_2458333, EPI_ISL_2458334, EPI_ISL_2458335, EPI_ISL_2458336, EPI_ISL_2458337, EPI_ISL_2458338, EPI_ISL_2458339, EPI_ISL_2458340, EPI_ISL_2458341, EPI_ISL_2458342, EPI_ISL_2458343, EPI_ISL_2458344, EPI_ISL_2458345, EPI_ISL_2458346, EPI_ISL_2458347, EPI_ISL_2458348, EPI_ISL_2458349, EPI_ISL_2458350, EPI_ISL_2458351, EPI_ISL_2458352, EPI_ISL_2458353, EPI_ISL_2458354, EPI_ISL_2458355, EPI_ISL_2458356, EPI_ISL_2458357, EPI_ISL_2458358, EPI_ISL_2458359, EPI_ISL_2458360, EPI_ISL_2458361, EPI_ISL_2458362, EPI_ISL_2458363, EPI_ISL_2458364, EPI_ISL_2458365, EPI_ISL_2458366, EPI_ISL_2458367, EPI_ISL_2458368, EPI_ISL_2458369, EPI_ISL_2458370, EPI_ISL_2458371, EPI_ISL_2458372, EPI_ISL_2458373, EPI_ISL_2458374, EPI_ISL_2458375, EPI_ISL_2458376, EPI_ISL_2458377, EPI_ISL_2458378, EPI_ISL_2458379, EPI_ISL_2458380, EPI_ISL_2458381, EPI_ISL_2458382, EPI_ISL_2458383, EPI_ISL_2458384, EPI_ISL_2458385, EPI_ISL_2458386, EPI_ISL_2458387, EPI_ISL_2458388, EPI_ISL_2458389, EPI_ISL_2458390, EPI_ISL_2458391, EPI_ISL_2458392, EPI_ISL_2458393, EPI_ISL_2458394, EPI_ISL_2458395, EPI_ISL_2458396, EPI_ISL_2458397, EPI_ISL_2458398, EPI_ISL_2458399, EPI_ISL_2458400, EPI_ISL_2458401, EPI_ISL_2458402, EPI_ISL_2458403, EPI_ISL_2458404, EPI_ISL_2458405, EPI_ISL_2458406, EPI_ISL_2458407, EPI_ISL_2458408, EPI_ISL_2458409, EPI_ISL_2458410, EPI_ISL_2458411, EPI_ISL_2458412, EPI_ISL_2458413, EPI_ISL_2458414, EPI_ISL_2458415, EPI_ISL_2458416, EPI_ISL_2458417, EPI_ISL_2458418, EPI_ISL_2458419, EPI_ISL_2458420, EPI_ISL_2458421, EPI_ISL_2458422, EPI_ISL_2458423, EPI_ISL_2458424, EPI_ISL_2458425, EPI_ISL_2458426, EPI_ISL_2458427, EPI_ISL_2458428, EPI_ISL_2458429, EPI_ISL_2458430, EPI_ISL_2458431, EPI_ISL_2458432, EPI_ISL_2458433, EPI_ISL_2458434, EPI_ISL_2458435, EPI_ISL_2458436, EPI_ISL_2458437, EPI_ISL_2458438, EPI_ISL_2458439, EPI_ISL_2458440, EPI_ISL_2458441, EPI_ISL_2458442, EPI_ISL_2458443, EPI_ISL_2458444, EPI_ISL_2458445, EPI_ISL_2458446, EPI_ISL_2458447, EPI_ISL_2458448, EPI_ISL_2458449, EPI_ISL_2458450, EPI_ISL_2458451, EPI_ISL_2458452, EPI_ISL_2458453, EPI_ISL_2458454, EPI_ISL_2458455, EPI_ISL_2458456, EPI_ISL_2458457, EPI_ISL_2458458, EPI_ISL_2458459, EPI_ISL_2458460, EPI_ISL_2458461, EPI_ISL_2458462, EPI_ISL_2458463, EPI_ISL_2458487, EPI_ISL_2458488, EPI_ISL_2458489, EPI_ISL_2458490, EPI_ISL_2458491, EPI_ISL_2458492, EPI_ISL_2458493, EPI_ISL_2458494, EPI_ISL_2458495, EPI_ISL_2458496, EPI_ISL_2458497, EPI_ISL_2458498, EPI_ISL_2458499, EPI_ISL_2458500, EPI_ISL_2458501, EPI_ISL_2458502, EPI_ISL_2458503, EPI_ISL_2458504, EPI_ISL_2458505, EPI_ISL_2458506, EPI_ISL_2458507, EPI_ISL_2458508, EPI_ISL_2458509, EPI_ISL_2458510, EPI_ISL_2458511, EPI_ISL_2458512, EPI_ISL_2458513, EPI_ISL_2458514, EPI_ISL_2458515, EPI_ISL_2458516, EPI_ISL_2458517, EPI_ISL_2458518, EPI_ISL_2458519, EPI_ISL_2458520, EPI_ISL_2458521, EPI_ISL_2458522, EPI_ISL_2458523, EPI_ISL_2458524, EPI_ISL_2458525, EPI_ISL_2458526, EPI_ISL_2458527, EPI_ISL_2458528, EPI_ISL_2458529, EPI_ISL_2458530, EPI_ISL_2458531, EPI_ISL_2458532, EPI_ISL_2458533, EPI_ISL_2458534, EPI_ISL_2458535, EPI_ISL_2458536, EPI_ISL_2458537, EPI_ISL_2458538, EPI_ISL_2458539, EPI_ISL_2458540, EPI_ISL_2458541, EPI_ISL_2458542, EPI_ISL_2458543, EPI_ISL_2458544, EPI_ISL_2458545, EPI_ISL_2458546, EPI_ISL_2458547, EPI_ISL_2458548, EPI_ISL_2458549, EPI_ISL_2458550, EPI_ISL_2458551, EPI_ISL_2458552, EPI_ISL_2458553, EPI_ISL_2458554, EPI_ISL_2458555, EPI_ISL_2458556, EPI_ISL_2458557, EPI_ISL_2458558, EPI_ISL_2458559, EPI_ISL_2458560, EPI_ISL_2458561, EPI_ISL_2458562, EPI_ISL_2458563, EPI_ISL_2458564, EPI_ISL_2458565, EPI_ISL_2458566, EPI_ISL_2458567, EPI_ISL_2458568, EPI_ISL_2458569, EPI_ISL_2458570, EPI_ISL_2458571, EPI_ISL_2458572, EPI_ISL_2458573, EPI_ISL_2458574, EPI_ISL_2458575, EPI_ISL_2458576, EPI_ISL_2458577, EPI_ISL_2458578, EPI_ISL_2458579, EPI_ISL_2458580, EPI_ISL_2458581, EPI_ISL_2458582, EPI_ISL_2458583, EPI_ISL_2458584, EPI_ISL_2458585, EPI_ISL_2458586, EPI_ISL_2458587, EPI_ISL_2458588, EPI_ISL_2458589, EPI_ISL_2458590, EPI_ISL_2458591, EPI_ISL_2458592, EPI_ISL_2458593, EPI_ISL_2458594, EPI_ISL_2458595, EPI_ISL_2458596, EPI_ISL_2458597, EPI_ISL_2458598, EPI_ISL_2458599, EPI_ISL_2458600, EPI_ISL_2458601, EPI_ISL_2458602, EPI_ISL_2458603, EPI_ISL_2458604, EPI_ISL_2458605, EPI_ISL_2458606, EPI_ISL_2458607, EPI_ISL_2458608, EPI_ISL_2458609, EPI_ISL_2458610, EPI_ISL_2458611, EPI_ISL_2458612, EPI_ISL_2458613, EPI_ISL_2458614, EPI_ISL_2458615, EPI_ISL_2458616, EPI_ISL_2458617, EPI_ISL_2458618, EPI_ISL_2458619, EPI_ISL_2458620, EPI_ISL_2458621, EPI_ISL_2458622, EPI_ISL_2458623 | | | |
| see above | Utah Public Health Laboratory | Utah Public Health Laboratory | Erin L. Young, Kelly F. Oakeson, Tara Gallagher |
| EPI_ISL_2462067, EPI_ISL_2462068 | Ohio Department of Health Laboratory | Ohio Department of Health Laboratory | Holmes, Jennifer; Eric Brandt, Keoni Omura, Glen McGillivary, Caitlin McDonnell, Jade Mowery, Stephanie Mccracken, Tyler Payne, Kirtana Ramadugu, Erica Leasure, Brent Lee, Kelsey Florek, Heather Blankenship, Allison Black, Quanta Brown, and Tammy Bannerman |
| EPI_ISL_2462069, EPI_ISL_2462070, EPI_ISL_2462071, EPI_ISL_2462072, EPI_ISL_2462073, EPI_ISL_2462074, EPI_ISL_2462075, EPI_ISL_2462076, EPI_ISL_2462078, EPI_ISL_2462079, EPI_ISL_2462080, EPI_ISL_2462081, EPI_ISL_2462082, EPI_ISL_2462084, EPI_ISL_2462085, EPI_ISL_2462086, EPI_ISL_2462087, EPI_ISL_2462088, EPI_ISL_2462089, EPI_ISL_2462090, EPI_ISL_2462091, EPI_ISL_2462092, EPI_ISL_2462093, EPI_ISL_2462094, EPI_ISL_2462095, EPI_ISL_2462096, EPI_ISL_2462097, EPI_ISL_2462098, EPI_ISL_2462101, EPI_ISL_2462102, EPI_ISL_2462103, EPI_ISL_2462104, EPI_ISL_2462105, EPI_ISL_2462106, EPI_ISL_2462107, EPI_ISL_2462108, EPI_ISL_2462109, EPI_ISL_2462110, EPI_ISL_2462111, EPI_ISL_2462112, EPI_ISL_2462113, EPI_ISL_2462115, EPI_ISL_2462116, EPI_ISL_2462117, EPI_ISL_2462118, EPI_ISL_2462119, EPI_ISL_2462120, EPI_ISL_2462122, EPI_ISL_2462123, EPI_ISL_2462124, EPI_ISL_2462125, EPI_ISL_2462126, EPI_ISL_2462127, EPI_ISL_2462128, EPI_ISL_2462129, EPI_ISL_2462130, EPI_ISL_2462131, EPI_ISL_2462132, EPI_ISL_2462133, EPI_ISL_2462134, EPI_ISL_2462135, EPI_ISL_2462136, EPI_ISL_2462137, EPI_ISL_2462138, EPI_ISL_2462139, EPI_ISL_2462140, EPI_ISL_2462142, EPI_ISL_2462143, EPI_ISL_2462144, EPI_ISL_2462145, EPI_ISL_2462146, EPI_ISL_2462147, EPI_ISL_2462149, EPI_ISL_2462150, EPI_ISL_2462151, EPI_ISL_2462152, EPI_ISL_2462153, EPI_ISL_2462154, EPI_ISL_2462155, EPI_ISL_2462156, EPI_ISL_2462157, EPI_ISL_2462158, EPI_ISL_2462159, EPI_ISL_2462160, EPI_ISL_2462161, EPI_ISL_2462163, EPI_ISL_2462164, EPI_ISL_2462165, EPI_ISL_2462166, EPI_ISL_2462167, EPI_ISL_2462168, EPI_ISL_2462169, EPI_ISL_2462170, EPI_ISL_2462171, EPI_ISL_2462172, EPI_ISL_2462173, EPI_ISL_2462175, EPI_ISL_2462176, EPI_ISL_2462177, EPI_ISL_2462179, EPI_ISL_2462180, EPI_ISL_2462181, EPI_ISL_2462182, EPI_ISL_2462183, EPI_ISL_2462184, EPI_ISL_2462185, EPI_ISL_2462186, EPI_ISL_2462187, EPI_ISL_2462188, EPI_ISL_2462190, EPI_ISL_2462191, EPI_ISL_2462192, EPI_ISL_2462193, EPI_ISL_2462194, EPI_ISL_2462195, EPI_ISL_2462196, EPI_ISL_2462198, EPI_ISL_2462199, EPI_ISL_2462201, EPI_ISL_2462202, EPI_ISL_2462203, EPI_ISL_2462204, EPI_ISL_2462205, EPI_ISL_2462206, EPI_ISL_2462207, EPI_ISL_2462208, EPI_ISL_2462209, EPI_ISL_2462210, EPI_ISL_2462211, EPI_ISL_2462212, EPI_ISL_2462213, EPI_ISL_2462214, EPI_ISL_2462215, EPI_ISL_2462216, EPI_ISL_2462217, EPI_ISL_2462218, EPI_ISL_2462219, EPI_ISL_2462220, EPI_ISL_2462221, EPI_ISL_2462222, EPI_ISL_2462223, EPI_ISL_2462224, EPI_ISL_2462225, EPI_ISL_2462226, EPI_ISL_2462227, EPI_ISL_2462228, EPI_ISL_2462229, EPI_ISL_2462230, EPI_ISL_2462231, EPI_ISL_2462232, EPI_ISL_2462233, EPI_ISL_2462234, EPI_ISL_2462235, EPI_ISL_2462236, EPI_ISL_2462237, EPI_ISL_2462238, EPI_ISL_2462239, EPI_ISL_2462240, EPI_ISL_2462241, EPI_ISL_2462242, EPI_ISL_2462243, EPI_ISL_2462244, EPI_ISL_2462245, EPI_ISL_2462246, EPI_ISL_2462247, EPI_ISL_2462248, EPI_ISL_2462249, EPI_ISL_2462250, EPI_ISL_2462251, EPI_ISL_2462252, EPI_ISL_2462253, EPI_ISL_2462254, EPI_ISL_2462255, EPI_ISL_2462256, EPI_ISL_2462257, EPI_ISL_2462258, EPI_ISL_2462259, EPI_ISL_2462260, EPI_ISL_2462261, EPI_ISL_2462262, EPI_ISL_2462263, EPI_ISL_2462264, EPI_ISL_2462265, EPI_ISL_2462266, EPI_ISL_2462267, EPI_ISL_2462268, EPI_ISL_2462269, EPI_ISL_2462270, EPI_ISL_2462271, EPI_ISL_2462272, EPI_ISL_2462273, EPI_ISL_2462274, EPI_ISL_2462277, EPI_ISL_2462278, EPI_ISL_2462279, EPI_ISL_2462280, EPI_ISL_2462281, EPI_ISL_2462282, EPI_ISL_2462283, EPI_ISL_2462284, EPI_ISL_2462285, EPI_ISL_2462286, EPI_ISL_2462287, EPI_ISL_2462288, EPI_ISL_2462289, EPI_ISL_2462290, EPI_ISL_2462291, EPI_ISL_2462292, EPI_ISL_2462293, EPI_ISL_2462294, EPI_ISL_2462295, EPI_ISL_2462296, EPI_ISL_2462297, EPI_ISL_2462298, EPI_ISL_2462299, EPI_ISL_2462300, EPI_ISL_2462301, EPI_ISL_2462302, EPI_ISL_2462303, EPI_ISL_2462304, EPI_ISL_2462305, EPI_ISL_2462306, EPI_ISL_2462307, EPI_ISL_2462308, EPI_ISL_2462309, EPI_ISL_2462310, EPI_ISL_2462311 | | | |
| see above | OHSU Lab Services Molecular Microbiology Lab | Oregon SARS-CoV-2 Genome Sequencing Center | Brendan L. O'Connell, Sonia Acharya, Cierra LaBlanc, Ruth V. Nichols, Alec J. Hirsch, Donna Hansel, Guang Fan, Xuan Qin, Daniel N. Streblow, William B. Messer, Andrew C. Adey, Benjamin N. Bimber, Brian J. O'Roak |
| EPI_ISL_2462312, | Ohio Department of Health Laboratory | Ohio Department of Health Laboratory | Holmes, Jennifer; Eric Brandt, Keoni Omura, Glen McGillivary, Caitlin McDonnell, Jade Mowery, Stephanie Mccracken, Tyler Payne, Kirtana Ramadugu, Erica Leasure, Brent Lee, Kelsey Florek, Heather Blankenship, Allison Black, Quanta Brown, and Tammy Bannerman |

| | | | |
|---|---|---|---|
| EPI_ISL_2462313, EPI_ISL_2462369, EPI_ISL_2462371, EPI_ISL_2462372, EPI_ISL_2462374, EPI_ISL_2462403 | | | |
| EPI_ISL_2462404 | SUNY Upstate Medical University | SUNY Upstate Medical University | Frank A Middleton |
| EPI_ISL_2462405, EPI_ISL_2462406, EPI_ISL_2462435, EPI_ISL_2462436, EPI_ISL_2462437, EPI_ISL_2462438, EPI_ISL_2462439 | Ohio Department of Health Laboratory | Ohio Department of Health Laboratory | Holmes, Jennifer; Eric Brandt, Keoni Omura, Glen McGillivary, Caitlin McDonnell, Jade Mowery, Stephanie Mccracken, Tyler Payne, Kirtana Ramadugu, Erica Leasure, Brent Lee, Kelsey Florek, Heather Blankenship, Allison Black, Quanta Brown, and Tammy Bannerman |
| EPI_ISL_2463001 | MT Public Health Laboratory | Centers for Disease Control and Prevention Division of Viral Diseases, Pathogen Discovery | Mili Sheth, Sarah Nobles, Jasmine Padilla, Mark Burroughs, Shoshona Le, Katie Dillon, Peter Cook, Clinton R. Paden, Dhwani Batra, Krista Queen, Kristen Knipe, Dakota Howard, Yvette Unoarumhi, Darlene Wagner, Matthew Schmerer, Ben L. Rambo-Martin, Kristine Lacek, Sam Shepard, Alison Laufer Halpin, Dave Wentworth, Vivien Dugan, Suxiang Tong, Justin Lee |
| EPI_ISL_2463002 | IL Dept. of Public Health Springfield Laboratory | Centers for Disease Control and Prevention Division of Viral Diseases, Pathogen Discovery | Mili Sheth, Sarah Nobles, Jasmine Padilla, Mark Burroughs, Shoshona Le, Katie Dillon, Peter Cook, Clinton R. Paden, Dhwani Batra, Krista Queen, Kristen Knipe, Dakota Howard, Yvette Unoarumhi, Darlene Wagner, Matthew Schmerer, Ben L. Rambo-Martin, Kristine Lacek, Sam Shepard, Alison Laufer Halpin, Dave Wentworth, Vivien Dugan, Suxiang Tong, Justin Lee |
| EPI_ISL_2463003 | "TX DSHS, Lab Services Section MC 1947" | Centers for Disease Control and Prevention Division of Viral Diseases, Pathogen Discovery | Mili Sheth, Sarah Nobles, Jasmine Padilla, Mark Burroughs, Shoshona Le, Katie Dillon, Peter Cook, Clinton R. Paden, Dhwani Batra, Krista Queen, Kristen Knipe, Dakota Howard, Yvette Unoarumhi, Darlene Wagner, Matthew Schmerer, Ben L. Rambo-Martin, Kristine Lacek, Sam Shepard, Alison Laufer Halpin, Dave Wentworth, Vivien Dugan, Suxiang Tong, Justin Lee |
| EPI_ISL_2463004 | IN State Department of Health Laboratory Services | Centers for Disease Control and Prevention Division of Viral Diseases, Pathogen Discovery | Mili Sheth, Sarah Nobles, Jasmine Padilla, Mark Burroughs, Shoshona Le, Katie Dillon, Peter Cook, Clinton R. Paden, Dhwani Batra, Krista Queen, Kristen Knipe, Dakota Howard, Yvette Unoarumhi, Darlene Wagner, Matthew Schmerer, Ben L. Rambo-Martin, Kristine Lacek, Sam Shepard, Alison Laufer Halpin, Dave Wentworth, Vivien Dugan, Suxiang Tong, Justin Lee |
| EPI_ISL_2463005 | RI State Health Laboratories | Centers for Disease Control and Prevention Division of Viral Diseases, Pathogen Discovery | Mili Sheth, Sarah Nobles, Jasmine Padilla, Mark Burroughs, Shoshona Le, Katie Dillon, Peter Cook, Clinton R. Paden, Dhwani Batra, Krista Queen, Kristen Knipe, Dakota Howard, Yvette Unoarumhi, Darlene Wagner, Matthew Schmerer, Ben L. Rambo-Martin, Kristine Lacek, Sam Shepard, Alison Laufer Halpin, Dave Wentworth, Vivien Dugan, Suxiang Tong, Justin Lee |
| EPI_ISL_2463006 | MD DOH Laboratories Administration | Centers for Disease Control and Prevention Division of Viral Diseases, Pathogen Discovery | Mili Sheth, Sarah Nobles, Jasmine Padilla, Mark Burroughs, Shoshona Le, Katie Dillon, Peter Cook, Clinton R. Paden, Dhwani Batra, Krista Queen, Kristen Knipe, Dakota Howard, Yvette Unoarumhi, Darlene Wagner, Matthew Schmerer, Ben L. Rambo-Martin, Kristine Lacek, Sam Shepard, Alison Laufer Halpin, Dave Wentworth, Vivien Dugan, Suxiang Tong, Justin Lee |
| EPI_ISL_2463007, EPI_ISL_2463008, EPI_ISL_2463009, EPI_ISL_2463010, EPI_ISL_2463011, EPI_ISL_2463012, EPI_ISL_2463013, EPI_ISL_2463014, EPI_ISL_2463015, EPI_ISL_2463016, EPI_ISL_2463017, EPI_ISL_2463018, EPI_ISL_2463019, EPI_ISL_2463020, EPI_ISL_2463021, EPI_ISL_2463022, EPI_ISL_2463023, EPI_ISL_2463024, EPI_ISL_2463025, EPI_ISL_2463026, EPI_ISL_2463027, EPI_ISL_2463028, EPI_ISL_2463029, EPI_ISL_2463030, EPI_ISL_2463031, EPI_ISL_2463032, EPI_ISL_2463033, EPI_ISL_2463034, EPI_ISL_2463035, EPI_ISL_2463036, EPI_ISL_2463037, EPI_ISL_2463038, EPI_ISL_2463039, EPI_ISL_2463040, EPI_ISL_2463041, EPI_ISL_2463042, EPI_ISL_2463043, EPI_ISL_2463044, EPI_ISL_2463045, EPI_ISL_2463046, EPI_ISL_2463047, EPI_ISL_2463048, EPI_ISL_2463049, EPI_ISL_2463050, EPI_ISL_2463051, EPI_ISL_2463052, EPI_ISL_2463053, EPI_ISL_2463054, EPI_ISL_2463055, EPI_ISL_2463056, EPI_ISL_2463057, EPI_ISL_2463058, EPI_ISL_2463059, EPI_ISL_2463060, EPI_ISL_2463061, EPI_ISL_2463062, EPI_ISL_2463063, EPI_ISL_2463064, EPI_ISL_2463065, EPI_ISL_2463066, EPI_ISL_2463067, EPI_ISL_2463068, EPI_ISL_2463069, EPI_ISL_2463070, EPI_ISL_2463071, EPI_ISL_2463072, EPI_ISL_2463073, EPI_ISL_2463074, EPI_ISL_2463075, EPI_ISL_2463076, EPI_ISL_2463077, EPI_ISL_2463078, EPI_ISL_2463079, EPI_ISL_2463080, EPI_ISL_2463081, EPI_ISL_2463082, EPI_ISL_2463083, EPI_ISL_2463084, EPI_ISL_2463085, EPI_ISL_2463086, EPI_ISL_2463087, EPI_ISL_2463088, EPI_ISL_2463089, EPI_ISL_2463090, EPI_ISL_2463091, EPI_ISL_2463092, EPI_ISL_2463093, EPI_ISL_2463094, EPI_ISL_2463095, EPI_ISL_2463096, EPI_ISL_2463097, EPI_ISL_2463098, EPI_ISL_2463099, EPI_ISL_2463100, EPI_ISL_2463101, EPI_ISL_2463102, EPI_ISL_2463103, EPI_ISL_2463104, EPI_ISL_2463105, EPI_ISL_2463106, EPI_ISL_2463107, EPI_ISL_2463108, EPI_ISL_2463109, EPI_ISL_2463110, EPI_ISL_2463111, EPI_ISL_2463112, EPI_ISL_2463113, EPI_ISL_2463114, EPI_ISL_2463115, EPI_ISL_2463116, EPI_ISL_2463117, EPI_ISL_2463118, EPI_ISL_2463119, EPI_ISL_2463120, EPI_ISL_2463121, EPI_ISL_2463122, EPI_ISL_2463123, EPI_ISL_2463124, EPI_ISL_2463125, EPI_ISL_2463126, EPI_ISL_2463127, EPI_ISL_2463128, EPI_ISL_2463129, EPI_ISL_2463130, EPI_ISL_2463131, EPI_ISL_2463132, EPI_ISL_2463133, EPI_ISL_2463134, EPI_ISL_2463135, EPI_ISL_2463136, EPI_ISL_2463137, EPI_ISL_2463138, EPI_ISL_2463139, EPI_ISL_2463140, EPI_ISL_2463141, EPI_ISL_2463142, EPI_ISL_2463143, EPI_ISL_2463144, EPI_ISL_2463145, EPI_ISL_2463146, EPI_ISL_2463147, EPI_ISL_2463148, EPI_ISL_2463149, EPI_ISL_2463150, EPI_ISL_2463151, EPI_ISL_2463152, EPI_ISL_2463153, EPI_ISL_2463154, EPI_ISL_2463155, EPI_ISL_2463156, EPI_ISL_2463157, EPI_ISL_2463158, EPI_ISL_2463159, EPI_ISL_2463160, EPI_ISL_2463161, EPI_ISL_2463162, EPI_ISL_2463163, EPI_ISL_2463164, EPI_ISL_2463165, EPI_ISL_2463166, EPI_ISL_2463167, EPI_ISL_2463168, EPI_ISL_2463169, EPI_ISL_2463170, EPI_ISL_2463171, EPI_ISL_2463172, EPI_ISL_2463173, EPI_ISL_2463174, EPI_ISL_2463175, EPI_ISL_2463176, EPI_ISL_2463177, EPI_ISL_2463178, EPI_ISL_2463179, EPI_ISL_2463180, EPI_ISL_2463181, EPI_ISL_2463182, EPI_ISL_2463183, EPI_ISL_2463184, EPI_ISL_2463185, EPI_ISL_2463186, EPI_ISL_2463187, EPI_ISL_2463188, EPI_ISL_2463189, EPI_ISL_2463190, EPI_ISL_2463191, EPI_ISL_2463192, EPI_ISL_2463193, EPI_ISL_2463194, EPI_ISL_2463195, EPI_ISL_2463196, EPI_ISL_2463197, EPI_ISL_2463198, EPI_ISL_2463199, EPI_ISL_2463200, EPI_ISL_2463201, EPI_ISL_2463202, EPI_ISL_2463203, EPI_ISL_2463204, EPI_ISL_2463205, EPI_ISL_2463206, EPI_ISL_2463207, EPI_ISL_2463208, EPI_ISL_2463209, EPI_ISL_2463210, EPI_ISL_2463211, EPI_ISL_2463212, EPI_ISL_2463213, EPI_ISL_2463214, EPI_ISL_2463215, EPI_ISL_2463216, EPI_ISL_2463217, EPI_ISL_2463218, EPI_ISL_2463219, EPI_ISL_2463220, EPI_ISL_2463221, EPI_ISL_2463222, EPI_ISL_2463223, EPI_ISL_2463224, EPI_ISL_2463225, EPI_ISL_2463226, EPI_ISL_2463227, EPI_ISL_2463228, EPI_ISL_2463229, EPI_ISL_2463230, EPI_ISL_2463231, EPI_ISL_2463232, EPI_ISL_2463233, EPI_ISL_2463234, EPI_ISL_2463235, EPI_ISL_2463236, EPI_ISL_2463237, EPI_ISL_2463238, EPI_ISL_2463239, EPI_ISL_2463240, EPI_ISL_2463241, EPI_ISL_2463242, EPI_ISL_2463243, EPI_ISL_2463244, EPI_ISL_2463245, EPI_ISL_2463246, EPI_ISL_2463247, EPI_ISL_2463248, EPI_ISL_2463249, EPI_ISL_2463250, EPI_ISL_2463251, EPI_ISL_2463252, EPI_ISL_2463253, EPI_ISL_2463254, EPI_ISL_2463255, EPI_ISL_2463256, EPI_ISL_2463257, EPI_ISL_2463258, EPI_ISL_2463259, EPI_ISL_2463260, EPI_ISL_2463261, EPI_ISL_2463262, EPI_ISL_2463263, EPI_ISL_2463264, EPI_ISL_2463265, EPI_ISL_2463266, EPI_ISL_2463267, EPI_ISL_2463268, EPI_ISL_2463269, EPI_ISL_2463270, EPI_ISL_2463271, EPI_ISL_2463272, EPI_ISL_2463273, EPI_ISL_2463274, EPI_ISL_2463275, EPI_ISL_2463276, EPI_ISL_2463277, EPI_ISL_2463278, EPI_ISL_2463279, EPI_ISL_2463280, EPI_ISL_2463281 | | | |
| see above | Quest Diagnostics Incorporated | Centers for Disease Control and Prevention Division of Viral Diseases, Pathogen Discovery | Dakota Howard, Dhwani Batra, Peter W. Cook, Kara Moser, Adrian Paskey, Jason Caravas, Benjamin Rambo-Martin, Shatavia Morrison, Christopher Gulvick, Scott Sammons, Yvette Unoarumhi, Darlene Wagner, Matthew Schmerer, S. H. Rosenthal, A. Gerasimova, R. M. Kagan, B. Anderson, M. Hua, Y. Liu, L.E. Bernstein, K.E. Livingston, A. Perez, I. A. Shlyakhter, R. V. Rolando, R. Owen, P. Tanpaiboon, F. Lacbawan, Clinton R. Paden, Duncan MacCannell |
| EPI_ISL_2463282, EPI_ISL_2463283, EPI_ISL_2463287 | Fulgent Genetics | Fulgent Genetics | Harry Gao, Mickey Li, John Gao, Joseph Fierro, Benafsh Sapra, Becky Tsai, Yan Meng, Doreen Ng, James Xie |
| EPI_ISL_2463288 | University of Wisconsin-Madison AIDS Vaccine Research Laboratories | University of Wisconsin-Madison AIDS Vaccine Research Laboratories | Gage Moreno, Katarina Braun, et al. AIDS Vaccine Research Laboratories |
| EPI_ISL_2463297 | NYU Langone Health | Departments of Pathology and Medicine, New York University School of Medicine | Adriana Heguy, Dacia Dimartino, Emily Guzman, Christian Marier, Peter Meyn, Sitharam Ramaswami, Gael Westby, Paul Zappile, Yutong Zhang, Paolo Cotzia, Guiqing Wang |
| EPI_ISL_2463299, EPI_ISL_2463761, EPI_ISL_2463836, EPI_ISL_2463838, EPI_ISL_2464021, EPI_ISL_2464022, EPI_ISL_2464023 | Fulgent Genetics | Fulgent Genetics | Harry Gao, Mickey Li, John Gao, Joseph Fierro, Benafsh Sapra, Becky Tsai, Yan Meng, Doreen Ng, James Xie |
| EPI_ISL_2464133, EPI_ISL_2464167, EPI_ISL_2464279, EPI_ISL_2464280, EPI_ISL_2464376, EPI_ISL_2464377, EPI_ISL_2464378, EPI_ISL_2464379, EPI_ISL_2464380, EPI_ISL_2464385, EPI_ISL_2464412, EPI_ISL_2464413, EPI_ISL_2464415, EPI_ISL_2464416, EPI_ISL_2464417, EPI_ISL_2464418 | | | |
| see above | Ohio Department of Health Laboratory | Ohio Department of Health Laboratory | Holmes, Jennifer; Eric Brandt, Keoni Omura, Glen McGillivary, Caitlin McDonnell, Jade Mowery, Stephanie Mccracken, Tyler Payne, Kirtana Ramadugu, Erica Leasure, Brent Lee, Kelsey Florek, Heather Blankenship, Allison Black, Quanta Brown, and Tammy Bannerman |
| EPI_ISL_2464565, EPI_ISL_2464566, EPI_ISL_2464568, EPI_ISL_2464569, EPI_ISL_2464570, EPI_ISL_2464571, EPI_ISL_2464573, EPI_ISL_2464574, EPI_ISL_2464575, EPI_ISL_2464576, EPI_ISL_2464577, EPI_ISL_2464578, EPI_ISL_2464579, EPI_ISL_2464581, EPI_ISL_2464582, EPI_ISL_2464583, EPI_ISL_2464584, EPI_ISL_2464585, EPI_ISL_2464586, EPI_ISL_2464587, EPI_ISL_2464588, EPI_ISL_2464589, EPI_ISL_2464590, EPI_ISL_2464591, EPI_ISL_2464592, EPI_ISL_2464593, EPI_ISL_2464594, EPI_ISL_2464595, EPI_ISL_2464596, EPI_ISL_2464598, EPI_ISL_2464599, EPI_ISL_2464600, EPI_ISL_2464601, EPI_ISL_2464602, EPI_ISL_2464604, EPI_ISL_2464605, EPI_ISL_2464606, EPI_ISL_2464607, EPI_ISL_2464608, EPI_ISL_2464609, EPI_ISL_2464610, EPI_ISL_2464611, EPI_ISL_2464613, EPI_ISL_2464614, EPI_ISL_2464615, EPI_ISL_2464617, EPI_ISL_2464618, EPI_ISL_2464620, EPI_ISL_2464621, EPI_ISL_2464622, EPI_ISL_2464623, EPI_ISL_2464625 | | | |
| see above | Maryland Genomics, Institute for Genome Sciences, University of Maryland School of Medicine | Maryland Genomics, Institute for Genome Sciences, University of Maryland School of Medicine | Tallon, Luke J; Sadzewicz, Lisa D; Humphrys, Mike; Ott, Sandra; Roussey, Holly; Mehta, Aditya; Vavikolanu, Kranthi; Fraser, Claire M; Ravel, Jacques |
| EPI_ISL_2465955, EPI_ISL_2465956, EPI_ISL_2465957, EPI_ISL_2465958, EPI_ISL_2465963, EPI_ISL_2465964, EPI_ISL_2465966, EPI_ISL_2465967, EPI_ISL_2465968, EPI_ISL_2465969, EPI_ISL_2465970, EPI_ISL_2465971, EPI_ISL_2465973, EPI_ISL_2465975, EPI_ISL_2465976, EPI_ISL_2465977, EPI_ISL_2465978, EPI_ISL_2465980, EPI_ISL_2465981, EPI_ISL_2465982, EPI_ISL_2465984, EPI_ISL_2465986, EPI_ISL_2465988, EPI_ISL_2465989, EPI_ISL_2465990, EPI_ISL_2465991, EPI_ISL_2465993, EPI_ISL_2465995, EPI_ISL_2465997, EPI_ISL_2466000, EPI_ISL_2466001, EPI_ISL_2466003, EPI_ISL_2466004, EPI_ISL_2466005, EPI_ISL_2466006, EPI_ISL_2466007, EPI_ISL_2466008, EPI_ISL_2466009, EPI_ISL_2466010, EPI_ISL_2466011, EPI_ISL_2466014, EPI_ISL_2466016, EPI_ISL_2466017, EPI_ISL_2466018, EPI_ISL_2466019, EPI_ISL_2466021, EPI_ISL_2466022, EPI_ISL_2466024, EPI_ISL_2466025, EPI_ISL_2466026, EPI_ISL_2466031, EPI_ISL_2466032, EPI_ISL_2466033, EPI_ISL_2466034, EPI_ISL_2466035, EPI_ISL_2466036 | | | |
| see above | Curative | New Mexico Department of Health Scientific Laboratory | Ellie Johnson, D'eldra Malone, Jennifer Benoit, Linda Salazar, Ratheesh Rajan, Keila Gutierrez, Anastacia Griego-Fisher |
| EPI_ISL_2466040 | Oregon State Public Health Laboratory | Oregon State Public Health Laboratory | Rafia Razzaque, Eugene Yeboah, Vanda Makris, Laura Tsaknaridis, John Fontana and Shane Sevey |
| EPI_ISL_2466041, EPI_ISL_2466042, EPI_ISL_2466043 | Department of General Services Division of Consolidated Laboratory Services, Virginia Division of Consolidated Laboratory Services Sequencing Submission Group | Department of General Services Division of Consolidated Laboratory Services, Virginia Division of Consolidated Laboratory Services Sequencing Submission Group | Fink,L. |
| EPI_ISL_2466046 | Respiratory Viruses Branch, Centers for Disease Control and Prevention | Respiratory Viruses Branch, Centers for Disease Control and Prevention | Howard,D., Batra,D., Cook,P.W., Moser,K., Paskey,A., Caravas,J., Rambo-Martin,B., Morrison,S., Gulvick,C., Sammons,S., Unoarumhi,Y., Schmerer,M., Roychoudhury,P., Xie,H., Shrestha,L., Nguyen,T.V., Bakhash,S.M., Lin,M., Baker,N.R., Perez,R., Ellis,S., Breit,N., Livingston,R.J., Huang,M.-L., Jerome,K.R., Mathias,P., Greninger,A., Paden,C.R. and MacCannell,D. |
| EPI_ISL_2466047, EPI_ISL_2466048, EPI_ISL_2466049 | Alaska State Virology Lab, Alaska State Public Health Labs | Alaska State Virology Lab, Alaska State Public Health Labs | DeRonde,S., House,E., Zidek,J., Redlinger,M., Smith,L., Chen,J. |
| EPI_ISL_2466468, EPI_ISL_2466469, EPI_ISL_2466470, EPI_ISL_2466471, EPI_ISL_2466472, EPI_ISL_2466473, EPI_ISL_2466474, EPI_ISL_2466475, EPI_ISL_2466476, EPI_ISL_2466477, EPI_ISL_2466478, EPI_ISL_2466480, EPI_ISL_2466481, EPI_ISL_2466482, EPI_ISL_2466483, EPI_ISL_2466484, EPI_ISL_2466485, EPI_ISL_2466486, EPI_ISL_2466487, EPI_ISL_2466488, EPI_ISL_2466489, EPI_ISL_2466492, EPI_ISL_2466493 | | | |
| see above | Wyoming Public Health Laboratory | Wyoming Public Health Laboratory | Jim Mildenberger, Taylor Fearing, Channing Weber, Ashley Norberg, Chayse Rowley, Marley Goetz, Brian Dominguez, Elliot Thomasson, Sam Britz, Cari Sloma, Robert Petit, and Rob Christensen |
| EPI_ISL_2466511, EPI_ISL_2466512, EPI_ISL_2466513, EPI_ISL_2466514, EPI_ISL_2466515, EPI_ISL_2466516, EPI_ISL_2466517, EPI_ISL_2466518, EPI_ISL_2466519, EPI_ISL_2466520, EPI_ISL_2466521, EPI_ISL_2466522, EPI_ISL_2466523, EPI_ISL_2466524 | | | |
| see above | Wexner Medical Center | The Ohio State University College of Medicine | Koenig,S., Seminetta,J. |
| EPI_ISL_2466525, EPI_ISL_2466526, EPI_ISL_2466527, EPI_ISL_2466529, EPI_ISL_2466530, EPI_ISL_2466531, EPI_ISL_2466532, EPI_ISL_2466533, EPI_ISL_2466534, EPI_ISL_2466537, EPI_ISL_2466538, EPI_ISL_2466539, EPI_ISL_2466540, EPI_ISL_2466541, EPI_ISL_2466542, EPI_ISL_2466543, EPI_ISL_2466544, EPI_ISL_2466545, EPI_ISL_2466546, EPI_ISL_2466547, EPI_ISL_2466548, EPI_ISL_2466549 | | | |
| see above | Wyoming Public Health Laboratory | Wyoming Public Health Laboratory | Jim Mildenberger, Taylor Fearing, Channing Weber, Ashley Norberg, Chayse Rowley, Marley Goetz, Brian Dominguez, Elliot Thomasson,Sam Britz, Cari Sloma, Robert Petit, and Rob Christensen |
| EPI_ISL_2466551 | Division of Viral Diseases, CDC Pathogen Discovery Team | Division of Viral Diseases, CDC Pathogen Discovery Team | Piantadosi,A., Babiker,A., Sheth,M., Nobles,S., Padilla,J.,Burroughs,M., Davis,M., Cook,P., Paden,C.R., Batra,D., Knipe,K.,Howard,D., Unoarumhi,Y., Schmerer,M., Rambo-Martin,B.L., Lacek,K.,Shepard,S., Laufer Halpin,A., Wentworth,D., Dugan,V. and Lee,J. |

| | | | |
|---|---|---|---|
| EPI_ISL_2466668, EPI_ISL_2466669, EPI_ISL_2466670 | Stony Brook Medicine | New York Genome Center | Michael Zody, Andre Corvelo, Dayna M. Oschwald, Samantha Fennessey, Tom Maniatis , Sarah A. Halbohn, Alaa B. Salaheldin, Sarah A. Malik, Josue M. Flores Acuna, Dimitri Gnatenko, Silvia G. Spitzer, Margaret M. McGovern, Eric D. Spitzer, Richard R. Kew, Kenneth R. Shroyer, Jiang Chen, Susan V. Donelan |
| EPI_ISL_2466681 | DOHMH PHL | New York City Public Health Laboratory | Jade Wang, et al. |
| EPI_ISL_2466682, EPI_ISL_2466683 | DOHMH Corona | New York City Public Health Laboratory | Jade Wang, et al. |
| EPI_ISL_2466684, EPI_ISL_2466685 | DOHMH Central Harlem | New York City Public Health Laboratory | Jade Wang, et al. |
| EPI_ISL_2466686 | DOHMH Jamaica | New York City Public Health Laboratory | Jade Wang, et al. |
| EPI_ISL_2466687 | DOHMH Chelsea | New York City Public Health Laboratory | Jade Wang, et al. |
| EPI_ISL_2466688 | DOHMH Corona | New York City Public Health Laboratory | Jade Wang, et al. |
| EPI_ISL_2466689 | DOHMH Jamaica | New York City Public Health Laboratory | Jade Wang, et al. |
| EPI_ISL_2466690 | DOHMH Corona | New York City Public Health Laboratory | Jade Wang, et al. |
| EPI_ISL_2466691, EPI_ISL_2466692, EPI_ISL_2466693, EPI_ISL_2466694, EPI_ISL_2466695, EPI_ISL_2466696 | Sherman Abrams Laboratory | New York City Public Health Laboratory | Jade Wang, et al. |
| EPI_ISL_2466697 | Department of Homeless Services | New York City Public Health Laboratory | Jade Wang, et al. |
| EPI_ISL_2466698, EPI_ISL_2466699 | Quest Diagnostics | New York City Public Health Laboratory | Jade Wang, et al. |
| EPI_ISL_2466700, EPI_ISL_2466701, EPI_ISL_2466702 | OCME Office Of Chief Medical Examiner | New York City Public Health Laboratory | Jade Wang, et al. |
| EPI_ISL_2466704, EPI_ISL_2466705, EPI_ISL_2466706, EPI_ISL_2466708, EPI_ISL_2466711 | The Jackson Laboratory | The Jackson Laboratory | Long J, Bergeron D, Renzette N, Adams M, Omerza G, Kelly K, Li L |
| EPI_ISL_2466736, EPI_ISL_2466737, EPI_ISL_2466738, EPI_ISL_2466739, EPI_ISL_2466740, EPI_ISL_2466741, EPI_ISL_2466742, EPI_ISL_2466743, EPI_ISL_2466744, EPI_ISL_2466745, EPI_ISL_2466746, EPI_ISL_2466747, EPI_ISL_2466748, EPI_ISL_2466749, EPI_ISL_2466750 | | | |
| see above | Nebraska Public Health Laboratory | NPHL COVID-19 Response Team | NPHL COVID-19 Response Team |
| EPI_ISL_2466752, EPI_ISL_2466753 | Kansas Health and Environmental Lab | Kansas Health and Environmental Lab | Mike Grose, Katherine Wiggins, Jonathan Barnell, Ben Olsen, and Phil Adam |
| EPI_ISL_2466754, EPI_ISL_2466755, EPI_ISL_2466756, EPI_ISL_2466757, EPI_ISL_2466758, EPI_ISL_2466759, EPI_ISL_2466760, EPI_ISL_2466761, EPI_ISL_2466762 | Wichita State University - Molecular Diagnostics Lab | Kansas Health and Environmental Lab | Mike Grose, Katherine Wiggins, Jonathan Barnell, Ben Olsen, and Phil Adam |
| EPI_ISL_2466763, EPI_ISL_2466764, EPI_ISL_2466765, EPI_ISL_2466766, EPI_ISL_2466767, EPI_ISL_2466768, EPI_ISL_2466769, EPI_ISL_2466770, EPI_ISL_2466771 | Kansas Health and Environmental Lab | Kansas Health and Environmental Lab | Mike Grose, Katherine Wiggins, Jonathan Barnell, Ben Olsen, and Phil Adam |
| EPI_ISL_2466831, EPI_ISL_2466832, EPI_ISL_2466833, EPI_ISL_2466834, EPI_ISL_2466835, EPI_ISL_2466836, EPI_ISL_2466837, EPI_ISL_2466838, EPI_ISL_2466839 | Wexner Medical Center | The Ohio State University College of Medicine | Koenig,S., Seminetta,J. |
| EPI_ISL_2466842, EPI_ISL_2466843, EPI_ISL_2466844, EPI_ISL_2466845, EPI_ISL_2466846, EPI_ISL_2466847, EPI_ISL_2466848, EPI_ISL_2466849, EPI_ISL_2466850, EPI_ISL_2466851, EPI_ISL_2466852, EPI_ISL_2466853, EPI_ISL_2466854, EPI_ISL_2466855, EPI_ISL_2466856, EPI_ISL_2466857, EPI_ISL_2466858, EPI_ISL_2466859, EPI_ISL_2466860, EPI_ISL_2466861, EPI_ISL_2466862, EPI_ISL_2466863, EPI_ISL_2466864, EPI_ISL_2466865, EPI_ISL_2466866, EPI_ISL_2466867, EPI_ISL_2466868, EPI_ISL_2466869, EPI_ISL_2466870, EPI_ISL_2466871, EPI_ISL_2466872, EPI_ISL_2466873, EPI_ISL_2466874, EPI_ISL_2466875, EPI_ISL_2466876, EPI_ISL_2466877, EPI_ISL_2466878, EPI_ISL_2466879, EPI_ISL_2466880, EPI_ISL_2466881, EPI_ISL_2466882, EPI_ISL_2466883, EPI_ISL_2466884, EPI_ISL_2466885, EPI_ISL_2466886, EPI_ISL_2466887, EPI_ISL_2466888, EPI_ISL_2466889, EPI_ISL_2466890, EPI_ISL_2466891, EPI_ISL_2466892, EPI_ISL_2466893, EPI_ISL_2466894, EPI_ISL_2466895, EPI_ISL_2466896, EPI_ISL_2466897, EPI_ISL_2466898, EPI_ISL_2466899, EPI_ISL_2466900, EPI_ISL_2466901, EPI_ISL_2466902, EPI_ISL_2466903, EPI_ISL_2466904, EPI_ISL_2466905, EPI_ISL_2466906, EPI_ISL_2466907, EPI_ISL_2466908, EPI_ISL_2466909, EPI_ISL_2466910, EPI_ISL_2466911, EPI_ISL_2466912, EPI_ISL_2466913, EPI_ISL_2466914, EPI_ISL_2466915, EPI_ISL_2466916, EPI_ISL_2466917, EPI_ISL_2466918, EPI_ISL_2466919, EPI_ISL_2466920, EPI_ISL_2466921, EPI_ISL_2466922, EPI_ISL_2466923, EPI_ISL_2466924, EPI_ISL_2466925, EPI_ISL_2466926, EPI_ISL_2466927, EPI_ISL_2466928, EPI_ISL_2466929, EPI_ISL_2466930, EPI_ISL_2466931, EPI_ISL_2466932, EPI_ISL_2466933, EPI_ISL_2466934, EPI_ISL_2466935, EPI_ISL_2466936, EPI_ISL_2466937, EPI_ISL_2466938, EPI_ISL_2466939, EPI_ISL_2466940, EPI_ISL_2466941, EPI_ISL_2466942, EPI_ISL_2466943, EPI_ISL_2466944, EPI_ISL_2466945, EPI_ISL_2466946, EPI_ISL_2466947, EPI_ISL_2466948, EPI_ISL_2466949, EPI_ISL_2466950, EPI_ISL_2466951, EPI_ISL_2466952, EPI_ISL_2466953, EPI_ISL_2466954, EPI_ISL_2466955, EPI_ISL_2466956, EPI_ISL_2466957, EPI_ISL_2466958, EPI_ISL_2466959, EPI_ISL_2466960, EPI_ISL_2466961, EPI_ISL_2466962, EPI_ISL_2466963, EPI_ISL_2466964, EPI_ISL_2466965, EPI_ISL_2466966, EPI_ISL_2466967, EPI_ISL_2466968, EPI_ISL_2466969, EPI_ISL_2466970, EPI_ISL_2466971, EPI_ISL_2466972, EPI_ISL_2466973, EPI_ISL_2466974, EPI_ISL_2466975, EPI_ISL_2466976, EPI_ISL_2466977, EPI_ISL_2466978, EPI_ISL_2466979, EPI_ISL_2466980, EPI_ISL_2466981, EPI_ISL_2466982, EPI_ISL_2466983, EPI_ISL_2466984, EPI_ISL_2466985, EPI_ISL_2466986, EPI_ISL_2466987, EPI_ISL_2466988, EPI_ISL_2466989, EPI_ISL_2466990, EPI_ISL_2466991, EPI_ISL_2466992, EPI_ISL_2466993, EPI_ISL_2466994, EPI_ISL_2466995, EPI_ISL_2466996, EPI_ISL_2466997, EPI_ISL_2466998, EPI_ISL_2466999, EPI_ISL_2467000, EPI_ISL_2467001, EPI_ISL_2467002, EPI_ISL_2467003, EPI_ISL_2467004, EPI_ISL_2467005, EPI_ISL_2467006, EPI_ISL_2467007, EPI_ISL_2467008, EPI_ISL_2467009, EPI_ISL_2467010, EPI_ISL_2467011, EPI_ISL_2467012, EPI_ISL_2467013, EPI_ISL_2467014, EPI_ISL_2467015, EPI_ISL_2467016, EPI_ISL_2467017, EPI_ISL_2467018, EPI_ISL_2467020, EPI_ISL_2467021, EPI_ISL_2467022, EPI_ISL_2467023, EPI_ISL_2467024, EPI_ISL_2467025, EPI_ISL_2467026, EPI_ISL_2467027, EPI_ISL_2467028, EPI_ISL_2467029, EPI_ISL_2467030, EPI_ISL_2467031, EPI_ISL_2467032, EPI_ISL_2467033, EPI_ISL_2467034, EPI_ISL_2467035, EPI_ISL_2467036, EPI_ISL_2467037, EPI_ISL_2467038, EPI_ISL_2467039, EPI_ISL_2467040, EPI_ISL_2467041, EPI_ISL_2467042, EPI_ISL_2467043, EPI_ISL_2467044, EPI_ISL_2467045, EPI_ISL_2467046, EPI_ISL_2467047, EPI_ISL_2467048, EPI_ISL_2467049, EPI_ISL_2467050, EPI_ISL_2467051, EPI_ISL_2467052, EPI_ISL_2467053, EPI_ISL_2467054, EPI_ISL_2467055, EPI_ISL_2467056, EPI_ISL_2467057, EPI_ISL_2467058, EPI_ISL_2467059, EPI_ISL_2467060, EPI_ISL_2467061, EPI_ISL_2467062, EPI_ISL_2467063, EPI_ISL_2467064, EPI_ISL_2467065, EPI_ISL_2467066, EPI_ISL_2467067, EPI_ISL_2467068, EPI_ISL_2467069, EPI_ISL_2467070, EPI_ISL_2467071, EPI_ISL_2467072, EPI_ISL_2467073, EPI_ISL_2467074 | | | |
| see above | Utah Public Health Laboratory | Utah Public Health Laboratory | Erin L. Young, Kelly F. Oakeson, Tara Gallagher |
| EPI_ISL_2467374, EPI_ISL_2467375, EPI_ISL_2467376, EPI_ISL_2467380, EPI_ISL_2467385, EPI_ISL_2467388, EPI_ISL_2467390, EPI_ISL_2467392, EPI_ISL_2467393, EPI_ISL_2467394, EPI_ISL_2467399, EPI_ISL_2467401, EPI_ISL_2467402, EPI_ISL_2467403, EPI_ISL_2467405, EPI_ISL_2467406, EPI_ISL_2467407, EPI_ISL_2467410, EPI_ISL_2467411, EPI_ISL_2467412, EPI_ISL_2467413, EPI_ISL_2467414, EPI_ISL_2467415, EPI_ISL_2467416, EPI_ISL_2467417, EPI_ISL_2467418, EPI_ISL_2467419, EPI_ISL_2467420, EPI_ISL_2467422, EPI_ISL_2467423, EPI_ISL_2467424, EPI_ISL_2467425, EPI_ISL_2467426, EPI_ISL_2467427, EPI_ISL_2467428, EPI_ISL_2467430, EPI_ISL_2467432, EPI_ISL_2467433, EPI_ISL_2467434, EPI_ISL_2467435, EPI_ISL_2467436, EPI_ISL_2467437, EPI_ISL_2467439, EPI_ISL_2467440, EPI_ISL_2467441, EPI_ISL_2467442, EPI_ISL_2467443, EPI_ISL_2467444, EPI_ISL_2467445, EPI_ISL_2467446, EPI_ISL_2467448, EPI_ISL_2467450, EPI_ISL_2467452, EPI_ISL_2467453, EPI_ISL_2467455, EPI_ISL_2467456, EPI_ISL_2467457, EPI_ISL_2467458, EPI_ISL_2467459, EPI_ISL_2467460, EPI_ISL_2467461, EPI_ISL_2467462, EPI_ISL_2467463, EPI_ISL_2467464, EPI_ISL_2467465, EPI_ISL_2467466, EPI_ISL_2467467, EPI_ISL_2467468, EPI_ISL_2467469, EPI_ISL_2467470, EPI_ISL_2467471, EPI_ISL_2467472, EPI_ISL_2467473, EPI_ISL_2467474, EPI_ISL_2467476, EPI_ISL_2467478, EPI_ISL_2467481, EPI_ISL_2467482, EPI_ISL_2467483, EPI_ISL_2467484, EPI_ISL_2467485, EPI_ISL_2467486, EPI_ISL_2467487, EPI_ISL_2467488, EPI_ISL_2467489, EPI_ISL_2467490, EPI_ISL_2467491, EPI_ISL_2467492, EPI_ISL_2467493, EPI_ISL_2467494, EPI_ISL_2467495, EPI_ISL_2467497, EPI_ISL_2467498, EPI_ISL_2467500, EPI_ISL_2467501, EPI_ISL_2467502, EPI_ISL_2467503, EPI_ISL_2467504, EPI_ISL_2467505, EPI_ISL_2467506, EPI_ISL_2467507, EPI_ISL_2467508, EPI_ISL_2467510, EPI_ISL_2467511, EPI_ISL_2467513, EPI_ISL_2467514, EPI_ISL_2467515, EPI_ISL_2467516 | | | |
| see above | Illinois Department of Public Health | Illinois Department of Public Health - Chicago Lab | Vineet K. Dhiman, Ira Heimler, Joel Price |
| EPI_ISL_2467520, EPI_ISL_2467521, EPI_ISL_2467522, EPI_ISL_2467523, EPI_ISL_2467524, EPI_ISL_2467525, EPI_ISL_2467526, EPI_ISL_2467527, EPI_ISL_2467528 | GA Department of Public Health | GA Department of Public Health | Stacy Reeves, Jonathan Edwards, Cynthia Dixey, Tonia Parrott, Aliyah Fields, Taylor Smith |
| EPI_ISL_2467782, EPI_ISL_2467783, EPI_ISL_2467784, EPI_ISL_2467785, EPI_ISL_2467786, EPI_ISL_2467787, EPI_ISL_2467788, EPI_ISL_2467790, EPI_ISL_2467791, EPI_ISL_2467792, EPI_ISL_2467793, EPI_ISL_2467794, EPI_ISL_2467796 | | | |
| see above | University of Wisconsin-Madison AIDS Vaccine Research Laboratories | University of Wisconsin-Madison AIDS Vaccine Research Laboratories | Gage Moreno, Katarina Braun, et al. AIDS Vaccine Research Laboratories |
| EPI_ISL_2467798, EPI_ISL_2467799, EPI_ISL_2467800, EPI_ISL_2467801, EPI_ISL_2467802, EPI_ISL_2467803, EPI_ISL_2467804, EPI_ISL_2467805, EPI_ISL_2467806, EPI_ISL_2467807, EPI_ISL_2467808, EPI_ISL_2467809, EPI_ISL_2467810, EPI_ISL_2467811, EPI_ISL_2467812, EPI_ISL_2467813, EPI_ISL_2467814, EPI_ISL_2467815, EPI_ISL_2467816, EPI_ISL_2467817, EPI_ISL_2467818, EPI_ISL_2467819, EPI_ISL_2467820, EPI_ISL_2467821, EPI_ISL_2467822, EPI_ISL_2467823, EPI_ISL_2467824, EPI_ISL_2467825, EPI_ISL_2467826, EPI_ISL_2467827, EPI_ISL_2467828, EPI_ISL_2467829, EPI_ISL_2467830, EPI_ISL_2467831, EPI_ISL_2467832, EPI_ISL_2467833 | | | |
| see above | Alaska State Virology Laboratory | Alaska State Virology Laboratory | Stephanie DeRonde, Elva House, Jacob Zidek, Lisa Smith, Ph.D., Jack Chen, Ph.D. |
| EPI_ISL_2467866, EPI_ISL_2467867, | Wexner Medical Center | The Ohio State University College of Medicine | Koenig,S., Seminetta,J. |

| | | | |
|---|---|---|---|
| EPI_ISL_2467868, EPI_ISL_2467869, EPI_ISL_2467870 | | | |
| EPI_ISL_2467872, EPI_ISL_2467873, EPI_ISL_2467874, EPI_ISL_2467875, EPI_ISL_2467876, EPI_ISL_2467877, EPI_ISL_2467878, EPI_ISL_2467879 | Rockford Ambulatory Surgery Center | BioInfoExperts LLC | Sarah E. Hagarty, David J Nolan, Rebecca Rose, Susanna L Lamers, Ed Yavitz |
| EPI_ISL_2467880 | WVDHHR - Office of Laboratory Services | Genomics and Discovery, Respiratory Viruses Branch, Division of Viral Diseases, Centers for Disease Control and Prevention | Ying Tao, Yan Li, Jing Zhang, Anna Kelleher, Brian Lynch, Krista Queen, Anna Uehara, Peter Cook, Han Jia Justin Ng, Rachel Marine, Clinton R. Paden, Dhwani Batra, Haibin Wang, Tara Coalter, Jasmine Padilla, Morgan Davis, Mili Sheth, Sarah Nobles, Mark Burroughs, Justin Lee, Adam Retchless, Suxiang Tong |
| EPI_ISL_2467881 | NC State Laboratory of Public Health | Genomics and Discovery, Respiratory Viruses Branch, Division of Viral Diseases, Centers for Disease Control and Prevention | Ying Tao, Yan Li, Jing Zhang, Anna Kelleher, Brian Lynch, Krista Queen, Anna Uehara, Peter Cook, Han Jia Justin Ng, Rachel Marine, Clinton R. Paden, Dhwani Batra, Haibin Wang, Tara Coalter, Jasmine Padilla, Morgan Davis, Mili Sheth, Sarah Nobles, Mark Burroughs, Justin Lee, Adam Retchless, Suxiang Tong |
| EPI_ISL_2467882, EPI_ISL_2467883 | AR Dept. of Health-PHL, Molecular Diagnostics | Genomics and Discovery, Respiratory Viruses Branch, Division of Viral Diseases, Centers for Disease Control and Prevention | Ying Tao, Yan Li, Jing Zhang, Anna Kelleher, Brian Lynch, Krista Queen, Anna Uehara, Peter Cook, Han Jia Justin Ng, Rachel Marine, Clinton R. Paden, Dhwani Batra, Haibin Wang, Tara Coalter, Jasmine Padilla, Morgan Davis, Mili Sheth, Sarah Nobles, Mark Burroughs, Justin Lee, Adam Retchless, Suxiang Tong |
| EPI_ISL_2467884, EPI_ISL_2467885, EPI_ISL_2467886 | OK Public Health Laboratory, Oklahoma State DOH | Genomics and Discovery, Respiratory Viruses Branch, Division of Viral Diseases, Centers for Disease Control and Prevention | Ying Tao, Yan Li, Jing Zhang, Anna Kelleher, Brian Lynch, Krista Queen, Anna Uehara, Peter Cook, Han Jia Justin Ng, Rachel Marine, Clinton R. Paden, Dhwani Batra, Haibin Wang, Tara Coalter, Jasmine Padilla, Morgan Davis, Mili Sheth, Sarah Nobles, Mark Burroughs, Justin Lee, Adam Retchless, Suxiang Tong |
| EPI_ISL_2467887, EPI_ISL_2467888, EPI_ISL_2467889, EPI_ISL_2467890 | GA Department of Public Health Laboratory | Genomics and Discovery, Respiratory Viruses Branch, Division of Viral Diseases, Centers for Disease Control and Prevention | Ying Tao, Yan Li, Jing Zhang, Anna Kelleher, Brian Lynch, Krista Queen, Anna Uehara, Peter Cook, Han Jia Justin Ng, Rachel Marine, Clinton R. Paden, Dhwani Batra, Haibin Wang, Tara Coalter, Jasmine Padilla, Morgan Davis, Mili Sheth, Sarah Nobles, Mark Burroughs, Justin Lee, Adam Retchless, Suxiang Tong |
| EPI_ISL_2467891, EPI_ISL_2467892, EPI_ISL_2467893, EPI_ISL_2467894, EPI_ISL_2467895, EPI_ISL_2467896, EPI_ISL_2467897, EPI_ISL_2467898, EPI_ISL_2467899, EPI_ISL_2467900, EPI_ISL_2467901, EPI_ISL_2467902, EPI_ISL_2467903, EPI_ISL_2467904 | | | |
| see above | PA Department of Health, Bureau of Laboratories | Genomics and Discovery, Respiratory Viruses Branch, Division of Viral Diseases, Centers for Disease Control and Prevention | Ying Tao, Yan Li, Jing Zhang, Anna Kelleher, Brian Lynch, Krista Queen, Anna Uehara, Peter Cook, Han Jia Justin Ng, Rachel Marine, Clinton R. Paden, Dhwani Batra, Haibin Wang, Tara Coalter, Jasmine Padilla, Morgan Davis, Mili Sheth, Sarah Nobles, Mark Burroughs, Justin Lee, Adam Retchless, Suxiang Tong |
| EPI_ISL_2472919, EPI_ISL_2472920, EPI_ISL_2472921, EPI_ISL_2472922, EPI_ISL_2472923, EPI_ISL_2472924, EPI_ISL_2472925, EPI_ISL_2472926, EPI_ISL_2472927, EPI_ISL_2472928, EPI_ISL_2472929, EPI_ISL_2472930, EPI_ISL_2472931, EPI_ISL_2472932, EPI_ISL_2472933, EPI_ISL_2472934, EPI_ISL_2472935, EPI_ISL_2472937, EPI_ISL_2472938, EPI_ISL_2472939, EPI_ISL_2472940, EPI_ISL_2472941, EPI_ISL_2472942, EPI_ISL_2472943, EPI_ISL_2472944, EPI_ISL_2472945, EPI_ISL_2472946, EPI_ISL_2472947, EPI_ISL_2472948, EPI_ISL_2472949, EPI_ISL_2472950, EPI_ISL_2472951, EPI_ISL_2472952, EPI_ISL_2472953, EPI_ISL_2472954, EPI_ISL_2472955, EPI_ISL_2472956, EPI_ISL_2472957, EPI_ISL_2472958, EPI_ISL_2472959, EPI_ISL_2472960, EPI_ISL_2472961, EPI_ISL_2472962, EPI_ISL_2472964, EPI_ISL_2472965, EPI_ISL_2472966, EPI_ISL_2472967, EPI_ISL_2472968, EPI_ISL_2472969, EPI_ISL_2472970, EPI_ISL_2472971, EPI_ISL_2472972, EPI_ISL_2472973, EPI_ISL_2472974, EPI_ISL_2472975, EPI_ISL_2472976, EPI_ISL_2472977, EPI_ISL_2472978, EPI_ISL_2472979, EPI_ISL_2472980, EPI_ISL_2472981, EPI_ISL_2472982, EPI_ISL_2472983, EPI_ISL_2472984, EPI_ISL_2472986, EPI_ISL_2472987, EPI_ISL_2472988, EPI_ISL_2472989, EPI_ISL_2472990, EPI_ISL_2472991, EPI_ISL_2472992, EPI_ISL_2472993, EPI_ISL_2472994, EPI_ISL_2472995, EPI_ISL_2472996, EPI_ISL_2472997, EPI_ISL_2472998, EPI_ISL_2472999, EPI_ISL_2473000, EPI_ISL_2473001, EPI_ISL_2473002, EPI_ISL_2473003, EPI_ISL_2473004, EPI_ISL_2473005, EPI_ISL_2473006, EPI_ISL_2473007, EPI_ISL_2473008, EPI_ISL_2473009, EPI_ISL_2473010, EPI_ISL_2473011, EPI_ISL_2473012, EPI_ISL_2473013, EPI_ISL_2473015, EPI_ISL_2473016, EPI_ISL_2473017, EPI_ISL_2473018, EPI_ISL_2473019, EPI_ISL_2473020, EPI_ISL_2473021, EPI_ISL_2473022, EPI_ISL_2473023, EPI_ISL_2473024, EPI_ISL_2473025, EPI_ISL_2473026, EPI_ISL_2473027, EPI_ISL_2473028, EPI_ISL_2473029, EPI_ISL_2473030, EPI_ISL_2473031, EPI_ISL_2473032, EPI_ISL_2473033, EPI_ISL_2473034, EPI_ISL_2473035, EPI_ISL_2473036, EPI_ISL_2473037, EPI_ISL_2473038, EPI_ISL_2473039, EPI_ISL_2473040, EPI_ISL_2473041, EPI_ISL_2473042, EPI_ISL_2473043, EPI_ISL_2473044, EPI_ISL_2473045, EPI_ISL_2473046, EPI_ISL_2473047, EPI_ISL_2473048, EPI_ISL_2473049, EPI_ISL_2473050, EPI_ISL_2473051, EPI_ISL_2473052, EPI_ISL_2473053, EPI_ISL_2473054, EPI_ISL_2473055, EPI_ISL_2473056, EPI_ISL_2473057, EPI_ISL_2473058, EPI_ISL_2473059, EPI_ISL_2473060, EPI_ISL_2473061, EPI_ISL_2473062, EPI_ISL_2473063, EPI_ISL_2473064, EPI_ISL_2473065, EPI_ISL_2473066, EPI_ISL_2473067, EPI_ISL_2473068, EPI_ISL_2473069, EPI_ISL_2473070, EPI_ISL_2473071, EPI_ISL_2473072, EPI_ISL_2473073, EPI_ISL_2473074, EPI_ISL_2473075, EPI_ISL_2473076, EPI_ISL_2473077, EPI_ISL_2473078, EPI_ISL_2473079, EPI_ISL_2473080, EPI_ISL_2473081, EPI_ISL_2473082, EPI_ISL_2473083, EPI_ISL_2473084, EPI_ISL_2473085, EPI_ISL_2473086, EPI_ISL_2473087, EPI_ISL_2473088, EPI_ISL_2473090, EPI_ISL_2473091, EPI_ISL_2473092, EPI_ISL_2473093, EPI_ISL_2473094, EPI_ISL_2473095, EPI_ISL_2473096, EPI_ISL_2473097, EPI_ISL_2473098, EPI_ISL_2473099, EPI_ISL_2473100, EPI_ISL_2473101, EPI_ISL_2473102, EPI_ISL_2473103, EPI_ISL_2473104, EPI_ISL_2473105, EPI_ISL_2473106, EPI_ISL_2473107, EPI_ISL_2473108, EPI_ISL_2473109, EPI_ISL_2473110, EPI_ISL_2473111, EPI_ISL_2473112, EPI_ISL_2473113, EPI_ISL_2473114, EPI_ISL_2473115, EPI_ISL_2473116, EPI_ISL_2473117, EPI_ISL_2473118, EPI_ISL_2473119, EPI_ISL_2473120, EPI_ISL_2473121, EPI_ISL_2473122, EPI_ISL_2473123, EPI_ISL_2473124, EPI_ISL_2473125, EPI_ISL_2473126, EPI_ISL_2473127, EPI_ISL_2473128, EPI_ISL_2473129, EPI_ISL_2473130, EPI_ISL_2473131, EPI_ISL_2473132, EPI_ISL_2473133, EPI_ISL_2473134, EPI_ISL_2473135, EPI_ISL_2473136, EPI_ISL_2473137, EPI_ISL_2473138, EPI_ISL_2473139, EPI_ISL_2473140, EPI_ISL_2473141, EPI_ISL_2473142, EPI_ISL_2473143, EPI_ISL_2473144, EPI_ISL_2473145, EPI_ISL_2473146, EPI_ISL_2473147, EPI_ISL_2473148, EPI_ISL_2473150, EPI_ISL_2473151, EPI_ISL_2473152, EPI_ISL_2473153, EPI_ISL_2473154, EPI_ISL_2473155, EPI_ISL_2473156, EPI_ISL_2473157, EPI_ISL_2473158, EPI_ISL_2473159, EPI_ISL_2473160, EPI_ISL_2473161, EPI_ISL_2473162, EPI_ISL_2473163, EPI_ISL_2473164, EPI_ISL_2473165, EPI_ISL_2473166, EPI_ISL_2473167, EPI_ISL_2473168, EPI_ISL_2473169, EPI_ISL_2473170, EPI_ISL_2473171, EPI_ISL_2473172, EPI_ISL_2473173, EPI_ISL_2473174, EPI_ISL_2473175, EPI_ISL_2473176, EPI_ISL_2473177, EPI_ISL_2473178, EPI_ISL_2473179, EPI_ISL_2473180, EPI_ISL_2473181, EPI_ISL_2473182, EPI_ISL_2473183, EPI_ISL_2473184, EPI_ISL_2473185, EPI_ISL_2473186, EPI_ISL_2473187, EPI_ISL_2473188, EPI_ISL_2473189, EPI_ISL_2473190, EPI_ISL_2473191, EPI_ISL_2473192, EPI_ISL_2473193, EPI_ISL_2473194, EPI_ISL_2473195, EPI_ISL_2473196, EPI_ISL_2473197, EPI_ISL_2473198, EPI_ISL_2473199, EPI_ISL_2473200, EPI_ISL_2473201, EPI_ISL_2473202, EPI_ISL_2473203, EPI_ISL_2473204, EPI_ISL_2473205, EPI_ISL_2473206, EPI_ISL_2473207, EPI_ISL_2473208, EPI_ISL_2473209, EPI_ISL_2473210, EPI_ISL_2473211, EPI_ISL_2473212, EPI_ISL_2473213, EPI_ISL_2473214, EPI_ISL_2473215, EPI_ISL_2473216, EPI_ISL_2473217, EPI_ISL_2473218, EPI_ISL_2473219, EPI_ISL_2473220, EPI_ISL_2473221, EPI_ISL_2473222, EPI_ISL_2473223, EPI_ISL_2473224, EPI_ISL_2473225, EPI_ISL_2473226, EPI_ISL_2473227, EPI_ISL_2473228, EPI_ISL_2473229, EPI_ISL_2473230, EPI_ISL_2473231, EPI_ISL_2473232, EPI_ISL_2473233, EPI_ISL_2473234, EPI_ISL_2473235, EPI_ISL_2473236, EPI_ISL_2473237, EPI_ISL_2473238, EPI_ISL_2473239, EPI_ISL_2473240, EPI_ISL_2473241, EPI_ISL_2473242, EPI_ISL_2473243, EPI_ISL_2473244, EPI_ISL_2473245, EPI_ISL_2473246, EPI_ISL_2473247, EPI_ISL_2473248, EPI_ISL_2473249, EPI_ISL_2473250, EPI_ISL_2473251, EPI_ISL_2473252, EPI_ISL_2473253, EPI_ISL_2473254, EPI_ISL_2473255, EPI_ISL_2473256, EPI_ISL_2473257, EPI_ISL_2473258, EPI_ISL_2473259, EPI_ISL_2473260, EPI_ISL_2473261, EPI_ISL_2473262, EPI_ISL_2473263, EPI_ISL_2473264, EPI_ISL_2473265, EPI_ISL_2473266, EPI_ISL_2473267, EPI_ISL_2473268, EPI_ISL_2473269, EPI_ISL_2473270, EPI_ISL_2473271, EPI_ISL_2473272, EPI_ISL_2473273, EPI_ISL_2473274, EPI_ISL_2473275, EPI_ISL_2473276, EPI_ISL_2473277, EPI_ISL_2473278, EPI_ISL_2473279, EPI_ISL_2473280, EPI_ISL_2473281, EPI_ISL_2473282, EPI_ISL_2473283, EPI_ISL_2473284, EPI_ISL_2473285, EPI_ISL_2473286, EPI_ISL_2473287, EPI_ISL_2473288, EPI_ISL_2473289, EPI_ISL_2473290, EPI_ISL_2473291, EPI_ISL_2473292, EPI_ISL_2473293, EPI_ISL_2473294, EPI_ISL_2473295, EPI_ISL_2473296, EPI_ISL_2473297, EPI_ISL_2473298, EPI_ISL_2473299, EPI_ISL_2473300, EPI_ISL_2473301, EPI_ISL_2473302, EPI_ISL_2473303, EPI_ISL_2473304, EPI_ISL_2473305, EPI_ISL_2473306, EPI_ISL_2473307, EPI_ISL_2473308, EPI_ISL_2473309, EPI_ISL_2473310, EPI_ISL_2473311, EPI_ISL_2473312, EPI_ISL_2473313, EPI_ISL_2473314, EPI_ISL_2473315, EPI_ISL_2473316, EPI_ISL_2473317, EPI_ISL_2473318, EPI_ISL_2473319, EPI_ISL_2473320, EPI_ISL_2473321, EPI_ISL_2473322, EPI_ISL_2473323, EPI_ISL_2473325, EPI_ISL_2473327, EPI_ISL_2473328, EPI_ISL_2473330, EPI_ISL_2473331, EPI_ISL_2473332, EPI_ISL_2473333, EPI_ISL_2473334, EPI_ISL_2473335, EPI_ISL_2473336, EPI_ISL_2473337, EPI_ISL_2473338, EPI_ISL_2473339, EPI_ISL_2473340, EPI_ISL_2473341, EPI_ISL_2473342, EPI_ISL_2473343, EPI_ISL_2473344, EPI_ISL_2473345, EPI_ISL_2473346, EPI_ISL_2473347, EPI_ISL_2473348, EPI_ISL_2473349, EPI_ISL_2473350, EPI_ISL_2473351, EPI_ISL_2473352, EPI_ISL_2473353, EPI_ISL_2473354, EPI_ISL_2473355, EPI_ISL_2473356, EPI_ISL_2473357, EPI_ISL_2473358, EPI_ISL_2473359, EPI_ISL_2473360, EPI_ISL_2473361, EPI_ISL_2473362, EPI_ISL_2473363, EPI_ISL_2473364, EPI_ISL_2473365, EPI_ISL_2473366, EPI_ISL_2473367, EPI_ISL_2473368, EPI_ISL_2473369, EPI_ISL_2473370, EPI_ISL_2473371, EPI_ISL_2473372, EPI_ISL_2473373, EPI_ISL_2473374, EPI_ISL_2473375, EPI_ISL_2473376, EPI_ISL_2473377, EPI_ISL_2473378, EPI_ISL_2473379, EPI_ISL_2473380, EPI_ISL_2473381, EPI_ISL_2473382, EPI_ISL_2473383, EPI_ISL_2473384, EPI_ISL_2473385, EPI_ISL_2473386, EPI_ISL_2473387, EPI_ISL_2473388, EPI_ISL_2473389, EPI_ISL_2473390, EPI_ISL_2473391, EPI_ISL_2473392, EPI_ISL_2473393, EPI_ISL_2473394, EPI_ISL_2473395, EPI_ISL_2473396, EPI_ISL_2473397, EPI_ISL_2473398, EPI_ISL_2473399, EPI_ISL_2473400, EPI_ISL_2473401, EPI_ISL_2473402 | | | |
| see above | State Testing Facility | Altius Institute for Biomedical Research | Daniel Bates, Rebecca Bruders, Michael Buckley, Mark Frerker, Amanda Gale, Clem Green, Muhammad Halimun, Kneshay Harper, Matt Hartman, Alex Isner, Audra Johnson, Jessica Kunder, Lauren Mitchell, Jemma Nelson, Alex Nguyen, Sofia Olsson, Sadie Patraw, Tobias Ragoczy, Joshua Richards, Jean Robinston, Jacob Rodriguez, John Stamatoyannopoulos, Eric Thorland, Julia Wald |
| EPI_ISL_2473403 | ResCare Washington | Altius Institute for Biomedical Research | Daniel Bates, Rebecca Bruders, Michael Buckley, Mark Frerker, Amanda Gale, Clem Green, Muhammad Halimun, Kneshay Harper, Matt Hartman, Alex Isner, Audra Johnson, Jessica Kunder, Lauren Mitchell, Jemma Nelson, Alex Nguyen, Sofia Olsson, Sadie Patraw, Tobias Ragoczy, Joshua Richards, Jean Robinston, Jacob Rodriguez, John Stamatoyannopoulos, Eric Thorland, Julia Wald |
| EPI_ISL_2473404, EPI_ISL_2473405, EPI_ISL_2473406, EPI_ISL_2473407, EPI_ISL_2473408, EPI_ISL_2473409, EPI_ISL_2473410, EPI_ISL_2473411, EPI_ISL_2473412, EPI_ISL_2473413, EPI_ISL_2473414, EPI_ISL_2473415, EPI_ISL_2473416, EPI_ISL_2473417, EPI_ISL_2473418, EPI_ISL_2473419, EPI_ISL_2473420, EPI_ISL_2473421, EPI_ISL_2473422, EPI_ISL_2473423, EPI_ISL_2473424, EPI_ISL_2473425, EPI_ISL_2473426, EPI_ISL_2473427, EPI_ISL_2473428, EPI_ISL_2473429, EPI_ISL_2473430, EPI_ISL_2473431, EPI_ISL_2473432, EPI_ISL_2473433, EPI_ISL_2473434, EPI_ISL_2473435, EPI_ISL_2473436, EPI_ISL_2473437, EPI_ISL_2473438, EPI_ISL_2473439, EPI_ISL_2473440, EPI_ISL_2473441, EPI_ISL_2473442, EPI_ISL_2473443, EPI_ISL_2473444, EPI_ISL_2473445, EPI_ISL_2473446, EPI_ISL_2473447, EPI_ISL_2473448, EPI_ISL_2473449, EPI_ISL_2473450, EPI_ISL_2473451, EPI_ISL_2473452, EPI_ISL_2473453, EPI_ISL_2473454, EPI_ISL_2473455, EPI_ISL_2473456, EPI_ISL_2473457, EPI_ISL_2473458, EPI_ISL_2473459, EPI_ISL_2473460, EPI_ISL_2473461, EPI_ISL_2473462, EPI_ISL_2473463, EPI_ISL_2473464, EPI_ISL_2473465, EPI_ISL_2473466, EPI_ISL_2473467, EPI_ISL_2473468, EPI_ISL_2473469, EPI_ISL_2473470, EPI_ISL_2473471, EPI_ISL_2473472, EPI_ISL_2473473, EPI_ISL_2473474, EPI_ISL_2473475, EPI_ISL_2473476, EPI_ISL_2473477, EPI_ISL_2473478, EPI_ISL_2473479, EPI_ISL_2473480, EPI_ISL_2473481, EPI_ISL_2473482, EPI_ISL_2473483, EPI_ISL_2473484, EPI_ISL_2473485, EPI_ISL_2473486, EPI_ISL_2473487, EPI_ISL_2473488, EPI_ISL_2473489, EPI_ISL_2473490, EPI_ISL_2473491, EPI_ISL_2473492, EPI_ISL_2473493, EPI_ISL_2473494, EPI_ISL_2473495, EPI_ISL_2473496, EPI_ISL_2473497, EPI_ISL_2473498, EPI_ISL_2473499, EPI_ISL_2473500, EPI_ISL_2473501, EPI_ISL_2473502, EPI_ISL_2473503, EPI_ISL_2473504, EPI_ISL_2473505, EPI_ISL_2473506, EPI_ISL_2473507, EPI_ISL_2473509, EPI_ISL_2473510, EPI_ISL_2473511, EPI_ISL_2473512, EPI_ISL_2473513, EPI_ISL_2473514, EPI_ISL_2473515, EPI_ISL_2473516, EPI_ISL_2473517, EPI_ISL_2473518, EPI_ISL_2473519, EPI_ISL_2473520, EPI_ISL_2473521, EPI_ISL_2473522, EPI_ISL_2473523, EPI_ISL_2473524, EPI_ISL_2473525, EPI_ISL_2473526, EPI_ISL_2473527, EPI_ISL_2473528, EPI_ISL_2473529, EPI_ISL_2473530, EPI_ISL_2473531, EPI_ISL_2473532, EPI_ISL_2473533, EPI_ISL_2473534, EPI_ISL_2473535, EPI_ISL_2473536, EPI_ISL_2473537, EPI_ISL_2473538, EPI_ISL_2473539, EPI_ISL_2473540, EPI_ISL_2473541, EPI_ISL_2473542, EPI_ISL_2473543, EPI_ISL_2473544, EPI_ISL_2473545, EPI_ISL_2473546, EPI_ISL_2473547, EPI_ISL_2473548, EPI_ISL_2473549, EPI_ISL_2473550, EPI_ISL_2473551, EPI_ISL_2473552, EPI_ISL_2473553, EPI_ISL_2473554, EPI_ISL_2473555, EPI_ISL_2473556, EPI_ISL_2473557, EPI_ISL_2473558, EPI_ISL_2473559, EPI_ISL_2473560, EPI_ISL_2473561, EPI_ISL_2473562, EPI_ISL_2473563, EPI_ISL_2473564, EPI_ISL_2473565, EPI_ISL_2473566, EPI_ISL_2473567, EPI_ISL_2473568, EPI_ISL_2473569, EPI_ISL_2473570, EPI_ISL_2473571, EPI_ISL_2473572, EPI_ISL_2473573, EPI_ISL_2473574, EPI_ISL_2473575, EPI_ISL_2473577, EPI_ISL_2473578, EPI_ISL_2473579, EPI_ISL_2473580, EPI_ISL_2473581, EPI_ISL_2473582, EPI_ISL_2473583, EPI_ISL_2473584, EPI_ISL_2473585, EPI_ISL_2473586, EPI_ISL_2473587, EPI_ISL_2473588, EPI_ISL_2473589, EPI_ISL_2473590, EPI_ISL_2473591, EPI_ISL_2473592, EPI_ISL_2473593, EPI_ISL_2473594, EPI_ISL_2473595, EPI_ISL_2473596, EPI_ISL_2473597, EPI_ISL_2473598, EPI_ISL_2473599, EPI_ISL_2473600, EPI_ISL_2473601, EPI_ISL_2473602, EPI_ISL_2473603, EPI_ISL_2473604, EPI_ISL_2473605, EPI_ISL_2473606, EPI_ISL_2473607, EPI_ISL_2473608, EPI_ISL_2473609, EPI_ISL_2473610, EPI_ISL_2473611, EPI_ISL_2473612, EPI_ISL_2473613, EPI_ISL_2473614, EPI_ISL_2473615, EPI_ISL_2473616, EPI_ISL_2473617, EPI_ISL_2473618, EPI_ISL_2473619, EPI_ISL_2473620, EPI_ISL_2473621, EPI_ISL_2473622, EPI_ISL_2473623, EPI_ISL_2473624, EPI_ISL_2473625, EPI_ISL_2473626, EPI_ISL_2473627, EPI_ISL_2473628, EPI_ISL_2473629, EPI_ISL_2473630, EPI_ISL_2473631, EPI_ISL_2473632, EPI_ISL_2473633, EPI_ISL_2473634, EPI_ISL_2473635, EPI_ISL_2473636, EPI_ISL_2473638, EPI_ISL_2473639, EPI_ISL_2473640, EPI_ISL_2473641, EPI_ISL_2473642, EPI_ISL_2473643, EPI_ISL_2473644, EPI_ISL_2473645, EPI_ISL_2473646, EPI_ISL_2473647, EPI_ISL_2473648, EPI_ISL_2473649, EPI_ISL_2473650, EPI_ISL_2473651, EPI_ISL_2473652, EPI_ISL_2473653, EPI_ISL_2473654 | | | |
| see above | State Testing Facility | Altius Institute for Biomedical Research | Daniel Bates, Rebecca Bruders, Michael Buckley, Mark Frerker, Amanda Gale, Clem Green, Muhammad Halimun, Kneshay Harper, Matt Hartman, Alex Isner, Audra Johnson, Jessica Kunder, Lauren Mitchell, Jemma Nelson, Alex Nguyen, Sofia Olsson, Sadie Patraw, Tobias Ragoczy, Joshua Richards, Jean Robinston, Jacob Rodriguez, John Stamatoyannopoulos, Eric Thorland, Julia Wald |
| EPI_ISL_2473655, EPI_ISL_2473656, EPI_ISL_2473657, EPI_ISL_2473658, EPI_ISL_2473659, EPI_ISL_2473660, EPI_ISL_2473661, EPI_ISL_2473662, EPI_ISL_2473663, EPI_ISL_2473664, EPI_ISL_2473665, EPI_ISL_2473666, EPI_ISL_2473667, EPI_ISL_2473668, EPI_ISL_2473669, EPI_ISL_2473670, EPI_ISL_2473671, EPI_ISL_2473672, EPI_ISL_2473673, EPI_ISL_2473674, EPI_ISL_2473675, EPI_ISL_2473676, EPI_ISL_2473677, EPI_ISL_2473678, EPI_ISL_2473679 | | | |
| see above | Oregon State Public Health Laboratory | Oregon State Public Health Laboratory | Rafia Razzaque, Eugene Yeboah, Vanda Makris, Laura Tsaknaridis, John Fontana and Shane Sevey |
| EPI_ISL_2473680, EPI_ISL_2473681, EPI_ISL_2473682, EPI_ISL_2473683, EPI_ISL_2473684, EPI_ISL_2473685, EPI_ISL_2473686, EPI_ISL_2473687, EPI_ISL_2473688, EPI_ISL_2473689 | Alaska State Virology Laboratory | Alaska State Virology Laboratory | Stephanie DeRonde, Elva House, Jacob Zidek, Lisa Smith, Ph.D., Jack Chen, Ph.D. |
| EPI_ISL_2473836, EPI_ISL_2473837, EPI_ISL_2473838, EPI_ISL_2473839, EPI_ISL_2473840, EPI_ISL_2473841, EPI_ISL_2473842, EPI_ISL_2473843, EPI_ISL_2473844, EPI_ISL_2473845, EPI_ISL_2473846, EPI_ISL_2473847, EPI_ISL_2473848, EPI_ISL_2473849, EPI_ISL_2473850, EPI_ISL_2473851, EPI_ISL_2473852, EPI_ISL_2473853, EPI_ISL_2473854, EPI_ISL_2473855, EPI_ISL_2473856, EPI_ISL_2473857, EPI_ISL_2473858, EPI_ISL_2473859, EPI_ISL_2473860, EPI_ISL_2473861, EPI_ISL_2473862, EPI_ISL_2473863, EPI_ISL_2473864, EPI_ISL_2473865, EPI_ISL_2473866, EPI_ISL_2473867, EPI_ISL_2473868, EPI_ISL_2473869, EPI_ISL_2473870, EPI_ISL_2473871, EPI_ISL_2473872, EPI_ISL_2473873, EPI_ISL_2473874, EPI_ISL_2473875, EPI_ISL_2473876, EPI_ISL_2473877, EPI_ISL_2473878, EPI_ISL_2473879, EPI_ISL_2473880, EPI_ISL_2473881, EPI_ISL_2473882, EPI_ISL_2473883, EPI_ISL_2473884, EPI_ISL_2473885, EPI_ISL_2473886, EPI_ISL_2473887, EPI_ISL_2473888, EPI_ISL_2473889, EPI_ISL_2473890, EPI_ISL_2473891, EPI_ISL_2473892, EPI_ISL_2473893, EPI_ISL_2473894, EPI_ISL_2473895, EPI_ISL_2473896, EPI_ISL_2473897, EPI_ISL_2473898, EPI_ISL_2473899, EPI_ISL_2473900, EPI_ISL_2473901, EPI_ISL_2473902, EPI_ISL_2473903, EPI_ISL_2473904, EPI_ISL_2473905, EPI_ISL_2473906, EPI_ISL_2473907, EPI_ISL_2473908, EPI_ISL_2473909, EPI_ISL_2473910, EPI_ISL_2473911, EPI_ISL_2473912, EPI_ISL_2473913, EPI_ISL_2473914, EPI_ISL_2473915, EPI_ISL_2473916, EPI_ISL_2473917, EPI_ISL_2473918, EPI_ISL_2473919, EPI_ISL_2473920, EPI_ISL_2473921, EPI_ISL_2473922, EPI_ISL_2473923, EPI_ISL_2473924, EPI_ISL_2473925, EPI_ISL_2473926, EPI_ISL_2473927, EPI_ISL_2473928, EPI_ISL_2473929, EPI_ISL_2473930, EPI_ISL_2473931, EPI_ISL_2473932, EPI_ISL_2473933, EPI_ISL_2473934, EPI_ISL_2473935, EPI_ISL_2473936, EPI_ISL_2473937, EPI_ISL_2473938, EPI_ISL_2473939, EPI_ISL_2473940, EPI_ISL_2473941, EPI_ISL_2473942, EPI_ISL_2473943, EPI_ISL_2473944, EPI_ISL_2473945, EPI_ISL_2473946, EPI_ISL_2473947, EPI_ISL_2473948, EPI_ISL_2473949, EPI_ISL_2473950, EPI_ISL_2473951, EPI_ISL_2473952, EPI_ISL_2473953, EPI_ISL_2473954, EPI_ISL_2473955, EPI_ISL_2473956, EPI_ISL_2473957, EPI_ISL_2473958, EPI_ISL_2473959, EPI_ISL_2473960, EPI_ISL_2473961, EPI_ISL_2473962, EPI_ISL_2473963, EPI_ISL_2473964, EPI_ISL_2473965, EPI_ISL_2473966, EPI_ISL_2473967, EPI_ISL_2473968, EPI_ISL_2473969, EPI_ISL_2473970, EPI_ISL_2473971, EPI_ISL_2473972, EPI_ISL_2473973, EPI_ISL_2473974, EPI_ISL_2473975, EPI_ISL_2473976, EPI_ISL_2473977, EPI_ISL_2473978, EPI_ISL_2473979, EPI_ISL_2473980, EPI_ISL_2473981, EPI_ISL_2473982, EPI_ISL_2473983, EPI_ISL_2473984, EPI_ISL_2473985, EPI_ISL_2473986, EPI_ISL_2473987, EPI_ISL_2473988, EPI_ISL_2473989, EPI_ISL_2473990, EPI_ISL_2473991, EPI_ISL_2473992, EPI_ISL_2473993, EPI_ISL_2473994, EPI_ISL_2473995, EPI_ISL_2473996, EPI_ISL_2473997, EPI_ISL_2473998, EPI_ISL_2473999, EPI_ISL_2474000, EPI_ISL_2474001, EPI_ISL_2474002, EPI_ISL_2474003, EPI_ISL_2474004, EPI_ISL_2474005, EPI_ISL_2474006, EPI_ISL_2474007, EPI_ISL_2474008, EPI_ISL_2474009, EPI_ISL_2474010, EPI_ISL_2474011, EPI_ISL_2474012, EPI_ISL_2474013, EPI_ISL_2474014, EPI_ISL_2474015, EPI_ISL_2474016, EPI_ISL_2474017, EPI_ISL_2474018, EPI_ISL_2474019, EPI_ISL_2474020, EPI_ISL_2474021, EPI_ISL_2474022, EPI_ISL_2474023, EPI_ISL_2474024, EPI_ISL_2474025, EPI_ISL_2474026, EPI_ISL_2474027, EPI_ISL_2474028, EPI_ISL_2474029, EPI_ISL_2474030, EPI_ISL_2474031, EPI_ISL_2474032, EPI_ISL_2474033, EPI_ISL_2474034, EPI_ISL_2474035, EPI_ISL_2474036, EPI_ISL_2474037, EPI_ISL_2474038, EPI_ISL_2474039, EPI_ISL_2474040, EPI_ISL_2474041, EPI_ISL_2474042, EPI_ISL_2474043, EPI_ISL_2474044, EPI_ISL_2474045, EPI_ISL_2474046, EPI_ISL_2474047, EPI_ISL_2474048, EPI_ISL_2474049, EPI_ISL_2474050, EPI_ISL_2474051, EPI_ISL_2474052, EPI_ISL_2474053, EPI_ISL_2474054, EPI_ISL_2474055, EPI_ISL_2474056, EPI_ISL_2474057, EPI_ISL_2474058, EPI_ISL_2474059, EPI_ISL_2474060, EPI_ISL_2474061, EPI_ISL_2474062, EPI_ISL_2474063, EPI_ISL_2474064, EPI_ISL_2474065, EPI_ISL_2474066, EPI_ISL_2474067, EPI_ISL_2474068, EPI_ISL_2474069, EPI_ISL_2474070, EPI_ISL_2474071, EPI_ISL_2474072, EPI_ISL_2474073, EPI_ISL_2474074, EPI_ISL_2474075, EPI_ISL_2474076, EPI_ISL_2474077, EPI_ISL_2474078, EPI_ISL_2474079, EPI_ISL_2474080, EPI_ISL_2474081, EPI_ISL_2474082, EPI_ISL_2474083, EPI_ISL_2474084, EPI_ISL_2474085, EPI_ISL_2474086, EPI_ISL_2474087, EPI_ISL_2474088, EPI_ISL_2474089, EPI_ISL_2474090, EPI_ISL_2474091, EPI_ISL_2474092, EPI_ISL_2474093, EPI_ISL_2474094, EPI_ISL_2474095, EPI_ISL_2474096, EPI_ISL_2474097, EPI_ISL_2474098, EPI_ISL_2474099, EPI_ISL_2474100, EPI_ISL_2474101, EPI_ISL_2474102, EPI_ISL_2474103, EPI_ISL_2474104, EPI_ISL_2474105, EPI_ISL_2474106, EPI_ISL_2474107, EPI_ISL_2474108, EPI_ISL_2474109, EPI_ISL_2474110, EPI_ISL_2474111, EPI_ISL_2474112, EPI_ISL_2474113, EPI_ISL_2474114, EPI_ISL_2474115, EPI_ISL_2474116, EPI_ISL_2474117, EPI_ISL_2474118, EPI_ISL_2474119, EPI_ISL_2474120, EPI_ISL_2474121, EPI_ISL_2474122, EPI_ISL_2474123, EPI_ISL_2474124, EPI_ISL_2474125, EPI_ISL_2474126, EPI_ISL_2474127, EPI_ISL_2474128, EPI_ISL_2474129, EPI_ISL_2474130, EPI_ISL_2474131, EPI_ISL_2474132, EPI_ISL_2474133, EPI_ISL_2474134, EPI_ISL_2474135, EPI_ISL_2474136, EPI_ISL_2474137, EPI_ISL_2474138, EPI_ISL_2474139, EPI_ISL_2474140, EPI_ISL_2474141, EPI_ISL_2474142, EPI_ISL_2474143, EPI_ISL_2474144, EPI_ISL_2474145, EPI_ISL_2474146, EPI_ISL_2474147, EPI_ISL_2474148, EPI_ISL_2474149, EPI_ISL_2474150, EPI_ISL_2474151, EPI_ISL_2474152, EPI_ISL_2474153, EPI_ISL_2474154, EPI_ISL_2474155, EPI_ISL_2474156, EPI_ISL_2474157, EPI_ISL_2474158, EPI_ISL_2474159, EPI_ISL_2474160, EPI_ISL_2474161, EPI_ISL_2474162, EPI_ISL_2474163, EPI_ISL_2474164, EPI_ISL_2474165, EPI_ISL_2474166, EPI_ISL_2474167, EPI_ISL_2474168, EPI_ISL_2474169, EPI_ISL_2474170, EPI_ISL_2474171, EPI_ISL_2474172, EPI_ISL_2474173, EPI_ISL_2474174, EPI_ISL_2474175, EPI_ISL_2474176, EPI_ISL_2474177 | | | |
| see above | Reditus Laboratories | Reditus Laboratories | Joshua J. Geltz, Ph.D., Robert M. Sgambelluri, Ph.D., Rex Dyer, Ph.D., Cassy Philips, M.S., Alexa Eichelberger, M.S. |
| EPI_ISL_2476348, EPI_ISL_2476349, EPI_ISL_2476350, EPI_ISL_2476351, EPI_ISL_2476352, EPI_ISL_2476353, EPI_ISL_2476354, EPI_ISL_2476355, EPI_ISL_2476356, EPI_ISL_2476357, EPI_ISL_2476358, EPI_ISL_2476359, EPI_ISL_2476360, EPI_ISL_2476361, EPI_ISL_2476362, EPI_ISL_2476363, EPI_ISL_2476364, EPI_ISL_2476365, EPI_ISL_2476366, EPI_ISL_2476367, EPI_ISL_2476368, EPI_ISL_2476369, | | | |

| | | | |
|---|---|---|---|
| EPI_ISL_2476370, EPI_ISL_2476371, EPI_ISL_2476372 | | | |
| see above | University of Chicago Medicine | RIPHL at Rush University Medical Center | Stefan Green, Kevin Kunstman, Max Kolton, Marieta Hyde, Laura Furtado, Felix Araujo Perez, Lauren Megger |
| EPI_ISL_2476373, EPI_ISL_2476374, EPI_ISL_2476375, EPI_ISL_2476376, EPI_ISL_2476377, EPI_ISL_2476378, EPI_ISL_2476379, EPI_ISL_2476380, EPI_ISL_2476381, EPI_ISL_2476382, EPI_ISL_2476383, EPI_ISL_2476384, EPI_ISL_2476385, EPI_ISL_2476386, EPI_ISL_2476387, EPI_ISL_2476388 | | | |
| see above | John H Stroger Jr Hospital of Cook County | RIPHL at Rush University Medical Center | Stefan Green, Kevin Kunstman, Max Kolton, Marieta Hyde, Laura Furtado, Felix Araujo Perez, Ah-Young Sung, Sharon Welbel |
| EPI_ISL_2476389, EPI_ISL_2476390, EPI_ISL_2476391, EPI_ISL_2476392, EPI_ISL_2476393, EPI_ISL_2476394, EPI_ISL_2476395, EPI_ISL_2476396, EPI_ISL_2476397, EPI_ISL_2476398, EPI_ISL_2476399, EPI_ISL_2476400 | | | |
| see above | University of Illinois at Chicago | RIPHL at Rush University Medical Center | Stefan Green, Kevin Kunstman, Max Kolton, Marieta Hyde, Laura Furtado, Felix Araujo Perez, Julie Bentea, Omar Perez |
| EPI_ISL_2476409, EPI_ISL_2476410, EPI_ISL_2476411, EPI_ISL_2476412 | LESP Ciudad de Mexico | Instituto de Diagnostico y Referencia Epidemiologicos (INDRE) | Claudia Wong-Arambula, Abril Rodriguez-Maldonado, Vanessa Rivero-Arredondo, Ariadna Medina-Benitez, Joaquin Quiroz-Mercado, Sergio Rangel-Guerrero, Natividad Cruz-Ortiz, Tatiana Nunez-Garcia, Gisela Barrera-Badillo, Lucia Hernandez-Rivas, Irma Lopez-Martinez, Ernesto Ramirez-Gonzalez. |
| EPI_ISL_2476413 | LESP Guanajuato | Instituto de Diagnostico y Referencia Epidemiologicos (INDRE) | Claudia Wong-Arambula, Abril Rodriguez-Maldonado, Vanessa Rivero-Arredondo, Ariadna Medina-Benitez, Joaquin Quiroz-Mercado, Sergio Rangel-Guerrero, Natividad Cruz-Ortiz, Tatiana Nunez-Garcia, Gisela Barrera-Badillo, Lucia Hernandez-Rivas, Irma Lopez-Martinez, Ernesto Ramirez-Gonzalez. |
| EPI_ISL_2476414 | LESP Yucatan | Instituto de Diagnostico y Referencia Epidemiologicos (INDRE) | Claudia Wong-Arambula, Abril Rodriguez-Maldonado, Vanessa Rivero-Arredondo, Ariadna Medina-Benitez, Joaquin Quiroz-Mercado, Sergio Rangel-Guerrero, Natividad Cruz-Ortiz, Tatiana Nunez-Garcia, Gisela Barrera-Badillo, Lucia Hernandez-Rivas, Irma Lopez-Martinez, Ernesto Ramirez-Gonzalez. |
| EPI_ISL_2476415 | LESP Guanajuato | Instituto de Diagnostico y Referencia Epidemiologicos (INDRE) | Claudia Wong-Arambula, Abril Rodriguez-Maldonado, Vanessa Rivero-Arredondo, Ariadna Medina-Benitez, Joaquin Quiroz-Mercado, Sergio Rangel-Guerrero, Natividad Cruz-Ortiz, Tatiana Nunez-Garcia, Gisela Barrera-Badillo, Lucia Hernandez-Rivas, Irma Lopez-Martinez, Ernesto Ramirez-Gonzalez. |
| EPI_ISL_2476416 | LESP Tamaulipas | Instituto de Diagnostico y Referencia Epidemiologicos (INDRE) | Claudia Wong-Arambula, Abril Rodriguez-Maldonado, Vanessa Rivero-Arredondo, Ariadna Medina-Benitez, Joaquin Quiroz-Mercado, Sergio Rangel-Guerrero, Natividad Cruz-Ortiz, Tatiana Nunez-Garcia, Gisela Barrera-Badillo, Lucia Hernandez-Rivas, Irma Lopez-Martinez, Ernesto Ramirez-Gonzalez. |
| EPI_ISL_2476417 | LESP Zacatecas | Instituto de Diagnostico y Referencia Epidemiologicos (INDRE) | Claudia Wong-Arambula, Abril Rodriguez-Maldonado, Vanessa Rivero-Arredondo, Ariadna Medina-Benitez, Joaquin Quiroz-Mercado, Sergio Rangel-Guerrero, Natividad Cruz-Ortiz, Tatiana Nunez-Garcia, Gisela Barrera-Badillo, Lucia Hernandez-Rivas, Irma Lopez-Martinez, Ernesto Ramirez-Gonzalez. |
| EPI_ISL_2476418, EPI_ISL_2476419, EPI_ISL_2476420, EPI_ISL_2476421, EPI_ISL_2476422 | LESP Guanajuato | Instituto de Diagnostico y Referencia Epidemiologicos (INDRE) | Claudia Wong-Arambula, Abril Rodriguez-Maldonado, Vanessa Rivero-Arredondo, Ariadna Medina-Benitez, Joaquin Quiroz-Mercado, Sergio Rangel-Guerrero, Natividad Cruz-Ortiz, Tatiana Nunez-Garcia, Gisela Barrera-Badillo, Lucia Hernandez-Rivas, Irma Lopez-Martinez, Ernesto Ramirez-Gonzalez. |
| EPI_ISL_2476423 | LESP Ciudad de Mexico | Instituto de Diagnostico y Referencia Epidemiologicos (INDRE) | Claudia Wong-Arambula, Abril Rodriguez-Maldonado, Vanessa Rivero-Arredondo, Ariadna Medina-Benitez, Joaquin Quiroz-Mercado, Sergio Rangel-Guerrero, Natividad Cruz-Ortiz, Tatiana Nunez-Garcia, Gisela Barrera-Badillo, Lucia Hernandez-Rivas, Irma Lopez-Martinez, Ernesto Ramirez-Gonzalez. |
| EPI_ISL_2476424 | LESP Guanajuato | Instituto de Diagnostico y Referencia Epidemiologicos (INDRE) | Claudia Wong-Arambula, Abril Rodriguez-Maldonado, Vanessa Rivero-Arredondo, Ariadna Medina-Benitez, Joaquin Quiroz-Mercado, Sergio Rangel-Guerrero, Natividad Cruz-Ortiz, Tatiana Nunez-Garcia, Gisela Barrera-Badillo, Lucia Hernandez-Rivas, Irma Lopez-Martinez, Ernesto Ramirez-Gonzalez. |
| EPI_ISL_2476425 | LESP Tamaulipas | Instituto de Diagnostico y Referencia Epidemiologicos (INDRE) | Claudia Wong-Arambula, Abril Rodriguez-Maldonado, Vanessa Rivero-Arredondo, Ariadna Medina-Benitez, Joaquin Quiroz-Mercado, Sergio Rangel-Guerrero, Natividad Cruz-Ortiz, Tatiana Nunez-Garcia, Gisela Barrera-Badillo, Lucia Hernandez-Rivas, Irma Lopez-Martinez, Ernesto Ramirez-Gonzalez. |
| EPI_ISL_2476426 | LESP Guanajuato | Instituto de Diagnostico y Referencia Epidemiologicos (INDRE) | Claudia Wong-Arambula, Abril Rodriguez-Maldonado, Vanessa Rivero-Arredondo, Ariadna Medina-Benitez, Joaquin Quiroz-Mercado, Sergio Rangel-Guerrero, Natividad Cruz-Ortiz, Tatiana Nunez-Garcia, Gisela Barrera-Badillo, Lucia Hernandez-Rivas, Irma Lopez-Martinez, Ernesto Ramirez-Gonzalez. |
| EPI_ISL_2476427, EPI_ISL_2476428, EPI_ISL_2476429 | LESP Ciudad de Mexico | Instituto de Diagnostico y Referencia Epidemiologicos (INDRE) | Claudia Wong-Arambula, Abril Rodriguez-Maldonado, Vanessa Rivero-Arredondo, Ariadna Medina-Benitez, Joaquin Quiroz-Mercado, Sergio Rangel-Guerrero, Natividad Cruz-Ortiz, Tatiana Nunez-Garcia, Gisela Barrera-Badillo, Lucia Hernandez-Rivas, Irma Lopez-Martinez, Ernesto Ramirez-Gonzalez. |
| EPI_ISL_2476430 | LESP Oaxaca | Instituto de Diagnostico y Referencia Epidemiologicos (INDRE) | Claudia Wong-Arambula, Abril Rodriguez-Maldonado, Vanessa Rivero-Arredondo, Ariadna Medina-Benitez, Joaquin Quiroz-Mercado, Sergio Rangel-Guerrero, Natividad Cruz-Ortiz, Tatiana Nunez-Garcia, Gisela Barrera-Badillo, Lucia Hernandez-Rivas, Irma Lopez-Martinez, Ernesto Ramirez-Gonzalez. |
| EPI_ISL_2476431, EPI_ISL_2476432, EPI_ISL_2476433, EPI_ISL_2476434, EPI_ISL_2476435, EPI_ISL_2476436 | LESP Tamaulipas | Instituto de Diagnostico y Referencia Epidemiologicos (INDRE) | Claudia Wong-Arambula, Abril Rodriguez-Maldonado, Vanessa Rivero-Arredondo, Ariadna Medina-Benitez, Joaquin Quiroz-Mercado, Sergio Rangel-Guerrero, Natividad Cruz-Ortiz, Tatiana Nunez-Garcia, Gisela Barrera-Badillo, Lucia Hernandez-Rivas, Irma Lopez-Martinez, Ernesto Ramirez-Gonzalez. |
| EPI_ISL_2476437 | LESP Sinaloa | Instituto de Diagnostico y Referencia Epidemiologicos (INDRE) | Claudia Wong-Arambula, Abril Rodriguez-Maldonado, Vanessa Rivero-Arredondo, Ariadna Medina-Benitez, Joaquin Quiroz-Mercado, Sergio Rangel-Guerrero, Natividad Cruz-Ortiz, Tatiana Nunez-Garcia, Gisela Barrera-Badillo, Lucia Hernandez-Rivas, Irma Lopez-Martinez, Ernesto Ramirez-Gonzalez. |
| EPI_ISL_2476438 | LESP Colima | Instituto de Diagnostico y Referencia Epidemiologicos (INDRE) | Claudia Wong-Arambula, Abril Rodriguez-Maldonado, Vanessa Rivero-Arredondo, Ariadna Medina-Benitez, Joaquin Quiroz-Mercado, Sergio Rangel-Guerrero, Natividad Cruz-Ortiz, Tatiana Nunez-Garcia, Gisela Barrera-Badillo, Lucia Hernandez-Rivas, Irma Lopez-Martinez, Ernesto Ramirez-Gonzalez. |
| EPI_ISL_2476439 | LESP Baja California | Instituto de Diagnostico y Referencia Epidemiologicos (INDRE) | Claudia Wong-Arambula, Abril Rodriguez-Maldonado, Vanessa Rivero-Arredondo, Ariadna Medina-Benitez, Joaquin Quiroz-Mercado, Sergio Rangel-Guerrero, Natividad Cruz-Ortiz, Tatiana Nunez-Garcia, Gisela Barrera-Badillo, Lucia Hernandez-Rivas, Irma Lopez-Martinez, Ernesto Ramirez-Gonzalez. |
| EPI_ISL_2476440 | LESP Morelos | Instituto de Diagnostico y Referencia Epidemiologicos (INDRE) | Claudia Wong-Arambula, Abril Rodriguez-Maldonado, Vanessa Rivero-Arredondo, Ariadna Medina-Benitez, Joaquin Quiroz-Mercado, Sergio Rangel-Guerrero, Natividad Cruz-Ortiz, Tatiana Nunez-Garcia, Gisela Barrera-Badillo, Lucia Hernandez-Rivas, Irma Lopez-Martinez, Ernesto Ramirez-Gonzalez. |
| EPI_ISL_2476441 | LESP Chihuahua | Instituto de Diagnostico y Referencia Epidemiologicos (INDRE) | Claudia Wong-Arambula, Abril Rodriguez-Maldonado, Vanessa Rivero-Arredondo, Ariadna Medina-Benitez, Joaquin Quiroz-Mercado, Sergio Rangel-Guerrero, Natividad Cruz-Ortiz, Tatiana Nunez-Garcia, Gisela Barrera-Badillo, Lucia Hernandez-Rivas, Irma Lopez-Martinez, Ernesto Ramirez-Gonzalez. |
| EPI_ISL_2476442 | LESP Campeche | Instituto de Diagnostico y Referencia Epidemiologicos (INDRE) | Claudia Wong-Arambula, Abril Rodriguez-Maldonado, Vanessa Rivero-Arredondo, Ariadna Medina-Benitez, Joaquin Quiroz-Mercado, Sergio Rangel-Guerrero, Natividad Cruz-Ortiz, Tatiana Nunez-Garcia, Gisela Barrera-Badillo, Lucia Hernandez-Rivas, Irma Lopez-Martinez, Ernesto Ramirez-Gonzalez. |
| EPI_ISL_2476443 | LESP Baja California | Instituto de Diagnostico y Referencia Epidemiologicos (INDRE) | Claudia Wong-Arambula, Abril Rodriguez-Maldonado, Vanessa Rivero-Arredondo, Ariadna Medina-Benitez, Joaquin Quiroz-Mercado, Sergio Rangel-Guerrero, Natividad Cruz-Ortiz, Tatiana Nunez-Garcia, Gisela Barrera-Badillo, Lucia Hernandez-Rivas, Irma Lopez-Martinez, Ernesto Ramirez-Gonzalez. |
| EPI_ISL_2476444 | LESP Sinaloa | Instituto de Diagnostico y Referencia Epidemiologicos (INDRE) | Claudia Wong-Arambula, Abril Rodriguez-Maldonado, Vanessa Rivero-Arredondo, Ariadna Medina-Benitez, Joaquin Quiroz-Mercado, Sergio Rangel-Guerrero, Natividad Cruz-Ortiz, Tatiana Nunez-Garcia, Gisela Barrera-Badillo, Lucia Hernandez-Rivas, Irma Lopez-Martinez, Ernesto Ramirez-Gonzalez. |
| EPI_ISL_2477504 | Stanford Health Care | Stanford University School of Medicine, Clinical Virology Laboratory | Daniel Solis, Mamdouh Sibai, Bernadette Troung, Becky Jiang, Michelle Verghese, ChunHong Huang, James Zehnder, Malaya K. Sahoo, and Benjamin A. Pinsky |
| EPI_ISL_2479502 | Ohio Department of Health Laboratory | Ohio Department of Health Laboratory | Holmes, Jennifer; Eric Brandt, Keoni Omura, Glen McGillivary, Caitlin McDonnell, Jade Mowery, Stephanie Mccracken, Tyler Payne, Kirtana Ramadugu, Erica Leasure, Brent Lee, Kelsey Florek, Heather Blankenship, Allison Black, Quanta Brown, and Tammy Bannerman |
| EPI_ISL_2479877, EPI_ISL_2479878, EPI_ISL_2479879, EPI_ISL_2479880, EPI_ISL_2479881 | LESP Queretaro | Instituto de Diagnostico y Referencia Epidemiologicos (INDRE) | Claudia Wong-Arambula, Abril Rodriguez-Maldonado, Vanessa Rivero-Arredondo, Ariadna Medina-Benitez, Joaquin Quiroz-Mercado, Sergio Rangel-Guerrero, Natividad Cruz-Ortiz, Tatiana Nunez-Garcia, Gisela Barrera-Badillo, Lucia Hernandez-Rivas, Irma Lopez-Martinez, Ernesto Ramirez-Gonzalez. |
| EPI_ISL_2479882, EPI_ISL_2479883 | LESP Ciudad de Mexico | Instituto de Diagnostico y Referencia Epidemiologicos (INDRE) | Claudia Wong-Arambula, Abril Rodriguez-Maldonado, Vanessa Rivero-Arredondo, Ariadna Medina-Benitez, Joaquin Quiroz-Mercado, Sergio Rangel-Guerrero, Natividad Cruz-Ortiz, Tatiana Nunez-Garcia, Gisela Barrera-Badillo, Lucia Hernandez-Rivas, Irma Lopez-Martinez, Ernesto Ramirez-Gonzalez. |
| EPI_ISL_2479884 | LESP Baja California | Instituto de Diagnostico y Referencia Epidemiologicos (INDRE) | Claudia Wong-Arambula, Abril Rodriguez-Maldonado, Vanessa Rivero-Arredondo, Ariadna Medina-Benitez, Joaquin Quiroz-Mercado, Sergio Rangel-Guerrero, Natividad Cruz-Ortiz, Tatiana Nunez-Garcia, Gisela Barrera-Badillo, Lucia Hernandez-Rivas, Irma Lopez-Martinez, Ernesto Ramirez-Gonzalez. |
| EPI_ISL_2479885 | LESP Chiapas | Instituto de Diagnostico y Referencia Epidemiologicos (INDRE) | Claudia Wong-Arambula, Abril Rodriguez-Maldonado, Vanessa Rivero-Arredondo, Ariadna Medina-Benitez, Joaquin Quiroz-Mercado, Sergio Rangel-Guerrero, Natividad Cruz-Ortiz, Tatiana Nunez-Garcia, Gisela Barrera-Badillo, Lucia Hernandez-Rivas, Irma Lopez-Martinez, Ernesto Ramirez-Gonzalez. |
| EPI_ISL_2479886, EPI_ISL_2479887 | LESP Yucatan | Instituto de Diagnostico y Referencia Epidemiologicos (INDRE) | Claudia Wong-Arambula, Abril Rodriguez-Maldonado, Vanessa Rivero-Arredondo, Ariadna Medina-Benitez, Joaquin Quiroz-Mercado, Sergio Rangel-Guerrero, Natividad Cruz-Ortiz, Tatiana Nunez-Garcia, Gisela Barrera-Badillo, Lucia Hernandez-Rivas, Irma Lopez-Martinez, Ernesto Ramirez-Gonzalez. |
| EPI_ISL_2479888 | LESP Baja California | Instituto de Diagnostico y Referencia Epidemiologicos (INDRE) | Claudia Wong-Arambula, Abril Rodriguez-Maldonado, Vanessa Rivero-Arredondo, Ariadna Medina-Benitez, Joaquin Quiroz-Mercado, Sergio Rangel-Guerrero, Natividad Cruz-Ortiz, Tatiana Nunez-Garcia, Gisela Barrera-Badillo, Lucia Hernandez-Rivas, Irma Lopez-Martinez, Ernesto Ramirez-Gonzalez. |
| EPI_ISL_2479889, EPI_ISL_2479890, EPI_ISL_2479891 | LESP Quintana Roo | Instituto de Diagnostico y Referencia Epidemiologicos (INDRE) | Claudia Wong-Arambula, Abril Rodriguez-Maldonado, Vanessa Rivero-Arredondo, Ariadna Medina-Benitez, Joaquin Quiroz-Mercado, Sergio Rangel-Guerrero, Natividad Cruz-Ortiz, Tatiana Nunez-Garcia, Gisela Barrera-Badillo, Lucia Hernandez-Rivas, Irma Lopez-Martinez, Ernesto Ramirez-Gonzalez. |
| EPI_ISL_2479892 | LESP Colima | Instituto de Diagnostico y Referencia Epidemiologicos (INDRE) | Claudia Wong-Arambula, Abril Rodriguez-Maldonado, Vanessa Rivero-Arredondo, Ariadna Medina-Benitez, Joaquin Quiroz-Mercado, Sergio Rangel-Guerrero, Natividad Cruz-Ortiz, Tatiana Nunez-Garcia, Gisela Barrera-Badillo, Lucia Hernandez-Rivas, Irma Lopez-Martinez, Ernesto Ramirez-Gonzalez. |
| EPI_ISL_2479893 | LESP Queretaro | Instituto de Diagnostico y Referencia Epidemiologicos (INDRE) | Claudia Wong-Arambula, Abril Rodriguez-Maldonado, Vanessa Rivero-Arredondo, Ariadna Medina-Benitez, Joaquin Quiroz-Mercado, Sergio Rangel-Guerrero, Natividad Cruz-Ortiz, Tatiana Nunez-Garcia, Gisela Barrera-Badillo, Lucia Hernandez-Rivas, Irma Lopez-Martinez, Ernesto Ramirez-Gonzalez. |
| EPI_ISL_2479894, EPI_ISL_2479895, EPI_ISL_2479896, EPI_ISL_2479897, EPI_ISL_2479898 | LESP Nuevo Leon | Instituto de Diagnostico y Referencia Epidemiologicos (INDRE) | Claudia Wong-Arambula, Abril Rodriguez-Maldonado, Vanessa Rivero-Arredondo, Ariadna Medina-Benitez, Joaquin Quiroz-Mercado, Sergio Rangel-Guerrero, Natividad Cruz-Ortiz, Tatiana Nunez-Garcia, Gisela Barrera-Badillo, Lucia Hernandez-Rivas, Irma Lopez-Martinez, Ernesto Ramirez-Gonzalez. |
| EPI_ISL_2479899 | LESP Guanajuato | Instituto de Diagnostico y Referencia Epidemiologicos (INDRE) | Claudia Wong-Arambula, Abril Rodriguez-Maldonado, Vanessa Rivero-Arredondo, Ariadna Medina-Benitez, Joaquin Quiroz-Mercado, Sergio Rangel-Guerrero, Natividad Cruz-Ortiz, Tatiana Nunez-Garcia, Gisela Barrera-Badillo, Lucia Hernandez-Rivas, Irma Lopez-Martinez, Ernesto Ramirez-Gonzalez. |

| | | | |
|---|---|---|---|
| EPI_ISL_2479900 | LESP Morelos | Instituto de Diagnostico y Referencia Epidemiologicos (INDRE) | Claudia Wong-Arambula, Abril Rodriguez-Maldonado, Vanessa Rivero-Arredondo, Ariadna Medina-Benitez, Joaquin Quiroz-Mercado, Sergio Rangel-Guerrero, Natividad Cruz-Ortiz, Tatiana Nunez-Garcia, Gisela Barrera-Badillo, Lucia Hernandez-Rivas, Irma Lopez-Martinez, Ernesto Ramirez-Gonzalez. |
| EPI_ISL_2479901 | LESP Hidalgo | Instituto de Diagnostico y Referencia Epidemiologicos (INDRE) | Claudia Wong-Arambula, Abril Rodriguez-Maldonado, Vanessa Rivero-Arredondo, Ariadna Medina-Benitez, Joaquin Quiroz-Mercado, Sergio Rangel-Guerrero, Natividad Cruz-Ortiz, Tatiana Nunez-Garcia, Gisela Barrera-Badillo, Lucia Hernandez-Rivas, Irma Lopez-Martinez, Ernesto Ramirez-Gonzalez. |
| EPI_ISL_2479902 | LESP Nuevo Leon | Instituto de Diagnostico y Referencia Epidemiologicos (INDRE) | Claudia Wong-Arambula, Abril Rodriguez-Maldonado, Vanessa Rivero-Arredondo, Ariadna Medina-Benitez, Joaquin Quiroz-Mercado, Sergio Rangel-Guerrero, Natividad Cruz-Ortiz, Tatiana Nunez-Garcia, Gisela Barrera-Badillo, Lucia Hernandez-Rivas, Irma Lopez-Martinez, Ernesto Ramirez-Gonzalez. |
| EPI_ISL_2479903, EPI_ISL_2479904, EPI_ISL_2479905 | LESP Puebla | Instituto de Diagnostico y Referencia Epidemiologicos (INDRE) | Claudia Wong-Arambula, Abril Rodriguez-Maldonado, Vanessa Rivero-Arredondo, Ariadna Medina-Benitez, Joaquin Quiroz-Mercado, Sergio Rangel-Guerrero, Natividad Cruz-Ortiz, Tatiana Nunez-Garcia, Gisela Barrera-Badillo, Lucia Hernandez-Rivas, Irma Lopez-Martinez, Ernesto Ramirez-Gonzalez. |
| EPI_ISL_2479906 | LESP Tamaulipas | Instituto de Diagnostico y Referencia Epidemiologicos (INDRE) | Claudia Wong-Arambula, Abril Rodriguez-Maldonado, Vanessa Rivero-Arredondo, Ariadna Medina-Benitez, Joaquin Quiroz-Mercado, Sergio Rangel-Guerrero, Natividad Cruz-Ortiz, Tatiana Nunez-Garcia, Gisela Barrera-Badillo, Lucia Hernandez-Rivas, Irma Lopez-Martinez, Ernesto Ramirez-Gonzalez. |
| EPI_ISL_2479907 | LESP Sonora | Instituto de Diagnostico y Referencia Epidemiologicos (INDRE) | Claudia Wong-Arambula, Abril Rodriguez-Maldonado, Vanessa Rivero-Arredondo, Ariadna Medina-Benitez, Joaquin Quiroz-Mercado, Sergio Rangel-Guerrero, Natividad Cruz-Ortiz, Tatiana Nunez-Garcia, Gisela Barrera-Badillo, Lucia Hernandez-Rivas, Irma Lopez-Martinez, Ernesto Ramirez-Gonzalez. |
| EPI_ISL_2479908 | LESP Sinaloa | Instituto de Diagnostico y Referencia Epidemiologicos (INDRE) | Claudia Wong-Arambula, Abril Rodriguez-Maldonado, Vanessa Rivero-Arredondo, Ariadna Medina-Benitez, Joaquin Quiroz-Mercado, Sergio Rangel-Guerrero, Natividad Cruz-Ortiz, Tatiana Nunez-Garcia, Gisela Barrera-Badillo, Lucia Hernandez-Rivas, Irma Lopez-Martinez, Ernesto Ramirez-Gonzalez. |
| EPI_ISL_2479909, EPI_ISL_2479910 | LESP Colima | Instituto de Diagnostico y Referencia Epidemiologicos (INDRE) | Claudia Wong-Arambula, Abril Rodriguez-Maldonado, Vanessa Rivero-Arredondo, Ariadna Medina-Benitez, Joaquin Quiroz-Mercado, Sergio Rangel-Guerrero, Natividad Cruz-Ortiz, Tatiana Nunez-Garcia, Gisela Barrera-Badillo, Lucia Hernandez-Rivas, Irma Lopez-Martinez, Ernesto Ramirez-Gonzalez. |
| EPI_ISL_2479911, EPI_ISL_2479912 | LESP Hidalgo | Instituto de Diagnostico y Referencia Epidemiologicos (INDRE) | Claudia Wong-Arambula, Abril Rodriguez-Maldonado, Vanessa Rivero-Arredondo, Ariadna Medina-Benitez, Joaquin Quiroz-Mercado, Sergio Rangel-Guerrero, Natividad Cruz-Ortiz, Tatiana Nunez-Garcia, Gisela Barrera-Badillo, Lucia Hernandez-Rivas, Irma Lopez-Martinez, Ernesto Ramirez-Gonzalez. |
| EPI_ISL_2479913 | LESP Queretaro | Instituto de Diagnostico y Referencia Epidemiologicos (INDRE) | Claudia Wong-Arambula, Abril Rodriguez-Maldonado, Vanessa Rivero-Arredondo, Ariadna Medina-Benitez, Joaquin Quiroz-Mercado, Sergio Rangel-Guerrero, Natividad Cruz-Ortiz, Tatiana Nunez-Garcia, Gisela Barrera-Badillo, Lucia Hernandez-Rivas, Irma Lopez-Martinez, Ernesto Ramirez-Gonzalez. |
| EPI_ISL_2479914, EPI_ISL_2479915 | LESP Quintana Roo | Instituto de Diagnostico y Referencia Epidemiologicos (INDRE) | Claudia Wong-Arambula, Abril Rodriguez-Maldonado, Vanessa Rivero-Arredondo, Ariadna Medina-Benitez, Joaquin Quiroz-Mercado, Sergio Rangel-Guerrero, Natividad Cruz-Ortiz, Tatiana Nunez-Garcia, Gisela Barrera-Badillo, Lucia Hernandez-Rivas, Irma Lopez-Martinez, Ernesto Ramirez-Gonzalez. |
| EPI_ISL_2479916 | LESP Jalisco | Instituto de Diagnostico y Referencia Epidemiologicos (INDRE) | Claudia Wong-Arambula, Abril Rodriguez-Maldonado, Vanessa Rivero-Arredondo, Ariadna Medina-Benitez, Joaquin Quiroz-Mercado, Sergio Rangel-Guerrero, Natividad Cruz-Ortiz, Tatiana Nunez-Garcia, Gisela Barrera-Badillo, Lucia Hernandez-Rivas, Irma Lopez-Martinez, Ernesto Ramirez-Gonzalez. |
| EPI_ISL_2479917, EPI_ISL_2479918 | LESP Yucatan | Instituto de Diagnostico y Referencia Epidemiologicos (INDRE) | Claudia Wong-Arambula, Abril Rodriguez-Maldonado, Vanessa Rivero-Arredondo, Ariadna Medina-Benitez, Joaquin Quiroz-Mercado, Sergio Rangel-Guerrero, Natividad Cruz-Ortiz, Tatiana Nunez-Garcia, Gisela Barrera-Badillo, Lucia Hernandez-Rivas, Irma Lopez-Martinez, Ernesto Ramirez-Gonzalez. |
| EPI_ISL_2479948, EPI_ISL_2479949, EPI_ISL_2479950, EPI_ISL_2479986 | Ohio Department of Health Laboratory | Ohio Department of Health Laboratory | Holmes, Jennifer; Eric Brandt, Keoni Omura, Glen McGillivary, Caitlin McDonnell, Jade Mowery, Stephanie Mccracken, Tyler Payne, Kirtana Ramadugu, Erica Leasure, Brent Lee, Kelsey Florek, Heather Blankenship, Allison Black, Quanta Brown, and Tammy Bannerman |
| EPI_ISL_2479997, EPI_ISL_2479998, EPI_ISL_2479999, EPI_ISL_2480000, EPI_ISL_2480001, EPI_ISL_2480002, EPI_ISL_2480004, EPI_ISL_2480005, EPI_ISL_2480006, EPI_ISL_2480007, EPI_ISL_2480008, EPI_ISL_2480009, EPI_ISL_2480010, EPI_ISL_2480011, EPI_ISL_2480012, EPI_ISL_2480013, EPI_ISL_2480017, EPI_ISL_2480018, EPI_ISL_2480019, EPI_ISL_2480020, EPI_ISL_2480021, EPI_ISL_2480022, EPI_ISL_2480023, EPI_ISL_2480024, EPI_ISL_2480025, EPI_ISL_2480026, EPI_ISL_2480027, EPI_ISL_2480028, EPI_ISL_2480029, EPI_ISL_2480032, EPI_ISL_2480033, EPI_ISL_2480034, EPI_ISL_2480036, EPI_ISL_2480037, EPI_ISL_2480038, EPI_ISL_2480039, EPI_ISL_2480040, EPI_ISL_2480041, EPI_ISL_2480042, EPI_ISL_2480044, EPI_ISL_2480045, EPI_ISL_2480046, EPI_ISL_2480047, EPI_ISL_2480048, EPI_ISL_2480049, EPI_ISL_2480050, EPI_ISL_2480053, EPI_ISL_2480055, EPI_ISL_2480056, EPI_ISL_2480057, EPI_ISL_2480058, EPI_ISL_2480059, EPI_ISL_2480060, EPI_ISL_2480061, EPI_ISL_2480062, EPI_ISL_2480063, EPI_ISL_2480064, EPI_ISL_2480065, EPI_ISL_2480066, EPI_ISL_2480067, EPI_ISL_2480068, EPI_ISL_2480069, EPI_ISL_2480070, EPI_ISL_2480071, EPI_ISL_2480072, EPI_ISL_2480073, EPI_ISL_2480074, EPI_ISL_2480075, EPI_ISL_2480076, EPI_ISL_2480077, EPI_ISL_2480078, EPI_ISL_2480079, EPI_ISL_2480080, EPI_ISL_2480081, EPI_ISL_2480082, EPI_ISL_2480083, EPI_ISL_2480084, EPI_ISL_2480085, EPI_ISL_2480086, EPI_ISL_2480087, EPI_ISL_2480088, EPI_ISL_2480089, EPI_ISL_2480090, EPI_ISL_2480091, EPI_ISL_2480092, EPI_ISL_2480093, EPI_ISL_2480094, EPI_ISL_2480095, EPI_ISL_2480096, EPI_ISL_2480097, EPI_ISL_2480099, EPI_ISL_2480100, EPI_ISL_2480101, EPI_ISL_2480102, EPI_ISL_2480103, EPI_ISL_2480104, EPI_ISL_2480105, EPI_ISL_2480106, EPI_ISL_2480107, EPI_ISL_2480108, EPI_ISL_2480109, EPI_ISL_2480110, EPI_ISL_2480111, EPI_ISL_2480112, EPI_ISL_2480114, EPI_ISL_2480115, EPI_ISL_2480116, EPI_ISL_2480117, EPI_ISL_2480118, EPI_ISL_2480119, EPI_ISL_2480120, EPI_ISL_2480121, EPI_ISL_2480122, EPI_ISL_2480123, EPI_ISL_2480124, EPI_ISL_2480125, EPI_ISL_2480126, EPI_ISL_2480127, EPI_ISL_2480128, EPI_ISL_2480129, EPI_ISL_2480130, EPI_ISL_2480131, EPI_ISL_2480132, EPI_ISL_2480133, EPI_ISL_2480134, EPI_ISL_2480135, EPI_ISL_2480136, EPI_ISL_2480137, EPI_ISL_2480138, EPI_ISL_2480139, EPI_ISL_2480140, EPI_ISL_2480141, EPI_ISL_2480142, EPI_ISL_2480143, EPI_ISL_2480144, EPI_ISL_2480145, EPI_ISL_2480146, EPI_ISL_2480147, EPI_ISL_2480148, EPI_ISL_2480149, EPI_ISL_2480150, EPI_ISL_2480151, EPI_ISL_2480152, EPI_ISL_2480153, EPI_ISL_2480154, EPI_ISL_2480155, EPI_ISL_2480156, EPI_ISL_2480157, EPI_ISL_2480158, EPI_ISL_2480159, EPI_ISL_2480160, EPI_ISL_2480161, EPI_ISL_2480162, EPI_ISL_2480165, EPI_ISL_2480166, EPI_ISL_2480167, EPI_ISL_2480168, EPI_ISL_2480169, EPI_ISL_2480170, EPI_ISL_2480171, EPI_ISL_2480172, EPI_ISL_2480173, EPI_ISL_2480175, EPI_ISL_2480176, EPI_ISL_2480177, EPI_ISL_2480178, EPI_ISL_2480179, EPI_ISL_2480180, EPI_ISL_2480181, EPI_ISL_2480182, EPI_ISL_2480185, EPI_ISL_2480186, EPI_ISL_2480187, EPI_ISL_2480188, EPI_ISL_2480189, EPI_ISL_2480190, EPI_ISL_2480191, EPI_ISL_2480192, EPI_ISL_2480193, EPI_ISL_2480194, EPI_ISL_2480195, EPI_ISL_2480196 | | | |
| see above | Fulgent Genetics | Centers for Disease Control and Prevention Division of Viral Diseases, Pathogen Discovery | Dakota Howard, Dhwani Batra, Peter W. Cook, Kara Moser, Adrian Paskey, Jason Caravas, Benjamin Rambo-Martin, Shatavia Morrison, Christopher Gulvick, Scott Sammons, Yvette Unoarumhi, Darlene Wagner, Matthew Schmerer, Harry Gao, Mickey Li, John Gao, Joseph Fierro, Benafsh Sapra, Becky Tsai, Yan Meng, Doreen Ng, James Xie, Clinton R. Paden, Duncan MacCannell |
| EPI_ISL_2480197, EPI_ISL_2480198, EPI_ISL_2480199, EPI_ISL_2480200, EPI_ISL_2480201, EPI_ISL_2480202, EPI_ISL_2480203, EPI_ISL_2480204, EPI_ISL_2480205, EPI_ISL_2480206, EPI_ISL_2480207, EPI_ISL_2480208, EPI_ISL_2480209, EPI_ISL_2480210, EPI_ISL_2480211, EPI_ISL_2480212, EPI_ISL_2480213, EPI_ISL_2480214, EPI_ISL_2480215, EPI_ISL_2480216, EPI_ISL_2480217, EPI_ISL_2480218, EPI_ISL_2480219, EPI_ISL_2480220, EPI_ISL_2480221, EPI_ISL_2480222, EPI_ISL_2480223, EPI_ISL_2480224, EPI_ISL_2480225, EPI_ISL_2480226, EPI_ISL_2480227, EPI_ISL_2480228, EPI_ISL_2480229, EPI_ISL_2480230, EPI_ISL_2480231, EPI_ISL_2480232, EPI_ISL_2480233, EPI_ISL_2480234, EPI_ISL_2480235, EPI_ISL_2480236, EPI_ISL_2480237, EPI_ISL_2480238, EPI_ISL_2480239, EPI_ISL_2480240, EPI_ISL_2480241, EPI_ISL_2480242, EPI_ISL_2480243, EPI_ISL_2480244, EPI_ISL_2480245, EPI_ISL_2480246, EPI_ISL_2480247, EPI_ISL_2480248, EPI_ISL_2480249, EPI_ISL_2480250, EPI_ISL_2480251, EPI_ISL_2480252, EPI_ISL_2480253, EPI_ISL_2480254, EPI_ISL_2480255, EPI_ISL_2480256, EPI_ISL_2480257, EPI_ISL_2480258, EPI_ISL_2480259, EPI_ISL_2480260, EPI_ISL_2480261, EPI_ISL_2480262, EPI_ISL_2480263, EPI_ISL_2480264, EPI_ISL_2480265, EPI_ISL_2480266, EPI_ISL_2480267, EPI_ISL_2480268, EPI_ISL_2480269, EPI_ISL_2480270, EPI_ISL_2480271, EPI_ISL_2480272, EPI_ISL_2480274 | | | |
| see above | Laboratory Corporation of America | Centers for Disease Control and Prevention Division of Viral Diseases, Pathogen Discovery | Dakota Howard, Dhwani Batra, Peter W. Cook, Kara Moser, Adrian Paskey, Jason Caravas, Benjamin Rambo-Martin, Shatavia Morrison, Christopher Gulvick, Scott Sammons, Yvette Unoarumhi, Darlene Wagner, Matthew Schmerer, Minoo Agarwal, Eyad Almasri, Debbie Boles, Ayla Burns, Nuthawin Charoensri, Oren Cohen, Susan Countryman, Mary Ann Cristobal, Bobbi Croy, Suzanne Dale, Hrushikesh Deshmukh, Amanda Douglas, Vincent Drouillon, Marcia Eisenberg, Howard Engler, Rama Ghatti, Prashant Gupta, Susan Hicks, Jake Humphrey, Lax Iyer, Lisa Pfefferle, Manoj Jain, Matthew Robinson, Mohan Kolli, Brian Krueger, Tim Kuphal, Stanley Letovsky, Michael Levandoski, Craig Lukasik, Jonathan Meltzer, Brian Norvell, Mindy Nye, Scott Parker, Christos Petropoulos, John Pruitt, Steven Ragan, Scott Ryan, Mike Sapeta, Jana Schroth, Suresh Babu Selvaraju, Goran Stevovic, Amanda Suchanek, Andrea Throop, Lyndon Tilson, Thomas Urban, Joe Voshell, Kimberly Wagner, Jonathan Williams, Mary Williamson, Qian Zeng, Tricia Zwiefelhofer, Clinton R. Paden, Duncan MacCannell |
| EPI_ISL_2480275 | Curative | New Mexico Department of Health Scientific Laboratory | Ellie Johnson, D'eldra Malone, Jennifer Benoit, Linda Salazar, Ratheesh Rajan, Keila Gutierrez, Anastacia Griego-Fisher |
| EPI_ISL_2480277 | Laboratory Corporation of America | Centers for Disease Control and Prevention Division of Viral Diseases, Pathogen Discovery | Dakota Howard, Dhwani Batra, Peter W. Cook, Kara Moser, Adrian Paskey, Jason Caravas, Benjamin Rambo-Martin, Shatavia Morrison, Christopher Gulvick, Scott Sammons, Yvette Unoarumhi, Darlene Wagner, Matthew Schmerer, Minoo Agarwal, Eyad Almasri, Debbie Boles, Ayla Burns, Nuthawin Charoensri, Oren Cohen, Susan Countryman, Mary Ann Cristobal, Bobbi Croy, Suzanne Dale, Hrushikesh Deshmukh, Amanda Douglas, Vincent Drouillon, Marcia Eisenberg, Howard Engler, Rama Ghatti, Prashant Gupta, Susan Hicks, Jake Humphrey, Lax Iyer, Lisa Pfefferle, Manoj Jain, Matthew Robinson, Mohan Kolli, Brian Krueger, Tim Kuphal, Stanley Letovsky, Michael Levandoski, Craig Lukasik, Jonathan Meltzer, Brian Norvell, Mindy Nye, Scott Parker, Christos Petropoulos, John Pruitt, Steven Ragan, Scott Ryan, Mike Sapeta, Jana Schroth, Suresh Babu Selvaraju, Goran Stevovic, Amanda Suchanek, Andrea Throop, Lyndon Tilson, Thomas Urban, Joe Voshell, Kimberly Wagner, Jonathan Williams, Mary Williamson, Qian Zeng, Tricia Zwiefelhofer, Clinton R. Paden, Duncan MacCannell |
| EPI_ISL_2480278 | Curative | New Mexico Department of Health Scientific Laboratory | Ellie Johnson, D'eldra Malone, Jennifer Benoit, Linda Salazar, Ratheesh Rajan, Keila Gutierrez, Anastacia Griego-Fisher |
| EPI_ISL_2480279 | Laboratory Corporation of America | Centers for Disease Control and Prevention Division of Viral Diseases, Pathogen Discovery | Dakota Howard, Dhwani Batra, Peter W. Cook, Kara Moser, Adrian Paskey, Jason Caravas, Benjamin Rambo-Martin, Shatavia Morrison, Christopher Gulvick, Scott Sammons, Yvette Unoarumhi, Darlene Wagner, Matthew Schmerer, Minoo Agarwal, Eyad Almasri, Debbie Boles, Ayla Burns, Nuthawin Charoensri, Oren Cohen, Susan Countryman, Mary Ann Cristobal, Bobbi Croy, Suzanne Dale, Hrushikesh Deshmukh, Amanda Douglas, Vincent Drouillon, Marcia Eisenberg, Howard Engler, Rama Ghatti, Prashant Gupta, Susan Hicks, Jake Humphrey, Lax Iyer, Lisa Pfefferle, Manoj Jain, Matthew Robinson, Mohan Kolli, Brian Krueger, Tim Kuphal, Stanley Letovsky, Michael Levandoski, Craig Lukasik, Jonathan Meltzer, Brian Norvell, Mindy Nye, Scott Parker, Christos Petropoulos, John Pruitt, Steven Ragan, Scott Ryan, Mike Sapeta, Jana Schroth, Suresh Babu Selvaraju, Goran Stevovic, Amanda Suchanek, Andrea Throop, Lyndon Tilson, Thomas Urban, Joe Voshell, Kimberly Wagner, Jonathan Williams, Mary Williamson, Qian Zeng, Tricia Zwiefelhofer, Clinton R. Paden, Duncan MacCannell |
| EPI_ISL_2480280 | Curative | New Mexico Department of Health Scientific Laboratory | Ellie Johnson, D'eldra Malone, Jennifer Benoit, Linda Salazar, Ratheesh Rajan, Keila Gutierrez, Anastacia Griego-Fisher |
| EPI_ISL_2480281, EPI_ISL_2480283, EPI_ISL_2480284 | Laboratory Corporation of America | Centers for Disease Control and Prevention Division of Viral Diseases, Pathogen Discovery | Dakota Howard, Dhwani Batra, Peter W. Cook, Kara Moser, Adrian Paskey, Jason Caravas, Benjamin Rambo-Martin, Shatavia Morrison, Christopher Gulvick, Scott Sammons, Yvette Unoarumhi, Darlene Wagner, Matthew Schmerer, Minoo Agarwal, Eyad Almasri, Debbie Boles, Ayla Burns, Nuthawin Charoensri, Oren Cohen, Susan Countryman, Mary Ann Cristobal, Bobbi Croy, Suzanne Dale, Hrushikesh Deshmukh, Amanda Douglas, Vincent Drouillon, Marcia Eisenberg, Howard Engler, Rama Ghatti, Prashant Gupta, Susan Hicks, Jake Humphrey, Lax Iyer, Lisa Pfefferle, Manoj Jain, Matthew Robinson, Mohan Kolli, Brian Krueger, Tim Kuphal, Stanley Letovsky, Michael Levandoski, Craig Lukasik, Jonathan Meltzer, Brian Norvell, Mindy Nye, Scott Parker, Christos Petropoulos, John Pruitt, Steven Ragan, Scott Ryan, Mike Sapeta, Jana Schroth, Suresh Babu Selvaraju, Goran Stevovic, Amanda Suchanek, Andrea Throop, Lyndon Tilson, Thomas Urban, Joe Voshell, Kimberly Wagner, Jonathan Williams, Mary Williamson, Qian Zeng, Tricia Zwiefelhofer, Clinton R. Paden, Duncan MacCannell |
| EPI_ISL_2480285 | Curative | New Mexico Department of Health Scientific Laboratory | Ellie Johnson, D'eldra Malone, Jennifer Benoit, Linda Salazar, Ratheesh Rajan, Keila Gutierrez, Anastacia Griego-Fisher |
| EPI_ISL_2480286, EPI_ISL_2480288, EPI_ISL_2480289, EPI_ISL_2480291 | Laboratory Corporation of America | Centers for Disease Control and Prevention Division of Viral Diseases, Pathogen Discovery | Dakota Howard, Dhwani Batra, Peter W. Cook, Kara Moser, Adrian Paskey, Jason Caravas, Benjamin Rambo-Martin, Shatavia Morrison, Christopher Gulvick, Scott Sammons, Yvette Unoarumhi, Darlene Wagner, Matthew Schmerer, Minoo Agarwal, Eyad Almasri, Debbie Boles, Ayla Burns, Nuthawin Charoensri, Oren Cohen, Susan Countryman, Mary Ann Cristobal, Bobbi Croy, Suzanne Dale, Hrushikesh Deshmukh, Amanda Douglas, Vincent Drouillon, Marcia Eisenberg, Howard Engler, Rama Ghatti, Prashant Gupta, Susan Hicks, Jake Humphrey, Lax Iyer, Lisa Pfefferle, Manoj Jain, Matthew Robinson, Mohan Kolli, Brian Krueger, Tim Kuphal, Stanley Letovsky, Michael Levandoski, Craig Lukasik, Jonathan Meltzer, Brian Norvell, Mindy Nye, Scott Parker, Christos Petropoulos, John Pruitt, Steven Ragan, Scott Ryan, Mike Sapeta, Jana Schroth, Suresh Babu Selvaraju, Goran Stevovic, Amanda Suchanek, Andrea Throop, Lyndon Tilson, Thomas Urban, Joe Voshell, Kimberly Wagner, Jonathan Williams, Mary Williamson, Qian Zeng, Tricia Zwiefelhofer, Clinton R. Paden, Duncan MacCannell |
| EPI_ISL_2480292 | Curative | New Mexico Department of Health Scientific Laboratory | Ellie Johnson, D'eldra Malone, Jennifer Benoit, Linda Salazar, Ratheesh Rajan, Keila Gutierrez, Anastacia Griego-Fisher |
| EPI_ISL_2480293 | Laboratory Corporation of America | Centers for Disease Control and Prevention Division of Viral Diseases, Pathogen Discovery | Dakota Howard, Dhwani Batra, Peter W. Cook, Kara Moser, Adrian Paskey, Jason Caravas, Benjamin Rambo-Martin, Shatavia Morrison, Christopher Gulvick, Scott Sammons, Yvette Unoarumhi, Darlene Wagner, Matthew Schmerer, Minoo Agarwal, Eyad Almasri, Debbie Boles, Ayla Burns, Nuthawin Charoensri, Oren Cohen, Susan Countryman, Mary Ann Cristobal, Bobbi Croy, Suzanne Dale, Hrushikesh Deshmukh, Amanda Douglas, Vincent Drouillon, Marcia Eisenberg, Howard Engler, Rama Ghatti, Prashant Gupta, Susan Hicks, Jake Humphrey, Lax Iyer, Lisa Pfefferle, Manoj Jain, Matthew Robinson, Mohan Kolli, Brian Krueger, Tim Kuphal, Stanley Letovsky, Michael Levandoski, Craig Lukasik, Jonathan Meltzer, Brian Norvell, Mindy Nye, Scott Parker, Christos Petropoulos, John Pruitt, Steven Ragan, Scott Ryan, Mike Sapeta, Jana Schroth, Suresh Babu Selvaraju, Goran Stevovic, Amanda Suchanek, Andrea Throop, Lyndon Tilson, Thomas Urban, Joe Voshell, Kimberly Wagner, Jonathan Williams, Mary Williamson, Qian Zeng, Tricia Zwiefelhofer, Clinton R. Paden, Duncan MacCannell |
| EPI_ISL_2480294 | Curative | New Mexico Department of Health Scientific Laboratory | Ellie Johnson, D'eldra Malone, Jennifer Benoit, Linda Salazar, Ratheesh Rajan, Keila Gutierrez, Anastacia Griego-Fisher |
| EPI_ISL_2480295, EPI_ISL_2480296, EPI_ISL_2480298 | Laboratory Corporation of America | Centers for Disease Control and Prevention Division of Viral Diseases, Pathogen Discovery | Dakota Howard, Dhwani Batra, Peter W. Cook, Kara Moser, Adrian Paskey, Jason Caravas, Benjamin Rambo-Martin, Shatavia Morrison, Christopher Gulvick, Scott Sammons, Yvette Unoarumhi, Darlene Wagner, Matthew Schmerer, Minoo Agarwal, Eyad Almasri, Debbie Boles, Ayla Burns, Nuthawin Charoensri, Oren Cohen, Susan Countryman, Mary Ann Cristobal, Bobbi Croy, Suzanne Dale, Hrushikesh Deshmukh, Amanda Douglas, Vincent Drouillon, Marcia Eisenberg, Howard Engler, Rama Ghatti, Prashant Gupta, Susan Hicks, Jake Humphrey, Lax Iyer, Lisa Pfefferle, Manoj Jain, Matthew Robinson, Mohan Kolli, Brian Krueger, Tim Kuphal, Stanley Letovsky, Michael Levandoski, Craig Lukasik, Jonathan Meltzer, Brian Norvell, Mindy Nye, Scott Parker, Christos Petropoulos, John Pruitt, Steven Ragan, Scott Ryan, Mike Sapeta, Jana Schroth, Suresh Babu Selvaraju, Goran Stevovic, Amanda Suchanek, Andrea Throop, Lyndon Tilson, Thomas Urban, Joe Voshell, Kimberly Wagner, Jonathan Williams, Mary Williamson, Qian Zeng, Tricia Zwiefelhofer, Clinton R. Paden, Duncan MacCannell |
| EPI_ISL_2480299 | Curative | New Mexico Department of Health Scientific Laboratory | Ellie Johnson, D'eldra Malone, Jennifer Benoit, Linda Salazar, Ratheesh Rajan, Keila Gutierrez, Anastacia Griego-Fisher |
| EPI_ISL_2480300 | Laboratory Corporation of America | Centers for Disease Control and Prevention Division of Viral Diseases, Pathogen Discovery | Dakota Howard, Dhwani Batra, Peter W. Cook, Kara Moser, Adrian Paskey, Jason Caravas, Benjamin Rambo-Martin, Shatavia Morrison, Christopher Gulvick, Scott Sammons, Yvette Unoarumhi, Darlene Wagner, Matthew Schmerer, Minoo Agarwal, Eyad Almasri, Debbie Boles, Ayla Burns, Nuthawin Charoensri, Oren Cohen, Susan Countryman, Mary Ann Cristobal, Bobbi Croy, Suzanne Dale, Hrushikesh Deshmukh, Amanda Douglas, Vincent Drouillon, Marcia Eisenberg, Howard Engler, Rama Ghatti, Prashant Gupta, Susan Hicks, Jake Humphrey, Lax Iyer, Lisa Pfefferle, Manoj Jain, Matthew Robinson, Mohan Kolli, Brian Krueger, Tim Kuphal, Stanley Letovsky, Michael Levandoski, Craig Lukasik, Jonathan Meltzer, Brian Norvell, Mindy Nye, Scott Parker, Christos Petropoulos, John Pruitt, Steven Ragan, Scott Ryan, Mike Sapeta, Jana Schroth, Suresh Babu Selvaraju, Goran Stevovic, Amanda Suchanek, Andrea Throop, Lyndon Tilson, Thomas Urban, Joe Voshell, Kimberly Wagner, Jonathan Williams, Mary Williamson, Qian Zeng, Tricia Zwiefelhofer, Clinton R. Paden, Duncan MacCannell |
| EPI_ISL_2480301 | Curative | New Mexico Department of Health Scientific | Ellie Johnson, D'eldra Malone, Jennifer Benoit, Linda Salazar, Ratheesh Rajan, Keila Gutierrez, Anastacia Griego-Fisher |

| | | | |
|---|---|---|---|
| | | Laboratory | |
| EPI_ISL_2480302, EPI_ISL_2480303 | Laboratory Corporation of America | Centers for Disease Control and Prevention Division of Viral Diseases, Pathogen Discovery | Dakota Howard, Dhwani Batra, Peter W. Cook, Kara Moser, Adrian Paskey, Jason Caravas, Benjamin Rambo-Martin, Shatavia Morrison, Christopher Gulvick, Scott Sammons, Yvette Unoarumhi, Darlene Wagner, Matthew Schmerer, Minoo Agarwal, Eyad Almasri, Debbie Boles, Ayla Burns, Nuthawin Charoensri, Oren Cohen, Susan Countryman, Mary Ann Cristobal, Bobbi Croy, Suzanne Dale, Hrushikesh Deshmukh, Amanda Douglas, Vincent Drouillon, Marcia Eisenberg, Howard Engler, Rama Ghatti, Prashant Gupta, Susan Hicks, Jake Humphrey, Lax Iyer, Lisa Pfefferle, Manoj Jain, Matthew Robinson, Mohan Kolli, Brian Krueger, Tim Kuphal, Stanley Letovsky, Michael Levandoski, Craig Lukasik, Jonathan Meltzer, Brian Norvell, Mindy Nye, Scott Parker, Christos Petropoulos, John Pruitt, Steven Ragan, Scott Ryan, Mike Sapeta, Jana Schroth, Suresh Babu Selvaraju, Goran Stevovic, Amanda Suchanek, Andrea Throop, Lyndon Tilson, Thomas Urban, Joe Voshell, Kimberly Wagner, Jonathan Williams, Mary Williamson, Qian Zeng, Tricia Zwiefelhofer, Clinton R. Paden, Duncan MacCannell |
| EPI_ISL_2480304 | Curative | New Mexico Department of Health Scientific Laboratory | Ellie Johnson, D'eldra Malone, Jennifer Benoit, Linda Salazar, Ratheesh Rajan, Keila Gutierrez, Anastacia Griego-Fisher |
| EPI_ISL_2480305, EPI_ISL_2480307 | Laboratory Corporation of America | Centers for Disease Control and Prevention Division of Viral Diseases, Pathogen Discovery | Dakota Howard, Dhwani Batra, Peter W. Cook, Kara Moser, Adrian Paskey, Jason Caravas, Benjamin Rambo-Martin, Shatavia Morrison, Christopher Gulvick, Scott Sammons, Yvette Unoarumhi, Darlene Wagner, Matthew Schmerer, Minoo Agarwal, Eyad Almasri, Debbie Boles, Ayla Burns, Nuthawin Charoensri, Oren Cohen, Susan Countryman, Mary Ann Cristobal, Bobbi Croy, Suzanne Dale, Hrushikesh Deshmukh, Amanda Douglas, Vincent Drouillon, Marcia Eisenberg, Howard Engler, Rama Ghatti, Prashant Gupta, Susan Hicks, Jake Humphrey, Lax Iyer, Lisa Pfefferle, Manoj Jain, Matthew Robinson, Mohan Kolli, Brian Krueger, Tim Kuphal, Stanley Letovsky, Michael Levandoski, Craig Lukasik, Jonathan Meltzer, Brian Norvell, Mindy Nye, Scott Parker, Christos Petropoulos, John Pruitt, Steven Ragan, Scott Ryan, Mike Sapeta, Jana Schroth, Suresh Babu Selvaraju, Goran Stevovic, Amanda Suchanek, Andrea Throop, Lyndon Tilson, Thomas Urban, Joe Voshell, Kimberly Wagner, Jonathan Williams, Mary Williamson, Qian Zeng, Tricia Zwiefelhofer, Clinton R. Paden, Duncan MacCannell |
| EPI_ISL_2480308 | Curative | New Mexico Department of Health Scientific Laboratory | Ellie Johnson, D'eldra Malone, Jennifer Benoit, Linda Salazar, Ratheesh Rajan, Keila Gutierrez, Anastacia Griego-Fisher |
| EPI_ISL_2480309, EPI_ISL_2480311, EPI_ISL_2480312, EPI_ISL_2480314 | Laboratory Corporation of America | Centers for Disease Control and Prevention Division of Viral Diseases, Pathogen Discovery | Dakota Howard, Dhwani Batra, Peter W. Cook, Kara Moser, Adrian Paskey, Jason Caravas, Benjamin Rambo-Martin, Shatavia Morrison, Christopher Gulvick, Scott Sammons, Yvette Unoarumhi, Darlene Wagner, Matthew Schmerer, Minoo Agarwal, Eyad Almasri, Debbie Boles, Ayla Burns, Nuthawin Charoensri, Oren Cohen, Susan Countryman, Mary Ann Cristobal, Bobbi Croy, Suzanne Dale, Hrushikesh Deshmukh, Amanda Douglas, Vincent Drouillon, Marcia Eisenberg, Howard Engler, Rama Ghatti, Prashant Gupta, Susan Hicks, Jake Humphrey, Lax Iyer, Lisa Pfefferle, Manoj Jain, Matthew Robinson, Mohan Kolli, Brian Krueger, Tim Kuphal, Stanley Letovsky, Michael Levandoski, Craig Lukasik, Jonathan Meltzer, Brian Norvell, Mindy Nye, Scott Parker, Christos Petropoulos, John Pruitt, Steven Ragan, Scott Ryan, Mike Sapeta, Jana Schroth, Suresh Babu Selvaraju, Goran Stevovic, Amanda Suchanek, Andrea Throop, Lyndon Tilson, Thomas Urban, Joe Voshell, Kimberly Wagner, Jonathan Williams, Mary Williamson, Qian Zeng, Tricia Zwiefelhofer, Clinton R. Paden, Duncan MacCannell |
| EPI_ISL_2480317 | Curative | New Mexico Department of Health Scientific Laboratory | Ellie Johnson, D'eldra Malone, Jennifer Benoit, Linda Salazar, Ratheesh Rajan, Keila Gutierrez, Anastacia Griego-Fisher |
| EPI_ISL_2480318, EPI_ISL_2480320, EPI_ISL_2480321 | Laboratory Corporation of America | Centers for Disease Control and Prevention Division of Viral Diseases, Pathogen Discovery | Dakota Howard, Dhwani Batra, Peter W. Cook, Kara Moser, Adrian Paskey, Jason Caravas, Benjamin Rambo-Martin, Shatavia Morrison, Christopher Gulvick, Scott Sammons, Yvette Unoarumhi, Darlene Wagner, Matthew Schmerer, Minoo Agarwal, Eyad Almasri, Debbie Boles, Ayla Burns, Nuthawin Charoensri, Oren Cohen, Susan Countryman, Mary Ann Cristobal, Bobbi Croy, Suzanne Dale, Hrushikesh Deshmukh, Amanda Douglas, Vincent Drouillon, Marcia Eisenberg, Howard Engler, Rama Ghatti, Prashant Gupta, Susan Hicks, Jake Humphrey, Lax Iyer, Lisa Pfefferle, Manoj Jain, Matthew Robinson, Mohan Kolli, Brian Krueger, Tim Kuphal, Stanley Letovsky, Michael Levandoski, Craig Lukasik, Jonathan Meltzer, Brian Norvell, Mindy Nye, Scott Parker, Christos Petropoulos, John Pruitt, Steven Ragan, Scott Ryan, Mike Sapeta, Jana Schroth, Suresh Babu Selvaraju, Goran Stevovic, Amanda Suchanek, Andrea Throop, Lyndon Tilson, Thomas Urban, Joe Voshell, Kimberly Wagner, Jonathan Williams, Mary Williamson, Qian Zeng, Tricia Zwiefelhofer, Clinton R. Paden, Duncan MacCannell |
| EPI_ISL_2480322 | Curative | New Mexico Department of Health Scientific Laboratory | Ellie Johnson, D'eldra Malone, Jennifer Benoit, Linda Salazar, Ratheesh Rajan, Keila Gutierrez, Anastacia Griego-Fisher |
| EPI_ISL_2480323 | Laboratory Corporation of America | Centers for Disease Control and Prevention Division of Viral Diseases, Pathogen Discovery | Dakota Howard, Dhwani Batra, Peter W. Cook, Kara Moser, Adrian Paskey, Jason Caravas, Benjamin Rambo-Martin, Shatavia Morrison, Christopher Gulvick, Scott Sammons, Yvette Unoarumhi, Darlene Wagner, Matthew Schmerer, Minoo Agarwal, Eyad Almasri, Debbie Boles, Ayla Burns, Nuthawin Charoensri, Oren Cohen, Susan Countryman, Mary Ann Cristobal, Bobbi Croy, Suzanne Dale, Hrushikesh Deshmukh, Amanda Douglas, Vincent Drouillon, Marcia Eisenberg, Howard Engler, Rama Ghatti, Prashant Gupta, Susan Hicks, Jake Humphrey, Lax Iyer, Lisa Pfefferle, Manoj Jain, Matthew Robinson, Mohan Kolli, Brian Krueger, Tim Kuphal, Stanley Letovsky, Michael Levandoski, Craig Lukasik, Jonathan Meltzer, Brian Norvell, Mindy Nye, Scott Parker, Christos Petropoulos, John Pruitt, Steven Ragan, Scott Ryan, Mike Sapeta, Jana Schroth, Suresh Babu Selvaraju, Goran Stevovic, Amanda Suchanek, Andrea Throop, Lyndon Tilson, Thomas Urban, Joe Voshell, Kimberly Wagner, Jonathan Williams, Mary Williamson, Qian Zeng, Tricia Zwiefelhofer, Clinton R. Paden, Duncan MacCannell |
| EPI_ISL_2480324 | Curative | New Mexico Department of Health Scientific Laboratory | Ellie Johnson, D'eldra Malone, Jennifer Benoit, Linda Salazar, Ratheesh Rajan, Keila Gutierrez, Anastacia Griego-Fisher |
| EPI_ISL_2480325 | Laboratory Corporation of America | Centers for Disease Control and Prevention Division of Viral Diseases, Pathogen Discovery | Dakota Howard, Dhwani Batra, Peter W. Cook, Kara Moser, Adrian Paskey, Jason Caravas, Benjamin Rambo-Martin, Shatavia Morrison, Christopher Gulvick, Scott Sammons, Yvette Unoarumhi, Darlene Wagner, Matthew Schmerer, Minoo Agarwal, Eyad Almasri, Debbie Boles, Ayla Burns, Nuthawin Charoensri, Oren Cohen, Susan Countryman, Mary Ann Cristobal, Bobbi Croy, Suzanne Dale, Hrushikesh Deshmukh, Amanda Douglas, Vincent Drouillon, Marcia Eisenberg, Howard Engler, Rama Ghatti, Prashant Gupta, Susan Hicks, Jake Humphrey, Lax Iyer, Lisa Pfefferle, Manoj Jain, Matthew Robinson, Mohan Kolli, Brian Krueger, Tim Kuphal, Stanley Letovsky, Michael Levandoski, Craig Lukasik, Jonathan Meltzer, Brian Norvell, Mindy Nye, Scott Parker, Christos Petropoulos, John Pruitt, Steven Ragan, Scott Ryan, Mike Sapeta, Jana Schroth, Suresh Babu Selvaraju, Goran Stevovic, Amanda Suchanek, Andrea Throop, Lyndon Tilson, Thomas Urban, Joe Voshell, Kimberly Wagner, Jonathan Williams, Mary Williamson, Qian Zeng, Tricia Zwiefelhofer, Clinton R. Paden, Duncan MacCannell |
| EPI_ISL_2480326 | Curative | New Mexico Department of Health Scientific Laboratory | Ellie Johnson, D'eldra Malone, Jennifer Benoit, Linda Salazar, Ratheesh Rajan, Keila Gutierrez, Anastacia Griego-Fisher |
| EPI_ISL_2480327, EPI_ISL_2480328 | Laboratory Corporation of America | Centers for Disease Control and Prevention Division of Viral Diseases, Pathogen Discovery | Dakota Howard, Dhwani Batra, Peter W. Cook, Kara Moser, Adrian Paskey, Jason Caravas, Benjamin Rambo-Martin, Shatavia Morrison, Christopher Gulvick, Scott Sammons, Yvette Unoarumhi, Darlene Wagner, Matthew Schmerer, Minoo Agarwal, Eyad Almasri, Debbie Boles, Ayla Burns, Nuthawin Charoensri, Oren Cohen, Susan Countryman, Mary Ann Cristobal, Bobbi Croy, Suzanne Dale, Hrushikesh Deshmukh, Amanda Douglas, Vincent Drouillon, Marcia Eisenberg, Howard Engler, Rama Ghatti, Prashant Gupta, Susan Hicks, Jake Humphrey, Lax Iyer, Lisa Pfefferle, Manoj Jain, Matthew Robinson, Mohan Kolli, Brian Krueger, Tim Kuphal, Stanley Letovsky, Michael Levandoski, Craig Lukasik, Jonathan Meltzer, Brian Norvell, Mindy Nye, Scott Parker, Christos Petropoulos, John Pruitt, Steven Ragan, Scott Ryan, Mike Sapeta, Jana Schroth, Suresh Babu Selvaraju, Goran Stevovic, Amanda Suchanek, Andrea Throop, Lyndon Tilson, Thomas Urban, Joe Voshell, Kimberly Wagner, Jonathan Williams, Mary Williamson, Qian Zeng, Tricia Zwiefelhofer, Clinton R. Paden, Duncan MacCannell |
| EPI_ISL_2480329 | Curative | New Mexico Department of Health Scientific Laboratory | Ellie Johnson, D'eldra Malone, Jennifer Benoit, Linda Salazar, Ratheesh Rajan, Keila Gutierrez, Anastacia Griego-Fisher |
| EPI_ISL_2480330 | Laboratory Corporation of America | Centers for Disease Control and Prevention Division of Viral Diseases, Pathogen Discovery | Dakota Howard, Dhwani Batra, Peter W. Cook, Kara Moser, Adrian Paskey, Jason Caravas, Benjamin Rambo-Martin, Shatavia Morrison, Christopher Gulvick, Scott Sammons, Yvette Unoarumhi, Darlene Wagner, Matthew Schmerer, Minoo Agarwal, Eyad Almasri, Debbie Boles, Ayla Burns, Nuthawin Charoensri, Oren Cohen, Susan Countryman, Mary Ann Cristobal, Bobbi Croy, Suzanne Dale, Hrushikesh Deshmukh, Amanda Douglas, Vincent Drouillon, Marcia Eisenberg, Howard Engler, Rama Ghatti, Prashant Gupta, Susan Hicks, Jake Humphrey, Lax Iyer, Lisa Pfefferle, Manoj Jain, Matthew Robinson, Mohan Kolli, Brian Krueger, Tim Kuphal, Stanley Letovsky, Michael Levandoski, Craig Lukasik, Jonathan Meltzer, Brian Norvell, Mindy Nye, Scott Parker, Christos Petropoulos, John Pruitt, Steven Ragan, Scott Ryan, Mike Sapeta, Jana Schroth, Suresh Babu Selvaraju, Goran Stevovic, Amanda Suchanek, Andrea Throop, Lyndon Tilson, Thomas Urban, Joe Voshell, Kimberly Wagner, Jonathan Williams, Mary Williamson, Qian Zeng, Tricia Zwiefelhofer, Clinton R. Paden, Duncan MacCannell |
| EPI_ISL_2480331 | Curative | New Mexico Department of Health Scientific Laboratory | Ellie Johnson, D'eldra Malone, Jennifer Benoit, Linda Salazar, Ratheesh Rajan, Keila Gutierrez, Anastacia Griego-Fisher |
| EPI_ISL_2480332 | Laboratory Corporation of America | Centers for Disease Control and Prevention Division of Viral Diseases, Pathogen Discovery | Dakota Howard, Dhwani Batra, Peter W. Cook, Kara Moser, Adrian Paskey, Jason Caravas, Benjamin Rambo-Martin, Shatavia Morrison, Christopher Gulvick, Scott Sammons, Yvette Unoarumhi, Darlene Wagner, Matthew Schmerer, Minoo Agarwal, Eyad Almasri, Debbie Boles, Ayla Burns, Nuthawin Charoensri, Oren Cohen, Susan Countryman, Mary Ann Cristobal, Bobbi Croy, Suzanne Dale, Hrushikesh Deshmukh, Amanda Douglas, Vincent Drouillon, Marcia Eisenberg, Howard Engler, Rama Ghatti, Prashant Gupta, Susan Hicks, Jake Humphrey, Lax Iyer, Lisa Pfefferle, Manoj Jain, Matthew Robinson, Mohan Kolli, Brian Krueger, Tim Kuphal, Stanley Letovsky, Michael Levandoski, Craig Lukasik, Jonathan Meltzer, Brian Norvell, Mindy Nye, Scott Parker, Christos Petropoulos, John Pruitt, Steven Ragan, Scott Ryan, Mike Sapeta, Jana Schroth, Suresh Babu Selvaraju, Goran Stevovic, Amanda Suchanek, Andrea Throop, Lyndon Tilson, Thomas Urban, Joe Voshell, Kimberly Wagner, Jonathan Williams, Mary Williamson, Qian Zeng, Tricia Zwiefelhofer, Clinton R. Paden, Duncan MacCannell |
| EPI_ISL_2480333 | Curative | New Mexico Department of Health Scientific Laboratory | Ellie Johnson, D'eldra Malone, Jennifer Benoit, Linda Salazar, Ratheesh Rajan, Keila Gutierrez, Anastacia Griego-Fisher |
| EPI_ISL_2480334, EPI_ISL_2480335, EPI_ISL_2480337 | Laboratory Corporation of America | Centers for Disease Control and Prevention Division of Viral Diseases, Pathogen Discovery | Dakota Howard, Dhwani Batra, Peter W. Cook, Kara Moser, Adrian Paskey, Jason Caravas, Benjamin Rambo-Martin, Shatavia Morrison, Christopher Gulvick, Scott Sammons, Yvette Unoarumhi, Darlene Wagner, Matthew Schmerer, Minoo Agarwal, Eyad Almasri, Debbie Boles, Ayla Burns, Nuthawin Charoensri, Oren Cohen, Susan Countryman, Mary Ann Cristobal, Bobbi Croy, Suzanne Dale, Hrushikesh Deshmukh, Amanda Douglas, Vincent Drouillon, Marcia Eisenberg, Howard Engler, Rama Ghatti, Prashant Gupta, Susan Hicks, Jake Humphrey, Lax Iyer, Lisa Pfefferle, Manoj Jain, Matthew Robinson, Mohan Kolli, Brian Krueger, Tim Kuphal, Stanley Letovsky, Michael Levandoski, Craig Lukasik, Jonathan Meltzer, Brian Norvell, Mindy Nye, Scott Parker, Christos Petropoulos, John Pruitt, Steven Ragan, Scott Ryan, Mike Sapeta, Jana Schroth, Suresh Babu Selvaraju, Goran Stevovic, Amanda Suchanek, Andrea Throop, Lyndon Tilson, Thomas Urban, Joe Voshell, Kimberly Wagner, Jonathan Williams, Mary Williamson, Qian Zeng, Tricia Zwiefelhofer, Clinton R. Paden, Duncan MacCannell |
| EPI_ISL_2480338 | Curative | New Mexico Department of Health Scientific Laboratory | Ellie Johnson, D'eldra Malone, Jennifer Benoit, Linda Salazar, Ratheesh Rajan, Keila Gutierrez, Anastacia Griego-Fisher |
| EPI_ISL_2480339, EPI_ISL_2480340, EPI_ISL_2480342, EPI_ISL_2480344, EPI_ISL_2480345, EPI_ISL_2480347, EPI_ISL_2480349 | Laboratory Corporation of America | Centers for Disease Control and Prevention Division of Viral Diseases, Pathogen Discovery | Dakota Howard, Dhwani Batra, Peter W. Cook, Kara Moser, Adrian Paskey, Jason Caravas, Benjamin Rambo-Martin, Shatavia Morrison, Christopher Gulvick, Scott Sammons, Yvette Unoarumhi, Darlene Wagner, Matthew Schmerer, Minoo Agarwal, Eyad Almasri, Debbie Boles, Ayla Burns, Nuthawin Charoensri, Oren Cohen, Susan Countryman, Mary Ann Cristobal, Bobbi Croy, Suzanne Dale, Hrushikesh Deshmukh, Amanda Douglas, Vincent Drouillon, Marcia Eisenberg, Howard Engler, Rama Ghatti, Prashant Gupta, Susan Hicks, Jake Humphrey, Lax Iyer, Lisa Pfefferle, Manoj Jain, Matthew Robinson, Mohan Kolli, Brian Krueger, Tim Kuphal, Stanley Letovsky, Michael Levandoski, Craig Lukasik, Jonathan Meltzer, Brian Norvell, Mindy Nye, Scott Parker, Christos Petropoulos, John Pruitt, Steven Ragan, Scott Ryan, Mike Sapeta, Jana Schroth, Suresh Babu Selvaraju, Goran Stevovic, Amanda Suchanek, Andrea Throop, Lyndon Tilson, Thomas Urban, Joe Voshell, Kimberly Wagner, Jonathan Williams, Mary Williamson, Qian Zeng, Tricia Zwiefelhofer, Clinton R. Paden, Duncan MacCannell |
| EPI_ISL_2480350 | Curative | New Mexico Department of Health Scientific Laboratory | Ellie Johnson, D'eldra Malone, Jennifer Benoit, Linda Salazar, Ratheesh Rajan, Keila Gutierrez, Anastacia Griego-Fisher |
| EPI_ISL_2480351, EPI_ISL_2480352, EPI_ISL_2480354 | Laboratory Corporation of America | Centers for Disease Control and Prevention Division of Viral Diseases, Pathogen Discovery | Dakota Howard, Dhwani Batra, Peter W. Cook, Kara Moser, Adrian Paskey, Jason Caravas, Benjamin Rambo-Martin, Shatavia Morrison, Christopher Gulvick, Scott Sammons, Yvette Unoarumhi, Darlene Wagner, Matthew Schmerer, Minoo Agarwal, Eyad Almasri, Debbie Boles, Ayla Burns, Nuthawin Charoensri, Oren Cohen, Susan Countryman, Mary Ann Cristobal, Bobbi Croy, Suzanne Dale, Hrushikesh Deshmukh, Amanda Douglas, Vincent Drouillon, Marcia Eisenberg, Howard Engler, Rama Ghatti, Prashant Gupta, Susan Hicks, Jake Humphrey, Lax Iyer, Lisa Pfefferle, Manoj Jain, Matthew Robinson, Mohan Kolli, Brian Krueger, Tim Kuphal, Stanley Letovsky, Michael Levandoski, Craig Lukasik, Jonathan Meltzer, Brian Norvell, Mindy Nye, Scott Parker, Christos Petropoulos, John Pruitt, Steven Ragan, Scott Ryan, Mike Sapeta, Jana Schroth, Suresh Babu Selvaraju, Goran Stevovic, Amanda Suchanek, Andrea Throop, Lyndon Tilson, Thomas Urban, Joe Voshell, Kimberly Wagner, Jonathan Williams, Mary Williamson, Qian Zeng, Tricia Zwiefelhofer, Clinton R. Paden, Duncan MacCannell |
| EPI_ISL_2480355 | Curative | New Mexico Department of Health Scientific Laboratory | Ellie Johnson, D'eldra Malone, Jennifer Benoit, Linda Salazar, Ratheesh Rajan, Keila Gutierrez, Anastacia Griego-Fisher |
| EPI_ISL_2480358 | Laboratory Corporation of America | Centers for Disease Control and Prevention Division of Viral Diseases, Pathogen Discovery | Dakota Howard, Dhwani Batra, Peter W. Cook, Kara Moser, Adrian Paskey, Jason Caravas, Benjamin Rambo-Martin, Shatavia Morrison, Christopher Gulvick, Scott Sammons, Yvette Unoarumhi, Darlene Wagner, Matthew Schmerer, Minoo Agarwal, Eyad Almasri, Debbie Boles, Ayla Burns, Nuthawin Charoensri, Oren Cohen, Susan Countryman, Mary Ann Cristobal, Bobbi Croy, Suzanne Dale, Hrushikesh Deshmukh, Amanda Douglas, Vincent Drouillon, Marcia Eisenberg, Howard Engler, Rama Ghatti, Prashant Gupta, Susan Hicks, Jake Humphrey, Lax Iyer, Lisa Pfefferle, Manoj Jain, Matthew Robinson, Mohan Kolli, Brian Krueger, Tim Kuphal, Stanley Letovsky, Michael Levandoski, Craig Lukasik, Jonathan Meltzer, Brian Norvell, Mindy Nye, Scott Parker, Christos Petropoulos, John Pruitt, Steven Ragan, Scott Ryan, Mike Sapeta, Jana Schroth, Suresh Babu Selvaraju, Goran Stevovic, Amanda Suchanek, Andrea Throop, Lyndon Tilson, Thomas Urban, Joe Voshell, Kimberly Wagner, Jonathan Williams, Mary Williamson, Qian Zeng, Tricia Zwiefelhofer, Clinton R. Paden, Duncan MacCannell |
| EPI_ISL_2480359 | Curative | New Mexico Department of Health Scientific Laboratory | Ellie Johnson, D'eldra Malone, Jennifer Benoit, Linda Salazar, Ratheesh Rajan, Keila Gutierrez, Anastacia Griego-Fisher |
| EPI_ISL_2480360, EPI_ISL_2480361, EPI_ISL_2480363, EPI_ISL_2480364, EPI_ISL_2480366, EPI_ISL_2480368, EPI_ISL_2480369, EPI_ISL_2480371 | Laboratory Corporation of America | Centers for Disease Control and Prevention Division of Viral Diseases, Pathogen Discovery | Dakota Howard, Dhwani Batra, Peter W. Cook, Kara Moser, Adrian Paskey, Jason Caravas, Benjamin Rambo-Martin, Shatavia Morrison, Christopher Gulvick, Scott Sammons, Yvette Unoarumhi, Darlene Wagner, Matthew Schmerer, Minoo Agarwal, Eyad Almasri, Debbie Boles, Ayla Burns, Nuthawin Charoensri, Oren Cohen, Susan Countryman, Mary Ann Cristobal, Bobbi Croy, Suzanne Dale, Hrushikesh Deshmukh, Amanda Douglas, Vincent Drouillon, Marcia Eisenberg, Howard Engler, Rama Ghatti, Prashant Gupta, Susan Hicks, Jake Humphrey, Lax Iyer, Lisa Pfefferle, Manoj Jain, Matthew Robinson, Mohan Kolli, Brian Krueger, Tim Kuphal, Stanley Letovsky, Michael Levandoski, Craig Lukasik, Jonathan Meltzer, Brian Norvell, Mindy Nye, Scott Parker, Christos Petropoulos, John Pruitt, Steven Ragan, Scott Ryan, Mike Sapeta, Jana Schroth, Suresh Babu Selvaraju, Goran Stevovic, Amanda Suchanek, Andrea Throop, Lyndon Tilson, Thomas Urban, Joe Voshell, Kimberly Wagner, Jonathan Williams, Mary Williamson, Qian Zeng, Tricia Zwiefelhofer, Clinton R. Paden, Duncan MacCannell |
| EPI_ISL_2480372 | Curative | New Mexico Department of Health Scientific Laboratory | Ellie Johnson, D'eldra Malone, Jennifer Benoit, Linda Salazar, Ratheesh Rajan, Keila Gutierrez, Anastacia Griego-Fisher |
| EPI_ISL_2480375 | Laboratory Corporation of America | Centers for Disease Control and Prevention Division of Viral Diseases, Pathogen Discovery | Dakota Howard, Dhwani Batra, Peter W. Cook, Kara Moser, Adrian Paskey, Jason Caravas, Benjamin Rambo-Martin, Shatavia Morrison, Christopher Gulvick, Scott Sammons, Yvette Unoarumhi, Darlene Wagner, Matthew Schmerer, Minoo Agarwal, Eyad Almasri, Debbie Boles, Ayla Burns, Nuthawin Charoensri, Oren Cohen, Susan Countryman, Mary Ann Cristobal, Bobbi Croy, Suzanne Dale, Hrushikesh Deshmukh, Amanda Douglas, Vincent Drouillon, Marcia Eisenberg, Howard Engler, Rama Ghatti, Prashant Gupta, Susan Hicks, Jake Humphrey, Lax Iyer, Lisa Pfefferle, Manoj Jain, Matthew Robinson, Mohan Kolli, Brian Krueger, Tim Kuphal, Stanley Letovsky, Michael Levandoski, Craig Lukasik, Jonathan Meltzer, Brian Norvell, Mindy Nye, Scott Parker, Christos Petropoulos, John Pruitt, Steven Ragan, Scott Ryan, Mike Sapeta, Jana Schroth, Suresh Babu Selvaraju, Goran Stevovic, Amanda Suchanek, Andrea Throop, Lyndon Tilson, Thomas Urban, Joe Voshell, Kimberly Wagner, Jonathan Williams, Mary Williamson, Qian Zeng, Tricia Zwiefelhofer, Clinton R. Paden, Duncan MacCannell |
| EPI_ISL_2480376 | Curative | New Mexico Department of Health Scientific | Ellie Johnson, D'eldra Malone, Jennifer Benoit, Linda Salazar, Ratheesh Rajan, Keila Gutierrez, Anastacia Griego-Fisher |

| | | | |
|---|---|---|---|
| | | Laboratory | |
| EPI_ISL_2480377, EPI_ISL_2480378, EPI_ISL_2480380 | Laboratory Corporation of America | Centers for Disease Control and Prevention Division of Viral Diseases, Pathogen Discovery | Dakota Howard, Dhwani Batra, Peter W. Cook, Kara Moser, Adrian Paskey, Jason Caravas, Benjamin Rambo-Martin, Shatavia Morrison, Christopher Gulvick, Scott Sammons, Yvette Unoarumhi, Darlene Wagner, Matthew Schmerer, Minoo Agarwal, Eyad Almasri, Debbie Boles, Ayla Burns, Nuthawin Charoensri, Oren Cohen, Susan Countryman, Mary Ann Cristobal, Bobbi Croy, Suzanne Dale, Hrushikesh Deshmukh, Amanda Douglas, Vincent Drouillon, Marcia Eisenberg, Howard Engler, Rama Ghatti, Prashant Gupta, Susan Hicks, Jake Humphrey, Lax Iyer, Lisa Pfefferle, Manoj Jain, Matthew Robinson, Mohan Kolli, Brian Krueger, Tim Kuphal, Stanley Letovsky, Michael Levandoski, Craig Lukasik, Jonathan Meltzer, Brian Norvell, Mindy Nye, Scott Parker, Christos Petropoulos, John Pruitt, Steven Ragan, Scott Ryan, Mike Sapeta, Jana Schroth, Suresh Babu Selvaraju, Goran Stevovic, Amanda Suchanek, Andrea Throop, Lyndon Tilson, Thomas Urban, Joe Voshell, Kimberly Wagner, Jonathan Williams, Mary Williamson, Qian Zeng, Tricia Zwiefelhofer, Clinton R. Paden, Duncan MacCannell |
| EPI_ISL_2480381 | Curative | New Mexico Department of Health Scientific Laboratory | Ellie Johnson, D'eldra Malone, Jennifer Benoit, Linda Salazar, Ratheesh Rajan, Keila Gutierrez, Anastacia Griego-Fisher |
| EPI_ISL_2480382, EPI_ISL_2480384 | Laboratory Corporation of America | Centers for Disease Control and Prevention Division of Viral Diseases, Pathogen Discovery | Dakota Howard, Dhwani Batra, Peter W. Cook, Kara Moser, Adrian Paskey, Jason Caravas, Benjamin Rambo-Martin, Shatavia Morrison, Christopher Gulvick, Scott Sammons, Yvette Unoarumhi, Darlene Wagner, Matthew Schmerer, Minoo Agarwal, Eyad Almasri, Debbie Boles, Ayla Burns, Nuthawin Charoensri, Oren Cohen, Susan Countryman, Mary Ann Cristobal, Bobbi Croy, Suzanne Dale, Hrushikesh Deshmukh, Amanda Douglas, Vincent Drouillon, Marcia Eisenberg, Howard Engler, Rama Ghatti, Prashant Gupta, Susan Hicks, Jake Humphrey, Lax Iyer, Lisa Pfefferle, Manoj Jain, Matthew Robinson, Mohan Kolli, Brian Krueger, Tim Kuphal, Stanley Letovsky, Michael Levandoski, Craig Lukasik, Jonathan Meltzer, Brian Norvell, Mindy Nye, Scott Parker, Christos Petropoulos, John Pruitt, Steven Ragan, Scott Ryan, Mike Sapeta, Jana Schroth, Suresh Babu Selvaraju, Goran Stevovic, Amanda Suchanek, Andrea Throop, Lyndon Tilson, Thomas Urban, Joe Voshell, Kimberly Wagner, Jonathan Williams, Mary Williamson, Qian Zeng, Tricia Zwiefelhofer, Clinton R. Paden, Duncan MacCannell |
| EPI_ISL_2480385 | Curative | New Mexico Department of Health Scientific Laboratory | Ellie Johnson, D'eldra Malone, Jennifer Benoit, Linda Salazar, Ratheesh Rajan, Keila Gutierrez, Anastacia Griego-Fisher |
| EPI_ISL_2480386, EPI_ISL_2480388, EPI_ISL_2480389 | Laboratory Corporation of America | Centers for Disease Control and Prevention Division of Viral Diseases, Pathogen Discovery | Dakota Howard, Dhwani Batra, Peter W. Cook, Kara Moser, Adrian Paskey, Jason Caravas, Benjamin Rambo-Martin, Shatavia Morrison, Christopher Gulvick, Scott Sammons, Yvette Unoarumhi, Darlene Wagner, Matthew Schmerer, Minoo Agarwal, Eyad Almasri, Debbie Boles, Ayla Burns, Nuthawin Charoensri, Oren Cohen, Susan Countryman, Mary Ann Cristobal, Bobbi Croy, Suzanne Dale, Hrushikesh Deshmukh, Amanda Douglas, Vincent Drouillon, Marcia Eisenberg, Howard Engler, Rama Ghatti, Prashant Gupta, Susan Hicks, Jake Humphrey, Lax Iyer, Lisa Pfefferle, Manoj Jain, Matthew Robinson, Mohan Kolli, Brian Krueger, Tim Kuphal, Stanley Letovsky, Michael Levandoski, Craig Lukasik, Jonathan Meltzer, Brian Norvell, Mindy Nye, Scott Parker, Christos Petropoulos, John Pruitt, Steven Ragan, Scott Ryan, Mike Sapeta, Jana Schroth, Suresh Babu Selvaraju, Goran Stevovic, Amanda Suchanek, Andrea Throop, Lyndon Tilson, Thomas Urban, Joe Voshell, Kimberly Wagner, Jonathan Williams, Mary Williamson, Qian Zeng, Tricia Zwiefelhofer, Clinton R. Paden, Duncan MacCannell |
| EPI_ISL_2480390 | Curative | New Mexico Department of Health Scientific Laboratory | Ellie Johnson, D'eldra Malone, Jennifer Benoit, Linda Salazar, Ratheesh Rajan, Keila Gutierrez, Anastacia Griego-Fisher |
| EPI_ISL_2480391, EPI_ISL_2480393, EPI_ISL_2480394 | Laboratory Corporation of America | Centers for Disease Control and Prevention Division of Viral Diseases, Pathogen Discovery | Dakota Howard, Dhwani Batra, Peter W. Cook, Kara Moser, Adrian Paskey, Jason Caravas, Benjamin Rambo-Martin, Shatavia Morrison, Christopher Gulvick, Scott Sammons, Yvette Unoarumhi, Darlene Wagner, Matthew Schmerer, Minoo Agarwal, Eyad Almasri, Debbie Boles, Ayla Burns, Nuthawin Charoensri, Oren Cohen, Susan Countryman, Mary Ann Cristobal, Bobbi Croy, Suzanne Dale, Hrushikesh Deshmukh, Amanda Douglas, Vincent Drouillon, Marcia Eisenberg, Howard Engler, Rama Ghatti, Prashant Gupta, Susan Hicks, Jake Humphrey, Lax Iyer, Lisa Pfefferle, Manoj Jain, Matthew Robinson, Mohan Kolli, Brian Krueger, Tim Kuphal, Stanley Letovsky, Michael Levandoski, Craig Lukasik, Jonathan Meltzer, Brian Norvell, Mindy Nye, Scott Parker, Christos Petropoulos, John Pruitt, Steven Ragan, Scott Ryan, Mike Sapeta, Jana Schroth, Suresh Babu Selvaraju, Goran Stevovic, Amanda Suchanek, Andrea Throop, Lyndon Tilson, Thomas Urban, Joe Voshell, Kimberly Wagner, Jonathan Williams, Mary Williamson, Qian Zeng, Tricia Zwiefelhofer, Clinton R. Paden, Duncan MacCannell |
| EPI_ISL_2480395 | Curative | New Mexico Department of Health Scientific Laboratory | Ellie Johnson, D'eldra Malone, Jennifer Benoit, Linda Salazar, Ratheesh Rajan, Keila Gutierrez, Anastacia Griego-Fisher |
| EPI_ISL_2480396, EPI_ISL_2480400, EPI_ISL_2480402, EPI_ISL_2480404, EPI_ISL_2480405, EPI_ISL_2480407 | Laboratory Corporation of America | Centers for Disease Control and Prevention Division of Viral Diseases, Pathogen Discovery | Dakota Howard, Dhwani Batra, Peter W. Cook, Kara Moser, Adrian Paskey, Jason Caravas, Benjamin Rambo-Martin, Shatavia Morrison, Christopher Gulvick, Scott Sammons, Yvette Unoarumhi, Darlene Wagner, Matthew Schmerer, Minoo Agarwal, Eyad Almasri, Debbie Boles, Ayla Burns, Nuthawin Charoensri, Oren Cohen, Susan Countryman, Mary Ann Cristobal, Bobbi Croy, Suzanne Dale, Hrushikesh Deshmukh, Amanda Douglas, Vincent Drouillon, Marcia Eisenberg, Howard Engler, Rama Ghatti, Prashant Gupta, Susan Hicks, Jake Humphrey, Lax Iyer, Lisa Pfefferle, Manoj Jain, Matthew Robinson, Mohan Kolli, Brian Krueger, Tim Kuphal, Stanley Letovsky, Michael Levandoski, Craig Lukasik, Jonathan Meltzer, Brian Norvell, Mindy Nye, Scott Parker, Christos Petropoulos, John Pruitt, Steven Ragan, Scott Ryan, Mike Sapeta, Jana Schroth, Suresh Babu Selvaraju, Goran Stevovic, Amanda Suchanek, Andrea Throop, Lyndon Tilson, Thomas Urban, Joe Voshell, Kimberly Wagner, Jonathan Williams, Mary Williamson, Qian Zeng, Tricia Zwiefelhofer, Clinton R. Paden, Duncan MacCannell |
| EPI_ISL_2480408 | Curative | New Mexico Department of Health Scientific Laboratory | Ellie Johnson, D'eldra Malone, Jennifer Benoit, Linda Salazar, Ratheesh Rajan, Keila Gutierrez, Anastacia Griego-Fisher |
| EPI_ISL_2480409 | Laboratory Corporation of America | Centers for Disease Control and Prevention Division of Viral Diseases, Pathogen Discovery | Dakota Howard, Dhwani Batra, Peter W. Cook, Kara Moser, Adrian Paskey, Jason Caravas, Benjamin Rambo-Martin, Shatavia Morrison, Christopher Gulvick, Scott Sammons, Yvette Unoarumhi, Darlene Wagner, Matthew Schmerer, Minoo Agarwal, Eyad Almasri, Debbie Boles, Ayla Burns, Nuthawin Charoensri, Oren Cohen, Susan Countryman, Mary Ann Cristobal, Bobbi Croy, Suzanne Dale, Hrushikesh Deshmukh, Amanda Douglas, Vincent Drouillon, Marcia Eisenberg, Howard Engler, Rama Ghatti, Prashant Gupta, Susan Hicks, Jake Humphrey, Lax Iyer, Lisa Pfefferle, Manoj Jain, Matthew Robinson, Mohan Kolli, Brian Krueger, Tim Kuphal, Stanley Letovsky, Michael Levandoski, Craig Lukasik, Jonathan Meltzer, Brian Norvell, Mindy Nye, Scott Parker, Christos Petropoulos, John Pruitt, Steven Ragan, Scott Ryan, Mike Sapeta, Jana Schroth, Suresh Babu Selvaraju, Goran Stevovic, Amanda Suchanek, Andrea Throop, Lyndon Tilson, Thomas Urban, Joe Voshell, Kimberly Wagner, Jonathan Williams, Mary Williamson, Qian Zeng, Tricia Zwiefelhofer, Clinton R. Paden, Duncan MacCannell |
| EPI_ISL_2480410 | Curative | New Mexico Department of Health Scientific Laboratory | Ellie Johnson, D'eldra Malone, Jennifer Benoit, Linda Salazar, Ratheesh Rajan, Keila Gutierrez, Anastacia Griego-Fisher |
| EPI_ISL_2480411, EPI_ISL_2480412 | Laboratory Corporation of America | Centers for Disease Control and Prevention Division of Viral Diseases, Pathogen Discovery | Dakota Howard, Dhwani Batra, Peter W. Cook, Kara Moser, Adrian Paskey, Jason Caravas, Benjamin Rambo-Martin, Shatavia Morrison, Christopher Gulvick, Scott Sammons, Yvette Unoarumhi, Darlene Wagner, Matthew Schmerer, Minoo Agarwal, Eyad Almasri, Debbie Boles, Ayla Burns, Nuthawin Charoensri, Oren Cohen, Susan Countryman, Mary Ann Cristobal, Bobbi Croy, Suzanne Dale, Hrushikesh Deshmukh, Amanda Douglas, Vincent Drouillon, Marcia Eisenberg, Howard Engler, Rama Ghatti, Prashant Gupta, Susan Hicks, Jake Humphrey, Lax Iyer, Lisa Pfefferle, Manoj Jain, Matthew Robinson, Mohan Kolli, Brian Krueger, Tim Kuphal, Stanley Letovsky, Michael Levandoski, Craig Lukasik, Jonathan Meltzer, Brian Norvell, Mindy Nye, Scott Parker, Christos Petropoulos, John Pruitt, Steven Ragan, Scott Ryan, Mike Sapeta, Jana Schroth, Suresh Babu Selvaraju, Goran Stevovic, Amanda Suchanek, Andrea Throop, Lyndon Tilson, Thomas Urban, Joe Voshell, Kimberly Wagner, Jonathan Williams, Mary Williamson, Qian Zeng, Tricia Zwiefelhofer, Clinton R. Paden, Duncan MacCannell |
| EPI_ISL_2480413 | Curative | New Mexico Department of Health Scientific Laboratory | Ellie Johnson, D'eldra Malone, Jennifer Benoit, Linda Salazar, Ratheesh Rajan, Keila Gutierrez, Anastacia Griego-Fisher |
| EPI_ISL_2480414 | Laboratory Corporation of America | Centers for Disease Control and Prevention Division of Viral Diseases, Pathogen Discovery | Dakota Howard, Dhwani Batra, Peter W. Cook, Kara Moser, Adrian Paskey, Jason Caravas, Benjamin Rambo-Martin, Shatavia Morrison, Christopher Gulvick, Scott Sammons, Yvette Unoarumhi, Darlene Wagner, Matthew Schmerer, Minoo Agarwal, Eyad Almasri, Debbie Boles, Ayla Burns, Nuthawin Charoensri, Oren Cohen, Susan Countryman, Mary Ann Cristobal, Bobbi Croy, Suzanne Dale, Hrushikesh Deshmukh, Amanda Douglas, Vincent Drouillon, Marcia Eisenberg, Howard Engler, Rama Ghatti, Prashant Gupta, Susan Hicks, Jake Humphrey, Lax Iyer, Lisa Pfefferle, Manoj Jain, Matthew Robinson, Mohan Kolli, Brian Krueger, Tim Kuphal, Stanley Letovsky, Michael Levandoski, Craig Lukasik, Jonathan Meltzer, Brian Norvell, Mindy Nye, Scott Parker, Christos Petropoulos, John Pruitt, Steven Ragan, Scott Ryan, Mike Sapeta, Jana Schroth, Suresh Babu Selvaraju, Goran Stevovic, Amanda Suchanek, Andrea Throop, Lyndon Tilson, Thomas Urban, Joe Voshell, Kimberly Wagner, Jonathan Williams, Mary Williamson, Qian Zeng, Tricia Zwiefelhofer, Clinton R. Paden, Duncan MacCannell |
| EPI_ISL_2480415 | Curative | New Mexico Department of Health Scientific Laboratory | Ellie Johnson, D'eldra Malone, Jennifer Benoit, Linda Salazar, Ratheesh Rajan, Keila Gutierrez, Anastacia Griego-Fisher |
| EPI_ISL_2480416 | Laboratory Corporation of America | Centers for Disease Control and Prevention Division of Viral Diseases, Pathogen Discovery | Dakota Howard, Dhwani Batra, Peter W. Cook, Kara Moser, Adrian Paskey, Jason Caravas, Benjamin Rambo-Martin, Shatavia Morrison, Christopher Gulvick, Scott Sammons, Yvette Unoarumhi, Darlene Wagner, Matthew Schmerer, Minoo Agarwal, Eyad Almasri, Debbie Boles, Ayla Burns, Nuthawin Charoensri, Oren Cohen, Susan Countryman, Mary Ann Cristobal, Bobbi Croy, Suzanne Dale, Hrushikesh Deshmukh, Amanda Douglas, Vincent Drouillon, Marcia Eisenberg, Howard Engler, Rama Ghatti, Prashant Gupta, Susan Hicks, Jake Humphrey, Lax Iyer, Lisa Pfefferle, Manoj Jain, Matthew Robinson, Mohan Kolli, Brian Krueger, Tim Kuphal, Stanley Letovsky, Michael Levandoski, Craig Lukasik, Jonathan Meltzer, Brian Norvell, Mindy Nye, Scott Parker, Christos Petropoulos, John Pruitt, Steven Ragan, Scott Ryan, Mike Sapeta, Jana Schroth, Suresh Babu Selvaraju, Goran Stevovic, Amanda Suchanek, Andrea Throop, Lyndon Tilson, Thomas Urban, Joe Voshell, Kimberly Wagner, Jonathan Williams, Mary Williamson, Qian Zeng, Tricia Zwiefelhofer, Clinton R. Paden, Duncan MacCannell |
| EPI_ISL_2480417 | Curative | New Mexico Department of Health Scientific Laboratory | Ellie Johnson, D'eldra Malone, Jennifer Benoit, Linda Salazar, Ratheesh Rajan, Keila Gutierrez, Anastacia Griego-Fisher |
| EPI_ISL_2480418, EPI_ISL_2480419, EPI_ISL_2480421, EPI_ISL_2480423 | Laboratory Corporation of America | Centers for Disease Control and Prevention Division of Viral Diseases, Pathogen Discovery | Dakota Howard, Dhwani Batra, Peter W. Cook, Kara Moser, Adrian Paskey, Jason Caravas, Benjamin Rambo-Martin, Shatavia Morrison, Christopher Gulvick, Scott Sammons, Yvette Unoarumhi, Darlene Wagner, Matthew Schmerer, Minoo Agarwal, Eyad Almasri, Debbie Boles, Ayla Burns, Nuthawin Charoensri, Oren Cohen, Susan Countryman, Mary Ann Cristobal, Bobbi Croy, Suzanne Dale, Hrushikesh Deshmukh, Amanda Douglas, Vincent Drouillon, Marcia Eisenberg, Howard Engler, Rama Ghatti, Prashant Gupta, Susan Hicks, Jake Humphrey, Lax Iyer, Lisa Pfefferle, Manoj Jain, Matthew Robinson, Mohan Kolli, Brian Krueger, Tim Kuphal, Stanley Letovsky, Michael Levandoski, Craig Lukasik, Jonathan Meltzer, Brian Norvell, Mindy Nye, Scott Parker, Christos Petropoulos, John Pruitt, Steven Ragan, Scott Ryan, Mike Sapeta, Jana Schroth, Suresh Babu Selvaraju, Goran Stevovic, Amanda Suchanek, Andrea Throop, Lyndon Tilson, Thomas Urban, Joe Voshell, Kimberly Wagner, Jonathan Williams, Mary Williamson, Qian Zeng, Tricia Zwiefelhofer, Clinton R. Paden, Duncan MacCannell |
| EPI_ISL_2480424 | Curative | New Mexico Department of Health Scientific Laboratory | Ellie Johnson, D'eldra Malone, Jennifer Benoit, Linda Salazar, Ratheesh Rajan, Keila Gutierrez, Anastacia Griego-Fisher |
| EPI_ISL_2480425, EPI_ISL_2480427, EPI_ISL_2480428, EPI_ISL_2480430, EPI_ISL_2480432, EPI_ISL_2480434, EPI_ISL_2480435, EPI_ISL_2480437, EPI_ISL_2480438 | Laboratory Corporation of America | Centers for Disease Control and Prevention Division of Viral Diseases, Pathogen Discovery | Dakota Howard, Dhwani Batra, Peter W. Cook, Kara Moser, Adrian Paskey, Jason Caravas, Benjamin Rambo-Martin, Shatavia Morrison, Christopher Gulvick, Scott Sammons, Yvette Unoarumhi, Darlene Wagner, Matthew Schmerer, Minoo Agarwal, Eyad Almasri, Debbie Boles, Ayla Burns, Nuthawin Charoensri, Oren Cohen, Susan Countryman, Mary Ann Cristobal, Bobbi Croy, Suzanne Dale, Hrushikesh Deshmukh, Amanda Douglas, Vincent Drouillon, Marcia Eisenberg, Howard Engler, Rama Ghatti, Prashant Gupta, Susan Hicks, Jake Humphrey, Lax Iyer, Lisa Pfefferle, Manoj Jain, Matthew Robinson, Mohan Kolli, Brian Krueger, Tim Kuphal, Stanley Letovsky, Michael Levandoski, Craig Lukasik, Jonathan Meltzer, Brian Norvell, Mindy Nye, Scott Parker, Christos Petropoulos, John Pruitt, Steven Ragan, Scott Ryan, Mike Sapeta, Jana Schroth, Suresh Babu Selvaraju, Goran Stevovic, Amanda Suchanek, Andrea Throop, Lyndon Tilson, Thomas Urban, Joe Voshell, Kimberly Wagner, Jonathan Williams, Mary Williamson, Qian Zeng, Tricia Zwiefelhofer, Clinton R. Paden, Duncan MacCannell |
| EPI_ISL_2480439 | Curative | New Mexico Department of Health Scientific Laboratory | Ellie Johnson, D'eldra Malone, Jennifer Benoit, Linda Salazar, Ratheesh Rajan, Keila Gutierrez, Anastacia Griego-Fisher |
| EPI_ISL_2480440, EPI_ISL_2480442, EPI_ISL_2480444, EPI_ISL_2480445 | Laboratory Corporation of America | Centers for Disease Control and Prevention Division of Viral Diseases, Pathogen Discovery | Dakota Howard, Dhwani Batra, Peter W. Cook, Kara Moser, Adrian Paskey, Jason Caravas, Benjamin Rambo-Martin, Shatavia Morrison, Christopher Gulvick, Scott Sammons, Yvette Unoarumhi, Darlene Wagner, Matthew Schmerer, Minoo Agarwal, Eyad Almasri, Debbie Boles, Ayla Burns, Nuthawin Charoensri, Oren Cohen, Susan Countryman, Mary Ann Cristobal, Bobbi Croy, Suzanne Dale, Hrushikesh Deshmukh, Amanda Douglas, Vincent Drouillon, Marcia Eisenberg, Howard Engler, Rama Ghatti, Prashant Gupta, Susan Hicks, Jake Humphrey, Lax Iyer, Lisa Pfefferle, Manoj Jain, Matthew Robinson, Mohan Kolli, Brian Krueger, Tim Kuphal, Stanley Letovsky, Michael Levandoski, Craig Lukasik, Jonathan Meltzer, Brian Norvell, Mindy Nye, Scott Parker, Christos Petropoulos, John Pruitt, Steven Ragan, Scott Ryan, Mike Sapeta, Jana Schroth, Suresh Babu Selvaraju, Goran Stevovic, Amanda Suchanek, Andrea Throop, Lyndon Tilson, Thomas Urban, Joe Voshell, Kimberly Wagner, Jonathan Williams, Mary Williamson, Qian Zeng, Tricia Zwiefelhofer, Clinton R. Paden, Duncan MacCannell |
| EPI_ISL_2480446 | Curative | New Mexico Department of Health Scientific Laboratory | Ellie Johnson, D'eldra Malone, Jennifer Benoit, Linda Salazar, Ratheesh Rajan, Keila Gutierrez, Anastacia Griego-Fisher |
| EPI_ISL_2480447, EPI_ISL_2480449, EPI_ISL_2480451, EPI_ISL_2480452 | Laboratory Corporation of America | Centers for Disease Control and Prevention Division of Viral Diseases, Pathogen Discovery | Dakota Howard, Dhwani Batra, Peter W. Cook, Kara Moser, Adrian Paskey, Jason Caravas, Benjamin Rambo-Martin, Shatavia Morrison, Christopher Gulvick, Scott Sammons, Yvette Unoarumhi, Darlene Wagner, Matthew Schmerer, Minoo Agarwal, Eyad Almasri, Debbie Boles, Ayla Burns, Nuthawin Charoensri, Oren Cohen, Susan Countryman, Mary Ann Cristobal, Bobbi Croy, Suzanne Dale, Hrushikesh Deshmukh, Amanda Douglas, Vincent Drouillon, Marcia Eisenberg, Howard Engler, Rama Ghatti, Prashant Gupta, Susan Hicks, Jake Humphrey, Lax Iyer, Lisa Pfefferle, Manoj Jain, Matthew Robinson, Mohan Kolli, Brian Krueger, Tim Kuphal, Stanley Letovsky, Michael Levandoski, Craig Lukasik, Jonathan Meltzer, Brian Norvell, Mindy Nye, Scott Parker, Christos Petropoulos, John Pruitt, Steven Ragan, Scott Ryan, Mike Sapeta, Jana Schroth, Suresh Babu Selvaraju, Goran Stevovic, Amanda Suchanek, Andrea Throop, Lyndon Tilson, Thomas Urban, Joe Voshell, Kimberly Wagner, Jonathan Williams, Mary Williamson, Qian Zeng, Tricia Zwiefelhofer, Clinton R. Paden, Duncan MacCannell |
| EPI_ISL_2480453 | Curative | New Mexico Department of Health Scientific Laboratory | Ellie Johnson, D'eldra Malone, Jennifer Benoit, Linda Salazar, Ratheesh Rajan, Keila Gutierrez, Anastacia Griego-Fisher |
| EPI_ISL_2480454 | Laboratory Corporation of America | Centers for Disease Control and Prevention Division of Viral Diseases, Pathogen Discovery | Dakota Howard, Dhwani Batra, Peter W. Cook, Kara Moser, Adrian Paskey, Jason Caravas, Benjamin Rambo-Martin, Shatavia Morrison, Christopher Gulvick, Scott Sammons, Yvette Unoarumhi, Darlene Wagner, Matthew Schmerer, Minoo Agarwal, Eyad Almasri, Debbie Boles, Ayla Burns, Nuthawin Charoensri, Oren Cohen, Susan Countryman, Mary Ann Cristobal, Bobbi Croy, Suzanne Dale, Hrushikesh Deshmukh, Amanda Douglas, Vincent Drouillon, Marcia Eisenberg, Howard Engler, Rama Ghatti, Prashant Gupta, Susan Hicks, Jake Humphrey, Lax Iyer, Lisa Pfefferle, Manoj Jain, Matthew Robinson, Mohan Kolli, Brian Krueger, Tim Kuphal, Stanley Letovsky, Michael Levandoski, Craig Lukasik, Jonathan Meltzer, Brian Norvell, Mindy Nye, Scott Parker, Christos Petropoulos, John Pruitt, Steven Ragan, Scott Ryan, Mike Sapeta, Jana Schroth, Suresh Babu Selvaraju, Goran Stevovic, Amanda Suchanek, Andrea Throop, Lyndon Tilson, Thomas Urban, Joe Voshell, Kimberly Wagner, Jonathan Williams, Mary Williamson, Qian Zeng, Tricia Zwiefelhofer, Clinton R. Paden, Duncan MacCannell |
| EPI_ISL_2480455 | Curative | New Mexico Department of Health Scientific Laboratory | Ellie Johnson, D'eldra Malone, Jennifer Benoit, Linda Salazar, Ratheesh Rajan, Keila Gutierrez, Anastacia Griego-Fisher |
| EPI_ISL_2480456, EPI_ISL_2480457 | Laboratory Corporation of America | Centers for Disease Control and Prevention Division of Viral Diseases, Pathogen Discovery | Dakota Howard, Dhwani Batra, Peter W. Cook, Kara Moser, Adrian Paskey, Jason Caravas, Benjamin Rambo-Martin, Shatavia Morrison, Christopher Gulvick, Scott Sammons, Yvette Unoarumhi, Darlene Wagner, Matthew Schmerer, Minoo Agarwal, Eyad Almasri, Debbie Boles, Ayla Burns, Nuthawin Charoensri, Oren Cohen, Susan Countryman, Mary Ann Cristobal, Bobbi Croy, Suzanne Dale, Hrushikesh Deshmukh, Amanda Douglas, Vincent Drouillon, Marcia Eisenberg, Howard Engler, Rama Ghatti, Prashant Gupta, Susan Hicks, Jake Humphrey, Lax Iyer, Lisa Pfefferle, Manoj Jain, Matthew Robinson, Mohan Kolli, Brian Krueger, Tim Kuphal, Stanley Letovsky, Michael Levandoski, Craig Lukasik, Jonathan Meltzer, Brian Norvell, Mindy Nye, Scott Parker, Christos Petropoulos, John Pruitt, Steven Ragan, Scott Ryan, Mike Sapeta, Jana Schroth, Suresh Babu Selvaraju, Goran Stevovic, Amanda Suchanek, Andrea Throop, Lyndon Tilson, Thomas Urban, Joe Voshell, Kimberly Wagner, Jonathan Williams, Mary Williamson, Qian Zeng, Tricia Zwiefelhofer, Clinton R. Paden, Duncan MacCannell |

| | | | |
|---|---|---|---|
| EPI_ISL_2480458 | Curative | New Mexico Department of Health Scientific Laboratory | Ellie Johnson, D'eldra Malone, Jennifer Benoit, Linda Salazar, Ratheesh Rajan, Keila Gutierrez, Anastacia Griego-Fisher |
| EPI_ISL_2480459 | Laboratory Corporation of America | Centers for Disease Control and Prevention Division of Viral Diseases, Pathogen Discovery | Dakota Howard, Dhwani Batra, Peter W. Cook, Kara Moser, Adrian Paskey, Jason Caravas, Benjamin Rambo-Martin, Shatavia Morrison, Christopher Gulvick, Scott Sammons, Yvette Unoarumhi, Darlene Wagner, Matthew Schmerer, Minoo Agarwal, Eyad Almasri, Debbie Boles, Ayla Burns, Nuthawin Charoensri, Oren Cohen, Susan Countryman, Mary Ann Cristobal, Bobbi Croy, Suzanne Dale, Hrushikesh Deshmukh, Amanda Douglas, Vincent Drouillon, Marcia Eisenberg, Howard Engler, Rama Ghatti, Prashant Gupta, Susan Hicks, Jake Humphrey, Lax Iyer, Lisa Pfefferle, Manoj Jain, Matthew Robinson, Mohan Kolli, Brian Krueger, Tim Kuphal, Stanley Letovsky, Michael Levandoski, Craig Lukasik, Jonathan Meltzer, Brian Norvell, Mindy Nye, Scott Parker, Christos Petropoulos, John Pruitt, Steven Ragan, Scott Ryan, Mike Sapeta, Jana Schroth, Suresh Babu Selvaraju, Goran Stevovic, Amanda Suchanek, Andrea Throop, Lyndon Tilson, Thomas Urban, Joe Voshell, Kimberly Wagner, Jonathan Williams, Mary Williamson, Qian Zeng, Tricia Zwiefelhofer, Clinton R. Paden, Duncan MacCannell |
| EPI_ISL_2480460 | Curative | New Mexico Department of Health Scientific Laboratory | Ellie Johnson, D'eldra Malone, Jennifer Benoit, Linda Salazar, Ratheesh Rajan, Keila Gutierrez, Anastacia Griego-Fisher |
| EPI_ISL_2480461 | Laboratory Corporation of America | Centers for Disease Control and Prevention Division of Viral Diseases, Pathogen Discovery | Dakota Howard, Dhwani Batra, Peter W. Cook, Kara Moser, Adrian Paskey, Jason Caravas, Benjamin Rambo-Martin, Shatavia Morrison, Christopher Gulvick, Scott Sammons, Yvette Unoarumhi, Darlene Wagner, Matthew Schmerer, Minoo Agarwal, Eyad Almasri, Debbie Boles, Ayla Burns, Nuthawin Charoensri, Oren Cohen, Susan Countryman, Mary Ann Cristobal, Bobbi Croy, Suzanne Dale, Hrushikesh Deshmukh, Amanda Douglas, Vincent Drouillon, Marcia Eisenberg, Howard Engler, Rama Ghatti, Prashant Gupta, Susan Hicks, Jake Humphrey, Lax Iyer, Lisa Pfefferle, Manoj Jain, Matthew Robinson, Mohan Kolli, Brian Krueger, Tim Kuphal, Stanley Letovsky, Michael Levandoski, Craig Lukasik, Jonathan Meltzer, Brian Norvell, Mindy Nye, Scott Parker, Christos Petropoulos, John Pruitt, Steven Ragan, Scott Ryan, Mike Sapeta, Jana Schroth, Suresh Babu Selvaraju, Goran Stevovic, Amanda Suchanek, Andrea Throop, Lyndon Tilson, Thomas Urban, Joe Voshell, Kimberly Wagner, Jonathan Williams, Mary Williamson, Qian Zeng, Tricia Zwiefelhofer, Clinton R. Paden, Duncan MacCannell |
| EPI_ISL_2480462 | Curative | New Mexico Department of Health Scientific Laboratory | Ellie Johnson, D'eldra Malone, Jennifer Benoit, Linda Salazar, Ratheesh Rajan, Keila Gutierrez, Anastacia Griego-Fisher |
| EPI_ISL_2480463, EPI_ISL_2480464 | Laboratory Corporation of America | Centers for Disease Control and Prevention Division of Viral Diseases, Pathogen Discovery | Dakota Howard, Dhwani Batra, Peter W. Cook, Kara Moser, Adrian Paskey, Jason Caravas, Benjamin Rambo-Martin, Shatavia Morrison, Christopher Gulvick, Scott Sammons, Yvette Unoarumhi, Darlene Wagner, Matthew Schmerer, Minoo Agarwal, Eyad Almasri, Debbie Boles, Ayla Burns, Nuthawin Charoensri, Oren Cohen, Susan Countryman, Mary Ann Cristobal, Bobbi Croy, Suzanne Dale, Hrushikesh Deshmukh, Amanda Douglas, Vincent Drouillon, Marcia Eisenberg, Howard Engler, Rama Ghatti, Prashant Gupta, Susan Hicks, Jake Humphrey, Lax Iyer, Lisa Pfefferle, Manoj Jain, Matthew Robinson, Mohan Kolli, Brian Krueger, Tim Kuphal, Stanley Letovsky, Michael Levandoski, Craig Lukasik, Jonathan Meltzer, Brian Norvell, Mindy Nye, Scott Parker, Christos Petropoulos, John Pruitt, Steven Ragan, Scott Ryan, Mike Sapeta, Jana Schroth, Suresh Babu Selvaraju, Goran Stevovic, Amanda Suchanek, Andrea Throop, Lyndon Tilson, Thomas Urban, Joe Voshell, Kimberly Wagner, Jonathan Williams, Mary Williamson, Qian Zeng, Tricia Zwiefelhofer, Clinton R. Paden, Duncan MacCannell |
| EPI_ISL_2480465 | Curative | New Mexico Department of Health Scientific Laboratory | Ellie Johnson, D'eldra Malone, Jennifer Benoit, Linda Salazar, Ratheesh Rajan, Keila Gutierrez, Anastacia Griego-Fisher |
| EPI_ISL_2480466 | Laboratory Corporation of America | Centers for Disease Control and Prevention Division of Viral Diseases, Pathogen Discovery | Dakota Howard, Dhwani Batra, Peter W. Cook, Kara Moser, Adrian Paskey, Jason Caravas, Benjamin Rambo-Martin, Shatavia Morrison, Christopher Gulvick, Scott Sammons, Yvette Unoarumhi, Darlene Wagner, Matthew Schmerer, Minoo Agarwal, Eyad Almasri, Debbie Boles, Ayla Burns, Nuthawin Charoensri, Oren Cohen, Susan Countryman, Mary Ann Cristobal, Bobbi Croy, Suzanne Dale, Hrushikesh Deshmukh, Amanda Douglas, Vincent Drouillon, Marcia Eisenberg, Howard Engler, Rama Ghatti, Prashant Gupta, Susan Hicks, Jake Humphrey, Lax Iyer, Lisa Pfefferle, Manoj Jain, Matthew Robinson, Mohan Kolli, Brian Krueger, Tim Kuphal, Stanley Letovsky, Michael Levandoski, Craig Lukasik, Jonathan Meltzer, Brian Norvell, Mindy Nye, Scott Parker, Christos Petropoulos, John Pruitt, Steven Ragan, Scott Ryan, Mike Sapeta, Jana Schroth, Suresh Babu Selvaraju, Goran Stevovic, Amanda Suchanek, Andrea Throop, Lyndon Tilson, Thomas Urban, Joe Voshell, Kimberly Wagner, Jonathan Williams, Mary Williamson, Qian Zeng, Tricia Zwiefelhofer, Clinton R. Paden, Duncan MacCannell |
| EPI_ISL_2480467 | Curative | New Mexico Department of Health Scientific Laboratory | Ellie Johnson, D'eldra Malone, Jennifer Benoit, Linda Salazar, Ratheesh Rajan, Keila Gutierrez, Anastacia Griego-Fisher |
| EPI_ISL_2480468 | Laboratory Corporation of America | Centers for Disease Control and Prevention Division of Viral Diseases, Pathogen Discovery | Dakota Howard, Dhwani Batra, Peter W. Cook, Kara Moser, Adrian Paskey, Jason Caravas, Benjamin Rambo-Martin, Shatavia Morrison, Christopher Gulvick, Scott Sammons, Yvette Unoarumhi, Darlene Wagner, Matthew Schmerer, Minoo Agarwal, Eyad Almasri, Debbie Boles, Ayla Burns, Nuthawin Charoensri, Oren Cohen, Susan Countryman, Mary Ann Cristobal, Bobbi Croy, Suzanne Dale, Hrushikesh Deshmukh, Amanda Douglas, Vincent Drouillon, Marcia Eisenberg, Howard Engler, Rama Ghatti, Prashant Gupta, Susan Hicks, Jake Humphrey, Lax Iyer, Lisa Pfefferle, Manoj Jain, Matthew Robinson, Mohan Kolli, Brian Krueger, Tim Kuphal, Stanley Letovsky, Michael Levandoski, Craig Lukasik, Jonathan Meltzer, Brian Norvell, Mindy Nye, Scott Parker, Christos Petropoulos, John Pruitt, Steven Ragan, Scott Ryan, Mike Sapeta, Jana Schroth, Suresh Babu Selvaraju, Goran Stevovic, Amanda Suchanek, Andrea Throop, Lyndon Tilson, Thomas Urban, Joe Voshell, Kimberly Wagner, Jonathan Williams, Mary Williamson, Qian Zeng, Tricia Zwiefelhofer, Clinton R. Paden, Duncan MacCannell |
| EPI_ISL_2480469 | Curative | New Mexico Department of Health Scientific Laboratory | Ellie Johnson, D'eldra Malone, Jennifer Benoit, Linda Salazar, Ratheesh Rajan, Keila Gutierrez, Anastacia Griego-Fisher |

EPI_ISL_2480470, EPI_ISL_2480471, EPI_ISL_2480473, EPI_ISL_2480474, EPI_ISL_2480476, EPI_ISL_2480478, EPI_ISL_2480480, EPI_ISL_2480481, EPI_ISL_2480482, EPI_ISL_2480483, EPI_ISL_2480484, EPI_ISL_2480485, EPI_ISL_2480486, EPI_ISL_2480487, EPI_ISL_2480488, EPI_ISL_2480489, EPI_ISL_2480490, EPI_ISL_2480491, EPI_ISL_2480492, EPI_ISL_2480493, EPI_ISL_2480494, EPI_ISL_2480495,
EPI_ISL_2480496, EPI_ISL_2480497, EPI_ISL_2480498, EPI_ISL_2480499, EPI_ISL_2480500, EPI_ISL_2480501, EPI_ISL_2480502, EPI_ISL_2480503, EPI_ISL_2480504, EPI_ISL_2480505, EPI_ISL_2480506, EPI_ISL_2480507, EPI_ISL_2480508, EPI_ISL_2480509, EPI_ISL_2480510, EPI_ISL_2480511, EPI_ISL_2480512, EPI_ISL_2480513, EPI_ISL_2480514, EPI_ISL_2480515, EPI_ISL_2480516, EPI_ISL_2480517,
EPI_ISL_2480518, EPI_ISL_2480519, EPI_ISL_2480520, EPI_ISL_2480521, EPI_ISL_2480522, EPI_ISL_2480523, EPI_ISL_2480524, EPI_ISL_2480525, EPI_ISL_2480526, EPI_ISL_2480527, EPI_ISL_2480528, EPI_ISL_2480529, EPI_ISL_2480530, EPI_ISL_2480531, EPI_ISL_2480532, EPI_ISL_2480533, EPI_ISL_2480534, EPI_ISL_2480535, EPI_ISL_2480536, EPI_ISL_2480537, EPI_ISL_2480538, EPI_ISL_2480539,
EPI_ISL_2480540, EPI_ISL_2480541, EPI_ISL_2480542, EPI_ISL_2480543, EPI_ISL_2480544, EPI_ISL_2480545, EPI_ISL_2480546, EPI_ISL_2480547, EPI_ISL_2480548, EPI_ISL_2480549, EPI_ISL_2480550, EPI_ISL_2480551, EPI_ISL_2480552, EPI_ISL_2480553, EPI_ISL_2480554, EPI_ISL_2480555, EPI_ISL_2480556, EPI_ISL_2480557, EPI_ISL_2480558, EPI_ISL_2480560, EPI_ISL_2480561, EPI_ISL_2480562,
EPI_ISL_2480563, EPI_ISL_2480564, EPI_ISL_2480565, EPI_ISL_2480566, EPI_ISL_2480567, EPI_ISL_2480568, EPI_ISL_2480569, EPI_ISL_2480570, EPI_ISL_2480572, EPI_ISL_2480573, EPI_ISL_2480574, EPI_ISL_2480575, EPI_ISL_2480576, EPI_ISL_2480577, EPI_ISL_2480579, EPI_ISL_2480580, EPI_ISL_2480581, EPI_ISL_2480582, EPI_ISL_2480583, EPI_ISL_2480584, EPI_ISL_2480585, EPI_ISL_2480586,
EPI_ISL_2480588, EPI_ISL_2480589, EPI_ISL_2480590, EPI_ISL_2480591, EPI_ISL_2480592, EPI_ISL_2480593, EPI_ISL_2480594, EPI_ISL_2480595, EPI_ISL_2480596, EPI_ISL_2480597, EPI_ISL_2480598, EPI_ISL_2480599, EPI_ISL_2480600, EPI_ISL_2480601, EPI_ISL_2480602, EPI_ISL_2480603, EPI_ISL_2480604, EPI_ISL_2480605, EPI_ISL_2480606, EPI_ISL_2480607, EPI_ISL_2480608, EPI_ISL_2480609,
EPI_ISL_2480610, EPI_ISL_2480611, EPI_ISL_2480612, EPI_ISL_2480613, EPI_ISL_2480614, EPI_ISL_2480615, EPI_ISL_2480616, EPI_ISL_2480617, EPI_ISL_2480618, EPI_ISL_2480619, EPI_ISL_2480620, EPI_ISL_2480621, EPI_ISL_2480622, EPI_ISL_2480623, EPI_ISL_2480624, EPI_ISL_2480625, EPI_ISL_2480626, EPI_ISL_2480627, EPI_ISL_2480628, EPI_ISL_2480629, EPI_ISL_2480630, EPI_ISL_2480631,
EPI_ISL_2480632, EPI_ISL_2480633, EPI_ISL_2480634, EPI_ISL_2480635, EPI_ISL_2480636, EPI_ISL_2480637, EPI_ISL_2480638, EPI_ISL_2480639, EPI_ISL_2480641, EPI_ISL_2480642, EPI_ISL_2480644, EPI_ISL_2480645, EPI_ISL_2480646, EPI_ISL_2480648, EPI_ISL_2480650, EPI_ISL_2480651, EPI_ISL_2480654, EPI_ISL_2480655, EPI_ISL_2480656, EPI_ISL_2480657, EPI_ISL_2480658, EPI_ISL_2480659,
EPI_ISL_2480660, EPI_ISL_2480661, EPI_ISL_2480662, EPI_ISL_2480663, EPI_ISL_2480664, EPI_ISL_2480665, EPI_ISL_2480667, EPI_ISL_2480668, EPI_ISL_2480669, EPI_ISL_2480670, EPI_ISL_2480671, EPI_ISL_2480672, EPI_ISL_2480673, EPI_ISL_2480674, EPI_ISL_2480675, EPI_ISL_2480676, EPI_ISL_2480677, EPI_ISL_2480678, EPI_ISL_2480679, EPI_ISL_2480680, EPI_ISL_2480681, EPI_ISL_2480682,
EPI_ISL_2480683, EPI_ISL_2480684, EPI_ISL_2480685, EPI_ISL_2480686, EPI_ISL_2480687, EPI_ISL_2480688, EPI_ISL_2480689, EPI_ISL_2480690, EPI_ISL_2480691, EPI_ISL_2480692, EPI_ISL_2480693, EPI_ISL_2480694, EPI_ISL_2480695, EPI_ISL_2480696, EPI_ISL_2480697, EPI_ISL_2480698, EPI_ISL_2480699, EPI_ISL_2480700, EPI_ISL_2480701, EPI_ISL_2480702, EPI_ISL_2480703, EPI_ISL_2480704,
EPI_ISL_2480705, EPI_ISL_2480706, EPI_ISL_2480707, EPI_ISL_2480708, EPI_ISL_2480709, EPI_ISL_2480710, EPI_ISL_2480711, EPI_ISL_2480712, EPI_ISL_2480714, EPI_ISL_2480715, EPI_ISL_2480716, EPI_ISL_2480717, EPI_ISL_2480718, EPI_ISL_2480719, EPI_ISL_2480720, EPI_ISL_2480721, EPI_ISL_2480722, EPI_ISL_2480723, EPI_ISL_2480724, EPI_ISL_2480725, EPI_ISL_2480726, EPI_ISL_2480727,
EPI_ISL_2480728, EPI_ISL_2480729, EPI_ISL_2480730, EPI_ISL_2480731, EPI_ISL_2480732, EPI_ISL_2480733, EPI_ISL_2480734, EPI_ISL_2480735, EPI_ISL_2480736, EPI_ISL_2480737, EPI_ISL_2480738, EPI_ISL_2480739, EPI_ISL_2480740, EPI_ISL_2480741, EPI_ISL_2480742, EPI_ISL_2480743, EPI_ISL_2480744, EPI_ISL_2480745, EPI_ISL_2480746, EPI_ISL_2480747, EPI_ISL_2480748, EPI_ISL_2480749,
EPI_ISL_2480750, EPI_ISL_2480751, EPI_ISL_2480752, EPI_ISL_2480753, EPI_ISL_2480754, EPI_ISL_2480755, EPI_ISL_2480756, EPI_ISL_2480757, EPI_ISL_2480758, EPI_ISL_2480759, EPI_ISL_2480760, EPI_ISL_2480761, EPI_ISL_2480762, EPI_ISL_2480763, EPI_ISL_2480764, EPI_ISL_2480765, EPI_ISL_2480766, EPI_ISL_2480767, EPI_ISL_2480768, EPI_ISL_2480769, EPI_ISL_2480770, EPI_ISL_2480772,
EPI_ISL_2480773, EPI_ISL_2480774, EPI_ISL_2480775, EPI_ISL_2480776, EPI_ISL_2480777, EPI_ISL_2480778, EPI_ISL_2480779, EPI_ISL_2480780, EPI_ISL_2480781, EPI_ISL_2480782, EPI_ISL_2480783, EPI_ISL_2480784, EPI_ISL_2480785, EPI_ISL_2480786, EPI_ISL_2480787, EPI_ISL_2480788, EPI_ISL_2480789, EPI_ISL_2480790, EPI_ISL_2480791, EPI_ISL_2480792, EPI_ISL_2480793, EPI_ISL_2480794,
EPI_ISL_2480795, EPI_ISL_2480796, EPI_ISL_2480797, EPI_ISL_2480798, EPI_ISL_2480799, EPI_ISL_2480800, EPI_ISL_2480801, EPI_ISL_2480802, EPI_ISL_2480803, EPI_ISL_2480804, EPI_ISL_2480805, EPI_ISL_2480806, EPI_ISL_2480807, EPI_ISL_2480808, EPI_ISL_2480809, EPI_ISL_2480810, EPI_ISL_2480811, EPI_ISL_2480812, EPI_ISL_2480813, EPI_ISL_2480814, EPI_ISL_2480815, EPI_ISL_2480816,
EPI_ISL_2480818, EPI_ISL_2480819, EPI_ISL_2480820, EPI_ISL_2480821, EPI_ISL_2480822, EPI_ISL_2480823, EPI_ISL_2480824, EPI_ISL_2480825, EPI_ISL_2480826, EPI_ISL_2480827, EPI_ISL_2480828, EPI_ISL_2480829, EPI_ISL_2480830, EPI_ISL_2480831, EPI_ISL_2480832, EPI_ISL_2480833, EPI_ISL_2480835, EPI_ISL_2480836, EPI_ISL_2480837, EPI_ISL_2480838, EPI_ISL_2480839, EPI_ISL_2480840,
EPI_ISL_2480841, EPI_ISL_2480842, EPI_ISL_2480843, EPI_ISL_2480845, EPI_ISL_2480846, EPI_ISL_2480847, EPI_ISL_2480848, EPI_ISL_2480849, EPI_ISL_2480850, EPI_ISL_2480851, EPI_ISL_2480852, EPI_ISL_2480854, EPI_ISL_2480855, EPI_ISL_2480856, EPI_ISL_2480857, EPI_ISL_2480858, EPI_ISL_2480859, EPI_ISL_2480860, EPI_ISL_2480861, EPI_ISL_2480862, EPI_ISL_2480863, EPI_ISL_2480864,
EPI_ISL_2480865, EPI_ISL_2480866, EPI_ISL_2480867, EPI_ISL_2480868, EPI_ISL_2480869, EPI_ISL_2480870, EPI_ISL_2480871, EPI_ISL_2480872, EPI_ISL_2480873, EPI_ISL_2480874, EPI_ISL_2480875, EPI_ISL_2480876, EPI_ISL_2480877, EPI_ISL_2480878, EPI_ISL_2480879, EPI_ISL_2480880, EPI_ISL_2480881, EPI_ISL_2480882, EPI_ISL_2480883, EPI_ISL_2480884, EPI_ISL_2480885, EPI_ISL_2480886,
EPI_ISL_2480887, EPI_ISL_2480888, EPI_ISL_2480889, EPI_ISL_2480890, EPI_ISL_2480891, EPI_ISL_2480892, EPI_ISL_2480893, EPI_ISL_2480894, EPI_ISL_2480895, EPI_ISL_2480896, EPI_ISL_2480897, EPI_ISL_2480898, EPI_ISL_2480899, EPI_ISL_2480900, EPI_ISL_2480901, EPI_ISL_2480902, EPI_ISL_2480903, EPI_ISL_2480904, EPI_ISL_2480905, EPI_ISL_2480907, EPI_ISL_2480908, EPI_ISL_2480909,
EPI_ISL_2480910, EPI_ISL_2480911, EPI_ISL_2480912, EPI_ISL_2480914, EPI_ISL_2480915, EPI_ISL_2480916, EPI_ISL_2480917, EPI_ISL_2480918, EPI_ISL_2480919, EPI_ISL_2480920, EPI_ISL_2480921, EPI_ISL_2480922, EPI_ISL_2480923, EPI_ISL_2480924, EPI_ISL_2480926, EPI_ISL_2480927, EPI_ISL_2480928, EPI_ISL_2480929, EPI_ISL_2480930, EPI_ISL_2480931, EPI_ISL_2480932,
EPI_ISL_2480933, EPI_ISL_2480934, EPI_ISL_2480935, EPI_ISL_2480936, EPI_ISL_2480937, EPI_ISL_2480938, EPI_ISL_2480939, EPI_ISL_2480941, EPI_ISL_2480942, EPI_ISL_2480943, EPI_ISL_2480944, EPI_ISL_2480945, EPI_ISL_2480946, EPI_ISL_2480947, EPI_ISL_2480948, EPI_ISL_2480949, EPI_ISL_2480950, EPI_ISL_2480951, EPI_ISL_2480952, EPI_ISL_2480953, EPI_ISL_2480954, EPI_ISL_2480955,
EPI_ISL_2480956, EPI_ISL_2480957, EPI_ISL_2480958, EPI_ISL_2480959, EPI_ISL_2480960, EPI_ISL_2480961, EPI_ISL_2480962, EPI_ISL_2480963, EPI_ISL_2480964, EPI_ISL_2480965, EPI_ISL_2480966, EPI_ISL_2480967, EPI_ISL_2480968, EPI_ISL_2480969, EPI_ISL_2480970, EPI_ISL_2480971, EPI_ISL_2480972, EPI_ISL_2480973, EPI_ISL_2480974, EPI_ISL_2480976, EPI_ISL_2480977,
EPI_ISL_2480980, EPI_ISL_2480981, EPI_ISL_2480982, EPI_ISL_2480983, EPI_ISL_2480984, EPI_ISL_2480985, EPI_ISL_2480986, EPI_ISL_2480987, EPI_ISL_2480988, EPI_ISL_2480989, EPI_ISL_2480990, EPI_ISL_2480992, EPI_ISL_2480993, EPI_ISL_2480994, EPI_ISL_2480995, EPI_ISL_2480996, EPI_ISL_2480997, EPI_ISL_2480998, EPI_ISL_2480999, EPI_ISL_2481000, EPI_ISL_2481001, EPI_ISL_2481002,
EPI_ISL_2481003, EPI_ISL_2481005, EPI_ISL_2481006, EPI_ISL_2481007, EPI_ISL_2481008, EPI_ISL_2481009, EPI_ISL_2481011, EPI_ISL_2481013, EPI_ISL_2481014, EPI_ISL_2481015, EPI_ISL_2481016, EPI_ISL_2481017, EPI_ISL_2481018, EPI_ISL_2481019, EPI_ISL_2481020, EPI_ISL_2481021, EPI_ISL_2481022, EPI_ISL_2481023, EPI_ISL_2481024, EPI_ISL_2481025, EPI_ISL_2481026, EPI_ISL_2481027,
EPI_ISL_2481028, EPI_ISL_2481030, EPI_ISL_2481031, EPI_ISL_2481032, EPI_ISL_2481033, EPI_ISL_2481034, EPI_ISL_2481035, EPI_ISL_2481036, EPI_ISL_2481037, EPI_ISL_2481038, EPI_ISL_2481039, EPI_ISL_2481040, EPI_ISL_2481041, EPI_ISL_2481043, EPI_ISL_2481044, EPI_ISL_2481045, EPI_ISL_2481046, EPI_ISL_2481047, EPI_ISL_2481048, EPI_ISL_2481049, EPI_ISL_2481051, EPI_ISL_2481052,
EPI_ISL_2481053, EPI_ISL_2481054, EPI_ISL_2481055, EPI_ISL_2481056, EPI_ISL_2481057, EPI_ISL_2481058, EPI_ISL_2481059, EPI_ISL_2481060, EPI_ISL_2481061, EPI_ISL_2481062, EPI_ISL_2481063, EPI_ISL_2481064, EPI_ISL_2481066, EPI_ISL_2481067, EPI_ISL_2481068, EPI_ISL_2481069, EPI_ISL_2481070, EPI_ISL_2481072, EPI_ISL_2481073, EPI_ISL_2481074, EPI_ISL_2481076, EPI_ISL_2481077,
EPI_ISL_2481078, EPI_ISL_2481079, EPI_ISL_2481080, EPI_ISL_2481081, EPI_ISL_2481082, EPI_ISL_2481083, EPI_ISL_2481084, EPI_ISL_2481085, EPI_ISL_2481086, EPI_ISL_2481087, EPI_ISL_2481088, EPI_ISL_2481089, EPI_ISL_2481090, EPI_ISL_2481091, EPI_ISL_2481092, EPI_ISL_2481093, EPI_ISL_2481094, EPI_ISL_2481095, EPI_ISL_2481096, EPI_ISL_2481097, EPI_ISL_2481098, EPI_ISL_2481099,
EPI_ISL_2481100, EPI_ISL_2481101, EPI_ISL_2481102, EPI_ISL_2481103, EPI_ISL_2481104, EPI_ISL_2481105, EPI_ISL_2481106, EPI_ISL_2481107, EPI_ISL_2481108, EPI_ISL_2481109, EPI_ISL_2481110, EPI_ISL_2481111, EPI_ISL_2481113, EPI_ISL_2481114, EPI_ISL_2481115, EPI_ISL_2481116, EPI_ISL_2481117, EPI_ISL_2481118, EPI_ISL_2481119, EPI_ISL_2481120, EPI_ISL_2481121, EPI_ISL_2481122,
EPI_ISL_2481123, EPI_ISL_2481124, EPI_ISL_2481125, EPI_ISL_2481126, EPI_ISL_2481127, EPI_ISL_2481128, EPI_ISL_2481129, EPI_ISL_2481130, EPI_ISL_2481132, EPI_ISL_2481133, EPI_ISL_2481134, EPI_ISL_2481135, EPI_ISL_2481136, EPI_ISL_2481137, EPI_ISL_2481138, EPI_ISL_2481139, EPI_ISL_2481140, EPI_ISL_2481141, EPI_ISL_2481142, EPI_ISL_2481143, EPI_ISL_2481144, EPI_ISL_2481145,
EPI_ISL_2481146, EPI_ISL_2481147, EPI_ISL_2481148, EPI_ISL_2481149, EPI_ISL_2481150, EPI_ISL_2481151, EPI_ISL_2481152, EPI_ISL_2481153, EPI_ISL_2481154, EPI_ISL_2481155, EPI_ISL_2481156, EPI_ISL_2481157, EPI_ISL_2481158, EPI_ISL_2481160, EPI_ISL_2481161, EPI_ISL_2481162, EPI_ISL_2481163, EPI_ISL_2481165, EPI_ISL_2481166, EPI_ISL_2481167, EPI_ISL_2481168, EPI_ISL_2481169,
EPI_ISL_2481170, EPI_ISL_2481171, EPI_ISL_2481172, EPI_ISL_2481173, EPI_ISL_2481174, EPI_ISL_2481175, EPI_ISL_2481176, EPI_ISL_2481178, EPI_ISL_2481179, EPI_ISL_2481180, EPI_ISL_2481181, EPI_ISL_2481182, EPI_ISL_2481183, EPI_ISL_2481184, EPI_ISL_2481185, EPI_ISL_2481186, EPI_ISL_2481188, EPI_ISL_2481189, EPI_ISL_2481190, EPI_ISL_2481191, EPI_ISL_2481192,
EPI_ISL_2481195, EPI_ISL_2481196, EPI_ISL_2481197, EPI_ISL_2481198, EPI_ISL_2481199, EPI_ISL_2481200, EPI_ISL_2481201, EPI_ISL_2481203, EPI_ISL_2481205, EPI_ISL_2481206, EPI_ISL_2481207, EPI_ISL_2481208, EPI_ISL_2481209, EPI_ISL_2481210, EPI_ISL_2481212, EPI_ISL_2481213, EPI_ISL_2481214, EPI_ISL_2481215, EPI_ISL_2481217, EPI_ISL_2481218, EPI_ISL_2481219, EPI_ISL_2481220,
EPI_ISL_2481221, EPI_ISL_2481222, EPI_ISL_2481223, EPI_ISL_2481224, EPI_ISL_2481225, EPI_ISL_2481226, EPI_ISL_2481227, EPI_ISL_2481228, EPI_ISL_2481229, EPI_ISL_2481230, EPI_ISL_2481232, EPI_ISL_2481233, EPI_ISL_2481234, EPI_ISL_2481235, EPI_ISL_2481236, EPI_ISL_2481237, EPI_ISL_2481238, EPI_ISL_2481239, EPI_ISL_2481240, EPI_ISL_2481241, EPI_ISL_2481242, EPI_ISL_2481243,
EPI_ISL_2481244, EPI_ISL_2481245, EPI_ISL_2481246, EPI_ISL_2481247, EPI_ISL_2481248, EPI_ISL_2481249, EPI_ISL_2481250, EPI_ISL_2481251, EPI_ISL_2481252, EPI_ISL_2481253, EPI_ISL_2481254, EPI_ISL_2481255, EPI_ISL_2481256, EPI_ISL_2481257, EPI_ISL_2481258, EPI_ISL_2481259, EPI_ISL_2481260, EPI_ISL_2481261, EPI_ISL_2481262, EPI_ISL_2481263, EPI_ISL_2481264, EPI_ISL_2481265,
EPI_ISL_2481267, EPI_ISL_2481269, EPI_ISL_2481270, EPI_ISL_2481271, EPI_ISL_2481272, EPI_ISL_2481273, EPI_ISL_2481274, EPI_ISL_2481275, EPI_ISL_2481276, EPI_ISL_2481277, EPI_ISL_2481278, EPI_ISL_2481279, EPI_ISL_2481280, EPI_ISL_2481281, EPI_ISL_2481282, EPI_ISL_2481283, EPI_ISL_2481284, EPI_ISL_2481285, EPI_ISL_2481287, EPI_ISL_2481288, EPI_ISL_2481289, EPI_ISL_2481290,
EPI_ISL_2481291, EPI_ISL_2481292, EPI_ISL_2481293, EPI_ISL_2481294, EPI_ISL_2481295, EPI_ISL_2481296, EPI_ISL_2481298, EPI_ISL_2481299, EPI_ISL_2481301, EPI_ISL_2481302, EPI_ISL_2481303, EPI_ISL_2481304, EPI_ISL_2481305, EPI_ISL_2481306, EPI_ISL_2481307, EPI_ISL_2481308, EPI_ISL_2481310, EPI_ISL_2481312, EPI_ISL_2481313, EPI_ISL_2481314, EPI_ISL_2481315, EPI_ISL_2481316,
EPI_ISL_2481317, EPI_ISL_2481318, EPI_ISL_2481319, EPI_ISL_2481322, EPI_ISL_2481325, EPI_ISL_2481328, EPI_ISL_2481331, EPI_ISL_2481337, EPI_ISL_2481340, EPI_ISL_2481344, EPI_ISL_2481347, EPI_ISL_2481350, EPI_ISL_2481353, EPI_ISL_2481362, EPI_ISL_2481365, EPI_ISL_2481369, EPI_ISL_2481372, EPI_ISL_2481375, EPI_ISL_2481377, EPI_ISL_2481383, EPI_ISL_2481386, EPI_ISL_2481388,
EPI_ISL_2481392, EPI_ISL_2481395, EPI_ISL_2481401, EPI_ISL_2481404, EPI_ISL_2481407, EPI_ISL_2481410, EPI_ISL_2481414, EPI_ISL_2481416, EPI_ISL_2481420, EPI_ISL_2481423, EPI_ISL_2481426, EPI_ISL_2481429, EPI_ISL_2481432, EPI_ISL_2481436, EPI_ISL_2481439, EPI_ISL_2481440, EPI_ISL_2481441, EPI_ISL_2481442, EPI_ISL_2481443, EPI_ISL_2481444, EPI_ISL_2481445, EPI_ISL_2481446,
EPI_ISL_2481447, EPI_ISL_2481448, EPI_ISL_2481449, EPI_ISL_2481450, EPI_ISL_2481451, EPI_ISL_2481452, EPI_ISL_2481453, EPI_ISL_2481454, EPI_ISL_2481455, EPI_ISL_2481456, EPI_ISL_2481457, EPI_ISL_2481458, EPI_ISL_2481460, EPI_ISL_2481461, EPI_ISL_2481462, EPI_ISL_2481463, EPI_ISL_2481464, EPI_ISL_2481465, EPI_ISL_2481466, EPI_ISL_2481468, EPI_ISL_2481469,
EPI_ISL_2481470, EPI_ISL_2481472, EPI_ISL_2481473, EPI_ISL_2481474, EPI_ISL_2481475, EPI_ISL_2481476, EPI_ISL_2481477, EPI_ISL_2481478, EPI_ISL_2481479, EPI_ISL_2481480, EPI_ISL_2481481, EPI_ISL_2481482, EPI_ISL_2481483, EPI_ISL_2481484, EPI_ISL_2481485, EPI_ISL_2481486, EPI_ISL_2481487, EPI_ISL_2481489, EPI_ISL_2481490, EPI_ISL_2481491, EPI_ISL_2481492, EPI_ISL_2481493,
EPI_ISL_2481494, EPI_ISL_2481495, EPI_ISL_2481496, EPI_ISL_2481497, EPI_ISL_2481498, EPI_ISL_2481499, EPI_ISL_2481500, EPI_ISL_2481501, EPI_ISL_2481502, EPI_ISL_2481503, EPI_ISL_2481504, EPI_ISL_2481505, EPI_ISL_2481506, EPI_ISL_2481507, EPI_ISL_2481508, EPI_ISL_2481509, EPI_ISL_2481510, EPI_ISL_2481511, EPI_ISL_2481512, EPI_ISL_2481514, EPI_ISL_2481515,
EPI_ISL_2481516, EPI_ISL_2481518, EPI_ISL_2481520, EPI_ISL_2481521, EPI_ISL_2481522, EPI_ISL_2481523, EPI_ISL_2481525, EPI_ISL_2481526, EPI_ISL_2481527, EPI_ISL_2481528, EPI_ISL_2481529, EPI_ISL_2481530, EPI_ISL_2481531, EPI_ISL_2481532, EPI_ISL_2481534, EPI_ISL_2481535, EPI_ISL_2481536, EPI_ISL_2481537, EPI_ISL_2481538, EPI_ISL_2481539, EPI_ISL_2481540, EPI_ISL_2481541,
EPI_ISL_2481544, EPI_ISL_2481545, EPI_ISL_2481546, EPI_ISL_2481547, EPI_ISL_2481549, EPI_ISL_2481551, EPI_ISL_2481552, EPI_ISL_2481553, EPI_ISL_2481555, EPI_ISL_2481556, EPI_ISL_2481557, EPI_ISL_2481558, EPI_ISL_2481559, EPI_ISL_2481560, EPI_ISL_2481561, EPI_ISL_2481562, EPI_ISL_2481563, EPI_ISL_2481565, EPI_ISL_2481566, EPI_ISL_2481567,
EPI_ISL_2481568, EPI_ISL_2481570, EPI_ISL_2481571, EPI_ISL_2481572, EPI_ISL_2481573, EPI_ISL_2481574, EPI_ISL_2481575, EPI_ISL_2481576, EPI_ISL_2481577, EPI_ISL_2481579, EPI_ISL_2481580, EPI_ISL_2481581, EPI_ISL_2481583, EPI_ISL_2481584, EPI_ISL_2481585, EPI_ISL_2481586, EPI_ISL_2481587, EPI_ISL_2481588, EPI_ISL_2481589, EPI_ISL_2481590, EPI_ISL_2481591,
EPI_ISL_2481592, EPI_ISL_2481593, EPI_ISL_2481594, EPI_ISL_2481596, EPI_ISL_2481597, EPI_ISL_2481598, EPI_ISL_2481599, EPI_ISL_2481600, EPI_ISL_2481601, EPI_ISL_2481602, EPI_ISL_2481603, EPI_ISL_2481604, EPI_ISL_2481605, EPI_ISL_2481606, EPI_ISL_2481607, EPI_ISL_2481608, EPI_ISL_2481609, EPI_ISL_2481610, EPI_ISL_2481611, EPI_ISL_2481612, EPI_ISL_2481614, EPI_ISL_2481615,
EPI_ISL_2481616, EPI_ISL_2481617, EPI_ISL_2481618, EPI_ISL_2481619, EPI_ISL_2481620, EPI_ISL_2481621, EPI_ISL_2481622, EPI_ISL_2481624, EPI_ISL_2481625, EPI_ISL_2481626, EPI_ISL_2481627, EPI_ISL_2481628, EPI_ISL_2481629, EPI_ISL_2481630, EPI_ISL_2481631, EPI_ISL_2481632, EPI_ISL_2481633, EPI_ISL_2481634, EPI_ISL_2481635, EPI_ISL_2481636, EPI_ISL_2481637,
EPI_ISL_2481638, EPI_ISL_2481639, EPI_ISL_2481640, EPI_ISL_2481641, EPI_ISL_2481642, EPI_ISL_2481643, EPI_ISL_2481644, EPI_ISL_2481645, EPI_ISL_2481646, EPI_ISL_2481647, EPI_ISL_2481649, EPI_ISL_2481650, EPI_ISL_2481651, EPI_ISL_2481652, EPI_ISL_2481653, EPI_ISL_2481654, EPI_ISL_2481655, EPI_ISL_2481656, EPI_ISL_2481657, EPI_ISL_2481658, EPI_ISL_2481659, EPI_ISL_2481660,
EPI_ISL_2481661, EPI_ISL_2481662, EPI_ISL_2481663, EPI_ISL_2481664, EPI_ISL_2481665, EPI_ISL_2481666, EPI_ISL_2481667, EPI_ISL_2481668, EPI_ISL_2481669, EPI_ISL_2481671, EPI_ISL_2481672, EPI_ISL_2481673, EPI_ISL_2481674, EPI_ISL_2481675, EPI_ISL_2481676, EPI_ISL_2481677, EPI_ISL_2481678, EPI_ISL_2481679, EPI_ISL_2481680, EPI_ISL_2481681, EPI_ISL_2481682, EPI_ISL_2481683,
EPI_ISL_2481684, EPI_ISL_2481685, EPI_ISL_2481686, EPI_ISL_2481687, EPI_ISL_2481688, EPI_ISL_2481689, EPI_ISL_2481690, EPI_ISL_2481691, EPI_ISL_2481692, EPI_ISL_2481693, EPI_ISL_2481694, EPI_ISL_2481695, EPI_ISL_2481696, EPI_ISL_2481697, EPI_ISL_2481698, EPI_ISL_2481699, EPI_ISL_2481700, EPI_ISL_2481701, EPI_ISL_2481702, EPI_ISL_2481703, EPI_ISL_2481704, EPI_ISL_2481706,
EPI_ISL_2481707, EPI_ISL_2481708, EPI_ISL_2481709, EPI_ISL_2481710, EPI_ISL_2481711, EPI_ISL_2481712, EPI_ISL_2481713, EPI_ISL_2481714, EPI_ISL_2481716, EPI_ISL_2481717, EPI_ISL_2481718, EPI_ISL_2481719, EPI_ISL_2481720, EPI_ISL_2481721, EPI_ISL_2481722, EPI_ISL_2481723, EPI_ISL_2481724, EPI_ISL_2481725, EPI_ISL_2481726, EPI_ISL_2481727, EPI_ISL_2481728, EPI_ISL_2481729,
EPI_ISL_2481730, EPI_ISL_2481732, EPI_ISL_2481733, EPI_ISL_2481735, EPI_ISL_2481736, EPI_ISL_2481737, EPI_ISL_2481738, EPI_ISL_2481739, EPI_ISL_2481740, EPI_ISL_2481741, EPI_ISL_2481742, EPI_ISL_2481743, EPI_ISL_2481744, EPI_ISL_2481745, EPI_ISL_2481747, EPI_ISL_2481748, EPI_ISL_2481749, EPI_ISL_2481750, EPI_ISL_2481751, EPI_ISL_2481753, EPI_ISL_2481754,
EPI_ISL_2481755, EPI_ISL_2481756, EPI_ISL_2481757, EPI_ISL_2481758, EPI_ISL_2481759, EPI_ISL_2481761, EPI_ISL_2481762, EPI_ISL_2481763, EPI_ISL_2481764, EPI_ISL_2481765, EPI_ISL_2481767, EPI_ISL_2481768, EPI_ISL_2481769, EPI_ISL_2481770, EPI_ISL_2481771, EPI_ISL_2481772, EPI_ISL_2481773, EPI_ISL_2481774, EPI_ISL_2481775, EPI_ISL_2481776, EPI_ISL_2481778, EPI_ISL_2481779,
EPI_ISL_2481780, EPI_ISL_2481781, EPI_ISL_2481782, EPI_ISL_2481783, EPI_ISL_2481784, EPI_ISL_2481785, EPI_ISL_2481786, EPI_ISL_2481787, EPI_ISL_2481788, EPI_ISL_2481789, EPI_ISL_2481791, EPI_ISL_2481793, EPI_ISL_2481794, EPI_ISL_2481795, EPI_ISL_2481796, EPI_ISL_2481797, EPI_ISL_2481799, EPI_ISL_2481800, EPI_ISL_2481801, EPI_ISL_2481802, EPI_ISL_2481803, EPI_ISL_2481804,
EPI_ISL_2481805, EPI_ISL_2481806, EPI_ISL_2481807, EPI_ISL_2481808, EPI_ISL_2481809, EPI_ISL_2481810, EPI_ISL_2481811, EPI_ISL_2481812, EPI_ISL_2481813, EPI_ISL_2481814, EPI_ISL_2481815, EPI_ISL_2481816, EPI_ISL_2481817, EPI_ISL_2481818, EPI_ISL_2481819, EPI_ISL_2481820, EPI_ISL_2481821, EPI_ISL_2481822, EPI_ISL_2481823, EPI_ISL_2481824, EPI_ISL_2481825, EPI_ISL_2481827,
EPI_ISL_2481828, EPI_ISL_2481829, EPI_ISL_2481831, EPI_ISL_2481832, EPI_ISL_2481833, EPI_ISL_2481834, EPI_ISL_2481835, EPI_ISL_2481836, EPI_ISL_2481837, EPI_ISL_2481838, EPI_ISL_2481839, EPI_ISL_2481840, EPI_ISL_2481841, EPI_ISL_2481842, EPI_ISL_2481843, EPI_ISL_2481844, EPI_ISL_2481845, EPI_ISL_2481846, EPI_ISL_2481847, EPI_ISL_2481848, EPI_ISL_2481849, EPI_ISL_2481850,
EPI_ISL_2481851, EPI_ISL_2481852, EPI_ISL_2481853, EPI_ISL_2481854, EPI_ISL_2481855, EPI_ISL_2481856, EPI_ISL_2481857, EPI_ISL_2481858, EPI_ISL_2481859, EPI_ISL_2481860, EPI_ISL_2481861, EPI_ISL_2481862, EPI_ISL_2481863, EPI_ISL_2481865, EPI_ISL_2481866, EPI_ISL_2481867, EPI_ISL_2481868, EPI_ISL_2481869, EPI_ISL_2481871, EPI_ISL_2481872, EPI_ISL_2481873, EPI_ISL_2481874,
EPI_ISL_2481875, EPI_ISL_2481876, EPI_ISL_2481877, EPI_ISL_2481879, EPI_ISL_2481880, EPI_ISL_2481881, EPI_ISL_2481882, EPI_ISL_2481883, EPI_ISL_2481884, EPI_ISL_2481885, EPI_ISL_2481886, EPI_ISL_2481887, EPI_ISL_2481888, EPI_ISL_2481889, EPI_ISL_2481890, EPI_ISL_2481891, EPI_ISL_2481892, EPI_ISL_2481893, EPI_ISL_2481894, EPI_ISL_2481895, EPI_ISL_2481896, EPI_ISL_2481897,
EPI_ISL_2481898, EPI_ISL_2481899, EPI_ISL_2481900, EPI_ISL_2481901, EPI_ISL_2481902, EPI_ISL_2481903, EPI_ISL_2481904, EPI_ISL_2481905, EPI_ISL_2481906, EPI_ISL_2481907, EPI_ISL_2481908, EPI_ISL_2481909, EPI_ISL_2481910, EPI_ISL_2481911, EPI_ISL_2481912, EPI_ISL_2481913, EPI_ISL_2481914, EPI_ISL_2481916, EPI_ISL_2481917, EPI_ISL_2481918, EPI_ISL_2481919,
EPI_ISL_2481920, EPI_ISL_2481921, EPI_ISL_2481923, EPI_ISL_2481924, EPI_ISL_2481925, EPI_ISL_2481927, EPI_ISL_2481928, EPI_ISL_2481929, EPI_ISL_2481930, EPI_ISL_2481931, EPI_ISL_2481932, EPI_ISL_2481933, EPI_ISL_2481934, EPI_ISL_2481935, EPI_ISL_2481936, EPI_ISL_2481937, EPI_ISL_2481938, EPI_ISL_2481939, EPI_ISL_2481940, EPI_ISL_2481941, EPI_ISL_2481942, EPI_ISL_2481943,
EPI_ISL_2481944, EPI_ISL_2481946, EPI_ISL_2481947, EPI_ISL_2481949, EPI_ISL_2481950, EPI_ISL_2481952, EPI_ISL_2481953, EPI_ISL_2481954, EPI_ISL_2481955, EPI_ISL_2481956, EPI_ISL_2481957, EPI_ISL_2481958, EPI_ISL_2481959, EPI_ISL_2481960, EPI_ISL_2481961, EPI_ISL_2481962

| | | | |
|---|---|---|---|
| see above | Laboratory Corporation of America | Centers for Disease Control and Prevention Division of Viral Diseases, Pathogen Discovery | Dakota Howard, Dhwani Batra, Peter W. Cook, Kara Moser, Adrian Paskey, Jason Caravas, Benjamin Rambo-Martin, Shatavia Morrison, Christopher Gulvick, Scott Sammons, Yvette Unoarumhi, Darlene Wagner, Matthew Schmerer, Minoo Agarwal, Eyad Almasri, Debbie Boles, Ayla Burns, Nuthawin Charoensri, Oren Cohen, Susan Countryman, Mary Ann Cristobal, Bobbi Croy, Suzanne Dale, Hrushikesh Deshmukh, Amanda Douglas, Vincent Drouillon, Marcia Eisenberg, Howard Engler, Rama Ghatti, Prashant Gupta, Susan Hicks, Jake Humphrey, Lax Iyer, Lisa Pfefferle, Manoj Jain, Matthew Robinson, Mohan Kolli, Brian Krueger, Tim Kuphal, Stanley Letovsky, Michael Levandoski, Craig Lukasik, Jonathan Meltzer, Brian Norvell, Mindy Nye, Scott Parker, Christos Petropoulos, John Pruitt, Steven Ragan, Scott Ryan, Mike Sapeta, Jana Schroth, Suresh Babu Selvaraju, Goran Stevovic, Amanda Suchanek, Andrea Throop, Lyndon Tilson, Thomas Urban, Joe Voshell, Kimberly Wagner, Jonathan Williams, Mary Williamson, Qian Zeng, Tricia Zwiefelhofer, Clinton R. Paden, Duncan MacCannell |
| EPI_ISL_2481963 | Ohio Department of Health Laboratory | Ohio Department of Health Laboratory | Holmes, Jennifer; Eric Brandt, Keoni Omura, Glen McGillivary, Caitlin McDonnell, Jade Mowery, Stephanie Mccracken, Tyler Payne, Kirtana Ramadugu, Erica Leasure, Brent Lee, Kelsey Florek, Heather Blankenship, Allison Black, Quanta Brown, and Tammy Bannerman |

EPI_ISL_2481964, EPI_ISL_2481965, EPI_ISL_2481966, EPI_ISL_2481967, EPI_ISL_2481968, EPI_ISL_2481969, EPI_ISL_2481970, EPI_ISL_2481972, EPI_ISL_2481973, EPI_ISL_2481974, EPI_ISL_2481975, EPI_ISL_2481977, EPI_ISL_2481978, EPI_ISL_2481979, EPI_ISL_2481980, EPI_ISL_2481981, EPI_ISL_2481982, EPI_ISL_2481983, EPI_ISL_2481984, EPI_ISL_2481985, EPI_ISL_2481986, EPI_ISL_2481987,
EPI_ISL_2481988, EPI_ISL_2481989, EPI_ISL_2481990, EPI_ISL_2481991, EPI_ISL_2481992, EPI_ISL_2481993, EPI_ISL_2481994, EPI_ISL_2481995, EPI_ISL_2481996, EPI_ISL_2481997, EPI_ISL_2481998, EPI_ISL_2481999, EPI_ISL_2482000, EPI_ISL_2482001, EPI_ISL_2482002, EPI_ISL_2482003, EPI_ISL_2482005, EPI_ISL_2482006, EPI_ISL_2482007, EPI_ISL_2482008, EPI_ISL_2482009, EPI_ISL_2482010,
EPI_ISL_2482011, EPI_ISL_2482012, EPI_ISL_2482014, EPI_ISL_2482015, EPI_ISL_2482016, EPI_ISL_2482017, EPI_ISL_2482018, EPI_ISL_2482019, EPI_ISL_2482020, EPI_ISL_2482021, EPI_ISL_2482022, EPI_ISL_2482023, EPI_ISL_2482024, EPI_ISL_2482025, EPI_ISL_2482026, EPI_ISL_2482027, EPI_ISL_2482028, EPI_ISL_2482029, EPI_ISL_2482031, EPI_ISL_2482032, EPI_ISL_2482034, EPI_ISL_2482035,
EPI_ISL_2482036, EPI_ISL_2482037, EPI_ISL_2482038, EPI_ISL_2482039, EPI_ISL_2482040, EPI_ISL_2482041, EPI_ISL_2482042, EPI_ISL_2482044, EPI_ISL_2482045, EPI_ISL_2482046, EPI_ISL_2482047, EPI_ISL_2482048, EPI_ISL_2482049, EPI_ISL_2482050, EPI_ISL_2482051, EPI_ISL_2482052, EPI_ISL_2482053, EPI_ISL_2482054, EPI_ISL_2482055, EPI_ISL_2482056, EPI_ISL_2482057, EPI_ISL_2482058,
EPI_ISL_2482059, EPI_ISL_2482060, EPI_ISL_2482061, EPI_ISL_2482062, EPI_ISL_2482063, EPI_ISL_2482064, EPI_ISL_2482065, EPI_ISL_2482066, EPI_ISL_2482067, EPI_ISL_2482068, EPI_ISL_2482069, EPI_ISL_2482070, EPI_ISL_2482071, EPI_ISL_2482072, EPI_ISL_2482073, EPI_ISL_2482074, EPI_ISL_2482075, EPI_ISL_2482076, EPI_ISL_2482077, EPI_ISL_2482078, EPI_ISL_2482079, EPI_ISL_2482080,
EPI_ISL_2482081, EPI_ISL_2482082, EPI_ISL_2482083, EPI_ISL_2482084, EPI_ISL_2482085, EPI_ISL_2482086, EPI_ISL_2482087, EPI_ISL_2482088, EPI_ISL_2482089, EPI_ISL_2482090, EPI_ISL_2482091, EPI_ISL_2482092, EPI_ISL_2482093, EPI_ISL_2482094, EPI_ISL_2482095, EPI_ISL_2482096, EPI_ISL_2482097, EPI_ISL_2482098, EPI_ISL_2482099, EPI_ISL_2482100, EPI_ISL_2482101, EPI_ISL_2482102,

| | | | |
|---|---|---|---|
| EPI_ISL_2482103, EPI_ISL_2482104, EPI_ISL_2482105, EPI_ISL_2482106, EPI_ISL_2482107, EPI_ISL_2482108, EPI_ISL_2482109, EPI_ISL_2482110, EPI_ISL_2482111, EPI_ISL_2482112, EPI_ISL_2482113, EPI_ISL_2482114, EPI_ISL_2482115, EPI_ISL_2482116, EPI_ISL_2482117, EPI_ISL_2482118, EPI_ISL_2482119, EPI_ISL_2482120, EPI_ISL_2482121, EPI_ISL_2482122, EPI_ISL_2482123, EPI_ISL_2482124, EPI_ISL_2482125, EPI_ISL_2482126, EPI_ISL_2482127, EPI_ISL_2482129, EPI_ISL_2482130, EPI_ISL_2482131, EPI_ISL_2482132, EPI_ISL_2482133, EPI_ISL_2482134, EPI_ISL_2482135, EPI_ISL_2482136, EPI_ISL_2482137, EPI_ISL_2482138, EPI_ISL_2482139, EPI_ISL_2482140, EPI_ISL_2482141, EPI_ISL_2482142, EPI_ISL_2482143, EPI_ISL_2482145, EPI_ISL_2482146, EPI_ISL_2482147, EPI_ISL_2482148, EPI_ISL_2482149, EPI_ISL_2482150, EPI_ISL_2482151, EPI_ISL_2482152, EPI_ISL_2482153, EPI_ISL_2482154, EPI_ISL_2482155, EPI_ISL_2482156, EPI_ISL_2482157, EPI_ISL_2482158, EPI_ISL_2482159, EPI_ISL_2482160, EPI_ISL_2482161, EPI_ISL_2482162, EPI_ISL_2482163, EPI_ISL_2482164, EPI_ISL_2482165, EPI_ISL_2482166, EPI_ISL_2482167, EPI_ISL_2482168, EPI_ISL_2482169, EPI_ISL_2482170, EPI_ISL_2482171, EPI_ISL_2482172, EPI_ISL_2482173, EPI_ISL_2482174, EPI_ISL_2482175, EPI_ISL_2482176, EPI_ISL_2482177, EPI_ISL_2482178, EPI_ISL_2482179, EPI_ISL_2482180, EPI_ISL_2482181, EPI_ISL_2482182, EPI_ISL_2482183, EPI_ISL_2482184, EPI_ISL_2482185, EPI_ISL_2482187, EPI_ISL_2482188, EPI_ISL_2482189, EPI_ISL_2482190, EPI_ISL_2482191, EPI_ISL_2482192, EPI_ISL_2482193, EPI_ISL_2482194, EPI_ISL_2482195, EPI_ISL_2482196, EPI_ISL_2482198, EPI_ISL_2482199, EPI_ISL_2482200, EPI_ISL_2482201, EPI_ISL_2482202, EPI_ISL_2482203, EPI_ISL_2482204, EPI_ISL_2482205, EPI_ISL_2482206, EPI_ISL_2482207, EPI_ISL_2482208, EPI_ISL_2482209, EPI_ISL_2482210, EPI_ISL_2482211, EPI_ISL_2482212, EPI_ISL_2482214, EPI_ISL_2482215, EPI_ISL_2482216, EPI_ISL_2482217, EPI_ISL_2482218, EPI_ISL_2482219, EPI_ISL_2482220, EPI_ISL_2482221, EPI_ISL_2482222, EPI_ISL_2482223, EPI_ISL_2482224, EPI_ISL_2482225, EPI_ISL_2482226, EPI_ISL_2482227, EPI_ISL_2482228, EPI_ISL_2482229, EPI_ISL_2482230, EPI_ISL_2482231, EPI_ISL_2482232, EPI_ISL_2482233, EPI_ISL_2482234, EPI_ISL_2482235, EPI_ISL_2482236, EPI_ISL_2482237, EPI_ISL_2482238, EPI_ISL_2482239, EPI_ISL_2482240, EPI_ISL_2482241, EPI_ISL_2482242, EPI_ISL_2482243, EPI_ISL_2482244, EPI_ISL_2482245, EPI_ISL_2482246, EPI_ISL_2482247, EPI_ISL_2482248, EPI_ISL_2482249, EPI_ISL_2482250, EPI_ISL_2482251, EPI_ISL_2482252, EPI_ISL_2482253, EPI_ISL_2482254, EPI_ISL_2482255, EPI_ISL_2482256, EPI_ISL_2482257, EPI_ISL_2482258, EPI_ISL_2482259, EPI_ISL_2482260, EPI_ISL_2482261, EPI_ISL_2482262, EPI_ISL_2482263, EPI_ISL_2482265, EPI_ISL_2482266, EPI_ISL_2482267, EPI_ISL_2482268, EPI_ISL_2482269, EPI_ISL_2482270, EPI_ISL_2482271, EPI_ISL_2482272, EPI_ISL_2482273, EPI_ISL_2482274, EPI_ISL_2482275, EPI_ISL_2482276, EPI_ISL_2482277, EPI_ISL_2482278, EPI_ISL_2482279 | | | |
| see above | Laboratory Corporation of America | Centers for Disease Control and Prevention Division of Viral Diseases, Pathogen Discovery | Dakota Howard, Dhwani Batra, Peter W. Cook, Kara Moser, Adrian Paskey, Jason Caravas, Benjamin Rambo-Martin, Shatavia Morrison, Christopher Gulvick, Scott Sammons, Yvette Unoarumhi, Darlene Wagner, Matthew Schmerer, Minoo Agarwal, Eyad Almasri, Debbie Boles, Ayla Burns, Nuthawin Charoensri, Oren Cohen, Susan Countryman, Mary Ann Cristobal, Bobbi Croy, Suzanne Dale, Hrushikesh Deshmukh, Amanda Douglas, Vincent Drouillon, Marcia Eisenberg, Howard Engler, Rama Ghatti, Prashant Gupta, Susan Hicks, Jake Humphrey, Lax Iyer, Lisa Pfefferle, Manoj Jain, Matthew Robinson, Mohan Kolli, Brian Krueger, Tim Kuphal, Stanley Letovsky, Michael Levandoski, Craig Lukasik, Jonathan Meltzer, Brian Norvell, Mindy Nye, Scott Parker, Christos Petropoulos, John Pruitt, Steven Ragan, Scott Ryan, Mike Sapeta, Jana Schroth, Suresh Babu Selvaraju, Goran Stevovic, Amanda Suchanek, Andrea Throop, Lyndon Tilson, Thomas Urban, Joe Voshell, Kimberly Wagner, Jonathan Williams, Mary Williamson, Qian Zeng, Tricia Zwiefelhofer, Clinton R. Paden, Duncan MacCannell |
| EPI_ISL_2482280, EPI_ISL_2482281, EPI_ISL_2482282, EPI_ISL_2482283, EPI_ISL_2482284, EPI_ISL_2482285, EPI_ISL_2482286, EPI_ISL_2482287, EPI_ISL_2482288, EPI_ISL_2482289, EPI_ISL_2482290, EPI_ISL_2482291, EPI_ISL_2482292, EPI_ISL_2482293, EPI_ISL_2482294, EPI_ISL_2482295, EPI_ISL_2482296, EPI_ISL_2482297, EPI_ISL_2482298, EPI_ISL_2482299, EPI_ISL_2482300, EPI_ISL_2482301 | | | |
| see above | Aegis Sciences Corporation | Centers for Disease Control and Prevention Division of Viral Diseases, Pathogen Discovery | Dakota Howard, Dhwani Batra, Peter W. Cook, Kara Moser, Adrian Paskey, Jason Caravas, Benjamin Rambo-Martin, Shatavia Morrison, Christopher Gulvick, Scott Sammons, Yvette Unoarumhi, Darlene Wagner, Matthew Schmerer, Cyndi Clark, Patrick Campbell, Rob Case, Vikramsinha Ghorpade, Holly Houdeshell, Ola Kvalvaag, Dillon Nall, Ethan Sanders, Alec Vest, Shaun Westlund, Matthew Hardison, Clinton R. Paden, Duncan MacCannell |
| EPI_ISL_2482302 | Ohio Department of Health Laboratory | Ohio Department of Health Laboratory | Holmes, Jennifer; Eric Brandt, Keoni Omura, Glen McGillivary, Caitlin McDonnell, Jade Mowery, Stephanie Mccracken, Tyler Payne, Kirtana Ramadugu, Erica Leasure, Brent Lee, Kelsey Florek, Heather Blankenship, Allison Black, Quanta Brown, and Tammy Bannerman |
| EPI_ISL_2482303, EPI_ISL_2482304, EPI_ISL_2482305, EPI_ISL_2482307, EPI_ISL_2482308, EPI_ISL_2482309, EPI_ISL_2482310, EPI_ISL_2482311, EPI_ISL_2482312, EPI_ISL_2482313, EPI_ISL_2482314, EPI_ISL_2482315, EPI_ISL_2482316, EPI_ISL_2482317, EPI_ISL_2482318, EPI_ISL_2482319, EPI_ISL_2482320, EPI_ISL_2482321, EPI_ISL_2482322, EPI_ISL_2482323, EPI_ISL_2482324, EPI_ISL_2482325, EPI_ISL_2482326, EPI_ISL_2482327, EPI_ISL_2482328, EPI_ISL_2482329, EPI_ISL_2482330, EPI_ISL_2482331, EPI_ISL_2482332, EPI_ISL_2482333, EPI_ISL_2482334, EPI_ISL_2482335, EPI_ISL_2482336, EPI_ISL_2482337, EPI_ISL_2482338, EPI_ISL_2482339, EPI_ISL_2482340, EPI_ISL_2482341, EPI_ISL_2482342, EPI_ISL_2482343, EPI_ISL_2482344, EPI_ISL_2482345, EPI_ISL_2482346, EPI_ISL_2482347, EPI_ISL_2482348, EPI_ISL_2482349, EPI_ISL_2482350, EPI_ISL_2482352, EPI_ISL_2482353, EPI_ISL_2482354, EPI_ISL_2482355, EPI_ISL_2482356, EPI_ISL_2482357, EPI_ISL_2482358, EPI_ISL_2482359, EPI_ISL_2482360, EPI_ISL_2482361, EPI_ISL_2482362, EPI_ISL_2482363, EPI_ISL_2482364, EPI_ISL_2482365, EPI_ISL_2482366, EPI_ISL_2482367, EPI_ISL_2482368, EPI_ISL_2482369, EPI_ISL_2482370, EPI_ISL_2482371, EPI_ISL_2482372, EPI_ISL_2482373, EPI_ISL_2482374, EPI_ISL_2482375, EPI_ISL_2482376, EPI_ISL_2482377, EPI_ISL_2482378, EPI_ISL_2482379, EPI_ISL_2482380, EPI_ISL_2482381, EPI_ISL_2482382, EPI_ISL_2482383, EPI_ISL_2482384, EPI_ISL_2482385, EPI_ISL_2482386, EPI_ISL_2482387, EPI_ISL_2482388, EPI_ISL_2482389, EPI_ISL_2482390, EPI_ISL_2482391, EPI_ISL_2482392, EPI_ISL_2482393, EPI_ISL_2482394, EPI_ISL_2482395, EPI_ISL_2482396, EPI_ISL_2482397, EPI_ISL_2482398, EPI_ISL_2482399, EPI_ISL_2482400, EPI_ISL_2482401, EPI_ISL_2482402, EPI_ISL_2482403, EPI_ISL_2482404, EPI_ISL_2482405, EPI_ISL_2482406, EPI_ISL_2482407, EPI_ISL_2482408, EPI_ISL_2482409, EPI_ISL_2482410, EPI_ISL_2482411, EPI_ISL_2482412, EPI_ISL_2482413, EPI_ISL_2482414, EPI_ISL_2482415, EPI_ISL_2482416, EPI_ISL_2482417, EPI_ISL_2482418, EPI_ISL_2482419, EPI_ISL_2482420, EPI_ISL_2482421, EPI_ISL_2482422, EPI_ISL_2482423, EPI_ISL_2482424, EPI_ISL_2482425, EPI_ISL_2482426, EPI_ISL_2482427, EPI_ISL_2482428, EPI_ISL_2482429, EPI_ISL_2482430, EPI_ISL_2482431, EPI_ISL_2482432 | | | |
| see above | Aegis Sciences Corporation | Centers for Disease Control and Prevention Division of Viral Diseases, Pathogen Discovery | Dakota Howard, Dhwani Batra, Peter W. Cook, Kara Moser, Adrian Paskey, Jason Caravas, Benjamin Rambo-Martin, Shatavia Morrison, Christopher Gulvick, Scott Sammons, Yvette Unoarumhi, Darlene Wagner, Matthew Schmerer, Cyndi Clark, Patrick Campbell, Rob Case, Vikramsinha Ghorpade, Holly Houdeshell, Ola Kvalvaag, Dillon Nall, Ethan Sanders, Alec Vest, Shaun Westlund, Matthew Hardison, Clinton R. Paden, Duncan MacCannell |
| EPI_ISL_2483069, EPI_ISL_2483070, EPI_ISL_2483071 | Wyoming Public Health Laboratory | Wyoming Public Health Laboratory | Jim Mildenberger, Wanda Manley, Noah Hull, Taylor Fearing, Lynette Gumbleton, Channing Weber, Ashley Norberg, Chayse Rowley, Marley Goetz, Brian Dominguez, Elliot Thomasson, Sam Britz, Cari Sloma, and Rob Christensen |
| EPI_ISL_2483329, EPI_ISL_2483330, EPI_ISL_2483331, EPI_ISL_2483332, EPI_ISL_2483333, EPI_ISL_2483334, EPI_ISL_2483335, EPI_ISL_2483336, EPI_ISL_2483337, EPI_ISL_2483338, EPI_ISL_2483339, EPI_ISL_2483340, EPI_ISL_2483341, EPI_ISL_2483342, EPI_ISL_2483343, EPI_ISL_2483344, EPI_ISL_2483345, EPI_ISL_2483346, EPI_ISL_2483347, EPI_ISL_2483348, EPI_ISL_2483349, EPI_ISL_2483350, EPI_ISL_2483351, EPI_ISL_2483352, EPI_ISL_2483353, EPI_ISL_2483354, EPI_ISL_2483355, EPI_ISL_2483356, EPI_ISL_2483357, EPI_ISL_2483358, EPI_ISL_2483359, EPI_ISL_2483360, EPI_ISL_2483361, EPI_ISL_2483362, EPI_ISL_2483363, EPI_ISL_2483364, EPI_ISL_2483365, EPI_ISL_2483366, EPI_ISL_2483367, EPI_ISL_2483368, EPI_ISL_2483369, EPI_ISL_2483370, EPI_ISL_2483371, EPI_ISL_2483372, EPI_ISL_2483373, EPI_ISL_2483374, EPI_ISL_2483375, EPI_ISL_2483376, EPI_ISL_2483377, EPI_ISL_2483378, EPI_ISL_2483379, EPI_ISL_2483380, EPI_ISL_2483381, EPI_ISL_2483382, EPI_ISL_2483383, EPI_ISL_2483384, EPI_ISL_2483385, EPI_ISL_2483386, EPI_ISL_2483387, EPI_ISL_2483388, EPI_ISL_2483389, EPI_ISL_2483390, EPI_ISL_2483391, EPI_ISL_2483392, EPI_ISL_2483393, EPI_ISL_2483394, EPI_ISL_2483395, EPI_ISL_2483396, EPI_ISL_2483397, EPI_ISL_2483398, EPI_ISL_2483399, EPI_ISL_2483400, EPI_ISL_2483401, EPI_ISL_2483402, EPI_ISL_2483403, EPI_ISL_2483404, EPI_ISL_2483405, EPI_ISL_2483406, EPI_ISL_2483407, EPI_ISL_2483408, EPI_ISL_2483409, EPI_ISL_2483410, EPI_ISL_2483411, EPI_ISL_2483412, EPI_ISL_2483413, EPI_ISL_2483414, EPI_ISL_2483415, EPI_ISL_2483416, EPI_ISL_2483417, EPI_ISL_2483418, EPI_ISL_2483419, EPI_ISL_2483420, EPI_ISL_2483421, EPI_ISL_2483422, EPI_ISL_2483423, EPI_ISL_2483424, EPI_ISL_2483425, EPI_ISL_2483426, EPI_ISL_2483427, EPI_ISL_2483428, EPI_ISL_2483429, EPI_ISL_2483430, EPI_ISL_2483431, EPI_ISL_2483432, EPI_ISL_2483433, EPI_ISL_2483434, EPI_ISL_2483435, EPI_ISL_2483436, EPI_ISL_2483437, EPI_ISL_2483438, EPI_ISL_2483439, EPI_ISL_2483440, EPI_ISL_2483441, EPI_ISL_2483442, EPI_ISL_2483443, EPI_ISL_2483444, EPI_ISL_2483445, EPI_ISL_2483446, EPI_ISL_2483447, EPI_ISL_2483448, EPI_ISL_2483449, EPI_ISL_2483450, EPI_ISL_2483451, EPI_ISL_2483452, EPI_ISL_2483453, EPI_ISL_2483454, EPI_ISL_2483455, EPI_ISL_2483456, EPI_ISL_2483457, EPI_ISL_2483458, EPI_ISL_2483460, EPI_ISL_2483461, EPI_ISL_2483462, EPI_ISL_2483464, EPI_ISL_2483465 | | | |
| see above | Hackensack Medical Center | New York Genome Center | Michael Zody, Andre Corvelo, Dayna M. Oschwald, Samantha Fennessey, Tom Maniatis, Liang Chen, Jose Mediavilla, Marcus Cunningham, Kaelea Composto, Kar Chow, David Perlin, Barry Kreiswirth |
| EPI_ISL_2483773, EPI_ISL_2483774, EPI_ISL_2483775, EPI_ISL_2483776, EPI_ISL_2483777, EPI_ISL_2483778, EPI_ISL_2483779, EPI_ISL_2483780, EPI_ISL_2483781, EPI_ISL_2483782, EPI_ISL_2483783, EPI_ISL_2483784, EPI_ISL_2483785, EPI_ISL_2483787, EPI_ISL_2483789, EPI_ISL_2483790, EPI_ISL_2483791, EPI_ISL_2483792, EPI_ISL_2483794, EPI_ISL_2483795, EPI_ISL_2483797, EPI_ISL_2483799, EPI_ISL_2483800, EPI_ISL_2483801, EPI_ISL_2483803, EPI_ISL_2483804, EPI_ISL_2483805, EPI_ISL_2483807, EPI_ISL_2483808, EPI_ISL_2483809 | | | |
| see above | Massachusetts State Public Health Laboratory | Massachusetts State Public Health Laboratory | Andrew Lang, Timelia Fink, Glen Gallagher, Sandra Smole |
| EPI_ISL_2484038, EPI_ISL_2484040, EPI_ISL_2484041, EPI_ISL_2484042, EPI_ISL_2484043, EPI_ISL_2484044, EPI_ISL_2484045, EPI_ISL_2484046, EPI_ISL_2484048, EPI_ISL_2484050, EPI_ISL_2484051, EPI_ISL_2484052, EPI_ISL_2484054, EPI_ISL_2484055, EPI_ISL_2484058, EPI_ISL_2484059, EPI_ISL_2484060, EPI_ISL_2484061, EPI_ISL_2484062, EPI_ISL_2484064, EPI_ISL_2484065, EPI_ISL_2484066, EPI_ISL_2484068, EPI_ISL_2484070, EPI_ISL_2484071, EPI_ISL_2484072, EPI_ISL_2484073, EPI_ISL_2484076, EPI_ISL_2484077, EPI_ISL_2484079, EPI_ISL_2484080, EPI_ISL_2484081, EPI_ISL_2484082, EPI_ISL_2484083, EPI_ISL_2484084, EPI_ISL_2484085, EPI_ISL_2484087, EPI_ISL_2484089, EPI_ISL_2484090, EPI_ISL_2484092, EPI_ISL_2484093, EPI_ISL_2484095, EPI_ISL_2484096, EPI_ISL_2484100, EPI_ISL_2484101, EPI_ISL_2484102, EPI_ISL_2484104, EPI_ISL_2484105, EPI_ISL_2484108, EPI_ISL_2484109, EPI_ISL_2484110, EPI_ISL_2484111, EPI_ISL_2484112, EPI_ISL_2484113, EPI_ISL_2484114, EPI_ISL_2484115, EPI_ISL_2484116, EPI_ISL_2484117, EPI_ISL_2484118, EPI_ISL_2484119, EPI_ISL_2484120, EPI_ISL_2484121, EPI_ISL_2484122, EPI_ISL_2484124, EPI_ISL_2484126, EPI_ISL_2484127, EPI_ISL_2484130, EPI_ISL_2484132, EPI_ISL_2484133, EPI_ISL_2484134, EPI_ISL_2484135, EPI_ISL_2484136, EPI_ISL_2484137 | | | |
| see above | Arizona State University | Arizona State University | Peter T. Skidmore, LaRinda A. Holland, Matthew F. Smith, Rabia Maqsood, Nicholas J. Mellor, Joy M. Blain, Valerie Harris, Joshua LaBaer, Vel Murugan, Efrem S. Lim |
| EPI_ISL_2484154, EPI_ISL_2484156, EPI_ISL_2484157, EPI_ISL_2484158, EPI_ISL_2484160, EPI_ISL_2484163 | Ohio Department of Health Laboratory | Ohio Department of Health Laboratory | Holmes, Jennifer; Eric Brandt, Keoni Omura, Glen McGillivary, Caitlin McDonnell, Jade Mowery, Stephanie Mccracken, Tyler Payne, Kirtana Ramadugu, Erica Leasure, Brent Lee, Kelsey Florek, Heather Blankenship, Allison Black, Quanta Brown, and Tammy Bannerman |
| EPI_ISL_2484164 | LESP Chihuahua | Instituto de Diagnostico y Referencia Epidemiologicos (INDRE) | Claudia Wong-Arambula, Abril Rodriguez-Maldonado, Vanessa Rivero-Arredondo, Ariadna Medina-Benitez, Joaquin Quiroz-Mercado, Sergio Rangel-Guerrero, Natividad Cruz-Ortiz, Tatiana Nunez-Garcia, Gisela Barrera-Badillo, Lucia Hernandez-Rivas, Irma Lopez-Martinez, Ernesto Ramirez-Gonzalez. |
| EPI_ISL_2484165 | LESP Yucatan | Instituto de Diagnostico y Referencia Epidemiologicos (INDRE) | Claudia Wong-Arambula, Abril Rodriguez-Maldonado, Vanessa Rivero-Arredondo, Ariadna Medina-Benitez, Joaquin Quiroz-Mercado, Sergio Rangel-Guerrero, Natividad Cruz-Ortiz, Tatiana Nunez-Garcia, Gisela Barrera-Badillo, Lucia Hernandez-Rivas, Irma Lopez-Martinez, Ernesto Ramirez-Gonzalez. |
| EPI_ISL_2484166 | LESP Zacatecas | Instituto de Diagnostico y Referencia Epidemiologicos (INDRE) | Claudia Wong-Arambula, Abril Rodriguez-Maldonado, Vanessa Rivero-Arredondo, Ariadna Medina-Benitez, Joaquin Quiroz-Mercado, Sergio Rangel-Guerrero, Natividad Cruz-Ortiz, Tatiana Nunez-Garcia, Gisela Barrera-Badillo, Lucia Hernandez-Rivas, Irma Lopez-Martinez, Ernesto Ramirez-Gonzalez. |
| EPI_ISL_2484168, EPI_ISL_2484169, EPI_ISL_2484170, EPI_ISL_2484171, EPI_ISL_2484172, EPI_ISL_2484175, EPI_ISL_2484176, EPI_ISL_2484177, EPI_ISL_2484178, EPI_ISL_2484179, EPI_ISL_2484180, EPI_ISL_2484181, EPI_ISL_2484182, EPI_ISL_2484183, EPI_ISL_2484184, EPI_ISL_2484185, EPI_ISL_2484186, EPI_ISL_2484187, EPI_ISL_2484188, EPI_ISL_2484189, EPI_ISL_2484190, EPI_ISL_2484191, EPI_ISL_2484192, EPI_ISL_2484193, EPI_ISL_2484194, EPI_ISL_2484195, EPI_ISL_2484196, EPI_ISL_2484197, EPI_ISL_2484198, EPI_ISL_2484199, EPI_ISL_2484200, EPI_ISL_2484201, EPI_ISL_2484202, EPI_ISL_2484203, EPI_ISL_2484204, EPI_ISL_2484205, EPI_ISL_2484206, EPI_ISL_2484207, EPI_ISL_2484208 | | | |
| see above | Tampa General Hospital Esoteric Lab | Tampa General Hospital Esoteric Research & Development Lab | Amorce Lima, Jon Faughn, Deanna Becker, Suzane Silbert |
| EPI_ISL_2484424 | Ohio Department of Health Laboratory | Ohio Department of Health Laboratory | Holmes, Jennifer; Eric Brandt, Keoni Omura, Glen McGillivary, Caitlin McDonnell, Jade Mowery, Stephanie Mccracken, Tyler Payne, Kirtana Ramadugu, Erica Leasure, Brent Lee, Kelsey Florek, Heather Blankenship, Allison Black, Quanta Brown, and Tammy Bannerman |
| EPI_ISL_2484683, EPI_ISL_2484684 | Arkansas Department of Health | Center for Global Health, University of New Mexico Health Sciences Center | Daryl Domman, Kurt Schwalm, Valerie Morley, Atul Kothari, Joshua L. Kennedy, Darrell Dinwiddie |
| EPI_ISL_2484685, EPI_ISL_2484686, EPI_ISL_2484687 | Baptist Health Medical Center | Center for Global Health, University of New Mexico Health Sciences Center | Daryl Domman, Kurt Schwalm, Valerie Morley, Amanda Novack, Dirk Haselow, Joshua L. Kennedy, Darrell Dinwiddie |
| EPI_ISL_2484688 | Arkansas Children's Hospital | Center for Global Health, University of New Mexico Health Sciences Center | Daryl Domman, Kurt Schwalm, Valerie Morley, Amanda Novack, Dirk Haselow, Joshua L. Kennedy, Darrell Dinwiddie |
| EPI_ISL_2484689, EPI_ISL_2484690, EPI_ISL_2484691 | Arkansas Department of Health | Center for Global Health, University of New Mexico Health Sciences Center | Daryl Domman, Kurt Schwalm, Valerie Morley, Atul Kothari, Joshua L. Kennedy, Darrell Dinwiddie |
| EPI_ISL_2484692 | Baptist Health Medical Center | Center for Global Health, University of New Mexico Health Sciences Center | Daryl Domman, Kurt Schwalm, Valerie Morley, Amanda Novack, Dirk Haselow, Joshua L. Kennedy, Darrell Dinwiddie |
| EPI_ISL_2484693, EPI_ISL_2484694, EPI_ISL_2484695, EPI_ISL_2484696, EPI_ISL_2484697, EPI_ISL_2484698 | Arkansas Children's Hospital | Center for Global Health, University of New Mexico Health Sciences Center | Daryl Domman, Kurt Schwalm, Valerie Morley, Amanda Novack, Dirk Haselow, Joshua L. Kennedy, Darrell Dinwiddie |
| EPI_ISL_2484699 | Arkansas Department of Health | Center for Global Health, University of New Mexico Health Sciences Center | Daryl Domman, Kurt Schwalm, Valerie Morley, Atul Kothari, Joshua L. Kennedy, Darrell Dinwiddie |
| EPI_ISL_2484700 | Baptist Health Medical Center | Center for Global Health, University of New Mexico Health Sciences Center | Daryl Domman, Kurt Schwalm, Valerie Morley, Amanda Novack, Dirk Haselow, Joshua L. Kennedy, Darrell Dinwiddie |
| EPI_ISL_2484701, EPI_ISL_2484702, EPI_ISL_2484703, EPI_ISL_2484704 | Arkansas Children's Hospital | Center for Global Health, University of New Mexico Health Sciences Center | Daryl Domman, Kurt Schwalm, Valerie Morley, Amanda Novack, Dirk Haselow, Joshua L. Kennedy, Darrell Dinwiddie |
| EPI_ISL_2484705, EPI_ISL_2484706, EPI_ISL_2484707, EPI_ISL_2484708, EPI_ISL_2484709, EPI_ISL_2484710, EPI_ISL_2484711, EPI_ISL_2484712, | Baptist Health Medical Center | Center for Global Health, University of New Mexico Health Sciences Center | Daryl Domman, Kurt Schwalm, Valerie Morley, Amanda Novack, Dirk Haselow, Joshua L. Kennedy, Darrell Dinwiddie |

| | | | |
|---|---|---|---|
| EPI_ISL_2484713, EPI_ISL_2484714 | | | |
| EPI_ISL_2484715, EPI_ISL_2484716, EPI_ISL_2484717, EPI_ISL_2484718 | Arkansas Department of Health | Center for Global Health, University of New Mexico Health Sciences Center | Daryl Domman, Kurt Schwalm, Valerie Morley, Atul Kothari, Joshua L. Kennedy, Darrell Dinwiddie |
| EPI_ISL_2484719, EPI_ISL_2484720, EPI_ISL_2484721, EPI_ISL_2484722, EPI_ISL_2484723, EPI_ISL_2484724, EPI_ISL_2484725, EPI_ISL_2484726, EPI_ISL_2484727, EPI_ISL_2484728, EPI_ISL_2484729, EPI_ISL_2484730, EPI_ISL_2484731, EPI_ISL_2484732, EPI_ISL_2484733, EPI_ISL_2484734 | | | |
| see above | Arkansas Children's Hospital | Center for Global Health, University of New Mexico Health Sciences Center | Daryl Domman, Kurt Schwalm, Valerie Morley, Amanda Novack, Dirk Haselow, Joshua L. Kennedy, Darrell Dinwiddie |
| EPI_ISL_2484739 | Baptist Health Medical Center | Center for Global Health, University of New Mexico Health Sciences Center | Daryl Domman, Kurt Schwalm, Valerie Morley, Amanda Novack, Dirk Haselow, Joshua L. Kennedy, Darrell Dinwiddie |
| EPI_ISL_2484740 | Arkansas Department of Health | Center for Global Health, University of New Mexico Health Sciences Center | Daryl Domman, Kurt Schwalm, Valerie Morley, Atul Kothari, Joshua L. Kennedy, Darrell Dinwiddie |
| EPI_ISL_2484741, EPI_ISL_2484742 | Arkansas Children's Hospital | Center for Global Health, University of New Mexico Health Sciences Center | Daryl Domman, Kurt Schwalm, Valerie Morley, Amanda Novack, Dirk Haselow, Joshua L. Kennedy, Darrell Dinwiddie |
| EPI_ISL_2484743 | Baptist Health Medical Center | Center for Global Health, University of New Mexico Health Sciences Center | Daryl Domman, Kurt Schwalm, Valerie Morley, Amanda Novack, Dirk Haselow, Joshua L. Kennedy, Darrell Dinwiddie |
| EPI_ISL_2484744, EPI_ISL_2484745, EPI_ISL_2484746, EPI_ISL_2484747 | Arkansas Children's Hospital | Center for Global Health, University of New Mexico Health Sciences Center | Daryl Domman, Kurt Schwalm, Valerie Morley, Amanda Novack, Dirk Haselow, Joshua L. Kennedy, Darrell Dinwiddie |
| EPI_ISL_2484748, EPI_ISL_2484749, EPI_ISL_2484750, EPI_ISL_2484751 | Arkansas Department of Health | Center for Global Health, University of New Mexico Health Sciences Center | Daryl Domman, Kurt Schwalm, Valerie Morley, Atul Kothari, Joshua L. Kennedy, Darrell Dinwiddie |
| EPI_ISL_2484752 | Arkansas Children's Hospital | Center for Global Health, University of New Mexico Health Sciences Center | Daryl Domman, Kurt Schwalm, Valerie Morley, Amanda Novack, Dirk Haselow, Joshua L. Kennedy, Darrell Dinwiddie |
| EPI_ISL_2484753 | Baptist Health Medical Center | Center for Global Health, University of New Mexico Health Sciences Center | Daryl Domman, Kurt Schwalm, Valerie Morley, Amanda Novack, Dirk Haselow, Joshua L. Kennedy, Darrell Dinwiddie |
| EPI_ISL_2484754, EPI_ISL_2484755 | Arkansas Children's Hospital | Center for Global Health, University of New Mexico Health Sciences Center | Daryl Domman, Kurt Schwalm, Valerie Morley, Amanda Novack, Dirk Haselow, Joshua L. Kennedy, Darrell Dinwiddie |
| EPI_ISL_2484756, EPI_ISL_2484757 | Baptist Health Medical Center | Center for Global Health, University of New Mexico Health Sciences Center | Daryl Domman, Kurt Schwalm, Valerie Morley, Amanda Novack, Dirk Haselow, Joshua L. Kennedy, Darrell Dinwiddie |
| EPI_ISL_2484758 | Arkansas Children's Hospital | Center for Global Health, University of New Mexico Health Sciences Center | Daryl Domman, Kurt Schwalm, Valerie Morley, Amanda Novack, Dirk Haselow, Joshua L. Kennedy, Darrell Dinwiddie |
| EPI_ISL_2484801, EPI_ISL_2484802 | TriCore Reference Laboratories | Center for Global Health, University of New Mexico Health Sciences Center | Daryl Domman, Kurt Schwalm, Valerie Morley, Cecilia Thompson, Kendra Pesko, Karissa Culbreath, Darrell Dinwiddie |
| EPI_ISL_2484852, EPI_ISL_2484853, EPI_ISL_2484854, EPI_ISL_2484855, EPI_ISL_2484856, EPI_ISL_2484857, EPI_ISL_2484858, EPI_ISL_2484859, EPI_ISL_2484860, EPI_ISL_2484861, EPI_ISL_2484862, EPI_ISL_2484863, EPI_ISL_2484864, EPI_ISL_2484865, EPI_ISL_2484866, EPI_ISL_2484867, EPI_ISL_2484868, EPI_ISL_2484869, EPI_ISL_2484870, EPI_ISL_2484871, EPI_ISL_2484872, EPI_ISL_2484873, EPI_ISL_2484874, EPI_ISL_2484875, EPI_ISL_2484876, EPI_ISL_2484877, EPI_ISL_2484878, EPI_ISL_2484879, EPI_ISL_2484880, EPI_ISL_2484881, EPI_ISL_2484882, EPI_ISL_2484886, EPI_ISL_2484887, EPI_ISL_2484888, EPI_ISL_2484889, EPI_ISL_2484890, EPI_ISL_2484891, EPI_ISL_2484892 | | | |
| see above | Southern Nevada Public Health Laboratory | Southern Nevada Public Health Laboratory | Michael Picker |
| EPI_ISL_2484901, EPI_ISL_2484905, EPI_ISL_2484906, EPI_ISL_2484916, EPI_ISL_2484917, EPI_ISL_2484918, EPI_ISL_2484920, EPI_ISL_2484923, EPI_ISL_2484924, EPI_ISL_2484925, EPI_ISL_2484926, EPI_ISL_2484927, EPI_ISL_2484928 | | | |
| see above | Wyoming Public Health Laboratory | Wyoming Public Health Laboratory | Jim Mildenberger, Taylor Fearing, Channing Weber, Ashley Norberg, Chayse Rowley, Marley Goetz, Brian Dominguez, Elliot Thomasson, Brittany Oher, Sam Britz, Cari Sloma, and Rob Christensen |
| EPI_ISL_2484933, EPI_ISL_2484934, EPI_ISL_2484935, EPI_ISL_2484936, EPI_ISL_2484937, EPI_ISL_2484939, EPI_ISL_2484940, EPI_ISL_2484941, EPI_ISL_2484942, EPI_ISL_2484943, EPI_ISL_2484944, EPI_ISL_2484945, EPI_ISL_2484946, EPI_ISL_2484947, EPI_ISL_2484948, EPI_ISL_2484949, EPI_ISL_2484950, EPI_ISL_2484951, EPI_ISL_2484952, EPI_ISL_2484953, EPI_ISL_2484954, EPI_ISL_2484955, EPI_ISL_2484956, EPI_ISL_2484957, EPI_ISL_2484958, EPI_ISL_2484959 | | | |
| see above | Massachusetts State Public Health Laboratory | Massachusetts State Public Health Laboratory | Andrew Lang, Timelia Fink, Glen Gallagher, Sandra Smole |
| EPI_ISL_2485256, EPI_ISL_2486194, EPI_ISL_2487260, EPI_ISL_2488708, EPI_ISL_2488709 | Ohio Department of Health Laboratory | Ohio Department of Health Laboratory | Holmes, Jennifer; Eric Brandt, Keoni Omura, Glen McGillivary, Caitlin McDonnell, Jade Mowery, Stephanie Mccracken, Tyler Payne, Kirtana Ramadugu, Erica Leasure, Brent Lee, Kelsey Florek, Heather Blankenship, Allison Black, Quanta Brown, and Tammy Bannerman |
| EPI_ISL_2488756 | NCSLPH | NCSLPH | Chase K, Miller MC, Greene S, Glover W |
| EPI_ISL_2488759 | University of Wisconsin-Madison AIDS Vaccine Research Laboratories | University of Wisconsin-Madison AIDS Vaccine Research Laboratories | Gage Moreno, Katarina Braun, et al. AIDS Vaccine Research Laboratories |
| EPI_ISL_2488820, EPI_ISL_2488821, EPI_ISL_2488822, EPI_ISL_2488823, EPI_ISL_2488824, EPI_ISL_2488825, EPI_ISL_2488826, EPI_ISL_2488827, EPI_ISL_2488828, EPI_ISL_2488829, EPI_ISL_2488830, EPI_ISL_2488831, EPI_ISL_2488832, EPI_ISL_2488833, EPI_ISL_2488834, EPI_ISL_2488835, EPI_ISL_2488836, EPI_ISL_2488837, EPI_ISL_2488838, EPI_ISL_2488839, EPI_ISL_2488840, EPI_ISL_2488841, EPI_ISL_2488842, EPI_ISL_2488843, EPI_ISL_2488844, EPI_ISL_2488845, EPI_ISL_2488846, EPI_ISL_2488847, EPI_ISL_2488848, EPI_ISL_2488849, EPI_ISL_2488850, EPI_ISL_2488851, EPI_ISL_2488852, EPI_ISL_2488853, EPI_ISL_2488854, EPI_ISL_2488855, EPI_ISL_2488856, EPI_ISL_2488857, EPI_ISL_2488858, EPI_ISL_2488859, EPI_ISL_2488860, EPI_ISL_2488861 | | | |
| see above | Quest Diagnostics Incorporated | Centers for Disease Control and Prevention Division of Viral Diseases, Pathogen Discovery | Dakota Howard, Dhwani Batra, Peter W. Cook, Kara Moser, Adrian Paskey, Jason Caravas, Benjamin Rambo-Martin, Shatavia Morrison, Christopher Gulvick, Scott Sammons, Yvette Unoarumhi, Darlene Wagner, Matthew Schmerer, S. H. Rosenthal, A. Gerasimova, R. M. Kagan, B. Anderson, M. Hua, Y. Liu, L.E. Bernstein, K.E. Livingston, A. Perez, I. A. Shlyakhter, R. V. Rolando, R. Owen, P. Tanpaiboon, F. Lacbawan, Clinton R. Paden, Duncan MacCannell |
| EPI_ISL_2488862, EPI_ISL_2488863, EPI_ISL_2488864, EPI_ISL_2488865, EPI_ISL_2488866, EPI_ISL_2488867, EPI_ISL_2488868, EPI_ISL_2488869, EPI_ISL_2488870, EPI_ISL_2488871, EPI_ISL_2488872, EPI_ISL_2488873, EPI_ISL_2488874, EPI_ISL_2488875, EPI_ISL_2488876, EPI_ISL_2488877, EPI_ISL_2488878, EPI_ISL_2488879, EPI_ISL_2488880, EPI_ISL_2488881, EPI_ISL_2488882, EPI_ISL_2488883, EPI_ISL_2488884, EPI_ISL_2488885, EPI_ISL_2488886, EPI_ISL_2488887, EPI_ISL_2488888, EPI_ISL_2488889, EPI_ISL_2488890, EPI_ISL_2488891, EPI_ISL_2488892, EPI_ISL_2488893, EPI_ISL_2488894, EPI_ISL_2488895, EPI_ISL_2488896, EPI_ISL_2488897, EPI_ISL_2488898, EPI_ISL_2488899, EPI_ISL_2488900, EPI_ISL_2488901, EPI_ISL_2488902, EPI_ISL_2488903, EPI_ISL_2488904, EPI_ISL_2488905, EPI_ISL_2488906, EPI_ISL_2488907, EPI_ISL_2488908, EPI_ISL_2488909, EPI_ISL_2488910, EPI_ISL_2488911, EPI_ISL_2488912, EPI_ISL_2488913, EPI_ISL_2488914, EPI_ISL_2488915, EPI_ISL_2488916, EPI_ISL_2488917, EPI_ISL_2488918, EPI_ISL_2488919, EPI_ISL_2488920, EPI_ISL_2488921, EPI_ISL_2488922, EPI_ISL_2488923, EPI_ISL_2488924, EPI_ISL_2488925, EPI_ISL_2488926, EPI_ISL_2488927, EPI_ISL_2488928, EPI_ISL_2488929, EPI_ISL_2488930, EPI_ISL_2488931, EPI_ISL_2488932, EPI_ISL_2488933, EPI_ISL_2488934, EPI_ISL_2488935, EPI_ISL_2488936, EPI_ISL_2488937, EPI_ISL_2488938, EPI_ISL_2488939, EPI_ISL_2488940, EPI_ISL_2488941, EPI_ISL_2488942, EPI_ISL_2488943, EPI_ISL_2488944, EPI_ISL_2488945, EPI_ISL_2488946, EPI_ISL_2488947, EPI_ISL_2488948, EPI_ISL_2488949, EPI_ISL_2488950, EPI_ISL_2488951, EPI_ISL_2488952, EPI_ISL_2488953, EPI_ISL_2488954, EPI_ISL_2488955, EPI_ISL_2488956, EPI_ISL_2488957, EPI_ISL_2488958, EPI_ISL_2488959, EPI_ISL_2488960, EPI_ISL_2488961, EPI_ISL_2488962, EPI_ISL_2488963, EPI_ISL_2488964, EPI_ISL_2488965, EPI_ISL_2488966, EPI_ISL_2488967, EPI_ISL_2488968, EPI_ISL_2488969, EPI_ISL_2488970, EPI_ISL_2488971, EPI_ISL_2488972, EPI_ISL_2488973, EPI_ISL_2488974, EPI_ISL_2488975, EPI_ISL_2488976, EPI_ISL_2488977, EPI_ISL_2488978, EPI_ISL_2488979, EPI_ISL_2488980, EPI_ISL_2488981, EPI_ISL_2488982, EPI_ISL_2488983, EPI_ISL_2488984, EPI_ISL_2488985, EPI_ISL_2488986, EPI_ISL_2488987, EPI_ISL_2488988, EPI_ISL_2488989, EPI_ISL_2488990, EPI_ISL_2488991, EPI_ISL_2488992, EPI_ISL_2488993, EPI_ISL_2488994, EPI_ISL_2488995, EPI_ISL_2488996, EPI_ISL_2488997, EPI_ISL_2488998, EPI_ISL_2488999, EPI_ISL_2489000, EPI_ISL_2489001, EPI_ISL_2489002, EPI_ISL_2489003, EPI_ISL_2489004, EPI_ISL_2489005, EPI_ISL_2489006, EPI_ISL_2489007, EPI_ISL_2489008, EPI_ISL_2489009, EPI_ISL_2489010, EPI_ISL_2489011, EPI_ISL_2489012, EPI_ISL_2489013, EPI_ISL_2489014, EPI_ISL_2489015, EPI_ISL_2489016, EPI_ISL_2489017, EPI_ISL_2489018, EPI_ISL_2489019, EPI_ISL_2489020, EPI_ISL_2489021, EPI_ISL_2489022, EPI_ISL_2489023, EPI_ISL_2489024, EPI_ISL_2489025, EPI_ISL_2489026, EPI_ISL_2489027, EPI_ISL_2489028, EPI_ISL_2489029, EPI_ISL_2489030, EPI_ISL_2489031, EPI_ISL_2489032, EPI_ISL_2489033, EPI_ISL_2489034, EPI_ISL_2489035, EPI_ISL_2489036, EPI_ISL_2489037, EPI_ISL_2489038, EPI_ISL_2489039, EPI_ISL_2489040, EPI_ISL_2489041, EPI_ISL_2489042, EPI_ISL_2489043, EPI_ISL_2489044, EPI_ISL_2489045, EPI_ISL_2489046, EPI_ISL_2489047, EPI_ISL_2489048, EPI_ISL_2489049, EPI_ISL_2489050, EPI_ISL_2489051, EPI_ISL_2489052, EPI_ISL_2489053, EPI_ISL_2489054, EPI_ISL_2489055, EPI_ISL_2489056, EPI_ISL_2489057, EPI_ISL_2489058, EPI_ISL_2489059, EPI_ISL_2489060, EPI_ISL_2489061 | | | |
| see above | Mako Medical | Centers for Disease Control and Prevention Division of Viral Diseases, Pathogen Discovery | Dakota Howard, Dhwani Batra, Peter W. Cook, Kara Moser, Adrian Paskey, Jason Caravas, Benjamin Rambo-Martin, Shatavia Morrison, Christopher Gulvick, Scott Sammons, Yvette Unoarumhi, Darlene Wagner, Matthew Schmerer,Matthew Tugwell, Lauren Moon, Clinton R. Paden, Duncan MacCannell |
| EPI_ISL_2489062, EPI_ISL_2489063, EPI_ISL_2489064, EPI_ISL_2489065, EPI_ISL_2489066, EPI_ISL_2489068, EPI_ISL_2489069, EPI_ISL_2489070, EPI_ISL_2489071, EPI_ISL_2489072, EPI_ISL_2489073, EPI_ISL_2489074, EPI_ISL_2489075, EPI_ISL_2489076, EPI_ISL_2489078, EPI_ISL_2489079, EPI_ISL_2489080, EPI_ISL_2489081, EPI_ISL_2489083, EPI_ISL_2489084, EPI_ISL_2489085, EPI_ISL_2489086, EPI_ISL_2489087, EPI_ISL_2489088, EPI_ISL_2489089, EPI_ISL_2489090, EPI_ISL_2489091, EPI_ISL_2489092, EPI_ISL_2489093, EPI_ISL_2489094, EPI_ISL_2489095, EPI_ISL_2489096, EPI_ISL_2489097, EPI_ISL_2489098, EPI_ISL_2489099, EPI_ISL_2489100, EPI_ISL_2489101, EPI_ISL_2489102, EPI_ISL_2489104, EPI_ISL_2489105, EPI_ISL_2489106, EPI_ISL_2489107, EPI_ISL_2489108, EPI_ISL_2489109, EPI_ISL_2489110, EPI_ISL_2489111, EPI_ISL_2489112, EPI_ISL_2489113, EPI_ISL_2489114, EPI_ISL_2489116, EPI_ISL_2489119, EPI_ISL_2489120, EPI_ISL_2489121, EPI_ISL_2489122, EPI_ISL_2489124, EPI_ISL_2489125, EPI_ISL_2489126, EPI_ISL_2489127, EPI_ISL_2489128, EPI_ISL_2489129, EPI_ISL_2489130, EPI_ISL_2489131, EPI_ISL_2489132, EPI_ISL_2489133, EPI_ISL_2489134, EPI_ISL_2489135, EPI_ISL_2489136, EPI_ISL_2489137, EPI_ISL_2489139, EPI_ISL_2489140, EPI_ISL_2489141, EPI_ISL_2489142, EPI_ISL_2489143, EPI_ISL_2489144, EPI_ISL_2489145, EPI_ISL_2489146, EPI_ISL_2489148, EPI_ISL_2489149, EPI_ISL_2489150, EPI_ISL_2489151, EPI_ISL_2489152, EPI_ISL_2489153, EPI_ISL_2489154, EPI_ISL_2489155, EPI_ISL_2489156, EPI_ISL_2489157, EPI_ISL_2489158, EPI_ISL_2489159, EPI_ISL_2489160, EPI_ISL_2489162, EPI_ISL_2489163, EPI_ISL_2489164, EPI_ISL_2489165, EPI_ISL_2489166, EPI_ISL_2489167, EPI_ISL_2489169, EPI_ISL_2489170, EPI_ISL_2489171, EPI_ISL_2489172, EPI_ISL_2489173, EPI_ISL_2489174, EPI_ISL_2489176, EPI_ISL_2489177, EPI_ISL_2489178, EPI_ISL_2489179, EPI_ISL_2489180, EPI_ISL_2489181, EPI_ISL_2489183, EPI_ISL_2489184, EPI_ISL_2489185, EPI_ISL_2489186, EPI_ISL_2489187, EPI_ISL_2489188, EPI_ISL_2489189, EPI_ISL_2489190, EPI_ISL_2489191, EPI_ISL_2489192, EPI_ISL_2489193, EPI_ISL_2489194, EPI_ISL_2489195, EPI_ISL_2489196, EPI_ISL_2489197, EPI_ISL_2489198, EPI_ISL_2489199, EPI_ISL_2489200, EPI_ISL_2489201, EPI_ISL_2489202, EPI_ISL_2489204, EPI_ISL_2489205, EPI_ISL_2489206, EPI_ISL_2489207, EPI_ISL_2489208, EPI_ISL_2489209, EPI_ISL_2489210, EPI_ISL_2489211, EPI_ISL_2489212, EPI_ISL_2489213, EPI_ISL_2489214, EPI_ISL_2489215, EPI_ISL_2489216, EPI_ISL_2489217, EPI_ISL_2489218, EPI_ISL_2489219, EPI_ISL_2489220, EPI_ISL_2489221, EPI_ISL_2489222, EPI_ISL_2489223, EPI_ISL_2489224, EPI_ISL_2489225, EPI_ISL_2489226, EPI_ISL_2489228, EPI_ISL_2489229, EPI_ISL_2489230, EPI_ISL_2489232, EPI_ISL_2489233, EPI_ISL_2489234, EPI_ISL_2489235, EPI_ISL_2489236, EPI_ISL_2489237, EPI_ISL_2489238, EPI_ISL_2489239, EPI_ISL_2489240, EPI_ISL_2489241, EPI_ISL_2489242, EPI_ISL_2489243, EPI_ISL_2489244, EPI_ISL_2489245, EPI_ISL_2489246, EPI_ISL_2489247, EPI_ISL_2489248, EPI_ISL_2489249, EPI_ISL_2489250, EPI_ISL_2489251, EPI_ISL_2489252, EPI_ISL_2489253, EPI_ISL_2489254, EPI_ISL_2489255, EPI_ISL_2489256, EPI_ISL_2489258, EPI_ISL_2489259, EPI_ISL_2489260, EPI_ISL_2489261, EPI_ISL_2489262, EPI_ISL_2489263, EPI_ISL_2489264, EPI_ISL_2489265, EPI_ISL_2489266, EPI_ISL_2489267, EPI_ISL_2489268, EPI_ISL_2489269, EPI_ISL_2489270, EPI_ISL_2489271, EPI_ISL_2489272, EPI_ISL_2489273, EPI_ISL_2489274, EPI_ISL_2489275, EPI_ISL_2489276, EPI_ISL_2489277, EPI_ISL_2489278, EPI_ISL_2489279, EPI_ISL_2489280, EPI_ISL_2489281, EPI_ISL_2489282, EPI_ISL_2489283, EPI_ISL_2489284, EPI_ISL_2489285, EPI_ISL_2489286, EPI_ISL_2489287, EPI_ISL_2489288, EPI_ISL_2489289, EPI_ISL_2489290, EPI_ISL_2489292, EPI_ISL_2489293, EPI_ISL_2489294, EPI_ISL_2489295, EPI_ISL_2489296, EPI_ISL_2489297, EPI_ISL_2489298, EPI_ISL_2489299, EPI_ISL_2489300, EPI_ISL_2489301, EPI_ISL_2489302, EPI_ISL_2489303, EPI_ISL_2489304, EPI_ISL_2489305, EPI_ISL_2489306, EPI_ISL_2489307, EPI_ISL_2489308, EPI_ISL_2489309, EPI_ISL_2489310, EPI_ISL_2489311, EPI_ISL_2489312, EPI_ISL_2489313, EPI_ISL_2489314, EPI_ISL_2489315, EPI_ISL_2489316, EPI_ISL_2489317, EPI_ISL_2489318, EPI_ISL_2489319, EPI_ISL_2489320, EPI_ISL_2489321, EPI_ISL_2489322, EPI_ISL_2489323, EPI_ISL_2489324, EPI_ISL_2489325, EPI_ISL_2489326, EPI_ISL_2489327, EPI_ISL_2489328, EPI_ISL_2489329, EPI_ISL_2489330, EPI_ISL_2489331, EPI_ISL_2489332, EPI_ISL_2489333, EPI_ISL_2489334, EPI_ISL_2489335, EPI_ISL_2489336, EPI_ISL_2489337, EPI_ISL_2489338, EPI_ISL_2489339, EPI_ISL_2489340, EPI_ISL_2489341, EPI_ISL_2489342, EPI_ISL_2489343, EPI_ISL_2489344, EPI_ISL_2489345, EPI_ISL_2489346, EPI_ISL_2489347, EPI_ISL_2489348, EPI_ISL_2489349, EPI_ISL_2489350, EPI_ISL_2489351, EPI_ISL_2489352, EPI_ISL_2489353, EPI_ISL_2489354, EPI_ISL_2489355, EPI_ISL_2489356, EPI_ISL_2489358, EPI_ISL_2489359, EPI_ISL_2489360, EPI_ISL_2489361, EPI_ISL_2489362, EPI_ISL_2489363, EPI_ISL_2489364, EPI_ISL_2489365, EPI_ISL_2489366, EPI_ISL_2489367, EPI_ISL_2489368, EPI_ISL_2489369, EPI_ISL_2489370, EPI_ISL_2489371, EPI_ISL_2489372, EPI_ISL_2489373, EPI_ISL_2489374, EPI_ISL_2489375, EPI_ISL_2489376, EPI_ISL_2489377, EPI_ISL_2489378, EPI_ISL_2489379, EPI_ISL_2489380, EPI_ISL_2489381, EPI_ISL_2489382, EPI_ISL_2489383, EPI_ISL_2489384, EPI_ISL_2489385, EPI_ISL_2489387, EPI_ISL_2489388, EPI_ISL_2489389, EPI_ISL_2489391, EPI_ISL_2489392, EPI_ISL_2489393, EPI_ISL_2489394, EPI_ISL_2489395, EPI_ISL_2489396, EPI_ISL_2489397, EPI_ISL_2489398, EPI_ISL_2489399, EPI_ISL_2489400, EPI_ISL_2489401, EPI_ISL_2489402, EPI_ISL_2489403, EPI_ISL_2489404, EPI_ISL_2489405, EPI_ISL_2489406, EPI_ISL_2489407, EPI_ISL_2489408, EPI_ISL_2489409, EPI_ISL_2489410, EPI_ISL_2489411, EPI_ISL_2489412, EPI_ISL_2489413, EPI_ISL_2489414, EPI_ISL_2489415, EPI_ISL_2489416, EPI_ISL_2489417, EPI_ISL_2489418, EPI_ISL_2489419, EPI_ISL_2489420, EPI_ISL_2489421, EPI_ISL_2489422, EPI_ISL_2489424, EPI_ISL_2489425, EPI_ISL_2489426, EPI_ISL_2489427, EPI_ISL_2489428, EPI_ISL_2489429, EPI_ISL_2489430, EPI_ISL_2489431, EPI_ISL_2489432, EPI_ISL_2489433, EPI_ISL_2489434, EPI_ISL_2489435, EPI_ISL_2489436, EPI_ISL_2489437, EPI_ISL_2489438, EPI_ISL_2489439, EPI_ISL_2489440, EPI_ISL_2489441, EPI_ISL_2489442, EPI_ISL_2489443, EPI_ISL_2489444, EPI_ISL_2489445, EPI_ISL_2489446, EPI_ISL_2489447, EPI_ISL_2489448, EPI_ISL_2489449, EPI_ISL_2489450, EPI_ISL_2489451, EPI_ISL_2489452, EPI_ISL_2489453, EPI_ISL_2489454, EPI_ISL_2489455, EPI_ISL_2489456, EPI_ISL_2489457, EPI_ISL_2489458, EPI_ISL_2489459, EPI_ISL_2489460, EPI_ISL_2489461, EPI_ISL_2489462, EPI_ISL_2489463, EPI_ISL_2489464, EPI_ISL_2489465, EPI_ISL_2489466, EPI_ISL_2489467, EPI_ISL_2489468, EPI_ISL_2489469, EPI_ISL_2489470, EPI_ISL_2489471, EPI_ISL_2489472, EPI_ISL_2489473, EPI_ISL_2489474, EPI_ISL_2489475, EPI_ISL_2489477, EPI_ISL_2489478, EPI_ISL_2489479, EPI_ISL_2489480, EPI_ISL_2489481, EPI_ISL_2489482, EPI_ISL_2489483, EPI_ISL_2489484, EPI_ISL_2489485, EPI_ISL_2489486, EPI_ISL_2489487, EPI_ISL_2489488, EPI_ISL_2489489, EPI_ISL_2489490, EPI_ISL_2489491, EPI_ISL_2489492, EPI_ISL_2489493, EPI_ISL_2489494, EPI_ISL_2489495, EPI_ISL_2489496, EPI_ISL_2489497, EPI_ISL_2489498, EPI_ISL_2489499, EPI_ISL_2489500, EPI_ISL_2489501, EPI_ISL_2489502, EPI_ISL_2489503, EPI_ISL_2489504, EPI_ISL_2489505, EPI_ISL_2489506, EPI_ISL_2489507, EPI_ISL_2489508, EPI_ISL_2489509, EPI_ISL_2489510, EPI_ISL_2489511, EPI_ISL_2489513, EPI_ISL_2489514, EPI_ISL_2489515, EPI_ISL_2489516, EPI_ISL_2489517, EPI_ISL_2489518, EPI_ISL_2489519, EPI_ISL_2489520, EPI_ISL_2489521, EPI_ISL_2489522, EPI_ISL_2489523, EPI_ISL_2489524, EPI_ISL_2489525, EPI_ISL_2489526, EPI_ISL_2489528, EPI_ISL_2489529, EPI_ISL_2489530, EPI_ISL_2489532, EPI_ISL_2489533, EPI_ISL_2489535, EPI_ISL_2489536, EPI_ISL_2489538, EPI_ISL_2489539, EPI_ISL_2489541, EPI_ISL_2489544, EPI_ISL_2489545, EPI_ISL_2489546, EPI_ISL_2489548, EPI_ISL_2489550, EPI_ISL_2489551, EPI_ISL_2489553, EPI_ISL_2489554, EPI_ISL_2489556, EPI_ISL_2489557, EPI_ISL_2489559, EPI_ISL_2489560, EPI_ISL_2489561, EPI_ISL_2489563, EPI_ISL_2489564, EPI_ISL_2489566, EPI_ISL_2489568, EPI_ISL_2489569, EPI_ISL_2489571, EPI_ISL_2489572, EPI_ISL_2489574, EPI_ISL_2489575, EPI_ISL_2489576, EPI_ISL_2489578, EPI_ISL_2489581, EPI_ISL_2489582, EPI_ISL_2489583, EPI_ISL_2489584, EPI_ISL_2489585, EPI_ISL_2489586, EPI_ISL_2489587, EPI_ISL_2489588, EPI_ISL_2489589, EPI_ISL_2489590, EPI_ISL_2489591, EPI_ISL_2489592, EPI_ISL_2489593, EPI_ISL_2489594, EPI_ISL_2489595, EPI_ISL_2489596, EPI_ISL_2489597, EPI_ISL_2489598, EPI_ISL_2489599, EPI_ISL_2489600, EPI_ISL_2489601, EPI_ISL_2489602, EPI_ISL_2489603, EPI_ISL_2489604, EPI_ISL_2489605, EPI_ISL_2489606, EPI_ISL_2489607, EPI_ISL_2489608, EPI_ISL_2489609, EPI_ISL_2489610, EPI_ISL_2489611, EPI_ISL_2489612, EPI_ISL_2489613, EPI_ISL_2489614, EPI_ISL_2489615, EPI_ISL_2489616, EPI_ISL_2489617, EPI_ISL_2489618, EPI_ISL_2489619, EPI_ISL_2489620, EPI_ISL_2489621, EPI_ISL_2489622, EPI_ISL_2489623, EPI_ISL_2489624, EPI_ISL_2489625, EPI_ISL_2489626, EPI_ISL_2489627, EPI_ISL_2489628, EPI_ISL_2489629, EPI_ISL_2489630, EPI_ISL_2489631, EPI_ISL_2489632, EPI_ISL_2489633, EPI_ISL_2489634, | | | |

| | | | |
|---|---|---|---|
| EPI_ISL_2489635, EPI_ISL_2489636, EPI_ISL_2489637, EPI_ISL_2489638, EPI_ISL_2489639, EPI_ISL_2489640, EPI_ISL_2489641, EPI_ISL_2489642, EPI_ISL_2489643, EPI_ISL_2489644, EPI_ISL_2489645, EPI_ISL_2489646, EPI_ISL_2489647, EPI_ISL_2489648, EPI_ISL_2489649, EPI_ISL_2489650, EPI_ISL_2489651, EPI_ISL_2489652, EPI_ISL_2489653, EPI_ISL_2489654, EPI_ISL_2489655, EPI_ISL_2489656, EPI_ISL_2489657, EPI_ISL_2489658, EPI_ISL_2489659, EPI_ISL_2489660, EPI_ISL_2489661, EPI_ISL_2489662, EPI_ISL_2489663, EPI_ISL_2489664, EPI_ISL_2489666, EPI_ISL_2489667, EPI_ISL_2489668, EPI_ISL_2489669, EPI_ISL_2489670, EPI_ISL_2489671, EPI_ISL_2489672, EPI_ISL_2489673, EPI_ISL_2489674, EPI_ISL_2489675, EPI_ISL_2489676, EPI_ISL_2489677, EPI_ISL_2489678, EPI_ISL_2489679, EPI_ISL_2489680, EPI_ISL_2489681, EPI_ISL_2489682, EPI_ISL_2489683, EPI_ISL_2489684, EPI_ISL_2489685, EPI_ISL_2489686, EPI_ISL_2489687, EPI_ISL_2489688, EPI_ISL_2489689, EPI_ISL_2489690, EPI_ISL_2489691, EPI_ISL_2489692, EPI_ISL_2489693, EPI_ISL_2489694, EPI_ISL_2489696, EPI_ISL_2489697, EPI_ISL_2489699, EPI_ISL_2489700, EPI_ISL_2489701, EPI_ISL_2489702, EPI_ISL_2489703, EPI_ISL_2489704, EPI_ISL_2489705, EPI_ISL_2489706, EPI_ISL_2489707, EPI_ISL_2489708, EPI_ISL_2489709, EPI_ISL_2489710, EPI_ISL_2489711, EPI_ISL_2489712, EPI_ISL_2489713, EPI_ISL_2489714, EPI_ISL_2489715, EPI_ISL_2489716, EPI_ISL_2489717, EPI_ISL_2489718, EPI_ISL_2489719, EPI_ISL_2489720, EPI_ISL_2489721, EPI_ISL_2489722, EPI_ISL_2489723, EPI_ISL_2489724, EPI_ISL_2489725, EPI_ISL_2489726, EPI_ISL_2489727, EPI_ISL_2489728, EPI_ISL_2489729, EPI_ISL_2489730, EPI_ISL_2489731, EPI_ISL_2489732, EPI_ISL_2489733, EPI_ISL_2489734, EPI_ISL_2489735, EPI_ISL_2489736, EPI_ISL_2489737, EPI_ISL_2489738, EPI_ISL_2489739, EPI_ISL_2489740, EPI_ISL_2489741 | | | |
| see above | Aegis Sciences Corporation | Centers for Disease Control and Prevention Division of Viral Diseases, Pathogen Discovery | Dakota Howard, Dhwani Batra, Peter W. Cook, Kara Moser, Adrian Paskey, Jason Caravas, Benjamin Rambo-Martin, Shatavia Morrison, Christopher Gulvick, Scott Sammons, Yvette Unoarumhi, Darlene Wagner, Matthew Schmerer, Cyndi Clark, Patrick Campbell, Rob Case, Vikramsinha Ghorpade, Holly Houdeshell, Ola Kvalvaag, Dillon Nall, Ethan Sanders, Alec Vest, Shaun Westlund, Matthew Hardison, Clinton R. Paden, Duncan MacCannell |
| EPI_ISL_2489742 | Wyoming Public Health Laboratory | Wyoming Public Health Laboratory | Jim Mildenberger, Wanda Manley, Noah Hull, Taylor Fearing, Lynette Gumbleton, Channing Weber, Ashley Norberg, Chayse Rowley, Marley Goetz, Brian Dominguez, Elliot Thomasson, Sam Britz, Cari Sloma, and Rob Christensen |
| EPI_ISL_2489743, EPI_ISL_2489744, EPI_ISL_2489745, EPI_ISL_2489746, EPI_ISL_2489747, EPI_ISL_2489748, EPI_ISL_2489749, EPI_ISL_2489750, EPI_ISL_2489751, EPI_ISL_2489752, EPI_ISL_2489753, EPI_ISL_2489754, EPI_ISL_2489755, EPI_ISL_2489756, EPI_ISL_2489757, EPI_ISL_2489758, EPI_ISL_2489759, EPI_ISL_2489760, EPI_ISL_2489761, EPI_ISL_2489762, EPI_ISL_2489763, EPI_ISL_2489764, EPI_ISL_2489765, EPI_ISL_2489766, EPI_ISL_2489768 | | | |
| see above | Aegis Sciences Corporation | Centers for Disease Control and Prevention Division of Viral Diseases, Pathogen Discovery | Dakota Howard, Dhwani Batra, Peter W. Cook, Kara Moser, Adrian Paskey, Jason Caravas, Benjamin Rambo-Martin, Shatavia Morrison, Christopher Gulvick, Scott Sammons, Yvette Unoarumhi, Darlene Wagner, Matthew Schmerer, Cyndi Clark, Patrick Campbell, Rob Case, Vikramsinha Ghorpade, Holly Houdeshell, Ola Kvalvaag, Dillon Nall, Ethan Sanders, Alec Vest, Shaun Westlund, Matthew Hardison, Clinton R. Paden, Duncan MacCannell |
| EPI_ISL_2489769, EPI_ISL_2489770, EPI_ISL_2489771 | State Testing Facility | Altius Institute for Biomedical Research | Daniel Bates, Rebecca Bruders, Michael Buckley, Mark Frerker, Amanda Gale, Clem Green, Muhammad Halimun, Kneshay Harper, Matt Hartman, Alex Isner, Audra Johnson, Jessica Kunder, Lauren Mitchell, Jemma Nelson, Alex Nguyen, Sofia Olsson, Sadie Patraw, Tobias Ragoczy, Joshua Richards, Jean Robinston, Jacob Rodriguez, John Stamatoyannopoulos, Eric Thorland, Julia Wald |
| EPI_ISL_2489772 | Aegis Sciences Corporation | Centers for Disease Control and Prevention Division of Viral Diseases, Pathogen Discovery | Dakota Howard, Dhwani Batra, Peter W. Cook, Kara Moser, Adrian Paskey, Jason Caravas, Benjamin Rambo-Martin, Shatavia Morrison, Christopher Gulvick, Scott Sammons, Yvette Unoarumhi, Darlene Wagner, Matthew Schmerer, Cyndi Clark, Patrick Campbell, Rob Case, Vikramsinha Ghorpade, Holly Houdeshell, Ola Kvalvaag, Dillon Nall, Ethan Sanders, Alec Vest, Shaun Westlund, Matthew Hardison, Clinton R. Paden, Duncan MacCannell |
| EPI_ISL_2489773, EPI_ISL_2489774 | State Testing Facility | Altius Institute for Biomedical Research | Daniel Bates, Rebecca Bruders, Michael Buckley, Mark Frerker, Amanda Gale, Clem Green, Muhammad Halimun, Kneshay Harper, Matt Hartman, Alex Isner, Audra Johnson, Jessica Kunder, Lauren Mitchell, Jemma Nelson, Alex Nguyen, Sofia Olsson, Sadie Patraw, Tobias Ragoczy, Joshua Richards, Jean Robinston, Jacob Rodriguez, John Stamatoyannopoulos, Eric Thorland, Julia Wald |
| EPI_ISL_2489775 | Aegis Sciences Corporation | Centers for Disease Control and Prevention Division of Viral Diseases, Pathogen Discovery | Dakota Howard, Dhwani Batra, Peter W. Cook, Kara Moser, Adrian Paskey, Jason Caravas, Benjamin Rambo-Martin, Shatavia Morrison, Christopher Gulvick, Scott Sammons, Yvette Unoarumhi, Darlene Wagner, Matthew Schmerer, Cyndi Clark, Patrick Campbell, Rob Case, Vikramsinha Ghorpade, Holly Houdeshell, Ola Kvalvaag, Dillon Nall, Ethan Sanders, Alec Vest, Shaun Westlund, Matthew Hardison, Clinton R. Paden, Duncan MacCannell |
| EPI_ISL_2489776, EPI_ISL_2489777 | State Testing Facility | Altius Institute for Biomedical Research | Daniel Bates, Rebecca Bruders, Michael Buckley, Mark Frerker, Amanda Gale, Clem Green, Muhammad Halimun, Kneshay Harper, Matt Hartman, Alex Isner, Audra Johnson, Jessica Kunder, Lauren Mitchell, Jemma Nelson, Alex Nguyen, Sofia Olsson, Sadie Patraw, Tobias Ragoczy, Joshua Richards, Jean Robinston, Jacob Rodriguez, John Stamatoyannopoulos, Eric Thorland, Julia Wald |
| EPI_ISL_2489778 | Aegis Sciences Corporation | Centers for Disease Control and Prevention Division of Viral Diseases, Pathogen Discovery | Dakota Howard, Dhwani Batra, Peter W. Cook, Kara Moser, Adrian Paskey, Jason Caravas, Benjamin Rambo-Martin, Shatavia Morrison, Christopher Gulvick, Scott Sammons, Yvette Unoarumhi, Darlene Wagner, Matthew Schmerer, Cyndi Clark, Patrick Campbell, Rob Case, Vikramsinha Ghorpade, Holly Houdeshell, Ola Kvalvaag, Dillon Nall, Ethan Sanders, Alec Vest, Shaun Westlund, Matthew Hardison, Clinton R. Paden, Duncan MacCannell |
| EPI_ISL_2489779 | State Testing Facility | Altius Institute for Biomedical Research | Daniel Bates, Rebecca Bruders, Michael Buckley, Mark Frerker, Amanda Gale, Clem Green, Muhammad Halimun, Kneshay Harper, Matt Hartman, Alex Isner, Audra Johnson, Jessica Kunder, Lauren Mitchell, Jemma Nelson, Alex Nguyen, Sofia Olsson, Sadie Patraw, Tobias Ragoczy, Joshua Richards, Jean Robinston, Jacob Rodriguez, John Stamatoyannopoulos, Eric Thorland, Julia Wald |
| EPI_ISL_2489780 | Aegis Sciences Corporation | Centers for Disease Control and Prevention Division of Viral Diseases, Pathogen Discovery | Dakota Howard, Dhwani Batra, Peter W. Cook, Kara Moser, Adrian Paskey, Jason Caravas, Benjamin Rambo-Martin, Shatavia Morrison, Christopher Gulvick, Scott Sammons, Yvette Unoarumhi, Darlene Wagner, Matthew Schmerer, Cyndi Clark, Patrick Campbell, Rob Case, Vikramsinha Ghorpade, Holly Houdeshell, Ola Kvalvaag, Dillon Nall, Ethan Sanders, Alec Vest, Shaun Westlund, Matthew Hardison, Clinton R. Paden, Duncan MacCannell |
| EPI_ISL_2489781 | State Testing Facility | Altius Institute for Biomedical Research | Daniel Bates, Rebecca Bruders, Michael Buckley, Mark Frerker, Amanda Gale, Clem Green, Muhammad Halimun, Kneshay Harper, Matt Hartman, Alex Isner, Audra Johnson, Jessica Kunder, Lauren Mitchell, Jemma Nelson, Alex Nguyen, Sofia Olsson, Sadie Patraw, Tobias Ragoczy, Joshua Richards, Jean Robinston, Jacob Rodriguez, John Stamatoyannopoulos, Eric Thorland, Julia Wald |
| EPI_ISL_2489783 | Aegis Sciences Corporation | Centers for Disease Control and Prevention Division of Viral Diseases, Pathogen Discovery | Dakota Howard, Dhwani Batra, Peter W. Cook, Kara Moser, Adrian Paskey, Jason Caravas, Benjamin Rambo-Martin, Shatavia Morrison, Christopher Gulvick, Scott Sammons, Yvette Unoarumhi, Darlene Wagner, Matthew Schmerer, Cyndi Clark, Patrick Campbell, Rob Case, Vikramsinha Ghorpade, Holly Houdeshell, Ola Kvalvaag, Dillon Nall, Ethan Sanders, Alec Vest, Shaun Westlund, Matthew Hardison, Clinton R. Paden, Duncan MacCannell |
| EPI_ISL_2489784, EPI_ISL_2489785 | State Testing Facility | Altius Institute for Biomedical Research | Daniel Bates, Rebecca Bruders, Michael Buckley, Mark Frerker, Amanda Gale, Clem Green, Muhammad Halimun, Kneshay Harper, Matt Hartman, Alex Isner, Audra Johnson, Jessica Kunder, Lauren Mitchell, Jemma Nelson, Alex Nguyen, Sofia Olsson, Sadie Patraw, Tobias Ragoczy, Joshua Richards, Jean Robinston, Jacob Rodriguez, John Stamatoyannopoulos, Eric Thorland, Julia Wald |
| EPI_ISL_2489786 | Aegis Sciences Corporation | Centers for Disease Control and Prevention Division of Viral Diseases, Pathogen Discovery | Dakota Howard, Dhwani Batra, Peter W. Cook, Kara Moser, Adrian Paskey, Jason Caravas, Benjamin Rambo-Martin, Shatavia Morrison, Christopher Gulvick, Scott Sammons, Yvette Unoarumhi, Darlene Wagner, Matthew Schmerer, Cyndi Clark, Patrick Campbell, Rob Case, Vikramsinha Ghorpade, Holly Houdeshell, Ola Kvalvaag, Dillon Nall, Ethan Sanders, Alec Vest, Shaun Westlund, Matthew Hardison, Clinton R. Paden, Duncan MacCannell |
| EPI_ISL_2489787 | State Testing Facility | Altius Institute for Biomedical Research | Daniel Bates, Rebecca Bruders, Michael Buckley, Mark Frerker, Amanda Gale, Clem Green, Muhammad Halimun, Kneshay Harper, Matt Hartman, Alex Isner, Audra Johnson, Jessica Kunder, Lauren Mitchell, Jemma Nelson, Alex Nguyen, Sofia Olsson, Sadie Patraw, Tobias Ragoczy, Joshua Richards, Jean Robinston, Jacob Rodriguez, John Stamatoyannopoulos, Eric Thorland, Julia Wald |
| EPI_ISL_2489788 | Aegis Sciences Corporation | Centers for Disease Control and Prevention Division of Viral Diseases, Pathogen Discovery | Dakota Howard, Dhwani Batra, Peter W. Cook, Kara Moser, Adrian Paskey, Jason Caravas, Benjamin Rambo-Martin, Shatavia Morrison, Christopher Gulvick, Scott Sammons, Yvette Unoarumhi, Darlene Wagner, Matthew Schmerer, Cyndi Clark, Patrick Campbell, Rob Case, Vikramsinha Ghorpade, Holly Houdeshell, Ola Kvalvaag, Dillon Nall, Ethan Sanders, Alec Vest, Shaun Westlund, Matthew Hardison, Clinton R. Paden, Duncan MacCannell |
| EPI_ISL_2489789, EPI_ISL_2489790 | State Testing Facility | Altius Institute for Biomedical Research | Daniel Bates, Rebecca Bruders, Michael Buckley, Mark Frerker, Amanda Gale, Clem Green, Muhammad Halimun, Kneshay Harper, Matt Hartman, Alex Isner, Audra Johnson, Jessica Kunder, Lauren Mitchell, Jemma Nelson, Alex Nguyen, Sofia Olsson, Sadie Patraw, Tobias Ragoczy, Joshua Richards, Jean Robinston, Jacob Rodriguez, John Stamatoyannopoulos, Eric Thorland, Julia Wald |
| EPI_ISL_2489791 | Aegis Sciences Corporation | Centers for Disease Control and Prevention Division of Viral Diseases, Pathogen Discovery | Dakota Howard, Dhwani Batra, Peter W. Cook, Kara Moser, Adrian Paskey, Jason Caravas, Benjamin Rambo-Martin, Shatavia Morrison, Christopher Gulvick, Scott Sammons, Yvette Unoarumhi, Darlene Wagner, Matthew Schmerer, Cyndi Clark, Patrick Campbell, Rob Case, Vikramsinha Ghorpade, Holly Houdeshell, Ola Kvalvaag, Dillon Nall, Ethan Sanders, Alec Vest, Shaun Westlund, Matthew Hardison, Clinton R. Paden, Duncan MacCannell |
| EPI_ISL_2489792, EPI_ISL_2489793 | State Testing Facility | Altius Institute for Biomedical Research | Daniel Bates, Rebecca Bruders, Michael Buckley, Mark Frerker, Amanda Gale, Clem Green, Muhammad Halimun, Kneshay Harper, Matt Hartman, Alex Isner, Audra Johnson, Jessica Kunder, Lauren Mitchell, Jemma Nelson, Alex Nguyen, Sofia Olsson, Sadie Patraw, Tobias Ragoczy, Joshua Richards, Jean Robinston, Jacob Rodriguez, John Stamatoyannopoulos, Eric Thorland, Julia Wald |
| EPI_ISL_2489794 | Aegis Sciences Corporation | Centers for Disease Control and Prevention Division of Viral Diseases, Pathogen Discovery | Dakota Howard, Dhwani Batra, Peter W. Cook, Kara Moser, Adrian Paskey, Jason Caravas, Benjamin Rambo-Martin, Shatavia Morrison, Christopher Gulvick, Scott Sammons, Yvette Unoarumhi, Darlene Wagner, Matthew Schmerer, Cyndi Clark, Patrick Campbell, Rob Case, Vikramsinha Ghorpade, Holly Houdeshell, Ola Kvalvaag, Dillon Nall, Ethan Sanders, Alec Vest, Shaun Westlund, Matthew Hardison, Clinton R. Paden, Duncan MacCannell |
| EPI_ISL_2489795 | State Testing Facility | Altius Institute for Biomedical Research | Daniel Bates, Rebecca Bruders, Michael Buckley, Mark Frerker, Amanda Gale, Clem Green, Muhammad Halimun, Kneshay Harper, Matt Hartman, Alex Isner, Audra Johnson, Jessica Kunder, Lauren Mitchell, Jemma Nelson, Alex Nguyen, Sofia Olsson, Sadie Patraw, Tobias Ragoczy, Joshua Richards, Jean Robinston, Jacob Rodriguez, John Stamatoyannopoulos, Eric Thorland, Julia Wald |
| EPI_ISL_2489796 | Aegis Sciences Corporation | Centers for Disease Control and Prevention Division of Viral Diseases, Pathogen Discovery | Dakota Howard, Dhwani Batra, Peter W. Cook, Kara Moser, Adrian Paskey, Jason Caravas, Benjamin Rambo-Martin, Shatavia Morrison, Christopher Gulvick, Scott Sammons, Yvette Unoarumhi, Darlene Wagner, Matthew Schmerer, Cyndi Clark, Patrick Campbell, Rob Case, Vikramsinha Ghorpade, Holly Houdeshell, Ola Kvalvaag, Dillon Nall, Ethan Sanders, Alec Vest, Shaun Westlund, Matthew Hardison, Clinton R. Paden, Duncan MacCannell |
| EPI_ISL_2489797, EPI_ISL_2489798 | State Testing Facility | Altius Institute for Biomedical Research | Daniel Bates, Rebecca Bruders, Michael Buckley, Mark Frerker, Amanda Gale, Clem Green, Muhammad Halimun, Kneshay Harper, Matt Hartman, Alex Isner, Audra Johnson, Jessica Kunder, Lauren Mitchell, Jemma Nelson, Alex Nguyen, Sofia Olsson, Sadie Patraw, Tobias Ragoczy, Joshua Richards, Jean Robinston, Jacob Rodriguez, John Stamatoyannopoulos, Eric Thorland, Julia Wald |
| EPI_ISL_2489799 | Aegis Sciences Corporation | Centers for Disease Control and Prevention Division of Viral Diseases, Pathogen Discovery | Dakota Howard, Dhwani Batra, Peter W. Cook, Kara Moser, Adrian Paskey, Jason Caravas, Benjamin Rambo-Martin, Shatavia Morrison, Christopher Gulvick, Scott Sammons, Yvette Unoarumhi, Darlene Wagner, Matthew Schmerer, Cyndi Clark, Patrick Campbell, Rob Case, Vikramsinha Ghorpade, Holly Houdeshell, Ola Kvalvaag, Dillon Nall, Ethan Sanders, Alec Vest, Shaun Westlund, Matthew Hardison, Clinton R. Paden, Duncan MacCannell |
| EPI_ISL_2489800, EPI_ISL_2489801 | State Testing Facility | Altius Institute for Biomedical Research | Daniel Bates, Rebecca Bruders, Michael Buckley, Mark Frerker, Amanda Gale, Clem Green, Muhammad Halimun, Kneshay Harper, Matt Hartman, Alex Isner, Audra Johnson, Jessica Kunder, Lauren Mitchell, Jemma Nelson, Alex Nguyen, Sofia Olsson, Sadie Patraw, Tobias Ragoczy, Joshua Richards, Jean Robinston, Jacob Rodriguez, John Stamatoyannopoulos, Eric Thorland, Julia Wald |
| EPI_ISL_2489802 | Aegis Sciences Corporation | Centers for Disease Control and Prevention Division of Viral Diseases, Pathogen Discovery | Dakota Howard, Dhwani Batra, Peter W. Cook, Kara Moser, Adrian Paskey, Jason Caravas, Benjamin Rambo-Martin, Shatavia Morrison, Christopher Gulvick, Scott Sammons, Yvette Unoarumhi, Darlene Wagner, Matthew Schmerer, Cyndi Clark, Patrick Campbell, Rob Case, Vikramsinha Ghorpade, Holly Houdeshell, Ola Kvalvaag, Dillon Nall, Ethan Sanders, Alec Vest, Shaun Westlund, Matthew Hardison, Clinton R. Paden, Duncan MacCannell |
| EPI_ISL_2489803, EPI_ISL_2489804 | State Testing Facility | Altius Institute for Biomedical Research | Daniel Bates, Rebecca Bruders, Michael Buckley, Mark Frerker, Amanda Gale, Clem Green, Muhammad Halimun, Kneshay Harper, Matt Hartman, Alex Isner, Audra Johnson, Jessica Kunder, Lauren Mitchell, Jemma Nelson, Alex Nguyen, Sofia Olsson, Sadie Patraw, Tobias Ragoczy, Joshua Richards, Jean Robinston, Jacob Rodriguez, John Stamatoyannopoulos, Eric Thorland, Julia Wald |
| EPI_ISL_2489806, EPI_ISL_2489807, EPI_ISL_2489808, EPI_ISL_2489809, EPI_ISL_2489810, EPI_ISL_2489811, EPI_ISL_2489812, EPI_ISL_2489814, EPI_ISL_2489815, EPI_ISL_2489816, EPI_ISL_2489817, EPI_ISL_2489818, EPI_ISL_2489819, EPI_ISL_2489820, EPI_ISL_2489821, EPI_ISL_2489822, EPI_ISL_2489823, EPI_ISL_2489824, EPI_ISL_2489825, EPI_ISL_2489826, EPI_ISL_2489827, EPI_ISL_2489828, EPI_ISL_2489829, EPI_ISL_2489830, EPI_ISL_2489831, EPI_ISL_2489832, EPI_ISL_2489833, EPI_ISL_2489834, EPI_ISL_2489835, EPI_ISL_2489836, EPI_ISL_2489837, EPI_ISL_2489838, EPI_ISL_2489839, EPI_ISL_2489840, EPI_ISL_2489841, EPI_ISL_2489842, EPI_ISL_2489843, EPI_ISL_2489844, EPI_ISL_2489845, EPI_ISL_2489846, EPI_ISL_2489847, EPI_ISL_2489848, EPI_ISL_2489849, EPI_ISL_2489850, EPI_ISL_2489851, EPI_ISL_2489852, EPI_ISL_2489853, EPI_ISL_2489854, EPI_ISL_2489855, EPI_ISL_2489856, EPI_ISL_2489857, EPI_ISL_2489858, EPI_ISL_2489859, EPI_ISL_2489860, EPI_ISL_2489861, EPI_ISL_2489862, EPI_ISL_2489863, EPI_ISL_2489864, EPI_ISL_2489865, EPI_ISL_2489866, EPI_ISL_2489867, EPI_ISL_2489868, EPI_ISL_2489870, EPI_ISL_2489871, EPI_ISL_2489872, EPI_ISL_2489873, EPI_ISL_2489874, EPI_ISL_2489875, EPI_ISL_2489876, EPI_ISL_2489878, EPI_ISL_2489879, EPI_ISL_2489880, EPI_ISL_2489881, EPI_ISL_2489882, EPI_ISL_2489883, EPI_ISL_2489884, EPI_ISL_2489885, EPI_ISL_2489886, EPI_ISL_2489887 | | | |
| see above | Aegis Sciences Corporation | Centers for Disease Control and Prevention Division of Viral Diseases, Pathogen Discovery | Dakota Howard, Dhwani Batra, Peter W. Cook, Kara Moser, Adrian Paskey, Jason Caravas, Benjamin Rambo-Martin, Shatavia Morrison, Christopher Gulvick, Scott Sammons, Yvette Unoarumhi, Darlene Wagner, Matthew Schmerer, Cyndi Clark, Patrick Campbell, Rob Case, Vikramsinha Ghorpade, Holly Houdeshell, Ola Kvalvaag, Dillon Nall, Ethan Sanders, Alec Vest, Shaun Westlund, Matthew Hardison, Clinton R. Paden, Duncan MacCannell |
| EPI_ISL_2489888 | Laboratory Corporation of America | Centers for Disease Control and Prevention Division of Viral Diseases, Pathogen Discovery | Dakota Howard, Dhwani Batra, Peter W. Cook, Kara Moser, Adrian Paskey, Jason Caravas, Benjamin Rambo-Martin, Shatavia Morrison, Christopher Gulvick, Scott Sammons, Yvette Unoarumhi, Darlene Wagner, Matthew Schmerer, Minoo Agarwal, Eyad Almasri, Debbie Boles, Ayla Burns, Nuthawin Charoensri, Oren Cohen, Susan Countryman, Mary Ann Cristobal, Bobbi Croy, Suzanne Dale, Hrushikesh Deshmukh, Amanda Douglas, Vincent Drouillon, Marcia Eisenberg, Howard Engler, Rama Ghatti, Prashant Gupta, Susan Hicks, Jake Humphrey, Lax Iyer, Lisa Pfefferle, Manoj Jain, Matthew Robinson, Mohan Kolli, Brian Krueger, Tim Kuphal, Stanley Letovsky, Michael Levandoski, Craig Lukasik, Jonathan Meltzer, Brian Norvell, Mindy Nye, Scott Parker, Christos Petropoulos, John Pruitt, Steven Ragan, Scott Ryan, Mike Sapeta, Jana Schroth, Suresh Babu Selvaraju, Goran Stevovic, Amanda Suchanek, Andrea Throop, Lyndon Tilson, Thomas Urban, Joe Voshell, Kimberly Wagner, Jonathan Williams, Mary Williamson, Qian Zeng, Tricia Zwiefelhofer, Clinton R. Paden, |
| EPI_ISL_2489889, EPI_ISL_2489891, EPI_ISL_2489893, EPI_ISL_2489894, EPI_ISL_2489895, EPI_ISL_2489896, EPI_ISL_2489897, EPI_ISL_2489899, EPI_ISL_2489900, EPI_ISL_2489901, EPI_ISL_2489902, EPI_ISL_2489903, EPI_ISL_2489904, EPI_ISL_2489905, EPI_ISL_2489906, EPI_ISL_2489907, EPI_ISL_2489909, EPI_ISL_2489910, EPI_ISL_2489911, EPI_ISL_2489912, EPI_ISL_2489913, EPI_ISL_2489916, EPI_ISL_2489917, EPI_ISL_2489918, EPI_ISL_2489919, EPI_ISL_2489920, EPI_ISL_2489921, EPI_ISL_2489923, EPI_ISL_2489924, EPI_ISL_2489925, EPI_ISL_2489926, EPI_ISL_2489931, EPI_ISL_2489932, EPI_ISL_2489933, EPI_ISL_2489934, EPI_ISL_2489935, EPI_ISL_2489936, EPI_ISL_2489937, EPI_ISL_2489938, EPI_ISL_2489939, EPI_ISL_2489940, EPI_ISL_2489941, EPI_ISL_2489942, EPI_ISL_2489943, EPI_ISL_2489944, EPI_ISL_2489945, EPI_ISL_2489946, EPI_ISL_2489947, EPI_ISL_2489948, EPI_ISL_2489950, EPI_ISL_2489951, EPI_ISL_2489952, EPI_ISL_2489953, EPI_ISL_2489955, EPI_ISL_2489956 | | | |
| see above | North Dakota Department of Health, Public Health Laboratory | North Dakota Department of Health, Public Health Laboratory | Lisa Wingerter |
| EPI_ISL_2489957, EPI_ISL_2489958, | Seattle Flu Study | Seattle Flu Study | Deborah A. Nickerson, Chris D. Frazar, Jover Lee, Benjamin Pelle, Erica Ryke, Matthew Richardson, Amanda Adler, Elisabeth Brandstetter, Peter D. Han, Kairsten Fay, Misja Ilcisin, Kirsten Lacombe, Thomas R. Sibley, Melissa Truong, Caitlin R. Wolf, Karen Cowgill, Stephanie Schrag, Jeff Duchin, Michael Boeckh, Janet A. Englund, Michael Famulare, Barry R. Lutz, Mark J. Rieder, Lea M. Starita, Matthew Thompson, Helen Y. Chu, Trevor Bedford, Jay Shendure |

| | | | |
|---|---|---|---|
| EPI_ISL_2489959, EPI_ISL_2489960, EPI_ISL_2489961 | | | |
| EPI_ISL_2489962, EPI_ISL_2489963 | Seattle Flu Study | Seattle Flu Study | Deborah A. Nickerson, Chris D. Frazar, Jover Lee, Benjamin Pelle, Erica Ryke, Matthew Richardson, Amanda Adler, Elisabeth Brandstetter, Peter D. Han, Kairsten Fay, Misja Ilcisin, Kirsten Lacombe, Thomas R. Sibley, Melissa Truong, Caitlin R. Wolf, Michael Boeckh, Janet A. Englund, Michael Famulare, Barry R. Lutz, Mark J. Rieder, Lea M. Starita, Matthew Thompson, Jay Shendure, Trevor Bedford, Helen Y. Chu |
| EPI_ISL_2489964, EPI_ISL_2489965, EPI_ISL_2489966, EPI_ISL_2489967 | Central Washington Hosptial | Seattle Flu Study | Deborah A. Nickerson, Chris D. Frazar, Jover Lee, Benjamin Pelle, Erica Ryke, Matthew Richardson, Amanda Adler, Elisabeth Brandstetter, Peter D. Han, Kairsten Fay, Misja Ilcisin, Kirsten Lacombe, Thomas R. Sibley, Melissa Truong, Caitlin R. Wolf, Romesh Gautom, Geoff Melly, Brian Hiatt, Philip Dykema, Scott Lindquist, Michael Boeckh, Janet A. Englund, Michael Famulare, Barry R. Lutz, Mark J. Rieder, Lea M. Starita, Matthew Thompson, Helen Y. Chu, Jay Shendure, Trevor Bedford |
| EPI_ISL_2489969, EPI_ISL_2489970, EPI_ISL_2489971 | Seattle Flu Study | Seattle Flu Study | Deborah A. Nickerson, Chris D. Frazar, Jover Lee, Benjamin Pelle, Erica Ryke, Matthew Richardson, Amanda Adler, Elisabeth Brandstetter, Peter D. Han, Kairsten Fay, Misja Ilcisin, Kirsten Lacombe, Thomas R. Sibley, Melissa Truong, Caitlin R. Wolf, Karen Cowgill, Stephanie Schrag, Jeff Duchin, Michael Boeckh, Janet A. Englund, Michael Famulare, Barry R. Lutz, Mark J. Rieder, Lea M. Starita, Matthew Thompson, Helen Y. Chu, Trevor Bedford, Jay Shendure |
| EPI_ISL_2489972 | Seattle Flu Study | Seattle Flu Study | Deborah A. Nickerson, Chris D. Frazar, Jover Lee, Benjamin Pelle, Erica Ryke, Matthew Richardson, Amanda Adler, Elisabeth Brandstetter, Peter D. Han, Kairsten Fay, Misja Ilcisin, Kirsten Lacombe, Thomas R. Sibley, Melissa Truong, Caitlin R. Wolf, Michael Boeckh, Janet A. Englund, Michael Famulare, Barry R. Lutz, Mark J. Rieder, Lea M. Starita, Matthew Thompson, Jay Shendure, Trevor Bedford, Helen Y. Chu |
| EPI_ISL_2489973 | Seattle Flu Study | Seattle Flu Study | Deborah A. Nickerson, Chris D. Frazar, Jover Lee, Benjamin Pelle, Erica Ryke, Matthew Richardson, Amanda Adler, Elisabeth Brandstetter, Peter D. Han, Kairsten Fay, Misja Ilcisin, Kirsten Lacombe, Thomas R. Sibley, Melissa Truong, Caitlin R. Wolf, Karen Cowgill, Stephanie Schrag, Jeff Duchin, Michael Boeckh, Janet A. Englund, Michael Famulare, Barry R. Lutz, Mark J. Rieder, Lea M. Starita, Matthew Thompson, Helen Y. Chu, Trevor Bedford, Jay Shendure |
| EPI_ISL_2489974 | Seattle Flu Study | Seattle Flu Study | Deborah A. Nickerson, Chris D. Frazar, Jover Lee, Benjamin Pelle, Erica Ryke, Matthew Richardson, Amanda Adler, Elisabeth Brandstetter, Peter D. Han, Kairsten Fay, Misja Ilcisin, Kirsten Lacombe, Thomas R. Sibley, Melissa Truong, Caitlin R. Wolf, Michael Boeckh, Janet A. Englund, Michael Famulare, Barry R. Lutz, Mark J. Rieder, Lea M. Starita, Matthew Thompson, Jay Shendure, Trevor Bedford, Helen Y. Chu |
| EPI_ISL_2489975, EPI_ISL_2489976, EPI_ISL_2489977, EPI_ISL_2489978, EPI_ISL_2489979, EPI_ISL_2489980, EPI_ISL_2489981, EPI_ISL_2489982, EPI_ISL_2489983, EPI_ISL_2489984, EPI_ISL_2489985, EPI_ISL_2489986, EPI_ISL_2489987, EPI_ISL_2489988, EPI_ISL_2489989, EPI_ISL_2489990, EPI_ISL_2489991, EPI_ISL_2489992, EPI_ISL_2489993, EPI_ISL_2489994, EPI_ISL_2489995, EPI_ISL_2489996, EPI_ISL_2489997 | | | |
| see above | Seattle Flu Study | Seattle Flu Study | Deborah A. Nickerson, Chris D. Frazar, Jover Lee, Benjamin Pelle, Erica Ryke, Matthew Richardson, Amanda Adler, Elisabeth Brandstetter, Peter D. Han, Kairsten Fay, Misja Ilcisin, Kirsten Lacombe, Thomas R. Sibley, Melissa Truong, Caitlin R. Wolf, Karen Cowgill, Stephanie Schrag, Jeff Duchin, Michael Boeckh, Janet A. Englund, Michael Famulare, Barry R. Lutz, Mark J. Rieder, Lea M. Starita, Matthew Thompson, Helen Y. Chu, Trevor Bedford, Jay Shendure |
| EPI_ISL_2489998 | Seattle Flu Study | Seattle Flu Study | Deborah A. Nickerson, Chris D. Frazar, Jover Lee, Benjamin Pelle, Erica Ryke, Matthew Richardson, Amanda Adler, Elisabeth Brandstetter, Peter D. Han, Kairsten Fay, Misja Ilcisin, Kirsten Lacombe, Thomas R. Sibley, Melissa Truong, Caitlin R. Wolf, Michael Boeckh, Janet A. Englund, Michael Famulare, Barry R. Lutz, Mark J. Rieder, Lea M. Starita, Matthew Thompson, Jay Shendure, Trevor Bedford, Helen Y. Chu |
| EPI_ISL_2490000 | Seattle Flu Study | Seattle Flu Study | Deborah A. Nickerson, Chris D. Frazar, Jover Lee, Benjamin Pelle, Erica Ryke, Matthew Richardson, Amanda Adler, Elisabeth Brandstetter, Peter D. Han, Kairsten Fay, Misja Ilcisin, Kirsten Lacombe, Thomas R. Sibley, Melissa Truong, Caitlin R. Wolf, Karen Cowgill, Stephanie Schrag, Jeff Duchin, Michael Boeckh, Janet A. Englund, Michael Famulare, Barry R. Lutz, Mark J. Rieder, Lea M. Starita, Matthew Thompson, Helen Y. Chu, Trevor Bedford, Jay Shendure |
| EPI_ISL_2490001 | Seattle Flu Study | Seattle Flu Study | Deborah A. Nickerson, Chris D. Frazar, Jover Lee, Benjamin Pelle, Erica Ryke, Matthew Richardson, Amanda Adler, Elisabeth Brandstetter, Peter D. Han, Kairsten Fay, Misja Ilcisin, Kirsten Lacombe, Thomas R. Sibley, Melissa Truong, Caitlin R. Wolf, Michael Boeckh, Janet A. Englund, Michael Famulare, Barry R. Lutz, Mark J. Rieder, Lea M. Starita, Matthew Thompson, Jay Shendure, Trevor Bedford, Helen Y. Chu |
| EPI_ISL_2490002 | Seattle Flu Study | Seattle Flu Study | Deborah A. Nickerson, Chris D. Frazar, Jover Lee, Benjamin Pelle, Erica Ryke, Matthew Richardson, Amanda Adler, Elisabeth Brandstetter, Peter D. Han, Kairsten Fay, Misja Ilcisin, Kirsten Lacombe, Thomas R. Sibley, Melissa Truong, Caitlin R. Wolf, Karen Cowgill, Stephanie Schrag, Jeff Duchin, Michael Boeckh, Janet A. Englund, Michael Famulare, Barry R. Lutz, Mark J. Rieder, Lea M. Starita, Matthew Thompson, Helen Y. Chu, Trevor Bedford, Jay Shendure |
| EPI_ISL_2490003, EPI_ISL_2490004 | Seattle Flu Study | Seattle Flu Study | Deborah A. Nickerson, Chris D. Frazar, Jover Lee, Benjamin Pelle, Erica Ryke, Matthew Richardson, Amanda Adler, Elisabeth Brandstetter, Peter D. Han, Kairsten Fay, Misja Ilcisin, Kirsten Lacombe, Thomas R. Sibley, Melissa Truong, Caitlin R. Wolf, Michael Boeckh, Janet A. Englund, Michael Famulare, Barry R. Lutz, Mark J. Rieder, Lea M. Starita, Matthew Thompson, Jay Shendure, Trevor Bedford, Helen Y. Chu |
| EPI_ISL_2490005, EPI_ISL_2490006, EPI_ISL_2490007, EPI_ISL_2490008, EPI_ISL_2490009, EPI_ISL_2490010, EPI_ISL_2490012 | Seattle Flu Study | Seattle Flu Study | Deborah A. Nickerson, Chris D. Frazar, Jover Lee, Benjamin Pelle, Erica Ryke, Matthew Richardson, Amanda Adler, Elisabeth Brandstetter, Peter D. Han, Kairsten Fay, Misja Ilcisin, Kirsten Lacombe, Thomas R. Sibley, Melissa Truong, Caitlin R. Wolf, Karen Cowgill, Stephanie Schrag, Jeff Duchin, Michael Boeckh, Janet A. Englund, Michael Famulare, Barry R. Lutz, Mark J. Rieder, Lea M. Starita, Matthew Thompson, Helen Y. Chu, Trevor Bedford, Jay Shendure |
| EPI_ISL_2490013, EPI_ISL_2490014 | Seattle Flu Study | Seattle Flu Study | Deborah A. Nickerson, Chris D. Frazar, Jover Lee, Benjamin Pelle, Erica Ryke, Matthew Richardson, Amanda Adler, Elisabeth Brandstetter, Peter D. Han, Kairsten Fay, Misja Ilcisin, Kirsten Lacombe, Thomas R. Sibley, Melissa Truong, Caitlin R. Wolf, Michael Boeckh, Janet A. Englund, Michael Famulare, Barry R. Lutz, Mark J. Rieder, Lea M. Starita, Matthew Thompson, Jay Shendure, Trevor Bedford, Helen Y. Chu |
| EPI_ISL_2490015, EPI_ISL_2490016, EPI_ISL_2490017 | Seattle Flu Study | Seattle Flu Study | Deborah A. Nickerson, Chris D. Frazar, Jover Lee, Benjamin Pelle, Erica Ryke, Matthew Richardson, Amanda Adler, Elisabeth Brandstetter, Peter D. Han, Kairsten Fay, Misja Ilcisin, Kirsten Lacombe, Thomas R. Sibley, Melissa Truong, Caitlin R. Wolf, Karen Cowgill, Stephanie Schrag, Jeff Duchin, Michael Boeckh, Janet A. Englund, Michael Famulare, Barry R. Lutz, Mark J. Rieder, Lea M. Starita, Matthew Thompson, Helen Y. Chu, Trevor Bedford, Jay Shendure |
| EPI_ISL_2490018 | Seattle Flu Study | Seattle Flu Study | Deborah A. Nickerson, Chris D. Frazar, Jover Lee, Benjamin Pelle, Erica Ryke, Matthew Richardson, Amanda Adler, Elisabeth Brandstetter, Peter D. Han, Kairsten Fay, Misja Ilcisin, Kirsten Lacombe, Thomas R. Sibley, Melissa Truong, Caitlin R. Wolf, Michael Boeckh, Janet A. Englund, Michael Famulare, Barry R. Lutz, Mark J. Rieder, Lea M. Starita, Matthew Thompson, Jay Shendure, Trevor Bedford, Helen Y. Chu |
| EPI_ISL_2490019, EPI_ISL_2490020, EPI_ISL_2490021, EPI_ISL_2490023, EPI_ISL_2490024, EPI_ISL_2490025, EPI_ISL_2490026, EPI_ISL_2490027, EPI_ISL_2490028, EPI_ISL_2490029, EPI_ISL_2490030, EPI_ISL_2490031, EPI_ISL_2490033, EPI_ISL_2490034, EPI_ISL_2490035, EPI_ISL_2490036, EPI_ISL_2490037, EPI_ISL_2490038, EPI_ISL_2490039, EPI_ISL_2490040 | | | |
| see above | Seattle Flu Study | Seattle Flu Study | Deborah A. Nickerson, Chris D. Frazar, Jover Lee, Benjamin Pelle, Erica Ryke, Matthew Richardson, Amanda Adler, Elisabeth Brandstetter, Peter D. Han, Kairsten Fay, Misja Ilcisin, Kirsten Lacombe, Thomas R. Sibley, Melissa Truong, Caitlin R. Wolf, Karen Cowgill, Stephanie Schrag, Jeff Duchin, Michael Boeckh, Janet A. Englund, Michael Famulare, Barry R. Lutz, Mark J. Rieder, Lea M. Starita, Matthew Thompson, Helen Y. Chu, Trevor Bedford, Jay Shendure |
| EPI_ISL_2490041 | Seattle Flu Study | Seattle Flu Study | Deborah A. Nickerson, Chris D. Frazar, Jover Lee, Benjamin Pelle, Erica Ryke, Matthew Richardson, Amanda Adler, Elisabeth Brandstetter, Peter D. Han, Kairsten Fay, Misja Ilcisin, Kirsten Lacombe, Thomas R. Sibley, Melissa Truong, Caitlin R. Wolf, Michael Boeckh, Janet A. Englund, Michael Famulare, Barry R. Lutz, Mark J. Rieder, Lea M. Starita, Matthew Thompson, Jay Shendure, Trevor Bedford, Helen Y. Chu |
| EPI_ISL_2490043, EPI_ISL_2490044, EPI_ISL_2490045, EPI_ISL_2490046 | Seattle Flu Study | Seattle Flu Study | Deborah A. Nickerson, Chris D. Frazar, Jover Lee, Benjamin Pelle, Erica Ryke, Matthew Richardson, Amanda Adler, Elisabeth Brandstetter, Peter D. Han, Kairsten Fay, Misja Ilcisin, Kirsten Lacombe, Thomas R. Sibley, Melissa Truong, Caitlin R. Wolf, Karen Cowgill, Stephanie Schrag, Jeff Duchin, Michael Boeckh, Janet A. Englund, Michael Famulare, Barry R. Lutz, Mark J. Rieder, Lea M. Starita, Matthew Thompson, Helen Y. Chu, Trevor Bedford, Jay Shendure |
| EPI_ISL_2490047 | Seattle Flu Study | Seattle Flu Study | Deborah A. Nickerson, Chris D. Frazar, Jover Lee, Benjamin Pelle, Erica Ryke, Matthew Richardson, Amanda Adler, Elisabeth Brandstetter, Peter D. Han, Kairsten Fay, Misja Ilcisin, Kirsten Lacombe, Thomas R. Sibley, Melissa Truong, Caitlin R. Wolf, Michael Boeckh, Janet A. Englund, Michael Famulare, Barry R. Lutz, Mark J. Rieder, Lea M. Starita, Matthew Thompson, Jay Shendure, Trevor Bedford, Helen Y. Chu |
| EPI_ISL_2490048, EPI_ISL_2490049, EPI_ISL_2490050, EPI_ISL_2490051 | Seattle Flu Study | Seattle Flu Study | Deborah A. Nickerson, Chris D. Frazar, Jover Lee, Benjamin Pelle, Erica Ryke, Matthew Richardson, Amanda Adler, Elisabeth Brandstetter, Peter D. Han, Kairsten Fay, Misja Ilcisin, Kirsten Lacombe, Thomas R. Sibley, Melissa Truong, Caitlin R. Wolf, Karen Cowgill, Stephanie Schrag, Jeff Duchin, Michael Boeckh, Janet A. Englund, Michael Famulare, Barry R. Lutz, Mark J. Rieder, Lea M. Starita, Matthew Thompson, Helen Y. Chu, Trevor Bedford, Jay Shendure |
| EPI_ISL_2490052, EPI_ISL_2490053, EPI_ISL_2490054, EPI_ISL_2490055, EPI_ISL_2490056, EPI_ISL_2490057, EPI_ISL_2490058 | Seattle Flu Study | Seattle Flu Study | Deborah A. Nickerson, Chris D. Frazar, Jover Lee, Benjamin Pelle, Erica Ryke, Matthew Richardson, Amanda Adler, Elisabeth Brandstetter, Peter D. Han, Kairsten Fay, Misja Ilcisin, Kirsten Lacombe, Thomas R. Sibley, Melissa Truong, Caitlin R. Wolf, Michael Boeckh, Janet A. Englund, Michael Famulare, Barry R. Lutz, Mark J. Rieder, Lea M. Starita, Matthew Thompson, Jay Shendure, Trevor Bedford, Helen Y. Chu |
| EPI_ISL_2490059, EPI_ISL_2490060 | Seattle Flu Study | Seattle Flu Study | Deborah A. Nickerson, Chris D. Frazar, Jover Lee, Benjamin Pelle, Erica Ryke, Matthew Richardson, Amanda Adler, Elisabeth Brandstetter, Peter D. Han, Kairsten Fay, Misja Ilcisin, Kirsten Lacombe, Thomas R. Sibley, Melissa Truong, Caitlin R. Wolf, Karen Cowgill, Stephanie Schrag, Jeff Duchin, Michael Boeckh, Janet A. Englund, Michael Famulare, Barry R. Lutz, Mark J. Rieder, Lea M. Starita, Matthew Thompson, Helen Y. Chu, Trevor Bedford, Jay Shendure |
| EPI_ISL_2490061, EPI_ISL_2490062 | Seattle Flu Study | Seattle Flu Study | Deborah A. Nickerson, Chris D. Frazar, Jover Lee, Benjamin Pelle, Erica Ryke, Matthew Richardson, Amanda Adler, Elisabeth Brandstetter, Peter D. Han, Kairsten Fay, Misja Ilcisin, Kirsten Lacombe, Thomas R. Sibley, Melissa Truong, Caitlin R. Wolf, Michael Boeckh, Janet A. Englund, Michael Famulare, Barry R. Lutz, Mark J. Rieder, Lea M. Starita, Matthew Thompson, Jay Shendure, Trevor Bedford, Helen Y. Chu |
| EPI_ISL_2490063 | Seattle Flu Study | Seattle Flu Study | Deborah A. Nickerson, Chris D. Frazar, Jover Lee, Benjamin Pelle, Erica Ryke, Matthew Richardson, Amanda Adler, Elisabeth Brandstetter, Peter D. Han, Kairsten Fay, Misja Ilcisin, Kirsten Lacombe, Thomas R. Sibley, Melissa Truong, Caitlin R. Wolf, Karen Cowgill, Stephanie Schrag, Jeff Duchin, Michael Boeckh, Janet A. Englund, Michael Famulare, Barry R. Lutz, Mark J. Rieder, Lea M. Starita, Matthew Thompson, Helen Y. Chu, Trevor Bedford, Jay Shendure |
| EPI_ISL_2490064, EPI_ISL_2490065 | Seattle Flu Study | Seattle Flu Study | Deborah A. Nickerson, Chris D. Frazar, Jover Lee, Benjamin Pelle, Erica Ryke, Matthew Richardson, Amanda Adler, Elisabeth Brandstetter, Peter D. Han, Kairsten Fay, Misja Ilcisin, Kirsten Lacombe, Thomas R. Sibley, Melissa Truong, Caitlin R. Wolf, Michael Boeckh, Janet A. Englund, Michael Famulare, Barry R. Lutz, Mark J. Rieder, Lea M. Starita, Matthew Thompson, Jay Shendure, Trevor Bedford, Helen Y. Chu |
| EPI_ISL_2490066 | Seattle Flu Study | Seattle Flu Study | Deborah A. Nickerson, Chris D. Frazar, Jover Lee, Benjamin Pelle, Erica Ryke, Matthew Richardson, Amanda Adler, Elisabeth Brandstetter, Peter D. Han, Kairsten Fay, Misja Ilcisin, Kirsten Lacombe, Thomas R. Sibley, Melissa Truong, Caitlin R. Wolf, Karen Cowgill, Stephanie Schrag, Jeff Duchin, Michael Boeckh, Janet A. Englund, Michael Famulare, Barry R. Lutz, Mark J. Rieder, Lea M. Starita, Matthew Thompson, Helen Y. Chu, Trevor Bedford, Jay Shendure |
| EPI_ISL_2490067, EPI_ISL_2490068 | Seattle Flu Study | Seattle Flu Study | Deborah A. Nickerson, Chris D. Frazar, Jover Lee, Benjamin Pelle, Erica Ryke, Matthew Richardson, Amanda Adler, Elisabeth Brandstetter, Peter D. Han, Kairsten Fay, Misja Ilcisin, Kirsten Lacombe, Thomas R. Sibley, Melissa Truong, Caitlin R. Wolf, Michael Boeckh, Janet A. Englund, Michael Famulare, Barry R. Lutz, Mark J. Rieder, Lea M. Starita, Matthew Thompson, Jay Shendure, Trevor Bedford, Helen Y. Chu |
| EPI_ISL_2490070 | Seattle Flu Study | Seattle Flu Study | Deborah A. Nickerson, Chris D. Frazar, Jover Lee, Benjamin Pelle, Erica Ryke, Matthew Richardson, Amanda Adler, Elisabeth Brandstetter, Peter D. Han, Kairsten Fay, Misja Ilcisin, Kirsten Lacombe, Thomas R. Sibley, Melissa Truong, Caitlin R. Wolf, Karen Cowgill, Stephanie Schrag, Jeff Duchin, Michael Boeckh, Janet A. Englund, Michael Famulare, Barry R. Lutz, Mark J. Rieder, Lea M. Starita, Matthew Thompson, Helen Y. Chu, Trevor Bedford, Jay Shendure |
| EPI_ISL_2490071, EPI_ISL_2490072, EPI_ISL_2490073, EPI_ISL_2490074, EPI_ISL_2490075, EPI_ISL_2490076 | Seattle Flu Study | Seattle Flu Study | Deborah A. Nickerson, Chris D. Frazar, Jover Lee, Benjamin Pelle, Erica Ryke, Matthew Richardson, Amanda Adler, Elisabeth Brandstetter, Peter D. Han, Kairsten Fay, Misja Ilcisin, Kirsten Lacombe, Thomas R. Sibley, Melissa Truong, Caitlin R. Wolf, Michael Boeckh, Janet A. Englund, Michael Famulare, Barry R. Lutz, Mark J. Rieder, Lea M. Starita, Matthew Thompson, Jay Shendure, Trevor Bedford, Helen Y. Chu |
| EPI_ISL_2490077, EPI_ISL_2490078 | Seattle Flu Study | Seattle Flu Study | Deborah A. Nickerson, Chris D. Frazar, Jover Lee, Benjamin Pelle, Erica Ryke, Matthew Richardson, Amanda Adler, Elisabeth Brandstetter, Peter D. Han, Kairsten Fay, Misja Ilcisin, Kirsten Lacombe, Thomas R. Sibley, Melissa Truong, Caitlin R. Wolf, Karen Cowgill, Stephanie Schrag, Jeff Duchin, Michael Boeckh, Janet A. Englund, Michael Famulare, Barry R. Lutz, Mark J. Rieder, Lea M. Starita, Matthew Thompson, Helen Y. Chu, Trevor Bedford, Jay Shendure |
| EPI_ISL_2490079, EPI_ISL_2490080 | Seattle Flu Study | Seattle Flu Study | Deborah A. Nickerson, Chris D. Frazar, Jover Lee, Benjamin Pelle, Erica Ryke, Matthew Richardson, Amanda Adler, Elisabeth Brandstetter, Peter D. Han, Kairsten Fay, Misja Ilcisin, Kirsten Lacombe, Thomas R. Sibley, Melissa Truong, Caitlin R. Wolf, Michael Boeckh, Janet A. Englund, Michael Famulare, Barry R. Lutz, Mark J. Rieder, Lea M. Starita, Matthew Thompson, Jay Shendure, Trevor Bedford, Helen Y. Chu |
| EPI_ISL_2490081, EPI_ISL_2490082, EPI_ISL_2490083, | Atlas Genomics | Seattle Flu Study | Deborah A. Nickerson, Chris D. Frazar, Jover Lee, Benjamin Pelle, Erica Ryke, Matthew Richardson, Amanda Adler, Elisabeth Brandstetter, Peter D. Han, Kairsten Fay, Misja Ilcisin, Kirsten Lacombe, Thomas R. Sibley, Melissa Truong, Caitlin R. Wolf, Romesh Gautom, Geoff Melly, Brian Hiatt, Philip Dykema, Scott Lindquist, Michael Boeckh, Janet A. Englund, Michael Famulare, Barry R. Lutz, Mark J. Rieder, Lea M. Starita, Matthew Thompson, Helen Y. Chu, Jay Shendure, Trevor Bedford |

| | | | |
|---|---|---|---|
| EPI_ISL_2490084, EPI_ISL_2490085, EPI_ISL_2490086, EPI_ISL_2490087 | | | |
| EPI_ISL_2490088, EPI_ISL_2490089, EPI_ISL_2490090, EPI_ISL_2490091, EPI_ISL_2490092, EPI_ISL_2490093, EPI_ISL_2490094 | Seattle Flu Study | Seattle Flu Study | Deborah A. Nickerson, Chris D. Frazar, Jover Lee, Benjamin Pelle, Erica Ryke, Matthew Richardson, Amanda Adler, Elisabeth Brandstetter, Peter D. Han, Kairsten Fay, Misja Ilcisin, Kirsten Lacombe, Thomas R. Sibley, Melissa Truong, Caitlin R. Wolf, Karen Cowgill, Stephanie Schrag, Jeff Duchin, Michael Boeckh, Janet A. Englund, Michael Famulare, Barry R. Lutz, Mark J. Rieder, Lea M. Starita, Matthew Thompson, Helen Y. Chu, Trevor Bedford, Jay Shendure |
| EPI_ISL_2490095, EPI_ISL_2490096, EPI_ISL_2490097, EPI_ISL_2490098, EPI_ISL_2490099, EPI_ISL_2490100, EPI_ISL_2490101, EPI_ISL_2490102, EPI_ISL_2490103, EPI_ISL_2490105, EPI_ISL_2490106, EPI_ISL_2490107, EPI_ISL_2490108, EPI_ISL_2490109, EPI_ISL_2490110, EPI_ISL_2490111, EPI_ISL_2490112, EPI_ISL_2490113, EPI_ISL_2490114, EPI_ISL_2490115, EPI_ISL_2490116, EPI_ISL_2490117, EPI_ISL_2490118, EPI_ISL_2490119, EPI_ISL_2490120, EPI_ISL_2490121, EPI_ISL_2490123, EPI_ISL_2490125, EPI_ISL_2490126, EPI_ISL_2490127, EPI_ISL_2490128, EPI_ISL_2490129, EPI_ISL_2490130, EPI_ISL_2490132, EPI_ISL_2490133, EPI_ISL_2490134, EPI_ISL_2490135, EPI_ISL_2490136, EPI_ISL_2490137, EPI_ISL_2490138, EPI_ISL_2490139, EPI_ISL_2490141, EPI_ISL_2490143, EPI_ISL_2490144, EPI_ISL_2490145, EPI_ISL_2490146, EPI_ISL_2490147, EPI_ISL_2490148, EPI_ISL_2490149, EPI_ISL_2490150, EPI_ISL_2490151, EPI_ISL_2490152, EPI_ISL_2490153, EPI_ISL_2490154, EPI_ISL_2490155, EPI_ISL_2490156, EPI_ISL_2490157, EPI_ISL_2490158, EPI_ISL_2490159, EPI_ISL_2490160, EPI_ISL_2490161, EPI_ISL_2490163, EPI_ISL_2490164, EPI_ISL_2490165, EPI_ISL_2490166, EPI_ISL_2490167, EPI_ISL_2490168, EPI_ISL_2490169, EPI_ISL_2490170, EPI_ISL_2490171, EPI_ISL_2490172, EPI_ISL_2490173, EPI_ISL_2490174, EPI_ISL_2490175, EPI_ISL_2490176, EPI_ISL_2490177, EPI_ISL_2490179, EPI_ISL_2490180, EPI_ISL_2490181, EPI_ISL_2490182, EPI_ISL_2490183, EPI_ISL_2490184, EPI_ISL_2490185, EPI_ISL_2490186, EPI_ISL_2490187, EPI_ISL_2490188, EPI_ISL_2490189, EPI_ISL_2490190, EPI_ISL_2490191, EPI_ISL_2490192, EPI_ISL_2490193, EPI_ISL_2490194, EPI_ISL_2490195, EPI_ISL_2490196, EPI_ISL_2490197, EPI_ISL_2490198, EPI_ISL_2490199, EPI_ISL_2490200, EPI_ISL_2490201, EPI_ISL_2490203, EPI_ISL_2490204, EPI_ISL_2490205, EPI_ISL_2490206, EPI_ISL_2490207, EPI_ISL_2490208, EPI_ISL_2490209, EPI_ISL_2490210, EPI_ISL_2490212, EPI_ISL_2490213, EPI_ISL_2490214, EPI_ISL_2490217, EPI_ISL_2490218, EPI_ISL_2490221, EPI_ISL_2490222, EPI_ISL_2490223, EPI_ISL_2490224, EPI_ISL_2490225, EPI_ISL_2490226, EPI_ISL_2490227, EPI_ISL_2490228, EPI_ISL_2490229, EPI_ISL_2490230, EPI_ISL_2490231, EPI_ISL_2490232, EPI_ISL_2490233, EPI_ISL_2490234, EPI_ISL_2490235, EPI_ISL_2490236, EPI_ISL_2490237, EPI_ISL_2490238, EPI_ISL_2490240, EPI_ISL_2490241, EPI_ISL_2490242, EPI_ISL_2490243, EPI_ISL_2490244, EPI_ISL_2490245, EPI_ISL_2490246, EPI_ISL_2490249, EPI_ISL_2490250, EPI_ISL_2490251, EPI_ISL_2490253, EPI_ISL_2490254, EPI_ISL_2490255, EPI_ISL_2490256, EPI_ISL_2490257, EPI_ISL_2490258, EPI_ISL_2490259, EPI_ISL_2490260, EPI_ISL_2490261, EPI_ISL_2490262, EPI_ISL_2490263, EPI_ISL_2490264, EPI_ISL_2490265 | | | |
| see above | Northwest Laboratory | Seattle Flu Study | Deborah A. Nickerson, Chris D. Frazar, Jover Lee, Benjamin Pelle, Erica Ryke, Matthew Richardson, Amanda Adler, Elisabeth Brandstetter, Peter D. Han, Kairsten Fay, Misja Ilcisin, Kirsten Lacombe, Thomas R. Sibley, Melissa Truong, Caitlin R. Wolf, Romesh Gautom, Geoff Melly, Brian Hiatt, Philip Dykema, Scott Lindquist, Michael Boeckh, Janet A. Englund, Michael Famulare, Barry R. Lutz, Mark J. Rieder, Lea M. Starita, Matthew Thompson, Helen Y. Chu, Jay Shendure, Trevor Bedford |
| EPI_ISL_2490266, EPI_ISL_2490267, EPI_ISL_2490269, EPI_ISL_2490270, EPI_ISL_2490271, EPI_ISL_2490272, EPI_ISL_2490273, EPI_ISL_2490274, EPI_ISL_2490275, EPI_ISL_2490276, EPI_ISL_2490277, EPI_ISL_2490278, EPI_ISL_2490279, EPI_ISL_2490280, EPI_ISL_2490281, EPI_ISL_2490282, EPI_ISL_2490283, EPI_ISL_2490284, EPI_ISL_2490285, EPI_ISL_2490286, EPI_ISL_2490288, EPI_ISL_2490289, EPI_ISL_2490290, EPI_ISL_2490291, EPI_ISL_2490292 | | | |
| see above | Incyte Diagnostics Spokane | Seattle Flu Study | Deborah A. Nickerson, Chris D. Frazar, Jover Lee, Benjamin Pelle, Erica Ryke, Matthew Richardson, Amanda Adler, Elisabeth Brandstetter, Peter D. Han, Kairsten Fay, Misja Ilcisin, Kirsten Lacombe, Thomas R. Sibley, Melissa Truong, Caitlin R. Wolf, Romesh Gautom, Geoff Melly, Brian Hiatt, Philip Dykema, Scott Lindquist, Michael Boeckh, Janet A. Englund, Michael Famulare, Barry R. Lutz, Mark J. Rieder, Lea M. Starita, Matthew Thompson, Helen Y. Chu, Jay Shendure, Trevor Bedford |
| EPI_ISL_2490293, EPI_ISL_2490294, EPI_ISL_2490295 | Seattle Flu Study | Seattle Flu Study | Deborah A. Nickerson, Chris D. Frazar, Jover Lee, Benjamin Pelle, Erica Ryke, Matthew Richardson, Amanda Adler, Elisabeth Brandstetter, Peter D. Han, Kairsten Fay, Misja Ilcisin, Kirsten Lacombe, Thomas R. Sibley, Melissa Truong, Caitlin R. Wolf, Michael Boeckh, Janet A. Englund, Michael Famulare, Barry R. Lutz, Mark J. Rieder, Lea M. Starita, Matthew Thompson, Jay Shendure, Trevor Bedford, Helen Y. Chu |
| EPI_ISL_2490310 | Unidad de Investigación Biomédica de Zacatecas (UIBZ) | Centro de Investigación en Enfermedades Infecciosas (CIENI), Instituto Nacional de Enfermedades Respiratorias (INER) | Consorcio Mexicano de Vigilancia Genómica (CoViGen-Mex). Authors (in alphabetical order): Julio Elias Alvarado-Yaah, Carlos F. Arias, Santiago Ávila-Ríos, Eduardo Becerril-Vargas, Víctor Hugo Borja-Aburto, Celia Boukadida, Cristóbal Cháidez-Quiróz, Juan Bautista Chale-Dzul, Ricardo Ciria Merce, Andreu Comas-García, Célida Duque Molina, Julissa Enciso-Ibarra, José Antonio Enciso-Moreno, Gloria Elena Espinosa-Ayala, Fernando Fontove-Herrera, Daniel Fregoso-Rueda, Victor Eduardo Garcia-Arias, Alejandra Garcia-Gasca, Bruno Gomez-Gil, Jean Pierre González, Irvin González-López, Concepción Grajales-Muñiz, Ricardo Grande, Rosa María Gutierrez Rios, Alejandra Hernández-Terán, Alfredo Herrera-Estrella, Carla Ivón Herrera-Najera, Pavel Isa, Daniel Lira Morales, Susana Lopez, Antonio Loza Román, Brenda Irasema Maldonado-Meza, Bernardo Martínez-Miguel, José Arturo Martínez-Orozco, Célida Martínez- Rodríguez, Margarita Matías-Florentino, Fidencio Mejía-Nepomuceno, María Guadalupe de Jesús Mireles-Rivera, Gloria María Molina-Salinas, Hector Montoya-Fuentes, , Mario Mújica-Sánchez, José Esteban Muñoz-Medina, José de Jesús Nuñez-Contreras, Alicia Ocaña-Mondragón, Luis Alberto Ochoa-Carrera, Hector Esteban Paz-Juárez, Marissa Perez-Garcia, Francisco Pulido, Helen Haydee Fernanda Ramírez-Plascencia, Jorge Salas-Hernández, Angel Gustavo Salas-Lais, Alejandro Sanchez-Flores, Clara Esperanza Santacruz-Tinoco, María Guadalupe Santiago-Mauricio, Selene Zárate, Nelly Sélem-Mojica, Blanca Taboada, Gloria Vazquez, Joel Armando Vázquez-Pérez. |
| EPI_ISL_2490311 | Unidad de Investigación Médica de Yucatán (UIMY) | Centro de Investigación en Enfermedades Infecciosas (CIENI), Instituto Nacional de Enfermedades Respiratorias (INER) | Consorcio Mexicano de Vigilancia Genómica (CoViGen-Mex). Authors (in alphabetical order): Julio Elias Alvarado-Yaah, Carlos F. Arias, Santiago Ávila-Ríos, Eduardo Becerril-Vargas, Víctor Hugo Borja-Aburto, Celia Boukadida, Cristóbal Cháidez-Quiróz, Juan Bautista Chale-Dzul, Ricardo Ciria Merce, Andreu Comas-García, Célida Duque Molina, Julissa Enciso-Ibarra, José Antonio Enciso-Moreno, Gloria Elena Espinosa-Ayala, Fernando Fontove-Herrera, Daniel Fregoso-Rueda, Victor Eduardo Garcia-Arias, Alejandra Garcia-Gasca, Bruno Gomez-Gil, Jean Pierre González, Irvin González-López, Concepción Grajales-Muñiz, Ricardo Grande, Rosa María Gutierrez Rios, Alejandra Hernández-Terán, Alfredo Herrera-Estrella, Carla Ivón Herrera-Najera, Pavel Isa, Daniel Lira Morales, Susana Lopez, Antonio Loza Román, Brenda Irasema Maldonado-Meza, Bernardo Martínez-Miguel, José Arturo Martínez-Orozco, Célida Martínez- Rodríguez, Margarita Matías-Florentino, Fidencio Mejía-Nepomuceno, María Guadalupe de Jesús Mireles-Rivera, Gloria María Molina-Salinas, Hector Montoya-Fuentes, , Mario Mújica-Sánchez, José Esteban Muñoz-Medina, José de Jesús Nuñez-Contreras, Alicia Ocaña-Mondragón, Luis Alberto Ochoa-Carrera, Hector Esteban Paz-Juárez, Marissa Perez-Garcia, Francisco Pulido, Helen Haydee Fernanda Ramírez-Plascencia, Jorge Salas-Hernández, Angel Gustavo Salas-Lais, Alejandro Sanchez-Flores, Clara Esperanza Santacruz-Tinoco, María Guadalupe Santiago-Mauricio, Selene Zárate, Nelly Sélem-Mojica, Blanca Taboada, Gloria Vazquez, Joel Armando Vázquez-Pérez. |
| EPI_ISL_2490312, EPI_ISL_2490313, EPI_ISL_2490314, EPI_ISL_2490315, EPI_ISL_2490316, EPI_ISL_2490317, EPI_ISL_2490318, EPI_ISL_2490319, EPI_ISL_2490320, EPI_ISL_2490321, EPI_ISL_2490322, EPI_ISL_2490323, EPI_ISL_2490324, EPI_ISL_2490325, EPI_ISL_2490326 | | | |
| see above | Centro de Investigación Biomédica del Noreste (CIBIN) | Centro de Investigación en Enfermedades Infecciosas (CIENI), Instituto Nacional de Enfermedades Respiratorias (INER) | Consorcio Mexicano de Vigilancia Genómica (CoViGen-Mex). Authors (in alphabetical order): Julio Elias Alvarado-Yaah, Carlos F. Arias, Santiago Ávila-Ríos, Eduardo Becerril-Vargas, Víctor Hugo Borja-Aburto, Celia Boukadida, Cristóbal Cháidez-Quiróz, Juan Bautista Chale-Dzul, Ricardo Ciria Merce, Andreu Comas-García, Célida Duque Molina, Julissa Enciso-Ibarra, José Antonio Enciso-Moreno, Gloria Elena Espinosa-Ayala, Fernando Fontove-Herrera, Daniel Fregoso-Rueda, Victor Eduardo Garcia-Arias, Alejandra Garcia-Gasca, Bruno Gomez-Gil, Jean Pierre González, Irvin González-López, Concepción Grajales-Muñiz, Ricardo Grande, Rosa María Gutierrez Rios, Alejandra Hernández-Terán, Alfredo Herrera-Estrella, Carla Ivón Herrera-Najera, Pavel Isa, Daniel Lira Morales, Susana Lopez, Antonio Loza Román, Brenda Irasema Maldonado-Meza, Bernardo Martínez-Miguel, José Arturo Martínez-Orozco, Célida Martínez- Rodríguez, Margarita Matías-Florentino, Fidencio Mejía-Nepomuceno, María Guadalupe de Jesús Mireles-Rivera, Gloria María Molina-Salinas, Hector Montoya-Fuentes, , Mario Mújica-Sánchez, José Esteban Muñoz-Medina, José de Jesús Nuñez-Contreras, Alicia Ocaña-Mondragón, Luis Alberto Ochoa-Carrera, Hector Esteban Paz-Juárez, Marissa Perez-Garcia, Francisco Pulido, Helen Haydee Fernanda Ramírez-Plascencia, Jorge Salas-Hernández, Angel Gustavo Salas-Lais, Alejandro Sanchez-Flores, Clara Esperanza Santacruz-Tinoco, María Guadalupe Santiago-Mauricio, Selene Zárate, Nelly Sélem-Mojica, Blanca Taboada, Gloria Vazquez, Joel Armando Vázquez-Pérez. |
| EPI_ISL_2490327, EPI_ISL_2490328, EPI_ISL_2490329, EPI_ISL_2490330 | Unidad de Investigación Médica de Yucatán (UIMY) | Centro de Investigación en Enfermedades Infecciosas (CIENI), Instituto Nacional de Enfermedades Respiratorias (INER) | Consorcio Mexicano de Vigilancia Genómica (CoViGen-Mex). Authors (in alphabetical order): Julio Elias Alvarado-Yaah, Carlos F. Arias, Santiago Ávila-Ríos, Eduardo Becerril-Vargas, Víctor Hugo Borja-Aburto, Celia Boukadida, Cristóbal Cháidez-Quiróz, Juan Bautista Chale-Dzul, Ricardo Ciria Merce, Andreu Comas-García, Célida Duque Molina, Julissa Enciso-Ibarra, José Antonio Enciso-Moreno, Gloria Elena Espinosa-Ayala, Fernando Fontove-Herrera, Daniel Fregoso-Rueda, Victor Eduardo Garcia-Arias, Alejandra Garcia-Gasca, Bruno Gomez-Gil, Jean Pierre González, Irvin González-López, Concepción Grajales-Muñiz, Ricardo Grande, Rosa María Gutierrez Rios, Alejandra Hernández-Terán, Alfredo Herrera-Estrella, Carla Ivón Herrera-Najera, Pavel Isa, Daniel Lira Morales, Susana Lopez, Antonio Loza Román, Brenda Irasema Maldonado-Meza, Bernardo Martínez-Miguel, José Arturo Martínez-Orozco, Célida Martínez- Rodríguez, Margarita Matías-Florentino, Fidencio Mejía-Nepomuceno, María Guadalupe de Jesús Mireles-Rivera, Gloria María Molina-Salinas, Hector Montoya-Fuentes, , Mario Mújica-Sánchez, José Esteban Muñoz-Medina, José de Jesús Nuñez-Contreras, Alicia Ocaña-Mondragón, Luis Alberto Ochoa-Carrera, Hector Esteban Paz-Juárez, Marissa Perez-Garcia, Francisco Pulido, Helen Haydee Fernanda Ramírez-Plascencia, Jorge Salas-Hernández, Angel Gustavo Salas-Lais, Alejandro Sanchez-Flores, Clara Esperanza Santacruz-Tinoco, María Guadalupe Santiago-Mauricio, Selene Zárate, Nelly Sélem-Mojica, Blanca Taboada, Gloria Vazquez, Joel Armando Vázquez-Pérez. |
| EPI_ISL_2490331, EPI_ISL_2490332 | Centro de Investigación Biomédica de Occidente (CIBO) | Centro de Investigación en Enfermedades Infecciosas (CIENI), Instituto Nacional de Enfermedades Respiratorias (INER) | Consorcio Mexicano de Vigilancia Genómica (CoViGen-Mex). Authors (in alphabetical order): Julio Elias Alvarado-Yaah, Carlos F. Arias, Santiago Ávila-Ríos, Eduardo Becerril-Vargas, Víctor Hugo Borja-Aburto, Celia Boukadida, Cristóbal Cháidez-Quiróz, Juan Bautista Chale-Dzul, Ricardo Ciria Merce, Andreu Comas-García, Célida Duque Molina, Julissa Enciso-Ibarra, José Antonio Enciso-Moreno, Gloria Elena Espinosa-Ayala, Fernando Fontove-Herrera, Daniel Fregoso-Rueda, Victor Eduardo Garcia-Arias, Alejandra Garcia-Gasca, Bruno Gomez-Gil, Jean Pierre González, Irvin González-López, Concepción Grajales-Muñiz, Ricardo Grande, Rosa María Gutierrez Rios, Alejandra Hernández-Terán, Alfredo Herrera-Estrella, Carla Ivón Herrera-Najera, Pavel Isa, Daniel Lira Morales, Susana Lopez, Antonio Loza Román, Brenda Irasema Maldonado-Meza, Bernardo Martínez-Miguel, José Arturo Martínez-Orozco, Célida Martínez- Rodríguez, Margarita Matías-Florentino, Fidencio Mejía-Nepomuceno, María Guadalupe de Jesús Mireles-Rivera, Gloria María Molina-Salinas, Hector Montoya-Fuentes, , Mario Mújica-Sánchez, José Esteban Muñoz-Medina, José de Jesús Nuñez-Contreras, Alicia Ocaña-Mondragón, Luis Alberto Ochoa-Carrera, Hector Esteban Paz-Juárez, Marissa Perez-Garcia, Francisco Pulido, Helen Haydee Fernanda Ramírez-Plascencia, Jorge Salas-Hernández, Angel Gustavo Salas-Lais, Alejandro Sanchez-Flores, Clara Esperanza Santacruz-Tinoco, María Guadalupe Santiago-Mauricio, Selene Zárate, Nelly Sélem-Mojica, Blanca Taboada, Gloria Vazquez, Joel Armando Vázquez-Pérez. |
| EPI_ISL_2490333 | Unidad de Investigación Biomédica de Zacatecas (UIBZ) | Centro de Investigación en Enfermedades Infecciosas (CIENI), Instituto Nacional de Enfermedades Respiratorias (INER) | Consorcio Mexicano de Vigilancia Genómica (CoViGen-Mex). Authors (in alphabetical order): Julio Elias Alvarado-Yaah, Carlos F. Arias, Santiago Ávila-Ríos, Eduardo Becerril-Vargas, Víctor Hugo Borja-Aburto, Celia Boukadida, Cristóbal Cháidez-Quiróz, Juan Bautista Chale-Dzul, Ricardo Ciria Merce, Andreu Comas-García, Célida Duque Molina, Julissa Enciso-Ibarra, José Antonio Enciso-Moreno, Gloria Elena Espinosa-Ayala, Fernando Fontove-Herrera, Daniel Fregoso-Rueda, Victor Eduardo Garcia-Arias, Alejandra Garcia-Gasca, Bruno Gomez-Gil, Jean Pierre González, Irvin González-López, Concepción Grajales-Muñiz, Ricardo Grande, Rosa María Gutierrez Rios, Alejandra Hernández-Terán, Alfredo Herrera-Estrella, Carla Ivón Herrera-Najera, Pavel Isa, Daniel Lira Morales, Susana Lopez, Antonio Loza Román, Brenda Irasema Maldonado-Meza, Bernardo Martínez-Miguel, José Arturo Martínez-Orozco, Célida Martínez- Rodríguez, Margarita Matías-Florentino, Fidencio Mejía-Nepomuceno, María Guadalupe de Jesús Mireles-Rivera, Gloria María Molina-Salinas, Hector Montoya-Fuentes, , Mario Mújica-Sánchez, José Esteban Muñoz-Medina, José de Jesús Nuñez-Contreras, Alicia Ocaña-Mondragón, Luis Alberto Ochoa-Carrera, Hector Esteban Paz-Juárez, Marissa Perez-Garcia, Francisco Pulido, Helen Haydee Fernanda Ramírez-Plascencia, Jorge Salas-Hernández, Angel Gustavo Salas-Lais, Alejandro Sanchez-Flores, Clara Esperanza Santacruz-Tinoco, María Guadalupe Santiago-Mauricio, Selene Zárate, Nelly Sélem-Mojica, Blanca Taboada, Gloria Vazquez, Joel Armando Vázquez-Pérez. |
| EPI_ISL_2490334 | Centro de Investigación Biomédica del Noreste (CIBIN) | Centro de Investigación en Enfermedades Infecciosas (CIENI), Instituto Nacional de Enfermedades Respiratorias (INER) | Consorcio Mexicano de Vigilancia Genómica (CoViGen-Mex). Authors (in alphabetical order): Julio Elias Alvarado-Yaah, Carlos F. Arias, Santiago Ávila-Ríos, Eduardo Becerril-Vargas, Víctor Hugo Borja-Aburto, Celia Boukadida, Cristóbal Cháidez-Quiróz, Juan Bautista Chale-Dzul, Ricardo Ciria Merce, Andreu Comas-García, Célida Duque Molina, Julissa Enciso-Ibarra, José Antonio Enciso-Moreno, Gloria Elena Espinosa-Ayala, Fernando Fontove-Herrera, Daniel Fregoso-Rueda, Victor Eduardo Garcia-Arias, Alejandra Garcia-Gasca, Bruno Gomez-Gil, Jean Pierre González, Irvin González-López, Concepción Grajales-Muñiz, Ricardo Grande, Rosa María Gutierrez Rios, Alejandra Hernández-Terán, Alfredo Herrera-Estrella, Carla Ivón Herrera-Najera, Pavel Isa, Daniel Lira Morales, Susana Lopez, Antonio Loza Román, Brenda Irasema Maldonado-Meza, Bernardo Martínez-Miguel, José Arturo Martínez-Orozco, Célida Martínez- Rodríguez, Margarita Matías-Florentino, Fidencio Mejía-Nepomuceno, María Guadalupe de Jesús Mireles-Rivera, Gloria María Molina-Salinas, Hector Montoya-Fuentes, , Mario Mújica-Sánchez, José Esteban Muñoz-Medina, José de Jesús Nuñez-Contreras, Alicia Ocaña-Mondragón, Luis Alberto Ochoa-Carrera, Hector Esteban Paz-Juárez, Marissa Perez-Garcia, Francisco Pulido, Helen Haydee Fernanda Ramírez-Plascencia, Jorge Salas-Hernández, Angel Gustavo Salas-Lais, Alejandro Sanchez-Flores, Clara Esperanza Santacruz-Tinoco, María Guadalupe Santiago-Mauricio, Selene Zárate, Nelly Sélem-Mojica, Blanca Taboada, Gloria Vazquez, Joel Armando Vázquez-Pérez. |
| EPI_ISL_2490335 | Unidad de Investigación Biomédica de Zacatecas (UIBZ) | Centro de Investigación en Enfermedades Infecciosas (CIENI), Instituto Nacional de Enfermedades Respiratorias (INER) | Consorcio Mexicano de Vigilancia Genómica (CoViGen-Mex). Authors (in alphabetical order): Julio Elias Alvarado-Yaah, Carlos F. Arias, Santiago Ávila-Ríos, Eduardo Becerril-Vargas, Víctor Hugo Borja-Aburto, Celia Boukadida, Cristóbal Cháidez-Quiróz, Juan Bautista Chale-Dzul, Ricardo Ciria Merce, Andreu Comas-García, Célida Duque Molina, Julissa Enciso-Ibarra, José Antonio Enciso-Moreno, Gloria Elena Espinosa-Ayala, Fernando Fontove-Herrera, Daniel Fregoso-Rueda, Victor Eduardo Garcia-Arias, Alejandra Garcia-Gasca, Bruno Gomez-Gil, Jean Pierre González, Irvin González-López, Concepción Grajales-Muñiz, Ricardo Grande, Rosa María Gutierrez Rios, Alejandra Hernández-Terán, Alfredo Herrera-Estrella, Carla Ivón Herrera-Najera, Pavel Isa, Daniel Lira Morales, Susana Lopez, Antonio Loza Román, Brenda Irasema Maldonado-Meza, Bernardo Martínez-Miguel, José Arturo Martínez-Orozco, Célida Martínez- Rodríguez, Margarita Matías-Florentino, Fidencio Mejía-Nepomuceno, María Guadalupe de Jesús Mireles-Rivera, Gloria María Molina-Salinas, Hector Montoya-Fuentes, , Mario Mújica-Sánchez, José Esteban Muñoz-Medina, José de Jesús Nuñez-Contreras, Alicia Ocaña-Mondragón, Luis Alberto Ochoa-Carrera, Hector Esteban Paz-Juárez, Marissa Perez-Garcia, Francisco Pulido, Helen Haydee Fernanda Ramírez-Plascencia, Jorge Salas-Hernández, Angel Gustavo Salas-Lais, Alejandro Sanchez-Flores, Clara Esperanza Santacruz-Tinoco, María Guadalupe Santiago-Mauricio, Selene Zárate, Nelly Sélem-Mojica, Blanca Taboada, Gloria Vazquez, Joel Armando Vázquez-Pérez. |
| EPI_ISL_2490336 | Laboratorio Central de Epidemiología (LCE) | Centro de Investigación en Enfermedades Infecciosas (CIENI), Instituto Nacional de Enfermedades Respiratorias (INER) | Consorcio Mexicano de Vigilancia Genómica (CoViGen-Mex). Authors (in alphabetical order): Julio Elias Alvarado-Yaah, Carlos F. Arias, Santiago Ávila-Ríos, Eduardo Becerril-Vargas, Víctor Hugo Borja-Aburto, Celia Boukadida, Cristóbal Cháidez-Quiróz, Juan Bautista Chale-Dzul, Ricardo Ciria Merce, Andreu Comas-García, Célida Duque Molina, Julissa Enciso-Ibarra, José Antonio Enciso-Moreno, Gloria Elena Espinosa-Ayala, Fernando Fontove-Herrera, Daniel Fregoso-Rueda, Victor Eduardo Garcia-Arias, Alejandra Garcia-Gasca, Bruno Gomez-Gil, Jean Pierre González, Irvin González-López, Concepción Grajales-Muñiz, Ricardo Grande, Rosa María Gutierrez Rios, Alejandra Hernández-Terán, Alfredo Herrera-Estrella, Carla Ivón Herrera-Najera, Pavel Isa, Daniel Lira Morales, Susana Lopez, Antonio Loza Román, Brenda Irasema Maldonado-Meza, Bernardo Martínez-Miguel, José Arturo Martínez-Orozco, Célida Martínez- Rodríguez, Margarita Matías-Florentino, Fidencio Mejía-Nepomuceno, María Guadalupe de Jesús Mireles-Rivera, Gloria María Molina-Salinas, Hector Montoya-Fuentes, , Mario Mújica-Sánchez, José Esteban Muñoz-Medina, José de Jesús Nuñez-Contreras, Alicia Ocaña-Mondragón, Luis Alberto Ochoa-Carrera, Hector Esteban Paz-Juárez, Marissa Perez-Garcia, Francisco Pulido, Helen Haydee Fernanda Ramírez-Plascencia, Jorge Salas-Hernández, Angel Gustavo Salas-Lais, Alejandro Sanchez-Flores, Clara Esperanza Santacruz-Tinoco, María Guadalupe Santiago-Mauricio, Selene Zárate, Nelly Sélem-Mojica, Blanca Taboada, Gloria Vazquez, Joel Armando Vázquez-Pérez. |
| EPI_ISL_2490337, EPI_ISL_2490338, EPI_ISL_2490339, EPI_ISL_2490340, EPI_ISL_2490341, EPI_ISL_2490342 | Centro de Investigación Biomédica del Noreste (CIBIN) | Centro de Investigación en Enfermedades Infecciosas (CIENI), Instituto Nacional de Enfermedades Respiratorias (INER) | Consorcio Mexicano de Vigilancia Genómica (CoViGen-Mex). Authors (in alphabetical order): Julio Elias Alvarado-Yaah, Carlos F. Arias, Santiago Ávila-Ríos, Eduardo Becerril-Vargas, Víctor Hugo Borja-Aburto, Celia Boukadida, Cristóbal Cháidez-Quiróz, Juan Bautista Chale-Dzul, Ricardo Ciria Merce, Andreu Comas-García, Célida Duque Molina, Julissa Enciso-Ibarra, José Antonio Enciso-Moreno, Gloria Elena Espinosa-Ayala, Fernando Fontove-Herrera, Daniel Fregoso-Rueda, Victor Eduardo Garcia-Arias, Alejandra Garcia-Gasca, Bruno Gomez-Gil, Jean Pierre González, Irvin González-López, Concepción Grajales-Muñiz, Ricardo Grande, Rosa María Gutierrez Rios, Alejandra Hernández-Terán, Alfredo Herrera-Estrella, Carla Ivón Herrera-Najera, Pavel Isa, Daniel Lira Morales, Susana Lopez, Antonio Loza Román, Brenda Irasema Maldonado-Meza, Bernardo Martínez-Miguel, José Arturo Martínez-Orozco, Célida Martínez- Rodríguez, Margarita Matías-Florentino, Fidencio Mejía-Nepomuceno, María Guadalupe de Jesús Mireles-Rivera, Gloria María Molina-Salinas, Hector Montoya-Fuentes, , Mario Mújica-Sánchez, José Esteban Muñoz-Medina, José de Jesús Nuñez-Contreras, Alicia Ocaña-Mondragón, Luis Alberto Ochoa-Carrera, Hector Esteban Paz-Juárez, Marissa Perez-Garcia, Francisco Pulido, Helen Haydee Fernanda Ramírez-Plascencia, Jorge Salas-Hernández, Angel Gustavo Salas-Lais, Alejandro Sanchez-Flores, Clara Esperanza Santacruz-Tinoco, María Guadalupe Santiago-Mauricio, Selene Zárate, Nelly Sélem-Mojica, Blanca Taboada, Gloria Vazquez, Joel Armando Vázquez-Pérez. |
| EPI_ISL_2490343, EPI_ISL_2490344, EPI_ISL_2490345, EPI_ISL_2490346, EPI_ISL_2490347 | Unidad de Investigación Médica de Yucatán (UIMY) | Centro de Investigación en Enfermedades Infecciosas (CIENI), Instituto Nacional de Enfermedades Respiratorias (INER) | Consorcio Mexicano de Vigilancia Genómica (CoViGen-Mex). Authors (in alphabetical order): Julio Elias Alvarado-Yaah, Carlos F. Arias, Santiago Ávila-Ríos, Eduardo Becerril-Vargas, Víctor Hugo Borja-Aburto, Celia Boukadida, Cristóbal Cháidez-Quiróz, Juan Bautista Chale-Dzul, Ricardo Ciria Merce, Andreu Comas-García, Célida Duque Molina, Julissa Enciso-Ibarra, José Antonio Enciso-Moreno, Gloria Elena Espinosa-Ayala, Fernando Fontove-Herrera, Daniel Fregoso-Rueda, Victor Eduardo Garcia-Arias, Alejandra Garcia-Gasca, Bruno Gomez-Gil, Jean Pierre González, Irvin González-López, Concepción Grajales-Muñiz, Ricardo Grande, Rosa María Gutierrez Rios, Alejandra Hernández-Terán, Alfredo Herrera-Estrella, Carla Ivón Herrera-Najera, Pavel Isa, Daniel Lira Morales, Susana Lopez, Antonio Loza Román, Brenda Irasema Maldonado-Meza, Bernardo Martínez-Miguel, José Arturo Martínez-Orozco, Célida Martínez- Rodríguez, Margarita Matías-Florentino, Fidencio Mejía-Nepomuceno, María Guadalupe de Jesús Mireles-Rivera, Gloria María Molina-Salinas, Hector Montoya-Fuentes, , Mario Mújica-Sánchez, José Esteban Muñoz-Medina, José de Jesús Nuñez-Contreras, Alicia Ocaña-Mondragón, Luis Alberto Ochoa-Carrera, Hector Esteban Paz-Juárez, Marissa Perez-Garcia, Francisco Pulido, Helen Haydee Fernanda Ramírez-Plascencia, Jorge Salas-Hernández, Angel Gustavo Salas-Lais, Alejandro Sanchez-Flores, Clara Esperanza Santacruz-Tinoco, María Guadalupe Santiago-Mauricio, Selene Zárate, Nelly Sélem-Mojica, Blanca Taboada, Gloria Vazquez, Joel Armando Vázquez-Pérez. |
| EPI_ISL_2490348 | Unidad de Investigación Biomédica de Zacatecas (UIBZ) | Centro de Investigación en Enfermedades Infecciosas (CIENI), Instituto Nacional de Enfermedades Respiratorias (INER) | Consorcio Mexicano de Vigilancia Genómica (CoViGen-Mex). Authors (in alphabetical order): Julio Elias Alvarado-Yaah, Carlos F. Arias, Santiago Ávila-Ríos, Eduardo Becerril-Vargas, Víctor Hugo Borja-Aburto, Celia Boukadida, Cristóbal Cháidez-Quiróz, Juan Bautista Chale-Dzul, Ricardo Ciria Merce, Andreu Comas-García, Célida Duque Molina, Julissa Enciso-Ibarra, José Antonio Enciso-Moreno, Gloria Elena Espinosa-Ayala, Fernando Fontove-Herrera, Daniel Fregoso-Rueda, Victor Eduardo Garcia-Arias, Alejandra Garcia-Gasca, Bruno Gomez-Gil, Jean Pierre González, Irvin González-López, Concepción Grajales-Muñiz, Ricardo Grande, Rosa María Gutierrez Rios, Alejandra Hernández-Terán, Alfredo Herrera-Estrella, Carla Ivón Herrera-Najera, Pavel Isa, Daniel Lira Morales, Susana Lopez, Antonio Loza Román, Brenda Irasema Maldonado-Meza, Bernardo Martínez-Miguel, José Arturo Martínez-Orozco, Célida Martínez- Rodríguez, Margarita Matías-Florentino, Fidencio Mejía-Nepomuceno, María Guadalupe de Jesús Mireles-Rivera, Gloria María Molina-Salinas, Hector Montoya-Fuentes, , Mario Mújica-Sánchez, José Esteban Muñoz-Medina, José de Jesús Nuñez-Contreras, Alicia Ocaña-Mondragón, Luis Alberto Ochoa-Carrera, Hector Esteban Paz-Juárez, Marissa Perez-Garcia, Francisco Pulido, Helen Haydee Fernanda Ramírez-Plascencia, Jorge Salas-Hernández, Angel Gustavo Salas-Lais, Alejandro Sanchez-Flores, Clara Esperanza Santacruz-Tinoco, María Guadalupe Santiago-Mauricio, Selene Zárate, Nelly Sélem-Mojica, Blanca Taboada, Gloria Vazquez, Joel Armando Vázquez-Pérez. |
| EPI_ISL_2490349, | Laboratorio Central de Epidemiología | Centro de Investigación en Enfermedades | Consorcio Mexicano de Vigilancia Genómica (CoViGen-Mex). Authors (in alphabetical order): Julio Elias Alvarado-Yaah, Carlos F. Arias, Santiago Ávila-Ríos, Eduardo Becerril-Vargas, Víctor Hugo Borja-Aburto, Celia Boukadida, Cristóbal Cháidez-Quiróz, Juan Bautista Chale-Dzul, Ricardo |

| | | | |
|---|---|---|---|
| EPI_ISL_2490350, EPI_ISL_2490351, EPI_ISL_2490352, EPI_ISL_2490353, EPI_ISL_2490354, EPI_ISL_2490355 | (LCE) | Infecciosas (CIENI), Instituto Nacional de Enfermedades Respiratorias (INER) | Ciria Merce, Andreu Comas-García, Célida Duque Molina, Julissa Enciso-Ibarra, José Antonio Enciso-Moreno, Gloria Elena Espinosa-Ayala, Fernando Fontove-Herrera, Daniel Fregoso-Rueda, Victor Eduardo Garcia-Arias, Alejandra Garcia-Gasca, Bruno Gomez-Gil, Jean Pierre González, Irvin González-López, Concepción Grajales-Muñiz, Ricardo Grande, Rosa María Gutierrez Rios, Alejandra Hernández-Terán, Alfredo Herrera-Estrella, Carla Ivón Herrera-Najera, Pavel Isa, Daniel Lira Morales, Susana Lopez, Antonio Loza Román, Brenda Irasema Maldonado-Meza, Bernardo Martínez-Miguel, José Arturo Martínez-Orozco, Célida Martínez- Rodríguez, Margarita Matías-Florentino, Fidencio Mejía-Nepomuceno, María Guadalupe de Jesús Mireles-Rivera, Gloria María Molina-Salinas, Hector Montoya-Fuentes, , Mario Mújica-Sánchez, José Esteban Muñoz-Medina, José de Jesús Nuñez-Contreras, Alicia Ocaña-Mondragón, Luis Alberto Ochoa-Carrera, Hector Esteban Paz-Juárez, Marissa Perez-Garcia, Francisco Pulido, Helen Haydee Fernanda Ramírez-Plascencia, Jorge Salas-Hernández, Angel Gustavo Salas-Lais, Alejandro Sanchez-Flores, Clara Esperanza Santacruz-Tinoco, María Guadalupe Santiago-Mauricio, Selene Zárate, Nelly Sélem-Mojica, Blanca Taboada, Gloria Vazquez, Joel Armando Vázquez-Pérez. |
| EPI_ISL_2490356 | Centro de Investigación Biomédica del Noreste (CIBIN) | Centro de Investigación en Enfermedades Infecciosas (CIENI), Instituto Nacional de Enfermedades Respiratorias (INER) | Consorcio Mexicano de Vigilancia Genómica (CoViGen-Mex). Authors (in alphabetical order): Julio Elias Alvarado-Yaah, Carlos F. Arias, Santiago Ávila-Ríos, Eduardo Becerril-Vargas, Víctor Hugo Borja-Aburto, Celia Boukadida, Cristóbal Cháidez-Quiróz, Juan Bautista Chale-Dzul, Ricardo Ciria Merce, Andreu Comas-García, Célida Duque Molina, Julissa Enciso-Ibarra, José Antonio Enciso-Moreno, Gloria Elena Espinosa-Ayala, Fernando Fontove-Herrera, Daniel Fregoso-Rueda, Victor Eduardo Garcia-Arias, Alejandra Garcia-Gasca, Bruno Gomez-Gil, Jean Pierre González, Irvin González-López, Concepción Grajales-Muñiz, Ricardo Grande, Rosa María Gutierrez Rios, Alejandra Hernández-Terán, Alfredo Herrera-Estrella, Carla Ivón Herrera-Najera, Pavel Isa, Daniel Lira Morales, Susana Lopez, Antonio Loza Román, Brenda Irasema Maldonado-Meza, Bernardo Martínez-Miguel, José Arturo Martínez-Orozco, Célida Martínez- Rodríguez, Margarita Matías-Florentino, Fidencio Mejía-Nepomuceno, María Guadalupe de Jesús Mireles-Rivera, Gloria María Molina-Salinas, Hector Montoya-Fuentes, , Mario Mújica-Sánchez, José Esteban Muñoz-Medina, José de Jesús Nuñez-Contreras, Alicia Ocaña-Mondragón, Luis Alberto Ochoa-Carrera, Hector Esteban Paz-Juárez, Marissa Perez-Garcia, Francisco Pulido, Helen Haydee Fernanda Ramírez-Plascencia, Jorge Salas-Hernández, Angel Gustavo Salas-Lais, Alejandro Sanchez-Flores, Clara Esperanza Santacruz-Tinoco, María Guadalupe Santiago-Mauricio, Selene Zárate, Nelly Sélem-Mojica, Blanca Taboada, Gloria Vazquez, Joel Armando Vázquez-Pérez. |
| EPI_ISL_2490357 | Unidad de Investigación Médica de Yucatán (UIMY) | Centro de Investigación en Enfermedades Infecciosas (CIENI), Instituto Nacional de Enfermedades Respiratorias (INER) | Consorcio Mexicano de Vigilancia Genómica (CoViGen-Mex). Authors (in alphabetical order): Julio Elias Alvarado-Yaah, Carlos F. Arias, Santiago Ávila-Ríos, Eduardo Becerril-Vargas, Víctor Hugo Borja-Aburto, Celia Boukadida, Cristóbal Cháidez-Quiróz, Juan Bautista Chale-Dzul, Ricardo Ciria Merce, Andreu Comas-García, Célida Duque Molina, Julissa Enciso-Ibarra, José Antonio Enciso-Moreno, Gloria Elena Espinosa-Ayala, Fernando Fontove-Herrera, Daniel Fregoso-Rueda, Victor Eduardo Garcia-Arias, Alejandra Garcia-Gasca, Bruno Gomez-Gil, Jean Pierre González, Irvin González-López, Concepción Grajales-Muñiz, Ricardo Grande, Rosa María Gutierrez Rios, Alejandra Hernández-Terán, Alfredo Herrera-Estrella, Carla Ivón Herrera-Najera, Pavel Isa, Daniel Lira Morales, Susana Lopez, Antonio Loza Román, Brenda Irasema Maldonado-Meza, Bernardo Martínez-Miguel, José Arturo Martínez-Orozco, Célida Martínez- Rodríguez, Margarita Matías-Florentino, Fidencio Mejía-Nepomuceno, María Guadalupe de Jesús Mireles-Rivera, Gloria María Molina-Salinas, Hector Montoya-Fuentes, , Mario Mújica-Sánchez, José Esteban Muñoz-Medina, José de Jesús Nuñez-Contreras, Alicia Ocaña-Mondragón, Luis Alberto Ochoa-Carrera, Hector Esteban Paz-Juárez, Marissa Perez-Garcia, Francisco Pulido, Helen Haydee Fernanda Ramírez-Plascencia, Jorge Salas-Hernández, Angel Gustavo Salas-Lais, Alejandro Sanchez-Flores, Clara Esperanza Santacruz-Tinoco, María Guadalupe Santiago-Mauricio, Selene Zárate, Nelly Sélem-Mojica, Blanca Taboada, Gloria Vazquez, Joel Armando Vázquez-Pérez. |
| EPI_ISL_2490358 | Unidad de Investigación Biomédica de Zacatecas (UIBZ) | Centro de Investigación en Enfermedades Infecciosas (CIENI), Instituto Nacional de Enfermedades Respiratorias (INER) | Consorcio Mexicano de Vigilancia Genómica (CoViGen-Mex). Authors (in alphabetical order): Julio Elias Alvarado-Yaah, Carlos F. Arias, Santiago Ávila-Ríos, Eduardo Becerril-Vargas, Víctor Hugo Borja-Aburto, Celia Boukadida, Cristóbal Cháidez-Quiróz, Juan Bautista Chale-Dzul, Ricardo Ciria Merce, Andreu Comas-García, Célida Duque Molina, Julissa Enciso-Ibarra, José Antonio Enciso-Moreno, Gloria Elena Espinosa-Ayala, Fernando Fontove-Herrera, Daniel Fregoso-Rueda, Victor Eduardo Garcia-Arias, Alejandra Garcia-Gasca, Bruno Gomez-Gil, Jean Pierre González, Irvin González-López, Concepción Grajales-Muñiz, Ricardo Grande, Rosa María Gutierrez Rios, Alejandra Hernández-Terán, Alfredo Herrera-Estrella, Carla Ivón Herrera-Najera, Pavel Isa, Daniel Lira Morales, Susana Lopez, Antonio Loza Román, Brenda Irasema Maldonado-Meza, Bernardo Martínez-Miguel, José Arturo Martínez-Orozco, Célida Martínez- Rodríguez, Margarita Matías-Florentino, Fidencio Mejía-Nepomuceno, María Guadalupe de Jesús Mireles-Rivera, Gloria María Molina-Salinas, Hector Montoya-Fuentes, , Mario Mújica-Sánchez, José Esteban Muñoz-Medina, José de Jesús Nuñez-Contreras, Alicia Ocaña-Mondragón, Luis Alberto Ochoa-Carrera, Hector Esteban Paz-Juárez, Marissa Perez-Garcia, Francisco Pulido, Helen Haydee Fernanda Ramírez-Plascencia, Jorge Salas-Hernández, Angel Gustavo Salas-Lais, Alejandro Sanchez-Flores, Clara Esperanza Santacruz-Tinoco, María Guadalupe Santiago-Mauricio, Selene Zárate, Nelly Sélem-Mojica, Blanca Taboada, Gloria Vazquez, Joel Armando Vázquez-Pérez. |
| EPI_ISL_2490359, EPI_ISL_2490360, EPI_ISL_2490361, EPI_ISL_2490362, EPI_ISL_2490363, EPI_ISL_2490364, EPI_ISL_2490365, EPI_ISL_2490366 | Centro de Investigación Biomédica del Noreste (CIBIN) | Centro de Investigación en Enfermedades Infecciosas (CIENI), Instituto Nacional de Enfermedades Respiratorias (INER) | Consorcio Mexicano de Vigilancia Genómica (CoViGen-Mex). Authors (in alphabetical order): Julio Elias Alvarado-Yaah, Carlos F. Arias, Santiago Ávila-Ríos, Eduardo Becerril-Vargas, Víctor Hugo Borja-Aburto, Celia Boukadida, Cristóbal Cháidez-Quiróz, Juan Bautista Chale-Dzul, Ricardo Ciria Merce, Andreu Comas-García, Célida Duque Molina, Julissa Enciso-Ibarra, José Antonio Enciso-Moreno, Gloria Elena Espinosa-Ayala, Fernando Fontove-Herrera, Daniel Fregoso-Rueda, Victor Eduardo Garcia-Arias, Alejandra Garcia-Gasca, Bruno Gomez-Gil, Jean Pierre González, Irvin González-López, Concepción Grajales-Muñiz, Ricardo Grande, Rosa María Gutierrez Rios, Alejandra Hernández-Terán, Alfredo Herrera-Estrella, Carla Ivón Herrera-Najera, Pavel Isa, Daniel Lira Morales, Susana Lopez, Antonio Loza Román, Brenda Irasema Maldonado-Meza, Bernardo Martínez-Miguel, José Arturo Martínez-Orozco, Célida Martínez- Rodríguez, Margarita Matías-Florentino, Fidencio Mejía-Nepomuceno, María Guadalupe de Jesús Mireles-Rivera, Gloria María Molina-Salinas, Hector Montoya-Fuentes, , Mario Mújica-Sánchez, José Esteban Muñoz-Medina, José de Jesús Nuñez-Contreras, Alicia Ocaña-Mondragón, Luis Alberto Ochoa-Carrera, Hector Esteban Paz-Juárez, Marissa Perez-Garcia, Francisco Pulido, Helen Haydee Fernanda Ramírez-Plascencia, Jorge Salas-Hernández, Angel Gustavo Salas-Lais, Alejandro Sanchez-Flores, Clara Esperanza Santacruz-Tinoco, María Guadalupe Santiago-Mauricio, Selene Zárate, Nelly Sélem-Mojica, Blanca Taboada, Gloria Vazquez, Joel Armando Vázquez-Pérez. |
| EPI_ISL_2490367, EPI_ISL_2490368, EPI_ISL_2490369, EPI_ISL_2490370, EPI_ISL_2490371, EPI_ISL_2490372, EPI_ISL_2490373, EPI_ISL_2490374, EPI_ISL_2490375, EPI_ISL_2490376, EPI_ISL_2490377, EPI_ISL_2490378, EPI_ISL_2490379, EPI_ISL_2490380, EPI_ISL_2490381 | | | |
| see above | Unidad de Investigación Médica de Yucatán (UIMY) | Centro de Investigación en Enfermedades Infecciosas (CIENI), Instituto Nacional de Enfermedades Respiratorias (INER) | Consorcio Mexicano de Vigilancia Genómica (CoViGen-Mex). Authors (in alphabetical order): Julio Elias Alvarado-Yaah, Carlos F. Arias, Santiago Ávila-Ríos, Eduardo Becerril-Vargas, Víctor Hugo Borja-Aburto, Celia Boukadida, Cristóbal Cháidez-Quiróz, Juan Bautista Chale-Dzul, Ricardo Ciria Merce, Andreu Comas-García, Célida Duque Molina, Julissa Enciso-Ibarra, José Antonio Enciso-Moreno, Gloria Elena Espinosa-Ayala, Fernando Fontove-Herrera, Daniel Fregoso-Rueda, Victor Eduardo Garcia-Arias, Alejandra Garcia-Gasca, Bruno Gomez-Gil, Jean Pierre González, Irvin González-López, Concepción Grajales-Muñiz, Ricardo Grande, Rosa María Gutierrez Rios, Alejandra Hernández-Terán, Alfredo Herrera-Estrella, Carla Ivón Herrera-Najera, Pavel Isa, Daniel Lira Morales, Susana Lopez, Antonio Loza Román, Brenda Irasema Maldonado-Meza, Bernardo Martínez-Miguel, José Arturo Martínez-Orozco, Célida Martínez- Rodríguez, Margarita Matías-Florentino, Fidencio Mejía-Nepomuceno, María Guadalupe de Jesús Mireles-Rivera, Gloria María Molina-Salinas, Hector Montoya-Fuentes, , Mario Mújica-Sánchez, José Esteban Muñoz-Medina, José de Jesús Nuñez-Contreras, Alicia Ocaña-Mondragón, Luis Alberto Ochoa-Carrera, Hector Esteban Paz-Juárez, Marissa Perez-Garcia, Francisco Pulido, Helen Haydee Fernanda Ramírez-Plascencia, Jorge Salas-Hernández, Angel Gustavo Salas-Lais, Alejandro Sanchez-Flores, Clara Esperanza Santacruz-Tinoco, María Guadalupe Santiago-Mauricio, Selene Zárate, Nelly Sélem-Mojica, Blanca Taboada, Gloria Vazquez, Joel Armando Vázquez-Pérez. |
| EPI_ISL_2490382, EPI_ISL_2490383, EPI_ISL_2490384 | Centro de Investigación Biomédica de Occidente (CIBO) | Centro de Investigación en Enfermedades Infecciosas (CIENI), Instituto Nacional de Enfermedades Respiratorias (INER) | Consorcio Mexicano de Vigilancia Genómica (CoViGen-Mex). Authors (in alphabetical order): Julio Elias Alvarado-Yaah, Carlos F. Arias, Santiago Ávila-Ríos, Eduardo Becerril-Vargas, Víctor Hugo Borja-Aburto, Celia Boukadida, Cristóbal Cháidez-Quiróz, Juan Bautista Chale-Dzul, Ricardo Ciria Merce, Andreu Comas-García, Célida Duque Molina, Julissa Enciso-Ibarra, José Antonio Enciso-Moreno, Gloria Elena Espinosa-Ayala, Fernando Fontove-Herrera, Daniel Fregoso-Rueda, Victor Eduardo Garcia-Arias, Alejandra Garcia-Gasca, Bruno Gomez-Gil, Jean Pierre González, Irvin González-López, Concepción Grajales-Muñiz, Ricardo Grande, Rosa María Gutierrez Rios, Alejandra Hernández-Terán, Alfredo Herrera-Estrella, Carla Ivón Herrera-Najera, Pavel Isa, Daniel Lira Morales, Susana Lopez, Antonio Loza Román, Brenda Irasema Maldonado-Meza, Bernardo Martínez-Miguel, José Arturo Martínez-Orozco, Célida Martínez- Rodríguez, Margarita Matías-Florentino, Fidencio Mejía-Nepomuceno, María Guadalupe de Jesús Mireles-Rivera, Gloria María Molina-Salinas, Hector Montoya-Fuentes, , Mario Mújica-Sánchez, José Esteban Muñoz-Medina, José de Jesús Nuñez-Contreras, Alicia Ocaña-Mondragón, Luis Alberto Ochoa-Carrera, Hector Esteban Paz-Juárez, Marissa Perez-Garcia, Francisco Pulido, Helen Haydee Fernanda Ramírez-Plascencia, Jorge Salas-Hernández, Angel Gustavo Salas-Lais, Alejandro Sanchez-Flores, Clara Esperanza Santacruz-Tinoco, María Guadalupe Santiago-Mauricio, Selene Zárate, Nelly Sélem-Mojica, Blanca Taboada, Gloria Vazquez, Joel Armando Vázquez-Pérez. |
| EPI_ISL_2490385, EPI_ISL_2490386 | Unidad de Investigación Biomédica de Zacatecas (UIBZ) | Centro de Investigación en Enfermedades Infecciosas (CIENI), Instituto Nacional de Enfermedades Respiratorias (INER) | Consorcio Mexicano de Vigilancia Genómica (CoViGen-Mex). Authors (in alphabetical order): Julio Elias Alvarado-Yaah, Carlos F. Arias, Santiago Ávila-Ríos, Eduardo Becerril-Vargas, Víctor Hugo Borja-Aburto, Celia Boukadida, Cristóbal Cháidez-Quiróz, Juan Bautista Chale-Dzul, Ricardo Ciria Merce, Andreu Comas-García, Célida Duque Molina, Julissa Enciso-Ibarra, José Antonio Enciso-Moreno, Gloria Elena Espinosa-Ayala, Fernando Fontove-Herrera, Daniel Fregoso-Rueda, Victor Eduardo Garcia-Arias, Alejandra Garcia-Gasca, Bruno Gomez-Gil, Jean Pierre González, Irvin González-López, Concepción Grajales-Muñiz, Ricardo Grande, Rosa María Gutierrez Rios, Alejandra Hernández-Terán, Alfredo Herrera-Estrella, Carla Ivón Herrera-Najera, Pavel Isa, Daniel Lira Morales, Susana Lopez, Antonio Loza Román, Brenda Irasema Maldonado-Meza, Bernardo Martínez-Miguel, José Arturo Martínez-Orozco, Célida Martínez- Rodríguez, Margarita Matías-Florentino, Fidencio Mejía-Nepomuceno, María Guadalupe de Jesús Mireles-Rivera, Gloria María Molina-Salinas, Hector Montoya-Fuentes, , Mario Mújica-Sánchez, José Esteban Muñoz-Medina, José de Jesús Nuñez-Contreras, Alicia Ocaña-Mondragón, Luis Alberto Ochoa-Carrera, Hector Esteban Paz-Juárez, Marissa Perez-Garcia, Francisco Pulido, Helen Haydee Fernanda Ramírez-Plascencia, Jorge Salas-Hernández, Angel Gustavo Salas-Lais, Alejandro Sanchez-Flores, Clara Esperanza Santacruz-Tinoco, María Guadalupe Santiago-Mauricio, Selene Zárate, Nelly Sélem-Mojica, Blanca Taboada, Gloria Vazquez, Joel Armando Vázquez-Pérez. |
| EPI_ISL_2490387, EPI_ISL_2490388, EPI_ISL_2490389, EPI_ISL_2490390, EPI_ISL_2490391, EPI_ISL_2490392, EPI_ISL_2490393 | Laboratorio Central de Epidemiología (LCE) | Centro de Investigación en Enfermedades Infecciosas (CIENI), Instituto Nacional de Enfermedades Respiratorias (INER) | Consorcio Mexicano de Vigilancia Genómica (CoViGen-Mex). Authors (in alphabetical order): Julio Elias Alvarado-Yaah, Carlos F. Arias, Santiago Ávila-Ríos, Eduardo Becerril-Vargas, Víctor Hugo Borja-Aburto, Celia Boukadida, Cristóbal Cháidez-Quiróz, Juan Bautista Chale-Dzul, Ricardo Ciria Merce, Andreu Comas-García, Célida Duque Molina, Julissa Enciso-Ibarra, José Antonio Enciso-Moreno, Gloria Elena Espinosa-Ayala, Fernando Fontove-Herrera, Daniel Fregoso-Rueda, Victor Eduardo Garcia-Arias, Alejandra Garcia-Gasca, Bruno Gomez-Gil, Jean Pierre González, Irvin González-López, Concepción Grajales-Muñiz, Ricardo Grande, Rosa María Gutierrez Rios, Alejandra Hernández-Terán, Alfredo Herrera-Estrella, Carla Ivón Herrera-Najera, Pavel Isa, Daniel Lira Morales, Susana Lopez, Antonio Loza Román, Brenda Irasema Maldonado-Meza, Bernardo Martínez-Miguel, José Arturo Martínez-Orozco, Célida Martínez- Rodríguez, Margarita Matías-Florentino, Fidencio Mejía-Nepomuceno, María Guadalupe de Jesús Mireles-Rivera, Gloria María Molina-Salinas, Hector Montoya-Fuentes, , Mario Mújica-Sánchez, José Esteban Muñoz-Medina, José de Jesús Nuñez-Contreras, Alicia Ocaña-Mondragón, Luis Alberto Ochoa-Carrera, Hector Esteban Paz-Juárez, Marissa Perez-Garcia, Francisco Pulido, Helen Haydee Fernanda Ramírez-Plascencia, Jorge Salas-Hernández, Angel Gustavo Salas-Lais, Alejandro Sanchez-Flores, Clara Esperanza Santacruz-Tinoco, María Guadalupe Santiago-Mauricio, Selene Zárate, Nelly Sélem-Mojica, Blanca Taboada, Gloria Vazquez, Joel Armando Vázquez-Pérez. |
| EPI_ISL_2490394 | Centro de Investigación Biomédica del Noreste (CIBIN) | Centro de Investigación en Enfermedades Infecciosas (CIENI), Instituto Nacional de Enfermedades Respiratorias (INER) | Consorcio Mexicano de Vigilancia Genómica (CoViGen-Mex). Authors (in alphabetical order): Julio Elias Alvarado-Yaah, Carlos F. Arias, Santiago Ávila-Ríos, Eduardo Becerril-Vargas, Víctor Hugo Borja-Aburto, Celia Boukadida, Cristóbal Cháidez-Quiróz, Juan Bautista Chale-Dzul, Ricardo Ciria Merce, Andreu Comas-García, Célida Duque Molina, Julissa Enciso-Ibarra, José Antonio Enciso-Moreno, Gloria Elena Espinosa-Ayala, Fernando Fontove-Herrera, Daniel Fregoso-Rueda, Victor Eduardo Garcia-Arias, Alejandra Garcia-Gasca, Bruno Gomez-Gil, Jean Pierre González, Irvin González-López, Concepción Grajales-Muñiz, Ricardo Grande, Rosa María Gutierrez Rios, Alejandra Hernández-Terán, Alfredo Herrera-Estrella, Carla Ivón Herrera-Najera, Pavel Isa, Daniel Lira Morales, Susana Lopez, Antonio Loza Román, Brenda Irasema Maldonado-Meza, Bernardo Martínez-Miguel, José Arturo Martínez-Orozco, Célida Martínez- Rodríguez, Margarita Matías-Florentino, Fidencio Mejía-Nepomuceno, María Guadalupe de Jesús Mireles-Rivera, Gloria María Molina-Salinas, Hector Montoya-Fuentes, , Mario Mújica-Sánchez, José Esteban Muñoz-Medina, José de Jesús Nuñez-Contreras, Alicia Ocaña-Mondragón, Luis Alberto Ochoa-Carrera, Hector Esteban Paz-Juárez, Marissa Perez-Garcia, Francisco Pulido, Helen Haydee Fernanda Ramírez-Plascencia, Jorge Salas-Hernández, Angel Gustavo Salas-Lais, Alejandro Sanchez-Flores, Clara Esperanza Santacruz-Tinoco, María Guadalupe Santiago-Mauricio, Selene Zárate, Nelly Sélem-Mojica, Blanca Taboada, Gloria Vazquez, Joel Armando Vázquez-Pérez. |
| EPI_ISL_2490395 | Centro de Investigación Biomédica de Occidente (CIBO) | Centro de Investigación en Enfermedades Infecciosas (CIENI), Instituto Nacional de Enfermedades Respiratorias (INER) | Consorcio Mexicano de Vigilancia Genómica (CoViGen-Mex). Authors (in alphabetical order): Julio Elias Alvarado-Yaah, Carlos F. Arias, Santiago Ávila-Ríos, Eduardo Becerril-Vargas, Víctor Hugo Borja-Aburto, Celia Boukadida, Cristóbal Cháidez-Quiróz, Juan Bautista Chale-Dzul, Ricardo Ciria Merce, Andreu Comas-García, Célida Duque Molina, Julissa Enciso-Ibarra, José Antonio Enciso-Moreno, Gloria Elena Espinosa-Ayala, Fernando Fontove-Herrera, Daniel Fregoso-Rueda, Victor Eduardo Garcia-Arias, Alejandra Garcia-Gasca, Bruno Gomez-Gil, Jean Pierre González, Irvin González-López, Concepción Grajales-Muñiz, Ricardo Grande, Rosa María Gutierrez Rios, Alejandra Hernández-Terán, Alfredo Herrera-Estrella, Carla Ivón Herrera-Najera, Pavel Isa, Daniel Lira Morales, Susana Lopez, Antonio Loza Román, Brenda Irasema Maldonado-Meza, Bernardo Martínez-Miguel, José Arturo Martínez-Orozco, Célida Martínez- Rodríguez, Margarita Matías-Florentino, Fidencio Mejía-Nepomuceno, María Guadalupe de Jesús Mireles-Rivera, Gloria María Molina-Salinas, Hector Montoya-Fuentes, , Mario Mújica-Sánchez, José Esteban Muñoz-Medina, José de Jesús Nuñez-Contreras, Alicia Ocaña-Mondragón, Luis Alberto Ochoa-Carrera, Hector Esteban Paz-Juárez, Marissa Perez-Garcia, Francisco Pulido, Helen Haydee Fernanda Ramírez-Plascencia, Jorge Salas-Hernández, Angel Gustavo Salas-Lais, Alejandro Sanchez-Flores, Clara Esperanza Santacruz-Tinoco, María Guadalupe Santiago-Mauricio, Selene Zárate, Nelly Sélem-Mojica, Blanca Taboada, Gloria Vazquez, Joel Armando Vázquez-Pérez. |
| EPI_ISL_2490396 | Laboratorio Central de Epidemiología (LCE) | Centro de Investigación en Enfermedades Infecciosas (CIENI), Instituto Nacional de Enfermedades Respiratorias (INER) | Consorcio Mexicano de Vigilancia Genómica (CoViGen-Mex). Authors (in alphabetical order): Julio Elias Alvarado-Yaah, Carlos F. Arias, Santiago Ávila-Ríos, Eduardo Becerril-Vargas, Víctor Hugo Borja-Aburto, Celia Boukadida, Cristóbal Cháidez-Quiróz, Juan Bautista Chale-Dzul, Ricardo Ciria Merce, Andreu Comas-García, Célida Duque Molina, Julissa Enciso-Ibarra, José Antonio Enciso-Moreno, Gloria Elena Espinosa-Ayala, Fernando Fontove-Herrera, Daniel Fregoso-Rueda, Victor Eduardo Garcia-Arias, Alejandra Garcia-Gasca, Bruno Gomez-Gil, Jean Pierre González, Irvin González-López, Concepción Grajales-Muñiz, Ricardo Grande, Rosa María Gutierrez Rios, Alejandra Hernández-Terán, Alfredo Herrera-Estrella, Carla Ivón Herrera-Najera, Pavel Isa, Daniel Lira Morales, Susana Lopez, Antonio Loza Román, Brenda Irasema Maldonado-Meza, Bernardo Martínez-Miguel, José Arturo Martínez-Orozco, Célida Martínez- Rodríguez, Margarita Matías-Florentino, Fidencio Mejía-Nepomuceno, María Guadalupe de Jesús Mireles-Rivera, Gloria María Molina-Salinas, Hector Montoya-Fuentes, , Mario Mújica-Sánchez, José Esteban Muñoz-Medina, José de Jesús Nuñez-Contreras, Alicia Ocaña-Mondragón, Luis Alberto Ochoa-Carrera, Hector Esteban Paz-Juárez, Marissa Perez-Garcia, Francisco Pulido, Helen Haydee Fernanda Ramírez-Plascencia, Jorge Salas-Hernández, Angel Gustavo Salas-Lais, Alejandro Sanchez-Flores, Clara Esperanza Santacruz-Tinoco, María Guadalupe Santiago-Mauricio, Selene Zárate, Nelly Sélem-Mojica, Blanca Taboada, Gloria Vazquez, Joel Armando Vázquez-Pérez. |
| EPI_ISL_2490397, EPI_ISL_2490398, EPI_ISL_2490399, EPI_ISL_2490400 | Centro de Investigación Biomédica de Occidente (CIBO) | Centro de Investigación en Enfermedades Infecciosas (CIENI), Instituto Nacional de Enfermedades Respiratorias (INER) | Consorcio Mexicano de Vigilancia Genómica (CoViGen-Mex). Authors (in alphabetical order): Julio Elias Alvarado-Yaah, Carlos F. Arias, Santiago Ávila-Ríos, Eduardo Becerril-Vargas, Víctor Hugo Borja-Aburto, Celia Boukadida, Cristóbal Cháidez-Quiróz, Juan Bautista Chale-Dzul, Ricardo Ciria Merce, Andreu Comas-García, Célida Duque Molina, Julissa Enciso-Ibarra, José Antonio Enciso-Moreno, Gloria Elena Espinosa-Ayala, Fernando Fontove-Herrera, Daniel Fregoso-Rueda, Victor Eduardo Garcia-Arias, Alejandra Garcia-Gasca, Bruno Gomez-Gil, Jean Pierre González, Irvin González-López, Concepción Grajales-Muñiz, Ricardo Grande, Rosa María Gutierrez Rios, Alejandra Hernández-Terán, Alfredo Herrera-Estrella, Carla Ivón Herrera-Najera, Pavel Isa, Daniel Lira Morales, Susana Lopez, Antonio Loza Román, Brenda Irasema Maldonado-Meza, Bernardo Martínez-Miguel, José Arturo Martínez-Orozco, Célida Martínez- Rodríguez, Margarita Matías-Florentino, Fidencio Mejía-Nepomuceno, María Guadalupe de Jesús Mireles-Rivera, Gloria María Molina-Salinas, Hector Montoya-Fuentes, , Mario Mújica-Sánchez, José Esteban Muñoz-Medina, José de Jesús Nuñez-Contreras, Alicia Ocaña-Mondragón, Luis Alberto Ochoa-Carrera, Hector Esteban Paz-Juárez, Marissa Perez-Garcia, Francisco Pulido, Helen Haydee Fernanda Ramírez-Plascencia, Jorge Salas-Hernández, Angel Gustavo Salas-Lais, Alejandro Sanchez-Flores, Clara Esperanza Santacruz-Tinoco, María Guadalupe Santiago-Mauricio, Selene Zárate, Nelly Sélem-Mojica, Blanca Taboada, Gloria Vazquez, Joel Armando Vázquez-Pérez. |
| EPI_ISL_2490401 | Unidad de Investigación Biomédica de Zacatecas (UIBZ) | Centro de Investigación en Enfermedades Infecciosas (CIENI), Instituto Nacional de Enfermedades Respiratorias (INER) | Consorcio Mexicano de Vigilancia Genómica (CoViGen-Mex). Authors (in alphabetical order): Julio Elias Alvarado-Yaah, Carlos F. Arias, Santiago Ávila-Ríos, Eduardo Becerril-Vargas, Víctor Hugo Borja-Aburto, Celia Boukadida, Cristóbal Cháidez-Quiróz, Juan Bautista Chale-Dzul, Ricardo Ciria Merce, Andreu Comas-García, Célida Duque Molina, Julissa Enciso-Ibarra, José Antonio Enciso-Moreno, Gloria Elena Espinosa-Ayala, Fernando Fontove-Herrera, Daniel Fregoso-Rueda, Victor Eduardo Garcia-Arias, Alejandra Garcia-Gasca, Bruno Gomez-Gil, Jean Pierre González, Irvin González-López, Concepción Grajales-Muñiz, Ricardo Grande, Rosa María Gutierrez Rios, Alejandra Hernández-Terán, Alfredo Herrera-Estrella, Carla Ivón Herrera-Najera, Pavel Isa, Daniel Lira Morales, Susana Lopez, Antonio Loza Román, Brenda Irasema Maldonado-Meza, Bernardo Martínez-Miguel, José Arturo Martínez-Orozco, Célida Martínez- Rodríguez, Margarita Matías-Florentino, Fidencio Mejía-Nepomuceno, María Guadalupe de Jesús Mireles-Rivera, Gloria María Molina-Salinas, Hector Montoya-Fuentes, , Mario Mújica-Sánchez, José Esteban Muñoz-Medina, José de Jesús Nuñez-Contreras, Alicia Ocaña-Mondragón, Luis Alberto Ochoa-Carrera, Hector Esteban Paz-Juárez, Marissa Perez-Garcia, Francisco Pulido, Helen Haydee Fernanda Ramírez-Plascencia, Jorge Salas-Hernández, Angel Gustavo Salas-Lais, Alejandro Sanchez-Flores, Clara Esperanza Santacruz-Tinoco, María Guadalupe Santiago-Mauricio, Selene Zárate, Nelly Sélem-Mojica, Blanca Taboada, Gloria Vazquez, Joel Armando Vázquez-Pérez. |
| EPI_ISL_2490402, EPI_ISL_2490403 | Laboratorio Central de Epidemiología (LCE) | Centro de Investigación en Enfermedades Infecciosas (CIENI), Instituto Nacional de Enfermedades Respiratorias (INER) | Consorcio Mexicano de Vigilancia Genómica (CoViGen-Mex). Authors (in alphabetical order): Julio Elias Alvarado-Yaah, Carlos F. Arias, Santiago Ávila-Ríos, Eduardo Becerril-Vargas, Víctor Hugo Borja-Aburto, Celia Boukadida, Cristóbal Cháidez-Quiróz, Juan Bautista Chale-Dzul, Ricardo Ciria Merce, Andreu Comas-García, Célida Duque Molina, Julissa Enciso-Ibarra, José Antonio Enciso-Moreno, Gloria Elena Espinosa-Ayala, Fernando Fontove-Herrera, Daniel Fregoso-Rueda, Victor Eduardo Garcia-Arias, Alejandra Garcia-Gasca, Bruno Gomez-Gil, Jean Pierre González, Irvin González-López, Concepción Grajales-Muñiz, Ricardo Grande, Rosa María Gutierrez Rios, Alejandra Hernández-Terán, Alfredo Herrera-Estrella, Carla Ivón Herrera-Najera, Pavel Isa, Daniel Lira Morales, Susana Lopez, Antonio Loza Román, Brenda Irasema Maldonado-Meza, Bernardo Martínez-Miguel, José Arturo Martínez-Orozco, Célida Martínez- Rodríguez, Margarita Matías-Florentino, Fidencio Mejía-Nepomuceno, María Guadalupe de Jesús Mireles-Rivera, Gloria María Molina-Salinas, Hector Montoya-Fuentes, , Mario Mújica-Sánchez, José Esteban Muñoz-Medina, José de Jesús Nuñez-Contreras, Alicia Ocaña-Mondragón, Luis Alberto Ochoa-Carrera, Hector Esteban Paz-Juárez, Marissa Perez-Garcia, Francisco Pulido, Helen Haydee Fernanda Ramírez-Plascencia, Jorge Salas-Hernández, Angel Gustavo Salas-Lais, Alejandro Sanchez-Flores, Clara Esperanza Santacruz-Tinoco, María Guadalupe Santiago-Mauricio, Selene Zárate, Nelly Sélem-Mojica, Blanca Taboada, Gloria Vazquez, Joel Armando Vázquez-Pérez. |
| EPI_ISL_2490404 | Centro de Investigación Biomédica de Occidente (CIBO) | Centro de Investigación en Enfermedades Infecciosas (CIENI), Instituto Nacional de Enfermedades Respiratorias (INER) | Consorcio Mexicano de Vigilancia Genómica (CoViGen-Mex). Authors (in alphabetical order): Julio Elias Alvarado-Yaah, Carlos F. Arias, Santiago Ávila-Ríos, Eduardo Becerril-Vargas, Víctor Hugo Borja-Aburto, Celia Boukadida, Cristóbal Cháidez-Quiróz, Juan Bautista Chale-Dzul, Ricardo Ciria Merce, Andreu Comas-García, Célida Duque Molina, Julissa Enciso-Ibarra, José Antonio Enciso-Moreno, Gloria Elena Espinosa-Ayala, Fernando Fontove-Herrera, Daniel Fregoso-Rueda, Victor Eduardo Garcia-Arias, Alejandra Garcia-Gasca, Bruno Gomez-Gil, Jean Pierre González, Irvin González-López, Concepción Grajales-Muñiz, Ricardo Grande, Rosa María Gutierrez Rios, Alejandra Hernández-Terán, Alfredo Herrera-Estrella, Carla Ivón Herrera-Najera, Pavel Isa, Daniel Lira Morales, Susana Lopez, Antonio Loza Román, Brenda Irasema Maldonado-Meza, Bernardo Martínez-Miguel, José Arturo Martínez-Orozco, Célida Martínez- Rodríguez, Margarita Matías-Florentino, Fidencio Mejía-Nepomuceno, María Guadalupe de Jesús Mireles-Rivera, Gloria María Molina-Salinas, Hector Montoya-Fuentes, , Mario Mújica-Sánchez, José Esteban Muñoz-Medina, José de Jesús Nuñez-Contreras, Alicia Ocaña-Mondragón, Luis Alberto Ochoa-Carrera, Hector Esteban Paz-Juárez, Marissa Perez-Garcia, Francisco Pulido, Helen Haydee Fernanda Ramírez-Plascencia, Jorge Salas-Hernández, Angel Gustavo Salas-Lais, Alejandro Sanchez-Flores, Clara Esperanza Santacruz-Tinoco, María Guadalupe Santiago-Mauricio, Selene Zárate, Nelly Sélem-Mojica, Blanca Taboada, Gloria Vazquez, Joel Armando Vázquez-Pérez. |
| EPI_ISL_2490405, EPI_ISL_2490406 | Centro de Investigación Biomédica del Noreste (CIBIN) | Centro de Investigación en Enfermedades Infecciosas (CIENI), Instituto Nacional de | Consorcio Mexicano de Vigilancia Genómica (CoViGen-Mex). Authors (in alphabetical order): Julio Elias Alvarado-Yaah, Carlos F. Arias, Santiago Ávila-Ríos, Eduardo Becerril-Vargas, Víctor Hugo Borja-Aburto, Celia Boukadida, Cristóbal Cháidez-Quiróz, Juan Bautista Chale-Dzul, Ricardo Ciria Merce, Andreu Comas-García, Célida Duque Molina, Julissa Enciso-Ibarra, José Antonio Enciso-Moreno, Gloria Elena Espinosa-Ayala, Fernando Fontove-Herrera, Daniel Fregoso-Rueda, Victor Eduardo Garcia-Arias, Alejandra Garcia-Gasca, Bruno Gomez-Gil, Jean Pierre González, Irvin |

| | | | |
|---|---|---|---|
| | | Enfermedades Respiratorias (INER) | González-López, Concepción Grajales-Muñiz, Ricardo Grande, Rosa María Gutierrez Rios, Alejandra Hernández-Terán, Alfredo Herrera-Estrella, Carla Ivón Herrera-Najera, Pavel Isa, Daniel Lira Morales, Susana Lopez, Antonio Loza Román, Brenda Irasema Maldonado-Meza, Bernardo Martínez-Miguel, José Arturo Martínez-Orozco, Célida Martínez- Rodríguez, Margarita Matías-Florentino, Fidencio Mejía-Nepomuceno, María Guadalupe de Jesús Mireles-Rivera, Gloria María Molina-Salinas, Hector Montoya-Fuentes, , Mario Mújica-Sánchez, José Esteban Muñoz-Medina, José de Jesús Nuñez-Contreras, Alicia Ocaña-Mondragón, Luis Alberto Ochoa-Carrera, Hector Esteban Paz-Juárez, Marissa Perez-Garcia, Francisco Pulido, Helen Haydee Fernanda Ramírez-Plascencia, Jorge Salas-Hernández, Angel Gustavo Salas-Lais, Alejandro Sanchez-Flores, Clara Esperanza Santacruz-Tinoco, María Guadalupe Santiago-Mauricio, Selene Zárate, Nelly Sélem-Mojica, Blanca Taboada, Gloria Vazquez, Joel Armando Vázquez-Pérez. |
| EPI_ISL_2490407, EPI_ISL_2490408 | Laboratorio Central de Epidemiología (LCE) | Centro de Investigación en Enfermedades Infecciosas (CIENI), Instituto Nacional de Enfermedades Respiratorias (INER) | Consorcio Mexicano de Vigilancia Genómica (CoViGen-Mex). Authors (in alphabetical order): Julio Elias Alvarado-Yaah, Carlos F. Arias, Santiago Ávila-Ríos, Eduardo Becerril-Vargas, Víctor Hugo Borja-Aburto, Celia Boukadida, Cristóbal Cháidez-Quiróz, Juan Bautista Chale-Dzul, Ricardo Ciria Merce, Andreu Comas-García, Célida Duque Molina, Julissa Enciso-Ibarra, José Antonio Enciso-Moreno, Gloria Elena Espinosa-Ayala, Fernando Fontove-Herrera, Daniel Fregoso-Rueda, Victor Eduardo Garcia-Arias, Alejandra Garcia-Gasca, Bruno Gomez-Gil, Jean Pierre González, Irvin González-López, Concepción Grajales-Muñiz, Ricardo Grande, Rosa María Gutierrez Rios, Alejandra Hernández-Terán, Alfredo Herrera-Estrella, Carla Ivón Herrera-Najera, Pavel Isa, Daniel Lira Morales, Susana Lopez, Antonio Loza Román, Brenda Irasema Maldonado-Meza, Bernardo Martínez-Miguel, José Arturo Martínez-Orozco, Célida Martínez- Rodríguez, Margarita Matías-Florentino, Fidencio Mejía-Nepomuceno, María Guadalupe de Jesús Mireles-Rivera, Gloria María Molina-Salinas, Hector Montoya-Fuentes, , Mario Mújica-Sánchez, José Esteban Muñoz-Medina, José de Jesús Nuñez-Contreras, Alicia Ocaña-Mondragón, Luis Alberto Ochoa-Carrera, Hector Esteban Paz-Juárez, Marissa Perez-Garcia, Francisco Pulido, Helen Haydee Fernanda Ramírez-Plascencia, Jorge Salas-Hernández, Angel Gustavo Salas-Lais, Alejandro Sanchez-Flores, Clara Esperanza Santacruz-Tinoco, María Guadalupe Santiago-Mauricio, Selene Zárate, Nelly Sélem-Mojica, Blanca Taboada, Gloria Vazquez, Joel Armando Vázquez-Pérez. |
| EPI_ISL_2490409, EPI_ISL_2490410, EPI_ISL_2490411 | Centro de Investigación Biomédica del Noreste (CIBIN) | Centro de Investigación en Enfermedades Infecciosas (CIENI), Instituto Nacional de Enfermedades Respiratorias (INER) | Consorcio Mexicano de Vigilancia Genómica (CoViGen-Mex). Authors (in alphabetical order): Julio Elias Alvarado-Yaah, Carlos F. Arias, Santiago Ávila-Ríos, Eduardo Becerril-Vargas, Víctor Hugo Borja-Aburto, Celia Boukadida, Cristóbal Cháidez-Quiróz, Juan Bautista Chale-Dzul, Ricardo Ciria Merce, Andreu Comas-García, Célida Duque Molina, Julissa Enciso-Ibarra, José Antonio Enciso-Moreno, Gloria Elena Espinosa-Ayala, Fernando Fontove-Herrera, Daniel Fregoso-Rueda, Victor Eduardo Garcia-Arias, Alejandra Garcia-Gasca, Bruno Gomez-Gil, Jean Pierre González, Irvin González-López, Concepción Grajales-Muñiz, Ricardo Grande, Rosa María Gutierrez Rios, Alejandra Hernández-Terán, Alfredo Herrera-Estrella, Carla Ivón Herrera-Najera, Pavel Isa, Daniel Lira Morales, Susana Lopez, Antonio Loza Román, Brenda Irasema Maldonado-Meza, Bernardo Martínez-Miguel, José Arturo Martínez-Orozco, Célida Martínez- Rodríguez, Margarita Matías-Florentino, Fidencio Mejía-Nepomuceno, María Guadalupe de Jesús Mireles-Rivera, Gloria María Molina-Salinas, Hector Montoya-Fuentes, , Mario Mújica-Sánchez, José Esteban Muñoz-Medina, José de Jesús Nuñez-Contreras, Alicia Ocaña-Mondragón, Luis Alberto Ochoa-Carrera, Hector Esteban Paz-Juárez, Marissa Perez-Garcia, Francisco Pulido, Helen Haydee Fernanda Ramírez-Plascencia, Jorge Salas-Hernández, Angel Gustavo Salas-Lais, Alejandro Sanchez-Flores, Clara Esperanza Santacruz-Tinoco, María Guadalupe Santiago-Mauricio, Selene Zárate, Nelly Sélem-Mojica, Blanca Taboada, Gloria Vazquez, Joel Armando Vázquez-Pérez. |
| EPI_ISL_2490412, EPI_ISL_2490413, EPI_ISL_2490414, EPI_ISL_2490415, EPI_ISL_2490416, EPI_ISL_2490417 | Laboratorio Central de Epidemiología (LCE) | Centro de Investigación en Enfermedades Infecciosas (CIENI), Instituto Nacional de Enfermedades Respiratorias (INER) | Consorcio Mexicano de Vigilancia Genómica (CoViGen-Mex). Authors (in alphabetical order): Julio Elias Alvarado-Yaah, Carlos F. Arias, Santiago Ávila-Ríos, Eduardo Becerril-Vargas, Víctor Hugo Borja-Aburto, Celia Boukadida, Cristóbal Cháidez-Quiróz, Juan Bautista Chale-Dzul, Ricardo Ciria Merce, Andreu Comas-García, Célida Duque Molina, Julissa Enciso-Ibarra, José Antonio Enciso-Moreno, Gloria Elena Espinosa-Ayala, Fernando Fontove-Herrera, Daniel Fregoso-Rueda, Victor Eduardo Garcia-Arias, Alejandra Garcia-Gasca, Bruno Gomez-Gil, Jean Pierre González, Irvin González-López, Concepción Grajales-Muñiz, Ricardo Grande, Rosa María Gutierrez Rios, Alejandra Hernández-Terán, Alfredo Herrera-Estrella, Carla Ivón Herrera-Najera, Pavel Isa, Daniel Lira Morales, Susana Lopez, Antonio Loza Román, Brenda Irasema Maldonado-Meza, Bernardo Martínez-Miguel, José Arturo Martínez-Orozco, Célida Martínez- Rodríguez, Margarita Matías-Florentino, Fidencio Mejía-Nepomuceno, María Guadalupe de Jesús Mireles-Rivera, Gloria María Molina-Salinas, Hector Montoya-Fuentes, , Mario Mújica-Sánchez, José Esteban Muñoz-Medina, José de Jesús Nuñez-Contreras, Alicia Ocaña-Mondragón, Luis Alberto Ochoa-Carrera, Hector Esteban Paz-Juárez, Marissa Perez-Garcia, Francisco Pulido, Helen Haydee Fernanda Ramírez-Plascencia, Jorge Salas-Hernández, Angel Gustavo Salas-Lais, Alejandro Sanchez-Flores, Clara Esperanza Santacruz-Tinoco, María Guadalupe Santiago-Mauricio, Selene Zárate, Nelly Sélem-Mojica, Blanca Taboada, Gloria Vazquez, Joel Armando Vázquez-Pérez. |
| EPI_ISL_2490418, EPI_ISL_2490419, EPI_ISL_2490420, EPI_ISL_2490421, EPI_ISL_2490422 | Unidad de Investigación Médica de Yucatán (UIMY) | Centro de Investigación en Enfermedades Infecciosas (CIENI), Instituto Nacional de Enfermedades Respiratorias (INER) | Consorcio Mexicano de Vigilancia Genómica (CoViGen-Mex). Authors (in alphabetical order): Julio Elias Alvarado-Yaah, Carlos F. Arias, Santiago Ávila-Ríos, Eduardo Becerril-Vargas, Víctor Hugo Borja-Aburto, Celia Boukadida, Cristóbal Cháidez-Quiróz, Juan Bautista Chale-Dzul, Ricardo Ciria Merce, Andreu Comas-García, Célida Duque Molina, Julissa Enciso-Ibarra, José Antonio Enciso-Moreno, Gloria Elena Espinosa-Ayala, Fernando Fontove-Herrera, Daniel Fregoso-Rueda, Victor Eduardo Garcia-Arias, Alejandra Garcia-Gasca, Bruno Gomez-Gil, Jean Pierre González, Irvin González-López, Concepción Grajales-Muñiz, Ricardo Grande, Rosa María Gutierrez Rios, Alejandra Hernández-Terán, Alfredo Herrera-Estrella, Carla Ivón Herrera-Najera, Pavel Isa, Daniel Lira Morales, Susana Lopez, Antonio Loza Román, Brenda Irasema Maldonado-Meza, Bernardo Martínez-Miguel, José Arturo Martínez-Orozco, Célida Martínez- Rodríguez, Margarita Matías-Florentino, Fidencio Mejía-Nepomuceno, María Guadalupe de Jesús Mireles-Rivera, Gloria María Molina-Salinas, Hector Montoya-Fuentes, , Mario Mújica-Sánchez, José Esteban Muñoz-Medina, José de Jesús Nuñez-Contreras, Alicia Ocaña-Mondragón, Luis Alberto Ochoa-Carrera, Hector Esteban Paz-Juárez, Marissa Perez-Garcia, Francisco Pulido, Helen Haydee Fernanda Ramírez-Plascencia, Jorge Salas-Hernández, Angel Gustavo Salas-Lais, Alejandro Sanchez-Flores, Clara Esperanza Santacruz-Tinoco, María Guadalupe Santiago-Mauricio, Selene Zárate, Nelly Sélem-Mojica, Blanca Taboada, Gloria Vazquez, Joel Armando Vázquez-Pérez. |
| EPI_ISL_2490423 | Laboratorio Central de Epidemiología (LCE) | Centro de Investigación en Enfermedades Infecciosas (CIENI), Instituto Nacional de Enfermedades Respiratorias (INER) | Consorcio Mexicano de Vigilancia Genómica (CoViGen-Mex). Authors (in alphabetical order): Julio Elias Alvarado-Yaah, Carlos F. Arias, Santiago Ávila-Ríos, Eduardo Becerril-Vargas, Víctor Hugo Borja-Aburto, Celia Boukadida, Cristóbal Cháidez-Quiróz, Juan Bautista Chale-Dzul, Ricardo Ciria Merce, Andreu Comas-García, Célida Duque Molina, Julissa Enciso-Ibarra, José Antonio Enciso-Moreno, Gloria Elena Espinosa-Ayala, Fernando Fontove-Herrera, Daniel Fregoso-Rueda, Victor Eduardo Garcia-Arias, Alejandra Garcia-Gasca, Bruno Gomez-Gil, Jean Pierre González, Irvin González-López, Concepción Grajales-Muñiz, Ricardo Grande, Rosa María Gutierrez Rios, Alejandra Hernández-Terán, Alfredo Herrera-Estrella, Carla Ivón Herrera-Najera, Pavel Isa, Daniel Lira Morales, Susana Lopez, Antonio Loza Román, Brenda Irasema Maldonado-Meza, Bernardo Martínez-Miguel, José Arturo Martínez-Orozco, Célida Martínez- Rodríguez, Margarita Matías-Florentino, Fidencio Mejía-Nepomuceno, María Guadalupe de Jesús Mireles-Rivera, Gloria María Molina-Salinas, Hector Montoya-Fuentes, , Mario Mújica-Sánchez, José Esteban Muñoz-Medina, José de Jesús Nuñez-Contreras, Alicia Ocaña-Mondragón, Luis Alberto Ochoa-Carrera, Hector Esteban Paz-Juárez, Marissa Perez-Garcia, Francisco Pulido, Helen Haydee Fernanda Ramírez-Plascencia, Jorge Salas-Hernández, Angel Gustavo Salas-Lais, Alejandro Sanchez-Flores, Clara Esperanza Santacruz-Tinoco, María Guadalupe Santiago-Mauricio, Selene Zárate, Nelly Sélem-Mojica, Blanca Taboada, Gloria Vazquez, Joel Armando Vázquez-Pérez. |
| EPI_ISL_2490424 | Unidad de Investigación Biomédica de Zacatecas (UIBZ) | Centro de Investigación en Enfermedades Infecciosas (CIENI), Instituto Nacional de Enfermedades Respiratorias (INER) | Consorcio Mexicano de Vigilancia Genómica (CoViGen-Mex). Authors (in alphabetical order): Julio Elias Alvarado-Yaah, Carlos F. Arias, Santiago Ávila-Ríos, Eduardo Becerril-Vargas, Víctor Hugo Borja-Aburto, Celia Boukadida, Cristóbal Cháidez-Quiróz, Juan Bautista Chale-Dzul, Ricardo Ciria Merce, Andreu Comas-García, Célida Duque Molina, Julissa Enciso-Ibarra, José Antonio Enciso-Moreno, Gloria Elena Espinosa-Ayala, Fernando Fontove-Herrera, Daniel Fregoso-Rueda, Victor Eduardo Garcia-Arias, Alejandra Garcia-Gasca, Bruno Gomez-Gil, Jean Pierre González, Irvin González-López, Concepción Grajales-Muñiz, Ricardo Grande, Rosa María Gutierrez Rios, Alejandra Hernández-Terán, Alfredo Herrera-Estrella, Carla Ivón Herrera-Najera, Pavel Isa, Daniel Lira Morales, Susana Lopez, Antonio Loza Román, Brenda Irasema Maldonado-Meza, Bernardo Martínez-Miguel, José Arturo Martínez-Orozco, Célida Martínez- Rodríguez, Margarita Matías-Florentino, Fidencio Mejía-Nepomuceno, María Guadalupe de Jesús Mireles-Rivera, Gloria María Molina-Salinas, Hector Montoya-Fuentes, , Mario Mújica-Sánchez, José Esteban Muñoz-Medina, José de Jesús Nuñez-Contreras, Alicia Ocaña-Mondragón, Luis Alberto Ochoa-Carrera, Hector Esteban Paz-Juárez, Marissa Perez-Garcia, Francisco Pulido, Helen Haydee Fernanda Ramírez-Plascencia, Jorge Salas-Hernández, Angel Gustavo Salas-Lais, Alejandro Sanchez-Flores, Clara Esperanza Santacruz-Tinoco, María Guadalupe Santiago-Mauricio, Selene Zárate, Nelly Sélem-Mojica, Blanca Taboada, Gloria Vazquez, Joel Armando Vázquez-Pérez. |
| EPI_ISL_2490425 | Laboratorio Central de Epidemiología (LCE) | Centro de Investigación en Enfermedades Infecciosas (CIENI), Instituto Nacional de Enfermedades Respiratorias (INER) | Consorcio Mexicano de Vigilancia Genómica (CoViGen-Mex). Authors (in alphabetical order): Julio Elias Alvarado-Yaah, Carlos F. Arias, Santiago Ávila-Ríos, Eduardo Becerril-Vargas, Víctor Hugo Borja-Aburto, Celia Boukadida, Cristóbal Cháidez-Quiróz, Juan Bautista Chale-Dzul, Ricardo Ciria Merce, Andreu Comas-García, Célida Duque Molina, Julissa Enciso-Ibarra, José Antonio Enciso-Moreno, Gloria Elena Espinosa-Ayala, Fernando Fontove-Herrera, Daniel Fregoso-Rueda, Victor Eduardo Garcia-Arias, Alejandra Garcia-Gasca, Bruno Gomez-Gil, Jean Pierre González, Irvin González-López, Concepción Grajales-Muñiz, Ricardo Grande, Rosa María Gutierrez Rios, Alejandra Hernández-Terán, Alfredo Herrera-Estrella, Carla Ivón Herrera-Najera, Pavel Isa, Daniel Lira Morales, Susana Lopez, Antonio Loza Román, Brenda Irasema Maldonado-Meza, Bernardo Martínez-Miguel, José Arturo Martínez-Orozco, Célida Martínez- Rodríguez, Margarita Matías-Florentino, Fidencio Mejía-Nepomuceno, María Guadalupe de Jesús Mireles-Rivera, Gloria María Molina-Salinas, Hector Montoya-Fuentes, , Mario Mújica-Sánchez, José Esteban Muñoz-Medina, José de Jesús Nuñez-Contreras, Alicia Ocaña-Mondragón, Luis Alberto Ochoa-Carrera, Hector Esteban Paz-Juárez, Marissa Perez-Garcia, Francisco Pulido, Helen Haydee Fernanda Ramírez-Plascencia, Jorge Salas-Hernández, Angel Gustavo Salas-Lais, Alejandro Sanchez-Flores, Clara Esperanza Santacruz-Tinoco, María Guadalupe Santiago-Mauricio, Selene Zárate, Nelly Sélem-Mojica, Blanca Taboada, Gloria Vazquez, Joel Armando Vázquez-Pérez. |
| EPI_ISL_2490426, EPI_ISL_2490427, EPI_ISL_2490428, EPI_ISL_2490429, EPI_ISL_2490430, EPI_ISL_2490431, EPI_ISL_2490432 | Centro de Investigación Biomédica del Noreste (CIBIN) | Centro de Investigación en Enfermedades Infecciosas (CIENI), Instituto Nacional de Enfermedades Respiratorias (INER) | Consorcio Mexicano de Vigilancia Genómica (CoViGen-Mex). Authors (in alphabetical order): Julio Elias Alvarado-Yaah, Carlos F. Arias, Santiago Ávila-Ríos, Eduardo Becerril-Vargas, Víctor Hugo Borja-Aburto, Celia Boukadida, Cristóbal Cháidez-Quiróz, Juan Bautista Chale-Dzul, Ricardo Ciria Merce, Andreu Comas-García, Célida Duque Molina, Julissa Enciso-Ibarra, José Antonio Enciso-Moreno, Gloria Elena Espinosa-Ayala, Fernando Fontove-Herrera, Daniel Fregoso-Rueda, Victor Eduardo Garcia-Arias, Alejandra Garcia-Gasca, Bruno Gomez-Gil, Jean Pierre González, Irvin González-López, Concepción Grajales-Muñiz, Ricardo Grande, Rosa María Gutierrez Rios, Alejandra Hernández-Terán, Alfredo Herrera-Estrella, Carla Ivón Herrera-Najera, Pavel Isa, Daniel Lira Morales, Susana Lopez, Antonio Loza Román, Brenda Irasema Maldonado-Meza, Bernardo Martínez-Miguel, José Arturo Martínez-Orozco, Célida Martínez- Rodríguez, Margarita Matías-Florentino, Fidencio Mejía-Nepomuceno, María Guadalupe de Jesús Mireles-Rivera, Gloria María Molina-Salinas, Hector Montoya-Fuentes, , Mario Mújica-Sánchez, José Esteban Muñoz-Medina, José de Jesús Nuñez-Contreras, Alicia Ocaña-Mondragón, Luis Alberto Ochoa-Carrera, Hector Esteban Paz-Juárez, Marissa Perez-Garcia, Francisco Pulido, Helen Haydee Fernanda Ramírez-Plascencia, Jorge Salas-Hernández, Angel Gustavo Salas-Lais, Alejandro Sanchez-Flores, Clara Esperanza Santacruz-Tinoco, María Guadalupe Santiago-Mauricio, Selene Zárate, Nelly Sélem-Mojica, Blanca Taboada, Gloria Vazquez, Joel Armando Vázquez-Pérez. |
| EPI_ISL_2490433, EPI_ISL_2490434, EPI_ISL_2490435, EPI_ISL_2490436, EPI_ISL_2490437, EPI_ISL_2490438 | Laboratorio Central de Epidemiología (LCE) | Centro de Investigación en Enfermedades Infecciosas (CIENI), Instituto Nacional de Enfermedades Respiratorias (INER) | Consorcio Mexicano de Vigilancia Genómica (CoViGen-Mex). Authors (in alphabetical order): Julio Elias Alvarado-Yaah, Carlos F. Arias, Santiago Ávila-Ríos, Eduardo Becerril-Vargas, Víctor Hugo Borja-Aburto, Celia Boukadida, Cristóbal Cháidez-Quiróz, Juan Bautista Chale-Dzul, Ricardo Ciria Merce, Andreu Comas-García, Célida Duque Molina, Julissa Enciso-Ibarra, José Antonio Enciso-Moreno, Gloria Elena Espinosa-Ayala, Fernando Fontove-Herrera, Daniel Fregoso-Rueda, Victor Eduardo Garcia-Arias, Alejandra Garcia-Gasca, Bruno Gomez-Gil, Jean Pierre González, Irvin González-López, Concepción Grajales-Muñiz, Ricardo Grande, Rosa María Gutierrez Rios, Alejandra Hernández-Terán, Alfredo Herrera-Estrella, Carla Ivón Herrera-Najera, Pavel Isa, Daniel Lira Morales, Susana Lopez, Antonio Loza Román, Brenda Irasema Maldonado-Meza, Bernardo Martínez-Miguel, José Arturo Martínez-Orozco, Célida Martínez- Rodríguez, Margarita Matías-Florentino, Fidencio Mejía-Nepomuceno, María Guadalupe de Jesús Mireles-Rivera, Gloria María Molina-Salinas, Hector Montoya-Fuentes, , Mario Mújica-Sánchez, José Esteban Muñoz-Medina, José de Jesús Nuñez-Contreras, Alicia Ocaña-Mondragón, Luis Alberto Ochoa-Carrera, Hector Esteban Paz-Juárez, Marissa Perez-Garcia, Francisco Pulido, Helen Haydee Fernanda Ramírez-Plascencia, Jorge Salas-Hernández, Angel Gustavo Salas-Lais, Alejandro Sanchez-Flores, Clara Esperanza Santacruz-Tinoco, María Guadalupe Santiago-Mauricio, Selene Zárate, Nelly Sélem-Mojica, Blanca Taboada, Gloria Vazquez, Joel Armando Vázquez-Pérez. |
| EPI_ISL_2490439, EPI_ISL_2490440, EPI_ISL_2490441, EPI_ISL_2490442, EPI_ISL_2490443, EPI_ISL_2490444, EPI_ISL_2490445, EPI_ISL_2490446, EPI_ISL_2490447, EPI_ISL_2490448, EPI_ISL_2490449, EPI_ISL_2490450, EPI_ISL_2490451, EPI_ISL_2490452, EPI_ISL_2490453, EPI_ISL_2490454, EPI_ISL_2490455, EPI_ISL_2490456, EPI_ISL_2490457, EPI_ISL_2490458, EPI_ISL_2490459, EPI_ISL_2490460, EPI_ISL_2490461, EPI_ISL_2490462, EPI_ISL_2490463, EPI_ISL_2490464, EPI_ISL_2490465 | | | |
| see above | Centro de Investigación Biomédica del Noreste (CIBIN) | Centro de Investigación en Enfermedades Infecciosas (CIENI), Instituto Nacional de Enfermedades Respiratorias (INER) | Consorcio Mexicano de Vigilancia Genómica (CoViGen-Mex). Authors (in alphabetical order): Julio Elias Alvarado-Yaah, Carlos F. Arias, Santiago Ávila-Ríos, Eduardo Becerril-Vargas, Víctor Hugo Borja-Aburto, Celia Boukadida, Cristóbal Cháidez-Quiróz, Juan Bautista Chale-Dzul, Ricardo Ciria Merce, Andreu Comas-García, Célida Duque Molina, Julissa Enciso-Ibarra, José Antonio Enciso-Moreno, Gloria Elena Espinosa-Ayala, Fernando Fontove-Herrera, Daniel Fregoso-Rueda, Victor Eduardo Garcia-Arias, Alejandra Garcia-Gasca, Bruno Gomez-Gil, Jean Pierre González, Irvin González-López, Concepción Grajales-Muñiz, Ricardo Grande, Rosa María Gutierrez Rios, Alejandra Hernández-Terán, Alfredo Herrera-Estrella, Carla Ivón Herrera-Najera, Pavel Isa, Daniel Lira Morales, Susana Lopez, Antonio Loza Román, Brenda Irasema Maldonado-Meza, Bernardo Martínez-Miguel, José Arturo Martínez-Orozco, Célida Martínez- Rodríguez, Margarita Matías-Florentino, Fidencio Mejía-Nepomuceno, María Guadalupe de Jesús Mireles-Rivera, Gloria María Molina-Salinas, Hector Montoya-Fuentes, , Mario Mújica-Sánchez, José Esteban Muñoz-Medina, José de Jesús Nuñez-Contreras, Alicia Ocaña-Mondragón, Luis Alberto Ochoa-Carrera, Hector Esteban Paz-Juárez, Marissa Perez-Garcia, Francisco Pulido, Helen Haydee Fernanda Ramírez-Plascencia, Jorge Salas-Hernández, Angel Gustavo Salas-Lais, Alejandro Sanchez-Flores, Clara Esperanza Santacruz-Tinoco, María Guadalupe Santiago-Mauricio, Selene Zárate, Nelly Sélem-Mojica, Blanca Taboada, Gloria Vazquez, Joel Armando Vázquez-Pérez. |
| EPI_ISL_2490466, EPI_ISL_2490467, EPI_ISL_2490468, EPI_ISL_2490469, EPI_ISL_2490470, EPI_ISL_2490471, EPI_ISL_2490472, EPI_ISL_2490473, EPI_ISL_2490474, EPI_ISL_2490475, EPI_ISL_2490476, EPI_ISL_2490477, EPI_ISL_2490478, EPI_ISL_2490479, EPI_ISL_2490480, EPI_ISL_2490481, EPI_ISL_2490482, EPI_ISL_2490483, EPI_ISL_2490484, EPI_ISL_2490485, EPI_ISL_2490486, EPI_ISL_2490487, EPI_ISL_2490488, EPI_ISL_2490489, EPI_ISL_2490490, EPI_ISL_2490491, EPI_ISL_2490492 | | | |
| see above | Unidad de Investigación Médica de Yucatán (UIMY) | Centro de Investigación en Enfermedades Infecciosas (CIENI), Instituto Nacional de Enfermedades Respiratorias (INER) | Consorcio Mexicano de Vigilancia Genómica (CoViGen-Mex). Authors (in alphabetical order): Julio Elias Alvarado-Yaah, Carlos F. Arias, Santiago Ávila-Ríos, Eduardo Becerril-Vargas, Víctor Hugo Borja-Aburto, Celia Boukadida, Cristóbal Cháidez-Quiróz, Juan Bautista Chale-Dzul, Ricardo Ciria Merce, Andreu Comas-García, Célida Duque Molina, Julissa Enciso-Ibarra, José Antonio Enciso-Moreno, Gloria Elena Espinosa-Ayala, Fernando Fontove-Herrera, Daniel Fregoso-Rueda, Victor Eduardo Garcia-Arias, Alejandra Garcia-Gasca, Bruno Gomez-Gil, Jean Pierre González, Irvin González-López, Concepción Grajales-Muñiz, Ricardo Grande, Rosa María Gutierrez Rios, Alejandra Hernández-Terán, Alfredo Herrera-Estrella, Carla Ivón Herrera-Najera, Pavel Isa, Daniel Lira Morales, Susana Lopez, Antonio Loza Román, Brenda Irasema Maldonado-Meza, Bernardo Martínez-Miguel, José Arturo Martínez-Orozco, Célida Martínez- Rodríguez, Margarita Matías-Florentino, Fidencio Mejía-Nepomuceno, María Guadalupe de Jesús Mireles-Rivera, Gloria María Molina-Salinas, Hector Montoya-Fuentes, , Mario Mújica-Sánchez, José Esteban Muñoz-Medina, José de Jesús Nuñez-Contreras, Alicia Ocaña-Mondragón, Luis Alberto Ochoa-Carrera, Hector Esteban Paz-Juárez, Marissa Perez-Garcia, Francisco Pulido, Helen Haydee Fernanda Ramírez-Plascencia, Jorge Salas-Hernández, Angel Gustavo Salas-Lais, Alejandro Sanchez-Flores, Clara Esperanza Santacruz-Tinoco, María Guadalupe Santiago-Mauricio, Selene Zárate, Nelly Sélem-Mojica, Blanca Taboada, Gloria Vazquez, Joel Armando Vázquez-Pérez. |
| EPI_ISL_2490493, EPI_ISL_2490494, EPI_ISL_2490495, EPI_ISL_2490496, EPI_ISL_2490497, EPI_ISL_2490498, EPI_ISL_2490499 | Centro de Investigación Biomédica de Occidente (CIBO) | Centro de Investigación en Enfermedades Infecciosas (CIENI), Instituto Nacional de Enfermedades Respiratorias (INER) | Consorcio Mexicano de Vigilancia Genómica (CoViGen-Mex). Authors (in alphabetical order): Julio Elias Alvarado-Yaah, Carlos F. Arias, Santiago Ávila-Ríos, Eduardo Becerril-Vargas, Víctor Hugo Borja-Aburto, Celia Boukadida, Cristóbal Cháidez-Quiróz, Juan Bautista Chale-Dzul, Ricardo Ciria Merce, Andreu Comas-García, Célida Duque Molina, Julissa Enciso-Ibarra, José Antonio Enciso-Moreno, Gloria Elena Espinosa-Ayala, Fernando Fontove-Herrera, Daniel Fregoso-Rueda, Victor Eduardo Garcia-Arias, Alejandra Garcia-Gasca, Bruno Gomez-Gil, Jean Pierre González, Irvin González-López, Concepción Grajales-Muñiz, Ricardo Grande, Rosa María Gutierrez Rios, Alejandra Hernández-Terán, Alfredo Herrera-Estrella, Carla Ivón Herrera-Najera, Pavel Isa, Daniel Lira Morales, Susana Lopez, Antonio Loza Román, Brenda Irasema Maldonado-Meza, Bernardo Martínez-Miguel, José Arturo Martínez-Orozco, Célida Martínez- Rodríguez, Margarita Matías-Florentino, Fidencio Mejía-Nepomuceno, María Guadalupe de Jesús Mireles-Rivera, Gloria María Molina-Salinas, Hector Montoya-Fuentes, , Mario Mújica-Sánchez, José Esteban Muñoz-Medina, José de Jesús Nuñez-Contreras, Alicia Ocaña-Mondragón, Luis Alberto Ochoa-Carrera, Hector Esteban Paz-Juárez, Marissa Perez-Garcia, Francisco Pulido, Helen Haydee Fernanda Ramírez-Plascencia, Jorge Salas-Hernández, Angel Gustavo Salas-Lais, Alejandro Sanchez-Flores, Clara Esperanza Santacruz-Tinoco, María Guadalupe Santiago-Mauricio, Selene Zárate, Nelly Sélem-Mojica, Blanca Taboada, Gloria Vazquez, Joel Armando Vázquez-Pérez. |
| EPI_ISL_2490500, EPI_ISL_2490501, EPI_ISL_2490502, EPI_ISL_2490503, EPI_ISL_2490504, EPI_ISL_2490505, EPI_ISL_2490506, EPI_ISL_2490507, EPI_ISL_2490508, EPI_ISL_2490509, EPI_ISL_2490510, EPI_ISL_2490511, EPI_ISL_2490512, EPI_ISL_2490513, EPI_ISL_2490514, EPI_ISL_2490515, EPI_ISL_2490516, EPI_ISL_2490517, EPI_ISL_2490518, EPI_ISL_2490519, EPI_ISL_2490520, EPI_ISL_2490521, EPI_ISL_2490522, EPI_ISL_2490523, EPI_ISL_2490524, EPI_ISL_2490525, EPI_ISL_2490526, EPI_ISL_2490527, EPI_ISL_2490528, EPI_ISL_2490529, EPI_ISL_2490530, EPI_ISL_2490531, EPI_ISL_2490532, EPI_ISL_2490533, EPI_ISL_2490535, EPI_ISL_2490536, EPI_ISL_2490537, EPI_ISL_2490538, EPI_ISL_2490539, EPI_ISL_2490540, EPI_ISL_2490541 | | | |
| see above | Unidad de Investigación Biomédica de Zacatecas (UIBZ) | Centro de Investigación en Enfermedades Infecciosas (CIENI), Instituto Nacional de Enfermedades Respiratorias (INER) | Consorcio Mexicano de Vigilancia Genómica (CoViGen-Mex). Authors (in alphabetical order): Julio Elias Alvarado-Yaah, Carlos F. Arias, Santiago Ávila-Ríos, Eduardo Becerril-Vargas, Víctor Hugo Borja-Aburto, Celia Boukadida, Cristóbal Cháidez-Quiróz, Juan Bautista Chale-Dzul, Ricardo Ciria Merce, Andreu Comas-García, Célida Duque Molina, Julissa Enciso-Ibarra, José Antonio Enciso-Moreno, Gloria Elena Espinosa-Ayala, Fernando Fontove-Herrera, Daniel Fregoso-Rueda, Victor Eduardo Garcia-Arias, Alejandra Garcia-Gasca, Bruno Gomez-Gil, Jean Pierre González, Irvin González-López, Concepción Grajales-Muñiz, Ricardo Grande, Rosa María Gutierrez Rios, Alejandra Hernández-Terán, Alfredo Herrera-Estrella, Carla Ivón Herrera-Najera, Pavel Isa, Daniel Lira Morales, Susana Lopez, Antonio Loza Román, Brenda Irasema Maldonado-Meza, Bernardo Martínez-Miguel, José Arturo Martínez-Orozco, Célida Martínez- Rodríguez, Margarita Matías-Florentino, Fidencio Mejía-Nepomuceno, María Guadalupe de Jesús Mireles-Rivera, Gloria María Molina-Salinas, Hector Montoya-Fuentes, , Mario Mújica-Sánchez, José Esteban Muñoz-Medina, José de Jesús Nuñez-Contreras, Alicia Ocaña-Mondragón, Luis Alberto Ochoa-Carrera, Hector Esteban Paz-Juárez, Marissa Perez-Garcia, Francisco Pulido, Helen Haydee Fernanda Ramírez-Plascencia, Jorge Salas-Hernández, Angel Gustavo Salas-Lais, Alejandro Sanchez-Flores, Clara Esperanza Santacruz-Tinoco, María Guadalupe Santiago-Mauricio, Selene Zárate, Nelly Sélem-Mojica, Blanca Taboada, Gloria Vazquez, Joel Armando Vázquez-Pérez. |
| EPI_ISL_2490542, EPI_ISL_2490543, EPI_ISL_2490544, EPI_ISL_2490545, EPI_ISL_2490546, EPI_ISL_2490547, EPI_ISL_2490548, EPI_ISL_2490549, EPI_ISL_2490550, EPI_ISL_2490551, EPI_ISL_2490552, EPI_ISL_2490553, EPI_ISL_2490554, EPI_ISL_2490555, EPI_ISL_2490556, EPI_ISL_2490557, EPI_ISL_2490558, EPI_ISL_2490559, EPI_ISL_2490560, EPI_ISL_2490561, EPI_ISL_2490562, EPI_ISL_2490563, EPI_ISL_2490564, EPI_ISL_2490565, EPI_ISL_2490566, EPI_ISL_2490567, EPI_ISL_2490568, EPI_ISL_2490569, EPI_ISL_2490570, EPI_ISL_2490571, EPI_ISL_2490572, EPI_ISL_2490573, EPI_ISL_2490574, EPI_ISL_2490575, EPI_ISL_2490576, EPI_ISL_2490577, EPI_ISL_2490578, EPI_ISL_2490579, EPI_ISL_2490580, EPI_ISL_2490581, EPI_ISL_2490582, EPI_ISL_2490583, EPI_ISL_2490584 | | | |
| see above | Laboratorio Central de Epidemiología (LCE) | Centro de Investigación en Enfermedades Infecciosas (CIENI), Instituto Nacional de Enfermedades Respiratorias (INER) | Consorcio Mexicano de Vigilancia Genómica (CoViGen-Mex). Authors (in alphabetical order): Julio Elias Alvarado-Yaah, Carlos F. Arias, Santiago Ávila-Ríos, Eduardo Becerril-Vargas, Víctor Hugo Borja-Aburto, Celia Boukadida, Cristóbal Cháidez-Quiróz, Juan Bautista Chale-Dzul, Ricardo Ciria Merce, Andreu Comas-García, Célida Duque Molina, Julissa Enciso-Ibarra, José Antonio Enciso-Moreno, Gloria Elena Espinosa-Ayala, Fernando Fontove-Herrera, Daniel Fregoso-Rueda, Victor Eduardo Garcia-Arias, Alejandra Garcia-Gasca, Bruno Gomez-Gil, Jean Pierre González, Irvin González-López, Concepción Grajales-Muñiz, Ricardo Grande, Rosa María Gutierrez Rios, Alejandra Hernández-Terán, Alfredo Herrera-Estrella, Carla Ivón Herrera-Najera, Pavel Isa, Daniel Lira Morales, Susana Lopez, Antonio Loza Román, Brenda Irasema Maldonado-Meza, Bernardo Martínez-Miguel, José Arturo Martínez-Orozco, Célida Martínez- Rodríguez, Margarita Matías-Florentino, Fidencio Mejía-Nepomuceno, María Guadalupe de Jesús Mireles-Rivera, Gloria María Molina-Salinas, Hector Montoya-Fuentes, , Mario Mújica-Sánchez, José Esteban Muñoz-Medina, José de Jesús Nuñez-Contreras, Alicia Ocaña-Mondragón, Luis Alberto Ochoa-Carrera, Hector Esteban Paz-Juárez, Marissa Perez-Garcia, Francisco Pulido, Helen Haydee Fernanda Ramírez-Plascencia, Jorge Salas-Hernández, Angel Gustavo Salas-Lais, Alejandro Sanchez-Flores, Clara Esperanza Santacruz-Tinoco, María Guadalupe Santiago-Mauricio, Selene Zárate, Nelly Sélem-Mojica, Blanca Taboada, Gloria Vazquez, Joel Armando Vázquez-Pérez. |
| EPI_ISL_2490585, EPI_ISL_2490586, EPI_ISL_2490587, EPI_ISL_2490588, EPI_ISL_2490589, EPI_ISL_2490590, EPI_ISL_2490591, EPI_ISL_2490592, EPI_ISL_2490593, EPI_ISL_2490594, EPI_ISL_2490595, EPI_ISL_2490596, EPI_ISL_2490597, EPI_ISL_2490598, EPI_ISL_2490599, EPI_ISL_2490600, EPI_ISL_2490601, EPI_ISL_2490602, EPI_ISL_2490603, EPI_ISL_2490604, EPI_ISL_2490605, EPI_ISL_2490606, EPI_ISL_2490607, EPI_ISL_2490608 | | | |
| see above | Instituto Nacional de Enfermedades | Centro de Investigación en Enfermedades | Consorcio Mexicano de Vigilancia Genómica (CoViGen-Mex). Authors (in alphabetical order): Julio Elias Alvarado-Yaah, Carlos F. Arias, Santiago Ávila-Ríos, Eduardo Becerril-Vargas, Víctor Hugo Borja-Aburto, Celia Boukadida, Cristóbal Cháidez-Quiróz, Juan Bautista Chale-Dzul, Ricardo |

| | | | |
|---|---|---|---|
| | Respiratorias (INER) | Infecciosas (CIENI), Instituto Nacional de Enfermedades Respiratorias (INER) | Ciria Merce, Andreu Comas-García, Célida Duque Molina, Julissa Enciso-Ibarra, José Antonio Enciso-Moreno, Gloria Elena Espinosa-Ayala, Fernando Fontove-Herrera, Daniel Fregoso-Rueda, Victor Eduardo Garcia-Arias, Alejandra Garcia-Gasca, Bruno Gomez-Gil, Jean Pierre González, Irvin González-López, Concepción Grajales-Muñiz, Ricardo Grande, Rosa María Gutierrez Rios, Alejandra Hernández-Terán, Alfredo Herrera-Estrella, Carla Ivón Herrera-Najera, Pavel Isa, Daniel Lira Morales, Susana Lopez, Antonio Loza Román, Brenda Irasema Maldonado-Meza, Bernardo Martínez-Miguel, José Arturo Martínez-Orozco, Célida Martínez- Rodríguez, Margarita Matías-Florentino, Fidencio Mejía-Nepomuceno, María Guadalupe de Jesús Mireles-Rivera, Gloria María Molina-Salinas, Hector Montoya-Fuentes, , Mario Mújica-Sánchez, José Esteban Muñoz-Medina, José de Jesús Nuñez-Contreras, Alicia Ocaña-Mondragón, Luis Alberto Ochoa-Carrera, Hector Esteban Paz-Juárez, Marissa Perez-Garcia, Francisco Pulido, Helen Haydee Fernanda Ramírez-Plascencia, Jorge Salas-Hernández, Angel Gustavo Salas-Lais, Alejandro Sanchez-Flores, Clara Esperanza Santacruz-Tinoco, María Guadalupe Santiago-Mauricio, Selene Zárate, Nelly Sélem-Mojica, Blanca Taboada, Gloria Vazquez, Joel Armando Vázquez-Pérez. |
| EPI_ISL_2490634 | Ohio Department of Health Laboratory | Ohio Department of Health Laboratory | Holmes, Jennifer; Eric Brandt, Keoni Omura, Glen McGillivary, Caitlin McDonnell, Jade Mowery, Stephanie Mccracken, Tyler Payne, Kirtana Ramadugu, Erica Leasure, Brent Lee, Kelsey Florek, Heather Blankenship, Allison Black, Quanta Brown, and Tammy Bannerman |
| EPI_ISL_2490916, EPI_ISL_2490920, EPI_ISL_2490931, EPI_ISL_2490939, EPI_ISL_2490957, EPI_ISL_2490961, EPI_ISL_2490965, EPI_ISL_2490968, EPI_ISL_2490975, EPI_ISL_2490983, EPI_ISL_2490995, EPI_ISL_2490997, EPI_ISL_2490999, EPI_ISL_2491003, EPI_ISL_2491010, EPI_ISL_2491020, EPI_ISL_2491032, EPI_ISL_2491039, EPI_ISL_2491043, EPI_ISL_2491051, EPI_ISL_2491062, EPI_ISL_2491070, EPI_ISL_2491077, EPI_ISL_2491100, EPI_ISL_2491107, EPI_ISL_2491111, EPI_ISL_2491118, EPI_ISL_2491122, EPI_ISL_2491130, EPI_ISL_2491142, EPI_ISL_2491146, EPI_ISL_2491148, EPI_ISL_2491150, EPI_ISL_2491151, EPI_ISL_2491153, EPI_ISL_2491155, EPI_ISL_2491156, EPI_ISL_2491158, EPI_ISL_2491159, EPI_ISL_2491160, EPI_ISL_2491162, EPI_ISL_2491165, EPI_ISL_2491170, EPI_ISL_2491172, EPI_ISL_2491176, EPI_ISL_2491177, EPI_ISL_2491178, EPI_ISL_2491182, EPI_ISL_2491183, EPI_ISL_2491186, EPI_ISL_2491189, EPI_ISL_2491202, EPI_ISL_2491209, EPI_ISL_2491212, EPI_ISL_2491215, EPI_ISL_2491218, EPI_ISL_2491219, EPI_ISL_2491225, EPI_ISL_2491228, EPI_ISL_2491231, EPI_ISL_2491233, EPI_ISL_2491241, EPI_ISL_2491243, EPI_ISL_2491244, EPI_ISL_2491245, EPI_ISL_2491247, EPI_ISL_2491248, EPI_ISL_2491249, EPI_ISL_2491257, EPI_ISL_2491259, EPI_ISL_2491260, EPI_ISL_2491261, EPI_ISL_2491262, EPI_ISL_2491263, EPI_ISL_2491265, EPI_ISL_2491269, EPI_ISL_2491270, EPI_ISL_2491273, EPI_ISL_2491275, EPI_ISL_2491282, EPI_ISL_2491284, EPI_ISL_2491285, EPI_ISL_2491287, EPI_ISL_2491289, EPI_ISL_2491292, EPI_ISL_2491294, EPI_ISL_2491296, EPI_ISL_2491300, EPI_ISL_2491306, EPI_ISL_2491307, EPI_ISL_2491309, EPI_ISL_2491311, EPI_ISL_2491314, EPI_ISL_2491315, EPI_ISL_2491317, EPI_ISL_2491318, EPI_ISL_2491322, EPI_ISL_2491323, EPI_ISL_2491325, EPI_ISL_2491327, EPI_ISL_2491329 | | | |
| see above | TGen North | TGen North | "Jolene Bowers, Heather Centner, Chris French, Hayley Yaglom, Ashlyn Pfeiffer, Darrin Lemmer, Dave Engelthaler, The Arizona COVID Genomics Union (ACGU)" |
| EPI_ISL_2491369, EPI_ISL_2491376 | Ohio Department of Health Laboratory | Ohio Department of Health Laboratory | Holmes, Jennifer; Eric Brandt, Keoni Omura, Glen McGillivary, Caitlin McDonnell, Jade Mowery, Stephanie Mccracken, Tyler Payne, Kirtana Ramadugu, Erica Leasure, Brent Lee, Kelsey Florek, Heather Blankenship, Allison Black, Quanta Brown, and Tammy Bannerman |
| EPI_ISL_2491598, EPI_ISL_2491599 | Maryland Genomics, Institute for Genome Sciences, University of Maryland School of Medicine | Maryland Genomics, Institute for Genome Sciences, University of Maryland School of Medicine | Tallon, Luke J; Sadzewicz, Lisa D; Humphrys, Mike; Ott, Sandra; Roussey, Holly; Mehta, Aditya; Vavikolanu, Kranthi; Fraser, Claire M; Ravel, Jacques |
| EPI_ISL_2491672, EPI_ISL_2492328, EPI_ISL_2492358 | Ohio Department of Health Laboratory | Ohio Department of Health Laboratory | Holmes, Jennifer; Eric Brandt, Keoni Omura, Glen McGillivary, Caitlin McDonnell, Jade Mowery, Stephanie Mccracken, Tyler Payne, Kirtana Ramadugu, Erica Leasure, Brent Lee, Kelsey Florek, Heather Blankenship, Allison Black, Quanta Brown, and Tammy Bannerman |
| EPI_ISL_2492441, EPI_ISL_2492442, EPI_ISL_2492443, EPI_ISL_2492444, EPI_ISL_2492445, EPI_ISL_2492446, EPI_ISL_2492447, EPI_ISL_2492448, EPI_ISL_2492449, EPI_ISL_2492450, EPI_ISL_2492451, EPI_ISL_2492452, EPI_ISL_2492453, EPI_ISL_2492454, EPI_ISL_2492455, EPI_ISL_2492456, EPI_ISL_2492457, EPI_ISL_2492458, EPI_ISL_2492459, EPI_ISL_2492460, EPI_ISL_2492461, EPI_ISL_2492462, EPI_ISL_2492463 | | | |
| see above | LESP Queretaro | Instituto de Diagnostico y Referencia Epidemiologicos (INDRE) | Claudia Wong-Arambula, Abril Rodriguez-Maldonado, Vanessa Rivero-Arredondo, Ariadna Medina-Benitez, Joaquin Quiroz-Mercado, Sergio Rangel-Guerrero, Natividad Cruz-Ortiz, Tatiana Nunez-Garcia, Gisela Barrera-Badillo, Lucia Hernandez-Rivas, Irma Lopez-Martinez, Ernesto Ramirez-Gonzalez. |
| EPI_ISL_2492464 | LESP Nuevo Leon | Instituto de Diagnostico y Referencia Epidemiologicos (INDRE) | Claudia Wong-Arambula, Abril Rodriguez-Maldonado, Vanessa Rivero-Arredondo, Ariadna Medina-Benitez, Joaquin Quiroz-Mercado, Sergio Rangel-Guerrero, Natividad Cruz-Ortiz, Tatiana Nunez-Garcia, Gisela Barrera-Badillo, Lucia Hernandez-Rivas, Irma Lopez-Martinez, Ernesto Ramirez-Gonzalez. |
| EPI_ISL_2492465 | LESP Guanajuato | Instituto de Diagnostico y Referencia Epidemiologicos (INDRE) | Claudia Wong-Arambula, Abril Rodriguez-Maldonado, Vanessa Rivero-Arredondo, Ariadna Medina-Benitez, Joaquin Quiroz-Mercado, Sergio Rangel-Guerrero, Natividad Cruz-Ortiz, Tatiana Nunez-Garcia, Gisela Barrera-Badillo, Lucia Hernandez-Rivas, Irma Lopez-Martinez, Ernesto Ramirez-Gonzalez. |
| EPI_ISL_2492466 | LESP Ciudad de Mexico | Instituto de Diagnostico y Referencia Epidemiologicos (INDRE) | Claudia Wong-Arambula, Abril Rodriguez-Maldonado, Vanessa Rivero-Arredondo, Ariadna Medina-Benitez, Joaquin Quiroz-Mercado, Sergio Rangel-Guerrero, Natividad Cruz-Ortiz, Tatiana Nunez-Garcia, Gisela Barrera-Badillo, Lucia Hernandez-Rivas, Irma Lopez-Martinez, Ernesto Ramirez-Gonzalez. |
| EPI_ISL_2492467 | LESP Nuevo Leon | Instituto de Diagnostico y Referencia Epidemiologicos (INDRE) | Claudia Wong-Arambula, Abril Rodriguez-Maldonado, Vanessa Rivero-Arredondo, Ariadna Medina-Benitez, Joaquin Quiroz-Mercado, Sergio Rangel-Guerrero, Natividad Cruz-Ortiz, Tatiana Nunez-Garcia, Gisela Barrera-Badillo, Lucia Hernandez-Rivas, Irma Lopez-Martinez, Ernesto Ramirez-Gonzalez. |
| EPI_ISL_2492468, EPI_ISL_2492469, EPI_ISL_2492470 | LESP Queretaro | Instituto de Diagnostico y Referencia Epidemiologicos (INDRE) | Claudia Wong-Arambula, Abril Rodriguez-Maldonado, Vanessa Rivero-Arredondo, Ariadna Medina-Benitez, Joaquin Quiroz-Mercado, Sergio Rangel-Guerrero, Natividad Cruz-Ortiz, Tatiana Nunez-Garcia, Gisela Barrera-Badillo, Lucia Hernandez-Rivas, Irma Lopez-Martinez, Ernesto Ramirez-Gonzalez. |
| EPI_ISL_2492471 | LESP Guanajuato | Instituto de Diagnostico y Referencia Epidemiologicos (INDRE) | Claudia Wong-Arambula, Abril Rodriguez-Maldonado, Vanessa Rivero-Arredondo, Ariadna Medina-Benitez, Joaquin Quiroz-Mercado, Sergio Rangel-Guerrero, Natividad Cruz-Ortiz, Tatiana Nunez-Garcia, Gisela Barrera-Badillo, Lucia Hernandez-Rivas, Irma Lopez-Martinez, Ernesto Ramirez-Gonzalez. |
| EPI_ISL_2492472 | LESP Oaxaca | Instituto de Diagnostico y Referencia Epidemiologicos (INDRE) | Claudia Wong-Arambula, Abril Rodriguez-Maldonado, Vanessa Rivero-Arredondo, Ariadna Medina-Benitez, Joaquin Quiroz-Mercado, Sergio Rangel-Guerrero, Natividad Cruz-Ortiz, Tatiana Nunez-Garcia, Gisela Barrera-Badillo, Lucia Hernandez-Rivas, Irma Lopez-Martinez, Ernesto Ramirez-Gonzalez. |
| EPI_ISL_2492473 | LESP San Luis Potosi | Instituto de Diagnostico y Referencia Epidemiologicos (INDRE) | Claudia Wong-Arambula, Abril Rodriguez-Maldonado, Vanessa Rivero-Arredondo, Ariadna Medina-Benitez, Joaquin Quiroz-Mercado, Sergio Rangel-Guerrero, Natividad Cruz-Ortiz, Tatiana Nunez-Garcia, Gisela Barrera-Badillo, Lucia Hernandez-Rivas, Irma Lopez-Martinez, Ernesto Ramirez-Gonzalez. |
| EPI_ISL_2492474 | LESP Queretaro | Instituto de Diagnostico y Referencia Epidemiologicos (INDRE) | Claudia Wong-Arambula, Abril Rodriguez-Maldonado, Vanessa Rivero-Arredondo, Ariadna Medina-Benitez, Joaquin Quiroz-Mercado, Sergio Rangel-Guerrero, Natividad Cruz-Ortiz, Tatiana Nunez-Garcia, Gisela Barrera-Badillo, Lucia Hernandez-Rivas, Irma Lopez-Martinez, Ernesto Ramirez-Gonzalez. |
| EPI_ISL_2492475, EPI_ISL_2492476, EPI_ISL_2492477 | LESP Nuevo Leon | Instituto de Diagnostico y Referencia Epidemiologicos (INDRE) | Claudia Wong-Arambula, Abril Rodriguez-Maldonado, Vanessa Rivero-Arredondo, Ariadna Medina-Benitez, Joaquin Quiroz-Mercado, Sergio Rangel-Guerrero, Natividad Cruz-Ortiz, Tatiana Nunez-Garcia, Gisela Barrera-Badillo, Lucia Hernandez-Rivas, Irma Lopez-Martinez, Ernesto Ramirez-Gonzalez. |
| EPI_ISL_2492478, EPI_ISL_2492479, EPI_ISL_2492480, EPI_ISL_2492481, EPI_ISL_2492482, EPI_ISL_2492483 | LESP Guanajuato | Instituto de Diagnostico y Referencia Epidemiologicos (INDRE) | Claudia Wong-Arambula, Abril Rodriguez-Maldonado, Vanessa Rivero-Arredondo, Ariadna Medina-Benitez, Joaquin Quiroz-Mercado, Sergio Rangel-Guerrero, Natividad Cruz-Ortiz, Tatiana Nunez-Garcia, Gisela Barrera-Badillo, Lucia Hernandez-Rivas, Irma Lopez-Martinez, Ernesto Ramirez-Gonzalez. |
| EPI_ISL_2492484 | LESP Ciudad de Mexico | Instituto de Diagnostico y Referencia Epidemiologicos (INDRE) | Claudia Wong-Arambula, Abril Rodriguez-Maldonado, Vanessa Rivero-Arredondo, Ariadna Medina-Benitez, Joaquin Quiroz-Mercado, Sergio Rangel-Guerrero, Natividad Cruz-Ortiz, Tatiana Nunez-Garcia, Gisela Barrera-Badillo, Lucia Hernandez-Rivas, Irma Lopez-Martinez, Ernesto Ramirez-Gonzalez. |
| EPI_ISL_2492485 | LESP Guanajuato | Instituto de Diagnostico y Referencia Epidemiologicos (INDRE) | Claudia Wong-Arambula, Abril Rodriguez-Maldonado, Vanessa Rivero-Arredondo, Ariadna Medina-Benitez, Joaquin Quiroz-Mercado, Sergio Rangel-Guerrero, Natividad Cruz-Ortiz, Tatiana Nunez-Garcia, Gisela Barrera-Badillo, Lucia Hernandez-Rivas, Irma Lopez-Martinez, Ernesto Ramirez-Gonzalez. |
| EPI_ISL_2492486 | LESP Nuevo Leon | Instituto de Diagnostico y Referencia Epidemiologicos (INDRE) | Claudia Wong-Arambula, Abril Rodriguez-Maldonado, Vanessa Rivero-Arredondo, Ariadna Medina-Benitez, Joaquin Quiroz-Mercado, Sergio Rangel-Guerrero, Natividad Cruz-Ortiz, Tatiana Nunez-Garcia, Gisela Barrera-Badillo, Lucia Hernandez-Rivas, Irma Lopez-Martinez, Ernesto Ramirez-Gonzalez. |
| EPI_ISL_2492488 | LESP Puebla | Instituto de Diagnostico y Referencia Epidemiologicos (INDRE) | Claudia Wong-Arambula, Abril Rodriguez-Maldonado, Vanessa Rivero-Arredondo, Ariadna Medina-Benitez, Joaquin Quiroz-Mercado, Sergio Rangel-Guerrero, Natividad Cruz-Ortiz, Tatiana Nunez-Garcia, Gisela Barrera-Badillo, Lucia Hernandez-Rivas, Irma Lopez-Martinez, Ernesto Ramirez-Gonzalez. |
| EPI_ISL_2492489 | LESP Chiapas | Instituto de Diagnostico y Referencia Epidemiologicos (INDRE) | Claudia Wong-Arambula, Abril Rodriguez-Maldonado, Vanessa Rivero-Arredondo, Ariadna Medina-Benitez, Joaquin Quiroz-Mercado, Sergio Rangel-Guerrero, Natividad Cruz-Ortiz, Tatiana Nunez-Garcia, Gisela Barrera-Badillo, Lucia Hernandez-Rivas, Irma Lopez-Martinez, Ernesto Ramirez-Gonzalez. |
| EPI_ISL_2492490, EPI_ISL_2492491 | LESP Ciudad de Mexico | Instituto de Diagnostico y Referencia Epidemiologicos (INDRE) | Claudia Wong-Arambula, Abril Rodriguez-Maldonado, Vanessa Rivero-Arredondo, Ariadna Medina-Benitez, Joaquin Quiroz-Mercado, Sergio Rangel-Guerrero, Natividad Cruz-Ortiz, Tatiana Nunez-Garcia, Gisela Barrera-Badillo, Lucia Hernandez-Rivas, Irma Lopez-Martinez, Ernesto Ramirez-Gonzalez. |
| EPI_ISL_2492492, EPI_ISL_2492493 | LESP Tamaulipas | Instituto de Diagnostico y Referencia Epidemiologicos (INDRE) | Claudia Wong-Arambula, Abril Rodriguez-Maldonado, Vanessa Rivero-Arredondo, Ariadna Medina-Benitez, Joaquin Quiroz-Mercado, Sergio Rangel-Guerrero, Natividad Cruz-Ortiz, Tatiana Nunez-Garcia, Gisela Barrera-Badillo, Lucia Hernandez-Rivas, Irma Lopez-Martinez, Ernesto Ramirez-Gonzalez. |
| EPI_ISL_2492494 | LESP Chiapas | Instituto de Diagnostico y Referencia Epidemiologicos (INDRE) | Claudia Wong-Arambula, Abril Rodriguez-Maldonado, Vanessa Rivero-Arredondo, Ariadna Medina-Benitez, Joaquin Quiroz-Mercado, Sergio Rangel-Guerrero, Natividad Cruz-Ortiz, Tatiana Nunez-Garcia, Gisela Barrera-Badillo, Lucia Hernandez-Rivas, Irma Lopez-Martinez, Ernesto Ramirez-Gonzalez. |
| EPI_ISL_2492495, EPI_ISL_2492496 | LESP Quintana Roo | Instituto de Diagnostico y Referencia Epidemiologicos (INDRE) | Claudia Wong-Arambula, Abril Rodriguez-Maldonado, Vanessa Rivero-Arredondo, Ariadna Medina-Benitez, Joaquin Quiroz-Mercado, Sergio Rangel-Guerrero, Natividad Cruz-Ortiz, Tatiana Nunez-Garcia, Gisela Barrera-Badillo, Lucia Hernandez-Rivas, Irma Lopez-Martinez, Ernesto Ramirez-Gonzalez. |
| EPI_ISL_2492497, EPI_ISL_2492498 | LESP Queretaro | Instituto de Diagnostico y Referencia Epidemiologicos (INDRE) | Claudia Wong-Arambula, Abril Rodriguez-Maldonado, Vanessa Rivero-Arredondo, Ariadna Medina-Benitez, Joaquin Quiroz-Mercado, Sergio Rangel-Guerrero, Natividad Cruz-Ortiz, Tatiana Nunez-Garcia, Gisela Barrera-Badillo, Lucia Hernandez-Rivas, Irma Lopez-Martinez, Ernesto Ramirez-Gonzalez. |
| EPI_ISL_2494632, EPI_ISL_2494633, EPI_ISL_2494634, EPI_ISL_2494635, EPI_ISL_2494637, EPI_ISL_2494638, EPI_ISL_2494640, EPI_ISL_2494642, EPI_ISL_2494643, EPI_ISL_2494644, EPI_ISL_2494645, EPI_ISL_2494647, EPI_ISL_2494651, EPI_ISL_2494652, EPI_ISL_2494653, EPI_ISL_2494654, EPI_ISL_2494655, EPI_ISL_2494656, EPI_ISL_2494657, EPI_ISL_2494658, EPI_ISL_2494659, EPI_ISL_2494660, EPI_ISL_2494661, EPI_ISL_2494662, EPI_ISL_2494663, EPI_ISL_2494664, EPI_ISL_2494665, EPI_ISL_2494666, EPI_ISL_2494667, EPI_ISL_2494668, EPI_ISL_2494669, EPI_ISL_2494670, EPI_ISL_2494671, EPI_ISL_2494672, EPI_ISL_2494673, EPI_ISL_2494674, EPI_ISL_2494676, EPI_ISL_2494677, EPI_ISL_2494679, EPI_ISL_2494680, EPI_ISL_2494681, EPI_ISL_2494682, EPI_ISL_2494683, EPI_ISL_2494686, EPI_ISL_2494688, EPI_ISL_2494689, EPI_ISL_2494691, EPI_ISL_2494695, EPI_ISL_2494697, EPI_ISL_2494699, EPI_ISL_2494700, EPI_ISL_2494701, EPI_ISL_2494702, EPI_ISL_2494703, EPI_ISL_2494704, EPI_ISL_2494706, EPI_ISL_2494709 | | | |
| see above | UCSC Genomics Institute | UCSC Genomics Institute | Mark Akeson, Ikenna Anigbogu, Eric Beraut, Isabel Bjork, Molly Cassatt-Johnstone, Terren Chang, Russell Corbett-Detig, Namrita Dhillon, Maximilian Haeussler, David Haussler, Angie Hinrichs, Miten Jain, Joshua Kapp, A. Marm Kilpatrick, Jakob McBroome, Hugh Olsen, Jeremy Sanford, Beth Shapiro, Michael Stone, Bryan Thornlow, Yatish Turakhia, Ciara Wanket |
| EPI_ISL_2495922, EPI_ISL_2495923 | LESP Queretaro | Instituto de Diagnostico y Referencia Epidemiologicos (INDRE) | Claudia Wong-Arambula, Abril Rodriguez-Maldonado, Vanessa Rivero-Arredondo, Ariadna Medina-Benitez, Joaquin Quiroz-Mercado, Sergio Rangel-Guerrero, Natividad Cruz-Ortiz, Tatiana Nunez-Garcia, Gisela Barrera-Badillo, Lucia Hernandez-Rivas, Irma Lopez-Martinez, Ernesto Ramirez-Gonzalez. |
| EPI_ISL_2495924 | LESP Aguascalientes | Instituto de Diagnostico y Referencia Epidemiologicos (INDRE) | Claudia Wong-Arambula, Abril Rodriguez-Maldonado, Vanessa Rivero-Arredondo, Ariadna Medina-Benitez, Joaquin Quiroz-Mercado, Sergio Rangel-Guerrero, Natividad Cruz-Ortiz, Tatiana Nunez-Garcia, Gisela Barrera-Badillo, Lucia Hernandez-Rivas, Irma Lopez-Martinez, Ernesto Ramirez-Gonzalez. |
| EPI_ISL_2495925, EPI_ISL_2495926, EPI_ISL_2495927, EPI_ISL_2495928 | LESP Queretaro | Instituto de Diagnostico y Referencia Epidemiologicos (INDRE) | Claudia Wong-Arambula, Abril Rodriguez-Maldonado, Vanessa Rivero-Arredondo, Ariadna Medina-Benitez, Joaquin Quiroz-Mercado, Sergio Rangel-Guerrero, Natividad Cruz-Ortiz, Tatiana Nunez-Garcia, Gisela Barrera-Badillo, Lucia Hernandez-Rivas, Irma Lopez-Martinez, Ernesto Ramirez-Gonzalez. |
| EPI_ISL_2495929 | LESP Puebla | Instituto de Diagnostico y Referencia Epidemiologicos (INDRE) | Claudia Wong-Arambula, Abril Rodriguez-Maldonado, Vanessa Rivero-Arredondo, Ariadna Medina-Benitez, Joaquin Quiroz-Mercado, Sergio Rangel-Guerrero, Natividad Cruz-Ortiz, Tatiana Nunez-Garcia, Gisela Barrera-Badillo, Lucia Hernandez-Rivas, Irma Lopez-Martinez, Ernesto Ramirez-Gonzalez. |
| EPI_ISL_2495930 | LESP Jalisco | Instituto de Diagnostico y Referencia Epidemiologicos (INDRE) | Claudia Wong-Arambula, Abril Rodriguez-Maldonado, Vanessa Rivero-Arredondo, Ariadna Medina-Benitez, Joaquin Quiroz-Mercado, Sergio Rangel-Guerrero, Natividad Cruz-Ortiz, Tatiana Nunez-Garcia, Gisela Barrera-Badillo, Lucia Hernandez-Rivas, Irma Lopez-Martinez, Ernesto Ramirez-Gonzalez. |
| EPI_ISL_2495931, EPI_ISL_2495932 | LESP Queretaro | Instituto de Diagnostico y Referencia Epidemiologicos (INDRE) | Claudia Wong-Arambula, Abril Rodriguez-Maldonado, Vanessa Rivero-Arredondo, Ariadna Medina-Benitez, Joaquin Quiroz-Mercado, Sergio Rangel-Guerrero, Natividad Cruz-Ortiz, Tatiana Nunez-Garcia, Gisela Barrera-Badillo, Lucia Hernandez-Rivas, Irma Lopez-Martinez, Ernesto Ramirez-Gonzalez. |
| EPI_ISL_2495933 | LESP Morelos | Instituto de Diagnostico y Referencia Epidemiologicos (INDRE) | Claudia Wong-Arambula, Abril Rodriguez-Maldonado, Vanessa Rivero-Arredondo, Ariadna Medina-Benitez, Joaquin Quiroz-Mercado, Sergio Rangel-Guerrero, Natividad Cruz-Ortiz, Tatiana Nunez-Garcia, Gisela Barrera-Badillo, Lucia Hernandez-Rivas, Irma Lopez-Martinez, Ernesto Ramirez-Gonzalez. |

| | | | |
|---|---|---|---|
| EPI_ISL_2495934 | LESP Nuevo Leon | Instituto de Diagnostico y Referencia Epidemiologicos (INDRE) | Claudia Wong-Arambula, Abril Rodriguez-Maldonado, Vanessa Rivero-Arredondo, Ariadna Medina-Benitez, Joaquin Quiroz-Mercado, Sergio Rangel-Guerrero, Natividad Cruz-Ortiz, Tatiana Nunez-Garcia, Gisela Barrera-Badillo, Lucia Hernandez-Rivas, Irma Lopez-Martinez, Ernesto Ramirez-Gonzalez. |
| EPI_ISL_2495935 | LESP Morelos | Instituto de Diagnostico y Referencia Epidemiologicos (INDRE) | Claudia Wong-Arambula, Abril Rodriguez-Maldonado, Vanessa Rivero-Arredondo, Ariadna Medina-Benitez, Joaquin Quiroz-Mercado, Sergio Rangel-Guerrero, Natividad Cruz-Ortiz, Tatiana Nunez-Garcia, Gisela Barrera-Badillo, Lucia Hernandez-Rivas, Irma Lopez-Martinez, Ernesto Ramirez-Gonzalez. |
| EPI_ISL_2495936 | LESP Michoacan | Instituto de Diagnostico y Referencia Epidemiologicos (INDRE) | Claudia Wong-Arambula, Abril Rodriguez-Maldonado, Vanessa Rivero-Arredondo, Ariadna Medina-Benitez, Joaquin Quiroz-Mercado, Sergio Rangel-Guerrero, Natividad Cruz-Ortiz, Tatiana Nunez-Garcia, Gisela Barrera-Badillo, Lucia Hernandez-Rivas, Irma Lopez-Martinez, Ernesto Ramirez-Gonzalez. |
| EPI_ISL_2495937, EPI_ISL_2495938, EPI_ISL_2495939, EPI_ISL_2495940, EPI_ISL_2495941, EPI_ISL_2495942, EPI_ISL_2495943, EPI_ISL_2495944, EPI_ISL_2495945, EPI_ISL_2495946, EPI_ISL_2495947, EPI_ISL_2495948, EPI_ISL_2495949, EPI_ISL_2495950, EPI_ISL_2495951, EPI_ISL_2495952, EPI_ISL_2495953, EPI_ISL_2495954, EPI_ISL_2495955, EPI_ISL_2495956, EPI_ISL_2495957, EPI_ISL_2495958, EPI_ISL_2495959, EPI_ISL_2495960, EPI_ISL_2495961, EPI_ISL_2495962 | | | |
| see above | LESP Queretaro | Instituto de Diagnostico y Referencia Epidemiologicos (INDRE) | Claudia Wong-Arambula, Abril Rodriguez-Maldonado, Vanessa Rivero-Arredondo, Ariadna Medina-Benitez, Joaquin Quiroz-Mercado, Sergio Rangel-Guerrero, Natividad Cruz-Ortiz, Tatiana Nunez-Garcia, Gisela Barrera-Badillo, Lucia Hernandez-Rivas, Irma Lopez-Martinez, Ernesto Ramirez-Gonzalez. |
| EPI_ISL_2495963 | LESP Nuevo Leon | Instituto de Diagnostico y Referencia Epidemiologicos (INDRE) | Claudia Wong-Arambula, Abril Rodriguez-Maldonado, Vanessa Rivero-Arredondo, Ariadna Medina-Benitez, Joaquin Quiroz-Mercado, Sergio Rangel-Guerrero, Natividad Cruz-Ortiz, Tatiana Nunez-Garcia, Gisela Barrera-Badillo, Lucia Hernandez-Rivas, Irma Lopez-Martinez, Ernesto Ramirez-Gonzalez. |
| EPI_ISL_2495964 | LESP Guanajuato | Instituto de Diagnostico y Referencia Epidemiologicos (INDRE) | Claudia Wong-Arambula, Abril Rodriguez-Maldonado, Vanessa Rivero-Arredondo, Ariadna Medina-Benitez, Joaquin Quiroz-Mercado, Sergio Rangel-Guerrero, Natividad Cruz-Ortiz, Tatiana Nunez-Garcia, Gisela Barrera-Badillo, Lucia Hernandez-Rivas, Irma Lopez-Martinez, Ernesto Ramirez-Gonzalez. |
| EPI_ISL_2495965 | LESP Yucatan | Instituto de Diagnostico y Referencia Epidemiologicos (INDRE) | Claudia Wong-Arambula, Abril Rodriguez-Maldonado, Vanessa Rivero-Arredondo, Ariadna Medina-Benitez, Joaquin Quiroz-Mercado, Sergio Rangel-Guerrero, Natividad Cruz-Ortiz, Tatiana Nunez-Garcia, Gisela Barrera-Badillo, Lucia Hernandez-Rivas, Irma Lopez-Martinez, Ernesto Ramirez-Gonzalez. |
| EPI_ISL_2495966 | LESP Quintana Roo | Instituto de Diagnostico y Referencia Epidemiologicos (INDRE) | Claudia Wong-Arambula, Abril Rodriguez-Maldonado, Vanessa Rivero-Arredondo, Ariadna Medina-Benitez, Joaquin Quiroz-Mercado, Sergio Rangel-Guerrero, Natividad Cruz-Ortiz, Tatiana Nunez-Garcia, Gisela Barrera-Badillo, Lucia Hernandez-Rivas, Irma Lopez-Martinez, Ernesto Ramirez-Gonzalez. |
| EPI_ISL_2495967, EPI_ISL_2495968, EPI_ISL_2495969 | LESP Queretaro | Instituto de Diagnostico y Referencia Epidemiologicos (INDRE) | Claudia Wong-Arambula, Abril Rodriguez-Maldonado, Vanessa Rivero-Arredondo, Ariadna Medina-Benitez, Joaquin Quiroz-Mercado, Sergio Rangel-Guerrero, Natividad Cruz-Ortiz, Tatiana Nunez-Garcia, Gisela Barrera-Badillo, Lucia Hernandez-Rivas, Irma Lopez-Martinez, Ernesto Ramirez-Gonzalez. |
| EPI_ISL_2495970 | LESP Michoacan | Instituto de Diagnostico y Referencia Epidemiologicos (INDRE) | Claudia Wong-Arambula, Abril Rodriguez-Maldonado, Vanessa Rivero-Arredondo, Ariadna Medina-Benitez, Joaquin Quiroz-Mercado, Sergio Rangel-Guerrero, Natividad Cruz-Ortiz, Tatiana Nunez-Garcia, Gisela Barrera-Badillo, Lucia Hernandez-Rivas, Irma Lopez-Martinez, Ernesto Ramirez-Gonzalez. |
| EPI_ISL_2495971, EPI_ISL_2495972, EPI_ISL_2495973, EPI_ISL_2495974, EPI_ISL_2495975 | LESP Zacatecas | Instituto de Diagnostico y Referencia Epidemiologicos (INDRE) | Claudia Wong-Arambula, Abril Rodriguez-Maldonado, Vanessa Rivero-Arredondo, Ariadna Medina-Benitez, Joaquin Quiroz-Mercado, Sergio Rangel-Guerrero, Natividad Cruz-Ortiz, Tatiana Nunez-Garcia, Gisela Barrera-Badillo, Lucia Hernandez-Rivas, Irma Lopez-Martinez, Ernesto Ramirez-Gonzalez. |
| EPI_ISL_2495976 | LESP Hidalgo | Instituto de Diagnostico y Referencia Epidemiologicos (INDRE) | Claudia Wong-Arambula, Abril Rodriguez-Maldonado, Vanessa Rivero-Arredondo, Ariadna Medina-Benitez, Joaquin Quiroz-Mercado, Sergio Rangel-Guerrero, Natividad Cruz-Ortiz, Tatiana Nunez-Garcia, Gisela Barrera-Badillo, Lucia Hernandez-Rivas, Irma Lopez-Martinez, Ernesto Ramirez-Gonzalez. |
| EPI_ISL_2495977, EPI_ISL_2495978 | LESP Queretaro | Instituto de Diagnostico y Referencia Epidemiologicos (INDRE) | Claudia Wong-Arambula, Abril Rodriguez-Maldonado, Vanessa Rivero-Arredondo, Ariadna Medina-Benitez, Joaquin Quiroz-Mercado, Sergio Rangel-Guerrero, Natividad Cruz-Ortiz, Tatiana Nunez-Garcia, Gisela Barrera-Badillo, Lucia Hernandez-Rivas, Irma Lopez-Martinez, Ernesto Ramirez-Gonzalez. |
| EPI_ISL_2495979 | LESP Tabasco | Instituto de Diagnostico y Referencia Epidemiologicos (INDRE) | Claudia Wong-Arambula, Abril Rodriguez-Maldonado, Vanessa Rivero-Arredondo, Ariadna Medina-Benitez, Joaquin Quiroz-Mercado, Sergio Rangel-Guerrero, Natividad Cruz-Ortiz, Tatiana Nunez-Garcia, Gisela Barrera-Badillo, Lucia Hernandez-Rivas, Irma Lopez-Martinez, Ernesto Ramirez-Gonzalez. |
| EPI_ISL_2495980, EPI_ISL_2495981 | LESP Nuevo Leon | Instituto de Diagnostico y Referencia Epidemiologicos (INDRE) | Claudia Wong-Arambula, Abril Rodriguez-Maldonado, Vanessa Rivero-Arredondo, Ariadna Medina-Benitez, Joaquin Quiroz-Mercado, Sergio Rangel-Guerrero, Natividad Cruz-Ortiz, Tatiana Nunez-Garcia, Gisela Barrera-Badillo, Lucia Hernandez-Rivas, Irma Lopez-Martinez, Ernesto Ramirez-Gonzalez. |
| EPI_ISL_2495982, EPI_ISL_2495983, EPI_ISL_2495984, EPI_ISL_2495985, EPI_ISL_2495986, EPI_ISL_2495987 | LESP Guanajuato | Instituto de Diagnostico y Referencia Epidemiologicos (INDRE) | Claudia Wong-Arambula, Abril Rodriguez-Maldonado, Vanessa Rivero-Arredondo, Ariadna Medina-Benitez, Joaquin Quiroz-Mercado, Sergio Rangel-Guerrero, Natividad Cruz-Ortiz, Tatiana Nunez-Garcia, Gisela Barrera-Badillo, Lucia Hernandez-Rivas, Irma Lopez-Martinez, Ernesto Ramirez-Gonzalez. |
| EPI_ISL_2495988 | LESP Guerrero | Instituto de Diagnostico y Referencia Epidemiologicos (INDRE) | Claudia Wong-Arambula, Abril Rodriguez-Maldonado, Vanessa Rivero-Arredondo, Ariadna Medina-Benitez, Joaquin Quiroz-Mercado, Sergio Rangel-Guerrero, Natividad Cruz-Ortiz, Tatiana Nunez-Garcia, Gisela Barrera-Badillo, Lucia Hernandez-Rivas, Irma Lopez-Martinez, Ernesto Ramirez-Gonzalez. |
| EPI_ISL_2495989, EPI_ISL_2495990, EPI_ISL_2495991 | LESP Guanajuato | Instituto de Diagnostico y Referencia Epidemiologicos (INDRE) | Claudia Wong-Arambula, Abril Rodriguez-Maldonado, Vanessa Rivero-Arredondo, Ariadna Medina-Benitez, Joaquin Quiroz-Mercado, Sergio Rangel-Guerrero, Natividad Cruz-Ortiz, Tatiana Nunez-Garcia, Gisela Barrera-Badillo, Lucia Hernandez-Rivas, Irma Lopez-Martinez, Ernesto Ramirez-Gonzalez. |
| EPI_ISL_2495992 | LESP Morelos | Instituto de Diagnostico y Referencia Epidemiologicos (INDRE) | Claudia Wong-Arambula, Abril Rodriguez-Maldonado, Vanessa Rivero-Arredondo, Ariadna Medina-Benitez, Joaquin Quiroz-Mercado, Sergio Rangel-Guerrero, Natividad Cruz-Ortiz, Tatiana Nunez-Garcia, Gisela Barrera-Badillo, Lucia Hernandez-Rivas, Irma Lopez-Martinez, Ernesto Ramirez-Gonzalez. |
| EPI_ISL_2495993 | LESP Campeche | Instituto de Diagnostico y Referencia Epidemiologicos (INDRE) | Claudia Wong-Arambula, Abril Rodriguez-Maldonado, Vanessa Rivero-Arredondo, Ariadna Medina-Benitez, Joaquin Quiroz-Mercado, Sergio Rangel-Guerrero, Natividad Cruz-Ortiz, Tatiana Nunez-Garcia, Gisela Barrera-Badillo, Lucia Hernandez-Rivas, Irma Lopez-Martinez, Ernesto Ramirez-Gonzalez. |
| EPI_ISL_2495994, EPI_ISL_2495995, EPI_ISL_2495996 | LESP Morelos | Instituto de Diagnostico y Referencia Epidemiologicos (INDRE) | Claudia Wong-Arambula, Abril Rodriguez-Maldonado, Vanessa Rivero-Arredondo, Ariadna Medina-Benitez, Joaquin Quiroz-Mercado, Sergio Rangel-Guerrero, Natividad Cruz-Ortiz, Tatiana Nunez-Garcia, Gisela Barrera-Badillo, Lucia Hernandez-Rivas, Irma Lopez-Martinez, Ernesto Ramirez-Gonzalez. |
| EPI_ISL_2495997 | LESP Guanajuato | Instituto de Diagnostico y Referencia Epidemiologicos (INDRE) | Claudia Wong-Arambula, Abril Rodriguez-Maldonado, Vanessa Rivero-Arredondo, Ariadna Medina-Benitez, Joaquin Quiroz-Mercado, Sergio Rangel-Guerrero, Natividad Cruz-Ortiz, Tatiana Nunez-Garcia, Gisela Barrera-Badillo, Lucia Hernandez-Rivas, Irma Lopez-Martinez, Ernesto Ramirez-Gonzalez. |
| EPI_ISL_2495998 | LESP Nuevo Leon | Instituto de Diagnostico y Referencia Epidemiologicos (INDRE) | Claudia Wong-Arambula, Abril Rodriguez-Maldonado, Vanessa Rivero-Arredondo, Ariadna Medina-Benitez, Joaquin Quiroz-Mercado, Sergio Rangel-Guerrero, Natividad Cruz-Ortiz, Tatiana Nunez-Garcia, Gisela Barrera-Badillo, Lucia Hernandez-Rivas, Irma Lopez-Martinez, Ernesto Ramirez-Gonzalez. |
| EPI_ISL_2495999, EPI_ISL_2496000 | LESP Guanajuato | Instituto de Diagnostico y Referencia Epidemiologicos (INDRE) | Claudia Wong-Arambula, Abril Rodriguez-Maldonado, Vanessa Rivero-Arredondo, Ariadna Medina-Benitez, Joaquin Quiroz-Mercado, Sergio Rangel-Guerrero, Natividad Cruz-Ortiz, Tatiana Nunez-Garcia, Gisela Barrera-Badillo, Lucia Hernandez-Rivas, Irma Lopez-Martinez, Ernesto Ramirez-Gonzalez. |
| EPI_ISL_2496001 | LESP Puebla | Instituto de Diagnostico y Referencia Epidemiologicos (INDRE) | Claudia Wong-Arambula, Abril Rodriguez-Maldonado, Vanessa Rivero-Arredondo, Ariadna Medina-Benitez, Joaquin Quiroz-Mercado, Sergio Rangel-Guerrero, Natividad Cruz-Ortiz, Tatiana Nunez-Garcia, Gisela Barrera-Badillo, Lucia Hernandez-Rivas, Irma Lopez-Martinez, Ernesto Ramirez-Gonzalez. |
| EPI_ISL_2496002 | LESP Guanajuato | Instituto de Diagnostico y Referencia Epidemiologicos (INDRE) | Claudia Wong-Arambula, Abril Rodriguez-Maldonado, Vanessa Rivero-Arredondo, Ariadna Medina-Benitez, Joaquin Quiroz-Mercado, Sergio Rangel-Guerrero, Natividad Cruz-Ortiz, Tatiana Nunez-Garcia, Gisela Barrera-Badillo, Lucia Hernandez-Rivas, Irma Lopez-Martinez, Ernesto Ramirez-Gonzalez. |
| EPI_ISL_2496003 | LESP Chihuahua | Instituto de Diagnostico y Referencia Epidemiologicos (INDRE) | Claudia Wong-Arambula, Abril Rodriguez-Maldonado, Vanessa Rivero-Arredondo, Ariadna Medina-Benitez, Joaquin Quiroz-Mercado, Sergio Rangel-Guerrero, Natividad Cruz-Ortiz, Tatiana Nunez-Garcia, Gisela Barrera-Badillo, Lucia Hernandez-Rivas, Irma Lopez-Martinez, Ernesto Ramirez-Gonzalez. |
| EPI_ISL_2496004 | LESP Tamaulipas | Instituto de Diagnostico y Referencia Epidemiologicos (INDRE) | Claudia Wong-Arambula, Abril Rodriguez-Maldonado, Vanessa Rivero-Arredondo, Ariadna Medina-Benitez, Joaquin Quiroz-Mercado, Sergio Rangel-Guerrero, Natividad Cruz-Ortiz, Tatiana Nunez-Garcia, Gisela Barrera-Badillo, Lucia Hernandez-Rivas, Irma Lopez-Martinez, Ernesto Ramirez-Gonzalez. |
| EPI_ISL_2496005 | LESP Campeche | Instituto de Diagnostico y Referencia Epidemiologicos (INDRE) | Claudia Wong-Arambula, Abril Rodriguez-Maldonado, Vanessa Rivero-Arredondo, Ariadna Medina-Benitez, Joaquin Quiroz-Mercado, Sergio Rangel-Guerrero, Natividad Cruz-Ortiz, Tatiana Nunez-Garcia, Gisela Barrera-Badillo, Lucia Hernandez-Rivas, Irma Lopez-Martinez, Ernesto Ramirez-Gonzalez. |
| EPI_ISL_2496006, EPI_ISL_2496007, EPI_ISL_2496008, EPI_ISL_2496009, EPI_ISL_2496010, EPI_ISL_2496011, EPI_ISL_2496012 | LESP Michoacan | Instituto de Diagnostico y Referencia Epidemiologicos (INDRE) | Claudia Wong-Arambula, Abril Rodriguez-Maldonado, Vanessa Rivero-Arredondo, Ariadna Medina-Benitez, Joaquin Quiroz-Mercado, Sergio Rangel-Guerrero, Natividad Cruz-Ortiz, Tatiana Nunez-Garcia, Gisela Barrera-Badillo, Lucia Hernandez-Rivas, Irma Lopez-Martinez, Ernesto Ramirez-Gonzalez. |
| EPI_ISL_2496013, EPI_ISL_2496014 | LESP Sinaloa | Instituto de Diagnostico y Referencia Epidemiologicos (INDRE) | Claudia Wong-Arambula, Abril Rodriguez-Maldonado, Vanessa Rivero-Arredondo, Ariadna Medina-Benitez, Joaquin Quiroz-Mercado, Sergio Rangel-Guerrero, Natividad Cruz-Ortiz, Tatiana Nunez-Garcia, Gisela Barrera-Badillo, Lucia Hernandez-Rivas, Irma Lopez-Martinez, Ernesto Ramirez-Gonzalez. |
| EPI_ISL_2496015, EPI_ISL_2496016, EPI_ISL_2496017, EPI_ISL_2496018, EPI_ISL_2496019 | LESP Ciudad de Mexico | Instituto de Diagnostico y Referencia Epidemiologicos (INDRE) | Claudia Wong-Arambula, Abril Rodriguez-Maldonado, Vanessa Rivero-Arredondo, Ariadna Medina-Benitez, Joaquin Quiroz-Mercado, Sergio Rangel-Guerrero, Natividad Cruz-Ortiz, Tatiana Nunez-Garcia, Gisela Barrera-Badillo, Lucia Hernandez-Rivas, Irma Lopez-Martinez, Ernesto Ramirez-Gonzalez. |
| EPI_ISL_2496020, EPI_ISL_2496021, EPI_ISL_2496022 | LESP Queretaro | Instituto de Diagnostico y Referencia Epidemiologicos (INDRE) | Claudia Wong-Arambula, Abril Rodriguez-Maldonado, Vanessa Rivero-Arredondo, Ariadna Medina-Benitez, Joaquin Quiroz-Mercado, Sergio Rangel-Guerrero, Natividad Cruz-Ortiz, Tatiana Nunez-Garcia, Gisela Barrera-Badillo, Lucia Hernandez-Rivas, Irma Lopez-Martinez, Ernesto Ramirez-Gonzalez. |
| EPI_ISL_2496023, EPI_ISL_2496024 | LESP Jalisco | Instituto de Diagnostico y Referencia Epidemiologicos (INDRE) | Claudia Wong-Arambula, Abril Rodriguez-Maldonado, Vanessa Rivero-Arredondo, Ariadna Medina-Benitez, Joaquin Quiroz-Mercado, Sergio Rangel-Guerrero, Natividad Cruz-Ortiz, Tatiana Nunez-Garcia, Gisela Barrera-Badillo, Lucia Hernandez-Rivas, Irma Lopez-Martinez, Ernesto Ramirez-Gonzalez. |
| EPI_ISL_2496025 | LESP Quintana Roo | Instituto de Diagnostico y Referencia Epidemiologicos (INDRE) | Claudia Wong-Arambula, Abril Rodriguez-Maldonado, Vanessa Rivero-Arredondo, Ariadna Medina-Benitez, Joaquin Quiroz-Mercado, Sergio Rangel-Guerrero, Natividad Cruz-Ortiz, Tatiana Nunez-Garcia, Gisela Barrera-Badillo, Lucia Hernandez-Rivas, Irma Lopez-Martinez, Ernesto Ramirez-Gonzalez. |
| EPI_ISL_2496026, EPI_ISL_2496027, EPI_ISL_2496028, EPI_ISL_2496029, EPI_ISL_2496030, EPI_ISL_2496031, EPI_ISL_2496032, EPI_ISL_2496033, EPI_ISL_2496034, EPI_ISL_2496035, EPI_ISL_2496036, EPI_ISL_2496037, EPI_ISL_2496038, EPI_ISL_2496039, EPI_ISL_2496040, EPI_ISL_2496041, EPI_ISL_2496042, EPI_ISL_2496043, EPI_ISL_2496044, EPI_ISL_2496045, EPI_ISL_2496046, EPI_ISL_2496047, EPI_ISL_2496048, EPI_ISL_2496049, EPI_ISL_2496050, EPI_ISL_2496051, EPI_ISL_2496052, EPI_ISL_2496053, EPI_ISL_2496054, EPI_ISL_2496055, EPI_ISL_2496056, EPI_ISL_2496057, EPI_ISL_2496058, EPI_ISL_2496059, EPI_ISL_2496060, EPI_ISL_2496061, EPI_ISL_2496062, EPI_ISL_2496063, EPI_ISL_2496064, EPI_ISL_2496065, EPI_ISL_2496066, EPI_ISL_2496067, EPI_ISL_2496068, EPI_ISL_2496069, EPI_ISL_2496070, EPI_ISL_2496071, EPI_ISL_2496072, EPI_ISL_2496090, EPI_ISL_2496091, EPI_ISL_2496092 | | | |

| | | | |
|---|---|---|---|
| see above | North Dakota Department of Health, Public Health Laboratory | North Dakota Department of Health, Public Health Laboratory | Lisa Wingerter |

EPI_ISL_2496100, EPI_ISL_2496101, EPI_ISL_2496102, EPI_ISL_2496103, EPI_ISL_2496104, EPI_ISL_2496105, EPI_ISL_2496106, EPI_ISL_2496107, EPI_ISL_2496108, EPI_ISL_2496109, EPI_ISL_2496110, EPI_ISL_2496111, EPI_ISL_2496112, EPI_ISL_2496113, EPI_ISL_2496114, EPI_ISL_2496115, EPI_ISL_2496116, EPI_ISL_2496117, EPI_ISL_2496118, EPI_ISL_2496119, EPI_ISL_2496120, EPI_ISL_2496121, EPI_ISL_2496122, EPI_ISL_2496123, EPI_ISL_2496124, EPI_ISL_2496125, EPI_ISL_2496126, EPI_ISL_2496127, EPI_ISL_2496128, EPI_ISL_2496129, EPI_ISL_2496130, EPI_ISL_2496131, EPI_ISL_2496132, EPI_ISL_2496133, EPI_ISL_2496134, EPI_ISL_2496135, EPI_ISL_2496136, EPI_ISL_2496137, EPI_ISL_2496138, EPI_ISL_2496139, EPI_ISL_2496140, EPI_ISL_2496141, EPI_ISL_2496142, EPI_ISL_2496143, EPI_ISL_2496144, EPI_ISL_2496145, EPI_ISL_2496146, EPI_ISL_2496147, EPI_ISL_2496148, EPI_ISL_2496149, EPI_ISL_2496150, EPI_ISL_2496151, EPI_ISL_2496152, EPI_ISL_2496153, EPI_ISL_2496154, EPI_ISL_2496155, EPI_ISL_2496156, EPI_ISL_2496157, EPI_ISL_2496158, EPI_ISL_2496159, EPI_ISL_2496160, EPI_ISL_2496161, EPI_ISL_2496162, EPI_ISL_2496186, EPI_ISL_2496187, EPI_ISL_2496188, EPI_ISL_2496189, EPI_ISL_2496190, EPI_ISL_2496191, EPI_ISL_2496192, EPI_ISL_2496193, EPI_ISL_2496194, EPI_ISL_2496195, EPI_ISL_2496196, EPI_ISL_2496197, EPI_ISL_2496198, EPI_ISL_2496199, EPI_ISL_2496200, EPI_ISL_2496201, EPI_ISL_2496202, EPI_ISL_2496203, EPI_ISL_2496204, EPI_ISL_2496205, EPI_ISL_2496206, EPI_ISL_2496207, EPI_ISL_2496208, EPI_ISL_2496209, EPI_ISL_2496210, EPI_ISL_2496211, EPI_ISL_2496212, EPI_ISL_2496213, EPI_ISL_2496214, EPI_ISL_2496215, EPI_ISL_2496216, EPI_ISL_2496217, EPI_ISL_2496218, EPI_ISL_2496219, EPI_ISL_2496220, EPI_ISL_2496221, EPI_ISL_2496222, EPI_ISL_2496223, EPI_ISL_2496224, EPI_ISL_2496225, EPI_ISL_2496226, EPI_ISL_2496227, EPI_ISL_2496228, EPI_ISL_2496229, EPI_ISL_2496230, EPI_ISL_2496231, EPI_ISL_2496232, EPI_ISL_2496233, EPI_ISL_2496234, EPI_ISL_2496235, EPI_ISL_2496236, EPI_ISL_2496237, EPI_ISL_2496238, EPI_ISL_2496239, EPI_ISL_2496240, EPI_ISL_2496241, EPI_ISL_2496242, EPI_ISL_2496243, EPI_ISL_2496244, EPI_ISL_2496245, EPI_ISL_2496246, EPI_ISL_2496247, EPI_ISL_2496248, EPI_ISL_2496249, EPI_ISL_2496250, EPI_ISL_2496251, EPI_ISL_2496252, EPI_ISL_2496253, EPI_ISL_2496254, EPI_ISL_2496255, EPI_ISL_2496256, EPI_ISL_2496257, EPI_ISL_2496258, EPI_ISL_2496259, EPI_ISL_2496260, EPI_ISL_2496261, EPI_ISL_2496262, EPI_ISL_2496263, EPI_ISL_2496264, EPI_ISL_2496265, EPI_ISL_2496266, EPI_ISL_2496267, EPI_ISL_2496268, EPI_ISL_2496269, EPI_ISL_2496270, EPI_ISL_2496271, EPI_ISL_2496272, EPI_ISL_2496273, EPI_ISL_2496274, EPI_ISL_2496275, EPI_ISL_2496276, EPI_ISL_2496277, EPI_ISL_2496278, EPI_ISL_2496279, EPI_ISL_2496280, EPI_ISL_2496281, EPI_ISL_2496282, EPI_ISL_2496283, EPI_ISL_2496284, EPI_ISL_2496285, EPI_ISL_2496286, EPI_ISL_2496287, EPI_ISL_2496288, EPI_ISL_2496289, EPI_ISL_2496290, EPI_ISL_2496291, EPI_ISL_2496292, EPI_ISL_2496293, EPI_ISL_2496294, EPI_ISL_2496295, EPI_ISL_2496296, EPI_ISL_2496297, EPI_ISL_2496298, EPI_ISL_2496299, EPI_ISL_2496300, EPI_ISL_2496301, EPI_ISL_2496302, EPI_ISL_2496303, EPI_ISL_2496304, EPI_ISL_2496305, EPI_ISL_2496306, EPI_ISL_2496307, EPI_ISL_2496308, EPI_ISL_2496309, EPI_ISL_2496310, EPI_ISL_2496311, EPI_ISL_2496312, EPI_ISL_2496313, EPI_ISL_2496314, EPI_ISL_2496315, EPI_ISL_2496316, EPI_ISL_2496317, EPI_ISL_2496318, EPI_ISL_2496319, EPI_ISL_2496320, EPI_ISL_2496321, EPI_ISL_2496322, EPI_ISL_2496323, EPI_ISL_2496324, EPI_ISL_2496325, EPI_ISL_2496326, EPI_ISL_2496327, EPI_ISL_2496328, EPI_ISL_2496329, EPI_ISL_2496330, EPI_ISL_2496331, EPI_ISL_2496332, EPI_ISL_2496333, EPI_ISL_2496334, EPI_ISL_2496335, EPI_ISL_2496336, EPI_ISL_2496337, EPI_ISL_2496338, EPI_ISL_2496339, EPI_ISL_2496340, EPI_ISL_2496341, EPI_ISL_2496342, EPI_ISL_2496343, EPI_ISL_2496344, EPI_ISL_2496345, EPI_ISL_2496346, EPI_ISL_2496347, EPI_ISL_2496348, EPI_ISL_2496349, EPI_ISL_2496350, EPI_ISL_2496351, EPI_ISL_2496352, EPI_ISL_2496353, EPI_ISL_2496354, EPI_ISL_2496355, EPI_ISL_2496356, EPI_ISL_2496357, EPI_ISL_2496358, EPI_ISL_2496359, EPI_ISL_2496360, EPI_ISL_2496361, EPI_ISL_2496362, EPI_ISL_2496363, EPI_ISL_2496364, EPI_ISL_2496365, EPI_ISL_2496366, EPI_ISL_2496367, EPI_ISL_2496368, EPI_ISL_2496369, EPI_ISL_2496370, EPI_ISL_2496371, EPI_ISL_2496372, EPI_ISL_2496373, EPI_ISL_2496374, EPI_ISL_2496375, EPI_ISL_2496376, EPI_ISL_2496377, EPI_ISL_2496378, EPI_ISL_2496391, EPI_ISL_2496392, EPI_ISL_2496393, EPI_ISL_2496394, EPI_ISL_2496395, EPI_ISL_2496396, EPI_ISL_2496397, EPI_ISL_2496398, EPI_ISL_2496399, EPI_ISL_2496400, EPI_ISL_2496401, EPI_ISL_2496402, EPI_ISL_2496403, EPI_ISL_2496404, EPI_ISL_2496405, EPI_ISL_2496406, EPI_ISL_2496407, EPI_ISL_2496408, EPI_ISL_2496409, EPI_ISL_2496410, EPI_ISL_2496411, EPI_ISL_2496412, EPI_ISL_2496413, EPI_ISL_2496414, EPI_ISL_2496415, EPI_ISL_2496416, EPI_ISL_2496417, EPI_ISL_2496418, EPI_ISL_2496419, EPI_ISL_2496420, EPI_ISL_2496421, EPI_ISL_2496422, EPI_ISL_2496423, EPI_ISL_2496424, EPI_ISL_2496425, EPI_ISL_2496426, EPI_ISL_2496427, EPI_ISL_2496428, EPI_ISL_2496429, EPI_ISL_2496430, EPI_ISL_2496431, EPI_ISL_2496432, EPI_ISL_2496433, EPI_ISL_2496434, EPI_ISL_2496435, EPI_ISL_2496436, EPI_ISL_2496437, EPI_ISL_2496438, EPI_ISL_2496439, EPI_ISL_2496440, EPI_ISL_2496441, EPI_ISL_2496442, EPI_ISL_2496443, EPI_ISL_2496444, EPI_ISL_2496445, EPI_ISL_2496446, EPI_ISL_2496447, EPI_ISL_2496448, EPI_ISL_2496449, EPI_ISL_2496450, EPI_ISL_2496451, EPI_ISL_2496452, EPI_ISL_2496453, EPI_ISL_2496454, EPI_ISL_2496455, EPI_ISL_2496456, EPI_ISL_2496457, EPI_ISL_2496458, EPI_ISL_2496459, EPI_ISL_2496460, EPI_ISL_2496461, EPI_ISL_2496462, EPI_ISL_2496463, EPI_ISL_2496464, EPI_ISL_2496465, EPI_ISL_2496466, EPI_ISL_2496467, EPI_ISL_2496468, EPI_ISL_2496469, EPI_ISL_2496470, EPI_ISL_2496471, EPI_ISL_2496472, EPI_ISL_2496473, EPI_ISL_2496474, EPI_ISL_2496475, EPI_ISL_2496476, EPI_ISL_2496477, EPI_ISL_2496478, EPI_ISL_2496479, EPI_ISL_2496480, EPI_ISL_2496481, EPI_ISL_2496482, EPI_ISL_2496483, EPI_ISL_2496484, EPI_ISL_2496485, EPI_ISL_2496486, EPI_ISL_2496487, EPI_ISL_2496488, EPI_ISL_2496489, EPI_ISL_2496490, EPI_ISL_2496491, EPI_ISL_2496492, EPI_ISL_2496493, EPI_ISL_2496494, EPI_ISL_2496495, EPI_ISL_2496496, EPI_ISL_2496497, EPI_ISL_2496498, EPI_ISL_2496499, EPI_ISL_2496500, EPI_ISL_2496501, EPI_ISL_2496502, EPI_ISL_2496503, EPI_ISL_2496504, EPI_ISL_2496505, EPI_ISL_2496506, EPI_ISL_2496507, EPI_ISL_2496508, EPI_ISL_2496509, EPI_ISL_2496510, EPI_ISL_2496511, EPI_ISL_2496512, EPI_ISL_2496513, EPI_ISL_2496514, EPI_ISL_2496515, EPI_ISL_2496516, EPI_ISL_2496517, EPI_ISL_2496518, EPI_ISL_2496519, EPI_ISL_2496520, EPI_ISL_2496521, EPI_ISL_2496522, EPI_ISL_2496523, EPI_ISL_2496524, EPI_ISL_2496525, EPI_ISL_2496526, EPI_ISL_2496527, EPI_ISL_2496528, EPI_ISL_2496529, EPI_ISL_2496530, EPI_ISL_2496531, EPI_ISL_2496532, EPI_ISL_2496533, EPI_ISL_2496534, EPI_ISL_2496535, EPI_ISL_2496536, EPI_ISL_2496537, EPI_ISL_2496538, EPI_ISL_2496539, EPI_ISL_2496540, EPI_ISL_2496541, EPI_ISL_2496542, EPI_ISL_2496543, EPI_ISL_2496544, EPI_ISL_2496545, EPI_ISL_2496546, EPI_ISL_2496547, EPI_ISL_2496548, EPI_ISL_2496549, EPI_ISL_2496550, EPI_ISL_2496551, EPI_ISL_2496552, EPI_ISL_2496553, EPI_ISL_2496554, EPI_ISL_2496555, EPI_ISL_2496556, EPI_ISL_2496557, EPI_ISL_2496558, EPI_ISL_2496559, EPI_ISL_2496560, EPI_ISL_2496561, EPI_ISL_2496562, EPI_ISL_2496563, EPI_ISL_2496564, EPI_ISL_2496565, EPI_ISL_2496566, EPI_ISL_2496567, EPI_ISL_2496568, EPI_ISL_2496569, EPI_ISL_2496570, EPI_ISL_2496571, EPI_ISL_2496572, EPI_ISL_2496573, EPI_ISL_2496574, EPI_ISL_2496575, EPI_ISL_2496576, EPI_ISL_2496577, EPI_ISL_2496578, EPI_ISL_2496579, EPI_ISL_2496580, EPI_ISL_2496581, EPI_ISL_2496582, EPI_ISL_2496583, EPI_ISL_2496584, EPI_ISL_2496585, EPI_ISL_2496586, EPI_ISL_2496587, EPI_ISL_2496588, EPI_ISL_2496589, EPI_ISL_2496590, EPI_ISL_2496591, EPI_ISL_2496592, EPI_ISL_2496593, EPI_ISL_2496594, EPI_ISL_2496595, EPI_ISL_2496596, EPI_ISL_2496597, EPI_ISL_2496598, EPI_ISL_2496599, EPI_ISL_2496600, EPI_ISL_2496601, EPI_ISL_2496602, EPI_ISL_2496603, EPI_ISL_2496604, EPI_ISL_2496605, EPI_ISL_2496606, EPI_ISL_2496607, EPI_ISL_2496608, EPI_ISL_2496609, EPI_ISL_2496610, EPI_ISL_2496611, EPI_ISL_2496612, EPI_ISL_2496613, EPI_ISL_2496614, EPI_ISL_2496615, EPI_ISL_2496616, EPI_ISL_2496617, EPI_ISL_2496618, EPI_ISL_2496619, EPI_ISL_2496620, EPI_ISL_2496621, EPI_ISL_2496622, EPI_ISL_2496623, EPI_ISL_2496624, EPI_ISL_2496625, EPI_ISL_2496626, EPI_ISL_2496627, EPI_ISL_2496628, EPI_ISL_2496629, EPI_ISL_2496630, EPI_ISL_2496631, EPI_ISL_2496632, EPI_ISL_2496633, EPI_ISL_2496634, EPI_ISL_2496635, EPI_ISL_2496636, EPI_ISL_2496637, EPI_ISL_2496638, EPI_ISL_2496639, EPI_ISL_2496640, EPI_ISL_2496641, EPI_ISL_2496642, EPI_ISL_2496643, EPI_ISL_2496644, EPI_ISL_2496645, EPI_ISL_2496646, EPI_ISL_2496647, EPI_ISL_2496648, EPI_ISL_2496649, EPI_ISL_2496650, EPI_ISL_2496651, EPI_ISL_2496652, EPI_ISL_2496653, EPI_ISL_2496654, EPI_ISL_2496655, EPI_ISL_2496656, EPI_ISL_2496657, EPI_ISL_2496658, EPI_ISL_2496659, EPI_ISL_2496660, EPI_ISL_2496661, EPI_ISL_2496662, EPI_ISL_2496663, EPI_ISL_2496664, EPI_ISL_2496665, EPI_ISL_2496666, EPI_ISL_2496667, EPI_ISL_2496668, EPI_ISL_2496669, EPI_ISL_2496670, EPI_ISL_2496671, EPI_ISL_2496672, EPI_ISL_2496673, EPI_ISL_2496674, EPI_ISL_2496675, EPI_ISL_2496676, EPI_ISL_2496677, EPI_ISL_2496678, EPI_ISL_2496679, EPI_ISL_2496680, EPI_ISL_2496681, EPI_ISL_2496682, EPI_ISL_2496683, EPI_ISL_2496684, EPI_ISL_2496685, EPI_ISL_2496686, EPI_ISL_2496687, EPI_ISL_2496688, EPI_ISL_2496689, EPI_ISL_2496690, EPI_ISL_2496691, EPI_ISL_2496692, EPI_ISL_2496693, EPI_ISL_2496694, EPI_ISL_2496695, EPI_ISL_2496696, EPI_ISL_2496697, EPI_ISL_2496698, EPI_ISL_2496701, EPI_ISL_2496702, EPI_ISL_2496703, EPI_ISL_2496704, EPI_ISL_2496705, EPI_ISL_2496706, EPI_ISL_2496707, EPI_ISL_2496708, EPI_ISL_2496709, EPI_ISL_2496710, EPI_ISL_2496711, EPI_ISL_2496712, EPI_ISL_2496713, EPI_ISL_2496714, EPI_ISL_2496715, EPI_ISL_2496716, EPI_ISL_2496717, EPI_ISL_2496718, EPI_ISL_2496719, EPI_ISL_2496720, EPI_ISL_2496721, EPI_ISL_2496722, EPI_ISL_2496723, EPI_ISL_2496724, EPI_ISL_2496725, EPI_ISL_2496726, EPI_ISL_2496727, EPI_ISL_2496728, EPI_ISL_2496729, EPI_ISL_2496730, EPI_ISL_2496731, EPI_ISL_2496732, EPI_ISL_2496733, EPI_ISL_2496734, EPI_ISL_2496735, EPI_ISL_2496736, EPI_ISL_2496737, EPI_ISL_2496738, EPI_ISL_2496739, EPI_ISL_2496740, EPI_ISL_2496741, EPI_ISL_2496742, EPI_ISL_2496743, EPI_ISL_2496744, EPI_ISL_2496745, EPI_ISL_2496746, EPI_ISL_2496747, EPI_ISL_2496748, EPI_ISL_2496749, EPI_ISL_2496750, EPI_ISL_2496751, EPI_ISL_2496752, EPI_ISL_2496753, EPI_ISL_2496754, EPI_ISL_2496755, EPI_ISL_2496756, EPI_ISL_2496757, EPI_ISL_2496758, EPI_ISL_2496759, EPI_ISL_2496760, EPI_ISL_2496761, EPI_ISL_2496762, EPI_ISL_2496763, EPI_ISL_2496764, EPI_ISL_2496765, EPI_ISL_2496766, EPI_ISL_2496767, EPI_ISL_2496768, EPI_ISL_2496769

| | | | |
|---|---|---|---|
| see above | Utah Public Health Laboratory | Utah Public Health Laboratory | Erin L. Young, Kelly F. Oakeson, Tara Gallagher |
| EPI_ISL_2496770, EPI_ISL_2496771, EPI_ISL_2496772, EPI_ISL_2496773, EPI_ISL_2496774, EPI_ISL_2496775 | CA DPH Viral and Rickettsial Disease Laboratory | Chan-Zuckerberg Biohub | CZB Cliahub Consortium |
| EPI_ISL_2496776, EPI_ISL_2496777 | Santa Clara County Public Health Laboratory | Chan-Zuckerberg Biohub | CZB Cliahub Consortium |
| EPI_ISL_2496778, EPI_ISL_2496779, EPI_ISL_2496780, EPI_ISL_2496781, EPI_ISL_2496782, EPI_ISL_2496783 | CA DPH Viral and Rickettsial Disease Laboratory | Chan-Zuckerberg Biohub | CZB Cliahub Consortium |
| EPI_ISL_2496784 | Contra Costa County Public Health Lab | Chan-Zuckerberg Biohub | CZB Cliahub Consortium |
| EPI_ISL_2496785, EPI_ISL_2496786 | CA DPH Viral and Rickettsial Disease Laboratory | Chan-Zuckerberg Biohub | CZB Cliahub Consortium |
| EPI_ISL_2496787, EPI_ISL_2496788 | Santa Clara County Public Health Laboratory | Chan-Zuckerberg Biohub | CZB Cliahub Consortium |
| EPI_ISL_2496789, EPI_ISL_2496790, EPI_ISL_2496791, EPI_ISL_2496792 | CA DPH Viral and Rickettsial Disease Laboratory | Chan-Zuckerberg Biohub | CZB Cliahub Consortium |
| EPI_ISL_2496793 | Contra Costa County Public Health Lab | Chan-Zuckerberg Biohub | CZB Cliahub Consortium |
| EPI_ISL_2496794 | CA DPH Viral and Rickettsial Disease Laboratory | Chan-Zuckerberg Biohub | CZB Cliahub Consortium |
| EPI_ISL_2496795 | Contra Costa County Public Health Lab | Chan-Zuckerberg Biohub | CZB Cliahub Consortium |
| EPI_ISL_2496796, EPI_ISL_2496797, EPI_ISL_2496798, EPI_ISL_2496799 | CA DPH Viral and Rickettsial Disease Laboratory | Chan-Zuckerberg Biohub | CZB Cliahub Consortium |
| EPI_ISL_2496800 | Contra Costa County Public Health Lab | Chan-Zuckerberg Biohub | CZB Cliahub Consortium |
| EPI_ISL_2496801, EPI_ISL_2496802, EPI_ISL_2496803, EPI_ISL_2496804 | CA DPH Viral and Rickettsial Disease Laboratory | Chan-Zuckerberg Biohub | CZB Cliahub Consortium |
| EPI_ISL_2496805 | Contra Costa County Public Health Lab | Chan-Zuckerberg Biohub | CZB Cliahub Consortium |
| EPI_ISL_2496806, EPI_ISL_2496807, EPI_ISL_2496808, EPI_ISL_2496809 | CA DPH Viral and Rickettsial Disease Laboratory | Chan-Zuckerberg Biohub | CZB Cliahub Consortium |
| EPI_ISL_2496810 | Santa Clara County Public Health Laboratory | Chan-Zuckerberg Biohub | CZB Cliahub Consortium |
| EPI_ISL_2496811 | CA DPH Viral and Rickettsial Disease Laboratory | Chan-Zuckerberg Biohub | CZB Cliahub Consortium |
| EPI_ISL_2496812 | Contra Costa County Public Health Lab | Chan-Zuckerberg Biohub | CZB Cliahub Consortium |
| EPI_ISL_2496813 | CA DPH Viral and Rickettsial Disease Laboratory | Chan-Zuckerberg Biohub | CZB Cliahub Consortium |
| EPI_ISL_2496814 | Contra Costa County Public Health Lab | Chan-Zuckerberg Biohub | CZB Cliahub Consortium |
| EPI_ISL_2496815, EPI_ISL_2496816, EPI_ISL_2496817, EPI_ISL_2496818, EPI_ISL_2496819, EPI_ISL_2496820 | CA DPH Viral and Rickettsial Disease Laboratory | Chan-Zuckerberg Biohub | CZB Cliahub Consortium |
| EPI_ISL_2496821 | Contra Costa County Public Health Lab | Chan-Zuckerberg Biohub | CZB Cliahub Consortium |
| EPI_ISL_2496822, EPI_ISL_2496823, EPI_ISL_2496824, EPI_ISL_2496825, EPI_ISL_2496826 | CA DPH Viral and Rickettsial Disease Laboratory | Chan-Zuckerberg Biohub | CZB Cliahub Consortium |
| EPI_ISL_2496827, EPI_ISL_2496828, EPI_ISL_2496829 | Contra Costa County Public Health Lab | Chan-Zuckerberg Biohub | CZB Cliahub Consortium |
| EPI_ISL_2496830, EPI_ISL_2496831 | CA DPH Viral and Rickettsial Disease Laboratory | Chan-Zuckerberg Biohub | CZB Cliahub Consortium |

| | | | |
|---|---|---|---|
| EPI_ISL_2496832 | Contra Costa County Public Health Lab | Chan-Zuckerberg Biohub | CZB Cliahub Consortium |
| EPI_ISL_2496833 | CA DPH Viral and Rickettsial Disease Laboratory | Chan-Zuckerberg Biohub | CZB Cliahub Consortium |
| EPI_ISL_2496834 | Contra Costa County Public Health Lab | Chan-Zuckerberg Biohub | CZB Cliahub Consortium |
| EPI_ISL_2496835, EPI_ISL_2496836, EPI_ISL_2496837 | CA DPH Viral and Rickettsial Disease Laboratory | Chan-Zuckerberg Biohub | CZB Cliahub Consortium |
| EPI_ISL_2496838 | Santa Clara County Public Health Laboratory | Chan-Zuckerberg Biohub | CZB Cliahub Consortium |
| EPI_ISL_2496839, EPI_ISL_2496840, EPI_ISL_2496841, EPI_ISL_2496842 | CA DPH Viral and Rickettsial Disease Laboratory | Chan-Zuckerberg Biohub | CZB Cliahub Consortium |
| EPI_ISL_2496843 | Contra Costa County Public Health Lab | Chan-Zuckerberg Biohub | CZB Cliahub Consortium |
| EPI_ISL_2496844 | CA DPH Viral and Rickettsial Disease Laboratory | Chan-Zuckerberg Biohub | CZB Cliahub Consortium |
| EPI_ISL_2496845 | Santa Clara County Public Health Laboratory | Chan-Zuckerberg Biohub | CZB Cliahub Consortium |
| EPI_ISL_2496846 | CA DPH Viral and Rickettsial Disease Laboratory | Chan-Zuckerberg Biohub | CZB Cliahub Consortium |
| EPI_ISL_2496847 | Contra Costa County Public Health Lab | Chan-Zuckerberg Biohub | CZB Cliahub Consortium |
| EPI_ISL_2496848, EPI_ISL_2496849, EPI_ISL_2496850, EPI_ISL_2496851 | CA DPH Viral and Rickettsial Disease Laboratory | Chan-Zuckerberg Biohub | CZB Cliahub Consortium |
| EPI_ISL_2496852, EPI_ISL_2496853 | Contra Costa County Public Health Lab | Chan-Zuckerberg Biohub | CZB Cliahub Consortium |
| EPI_ISL_2496854 | CA DPH Viral and Rickettsial Disease Laboratory | Chan-Zuckerberg Biohub | CZB Cliahub Consortium |
| EPI_ISL_2496855 | Contra Costa County Public Health Lab | Chan-Zuckerberg Biohub | CZB Cliahub Consortium |
| EPI_ISL_2496856, EPI_ISL_2496857 | CA DPH Viral and Rickettsial Disease Laboratory | Chan-Zuckerberg Biohub | CZB Cliahub Consortium |
| EPI_ISL_2496858 | Contra Costa County Public Health Lab | Chan-Zuckerberg Biohub | CZB Cliahub Consortium |
| EPI_ISL_2496859, EPI_ISL_2496860, EPI_ISL_2496861, EPI_ISL_2496862, EPI_ISL_2496863 | CA DPH Viral and Rickettsial Disease Laboratory | Chan-Zuckerberg Biohub | CZB Cliahub Consortium |
| EPI_ISL_2496864 | Contra Costa County Public Health Lab | Chan-Zuckerberg Biohub | CZB Cliahub Consortium |
| EPI_ISL_2496865 | CA DPH Viral and Rickettsial Disease Laboratory | Chan-Zuckerberg Biohub | CZB Cliahub Consortium |
| EPI_ISL_2496866, EPI_ISL_2496867 | Contra Costa County Public Health Lab | Chan-Zuckerberg Biohub | CZB Cliahub Consortium |
| EPI_ISL_2496868, EPI_ISL_2496869, EPI_ISL_2496870, EPI_ISL_2496871 | CA DPH Viral and Rickettsial Disease Laboratory | Chan-Zuckerberg Biohub | CZB Cliahub Consortium |
| EPI_ISL_2496872 | Contra Costa County Public Health Lab | Chan-Zuckerberg Biohub | CZB Cliahub Consortium |
| EPI_ISL_2496873, EPI_ISL_2496874, EPI_ISL_2496875, EPI_ISL_2496876 | CA DPH Viral and Rickettsial Disease Laboratory | Chan-Zuckerberg Biohub | CZB Cliahub Consortium |
| EPI_ISL_2496877, EPI_ISL_2496878, EPI_ISL_2496879 | Contra Costa County Public Health Lab | Chan-Zuckerberg Biohub | CZB Cliahub Consortium |
| EPI_ISL_2496880 | CA DPH Viral and Rickettsial Disease Laboratory | Chan-Zuckerberg Biohub | CZB Cliahub Consortium |
| EPI_ISL_2496881 | Contra Costa County Public Health Lab | Chan-Zuckerberg Biohub | CZB Cliahub Consortium |
| EPI_ISL_2496882, EPI_ISL_2496883 | Orange County Public Health Lab | Chan-Zuckerberg Biohub | CZB Cliahub Consortium |
| EPI_ISL_2496884 | CA DPH Viral and Rickettsial Disease Laboratory | Chan-Zuckerberg Biohub | CZB Cliahub Consortium |
| EPI_ISL_2496885, EPI_ISL_2496886, EPI_ISL_2496887 | Contra Costa County Public Health Lab | Chan-Zuckerberg Biohub | CZB Cliahub Consortium |
| EPI_ISL_2496888 | Orange County Public Health Lab | Chan-Zuckerberg Biohub | CZB Cliahub Consortium |
| EPI_ISL_2496889, EPI_ISL_2496890, EPI_ISL_2496891 | CA DPH Viral and Rickettsial Disease Laboratory | Chan-Zuckerberg Biohub | CZB Cliahub Consortium |
| EPI_ISL_2496892 | Santa Clara County Public Health Laboratory | Chan-Zuckerberg Biohub | CZB Cliahub Consortium |
| EPI_ISL_2496893, EPI_ISL_2496898, EPI_ISL_2496899, EPI_ISL_2496900, EPI_ISL_2496901, EPI_ISL_2496902, EPI_ISL_2496903 | CA DPH Viral and Rickettsial Disease Laboratory | Chan-Zuckerberg Biohub | CZB Cliahub Consortium |
| EPI_ISL_2496904 | Contra Costa County Public Health Lab | Chan-Zuckerberg Biohub | CZB Cliahub Consortium |
| EPI_ISL_2496905, EPI_ISL_2496906, EPI_ISL_2496907, EPI_ISL_2496908, EPI_ISL_2496909 | CA DPH Viral and Rickettsial Disease Laboratory | Chan-Zuckerberg Biohub | CZB Cliahub Consortium |
| EPI_ISL_2496910 | Contra Costa County Public Health Lab | Chan-Zuckerberg Biohub | CZB Cliahub Consortium |
| EPI_ISL_2496911 | CA DPH Viral and Rickettsial Disease Laboratory | Chan-Zuckerberg Biohub | CZB Cliahub Consortium |
| EPI_ISL_2496912 | Contra Costa County Public Health Lab | Chan-Zuckerberg Biohub | CZB Cliahub Consortium |
| EPI_ISL_2496913 | CA DPH Viral and Rickettsial Disease Laboratory | Chan-Zuckerberg Biohub | CZB Cliahub Consortium |
| EPI_ISL_2496914 | Contra Costa County Public Health Lab | Chan-Zuckerberg Biohub | CZB Cliahub Consortium |

| | | | |
|---|---|---|---|
| EPI_ISL_2496915 | CA DPH Viral and Rickettsial Disease Laboratory | Chan-Zuckerberg Biohub | CZB Cliahub Consortium |
| EPI_ISL_2496916, EPI_ISL_2496917 | Contra Costa County Public Health Lab | Chan-Zuckerberg Biohub | CZB Cliahub Consortium |
| EPI_ISL_2496918 | CA DPH Viral and Rickettsial Disease Laboratory | Chan-Zuckerberg Biohub | CZB Cliahub Consortium |
| EPI_ISL_2496919, EPI_ISL_2496920, EPI_ISL_2496921 | Contra Costa County Public Health Lab | Chan-Zuckerberg Biohub | CZB Cliahub Consortium |
| EPI_ISL_2496922 | CA DPH Viral and Rickettsial Disease Laboratory | Chan-Zuckerberg Biohub | CZB Cliahub Consortium |
| EPI_ISL_2496923 | Santa Clara County Public Health Laboratory | Chan-Zuckerberg Biohub | CZB Cliahub Consortium |
| EPI_ISL_2496924, EPI_ISL_2496925 | Contra Costa County Public Health Lab | Chan-Zuckerberg Biohub | CZB Cliahub Consortium |
| EPI_ISL_2496926, EPI_ISL_2496927, EPI_ISL_2496928, EPI_ISL_2496929, EPI_ISL_2496930 | CA DPH Viral and Rickettsial Disease Laboratory | Chan-Zuckerberg Biohub | CZB Cliahub Consortium |
| EPI_ISL_2496931 | Santa Clara County Public Health Laboratory | Chan-Zuckerberg Biohub | CZB Cliahub Consortium |
| EPI_ISL_2496932, EPI_ISL_2496933 | Contra Costa County Public Health Lab | Chan-Zuckerberg Biohub | CZB Cliahub Consortium |
| EPI_ISL_2496934 | CA DPH Viral and Rickettsial Disease Laboratory | Chan-Zuckerberg Biohub | CZB Cliahub Consortium |
| EPI_ISL_2496935, EPI_ISL_2496936 | Contra Costa County Public Health Lab | Chan-Zuckerberg Biohub | CZB Cliahub Consortium |
| EPI_ISL_2496937 | CA DPH Viral and Rickettsial Disease Laboratory | Chan-Zuckerberg Biohub | CZB Cliahub Consortium |
| EPI_ISL_2496938, EPI_ISL_2496939 | Contra Costa County Public Health Lab | Chan-Zuckerberg Biohub | CZB Cliahub Consortium |
| EPI_ISL_2496940 | CA DPH Viral and Rickettsial Disease Laboratory | Chan-Zuckerberg Biohub | CZB Cliahub Consortium |
| EPI_ISL_2496941 | Santa Clara County Public Health Laboratory | Chan-Zuckerberg Biohub | CZB Cliahub Consortium |
| EPI_ISL_2496942, EPI_ISL_2496943 | Contra Costa County Public Health Lab | Chan-Zuckerberg Biohub | CZB Cliahub Consortium |
| EPI_ISL_2496944 | CA DPH Viral and Rickettsial Disease Laboratory | Chan-Zuckerberg Biohub | CZB Cliahub Consortium |
| EPI_ISL_2496945 | Contra Costa County Public Health Lab | Chan-Zuckerberg Biohub | CZB Cliahub Consortium |
| EPI_ISL_2496946 | CA DPH Viral and Rickettsial Disease Laboratory | Chan-Zuckerberg Biohub | CZB Cliahub Consortium |
| EPI_ISL_2496947 | Contra Costa County Public Health Lab | Chan-Zuckerberg Biohub | CZB Cliahub Consortium |
| EPI_ISL_2496948, EPI_ISL_2496949, EPI_ISL_2496950, EPI_ISL_2496951, EPI_ISL_2496952, EPI_ISL_2496953, EPI_ISL_2496954, EPI_ISL_2496955 | CA DPH Viral and Rickettsial Disease Laboratory | Chan-Zuckerberg Biohub | CZB Cliahub Consortium |
| EPI_ISL_2496956 | Contra Costa County Public Health Lab | Chan-Zuckerberg Biohub | CZB Cliahub Consortium |
| EPI_ISL_2496957, EPI_ISL_2496958, EPI_ISL_2496959 | CA DPH Viral and Rickettsial Disease Laboratory | Chan-Zuckerberg Biohub | CZB Cliahub Consortium |
| EPI_ISL_2496960 | Contra Costa County Public Health Lab | Chan-Zuckerberg Biohub | CZB Cliahub Consortium |
| EPI_ISL_2496961, EPI_ISL_2496962, EPI_ISL_2496963, EPI_ISL_2496964 | CA DPH Viral and Rickettsial Disease Laboratory | Chan-Zuckerberg Biohub | CZB Cliahub Consortium |
| EPI_ISL_2496965 | Contra Costa County Public Health Lab | Chan-Zuckerberg Biohub | CZB Cliahub Consortium |
| EPI_ISL_2496966, EPI_ISL_2496967 | CA DPH Viral and Rickettsial Disease Laboratory | Chan-Zuckerberg Biohub | CZB Cliahub Consortium |
| EPI_ISL_2496968, EPI_ISL_2496969 | Contra Costa County Public Health Lab | Chan-Zuckerberg Biohub | CZB Cliahub Consortium |
| EPI_ISL_2496970 | CA DPH Viral and Rickettsial Disease Laboratory | Chan-Zuckerberg Biohub | CZB Cliahub Consortium |
| EPI_ISL_2496971, EPI_ISL_2496972, EPI_ISL_2496973 | Contra Costa County Public Health Lab | Chan-Zuckerberg Biohub | CZB Cliahub Consortium |
| EPI_ISL_2496974 | CA DPH Viral and Rickettsial Disease Laboratory | Chan-Zuckerberg Biohub | CZB Cliahub Consortium |
| EPI_ISL_2496975 | Contra Costa County Public Health Lab | Chan-Zuckerberg Biohub | CZB Cliahub Consortium |
| EPI_ISL_2496976 | Santa Clara County Public Health Laboratory | Chan-Zuckerberg Biohub | CZB Cliahub Consortium |
| EPI_ISL_2496977 | Contra Costa County Public Health Lab | Chan-Zuckerberg Biohub | CZB Cliahub Consortium |
| EPI_ISL_2496978 | CA DPH Viral and Rickettsial Disease Laboratory | Chan-Zuckerberg Biohub | CZB Cliahub Consortium |
| EPI_ISL_2496979, EPI_ISL_2496980, EPI_ISL_2496981 | Contra Costa County Public Health Lab | Chan-Zuckerberg Biohub | CZB Cliahub Consortium |
| EPI_ISL_2496982, EPI_ISL_2496983, EPI_ISL_2496984, EPI_ISL_2496985, EPI_ISL_2496986 | CA DPH Viral and Rickettsial Disease Laboratory | Chan-Zuckerberg Biohub | CZB Cliahub Consortium |
| EPI_ISL_2496987 | Contra Costa County Public Health Lab | Chan-Zuckerberg Biohub | CZB Cliahub Consortium |
| EPI_ISL_2496988 | CA DPH Viral and Rickettsial Disease Laboratory | Chan-Zuckerberg Biohub | CZB Cliahub Consortium |
| EPI_ISL_2496989, EPI_ISL_2496990 | Contra Costa County Public Health Lab | Chan-Zuckerberg Biohub | CZB Cliahub Consortium |

| | | | |
|---|---|---|---|
| EPI_ISL_2496991 | CA DPH Viral and Rickettsial Disease Laboratory | Chan-Zuckerberg Biohub | CZB Cliahub Consortium |
| EPI_ISL_2496992 | Contra Costa County Public Health Lab | Chan-Zuckerberg Biohub | CZB Cliahub Consortium |
| EPI_ISL_2496993, EPI_ISL_2496994 | CA DPH Viral and Rickettsial Disease Laboratory | Chan-Zuckerberg Biohub | CZB Cliahub Consortium |
| EPI_ISL_2496995, EPI_ISL_2496996 | Contra Costa County Public Health Lab | Chan-Zuckerberg Biohub | CZB Cliahub Consortium |
| EPI_ISL_2496997, EPI_ISL_2496998 | CA DPH Viral and Rickettsial Disease Laboratory | Chan-Zuckerberg Biohub | CZB Cliahub Consortium |
| EPI_ISL_2496999, EPI_ISL_2497000 | Contra Costa County Public Health Lab | Chan-Zuckerberg Biohub | CZB Cliahub Consortium |
| EPI_ISL_2497001, EPI_ISL_2497002, EPI_ISL_2497003, EPI_ISL_2497004, EPI_ISL_2497005, EPI_ISL_2497006, EPI_ISL_2497007, EPI_ISL_2497009, EPI_ISL_2497010, EPI_ISL_2497011, EPI_ISL_2497012, EPI_ISL_2497013, EPI_ISL_2497014, EPI_ISL_2497015, EPI_ISL_2497016, EPI_ISL_2497017, EPI_ISL_2497018, EPI_ISL_2497020, EPI_ISL_2497021, EPI_ISL_2497022, EPI_ISL_2497023, EPI_ISL_2497024, EPI_ISL_2497025, EPI_ISL_2497026, EPI_ISL_2497027, EPI_ISL_2497028, EPI_ISL_2497029, EPI_ISL_2497030, EPI_ISL_2497031, EPI_ISL_2497032, EPI_ISL_2497033, EPI_ISL_2497034, EPI_ISL_2497035, EPI_ISL_2497036, EPI_ISL_2497037, EPI_ISL_2497038, EPI_ISL_2497039, EPI_ISL_2497040, EPI_ISL_2497041, EPI_ISL_2497042, EPI_ISL_2497043, EPI_ISL_2497044, EPI_ISL_2497045, EPI_ISL_2497046, EPI_ISL_2497047, EPI_ISL_2497048, EPI_ISL_2497049, EPI_ISL_2497050, EPI_ISL_2497051, EPI_ISL_2497052, EPI_ISL_2497053, EPI_ISL_2497054, EPI_ISL_2497055, EPI_ISL_2497056, EPI_ISL_2497057, EPI_ISL_2497058, EPI_ISL_2497059, EPI_ISL_2497060, EPI_ISL_2497061, EPI_ISL_2497062, EPI_ISL_2497063, EPI_ISL_2497064, EPI_ISL_2497065, EPI_ISL_2497066, EPI_ISL_2497067, EPI_ISL_2497068, EPI_ISL_2497069, EPI_ISL_2497070, EPI_ISL_2497071, EPI_ISL_2497072, EPI_ISL_2497073, EPI_ISL_2497074, EPI_ISL_2497075, EPI_ISL_2497076, EPI_ISL_2497077, EPI_ISL_2497078, EPI_ISL_2497079, EPI_ISL_2497080, EPI_ISL_2497081, EPI_ISL_2497082, EPI_ISL_2497083, EPI_ISL_2497084, EPI_ISL_2497085, EPI_ISL_2497086, EPI_ISL_2497087, EPI_ISL_2497088, EPI_ISL_2497089, EPI_ISL_2497090, EPI_ISL_2497091, EPI_ISL_2497092, EPI_ISL_2497093, EPI_ISL_2497094, EPI_ISL_2497095, EPI_ISL_2497096, EPI_ISL_2497097, EPI_ISL_2497098, EPI_ISL_2497099, EPI_ISL_2497100, EPI_ISL_2497101, EPI_ISL_2497102, EPI_ISL_2497103, EPI_ISL_2497104, EPI_ISL_2497105, EPI_ISL_2497106, EPI_ISL_2497107, EPI_ISL_2497108, EPI_ISL_2497109, EPI_ISL_2497110, EPI_ISL_2497111, EPI_ISL_2497112, EPI_ISL_2497113, EPI_ISL_2497114, EPI_ISL_2497116, EPI_ISL_2497117, EPI_ISL_2497118, EPI_ISL_2497119, EPI_ISL_2497120, EPI_ISL_2497121, EPI_ISL_2497122, EPI_ISL_2497123, EPI_ISL_2497124, EPI_ISL_2497125, EPI_ISL_2497126, EPI_ISL_2497127, EPI_ISL_2497128, EPI_ISL_2497129, EPI_ISL_2497130, EPI_ISL_2497131, EPI_ISL_2497132, EPI_ISL_2497133, EPI_ISL_2497134, EPI_ISL_2497135, EPI_ISL_2497136, EPI_ISL_2497137, EPI_ISL_2497138, EPI_ISL_2497139, EPI_ISL_2497140, EPI_ISL_2497141, EPI_ISL_2497142, EPI_ISL_2497144, EPI_ISL_2497145, EPI_ISL_2497146, EPI_ISL_2497147, EPI_ISL_2497148, EPI_ISL_2497149, EPI_ISL_2497150, EPI_ISL_2497151, EPI_ISL_2497152, EPI_ISL_2497153, EPI_ISL_2497154, EPI_ISL_2497155, EPI_ISL_2497156, EPI_ISL_2497157, EPI_ISL_2497158, EPI_ISL_2497159, EPI_ISL_2497160, EPI_ISL_2497161, EPI_ISL_2497162, EPI_ISL_2497163, EPI_ISL_2497164, EPI_ISL_2497165, EPI_ISL_2497166, EPI_ISL_2497167, EPI_ISL_2497169, EPI_ISL_2497170, EPI_ISL_2497171, EPI_ISL_2497172, EPI_ISL_2497173, EPI_ISL_2497174, EPI_ISL_2497175, EPI_ISL_2497176, EPI_ISL_2497177, EPI_ISL_2497178, EPI_ISL_2497179, EPI_ISL_2497180, EPI_ISL_2497181, EPI_ISL_2497182, EPI_ISL_2497183, EPI_ISL_2497184 | | | |
| see above | Cayuga Medical Center | Cornell Covid-19 Testing Laboratory | Patrick Mitchell, Leonardo Caserta, Brittany Cronk, Renee Anderson, Melissa Laverack, Roopa Venugopalan, Elizabeth Plocharczyk, Diego Diel |
| EPI_ISL_2497185, EPI_ISL_2497186, EPI_ISL_2497187, EPI_ISL_2497188, EPI_ISL_2497189, EPI_ISL_2497190, EPI_ISL_2497191, EPI_ISL_2497192, EPI_ISL_2497193, EPI_ISL_2497194, EPI_ISL_2497195, EPI_ISL_2497196, EPI_ISL_2497198, EPI_ISL_2497199, EPI_ISL_2497200, EPI_ISL_2497201, EPI_ISL_2497202, EPI_ISL_2497203, EPI_ISL_2497205, EPI_ISL_2497206, EPI_ISL_2497207, EPI_ISL_2497208, EPI_ISL_2497209, EPI_ISL_2497210, EPI_ISL_2497211, EPI_ISL_2497212, EPI_ISL_2497213, EPI_ISL_2497214, EPI_ISL_2497215, EPI_ISL_2497216, EPI_ISL_2497217, EPI_ISL_2497218, EPI_ISL_2497219, EPI_ISL_2497220, EPI_ISL_2497221, EPI_ISL_2497222, EPI_ISL_2497223, EPI_ISL_2497224, EPI_ISL_2497225, EPI_ISL_2497226, EPI_ISL_2497227, EPI_ISL_2497228, EPI_ISL_2497229, EPI_ISL_2497230, EPI_ISL_2497231, EPI_ISL_2497232, EPI_ISL_2497233, EPI_ISL_2497234, EPI_ISL_2497235, EPI_ISL_2497236, EPI_ISL_2497237, EPI_ISL_2497238, EPI_ISL_2497239, EPI_ISL_2497240, EPI_ISL_2497241, EPI_ISL_2497242, EPI_ISL_2497243, EPI_ISL_2497244, EPI_ISL_2497245, EPI_ISL_2497246, EPI_ISL_2497247, EPI_ISL_2497248, EPI_ISL_2497249, EPI_ISL_2497250, EPI_ISL_2497251, EPI_ISL_2497252, EPI_ISL_2497253, EPI_ISL_2497254, EPI_ISL_2497255, EPI_ISL_2497256, EPI_ISL_2497257, EPI_ISL_2497258, EPI_ISL_2497259, EPI_ISL_2497260, EPI_ISL_2497261, EPI_ISL_2497262, EPI_ISL_2497263, EPI_ISL_2497264, EPI_ISL_2497265, EPI_ISL_2497266, EPI_ISL_2497267, EPI_ISL_2497268, EPI_ISL_2497269, EPI_ISL_2497270, EPI_ISL_2497271, EPI_ISL_2497272, EPI_ISL_2497273, EPI_ISL_2497275, EPI_ISL_2497276, EPI_ISL_2497277, EPI_ISL_2497279, EPI_ISL_2497281, EPI_ISL_2497282, EPI_ISL_2497283, EPI_ISL_2497285, EPI_ISL_2497286, EPI_ISL_2497287, EPI_ISL_2497288, EPI_ISL_2497289, EPI_ISL_2497290, EPI_ISL_2497291, EPI_ISL_2497292, EPI_ISL_2497293, EPI_ISL_2497294, EPI_ISL_2497295, EPI_ISL_2497296, EPI_ISL_2497297, EPI_ISL_2497298, EPI_ISL_2497299, EPI_ISL_2497300, EPI_ISL_2497301, EPI_ISL_2497302, EPI_ISL_2497303, EPI_ISL_2497304, EPI_ISL_2497305, EPI_ISL_2497306, EPI_ISL_2497307, EPI_ISL_2497308, EPI_ISL_2497309, EPI_ISL_2497310, EPI_ISL_2497311, EPI_ISL_2497312, EPI_ISL_2497313, EPI_ISL_2497314, EPI_ISL_2497315, EPI_ISL_2497316, EPI_ISL_2497317, EPI_ISL_2497318, EPI_ISL_2497319, EPI_ISL_2497320, EPI_ISL_2497321, EPI_ISL_2497322, EPI_ISL_2497323, EPI_ISL_2497324, EPI_ISL_2497325, EPI_ISL_2497326, EPI_ISL_2497327, EPI_ISL_2497328, EPI_ISL_2497329, EPI_ISL_2497330, EPI_ISL_2497331, EPI_ISL_2497332, EPI_ISL_2497333, EPI_ISL_2497334, EPI_ISL_2497335, EPI_ISL_2497336, EPI_ISL_2497337, EPI_ISL_2497338, EPI_ISL_2497339, EPI_ISL_2497340, EPI_ISL_2497341, EPI_ISL_2497342, EPI_ISL_2497343, EPI_ISL_2497344, EPI_ISL_2497345, EPI_ISL_2497346, EPI_ISL_2497347, EPI_ISL_2497348, EPI_ISL_2497349, EPI_ISL_2497350, EPI_ISL_2497351, EPI_ISL_2497352, EPI_ISL_2497353, EPI_ISL_2497354, EPI_ISL_2497355, EPI_ISL_2497356, EPI_ISL_2497357, EPI_ISL_2497358, EPI_ISL_2497359, EPI_ISL_2497360, EPI_ISL_2497361, EPI_ISL_2497362, EPI_ISL_2497363, EPI_ISL_2497364, EPI_ISL_2497365, EPI_ISL_2497366, EPI_ISL_2497368, EPI_ISL_2497369, EPI_ISL_2497370, EPI_ISL_2497371, EPI_ISL_2497372, EPI_ISL_2497373, EPI_ISL_2497374, EPI_ISL_2497375, EPI_ISL_2497376, EPI_ISL_2497377, EPI_ISL_2497378, EPI_ISL_2497379, EPI_ISL_2497380, EPI_ISL_2497381, EPI_ISL_2497382, EPI_ISL_2497383, EPI_ISL_2497385, EPI_ISL_2497386, EPI_ISL_2497387, EPI_ISL_2497388, EPI_ISL_2497389, EPI_ISL_2497390, EPI_ISL_2497391, EPI_ISL_2497392, EPI_ISL_2497393, EPI_ISL_2497394, EPI_ISL_2497395, EPI_ISL_2497396, EPI_ISL_2497397, EPI_ISL_2497398, EPI_ISL_2497399, EPI_ISL_2497400, EPI_ISL_2497401, EPI_ISL_2497402, EPI_ISL_2497403, EPI_ISL_2497404, EPI_ISL_2497405, EPI_ISL_2497407, EPI_ISL_2497408, EPI_ISL_2497409, EPI_ISL_2497410, EPI_ISL_2497411, EPI_ISL_2497412, EPI_ISL_2497413, EPI_ISL_2497414, EPI_ISL_2497415, EPI_ISL_2497416, EPI_ISL_2497417, EPI_ISL_2497418, EPI_ISL_2497419, EPI_ISL_2497420, EPI_ISL_2497421, EPI_ISL_2497422, EPI_ISL_2497423, EPI_ISL_2497424, EPI_ISL_2497425, EPI_ISL_2497426, EPI_ISL_2497427, EPI_ISL_2497428, EPI_ISL_2497429, EPI_ISL_2497430, EPI_ISL_2497431, EPI_ISL_2497432 | | | |
| see above | UW Virology Lab | UW Virology Lab | Pavitra Roychoudhury, Hong Xie, Lasata Shrestha, Tien V. Nguyen, Shah Mohamed Bakhash, Michelle Lin, Noah R. Baker, Ricardo Perez, Sean Ellis, Nathan Breit, Robert J. Livingston, Meei-Li Huang, Keith R Jerome, Patrick Mathias, Alexander Greninger |
| EPI_ISL_2497544 | CA DPH Viral and Rickettsial Disease Laboratory | Chan-Zuckerberg Biohub | CZB Cliahub Consortium |
| EPI_ISL_2497545 | County of San Luis Obispo Public Health Laboratory | Chan-Zuckerberg Biohub | CZB Cliahub Consortium |
| EPI_ISL_2497546 | CA DPH Viral and Rickettsial Disease Laboratory | Chan-Zuckerberg Biohub | CZB Cliahub Consortium |
| EPI_ISL_2497547 | County of San Luis Obispo Public Health Laboratory | Chan-Zuckerberg Biohub | CZB Cliahub Consortium |
| EPI_ISL_2497548 | CA DPH Viral and Rickettsial Disease Laboratory | Chan-Zuckerberg Biohub | CZB Cliahub Consortium |
| EPI_ISL_2497549 | Humboldt County Public Health Laboratory | Chan-Zuckerberg Biohub | CZB Cliahub Consortium |
| EPI_ISL_2497550, EPI_ISL_2497551, EPI_ISL_2497552, EPI_ISL_2497553, EPI_ISL_2497554 | County of San Luis Obispo Public Health Laboratory | Chan-Zuckerberg Biohub | CZB Cliahub Consortium |
| EPI_ISL_2497555 | Contra Costa County Public Health Lab | Chan-Zuckerberg Biohub | CZB Cliahub Consortium |
| EPI_ISL_2497556, EPI_ISL_2497557 | CA DPH Viral and Rickettsial Disease Laboratory | Chan-Zuckerberg Biohub | CZB Cliahub Consortium |
| EPI_ISL_2497558, EPI_ISL_2497559 | Contra Costa County Public Health Lab | Chan-Zuckerberg Biohub | CZB Cliahub Consortium |
| EPI_ISL_2497560, EPI_ISL_2497561 | CA DPH Viral and Rickettsial Disease Laboratory | Chan-Zuckerberg Biohub | CZB Cliahub Consortium |
| EPI_ISL_2497562 | Contra Costa County Public Health Lab | Chan-Zuckerberg Biohub | CZB Cliahub Consortium |
| EPI_ISL_2497563, EPI_ISL_2497564 | CA DPH Viral and Rickettsial Disease Laboratory | Chan-Zuckerberg Biohub | CZB Cliahub Consortium |
| EPI_ISL_2497565, EPI_ISL_2497566 | Contra Costa County Public Health Lab | Chan-Zuckerberg Biohub | CZB Cliahub Consortium |
| EPI_ISL_2497567 | Humboldt County Public Health Laboratory | Chan-Zuckerberg Biohub | CZB Cliahub Consortium |
| EPI_ISL_2497568 | CA DPH Viral and Rickettsial Disease Laboratory | Chan-Zuckerberg Biohub | CZB Cliahub Consortium |
| EPI_ISL_2497569 | Contra Costa County Public Health Lab | Chan-Zuckerberg Biohub | CZB Cliahub Consortium |
| EPI_ISL_2497570 | CA DPH Viral and Rickettsial Disease Laboratory | Chan-Zuckerberg Biohub | CZB Cliahub Consortium |
| EPI_ISL_2497571, EPI_ISL_2497572 | Contra Costa County Public Health Lab | Chan-Zuckerberg Biohub | CZB Cliahub Consortium |
| EPI_ISL_2497573, EPI_ISL_2497574 | County of San Luis Obispo Public Health Laboratory | Chan-Zuckerberg Biohub | CZB Cliahub Consortium |
| EPI_ISL_2497575 | CA DPH Viral and Rickettsial Disease Laboratory | Chan-Zuckerberg Biohub | CZB Cliahub Consortium |
| EPI_ISL_2497576, EPI_ISL_2497577, EPI_ISL_2497578, EPI_ISL_2497579 | Contra Costa County Public Health Lab | Chan-Zuckerberg Biohub | CZB Cliahub Consortium |
| EPI_ISL_2497580 | CA DPH Viral and Rickettsial Disease Laboratory | Chan-Zuckerberg Biohub | CZB Cliahub Consortium |
| EPI_ISL_2497581 | Contra Costa County Public Health Lab | Chan-Zuckerberg Biohub | CZB Cliahub Consortium |
| EPI_ISL_2497582, EPI_ISL_2497583 | CA DPH Viral and Rickettsial Disease Laboratory | Chan-Zuckerberg Biohub | CZB Cliahub Consortium |
| EPI_ISL_2497584 | Contra Costa County Public Health Lab | Chan-Zuckerberg Biohub | CZB Cliahub Consortium |
| EPI_ISL_2497585 | CA DPH Viral and Rickettsial Disease Laboratory | Chan-Zuckerberg Biohub | CZB Cliahub Consortium |
| EPI_ISL_2497586, EPI_ISL_2497587 | Contra Costa County Public Health Lab | Chan-Zuckerberg Biohub | CZB Cliahub Consortium |
| EPI_ISL_2497588 | CA DPH Viral and Rickettsial Disease Laboratory | Chan-Zuckerberg Biohub | CZB Cliahub Consortium |
| EPI_ISL_2497589, EPI_ISL_2497590, | Contra Costa County Public Health Lab | Chan-Zuckerberg Biohub | CZB Cliahub Consortium |

| | | | |
|---|---|---|---|
| EPI_ISL_2497591, EPI_ISL_2497592, EPI_ISL_2497593 | | | |
| EPI_ISL_2497594 | CA DPH Viral and Rickettsial Disease Laboratory | Chan-Zuckerberg Biohub | CZB Cliahub Consortium |
| EPI_ISL_2497595, EPI_ISL_2497596 | Contra Costa County Public Health Lab | Chan-Zuckerberg Biohub | CZB Cliahub Consortium |
| EPI_ISL_2497597, EPI_ISL_2497598 | CA DPH Viral and Rickettsial Disease Laboratory | Chan-Zuckerberg Biohub | CZB Cliahub Consortium |
| EPI_ISL_2497599 | Contra Costa County Public Health Lab | Chan-Zuckerberg Biohub | CZB Cliahub Consortium |
| EPI_ISL_2497600, EPI_ISL_2497601 | CA DPH Viral and Rickettsial Disease Laboratory | Chan-Zuckerberg Biohub | CZB Cliahub Consortium |
| EPI_ISL_2497602, EPI_ISL_2497603, EPI_ISL_2497604, EPI_ISL_2497605 | Contra Costa County Public Health Lab | Chan-Zuckerberg Biohub | CZB Cliahub Consortium |
| EPI_ISL_2497606 | CA DPH Viral and Rickettsial Disease Laboratory | Chan-Zuckerberg Biohub | CZB Cliahub Consortium |
| EPI_ISL_2497607 | Contra Costa County Public Health Lab | Chan-Zuckerberg Biohub | CZB Cliahub Consortium |
| EPI_ISL_2497608, EPI_ISL_2497609, EPI_ISL_2497610 | CA DPH Viral and Rickettsial Disease Laboratory | Chan-Zuckerberg Biohub | CZB Cliahub Consortium |
| EPI_ISL_2497611, EPI_ISL_2497612 | Contra Costa County Public Health Lab | Chan-Zuckerberg Biohub | CZB Cliahub Consortium |
| EPI_ISL_2497613 | CA DPH Viral and Rickettsial Disease Laboratory | Chan-Zuckerberg Biohub | CZB Cliahub Consortium |
| EPI_ISL_2497614 | Contra Costa County Public Health Lab | Chan-Zuckerberg Biohub | CZB Cliahub Consortium |
| EPI_ISL_2497615, EPI_ISL_2497616, EPI_ISL_2497617 | CA DPH Viral and Rickettsial Disease Laboratory | Chan-Zuckerberg Biohub | CZB Cliahub Consortium |
| EPI_ISL_2497618 | Contra Costa County Public Health Lab | Chan-Zuckerberg Biohub | CZB Cliahub Consortium |
| EPI_ISL_2497619 | County of San Luis Obispo Public Health Laboratory | Chan-Zuckerberg Biohub | CZB Cliahub Consortium |
| EPI_ISL_2497620 | CA DPH Viral and Rickettsial Disease Laboratory | Chan-Zuckerberg Biohub | CZB Cliahub Consortium |
| EPI_ISL_2497621 | County of San Luis Obispo Public Health Laboratory | Chan-Zuckerberg Biohub | CZB Cliahub Consortium |
| EPI_ISL_2497622 | Humboldt County Public Health Laboratory | Chan-Zuckerberg Biohub | CZB Cliahub Consortium |
| EPI_ISL_2497623, EPI_ISL_2497624 | CA DPH Viral and Rickettsial Disease Laboratory | Chan-Zuckerberg Biohub | CZB Cliahub Consortium |
| EPI_ISL_2497625 | Humboldt County Public Health Laboratory | Chan-Zuckerberg Biohub | CZB Cliahub Consortium |
| EPI_ISL_2497626, EPI_ISL_2497627 | CA DPH Viral and Rickettsial Disease Laboratory | Chan-Zuckerberg Biohub | CZB Cliahub Consortium |
| EPI_ISL_2497628 | Humboldt County Public Health Laboratory | Chan-Zuckerberg Biohub | CZB Cliahub Consortium |
| EPI_ISL_2497629 | Contra Costa County Public Health Lab | Chan-Zuckerberg Biohub | CZB Cliahub Consortium |
| EPI_ISL_2497630, EPI_ISL_2497631, EPI_ISL_2497632 | CA DPH Viral and Rickettsial Disease Laboratory | Chan-Zuckerberg Biohub | CZB Cliahub Consortium |
| EPI_ISL_2497633 | Humboldt County Public Health Laboratory | Chan-Zuckerberg Biohub | CZB Cliahub Consortium |
| EPI_ISL_2497634 | CA DPH Viral and Rickettsial Disease Laboratory | Chan-Zuckerberg Biohub | CZB Cliahub Consortium |
| EPI_ISL_2497635 | Contra Costa County Public Health Lab | Chan-Zuckerberg Biohub | CZB Cliahub Consortium |
| EPI_ISL_2497636, EPI_ISL_2497637 | CA DPH Viral and Rickettsial Disease Laboratory | Chan-Zuckerberg Biohub | CZB Cliahub Consortium |
| EPI_ISL_2497638, EPI_ISL_2497639 | Contra Costa County Public Health Lab | Chan-Zuckerberg Biohub | CZB Cliahub Consortium |
| EPI_ISL_2497640 | CA DPH Viral and Rickettsial Disease Laboratory | Chan-Zuckerberg Biohub | CZB Cliahub Consortium |
| EPI_ISL_2497641 | Contra Costa County Public Health Lab | Chan-Zuckerberg Biohub | CZB Cliahub Consortium |
| EPI_ISL_2497642, EPI_ISL_2497643 | County of San Luis Obispo Public Health Laboratory | Chan-Zuckerberg Biohub | CZB Cliahub Consortium |
| EPI_ISL_2497644 | Contra Costa County Public Health Lab | Chan-Zuckerberg Biohub | CZB Cliahub Consortium |
| EPI_ISL_2497645, EPI_ISL_2497646, EPI_ISL_2497648, EPI_ISL_2497649, EPI_ISL_2497650, EPI_ISL_2497651, EPI_ISL_2497652 | CA DPH Viral and Rickettsial Disease Laboratory | Chan-Zuckerberg Biohub | CZB Cliahub Consortium |
| EPI_ISL_2497653 | Humboldt County Public Health Laboratory | Chan-Zuckerberg Biohub | CZB Cliahub Consortium |
| EPI_ISL_2497654, EPI_ISL_2497655, EPI_ISL_2497656 | CA DPH Viral and Rickettsial Disease Laboratory | Chan-Zuckerberg Biohub | CZB Cliahub Consortium |
| EPI_ISL_2497657 | Humboldt County Public Health Laboratory | Chan-Zuckerberg Biohub | CZB Cliahub Consortium |
| EPI_ISL_2497658 | CA DPH Viral and Rickettsial Disease Laboratory | Chan-Zuckerberg Biohub | CZB Cliahub Consortium |
| EPI_ISL_2497659 | Contra Costa County Public Health Lab | Chan-Zuckerberg Biohub | CZB Cliahub Consortium |
| EPI_ISL_2497660 | Humboldt County Public Health Laboratory | Chan-Zuckerberg Biohub | CZB Cliahub Consortium |
| EPI_ISL_2497661 | CA DPH Viral and Rickettsial Disease Laboratory | Chan-Zuckerberg Biohub | CZB Cliahub Consortium |
| EPI_ISL_2497662, EPI_ISL_2497663, EPI_ISL_2497664 | Humboldt County Public Health Laboratory | Chan-Zuckerberg Biohub | CZB Cliahub Consortium |
| EPI_ISL_2497665, | CA DPH Viral and Rickettsial Disease | Chan-Zuckerberg Biohub | CZB Cliahub Consortium |

| | | | |
|---|---|---|---|
| EPI_ISL_2497666 | Laboratory | | |
| EPI_ISL_2497667 | Contra Costa County Public Health Lab | Chan-Zuckerberg Biohub | CZB Cliahub Consortium |
| EPI_ISL_2497668 | CA DPH Viral and Rickettsial Disease Laboratory | Chan-Zuckerberg Biohub | CZB Cliahub Consortium |
| EPI_ISL_2497669 | Humboldt County Public Health Laboratory | Chan-Zuckerberg Biohub | CZB Cliahub Consortium |
| EPI_ISL_2497670, EPI_ISL_2497671, EPI_ISL_2497672, EPI_ISL_2497673, EPI_ISL_2497674 | CA DPH Viral and Rickettsial Disease Laboratory | Chan-Zuckerberg Biohub | CZB Cliahub Consortium |
| EPI_ISL_2497675 | Contra Costa County Public Health Lab | Chan-Zuckerberg Biohub | CZB Cliahub Consortium |
| EPI_ISL_2497676 | County of San Luis Obispo Public Health Laboratory | Chan-Zuckerberg Biohub | CZB Cliahub Consortium |
| EPI_ISL_2497677, EPI_ISL_2497678, EPI_ISL_2497679 | Contra Costa County Public Health Lab | Chan-Zuckerberg Biohub | CZB Cliahub Consortium |
| EPI_ISL_2497680 | County of San Luis Obispo Public Health Laboratory | Chan-Zuckerberg Biohub | CZB Cliahub Consortium |
| EPI_ISL_2497681 | Contra Costa County Public Health Lab | Chan-Zuckerberg Biohub | CZB Cliahub Consortium |
| EPI_ISL_2497682, EPI_ISL_2497683 | County of San Luis Obispo Public Health Laboratory | Chan-Zuckerberg Biohub | CZB Cliahub Consortium |
| EPI_ISL_2497684 | Contra Costa County Public Health Lab | Chan-Zuckerberg Biohub | CZB Cliahub Consortium |
| EPI_ISL_2497685 | CA DPH Viral and Rickettsial Disease Laboratory | Chan-Zuckerberg Biohub | CZB Cliahub Consortium |
| EPI_ISL_2497686 | Humboldt County Public Health Laboratory | Chan-Zuckerberg Biohub | CZB Cliahub Consortium |
| EPI_ISL_2497687, EPI_ISL_2497688 | Contra Costa County Public Health Lab | Chan-Zuckerberg Biohub | CZB Cliahub Consortium |
| EPI_ISL_2497689 | CA DPH Viral and Rickettsial Disease Laboratory | Chan-Zuckerberg Biohub | CZB Cliahub Consortium |
| EPI_ISL_2497690, EPI_ISL_2497691 | Humboldt County Public Health Laboratory | Chan-Zuckerberg Biohub | CZB Cliahub Consortium |
| EPI_ISL_2497692 | Contra Costa County Public Health Lab | Chan-Zuckerberg Biohub | CZB Cliahub Consortium |
| EPI_ISL_2497693 | Humboldt County Public Health Laboratory | Chan-Zuckerberg Biohub | CZB Cliahub Consortium |
| EPI_ISL_2497694 | CA DPH Viral and Rickettsial Disease Laboratory | Chan-Zuckerberg Biohub | CZB Cliahub Consortium |
| EPI_ISL_2497695, EPI_ISL_2497696 | County of San Luis Obispo Public Health Laboratory | Chan-Zuckerberg Biohub | CZB Cliahub Consortium |
| EPI_ISL_2497697 | Humboldt County Public Health Laboratory | Chan-Zuckerberg Biohub | CZB Cliahub Consortium |
| EPI_ISL_2497698 | Contra Costa County Public Health Lab | Chan-Zuckerberg Biohub | CZB Cliahub Consortium |
| EPI_ISL_2497699 | County of San Luis Obispo Public Health Laboratory | Chan-Zuckerberg Biohub | CZB Cliahub Consortium |
| EPI_ISL_2497700 | Contra Costa County Public Health Lab | Chan-Zuckerberg Biohub | CZB Cliahub Consortium |
| EPI_ISL_2497701, EPI_ISL_2497702, EPI_ISL_2497703 | County of San Luis Obispo Public Health Laboratory | Chan-Zuckerberg Biohub | CZB Cliahub Consortium |
| EPI_ISL_2497704 | Contra Costa County Public Health Lab | Chan-Zuckerberg Biohub | CZB Cliahub Consortium |
| EPI_ISL_2497705, EPI_ISL_2497706, EPI_ISL_2497707 | County of San Luis Obispo Public Health Laboratory | Chan-Zuckerberg Biohub | CZB Cliahub Consortium |
| EPI_ISL_2497708, EPI_ISL_2497709 | Contra Costa County Public Health Lab | Chan-Zuckerberg Biohub | CZB Cliahub Consortium |
| EPI_ISL_2497710 | CA DPH Viral and Rickettsial Disease Laboratory | Chan-Zuckerberg Biohub | CZB Cliahub Consortium |
| EPI_ISL_2497711, EPI_ISL_2497712, EPI_ISL_2497713 | County of San Luis Obispo Public Health Laboratory | Chan-Zuckerberg Biohub | CZB Cliahub Consortium |
| EPI_ISL_2497714 | Contra Costa County Public Health Lab | Chan-Zuckerberg Biohub | CZB Cliahub Consortium |
| EPI_ISL_2497715, EPI_ISL_2497716, EPI_ISL_2497717, EPI_ISL_2497718 | County of San Luis Obispo Public Health Laboratory | Chan-Zuckerberg Biohub | CZB Cliahub Consortium |
| EPI_ISL_2497719 | Contra Costa County Public Health Lab | Chan-Zuckerberg Biohub | CZB Cliahub Consortium |
| EPI_ISL_2497720 | County of San Luis Obispo Public Health Laboratory | Chan-Zuckerberg Biohub | CZB Cliahub Consortium |
| EPI_ISL_2497721 | Contra Costa County Public Health Lab | Chan-Zuckerberg Biohub | CZB Cliahub Consortium |
| EPI_ISL_2497722, EPI_ISL_2497723 | County of San Luis Obispo Public Health Laboratory | Chan-Zuckerberg Biohub | CZB Cliahub Consortium |
| EPI_ISL_2497724 | Contra Costa County Public Health Lab | Chan-Zuckerberg Biohub | CZB Cliahub Consortium |
| EPI_ISL_2497725, EPI_ISL_2497726, EPI_ISL_2497727, EPI_ISL_2497728, EPI_ISL_2497729, EPI_ISL_2497730, EPI_ISL_2497731, EPI_ISL_2497732 | County of San Luis Obispo Public Health Laboratory | Chan-Zuckerberg Biohub | CZB Cliahub Consortium |
| EPI_ISL_2497733, EPI_ISL_2497734 | Contra Costa County Public Health Lab | Chan-Zuckerberg Biohub | CZB Cliahub Consortium |
| EPI_ISL_2497735, EPI_ISL_2497736 | County of San Luis Obispo Public Health Laboratory | Chan-Zuckerberg Biohub | CZB Cliahub Consortium |
| EPI_ISL_2497737 | CA DPH Viral and Rickettsial Disease Laboratory | Chan-Zuckerberg Biohub | CZB Cliahub Consortium |
| EPI_ISL_2497738, EPI_ISL_2497739, EPI_ISL_2497740, EPI_ISL_2497741, EPI_ISL_2497742 | County of San Luis Obispo Public Health Laboratory | Chan-Zuckerberg Biohub | CZB Cliahub Consortium |

| | | | |
|---|---|---|---|
| EPI_ISL_2497743 | Contra Costa County Public Health Lab | Chan-Zuckerberg Biohub | CZB Cliahub Consortium |
| EPI_ISL_2497744, EPI_ISL_2497745, EPI_ISL_2497746, EPI_ISL_2497747, EPI_ISL_2497748 | County of San Luis Obispo Public Health Laboratory | Chan-Zuckerberg Biohub | CZB Cliahub Consortium |
| EPI_ISL_2497749 | Contra Costa County Public Health Lab | Chan-Zuckerberg Biohub | CZB Cliahub Consortium |
| EPI_ISL_2497750, EPI_ISL_2497751 | County of San Luis Obispo Public Health Laboratory | Chan-Zuckerberg Biohub | CZB Cliahub Consortium |
| EPI_ISL_2497752 | Contra Costa County Public Health Lab | Chan-Zuckerberg Biohub | CZB Cliahub Consortium |
| EPI_ISL_2497753, EPI_ISL_2497754, EPI_ISL_2497755 | County of San Luis Obispo Public Health Laboratory | Chan-Zuckerberg Biohub | CZB Cliahub Consortium |
| EPI_ISL_2497756, EPI_ISL_2497757 | Contra Costa County Public Health Lab | Chan-Zuckerberg Biohub | CZB Cliahub Consortium |
| EPI_ISL_2497758 | County of San Luis Obispo Public Health Laboratory | Chan-Zuckerberg Biohub | CZB Cliahub Consortium |
| EPI_ISL_2497759 | Contra Costa County Public Health Lab | Chan-Zuckerberg Biohub | CZB Cliahub Consortium |
| EPI_ISL_2497760, EPI_ISL_2497761 | County of San Luis Obispo Public Health Laboratory | Chan-Zuckerberg Biohub | CZB Cliahub Consortium |
| EPI_ISL_2497762 | Contra Costa County Public Health Lab | Chan-Zuckerberg Biohub | CZB Cliahub Consortium |
| EPI_ISL_2497763, EPI_ISL_2497764, EPI_ISL_2497765, EPI_ISL_2497766, EPI_ISL_2497767 | County of San Luis Obispo Public Health Laboratory | Chan-Zuckerberg Biohub | CZB Cliahub Consortium |
| EPI_ISL_2497768 | CA DPH Viral and Rickettsial Disease Laboratory | Chan-Zuckerberg Biohub | CZB Cliahub Consortium |
| EPI_ISL_2497769, EPI_ISL_2497770, EPI_ISL_2497771, EPI_ISL_2497772, EPI_ISL_2497773 | County of San Luis Obispo Public Health Laboratory | Chan-Zuckerberg Biohub | CZB Cliahub Consortium |
| EPI_ISL_2497774 | Contra Costa County Public Health Lab | Chan-Zuckerberg Biohub | CZB Cliahub Consortium |
| EPI_ISL_2497775, EPI_ISL_2497776, EPI_ISL_2497777, EPI_ISL_2497778, EPI_ISL_2497779, EPI_ISL_2497780, EPI_ISL_2497781, EPI_ISL_2497782 | County of San Luis Obispo Public Health Laboratory | Chan-Zuckerberg Biohub | CZB Cliahub Consortium |
| EPI_ISL_2497783 | Contra Costa County Public Health Lab | Chan-Zuckerberg Biohub | CZB Cliahub Consortium |
| EPI_ISL_2497784, EPI_ISL_2497785, EPI_ISL_2497786, EPI_ISL_2497787 | County of San Luis Obispo Public Health Laboratory | Chan-Zuckerberg Biohub | CZB Cliahub Consortium |
| EPI_ISL_2497788 | Contra Costa County Public Health Lab | Chan-Zuckerberg Biohub | CZB Cliahub Consortium |
| EPI_ISL_2497789, EPI_ISL_2497790, EPI_ISL_2497791, EPI_ISL_2497792 | County of San Luis Obispo Public Health Laboratory | Chan-Zuckerberg Biohub | CZB Cliahub Consortium |
| EPI_ISL_2497793, EPI_ISL_2497794 | Contra Costa County Public Health Lab | Chan-Zuckerberg Biohub | CZB Cliahub Consortium |
| EPI_ISL_2497795, EPI_ISL_2497796, EPI_ISL_2497797 | County of San Luis Obispo Public Health Laboratory | Chan-Zuckerberg Biohub | CZB Cliahub Consortium |
| EPI_ISL_2497798 | Contra Costa County Public Health Lab | Chan-Zuckerberg Biohub | CZB Cliahub Consortium |
| EPI_ISL_2497799, EPI_ISL_2497800 | County of San Luis Obispo Public Health Laboratory | Chan-Zuckerberg Biohub | CZB Cliahub Consortium |
| EPI_ISL_2497801 | Contra Costa County Public Health Lab | Chan-Zuckerberg Biohub | CZB Cliahub Consortium |
| EPI_ISL_2497802, EPI_ISL_2497803, EPI_ISL_2497804, EPI_ISL_2497805, EPI_ISL_2497806, EPI_ISL_2497807, EPI_ISL_2497808, EPI_ISL_2497809, EPI_ISL_2497810 | County of San Luis Obispo Public Health Laboratory | Chan-Zuckerberg Biohub | CZB Cliahub Consortium |
| EPI_ISL_2497811, EPI_ISL_2497812, EPI_ISL_2497813 | Contra Costa County Public Health Lab | Chan-Zuckerberg Biohub | CZB Cliahub Consortium |
| EPI_ISL_2497814, EPI_ISL_2497815 | County of San Luis Obispo Public Health Laboratory | Chan-Zuckerberg Biohub | CZB Cliahub Consortium |
| EPI_ISL_2497816 | Contra Costa County Public Health Lab | Chan-Zuckerberg Biohub | CZB Cliahub Consortium |
| EPI_ISL_2497817, EPI_ISL_2497818, EPI_ISL_2497819, EPI_ISL_2497820 | County of San Luis Obispo Public Health Laboratory | Chan-Zuckerberg Biohub | CZB Cliahub Consortium |
| EPI_ISL_2497821, EPI_ISL_2497822 | Contra Costa County Public Health Lab | Chan-Zuckerberg Biohub | CZB Cliahub Consortium |
| EPI_ISL_2497823, EPI_ISL_2497824, EPI_ISL_2497825, EPI_ISL_2497826 | County of San Luis Obispo Public Health Laboratory | Chan-Zuckerberg Biohub | CZB Cliahub Consortium |
| EPI_ISL_2497827 | Contra Costa County Public Health Lab | Chan-Zuckerberg Biohub | CZB Cliahub Consortium |
| EPI_ISL_2497828, EPI_ISL_2497829, EPI_ISL_2497830, EPI_ISL_2497831, EPI_ISL_2497832, EPI_ISL_2497833, EPI_ISL_2497834 | County of San Luis Obispo Public Health Laboratory | Chan-Zuckerberg Biohub | CZB Cliahub Consortium |
| EPI_ISL_2497835 | Contra Costa County Public Health Lab | Chan-Zuckerberg Biohub | CZB Cliahub Consortium |

| | | | |
|---|---|---|---|
| EPI_ISL_2497836 | County of San Luis Obispo Public Health Laboratory | Chan-Zuckerberg Biohub | CZB Cliahub Consortium |
| EPI_ISL_2497837 | Contra Costa County Public Health Lab | Chan-Zuckerberg Biohub | CZB Cliahub Consortium |
| EPI_ISL_2497838, EPI_ISL_2497839, EPI_ISL_2497840 | County of San Luis Obispo Public Health Laboratory | Chan-Zuckerberg Biohub | CZB Cliahub Consortium |
| EPI_ISL_2497841 | Contra Costa County Public Health Lab | Chan-Zuckerberg Biohub | CZB Cliahub Consortium |
| EPI_ISL_2497842 | County of San Luis Obispo Public Health Laboratory | Chan-Zuckerberg Biohub | CZB Cliahub Consortium |
| EPI_ISL_2497843 | Contra Costa County Public Health Lab | Chan-Zuckerberg Biohub | CZB Cliahub Consortium |
| EPI_ISL_2497844 | County of San Luis Obispo Public Health Laboratory | Chan-Zuckerberg Biohub | CZB Cliahub Consortium |
| EPI_ISL_2497845 | CA DPH Viral and Rickettsial Disease Laboratory | Chan-Zuckerberg Biohub | CZB Cliahub Consortium |
| EPI_ISL_2497846 | County of San Luis Obispo Public Health Laboratory | Chan-Zuckerberg Biohub | CZB Cliahub Consortium |
| EPI_ISL_2497848, EPI_ISL_2497866, EPI_ISL_2497868 | Ohio Department of Health Laboratory | Ohio Department of Health Laboratory | Holmes, Jennifer; Eric Brandt, Keoni Omura, Glen McGillivary, Caitlin McDonnell, Jade Mowery, Stephanie Mccracken, Tyler Payne, Kirtana Ramadugu, Erica Leasure, Brent Lee, Kelsey Florek, Heather Blankenship, Allison Black, Quanta Brown, and Tammy Bannerman |
| EPI_ISL_2497883 | MS Public Health Laboratory | University of Mississippi Medical Center, Molecular and Genomics Core Facility | Ashley C. Johnson, Ithiel J. Frame, Krishna K. Ayyalasomayajula, Michael R. Garrett, D. Ashley Robinson |
| EPI_ISL_2497885, EPI_ISL_2497886, EPI_ISL_2497888, EPI_ISL_2497889, EPI_ISL_2497892, EPI_ISL_2497893, EPI_ISL_2497894, EPI_ISL_2497895 | University of Mississippi Medical Center, Department of Pathology | University of Mississippi Medical Center, Molecular and Genomics Core Facility | Ashley C. Johnson, Ithiel J. Frame, Krishna K. Ayyalasomayajula, Michael R. Garrett, D. Ashley Robinson |
| EPI_ISL_2497897, EPI_ISL_2497907 | Ohio Department of Health Laboratory | Ohio Department of Health Laboratory | Holmes, Jennifer; Eric Brandt, Keoni Omura, Glen McGillivary, Caitlin McDonnell, Jade Mowery, Stephanie Mccracken, Tyler Payne, Kirtana Ramadugu, Erica Leasure, Brent Lee, Kelsey Florek, Heather Blankenship, Allison Black, Quanta Brown, and Tammy Bannerman |
| EPI_ISL_2497908, EPI_ISL_2497909, EPI_ISL_2497910, EPI_ISL_2497911, EPI_ISL_2497912, EPI_ISL_2497914, EPI_ISL_2497915, EPI_ISL_2497916, EPI_ISL_2497918, EPI_ISL_2497920, EPI_ISL_2497921 | | | |
| see above | Wichita State University - Molecular Diagnostics Lab | Kansas Health and Environmental Lab | Mike Grose, Katherine Wiggins, Jonathan Barnell, Ben Olsen, and Phil Adam |
| EPI_ISL_2497924 | Ohio Department of Health Laboratory | Ohio Department of Health Laboratory | Holmes, Jennifer; Eric Brandt, Keoni Omura, Glen McGillivary, Caitlin McDonnell, Jade Mowery, Stephanie Mccracken, Tyler Payne, Kirtana Ramadugu, Erica Leasure, Brent Lee, Kelsey Florek, Heather Blankenship, Allison Black, Quanta Brown, and Tammy Bannerman |
| EPI_ISL_2497925, EPI_ISL_2497926, EPI_ISL_2497927, EPI_ISL_2497928, EPI_ISL_2497929, EPI_ISL_2497930, EPI_ISL_2497931, EPI_ISL_2497932, EPI_ISL_2497933, EPI_ISL_2497934, EPI_ISL_2497935, EPI_ISL_2497936, EPI_ISL_2497937, EPI_ISL_2497938, EPI_ISL_2497939, EPI_ISL_2497940 | | | |
| see above | UW Virology Lab | UW Virology Lab | Pavitra Roychoudhury, Hong Xie, Lasata Shrestha, Tien V. Nguyen, Shah Mohamed Bakhash, Michelle Lin, Noah R. Baker, Ricardo Perez, Sean Ellis, Nathan Breit, Robert J. Livingston, Meei-Li Huang, Keith R Jerome, Patrick Mathias, Alexander Greninger |
| EPI_ISL_2499752, EPI_ISL_2499753, EPI_ISL_2499754, EPI_ISL_2499755, EPI_ISL_2499756, EPI_ISL_2499778 | Alaska State Virology Laboratory | Alaska State Virology Laboratory | Stephanie DeRonde, Elva House, Jacob Zidek, Lisa Smith, Ph.D., Jack Chen, Ph.D. |
| EPI_ISL_2499788, EPI_ISL_2499789, EPI_ISL_2499790, EPI_ISL_2499791, EPI_ISL_2499792, EPI_ISL_2499793, EPI_ISL_2499794, EPI_ISL_2499795, EPI_ISL_2499796, EPI_ISL_2499797, EPI_ISL_2499798, EPI_ISL_2499799, EPI_ISL_2499800, EPI_ISL_2499801, EPI_ISL_2499802, EPI_ISL_2499803, EPI_ISL_2499805, EPI_ISL_2499806, EPI_ISL_2499807, EPI_ISL_2499808, EPI_ISL_2499809, EPI_ISL_2499810, EPI_ISL_2499811, EPI_ISL_2499812, EPI_ISL_2499813, EPI_ISL_2499814, EPI_ISL_2499815 | | | |
| see above | Wyoming Public Health Laboratory | Wyoming Public Health Laboratory | Jim Mildenberger, Taylor Fearing, Channing Weber, Ashley Norberg, Chayse Rowley, Marley Goetz, Brian Dominguez, Elliot Thomasson, Sam Britz, Cari Sloma, Robert Petit, and Rob Christensen |
| EPI_ISL_2499828, EPI_ISL_2499829, EPI_ISL_2499832, EPI_ISL_2499834, EPI_ISL_2499835, EPI_ISL_2499836, EPI_ISL_2499838, EPI_ISL_2499839, EPI_ISL_2499840, EPI_ISL_2499842 | Arizona State Public Health Laboratory | Arizona State Public Health Laboratory | Trung Huynh, Jessica Escobar, Katherine Fullerton, Nobuko Fukushima, Stacy White, Linda Getsinger, Victor Waddell |
| EPI_ISL_2499849, EPI_ISL_2499850, EPI_ISL_2499851, EPI_ISL_2499852 | NCSLPH | NCSLPH | Chase K, Miller MC, Greene S, Glover W |
| EPI_ISL_2499853, EPI_ISL_2499854, EPI_ISL_2499855, EPI_ISL_2499856, EPI_ISL_2499857, EPI_ISL_2499858, EPI_ISL_2499859, EPI_ISL_2499860, EPI_ISL_2499861 | DC Public Health Lab/ Dept. of Forensic Sciences | DC Public Health Lab/ Dept. of Forensic Sciences | Janis Doss, Scott Nguyen, Elizabeth Zelaya, Sarah Scott, Connie Maza, Monica Mann, Brittany Hamilton, David Payne, Jocelyn Hauser |
| EPI_ISL_2499918, EPI_ISL_2499919, EPI_ISL_2499920, EPI_ISL_2499921, EPI_ISL_2499922, EPI_ISL_2499923, EPI_ISL_2499925, EPI_ISL_2499926, EPI_ISL_2499927, EPI_ISL_2499928, EPI_ISL_2499929, EPI_ISL_2499930 | | | |
| see above | Alaska State Virology Laboratory | Alaska State Virology Laboratory | Stephanie DeRonde, Elva House, Jacob Zidek, Lisa Smith, Ph.D., Jack Chen, Ph.D. |
| EPI_ISL_2499934, EPI_ISL_2499935 | Alaska State Virology Laboratory | Alaska State Virology Laboratory | Stephanie DeRonde, Elva House, Lisa Smith, Ph.D., Jack Chen, Ph.D. |
| EPI_ISL_2499936 | AL Dept. of Public Health Bureau of Clinical Laboratories | Genomics and Discovery, Respiratory Viruses Branch, Division of Viral Diseases, Centers for Disease Control and Prevention | Ying Tao, Yan Li, Jing Zhang, Anna Kelleher, Brian Lynch, Krista Queen, Anna Uehara, Peter Cook, Han Jia Justin Ng, Rachel Marine, Clinton R. Paden, Dhwani Batra, Haibin Wang, Tara Coalter, Jasmine Padilla, Morgan Davis, Mili Sheth, Sarah Nobles, Mark Burroughs, Justin Lee, Adam Retchless, Suxiang Tong |
| EPI_ISL_2499937, EPI_ISL_2499938, EPI_ISL_2499939, EPI_ISL_2499940, EPI_ISL_2499941 | Alaska State Virology Laboratory | Alaska State Virology Laboratory | Stephanie DeRonde, Elva House, Jacob Zidek, Lisa Smith, Ph.D., Jack Chen, Ph.D. |
| EPI_ISL_2499942, EPI_ISL_2499943, EPI_ISL_2499944, EPI_ISL_2499945, EPI_ISL_2499946, EPI_ISL_2499947 | Wichita State University - Molecular Diagnostics Lab | Kansas Health and Environmental Lab | Mike Grose, Katherine Wiggins, Jonathan Barnell, Ben Olsen, and Phil Adam |
| EPI_ISL_2499948, EPI_ISL_2499949, EPI_ISL_2499950, EPI_ISL_2499951, EPI_ISL_2499952, EPI_ISL_2499953 | Kansas Health and Environmental Lab | Kansas Health and Environmental Lab | Mike Grose, Katherine Wiggins, Jonathan Barnell, Ben Olsen, and Phil Adam |
| EPI_ISL_2499954, EPI_ISL_2499955 | Wichita State University - Molecular Diagnostics Lab | Kansas Health and Environmental Lab | Mike Grose, Katherine Wiggins, Jonathan Barnell, Ben Olsen, and Phil Adam |
| EPI_ISL_2499956 | Alaska State Virology Laboratory | Alaska State Virology Laboratory | Stephanie DeRonde, Elva House, Lisa Smith, Ph.D., Jack Chen, Ph.D. |

| | | | |
|---|---|---|---|
| EPI_ISL_2499957, EPI_ISL_2499958, EPI_ISL_2499959 | Alaska State Virology Laboratory | Alaska State Virology Laboratory | Stephanie DeRonde, Elva House, Jacob Zidek, Lisa Smith, Ph.D., Jack Chen, Ph.D. |
| EPI_ISL_2499966 | LESP Sinaloa | Instituto de Diagnostico y Referencia Epidemiologicos (INDRE) | Claudia Wong-Arambula, Abril Rodriguez-Maldonado, Vanessa Rivero-Arredondo, Ariadna Medina-Benitez, Joaquin Quiroz-Mercado, Sergio Rangel-Guerrero, Natividad Cruz-Ortiz, Tatiana Nunez-Garcia, Gisela Barrera-Badillo, Lucia Hernandez-Rivas, Irma Lopez-Martinez, Ernesto Ramirez-Gonzalez. |
| EPI_ISL_2499967 | LESP Michoacan | Instituto de Diagnostico y Referencia Epidemiologicos (INDRE) | Claudia Wong-Arambula, Abril Rodriguez-Maldonado, Vanessa Rivero-Arredondo, Ariadna Medina-Benitez, Joaquin Quiroz-Mercado, Sergio Rangel-Guerrero, Natividad Cruz-Ortiz, Tatiana Nunez-Garcia, Gisela Barrera-Badillo, Lucia Hernandez-Rivas, Irma Lopez-Martinez, Ernesto Ramirez-Gonzalez. |
| EPI_ISL_2499969 | University of Wisconsin-Madison AIDS Vaccine Research Laboratories | University of Wisconsin-Madison AIDS Vaccine Research Laboratories | Gage Moreno, Katarina Braun, et al. AIDS Vaccine Research Laboratories |
| EPI_ISL_2500024 | CA DPH Viral and Rickettsial Disease Laboratory | Chan-Zuckerberg Biohub | CZB Cliahub Consortium |
| EPI_ISL_2500025 | Humboldt County Public Health Laboratory | Chan-Zuckerberg Biohub | CZB Cliahub Consortium |
| EPI_ISL_2500026 | CA DPH Viral and Rickettsial Disease Laboratory | Chan-Zuckerberg Biohub | CZB Cliahub Consortium |
| EPI_ISL_2500027 | County of San Luis Obispo Public Health Laboratory | Chan-Zuckerberg Biohub | CZB Cliahub Consortium |
| EPI_ISL_2500028 | CA DPH Viral and Rickettsial Disease Laboratory | Chan-Zuckerberg Biohub | CZB Cliahub Consortium |
| EPI_ISL_2500029 | Orange County Public Health Lab | Chan-Zuckerberg Biohub | CZB Cliahub Consortium |
| EPI_ISL_2500030 | CA DPH Viral and Rickettsial Disease Laboratory | Chan-Zuckerberg Biohub | CZB Cliahub Consortium |
| EPI_ISL_2500031, EPI_ISL_2500032 | Humboldt County Public Health Laboratory | Chan-Zuckerberg Biohub | CZB Cliahub Consortium |
| EPI_ISL_2500033, EPI_ISL_2500034 | Santa Clara County Public Health Laboratory | Chan-Zuckerberg Biohub | CZB Cliahub Consortium |
| EPI_ISL_2500035, EPI_ISL_2500036, EPI_ISL_2500037, EPI_ISL_2500038, EPI_ISL_2500039 | CA DPH Viral and Rickettsial Disease Laboratory | Chan-Zuckerberg Biohub | CZB Cliahub Consortium |
| EPI_ISL_2500040, EPI_ISL_2500041 | County of San Luis Obispo Public Health Laboratory | Chan-Zuckerberg Biohub | CZB Cliahub Consortium |
| EPI_ISL_2500042 | CA DPH Viral and Rickettsial Disease Laboratory | Chan-Zuckerberg Biohub | CZB Cliahub Consortium |
| EPI_ISL_2500043 | Humboldt County Public Health Laboratory | Chan-Zuckerberg Biohub | CZB Cliahub Consortium |
| EPI_ISL_2500044 | CA DPH Viral and Rickettsial Disease Laboratory | Chan-Zuckerberg Biohub | CZB Cliahub Consortium |
| EPI_ISL_2500045, EPI_ISL_2500046 | County of San Luis Obispo Public Health Laboratory | Chan-Zuckerberg Biohub | CZB Cliahub Consortium |
| EPI_ISL_2500047 | CA DPH Viral and Rickettsial Disease Laboratory | Chan-Zuckerberg Biohub | CZB Cliahub Consortium |
| EPI_ISL_2500048, EPI_ISL_2500049 | Humboldt County Public Health Laboratory | Chan-Zuckerberg Biohub | CZB Cliahub Consortium |
| EPI_ISL_2500050, EPI_ISL_2500051, EPI_ISL_2500052 | CA DPH Viral and Rickettsial Disease Laboratory | Chan-Zuckerberg Biohub | CZB Cliahub Consortium |
| EPI_ISL_2500053 | Santa Clara County Public Health Laboratory | Chan-Zuckerberg Biohub | CZB Cliahub Consortium |
| EPI_ISL_2500054 | Humboldt County Public Health Laboratory | Chan-Zuckerberg Biohub | CZB Cliahub Consortium |
| EPI_ISL_2500055, EPI_ISL_2500056 | County of San Luis Obispo Public Health Laboratory | Chan-Zuckerberg Biohub | CZB Cliahub Consortium |
| EPI_ISL_2500057 | CA DPH Viral and Rickettsial Disease Laboratory | Chan-Zuckerberg Biohub | CZB Cliahub Consortium |
| EPI_ISL_2500058 | Humboldt County Public Health Laboratory | Chan-Zuckerberg Biohub | CZB Cliahub Consortium |
| EPI_ISL_2500059 | Orange County Public Health Lab | Chan-Zuckerberg Biohub | CZB Cliahub Consortium |
| EPI_ISL_2500060, EPI_ISL_2500061 | Humboldt County Public Health Laboratory | Chan-Zuckerberg Biohub | CZB Cliahub Consortium |
| EPI_ISL_2500062 | CA DPH Viral and Rickettsial Disease Laboratory | Chan-Zuckerberg Biohub | CZB Cliahub Consortium |
| EPI_ISL_2500063 | Humboldt County Public Health Laboratory | Chan-Zuckerberg Biohub | CZB Cliahub Consortium |
| EPI_ISL_2500064 | Santa Clara County Public Health Laboratory | Chan-Zuckerberg Biohub | CZB Cliahub Consortium |
| EPI_ISL_2500065, EPI_ISL_2500066 | County of San Luis Obispo Public Health Laboratory | Chan-Zuckerberg Biohub | CZB Cliahub Consortium |
| EPI_ISL_2500067, EPI_ISL_2500068 | CA DPH Viral and Rickettsial Disease Laboratory | Chan-Zuckerberg Biohub | CZB Cliahub Consortium |
| EPI_ISL_2500069 | Humboldt County Public Health Laboratory | Chan-Zuckerberg Biohub | CZB Cliahub Consortium |
| EPI_ISL_2500070 | CA DPH Viral and Rickettsial Disease Laboratory | Chan-Zuckerberg Biohub | CZB Cliahub Consortium |
| EPI_ISL_2500071 | County of San Luis Obispo Public Health Laboratory | Chan-Zuckerberg Biohub | CZB Cliahub Consortium |
| EPI_ISL_2500072, EPI_ISL_2500073 | CA DPH Viral and Rickettsial Disease Laboratory | Chan-Zuckerberg Biohub | CZB Cliahub Consortium |
| EPI_ISL_2500074 | Humboldt County Public Health Laboratory | Chan-Zuckerberg Biohub | CZB Cliahub Consortium |
| EPI_ISL_2500075 | CA DPH Viral and Rickettsial Disease Laboratory | Chan-Zuckerberg Biohub | CZB Cliahub Consortium |
| EPI_ISL_2500076 | Santa Clara County Public Health Laboratory | Chan-Zuckerberg Biohub | CZB Cliahub Consortium |
| EPI_ISL_2500077, EPI_ISL_2500078 | Humboldt County Public Health Laboratory | Chan-Zuckerberg Biohub | CZB Cliahub Consortium |
| EPI_ISL_2500079, EPI_ISL_2500080, EPI_ISL_2500081, EPI_ISL_2500082, | CA DPH Viral and Rickettsial Disease Laboratory | Chan-Zuckerberg Biohub | CZB Cliahub Consortium |

| | | | |
|---|---|---|---|
| EPI_ISL_2500083, EPI_ISL_2500084 | | | |
| EPI_ISL_2500085 | Santa Clara County Public Health Laboratory | Chan-Zuckerberg Biohub | CZB Cliahub Consortium |
| EPI_ISL_2500086 | CA DPH Viral and Rickettsial Disease Laboratory | Chan-Zuckerberg Biohub | CZB Cliahub Consortium |
| EPI_ISL_2500087, EPI_ISL_2500088 | County of San Luis Obispo Public Health Laboratory | Chan-Zuckerberg Biohub | CZB Cliahub Consortium |
| EPI_ISL_2500089, EPI_ISL_2500090, EPI_ISL_2500093, EPI_ISL_2500094 | CA DPH Viral and Rickettsial Disease Laboratory | Chan-Zuckerberg Biohub | CZB Cliahub Consortium |
| EPI_ISL_2500095 | County of San Luis Obispo Public Health Laboratory | Chan-Zuckerberg Biohub | CZB Cliahub Consortium |
| EPI_ISL_2500096, EPI_ISL_2500097, EPI_ISL_2500098, EPI_ISL_2500099 | CA DPH Viral and Rickettsial Disease Laboratory | Chan-Zuckerberg Biohub | CZB Cliahub Consortium |
| EPI_ISL_2500100 | Humboldt County Public Health Laboratory | Chan-Zuckerberg Biohub | CZB Cliahub Consortium |
| EPI_ISL_2500101 | County of San Luis Obispo Public Health Laboratory | Chan-Zuckerberg Biohub | CZB Cliahub Consortium |
| EPI_ISL_2500102 | CA DPH Viral and Rickettsial Disease Laboratory | Chan-Zuckerberg Biohub | CZB Cliahub Consortium |
| EPI_ISL_2500103 | County of San Luis Obispo Public Health Laboratory | Chan-Zuckerberg Biohub | CZB Cliahub Consortium |
| EPI_ISL_2500104 | Orange County Public Health Lab | Chan-Zuckerberg Biohub | CZB Cliahub Consortium |
| EPI_ISL_2500105 | County of San Luis Obispo Public Health Laboratory | Chan-Zuckerberg Biohub | CZB Cliahub Consortium |
| EPI_ISL_2500106 | CA DPH Viral and Rickettsial Disease Laboratory | Chan-Zuckerberg Biohub | CZB Cliahub Consortium |
| EPI_ISL_2500107 | County of San Luis Obispo Public Health Laboratory | Chan-Zuckerberg Biohub | CZB Cliahub Consortium |
| EPI_ISL_2500108, EPI_ISL_2500109 | Humboldt County Public Health Laboratory | Chan-Zuckerberg Biohub | CZB Cliahub Consortium |
| EPI_ISL_2500110, EPI_ISL_2500111, EPI_ISL_2500112, EPI_ISL_2500113 | CA DPH Viral and Rickettsial Disease Laboratory | Chan-Zuckerberg Biohub | CZB Cliahub Consortium |
| EPI_ISL_2500114 | County of San Luis Obispo Public Health Laboratory | Chan-Zuckerberg Biohub | CZB Cliahub Consortium |
| EPI_ISL_2500115 | Santa Clara County Public Health Laboratory | Chan-Zuckerberg Biohub | CZB Cliahub Consortium |
| EPI_ISL_2500116 | Humboldt County Public Health Laboratory | Chan-Zuckerberg Biohub | CZB Cliahub Consortium |
| EPI_ISL_2500117 | County of San Luis Obispo Public Health Laboratory | Chan-Zuckerberg Biohub | CZB Cliahub Consortium |
| EPI_ISL_2500118 | CA DPH Viral and Rickettsial Disease Laboratory | Chan-Zuckerberg Biohub | CZB Cliahub Consortium |
| EPI_ISL_2500119 | County of San Luis Obispo Public Health Laboratory | Chan-Zuckerberg Biohub | CZB Cliahub Consortium |
| EPI_ISL_2500120, EPI_ISL_2500121 | CA DPH Viral and Rickettsial Disease Laboratory | Chan-Zuckerberg Biohub | CZB Cliahub Consortium |
| EPI_ISL_2500122 | Humboldt County Public Health Laboratory | Chan-Zuckerberg Biohub | CZB Cliahub Consortium |
| EPI_ISL_2500123, EPI_ISL_2500124 | County of San Luis Obispo Public Health Laboratory | Chan-Zuckerberg Biohub | CZB Cliahub Consortium |
| EPI_ISL_2500125 | Humboldt County Public Health Laboratory | Chan-Zuckerberg Biohub | CZB Cliahub Consortium |
| EPI_ISL_2500126 | CA DPH Viral and Rickettsial Disease Laboratory | Chan-Zuckerberg Biohub | CZB Cliahub Consortium |
| EPI_ISL_2500127 | Santa Clara County Public Health Laboratory | Chan-Zuckerberg Biohub | CZB Cliahub Consortium |
| EPI_ISL_2500128 | County of San Luis Obispo Public Health Laboratory | Chan-Zuckerberg Biohub | CZB Cliahub Consortium |
| EPI_ISL_2500129 | Santa Clara County Public Health Laboratory | Chan-Zuckerberg Biohub | CZB Cliahub Consortium |
| EPI_ISL_2500130 | Orange County Public Health Lab | Chan-Zuckerberg Biohub | CZB Cliahub Consortium |
| EPI_ISL_2500131 | Santa Clara County Public Health Laboratory | Chan-Zuckerberg Biohub | CZB Cliahub Consortium |
| EPI_ISL_2500132, EPI_ISL_2500133 | Humboldt County Public Health Laboratory | Chan-Zuckerberg Biohub | CZB Cliahub Consortium |
| EPI_ISL_2500134 | Santa Clara County Public Health Laboratory | Chan-Zuckerberg Biohub | CZB Cliahub Consortium |
| EPI_ISL_2500135 | CA DPH Viral and Rickettsial Disease Laboratory | Chan-Zuckerberg Biohub | CZB Cliahub Consortium |
| EPI_ISL_2500136 | Santa Clara County Public Health Laboratory | Chan-Zuckerberg Biohub | CZB Cliahub Consortium |
| EPI_ISL_2500137, EPI_ISL_2500138, EPI_ISL_2500139, EPI_ISL_2500140 | CA DPH Viral and Rickettsial Disease Laboratory | Chan-Zuckerberg Biohub | CZB Cliahub Consortium |
| EPI_ISL_2500141 | County of San Luis Obispo Public Health Laboratory | Chan-Zuckerberg Biohub | CZB Cliahub Consortium |
| EPI_ISL_2500142 | Humboldt County Public Health Laboratory | Chan-Zuckerberg Biohub | CZB Cliahub Consortium |
| EPI_ISL_2500143, EPI_ISL_2500144, EPI_ISL_2500145, EPI_ISL_2500146, EPI_ISL_2500147 | CA DPH Viral and Rickettsial Disease Laboratory | Chan-Zuckerberg Biohub | CZB Cliahub Consortium |
| EPI_ISL_2500148 | County of San Luis Obispo Public Health | Chan-Zuckerberg Biohub | CZB Cliahub Consortium |

| | | | |
|---|---|---|---|
| | Laboratory | | |
| EPI_ISL_2500149 | CA DPH Viral and Rickettsial Disease Laboratory | Chan-Zuckerberg Biohub | CZB Cliahub Consortium |
| EPI_ISL_2500150, EPI_ISL_2500151 | Humboldt County Public Health Laboratory | Chan-Zuckerberg Biohub | CZB Cliahub Consortium |
| EPI_ISL_2500152, EPI_ISL_2500153 | CA DPH Viral and Rickettsial Disease Laboratory | Chan-Zuckerberg Biohub | CZB Cliahub Consortium |
| EPI_ISL_2500154, EPI_ISL_2500155, EPI_ISL_2500156 | Santa Clara County Public Health Laboratory | Chan-Zuckerberg Biohub | CZB Cliahub Consortium |
| EPI_ISL_2500157 | Humboldt County Public Health Laboratory | Chan-Zuckerberg Biohub | CZB Cliahub Consortium |
| EPI_ISL_2500158 | CA DPH Viral and Rickettsial Disease Laboratory | Chan-Zuckerberg Biohub | CZB Cliahub Consortium |
| EPI_ISL_2500159 | Humboldt County Public Health Laboratory | Chan-Zuckerberg Biohub | CZB Cliahub Consortium |
| EPI_ISL_2500160 | CA DPH Viral and Rickettsial Disease Laboratory | Chan-Zuckerberg Biohub | CZB Cliahub Consortium |
| EPI_ISL_2500161 | County of San Luis Obispo Public Health Laboratory | Chan-Zuckerberg Biohub | CZB Cliahub Consortium |
| EPI_ISL_2500162 | Santa Clara County Public Health Laboratory | Chan-Zuckerberg Biohub | CZB Cliahub Consortium |
| EPI_ISL_2500163, EPI_ISL_2500164 | CA DPH Viral and Rickettsial Disease Laboratory | Chan-Zuckerberg Biohub | CZB Cliahub Consortium |
| EPI_ISL_2500165 | County of San Luis Obispo Public Health Laboratory | Chan-Zuckerberg Biohub | CZB Cliahub Consortium |
| EPI_ISL_2500166 | CA DPH Viral and Rickettsial Disease Laboratory | Chan-Zuckerberg Biohub | CZB Cliahub Consortium |
| EPI_ISL_2500167 | County of San Luis Obispo Public Health Laboratory | Chan-Zuckerberg Biohub | CZB Cliahub Consortium |
| EPI_ISL_2500168, EPI_ISL_2500169, EPI_ISL_2500170 | CA DPH Viral and Rickettsial Disease Laboratory | Chan-Zuckerberg Biohub | CZB Cliahub Consortium |
| EPI_ISL_2500171 | Santa Clara County Public Health Laboratory | Chan-Zuckerberg Biohub | CZB Cliahub Consortium |
| EPI_ISL_2500172 | County of San Luis Obispo Public Health Laboratory | Chan-Zuckerberg Biohub | CZB Cliahub Consortium |
| EPI_ISL_2500173 | Santa Clara County Public Health Laboratory | Chan-Zuckerberg Biohub | CZB Cliahub Consortium |
| EPI_ISL_2500174 | CA DPH Viral and Rickettsial Disease Laboratory | Chan-Zuckerberg Biohub | CZB Cliahub Consortium |
| EPI_ISL_2500176 | County of San Luis Obispo Public Health Laboratory | Chan-Zuckerberg Biohub | CZB Cliahub Consortium |
| EPI_ISL_2500177, EPI_ISL_2500178, EPI_ISL_2500179, EPI_ISL_2500180, EPI_ISL_2500181 | CA DPH Viral and Rickettsial Disease Laboratory | Chan-Zuckerberg Biohub | CZB Cliahub Consortium |
| EPI_ISL_2500182 | Humboldt County Public Health Laboratory | Chan-Zuckerberg Biohub | CZB Cliahub Consortium |
| EPI_ISL_2500183 | Santa Clara County Public Health Laboratory | Chan-Zuckerberg Biohub | CZB Cliahub Consortium |
| EPI_ISL_2500184 | CA DPH Viral and Rickettsial Disease Laboratory | Chan-Zuckerberg Biohub | CZB Cliahub Consortium |
| EPI_ISL_2500185 | Humboldt County Public Health Laboratory | Chan-Zuckerberg Biohub | CZB Cliahub Consortium |
| EPI_ISL_2500186 | CA DPH Viral and Rickettsial Disease Laboratory | Chan-Zuckerberg Biohub | CZB Cliahub Consortium |
| EPI_ISL_2500187, EPI_ISL_2500188, EPI_ISL_2500189 | Humboldt County Public Health Laboratory | Chan-Zuckerberg Biohub | CZB Cliahub Consortium |
| EPI_ISL_2500190 | CA DPH Viral and Rickettsial Disease Laboratory | Chan-Zuckerberg Biohub | CZB Cliahub Consortium |
| EPI_ISL_2500191 | County of San Luis Obispo Public Health Laboratory | Chan-Zuckerberg Biohub | CZB Cliahub Consortium |
| EPI_ISL_2500192 | Santa Clara County Public Health Laboratory | Chan-Zuckerberg Biohub | CZB Cliahub Consortium |
| EPI_ISL_2500193, EPI_ISL_2500194, EPI_ISL_2500195 | CA DPH Viral and Rickettsial Disease Laboratory | Chan-Zuckerberg Biohub | CZB Cliahub Consortium |
| EPI_ISL_2500196 | Humboldt County Public Health Laboratory | Chan-Zuckerberg Biohub | CZB Cliahub Consortium |
| EPI_ISL_2500197, EPI_ISL_2500198 | County of San Luis Obispo Public Health Laboratory | Chan-Zuckerberg Biohub | CZB Cliahub Consortium |
| EPI_ISL_2500200 | Santa Clara County Public Health Laboratory | Chan-Zuckerberg Biohub | CZB Cliahub Consortium |
| EPI_ISL_2500201, EPI_ISL_2500202 | Humboldt County Public Health Laboratory | Chan-Zuckerberg Biohub | CZB Cliahub Consortium |
| EPI_ISL_2500203, EPI_ISL_2500204, EPI_ISL_2500205 | CA DPH Viral and Rickettsial Disease Laboratory | Chan-Zuckerberg Biohub | CZB Cliahub Consortium |
| EPI_ISL_2500206 | Humboldt County Public Health Laboratory | Chan-Zuckerberg Biohub | CZB Cliahub Consortium |
| EPI_ISL_2500207 | Santa Clara County Public Health Laboratory | Chan-Zuckerberg Biohub | CZB Cliahub Consortium |
| EPI_ISL_2500208, EPI_ISL_2500209, EPI_ISL_2500210 | CA DPH Viral and Rickettsial Disease Laboratory | Chan-Zuckerberg Biohub | CZB Cliahub Consortium |
| EPI_ISL_2500211 | Orange County Public Health Lab | Chan-Zuckerberg Biohub | CZB Cliahub Consortium |
| EPI_ISL_2500212 | CA DPH Viral and Rickettsial Disease Laboratory | Chan-Zuckerberg Biohub | CZB Cliahub Consortium |

| | | | |
|---|---|---|---|
| EPI_ISL_2500213, EPI_ISL_2500214 | Humboldt County Public Health Laboratory | Chan-Zuckerberg Biohub | CZB Cliahub Consortium |
| EPI_ISL_2500215 | County of San Luis Obispo Public Health Laboratory | Chan-Zuckerberg Biohub | CZB Cliahub Consortium |
| EPI_ISL_2500216 | CA DPH Viral and Rickettsial Disease Laboratory | Chan-Zuckerberg Biohub | CZB Cliahub Consortium |
| EPI_ISL_2500306, EPI_ISL_2500307, EPI_ISL_2500308, EPI_ISL_2500309, EPI_ISL_2500311, EPI_ISL_2500312, EPI_ISL_2500313, EPI_ISL_2500314, EPI_ISL_2500315, EPI_ISL_2500316, EPI_ISL_2500317, EPI_ISL_2500318, EPI_ISL_2500319, EPI_ISL_2500320, EPI_ISL_2500321, EPI_ISL_2500323, EPI_ISL_2500324, EPI_ISL_2500326, EPI_ISL_2500327, EPI_ISL_2500340, EPI_ISL_2500343, EPI_ISL_2500344, EPI_ISL_2500345, EPI_ISL_2500348, EPI_ISL_2500354, EPI_ISL_2500358, EPI_ISL_2500360 | | | |
| see above | City of Milwaukee Health Department Laboratory | City of Milwaukee Health Department Laboratory | Sanjib Bhattacharyya, Manjeet Khubbar, Amy Bauer, Jennifer Lentz, Samantha Scott |
| EPI_ISL_2500371, EPI_ISL_2500372, EPI_ISL_2500373, EPI_ISL_2500374, EPI_ISL_2500375, EPI_ISL_2500377, EPI_ISL_2500378, EPI_ISL_2500379, EPI_ISL_2500380, EPI_ISL_2500381, EPI_ISL_2500382, EPI_ISL_2500383, EPI_ISL_2500384, EPI_ISL_2500385, EPI_ISL_2500386, EPI_ISL_2500387, EPI_ISL_2500388, EPI_ISL_2500389, EPI_ISL_2500390, EPI_ISL_2500391, EPI_ISL_2500392, EPI_ISL_2500393, EPI_ISL_2500394, EPI_ISL_2500395, EPI_ISL_2500396, EPI_ISL_2500397, EPI_ISL_2500398, EPI_ISL_2500399, EPI_ISL_2500400, EPI_ISL_2500401, EPI_ISL_2500402, EPI_ISL_2500403, EPI_ISL_2500404, EPI_ISL_2500405, EPI_ISL_2500406, EPI_ISL_2500407, EPI_ISL_2500408, EPI_ISL_2500409, EPI_ISL_2500410, EPI_ISL_2500411, EPI_ISL_2500412, EPI_ISL_2500413, EPI_ISL_2500414, EPI_ISL_2500415, EPI_ISL_2500416, EPI_ISL_2500417, EPI_ISL_2500418, EPI_ISL_2500419, EPI_ISL_2500420, EPI_ISL_2500421 | | | |
| see above | IVY3 Central Lab, Vanderbilt University Medical Center | Lauring Lab, University of Michigan, Department of Microbiology and Immunology | Valesano |
| EPI_ISL_2500422, EPI_ISL_2500423, EPI_ISL_2500424, EPI_ISL_2500427, EPI_ISL_2500428, EPI_ISL_2500429, EPI_ISL_2500430, EPI_ISL_2500431, EPI_ISL_2500433, EPI_ISL_2500434, EPI_ISL_2500436, EPI_ISL_2500437, EPI_ISL_2500438 | | | |
| see above | CT Department of Public Health | CT Department of Public Health | Claire_Pearson,Tu_N_Nguyen |
| EPI_ISL_2500439, EPI_ISL_2500440, EPI_ISL_2500441, EPI_ISL_2500442, EPI_ISL_2500443, EPI_ISL_2500444, EPI_ISL_2500445, EPI_ISL_2500446, EPI_ISL_2500447, EPI_ISL_2500448, EPI_ISL_2500449, EPI_ISL_2500450, EPI_ISL_2500451, EPI_ISL_2500452, EPI_ISL_2500453, EPI_ISL_2500454, EPI_ISL_2500455, EPI_ISL_2500456, EPI_ISL_2500457, EPI_ISL_2500458, EPI_ISL_2500459, EPI_ISL_2500460, EPI_ISL_2500461, EPI_ISL_2500462, EPI_ISL_2500463 | | | |
| see above | Hackensack Medical Center | New York Genome Center | Michael Zody, Andre Corvelo, Dayna M. Oschwald, Samantha Fennessey, Tom Maniatis, Liang Chen, Jose Mediavilla, Marcus Cunningham, Kaelea Composto, Kar Chow, David Perlin, Barry Kreiswirth |
| EPI_ISL_2500544, EPI_ISL_2500545, EPI_ISL_2500546, EPI_ISL_2500547, EPI_ISL_2500548, EPI_ISL_2500549, EPI_ISL_2500550, EPI_ISL_2500551, EPI_ISL_2500553, EPI_ISL_2500554, EPI_ISL_2500555, EPI_ISL_2500556, EPI_ISL_2500557, EPI_ISL_2500559, EPI_ISL_2500560, EPI_ISL_2500561, EPI_ISL_2500562, EPI_ISL_2500563, EPI_ISL_2500564, EPI_ISL_2500565, EPI_ISL_2500566, EPI_ISL_2500567, EPI_ISL_2500568, EPI_ISL_2500569, EPI_ISL_2500570, EPI_ISL_2500571, EPI_ISL_2500572, EPI_ISL_2500573, EPI_ISL_2500574, EPI_ISL_2500575, EPI_ISL_2500576, EPI_ISL_2500577, EPI_ISL_2500578, EPI_ISL_2500579, EPI_ISL_2500580, EPI_ISL_2500581, EPI_ISL_2500582, EPI_ISL_2500583, EPI_ISL_2500584, EPI_ISL_2500585, EPI_ISL_2500586, EPI_ISL_2500587, EPI_ISL_2500588, EPI_ISL_2500589, EPI_ISL_2500590, EPI_ISL_2500591, EPI_ISL_2500592, EPI_ISL_2500593, EPI_ISL_2500594, EPI_ISL_2500595, EPI_ISL_2500596, EPI_ISL_2500598, EPI_ISL_2500599, EPI_ISL_2500600, EPI_ISL_2500601, EPI_ISL_2500602, EPI_ISL_2500603, EPI_ISL_2500604, EPI_ISL_2500605, EPI_ISL_2500606, EPI_ISL_2500607, EPI_ISL_2500608, EPI_ISL_2500609, EPI_ISL_2500610, EPI_ISL_2500611, EPI_ISL_2500612, EPI_ISL_2500613, EPI_ISL_2500614, EPI_ISL_2500615, EPI_ISL_2500616, EPI_ISL_2500617, EPI_ISL_2500618, EPI_ISL_2500619, EPI_ISL_2500620, EPI_ISL_2500622, EPI_ISL_2500623, EPI_ISL_2500624, EPI_ISL_2500625, EPI_ISL_2500626, EPI_ISL_2500629, EPI_ISL_2500631, EPI_ISL_2500632, EPI_ISL_2500633, EPI_ISL_2500635, EPI_ISL_2500636, EPI_ISL_2500638, EPI_ISL_2500639, EPI_ISL_2500640, EPI_ISL_2500641, EPI_ISL_2500642, EPI_ISL_2500643, EPI_ISL_2500644, EPI_ISL_2500645, EPI_ISL_2500646, EPI_ISL_2500647, EPI_ISL_2500648, EPI_ISL_2500649, EPI_ISL_2500650, EPI_ISL_2500651, EPI_ISL_2500652, EPI_ISL_2500653, EPI_ISL_2500654, EPI_ISL_2500655, EPI_ISL_2500656, EPI_ISL_2500657, EPI_ISL_2500659, EPI_ISL_2500660, EPI_ISL_2500662, EPI_ISL_2500663, EPI_ISL_2500664, EPI_ISL_2500665, EPI_ISL_2500666, EPI_ISL_2500667, EPI_ISL_2500668, EPI_ISL_2500669, EPI_ISL_2500670, EPI_ISL_2500671, EPI_ISL_2500672, EPI_ISL_2500673, EPI_ISL_2500674, EPI_ISL_2500675, EPI_ISL_2500676, EPI_ISL_2500677, EPI_ISL_2500679, EPI_ISL_2500680, EPI_ISL_2500681, EPI_ISL_2500682, EPI_ISL_2500684, EPI_ISL_2500685, EPI_ISL_2500687, EPI_ISL_2500688, EPI_ISL_2500689, EPI_ISL_2500690, EPI_ISL_2500691, EPI_ISL_2500692, EPI_ISL_2500693, EPI_ISL_2500694, EPI_ISL_2500695, EPI_ISL_2500698, EPI_ISL_2500699, EPI_ISL_2500700, EPI_ISL_2500701, EPI_ISL_2500702, EPI_ISL_2500703, EPI_ISL_2500704, EPI_ISL_2500705, EPI_ISL_2500706, EPI_ISL_2500707, EPI_ISL_2500708, EPI_ISL_2500709, EPI_ISL_2500710, EPI_ISL_2500711, EPI_ISL_2500712, EPI_ISL_2500713, EPI_ISL_2500714, EPI_ISL_2500716, EPI_ISL_2500717, EPI_ISL_2500718, EPI_ISL_2500719, EPI_ISL_2500720, EPI_ISL_2500721, EPI_ISL_2500722, EPI_ISL_2500723, EPI_ISL_2500724, EPI_ISL_2500725, EPI_ISL_2500726, EPI_ISL_2500727, EPI_ISL_2500728, EPI_ISL_2500729, EPI_ISL_2500730, EPI_ISL_2500731, EPI_ISL_2500732, EPI_ISL_2500733, EPI_ISL_2500734, EPI_ISL_2500735, EPI_ISL_2500736, EPI_ISL_2500737, EPI_ISL_2500738, EPI_ISL_2500739, EPI_ISL_2500740, EPI_ISL_2500741, EPI_ISL_2500742, EPI_ISL_2500743, EPI_ISL_2500744, EPI_ISL_2500745, EPI_ISL_2500747, EPI_ISL_2500748, EPI_ISL_2500750, EPI_ISL_2500751, EPI_ISL_2500752, EPI_ISL_2500753, EPI_ISL_2500754, EPI_ISL_2500755, EPI_ISL_2500756, EPI_ISL_2500757, EPI_ISL_2500759, EPI_ISL_2500763, EPI_ISL_2500765, EPI_ISL_2500766, EPI_ISL_2500767, EPI_ISL_2500768, EPI_ISL_2500769, EPI_ISL_2500770, EPI_ISL_2500771, EPI_ISL_2500772, EPI_ISL_2500773, EPI_ISL_2500775, EPI_ISL_2500776, EPI_ISL_2500777, EPI_ISL_2500778, EPI_ISL_2500779, EPI_ISL_2500780, EPI_ISL_2500781, EPI_ISL_2500782, EPI_ISL_2500784, EPI_ISL_2500786, EPI_ISL_2500787, EPI_ISL_2500788, EPI_ISL_2500789, EPI_ISL_2500790, EPI_ISL_2500791, EPI_ISL_2500792, EPI_ISL_2500793, EPI_ISL_2500794, EPI_ISL_2500796, EPI_ISL_2500797, EPI_ISL_2500798, EPI_ISL_2500799, EPI_ISL_2500800, EPI_ISL_2500801, EPI_ISL_2500802, EPI_ISL_2500803, EPI_ISL_2500804, EPI_ISL_2500805, EPI_ISL_2500806, EPI_ISL_2500807, EPI_ISL_2500810, EPI_ISL_2500812, EPI_ISL_2500813, EPI_ISL_2500814, EPI_ISL_2500815, EPI_ISL_2500816, EPI_ISL_2500817, EPI_ISL_2500818, EPI_ISL_2500819, EPI_ISL_2500820, EPI_ISL_2500821, EPI_ISL_2500822, EPI_ISL_2500823, EPI_ISL_2500824, EPI_ISL_2500825, EPI_ISL_2500826, EPI_ISL_2500827, EPI_ISL_2500828, EPI_ISL_2500829, EPI_ISL_2500830, EPI_ISL_2500831, EPI_ISL_2500832, EPI_ISL_2500833, EPI_ISL_2500834, EPI_ISL_2500835, EPI_ISL_2500836, EPI_ISL_2500837, EPI_ISL_2500838, EPI_ISL_2500839, EPI_ISL_2500840, EPI_ISL_2500842, EPI_ISL_2500843, EPI_ISL_2500844, EPI_ISL_2500845, EPI_ISL_2500846, EPI_ISL_2500847, EPI_ISL_2500848 | | | |
| see above | Clinical Microbiology Laboratory, NewYork Presbyterian Hospital/Columbia University Irving Medical Center | Uhlemann Laboratory, Columbia University Irving Medical Center | Medini K. Annavajhala, Anne Kelley, Maya Tagliavia, Anne-Catrin Uhlemann |
| EPI_ISL_2500988, EPI_ISL_2500989, EPI_ISL_2500990, EPI_ISL_2500991, EPI_ISL_2500992, EPI_ISL_2500993, EPI_ISL_2500994, EPI_ISL_2500995, EPI_ISL_2500996, EPI_ISL_2500997, EPI_ISL_2500998, EPI_ISL_2500999, EPI_ISL_2501000, EPI_ISL_2501001, EPI_ISL_2501002, EPI_ISL_2501003, EPI_ISL_2501004, EPI_ISL_2501005, EPI_ISL_2501006, EPI_ISL_2501007, EPI_ISL_2501008, EPI_ISL_2501009, EPI_ISL_2501010, EPI_ISL_2501011, EPI_ISL_2501012, EPI_ISL_2501013, EPI_ISL_2501014, EPI_ISL_2501015, EPI_ISL_2501016, EPI_ISL_2501017, EPI_ISL_2501018, EPI_ISL_2501019 | | | |
| see above | Gundersen Clinical Microbiology Laboratory | Kabara Cancer Research Institute | Craig S. Richmond, Paraic A. Kenny |
| EPI_ISL_2501096 | Allina Health Laboratory | Minnesota Department of Health, Public Health Laboratory | Alexandra Lorentz, Jacob Garfin, Matt Plumb, and Xiong Wang |
| EPI_ISL_2501097, EPI_ISL_2501098 | M Health Fairview | Minnesota Department of Health, Public Health Laboratory | Alexandra Lorentz, Jacob Garfin, Matt Plumb, and Xiong Wang |
| EPI_ISL_2501099 | Allina Health Laboratory | Minnesota Department of Health, Public Health Laboratory | Alexandra Lorentz, Jacob Garfin, Matt Plumb, and Xiong Wang |
| EPI_ISL_2501100 | M Health Fairview | Minnesota Department of Health, Public Health Laboratory | Alexandra Lorentz, Jacob Garfin, Matt Plumb, and Xiong Wang |
| EPI_ISL_2501101 | Hennepin County Medical Center | Minnesota Department of Health, Public Health Laboratory | Alexandra Lorentz, Jacob Garfin, Matt Plumb, and Xiong Wang |
| EPI_ISL_2501102 | Regions Hospital | Minnesota Department of Health, Public Health Laboratory | Alexandra Lorentz, Jacob Garfin, Matt Plumb, and Xiong Wang |
| EPI_ISL_2501103, EPI_ISL_2501104, EPI_ISL_2501105 | M Health Fairview | Minnesota Department of Health, Public Health Laboratory | Alexandra Lorentz, Jacob Garfin, Matt Plumb, and Xiong Wang |
| EPI_ISL_2501106 | Regions Hospital | Minnesota Department of Health, Public Health Laboratory | Alexandra Lorentz, Jacob Garfin, Matt Plumb, and Xiong Wang |
| EPI_ISL_2501107 | Premier Medical Lab | Minnesota Department of Health, Public Health Laboratory | Alexandra Lorentz, Jacob Garfin, Matt Plumb, and Xiong Wang |
| EPI_ISL_2501108 | M Health Fairview | Minnesota Department of Health, Public Health Laboratory | Alexandra Lorentz, Jacob Garfin, Matt Plumb, and Xiong Wang |
| EPI_ISL_2501109 | Allina Health Laboratory | Minnesota Department of Health, Public Health Laboratory | Alexandra Lorentz, Jacob Garfin, Matt Plumb, and Xiong Wang |
| EPI_ISL_2501110 | Maple Grove Hospital | Minnesota Department of Health, Public Health Laboratory | Alexandra Lorentz, Jacob Garfin, Matt Plumb, and Xiong Wang |
| EPI_ISL_2501111 | M Health Fairview | Minnesota Department of Health, Public Health Laboratory | Alexandra Lorentz, Jacob Garfin, Matt Plumb, and Xiong Wang |
| EPI_ISL_2501112 | Hennepin County Medical Center | Minnesota Department of Health, Public Health Laboratory | Alexandra Lorentz, Jacob Garfin, Matt Plumb, and Xiong Wang |
| EPI_ISL_2501113 | M Health Fairview | Minnesota Department of Health, Public Health Laboratory | Alexandra Lorentz, Jacob Garfin, Matt Plumb, and Xiong Wang |
| EPI_ISL_2501114, EPI_ISL_2501115 | Hennepin County Medical Center | Minnesota Department of Health, Public Health Laboratory | Alexandra Lorentz, Jacob Garfin, Matt Plumb, and Xiong Wang |
| EPI_ISL_2501116, EPI_ISL_2501117 | Minnesota Department of Health, Public Health Laboratory | Minnesota Department of Health, Public Health Laboratory | Alexandra Lorentz, Jacob Garfin, Matt Plumb, and Xiong Wang |
| EPI_ISL_2501118 | Mayo Clinic & Mayo Clinic Laboratories | Minnesota Department of Health, Public Health Laboratory | Alexandra Lorentz, Jacob Garfin, Matt Plumb, and Xiong Wang |
| EPI_ISL_2501119 | Maple Grove Hospital | Minnesota Department of Health, Public Health Laboratory | Alexandra Lorentz, Jacob Garfin, Matt Plumb, and Xiong Wang |
| EPI_ISL_2501120, EPI_ISL_2501121, EPI_ISL_2501122, EPI_ISL_2501123, EPI_ISL_2501124, EPI_ISL_2501125, EPI_ISL_2501126, EPI_ISL_2501127 | M Health Fairview | Minnesota Department of Health, Public Health Laboratory | Alexandra Lorentz, Jacob Garfin, Matt Plumb, and Xiong Wang |
| EPI_ISL_2501128 | Minnesota Department of Health, Public Health Laboratory | Minnesota Department of Health, Public Health Laboratory | Alexandra Lorentz, Jacob Garfin, Matt Plumb, and Xiong Wang |
| EPI_ISL_2501129, EPI_ISL_2501130, EPI_ISL_2501131, EPI_ISL_2501132, | M Health Fairview | Minnesota Department of Health, Public Health Laboratory | Alexandra Lorentz, Jacob Garfin, Matt Plumb, and Xiong Wang |

| | | | |
|---|---|---|---|
| EPI_ISL_2501133, EPI_ISL_2501134 | | | |
| EPI_ISL_2501145, EPI_ISL_2501146 | Mayo Clinic & Mayo Clinic Laboratories | Minnesota Department of Health, Public Health Laboratory | Alexandra Lorentz, Jacob Garfin, Matt Plumb, and Xiong Wang |
| EPI_ISL_2501147, EPI_ISL_2501148 | Allina Health Laboratory | Minnesota Department of Health, Public Health Laboratory | Alexandra Lorentz, Jacob Garfin, Matt Plumb, and Xiong Wang |
| EPI_ISL_2501149, EPI_ISL_2501150, EPI_ISL_2501151 | M Health Fairview | Minnesota Department of Health, Public Health Laboratory | Alexandra Lorentz, Jacob Garfin, Matt Plumb, and Xiong Wang |
| EPI_ISL_2501152 | Minnesota Department of Health, Public Health Laboratory | Minnesota Department of Health, Public Health Laboratory | Alexandra Lorentz, Jacob Garfin, Matt Plumb, and Xiong Wang |
| EPI_ISL_2501153, EPI_ISL_2501154 | Florida Bureau of Public Health Laboratories | Florida Bureau of Public Health Laboratories | Sarah Schmedes, Jason Blanton |
| EPI_ISL_2501156, EPI_ISL_2501157, EPI_ISL_2501158, EPI_ISL_2501159, EPI_ISL_2501160, EPI_ISL_2501161, EPI_ISL_2501162, EPI_ISL_2501163, EPI_ISL_2501164, EPI_ISL_2501165, EPI_ISL_2501166, EPI_ISL_2501167, EPI_ISL_2501168, EPI_ISL_2501169, EPI_ISL_2501170, EPI_ISL_2501171, EPI_ISL_2501172, EPI_ISL_2501173, EPI_ISL_2501174, EPI_ISL_2501175, EPI_ISL_2501176, EPI_ISL_2501177, EPI_ISL_2501178, EPI_ISL_2501179, EPI_ISL_2501180, EPI_ISL_2501181, EPI_ISL_2501182, EPI_ISL_2501183, EPI_ISL_2501184, EPI_ISL_2501185, EPI_ISL_2501186, EPI_ISL_2501187, EPI_ISL_2501190, EPI_ISL_2501191, EPI_ISL_2501192, EPI_ISL_2501193, EPI_ISL_2501194, EPI_ISL_2501195, EPI_ISL_2501196, EPI_ISL_2501197, EPI_ISL_2501198, EPI_ISL_2501199, EPI_ISL_2501200, EPI_ISL_2501201, EPI_ISL_2501202, EPI_ISL_2501203, EPI_ISL_2501204, EPI_ISL_2501205, EPI_ISL_2501206, EPI_ISL_2501207, EPI_ISL_2501208, EPI_ISL_2501209, EPI_ISL_2501210, EPI_ISL_2501211, EPI_ISL_2501212, EPI_ISL_2501213, EPI_ISL_2501214, EPI_ISL_2501215, EPI_ISL_2501216, EPI_ISL_2501217, EPI_ISL_2501218, EPI_ISL_2501219, EPI_ISL_2501220, EPI_ISL_2501221, EPI_ISL_2501222 | | | |
| see above | Maine HETL | Health and Environmental Testing Laboratory | Matluk,N., Dewey,H., Martha,J., Meak,S., Grieser,H.,Barter,M., Lynch,R., Munger,H. and Tewhey,R. |
| EPI_ISL_2501396, EPI_ISL_2501397, EPI_ISL_2501398, EPI_ISL_2501399, EPI_ISL_2501400, EPI_ISL_2501401, EPI_ISL_2501402, EPI_ISL_2501403, EPI_ISL_2501404, EPI_ISL_2501405, EPI_ISL_2501406, EPI_ISL_2501407, EPI_ISL_2501408, EPI_ISL_2501409, EPI_ISL_2501410, EPI_ISL_2501411, EPI_ISL_2501412, EPI_ISL_2501413, EPI_ISL_2501414, EPI_ISL_2501415, EPI_ISL_2501416, EPI_ISL_2501417, EPI_ISL_2501418, EPI_ISL_2501419, EPI_ISL_2501420, EPI_ISL_2501421, EPI_ISL_2501422, EPI_ISL_2501423, EPI_ISL_2501424, EPI_ISL_2501425, EPI_ISL_2501426, EPI_ISL_2501427, EPI_ISL_2501428, EPI_ISL_2501429, EPI_ISL_2501430, EPI_ISL_2501431, EPI_ISL_2501432, EPI_ISL_2501433, EPI_ISL_2501434, EPI_ISL_2501435, EPI_ISL_2501436, EPI_ISL_2501437, EPI_ISL_2501438, EPI_ISL_2501439, EPI_ISL_2501440, EPI_ISL_2501441, EPI_ISL_2501442, EPI_ISL_2501443, EPI_ISL_2501444, EPI_ISL_2501445, EPI_ISL_2501446, EPI_ISL_2501447, EPI_ISL_2501448, EPI_ISL_2501449, EPI_ISL_2501450, EPI_ISL_2501451, EPI_ISL_2501452, EPI_ISL_2501453, EPI_ISL_2501454, EPI_ISL_2501455, EPI_ISL_2501456, EPI_ISL_2501457, EPI_ISL_2501458, EPI_ISL_2501459, EPI_ISL_2501460, EPI_ISL_2501461, EPI_ISL_2501462, EPI_ISL_2501463, EPI_ISL_2501464, EPI_ISL_2501465, EPI_ISL_2501466, EPI_ISL_2501467, EPI_ISL_2501468, EPI_ISL_2501472, EPI_ISL_2501473, EPI_ISL_2501474, EPI_ISL_2501475, EPI_ISL_2501476, EPI_ISL_2501477, EPI_ISL_2501478, EPI_ISL_2501479, EPI_ISL_2501480, EPI_ISL_2501481, EPI_ISL_2501482, EPI_ISL_2501483, EPI_ISL_2501484, EPI_ISL_2501485, EPI_ISL_2501486, EPI_ISL_2501487, EPI_ISL_2501488, EPI_ISL_2501489, EPI_ISL_2501490, EPI_ISL_2501491, EPI_ISL_2501492, EPI_ISL_2501493, EPI_ISL_2501494, EPI_ISL_2501495, EPI_ISL_2501496, EPI_ISL_2501497, EPI_ISL_2501498, EPI_ISL_2501499, EPI_ISL_2501500, EPI_ISL_2501501, EPI_ISL_2501502, EPI_ISL_2501503, EPI_ISL_2501504, EPI_ISL_2501505, EPI_ISL_2501506, EPI_ISL_2501507, EPI_ISL_2501508, EPI_ISL_2501509, EPI_ISL_2501510, EPI_ISL_2501511, EPI_ISL_2501512, EPI_ISL_2501513, EPI_ISL_2501514, EPI_ISL_2501515, EPI_ISL_2501516, EPI_ISL_2501517, EPI_ISL_2501518, EPI_ISL_2501519, EPI_ISL_2501520, EPI_ISL_2501521, EPI_ISL_2501522, EPI_ISL_2501523, EPI_ISL_2501524, EPI_ISL_2501525 | | | |
| see above | UW Virology Lab | UW Virology Lab | Pavitra Roychoudhury, Hong Xie, Lasata Shrestha, Tien V. Nguyen, Shah Mohamed Bakhash, Michelle Lin, Noah R. Baker, Ricardo Perez, Sean Ellis, Nathan Breit, Robert J. Livingston, Meei-Li Huang, Keith R Jerome, Patrick Mathias, Alexander Greninger |
| EPI_ISL_2501535, EPI_ISL_2501536, EPI_ISL_2501537, EPI_ISL_2501539, EPI_ISL_2501540, EPI_ISL_2501541 | CT Department of Public Health | CT Department of Public Health | Claire_Pearson,Tu_N_Nguyen |
| EPI_ISL_2501542, EPI_ISL_2501543, EPI_ISL_2501544, EPI_ISL_2501549 | Maine HETL | Health and Environmental Testing Laboratory | Matluk,N., Dewey,H., Martha,J., Meak,S., Grieser,H.,Barter,M., Lynch,R., Munger,H. and Tewhey,R. |
| EPI_ISL_2501550, EPI_ISL_2501551, EPI_ISL_2501552, EPI_ISL_2501553, EPI_ISL_2501554, EPI_ISL_2501555, EPI_ISL_2501556, EPI_ISL_2501557, EPI_ISL_2501558, EPI_ISL_2501559, EPI_ISL_2501560, EPI_ISL_2501561, EPI_ISL_2501562, EPI_ISL_2501563, EPI_ISL_2501564, EPI_ISL_2501565, EPI_ISL_2501566, EPI_ISL_2501567, EPI_ISL_2501568, EPI_ISL_2501569, EPI_ISL_2501570, EPI_ISL_2501571, EPI_ISL_2501572, EPI_ISL_2501573, EPI_ISL_2501574, EPI_ISL_2501575, EPI_ISL_2501576, EPI_ISL_2501577, EPI_ISL_2501578, EPI_ISL_2501579, EPI_ISL_2501580, EPI_ISL_2501581, EPI_ISL_2501582, EPI_ISL_2501583, EPI_ISL_2501584, EPI_ISL_2501585, EPI_ISL_2501586, EPI_ISL_2501587, EPI_ISL_2501588, EPI_ISL_2501589, EPI_ISL_2501590, EPI_ISL_2501591, EPI_ISL_2501592, EPI_ISL_2501593, EPI_ISL_2501594, EPI_ISL_2501595, EPI_ISL_2501596, EPI_ISL_2501597, EPI_ISL_2501598, EPI_ISL_2501599, EPI_ISL_2501600, EPI_ISL_2501601, EPI_ISL_2501602, EPI_ISL_2501603, EPI_ISL_2501604, EPI_ISL_2501605, EPI_ISL_2501617, EPI_ISL_2501618, EPI_ISL_2501619, EPI_ISL_2501620, EPI_ISL_2501621, EPI_ISL_2501622, EPI_ISL_2501623, EPI_ISL_2501624 | | | |
| see above | Texas Children's Hospital | Texas Children's Microbiome Center | Ruth Ann Luna, Jennifer K. Spinler, James Dunn, James Versalovic, Ila Singh |
| EPI_ISL_2501625, EPI_ISL_2501626, EPI_ISL_2501627, EPI_ISL_2501628, EPI_ISL_2501629, EPI_ISL_2501630, EPI_ISL_2501631, EPI_ISL_2501632, EPI_ISL_2501633, EPI_ISL_2501634, EPI_ISL_2501635, EPI_ISL_2501636, EPI_ISL_2501637, EPI_ISL_2501638, EPI_ISL_2501639, EPI_ISL_2501640, EPI_ISL_2501641, EPI_ISL_2501642, EPI_ISL_2501643, EPI_ISL_2501644, EPI_ISL_2501645, EPI_ISL_2501646, EPI_ISL_2501647, EPI_ISL_2501648, EPI_ISL_2501650, EPI_ISL_2501651, EPI_ISL_2501652, EPI_ISL_2501653, EPI_ISL_2501654, EPI_ISL_2501655, EPI_ISL_2501656, EPI_ISL_2501657, EPI_ISL_2501658, EPI_ISL_2501659, EPI_ISL_2501660, EPI_ISL_2501661, EPI_ISL_2501662 | | | |
| see above | GA Department of Public Health Laboratory | Genomics and Discovery, Respiratory Viruses Branch, Division of Viral Diseases, Centers for Disease Control and Prevention | Ying Tao, Yan Li, Jing Zhang, Anna Kelleher, Brian Lynch, Krista Queen, Anna Uehara, Peter Cook, Han Jia Justin Ng, Rachel Marine, Clinton R. Paden, Dhwani Batra, Haibin Wang, Tara Coalter, Jasmine Padilla, Morgan Davis, Mili Sheth, Sarah Nobles, Mark Burroughs, Justin Lee, Adam Retchless, Suxiang Tong |
| EPI_ISL_2501663, EPI_ISL_2501666, EPI_ISL_2501667, EPI_ISL_2501668, EPI_ISL_2501669, EPI_ISL_2501672, EPI_ISL_2501673, EPI_ISL_2501674, EPI_ISL_2501675, EPI_ISL_2501676, EPI_ISL_2501677, EPI_ISL_2501678, EPI_ISL_2501679, EPI_ISL_2501680, EPI_ISL_2501681, EPI_ISL_2501682, EPI_ISL_2501683, EPI_ISL_2501684, EPI_ISL_2501685, EPI_ISL_2501686, EPI_ISL_2501687, EPI_ISL_2501688, EPI_ISL_2501689, EPI_ISL_2501690, EPI_ISL_2501691, EPI_ISL_2501692, EPI_ISL_2501693, EPI_ISL_2501694, EPI_ISL_2501695, EPI_ISL_2501696, EPI_ISL_2501697, EPI_ISL_2501703, EPI_ISL_2501704, EPI_ISL_2501705, EPI_ISL_2501706, EPI_ISL_2501719, EPI_ISL_2501721, EPI_ISL_2501722, EPI_ISL_2501724, EPI_ISL_2501725, EPI_ISL_2501726, EPI_ISL_2501727 | | | |
| see above | Yale Clinical Virology Lab | Grubaugh Lab - Yale School of Public Health | Joseph Fauver, Mallery Breban, Isabel Ott, Tara Alpert, Mary Petrone, Anderson Brito, Chantal Vogels, Annie Watkins, Chaney Kalinich, Jessica Rothman, Marie L. Landry, Nathan Grubaugh |
| EPI_ISL_2501731, EPI_ISL_2501732, EPI_ISL_2501733, EPI_ISL_2501734, EPI_ISL_2501735, EPI_ISL_2501736, EPI_ISL_2501737, EPI_ISL_2501738 | Premier Laboratory Services | BioInfoExperts LLC | Cheryl Metzler, Valerie Murray, Rebecca Rose, David Nolan, Susanna Lamers |
| EPI_ISL_2501739, EPI_ISL_2501740, EPI_ISL_2501741, EPI_ISL_2501742, EPI_ISL_2501743, EPI_ISL_2501744, EPI_ISL_2501745, EPI_ISL_2501746, EPI_ISL_2501747 | TXDSHS | TXDSHS | Rashmi Tuladhar, Bonnie Oh, Jenny Zhang, Maliha Rahman, Mayela Pedrueza, Anita Pokharel, Karen Bobier, Lorraine Rodriguez, Myong Koag, Chun Wang, Rachel Lee, Grace Kubin |
| EPI_ISL_2501748, EPI_ISL_2501749, EPI_ISL_2501750, EPI_ISL_2501751, EPI_ISL_2501752, EPI_ISL_2501753, EPI_ISL_2501754, EPI_ISL_2501755, EPI_ISL_2501756, EPI_ISL_2501757, EPI_ISL_2501758, EPI_ISL_2501759, EPI_ISL_2501760, EPI_ISL_2501761, EPI_ISL_2501762, EPI_ISL_2501763, EPI_ISL_2501764, EPI_ISL_2501765, EPI_ISL_2501766, EPI_ISL_2501767, EPI_ISL_2501768, EPI_ISL_2501769, EPI_ISL_2501770, EPI_ISL_2501771, EPI_ISL_2501772, EPI_ISL_2501773, EPI_ISL_2501774, EPI_ISL_2501775, EPI_ISL_2501776, EPI_ISL_2501777, EPI_ISL_2501778, EPI_ISL_2501779, EPI_ISL_2501780, EPI_ISL_2501781, EPI_ISL_2501782, EPI_ISL_2501783, EPI_ISL_2501784, EPI_ISL_2501785, EPI_ISL_2501786, EPI_ISL_2501787, EPI_ISL_2501788, EPI_ISL_2501789, EPI_ISL_2501790, EPI_ISL_2501791, EPI_ISL_2501792, EPI_ISL_2501793, EPI_ISL_2501794, EPI_ISL_2501795, EPI_ISL_2501796, EPI_ISL_2501797, EPI_ISL_2501798, EPI_ISL_2501799, EPI_ISL_2501800, EPI_ISL_2501801, EPI_ISL_2501802, EPI_ISL_2501803, EPI_ISL_2501804, EPI_ISL_2501805, EPI_ISL_2501806, EPI_ISL_2501807, EPI_ISL_2501808, EPI_ISL_2501809, EPI_ISL_2501810, EPI_ISL_2501811, EPI_ISL_2501812, EPI_ISL_2501813, EPI_ISL_2501814, EPI_ISL_2501815, EPI_ISL_2501816, EPI_ISL_2501817, EPI_ISL_2501818, EPI_ISL_2501819, EPI_ISL_2501820, EPI_ISL_2501821, EPI_ISL_2501822, EPI_ISL_2501831, EPI_ISL_2501832, EPI_ISL_2501833, EPI_ISL_2501834, EPI_ISL_2501835, EPI_ISL_2501836, EPI_ISL_2501837, EPI_ISL_2501838, EPI_ISL_2501839, EPI_ISL_2501840, EPI_ISL_2501841, EPI_ISL_2501842, EPI_ISL_2501843, EPI_ISL_2501844, EPI_ISL_2501845, EPI_ISL_2501846, EPI_ISL_2501847, EPI_ISL_2501848, EPI_ISL_2501849, EPI_ISL_2501850, EPI_ISL_2501851, EPI_ISL_2501852, EPI_ISL_2501853 | | | |
| see above | Maryland Genomics, Institute for Genome Sciences, University of Maryland School of Medicine | Maryland Genomics, Institute for Genome Sciences, University of Maryland School of Medicine | Tallon, Luke J; Sadzewicz, Lisa D; Humphrys, Mike; Ott, Sandra; Roussey, Holly; Santana-Cruz, Ivette; Vavikolanu, Kranthi; Lim, Jonathan;Hazen, Tracy;Fraser, Claire M; Ravel, Jacques |
| EPI_ISL_2501867, EPI_ISL_2501868, EPI_ISL_2501869, EPI_ISL_2501870, EPI_ISL_2501871, EPI_ISL_2501872, EPI_ISL_2501873, EPI_ISL_2501874, EPI_ISL_2501875, EPI_ISL_2501876, EPI_ISL_2501877, EPI_ISL_2501878, EPI_ISL_2501879, EPI_ISL_2501880, EPI_ISL_2501881, EPI_ISL_2501882, EPI_ISL_2501883, EPI_ISL_2501884, EPI_ISL_2501885, EPI_ISL_2501886, EPI_ISL_2501887, EPI_ISL_2501888, EPI_ISL_2501889, EPI_ISL_2501890, EPI_ISL_2501891, EPI_ISL_2501892, EPI_ISL_2501893, EPI_ISL_2501894, EPI_ISL_2501895, EPI_ISL_2501896, EPI_ISL_2501897, EPI_ISL_2501898, EPI_ISL_2501899, EPI_ISL_2501900, EPI_ISL_2501901, EPI_ISL_2501902, EPI_ISL_2501903, EPI_ISL_2501904, EPI_ISL_2501905, EPI_ISL_2501906, EPI_ISL_2501907, EPI_ISL_2501908, EPI_ISL_2501909, EPI_ISL_2501910, EPI_ISL_2501911, EPI_ISL_2501912, EPI_ISL_2501913, EPI_ISL_2501914, EPI_ISL_2501915, EPI_ISL_2501916, EPI_ISL_2501917, EPI_ISL_2501918, EPI_ISL_2501919, EPI_ISL_2501920, EPI_ISL_2501921, EPI_ISL_2501926, EPI_ISL_2501927, EPI_ISL_2501928, EPI_ISL_2501929, EPI_ISL_2501930, EPI_ISL_2501931, EPI_ISL_2501932, EPI_ISL_2501933, EPI_ISL_2501934, EPI_ISL_2501935, EPI_ISL_2501936, EPI_ISL_2501937, EPI_ISL_2501938, EPI_ISL_2501939, EPI_ISL_2501940, EPI_ISL_2501941, EPI_ISL_2501942, EPI_ISL_2501943, EPI_ISL_2501944, EPI_ISL_2501945, EPI_ISL_2501946, EPI_ISL_2501947, EPI_ISL_2501948 | | | |
| see above | Missouri State Public Health Laboratory | Missouri State Public Health Laboratory | Matthew Sinn, Joshua Barry, Ashley New |
| EPI_ISL_2501949, EPI_ISL_2501950, EPI_ISL_2501951, EPI_ISL_2501952, EPI_ISL_2501957, EPI_ISL_2501958, EPI_ISL_2501959, EPI_ISL_2501960, EPI_ISL_2501961, EPI_ISL_2501962, EPI_ISL_2501963, EPI_ISL_2501964, EPI_ISL_2501965, EPI_ISL_2501966, EPI_ISL_2501967, EPI_ISL_2501968, EPI_ISL_2501969, EPI_ISL_2501970, EPI_ISL_2501971, EPI_ISL_2501972, EPI_ISL_2501973, EPI_ISL_2501974, EPI_ISL_2501975, EPI_ISL_2501976, EPI_ISL_2501977, EPI_ISL_2501978, EPI_ISL_2501979, EPI_ISL_2501980, EPI_ISL_2501981, EPI_ISL_2501982, EPI_ISL_2501983, EPI_ISL_2501984, EPI_ISL_2501985, EPI_ISL_2501986 | | | |
| see above | Wyoming Public Health Laboratory | Wyoming Public Health Laboratory | Jim Mildenberger, Taylor Fearing, Channing Weber, Ashley Norberg, Chayse Rowley, Marley Goetz, Brian Dominguez, Elliot Thomasson, Sam Britz, Cari Sloma, Robert Petit, and Rob Christensen |
| EPI_ISL_2501987, EPI_ISL_2501988, EPI_ISL_2502003 | City of Milwaukee Health Department Laboratory | City of Milwaukee Health Department Laboratory | Manjeet Khubbar, Amy Bauer, Jennifer Lentz, Samantha Scott, Sanjib Bhattacharyya |
| EPI_ISL_2502004, EPI_ISL_2502005, EPI_ISL_2502006, EPI_ISL_2502007, EPI_ISL_2502008, EPI_ISL_2502009 | Alaska State Virology Laboratory | Alaska State Virology Laboratory | Stephanie DeRonde, Elva House, Jacob Zidek, Lisa Smith, Ph.D., Jack Chen, Ph.D. |
| EPI_ISL_2502010 | Alaska State Virology Laboratory | Alaska State Virology Laboratory | Stephanie DeRonde, Elva House, Lisa Smith, Ph.D., Jack Chen, Ph.D. |
| EPI_ISL_2502012, EPI_ISL_2502013, EPI_ISL_2502014 | Alaska State Virology Laboratory | Alaska State Virology Laboratory | Stephanie DeRonde, Elva House, Jacob Zidek, Lisa Smith, Ph.D., Jack Chen, Ph.D. |
| EPI_ISL_2502578 | DPH, Massachusetts State Public Health Lab | DPH, Massachusetts State Public Health Lab | Lang,A.S., Fink,T., Gallagher,G.R. and Smole,S.C. |
| EPI_ISL_2502579 | Alaska State Virology Lab, Alaska State | Alaska State Virology Lab, Alaska State | DeRonde,S., House,E., Zidek,J., Redlinger,M., Smith,L. and Chen,J. |

| | | | |
|---|---|---|---|
| | Public Health Labs | Public Health Labs | |
| EPI_ISL_2502591, EPI_ISL_2502592, EPI_ISL_2502593, EPI_ISL_2502594, EPI_ISL_2502595 | Operation Infectious Diseases, Naval Health Research Center | Operation Infectious Diseases, Naval Health Research Center | Pan,R.W., Underwood,R.S., Balansay-Ames,M.S., Myers,C.A. |
| EPI_ISL_2502596, EPI_ISL_2502597, EPI_ISL_2502598, EPI_ISL_2502599, EPI_ISL_2502600, EPI_ISL_2502601, EPI_ISL_2502602, EPI_ISL_2502603, EPI_ISL_2502604, EPI_ISL_2502605, EPI_ISL_2502606, EPI_ISL_2502607, EPI_ISL_2502608, EPI_ISL_2502609, EPI_ISL_2502610, EPI_ISL_2502611, EPI_ISL_2502612, EPI_ISL_2502613, EPI_ISL_2502614 | | | |
| see above | Operation Infectious Diseases, Naval Health Research Center | Operation Infectious Diseases, Naval Health Research Center | Pan,R.W., Underwood,R.S., Balansay-Ames,M.S., Myers,C.A., Fierro,M., Kriner,P., Iniguez-Stevens,E., Moser,K., Phippard,A.E. |
| EPI_ISL_2502766, EPI_ISL_2502767, EPI_ISL_2502768, EPI_ISL_2502769, EPI_ISL_2502770, EPI_ISL_2502771, EPI_ISL_2502772, EPI_ISL_2502773, EPI_ISL_2502774, EPI_ISL_2502775, EPI_ISL_2502776, EPI_ISL_2502777, EPI_ISL_2502778, EPI_ISL_2502779, EPI_ISL_2502780, EPI_ISL_2502781, EPI_ISL_2502782, EPI_ISL_2502783, EPI_ISL_2502784, EPI_ISL_2502785, EPI_ISL_2502786, EPI_ISL_2502787, EPI_ISL_2502788, EPI_ISL_2502789, EPI_ISL_2502790, EPI_ISL_2502791, EPI_ISL_2502792, EPI_ISL_2502793, EPI_ISL_2502794, EPI_ISL_2502795, EPI_ISL_2502796, EPI_ISL_2502797, EPI_ISL_2502798, EPI_ISL_2502799, EPI_ISL_2502800, EPI_ISL_2502801, EPI_ISL_2502802, EPI_ISL_2502803, EPI_ISL_2502804, EPI_ISL_2502805, EPI_ISL_2502806, EPI_ISL_2502807, EPI_ISL_2502808, EPI_ISL_2502809, EPI_ISL_2502810, EPI_ISL_2502811, EPI_ISL_2502812, EPI_ISL_2502813, EPI_ISL_2502814, EPI_ISL_2502815, EPI_ISL_2502816, EPI_ISL_2502817, EPI_ISL_2502818, EPI_ISL_2502819, EPI_ISL_2502820, EPI_ISL_2502821, EPI_ISL_2502822, EPI_ISL_2502823, EPI_ISL_2502824, EPI_ISL_2502825, EPI_ISL_2502826, EPI_ISL_2502827, EPI_ISL_2502828, EPI_ISL_2502829 | | | |
| see above | UW Virology Lab | UW Virology Lab | Pavitra Roychoudhury, Hong Xie, Lasata Shrestha, Shah Mohamed Bakhash, Margaret Mills, Noah Baker, Sean Ellis, Meei-Li Huang, Keith R Jerome, Alexander Greninger |
| EPI_ISL_2502830, EPI_ISL_2502831, EPI_ISL_2502832, EPI_ISL_2502833, EPI_ISL_2502834, EPI_ISL_2502835, EPI_ISL_2502837, EPI_ISL_2502839, EPI_ISL_2502840, EPI_ISL_2502841, EPI_ISL_2502842, EPI_ISL_2502843, EPI_ISL_2502844, EPI_ISL_2502845, EPI_ISL_2502846, EPI_ISL_2502847, EPI_ISL_2502848, EPI_ISL_2502849, EPI_ISL_2502850, EPI_ISL_2502851, EPI_ISL_2502852, EPI_ISL_2502853, EPI_ISL_2502854, EPI_ISL_2502855, EPI_ISL_2502856, EPI_ISL_2502857, EPI_ISL_2502858, EPI_ISL_2502859, EPI_ISL_2502860, EPI_ISL_2502861, EPI_ISL_2502862, EPI_ISL_2502863, EPI_ISL_2502864, EPI_ISL_2502865, EPI_ISL_2502867, EPI_ISL_2502868, EPI_ISL_2502869, EPI_ISL_2502870, EPI_ISL_2502871, EPI_ISL_2502872, EPI_ISL_2502873, EPI_ISL_2502874, EPI_ISL_2502875, EPI_ISL_2502876, EPI_ISL_2502877, EPI_ISL_2502878, EPI_ISL_2502879, EPI_ISL_2502880, EPI_ISL_2502881, EPI_ISL_2502882, EPI_ISL_2502883, EPI_ISL_2502884, EPI_ISL_2502885, EPI_ISL_2502886, EPI_ISL_2502887, EPI_ISL_2502888, EPI_ISL_2502889, EPI_ISL_2502890, EPI_ISL_2502891, EPI_ISL_2502892, EPI_ISL_2502893, EPI_ISL_2502894, EPI_ISL_2502895, EPI_ISL_2502896, EPI_ISL_2502897, EPI_ISL_2502898, EPI_ISL_2502899, EPI_ISL_2502900, EPI_ISL_2502901, EPI_ISL_2502902, EPI_ISL_2502903, EPI_ISL_2502904, EPI_ISL_2502905, EPI_ISL_2502906, EPI_ISL_2502907, EPI_ISL_2502908, EPI_ISL_2502909, EPI_ISL_2502910, EPI_ISL_2502911, EPI_ISL_2502912, EPI_ISL_2502913, EPI_ISL_2502914, EPI_ISL_2502915, EPI_ISL_2502916, EPI_ISL_2502917, EPI_ISL_2502918, EPI_ISL_2502919, EPI_ISL_2502920, EPI_ISL_2502921, EPI_ISL_2502922, EPI_ISL_2502923, EPI_ISL_2502924, EPI_ISL_2502925, EPI_ISL_2502926, EPI_ISL_2502927, EPI_ISL_2502928, EPI_ISL_2502929, EPI_ISL_2502930, EPI_ISL_2502931, EPI_ISL_2502932, EPI_ISL_2502933, EPI_ISL_2502934, EPI_ISL_2502935, EPI_ISL_2502937, EPI_ISL_2502938, EPI_ISL_2502939, EPI_ISL_2502940, EPI_ISL_2502941, EPI_ISL_2502942, EPI_ISL_2502943, EPI_ISL_2502944, EPI_ISL_2502945, EPI_ISL_2502946, EPI_ISL_2502947, EPI_ISL_2502948, EPI_ISL_2502949, EPI_ISL_2502950, EPI_ISL_2502951, EPI_ISL_2502952, EPI_ISL_2502954, EPI_ISL_2502955, EPI_ISL_2502956, EPI_ISL_2502957, EPI_ISL_2502959, EPI_ISL_2502960, EPI_ISL_2502961, EPI_ISL_2502962, EPI_ISL_2502963, EPI_ISL_2502964, EPI_ISL_2502965, EPI_ISL_2502966, EPI_ISL_2502967, EPI_ISL_2502968, EPI_ISL_2502969, EPI_ISL_2502970, EPI_ISL_2502972, EPI_ISL_2502973, EPI_ISL_2502974, EPI_ISL_2502975, EPI_ISL_2502976, EPI_ISL_2502977, EPI_ISL_2502978, EPI_ISL_2502979, EPI_ISL_2502980, EPI_ISL_2502981, EPI_ISL_2502982, EPI_ISL_2502983, EPI_ISL_2502984, EPI_ISL_2502985, EPI_ISL_2502986, EPI_ISL_2502987, EPI_ISL_2502988, EPI_ISL_2502989, EPI_ISL_2502990, EPI_ISL_2502991, EPI_ISL_2502992, EPI_ISL_2502993, EPI_ISL_2502994, EPI_ISL_2502995, EPI_ISL_2502996, EPI_ISL_2502997, EPI_ISL_2502998, EPI_ISL_2502999, EPI_ISL_2503000, EPI_ISL_2503001, EPI_ISL_2503002, EPI_ISL_2503003, EPI_ISL_2503004, EPI_ISL_2503005, EPI_ISL_2503006, EPI_ISL_2503007, EPI_ISL_2503008, EPI_ISL_2503009, EPI_ISL_2503010, EPI_ISL_2503011, EPI_ISL_2503012, EPI_ISL_2503013, EPI_ISL_2503014, EPI_ISL_2503015, EPI_ISL_2503016, EPI_ISL_2503017, EPI_ISL_2503018, EPI_ISL_2503019, EPI_ISL_2503020, EPI_ISL_2503021, EPI_ISL_2503022, EPI_ISL_2503023, EPI_ISL_2503024, EPI_ISL_2503025, EPI_ISL_2503026, EPI_ISL_2503028, EPI_ISL_2503029, EPI_ISL_2503030, EPI_ISL_2503031, EPI_ISL_2503032, EPI_ISL_2503033, EPI_ISL_2503034, EPI_ISL_2503035, EPI_ISL_2503036, EPI_ISL_2503037, EPI_ISL_2503038, EPI_ISL_2503039, EPI_ISL_2503040, EPI_ISL_2503041, EPI_ISL_2503042, EPI_ISL_2503043, EPI_ISL_2503044, EPI_ISL_2503045, EPI_ISL_2503046, EPI_ISL_2503047, EPI_ISL_2503048, EPI_ISL_2503049, EPI_ISL_2503050, EPI_ISL_2503051, EPI_ISL_2503052, EPI_ISL_2503054, EPI_ISL_2503055, EPI_ISL_2503056, EPI_ISL_2503057, EPI_ISL_2503058, EPI_ISL_2503059, EPI_ISL_2503060, EPI_ISL_2503061, EPI_ISL_2503062, EPI_ISL_2503063, EPI_ISL_2503064, EPI_ISL_2503065, EPI_ISL_2503066, EPI_ISL_2503067, EPI_ISL_2503068, EPI_ISL_2503069, EPI_ISL_2503070, EPI_ISL_2503071, EPI_ISL_2503072, EPI_ISL_2503073, EPI_ISL_2503074, EPI_ISL_2503075, EPI_ISL_2503076, EPI_ISL_2503077, EPI_ISL_2503078, EPI_ISL_2503079, EPI_ISL_2503080, EPI_ISL_2503081, EPI_ISL_2503082, EPI_ISL_2503083, EPI_ISL_2503084, EPI_ISL_2503086, EPI_ISL_2503087, EPI_ISL_2503088, EPI_ISL_2503089, EPI_ISL_2503090, EPI_ISL_2503091, EPI_ISL_2503092, EPI_ISL_2503093, EPI_ISL_2503094, EPI_ISL_2503095, EPI_ISL_2503096, EPI_ISL_2503097, EPI_ISL_2503098, EPI_ISL_2503099, EPI_ISL_2503100, EPI_ISL_2503101, EPI_ISL_2503102, EPI_ISL_2503103, EPI_ISL_2503104, EPI_ISL_2503105, EPI_ISL_2503106, EPI_ISL_2503107, EPI_ISL_2503108, EPI_ISL_2503109, EPI_ISL_2503110, EPI_ISL_2503111, EPI_ISL_2503112, EPI_ISL_2503113, EPI_ISL_2503114, EPI_ISL_2503115, EPI_ISL_2503116, EPI_ISL_2503117, EPI_ISL_2503118, EPI_ISL_2503119, EPI_ISL_2503120, EPI_ISL_2503121, EPI_ISL_2503123, EPI_ISL_2503124, EPI_ISL_2503125, EPI_ISL_2503126 | | | |
| see above | UW Virology Lab | UW Virology Lab | Pavitra Roychoudhury, Hong Xie, Lasata Shrestha, Shah Mohamed Bakhash, Tien V. Nguyen, Noah R. Baker, Sean Ellis, Meei-Li Huang, Keith R Jerome, Alexander Greninger |
| EPI_ISL_2503127, EPI_ISL_2503128, EPI_ISL_2503129, EPI_ISL_2503130, EPI_ISL_2503131, EPI_ISL_2503132, EPI_ISL_2503133 | UW Virology Lab | UW Virology Lab | Pavitra Roychoudhury, Hong Xie, Lasata Shrestha, Tien V. Nguyen, Shah Mohamed Bakhash, Michelle Lin, Noah R. Baker, Sean Ellis, Meei-Li Huang, Keith R Jerome, Alexander Greninger |
| EPI_ISL_2503977, EPI_ISL_2503978, EPI_ISL_2503979, EPI_ISL_2503980, EPI_ISL_2503981, EPI_ISL_2503982, EPI_ISL_2503983, EPI_ISL_2503984, EPI_ISL_2503985, EPI_ISL_2503986, EPI_ISL_2503987, EPI_ISL_2503988, EPI_ISL_2503989, EPI_ISL_2503990, EPI_ISL_2503991, EPI_ISL_2503992, EPI_ISL_2503993, EPI_ISL_2503994, EPI_ISL_2503995, EPI_ISL_2503996, EPI_ISL_2503997, EPI_ISL_2503998, EPI_ISL_2503999, EPI_ISL_2504000, EPI_ISL_2504001, EPI_ISL_2504002, EPI_ISL_2504003, EPI_ISL_2504004, EPI_ISL_2504005, EPI_ISL_2504006, EPI_ISL_2504007, EPI_ISL_2504008, EPI_ISL_2504009, EPI_ISL_2504010, EPI_ISL_2504011, EPI_ISL_2504012, EPI_ISL_2504013 | | | |
| see above | Alaska State Virology Laboratory | Alaska State Virology Laboratory | Stephanie DeRonde, Elva House, Jacob Zidek, Lisa Smith, Ph.D., Jack Chen, Ph.D. |
| EPI_ISL_2504181, EPI_ISL_2504182, EPI_ISL_2504183, EPI_ISL_2504184, EPI_ISL_2504185, EPI_ISL_2504186, EPI_ISL_2504187, EPI_ISL_2504188, EPI_ISL_2504189 | Swedish Medical Center, Seattle, WA, USA | Institute for Systems Biology | Jason D. Goldman, Chris Lausted, Kai Wang, Jared C. Roach, James R. Heath, Julie A. Wallick |
| EPI_ISL_2507105, EPI_ISL_2507113, EPI_ISL_2507117, EPI_ISL_2507126, EPI_ISL_2507130, EPI_ISL_2507131, EPI_ISL_2507135, EPI_ISL_2507156, EPI_ISL_2507162, EPI_ISL_2507163, EPI_ISL_2507167, EPI_ISL_2507195, EPI_ISL_2507204, EPI_ISL_2507225, EPI_ISL_2507229, EPI_ISL_2507231, EPI_ISL_2507239, EPI_ISL_2507240, EPI_ISL_2507242, EPI_ISL_2507250, EPI_ISL_2507254, EPI_ISL_2507296, EPI_ISL_2507303, EPI_ISL_2507317, EPI_ISL_2507326, EPI_ISL_2507335, EPI_ISL_2507358 | | | |
| see above | Curative Labs | Curative Labs | Mikhail Hanewich-Hollatz, J.Cesar Ignacio-Espinoza, Elias L. Salfati, Nabjot Sandhu, George Way, Zhiyi Xie, Eugenia Khorosheva, Sophia Quasem, Janet Chen, Vladimir Slepnev |
| EPI_ISL_2508349, EPI_ISL_2508350, EPI_ISL_2508352, EPI_ISL_2508353, EPI_ISL_2508354, EPI_ISL_2508356, EPI_ISL_2508357, EPI_ISL_2508358, EPI_ISL_2508359, EPI_ISL_2508360, EPI_ISL_2508361, EPI_ISL_2508362, EPI_ISL_2508363, EPI_ISL_2508364, EPI_ISL_2508365, EPI_ISL_2508366, EPI_ISL_2508367, EPI_ISL_2508369 | | | |
| see above | DeRisi Lab, University of California, San Francisco | DeRisi Lab, University of California, San Francisco | Sabrina Mann, Anthea Mitchell, Jamin Liu, Sara Sunshine, Matthew Laurie, Genay Pilarowski, Amy Kistler, Manu Vanaerschot, Patrick Ayscue, Lucy Li, Eric Chow, IDseq Team, James Peng, Carina Marquez, Luis Rubio, Gabriel Chamie, Diane Jones, Jon Jacobo, Susana Rojas, Susy Rojas, Valerie Tulier-Laiwa, Douglas Black, Jackie Martinez, Maya Petersen, Diane Havlir, and Joseph DeRisi |
| EPI_ISL_2508397, EPI_ISL_2508399, EPI_ISL_2508400, EPI_ISL_2508401, EPI_ISL_2508402, EPI_ISL_2508403, EPI_ISL_2508404, EPI_ISL_2508405, EPI_ISL_2508406, EPI_ISL_2508407, EPI_ISL_2508408, EPI_ISL_2508410, EPI_ISL_2508411, EPI_ISL_2508412, EPI_ISL_2508413, EPI_ISL_2508414, EPI_ISL_2508416, EPI_ISL_2508418, EPI_ISL_2508419, EPI_ISL_2508420, EPI_ISL_2508421, EPI_ISL_2508422, EPI_ISL_2508423, EPI_ISL_2508424, EPI_ISL_2508425, EPI_ISL_2508426, EPI_ISL_2508427, EPI_ISL_2508428, EPI_ISL_2508429, EPI_ISL_2508430, EPI_ISL_2508431, EPI_ISL_2508433, EPI_ISL_2508434, EPI_ISL_2508435, EPI_ISL_2508436, EPI_ISL_2508437, EPI_ISL_2508438, EPI_ISL_2508439, EPI_ISL_2508440, EPI_ISL_2508441, EPI_ISL_2508442, EPI_ISL_2508444, EPI_ISL_2508445, EPI_ISL_2508446, EPI_ISL_2508447, EPI_ISL_2508449, EPI_ISL_2508450, EPI_ISL_2508451, EPI_ISL_2508452, EPI_ISL_2508453, EPI_ISL_2508454, EPI_ISL_2508455, EPI_ISL_2508456, EPI_ISL_2508457, EPI_ISL_2508458, EPI_ISL_2508459, EPI_ISL_2508460, EPI_ISL_2508461, EPI_ISL_2508462, EPI_ISL_2508463, EPI_ISL_2508464, EPI_ISL_2508465, EPI_ISL_2508466, EPI_ISL_2508467, EPI_ISL_2508468, EPI_ISL_2508469, EPI_ISL_2508470, EPI_ISL_2508471, EPI_ISL_2508472, EPI_ISL_2508473, EPI_ISL_2508474, EPI_ISL_2508475, EPI_ISL_2508476, EPI_ISL_2508477, EPI_ISL_2508478, EPI_ISL_2508479, EPI_ISL_2508480, EPI_ISL_2508481, EPI_ISL_2508482, EPI_ISL_2508483, EPI_ISL_2508484, EPI_ISL_2508486, EPI_ISL_2508487, EPI_ISL_2508488, EPI_ISL_2508489, EPI_ISL_2508490, EPI_ISL_2508491, EPI_ISL_2508492, EPI_ISL_2508494, EPI_ISL_2508495, EPI_ISL_2508496, EPI_ISL_2508497, EPI_ISL_2508498, EPI_ISL_2508499, EPI_ISL_2508500, EPI_ISL_2508501, EPI_ISL_2508502, EPI_ISL_2508503, EPI_ISL_2508504, EPI_ISL_2508507, EPI_ISL_2508508, EPI_ISL_2508509, EPI_ISL_2508510, EPI_ISL_2508511, EPI_ISL_2508512, EPI_ISL_2508513, EPI_ISL_2508514, EPI_ISL_2508515, EPI_ISL_2508516, EPI_ISL_2508517, EPI_ISL_2508518, EPI_ISL_2508519, EPI_ISL_2508520, EPI_ISL_2508522, EPI_ISL_2508523, EPI_ISL_2508524, EPI_ISL_2508525, EPI_ISL_2508526, EPI_ISL_2508527, EPI_ISL_2508529, EPI_ISL_2508530, EPI_ISL_2508531, EPI_ISL_2508532, EPI_ISL_2508533, EPI_ISL_2508534, EPI_ISL_2508535, EPI_ISL_2508536, EPI_ISL_2508537, EPI_ISL_2508538, EPI_ISL_2508539, EPI_ISL_2508540, EPI_ISL_2508541, EPI_ISL_2508542, EPI_ISL_2508543, EPI_ISL_2508544, EPI_ISL_2508545, EPI_ISL_2508546 | | | |
| see above | Microbiology Division, SC DHEC | Microbiology Division, SC DHEC | Freeman,J., Flores,H. |
| EPI_ISL_2509338, EPI_ISL_2509339, EPI_ISL_2509340, EPI_ISL_2509341, EPI_ISL_2509342 | Public Health Laboratory - Infectious Disease Lab, Minnesota Department of Health Infectious Disease Laboratory Submission Group | Public Health Laboratory - Infectious Disease Lab, Minnesota Department of Health Infectious Disease Laboratory Submission Group | Lorentz,A., Garfin,J., Plumb,M., Wang,X. |
| EPI_ISL_2509960 | VGTI/ONPRC, Oregon Health & Science University | VGTI/ONPRC, Oregon Health & Science University | Bimber,B.N. |
| EPI_ISL_2514921 | CEIRS Data Processing and Coordinating Center, St. Jude Center of Excellence for Influenza Research and Surveillance (CEIRS) | St. Jude Children's Research Hospital | Jeevan,T., Truong,M., Patel,A., Fabrizio,T., Allay,J., Webb,A. and Webby,R. |
| EPI_ISL_2516977, EPI_ISL_2516978, EPI_ISL_2516979, EPI_ISL_2516980, EPI_ISL_2516981, EPI_ISL_2516982, EPI_ISL_2516983, EPI_ISL_2516984, EPI_ISL_2516985, EPI_ISL_2516986, EPI_ISL_2516987, EPI_ISL_2516988, EPI_ISL_2516989, EPI_ISL_2516990, EPI_ISL_2516991, EPI_ISL_2516992, EPI_ISL_2516993, EPI_ISL_2516994, EPI_ISL_2516995, EPI_ISL_2516996, EPI_ISL_2516997, EPI_ISL_2516998, EPI_ISL_2516999, EPI_ISL_2517000, EPI_ISL_2517001, EPI_ISL_2517002, EPI_ISL_2517003, EPI_ISL_2517004, EPI_ISL_2517005, EPI_ISL_2517006, EPI_ISL_2517007, EPI_ISL_2517008, EPI_ISL_2517009, EPI_ISL_2517010, EPI_ISL_2517011, EPI_ISL_2517012, EPI_ISL_2517013, EPI_ISL_2517014, EPI_ISL_2517015, EPI_ISL_2517016, EPI_ISL_2517017, EPI_ISL_2517018, EPI_ISL_2517019, EPI_ISL_2517020, EPI_ISL_2517021, EPI_ISL_2517022, EPI_ISL_2517023, EPI_ISL_2517024, EPI_ISL_2517025, EPI_ISL_2517026, EPI_ISL_2517027, EPI_ISL_2517028, EPI_ISL_2517029, EPI_ISL_2517030, EPI_ISL_2517031, EPI_ISL_2517032, EPI_ISL_2517033, EPI_ISL_2517034, EPI_ISL_2517035, EPI_ISL_2517036, EPI_ISL_2517037, EPI_ISL_2517038, EPI_ISL_2517039, EPI_ISL_2517040, EPI_ISL_2517041, EPI_ISL_2517042, EPI_ISL_2517043 | | | |
| see above | Molecular Microbiology, Washington University of St. Louis | Molecular Microbiology, Washington University of St. Louis | Wang,D., Handley,S. |
| EPI_ISL_2521311, EPI_ISL_2521326, EPI_ISL_2521327, EPI_ISL_2521382, EPI_ISL_2521597, EPI_ISL_2521599, EPI_ISL_2521736, EPI_ISL_2521747, EPI_ISL_2521750 | Ohio Department of Health Laboratory | Ohio Department of Health Laboratory | Holmes, Jennifer; Eric Brandt, Keoni Omura, Glen McGillivary, Caitlin McDonnell, Jade Mowery, Stephanie Mccracken, Tyler Payne, Kirtana Ramadugu, Erica Leasure, Brent Lee, Kelsey Florek, Heather Blankenship, Allison Black, Quanta Brown, and Tammy Bannerman |
| EPI_ISL_2522093 | DeRisi Lab, University of California, San Francisco | DeRisi Lab, University of California, San Francisco | Sabrina Mann, Anthea Mitchell, Jamin Liu, Sara Sunshine, Matthew Laurie, Genay Pilarowski, Amy Kistler, Manu Vanaerschot, Patrick Ayscue, Lucy Li, Eric Chow, IDseq Team, James Peng, Carina Marquez, Luis Rubio, Gabriel Chamie, Diane Jones, Jon Jacobo, Susana Rojas, Susy Rojas, Valerie Tulier-Laiwa, Douglas Black, Jackie Martinez, Maya Petersen, Diane Havlir, and Joseph DeRisi |
| EPI_ISL_2522462, EPI_ISL_2523509, EPI_ISL_2524010, EPI_ISL_2524335, EPI_ISL_2524848 | Ohio Department of Health Laboratory | Ohio Department of Health Laboratory | Holmes, Jennifer; Eric Brandt, Keoni Omura, Glen McGillivary, Caitlin McDonnell, Jade Mowery, Stephanie Mccracken, Tyler Payne, Kirtana Ramadugu, Erica Leasure, Brent Lee, Kelsey Florek, Heather Blankenship, Allison Black, Quanta Brown, and Tammy Bannerman |
| EPI_ISL_2524952, EPI_ISL_2524954, EPI_ISL_2524956, EPI_ISL_2524958, EPI_ISL_2524960, EPI_ISL_2524962, EPI_ISL_2524964, EPI_ISL_2524966, EPI_ISL_2524968, EPI_ISL_2524970, EPI_ISL_2524973, EPI_ISL_2524975, EPI_ISL_2524978, EPI_ISL_2524980, EPI_ISL_2524983, EPI_ISL_2524985, EPI_ISL_2524987, EPI_ISL_2524989, EPI_ISL_2524991, EPI_ISL_2524993, EPI_ISL_2524996, EPI_ISL_2524999, EPI_ISL_2525001, EPI_ISL_2525003, EPI_ISL_2525005, EPI_ISL_2525007, EPI_ISL_2525009, EPI_ISL_2525011, EPI_ISL_2525014, EPI_ISL_2525016, EPI_ISL_2525019, EPI_ISL_2525021, EPI_ISL_2525023, EPI_ISL_2525025, EPI_ISL_2525028, EPI_ISL_2525030, EPI_ISL_2525032, EPI_ISL_2525034, EPI_ISL_2525036, EPI_ISL_2525038, EPI_ISL_2525041, EPI_ISL_2525044, EPI_ISL_2525046, EPI_ISL_2525048, EPI_ISL_2525051, EPI_ISL_2525053, EPI_ISL_2525055, EPI_ISL_2525057, EPI_ISL_2525059, EPI_ISL_2525062, EPI_ISL_2525064, EPI_ISL_2525066, EPI_ISL_2525068, EPI_ISL_2525070, EPI_ISL_2525072, EPI_ISL_2525074, EPI_ISL_2525076, EPI_ISL_2525078, EPI_ISL_2525081, EPI_ISL_2525084, EPI_ISL_2525086, EPI_ISL_2525088, EPI_ISL_2525091, EPI_ISL_2525093, EPI_ISL_2525095, EPI_ISL_2525097, EPI_ISL_2525100, EPI_ISL_2525102, EPI_ISL_2525105, EPI_ISL_2525107, EPI_ISL_2525109, EPI_ISL_2525111, EPI_ISL_2525113, EPI_ISL_2525116, EPI_ISL_2525118, EPI_ISL_2525120, EPI_ISL_2525122, EPI_ISL_2525125, EPI_ISL_2525127, EPI_ISL_2525129, EPI_ISL_2525131, EPI_ISL_2525133, EPI_ISL_2525135, EPI_ISL_2525137, EPI_ISL_2525139, EPI_ISL_2525142, EPI_ISL_2525145, EPI_ISL_2525147, EPI_ISL_2525150, EPI_ISL_2525152, EPI_ISL_2525154, EPI_ISL_2525156, EPI_ISL_2525158, EPI_ISL_2525160, EPI_ISL_2525162, EPI_ISL_2525164, EPI_ISL_2525167, EPI_ISL_2525170, EPI_ISL_2525172, EPI_ISL_2525174, EPI_ISL_2525176, EPI_ISL_2525178, EPI_ISL_2525181, EPI_ISL_2525183, EPI_ISL_2525185, EPI_ISL_2525187, EPI_ISL_2525190, EPI_ISL_2525192, EPI_ISL_2525194, EPI_ISL_2525196, | | | |

EPI_ISL_2525198, EPI_ISL_2525200, EPI_ISL_2525202, EPI_ISL_2525204, EPI_ISL_2525206, EPI_ISL_2525208, EPI_ISL_2525212, EPI_ISL_2525214, EPI_ISL_2525216, EPI_ISL_2525218, EPI_ISL_2525220, EPI_ISL_2525222, EPI_ISL_2525225, EPI_ISL_2525227, EPI_ISL_2525229, EPI_ISL_2525232, EPI_ISL_2525235, EPI_ISL_2525237, EPI_ISL_2525239, EPI_ISL_2525241, EPI_ISL_2525243, EPI_ISL_2525246, EPI_ISL_2525248, EPI_ISL_2525250, EPI_ISL_2525252, EPI_ISL_2525255, EPI_ISL_2525257, EPI_ISL_2525259, EPI_ISL_2525261, EPI_ISL_2525263, EPI_ISL_2525265, EPI_ISL_2525267, EPI_ISL_2525269, EPI_ISL_2525272, EPI_ISL_2525274, EPI_ISL_2525277, EPI_ISL_2525281, EPI_ISL_2525284, EPI_ISL_2525286, EPI_ISL_2525288, EPI_ISL_2525290, EPI_ISL_2525292, EPI_ISL_2525295, EPI_ISL_2525297, EPI_ISL_2525299, EPI_ISL_2525301, EPI_ISL_2525304, EPI_ISL_2525306, EPI_ISL_2525308, EPI_ISL_2525310, EPI_ISL_2525313, EPI_ISL_2525316, EPI_ISL_2525318, EPI_ISL_2525320, EPI_ISL_2525322, EPI_ISL_2525324, EPI_ISL_2525326, EPI_ISL_2525328, EPI_ISL_2525331, EPI_ISL_2525333, EPI_ISL_2525335, EPI_ISL_2525338, EPI_ISL_2525340, EPI_ISL_2525342, EPI_ISL_2525344, EPI_ISL_2525346, EPI_ISL_2525348, EPI_ISL_2525351, EPI_ISL_2525353, EPI_ISL_2525355, EPI_ISL_2525358, EPI_ISL_2525360, EPI_ISL_2525363, EPI_ISL_2525365, EPI_ISL_2525367, EPI_ISL_2525369

| | | | |
|---|---|---|---|
| see above | Mako Medical | Centers for Disease Control and Prevention Division of Viral Diseases, Pathogen Discovery | Dakota Howard, Dhwani Batra, Peter W. Cook, Kara Moser, Adrian Paskey, Jason Caravas, Benjamin Rambo-Martin, Shatavia Morrison, Christopher Gulvick, Scott Sammons, Yvette Unoarumhi, Darlene Wagner, Matthew Schmerer,Matthew Tugwell, Lauren Moon, Clinton R. Paden, Duncan MacCannell |

EPI_ISL_2525430, EPI_ISL_2525433, EPI_ISL_2525435, EPI_ISL_2525437, EPI_ISL_2525439, EPI_ISL_2525441, EPI_ISL_2525443, EPI_ISL_2525446, EPI_ISL_2525448, EPI_ISL_2525450, EPI_ISL_2525452, EPI_ISL_2525454, EPI_ISL_2525457, EPI_ISL_2525459, EPI_ISL_2525461, EPI_ISL_2525464, EPI_ISL_2525466, EPI_ISL_2525468, EPI_ISL_2525470, EPI_ISL_2525472, EPI_ISL_2525474, EPI_ISL_2525476, EPI_ISL_2525478, EPI_ISL_2525480, EPI_ISL_2525482, EPI_ISL_2525484, EPI_ISL_2525488, EPI_ISL_2525490, EPI_ISL_2525492, EPI_ISL_2525494, EPI_ISL_2525497, EPI_ISL_2525499, EPI_ISL_2525501, EPI_ISL_2525503, EPI_ISL_2525506, EPI_ISL_2525509, EPI_ISL_2525511, EPI_ISL_2525513, EPI_ISL_2525515, EPI_ISL_2525517, EPI_ISL_2525519, EPI_ISL_2525521, EPI_ISL_2525523, EPI_ISL_2525525, EPI_ISL_2525527, EPI_ISL_2525529, EPI_ISL_2525531, EPI_ISL_2525533, EPI_ISL_2525536, EPI_ISL_2525538, EPI_ISL_2525540, EPI_ISL_2525542, EPI_ISL_2525544, EPI_ISL_2525546, EPI_ISL_2525551, EPI_ISL_2525553, EPI_ISL_2525555, EPI_ISL_2525557, EPI_ISL_2525560, EPI_ISL_2525562, EPI_ISL_2525566, EPI_ISL_2525568, EPI_ISL_2525570, EPI_ISL_2525572, EPI_ISL_2525574, EPI_ISL_2525576, EPI_ISL_2525579, EPI_ISL_2525581, EPI_ISL_2525583, EPI_ISL_2525585, EPI_ISL_2525587, EPI_ISL_2525592, EPI_ISL_2525594, EPI_ISL_2525596, EPI_ISL_2525599, EPI_ISL_2525601, EPI_ISL_2525602, EPI_ISL_2525605, EPI_ISL_2525607, EPI_ISL_2525609, EPI_ISL_2525611, EPI_ISL_2525613, EPI_ISL_2525615, EPI_ISL_2525617, EPI_ISL_2525619, EPI_ISL_2525621, EPI_ISL_2525623, EPI_ISL_2525625, EPI_ISL_2525631, EPI_ISL_2525633, EPI_ISL_2525635, EPI_ISL_2525637, EPI_ISL_2525640, EPI_ISL_2525642, EPI_ISL_2525644, EPI_ISL_2525646, EPI_ISL_2525648, EPI_ISL_2525650, EPI_ISL_2525652, EPI_ISL_2525654, EPI_ISL_2525656, EPI_ISL_2525658, EPI_ISL_2525660, EPI_ISL_2525662, EPI_ISL_2525665, EPI_ISL_2525667, EPI_ISL_2525669, EPI_ISL_2525671, EPI_ISL_2525674, EPI_ISL_2525676, EPI_ISL_2525678, EPI_ISL_2525679, EPI_ISL_2525680, EPI_ISL_2525681, EPI_ISL_2525683, EPI_ISL_2525685, EPI_ISL_2525687, EPI_ISL_2525689, EPI_ISL_2525691, EPI_ISL_2525693, EPI_ISL_2525695, EPI_ISL_2525698, EPI_ISL_2525700, EPI_ISL_2525702, EPI_ISL_2525704, EPI_ISL_2525706, EPI_ISL_2525708, EPI_ISL_2525710, EPI_ISL_2525712, EPI_ISL_2525714, EPI_ISL_2525716, EPI_ISL_2525718, EPI_ISL_2525720, EPI_ISL_2525725, EPI_ISL_2525727, EPI_ISL_2525729, EPI_ISL_2525731, EPI_ISL_2525733, EPI_ISL_2525735, EPI_ISL_2525737, EPI_ISL_2525739, EPI_ISL_2525741, EPI_ISL_2525744, EPI_ISL_2525746, EPI_ISL_2525748, EPI_ISL_2525750, EPI_ISL_2525752, EPI_ISL_2525754, EPI_ISL_2525756, EPI_ISL_2525759, EPI_ISL_2525761, EPI_ISL_2525763, EPI_ISL_2525767, EPI_ISL_2525769, EPI_ISL_2525771, EPI_ISL_2525776, EPI_ISL_2525778, EPI_ISL_2525780, EPI_ISL_2525785, EPI_ISL_2525787, EPI_ISL_2525789, EPI_ISL_2525791, EPI_ISL_2525795, EPI_ISL_2525797, EPI_ISL_2525799, EPI_ISL_2525801, EPI_ISL_2525803, EPI_ISL_2525805, EPI_ISL_2525807, EPI_ISL_2525809, EPI_ISL_2525811, EPI_ISL_2525814, EPI_ISL_2525816, EPI_ISL_2525818, EPI_ISL_2525825, EPI_ISL_2525827, EPI_ISL_2525829, EPI_ISL_2525831, EPI_ISL_2525834, EPI_ISL_2525836, EPI_ISL_2525838, EPI_ISL_2525845, EPI_ISL_2525848, EPI_ISL_2525850, EPI_ISL_2525852, EPI_ISL_2525856, EPI_ISL_2525858, EPI_ISL_2525861, EPI_ISL_2525863, EPI_ISL_2525865, EPI_ISL_2525867, EPI_ISL_2525869, EPI_ISL_2525871, EPI_ISL_2525873, EPI_ISL_2525875, EPI_ISL_2525877, EPI_ISL_2525879, EPI_ISL_2525881, EPI_ISL_2525883, EPI_ISL_2525885, EPI_ISL_2525888, EPI_ISL_2525890, EPI_ISL_2525892, EPI_ISL_2525895, EPI_ISL_2525897, EPI_ISL_2525899, EPI_ISL_2525901, EPI_ISL_2525903, EPI_ISL_2525905, EPI_ISL_2525907, EPI_ISL_2525910, EPI_ISL_2525912, EPI_ISL_2525914, EPI_ISL_2525916, EPI_ISL_2525921, EPI_ISL_2525923, EPI_ISL_2525925, EPI_ISL_2525927, EPI_ISL_2525931, EPI_ISL_2525933, EPI_ISL_2525935, EPI_ISL_2525937, EPI_ISL_2525940, EPI_ISL_2525942, EPI_ISL_2525944, EPI_ISL_2525946, EPI_ISL_2525948, EPI_ISL_2525951, EPI_ISL_2525953, EPI_ISL_2525955, EPI_ISL_2525960, EPI_ISL_2525962, EPI_ISL_2525964, EPI_ISL_2525966, EPI_ISL_2525968, EPI_ISL_2525970, EPI_ISL_2525973, EPI_ISL_2525975, EPI_ISL_2525977, EPI_ISL_2525980, EPI_ISL_2525982, EPI_ISL_2525987, EPI_ISL_2525989, EPI_ISL_2525991, EPI_ISL_2525993, EPI_ISL_2525995, EPI_ISL_2525998, EPI_ISL_2526000, EPI_ISL_2526002, EPI_ISL_2526004, EPI_ISL_2526006, EPI_ISL_2526008, EPI_ISL_2526010, EPI_ISL_2526012, EPI_ISL_2526015, EPI_ISL_2526017, EPI_ISL_2526019, EPI_ISL_2526022, EPI_ISL_2526024, EPI_ISL_2526026, EPI_ISL_2526028, EPI_ISL_2526031, EPI_ISL_2526033, EPI_ISL_2526035, EPI_ISL_2526038, EPI_ISL_2526040, EPI_ISL_2526042, EPI_ISL_2526044, EPI_ISL_2526046, EPI_ISL_2526048, EPI_ISL_2526050, EPI_ISL_2526053, EPI_ISL_2526055, EPI_ISL_2526057, EPI_ISL_2526063, EPI_ISL_2526065, EPI_ISL_2526067, EPI_ISL_2526069, EPI_ISL_2526071, EPI_ISL_2526689, EPI_ISL_2526691

| | | | |
|---|---|---|---|
| see above | Fulgent Genetics | Centers for Disease Control and Prevention Division of Viral Diseases, Pathogen Discovery | Dakota Howard, Dhwani Batra, Peter W. Cook, Kara Moser, Adrian Paskey, Jason Caravas, Benjamin Rambo-Martin, Shatavia Morrison, Christopher Gulvick, Scott Sammons, Yvette Unoarumhi, Darlene Wagner, Matthew Schmerer, Harry Gao, Mickey Li, John Gao, Joseph Fierro, Benafsh Sapra, Becky Tsai, Yan Meng, Doreen Ng, James Xie, Clinton R. Paden, Duncan MacCannell |
| EPI_ISL_2526703 | Laboratory Corporation of America | Centers for Disease Control and Prevention Division of Viral Diseases, Pathogen Discovery | Dakota Howard, Dhwani Batra, Peter W. Cook, Kara Moser, Adrian Paskey, Jason Caravas, Benjamin Rambo-Martin, Shatavia Morrison, Christopher Gulvick, Scott Sammons, Yvette Unoarumhi, Darlene Wagner, Matthew Schmerer, Minoo Agarwal, Eyad Almasri, Debbie Boles, Ayla Burns, Nuthawin Charoensri, Oren Cohen, Susan Countryman, Mary Ann Cristobal, Bobbi Croy, Suzanne Dale, Hrushikesh Deshmukh, Amanda Douglas, Vincent Drouillon, Marcia Eisenberg, Howard Engler, Rama Ghatti, Prashant Gupta, Susan Hicks, Jake Humphrey, Lax Iyer, Lisa Pfefferle, Manoj Jain, Matthew Robinson, Mohan Kolli, Brian Krueger, Tim Kuphal, Stanley Letovsky, Michael Levandoski, Craig Lukasik, Jonathan Meltzer, Brian Norvell, Mindy Nye, Scott Parker, Christos Petropoulos, John Pruitt, Steven Ragan, Scott Ryan, Mike Sapeta, Jana Schroth, Suresh Babu Selvaraju, Goran Stevovic, Amanda Suchanek, Andrea Throop, Lyndon Tilson, Thomas Urban, Joe Voshell, Kimberly Wagner, Jonathan Williams, Mary Williamson, Qian Zeng, Tricia Zwiefelhofer, Clinton R. Paden, Duncan MacCannell |

EPI_ISL_2526714, EPI_ISL_2526716, EPI_ISL_2526718, EPI_ISL_2526720, EPI_ISL_2526722, EPI_ISL_2526724, EPI_ISL_2526726, EPI_ISL_2526728, EPI_ISL_2526730, EPI_ISL_2526732, EPI_ISL_2526734

| | | | |
|---|---|---|---|
| see above | Aegis Sciences Corporation | Centers for Disease Control and Prevention Division of Viral Diseases, Pathogen Discovery | Dakota Howard, Dhwani Batra, Peter W. Cook, Kara Moser, Adrian Paskey, Jason Caravas, Benjamin Rambo-Martin, Shatavia Morrison, Christopher Gulvick, Scott Sammons, Yvette Unoarumhi, Darlene Wagner, Matthew Schmerer, Cyndi Clark, Patrick Campbell, Rob Case, Vikramsinha Ghorpade, Holly Houdeshell, Ola Kvalvaag, Dillon Nall, Ethan Sanders, Alec Vest, Shaun Westlund, Matthew Hardison, Clinton R. Paden, Duncan MacCannell |

EPI_ISL_2526746, EPI_ISL_2526748, EPI_ISL_2526751, EPI_ISL_2526753, EPI_ISL_2526755, EPI_ISL_2526757, EPI_ISL_2526759, EPI_ISL_2526761, EPI_ISL_2526763, EPI_ISL_2526766, EPI_ISL_2526768, EPI_ISL_2526770, EPI_ISL_2526772, EPI_ISL_2526774, EPI_ISL_2526777, EPI_ISL_2526779, EPI_ISL_2526781, EPI_ISL_2526783, EPI_ISL_2526785, EPI_ISL_2526788, EPI_ISL_2526790, EPI_ISL_2526792, EPI_ISL_2526794, EPI_ISL_2526796, EPI_ISL_2526798, EPI_ISL_2526800, EPI_ISL_2526803, EPI_ISL_2526805, EPI_ISL_2526807, EPI_ISL_2526809, EPI_ISL_2526811, EPI_ISL_2526814, EPI_ISL_2526816, EPI_ISL_2526818, EPI_ISL_2526820, EPI_ISL_2526822, EPI_ISL_2526825, EPI_ISL_2526827, EPI_ISL_2526829, EPI_ISL_2526831, EPI_ISL_2526833, EPI_ISL_2526835, EPI_ISL_2526837, EPI_ISL_2526840, EPI_ISL_2526842, EPI_ISL_2526844, EPI_ISL_2526846, EPI_ISL_2526849, EPI_ISL_2526851, EPI_ISL_2526853, EPI_ISL_2526855, EPI_ISL_2526857, EPI_ISL_2526859, EPI_ISL_2526861, EPI_ISL_2526863, EPI_ISL_2526865, EPI_ISL_2526867, EPI_ISL_2526869, EPI_ISL_2526871, EPI_ISL_2526873, EPI_ISL_2526875, EPI_ISL_2526877, EPI_ISL_2526879, EPI_ISL_2526881, EPI_ISL_2526884, EPI_ISL_2526886, EPI_ISL_2526888, EPI_ISL_2526890, EPI_ISL_2526892, EPI_ISL_2526894, EPI_ISL_2526896, EPI_ISL_2526898, EPI_ISL_2526900, EPI_ISL_2526902, EPI_ISL_2526904, EPI_ISL_2526906, EPI_ISL_2526909, EPI_ISL_2526911, EPI_ISL_2526913, EPI_ISL_2526915, EPI_ISL_2526917, EPI_ISL_2526919, EPI_ISL_2526922

| | | | |
|---|---|---|---|
| see above | Quest Diagnostics Incorporated | Centers for Disease Control and Prevention Division of Viral Diseases, Pathogen Discovery | Dakota Howard, Dhwani Batra, Peter W. Cook, Kara Moser, Adrian Paskey, Jason Caravas, Benjamin Rambo-Martin, Shatavia Morrison, Christopher Gulvick, Scott Sammons, Yvette Unoarumhi, Darlene Wagner, Matthew Schmerer, S. H. Rosenthal, A. Gerasimova, R. M. Kagan, B. Anderson, M. Hua, Y. Liu, L.E. Bernstein, K.E. Livingston, A. Perez, I. A. Shlyakhter, R. V. Rolando, R. Owen, P. Tanpaiboon, F. Lacbawan, Clinton R. Paden, Duncan MacCannell |
| EPI_ISL_2526923 | Ohio Department of Health Laboratory | Ohio Department of Health Laboratory | Holmes, Jennifer; Eric Brandt, Keoni Omura, Glen McGillivary, Caitlin McDonnell, Jade Mowery, Stephanie Mccracken, Tyler Payne, Kirtana Ramadugu, Erica Leasure, Brent Lee, Kelsey Florek, Heather Blankenship, Allison Black, Quanta Brown, and Tammy Bannerman |

EPI_ISL_2526925, EPI_ISL_2526927, EPI_ISL_2526929, EPI_ISL_2526931, EPI_ISL_2526933, EPI_ISL_2526935, EPI_ISL_2526938, EPI_ISL_2526940, EPI_ISL_2526942, EPI_ISL_2526944, EPI_ISL_2526946, EPI_ISL_2526948, EPI_ISL_2526950, EPI_ISL_2526952, EPI_ISL_2526954, EPI_ISL_2526956, EPI_ISL_2526959, EPI_ISL_2526961, EPI_ISL_2526963, EPI_ISL_2526965, EPI_ISL_2526967, EPI_ISL_2526969, EPI_ISL_2526972, EPI_ISL_2526974, EPI_ISL_2526976, EPI_ISL_2526978, EPI_ISL_2526980, EPI_ISL_2526983, EPI_ISL_2526985, EPI_ISL_2526987, EPI_ISL_2526989, EPI_ISL_2526991, EPI_ISL_2526993, EPI_ISL_2526994, EPI_ISL_2526995, EPI_ISL_2526996, EPI_ISL_2526998, EPI_ISL_2527001, EPI_ISL_2527003, EPI_ISL_2527005, EPI_ISL_2527007, EPI_ISL_2527009, EPI_ISL_2527012, EPI_ISL_2527014, EPI_ISL_2527016, EPI_ISL_2527017, EPI_ISL_2527020, EPI_ISL_2527022, EPI_ISL_2527024, EPI_ISL_2527026, EPI_ISL_2527028, EPI_ISL_2527030, EPI_ISL_2527032, EPI_ISL_2527034, EPI_ISL_2527036, EPI_ISL_2527038, EPI_ISL_2527040, EPI_ISL_2527043

| | | | |
|---|---|---|---|
| see above | Quest Diagnostics Incorporated | Centers for Disease Control and Prevention Division of Viral Diseases, Pathogen Discovery | Dakota Howard, Dhwani Batra, Peter W. Cook, Kara Moser, Adrian Paskey, Jason Caravas, Benjamin Rambo-Martin, Shatavia Morrison, Christopher Gulvick, Scott Sammons, Yvette Unoarumhi, Darlene Wagner, Matthew Schmerer, S. H. Rosenthal, A. Gerasimova, R. M. Kagan, B. Anderson, M. Hua, Y. Liu, L.E. Bernstein, K.E. Livingston, A. Perez, I. A. Shlyakhter, R. V. Rolando, R. Owen, P. Tanpaiboon, F. Lacbawan, Clinton R. Paden, Duncan MacCannell |
| EPI_ISL_2527432 | Ohio Department of Health Laboratory | Ohio Department of Health Laboratory | Holmes, Jennifer; Eric Brandt, Keoni Omura, Glen McGillivary, Caitlin McDonnell, Jade Mowery, Stephanie Mccracken, Tyler Payne, Kirtana Ramadugu, Erica Leasure, Brent Lee, Kelsey Florek, Heather Blankenship, Allison Black, Quanta Brown, and Tammy Bannerman |

EPI_ISL_2527563, EPI_ISL_2527565, EPI_ISL_2527567, EPI_ISL_2527570, EPI_ISL_2527573, EPI_ISL_2527575, EPI_ISL_2527577, EPI_ISL_2527581, EPI_ISL_2527584, EPI_ISL_2527586, EPI_ISL_2527588, EPI_ISL_2527590, EPI_ISL_2527592, EPI_ISL_2527594, EPI_ISL_2527597, EPI_ISL_2527599, EPI_ISL_2527601, EPI_ISL_2527603, EPI_ISL_2527605, EPI_ISL_2527608, EPI_ISL_2527610, EPI_ISL_2527612, EPI_ISL_2527614, EPI_ISL_2527616, EPI_ISL_2527618, EPI_ISL_2527620, EPI_ISL_2527622, EPI_ISL_2527624, EPI_ISL_2527629, EPI_ISL_2527632, EPI_ISL_2527634, EPI_ISL_2527636, EPI_ISL_2527638, EPI_ISL_2527641, EPI_ISL_2527643, EPI_ISL_2527645, EPI_ISL_2527650, EPI_ISL_2527652, EPI_ISL_2527654, EPI_ISL_2527656, EPI_ISL_2527658, EPI_ISL_2527660, EPI_ISL_2527663, EPI_ISL_2527665, EPI_ISL_2527669, EPI_ISL_2527672, EPI_ISL_2527674, EPI_ISL_2527676, EPI_ISL_2527678, EPI_ISL_2527680, EPI_ISL_2527682, EPI_ISL_2527685, EPI_ISL_2527687, EPI_ISL_2527688, EPI_ISL_2527693, EPI_ISL_2527695, EPI_ISL_2527697, EPI_ISL_2527700, EPI_ISL_2527702, EPI_ISL_2527704, EPI_ISL_2527708, EPI_ISL_2527712, EPI_ISL_2527714, EPI_ISL_2527716, EPI_ISL_2527719, EPI_ISL_2527721, EPI_ISL_2527723, EPI_ISL_2527727, EPI_ISL_2527731, EPI_ISL_2527733, EPI_ISL_2527735, EPI_ISL_2527739, EPI_ISL_2527741, EPI_ISL_2527743, EPI_ISL_2527745, EPI_ISL_2527747, EPI_ISL_2527749, EPI_ISL_2527751, EPI_ISL_2527753, EPI_ISL_2527755, EPI_ISL_2527757, EPI_ISL_2527760, EPI_ISL_2527762, EPI_ISL_2527764, EPI_ISL_2527768, EPI_ISL_2527772, EPI_ISL_2527774, EPI_ISL_2527775, EPI_ISL_2527777, EPI_ISL_2527779, EPI_ISL_2527781, EPI_ISL_2527784, EPI_ISL_2527786, EPI_ISL_2527788, EPI_ISL_2527790, EPI_ISL_2527792, EPI_ISL_2527794, EPI_ISL_2527796, EPI_ISL_2527798, EPI_ISL_2527801, EPI_ISL_2527803, EPI_ISL_2527805, EPI_ISL_2527807, EPI_ISL_2527809, EPI_ISL_2527811, EPI_ISL_2527813, EPI_ISL_2527816, EPI_ISL_2527818, EPI_ISL_2527820, EPI_ISL_2527823, EPI_ISL_2527825, EPI_ISL_2527827, EPI_ISL_2527831, EPI_ISL_2527833, EPI_ISL_2527835, EPI_ISL_2527837, EPI_ISL_2527841

| | | | |
|---|---|---|---|
| see above | Fulgent Genetics | Centers for Disease Control and Prevention Division of Viral Diseases, Pathogen Discovery | Dakota Howard, Dhwani Batra, Peter W. Cook, Kara Moser, Adrian Paskey, Jason Caravas, Benjamin Rambo-Martin, Shatavia Morrison, Christopher Gulvick, Scott Sammons, Yvette Unoarumhi, Darlene Wagner, Matthew Schmerer, Harry Gao, Mickey Li, John Gao, Joseph Fierro, Benafsh Sapra, Becky Tsai, Yan Meng, Doreen Ng, James Xie, Clinton R. Paden, Duncan MacCannell |

EPI_ISL_2527872, EPI_ISL_2527874, EPI_ISL_2527876, EPI_ISL_2527878, EPI_ISL_2527880, EPI_ISL_2527882, EPI_ISL_2527884, EPI_ISL_2527886, EPI_ISL_2527889, EPI_ISL_2527891, EPI_ISL_2527893, EPI_ISL_2527895, EPI_ISL_2527899, EPI_ISL_2527901, EPI_ISL_2527904, EPI_ISL_2527906, EPI_ISL_2527909, EPI_ISL_2527911, EPI_ISL_2527913, EPI_ISL_2527915, EPI_ISL_2527917, EPI_ISL_2527920, EPI_ISL_2527922, EPI_ISL_2527924, EPI_ISL_2527927, EPI_ISL_2527929, EPI_ISL_2527931, EPI_ISL_2527933, EPI_ISL_2527935, EPI_ISL_2527937, EPI_ISL_2527939, EPI_ISL_2527942, EPI_ISL_2527944, EPI_ISL_2527946, EPI_ISL_2527948, EPI_ISL_2527950, EPI_ISL_2527952, EPI_ISL_2527954, EPI_ISL_2527956, EPI_ISL_2527959, EPI_ISL_2527961, EPI_ISL_2527963, EPI_ISL_2527965, EPI_ISL_2527967, EPI_ISL_2527969, EPI_ISL_2527974, EPI_ISL_2527976, EPI_ISL_2527978, EPI_ISL_2527981, EPI_ISL_2527983, EPI_ISL_2527985, EPI_ISL_2527988, EPI_ISL_2527990, EPI_ISL_2527992, EPI_ISL_2527995

| | | | |
|---|---|---|---|
| see above | Aegis Sciences Corporation | Centers for Disease Control and Prevention Division of Viral Diseases, Pathogen Discovery | Dakota Howard, Dhwani Batra, Peter W. Cook, Kara Moser, Adrian Paskey, Jason Caravas, Benjamin Rambo-Martin, Shatavia Morrison, Christopher Gulvick, Scott Sammons, Yvette Unoarumhi, Darlene Wagner, Matthew Schmerer, Cyndi Clark, Patrick Campbell, Rob Case, Vikramsinha Ghorpade, Holly Houdeshell, Ola Kvalvaag, Dillon Nall, Ethan Sanders, Alec Vest, Shaun Westlund, Matthew Hardison, Clinton R. Paden, Duncan MacCannell |
| EPI_ISL_2527996 | Ohio Department of Health Laboratory | Ohio Department of Health Laboratory | Holmes, Jennifer; Eric Brandt, Keoni Omura, Glen McGillivary, Caitlin McDonnell, Jade Mowery, Stephanie Mccracken, Tyler Payne, Kirtana Ramadugu, Erica Leasure, Brent Lee, Kelsey Florek, Heather Blankenship, Allison Black, Quanta Brown, and Tammy Bannerman |

EPI_ISL_2527998, EPI_ISL_2528000, EPI_ISL_2528002, EPI_ISL_2528004, EPI_ISL_2528006, EPI_ISL_2528008, EPI_ISL_2528010, EPI_ISL_2528012, EPI_ISL_2528014, EPI_ISL_2528016, EPI_ISL_2528018, EPI_ISL_2528019, EPI_ISL_2528021, EPI_ISL_2528023, EPI_ISL_2528025, EPI_ISL_2528028, EPI_ISL_2528030, EPI_ISL_2528032, EPI_ISL_2528034, EPI_ISL_2528037, EPI_ISL_2528039, EPI_ISL_2528041, EPI_ISL_2528043, EPI_ISL_2528045, EPI_ISL_2528047, EPI_ISL_2528049, EPI_ISL_2528052, EPI_ISL_2528054, EPI_ISL_2528056, EPI_ISL_2528058, EPI_ISL_2528060, EPI_ISL_2528062, EPI_ISL_2528063, EPI_ISL_2528065, EPI_ISL_2528067, EPI_ISL_2528069, EPI_ISL_2528072, EPI_ISL_2528074, EPI_ISL_2528076, EPI_ISL_2528078, EPI_ISL_2528081, EPI_ISL_2528083, EPI_ISL_2528085, EPI_ISL_2528087, EPI_ISL_2528089, EPI_ISL_2528091, EPI_ISL_2528093, EPI_ISL_2528096, EPI_ISL_2528098, EPI_ISL_2528100, EPI_ISL_2528102, EPI_ISL_2528104, EPI_ISL_2528106, EPI_ISL_2528109, EPI_ISL_2528111, EPI_ISL_2528114, EPI_ISL_2528116, EPI_ISL_2528118, EPI_ISL_2528120, EPI_ISL_2528121, EPI_ISL_2528123, EPI_ISL_2528125, EPI_ISL_2528128, EPI_ISL_2528130, EPI_ISL_2528132, EPI_ISL_2528135, EPI_ISL_2528137, EPI_ISL_2528139, EPI_ISL_2528141, EPI_ISL_2528143, EPI_ISL_2528146, EPI_ISL_2528148, EPI_ISL_2528151, EPI_ISL_2528153, EPI_ISL_2528155, EPI_ISL_2528157, EPI_ISL_2528159, EPI_ISL_2528161, EPI_ISL_2528163, EPI_ISL_2528165, EPI_ISL_2528167, EPI_ISL_2528169, EPI_ISL_2528171, EPI_ISL_2528173, EPI_ISL_2528175, EPI_ISL_2528177, EPI_ISL_2528179, EPI_ISL_2528182, EPI_ISL_2528184, EPI_ISL_2528186, EPI_ISL_2528188, EPI_ISL_2528191, EPI_ISL_2528193, EPI_ISL_2528195, EPI_ISL_2528197, EPI_ISL_2528199, EPI_ISL_2528201, EPI_ISL_2528203, EPI_ISL_2528205, EPI_ISL_2528208, EPI_ISL_2528210, EPI_ISL_2528212, EPI_ISL_2528218, EPI_ISL_2528220, EPI_ISL_2528222, EPI_ISL_2528224, EPI_ISL_2528226, EPI_ISL_2528228, EPI_ISL_2528230, EPI_ISL_2528233, EPI_ISL_2528235, EPI_ISL_2528237, EPI_ISL_2528239, EPI_ISL_2528241, EPI_ISL_2528243, EPI_ISL_2528245, EPI_ISL_2528247, EPI_ISL_2528249, EPI_ISL_2528252, EPI_ISL_2528256, EPI_ISL_2528257, EPI_ISL_2528260, EPI_ISL_2528263, EPI_ISL_2528265, EPI_ISL_2528267, EPI_ISL_2528269, EPI_ISL_2528271, EPI_ISL_2528274, EPI_ISL_2528276, EPI_ISL_2528278, EPI_ISL_2528280, EPI_ISL_2528283, EPI_ISL_2528285, EPI_ISL_2528287, EPI_ISL_2528289, EPI_ISL_2528291, EPI_ISL_2528293, EPI_ISL_2528295, EPI_ISL_2528297, EPI_ISL_2528299, EPI_ISL_2528301, EPI_ISL_2528303, EPI_ISL_2528306, EPI_ISL_2528308, EPI_ISL_2528310, EPI_ISL_2528312, EPI_ISL_2528314, EPI_ISL_2528316, EPI_ISL_2528318, EPI_ISL_2528320, EPI_ISL_2528322, EPI_ISL_2528324, EPI_ISL_2528326, EPI_ISL_2528328, EPI_ISL_2528330, EPI_ISL_2528333, EPI_ISL_2528335, EPI_ISL_2528337, EPI_ISL_2528339, EPI_ISL_2528341, EPI_ISL_2528343, EPI_ISL_2528345, EPI_ISL_2528347, EPI_ISL_2528350, EPI_ISL_2528352, EPI_ISL_2528354, EPI_ISL_2528356, EPI_ISL_2528360, EPI_ISL_2528363, EPI_ISL_2528365, EPI_ISL_2528367, EPI_ISL_2528370, EPI_ISL_2528372, EPI_ISL_2528374, EPI_ISL_2528376, EPI_ISL_2528378, EPI_ISL_2528380, EPI_ISL_2528383, EPI_ISL_2528385, EPI_ISL_2528387, EPI_ISL_2528390, EPI_ISL_2528392, EPI_ISL_2528394, EPI_ISL_2528396, EPI_ISL_2528398, EPI_ISL_2528400, EPI_ISL_2528401, EPI_ISL_2528402, EPI_ISL_2528404, EPI_ISL_2528406, EPI_ISL_2528408, EPI_ISL_2528410, EPI_ISL_2528413, EPI_ISL_2528415, EPI_ISL_2528417, EPI_ISL_2528419, EPI_ISL_2528421, EPI_ISL_2528423, EPI_ISL_2528425, EPI_ISL_2528428, EPI_ISL_2528430, EPI_ISL_2528432, EPI_ISL_2528434, EPI_ISL_2528436, EPI_ISL_2528438, EPI_ISL_2528440, EPI_ISL_2528442, EPI_ISL_2528444, EPI_ISL_2528446, EPI_ISL_2528449, EPI_ISL_2528451, EPI_ISL_2528453, EPI_ISL_2528455, EPI_ISL_2528458, EPI_ISL_2528460, EPI_ISL_2528462, EPI_ISL_2528464, EPI_ISL_2528466, EPI_ISL_2528468, EPI_ISL_2528470, EPI_ISL_2528472, EPI_ISL_2528475, EPI_ISL_2528479, EPI_ISL_2528482, EPI_ISL_2528484, EPI_ISL_2528486, EPI_ISL_2528488, EPI_ISL_2528490, EPI_ISL_2528492, EPI_ISL_2528494, EPI_ISL_2528496, EPI_ISL_2528498, EPI_ISL_2528500, EPI_ISL_2528502, EPI_ISL_2528504, EPI_ISL_2528506, EPI_ISL_2528508, EPI_ISL_2528510, EPI_ISL_2528513, EPI_ISL_2528517, EPI_ISL_2528519, EPI_ISL_2528521, EPI_ISL_2528523, EPI_ISL_2528525, EPI_ISL_2528528, EPI_ISL_2528530, EPI_ISL_2528532, EPI_ISL_2528534, EPI_ISL_2528537, EPI_ISL_2528539, EPI_ISL_2528541, EPI_ISL_2528544, EPI_ISL_2528546, EPI_ISL_2528548, EPI_ISL_2528550, EPI_ISL_2528552, EPI_ISL_2528554, EPI_ISL_2528556, EPI_ISL_2528559, EPI_ISL_2528561, EPI_ISL_2528563, EPI_ISL_2528565, EPI_ISL_2528568, EPI_ISL_2528570, EPI_ISL_2528572, EPI_ISL_2528574, EPI_ISL_2528576, EPI_ISL_2528578, EPI_ISL_2528580, EPI_ISL_2528582, EPI_ISL_2528585, EPI_ISL_2528587, EPI_ISL_2528589, EPI_ISL_2528591, EPI_ISL_2528594, EPI_ISL_2528598, EPI_ISL_2528601, EPI_ISL_2528603, EPI_ISL_2528605, EPI_ISL_2528608, EPI_ISL_2528610, EPI_ISL_2528612, EPI_ISL_2528614, EPI_ISL_2528616, EPI_ISL_2528619

| | | | |
|---|---|---|---|
| see above | Aegis Sciences Corporation | Centers for Disease Control and Prevention Division of Viral Diseases, Pathogen Discovery | Dakota Howard, Dhwani Batra, Peter W. Cook, Kara Moser, Adrian Paskey, Jason Caravas, Benjamin Rambo-Martin, Shatavia Morrison, Christopher Gulvick, Scott Sammons, Yvette Unoarumhi, Darlene Wagner, Matthew Schmerer, Cyndi Clark, Patrick Campbell, Rob Case, Vikramsinha Ghorpade, Holly Houdeshell, Ola Kvalvaag, Dillon Nall, Ethan Sanders, Alec Vest, Shaun Westlund, Matthew Hardison, Clinton R. Paden, Duncan MacCannell |
| EPI_ISL_2528620 | Ohio Department of Health Laboratory | Ohio Department of Health Laboratory | Holmes, Jennifer; Eric Brandt, Keoni Omura, Glen McGillivary, Caitlin McDonnell, Jade Mowery, Stephanie Mccracken, Tyler Payne, Kirtana Ramadugu, Erica Leasure, Brent Lee, Kelsey Florek, Heather Blankenship, Allison Black, Quanta Brown, and Tammy Bannerman |

EPI_ISL_2528622, EPI_ISL_2528624, EPI_ISL_2528626, EPI_ISL_2528629, EPI_ISL_2528633, EPI_ISL_2528635, EPI_ISL_2528637, EPI_ISL_2528640, EPI_ISL_2528642, EPI_ISL_2528644, EPI_ISL_2528646, EPI_ISL_2528647, EPI_ISL_2528650, EPI_ISL_2528652, EPI_ISL_2528654, EPI_ISL_2528656, EPI_ISL_2528658, EPI_ISL_2528659, EPI_ISL_2528662, EPI_ISL_2528664, EPI_ISL_2528666, EPI_ISL_2528668, EPI_ISL_2528670, EPI_ISL_2528673, EPI_ISL_2528675, EPI_ISL_2528677, EPI_ISL_2528679, EPI_ISL_2528682, EPI_ISL_2528684, EPI_ISL_2528686, EPI_ISL_2528688, EPI_ISL_2528690, EPI_ISL_2528692, EPI_ISL_2528694, EPI_ISL_2528697, EPI_ISL_2528699, EPI_ISL_2528701, EPI_ISL_2528703, EPI_ISL_2528705, EPI_ISL_2528707, EPI_ISL_2528709, EPI_ISL_2528711, EPI_ISL_2528714, EPI_ISL_2528716, EPI_ISL_2528718, EPI_ISL_2528720, EPI_ISL_2528722, EPI_ISL_2528725, EPI_ISL_2528727, EPI_ISL_2528732, EPI_ISL_2528734, EPI_ISL_2528737, EPI_ISL_2528739, EPI_ISL_2528741, EPI_ISL_2528743, EPI_ISL_2528745, EPI_ISL_2528747, EPI_ISL_2528749, EPI_ISL_2528751, EPI_ISL_2528753, EPI_ISL_2528755, EPI_ISL_2528760, EPI_ISL_2528761, EPI_ISL_2528764, EPI_ISL_2528766, EPI_ISL_2528768, EPI_ISL_2528770, EPI_ISL_2528772, EPI_ISL_2528774, EPI_ISL_2528776, EPI_ISL_2528779, EPI_ISL_2528781, EPI_ISL_2528783, EPI_ISL_2528785, EPI_ISL_2528787, EPI_ISL_2528789, EPI_ISL_2528791, EPI_ISL_2528793, EPI_ISL_2528795, EPI_ISL_2528797, EPI_ISL_2528800, EPI_ISL_2528802, EPI_ISL_2528804, EPI_ISL_2528806, EPI_ISL_2528808, EPI_ISL_2528811, EPI_ISL_2528813, EPI_ISL_2528815, EPI_ISL_2528817, EPI_ISL_2528819, EPI_ISL_2528821, EPI_ISL_2528823, EPI_ISL_2528825, EPI_ISL_2528828, EPI_ISL_2528830, EPI_ISL_2528832, EPI_ISL_2528834, EPI_ISL_2528836, EPI_ISL_2528838, EPI_ISL_2528841, EPI_ISL_2528843, EPI_ISL_2528845, EPI_ISL_2528847, EPI_ISL_2528849, EPI_ISL_2528851, EPI_ISL_2528854, EPI_ISL_2528856, EPI_ISL_2528858, EPI_ISL_2528860, EPI_ISL_2528862, EPI_ISL_2528864, EPI_ISL_2528868, EPI_ISL_2528871, EPI_ISL_2528873, EPI_ISL_2528875, EPI_ISL_2528879, EPI_ISL_2528880, EPI_ISL_2528883, EPI_ISL_2528885, EPI_ISL_2528887, EPI_ISL_2528889, EPI_ISL_2528891, EPI_ISL_2528893, EPI_ISL_2528896, EPI_ISL_2528898, EPI_ISL_2528900, EPI_ISL_2528902, EPI_ISL_2528904, EPI_ISL_2528907, EPI_ISL_2528909, EPI_ISL_2528911, EPI_ISL_2528913, EPI_ISL_2528916, EPI_ISL_2528918, EPI_ISL_2528920, EPI_ISL_2528923, EPI_ISL_2528925, EPI_ISL_2528927, EPI_ISL_2528929, EPI_ISL_2528931, EPI_ISL_2528933, EPI_ISL_2528935, EPI_ISL_2528937, EPI_ISL_2528939, EPI_ISL_2528941, EPI_ISL_2528944, EPI_ISL_2528949, EPI_ISL_2528951, EPI_ISL_2528953, EPI_ISL_2528955, EPI_ISL_2528957, EPI_ISL_2528959, EPI_ISL_2528961, EPI_ISL_2528963, EPI_ISL_2528965, EPI_ISL_2528968, EPI_ISL_2528970, EPI_ISL_2528971, EPI_ISL_2528974, EPI_ISL_2528976, EPI_ISL_2528978, EPI_ISL_2528980, EPI_ISL_2528983, EPI_ISL_2528985, EPI_ISL_2528987, EPI_ISL_2528990, EPI_ISL_2528992, EPI_ISL_2528994, EPI_ISL_2528996, EPI_ISL_2528999, EPI_ISL_2529001, EPI_ISL_2529003, EPI_ISL_2529005, EPI_ISL_2529007, EPI_ISL_2529009, EPI_ISL_2529011, EPI_ISL_2529013, EPI_ISL_2529015, EPI_ISL_2529017, EPI_ISL_2529020, EPI_ISL_2529022

| | | | |
|---|---|---|---|
| see above | Aegis Sciences Corporation | Centers for Disease Control and Prevention Division of Viral Diseases, Pathogen Discovery | Dakota Howard, Dhwani Batra, Peter W. Cook, Kara Moser, Adrian Paskey, Jason Caravas, Benjamin Rambo-Martin, Shatavia Morrison, Christopher Gulvick, Scott Sammons, Yvette Unoarumhi, Darlene Wagner, Matthew Schmerer, Cyndi Clark, Patrick Campbell, Rob Case, Vikramsinha Ghorpade, Holly Houdeshell, Ola Kvalvaag, Dillon Nall, Ethan Sanders, Alec Vest, Shaun Westlund, Matthew Hardison, Clinton R. Paden, Duncan MacCannell |

EPI_ISL_2529075, EPI_ISL_2529077, EPI_ISL_2529080, EPI_ISL_2529083, EPI_ISL_2529085, EPI_ISL_2529087, EPI_ISL_2529089, EPI_ISL_2529091, EPI_ISL_2529093, EPI_ISL_2529095, EPI_ISL_2529097, EPI_ISL_2529099, EPI_ISL_2529102, EPI_ISL_2529104, EPI_ISL_2529106, EPI_ISL_2529108, EPI_ISL_2529111, EPI_ISL_2529113, EPI_ISL_2529115, EPI_ISL_2529117, EPI_ISL_2529119, EPI_ISL_2529121, EPI_ISL_2529123, EPI_ISL_2529125, EPI_ISL_2529127, EPI_ISL_2529129, EPI_ISL_2529131, EPI_ISL_2529134, EPI_ISL_2529136, EPI_ISL_2529138, EPI_ISL_2529140, EPI_ISL_2529142, EPI_ISL_2529144, EPI_ISL_2529146, EPI_ISL_2529148, EPI_ISL_2529150, EPI_ISL_2529152, EPI_ISL_2529155, EPI_ISL_2529157, EPI_ISL_2529159, EPI_ISL_2529161, EPI_ISL_2529163, EPI_ISL_2529166, EPI_ISL_2529168, EPI_ISL_2529170, EPI_ISL_2529172, EPI_ISL_2529174, EPI_ISL_2529176, EPI_ISL_2529178, EPI_ISL_2529181, EPI_ISL_2529183, EPI_ISL_2529185, EPI_ISL_2529187, EPI_ISL_2529190, EPI_ISL_2529191, EPI_ISL_2529193, EPI_ISL_2529195, EPI_ISL_2529197, EPI_ISL_2529200, EPI_ISL_2529202, EPI_ISL_2529204, EPI_ISL_2529206, EPI_ISL_2529208, EPI_ISL_2529210, EPI_ISL_2529212, EPI_ISL_2529214, EPI_ISL_2529216, EPI_ISL_2529218, EPI_ISL_2529219, EPI_ISL_2529221, EPI_ISL_2529223, EPI_ISL_2529225, EPI_ISL_2529228, EPI_ISL_2529230, EPI_ISL_2529232, EPI_ISL_2529234, EPI_ISL_2529236, EPI_ISL_2529238, EPI_ISL_2529240, EPI_ISL_2529242, EPI_ISL_2529244, EPI_ISL_2529246, EPI_ISL_2529249, EPI_ISL_2529251, EPI_ISL_2529254, EPI_ISL_2529256, EPI_ISL_2529258, EPI_ISL_2529260, EPI_ISL_2529262, EPI_ISL_2529265, EPI_ISL_2529267, EPI_ISL_2529269, EPI_ISL_2529271, EPI_ISL_2529274, EPI_ISL_2529276, EPI_ISL_2529278, EPI_ISL_2529281, EPI_ISL_2529283, EPI_ISL_2529286, EPI_ISL_2529288, EPI_ISL_2529290, EPI_ISL_2529291, EPI_ISL_2529292, EPI_ISL_2529293, EPI_ISL_2529295, EPI_ISL_2529297, EPI_ISL_2529299, EPI_ISL_2529301, EPI_ISL_2529303, EPI_ISL_2529306, EPI_ISL_2529308, EPI_ISL_2529310, EPI_ISL_2529312, EPI_ISL_2529314, EPI_ISL_2529316, EPI_ISL_2529318, EPI_ISL_2529320, EPI_ISL_2529323, EPI_ISL_2529325, EPI_ISL_2529327, EPI_ISL_2529329, EPI_ISL_2529331, EPI_ISL_2529333, EPI_ISL_2529335, EPI_ISL_2529338, EPI_ISL_2529340, EPI_ISL_2529342, EPI_ISL_2529344, EPI_ISL_2529347

| | | | |
|---|---|---|---|
| see above | Quest Diagnostics Incorporated | Centers for Disease Control and Prevention Division of Viral Diseases, Pathogen Discovery | Dakota Howard, Dhwani Batra, Peter W. Cook, Kara Moser, Adrian Paskey, Jason Caravas, Benjamin Rambo-Martin, Shatavia Morrison, Christopher Gulvick, Scott Sammons, Yvette Unoarumhi, Darlene Wagner, Matthew Schmerer, S. H. Rosenthal, A. Gerasimova, R. M. Kagan, B. Anderson, M. Hua, Y. Liu, L.E. Bernstein, K.E. Livingston, A. Perez, I. A. Shlyakhter, R. V. Rolando, R. Owen, P. Tanpaiboon, F. Lacbawan, Clinton R. Paden, Duncan MacCannell |

| | | | |
|---|---|---|---|
| EPI_ISL_2529850, EPI_ISL_2529852, EPI_ISL_2529854, EPI_ISL_2529856, EPI_ISL_2529858, EPI_ISL_2529861 | Baylor College of Medicine/ GCID | Baylor College of Medicine/ GCID | Timothy Farinholt, Qing Meng, Qinchang Meng, Vipin Menon, Xiang Qin, Harsha Doddapaneni, Paige Farinholt, Charu Agrawal, Donna Muzny, Pedro A. Piedra, Richard A. Gibbs, Joseph Pertosino |
| EPI_ISL_2530084, EPI_ISL_2530579, EPI_ISL_2530928 | Ohio Department of Health Laboratory | Ohio Department of Health Laboratory | Holmes, Jennifer; Eric Brandt, Keoni Omura, Glen McGillivary, Caitlin McDonnell, Jade Mowery, Stephanie Mccracken, Tyler Payne, Kirtana Ramadugu, Erica Leasure, Brent Lee, Kelsey Florek, Heather Blankenship, Allison Black, Quanta Brown, and Tammy Bannerman |
| EPI_ISL_2530930, EPI_ISL_2530932, EPI_ISL_2530934, EPI_ISL_2530936, EPI_ISL_2530938, EPI_ISL_2530940, EPI_ISL_2530942, EPI_ISL_2530944, EPI_ISL_2530946, EPI_ISL_2530948, EPI_ISL_2530950, EPI_ISL_2530953, EPI_ISL_2530955, EPI_ISL_2530957, EPI_ISL_2530959, EPI_ISL_2530961, EPI_ISL_2530964, EPI_ISL_2530966, EPI_ISL_2530968, EPI_ISL_2530970, EPI_ISL_2530972, EPI_ISL_2530974, EPI_ISL_2530977, EPI_ISL_2530981, EPI_ISL_2530983, EPI_ISL_2530985, EPI_ISL_2530988, EPI_ISL_2530989, EPI_ISL_2530993, EPI_ISL_2530995, EPI_ISL_2530997, EPI_ISL_2531000, EPI_ISL_2531002, EPI_ISL_2531004, EPI_ISL_2531006, EPI_ISL_2531008, EPI_ISL_2531010, EPI_ISL_2531012, EPI_ISL_2531014, EPI_ISL_2531017, EPI_ISL_2531021, EPI_ISL_2531023, EPI_ISL_2531027, EPI_ISL_2531029, EPI_ISL_2531030, EPI_ISL_2531032, EPI_ISL_2531033, EPI_ISL_2531034, EPI_ISL_2531036, EPI_ISL_2531038, EPI_ISL_2531040, EPI_ISL_2531042, EPI_ISL_2531044, EPI_ISL_2531047, EPI_ISL_2531049, EPI_ISL_2531051, EPI_ISL_2531053, EPI_ISL_2531057, EPI_ISL_2531059, EPI_ISL_2531061, EPI_ISL_2531063, EPI_ISL_2531065, EPI_ISL_2531068, EPI_ISL_2531070, EPI_ISL_2531072, EPI_ISL_2531074, EPI_ISL_2531076, EPI_ISL_2531078, EPI_ISL_2531080, EPI_ISL_2531083, EPI_ISL_2531085, EPI_ISL_2531087, EPI_ISL_2531089, EPI_ISL_2531091, EPI_ISL_2531093 | | | |
| see above | Aegis Sciences Corporation | Centers for Disease Control and Prevention Division of Viral Diseases, Pathogen Discovery | Dakota Howard, Dhwani Batra, Peter W. Cook, Kara Moser, Adrian Paskey, Jason Caravas, Benjamin Rambo-Martin, Shatavia Morrison, Christopher Gulvick, Scott Sammons, Yvette Unoarumhi, Darlene Wagner, Matthew Schmerer, Cyndi Clark, Patrick Campbell, Rob Case, Vikramsinha Ghorpade, Holly Houdeshell, Ola Kvalvaag, Dillon Nall, Ethan Sanders, Alec Vest, Shaun Westlund, Matthew Hardison, Clinton R. Paden, Duncan MacCannell |
| EPI_ISL_2531308 | Ohio Department of Health Laboratory | Ohio Department of Health Laboratory | Holmes, Jennifer; Eric Brandt, Keoni Omura, Glen McGillivary, Caitlin McDonnell, Jade Mowery, Stephanie Mccracken, Tyler Payne, Kirtana Ramadugu, Erica Leasure, Brent Lee, Kelsey Florek, Heather Blankenship, Allison Black, Quanta Brown, and Tammy Bannerman |
| EPI_ISL_2532642, EPI_ISL_2532643, EPI_ISL_2532646, EPI_ISL_2532647, EPI_ISL_2532648, EPI_ISL_2532649, EPI_ISL_2532650, EPI_ISL_2532651, EPI_ISL_2532652, EPI_ISL_2532653, EPI_ISL_2532654, EPI_ISL_2532655, EPI_ISL_2532656, EPI_ISL_2532657, EPI_ISL_2532658, EPI_ISL_2532659, EPI_ISL_2532660, EPI_ISL_2532661, EPI_ISL_2532662, EPI_ISL_2532663, EPI_ISL_2532664, EPI_ISL_2532665, EPI_ISL_2532666, EPI_ISL_2532667, EPI_ISL_2532668, EPI_ISL_2532669, EPI_ISL_2532670, EPI_ISL_2532671, EPI_ISL_2532677, EPI_ISL_2532678, EPI_ISL_2532679, EPI_ISL_2532680, EPI_ISL_2532683, EPI_ISL_2532688, EPI_ISL_2532689, EPI_ISL_2532692, EPI_ISL_2532693, EPI_ISL_2532694, EPI_ISL_2532695, EPI_ISL_2532697, EPI_ISL_2532699, EPI_ISL_2532701, EPI_ISL_2532703, EPI_ISL_2532704, EPI_ISL_2532705, EPI_ISL_2532707, EPI_ISL_2532708, EPI_ISL_2532709, EPI_ISL_2532710, EPI_ISL_2532711, EPI_ISL_2532712, EPI_ISL_2532713, EPI_ISL_2532715, EPI_ISL_2532716, EPI_ISL_2532717, EPI_ISL_2532718, EPI_ISL_2532719, EPI_ISL_2532720, EPI_ISL_2532722, EPI_ISL_2532723, EPI_ISL_2532724, EPI_ISL_2532725, EPI_ISL_2532726 | | | |
| see above | Wyoming Public Health Laboratory | Wyoming Public Health Laboratory | Jim Mildenberger, Taylor Fearing, Channing Weber, Ashley Norberg, Chayse Rowley, Marley Goetz, Brian Dominguez, Elliot Thomasson, Sam Britz, Cari Sloma, Robert Petit, and Rob Christensen |
| EPI_ISL_2533751 | VA Palo Alto Health Care System | Genomics and Discovery, Respiratory Viruses Branch, Division of Viral Diseases, Centers for Disease Control and Prevention | Yan Li, Jing Zhang, Anna Kelleher, Ying Tao, Brian Lynch, Krista Queen, Anna Uehara, Peter Cook, Han Jia Justin Ng, Rachel Marine, Clinton R. Paden, Dhwani Batra, Haibin Wang, Tara Coalter, Jasmine Padilla, Morgan Davis, Mili Sheth, Sarah Nobles, Mark Burroughs, Justin Lee, Adam Retchless, Suxiang Tong |
| EPI_ISL_2533752, EPI_ISL_2533753, EPI_ISL_2533754, EPI_ISL_2533755, EPI_ISL_2533756, EPI_ISL_2533757 | Michael E. Debakey VA Medical Center | Genomics and Discovery, Respiratory Viruses Branch, Division of Viral Diseases, Centers for Disease Control and Prevention | Yan Li, Jing Zhang, Anna Kelleher, Ying Tao, Brian Lynch, Krista Queen, Anna Uehara, Peter Cook, Han Jia Justin Ng, Rachel Marine, Clinton R. Paden, Dhwani Batra, Haibin Wang, Tara Coalter, Jasmine Padilla, Morgan Davis, Mili Sheth, Sarah Nobles, Mark Burroughs, Justin Lee, Adam Retchless, Suxiang Tong |
| EPI_ISL_2533758 | WLA VAMC | Genomics and Discovery, Respiratory Viruses Branch, Division of Viral Diseases, Centers for Disease Control and Prevention | Yan Li, Jing Zhang, Anna Kelleher, Ying Tao, Brian Lynch, Krista Queen, Anna Uehara, Peter Cook, Han Jia Justin Ng, Rachel Marine, Clinton R. Paden, Dhwani Batra, Haibin Wang, Tara Coalter, Jasmine Padilla, Morgan Davis, Mili Sheth, Sarah Nobles, Mark Burroughs, Justin Lee, Adam Retchless, Suxiang Tong |
| EPI_ISL_2533759, EPI_ISL_2533760, EPI_ISL_2533761, EPI_ISL_2533762 | LESP Chihuahua | Instituto de Diagnostico y Referencia Epidemiologicos (INDRE) | Claudia Wong-Arambula, Abril Rodriguez-Maldonado, Vanessa Rivero-Arredondo, Ariadna Medina-Benitez, Joaquin Quiroz-Mercado, Sergio Rangel-Guerrero, Natividad Cruz-Ortiz, Tatiana Nunez-Garcia, Gisela Barrera-Badillo, Lucia Hernandez-Rivas, Irma Lopez-Martinez, Ernesto Ramirez-Gonzalez. |
| EPI_ISL_2533763 | LESP Coahuila | Instituto de Diagnostico y Referencia Epidemiologicos (INDRE) | Claudia Wong-Arambula, Abril Rodriguez-Maldonado, Vanessa Rivero-Arredondo, Ariadna Medina-Benitez, Joaquin Quiroz-Mercado, Sergio Rangel-Guerrero, Natividad Cruz-Ortiz, Tatiana Nunez-Garcia, Gisela Barrera-Badillo, Lucia Hernandez-Rivas, Irma Lopez-Martinez, Ernesto Ramirez-Gonzalez. |
| EPI_ISL_2533764, EPI_ISL_2533765, EPI_ISL_2533766 | LESP Queretaro | Instituto de Diagnostico y Referencia Epidemiologicos (INDRE) | Claudia Wong-Arambula, Abril Rodriguez-Maldonado, Vanessa Rivero-Arredondo, Ariadna Medina-Benitez, Joaquin Quiroz-Mercado, Sergio Rangel-Guerrero, Natividad Cruz-Ortiz, Tatiana Nunez-Garcia, Gisela Barrera-Badillo, Lucia Hernandez-Rivas, Irma Lopez-Martinez, Ernesto Ramirez-Gonzalez. |
| EPI_ISL_2533767 | LESP Jalisco | Instituto de Diagnostico y Referencia Epidemiologicos (INDRE) | Claudia Wong-Arambula, Abril Rodriguez-Maldonado, Vanessa Rivero-Arredondo, Ariadna Medina-Benitez, Joaquin Quiroz-Mercado, Sergio Rangel-Guerrero, Natividad Cruz-Ortiz, Tatiana Nunez-Garcia, Gisela Barrera-Badillo, Lucia Hernandez-Rivas, Irma Lopez-Martinez, Ernesto Ramirez-Gonzalez. |
| EPI_ISL_2533768, EPI_ISL_2533769 | LESP Tabasco | Instituto de Diagnostico y Referencia Epidemiologicos (INDRE) | Claudia Wong-Arambula, Abril Rodriguez-Maldonado, Vanessa Rivero-Arredondo, Ariadna Medina-Benitez, Joaquin Quiroz-Mercado, Sergio Rangel-Guerrero, Natividad Cruz-Ortiz, Tatiana Nunez-Garcia, Gisela Barrera-Badillo, Lucia Hernandez-Rivas, Irma Lopez-Martinez, Ernesto Ramirez-Gonzalez. |
| EPI_ISL_2533770, EPI_ISL_2533771, EPI_ISL_2533772, EPI_ISL_2533773, EPI_ISL_2533774, EPI_ISL_2533775, EPI_ISL_2533776, EPI_ISL_2533777, EPI_ISL_2533778, EPI_ISL_2533779, EPI_ISL_2533780, EPI_ISL_2533781, EPI_ISL_2533782, EPI_ISL_2533783, EPI_ISL_2533784, EPI_ISL_2533785, EPI_ISL_2533786, EPI_ISL_2533787, EPI_ISL_2533788, EPI_ISL_2533789, EPI_ISL_2533790, EPI_ISL_2533791, EPI_ISL_2533792, EPI_ISL_2533793, EPI_ISL_2533794, EPI_ISL_2533795, EPI_ISL_2533796 | | | |
| see above | LESP Yucatan | Instituto de Diagnostico y Referencia Epidemiologicos (INDRE) | Claudia Wong-Arambula, Abril Rodriguez-Maldonado, Vanessa Rivero-Arredondo, Ariadna Medina-Benitez, Joaquin Quiroz-Mercado, Sergio Rangel-Guerrero, Natividad Cruz-Ortiz, Tatiana Nunez-Garcia, Gisela Barrera-Badillo, Lucia Hernandez-Rivas, Irma Lopez-Martinez, Ernesto Ramirez-Gonzalez. |
| EPI_ISL_2533797, EPI_ISL_2533798, EPI_ISL_2533799, EPI_ISL_2533800, EPI_ISL_2533801 | LESP Tamaulipas | Instituto de Diagnostico y Referencia Epidemiologicos (INDRE) | Claudia Wong-Arambula, Abril Rodriguez-Maldonado, Vanessa Rivero-Arredondo, Ariadna Medina-Benitez, Joaquin Quiroz-Mercado, Sergio Rangel-Guerrero, Natividad Cruz-Ortiz, Tatiana Nunez-Garcia, Gisela Barrera-Badillo, Lucia Hernandez-Rivas, Irma Lopez-Martinez, Ernesto Ramirez-Gonzalez. |
| EPI_ISL_2533802 | Lesp Jalisco /Unidad de Patologia | Instituto de Diagnostico y Referencia Epidemiologicos (INDRE) | Claudia Wong-Arambula, Abril Rodriguez-Maldonado, Vanessa Rivero-Arredondo, Ariadna Medina-Benitez, Joaquin Quiroz-Mercado, Sergio Rangel-Guerrero, Natividad Cruz-Ortiz, Tatiana Nunez-Garcia, Gisela Barrera-Badillo, Lucia Hernandez-Rivas, Irma Lopez-Martinez, Ernesto Ramirez-Gonzalez. |
| EPI_ISL_2533803 | LESP Coahuila | Instituto de Diagnostico y Referencia Epidemiologicos (INDRE) | Claudia Wong-Arambula, Abril Rodriguez-Maldonado, Vanessa Rivero-Arredondo, Ariadna Medina-Benitez, Joaquin Quiroz-Mercado, Sergio Rangel-Guerrero, Natividad Cruz-Ortiz, Tatiana Nunez-Garcia, Gisela Barrera-Badillo, Lucia Hernandez-Rivas, Irma Lopez-Martinez, Ernesto Ramirez-Gonzalez. |
| EPI_ISL_2533804, EPI_ISL_2533805 | LESP Quintana Roo | Instituto de Diagnostico y Referencia Epidemiologicos (INDRE) | Claudia Wong-Arambula, Abril Rodriguez-Maldonado, Vanessa Rivero-Arredondo, Ariadna Medina-Benitez, Joaquin Quiroz-Mercado, Sergio Rangel-Guerrero, Natividad Cruz-Ortiz, Tatiana Nunez-Garcia, Gisela Barrera-Badillo, Lucia Hernandez-Rivas, Irma Lopez-Martinez, Ernesto Ramirez-Gonzalez. |
| EPI_ISL_2533806 | LESP San Luis Potosi | Instituto de Diagnostico y Referencia Epidemiologicos (INDRE) | Claudia Wong-Arambula, Abril Rodriguez-Maldonado, Vanessa Rivero-Arredondo, Ariadna Medina-Benitez, Joaquin Quiroz-Mercado, Sergio Rangel-Guerrero, Natividad Cruz-Ortiz, Tatiana Nunez-Garcia, Gisela Barrera-Badillo, Lucia Hernandez-Rivas, Irma Lopez-Martinez, Ernesto Ramirez-Gonzalez. |
| EPI_ISL_2533807, EPI_ISL_2533808 | LESP Ciudad de Mexico | Instituto de Diagnostico y Referencia Epidemiologicos (INDRE) | Claudia Wong-Arambula, Abril Rodriguez-Maldonado, Vanessa Rivero-Arredondo, Ariadna Medina-Benitez, Joaquin Quiroz-Mercado, Sergio Rangel-Guerrero, Natividad Cruz-Ortiz, Tatiana Nunez-Garcia, Gisela Barrera-Badillo, Lucia Hernandez-Rivas, Irma Lopez-Martinez, Ernesto Ramirez-Gonzalez. |
| EPI_ISL_2533809 | LESP Estado de Mexico | Instituto de Diagnostico y Referencia Epidemiologicos (INDRE) | Claudia Wong-Arambula, Abril Rodriguez-Maldonado, Vanessa Rivero-Arredondo, Ariadna Medina-Benitez, Joaquin Quiroz-Mercado, Sergio Rangel-Guerrero, Natividad Cruz-Ortiz, Tatiana Nunez-Garcia, Gisela Barrera-Badillo, Lucia Hernandez-Rivas, Irma Lopez-Martinez, Ernesto Ramirez-Gonzalez. |
| EPI_ISL_2533810, EPI_ISL_2533811, EPI_ISL_2533812, EPI_ISL_2533813, EPI_ISL_2533814, EPI_ISL_2533815 | LESP Ciudad de Mexico | Instituto de Diagnostico y Referencia Epidemiologicos (INDRE) | Claudia Wong-Arambula, Abril Rodriguez-Maldonado, Vanessa Rivero-Arredondo, Ariadna Medina-Benitez, Joaquin Quiroz-Mercado, Sergio Rangel-Guerrero, Natividad Cruz-Ortiz, Tatiana Nunez-Garcia, Gisela Barrera-Badillo, Lucia Hernandez-Rivas, Irma Lopez-Martinez, Ernesto Ramirez-Gonzalez. |
| EPI_ISL_2533816 | LESP Campeche | Instituto de Diagnostico y Referencia Epidemiologicos (INDRE) | Claudia Wong-Arambula, Abril Rodriguez-Maldonado, Vanessa Rivero-Arredondo, Ariadna Medina-Benitez, Joaquin Quiroz-Mercado, Sergio Rangel-Guerrero, Natividad Cruz-Ortiz, Tatiana Nunez-Garcia, Gisela Barrera-Badillo, Lucia Hernandez-Rivas, Irma Lopez-Martinez, Ernesto Ramirez-Gonzalez. |
| EPI_ISL_2533817 | LESP Ciudad de Mexico | Instituto de Diagnostico y Referencia Epidemiologicos (INDRE) | Claudia Wong-Arambula, Abril Rodriguez-Maldonado, Vanessa Rivero-Arredondo, Ariadna Medina-Benitez, Joaquin Quiroz-Mercado, Sergio Rangel-Guerrero, Natividad Cruz-Ortiz, Tatiana Nunez-Garcia, Gisela Barrera-Badillo, Lucia Hernandez-Rivas, Irma Lopez-Martinez, Ernesto Ramirez-Gonzalez. |
| EPI_ISL_2533818 | LESP Baja California Sur | Instituto de Diagnostico y Referencia Epidemiologicos (INDRE) | Claudia Wong-Arambula, Abril Rodriguez-Maldonado, Vanessa Rivero-Arredondo, Ariadna Medina-Benitez, Joaquin Quiroz-Mercado, Sergio Rangel-Guerrero, Natividad Cruz-Ortiz, Tatiana Nunez-Garcia, Gisela Barrera-Badillo, Lucia Hernandez-Rivas, Irma Lopez-Martinez, Ernesto Ramirez-Gonzalez. |
| EPI_ISL_2533819 | LESP Yucatan | Instituto de Diagnostico y Referencia Epidemiologicos (INDRE) | Claudia Wong-Arambula, Abril Rodriguez-Maldonado, Vanessa Rivero-Arredondo, Ariadna Medina-Benitez, Joaquin Quiroz-Mercado, Sergio Rangel-Guerrero, Natividad Cruz-Ortiz, Tatiana Nunez-Garcia, Gisela Barrera-Badillo, Lucia Hernandez-Rivas, Irma Lopez-Martinez, Ernesto Ramirez-Gonzalez. |
| EPI_ISL_2533820 | LESP Quintana Roo | Instituto de Diagnostico y Referencia Epidemiologicos (INDRE) | Claudia Wong-Arambula, Abril Rodriguez-Maldonado, Vanessa Rivero-Arredondo, Ariadna Medina-Benitez, Joaquin Quiroz-Mercado, Sergio Rangel-Guerrero, Natividad Cruz-Ortiz, Tatiana Nunez-Garcia, Gisela Barrera-Badillo, Lucia Hernandez-Rivas, Irma Lopez-Martinez, Ernesto Ramirez-Gonzalez. |
| EPI_ISL_2533821 | LESP Ciudad de México /DIAGNOMOL | Instituto de Diagnostico y Referencia Epidemiologicos (INDRE) | Claudia Wong-Arambula, Abril Rodriguez-Maldonado, Vanessa Rivero-Arredondo, Ariadna Medina-Benitez, Joaquin Quiroz-Mercado, Sergio Rangel-Guerrero, Natividad Cruz-Ortiz, Tatiana Nunez-Garcia, Gisela Barrera-Badillo, Lucia Hernandez-Rivas, Irma Lopez-Martinez, Ernesto Ramirez-Gonzalez. |
| EPI_ISL_2533822 | LESP Queretaro | Instituto de Diagnostico y Referencia Epidemiologicos (INDRE) | Claudia Wong-Arambula, Abril Rodriguez-Maldonado, Vanessa Rivero-Arredondo, Ariadna Medina-Benitez, Joaquin Quiroz-Mercado, Sergio Rangel-Guerrero, Natividad Cruz-Ortiz, Tatiana Nunez-Garcia, Gisela Barrera-Badillo, Lucia Hernandez-Rivas, Irma Lopez-Martinez, Ernesto Ramirez-Gonzalez. |
| EPI_ISL_2533823 | LESP Sinaloa | Instituto de Diagnostico y Referencia Epidemiologicos (INDRE) | Claudia Wong-Arambula, Abril Rodriguez-Maldonado, Vanessa Rivero-Arredondo, Ariadna Medina-Benitez, Joaquin Quiroz-Mercado, Sergio Rangel-Guerrero, Natividad Cruz-Ortiz, Tatiana Nunez-Garcia, Gisela Barrera-Badillo, Lucia Hernandez-Rivas, Irma Lopez-Martinez, Ernesto Ramirez-Gonzalez. |
| EPI_ISL_2533824 | LESP Ciudad de Mexico | Instituto de Diagnostico y Referencia Epidemiologicos (INDRE) | Claudia Wong-Arambula, Abril Rodriguez-Maldonado, Vanessa Rivero-Arredondo, Ariadna Medina-Benitez, Joaquin Quiroz-Mercado, Sergio Rangel-Guerrero, Natividad Cruz-Ortiz, Tatiana Nunez-Garcia, Gisela Barrera-Badillo, Lucia Hernandez-Rivas, Irma Lopez-Martinez, Ernesto Ramirez-Gonzalez. |
| EPI_ISL_2533884, EPI_ISL_2533885, EPI_ISL_2533886, EPI_ISL_2533887, | Naval Infectious Diseases Diagnostic Laboratory | Naval Medical Research Center Biological Defense Research Directorate | Logan J. Voegtly, Catherine E. Arnold, Bishwo N. Adhikari, Francisco Malagon, Andrea E. Luquette, Gregory K. Rice, Andrew J. Bennett, Kyle A. Long, Lindsay A. Glang, Michael V. Deschenes, Megan A. Schilling, Victor A. Sugiharto, Regina Z. Cer, Kimberly A. Bishop-Lilly |

| | | | |
|---|---|---|---|
| EPI_ISL_2533888, EPI_ISL_2533889 | | | |
| EPI_ISL_2533890, EPI_ISL_2533891, EPI_ISL_2533892, EPI_ISL_2533893, EPI_ISL_2533894, EPI_ISL_2533895, EPI_ISL_2533896, EPI_ISL_2533897, EPI_ISL_2533898, EPI_ISL_2533899, EPI_ISL_2533900, EPI_ISL_2533901, EPI_ISL_2533902 | | | |
| see above | Montana Public Health Laboratory | Montana Public Health Laboratory | Joy Ritter, Michelle Mozer, Carrie Biskupiak, Deborah Gibson, Michael Dills |
| EPI_ISL_2533903 | Imágenes Diagnósticas de Mazatlán | Microbial Genomics Laboratory | Consorcio Mexicano de Vigilancia Genómica (CoViGen-Mex). Authors (in alphabetical order): Julio Elias Alvarado-Yaah, Carlos F. Arias, Santiago Ávila-Ríos, Eduardo Becerril-Vargas, Víctor Hugo Borja-Aburto, Celia Boukadida, Cristóbal Cháidez-Quiróz, Juan Bautista Chale-Dzul, Ricardo Ciria Merce, Andreu Comas-García, Célida Duque Molina, Julissa Enciso-Ibarra, José Antonio Enciso-Moreno, Gloria Elena Espinosa-Ayala, Fernando Fontove-Herrera, Daniel Fregoso-Rueda, Victor Eduardo García-Arias, Alejandra García-Gasca, Bruno Gómez-Gil, Jean Pierre González, Irvin González-López, Concepción Grajales-Muñiz, Ricardo Grande, Rosa María Gutiérrez Rios, Jesús Hernández, Alejandra Hernández-Terán, Alfredo Herrera-Estrella, Carla Ivón Herrera-Najera, Pavel Isa, Verónica Mata-Haro, Daniel Lira Morales, Susana Lopez, Antonio Loza Román, Brenda Irasema Maldonado-Meza, Bernardo Martínez-Miguel, José Arturo Martínez-Orozco, Célida Martínez- Rodríguez, Margarita Matías-Florentino, Fidencio Mejía-Nepomuceno, María Guadalupe de Jesús Mireles-Rivera, Gloria María Molina-Salinas, Hector Montoya-Fuentes, , Mario Mújica-Sánchez, José Esteban Muñoz-Medina, José de Jesús Nuñez-Contreras, Alicia Ocaña-Mondragón, Luis Alberto Ochoa-Carrera, Hector Esteban Paz-Juárez, Marissa Perez-Garcia, Francisco Pulido, Helen Haydee Fernanda Ramírez-Plascencia, Jorge Salas-Hernández, Angel Gustavo Salas-Lais, Alejandro Sánchez-Flores, Clara Esperanza Santacruz-Tinoco, María Guadalupe Santiago-Mauricio, Selene Zárate, Nelly Sélem-Mojica, Blanca Taboada, Gloria Vazquez, Joel Armando Vázquez-Pérez. |
| EPI_ISL_2533904 | Hospital Sharp | Microbial Genomics Laboratory | Consorcio Mexicano de Vigilancia Genómica (CoViGen-Mex). Authors (in alphabetical order): Julio Elias Alvarado-Yaah, Carlos F. Arias, Santiago Ávila-Ríos, Eduardo Becerril-Vargas, Víctor Hugo Borja-Aburto, Celia Boukadida, Cristóbal Cháidez-Quiróz, Juan Bautista Chale-Dzul, Ricardo Ciria Merce, Andreu Comas-García, Célida Duque Molina, Julissa Enciso-Ibarra, José Antonio Enciso-Moreno, Gloria Elena Espinosa-Ayala, Fernando Fontove-Herrera, Daniel Fregoso-Rueda, Victor Eduardo García-Arias, Alejandra García-Gasca, Bruno Gómez-Gil, Jean Pierre González, Irvin González-López, Concepción Grajales-Muñiz, Ricardo Grande, Rosa María Gutiérrez Rios, Jesús Hernández, Alejandra Hernández-Terán, Alfredo Herrera-Estrella, Carla Ivón Herrera-Najera, Pavel Isa, Verónica Mata-Haro, Daniel Lira Morales, Susana Lopez, Antonio Loza Román, Brenda Irasema Maldonado-Meza, Bernardo Martínez-Miguel, José Arturo Martínez-Orozco, Célida Martínez- Rodríguez, Margarita Matías-Florentino, Fidencio Mejía-Nepomuceno, María Guadalupe de Jesús Mireles-Rivera, Gloria María Molina-Salinas, Hector Montoya-Fuentes, , Mario Mújica-Sánchez, José Esteban Muñoz-Medina, José de Jesús Nuñez-Contreras, Alicia Ocaña-Mondragón, Luis Alberto Ochoa-Carrera, Hector Esteban Paz-Juárez, Marissa Perez-Garcia, Francisco Pulido, Helen Haydee Fernanda Ramírez-Plascencia, Jorge Salas-Hernández, Angel Gustavo Salas-Lais, Alejandro Sánchez-Flores, Clara Esperanza Santacruz-Tinoco, María Guadalupe Santiago-Mauricio, Selene Zárate, Nelly Sélem-Mojica, Blanca Taboada, Gloria Vazquez, Joel Armando Vázquez-Pérez. |
| EPI_ISL_2533905 | Hospital Margarita Maza de Juárez | Microbial Genomics Laboratory | Consorcio Mexicano de Vigilancia Genómica (CoViGen-Mex). Authors (in alphabetical order): Julio Elias Alvarado-Yaah, Carlos F. Arias, Santiago Ávila-Ríos, Eduardo Becerril-Vargas, Víctor Hugo Borja-Aburto, Celia Boukadida, Cristóbal Cháidez-Quiróz, Juan Bautista Chale-Dzul, Ricardo Ciria Merce, Andreu Comas-García, Célida Duque Molina, Julissa Enciso-Ibarra, José Antonio Enciso-Moreno, Gloria Elena Espinosa-Ayala, Fernando Fontove-Herrera, Daniel Fregoso-Rueda, Victor Eduardo García-Arias, Alejandra García-Gasca, Bruno Gómez-Gil, Jean Pierre González, Irvin González-López, Concepción Grajales-Muñiz, Ricardo Grande, Rosa María Gutiérrez Rios, Jesús Hernández, Alejandra Hernández-Terán, Alfredo Herrera-Estrella, Carla Ivón Herrera-Najera, Pavel Isa, Verónica Mata-Haro, Daniel Lira Morales, Susana Lopez, Antonio Loza Román, Brenda Irasema Maldonado-Meza, Bernardo Martínez-Miguel, José Arturo Martínez-Orozco, Célida Martínez- Rodríguez, Margarita Matías-Florentino, Fidencio Mejía-Nepomuceno, María Guadalupe de Jesús Mireles-Rivera, Gloria María Molina-Salinas, Hector Montoya-Fuentes, , Mario Mújica-Sánchez, José Esteban Muñoz-Medina, José de Jesús Nuñez-Contreras, Alicia Ocaña-Mondragón, Luis Alberto Ochoa-Carrera, Hector Esteban Paz-Juárez, Marissa Perez-Garcia, Francisco Pulido, Helen Haydee Fernanda Ramírez-Plascencia, Jorge Salas-Hernández, Angel Gustavo Salas-Lais, Alejandro Sánchez-Flores, Clara Esperanza Santacruz-Tinoco, María Guadalupe Santiago-Mauricio, Selene Zárate, Nelly Sélem-Mojica, Blanca Taboada, Gloria Vazquez, Joel Armando Vázquez-Pérez. |
| EPI_ISL_2533906 | Imágenes Diagnósticas de Mazatlán | Microbial Genomics Laboratory | Consorcio Mexicano de Vigilancia Genómica (CoViGen-Mex). Authors (in alphabetical order): Julio Elias Alvarado-Yaah, Carlos F. Arias, Santiago Ávila-Ríos, Eduardo Becerril-Vargas, Víctor Hugo Borja-Aburto, Celia Boukadida, Cristóbal Cháidez-Quiróz, Juan Bautista Chale-Dzul, Ricardo Ciria Merce, Andreu Comas-García, Célida Duque Molina, Julissa Enciso-Ibarra, José Antonio Enciso-Moreno, Gloria Elena Espinosa-Ayala, Fernando Fontove-Herrera, Daniel Fregoso-Rueda, Victor Eduardo García-Arias, Alejandra García-Gasca, Bruno Gómez-Gil, Jean Pierre González, Irvin González-López, Concepción Grajales-Muñiz, Ricardo Grande, Rosa María Gutiérrez Rios, Jesús Hernández, Alejandra Hernández-Terán, Alfredo Herrera-Estrella, Carla Ivón Herrera-Najera, Pavel Isa, Verónica Mata-Haro, Daniel Lira Morales, Susana Lopez, Antonio Loza Román, Brenda Irasema Maldonado-Meza, Bernardo Martínez-Miguel, José Arturo Martínez-Orozco, Célida Martínez- Rodríguez, Margarita Matías-Florentino, Fidencio Mejía-Nepomuceno, María Guadalupe de Jesús Mireles-Rivera, Gloria María Molina-Salinas, Hector Montoya-Fuentes, , Mario Mújica-Sánchez, José Esteban Muñoz-Medina, José de Jesús Nuñez-Contreras, Alicia Ocaña-Mondragón, Luis Alberto Ochoa-Carrera, Hector Esteban Paz-Juárez, Marissa Perez-Garcia, Francisco Pulido, Helen Haydee Fernanda Ramírez-Plascencia, Jorge Salas-Hernández, Angel Gustavo Salas-Lais, Alejandro Sánchez-Flores, Clara Esperanza Santacruz-Tinoco, María Guadalupe Santiago-Mauricio, Selene Zárate, Nelly Sélem-Mojica, Blanca Taboada, Gloria Vazquez, Joel Armando Vázquez-Pérez. |
| EPI_ISL_2533907 | Hospital Margarita Maza de Juárez | Microbial Genomics Laboratory | Consorcio Mexicano de Vigilancia Genómica (CoViGen-Mex). Authors (in alphabetical order): Julio Elias Alvarado-Yaah, Carlos F. Arias, Santiago Ávila-Ríos, Eduardo Becerril-Vargas, Víctor Hugo Borja-Aburto, Celia Boukadida, Cristóbal Cháidez-Quiróz, Juan Bautista Chale-Dzul, Ricardo Ciria Merce, Andreu Comas-García, Célida Duque Molina, Julissa Enciso-Ibarra, José Antonio Enciso-Moreno, Gloria Elena Espinosa-Ayala, Fernando Fontove-Herrera, Daniel Fregoso-Rueda, Victor Eduardo García-Arias, Alejandra García-Gasca, Bruno Gómez-Gil, Jean Pierre González, Irvin González-López, Concepción Grajales-Muñiz, Ricardo Grande, Rosa María Gutiérrez Rios, Jesús Hernández, Alejandra Hernández-Terán, Alfredo Herrera-Estrella, Carla Ivón Herrera-Najera, Pavel Isa, Verónica Mata-Haro, Daniel Lira Morales, Susana Lopez, Antonio Loza Román, Brenda Irasema Maldonado-Meza, Bernardo Martínez-Miguel, José Arturo Martínez-Orozco, Célida Martínez- Rodríguez, Margarita Matías-Florentino, Fidencio Mejía-Nepomuceno, María Guadalupe de Jesús Mireles-Rivera, Gloria María Molina-Salinas, Hector Montoya-Fuentes, , Mario Mújica-Sánchez, José Esteban Muñoz-Medina, José de Jesús Nuñez-Contreras, Alicia Ocaña-Mondragón, Luis Alberto Ochoa-Carrera, Hector Esteban Paz-Juárez, Marissa Perez-Garcia, Francisco Pulido, Helen Haydee Fernanda Ramírez-Plascencia, Jorge Salas-Hernández, Angel Gustavo Salas-Lais, Alejandro Sánchez-Flores, Clara Esperanza Santacruz-Tinoco, María Guadalupe Santiago-Mauricio, Selene Zárate, Nelly Sélem-Mojica, Blanca Taboada, Gloria Vazquez, Joel Armando Vázquez-Pérez. |
| EPI_ISL_2533908, EPI_ISL_2533909, EPI_ISL_2533910, EPI_ISL_2533911 | Hospital Sharp | Microbial Genomics Laboratory | Consorcio Mexicano de Vigilancia Genómica (CoViGen-Mex). Authors (in alphabetical order): Julio Elias Alvarado-Yaah, Carlos F. Arias, Santiago Ávila-Ríos, Eduardo Becerril-Vargas, Víctor Hugo Borja-Aburto, Celia Boukadida, Cristóbal Cháidez-Quiróz, Juan Bautista Chale-Dzul, Ricardo Ciria Merce, Andreu Comas-García, Célida Duque Molina, Julissa Enciso-Ibarra, José Antonio Enciso-Moreno, Gloria Elena Espinosa-Ayala, Fernando Fontove-Herrera, Daniel Fregoso-Rueda, Victor Eduardo García-Arias, Alejandra García-Gasca, Bruno Gómez-Gil, Jean Pierre González, Irvin González-López, Concepción Grajales-Muñiz, Ricardo Grande, Rosa María Gutiérrez Rios, Jesús Hernández, Alejandra Hernández-Terán, Alfredo Herrera-Estrella, Carla Ivón Herrera-Najera, Pavel Isa, Verónica Mata-Haro, Daniel Lira Morales, Susana Lopez, Antonio Loza Román, Brenda Irasema Maldonado-Meza, Bernardo Martínez-Miguel, José Arturo Martínez-Orozco, Célida Martínez- Rodríguez, Margarita Matías-Florentino, Fidencio Mejía-Nepomuceno, María Guadalupe de Jesús Mireles-Rivera, Gloria María Molina-Salinas, Hector Montoya-Fuentes, , Mario Mújica-Sánchez, José Esteban Muñoz-Medina, José de Jesús Nuñez-Contreras, Alicia Ocaña-Mondragón, Luis Alberto Ochoa-Carrera, Hector Esteban Paz-Juárez, Marissa Perez-Garcia, Francisco Pulido, Helen Haydee Fernanda Ramírez-Plascencia, Jorge Salas-Hernández, Angel Gustavo Salas-Lais, Alejandro Sánchez-Flores, Clara Esperanza Santacruz-Tinoco, María Guadalupe Santiago-Mauricio, Selene Zárate, Nelly Sélem-Mojica, Blanca Taboada, Gloria Vazquez, Joel Armando Vázquez-Pérez. |
| EPI_ISL_2533913 | Imágenes Diagnósticas de Mazatlán | Microbial Genomics Laboratory | Consorcio Mexicano de Vigilancia Genómica (CoViGen-Mex). Authors (in alphabetical order): Julio Elias Alvarado-Yaah, Carlos F. Arias, Santiago Ávila-Ríos, Eduardo Becerril-Vargas, Víctor Hugo Borja-Aburto, Celia Boukadida, Cristóbal Cháidez-Quiróz, Juan Bautista Chale-Dzul, Ricardo Ciria Merce, Andreu Comas-García, Célida Duque Molina, Julissa Enciso-Ibarra, José Antonio Enciso-Moreno, Gloria Elena Espinosa-Ayala, Fernando Fontove-Herrera, Daniel Fregoso-Rueda, Victor Eduardo García-Arias, Alejandra García-Gasca, Bruno Gómez-Gil, Jean Pierre González, Irvin González-López, Concepción Grajales-Muñiz, Ricardo Grande, Rosa María Gutiérrez Rios, Jesús Hernández, Alejandra Hernández-Terán, Alfredo Herrera-Estrella, Carla Ivón Herrera-Najera, Pavel Isa, Verónica Mata-Haro, Daniel Lira Morales, Susana Lopez, Antonio Loza Román, Brenda Irasema Maldonado-Meza, Bernardo Martínez-Miguel, José Arturo Martínez-Orozco, Célida Martínez- Rodríguez, Margarita Matías-Florentino, Fidencio Mejía-Nepomuceno, María Guadalupe de Jesús Mireles-Rivera, Gloria María Molina-Salinas, Hector Montoya-Fuentes, , Mario Mújica-Sánchez, José Esteban Muñoz-Medina, José de Jesús Nuñez-Contreras, Alicia Ocaña-Mondragón, Luis Alberto Ochoa-Carrera, Hector Esteban Paz-Juárez, Marissa Perez-Garcia, Francisco Pulido, Helen Haydee Fernanda Ramírez-Plascencia, Jorge Salas-Hernández, Angel Gustavo Salas-Lais, Alejandro Sánchez-Flores, Clara Esperanza Santacruz-Tinoco, María Guadalupe Santiago-Mauricio, Selene Zárate, Nelly Sélem-Mojica, Blanca Taboada, Gloria Vazquez, Joel Armando Vázquez-Pérez. |
| EPI_ISL_2533914, EPI_ISL_2533915, EPI_ISL_2533916 | Hospital Sharp | Microbial Genomics Laboratory | Consorcio Mexicano de Vigilancia Genómica (CoViGen-Mex). Authors (in alphabetical order): Julio Elias Alvarado-Yaah, Carlos F. Arias, Santiago Ávila-Ríos, Eduardo Becerril-Vargas, Víctor Hugo Borja-Aburto, Celia Boukadida, Cristóbal Cháidez-Quiróz, Juan Bautista Chale-Dzul, Ricardo Ciria Merce, Andreu Comas-García, Célida Duque Molina, Julissa Enciso-Ibarra, José Antonio Enciso-Moreno, Gloria Elena Espinosa-Ayala, Fernando Fontove-Herrera, Daniel Fregoso-Rueda, Victor Eduardo García-Arias, Alejandra García-Gasca, Bruno Gómez-Gil, Jean Pierre González, Irvin González-López, Concepción Grajales-Muñiz, Ricardo Grande, Rosa María Gutiérrez Rios, Jesús Hernández, Alejandra Hernández-Terán, Alfredo Herrera-Estrella, Carla Ivón Herrera-Najera, Pavel Isa, Verónica Mata-Haro, Daniel Lira Morales, Susana Lopez, Antonio Loza Román, Brenda Irasema Maldonado-Meza, Bernardo Martínez-Miguel, José Arturo Martínez-Orozco, Célida Martínez- Rodríguez, Margarita Matías-Florentino, Fidencio Mejía-Nepomuceno, María Guadalupe de Jesús Mireles-Rivera, Gloria María Molina-Salinas, Hector Montoya-Fuentes, , Mario Mújica-Sánchez, José Esteban Muñoz-Medina, José de Jesús Nuñez-Contreras, Alicia Ocaña-Mondragón, Luis Alberto Ochoa-Carrera, Hector Esteban Paz-Juárez, Marissa Perez-Garcia, Francisco Pulido, Helen Haydee Fernanda Ramírez-Plascencia, Jorge Salas-Hernández, Angel Gustavo Salas-Lais, Alejandro Sánchez-Flores, Clara Esperanza Santacruz-Tinoco, María Guadalupe Santiago-Mauricio, Selene Zárate, Nelly Sélem-Mojica, Blanca Taboada, Gloria Vazquez, Joel Armando Vázquez-Pérez. |
| EPI_ISL_2533917 | Hospital Margarita Maza de Juárez | Microbial Genomics Laboratory | Consorcio Mexicano de Vigilancia Genómica (CoViGen-Mex). Authors (in alphabetical order): Julio Elias Alvarado-Yaah, Carlos F. Arias, Santiago Ávila-Ríos, Eduardo Becerril-Vargas, Víctor Hugo Borja-Aburto, Celia Boukadida, Cristóbal Cháidez-Quiróz, Juan Bautista Chale-Dzul, Ricardo Ciria Merce, Andreu Comas-García, Célida Duque Molina, Julissa Enciso-Ibarra, José Antonio Enciso-Moreno, Gloria Elena Espinosa-Ayala, Fernando Fontove-Herrera, Daniel Fregoso-Rueda, Victor Eduardo García-Arias, Alejandra García-Gasca, Bruno Gómez-Gil, Jean Pierre González, Irvin González-López, Concepción Grajales-Muñiz, Ricardo Grande, Rosa María Gutiérrez Rios, Jesús Hernández, Alejandra Hernández-Terán, Alfredo Herrera-Estrella, Carla Ivón Herrera-Najera, Pavel Isa, Verónica Mata-Haro, Daniel Lira Morales, Susana Lopez, Antonio Loza Román, Brenda Irasema Maldonado-Meza, Bernardo Martínez-Miguel, José Arturo Martínez-Orozco, Célida Martínez- Rodríguez, Margarita Matías-Florentino, Fidencio Mejía-Nepomuceno, María Guadalupe de Jesús Mireles-Rivera, Gloria María Molina-Salinas, Hector Montoya-Fuentes, , Mario Mújica-Sánchez, José Esteban Muñoz-Medina, José de Jesús Nuñez-Contreras, Alicia Ocaña-Mondragón, Luis Alberto Ochoa-Carrera, Hector Esteban Paz-Juárez, Marissa Perez-Garcia, Francisco Pulido, Helen Haydee Fernanda Ramírez-Plascencia, Jorge Salas-Hernández, Angel Gustavo Salas-Lais, Alejandro Sánchez-Flores, Clara Esperanza Santacruz-Tinoco, María Guadalupe Santiago-Mauricio, Selene Zárate, Nelly Sélem-Mojica, Blanca Taboada, Gloria Vazquez, Joel Armando Vázquez-Pérez. |
| EPI_ISL_2533918 | Imágenes Diagnósticas de Mazatlán | Microbial Genomics Laboratory | Consorcio Mexicano de Vigilancia Genómica (CoViGen-Mex). Authors (in alphabetical order): Julio Elias Alvarado-Yaah, Carlos F. Arias, Santiago Ávila-Ríos, Eduardo Becerril-Vargas, Víctor Hugo Borja-Aburto, Celia Boukadida, Cristóbal Cháidez-Quiróz, Juan Bautista Chale-Dzul, Ricardo Ciria Merce, Andreu Comas-García, Célida Duque Molina, Julissa Enciso-Ibarra, José Antonio Enciso-Moreno, Gloria Elena Espinosa-Ayala, Fernando Fontove-Herrera, Daniel Fregoso-Rueda, Victor Eduardo García-Arias, Alejandra García-Gasca, Bruno Gómez-Gil, Jean Pierre González, Irvin González-López, Concepción Grajales-Muñiz, Ricardo Grande, Rosa María Gutiérrez Rios, Jesús Hernández, Alejandra Hernández-Terán, Alfredo Herrera-Estrella, Carla Ivón Herrera-Najera, Pavel Isa, Verónica Mata-Haro, Daniel Lira Morales, Susana Lopez, Antonio Loza Román, Brenda Irasema Maldonado-Meza, Bernardo Martínez-Miguel, José Arturo Martínez-Orozco, Célida Martínez- Rodríguez, Margarita Matías-Florentino, Fidencio Mejía-Nepomuceno, María Guadalupe de Jesús Mireles-Rivera, Gloria María Molina-Salinas, Hector Montoya-Fuentes, , Mario Mújica-Sánchez, José Esteban Muñoz-Medina, José de Jesús Nuñez-Contreras, Alicia Ocaña-Mondragón, Luis Alberto Ochoa-Carrera, Hector Esteban Paz-Juárez, Marissa Perez-Garcia, Francisco Pulido, Helen Haydee Fernanda Ramírez-Plascencia, Jorge Salas-Hernández, Angel Gustavo Salas-Lais, Alejandro Sánchez-Flores, Clara Esperanza Santacruz-Tinoco, María Guadalupe Santiago-Mauricio, Selene Zárate, Nelly Sélem-Mojica, Blanca Taboada, Gloria Vazquez, Joel Armando Vázquez-Pérez. |
| EPI_ISL_2533919, EPI_ISL_2533920, EPI_ISL_2533921, EPI_ISL_2533922, EPI_ISL_2533923, EPI_ISL_2533924 | Hospital Sharp | Microbial Genomics Laboratory | Consorcio Mexicano de Vigilancia Genómica (CoViGen-Mex). Authors (in alphabetical order): Julio Elias Alvarado-Yaah, Carlos F. Arias, Santiago Ávila-Ríos, Eduardo Becerril-Vargas, Víctor Hugo Borja-Aburto, Celia Boukadida, Cristóbal Cháidez-Quiróz, Juan Bautista Chale-Dzul, Ricardo Ciria Merce, Andreu Comas-García, Célida Duque Molina, Julissa Enciso-Ibarra, José Antonio Enciso-Moreno, Gloria Elena Espinosa-Ayala, Fernando Fontove-Herrera, Daniel Fregoso-Rueda, Victor Eduardo García-Arias, Alejandra García-Gasca, Bruno Gómez-Gil, Jean Pierre González, Irvin González-López, Concepción Grajales-Muñiz, Ricardo Grande, Rosa María Gutiérrez Rios, Jesús Hernández, Alejandra Hernández-Terán, Alfredo Herrera-Estrella, Carla Ivón Herrera-Najera, Pavel Isa, Verónica Mata-Haro, Daniel Lira Morales, Susana Lopez, Antonio Loza Román, Brenda Irasema Maldonado-Meza, Bernardo Martínez-Miguel, José Arturo Martínez-Orozco, Célida Martínez- Rodríguez, Margarita Matías-Florentino, Fidencio Mejía-Nepomuceno, María Guadalupe de Jesús Mireles-Rivera, Gloria María Molina-Salinas, Hector Montoya-Fuentes, , Mario Mújica-Sánchez, José Esteban Muñoz-Medina, José de Jesús Nuñez-Contreras, Alicia Ocaña-Mondragón, Luis Alberto Ochoa-Carrera, Hector Esteban Paz-Juárez, Marissa Perez-Garcia, Francisco Pulido, Helen Haydee Fernanda Ramírez-Plascencia, Jorge Salas-Hernández, Angel Gustavo Salas-Lais, Alejandro Sánchez-Flores, Clara Esperanza Santacruz-Tinoco, María Guadalupe Santiago-Mauricio, Selene Zárate, Nelly Sélem-Mojica, Blanca Taboada, Gloria Vazquez, Joel Armando Vázquez-Pérez. |
| EPI_ISL_2533925 | Imágenes Diagnósticas de Mazatlán | Microbial Genomics Laboratory | Consorcio Mexicano de Vigilancia Genómica (CoViGen-Mex). Authors (in alphabetical order): Julio Elias Alvarado-Yaah, Carlos F. Arias, Santiago Ávila-Ríos, Eduardo Becerril-Vargas, Víctor Hugo Borja-Aburto, Celia Boukadida, Cristóbal Cháidez-Quiróz, Juan Bautista Chale-Dzul, Ricardo Ciria Merce, Andreu Comas-García, Célida Duque Molina, Julissa Enciso-Ibarra, José Antonio Enciso-Moreno, Gloria Elena Espinosa-Ayala, Fernando Fontove-Herrera, Daniel Fregoso-Rueda, Victor Eduardo García-Arias, Alejandra García-Gasca, Bruno Gómez-Gil, Jean Pierre González, Irvin González-López, Concepción Grajales-Muñiz, Ricardo Grande, Rosa María Gutiérrez Rios, Jesús Hernández, Alejandra Hernández-Terán, Alfredo Herrera-Estrella, Carla Ivón Herrera-Najera, Pavel Isa, Verónica Mata-Haro, Daniel Lira Morales, Susana Lopez, Antonio Loza Román, Brenda Irasema Maldonado-Meza, Bernardo Martínez-Miguel, José Arturo Martínez-Orozco, Célida Martínez- Rodríguez, Margarita Matías-Florentino, Fidencio Mejía-Nepomuceno, María Guadalupe de Jesús Mireles-Rivera, Gloria María Molina-Salinas, Hector Montoya-Fuentes, , Mario Mújica-Sánchez, José Esteban Muñoz-Medina, José de Jesús Nuñez-Contreras, Alicia Ocaña-Mondragón, Luis Alberto Ochoa-Carrera, Hector Esteban Paz-Juárez, Marissa Perez-Garcia, Francisco Pulido, Helen Haydee Fernanda Ramírez-Plascencia, Jorge Salas-Hernández, Angel Gustavo Salas-Lais, Alejandro Sánchez-Flores, Clara Esperanza Santacruz-Tinoco, María Guadalupe Santiago-Mauricio, Selene Zárate, Nelly Sélem-Mojica, Blanca Taboada, Gloria Vazquez, Joel Armando Vázquez-Pérez. |
| EPI_ISL_2533926, EPI_ISL_2533927 | Hospital Sharp | Microbial Genomics Laboratory | Consorcio Mexicano de Vigilancia Genómica (CoViGen-Mex). Authors (in alphabetical order): Julio Elias Alvarado-Yaah, Carlos F. Arias, Santiago Ávila-Ríos, Eduardo Becerril-Vargas, Víctor Hugo Borja-Aburto, Celia Boukadida, Cristóbal Cháidez-Quiróz, Juan Bautista Chale-Dzul, Ricardo Ciria Merce, Andreu Comas-García, Célida Duque Molina, Julissa Enciso-Ibarra, José Antonio Enciso-Moreno, Gloria Elena Espinosa-Ayala, Fernando Fontove-Herrera, Daniel Fregoso-Rueda, Victor Eduardo García-Arias, Alejandra García-Gasca, Bruno Gómez-Gil, Jean Pierre González, Irvin González-López, Concepción Grajales-Muñiz, Ricardo Grande, Rosa María Gutiérrez Rios, Jesús Hernández, Alejandra Hernández-Terán, Alfredo Herrera-Estrella, Carla Ivón Herrera-Najera, Pavel Isa, Verónica Mata-Haro, Daniel Lira Morales, Susana Lopez, Antonio Loza Román, Brenda Irasema Maldonado-Meza, Bernardo Martínez-Miguel, José Arturo Martínez-Orozco, Célida Martínez- Rodríguez, Margarita Matías-Florentino, Fidencio Mejía-Nepomuceno, María Guadalupe de Jesús Mireles-Rivera, Gloria María Molina-Salinas, Hector Montoya-Fuentes, , Mario Mújica-Sánchez, José Esteban Muñoz-Medina, José de Jesús Nuñez-Contreras, Alicia Ocaña-Mondragón, Luis Alberto Ochoa-Carrera, Hector Esteban Paz-Juárez, Marissa Perez-Garcia, Francisco Pulido, Helen Haydee Fernanda Ramírez-Plascencia, Jorge Salas-Hernández, Angel Gustavo Salas-Lais, Alejandro Sánchez-Flores, Clara Esperanza Santacruz-Tinoco, María Guadalupe Santiago-Mauricio, Selene Zárate, Nelly Sélem-Mojica, Blanca Taboada, Gloria Vazquez, Joel Armando Vázquez-Pérez. |
| EPI_ISL_2533928, EPI_ISL_2533929 | Imágenes Diagnósticas de Mazatlán | Microbial Genomics Laboratory | Consorcio Mexicano de Vigilancia Genómica (CoViGen-Mex). Authors (in alphabetical order): Julio Elias Alvarado-Yaah, Carlos F. Arias, Santiago Ávila-Ríos, Eduardo Becerril-Vargas, Víctor Hugo Borja-Aburto, Celia Boukadida, Cristóbal Cháidez-Quiróz, Juan Bautista Chale-Dzul, Ricardo Ciria Merce, Andreu Comas-García, Célida Duque Molina, Julissa Enciso-Ibarra, José Antonio Enciso-Moreno, Gloria Elena Espinosa-Ayala, Fernando Fontove-Herrera, Daniel Fregoso-Rueda, Victor Eduardo García-Arias, Alejandra García-Gasca, Bruno Gómez-Gil, Jean Pierre González, Irvin González-López, Concepción Grajales-Muñiz, Ricardo Grande, Rosa María Gutiérrez Rios, Jesús Hernández, Alejandra Hernández-Terán, Alfredo Herrera-Estrella, Carla Ivón Herrera-Najera, Pavel Isa, Verónica Mata-Haro, Daniel Lira Morales, Susana Lopez, Antonio Loza Román, Brenda Irasema Maldonado-Meza, Bernardo Martínez-Miguel, José Arturo Martínez-Orozco, Célida Martínez- Rodríguez, Margarita Matías-Florentino, Fidencio Mejía-Nepomuceno, María Guadalupe de Jesús Mireles-Rivera, Gloria María Molina-Salinas, Hector Montoya-Fuentes, , Mario Mújica-Sánchez, José Esteban Muñoz-Medina, José de Jesús Nuñez-Contreras, Alicia Ocaña-Mondragón, Luis Alberto Ochoa-Carrera, Hector Esteban Paz-Juárez, Marissa Perez-Garcia, Francisco Pulido, Helen Haydee Fernanda Ramírez-Plascencia, Jorge Salas-Hernández, Angel Gustavo Salas-Lais, Alejandro Sánchez-Flores, Clara Esperanza Santacruz-Tinoco, María Guadalupe Santiago-Mauricio, Selene Zárate, Nelly Sélem-Mojica, Blanca Taboada, Gloria Vazquez, Joel Armando Vázquez-Pérez. |
| EPI_ISL_2533930, EPI_ISL_2533931, EPI_ISL_2533932 | CIAD-Hermosillo | Immunology Laboratory | Consorcio Mexicano de Vigilancia Genómica (CoViGen-Mex). Authors (in alphabetical order): Julio Elias Alvarado-Yaah, Carlos F. Arias, Santiago Ávila-Ríos, Eduardo Becerril-Vargas, Víctor Hugo Borja-Aburto, Celia Boukadida, Cristóbal Cháidez-Quiróz, Juan Bautista Chale-Dzul, Ricardo Ciria Merce, Andreu Comas-García, Célida Duque Molina, Julissa Enciso-Ibarra, José Antonio Enciso-Moreno, Gloria Elena Espinosa-Ayala, Fernando Fontove-Herrera, Daniel Fregoso-Rueda, Victor Eduardo García-Arias, Alejandra García-Gasca, Bruno Gómez-Gil, Jean Pierre González, Irvin González-López, Concepción Grajales-Muñiz, Ricardo Grande, Rosa María Gutiérrez Rios, Jesús Hernández, Alejandra Hernández-Terán, Alfredo Herrera-Estrella, Carla Ivón Herrera-Najera, Pavel Isa, Verónica Mata-Haro, Daniel Lira Morales, Susana Lopez, Antonio Loza Román, Brenda Irasema Maldonado-Meza, Bernardo Martínez-Miguel, José Arturo Martínez-Orozco, Célida Martínez- Rodríguez, Margarita Matías-Florentino, Fidencio Mejía-Nepomuceno, María Guadalupe de Jesús Mireles-Rivera, Gloria María Molina-Salinas, Hector Montoya-Fuentes, , Mario Mújica-Sánchez, José Esteban Muñoz-Medina, José de Jesús Nuñez-Contreras, Alicia Ocaña-Mondragón, Luis Alberto Ochoa-Carrera, Hector Esteban Paz-Juárez, Marissa Perez-Garcia, Francisco Pulido, Helen Haydee Fernanda Ramírez-Plascencia, Jorge Salas-Hernández, Angel Gustavo Salas-Lais, Alejandro Sánchez-Flores, Clara Esperanza Santacruz-Tinoco, María Guadalupe Santiago-Mauricio, Selene Zárate, Nelly Sélem-Mojica, Blanca Taboada, Gloria Vazquez, Joel Armando Vázquez-Pérez. |
| EPI_ISL_2533933, EPI_ISL_2533934, EPI_ISL_2533935 | Servicio Diagnóstico Clínico | Laboratorio Nacional-LANIIA-CIAD | Consorcio Mexicano de Vigilancia Genómica (CoViGen-Mex). Authors (in alphabetical order): Julio Elias Alvarado-Yaah, Carlos F. Arias, Santiago Ávila-Ríos, Eduardo Becerril-Vargas, Víctor Hugo Borja-Aburto, Celia Boukadida, Cristóbal Cháidez-Quiróz, Juan Bautista Chale-Dzul, Ricardo Ciria Merce, Andreu Comas-García, Célida Duque Molina, Julissa Enciso-Ibarra, José Antonio Enciso-Moreno, Gloria Elena Espinosa-Ayala, Fernando Fontove-Herrera, Daniel Fregoso-Rueda, Victor Eduardo García-Arias, Alejandra García-Gasca, Bruno Gómez-Gil, Jean Pierre González, Irvin González-López, Concepción Grajales-Muñiz, Ricardo Grande, Rosa María Gutiérrez Rios, Jesús Hernández, Alejandra Hernández-Terán, Alfredo Herrera-Estrella, Carla Ivón Herrera-Najera, Pavel Isa, Verónica Mata-Haro, Daniel Lira Morales, Susana Lopez, Antonio Loza Román, Brenda Irasema Maldonado-Meza, Bernardo Martínez-Miguel, José Arturo Martínez-Orozco, Célida Martínez- Rodríguez, Margarita Matías-Florentino, Fidencio Mejía-Nepomuceno, María Guadalupe de Jesús Mireles-Rivera, Gloria María Molina-Salinas, Hector Montoya-Fuentes, , Mario Mújica-Sánchez, José Esteban Muñoz-Medina, José de Jesús Nuñez-Contreras, Alicia Ocaña-Mondragón, Luis Alberto Ochoa-Carrera, Hector Esteban Paz-Juárez, Marissa Perez-Garcia, Francisco Pulido, Helen Haydee Fernanda Ramírez-Plascencia, Jorge Salas-Hernández, Angel Gustavo Salas-Lais, Alejandro Sánchez-Flores, Clara Esperanza Santacruz-Tinoco, María Guadalupe Santiago-Mauricio, Selene Zárate, Nelly Sélem-Mojica, Blanca Taboada, Gloria Vazquez, Joel Armando Vázquez-Pérez. |
| EPI_ISL_2533936 | Hospital Fátima S.A. de C.V. | Laboratorio Nacional-LANIIA-CIAD | Consorcio Mexicano de Vigilancia Genómica (CoViGen-Mex). Authors (in alphabetical order): Julio Elias Alvarado-Yaah, Carlos F. Arias, Santiago Ávila-Ríos, Eduardo Becerril-Vargas, Víctor Hugo Borja-Aburto, Celia Boukadida, Cristóbal Cháidez-Quiróz, Juan Bautista Chale-Dzul, Ricardo |

| | | | |
|---|---|---|---|
| | | | Ciria Merce, Andreu Comas-García, Célida Duque Molina, Julissa Enciso-Ibarra, José Antonio Enciso-Moreno, Gloria Elena Espinosa-Ayala, Fernando Fontove-Herrera, Daniel Fregoso-Rueda, Victor Eduardo García-Arias, Alejandra García-Gasca, Bruno Gómez-Gil, Jean Pierre González, Irvin González-López, Concepción Grajales-Muñiz, Ricardo Grande, Rosa María Gutiérrez Rios, Jesús Hernández, Alejandra Hernández-Terán, Alfredo Herrera-Estrella, Carla Ivón Herrera-Najera, Pavel Isa, Verónica Mata-Haro, Daniel Lira Morales, Susana Lopez, Antonio Loza Román, Brenda Irasema Maldonado-Meza, Bernardo Martínez-Miguel, José Arturo Martínez-Orozco, Célida Martínez- Rodríguez, Margarita Matías-Florentino, Fidencio Mejía-Nepomuceno, María Guadalupe de Jesús Mireles-Rivera, Gloria María Molina-Salinas, Hector Montoya-Fuentes, , Mario Mújica-Sánchez, José Esteban Muñoz-Medina, José de Jesús Nuñez-Contreras, Alicia Ocaña-Mondragón, Luis Alberto Ochoa-Carrera, Hector Esteban Paz-Juárez, Marissa Perez-Garcia, Francisco Pulido, Helen Haydee Fernanda Ramírez-Plascencia, Jorge Salas-Hernández, Angel Gustavo Salas-Lais, Alejandro Sánchez-Flores, Clara Esperanza Santacruz-Tinoco, María Guadalupe Santiago-Mauricio, Selene Zárate, Nelly Sélem-Mojica, Blanca Taboada, Gloria Vazquez, Joel Armando Vázquez-Pérez. |
| EPI_ISL_2533954, EPI_ISL_2533955, EPI_ISL_2533956, EPI_ISL_2533957, EPI_ISL_2533958, EPI_ISL_2533959, EPI_ISL_2533960, EPI_ISL_2533961, EPI_ISL_2533962, EPI_ISL_2533963, EPI_ISL_2533964, EPI_ISL_2533965, EPI_ISL_2533966, EPI_ISL_2533967, EPI_ISL_2533968, EPI_ISL_2533969, EPI_ISL_2533970, EPI_ISL_2533971, EPI_ISL_2533972, EPI_ISL_2533973, EPI_ISL_2533974, EPI_ISL_2533975, EPI_ISL_2533976, EPI_ISL_2533977, EPI_ISL_2533978, EPI_ISL_2533979, EPI_ISL_2533980, EPI_ISL_2533981, EPI_ISL_2533982, EPI_ISL_2533983, EPI_ISL_2533984, EPI_ISL_2533985, EPI_ISL_2533986, EPI_ISL_2533987, EPI_ISL_2533988, EPI_ISL_2533989, EPI_ISL_2533990, EPI_ISL_2533991, EPI_ISL_2533992, EPI_ISL_2533993, EPI_ISL_2533994, EPI_ISL_2533995, EPI_ISL_2533996, EPI_ISL_2533997, EPI_ISL_2533998, EPI_ISL_2533999, EPI_ISL_2534000, EPI_ISL_2534001, EPI_ISL_2534002, EPI_ISL_2534003, EPI_ISL_2534004, EPI_ISL_2534005, EPI_ISL_2534006, EPI_ISL_2534007, EPI_ISL_2534008, EPI_ISL_2534009, EPI_ISL_2534010, EPI_ISL_2534011, EPI_ISL_2534012, EPI_ISL_2534013, EPI_ISL_2534014, EPI_ISL_2534015, EPI_ISL_2534016, EPI_ISL_2534017, EPI_ISL_2534018, EPI_ISL_2534019, EPI_ISL_2534020, EPI_ISL_2534021, EPI_ISL_2534022 | | | |
| see above | Kaiser Permanente Washington Health Research Institute | Genomics and Discovery, Respiratory Viruses Branch, Division of Viral Diseases, Centers for Disease Control and Prevention | Yan Li, Jing Zhang, Anna Kelleher, Ying Tao, Brian Lynch, Krista Queen, Anna Uehara, Peter Cook, Han Jia Justin Ng, Rachel Marine, Clinton R. Paden, Dhwani Batra, Haibin Wang, Tara Coalter, Jasmine Padilla, Morgan Davis, Mili Sheth, Sarah Nobles, Mark Burroughs, Justin Lee, Adam Retchless, Suxiang Tong |
| EPI_ISL_2534083, EPI_ISL_2534086, EPI_ISL_2534087, EPI_ISL_2534088, EPI_ISL_2534089, EPI_ISL_2534090, EPI_ISL_2534091, EPI_ISL_2534092, EPI_ISL_2534093, EPI_ISL_2534094, EPI_ISL_2534095, EPI_ISL_2534096, EPI_ISL_2534097, EPI_ISL_2534098, EPI_ISL_2534102, EPI_ISL_2534103, EPI_ISL_2534104, EPI_ISL_2534105, EPI_ISL_2534107, EPI_ISL_2534108, EPI_ISL_2534109, EPI_ISL_2534110, EPI_ISL_2534112, EPI_ISL_2534113, EPI_ISL_2534114, EPI_ISL_2534115, EPI_ISL_2534116, EPI_ISL_2534117, EPI_ISL_2534118, EPI_ISL_2534119, EPI_ISL_2534120, EPI_ISL_2534121, EPI_ISL_2534122, EPI_ISL_2534123, EPI_ISL_2534124 | | | |
| see above | Maryland Genomics, Institute for Genome Sciences, University of Maryland School of Medicine | Maryland Genomics, Institute for Genome Sciences, University of Maryland School of Medicine | Tallon, Luke J; Sadzewicz, Lisa D; Humphrys, Mike; Ott, Sandra; Roussey, Holly; Santana-Cruz, Ivette; Vavikolanu, Kranthi; Lim, Jonathan;Hazen, Tracy;Fraser, Claire M; Ravel, Jacques |
| EPI_ISL_2534148, EPI_ISL_2534150, EPI_ISL_2534151, EPI_ISL_2534152, EPI_ISL_2534154, EPI_ISL_2534156, EPI_ISL_2534160, EPI_ISL_2534162, EPI_ISL_2534164, EPI_ISL_2534167, EPI_ISL_2534168, EPI_ISL_2534170, EPI_ISL_2534171, EPI_ISL_2534173, EPI_ISL_2534176, EPI_ISL_2534177, EPI_ISL_2534182, EPI_ISL_2534183, EPI_ISL_2534184, EPI_ISL_2534188, EPI_ISL_2534189, EPI_ISL_2534190, EPI_ISL_2534191, EPI_ISL_2534192, EPI_ISL_2534193, EPI_ISL_2534196, EPI_ISL_2534197, EPI_ISL_2534199, EPI_ISL_2534203 | | | |
| see above | INNO Diagnostics Reference Laboratory | RCMI-Center for Research Resources, Ponce Research Institute | Vanessa Rivera-Amill, Andrea Arias-García, Raphael Sánchez-Torres |
| EPI_ISL_2534219, EPI_ISL_2534220, EPI_ISL_2534221, EPI_ISL_2534222, EPI_ISL_2534223 | Wichita State University- Molecular Diagnostic Lab | Kansas Health and Environmental Lab | Katherine Wiggins, Mike Grose, Jonathan Barnell, Ben Olsen, and Phil Adam |
| EPI_ISL_2534224, EPI_ISL_2534225, EPI_ISL_2534226, EPI_ISL_2534228 | Cytocheck Laboratory | Kansas Health and Environmental Lab | Katherine Wiggins, Mike Grose, Jonathan Barnell, Ben Olsen, and Phil Adam |
| EPI_ISL_2534229, EPI_ISL_2534230, EPI_ISL_2534231, EPI_ISL_2534234, EPI_ISL_2534235 | Kansas Health and Environmental Lab | Kansas Health and Environmental Lab | Katherine Wiggins, Mike Grose, Jonathan Barnell, Ben Olsen, and Phil Adam |
| EPI_ISL_2534236, EPI_ISL_2534237 | Clinical Reference Laboratory | Kansas Health and Environmental Lab | Mike Grose, Katherine Wiggins, Jonathan Barnell, Ben Olsen, and Phil Adam |
| EPI_ISL_2534238, EPI_ISL_2534239, EPI_ISL_2534241, EPI_ISL_2534242, EPI_ISL_2534243, EPI_ISL_2534244, EPI_ISL_2534245, EPI_ISL_2534246, EPI_ISL_2534247, EPI_ISL_2534248, EPI_ISL_2534250 | | | |
| see above | Kansas Health and Environmental Lab | Kansas Health and Environmental Lab | Mike Grose, Katherine Wiggins, Jonathan Barnell, Ben Olsen, and Phil Adam |
| EPI_ISL_2534251, EPI_ISL_2534252, EPI_ISL_2534253, EPI_ISL_2534254, EPI_ISL_2534255, EPI_ISL_2534256, EPI_ISL_2534257, EPI_ISL_2534258, EPI_ISL_2534259, EPI_ISL_2534260, EPI_ISL_2534261, EPI_ISL_2534262, EPI_ISL_2534263 | | | |
| see above | Wichita State University - Molecular Diagnostics Lab | Kansas Health and Environmental Lab | Mike Grose, Katherine Wiggins, Jonathan Barnell, Ben Olsen, and Phil Adam |
| EPI_ISL_2534264, EPI_ISL_2534265, EPI_ISL_2534266, EPI_ISL_2534267, EPI_ISL_2534268, EPI_ISL_2534269, EPI_ISL_2534270, EPI_ISL_2534271, EPI_ISL_2534272, EPI_ISL_2534273, EPI_ISL_2534274, EPI_ISL_2534275, EPI_ISL_2534276, EPI_ISL_2534277, EPI_ISL_2534278, EPI_ISL_2534279, EPI_ISL_2534280, EPI_ISL_2534281, EPI_ISL_2534282, EPI_ISL_2534283, EPI_ISL_2534284, EPI_ISL_2534285, EPI_ISL_2534286, EPI_ISL_2534287, EPI_ISL_2534288, EPI_ISL_2534290, EPI_ISL_2534291, EPI_ISL_2534292, EPI_ISL_2534293, EPI_ISL_2534294, EPI_ISL_2534295, EPI_ISL_2534296, EPI_ISL_2534297, EPI_ISL_2534298, EPI_ISL_2534299, EPI_ISL_2534300, EPI_ISL_2534301, EPI_ISL_2534302, EPI_ISL_2534303, EPI_ISL_2534304, EPI_ISL_2534305, EPI_ISL_2534306, EPI_ISL_2534307, EPI_ISL_2534308 | | | |
| see above | Department of Pathology, The Medical College of Wisconsin, Inc. | Center for Infection and Immunity, Columbia University | Nischay Mishra, Mitali Mishra, James Ng, Komal Jain, W. Ian Lipkin, Blake W. Buchan |
| EPI_ISL_2534309, EPI_ISL_2534310, EPI_ISL_2534311, EPI_ISL_2534312, EPI_ISL_2534313, EPI_ISL_2534314, EPI_ISL_2534315, EPI_ISL_2534316 | The Ohio State University Applied Microbiology Services Laboratory | The Ohio State University Applied Microbiology Services Laboratory | Seth A. Faith PhD |
| EPI_ISL_2534318 | AR Dept. of Health-PHL, Molecular Diagnostics | Genomics and Discovery, Respiratory Viruses Branch, Division of Viral Diseases, Centers for Disease Control and Prevention | Yan Li, Jing Zhang, Anna Kelleher, Ying Tao, Brian Lynch, Krista Queen, Anna Uehara, Peter Cook, Han Jia Justin Ng, Rachel Marine, Clinton R. Paden, Dhwani Batra, Haibin Wang, Tara Coalter, Jasmine Padilla, Morgan Davis, Mili Sheth, Sarah Nobles, Mark Burroughs, Justin Lee, Adam Retchless, Suxiang Tong |
| EPI_ISL_2534319, EPI_ISL_2534322, EPI_ISL_2534323 | PA Department of Health, Bureau of Laboratories | Genomics and Discovery, Respiratory Viruses Branch, Division of Viral Diseases, Centers for Disease Control and Prevention | Yan Li, Jing Zhang, Anna Kelleher, Ying Tao, Brian Lynch, Krista Queen, Anna Uehara, Peter Cook, Han Jia Justin Ng, Rachel Marine, Clinton R. Paden, Dhwani Batra, Haibin Wang, Tara Coalter, Jasmine Padilla, Morgan Davis, Mili Sheth, Sarah Nobles, Mark Burroughs, Justin Lee, Adam Retchless, Suxiang Tong |
| EPI_ISL_2534329, EPI_ISL_2534330, EPI_ISL_2534331, EPI_ISL_2534332, EPI_ISL_2534333, EPI_ISL_2534334, EPI_ISL_2534335, EPI_ISL_2534336, EPI_ISL_2534337 | IVY3 Central Lab, Vanderbilt University Medical Center | Lauring Lab, University of Michigan, Department of Microbiology and Immunology | Valesano |
| EPI_ISL_2534342, EPI_ISL_2534343, EPI_ISL_2534344, EPI_ISL_2534345 | Maryland Genomics, Institute for Genome Sciences, University of Maryland School of Medicine | Maryland Genomics, Institute for Genome Sciences, University of Maryland School of Medicine | Tallon, Luke J; Sadzewicz, Lisa D; Humphrys, Mike; Ott, Sandra; Roussey, Holly; Santana-Cruz, Ivette; Vavikolanu, Kranthi; Lim, Jonathan;Hazen, Tracy;Fraser, Claire M; Ravel, Jacques |
| EPI_ISL_2534347, EPI_ISL_2534348, EPI_ISL_2534349, EPI_ISL_2534350, EPI_ISL_2534351, EPI_ISL_2534352, EPI_ISL_2534353, EPI_ISL_2534354, EPI_ISL_2534355, EPI_ISL_2534356, EPI_ISL_2534357, EPI_ISL_2534358, EPI_ISL_2534359, EPI_ISL_2534360, EPI_ISL_2534361, EPI_ISL_2534362, EPI_ISL_2534363, EPI_ISL_2534364, EPI_ISL_2534365, EPI_ISL_2534366, EPI_ISL_2534367, EPI_ISL_2534368, EPI_ISL_2534369, EPI_ISL_2534370, EPI_ISL_2534371, EPI_ISL_2534372, EPI_ISL_2534373, EPI_ISL_2534374, EPI_ISL_2534375 | | | |
| see above | Nebraska Public Health Laboratory | NPHL COVID-19 Response Team | NPHL COVID-19 Response Team |
| EPI_ISL_2534492, EPI_ISL_2534493, EPI_ISL_2534494, EPI_ISL_2534496, EPI_ISL_2534498, EPI_ISL_2534499, EPI_ISL_2534500, EPI_ISL_2534501, EPI_ISL_2534503, EPI_ISL_2534505, EPI_ISL_2534507, EPI_ISL_2534508, EPI_ISL_2534509, EPI_ISL_2534510, EPI_ISL_2534511, EPI_ISL_2534512, EPI_ISL_2534513, EPI_ISL_2534514, EPI_ISL_2534516, EPI_ISL_2534517 | | | |
| see above | Kansas Health and Environmental Lab | Kansas Health and Environmental Lab | Katherine Wiggins, Mike Grose, Jonathan Barnell, Ben Olsen, and Phil Adam |
| EPI_ISL_2534519, EPI_ISL_2534520, EPI_ISL_2534521, EPI_ISL_2534522, EPI_ISL_2534523, EPI_ISL_2534524, EPI_ISL_2534525, EPI_ISL_2534526, EPI_ISL_2534527, EPI_ISL_2534528, EPI_ISL_2534529, EPI_ISL_2534530, EPI_ISL_2534531, EPI_ISL_2534532 | | | |
| see above | Quest Diagnostics Incorporated | Centers for Disease Control and Prevention Division of Viral Diseases, Pathogen Discovery | Dakota Howard, Dhwani Batra, Peter W. Cook, Kara Moser, Adrian Paskey, Jason Caravas, Benjamin Rambo-Martin, Shatavia Morrison, Christopher Gulvick, Scott Sammons, Yvette Unoarumhi, Darlene Wagner, Matthew Schmerer, S. H. Rosenthal, A. Gerasimova, R. M. Kagan, B. Anderson, M. Hua, Y. Liu, L.E. Bernstein, K.E. Livingston, A. Perez, I. A. Shlyakhter, R. V. Rolando, R. Owen, P. Tanpaiboon, F. Lacbawan, Clinton R. Paden, Duncan MacCannell |
| EPI_ISL_2534533, EPI_ISL_2534534, EPI_ISL_2534535, EPI_ISL_2534536, EPI_ISL_2534537, EPI_ISL_2534538, EPI_ISL_2534539, EPI_ISL_2534540, EPI_ISL_2534541, EPI_ISL_2534543, EPI_ISL_2534544, EPI_ISL_2534545, EPI_ISL_2534546, EPI_ISL_2534547, EPI_ISL_2534548, EPI_ISL_2534549, EPI_ISL_2534550, EPI_ISL_2534551, EPI_ISL_2534552 | | | |
| see above | Aegis Sciences Corporation | Centers for Disease Control and Prevention Division of Viral Diseases, Pathogen Discovery | Dakota Howard, Dhwani Batra, Peter W. Cook, Kara Moser, Adrian Paskey, Jason Caravas, Benjamin Rambo-Martin, Shatavia Morrison, Christopher Gulvick, Scott Sammons, Yvette Unoarumhi, Darlene Wagner, Matthew Schmerer, Cyndi Clark, Patrick Campbell, Rob Case, Vikramsinha Ghorpade, Holly Houdeshell, Ola Kvalvaag, Dillon Nall, Ethan Sanders, Alec Vest, Shaun Westlund, Matthew Hardison, Clinton R. Paden, Duncan MacCannell |
| EPI_ISL_2534553, EPI_ISL_2534554, EPI_ISL_2534555, EPI_ISL_2534556, EPI_ISL_2534557, EPI_ISL_2534558, EPI_ISL_2534559, EPI_ISL_2534560, EPI_ISL_2534561, EPI_ISL_2534562, EPI_ISL_2534563, EPI_ISL_2534564, EPI_ISL_2534565 | | | |
| see above | Quest Diagnostics Incorporated | Centers for Disease Control and Prevention Division of Viral Diseases, Pathogen Discovery | Dakota Howard, Dhwani Batra, Peter W. Cook, Kara Moser, Adrian Paskey, Jason Caravas, Benjamin Rambo-Martin, Shatavia Morrison, Christopher Gulvick, Scott Sammons, Yvette Unoarumhi, Darlene Wagner, Matthew Schmerer, S. H. Rosenthal, A. Gerasimova, R. M. Kagan, B. Anderson, M. Hua, Y. Liu, L.E. Bernstein, K.E. Livingston, A. Perez, I. A. Shlyakhter, R. V. Rolando, R. Owen, P. Tanpaiboon, F. Lacbawan, Clinton R. Paden, Duncan MacCannell |
| EPI_ISL_2535839, EPI_ISL_2535840, EPI_ISL_2535841, EPI_ISL_2535842, EPI_ISL_2535843, EPI_ISL_2535844, EPI_ISL_2535845, EPI_ISL_2535846, EPI_ISL_2535847, EPI_ISL_2535848, EPI_ISL_2535849, EPI_ISL_2535850, EPI_ISL_2535851, EPI_ISL_2535852, EPI_ISL_2535853, EPI_ISL_2535854, EPI_ISL_2535855, EPI_ISL_2535856, EPI_ISL_2535857, EPI_ISL_2535858, EPI_ISL_2535859, EPI_ISL_2535860, EPI_ISL_2535861, EPI_ISL_2535862, EPI_ISL_2535863, EPI_ISL_2535864, EPI_ISL_2535865, EPI_ISL_2535867, EPI_ISL_2535868, EPI_ISL_2535869, EPI_ISL_2535870, EPI_ISL_2535871, EPI_ISL_2535872, EPI_ISL_2535873, EPI_ISL_2535874, EPI_ISL_2535875, EPI_ISL_2535876, EPI_ISL_2535877, EPI_ISL_2535878, EPI_ISL_2535879, EPI_ISL_2535880, EPI_ISL_2535881, EPI_ISL_2535882, EPI_ISL_2535883, EPI_ISL_2535884, EPI_ISL_2535885, EPI_ISL_2535886, EPI_ISL_2535887, EPI_ISL_2535888, EPI_ISL_2535889, EPI_ISL_2535890, EPI_ISL_2535891, EPI_ISL_2535892, EPI_ISL_2535893, EPI_ISL_2535894, EPI_ISL_2535895, EPI_ISL_2535896, EPI_ISL_2535897, EPI_ISL_2535898, EPI_ISL_2535899, EPI_ISL_2535900, EPI_ISL_2535901, EPI_ISL_2535902, EPI_ISL_2535903, EPI_ISL_2535904, EPI_ISL_2535905, EPI_ISL_2535906, EPI_ISL_2535907, EPI_ISL_2535909, EPI_ISL_2535910, EPI_ISL_2535911, EPI_ISL_2535912, EPI_ISL_2535913, EPI_ISL_2535914, EPI_ISL_2535915, EPI_ISL_2535916, EPI_ISL_2535917, EPI_ISL_2535918, EPI_ISL_2535919, EPI_ISL_2535920, EPI_ISL_2535921, EPI_ISL_2535922, EPI_ISL_2535923, EPI_ISL_2535924, EPI_ISL_2535925, EPI_ISL_2535926, EPI_ISL_2535927, EPI_ISL_2535928, EPI_ISL_2535929, EPI_ISL_2535930, EPI_ISL_2535931, EPI_ISL_2535932, EPI_ISL_2535933, EPI_ISL_2535934, EPI_ISL_2535935, EPI_ISL_2535936, EPI_ISL_2535937, EPI_ISL_2535938, EPI_ISL_2535939, EPI_ISL_2535940, EPI_ISL_2535941, EPI_ISL_2535942, EPI_ISL_2535943, EPI_ISL_2535944, EPI_ISL_2535945, EPI_ISL_2535946, EPI_ISL_2535947, EPI_ISL_2535948, EPI_ISL_2535949, EPI_ISL_2535950, EPI_ISL_2535951, EPI_ISL_2535952, EPI_ISL_2535953, EPI_ISL_2535954, EPI_ISL_2535955, EPI_ISL_2535956, EPI_ISL_2535957, EPI_ISL_2535958, EPI_ISL_2535959, EPI_ISL_2535960, EPI_ISL_2535961, EPI_ISL_2535962, EPI_ISL_2535963, EPI_ISL_2535964, EPI_ISL_2535965, EPI_ISL_2535966, EPI_ISL_2535967, EPI_ISL_2535968, EPI_ISL_2535969, EPI_ISL_2535970, EPI_ISL_2535972, EPI_ISL_2535973, | | | |

| | | | |
|---|---|---|---|
| EPI_ISL_2535974, EPI_ISL_2535975, EPI_ISL_2535976, EPI_ISL_2535977, EPI_ISL_2535978, EPI_ISL_2535979, EPI_ISL_2535980, EPI_ISL_2535981, EPI_ISL_2535982, EPI_ISL_2535983, EPI_ISL_2535984, EPI_ISL_2535985, EPI_ISL_2535986, EPI_ISL_2535987, EPI_ISL_2535988, EPI_ISL_2535989, EPI_ISL_2535990, EPI_ISL_2535991, EPI_ISL_2535992, EPI_ISL_2535993, EPI_ISL_2535994, EPI_ISL_2535995, EPI_ISL_2535996, EPI_ISL_2535997, EPI_ISL_2535998, EPI_ISL_2535999, EPI_ISL_2536000, EPI_ISL_2536001, EPI_ISL_2536002, EPI_ISL_2536003, EPI_ISL_2536004, EPI_ISL_2536005, EPI_ISL_2536006, EPI_ISL_2536007, EPI_ISL_2536008, EPI_ISL_2536010, EPI_ISL_2536011, EPI_ISL_2536012, EPI_ISL_2536013, EPI_ISL_2536014, EPI_ISL_2536015, EPI_ISL_2536016, EPI_ISL_2536017, EPI_ISL_2536018, EPI_ISL_2536019, EPI_ISL_2536020, EPI_ISL_2536021, EPI_ISL_2536022, EPI_ISL_2536023, EPI_ISL_2536024, EPI_ISL_2536025, EPI_ISL_2536026, EPI_ISL_2536027, EPI_ISL_2536028, EPI_ISL_2536030, EPI_ISL_2536031, EPI_ISL_2536032, EPI_ISL_2536033, EPI_ISL_2536034, EPI_ISL_2536035, EPI_ISL_2536036, EPI_ISL_2536037, EPI_ISL_2536038, EPI_ISL_2536039 | | | |
| see above | Gravity Diagnostics, LLC | Gravity Diagnostics, LLC | Gravity Diagnostics |
| EPI_ISL_2536161, EPI_ISL_2536163, EPI_ISL_2536164, EPI_ISL_2536167, EPI_ISL_2536169, EPI_ISL_2536170, EPI_ISL_2536171, EPI_ISL_2536174, EPI_ISL_2536176, EPI_ISL_2536177, EPI_ISL_2536178, EPI_ISL_2536179, EPI_ISL_2536180 | | | |
| see above | Wyoming Public Health Laboratory | Wyoming Public Health Laboratory | Jim Mildenberger, Taylor Fearing, Channing Weber, Ashley Norberg, Chayse Rowley, Marley Goetz, Brian Dominguez, Elliot Thomasson, Brittany Oher, Sam Britz, Cari Sloma, and Rob Christensen |
| EPI_ISL_2536182, EPI_ISL_2536183, EPI_ISL_2536184, EPI_ISL_2536185, EPI_ISL_2536186, EPI_ISL_2536187, EPI_ISL_2536188, EPI_ISL_2536189, EPI_ISL_2536190, EPI_ISL_2536191, EPI_ISL_2536192, EPI_ISL_2536193, EPI_ISL_2536194, EPI_ISL_2536195, EPI_ISL_2536196 | | | |
| see above | GA Department of Public Health | GA Department of Public Health | Stacy Reeves, Jonathan Edwards, Cynthia Dixey, Tonia Parrott, Aliyah Fields, Taylor Smith |
| EPI_ISL_2536197, EPI_ISL_2536198 | California Institute of Technology | Chan-Zuckerberg Biohub | CZB Cliahub Consortium |
| EPI_ISL_2536375, EPI_ISL_2536376, EPI_ISL_2536377, EPI_ISL_2536378, EPI_ISL_2536379, EPI_ISL_2536380, EPI_ISL_2536381, EPI_ISL_2536383, EPI_ISL_2536384, EPI_ISL_2536385, EPI_ISL_2536386, EPI_ISL_2536387, EPI_ISL_2536388, EPI_ISL_2536389, EPI_ISL_2536390, EPI_ISL_2536391, EPI_ISL_2536392, EPI_ISL_2536393, EPI_ISL_2536394, EPI_ISL_2536395, EPI_ISL_2536396, EPI_ISL_2536397, EPI_ISL_2536398, EPI_ISL_2536399, EPI_ISL_2536400, EPI_ISL_2536401, EPI_ISL_2536402, EPI_ISL_2536403, EPI_ISL_2536404, EPI_ISL_2536406, EPI_ISL_2536407, EPI_ISL_2536408, EPI_ISL_2536409, EPI_ISL_2536410, EPI_ISL_2536411, EPI_ISL_2536412, EPI_ISL_2536413, EPI_ISL_2536414, EPI_ISL_2536415, EPI_ISL_2536416, EPI_ISL_2536417, EPI_ISL_2536418, EPI_ISL_2536419, EPI_ISL_2536421, EPI_ISL_2536422, EPI_ISL_2536423, EPI_ISL_2536424, EPI_ISL_2536425, EPI_ISL_2536426, EPI_ISL_2536427, EPI_ISL_2536428, EPI_ISL_2536429, EPI_ISL_2536430, EPI_ISL_2536431, EPI_ISL_2536432, EPI_ISL_2536433, EPI_ISL_2536434, EPI_ISL_2536435, EPI_ISL_2536436, EPI_ISL_2536437, EPI_ISL_2536438, EPI_ISL_2536439, EPI_ISL_2536440, EPI_ISL_2536441, EPI_ISL_2536442, EPI_ISL_2536443, EPI_ISL_2536444, EPI_ISL_2536445, EPI_ISL_2536446, EPI_ISL_2536447, EPI_ISL_2536448, EPI_ISL_2536449, EPI_ISL_2536450, EPI_ISL_2536451, EPI_ISL_2536452, EPI_ISL_2536453, EPI_ISL_2536454, EPI_ISL_2536455, EPI_ISL_2536456, EPI_ISL_2536457, EPI_ISL_2536458, EPI_ISL_2536459, EPI_ISL_2536460, EPI_ISL_2536461, EPI_ISL_2536462, EPI_ISL_2536463, EPI_ISL_2536464, EPI_ISL_2536465, EPI_ISL_2536466, EPI_ISL_2536467, EPI_ISL_2536468, EPI_ISL_2536469, EPI_ISL_2536470, EPI_ISL_2536472, EPI_ISL_2536473, EPI_ISL_2536474, EPI_ISL_2536475, EPI_ISL_2536476, EPI_ISL_2536477, EPI_ISL_2536478, EPI_ISL_2536479, EPI_ISL_2536480, EPI_ISL_2536481, EPI_ISL_2536482, EPI_ISL_2536483, EPI_ISL_2536484, EPI_ISL_2536485, EPI_ISL_2536486, EPI_ISL_2536487, EPI_ISL_2536488, EPI_ISL_2536489, EPI_ISL_2536490, EPI_ISL_2536491, EPI_ISL_2536492, EPI_ISL_2536493, EPI_ISL_2536494, EPI_ISL_2536495, EPI_ISL_2536496, EPI_ISL_2536497, EPI_ISL_2536499, EPI_ISL_2536500, EPI_ISL_2536502, EPI_ISL_2536503, EPI_ISL_2536504, EPI_ISL_2536505, EPI_ISL_2536506, EPI_ISL_2536507 | | | |
| see above | Incyte Diagnostics Spokane | Seattle Flu Study | Deborah A. Nickerson, Chris D. Frazar, Jover Lee, Benjamin Pelle, Erica Ryke, Matthew Richardson, Amanda Adler, Elisabeth Brandstetter, Peter D. Han, Kairsten Fay, Misja Ilcisin, Kirsten Lacombe, Thomas R. Sibley, Melissa Truong, Caitlin R. Wolf, Romesh Gautom, Geoff Melly, Brian Hiatt, Philip Dykema, Scott Lindquist, Michael Boeckh, Janet A. Englund, Michael Famulare, Barry R. Lutz, Mark J. Rieder, Lea M. Starita, Matthew Thompson, Helen Y. Chu, Jay Shendure, Trevor Bedford |
| EPI_ISL_2536508, EPI_ISL_2536510, EPI_ISL_2536511, EPI_ISL_2536512, EPI_ISL_2536513, EPI_ISL_2536514, EPI_ISL_2536515, EPI_ISL_2536516, EPI_ISL_2536517, EPI_ISL_2536518, EPI_ISL_2536519, EPI_ISL_2536520, EPI_ISL_2536521, EPI_ISL_2536522, EPI_ISL_2536523, EPI_ISL_2536524, EPI_ISL_2536525, EPI_ISL_2536526, EPI_ISL_2536527, EPI_ISL_2536528, EPI_ISL_2536529, EPI_ISL_2536530, EPI_ISL_2536532, EPI_ISL_2536533, EPI_ISL_2536534, EPI_ISL_2536535, EPI_ISL_2536536, EPI_ISL_2536537, EPI_ISL_2536538, EPI_ISL_2536540, EPI_ISL_2536541 | | | |
| see above | Interpath Laboratory | Seattle Flu Study | Deborah A. Nickerson, Chris D. Frazar, Jover Lee, Benjamin Pelle, Erica Ryke, Matthew Richardson, Amanda Adler, Elisabeth Brandstetter, Peter D. Han, Kairsten Fay, Misja Ilcisin, Kirsten Lacombe, Thomas R. Sibley, Melissa Truong, Caitlin R. Wolf, Romesh Gautom, Geoff Melly, Brian Hiatt, Philip Dykema, Scott Lindquist, Michael Boeckh, Janet A. Englund, Michael Famulare, Barry R. Lutz, Mark J. Rieder, Lea M. Starita, Matthew Thompson, Helen Y. Chu, Jay Shendure, Trevor Bedford |
| EPI_ISL_2536542, EPI_ISL_2536543, EPI_ISL_2536544, EPI_ISL_2536545, EPI_ISL_2536550, EPI_ISL_2536551, EPI_ISL_2536552, EPI_ISL_2536553, EPI_ISL_2536554, EPI_ISL_2536555, EPI_ISL_2536557, EPI_ISL_2536558, EPI_ISL_2536559, EPI_ISL_2536560, EPI_ISL_2536561, EPI_ISL_2536562, EPI_ISL_2536563, EPI_ISL_2536564, EPI_ISL_2536565, EPI_ISL_2536566, EPI_ISL_2536567, EPI_ISL_2536568, EPI_ISL_2536569, EPI_ISL_2536570, EPI_ISL_2536571, EPI_ISL_2536572, EPI_ISL_2536573, EPI_ISL_2536574, EPI_ISL_2536576, EPI_ISL_2536577, EPI_ISL_2536578, EPI_ISL_2536579, EPI_ISL_2536580, EPI_ISL_2536581, EPI_ISL_2536582, EPI_ISL_2536583, EPI_ISL_2536584, EPI_ISL_2536585, EPI_ISL_2536586, EPI_ISL_2536587, EPI_ISL_2536588, EPI_ISL_2536589, EPI_ISL_2536590, EPI_ISL_2536591, EPI_ISL_2536592, EPI_ISL_2536593, EPI_ISL_2536594, EPI_ISL_2536595, EPI_ISL_2536597, EPI_ISL_2536598, EPI_ISL_2536599, EPI_ISL_2536600, EPI_ISL_2536601, EPI_ISL_2536602, EPI_ISL_2536603, EPI_ISL_2536604, EPI_ISL_2536605, EPI_ISL_2536606, EPI_ISL_2536607, EPI_ISL_2536608, EPI_ISL_2536609, EPI_ISL_2536610, EPI_ISL_2536611, EPI_ISL_2536612, EPI_ISL_2536613, EPI_ISL_2536614, EPI_ISL_2536616, EPI_ISL_2536617, EPI_ISL_2536618, EPI_ISL_2536619, EPI_ISL_2536620, EPI_ISL_2536621, EPI_ISL_2536622, EPI_ISL_2536623, EPI_ISL_2536624, EPI_ISL_2536625, EPI_ISL_2536626 | | | |
| see above | Northwest Laboratory | Seattle Flu Study | Deborah A. Nickerson, Chris D. Frazar, Jover Lee, Benjamin Pelle, Erica Ryke, Matthew Richardson, Amanda Adler, Elisabeth Brandstetter, Peter D. Han, Kairsten Fay, Misja Ilcisin, Kirsten Lacombe, Thomas R. Sibley, Melissa Truong, Caitlin R. Wolf, Romesh Gautom, Geoff Melly, Brian Hiatt, Philip Dykema, Scott Lindquist, Michael Boeckh, Janet A. Englund, Michael Famulare, Barry R. Lutz, Mark J. Rieder, Lea M. Starita, Matthew Thompson, Helen Y. Chu, Jay Shendure, Trevor Bedford |
| EPI_ISL_2536627, EPI_ISL_2536628, EPI_ISL_2536629, EPI_ISL_2536630, EPI_ISL_2536631, EPI_ISL_2536632, EPI_ISL_2536633, EPI_ISL_2536634, EPI_ISL_2536635, EPI_ISL_2536636, EPI_ISL_2536637, EPI_ISL_2536638, EPI_ISL_2536639, EPI_ISL_2536640, EPI_ISL_2536641, EPI_ISL_2536642, EPI_ISL_2536643, EPI_ISL_2536644, EPI_ISL_2536646, EPI_ISL_2536647, EPI_ISL_2536648 | | | |
| see above | Overlake Hospital | Seattle Flu Study | Deborah A. Nickerson, Chris D. Frazar, Jover Lee, Benjamin Pelle, Erica Ryke, Matthew Richardson, Amanda Adler, Elisabeth Brandstetter, Peter D. Han, Kairsten Fay, Misja Ilcisin, Kirsten Lacombe, Thomas R. Sibley, Melissa Truong, Caitlin R. Wolf, Romesh Gautom, Geoff Melly, Brian Hiatt, Philip Dykema, Scott Lindquist, Michael Boeckh, Janet A. Englund, Michael Famulare, Barry R. Lutz, Mark J. Rieder, Lea M. Starita, Matthew Thompson, Helen Y. Chu, Jay Shendure, Trevor Bedford |
| EPI_ISL_2536651, EPI_ISL_2536652, EPI_ISL_2536654, EPI_ISL_2536655, EPI_ISL_2536656, EPI_ISL_2536657, EPI_ISL_2536658, EPI_ISL_2536659, EPI_ISL_2536660, EPI_ISL_2536661, EPI_ISL_2536662, EPI_ISL_2536663, EPI_ISL_2536664, EPI_ISL_2536665, EPI_ISL_2536666, EPI_ISL_2536667, EPI_ISL_2536668, EPI_ISL_2536669, EPI_ISL_2536670, EPI_ISL_2536671, EPI_ISL_2536672, EPI_ISL_2536673, EPI_ISL_2536674, EPI_ISL_2536675, EPI_ISL_2536677, EPI_ISL_2536678, EPI_ISL_2536679, EPI_ISL_2536680, EPI_ISL_2536683, EPI_ISL_2536684, EPI_ISL_2536685, EPI_ISL_2536687, EPI_ISL_2536688, EPI_ISL_2536689, EPI_ISL_2536690, EPI_ISL_2536691, EPI_ISL_2536693, EPI_ISL_2536694, EPI_ISL_2536696, EPI_ISL_2536697, EPI_ISL_2536698, EPI_ISL_2536699, EPI_ISL_2536700 | | | |
| see above | Seattle Flu Study | Seattle Flu Study | Deborah A. Nickerson, Chris D. Frazar, Jover Lee, Benjamin Pelle, Erica Ryke, Matthew Richardson, Amanda Adler, Elisabeth Brandstetter, Peter D. Han, Kairsten Fay, Misja Ilcisin, Kirsten Lacombe, Thomas R. Sibley, Melissa Truong, Caitlin R. Wolf, Michael Boeckh, Janet A. Englund, Michael Famulare, Barry R. Lutz, Mark J. Rieder, Lea M. Starita, Matthew Thompson, Jay Shendure, Trevor Bedford, Helen Y. Chu |
| EPI_ISL_2536893, EPI_ISL_2536895, EPI_ISL_2536896, EPI_ISL_2536897, EPI_ISL_2536898, EPI_ISL_2536899, EPI_ISL_2536900, EPI_ISL_2536901, EPI_ISL_2536902, EPI_ISL_2536903, EPI_ISL_2536904 | | | |
| see above | Minnesota Department of Health, Public Health Laboratory | Minnesota Department of Health, Public Health Laboratory | Alexandra Lorentz, Jacob Garfin, Matt Plumb, and Xiong Wang |
| EPI_ISL_2536905, EPI_ISL_2536906 | Centracare Laboratory Services | Minnesota Department of Health, Public Health Laboratory | Alexandra Lorentz, Jacob Garfin, Matt Plumb, and Xiong Wang |
| EPI_ISL_2536907, EPI_ISL_2536908, EPI_ISL_2536909, EPI_ISL_2536910, EPI_ISL_2536911, EPI_ISL_2536912 | HealthPartners Central Lab | Minnesota Department of Health, Public Health Laboratory | Alexandra Lorentz, Jacob Garfin, Matt Plumb, and Xiong Wang |
| EPI_ISL_2536913, EPI_ISL_2536914, EPI_ISL_2536915, EPI_ISL_2536916, EPI_ISL_2536917, EPI_ISL_2536918, EPI_ISL_2536919, EPI_ISL_2536920, EPI_ISL_2536921, EPI_ISL_2536922, EPI_ISL_2536923, EPI_ISL_2536924, EPI_ISL_2536925, EPI_ISL_2536926, EPI_ISL_2536927, EPI_ISL_2536928, EPI_ISL_2536929, EPI_ISL_2536930, EPI_ISL_2536931, EPI_ISL_2536932, EPI_ISL_2536933, EPI_ISL_2536934, EPI_ISL_2536935, EPI_ISL_2536936, EPI_ISL_2536937, EPI_ISL_2536938, EPI_ISL_2536939, EPI_ISL_2536940, EPI_ISL_2536941, EPI_ISL_2536942, EPI_ISL_2536943, EPI_ISL_2536944, EPI_ISL_2536945, EPI_ISL_2536946, EPI_ISL_2536947, EPI_ISL_2536948, EPI_ISL_2536949, EPI_ISL_2536950, EPI_ISL_2536951, EPI_ISL_2536952, EPI_ISL_2536953 | | | |
| see above | Centracare Laboratory Services | Minnesota Department of Health, Public Health Laboratory | Alexandra Lorentz, Jacob Garfin, Matt Plumb, and Xiong Wang |
| EPI_ISL_2536955, EPI_ISL_2536956, EPI_ISL_2536957, EPI_ISL_2536958, EPI_ISL_2536959, EPI_ISL_2536960, EPI_ISL_2536961, EPI_ISL_2536962, EPI_ISL_2536963, EPI_ISL_2536964, EPI_ISL_2536965, EPI_ISL_2536966, EPI_ISL_2536967, EPI_ISL_2536968, EPI_ISL_2536969, EPI_ISL_2536970, EPI_ISL_2536971, EPI_ISL_2536972, EPI_ISL_2536973, EPI_ISL_2536974, EPI_ISL_2536975, EPI_ISL_2536976, EPI_ISL_2536977, EPI_ISL_2536978, EPI_ISL_2536979, EPI_ISL_2536980, EPI_ISL_2536981, EPI_ISL_2536982, EPI_ISL_2536983, EPI_ISL_2536984, EPI_ISL_2536985, EPI_ISL_2536986, EPI_ISL_2536987, EPI_ISL_2536988, EPI_ISL_2536989, EPI_ISL_2536990, EPI_ISL_2536991, EPI_ISL_2536992, EPI_ISL_2536993, EPI_ISL_2536994, EPI_ISL_2536995, EPI_ISL_2536997, EPI_ISL_2536998, EPI_ISL_2536999, EPI_ISL_2537000, EPI_ISL_2537001, EPI_ISL_2537002, EPI_ISL_2537003, EPI_ISL_2537004, EPI_ISL_2537005, EPI_ISL_2537007, EPI_ISL_2537009, EPI_ISL_2537010, EPI_ISL_2537011, EPI_ISL_2537012, EPI_ISL_2537013, EPI_ISL_2537014, EPI_ISL_2537015, EPI_ISL_2537016, EPI_ISL_2537017, EPI_ISL_2537018, EPI_ISL_2537019, EPI_ISL_2537020, EPI_ISL_2537021, EPI_ISL_2537022, EPI_ISL_2537023, EPI_ISL_2537024, EPI_ISL_2537025, EPI_ISL_2537026, EPI_ISL_2537027, EPI_ISL_2537028, EPI_ISL_2537029, EPI_ISL_2537030, EPI_ISL_2537031, EPI_ISL_2537032, EPI_ISL_2537033, EPI_ISL_2537034, EPI_ISL_2537035, EPI_ISL_2537036, EPI_ISL_2537037, EPI_ISL_2537038, EPI_ISL_2537039, EPI_ISL_2537040, EPI_ISL_2537041, EPI_ISL_2537042, EPI_ISL_2537043, EPI_ISL_2537044, EPI_ISL_2537045, EPI_ISL_2537046, EPI_ISL_2537047, EPI_ISL_2537048, EPI_ISL_2537049, EPI_ISL_2537050, EPI_ISL_2537051, EPI_ISL_2537052, EPI_ISL_2537053, EPI_ISL_2537054, EPI_ISL_2537055, EPI_ISL_2537056, EPI_ISL_2537057, EPI_ISL_2537058, EPI_ISL_2537059, EPI_ISL_2537060, EPI_ISL_2537061, EPI_ISL_2537062, EPI_ISL_2537063, EPI_ISL_2537064, EPI_ISL_2537065, EPI_ISL_2537066, EPI_ISL_2537067, EPI_ISL_2537068, EPI_ISL_2537069, EPI_ISL_2537070, EPI_ISL_2537071, EPI_ISL_2537072, EPI_ISL_2537073, EPI_ISL_2537074, EPI_ISL_2537075, EPI_ISL_2537076, EPI_ISL_2537077 | | | |
| see above | Arizona State Public Health Laboratory | Arizona State Public Health Laboratory | Trung Huynh, Jessica Escobar, Katherine Fullerton, Nobuko Fukushima, Stacy White, Linda Getsinger, Victor Waddell |
| EPI_ISL_2537079, EPI_ISL_2537080, EPI_ISL_2537081, EPI_ISL_2537082, EPI_ISL_2537083, EPI_ISL_2537084, EPI_ISL_2537085, EPI_ISL_2537086, EPI_ISL_2537087, EPI_ISL_2537088, EPI_ISL_2537089, EPI_ISL_2537090, EPI_ISL_2537091, EPI_ISL_2537092, EPI_ISL_2537093, EPI_ISL_2537094, EPI_ISL_2537095, EPI_ISL_2537096, EPI_ISL_2537097, EPI_ISL_2537098, EPI_ISL_2537099, EPI_ISL_2537100, EPI_ISL_2537101, EPI_ISL_2537102, EPI_ISL_2537103, EPI_ISL_2537104, EPI_ISL_2537105, EPI_ISL_2537106, EPI_ISL_2537107, EPI_ISL_2537108, EPI_ISL_2537109, EPI_ISL_2537110, EPI_ISL_2537112, EPI_ISL_2537113, EPI_ISL_2537114, EPI_ISL_2537115, EPI_ISL_2537116, EPI_ISL_2537117, EPI_ISL_2537118, EPI_ISL_2537119, EPI_ISL_2537120, EPI_ISL_2537121, EPI_ISL_2537122, EPI_ISL_2537123, EPI_ISL_2537124, EPI_ISL_2537125, EPI_ISL_2537126, EPI_ISL_2537127, EPI_ISL_2537128, EPI_ISL_2537129, EPI_ISL_2537130, EPI_ISL_2537131, EPI_ISL_2537132, EPI_ISL_2537133, EPI_ISL_2537134, EPI_ISL_2537135, EPI_ISL_2537136, EPI_ISL_2537137, EPI_ISL_2537138, EPI_ISL_2537139, EPI_ISL_2537140, EPI_ISL_2537141, EPI_ISL_2537142, EPI_ISL_2537143, EPI_ISL_2537144, EPI_ISL_2537145, EPI_ISL_2537146, EPI_ISL_2537147, EPI_ISL_2537148, EPI_ISL_2537149, EPI_ISL_2537150, EPI_ISL_2537151, EPI_ISL_2537152, EPI_ISL_2537153, EPI_ISL_2537154, EPI_ISL_2537155, EPI_ISL_2537156, EPI_ISL_2537157, EPI_ISL_2537158, EPI_ISL_2537159, EPI_ISL_2537160, EPI_ISL_2537161, EPI_ISL_2537162, EPI_ISL_2537163, EPI_ISL_2537164, EPI_ISL_2537166, EPI_ISL_2537167, EPI_ISL_2537169, EPI_ISL_2537170, EPI_ISL_2537171, EPI_ISL_2537172, EPI_ISL_2537174, EPI_ISL_2537175, EPI_ISL_2537176, EPI_ISL_2537177, EPI_ISL_2537178, EPI_ISL_2537179, EPI_ISL_2537180, EPI_ISL_2537183, EPI_ISL_2537184, EPI_ISL_2537185, EPI_ISL_2537186, EPI_ISL_2537187, EPI_ISL_2537188, EPI_ISL_2537189, EPI_ISL_2537190, EPI_ISL_2537191, EPI_ISL_2537192, EPI_ISL_2537193, EPI_ISL_2537194, EPI_ISL_2537195, EPI_ISL_2537196, EPI_ISL_2537197, EPI_ISL_2537198, EPI_ISL_2537199, EPI_ISL_2537200, EPI_ISL_2537201, EPI_ISL_2537202, EPI_ISL_2537203, EPI_ISL_2537204, EPI_ISL_2537205, EPI_ISL_2537206, EPI_ISL_2537207, EPI_ISL_2537208, EPI_ISL_2537209, EPI_ISL_2537210, EPI_ISL_2537211, EPI_ISL_2537212, EPI_ISL_2537213, EPI_ISL_2537214, EPI_ISL_2537215, EPI_ISL_2537217, EPI_ISL_2537218, EPI_ISL_2537219, EPI_ISL_2537220, EPI_ISL_2537221, EPI_ISL_2537222, EPI_ISL_2537223, EPI_ISL_2537224, EPI_ISL_2537225, EPI_ISL_2537226, EPI_ISL_2537227, EPI_ISL_2537228, EPI_ISL_2537229, EPI_ISL_2537230, EPI_ISL_2537231, EPI_ISL_2537232, EPI_ISL_2537233, EPI_ISL_2537234, EPI_ISL_2537235, EPI_ISL_2537236, EPI_ISL_2537237, EPI_ISL_2537238, EPI_ISL_2537239, EPI_ISL_2537240, EPI_ISL_2537241, EPI_ISL_2537242, EPI_ISL_2537244, EPI_ISL_2537245, EPI_ISL_2537246, EPI_ISL_2537247, EPI_ISL_2537248, EPI_ISL_2537249, EPI_ISL_2537250, EPI_ISL_2537251, EPI_ISL_2537252, EPI_ISL_2537253, EPI_ISL_2537254, EPI_ISL_2537255, EPI_ISL_2537256, EPI_ISL_2537257, EPI_ISL_2537258, EPI_ISL_2537259, EPI_ISL_2537260, EPI_ISL_2537261, EPI_ISL_2537262, EPI_ISL_2537263, EPI_ISL_2537264, EPI_ISL_2537265, EPI_ISL_2537266, EPI_ISL_2537267, EPI_ISL_2537268, EPI_ISL_2537269, EPI_ISL_2537270, EPI_ISL_2537271, EPI_ISL_2537273, EPI_ISL_2537274, EPI_ISL_2537275, EPI_ISL_2537276, EPI_ISL_2537277 | | | |
| see above | Gravity Diagnostics, LLC | Gravity Diagnostics, LLC | Gravity Diagnostics |
| EPI_ISL_2537355, EPI_ISL_2537356, EPI_ISL_2537357, EPI_ISL_2537358, EPI_ISL_2537359, EPI_ISL_2537360, EPI_ISL_2537361, EPI_ISL_2537362, EPI_ISL_2537363, EPI_ISL_2537364, EPI_ISL_2537365, EPI_ISL_2537366, EPI_ISL_2537367, EPI_ISL_2537368, EPI_ISL_2537369, EPI_ISL_2537370, EPI_ISL_2537371, EPI_ISL_2537372, EPI_ISL_2537373, EPI_ISL_2537374, EPI_ISL_2537375, EPI_ISL_2537376, EPI_ISL_2537377, EPI_ISL_2537378, EPI_ISL_2537379, EPI_ISL_2537380, EPI_ISL_2537381, EPI_ISL_2537382, EPI_ISL_2537383, EPI_ISL_2537384, EPI_ISL_2537385 | | | |
| see above | Montana Public Health Laboratory | Montana Public Health Laboratory | Joy Ritter, Michelle Mozer, Carrie Biskupiak, Deborah Gibson, Michael Dills |
| EPI_ISL_2537388, EPI_ISL_2537389, EPI_ISL_2537390, EPI_ISL_2537391, EPI_ISL_2537392, EPI_ISL_2537393, EPI_ISL_2537394, EPI_ISL_2537395, EPI_ISL_2537396, EPI_ISL_2537397, EPI_ISL_2537398, EPI_ISL_2537399, EPI_ISL_2537400, EPI_ISL_2537401, EPI_ISL_2537402, EPI_ISL_2537403, EPI_ISL_2537404, EPI_ISL_2537405, EPI_ISL_2537406, EPI_ISL_2537407, EPI_ISL_2537408, EPI_ISL_2537409, EPI_ISL_2537410, EPI_ISL_2537411, EPI_ISL_2537412, EPI_ISL_2537413, EPI_ISL_2537414, EPI_ISL_2537415, EPI_ISL_2537416, EPI_ISL_2537417, EPI_ISL_2537418, EPI_ISL_2537419, EPI_ISL_2537420, EPI_ISL_2537421, EPI_ISL_2537422, EPI_ISL_2537423, EPI_ISL_2537424, EPI_ISL_2537425, EPI_ISL_2537426, EPI_ISL_2537427, EPI_ISL_2537428, EPI_ISL_2537429, EPI_ISL_2537430, EPI_ISL_2537431, EPI_ISL_2537432, EPI_ISL_2537433, EPI_ISL_2537434, EPI_ISL_2537435, EPI_ISL_2537436, EPI_ISL_2537437, EPI_ISL_2537438, EPI_ISL_2537439, EPI_ISL_2537440, EPI_ISL_2537441, EPI_ISL_2537442, EPI_ISL_2537443, EPI_ISL_2537444, EPI_ISL_2537445, EPI_ISL_2537446, EPI_ISL_2537447, EPI_ISL_2537449, EPI_ISL_2537450, EPI_ISL_2537451, EPI_ISL_2537452, EPI_ISL_2537453, EPI_ISL_2537454, EPI_ISL_2537455, EPI_ISL_2537456, EPI_ISL_2537457, EPI_ISL_2537458, EPI_ISL_2537459, EPI_ISL_2537460, EPI_ISL_2537461, EPI_ISL_2537462, EPI_ISL_2537463, EPI_ISL_2537464, EPI_ISL_2537465, EPI_ISL_2537466, EPI_ISL_2537467, EPI_ISL_2537468, EPI_ISL_2537469, EPI_ISL_2537470, EPI_ISL_2537471, EPI_ISL_2537472, EPI_ISL_2537473, EPI_ISL_2537474, EPI_ISL_2537475, EPI_ISL_2537478, EPI_ISL_2537479 | | | |
| see above | California Department of Public Health | California Department of Public Health | CDPH IDLB COVIDNet et al |
| EPI_ISL_2537484 | Respiratory Viruses Branch, Centers for Disease Control and Prevention | Respiratory Viruses Branch, Centers for Disease Control and Prevention | Howard,D., Batra,D., Cook,P.W., Moser,K., Paskey,A., Caravas,J.,Rambo-Martin,B., Morrison,S., Gulvick,C., Sammons,S., Unoarumhi,Y.,Schmerer,M., Roychoudhury,P., Xie,H., Shrestha,L., Nguyen,T.V.,Bakhash,S.M., Lin,M., Baker,N.R., Perez,R., Ellis,S., Breit,N.,Livingston,R.J., Huang,M.-L., Jerome,K.R., Mathias,P.,Greninger,A., Paden,C.R. and MacCannell,D. |
| EPI_ISL_2543920, EPI_ISL_2544284, EPI_ISL_2544285, EPI_ISL_2544381, EPI_ISL_2544495, EPI_ISL_2544497, EPI_ISL_2544502, EPI_ISL_2544504, EPI_ISL_2544505, EPI_ISL_2544511, EPI_ISL_2544527, EPI_ISL_2544530, EPI_ISL_2544531 | | | |
| see above | Ohio Department of Health Laboratory | Ohio Department of Health Laboratory | Holmes, Jennifer; Eric Brandt, Keoni Omura, Glen McGillivary, Caitlin McDonnell, Jade Mowery, Stephanie Mccracken, Tyler Payne, Kirtana Ramadugu, Erica Leasure, Brent Lee, Kelsey Florek, Heather Blankenship, Allison Black, Quanta Brown, and Tammy Bannerman |
| EPI_ISL_2544533, EPI_ISL_2544534, EPI_ISL_2544535, EPI_ISL_2544537, EPI_ISL_2544538, EPI_ISL_2544539, EPI_ISL_2544541, EPI_ISL_2544542, EPI_ISL_2544543, EPI_ISL_2544544, EPI_ISL_2544545, EPI_ISL_2544546, EPI_ISL_2544548, EPI_ISL_2544549, EPI_ISL_2544551, EPI_ISL_2544552, EPI_ISL_2544554, EPI_ISL_2544555, EPI_ISL_2544556, EPI_ISL_2544559, EPI_ISL_2544560, EPI_ISL_2544561, EPI_ISL_2544562, EPI_ISL_2544563, EPI_ISL_2544564, EPI_ISL_2544567, EPI_ISL_2544569, EPI_ISL_2544570, EPI_ISL_2544572, EPI_ISL_2544573, EPI_ISL_2544574, EPI_ISL_2544577, EPI_ISL_2544578, EPI_ISL_2544579, EPI_ISL_2544580, EPI_ISL_2544581, EPI_ISL_2544582, EPI_ISL_2544583, EPI_ISL_2544584, EPI_ISL_2544585, EPI_ISL_2544586, EPI_ISL_2544587, EPI_ISL_2544588, EPI_ISL_2544589, EPI_ISL_2544590, EPI_ISL_2544592, EPI_ISL_2544593, EPI_ISL_2544594, EPI_ISL_2544595, EPI_ISL_2544596, EPI_ISL_2544597, EPI_ISL_2544598, EPI_ISL_2544599, EPI_ISL_2544600, EPI_ISL_2544601, EPI_ISL_2544602, EPI_ISL_2544603, EPI_ISL_2544604, EPI_ISL_2544605, EPI_ISL_2544606, EPI_ISL_2544607, EPI_ISL_2544608, EPI_ISL_2544609, EPI_ISL_2544610, EPI_ISL_2544611, EPI_ISL_2544612, EPI_ISL_2544613, EPI_ISL_2544614, EPI_ISL_2544615, EPI_ISL_2544616, EPI_ISL_2544617, EPI_ISL_2544618, EPI_ISL_2544619, EPI_ISL_2544620, EPI_ISL_2544621, EPI_ISL_2544623, EPI_ISL_2544624, EPI_ISL_2544625, EPI_ISL_2544626, EPI_ISL_2544627, EPI_ISL_2544628, EPI_ISL_2544629, EPI_ISL_2544630, EPI_ISL_2544631, EPI_ISL_2544632, EPI_ISL_2544633, EPI_ISL_2544635, EPI_ISL_2544636, EPI_ISL_2544637, EPI_ISL_2544638, EPI_ISL_2544640, EPI_ISL_2544642, EPI_ISL_2544643, EPI_ISL_2544644, EPI_ISL_2544646, EPI_ISL_2544647, EPI_ISL_2544649, EPI_ISL_2544650, EPI_ISL_2544651, EPI_ISL_2544652, EPI_ISL_2544653, EPI_ISL_2544654, EPI_ISL_2544655, EPI_ISL_2544656, EPI_ISL_2544657, EPI_ISL_2544658, EPI_ISL_2544659, EPI_ISL_2544660, EPI_ISL_2544661, EPI_ISL_2544662, EPI_ISL_2544663, EPI_ISL_2544664, EPI_ISL_2544665, EPI_ISL_2544666, EPI_ISL_2544667, EPI_ISL_2544669, EPI_ISL_2544670, EPI_ISL_2544671, EPI_ISL_2544674, EPI_ISL_2544676 | | | |
| see above | IN State Department of Health Laboratory Services | IN State Department of Health Laboratory Services | Cassandra Campion, Jamie Yeadon, Brian Pope, Lixia Liu, Kyle Brownlee, Melissa Hindenlang, Mark Glazier |
| EPI_ISL_2544677, EPI_ISL_2544680, EPI_ISL_2544699, EPI_ISL_2544742 | Ohio Department of Health Laboratory | Ohio Department of Health Laboratory | Holmes, Jennifer; Eric Brandt, Keoni Omura, Glen McGillivary, Caitlin McDonnell, Jade Mowery, Stephanie Mccracken, Tyler Payne, Kirtana Ramadugu, Erica Leasure, Brent Lee, Kelsey Florek, Heather Blankenship, Allison Black, Quanta Brown, and Tammy Bannerman |
| EPI_ISL_2544750, EPI_ISL_2544751 | Centracare Laboratory Services | Minnesota Department of Health, Public Health Laboratory | Alexandra Lorentz, Jacob Garfin, Matt Plumb, and Xiong Wang |

| | | | |
|---|---|---|---|
| EPI_ISL_2544759, EPI_ISL_2544760 | PA Department of Health, Bureau of Laboratories | Genomics and Discovery, Respiratory Viruses Branch, Division of Viral Diseases, Centers for Disease Control and Prevention | Ying Tao, Yan Li, Jing Zhang, Anna Kelleher, Brian Lynch, Krista Queen, Anna Uehara, Peter Cook, Han Jia Justin Ng, Rachel Marine, Clinton R. Paden, Dhwani Batra, Haibin Wang, Tara Coalter, Jasmine Padilla, Morgan Davis, Mili Sheth, Sarah Nobles, Mark Burroughs, Justin Lee, Adam Retchless, Suxiang Tong |
| EPI_ISL_2544761, EPI_ISL_2544762, EPI_ISL_2544763, EPI_ISL_2544764, EPI_ISL_2544765 | University of Michigan Clinical Microbiology Laboratory | Lauring Lab, University of Michigan, Department of Microbiology and Immunology | Gilbert |
| EPI_ISL_2544766, EPI_ISL_2544767, EPI_ISL_2544768, EPI_ISL_2544769, EPI_ISL_2544770, EPI_ISL_2544771, EPI_ISL_2544772, EPI_ISL_2544773, EPI_ISL_2544774, EPI_ISL_2544775, EPI_ISL_2544776, EPI_ISL_2544777, EPI_ISL_2544778, EPI_ISL_2544779, EPI_ISL_2544780, EPI_ISL_2544781, EPI_ISL_2544782, EPI_ISL_2544783, EPI_ISL_2544784, EPI_ISL_2544785, EPI_ISL_2544786, EPI_ISL_2544787, EPI_ISL_2544788, EPI_ISL_2544789, EPI_ISL_2544790, EPI_ISL_2544791, EPI_ISL_2544792, EPI_ISL_2544793, EPI_ISL_2544794, EPI_ISL_2544795, EPI_ISL_2544796, EPI_ISL_2544797, EPI_ISL_2544798, EPI_ISL_2544799 | | | |
| see above | MD PHL | MD PHL | Maryland Department of Health Laboratories Administration |
| EPI_ISL_2544913, EPI_ISL_2544914, EPI_ISL_2544915, EPI_ISL_2544916, EPI_ISL_2544917, EPI_ISL_2544918, EPI_ISL_2544919, EPI_ISL_2544920, EPI_ISL_2544921, EPI_ISL_2544922, EPI_ISL_2544923, EPI_ISL_2544925, EPI_ISL_2544926, EPI_ISL_2544927, EPI_ISL_2544928, EPI_ISL_2544929, EPI_ISL_2544930, EPI_ISL_2544931, EPI_ISL_2544932, EPI_ISL_2544933, EPI_ISL_2544934, EPI_ISL_2544935, EPI_ISL_2544936, EPI_ISL_2544937, EPI_ISL_2544938, EPI_ISL_2544940, EPI_ISL_2544941, EPI_ISL_2544942, EPI_ISL_2544943, EPI_ISL_2544944, EPI_ISL_2544945, EPI_ISL_2544946, EPI_ISL_2544947, EPI_ISL_2544948, EPI_ISL_2544949, EPI_ISL_2544950, EPI_ISL_2544951, EPI_ISL_2544952, EPI_ISL_2544953, EPI_ISL_2544955, EPI_ISL_2544956, EPI_ISL_2544957, EPI_ISL_2544958, EPI_ISL_2544959, EPI_ISL_2544960, EPI_ISL_2544961, EPI_ISL_2544962, EPI_ISL_2544963, EPI_ISL_2544964, EPI_ISL_2544965, EPI_ISL_2544966, EPI_ISL_2544967, EPI_ISL_2544968, EPI_ISL_2544969, EPI_ISL_2544970, EPI_ISL_2544971, EPI_ISL_2544972, EPI_ISL_2544973, EPI_ISL_2544974, EPI_ISL_2544975, EPI_ISL_2544976, EPI_ISL_2544977, EPI_ISL_2544978, EPI_ISL_2544979, EPI_ISL_2544980, EPI_ISL_2544981, EPI_ISL_2544982, EPI_ISL_2544983, EPI_ISL_2544984, EPI_ISL_2544985, EPI_ISL_2544986, EPI_ISL_2544987, EPI_ISL_2544988, EPI_ISL_2544990, EPI_ISL_2544991, EPI_ISL_2544992, EPI_ISL_2544993, EPI_ISL_2544994, EPI_ISL_2544995, EPI_ISL_2544996, EPI_ISL_2544997, EPI_ISL_2544998, EPI_ISL_2544999, EPI_ISL_2545000, EPI_ISL_2545001, EPI_ISL_2545002, EPI_ISL_2545003, EPI_ISL_2545004, EPI_ISL_2545005, EPI_ISL_2545006, EPI_ISL_2545007, EPI_ISL_2545008, EPI_ISL_2545009, EPI_ISL_2545010, EPI_ISL_2545011, EPI_ISL_2545012, EPI_ISL_2545014, EPI_ISL_2545015, EPI_ISL_2545016, EPI_ISL_2545017, EPI_ISL_2545018, EPI_ISL_2545019, EPI_ISL_2545020, EPI_ISL_2545021, EPI_ISL_2545022, EPI_ISL_2545023, EPI_ISL_2545024, EPI_ISL_2545025, EPI_ISL_2545026, EPI_ISL_2545027, EPI_ISL_2545028, EPI_ISL_2545029, EPI_ISL_2545030, EPI_ISL_2545031 | | | |
| see above | UW Virology Lab | UW Virology Lab | Pavitra Roychoudhury, Hong Xie, Lasata Shrestha, Shah Mohamed Bakhash, Tien V. Nguyen, Noah R. Baker, Sean Ellis, Meei-Li Huang, Keith R Jerome, Alexander Greninger |
| EPI_ISL_2545032 | University of Michigan Clinical Microbiology Laboratory | Lauring Lab, University of Michigan, Department of Microbiology and Immunology | Gilbert |
| EPI_ISL_2545034, EPI_ISL_2545035, EPI_ISL_2545036, EPI_ISL_2545037, EPI_ISL_2545038, EPI_ISL_2545039, EPI_ISL_2545040, EPI_ISL_2545041, EPI_ISL_2545042, EPI_ISL_2545043, EPI_ISL_2545044, EPI_ISL_2545045, EPI_ISL_2545046, EPI_ISL_2545047, EPI_ISL_2545048, EPI_ISL_2545049, EPI_ISL_2545050, EPI_ISL_2545051, EPI_ISL_2545052, EPI_ISL_2545053, EPI_ISL_2545054, EPI_ISL_2545055, EPI_ISL_2545056, EPI_ISL_2545057, EPI_ISL_2545058, EPI_ISL_2545059, EPI_ISL_2545060, EPI_ISL_2545062, EPI_ISL_2545063, EPI_ISL_2545064, EPI_ISL_2545065, EPI_ISL_2545067, EPI_ISL_2545068, EPI_ISL_2545069, EPI_ISL_2545070, EPI_ISL_2545071, EPI_ISL_2545072, EPI_ISL_2545073, EPI_ISL_2545075, EPI_ISL_2545076, EPI_ISL_2545077, EPI_ISL_2545078, EPI_ISL_2545079, EPI_ISL_2545080, EPI_ISL_2545081, EPI_ISL_2545082, EPI_ISL_2545083, EPI_ISL_2545084, EPI_ISL_2545085, EPI_ISL_2545086, EPI_ISL_2545087, EPI_ISL_2545088, EPI_ISL_2545089, EPI_ISL_2545090, EPI_ISL_2545091, EPI_ISL_2545092, EPI_ISL_2545093, EPI_ISL_2545094, EPI_ISL_2545095, EPI_ISL_2545096, EPI_ISL_2545097, EPI_ISL_2545098, EPI_ISL_2545099, EPI_ISL_2545100, EPI_ISL_2545101, EPI_ISL_2545102, EPI_ISL_2545103, EPI_ISL_2545104, EPI_ISL_2545105, EPI_ISL_2545106, EPI_ISL_2545107, EPI_ISL_2545108, EPI_ISL_2545109, EPI_ISL_2545111, EPI_ISL_2545112, EPI_ISL_2545113, EPI_ISL_2545114, EPI_ISL_2545115, EPI_ISL_2545117, EPI_ISL_2545118, EPI_ISL_2545119, EPI_ISL_2545120, EPI_ISL_2545121, EPI_ISL_2545122, EPI_ISL_2545123, EPI_ISL_2545125, EPI_ISL_2545126, EPI_ISL_2545127, EPI_ISL_2545128, EPI_ISL_2545129, EPI_ISL_2545130, EPI_ISL_2545131, EPI_ISL_2545132, EPI_ISL_2545133, EPI_ISL_2545134, EPI_ISL_2545135, EPI_ISL_2545137, EPI_ISL_2545138, EPI_ISL_2545139, EPI_ISL_2545140, EPI_ISL_2545141, EPI_ISL_2545142, EPI_ISL_2545143, EPI_ISL_2545144, EPI_ISL_2545145, EPI_ISL_2545146, EPI_ISL_2545148, EPI_ISL_2545149, EPI_ISL_2545150, EPI_ISL_2545151, EPI_ISL_2545152, EPI_ISL_2545153, EPI_ISL_2545154, EPI_ISL_2545155, EPI_ISL_2545156, EPI_ISL_2545157, EPI_ISL_2545158, EPI_ISL_2545159, EPI_ISL_2545160, EPI_ISL_2545161, EPI_ISL_2545162, EPI_ISL_2545163, EPI_ISL_2545164, EPI_ISL_2545165, EPI_ISL_2545166, EPI_ISL_2545167, EPI_ISL_2545168, EPI_ISL_2545169, EPI_ISL_2545170, EPI_ISL_2545171, EPI_ISL_2545172, EPI_ISL_2545173, EPI_ISL_2545174, EPI_ISL_2545175, EPI_ISL_2545176, EPI_ISL_2545177, EPI_ISL_2545178, EPI_ISL_2545179, EPI_ISL_2545180, EPI_ISL_2545181, EPI_ISL_2545182, EPI_ISL_2545183, EPI_ISL_2545184 | | | |
| see above | UW Virology Lab | UW Virology Lab | Pavitra Roychoudhury, Hong Xie, Lasata Shrestha, Shah Mohamed Bakhash, Tien V. Nguyen, Noah R. Baker, Sean Ellis, Meei-Li Huang, Keith R Jerome, Alexander Greninger |
| EPI_ISL_2545422, EPI_ISL_2545423, EPI_ISL_2545424, EPI_ISL_2545427, EPI_ISL_2545431, EPI_ISL_2545432, EPI_ISL_2545433, EPI_ISL_2545434 | Kansas Health and Environmental Lab | Kansas Health and Environmental Lab | Katherine Wiggins, Mike Grose, Jonathan Barnell, Ben Olsen, and Phil Adam |
| EPI_ISL_2545585 | GA Department of Public Health Laboratory | Genomics and Discovery, Respiratory Viruses Branch, Division of Viral Diseases, Centers for Disease Control and Prevention | Yan Li, Jing Zhang, Anna Kelleher, Ying Tao, Brian Lynch, Krista Queen, Anna Uehara, Peter Cook, Han Jia Justin Ng, Rachel Marine, Clinton R. Paden, Dhwani Batra, Haibin Wang, Tara Coalter, Jasmine Padilla, Morgan Davis, Mili Sheth, Sarah Nobles, Mark Burroughs, Justin Lee, Adam Retchless, Suxiang Tong |
| EPI_ISL_2545586 | PA Department of Health, Bureau of Laboratories | Genomics and Discovery, Respiratory Viruses Branch, Division of Viral Diseases, Centers for Disease Control and Prevention | Yan Li, Jing Zhang, Anna Kelleher, Ying Tao, Brian Lynch, Krista Queen, Anna Uehara, Peter Cook, Han Jia Justin Ng, Rachel Marine, Clinton R. Paden, Dhwani Batra, Haibin Wang, Tara Coalter, Jasmine Padilla, Morgan Davis, Mili Sheth, Sarah Nobles, Mark Burroughs, Justin Lee, Adam Retchless, Suxiang Tong |
| EPI_ISL_2545667 | Tripler Army Medical Center | Tripler Army Medical Center | Rachael L. Downham, Jonathan A. D'Ambrozio, Kendra N. Carter, Lindsey L. O'Neal, Zakariae Bram, and Catherine F. Uyehara |
| EPI_ISL_2545668, EPI_ISL_2545669, EPI_ISL_2545670, EPI_ISL_2545671, EPI_ISL_2545672, EPI_ISL_2545673, EPI_ISL_2545674, EPI_ISL_2545675, EPI_ISL_2545676, EPI_ISL_2545677, EPI_ISL_2545678, EPI_ISL_2545679 | | | |
| see above | Kaiser Permanente Washington Health Research Institute | Genomics and Discovery, Respiratory Viruses Branch, Division of Viral Diseases, Centers for Disease Control and Prevention | Yan Li, Jing Zhang, Anna Kelleher, Ying Tao, Brian Lynch, Krista Queen, Anna Uehara, Peter Cook, Han Jia Justin Ng, Rachel Marine, Clinton R. Paden, Dhwani Batra, Haibin Wang, Tara Coalter, Jasmine Padilla, Morgan Davis, Mili Sheth, Sarah Nobles, Mark Burroughs, Justin Lee, Adam Retchless, Suxiang Tong |
| EPI_ISL_2545680, EPI_ISL_2545681 | LESP Tamaulipas | Instituto de Diagnostico y Referencia Epidemiologicos (INDRE) | Claudia Wong-Arambula, Abril Rodriguez-Maldonado, Vanessa Rivero-Arredondo, Ariadna Medina-Benitez, Joaquin Quiroz-Mercado, Sergio Rangel-Guerrero, Natividad Cruz-Ortiz, Tatiana Nunez-Garcia, Gisela Barrera-Badillo, Lucia Hernandez-Rivas, Irma Lopez-Martinez, Ernesto Ramirez-Gonzalez. |
| EPI_ISL_2545682 | LESP Queretaro | Instituto de Diagnostico y Referencia Epidemiologicos (INDRE) | Claudia Wong-Arambula, Abril Rodriguez-Maldonado, Vanessa Rivero-Arredondo, Ariadna Medina-Benitez, Joaquin Quiroz-Mercado, Sergio Rangel-Guerrero, Natividad Cruz-Ortiz, Tatiana Nunez-Garcia, Gisela Barrera-Badillo, Lucia Hernandez-Rivas, Irma Lopez-Martinez, Ernesto Ramirez-Gonzalez. |
| EPI_ISL_2545683 | LESP Jalisco | Instituto de Diagnostico y Referencia Epidemiologicos (INDRE) | Claudia Wong-Arambula, Abril Rodriguez-Maldonado, Vanessa Rivero-Arredondo, Ariadna Medina-Benitez, Joaquin Quiroz-Mercado, Sergio Rangel-Guerrero, Natividad Cruz-Ortiz, Tatiana Nunez-Garcia, Gisela Barrera-Badillo, Lucia Hernandez-Rivas, Irma Lopez-Martinez, Ernesto Ramirez-Gonzalez. |
| EPI_ISL_2545684, EPI_ISL_2545685, EPI_ISL_2545686, EPI_ISL_2545687, EPI_ISL_2545688 | LESP Yucatan | Instituto de Diagnostico y Referencia Epidemiologicos (INDRE) | Claudia Wong-Arambula, Abril Rodriguez-Maldonado, Vanessa Rivero-Arredondo, Ariadna Medina-Benitez, Joaquin Quiroz-Mercado, Sergio Rangel-Guerrero, Natividad Cruz-Ortiz, Tatiana Nunez-Garcia, Gisela Barrera-Badillo, Lucia Hernandez-Rivas, Irma Lopez-Martinez, Ernesto Ramirez-Gonzalez. |
| EPI_ISL_2545689, EPI_ISL_2545690, EPI_ISL_2545691 | LESP Tamaulipas | Instituto de Diagnostico y Referencia Epidemiologicos (INDRE) | Claudia Wong-Arambula, Abril Rodriguez-Maldonado, Vanessa Rivero-Arredondo, Ariadna Medina-Benitez, Joaquin Quiroz-Mercado, Sergio Rangel-Guerrero, Natividad Cruz-Ortiz, Tatiana Nunez-Garcia, Gisela Barrera-Badillo, Lucia Hernandez-Rivas, Irma Lopez-Martinez, Ernesto Ramirez-Gonzalez. |
| EPI_ISL_2545692 | LESP Sinaloa | Instituto de Diagnostico y Referencia Epidemiologicos (INDRE) | Claudia Wong-Arambula, Abril Rodriguez-Maldonado, Vanessa Rivero-Arredondo, Ariadna Medina-Benitez, Joaquin Quiroz-Mercado, Sergio Rangel-Guerrero, Natividad Cruz-Ortiz, Tatiana Nunez-Garcia, Gisela Barrera-Badillo, Lucia Hernandez-Rivas, Irma Lopez-Martinez, Ernesto Ramirez-Gonzalez. |
| EPI_ISL_2545693, EPI_ISL_2545694 | LESP Quintana Roo | Instituto de Diagnostico y Referencia Epidemiologicos (INDRE) | Claudia Wong-Arambula, Abril Rodriguez-Maldonado, Vanessa Rivero-Arredondo, Ariadna Medina-Benitez, Joaquin Quiroz-Mercado, Sergio Rangel-Guerrero, Natividad Cruz-Ortiz, Tatiana Nunez-Garcia, Gisela Barrera-Badillo, Lucia Hernandez-Rivas, Irma Lopez-Martinez, Ernesto Ramirez-Gonzalez. |
| EPI_ISL_2545695, EPI_ISL_2545696, EPI_ISL_2545697 | LESP Ciudad de Mexico | Instituto de Diagnostico y Referencia Epidemiologicos (INDRE) | Claudia Wong-Arambula, Abril Rodriguez-Maldonado, Vanessa Rivero-Arredondo, Ariadna Medina-Benitez, Joaquin Quiroz-Mercado, Sergio Rangel-Guerrero, Natividad Cruz-Ortiz, Tatiana Nunez-Garcia, Gisela Barrera-Badillo, Lucia Hernandez-Rivas, Irma Lopez-Martinez, Ernesto Ramirez-Gonzalez. |
| EPI_ISL_2545698 | LESP Estado de Mexico | Instituto de Diagnostico y Referencia Epidemiologicos (INDRE) | Claudia Wong-Arambula, Abril Rodriguez-Maldonado, Vanessa Rivero-Arredondo, Ariadna Medina-Benitez, Joaquin Quiroz-Mercado, Sergio Rangel-Guerrero, Natividad Cruz-Ortiz, Tatiana Nunez-Garcia, Gisela Barrera-Badillo, Lucia Hernandez-Rivas, Irma Lopez-Martinez, Ernesto Ramirez-Gonzalez. |
| EPI_ISL_2545699 | LESP Campeche | Instituto de Diagnostico y Referencia Epidemiologicos (INDRE) | Claudia Wong-Arambula, Abril Rodriguez-Maldonado, Vanessa Rivero-Arredondo, Ariadna Medina-Benitez, Joaquin Quiroz-Mercado, Sergio Rangel-Guerrero, Natividad Cruz-Ortiz, Tatiana Nunez-Garcia, Gisela Barrera-Badillo, Lucia Hernandez-Rivas, Irma Lopez-Martinez, Ernesto Ramirez-Gonzalez. |
| EPI_ISL_2545700, EPI_ISL_2545701 | LESP Queretaro | Instituto de Diagnostico y Referencia Epidemiologicos (INDRE) | Claudia Wong-Arambula, Abril Rodriguez-Maldonado, Vanessa Rivero-Arredondo, Ariadna Medina-Benitez, Joaquin Quiroz-Mercado, Sergio Rangel-Guerrero, Natividad Cruz-Ortiz, Tatiana Nunez-Garcia, Gisela Barrera-Badillo, Lucia Hernandez-Rivas, Irma Lopez-Martinez, Ernesto Ramirez-Gonzalez. |
| EPI_ISL_2545702 | LESP Michoacan | Instituto de Diagnostico y Referencia Epidemiologicos (INDRE) | Claudia Wong-Arambula, Abril Rodriguez-Maldonado, Vanessa Rivero-Arredondo, Ariadna Medina-Benitez, Joaquin Quiroz-Mercado, Sergio Rangel-Guerrero, Natividad Cruz-Ortiz, Tatiana Nunez-Garcia, Gisela Barrera-Badillo, Lucia Hernandez-Rivas, Irma Lopez-Martinez, Ernesto Ramirez-Gonzalez. |
| EPI_ISL_2545703, EPI_ISL_2545704 | LESP Sinaloa | Instituto de Diagnostico y Referencia Epidemiologicos (INDRE) | Claudia Wong-Arambula, Abril Rodriguez-Maldonado, Vanessa Rivero-Arredondo, Ariadna Medina-Benitez, Joaquin Quiroz-Mercado, Sergio Rangel-Guerrero, Natividad Cruz-Ortiz, Tatiana Nunez-Garcia, Gisela Barrera-Badillo, Lucia Hernandez-Rivas, Irma Lopez-Martinez, Ernesto Ramirez-Gonzalez. |
| EPI_ISL_2545706 | LESP Jalisco | Instituto de Diagnostico y Referencia Epidemiologicos (INDRE) | Claudia Wong-Arambula, Abril Rodriguez-Maldonado, Vanessa Rivero-Arredondo, Ariadna Medina-Benitez, Joaquin Quiroz-Mercado, Sergio Rangel-Guerrero, Natividad Cruz-Ortiz, Tatiana Nunez-Garcia, Gisela Barrera-Badillo, Lucia Hernandez-Rivas, Irma Lopez-Martinez, Ernesto Ramirez-Gonzalez. |
| EPI_ISL_2545707, EPI_ISL_2545708, EPI_ISL_2545709 | LESP Chihuahua | Instituto de Diagnostico y Referencia Epidemiologicos (INDRE) | Claudia Wong-Arambula, Abril Rodriguez-Maldonado, Vanessa Rivero-Arredondo, Ariadna Medina-Benitez, Joaquin Quiroz-Mercado, Sergio Rangel-Guerrero, Natividad Cruz-Ortiz, Tatiana Nunez-Garcia, Gisela Barrera-Badillo, Lucia Hernandez-Rivas, Irma Lopez-Martinez, Ernesto Ramirez-Gonzalez. |
| EPI_ISL_2545710, EPI_ISL_2545711 | LESP Veracruz | Instituto de Diagnostico y Referencia Epidemiologicos (INDRE) | Claudia Wong-Arambula, Abril Rodriguez-Maldonado, Vanessa Rivero-Arredondo, Ariadna Medina-Benitez, Joaquin Quiroz-Mercado, Sergio Rangel-Guerrero, Natividad Cruz-Ortiz, Tatiana Nunez-Garcia, Gisela Barrera-Badillo, Lucia Hernandez-Rivas, Irma Lopez-Martinez, Ernesto Ramirez-Gonzalez. |
| EPI_ISL_2545712 | LESP Tamaulipas | Instituto de Diagnostico y Referencia Epidemiologicos (INDRE) | Claudia Wong-Arambula, Abril Rodriguez-Maldonado, Vanessa Rivero-Arredondo, Ariadna Medina-Benitez, Joaquin Quiroz-Mercado, Sergio Rangel-Guerrero, Natividad Cruz-Ortiz, Tatiana Nunez-Garcia, Gisela Barrera-Badillo, Lucia Hernandez-Rivas, Irma Lopez-Martinez, Ernesto Ramirez-Gonzalez. |
| EPI_ISL_2545713, EPI_ISL_2545714, EPI_ISL_2545715, EPI_ISL_2545716, EPI_ISL_2545717, EPI_ISL_2545718, EPI_ISL_2545719, EPI_ISL_2545720 | LESP Queretaro | Instituto de Diagnostico y Referencia Epidemiologicos (INDRE) | Claudia Wong-Arambula, Abril Rodriguez-Maldonado, Vanessa Rivero-Arredondo, Ariadna Medina-Benitez, Joaquin Quiroz-Mercado, Sergio Rangel-Guerrero, Natividad Cruz-Ortiz, Tatiana Nunez-Garcia, Gisela Barrera-Badillo, Lucia Hernandez-Rivas, Irma Lopez-Martinez, Ernesto Ramirez-Gonzalez. |

| | | | |
|---|---|---|---|
| EPI_ISL_2545721 | LESP Jalisco | Instituto de Diagnostico y Referencia Epidemiologicos (INDRE) | Claudia Wong-Arambula, Abril Rodriguez-Maldonado, Vanessa Rivero-Arredondo, Ariadna Medina-Benitez, Joaquin Quiroz-Mercado, Sergio Rangel-Guerrero, Natividad Cruz-Ortiz, Tatiana Nunez-Garcia, Gisela Barrera-Badillo, Lucia Hernandez-Rivas, Irma Lopez-Martinez, Ernesto Ramirez-Gonzalez. |
| EPI_ISL_2545722 | LESP Ciudad de Mexico | Instituto de Diagnostico y Referencia Epidemiologicos (INDRE) | Claudia Wong-Arambula, Abril Rodriguez-Maldonado, Vanessa Rivero-Arredondo, Ariadna Medina-Benitez, Joaquin Quiroz-Mercado, Sergio Rangel-Guerrero, Natividad Cruz-Ortiz, Tatiana Nunez-Garcia, Gisela Barrera-Badillo, Lucia Hernandez-Rivas, Irma Lopez-Martinez, Ernesto Ramirez-Gonzalez. |
| EPI_ISL_2545723 | LESP Estado de Mexico | Instituto de Diagnostico y Referencia Epidemiologicos (INDRE) | Claudia Wong-Arambula, Abril Rodriguez-Maldonado, Vanessa Rivero-Arredondo, Ariadna Medina-Benitez, Joaquin Quiroz-Mercado, Sergio Rangel-Guerrero, Natividad Cruz-Ortiz, Tatiana Nunez-Garcia, Gisela Barrera-Badillo, Lucia Hernandez-Rivas, Irma Lopez-Martinez, Ernesto Ramirez-Gonzalez. |
| EPI_ISL_2545724, EPI_ISL_2545725, EPI_ISL_2545726, EPI_ISL_2545727 | LESP Campeche | Instituto de Diagnostico y Referencia Epidemiologicos (INDRE) | Claudia Wong-Arambula, Abril Rodriguez-Maldonado, Vanessa Rivero-Arredondo, Ariadna Medina-Benitez, Joaquin Quiroz-Mercado, Sergio Rangel-Guerrero, Natividad Cruz-Ortiz, Tatiana Nunez-Garcia, Gisela Barrera-Badillo, Lucia Hernandez-Rivas, Irma Lopez-Martinez, Ernesto Ramirez-Gonzalez. |
| EPI_ISL_2545728 | LESP Coahuila | Instituto de Diagnostico y Referencia Epidemiologicos (INDRE) | Claudia Wong-Arambula, Abril Rodriguez-Maldonado, Vanessa Rivero-Arredondo, Ariadna Medina-Benitez, Joaquin Quiroz-Mercado, Sergio Rangel-Guerrero, Natividad Cruz-Ortiz, Tatiana Nunez-Garcia, Gisela Barrera-Badillo, Lucia Hernandez-Rivas, Irma Lopez-Martinez, Ernesto Ramirez-Gonzalez. |
| EPI_ISL_2545729, EPI_ISL_2545730 | LESP Baja California Sur | Instituto de Diagnostico y Referencia Epidemiologicos (INDRE) | Claudia Wong-Arambula, Abril Rodriguez-Maldonado, Vanessa Rivero-Arredondo, Ariadna Medina-Benitez, Joaquin Quiroz-Mercado, Sergio Rangel-Guerrero, Natividad Cruz-Ortiz, Tatiana Nunez-Garcia, Gisela Barrera-Badillo, Lucia Hernandez-Rivas, Irma Lopez-Martinez, Ernesto Ramirez-Gonzalez. |
| EPI_ISL_2545731, EPI_ISL_2545732 | LESP Queretaro | Instituto de Diagnostico y Referencia Epidemiologicos (INDRE) | Claudia Wong-Arambula, Abril Rodriguez-Maldonado, Vanessa Rivero-Arredondo, Ariadna Medina-Benitez, Joaquin Quiroz-Mercado, Sergio Rangel-Guerrero, Natividad Cruz-Ortiz, Tatiana Nunez-Garcia, Gisela Barrera-Badillo, Lucia Hernandez-Rivas, Irma Lopez-Martinez, Ernesto Ramirez-Gonzalez. |
| EPI_ISL_2545733, EPI_ISL_2545735, EPI_ISL_2545736, EPI_ISL_2545737, EPI_ISL_2545738, EPI_ISL_2545739, EPI_ISL_2545740, EPI_ISL_2545741, EPI_ISL_2545742, EPI_ISL_2545743, EPI_ISL_2545744, EPI_ISL_2545745, EPI_ISL_2545746, EPI_ISL_2545747, EPI_ISL_2545748, EPI_ISL_2545749, EPI_ISL_2545750, EPI_ISL_2545751, EPI_ISL_2545752, EPI_ISL_2545753, EPI_ISL_2545754, EPI_ISL_2545755, EPI_ISL_2545756, EPI_ISL_2545757, EPI_ISL_2545758, EPI_ISL_2545759, EPI_ISL_2545760, EPI_ISL_2545761, EPI_ISL_2545762, EPI_ISL_2545763, EPI_ISL_2545764, EPI_ISL_2545765, EPI_ISL_2545766, EPI_ISL_2545767, EPI_ISL_2545768, EPI_ISL_2545769, EPI_ISL_2545770, EPI_ISL_2545771, EPI_ISL_2545772, EPI_ISL_2545773, EPI_ISL_2545774, EPI_ISL_2545775, EPI_ISL_2545776, EPI_ISL_2545777, EPI_ISL_2545778, EPI_ISL_2545779, EPI_ISL_2545780, EPI_ISL_2545781, EPI_ISL_2545782, EPI_ISL_2545783, EPI_ISL_2545784, EPI_ISL_2545785, EPI_ISL_2545786, EPI_ISL_2545787, EPI_ISL_2545788, EPI_ISL_2545789, EPI_ISL_2545790, EPI_ISL_2545791, EPI_ISL_2545792, EPI_ISL_2545793, EPI_ISL_2545794, EPI_ISL_2545795, EPI_ISL_2545796, EPI_ISL_2545797, EPI_ISL_2545798, EPI_ISL_2545799, EPI_ISL_2545800, EPI_ISL_2545801, EPI_ISL_2545802, EPI_ISL_2545803 | | | |
| see above | LESP Yucatan | Instituto de Diagnostico y Referencia Epidemiologicos (INDRE) | Claudia Wong-Arambula, Abril Rodriguez-Maldonado, Vanessa Rivero-Arredondo, Ariadna Medina-Benitez, Joaquin Quiroz-Mercado, Sergio Rangel-Guerrero, Natividad Cruz-Ortiz, Tatiana Nunez-Garcia, Gisela Barrera-Badillo, Lucia Hernandez-Rivas, Irma Lopez-Martinez, Ernesto Ramirez-Gonzalez. |
| EPI_ISL_2545804 | LESP Michoacan | Instituto de Diagnostico y Referencia Epidemiologicos (INDRE) | Claudia Wong-Arambula, Abril Rodriguez-Maldonado, Vanessa Rivero-Arredondo, Ariadna Medina-Benitez, Joaquin Quiroz-Mercado, Sergio Rangel-Guerrero, Natividad Cruz-Ortiz, Tatiana Nunez-Garcia, Gisela Barrera-Badillo, Lucia Hernandez-Rivas, Irma Lopez-Martinez, Ernesto Ramirez-Gonzalez. |
| EPI_ISL_2545805 | LESP Durango | Instituto de Diagnostico y Referencia Epidemiologicos (INDRE) | Claudia Wong-Arambula, Abril Rodriguez-Maldonado, Vanessa Rivero-Arredondo, Ariadna Medina-Benitez, Joaquin Quiroz-Mercado, Sergio Rangel-Guerrero, Natividad Cruz-Ortiz, Tatiana Nunez-Garcia, Gisela Barrera-Badillo, Lucia Hernandez-Rivas, Irma Lopez-Martinez, Ernesto Ramirez-Gonzalez. |
| EPI_ISL_2545806 | LESP Campeche | Instituto de Diagnostico y Referencia Epidemiologicos (INDRE) | Claudia Wong-Arambula, Abril Rodriguez-Maldonado, Vanessa Rivero-Arredondo, Ariadna Medina-Benitez, Joaquin Quiroz-Mercado, Sergio Rangel-Guerrero, Natividad Cruz-Ortiz, Tatiana Nunez-Garcia, Gisela Barrera-Badillo, Lucia Hernandez-Rivas, Irma Lopez-Martinez, Ernesto Ramirez-Gonzalez. |
| EPI_ISL_2545807 | LESP Baja California | Instituto de Diagnostico y Referencia Epidemiologicos (INDRE) | Claudia Wong-Arambula, Abril Rodriguez-Maldonado, Vanessa Rivero-Arredondo, Ariadna Medina-Benitez, Joaquin Quiroz-Mercado, Sergio Rangel-Guerrero, Natividad Cruz-Ortiz, Tatiana Nunez-Garcia, Gisela Barrera-Badillo, Lucia Hernandez-Rivas, Irma Lopez-Martinez, Ernesto Ramirez-Gonzalez. |
| EPI_ISL_2545808, EPI_ISL_2545809, EPI_ISL_2545810, EPI_ISL_2545811 | LESP Quintana Roo | Instituto de Diagnostico y Referencia Epidemiologicos (INDRE) | Claudia Wong-Arambula, Abril Rodriguez-Maldonado, Vanessa Rivero-Arredondo, Ariadna Medina-Benitez, Joaquin Quiroz-Mercado, Sergio Rangel-Guerrero, Natividad Cruz-Ortiz, Tatiana Nunez-Garcia, Gisela Barrera-Badillo, Lucia Hernandez-Rivas, Irma Lopez-Martinez, Ernesto Ramirez-Gonzalez. |
| EPI_ISL_2545812 | LESP Sonora | Instituto de Diagnostico y Referencia Epidemiologicos (INDRE) | Claudia Wong-Arambula, Abril Rodriguez-Maldonado, Vanessa Rivero-Arredondo, Ariadna Medina-Benitez, Joaquin Quiroz-Mercado, Sergio Rangel-Guerrero, Natividad Cruz-Ortiz, Tatiana Nunez-Garcia, Gisela Barrera-Badillo, Lucia Hernandez-Rivas, Irma Lopez-Martinez, Ernesto Ramirez-Gonzalez. |
| EPI_ISL_2545813, EPI_ISL_2545814 | LESP San Luis Potosi | Instituto de Diagnostico y Referencia Epidemiologicos (INDRE) | Claudia Wong-Arambula, Abril Rodriguez-Maldonado, Vanessa Rivero-Arredondo, Ariadna Medina-Benitez, Joaquin Quiroz-Mercado, Sergio Rangel-Guerrero, Natividad Cruz-Ortiz, Tatiana Nunez-Garcia, Gisela Barrera-Badillo, Lucia Hernandez-Rivas, Irma Lopez-Martinez, Ernesto Ramirez-Gonzalez. |
| EPI_ISL_2545815, EPI_ISL_2545816 | LESP Ciudad de Mexico | Instituto de Diagnostico y Referencia Epidemiologicos (INDRE) | Claudia Wong-Arambula, Abril Rodriguez-Maldonado, Vanessa Rivero-Arredondo, Ariadna Medina-Benitez, Joaquin Quiroz-Mercado, Sergio Rangel-Guerrero, Natividad Cruz-Ortiz, Tatiana Nunez-Garcia, Gisela Barrera-Badillo, Lucia Hernandez-Rivas, Irma Lopez-Martinez, Ernesto Ramirez-Gonzalez. |
| EPI_ISL_2545817 | LESP Estado de Mexico | Instituto de Diagnostico y Referencia Epidemiologicos (INDRE) | Claudia Wong-Arambula, Abril Rodriguez-Maldonado, Vanessa Rivero-Arredondo, Ariadna Medina-Benitez, Joaquin Quiroz-Mercado, Sergio Rangel-Guerrero, Natividad Cruz-Ortiz, Tatiana Nunez-Garcia, Gisela Barrera-Badillo, Lucia Hernandez-Rivas, Irma Lopez-Martinez, Ernesto Ramirez-Gonzalez. |
| EPI_ISL_2545818 | LESP Ciudad de Mexico | Instituto de Diagnostico y Referencia Epidemiologicos (INDRE) | Claudia Wong-Arambula, Abril Rodriguez-Maldonado, Vanessa Rivero-Arredondo, Ariadna Medina-Benitez, Joaquin Quiroz-Mercado, Sergio Rangel-Guerrero, Natividad Cruz-Ortiz, Tatiana Nunez-Garcia, Gisela Barrera-Badillo, Lucia Hernandez-Rivas, Irma Lopez-Martinez, Ernesto Ramirez-Gonzalez. |
| EPI_ISL_2545819 | LESP San Luis Potosi | Instituto de Diagnostico y Referencia Epidemiologicos (INDRE) | Claudia Wong-Arambula, Abril Rodriguez-Maldonado, Vanessa Rivero-Arredondo, Ariadna Medina-Benitez, Joaquin Quiroz-Mercado, Sergio Rangel-Guerrero, Natividad Cruz-Ortiz, Tatiana Nunez-Garcia, Gisela Barrera-Badillo, Lucia Hernandez-Rivas, Irma Lopez-Martinez, Ernesto Ramirez-Gonzalez. |
| EPI_ISL_2547287 | CEIRS Data Processing and Coordinating Center, St. Jude Center of Excellence for Influenza Research and Surveillance (CEIRS) | CEIRS Data Processing and Coordinating Center, St. Jude Center of Excellence for Influenza Research and Surveillance (CEIRS) | Jeevan,T., Truong,M., Patel,A., Fabrizio,T., Allay,J., Webb,A. and Webby,R. |
| EPI_ISL_2547288, EPI_ISL_2547289, EPI_ISL_2547290, EPI_ISL_2547291, EPI_ISL_2547292, EPI_ISL_2547293, EPI_ISL_2547294, EPI_ISL_2547295, EPI_ISL_2547296, EPI_ISL_2547297, EPI_ISL_2547298, EPI_ISL_2547299, EPI_ISL_2547300, EPI_ISL_2547301, EPI_ISL_2547302, EPI_ISL_2547303, EPI_ISL_2547304, EPI_ISL_2547305, EPI_ISL_2547306, EPI_ISL_2547307, EPI_ISL_2547308, EPI_ISL_2547309, EPI_ISL_2547310, EPI_ISL_2547311, EPI_ISL_2547312, EPI_ISL_2547313, EPI_ISL_2547314, EPI_ISL_2547315, EPI_ISL_2547316, EPI_ISL_2547317, EPI_ISL_2547318, EPI_ISL_2547319, EPI_ISL_2547320, EPI_ISL_2547321, EPI_ISL_2547322, EPI_ISL_2547323, EPI_ISL_2547324, EPI_ISL_2547325, EPI_ISL_2547326, EPI_ISL_2547327, EPI_ISL_2547328, EPI_ISL_2547329, EPI_ISL_2547330, EPI_ISL_2547331, EPI_ISL_2547332, EPI_ISL_2547333, EPI_ISL_2547334, EPI_ISL_2547335 | | | |
| see above | UHTL, University Hospitals | UHTL, University Hospitals | Sadri,N., Alouani,D., Song,X. |
| EPI_ISL_2549326, EPI_ISL_2549327, EPI_ISL_2549329, EPI_ISL_2549330, EPI_ISL_2549331, EPI_ISL_2549332, EPI_ISL_2549333, EPI_ISL_2549334, EPI_ISL_2549335, EPI_ISL_2549336, EPI_ISL_2549337, EPI_ISL_2549338, EPI_ISL_2549339, EPI_ISL_2549340, EPI_ISL_2549341, EPI_ISL_2549342, EPI_ISL_2549343, EPI_ISL_2549344, EPI_ISL_2549345, EPI_ISL_2549346, EPI_ISL_2549347, EPI_ISL_2549348, EPI_ISL_2549349, EPI_ISL_2549350, EPI_ISL_2549351, EPI_ISL_2549352, EPI_ISL_2549353, EPI_ISL_2549354, EPI_ISL_2549355, EPI_ISL_2549356, EPI_ISL_2549357, EPI_ISL_2549358, EPI_ISL_2549359, EPI_ISL_2549360, EPI_ISL_2549361, EPI_ISL_2549362, EPI_ISL_2549363, EPI_ISL_2549364, EPI_ISL_2549365, EPI_ISL_2549366, EPI_ISL_2549367, EPI_ISL_2549368, EPI_ISL_2549369, EPI_ISL_2549370, EPI_ISL_2549371, EPI_ISL_2549372, EPI_ISL_2549373, EPI_ISL_2549374, EPI_ISL_2549375, EPI_ISL_2549376, EPI_ISL_2549377, EPI_ISL_2549378, EPI_ISL_2549379, EPI_ISL_2549380, EPI_ISL_2549381, EPI_ISL_2549382, EPI_ISL_2549383, EPI_ISL_2549384, EPI_ISL_2549385, EPI_ISL_2549386, EPI_ISL_2549387, EPI_ISL_2549388, EPI_ISL_2549389, EPI_ISL_2549390, EPI_ISL_2549391, EPI_ISL_2549392, EPI_ISL_2549393, EPI_ISL_2549394, EPI_ISL_2549395, EPI_ISL_2549396, EPI_ISL_2549397, EPI_ISL_2549398, EPI_ISL_2549399, EPI_ISL_2549400, EPI_ISL_2549401, EPI_ISL_2549402, EPI_ISL_2549403, EPI_ISL_2549404, EPI_ISL_2549406, EPI_ISL_2549407, EPI_ISL_2549408, EPI_ISL_2549409, EPI_ISL_2549410, EPI_ISL_2549411, EPI_ISL_2549412, EPI_ISL_2549413, EPI_ISL_2549414, EPI_ISL_2549415, EPI_ISL_2549416, EPI_ISL_2549417, EPI_ISL_2549418, EPI_ISL_2549419, EPI_ISL_2549420, EPI_ISL_2549421, EPI_ISL_2549422, EPI_ISL_2549423, EPI_ISL_2549424, EPI_ISL_2549425, EPI_ISL_2549426, EPI_ISL_2549427, EPI_ISL_2549428, EPI_ISL_2549429, EPI_ISL_2549430, EPI_ISL_2549431, EPI_ISL_2549432, EPI_ISL_2549433, EPI_ISL_2549434, EPI_ISL_2549435, EPI_ISL_2549436, EPI_ISL_2549437, EPI_ISL_2549438, EPI_ISL_2549439, EPI_ISL_2549440, EPI_ISL_2549441, EPI_ISL_2549442, EPI_ISL_2549443, EPI_ISL_2549444, EPI_ISL_2549445, EPI_ISL_2549446, EPI_ISL_2549447, EPI_ISL_2549448, EPI_ISL_2549449, EPI_ISL_2549450, EPI_ISL_2549451, EPI_ISL_2549452, EPI_ISL_2549453, EPI_ISL_2549454, EPI_ISL_2549455, EPI_ISL_2549456, EPI_ISL_2549457, EPI_ISL_2549458, EPI_ISL_2549459, EPI_ISL_2549460, EPI_ISL_2549461, EPI_ISL_2549462, EPI_ISL_2549463, EPI_ISL_2549464, EPI_ISL_2549465, EPI_ISL_2549466, EPI_ISL_2549467, EPI_ISL_2549468, EPI_ISL_2549469, EPI_ISL_2549470, EPI_ISL_2549471, EPI_ISL_2549472, EPI_ISL_2549473, EPI_ISL_2549474, EPI_ISL_2549475, EPI_ISL_2549476, EPI_ISL_2549477, EPI_ISL_2549478, EPI_ISL_2549479, EPI_ISL_2549480, EPI_ISL_2549481, EPI_ISL_2549482, EPI_ISL_2549483, EPI_ISL_2549484, EPI_ISL_2549485, EPI_ISL_2549486, EPI_ISL_2549487, EPI_ISL_2549488, EPI_ISL_2549489, EPI_ISL_2549490, EPI_ISL_2549491, EPI_ISL_2549492, EPI_ISL_2549493, EPI_ISL_2549494, EPI_ISL_2549495, EPI_ISL_2549496, EPI_ISL_2549497, EPI_ISL_2549498, EPI_ISL_2549499, EPI_ISL_2549500, EPI_ISL_2549501, EPI_ISL_2549502, EPI_ISL_2549503, EPI_ISL_2549504, EPI_ISL_2549505, EPI_ISL_2549506, EPI_ISL_2549507, EPI_ISL_2549508, EPI_ISL_2549509, EPI_ISL_2549510, EPI_ISL_2549511, EPI_ISL_2549512, EPI_ISL_2549513, EPI_ISL_2549514, EPI_ISL_2549515, EPI_ISL_2549516, EPI_ISL_2549517, EPI_ISL_2549518, EPI_ISL_2549519, EPI_ISL_2549520, EPI_ISL_2549521, EPI_ISL_2549522, EPI_ISL_2549523, EPI_ISL_2549524, EPI_ISL_2549525, EPI_ISL_2549526, EPI_ISL_2549527, EPI_ISL_2549528, EPI_ISL_2549529, EPI_ISL_2549530, EPI_ISL_2549531, EPI_ISL_2549532, EPI_ISL_2549533, EPI_ISL_2549534, EPI_ISL_2549535, EPI_ISL_2549536, EPI_ISL_2549537, EPI_ISL_2549538, EPI_ISL_2549540, EPI_ISL_2549541, EPI_ISL_2549542, EPI_ISL_2549543, EPI_ISL_2549544, EPI_ISL_2549545, EPI_ISL_2549546, EPI_ISL_2549547, EPI_ISL_2549548, EPI_ISL_2549549, EPI_ISL_2549550, EPI_ISL_2549551, EPI_ISL_2549552, EPI_ISL_2549553, EPI_ISL_2549554, EPI_ISL_2549555, EPI_ISL_2549556, EPI_ISL_2549557, EPI_ISL_2549558, EPI_ISL_2549559, EPI_ISL_2549560, EPI_ISL_2549561 | | | |
| see above | UW Virology Lab | UW Virology Lab | Pavitra Roychoudhury, Hong Xie, Lasata Shrestha, Tien V. Nguyen, Shah Mohamed Bakhash, Michelle Lin, Noah R. Baker, Ricardo Perez, Sean Ellis, Nathan Breit, Robert J. Livingston, Meei-Li Huang, Keith R Jerome, Patrick Mathias, Alexander Greninger |
| EPI_ISL_2549577, EPI_ISL_2549579, EPI_ISL_2549580, EPI_ISL_2549581, EPI_ISL_2549582, EPI_ISL_2549584, EPI_ISL_2549585, EPI_ISL_2549586, EPI_ISL_2549587, EPI_ISL_2549588, EPI_ISL_2549589, EPI_ISL_2549590, EPI_ISL_2549591, EPI_ISL_2549592, EPI_ISL_2549593, EPI_ISL_2549594, EPI_ISL_2549595, EPI_ISL_2549596, EPI_ISL_2549597, EPI_ISL_2549598, EPI_ISL_2549599, EPI_ISL_2549600, EPI_ISL_2549604, EPI_ISL_2549605, EPI_ISL_2549608, EPI_ISL_2549609, EPI_ISL_2549610, EPI_ISL_2549611, EPI_ISL_2549612, EPI_ISL_2549613, EPI_ISL_2549615, EPI_ISL_2549616, EPI_ISL_2549617, EPI_ISL_2549618, EPI_ISL_2549619, EPI_ISL_2549622, EPI_ISL_2549624, EPI_ISL_2549626, EPI_ISL_2549627, EPI_ISL_2549628, EPI_ISL_2549629, EPI_ISL_2549631, EPI_ISL_2549632, EPI_ISL_2549633, EPI_ISL_2549635, EPI_ISL_2549637, EPI_ISL_2549638, EPI_ISL_2549639, EPI_ISL_2549641, EPI_ISL_2549644, EPI_ISL_2549645, EPI_ISL_2549647, EPI_ISL_2549648, EPI_ISL_2549649, EPI_ISL_2549650, EPI_ISL_2549651, EPI_ISL_2549652, EPI_ISL_2549653, EPI_ISL_2549654, EPI_ISL_2549655, EPI_ISL_2549656, EPI_ISL_2549657, EPI_ISL_2549658, EPI_ISL_2549659, EPI_ISL_2549660, EPI_ISL_2549662, EPI_ISL_2549663, EPI_ISL_2549664, EPI_ISL_2549667, EPI_ISL_2549668, EPI_ISL_2549669, EPI_ISL_2549670, EPI_ISL_2549672, EPI_ISL_2549674, EPI_ISL_2549675, EPI_ISL_2549676, EPI_ISL_2549677, EPI_ISL_2549679, EPI_ISL_2549682, EPI_ISL_2549683, EPI_ISL_2549684, EPI_ISL_2549685, EPI_ISL_2549686, EPI_ISL_2549688, EPI_ISL_2549689, EPI_ISL_2549692, EPI_ISL_2549694, EPI_ISL_2549695, EPI_ISL_2549696, EPI_ISL_2549697, EPI_ISL_2549699, EPI_ISL_2549700, EPI_ISL_2549701, EPI_ISL_2549703, EPI_ISL_2549704, EPI_ISL_2549705, EPI_ISL_2549706, EPI_ISL_2549709, EPI_ISL_2549711, EPI_ISL_2549712, EPI_ISL_2549714, EPI_ISL_2549715, EPI_ISL_2549716, EPI_ISL_2549717, EPI_ISL_2549718, EPI_ISL_2549719, EPI_ISL_2549720, EPI_ISL_2549721, EPI_ISL_2549725, EPI_ISL_2549726, EPI_ISL_2549727, EPI_ISL_2549728, EPI_ISL_2549732, EPI_ISL_2549733, EPI_ISL_2549734, EPI_ISL_2549735, EPI_ISL_2549736, EPI_ISL_2549737, EPI_ISL_2549739, EPI_ISL_2549740, EPI_ISL_2549742, EPI_ISL_2549743, EPI_ISL_2549744, EPI_ISL_2549745, EPI_ISL_2549746, EPI_ISL_2549747, EPI_ISL_2549748, EPI_ISL_2549749, EPI_ISL_2549750, EPI_ISL_2549751, EPI_ISL_2549752, EPI_ISL_2549753, EPI_ISL_2549754, EPI_ISL_2549755, EPI_ISL_2549756, EPI_ISL_2549757, EPI_ISL_2549758, EPI_ISL_2549759, EPI_ISL_2549760, EPI_ISL_2549761, EPI_ISL_2549762, EPI_ISL_2549763, EPI_ISL_2549764, EPI_ISL_2549765, EPI_ISL_2549766, EPI_ISL_2549767, EPI_ISL_2549768, EPI_ISL_2549769, EPI_ISL_2549770, EPI_ISL_2549772, EPI_ISL_2549773, EPI_ISL_2549775, EPI_ISL_2549777, EPI_ISL_2549778, EPI_ISL_2549779, EPI_ISL_2549781, EPI_ISL_2549784, EPI_ISL_2549785, EPI_ISL_2549786, EPI_ISL_2549787, EPI_ISL_2549788, EPI_ISL_2549789, EPI_ISL_2549790, EPI_ISL_2549791, EPI_ISL_2549792, EPI_ISL_2549793, EPI_ISL_2549794, EPI_ISL_2549795, EPI_ISL_2549796, EPI_ISL_2549798, EPI_ISL_2549799, EPI_ISL_2549800, EPI_ISL_2549801, EPI_ISL_2549802, EPI_ISL_2549803, EPI_ISL_2549805, EPI_ISL_2549807, EPI_ISL_2549808, EPI_ISL_2549809, EPI_ISL_2549811, EPI_ISL_2549812, EPI_ISL_2549813, EPI_ISL_2549814, EPI_ISL_2549815, EPI_ISL_2549816, EPI_ISL_2549817, EPI_ISL_2549818, EPI_ISL_2549819, EPI_ISL_2549820, EPI_ISL_2549821, EPI_ISL_2549823, EPI_ISL_2549826, EPI_ISL_2549827, EPI_ISL_2549828, EPI_ISL_2549829, EPI_ISL_2549830, EPI_ISL_2549831, EPI_ISL_2549832, EPI_ISL_2549833, EPI_ISL_2549835, EPI_ISL_2549836, EPI_ISL_2549837, EPI_ISL_2549839, EPI_ISL_2549841, EPI_ISL_2549842, EPI_ISL_2549843, EPI_ISL_2549844, EPI_ISL_2549845, EPI_ISL_2549846, EPI_ISL_2549847, EPI_ISL_2549848, EPI_ISL_2549849, EPI_ISL_2549850, EPI_ISL_2549852, EPI_ISL_2549854, EPI_ISL_2549855, EPI_ISL_2549856, EPI_ISL_2549857, EPI_ISL_2549858, EPI_ISL_2549860, EPI_ISL_2549861, EPI_ISL_2549862, EPI_ISL_2549864, EPI_ISL_2549866, EPI_ISL_2549867, EPI_ISL_2549868, EPI_ISL_2549869, EPI_ISL_2549870, EPI_ISL_2549871, EPI_ISL_2549872, EPI_ISL_2549874, EPI_ISL_2549875, EPI_ISL_2549876, EPI_ISL_2549877, EPI_ISL_2549879, EPI_ISL_2549881, EPI_ISL_2549882, EPI_ISL_2549883, EPI_ISL_2549885, EPI_ISL_2549886, EPI_ISL_2549887, EPI_ISL_2549888, EPI_ISL_2549889, EPI_ISL_2549891, EPI_ISL_2549892, EPI_ISL_2549893, EPI_ISL_2549895, EPI_ISL_2549897, EPI_ISL_2549898, EPI_ISL_2549899, EPI_ISL_2549903, EPI_ISL_2549904, EPI_ISL_2549905, EPI_ISL_2549906, EPI_ISL_2549907, EPI_ISL_2549908, EPI_ISL_2549909, EPI_ISL_2549910, EPI_ISL_2549911, EPI_ISL_2549912, EPI_ISL_2549913, EPI_ISL_2549915, EPI_ISL_2549916, EPI_ISL_2549918, EPI_ISL_2549919, EPI_ISL_2549922, EPI_ISL_2549923, EPI_ISL_2549924, EPI_ISL_2549925, EPI_ISL_2549928, EPI_ISL_2549930, EPI_ISL_2549931, EPI_ISL_2549932, EPI_ISL_2549933, EPI_ISL_2549934, EPI_ISL_2549935, EPI_ISL_2549936, EPI_ISL_2549937, EPI_ISL_2549938, EPI_ISL_2549939, EPI_ISL_2549940, EPI_ISL_2549941, EPI_ISL_2549942, EPI_ISL_2549943, EPI_ISL_2549944, EPI_ISL_2549948, EPI_ISL_2549949, EPI_ISL_2549950, EPI_ISL_2549951, EPI_ISL_2549952, EPI_ISL_2549953, EPI_ISL_2549954, EPI_ISL_2549955, EPI_ISL_2549956, EPI_ISL_2549957, EPI_ISL_2549958, EPI_ISL_2549959, EPI_ISL_2549965, EPI_ISL_2549966, EPI_ISL_2549969, EPI_ISL_2549970, EPI_ISL_2549971, EPI_ISL_2549975, EPI_ISL_2549977, EPI_ISL_2549982, EPI_ISL_2549984, EPI_ISL_2549986, EPI_ISL_2549988, EPI_ISL_2549991, EPI_ISL_2549992, EPI_ISL_2549993, EPI_ISL_2549998, EPI_ISL_2549999, EPI_ISL_2550000, EPI_ISL_2550002, EPI_ISL_2550008, EPI_ISL_2550009, EPI_ISL_2550010, EPI_ISL_2550011, EPI_ISL_2550012, EPI_ISL_2550013, EPI_ISL_2550014, EPI_ISL_2550015, EPI_ISL_2550016, EPI_ISL_2550017, EPI_ISL_2550018, EPI_ISL_2550019, EPI_ISL_2550020, EPI_ISL_2550021, EPI_ISL_2550022, EPI_ISL_2550023, EPI_ISL_2550024, EPI_ISL_2550025, EPI_ISL_2550028, EPI_ISL_2550029, EPI_ISL_2550030, EPI_ISL_2550031, EPI_ISL_2550034, EPI_ISL_2550039, EPI_ISL_2550042, EPI_ISL_2550043, EPI_ISL_2550048, EPI_ISL_2550049, EPI_ISL_2550052, EPI_ISL_2550055, EPI_ISL_2550056, EPI_ISL_2550058, EPI_ISL_2550059, EPI_ISL_2550062, EPI_ISL_2550063, EPI_ISL_2550065, EPI_ISL_2550066, EPI_ISL_2550067, EPI_ISL_2550068, EPI_ISL_2550069, EPI_ISL_2550070, EPI_ISL_2550071, EPI_ISL_2550072, EPI_ISL_2550074, EPI_ISL_2550075, EPI_ISL_2550076, EPI_ISL_2550077, EPI_ISL_2550078, EPI_ISL_2550079, EPI_ISL_2550080, EPI_ISL_2550081, EPI_ISL_2550082, EPI_ISL_2550084, EPI_ISL_2550085, EPI_ISL_2550087, EPI_ISL_2550088, EPI_ISL_2550089, EPI_ISL_2550090, EPI_ISL_2550092, EPI_ISL_2550093, EPI_ISL_2550094, EPI_ISL_2550095, EPI_ISL_2550097, EPI_ISL_2550098, EPI_ISL_2550099, EPI_ISL_2550100, EPI_ISL_2550101, EPI_ISL_2550102, EPI_ISL_2550103, EPI_ISL_2550104, EPI_ISL_2550105, EPI_ISL_2550106, EPI_ISL_2550107, EPI_ISL_2550108, EPI_ISL_2550109, EPI_ISL_2550110, EPI_ISL_2550112, EPI_ISL_2550113, EPI_ISL_2550114, EPI_ISL_2550115, EPI_ISL_2550116, EPI_ISL_2550117, EPI_ISL_2550118, EPI_ISL_2550119, EPI_ISL_2550120, EPI_ISL_2550121, EPI_ISL_2550122, EPI_ISL_2550123, EPI_ISL_2550124, EPI_ISL_2550125, EPI_ISL_2550126, EPI_ISL_2550128, EPI_ISL_2550130, EPI_ISL_2550132, EPI_ISL_2550133, EPI_ISL_2550134, EPI_ISL_2550136, EPI_ISL_2550137, EPI_ISL_2550139, EPI_ISL_2550140, EPI_ISL_2550142, EPI_ISL_2550143, EPI_ISL_2550145, EPI_ISL_2550146, EPI_ISL_2550147, EPI_ISL_2550148, EPI_ISL_2550150, EPI_ISL_2550151, EPI_ISL_2550152, EPI_ISL_2550153, EPI_ISL_2550155, EPI_ISL_2550156, EPI_ISL_2550157, EPI_ISL_2550158, EPI_ISL_2550159, EPI_ISL_2550160, EPI_ISL_2550161, EPI_ISL_2550162, EPI_ISL_2550163, EPI_ISL_2550164, EPI_ISL_2550165, EPI_ISL_2550166, EPI_ISL_2550168, EPI_ISL_2550169, EPI_ISL_2550170, EPI_ISL_2550171, EPI_ISL_2550172, EPI_ISL_2550173, EPI_ISL_2550174, EPI_ISL_2550175, EPI_ISL_2550176, EPI_ISL_2550177, EPI_ISL_2550178, EPI_ISL_2550179, EPI_ISL_2550180, EPI_ISL_2550181, EPI_ISL_2550184, EPI_ISL_2550185, EPI_ISL_2550186, EPI_ISL_2550187, EPI_ISL_2550188, EPI_ISL_2550190, EPI_ISL_2550191, EPI_ISL_2550192, EPI_ISL_2550194, EPI_ISL_2550195, EPI_ISL_2550196, EPI_ISL_2550197, EPI_ISL_2550198, EPI_ISL_2550199, EPI_ISL_2550200, EPI_ISL_2550201, EPI_ISL_2550206, EPI_ISL_2550207, EPI_ISL_2550208, EPI_ISL_2550210, EPI_ISL_2550211, EPI_ISL_2550212, EPI_ISL_2550214, EPI_ISL_2550217, EPI_ISL_2550219, EPI_ISL_2550222, EPI_ISL_2550223, EPI_ISL_2550225, EPI_ISL_2550226, EPI_ISL_2550228, EPI_ISL_2550230, EPI_ISL_2550231, EPI_ISL_2550232, EPI_ISL_2550234, EPI_ISL_2550235, EPI_ISL_2550236, EPI_ISL_2550237, EPI_ISL_2550238, EPI_ISL_2550239, EPI_ISL_2550240, EPI_ISL_2550241, EPI_ISL_2550242, EPI_ISL_2550243, EPI_ISL_2550245, EPI_ISL_2550246, EPI_ISL_2550247, EPI_ISL_2550248, EPI_ISL_2550252, EPI_ISL_2550253, EPI_ISL_2550255, EPI_ISL_2550256, EPI_ISL_2550257, EPI_ISL_2550258, EPI_ISL_2550259, EPI_ISL_2550260, EPI_ISL_2550261, EPI_ISL_2550262, EPI_ISL_2550265, EPI_ISL_2550266, EPI_ISL_2550267, EPI_ISL_2550268, EPI_ISL_2550269, EPI_ISL_2550270, EPI_ISL_2550271, EPI_ISL_2550273, EPI_ISL_2550275, EPI_ISL_2550276, EPI_ISL_2550278, EPI_ISL_2550280, EPI_ISL_2550281, EPI_ISL_2550282, EPI_ISL_2550284, EPI_ISL_2550286, EPI_ISL_2550289, EPI_ISL_2550290, EPI_ISL_2550291, EPI_ISL_2550292, EPI_ISL_2550294, EPI_ISL_2550295, EPI_ISL_2550296, EPI_ISL_2550297, EPI_ISL_2550300, EPI_ISL_2550301, EPI_ISL_2550302, EPI_ISL_2550303, EPI_ISL_2550304, EPI_ISL_2550305, EPI_ISL_2550306, EPI_ISL_2550307, EPI_ISL_2550309, EPI_ISL_2550310, EPI_ISL_2550311, EPI_ISL_2550312, EPI_ISL_2550315, EPI_ISL_2550317, EPI_ISL_2550318, EPI_ISL_2550319, EPI_ISL_2550320, EPI_ISL_2550321, EPI_ISL_2550322, EPI_ISL_2550323, EPI_ISL_2550324, EPI_ISL_2550326, EPI_ISL_2550327, EPI_ISL_2550328, EPI_ISL_2550329, EPI_ISL_2550330, EPI_ISL_2550331, EPI_ISL_2550332, EPI_ISL_2550333, EPI_ISL_2550334, EPI_ISL_2550335, EPI_ISL_2550336, EPI_ISL_2550337, EPI_ISL_2550338, EPI_ISL_2550339, EPI_ISL_2550340, EPI_ISL_2550342, EPI_ISL_2550343, EPI_ISL_2550344, EPI_ISL_2550345, EPI_ISL_2550346, EPI_ISL_2550347, EPI_ISL_2550349, EPI_ISL_2550350, EPI_ISL_2550351, EPI_ISL_2550352, EPI_ISL_2550353, EPI_ISL_2550354, EPI_ISL_2550355, EPI_ISL_2550357, EPI_ISL_2550358, EPI_ISL_2550360, EPI_ISL_2550361, EPI_ISL_2550362, EPI_ISL_2550363, EPI_ISL_2550364, EPI_ISL_2550365, EPI_ISL_2550366, EPI_ISL_2550367, EPI_ISL_2550369, EPI_ISL_2550370, EPI_ISL_2550371, EPI_ISL_2550372, EPI_ISL_2550373, EPI_ISL_2550374, EPI_ISL_2550375, EPI_ISL_2550376, EPI_ISL_2550379, EPI_ISL_2550380, EPI_ISL_2550381, EPI_ISL_2550382, EPI_ISL_2550383, EPI_ISL_2550384, EPI_ISL_2550385, EPI_ISL_2550387, EPI_ISL_2550388, EPI_ISL_2550391, EPI_ISL_2550392, EPI_ISL_2550393, EPI_ISL_2550394, EPI_ISL_2550395, EPI_ISL_2550396, EPI_ISL_2550399, EPI_ISL_2550400, EPI_ISL_2550402, EPI_ISL_2550403, EPI_ISL_2550404, EPI_ISL_2550405, EPI_ISL_2550406, EPI_ISL_2550407, EPI_ISL_2550408, EPI_ISL_2550412, EPI_ISL_2550414, EPI_ISL_2550415, EPI_ISL_2550416, EPI_ISL_2550417, EPI_ISL_2550418, EPI_ISL_2550419, EPI_ISL_2550420, EPI_ISL_2550422, EPI_ISL_2550425, EPI_ISL_2550426, EPI_ISL_2550427, EPI_ISL_2550428, EPI_ISL_2550429, EPI_ISL_2550430, EPI_ISL_2550432, EPI_ISL_2550433, EPI_ISL_2550434, EPI_ISL_2550435, EPI_ISL_2550436, EPI_ISL_2550437, EPI_ISL_2550439, EPI_ISL_2550441, EPI_ISL_2550442, EPI_ISL_2550443, EPI_ISL_2550444, EPI_ISL_2550445, EPI_ISL_2550446, EPI_ISL_2550447, EPI_ISL_2550448, EPI_ISL_2550449, EPI_ISL_2550451, EPI_ISL_2550452, EPI_ISL_2550454, EPI_ISL_2550455, EPI_ISL_2550456, EPI_ISL_2550457, EPI_ISL_2550458, EPI_ISL_2550459, EPI_ISL_2550461, EPI_ISL_2550462, EPI_ISL_2550463, EPI_ISL_2550464, EPI_ISL_2550465, EPI_ISL_2550466, EPI_ISL_2550467, EPI_ISL_2550468, EPI_ISL_2550469, EPI_ISL_2550470, EPI_ISL_2550471, EPI_ISL_2550472, EPI_ISL_2550473, EPI_ISL_2550474, EPI_ISL_2550475, EPI_ISL_2550476, EPI_ISL_2550478, EPI_ISL_2550479, EPI_ISL_2550481, EPI_ISL_2550482, EPI_ISL_2550483, EPI_ISL_2550484, EPI_ISL_2550485, EPI_ISL_2550486, EPI_ISL_2550487, EPI_ISL_2550488, EPI_ISL_2550490, EPI_ISL_2550491, EPI_ISL_2550492, EPI_ISL_2550493, EPI_ISL_2550494, EPI_ISL_2550495, EPI_ISL_2550496, EPI_ISL_2550497, EPI_ISL_2550498, EPI_ISL_2550499, EPI_ISL_2550500, EPI_ISL_2550502, EPI_ISL_2550503, EPI_ISL_2550504, EPI_ISL_2550505, EPI_ISL_2550506, EPI_ISL_2550508, EPI_ISL_2550509, EPI_ISL_2550510, EPI_ISL_2550511, EPI_ISL_2550516, EPI_ISL_2550517, EPI_ISL_2550518, EPI_ISL_2550519 | | | |

| | | | |
|---|---|---|---|
| see above | Colorado Department of Public Health and Environment | Colorado Department of Public Health and Environment | Laura Bankers, Molly C. Hetherington-Rauth, Diana Ir, Alexandria Rossheim, Shannon R. Matzinger, Sarah Elizabeth Totten, Emily A. Travanty |
| EPI_ISL_2550708, EPI_ISL_2550709, EPI_ISL_2550710, EPI_ISL_2550711, EPI_ISL_2550712, EPI_ISL_2550713 | GA Department of Public Health | GA Department of Public Health | Stacy Reeves, Jonathan Edwards, Cynthia Dixey, Tonia Parrott, Aliyah Fields, Taylor Smith |
| EPI_ISL_2550734, EPI_ISL_2550735, EPI_ISL_2550736, EPI_ISL_2550737, EPI_ISL_2550738, EPI_ISL_2550739, EPI_ISL_2550740, EPI_ISL_2550741, EPI_ISL_2550742, EPI_ISL_2550743, EPI_ISL_2550744, EPI_ISL_2550745, EPI_ISL_2550746, EPI_ISL_2550747, EPI_ISL_2550748, EPI_ISL_2550749, EPI_ISL_2550751, EPI_ISL_2550752, EPI_ISL_2550753, EPI_ISL_2550754, EPI_ISL_2550755, EPI_ISL_2550756, EPI_ISL_2550757 | | | |
| see above | Salud Digna | Instituto Nacional de Medicina Genomica | Hidalgo-Miranda A, Cedro-Tanda A, Mendoza-Vargas A, Reyes-Grajeda JP, Abraham Campos-Romero, Moreno-Camacho José Luis, Rodriguez-Gallegos Jorge, Luna-Ruiz Marco, Gonzalez-Barrera D, Rangel-DeLeon D, Munguia-Garza P, Ramirez-Vega O, Escobar-Arrazola, M, Herrera-Montalvo LA. |
| EPI_ISL_2550760, EPI_ISL_2550761, EPI_ISL_2550762, EPI_ISL_2550763, EPI_ISL_2550764, EPI_ISL_2550765, EPI_ISL_2550766, EPI_ISL_2550767, EPI_ISL_2550768, EPI_ISL_2550769, EPI_ISL_2550770, EPI_ISL_2550771, EPI_ISL_2550772, EPI_ISL_2550773, EPI_ISL_2550774, EPI_ISL_2550775, EPI_ISL_2550776, EPI_ISL_2550777, EPI_ISL_2550778, EPI_ISL_2550779, EPI_ISL_2550780, EPI_ISL_2550781, EPI_ISL_2550782, EPI_ISL_2550783, EPI_ISL_2550784, EPI_ISL_2550785, EPI_ISL_2550786, EPI_ISL_2550787, EPI_ISL_2550788, EPI_ISL_2550789, EPI_ISL_2550790, EPI_ISL_2550791, EPI_ISL_2550792, EPI_ISL_2550793, EPI_ISL_2550794, EPI_ISL_2550795 | | | |
| see above | Maryland Public Health Laboratory (MD PHL) | Maryland Public Health Laboratory (MD PHL) | Maryland Department of Health Laboratories Administration |
| EPI_ISL_2550796, EPI_ISL_2550797, EPI_ISL_2550798, EPI_ISL_2550800, EPI_ISL_2550801, EPI_ISL_2550802, EPI_ISL_2550803, EPI_ISL_2550804, EPI_ISL_2550805, EPI_ISL_2550806, EPI_ISL_2550807, EPI_ISL_2550808, EPI_ISL_2550809, EPI_ISL_2550810, EPI_ISL_2550811, EPI_ISL_2550812, EPI_ISL_2550813, EPI_ISL_2550814 | | | |
| see above | Grady Memorial Hospital, Clinical Microbiology Laboratory | Piantadosi Lab, Emory Department of Pathology | Yun F (Wayne) Wang, Dara Khosravi, Anne Piantadosi |
| EPI_ISL_2550815, EPI_ISL_2550816, EPI_ISL_2550817, EPI_ISL_2550818, EPI_ISL_2550819, EPI_ISL_2550820, EPI_ISL_2550821, EPI_ISL_2550822 | Emory Molecular Diagnostics Laboratory, Emory Healthcare | Piantadosi Lab, Emory Department of Pathology | Ahmed Babiker, Dara Khosravi, Anne Piantadosi |
| EPI_ISL_2550823, EPI_ISL_2550824, EPI_ISL_2550826, EPI_ISL_2550827, EPI_ISL_2550828, EPI_ISL_2550829, EPI_ISL_2550830, EPI_ISL_2550831, EPI_ISL_2550832, EPI_ISL_2550833, EPI_ISL_2550834, EPI_ISL_2550835, EPI_ISL_2550836, EPI_ISL_2550838, EPI_ISL_2550840, EPI_ISL_2550841, EPI_ISL_2550842, EPI_ISL_2550844, EPI_ISL_2550845 | | | |
| see above | Wyoming Public Health Laboratory | Wyoming Public Health Laboratory | Jim Mildenberger, Taylor Fearing, Channing Weber, Ashley Norberg, Chayse Rowley, Marley Goetz, Brian Dominguez, Elliot Thomasson, Sam Britz, Cari Sloma, Robert Petit, and Rob Christensen |
| EPI_ISL_2550846 | IVY3 Central Lab, Vanderbilt University Medical Center | Lauring Lab, University of Michigan, Department of Microbiology and Immunology | Gilbert |
| EPI_ISL_2551157, EPI_ISL_2551159, EPI_ISL_2551160, EPI_ISL_2551161, EPI_ISL_2551162, EPI_ISL_2551163, EPI_ISL_2551166, EPI_ISL_2551178, EPI_ISL_2551182 | Wyoming Public Health Laboratory | Wyoming Public Health Laboratory | Jim Mildenberger, Taylor Fearing, Channing Weber, Ashley Norberg, Chayse Rowley, Marley Goetz, Brian Dominguez, Elliot Thomasson, Sam Britz, Cari Sloma, Robert Petit, and Rob Christensen |
| EPI_ISL_2551184 | Providence Oregon Regional Laboratories | Providence St. Joseph Health Molecular Genomics Laboratory | Alexa K Dowdell, Brian D Piening, Fred L Robinson, Carlo B Bifulco, Mary Campbell, Rogan Rattray |
| EPI_ISL_2551188, EPI_ISL_2551192, EPI_ISL_2551195, EPI_ISL_2551196, EPI_ISL_2551198, EPI_ISL_2551201, EPI_ISL_2551202, EPI_ISL_2551203, EPI_ISL_2551209, EPI_ISL_2551211, EPI_ISL_2551212, EPI_ISL_2551213, EPI_ISL_2551214, EPI_ISL_2551215, EPI_ISL_2551216, EPI_ISL_2551217, EPI_ISL_2551219, EPI_ISL_2551220, EPI_ISL_2551221 | | | |
| see above | Providence Saint Joseph Medical Center - Burbank | Providence St. Joseph Health Molecular Genomics Laboratory | Alexa K Dowdell, Brian D Piening, Fred L Robinson, Carlo B Bifulco, Mary Campbell, Rogan Rattray |
| EPI_ISL_2551223, EPI_ISL_2551224, EPI_ISL_2551225, EPI_ISL_2551227, EPI_ISL_2551230, EPI_ISL_2551231, EPI_ISL_2551232, EPI_ISL_2551233, EPI_ISL_2551234, EPI_ISL_2551235, EPI_ISL_2551237, EPI_ISL_2551238, EPI_ISL_2551240, EPI_ISL_2551241, EPI_ISL_2551243, EPI_ISL_2551244, EPI_ISL_2551245, EPI_ISL_2551246, EPI_ISL_2551247, EPI_ISL_2551249, EPI_ISL_2551251 | | | |
| see above | Providence St. Joseph Health Orange - Chapman | Providence St. Joseph Health Molecular Genomics Laboratory | Alexa K Dowdell, Brian D Piening, Fred L Robinson, Carlo B Bifulco, Mary Campbell, Rogan Rattray |
| EPI_ISL_2551254, EPI_ISL_2551255, EPI_ISL_2551256, EPI_ISL_2551257, EPI_ISL_2551262, EPI_ISL_2551266, EPI_ISL_2551267, EPI_ISL_2551269, EPI_ISL_2551274, EPI_ISL_2551276, EPI_ISL_2551278, EPI_ISL_2551279, EPI_ISL_2551280, EPI_ISL_2551281, EPI_ISL_2551285, EPI_ISL_2551286 | | | |
| see above | Providence Regional Medical Center Everett | Providence St. Joseph Health Molecular Genomics Laboratory | Alexa K Dowdell, Brian D Piening, Fred L Robinson, Carlo B Bifulco, Mary Campbell, Rogan Rattray |
| EPI_ISL_2551288, EPI_ISL_2551289, EPI_ISL_2551290, EPI_ISL_2551292, EPI_ISL_2551294, EPI_ISL_2551295, EPI_ISL_2551296, EPI_ISL_2551297, EPI_ISL_2551299, EPI_ISL_2551300, EPI_ISL_2551302, EPI_ISL_2551303, EPI_ISL_2551305, EPI_ISL_2551307, EPI_ISL_2551308, EPI_ISL_2551309, EPI_ISL_2551310, EPI_ISL_2551311, EPI_ISL_2551314, EPI_ISL_2551315, EPI_ISL_2551316, EPI_ISL_2551319, EPI_ISL_2551320, EPI_ISL_2551322, EPI_ISL_2551325, EPI_ISL_2551326, EPI_ISL_2551327, EPI_ISL_2551330, EPI_ISL_2551331, EPI_ISL_2551332, EPI_ISL_2551334, EPI_ISL_2551336, EPI_ISL_2551337, EPI_ISL_2551340, EPI_ISL_2551341, EPI_ISL_2551342, EPI_ISL_2551343, EPI_ISL_2551344, EPI_ISL_2551345, EPI_ISL_2551347, EPI_ISL_2551348, EPI_ISL_2551349, EPI_ISL_2551351, EPI_ISL_2551353, EPI_ISL_2551354, EPI_ISL_2551356, EPI_ISL_2551357, EPI_ISL_2551358, EPI_ISL_2551359, EPI_ISL_2551360, EPI_ISL_2551361, EPI_ISL_2551362, EPI_ISL_2551363, EPI_ISL_2551364, EPI_ISL_2551365, EPI_ISL_2551366, EPI_ISL_2551368, EPI_ISL_2551370, EPI_ISL_2551371, EPI_ISL_2551372, EPI_ISL_2551373, EPI_ISL_2551374, EPI_ISL_2551375, EPI_ISL_2551376, EPI_ISL_2551378, EPI_ISL_2551379 | | | |
| see above | Providence Oregon Regional Laboratories | Providence St. Joseph Health Molecular Genomics Laboratory | Alexa K Dowdell, Brian D Piening, Fred L Robinson, Carlo B Bifulco, Mary Campbell, Rogan Rattray |
| EPI_ISL_2551380, EPI_ISL_2551381, EPI_ISL_2551385, EPI_ISL_2551387, EPI_ISL_2551389, EPI_ISL_2551390, EPI_ISL_2551391, EPI_ISL_2551392, EPI_ISL_2551393, EPI_ISL_2551395, EPI_ISL_2551396, EPI_ISL_2551397, EPI_ISL_2551398, EPI_ISL_2551400, EPI_ISL_2551401, EPI_ISL_2551402, EPI_ISL_2551403, EPI_ISL_2551404, EPI_ISL_2551405, EPI_ISL_2551406, EPI_ISL_2551407, EPI_ISL_2551408, EPI_ISL_2551409, EPI_ISL_2551410, EPI_ISL_2551411, EPI_ISL_2551413, EPI_ISL_2551414, EPI_ISL_2551415, EPI_ISL_2551416, EPI_ISL_2551419, EPI_ISL_2551420 | | | |
| see above | Providence Regional Medical Center Everett | Providence St. Joseph Health Molecular Genomics Laboratory | Alexa K Dowdell, Brian D Piening, Fred L Robinson, Carlo B Bifulco, Mary Campbell, Rogan Rattray |
| EPI_ISL_2551422 | Providence Oregon Regional Laboratories | Providence St. Joseph Health Molecular Genomics Laboratory | Alexa K Dowdell, Brian D Piening, Fred L Robinson, Carlo B Bifulco, Mary Campbell, Rogan Rattray |
| EPI_ISL_2551423, EPI_ISL_2551426, EPI_ISL_2551427, EPI_ISL_2551428, EPI_ISL_2551429, EPI_ISL_2551432, EPI_ISL_2551433, EPI_ISL_2551434, EPI_ISL_2551436, EPI_ISL_2551437, EPI_ISL_2551438, EPI_ISL_2551439, EPI_ISL_2551440, EPI_ISL_2551441, EPI_ISL_2551442, EPI_ISL_2551443, EPI_ISL_2551445, EPI_ISL_2551447, EPI_ISL_2551448, EPI_ISL_2551449, EPI_ISL_2551452, EPI_ISL_2551453, EPI_ISL_2551454, EPI_ISL_2551455, EPI_ISL_2551457, EPI_ISL_2551458, EPI_ISL_2551459, EPI_ISL_2551460 | | | |
| see above | Providence Regional Medical Center Everett | Providence St. Joseph Health Molecular Genomics Laboratory | Alexa K Dowdell, Brian D Piening, Fred L Robinson, Carlo B Bifulco, Mary Campbell, Rogan Rattray |
| EPI_ISL_2551462, EPI_ISL_2551463, EPI_ISL_2551465, EPI_ISL_2551466, EPI_ISL_2551467, EPI_ISL_2551468, EPI_ISL_2551469, EPI_ISL_2551470, EPI_ISL_2551471, EPI_ISL_2551475, EPI_ISL_2551476, EPI_ISL_2551477, EPI_ISL_2551478, EPI_ISL_2551480, EPI_ISL_2551481, EPI_ISL_2551482, EPI_ISL_2551483, EPI_ISL_2551484, EPI_ISL_2551485, EPI_ISL_2551486, EPI_ISL_2551487, EPI_ISL_2551490, EPI_ISL_2551491, EPI_ISL_2551492, EPI_ISL_2551493, EPI_ISL_2551495, EPI_ISL_2551499, EPI_ISL_2551500, EPI_ISL_2551501, EPI_ISL_2551505, EPI_ISL_2551506, EPI_ISL_2551508, EPI_ISL_2551510, EPI_ISL_2551511, EPI_ISL_2551512, EPI_ISL_2551513, EPI_ISL_2551514, EPI_ISL_2551515 | | | |
| see above | Providence Oregon Regional Laboratories | Providence St. Joseph Health Molecular Genomics Laboratory | Alexa K Dowdell, Brian D Piening, Fred L Robinson, Carlo B Bifulco, Mary Campbell, Rogan Rattray |
| EPI_ISL_2551518 | Kaiser Permanente Washington Health Research Institute | Genomics and Discovery, Respiratory Viruses Branch, Division of Viral Diseases, Centers for Disease Control and Prevention | Yan Li, Jing Zhang, Anna Kelleher, Ying Tao, Brian Lynch, Krista Queen, Anna Uehara, Peter Cook, Han Jia Justin Ng, Rachel Marine, Clinton R. Paden, Dhwani Batra, Haibin Wang, Tara Coalter, Jasmine Padilla, Morgan Davis, Mili Sheth, Sarah Nobles, Mark Burroughs, Justin Lee, Adam Retchless, Suxiang Tong |
| EPI_ISL_2551537, EPI_ISL_2551541, EPI_ISL_2551543, EPI_ISL_2551544, EPI_ISL_2551545, EPI_ISL_2551546, EPI_ISL_2551547, EPI_ISL_2551548, EPI_ISL_2551549, EPI_ISL_2551550, EPI_ISL_2551551, EPI_ISL_2551552, EPI_ISL_2551553, EPI_ISL_2551554, EPI_ISL_2551556, EPI_ISL_2551557, EPI_ISL_2551558, EPI_ISL_2551559, EPI_ISL_2551560, EPI_ISL_2551561, EPI_ISL_2551562, EPI_ISL_2551563, EPI_ISL_2551564, EPI_ISL_2551566, EPI_ISL_2551567, EPI_ISL_2551568, EPI_ISL_2551569, EPI_ISL_2551570, EPI_ISL_2551571, EPI_ISL_2551572, EPI_ISL_2551573, EPI_ISL_2551574, EPI_ISL_2551575, EPI_ISL_2551576, EPI_ISL_2551577, EPI_ISL_2551578, EPI_ISL_2551579, EPI_ISL_2551581, EPI_ISL_2554083, EPI_ISL_2554084, EPI_ISL_2554085, EPI_ISL_2554086, EPI_ISL_2554087, EPI_ISL_2554088, EPI_ISL_2554089, EPI_ISL_2554090, EPI_ISL_2554091, EPI_ISL_2554092, EPI_ISL_2554093, EPI_ISL_2554094, EPI_ISL_2554095, EPI_ISL_2554096, EPI_ISL_2554097, EPI_ISL_2554098, EPI_ISL_2554099, EPI_ISL_2554100, EPI_ISL_2554101, EPI_ISL_2554102, EPI_ISL_2554103, EPI_ISL_2554104, EPI_ISL_2554105, EPI_ISL_2554106, EPI_ISL_2554107, EPI_ISL_2554108, EPI_ISL_2554109, EPI_ISL_2554110 | | | |
| see above | Wyoming Public Health Laboratory | Wyoming Public Health Laboratory | Jim Mildenberger, Taylor Fearing, Channing Weber, Ashley Norberg, Chayse Rowley, Marley Goetz, Brian Dominguez, Elliot Thomasson, Sam Britz, Cari Sloma, Robert Petit, and Rob Christensen |
| EPI_ISL_2555533, EPI_ISL_2555535, EPI_ISL_2555539, EPI_ISL_2555540, EPI_ISL_2556114, EPI_ISL_2556116, EPI_ISL_2556118, EPI_ISL_2556120, EPI_ISL_2557034 | Ohio Department of Health Laboratory | Ohio Department of Health Laboratory | Holmes, Jennifer; Eric Brandt, Keoni Omura, Glen McGillivary, Caitlin McDonnell, Jade Mowery, Stephanie Mccracken, Tyler Payne, Kirtana Ramadugu, Erica Leasure, Brent Lee, Kelsey Florek, Heather Blankenship, Allison Black, Quanta Brown, and Tammy Bannerman |
| EPI_ISL_2557035, EPI_ISL_2557036, EPI_ISL_2557037, EPI_ISL_2557038, EPI_ISL_2557039, EPI_ISL_2557040, EPI_ISL_2557041, EPI_ISL_2557042, EPI_ISL_2557043, EPI_ISL_2557044, EPI_ISL_2557045, EPI_ISL_2557046, EPI_ISL_2557047, EPI_ISL_2557048, EPI_ISL_2557049, EPI_ISL_2557050, EPI_ISL_2557051, EPI_ISL_2557052, EPI_ISL_2557053, EPI_ISL_2557054, EPI_ISL_2557055, EPI_ISL_2557056, EPI_ISL_2557057, EPI_ISL_2557058, EPI_ISL_2557059, EPI_ISL_2557060, EPI_ISL_2557061, EPI_ISL_2557062, EPI_ISL_2557063, EPI_ISL_2557064, EPI_ISL_2557066, EPI_ISL_2557067, EPI_ISL_2557068, EPI_ISL_2557069, EPI_ISL_2557070, EPI_ISL_2557071, EPI_ISL_2557072, EPI_ISL_2557073, EPI_ISL_2557074, EPI_ISL_2557075, EPI_ISL_2557077, EPI_ISL_2557078, EPI_ISL_2557079, EPI_ISL_2557080, EPI_ISL_2557081, EPI_ISL_2557082 | | | |
| see above | Washington State Department of Health Public Health Laboratories | Washington State Department of Health Public Health Laboratories | Drew MacKellar, Philip Dykema, Denny Russell, Joenice Gonzalez, Hannah Gray, Geoff Melly, Vanessa De Los Santos, Darren Lucas, JohnAric Peterson, Avi Singh, Rebecca Cao |
| EPI_ISL_2557083 | Ohio Department of Health Laboratory | Ohio Department of Health Laboratory | Holmes, Jennifer; Eric Brandt, Keoni Omura, Glen McGillivary, Caitlin McDonnell, Jade Mowery, Stephanie Mccracken, Tyler Payne, Kirtana Ramadugu, Erica Leasure, Brent Lee, Kelsey Florek, Heather Blankenship, Allison Black, Quanta Brown, and Tammy Bannerman |
| EPI_ISL_2557087, EPI_ISL_2557089, EPI_ISL_2557090, EPI_ISL_2557091, EPI_ISL_2557092, EPI_ISL_2557093, EPI_ISL_2557094, EPI_ISL_2557095, EPI_ISL_2557096, EPI_ISL_2557097, EPI_ISL_2557098, EPI_ISL_2557099, EPI_ISL_2557100, EPI_ISL_2557101, EPI_ISL_2557102, EPI_ISL_2557103, EPI_ISL_2557104, EPI_ISL_2557105, EPI_ISL_2557106, EPI_ISL_2557107, EPI_ISL_2557108, EPI_ISL_2557109, EPI_ISL_2557110, EPI_ISL_2557111, EPI_ISL_2557112, EPI_ISL_2557113, EPI_ISL_2557115, EPI_ISL_2557117, EPI_ISL_2557118, EPI_ISL_2557119, EPI_ISL_2557120, EPI_ISL_2557121, EPI_ISL_2557122, EPI_ISL_2557123, EPI_ISL_2557124, EPI_ISL_2557125, EPI_ISL_2557126, EPI_ISL_2557127, EPI_ISL_2557128, EPI_ISL_2557129, EPI_ISL_2557130, EPI_ISL_2557131, EPI_ISL_2557132, EPI_ISL_2557133, | | | |

| | | | |
|---|---|---|---|
| EPI_ISL_2557134, EPI_ISL_2557135, EPI_ISL_2557136, EPI_ISL_2557137, EPI_ISL_2557138, EPI_ISL_2557139, EPI_ISL_2557140, EPI_ISL_2557141, EPI_ISL_2557142, EPI_ISL_2557143, EPI_ISL_2557144, EPI_ISL_2557145, EPI_ISL_2557146, EPI_ISL_2557147, EPI_ISL_2557148, EPI_ISL_2557149, EPI_ISL_2557150, EPI_ISL_2557151, EPI_ISL_2557152, EPI_ISL_2557153, EPI_ISL_2557155, EPI_ISL_2557156, EPI_ISL_2557157, EPI_ISL_2557158 | | | |
| see above | NJDOH, Public Health and Environmental Laboratories | NJ_PHEL | Lindsey Bodnar, Shiv K. Verma, Ryan Pachucki, Chelsea San Filippo, Dana Woell, Allison Roder, Byeong Jeong |
| EPI_ISL_2557212, EPI_ISL_2557213, EPI_ISL_2557214, EPI_ISL_2557215, EPI_ISL_2557216, EPI_ISL_2557217, EPI_ISL_2557218, EPI_ISL_2557219, EPI_ISL_2557220, EPI_ISL_2557221, EPI_ISL_2557222, EPI_ISL_2557223, EPI_ISL_2557224, EPI_ISL_2557226, EPI_ISL_2557228, EPI_ISL_2557232, EPI_ISL_2557234, EPI_ISL_2557235, EPI_ISL_2557236, EPI_ISL_2557237, EPI_ISL_2557238, EPI_ISL_2557239, EPI_ISL_2557240, EPI_ISL_2557241, EPI_ISL_2557242 | | | |
| see above | IN State Department of Health Laboratory Services | IN State Department of Health Laboratory Services | Cassandra Campion, Jamie Yeadon, Brian Pope, Lixia Liu, Kyle Brownlee, Melissa Hindenlang, Mark Glazier |
| EPI_ISL_2557243 | Ohio Department of Health Laboratory | Ohio Department of Health Laboratory | Holmes, Jennifer; Eric Brandt, Keoni Omura, Glen McGillivary, Caitlin McDonnell, Jade Mowery, Stephanie Mccracken, Tyler Payne, Kirtana Ramadugu, Erica Leasure, Brent Lee, Kelsey Florek, Heather Blankenship, Allison Black, Quanta Brown, and Tammy Bannerman |
| EPI_ISL_2557244, EPI_ISL_2557245, EPI_ISL_2557246 | Kansas Health and Environmental Lab | Kansas Health and Environmental Lab | Katherine Wiggins, Mike Grose, Jonathan Barnell, Ben Olsen, and Phil Adam |
| EPI_ISL_2557247, EPI_ISL_2557249, EPI_ISL_2557305, EPI_ISL_2557306, EPI_ISL_2557307, EPI_ISL_2557308 | Ohio Department of Health Laboratory | Ohio Department of Health Laboratory | Holmes, Jennifer; Eric Brandt, Keoni Omura, Glen McGillivary, Caitlin McDonnell, Jade Mowery, Stephanie Mccracken, Tyler Payne, Kirtana Ramadugu, Erica Leasure, Brent Lee, Kelsey Florek, Heather Blankenship, Allison Black, Quanta Brown, and Tammy Bannerman |
| EPI_ISL_2557431, EPI_ISL_2557432, EPI_ISL_2557433, EPI_ISL_2557435, EPI_ISL_2557436, EPI_ISL_2557437, EPI_ISL_2557438, EPI_ISL_2557439, EPI_ISL_2557441, EPI_ISL_2557442, EPI_ISL_2557443, EPI_ISL_2557444, EPI_ISL_2557445, EPI_ISL_2557446, EPI_ISL_2557448, EPI_ISL_2557449, EPI_ISL_2557450, EPI_ISL_2557451, EPI_ISL_2557453, EPI_ISL_2557454, EPI_ISL_2557455, EPI_ISL_2557456, EPI_ISL_2557457, EPI_ISL_2557458, EPI_ISL_2557460, EPI_ISL_2557461, EPI_ISL_2557462, EPI_ISL_2557463, EPI_ISL_2557465, EPI_ISL_2557466, EPI_ISL_2557467, EPI_ISL_2557468, EPI_ISL_2557469, EPI_ISL_2557470, EPI_ISL_2557471, EPI_ISL_2557472, EPI_ISL_2557473, EPI_ISL_2557474, EPI_ISL_2557475, EPI_ISL_2557477, EPI_ISL_2557479, EPI_ISL_2557480, EPI_ISL_2557481, EPI_ISL_2557482, EPI_ISL_2557483, EPI_ISL_2557485, EPI_ISL_2557486, EPI_ISL_2557487, EPI_ISL_2557488, EPI_ISL_2557489, EPI_ISL_2557490, EPI_ISL_2557492, EPI_ISL_2557493, EPI_ISL_2557494, EPI_ISL_2557495, EPI_ISL_2557496, EPI_ISL_2557498, EPI_ISL_2557499, EPI_ISL_2557500, EPI_ISL_2557501, EPI_ISL_2557502, EPI_ISL_2557503, EPI_ISL_2557505, EPI_ISL_2557506, EPI_ISL_2557507, EPI_ISL_2557508, EPI_ISL_2557509, EPI_ISL_2557510, EPI_ISL_2557512, EPI_ISL_2557513, EPI_ISL_2557514, EPI_ISL_2557515, EPI_ISL_2557516, EPI_ISL_2557517, EPI_ISL_2557518, EPI_ISL_2557519, EPI_ISL_2557520, EPI_ISL_2557521, EPI_ISL_2557523, EPI_ISL_2557524, EPI_ISL_2557525, EPI_ISL_2557526, EPI_ISL_2557527, EPI_ISL_2557528, EPI_ISL_2557529, EPI_ISL_2557530, EPI_ISL_2557531, EPI_ISL_2557532, EPI_ISL_2557533, EPI_ISL_2557534, EPI_ISL_2557537, EPI_ISL_2557538, EPI_ISL_2557539, EPI_ISL_2557540, EPI_ISL_2557541, EPI_ISL_2557542, EPI_ISL_2557543, EPI_ISL_2557544, EPI_ISL_2557547, EPI_ISL_2557548, EPI_ISL_2557549, EPI_ISL_2557551, EPI_ISL_2557552, EPI_ISL_2557553, EPI_ISL_2557554, EPI_ISL_2557555, EPI_ISL_2557556, EPI_ISL_2557558, EPI_ISL_2557559, EPI_ISL_2557560, EPI_ISL_2557561, EPI_ISL_2557562, EPI_ISL_2557563, EPI_ISL_2557564, EPI_ISL_2557565, EPI_ISL_2557566, EPI_ISL_2557568, EPI_ISL_2557569, EPI_ISL_2557570, EPI_ISL_2557571, EPI_ISL_2557572, EPI_ISL_2557573, EPI_ISL_2557574, EPI_ISL_2557575, EPI_ISL_2557576, EPI_ISL_2557578, EPI_ISL_2557579, EPI_ISL_2557580, EPI_ISL_2557581, EPI_ISL_2557582, EPI_ISL_2557583, EPI_ISL_2557584, EPI_ISL_2557585, EPI_ISL_2557586, EPI_ISL_2557588, EPI_ISL_2557589, EPI_ISL_2557590, EPI_ISL_2557591, EPI_ISL_2557592, EPI_ISL_2557593, EPI_ISL_2557594, EPI_ISL_2557595, EPI_ISL_2557596, EPI_ISL_2557598, EPI_ISL_2557600, EPI_ISL_2557601, EPI_ISL_2557602, EPI_ISL_2557603, EPI_ISL_2557604, EPI_ISL_2557605, EPI_ISL_2557606, EPI_ISL_2557608, EPI_ISL_2557609, EPI_ISL_2557610, EPI_ISL_2557611, EPI_ISL_2557612, EPI_ISL_2557613, EPI_ISL_2557614, EPI_ISL_2557615, EPI_ISL_2557616, EPI_ISL_2557618, EPI_ISL_2557619, EPI_ISL_2557620, EPI_ISL_2557621, EPI_ISL_2557622, EPI_ISL_2557623, EPI_ISL_2557624, EPI_ISL_2557625, EPI_ISL_2557626, EPI_ISL_2557627, EPI_ISL_2557629 | | | |
| see above | Infinity Biologix | Centers for Disease Control and Prevention Division of Viral Diseases, Pathogen Discovery | Dakota Howard, Dhwani Batra, Peter W. Cook, Kara Moser, Adrian Paskey, Jason Caravas, Benjamin Rambo-Martin, Shatavia Morrison, Christopher Gulvick, Scott Sammons, Yvette Unoarumhi, Darlene Wagner, Matthew Schmerer, Christian Bixby, Yihe Wang, Jonathan Schultz, Chirayu Goswami, Russ Hager, Robin Grimwood, Clinton R. Paden, Duncan MacCannell |
| EPI_ISL_2557644 | Ohio Department of Health Laboratory | Ohio Department of Health Laboratory | Holmes, Jennifer; Eric Brandt, Keoni Omura, Glen McGillivary, Caitlin McDonnell, Jade Mowery, Stephanie Mccracken, Tyler Payne, Kirtana Ramadugu, Erica Leasure, Brent Lee, Kelsey Florek, Heather Blankenship, Allison Black, Quanta Brown, and Tammy Bannerman |
| EPI_ISL_2557747, EPI_ISL_2557748, EPI_ISL_2557749, EPI_ISL_2557750, EPI_ISL_2557751, EPI_ISL_2557752, EPI_ISL_2557753, EPI_ISL_2557754, EPI_ISL_2557755, EPI_ISL_2557756, EPI_ISL_2557757, EPI_ISL_2557758, EPI_ISL_2557759, EPI_ISL_2557760, EPI_ISL_2557761, EPI_ISL_2557763, EPI_ISL_2557764, EPI_ISL_2557765, EPI_ISL_2557767, EPI_ISL_2557770, EPI_ISL_2557771, EPI_ISL_2557772, EPI_ISL_2557773, EPI_ISL_2557774, EPI_ISL_2557777, EPI_ISL_2557778, EPI_ISL_2557779, EPI_ISL_2557780, EPI_ISL_2557781, EPI_ISL_2557782, EPI_ISL_2557783, EPI_ISL_2557785, EPI_ISL_2557786, EPI_ISL_2557787, EPI_ISL_2557788, EPI_ISL_2557789, EPI_ISL_2557790, EPI_ISL_2557791, EPI_ISL_2557792, EPI_ISL_2557793, EPI_ISL_2557794, EPI_ISL_2557795, EPI_ISL_2557796, EPI_ISL_2557797, EPI_ISL_2557798, EPI_ISL_2557799, EPI_ISL_2557800, EPI_ISL_2557801, EPI_ISL_2557802, EPI_ISL_2557803, EPI_ISL_2557804, EPI_ISL_2557805, EPI_ISL_2557806, EPI_ISL_2557807, EPI_ISL_2557808, EPI_ISL_2557809, EPI_ISL_2557810, EPI_ISL_2557811, EPI_ISL_2557812, EPI_ISL_2557813, EPI_ISL_2557814, EPI_ISL_2557815, EPI_ISL_2557816, EPI_ISL_2557817, EPI_ISL_2557819, EPI_ISL_2557820, EPI_ISL_2557821, EPI_ISL_2557822, EPI_ISL_2557823 | | | |
| see above | Michigan Department of Health and Human Services, Bureau of Laboratories | Michigan Department of Health and Human Services, Bureau of Laboratories | Blankenship HM, Riner D, Soehnlen MK |
| EPI_ISL_2557866, EPI_ISL_2557867, EPI_ISL_2557868 | Oregon State University TRACE Project | OSU Center for Genome Research and Biocomputing | Oregon State University TRACE Project |
| EPI_ISL_2557914, EPI_ISL_2557915, EPI_ISL_2557916, EPI_ISL_2557917, EPI_ISL_2557918, EPI_ISL_2557919, EPI_ISL_2557920, EPI_ISL_2557921, EPI_ISL_2557922, EPI_ISL_2557923, EPI_ISL_2557924, EPI_ISL_2557925, EPI_ISL_2557926, EPI_ISL_2557927, EPI_ISL_2557928, EPI_ISL_2557929, EPI_ISL_2557930, EPI_ISL_2557931, EPI_ISL_2557932, EPI_ISL_2557933, EPI_ISL_2557934, EPI_ISL_2557935, EPI_ISL_2557936, EPI_ISL_2557937, EPI_ISL_2557938, EPI_ISL_2557939, EPI_ISL_2557940, EPI_ISL_2557941 | | | |
| see above | Oregon State Public Health Laboratory | Oregon State Public Health Laboratory | Rafia Razzaque, Eugene Yeboah, Vanda Makris, Laura Tsaknaridis, John Fontana and Shane Sevey |
| EPI_ISL_2558047, EPI_ISL_2558048, EPI_ISL_2558049, EPI_ISL_2558050, EPI_ISL_2558051, EPI_ISL_2558064 | Ohio Department of Health Laboratory | Ohio Department of Health Laboratory | Holmes, Jennifer; Eric Brandt, Keoni Omura, Glen McGillivary, Caitlin McDonnell, Jade Mowery, Stephanie Mccracken, Tyler Payne, Kirtana Ramadugu, Erica Leasure, Brent Lee, Kelsey Florek, Heather Blankenship, Allison Black, Quanta Brown, and Tammy Bannerman |
| EPI_ISL_2558085, EPI_ISL_2558086, EPI_ISL_2558087, EPI_ISL_2558088, EPI_ISL_2558089, EPI_ISL_2558090, EPI_ISL_2558091, EPI_ISL_2558092, EPI_ISL_2558093, EPI_ISL_2558094, EPI_ISL_2558095, EPI_ISL_2558096, EPI_ISL_2558097, EPI_ISL_2558098, EPI_ISL_2558099, EPI_ISL_2558100, EPI_ISL_2558101, EPI_ISL_2558102, EPI_ISL_2558103, EPI_ISL_2558104, EPI_ISL_2558105, EPI_ISL_2558106, EPI_ISL_2558107, EPI_ISL_2558108, EPI_ISL_2558109, EPI_ISL_2558110, EPI_ISL_2558111, EPI_ISL_2558112, EPI_ISL_2558114, EPI_ISL_2558115, EPI_ISL_2558116, EPI_ISL_2558117, EPI_ISL_2558118, EPI_ISL_2558119, EPI_ISL_2558120, EPI_ISL_2558121, EPI_ISL_2558122, EPI_ISL_2558123, EPI_ISL_2558124, EPI_ISL_2558125, EPI_ISL_2558126, EPI_ISL_2558127, EPI_ISL_2558128, EPI_ISL_2558129, EPI_ISL_2558130, EPI_ISL_2558131, EPI_ISL_2558132, EPI_ISL_2558133, EPI_ISL_2558134, EPI_ISL_2558135, EPI_ISL_2558136, EPI_ISL_2558137, EPI_ISL_2558139, EPI_ISL_2558140, EPI_ISL_2558141, EPI_ISL_2558142, EPI_ISL_2558143, EPI_ISL_2558144, EPI_ISL_2558145, EPI_ISL_2558146, EPI_ISL_2558147, EPI_ISL_2558148, EPI_ISL_2558149, EPI_ISL_2558150, EPI_ISL_2558151, EPI_ISL_2558152, EPI_ISL_2558153, EPI_ISL_2558154, EPI_ISL_2558155, EPI_ISL_2558156, EPI_ISL_2558157, EPI_ISL_2558158, EPI_ISL_2558159, EPI_ISL_2558160, EPI_ISL_2558161, EPI_ISL_2558162, EPI_ISL_2558163, EPI_ISL_2558164, EPI_ISL_2558165, EPI_ISL_2558166, EPI_ISL_2558167, EPI_ISL_2558168, EPI_ISL_2558169, EPI_ISL_2558170, EPI_ISL_2558171, EPI_ISL_2558172, EPI_ISL_2558173, EPI_ISL_2558174, EPI_ISL_2558175, EPI_ISL_2558176, EPI_ISL_2558177, EPI_ISL_2558178, EPI_ISL_2558179, EPI_ISL_2558180, EPI_ISL_2558181, EPI_ISL_2558182, EPI_ISL_2558183, EPI_ISL_2558184, EPI_ISL_2558185, EPI_ISL_2558186, EPI_ISL_2558187, EPI_ISL_2558188, EPI_ISL_2558189, EPI_ISL_2558190, EPI_ISL_2558191, EPI_ISL_2558192, EPI_ISL_2558193, EPI_ISL_2558194, EPI_ISL_2558195, EPI_ISL_2558196, EPI_ISL_2558197, EPI_ISL_2558198, EPI_ISL_2558199, EPI_ISL_2558200, EPI_ISL_2558201, EPI_ISL_2558202, EPI_ISL_2558203, EPI_ISL_2558204, EPI_ISL_2558205, EPI_ISL_2558206 | | | |
| see above | Wisconsin State Laboratory of Hygiene Communicable Disease Division | Wisconsin State Laboratory of Hygiene Communicable Disease Division | Abigail C. Shockey, Alicia J. Mooney, Erika M. Hanson, Tonya Danz, Richard Griesser, Sara Wagner, Kelsey R. Florek |
| EPI_ISL_2558207 | Ohio Department of Health Laboratory | Ohio Department of Health Laboratory | Holmes, Jennifer; Eric Brandt, Keoni Omura, Glen McGillivary, Caitlin McDonnell, Jade Mowery, Stephanie Mccracken, Tyler Payne, Kirtana Ramadugu, Erica Leasure, Brent Lee, Kelsey Florek, Heather Blankenship, Allison Black, Quanta Brown, and Tammy Bannerman |
| EPI_ISL_2558208, EPI_ISL_2558210, EPI_ISL_2558211, EPI_ISL_2558212, EPI_ISL_2558213, EPI_ISL_2558214, EPI_ISL_2558215, EPI_ISL_2558216, EPI_ISL_2558217, EPI_ISL_2558218, EPI_ISL_2558219, EPI_ISL_2558220, EPI_ISL_2558221, EPI_ISL_2558222, EPI_ISL_2558223, EPI_ISL_2558224, EPI_ISL_2558225, EPI_ISL_2558226, EPI_ISL_2558227, EPI_ISL_2558228, EPI_ISL_2558229, EPI_ISL_2558230, EPI_ISL_2558231, EPI_ISL_2558232, EPI_ISL_2558233, EPI_ISL_2558234, EPI_ISL_2558235, EPI_ISL_2558236, EPI_ISL_2558237, EPI_ISL_2558238, EPI_ISL_2558239, EPI_ISL_2558240, EPI_ISL_2558241, EPI_ISL_2558242, EPI_ISL_2558243, EPI_ISL_2558245, EPI_ISL_2558246, EPI_ISL_2558247, EPI_ISL_2558248, EPI_ISL_2558249, EPI_ISL_2558250, EPI_ISL_2558251, EPI_ISL_2558252, EPI_ISL_2558253, EPI_ISL_2558254, EPI_ISL_2558255, EPI_ISL_2558256, EPI_ISL_2558257, EPI_ISL_2558258, EPI_ISL_2558260, EPI_ISL_2558261, EPI_ISL_2558262, EPI_ISL_2558263, EPI_ISL_2558264, EPI_ISL_2558265, EPI_ISL_2558266, EPI_ISL_2558267, EPI_ISL_2558268, EPI_ISL_2558269, EPI_ISL_2558270, EPI_ISL_2558271, EPI_ISL_2558272, EPI_ISL_2558273, EPI_ISL_2558274, EPI_ISL_2558275, EPI_ISL_2558276, EPI_ISL_2558277, EPI_ISL_2558278, EPI_ISL_2558279, EPI_ISL_2558280, EPI_ISL_2558281, EPI_ISL_2558282, EPI_ISL_2558283, EPI_ISL_2558284, EPI_ISL_2558285, EPI_ISL_2558287, EPI_ISL_2558289, EPI_ISL_2558291, EPI_ISL_2558292, EPI_ISL_2558293, EPI_ISL_2558295, EPI_ISL_2558297, EPI_ISL_2558298, EPI_ISL_2558299, EPI_ISL_2558301, EPI_ISL_2558304, EPI_ISL_2558305, EPI_ISL_2558307, EPI_ISL_2558309, EPI_ISL_2558310, EPI_ISL_2558311, EPI_ISL_2558312, EPI_ISL_2558313, EPI_ISL_2558314, EPI_ISL_2558315, EPI_ISL_2558316, EPI_ISL_2558317, EPI_ISL_2558320, EPI_ISL_2558321, EPI_ISL_2558322, EPI_ISL_2558323, EPI_ISL_2558324, EPI_ISL_2558325, EPI_ISL_2558326, EPI_ISL_2558327, EPI_ISL_2558328, EPI_ISL_2558329, EPI_ISL_2558330, EPI_ISL_2558331, EPI_ISL_2558332, EPI_ISL_2558333, EPI_ISL_2558334, EPI_ISL_2558335, EPI_ISL_2558336, EPI_ISL_2558337, EPI_ISL_2558338, EPI_ISL_2558339, EPI_ISL_2558340, EPI_ISL_2558341, EPI_ISL_2558342, EPI_ISL_2558343, EPI_ISL_2558344, EPI_ISL_2558345, EPI_ISL_2558346, EPI_ISL_2558347, EPI_ISL_2558348, EPI_ISL_2558349, EPI_ISL_2558350, EPI_ISL_2558351, EPI_ISL_2558352, EPI_ISL_2558353, EPI_ISL_2558355, EPI_ISL_2558356, EPI_ISL_2558357, EPI_ISL_2558358, EPI_ISL_2558359, EPI_ISL_2558360, EPI_ISL_2558361, EPI_ISL_2558362, EPI_ISL_2558363, EPI_ISL_2558364, EPI_ISL_2558365, EPI_ISL_2558366, EPI_ISL_2558367, EPI_ISL_2558368, EPI_ISL_2558369, EPI_ISL_2558370, EPI_ISL_2558371, EPI_ISL_2558372, EPI_ISL_2558373, EPI_ISL_2558374, EPI_ISL_2558375, EPI_ISL_2558376, EPI_ISL_2558377, EPI_ISL_2558378, EPI_ISL_2558379, EPI_ISL_2558380, EPI_ISL_2558381, EPI_ISL_2558382, EPI_ISL_2558383, EPI_ISL_2558384, EPI_ISL_2558385, EPI_ISL_2558386, EPI_ISL_2558387, EPI_ISL_2558388, EPI_ISL_2558389, EPI_ISL_2558390, EPI_ISL_2558391, EPI_ISL_2558392, EPI_ISL_2558393, EPI_ISL_2558394, EPI_ISL_2558395, EPI_ISL_2558396, EPI_ISL_2558397, EPI_ISL_2558398, EPI_ISL_2558401, EPI_ISL_2558402, EPI_ISL_2558403, EPI_ISL_2558404, EPI_ISL_2558405, EPI_ISL_2558406, EPI_ISL_2558407, EPI_ISL_2558408, EPI_ISL_2558409, EPI_ISL_2558410, EPI_ISL_2558412, EPI_ISL_2558413, EPI_ISL_2558414, EPI_ISL_2558415, EPI_ISL_2558416, EPI_ISL_2558417, EPI_ISL_2558418, EPI_ISL_2558419, EPI_ISL_2558420, EPI_ISL_2558421, EPI_ISL_2558422, EPI_ISL_2558423, EPI_ISL_2558424, EPI_ISL_2558426, EPI_ISL_2558427, EPI_ISL_2558428, EPI_ISL_2558429, EPI_ISL_2558430, EPI_ISL_2558431, EPI_ISL_2558432, EPI_ISL_2558433, EPI_ISL_2558434, EPI_ISL_2558435, EPI_ISL_2558436, EPI_ISL_2558437, EPI_ISL_2558438, EPI_ISL_2558439, EPI_ISL_2558440, EPI_ISL_2558441, EPI_ISL_2558442, EPI_ISL_2558443, EPI_ISL_2558444, EPI_ISL_2558445, EPI_ISL_2558446, EPI_ISL_2558447, EPI_ISL_2558448, EPI_ISL_2558449, EPI_ISL_2558450, EPI_ISL_2558451, EPI_ISL_2558452, EPI_ISL_2558453, EPI_ISL_2558454, EPI_ISL_2558456, EPI_ISL_2558457, EPI_ISL_2558458, EPI_ISL_2558459, EPI_ISL_2558460, EPI_ISL_2558461, EPI_ISL_2558462, EPI_ISL_2558463, EPI_ISL_2558464, EPI_ISL_2558465, EPI_ISL_2558466, EPI_ISL_2558467, EPI_ISL_2558468, EPI_ISL_2558469, EPI_ISL_2558470, EPI_ISL_2558471, EPI_ISL_2558473, EPI_ISL_2558474, EPI_ISL_2558475, EPI_ISL_2558476, EPI_ISL_2558477, EPI_ISL_2558478, EPI_ISL_2558479, EPI_ISL_2558480, EPI_ISL_2558481, EPI_ISL_2558482, EPI_ISL_2558483, EPI_ISL_2558484, EPI_ISL_2558485, EPI_ISL_2558486, EPI_ISL_2558487, EPI_ISL_2558488, EPI_ISL_2558489, EPI_ISL_2558490, EPI_ISL_2558491, EPI_ISL_2558492, EPI_ISL_2558493, EPI_ISL_2558494, EPI_ISL_2558495, EPI_ISL_2558496, EPI_ISL_2558497, EPI_ISL_2558498, EPI_ISL_2558499, EPI_ISL_2558500, EPI_ISL_2558501, EPI_ISL_2558502, EPI_ISL_2558503, EPI_ISL_2558504, EPI_ISL_2558505, EPI_ISL_2558506, EPI_ISL_2558507, EPI_ISL_2558508, EPI_ISL_2558509, EPI_ISL_2558510, EPI_ISL_2558511, EPI_ISL_2558512, EPI_ISL_2558513, EPI_ISL_2558514, EPI_ISL_2558515, EPI_ISL_2558516, EPI_ISL_2558517, EPI_ISL_2558518, EPI_ISL_2558519, EPI_ISL_2558520, EPI_ISL_2558521, EPI_ISL_2558522, EPI_ISL_2558524, EPI_ISL_2558525, EPI_ISL_2558526, EPI_ISL_2558527, EPI_ISL_2558528, EPI_ISL_2558529, EPI_ISL_2558530, EPI_ISL_2558532, EPI_ISL_2558533, EPI_ISL_2558534, EPI_ISL_2558535, EPI_ISL_2558536, EPI_ISL_2558537, EPI_ISL_2558538, EPI_ISL_2558539, EPI_ISL_2558540, EPI_ISL_2558541, EPI_ISL_2558542, EPI_ISL_2558543, EPI_ISL_2558544, EPI_ISL_2558545, EPI_ISL_2558546, EPI_ISL_2558547, EPI_ISL_2558548, EPI_ISL_2558549, EPI_ISL_2558550, EPI_ISL_2558553, EPI_ISL_2558554, EPI_ISL_2558555, EPI_ISL_2558556, EPI_ISL_2558557, EPI_ISL_2558558, EPI_ISL_2558559, EPI_ISL_2558560, EPI_ISL_2558561, EPI_ISL_2558562, EPI_ISL_2558563, EPI_ISL_2558564, EPI_ISL_2558565, EPI_ISL_2558566, EPI_ISL_2558567, EPI_ISL_2558568, EPI_ISL_2558569, EPI_ISL_2558570, EPI_ISL_2558571, EPI_ISL_2558572, EPI_ISL_2558573, EPI_ISL_2558574, EPI_ISL_2558575, EPI_ISL_2558576, EPI_ISL_2558577, EPI_ISL_2558578, EPI_ISL_2558579, EPI_ISL_2558580, EPI_ISL_2558581, EPI_ISL_2558582, EPI_ISL_2558583, EPI_ISL_2558584, EPI_ISL_2558585, EPI_ISL_2558586, EPI_ISL_2558587, EPI_ISL_2558588, EPI_ISL_2558589, EPI_ISL_2558590, EPI_ISL_2558591, EPI_ISL_2558592, EPI_ISL_2558593, EPI_ISL_2558594, EPI_ISL_2558595, EPI_ISL_2558596, EPI_ISL_2558597, EPI_ISL_2558598, EPI_ISL_2558599, EPI_ISL_2558600, EPI_ISL_2558601, EPI_ISL_2558602, EPI_ISL_2558603, EPI_ISL_2558604, EPI_ISL_2558605, EPI_ISL_2558606, EPI_ISL_2558607, EPI_ISL_2558609, EPI_ISL_2558610, EPI_ISL_2558612, EPI_ISL_2558613, EPI_ISL_2558614, EPI_ISL_2558615, EPI_ISL_2558616, EPI_ISL_2558618, EPI_ISL_2558619, EPI_ISL_2558620, EPI_ISL_2558621, EPI_ISL_2558622, EPI_ISL_2558623, EPI_ISL_2558624, EPI_ISL_2558625, EPI_ISL_2558626, EPI_ISL_2558627, EPI_ISL_2558628, EPI_ISL_2558629, EPI_ISL_2558630, EPI_ISL_2558631, EPI_ISL_2558632, EPI_ISL_2558633, EPI_ISL_2558634, EPI_ISL_2558635, EPI_ISL_2558636, EPI_ISL_2558637, EPI_ISL_2558638, EPI_ISL_2558639, EPI_ISL_2558640, EPI_ISL_2558641, EPI_ISL_2558642, EPI_ISL_2558644, EPI_ISL_2558645, EPI_ISL_2558646, EPI_ISL_2558647, EPI_ISL_2558648, EPI_ISL_2558649, EPI_ISL_2558650, EPI_ISL_2558651, EPI_ISL_2558653, EPI_ISL_2558654, EPI_ISL_2558655, EPI_ISL_2558656, EPI_ISL_2558657, EPI_ISL_2558658, EPI_ISL_2558659, EPI_ISL_2558660, EPI_ISL_2558661, EPI_ISL_2558662, EPI_ISL_2558663, EPI_ISL_2558664, EPI_ISL_2558665, EPI_ISL_2558666, EPI_ISL_2558667, EPI_ISL_2558668, EPI_ISL_2558669, EPI_ISL_2558670, EPI_ISL_2558671, EPI_ISL_2558672, EPI_ISL_2558673, EPI_ISL_2558674, EPI_ISL_2558675 | | | |
| see above | Wisconsin State Laboratory of Hygiene Communicable Disease Division | Wisconsin State Laboratory of Hygiene Communicable Disease Division | Abigail C. Shockey, Alicia J. Mooney, Erika M. Hanson, Tonya Danz, Richard Griesser, Sara Wagner, Kelsey R. Florek |
| EPI_ISL_2558676, EPI_ISL_2558677, EPI_ISL_2558678, EPI_ISL_2558679, EPI_ISL_2558680, EPI_ISL_2558681, EPI_ISL_2558682, EPI_ISL_2558683, EPI_ISL_2558684, EPI_ISL_2558685, EPI_ISL_2558686, EPI_ISL_2558687, EPI_ISL_2558688, EPI_ISL_2558689, EPI_ISL_2558690, EPI_ISL_2558691, EPI_ISL_2558692, EPI_ISL_2558693, EPI_ISL_2558694 | | | |
| see above | Kansas Health and Environmental Lab | Kansas Health and Environmental Lab | Katherine Wiggins, Mike Grose, Jonathan Barnell, Ben Olsen, and Phil Adam |
| EPI_ISL_2558695, EPI_ISL_2558696, EPI_ISL_2558697, EPI_ISL_2558698 | Wichita State University Molecular Diagnotic Lab | Kansas Health and Environmental Lab | Katherine Wiggins, Mike Grose, Jonathan Barnell, Ben Olsen, and Phil Adam |
| EPI_ISL_2558700, EPI_ISL_2558702, EPI_ISL_2558703, EPI_ISL_2558704 | Kansas Health and Environmental Lab | Kansas Health and Environmental Lab | Katherine Wiggins, Mike Grose, Jonathan Barnell, Ben Olsen, and Phil Adam |
| EPI_ISL_2559101, EPI_ISL_2559104, EPI_ISL_2559106, EPI_ISL_2559108, EPI_ISL_2559110, EPI_ISL_2559112, EPI_ISL_2559115, EPI_ISL_2559117, EPI_ISL_2559119, EPI_ISL_2559122, EPI_ISL_2559127, EPI_ISL_2559129, EPI_ISL_2559131, EPI_ISL_2559133, EPI_ISL_2559135, EPI_ISL_2559137, EPI_ISL_2559139, EPI_ISL_2559142, EPI_ISL_2559144, EPI_ISL_2559146, EPI_ISL_2559149, EPI_ISL_2559151 | | | |
| see above | Wyoming Public Health Laboratory | Wyoming Public Health Laboratory | Jim Mildenberger, Taylor Fearing, Channing Weber, Ashley Norberg, Chayse Rowley, Marley Goetz, Brian Dominguez, Elliot Thomasson, Sam Britz, Cari Sloma, Robert Petit, and Rob Christensen |
| EPI_ISL_2559261 | LESP Jalisco | Instituto de Diagnostico y Referencia Epidemiologicos (INDRE) | Claudia Wong-Arambula, Abril Rodriguez-Maldonado, Vanessa Rivero-Arredondo, Ariadna Medina-Benitez, Joaquin Quiroz-Mercado, Sergio Rangel-Guerrero, Natividad Cruz-Ortiz, Tatiana Nunez-Garcia, Gisela Barrera-Badillo, Lucia Hernandez-Rivas, Irma Lopez-Martinez, Ernesto Ramirez-Gonzalez. |
| EPI_ISL_2559263, EPI_ISL_2559265, EPI_ISL_2559267, EPI_ISL_2559269, EPI_ISL_2559271, | LESP Chihuahua | Instituto de Diagnostico y Referencia Epidemiologicos (INDRE) | Claudia Wong-Arambula, Abril Rodriguez-Maldonado, Vanessa Rivero-Arredondo, Ariadna Medina-Benitez, Joaquin Quiroz-Mercado, Sergio Rangel-Guerrero, Natividad Cruz-Ortiz, Tatiana Nunez-Garcia, Gisela Barrera-Badillo, Lucia Hernandez-Rivas, Irma Lopez-Martinez, Ernesto Ramirez-Gonzalez. |

| | | | |
|---|---|---|---|
| EPI_ISL_2559273, EPI_ISL_2559275 | | | |
| EPI_ISL_2559277, EPI_ISL_2559279 | LESP Veracruz | Instituto de Diagnostico y Referencia Epidemiologicos (INDRE) | Claudia Wong-Arambula, Abril Rodriguez-Maldonado, Vanessa Rivero-Arredondo, Ariadna Medina-Benitez, Joaquin Quiroz-Mercado, Sergio Rangel-Guerrero, Natividad Cruz-Ortiz, Tatiana Nunez-Garcia, Gisela Barrera-Badillo, Lucia Hernandez-Rivas, Irma Lopez-Martinez, Ernesto Ramirez-Gonzalez. |
| EPI_ISL_2559281 | LESP Tamaulipas | Instituto de Diagnostico y Referencia Epidemiologicos (INDRE) | Claudia Wong-Arambula, Abril Rodriguez-Maldonado, Vanessa Rivero-Arredondo, Ariadna Medina-Benitez, Joaquin Quiroz-Mercado, Sergio Rangel-Guerrero, Natividad Cruz-Ortiz, Tatiana Nunez-Garcia, Gisela Barrera-Badillo, Lucia Hernandez-Rivas, Irma Lopez-Martinez, Ernesto Ramirez-Gonzalez. |
| EPI_ISL_2559283, EPI_ISL_2559285, EPI_ISL_2559287, EPI_ISL_2559289, EPI_ISL_2559291, EPI_ISL_2559293, EPI_ISL_2559295, EPI_ISL_2559297, EPI_ISL_2559299, EPI_ISL_2559302, EPI_ISL_2559303, EPI_ISL_2559305, EPI_ISL_2559307, EPI_ISL_2559309, EPI_ISL_2559311, EPI_ISL_2559314, EPI_ISL_2559316, EPI_ISL_2559317, EPI_ISL_2559319, EPI_ISL_2559321, EPI_ISL_2559323, EPI_ISL_2559325, EPI_ISL_2559327, EPI_ISL_2559329, EPI_ISL_2559331, EPI_ISL_2559333, EPI_ISL_2559336, EPI_ISL_2559338, EPI_ISL_2559340, EPI_ISL_2559342, EPI_ISL_2559343, EPI_ISL_2559346, EPI_ISL_2559347, EPI_ISL_2559350, EPI_ISL_2559352 | | | |
| see above | LESP Queretaro | Instituto de Diagnostico y Referencia Epidemiologicos (INDRE) | Claudia Wong-Arambula, Abril Rodriguez-Maldonado, Vanessa Rivero-Arredondo, Ariadna Medina-Benitez, Joaquin Quiroz-Mercado, Sergio Rangel-Guerrero, Natividad Cruz-Ortiz, Tatiana Nunez-Garcia, Gisela Barrera-Badillo, Lucia Hernandez-Rivas, Irma Lopez-Martinez, Ernesto Ramirez-Gonzalez. |
| EPI_ISL_2559354 | LESP Morelos | Instituto de Diagnostico y Referencia Epidemiologicos (INDRE) | Claudia Wong-Arambula, Abril Rodriguez-Maldonado, Vanessa Rivero-Arredondo, Ariadna Medina-Benitez, Joaquin Quiroz-Mercado, Sergio Rangel-Guerrero, Natividad Cruz-Ortiz, Tatiana Nunez-Garcia, Gisela Barrera-Badillo, Lucia Hernandez-Rivas, Irma Lopez-Martinez, Ernesto Ramirez-Gonzalez. |
| EPI_ISL_2559356, EPI_ISL_2559358 | LESP Chihuahua | Instituto de Diagnostico y Referencia Epidemiologicos (INDRE) | Claudia Wong-Arambula, Abril Rodriguez-Maldonado, Vanessa Rivero-Arredondo, Ariadna Medina-Benitez, Joaquin Quiroz-Mercado, Sergio Rangel-Guerrero, Natividad Cruz-Ortiz, Tatiana Nunez-Garcia, Gisela Barrera-Badillo, Lucia Hernandez-Rivas, Irma Lopez-Martinez, Ernesto Ramirez-Gonzalez. |
| EPI_ISL_2559361 | LESP Campeche | Instituto de Diagnostico y Referencia Epidemiologicos (INDRE) | Claudia Wong-Arambula, Abril Rodriguez-Maldonado, Vanessa Rivero-Arredondo, Ariadna Medina-Benitez, Joaquin Quiroz-Mercado, Sergio Rangel-Guerrero, Natividad Cruz-Ortiz, Tatiana Nunez-Garcia, Gisela Barrera-Badillo, Lucia Hernandez-Rivas, Irma Lopez-Martinez, Ernesto Ramirez-Gonzalez. |
| EPI_ISL_2559363, EPI_ISL_2559365 | LESP Puebla | Instituto de Diagnostico y Referencia Epidemiologicos (INDRE) | Claudia Wong-Arambula, Abril Rodriguez-Maldonado, Vanessa Rivero-Arredondo, Ariadna Medina-Benitez, Joaquin Quiroz-Mercado, Sergio Rangel-Guerrero, Natividad Cruz-Ortiz, Tatiana Nunez-Garcia, Gisela Barrera-Badillo, Lucia Hernandez-Rivas, Irma Lopez-Martinez, Ernesto Ramirez-Gonzalez. |
| EPI_ISL_2559367, EPI_ISL_2559369, EPI_ISL_2559371, EPI_ISL_2559373 | LESP Queretaro | Instituto de Diagnostico y Referencia Epidemiologicos (INDRE) | Claudia Wong-Arambula, Abril Rodriguez-Maldonado, Vanessa Rivero-Arredondo, Ariadna Medina-Benitez, Joaquin Quiroz-Mercado, Sergio Rangel-Guerrero, Natividad Cruz-Ortiz, Tatiana Nunez-Garcia, Gisela Barrera-Badillo, Lucia Hernandez-Rivas, Irma Lopez-Martinez, Ernesto Ramirez-Gonzalez. |
| EPI_ISL_2559375, EPI_ISL_2559377 | LESP Tabasco | Instituto de Diagnostico y Referencia Epidemiologicos (INDRE) | Claudia Wong-Arambula, Abril Rodriguez-Maldonado, Vanessa Rivero-Arredondo, Ariadna Medina-Benitez, Joaquin Quiroz-Mercado, Sergio Rangel-Guerrero, Natividad Cruz-Ortiz, Tatiana Nunez-Garcia, Gisela Barrera-Badillo, Lucia Hernandez-Rivas, Irma Lopez-Martinez, Ernesto Ramirez-Gonzalez. |
| EPI_ISL_2559379 | LESP Veracruz | Instituto de Diagnostico y Referencia Epidemiologicos (INDRE) | Claudia Wong-Arambula, Abril Rodriguez-Maldonado, Vanessa Rivero-Arredondo, Ariadna Medina-Benitez, Joaquin Quiroz-Mercado, Sergio Rangel-Guerrero, Natividad Cruz-Ortiz, Tatiana Nunez-Garcia, Gisela Barrera-Badillo, Lucia Hernandez-Rivas, Irma Lopez-Martinez, Ernesto Ramirez-Gonzalez. |
| EPI_ISL_2559381, EPI_ISL_2559384, EPI_ISL_2559386, EPI_ISL_2559388, EPI_ISL_2559389, EPI_ISL_2559391, EPI_ISL_2559393, EPI_ISL_2559395, EPI_ISL_2559397, EPI_ISL_2559399, EPI_ISL_2559402, EPI_ISL_2559404, EPI_ISL_2559406, EPI_ISL_2559408, EPI_ISL_2559410, EPI_ISL_2559412, EPI_ISL_2559414, EPI_ISL_2559416, EPI_ISL_2559419, EPI_ISL_2559421, EPI_ISL_2559423 | | | |
| see above | LESP Yucatan | Instituto de Diagnostico y Referencia Epidemiologicos (INDRE) | Claudia Wong-Arambula, Abril Rodriguez-Maldonado, Vanessa Rivero-Arredondo, Ariadna Medina-Benitez, Joaquin Quiroz-Mercado, Sergio Rangel-Guerrero, Natividad Cruz-Ortiz, Tatiana Nunez-Garcia, Gisela Barrera-Badillo, Lucia Hernandez-Rivas, Irma Lopez-Martinez, Ernesto Ramirez-Gonzalez. |
| EPI_ISL_2559425, EPI_ISL_2559427, EPI_ISL_2559429, EPI_ISL_2559431, EPI_ISL_2559433, EPI_ISL_2559435, EPI_ISL_2559437, EPI_ISL_2559439, EPI_ISL_2559442, EPI_ISL_2559444, EPI_ISL_2559446 | | | |
| see above | LESP Michoacan | Instituto de Diagnostico y Referencia Epidemiologicos (INDRE) | Claudia Wong-Arambula, Abril Rodriguez-Maldonado, Vanessa Rivero-Arredondo, Ariadna Medina-Benitez, Joaquin Quiroz-Mercado, Sergio Rangel-Guerrero, Natividad Cruz-Ortiz, Tatiana Nunez-Garcia, Gisela Barrera-Badillo, Lucia Hernandez-Rivas, Irma Lopez-Martinez, Ernesto Ramirez-Gonzalez. |
| EPI_ISL_2559448 | LESP Durango | Instituto de Diagnostico y Referencia Epidemiologicos (INDRE) | Claudia Wong-Arambula, Abril Rodriguez-Maldonado, Vanessa Rivero-Arredondo, Ariadna Medina-Benitez, Joaquin Quiroz-Mercado, Sergio Rangel-Guerrero, Natividad Cruz-Ortiz, Tatiana Nunez-Garcia, Gisela Barrera-Badillo, Lucia Hernandez-Rivas, Irma Lopez-Martinez, Ernesto Ramirez-Gonzalez. |
| EPI_ISL_2559450 | LESP Sinaloa | Instituto de Diagnostico y Referencia Epidemiologicos (INDRE) | Claudia Wong-Arambula, Abril Rodriguez-Maldonado, Vanessa Rivero-Arredondo, Ariadna Medina-Benitez, Joaquin Quiroz-Mercado, Sergio Rangel-Guerrero, Natividad Cruz-Ortiz, Tatiana Nunez-Garcia, Gisela Barrera-Badillo, Lucia Hernandez-Rivas, Irma Lopez-Martinez, Ernesto Ramirez-Gonzalez. |
| EPI_ISL_2559452 | LESP Colima | Instituto de Diagnostico y Referencia Epidemiologicos (INDRE) | Claudia Wong-Arambula, Abril Rodriguez-Maldonado, Vanessa Rivero-Arredondo, Ariadna Medina-Benitez, Joaquin Quiroz-Mercado, Sergio Rangel-Guerrero, Natividad Cruz-Ortiz, Tatiana Nunez-Garcia, Gisela Barrera-Badillo, Lucia Hernandez-Rivas, Irma Lopez-Martinez, Ernesto Ramirez-Gonzalez. |
| EPI_ISL_2559454 | LESP San Luis Potosi | Instituto de Diagnostico y Referencia Epidemiologicos (INDRE) | Claudia Wong-Arambula, Abril Rodriguez-Maldonado, Vanessa Rivero-Arredondo, Ariadna Medina-Benitez, Joaquin Quiroz-Mercado, Sergio Rangel-Guerrero, Natividad Cruz-Ortiz, Tatiana Nunez-Garcia, Gisela Barrera-Badillo, Lucia Hernandez-Rivas, Irma Lopez-Martinez, Ernesto Ramirez-Gonzalez. |
| EPI_ISL_2559456, EPI_ISL_2559458 | LESP Ciudad de Mexico | Instituto de Diagnostico y Referencia Epidemiologicos (INDRE) | Claudia Wong-Arambula, Abril Rodriguez-Maldonado, Vanessa Rivero-Arredondo, Ariadna Medina-Benitez, Joaquin Quiroz-Mercado, Sergio Rangel-Guerrero, Natividad Cruz-Ortiz, Tatiana Nunez-Garcia, Gisela Barrera-Badillo, Lucia Hernandez-Rivas, Irma Lopez-Martinez, Ernesto Ramirez-Gonzalez. |
| EPI_ISL_2559460 | LESP Estado de Mexico | Instituto de Diagnostico y Referencia Epidemiologicos (INDRE) | Claudia Wong-Arambula, Abril Rodriguez-Maldonado, Vanessa Rivero-Arredondo, Ariadna Medina-Benitez, Joaquin Quiroz-Mercado, Sergio Rangel-Guerrero, Natividad Cruz-Ortiz, Tatiana Nunez-Garcia, Gisela Barrera-Badillo, Lucia Hernandez-Rivas, Irma Lopez-Martinez, Ernesto Ramirez-Gonzalez. |
| EPI_ISL_2559463, EPI_ISL_2559465, EPI_ISL_2559467 | LESP Ciudad de Mexico | Instituto de Diagnostico y Referencia Epidemiologicos (INDRE) | Claudia Wong-Arambula, Abril Rodriguez-Maldonado, Vanessa Rivero-Arredondo, Ariadna Medina-Benitez, Joaquin Quiroz-Mercado, Sergio Rangel-Guerrero, Natividad Cruz-Ortiz, Tatiana Nunez-Garcia, Gisela Barrera-Badillo, Lucia Hernandez-Rivas, Irma Lopez-Martinez, Ernesto Ramirez-Gonzalez. |
| EPI_ISL_2559469 | LESP Estado de Mexico | Instituto de Diagnostico y Referencia Epidemiologicos (INDRE) | Claudia Wong-Arambula, Abril Rodriguez-Maldonado, Vanessa Rivero-Arredondo, Ariadna Medina-Benitez, Joaquin Quiroz-Mercado, Sergio Rangel-Guerrero, Natividad Cruz-Ortiz, Tatiana Nunez-Garcia, Gisela Barrera-Badillo, Lucia Hernandez-Rivas, Irma Lopez-Martinez, Ernesto Ramirez-Gonzalez. |
| EPI_ISL_2559471, EPI_ISL_2559472, EPI_ISL_2559473, EPI_ISL_2559475, EPI_ISL_2559477, EPI_ISL_2559479 | LESP Ciudad de Mexico | Instituto de Diagnostico y Referencia Epidemiologicos (INDRE) | Claudia Wong-Arambula, Abril Rodriguez-Maldonado, Vanessa Rivero-Arredondo, Ariadna Medina-Benitez, Joaquin Quiroz-Mercado, Sergio Rangel-Guerrero, Natividad Cruz-Ortiz, Tatiana Nunez-Garcia, Gisela Barrera-Badillo, Lucia Hernandez-Rivas, Irma Lopez-Martinez, Ernesto Ramirez-Gonzalez. |
| EPI_ISL_2560418, EPI_ISL_2560420, EPI_ISL_2560424, EPI_ISL_2560426, EPI_ISL_2560429, EPI_ISL_2560431, EPI_ISL_2560433, EPI_ISL_2560439, EPI_ISL_2560441, EPI_ISL_2560443, EPI_ISL_2560446, EPI_ISL_2560448, EPI_ISL_2560450, EPI_ISL_2560453, EPI_ISL_2560457, EPI_ISL_2560460, EPI_ISL_2560462, EPI_ISL_2560465, EPI_ISL_2560467, EPI_ISL_2560469, EPI_ISL_2560472, EPI_ISL_2560474, EPI_ISL_2560477, EPI_ISL_2560479, EPI_ISL_2560482, EPI_ISL_2560484, EPI_ISL_2560486, EPI_ISL_2560491 | | | |
| see above | Wyoming Public Health Laboratory | Wyoming Public Health Laboratory | Jim Mildenberger, Taylor Fearing, Channing Weber, Ashley Norberg, Chayse Rowley, Marley Goetz, Brian Dominguez, Elliot Thomasson, Sam Britz, Cari Sloma, Robert Petit, and Rob Christensen |
| EPI_ISL_2561897 | Willis-Knighton Medical Center Hospital Laboratory | LSUHS Emerging Viral Threat Laboratory | Lorie M. Atkins, April N. Johnson, Gregory L. Ware, Alexander Mijalis, Jeremy P. Kamil, Jennifer L. Carroll, Maarten Van Diest, Rona S. Scott, Andrew D. Yurochko, Christopher G. Kevil, John A. Vanchiere, Joseph A. Bocchini |
| EPI_ISL_2561898 | LSUHS Emerging Viral Threat Laboratory | LSUHS Emerging Viral Threat Laboratory | Gregory L. Ware, Alexander Mijalis, Jeremy P. Kamil, Jennifer L. Carroll, Maarten Van Diest, Rona S. Scott, Andrew D. Yurochko, Christopher G. Kevil, John A. Vanchiere |
| EPI_ISL_2561903 | Willis-Knighton Medical Center Hospital Laboratory | LSUHS Emerging Viral Threat Laboratory | Lorie M. Atkins, April N. Johnson, Gregory L. Ware, Alexander Mijalis, Jeremy P. Kamil, Jennifer L. Carroll, Maarten Van Diest, Rona S. Scott, Andrew D. Yurochko, Christopher G. Kevil, John A. Vanchiere, Joseph A. Bocchini |
| EPI_ISL_2561909 | LSUHS Emerging Viral Threat Laboratory | LSUHS Emerging Viral Threat Laboratory | Gregory L. Ware, Alexander Mijalis, Jeremy P. Kamil, Jennifer L. Carroll, Maarten Van Diest, Rona S. Scott, Andrew D. Yurochko, Christopher G. Kevil, John A. Vanchiere |
| EPI_ISL_2561910, EPI_ISL_2561911, EPI_ISL_2561912, EPI_ISL_2561913 | Willis-Knighton Medical Center Hospital Laboratory | LSUHS Emerging Viral Threat Laboratory | Lorie M. Atkins, April N. Johnson, Gregory L. Ware, Alexander Mijalis, Jeremy P. Kamil, Jennifer L. Carroll, Maarten Van Diest, Rona S. Scott, Andrew D. Yurochko, Christopher G. Kevil, John A. Vanchiere, Joseph A. Bocchini |
| EPI_ISL_2561914 | LSUHS Emerging Viral Threat Laboratory | LSUHS Emerging Viral Threat Laboratory | Gregory L. Ware, Alexander Mijalis, Jeremy P. Kamil, Jennifer L. Carroll, Maarten Van Diest, Rona S. Scott, Andrew D. Yurochko, Christopher G. Kevil, John A. Vanchiere |
| EPI_ISL_2561915, EPI_ISL_2561916, EPI_ISL_2561917, EPI_ISL_2561919, EPI_ISL_2561920 | Willis-Knighton Medical Center Hospital Laboratory | LSUHS Emerging Viral Threat Laboratory | Lorie M. Atkins, April N. Johnson, Gregory L. Ware, Alexander Mijalis, Jeremy P. Kamil, Jennifer L. Carroll, Maarten Van Diest, Rona S. Scott, Andrew D. Yurochko, Christopher G. Kevil, John A. Vanchiere, Joseph A. Bocchini |
| EPI_ISL_2561921, EPI_ISL_2561922 | LSUHS Emerging Viral Threat Laboratory | LSUHS Emerging Viral Threat Laboratory | Alexander Mijalis, Jeremy P. Kamil, Jennifer L. Carroll, Maarten Van Diest, Rona S. Scott, Gregory L. Ware, Andrew D. Yurochko, Christopher G. Kevil, John A. Vanchiere |
| EPI_ISL_2561923, EPI_ISL_2561924, EPI_ISL_2561925, EPI_ISL_2561926, EPI_ISL_2561927, EPI_ISL_2561930, EPI_ISL_2561931 | Willis-Knighton Medical Center Hospital Laboratory | LSUHS Emerging Viral Threat Laboratory | Lorie M. Atkins, April N. Johnson, Gregory L. Ware, Alexander Mijalis, Jeremy P. Kamil, Jennifer L. Carroll, Maarten Van Diest, Rona S. Scott, Andrew D. Yurochko, Christopher G. Kevil, John A. Vanchiere, Joseph A. Bocchini |
| EPI_ISL_2561933 | LSUHS Emerging Viral Threat Laboratory | LSUHS Emerging Viral Threat Laboratory | Alexander Mijalis, Jeremy P. Kamil, Jennifer L. Carroll, Maarten Van Diest, Rona S. Scott, Gregory L. Ware, Andrew D. Yurochko, Christopher G. Kevil, John A. Vanchiere |
| EPI_ISL_2561934 | Willis-Knighton Medical Center Hospital Laboratory | LSUHS Emerging Viral Threat Laboratory | Lorie M. Atkins, April N. Johnson, Gregory L. Ware, Alexander Mijalis, Jeremy P. Kamil, Jennifer L. Carroll, Maarten Van Diest, Rona S. Scott, Andrew D. Yurochko, Christopher G. Kevil, John A. Vanchiere, Joseph A. Bocchini |
| EPI_ISL_2561936 | LSUHS Emerging Viral Threat Laboratory | LSUHS Emerging Viral Threat Laboratory | Alexander Mijalis, Jeremy P. Kamil, Jennifer L. Carroll, Maarten Van Diest, Rona S. Scott, Gregory L. Ware, Andrew D. Yurochko, Christopher G. Kevil, John A. Vanchiere |
| EPI_ISL_2561937 | Willis-Knighton Medical Center Hospital | LSUHS Emerging Viral Threat Laboratory | Lorie M. Atkins, April N. Johnson, Gregory L. Ware, Alexander Mijalis, Jeremy P. Kamil, Jennifer L. Carroll, Maarten Van Diest, Rona S. Scott, Andrew D. Yurochko, Christopher G. Kevil, John A. Vanchiere, Joseph A. Bocchini |

| | | | |
|---|---|---|---|
| | Laboratory | | |
| EPI_ISL_2562121, EPI_ISL_2562122, EPI_ISL_2562123, EPI_ISL_2562124, EPI_ISL_2562125 | The Jackson Laboratory | The Jackson Laboratory | Long J,Bergeron D, Renzette N, Adams M, Omerza G, Kelly K, Li L |
| EPI_ISL_2562214, EPI_ISL_2562218, EPI_ISL_2562221, EPI_ISL_2562228, EPI_ISL_2562242, EPI_ISL_2562243, EPI_ISL_2562316, EPI_ISL_2562322, EPI_ISL_2562325, EPI_ISL_2562329, EPI_ISL_2562330, EPI_ISL_2562337, EPI_ISL_2562341, EPI_ISL_2562346, EPI_ISL_2562358, EPI_ISL_2562407, EPI_ISL_2562424, EPI_ISL_2562425, EPI_ISL_2562433, EPI_ISL_2562438, EPI_ISL_2562449, EPI_ISL_2562457, EPI_ISL_2562462, EPI_ISL_2562477, EPI_ISL_2562489, EPI_ISL_2562495, EPI_ISL_2562502, EPI_ISL_2562517, EPI_ISL_2562537, EPI_ISL_2562675, EPI_ISL_2562676, EPI_ISL_2562677, EPI_ISL_2562678, EPI_ISL_2562680, EPI_ISL_2562681, EPI_ISL_2562695, EPI_ISL_2562706, EPI_ISL_2562707, EPI_ISL_2562708, EPI_ISL_2562709, EPI_ISL_2562711, EPI_ISL_2562712, EPI_ISL_2562721, EPI_ISL_2562722, EPI_ISL_2562723 | | | |
| see above | Creighton University Medical Center | Creighton COVID Consortium | Holly Stessman, Michael Belshan, Richard Goering, Cynthia Watson, Jake Siedlik |
| EPI_ISL_2562727, EPI_ISL_2562728 | NC State Laboratory of Public Health | Genomics and Discovery, Respiratory Viruses Branch, Division of Viral Diseases, Centers for Disease Control and Prevention | Yan Li, Jing Zhang, Anna Kelleher, Ying Tao, Brian Lynch, Krista Queen, Anna Uehara, Peter Cook, Han Jia Justin Ng, Rachel Marine, Clinton R. Paden, Dhwani Batra, Haibin Wang, Tara Coalter, Jasmine Padilla, Morgan Davis, Mili Sheth, Sarah Nobles, Mark Burroughs, Justin Lee, Adam Retchless, Suxiang Tong |
| EPI_ISL_2562729, EPI_ISL_2562730, EPI_ISL_2562731 | AR Dept. of Health-PHL, Molecular Diagnostics | Genomics and Discovery, Respiratory Viruses Branch, Division of Viral Diseases, Centers for Disease Control and Prevention | Yan Li, Jing Zhang, Anna Kelleher, Ying Tao, Brian Lynch, Krista Queen, Anna Uehara, Peter Cook, Han Jia Justin Ng, Rachel Marine, Clinton R. Paden, Dhwani Batra, Haibin Wang, Tara Coalter, Jasmine Padilla, Morgan Davis, Mili Sheth, Sarah Nobles, Mark Burroughs, Justin Lee, Adam Retchless, Suxiang Tong |
| EPI_ISL_2562732, EPI_ISL_2562733 | NM Dept. Health, Scientific Laboratory Division | Genomics and Discovery, Respiratory Viruses Branch, Division of Viral Diseases, Centers for Disease Control and Prevention | Yan Li, Jing Zhang, Anna Kelleher, Ying Tao, Brian Lynch, Krista Queen, Anna Uehara, Peter Cook, Han Jia Justin Ng, Rachel Marine, Clinton R. Paden, Dhwani Batra, Haibin Wang, Tara Coalter, Jasmine Padilla, Morgan Davis, Mili Sheth, Sarah Nobles, Mark Burroughs, Justin Lee, Adam Retchless, Suxiang Tong |
| EPI_ISL_2562734 | LA Office of Public Health Laboratories | Genomics and Discovery, Respiratory Viruses Branch, Division of Viral Diseases, Centers for Disease Control and Prevention | Yan Li, Jing Zhang, Anna Kelleher, Ying Tao, Brian Lynch, Krista Queen, Anna Uehara, Peter Cook, Han Jia Justin Ng, Rachel Marine, Clinton R. Paden, Dhwani Batra, Haibin Wang, Tara Coalter, Jasmine Padilla, Morgan Davis, Mili Sheth, Sarah Nobles, Mark Burroughs, Justin Lee, Adam Retchless, Suxiang Tong |
| EPI_ISL_2562735, EPI_ISL_2562736 | OK Public Health Laboratory, Oklahoma State DOH | Genomics and Discovery, Respiratory Viruses Branch, Division of Viral Diseases, Centers for Disease Control and Prevention | Yan Li, Jing Zhang, Anna Kelleher, Ying Tao, Brian Lynch, Krista Queen, Anna Uehara, Peter Cook, Han Jia Justin Ng, Rachel Marine, Clinton R. Paden, Dhwani Batra, Haibin Wang, Tara Coalter, Jasmine Padilla, Morgan Davis, Mili Sheth, Sarah Nobles, Mark Burroughs, Justin Lee, Adam Retchless, Suxiang Tong |
| EPI_ISL_2562737, EPI_ISL_2562738, EPI_ISL_2562739, EPI_ISL_2562740, EPI_ISL_2562741, EPI_ISL_2562742, EPI_ISL_2562745, EPI_ISL_2562746, EPI_ISL_2562747, EPI_ISL_2562748, EPI_ISL_2562749, EPI_ISL_2562750, EPI_ISL_2562751, EPI_ISL_2562752, EPI_ISL_2562753, EPI_ISL_2562754, EPI_ISL_2562755, EPI_ISL_2562756, EPI_ISL_2562757, EPI_ISL_2562758, EPI_ISL_2562760, EPI_ISL_2562761, EPI_ISL_2562762, EPI_ISL_2562763, EPI_ISL_2562764, EPI_ISL_2562765, EPI_ISL_2562766, EPI_ISL_2562767, EPI_ISL_2562768, EPI_ISL_2562769, EPI_ISL_2562770, EPI_ISL_2562771, EPI_ISL_2562772, EPI_ISL_2562773, EPI_ISL_2562774, EPI_ISL_2562775, EPI_ISL_2562776, EPI_ISL_2562777, EPI_ISL_2562778, EPI_ISL_2562779, EPI_ISL_2562780, EPI_ISL_2562781, EPI_ISL_2562782, EPI_ISL_2562783, EPI_ISL_2562784, EPI_ISL_2562785, EPI_ISL_2562786, EPI_ISL_2562787, EPI_ISL_2562788, EPI_ISL_2562789, EPI_ISL_2562790, EPI_ISL_2562791, EPI_ISL_2562792, EPI_ISL_2562793, EPI_ISL_2562794, EPI_ISL_2562795 | | | |
| see above | TriCore Reference Laboratories | Center for Global Health, University of New Mexico Health Sciences Center | Daryl Domman, Kurt Schwalm, Valerie Morley, Cecilia Thompson, Kendra Pesko, Karissa Culbreath, Darrell Dinwiddie |
| EPI_ISL_2565057, EPI_ISL_2565058, EPI_ISL_2565059, EPI_ISL_2565060, EPI_ISL_2565061, EPI_ISL_2565062, EPI_ISL_2565063, EPI_ISL_2565064, EPI_ISL_2565065 | GA Department of Public Health | GA Department of Public Health | Stacy Reeves, Jonathan Edwards, Cynthia Dixey, Tonia Parrott, Aliyah Fields, Taylor Smith |
| EPI_ISL_2565759, EPI_ISL_2565761, EPI_ISL_2565762, EPI_ISL_2565763, EPI_ISL_2565764, EPI_ISL_2565765, EPI_ISL_2565766, EPI_ISL_2565767, EPI_ISL_2565768, EPI_ISL_2565769, EPI_ISL_2565770, EPI_ISL_2565771, EPI_ISL_2565772, EPI_ISL_2565773, EPI_ISL_2565774, EPI_ISL_2565775, EPI_ISL_2565776, EPI_ISL_2565777, EPI_ISL_2565778, EPI_ISL_2565779, EPI_ISL_2565780, EPI_ISL_2565781, EPI_ISL_2565782, EPI_ISL_2565783, EPI_ISL_2565784, EPI_ISL_2565785, EPI_ISL_2565786, EPI_ISL_2565787, EPI_ISL_2565788, EPI_ISL_2565789, EPI_ISL_2565791, EPI_ISL_2565792, EPI_ISL_2565793, EPI_ISL_2565794, EPI_ISL_2565795, EPI_ISL_2565796, EPI_ISL_2565797, EPI_ISL_2565798, EPI_ISL_2565799 | | | |
| see above | TXDSHS | TXDSHS | Rashmi Tuladhar, Bonnie Oh, Jenny Zhang, Maliha Rahman, Mayela Pedrueza, Anita Pokharel, Karen Bobier, Lorraine Rodriguez, Myong Koag, Chun Wang, Rachel Lee, Grace Kubin |
| EPI_ISL_2565800, EPI_ISL_2565801, EPI_ISL_2565802, EPI_ISL_2565803, EPI_ISL_2565804, EPI_ISL_2565805, EPI_ISL_2565806, EPI_ISL_2565807, EPI_ISL_2565808, EPI_ISL_2565809, EPI_ISL_2565810, EPI_ISL_2565811, EPI_ISL_2565812, EPI_ISL_2565813, EPI_ISL_2565814, EPI_ISL_2565815, EPI_ISL_2565816, EPI_ISL_2565817, EPI_ISL_2565818, EPI_ISL_2565819 | | | |
| see above | Montana Public Health Laboratory | Montana Public Health Laboratory | Joy Ritter, Michelle Mozer, Carrie Biskupiak, Deborah Gibson, Michael Dills |
| EPI_ISL_2565820, EPI_ISL_2565821, EPI_ISL_2565822, EPI_ISL_2565823, EPI_ISL_2565824, EPI_ISL_2565825, EPI_ISL_2565826, EPI_ISL_2565827, EPI_ISL_2565828, EPI_ISL_2565829, EPI_ISL_2565830, EPI_ISL_2565831 | | | |
| see above | Swedish Medical Center, Seattle, WA, USA | Institute for Systems Biology | Jason D. Goldman, Chris Lausted, Kai Wang, Jared C. Roach, James R. Heath, Julie A. Wallick |
| EPI_ISL_2565832, EPI_ISL_2565833, EPI_ISL_2565834, EPI_ISL_2565835, EPI_ISL_2565836, EPI_ISL_2565837, EPI_ISL_2565838, EPI_ISL_2565839, EPI_ISL_2565840, EPI_ISL_2565841, EPI_ISL_2565842, EPI_ISL_2565843, EPI_ISL_2565844, EPI_ISL_2565845, EPI_ISL_2565846, EPI_ISL_2565847, EPI_ISL_2565848, EPI_ISL_2565849, EPI_ISL_2565850, EPI_ISL_2565851, EPI_ISL_2565852, EPI_ISL_2565853, EPI_ISL_2565854, EPI_ISL_2565856, EPI_ISL_2565857, EPI_ISL_2565858 | | | |
| see above | Kansas Health and Environmental Lab | Kansas Health and Environmental Lab | Mike Grose, Jonathan Barnell, Ben Olsen, and Phil Adam |
| EPI_ISL_2565862 | Minnesota Department of Health, Public Health Laboratory | Minnesota Department of Health, Public Health Laboratory | Alexandra Lorentz, Jacob Garfin, Matt Plumb, and Xiong Wang |
| EPI_ISL_2565863 | Regions Hospital | Minnesota Department of Health, Public Health Laboratory | Alexandra Lorentz, Jacob Garfin, Matt Plumb, and Xiong Wang |
| EPI_ISL_2565864, EPI_ISL_2565865, EPI_ISL_2565866, EPI_ISL_2565867, EPI_ISL_2565868, EPI_ISL_2565869 | Minnesota Department of Health, Public Health Laboratory | Minnesota Department of Health, Public Health Laboratory | Alexandra Lorentz, Jacob Garfin, Matt Plumb, and Xiong Wang |
| EPI_ISL_2565870, EPI_ISL_2565871, EPI_ISL_2565872, EPI_ISL_2565873, EPI_ISL_2565874, EPI_ISL_2565875 | Hennepin County Medical Center | Minnesota Department of Health, Public Health Laboratory | Alexandra Lorentz, Jacob Garfin, Matt Plumb, and Xiong Wang |
| EPI_ISL_2565876, EPI_ISL_2565877, EPI_ISL_2565878, EPI_ISL_2565879, EPI_ISL_2565880, EPI_ISL_2565881 | Minnesota Department of Health, Public Health Laboratory | Minnesota Department of Health, Public Health Laboratory | Alexandra Lorentz, Jacob Garfin, Matt Plumb, and Xiong Wang |
| EPI_ISL_2565882, EPI_ISL_2565883, EPI_ISL_2565884, EPI_ISL_2565885 | Premier Medical Lab | Minnesota Department of Health, Public Health Laboratory | Alexandra Lorentz, Jacob Garfin, Matt Plumb, and Xiong Wang |
| EPI_ISL_2565900, EPI_ISL_2565901, EPI_ISL_2565902, EPI_ISL_2565903, EPI_ISL_2565905, EPI_ISL_2565907, EPI_ISL_2565908, EPI_ISL_2565909, EPI_ISL_2565910, EPI_ISL_2565911, EPI_ISL_2565912, EPI_ISL_2565913, EPI_ISL_2565914, EPI_ISL_2565915, EPI_ISL_2565916, EPI_ISL_2565917, EPI_ISL_2565918, EPI_ISL_2565919, EPI_ISL_2565920, EPI_ISL_2565922, EPI_ISL_2565923, EPI_ISL_2565924, EPI_ISL_2565925, EPI_ISL_2565926, EPI_ISL_2565927, EPI_ISL_2565928, EPI_ISL_2565929, EPI_ISL_2565932, EPI_ISL_2565933, EPI_ISL_2565934, EPI_ISL_2565935, EPI_ISL_2565936, EPI_ISL_2565938, EPI_ISL_2565939, EPI_ISL_2565942, EPI_ISL_2565943, EPI_ISL_2565944, EPI_ISL_2565945, EPI_ISL_2565946, EPI_ISL_2565947, EPI_ISL_2565948, EPI_ISL_2565949, EPI_ISL_2565950, EPI_ISL_2565951, EPI_ISL_2565952, EPI_ISL_2565953, EPI_ISL_2565955, EPI_ISL_2565956, EPI_ISL_2565957, EPI_ISL_2565958, EPI_ISL_2565960, EPI_ISL_2565961, EPI_ISL_2565962, EPI_ISL_2565963, EPI_ISL_2565964, EPI_ISL_2565965, EPI_ISL_2565966, EPI_ISL_2565967 | | | |
| see above | North Dakota Department of Health, Public Health Laboratory | North Dakota Department of Health, Public Health Laboratory | Lisa Wingerter |
| EPI_ISL_2566083, EPI_ISL_2566084, EPI_ISL_2566085, EPI_ISL_2566086, EPI_ISL_2566087, EPI_ISL_2566089, EPI_ISL_2566090, EPI_ISL_2566091, EPI_ISL_2566092, EPI_ISL_2566093, EPI_ISL_2566094, EPI_ISL_2566095, EPI_ISL_2566096, EPI_ISL_2566097, EPI_ISL_2566098, EPI_ISL_2566099, EPI_ISL_2566100, EPI_ISL_2566101, EPI_ISL_2566102, EPI_ISL_2566103, EPI_ISL_2566104, EPI_ISL_2566105, EPI_ISL_2566106, EPI_ISL_2566107, EPI_ISL_2566108, EPI_ISL_2566109, EPI_ISL_2566110 | | | |
| see above | Wyoming Public Health Laboratory | Wyoming Public Health Laboratory | Jim Mildenberger, Taylor Fearing, Channing Weber, Ashley Norberg, Chayse Rowley, Marley Goetz, Brian Dominguez, Elliot Thomasson, Sam Britz, Cari Sloma, Robert Petit, and Rob Christensen |
| EPI_ISL_2566111, EPI_ISL_2566112, EPI_ISL_2566113, EPI_ISL_2566114, EPI_ISL_2566115, EPI_ISL_2566116, EPI_ISL_2566117, EPI_ISL_2566118, EPI_ISL_2566120 | Washington State Department of Health Public Health Laboratories | Washington State Department of Health Public Health Laboratories | Drew MacKellar, Philip Dykema, Denny Russell, Joenice Gonzalez, Hannah Gray, Geoff Melly, Vanessa De Los Santos, Darren Lucas, JohnAric Peterson, Avi Singh, Rebecca Cao |
| EPI_ISL_2566121, EPI_ISL_2566122 | Nevada State Public Health Laboratory | Nevada State Public Health Laboratory | Andrew Gorzalski, Mark Pandori |
| EPI_ISL_2566243, EPI_ISL_2566244, EPI_ISL_2566245, EPI_ISL_2566246, EPI_ISL_2566247, EPI_ISL_2566248, EPI_ISL_2566249, EPI_ISL_2566250, EPI_ISL_2566251, EPI_ISL_2566252, EPI_ISL_2566253, EPI_ISL_2566254, EPI_ISL_2566255, EPI_ISL_2566256, EPI_ISL_2566257, EPI_ISL_2566258, EPI_ISL_2566259, EPI_ISL_2566260, EPI_ISL_2566261, EPI_ISL_2566262, EPI_ISL_2566263, EPI_ISL_2566264, | | | |

| | | | |
|---|---|---|---|
| EPI_ISL_2566265, EPI_ISL_2566266, EPI_ISL_2566267, EPI_ISL_2566268, EPI_ISL_2566269, EPI_ISL_2566271, EPI_ISL_2566272, EPI_ISL_2566273, EPI_ISL_2566274, EPI_ISL_2566275, EPI_ISL_2566276, EPI_ISL_2566277, EPI_ISL_2566278, EPI_ISL_2566279, EPI_ISL_2566280, EPI_ISL_2566281, EPI_ISL_2566282, EPI_ISL_2566283, EPI_ISL_2566284, EPI_ISL_2566285, EPI_ISL_2566286, EPI_ISL_2566287, EPI_ISL_2566288, EPI_ISL_2566289, EPI_ISL_2566290, EPI_ISL_2566291, EPI_ISL_2566292, EPI_ISL_2566293, EPI_ISL_2566294, EPI_ISL_2566295, EPI_ISL_2566296, EPI_ISL_2566297, EPI_ISL_2566298, EPI_ISL_2566299, EPI_ISL_2566300, EPI_ISL_2566301, EPI_ISL_2566302, EPI_ISL_2566303 | | | |
| see above | Wyoming Public Health Laboratory | Wyoming Public Health Laboratory | Jim Mildenberger, Taylor Fearing, Channing Weber, Ashley Norberg, Chayse Rowley, Marley Goetz, Brian Dominguez, Elliot Thomasson, Sam Britz, Cari Sloma, Robert Petit, and Rob Christensen |
| EPI_ISL_2566304, EPI_ISL_2566305 | Utah Public Health Laboratory, Utah Public Health Laboratory Infectious Disease submission group | Utah Public Health Laboratory, Utah Public Health Laboratory Infectious Disease submission group | Young,E.L., Oakeson,K.F., Gallagher,T., Linares-Perdomo,O. |
| EPI_ISL_2566307, EPI_ISL_2566308, EPI_ISL_2566310, EPI_ISL_2566312, EPI_ISL_2566313 | Utah Public Health Laboratory, Utah Public Health Laboratory Infectious Disease submission group | Utah Public Health Laboratory, Utah Public Health Laboratory Infectious Disease submission group | Gallagher,T., Young,E.L., Oakeson,K.F. |
| EPI_ISL_2566314, EPI_ISL_2566315, EPI_ISL_2566316, EPI_ISL_2566317, EPI_ISL_2566318, EPI_ISL_2566319, EPI_ISL_2566320, EPI_ISL_2566321, EPI_ISL_2566322, EPI_ISL_2566323, EPI_ISL_2566324, EPI_ISL_2566325, EPI_ISL_2566326, EPI_ISL_2566328, EPI_ISL_2566329, EPI_ISL_2566330, EPI_ISL_2566331, EPI_ISL_2566332, EPI_ISL_2566333, EPI_ISL_2566334, EPI_ISL_2566335, EPI_ISL_2566336, EPI_ISL_2566337, EPI_ISL_2566338, EPI_ISL_2566339, EPI_ISL_2566340, EPI_ISL_2566341 | | | |
| see above | Wyoming Public Health Laboratory | Wyoming Public Health Laboratory | Jim Mildenberger, Taylor Fearing, Channing Weber, Ashley Norberg, Chayse Rowley, Marley Goetz, Brian Dominguez, Elliot Thomasson, Sam Britz, Cari Sloma, Robert Petit, and Rob Christensen |
| EPI_ISL_2566342, EPI_ISL_2566343, EPI_ISL_2566344, EPI_ISL_2566345, EPI_ISL_2566346, EPI_ISL_2566347, EPI_ISL_2566348, EPI_ISL_2566349, EPI_ISL_2566350, EPI_ISL_2566351, EPI_ISL_2566352, EPI_ISL_2566353 | | | |
| see above | Operation Infectious Diseases, Naval Health Research Center | Operation Infectious Diseases, Naval Health Research Center | Pan,R.W., Underwood,R.S., Balansay-Ames,M.S., Myers,C.A. |
| EPI_ISL_2566354, EPI_ISL_2566355, EPI_ISL_2566356, EPI_ISL_2566357, EPI_ISL_2566358, EPI_ISL_2566359, EPI_ISL_2566360, EPI_ISL_2566361, EPI_ISL_2566362, EPI_ISL_2566363, EPI_ISL_2566364, EPI_ISL_2566365, EPI_ISL_2566366, EPI_ISL_2566367, EPI_ISL_2566368, EPI_ISL_2566369, EPI_ISL_2566370, EPI_ISL_2566371, EPI_ISL_2566372, EPI_ISL_2566373, EPI_ISL_2566374, EPI_ISL_2566375, EPI_ISL_2566376, EPI_ISL_2566378, EPI_ISL_2566379 | | | |
| see above | Operation Infectious Diseases, Naval Health Research Center | Operation Infectious Diseases, Naval Health Research Center | Pan,R.W., Underwood,R.S., Balansay-Ames,M.S., Myers,C.A., Fierro,M., Kriner,P., Iniguez-Stevens,E., Moser,K., Phippard,A.E. |
| EPI_ISL_2567077, EPI_ISL_2567078, EPI_ISL_2567079, EPI_ISL_2567080, EPI_ISL_2567081, EPI_ISL_2567083, EPI_ISL_2567084, EPI_ISL_2567085, EPI_ISL_2567086, EPI_ISL_2567087, EPI_ISL_2567089, EPI_ISL_2567090, EPI_ISL_2567092, EPI_ISL_2567093, EPI_ISL_2567095, EPI_ISL_2567096, EPI_ISL_2567098, EPI_ISL_2567099, EPI_ISL_2567101, EPI_ISL_2567102, EPI_ISL_2567103, EPI_ISL_2567105, EPI_ISL_2567106, EPI_ISL_2567108, EPI_ISL_2567109, EPI_ISL_2567110, EPI_ISL_2567111, EPI_ISL_2567112, EPI_ISL_2567113, EPI_ISL_2567114, EPI_ISL_2567116, EPI_ISL_2567117, EPI_ISL_2567118, EPI_ISL_2567119, EPI_ISL_2567120, EPI_ISL_2567122, EPI_ISL_2567123, EPI_ISL_2567125, EPI_ISL_2567126, EPI_ISL_2567127, EPI_ISL_2567128, EPI_ISL_2567129, EPI_ISL_2567130, EPI_ISL_2567131, EPI_ISL_2567132, EPI_ISL_2567134, EPI_ISL_2567135, EPI_ISL_2567136, EPI_ISL_2567137, EPI_ISL_2567138, EPI_ISL_2567139, EPI_ISL_2567140, EPI_ISL_2567141, EPI_ISL_2567143, EPI_ISL_2567145, EPI_ISL_2567147, EPI_ISL_2567148, EPI_ISL_2567149, EPI_ISL_2567150, EPI_ISL_2567152, EPI_ISL_2567153, EPI_ISL_2567154, EPI_ISL_2567155, EPI_ISL_2567156, EPI_ISL_2567157, EPI_ISL_2567158, EPI_ISL_2567160, EPI_ISL_2567161, EPI_ISL_2567163, EPI_ISL_2567164, EPI_ISL_2567165, EPI_ISL_2567168, EPI_ISL_2567169, EPI_ISL_2567172, EPI_ISL_2567173, EPI_ISL_2567176, EPI_ISL_2567177, EPI_ISL_2567178, EPI_ISL_2567179, EPI_ISL_2567181, EPI_ISL_2567182, EPI_ISL_2567183, EPI_ISL_2567184, EPI_ISL_2567185, EPI_ISL_2567187, EPI_ISL_2567188, EPI_ISL_2567190, EPI_ISL_2567191, EPI_ISL_2567192, EPI_ISL_2567193, EPI_ISL_2567195, EPI_ISL_2567198, EPI_ISL_2567200, EPI_ISL_2567201, EPI_ISL_2567202, EPI_ISL_2567203, EPI_ISL_2567204, EPI_ISL_2567205, EPI_ISL_2567206, EPI_ISL_2567207, EPI_ISL_2567208, EPI_ISL_2567209, EPI_ISL_2567210, EPI_ISL_2567211, EPI_ISL_2567213, EPI_ISL_2567214, EPI_ISL_2567215, EPI_ISL_2567216, EPI_ISL_2567217, EPI_ISL_2567220, EPI_ISL_2567222, EPI_ISL_2567223, EPI_ISL_2567224, EPI_ISL_2567225, EPI_ISL_2567226, EPI_ISL_2567227, EPI_ISL_2567228, EPI_ISL_2567229, EPI_ISL_2567230, EPI_ISL_2567232, EPI_ISL_2567233, EPI_ISL_2567234, EPI_ISL_2567235, EPI_ISL_2567237, EPI_ISL_2567240, EPI_ISL_2567243, EPI_ISL_2567244, EPI_ISL_2567245, EPI_ISL_2567246, EPI_ISL_2567247, EPI_ISL_2567248, EPI_ISL_2567249, EPI_ISL_2567250, EPI_ISL_2567251, EPI_ISL_2567256, EPI_ISL_2567258, EPI_ISL_2567259, EPI_ISL_2567260, EPI_ISL_2567262, EPI_ISL_2567263 | | | |
| see above | Florida Bureau of Public Health Laboratories | Florida Bureau of Public Health Laboratories | Sarah Schmedes, Jason Blanton |
| EPI_ISL_2574006, EPI_ISL_2574008, EPI_ISL_2574009, EPI_ISL_2574010, EPI_ISL_2574011, EPI_ISL_2574012, EPI_ISL_2574013, EPI_ISL_2574014, EPI_ISL_2574015, EPI_ISL_2574016, EPI_ISL_2574017, EPI_ISL_2574018, EPI_ISL_2574019, EPI_ISL_2574020, EPI_ISL_2574021, EPI_ISL_2574022, EPI_ISL_2574023, EPI_ISL_2574024, EPI_ISL_2574025, EPI_ISL_2574026, EPI_ISL_2574027, EPI_ISL_2574028, EPI_ISL_2574029, EPI_ISL_2574030, EPI_ISL_2574031, EPI_ISL_2574032, EPI_ISL_2574033, EPI_ISL_2574034, EPI_ISL_2574035, EPI_ISL_2574036, EPI_ISL_2574037, EPI_ISL_2574038, EPI_ISL_2574039, EPI_ISL_2574040, EPI_ISL_2574041, EPI_ISL_2574042, EPI_ISL_2574043, EPI_ISL_2574044, EPI_ISL_2574045, EPI_ISL_2574046, EPI_ISL_2574047, EPI_ISL_2574048, EPI_ISL_2574049, EPI_ISL_2574050, EPI_ISL_2574051, EPI_ISL_2574052 | | | |
| see above | New Mexico Department of Health Scientific Laboratory | New Mexico Department of Health Scientific Laboratory | Ellie Johnson, D'eldra Malone, Jennifer Benoit, Ratheesh Rajan, Linda Salazar, Keila Gutierrez, Anastacia Griego-Fisher |
| EPI_ISL_2574054 | Kansas Health and Environmental Lab | Kansas Health and Environmental Lab | Katherine Wiggins, Mike Grose, Jonathan Barnell, Ben Olsen, and Phil Adam |
| EPI_ISL_2576912, EPI_ISL_2576913, EPI_ISL_2576914, EPI_ISL_2576915, EPI_ISL_2576916, EPI_ISL_2576917, EPI_ISL_2576919, EPI_ISL_2576920, EPI_ISL_2576921, EPI_ISL_2576922, EPI_ISL_2576923, EPI_ISL_2576924, EPI_ISL_2576925, EPI_ISL_2576926, EPI_ISL_2576927, EPI_ISL_2576928, EPI_ISL_2576929, EPI_ISL_2576930, EPI_ISL_2576931, EPI_ISL_2576932, EPI_ISL_2576933, EPI_ISL_2576934, EPI_ISL_2576935, EPI_ISL_2576936, EPI_ISL_2576937 | | | |
| see above | Sonic Reference Laboratory | Sonic Reference Laboratory | "Robert Moller, Bijaya Dhakal, Krishna Maddula, John Woody, Cora Lahey, Ellen Pizarek" |
| EPI_ISL_2578646 | Ohio Department of Health Laboratory | Ohio Department of Health Laboratory | Holmes, Jennifer; Eric Brandt, Keoni Omura, Glen McGillivary, Caitlin McDonnell, Jade Mowery, Stephanie Mccracken, Tyler Payne, Kirtana Ramadugu, Erica Leasure, Brent Lee, Kelsey Florek, Heather Blankenship, Allison Black, Quanta Brown, and Tammy Bannerman |
| EPI_ISL_2578647, EPI_ISL_2578648, EPI_ISL_2578649, EPI_ISL_2578650, EPI_ISL_2578651, EPI_ISL_2578652, EPI_ISL_2578653, EPI_ISL_2578654 | University of Wisconsin-Madison AIDS Vaccine Research Laboratories | University of Wisconsin-Madison AIDS Vaccine Research Laboratories | Gage Moreno, Katarina Braun, et al. AIDS Vaccine Research Laboratories |
| EPI_ISL_2578655, EPI_ISL_2579148, EPI_ISL_2582297, EPI_ISL_2583261, EPI_ISL_2583564 | Ohio Department of Health Laboratory | Ohio Department of Health Laboratory | Holmes, Jennifer; Eric Brandt, Keoni Omura, Glen McGillivary, Caitlin McDonnell, Jade Mowery, Stephanie Mccracken, Tyler Payne, Kirtana Ramadugu, Erica Leasure, Brent Lee, Kelsey Florek, Heather Blankenship, Allison Black, Quanta Brown, and Tammy Bannerman |
| EPI_ISL_2586500, EPI_ISL_2586504, EPI_ISL_2586508, EPI_ISL_2586513, EPI_ISL_2586520, EPI_ISL_2586526, EPI_ISL_2586533, EPI_ISL_2586538, EPI_ISL_2586544, EPI_ISL_2586556, EPI_ISL_2586563, EPI_ISL_2586568 | | | |
| see above | Sonic Reference Laboratory | Sonic Reference Laboratory | "Robert Moller, Bijaya Dhakal, Krishna Maddula, John Woody, Cora Lahey, Ellen Pizarek" |
| EPI_ISL_2588601, EPI_ISL_2588622, EPI_ISL_2588623, EPI_ISL_2588624, EPI_ISL_2588625, EPI_ISL_2588635, EPI_ISL_2588637, EPI_ISL_2588638, EPI_ISL_2588639, EPI_ISL_2588640, EPI_ISL_2588646 | | | |
| see above | Ohio Department of Health Laboratory | Ohio Department of Health Laboratory | Holmes, Jennifer; Eric Brandt, Keoni Omura, Glen McGillivary, Caitlin McDonnell, Jade Mowery, Stephanie Mccracken, Tyler Payne, Kirtana Ramadugu, Erica Leasure, Brent Lee, Kelsey Florek, Heather Blankenship, Allison Black, Quanta Brown, and Tammy Bannerman |
| EPI_ISL_2594104, EPI_ISL_2594108, EPI_ISL_2594112, EPI_ISL_2594117, EPI_ISL_2594122, EPI_ISL_2594127, EPI_ISL_2594132, EPI_ISL_2594137, EPI_ISL_2594141, EPI_ISL_2594146, EPI_ISL_2594150, EPI_ISL_2594154, EPI_ISL_2594159, EPI_ISL_2594163, EPI_ISL_2594168, EPI_ISL_2594173, EPI_ISL_2594178, EPI_ISL_2594182, EPI_ISL_2594187, EPI_ISL_2594192, EPI_ISL_2594198, EPI_ISL_2594203, EPI_ISL_2594206, EPI_ISL_2594215, EPI_ISL_2594220, EPI_ISL_2594224, EPI_ISL_2594228, EPI_ISL_2594233, EPI_ISL_2594237, EPI_ISL_2594241, EPI_ISL_2594247, EPI_ISL_2594252, EPI_ISL_2594257, EPI_ISL_2594262, EPI_ISL_2594276, EPI_ISL_2594281, EPI_ISL_2594286, EPI_ISL_2594291, EPI_ISL_2594296, EPI_ISL_2594301, EPI_ISL_2594305, EPI_ISL_2594315, EPI_ISL_2594319, EPI_ISL_2594324, EPI_ISL_2594328, EPI_ISL_2594333, EPI_ISL_2594338, EPI_ISL_2594343, EPI_ISL_2594348, EPI_ISL_2594352, EPI_ISL_2594356, EPI_ISL_2594361, EPI_ISL_2594366, EPI_ISL_2594370, EPI_ISL_2594373, EPI_ISL_2594378, EPI_ISL_2594383, EPI_ISL_2594387, EPI_ISL_2594392, EPI_ISL_2594398, EPI_ISL_2594402, EPI_ISL_2594407, EPI_ISL_2594411, EPI_ISL_2594416, EPI_ISL_2594420, EPI_ISL_2594435, EPI_ISL_2594444, EPI_ISL_2594450, EPI_ISL_2594460, EPI_ISL_2594465, EPI_ISL_2594475, EPI_ISL_2594481, EPI_ISL_2594486, EPI_ISL_2594491, EPI_ISL_2594501, EPI_ISL_2594504, EPI_ISL_2594513, EPI_ISL_2594518, EPI_ISL_2594529, EPI_ISL_2594533, EPI_ISL_2594539, EPI_ISL_2594543, EPI_ISL_2594553, EPI_ISL_2594556, EPI_ISL_2594565, EPI_ISL_2594569, EPI_ISL_2594574, EPI_ISL_2594583, EPI_ISL_2594588, EPI_ISL_2594592, EPI_ISL_2594601, EPI_ISL_2594605, EPI_ISL_2594610, EPI_ISL_2594620, EPI_ISL_2594638, EPI_ISL_2594642, EPI_ISL_2594646, EPI_ISL_2594651, EPI_ISL_2594655, EPI_ISL_2594660, EPI_ISL_2594664, EPI_ISL_2594669, EPI_ISL_2594671, EPI_ISL_2594674, EPI_ISL_2594676, EPI_ISL_2594679, EPI_ISL_2594681, EPI_ISL_2594683, EPI_ISL_2594687, EPI_ISL_2594690, EPI_ISL_2594692, EPI_ISL_2594694, EPI_ISL_2594697, EPI_ISL_2594699, EPI_ISL_2594701, EPI_ISL_2594704, EPI_ISL_2594706, EPI_ISL_2594708, EPI_ISL_2594710, EPI_ISL_2594712, EPI_ISL_2594715, EPI_ISL_2594720, EPI_ISL_2594722, EPI_ISL_2594724, EPI_ISL_2594726, EPI_ISL_2594729, EPI_ISL_2594731, EPI_ISL_2594733, EPI_ISL_2594735, EPI_ISL_2594737, EPI_ISL_2594740, EPI_ISL_2594742, EPI_ISL_2594744, EPI_ISL_2594747, EPI_ISL_2594749, EPI_ISL_2594751, EPI_ISL_2594753, EPI_ISL_2594756, EPI_ISL_2594758, EPI_ISL_2594760, EPI_ISL_2594763, EPI_ISL_2594765, EPI_ISL_2594767, EPI_ISL_2594769, EPI_ISL_2594772, EPI_ISL_2594774, EPI_ISL_2594776, EPI_ISL_2594778, EPI_ISL_2594781, EPI_ISL_2594783, EPI_ISL_2594785, EPI_ISL_2594788, EPI_ISL_2594790, EPI_ISL_2594792, EPI_ISL_2594795, EPI_ISL_2594797, EPI_ISL_2594799, EPI_ISL_2594802, EPI_ISL_2594803, EPI_ISL_2594805, EPI_ISL_2594808, EPI_ISL_2594810, EPI_ISL_2594814, EPI_ISL_2594816, EPI_ISL_2594818, EPI_ISL_2594820, EPI_ISL_2594822, EPI_ISL_2594824, EPI_ISL_2594827, EPI_ISL_2594829, EPI_ISL_2594834, EPI_ISL_2594843, EPI_ISL_2594845, EPI_ISL_2594854, EPI_ISL_2594856, EPI_ISL_2594860, EPI_ISL_2594862, EPI_ISL_2594865, EPI_ISL_2594867, EPI_ISL_2594869, EPI_ISL_2594872, EPI_ISL_2594874, EPI_ISL_2594875, EPI_ISL_2594877, EPI_ISL_2594880, EPI_ISL_2594882, EPI_ISL_2594886, EPI_ISL_2594888, EPI_ISL_2594891, EPI_ISL_2594895, EPI_ISL_2594902, EPI_ISL_2594904, EPI_ISL_2594909, EPI_ISL_2594912, EPI_ISL_2594916, EPI_ISL_2594918, EPI_ISL_2594921, EPI_ISL_2594923, EPI_ISL_2594927, EPI_ISL_2594930 | | | |
| see above | Wyoming Public Health Laboratory | Wyoming Public Health Laboratory | Jim Mildenberger, Taylor Fearing, Channing Weber, Ashley Norberg, Chayse Rowley, Marley Goetz, Brian Dominguez, Elliot Thomasson, Sam Britz, Cari Sloma, Robert Petit, and Rob Christensen |
| EPI_ISL_2595727, EPI_ISL_2595728, EPI_ISL_2595729, EPI_ISL_2595730, EPI_ISL_2595731, EPI_ISL_2595732, EPI_ISL_2595733, EPI_ISL_2595734, EPI_ISL_2595735, EPI_ISL_2595736, EPI_ISL_2595737, EPI_ISL_2595738, EPI_ISL_2595739, EPI_ISL_2595740, EPI_ISL_2595741 | | | |
| see above | Nebraska Public Health Laboratory | NPHL COVID-19 Response Team | NPHL COVID-19 Response Team |
| EPI_ISL_2597056 | Ohio Department of Health Laboratory | Ohio Department of Health Laboratory | Holmes, Jennifer; Eric Brandt, Keoni Omura, Glen McGillivary, Caitlin McDonnell, Jade Mowery, Stephanie Mccracken, Tyler Payne, Kirtana Ramadugu, Erica Leasure, Brent Lee, Kelsey Florek, Heather Blankenship, Allison Black, Quanta Brown, and Tammy Bannerman |
| EPI_ISL_2597313, EPI_ISL_2597314, EPI_ISL_2597315, EPI_ISL_2597316, EPI_ISL_2597317, EPI_ISL_2597318, EPI_ISL_2597319, EPI_ISL_2597320, EPI_ISL_2597321, EPI_ISL_2597322, EPI_ISL_2597323, EPI_ISL_2597324, EPI_ISL_2597325, EPI_ISL_2597326, EPI_ISL_2597327, EPI_ISL_2597328, EPI_ISL_2597329, EPI_ISL_2597330, EPI_ISL_2597331, EPI_ISL_2597332, EPI_ISL_2597333, EPI_ISL_2597334, EPI_ISL_2597335, EPI_ISL_2597336, EPI_ISL_2597337, EPI_ISL_2597338, EPI_ISL_2597339, EPI_ISL_2597340, EPI_ISL_2597341, EPI_ISL_2597342, EPI_ISL_2597343, EPI_ISL_2597344, EPI_ISL_2597345, EPI_ISL_2597346, EPI_ISL_2597347, EPI_ISL_2597348, EPI_ISL_2597349, EPI_ISL_2597350, EPI_ISL_2597351, EPI_ISL_2597352, EPI_ISL_2597353, EPI_ISL_2597354, EPI_ISL_2597355, EPI_ISL_2597356, EPI_ISL_2597357, EPI_ISL_2597358, EPI_ISL_2597359, EPI_ISL_2597360, EPI_ISL_2597361, EPI_ISL_2597362, EPI_ISL_2597363, EPI_ISL_2597364, EPI_ISL_2597365, EPI_ISL_2597366, EPI_ISL_2597367, EPI_ISL_2597368, EPI_ISL_2597369, EPI_ISL_2597370, EPI_ISL_2597371, EPI_ISL_2597372, EPI_ISL_2597373, EPI_ISL_2597374, EPI_ISL_2597375, EPI_ISL_2597376, EPI_ISL_2597377, EPI_ISL_2597378, EPI_ISL_2597379, EPI_ISL_2597380, EPI_ISL_2597381, EPI_ISL_2597382, EPI_ISL_2597383, EPI_ISL_2597384, EPI_ISL_2597385, EPI_ISL_2597386, EPI_ISL_2597387, EPI_ISL_2597388, EPI_ISL_2597389, EPI_ISL_2597390, EPI_ISL_2597391, EPI_ISL_2597392, EPI_ISL_2597393, EPI_ISL_2597394, EPI_ISL_2597395, EPI_ISL_2597396, EPI_ISL_2597397, EPI_ISL_2597398, EPI_ISL_2597399, EPI_ISL_2597400, EPI_ISL_2597401, EPI_ISL_2597402, EPI_ISL_2597403, EPI_ISL_2597404, EPI_ISL_2597405, EPI_ISL_2597406, EPI_ISL_2597407, EPI_ISL_2597408, EPI_ISL_2597409, EPI_ISL_2597410, EPI_ISL_2597411, EPI_ISL_2597412, EPI_ISL_2597413, EPI_ISL_2597414, EPI_ISL_2597415, EPI_ISL_2597416, EPI_ISL_2597417, EPI_ISL_2597418, EPI_ISL_2597419, EPI_ISL_2597420, EPI_ISL_2597421, EPI_ISL_2597422, EPI_ISL_2597423, EPI_ISL_2597424, EPI_ISL_2597425, EPI_ISL_2597426, EPI_ISL_2597427, EPI_ISL_2597428, EPI_ISL_2597429, EPI_ISL_2597430, EPI_ISL_2597431, EPI_ISL_2597432, EPI_ISL_2597433, EPI_ISL_2597434, EPI_ISL_2597435, EPI_ISL_2597436, EPI_ISL_2597437, EPI_ISL_2597438, EPI_ISL_2597439, EPI_ISL_2597440, EPI_ISL_2597441, EPI_ISL_2597442, EPI_ISL_2597443, EPI_ISL_2597444, EPI_ISL_2597445, EPI_ISL_2597446, EPI_ISL_2597447, EPI_ISL_2597448, EPI_ISL_2597449, EPI_ISL_2597450, EPI_ISL_2597451, EPI_ISL_2597452, EPI_ISL_2597453, EPI_ISL_2597454, EPI_ISL_2597455, EPI_ISL_2597456, EPI_ISL_2597457, EPI_ISL_2597458, EPI_ISL_2597459, EPI_ISL_2597460, EPI_ISL_2597461, EPI_ISL_2597462, EPI_ISL_2597463, EPI_ISL_2597464, EPI_ISL_2597465, EPI_ISL_2597466, EPI_ISL_2597467, EPI_ISL_2597468, EPI_ISL_2597469, EPI_ISL_2597470, EPI_ISL_2597471, EPI_ISL_2597472, EPI_ISL_2597473, EPI_ISL_2597474, EPI_ISL_2597475, EPI_ISL_2597476, EPI_ISL_2597477, EPI_ISL_2597478, EPI_ISL_2597479, EPI_ISL_2597480, EPI_ISL_2597481, EPI_ISL_2597482, EPI_ISL_2597483, EPI_ISL_2597484, EPI_ISL_2597485, EPI_ISL_2597486, EPI_ISL_2597487, EPI_ISL_2597488, EPI_ISL_2597489, EPI_ISL_2597490, EPI_ISL_2597491, EPI_ISL_2597492, EPI_ISL_2597493, EPI_ISL_2597494, EPI_ISL_2597495, EPI_ISL_2597496, EPI_ISL_2597497, EPI_ISL_2597498, EPI_ISL_2597499, EPI_ISL_2597500, EPI_ISL_2597501, EPI_ISL_2597502, EPI_ISL_2597503, EPI_ISL_2597504, EPI_ISL_2597505, EPI_ISL_2597506, EPI_ISL_2597507, EPI_ISL_2597508, EPI_ISL_2597509, EPI_ISL_2597510, EPI_ISL_2597511, EPI_ISL_2597512, EPI_ISL_2597513, EPI_ISL_2597514, EPI_ISL_2597515, EPI_ISL_2597516, EPI_ISL_2597517, EPI_ISL_2597518, EPI_ISL_2597519, EPI_ISL_2597520, EPI_ISL_2597521, EPI_ISL_2597522, EPI_ISL_2597523, EPI_ISL_2597524, EPI_ISL_2597525, EPI_ISL_2597526, EPI_ISL_2597527, EPI_ISL_2597528, EPI_ISL_2597529, EPI_ISL_2597530, EPI_ISL_2597531, EPI_ISL_2597532, EPI_ISL_2597533, EPI_ISL_2597534, EPI_ISL_2597535, EPI_ISL_2597536, EPI_ISL_2597537, EPI_ISL_2597538, EPI_ISL_2597539, EPI_ISL_2597540, EPI_ISL_2597541, EPI_ISL_2597542, EPI_ISL_2597543, EPI_ISL_2597544, EPI_ISL_2597545, EPI_ISL_2597546, EPI_ISL_2597547, EPI_ISL_2597548, EPI_ISL_2597549, EPI_ISL_2597550, EPI_ISL_2597551, EPI_ISL_2597552, EPI_ISL_2597553, EPI_ISL_2597554, EPI_ISL_2597555, EPI_ISL_2597556, EPI_ISL_2597557, EPI_ISL_2597558, EPI_ISL_2597559, EPI_ISL_2597560, EPI_ISL_2597561, EPI_ISL_2597562, EPI_ISL_2597563, EPI_ISL_2597564, EPI_ISL_2597565, EPI_ISL_2597566, EPI_ISL_2597567, EPI_ISL_2597568, EPI_ISL_2597569, EPI_ISL_2597570, EPI_ISL_2597571, EPI_ISL_2597572, EPI_ISL_2597573, EPI_ISL_2597574, EPI_ISL_2597575, EPI_ISL_2597576, EPI_ISL_2597577, EPI_ISL_2597578, EPI_ISL_2597579, EPI_ISL_2597580, EPI_ISL_2597581, EPI_ISL_2597582, EPI_ISL_2597583, EPI_ISL_2597584, EPI_ISL_2597585, EPI_ISL_2597586, EPI_ISL_2597587, EPI_ISL_2597588, EPI_ISL_2597589, EPI_ISL_2597590, EPI_ISL_2597591, EPI_ISL_2597592, EPI_ISL_2597593, EPI_ISL_2597594, EPI_ISL_2597595, EPI_ISL_2597596, EPI_ISL_2597597, EPI_ISL_2597598, EPI_ISL_2597599, EPI_ISL_2597600, EPI_ISL_2597601, EPI_ISL_2597602, EPI_ISL_2597603, EPI_ISL_2597604, EPI_ISL_2597605, EPI_ISL_2597606, EPI_ISL_2597607, EPI_ISL_2597608, EPI_ISL_2597609, EPI_ISL_2597610, EPI_ISL_2597611, EPI_ISL_2597612, EPI_ISL_2597613, EPI_ISL_2597614, EPI_ISL_2597615, EPI_ISL_2597616, EPI_ISL_2597617, EPI_ISL_2597618, EPI_ISL_2597619, EPI_ISL_2597620, EPI_ISL_2597621, EPI_ISL_2597622, EPI_ISL_2597623, EPI_ISL_2597624, EPI_ISL_2597625, EPI_ISL_2597626, EPI_ISL_2597627, EPI_ISL_2597628, EPI_ISL_2597629, EPI_ISL_2597630, EPI_ISL_2597631, EPI_ISL_2597632, EPI_ISL_2597633, EPI_ISL_2597634, EPI_ISL_2597635, EPI_ISL_2597636, EPI_ISL_2597637, EPI_ISL_2597638, EPI_ISL_2597639, EPI_ISL_2597640, EPI_ISL_2597641, EPI_ISL_2597642, EPI_ISL_2597643, EPI_ISL_2597644, EPI_ISL_2597645, EPI_ISL_2597646, EPI_ISL_2597647, EPI_ISL_2597648, EPI_ISL_2597649, EPI_ISL_2597650, EPI_ISL_2597651, EPI_ISL_2597652, EPI_ISL_2597653, EPI_ISL_2597654, EPI_ISL_2597655, EPI_ISL_2597656, EPI_ISL_2597657, EPI_ISL_2597658, EPI_ISL_2597659, EPI_ISL_2597660, EPI_ISL_2597661, EPI_ISL_2597662, EPI_ISL_2597663, EPI_ISL_2597664, EPI_ISL_2597665, EPI_ISL_2597666, EPI_ISL_2597667, EPI_ISL_2597668, EPI_ISL_2597669, EPI_ISL_2597670, EPI_ISL_2597671, EPI_ISL_2597672, EPI_ISL_2597673, EPI_ISL_2597674, EPI_ISL_2597675, EPI_ISL_2597676, EPI_ISL_2597677, EPI_ISL_2597678, EPI_ISL_2597679, EPI_ISL_2597680, EPI_ISL_2597681, EPI_ISL_2597682, EPI_ISL_2597683, EPI_ISL_2597684, EPI_ISL_2597685 | | | |
| see above | Hospital of the University of Pennsylvania Molecular Pathology Lab | Bushman Lab - University of Pennsylvania | John K. Everett, Kyle Rodino, Shantan Reddy, Aoife M. Roche, Young Hwang, Scott Sherrill-Mix, Samantha A. Whiteside, Jevon Graham-Wooten, Layla A. Khatib, Ayannah S. Fitzgerald, Arupa Ganguly, Mike Feldman, Brendan Kelly, Ronald G. Collman and Frederic Bushan |
| EPI_ISL_2597686 | Ohio Department of Health Laboratory | Ohio Department of Health Laboratory | Holmes, Jennifer; Eric Brandt, Keoni Omura, Glen McGillivary, Caitlin McDonnell, Jade Mowery, Stephanie Mccracken, Tyler Payne, Kirtana Ramadugu, Erica Leasure, Brent Lee, Kelsey Florek, Heather Blankenship, Allison Black, Quanta Brown, and Tammy Bannerman |
| EPI_ISL_2597687, EPI_ISL_2597688, EPI_ISL_2597689, EPI_ISL_2597690, EPI_ISL_2597691, EPI_ISL_2597692, EPI_ISL_2597693, EPI_ISL_2597694, EPI_ISL_2597695, EPI_ISL_2597696, EPI_ISL_2597697, EPI_ISL_2597698, EPI_ISL_2597699, EPI_ISL_2597700, EPI_ISL_2597701, EPI_ISL_2597702, EPI_ISL_2597703, EPI_ISL_2597704, EPI_ISL_2597705, EPI_ISL_2597706, EPI_ISL_2597707, EPI_ISL_2597708 | | | |
| see above | Hospital of the University of Pennsylvania Molecular Pathology Lab | Bushman Lab - University of Pennsylvania | John K. Everett, Kyle Rodino, Shantan Reddy, Aoife M. Roche, Young Hwang, Scott Sherrill-Mix, Samantha A. Whiteside, Jevon Graham-Wooten, Layla A. Khatib, Ayannah S. Fitzgerald, Arupa Ganguly, Mike Feldman, Brendan Kelly, Ronald G. Collman and Frederic Bushan |
| EPI_ISL_2597709, EPI_ISL_2597713, EPI_ISL_2597714, EPI_ISL_2597715, EPI_ISL_2597716, EPI_ISL_2597718, EPI_ISL_2597719, EPI_ISL_2597721, EPI_ISL_2597723, EPI_ISL_2597724, EPI_ISL_2597725, EPI_ISL_2597726, EPI_ISL_2597727, EPI_ISL_2597728, EPI_ISL_2597729, EPI_ISL_2597730, EPI_ISL_2597731, EPI_ISL_2597732, EPI_ISL_2597733, EPI_ISL_2597734, EPI_ISL_2597735, EPI_ISL_2597736, | | | |

| | | | |
|---|---|---|---|
| EPI_ISL_2597737, EPI_ISL_2597738, EPI_ISL_2597739, EPI_ISL_2597740, EPI_ISL_2597741, EPI_ISL_2597742, EPI_ISL_2597743, EPI_ISL_2597744, EPI_ISL_2597745, EPI_ISL_2597746, EPI_ISL_2597747, EPI_ISL_2597748, EPI_ISL_2597749, EPI_ISL_2597750, EPI_ISL_2597751, EPI_ISL_2597752, EPI_ISL_2597753, EPI_ISL_2597754, EPI_ISL_2597755, EPI_ISL_2597756, EPI_ISL_2597757, EPI_ISL_2597758, EPI_ISL_2597759, EPI_ISL_2597760, EPI_ISL_2597761, EPI_ISL_2597762, EPI_ISL_2597763, EPI_ISL_2597764, EPI_ISL_2597765, EPI_ISL_2597766, EPI_ISL_2597767, EPI_ISL_2597768, EPI_ISL_2597769, EPI_ISL_2597770, EPI_ISL_2597771, EPI_ISL_2597772, EPI_ISL_2597773, EPI_ISL_2597774, EPI_ISL_2597775, EPI_ISL_2597776, EPI_ISL_2597777, EPI_ISL_2597780, EPI_ISL_2597781, EPI_ISL_2597782, EPI_ISL_2597784, EPI_ISL_2597786, EPI_ISL_2597787, EPI_ISL_2597788, EPI_ISL_2597789, EPI_ISL_2597790, EPI_ISL_2597791, EPI_ISL_2597794, EPI_ISL_2597796, EPI_ISL_2597797, EPI_ISL_2597798, EPI_ISL_2597799, EPI_ISL_2597800, EPI_ISL_2597801 | | | |
| see above | Arizona State Public Health Laboratory | Arizona State Public Health Laboratory | Trung Huynh, Jessica Escobar, Katherine Fullerton, Nobuko Fukushima, Matthew Contursi, Stacy White, Linda Getsinger, Victor Waddell |
| EPI_ISL_2597901, EPI_ISL_2597902, EPI_ISL_2597903, EPI_ISL_2597904, EPI_ISL_2597905, EPI_ISL_2597906, EPI_ISL_2597907, EPI_ISL_2597908, EPI_ISL_2597909, EPI_ISL_2597910, EPI_ISL_2597911, EPI_ISL_2597912, EPI_ISL_2597913, EPI_ISL_2597914, EPI_ISL_2597915, EPI_ISL_2597916, EPI_ISL_2597917, EPI_ISL_2597918, EPI_ISL_2597919, EPI_ISL_2597920, EPI_ISL_2597921, EPI_ISL_2597922, EPI_ISL_2597923, EPI_ISL_2597924, EPI_ISL_2597925, EPI_ISL_2597926, EPI_ISL_2597927, EPI_ISL_2597928, EPI_ISL_2597929, EPI_ISL_2597930, EPI_ISL_2597931, EPI_ISL_2597932, EPI_ISL_2597933, EPI_ISL_2597934, EPI_ISL_2597935, EPI_ISL_2597936, EPI_ISL_2597937, EPI_ISL_2597938, EPI_ISL_2597939, EPI_ISL_2597940, EPI_ISL_2597941, EPI_ISL_2597942, EPI_ISL_2597943, EPI_ISL_2597944, EPI_ISL_2597945, EPI_ISL_2597946, EPI_ISL_2597947, EPI_ISL_2597948, EPI_ISL_2597949, EPI_ISL_2597950, EPI_ISL_2597951, EPI_ISL_2597952, EPI_ISL_2597953, EPI_ISL_2597954, EPI_ISL_2597955 | | | |
| see above | Nebraska Public Health Laboratory | NPHL COVID-19 Response Team | NPHL COVID-19 Response Team |
| EPI_ISL_2597975, EPI_ISL_2597976 | Ohio Department of Health Laboratory | Ohio Department of Health Laboratory | Holmes, Jennifer; Eric Brandt, Keoni Omura, Glen McGillivary, Caitlin McDonnell, Jade Mowery, Stephanie Mccracken, Tyler Payne, Kirtana Ramadugu, Erica Leasure, Brent Lee, Kelsey Florek, Heather Blankenship, Allison Black, Quanta Brown, and Tammy Bannerman |
| EPI_ISL_2597977, EPI_ISL_2597978, EPI_ISL_2597979 | John H Stroger Jr Hospital of Cook County | RIPHL at Rush University Medical Center | Stefan Green, Kevin Kunstman, Max Kolton, Marieta Hyde, Laura Furtado, Felix Araujo Perez, Ah-Young Sung, Sharon Welbel |
| EPI_ISL_2597980 | Ohio Department of Health Laboratory | Ohio Department of Health Laboratory | Holmes, Jennifer; Eric Brandt, Keoni Omura, Glen McGillivary, Caitlin McDonnell, Jade Mowery, Stephanie Mccracken, Tyler Payne, Kirtana Ramadugu, Erica Leasure, Brent Lee, Kelsey Florek, Heather Blankenship, Allison Black, Quanta Brown, and Tammy Bannerman |
| EPI_ISL_2597981, EPI_ISL_2597982, EPI_ISL_2597983, EPI_ISL_2597984, EPI_ISL_2597985, EPI_ISL_2597986, EPI_ISL_2597987, EPI_ISL_2597988, EPI_ISL_2597989, EPI_ISL_2597990, EPI_ISL_2597991, EPI_ISL_2597992 | | | |
| see above | Atlanta VA Medical Center | Genomics and Discovery, Respiratory Viruses Branch, Division of Viral Diseases, Centers for Disease Control and Prevention | Yan Li, Jing Zhang, Anna Kelleher, Ying Tao, Brian Lynch, Krista Queen, Anna Uehara, Peter Cook, Han Jia Justin Ng, Rachel Marine, Clinton R. Paden, Dhwani Batra, Haibin Wang, Tara Coalter, Jasmine Padilla, Morgan Davis, Mili Sheth, Sarah Nobles, Mark Burroughs, Justin Lee, Adam Retchless, Suxiang Tong |
| EPI_ISL_2597994 | Ohio Department of Health Laboratory | Ohio Department of Health Laboratory | Holmes, Jennifer; Eric Brandt, Keoni Omura, Glen McGillivary, Caitlin McDonnell, Jade Mowery, Stephanie Mccracken, Tyler Payne, Kirtana Ramadugu, Erica Leasure, Brent Lee, Kelsey Florek, Heather Blankenship, Allison Black, Quanta Brown, and Tammy Bannerman |
| EPI_ISL_2597995 | LESP Ciudad de Mexico | Instituto de Diagnostico y Referencia Epidemiologicos (INDRE) | Claudia Wong-Arambula, Abril Rodriguez-Maldonado, Vanessa Rivero-Arredondo, Ariadna Medina-Benitez, Joaquin Quiroz-Mercado, Sergio Rangel-Guerrero, Natividad Cruz-Ortiz, Tatiana Nunez-Garcia, Gisela Barrera-Badillo, Lucia Hernandez-Rivas, Irma Lopez-Martinez, Ernesto Ramirez-Gonzalez. |
| EPI_ISL_2597996 | LESP Puebla | Instituto de Diagnostico y Referencia Epidemiologicos (INDRE) | Claudia Wong-Arambula, Abril Rodriguez-Maldonado, Vanessa Rivero-Arredondo, Ariadna Medina-Benitez, Joaquin Quiroz-Mercado, Sergio Rangel-Guerrero, Natividad Cruz-Ortiz, Tatiana Nunez-Garcia, Gisela Barrera-Badillo, Lucia Hernandez-Rivas, Irma Lopez-Martinez, Ernesto Ramirez-Gonzalez. |
| EPI_ISL_2597997, EPI_ISL_2597998, EPI_ISL_2597999, EPI_ISL_2598000, EPI_ISL_2598001 | LESP Queretaro | Instituto de Diagnostico y Referencia Epidemiologicos (INDRE) | Claudia Wong-Arambula, Abril Rodriguez-Maldonado, Vanessa Rivero-Arredondo, Ariadna Medina-Benitez, Joaquin Quiroz-Mercado, Sergio Rangel-Guerrero, Natividad Cruz-Ortiz, Tatiana Nunez-Garcia, Gisela Barrera-Badillo, Lucia Hernandez-Rivas, Irma Lopez-Martinez, Ernesto Ramirez-Gonzalez. |
| EPI_ISL_2598002 | LESP Tamaulipas | Instituto de Diagnostico y Referencia Epidemiologicos (INDRE) | Claudia Wong-Arambula, Abril Rodriguez-Maldonado, Vanessa Rivero-Arredondo, Ariadna Medina-Benitez, Joaquin Quiroz-Mercado, Sergio Rangel-Guerrero, Natividad Cruz-Ortiz, Tatiana Nunez-Garcia, Gisela Barrera-Badillo, Lucia Hernandez-Rivas, Irma Lopez-Martinez, Ernesto Ramirez-Gonzalez. |
| EPI_ISL_2598003 | LESP Coahuila | Instituto de Diagnostico y Referencia Epidemiologicos (INDRE) | Claudia Wong-Arambula, Abril Rodriguez-Maldonado, Vanessa Rivero-Arredondo, Ariadna Medina-Benitez, Joaquin Quiroz-Mercado, Sergio Rangel-Guerrero, Natividad Cruz-Ortiz, Tatiana Nunez-Garcia, Gisela Barrera-Badillo, Lucia Hernandez-Rivas, Irma Lopez-Martinez, Ernesto Ramirez-Gonzalez. |
| EPI_ISL_2598004, EPI_ISL_2598005, EPI_ISL_2598006, EPI_ISL_2598007, EPI_ISL_2598008, EPI_ISL_2598009, EPI_ISL_2598010, EPI_ISL_2598011, EPI_ISL_2598012, EPI_ISL_2598013, EPI_ISL_2598014, EPI_ISL_2598015 | | | |
| see above | LESP Ciudad de Mexico | Instituto de Diagnostico y Referencia Epidemiologicos (INDRE) | Claudia Wong-Arambula, Abril Rodriguez-Maldonado, Vanessa Rivero-Arredondo, Ariadna Medina-Benitez, Joaquin Quiroz-Mercado, Sergio Rangel-Guerrero, Natividad Cruz-Ortiz, Tatiana Nunez-Garcia, Gisela Barrera-Badillo, Lucia Hernandez-Rivas, Irma Lopez-Martinez, Ernesto Ramirez-Gonzalez. |
| EPI_ISL_2598016 | Hackensack Medical Center | New York Genome Center | Michael Zody, Andre Corvelo, Dayna M. Oschwald, Samantha Fennessey, Tom Maniatis, Liang Chen, Jose Mediavilla, Marcus Cunningham, Kaelea Composto, Kar Chow, David Perlin, Barry Kreiswirth |
| EPI_ISL_2598017, EPI_ISL_2598018, EPI_ISL_2598019, EPI_ISL_2598020, EPI_ISL_2598021, EPI_ISL_2598022, EPI_ISL_2598023, EPI_ISL_2598024 | MD PHL | MD PHL | Maryland Department of Health Laboratories Administration |
| EPI_ISL_2598025 | Ohio Department of Health Laboratory | Ohio Department of Health Laboratory | Holmes, Jennifer; Eric Brandt, Keoni Omura, Glen McGillivary, Caitlin McDonnell, Jade Mowery, Stephanie Mccracken, Tyler Payne, Kirtana Ramadugu, Erica Leasure, Brent Lee, Kelsey Florek, Heather Blankenship, Allison Black, Quanta Brown, and Tammy Bannerman |
| EPI_ISL_2598026, EPI_ISL_2598027, EPI_ISL_2598028, EPI_ISL_2598029, EPI_ISL_2598030, EPI_ISL_2598031, EPI_ISL_2598032, EPI_ISL_2598033, EPI_ISL_2598034, EPI_ISL_2598035, EPI_ISL_2598036, EPI_ISL_2598037, EPI_ISL_2598038, EPI_ISL_2598039, EPI_ISL_2598040, EPI_ISL_2598041, EPI_ISL_2598042, EPI_ISL_2598043, EPI_ISL_2598044, EPI_ISL_2598045, EPI_ISL_2598046, EPI_ISL_2598047, EPI_ISL_2598048, EPI_ISL_2598049, EPI_ISL_2598050, EPI_ISL_2598051, EPI_ISL_2598052, EPI_ISL_2598053, EPI_ISL_2598054, EPI_ISL_2598055, EPI_ISL_2598056, EPI_ISL_2598057, EPI_ISL_2598058, EPI_ISL_2598059, EPI_ISL_2598060, EPI_ISL_2598061, EPI_ISL_2598062, EPI_ISL_2598063, EPI_ISL_2598064, EPI_ISL_2598065, EPI_ISL_2598066, EPI_ISL_2598067, EPI_ISL_2598068, EPI_ISL_2598069, EPI_ISL_2598070, EPI_ISL_2598071, EPI_ISL_2598072, EPI_ISL_2598073, EPI_ISL_2598074, EPI_ISL_2598075, EPI_ISL_2598076, EPI_ISL_2598077, EPI_ISL_2598078, EPI_ISL_2598079, EPI_ISL_2598080, EPI_ISL_2598081, EPI_ISL_2598082, EPI_ISL_2598083, EPI_ISL_2598084 | | | |
| see above | MD PHL | MD PHL | Maryland Department of Health Laboratories Administration |
| EPI_ISL_2598085, EPI_ISL_2598086 | Ohio Department of Health Laboratory | Ohio Department of Health Laboratory | Holmes, Jennifer; Eric Brandt, Keoni Omura, Glen McGillivary, Caitlin McDonnell, Jade Mowery, Stephanie Mccracken, Tyler Payne, Kirtana Ramadugu, Erica Leasure, Brent Lee, Kelsey Florek, Heather Blankenship, Allison Black, Quanta Brown, and Tammy Bannerman |
| EPI_ISL_2598087, EPI_ISL_2598088, EPI_ISL_2598089, EPI_ISL_2598090, EPI_ISL_2598091, EPI_ISL_2598092, EPI_ISL_2598093, EPI_ISL_2598094, EPI_ISL_2598095, EPI_ISL_2598096, EPI_ISL_2598097, EPI_ISL_2598098, EPI_ISL_2598099, EPI_ISL_2598100, EPI_ISL_2598101, EPI_ISL_2598102, EPI_ISL_2598103, EPI_ISL_2598104, EPI_ISL_2598105, EPI_ISL_2598107, EPI_ISL_2598108, EPI_ISL_2598109, EPI_ISL_2598110, EPI_ISL_2598111, EPI_ISL_2598112, EPI_ISL_2598113, EPI_ISL_2598114, EPI_ISL_2598115, EPI_ISL_2598116, EPI_ISL_2598117, EPI_ISL_2598118 | | | |
| see above | TriCore Reference Laboratories | Center for Global Health, University of New Mexico Health Sciences Center | Daryl Domman, Kurt Schwalm, Valerie Morley, Cecilia Thompson, Kendra Pesko, Karissa Culbreath, Darrell Dinwiddie |
| EPI_ISL_2598120, EPI_ISL_2598121, EPI_ISL_2598122, EPI_ISL_2598123, EPI_ISL_2598124, EPI_ISL_2598125, EPI_ISL_2598126, EPI_ISL_2598127, EPI_ISL_2598128, EPI_ISL_2598129, EPI_ISL_2598130, EPI_ISL_2598131, EPI_ISL_2598132, EPI_ISL_2598133, EPI_ISL_2598134 | | | |
| see above | Idaho Bureau of Laboratories | Center for Global Health, University of New Mexico Health Sciences Center | Daryl Domman, Kurt Schwalm, Valerie Morley, Matthew Burns, Robert Voermans, Christopher Ball, Darrell Dinwiddie |
| EPI_ISL_2598135, EPI_ISL_2598136, EPI_ISL_2598137, EPI_ISL_2598138 | New Mexico Department of Health Scientific Laboratory | Center for Global Health, University of New Mexico Health Sciences Center | Daryl Domman, Kurt Schwalm, Twila Kunde, Joseph Hicks, Anastacia Griego, Michael Edwards, Valerie Morley, Darrell Dinwiddie |
| EPI_ISL_2598140, EPI_ISL_2598141, EPI_ISL_2598142, EPI_ISL_2598143, EPI_ISL_2598144, EPI_ISL_2598145, EPI_ISL_2598146, EPI_ISL_2598147, EPI_ISL_2598148, EPI_ISL_2598149, EPI_ISL_2598150, EPI_ISL_2598151, EPI_ISL_2598152, EPI_ISL_2598153, EPI_ISL_2598154 | | | |
| see above | Minnesota Department of Health, Public Health Laboratory | Minnesota Department of Health, Public Health Laboratory | Alexandra Lorentz, Jacob Garfin, Matt Plumb, and Xiong Wang |
| EPI_ISL_2598155 | Centracare Laboratory Services | Minnesota Department of Health, Public Health Laboratory | Alexandra Lorentz, Jacob Garfin, Matt Plumb, and Xiong Wang |
| EPI_ISL_2598156, EPI_ISL_2598157, EPI_ISL_2598158, EPI_ISL_2598159, EPI_ISL_2598160 | Minnesota Department of Health, Public Health Laboratory | Minnesota Department of Health, Public Health Laboratory | Alexandra Lorentz, Jacob Garfin, Matt Plumb, and Xiong Wang |
| EPI_ISL_2598161 | Ohio Department of Health Laboratory | Ohio Department of Health Laboratory | Holmes, Jennifer; Eric Brandt, Keoni Omura, Glen McGillivary, Caitlin McDonnell, Jade Mowery, Stephanie Mccracken, Tyler Payne, Kirtana Ramadugu, Erica Leasure, Brent Lee, Kelsey Florek, Heather Blankenship, Allison Black, Quanta Brown, and Tammy Bannerman |
| EPI_ISL_2598162, EPI_ISL_2598163, EPI_ISL_2598164, EPI_ISL_2598165, EPI_ISL_2598167, EPI_ISL_2598168, EPI_ISL_2598169, EPI_ISL_2598171, EPI_ISL_2598172, EPI_ISL_2598173, EPI_ISL_2598174, EPI_ISL_2598175, EPI_ISL_2598176, EPI_ISL_2598177, EPI_ISL_2598179, EPI_ISL_2598180, EPI_ISL_2598182, EPI_ISL_2598183, EPI_ISL_2598184, EPI_ISL_2598185, EPI_ISL_2598186, EPI_ISL_2598187, EPI_ISL_2598188, EPI_ISL_2598189, EPI_ISL_2598191, EPI_ISL_2598192, EPI_ISL_2598193, EPI_ISL_2598194, EPI_ISL_2598195, EPI_ISL_2598196, EPI_ISL_2598197, EPI_ISL_2598198, EPI_ISL_2598199, EPI_ISL_2598200, EPI_ISL_2598202, EPI_ISL_2598203, EPI_ISL_2598204, EPI_ISL_2598205, EPI_ISL_2598206, EPI_ISL_2598207, EPI_ISL_2598208, EPI_ISL_2598209, EPI_ISL_2598210, EPI_ISL_2598211, EPI_ISL_2598212, EPI_ISL_2598213, EPI_ISL_2598215, EPI_ISL_2598216, EPI_ISL_2598217, EPI_ISL_2598218, EPI_ISL_2598220, EPI_ISL_2598221, EPI_ISL_2598222, EPI_ISL_2598223, EPI_ISL_2598224, EPI_ISL_2598225, EPI_ISL_2598227, EPI_ISL_2598228, EPI_ISL_2598229, EPI_ISL_2598230, EPI_ISL_2598231, EPI_ISL_2598232, EPI_ISL_2598233, EPI_ISL_2598234, EPI_ISL_2598235, EPI_ISL_2598237, EPI_ISL_2598238, EPI_ISL_2598239, EPI_ISL_2598240, EPI_ISL_2598242, EPI_ISL_2598243, EPI_ISL_2598244, EPI_ISL_2598245, EPI_ISL_2598246, EPI_ISL_2598247 | | | |
| see above | Illinois Department of Public Health - Springfield Lab | Illinois Department of Public Health - Springfield Lab | Bryan Sim, Gordon McCall |
| EPI_ISL_2598248 | Ohio Department of Health Laboratory | Ohio Department of Health Laboratory | Holmes, Jennifer; Eric Brandt, Keoni Omura, Glen McGillivary, Caitlin McDonnell, Jade Mowery, Stephanie Mccracken, Tyler Payne, Kirtana Ramadugu, Erica Leasure, Brent Lee, Kelsey Florek, Heather Blankenship, Allison Black, Quanta Brown, and Tammy Bannerman |
| EPI_ISL_2598250, EPI_ISL_2598252, EPI_ISL_2598253, EPI_ISL_2598254, EPI_ISL_2598256, EPI_ISL_2598257, EPI_ISL_2598258, EPI_ISL_2598259, EPI_ISL_2598260, EPI_ISL_2598261, EPI_ISL_2598262, EPI_ISL_2598263, EPI_ISL_2598264, EPI_ISL_2598266, EPI_ISL_2598267, EPI_ISL_2598268 | | | |
| see above | University of Michigan Clinical Microbiology Laboratory | Lauring Lab, University of Michigan, Department of Microbiology and Immunology | Valesano |
| EPI_ISL_2598269, EPI_ISL_2598270, EPI_ISL_2598272, EPI_ISL_2598273, EPI_ISL_2598274, EPI_ISL_2598275, EPI_ISL_2598277, EPI_ISL_2598278, EPI_ISL_2598279, EPI_ISL_2598280, EPI_ISL_2598283, EPI_ISL_2598284, EPI_ISL_2598285, EPI_ISL_2598286, EPI_ISL_2598287, EPI_ISL_2598288, EPI_ISL_2598289, EPI_ISL_2598291, EPI_ISL_2598292, EPI_ISL_2598293, EPI_ISL_2598294 | | | |
| see above | IVY3 Central Lab, Vanderbilt University Medical Center | Lauring Lab, University of Michigan, Department of Microbiology and Immunology | Valesano |
| EPI_ISL_2598402, EPI_ISL_2598403, EPI_ISL_2598404, EPI_ISL_2598405, EPI_ISL_2598406, EPI_ISL_2598407, EPI_ISL_2598408, EPI_ISL_2598409, EPI_ISL_2598410, EPI_ISL_2598411, EPI_ISL_2598412, EPI_ISL_2598413, EPI_ISL_2598414, EPI_ISL_2598415, EPI_ISL_2598416, EPI_ISL_2598417, EPI_ISL_2598418, EPI_ISL_2598419, EPI_ISL_2598420, EPI_ISL_2598421, EPI_ISL_2598422, EPI_ISL_2598423, EPI_ISL_2598424, EPI_ISL_2598425, EPI_ISL_2598426, EPI_ISL_2598427, EPI_ISL_2598428, EPI_ISL_2598429, EPI_ISL_2598430, EPI_ISL_2598431, EPI_ISL_2598432, EPI_ISL_2598433, EPI_ISL_2598434, EPI_ISL_2598435, EPI_ISL_2598436, EPI_ISL_2598438, EPI_ISL_2598439, EPI_ISL_2598440, EPI_ISL_2598442, EPI_ISL_2598443, EPI_ISL_2598444, EPI_ISL_2598445, EPI_ISL_2598446, EPI_ISL_2598447, EPI_ISL_2598448, EPI_ISL_2598449, EPI_ISL_2598450, EPI_ISL_2598451, EPI_ISL_2598452, EPI_ISL_2598453, EPI_ISL_2598454, EPI_ISL_2598455, EPI_ISL_2598456, EPI_ISL_2598457, EPI_ISL_2598458, EPI_ISL_2598459, EPI_ISL_2598460, EPI_ISL_2598461, EPI_ISL_2598462, EPI_ISL_2598463, EPI_ISL_2598464, EPI_ISL_2598465, EPI_ISL_2598466, EPI_ISL_2598467, EPI_ISL_2598468, EPI_ISL_2598469, EPI_ISL_2598470, EPI_ISL_2598471, EPI_ISL_2598472, EPI_ISL_2598473, EPI_ISL_2598474, EPI_ISL_2598475, EPI_ISL_2598476, EPI_ISL_2598477, EPI_ISL_2598478, EPI_ISL_2598479, EPI_ISL_2598480, EPI_ISL_2598482, EPI_ISL_2598483, EPI_ISL_2598484, EPI_ISL_2598485, EPI_ISL_2598486, EPI_ISL_2598487, EPI_ISL_2598488, EPI_ISL_2598489, EPI_ISL_2598490, EPI_ISL_2598491, EPI_ISL_2598492, EPI_ISL_2598493 | | | |

| | | | |
|---|---|---|---|
| see above | Maryland Genomics, Institute for Genome Sciences, University of Maryland School of Medicine | Maryland Genomics, Institute for Genome Sciences, University of Maryland School of Medicine | Tallon, Luke J; Sadzewicz, Lisa D; Humphrys, Mike; Ott, Sandra; Roussey, Holly; Santana-Cruz, Ivette; Vavikolanu, Kranthi; Lim, Jonathan;Hazen, Tracy;Fraser, Claire M; Ravel, Jacques |
| EPI_ISL_2598674, EPI_ISL_2598675, EPI_ISL_2598676, EPI_ISL_2598678, EPI_ISL_2598679, EPI_ISL_2598680, EPI_ISL_2598681, EPI_ISL_2598683, EPI_ISL_2598684, EPI_ISL_2598685, EPI_ISL_2598686, EPI_ISL_2598688, EPI_ISL_2598689, EPI_ISL_2598691 | | | |
| see above | Yale Clinical Virology Lab | Grubaugh Lab - Yale School of Public Health | Tara Alpert, Joseph Fauver, Anderson Brito, Mallery Breban, Anne Wyllie, Chantal Vogels, Mary Petrone, Chaney Kalinich, Isabel Ott, Nathan Grubaugh |
| EPI_ISL_2598692 | Grubaugh Lab - Yale School of Public Health | Grubaugh Lab - Yale School of Public Health | Tara Alpert, Joseph Fauver, Anderson Brito, Mallery Breban, Anne Wyllie, Chantal Vogels, Mary Petrone, Chaney Kalinich, Isabel Ott, Nathan Grubaugh |
| EPI_ISL_2598693, EPI_ISL_2598694, EPI_ISL_2598695, EPI_ISL_2598696, EPI_ISL_2598697 | Yale Clinical Virology Lab | Grubaugh Lab - Yale School of Public Health | Tara Alpert, Joseph Fauver, Anderson Brito, Mallery Breban, Anne Wyllie, Chantal Vogels, Mary Petrone, Chaney Kalinich, Isabel Ott, Nathan Grubaugh |
| EPI_ISL_2598700, EPI_ISL_2598701, EPI_ISL_2598702, EPI_ISL_2598703, EPI_ISL_2598704, EPI_ISL_2598705, EPI_ISL_2598706, EPI_ISL_2598707, EPI_ISL_2598708, EPI_ISL_2598709, EPI_ISL_2598710, EPI_ISL_2598711, EPI_ISL_2598712, EPI_ISL_2598713, EPI_ISL_2598714, EPI_ISL_2598715, EPI_ISL_2598716, EPI_ISL_2598717, EPI_ISL_2598718, EPI_ISL_2598719, EPI_ISL_2598720, EPI_ISL_2598722, EPI_ISL_2598723, EPI_ISL_2598724, EPI_ISL_2598726, EPI_ISL_2598728, EPI_ISL_2598730, EPI_ISL_2598731, EPI_ISL_2598733, EPI_ISL_2598734, EPI_ISL_2598735, EPI_ISL_2598736, EPI_ISL_2598737, EPI_ISL_2598738, EPI_ISL_2598740, EPI_ISL_2598741, EPI_ISL_2598742, EPI_ISL_2598743, EPI_ISL_2598745, EPI_ISL_2598747, EPI_ISL_2598748, EPI_ISL_2598749, EPI_ISL_2598750, EPI_ISL_2598751, EPI_ISL_2598753, EPI_ISL_2598754, EPI_ISL_2598755, EPI_ISL_2598756, EPI_ISL_2598757, EPI_ISL_2598759, EPI_ISL_2598760, EPI_ISL_2598761, EPI_ISL_2598762, EPI_ISL_2598763, EPI_ISL_2598764, EPI_ISL_2598765, EPI_ISL_2598766, EPI_ISL_2598767, EPI_ISL_2598768, EPI_ISL_2598769, EPI_ISL_2598771, EPI_ISL_2598772, EPI_ISL_2598773, EPI_ISL_2598774, EPI_ISL_2598775, EPI_ISL_2598776, EPI_ISL_2598777, EPI_ISL_2598778, EPI_ISL_2598779, EPI_ISL_2598780, EPI_ISL_2598781, EPI_ISL_2598782, EPI_ISL_2598783, EPI_ISL_2598784, EPI_ISL_2598785, EPI_ISL_2598786, EPI_ISL_2598787, EPI_ISL_2598788, EPI_ISL_2598789, EPI_ISL_2598790, EPI_ISL_2598791, EPI_ISL_2598792, EPI_ISL_2598793, EPI_ISL_2598795, EPI_ISL_2598796, EPI_ISL_2598797, EPI_ISL_2598798, EPI_ISL_2598799, EPI_ISL_2598800, EPI_ISL_2598801, EPI_ISL_2598802, EPI_ISL_2598803, EPI_ISL_2598804, EPI_ISL_2598805, EPI_ISL_2598806, EPI_ISL_2598807, EPI_ISL_2598808, EPI_ISL_2598809, EPI_ISL_2598810, EPI_ISL_2598812, EPI_ISL_2598813, EPI_ISL_2598814, EPI_ISL_2598815, EPI_ISL_2598816, EPI_ISL_2598817, EPI_ISL_2598819, EPI_ISL_2598820, EPI_ISL_2598821, EPI_ISL_2598822, EPI_ISL_2598823, EPI_ISL_2598824, EPI_ISL_2598825, EPI_ISL_2598826, EPI_ISL_2598827, EPI_ISL_2598828, EPI_ISL_2598829, EPI_ISL_2598830, EPI_ISL_2598831, EPI_ISL_2598832, EPI_ISL_2598833, EPI_ISL_2598835, EPI_ISL_2598836, EPI_ISL_2598837, EPI_ISL_2598838 | | | |
| see above | Fulgent Genetics | Centers for Disease Control and Prevention Division of Viral Diseases, Pathogen Discovery | Dakota Howard, Dhwani Batra, Peter W. Cook, Kara Moser, Adrian Paskey, Jason Caravas, Benjamin Rambo-Martin, Shatavia Morrison, Christopher Gulvick, Scott Sammons, Yvette Unoarumhi, Darlene Wagner, Matthew Schmerer, Harry Gao, Mickey Li, John Gao, Joseph Fierro, Benafsh Sapra, Becky Tsai, Yan Meng, Doreen Ng, James Xie, Clinton R. Paden, Duncan MacCannell |
| EPI_ISL_2598839, EPI_ISL_2598840, EPI_ISL_2598841, EPI_ISL_2598842, EPI_ISL_2598843, EPI_ISL_2598844, EPI_ISL_2598845, EPI_ISL_2598846, EPI_ISL_2598847, EPI_ISL_2598848, EPI_ISL_2598849, EPI_ISL_2598850, EPI_ISL_2598851, EPI_ISL_2598852, EPI_ISL_2598853, EPI_ISL_2598854, EPI_ISL_2598855, EPI_ISL_2598856, EPI_ISL_2598857, EPI_ISL_2598858, EPI_ISL_2598859, EPI_ISL_2598860, EPI_ISL_2598861, EPI_ISL_2598862, EPI_ISL_2598863, EPI_ISL_2598864, EPI_ISL_2598865, EPI_ISL_2598866, EPI_ISL_2598867, EPI_ISL_2598868, EPI_ISL_2598869, EPI_ISL_2598870, EPI_ISL_2598871, EPI_ISL_2598872, EPI_ISL_2598873, EPI_ISL_2598874, EPI_ISL_2598875, EPI_ISL_2598876, EPI_ISL_2598877, EPI_ISL_2598878, EPI_ISL_2598879, EPI_ISL_2598880, EPI_ISL_2598881, EPI_ISL_2598882, EPI_ISL_2598883, EPI_ISL_2598884, EPI_ISL_2598885, EPI_ISL_2598886, EPI_ISL_2598887, EPI_ISL_2598888, EPI_ISL_2598889, EPI_ISL_2598890, EPI_ISL_2598891, EPI_ISL_2598892, EPI_ISL_2598893, EPI_ISL_2598894, EPI_ISL_2598895, EPI_ISL_2598896, EPI_ISL_2598897, EPI_ISL_2598898, EPI_ISL_2598899, EPI_ISL_2598900, EPI_ISL_2598901, EPI_ISL_2598902, EPI_ISL_2598903, EPI_ISL_2598904, EPI_ISL_2598905, EPI_ISL_2598906, EPI_ISL_2598907, EPI_ISL_2598908, EPI_ISL_2598909, EPI_ISL_2598910, EPI_ISL_2598911, EPI_ISL_2598912, EPI_ISL_2598913, EPI_ISL_2598914, EPI_ISL_2598915, EPI_ISL_2598916, EPI_ISL_2598917, EPI_ISL_2598918, EPI_ISL_2598919, EPI_ISL_2598920, EPI_ISL_2598921, EPI_ISL_2598922, EPI_ISL_2598923, EPI_ISL_2598924, EPI_ISL_2598925, EPI_ISL_2598926, EPI_ISL_2598927, EPI_ISL_2598928, EPI_ISL_2598929, EPI_ISL_2598930, EPI_ISL_2598931, EPI_ISL_2598932, EPI_ISL_2598933, EPI_ISL_2598934, EPI_ISL_2598935, EPI_ISL_2598936, EPI_ISL_2598937, EPI_ISL_2598938, EPI_ISL_2598939, EPI_ISL_2598940, EPI_ISL_2598941, EPI_ISL_2598942, EPI_ISL_2598943, EPI_ISL_2598944, EPI_ISL_2598945, EPI_ISL_2598946, EPI_ISL_2598947, EPI_ISL_2598948, EPI_ISL_2598949, EPI_ISL_2598950, EPI_ISL_2598951, EPI_ISL_2598952, EPI_ISL_2598953, EPI_ISL_2598954, EPI_ISL_2598955, EPI_ISL_2598956, EPI_ISL_2598957, EPI_ISL_2598958, EPI_ISL_2598959, EPI_ISL_2598960, EPI_ISL_2598961, EPI_ISL_2598962, EPI_ISL_2598963, EPI_ISL_2598964, EPI_ISL_2598965, EPI_ISL_2598966, EPI_ISL_2598967, EPI_ISL_2598968, EPI_ISL_2598969, EPI_ISL_2598970, EPI_ISL_2598971, EPI_ISL_2598972, EPI_ISL_2598973, EPI_ISL_2598974, EPI_ISL_2598975, EPI_ISL_2598976, EPI_ISL_2598977, EPI_ISL_2598978, EPI_ISL_2598979, EPI_ISL_2598980, EPI_ISL_2598981, EPI_ISL_2598982, EPI_ISL_2598983, EPI_ISL_2598984, EPI_ISL_2598985, EPI_ISL_2598986, EPI_ISL_2598987, EPI_ISL_2598988, EPI_ISL_2598989, EPI_ISL_2598990, EPI_ISL_2598991, EPI_ISL_2598992, EPI_ISL_2598993, EPI_ISL_2598994, EPI_ISL_2598995, EPI_ISL_2598996, EPI_ISL_2598997, EPI_ISL_2598998, EPI_ISL_2598999, EPI_ISL_2599000, EPI_ISL_2599001, EPI_ISL_2599002, EPI_ISL_2599003, EPI_ISL_2599004, EPI_ISL_2599005, EPI_ISL_2599006, EPI_ISL_2599007, EPI_ISL_2599008, EPI_ISL_2599009, EPI_ISL_2599010, EPI_ISL_2599011, EPI_ISL_2599012, EPI_ISL_2599013, EPI_ISL_2599014, EPI_ISL_2599015, EPI_ISL_2599016, EPI_ISL_2599017, EPI_ISL_2599018, EPI_ISL_2599019, EPI_ISL_2599021, EPI_ISL_2599022, EPI_ISL_2599023, EPI_ISL_2599024, EPI_ISL_2599025, EPI_ISL_2599026, EPI_ISL_2599027, EPI_ISL_2599028, EPI_ISL_2599029, EPI_ISL_2599030, EPI_ISL_2599031, EPI_ISL_2599032, EPI_ISL_2599033, EPI_ISL_2599034, EPI_ISL_2599035, EPI_ISL_2599036, EPI_ISL_2599037, EPI_ISL_2599038, EPI_ISL_2599039, EPI_ISL_2599040, EPI_ISL_2599041, EPI_ISL_2599042, EPI_ISL_2599043, EPI_ISL_2599044, EPI_ISL_2599045, EPI_ISL_2599046, EPI_ISL_2599047, EPI_ISL_2599048, EPI_ISL_2599049, EPI_ISL_2599050, EPI_ISL_2599051, EPI_ISL_2599052, EPI_ISL_2599053, EPI_ISL_2599054, EPI_ISL_2599055, EPI_ISL_2599056, EPI_ISL_2599057, EPI_ISL_2599058, EPI_ISL_2599059, EPI_ISL_2599060, EPI_ISL_2599061, EPI_ISL_2599062, EPI_ISL_2599063, EPI_ISL_2599064, EPI_ISL_2599065, EPI_ISL_2599066, EPI_ISL_2599067, EPI_ISL_2599068, EPI_ISL_2599069, EPI_ISL_2599070, EPI_ISL_2599071, EPI_ISL_2599072, EPI_ISL_2599073, EPI_ISL_2599074, EPI_ISL_2599075, EPI_ISL_2599076, EPI_ISL_2599077, EPI_ISL_2599078, EPI_ISL_2599079, EPI_ISL_2599080, EPI_ISL_2599081, EPI_ISL_2599082, EPI_ISL_2599083, EPI_ISL_2599084, EPI_ISL_2599085, EPI_ISL_2599086, EPI_ISL_2599087, EPI_ISL_2599088, EPI_ISL_2599089, EPI_ISL_2599090, EPI_ISL_2599091, EPI_ISL_2599092, EPI_ISL_2599093, EPI_ISL_2599094, EPI_ISL_2599095, EPI_ISL_2599096, EPI_ISL_2599097, EPI_ISL_2599098, EPI_ISL_2599099, EPI_ISL_2599100, EPI_ISL_2599101, EPI_ISL_2599102, EPI_ISL_2599103, EPI_ISL_2599104, EPI_ISL_2599105, EPI_ISL_2599106, EPI_ISL_2599107, EPI_ISL_2599108, EPI_ISL_2599109, EPI_ISL_2599110, EPI_ISL_2599111, EPI_ISL_2599112, EPI_ISL_2599113, EPI_ISL_2599114, EPI_ISL_2599115, EPI_ISL_2599116, EPI_ISL_2599117, EPI_ISL_2599118, EPI_ISL_2599119, EPI_ISL_2599120, EPI_ISL_2599121, EPI_ISL_2599122, EPI_ISL_2599123, EPI_ISL_2599124, EPI_ISL_2599125, EPI_ISL_2599126, EPI_ISL_2599127, EPI_ISL_2599128, EPI_ISL_2599129, EPI_ISL_2599130, EPI_ISL_2599131, EPI_ISL_2599133, EPI_ISL_2599134, EPI_ISL_2599135, EPI_ISL_2599136, EPI_ISL_2599137, EPI_ISL_2599138, EPI_ISL_2599139, EPI_ISL_2599140, EPI_ISL_2599141, EPI_ISL_2599142, EPI_ISL_2599143, EPI_ISL_2599144, EPI_ISL_2599145, EPI_ISL_2599146, EPI_ISL_2599147, EPI_ISL_2599148, EPI_ISL_2599149, EPI_ISL_2599150, EPI_ISL_2599151, EPI_ISL_2599152, EPI_ISL_2599153, EPI_ISL_2599154, EPI_ISL_2599155, EPI_ISL_2599156, EPI_ISL_2599157, EPI_ISL_2599158, EPI_ISL_2599159, EPI_ISL_2599160, EPI_ISL_2599161, EPI_ISL_2599162, EPI_ISL_2599163, EPI_ISL_2599164, EPI_ISL_2599165, EPI_ISL_2599166, EPI_ISL_2599167, EPI_ISL_2599168, EPI_ISL_2599169, EPI_ISL_2599170, EPI_ISL_2599171, EPI_ISL_2599172, EPI_ISL_2599173, EPI_ISL_2599174, EPI_ISL_2599175, EPI_ISL_2599176, EPI_ISL_2599177, EPI_ISL_2599178, EPI_ISL_2599179, EPI_ISL_2599180, EPI_ISL_2599181, EPI_ISL_2599182, EPI_ISL_2599183, EPI_ISL_2599184, EPI_ISL_2599185, EPI_ISL_2599186, EPI_ISL_2599187, EPI_ISL_2599188, EPI_ISL_2599189, EPI_ISL_2599190, EPI_ISL_2599191, EPI_ISL_2599192, EPI_ISL_2599193, EPI_ISL_2599194, EPI_ISL_2599195, EPI_ISL_2599196, EPI_ISL_2599197, EPI_ISL_2599198, EPI_ISL_2599199, EPI_ISL_2599200, EPI_ISL_2599201, EPI_ISL_2599202, EPI_ISL_2599203, EPI_ISL_2599204, EPI_ISL_2599205, EPI_ISL_2599206, EPI_ISL_2599207, EPI_ISL_2599208, EPI_ISL_2599209, EPI_ISL_2599210, EPI_ISL_2599211, EPI_ISL_2599212, EPI_ISL_2599213, EPI_ISL_2599214, EPI_ISL_2599215, EPI_ISL_2599216, EPI_ISL_2599217, EPI_ISL_2599218, EPI_ISL_2599219, EPI_ISL_2599220, EPI_ISL_2599221, EPI_ISL_2599222, EPI_ISL_2599223, EPI_ISL_2599224, EPI_ISL_2599225, EPI_ISL_2599226, EPI_ISL_2599227, EPI_ISL_2599228, EPI_ISL_2599229, EPI_ISL_2599230, EPI_ISL_2599231, EPI_ISL_2599232, EPI_ISL_2599233, EPI_ISL_2599234, EPI_ISL_2599235, EPI_ISL_2599236, EPI_ISL_2599237, EPI_ISL_2599238, EPI_ISL_2599239, EPI_ISL_2599240, EPI_ISL_2599241, EPI_ISL_2599242, EPI_ISL_2599243, EPI_ISL_2599244, EPI_ISL_2599245, EPI_ISL_2599246, EPI_ISL_2599247, EPI_ISL_2599248, EPI_ISL_2599249, EPI_ISL_2599250, EPI_ISL_2599251, EPI_ISL_2599252, EPI_ISL_2599253, EPI_ISL_2599254, EPI_ISL_2599255, EPI_ISL_2599256, EPI_ISL_2599257, EPI_ISL_2599258, EPI_ISL_2599259, EPI_ISL_2599260, EPI_ISL_2599261, EPI_ISL_2599262, EPI_ISL_2599263, EPI_ISL_2599264, EPI_ISL_2599265, EPI_ISL_2599266, EPI_ISL_2599267, EPI_ISL_2599268, EPI_ISL_2599269, EPI_ISL_2599270, EPI_ISL_2599271, EPI_ISL_2599272, EPI_ISL_2599273, EPI_ISL_2599274, EPI_ISL_2599275, EPI_ISL_2599276, EPI_ISL_2599277, EPI_ISL_2599278, EPI_ISL_2599279, EPI_ISL_2599280, EPI_ISL_2599281, EPI_ISL_2599282, EPI_ISL_2599283, EPI_ISL_2599284, EPI_ISL_2599285, EPI_ISL_2599286, EPI_ISL_2599287, EPI_ISL_2599288, EPI_ISL_2599289, EPI_ISL_2599290, EPI_ISL_2599291, EPI_ISL_2599292, EPI_ISL_2599293, EPI_ISL_2599294, EPI_ISL_2599295, EPI_ISL_2599296, EPI_ISL_2599297, EPI_ISL_2599298, EPI_ISL_2599299, EPI_ISL_2599300, EPI_ISL_2599301, EPI_ISL_2599302, EPI_ISL_2599303, EPI_ISL_2599304, EPI_ISL_2599305, EPI_ISL_2599306, EPI_ISL_2599307, EPI_ISL_2599308, EPI_ISL_2599309, EPI_ISL_2599310, EPI_ISL_2599311, EPI_ISL_2599312, EPI_ISL_2599313, EPI_ISL_2599314, EPI_ISL_2599315, EPI_ISL_2599316, EPI_ISL_2599317, EPI_ISL_2599318, EPI_ISL_2599319, EPI_ISL_2599320, EPI_ISL_2599321, EPI_ISL_2599322, EPI_ISL_2599323, EPI_ISL_2599324, EPI_ISL_2599325, EPI_ISL_2599326, EPI_ISL_2599327, EPI_ISL_2599328, EPI_ISL_2599329, EPI_ISL_2599330, EPI_ISL_2599331, EPI_ISL_2599332, EPI_ISL_2599333, EPI_ISL_2599334, EPI_ISL_2599335, EPI_ISL_2599336, EPI_ISL_2599337, EPI_ISL_2599338, EPI_ISL_2599339, EPI_ISL_2599340, EPI_ISL_2599341, EPI_ISL_2599342, EPI_ISL_2599343, EPI_ISL_2599344, EPI_ISL_2599345, EPI_ISL_2599346, EPI_ISL_2599347, EPI_ISL_2599348, EPI_ISL_2599349, EPI_ISL_2599350, EPI_ISL_2599351, EPI_ISL_2599352, EPI_ISL_2599353, EPI_ISL_2599354, EPI_ISL_2599355, EPI_ISL_2599356, EPI_ISL_2599357, EPI_ISL_2599358, EPI_ISL_2599359, EPI_ISL_2599360, EPI_ISL_2599361, EPI_ISL_2599362, EPI_ISL_2599363, EPI_ISL_2599364, EPI_ISL_2599365, EPI_ISL_2599366, EPI_ISL_2599367, EPI_ISL_2599368, EPI_ISL_2599369, EPI_ISL_2599370, EPI_ISL_2599371, EPI_ISL_2599372, EPI_ISL_2599373, EPI_ISL_2599374, EPI_ISL_2599375, EPI_ISL_2599376, EPI_ISL_2599377, EPI_ISL_2599378, EPI_ISL_2599379, EPI_ISL_2599380, EPI_ISL_2599381, EPI_ISL_2599382, EPI_ISL_2599383, EPI_ISL_2599384, EPI_ISL_2599385, EPI_ISL_2599386, EPI_ISL_2599387, EPI_ISL_2599388, EPI_ISL_2599389, EPI_ISL_2599390, EPI_ISL_2599391, EPI_ISL_2599392, EPI_ISL_2599393, EPI_ISL_2599394, EPI_ISL_2599395, EPI_ISL_2599396, EPI_ISL_2599397, EPI_ISL_2599398, EPI_ISL_2599399, EPI_ISL_2599400, EPI_ISL_2599401, EPI_ISL_2599402, EPI_ISL_2599403, EPI_ISL_2599404, EPI_ISL_2599405, EPI_ISL_2599406, EPI_ISL_2599407, EPI_ISL_2599408, EPI_ISL_2599409, EPI_ISL_2599410, EPI_ISL_2599411, EPI_ISL_2599412, EPI_ISL_2599413, EPI_ISL_2599414, EPI_ISL_2599415, EPI_ISL_2599416, EPI_ISL_2599417, EPI_ISL_2599418, EPI_ISL_2599419, EPI_ISL_2599420, EPI_ISL_2599421, EPI_ISL_2599422, EPI_ISL_2599423, EPI_ISL_2599424, EPI_ISL_2599426, EPI_ISL_2599427, EPI_ISL_2599428, EPI_ISL_2599429, EPI_ISL_2599430, EPI_ISL_2599431, EPI_ISL_2599432, EPI_ISL_2599433, EPI_ISL_2599434, EPI_ISL_2599436, EPI_ISL_2599437, EPI_ISL_2599438, EPI_ISL_2599439, EPI_ISL_2599440, EPI_ISL_2599441, EPI_ISL_2599442, EPI_ISL_2599443, EPI_ISL_2599444, EPI_ISL_2599445, EPI_ISL_2599446, EPI_ISL_2599447, EPI_ISL_2599448, EPI_ISL_2599449, EPI_ISL_2599450, EPI_ISL_2599451, EPI_ISL_2599452, EPI_ISL_2599453, EPI_ISL_2599454, EPI_ISL_2599455, EPI_ISL_2599456, EPI_ISL_2599457, EPI_ISL_2599458, EPI_ISL_2599459, EPI_ISL_2599460, EPI_ISL_2599461, EPI_ISL_2599462, EPI_ISL_2599463, EPI_ISL_2599464, EPI_ISL_2599465, EPI_ISL_2599466, EPI_ISL_2599467, EPI_ISL_2599468, EPI_ISL_2599469, EPI_ISL_2599470, EPI_ISL_2599471, EPI_ISL_2599472, EPI_ISL_2599473, EPI_ISL_2599474, EPI_ISL_2599475, EPI_ISL_2599476, EPI_ISL_2599477, EPI_ISL_2599478, EPI_ISL_2599479, EPI_ISL_2599480, EPI_ISL_2599481, EPI_ISL_2599482, EPI_ISL_2599483, EPI_ISL_2599484, EPI_ISL_2599485, EPI_ISL_2599486, EPI_ISL_2599487, EPI_ISL_2599488, EPI_ISL_2599489, EPI_ISL_2599490, EPI_ISL_2599491, EPI_ISL_2599492, EPI_ISL_2599493, EPI_ISL_2599494, EPI_ISL_2599495, EPI_ISL_2599496, EPI_ISL_2599497, EPI_ISL_2599498, EPI_ISL_2599499, EPI_ISL_2599500, EPI_ISL_2599501, EPI_ISL_2599502, EPI_ISL_2599503, EPI_ISL_2599504, EPI_ISL_2599506, EPI_ISL_2599507, EPI_ISL_2599508, EPI_ISL_2599509, EPI_ISL_2599510, EPI_ISL_2599511, EPI_ISL_2599512, EPI_ISL_2599513, EPI_ISL_2599514, EPI_ISL_2599515, EPI_ISL_2599516, EPI_ISL_2599517, EPI_ISL_2599518, EPI_ISL_2599519, EPI_ISL_2599520, EPI_ISL_2599521, EPI_ISL_2599522, EPI_ISL_2599523, EPI_ISL_2599524, EPI_ISL_2599525, EPI_ISL_2599526, EPI_ISL_2599527, EPI_ISL_2599528, EPI_ISL_2599529, EPI_ISL_2599530, EPI_ISL_2599531, EPI_ISL_2599533, EPI_ISL_2599534, EPI_ISL_2599535, EPI_ISL_2599536, EPI_ISL_2599537, EPI_ISL_2599538, EPI_ISL_2599539, EPI_ISL_2599540, EPI_ISL_2599541, EPI_ISL_2599542, EPI_ISL_2599543, EPI_ISL_2599544, EPI_ISL_2599545, EPI_ISL_2599546, EPI_ISL_2599547, EPI_ISL_2599548, EPI_ISL_2599549, EPI_ISL_2599550, EPI_ISL_2599551, EPI_ISL_2599552, EPI_ISL_2599553, EPI_ISL_2599554, EPI_ISL_2599555, EPI_ISL_2599556, EPI_ISL_2599557, EPI_ISL_2599558, EPI_ISL_2599559, EPI_ISL_2599560, EPI_ISL_2599561, EPI_ISL_2599562, EPI_ISL_2599563, EPI_ISL_2599564, EPI_ISL_2599565, EPI_ISL_2599566, EPI_ISL_2599567, EPI_ISL_2599568, EPI_ISL_2599569, EPI_ISL_2599570, EPI_ISL_2599571, EPI_ISL_2599573, EPI_ISL_2599574, EPI_ISL_2599575, EPI_ISL_2599576, EPI_ISL_2599577, EPI_ISL_2599578, EPI_ISL_2599579, EPI_ISL_2599580, EPI_ISL_2599581, EPI_ISL_2599582, EPI_ISL_2599583, EPI_ISL_2599584, EPI_ISL_2599585, EPI_ISL_2599586, EPI_ISL_2599587, EPI_ISL_2599588, EPI_ISL_2599589, EPI_ISL_2599590, EPI_ISL_2599591, EPI_ISL_2599592, EPI_ISL_2599593, EPI_ISL_2599594, EPI_ISL_2599595, EPI_ISL_2599596, EPI_ISL_2599597, EPI_ISL_2599598, EPI_ISL_2599599, EPI_ISL_2599600, EPI_ISL_2599601, EPI_ISL_2599602, EPI_ISL_2599603, EPI_ISL_2599604, EPI_ISL_2599605, EPI_ISL_2599606, EPI_ISL_2599607, EPI_ISL_2599608, EPI_ISL_2599609, EPI_ISL_2599610, EPI_ISL_2599611, EPI_ISL_2599612, EPI_ISL_2599613, EPI_ISL_2599614, EPI_ISL_2599615, EPI_ISL_2599616, EPI_ISL_2599618, EPI_ISL_2599619, EPI_ISL_2599620, EPI_ISL_2599621, EPI_ISL_2599622, EPI_ISL_2599623, EPI_ISL_2599624, EPI_ISL_2599625, EPI_ISL_2599626, EPI_ISL_2599627, EPI_ISL_2599628, EPI_ISL_2599629, EPI_ISL_2599630 | | | |
| see above | Helix/Illumina | Centers for Disease Control and Prevention Division of Viral Diseases, Pathogen Discovery | Dakota Howard, Dhwani Batra, Peter W. Cook, Kara Moser, Adrian Paskey, Jason Caravas, Benjamin Rambo-Martin, Shatavia Morrison, Christopher Gulvick, Scott Sammons, Yvette Unoarumhi, Darlene Wagner, Matthew Schmerer, Eileen de Feo, Jan Antico, Christine Tran, Matthew Tolentino, Shannon Wickline, Kim Gietzen, Brad Sickler, Jingtao Liu, Eric Allen, Phil Febbo, Nicole L. Washington, Simon White, Geraint Levan, Kelly Schiabor Barrett, Elizabeth Cirulli, Alexandre Bolze, Ary Ascencio, Charlotte Rivera-Garcia, Ryan Cho, Jason Nguyen, Sherry Wang, Jimmy Ramirez, Tyler Cassens, Efren Sandoval, Magnus Isaksson, William Lee, David Becker, Marc Laurent, James Lu, Clinton R. Paden, Duncan MacCannell |
| EPI_ISL_2599631, EPI_ISL_2599632, EPI_ISL_2599633, EPI_ISL_2599634, EPI_ISL_2599635, EPI_ISL_2599636, EPI_ISL_2599637, EPI_ISL_2599638, EPI_ISL_2599639, EPI_ISL_2599640, EPI_ISL_2599641, EPI_ISL_2599642, EPI_ISL_2599643, EPI_ISL_2599644, EPI_ISL_2599645, EPI_ISL_2599646, EPI_ISL_2599647, EPI_ISL_2599648, EPI_ISL_2599649, EPI_ISL_2599650, EPI_ISL_2599651, EPI_ISL_2599652, EPI_ISL_2599653, EPI_ISL_2599654, EPI_ISL_2599655, EPI_ISL_2599656, EPI_ISL_2599657, EPI_ISL_2599658, EPI_ISL_2599659, EPI_ISL_2599660, EPI_ISL_2599661, EPI_ISL_2599662, EPI_ISL_2599663, EPI_ISL_2599664, EPI_ISL_2599665, EPI_ISL_2599666, EPI_ISL_2599667, EPI_ISL_2599668, EPI_ISL_2599669, EPI_ISL_2599670, EPI_ISL_2599671, EPI_ISL_2599672, EPI_ISL_2599673, EPI_ISL_2599674, EPI_ISL_2599675, EPI_ISL_2599676, EPI_ISL_2599677, EPI_ISL_2599678, EPI_ISL_2599679, EPI_ISL_2599680, EPI_ISL_2599681, EPI_ISL_2599682, EPI_ISL_2599683, EPI_ISL_2599684, EPI_ISL_2599685, EPI_ISL_2599686, EPI_ISL_2599687, EPI_ISL_2599688, EPI_ISL_2599689, EPI_ISL_2599690, EPI_ISL_2599691, EPI_ISL_2599692, EPI_ISL_2599693, EPI_ISL_2599694, EPI_ISL_2599695, EPI_ISL_2599696, EPI_ISL_2599697, EPI_ISL_2599698, EPI_ISL_2599699, EPI_ISL_2599700, EPI_ISL_2599701, EPI_ISL_2599702, EPI_ISL_2599703, EPI_ISL_2599704, EPI_ISL_2599705, EPI_ISL_2599706, EPI_ISL_2599707, EPI_ISL_2599708, EPI_ISL_2599709, EPI_ISL_2599710, EPI_ISL_2599711, EPI_ISL_2599712, EPI_ISL_2599713, EPI_ISL_2599714, EPI_ISL_2599715, EPI_ISL_2599716, EPI_ISL_2599717, EPI_ISL_2599718, EPI_ISL_2599719, EPI_ISL_2599720, EPI_ISL_2599721, EPI_ISL_2599722, EPI_ISL_2599723, EPI_ISL_2599724, EPI_ISL_2599725, EPI_ISL_2599726, EPI_ISL_2599727, EPI_ISL_2599728, EPI_ISL_2599729, EPI_ISL_2599730, EPI_ISL_2599731, EPI_ISL_2599732, EPI_ISL_2599733, EPI_ISL_2599734, EPI_ISL_2599735, EPI_ISL_2599736, EPI_ISL_2599737, EPI_ISL_2599738, EPI_ISL_2599739, EPI_ISL_2599740, EPI_ISL_2599741, EPI_ISL_2599742, EPI_ISL_2599743, EPI_ISL_2599744, EPI_ISL_2599745, EPI_ISL_2599746, EPI_ISL_2599747, EPI_ISL_2599748, EPI_ISL_2599749, EPI_ISL_2599750, EPI_ISL_2599751, EPI_ISL_2599752, EPI_ISL_2599753, EPI_ISL_2599754, EPI_ISL_2599755, EPI_ISL_2599756, EPI_ISL_2599757, EPI_ISL_2599758, EPI_ISL_2599759, EPI_ISL_2599760, EPI_ISL_2599761, EPI_ISL_2599762, EPI_ISL_2599763, EPI_ISL_2599764, EPI_ISL_2599765, EPI_ISL_2599766, EPI_ISL_2599767, EPI_ISL_2599768, EPI_ISL_2599769, EPI_ISL_2599771, EPI_ISL_2599773, EPI_ISL_2599776, EPI_ISL_2599779, EPI_ISL_2599782, EPI_ISL_2599785, EPI_ISL_2599788, EPI_ISL_2599791, EPI_ISL_2599794, EPI_ISL_2599796, EPI_ISL_2599799, EPI_ISL_2599801, EPI_ISL_2599804, EPI_ISL_2599807, EPI_ISL_2599810, EPI_ISL_2599813, EPI_ISL_2599816, EPI_ISL_2599819, EPI_ISL_2599821, EPI_ISL_2599824, EPI_ISL_2599827, EPI_ISL_2599830, EPI_ISL_2599831, EPI_ISL_2599832, EPI_ISL_2599833, EPI_ISL_2599834, EPI_ISL_2599835, EPI_ISL_2599836, EPI_ISL_2599837, EPI_ISL_2599838, EPI_ISL_2599839, EPI_ISL_2599840, EPI_ISL_2599841, EPI_ISL_2599842, EPI_ISL_2599843, EPI_ISL_2599844, EPI_ISL_2599845, EPI_ISL_2599846, EPI_ISL_2599847, EPI_ISL_2599848, EPI_ISL_2599849, EPI_ISL_2599850, EPI_ISL_2599851, EPI_ISL_2599852, EPI_ISL_2599853, EPI_ISL_2599854, EPI_ISL_2599855, EPI_ISL_2599856, EPI_ISL_2599857, EPI_ISL_2599858, EPI_ISL_2599859, EPI_ISL_2599860, EPI_ISL_2599861, EPI_ISL_2599862, EPI_ISL_2599863, EPI_ISL_2599864, EPI_ISL_2599865, EPI_ISL_2599866, EPI_ISL_2599867, EPI_ISL_2599868, EPI_ISL_2599869, EPI_ISL_2599870, EPI_ISL_2599871, EPI_ISL_2599872, EPI_ISL_2599873, EPI_ISL_2599874, EPI_ISL_2599875, EPI_ISL_2599876, EPI_ISL_2599877, EPI_ISL_2599878, EPI_ISL_2599879, EPI_ISL_2599880, EPI_ISL_2599881, EPI_ISL_2599882, EPI_ISL_2599883, EPI_ISL_2599884, EPI_ISL_2599885, EPI_ISL_2599886, EPI_ISL_2599887, EPI_ISL_2599888, EPI_ISL_2599889, EPI_ISL_2599890, EPI_ISL_2599891, EPI_ISL_2599892, EPI_ISL_2599893, EPI_ISL_2599894, EPI_ISL_2599895, EPI_ISL_2599896, EPI_ISL_2599897, EPI_ISL_2599898, EPI_ISL_2599899, EPI_ISL_2599900, EPI_ISL_2599901, EPI_ISL_2599902, EPI_ISL_2599903, EPI_ISL_2599904, EPI_ISL_2599905, EPI_ISL_2599906, EPI_ISL_2599907, EPI_ISL_2599908, EPI_ISL_2599909, EPI_ISL_2599910, EPI_ISL_2599911, EPI_ISL_2599912, EPI_ISL_2599913, EPI_ISL_2599914, EPI_ISL_2599915, EPI_ISL_2599916, EPI_ISL_2599917, EPI_ISL_2599918, EPI_ISL_2599919, EPI_ISL_2599920, EPI_ISL_2599921 | | | |
| see above | Quest Diagnostics Incorporated | Centers for Disease Control and Prevention Division of Viral Diseases, Pathogen Discovery | Dakota Howard, Dhwani Batra, Peter W. Cook, Kara Moser, Adrian Paskey, Jason Caravas, Benjamin Rambo-Martin, Shatavia Morrison, Christopher Gulvick, Scott Sammons, Yvette Unoarumhi, Darlene Wagner, Matthew Schmerer, S. H. Rosenthal, A. Gerasimova, R. M. Kagan, B. Anderson, M. Hua, Y. Liu, L.E. Bernstein, K.E. Livingston, A. Perez, I. A. Shlyakhter, R. V. Rolando, R. Owen, P. Tanpaiboon, F. Lacbawan, Clinton R. Paden, Duncan MacCannell |
| EPI_ISL_2599922, EPI_ISL_2599923, EPI_ISL_2599924 | Fulgent Genetics | Centers for Disease Control and Prevention Division of Viral Diseases, Pathogen Discovery | Dakota Howard, Dhwani Batra, Peter W. Cook, Kara Moser, Adrian Paskey, Jason Caravas, Benjamin Rambo-Martin, Shatavia Morrison, Christopher Gulvick, Scott Sammons, Yvette Unoarumhi, Darlene Wagner, Matthew Schmerer, Harry Gao, Mickey Li, John Gao, Joseph Fierro, Benafsh Sapra, Becky Tsai, Yan Meng, Doreen Ng, James Xie, Clinton R. Paden, Duncan MacCannell |
| EPI_ISL_2599925, EPI_ISL_2599926, EPI_ISL_2599927, EPI_ISL_2599928, EPI_ISL_2599929, EPI_ISL_2599930, EPI_ISL_2599931, EPI_ISL_2599932, EPI_ISL_2599933, EPI_ISL_2599934, EPI_ISL_2599935, EPI_ISL_2599936, EPI_ISL_2599937, EPI_ISL_2599938, EPI_ISL_2599939, EPI_ISL_2599941, EPI_ISL_2599942, EPI_ISL_2599943, EPI_ISL_2599944, EPI_ISL_2599945, EPI_ISL_2599946, EPI_ISL_2599947, EPI_ISL_2599948, EPI_ISL_2599949, EPI_ISL_2599950, EPI_ISL_2599951, EPI_ISL_2599952, EPI_ISL_2599953, EPI_ISL_2599954, EPI_ISL_2599955, EPI_ISL_2599956, EPI_ISL_2599957, EPI_ISL_2599958, EPI_ISL_2599959, EPI_ISL_2599960, EPI_ISL_2599961, EPI_ISL_2599962, EPI_ISL_2599963, EPI_ISL_2599964, EPI_ISL_2599965, EPI_ISL_2599966, EPI_ISL_2599967, EPI_ISL_2599968, EPI_ISL_2599969, EPI_ISL_2599970, EPI_ISL_2599971, EPI_ISL_2599972, EPI_ISL_2599973, EPI_ISL_2599974, EPI_ISL_2599975, EPI_ISL_2599976, EPI_ISL_2599977, EPI_ISL_2599978, EPI_ISL_2599979, EPI_ISL_2599980, EPI_ISL_2599981, EPI_ISL_2599982, EPI_ISL_2599983, EPI_ISL_2599984, EPI_ISL_2599985, EPI_ISL_2599986, EPI_ISL_2599987, EPI_ISL_2599988, EPI_ISL_2599989, EPI_ISL_2599990, EPI_ISL_2599991, EPI_ISL_2599992, EPI_ISL_2599993, EPI_ISL_2599994, EPI_ISL_2599995, EPI_ISL_2599996, EPI_ISL_2599997, EPI_ISL_2599998, EPI_ISL_2599999, EPI_ISL_2600000, EPI_ISL_2600001, EPI_ISL_2600002, EPI_ISL_2600003, EPI_ISL_2600004, EPI_ISL_2600005, EPI_ISL_2600006, EPI_ISL_2600008, EPI_ISL_2600009, EPI_ISL_2600010, EPI_ISL_2600011, EPI_ISL_2600012, EPI_ISL_2600013, EPI_ISL_2600014, EPI_ISL_2600015, EPI_ISL_2600016, EPI_ISL_2600017, EPI_ISL_2600018, EPI_ISL_2600019, EPI_ISL_2600020, EPI_ISL_2600021, EPI_ISL_2600022, EPI_ISL_2600023, EPI_ISL_2600024, EPI_ISL_2600025, EPI_ISL_2600026, EPI_ISL_2600027, EPI_ISL_2600028, EPI_ISL_2600029, EPI_ISL_2600030, EPI_ISL_2600031, EPI_ISL_2600032, EPI_ISL_2600033, EPI_ISL_2600034, EPI_ISL_2600035, EPI_ISL_2600036, EPI_ISL_2600037, EPI_ISL_2600038, EPI_ISL_2600039, EPI_ISL_2600040, EPI_ISL_2600041, EPI_ISL_2600042, EPI_ISL_2600043, EPI_ISL_2600044, EPI_ISL_2600045, EPI_ISL_2600046, EPI_ISL_2600047, EPI_ISL_2600048, EPI_ISL_2600049, EPI_ISL_2600050, EPI_ISL_2600051, EPI_ISL_2600052, EPI_ISL_2600053, EPI_ISL_2600054, EPI_ISL_2600055, EPI_ISL_2600056, EPI_ISL_2600057, EPI_ISL_2600058, EPI_ISL_2600059, EPI_ISL_2600060, EPI_ISL_2600061, EPI_ISL_2600062, EPI_ISL_2600063, EPI_ISL_2600064, EPI_ISL_2600065, EPI_ISL_2600066, EPI_ISL_2600067, EPI_ISL_2600068, EPI_ISL_2600069, EPI_ISL_2600070, EPI_ISL_2600071, EPI_ISL_2600072, EPI_ISL_2600073, EPI_ISL_2600074, EPI_ISL_2600075, EPI_ISL_2600076, EPI_ISL_2600077, EPI_ISL_2600078, EPI_ISL_2600079, EPI_ISL_2600080, EPI_ISL_2600081, EPI_ISL_2600082, EPI_ISL_2600083, EPI_ISL_2600084, EPI_ISL_2600085, EPI_ISL_2600086, EPI_ISL_2600087, EPI_ISL_2600088, EPI_ISL_2600089, EPI_ISL_2600090, EPI_ISL_2600091, EPI_ISL_2600092, EPI_ISL_2600093, EPI_ISL_2600094, EPI_ISL_2600095, EPI_ISL_2600096, EPI_ISL_2600097, EPI_ISL_2600098, EPI_ISL_2600099, EPI_ISL_2600100, EPI_ISL_2600101, EPI_ISL_2600103, EPI_ISL_2600104, EPI_ISL_2600105, EPI_ISL_2600106, EPI_ISL_2600107, EPI_ISL_2600108, EPI_ISL_2600109, EPI_ISL_2600110, EPI_ISL_2600111, EPI_ISL_2600112, EPI_ISL_2600113, EPI_ISL_2600115, EPI_ISL_2600116, EPI_ISL_2600117, EPI_ISL_2600118, EPI_ISL_2600119, EPI_ISL_2600120, EPI_ISL_2600121, EPI_ISL_2600122, EPI_ISL_2600123, EPI_ISL_2600124, EPI_ISL_2600125, EPI_ISL_2600126, EPI_ISL_2600127, EPI_ISL_2600128, EPI_ISL_2600129, EPI_ISL_2600130, EPI_ISL_2600131, EPI_ISL_2600132, EPI_ISL_2600133, EPI_ISL_2600134, EPI_ISL_2600135, EPI_ISL_2600136, EPI_ISL_2600137, EPI_ISL_2600138, EPI_ISL_2600139, EPI_ISL_2600140, EPI_ISL_2600141, EPI_ISL_2600142, EPI_ISL_2600143, EPI_ISL_2600144, EPI_ISL_2600145, EPI_ISL_2600146, EPI_ISL_2600147, EPI_ISL_2600148, EPI_ISL_2600149, EPI_ISL_2600150, EPI_ISL_2600151, EPI_ISL_2600152, EPI_ISL_2600153, EPI_ISL_2600154, EPI_ISL_2600155, EPI_ISL_2600156, EPI_ISL_2600157, EPI_ISL_2600158, EPI_ISL_2600159, EPI_ISL_2600160, EPI_ISL_2600161, EPI_ISL_2600162, EPI_ISL_2600163, EPI_ISL_2600164, EPI_ISL_2600165, EPI_ISL_2600166, EPI_ISL_2600167, EPI_ISL_2600168, EPI_ISL_2600169, EPI_ISL_2600170, EPI_ISL_2600171, EPI_ISL_2600172, EPI_ISL_2600173, EPI_ISL_2600174, EPI_ISL_2600175, EPI_ISL_2600176, EPI_ISL_2600177, EPI_ISL_2600178, EPI_ISL_2600179, EPI_ISL_2600180, EPI_ISL_2600181, EPI_ISL_2600182, EPI_ISL_2600183, EPI_ISL_2600184, EPI_ISL_2600185, EPI_ISL_2600186, EPI_ISL_2600187, EPI_ISL_2600188, EPI_ISL_2600189, EPI_ISL_2600191, EPI_ISL_2600192, EPI_ISL_2600193, EPI_ISL_2600194, EPI_ISL_2600195, EPI_ISL_2600196, EPI_ISL_2600197, EPI_ISL_2600198, EPI_ISL_2600199, EPI_ISL_2600200, EPI_ISL_2600201, EPI_ISL_2600202, EPI_ISL_2600203, EPI_ISL_2600204, EPI_ISL_2600205, EPI_ISL_2600206, EPI_ISL_2600207, EPI_ISL_2600208, EPI_ISL_2600209, EPI_ISL_2600210, EPI_ISL_2600211, EPI_ISL_2600212, EPI_ISL_2600213, EPI_ISL_2600214, EPI_ISL_2600215, EPI_ISL_2600216, EPI_ISL_2600217, EPI_ISL_2600218, EPI_ISL_2600219, EPI_ISL_2600220, EPI_ISL_2600221, EPI_ISL_2600222, EPI_ISL_2600223, EPI_ISL_2600224, EPI_ISL_2600225, EPI_ISL_2600226, EPI_ISL_2600227, EPI_ISL_2600228, EPI_ISL_2600229, EPI_ISL_2600230, EPI_ISL_2600231, EPI_ISL_2600232, EPI_ISL_2600233, EPI_ISL_2600234, EPI_ISL_2600235, EPI_ISL_2600236, EPI_ISL_2600237, EPI_ISL_2600238, EPI_ISL_2600239, EPI_ISL_2600240, EPI_ISL_2600241, EPI_ISL_2600242, EPI_ISL_2600243, EPI_ISL_2600244, EPI_ISL_2600245, EPI_ISL_2600246, EPI_ISL_2600247, EPI_ISL_2600248, EPI_ISL_2600249, EPI_ISL_2600250, EPI_ISL_2600251, EPI_ISL_2600252, EPI_ISL_2600253, EPI_ISL_2600254, EPI_ISL_2600255, EPI_ISL_2600256, EPI_ISL_2600257, EPI_ISL_2600258, EPI_ISL_2600259, EPI_ISL_2600260, EPI_ISL_2600261, EPI_ISL_2600262, EPI_ISL_2600263, EPI_ISL_2600264, EPI_ISL_2600265, EPI_ISL_2600266, EPI_ISL_2600267, EPI_ISL_2600268, EPI_ISL_2600269, EPI_ISL_2600270, EPI_ISL_2600271, EPI_ISL_2600272, EPI_ISL_2600273, EPI_ISL_2600274, EPI_ISL_2600275, EPI_ISL_2600276, EPI_ISL_2600277, EPI_ISL_2600278, EPI_ISL_2600279, EPI_ISL_2600280, EPI_ISL_2600281, EPI_ISL_2600282, EPI_ISL_2600283, EPI_ISL_2600284, EPI_ISL_2600285, EPI_ISL_2600286, EPI_ISL_2600287, EPI_ISL_2600288, EPI_ISL_2600289, EPI_ISL_2600290, EPI_ISL_2600291, EPI_ISL_2600292, EPI_ISL_2600293, EPI_ISL_2600294, EPI_ISL_2600295, EPI_ISL_2600296, EPI_ISL_2600297, EPI_ISL_2600298, EPI_ISL_2600299, EPI_ISL_2600300, EPI_ISL_2600301, EPI_ISL_2600302, EPI_ISL_2600303, EPI_ISL_2600304, EPI_ISL_2600305, EPI_ISL_2600307, EPI_ISL_2600310, EPI_ISL_2600311, EPI_ISL_2600312 | | | |
| see above | Aegis Sciences Corporation | Centers for Disease Control and Prevention Division of Viral Diseases, Pathogen Discovery | Dakota Howard, Dhwani Batra, Peter W. Cook, Kara Moser, Adrian Paskey, Jason Caravas, Benjamin Rambo-Martin, Shatavia Morrison, Christopher Gulvick, Scott Sammons, Yvette Unoarumhi, Darlene Wagner, Matthew Schmerer, Cyndi Clark, Patrick Campbell, Rob Case, Vikramsinha Ghorpade, Holly Houdeshell, Ola Kvalvaag, Dillon Nall, Ethan Sanders, Alec Vest, Shaun Westlund, Matthew Hardison, Clinton R. Paden, Duncan MacCannell |
| EPI_ISL_2600313, EPI_ISL_2600314, EPI_ISL_2600315, EPI_ISL_2600316, EPI_ISL_2600317, EPI_ISL_2600318, EPI_ISL_2600319, EPI_ISL_2600320, EPI_ISL_2600321, EPI_ISL_2600322, EPI_ISL_2600323, EPI_ISL_2600324, EPI_ISL_2600325, EPI_ISL_2600326, EPI_ISL_2600327, EPI_ISL_2600328, EPI_ISL_2600329, EPI_ISL_2600330, EPI_ISL_2600331, EPI_ISL_2600332, EPI_ISL_2600333, EPI_ISL_2600334, EPI_ISL_2600335, EPI_ISL_2600336, EPI_ISL_2600337 | | | |
| see above | Infinity Biologix | Centers for Disease Control and Prevention Division of Viral Diseases, Pathogen Discovery | Dakota Howard, Dhwani Batra, Peter W. Cook, Kara Moser, Adrian Paskey, Jason Caravas, Benjamin Rambo-Martin, Shatavia Morrison, Christopher Gulvick, Scott Sammons, Yvette Unoarumhi, Darlene Wagner, Matthew Schmerer, Christian Bixby, Yihe Wang, Jonathan Schultz, Chirayu Goswami, Russ Hager, Robin Grimwood, Clinton R. Paden, Duncan MacCannell |

| | | | |
|---|---|---|---|
| EPI_ISL_2600374 | Tulare County Public Health Laboratory | Tulare County Public Health Laboratory | Denise L Lopez, Joshua Kootstra |
| EPI_ISL_2600380, EPI_ISL_2600383, EPI_ISL_2600388, EPI_ISL_2600391, EPI_ISL_2600396, EPI_ISL_2600397, EPI_ISL_2600402, EPI_ISL_2600403, EPI_ISL_2600405, EPI_ISL_2600407, EPI_ISL_2600408, EPI_ISL_2600411, EPI_ISL_2600415, EPI_ISL_2600416, EPI_ISL_2600417, EPI_ISL_2600421, EPI_ISL_2600422, EPI_ISL_2600423, EPI_ISL_2600424, EPI_ISL_2600433, EPI_ISL_2600439, EPI_ISL_2600440, EPI_ISL_2600441, EPI_ISL_2600442, EPI_ISL_2600443, EPI_ISL_2600444, EPI_ISL_2600445, EPI_ISL_2600446, EPI_ISL_2600447, EPI_ISL_2600449, EPI_ISL_2600450, EPI_ISL_2600451, EPI_ISL_2600452, EPI_ISL_2600457, EPI_ISL_2600461, EPI_ISL_2600463, EPI_ISL_2600464, EPI_ISL_2600465, EPI_ISL_2600467, EPI_ISL_2600468, EPI_ISL_2600473, EPI_ISL_2600474, EPI_ISL_2600482, EPI_ISL_2600484, EPI_ISL_2600485, EPI_ISL_2600486, EPI_ISL_2600487, EPI_ISL_2600488, EPI_ISL_2600489, EPI_ISL_2600494, EPI_ISL_2600495, EPI_ISL_2600496, EPI_ISL_2600502, EPI_ISL_2600503, EPI_ISL_2600506, EPI_ISL_2600509, EPI_ISL_2600512, EPI_ISL_2600513, EPI_ISL_2600515, EPI_ISL_2600516, EPI_ISL_2600517, EPI_ISL_2600518, EPI_ISL_2600519, EPI_ISL_2600522, EPI_ISL_2600524, EPI_ISL_2600525, EPI_ISL_2600527, EPI_ISL_2600528, EPI_ISL_2600533 | | | |
| see above | TGen North | TGen North | "Jolene Bowers, Heather Centner, Chris French, Hayley Yaglom, Ashlyn Pfeiffer, Darrin Lemmer, Dave Engelthaler, The Arizona COVID Genomics Union (ACGU)" |
| EPI_ISL_2600577, EPI_ISL_2600578, EPI_ISL_2600579, EPI_ISL_2600580, EPI_ISL_2600581, EPI_ISL_2600583, EPI_ISL_2600584, EPI_ISL_2600585, EPI_ISL_2600586, EPI_ISL_2600587, EPI_ISL_2600588, EPI_ISL_2600589, EPI_ISL_2600590, EPI_ISL_2600591, EPI_ISL_2600592, EPI_ISL_2600593, EPI_ISL_2600594, EPI_ISL_2600595, EPI_ISL_2600596, EPI_ISL_2600597, EPI_ISL_2600598, EPI_ISL_2600599, EPI_ISL_2600600, EPI_ISL_2600601, EPI_ISL_2600602, EPI_ISL_2600603, EPI_ISL_2600604, EPI_ISL_2600605, EPI_ISL_2600606, EPI_ISL_2600607, EPI_ISL_2600608, EPI_ISL_2600609, EPI_ISL_2600610, EPI_ISL_2600611, EPI_ISL_2600612, EPI_ISL_2600613, EPI_ISL_2600614, EPI_ISL_2600615, EPI_ISL_2600616, EPI_ISL_2600617, EPI_ISL_2600618, EPI_ISL_2600619, EPI_ISL_2600620, EPI_ISL_2600621, EPI_ISL_2600622, EPI_ISL_2600623, EPI_ISL_2600624, EPI_ISL_2600625, EPI_ISL_2600626, EPI_ISL_2600627, EPI_ISL_2600628, EPI_ISL_2600629, EPI_ISL_2600630, EPI_ISL_2600631, EPI_ISL_2600632, EPI_ISL_2600633, EPI_ISL_2600634, EPI_ISL_2600635, EPI_ISL_2600636, EPI_ISL_2600637, EPI_ISL_2600638, EPI_ISL_2600639, EPI_ISL_2600640, EPI_ISL_2600641, EPI_ISL_2600642, EPI_ISL_2600643, EPI_ISL_2600644, EPI_ISL_2600645, EPI_ISL_2600646, EPI_ISL_2600647, EPI_ISL_2600648, EPI_ISL_2600649, EPI_ISL_2600650, EPI_ISL_2600651, EPI_ISL_2600652, EPI_ISL_2600653, EPI_ISL_2600654, EPI_ISL_2600655, EPI_ISL_2600656, EPI_ISL_2600657, EPI_ISL_2600658, EPI_ISL_2600659, EPI_ISL_2600660, EPI_ISL_2600661, EPI_ISL_2600662, EPI_ISL_2600663, EPI_ISL_2600664, EPI_ISL_2600665, EPI_ISL_2600666, EPI_ISL_2600667, EPI_ISL_2600668, EPI_ISL_2600669, EPI_ISL_2600670, EPI_ISL_2600671, EPI_ISL_2600672, EPI_ISL_2600673, EPI_ISL_2600674, EPI_ISL_2600675, EPI_ISL_2600676, EPI_ISL_2600677, EPI_ISL_2600678, EPI_ISL_2600679, EPI_ISL_2600680, EPI_ISL_2600681, EPI_ISL_2600682, EPI_ISL_2600683, EPI_ISL_2600684, EPI_ISL_2600685, EPI_ISL_2600686, EPI_ISL_2600687, EPI_ISL_2600688, EPI_ISL_2600689, EPI_ISL_2600690, EPI_ISL_2600691, EPI_ISL_2600692, EPI_ISL_2600693, EPI_ISL_2600694, EPI_ISL_2600695, EPI_ISL_2600696, EPI_ISL_2600697, EPI_ISL_2600698, EPI_ISL_2600699, EPI_ISL_2600700, EPI_ISL_2600701, EPI_ISL_2600702, EPI_ISL_2600703, EPI_ISL_2600704, EPI_ISL_2600705, EPI_ISL_2600706, EPI_ISL_2600707, EPI_ISL_2600708, EPI_ISL_2600709, EPI_ISL_2600710, EPI_ISL_2600711, EPI_ISL_2600712, EPI_ISL_2600713, EPI_ISL_2600714, EPI_ISL_2600715, EPI_ISL_2600716, EPI_ISL_2600717, EPI_ISL_2600718, EPI_ISL_2600719, EPI_ISL_2600720, EPI_ISL_2600721, EPI_ISL_2600722, EPI_ISL_2600723, EPI_ISL_2600724, EPI_ISL_2600725, EPI_ISL_2600726, EPI_ISL_2600727, EPI_ISL_2600728, EPI_ISL_2600729, EPI_ISL_2600730, EPI_ISL_2600731, EPI_ISL_2600732 | | | |
| see above | University of Pittsburgh | Genomics and Discovery, Respiratory Viruses Branch, Division of Viral Diseases, Centers for Disease Control and Prevention | Yan Li, Jing Zhang, Anna Kelleher, Ying Tao, Brian Lynch, Krista Queen, Anna Uehara, Peter Cook, Han Jia Justin Ng, Rachel Marine, Clinton R. Paden, Dhwani Batra, Haibin Wang, Tara Coalter, Jasmine Padilla, Morgan Davis, Mili Sheth, Sarah Nobles, Mark Burroughs, Justin Lee, Adam Retchless, Suxiang Tong |
| EPI_ISL_2601739, EPI_ISL_2601740, EPI_ISL_2601741, EPI_ISL_2601742, EPI_ISL_2601743, EPI_ISL_2601744, EPI_ISL_2601745, EPI_ISL_2601746, EPI_ISL_2601747, EPI_ISL_2601748, EPI_ISL_2601749, EPI_ISL_2601750, EPI_ISL_2601751, EPI_ISL_2601752, EPI_ISL_2601753, EPI_ISL_2601754, EPI_ISL_2601755, EPI_ISL_2601756, EPI_ISL_2601757, EPI_ISL_2601758, EPI_ISL_2601759, EPI_ISL_2601760, EPI_ISL_2601761, EPI_ISL_2601762, EPI_ISL_2601763, EPI_ISL_2601764, EPI_ISL_2601765, EPI_ISL_2601766, EPI_ISL_2601767, EPI_ISL_2601768, EPI_ISL_2601769, EPI_ISL_2601770, EPI_ISL_2601771, EPI_ISL_2601772, EPI_ISL_2601773, EPI_ISL_2601774, EPI_ISL_2601775, EPI_ISL_2601776, EPI_ISL_2601777, EPI_ISL_2601778, EPI_ISL_2601779, EPI_ISL_2601780, EPI_ISL_2601781, EPI_ISL_2601782, EPI_ISL_2601783, EPI_ISL_2601784, EPI_ISL_2601785, EPI_ISL_2601786, EPI_ISL_2601787, EPI_ISL_2601789, EPI_ISL_2601790, EPI_ISL_2601792, EPI_ISL_2601793, EPI_ISL_2601794, EPI_ISL_2601795, EPI_ISL_2601796, EPI_ISL_2601797, EPI_ISL_2601798, EPI_ISL_2601799, EPI_ISL_2601800, EPI_ISL_2601801, EPI_ISL_2601802, EPI_ISL_2601803, EPI_ISL_2601804, EPI_ISL_2601805, EPI_ISL_2601806, EPI_ISL_2601807, EPI_ISL_2601808, EPI_ISL_2601810, EPI_ISL_2601811, EPI_ISL_2601812, EPI_ISL_2601813, EPI_ISL_2601814, EPI_ISL_2601815, EPI_ISL_2601816, EPI_ISL_2601817, EPI_ISL_2601818, EPI_ISL_2601819, EPI_ISL_2601820, EPI_ISL_2601821, EPI_ISL_2601822, EPI_ISL_2601823, EPI_ISL_2601824, EPI_ISL_2601825, EPI_ISL_2601826, EPI_ISL_2601827, EPI_ISL_2601828, EPI_ISL_2601829, EPI_ISL_2601830, EPI_ISL_2601831, EPI_ISL_2601832, EPI_ISL_2601833, EPI_ISL_2601834, EPI_ISL_2601835, EPI_ISL_2601836, EPI_ISL_2601837, EPI_ISL_2601838, EPI_ISL_2601839, EPI_ISL_2601840, EPI_ISL_2601841, EPI_ISL_2601842, EPI_ISL_2601843, EPI_ISL_2601844, EPI_ISL_2601845, EPI_ISL_2601846, EPI_ISL_2601847, EPI_ISL_2601848, EPI_ISL_2601849, EPI_ISL_2601850, EPI_ISL_2601851, EPI_ISL_2601852, EPI_ISL_2601853, EPI_ISL_2601854, EPI_ISL_2601855, EPI_ISL_2601856, EPI_ISL_2601857, EPI_ISL_2601858, EPI_ISL_2601859, EPI_ISL_2601860, EPI_ISL_2601861, EPI_ISL_2601862, EPI_ISL_2601863, EPI_ISL_2601864, EPI_ISL_2601865, EPI_ISL_2601866, EPI_ISL_2601867, EPI_ISL_2601868, EPI_ISL_2601869, EPI_ISL_2601870, EPI_ISL_2601872, EPI_ISL_2601873, EPI_ISL_2601874, EPI_ISL_2601875, EPI_ISL_2601876, EPI_ISL_2601877, EPI_ISL_2601878, EPI_ISL_2601880, EPI_ISL_2601881, EPI_ISL_2601882, EPI_ISL_2601883, EPI_ISL_2601884, EPI_ISL_2601885, EPI_ISL_2601886, EPI_ISL_2601887, EPI_ISL_2601888, EPI_ISL_2601889, EPI_ISL_2601890, EPI_ISL_2601891, EPI_ISL_2601892, EPI_ISL_2601893, EPI_ISL_2601894, EPI_ISL_2601895, EPI_ISL_2601896, EPI_ISL_2601897, EPI_ISL_2601898, EPI_ISL_2601899, EPI_ISL_2601900, EPI_ISL_2601901, EPI_ISL_2601902, EPI_ISL_2601903, EPI_ISL_2601904, EPI_ISL_2601905, EPI_ISL_2601907, EPI_ISL_2601908, EPI_ISL_2601909, EPI_ISL_2601910, EPI_ISL_2601911, EPI_ISL_2601912, EPI_ISL_2601913, EPI_ISL_2601914, EPI_ISL_2601915, EPI_ISL_2601916, EPI_ISL_2601917, EPI_ISL_2601918, EPI_ISL_2601919, EPI_ISL_2601920, EPI_ISL_2601921, EPI_ISL_2601922, EPI_ISL_2601923, EPI_ISL_2601925, EPI_ISL_2601926, EPI_ISL_2601927, EPI_ISL_2601928, EPI_ISL_2601929, EPI_ISL_2601930, EPI_ISL_2601931, EPI_ISL_2601932, EPI_ISL_2601933, EPI_ISL_2601934, EPI_ISL_2601935, EPI_ISL_2601936, EPI_ISL_2601937, EPI_ISL_2601938, EPI_ISL_2601939, EPI_ISL_2601981, EPI_ISL_2601982, EPI_ISL_2601983, EPI_ISL_2601984, EPI_ISL_2601985, EPI_ISL_2601986, EPI_ISL_2601987, EPI_ISL_2601988, EPI_ISL_2601989, EPI_ISL_2601990, EPI_ISL_2601991, EPI_ISL_2601992, EPI_ISL_2601993, EPI_ISL_2601994, EPI_ISL_2601995, EPI_ISL_2601996, EPI_ISL_2601997, EPI_ISL_2601998, EPI_ISL_2601999, EPI_ISL_2602000, EPI_ISL_2602001, EPI_ISL_2602002, EPI_ISL_2602003, EPI_ISL_2602004, EPI_ISL_2602005, EPI_ISL_2602006, EPI_ISL_2602007, EPI_ISL_2602008, EPI_ISL_2602009, EPI_ISL_2602010, EPI_ISL_2602011, EPI_ISL_2602012, EPI_ISL_2602013, EPI_ISL_2602014, EPI_ISL_2602015, EPI_ISL_2602016, EPI_ISL_2602017, EPI_ISL_2602018, EPI_ISL_2602019, EPI_ISL_2602020, EPI_ISL_2602021, EPI_ISL_2602022, EPI_ISL_2602023, EPI_ISL_2602024, EPI_ISL_2602025, EPI_ISL_2602026, EPI_ISL_2602027, EPI_ISL_2602028, EPI_ISL_2602029, EPI_ISL_2602030, EPI_ISL_2602031, EPI_ISL_2602032, EPI_ISL_2602033, EPI_ISL_2602034, EPI_ISL_2602035, EPI_ISL_2602036, EPI_ISL_2602037, EPI_ISL_2602038, EPI_ISL_2602039, EPI_ISL_2602040, EPI_ISL_2602041, EPI_ISL_2602042, EPI_ISL_2602043, EPI_ISL_2602044, EPI_ISL_2602045, EPI_ISL_2602047, EPI_ISL_2602048, EPI_ISL_2602049, EPI_ISL_2602050, EPI_ISL_2602051, EPI_ISL_2602052, EPI_ISL_2602053, EPI_ISL_2602054, EPI_ISL_2602055, EPI_ISL_2602056, EPI_ISL_2602057, EPI_ISL_2602058, EPI_ISL_2602059, EPI_ISL_2602060, EPI_ISL_2602061, EPI_ISL_2602062, EPI_ISL_2602063, EPI_ISL_2602064, EPI_ISL_2602065, EPI_ISL_2602066, EPI_ISL_2602067, EPI_ISL_2602068, EPI_ISL_2602069, EPI_ISL_2602070, EPI_ISL_2602071, EPI_ISL_2602072, EPI_ISL_2602073, EPI_ISL_2602074, EPI_ISL_2602075, EPI_ISL_2602076, EPI_ISL_2602077, EPI_ISL_2602078, EPI_ISL_2602079, EPI_ISL_2602080, EPI_ISL_2602082, EPI_ISL_2602084, EPI_ISL_2602085, EPI_ISL_2602086, EPI_ISL_2602087, EPI_ISL_2602088, EPI_ISL_2602089, EPI_ISL_2602090, EPI_ISL_2602091, EPI_ISL_2602092, EPI_ISL_2602093, EPI_ISL_2602094, EPI_ISL_2602095, EPI_ISL_2602096, EPI_ISL_2602097, EPI_ISL_2602099, EPI_ISL_2602100, EPI_ISL_2602101, EPI_ISL_2602102, EPI_ISL_2602103, EPI_ISL_2602104, EPI_ISL_2602105, EPI_ISL_2602106, EPI_ISL_2602107, EPI_ISL_2602108, EPI_ISL_2602109, EPI_ISL_2602110, EPI_ISL_2602111, EPI_ISL_2602112, EPI_ISL_2602113, EPI_ISL_2602114, EPI_ISL_2602115, EPI_ISL_2602116, EPI_ISL_2602117, EPI_ISL_2602118, EPI_ISL_2602119, EPI_ISL_2602120, EPI_ISL_2602121, EPI_ISL_2602123, EPI_ISL_2602124, EPI_ISL_2602125, EPI_ISL_2602126, EPI_ISL_2602127, EPI_ISL_2602128, EPI_ISL_2602129, EPI_ISL_2602130, EPI_ISL_2602131, EPI_ISL_2602132, EPI_ISL_2602133, EPI_ISL_2602134, EPI_ISL_2602135, EPI_ISL_2602136, EPI_ISL_2602137, EPI_ISL_2602138, EPI_ISL_2602139, EPI_ISL_2602140, EPI_ISL_2602142, EPI_ISL_2602143, EPI_ISL_2602144, EPI_ISL_2602145, EPI_ISL_2602146, EPI_ISL_2602147, EPI_ISL_2602148, EPI_ISL_2602149, EPI_ISL_2602150, EPI_ISL_2602151, EPI_ISL_2602152, EPI_ISL_2602153, EPI_ISL_2602154, EPI_ISL_2602155, EPI_ISL_2602156, EPI_ISL_2602157, EPI_ISL_2602158, EPI_ISL_2602159, EPI_ISL_2602160, EPI_ISL_2602161, EPI_ISL_2602162, EPI_ISL_2602163, EPI_ISL_2602164, EPI_ISL_2602165, EPI_ISL_2602166, EPI_ISL_2602167, EPI_ISL_2602168, EPI_ISL_2602169, EPI_ISL_2602170, EPI_ISL_2602171, EPI_ISL_2602172, EPI_ISL_2602173, EPI_ISL_2602174, EPI_ISL_2602175, EPI_ISL_2602176, EPI_ISL_2602177, EPI_ISL_2602178, EPI_ISL_2602179, EPI_ISL_2602180, EPI_ISL_2602181, EPI_ISL_2602182, EPI_ISL_2602183, EPI_ISL_2602184, EPI_ISL_2602185, EPI_ISL_2602186, EPI_ISL_2602187, EPI_ISL_2602188, EPI_ISL_2602189, EPI_ISL_2602190, EPI_ISL_2602191, EPI_ISL_2602192, EPI_ISL_2602193, EPI_ISL_2602194, EPI_ISL_2602195, EPI_ISL_2602196, EPI_ISL_2602197, EPI_ISL_2602198, EPI_ISL_2602199, EPI_ISL_2602200, EPI_ISL_2602201, EPI_ISL_2602202, EPI_ISL_2602203, EPI_ISL_2602204, EPI_ISL_2602205, EPI_ISL_2602207, EPI_ISL_2602208, EPI_ISL_2602209, EPI_ISL_2602211, EPI_ISL_2602212, EPI_ISL_2602213, EPI_ISL_2602214, EPI_ISL_2602215, EPI_ISL_2602216, EPI_ISL_2602217, EPI_ISL_2602218, EPI_ISL_2602219, EPI_ISL_2602220, EPI_ISL_2602221, EPI_ISL_2602222, EPI_ISL_2602223, EPI_ISL_2602224, EPI_ISL_2602225, EPI_ISL_2602226, EPI_ISL_2602227, EPI_ISL_2602228, EPI_ISL_2602229, EPI_ISL_2602230, EPI_ISL_2602231, EPI_ISL_2602232, EPI_ISL_2602234, EPI_ISL_2602235, EPI_ISL_2602236, EPI_ISL_2602237, EPI_ISL_2602238, EPI_ISL_2602239, EPI_ISL_2602240, EPI_ISL_2602241, EPI_ISL_2602242, EPI_ISL_2602243, EPI_ISL_2602244, EPI_ISL_2602245, EPI_ISL_2602246, EPI_ISL_2602247, EPI_ISL_2602248, EPI_ISL_2602249, EPI_ISL_2602250, EPI_ISL_2602251, EPI_ISL_2602252, EPI_ISL_2602253, EPI_ISL_2602254, EPI_ISL_2602255, EPI_ISL_2602256, EPI_ISL_2602257, EPI_ISL_2602259, EPI_ISL_2602260, EPI_ISL_2602261, EPI_ISL_2602262, EPI_ISL_2602263, EPI_ISL_2602264, EPI_ISL_2602265, EPI_ISL_2602266, EPI_ISL_2602267, EPI_ISL_2602268, EPI_ISL_2602269, EPI_ISL_2602270, EPI_ISL_2602271, EPI_ISL_2602272, EPI_ISL_2602273, EPI_ISL_2602274, EPI_ISL_2602275, EPI_ISL_2602276, EPI_ISL_2602277, EPI_ISL_2602278, EPI_ISL_2602279, EPI_ISL_2602280, EPI_ISL_2602281, EPI_ISL_2602282, EPI_ISL_2602283, EPI_ISL_2602284, EPI_ISL_2602285, EPI_ISL_2602286, EPI_ISL_2602287, EPI_ISL_2602288, EPI_ISL_2602289, EPI_ISL_2602290, EPI_ISL_2602291, EPI_ISL_2602292, EPI_ISL_2602293, EPI_ISL_2602294, EPI_ISL_2602295, EPI_ISL_2602296, EPI_ISL_2602297, EPI_ISL_2602298, EPI_ISL_2602299, EPI_ISL_2602300, EPI_ISL_2602301, EPI_ISL_2602302, EPI_ISL_2602303, EPI_ISL_2602304, EPI_ISL_2602305, EPI_ISL_2602306, EPI_ISL_2602307, EPI_ISL_2602308, EPI_ISL_2602309, EPI_ISL_2602310, EPI_ISL_2602311, EPI_ISL_2602312, EPI_ISL_2602313, EPI_ISL_2602314, EPI_ISL_2602315, EPI_ISL_2602316, EPI_ISL_2602317, EPI_ISL_2602318, EPI_ISL_2602319, EPI_ISL_2602320, EPI_ISL_2602321, EPI_ISL_2602322, EPI_ISL_2602323, EPI_ISL_2602324, EPI_ISL_2602325, EPI_ISL_2602326, EPI_ISL_2602327, EPI_ISL_2602328, EPI_ISL_2602329, EPI_ISL_2602330, EPI_ISL_2602331, EPI_ISL_2602332, EPI_ISL_2602333, EPI_ISL_2602334, EPI_ISL_2602335, EPI_ISL_2602337, EPI_ISL_2602338, EPI_ISL_2602339, EPI_ISL_2602340, EPI_ISL_2602341, EPI_ISL_2602342, EPI_ISL_2602343, EPI_ISL_2602344, EPI_ISL_2602345, EPI_ISL_2602346, EPI_ISL_2602347, EPI_ISL_2602348, EPI_ISL_2602349, EPI_ISL_2602350, EPI_ISL_2602351, EPI_ISL_2602352, EPI_ISL_2602353, EPI_ISL_2602354, EPI_ISL_2602355, EPI_ISL_2602356, EPI_ISL_2602357, EPI_ISL_2602358, EPI_ISL_2602359, EPI_ISL_2602360, EPI_ISL_2602361, EPI_ISL_2602362, EPI_ISL_2602364, EPI_ISL_2602365, EPI_ISL_2602366, EPI_ISL_2602367, EPI_ISL_2602368, EPI_ISL_2602369, EPI_ISL_2602370, EPI_ISL_2602371, EPI_ISL_2602372, EPI_ISL_2602373, EPI_ISL_2602374, EPI_ISL_2602375, EPI_ISL_2602376, EPI_ISL_2602377, EPI_ISL_2602378, EPI_ISL_2602379, EPI_ISL_2602380, EPI_ISL_2602381, EPI_ISL_2602382, EPI_ISL_2602383, EPI_ISL_2602384, EPI_ISL_2602385, EPI_ISL_2602386, EPI_ISL_2602387, EPI_ISL_2602388, EPI_ISL_2602389, EPI_ISL_2602390, EPI_ISL_2602391, EPI_ISL_2602392, EPI_ISL_2602393, EPI_ISL_2602394, EPI_ISL_2602395, EPI_ISL_2602396, EPI_ISL_2602397, EPI_ISL_2602398 | | | |
| see above | Gravity Diagnostics, LLC | Gravity Diagnostics, LLC | Gravity Diagnostics |
| EPI_ISL_2603122, EPI_ISL_2603123 | Utah Public Health Laboratory, Utah Public Health Laboratory Infectious Disease submission group | Utah Public Health Laboratory, Utah Public Health Laboratory Infectious Disease submission group | Young,E.L., Oakeson,K.F., Gallagher,T., Linares-Perdomo,O. |
| EPI_ISL_2603184 | University of Washington Medical Center, Seattle Flu Study | University of Washington Medical Center, Seattle Flu Study | Nickerson,D.A., Frazar,C.D., Lee,J., Pelle,B., Ryke,E., Richardson,M., Adler,A., Brandstetter,E., Han,P.D., Fay,K., Ilcisin,M., Lacombe,K., Sibley,T.R., Truong,M., Wolf,C.R., Cowgill,K., Schrag,S., Duchin,J., Boeckh,M., Englund,J.A., Famulare,M., Lutz,B.R., Rieder,M.J., Starita,L.M., Thompson,M., Chu,H.Y., Bedford,T., Shendure,J. |
| EPI_ISL_2603185, EPI_ISL_2603186, EPI_ISL_2603187, EPI_ISL_2603188, EPI_ISL_2603189, EPI_ISL_2603190, EPI_ISL_2603191 | University of Washington Medical Center, Seattle Flu Study | University of Washington Medical Center, Seattle Flu Study | Nickerson,D.A., Frazar,C.D., Lee,J., Pelle,B., Ryke,E., Richardson,M., Adler,A., Brandstetter,E., Han,P.D., Fay,K., Ilcisin,M., Lacombe,K., Sibley,T.R., Truong,M., Wolf,C.R., Boeckh,M., Englund,J.A., Famulare,M., Lutz,B.R., Rieder,M.J., Starita,L.M., Thompson,M., Shendure,J., Bedford,T., Chu,H.Y. |
| EPI_ISL_2603192, EPI_ISL_2603193 | University of Washington Medical Center, Seattle Flu Study | University of Washington Medical Center, Seattle Flu Study | Nickerson,D.A., Frazar,C.D., Lee,J., Pelle,B., Ryke,E., Richardson,M., Adler,A., Brandstetter,E., Han,P.D., Fay,K., Ilcisin,M., Lacombe,K., Sibley,T.R., Truong,M., Wolf,C.R., Gautom,R., Melly,G., Hiatt,B., Dykema,P., Lindquist,S., Boeckh,M., Englund,J.A., Famulare,M., Lutz,B.R., Rieder,M.J., Starita,L.M., Thompson,M., Chu,H.Y., Shendure,J., Bedford,T. |
| EPI_ISL_2603202, EPI_ISL_2603203, EPI_ISL_2603204, EPI_ISL_2603205, EPI_ISL_2603206, EPI_ISL_2603207, EPI_ISL_2603208, EPI_ISL_2603209, EPI_ISL_2603210, EPI_ISL_2603211, EPI_ISL_2603212, EPI_ISL_2603213, EPI_ISL_2603214, EPI_ISL_2603215, EPI_ISL_2603216, EPI_ISL_2603217, EPI_ISL_2603218, EPI_ISL_2603219, EPI_ISL_2603220, EPI_ISL_2603222, EPI_ISL_2603223, EPI_ISL_2603224, EPI_ISL_2603225, EPI_ISL_2603226, EPI_ISL_2603227, EPI_ISL_2603228, EPI_ISL_2603229, EPI_ISL_2603230, EPI_ISL_2603231, EPI_ISL_2603232, EPI_ISL_2603233, EPI_ISL_2603234, EPI_ISL_2603235, EPI_ISL_2603236 | | | |
| see above | Virginia Division of Consolidated Laboratory Services | Virginia Division of Consolidated Laboratory Services | Virginia DCLS |
| EPI_ISL_2603257, EPI_ISL_2603258, EPI_ISL_2603259, EPI_ISL_2603260, EPI_ISL_2603261, EPI_ISL_2603262, EPI_ISL_2603263, EPI_ISL_2603264, EPI_ISL_2603265, EPI_ISL_2603266, EPI_ISL_2603267, EPI_ISL_2603268, EPI_ISL_2603269, EPI_ISL_2603270, EPI_ISL_2603271, EPI_ISL_2603272, EPI_ISL_2603273 | | | |
| see above | Respiratory Viruses Branch, US Centers for Disease Control and Prevention | Respiratory Viruses Branch, US Centers for Disease Control and Prevention | Tao,Y., Li,Y., Zhang,J., Kelleher,A., Lynch,B., Queen,K., Uehara,A., Cook,P., Ng,H.J.J., Marine,R., Paden,C.R., Batra,D., Wang,H., Coalter,T., Padilla,J., Davis,M., Sheth,M., Nobles,S., Burroughs,M., Lee,J., Retchless,A., Tong,S. |
| EPI_ISL_2603274, EPI_ISL_2603275, EPI_ISL_2603276, EPI_ISL_2603277, EPI_ISL_2603278, EPI_ISL_2603280, EPI_ISL_2603281, EPI_ISL_2603282, EPI_ISL_2603283, EPI_ISL_2603284, EPI_ISL_2603285, EPI_ISL_2603286, EPI_ISL_2603287, EPI_ISL_2603288, EPI_ISL_2603289, EPI_ISL_2603290, EPI_ISL_2603291, EPI_ISL_2603292, EPI_ISL_2603293 | | | |
| see above | Wyoming Public Health Laboratory | Wyoming Public Health Laboratory | Jim Mildenberger, Taylor Fearing, Channing Weber, Ashley Norberg, Chayse Rowley, Marley Goetz, Brian Dominguez, Elliot Thomasson, Sam Britz, Cari Sloma, Robert Petit, and Rob Christensen |
| EPI_ISL_2603297, EPI_ISL_2603298, EPI_ISL_2603299, EPI_ISL_2603300, EPI_ISL_2603301, EPI_ISL_2603302, EPI_ISL_2603303, EPI_ISL_2603304, EPI_ISL_2603305, EPI_ISL_2603306, EPI_ISL_2603307, EPI_ISL_2603308, EPI_ISL_2603309, EPI_ISL_2603310, EPI_ISL_2603311, EPI_ISL_2603312, EPI_ISL_2603313, EPI_ISL_2603314, EPI_ISL_2603315, EPI_ISL_2603316, EPI_ISL_2603317, EPI_ISL_2603318, EPI_ISL_2603319, EPI_ISL_2603320, EPI_ISL_2603321, EPI_ISL_2603322, EPI_ISL_2603323, EPI_ISL_2603324, EPI_ISL_2603325, EPI_ISL_2603326, EPI_ISL_2603327, EPI_ISL_2603328, EPI_ISL_2603329, EPI_ISL_2603330, EPI_ISL_2603331, EPI_ISL_2603332, EPI_ISL_2603333, EPI_ISL_2603334, EPI_ISL_2603335 | | | |
| see above | Rhode Island State Health Laboratory | Rhode Island State Health Laboratory | Ewa King, Richard C. Huard, Kristin Carpenter-Azevedo |
| EPI_ISL_2603336, EPI_ISL_2603337, EPI_ISL_2603338, EPI_ISL_2603339, EPI_ISL_2603340, EPI_ISL_2603341, EPI_ISL_2603342, EPI_ISL_2603343, EPI_ISL_2603344, EPI_ISL_2603345, EPI_ISL_2603346, EPI_ISL_2603347, EPI_ISL_2603348, EPI_ISL_2603349, EPI_ISL_2603350, EPI_ISL_2603351, EPI_ISL_2603352, EPI_ISL_2603353, EPI_ISL_2603354, EPI_ISL_2603355, EPI_ISL_2603356, EPI_ISL_2603357, EPI_ISL_2603358, EPI_ISL_2603359, EPI_ISL_2603360, EPI_ISL_2603361, EPI_ISL_2603362, EPI_ISL_2603363 | | | |
| see above | Virginia Division of Consolidated Laboratory Services | Virginia Division of Consolidated Laboratory Services | Virginia DCLS |
| EPI_ISL_2603365, EPI_ISL_2603366, EPI_ISL_2603367, EPI_ISL_2603368, EPI_ISL_2603369 | Pathology, University of Alabama at Birmingham | Pathology, University of Alabama at Birmingham | Moates,D., Hendrickson,R.C., Lefkowitz,E.J., Leal,S.M. |
| EPI_ISL_2603370, EPI_ISL_2603371, EPI_ISL_2603372, EPI_ISL_2603373, EPI_ISL_2603374, EPI_ISL_2603375, EPI_ISL_2603376, EPI_ISL_2603377, EPI_ISL_2603378, EPI_ISL_2603379, EPI_ISL_2603380, EPI_ISL_2603381, EPI_ISL_2603382, EPI_ISL_2603383, EPI_ISL_2603384, EPI_ISL_2603385, EPI_ISL_2603386, EPI_ISL_2603388, EPI_ISL_2603389, EPI_ISL_2603390, EPI_ISL_2603391, EPI_ISL_2603392, EPI_ISL_2603393, EPI_ISL_2603394, EPI_ISL_2603396, EPI_ISL_2603399, EPI_ISL_2603400, EPI_ISL_2603401, EPI_ISL_2603403, EPI_ISL_2603404, EPI_ISL_2603405, EPI_ISL_2603406, EPI_ISL_2603407, EPI_ISL_2603408 | | | |
| see above | Wyoming Public Health Laboratory | Wyoming Public Health Laboratory | Jim Mildenberger, Taylor Fearing, Channing Weber, Ashley Norberg, Chayse Rowley, Marley Goetz, Brian Dominguez, Elliot Thomasson, Sam Britz, Cari Sloma, Robert Petit, and Rob Christensen |
| EPI_ISL_2603592, EPI_ISL_2603593 | NIAID - IRF Frederick, NIH | NIAID - IRF Frederick, NIH | Georgia,S., Kocher,G. |
| EPI_ISL_2603595, EPI_ISL_2603596, EPI_ISL_2603598, EPI_ISL_2603599, EPI_ISL_2603600, EPI_ISL_2603601, EPI_ISL_2603602, EPI_ISL_2603603, EPI_ISL_2603604, EPI_ISL_2603605, EPI_ISL_2603606, EPI_ISL_2603607, EPI_ISL_2603608, EPI_ISL_2603609, EPI_ISL_2603610, EPI_ISL_2603611, EPI_ISL_2603612, EPI_ISL_2603613, EPI_ISL_2603614, EPI_ISL_2603615, EPI_ISL_2603616, EPI_ISL_2603617, EPI_ISL_2603618, EPI_ISL_2603619, EPI_ISL_2603620, EPI_ISL_2603621, EPI_ISL_2603622, EPI_ISL_2603623, EPI_ISL_2603624, EPI_ISL_2603625, EPI_ISL_2603626, EPI_ISL_2603627, EPI_ISL_2603628, EPI_ISL_2603629, EPI_ISL_2603630, EPI_ISL_2603631, EPI_ISL_2603632, EPI_ISL_2603633, EPI_ISL_2603634, EPI_ISL_2603635, EPI_ISL_2603637, EPI_ISL_2603638, EPI_ISL_2603639, EPI_ISL_2603640, EPI_ISL_2603641, EPI_ISL_2603642, EPI_ISL_2603643, EPI_ISL_2603644, EPI_ISL_2603645, EPI_ISL_2603646, EPI_ISL_2603647, EPI_ISL_2603648, EPI_ISL_2603649, EPI_ISL_2603650, EPI_ISL_2603651, EPI_ISL_2603652, EPI_ISL_2603653, EPI_ISL_2603654, EPI_ISL_2603655, EPI_ISL_2603656, EPI_ISL_2603657, EPI_ISL_2603658 | | | |
| see above | InDRE | Instituto Nacional de Medicina Genomica | Hidalgo-Miranda A, Mendoza-Vargas A, Reyes-Grajeda JP, Cedro-Tanda A, Gisela Barrera-Badillo, Irma Lopez-Martinez, Jose Ernesto Ramirez González, Gonzalez-Barrera D, Rangel-DeLeon D, Munguia-Garza P, Garcia-Cardenas FJ, Gonzalez-Woge MA, Herrera-Montalvo LA. |
| EPI_ISL_2603659, EPI_ISL_2603660, EPI_ISL_2603661, EPI_ISL_2603662, EPI_ISL_2603663, EPI_ISL_2603664, EPI_ISL_2603665, EPI_ISL_2603666, EPI_ISL_2603667, EPI_ISL_2603668, EPI_ISL_2603669, EPI_ISL_2603670, EPI_ISL_2603671, EPI_ISL_2603672, EPI_ISL_2603673, EPI_ISL_2603674, EPI_ISL_2603675, EPI_ISL_2603676, EPI_ISL_2603677, EPI_ISL_2603678, EPI_ISL_2603679, EPI_ISL_2603680, EPI_ISL_2603681, EPI_ISL_2603682, EPI_ISL_2603684, EPI_ISL_2603685, EPI_ISL_2603686, EPI_ISL_2603687, EPI_ISL_2603688, EPI_ISL_2603689, EPI_ISL_2603690, EPI_ISL_2603691, EPI_ISL_2603692, EPI_ISL_2603693, EPI_ISL_2603694, EPI_ISL_2603695, EPI_ISL_2603696, EPI_ISL_2603697, EPI_ISL_2603698, EPI_ISL_2603699, EPI_ISL_2603701, EPI_ISL_2603702, EPI_ISL_2603703, EPI_ISL_2603704, EPI_ISL_2603705, EPI_ISL_2603706, EPI_ISL_2603707, EPI_ISL_2603708, EPI_ISL_2603709, EPI_ISL_2603710, EPI_ISL_2603711, EPI_ISL_2603713, EPI_ISL_2603714, EPI_ISL_2603715, EPI_ISL_2603716, EPI_ISL_2603717, EPI_ISL_2603718, EPI_ISL_2603719, EPI_ISL_2603720, EPI_ISL_2603721, EPI_ISL_2603722, EPI_ISL_2603723, EPI_ISL_2603724, EPI_ISL_2603725, EPI_ISL_2603726, EPI_ISL_2603727, EPI_ISL_2603728, EPI_ISL_2603729, EPI_ISL_2603730, EPI_ISL_2603731, EPI_ISL_2603732, EPI_ISL_2603733, EPI_ISL_2603734, EPI_ISL_2603735, EPI_ISL_2603736, EPI_ISL_2603737, EPI_ISL_2603738, EPI_ISL_2603739, EPI_ISL_2603740, EPI_ISL_2603741, EPI_ISL_2603742, EPI_ISL_2603743, EPI_ISL_2603744, EPI_ISL_2603745, EPI_ISL_2603746, EPI_ISL_2603747, EPI_ISL_2603748, EPI_ISL_2603749, EPI_ISL_2603750, EPI_ISL_2603751, EPI_ISL_2603752, EPI_ISL_2603753, EPI_ISL_2603754, EPI_ISL_2603755, EPI_ISL_2603756, EPI_ISL_2603757, EPI_ISL_2603758, EPI_ISL_2603759, EPI_ISL_2603760, EPI_ISL_2603761, EPI_ISL_2603762, EPI_ISL_2603763, EPI_ISL_2603764, EPI_ISL_2603765, EPI_ISL_2603766, EPI_ISL_2603767, EPI_ISL_2603768, EPI_ISL_2603769, EPI_ISL_2603770, EPI_ISL_2603771, EPI_ISL_2603772, EPI_ISL_2603773, EPI_ISL_2603774, EPI_ISL_2603775, EPI_ISL_2603776, EPI_ISL_2603777, EPI_ISL_2603778, EPI_ISL_2603779, EPI_ISL_2603780, EPI_ISL_2603781, EPI_ISL_2603782, EPI_ISL_2603783, EPI_ISL_2603784, EPI_ISL_2603785, EPI_ISL_2603786, EPI_ISL_2603787, EPI_ISL_2603788, EPI_ISL_2603789, EPI_ISL_2603790, EPI_ISL_2603791, EPI_ISL_2603792, EPI_ISL_2603793, EPI_ISL_2603794, EPI_ISL_2603795, EPI_ISL_2603796, EPI_ISL_2603797, EPI_ISL_2603798, EPI_ISL_2603799, EPI_ISL_2603800, EPI_ISL_2603801, EPI_ISL_2603802, EPI_ISL_2603803, EPI_ISL_2603804, EPI_ISL_2603805, EPI_ISL_2603806, EPI_ISL_2603807, EPI_ISL_2603808, EPI_ISL_2603809, EPI_ISL_2603810, EPI_ISL_2603811, EPI_ISL_2603812, EPI_ISL_2603813, EPI_ISL_2603814, EPI_ISL_2603815 | | | |

| | | | |
|---|---|---|---|
| see above | Salud Digna | Instituto Nacional de Medicina Genomica | Hidalgo-Miranda A, Cedro-Tanda A, Mendoza-Vargas A, Reyes-Grajeda JP, Abraham Campos-Romero, Moreno-Camacho José Luis, Rodriguez-Gallegos Jorge, Luna-Ruiz Marco, Gonzalez-Barrera D, Rangel-DeLeon D, Munguia-Garza P, Ramirez-Vega O, Escobar-Arrazola, M, Herrera-Montalvo LA. |
| EPI_ISL_2603844, EPI_ISL_2603845, EPI_ISL_2603846, EPI_ISL_2603847, EPI_ISL_2603848, EPI_ISL_2603849, EPI_ISL_2603850, EPI_ISL_2603851, EPI_ISL_2603852, EPI_ISL_2603853 | Kansas Health and Environmental Lab | Kansas Health and Environmental Lab | Katherine Wiggins, Mike Grose, Jonathan Barnell, Ben Olsen, and Phil Adam |
| EPI_ISL_2603854 | Cytocheck Laboratory | Kansas Health and Environmental Lab | Katherine Wiggins, Mike Grose, Jonathan Barnell, Ben Olsen, and Phil Adam |
| EPI_ISL_2603858, EPI_ISL_2603859, EPI_ISL_2603860, EPI_ISL_2603861, EPI_ISL_2603862, EPI_ISL_2603863, EPI_ISL_2603864, EPI_ISL_2603865, EPI_ISL_2603866, EPI_ISL_2603867, EPI_ISL_2603868, EPI_ISL_2603869, EPI_ISL_2603870, EPI_ISL_2603871, EPI_ISL_2603872, EPI_ISL_2603873, EPI_ISL_2603874, EPI_ISL_2603875, EPI_ISL_2603876, EPI_ISL_2603877, EPI_ISL_2603878, EPI_ISL_2603879, EPI_ISL_2603880, EPI_ISL_2603881, EPI_ISL_2603882, EPI_ISL_2603883, EPI_ISL_2603884, EPI_ISL_2603885, EPI_ISL_2603886, EPI_ISL_2603887, EPI_ISL_2603888, EPI_ISL_2603889, EPI_ISL_2603890, EPI_ISL_2603891, EPI_ISL_2603892, EPI_ISL_2603893, EPI_ISL_2603894, EPI_ISL_2603895, EPI_ISL_2603896, EPI_ISL_2603897 | | | |
| see above | Rhode Island State Health Laboratory | Rhode Island State Health Laboratory | Ewa King, Richard C. Huard, Kristin Carpenter-Azevedo |
| EPI_ISL_2603900, EPI_ISL_2603901, EPI_ISL_2603902, EPI_ISL_2603903, EPI_ISL_2603904, EPI_ISL_2603905, EPI_ISL_2603906, EPI_ISL_2603907, EPI_ISL_2603908, EPI_ISL_2603909, EPI_ISL_2603910, EPI_ISL_2603911, EPI_ISL_2603912, EPI_ISL_2603913, EPI_ISL_2603914, EPI_ISL_2603915, EPI_ISL_2603916, EPI_ISL_2603917, EPI_ISL_2603918, EPI_ISL_2603919, EPI_ISL_2603920, EPI_ISL_2603921, EPI_ISL_2603922, EPI_ISL_2603923, EPI_ISL_2603924, EPI_ISL_2603925, EPI_ISL_2603926, EPI_ISL_2603927, EPI_ISL_2603928, EPI_ISL_2603929, EPI_ISL_2603930, EPI_ISL_2603931, EPI_ISL_2603932, EPI_ISL_2603933, EPI_ISL_2603934, EPI_ISL_2603935, EPI_ISL_2603936, EPI_ISL_2603937, EPI_ISL_2603938, EPI_ISL_2603939, EPI_ISL_2603940, EPI_ISL_2603941, EPI_ISL_2603942, EPI_ISL_2603943, EPI_ISL_2603944, EPI_ISL_2603945, EPI_ISL_2603946, EPI_ISL_2603947, EPI_ISL_2603948, EPI_ISL_2603949, EPI_ISL_2603950, EPI_ISL_2603951, EPI_ISL_2603952, EPI_ISL_2603953, EPI_ISL_2603954, EPI_ISL_2603955, EPI_ISL_2603956, EPI_ISL_2603957, EPI_ISL_2603958, EPI_ISL_2603959, EPI_ISL_2603960, EPI_ISL_2603961, EPI_ISL_2603962, EPI_ISL_2603963, EPI_ISL_2603964, EPI_ISL_2603965, EPI_ISL_2603966, EPI_ISL_2603967, EPI_ISL_2603968, EPI_ISL_2603969, EPI_ISL_2603970, EPI_ISL_2603971, EPI_ISL_2603972, EPI_ISL_2603973, EPI_ISL_2603974, EPI_ISL_2603975, EPI_ISL_2603976, EPI_ISL_2603977, EPI_ISL_2603978, EPI_ISL_2603979, EPI_ISL_2603980, EPI_ISL_2603981, EPI_ISL_2603982, EPI_ISL_2603983, EPI_ISL_2603984, EPI_ISL_2603985, EPI_ISL_2603986, EPI_ISL_2603987, EPI_ISL_2603988, EPI_ISL_2603989, EPI_ISL_2603990, EPI_ISL_2603991, EPI_ISL_2603992, EPI_ISL_2603993, EPI_ISL_2603994, EPI_ISL_2603995, EPI_ISL_2603996, EPI_ISL_2603997, EPI_ISL_2603998, EPI_ISL_2603999, EPI_ISL_2604000, EPI_ISL_2604001, EPI_ISL_2604002, EPI_ISL_2604003, EPI_ISL_2604004, EPI_ISL_2604005, EPI_ISL_2604006, EPI_ISL_2604007, EPI_ISL_2604008, EPI_ISL_2604009, EPI_ISL_2604010, EPI_ISL_2604011, EPI_ISL_2604012, EPI_ISL_2604013, EPI_ISL_2604014, EPI_ISL_2604015, EPI_ISL_2604016, EPI_ISL_2604017, EPI_ISL_2604018, EPI_ISL_2604019, EPI_ISL_2604020, EPI_ISL_2604021, EPI_ISL_2604022, EPI_ISL_2604023, EPI_ISL_2604024, EPI_ISL_2604025, EPI_ISL_2604026, EPI_ISL_2604027, EPI_ISL_2604028, EPI_ISL_2604029, EPI_ISL_2604030, EPI_ISL_2604031, EPI_ISL_2604032, EPI_ISL_2604035, EPI_ISL_2604036, EPI_ISL_2604037, EPI_ISL_2604038, EPI_ISL_2604039, EPI_ISL_2604040, EPI_ISL_2604041, EPI_ISL_2604042, EPI_ISL_2604043, EPI_ISL_2604044, EPI_ISL_2604045, EPI_ISL_2604046, EPI_ISL_2604047, EPI_ISL_2604048, EPI_ISL_2604049, EPI_ISL_2604050, EPI_ISL_2604051, EPI_ISL_2604052, EPI_ISL_2604053, EPI_ISL_2604054, EPI_ISL_2604055, EPI_ISL_2604056, EPI_ISL_2604057, EPI_ISL_2604058, EPI_ISL_2604059, EPI_ISL_2604060, EPI_ISL_2604061, EPI_ISL_2604062, EPI_ISL_2604063, EPI_ISL_2604064, EPI_ISL_2604065, EPI_ISL_2604066, EPI_ISL_2604067, EPI_ISL_2604068, EPI_ISL_2604069, EPI_ISL_2604070, EPI_ISL_2604071, EPI_ISL_2604072, EPI_ISL_2604073, EPI_ISL_2604074, EPI_ISL_2604075, EPI_ISL_2604076, EPI_ISL_2604077, EPI_ISL_2604078, EPI_ISL_2604079, EPI_ISL_2604080, EPI_ISL_2604081, EPI_ISL_2604082, EPI_ISL_2604083, EPI_ISL_2604084, EPI_ISL_2604085, EPI_ISL_2604086, EPI_ISL_2604087, EPI_ISL_2604088, EPI_ISL_2604089, EPI_ISL_2604090, EPI_ISL_2604091, EPI_ISL_2604092, EPI_ISL_2604093, EPI_ISL_2604094, EPI_ISL_2604095, EPI_ISL_2604096, EPI_ISL_2604097, EPI_ISL_2604098, EPI_ISL_2604099, EPI_ISL_2604100 | | | |
| see above | UW Virology Lab | UW Virology Lab | Pavitra Roychoudhury, Hong Xie, Lasata Shrestha, Shah Mohamed Bakhash, Margaret Mills, Noah Baker, Sean Ellis, Meei-Li Huang, Keith R Jerome, Alexander Greninger |
| EPI_ISL_2604106, EPI_ISL_2604108, EPI_ISL_2604110, EPI_ISL_2604111, EPI_ISL_2604113, EPI_ISL_2604115, EPI_ISL_2604117, EPI_ISL_2604118, EPI_ISL_2604119, EPI_ISL_2604120, EPI_ISL_2604121, EPI_ISL_2604122, EPI_ISL_2604123, EPI_ISL_2604125, EPI_ISL_2604126, EPI_ISL_2604127, EPI_ISL_2604128, EPI_ISL_2604129, EPI_ISL_2604130, EPI_ISL_2604131, EPI_ISL_2604132, EPI_ISL_2604133, EPI_ISL_2604135, EPI_ISL_2604136, EPI_ISL_2604137, EPI_ISL_2604139, EPI_ISL_2604142, EPI_ISL_2604144, EPI_ISL_2604148, EPI_ISL_2604149, EPI_ISL_2604151, EPI_ISL_2604153, EPI_ISL_2604156, EPI_ISL_2604157, EPI_ISL_2604158, EPI_ISL_2604159, EPI_ISL_2604160, EPI_ISL_2604161, EPI_ISL_2604163, EPI_ISL_2604164, EPI_ISL_2604165, EPI_ISL_2604166, EPI_ISL_2604167, EPI_ISL_2604168, EPI_ISL_2604169, EPI_ISL_2604170, EPI_ISL_2604171, EPI_ISL_2604172, EPI_ISL_2604173, EPI_ISL_2604174, EPI_ISL_2604176, EPI_ISL_2604177, EPI_ISL_2604178, EPI_ISL_2604179, EPI_ISL_2604180, EPI_ISL_2604181, EPI_ISL_2604182 | | | |
| see above | Arizona State University | Arizona State University | Peter T. Skidmore, LaRinda A. Holland, Matthew F. Smith, Rabia Maqsood, Nicholas J. Mellor, Joy M. Blain, Valerie Harris, Joshua LaBaer, Vel Murugan, Efrem S. Lim |
| EPI_ISL_2604198, EPI_ISL_2604199, EPI_ISL_2604200, EPI_ISL_2604201, EPI_ISL_2604202, EPI_ISL_2604203, EPI_ISL_2604204, EPI_ISL_2604205, EPI_ISL_2604206, EPI_ISL_2604207, EPI_ISL_2604208, EPI_ISL_2604209, EPI_ISL_2604210, EPI_ISL_2604211, EPI_ISL_2604212, EPI_ISL_2604213, EPI_ISL_2604214, EPI_ISL_2604215 | | | |
| see above | Oregon State Public Health Laboratory | Oregon State Public Health Laboratory | Rafia Razzaque, Eugene Yeboah, Vanda Makris, Laura Tsaknaridis, John Fontana and Shane Sevey |
| EPI_ISL_2604217, EPI_ISL_2604218, EPI_ISL_2604219, EPI_ISL_2604220, EPI_ISL_2604221, EPI_ISL_2604222, EPI_ISL_2604223, EPI_ISL_2604224, EPI_ISL_2604225, EPI_ISL_2604226, EPI_ISL_2604227, EPI_ISL_2604228, EPI_ISL_2604229, EPI_ISL_2604230, EPI_ISL_2604231, EPI_ISL_2604232, EPI_ISL_2604233, EPI_ISL_2604234, EPI_ISL_2604235, EPI_ISL_2604236, EPI_ISL_2604237, EPI_ISL_2604238, EPI_ISL_2604239, EPI_ISL_2604240 | | | |
| see above | Montana Public Health Laboratory | Montana Public Health Laboratory | Joy Ritter, Michelle Mozer, Carrie Biskupiak, Deborah Gibson, Michael Dills |
| EPI_ISL_2604241, EPI_ISL_2604242, EPI_ISL_2604243, EPI_ISL_2604244, EPI_ISL_2604245, EPI_ISL_2604246, EPI_ISL_2604247, EPI_ISL_2604248, EPI_ISL_2604249, EPI_ISL_2604250 | Infectious Diseases, Chan-Zuckerberg Biohub Covidtracker | Infectious Diseases, Chan-Zuckerberg Biohub Covidtracker | CZB Cliahub Consortium |
| EPI_ISL_2604782, EPI_ISL_2604783, EPI_ISL_2604784, EPI_ISL_2604785, EPI_ISL_2604786, EPI_ISL_2604787, EPI_ISL_2604788, EPI_ISL_2604789, EPI_ISL_2604790, EPI_ISL_2604791, EPI_ISL_2604792, EPI_ISL_2604793, EPI_ISL_2604794, EPI_ISL_2604795, EPI_ISL_2604796, EPI_ISL_2604797, EPI_ISL_2604798, EPI_ISL_2604799, EPI_ISL_2604800, EPI_ISL_2604801, EPI_ISL_2604802, EPI_ISL_2604803, EPI_ISL_2604804, EPI_ISL_2604805, EPI_ISL_2604806, EPI_ISL_2604807, EPI_ISL_2604808, EPI_ISL_2604809, EPI_ISL_2604810, EPI_ISL_2604811, EPI_ISL_2604812, EPI_ISL_2604813, EPI_ISL_2604814, EPI_ISL_2604815, EPI_ISL_2604816, EPI_ISL_2604817, EPI_ISL_2604818, EPI_ISL_2604819, EPI_ISL_2604820, EPI_ISL_2604821, EPI_ISL_2604823, EPI_ISL_2604824, EPI_ISL_2604825, EPI_ISL_2604826, EPI_ISL_2604827, EPI_ISL_2604828, EPI_ISL_2604829, EPI_ISL_2604830, EPI_ISL_2604831, EPI_ISL_2604832, EPI_ISL_2604833, EPI_ISL_2604834, EPI_ISL_2604835, EPI_ISL_2604836, EPI_ISL_2604837, EPI_ISL_2604838, EPI_ISL_2604839, EPI_ISL_2604840, EPI_ISL_2604841, EPI_ISL_2604842, EPI_ISL_2604843, EPI_ISL_2604844, EPI_ISL_2604845, EPI_ISL_2604846, EPI_ISL_2604847, EPI_ISL_2604848, EPI_ISL_2604849, EPI_ISL_2604850, EPI_ISL_2604851, EPI_ISL_2604852, EPI_ISL_2604853, EPI_ISL_2604854, EPI_ISL_2604855, EPI_ISL_2604856, EPI_ISL_2604857, EPI_ISL_2604858, EPI_ISL_2604859, EPI_ISL_2604860, EPI_ISL_2604861, EPI_ISL_2604862, EPI_ISL_2604863, EPI_ISL_2604864, EPI_ISL_2604865 | | | |
| see above | Rhode Island State Health Laboratories | Rhode Island State Health Laboratories | Ewa King, Richard C. Huard, Kristin Carpenter-Azevedo |
| EPI_ISL_2604866 | NIAID - IRF Frederick, NIH | NIAID - IRF Frederick, NIH | Georgia,S. and Kocher,G. |
| EPI_ISL_2611120, EPI_ISL_2611121, EPI_ISL_2611122, EPI_ISL_2611123, EPI_ISL_2611124, EPI_ISL_2611125, EPI_ISL_2611126, EPI_ISL_2611127, EPI_ISL_2611128, EPI_ISL_2611129, EPI_ISL_2611130, EPI_ISL_2611131, EPI_ISL_2611132, EPI_ISL_2611133, EPI_ISL_2611134, EPI_ISL_2611135, EPI_ISL_2611136, EPI_ISL_2611137, EPI_ISL_2611138, EPI_ISL_2611139, EPI_ISL_2611140, EPI_ISL_2611141, EPI_ISL_2611142, EPI_ISL_2611143, EPI_ISL_2611144, EPI_ISL_2611145, EPI_ISL_2611146, EPI_ISL_2611147, EPI_ISL_2611148, EPI_ISL_2611149, EPI_ISL_2611150, EPI_ISL_2611151, EPI_ISL_2611152, EPI_ISL_2611153, EPI_ISL_2611154, EPI_ISL_2611155, EPI_ISL_2611156, EPI_ISL_2611157, EPI_ISL_2611158, EPI_ISL_2611159, EPI_ISL_2611160, EPI_ISL_2611161, EPI_ISL_2611162, EPI_ISL_2611163, EPI_ISL_2611164, EPI_ISL_2611165, EPI_ISL_2611166, EPI_ISL_2611167, EPI_ISL_2611168, EPI_ISL_2611169, EPI_ISL_2611170, EPI_ISL_2611171, EPI_ISL_2611172, EPI_ISL_2611173, EPI_ISL_2611174, EPI_ISL_2611175, EPI_ISL_2611176, EPI_ISL_2611177, EPI_ISL_2611178, EPI_ISL_2611179, EPI_ISL_2611180, EPI_ISL_2611181, EPI_ISL_2611182, EPI_ISL_2611183, EPI_ISL_2611184, EPI_ISL_2611185, EPI_ISL_2611186, EPI_ISL_2611187, EPI_ISL_2611188, EPI_ISL_2611189, EPI_ISL_2611190, EPI_ISL_2611191, EPI_ISL_2611192, EPI_ISL_2611193, EPI_ISL_2611194, EPI_ISL_2611195, EPI_ISL_2611196, EPI_ISL_2611197, EPI_ISL_2611198, EPI_ISL_2611199, EPI_ISL_2611200, EPI_ISL_2611201, EPI_ISL_2611202, EPI_ISL_2611203, EPI_ISL_2611204, EPI_ISL_2611205, EPI_ISL_2611206, EPI_ISL_2611207, EPI_ISL_2611208, EPI_ISL_2611209, EPI_ISL_2611210, EPI_ISL_2611211, EPI_ISL_2611212, EPI_ISL_2611213, EPI_ISL_2611214, EPI_ISL_2611215, EPI_ISL_2611216, EPI_ISL_2611217, EPI_ISL_2611218, EPI_ISL_2611219, EPI_ISL_2611220, EPI_ISL_2611221, EPI_ISL_2611222, EPI_ISL_2611224, EPI_ISL_2611225, EPI_ISL_2611226, EPI_ISL_2611227, EPI_ISL_2611228, EPI_ISL_2611229, EPI_ISL_2611230, EPI_ISL_2611231, EPI_ISL_2611232, EPI_ISL_2611233, EPI_ISL_2611234, EPI_ISL_2611235, EPI_ISL_2611236, EPI_ISL_2611237, EPI_ISL_2611238, EPI_ISL_2611239, EPI_ISL_2611240, EPI_ISL_2611241, EPI_ISL_2611242, EPI_ISL_2611243, EPI_ISL_2611244, EPI_ISL_2611245, EPI_ISL_2611247, EPI_ISL_2611249, EPI_ISL_2611250, EPI_ISL_2611251, EPI_ISL_2611253, EPI_ISL_2611254, EPI_ISL_2611255, EPI_ISL_2611256, EPI_ISL_2611257, EPI_ISL_2611258, EPI_ISL_2611259, EPI_ISL_2611260, EPI_ISL_2611261, EPI_ISL_2611262, EPI_ISL_2611263, EPI_ISL_2611264, EPI_ISL_2611265, EPI_ISL_2611266, EPI_ISL_2611267, EPI_ISL_2611268, EPI_ISL_2611269, EPI_ISL_2611270, EPI_ISL_2611271, EPI_ISL_2611272, EPI_ISL_2611273, EPI_ISL_2611274, EPI_ISL_2611275, EPI_ISL_2611276, EPI_ISL_2611277, EPI_ISL_2611278, EPI_ISL_2611279, EPI_ISL_2611280 | | | |
| see above | Laboratory Corporation of America | Centers for Disease Control and Prevention Division of Viral Diseases, Pathogen Discovery | Dakota Howard, Dhwani Batra, Peter W. Cook, Kara Moser, Adrian Paskey, Jason Caravas, Benjamin Rambo-Martin, Shatavia Morrison, Christopher Gulvick, Scott Sammons, Yvette Unoarumhi, Darlene Wagner, Matthew Schmerer, Minoo Agarwal, Eyad Almasri, Debbie Boles, Ayla Burns, Nuthawin Charoensri, Oren Cohen, Susan Countryman, Mary Ann Cristobal, Bobbi Croy, Suzanne Dale, Hrushikesh Deshmukh, Amanda Douglas, Vincent Drouillon, Marcia Eisenberg, Howard Engler, Rama Ghatti, Prashant Gupta, Susan Hicks, Jake Humphrey, Lax Iyer, Lisa Pfefferle, Manoj Jain, Matthew Robinson, Mohan Kolli, Brian Krueger, Tim Kuphal, Stanley Letovsky, Michael Levandoski, Craig Lukasik, Jonathan Meltzer, Brian Norvell, Mindy Nye, Scott Parker, Christos Petropoulos, John Pruitt, Steven Ragan, Scott Ryan, Mike Sapeta, Jana Schroth, Suresh Babu Selvaraju, Goran Stevovic, Amanda Suchanek, Andrea Throop, Lyndon Tilson, Thomas Urban, Joe Voshell, Kimberly Wagner, Jonathan Williams, Mary Williamson, Qian Zeng, Tricia Zwiefelhofer, Clinton R. Paden, Duncan MacCannell |
| EPI_ISL_2611281 | Laboratory Corporation of America | Centers for Disease Control and Prevention Division of Viral Diseases, Pathogen Discovery | Dakota Howard, Dhwani Batra, Peter W. Cook, Kara Moser, Adrian Paskey, Jason Caravas, Benjamin Rambo-Martin, Shatavia Morrison, Christopher Gulvick, Scott Sammons, Yvette Unoarumhi, Darlene Wagner, Matthew Schmerer, Minoo Agarwal, Eyad Almasri, Debbie Boles, Ayla Burns, Nuthawin Charoensri, Oren Cohen, Susan Countryman, Mary Ann Cristobal, Bobbi Croy, Suzanne Dale, Hrushikesh Deshmukh, Amanda Douglas, Vincent Drouillon, Marcia Eisenberg, Howard Engler, Rama Ghatti, Prashant Gupta, Susan Hicks, Jake Humphrey, Lax Iyer, Lisa Pfefferle, Manoj Jain, Matthew Robinson, Mohan Kolli, Brian Krueger, Tim Kuphal, Stanley Letovsky, Michael Levandoski, Craig Lukasik, Jonathan Meltzer, Brian Norvell, Mindy Nye, Scott Parker, Christos Petropoulos, John Pruitt, Steven Ragan, Scott Ryan, Mike Sapeta, Jana Schroth, Suresh Babu Selvaraju, Goran Stevovic, Amanda Suchanek, Andrea Throop, Lyndon Tilson, Thomas Urban, Joe Voshell, Kimberly Wagner, Jonathan Williams, Mary Williamson, Qian Zeng, Tricia Zwiefelhofer, Clinton R. Paden, |
| EPI_ISL_2611282, EPI_ISL_2611283, EPI_ISL_2611284, EPI_ISL_2611285, EPI_ISL_2611286, EPI_ISL_2611287, EPI_ISL_2611288, EPI_ISL_2611289, EPI_ISL_2611290, EPI_ISL_2611291, EPI_ISL_2611292, EPI_ISL_2611293, EPI_ISL_2611294, EPI_ISL_2611295, EPI_ISL_2611296, EPI_ISL_2611297, EPI_ISL_2611298, EPI_ISL_2611299, EPI_ISL_2611300, EPI_ISL_2611301, EPI_ISL_2611302, EPI_ISL_2611303, EPI_ISL_2611304, EPI_ISL_2611305, EPI_ISL_2611306, EPI_ISL_2611307, EPI_ISL_2611308, EPI_ISL_2611309, EPI_ISL_2611311, EPI_ISL_2611312, EPI_ISL_2611313, EPI_ISL_2611314, EPI_ISL_2611315, EPI_ISL_2611316, EPI_ISL_2611317, EPI_ISL_2611318, EPI_ISL_2611319, EPI_ISL_2611320, EPI_ISL_2611321, EPI_ISL_2611322, EPI_ISL_2611323, EPI_ISL_2611324, EPI_ISL_2611325, EPI_ISL_2611326, EPI_ISL_2611327, EPI_ISL_2611328, EPI_ISL_2611329, EPI_ISL_2611330, EPI_ISL_2611331, EPI_ISL_2611332, EPI_ISL_2611333, EPI_ISL_2611334, EPI_ISL_2611335, EPI_ISL_2611336, EPI_ISL_2611337, EPI_ISL_2611338, EPI_ISL_2611339, EPI_ISL_2611340, EPI_ISL_2611341, EPI_ISL_2611343, EPI_ISL_2611344, EPI_ISL_2611345, EPI_ISL_2611346, EPI_ISL_2611347, EPI_ISL_2611348, EPI_ISL_2611349, EPI_ISL_2611350, EPI_ISL_2611351, EPI_ISL_2611352, EPI_ISL_2611353, EPI_ISL_2611354, EPI_ISL_2611355, EPI_ISL_2611356, EPI_ISL_2611358, EPI_ISL_2611359, EPI_ISL_2611360, EPI_ISL_2611361, EPI_ISL_2611362, EPI_ISL_2611363, EPI_ISL_2611364, EPI_ISL_2611365, EPI_ISL_2611366, EPI_ISL_2611367, EPI_ISL_2611368, EPI_ISL_2611369, EPI_ISL_2611370, EPI_ISL_2611371, EPI_ISL_2611372, EPI_ISL_2611373, EPI_ISL_2611374, EPI_ISL_2611375, EPI_ISL_2611376, EPI_ISL_2611377, EPI_ISL_2611378, EPI_ISL_2611379, EPI_ISL_2611380, EPI_ISL_2611381, EPI_ISL_2611382, EPI_ISL_2611383, EPI_ISL_2611384, EPI_ISL_2611385, EPI_ISL_2611386, EPI_ISL_2611387, EPI_ISL_2611388, EPI_ISL_2611389, EPI_ISL_2611390, EPI_ISL_2611391, EPI_ISL_2611392, EPI_ISL_2611393, EPI_ISL_2611394, EPI_ISL_2611395, EPI_ISL_2611396, EPI_ISL_2611397, EPI_ISL_2611398, EPI_ISL_2611399, EPI_ISL_2611400, EPI_ISL_2611401, EPI_ISL_2611402, EPI_ISL_2611403, EPI_ISL_2611404, EPI_ISL_2611405, EPI_ISL_2611406, EPI_ISL_2611407, EPI_ISL_2611408, EPI_ISL_2611409, EPI_ISL_2611410, EPI_ISL_2611411, EPI_ISL_2611412, EPI_ISL_2611413, EPI_ISL_2611414, EPI_ISL_2611415, EPI_ISL_2611416, EPI_ISL_2611417, EPI_ISL_2611418, EPI_ISL_2611419, EPI_ISL_2611420, EPI_ISL_2611421, EPI_ISL_2611422, EPI_ISL_2611423, EPI_ISL_2611424, EPI_ISL_2611425, EPI_ISL_2611426, EPI_ISL_2611427, EPI_ISL_2611428, EPI_ISL_2611429, EPI_ISL_2611430, EPI_ISL_2611432, EPI_ISL_2611433, EPI_ISL_2611434, EPI_ISL_2611435, EPI_ISL_2611436, EPI_ISL_2611437, EPI_ISL_2611438, EPI_ISL_2611439, EPI_ISL_2611440, EPI_ISL_2611441, EPI_ISL_2611442, EPI_ISL_2611443, EPI_ISL_2611444, EPI_ISL_2611445, EPI_ISL_2611446, EPI_ISL_2611447, EPI_ISL_2611448, EPI_ISL_2611449, EPI_ISL_2611451, EPI_ISL_2611452, EPI_ISL_2611454, EPI_ISL_2611455, EPI_ISL_2611456, EPI_ISL_2611457, EPI_ISL_2611458, EPI_ISL_2611459, EPI_ISL_2611460, EPI_ISL_2611461, EPI_ISL_2611462, EPI_ISL_2611463, EPI_ISL_2611464, EPI_ISL_2611465, EPI_ISL_2611466, EPI_ISL_2611467, EPI_ISL_2611468, EPI_ISL_2611469, EPI_ISL_2611470, EPI_ISL_2611471, EPI_ISL_2611472, EPI_ISL_2611473, EPI_ISL_2611474, EPI_ISL_2611475, EPI_ISL_2611476, EPI_ISL_2611477, EPI_ISL_2611478, EPI_ISL_2611479, EPI_ISL_2611480, EPI_ISL_2611481, EPI_ISL_2611482, EPI_ISL_2611483, EPI_ISL_2611484, EPI_ISL_2611485, EPI_ISL_2611486, EPI_ISL_2611487, EPI_ISL_2611488, EPI_ISL_2611489, EPI_ISL_2611490, EPI_ISL_2611491, EPI_ISL_2611492, EPI_ISL_2611493, EPI_ISL_2611494, EPI_ISL_2611495, EPI_ISL_2611496, EPI_ISL_2611498, EPI_ISL_2611499, EPI_ISL_2611500, EPI_ISL_2611501, EPI_ISL_2611502, EPI_ISL_2611503, EPI_ISL_2611504, EPI_ISL_2611505, EPI_ISL_2611506, EPI_ISL_2611507, EPI_ISL_2611508, EPI_ISL_2611509, EPI_ISL_2611510, EPI_ISL_2611511, EPI_ISL_2611512, EPI_ISL_2611513, EPI_ISL_2611514, EPI_ISL_2611515, EPI_ISL_2611516, EPI_ISL_2611517, EPI_ISL_2611518, EPI_ISL_2611519, EPI_ISL_2611520, EPI_ISL_2611521, EPI_ISL_2611522, EPI_ISL_2611523, EPI_ISL_2611524, EPI_ISL_2611526, EPI_ISL_2611527, EPI_ISL_2611528, EPI_ISL_2611529, EPI_ISL_2611530, EPI_ISL_2611531, EPI_ISL_2611532, EPI_ISL_2611533, EPI_ISL_2611534, EPI_ISL_2611535, EPI_ISL_2611536, EPI_ISL_2611537, EPI_ISL_2611538, EPI_ISL_2611539, EPI_ISL_2611540, EPI_ISL_2611541, EPI_ISL_2611542, EPI_ISL_2611543, EPI_ISL_2611545, EPI_ISL_2611546, EPI_ISL_2611547, EPI_ISL_2611548, EPI_ISL_2611549, EPI_ISL_2611550, EPI_ISL_2611551, EPI_ISL_2611552, EPI_ISL_2611553, EPI_ISL_2611554, EPI_ISL_2611555, EPI_ISL_2611556, EPI_ISL_2611558, EPI_ISL_2611559, EPI_ISL_2611560, EPI_ISL_2611561, EPI_ISL_2611562, EPI_ISL_2611563, EPI_ISL_2611564, EPI_ISL_2611565, EPI_ISL_2611566, EPI_ISL_2611567, EPI_ISL_2611568, EPI_ISL_2611569, EPI_ISL_2611570, EPI_ISL_2611571, EPI_ISL_2611572, EPI_ISL_2611573, EPI_ISL_2611574, EPI_ISL_2611575, EPI_ISL_2611576, EPI_ISL_2611577, EPI_ISL_2611578, EPI_ISL_2611579, EPI_ISL_2611580, EPI_ISL_2611581, EPI_ISL_2611582, EPI_ISL_2611583, EPI_ISL_2611584, EPI_ISL_2611585, EPI_ISL_2611586, EPI_ISL_2611587, EPI_ISL_2611588, EPI_ISL_2611589, EPI_ISL_2611590, EPI_ISL_2611591, EPI_ISL_2611592, EPI_ISL_2611593, EPI_ISL_2611594, EPI_ISL_2611595, EPI_ISL_2611596, EPI_ISL_2611597, EPI_ISL_2611599, EPI_ISL_2611600, EPI_ISL_2611601, EPI_ISL_2611602, EPI_ISL_2611603, EPI_ISL_2611604, EPI_ISL_2611605, EPI_ISL_2611607, EPI_ISL_2611608, EPI_ISL_2611609, EPI_ISL_2611610, EPI_ISL_2611611, EPI_ISL_2611612, EPI_ISL_2611613, EPI_ISL_2611614, EPI_ISL_2611615, EPI_ISL_2611616, EPI_ISL_2611617, EPI_ISL_2611618, EPI_ISL_2611619, EPI_ISL_2611620, EPI_ISL_2611621, EPI_ISL_2611622, EPI_ISL_2611623, EPI_ISL_2611624, EPI_ISL_2611625, EPI_ISL_2611626, EPI_ISL_2611627, EPI_ISL_2611628, EPI_ISL_2611629, EPI_ISL_2611630, EPI_ISL_2611631, EPI_ISL_2611632, EPI_ISL_2611633, EPI_ISL_2611634, EPI_ISL_2611635, EPI_ISL_2611636, EPI_ISL_2611637, EPI_ISL_2611638, EPI_ISL_2611640, EPI_ISL_2611641, EPI_ISL_2611642, EPI_ISL_2611643, EPI_ISL_2611644, EPI_ISL_2611645, EPI_ISL_2611646, EPI_ISL_2611647, EPI_ISL_2611648, EPI_ISL_2611649, EPI_ISL_2611650, EPI_ISL_2611651, EPI_ISL_2611652, EPI_ISL_2611653, EPI_ISL_2611654, EPI_ISL_2611656, EPI_ISL_2611657, EPI_ISL_2611658, EPI_ISL_2611659, EPI_ISL_2611660, EPI_ISL_2611663, EPI_ISL_2611666, EPI_ISL_2611669, EPI_ISL_2611672, EPI_ISL_2611675, EPI_ISL_2611676, EPI_ISL_2611677, EPI_ISL_2611678, EPI_ISL_2611679, EPI_ISL_2611680, EPI_ISL_2611681, EPI_ISL_2611682, EPI_ISL_2611683, EPI_ISL_2611684, EPI_ISL_2611686, EPI_ISL_2611688, EPI_ISL_2611689, EPI_ISL_2611690, EPI_ISL_2611691, EPI_ISL_2611692, EPI_ISL_2611693, EPI_ISL_2611694, EPI_ISL_2611695, EPI_ISL_2611696, EPI_ISL_2611697, EPI_ISL_2611698, EPI_ISL_2611699, EPI_ISL_2611700, EPI_ISL_2611703, EPI_ISL_2611704, EPI_ISL_2611705, EPI_ISL_2611706, EPI_ISL_2611707, EPI_ISL_2611708, EPI_ISL_2611709, EPI_ISL_2611710, EPI_ISL_2611711, EPI_ISL_2611712, EPI_ISL_2611713, EPI_ISL_2611714, EPI_ISL_2611715, EPI_ISL_2611716, EPI_ISL_2611718, EPI_ISL_2611719, EPI_ISL_2611720, EPI_ISL_2611721, EPI_ISL_2611722, EPI_ISL_2611723, EPI_ISL_2611724, EPI_ISL_2611725, EPI_ISL_2611726, EPI_ISL_2611727, EPI_ISL_2611728, EPI_ISL_2611729, EPI_ISL_2611730, EPI_ISL_2611731, EPI_ISL_2611732, EPI_ISL_2611733, EPI_ISL_2611734, EPI_ISL_2611735, EPI_ISL_2611736, EPI_ISL_2611738, EPI_ISL_2611739, EPI_ISL_2611740, EPI_ISL_2611741, EPI_ISL_2611742, EPI_ISL_2611743, EPI_ISL_2611744, EPI_ISL_2611745, EPI_ISL_2611746, EPI_ISL_2611747, EPI_ISL_2611748, EPI_ISL_2611749, EPI_ISL_2611750, EPI_ISL_2611751, EPI_ISL_2611752, EPI_ISL_2611753, EPI_ISL_2611754, EPI_ISL_2611755, EPI_ISL_2611756, EPI_ISL_2611757, EPI_ISL_2611758, EPI_ISL_2611760, EPI_ISL_2611761, EPI_ISL_2611762, EPI_ISL_2611763, EPI_ISL_2611764, EPI_ISL_2611765, EPI_ISL_2611766, EPI_ISL_2611767, EPI_ISL_2611768, EPI_ISL_2611769, EPI_ISL_2611770, EPI_ISL_2611771, EPI_ISL_2611772, EPI_ISL_2611773, EPI_ISL_2611774, EPI_ISL_2611775, EPI_ISL_2611776, EPI_ISL_2611777, EPI_ISL_2611778, EPI_ISL_2611779, EPI_ISL_2611780, EPI_ISL_2611781, EPI_ISL_2611782, EPI_ISL_2611783, EPI_ISL_2611784, EPI_ISL_2611785, EPI_ISL_2611787, EPI_ISL_2611788, EPI_ISL_2611789, EPI_ISL_2611790, EPI_ISL_2611791, EPI_ISL_2611792, EPI_ISL_2611793, EPI_ISL_2611794, EPI_ISL_2611795, EPI_ISL_2611796, EPI_ISL_2611797, EPI_ISL_2611798, EPI_ISL_2611799, EPI_ISL_2611800, EPI_ISL_2611801, EPI_ISL_2611802, EPI_ISL_2611803, EPI_ISL_2611804, EPI_ISL_2611806, EPI_ISL_2611807, EPI_ISL_2611808, EPI_ISL_2611809, EPI_ISL_2611811, EPI_ISL_2611812, EPI_ISL_2611813, EPI_ISL_2611814, EPI_ISL_2611815, EPI_ISL_2611816, EPI_ISL_2611818, EPI_ISL_2611819, EPI_ISL_2611820, EPI_ISL_2611821, EPI_ISL_2611822, EPI_ISL_2611823, EPI_ISL_2611824 | | | |
| see above | Laboratory Corporation of America | Centers for Disease Control and Prevention Division of Viral Diseases, Pathogen Discovery | Dakota Howard, Dhwani Batra, Peter W. Cook, Kara Moser, Adrian Paskey, Jason Caravas, Benjamin Rambo-Martin, Shatavia Morrison, Christopher Gulvick, Scott Sammons, Yvette Unoarumhi, Darlene Wagner, Matthew Schmerer, Minoo Agarwal, Eyad Almasri, Debbie Boles, Ayla Burns, Nuthawin Charoensri, Oren Cohen, Susan Countryman, Mary Ann Cristobal, Bobbi Croy, Suzanne Dale, Hrushikesh Deshmukh, Amanda Douglas, Vincent Drouillon, Marcia Eisenberg, Howard Engler, Rama Ghatti, Prashant Gupta, Susan Hicks, Jake Humphrey, Lax Iyer, Lisa Pfefferle, Manoj Jain, Matthew Robinson, Mohan Kolli, Brian Krueger, Tim Kuphal, Stanley Letovsky, Michael Levandoski, Craig Lukasik, Jonathan Meltzer, Brian Norvell, Mindy Nye, Scott Parker, Christos Petropoulos, John Pruitt, Steven Ragan, Scott Ryan, Mike Sapeta, Jana Schroth, Suresh Babu Selvaraju, Goran Stevovic, Amanda Suchanek, Andrea Throop, Lyndon Tilson, Thomas Urban, Joe Voshell, Kimberly Wagner, Jonathan Williams, Mary Williamson, Qian Zeng, Tricia Zwiefelhofer, Clinton R. Paden, Duncan MacCannell |
| EPI_ISL_2611825, EPI_ISL_2611826, EPI_ISL_2611827, EPI_ISL_2611828, EPI_ISL_2611829, EPI_ISL_2611830, EPI_ISL_2611831, EPI_ISL_2611832, EPI_ISL_2611833, EPI_ISL_2611834, EPI_ISL_2611835, EPI_ISL_2611836, EPI_ISL_2611837, EPI_ISL_2611838, EPI_ISL_2611839, EPI_ISL_2611840, EPI_ISL_2611841, EPI_ISL_2611842, EPI_ISL_2611843, EPI_ISL_2611844, EPI_ISL_2611845, EPI_ISL_2611846, EPI_ISL_2611847, EPI_ISL_2611848, EPI_ISL_2611849, EPI_ISL_2611850, EPI_ISL_2611851, EPI_ISL_2611852, EPI_ISL_2611853, EPI_ISL_2611854, EPI_ISL_2611855, EPI_ISL_2611856, EPI_ISL_2611857, EPI_ISL_2611858, EPI_ISL_2611859, EPI_ISL_2611860, EPI_ISL_2611861, EPI_ISL_2611862, EPI_ISL_2611863, EPI_ISL_2611864, EPI_ISL_2611865, EPI_ISL_2611866, EPI_ISL_2611867, EPI_ISL_2611868, EPI_ISL_2611869 | | | |

| | | | |
|---|---|---|---|
| see above | Quest Diagnostics Incorporated | Centers for Disease Control and Prevention Division of Viral Diseases, Pathogen Discovery | Dakota Howard, Dhwani Batra, Peter W. Cook, Kara Moser, Adrian Paskey, Jason Caravas, Benjamin Rambo-Martin, Shatavia Morrison, Christopher Gulvick, Scott Sammons, Yvette Unoarumhi, Darlene Wagner, Matthew Schmerer, S. H. Rosenthal, A. Gerasimova, R. M. Kagan, B. Anderson, M. Hua, Y. Liu, L.E. Bernstein, K.E. Livingston, A. Perez, I. A. Shlyakhter, R. V. Rolando, R. Owen, P. Tanpaiboon, F. Lacbawan, Clinton R. Paden, Duncan MacCannell |
| EPI_ISL_2612045, EPI_ISL_2612046, EPI_ISL_2612047, EPI_ISL_2612048, EPI_ISL_2612049, EPI_ISL_2612050, EPI_ISL_2612051, EPI_ISL_2612052, EPI_ISL_2612053, EPI_ISL_2612054, EPI_ISL_2612055, EPI_ISL_2612056, EPI_ISL_2612057, EPI_ISL_2612058, EPI_ISL_2612059, EPI_ISL_2612060, EPI_ISL_2612061 | | | |
| see above | Wyoming Public Health Laboratory | Wyoming Public Health Laboratory | Jim Mildenberger, Taylor Fearing, Channing Weber, Ashley Norberg, Chayse Rowley, Marley Goetz, Brian Dominguez, Elliot Thomasson, Sam Britz, Cari Sloma, Robert Petit, and Rob Christensen |
| EPI_ISL_2612139, EPI_ISL_2612141, EPI_ISL_2612143, EPI_ISL_2612144, EPI_ISL_2612145, EPI_ISL_2612146, EPI_ISL_2612147, EPI_ISL_2612149, EPI_ISL_2612150, EPI_ISL_2612151, EPI_ISL_2612153, EPI_ISL_2612154, EPI_ISL_2612155, EPI_ISL_2612156 | | | |
| see above | Wyoming Public Health Laboratory | Wyoming Public Health Laboratory | Jim Mildenberger, Taylor Fearing, Channing Weber, Ashley Norberg, Chayse Rowley, Marley Goetz, Brian Dominguez, Elliot Thomasson, Sam Britz, Cari Sloma, and Rob Christensen |
| EPI_ISL_2612158, EPI_ISL_2612159, EPI_ISL_2612160, EPI_ISL_2612161, EPI_ISL_2612162, EPI_ISL_2612163, EPI_ISL_2612164, EPI_ISL_2612165, EPI_ISL_2612166, EPI_ISL_2612167, EPI_ISL_2612168, EPI_ISL_2612169, EPI_ISL_2612170, EPI_ISL_2612171, EPI_ISL_2612172, EPI_ISL_2612173, EPI_ISL_2612174, EPI_ISL_2612175, EPI_ISL_2612176, EPI_ISL_2612177, EPI_ISL_2612178, EPI_ISL_2612179, EPI_ISL_2612180, EPI_ISL_2612183, EPI_ISL_2612184, EPI_ISL_2612185, EPI_ISL_2612186, EPI_ISL_2612187 | | | |
| see above | Kansas Health and Environmental Lab | Kansas Health and Environmental Lab | Mike Grose, Katherine Wiggins, Jonathan Barnell, Ben Olsen, and Phil Adam |
| EPI_ISL_2612188, EPI_ISL_2612190, EPI_ISL_2612191, EPI_ISL_2612192, EPI_ISL_2612193, EPI_ISL_2612194, EPI_ISL_2612195 | Wichita State University - Molecular Diagnostics Lab | Kansas Health and Environmental Lab | Mike Grose, Katherine Wiggins, Jonathan Barnell, Ben Olsen, and Phil Adam |
| EPI_ISL_2612278, EPI_ISL_2612279, EPI_ISL_2612280, EPI_ISL_2612281, EPI_ISL_2612282, EPI_ISL_2612283, EPI_ISL_2612284, EPI_ISL_2612285, EPI_ISL_2612286, EPI_ISL_2612287, EPI_ISL_2612288, EPI_ISL_2612289, EPI_ISL_2612290, EPI_ISL_2612291, EPI_ISL_2612292, EPI_ISL_2612293, EPI_ISL_2612294 | | | |
| see above | Kansas Health and Environmental Lab | Kansas Health and Environmental Lab | Mike Grose, Katherine Wiggins, Jonathan Barnell, Ben Olsen, and Phil Adam |
| EPI_ISL_2612295, EPI_ISL_2612296, EPI_ISL_2612297, EPI_ISL_2612298 | University of Michigan Clinical Microbiology Laboratory | Lauring Lab, University of Michigan, Department of Microbiology and Immunology | Valesano |
| EPI_ISL_2612299 | IVY3 Central Lab, Vanderbilt University Medical Center | Lauring Lab, University of Michigan, Department of Microbiology and Immunology | Valesano |
| EPI_ISL_2612455, EPI_ISL_2612456, EPI_ISL_2612457, EPI_ISL_2612458, EPI_ISL_2612459, EPI_ISL_2612460, EPI_ISL_2612461, EPI_ISL_2612462, EPI_ISL_2612463, EPI_ISL_2612464, EPI_ISL_2612465, EPI_ISL_2612466, EPI_ISL_2612467, EPI_ISL_2612468, EPI_ISL_2612469, EPI_ISL_2612470, EPI_ISL_2612471, EPI_ISL_2612472, EPI_ISL_2612473, EPI_ISL_2612474, EPI_ISL_2612475, EPI_ISL_2612476, EPI_ISL_2612477, EPI_ISL_2612478, EPI_ISL_2612479 | | | |
| see above | Montana Public Health Laboratory | Montana Public Health Laboratory | Joy Ritter, Michelle Mozer, Carrie Biskupiak, Deborah Gibson, Michael Dills |
| EPI_ISL_2612480, EPI_ISL_2612481, EPI_ISL_2612482, EPI_ISL_2612483, EPI_ISL_2612484, EPI_ISL_2612485, EPI_ISL_2612486, EPI_ISL_2612487, EPI_ISL_2612488, EPI_ISL_2612489, EPI_ISL_2612490, EPI_ISL_2612491, EPI_ISL_2612492, EPI_ISL_2612493, EPI_ISL_2612494, EPI_ISL_2612495, EPI_ISL_2612496, EPI_ISL_2612497, EPI_ISL_2612498, EPI_ISL_2612499, EPI_ISL_2612500 | | | |
| see above | ISDH | Quantigen Biosciences | Paul J. Childress |
| EPI_ISL_2612501, EPI_ISL_2612502, EPI_ISL_2612504, EPI_ISL_2612505, EPI_ISL_2612506, EPI_ISL_2612507, EPI_ISL_2612508 | Nevada State Public Health Laboratory | Nevada State Public Health Laboratory | Andrew Gorzalski, Mark Pandori |
| EPI_ISL_2612509, EPI_ISL_2612510, EPI_ISL_2612511, EPI_ISL_2612512, EPI_ISL_2612513, EPI_ISL_2612514, EPI_ISL_2612515, EPI_ISL_2612516, EPI_ISL_2612518, EPI_ISL_2612519 | Minnesota Department of Health, Public Health Laboratory | Minnesota Department of Health, Public Health Laboratory | Alexandra Lorentz, Jacob Garfin, Matt Plumb, and Xiong Wang |
| EPI_ISL_2612520, EPI_ISL_2612521 | Centracare Laboratory Services | Minnesota Department of Health, Public Health Laboratory | Alexandra Lorentz, Jacob Garfin, Matt Plumb, and Xiong Wang |
| EPI_ISL_2612522, EPI_ISL_2612523 | Minnesota Department of Health, Public Health Laboratory | Minnesota Department of Health, Public Health Laboratory | Alexandra Lorentz, Jacob Garfin, Matt Plumb, and Xiong Wang |
| EPI_ISL_2612524, EPI_ISL_2612525, EPI_ISL_2612526, EPI_ISL_2612527, EPI_ISL_2612528, EPI_ISL_2612529, EPI_ISL_2612530, EPI_ISL_2612531, EPI_ISL_2612532, EPI_ISL_2612533, EPI_ISL_2612534, EPI_ISL_2612535 | | | |
| see above | Centracare Laboratory Services | Minnesota Department of Health, Public Health Laboratory | Alexandra Lorentz, Jacob Garfin, Matt Plumb, and Xiong Wang |
| EPI_ISL_2612536, EPI_ISL_2612537 | Minnesota Department of Health, Public Health Laboratory | Minnesota Department of Health, Public Health Laboratory | Alexandra Lorentz, Jacob Garfin, Matt Plumb, and Xiong Wang |
| EPI_ISL_2612538, EPI_ISL_2612539, EPI_ISL_2612540, EPI_ISL_2612541, EPI_ISL_2612542, EPI_ISL_2612543, EPI_ISL_2612544, EPI_ISL_2612545, EPI_ISL_2612546, EPI_ISL_2612547, EPI_ISL_2612548, EPI_ISL_2612549 | | | |
| see above | Oregon State Public Health Laboratory | Oregon State Public Health Laboratory | Rafia Razzaque, Eugene Yeboah, Vanda Makris, Laura Tsaknaridis, John Fontana and Shane Sevey |
| EPI_ISL_2612550 | Lincoln Diagnostics | New York City Public Health Laboratory | Jade Wang, et al. |
| EPI_ISL_2612551 | Quest Diagnostics | New York City Public Health Laboratory | Jade Wang, et al. |
| EPI_ISL_2612552 | EastGene Laboratories | New York City Public Health Laboratory | Jade Wang, et al. |
| EPI_ISL_2612553, EPI_ISL_2612554, EPI_ISL_2612555, EPI_ISL_2612556 | DOHMH Corona | New York City Public Health Laboratory | Jade Wang, et al. |
| EPI_ISL_2612557, EPI_ISL_2612558, EPI_ISL_2612559 | DOHMH Jamaica | New York City Public Health Laboratory | Jade Wang, et al. |
| EPI_ISL_2612560, EPI_ISL_2612561 | DOHMH Riverside | New York City Public Health Laboratory | Jade Wang, et al. |
| EPI_ISL_2612562 | DOHMH Jamaica | New York City Public Health Laboratory | Jade Wang, et al. |
| EPI_ISL_2612563 | OCME Office Of Chief Medical Examiner | New York City Public Health Laboratory | Jade Wang, et al. |
| EPI_ISL_2613836, EPI_ISL_2613837, EPI_ISL_2613838, EPI_ISL_2613839, EPI_ISL_2613840, EPI_ISL_2613841, EPI_ISL_2613842, EPI_ISL_2613843, EPI_ISL_2613844, EPI_ISL_2613845, EPI_ISL_2613846, EPI_ISL_2613847, EPI_ISL_2613848, EPI_ISL_2613849, EPI_ISL_2613850, EPI_ISL_2613851, EPI_ISL_2613852, EPI_ISL_2613853, EPI_ISL_2613854, EPI_ISL_2613855, EPI_ISL_2613856, EPI_ISL_2613857, EPI_ISL_2613858, EPI_ISL_2613859, EPI_ISL_2613860, EPI_ISL_2613861, EPI_ISL_2613862, EPI_ISL_2613863, EPI_ISL_2613864, EPI_ISL_2613865, EPI_ISL_2613866, EPI_ISL_2613867, EPI_ISL_2613868, EPI_ISL_2613869, EPI_ISL_2613871, EPI_ISL_2613872, EPI_ISL_2613873, EPI_ISL_2613874, EPI_ISL_2613875, EPI_ISL_2613876, EPI_ISL_2613877, EPI_ISL_2613878, EPI_ISL_2613879, EPI_ISL_2613880, EPI_ISL_2613881, EPI_ISL_2613882, EPI_ISL_2613883, EPI_ISL_2613884, EPI_ISL_2613885, EPI_ISL_2613886, EPI_ISL_2613887, EPI_ISL_2613888, EPI_ISL_2613889, EPI_ISL_2613890, EPI_ISL_2613891, EPI_ISL_2613892, EPI_ISL_2613893, EPI_ISL_2613894, EPI_ISL_2613895, EPI_ISL_2613896, EPI_ISL_2613897, EPI_ISL_2613898, EPI_ISL_2613899, EPI_ISL_2613900, EPI_ISL_2613901, EPI_ISL_2613902, EPI_ISL_2613903, EPI_ISL_2613904, EPI_ISL_2613905, EPI_ISL_2613906, EPI_ISL_2613907, EPI_ISL_2613908, EPI_ISL_2613909, EPI_ISL_2613910, EPI_ISL_2613911, EPI_ISL_2613912, EPI_ISL_2613913, EPI_ISL_2613914, EPI_ISL_2613916, EPI_ISL_2613917, EPI_ISL_2613918, EPI_ISL_2613919, EPI_ISL_2613920, EPI_ISL_2613921, EPI_ISL_2613922, EPI_ISL_2613923, EPI_ISL_2613924, EPI_ISL_2613925, EPI_ISL_2613926, EPI_ISL_2613927, EPI_ISL_2613928, EPI_ISL_2613930, EPI_ISL_2613931, EPI_ISL_2613932, EPI_ISL_2613933, EPI_ISL_2613934, EPI_ISL_2613935, EPI_ISL_2613936, EPI_ISL_2613937, EPI_ISL_2613938, EPI_ISL_2613939, EPI_ISL_2613940, EPI_ISL_2613941, EPI_ISL_2613942, EPI_ISL_2613943, EPI_ISL_2613944, EPI_ISL_2613945, EPI_ISL_2613946, EPI_ISL_2613947, EPI_ISL_2613948, EPI_ISL_2613949, EPI_ISL_2613950, EPI_ISL_2613951, EPI_ISL_2613952, EPI_ISL_2613953, EPI_ISL_2613954, EPI_ISL_2613955, EPI_ISL_2613956, EPI_ISL_2613957, EPI_ISL_2613958, EPI_ISL_2613959, EPI_ISL_2613960, EPI_ISL_2613961, EPI_ISL_2613962, EPI_ISL_2613963, EPI_ISL_2613964, EPI_ISL_2613965, EPI_ISL_2613966, EPI_ISL_2613967, EPI_ISL_2613968, EPI_ISL_2613969, EPI_ISL_2613970, EPI_ISL_2613971, EPI_ISL_2613972, EPI_ISL_2613973, EPI_ISL_2613974, EPI_ISL_2613975, EPI_ISL_2613976, EPI_ISL_2613977, EPI_ISL_2613978, EPI_ISL_2613979, EPI_ISL_2613980, EPI_ISL_2613981, EPI_ISL_2613982, EPI_ISL_2613983, EPI_ISL_2613984, EPI_ISL_2613985, EPI_ISL_2613986, EPI_ISL_2613987, EPI_ISL_2613988, EPI_ISL_2613989, EPI_ISL_2613990, EPI_ISL_2613991, EPI_ISL_2613992, EPI_ISL_2613993, EPI_ISL_2613994, EPI_ISL_2613995, EPI_ISL_2613996, EPI_ISL_2613997, EPI_ISL_2613998, EPI_ISL_2613999, EPI_ISL_2614000, EPI_ISL_2614001, EPI_ISL_2614002, EPI_ISL_2614003, EPI_ISL_2614004, EPI_ISL_2614005, EPI_ISL_2614006, EPI_ISL_2614007, EPI_ISL_2614008, EPI_ISL_2614009, EPI_ISL_2614010, EPI_ISL_2614011, EPI_ISL_2614012, EPI_ISL_2614013, EPI_ISL_2614014, EPI_ISL_2614015, EPI_ISL_2614016, EPI_ISL_2614017, EPI_ISL_2614018, EPI_ISL_2614019, EPI_ISL_2614020, EPI_ISL_2614021, EPI_ISL_2614022, EPI_ISL_2614023, EPI_ISL_2614024, EPI_ISL_2614025, EPI_ISL_2614026, EPI_ISL_2614027, EPI_ISL_2614028, EPI_ISL_2614029, EPI_ISL_2614030, EPI_ISL_2614031, EPI_ISL_2614032, EPI_ISL_2614033, EPI_ISL_2614034, EPI_ISL_2614035, EPI_ISL_2614036, EPI_ISL_2614037, EPI_ISL_2614038, EPI_ISL_2614039, EPI_ISL_2614040, EPI_ISL_2614041, EPI_ISL_2614042, EPI_ISL_2614043, EPI_ISL_2614045, EPI_ISL_2614047, EPI_ISL_2614048, EPI_ISL_2614049, EPI_ISL_2614050, EPI_ISL_2614051, EPI_ISL_2614052, EPI_ISL_2614053, EPI_ISL_2614054, EPI_ISL_2614055, EPI_ISL_2614056, EPI_ISL_2614057, EPI_ISL_2614058, EPI_ISL_2614060, EPI_ISL_2614061, EPI_ISL_2614062, EPI_ISL_2614063, EPI_ISL_2614064, EPI_ISL_2614065, EPI_ISL_2614066, EPI_ISL_2614067, EPI_ISL_2614068, EPI_ISL_2614069 | | | |
| see above | Gravity Diagnostics, LLC | Gravity Diagnostics, LLC | Gravity Diagnostics |
| EPI_ISL_2614384 | County of San Luis Obispo Public Health Laboratory | County of San Luis Obispo Public Health Laboratory | Frances Sidhu, Katrina Erwin |
| EPI_ISL_2614398, EPI_ISL_2614399, EPI_ISL_2614401, EPI_ISL_2614402, EPI_ISL_2614403 | Lafene Health Center | Kansas State Veterinary Diagnostic Laboratory | Tyler Doerksen, Andreea Lu, Lance Noll, Elizabeth Porter, Emily Cox, Jianfa Bai, Jamie Henningson, Rachel Palinski |
| EPI_ISL_2615401, EPI_ISL_2615402 | Berkeley Medical Center | WVU and Marshall University Combined Genomics Core Facilities | James Denvir, Peter Stoilov, Peter Perrotta, Wesley Kimble, Ryan Percifield |

| | | | |
|---|---|---|---|
| EPI_ISL_2615403 | QLabs | WVU and Marshall University Combined Genomics Core Facilities | James Denvir, Peter Stoilov, Peter Perrotta, Wesley Kimble, Ryan Percifield |
| EPI_ISL_2615404 | J.W. Ruby Memorial Hospital | WVU and Marshall University Combined Genomics Core Facilities | James Denvir, Peter Stoilov, Peter Perrotta, Wesley Kimble, Ryan Percifield |
| EPI_ISL_2615405 | Berkeley Medical Center | WVU and Marshall University Combined Genomics Core Facilities | James Denvir, Peter Stoilov, Peter Perrotta, Wesley Kimble, Ryan Percifield |
| EPI_ISL_2615406, EPI_ISL_2615407, EPI_ISL_2615408 | QLabs | WVU and Marshall University Combined Genomics Core Facilities | James Denvir, Peter Stoilov, Peter Perrotta, Wesley Kimble, Ryan Percifield |
| EPI_ISL_2615409, EPI_ISL_2615410 | WV Labs | WVU and Marshall University Combined Genomics Core Facilities | James Denvir, Peter Stoilov, Peter Perrotta, Wesley Kimble, Ryan Percifield |
| EPI_ISL_2615411 | QLabs | WVU and Marshall University Combined Genomics Core Facilities | James Denvir, Peter Stoilov, Peter Perrotta, Wesley Kimble, Ryan Percifield |
| EPI_ISL_2615412, EPI_ISL_2615413, EPI_ISL_2615414 | WV Labs | WVU and Marshall University Combined Genomics Core Facilities | James Denvir, Peter Stoilov, Peter Perrotta, Wesley Kimble, Ryan Percifield |
| EPI_ISL_2615415, EPI_ISL_2615416 | J.W. Ruby Memorial Hospital | WVU and Marshall University Combined Genomics Core Facilities | James Denvir, Peter Stoilov, Peter Perrotta, Wesley Kimble, Ryan Percifield |
| EPI_ISL_2615417 | WV Labs | WVU and Marshall University Combined Genomics Core Facilities | James Denvir, Peter Stoilov, Peter Perrotta, Wesley Kimble, Ryan Percifield |
| EPI_ISL_2615418, EPI_ISL_2615419 | J.W. Ruby Memorial Hospital | WVU and Marshall University Combined Genomics Core Facilities | James Denvir, Peter Stoilov, Peter Perrotta, Wesley Kimble, Ryan Percifield |
| EPI_ISL_2615420 | WV Labs | WVU and Marshall University Combined Genomics Core Facilities | James Denvir, Peter Stoilov, Peter Perrotta, Wesley Kimble, Ryan Percifield |
| EPI_ISL_2615421 | QLabs | WVU and Marshall University Combined Genomics Core Facilities | James Denvir, Peter Stoilov, Peter Perrotta, Wesley Kimble, Ryan Percifield |
| EPI_ISL_2615422, EPI_ISL_2615423 | J.W. Ruby Memorial Hospital | WVU and Marshall University Combined Genomics Core Facilities | James Denvir, Peter Stoilov, Peter Perrotta, Wesley Kimble, Ryan Percifield |
| EPI_ISL_2615424, EPI_ISL_2615425 | Berkeley Medical Center | WVU and Marshall University Combined Genomics Core Facilities | James Denvir, Peter Stoilov, Peter Perrotta, Wesley Kimble, Ryan Percifield |
| EPI_ISL_2615426, EPI_ISL_2615427, EPI_ISL_2615428 | WV Labs | WVU and Marshall University Combined Genomics Core Facilities | James Denvir, Peter Stoilov, Peter Perrotta, Wesley Kimble, Ryan Percifield |
| EPI_ISL_2615429, EPI_ISL_2615430 | QLabs | WVU and Marshall University Combined Genomics Core Facilities | James Denvir, Peter Stoilov, Peter Perrotta, Wesley Kimble, Ryan Percifield |
| EPI_ISL_2615431, EPI_ISL_2615432, EPI_ISL_2615433 | J.W. Ruby Memorial Hospital | WVU and Marshall University Combined Genomics Core Facilities | James Denvir, Peter Stoilov, Peter Perrotta, Wesley Kimble, Ryan Percifield |
| EPI_ISL_2615434 | WV Labs | WVU and Marshall University Combined Genomics Core Facilities | James Denvir, Peter Stoilov, Peter Perrotta, Wesley Kimble, Ryan Percifield |
| EPI_ISL_2615435, EPI_ISL_2615436, EPI_ISL_2615437, EPI_ISL_2615438 | J.W. Ruby Memorial Hospital | WVU and Marshall University Combined Genomics Core Facilities | James Denvir, Peter Stoilov, Peter Perrotta, Wesley Kimble, Ryan Percifield |
| EPI_ISL_2615439 | WV Labs | WVU and Marshall University Combined Genomics Core Facilities | James Denvir, Peter Stoilov, Peter Perrotta, Wesley Kimble, Ryan Percifield |
| EPI_ISL_2615440 | J.W. Ruby Memorial Hospital | WVU and Marshall University Combined Genomics Core Facilities | James Denvir, Peter Stoilov, Peter Perrotta, Wesley Kimble, Ryan Percifield |
| EPI_ISL_2615441 | Mon Health Medical Center | WVU and Marshall University Combined Genomics Core Facilities | James Denvir, Peter Stoilov, Peter Perrotta, Wesley Kimble, Ryan Percifield |
| EPI_ISL_2615442 | J.W. Ruby Memorial Hospital | WVU and Marshall University Combined Genomics Core Facilities | James Denvir, Peter Stoilov, Peter Perrotta, Wesley Kimble, Ryan Percifield |
| EPI_ISL_2615443 | Preston Taylor Community Health Center | WVU and Marshall University Combined Genomics Core Facilities | James Denvir, Peter Stoilov, Peter Perrotta, Wesley Kimble, Ryan Percifield |
| EPI_ISL_2615444 | QLabs | WVU and Marshall University Combined Genomics Core Facilities | James Denvir, Peter Stoilov, Peter Perrotta, Wesley Kimble, Ryan Percifield |
| EPI_ISL_2615445, EPI_ISL_2615446 | J.W. Ruby Memorial Hospital | WVU and Marshall University Combined Genomics Core Facilities | James Denvir, Peter Stoilov, Peter Perrotta, Wesley Kimble, Ryan Percifield |
| EPI_ISL_2615447, EPI_ISL_2615448 | QLabs | WVU and Marshall University Combined Genomics Core Facilities | James Denvir, Peter Stoilov, Peter Perrotta, Wesley Kimble, Ryan Percifield |
| EPI_ISL_2615449, EPI_ISL_2615450 | WV Labs | WVU and Marshall University Combined Genomics Core Facilities | James Denvir, Peter Stoilov, Peter Perrotta, Wesley Kimble, Ryan Percifield |
| EPI_ISL_2615451 | Mon Health Medical Center | WVU and Marshall University Combined Genomics Core Facilities | James Denvir, Peter Stoilov, Peter Perrotta, Wesley Kimble, Ryan Percifield |
| EPI_ISL_2615452 | Berkeley Medical Center | WVU and Marshall University Combined Genomics Core Facilities | James Denvir, Peter Stoilov, Peter Perrotta, Wesley Kimble, Ryan Percifield |
| EPI_ISL_2615453 | J.W. Ruby Memorial Hospital | WVU and Marshall University Combined Genomics Core Facilities | James Denvir, Peter Stoilov, Peter Perrotta, Wesley Kimble, Ryan Percifield |
| EPI_ISL_2615454 | WV Labs | WVU and Marshall University Combined Genomics Core Facilities | James Denvir, Peter Stoilov, Peter Perrotta, Wesley Kimble, Ryan Percifield |
| EPI_ISL_2615455 | QLabs | WVU and Marshall University Combined Genomics Core Facilities | James Denvir, Peter Stoilov, Peter Perrotta, Wesley Kimble, Ryan Percifield |
| EPI_ISL_2615456, EPI_ISL_2615457, EPI_ISL_2615458 | J.W. Ruby Memorial Hospital | WVU and Marshall University Combined Genomics Core Facilities | James Denvir, Peter Stoilov, Peter Perrotta, Wesley Kimble, Ryan Percifield |
| EPI_ISL_2615459, EPI_ISL_2615460, EPI_ISL_2615461 | WV Labs | WVU and Marshall University Combined Genomics Core Facilities | James Denvir, Peter Stoilov, Peter Perrotta, Wesley Kimble, Ryan Percifield |
| EPI_ISL_2615462 | J.W. Ruby Memorial Hospital | WVU and Marshall University Combined Genomics Core Facilities | James Denvir, Peter Stoilov, Peter Perrotta, Wesley Kimble, Ryan Percifield |
| EPI_ISL_2615463 | Berkeley Medical Center | WVU and Marshall University Combined Genomics Core Facilities | James Denvir, Peter Stoilov, Peter Perrotta, Wesley Kimble, Ryan Percifield |
| EPI_ISL_2615464 | WV Labs | WVU and Marshall University Combined Genomics Core Facilities | James Denvir, Peter Stoilov, Peter Perrotta, Wesley Kimble, Ryan Percifield |
| EPI_ISL_2615465 | QLabs | WVU and Marshall University Combined Genomics Core Facilities | James Denvir, Peter Stoilov, Peter Perrotta, Wesley Kimble, Ryan Percifield |
| EPI_ISL_2615466 | WV Labs | WVU and Marshall University Combined Genomics Core Facilities | James Denvir, Peter Stoilov, Peter Perrotta, Wesley Kimble, Ryan Percifield |
| EPI_ISL_2615467 | QLabs | WVU and Marshall University Combined Genomics Core Facilities | James Denvir, Peter Stoilov, Peter Perrotta, Wesley Kimble, Ryan Percifield |
| EPI_ISL_2615468 | J.W. Ruby Memorial Hospital | WVU and Marshall University Combined Genomics Core Facilities | James Denvir, Peter Stoilov, Peter Perrotta, Wesley Kimble, Ryan Percifield |

| | | | |
|---|---|---|---|
| EPI_ISL_2615469 | QLabs | WVU and Marshall University Combined Genomics Core Facilities | James Denvir, Peter Stoilov, Peter Perrotta, Wesley Kimble, Ryan Percifield |
| EPI_ISL_2615470 | J.W. Ruby Memorial Hospital | WVU and Marshall University Combined Genomics Core Facilities | James Denvir, Peter Stoilov, Peter Perrotta, Wesley Kimble, Ryan Percifield |
| EPI_ISL_2615471 | QLabs | WVU and Marshall University Combined Genomics Core Facilities | James Denvir, Peter Stoilov, Peter Perrotta, Wesley Kimble, Ryan Percifield |
| EPI_ISL_2615472 | J.W. Ruby Memorial Hospital | WVU and Marshall University Combined Genomics Core Facilities | James Denvir, Peter Stoilov, Peter Perrotta, Wesley Kimble, Ryan Percifield |
| EPI_ISL_2615473, EPI_ISL_2615474, EPI_ISL_2615475 | QLabs | WVU and Marshall University Combined Genomics Core Facilities | James Denvir, Peter Stoilov, Peter Perrotta, Wesley Kimble, Ryan Percifield |
| EPI_ISL_2615476, EPI_ISL_2615477, EPI_ISL_2615478 | J.W. Ruby Memorial Hospital | WVU and Marshall University Combined Genomics Core Facilities | James Denvir, Peter Stoilov, Peter Perrotta, Wesley Kimble, Ryan Percifield |
| EPI_ISL_2615479, EPI_ISL_2615480, EPI_ISL_2615481 | WV Labs | WVU and Marshall University Combined Genomics Core Facilities | James Denvir, Peter Stoilov, Peter Perrotta, Wesley Kimble, Ryan Percifield |
| EPI_ISL_2615482, EPI_ISL_2615483 | J.W. Ruby Memorial Hospital | WVU and Marshall University Combined Genomics Core Facilities | James Denvir, Peter Stoilov, Peter Perrotta, Wesley Kimble, Ryan Percifield |
| EPI_ISL_2615484, EPI_ISL_2615485, EPI_ISL_2615486, EPI_ISL_2615487, EPI_ISL_2615488, EPI_ISL_2615489, EPI_ISL_2615490, EPI_ISL_2615491 | Berkeley Medical Center | WVU and Marshall University Combined Genomics Core Facilities | James Denvir, Peter Stoilov, Peter Perrotta, Wesley Kimble, Ryan Percifield |
| EPI_ISL_2615492, EPI_ISL_2615493, EPI_ISL_2615494, EPI_ISL_2615495, EPI_ISL_2615496, EPI_ISL_2615497, EPI_ISL_2615498, EPI_ISL_2615499, EPI_ISL_2615500, EPI_ISL_2615501, EPI_ISL_2615502, EPI_ISL_2615503, EPI_ISL_2615504, EPI_ISL_2615505, EPI_ISL_2615506 | | | |
| see above | QLabs | WVU and Marshall University Combined Genomics Core Facilities | James Denvir, Peter Stoilov, Peter Perrotta, Wesley Kimble, Ryan Percifield |
| EPI_ISL_2615507, EPI_ISL_2615508, EPI_ISL_2615509, EPI_ISL_2615510, EPI_ISL_2615511, EPI_ISL_2615512, EPI_ISL_2615513 | WV Labs | WVU and Marshall University Combined Genomics Core Facilities | James Denvir, Peter Stoilov, Peter Perrotta, Wesley Kimble, Ryan Percifield |
| EPI_ISL_2615514, EPI_ISL_2615515, EPI_ISL_2615516, EPI_ISL_2615517, EPI_ISL_2615518, EPI_ISL_2615519 | Wheeling Hospital | WVU and Marshall University Combined Genomics Core Facilities | James Denvir, Peter Stoilov, Peter Perrotta, Wesley Kimble, Ryan Percifield |
| EPI_ISL_2615520, EPI_ISL_2615521, EPI_ISL_2615522, EPI_ISL_2615523, EPI_ISL_2615524, EPI_ISL_2615525, EPI_ISL_2615526, EPI_ISL_2615527, EPI_ISL_2615528, EPI_ISL_2615529 | Berkeley Medical Center | WVU and Marshall University Combined Genomics Core Facilities | James Denvir, Peter Stoilov, Peter Perrotta, Wesley Kimble, Ryan Percifield |
| EPI_ISL_2615530, EPI_ISL_2615531 | QLabs | WVU and Marshall University Combined Genomics Core Facilities | James Denvir, Peter Stoilov, Peter Perrotta, Wesley Kimble, Ryan Percifield |
| EPI_ISL_2615532, EPI_ISL_2615533, EPI_ISL_2615534, EPI_ISL_2615535 | Berkeley Medical Center | WVU and Marshall University Combined Genomics Core Facilities | James Denvir, Peter Stoilov, Peter Perrotta, Wesley Kimble, Ryan Percifield |
| EPI_ISL_2615536, EPI_ISL_2615537, EPI_ISL_2615538, EPI_ISL_2615539, EPI_ISL_2615540, EPI_ISL_2615541, EPI_ISL_2615542 | United Hospital Center | WVU and Marshall University Combined Genomics Core Facilities | James Denvir, Peter Stoilov, Peter Perrotta, Wesley Kimble, Ryan Percifield |
| EPI_ISL_2615543, EPI_ISL_2615544, EPI_ISL_2615545, EPI_ISL_2615546, EPI_ISL_2615547, EPI_ISL_2615548, EPI_ISL_2615549, EPI_ISL_2615550, EPI_ISL_2615551, EPI_ISL_2615552, EPI_ISL_2615553, EPI_ISL_2615554, EPI_ISL_2615555, EPI_ISL_2615556, EPI_ISL_2615557, EPI_ISL_2615558, EPI_ISL_2615559, EPI_ISL_2615560, EPI_ISL_2615561, EPI_ISL_2615562, EPI_ISL_2615563, EPI_ISL_2615564, EPI_ISL_2615565, EPI_ISL_2615566, EPI_ISL_2615567, EPI_ISL_2615568, EPI_ISL_2615569, EPI_ISL_2615570, EPI_ISL_2615571 | | | |
| see above | QLabs | WVU and Marshall University Combined Genomics Core Facilities | James Denvir, Peter Stoilov, Peter Perrotta, Wesley Kimble, Ryan Percifield |
| EPI_ISL_2615572 | WVU Rapid Development Lab | WVU and Marshall University Combined Genomics Core Facilities | James Denvir, Peter Stoilov, Peter Perrotta, Wesley Kimble, Ryan Percifield |
| EPI_ISL_2615573, EPI_ISL_2615574 | WV Labs | WVU and Marshall University Combined Genomics Core Facilities | James Denvir, Peter Stoilov, Peter Perrotta, Wesley Kimble, Ryan Percifield |
| EPI_ISL_2615575, EPI_ISL_2615576 | QLabs | WVU and Marshall University Combined Genomics Core Facilities | James Denvir, Peter Stoilov, Peter Perrotta, Wesley Kimble, Ryan Percifield |
| EPI_ISL_2615577, EPI_ISL_2615578, EPI_ISL_2615579, EPI_ISL_2615580, EPI_ISL_2615581, EPI_ISL_2615582, EPI_ISL_2615583, EPI_ISL_2615584, EPI_ISL_2615585, EPI_ISL_2615586, EPI_ISL_2615587, EPI_ISL_2615588, EPI_ISL_2615589, EPI_ISL_2615590, EPI_ISL_2615591, EPI_ISL_2615592, EPI_ISL_2615593, EPI_ISL_2615594, EPI_ISL_2615595, EPI_ISL_2615596, EPI_ISL_2615597, EPI_ISL_2615598, EPI_ISL_2615599, EPI_ISL_2615600, EPI_ISL_2615601, EPI_ISL_2615602, EPI_ISL_2615603, EPI_ISL_2615604, EPI_ISL_2615605, EPI_ISL_2615606, EPI_ISL_2615607, EPI_ISL_2615608, EPI_ISL_2615609, EPI_ISL_2615610, EPI_ISL_2615611 | | | |
| see above | J.W. Ruby Memorial Hospital | WVU and Marshall University Combined Genomics Core Facilities | James Denvir, Peter Stoilov, Peter Perrotta, Wesley Kimble, Ryan Percifield |
| EPI_ISL_2615612 | Mon Health Medical Center | WVU and Marshall University Combined Genomics Core Facilities | James Denvir, Peter Stoilov, Peter Perrotta, Wesley Kimble, Ryan Percifield |
| EPI_ISL_2615613, EPI_ISL_2615614, EPI_ISL_2615615, EPI_ISL_2615616, EPI_ISL_2615617, EPI_ISL_2615618, EPI_ISL_2615619, EPI_ISL_2615620, EPI_ISL_2615621, EPI_ISL_2615622 | Berkeley Medical Center | WVU and Marshall University Combined Genomics Core Facilities | James Denvir, Peter Stoilov, Peter Perrotta, Wesley Kimble, Ryan Percifield |
| EPI_ISL_2615623, EPI_ISL_2615624, EPI_ISL_2615625, EPI_ISL_2615626, EPI_ISL_2615627, EPI_ISL_2615628, EPI_ISL_2615629, EPI_ISL_2615630, EPI_ISL_2615631, EPI_ISL_2615632, EPI_ISL_2615633, EPI_ISL_2615634, EPI_ISL_2615635, EPI_ISL_2615636, EPI_ISL_2615637, EPI_ISL_2615638, EPI_ISL_2615639 | | | |
| see above | J.W. Ruby Memorial Hospital | WVU and Marshall University Combined Genomics Core Facilities | James Denvir, Peter Stoilov, Peter Perrotta, Wesley Kimble, Ryan Percifield |
| EPI_ISL_2615640, EPI_ISL_2615641, EPI_ISL_2615642, | Berkeley Medical Center | WVU and Marshall University Combined Genomics Core Facilities | James Denvir, Peter Stoilov, Peter Perrotta, Wesley Kimble, Ryan Percifield |

| | | | |
|---|---|---|---|
| EPI_ISL_2615643 | | | |
| EPI_ISL_2615644, EPI_ISL_2615645, EPI_ISL_2615646, EPI_ISL_2615647, EPI_ISL_2615648, EPI_ISL_2615649, EPI_ISL_2615650, EPI_ISL_2615651, EPI_ISL_2615652, EPI_ISL_2615653, EPI_ISL_2615654, EPI_ISL_2615655, EPI_ISL_2615656, EPI_ISL_2615657, EPI_ISL_2615658, EPI_ISL_2615659, EPI_ISL_2615660, EPI_ISL_2615661, EPI_ISL_2615662, EPI_ISL_2615663, EPI_ISL_2615664, EPI_ISL_2615665, EPI_ISL_2615666, EPI_ISL_2615667, EPI_ISL_2615668, EPI_ISL_2615669, EPI_ISL_2615670, EPI_ISL_2615671, EPI_ISL_2615672 | | | |
| see above | QLabs | WVU and Marshall University Combined Genomics Core Facilities | James Denvir, Peter Stoilov, Peter Perrotta, Wesley Kimble, Ryan Percifield |
| EPI_ISL_2615673, EPI_ISL_2615674, EPI_ISL_2615675, EPI_ISL_2615676, EPI_ISL_2615677, EPI_ISL_2615678, EPI_ISL_2615679, EPI_ISL_2615680, EPI_ISL_2615681, EPI_ISL_2615682, EPI_ISL_2615683, EPI_ISL_2615684, EPI_ISL_2615685, EPI_ISL_2615686, EPI_ISL_2615687 | | | |
| see above | J.W. Ruby Memorial Hospital | WVU and Marshall University Combined Genomics Core Facilities | James Denvir, Peter Stoilov, Peter Perrotta, Wesley Kimble, Ryan Percifield |
| EPI_ISL_2615688, EPI_ISL_2615689 | Berkeley Medical Center | WVU and Marshall University Combined Genomics Core Facilities | James Denvir, Peter Stoilov, Peter Perrotta, Wesley Kimble, Ryan Percifield |
| EPI_ISL_2615690 | WV Labs | WVU and Marshall University Combined Genomics Core Facilities | James Denvir, Peter Stoilov, Peter Perrotta, Wesley Kimble, Ryan Percifield |
| EPI_ISL_2615691, EPI_ISL_2615692, EPI_ISL_2615693, EPI_ISL_2615694, EPI_ISL_2615695, EPI_ISL_2615696, EPI_ISL_2615697 | Berkeley Medical Center | WVU and Marshall University Combined Genomics Core Facilities | James Denvir, Peter Stoilov, Peter Perrotta, Wesley Kimble, Ryan Percifield |
| EPI_ISL_2615698 | Summersville Regional Medical Center | WVU and Marshall University Combined Genomics Core Facilities | James Denvir, Peter Stoilov, Peter Perrotta, Wesley Kimble, Ryan Percifield |
| EPI_ISL_2615699 | Wheeling Hospital | WVU and Marshall University Combined Genomics Core Facilities | James Denvir, Peter Stoilov, Peter Perrotta, Wesley Kimble, Ryan Percifield |
| EPI_ISL_2615700, EPI_ISL_2615701, EPI_ISL_2615702, EPI_ISL_2615703, EPI_ISL_2615704, EPI_ISL_2615705 | J.W. Ruby Memorial Hospital | WVU and Marshall University Combined Genomics Core Facilities | James Denvir, Peter Stoilov, Peter Perrotta, Wesley Kimble, Ryan Percifield |
| EPI_ISL_2615706 | Mon Health Medical Center | WVU and Marshall University Combined Genomics Core Facilities | James Denvir, Peter Stoilov, Peter Perrotta, Wesley Kimble, Ryan Percifield |
| EPI_ISL_2615707, EPI_ISL_2615708, EPI_ISL_2615709, EPI_ISL_2615710, EPI_ISL_2615711, EPI_ISL_2615712, EPI_ISL_2615713, EPI_ISL_2615714 | QLabs | WVU and Marshall University Combined Genomics Core Facilities | James Denvir, Peter Stoilov, Peter Perrotta, Wesley Kimble, Ryan Percifield |
| EPI_ISL_2615715, EPI_ISL_2615716, EPI_ISL_2615717, EPI_ISL_2615718, EPI_ISL_2615719, EPI_ISL_2615720 | Berkeley Medical Center | WVU and Marshall University Combined Genomics Core Facilities | James Denvir, Peter Stoilov, Peter Perrotta, Wesley Kimble, Ryan Percifield |
| EPI_ISL_2615721, EPI_ISL_2615722, EPI_ISL_2615723, EPI_ISL_2615724, EPI_ISL_2615725 | J.W. Ruby Memorial Hospital | WVU and Marshall University Combined Genomics Core Facilities | James Denvir, Peter Stoilov, Peter Perrotta, Wesley Kimble, Ryan Percifield |
| EPI_ISL_2615726, EPI_ISL_2615727, EPI_ISL_2615728 | QLabs | WVU and Marshall University Combined Genomics Core Facilities | James Denvir, Peter Stoilov, Peter Perrotta, Wesley Kimble, Ryan Percifield |
| EPI_ISL_2615729 | Preston Taylor Community Health Center | WVU and Marshall University Combined Genomics Core Facilities | James Denvir, Peter Stoilov, Peter Perrotta, Wesley Kimble, Ryan Percifield |
| EPI_ISL_2615730, EPI_ISL_2615731 | QLabs | WVU and Marshall University Combined Genomics Core Facilities | James Denvir, Peter Stoilov, Peter Perrotta, Wesley Kimble, Ryan Percifield |
| EPI_ISL_2615732 | Preston Taylor Community Health Center | WVU and Marshall University Combined Genomics Core Facilities | James Denvir, Peter Stoilov, Peter Perrotta, Wesley Kimble, Ryan Percifield |
| EPI_ISL_2615733, EPI_ISL_2615734 | Berkeley Medical Center | WVU and Marshall University Combined Genomics Core Facilities | James Denvir, Peter Stoilov, Peter Perrotta, Wesley Kimble, Ryan Percifield |
| EPI_ISL_2615735, EPI_ISL_2615736, EPI_ISL_2615737, EPI_ISL_2615738, EPI_ISL_2615739, EPI_ISL_2615740, EPI_ISL_2615741, EPI_ISL_2615742, EPI_ISL_2615743, EPI_ISL_2615744 | QLabs | WVU and Marshall University Combined Genomics Core Facilities | James Denvir, Peter Stoilov, Peter Perrotta, Wesley Kimble, Ryan Percifield |
| EPI_ISL_2615745, EPI_ISL_2615746 | WV Labs | WVU and Marshall University Combined Genomics Core Facilities | James Denvir, Peter Stoilov, Peter Perrotta, Wesley Kimble, Ryan Percifield |
| EPI_ISL_2615747, EPI_ISL_2615748, EPI_ISL_2615749, EPI_ISL_2615750 | Wheeling Hospital | WVU and Marshall University Combined Genomics Core Facilities | James Denvir, Peter Stoilov, Peter Perrotta, Wesley Kimble, Ryan Percifield |
| EPI_ISL_2615751, EPI_ISL_2615752, EPI_ISL_2615753, EPI_ISL_2615754, EPI_ISL_2615755 | Berkeley Medical Center | WVU and Marshall University Combined Genomics Core Facilities | James Denvir, Peter Stoilov, Peter Perrotta, Wesley Kimble, Ryan Percifield |
| EPI_ISL_2615756, EPI_ISL_2615757, EPI_ISL_2615758 | QLabs | WVU and Marshall University Combined Genomics Core Facilities | James Denvir, Peter Stoilov, Peter Perrotta, Wesley Kimble, Ryan Percifield |
| EPI_ISL_2615759, EPI_ISL_2615760, EPI_ISL_2615761, EPI_ISL_2615762, EPI_ISL_2615763, EPI_ISL_2615764, EPI_ISL_2615765 | Berkeley Medical Center | WVU and Marshall University Combined Genomics Core Facilities | James Denvir, Peter Stoilov, Peter Perrotta, Wesley Kimble, Ryan Percifield |
| EPI_ISL_2615766, EPI_ISL_2615767 | Hampshire Memorial Hospital | WVU and Marshall University Combined Genomics Core Facilities | James Denvir, Peter Stoilov, Peter Perrotta, Wesley Kimble, Ryan Percifield |
| EPI_ISL_2615768 | J.W. Ruby Memorial Hospital | WVU and Marshall University Combined Genomics Core Facilities | James Denvir, Peter Stoilov, Peter Perrotta, Wesley Kimble, Ryan Percifield |
| EPI_ISL_2615769, EPI_ISL_2615770, EPI_ISL_2615771, EPI_ISL_2615772, EPI_ISL_2615773, EPI_ISL_2615774, EPI_ISL_2615775, EPI_ISL_2615776, EPI_ISL_2615777, EPI_ISL_2615778, EPI_ISL_2615779, EPI_ISL_2615780 | | | |

| | | | |
|---|---|---|---|
| see above | QLabs | WVU and Marshall University Combined Genomics Core Facilities | James Denvir, Peter Stoilov, Peter Perrotta, Wesley Kimble, Ryan Percifield |
| EPI_ISL_2615781, EPI_ISL_2615782, EPI_ISL_2615783 | WVU Rapid Development Lab | WVU and Marshall University Combined Genomics Core Facilities | James Denvir, Peter Stoilov, Peter Perrotta, Wesley Kimble, Ryan Percifield |
| EPI_ISL_2615784, EPI_ISL_2615785, EPI_ISL_2615786, EPI_ISL_2615787, EPI_ISL_2615788, EPI_ISL_2615789, EPI_ISL_2615790, EPI_ISL_2615791, EPI_ISL_2615792, EPI_ISL_2615793, EPI_ISL_2615794, EPI_ISL_2615795, EPI_ISL_2615796, EPI_ISL_2615797, EPI_ISL_2615798, EPI_ISL_2615799, EPI_ISL_2615800, EPI_ISL_2615801, EPI_ISL_2615802, EPI_ISL_2615803, EPI_ISL_2615804, EPI_ISL_2615805, EPI_ISL_2615806, EPI_ISL_2615807, EPI_ISL_2615808, EPI_ISL_2615809, EPI_ISL_2615810, EPI_ISL_2615811, EPI_ISL_2615812, EPI_ISL_2615813, EPI_ISL_2615814, EPI_ISL_2615815, EPI_ISL_2615816, EPI_ISL_2615817, EPI_ISL_2615818, EPI_ISL_2615819, EPI_ISL_2615820, EPI_ISL_2615821, EPI_ISL_2615822, EPI_ISL_2615823, EPI_ISL_2615824 | | | |
| see above | J.W. Ruby Memorial Hospital | WVU and Marshall University Combined Genomics Core Facilities | James Denvir, Peter Stoilov, Peter Perrotta, Wesley Kimble, Ryan Percifield |
| EPI_ISL_2615825, EPI_ISL_2615826, EPI_ISL_2615827, EPI_ISL_2615828, EPI_ISL_2615829, EPI_ISL_2615830 | Berkeley Medical Center | WVU and Marshall University Combined Genomics Core Facilities | James Denvir, Peter Stoilov, Peter Perrotta, Wesley Kimble, Ryan Percifield |
| EPI_ISL_2615831, EPI_ISL_2615832, EPI_ISL_2615833, EPI_ISL_2615834, EPI_ISL_2615835, EPI_ISL_2615836, EPI_ISL_2615837, EPI_ISL_2615838, EPI_ISL_2615839, EPI_ISL_2615840, EPI_ISL_2615841 | | | |
| see above | J.W. Ruby Memorial Hospital | WVU and Marshall University Combined Genomics Core Facilities | James Denvir, Peter Stoilov, Peter Perrotta, Wesley Kimble, Ryan Percifield |
| EPI_ISL_2615842, EPI_ISL_2615843, EPI_ISL_2615844, EPI_ISL_2615845 | Berkeley Medical Center | WVU and Marshall University Combined Genomics Core Facilities | James Denvir, Peter Stoilov, Peter Perrotta, Wesley Kimble, Ryan Percifield |
| EPI_ISL_2615846, EPI_ISL_2615847, EPI_ISL_2615848, EPI_ISL_2615849, EPI_ISL_2615850 | QLabs | WVU and Marshall University Combined Genomics Core Facilities | James Denvir, Peter Stoilov, Peter Perrotta, Wesley Kimble, Ryan Percifield |
| EPI_ISL_2615851, EPI_ISL_2615852, EPI_ISL_2615853, EPI_ISL_2615854, EPI_ISL_2615855, EPI_ISL_2615856, EPI_ISL_2615857, EPI_ISL_2615858, EPI_ISL_2615859, EPI_ISL_2615860, EPI_ISL_2615861, EPI_ISL_2615862 | | | |
| see above | J.W. Ruby Memorial Hospital | WVU and Marshall University Combined Genomics Core Facilities | James Denvir, Peter Stoilov, Peter Perrotta, Wesley Kimble, Ryan Percifield |
| EPI_ISL_2615863, EPI_ISL_2615864, EPI_ISL_2615865, EPI_ISL_2615866, EPI_ISL_2615867, EPI_ISL_2615868 | Berkeley Medical Center | WVU and Marshall University Combined Genomics Core Facilities | James Denvir, Peter Stoilov, Peter Perrotta, Wesley Kimble, Ryan Percifield |
| EPI_ISL_2615869 | Mon Health Medical Center | WVU and Marshall University Combined Genomics Core Facilities | James Denvir, Peter Stoilov, Peter Perrotta, Wesley Kimble, Ryan Percifield |
| EPI_ISL_2615870, EPI_ISL_2615871 | Wheeling Hospital | WVU and Marshall University Combined Genomics Core Facilities | James Denvir, Peter Stoilov, Peter Perrotta, Wesley Kimble, Ryan Percifield |
| EPI_ISL_2615872, EPI_ISL_2615873, EPI_ISL_2615874, EPI_ISL_2615875, EPI_ISL_2615876 | J.W. Ruby Memorial Hospital | WVU and Marshall University Combined Genomics Core Facilities | James Denvir, Peter Stoilov, Peter Perrotta, Wesley Kimble, Ryan Percifield |
| EPI_ISL_2615877, EPI_ISL_2615878, EPI_ISL_2615879 | QLabs | WVU and Marshall University Combined Genomics Core Facilities | James Denvir, Peter Stoilov, Peter Perrotta, Wesley Kimble, Ryan Percifield |
| EPI_ISL_2615880 | Berkeley Medical Center | WVU and Marshall University Combined Genomics Core Facilities | James Denvir, Peter Stoilov, Peter Perrotta, Wesley Kimble, Ryan Percifield |
| EPI_ISL_2615881, EPI_ISL_2615882, EPI_ISL_2615883, EPI_ISL_2615884, EPI_ISL_2615885, EPI_ISL_2615886, EPI_ISL_2615887 | J.W. Ruby Memorial Hospital | WVU and Marshall University Combined Genomics Core Facilities | James Denvir, Peter Stoilov, Peter Perrotta, Wesley Kimble, Ryan Percifield |
| EPI_ISL_2615888 | QLabs | WVU and Marshall University Combined Genomics Core Facilities | James Denvir, Peter Stoilov, Peter Perrotta, Wesley Kimble, Ryan Percifield |
| EPI_ISL_2615889 | Berkeley Medical Center | WVU and Marshall University Combined Genomics Core Facilities | James Denvir, Peter Stoilov, Peter Perrotta, Wesley Kimble, Ryan Percifield |
| EPI_ISL_2615890, EPI_ISL_2615891 | J.W. Ruby Memorial Hospital | WVU and Marshall University Combined Genomics Core Facilities | James Denvir, Peter Stoilov, Peter Perrotta, Wesley Kimble, Ryan Percifield |
| EPI_ISL_2615892 | Berkeley Medical Center | WVU and Marshall University Combined Genomics Core Facilities | James Denvir, Peter Stoilov, Peter Perrotta, Wesley Kimble, Ryan Percifield |
| EPI_ISL_2615893, EPI_ISL_2615894 | J.W. Ruby Memorial Hospital | WVU and Marshall University Combined Genomics Core Facilities | James Denvir, Peter Stoilov, Peter Perrotta, Wesley Kimble, Ryan Percifield |
| EPI_ISL_2615917 | QLabs | WVU and Marshall University Combined Genomics Core Facilities | James Denvir, Peter Stoilov, Peter Perrotta, Wesley Kimble, Ryan Percifield |
| EPI_ISL_2615918, EPI_ISL_2615919 | J.W. Ruby Memorial Hospital | WVU and Marshall University Combined Genomics Core Facilities | James Denvir, Peter Stoilov, Peter Perrotta, Wesley Kimble, Ryan Percifield |
| EPI_ISL_2615920 | QLabs | WVU and Marshall University Combined Genomics Core Facilities | James Denvir, Peter Stoilov, Peter Perrotta, Wesley Kimble, Ryan Percifield |
| EPI_ISL_2615921, EPI_ISL_2615922, EPI_ISL_2615923, EPI_ISL_2615924, EPI_ISL_2615925, EPI_ISL_2615926 | J.W. Ruby Memorial Hospital | WVU and Marshall University Combined Genomics Core Facilities | James Denvir, Peter Stoilov, Peter Perrotta, Wesley Kimble, Ryan Percifield |
| EPI_ISL_2615927 | QLabs | WVU and Marshall University Combined Genomics Core Facilities | James Denvir, Peter Stoilov, Peter Perrotta, Wesley Kimble, Ryan Percifield |
| EPI_ISL_2615928, EPI_ISL_2615929 | Berkeley Medical Center | WVU and Marshall University Combined Genomics Core Facilities | James Denvir, Peter Stoilov, Peter Perrotta, Wesley Kimble, Ryan Percifield |
| EPI_ISL_2615930 | QLabs | WVU and Marshall University Combined Genomics Core Facilities | James Denvir, Peter Stoilov, Peter Perrotta, Wesley Kimble, Ryan Percifield |
| EPI_ISL_2615931 | WV Labs | WVU and Marshall University Combined Genomics Core Facilities | James Denvir, Peter Stoilov, Peter Perrotta, Wesley Kimble, Ryan Percifield |
| EPI_ISL_2615932, EPI_ISL_2615933, EPI_ISL_2615934, EPI_ISL_2615935, EPI_ISL_2615936, EPI_ISL_2615937, EPI_ISL_2615938, EPI_ISL_2615939, EPI_ISL_2615940, | QLabs | WVU and Marshall University Combined Genomics Core Facilities | James Denvir, Peter Stoilov, Peter Perrotta, Wesley Kimble, Ryan Percifield |

| | | | |
|---|---|---|---|
| EPI_ISL_2615941 | | | |
| EPI_ISL_2615942, EPI_ISL_2615943 | Berkeley Medical Center | WVU and Marshall University Combined Genomics Core Facilities | James Denvir, Peter Stoilov, Peter Perrotta, Wesley Kimble, Ryan Percifield |
| EPI_ISL_2615944, EPI_ISL_2615945, EPI_ISL_2615946 | QLabs | WVU and Marshall University Combined Genomics Core Facilities | James Denvir, Peter Stoilov, Peter Perrotta, Wesley Kimble, Ryan Percifield |
| EPI_ISL_2615947, EPI_ISL_2615948 | J.W. Ruby Memorial Hospital | WVU and Marshall University Combined Genomics Core Facilities | James Denvir, Peter Stoilov, Peter Perrotta, Wesley Kimble, Ryan Percifield |
| EPI_ISL_2615949 | Berkeley Medical Center | WVU and Marshall University Combined Genomics Core Facilities | James Denvir, Peter Stoilov, Peter Perrotta, Wesley Kimble, Ryan Percifield |
| EPI_ISL_2615950, EPI_ISL_2615951 | J.W. Ruby Memorial Hospital | WVU and Marshall University Combined Genomics Core Facilities | James Denvir, Peter Stoilov, Peter Perrotta, Wesley Kimble, Ryan Percifield |
| EPI_ISL_2615952 | Berkeley Medical Center | WVU and Marshall University Combined Genomics Core Facilities | James Denvir, Peter Stoilov, Peter Perrotta, Wesley Kimble, Ryan Percifield |
| EPI_ISL_2615953 | Raleigh General Hospital Microbiology Dept. | WVU and Marshall University Combined Genomics Core Facilities | James Denvir, Peter Stoilov, Peter Perrotta, Wesley Kimble, Ryan Percifield |
| EPI_ISL_2615954 | QLabs | WVU and Marshall University Combined Genomics Core Facilities | James Denvir, Peter Stoilov, Peter Perrotta, Wesley Kimble, Ryan Percifield |
| EPI_ISL_2615955, EPI_ISL_2615956 | J.W. Ruby Memorial Hospital | WVU and Marshall University Combined Genomics Core Facilities | James Denvir, Peter Stoilov, Peter Perrotta, Wesley Kimble, Ryan Percifield |
| EPI_ISL_2615957 | WV Labs | WVU and Marshall University Combined Genomics Core Facilities | James Denvir, Peter Stoilov, Peter Perrotta, Wesley Kimble, Ryan Percifield |
| EPI_ISL_2615958, EPI_ISL_2615959 | J.W. Ruby Memorial Hospital | WVU and Marshall University Combined Genomics Core Facilities | James Denvir, Peter Stoilov, Peter Perrotta, Wesley Kimble, Ryan Percifield |
| EPI_ISL_2615960 | Berkeley Medical Center | WVU and Marshall University Combined Genomics Core Facilities | James Denvir, Peter Stoilov, Peter Perrotta, Wesley Kimble, Ryan Percifield |
| EPI_ISL_2615961 | QLabs | WVU and Marshall University Combined Genomics Core Facilities | James Denvir, Peter Stoilov, Peter Perrotta, Wesley Kimble, Ryan Percifield |
| EPI_ISL_2615962 | WV Labs | WVU and Marshall University Combined Genomics Core Facilities | James Denvir, Peter Stoilov, Peter Perrotta, Wesley Kimble, Ryan Percifield |
| EPI_ISL_2615963, EPI_ISL_2615964, EPI_ISL_2615965 | Berkeley Medical Center | WVU and Marshall University Combined Genomics Core Facilities | James Denvir, Peter Stoilov, Peter Perrotta, Wesley Kimble, Ryan Percifield |
| EPI_ISL_2615966 | WV Labs | WVU and Marshall University Combined Genomics Core Facilities | James Denvir, Peter Stoilov, Peter Perrotta, Wesley Kimble, Ryan Percifield |
| EPI_ISL_2615967 | J.W. Ruby Memorial Hospital | WVU and Marshall University Combined Genomics Core Facilities | James Denvir, Peter Stoilov, Peter Perrotta, Wesley Kimble, Ryan Percifield |
| EPI_ISL_2615968, EPI_ISL_2615969 | Berkeley Medical Center | WVU and Marshall University Combined Genomics Core Facilities | James Denvir, Peter Stoilov, Peter Perrotta, Wesley Kimble, Ryan Percifield |
| EPI_ISL_2615970, EPI_ISL_2615971 | J.W. Ruby Memorial Hospital | WVU and Marshall University Combined Genomics Core Facilities | James Denvir, Peter Stoilov, Peter Perrotta, Wesley Kimble, Ryan Percifield |
| EPI_ISL_2615972 | Berkeley Medical Center | WVU and Marshall University Combined Genomics Core Facilities | James Denvir, Peter Stoilov, Peter Perrotta, Wesley Kimble, Ryan Percifield |
| EPI_ISL_2615973, EPI_ISL_2615974, EPI_ISL_2615975, EPI_ISL_2615976, EPI_ISL_2615977, EPI_ISL_2615978 | QLabs | WVU and Marshall University Combined Genomics Core Facilities | James Denvir, Peter Stoilov, Peter Perrotta, Wesley Kimble, Ryan Percifield |
| EPI_ISL_2615979, EPI_ISL_2615980, EPI_ISL_2615981, EPI_ISL_2615982, EPI_ISL_2615983, EPI_ISL_2615984 | J.W. Ruby Memorial Hospital | WVU and Marshall University Combined Genomics Core Facilities | James Denvir, Peter Stoilov, Peter Perrotta, Wesley Kimble, Ryan Percifield |
| EPI_ISL_2615985, EPI_ISL_2615986 | Summersville Regional Medical Center | WVU and Marshall University Combined Genomics Core Facilities | James Denvir, Peter Stoilov, Peter Perrotta, Wesley Kimble, Ryan Percifield |
| EPI_ISL_2615987 | QLabs | WVU and Marshall University Combined Genomics Core Facilities | James Denvir, Peter Stoilov, Peter Perrotta, Wesley Kimble, Ryan Percifield |
| EPI_ISL_2615988, EPI_ISL_2615989, EPI_ISL_2615990, EPI_ISL_2615991 | J.W. Ruby Memorial Hospital | WVU and Marshall University Combined Genomics Core Facilities | James Denvir, Peter Stoilov, Peter Perrotta, Wesley Kimble, Ryan Percifield |
| EPI_ISL_2615992 | Mon Health Medical Center | WVU and Marshall University Combined Genomics Core Facilities | James Denvir, Peter Stoilov, Peter Perrotta, Wesley Kimble, Ryan Percifield |
| EPI_ISL_2615993, EPI_ISL_2615994, EPI_ISL_2615995 | J.W. Ruby Memorial Hospital | WVU and Marshall University Combined Genomics Core Facilities | James Denvir, Peter Stoilov, Peter Perrotta, Wesley Kimble, Ryan Percifield |
| EPI_ISL_2615996, EPI_ISL_2615997 | QLabs | WVU and Marshall University Combined Genomics Core Facilities | James Denvir, Peter Stoilov, Peter Perrotta, Wesley Kimble, Ryan Percifield |
| EPI_ISL_2615998, EPI_ISL_2615999, EPI_ISL_2616000 | J.W. Ruby Memorial Hospital | WVU and Marshall University Combined Genomics Core Facilities | James Denvir, Peter Stoilov, Peter Perrotta, Wesley Kimble, Ryan Percifield |
| EPI_ISL_2616001, EPI_ISL_2616002, EPI_ISL_2616003, EPI_ISL_2616004, EPI_ISL_2616005, EPI_ISL_2616006, EPI_ISL_2616007, EPI_ISL_2616008, EPI_ISL_2616009, EPI_ISL_2616010, EPI_ISL_2616011, EPI_ISL_2616012, EPI_ISL_2616013, EPI_ISL_2616014 | | | |
| see above | QLabs | WVU and Marshall University Combined Genomics Core Facilities | James Denvir, Peter Stoilov, Peter Perrotta, Wesley Kimble, Ryan Percifield |
| EPI_ISL_2616015 | Berkeley Medical Center | WVU and Marshall University Combined Genomics Core Facilities | James Denvir, Peter Stoilov, Peter Perrotta, Wesley Kimble, Ryan Percifield |
| EPI_ISL_2616016, EPI_ISL_2616017 | QLabs | WVU and Marshall University Combined Genomics Core Facilities | James Denvir, Peter Stoilov, Peter Perrotta, Wesley Kimble, Ryan Percifield |
| EPI_ISL_2616018 | Berkeley Medical Center | WVU and Marshall University Combined Genomics Core Facilities | James Denvir, Peter Stoilov, Peter Perrotta, Wesley Kimble, Ryan Percifield |
| EPI_ISL_2616019, EPI_ISL_2616020, EPI_ISL_2616021, EPI_ISL_2616022, EPI_ISL_2616023, EPI_ISL_2616024, EPI_ISL_2616025, EPI_ISL_2616026, EPI_ISL_2616027, EPI_ISL_2616028, EPI_ISL_2616029, EPI_ISL_2616030 | | | |
| see above | QLabs | WVU and Marshall University Combined Genomics Core Facilities | James Denvir, Peter Stoilov, Peter Perrotta, Wesley Kimble, Ryan Percifield |
| EPI_ISL_2616031, EPI_ISL_2616032, EPI_ISL_2616033, EPI_ISL_2616034, EPI_ISL_2616035, EPI_ISL_2616036, EPI_ISL_2616037, EPI_ISL_2616038, | J.W. Ruby Memorial Hospital | WVU and Marshall University Combined Genomics Core Facilities | James Denvir, Peter Stoilov, Peter Perrotta, Wesley Kimble, Ryan Percifield |

| | | | |
|---|---|---|---|
| EPI_ISL_2616039, EPI_ISL_2616040 | | | |
| EPI_ISL_2616041 | Berkeley Medical Center | WVU and Marshall University Combined Genomics Core Facilities | James Denvir, Peter Stoilov, Peter Perrotta, Wesley Kimble, Ryan Percifield |
| EPI_ISL_2616042, EPI_ISL_2616043, EPI_ISL_2616044, EPI_ISL_2616045, EPI_ISL_2616046, EPI_ISL_2616047, EPI_ISL_2616048, EPI_ISL_2616049 | QLabs | WVU and Marshall University Combined Genomics Core Facilities | James Denvir, Peter Stoilov, Peter Perrotta, Wesley Kimble, Ryan Percifield |
| EPI_ISL_2616050, EPI_ISL_2616051, EPI_ISL_2616052, EPI_ISL_2616053, EPI_ISL_2616054, EPI_ISL_2616055, EPI_ISL_2616056 | J.W. Ruby Memorial Hospital | WVU and Marshall University Combined Genomics Core Facilities | James Denvir, Peter Stoilov, Peter Perrotta, Wesley Kimble, Ryan Percifield |
| EPI_ISL_2616057, EPI_ISL_2616058 | Berkeley Medical Center | WVU and Marshall University Combined Genomics Core Facilities | James Denvir, Peter Stoilov, Peter Perrotta, Wesley Kimble, Ryan Percifield |
| EPI_ISL_2616059, EPI_ISL_2616060, EPI_ISL_2616061 | J.W. Ruby Memorial Hospital | WVU and Marshall University Combined Genomics Core Facilities | James Denvir, Peter Stoilov, Peter Perrotta, Wesley Kimble, Ryan Percifield |
| EPI_ISL_2616062 | Mon Health Medical Center | WVU and Marshall University Combined Genomics Core Facilities | James Denvir, Peter Stoilov, Peter Perrotta, Wesley Kimble, Ryan Percifield |
| EPI_ISL_2616063, EPI_ISL_2616064, EPI_ISL_2616065, EPI_ISL_2616066, EPI_ISL_2616067 | QLabs | WVU and Marshall University Combined Genomics Core Facilities | James Denvir, Peter Stoilov, Peter Perrotta, Wesley Kimble, Ryan Percifield |
| EPI_ISL_2616068, EPI_ISL_2616069, EPI_ISL_2616070, EPI_ISL_2616071, EPI_ISL_2616072, EPI_ISL_2616073, EPI_ISL_2616074, EPI_ISL_2616075, EPI_ISL_2616076, EPI_ISL_2616077, EPI_ISL_2616078, EPI_ISL_2616079, EPI_ISL_2616080 | | | |
| see above | J.W. Ruby Memorial Hospital | WVU and Marshall University Combined Genomics Core Facilities | James Denvir, Peter Stoilov, Peter Perrotta, Wesley Kimble, Ryan Percifield |
| EPI_ISL_2616081 | WVU Rapid Development Lab | WVU and Marshall University Combined Genomics Core Facilities | James Denvir, Peter Stoilov, Peter Perrotta, Wesley Kimble, Ryan Percifield |
| EPI_ISL_2616082, EPI_ISL_2616083 | WV Labs | WVU and Marshall University Combined Genomics Core Facilities | James Denvir, Peter Stoilov, Peter Perrotta, Wesley Kimble, Ryan Percifield |
| EPI_ISL_2616084 | QLabs | WVU and Marshall University Combined Genomics Core Facilities | James Denvir, Peter Stoilov, Peter Perrotta, Wesley Kimble, Ryan Percifield |
| EPI_ISL_2616085 | WV Labs | WVU and Marshall University Combined Genomics Core Facilities | James Denvir, Peter Stoilov, Peter Perrotta, Wesley Kimble, Ryan Percifield |
| EPI_ISL_2616086, EPI_ISL_2616087, EPI_ISL_2616088 | QLabs | WVU and Marshall University Combined Genomics Core Facilities | James Denvir, Peter Stoilov, Peter Perrotta, Wesley Kimble, Ryan Percifield |
| EPI_ISL_2616089, EPI_ISL_2616090, EPI_ISL_2616091 | J.W. Ruby Memorial Hospital | WVU and Marshall University Combined Genomics Core Facilities | James Denvir, Peter Stoilov, Peter Perrotta, Wesley Kimble, Ryan Percifield |
| EPI_ISL_2616092, EPI_ISL_2616093, EPI_ISL_2616094 | Berkeley Medical Center | WVU and Marshall University Combined Genomics Core Facilities | James Denvir, Peter Stoilov, Peter Perrotta, Wesley Kimble, Ryan Percifield |
| EPI_ISL_2616095, EPI_ISL_2616096, EPI_ISL_2616097 | J.W. Ruby Memorial Hospital | WVU and Marshall University Combined Genomics Core Facilities | James Denvir, Peter Stoilov, Peter Perrotta, Wesley Kimble, Ryan Percifield |
| EPI_ISL_2616098, EPI_ISL_2616099, EPI_ISL_2616100, EPI_ISL_2616101, EPI_ISL_2616102 | QLabs | WVU and Marshall University Combined Genomics Core Facilities | James Denvir, Peter Stoilov, Peter Perrotta, Wesley Kimble, Ryan Percifield |
| EPI_ISL_2616103 | Berkeley Medical Center | WVU and Marshall University Combined Genomics Core Facilities | James Denvir, Peter Stoilov, Peter Perrotta, Wesley Kimble, Ryan Percifield |
| EPI_ISL_2616104 | Hampshire Memorial Hospital | WVU and Marshall University Combined Genomics Core Facilities | James Denvir, Peter Stoilov, Peter Perrotta, Wesley Kimble, Ryan Percifield |
| EPI_ISL_2616105, EPI_ISL_2616106 | QLabs | WVU and Marshall University Combined Genomics Core Facilities | James Denvir, Peter Stoilov, Peter Perrotta, Wesley Kimble, Ryan Percifield |
| EPI_ISL_2616107 | Berkeley Medical Center | WVU and Marshall University Combined Genomics Core Facilities | James Denvir, Peter Stoilov, Peter Perrotta, Wesley Kimble, Ryan Percifield |
| EPI_ISL_2616108, EPI_ISL_2616109 | J.W. Ruby Memorial Hospital | WVU and Marshall University Combined Genomics Core Facilities | James Denvir, Peter Stoilov, Peter Perrotta, Wesley Kimble, Ryan Percifield |
| EPI_ISL_2616110 | QLabs | WVU and Marshall University Combined Genomics Core Facilities | James Denvir, Peter Stoilov, Peter Perrotta, Wesley Kimble, Ryan Percifield |
| EPI_ISL_2616111, EPI_ISL_2616112, EPI_ISL_2616113, EPI_ISL_2616114, EPI_ISL_2616115, EPI_ISL_2616116, EPI_ISL_2616117, EPI_ISL_2616118, EPI_ISL_2616119, EPI_ISL_2616120, EPI_ISL_2616121, EPI_ISL_2616122, EPI_ISL_2616123, EPI_ISL_2616124, EPI_ISL_2616125, EPI_ISL_2616126, EPI_ISL_2616127, EPI_ISL_2616128, EPI_ISL_2616129, EPI_ISL_2616130, EPI_ISL_2616131, EPI_ISL_2616132, EPI_ISL_2616133, EPI_ISL_2616134, EPI_ISL_2616135, EPI_ISL_2616136, EPI_ISL_2616137, EPI_ISL_2616138, EPI_ISL_2616139, EPI_ISL_2616140, EPI_ISL_2616141, EPI_ISL_2616142, EPI_ISL_2616143, EPI_ISL_2616144, EPI_ISL_2616145, EPI_ISL_2616146, EPI_ISL_2616147, EPI_ISL_2616148, EPI_ISL_2616149, EPI_ISL_2616150, EPI_ISL_2616151, EPI_ISL_2616152, EPI_ISL_2616153, EPI_ISL_2616154, EPI_ISL_2616155, EPI_ISL_2616156, EPI_ISL_2616157, EPI_ISL_2616158, EPI_ISL_2616159, EPI_ISL_2616160, EPI_ISL_2616161, EPI_ISL_2616162, EPI_ISL_2616163, EPI_ISL_2616164, EPI_ISL_2616165, EPI_ISL_2616166, EPI_ISL_2616167, EPI_ISL_2616168, EPI_ISL_2616169, EPI_ISL_2616170, EPI_ISL_2616171, EPI_ISL_2616172, EPI_ISL_2616173, EPI_ISL_2616174, EPI_ISL_2616175, EPI_ISL_2616176, EPI_ISL_2616177, EPI_ISL_2616178, EPI_ISL_2616179, EPI_ISL_2616180, EPI_ISL_2616181, EPI_ISL_2616182, EPI_ISL_2616183, EPI_ISL_2616184, EPI_ISL_2616185, EPI_ISL_2616186, EPI_ISL_2616187, EPI_ISL_2616188, EPI_ISL_2616189, EPI_ISL_2616190 | | | |
| see above | J.W. Ruby Memorial Hospital | WVU and Marshall University Combined Genomics Core Facilities | James Denvir, Peter Stoilov, Peter Perrotta, Wesley Kimble, Ryan Percifield |
| EPI_ISL_2616522, EPI_ISL_2616523, EPI_ISL_2616524, EPI_ISL_2616525, EPI_ISL_2616526, EPI_ISL_2616527, EPI_ISL_2616528, EPI_ISL_2616529, EPI_ISL_2616530, EPI_ISL_2616531, EPI_ISL_2616532, EPI_ISL_2616533, EPI_ISL_2616534, EPI_ISL_2616535, EPI_ISL_2616536, EPI_ISL_2616537, EPI_ISL_2616538, EPI_ISL_2616539, EPI_ISL_2616540, EPI_ISL_2616541, EPI_ISL_2616542, EPI_ISL_2616543, EPI_ISL_2616544, EPI_ISL_2616545, EPI_ISL_2616546, EPI_ISL_2616547, EPI_ISL_2616548, EPI_ISL_2616549, EPI_ISL_2616550, EPI_ISL_2616551, EPI_ISL_2616552, EPI_ISL_2616553, EPI_ISL_2616554, EPI_ISL_2616555, EPI_ISL_2616556, EPI_ISL_2616557, EPI_ISL_2616558, EPI_ISL_2616559, EPI_ISL_2616560, EPI_ISL_2616561, EPI_ISL_2616562, EPI_ISL_2616563, EPI_ISL_2616564, EPI_ISL_2616565, EPI_ISL_2616566, EPI_ISL_2616567, EPI_ISL_2616568, EPI_ISL_2616569, EPI_ISL_2616570, EPI_ISL_2616571, EPI_ISL_2616572, EPI_ISL_2616573, EPI_ISL_2616574, EPI_ISL_2616575, EPI_ISL_2616576, EPI_ISL_2616577, EPI_ISL_2616578, EPI_ISL_2616579, EPI_ISL_2616580, EPI_ISL_2616581, EPI_ISL_2616582, EPI_ISL_2616583, EPI_ISL_2616584, EPI_ISL_2616585, EPI_ISL_2616586, EPI_ISL_2616587, EPI_ISL_2616588, EPI_ISL_2616589, EPI_ISL_2616590, EPI_ISL_2616591, EPI_ISL_2616592, EPI_ISL_2616593, EPI_ISL_2616594, EPI_ISL_2616595, EPI_ISL_2616596, EPI_ISL_2616597, EPI_ISL_2616598, EPI_ISL_2616599, EPI_ISL_2616600, EPI_ISL_2616601, EPI_ISL_2616602, EPI_ISL_2616603, EPI_ISL_2616604, EPI_ISL_2616605, EPI_ISL_2616606, EPI_ISL_2616607, EPI_ISL_2616608, EPI_ISL_2616609, EPI_ISL_2616610, EPI_ISL_2616611, EPI_ISL_2616612, EPI_ISL_2616613, EPI_ISL_2616614, EPI_ISL_2616615, EPI_ISL_2616616, EPI_ISL_2616617, EPI_ISL_2616618, EPI_ISL_2616619, EPI_ISL_2616620, EPI_ISL_2616621, EPI_ISL_2616622, EPI_ISL_2616653, EPI_ISL_2616654, EPI_ISL_2616655, EPI_ISL_2616656, EPI_ISL_2616657, EPI_ISL_2616658, EPI_ISL_2616659, EPI_ISL_2616660, EPI_ISL_2616661, EPI_ISL_2616662, EPI_ISL_2616663, EPI_ISL_2616664, EPI_ISL_2616665, EPI_ISL_2616666, EPI_ISL_2616667, EPI_ISL_2616668, EPI_ISL_2616669, EPI_ISL_2616670, EPI_ISL_2616671, EPI_ISL_2616672, EPI_ISL_2616673, EPI_ISL_2616674, EPI_ISL_2616675, EPI_ISL_2616676, EPI_ISL_2616677, EPI_ISL_2616678, EPI_ISL_2616679, EPI_ISL_2616680, EPI_ISL_2616681, EPI_ISL_2616682, EPI_ISL_2616683, EPI_ISL_2616684, EPI_ISL_2616685, EPI_ISL_2616686, EPI_ISL_2616687, EPI_ISL_2616688, EPI_ISL_2616689, EPI_ISL_2616690, EPI_ISL_2616691, EPI_ISL_2616692, EPI_ISL_2616693, EPI_ISL_2616694, EPI_ISL_2616695, EPI_ISL_2616696, EPI_ISL_2616697, EPI_ISL_2616698, EPI_ISL_2616699, EPI_ISL_2616700, EPI_ISL_2616701, EPI_ISL_2616702, EPI_ISL_2616703, EPI_ISL_2616704, EPI_ISL_2616705, EPI_ISL_2616706, EPI_ISL_2616707, EPI_ISL_2616708, EPI_ISL_2616709, EPI_ISL_2616710, EPI_ISL_2616711, EPI_ISL_2616712, EPI_ISL_2616713, EPI_ISL_2616714, EPI_ISL_2616715, EPI_ISL_2616716, EPI_ISL_2616717, EPI_ISL_2616718, EPI_ISL_2616719, EPI_ISL_2616720, EPI_ISL_2616721, EPI_ISL_2616722, EPI_ISL_2616723, EPI_ISL_2616724, EPI_ISL_2616725, EPI_ISL_2616726, EPI_ISL_2616727, EPI_ISL_2616728, EPI_ISL_2616729, EPI_ISL_2616730, EPI_ISL_2616731, EPI_ISL_2616732, EPI_ISL_2616733, EPI_ISL_2616734, EPI_ISL_2616735, EPI_ISL_2616736, EPI_ISL_2616737, EPI_ISL_2616738, EPI_ISL_2616739, EPI_ISL_2616740, EPI_ISL_2616741 | | | |
| see above | Wyoming Public Health Laboratory | Wyoming Public Health Laboratory | Jim Mildenberger, Taylor Fearing, Channing Weber, Ashley Norberg, Chayse Rowley, Marley Goetz, Brian Dominguez, Elliot Thomasson, Sam Britz, Cari Sloma, Robert Petit, and Rob Christensen |
| EPI_ISL_2616742, EPI_ISL_2616743, EPI_ISL_2616744, EPI_ISL_2616745, EPI_ISL_2616746, EPI_ISL_2616747, EPI_ISL_2616748, EPI_ISL_2616749, EPI_ISL_2616750, EPI_ISL_2616751, EPI_ISL_2616752, EPI_ISL_2616753, EPI_ISL_2616754, EPI_ISL_2616755, EPI_ISL_2616756, EPI_ISL_2616757, EPI_ISL_2616758, EPI_ISL_2616759, EPI_ISL_2616760, EPI_ISL_2616761, EPI_ISL_2616762, EPI_ISL_2616763, EPI_ISL_2616764, EPI_ISL_2616765, EPI_ISL_2616766, EPI_ISL_2616767, EPI_ISL_2616768, EPI_ISL_2616769, EPI_ISL_2616770, EPI_ISL_2616771, EPI_ISL_2616772, EPI_ISL_2616773, EPI_ISL_2616774, EPI_ISL_2616775, EPI_ISL_2616776, EPI_ISL_2616777, EPI_ISL_2616778, EPI_ISL_2616779, EPI_ISL_2616780, EPI_ISL_2616783 | | | |
| see above | Houston Health Dept. | Houston Health Dept. | Ryker Penn, Pamela Brown, Adolpho Lara, Yanlai Lai |
| EPI_ISL_2616784, EPI_ISL_2616785, EPI_ISL_2616786, EPI_ISL_2616787, EPI_ISL_2616788, EPI_ISL_2616789, EPI_ISL_2616790, EPI_ISL_2616791, EPI_ISL_2616792, EPI_ISL_2616793, EPI_ISL_2616794, EPI_ISL_2616795, EPI_ISL_2616796, EPI_ISL_2616797, EPI_ISL_2616798, EPI_ISL_2616799, EPI_ISL_2616800, EPI_ISL_2616801, EPI_ISL_2616802, EPI_ISL_2616803, EPI_ISL_2616804, EPI_ISL_2616805, EPI_ISL_2616806, EPI_ISL_2616807, EPI_ISL_2616808, EPI_ISL_2616809, EPI_ISL_2616810, EPI_ISL_2616811, EPI_ISL_2616812, EPI_ISL_2616813, EPI_ISL_2616814, EPI_ISL_2616815, EPI_ISL_2616816, EPI_ISL_2616817, EPI_ISL_2616818, EPI_ISL_2616819, EPI_ISL_2616820, EPI_ISL_2616821, EPI_ISL_2616822, EPI_ISL_2616823, EPI_ISL_2616824, EPI_ISL_2616825, EPI_ISL_2616826, EPI_ISL_2616827, EPI_ISL_2616828, EPI_ISL_2616829, EPI_ISL_2616830, EPI_ISL_2616831, EPI_ISL_2616832, EPI_ISL_2616833, EPI_ISL_2616834, EPI_ISL_2616835, EPI_ISL_2616836, EPI_ISL_2616837, EPI_ISL_2616838, EPI_ISL_2616839, EPI_ISL_2616840, EPI_ISL_2616841, EPI_ISL_2616842, EPI_ISL_2616843, EPI_ISL_2616844, EPI_ISL_2616845, EPI_ISL_2616846, EPI_ISL_2616847, EPI_ISL_2616848, EPI_ISL_2616849, EPI_ISL_2616850, EPI_ISL_2616851, EPI_ISL_2616852, EPI_ISL_2616853, EPI_ISL_2616854, EPI_ISL_2616855, EPI_ISL_2616856, EPI_ISL_2616857, EPI_ISL_2616858, EPI_ISL_2616859, EPI_ISL_2616860, EPI_ISL_2616861, EPI_ISL_2616862, EPI_ISL_2616863, EPI_ISL_2616864, EPI_ISL_2616865, EPI_ISL_2616866, EPI_ISL_2616867, EPI_ISL_2616868, EPI_ISL_2616869, EPI_ISL_2616870, EPI_ISL_2616871, EPI_ISL_2616872, EPI_ISL_2616873, EPI_ISL_2616874, EPI_ISL_2616875, EPI_ISL_2616876, EPI_ISL_2616877, EPI_ISL_2616878, EPI_ISL_2616879, EPI_ISL_2616880, EPI_ISL_2616881, EPI_ISL_2616882, EPI_ISL_2616883, EPI_ISL_2616884, EPI_ISL_2616885, EPI_ISL_2616886, EPI_ISL_2616887, EPI_ISL_2616888, EPI_ISL_2616889, EPI_ISL_2616890, EPI_ISL_2616891, EPI_ISL_2616892, EPI_ISL_2616893, EPI_ISL_2616894, EPI_ISL_2616895, EPI_ISL_2616896, EPI_ISL_2616897, EPI_ISL_2616898, EPI_ISL_2616899, EPI_ISL_2616900, EPI_ISL_2616901, EPI_ISL_2616902, EPI_ISL_2616903, EPI_ISL_2616904, EPI_ISL_2616905, EPI_ISL_2616906, EPI_ISL_2616907, EPI_ISL_2616908, EPI_ISL_2616909, EPI_ISL_2616910, EPI_ISL_2616911, EPI_ISL_2616912, EPI_ISL_2616913, EPI_ISL_2616914, EPI_ISL_2616915, EPI_ISL_2616916, EPI_ISL_2616917, EPI_ISL_2616918, EPI_ISL_2616919, EPI_ISL_2616920, EPI_ISL_2616921, EPI_ISL_2616922, EPI_ISL_2616923, EPI_ISL_2616924, EPI_ISL_2616925, EPI_ISL_2616926, EPI_ISL_2616927, EPI_ISL_2616928, EPI_ISL_2616929, EPI_ISL_2616930, EPI_ISL_2616931, EPI_ISL_2616932, EPI_ISL_2616933, EPI_ISL_2616934, EPI_ISL_2616935, EPI_ISL_2616936, EPI_ISL_2616937, | | | |

| | | | |
|---|---|---|---|
| EPI_ISL_2616938, EPI_ISL_2616939, EPI_ISL_2616940, EPI_ISL_2616941, EPI_ISL_2616942, EPI_ISL_2616943, EPI_ISL_2616944, EPI_ISL_2616945, EPI_ISL_2616946, EPI_ISL_2616947, EPI_ISL_2616948, EPI_ISL_2616949, EPI_ISL_2616950, EPI_ISL_2616951, EPI_ISL_2616952, EPI_ISL_2616953, EPI_ISL_2616954, EPI_ISL_2616955, EPI_ISL_2616956, EPI_ISL_2616957, EPI_ISL_2616958, EPI_ISL_2616959, EPI_ISL_2616960, EPI_ISL_2616961, EPI_ISL_2616962, EPI_ISL_2616963, EPI_ISL_2616964, EPI_ISL_2616965, EPI_ISL_2616966, EPI_ISL_2616967, EPI_ISL_2616968, EPI_ISL_2616969, EPI_ISL_2616970, EPI_ISL_2616971, EPI_ISL_2616972, EPI_ISL_2616973, EPI_ISL_2617004, EPI_ISL_2617005, EPI_ISL_2617006, EPI_ISL_2617007, EPI_ISL_2617008, EPI_ISL_2617009, EPI_ISL_2617010, EPI_ISL_2617011, EPI_ISL_2617012, EPI_ISL_2617013, EPI_ISL_2617014, EPI_ISL_2617015, EPI_ISL_2617016, EPI_ISL_2617017, EPI_ISL_2617018, EPI_ISL_2617019, EPI_ISL_2617020, EPI_ISL_2617021, EPI_ISL_2617022, EPI_ISL_2617023, EPI_ISL_2617024, EPI_ISL_2617025 | | | |
| see above | Salud Digna | Instituto Nacional de Medicina Genomica | Hidalgo-Miranda A, Cedro-Tanda A, Mendoza-Vargas A, Reyes-Grajeda JP, Abraham Campos-Romero, Moreno-Camacho José Luis, Rodriguez-Gallegos Jorge, Luna-Ruiz Marco, Gonzalez-Barrera D, Rangel-DeLeon D, Munguia-Garza P, Ramirez-Vega O, Escobar-Arrazola, M, Herrera-Montalvo LA. |
| EPI_ISL_2617026, EPI_ISL_2617027, EPI_ISL_2617028, EPI_ISL_2617029, EPI_ISL_2617030, EPI_ISL_2617031, EPI_ISL_2617032, EPI_ISL_2617033, EPI_ISL_2617034 | Salud Digna | Instituto Nacional de Medicina Genomica | Hidalgo-Miranda A, Cedro-Tanda A, Cisneros-Villanueva M, Orjuela-Rodríguez M, Mendoza-Vargas A, Reyes-Grajeda JP, Abraham Campos-Romero, Moreno-Camacho José Luis, Rodriguez-Gallegos Jorge, Luna-Ruiz Marco, Gonzalez-Barrera D, Rangel-DeLeon D, Munguia-Garza P, Ramirez-Vega O, Escobar-Arrazola, M, Herrera-Montalvo LA. |
| EPI_ISL_2617555, EPI_ISL_2617556, EPI_ISL_2617557, EPI_ISL_2617561, EPI_ISL_2617562, EPI_ISL_2617564, EPI_ISL_2617565, EPI_ISL_2617566, EPI_ISL_2617568, EPI_ISL_2617569, EPI_ISL_2617572, EPI_ISL_2617574, EPI_ISL_2617575, EPI_ISL_2617577, EPI_ISL_2617581, EPI_ISL_2617582, EPI_ISL_2617583 | | | |
| see above | City of Milwaukee Health Department Laboratory | City of Milwaukee Health Department Laboratory | Manjeet Khubbar, Amy Bauer, Jennifer Lentz, Samantha Scott, Sanjib Bhattacharyya |
| EPI_ISL_2617628, EPI_ISL_2617629, EPI_ISL_2617630 | UW Virology Lab | UW Virology Lab | Pavitra Roychoudhury, Hong Xie, Lasata Shrestha, Tien V. Nguyen, Shah Mohamed Bakhash, Michelle Lin, Noah R. Baker, Ricardo Perez, Sean Ellis, Nathan Breit, Robert J. Livingston, Meei-Li Huang, Keith R Jerome, Patrick Mathias, Alexander Greninger |
| EPI_ISL_2617631 | Omega Diagnostics at Mounes | Omega Diagnostics at Mounes | Cynthia Corley, Ashleigh McGrail, Latira Haynes-Jacob, Vivek Khare,MD |
| EPI_ISL_2617686, EPI_ISL_2617687, EPI_ISL_2617688, EPI_ISL_2617689, EPI_ISL_2617690, EPI_ISL_2617691, EPI_ISL_2617692, EPI_ISL_2617693, EPI_ISL_2617694, EPI_ISL_2617695, EPI_ISL_2617696, EPI_ISL_2617697, EPI_ISL_2617698, EPI_ISL_2617699, EPI_ISL_2617700, EPI_ISL_2617701, EPI_ISL_2617702, EPI_ISL_2617703, EPI_ISL_2617704, EPI_ISL_2617705, EPI_ISL_2617706, EPI_ISL_2617707, EPI_ISL_2617708, EPI_ISL_2617709, EPI_ISL_2617710, EPI_ISL_2617711, EPI_ISL_2617712, EPI_ISL_2617713, EPI_ISL_2617714, EPI_ISL_2617715, EPI_ISL_2617716, EPI_ISL_2617717, EPI_ISL_2617718, EPI_ISL_2617719, EPI_ISL_2617720, EPI_ISL_2617721, EPI_ISL_2617722, EPI_ISL_2617723, EPI_ISL_2617724, EPI_ISL_2617725, EPI_ISL_2617726, EPI_ISL_2617727, EPI_ISL_2617728, EPI_ISL_2617729, EPI_ISL_2617730, EPI_ISL_2617731, EPI_ISL_2617732, EPI_ISL_2617733, EPI_ISL_2617734, EPI_ISL_2617735, EPI_ISL_2617736, EPI_ISL_2617737, EPI_ISL_2617738, EPI_ISL_2617739, EPI_ISL_2617740, EPI_ISL_2617741, EPI_ISL_2617742, EPI_ISL_2617743, EPI_ISL_2617744, EPI_ISL_2617745, EPI_ISL_2617746, EPI_ISL_2617747, EPI_ISL_2617748, EPI_ISL_2617749, EPI_ISL_2617751, EPI_ISL_2617752, EPI_ISL_2617753, EPI_ISL_2617754, EPI_ISL_2617755, EPI_ISL_2617756, EPI_ISL_2617757, EPI_ISL_2617759, EPI_ISL_2617760, EPI_ISL_2617761, EPI_ISL_2617762, EPI_ISL_2617763, EPI_ISL_2617764, EPI_ISL_2617765, EPI_ISL_2617766, EPI_ISL_2617767, EPI_ISL_2617768, EPI_ISL_2617769, EPI_ISL_2617770, EPI_ISL_2617771, EPI_ISL_2617772, EPI_ISL_2617773, EPI_ISL_2617774, EPI_ISL_2617775, EPI_ISL_2617776, EPI_ISL_2617777, EPI_ISL_2617778, EPI_ISL_2617779, EPI_ISL_2617780, EPI_ISL_2617781, EPI_ISL_2617782, EPI_ISL_2617783, EPI_ISL_2617784, EPI_ISL_2617785, EPI_ISL_2617786, EPI_ISL_2617787, EPI_ISL_2617788, EPI_ISL_2617789, EPI_ISL_2617790, EPI_ISL_2617791, EPI_ISL_2617792, EPI_ISL_2617793, EPI_ISL_2617794, EPI_ISL_2617795, EPI_ISL_2617796, EPI_ISL_2617797, EPI_ISL_2617798, EPI_ISL_2617799, EPI_ISL_2617800, EPI_ISL_2617801, EPI_ISL_2617802, EPI_ISL_2617803, EPI_ISL_2617804, EPI_ISL_2617805, EPI_ISL_2617806, EPI_ISL_2617807, EPI_ISL_2617808, EPI_ISL_2617809, EPI_ISL_2617810, EPI_ISL_2617811, EPI_ISL_2617813, EPI_ISL_2617814, EPI_ISL_2617815, EPI_ISL_2617816, EPI_ISL_2617817, EPI_ISL_2617819, EPI_ISL_2617820, EPI_ISL_2617821, EPI_ISL_2617822, EPI_ISL_2617823, EPI_ISL_2617824, EPI_ISL_2617826 | | | |
| see above | Pathology and Laboratory Medicine Institute, Cleveland Clinic, Ohio, USA | Pathology and Laboratory Medicine Institute, Cleveland Clinic, Ohio, USA | Concetta Peck, Jennifer Starbuck, Joy Nakitandwe, Kristen McDonnell, David Plunkett, Zheng Jin Tu, Jay Brock, Yu-Wei Cheng, Gary Procop, Daniel Rhoads, Daniel H. Farkas, David Bosler |
| EPI_ISL_2617827, EPI_ISL_2617828, EPI_ISL_2617829, EPI_ISL_2617830, EPI_ISL_2617831, EPI_ISL_2617832, EPI_ISL_2617833, EPI_ISL_2617834, EPI_ISL_2617835, EPI_ISL_2617836, EPI_ISL_2617837, EPI_ISL_2617838, EPI_ISL_2617839, EPI_ISL_2617841, EPI_ISL_2617842, EPI_ISL_2617843, EPI_ISL_2617844, EPI_ISL_2617845, EPI_ISL_2617846, EPI_ISL_2617847, EPI_ISL_2617848, EPI_ISL_2617849, EPI_ISL_2617850, EPI_ISL_2617851, EPI_ISL_2617852, EPI_ISL_2617853 | | | |
| see above | USC Clinical Lab | Los Angeles County Public Health Laboratories | P. Hemarajata et al. |
| EPI_ISL_2617854 | Los Angeles County Public Health Laboratories | Los Angeles County Public Health Laboratories | P. Hemarajata et al. |
| EPI_ISL_2617855, EPI_ISL_2617856, EPI_ISL_2617857, EPI_ISL_2617858, EPI_ISL_2617859, EPI_ISL_2617860, EPI_ISL_2617861, EPI_ISL_2617862, EPI_ISL_2617863, EPI_ISL_2617864, EPI_ISL_2617865, EPI_ISL_2617866, EPI_ISL_2617867, EPI_ISL_2617868, EPI_ISL_2617869, EPI_ISL_2617870, EPI_ISL_2617871, EPI_ISL_2617872, EPI_ISL_2617873, EPI_ISL_2617874 | | | |
| see above | USC Clinical Lab | Los Angeles County Public Health Laboratories | P. Hemarajata et al. |
| EPI_ISL_2617942 | Pomona Valley Hospital Medical Center | Los Angeles County Public Health Laboratories | P. Hemarajata et al. |
| EPI_ISL_2617943, EPI_ISL_2617944, EPI_ISL_2617945, EPI_ISL_2617946, EPI_ISL_2617947, EPI_ISL_2617948, EPI_ISL_2617949, EPI_ISL_2617950, EPI_ISL_2617951, EPI_ISL_2617952, EPI_ISL_2617953, EPI_ISL_2617954, EPI_ISL_2617955, EPI_ISL_2617956, EPI_ISL_2617957, EPI_ISL_2617958, EPI_ISL_2617959 | | | |
| see above | UCLA Clinical Micro Lab | Los Angeles County Public Health Laboratories | P. Hemarajata et al. |
| EPI_ISL_2617961, EPI_ISL_2617962, EPI_ISL_2617964, EPI_ISL_2617965, EPI_ISL_2617966 | USC Clinical Lab | Los Angeles County Public Health Laboratories | P. Hemarajata et al. |
| EPI_ISL_2617969, EPI_ISL_2617970, EPI_ISL_2617971, EPI_ISL_2617973, EPI_ISL_2617974, EPI_ISL_2617975, EPI_ISL_2617976, EPI_ISL_2617977, EPI_ISL_2617978, EPI_ISL_2617979, EPI_ISL_2617980, EPI_ISL_2617981, EPI_ISL_2617982, EPI_ISL_2617983, EPI_ISL_2617984, EPI_ISL_2617985, EPI_ISL_2617986, EPI_ISL_2617987, EPI_ISL_2617988, EPI_ISL_2617989, EPI_ISL_2617990, EPI_ISL_2617991, EPI_ISL_2617992, EPI_ISL_2617993, EPI_ISL_2617994, EPI_ISL_2617995, EPI_ISL_2617996, EPI_ISL_2617997, EPI_ISL_2617998, EPI_ISL_2617999, EPI_ISL_2618000, EPI_ISL_2618001, EPI_ISL_2618002, EPI_ISL_2618003, EPI_ISL_2618004, EPI_ISL_2618005, EPI_ISL_2618006, EPI_ISL_2618007, EPI_ISL_2618008, EPI_ISL_2618009, EPI_ISL_2618010, EPI_ISL_2618012, EPI_ISL_2618013, EPI_ISL_2618014, EPI_ISL_2618015, EPI_ISL_2618016, EPI_ISL_2618017, EPI_ISL_2618018, EPI_ISL_2618019, EPI_ISL_2618021, EPI_ISL_2618022, EPI_ISL_2618023, EPI_ISL_2618024, EPI_ISL_2618025, EPI_ISL_2618026, EPI_ISL_2618027, EPI_ISL_2618028, EPI_ISL_2618029, EPI_ISL_2618031, EPI_ISL_2618032, EPI_ISL_2618033, EPI_ISL_2618034, EPI_ISL_2618035, EPI_ISL_2618036, EPI_ISL_2618037, EPI_ISL_2618038, EPI_ISL_2618039, EPI_ISL_2618040, EPI_ISL_2618041, EPI_ISL_2618042, EPI_ISL_2618043, EPI_ISL_2618044, EPI_ISL_2618045, EPI_ISL_2618046, EPI_ISL_2618047, EPI_ISL_2618048, EPI_ISL_2618049, EPI_ISL_2618050, EPI_ISL_2618051, EPI_ISL_2618052, EPI_ISL_2618053, EPI_ISL_2618054, EPI_ISL_2618055, EPI_ISL_2618056, EPI_ISL_2618057, EPI_ISL_2618058, EPI_ISL_2618059, EPI_ISL_2618060, EPI_ISL_2618061, EPI_ISL_2618062, EPI_ISL_2618063, EPI_ISL_2618064, EPI_ISL_2618065, EPI_ISL_2618066, EPI_ISL_2618067, EPI_ISL_2618068, EPI_ISL_2618069, EPI_ISL_2618070, EPI_ISL_2618071, EPI_ISL_2618072, EPI_ISL_2618073, EPI_ISL_2618074, EPI_ISL_2618075, EPI_ISL_2618076, EPI_ISL_2618077, EPI_ISL_2618078, EPI_ISL_2618079, EPI_ISL_2618080, EPI_ISL_2618081, EPI_ISL_2618082, EPI_ISL_2618083, EPI_ISL_2618084, EPI_ISL_2618085, EPI_ISL_2618086, EPI_ISL_2618087, EPI_ISL_2618088, EPI_ISL_2618089, EPI_ISL_2618090, EPI_ISL_2618091, EPI_ISL_2618092, EPI_ISL_2618094, EPI_ISL_2618095, EPI_ISL_2618096, EPI_ISL_2618097, EPI_ISL_2618098, EPI_ISL_2618099, EPI_ISL_2618100, EPI_ISL_2618101, EPI_ISL_2618102, EPI_ISL_2618103, EPI_ISL_2618104, EPI_ISL_2618105, EPI_ISL_2618106, EPI_ISL_2618107, EPI_ISL_2618108, EPI_ISL_2618109, EPI_ISL_2618110, EPI_ISL_2618111, EPI_ISL_2618112 | | | |
| see above | UW Virology Lab | UW Virology Lab | Pavitra Roychoudhury, Hong Xie, Lasata Shrestha, Tien V. Nguyen, Shah Mohamed Bakhash, Michelle Lin, Noah R. Baker, Ricardo Perez, Sean Ellis, Nathan Breit, Robert J. Livingston, Meei-Li Huang, Keith R Jerome, Patrick Mathias, Alexander Greninger |
| EPI_ISL_2618113, EPI_ISL_2618114, EPI_ISL_2618115, EPI_ISL_2618116, EPI_ISL_2618117, EPI_ISL_2618118, EPI_ISL_2618119, EPI_ISL_2618120, EPI_ISL_2618121, EPI_ISL_2618122, EPI_ISL_2618123, EPI_ISL_2618124, EPI_ISL_2618125, EPI_ISL_2618126, EPI_ISL_2618127, EPI_ISL_2618128, EPI_ISL_2618129, EPI_ISL_2618130, EPI_ISL_2618131, EPI_ISL_2618132, EPI_ISL_2618133, EPI_ISL_2618134, EPI_ISL_2618135, EPI_ISL_2618136, EPI_ISL_2618137, EPI_ISL_2618138, EPI_ISL_2618139, EPI_ISL_2618140, EPI_ISL_2618141, EPI_ISL_2618142, EPI_ISL_2618143, EPI_ISL_2618144, EPI_ISL_2618145, EPI_ISL_2618146, EPI_ISL_2618147, EPI_ISL_2618148, EPI_ISL_2618149, EPI_ISL_2618150, EPI_ISL_2618151, EPI_ISL_2618152, EPI_ISL_2618153, EPI_ISL_2618154, EPI_ISL_2618155, EPI_ISL_2618156, EPI_ISL_2618157, EPI_ISL_2618158, EPI_ISL_2618159, EPI_ISL_2618160, EPI_ISL_2618161, EPI_ISL_2618162, EPI_ISL_2618163, EPI_ISL_2618164, EPI_ISL_2618165, EPI_ISL_2618166, EPI_ISL_2618167, EPI_ISL_2618168, EPI_ISL_2618169, EPI_ISL_2618170, EPI_ISL_2618171, EPI_ISL_2618172, EPI_ISL_2618173, EPI_ISL_2618175, EPI_ISL_2618176, EPI_ISL_2618177, EPI_ISL_2618178, EPI_ISL_2618179, EPI_ISL_2618180, EPI_ISL_2618181, EPI_ISL_2618182, EPI_ISL_2618183, EPI_ISL_2618184, EPI_ISL_2618185, EPI_ISL_2618186, EPI_ISL_2618187, EPI_ISL_2618188, EPI_ISL_2618189, EPI_ISL_2618190, EPI_ISL_2618191, EPI_ISL_2618192, EPI_ISL_2618193, EPI_ISL_2618194, EPI_ISL_2618195, EPI_ISL_2618196, EPI_ISL_2618197, EPI_ISL_2618198, EPI_ISL_2618199, EPI_ISL_2618200, EPI_ISL_2618201, EPI_ISL_2618202, EPI_ISL_2618203, EPI_ISL_2618204, EPI_ISL_2618205, EPI_ISL_2618206, EPI_ISL_2618207, EPI_ISL_2618208, EPI_ISL_2618209, EPI_ISL_2618210, EPI_ISL_2618211, EPI_ISL_2618212, EPI_ISL_2618213, EPI_ISL_2618214, EPI_ISL_2618215, EPI_ISL_2618216, EPI_ISL_2618217, EPI_ISL_2618218, EPI_ISL_2618219, EPI_ISL_2618220, EPI_ISL_2618221, EPI_ISL_2618222, EPI_ISL_2618223, EPI_ISL_2618224, EPI_ISL_2618225, EPI_ISL_2618226, EPI_ISL_2618227, EPI_ISL_2618229, EPI_ISL_2618230, EPI_ISL_2618231, EPI_ISL_2618232, EPI_ISL_2618233, EPI_ISL_2618234, EPI_ISL_2618235, EPI_ISL_2618236, EPI_ISL_2618237, EPI_ISL_2618238, EPI_ISL_2618239, EPI_ISL_2618240, EPI_ISL_2618241, EPI_ISL_2618242, EPI_ISL_2618243, EPI_ISL_2618244, EPI_ISL_2618245, EPI_ISL_2618246, EPI_ISL_2618247, EPI_ISL_2618248, EPI_ISL_2618249, EPI_ISL_2618250, EPI_ISL_2618251, EPI_ISL_2618252, EPI_ISL_2618253, EPI_ISL_2618254, EPI_ISL_2618255, EPI_ISL_2618256, EPI_ISL_2618257, EPI_ISL_2618258, EPI_ISL_2618259, EPI_ISL_2618260, EPI_ISL_2618261, EPI_ISL_2618262, EPI_ISL_2618264, EPI_ISL_2618265, EPI_ISL_2618266, EPI_ISL_2618267, EPI_ISL_2618268, EPI_ISL_2618269, EPI_ISL_2618270, EPI_ISL_2618271, EPI_ISL_2618272, EPI_ISL_2618273, EPI_ISL_2618274, EPI_ISL_2618275, EPI_ISL_2618276, EPI_ISL_2618277, EPI_ISL_2618278, EPI_ISL_2618279, EPI_ISL_2618280, EPI_ISL_2618281, EPI_ISL_2618282, EPI_ISL_2618283, EPI_ISL_2618284, EPI_ISL_2618285, EPI_ISL_2618286, EPI_ISL_2618287, EPI_ISL_2618288, EPI_ISL_2618289, EPI_ISL_2618290, EPI_ISL_2618291, EPI_ISL_2618292, EPI_ISL_2618293, EPI_ISL_2618294, EPI_ISL_2618295, EPI_ISL_2618296, EPI_ISL_2618297, EPI_ISL_2618298, EPI_ISL_2618299, EPI_ISL_2618300, EPI_ISL_2618301, EPI_ISL_2618302, EPI_ISL_2618303, EPI_ISL_2618304, EPI_ISL_2618306, EPI_ISL_2618307, EPI_ISL_2618308, EPI_ISL_2618309, EPI_ISL_2618310, EPI_ISL_2618311, EPI_ISL_2618312, EPI_ISL_2618313, EPI_ISL_2618314, EPI_ISL_2618315, EPI_ISL_2618316, EPI_ISL_2618317, EPI_ISL_2618318, EPI_ISL_2618319, EPI_ISL_2618320, EPI_ISL_2618321, EPI_ISL_2618322, EPI_ISL_2618323, EPI_ISL_2618324, EPI_ISL_2618325, EPI_ISL_2618326, EPI_ISL_2618327, EPI_ISL_2618328, EPI_ISL_2618329, EPI_ISL_2618330, EPI_ISL_2618331, EPI_ISL_2618332, EPI_ISL_2618333, EPI_ISL_2618334, EPI_ISL_2618335, EPI_ISL_2618336, EPI_ISL_2618337, EPI_ISL_2618338, EPI_ISL_2618339, EPI_ISL_2618340, EPI_ISL_2618341, EPI_ISL_2618342, EPI_ISL_2618343, EPI_ISL_2618344, EPI_ISL_2618345, EPI_ISL_2618346, EPI_ISL_2618347, EPI_ISL_2618348, EPI_ISL_2618349, EPI_ISL_2618350, EPI_ISL_2618351, EPI_ISL_2618352, EPI_ISL_2618353, EPI_ISL_2618354, EPI_ISL_2618355, EPI_ISL_2618356, EPI_ISL_2618357, EPI_ISL_2618358, EPI_ISL_2618359, EPI_ISL_2618360, EPI_ISL_2618361, EPI_ISL_2618362, EPI_ISL_2618363, EPI_ISL_2618364, EPI_ISL_2618365, EPI_ISL_2618366, EPI_ISL_2618367, EPI_ISL_2618368, EPI_ISL_2618369, EPI_ISL_2618370, EPI_ISL_2618371, EPI_ISL_2618372, EPI_ISL_2618373, EPI_ISL_2618374, EPI_ISL_2618375, EPI_ISL_2618376, EPI_ISL_2618377, EPI_ISL_2618378, EPI_ISL_2618379, EPI_ISL_2618380, EPI_ISL_2618381, EPI_ISL_2618382, EPI_ISL_2618383, EPI_ISL_2618384, EPI_ISL_2618385, EPI_ISL_2618386, EPI_ISL_2618387, EPI_ISL_2618388, EPI_ISL_2618389, EPI_ISL_2618390, EPI_ISL_2618391, EPI_ISL_2618392, EPI_ISL_2618393, EPI_ISL_2618394, EPI_ISL_2618395, EPI_ISL_2618397, EPI_ISL_2618398, EPI_ISL_2618399, EPI_ISL_2618400, EPI_ISL_2618401, EPI_ISL_2618402, EPI_ISL_2618403, EPI_ISL_2618404, EPI_ISL_2618405, EPI_ISL_2618406, EPI_ISL_2618407, EPI_ISL_2618409, EPI_ISL_2618410, EPI_ISL_2618411, EPI_ISL_2618412, EPI_ISL_2618413, EPI_ISL_2618414, EPI_ISL_2618415, EPI_ISL_2618416, EPI_ISL_2618417, EPI_ISL_2618418, EPI_ISL_2618419, EPI_ISL_2618420, EPI_ISL_2618421, EPI_ISL_2618422, EPI_ISL_2618423, EPI_ISL_2618424, EPI_ISL_2618425, EPI_ISL_2618426, EPI_ISL_2618427, EPI_ISL_2618428, EPI_ISL_2618429, EPI_ISL_2618430, EPI_ISL_2618431, EPI_ISL_2618432, EPI_ISL_2618433, EPI_ISL_2618434, EPI_ISL_2618435, EPI_ISL_2618436, EPI_ISL_2618437, EPI_ISL_2618438, EPI_ISL_2618439, EPI_ISL_2618440, EPI_ISL_2618442, EPI_ISL_2618443, EPI_ISL_2618444, EPI_ISL_2618445, EPI_ISL_2618446, EPI_ISL_2618447, EPI_ISL_2618448, EPI_ISL_2618449, EPI_ISL_2618450, EPI_ISL_2618451, EPI_ISL_2618452, EPI_ISL_2618453, EPI_ISL_2618454, EPI_ISL_2618455, EPI_ISL_2618456, EPI_ISL_2618457, EPI_ISL_2618458, EPI_ISL_2618459, EPI_ISL_2618460, EPI_ISL_2618462, EPI_ISL_2618463, EPI_ISL_2618464, EPI_ISL_2618465, EPI_ISL_2618466, EPI_ISL_2618467, EPI_ISL_2618468, EPI_ISL_2618469, EPI_ISL_2618470, EPI_ISL_2618471, EPI_ISL_2618472, EPI_ISL_2618473, EPI_ISL_2618474, EPI_ISL_2618475, EPI_ISL_2618477, EPI_ISL_2618478, EPI_ISL_2618479, EPI_ISL_2618480, EPI_ISL_2618481, EPI_ISL_2618482, EPI_ISL_2618483, EPI_ISL_2618484, EPI_ISL_2618485, EPI_ISL_2618486, EPI_ISL_2618487, EPI_ISL_2618488, EPI_ISL_2618489, EPI_ISL_2618490, EPI_ISL_2618491, EPI_ISL_2618492, EPI_ISL_2618493, EPI_ISL_2618494, EPI_ISL_2618495, EPI_ISL_2618496, EPI_ISL_2618497, EPI_ISL_2618498, EPI_ISL_2618499, EPI_ISL_2618500, EPI_ISL_2618501, EPI_ISL_2618502, EPI_ISL_2618503, EPI_ISL_2618504, EPI_ISL_2618505, EPI_ISL_2618506, EPI_ISL_2618507, EPI_ISL_2618508, EPI_ISL_2618509, EPI_ISL_2618510, EPI_ISL_2618511, EPI_ISL_2618512, EPI_ISL_2618513, EPI_ISL_2618514, EPI_ISL_2618515, EPI_ISL_2618516, EPI_ISL_2618517, EPI_ISL_2618518, EPI_ISL_2618519, EPI_ISL_2618520, EPI_ISL_2618521, EPI_ISL_2618522, EPI_ISL_2618523, EPI_ISL_2618524, EPI_ISL_2618525, EPI_ISL_2618526, EPI_ISL_2618527, EPI_ISL_2618528, EPI_ISL_2618530, EPI_ISL_2618531, EPI_ISL_2618532, EPI_ISL_2618533, EPI_ISL_2618534, EPI_ISL_2618535, EPI_ISL_2618536, EPI_ISL_2618537, EPI_ISL_2618538, EPI_ISL_2618539, EPI_ISL_2618540, EPI_ISL_2618541, EPI_ISL_2618542, EPI_ISL_2618543, EPI_ISL_2618544, EPI_ISL_2618545, EPI_ISL_2618546, EPI_ISL_2618547, EPI_ISL_2618548, EPI_ISL_2618549, EPI_ISL_2618550, EPI_ISL_2618551, EPI_ISL_2618552, EPI_ISL_2618553, EPI_ISL_2618554, EPI_ISL_2618555, EPI_ISL_2618556, EPI_ISL_2618557, EPI_ISL_2618558, EPI_ISL_2618559, EPI_ISL_2618560, EPI_ISL_2618561, EPI_ISL_2618562, EPI_ISL_2618563, EPI_ISL_2618564, EPI_ISL_2618565, EPI_ISL_2618566, EPI_ISL_2618567, EPI_ISL_2618568, EPI_ISL_2618569, EPI_ISL_2618570, EPI_ISL_2618571, EPI_ISL_2618572, EPI_ISL_2618573, EPI_ISL_2618574, EPI_ISL_2618575, EPI_ISL_2618576, EPI_ISL_2618577, EPI_ISL_2618578, EPI_ISL_2618579, EPI_ISL_2618580, EPI_ISL_2618581, EPI_ISL_2618582, EPI_ISL_2618583, EPI_ISL_2618584, EPI_ISL_2618585, EPI_ISL_2618586, EPI_ISL_2618587, EPI_ISL_2618588, EPI_ISL_2618589, EPI_ISL_2618590, EPI_ISL_2618592, EPI_ISL_2618593, EPI_ISL_2618594, EPI_ISL_2618595, EPI_ISL_2618596, EPI_ISL_2618597, EPI_ISL_2618598, EPI_ISL_2618599, EPI_ISL_2618600, EPI_ISL_2618601, EPI_ISL_2618602, EPI_ISL_2618603, EPI_ISL_2618604, EPI_ISL_2618605, EPI_ISL_2618606, EPI_ISL_2618607, EPI_ISL_2618608, EPI_ISL_2618609, EPI_ISL_2618610, EPI_ISL_2618611, EPI_ISL_2618612, EPI_ISL_2618613, EPI_ISL_2618614, EPI_ISL_2618615, EPI_ISL_2618616, EPI_ISL_2618617, EPI_ISL_2618618, EPI_ISL_2618619, EPI_ISL_2618620, EPI_ISL_2618621, EPI_ISL_2618622, EPI_ISL_2618623, EPI_ISL_2618624, EPI_ISL_2618625, EPI_ISL_2618626, EPI_ISL_2618627, EPI_ISL_2618628, EPI_ISL_2618629, EPI_ISL_2618630, EPI_ISL_2618631, EPI_ISL_2618632, EPI_ISL_2618633, EPI_ISL_2618635, EPI_ISL_2618636, EPI_ISL_2618637, EPI_ISL_2618638, EPI_ISL_2618639, EPI_ISL_2618640, EPI_ISL_2618641, EPI_ISL_2618642, EPI_ISL_2618643, EPI_ISL_2618644, EPI_ISL_2618645, EPI_ISL_2618646, EPI_ISL_2618647, EPI_ISL_2618648, EPI_ISL_2618649, EPI_ISL_2618650, EPI_ISL_2618651, EPI_ISL_2618652, EPI_ISL_2618654, EPI_ISL_2618655, EPI_ISL_2618656, EPI_ISL_2618657, EPI_ISL_2618658, EPI_ISL_2618659, EPI_ISL_2618660, EPI_ISL_2618661, EPI_ISL_2618662, EPI_ISL_2618663, EPI_ISL_2618664, EPI_ISL_2618665, EPI_ISL_2618666, EPI_ISL_2618667, EPI_ISL_2618668, EPI_ISL_2618669, EPI_ISL_2618670, EPI_ISL_2618671, EPI_ISL_2618672, EPI_ISL_2618673, EPI_ISL_2618674, EPI_ISL_2618675, EPI_ISL_2618676, EPI_ISL_2618677, EPI_ISL_2618678, EPI_ISL_2618679, EPI_ISL_2618680, EPI_ISL_2618681, EPI_ISL_2618682, EPI_ISL_2618683, EPI_ISL_2618684, EPI_ISL_2618685, EPI_ISL_2618686, EPI_ISL_2618687, EPI_ISL_2618688, EPI_ISL_2618689, EPI_ISL_2618690, EPI_ISL_2618691, EPI_ISL_2618692, EPI_ISL_2618693, EPI_ISL_2618694, EPI_ISL_2618695, EPI_ISL_2618696, EPI_ISL_2618697, EPI_ISL_2618698, EPI_ISL_2618699, EPI_ISL_2618700, EPI_ISL_2618701, EPI_ISL_2618702, EPI_ISL_2618703, EPI_ISL_2618704, EPI_ISL_2618705, EPI_ISL_2618706, EPI_ISL_2618707, EPI_ISL_2618708, EPI_ISL_2618709, EPI_ISL_2618710, EPI_ISL_2618711, EPI_ISL_2618712, EPI_ISL_2618713, EPI_ISL_2618714, EPI_ISL_2618715, EPI_ISL_2618716, EPI_ISL_2618717, EPI_ISL_2618718, EPI_ISL_2618719, EPI_ISL_2618720, EPI_ISL_2618721, EPI_ISL_2618722, EPI_ISL_2618723, EPI_ISL_2618724, EPI_ISL_2618725, EPI_ISL_2618726, EPI_ISL_2618727, EPI_ISL_2618728, EPI_ISL_2618729, EPI_ISL_2618730, EPI_ISL_2618731, EPI_ISL_2618732, EPI_ISL_2618733, EPI_ISL_2618734, EPI_ISL_2618735, EPI_ISL_2618736, EPI_ISL_2618737, EPI_ISL_2618738, EPI_ISL_2618739 | | | |
| see above | UW Virology Lab | UW Virology Lab | Pavitra Roychoudhury, Hong Xie, Lasata Shrestha, Shah Mohamed Bakhash, Tien V. Nguyen, Noah R. Baker, Sean Ellis, Meei-Li Huang, Keith R Jerome, Alexander Greninger |
| EPI_ISL_2620746, EPI_ISL_2620747, EPI_ISL_2620748, EPI_ISL_2620749, EPI_ISL_2620750, EPI_ISL_2620751, EPI_ISL_2620752, EPI_ISL_2620753, EPI_ISL_2620754, EPI_ISL_2620755, EPI_ISL_2620756, EPI_ISL_2620757, EPI_ISL_2620758, EPI_ISL_2620759, EPI_ISL_2620760, EPI_ISL_2620761, EPI_ISL_2620762, EPI_ISL_2620763, EPI_ISL_2620764, EPI_ISL_2620765, EPI_ISL_2620766, EPI_ISL_2620767, EPI_ISL_2620768, EPI_ISL_2620769 | | | |
| see above | UCLA Clinical Micro Lab | Los Angeles County Public Health Laboratories | P. Hemarajata et al. |
| EPI_ISL_2620770, EPI_ISL_2620771, EPI_ISL_2620772, EPI_ISL_2620773, EPI_ISL_2620774 | USC Clinical Lab | Los Angeles County Public Health Laboratories | P. Hemarajata et al. |
| EPI_ISL_2620775 | Flow Diagnostics Lab | Los Angeles County Public Health Laboratories | P. Hemarajata et al. |
| EPI_ISL_2620776, EPI_ISL_2620777, | California Hospital Medical Center | Los Angeles County Public Health Laboratories | P. Hemarajata et al. |

| | | | |
|---|---|---|---|
| EPI_ISL_2620778 | | | |
| EPI_ISL_2620779 | Torrance Memorial Medical Center | Los Angeles County Public Health Laboratories | P. Hemarajata et al. |
| EPI_ISL_2620780, EPI_ISL_2620781, EPI_ISL_2620782 | Los Angeles County Public Health Laboratories | Los Angeles County Public Health Laboratories | P. Hemarajata et al. |
| EPI_ISL_2620783 | USC Clinical Lab | Los Angeles County Public Health Laboratories | P. Hemarajata et al. |
| EPI_ISL_2620784, EPI_ISL_2620785, EPI_ISL_2620786, EPI_ISL_2620787, EPI_ISL_2620788, EPI_ISL_2620789, EPI_ISL_2620790, EPI_ISL_2620791, EPI_ISL_2620792, EPI_ISL_2620793, EPI_ISL_2620794 | | | |
| see above | Los Angeles County Public Health Laboratories | Los Angeles County Public Health Laboratories | P. Hemarajata et al. |
| EPI_ISL_2620796, EPI_ISL_2620797, EPI_ISL_2620798, EPI_ISL_2620799 | USC Clinical Lab | Los Angeles County Public Health Laboratories | P. Hemarajata et al. |
| EPI_ISL_2620801 | Los Angeles County Public Health Laboratories | Los Angeles County Public Health Laboratories | P. Hemarajata et al. |
| EPI_ISL_2620802, EPI_ISL_2620803, EPI_ISL_2620804, EPI_ISL_2620805, EPI_ISL_2620806, EPI_ISL_2620807, EPI_ISL_2620808, EPI_ISL_2620809, EPI_ISL_2620810, EPI_ISL_2620811, EPI_ISL_2620812, EPI_ISL_2620813, EPI_ISL_2620814, EPI_ISL_2620815, EPI_ISL_2620816, EPI_ISL_2620817, EPI_ISL_2620818, EPI_ISL_2620819, EPI_ISL_2620820, EPI_ISL_2620821, EPI_ISL_2620822, EPI_ISL_2620823 | | | |
| see above | OmniPathology Laboratory | Los Angeles County Public Health Laboratories | P. Hemarajata et al. |
| EPI_ISL_2620824, EPI_ISL_2620825 | Los Angeles County Public Health Laboratories | Los Angeles County Public Health Laboratories | P. Hemarajata et al. |
| EPI_ISL_2620826 | Martin Luther King, Jr. Outpatient Center | Los Angeles County Public Health Laboratories | P. Hemarajata et al. |
| EPI_ISL_2620827, EPI_ISL_2620828, EPI_ISL_2620829 | Palmdale Regional Medical Center | Los Angeles County Public Health Laboratories | P. Hemarajata et al. |
| EPI_ISL_2620830, EPI_ISL_2620831, EPI_ISL_2620832, EPI_ISL_2620833, EPI_ISL_2620834, EPI_ISL_2620835, EPI_ISL_2620836, EPI_ISL_2620837, EPI_ISL_2620838, EPI_ISL_2620839, EPI_ISL_2620840, EPI_ISL_2620841, EPI_ISL_2620842, EPI_ISL_2620843, EPI_ISL_2620844, EPI_ISL_2620845, EPI_ISL_2620846, EPI_ISL_2620847, EPI_ISL_2620848, EPI_ISL_2620849 | | | |
| see above | OmniPathology Laboratory | Los Angeles County Public Health Laboratories | P. Hemarajata et al. |
| EPI_ISL_2620898, EPI_ISL_2620899, EPI_ISL_2620900 | Pathology and Laboratory Medicine Institute, Cleveland Clinic, Ohio, USA | Pathology and Laboratory Medicine Institute, Cleveland Clinic, Ohio, USA | Concetta Peck, Jennifer Starbuck, Joy Nakitandwe, Kristen McDonnell, David Plunkett, Zheng Jin Tu, Jay Brock, Yu-Wei Cheng, Gary Procop, Daniel Rhoads, Daniel H. Farkas, David Bosler |
| EPI_ISL_2621523, EPI_ISL_2621524, EPI_ISL_2621525, EPI_ISL_2621526, EPI_ISL_2621527, EPI_ISL_2621528, EPI_ISL_2621529, EPI_ISL_2621531, EPI_ISL_2621533, EPI_ISL_2621534, EPI_ISL_2621536, EPI_ISL_2621537, EPI_ISL_2621538, EPI_ISL_2621539, EPI_ISL_2621541, EPI_ISL_2621542, EPI_ISL_2621545, EPI_ISL_2621546, EPI_ISL_2621547 | | | |
| see above | Omega Diagnostics at Mounes | Omega Diagnostics at Mounes | Cynthia Corley, Ashleigh McGrail, Latira Haynes-Jacob, Vivek Khare, MD |
| EPI_ISL_2621549, EPI_ISL_2621550, EPI_ISL_2621551, EPI_ISL_2621552, EPI_ISL_2621554, EPI_ISL_2621555, EPI_ISL_2621556 | Genomics, Poplar Healthcare | Genomics, Poplar Healthcare | Truong,M. |
| EPI_ISL_2621557, EPI_ISL_2621558, EPI_ISL_2621559, EPI_ISL_2621560, EPI_ISL_2621561, EPI_ISL_2621562 | University of Washington Medical Center, Seattle Flu Study | University of Washington Medical Center, Seattle Flu Study | Nickerson,D.A., Frazar,C.D., Lee,J., Pelle,B., Ryke,E., Richardson,M., Adler,A., Brandstetter,E., Han,P.D., Fay,K., Ilcisin,M., Lacombe,K., Sibley,T.R., Truong,M., Wolf,C.R., Gautom,R., Melly,G., Hiatt,B., Dykema,P., Lindquist,S., Boeckh,M., Englund,J.A., Famulare,M., Lutz,B.R., Rieder,M.J., Starita,L.M., Thompson,M., Chu,H.Y., Shendure,J., Bedford,T. |
| EPI_ISL_2621563, EPI_ISL_2621564, EPI_ISL_2621565, EPI_ISL_2621566, EPI_ISL_2621567, EPI_ISL_2621568 | NIAID, Laboratory of Parasitic Diseases | NIAID, Laboratory of Parasitic Diseases | Roder,A.E., Banakis,S., Chung,M., Youn,J.-h., Palmore,T., Das,S., Nussenblatt,V., Ghedin,E. |
| EPI_ISL_2621575, EPI_ISL_2621576, EPI_ISL_2621577, EPI_ISL_2621578, EPI_ISL_2621579, EPI_ISL_2621580 | Laboratory, Gravity Diagnostics | Laboratory, Gravity Diagnostics | Diagnostics,G. |
| EPI_ISL_2621581, EPI_ISL_2621582 | CSL, Maryland Department of Health | CSL, Maryland Department of Health | Keller,E., Loomis,J., Moore,T.L., Low,K. |
| EPI_ISL_2621583, EPI_ISL_2621584, EPI_ISL_2621585, EPI_ISL_2621586, EPI_ISL_2621587, EPI_ISL_2621588, EPI_ISL_2621589, EPI_ISL_2621590, EPI_ISL_2621591, EPI_ISL_2621592, EPI_ISL_2621593, EPI_ISL_2621594, EPI_ISL_2621595, EPI_ISL_2621596, EPI_ISL_2621597, EPI_ISL_2621598, EPI_ISL_2621599, EPI_ISL_2621600, EPI_ISL_2621601, EPI_ISL_2621602, EPI_ISL_2621603, EPI_ISL_2621605, EPI_ISL_2621606, EPI_ISL_2621607, EPI_ISL_2621608, EPI_ISL_2621609, EPI_ISL_2621610, EPI_ISL_2621611, EPI_ISL_2621612, EPI_ISL_2621613, EPI_ISL_2621614, EPI_ISL_2621615, EPI_ISL_2621616, EPI_ISL_2621617 | | | |
| see above | Respiratory Viruses Branch, US Centers for Disease Control and Prevention | Respiratory Viruses Branch, US Centers for Disease Control and Prevention | Tao,Y., Li,Y., Zhang,J., Kelleher,A., Lynch,B., Queen,K., Uehara,A., Cook,P., Ng,H.J.J., Marine,R., Paden,C.R., Batra,D., Wang,H., Coalter,T., Padilla,J., Davis,M., Sheth,M., Nobles,S., Burroughs,M., Lee,J., Retchless,A., Tong,S. |
| EPI_ISL_2621618, EPI_ISL_2621619, EPI_ISL_2621620 | Laboratory, Gravity Diagnostics | Laboratory, Gravity Diagnostics | Diagnostics,G. |
| EPI_ISL_2621665, EPI_ISL_2621667, EPI_ISL_2621668, EPI_ISL_2621669, EPI_ISL_2621670, EPI_ISL_2621671, EPI_ISL_2621672, EPI_ISL_2621674 | Center for Genome Sciences, USAMRIID | Center for Genome Sciences, USAMRIID | Kugelman,J., Di Paola,N., Richardson,J. |
| EPI_ISL_2625634, EPI_ISL_2625636, EPI_ISL_2625639, EPI_ISL_2625640, EPI_ISL_2625732, EPI_ISL_2625733, EPI_ISL_2626188, EPI_ISL_2626435, EPI_ISL_2626436, EPI_ISL_2626437, EPI_ISL_2626438, EPI_ISL_2626439, EPI_ISL_2626440, EPI_ISL_2626442, EPI_ISL_2626449, EPI_ISL_2626450, EPI_ISL_2626457, EPI_ISL_2626472, EPI_ISL_2626473, EPI_ISL_2626474 | | | |
| see above | Ohio Department of Health Laboratory | Ohio Department of Health Laboratory | Holmes, Jennifer; Eric Brandt, Keoni Omura, Glen McGillivary, Caitlin McDonnell, Jade Mowery, Stephanie Mccracken, Tyler Payne, Kirtana Ramadugu, Erica Leasure, Brent Lee, Kelsey Florek, Heather Blankenship, Allison Black, Quanta Brown, and Tammy Bannerman |
| EPI_ISL_2626475, EPI_ISL_2626476, EPI_ISL_2626477, EPI_ISL_2626478, EPI_ISL_2626479, EPI_ISL_2626480, EPI_ISL_2626481, EPI_ISL_2626482, EPI_ISL_2626483, EPI_ISL_2626484 | Helix | Helix | Celina Andrade, Chelsea Andrews, David Becker, Tyler Cassens, Andrew dei Rossi, Justin Eagles, Magnus Isaksson, Marc Laurent, William Lee, Geraint Levan, James Lu, Shishi Luo, Henrique Machado, Jason Nguyen, Jimmy Ramirez, Efren Sandoval, Francisco Tanudajaja, Sherry Wang, Simon White, Dana Wyman |
| EPI_ISL_2626485, EPI_ISL_2626571, EPI_ISL_2626572 | Ohio Department of Health Laboratory | Ohio Department of Health Laboratory | Holmes, Jennifer; Eric Brandt, Keoni Omura, Glen McGillivary, Caitlin McDonnell, Jade Mowery, Stephanie Mccracken, Tyler Payne, Kirtana Ramadugu, Erica Leasure, Brent Lee, Kelsey Florek, Heather Blankenship, Allison Black, Quanta Brown, and Tammy Bannerman |
| EPI_ISL_2626592 | CT Department of Public Health | CT Department of Public Health | Claire_Pearson,Tu_N_Nguyen |
| EPI_ISL_2626621, EPI_ISL_2626622, EPI_ISL_2626623, EPI_ISL_2626624, EPI_ISL_2626625, EPI_ISL_2626626, EPI_ISL_2626627, EPI_ISL_2626628, EPI_ISL_2626629, EPI_ISL_2626630, EPI_ISL_2626631, EPI_ISL_2626632, EPI_ISL_2626633, EPI_ISL_2626634, EPI_ISL_2626635, EPI_ISL_2626636, EPI_ISL_2626637, EPI_ISL_2626638, EPI_ISL_2626639, EPI_ISL_2626640, EPI_ISL_2626641, EPI_ISL_2626642, EPI_ISL_2626643, EPI_ISL_2626644, EPI_ISL_2626645, EPI_ISL_2626646, EPI_ISL_2626647, EPI_ISL_2626648, EPI_ISL_2626649, EPI_ISL_2626650, EPI_ISL_2626651, EPI_ISL_2626652, EPI_ISL_2626653, EPI_ISL_2626654, EPI_ISL_2626655 | | | |
| see above | Tampa General Hospital Esoteric Lab | Tampa General Hospital Esoteric Research & | Jon Faughn, Deanna Becker, Dominic Uy, Vicki Healer, Amorce Lima, Suzane Silbert |

| | | | |
|---|---|---|---|
| | | Development Lab | |
| EPI_ISL_2626687, EPI_ISL_2626688, EPI_ISL_2626689, EPI_ISL_2626690, EPI_ISL_2626691, EPI_ISL_2626692, EPI_ISL_2626693, EPI_ISL_2626694, EPI_ISL_2626695, EPI_ISL_2626696, EPI_ISL_2626697, EPI_ISL_2626698, EPI_ISL_2626699, EPI_ISL_2626700, EPI_ISL_2626701, EPI_ISL_2626702, EPI_ISL_2626703, EPI_ISL_2626704, EPI_ISL_2626705, EPI_ISL_2626707, EPI_ISL_2626708, EPI_ISL_2626709, EPI_ISL_2626710, EPI_ISL_2626711, EPI_ISL_2626712, EPI_ISL_2626713, EPI_ISL_2626714, EPI_ISL_2626715, EPI_ISL_2626716, EPI_ISL_2626717, EPI_ISL_2626718, EPI_ISL_2626719, EPI_ISL_2626720, EPI_ISL_2626721, EPI_ISL_2626722, EPI_ISL_2626723, EPI_ISL_2626724, EPI_ISL_2626725, EPI_ISL_2626726, EPI_ISL_2626727, EPI_ISL_2626728, EPI_ISL_2626729, EPI_ISL_2626730, EPI_ISL_2626731, EPI_ISL_2626732, EPI_ISL_2626733, EPI_ISL_2626734, EPI_ISL_2626735, EPI_ISL_2626736, EPI_ISL_2626737, EPI_ISL_2626738, EPI_ISL_2626739, EPI_ISL_2626740, EPI_ISL_2626741, EPI_ISL_2626742, EPI_ISL_2626743, EPI_ISL_2626744, EPI_ISL_2626745, EPI_ISL_2626746, EPI_ISL_2626747, EPI_ISL_2626748, EPI_ISL_2626749, EPI_ISL_2626750, EPI_ISL_2626751, EPI_ISL_2626752, EPI_ISL_2626754, EPI_ISL_2626755 | | | |
| see above | Alaska State Virology Laboratory | Alaska State Virology Laboratory | Stephanie DeRonde, Elva House, Jacob Zidek, Lisa Smith, Ph.D., Jack Chen, Ph.D. |
| EPI_ISL_2626756, EPI_ISL_2626757, EPI_ISL_2626758, EPI_ISL_2626759, EPI_ISL_2626760, EPI_ISL_2626761, EPI_ISL_2626762, EPI_ISL_2626763 | University of Pittsburgh | Genomics and Discovery, Respiratory Viruses Branch, Division of Viral Diseases, Centers for Disease Control and Prevention | Yan Li, Jing Zhang, Anna Kelleher, Ying Tao, Brian Lynch, Krista Queen, Anna Uehara, Peter Cook, Han Jia Justin Ng, Rachel Marine, Clinton R. Paden, Dhwani Batra, Haibin Wang, Tara Coalter, Jasmine Padilla, Morgan Davis, Mili Sheth, Sarah Nobles, Mark Burroughs, Justin Lee, Adam Retchless, Suxiang Tong |
| EPI_ISL_2626770, EPI_ISL_2626771, EPI_ISL_2626772, EPI_ISL_2626773, EPI_ISL_2626774, EPI_ISL_2626775, EPI_ISL_2626776, EPI_ISL_2626777, EPI_ISL_2626778, EPI_ISL_2626779, EPI_ISL_2626780, EPI_ISL_2626781, EPI_ISL_2626782, EPI_ISL_2626783, EPI_ISL_2626784, EPI_ISL_2626785, EPI_ISL_2626786, EPI_ISL_2626787, EPI_ISL_2626788, EPI_ISL_2626789, EPI_ISL_2626790, EPI_ISL_2626791, EPI_ISL_2626792, EPI_ISL_2626793, EPI_ISL_2626794, EPI_ISL_2626795, EPI_ISL_2626796, EPI_ISL_2626797, EPI_ISL_2626798, EPI_ISL_2626799, EPI_ISL_2626800, EPI_ISL_2626801, EPI_ISL_2626802, EPI_ISL_2626803, EPI_ISL_2626804, EPI_ISL_2626805, EPI_ISL_2626806, EPI_ISL_2626807, EPI_ISL_2626808, EPI_ISL_2626809, EPI_ISL_2626810, EPI_ISL_2626811, EPI_ISL_2626812, EPI_ISL_2626813, EPI_ISL_2626814, EPI_ISL_2626815, EPI_ISL_2626816, EPI_ISL_2626817, EPI_ISL_2626818, EPI_ISL_2626819, EPI_ISL_2626820, EPI_ISL_2626821, EPI_ISL_2626822, EPI_ISL_2626823, EPI_ISL_2626824, EPI_ISL_2626825, EPI_ISL_2626826, EPI_ISL_2626827, EPI_ISL_2626828, EPI_ISL_2626829, EPI_ISL_2626831, EPI_ISL_2626832, EPI_ISL_2626833, EPI_ISL_2626834, EPI_ISL_2626835, EPI_ISL_2626836, EPI_ISL_2626837, EPI_ISL_2626838 | | | |
| see above | Baylor Scott & White-Temple | Baylor Scott & White-Temple | Ari Rao, Linden Morales, Kimberly Walker, Marcus Volz, Shelby Johnson |
| EPI_ISL_2626923 | MONTEFIORE MEDICAL CENTER LABORATORIES | Wadsworth Center, New York State Department of Health | Kirsten St. George, Daryl M. Lamson, Alexis Russell, Matthew Shudt, Melissa A Leisner, Jonathan Plitnick, Catharine Prussing, Navjot Singh, John Kelly, Erasmus Schneider, Erica Lasek-Nesselquist |
| EPI_ISL_2626924 | WESTCHESTER MEDICAL CENTER | Wadsworth Center, New York State Department of Health | Kirsten St. George, Daryl M. Lamson, Alexis Russell, Matthew Shudt, Melissa A Leisner, Jonathan Plitnick, Catharine Prussing, Navjot Singh, John Kelly, Erasmus Schneider, Erica Lasek-Nesselquist |
| EPI_ISL_2626925, EPI_ISL_2626926, EPI_ISL_2626927, EPI_ISL_2626928, EPI_ISL_2626929, EPI_ISL_2626930, EPI_ISL_2626931, EPI_ISL_2626932, EPI_ISL_2626933, EPI_ISL_2626934, EPI_ISL_2626935, EPI_ISL_2626936, EPI_ISL_2626937, EPI_ISL_2626938, EPI_ISL_2626939, EPI_ISL_2626940, EPI_ISL_2626941, EPI_ISL_2626942, EPI_ISL_2626943, EPI_ISL_2626944, EPI_ISL_2626945, EPI_ISL_2626946, EPI_ISL_2626947, EPI_ISL_2626948, EPI_ISL_2626949, EPI_ISL_2626950, EPI_ISL_2626951, EPI_ISL_2626952, EPI_ISL_2626953, EPI_ISL_2626954, EPI_ISL_2626955, EPI_ISL_2626956, EPI_ISL_2626957, EPI_ISL_2626958, EPI_ISL_2626959, EPI_ISL_2626960, EPI_ISL_2626961, EPI_ISL_2626962, EPI_ISL_2626963, EPI_ISL_2626964, EPI_ISL_2626965, EPI_ISL_2626966, EPI_ISL_2626967, EPI_ISL_2626968, EPI_ISL_2626969, EPI_ISL_2626970, EPI_ISL_2626971 | | | |
| see above | TEMPUS LABS INC | Wadsworth Center, New York State Department of Health | Kirsten St. George, Daryl M. Lamson, Alexis Russell, Matthew Shudt, Melissa A Leisner, Jonathan Plitnick, Catharine Prussing, Navjot Singh, John Kelly, Erasmus Schneider, Erica Lasek-Nesselquist |
| EPI_ISL_2626972 | PECONIC BAY MEDICAL CENTER LABORATORY | Wadsworth Center, New York State Department of Health | Kirsten St. George, Daryl M. Lamson, Alexis Russell, Matthew Shudt, Melissa A Leisner, Jonathan Plitnick, Catharine Prussing, Navjot Singh, John Kelly, Erasmus Schneider, Erica Lasek-Nesselquist |
| EPI_ISL_2626973, EPI_ISL_2626974, EPI_ISL_2626975, EPI_ISL_2626976, EPI_ISL_2626977, EPI_ISL_2626978, EPI_ISL_2626979, EPI_ISL_2626980, EPI_ISL_2626981, EPI_ISL_2626982, EPI_ISL_2626983, EPI_ISL_2626984, EPI_ISL_2626985, EPI_ISL_2626986, EPI_ISL_2626987, EPI_ISL_2626988, EPI_ISL_2626989, EPI_ISL_2626990, EPI_ISL_2626991, EPI_ISL_2626992 | | | |
| see above | URMC LABS | Wadsworth Center, New York State Department of Health | Kirsten St. George, Daryl M. Lamson, Alexis Russell, Matthew Shudt, Melissa A Leisner, Jonathan Plitnick, Catharine Prussing, Navjot Singh, John Kelly, Erasmus Schneider, Erica Lasek-Nesselquist |
| EPI_ISL_2626993, EPI_ISL_2626994, EPI_ISL_2626995, EPI_ISL_2626996, EPI_ISL_2626997, EPI_ISL_2626998 | MONTEFIORE MEDICAL CENTER LABORATORIES | Wadsworth Center, New York State Department of Health | Kirsten St. George, Daryl M. Lamson, Alexis Russell, Matthew Shudt, Melissa A Leisner, Jonathan Plitnick, Catharine Prussing, Navjot Singh, John Kelly, Erasmus Schneider, Erica Lasek-Nesselquist |
| EPI_ISL_2626999, EPI_ISL_2627000, EPI_ISL_2627001, EPI_ISL_2627002 | ALBANY MEDICAL CENTER | Wadsworth Center, New York State Department of Health | Kirsten St. George, Daryl M. Lamson, Alexis Russell, Matthew Shudt, Melissa A Leisner, Jonathan Plitnick, Catharine Prussing, Navjot Singh, John Kelly, Erasmus Schneider, Erica Lasek-Nesselquist |
| EPI_ISL_2627003, EPI_ISL_2627004, EPI_ISL_2627005, EPI_ISL_2627006 | Saratoga Hospital Laboratory | Wadsworth Center, New York State Department of Health | Kirsten St. George, Daryl M. Lamson, Alexis Russell, Matthew Shudt, Melissa A Leisner, Jonathan Plitnick, Catharine Prussing, Navjot Singh, John Kelly, Erasmus Schneider, Erica Lasek-Nesselquist |
| EPI_ISL_2627007, EPI_ISL_2627008, EPI_ISL_2627009, EPI_ISL_2627010 | URMC LABS | Wadsworth Center, New York State Department of Health | Kirsten St. George, Daryl M. Lamson, Alexis Russell, Matthew Shudt, Melissa A Leisner, Jonathan Plitnick, Catharine Prussing, Navjot Singh, John Kelly, Erasmus Schneider, Erica Lasek-Nesselquist |
| EPI_ISL_2627011 | WESTCHESTER MEDICAL CENTER | Wadsworth Center, New York State Department of Health | Kirsten St. George, Daryl M. Lamson, Alexis Russell, Matthew Shudt, Melissa A Leisner, Jonathan Plitnick, Catharine Prussing, Navjot Singh, John Kelly, Erasmus Schneider, Erica Lasek-Nesselquist |
| EPI_ISL_2627012, EPI_ISL_2627013, EPI_ISL_2627014, EPI_ISL_2627015, EPI_ISL_2627016, EPI_ISL_2627017, EPI_ISL_2627018, EPI_ISL_2627019, EPI_ISL_2627020, EPI_ISL_2627021, EPI_ISL_2627022, EPI_ISL_2627023, EPI_ISL_2627024, EPI_ISL_2627025, EPI_ISL_2627026, EPI_ISL_2627027, EPI_ISL_2627028, EPI_ISL_2627029, EPI_ISL_2627030, EPI_ISL_2627031, EPI_ISL_2627032, EPI_ISL_2627033, EPI_ISL_2627034, EPI_ISL_2627035, EPI_ISL_2627036, EPI_ISL_2627037, EPI_ISL_2627038, EPI_ISL_2627039, EPI_ISL_2627040, EPI_ISL_2627041, EPI_ISL_2627042, EPI_ISL_2627043, EPI_ISL_2627044, EPI_ISL_2627045, EPI_ISL_2627046, EPI_ISL_2627047, EPI_ISL_2627048, EPI_ISL_2627049, EPI_ISL_2627050, EPI_ISL_2627051, EPI_ISL_2627052, EPI_ISL_2627053, EPI_ISL_2627054, EPI_ISL_2627055, EPI_ISL_2627056, EPI_ISL_2627057, EPI_ISL_2627058, EPI_ISL_2627059, EPI_ISL_2627060, EPI_ISL_2627061, EPI_ISL_2627062, EPI_ISL_2627063, EPI_ISL_2627064, EPI_ISL_2627065, EPI_ISL_2627066, EPI_ISL_2627067, EPI_ISL_2627068, EPI_ISL_2627069, EPI_ISL_2627070, EPI_ISL_2627071, EPI_ISL_2627072, EPI_ISL_2627073, EPI_ISL_2627074, EPI_ISL_2627075, EPI_ISL_2627076, EPI_ISL_2627077, EPI_ISL_2627078, EPI_ISL_2627079, EPI_ISL_2627080, EPI_ISL_2627081, EPI_ISL_2627082, EPI_ISL_2627083, EPI_ISL_2627084, EPI_ISL_2627085, EPI_ISL_2627086, EPI_ISL_2627087, EPI_ISL_2627088, EPI_ISL_2627089, EPI_ISL_2627090, EPI_ISL_2627091, EPI_ISL_2627092, EPI_ISL_2627093, EPI_ISL_2627094, EPI_ISL_2627095, EPI_ISL_2627096, EPI_ISL_2627097, EPI_ISL_2627098, EPI_ISL_2627099, EPI_ISL_2627100, EPI_ISL_2627101, EPI_ISL_2627102, EPI_ISL_2627103, EPI_ISL_2627104, EPI_ISL_2627105, EPI_ISL_2627106, EPI_ISL_2627107, EPI_ISL_2627108, EPI_ISL_2627109, EPI_ISL_2627110, EPI_ISL_2627111, EPI_ISL_2627112, EPI_ISL_2627113, EPI_ISL_2627114 | | | |
| see above | URMC LABS | Wadsworth Center, New York State Department of Health | Kirsten St. George, Daryl M. Lamson, Alexis Russell, Matthew Shudt, Melissa A Leisner, Jonathan Plitnick, Catharine Prussing, Navjot Singh, John Kelly, Erasmus Schneider, Erica Lasek-Nesselquist |
| EPI_ISL_2627115, EPI_ISL_2627116, EPI_ISL_2627117, EPI_ISL_2627118, EPI_ISL_2627119, EPI_ISL_2627120, EPI_ISL_2627121, EPI_ISL_2627122, EPI_ISL_2627123, EPI_ISL_2627124, EPI_ISL_2627125, EPI_ISL_2627126, EPI_ISL_2627127, EPI_ISL_2627128, EPI_ISL_2627129, EPI_ISL_2627130, EPI_ISL_2627131, EPI_ISL_2627132, EPI_ISL_2627133, EPI_ISL_2627134 | | | |
| see above | TEMPUS LABS INC | Wadsworth Center, New York State Department of Health | Kirsten St. George, Daryl M. Lamson, Alexis Russell, Matthew Shudt, Melissa A Leisner, Jonathan Plitnick, Catharine Prussing, Navjot Singh, John Kelly, Erasmus Schneider, Erica Lasek-Nesselquist |
| EPI_ISL_2627135, EPI_ISL_2627136, EPI_ISL_2627137 | BOSTON HEART DIAGNOSTICS CORP | Wadsworth Center, New York State Department of Health | Kirsten St. George, Daryl M. Lamson, Alexis Russell, Matthew Shudt, Melissa A Leisner, Jonathan Plitnick, Catharine Prussing, Navjot Singh, John Kelly, Erasmus Schneider, Erica Lasek-Nesselquist |
| EPI_ISL_2627138, EPI_ISL_2627139, EPI_ISL_2627140, EPI_ISL_2627141 | WESTCHESTER MEDICAL CENTER | Wadsworth Center, New York State Department of Health | Kirsten St. George, Daryl M. Lamson, Alexis Russell, Matthew Shudt, Melissa A Leisner, Jonathan Plitnick, Catharine Prussing, Navjot Singh, John Kelly, Erasmus Schneider, Erica Lasek-Nesselquist |
| EPI_ISL_2627142 | SBH Health System | Wadsworth Center, New York State Department of Health | Kirsten St. George, Daryl M. Lamson, Alexis Russell, Matthew Shudt, Melissa A Leisner, Jonathan Plitnick, Catharine Prussing, Navjot Singh, John Kelly, Erasmus Schneider, Erica Lasek-Nesselquist |
| EPI_ISL_2627143, EPI_ISL_2627144, EPI_ISL_2627145, EPI_ISL_2627146, EPI_ISL_2627147 | MONTEFIORE MEDICAL CENTER LABORATORIES | Wadsworth Center, New York State Department of Health | Kirsten St. George, Daryl M. Lamson, Alexis Russell, Matthew Shudt, Melissa A Leisner, Jonathan Plitnick, Catharine Prussing, Navjot Singh, John Kelly, Erasmus Schneider, Erica Lasek-Nesselquist |
| EPI_ISL_2627148, EPI_ISL_2627149, EPI_ISL_2627150, EPI_ISL_2627151, EPI_ISL_2627152 | STONY BROOK UNIVERSITY HOSPITAL | Wadsworth Center, New York State Department of Health | Kirsten St. George, Daryl M. Lamson, Alexis Russell, Matthew Shudt, Melissa A Leisner, Jonathan Plitnick, Catharine Prussing, Navjot Singh, John Kelly, Erasmus Schneider, Erica Lasek-Nesselquist |
| EPI_ISL_2627153, EPI_ISL_2627154, EPI_ISL_2627155, EPI_ISL_2627156 | NORTHWELL HEALTH LABORATORIES | Wadsworth Center, New York State Department of Health | Kirsten St. George, Daryl M. Lamson, Alexis Russell, Matthew Shudt, Melissa A Leisner, Jonathan Plitnick, Catharine Prussing, Navjot Singh, John Kelly, Erasmus Schneider, Erica Lasek-Nesselquist |
| EPI_ISL_2627157, EPI_ISL_2627158, EPI_ISL_2627159, EPI_ISL_2627160, EPI_ISL_2627161, EPI_ISL_2627162, EPI_ISL_2627163 | ALBANY MEDICAL CENTER | Wadsworth Center, New York State Department of Health | Kirsten St. George, Daryl M. Lamson, Alexis Russell, Matthew Shudt, Melissa A Leisner, Jonathan Plitnick, Catharine Prussing, Navjot Singh, John Kelly, Erasmus Schneider, Erica Lasek-Nesselquist |
| EPI_ISL_2627164, EPI_ISL_2627165, EPI_ISL_2627166, EPI_ISL_2627167, EPI_ISL_2627168, EPI_ISL_2627169, EPI_ISL_2627170, EPI_ISL_2627171, EPI_ISL_2627172, EPI_ISL_2627173, EPI_ISL_2627174, EPI_ISL_2627175, EPI_ISL_2627176, EPI_ISL_2627177, EPI_ISL_2627178, EPI_ISL_2627179 | | | |
| see above | NORTHWELL HEALTH LABORATORIES | Wadsworth Center, New York State Department of Health | Kirsten St. George, Daryl M. Lamson, Alexis Russell, Matthew Shudt, Melissa A Leisner, Jonathan Plitnick, Catharine Prussing, Navjot Singh, John Kelly, Erasmus Schneider, Erica Lasek-Nesselquist |
| EPI_ISL_2627487 | NC State Laboratory of Public Health | Genomics and Discovery, Respiratory | Yan Li, Jing Zhang, Anna Kelleher, Ying Tao, Brian Lynch, Krista Queen, Anna Uehara, Peter Cook, Han Jia Justin Ng, Rachel Marine, Clinton R. Paden, Dhwani Batra, Haibin Wang, Tara Coalter, Jasmine Padilla, Morgan Davis, Mili Sheth, Sarah Nobles, Mark Burroughs, Justin Lee, Adam |

| | | | |
|---|---|---|---|
| | | Viruses Branch, Division of Viral Diseases, Centers for Disease Control and Prevention | Retchless, Suxiang Tong |
| EPI_ISL_2627488, EPI_ISL_2627489, EPI_ISL_2627490 | Kansas Health and Environmental Lab | Kansas Health and Environmental Lab | Katherine Wiggins, Mike Grose, Jonathan Barnell, Ben Olsen, and Phil Adam |
| EPI_ISL_2627765, EPI_ISL_2627766, EPI_ISL_2627767, EPI_ISL_2627768, EPI_ISL_2627769, EPI_ISL_2627770, EPI_ISL_2627771 | NCSLPH | NCSLPH | Chase K, Miller MC, Greene S, Glover W |
| EPI_ISL_2627907, EPI_ISL_2627908, EPI_ISL_2627910, EPI_ISL_2627911, EPI_ISL_2627912, EPI_ISL_2627913, EPI_ISL_2627914, EPI_ISL_2627915, EPI_ISL_2627916, EPI_ISL_2627917, EPI_ISL_2627918, EPI_ISL_2627919, EPI_ISL_2627920, EPI_ISL_2627921, EPI_ISL_2627922, EPI_ISL_2627925, EPI_ISL_2627926 | | | |
| see above | Kansas Health and Environmental Lab | Kansas Health and Environmental Lab | Mike Grose, Katherine Wiggins, Jonathan Barnell, Ben Olsen, and Phil Adam |
| EPI_ISL_2627929, EPI_ISL_2627930, EPI_ISL_2627931, EPI_ISL_2627932, EPI_ISL_2627933, EPI_ISL_2627934, EPI_ISL_2627935, EPI_ISL_2627936, EPI_ISL_2627937, EPI_ISL_2627938, EPI_ISL_2627939 | | | |
| see above | Wichita State University - Molecular Diagnostics Lab | Kansas Health and Environmental Lab | Mike Grose, Katherine Wiggins, Jonathan Barnell, Ben Olsen, and Phil Adam |
| EPI_ISL_2628006, EPI_ISL_2628007, EPI_ISL_2628008, EPI_ISL_2628009, EPI_ISL_2628010, EPI_ISL_2628011, EPI_ISL_2628012, EPI_ISL_2628013, EPI_ISL_2628014, EPI_ISL_2628015, EPI_ISL_2628016, EPI_ISL_2628017, EPI_ISL_2628018, EPI_ISL_2628019, EPI_ISL_2628020, EPI_ISL_2628021, EPI_ISL_2628022, EPI_ISL_2628023, EPI_ISL_2628024, EPI_ISL_2628025, EPI_ISL_2628026, EPI_ISL_2628027, EPI_ISL_2628028, EPI_ISL_2628029, EPI_ISL_2628030, EPI_ISL_2628031, EPI_ISL_2628032, EPI_ISL_2628033, EPI_ISL_2628034, EPI_ISL_2628035 | | | |
| see above | Montana Public Health Laboratory | Montana Public Health Laboratory | Joy Ritter, Michelle Mozer, Carrie Biskupiak, Deborah Gibson, Michael Dills |
| EPI_ISL_2628193 | Alaska State Virology Laboratory | Alaska State Virology Laboratory | Stephanie DeRonde, Elva House, Jacob Zidek, Lisa Smith, Ph.D., Jack Chen, Ph.D. |
| EPI_ISL_2628211, EPI_ISL_2628212, EPI_ISL_2628220, EPI_ISL_2628221, EPI_ISL_2628222, EPI_ISL_2628223, EPI_ISL_2628226, EPI_ISL_2628227, EPI_ISL_2628230, EPI_ISL_2628231, EPI_ISL_2628232, EPI_ISL_2628238, EPI_ISL_2628239, EPI_ISL_2628242, EPI_ISL_2628247, EPI_ISL_2628255, EPI_ISL_2628256, EPI_ISL_2628259, EPI_ISL_2628261, EPI_ISL_2628266 | | | |
| see above | Arizona State Public Health Laboratory | Arizona State Public Health Laboratory | Trung Huynh, Jessica Escobar, Katherine Fullerton, Nobuko Fukushima, Stacy White, Linda Getsinger, Victor Waddell |
| EPI_ISL_2628311, EPI_ISL_2628312, EPI_ISL_2628313, EPI_ISL_2628314, EPI_ISL_2628315, EPI_ISL_2628316, EPI_ISL_2628317, EPI_ISL_2628318, EPI_ISL_2628319, EPI_ISL_2628320, EPI_ISL_2628321, EPI_ISL_2628322, EPI_ISL_2628323, EPI_ISL_2628324, EPI_ISL_2628325, EPI_ISL_2628326, EPI_ISL_2628327, EPI_ISL_2628328, EPI_ISL_2628329, EPI_ISL_2628330, EPI_ISL_2628331, EPI_ISL_2628332, EPI_ISL_2628333, EPI_ISL_2628334, EPI_ISL_2628335, EPI_ISL_2628336, EPI_ISL_2628337, EPI_ISL_2628338, EPI_ISL_2628339, EPI_ISL_2628340, EPI_ISL_2628341, EPI_ISL_2628342, EPI_ISL_2628343, EPI_ISL_2628344, EPI_ISL_2628345, EPI_ISL_2628346, EPI_ISL_2628347, EPI_ISL_2628348, EPI_ISL_2628349, EPI_ISL_2628350, EPI_ISL_2628351, EPI_ISL_2628352, EPI_ISL_2628353, EPI_ISL_2628354, EPI_ISL_2628355 | | | |
| see above | San Diego County Public Health Laboratory | Andersen lab at Scripps Research | SEARCH Alliance San Diego with Ashleigh Murphy, Jovan Shephard, Brett Austin |
| EPI_ISL_2628356, EPI_ISL_2628357, EPI_ISL_2628359, EPI_ISL_2628360, EPI_ISL_2628361, EPI_ISL_2628362, EPI_ISL_2628363, EPI_ISL_2628364, EPI_ISL_2628365, EPI_ISL_2628366, EPI_ISL_2628367, EPI_ISL_2628368, EPI_ISL_2628369, EPI_ISL_2628370, EPI_ISL_2628371, EPI_ISL_2628372, EPI_ISL_2628373, EPI_ISL_2628374, EPI_ISL_2628375, EPI_ISL_2628376, EPI_ISL_2628377, EPI_ISL_2628378, EPI_ISL_2628379, EPI_ISL_2628380, EPI_ISL_2628381, EPI_ISL_2628382, EPI_ISL_2628383, EPI_ISL_2628384, EPI_ISL_2628385, EPI_ISL_2628386, EPI_ISL_2628389, EPI_ISL_2628390, EPI_ISL_2628391, EPI_ISL_2628392, EPI_ISL_2628393, EPI_ISL_2628394, EPI_ISL_2628395, EPI_ISL_2628397, EPI_ISL_2628398, EPI_ISL_2628399, EPI_ISL_2628400, EPI_ISL_2628401, EPI_ISL_2628402, EPI_ISL_2628403, EPI_ISL_2628404, EPI_ISL_2628405, EPI_ISL_2628406, EPI_ISL_2628407, EPI_ISL_2628408, EPI_ISL_2628409, EPI_ISL_2628410, EPI_ISL_2628411, EPI_ISL_2628412, EPI_ISL_2628413, EPI_ISL_2628414, EPI_ISL_2628415, EPI_ISL_2628416, EPI_ISL_2628417, EPI_ISL_2628418, EPI_ISL_2628419, EPI_ISL_2628420, EPI_ISL_2628421, EPI_ISL_2628422, EPI_ISL_2628423, EPI_ISL_2628424, EPI_ISL_2628425, EPI_ISL_2628426, EPI_ISL_2628427, EPI_ISL_2628428, EPI_ISL_2628429, EPI_ISL_2628430, EPI_ISL_2628431, EPI_ISL_2628432, EPI_ISL_2628433, EPI_ISL_2628434, EPI_ISL_2628435 | | | |
| see above | Sharp HealthCare Laboratory | Andersen lab at Scripps Research | SEARCH Alliance San Diego with Aaron Harding, Jacquelyn Berumen, Cathy Woerle, Liam McGinnis, Art Mendoza, Omid Bakhtar |
| EPI_ISL_2628436, EPI_ISL_2628437, EPI_ISL_2628438, EPI_ISL_2628440, EPI_ISL_2628441, EPI_ISL_2628442, EPI_ISL_2628443, EPI_ISL_2628444, EPI_ISL_2628445, EPI_ISL_2628446, EPI_ISL_2628448, EPI_ISL_2628449, EPI_ISL_2628450, EPI_ISL_2628451, EPI_ISL_2628452, EPI_ISL_2628453, EPI_ISL_2628454, EPI_ISL_2628455, EPI_ISL_2628456, EPI_ISL_2628458, EPI_ISL_2628459, EPI_ISL_2628460, EPI_ISL_2628461, EPI_ISL_2628462, EPI_ISL_2628463, EPI_ISL_2628464, EPI_ISL_2628465, EPI_ISL_2628466, EPI_ISL_2628467, EPI_ISL_2628468, EPI_ISL_2628469, EPI_ISL_2628470, EPI_ISL_2628471, EPI_ISL_2628472, EPI_ISL_2628474, EPI_ISL_2628475 | | | |
| see above | San Diego County Public Health Laboratory | Andersen lab at Scripps Research | SEARCH Alliance San Diego with Ashleigh Murphy, Jovan Shephard, Brett Austin |
| EPI_ISL_2628497, EPI_ISL_2628498, EPI_ISL_2628499, EPI_ISL_2628500, EPI_ISL_2628501, EPI_ISL_2628502, EPI_ISL_2628503, EPI_ISL_2628505, EPI_ISL_2628506 | Rady Children's Hospital - San Diego | Andersen lab at Scripps Research | SEARCH Alliance San Diego with Nanda Radamchar, David Dimmock, Linda Luo, Christina Clarke, Kathryn Bouic, Teresa Mueller, Denise Malicki |
| EPI_ISL_2628507, EPI_ISL_2628508, EPI_ISL_2628509, EPI_ISL_2628510, EPI_ISL_2628511 | San Diego County Public Health Laboratory | Andersen lab at Scripps Research | SEARCH Alliance San Diego with Ashleigh Murphy, Jovan Shephard, Brett Austin |
| EPI_ISL_2628512, EPI_ISL_2628513, EPI_ISL_2628514, EPI_ISL_2628515, EPI_ISL_2628516, EPI_ISL_2628517, EPI_ISL_2628518, EPI_ISL_2628519, EPI_ISL_2628520, EPI_ISL_2628521, EPI_ISL_2628522, EPI_ISL_2628523, EPI_ISL_2628524, EPI_ISL_2628525, EPI_ISL_2628526, EPI_ISL_2628527, EPI_ISL_2628528, EPI_ISL_2628529, EPI_ISL_2628530, EPI_ISL_2628531, EPI_ISL_2628532, EPI_ISL_2628533, EPI_ISL_2628534, EPI_ISL_2628535, EPI_ISL_2628536, EPI_ISL_2628537, EPI_ISL_2628538, EPI_ISL_2628539, EPI_ISL_2628540, EPI_ISL_2628541, EPI_ISL_2628542, EPI_ISL_2628543, EPI_ISL_2628544, EPI_ISL_2628545, EPI_ISL_2628546, EPI_ISL_2628547 | | | |
| see above | Sharp HealthCare Laboratory | Andersen lab at Scripps Research | SEARCH Alliance San Diego with Aaron Harding, Jacquelyn Berumen, Cathy Woerle, Liam McGinnis, Art Mendoza, Omid Bakhtar |
| EPI_ISL_2628548, EPI_ISL_2628549, EPI_ISL_2628550, EPI_ISL_2628551, EPI_ISL_2628552, EPI_ISL_2628553, EPI_ISL_2628554, EPI_ISL_2628555, EPI_ISL_2628556, EPI_ISL_2628557, EPI_ISL_2628558, EPI_ISL_2628559, EPI_ISL_2628560, EPI_ISL_2628561, EPI_ISL_2628562, EPI_ISL_2628563, EPI_ISL_2628564, EPI_ISL_2628565, EPI_ISL_2628566, EPI_ISL_2628567, EPI_ISL_2628568, EPI_ISL_2628569, EPI_ISL_2628570, EPI_ISL_2628571, EPI_ISL_2628572, EPI_ISL_2628573, EPI_ISL_2628574, EPI_ISL_2628575, EPI_ISL_2628576, EPI_ISL_2628577, EPI_ISL_2628578, EPI_ISL_2628579, EPI_ISL_2628580, EPI_ISL_2628581, EPI_ISL_2628582, EPI_ISL_2628583, EPI_ISL_2628584, EPI_ISL_2628585, EPI_ISL_2628586, EPI_ISL_2628587, EPI_ISL_2628588, EPI_ISL_2628589, EPI_ISL_2628590, EPI_ISL_2628591, EPI_ISL_2628592, EPI_ISL_2628593, EPI_ISL_2628594, EPI_ISL_2628595, EPI_ISL_2628596, EPI_ISL_2628597, EPI_ISL_2628598, EPI_ISL_2628599, EPI_ISL_2628600, EPI_ISL_2628601, EPI_ISL_2628602, EPI_ISL_2628603, EPI_ISL_2628604, EPI_ISL_2628606, EPI_ISL_2628608, EPI_ISL_2628609, EPI_ISL_2628610, EPI_ISL_2628611, EPI_ISL_2628612, EPI_ISL_2628613, EPI_ISL_2628614, EPI_ISL_2628615, EPI_ISL_2628618, EPI_ISL_2628619, EPI_ISL_2628623, EPI_ISL_2628627, EPI_ISL_2628630, EPI_ISL_2628634, EPI_ISL_2628639, EPI_ISL_2628643, EPI_ISL_2628647, EPI_ISL_2628650, EPI_ISL_2628654, EPI_ISL_2628658, EPI_ISL_2628661, EPI_ISL_2628665, EPI_ISL_2628669, EPI_ISL_2628673, EPI_ISL_2628677, EPI_ISL_2628684, EPI_ISL_2628689, EPI_ISL_2628692, EPI_ISL_2628695, EPI_ISL_2628699, EPI_ISL_2628703, EPI_ISL_2628707, EPI_ISL_2628711, EPI_ISL_2628715, EPI_ISL_2628719, EPI_ISL_2628723, EPI_ISL_2628731, EPI_ISL_2628739, EPI_ISL_2628742, EPI_ISL_2628745, EPI_ISL_2628760, EPI_ISL_2628768, EPI_ISL_2628771, EPI_ISL_2628786, EPI_ISL_2628790, EPI_ISL_2628793, EPI_ISL_2628804, EPI_ISL_2628812, EPI_ISL_2628815, EPI_ISL_2628819, EPI_ISL_2628833, EPI_ISL_2628837, EPI_ISL_2628844, EPI_ISL_2628855, EPI_ISL_2628859, EPI_ISL_2628870, EPI_ISL_2628901, EPI_ISL_2628904, EPI_ISL_2628908, EPI_ISL_2628911, EPI_ISL_2628918, EPI_ISL_2628922, EPI_ISL_2628926, EPI_ISL_2628929, EPI_ISL_2628933, EPI_ISL_2628937, EPI_ISL_2628940, EPI_ISL_2628944, EPI_ISL_2628948, EPI_ISL_2628951, EPI_ISL_2628955 | | | |
| see above | San Diego County Public Health Laboratory | Andersen lab at Scripps Research | SEARCH Alliance San Diego with Ashleigh Murphy, Jovan Shephard, Brett Austin |
| EPI_ISL_2629187, EPI_ISL_2629188, EPI_ISL_2629189, EPI_ISL_2629190, EPI_ISL_2629191, EPI_ISL_2629192, EPI_ISL_2629193, EPI_ISL_2629194, EPI_ISL_2629195, EPI_ISL_2629196, EPI_ISL_2629197, EPI_ISL_2629198, EPI_ISL_2629199, EPI_ISL_2629200, EPI_ISL_2629201, EPI_ISL_2629202, EPI_ISL_2629203, EPI_ISL_2629204, EPI_ISL_2629205, EPI_ISL_2629206, EPI_ISL_2629207, EPI_ISL_2629208, EPI_ISL_2629209, EPI_ISL_2629210, EPI_ISL_2629211, EPI_ISL_2629212, EPI_ISL_2629213, EPI_ISL_2629214, EPI_ISL_2629215, EPI_ISL_2629216, EPI_ISL_2629217, EPI_ISL_2629218, EPI_ISL_2629219, EPI_ISL_2629220, EPI_ISL_2629221, EPI_ISL_2629222 | | | |
| see above | Microbiology Division, South Carolina Department of Health and Environmental Control | Microbiology Division, South Carolina Department of Health and Environmental Control | Freeman,J.S., Flores,H.V. |
| EPI_ISL_2629584, EPI_ISL_2629585, EPI_ISL_2629586, EPI_ISL_2629587, EPI_ISL_2629588, EPI_ISL_2629589, EPI_ISL_2629590, EPI_ISL_2629591, EPI_ISL_2629592, EPI_ISL_2629593, EPI_ISL_2629594 | | | |
| see above | GA Department of Public Health | GA Department of Public Health | Stacy Reeves, Jonathan Edwards, Cynthia Dixey, Tonia Parrott, Aliyah Fields, Taylor Smith |
| EPI_ISL_2629861, EPI_ISL_2629867, EPI_ISL_2629875, EPI_ISL_2629877, EPI_ISL_2629878, EPI_ISL_2629880, EPI_ISL_2629881, EPI_ISL_2629882, EPI_ISL_2629883, EPI_ISL_2629884, EPI_ISL_2629885, EPI_ISL_2629886, EPI_ISL_2629888, EPI_ISL_2629889, EPI_ISL_2629890, EPI_ISL_2629891, EPI_ISL_2629892, EPI_ISL_2629893, EPI_ISL_2629894, EPI_ISL_2629895, EPI_ISL_2629897, EPI_ISL_2629899, EPI_ISL_2629900, EPI_ISL_2629902, EPI_ISL_2629904, EPI_ISL_2629905, EPI_ISL_2629908, EPI_ISL_2629909, EPI_ISL_2629910, EPI_ISL_2629912, EPI_ISL_2629913, EPI_ISL_2629915, EPI_ISL_2629916, EPI_ISL_2629917, EPI_ISL_2629918, EPI_ISL_2629919, EPI_ISL_2629920, EPI_ISL_2629921, EPI_ISL_2629923, EPI_ISL_2629924, EPI_ISL_2629925, EPI_ISL_2629926, EPI_ISL_2629928, EPI_ISL_2629929, EPI_ISL_2629930, EPI_ISL_2629931, EPI_ISL_2629932, EPI_ISL_2629933, EPI_ISL_2629934, EPI_ISL_2629935, EPI_ISL_2629936, EPI_ISL_2629937, EPI_ISL_2629938, EPI_ISL_2629939, EPI_ISL_2629940, EPI_ISL_2629941 | | | |
| see above | Wyoming Public Health Laboratory | Wyoming Public Health Laboratory | Jim Mildenberger, Taylor Fearing, Channing Weber, Ashley Norberg, Chayse Rowley, Marley Goetz, Brian Dominguez, Elliot Thomasson, Sam Britz, Cari Sloma, Robert Petit, and Rob Christensen |
| EPI_ISL_2631151, EPI_ISL_2631152, EPI_ISL_2631153, EPI_ISL_2631154, EPI_ISL_2631155, EPI_ISL_2631156, EPI_ISL_2631157, EPI_ISL_2631158, EPI_ISL_2631159, EPI_ISL_2631160, EPI_ISL_2631161, EPI_ISL_2631162, EPI_ISL_2631163, EPI_ISL_2631164, EPI_ISL_2631165, EPI_ISL_2631166, EPI_ISL_2631167, EPI_ISL_2631168, EPI_ISL_2631169, EPI_ISL_2631170, EPI_ISL_2631171, EPI_ISL_2631172, EPI_ISL_2631173, EPI_ISL_2631174, EPI_ISL_2631175, EPI_ISL_2631176, EPI_ISL_2631177, EPI_ISL_2631178, EPI_ISL_2631179, EPI_ISL_2631180, EPI_ISL_2631181, EPI_ISL_2631182, EPI_ISL_2631183, EPI_ISL_2631184, EPI_ISL_2631185, EPI_ISL_2631186, EPI_ISL_2631187 | | | |
| see above | Oregon State Public Health Laboratory | Oregon State Public Health Laboratory | Rafia Razzaque, Eugene Yeboah, Vanda Makris, Laura Tsaknaridis, John Fontana and Shane Sevey |
| EPI_ISL_2631192, EPI_ISL_2631193, EPI_ISL_2631194, EPI_ISL_2631197, EPI_ISL_2631198, EPI_ISL_2631201, EPI_ISL_2631202, EPI_ISL_2631203, EPI_ISL_2631204, EPI_ISL_2631205, EPI_ISL_2631206, EPI_ISL_2631207 | | | |
| see above | Santa Clara County Public Health Laboratory | Santa Clara County Public Health Laboratory | Santa Clara County Public Health Department |
| EPI_ISL_2631208, EPI_ISL_2631209, EPI_ISL_2631211, EPI_ISL_2631213, EPI_ISL_2631214, EPI_ISL_2631216, EPI_ISL_2631218 | Santa Clara Valley Medical Center | Santa Clara County Public Health Laboratory | Santa Clara County Public Health Department |
| EPI_ISL_2631221, EPI_ISL_2631222, EPI_ISL_2631223, EPI_ISL_2631226, EPI_ISL_2631229 | Worobey Lab, Department of Ecology and Evolutionary Biology, University of Arizona | Worobey Lab, Department of Ecology and Evolutionary Biology, University of Arizona | Grace Quirk, Brendan Larsen, Thomas Watts, David Baltrus, Michael Worobey |
| EPI_ISL_2631338, EPI_ISL_2631339, EPI_ISL_2631340, EPI_ISL_2631342, EPI_ISL_2631343, EPI_ISL_2631344, EPI_ISL_2631345, EPI_ISL_2631347, EPI_ISL_2631348, EPI_ISL_2631349, EPI_ISL_2631350, EPI_ISL_2631351, EPI_ISL_2631352, EPI_ISL_2631354, EPI_ISL_2631355, EPI_ISL_2631356, EPI_ISL_2631357, EPI_ISL_2631358, EPI_ISL_2631359, EPI_ISL_2631360, EPI_ISL_2631361, EPI_ISL_2631362, EPI_ISL_2631363, EPI_ISL_2631364, EPI_ISL_2631365, EPI_ISL_2631366, EPI_ISL_2631367, EPI_ISL_2631368, EPI_ISL_2631369, EPI_ISL_2631370, EPI_ISL_2631371, EPI_ISL_2631372, EPI_ISL_2631373, EPI_ISL_2631374, EPI_ISL_2631375, EPI_ISL_2631376, EPI_ISL_2631377, EPI_ISL_2631379, EPI_ISL_2631380, EPI_ISL_2631381, EPI_ISL_2631382, EPI_ISL_2631383, EPI_ISL_2631384, EPI_ISL_2631386, | | | |

| | | | |
|---|---|---|---|
| EPI_ISL_2631387, EPI_ISL_2631388, EPI_ISL_2631389, EPI_ISL_2631390, EPI_ISL_2631392, EPI_ISL_2631394, EPI_ISL_2631395, EPI_ISL_2631396, EPI_ISL_2631397, EPI_ISL_2631398, EPI_ISL_2631399, EPI_ISL_2631400, EPI_ISL_2631401, EPI_ISL_2631402, EPI_ISL_2631403, EPI_ISL_2631404, EPI_ISL_2631405, EPI_ISL_2631406, EPI_ISL_2631407, EPI_ISL_2632880, EPI_ISL_2632884, EPI_ISL_2632887, EPI_ISL_2632890, EPI_ISL_2632893, EPI_ISL_2632897, EPI_ISL_2632900, EPI_ISL_2632904, EPI_ISL_2632907, EPI_ISL_2632911, EPI_ISL_2632915, EPI_ISL_2632918, EPI_ISL_2632922, EPI_ISL_2632925, EPI_ISL_2632929, EPI_ISL_2632933, EPI_ISL_2632936, EPI_ISL_2632940, EPI_ISL_2632944, EPI_ISL_2632949, EPI_ISL_2632957, EPI_ISL_2632961, EPI_ISL_2632965, EPI_ISL_2632969, EPI_ISL_2632973, EPI_ISL_2632976, EPI_ISL_2632979, EPI_ISL_2632983, EPI_ISL_2632986, EPI_ISL_2632990, EPI_ISL_2632994, EPI_ISL_2632998, EPI_ISL_2633001, EPI_ISL_2633005, EPI_ISL_2633009, EPI_ISL_2633013, EPI_ISL_2633017, EPI_ISL_2633021, EPI_ISL_2633025, EPI_ISL_2633028, EPI_ISL_2633031, EPI_ISL_2633039, EPI_ISL_2633043, EPI_ISL_2633048, EPI_ISL_2633051, EPI_ISL_2633056, EPI_ISL_2633060, EPI_ISL_2633064, EPI_ISL_2633067, EPI_ISL_2633072, EPI_ISL_2633076, EPI_ISL_2633079, EPI_ISL_2633081, EPI_ISL_2633085, EPI_ISL_2633089, EPI_ISL_2633093, EPI_ISL_2633097, EPI_ISL_2633101, EPI_ISL_2633107, EPI_ISL_2633109, EPI_ISL_2633112, EPI_ISL_2633119, EPI_ISL_2633122, EPI_ISL_2633125, EPI_ISL_2633129, EPI_ISL_2633133, EPI_ISL_2633135, EPI_ISL_2633136, EPI_ISL_2633137, EPI_ISL_2633138, EPI_ISL_2633140, EPI_ISL_2633143, EPI_ISL_2633147, EPI_ISL_2633151, EPI_ISL_2633154, EPI_ISL_2633158, EPI_ISL_2633162, EPI_ISL_2633165, EPI_ISL_2633169, EPI_ISL_2633176, EPI_ISL_2633183, EPI_ISL_2633186, EPI_ISL_2633189, EPI_ISL_2633193, EPI_ISL_2633196, EPI_ISL_2633200, EPI_ISL_2633207, EPI_ISL_2633210, EPI_ISL_2633214, EPI_ISL_2633217, EPI_ISL_2633221, EPI_ISL_2633224, EPI_ISL_2633227, EPI_ISL_2633230, EPI_ISL_2633234, EPI_ISL_2633237, EPI_ISL_2633240, EPI_ISL_2633244, EPI_ISL_2633248, EPI_ISL_2633251, EPI_ISL_2633254, EPI_ISL_2633258, EPI_ISL_2633260, EPI_ISL_2633262, EPI_ISL_2633263, EPI_ISL_2633265, EPI_ISL_2633267, EPI_ISL_2633269, EPI_ISL_2633271, EPI_ISL_2633274, EPI_ISL_2633278, EPI_ISL_2633280, EPI_ISL_2633282, EPI_ISL_2633284, EPI_ISL_2633285, EPI_ISL_2633287, EPI_ISL_2633289, EPI_ISL_2633291, EPI_ISL_2633295, EPI_ISL_2633297, EPI_ISL_2633300, EPI_ISL_2633305, EPI_ISL_2633309, EPI_ISL_2633315, EPI_ISL_2633320, EPI_ISL_2633326, EPI_ISL_2633332, EPI_ISL_2633338, EPI_ISL_2633347, EPI_ISL_2633351, EPI_ISL_2633354, EPI_ISL_2633358, EPI_ISL_2633361, EPI_ISL_2633365, EPI_ISL_2633368, EPI_ISL_2633370, EPI_ISL_2633372, EPI_ISL_2633378, EPI_ISL_2633382, EPI_ISL_2633385, EPI_ISL_2633389, EPI_ISL_2633393, EPI_ISL_2633396, EPI_ISL_2633400, EPI_ISL_2633404, EPI_ISL_2633405, EPI_ISL_2633411, EPI_ISL_2633414, EPI_ISL_2633418, EPI_ISL_2633422, EPI_ISL_2633426, EPI_ISL_2633430, EPI_ISL_2633438, EPI_ISL_2633442, EPI_ISL_2633447, EPI_ISL_2633448, EPI_ISL_2633449, EPI_ISL_2633450, EPI_ISL_2633451, EPI_ISL_2633454, EPI_ISL_2633458, EPI_ISL_2633459, EPI_ISL_2633463, EPI_ISL_2633467, EPI_ISL_2633471, EPI_ISL_2633472, EPI_ISL_2633474, EPI_ISL_2633476, EPI_ISL_2633480, EPI_ISL_2633484, EPI_ISL_2633486, EPI_ISL_2633488, EPI_ISL_2633490, EPI_ISL_2633497, EPI_ISL_2633501, EPI_ISL_2633505, EPI_ISL_2633508, EPI_ISL_2633513, EPI_ISL_2633517, EPI_ISL_2633522, EPI_ISL_2633530, EPI_ISL_2633534, EPI_ISL_2633538, EPI_ISL_2633541, EPI_ISL_2633546, EPI_ISL_2633548, EPI_ISL_2633550, EPI_ISL_2633553, EPI_ISL_2633555, EPI_ISL_2633556, EPI_ISL_2633560, EPI_ISL_2633564, EPI_ISL_2633567, EPI_ISL_2633571, EPI_ISL_2633575, EPI_ISL_2633579, EPI_ISL_2633582, EPI_ISL_2633587, EPI_ISL_2633589, EPI_ISL_2633593, EPI_ISL_2633597, EPI_ISL_2633600, EPI_ISL_2633604, EPI_ISL_2633609, EPI_ISL_2633611, EPI_ISL_2633615, EPI_ISL_2633619, EPI_ISL_2633622, EPI_ISL_2633626, EPI_ISL_2633628, EPI_ISL_2633629, EPI_ISL_2633631, EPI_ISL_2633633, EPI_ISL_2633634, EPI_ISL_2633636, EPI_ISL_2633638, EPI_ISL_2633640, EPI_ISL_2633642, EPI_ISL_2633644, EPI_ISL_2633645, EPI_ISL_2633647, EPI_ISL_2633649, EPI_ISL_2633651, EPI_ISL_2633653, EPI_ISL_2633654, EPI_ISL_2633656, EPI_ISL_2633657, EPI_ISL_2633659, EPI_ISL_2633663, EPI_ISL_2633666, EPI_ISL_2633669, EPI_ISL_2633671, EPI_ISL_2633673, EPI_ISL_2633675, EPI_ISL_2633677, EPI_ISL_2633678, EPI_ISL_2633682, EPI_ISL_2633684, EPI_ISL_2633685, EPI_ISL_2633689, EPI_ISL_2633691, EPI_ISL_2633694, EPI_ISL_2633697, EPI_ISL_2633707, EPI_ISL_2633714, EPI_ISL_2633717, EPI_ISL_2633721, EPI_ISL_2633727, EPI_ISL_2633729, EPI_ISL_2633730, EPI_ISL_2633732, EPI_ISL_2633736, EPI_ISL_2633740, EPI_ISL_2633745, EPI_ISL_2633746, EPI_ISL_2633748, EPI_ISL_2633750, EPI_ISL_2633752, EPI_ISL_2633753, EPI_ISL_2633755, EPI_ISL_2633757, EPI_ISL_2633759, EPI_ISL_2633761, EPI_ISL_2633763, EPI_ISL_2633764, EPI_ISL_2633766, EPI_ISL_2633768, EPI_ISL_2633770, EPI_ISL_2633774, EPI_ISL_2633778, EPI_ISL_2633781, EPI_ISL_2633783, EPI_ISL_2633785, EPI_ISL_2633788, EPI_ISL_2633792, EPI_ISL_2633796, EPI_ISL_2633799, EPI_ISL_2633803, EPI_ISL_2633807, EPI_ISL_2633810, EPI_ISL_2633814, EPI_ISL_2633820, EPI_ISL_2633826, EPI_ISL_2633833, EPI_ISL_2633837, EPI_ISL_2633840, EPI_ISL_2633842, EPI_ISL_2633844, EPI_ISL_2633851, EPI_ISL_2633855, EPI_ISL_2633858, EPI_ISL_2633861, EPI_ISL_2633865, EPI_ISL_2633869, EPI_ISL_2633872, EPI_ISL_2633874, EPI_ISL_2633875, EPI_ISL_2633879, EPI_ISL_2633883, EPI_ISL_2633887, EPI_ISL_2633890, EPI_ISL_2633894, EPI_ISL_2633902, EPI_ISL_2633905, EPI_ISL_2633907, EPI_ISL_2633909, EPI_ISL_2633912, EPI_ISL_2633917, EPI_ISL_2633919, EPI_ISL_2633921, EPI_ISL_2633922, EPI_ISL_2633924, EPI_ISL_2633931, EPI_ISL_2633935, EPI_ISL_2633939, EPI_ISL_2633940, EPI_ISL_2633942, EPI_ISL_2633944, EPI_ISL_2633949, EPI_ISL_2633950, EPI_ISL_2633952, EPI_ISL_2633957, EPI_ISL_2633960, EPI_ISL_2633963, EPI_ISL_2633967, EPI_ISL_2633970, EPI_ISL_2633974, EPI_ISL_2633976, EPI_ISL_2633978, EPI_ISL_2633979, EPI_ISL_2633981, EPI_ISL_2633983, EPI_ISL_2633987, EPI_ISL_2633989, EPI_ISL_2633990, EPI_ISL_2633992, EPI_ISL_2633994, EPI_ISL_2633996, EPI_ISL_2633997, EPI_ISL_2633999, EPI_ISL_2634003, EPI_ISL_2634010, EPI_ISL_2634014, EPI_ISL_2634017, EPI_ISL_2634021, EPI_ISL_2634028, EPI_ISL_2634032, EPI_ISL_2634036, EPI_ISL_2634040, EPI_ISL_2634043, EPI_ISL_2634047, EPI_ISL_2634050, EPI_ISL_2634054, EPI_ISL_2634057, EPI_ISL_2634061, EPI_ISL_2634065, EPI_ISL_2634068, EPI_ISL_2634071, EPI_ISL_2634074, EPI_ISL_2634076, EPI_ISL_2634080, EPI_ISL_2634084, EPI_ISL_2634088, EPI_ISL_2634090, EPI_ISL_2634091, EPI_ISL_2634093, EPI_ISL_2634095, EPI_ISL_2634099, EPI_ISL_2634103, EPI_ISL_2634106, EPI_ISL_2634109, EPI_ISL_2634112, EPI_ISL_2634116, EPI_ISL_2634118, EPI_ISL_2634120, EPI_ISL_2634122, EPI_ISL_2634123, EPI_ISL_2634125, EPI_ISL_2634127, EPI_ISL_2634129, EPI_ISL_2634130, EPI_ISL_2634132, EPI_ISL_2634134, EPI_ISL_2634136, EPI_ISL_2634137, EPI_ISL_2634141, EPI_ISL_2634143, EPI_ISL_2634145, EPI_ISL_2634148, EPI_ISL_2634150, EPI_ISL_2634152, EPI_ISL_2634154, EPI_ISL_2634156, EPI_ISL_2634158, EPI_ISL_2634160, EPI_ISL_2634163, EPI_ISL_2634165, EPI_ISL_2634166, EPI_ISL_2634168, EPI_ISL_2634171, EPI_ISL_2634173, EPI_ISL_2634175, EPI_ISL_2634178, EPI_ISL_2634180, EPI_ISL_2634181, EPI_ISL_2634183, EPI_ISL_2634185, EPI_ISL_2634187, EPI_ISL_2634189, EPI_ISL_2634193, EPI_ISL_2634194, EPI_ISL_2634196, EPI_ISL_2634198, EPI_ISL_2634201, EPI_ISL_2634203, EPI_ISL_2634205, EPI_ISL_2634206, EPI_ISL_2634208, EPI_ISL_2634210, EPI_ISL_2634211, EPI_ISL_2634213, EPI_ISL_2634214, EPI_ISL_2634216, EPI_ISL_2634218, EPI_ISL_2634219, EPI_ISL_2634221, EPI_ISL_2634223, EPI_ISL_2634224, EPI_ISL_2634226, EPI_ISL_2634228, EPI_ISL_2634229, EPI_ISL_2634231, EPI_ISL_2634235, EPI_ISL_2634236, EPI_ISL_2634238, EPI_ISL_2634240, EPI_ISL_2634242, EPI_ISL_2634243, EPI_ISL_2634245, EPI_ISL_2634247, EPI_ISL_2634248, EPI_ISL_2634250, EPI_ISL_2634251, EPI_ISL_2634252, EPI_ISL_2634254, EPI_ISL_2634256, EPI_ISL_2634258, EPI_ISL_2634260, EPI_ISL_2634261, EPI_ISL_2634263, EPI_ISL_2634265, EPI_ISL_2634267, EPI_ISL_2634268, EPI_ISL_2634270, EPI_ISL_2634272, EPI_ISL_2634273, EPI_ISL_2634275, EPI_ISL_2634276, EPI_ISL_2634277, EPI_ISL_2634279, EPI_ISL_2634280, EPI_ISL_2634282, EPI_ISL_2634284, EPI_ISL_2634286, EPI_ISL_2634288, EPI_ISL_2634289, EPI_ISL_2634291, EPI_ISL_2634293, EPI_ISL_2634295, EPI_ISL_2634296, EPI_ISL_2634300, EPI_ISL_2634302, EPI_ISL_2634304, EPI_ISL_2634305, EPI_ISL_2634307, EPI_ISL_2634309, EPI_ISL_2634311, EPI_ISL_2634313, EPI_ISL_2634314, EPI_ISL_2634316, EPI_ISL_2634320, EPI_ISL_2634322, EPI_ISL_2634324, EPI_ISL_2634325, EPI_ISL_2634327, EPI_ISL_2634329, EPI_ISL_2634331, EPI_ISL_2634334, EPI_ISL_2634336, EPI_ISL_2634338, EPI_ISL_2634340, EPI_ISL_2634342, EPI_ISL_2634344, EPI_ISL_2634346, EPI_ISL_2634347, EPI_ISL_2634349, EPI_ISL_2634351, EPI_ISL_2634353, EPI_ISL_2634355, EPI_ISL_2634356, EPI_ISL_2634358, EPI_ISL_2634360, EPI_ISL_2634362, EPI_ISL_2634364, EPI_ISL_2634365, EPI_ISL_2634367, EPI_ISL_2634369, EPI_ISL_2634370, EPI_ISL_2634372, EPI_ISL_2634374, EPI_ISL_2634375, EPI_ISL_2634377, EPI_ISL_2634379, EPI_ISL_2634381, EPI_ISL_2634382, EPI_ISL_2634384, EPI_ISL_2634386, EPI_ISL_2634388, EPI_ISL_2634389, EPI_ISL_2634391, EPI_ISL_2634393, EPI_ISL_2634394, EPI_ISL_2634396, EPI_ISL_2634398, EPI_ISL_2634400, EPI_ISL_2634402, EPI_ISL_2634405, EPI_ISL_2634407, EPI_ISL_2634412, EPI_ISL_2634416, EPI_ISL_2634420, EPI_ISL_2634422, EPI_ISL_2634425, EPI_ISL_2634429, EPI_ISL_2634431, EPI_ISL_2634433, EPI_ISL_2634434, EPI_ISL_2634436, EPI_ISL_2634437, EPI_ISL_2634438, EPI_ISL_2634440, EPI_ISL_2634445, EPI_ISL_2634447, EPI_ISL_2634449, EPI_ISL_2634451, EPI_ISL_2634453, EPI_ISL_2634456, EPI_ISL_2634458, EPI_ISL_2634460, EPI_ISL_2634462, EPI_ISL_2634464, EPI_ISL_2634465, EPI_ISL_2634467, EPI_ISL_2634469, EPI_ISL_2634471, EPI_ISL_2634473, EPI_ISL_2634475, EPI_ISL_2634476, EPI_ISL_2634478, EPI_ISL_2634482, EPI_ISL_2634484, EPI_ISL_2634485, EPI_ISL_2634487, EPI_ISL_2634489, EPI_ISL_2634491, EPI_ISL_2634493, EPI_ISL_2634494, EPI_ISL_2634496, EPI_ISL_2634498, EPI_ISL_2634502, EPI_ISL_2634505, EPI_ISL_2634507, EPI_ISL_2634509, EPI_ISL_2634511, EPI_ISL_2634512, EPI_ISL_2634514, EPI_ISL_2634516, EPI_ISL_2634517, EPI_ISL_2634519, EPI_ISL_2634521, EPI_ISL_2634523, EPI_ISL_2634527, EPI_ISL_2634528, EPI_ISL_2634532, EPI_ISL_2634533, EPI_ISL_2634535, EPI_ISL_2634537, EPI_ISL_2634539, EPI_ISL_2634541, EPI_ISL_2634542, EPI_ISL_2634544, EPI_ISL_2634546, EPI_ISL_2634547, EPI_ISL_2634548, EPI_ISL_2634549, EPI_ISL_2634550, EPI_ISL_2634552, EPI_ISL_2634554, EPI_ISL_2634555, EPI_ISL_2634557, EPI_ISL_2634559, EPI_ISL_2634560, EPI_ISL_2634562, EPI_ISL_2634564, EPI_ISL_2634566, EPI_ISL_2634568, EPI_ISL_2634570, EPI_ISL_2634571, EPI_ISL_2634573, EPI_ISL_2634575, EPI_ISL_2634576, EPI_ISL_2634578, EPI_ISL_2634580, EPI_ISL_2634582, EPI_ISL_2634584, EPI_ISL_2634586, EPI_ISL_2634587, EPI_ISL_2634591, EPI_ISL_2634593, EPI_ISL_2634595, EPI_ISL_2634598, EPI_ISL_2634600, EPI_ISL_2634602, EPI_ISL_2634604, EPI_ISL_2634606, EPI_ISL_2634609, EPI_ISL_2634611, EPI_ISL_2634613, EPI_ISL_2634615, EPI_ISL_2634616, EPI_ISL_2634618, EPI_ISL_2634620, EPI_ISL_2634622, EPI_ISL_2634624, EPI_ISL_2634626, EPI_ISL_2634627, EPI_ISL_2634629, EPI_ISL_2634631, EPI_ISL_2634633, EPI_ISL_2634635, EPI_ISL_2634636, EPI_ISL_2634638, EPI_ISL_2634640, EPI_ISL_2634642, EPI_ISL_2634644, EPI_ISL_2634646, EPI_ISL_2634647, EPI_ISL_2634649, EPI_ISL_2634653, EPI_ISL_2634655, EPI_ISL_2634657, EPI_ISL_2634662, EPI_ISL_2634663, EPI_ISL_2634665, EPI_ISL_2634666, EPI_ISL_2634670, EPI_ISL_2634672, EPI_ISL_2634673, EPI_ISL_2634675, EPI_ISL_2634679, EPI_ISL_2634681, EPI_ISL_2634682, EPI_ISL_2634684, EPI_ISL_2634688, EPI_ISL_2634691, EPI_ISL_2634692, EPI_ISL_2634694, EPI_ISL_2634696, EPI_ISL_2634698, EPI_ISL_2634699, EPI_ISL_2634701, EPI_ISL_2634703, EPI_ISL_2634705, EPI_ISL_2634706, EPI_ISL_2634708, EPI_ISL_2634712, EPI_ISL_2634714, EPI_ISL_2634716, EPI_ISL_2634718, EPI_ISL_2634720, EPI_ISL_2634721, EPI_ISL_2634724, EPI_ISL_2634725, EPI_ISL_2634727, EPI_ISL_2634729, EPI_ISL_2634731, EPI_ISL_2634733, EPI_ISL_2634735, EPI_ISL_2634737, EPI_ISL_2634741, EPI_ISL_2634745, EPI_ISL_2634746, EPI_ISL_2634748, EPI_ISL_2634749, EPI_ISL_2634752, EPI_ISL_2634753, EPI_ISL_2634754, EPI_ISL_2634755, EPI_ISL_2634757, EPI_ISL_2634759, EPI_ISL_2634763, EPI_ISL_2634764, EPI_ISL_2634766, EPI_ISL_2634767, EPI_ISL_2634768, EPI_ISL_2634769, EPI_ISL_2634771, EPI_ISL_2634773, EPI_ISL_2634774, EPI_ISL_2634776, EPI_ISL_2634780, EPI_ISL_2634782, EPI_ISL_2634783, EPI_ISL_2634785, EPI_ISL_2634786, EPI_ISL_2634787, EPI_ISL_2634788, EPI_ISL_2634790, EPI_ISL_2634792, EPI_ISL_2634794, EPI_ISL_2634796, EPI_ISL_2634798, EPI_ISL_2634801, EPI_ISL_2634803, EPI_ISL_2634805, EPI_ISL_2634807, EPI_ISL_2634808, EPI_ISL_2634810, EPI_ISL_2634814, EPI_ISL_2634816, EPI_ISL_2634817, EPI_ISL_2634820, EPI_ISL_2634822, EPI_ISL_2634825, EPI_ISL_2634827, EPI_ISL_2634829, EPI_ISL_2634831, EPI_ISL_2634834, EPI_ISL_2634836, EPI_ISL_2634837, EPI_ISL_2634839, EPI_ISL_2634841, EPI_ISL_2634843, EPI_ISL_2634845, EPI_ISL_2634847, EPI_ISL_2634849, EPI_ISL_2634850, EPI_ISL_2634852, EPI_ISL_2634854, EPI_ISL_2634856, EPI_ISL_2634857, EPI_ISL_2634859, EPI_ISL_2634861, EPI_ISL_2634863, EPI_ISL_2634867, EPI_ISL_2634868, EPI_ISL_2634870, EPI_ISL_2634872, EPI_ISL_2634874, EPI_ISL_2634875, EPI_ISL_2634877, EPI_ISL_2634879, EPI_ISL_2634881, EPI_ISL_2634885, EPI_ISL_2634887, EPI_ISL_2634890, EPI_ISL_2634892, EPI_ISL_2634894, EPI_ISL_2634896, EPI_ISL_2634897, EPI_ISL_2634899, EPI_ISL_2634900, EPI_ISL_2634902, EPI_ISL_2634903, EPI_ISL_2634904, EPI_ISL_2634907, EPI_ISL_2634909, EPI_ISL_2634911, EPI_ISL_2634913, EPI_ISL_2634915, EPI_ISL_2634916, EPI_ISL_2634918, EPI_ISL_2634920, EPI_ISL_2634922, EPI_ISL_2634923, EPI_ISL_2634925, EPI_ISL_2634927, EPI_ISL_2634929, EPI_ISL_2634931, EPI_ISL_2634933, EPI_ISL_2634934, EPI_ISL_2634936, EPI_ISL_2634938, EPI_ISL_2634940, EPI_ISL_2634942, EPI_ISL_2634943, EPI_ISL_2634945, EPI_ISL_2634947, EPI_ISL_2634949, EPI_ISL_2634951, EPI_ISL_2634952, EPI_ISL_2634954, EPI_ISL_2634956, EPI_ISL_2634959, EPI_ISL_2634961, EPI_ISL_2634963, EPI_ISL_2634965, EPI_ISL_2634967, EPI_ISL_2634969, EPI_ISL_2634971, EPI_ISL_2634973 | | | |
| see above | Pandemic Response Lab - NYC | Pandemic Response Lab, R&D | Henry Lee, Michael Hammerling, Melissa Hopkins, Cybill del Castillo, Shinyoung Clair Kang, William Ward, Pradeep Bugga, Sol Rey, Dylan Law, Katharine Nelson, Haiping Hao, Jon Laurent |
| EPI_ISL_2635037, EPI_ISL_2635039, EPI_ISL_2635041, EPI_ISL_2635043, EPI_ISL_2635045, EPI_ISL_2635047, EPI_ISL_2635048, EPI_ISL_2635050, EPI_ISL_2635052, EPI_ISL_2635054, EPI_ISL_2635055, EPI_ISL_2635059, EPI_ISL_2635061, EPI_ISL_2635063, EPI_ISL_2635065, EPI_ISL_2635066, EPI_ISL_2635068, EPI_ISL_2635070, EPI_ISL_2635072, EPI_ISL_2635073, EPI_ISL_2635077, EPI_ISL_2635079, EPI_ISL_2635080, EPI_ISL_2635082, EPI_ISL_2635083, EPI_ISL_2635085, EPI_ISL_2635086, EPI_ISL_2635088, EPI_ISL_2635090, EPI_ISL_2635091, EPI_ISL_2635093, EPI_ISL_2635095, EPI_ISL_2635097, EPI_ISL_2635098, EPI_ISL_2635100, EPI_ISL_2635102, EPI_ISL_2635104, EPI_ISL_2635105, EPI_ISL_2635107, EPI_ISL_2635109, EPI_ISL_2635111, EPI_ISL_2635115, EPI_ISL_2635116, EPI_ISL_2635118, EPI_ISL_2635120, EPI_ISL_2635122, EPI_ISL_2635124, EPI_ISL_2635126, EPI_ISL_2635127, EPI_ISL_2635130, EPI_ISL_2635131, EPI_ISL_2635133, EPI_ISL_2635135, EPI_ISL_2635137, EPI_ISL_2635138, EPI_ISL_2635140, EPI_ISL_2635142, EPI_ISL_2635144, EPI_ISL_2635145, EPI_ISL_2635147, EPI_ISL_2635149, EPI_ISL_2635151, EPI_ISL_2635153, EPI_ISL_2635155, EPI_ISL_2635156, EPI_ISL_2635158 | | | |
| see above | Missouri State Public Health Laboratory | Missouri State Public Health Laboratory | Matthew Sinn, Joshua Barry, Ashley New |
| EPI_ISL_2642148, EPI_ISL_2642544, EPI_ISL_2642546, EPI_ISL_2642548, EPI_ISL_2644672, EPI_ISL_2644673, EPI_ISL_2644750, EPI_ISL_2644766, EPI_ISL_2644772, EPI_ISL_2644820, EPI_ISL_2644821 | | | |
| see above | Ohio Department of Health Laboratory | Ohio Department of Health Laboratory | Holmes, Jennifer; Eric Brandt, Keoni Omura, Glen McGillivary, Caitlin McDonnell, Jade Mowery, Stephanie Mccracken, Tyler Payne, Kirtana Ramadugu, Erica Leasure, Brent Lee, Kelsey Florek, Heather Blankenship, Allison Black, Quanta Brown, and Tammy Bannerman |
| EPI_ISL_2644834 | LESP HIDALGO | LESP HIDALGO GENE2LIFE | HERRERA-GOMEZ CONNIE ALEJANDRA; CASTILLO-ORTEGA LAURA SOFIA; GARCIA-REYES MELITO; VIGUERAS-MORALES YAJAIRA SULIM; GARCIA-GONZALEZ OCTAVIO; VILLAFUENTES-TELLEZ HECTOR ALBERTO |
| EPI_ISL_2644835, EPI_ISL_2644836 | LESP HIDALGO | LESP HIDALGO GENE2LIFE | HERREGA-GOMEZ CONNIE ALEJANDRA; CASTILLO-ORTEGA LAURA SOFIA; GARCIA-REYES MELITO; VIGUERAS-MORALES YAJAIRA SULIM; GARCIA-GONZALEZ OCTAVIO; VILLAFUENTES-TELLEZ HECTOR ALBERTO |
| EPI_ISL_2644837, EPI_ISL_2644838 | Ohio Department of Health Laboratory | Ohio Department of Health Laboratory | Holmes, Jennifer; Eric Brandt, Keoni Omura, Glen McGillivary, Caitlin McDonnell, Jade Mowery, Stephanie Mccracken, Tyler Payne, Kirtana Ramadugu, Erica Leasure, Brent Lee, Kelsey Florek, Heather Blankenship, Allison Black, Quanta Brown, and Tammy Bannerman |
| EPI_ISL_2645280 | IA State Hygienic Laboratory | Centers for Disease Control and Prevention Division of Viral Diseases, Pathogen Discovery | Mili Sheth, Sarah Nobles, Jasmine Padilla, Mark Burroughs, Shoshona Le, Katie Dillon, Peter Cook, Clinton R. Paden, Dhwani Batra, Krista Queen, Kristen Knipe, Dakota Howard, Yvette Unoarumhi, Darlene Wagner, Matthew Schmerer, Ben L. Rambo-Martin, Kristine Lacek, Sam Shepard, Alison Laufer Halpin, Dave Wentworth, Vivien Dugan, Suxiang Tong, Justin Lee |
| EPI_ISL_2645281, EPI_ISL_2645282, EPI_ISL_2645284, EPI_ISL_2645285 | SD Public Health Laboratory | Centers for Disease Control and Prevention Division of Viral Diseases, Pathogen Discovery | Mili Sheth, Sarah Nobles, Jasmine Padilla, Mark Burroughs, Shoshona Le, Katie Dillon, Peter Cook, Clinton R. Paden, Dhwani Batra, Krista Queen, Kristen Knipe, Dakota Howard, Yvette Unoarumhi, Darlene Wagner, Matthew Schmerer, Ben L. Rambo-Martin, Kristine Lacek, Sam Shepard, Alison Laufer Halpin, Dave Wentworth, Vivien Dugan, Suxiang Tong, Justin Lee |
| EPI_ISL_2645286, EPI_ISL_2645287 | DC Public Health Lab/ Dept. of Forensic Sciences | Centers for Disease Control and Prevention Division of Viral Diseases, Pathogen Discovery | Mili Sheth, Sarah Nobles, Jasmine Padilla, Mark Burroughs, Shoshona Le, Katie Dillon, Peter Cook, Clinton R. Paden, Dhwani Batra, Krista Queen, Kristen Knipe, Dakota Howard, Yvette Unoarumhi, Darlene Wagner, Matthew Schmerer, Ben L. Rambo-Martin, Kristine Lacek, Sam Shepard, Alison Laufer Halpin, Dave Wentworth, Vivien Dugan, Suxiang Tong, Justin Lee |
| EPI_ISL_2645288, EPI_ISL_2645289, EPI_ISL_2645290, EPI_ISL_2645291, EPI_ISL_2645292, EPI_ISL_2645293, EPI_ISL_2645294, EPI_ISL_2645295, EPI_ISL_2645296, EPI_ISL_2645297 | KS Health and Environmental Laboratories | Centers for Disease Control and Prevention Division of Viral Diseases, Pathogen Discovery | Mili Sheth, Sarah Nobles, Jasmine Padilla, Mark Burroughs, Shoshona Le, Katie Dillon, Peter Cook, Clinton R. Paden, Dhwani Batra, Krista Queen, Kristen Knipe, Dakota Howard, Yvette Unoarumhi, Darlene Wagner, Matthew Schmerer, Ben L. Rambo-Martin, Kristine Lacek, Sam Shepard, Alison Laufer Halpin, Dave Wentworth, Vivien Dugan, Suxiang Tong, Justin Lee |
| EPI_ISL_2645298, EPI_ISL_2645299, EPI_ISL_2645300, EPI_ISL_2645301, EPI_ISL_2645302, EPI_ISL_2645303, EPI_ISL_2645304, EPI_ISL_2645305, EPI_ISL_2645306, EPI_ISL_2645307, EPI_ISL_2645308 | | | |
| see above | "CO Dept. of Public Health and Environment, Lab Services Division" | Centers for Disease Control and Prevention Division of Viral Diseases, Pathogen Discovery | Mili Sheth, Sarah Nobles, Jasmine Padilla, Mark Burroughs, Shoshona Le, Katie Dillon, Peter Cook, Clinton R. Paden, Dhwani Batra, Krista Queen, Kristen Knipe, Dakota Howard, Yvette Unoarumhi, Darlene Wagner, Matthew Schmerer, Ben L. Rambo-Martin, Kristine Lacek, Sam Shepard, Alison Laufer Halpin, Dave Wentworth, Vivien Dugan, Suxiang Tong, Justin Lee |
| EPI_ISL_2645309, EPI_ISL_2645310, EPI_ISL_2645311, EPI_ISL_2645312, EPI_ISL_2645313, EPI_ISL_2645314, EPI_ISL_2645315, EPI_ISL_2645316, EPI_ISL_2645317, EPI_ISL_2645318, EPI_ISL_2645319, EPI_ISL_2645320 | | | |
| see above | SC Dept of Health and Env. Control-Bureau of Laboratories | Centers for Disease Control and Prevention Division of Viral Diseases, Pathogen Discovery | Mili Sheth, Sarah Nobles, Jasmine Padilla, Mark Burroughs, Shoshona Le, Katie Dillon, Peter Cook, Clinton R. Paden, Dhwani Batra, Krista Queen, Kristen Knipe, Dakota Howard, Yvette Unoarumhi, Darlene Wagner, Matthew Schmerer, Ben L. Rambo-Martin, Kristine Lacek, Sam Shepard, Alison Laufer Halpin, Dave Wentworth, Vivien Dugan, Suxiang Tong, Justin Lee |
| EPI_ISL_2645321 | "AZ SPHL, Arizona Department of Health Services" | Centers for Disease Control and Prevention Division of Viral Diseases, Pathogen Discovery | Mili Sheth, Sarah Nobles, Jasmine Padilla, Mark Burroughs, Shoshona Le, Katie Dillon, Peter Cook, Clinton R. Paden, Dhwani Batra, Krista Queen, Kristen Knipe, Dakota Howard, Yvette Unoarumhi, Darlene Wagner, Matthew Schmerer, Ben L. Rambo-Martin, Kristine Lacek, Sam Shepard, Alison Laufer Halpin, Dave Wentworth, Vivien Dugan, Suxiang Tong, Justin Lee |
| EPI_ISL_2645322, EPI_ISL_2645323, EPI_ISL_2645324, EPI_ISL_2645325, EPI_ISL_2645326, EPI_ISL_2645327, EPI_ISL_2645328, EPI_ISL_2645329, EPI_ISL_2645330, EPI_ISL_2645331, EPI_ISL_2645332, EPI_ISL_2645333, EPI_ISL_2645334 | | | |
| see above | OH Department of Health Laboratory | Centers for Disease Control and Prevention Division of Viral Diseases, Pathogen Discovery | Mili Sheth, Sarah Nobles, Jasmine Padilla, Mark Burroughs, Shoshona Le, Katie Dillon, Peter Cook, Clinton R. Paden, Dhwani Batra, Krista Queen, Kristen Knipe, Dakota Howard, Yvette Unoarumhi, Darlene Wagner, Matthew Schmerer, Ben L. Rambo-Martin, Kristine Lacek, Sam Shepard, Alison Laufer Halpin, Dave Wentworth, Vivien Dugan, Suxiang Tong, Justin Lee |
| EPI_ISL_2645335 | CT-Dr. Katherine A. Kelley State Public Health Lab | Centers for Disease Control and Prevention Division of Viral Diseases, Pathogen Discovery | Mili Sheth, Sarah Nobles, Jasmine Padilla, Mark Burroughs, Shoshona Le, Katie Dillon, Peter Cook, Clinton R. Paden, Dhwani Batra, Krista Queen, Kristen Knipe, Dakota Howard, Yvette Unoarumhi, Darlene Wagner, Matthew Schmerer, Ben L. Rambo-Martin, Kristine Lacek, Sam Shepard, Alison Laufer Halpin, Dave Wentworth, Vivien Dugan, Suxiang Tong, Justin Lee |
| EPI_ISL_2645336, EPI_ISL_2645337, EPI_ISL_2645338, EPI_ISL_2645339, EPI_ISL_2645340, EPI_ISL_2645341, EPI_ISL_2645342, EPI_ISL_2645343, EPI_ISL_2645344, EPI_ISL_2645345, EPI_ISL_2645346, EPI_ISL_2645347 | | | |
| see above | MI - Michigan Department of Health and Human Services - Bureau of Laboratories | Centers for Disease Control and Prevention Division of Viral Diseases, Pathogen Discovery | Mili Sheth, Sarah Nobles, Jasmine Padilla, Mark Burroughs, Shoshona Le, Katie Dillon, Peter Cook, Clinton R. Paden, Dhwani Batra, Krista Queen, Kristen Knipe, Dakota Howard, Yvette Unoarumhi, Darlene Wagner, Matthew Schmerer, Ben L. Rambo-Martin, Kristine Lacek, Sam Shepard, Alison Laufer Halpin, Dave Wentworth, Vivien Dugan, Suxiang Tong, Justin Lee |
| EPI_ISL_2645348, EPI_ISL_2645349, EPI_ISL_2645350, EPI_ISL_2645351, EPI_ISL_2645352, EPI_ISL_2645353, EPI_ISL_2645354, EPI_ISL_2645355, EPI_ISL_2645356, EPI_ISL_2645357, EPI_ISL_2645358, EPI_ISL_2645359, EPI_ISL_2645360, EPI_ISL_2645361, EPI_ISL_2645362, EPI_ISL_2645363, EPI_ISL_2645364 | | | |
| see above | RI State Health Laboratories | Centers for Disease Control and Prevention Division of Viral Diseases, Pathogen Discovery | Mili Sheth, Sarah Nobles, Jasmine Padilla, Mark Burroughs, Shoshona Le, Katie Dillon, Peter Cook, Clinton R. Paden, Dhwani Batra, Krista Queen, Kristen Knipe, Dakota Howard, Yvette Unoarumhi, Darlene Wagner, Matthew Schmerer, Ben L. Rambo-Martin, Kristine Lacek, Sam Shepard, Alison Laufer Halpin, Dave Wentworth, Vivien Dugan, Suxiang Tong, Justin Lee |
| EPI_ISL_2645365, EPI_ISL_2645366, EPI_ISL_2645367, | "PA Department of Health, Bureau of Laboratories" | Centers for Disease Control and Prevention Division of Viral Diseases, Pathogen Discovery | Mili Sheth, Sarah Nobles, Jasmine Padilla, Mark Burroughs, Shoshona Le, Katie Dillon, Peter Cook, Clinton R. Paden, Dhwani Batra, Krista Queen, Kristen Knipe, Dakota Howard, Yvette Unoarumhi, Darlene Wagner, Matthew Schmerer, Ben L. Rambo-Martin, Kristine Lacek, Sam Shepard, Alison Laufer Halpin, Dave Wentworth, Vivien Dugan, Suxiang Tong, Justin Lee |

| | | | |
|---|---|---|---|
| EPI_ISL_2645368, EPI_ISL_2645369, EPI_ISL_2645370, EPI_ISL_2645371, EPI_ISL_2645372 | | | |
| EPI_ISL_2645373, EPI_ISL_2645374 | NH Dept. of Health and Human Services Public Health Labs | Centers for Disease Control and Prevention Division of Viral Diseases, Pathogen Discovery | Mili Sheth, Sarah Nobles, Jasmine Padilla, Mark Burroughs, Shoshona Le, Katie Dillon, Peter Cook, Clinton R. Paden, Dhwani Batra, Krista Queen, Kristen Knipe, Dakota Howard, Yvette Unoarumhi, Darlene Wagner, Matthew Schmerer, Ben L. Rambo-Martin, Kristine Lacek, Sam Shepard, Alison Laufer Halpin, Dave Wentworth, Vivien Dugan, Suxiang Tong, Justin Lee |
| EPI_ISL_2645376, EPI_ISL_2645377, EPI_ISL_2645378, EPI_ISL_2645379 | Ohio Department of Health Laboratory | Ohio Department of Health Laboratory | Holmes, Jennifer; Eric Brandt, Keoni Omura, Glen McGillivary, Caitlin McDonnell, Jade Mowery, Stephanie Mccracken, Tyler Payne, Kirtana Ramadugu, Erica Leasure, Brent Lee, Kelsey Florek, Heather Blankenship, Allison Black, Quanta Brown, and Tammy Bannerman |
| EPI_ISL_2645742, EPI_ISL_2645743, EPI_ISL_2645744, EPI_ISL_2645745, EPI_ISL_2645746, EPI_ISL_2645747, EPI_ISL_2645748, EPI_ISL_2645749, EPI_ISL_2645750, EPI_ISL_2645751, EPI_ISL_2645752, EPI_ISL_2645753, EPI_ISL_2645754, EPI_ISL_2645755, EPI_ISL_2645756, EPI_ISL_2645757, EPI_ISL_2645758, EPI_ISL_2645759, EPI_ISL_2645760, EPI_ISL_2645761, EPI_ISL_2645762, EPI_ISL_2645763, EPI_ISL_2645764, EPI_ISL_2645765, EPI_ISL_2645766, EPI_ISL_2645767 | | | |
| see above | Nebraska Public Health Laboratory | NPHL COVID-19 Response Team | NPHL COVID-19 Response Team |
| EPI_ISL_2645768, EPI_ISL_2645769, EPI_ISL_2645770, EPI_ISL_2645771 | GA Department of Public Health | GA Department of Public Health | Stacy Reeves, Jonathan Edwards, Cynthia Dixey, Tonia Parrott, Aliyah Fields, Taylor Smith |
| EPI_ISL_2645772, EPI_ISL_2645773, EPI_ISL_2645774, EPI_ISL_2645775, EPI_ISL_2645776, EPI_ISL_2645777, EPI_ISL_2645778, EPI_ISL_2645779, EPI_ISL_2645780, EPI_ISL_2645781, EPI_ISL_2645782, EPI_ISL_2645783, EPI_ISL_2645784, EPI_ISL_2645785, EPI_ISL_2645786, EPI_ISL_2645787, EPI_ISL_2645788, EPI_ISL_2645789, EPI_ISL_2645790, EPI_ISL_2645791, EPI_ISL_2645792, EPI_ISL_2645793 | | | |
| see above | Nebraska Public Health Laboratory | NPHL COVID-19 Response Team | NPHL COVID-19 Response Team |
| EPI_ISL_2646121, EPI_ISL_2646122 | Tampa General Hospital Esoteric Lab | Tampa General Hospital Esoteric Research & Development Lab | Jon Faughn, Deanna Becker, Dominic Uy, Vicki Healer, Amorce Lima, Suzane Silbert |
| EPI_ISL_2646123, EPI_ISL_2646124, EPI_ISL_2646125, EPI_ISL_2646126, EPI_ISL_2646127, EPI_ISL_2646128, EPI_ISL_2646129, EPI_ISL_2646130, EPI_ISL_2646131, EPI_ISL_2646132, EPI_ISL_2646133, EPI_ISL_2646134, EPI_ISL_2646135, EPI_ISL_2646136, EPI_ISL_2646137, EPI_ISL_2646138, EPI_ISL_2646139 | | | |
| see above | Instituto Nacional de Medicina Genomica | Centro de Investigación en Enfermedades Infecciosas (CIENI), Instituto Nacional de Enfermedades Respiratorias (INER) | Matias-Florentino M, Perez-Garcia M, Hidalgo-Miranda A, Mendoza-Vargas A, Reyes-Grajeda JP, Cedro-Tanda A, Herrera-Montalvo LA, Reyes-Teran G, Avila-Rios S, Boukadida C |
| EPI_ISL_2646299, EPI_ISL_2646300, EPI_ISL_2646301 | Ohio Department of Health Laboratory | Ohio Department of Health Laboratory | Holmes, Jennifer; Eric Brandt, Keoni Omura, Glen McGillivary, Caitlin McDonnell, Jade Mowery, Stephanie Mccracken, Tyler Payne, Kirtana Ramadugu, Erica Leasure, Brent Lee, Kelsey Florek, Heather Blankenship, Allison Black, Quanta Brown, and Tammy Bannerman |
| EPI_ISL_2646319, EPI_ISL_2646320, EPI_ISL_2646321, EPI_ISL_2646322, EPI_ISL_2646323, EPI_ISL_2646324, EPI_ISL_2646325, EPI_ISL_2646326, EPI_ISL_2646327, EPI_ISL_2646328, EPI_ISL_2646329 | | | |
| see above | University of Mississippi Medical Center, Department of Pathology | University of Mississippi Medical Center, Molecular and Genomics Core Facility | Ashley C. Johnson, Wenjie Wu, Ithiel J. Frame, Krishna K. Ayyalasomayajula, Michael R. Garrett, D. Ashley Robinson |
| EPI_ISL_2646330, EPI_ISL_2646331, EPI_ISL_2646332, EPI_ISL_2646333, EPI_ISL_2646334, EPI_ISL_2646335, EPI_ISL_2646336, EPI_ISL_2646337, EPI_ISL_2646338, EPI_ISL_2646339, EPI_ISL_2646340, EPI_ISL_2646341, EPI_ISL_2646342, EPI_ISL_2646343, EPI_ISL_2646344, EPI_ISL_2646345, EPI_ISL_2646346, EPI_ISL_2646347, EPI_ISL_2646348, EPI_ISL_2646349, EPI_ISL_2646350, EPI_ISL_2646351, EPI_ISL_2646352 | | | |
| see above | MS Public Health Laboratory | University of Mississippi Medical Center, Molecular and Genomics Core Facility | Ashley C. Johnson, Wenjie Wu, Ithiel J. Frame, Krishna K. Ayyalasomayajula, Michael R. Garrett, D. Ashley Robinson |
| EPI_ISL_2646353, EPI_ISL_2646354, EPI_ISL_2646355, EPI_ISL_2646356, EPI_ISL_2646357, EPI_ISL_2646358, EPI_ISL_2646359, EPI_ISL_2646360 | Wexner Medical Center | The Ohio State University College of Medicine | Koenig,S., Seminetta,J. |
| EPI_ISL_2646361, EPI_ISL_2646363, EPI_ISL_2646364, EPI_ISL_2646365, EPI_ISL_2646366, EPI_ISL_2646368, EPI_ISL_2646370, EPI_ISL_2646371, EPI_ISL_2646373, EPI_ISL_2646374, EPI_ISL_2646375, EPI_ISL_2646376, EPI_ISL_2646377, EPI_ISL_2646378, EPI_ISL_2646381, EPI_ISL_2646382, EPI_ISL_2646384, EPI_ISL_2646386, EPI_ISL_2646388, EPI_ISL_2646389, EPI_ISL_2646390, EPI_ISL_2646392, EPI_ISL_2646393, EPI_ISL_2646394, EPI_ISL_2646395, EPI_ISL_2646396, EPI_ISL_2646397, EPI_ISL_2646399, EPI_ISL_2646400, EPI_ISL_2646402, EPI_ISL_2646403, EPI_ISL_2646404, EPI_ISL_2646405, EPI_ISL_2646406, EPI_ISL_2646408, EPI_ISL_2646409, EPI_ISL_2646410, EPI_ISL_2646412, EPI_ISL_2646413, EPI_ISL_2646416, EPI_ISL_2646417, EPI_ISL_2646418, EPI_ISL_2646419, EPI_ISL_2646420, EPI_ISL_2646421, EPI_ISL_2646422, EPI_ISL_2646423, EPI_ISL_2646424, EPI_ISL_2646425, EPI_ISL_2646426, EPI_ISL_2646427, EPI_ISL_2646428, EPI_ISL_2646429, EPI_ISL_2646430, EPI_ISL_2646431, EPI_ISL_2646432, EPI_ISL_2646433, EPI_ISL_2646435, EPI_ISL_2646436, EPI_ISL_2646437, EPI_ISL_2646438, EPI_ISL_2646439, EPI_ISL_2646440, EPI_ISL_2646443, EPI_ISL_2646444, EPI_ISL_2646445, EPI_ISL_2646446, EPI_ISL_2646447, EPI_ISL_2646448, EPI_ISL_2646449, EPI_ISL_2646450, EPI_ISL_2646451, EPI_ISL_2646452, EPI_ISL_2646453, EPI_ISL_2646454, EPI_ISL_2646455, EPI_ISL_2646456, EPI_ISL_2646457, EPI_ISL_2646458, EPI_ISL_2646459, EPI_ISL_2646460, EPI_ISL_2646461, EPI_ISL_2646462, EPI_ISL_2646463, EPI_ISL_2646464, EPI_ISL_2646465, EPI_ISL_2646466, EPI_ISL_2646467, EPI_ISL_2646468, EPI_ISL_2646469, EPI_ISL_2646470, EPI_ISL_2646471, EPI_ISL_2646472, EPI_ISL_2646473, EPI_ISL_2646474, EPI_ISL_2646475, EPI_ISL_2646476, EPI_ISL_2646477, EPI_ISL_2646478, EPI_ISL_2646479, EPI_ISL_2646480, EPI_ISL_2646481, EPI_ISL_2646482, EPI_ISL_2646483, EPI_ISL_2646484, EPI_ISL_2646485, EPI_ISL_2646486, EPI_ISL_2646488, EPI_ISL_2646489, EPI_ISL_2646490, EPI_ISL_2646491, EPI_ISL_2646492, EPI_ISL_2646494, EPI_ISL_2646495, EPI_ISL_2646498, EPI_ISL_2646499, EPI_ISL_2646500, EPI_ISL_2646501, EPI_ISL_2646502, EPI_ISL_2646503, EPI_ISL_2646504, EPI_ISL_2646505, EPI_ISL_2646506, EPI_ISL_2646507, EPI_ISL_2646508, EPI_ISL_2646509, EPI_ISL_2646510, EPI_ISL_2646511, EPI_ISL_2646512, EPI_ISL_2646513, EPI_ISL_2646514, EPI_ISL_2646515, EPI_ISL_2646516, EPI_ISL_2646517, EPI_ISL_2646518, EPI_ISL_2646519, EPI_ISL_2646520, EPI_ISL_2646521, EPI_ISL_2646522, EPI_ISL_2646523, EPI_ISL_2646524, EPI_ISL_2646525, EPI_ISL_2646526, EPI_ISL_2646527, EPI_ISL_2646528, EPI_ISL_2646529, EPI_ISL_2646530, EPI_ISL_2646531, EPI_ISL_2646532, EPI_ISL_2646533, EPI_ISL_2646534, EPI_ISL_2646535, EPI_ISL_2646536, EPI_ISL_2646537, EPI_ISL_2646538, EPI_ISL_2646539, EPI_ISL_2646540, EPI_ISL_2646541, EPI_ISL_2646542, EPI_ISL_2646543, EPI_ISL_2646544, EPI_ISL_2646545, EPI_ISL_2646546, EPI_ISL_2646547, EPI_ISL_2646548, EPI_ISL_2646549, EPI_ISL_2646550, EPI_ISL_2646551, EPI_ISL_2646552, EPI_ISL_2646553, EPI_ISL_2646554, EPI_ISL_2646555, EPI_ISL_2646556, EPI_ISL_2646557, EPI_ISL_2646558, EPI_ISL_2646559, EPI_ISL_2646560, EPI_ISL_2646561, EPI_ISL_2646562, EPI_ISL_2646563, EPI_ISL_2646564, EPI_ISL_2646565, EPI_ISL_2646566, EPI_ISL_2646567, EPI_ISL_2646568, EPI_ISL_2646569, EPI_ISL_2646570, EPI_ISL_2646571, EPI_ISL_2646572, EPI_ISL_2646573, EPI_ISL_2646574, EPI_ISL_2646575, EPI_ISL_2646576, EPI_ISL_2646577, EPI_ISL_2646578, EPI_ISL_2646579, EPI_ISL_2646580, EPI_ISL_2646581, EPI_ISL_2646582, EPI_ISL_2646583, EPI_ISL_2646584, EPI_ISL_2646585, EPI_ISL_2646586, EPI_ISL_2646587, EPI_ISL_2646588, EPI_ISL_2646589, EPI_ISL_2646590, EPI_ISL_2646591, EPI_ISL_2646592, EPI_ISL_2646593, EPI_ISL_2646594, EPI_ISL_2646595, EPI_ISL_2646596, EPI_ISL_2646597, EPI_ISL_2646598, EPI_ISL_2646599, EPI_ISL_2646600, EPI_ISL_2646601, EPI_ISL_2646602, EPI_ISL_2646603, EPI_ISL_2646604, EPI_ISL_2646605, EPI_ISL_2646606, EPI_ISL_2646607, EPI_ISL_2646608, EPI_ISL_2646609, EPI_ISL_2646610, EPI_ISL_2646611, EPI_ISL_2646612, EPI_ISL_2646613, EPI_ISL_2646614, EPI_ISL_2646615, EPI_ISL_2646616, EPI_ISL_2646617, EPI_ISL_2646618, EPI_ISL_2646619, EPI_ISL_2646620, EPI_ISL_2646621, EPI_ISL_2646622, EPI_ISL_2646623, EPI_ISL_2646624, EPI_ISL_2646625, EPI_ISL_2646626, EPI_ISL_2646627, EPI_ISL_2646628, EPI_ISL_2646629, EPI_ISL_2646630, EPI_ISL_2646631, EPI_ISL_2646632, EPI_ISL_2646633, EPI_ISL_2646634, EPI_ISL_2646635, EPI_ISL_2646636, EPI_ISL_2646637, EPI_ISL_2646638, EPI_ISL_2646639, EPI_ISL_2646640, EPI_ISL_2646641, EPI_ISL_2646642, EPI_ISL_2646643, EPI_ISL_2646644, EPI_ISL_2646645, EPI_ISL_2646646, EPI_ISL_2646647, EPI_ISL_2646648, EPI_ISL_2646649, EPI_ISL_2646650, EPI_ISL_2646651, EPI_ISL_2646652, EPI_ISL_2646653, EPI_ISL_2646654, EPI_ISL_2646655, EPI_ISL_2646656, EPI_ISL_2646657, EPI_ISL_2646658, EPI_ISL_2646659, EPI_ISL_2646660, EPI_ISL_2646661, EPI_ISL_2646662, EPI_ISL_2646663, EPI_ISL_2646664, EPI_ISL_2646665, EPI_ISL_2646666, EPI_ISL_2646667, EPI_ISL_2646668, EPI_ISL_2646669, EPI_ISL_2646670, EPI_ISL_2646671, EPI_ISL_2646672, EPI_ISL_2646673, EPI_ISL_2646674, EPI_ISL_2646675, EPI_ISL_2646676, EPI_ISL_2646677, EPI_ISL_2646678, EPI_ISL_2646679, EPI_ISL_2646680, EPI_ISL_2646681, EPI_ISL_2646683, EPI_ISL_2646684, EPI_ISL_2646685, EPI_ISL_2646686, EPI_ISL_2646687, EPI_ISL_2646688, EPI_ISL_2646689, EPI_ISL_2646690, EPI_ISL_2646691, EPI_ISL_2646692, EPI_ISL_2646693, EPI_ISL_2646694, EPI_ISL_2646695, EPI_ISL_2646696, EPI_ISL_2646697, EPI_ISL_2646698, EPI_ISL_2646699, EPI_ISL_2646700, EPI_ISL_2646701, EPI_ISL_2646702, EPI_ISL_2646703, EPI_ISL_2646704, EPI_ISL_2646705, EPI_ISL_2646706, EPI_ISL_2646707, EPI_ISL_2646708, EPI_ISL_2646709, EPI_ISL_2646710, EPI_ISL_2646711, EPI_ISL_2646712, EPI_ISL_2646713, EPI_ISL_2646714, EPI_ISL_2646715, EPI_ISL_2646716, EPI_ISL_2646717, EPI_ISL_2646718, EPI_ISL_2646719, EPI_ISL_2646720, EPI_ISL_2646721, EPI_ISL_2646722, EPI_ISL_2646723, EPI_ISL_2646724, EPI_ISL_2646725, EPI_ISL_2646726, EPI_ISL_2646727, EPI_ISL_2646728, EPI_ISL_2646729, EPI_ISL_2646730, EPI_ISL_2646731, EPI_ISL_2646732, EPI_ISL_2646733, EPI_ISL_2646734, EPI_ISL_2646735, EPI_ISL_2646736, EPI_ISL_2646737, EPI_ISL_2646738, EPI_ISL_2646739, EPI_ISL_2646740, EPI_ISL_2646741, EPI_ISL_2646742, EPI_ISL_2646743, EPI_ISL_2646744, EPI_ISL_2646745, EPI_ISL_2646746, EPI_ISL_2646747, EPI_ISL_2646748, EPI_ISL_2646749, EPI_ISL_2646750, EPI_ISL_2646751, EPI_ISL_2646752, EPI_ISL_2646753, EPI_ISL_2646754, EPI_ISL_2646755, EPI_ISL_2646756, EPI_ISL_2646757, EPI_ISL_2646758, EPI_ISL_2646759, EPI_ISL_2646760, EPI_ISL_2646761, EPI_ISL_2646762, EPI_ISL_2646763, EPI_ISL_2646764, EPI_ISL_2646765, EPI_ISL_2646766, EPI_ISL_2646767, EPI_ISL_2646768, EPI_ISL_2646769, EPI_ISL_2646770, EPI_ISL_2646771, EPI_ISL_2646772, EPI_ISL_2646773, EPI_ISL_2646774, EPI_ISL_2646775, EPI_ISL_2646776, EPI_ISL_2646777, EPI_ISL_2646778, EPI_ISL_2646779, EPI_ISL_2646780, EPI_ISL_2646781, EPI_ISL_2646782, EPI_ISL_2646783, EPI_ISL_2646784, EPI_ISL_2646785, EPI_ISL_2646786, EPI_ISL_2646787, EPI_ISL_2646788, EPI_ISL_2646789, EPI_ISL_2646790, EPI_ISL_2646791, EPI_ISL_2646792, EPI_ISL_2646793, EPI_ISL_2646794, EPI_ISL_2646795, EPI_ISL_2646796, EPI_ISL_2646797, EPI_ISL_2646798, EPI_ISL_2646799, EPI_ISL_2646800, EPI_ISL_2646801, EPI_ISL_2646802, EPI_ISL_2646803, EPI_ISL_2646804, EPI_ISL_2646805, EPI_ISL_2646806, EPI_ISL_2646807, EPI_ISL_2646808, EPI_ISL_2646809, EPI_ISL_2646810, EPI_ISL_2646811, EPI_ISL_2646812, EPI_ISL_2646813, EPI_ISL_2646814, EPI_ISL_2646815, EPI_ISL_2646816, EPI_ISL_2646817, EPI_ISL_2646818, EPI_ISL_2646819, EPI_ISL_2646821, EPI_ISL_2646822, EPI_ISL_2646823, EPI_ISL_2646824, EPI_ISL_2646825, EPI_ISL_2646826, EPI_ISL_2646827, EPI_ISL_2646828, EPI_ISL_2646829, EPI_ISL_2646830, EPI_ISL_2646831, EPI_ISL_2646832, EPI_ISL_2646833, EPI_ISL_2646834, EPI_ISL_2646835, EPI_ISL_2646836, EPI_ISL_2646837, EPI_ISL_2646838, EPI_ISL_2646839, EPI_ISL_2646840, EPI_ISL_2646841, EPI_ISL_2646842, EPI_ISL_2646843, EPI_ISL_2646844, EPI_ISL_2646845, EPI_ISL_2646846, EPI_ISL_2646847, EPI_ISL_2646848, EPI_ISL_2646849, EPI_ISL_2646850, EPI_ISL_2646852, EPI_ISL_2646853, EPI_ISL_2646854, EPI_ISL_2646855, EPI_ISL_2646856, EPI_ISL_2646857, EPI_ISL_2646858, EPI_ISL_2646859, EPI_ISL_2646860, EPI_ISL_2646861, EPI_ISL_2646862, EPI_ISL_2646863, EPI_ISL_2646864, EPI_ISL_2646865, EPI_ISL_2646866, EPI_ISL_2646867, EPI_ISL_2646868, EPI_ISL_2646869, EPI_ISL_2646870, EPI_ISL_2646871, EPI_ISL_2646872, EPI_ISL_2646873, EPI_ISL_2646874, EPI_ISL_2646875, EPI_ISL_2646876, EPI_ISL_2646877, EPI_ISL_2646878, EPI_ISL_2646879, EPI_ISL_2646880, EPI_ISL_2646881, EPI_ISL_2646882, EPI_ISL_2646883, EPI_ISL_2646884, EPI_ISL_2646885, EPI_ISL_2646886, EPI_ISL_2646887, EPI_ISL_2646888, EPI_ISL_2646889, EPI_ISL_2646890, EPI_ISL_2646891, EPI_ISL_2646892, EPI_ISL_2646893, EPI_ISL_2646894, EPI_ISL_2646895, EPI_ISL_2646896, EPI_ISL_2646897, EPI_ISL_2646898, EPI_ISL_2646899, EPI_ISL_2646900, EPI_ISL_2646901, EPI_ISL_2646902, EPI_ISL_2646903, EPI_ISL_2646904, EPI_ISL_2646905, EPI_ISL_2646906, EPI_ISL_2646907, EPI_ISL_2646908, EPI_ISL_2646909, EPI_ISL_2646910, EPI_ISL_2646911, EPI_ISL_2646912, EPI_ISL_2646913, EPI_ISL_2646914, EPI_ISL_2646915, EPI_ISL_2646916, EPI_ISL_2646917, EPI_ISL_2646918, EPI_ISL_2646919, EPI_ISL_2646920, EPI_ISL_2646921, EPI_ISL_2646922, EPI_ISL_2646923, EPI_ISL_2646924, EPI_ISL_2646925, EPI_ISL_2646926, EPI_ISL_2646927, EPI_ISL_2646928, EPI_ISL_2646929, EPI_ISL_2646930, EPI_ISL_2646931, EPI_ISL_2646932, EPI_ISL_2646933, EPI_ISL_2646934, EPI_ISL_2646935, EPI_ISL_2646936, EPI_ISL_2646937, EPI_ISL_2646938, EPI_ISL_2646939, EPI_ISL_2646940, EPI_ISL_2646941, EPI_ISL_2646942, EPI_ISL_2646943, EPI_ISL_2646944, EPI_ISL_2646945, EPI_ISL_2646946, EPI_ISL_2646947, EPI_ISL_2646948, EPI_ISL_2646949, EPI_ISL_2646950, EPI_ISL_2646951, EPI_ISL_2646952, EPI_ISL_2646953, EPI_ISL_2646954, EPI_ISL_2646955, EPI_ISL_2646956, EPI_ISL_2646957, EPI_ISL_2646958, EPI_ISL_2646959, EPI_ISL_2646960, EPI_ISL_2646961 | | | |
| see above | Mako Medical | Centers for Disease Control and Prevention Division of Viral Diseases, Pathogen Discovery | Dakota Howard, Dhwani Batra, Peter W. Cook, Kara Moser, Adrian Paskey, Jason Caravas, Benjamin Rambo-Martin, Shatavia Morrison, Christopher Gulvick, Scott Sammons, Yvette Unoarumhi, Darlene Wagner, Matthew Schmerer,Matthew Tugwell, Lauren Moon, Clinton R. Paden, Duncan MacCannell |
| EPI_ISL_2646962, EPI_ISL_2646963 | Montana Public Health Laboratory | Montana Public Health Laboratory | Joy Ritter, Michelle Mozer, Carrie Biskupiak, Deborah Gibson, Michael Dills |
| EPI_ISL_2646964 | Mako Medical | Centers for Disease Control and Prevention Division of Viral Diseases, Pathogen Discovery | Dakota Howard, Dhwani Batra, Peter W. Cook, Kara Moser, Adrian Paskey, Jason Caravas, Benjamin Rambo-Martin, Shatavia Morrison, Christopher Gulvick, Scott Sammons, Yvette Unoarumhi, Darlene Wagner, Matthew Schmerer,Matthew Tugwell, Lauren Moon, Clinton R. Paden, Duncan MacCannell |
| EPI_ISL_2646965, EPI_ISL_2646966 | Montana Public Health Laboratory | Montana Public Health Laboratory | Joy Ritter, Michelle Mozer, Carrie Biskupiak, Deborah Gibson, Michael Dills |
| EPI_ISL_2646967 | Mako Medical | Centers for Disease Control and Prevention Division of Viral Diseases, Pathogen Discovery | Dakota Howard, Dhwani Batra, Peter W. Cook, Kara Moser, Adrian Paskey, Jason Caravas, Benjamin Rambo-Martin, Shatavia Morrison, Christopher Gulvick, Scott Sammons, Yvette Unoarumhi, Darlene Wagner, Matthew Schmerer,Matthew Tugwell, Lauren Moon, Clinton R. Paden, Duncan MacCannell |
| EPI_ISL_2646968, EPI_ISL_2646969 | Montana Public Health Laboratory | Montana Public Health Laboratory | Joy Ritter, Michelle Mozer, Carrie Biskupiak, Deborah Gibson, Michael Dills |
| EPI_ISL_2646970 | Mako Medical | Centers for Disease Control and Prevention Division of Viral Diseases, Pathogen Discovery | Dakota Howard, Dhwani Batra, Peter W. Cook, Kara Moser, Adrian Paskey, Jason Caravas, Benjamin Rambo-Martin, Shatavia Morrison, Christopher Gulvick, Scott Sammons, Yvette Unoarumhi, Darlene Wagner, Matthew Schmerer,Matthew Tugwell, Lauren Moon, Clinton R. Paden, Duncan MacCannell |
| EPI_ISL_2646971, EPI_ISL_2646972 | Montana Public Health Laboratory | Montana Public Health Laboratory | Joy Ritter, Michelle Mozer, Carrie Biskupiak, Deborah Gibson, Michael Dills |
| EPI_ISL_2646973 | Mako Medical | Centers for Disease Control and Prevention Division of Viral Diseases, Pathogen Discovery | Dakota Howard, Dhwani Batra, Peter W. Cook, Kara Moser, Adrian Paskey, Jason Caravas, Benjamin Rambo-Martin, Shatavia Morrison, Christopher Gulvick, Scott Sammons, Yvette Unoarumhi, Darlene Wagner, Matthew Schmerer,Matthew Tugwell, Lauren Moon, Clinton R. Paden, Duncan MacCannell |
| EPI_ISL_2646974 | Montana Public Health Laboratory | Montana Public Health Laboratory | Joy Ritter, Michelle Mozer, Carrie Biskupiak, Deborah Gibson, Michael Dills |
| EPI_ISL_2646975 | Mako Medical | Centers for Disease Control and Prevention Division of Viral Diseases, Pathogen | Dakota Howard, Dhwani Batra, Peter W. Cook, Kara Moser, Adrian Paskey, Jason Caravas, Benjamin Rambo-Martin, Shatavia Morrison, Christopher Gulvick, Scott Sammons, Yvette Unoarumhi, Darlene Wagner, Matthew Schmerer,Matthew Tugwell, Lauren Moon, Clinton R. Paden, Duncan MacCannell |

| | | | |
|---|---|---|---|
| | | Discovery | |
| EPI_ISL_2646976, EPI_ISL_2646977 | Montana Public Health Laboratory | Montana Public Health Laboratory | Joy Ritter, Michelle Mozer, Carrie Biskupiak, Deborah Gibson, Michael Dills |
| EPI_ISL_2646978 | Mako Medical | Centers for Disease Control and Prevention Division of Viral Diseases, Pathogen Discovery | Dakota Howard, Dhwani Batra, Peter W. Cook, Kara Moser, Adrian Paskey, Jason Caravas, Benjamin Rambo-Martin, Shatavia Morrison, Christopher Gulvick, Scott Sammons, Yvette Unoarumhi, Darlene Wagner, Matthew Schmerer,Matthew Tugwell, Lauren Moon, Clinton R. Paden, Duncan MacCannell |
| EPI_ISL_2646979 | Montana Public Health Laboratory | Montana Public Health Laboratory | Joy Ritter, Michelle Mozer, Carrie Biskupiak, Deborah Gibson, Michael Dills |
| EPI_ISL_2646981 | Mako Medical | Centers for Disease Control and Prevention Division of Viral Diseases, Pathogen Discovery | Dakota Howard, Dhwani Batra, Peter W. Cook, Kara Moser, Adrian Paskey, Jason Caravas, Benjamin Rambo-Martin, Shatavia Morrison, Christopher Gulvick, Scott Sammons, Yvette Unoarumhi, Darlene Wagner, Matthew Schmerer,Matthew Tugwell, Lauren Moon, Clinton R. Paden, Duncan MacCannell |
| EPI_ISL_2646982 | Montana Public Health Laboratory | Montana Public Health Laboratory | Joy Ritter, Michelle Mozer, Carrie Biskupiak, Deborah Gibson, Michael Dills |
| EPI_ISL_2646983 | Mako Medical | Centers for Disease Control and Prevention Division of Viral Diseases, Pathogen Discovery | Dakota Howard, Dhwani Batra, Peter W. Cook, Kara Moser, Adrian Paskey, Jason Caravas, Benjamin Rambo-Martin, Shatavia Morrison, Christopher Gulvick, Scott Sammons, Yvette Unoarumhi, Darlene Wagner, Matthew Schmerer,Matthew Tugwell, Lauren Moon, Clinton R. Paden, Duncan MacCannell |
| EPI_ISL_2646984, EPI_ISL_2646985 | Montana Public Health Laboratory | Montana Public Health Laboratory | Joy Ritter, Michelle Mozer, Carrie Biskupiak, Deborah Gibson, Michael Dills |
| EPI_ISL_2646986 | Mako Medical | Centers for Disease Control and Prevention Division of Viral Diseases, Pathogen Discovery | Dakota Howard, Dhwani Batra, Peter W. Cook, Kara Moser, Adrian Paskey, Jason Caravas, Benjamin Rambo-Martin, Shatavia Morrison, Christopher Gulvick, Scott Sammons, Yvette Unoarumhi, Darlene Wagner, Matthew Schmerer,Matthew Tugwell, Lauren Moon, Clinton R. Paden, Duncan MacCannell |
| EPI_ISL_2646987, EPI_ISL_2646988 | Montana Public Health Laboratory | Montana Public Health Laboratory | Joy Ritter, Michelle Mozer, Carrie Biskupiak, Deborah Gibson, Michael Dills |
| EPI_ISL_2646989 | Mako Medical | Centers for Disease Control and Prevention Division of Viral Diseases, Pathogen Discovery | Dakota Howard, Dhwani Batra, Peter W. Cook, Kara Moser, Adrian Paskey, Jason Caravas, Benjamin Rambo-Martin, Shatavia Morrison, Christopher Gulvick, Scott Sammons, Yvette Unoarumhi, Darlene Wagner, Matthew Schmerer,Matthew Tugwell, Lauren Moon, Clinton R. Paden, Duncan MacCannell |
| EPI_ISL_2646990 | Montana Public Health Laboratory | Montana Public Health Laboratory | Joy Ritter, Michelle Mozer, Carrie Biskupiak, Deborah Gibson, Michael Dills |
| EPI_ISL_2646991 | Mako Medical | Centers for Disease Control and Prevention Division of Viral Diseases, Pathogen Discovery | Dakota Howard, Dhwani Batra, Peter W. Cook, Kara Moser, Adrian Paskey, Jason Caravas, Benjamin Rambo-Martin, Shatavia Morrison, Christopher Gulvick, Scott Sammons, Yvette Unoarumhi, Darlene Wagner, Matthew Schmerer,Matthew Tugwell, Lauren Moon, Clinton R. Paden, Duncan MacCannell |
| EPI_ISL_2646992, EPI_ISL_2646993 | Montana Public Health Laboratory | Montana Public Health Laboratory | Joy Ritter, Michelle Mozer, Carrie Biskupiak, Deborah Gibson, Michael Dills |
| EPI_ISL_2646994 | Mako Medical | Centers for Disease Control and Prevention Division of Viral Diseases, Pathogen Discovery | Dakota Howard, Dhwani Batra, Peter W. Cook, Kara Moser, Adrian Paskey, Jason Caravas, Benjamin Rambo-Martin, Shatavia Morrison, Christopher Gulvick, Scott Sammons, Yvette Unoarumhi, Darlene Wagner, Matthew Schmerer,Matthew Tugwell, Lauren Moon, Clinton R. Paden, Duncan MacCannell |
| EPI_ISL_2646995, EPI_ISL_2646996 | Montana Public Health Laboratory | Montana Public Health Laboratory | Joy Ritter, Michelle Mozer, Carrie Biskupiak, Deborah Gibson, Michael Dills |
| EPI_ISL_2646997 | Mako Medical | Centers for Disease Control and Prevention Division of Viral Diseases, Pathogen Discovery | Dakota Howard, Dhwani Batra, Peter W. Cook, Kara Moser, Adrian Paskey, Jason Caravas, Benjamin Rambo-Martin, Shatavia Morrison, Christopher Gulvick, Scott Sammons, Yvette Unoarumhi, Darlene Wagner, Matthew Schmerer,Matthew Tugwell, Lauren Moon, Clinton R. Paden, Duncan MacCannell |
| EPI_ISL_2646998, EPI_ISL_2646999 | Montana Public Health Laboratory | Montana Public Health Laboratory | Joy Ritter, Michelle Mozer, Carrie Biskupiak, Deborah Gibson, Michael Dills |
| EPI_ISL_2647000 | Mako Medical | Centers for Disease Control and Prevention Division of Viral Diseases, Pathogen Discovery | Dakota Howard, Dhwani Batra, Peter W. Cook, Kara Moser, Adrian Paskey, Jason Caravas, Benjamin Rambo-Martin, Shatavia Morrison, Christopher Gulvick, Scott Sammons, Yvette Unoarumhi, Darlene Wagner, Matthew Schmerer,Matthew Tugwell, Lauren Moon, Clinton R. Paden, Duncan MacCannell |
| EPI_ISL_2647001 | Montana Public Health Laboratory | Montana Public Health Laboratory | Joy Ritter, Michelle Mozer, Carrie Biskupiak, Deborah Gibson, Michael Dills |
| EPI_ISL_2647002, EPI_ISL_2647003, EPI_ISL_2647004, EPI_ISL_2647005, EPI_ISL_2647006, EPI_ISL_2647007, EPI_ISL_2647008, EPI_ISL_2647009, EPI_ISL_2647010, EPI_ISL_2647011, EPI_ISL_2647012, EPI_ISL_2647013, EPI_ISL_2647014, EPI_ISL_2647015, EPI_ISL_2647016, EPI_ISL_2647017, EPI_ISL_2647018, EPI_ISL_2647019, EPI_ISL_2647020, EPI_ISL_2647021, EPI_ISL_2647022, EPI_ISL_2647023, EPI_ISL_2647024, EPI_ISL_2647025, EPI_ISL_2647026, EPI_ISL_2647027, EPI_ISL_2647028, EPI_ISL_2647029, EPI_ISL_2647030, EPI_ISL_2647031, EPI_ISL_2647032, EPI_ISL_2647033, EPI_ISL_2647034, EPI_ISL_2647036, EPI_ISL_2647037, EPI_ISL_2647039, EPI_ISL_2647040, EPI_ISL_2647041, EPI_ISL_2647042, EPI_ISL_2647043, EPI_ISL_2647044, EPI_ISL_2647045, EPI_ISL_2647046, EPI_ISL_2647047, EPI_ISL_2647048, EPI_ISL_2647049, EPI_ISL_2647050, EPI_ISL_2647051, EPI_ISL_2647052, EPI_ISL_2647053, EPI_ISL_2647055, EPI_ISL_2647056, EPI_ISL_2647057, EPI_ISL_2647058, EPI_ISL_2647059, EPI_ISL_2647060, EPI_ISL_2647061, EPI_ISL_2647062, EPI_ISL_2647063, EPI_ISL_2647064, EPI_ISL_2647065, EPI_ISL_2647066, EPI_ISL_2647067, EPI_ISL_2647068, EPI_ISL_2647069, EPI_ISL_2647070, EPI_ISL_2647071, EPI_ISL_2647072, EPI_ISL_2647073, EPI_ISL_2647074, EPI_ISL_2647075, EPI_ISL_2647076, EPI_ISL_2647077, EPI_ISL_2647079, EPI_ISL_2647081, EPI_ISL_2647082, EPI_ISL_2647083, EPI_ISL_2647084, EPI_ISL_2647085, EPI_ISL_2647086, EPI_ISL_2647087, EPI_ISL_2647088, EPI_ISL_2647089, EPI_ISL_2647090, EPI_ISL_2647091, EPI_ISL_2647092, EPI_ISL_2647093, EPI_ISL_2647094, EPI_ISL_2647095, EPI_ISL_2647096, EPI_ISL_2647097, EPI_ISL_2647098, EPI_ISL_2647099, EPI_ISL_2647100, EPI_ISL_2647101, EPI_ISL_2647102, EPI_ISL_2647103, EPI_ISL_2647104, EPI_ISL_2647105, EPI_ISL_2647106, EPI_ISL_2647107, EPI_ISL_2647108, EPI_ISL_2647109, EPI_ISL_2647110, EPI_ISL_2647111, EPI_ISL_2647112, EPI_ISL_2647113, EPI_ISL_2647114, EPI_ISL_2647115, EPI_ISL_2647116, EPI_ISL_2647117, EPI_ISL_2647118, EPI_ISL_2647119, EPI_ISL_2647120, EPI_ISL_2647121, EPI_ISL_2647122, EPI_ISL_2647123, EPI_ISL_2647124, EPI_ISL_2647125, EPI_ISL_2647126, EPI_ISL_2647127, EPI_ISL_2647128, EPI_ISL_2647129, EPI_ISL_2647130, EPI_ISL_2647131, EPI_ISL_2647132, EPI_ISL_2647133, EPI_ISL_2647134, EPI_ISL_2647135, EPI_ISL_2647136, EPI_ISL_2647137, EPI_ISL_2647138, EPI_ISL_2647139, EPI_ISL_2647140, EPI_ISL_2647141, EPI_ISL_2647142, EPI_ISL_2647143, EPI_ISL_2647144, EPI_ISL_2647145, EPI_ISL_2647146, EPI_ISL_2647147, EPI_ISL_2647148, EPI_ISL_2647149, EPI_ISL_2647150, EPI_ISL_2647151, EPI_ISL_2647152, EPI_ISL_2647153, EPI_ISL_2647154, EPI_ISL_2647155, EPI_ISL_2647156, EPI_ISL_2647157, EPI_ISL_2647158, EPI_ISL_2647159, EPI_ISL_2647160, EPI_ISL_2647161, EPI_ISL_2647162, EPI_ISL_2647163, EPI_ISL_2647164, EPI_ISL_2647165, EPI_ISL_2647166, EPI_ISL_2647167, EPI_ISL_2647168, EPI_ISL_2647169, EPI_ISL_2647170, EPI_ISL_2647171, EPI_ISL_2647172, EPI_ISL_2647173, EPI_ISL_2647174, EPI_ISL_2647175, EPI_ISL_2647176, EPI_ISL_2647177, EPI_ISL_2647178, EPI_ISL_2647179, EPI_ISL_2647180, EPI_ISL_2647181, EPI_ISL_2647182, EPI_ISL_2647183, EPI_ISL_2647184, EPI_ISL_2647185, EPI_ISL_2647186, EPI_ISL_2647187, EPI_ISL_2647188, EPI_ISL_2647189, EPI_ISL_2647190, EPI_ISL_2647191, EPI_ISL_2647192, EPI_ISL_2647193, EPI_ISL_2647194, EPI_ISL_2647195, EPI_ISL_2647196, EPI_ISL_2647197, EPI_ISL_2647198, EPI_ISL_2647199, EPI_ISL_2647200, EPI_ISL_2647201, EPI_ISL_2647202, EPI_ISL_2647203, EPI_ISL_2647204, EPI_ISL_2647205, EPI_ISL_2647206, EPI_ISL_2647207, EPI_ISL_2647208, EPI_ISL_2647209, EPI_ISL_2647210, EPI_ISL_2647211, EPI_ISL_2647212, EPI_ISL_2647213, EPI_ISL_2647214, EPI_ISL_2647215, EPI_ISL_2647216, EPI_ISL_2647217, EPI_ISL_2647218, EPI_ISL_2647219, EPI_ISL_2647220, EPI_ISL_2647221, EPI_ISL_2647222, EPI_ISL_2647223, EPI_ISL_2647224, EPI_ISL_2647225, EPI_ISL_2647226, EPI_ISL_2647227, EPI_ISL_2647228, EPI_ISL_2647229, EPI_ISL_2647230, EPI_ISL_2647231, EPI_ISL_2647232, EPI_ISL_2647233, EPI_ISL_2647234, EPI_ISL_2647235, EPI_ISL_2647236, EPI_ISL_2647237, EPI_ISL_2647238, EPI_ISL_2647239, EPI_ISL_2647240, EPI_ISL_2647241, EPI_ISL_2647242, EPI_ISL_2647243, EPI_ISL_2647244, EPI_ISL_2647245, EPI_ISL_2647246, EPI_ISL_2647247, EPI_ISL_2647248, EPI_ISL_2647249, EPI_ISL_2647250, EPI_ISL_2647251, EPI_ISL_2647252, EPI_ISL_2647253, EPI_ISL_2647254, EPI_ISL_2647255, EPI_ISL_2647256, EPI_ISL_2647257, EPI_ISL_2647258, EPI_ISL_2647259, EPI_ISL_2647260, EPI_ISL_2647261, EPI_ISL_2647262, EPI_ISL_2647264, EPI_ISL_2647265, EPI_ISL_2647266, EPI_ISL_2647267, EPI_ISL_2647268, EPI_ISL_2647269, EPI_ISL_2647270, EPI_ISL_2647271, EPI_ISL_2647272, EPI_ISL_2647273, EPI_ISL_2647274, EPI_ISL_2647275, EPI_ISL_2647276, EPI_ISL_2647278, EPI_ISL_2647279, EPI_ISL_2647280, EPI_ISL_2647281, EPI_ISL_2647282, EPI_ISL_2647283, EPI_ISL_2647286, EPI_ISL_2647287, EPI_ISL_2647288, EPI_ISL_2647289, EPI_ISL_2647290, EPI_ISL_2647291, EPI_ISL_2647292, EPI_ISL_2647293, EPI_ISL_2647294, EPI_ISL_2647295, EPI_ISL_2647296, EPI_ISL_2647297, EPI_ISL_2647298, EPI_ISL_2647299, EPI_ISL_2647300, EPI_ISL_2647301, EPI_ISL_2647302, EPI_ISL_2647303, EPI_ISL_2647304, EPI_ISL_2647305, EPI_ISL_2647306, EPI_ISL_2647307, EPI_ISL_2647308, EPI_ISL_2647309, EPI_ISL_2647310, EPI_ISL_2647311, EPI_ISL_2647312, EPI_ISL_2647313, EPI_ISL_2647314, EPI_ISL_2647315, EPI_ISL_2647316, EPI_ISL_2647317, EPI_ISL_2647318, EPI_ISL_2647319, EPI_ISL_2647320, EPI_ISL_2647321, EPI_ISL_2647322, EPI_ISL_2647323, EPI_ISL_2647324, EPI_ISL_2647325, EPI_ISL_2647326, EPI_ISL_2647327, EPI_ISL_2647328, EPI_ISL_2647329, EPI_ISL_2647330, EPI_ISL_2647331, EPI_ISL_2647332, EPI_ISL_2647333, EPI_ISL_2647334, EPI_ISL_2647335, EPI_ISL_2647336, EPI_ISL_2647337, EPI_ISL_2647338, EPI_ISL_2647339, EPI_ISL_2647340, EPI_ISL_2647341, EPI_ISL_2647342, EPI_ISL_2647343, EPI_ISL_2647344, EPI_ISL_2647345, EPI_ISL_2647346, EPI_ISL_2647347, EPI_ISL_2647348, EPI_ISL_2647349, EPI_ISL_2647350, EPI_ISL_2647351, EPI_ISL_2647352, EPI_ISL_2647353, EPI_ISL_2647354, EPI_ISL_2647355, EPI_ISL_2647356, EPI_ISL_2647357, EPI_ISL_2647358, EPI_ISL_2647359, EPI_ISL_2647360, EPI_ISL_2647361, EPI_ISL_2647362, EPI_ISL_2647363, EPI_ISL_2647364, EPI_ISL_2647365, EPI_ISL_2647366, EPI_ISL_2647367, EPI_ISL_2647368, EPI_ISL_2647369, EPI_ISL_2647370, EPI_ISL_2647371, EPI_ISL_2647372, EPI_ISL_2647373, EPI_ISL_2647374, EPI_ISL_2647375, EPI_ISL_2647376, EPI_ISL_2647377, EPI_ISL_2647378, EPI_ISL_2647379, EPI_ISL_2647380, EPI_ISL_2647381, EPI_ISL_2647382, EPI_ISL_2647383, EPI_ISL_2647384, EPI_ISL_2647385, EPI_ISL_2647386, EPI_ISL_2647387, EPI_ISL_2647388, EPI_ISL_2647389, EPI_ISL_2647390, EPI_ISL_2647391, EPI_ISL_2647392, EPI_ISL_2647393, EPI_ISL_2647394, EPI_ISL_2647395, EPI_ISL_2647396, EPI_ISL_2647397, EPI_ISL_2647398, EPI_ISL_2647399, EPI_ISL_2647400, EPI_ISL_2647401, EPI_ISL_2647402, EPI_ISL_2647403, EPI_ISL_2647404, EPI_ISL_2647405, EPI_ISL_2647406, EPI_ISL_2647407, EPI_ISL_2647408, EPI_ISL_2647409, EPI_ISL_2647410, EPI_ISL_2647411, EPI_ISL_2647412, EPI_ISL_2647413, EPI_ISL_2647414, EPI_ISL_2647415, EPI_ISL_2647416, EPI_ISL_2647417, EPI_ISL_2647418, EPI_ISL_2647419, EPI_ISL_2647420, EPI_ISL_2647421, EPI_ISL_2647422, EPI_ISL_2647423, EPI_ISL_2647424, EPI_ISL_2647425, EPI_ISL_2647426, EPI_ISL_2647427, EPI_ISL_2647428, EPI_ISL_2647429, EPI_ISL_2647430, EPI_ISL_2647431, EPI_ISL_2647432, EPI_ISL_2647433, EPI_ISL_2647434, EPI_ISL_2647435, EPI_ISL_2647436, EPI_ISL_2647437, EPI_ISL_2647438, EPI_ISL_2647439, EPI_ISL_2647440, EPI_ISL_2647441, EPI_ISL_2647442, EPI_ISL_2647443, EPI_ISL_2647444, EPI_ISL_2647445, EPI_ISL_2647446, EPI_ISL_2647447, EPI_ISL_2647448, EPI_ISL_2647449, EPI_ISL_2647450, EPI_ISL_2647451, EPI_ISL_2647452, EPI_ISL_2647453, EPI_ISL_2647454, EPI_ISL_2647455, EPI_ISL_2647456, EPI_ISL_2647457, EPI_ISL_2647458, EPI_ISL_2647459, EPI_ISL_2647460, EPI_ISL_2647461, EPI_ISL_2647462, EPI_ISL_2647463, EPI_ISL_2647464, EPI_ISL_2647465, EPI_ISL_2647466, EPI_ISL_2647467, EPI_ISL_2647468, EPI_ISL_2647469, EPI_ISL_2647470, EPI_ISL_2647471, EPI_ISL_2647472, EPI_ISL_2647473, EPI_ISL_2647474, EPI_ISL_2647475, EPI_ISL_2647476, EPI_ISL_2647477, EPI_ISL_2647478, EPI_ISL_2647479, EPI_ISL_2647480, EPI_ISL_2647481, EPI_ISL_2647482, EPI_ISL_2647483, EPI_ISL_2647484, EPI_ISL_2647485, EPI_ISL_2647486, EPI_ISL_2647487, EPI_ISL_2647488, EPI_ISL_2647489, EPI_ISL_2647490, EPI_ISL_2647491, EPI_ISL_2647492, EPI_ISL_2647493, EPI_ISL_2647494, EPI_ISL_2647495, EPI_ISL_2647496, EPI_ISL_2647497, EPI_ISL_2647498, EPI_ISL_2647499 | | | |
| see above | Mako Medical | Centers for Disease Control and Prevention Division of Viral Diseases, Pathogen Discovery | Dakota Howard, Dhwani Batra, Peter W. Cook, Kara Moser, Adrian Paskey, Jason Caravas, Benjamin Rambo-Martin, Shatavia Morrison, Christopher Gulvick, Scott Sammons, Yvette Unoarumhi, Darlene Wagner, Matthew Schmerer,Matthew Tugwell, Lauren Moon, Clinton R. Paden, Duncan MacCannell |
| EPI_ISL_2647500 | Michael E. Debakey VA Medical Center | Genomics and Discovery, Respiratory Viruses Branch, Division of Viral Diseases, Centers for Disease Control and Prevention | Jing Zhang, Anna Kelleher, Ying Tao, Yan Li, Brian Lynch, Krista Queen, Anna Uehara, Peter Cook, Han Jia Justin Ng, Rachel Marine, Clinton R. Paden, Dhwani Batra, Haibin Wang, Tara Coalter, Jasmine Padilla, Morgan Davis, Mili Sheth, Sarah Nobles, Mark Burroughs, Justin Lee, Adam Retchless, Suxiang Tong |
| EPI_ISL_2647501 | Mako Medical | Centers for Disease Control and Prevention Division of Viral Diseases, Pathogen Discovery | Dakota Howard, Dhwani Batra, Peter W. Cook, Kara Moser, Adrian Paskey, Jason Caravas, Benjamin Rambo-Martin, Shatavia Morrison, Christopher Gulvick, Scott Sammons, Yvette Unoarumhi, Darlene Wagner, Matthew Schmerer,Matthew Tugwell, Lauren Moon, Clinton R. Paden, Duncan MacCannell |
| EPI_ISL_2647502, EPI_ISL_2647503 | Michael E. Debakey VA Medical Center | Genomics and Discovery, Respiratory Viruses Branch, Division of Viral Diseases, Centers for Disease Control and Prevention | Jing Zhang, Anna Kelleher, Ying Tao, Yan Li, Brian Lynch, Krista Queen, Anna Uehara, Peter Cook, Han Jia Justin Ng, Rachel Marine, Clinton R. Paden, Dhwani Batra, Haibin Wang, Tara Coalter, Jasmine Padilla, Morgan Davis, Mili Sheth, Sarah Nobles, Mark Burroughs, Justin Lee, Adam Retchless, Suxiang Tong |
| EPI_ISL_2647504 | Mako Medical | Centers for Disease Control and Prevention Division of Viral Diseases, Pathogen Discovery | Dakota Howard, Dhwani Batra, Peter W. Cook, Kara Moser, Adrian Paskey, Jason Caravas, Benjamin Rambo-Martin, Shatavia Morrison, Christopher Gulvick, Scott Sammons, Yvette Unoarumhi, Darlene Wagner, Matthew Schmerer,Matthew Tugwell, Lauren Moon, Clinton R. Paden, Duncan MacCannell |
| EPI_ISL_2647505 | Michael E. Debakey VA Medical Center | Genomics and Discovery, Respiratory Viruses Branch, Division of Viral Diseases, Centers for Disease Control and Prevention | Jing Zhang, Anna Kelleher, Ying Tao, Yan Li, Brian Lynch, Krista Queen, Anna Uehara, Peter Cook, Han Jia Justin Ng, Rachel Marine, Clinton R. Paden, Dhwani Batra, Haibin Wang, Tara Coalter, Jasmine Padilla, Morgan Davis, Mili Sheth, Sarah Nobles, Mark Burroughs, Justin Lee, Adam Retchless, Suxiang Tong |
| EPI_ISL_2647506, EPI_ISL_2647507, EPI_ISL_2647508, EPI_ISL_2647509, EPI_ISL_2647510, EPI_ISL_2647511, EPI_ISL_2647512, EPI_ISL_2647513, EPI_ISL_2647514, EPI_ISL_2647515, EPI_ISL_2647516, EPI_ISL_2647517, EPI_ISL_2647518, EPI_ISL_2647519, EPI_ISL_2647520, EPI_ISL_2647521, EPI_ISL_2647522, EPI_ISL_2647523, EPI_ISL_2647524, EPI_ISL_2647525, EPI_ISL_2647526, EPI_ISL_2647527, EPI_ISL_2647528, EPI_ISL_2647529, EPI_ISL_2647530, EPI_ISL_2647531, EPI_ISL_2647532, EPI_ISL_2647533, EPI_ISL_2647534, EPI_ISL_2647535, EPI_ISL_2647536, EPI_ISL_2647537, EPI_ISL_2647538, EPI_ISL_2647539, EPI_ISL_2647540, EPI_ISL_2647541, EPI_ISL_2647542, EPI_ISL_2647543, EPI_ISL_2647544, EPI_ISL_2647545, EPI_ISL_2647546, EPI_ISL_2647547, EPI_ISL_2647548, EPI_ISL_2647549, EPI_ISL_2647550, EPI_ISL_2647551, EPI_ISL_2647552, EPI_ISL_2647553, EPI_ISL_2647554, EPI_ISL_2647555, EPI_ISL_2647556, EPI_ISL_2647557, EPI_ISL_2647558, EPI_ISL_2647559, EPI_ISL_2647560, EPI_ISL_2647561, EPI_ISL_2647562, EPI_ISL_2647563, EPI_ISL_2647564, EPI_ISL_2647565, EPI_ISL_2647566, EPI_ISL_2647567, EPI_ISL_2647568, EPI_ISL_2647569, EPI_ISL_2647570, EPI_ISL_2647572, EPI_ISL_2647573, EPI_ISL_2647574, EPI_ISL_2647575, EPI_ISL_2647576, EPI_ISL_2647577, EPI_ISL_2647578, EPI_ISL_2647579, EPI_ISL_2647580, EPI_ISL_2647581, EPI_ISL_2647582, EPI_ISL_2647583, EPI_ISL_2647584, EPI_ISL_2647585, EPI_ISL_2647586, EPI_ISL_2647587, EPI_ISL_2647588, EPI_ISL_2647589, EPI_ISL_2647590, EPI_ISL_2647591, EPI_ISL_2647592, EPI_ISL_2647593, EPI_ISL_2647594, EPI_ISL_2647595, EPI_ISL_2647596, EPI_ISL_2647597, EPI_ISL_2647598, EPI_ISL_2647599, EPI_ISL_2647600, EPI_ISL_2647601, EPI_ISL_2647602, EPI_ISL_2647603, EPI_ISL_2647604, EPI_ISL_2647605, EPI_ISL_2647606, EPI_ISL_2647607, EPI_ISL_2647608, EPI_ISL_2647609, EPI_ISL_2647610, EPI_ISL_2647611, EPI_ISL_2647612, EPI_ISL_2647613, EPI_ISL_2647614, EPI_ISL_2647615, EPI_ISL_2647616, EPI_ISL_2647617, EPI_ISL_2647618, EPI_ISL_2647619, EPI_ISL_2647620, EPI_ISL_2647621, EPI_ISL_2647622, EPI_ISL_2647623, EPI_ISL_2647624, EPI_ISL_2647626, EPI_ISL_2647627, EPI_ISL_2647628, EPI_ISL_2647629, EPI_ISL_2647630, EPI_ISL_2647631, EPI_ISL_2647632, EPI_ISL_2647633, EPI_ISL_2647634, EPI_ISL_2647635, EPI_ISL_2647636, EPI_ISL_2647637, EPI_ISL_2647638, EPI_ISL_2647639, EPI_ISL_2647640, EPI_ISL_2647641, EPI_ISL_2647642, EPI_ISL_2647643, EPI_ISL_2647644, EPI_ISL_2647645, EPI_ISL_2647646, EPI_ISL_2647647, EPI_ISL_2647648, EPI_ISL_2647649, EPI_ISL_2647650, EPI_ISL_2647651, EPI_ISL_2647652, EPI_ISL_2647653, EPI_ISL_2647654, EPI_ISL_2647655, EPI_ISL_2647656, EPI_ISL_2647657, EPI_ISL_2647658, EPI_ISL_2647659, EPI_ISL_2647660, EPI_ISL_2647661, EPI_ISL_2647662, EPI_ISL_2647663, EPI_ISL_2647664, EPI_ISL_2647665, EPI_ISL_2647666, EPI_ISL_2647667, EPI_ISL_2647668, EPI_ISL_2647669, EPI_ISL_2647670, EPI_ISL_2647671, EPI_ISL_2647672, EPI_ISL_2647673, EPI_ISL_2647674, EPI_ISL_2647675, EPI_ISL_2647676, EPI_ISL_2647677, EPI_ISL_2647678, EPI_ISL_2647679, EPI_ISL_2647680, EPI_ISL_2647681, EPI_ISL_2647682, EPI_ISL_2647683, EPI_ISL_2647684, EPI_ISL_2647685, EPI_ISL_2647686, EPI_ISL_2647687, EPI_ISL_2647688, EPI_ISL_2647689, EPI_ISL_2647690, EPI_ISL_2647691, EPI_ISL_2647692, EPI_ISL_2647693, EPI_ISL_2647694, EPI_ISL_2647695, EPI_ISL_2647696, EPI_ISL_2647697, EPI_ISL_2647698, EPI_ISL_2647699, EPI_ISL_2647700, EPI_ISL_2647701, EPI_ISL_2647702, EPI_ISL_2647703, EPI_ISL_2647704, EPI_ISL_2647705, EPI_ISL_2647706, EPI_ISL_2647707, EPI_ISL_2647708, EPI_ISL_2647709, EPI_ISL_2647710, EPI_ISL_2647711, EPI_ISL_2647712, EPI_ISL_2647713, EPI_ISL_2647714, EPI_ISL_2647715, EPI_ISL_2647716, EPI_ISL_2647717, EPI_ISL_2647718, EPI_ISL_2647719, EPI_ISL_2647720, EPI_ISL_2647721, EPI_ISL_2647722, EPI_ISL_2647723, EPI_ISL_2647724, EPI_ISL_2647725, EPI_ISL_2647726, EPI_ISL_2647727, EPI_ISL_2647728 | | | |
| see above | Mako Medical | Centers for Disease Control and Prevention Division of Viral Diseases, Pathogen | Dakota Howard, Dhwani Batra, Peter W. Cook, Kara Moser, Adrian Paskey, Jason Caravas, Benjamin Rambo-Martin, Shatavia Morrison, Christopher Gulvick, Scott Sammons, Yvette Unoarumhi, Darlene Wagner, Matthew Schmerer,Matthew Tugwell, Lauren Moon, Clinton R. Paden, Duncan MacCannell |

| | | | |
|---|---|---|---|
| | | Discovery | |
| EPI_ISL_2647729, EPI_ISL_2647730 | Wyoming Public Health Laboratory | Wyoming Public Health Laboratory | Jim Mildenberger, Taylor Fearing, Channing Weber, Ashley Norberg, Chayse Rowley, Marley Goetz, Brian Dominguez, Elliot Thomasson, Sam Britz, Cari Sloma, Robert Petit, and Rob Christensen |
| EPI_ISL_2647731 | Mako Medical | Centers for Disease Control and Prevention Division of Viral Diseases, Pathogen Discovery | Dakota Howard, Dhwani Batra, Peter W. Cook, Kara Moser, Adrian Paskey, Jason Caravas, Benjamin Rambo-Martin, Shatavia Morrison, Christopher Gulvick, Scott Sammons, Yvette Unoarumhi, Darlene Wagner, Matthew Schmerer,Matthew Tugwell, Lauren Moon, Clinton R. Paden, Duncan MacCannell |
| EPI_ISL_2647732, EPI_ISL_2647733 | Wyoming Public Health Laboratory | Wyoming Public Health Laboratory | Jim Mildenberger, Taylor Fearing, Channing Weber, Ashley Norberg, Chayse Rowley, Marley Goetz, Brian Dominguez, Elliot Thomasson, Sam Britz, Cari Sloma, Robert Petit, and Rob Christensen |
| EPI_ISL_2647734 | Mako Medical | Centers for Disease Control and Prevention Division of Viral Diseases, Pathogen Discovery | Dakota Howard, Dhwani Batra, Peter W. Cook, Kara Moser, Adrian Paskey, Jason Caravas, Benjamin Rambo-Martin, Shatavia Morrison, Christopher Gulvick, Scott Sammons, Yvette Unoarumhi, Darlene Wagner, Matthew Schmerer,Matthew Tugwell, Lauren Moon, Clinton R. Paden, Duncan MacCannell |
| EPI_ISL_2647735, EPI_ISL_2647736 | Wyoming Public Health Laboratory | Wyoming Public Health Laboratory | Jim Mildenberger, Taylor Fearing, Channing Weber, Ashley Norberg, Chayse Rowley, Marley Goetz, Brian Dominguez, Elliot Thomasson, Sam Britz, Cari Sloma, Robert Petit, and Rob Christensen |
| EPI_ISL_2647737 | Mako Medical | Centers for Disease Control and Prevention Division of Viral Diseases, Pathogen Discovery | Dakota Howard, Dhwani Batra, Peter W. Cook, Kara Moser, Adrian Paskey, Jason Caravas, Benjamin Rambo-Martin, Shatavia Morrison, Christopher Gulvick, Scott Sammons, Yvette Unoarumhi, Darlene Wagner, Matthew Schmerer,Matthew Tugwell, Lauren Moon, Clinton R. Paden, Duncan MacCannell |
| EPI_ISL_2647738 | Wyoming Public Health Laboratory | Wyoming Public Health Laboratory | Jim Mildenberger, Taylor Fearing, Channing Weber, Ashley Norberg, Chayse Rowley, Marley Goetz, Brian Dominguez, Elliot Thomasson, Sam Britz, Cari Sloma, Robert Petit, and Rob Christensen |
| EPI_ISL_2647739 | Mako Medical | Centers for Disease Control and Prevention Division of Viral Diseases, Pathogen Discovery | Dakota Howard, Dhwani Batra, Peter W. Cook, Kara Moser, Adrian Paskey, Jason Caravas, Benjamin Rambo-Martin, Shatavia Morrison, Christopher Gulvick, Scott Sammons, Yvette Unoarumhi, Darlene Wagner, Matthew Schmerer,Matthew Tugwell, Lauren Moon, Clinton R. Paden, Duncan MacCannell |
| EPI_ISL_2647740, EPI_ISL_2647741 | Wyoming Public Health Laboratory | Wyoming Public Health Laboratory | Jim Mildenberger, Taylor Fearing, Channing Weber, Ashley Norberg, Chayse Rowley, Marley Goetz, Brian Dominguez, Elliot Thomasson, Sam Britz, Cari Sloma, Robert Petit, and Rob Christensen |
| EPI_ISL_2647742 | Mako Medical | Centers for Disease Control and Prevention Division of Viral Diseases, Pathogen Discovery | Dakota Howard, Dhwani Batra, Peter W. Cook, Kara Moser, Adrian Paskey, Jason Caravas, Benjamin Rambo-Martin, Shatavia Morrison, Christopher Gulvick, Scott Sammons, Yvette Unoarumhi, Darlene Wagner, Matthew Schmerer,Matthew Tugwell, Lauren Moon, Clinton R. Paden, Duncan MacCannell |
| EPI_ISL_2647743, EPI_ISL_2647744 | Wyoming Public Health Laboratory | Wyoming Public Health Laboratory | Jim Mildenberger, Taylor Fearing, Channing Weber, Ashley Norberg, Chayse Rowley, Marley Goetz, Brian Dominguez, Elliot Thomasson, Sam Britz, Cari Sloma, Robert Petit, and Rob Christensen |
| EPI_ISL_2647745 | Mako Medical | Centers for Disease Control and Prevention Division of Viral Diseases, Pathogen Discovery | Dakota Howard, Dhwani Batra, Peter W. Cook, Kara Moser, Adrian Paskey, Jason Caravas, Benjamin Rambo-Martin, Shatavia Morrison, Christopher Gulvick, Scott Sammons, Yvette Unoarumhi, Darlene Wagner, Matthew Schmerer,Matthew Tugwell, Lauren Moon, Clinton R. Paden, Duncan MacCannell |
| EPI_ISL_2647746, EPI_ISL_2647747 | Wyoming Public Health Laboratory | Wyoming Public Health Laboratory | Jim Mildenberger, Taylor Fearing, Channing Weber, Ashley Norberg, Chayse Rowley, Marley Goetz, Brian Dominguez, Elliot Thomasson, Sam Britz, Cari Sloma, Robert Petit, and Rob Christensen |
| EPI_ISL_2647748 | Mako Medical | Centers for Disease Control and Prevention Division of Viral Diseases, Pathogen Discovery | Dakota Howard, Dhwani Batra, Peter W. Cook, Kara Moser, Adrian Paskey, Jason Caravas, Benjamin Rambo-Martin, Shatavia Morrison, Christopher Gulvick, Scott Sammons, Yvette Unoarumhi, Darlene Wagner, Matthew Schmerer,Matthew Tugwell, Lauren Moon, Clinton R. Paden, Duncan MacCannell |
| EPI_ISL_2647749, EPI_ISL_2647750 | Wyoming Public Health Laboratory | Wyoming Public Health Laboratory | Jim Mildenberger, Taylor Fearing, Channing Weber, Ashley Norberg, Chayse Rowley, Marley Goetz, Brian Dominguez, Elliot Thomasson, Sam Britz, Cari Sloma, Robert Petit, and Rob Christensen |
| EPI_ISL_2647751 | Mako Medical | Centers for Disease Control and Prevention Division of Viral Diseases, Pathogen Discovery | Dakota Howard, Dhwani Batra, Peter W. Cook, Kara Moser, Adrian Paskey, Jason Caravas, Benjamin Rambo-Martin, Shatavia Morrison, Christopher Gulvick, Scott Sammons, Yvette Unoarumhi, Darlene Wagner, Matthew Schmerer,Matthew Tugwell, Lauren Moon, Clinton R. Paden, Duncan MacCannell |
| EPI_ISL_2647752 | Wyoming Public Health Laboratory | Wyoming Public Health Laboratory | Jim Mildenberger, Taylor Fearing, Channing Weber, Ashley Norberg, Chayse Rowley, Marley Goetz, Brian Dominguez, Elliot Thomasson, Sam Britz, Cari Sloma, Robert Petit, and Rob Christensen |
| EPI_ISL_2647753 | Mako Medical | Centers for Disease Control and Prevention Division of Viral Diseases, Pathogen Discovery | Dakota Howard, Dhwani Batra, Peter W. Cook, Kara Moser, Adrian Paskey, Jason Caravas, Benjamin Rambo-Martin, Shatavia Morrison, Christopher Gulvick, Scott Sammons, Yvette Unoarumhi, Darlene Wagner, Matthew Schmerer,Matthew Tugwell, Lauren Moon, Clinton R. Paden, Duncan MacCannell |
| EPI_ISL_2647754, EPI_ISL_2647755 | Wyoming Public Health Laboratory | Wyoming Public Health Laboratory | Jim Mildenberger, Taylor Fearing, Channing Weber, Ashley Norberg, Chayse Rowley, Marley Goetz, Brian Dominguez, Elliot Thomasson, Sam Britz, Cari Sloma, Robert Petit, and Rob Christensen |
| EPI_ISL_2647756 | Mako Medical | Centers for Disease Control and Prevention Division of Viral Diseases, Pathogen Discovery | Dakota Howard, Dhwani Batra, Peter W. Cook, Kara Moser, Adrian Paskey, Jason Caravas, Benjamin Rambo-Martin, Shatavia Morrison, Christopher Gulvick, Scott Sammons, Yvette Unoarumhi, Darlene Wagner, Matthew Schmerer,Matthew Tugwell, Lauren Moon, Clinton R. Paden, Duncan MacCannell |
| EPI_ISL_2647757, EPI_ISL_2647758 | Wyoming Public Health Laboratory | Wyoming Public Health Laboratory | Jim Mildenberger, Taylor Fearing, Channing Weber, Ashley Norberg, Chayse Rowley, Marley Goetz, Brian Dominguez, Elliot Thomasson, Sam Britz, Cari Sloma, Robert Petit, and Rob Christensen |
| EPI_ISL_2647759 | Mako Medical | Centers for Disease Control and Prevention Division of Viral Diseases, Pathogen Discovery | Dakota Howard, Dhwani Batra, Peter W. Cook, Kara Moser, Adrian Paskey, Jason Caravas, Benjamin Rambo-Martin, Shatavia Morrison, Christopher Gulvick, Scott Sammons, Yvette Unoarumhi, Darlene Wagner, Matthew Schmerer,Matthew Tugwell, Lauren Moon, Clinton R. Paden, Duncan MacCannell |
| EPI_ISL_2647760, EPI_ISL_2647761, EPI_ISL_2647763 | Wyoming Public Health Laboratory | Wyoming Public Health Laboratory | Jim Mildenberger, Taylor Fearing, Channing Weber, Ashley Norberg, Chayse Rowley, Marley Goetz, Brian Dominguez, Elliot Thomasson, Sam Britz, Cari Sloma, Robert Petit, and Rob Christensen |
| EPI_ISL_2647764 | Mako Medical | Centers for Disease Control and Prevention Division of Viral Diseases, Pathogen Discovery | Dakota Howard, Dhwani Batra, Peter W. Cook, Kara Moser, Adrian Paskey, Jason Caravas, Benjamin Rambo-Martin, Shatavia Morrison, Christopher Gulvick, Scott Sammons, Yvette Unoarumhi, Darlene Wagner, Matthew Schmerer,Matthew Tugwell, Lauren Moon, Clinton R. Paden, Duncan MacCannell |
| EPI_ISL_2647765, EPI_ISL_2647766 | Wyoming Public Health Laboratory | Wyoming Public Health Laboratory | Jim Mildenberger, Taylor Fearing, Channing Weber, Ashley Norberg, Chayse Rowley, Marley Goetz, Brian Dominguez, Elliot Thomasson, Sam Britz, Cari Sloma, Robert Petit, and Rob Christensen |
| EPI_ISL_2647767 | Mako Medical | Centers for Disease Control and Prevention Division of Viral Diseases, Pathogen Discovery | Dakota Howard, Dhwani Batra, Peter W. Cook, Kara Moser, Adrian Paskey, Jason Caravas, Benjamin Rambo-Martin, Shatavia Morrison, Christopher Gulvick, Scott Sammons, Yvette Unoarumhi, Darlene Wagner, Matthew Schmerer,Matthew Tugwell, Lauren Moon, Clinton R. Paden, Duncan MacCannell |
| EPI_ISL_2647768, EPI_ISL_2647769 | Wyoming Public Health Laboratory | Wyoming Public Health Laboratory | Jim Mildenberger, Taylor Fearing, Channing Weber, Ashley Norberg, Chayse Rowley, Marley Goetz, Brian Dominguez, Elliot Thomasson, Sam Britz, Cari Sloma, Robert Petit, and Rob Christensen |
| EPI_ISL_2647770 | Mako Medical | Centers for Disease Control and Prevention Division of Viral Diseases, Pathogen Discovery | Dakota Howard, Dhwani Batra, Peter W. Cook, Kara Moser, Adrian Paskey, Jason Caravas, Benjamin Rambo-Martin, Shatavia Morrison, Christopher Gulvick, Scott Sammons, Yvette Unoarumhi, Darlene Wagner, Matthew Schmerer,Matthew Tugwell, Lauren Moon, Clinton R. Paden, Duncan MacCannell |
| EPI_ISL_2647771, EPI_ISL_2647772 | Wyoming Public Health Laboratory | Wyoming Public Health Laboratory | Jim Mildenberger, Taylor Fearing, Channing Weber, Ashley Norberg, Chayse Rowley, Marley Goetz, Brian Dominguez, Elliot Thomasson, Sam Britz, Cari Sloma, Robert Petit, and Rob Christensen |
| EPI_ISL_2647773 | Mako Medical | Centers for Disease Control and Prevention Division of Viral Diseases, Pathogen Discovery | Dakota Howard, Dhwani Batra, Peter W. Cook, Kara Moser, Adrian Paskey, Jason Caravas, Benjamin Rambo-Martin, Shatavia Morrison, Christopher Gulvick, Scott Sammons, Yvette Unoarumhi, Darlene Wagner, Matthew Schmerer,Matthew Tugwell, Lauren Moon, Clinton R. Paden, Duncan MacCannell |
| EPI_ISL_2647774 | Wyoming Public Health Laboratory | Wyoming Public Health Laboratory | Jim Mildenberger, Taylor Fearing, Channing Weber, Ashley Norberg, Chayse Rowley, Marley Goetz, Brian Dominguez, Elliot Thomasson, Sam Britz, Cari Sloma, Robert Petit, and Rob Christensen |
| EPI_ISL_2647775 | Mako Medical | Centers for Disease Control and Prevention Division of Viral Diseases, Pathogen Discovery | Dakota Howard, Dhwani Batra, Peter W. Cook, Kara Moser, Adrian Paskey, Jason Caravas, Benjamin Rambo-Martin, Shatavia Morrison, Christopher Gulvick, Scott Sammons, Yvette Unoarumhi, Darlene Wagner, Matthew Schmerer,Matthew Tugwell, Lauren Moon, Clinton R. Paden, Duncan MacCannell |
| EPI_ISL_2647776 | Wyoming Public Health Laboratory | Wyoming Public Health Laboratory | Jim Mildenberger, Taylor Fearing, Channing Weber, Ashley Norberg, Chayse Rowley, Marley Goetz, Brian Dominguez, Elliot Thomasson, Sam Britz, Cari Sloma, Robert Petit, and Rob Christensen |
| EPI_ISL_2647777, EPI_ISL_2647778, EPI_ISL_2647779, EPI_ISL_2647780, EPI_ISL_2647781, EPI_ISL_2647782, EPI_ISL_2647783, EPI_ISL_2647784, EPI_ISL_2647785, EPI_ISL_2647786, EPI_ISL_2647787, EPI_ISL_2647788, EPI_ISL_2647789, EPI_ISL_2647790, EPI_ISL_2647791, EPI_ISL_2647792, EPI_ISL_2647793, EPI_ISL_2647794, EPI_ISL_2647795, EPI_ISL_2647797, EPI_ISL_2647798, EPI_ISL_2647799, EPI_ISL_2647800, EPI_ISL_2647801, EPI_ISL_2647802, EPI_ISL_2647803, EPI_ISL_2647804, EPI_ISL_2647805, EPI_ISL_2647806, EPI_ISL_2647807, EPI_ISL_2647808, EPI_ISL_2647809, EPI_ISL_2647810, EPI_ISL_2647811, EPI_ISL_2647812, EPI_ISL_2647813, EPI_ISL_2647814, EPI_ISL_2647815, EPI_ISL_2647816, EPI_ISL_2647817, EPI_ISL_2647818, EPI_ISL_2647820, EPI_ISL_2647821, EPI_ISL_2647822, EPI_ISL_2647823, EPI_ISL_2647824, EPI_ISL_2647825, EPI_ISL_2647826, EPI_ISL_2647827, EPI_ISL_2647828, EPI_ISL_2647829, EPI_ISL_2647831, EPI_ISL_2647832, EPI_ISL_2647833, EPI_ISL_2647834, EPI_ISL_2647835, EPI_ISL_2647836, EPI_ISL_2647837, EPI_ISL_2647838, EPI_ISL_2647839, EPI_ISL_2647840, EPI_ISL_2647841, EPI_ISL_2647842, EPI_ISL_2647843, EPI_ISL_2647844, EPI_ISL_2647845, EPI_ISL_2647846, EPI_ISL_2647847, EPI_ISL_2647848, EPI_ISL_2647849, EPI_ISL_2647850, EPI_ISL_2647851, EPI_ISL_2647852, EPI_ISL_2647853, EPI_ISL_2647854, EPI_ISL_2647855, EPI_ISL_2647856, EPI_ISL_2647857, EPI_ISL_2647858, EPI_ISL_2647859, EPI_ISL_2647860, EPI_ISL_2647861, EPI_ISL_2647862, EPI_ISL_2647863, EPI_ISL_2647864, EPI_ISL_2647865, EPI_ISL_2647866, EPI_ISL_2647867, EPI_ISL_2647868, EPI_ISL_2647869, EPI_ISL_2647870, EPI_ISL_2647871, EPI_ISL_2647872, EPI_ISL_2647873, EPI_ISL_2647874, EPI_ISL_2647875, EPI_ISL_2647876, EPI_ISL_2647877, EPI_ISL_2647878, EPI_ISL_2647879, EPI_ISL_2647880, EPI_ISL_2647881, EPI_ISL_2647882, EPI_ISL_2647883, EPI_ISL_2647884, EPI_ISL_2647885, EPI_ISL_2647886, EPI_ISL_2647887, EPI_ISL_2647888, EPI_ISL_2647889, EPI_ISL_2647890, EPI_ISL_2647891, EPI_ISL_2647892, EPI_ISL_2647893, EPI_ISL_2647894, EPI_ISL_2647895, EPI_ISL_2647896, EPI_ISL_2647897, EPI_ISL_2647898, EPI_ISL_2647899, EPI_ISL_2647900, EPI_ISL_2647901, EPI_ISL_2647902, EPI_ISL_2647903, EPI_ISL_2647904, EPI_ISL_2647905, EPI_ISL_2647906, EPI_ISL_2647907, EPI_ISL_2647908, EPI_ISL_2647909, EPI_ISL_2647910, EPI_ISL_2647911, EPI_ISL_2647912, EPI_ISL_2647913, EPI_ISL_2647914, EPI_ISL_2647915, EPI_ISL_2647916, EPI_ISL_2647917, EPI_ISL_2647918, EPI_ISL_2647919, EPI_ISL_2647921, EPI_ISL_2647922, EPI_ISL_2647923, EPI_ISL_2647924, EPI_ISL_2647926, EPI_ISL_2647927, EPI_ISL_2647929, EPI_ISL_2647930, EPI_ISL_2647931, EPI_ISL_2647932, EPI_ISL_2647933, EPI_ISL_2647934, EPI_ISL_2647935, EPI_ISL_2647936, EPI_ISL_2647937, EPI_ISL_2647938, EPI_ISL_2647939, EPI_ISL_2647940, EPI_ISL_2647941, EPI_ISL_2647942, EPI_ISL_2647943, EPI_ISL_2647944, EPI_ISL_2647945, EPI_ISL_2647946, EPI_ISL_2647947, EPI_ISL_2647948, EPI_ISL_2647949, EPI_ISL_2647950, EPI_ISL_2647951, EPI_ISL_2647952, EPI_ISL_2647953, EPI_ISL_2647954, EPI_ISL_2647955, EPI_ISL_2647956, EPI_ISL_2647957, EPI_ISL_2647958, EPI_ISL_2647959, EPI_ISL_2647960, EPI_ISL_2647961, EPI_ISL_2647962, EPI_ISL_2647963, EPI_ISL_2647964, EPI_ISL_2647965, EPI_ISL_2647966, EPI_ISL_2647968, EPI_ISL_2647969, EPI_ISL_2647970, EPI_ISL_2647971, EPI_ISL_2647972, EPI_ISL_2647973, EPI_ISL_2647974, EPI_ISL_2647975, EPI_ISL_2647976, EPI_ISL_2647977, EPI_ISL_2647978, EPI_ISL_2647979, EPI_ISL_2647980, EPI_ISL_2647981, EPI_ISL_2647982, EPI_ISL_2647983, EPI_ISL_2647984, EPI_ISL_2647985, EPI_ISL_2647986, EPI_ISL_2647987, EPI_ISL_2647988, EPI_ISL_2647989, EPI_ISL_2647990, EPI_ISL_2647991, EPI_ISL_2647992, EPI_ISL_2647993, EPI_ISL_2647994, EPI_ISL_2647995, EPI_ISL_2647996, EPI_ISL_2647997, EPI_ISL_2647998, EPI_ISL_2647999, EPI_ISL_2648000, EPI_ISL_2648001, EPI_ISL_2648002, EPI_ISL_2648003, EPI_ISL_2648004, EPI_ISL_2648005, EPI_ISL_2648006, EPI_ISL_2648007, EPI_ISL_2648008, EPI_ISL_2648009, EPI_ISL_2648010, EPI_ISL_2648011, EPI_ISL_2648012 | | | |
| see above | Mako Medical | Centers for Disease Control and Prevention Division of Viral Diseases, Pathogen Discovery | Dakota Howard, Dhwani Batra, Peter W. Cook, Kara Moser, Adrian Paskey, Jason Caravas, Benjamin Rambo-Martin, Shatavia Morrison, Christopher Gulvick, Scott Sammons, Yvette Unoarumhi, Darlene Wagner, Matthew Schmerer,Matthew Tugwell, Lauren Moon, Clinton R. Paden, Duncan MacCannell |

| | | | |
|---|---|---|---|
| EPI_ISL_2648013, EPI_ISL_2648014, EPI_ISL_2648015, EPI_ISL_2648016, EPI_ISL_2648017, EPI_ISL_2648019, EPI_ISL_2648020, EPI_ISL_2648021, EPI_ISL_2648022, EPI_ISL_2648023, EPI_ISL_2648024, EPI_ISL_2648025, EPI_ISL_2648026, EPI_ISL_2648027, EPI_ISL_2648028, EPI_ISL_2648029, EPI_ISL_2648030, EPI_ISL_2648031, EPI_ISL_2648032, EPI_ISL_2648033, EPI_ISL_2648034, EPI_ISL_2648035, EPI_ISL_2648036, EPI_ISL_2648037, EPI_ISL_2648038, EPI_ISL_2648039, EPI_ISL_2648040, EPI_ISL_2648041, EPI_ISL_2648042, EPI_ISL_2648043, EPI_ISL_2648044, EPI_ISL_2648046, EPI_ISL_2648047, EPI_ISL_2648048, EPI_ISL_2648049, EPI_ISL_2648051, EPI_ISL_2648053, EPI_ISL_2648055, EPI_ISL_2648056, EPI_ISL_2648057 | | | |
| see above | University of Florida Health Shands Hospital | Fusion Genomics Corporation | Shing H. Zhan, Sepideh M. Alamouti, Habib Daneshpajouh, Brian S. Kwok, Meng-Hsun Lee, Jaswinder Khattra, Herbert J. Houck, Kenneth H. Rand |
| EPI_ISL_2648110, EPI_ISL_2648111, EPI_ISL_2648112, EPI_ISL_2648113 | Instituto Nacional de Enfermedades Respiratorias (INER) | Centro de Investigación en Enfermedades Infecciosas (CIENI), Instituto Nacional de Enfermedades Respiratorias (INER) | Consorcio Mexicano de Vigilancia Genómica (CoViGen-Mex). Authors (in alphabetical order): Julio Elias Alvarado-Yaah, Carlos F. Arias, Santiago Ávila-Ríos, Eduardo Becerril-Vargas, Víctor Hugo Borja-Aburto, Celia Boukadida, Cristóbal Cháidez-Quiróz, Juan Bautista Chale-Dzul, Ricardo Ciria Merce, Andreu Comas-García, Célida Duque Molina, Julissa Enciso-Ibarra, José Antonio Enciso-Moreno, Gloria Elena Espinosa-Ayala, Fernando Fontove-Herrera, Daniel Fregoso-Rueda, Victor Eduardo Garcia-Arias, Alejandra Garcia-Gasca, Bruno Gomez-Gil, Jean Pierre González, Irvin González-López, Concepción Grajales-Muñiz, Ricardo Grande, Rosa María Gutierrez Rios, Alejandra Hernández-Terán, Alfredo Herrera-Estrella, Carla Ivón Herrera-Najera, Pavel Isa, Daniel Lira Morales, Susana Lopez, Antonio Loza Román, Brenda Irasema Maldonado-Meza, Bernardo Martínez-Miguel, José Arturo Martínez-Orozco, Célida Martínez- Rodríguez, Margarita Matías-Florentino, Fidencio Mejía-Nepomuceno, María Guadalupe de Jesús Mireles-Rivera, Gloria María Molina-Salinas, Hector Montoya-Fuentes, , Mario Mújica-Sánchez, José Esteban Muñoz-Medina, José de Jesús Nuñez-Contreras, Alicia Ocaña-Mondragón, Luis Alberto Ochoa-Carrera, Hector Esteban Paz-Juárez, Marissa Pérez-García, Francisco Pulido, Helen Haydee Fernanda Ramírez-Plascencia, Jorge Salas-Hernández, Angel Gustavo Salas-Lais, Alejandro Sanchez-Flores, Clara Esperanza Santacruz-Tinoco, María Guadalupe Santiago-Mauricio, Selene Zárate, Nelly Sélem-Mojica, Blanca Taboada, Gloria Vazquez, Joel Armando Vázquez-Pérez. |
| EPI_ISL_2648114 | Instituto Nacional de Cancerología | Centro de Investigación en Enfermedades Infecciosas (CIENI), Instituto Nacional de Enfermedades Respiratorias (INER) | Consorcio Mexicano de Vigilancia Genómica (CoViGen-Mex). Authors (in alphabetical order): Julio Elias Alvarado-Yaah, Carlos F. Arias, Santiago Ávila-Ríos, Eduardo Becerril-Vargas, Víctor Hugo Borja-Aburto, Celia Boukadida, Cristóbal Cháidez-Quiróz, Juan Bautista Chale-Dzul, Ricardo Ciria Merce, Andreu Comas-García, Célida Duque Molina, Julissa Enciso-Ibarra, José Antonio Enciso-Moreno, Gloria Elena Espinosa-Ayala, Fernando Fontove-Herrera, Daniel Fregoso-Rueda, Victor Eduardo Garcia-Arias, Alejandra Garcia-Gasca, Bruno Gomez-Gil, Jean Pierre González, Irvin González-López, Concepción Grajales-Muñiz, Ricardo Grande, Rosa María Gutierrez Rios, Alejandra Hernández-Terán, Alfredo Herrera-Estrella, Carla Ivón Herrera-Najera, Pavel Isa, Daniel Lira Morales, Susana Lopez, Antonio Loza Román, Brenda Irasema Maldonado-Meza, Bernardo Martínez-Miguel, José Arturo Martínez-Orozco, Célida Martínez- Rodríguez, Margarita Matías-Florentino, Fidencio Mejía-Nepomuceno, María Guadalupe de Jesús Mireles-Rivera, Gloria María Molina-Salinas, Hector Montoya-Fuentes, , Mario Mújica-Sánchez, José Esteban Muñoz-Medina, José de Jesús Nuñez-Contreras, Alicia Ocaña-Mondragón, Luis Alberto Ochoa-Carrera, Hector Esteban Paz-Juárez, Marissa Pérez-García, Francisco Pulido, Helen Haydee Fernanda Ramírez-Plascencia, Jorge Salas-Hernández, Angel Gustavo Salas-Lais, Alejandro Sanchez-Flores, Clara Esperanza Santacruz-Tinoco, María Guadalupe Santiago-Mauricio, Selene Zárate, Nelly Sélem-Mojica, Blanca Taboada, Gloria Vazquez, Joel Armando Vázquez-Pérez. |
| EPI_ISL_2648115, EPI_ISL_2648116, EPI_ISL_2648117, EPI_ISL_2648118 | Instituto Nacional de Enfermedades Respiratorias (INER) | Centro de Investigación en Enfermedades Infecciosas (CIENI), Instituto Nacional de Enfermedades Respiratorias (INER) | Consorcio Mexicano de Vigilancia Genómica (CoViGen-Mex). Authors (in alphabetical order): Julio Elias Alvarado-Yaah, Carlos F. Arias, Santiago Ávila-Ríos, Eduardo Becerril-Vargas, Víctor Hugo Borja-Aburto, Celia Boukadida, Cristóbal Cháidez-Quiróz, Juan Bautista Chale-Dzul, Ricardo Ciria Merce, Andreu Comas-García, Célida Duque Molina, Julissa Enciso-Ibarra, José Antonio Enciso-Moreno, Gloria Elena Espinosa-Ayala, Fernando Fontove-Herrera, Daniel Fregoso-Rueda, Victor Eduardo Garcia-Arias, Alejandra Garcia-Gasca, Bruno Gomez-Gil, Jean Pierre González, Irvin González-López, Concepción Grajales-Muñiz, Ricardo Grande, Rosa María Gutierrez Rios, Alejandra Hernández-Terán, Alfredo Herrera-Estrella, Carla Ivón Herrera-Najera, Pavel Isa, Daniel Lira Morales, Susana Lopez, Antonio Loza Román, Brenda Irasema Maldonado-Meza, Bernardo Martínez-Miguel, José Arturo Martínez-Orozco, Célida Martínez- Rodríguez, Margarita Matías-Florentino, Fidencio Mejía-Nepomuceno, María Guadalupe de Jesús Mireles-Rivera, Gloria María Molina-Salinas, Hector Montoya-Fuentes, , Mario Mújica-Sánchez, José Esteban Muñoz-Medina, José de Jesús Nuñez-Contreras, Alicia Ocaña-Mondragón, Luis Alberto Ochoa-Carrera, Hector Esteban Paz-Juárez, Marissa Pérez-García, Francisco Pulido, Helen Haydee Fernanda Ramírez-Plascencia, Jorge Salas-Hernández, Angel Gustavo Salas-Lais, Alejandro Sanchez-Flores, Clara Esperanza Santacruz-Tinoco, María Guadalupe Santiago-Mauricio, Selene Zárate, Nelly Sélem-Mojica, Blanca Taboada, Gloria Vazquez, Joel Armando Vázquez-Pérez. |
| EPI_ISL_2648119 | Instituto Nacional de Cancerología | Centro de Investigación en Enfermedades Infecciosas (CIENI), Instituto Nacional de Enfermedades Respiratorias (INER) | Consorcio Mexicano de Vigilancia Genómica (CoViGen-Mex). Authors (in alphabetical order): Julio Elias Alvarado-Yaah, Carlos F. Arias, Santiago Ávila-Ríos, Eduardo Becerril-Vargas, Víctor Hugo Borja-Aburto, Celia Boukadida, Cristóbal Cháidez-Quiróz, Juan Bautista Chale-Dzul, Ricardo Ciria Merce, Andreu Comas-García, Célida Duque Molina, Julissa Enciso-Ibarra, José Antonio Enciso-Moreno, Gloria Elena Espinosa-Ayala, Fernando Fontove-Herrera, Daniel Fregoso-Rueda, Victor Eduardo Garcia-Arias, Alejandra Garcia-Gasca, Bruno Gomez-Gil, Jean Pierre González, Irvin González-López, Concepción Grajales-Muñiz, Ricardo Grande, Rosa María Gutierrez Rios, Alejandra Hernández-Terán, Alfredo Herrera-Estrella, Carla Ivón Herrera-Najera, Pavel Isa, Daniel Lira Morales, Susana Lopez, Antonio Loza Román, Brenda Irasema Maldonado-Meza, Bernardo Martínez-Miguel, José Arturo Martínez-Orozco, Célida Martínez- Rodríguez, Margarita Matías-Florentino, Fidencio Mejía-Nepomuceno, María Guadalupe de Jesús Mireles-Rivera, Gloria María Molina-Salinas, Hector Montoya-Fuentes, , Mario Mújica-Sánchez, José Esteban Muñoz-Medina, José de Jesús Nuñez-Contreras, Alicia Ocaña-Mondragón, Luis Alberto Ochoa-Carrera, Hector Esteban Paz-Juárez, Marissa Pérez-García, Francisco Pulido, Helen Haydee Fernanda Ramírez-Plascencia, Jorge Salas-Hernández, Angel Gustavo Salas-Lais, Alejandro Sanchez-Flores, Clara Esperanza Santacruz-Tinoco, María Guadalupe Santiago-Mauricio, Selene Zárate, Nelly Sélem-Mojica, Blanca Taboada, Gloria Vazquez, Joel Armando Vázquez-Pérez. |
| EPI_ISL_2649555, EPI_ISL_2649557, EPI_ISL_2649558, EPI_ISL_2649560, EPI_ISL_2649561, EPI_ISL_2649563, EPI_ISL_2649564, EPI_ISL_2649565, EPI_ISL_2649567, EPI_ISL_2649569, EPI_ISL_2649570, EPI_ISL_2649572, EPI_ISL_2649573, EPI_ISL_2649574 | | | |
| see above | Wyoming Public Health Laboratory | Wyoming Public Health Laboratory | Jim Mildenberger, Taylor Fearing, Channing Weber, Ashley Norberg, Chayse Rowley, Marley Goetz, Brian Dominguez, Elliot Thomasson, Sam Britz, Cari Sloma, Robert Petit, and Rob Christensen |
| EPI_ISL_2649798, EPI_ISL_2649799, EPI_ISL_2649800, EPI_ISL_2649801, EPI_ISL_2649802, EPI_ISL_2649803, EPI_ISL_2649804, EPI_ISL_2649805, EPI_ISL_2649806, EPI_ISL_2649807, EPI_ISL_2649808, EPI_ISL_2649809, EPI_ISL_2649810, EPI_ISL_2649811, EPI_ISL_2649812, EPI_ISL_2649813, EPI_ISL_2649814, EPI_ISL_2649815, EPI_ISL_2649816, EPI_ISL_2649817, EPI_ISL_2649818, EPI_ISL_2649819, EPI_ISL_2649820, EPI_ISL_2649821, EPI_ISL_2649822, EPI_ISL_2649823, EPI_ISL_2649824, EPI_ISL_2649825, EPI_ISL_2649826, EPI_ISL_2649827 | | | |
| see above | Tampa General Hospital Esoteric Lab | Tampa General Hospital Esoteric Research & Development Lab | Jon Faughn, Deanna Becker, Dominic Uy, Vicki Healer, Amorce Lima, Suzane Silbert |
| EPI_ISL_2649830, EPI_ISL_2649831, EPI_ISL_2649832, EPI_ISL_2649833, EPI_ISL_2649834, EPI_ISL_2649836, EPI_ISL_2649837, EPI_ISL_2649838, EPI_ISL_2649839, EPI_ISL_2649840, EPI_ISL_2649841, EPI_ISL_2649842, EPI_ISL_2649843, EPI_ISL_2649844, EPI_ISL_2649845, EPI_ISL_2649847, EPI_ISL_2649848, EPI_ISL_2649850, EPI_ISL_2649851, EPI_ISL_2649852, EPI_ISL_2649853, EPI_ISL_2649854, EPI_ISL_2649855, EPI_ISL_2649856, EPI_ISL_2649857, EPI_ISL_2649858, EPI_ISL_2649859, EPI_ISL_2649860, EPI_ISL_2649861, EPI_ISL_2649862, EPI_ISL_2649863, EPI_ISL_2649865, EPI_ISL_2649866, EPI_ISL_2649867, EPI_ISL_2649868, EPI_ISL_2649869, EPI_ISL_2649870, EPI_ISL_2649871, EPI_ISL_2649872, EPI_ISL_2649873, EPI_ISL_2649874, EPI_ISL_2649875, EPI_ISL_2649876, EPI_ISL_2649877, EPI_ISL_2649879, EPI_ISL_2649880, EPI_ISL_2649881, EPI_ISL_2649882, EPI_ISL_2649883, EPI_ISL_2649884, EPI_ISL_2649885, EPI_ISL_2649886, EPI_ISL_2649887 | | | |
| see above | NJDOH, Public Health and Environmental Laboratories | NJ_PHEL | Lindsey Bodnar, Shiv K. Verma, Ryan Pachucki, Chelsea San Filippo, Jacquelyn Deverell, Dana Woell, Allison Roder, Byeong Jeong |
| EPI_ISL_2649901, EPI_ISL_2649902, EPI_ISL_2649903, EPI_ISL_2649904, EPI_ISL_2649905, EPI_ISL_2649906, EPI_ISL_2649907, EPI_ISL_2649908, EPI_ISL_2649909, EPI_ISL_2649910, EPI_ISL_2649911, EPI_ISL_2649912, EPI_ISL_2649913, EPI_ISL_2649914, EPI_ISL_2649915 | | | |
| see above | Aegis Sciences Corporation | Centers for Disease Control and Prevention Division of Viral Diseases, Pathogen Discovery | Dakota Howard, Dhwani Batra, Peter W. Cook, Kara Moser, Adrian Paskey, Jason Caravas, Benjamin Rambo-Martin, Shatavia Morrison, Christopher Gulvick, Scott Sammons, Yvette Unoarumhi, Darlene Wagner, Matthew Schmerer, Cyndi Clark, Patrick Campbell, Rob Case, Vikramsinha Ghorpade, Holly Houdeshell, Ola Kvalvaag, Dillon Nall, Ethan Sanders, Alec Vest, Shaun Westlund, Matthew Hardison, Clinton R. Paden, Duncan MacCannell |
| EPI_ISL_2649916, EPI_ISL_2649917, EPI_ISL_2649918, EPI_ISL_2649919, EPI_ISL_2649920, EPI_ISL_2649921, EPI_ISL_2649922, EPI_ISL_2649923, EPI_ISL_2649924, EPI_ISL_2649925, EPI_ISL_2649926, EPI_ISL_2649927, EPI_ISL_2649928, EPI_ISL_2649929, EPI_ISL_2649930, EPI_ISL_2649931, EPI_ISL_2649932, EPI_ISL_2649933, EPI_ISL_2649934, EPI_ISL_2649935, EPI_ISL_2649936, EPI_ISL_2649937, EPI_ISL_2649938, EPI_ISL_2649939, EPI_ISL_2649940, EPI_ISL_2649941, EPI_ISL_2649942, EPI_ISL_2649943, EPI_ISL_2649944, EPI_ISL_2649945, EPI_ISL_2649946, EPI_ISL_2649947 | | | |
| see above | Helix/Illumina | Centers for Disease Control and Prevention Division of Viral Diseases, Pathogen Discovery | Dakota Howard, Dhwani Batra, Peter W. Cook, Kara Moser, Adrian Paskey, Jason Caravas, Benjamin Rambo-Martin, Shatavia Morrison, Christopher Gulvick, Scott Sammons, Yvette Unoarumhi, Darlene Wagner, Matthew Schmerer, Eileen de Feo, Jan Antico, Christine Tran, Matthew Tolentino, Shannon Wickline, Kim Gietzen, Brad Sickler, Jingtao Liu, Eric Allen, Phil Febbo, Nicole L. Washington, Simon White, Geraint Levan, Kelly Schiabor Barrett, Elizabeth Cirulli, Alexandre Bolze, Ary Ascencio, Charlotte Rivera-Garcia, Ryan Cho, Jason Nguyen, Sherry Wang, Jimmy Ramirez, Tyler Cassens, Efren Sandoval, Magnus Isaksson, William Lee, David Becker, Marc Laurent, James Lu, Clinton R. Paden, Duncan MacCannell |
| EPI_ISL_2649948, EPI_ISL_2649949, EPI_ISL_2649950, EPI_ISL_2649951, EPI_ISL_2649952, EPI_ISL_2649953, EPI_ISL_2649954, EPI_ISL_2649955, EPI_ISL_2649956, EPI_ISL_2649957, EPI_ISL_2649958, EPI_ISL_2649959, EPI_ISL_2649960, EPI_ISL_2649961, EPI_ISL_2649962, EPI_ISL_2649963, EPI_ISL_2649964, EPI_ISL_2649965, EPI_ISL_2649966, EPI_ISL_2649967, EPI_ISL_2649968, EPI_ISL_2649969 | | | |
| see above | Quest Diagnostics Incorporated | Centers for Disease Control and Prevention Division of Viral Diseases, Pathogen Discovery | Dakota Howard, Dhwani Batra, Peter W. Cook, Kara Moser, Adrian Paskey, Jason Caravas, Benjamin Rambo-Martin, Shatavia Morrison, Christopher Gulvick, Scott Sammons, Yvette Unoarumhi, Darlene Wagner, Matthew Schmerer, S. H. Rosenthal, A. Gerasimova, R. M. Kagan, B. Anderson, M. Hua, Y. Liu, L.E. Bernstein, K.E. Livingston, A. Perez, I. A. Shlyakhter, R. V. Rolando, R. Owen, P. Tanpaiboon, F. Lacbawan, Clinton R. Paden, Duncan MacCannell |
| EPI_ISL_2649970, EPI_ISL_2649971, EPI_ISL_2649972, EPI_ISL_2649973, EPI_ISL_2649974, EPI_ISL_2649975 | Infinity Biologix | Centers for Disease Control and Prevention Division of Viral Diseases, Pathogen Discovery | Dakota Howard, Dhwani Batra, Peter W. Cook, Kara Moser, Adrian Paskey, Jason Caravas, Benjamin Rambo-Martin, Shatavia Morrison, Christopher Gulvick, Scott Sammons, Yvette Unoarumhi, Darlene Wagner, Matthew Schmerer, Christian Bixby, Yihe Wang, Jonathan Schultz, Chirayu Goswami, Russ Hager, Robin Grimwood, Clinton R. Paden, Duncan MacCannell |
| EPI_ISL_2649976, EPI_ISL_2649977, EPI_ISL_2649978, EPI_ISL_2649979, EPI_ISL_2649980, EPI_ISL_2649981, EPI_ISL_2649982, EPI_ISL_2649983, EPI_ISL_2649984, EPI_ISL_2649985, EPI_ISL_2649986, EPI_ISL_2649987, EPI_ISL_2649988, EPI_ISL_2649989, EPI_ISL_2649990 | | | |
| see above | Quest Diagnostics Incorporated | Centers for Disease Control and Prevention Division of Viral Diseases, Pathogen Discovery | Dakota Howard, Dhwani Batra, Peter W. Cook, Kara Moser, Adrian Paskey, Jason Caravas, Benjamin Rambo-Martin, Shatavia Morrison, Christopher Gulvick, Scott Sammons, Yvette Unoarumhi, Darlene Wagner, Matthew Schmerer, S. H. Rosenthal, A. Gerasimova, R. M. Kagan, B. Anderson, M. Hua, Y. Liu, L.E. Bernstein, K.E. Livingston, A. Perez, I. A. Shlyakhter, R. V. Rolando, R. Owen, P. Tanpaiboon, F. Lacbawan, Clinton R. Paden, Duncan MacCannell |
| EPI_ISL_2650029, EPI_ISL_2650030, EPI_ISL_2650031, EPI_ISL_2650034, EPI_ISL_2650036, EPI_ISL_2650040, EPI_ISL_2650044, EPI_ISL_2650048, EPI_ISL_2650049, EPI_ISL_2650051, EPI_ISL_2650053, EPI_ISL_2650054, EPI_ISL_2650056, EPI_ISL_2650069, EPI_ISL_2650072, EPI_ISL_2650078, EPI_ISL_2650081, EPI_ISL_2650088, EPI_ISL_2650092, EPI_ISL_2650096, EPI_ISL_2650097, EPI_ISL_2650098, EPI_ISL_2650099, EPI_ISL_2650100, EPI_ISL_2650101, EPI_ISL_2650102, EPI_ISL_2650103, EPI_ISL_2650104, EPI_ISL_2650105, EPI_ISL_2650106, EPI_ISL_2650107, EPI_ISL_2650108, EPI_ISL_2650109, EPI_ISL_2650110, EPI_ISL_2650111, EPI_ISL_2650112, EPI_ISL_2650113, EPI_ISL_2650114, EPI_ISL_2650115, EPI_ISL_2650116, EPI_ISL_2650117, EPI_ISL_2650118, EPI_ISL_2650119, EPI_ISL_2650120, EPI_ISL_2650121, EPI_ISL_2650122, EPI_ISL_2650123, EPI_ISL_2650124, EPI_ISL_2650125, EPI_ISL_2650126, EPI_ISL_2650127, EPI_ISL_2650128, EPI_ISL_2650129, EPI_ISL_2650130, EPI_ISL_2650132, EPI_ISL_2650133, EPI_ISL_2650134, EPI_ISL_2650135, EPI_ISL_2650136, EPI_ISL_2650137, EPI_ISL_2650138, EPI_ISL_2650139, EPI_ISL_2650140, EPI_ISL_2650141, EPI_ISL_2650142, EPI_ISL_2650143, EPI_ISL_2650144, EPI_ISL_2650145, EPI_ISL_2650146, EPI_ISL_2650147, EPI_ISL_2650148, EPI_ISL_2650149, EPI_ISL_2650150, EPI_ISL_2650151, EPI_ISL_2650152, EPI_ISL_2650154, EPI_ISL_2650156, EPI_ISL_2650157, EPI_ISL_2650158, EPI_ISL_2650159, EPI_ISL_2650160, EPI_ISL_2650161, EPI_ISL_2650162, EPI_ISL_2650163, EPI_ISL_2650164, EPI_ISL_2650165, EPI_ISL_2650166, EPI_ISL_2650167, EPI_ISL_2650168, EPI_ISL_2650169, EPI_ISL_2650170, EPI_ISL_2650171, EPI_ISL_2650172, EPI_ISL_2650173, EPI_ISL_2650175, EPI_ISL_2650176, EPI_ISL_2650178, EPI_ISL_2650179, EPI_ISL_2650180 | | | |
| see above | Arizona State Public Health Laboratory | Arizona State Public Health Laboratory | Trung Huynh, Jessica Escobar, Katherine Fullerton, Nobuko Fukushima, Matthew Contursi, Stacy White, Linda Getsinger, Victor Waddell |
| EPI_ISL_2650349, EPI_ISL_2650350, EPI_ISL_2650351, EPI_ISL_2650352, EPI_ISL_2650353, EPI_ISL_2650354, EPI_ISL_2650355, EPI_ISL_2650356, EPI_ISL_2650357, EPI_ISL_2650358, EPI_ISL_2650359, EPI_ISL_2650360, EPI_ISL_2650361, EPI_ISL_2650362, EPI_ISL_2650363, EPI_ISL_2650364, EPI_ISL_2650365, EPI_ISL_2650366, EPI_ISL_2650367, EPI_ISL_2650368, EPI_ISL_2650369, EPI_ISL_2650370, EPI_ISL_2650371, EPI_ISL_2650372, EPI_ISL_2650373, EPI_ISL_2650374, EPI_ISL_2650375, EPI_ISL_2650376, EPI_ISL_2650377, EPI_ISL_2650378, EPI_ISL_2650379, EPI_ISL_2650380, EPI_ISL_2650381, EPI_ISL_2650382, EPI_ISL_2650383, EPI_ISL_2650384, EPI_ISL_2650385, EPI_ISL_2650386, EPI_ISL_2650387, EPI_ISL_2650388, EPI_ISL_2650389, EPI_ISL_2650390, EPI_ISL_2650391, EPI_ISL_2650392, EPI_ISL_2650393, EPI_ISL_2650394, EPI_ISL_2650395, EPI_ISL_2650396, EPI_ISL_2650397, EPI_ISL_2650398, EPI_ISL_2650399, EPI_ISL_2650400, EPI_ISL_2650401, EPI_ISL_2650402, EPI_ISL_2650403, EPI_ISL_2650404, EPI_ISL_2650405, EPI_ISL_2650406, EPI_ISL_2650408, EPI_ISL_2650409, EPI_ISL_2650410, EPI_ISL_2650411, EPI_ISL_2650412, EPI_ISL_2650413, EPI_ISL_2650414, EPI_ISL_2650415, EPI_ISL_2650416, EPI_ISL_2650417, EPI_ISL_2650418, EPI_ISL_2650420, EPI_ISL_2650421, EPI_ISL_2650422, EPI_ISL_2650423, EPI_ISL_2650424, EPI_ISL_2650425, EPI_ISL_2650426, EPI_ISL_2650427, EPI_ISL_2650428, EPI_ISL_2650429 | | | |
| see above | Ochsner Health | Bioinfoexperts, LLC | Judy Crabtree, Amy Feehan, Rebecca Rose, David J. Nolan, Tessa LaFleur, Julia-Garcia-Diaz, Lucio Miele, Susanna L. Lamers |
| EPI_ISL_2650507, EPI_ISL_2650508, EPI_ISL_2650509, EPI_ISL_2650510, EPI_ISL_2650511, EPI_ISL_2650512, EPI_ISL_2650513, EPI_ISL_2650514, EPI_ISL_2650515, EPI_ISL_2650516, EPI_ISL_2650517, EPI_ISL_2650518, EPI_ISL_2650519, EPI_ISL_2650520, EPI_ISL_2650521, EPI_ISL_2650522 | | | |
| see above | University of Oregon COVID-19 MAP Laboratory | University of Oregon Genomics and Cell Characterization Core Facility (GC3F) | Douglas Turnbull, Ariana White, Jeff Bishop, Jason Carriere, Jason Sydes, Peter Batzel |
| EPI_ISL_2650540 | Kaiser San Jose | Santa Clara County Public Health Laboratory | Santa Clara County Public Health Department |
| EPI_ISL_2650541 | Kaiser Santa Clara | Santa Clara County Public Health Laboratory | Santa Clara County Public Health Department |
| EPI_ISL_2650542, EPI_ISL_2650543 | El Camino Hospital | Santa Clara County Public Health Laboratory | Santa Clara County Public Health Department |
| EPI_ISL_2650544, EPI_ISL_2650545 | PAMF Palo Alto | Santa Clara County Public Health Laboratory | Santa Clara County Public Health Department |
| EPI_ISL_2650550, EPI_ISL_2650552, EPI_ISL_2650553, EPI_ISL_2650555, EPI_ISL_2650557, EPI_ISL_2650560 | Santa Clara Valley Medical Center | Santa Clara County Public Health Laboratory | Santa Clara County Public Health Department |
| EPI_ISL_2650562 | PAMF Palo Alto | Santa Clara County Public Health Laboratory | Santa Clara County Public Health Department |
| EPI_ISL_2650567, EPI_ISL_2650568, EPI_ISL_2650569, EPI_ISL_2650571, EPI_ISL_2650572, EPI_ISL_2650573, EPI_ISL_2650574, EPI_ISL_2650576, EPI_ISL_2650577, EPI_ISL_2650578, EPI_ISL_2650579, EPI_ISL_2650580 | | | |
| see above | Santa Clara Valley Medical Center | Santa Clara County Public Health Laboratory | Santa Clara County Public Health Department |
| EPI_ISL_2650581 | Kaiser San Jose | Santa Clara County Public Health Laboratory | Santa Clara County Public Health Department |
| EPI_ISL_2650582, EPI_ISL_2650583 | Santa Clara Valley Medical Center | Santa Clara County Public Health Laboratory | Santa Clara County Public Health Department |

| | | | |
|---|---|---|---|
| EPI_ISL_2650584 | Kaiser San Jose | Santa Clara County Public Health Laboratory | Santa Clara County Public Health Department |
| EPI_ISL_2650585 | PAMF Palo Alto | Santa Clara County Public Health Laboratory | Santa Clara County Public Health Department |
| EPI_ISL_2651034, EPI_ISL_2651035, EPI_ISL_2651037, EPI_ISL_2651038, EPI_ISL_2651039, EPI_ISL_2651040, EPI_ISL_2651041, EPI_ISL_2651042, EPI_ISL_2651043, EPI_ISL_2651044, EPI_ISL_2651045, EPI_ISL_2651046, EPI_ISL_2651047, EPI_ISL_2651048, EPI_ISL_2651049, EPI_ISL_2651050, EPI_ISL_2651051, EPI_ISL_2651052, EPI_ISL_2651053, EPI_ISL_2651054, EPI_ISL_2651055, EPI_ISL_2651056, EPI_ISL_2651057, EPI_ISL_2651058, EPI_ISL_2651059, EPI_ISL_2651060, EPI_ISL_2651061, EPI_ISL_2651062, EPI_ISL_2651063, EPI_ISL_2651064, EPI_ISL_2651065, EPI_ISL_2651066, EPI_ISL_2651067, EPI_ISL_2651068, EPI_ISL_2651069, EPI_ISL_2651070, EPI_ISL_2651071, EPI_ISL_2651072, EPI_ISL_2651073, EPI_ISL_2651074, EPI_ISL_2651075, EPI_ISL_2651076, EPI_ISL_2651077, EPI_ISL_2651078, EPI_ISL_2651079, EPI_ISL_2651080, EPI_ISL_2651081, EPI_ISL_2651082, EPI_ISL_2651083, EPI_ISL_2651084, EPI_ISL_2651085, EPI_ISL_2651086, EPI_ISL_2651087, EPI_ISL_2651088, EPI_ISL_2651089, EPI_ISL_2651090, EPI_ISL_2651091, EPI_ISL_2651093, EPI_ISL_2651094, EPI_ISL_2651095, EPI_ISL_2651096, EPI_ISL_2651097, EPI_ISL_2651098, EPI_ISL_2651099, EPI_ISL_2651100, EPI_ISL_2651101, EPI_ISL_2651102, EPI_ISL_2651103, EPI_ISL_2651104, EPI_ISL_2651105, EPI_ISL_2651106, EPI_ISL_2651107, EPI_ISL_2651108, EPI_ISL_2651109, EPI_ISL_2651110, EPI_ISL_2651111, EPI_ISL_2651112, EPI_ISL_2651113, EPI_ISL_2651114 | | | |
| see above | Washington State Department of Health Public Health Laboratories | Washington State Department of Health Public Health Laboratories | Drew MacKellar, Philip Dykema, Denny Russell, Joenice Gonzalez, Hannah Gray, Geoff Melly, Vanessa De Los Santos, Darren Lucas, JohnAric Peterson, Avi Singh, Rebecca Cao |
| EPI_ISL_2651133, EPI_ISL_2651134, EPI_ISL_2651135, EPI_ISL_2651136, EPI_ISL_2651137, EPI_ISL_2651138, EPI_ISL_2651139, EPI_ISL_2651140, EPI_ISL_2651141, EPI_ISL_2651142, EPI_ISL_2651143, EPI_ISL_2651144, EPI_ISL_2651145, EPI_ISL_2651146, EPI_ISL_2651147, EPI_ISL_2651148, EPI_ISL_2651149, EPI_ISL_2651150, EPI_ISL_2651151, EPI_ISL_2651152, EPI_ISL_2651153, EPI_ISL_2651154, EPI_ISL_2651155, EPI_ISL_2651156, EPI_ISL_2651157, EPI_ISL_2651158 | | | |
| see above | Nebraska Public Health Laboratory | NPHL COVID-19 Response Team | NPHL COVID-19 Response Team |
| EPI_ISL_2652005 | Ohio Department of Health Laboratory | Ohio Department of Health Laboratory | Holmes, Jennifer; Eric Brandt, Keoni Omura, Glen McGillivary, Caitlin McDonnell, Jade Mowery, Stephanie Mccracken, Tyler Payne, Kirtana Ramadugu, Erica Leasure, Brent Lee, Kelsey Florek, Heather Blankenship, Allison Black, Quanta Brown, and Tammy Bannerman |
| EPI_ISL_2652006, EPI_ISL_2652007, EPI_ISL_2652008, EPI_ISL_2652009, EPI_ISL_2652010, EPI_ISL_2652011, EPI_ISL_2652012, EPI_ISL_2652013, EPI_ISL_2652014, EPI_ISL_2652015, EPI_ISL_2652016, EPI_ISL_2652017, EPI_ISL_2652018, EPI_ISL_2652019, EPI_ISL_2652020, EPI_ISL_2652021, EPI_ISL_2652023, EPI_ISL_2652024, EPI_ISL_2652025, EPI_ISL_2652026, EPI_ISL_2652027, EPI_ISL_2652029, EPI_ISL_2652030, EPI_ISL_2652031, EPI_ISL_2652032, EPI_ISL_2652033, EPI_ISL_2652034, EPI_ISL_2652035, EPI_ISL_2652036, EPI_ISL_2652037, EPI_ISL_2652038, EPI_ISL_2652039, EPI_ISL_2652040, EPI_ISL_2652041, EPI_ISL_2652042, EPI_ISL_2652043, EPI_ISL_2652044, EPI_ISL_2652045, EPI_ISL_2652046, EPI_ISL_2652047, EPI_ISL_2652048, EPI_ISL_2652050, EPI_ISL_2652053, EPI_ISL_2652056, EPI_ISL_2652060, EPI_ISL_2652062, EPI_ISL_2652065, EPI_ISL_2652067, EPI_ISL_2652068, EPI_ISL_2652069, EPI_ISL_2652070, EPI_ISL_2652071, EPI_ISL_2652072, EPI_ISL_2652073, EPI_ISL_2652074, EPI_ISL_2652075, EPI_ISL_2652076, EPI_ISL_2652077, EPI_ISL_2652078, EPI_ISL_2652079, EPI_ISL_2652080, EPI_ISL_2652081, EPI_ISL_2652082, EPI_ISL_2652083, EPI_ISL_2652084, EPI_ISL_2652085, EPI_ISL_2652086, EPI_ISL_2652087, EPI_ISL_2652088, EPI_ISL_2652089, EPI_ISL_2652090, EPI_ISL_2652091, EPI_ISL_2652093, EPI_ISL_2652095, EPI_ISL_2652096, EPI_ISL_2652097, EPI_ISL_2652098, EPI_ISL_2652099, EPI_ISL_2652100, EPI_ISL_2652101, EPI_ISL_2652102, EPI_ISL_2652103, EPI_ISL_2652104, EPI_ISL_2652106, EPI_ISL_2652109 | | | |
| see above | New Mexico Department of Health Scientific Laboratory | New Mexico Department of Health Scientific Laboratory | Ellie Johnson, D'eldra Malone, Jennifer Benoit, Ratheesh Rajan, Linda Salazar, Keila Gutierrez, Anastacia Griego-Fisher |
| EPI_ISL_2652110, EPI_ISL_2652111, EPI_ISL_2652112, EPI_ISL_2652113, EPI_ISL_2652150, EPI_ISL_2652153, EPI_ISL_2652156 | Ohio Department of Health Laboratory | Ohio Department of Health Laboratory | Holmes, Jennifer; Eric Brandt, Keoni Omura, Glen McGillivary, Caitlin McDonnell, Jade Mowery, Stephanie Mccracken, Tyler Payne, Kirtana Ramadugu, Erica Leasure, Brent Lee, Kelsey Florek, Heather Blankenship, Allison Black, Quanta Brown, and Tammy Bannerman |
| EPI_ISL_2652159 | OHSU Molecular Microbiology Laboratory | OHSU Molecular Microbiology Laboratory | Rebecca O'Gara |
| EPI_ISL_2652267, EPI_ISL_2652268, EPI_ISL_2652269, EPI_ISL_2652270, EPI_ISL_2652271, EPI_ISL_2652272, EPI_ISL_2652273, EPI_ISL_2652274, EPI_ISL_2652275, EPI_ISL_2652276, EPI_ISL_2652277, EPI_ISL_2652278, EPI_ISL_2652279, EPI_ISL_2652280, EPI_ISL_2652281, EPI_ISL_2652282, EPI_ISL_2652283, EPI_ISL_2652284, EPI_ISL_2652285, EPI_ISL_2652286, EPI_ISL_2652287, EPI_ISL_2652288, EPI_ISL_2652289, EPI_ISL_2652290, EPI_ISL_2652291, EPI_ISL_2652292, EPI_ISL_2652293, EPI_ISL_2652294, EPI_ISL_2652295, EPI_ISL_2652296, EPI_ISL_2652297, EPI_ISL_2652298, EPI_ISL_2652299, EPI_ISL_2652300, EPI_ISL_2652301, EPI_ISL_2652302, EPI_ISL_2652303, EPI_ISL_2652304, EPI_ISL_2652305, EPI_ISL_2652306, EPI_ISL_2652307, EPI_ISL_2652308, EPI_ISL_2652309, EPI_ISL_2652310, EPI_ISL_2652311, EPI_ISL_2652312, EPI_ISL_2652313, EPI_ISL_2652314, EPI_ISL_2652315, EPI_ISL_2652316, EPI_ISL_2652317, EPI_ISL_2652318, EPI_ISL_2652319, EPI_ISL_2652320, EPI_ISL_2652321, EPI_ISL_2652322, EPI_ISL_2652323, EPI_ISL_2652324, EPI_ISL_2652325, EPI_ISL_2652326, EPI_ISL_2652327, EPI_ISL_2652328, EPI_ISL_2652329, EPI_ISL_2652330, EPI_ISL_2652331, EPI_ISL_2652332, EPI_ISL_2652333, EPI_ISL_2652334, EPI_ISL_2652335, EPI_ISL_2652336, EPI_ISL_2652337, EPI_ISL_2652338, EPI_ISL_2652339, EPI_ISL_2652340, EPI_ISL_2652341, EPI_ISL_2652342, EPI_ISL_2652343, EPI_ISL_2652344, EPI_ISL_2652345, EPI_ISL_2652346, EPI_ISL_2652347, EPI_ISL_2652348, EPI_ISL_2652349, EPI_ISL_2652350, EPI_ISL_2652351, EPI_ISL_2652352, EPI_ISL_2652353, EPI_ISL_2652354, EPI_ISL_2652355, EPI_ISL_2652356, EPI_ISL_2652357, EPI_ISL_2652358, EPI_ISL_2652359, EPI_ISL_2652360, EPI_ISL_2652361, EPI_ISL_2652362, EPI_ISL_2652363, EPI_ISL_2652364, EPI_ISL_2652365, EPI_ISL_2652366, EPI_ISL_2652367, EPI_ISL_2652368, EPI_ISL_2652369, EPI_ISL_2652370, EPI_ISL_2652371, EPI_ISL_2652372, EPI_ISL_2652373, EPI_ISL_2652374, EPI_ISL_2652375, EPI_ISL_2652376, EPI_ISL_2652377, EPI_ISL_2652378, EPI_ISL_2652379, EPI_ISL_2652380, EPI_ISL_2652381, EPI_ISL_2652382, EPI_ISL_2652383, EPI_ISL_2652384, EPI_ISL_2652385, EPI_ISL_2652386, EPI_ISL_2652387, EPI_ISL_2652388, EPI_ISL_2652389, EPI_ISL_2652390, EPI_ISL_2652391, EPI_ISL_2652392, EPI_ISL_2652393, EPI_ISL_2652394, EPI_ISL_2652395, EPI_ISL_2652396, EPI_ISL_2652397, EPI_ISL_2652398, EPI_ISL_2652399, EPI_ISL_2652400, EPI_ISL_2652401, EPI_ISL_2652402, EPI_ISL_2652403, EPI_ISL_2652404, EPI_ISL_2652405, EPI_ISL_2652406, EPI_ISL_2652407, EPI_ISL_2652408, EPI_ISL_2652409, EPI_ISL_2652410, EPI_ISL_2652411, EPI_ISL_2652412, EPI_ISL_2652413, EPI_ISL_2652414, EPI_ISL_2652415, EPI_ISL_2652416, EPI_ISL_2652417, EPI_ISL_2652418, EPI_ISL_2652419, EPI_ISL_2652420, EPI_ISL_2652421, EPI_ISL_2652422, EPI_ISL_2652423, EPI_ISL_2652424, EPI_ISL_2652425, EPI_ISL_2652426, EPI_ISL_2652427, EPI_ISL_2652428, EPI_ISL_2652429, EPI_ISL_2652430, EPI_ISL_2652431, EPI_ISL_2652432, EPI_ISL_2652433, EPI_ISL_2652434, EPI_ISL_2652435, EPI_ISL_2652436, EPI_ISL_2652437, EPI_ISL_2652438, EPI_ISL_2652439, EPI_ISL_2652440, EPI_ISL_2652441, EPI_ISL_2652442, EPI_ISL_2652443, EPI_ISL_2652444, EPI_ISL_2652445, EPI_ISL_2652446, EPI_ISL_2652447, EPI_ISL_2652448, EPI_ISL_2652449, EPI_ISL_2652450, EPI_ISL_2652451, EPI_ISL_2652452, EPI_ISL_2652453, EPI_ISL_2652454, EPI_ISL_2652455, EPI_ISL_2652456, EPI_ISL_2652457, EPI_ISL_2652458, EPI_ISL_2652459, EPI_ISL_2652460, EPI_ISL_2652461, EPI_ISL_2652462, EPI_ISL_2652463, EPI_ISL_2652464, EPI_ISL_2652465, EPI_ISL_2652466, EPI_ISL_2652467, EPI_ISL_2652468, EPI_ISL_2652469, EPI_ISL_2652470, EPI_ISL_2652471, EPI_ISL_2652472, EPI_ISL_2652473, EPI_ISL_2652474, EPI_ISL_2652475, EPI_ISL_2652476, EPI_ISL_2652477, EPI_ISL_2652478 | | | |
| see above | Quest Diagnostics Incorporated | Centers for Disease Control and Prevention Division of Viral Diseases, Pathogen Discovery | Dakota Howard, Dhwani Batra, Peter W. Cook, Kara Moser, Adrian Paskey, Jason Caravas, Benjamin Rambo-Martin, Shatavia Morrison, Christopher Gulvick, Scott Sammons, Yvette Unoarumhi, Darlene Wagner, Matthew Schmerer, S. H. Rosenthal, A. Gerasimova, R. M. Kagan, B. Anderson, M. Hua, Y. Liu, L.E. Bernstein, K.E. Livingston, A. Perez, I. A. Shlyakhter, R. V. Rolando, R. Owen, P. Tanpaiboon, F. Lacbawan, Clinton R. Paden, Duncan MacCannell |
| EPI_ISL_2652479 | Ohio Department of Health Laboratory | Ohio Department of Health Laboratory | Holmes, Jennifer; Eric Brandt, Keoni Omura, Glen McGillivary, Caitlin McDonnell, Jade Mowery, Stephanie Mccracken, Tyler Payne, Kirtana Ramadugu, Erica Leasure, Brent Lee, Kelsey Florek, Heather Blankenship, Allison Black, Quanta Brown, and Tammy Bannerman |
| EPI_ISL_2652481, EPI_ISL_2652482, EPI_ISL_2652483, EPI_ISL_2652484, EPI_ISL_2652485, EPI_ISL_2652486 | Minnesota Department of Health, Public Health Laboratory | Minnesota Department of Health, Public Health Laboratory | Alexandra Lorentz, Jacob Garfin, Matt Plumb, and Xiong Wang |
| EPI_ISL_2652487 | M Health Fairview | Minnesota Department of Health, Public Health Laboratory | Alexandra Lorentz, Jacob Garfin, Matt Plumb, and Xiong Wang |
| EPI_ISL_2652488, EPI_ISL_2652489 | Minnesota Department of Health, Public Health Laboratory | Minnesota Department of Health, Public Health Laboratory | Alexandra Lorentz, Jacob Garfin, Matt Plumb, and Xiong Wang |
| EPI_ISL_2652490, EPI_ISL_2652491, EPI_ISL_2652492, EPI_ISL_2652493, EPI_ISL_2652494, EPI_ISL_2652495, EPI_ISL_2652496, EPI_ISL_2652497, EPI_ISL_2652498, EPI_ISL_2652499, EPI_ISL_2652500 | | | |
| see above | M Health Fairview | Minnesota Department of Health, Public Health Laboratory | Alexandra Lorentz, Jacob Garfin, Matt Plumb, and Xiong Wang |
| EPI_ISL_2652501, EPI_ISL_2652502 | Minnesota Department of Health, Public Health Laboratory | Minnesota Department of Health, Public Health Laboratory | Alexandra Lorentz, Jacob Garfin, Matt Plumb, and Xiong Wang |
| EPI_ISL_2652503 | Centracare Laboratory Services | Minnesota Department of Health, Public Health Laboratory | Alexandra Lorentz, Jacob Garfin, Matt Plumb, and Xiong Wang |
| EPI_ISL_2652504, EPI_ISL_2652505, EPI_ISL_2652506, EPI_ISL_2652509, EPI_ISL_2652510, EPI_ISL_2652511 | M Health Fairview | Minnesota Department of Health, Public Health Laboratory | Alexandra Lorentz, Jacob Garfin, Matt Plumb, and Xiong Wang |
| EPI_ISL_2652512, EPI_ISL_2652513, EPI_ISL_2652514, EPI_ISL_2652515, EPI_ISL_2652516, EPI_ISL_2652517, EPI_ISL_2652518, EPI_ISL_2652519, EPI_ISL_2652520, EPI_ISL_2652521, EPI_ISL_2652522, EPI_ISL_2652523, EPI_ISL_2652524, EPI_ISL_2652525, EPI_ISL_2652526, EPI_ISL_2652527, EPI_ISL_2652528, EPI_ISL_2652529, EPI_ISL_2652530, EPI_ISL_2652531, EPI_ISL_2652532, EPI_ISL_2652533, EPI_ISL_2652534, EPI_ISL_2652536, EPI_ISL_2652537, EPI_ISL_2652538, EPI_ISL_2652539, EPI_ISL_2652540, EPI_ISL_2652541, EPI_ISL_2652542, EPI_ISL_2652543, EPI_ISL_2652545, EPI_ISL_2652546, EPI_ISL_2652547, EPI_ISL_2652548, EPI_ISL_2652549, EPI_ISL_2652550, EPI_ISL_2652551, EPI_ISL_2652552, EPI_ISL_2652553, EPI_ISL_2652554, EPI_ISL_2652555, EPI_ISL_2652556, EPI_ISL_2652557, EPI_ISL_2652558, EPI_ISL_2652559, EPI_ISL_2652560, EPI_ISL_2652561, EPI_ISL_2652562, EPI_ISL_2652563, EPI_ISL_2652564, EPI_ISL_2652565, EPI_ISL_2652566, EPI_ISL_2652567, EPI_ISL_2652568, EPI_ISL_2652569, EPI_ISL_2652570, EPI_ISL_2652571, EPI_ISL_2652572, EPI_ISL_2652573, EPI_ISL_2652574, EPI_ISL_2652575, EPI_ISL_2652576, EPI_ISL_2652577, EPI_ISL_2652578, EPI_ISL_2652580, EPI_ISL_2652581, EPI_ISL_2652582, EPI_ISL_2652583, EPI_ISL_2652584, EPI_ISL_2652585, EPI_ISL_2652586, EPI_ISL_2652587, EPI_ISL_2652588, EPI_ISL_2652589, EPI_ISL_2652590, EPI_ISL_2652591, EPI_ISL_2652592, EPI_ISL_2652594, EPI_ISL_2652595, EPI_ISL_2652596, EPI_ISL_2652597, EPI_ISL_2652598, EPI_ISL_2652599, EPI_ISL_2652600, EPI_ISL_2652601, EPI_ISL_2652602, EPI_ISL_2652603, EPI_ISL_2652604, EPI_ISL_2652605, EPI_ISL_2652606, EPI_ISL_2652607, EPI_ISL_2652608, EPI_ISL_2652609, EPI_ISL_2652610, EPI_ISL_2652611, EPI_ISL_2652612, EPI_ISL_2652613, EPI_ISL_2652614, EPI_ISL_2652615, EPI_ISL_2652616, EPI_ISL_2652617, EPI_ISL_2652618, EPI_ISL_2652619, EPI_ISL_2652620, EPI_ISL_2652621, EPI_ISL_2652622, EPI_ISL_2652623, EPI_ISL_2652625, EPI_ISL_2652626, EPI_ISL_2652627, EPI_ISL_2652628, EPI_ISL_2652629, EPI_ISL_2652630, EPI_ISL_2652631, EPI_ISL_2652632, EPI_ISL_2652633, EPI_ISL_2652634, EPI_ISL_2652635, EPI_ISL_2652636, EPI_ISL_2652637, EPI_ISL_2652638, EPI_ISL_2652639, EPI_ISL_2652640, EPI_ISL_2652641, EPI_ISL_2652642, EPI_ISL_2652643, EPI_ISL_2652644, EPI_ISL_2652645, EPI_ISL_2652646, EPI_ISL_2652647, EPI_ISL_2652648, EPI_ISL_2652649, EPI_ISL_2652650, EPI_ISL_2652651, EPI_ISL_2652652, EPI_ISL_2652653, EPI_ISL_2652654, EPI_ISL_2652655, EPI_ISL_2652656, EPI_ISL_2652658, EPI_ISL_2652659, EPI_ISL_2652660, EPI_ISL_2652661, EPI_ISL_2652662, EPI_ISL_2652663, EPI_ISL_2652664, EPI_ISL_2652665, EPI_ISL_2652666, EPI_ISL_2652667, EPI_ISL_2652668, EPI_ISL_2652669, EPI_ISL_2652670, EPI_ISL_2652671, EPI_ISL_2652672, EPI_ISL_2652673, EPI_ISL_2652674, EPI_ISL_2652675, EPI_ISL_2652676, EPI_ISL_2652677, EPI_ISL_2652678, EPI_ISL_2652679, EPI_ISL_2652680, EPI_ISL_2652681, EPI_ISL_2652682, EPI_ISL_2652683, EPI_ISL_2652684, EPI_ISL_2652685, EPI_ISL_2652686, EPI_ISL_2652687, EPI_ISL_2652689, EPI_ISL_2652690, EPI_ISL_2652691, EPI_ISL_2652692, EPI_ISL_2652693, EPI_ISL_2652694, EPI_ISL_2652695, EPI_ISL_2652696, EPI_ISL_2652697, EPI_ISL_2652698, EPI_ISL_2652699, EPI_ISL_2652700, EPI_ISL_2652701, EPI_ISL_2652702, EPI_ISL_2652703, EPI_ISL_2652704, EPI_ISL_2652705, EPI_ISL_2652706, EPI_ISL_2652707, EPI_ISL_2652708, EPI_ISL_2652709, EPI_ISL_2652710, EPI_ISL_2652711, EPI_ISL_2652712, EPI_ISL_2652713, EPI_ISL_2652714, EPI_ISL_2652716, EPI_ISL_2652717, EPI_ISL_2652718, EPI_ISL_2652719, EPI_ISL_2652720, EPI_ISL_2652721, EPI_ISL_2652722, EPI_ISL_2652723, EPI_ISL_2652724, EPI_ISL_2652725, EPI_ISL_2652726, EPI_ISL_2652727, EPI_ISL_2652729, EPI_ISL_2652730, EPI_ISL_2652731, EPI_ISL_2652732, EPI_ISL_2652733, EPI_ISL_2652734, EPI_ISL_2652735, EPI_ISL_2652736, EPI_ISL_2652737, EPI_ISL_2652738 | | | |
| see above | Infinity Biologix | Centers for Disease Control and Prevention Division of Viral Diseases, Pathogen Discovery | Dakota Howard, Dhwani Batra, Peter W. Cook, Kara Moser, Adrian Paskey, Jason Caravas, Benjamin Rambo-Martin, Shatavia Morrison, Christopher Gulvick, Scott Sammons, Yvette Unoarumhi, Darlene Wagner, Matthew Schmerer, Christian Bixby, Yihe Wang, Jonathan Schultz, Chirayu Goswami, Russ Hager, Robin Grimwood, Clinton R. Paden, Duncan MacCannell |
| EPI_ISL_2652739 | OHSU-MM Lab | OHSU-MM Lab | Rebecca O'Gara |
| EPI_ISL_2652740, EPI_ISL_2652741, EPI_ISL_2652742, EPI_ISL_2652743, EPI_ISL_2652744, EPI_ISL_2652745, EPI_ISL_2652746, EPI_ISL_2652747, EPI_ISL_2652748, EPI_ISL_2652749, EPI_ISL_2652750, EPI_ISL_2652751, EPI_ISL_2652752, EPI_ISL_2652753, EPI_ISL_2652754, EPI_ISL_2652755, EPI_ISL_2652757, EPI_ISL_2652758, EPI_ISL_2652759, EPI_ISL_2652760, EPI_ISL_2652761, EPI_ISL_2652762, EPI_ISL_2652763, EPI_ISL_2652764, EPI_ISL_2652765, EPI_ISL_2652766, EPI_ISL_2652767, EPI_ISL_2652768, EPI_ISL_2652769, EPI_ISL_2652770, EPI_ISL_2652771, EPI_ISL_2652772, EPI_ISL_2652773, EPI_ISL_2652774, EPI_ISL_2652775, EPI_ISL_2652776, EPI_ISL_2652777, EPI_ISL_2652778, EPI_ISL_2652779, EPI_ISL_2652780, EPI_ISL_2652781, EPI_ISL_2652782, EPI_ISL_2652783, EPI_ISL_2652784, EPI_ISL_2652785, EPI_ISL_2652786, EPI_ISL_2652787, EPI_ISL_2652788, EPI_ISL_2652789, EPI_ISL_2652790, EPI_ISL_2652791, EPI_ISL_2652792, EPI_ISL_2652793, EPI_ISL_2652794, EPI_ISL_2652795, EPI_ISL_2652796, EPI_ISL_2652797, EPI_ISL_2652798, EPI_ISL_2652799, EPI_ISL_2652800, EPI_ISL_2652801, EPI_ISL_2652802, EPI_ISL_2652803, EPI_ISL_2652804, EPI_ISL_2652805, EPI_ISL_2652806, EPI_ISL_2652807, EPI_ISL_2652808, EPI_ISL_2652809, EPI_ISL_2652810, EPI_ISL_2652811, EPI_ISL_2652812, EPI_ISL_2652813, EPI_ISL_2652814, EPI_ISL_2652815, EPI_ISL_2652816, EPI_ISL_2652817, EPI_ISL_2652818, EPI_ISL_2652819, EPI_ISL_2652820, EPI_ISL_2652821, EPI_ISL_2652822, EPI_ISL_2652823, EPI_ISL_2652824, EPI_ISL_2652825, EPI_ISL_2652826, EPI_ISL_2652827, EPI_ISL_2652828, EPI_ISL_2652829, EPI_ISL_2652830, EPI_ISL_2652831, EPI_ISL_2652832, EPI_ISL_2652833, EPI_ISL_2652835, EPI_ISL_2652836, EPI_ISL_2652837, EPI_ISL_2652838, EPI_ISL_2652839, EPI_ISL_2652840, EPI_ISL_2652841, EPI_ISL_2652842, EPI_ISL_2652843, EPI_ISL_2652844, EPI_ISL_2652845, EPI_ISL_2652846, EPI_ISL_2652847, EPI_ISL_2652848, EPI_ISL_2652849, EPI_ISL_2652850, EPI_ISL_2652851, EPI_ISL_2652852, EPI_ISL_2652853, EPI_ISL_2652854, EPI_ISL_2652855, EPI_ISL_2652856, EPI_ISL_2652857, EPI_ISL_2652858, EPI_ISL_2652859, EPI_ISL_2652860, EPI_ISL_2652861, EPI_ISL_2652862, EPI_ISL_2652863, EPI_ISL_2652864, EPI_ISL_2652865, EPI_ISL_2652866, EPI_ISL_2652867, EPI_ISL_2652868, EPI_ISL_2652871, EPI_ISL_2652872, EPI_ISL_2652873, EPI_ISL_2652874, EPI_ISL_2652875, EPI_ISL_2652876, EPI_ISL_2652877, EPI_ISL_2652878, EPI_ISL_2652879, EPI_ISL_2652880, EPI_ISL_2652881, EPI_ISL_2652882, EPI_ISL_2652883, EPI_ISL_2652884, EPI_ISL_2652885, EPI_ISL_2652887, EPI_ISL_2652888, EPI_ISL_2652889, EPI_ISL_2652890, EPI_ISL_2652891, EPI_ISL_2652892, EPI_ISL_2652893, EPI_ISL_2652894, EPI_ISL_2652895, EPI_ISL_2652896, EPI_ISL_2652897, EPI_ISL_2652898, EPI_ISL_2652899, EPI_ISL_2652900, EPI_ISL_2652901, EPI_ISL_2652902, EPI_ISL_2652903, EPI_ISL_2652904, EPI_ISL_2652905, EPI_ISL_2652906, EPI_ISL_2652907, EPI_ISL_2652908, EPI_ISL_2652909, EPI_ISL_2652910, EPI_ISL_2652911, EPI_ISL_2652912, EPI_ISL_2652913, EPI_ISL_2652914, EPI_ISL_2652915, EPI_ISL_2652916, EPI_ISL_2652917, EPI_ISL_2652918, EPI_ISL_2652919, EPI_ISL_2652920, EPI_ISL_2652921, EPI_ISL_2652922, EPI_ISL_2652923, EPI_ISL_2652924, EPI_ISL_2652925, EPI_ISL_2652926, EPI_ISL_2652927, EPI_ISL_2652929, EPI_ISL_2652930, EPI_ISL_2652931, EPI_ISL_2652934, EPI_ISL_2652935, EPI_ISL_2652936, EPI_ISL_2652937, EPI_ISL_2652938, EPI_ISL_2652940, EPI_ISL_2652941, EPI_ISL_2652942, EPI_ISL_2652943, EPI_ISL_2652944, EPI_ISL_2652945, EPI_ISL_2652946, EPI_ISL_2652947, EPI_ISL_2652948, EPI_ISL_2652949, EPI_ISL_2652950, EPI_ISL_2652951, EPI_ISL_2652952, EPI_ISL_2652953, EPI_ISL_2652954, EPI_ISL_2652955, EPI_ISL_2652956, EPI_ISL_2652957, EPI_ISL_2652958, EPI_ISL_2652959, EPI_ISL_2652960, EPI_ISL_2652961, EPI_ISL_2652963, EPI_ISL_2652964, EPI_ISL_2652965, EPI_ISL_2652966, EPI_ISL_2652967, EPI_ISL_2652968, EPI_ISL_2652969, EPI_ISL_2652970, EPI_ISL_2652971, EPI_ISL_2652973, EPI_ISL_2652975 | | | |
| see above | Infinity Biologix | Centers for Disease Control and Prevention Division of Viral Diseases, Pathogen Discovery | Dakota Howard, Dhwani Batra, Peter W. Cook, Kara Moser, Adrian Paskey, Jason Caravas, Benjamin Rambo-Martin, Shatavia Morrison, Christopher Gulvick, Scott Sammons, Yvette Unoarumhi, Darlene Wagner, Matthew Schmerer, Christian Bixby, Yihe Wang, Jonathan Schultz, Chirayu Goswami, Russ Hager, Robin Grimwood, Clinton R. Paden, Duncan MacCannell |
| EPI_ISL_2652976 | Ohio Department of Health Laboratory | Ohio Department of Health Laboratory | Holmes, Jennifer; Eric Brandt, Keoni Omura, Glen McGillivary, Caitlin McDonnell, Jade Mowery, Stephanie Mccracken, Tyler Payne, Kirtana Ramadugu, Erica Leasure, Brent Lee, Kelsey Florek, Heather Blankenship, Allison Black, Quanta Brown, and Tammy Bannerman |
| EPI_ISL_2652977, EPI_ISL_2652978, EPI_ISL_2652979, EPI_ISL_2652980, EPI_ISL_2652981, EPI_ISL_2652982, EPI_ISL_2652983, EPI_ISL_2652984, EPI_ISL_2652985, EPI_ISL_2652986, EPI_ISL_2652987, EPI_ISL_2652989, EPI_ISL_2652990, EPI_ISL_2652991, EPI_ISL_2652992, EPI_ISL_2652993, EPI_ISL_2652994, EPI_ISL_2652995, EPI_ISL_2652996, EPI_ISL_2652997, EPI_ISL_2652999, EPI_ISL_2653000, EPI_ISL_2653001, EPI_ISL_2653002, EPI_ISL_2653003, EPI_ISL_2653004, EPI_ISL_2653005, EPI_ISL_2653006, EPI_ISL_2653007, EPI_ISL_2653008, EPI_ISL_2653009, EPI_ISL_2653011, EPI_ISL_2653012, EPI_ISL_2653013, EPI_ISL_2653014, EPI_ISL_2653015 | | | |
| see above | Infinity Biologix | Centers for Disease Control and Prevention Division of Viral Diseases, Pathogen | Dakota Howard, Dhwani Batra, Peter W. Cook, Kara Moser, Adrian Paskey, Jason Caravas, Benjamin Rambo-Martin, Shatavia Morrison, Christopher Gulvick, Scott Sammons, Yvette Unoarumhi, Darlene Wagner, Matthew Schmerer, Christian Bixby, Yihe Wang, Jonathan Schultz, Chirayu Goswami, Russ Hager, Robin Grimwood, Clinton R. Paden, Duncan MacCannell |

| | | | |
|---|---|---|---|
| | | Discovery | |
| EPI_ISL_2653016 | Hospital of the University of Pennsylvania Molecular Pathology Lab | Bushman Lab - University of Pennsylvania | John K. Everett, Kyle Rodino, Shantan Reddy, Aoife M. Roche, Young Hwang, Scott Sherrill-Mix, Samantha A. Whiteside, Jevon Graham-Wooten, Layla A. Khatib, Ayannah S. Fitzgerald, Arupa Ganguly, Mike Feldman, Brendan Kelly, Ronald G. Collman and Frederic Bushan |
| EPI_ISL_2653017 | Infinity Biologix | Centers for Disease Control and Prevention Division of Viral Diseases, Pathogen Discovery | Dakota Howard, Dhwani Batra, Peter W. Cook, Kara Moser, Adrian Paskey, Jason Caravas, Benjamin Rambo-Martin, Shatavia Morrison, Christopher Gulvick, Scott Sammons, Yvette Unoarumhi, Darlene Wagner, Matthew Schmerer, Christian Bixby, Yihe Wang, Jonathan Schultz, Chirayu Goswami, Russ Hager, Robin Grimwood, Clinton R. Paden, Duncan MacCannell |
| EPI_ISL_2653018, EPI_ISL_2653019, EPI_ISL_2653021, EPI_ISL_2653022 | Hospital of the University of Pennsylvania Molecular Pathology Lab | Bushman Lab - University of Pennsylvania | John K. Everett, Kyle Rodino, Shantan Reddy, Aoife M. Roche, Young Hwang, Scott Sherrill-Mix, Samantha A. Whiteside, Jevon Graham-Wooten, Layla A. Khatib, Ayannah S. Fitzgerald, Arupa Ganguly, Mike Feldman, Brendan Kelly, Ronald G. Collman and Frederic Bushan |
| EPI_ISL_2653023 | Infinity Biologix | Centers for Disease Control and Prevention Division of Viral Diseases, Pathogen Discovery | Dakota Howard, Dhwani Batra, Peter W. Cook, Kara Moser, Adrian Paskey, Jason Caravas, Benjamin Rambo-Martin, Shatavia Morrison, Christopher Gulvick, Scott Sammons, Yvette Unoarumhi, Darlene Wagner, Matthew Schmerer, Christian Bixby, Yihe Wang, Jonathan Schultz, Chirayu Goswami, Russ Hager, Robin Grimwood, Clinton R. Paden, Duncan MacCannell |
| EPI_ISL_2653024, EPI_ISL_2653025 | Hospital of the University of Pennsylvania Molecular Pathology Lab | Bushman Lab - University of Pennsylvania | John K. Everett, Kyle Rodino, Shantan Reddy, Aoife M. Roche, Young Hwang, Scott Sherrill-Mix, Samantha A. Whiteside, Jevon Graham-Wooten, Layla A. Khatib, Ayannah S. Fitzgerald, Arupa Ganguly, Mike Feldman, Brendan Kelly, Ronald G. Collman and Frederic Bushan |
| EPI_ISL_2653026 | Infinity Biologix | Centers for Disease Control and Prevention Division of Viral Diseases, Pathogen Discovery | Dakota Howard, Dhwani Batra, Peter W. Cook, Kara Moser, Adrian Paskey, Jason Caravas, Benjamin Rambo-Martin, Shatavia Morrison, Christopher Gulvick, Scott Sammons, Yvette Unoarumhi, Darlene Wagner, Matthew Schmerer, Christian Bixby, Yihe Wang, Jonathan Schultz, Chirayu Goswami, Russ Hager, Robin Grimwood, Clinton R. Paden, Duncan MacCannell |
| EPI_ISL_2653027 | Hospital of the University of Pennsylvania Molecular Pathology Lab | Bushman Lab - University of Pennsylvania | John K. Everett, Kyle Rodino, Shantan Reddy, Aoife M. Roche, Young Hwang, Scott Sherrill-Mix, Samantha A. Whiteside, Jevon Graham-Wooten, Layla A. Khatib, Ayannah S. Fitzgerald, Arupa Ganguly, Mike Feldman, Brendan Kelly, Ronald G. Collman and Frederic Bushan |
| EPI_ISL_2653028 | Infinity Biologix | Centers for Disease Control and Prevention Division of Viral Diseases, Pathogen Discovery | Dakota Howard, Dhwani Batra, Peter W. Cook, Kara Moser, Adrian Paskey, Jason Caravas, Benjamin Rambo-Martin, Shatavia Morrison, Christopher Gulvick, Scott Sammons, Yvette Unoarumhi, Darlene Wagner, Matthew Schmerer, Christian Bixby, Yihe Wang, Jonathan Schultz, Chirayu Goswami, Russ Hager, Robin Grimwood, Clinton R. Paden, Duncan MacCannell |
| EPI_ISL_2653029, EPI_ISL_2653030 | Hospital of the University of Pennsylvania Molecular Pathology Lab | Bushman Lab - University of Pennsylvania | John K. Everett, Kyle Rodino, Shantan Reddy, Aoife M. Roche, Young Hwang, Scott Sherrill-Mix, Samantha A. Whiteside, Jevon Graham-Wooten, Layla A. Khatib, Ayannah S. Fitzgerald, Arupa Ganguly, Mike Feldman, Brendan Kelly, Ronald G. Collman and Frederic Bushan |
| EPI_ISL_2653031 | Infinity Biologix | Centers for Disease Control and Prevention Division of Viral Diseases, Pathogen Discovery | Dakota Howard, Dhwani Batra, Peter W. Cook, Kara Moser, Adrian Paskey, Jason Caravas, Benjamin Rambo-Martin, Shatavia Morrison, Christopher Gulvick, Scott Sammons, Yvette Unoarumhi, Darlene Wagner, Matthew Schmerer, Christian Bixby, Yihe Wang, Jonathan Schultz, Chirayu Goswami, Russ Hager, Robin Grimwood, Clinton R. Paden, Duncan MacCannell |
| EPI_ISL_2653032, EPI_ISL_2653033, EPI_ISL_2653034 | Hospital of the University of Pennsylvania Molecular Pathology Lab | Bushman Lab - University of Pennsylvania | John K. Everett, Kyle Rodino, Shantan Reddy, Aoife M. Roche, Young Hwang, Scott Sherrill-Mix, Samantha A. Whiteside, Jevon Graham-Wooten, Layla A. Khatib, Ayannah S. Fitzgerald, Arupa Ganguly, Mike Feldman, Brendan Kelly, Ronald G. Collman and Frederic Bushan |
| EPI_ISL_2653035 | Infinity Biologix | Centers for Disease Control and Prevention Division of Viral Diseases, Pathogen Discovery | Dakota Howard, Dhwani Batra, Peter W. Cook, Kara Moser, Adrian Paskey, Jason Caravas, Benjamin Rambo-Martin, Shatavia Morrison, Christopher Gulvick, Scott Sammons, Yvette Unoarumhi, Darlene Wagner, Matthew Schmerer, Christian Bixby, Yihe Wang, Jonathan Schultz, Chirayu Goswami, Russ Hager, Robin Grimwood, Clinton R. Paden, Duncan MacCannell |
| EPI_ISL_2653036, EPI_ISL_2653037, EPI_ISL_2653039 | Hospital of the University of Pennsylvania Molecular Pathology Lab | Bushman Lab - University of Pennsylvania | John K. Everett, Kyle Rodino, Shantan Reddy, Aoife M. Roche, Young Hwang, Scott Sherrill-Mix, Samantha A. Whiteside, Jevon Graham-Wooten, Layla A. Khatib, Ayannah S. Fitzgerald, Arupa Ganguly, Mike Feldman, Brendan Kelly, Ronald G. Collman and Frederic Bushan |
| EPI_ISL_2653040 | Infinity Biologix | Centers for Disease Control and Prevention Division of Viral Diseases, Pathogen Discovery | Dakota Howard, Dhwani Batra, Peter W. Cook, Kara Moser, Adrian Paskey, Jason Caravas, Benjamin Rambo-Martin, Shatavia Morrison, Christopher Gulvick, Scott Sammons, Yvette Unoarumhi, Darlene Wagner, Matthew Schmerer, Christian Bixby, Yihe Wang, Jonathan Schultz, Chirayu Goswami, Russ Hager, Robin Grimwood, Clinton R. Paden, Duncan MacCannell |
| EPI_ISL_2653041, EPI_ISL_2653042 | Hospital of the University of Pennsylvania Molecular Pathology Lab | Bushman Lab - University of Pennsylvania | John K. Everett, Kyle Rodino, Shantan Reddy, Aoife M. Roche, Young Hwang, Scott Sherrill-Mix, Samantha A. Whiteside, Jevon Graham-Wooten, Layla A. Khatib, Ayannah S. Fitzgerald, Arupa Ganguly, Mike Feldman, Brendan Kelly, Ronald G. Collman and Frederic Bushan |
| EPI_ISL_2653043 | Infinity Biologix | Centers for Disease Control and Prevention Division of Viral Diseases, Pathogen Discovery | Dakota Howard, Dhwani Batra, Peter W. Cook, Kara Moser, Adrian Paskey, Jason Caravas, Benjamin Rambo-Martin, Shatavia Morrison, Christopher Gulvick, Scott Sammons, Yvette Unoarumhi, Darlene Wagner, Matthew Schmerer, Christian Bixby, Yihe Wang, Jonathan Schultz, Chirayu Goswami, Russ Hager, Robin Grimwood, Clinton R. Paden, Duncan MacCannell |
| EPI_ISL_2653044, EPI_ISL_2653045, EPI_ISL_2653046 | Hospital of the University of Pennsylvania Molecular Pathology Lab | Bushman Lab - University of Pennsylvania | John K. Everett, Kyle Rodino, Shantan Reddy, Aoife M. Roche, Young Hwang, Scott Sherrill-Mix, Samantha A. Whiteside, Jevon Graham-Wooten, Layla A. Khatib, Ayannah S. Fitzgerald, Arupa Ganguly, Mike Feldman, Brendan Kelly, Ronald G. Collman and Frederic Bushan |
| EPI_ISL_2653047, EPI_ISL_2653048, EPI_ISL_2653049, EPI_ISL_2653050, EPI_ISL_2653051, EPI_ISL_2653052, EPI_ISL_2653053, EPI_ISL_2653054, EPI_ISL_2653055, EPI_ISL_2653056, EPI_ISL_2653057, EPI_ISL_2653058, EPI_ISL_2653059, EPI_ISL_2653060, EPI_ISL_2653061, EPI_ISL_2653062, EPI_ISL_2653064, EPI_ISL_2653065, EPI_ISL_2653066, EPI_ISL_2653067 | | | |
| see above | Infinity Biologix | Centers for Disease Control and Prevention Division of Viral Diseases, Pathogen Discovery | Dakota Howard, Dhwani Batra, Peter W. Cook, Kara Moser, Adrian Paskey, Jason Caravas, Benjamin Rambo-Martin, Shatavia Morrison, Christopher Gulvick, Scott Sammons, Yvette Unoarumhi, Darlene Wagner, Matthew Schmerer, Christian Bixby, Yihe Wang, Jonathan Schultz, Chirayu Goswami, Russ Hager, Robin Grimwood, Clinton R. Paden, Duncan MacCannell |
| EPI_ISL_2653068, EPI_ISL_2653070, EPI_ISL_2653073, EPI_ISL_2653074 | Ohio Department of Health Laboratory | Ohio Department of Health Laboratory | Holmes, Jennifer; Eric Brandt, Keoni Omura, Glen McGillivary, Caitlin McDonnell, Jade Mowery, Stephanie Mccracken, Tyler Payne, Kirtana Ramadugu, Erica Leasure, Brent Lee, Kelsey Florek, Heather Blankenship, Allison Black, Quanta Brown, and Tammy Bannerman |
| EPI_ISL_2656117, EPI_ISL_2657575 | OHSU-MM Lab | OHSU-MM Lab | Rebecca O'Gara |
| EPI_ISL_2657576, EPI_ISL_2657577, EPI_ISL_2657578, EPI_ISL_2657579, EPI_ISL_2657581 | Ohio Department of Health Laboratory | Ohio Department of Health Laboratory | Holmes, Jennifer; Eric Brandt, Keoni Omura, Glen McGillivary, Caitlin McDonnell, Jade Mowery, Stephanie Mccracken, Tyler Payne, Kirtana Ramadugu, Erica Leasure, Brent Lee, Kelsey Florek, Heather Blankenship, Allison Black, Quanta Brown, and Tammy Bannerman |
| EPI_ISL_2657662 | Florida Bureau of Public Health Laboratories | Florida Bureau of Public Health Laboratories | Sarah Schmedes, Jason Blanton |
| EPI_ISL_2657663, EPI_ISL_2657664, EPI_ISL_2657665, EPI_ISL_2657666, EPI_ISL_2657667, EPI_ISL_2657668 | NCSLPH | NCSLPH | Chase K, Miller MC, Greene S, Glover W |
| EPI_ISL_2657670, EPI_ISL_2657672, EPI_ISL_2657673 | Tampa General Hospital Esoteric Lab | Tampa General Hospital Esoteric Research & Development Lab | Jon Faughn, Deanna Becker, Dominic Uy, Vicki Healer, Amorce Lima, Suzane Silbert |
| EPI_ISL_2657689, EPI_ISL_2657690, EPI_ISL_2657691, EPI_ISL_2657692, EPI_ISL_2657693, EPI_ISL_2657694, EPI_ISL_2657695 | Wexner Medical Center | The Ohio State University College of Medicine | Koenig,S., Seminetta,J. |
| EPI_ISL_2657761, EPI_ISL_2657762, EPI_ISL_2657763, EPI_ISL_2657764 | Kansas Health and Environmental Lab | Kansas Health and Environmental Lab | Katherine Wiggins, Mike Grose, Jonathan Barnell, Ben Olsen, and Phil Adam |
| EPI_ISL_2657765, EPI_ISL_2657766, EPI_ISL_2657767, EPI_ISL_2657768, EPI_ISL_2657769, EPI_ISL_2657856, EPI_ISL_2658057, EPI_ISL_2658149 | OHSU-MM Lab | OHSU-MM Lab | Rebecca O'Gara |
| EPI_ISL_2658150, EPI_ISL_2658151, EPI_ISL_2658152, EPI_ISL_2658153, EPI_ISL_2658154, EPI_ISL_2658155, EPI_ISL_2658156, EPI_ISL_2658157, EPI_ISL_2658158, EPI_ISL_2658159, EPI_ISL_2658160, EPI_ISL_2658161, EPI_ISL_2658162, EPI_ISL_2658163, EPI_ISL_2658164, EPI_ISL_2658165, EPI_ISL_2658166, EPI_ISL_2658167, EPI_ISL_2658168, EPI_ISL_2658169, EPI_ISL_2658170, EPI_ISL_2658171 | | | |
| see above | Kansas Health and Environmental Lab | Kansas Health and Environmental Lab | Mike Grose, Katherine Wiggins, Jonathan Barnell, Ben Olsen, and Phil Adam |
| EPI_ISL_2658173 | OHSU-MM Lab | OHSU-MM Lab | Rebecca O'Gara |

| | | | |
|---|---|---|---|
| EPI_ISL_2658286, EPI_ISL_2658287, EPI_ISL_2658288, EPI_ISL_2658289, EPI_ISL_2658290, EPI_ISL_2658291, EPI_ISL_2658292, EPI_ISL_2658293, EPI_ISL_2658294, EPI_ISL_2658295, EPI_ISL_2658296, EPI_ISL_2658297, EPI_ISL_2658298, EPI_ISL_2658299, EPI_ISL_2658300, EPI_ISL_2658301, EPI_ISL_2658302, EPI_ISL_2658304, EPI_ISL_2658305, EPI_ISL_2658306, EPI_ISL_2658307, EPI_ISL_2658308, EPI_ISL_2658309, EPI_ISL_2658310, EPI_ISL_2658311, EPI_ISL_2658312, EPI_ISL_2658313 | | | |
| see above | Wyoming Public Health Laboratory | Wyoming Public Health Laboratory | Jim Mildenberger, Taylor Fearing, Channing Weber, Ashley Norberg, Chayse Rowley, Marley Goetz, Brian Dominguez, Elliot Thomasson, Sam Britz, Cari Sloma, Robert Petit, and Rob Christensen |
| EPI_ISL_2658317 | Orion Laboratories | Orion Laboratories | Keisha Simoneaux, David Slaughter |
| EPI_ISL_2658453 | Michael E. Debakey VA Medical Center | Genomics and Discovery, Respiratory Viruses Branch, Division of Viral Diseases, Centers for Disease Control and Prevention | Jing Zhang, Anna Kelleher, Ying Tao, Yan Li, Brian Lynch, Krista Queen, Anna Uehara, Peter Cook, Han Jia Justin Ng, Rachel Marine, Clinton R. Paden, Dhwani Batra, Haibin Wang, Tara Coalter, Jasmine Padilla, Morgan Davis, Mili Sheth, Sarah Nobles, Mark Burroughs, Justin Lee, Adam Retchless, Suxiang Tong |
| EPI_ISL_2658514, EPI_ISL_2658515, EPI_ISL_2658516, EPI_ISL_2658517, EPI_ISL_2658518, EPI_ISL_2658519, EPI_ISL_2658520, EPI_ISL_2658521, EPI_ISL_2658522, EPI_ISL_2658523, EPI_ISL_2658524, EPI_ISL_2658525, EPI_ISL_2658526, EPI_ISL_2658527, EPI_ISL_2658529, EPI_ISL_2658531, EPI_ISL_2658532, EPI_ISL_2658533, EPI_ISL_2658534, EPI_ISL_2658535, EPI_ISL_2658536, EPI_ISL_2658537, EPI_ISL_2658538, EPI_ISL_2658539, EPI_ISL_2658540, EPI_ISL_2658541, EPI_ISL_2658542, EPI_ISL_2658543, EPI_ISL_2658544, EPI_ISL_2658545, EPI_ISL_2658546, EPI_ISL_2658547 | | | |
| see above | Minnesota Department of Health, Public Health Laboratory | Minnesota Department of Health, Public Health Laboratory | Alexandra Lorentz, Jacob Garfin, Matt Plumb, and Xiong Wang |
| EPI_ISL_2658548, EPI_ISL_2658549, EPI_ISL_2658550 | Premier Medical Lab | Minnesota Department of Health, Public Health Laboratory | Alexandra Lorentz, Jacob Garfin, Matt Plumb, and Xiong Wang |
| EPI_ISL_2658552, EPI_ISL_2658553, EPI_ISL_2658554, EPI_ISL_2658555, EPI_ISL_2658557, EPI_ISL_2658558, EPI_ISL_2658559, EPI_ISL_2658560, EPI_ISL_2658561, EPI_ISL_2658562, EPI_ISL_2658563, EPI_ISL_2658564, EPI_ISL_2658565, EPI_ISL_2658566, EPI_ISL_2658567 | | | |
| see above | Minnesota Department of Health, Public Health Laboratory | Minnesota Department of Health, Public Health Laboratory | Alexandra Lorentz, Jacob Garfin, Matt Plumb, and Xiong Wang |
| EPI_ISL_2658568, EPI_ISL_2658569 | Alameda County Public Health Lab | Chan-Zuckerberg Biohub | CZB Cliahub Consortium |
| EPI_ISL_2658570, EPI_ISL_2658572, EPI_ISL_2658573, EPI_ISL_2658574 | Santa Clara County Public Health Laboratory | Chan-Zuckerberg Biohub | CZB Cliahub Consortium |
| EPI_ISL_2658575 | Contra Costa County Public Health Lab | Chan-Zuckerberg Biohub | CZB Cliahub Consortium |
| EPI_ISL_2658576 | Santa Clara County Public Health Laboratory | Chan-Zuckerberg Biohub | CZB Cliahub Consortium |
| EPI_ISL_2658577 | Alameda County Public Health Lab | Chan-Zuckerberg Biohub | CZB Cliahub Consortium |
| EPI_ISL_2658578 | Santa Clara County Public Health Laboratory | Chan-Zuckerberg Biohub | CZB Cliahub Consortium |
| EPI_ISL_2658579 | Alameda County Public Health Lab | Chan-Zuckerberg Biohub | CZB Cliahub Consortium |
| EPI_ISL_2658580 | Orange County Public Health Lab | Chan-Zuckerberg Biohub | CZB Cliahub Consortium |
| EPI_ISL_2658582 | Humboldt County Public Health Laboratory | Chan-Zuckerberg Biohub | CZB Cliahub Consortium |
| EPI_ISL_2658583 | Alameda County Public Health Lab | Chan-Zuckerberg Biohub | CZB Cliahub Consortium |
| EPI_ISL_2658584, EPI_ISL_2658585 | Santa Clara County Public Health Laboratory | Chan-Zuckerberg Biohub | CZB Cliahub Consortium |
| EPI_ISL_2658587 | UCSF Clinical Microbiology Laboratory | Chan-Zuckerberg Biohub | CZB Cliahub Consortium |
| EPI_ISL_2658588, EPI_ISL_2658590, EPI_ISL_2658591, EPI_ISL_2658592 | Santa Clara County Public Health Laboratory | Chan-Zuckerberg Biohub | CZB Cliahub Consortium |
| EPI_ISL_2658594 | Contra Costa County Public Health Lab | Chan-Zuckerberg Biohub | CZB Cliahub Consortium |
| EPI_ISL_2658595, EPI_ISL_2658597 | CA DPH Viral and Rickettsial Disease Laboratory | Chan-Zuckerberg Biohub | CZB Cliahub Consortium |
| EPI_ISL_2658598 | Santa Clara County Public Health Laboratory | Chan-Zuckerberg Biohub | CZB Cliahub Consortium |
| EPI_ISL_2658599 | Alameda County Public Health Lab | Chan-Zuckerberg Biohub | CZB Cliahub Consortium |
| EPI_ISL_2658600 | Humboldt County Public Health Laboratory | Chan-Zuckerberg Biohub | CZB Cliahub Consortium |
| EPI_ISL_2658601 | County of San Luis Obispo Public Health Laboratory | Chan-Zuckerberg Biohub | CZB Cliahub Consortium |
| EPI_ISL_2658602 | Orange County Public Health Lab | Chan-Zuckerberg Biohub | CZB Cliahub Consortium |
| EPI_ISL_2658603 | Alameda County Public Health Lab | Chan-Zuckerberg Biohub | CZB Cliahub Consortium |
| EPI_ISL_2658604 | Humboldt County Public Health Laboratory | Chan-Zuckerberg Biohub | CZB Cliahub Consortium |
| EPI_ISL_2658605, EPI_ISL_2658606, EPI_ISL_2658607, EPI_ISL_2658608 | Santa Clara County Public Health Laboratory | Chan-Zuckerberg Biohub | CZB Cliahub Consortium |
| EPI_ISL_2658609 | Humboldt County Public Health Laboratory | Chan-Zuckerberg Biohub | CZB Cliahub Consortium |
| EPI_ISL_2658610 | Contra Costa County Public Health Lab | Chan-Zuckerberg Biohub | CZB Cliahub Consortium |
| EPI_ISL_2658611, EPI_ISL_2658612 | Santa Clara County Public Health Laboratory | Chan-Zuckerberg Biohub | CZB Cliahub Consortium |
| EPI_ISL_2658613 | County of San Luis Obispo Public Health Laboratory | Chan-Zuckerberg Biohub | CZB Cliahub Consortium |
| EPI_ISL_2658614 | Orange County Public Health Lab | Chan-Zuckerberg Biohub | CZB Cliahub Consortium |
| EPI_ISL_2658615 | CA DPH Viral and Rickettsial Disease Laboratory | Chan-Zuckerberg Biohub | CZB Cliahub Consortium |
| EPI_ISL_2658616 | Contra Costa County Public Health Lab | Chan-Zuckerberg Biohub | CZB Cliahub Consortium |
| EPI_ISL_2658618 | Alameda County Public Health Lab | Chan-Zuckerberg Biohub | CZB Cliahub Consortium |
| EPI_ISL_2658619 | Contra Costa County Public Health Lab | Chan-Zuckerberg Biohub | CZB Cliahub Consortium |
| EPI_ISL_2658620 | Alameda County Public Health Lab | Chan-Zuckerberg Biohub | CZB Cliahub Consortium |
| EPI_ISL_2658621 | CA DPH Viral and Rickettsial Disease Laboratory | Chan-Zuckerberg Biohub | CZB Cliahub Consortium |
| EPI_ISL_2658622, EPI_ISL_2658623, EPI_ISL_2658624 | Santa Clara County Public Health Laboratory | Chan-Zuckerberg Biohub | CZB Cliahub Consortium |
| EPI_ISL_2658625 | Orange County Public Health Lab | Chan-Zuckerberg Biohub | CZB Cliahub Consortium |
| EPI_ISL_2658626 | Alameda County Public Health Lab | Chan-Zuckerberg Biohub | CZB Cliahub Consortium |
| EPI_ISL_2658628 | Santa Clara County Public Health Laboratory | Chan-Zuckerberg Biohub | CZB Cliahub Consortium |
| EPI_ISL_2658631 | Alameda County Public Health Lab | Chan-Zuckerberg Biohub | CZB Cliahub Consortium |
| EPI_ISL_2658632 | County of San Luis Obispo Public Health Laboratory | Chan-Zuckerberg Biohub | CZB Cliahub Consortium |
| EPI_ISL_2658633 | Santa Clara County Public Health Laboratory | Chan-Zuckerberg Biohub | CZB Cliahub Consortium |
| EPI_ISL_2658636 | County of San Luis Obispo Public Health | Chan-Zuckerberg Biohub | CZB Cliahub Consortium |

| | | | |
|---|---|---|---|
| | Laboratory | | |
| EPI_ISL_2658638 | Alameda County Public Health Lab | Chan-Zuckerberg Biohub | CZB Cliahub Consortium |
| EPI_ISL_2658639 | Orange County Public Health Lab | Chan-Zuckerberg Biohub | CZB Cliahub Consortium |
| EPI_ISL_2658640, EPI_ISL_2658641, EPI_ISL_2658642 | Santa Clara County Public Health Laboratory | Chan-Zuckerberg Biohub | CZB Cliahub Consortium |
| EPI_ISL_2658643 | Orange County Public Health Lab | Chan-Zuckerberg Biohub | CZB Cliahub Consortium |
| EPI_ISL_2658644 | Contra Costa County Public Health Lab | Chan-Zuckerberg Biohub | CZB Cliahub Consortium |
| EPI_ISL_2658645 | Santa Clara County Public Health Laboratory | Chan-Zuckerberg Biohub | CZB Cliahub Consortium |
| EPI_ISL_2658646 | Humboldt County Public Health Laboratory | Chan-Zuckerberg Biohub | CZB Cliahub Consortium |
| EPI_ISL_2658648, EPI_ISL_2658650 | Santa Clara County Public Health Laboratory | Chan-Zuckerberg Biohub | CZB Cliahub Consortium |
| EPI_ISL_2658651 | Humboldt County Public Health Laboratory | Chan-Zuckerberg Biohub | CZB Cliahub Consortium |
| EPI_ISL_2658654 | Alameda County Public Health Lab | Chan-Zuckerberg Biohub | CZB Cliahub Consortium |
| EPI_ISL_2658655 | Santa Clara County Public Health Laboratory | Chan-Zuckerberg Biohub | CZB Cliahub Consortium |
| EPI_ISL_2658656 | Orange County Public Health Lab | Chan-Zuckerberg Biohub | CZB Cliahub Consortium |
| EPI_ISL_2658658 | Alameda County Public Health Lab | Chan-Zuckerberg Biohub | CZB Cliahub Consortium |
| EPI_ISL_2658659 | Orange County Public Health Lab | Chan-Zuckerberg Biohub | CZB Cliahub Consortium |
| EPI_ISL_2658660, EPI_ISL_2658661 | UCSF Clinical Microbiology Laboratory | Chan-Zuckerberg Biohub | CZB Cliahub Consortium |
| EPI_ISL_2658662 | Humboldt County Public Health Laboratory | Chan-Zuckerberg Biohub | CZB Cliahub Consortium |
| EPI_ISL_2658663 | Santa Clara County Public Health Laboratory | Chan-Zuckerberg Biohub | CZB Cliahub Consortium |
| EPI_ISL_2658664 | Alameda County Public Health Lab | Chan-Zuckerberg Biohub | CZB Cliahub Consortium |
| EPI_ISL_2658665 | CA DPH Viral and Rickettsial Disease Laboratory | Chan-Zuckerberg Biohub | CZB Cliahub Consortium |
| EPI_ISL_2658666 | Alameda County Public Health Lab | Chan-Zuckerberg Biohub | CZB Cliahub Consortium |
| EPI_ISL_2658668 | Humboldt County Public Health Laboratory | Chan-Zuckerberg Biohub | CZB Cliahub Consortium |
| EPI_ISL_2658670 | Madera County Department of Public Health | Chan-Zuckerberg Biohub | CZB Cliahub Consortium |
| EPI_ISL_2658671 | Alameda County Public Health Lab | Chan-Zuckerberg Biohub | CZB Cliahub Consortium |
| EPI_ISL_2658674 | Contra Costa County Public Health Lab | Chan-Zuckerberg Biohub | CZB Cliahub Consortium |
| EPI_ISL_2658675, EPI_ISL_2658676, EPI_ISL_2658677, EPI_ISL_2658679 | Santa Clara County Public Health Laboratory | Chan-Zuckerberg Biohub | CZB Cliahub Consortium |
| EPI_ISL_2658680, EPI_ISL_2658681 | CA DPH Viral and Rickettsial Disease Laboratory | Chan-Zuckerberg Biohub | CZB Cliahub Consortium |
| EPI_ISL_2658682 | Alameda County Public Health Lab | Chan-Zuckerberg Biohub | CZB Cliahub Consortium |
| EPI_ISL_2658683 | CA DPH Viral and Rickettsial Disease Laboratory | Chan-Zuckerberg Biohub | CZB Cliahub Consortium |
| EPI_ISL_2658684 | Santa Clara County Public Health Laboratory | Chan-Zuckerberg Biohub | CZB Cliahub Consortium |
| EPI_ISL_2658685 | CA DPH Viral and Rickettsial Disease Laboratory | Chan-Zuckerberg Biohub | CZB Cliahub Consortium |
| EPI_ISL_2658688 | Santa Clara County Public Health Laboratory | Chan-Zuckerberg Biohub | CZB Cliahub Consortium |
| EPI_ISL_2658690, EPI_ISL_2658693 | Alameda County Public Health Lab | Chan-Zuckerberg Biohub | CZB Cliahub Consortium |
| EPI_ISL_2658694 | Santa Clara County Public Health Laboratory | Chan-Zuckerberg Biohub | CZB Cliahub Consortium |
| EPI_ISL_2658698 | Contra Costa County Public Health Lab | Chan-Zuckerberg Biohub | CZB Cliahub Consortium |
| EPI_ISL_2658699, EPI_ISL_2658700 | Humboldt County Public Health Laboratory | Chan-Zuckerberg Biohub | CZB Cliahub Consortium |
| EPI_ISL_2658701 | CA DPH Viral and Rickettsial Disease Laboratory | Chan-Zuckerberg Biohub | CZB Cliahub Consortium |
| EPI_ISL_2658702 | Orange County Public Health Lab | Chan-Zuckerberg Biohub | CZB Cliahub Consortium |
| EPI_ISL_2658703 | Santa Clara County Public Health Laboratory | Chan-Zuckerberg Biohub | CZB Cliahub Consortium |
| EPI_ISL_2658704 | Humboldt County Public Health Laboratory | Chan-Zuckerberg Biohub | CZB Cliahub Consortium |
| EPI_ISL_2658705 | CA DPH Viral and Rickettsial Disease Laboratory | Chan-Zuckerberg Biohub | CZB Cliahub Consortium |
| EPI_ISL_2658707, EPI_ISL_2658708 | Contra Costa County Public Health Lab | Chan-Zuckerberg Biohub | CZB Cliahub Consortium |
| EPI_ISL_2658709 | Santa Clara County Public Health Laboratory | Chan-Zuckerberg Biohub | CZB Cliahub Consortium |
| EPI_ISL_2658710 | Humboldt County Public Health Laboratory | Chan-Zuckerberg Biohub | CZB Cliahub Consortium |
| EPI_ISL_2658711 | Santa Clara County Public Health Laboratory | Chan-Zuckerberg Biohub | CZB Cliahub Consortium |
| EPI_ISL_2658713 | Alameda County Public Health Lab | Chan-Zuckerberg Biohub | CZB Cliahub Consortium |
| EPI_ISL_2658714 | Santa Clara County Public Health Laboratory | Chan-Zuckerberg Biohub | CZB Cliahub Consortium |
| EPI_ISL_2658717 | Contra Costa County Public Health Lab | Chan-Zuckerberg Biohub | CZB Cliahub Consortium |
| EPI_ISL_2658718 | Orange County Public Health Lab | Chan-Zuckerberg Biohub | CZB Cliahub Consortium |
| EPI_ISL_2658719 | CA DPH Viral and Rickettsial Disease Laboratory | Chan-Zuckerberg Biohub | CZB Cliahub Consortium |
| EPI_ISL_2658721 | Orange County Public Health Lab | Chan-Zuckerberg Biohub | CZB Cliahub Consortium |
| EPI_ISL_2658722 | Contra Costa County Public Health Lab | Chan-Zuckerberg Biohub | CZB Cliahub Consortium |
| EPI_ISL_2658723, | Orange County Public Health Lab | Chan-Zuckerberg Biohub | CZB Cliahub Consortium |

| | | | |
|---|---|---|---|
| EPI_ISL_2658724 | | | |
| EPI_ISL_2658725, EPI_ISL_2658726, EPI_ISL_2658727, EPI_ISL_2658728 | Santa Clara County Public Health Laboratory | Chan-Zuckerberg Biohub | CZB Cliahub Consortium |
| EPI_ISL_2658729 | CA DPH Viral and Rickettsial Disease Laboratory | Chan-Zuckerberg Biohub | CZB Cliahub Consortium |
| EPI_ISL_2658730 | Orange County Public Health Lab | Chan-Zuckerberg Biohub | CZB Cliahub Consortium |
| EPI_ISL_2658731 | CA DPH Viral and Rickettsial Disease Laboratory | Chan-Zuckerberg Biohub | CZB Cliahub Consortium |
| EPI_ISL_2658732 | Humboldt County Public Health Laboratory | Chan-Zuckerberg Biohub | CZB Cliahub Consortium |
| EPI_ISL_2658733 | Santa Clara County Public Health Laboratory | Chan-Zuckerberg Biohub | CZB Cliahub Consortium |
| EPI_ISL_2658734 | Alameda County Public Health Lab | Chan-Zuckerberg Biohub | CZB Cliahub Consortium |
| EPI_ISL_2658735, EPI_ISL_2658736 | Santa Clara County Public Health Laboratory | Chan-Zuckerberg Biohub | CZB Cliahub Consortium |
| EPI_ISL_2658737 | Alameda County Public Health Lab | Chan-Zuckerberg Biohub | CZB Cliahub Consortium |
| EPI_ISL_2658738 | Humboldt County Public Health Laboratory | Chan-Zuckerberg Biohub | CZB Cliahub Consortium |
| EPI_ISL_2658740 | Orange County Public Health Lab | Chan-Zuckerberg Biohub | CZB Cliahub Consortium |
| EPI_ISL_2658741 | Contra Costa County Public Health Lab | Chan-Zuckerberg Biohub | CZB Cliahub Consortium |
| EPI_ISL_2658743, EPI_ISL_2658744 | Santa Clara County Public Health Laboratory | Chan-Zuckerberg Biohub | CZB Cliahub Consortium |
| EPI_ISL_2658745 | Madera County Department of Public Health | Chan-Zuckerberg Biohub | CZB Cliahub Consortium |
| EPI_ISL_2658746, EPI_ISL_2658747, EPI_ISL_2658748, EPI_ISL_2658749, EPI_ISL_2658750, EPI_ISL_2658751, EPI_ISL_2658752, EPI_ISL_2658753, EPI_ISL_2658754, EPI_ISL_2658755 | Wyoming Public Health Laboratory | Wyoming Public Health Laboratory | Jim Mildenberger, Wanda Manley, Noah Hull, Taylor Fearing, Lynette Gumbleton, Channing Weber, Ashley Norberg, Chayse Rowley, Marley Goetz, Brian Dominguez, Elliot Thomasson, Sam Britz, Cari Sloma, and Rob Christensen |
| EPI_ISL_2658946 | AR Dept. of Health-PHL, Molecular Diagnostics | Genomics and Discovery, Respiratory Viruses Branch, Division of Viral Diseases, Centers for Disease Control and Prevention | Jing Zhang, Anna Kelleher, Ying Tao, Yan Li, Brian Lynch, Krista Queen, Anna Uehara, Peter Cook, Han Jia Justin Ng, Rachel Marine, Clinton R. Paden, Dhwani Batra, Haibin Wang, Tara Coalter, Jasmine Padilla, Morgan Davis, Mili Sheth, Sarah Nobles, Mark Burroughs, Justin Lee, Adam Retchless, Suxiang Tong |
| EPI_ISL_2658947 | TN Division of Laboratory Services | Genomics and Discovery, Respiratory Viruses Branch, Division of Viral Diseases, Centers for Disease Control and Prevention | Jing Zhang, Anna Kelleher, Ying Tao, Yan Li, Brian Lynch, Krista Queen, Anna Uehara, Peter Cook, Han Jia Justin Ng, Rachel Marine, Clinton R. Paden, Dhwani Batra, Haibin Wang, Tara Coalter, Jasmine Padilla, Morgan Davis, Mili Sheth, Sarah Nobles, Mark Burroughs, Justin Lee, Adam Retchless, Suxiang Tong |
| EPI_ISL_2658948, EPI_ISL_2658949 | OK Public Health Laboratory, Oklahoma State DOH | Genomics and Discovery, Respiratory Viruses Branch, Division of Viral Diseases, Centers for Disease Control and Prevention | Jing Zhang, Anna Kelleher, Ying Tao, Yan Li, Brian Lynch, Krista Queen, Anna Uehara, Peter Cook, Han Jia Justin Ng, Rachel Marine, Clinton R. Paden, Dhwani Batra, Haibin Wang, Tara Coalter, Jasmine Padilla, Morgan Davis, Mili Sheth, Sarah Nobles, Mark Burroughs, Justin Lee, Adam Retchless, Suxiang Tong |
| EPI_ISL_2658951, EPI_ISL_2658952 | LESP San Luis Potosi | Instituto de Diagnostico y Referencia Epidemiologicos (INDRE) | Claudia Wong-Arambula, Abril Rodriguez-Maldonado, Vanessa Rivero-Arredondo, Ariadna Medina-Benitez, Joaquin Quiroz-Mercado, Sergio Rangel-Guerrero, Natividad Cruz-Ortiz, Tatiana Nunez-Garcia, Gisela Barrera-Badillo, Lucia Hernandez-Rivas, Irma Lopez-Martinez, Ernesto Ramirez-Gonzalez. |
| EPI_ISL_2658953 | LESP Nuevo Leon | Instituto de Diagnostico y Referencia Epidemiologicos (INDRE) | Claudia Wong-Arambula, Abril Rodriguez-Maldonado, Vanessa Rivero-Arredondo, Ariadna Medina-Benitez, Joaquin Quiroz-Mercado, Sergio Rangel-Guerrero, Natividad Cruz-Ortiz, Tatiana Nunez-Garcia, Gisela Barrera-Badillo, Lucia Hernandez-Rivas, Irma Lopez-Martinez, Ernesto Ramirez-Gonzalez. |
| EPI_ISL_2658955, EPI_ISL_2658956, EPI_ISL_2658957, EPI_ISL_2658958, EPI_ISL_2658959, EPI_ISL_2658960, EPI_ISL_2658962, EPI_ISL_2658963, EPI_ISL_2658965, EPI_ISL_2658966, EPI_ISL_2658967, EPI_ISL_2658968, EPI_ISL_2658969, EPI_ISL_2658970, EPI_ISL_2658971, EPI_ISL_2658972, EPI_ISL_2658973, EPI_ISL_2658974, EPI_ISL_2658976, EPI_ISL_2658977, EPI_ISL_2658978, EPI_ISL_2658979, EPI_ISL_2658980 | | | |
| see above | TXDSHS | TXDSHS | Rashmi Tuladhar, Bonnie Oh, Jenny Zhang, Maliha Rahman, Mayela Pedrueza, Anita Pokharel, Karen Bobier, Lorraine Rodriguez, Myong Koag, Chun Wang, Rachel Lee, Grace Kubin |
| EPI_ISL_2658981, EPI_ISL_2658982 | County of San Luis Obispo Public Health Laboratory | County of San Luis Obispo Public Health Laboratory | Katrina Erwin, Frances Sidhu |
| EPI_ISL_2658984 | Kaiser Santa Clara | Santa Clara County Public Health Laboratory | Santa Clara County Public Health Department |
| EPI_ISL_2658985, EPI_ISL_2658986, EPI_ISL_2658987, EPI_ISL_2658988, EPI_ISL_2658990, EPI_ISL_2658991, EPI_ISL_2658992, EPI_ISL_2658993, EPI_ISL_2658994, EPI_ISL_2658995, EPI_ISL_2658996, EPI_ISL_2658997, EPI_ISL_2658998, EPI_ISL_2658999, EPI_ISL_2659000, EPI_ISL_2659001, EPI_ISL_2659002, EPI_ISL_2659003, EPI_ISL_2659004, EPI_ISL_2659005, EPI_ISL_2659006, EPI_ISL_2659007, EPI_ISL_2659008, EPI_ISL_2659009, EPI_ISL_2659010, EPI_ISL_2659011, EPI_ISL_2659012, EPI_ISL_2659013, EPI_ISL_2659014, EPI_ISL_2659015, EPI_ISL_2659016, EPI_ISL_2659017, EPI_ISL_2659018, EPI_ISL_2659019, EPI_ISL_2659020, EPI_ISL_2659021, EPI_ISL_2659022, EPI_ISL_2659023, EPI_ISL_2659024, EPI_ISL_2659025, EPI_ISL_2659026, EPI_ISL_2659027, EPI_ISL_2659028, EPI_ISL_2659029, EPI_ISL_2659030, EPI_ISL_2659031, EPI_ISL_2659032, EPI_ISL_2659033, EPI_ISL_2659034, EPI_ISL_2659035, EPI_ISL_2659036, EPI_ISL_2659037, EPI_ISL_2659038, EPI_ISL_2659039, EPI_ISL_2659041, EPI_ISL_2659045, EPI_ISL_2659047, EPI_ISL_2659048, EPI_ISL_2659050 | | | |
| see above | Santa Clara Valley Medical Center | Santa Clara County Public Health Laboratory | Santa Clara County Public Health Department |
| EPI_ISL_2659052, EPI_ISL_2659054, EPI_ISL_2659055, EPI_ISL_2659056, EPI_ISL_2659057 | Santa Clara County Public Health Laboratory | Santa Clara County Public Health Laboratory | Santa Clara County Public Health Department |
| EPI_ISL_2659058, EPI_ISL_2659061, EPI_ISL_2659062 | Stanford Clinical Virology Laboratory | Santa Clara County Public Health Laboratory | Santa Clara County Public Health Department |
| EPI_ISL_2659065, EPI_ISL_2659068, EPI_ISL_2659069, EPI_ISL_2659070, EPI_ISL_2659072 | El Camino Hospital | Santa Clara County Public Health Laboratory | Santa Clara County Public Health Department |
| EPI_ISL_2659075, EPI_ISL_2659076, EPI_ISL_2659078, EPI_ISL_2659081, EPI_ISL_2659082, EPI_ISL_2659086, EPI_ISL_2659087, EPI_ISL_2659088, EPI_ISL_2659089, EPI_ISL_2659091, EPI_ISL_2659094, EPI_ISL_2659095, EPI_ISL_2659096, EPI_ISL_2659097, EPI_ISL_2659101, EPI_ISL_2659102 | | | |
| see above | Santa Clara Valley Medical Center | Santa Clara County Public Health Laboratory | Santa Clara County Public Health Department |
| EPI_ISL_2659105, EPI_ISL_2659106 | PAMF Palo Alto | Santa Clara County Public Health Laboratory | Santa Clara County Public Health Department |
| EPI_ISL_2659110, EPI_ISL_2659111, EPI_ISL_2659112, EPI_ISL_2659113, EPI_ISL_2659114, EPI_ISL_2659115 | Santa Clara Valley Medical Center | Santa Clara County Public Health Laboratory | Santa Clara County Public Health Department |
| EPI_ISL_2659116 | County of San Luis Obispo Public Health Laboratory | County of San Luis Obispo Public Health Laboratory | Katrina Erwin, Frances Sidhu |
| EPI_ISL_2659199 | Santa Clara County Public Health Laboratory | Chan-Zuckerberg Biohub | CZB Cliahub Consortium |
| EPI_ISL_2659200 | CA DPH Viral and Rickettsial Disease Laboratory | Chan-Zuckerberg Biohub | CZB Cliahub Consortium |
| EPI_ISL_2659201 | Santa Clara County Public Health Laboratory | Chan-Zuckerberg Biohub | CZB Cliahub Consortium |
| EPI_ISL_2659202 | Contra Costa County Public Health Lab | Chan-Zuckerberg Biohub | CZB Cliahub Consortium |
| EPI_ISL_2659203 | Madera County Department of Public | Chan-Zuckerberg Biohub | CZB Cliahub Consortium |

| | | | |
|---|---|---|---|
| | Health | | |
| EPI_ISL_2659204 | CA DPH Viral and Rickettsial Disease Laboratory | Chan-Zuckerberg Biohub | CZB Cliahub Consortium |
| EPI_ISL_2659205 | Humboldt County Public Health Laboratory | Chan-Zuckerberg Biohub | CZB Cliahub Consortium |
| EPI_ISL_2659206 | Orange County Public Health Lab | Chan-Zuckerberg Biohub | CZB Cliahub Consortium |
| EPI_ISL_2659207 | CA DPH Viral and Rickettsial Disease Laboratory | Chan-Zuckerberg Biohub | CZB Cliahub Consortium |
| EPI_ISL_2659208 | Santa Clara County Public Health Laboratory | Chan-Zuckerberg Biohub | CZB Cliahub Consortium |
| EPI_ISL_2659209 | Humboldt County Public Health Laboratory | Chan-Zuckerberg Biohub | CZB Cliahub Consortium |
| EPI_ISL_2659210 | Contra Costa County Public Health Lab | Chan-Zuckerberg Biohub | CZB Cliahub Consortium |
| EPI_ISL_2659211 | CA DPH Viral and Rickettsial Disease Laboratory | Chan-Zuckerberg Biohub | CZB Cliahub Consortium |
| EPI_ISL_2659212 | Contra Costa County Public Health Lab | Chan-Zuckerberg Biohub | CZB Cliahub Consortium |
| EPI_ISL_2659213 | Humboldt County Public Health Laboratory | Chan-Zuckerberg Biohub | CZB Cliahub Consortium |
| EPI_ISL_2659214 | Santa Clara County Public Health Laboratory | Chan-Zuckerberg Biohub | CZB Cliahub Consortium |
| EPI_ISL_2659215 | Alameda County Public Health Lab | Chan-Zuckerberg Biohub | CZB Cliahub Consortium |
| EPI_ISL_2659217 | Orange County Public Health Lab | Chan-Zuckerberg Biohub | CZB Cliahub Consortium |
| EPI_ISL_2659218, EPI_ISL_2659219 | Santa Clara County Public Health Laboratory | Chan-Zuckerberg Biohub | CZB Cliahub Consortium |
| EPI_ISL_2659220 | Alameda County Public Health Lab | Chan-Zuckerberg Biohub | CZB Cliahub Consortium |
| EPI_ISL_2659221, EPI_ISL_2659222, EPI_ISL_2659223 | Santa Clara County Public Health Laboratory | Chan-Zuckerberg Biohub | CZB Cliahub Consortium |
| EPI_ISL_2659224 | Alameda County Public Health Lab | Chan-Zuckerberg Biohub | CZB Cliahub Consortium |
| EPI_ISL_2659225 | Santa Clara County Public Health Laboratory | Chan-Zuckerberg Biohub | CZB Cliahub Consortium |
| EPI_ISL_2659226 | Contra Costa County Public Health Lab | Chan-Zuckerberg Biohub | CZB Cliahub Consortium |
| EPI_ISL_2659227, EPI_ISL_2659228 | Santa Clara County Public Health Laboratory | Chan-Zuckerberg Biohub | CZB Cliahub Consortium |
| EPI_ISL_2659229 | Alameda County Public Health Lab | Chan-Zuckerberg Biohub | CZB Cliahub Consortium |
| EPI_ISL_2659230 | Santa Clara County Public Health Laboratory | Chan-Zuckerberg Biohub | CZB Cliahub Consortium |
| EPI_ISL_2659231 | CA DPH Viral and Rickettsial Disease Laboratory | Chan-Zuckerberg Biohub | CZB Cliahub Consortium |
| EPI_ISL_2659232 | Alameda County Public Health Lab | Chan-Zuckerberg Biohub | CZB Cliahub Consortium |
| EPI_ISL_2659233 | CA DPH Viral and Rickettsial Disease Laboratory | Chan-Zuckerberg Biohub | CZB Cliahub Consortium |
| EPI_ISL_2659234 | Santa Clara County Public Health Laboratory | Chan-Zuckerberg Biohub | CZB Cliahub Consortium |
| EPI_ISL_2659235 | Alameda County Public Health Lab | Chan-Zuckerberg Biohub | CZB Cliahub Consortium |
| EPI_ISL_2659236, EPI_ISL_2659237, EPI_ISL_2659238, EPI_ISL_2659239 | Santa Clara County Public Health Laboratory | Chan-Zuckerberg Biohub | CZB Cliahub Consortium |
| EPI_ISL_2659240 | Orange County Public Health Lab | Chan-Zuckerberg Biohub | CZB Cliahub Consortium |
| EPI_ISL_2659241, EPI_ISL_2659242, EPI_ISL_2659243, EPI_ISL_2659244 | Santa Clara County Public Health Laboratory | Chan-Zuckerberg Biohub | CZB Cliahub Consortium |
| EPI_ISL_2659245 | Alameda County Public Health Lab | Chan-Zuckerberg Biohub | CZB Cliahub Consortium |
| EPI_ISL_2659246, EPI_ISL_2659247, EPI_ISL_2659248, EPI_ISL_2659249, EPI_ISL_2659250 | Santa Clara County Public Health Laboratory | Chan-Zuckerberg Biohub | CZB Cliahub Consortium |
| EPI_ISL_2659251 | Humboldt County Public Health Laboratory | Chan-Zuckerberg Biohub | CZB Cliahub Consortium |
| EPI_ISL_2659252 | County of San Luis Obispo Public Health Laboratory | Chan-Zuckerberg Biohub | CZB Cliahub Consortium |
| EPI_ISL_2659253, EPI_ISL_2659254 | Santa Clara County Public Health Laboratory | Chan-Zuckerberg Biohub | CZB Cliahub Consortium |
| EPI_ISL_2659255 | Contra Costa County Public Health Lab | Chan-Zuckerberg Biohub | CZB Cliahub Consortium |
| EPI_ISL_2659256 | Santa Clara County Public Health Laboratory | Chan-Zuckerberg Biohub | CZB Cliahub Consortium |
| EPI_ISL_2659257 | Contra Costa County Public Health Lab | Chan-Zuckerberg Biohub | CZB Cliahub Consortium |
| EPI_ISL_2659258 | CA DPH Viral and Rickettsial Disease Laboratory | Chan-Zuckerberg Biohub | CZB Cliahub Consortium |
| EPI_ISL_2659259, EPI_ISL_2659260 | Humboldt County Public Health Laboratory | Chan-Zuckerberg Biohub | CZB Cliahub Consortium |
| EPI_ISL_2659261 | Alameda County Public Health Lab | Chan-Zuckerberg Biohub | CZB Cliahub Consortium |
| EPI_ISL_2659262 | Orange County Public Health Lab | Chan-Zuckerberg Biohub | CZB Cliahub Consortium |
| EPI_ISL_2659263 | Santa Clara County Public Health Laboratory | Chan-Zuckerberg Biohub | CZB Cliahub Consortium |
| EPI_ISL_2659265 | Humboldt County Public Health Laboratory | Chan-Zuckerberg Biohub | CZB Cliahub Consortium |
| EPI_ISL_2659266, EPI_ISL_2659267 | Alameda County Public Health Lab | Chan-Zuckerberg Biohub | CZB Cliahub Consortium |
| EPI_ISL_2659268, EPI_ISL_2659269 | Santa Clara County Public Health Laboratory | Chan-Zuckerberg Biohub | CZB Cliahub Consortium |
| EPI_ISL_2659270 | Orange County Public Health Lab | Chan-Zuckerberg Biohub | CZB Cliahub Consortium |
| EPI_ISL_2659271 | Contra Costa County Public Health Lab | Chan-Zuckerberg Biohub | CZB Cliahub Consortium |
| EPI_ISL_2659272 | Alameda County Public Health Lab | Chan-Zuckerberg Biohub | CZB Cliahub Consortium |
| EPI_ISL_2659273, | Santa Clara County Public Health | Chan-Zuckerberg Biohub | CZB Cliahub Consortium |

| | | | |
|---|---|---|---|
| EPI_ISL_2659274, EPI_ISL_2659275 | Laboratory | | |
| EPI_ISL_2659276 | Orange County Public Health Lab | Chan-Zuckerberg Biohub | CZB Cliahub Consortium |
| EPI_ISL_2659277 | CA DPH Viral and Rickettsial Disease Laboratory | Chan-Zuckerberg Biohub | CZB Cliahub Consortium |
| EPI_ISL_2659278 | Orange County Public Health Lab | Chan-Zuckerberg Biohub | CZB Cliahub Consortium |
| EPI_ISL_2659279, EPI_ISL_2659280 | CA DPH Viral and Rickettsial Disease Laboratory | Chan-Zuckerberg Biohub | CZB Cliahub Consortium |
| EPI_ISL_2659281 | Santa Clara County Public Health Laboratory | Chan-Zuckerberg Biohub | CZB Cliahub Consortium |
| EPI_ISL_2659282, EPI_ISL_2659283 | CA DPH Viral and Rickettsial Disease Laboratory | Chan-Zuckerberg Biohub | CZB Cliahub Consortium |
| EPI_ISL_2659284, EPI_ISL_2659285, EPI_ISL_2659286 | Santa Clara County Public Health Laboratory | Chan-Zuckerberg Biohub | CZB Cliahub Consortium |
| EPI_ISL_2659287 | Orange County Public Health Lab | Chan-Zuckerberg Biohub | CZB Cliahub Consortium |
| EPI_ISL_2659288, EPI_ISL_2659290 | Santa Clara County Public Health Laboratory | Chan-Zuckerberg Biohub | CZB Cliahub Consortium |
| EPI_ISL_2659291 | Orange County Public Health Lab | Chan-Zuckerberg Biohub | CZB Cliahub Consortium |
| EPI_ISL_2659292, EPI_ISL_2659293 | Santa Clara County Public Health Laboratory | Chan-Zuckerberg Biohub | CZB Cliahub Consortium |
| EPI_ISL_2659294 | Contra Costa County Public Health Lab | Chan-Zuckerberg Biohub | CZB Cliahub Consortium |
| EPI_ISL_2659295 | San Joaquin County Public Health Lab | Chan-Zuckerberg Biohub | CZB Cliahub Consortium |
| EPI_ISL_2659296 | CA DPH Viral and Rickettsial Disease Laboratory | Chan-Zuckerberg Biohub | CZB Cliahub Consortium |
| EPI_ISL_2659297 | County of San Luis Obispo Public Health Laboratory | Chan-Zuckerberg Biohub | CZB Cliahub Consortium |
| EPI_ISL_2659298, EPI_ISL_2659299 | Santa Clara County Public Health Laboratory | Chan-Zuckerberg Biohub | CZB Cliahub Consortium |
| EPI_ISL_2659300 | CA DPH Viral and Rickettsial Disease Laboratory | Chan-Zuckerberg Biohub | CZB Cliahub Consortium |
| EPI_ISL_2659301, EPI_ISL_2659302, EPI_ISL_2659303 | Santa Clara County Public Health Laboratory | Chan-Zuckerberg Biohub | CZB Cliahub Consortium |
| EPI_ISL_2659304 | Orange County Public Health Lab | Chan-Zuckerberg Biohub | CZB Cliahub Consortium |
| EPI_ISL_2659305, EPI_ISL_2659306 | CA DPH Viral and Rickettsial Disease Laboratory | Chan-Zuckerberg Biohub | CZB Cliahub Consortium |
| EPI_ISL_2659307 | Santa Clara County Public Health Laboratory | Chan-Zuckerberg Biohub | CZB Cliahub Consortium |
| EPI_ISL_2659308, EPI_ISL_2659309, EPI_ISL_2659310 | Humboldt County Public Health Laboratory | Chan-Zuckerberg Biohub | CZB Cliahub Consortium |
| EPI_ISL_2659312 | Contra Costa County Public Health Lab | Chan-Zuckerberg Biohub | CZB Cliahub Consortium |
| EPI_ISL_2659313, EPI_ISL_2659314, EPI_ISL_2659315 | Santa Clara County Public Health Laboratory | Chan-Zuckerberg Biohub | CZB Cliahub Consortium |
| EPI_ISL_2659316 | Humboldt County Public Health Laboratory | Chan-Zuckerberg Biohub | CZB Cliahub Consortium |
| EPI_ISL_2659317 | Santa Clara County Public Health Laboratory | Chan-Zuckerberg Biohub | CZB Cliahub Consortium |
| EPI_ISL_2659318 | CA DPH Viral and Rickettsial Disease Laboratory | Chan-Zuckerberg Biohub | CZB Cliahub Consortium |
| EPI_ISL_2659319 | Humboldt County Public Health Laboratory | Chan-Zuckerberg Biohub | CZB Cliahub Consortium |
| EPI_ISL_2659320, EPI_ISL_2659321 | Santa Clara County Public Health Laboratory | Chan-Zuckerberg Biohub | CZB Cliahub Consortium |
| EPI_ISL_2659322 | CA DPH Viral and Rickettsial Disease Laboratory | Chan-Zuckerberg Biohub | CZB Cliahub Consortium |
| EPI_ISL_2659323 | Santa Clara County Public Health Laboratory | Chan-Zuckerberg Biohub | CZB Cliahub Consortium |
| EPI_ISL_2659324, EPI_ISL_2659325 | Alameda County Public Health Lab | Chan-Zuckerberg Biohub | CZB Cliahub Consortium |
| EPI_ISL_2659326, EPI_ISL_2659327 | Santa Clara County Public Health Laboratory | Chan-Zuckerberg Biohub | CZB Cliahub Consortium |
| EPI_ISL_2659328 | Alameda County Public Health Lab | Chan-Zuckerberg Biohub | CZB Cliahub Consortium |
| EPI_ISL_2659329 | Santa Clara County Public Health Laboratory | Chan-Zuckerberg Biohub | CZB Cliahub Consortium |
| EPI_ISL_2659330 | Orange County Public Health Lab | Chan-Zuckerberg Biohub | CZB Cliahub Consortium |
| EPI_ISL_2659331, EPI_ISL_2659332 | Alameda County Public Health Lab | Chan-Zuckerberg Biohub | CZB Cliahub Consortium |
| EPI_ISL_2659333, EPI_ISL_2659334 | CA DPH Viral and Rickettsial Disease Laboratory | Chan-Zuckerberg Biohub | CZB Cliahub Consortium |
| EPI_ISL_2659335 | Santa Clara County Public Health Laboratory | Chan-Zuckerberg Biohub | CZB Cliahub Consortium |
| EPI_ISL_2659336 | Humboldt County Public Health Laboratory | Chan-Zuckerberg Biohub | CZB Cliahub Consortium |
| EPI_ISL_2659337 | County of San Luis Obispo Public Health Laboratory | Chan-Zuckerberg Biohub | CZB Cliahub Consortium |
| EPI_ISL_2659338 | Humboldt County Public Health Laboratory | Chan-Zuckerberg Biohub | CZB Cliahub Consortium |
| EPI_ISL_2659339 | Contra Costa County Public Health Lab | Chan-Zuckerberg Biohub | CZB Cliahub Consortium |
| EPI_ISL_2659340 | Santa Clara County Public Health Laboratory | Chan-Zuckerberg Biohub | CZB Cliahub Consortium |
| EPI_ISL_2659341 | Orange County Public Health Lab | Chan-Zuckerberg Biohub | CZB Cliahub Consortium |
| EPI_ISL_2659342 | Santa Clara County Public Health Laboratory | Chan-Zuckerberg Biohub | CZB Cliahub Consortium |
| EPI_ISL_2659343, EPI_ISL_2659344 | Contra Costa County Public Health Lab | Chan-Zuckerberg Biohub | CZB Cliahub Consortium |
| EPI_ISL_2659345 | Santa Clara County Public Health | Chan-Zuckerberg Biohub | CZB Cliahub Consortium |

| | | | |
|---|---|---|---|
| | Laboratory | | |
| EPI_ISL_2659346 | Humboldt County Public Health Laboratory | Chan-Zuckerberg Biohub | CZB Cliahub Consortium |
| EPI_ISL_2659347 | Contra Costa County Public Health Lab | Chan-Zuckerberg Biohub | CZB Cliahub Consortium |
| EPI_ISL_2659348, EPI_ISL_2659349 | CA DPH Viral and Rickettsial Disease Laboratory | Chan-Zuckerberg Biohub | CZB Cliahub Consortium |
| EPI_ISL_2659350, EPI_ISL_2659351, EPI_ISL_2659352 | Santa Clara County Public Health Laboratory | Chan-Zuckerberg Biohub | CZB Cliahub Consortium |
| EPI_ISL_2659353 | Contra Costa County Public Health Lab | Chan-Zuckerberg Biohub | CZB Cliahub Consortium |
| EPI_ISL_2659354 | Santa Clara County Public Health Laboratory | Chan-Zuckerberg Biohub | CZB Cliahub Consortium |
| EPI_ISL_2659355, EPI_ISL_2659356 | Alameda County Public Health Lab | Chan-Zuckerberg Biohub | CZB Cliahub Consortium |
| EPI_ISL_2659357 | Orange County Public Health Lab | Chan-Zuckerberg Biohub | CZB Cliahub Consortium |
| EPI_ISL_2659358 | Santa Clara County Public Health Laboratory | Chan-Zuckerberg Biohub | CZB Cliahub Consortium |
| EPI_ISL_2659359 | Humboldt County Public Health Laboratory | Chan-Zuckerberg Biohub | CZB Cliahub Consortium |
| EPI_ISL_2659360 | Santa Clara County Public Health Laboratory | Chan-Zuckerberg Biohub | CZB Cliahub Consortium |
| EPI_ISL_2659361 | CA DPH Viral and Rickettsial Disease Laboratory | Chan-Zuckerberg Biohub | CZB Cliahub Consortium |
| EPI_ISL_2659362 | Humboldt County Public Health Laboratory | Chan-Zuckerberg Biohub | CZB Cliahub Consortium |
| EPI_ISL_2659363, EPI_ISL_2659364 | Santa Clara County Public Health Laboratory | Chan-Zuckerberg Biohub | CZB Cliahub Consortium |
| EPI_ISL_2659365 | Madera County Department of Public Health | Chan-Zuckerberg Biohub | CZB Cliahub Consortium |
| EPI_ISL_2659366, EPI_ISL_2659367, EPI_ISL_2659368 | Santa Clara County Public Health Laboratory | Chan-Zuckerberg Biohub | CZB Cliahub Consortium |
| EPI_ISL_2659369 | CA DPH Viral and Rickettsial Disease Laboratory | Chan-Zuckerberg Biohub | CZB Cliahub Consortium |
| EPI_ISL_2659370 | Santa Clara County Public Health Laboratory | Chan-Zuckerberg Biohub | CZB Cliahub Consortium |
| EPI_ISL_2659371 | Humboldt County Public Health Laboratory | Chan-Zuckerberg Biohub | CZB Cliahub Consortium |
| EPI_ISL_2659372 | Alameda County Public Health Lab | Chan-Zuckerberg Biohub | CZB Cliahub Consortium |
| EPI_ISL_2659373, EPI_ISL_2659374, EPI_ISL_2659375, EPI_ISL_2659376 | Santa Clara County Public Health Laboratory | Chan-Zuckerberg Biohub | CZB Cliahub Consortium |
| EPI_ISL_2659377 | Alameda County Public Health Lab | Chan-Zuckerberg Biohub | CZB Cliahub Consortium |
| EPI_ISL_2659378, EPI_ISL_2659379 | Santa Clara County Public Health Laboratory | Chan-Zuckerberg Biohub | CZB Cliahub Consortium |
| EPI_ISL_2659380 | Humboldt County Public Health Laboratory | Chan-Zuckerberg Biohub | CZB Cliahub Consortium |
| EPI_ISL_2659381, EPI_ISL_2659382, EPI_ISL_2659383 | Santa Clara County Public Health Laboratory | Chan-Zuckerberg Biohub | CZB Cliahub Consortium |
| EPI_ISL_2659384, EPI_ISL_2659385 | Humboldt County Public Health Laboratory | Chan-Zuckerberg Biohub | CZB Cliahub Consortium |
| EPI_ISL_2659386 | Contra Costa County Public Health Lab | Chan-Zuckerberg Biohub | CZB Cliahub Consortium |
| EPI_ISL_2659387 | CA DPH Viral and Rickettsial Disease Laboratory | Chan-Zuckerberg Biohub | CZB Cliahub Consortium |
| EPI_ISL_2659388, EPI_ISL_2659389 | Santa Clara County Public Health Laboratory | Chan-Zuckerberg Biohub | CZB Cliahub Consortium |
| EPI_ISL_2659390 | Alameda County Public Health Lab | Chan-Zuckerberg Biohub | CZB Cliahub Consortium |
| EPI_ISL_2659391 | Santa Clara County Public Health Laboratory | Chan-Zuckerberg Biohub | CZB Cliahub Consortium |
| EPI_ISL_2659392 | CA DPH Viral and Rickettsial Disease Laboratory | Chan-Zuckerberg Biohub | CZB Cliahub Consortium |
| EPI_ISL_2659393 | County of San Luis Obispo Public Health Laboratory | Chan-Zuckerberg Biohub | CZB Cliahub Consortium |
| EPI_ISL_2659394, EPI_ISL_2659395 | Santa Clara County Public Health Laboratory | Chan-Zuckerberg Biohub | CZB Cliahub Consortium |
| EPI_ISL_2659396 | Contra Costa County Public Health Lab | Chan-Zuckerberg Biohub | CZB Cliahub Consortium |
| EPI_ISL_2659397 | Santa Clara County Public Health Laboratory | Chan-Zuckerberg Biohub | CZB Cliahub Consortium |
| EPI_ISL_2659398 | Orange County Public Health Lab | Chan-Zuckerberg Biohub | CZB Cliahub Consortium |
| EPI_ISL_2659399 | Santa Clara County Public Health Laboratory | Chan-Zuckerberg Biohub | CZB Cliahub Consortium |
| EPI_ISL_2659400 | CA DPH Viral and Rickettsial Disease Laboratory | Chan-Zuckerberg Biohub | CZB Cliahub Consortium |
| EPI_ISL_2659401, EPI_ISL_2659402 | Santa Clara County Public Health Laboratory | Chan-Zuckerberg Biohub | CZB Cliahub Consortium |
| EPI_ISL_2659403 | Humboldt County Public Health Laboratory | Chan-Zuckerberg Biohub | CZB Cliahub Consortium |
| EPI_ISL_2659404 | Alameda County Public Health Lab | Chan-Zuckerberg Biohub | CZB Cliahub Consortium |
| EPI_ISL_2659405 | Humboldt County Public Health Laboratory | Chan-Zuckerberg Biohub | CZB Cliahub Consortium |
| EPI_ISL_2659406 | Santa Clara County Public Health Laboratory | Chan-Zuckerberg Biohub | CZB Cliahub Consortium |
| EPI_ISL_2659407 | Alameda County Public Health Lab | Chan-Zuckerberg Biohub | CZB Cliahub Consortium |
| EPI_ISL_2659408 | Humboldt County Public Health Laboratory | Chan-Zuckerberg Biohub | CZB Cliahub Consortium |
| EPI_ISL_2659409 | Contra Costa County Public Health Lab | Chan-Zuckerberg Biohub | CZB Cliahub Consortium |
| EPI_ISL_2659410, | Santa Clara County Public Health | Chan-Zuckerberg Biohub | CZB Cliahub Consortium |

| | | | |
|---|---|---|---|
| EPI_ISL_2659411 | Laboratory | | |
| EPI_ISL_2659412 | Contra Costa County Public Health Lab | Chan-Zuckerberg Biohub | CZB Cliahub Consortium |
| EPI_ISL_2659413 | Santa Clara County Public Health Laboratory | Chan-Zuckerberg Biohub | CZB Cliahub Consortium |
| EPI_ISL_2659414 | Humboldt County Public Health Laboratory | Chan-Zuckerberg Biohub | CZB Cliahub Consortium |
| EPI_ISL_2659415 | Santa Clara County Public Health Laboratory | Chan-Zuckerberg Biohub | CZB Cliahub Consortium |
| EPI_ISL_2659416, EPI_ISL_2659417 | Alameda County Public Health Lab | Chan-Zuckerberg Biohub | CZB Cliahub Consortium |
| EPI_ISL_2659418 | Orange County Public Health Lab | Chan-Zuckerberg Biohub | CZB Cliahub Consortium |
| EPI_ISL_2659419 | Alameda County Public Health Lab | Chan-Zuckerberg Biohub | CZB Cliahub Consortium |
| EPI_ISL_2659420 | Santa Clara County Public Health Laboratory | Chan-Zuckerberg Biohub | CZB Cliahub Consortium |
| EPI_ISL_2659421 | Humboldt County Public Health Laboratory | Chan-Zuckerberg Biohub | CZB Cliahub Consortium |
| EPI_ISL_2659422 | Santa Clara County Public Health Laboratory | Chan-Zuckerberg Biohub | CZB Cliahub Consortium |
| EPI_ISL_2659423 | Alameda County Public Health Lab | Chan-Zuckerberg Biohub | CZB Cliahub Consortium |
| EPI_ISL_2659424, EPI_ISL_2659425, EPI_ISL_2659426, EPI_ISL_2659427 | Santa Clara County Public Health Laboratory | Chan-Zuckerberg Biohub | CZB Cliahub Consortium |
| EPI_ISL_2659428 | Alameda County Public Health Lab | Chan-Zuckerberg Biohub | CZB Cliahub Consortium |
| EPI_ISL_2659429 | Santa Clara County Public Health Laboratory | Chan-Zuckerberg Biohub | CZB Cliahub Consortium |
| EPI_ISL_2659430 | Orange County Public Health Lab | Chan-Zuckerberg Biohub | CZB Cliahub Consortium |
| EPI_ISL_2659432, EPI_ISL_2659433 | Santa Clara County Public Health Laboratory | Chan-Zuckerberg Biohub | CZB Cliahub Consortium |
| EPI_ISL_2659434 | Contra Costa County Public Health Lab | Chan-Zuckerberg Biohub | CZB Cliahub Consortium |
| EPI_ISL_2659436, EPI_ISL_2659437, EPI_ISL_2659438, EPI_ISL_2659439, EPI_ISL_2659440, EPI_ISL_2659441, EPI_ISL_2659442, EPI_ISL_2659443, EPI_ISL_2659444 | Santa Clara County Public Health Laboratory | Chan-Zuckerberg Biohub | CZB Cliahub Consortium |
| EPI_ISL_2659445 | CA DPH Viral and Rickettsial Disease Laboratory | Chan-Zuckerberg Biohub | CZB Cliahub Consortium |
| EPI_ISL_2659446, EPI_ISL_2659447, EPI_ISL_2659448 | Santa Clara County Public Health Laboratory | Chan-Zuckerberg Biohub | CZB Cliahub Consortium |
| EPI_ISL_2659449, EPI_ISL_2659450 | Alameda County Public Health Lab | Chan-Zuckerberg Biohub | CZB Cliahub Consortium |
| EPI_ISL_2659451 | Santa Clara County Public Health Laboratory | Chan-Zuckerberg Biohub | CZB Cliahub Consortium |
| EPI_ISL_2659452 | CA DPH Viral and Rickettsial Disease Laboratory | Chan-Zuckerberg Biohub | CZB Cliahub Consortium |
| EPI_ISL_2659453, EPI_ISL_2659454 | Santa Clara County Public Health Laboratory | Chan-Zuckerberg Biohub | CZB Cliahub Consortium |
| EPI_ISL_2659455 | Alameda County Public Health Lab | Chan-Zuckerberg Biohub | CZB Cliahub Consortium |
| EPI_ISL_2659456 | Santa Clara County Public Health Laboratory | Chan-Zuckerberg Biohub | CZB Cliahub Consortium |
| EPI_ISL_2659457 | CA DPH Viral and Rickettsial Disease Laboratory | Chan-Zuckerberg Biohub | CZB Cliahub Consortium |
| EPI_ISL_2659458, EPI_ISL_2659459, EPI_ISL_2659460 | Santa Clara County Public Health Laboratory | Chan-Zuckerberg Biohub | CZB Cliahub Consortium |
| EPI_ISL_2659461 | Humboldt County Public Health Laboratory | Chan-Zuckerberg Biohub | CZB Cliahub Consortium |
| EPI_ISL_2659462 | Orange County Public Health Lab | Chan-Zuckerberg Biohub | CZB Cliahub Consortium |
| EPI_ISL_2659463 | Santa Clara County Public Health Laboratory | Chan-Zuckerberg Biohub | CZB Cliahub Consortium |
| EPI_ISL_2659464 | CA DPH Viral and Rickettsial Disease Laboratory | Chan-Zuckerberg Biohub | CZB Cliahub Consortium |
| EPI_ISL_2659465 | Ventura County Public Health Lab | Chan-Zuckerberg Biohub | CZB Cliahub Consortium |
| EPI_ISL_2659466 | Santa Clara County Public Health Laboratory | Chan-Zuckerberg Biohub | CZB Cliahub Consortium |
| EPI_ISL_2659467 | Contra Costa County Public Health Lab | Chan-Zuckerberg Biohub | CZB Cliahub Consortium |
| EPI_ISL_2659468 | UCSF Clinical Microbiology Laboratory | Chan-Zuckerberg Biohub | CZB Cliahub Consortium |
| EPI_ISL_2659469 | Contra Costa County Public Health Lab | Chan-Zuckerberg Biohub | CZB Cliahub Consortium |
| EPI_ISL_2659471, EPI_ISL_2659472, EPI_ISL_2659473 | Santa Clara County Public Health Laboratory | Chan-Zuckerberg Biohub | CZB Cliahub Consortium |
| EPI_ISL_2659474 | Orange County Public Health Lab | Chan-Zuckerberg Biohub | CZB Cliahub Consortium |
| EPI_ISL_2659475, EPI_ISL_2659476 | Santa Clara County Public Health Laboratory | Chan-Zuckerberg Biohub | CZB Cliahub Consortium |
| EPI_ISL_2659477 | CA DPH Viral and Rickettsial Disease Laboratory | Chan-Zuckerberg Biohub | CZB Cliahub Consortium |
| EPI_ISL_2659478, EPI_ISL_2659479 | Santa Clara County Public Health Laboratory | Chan-Zuckerberg Biohub | CZB Cliahub Consortium |
| EPI_ISL_2659480 | Alameda County Public Health Lab | Chan-Zuckerberg Biohub | CZB Cliahub Consortium |
| EPI_ISL_2659481 | CA DPH Viral and Rickettsial Disease Laboratory | Chan-Zuckerberg Biohub | CZB Cliahub Consortium |
| EPI_ISL_2659482 | Alameda County Public Health Lab | Chan-Zuckerberg Biohub | CZB Cliahub Consortium |
| EPI_ISL_2659483 | Santa Clara County Public Health Laboratory | Chan-Zuckerberg Biohub | CZB Cliahub Consortium |

| | | | |
|---|---|---|---|
| EPI_ISL_2659484 | Alameda County Public Health Lab | Chan-Zuckerberg Biohub | CZB Cliahub Consortium |
| EPI_ISL_2659485 | Santa Clara County Public Health Laboratory | Chan-Zuckerberg Biohub | CZB Cliahub Consortium |
| EPI_ISL_2659486, EPI_ISL_2659487 | Humboldt County Public Health Laboratory | Chan-Zuckerberg Biohub | CZB Cliahub Consortium |
| EPI_ISL_2659488 | Orange County Public Health Lab | Chan-Zuckerberg Biohub | CZB Cliahub Consortium |
| EPI_ISL_2659489, EPI_ISL_2659491 | Santa Clara County Public Health Laboratory | Chan-Zuckerberg Biohub | CZB Cliahub Consortium |
| EPI_ISL_2659492 | Humboldt County Public Health Laboratory | Chan-Zuckerberg Biohub | CZB Cliahub Consortium |
| EPI_ISL_2659493, EPI_ISL_2659494 | Santa Clara County Public Health Laboratory | Chan-Zuckerberg Biohub | CZB Cliahub Consortium |
| EPI_ISL_2659495 | CA DPH Viral and Rickettsial Disease Laboratory | Chan-Zuckerberg Biohub | CZB Cliahub Consortium |
| EPI_ISL_2659496 | Santa Clara County Public Health Laboratory | Chan-Zuckerberg Biohub | CZB Cliahub Consortium |
| EPI_ISL_2659497 | Contra Costa County Public Health Lab | Chan-Zuckerberg Biohub | CZB Cliahub Consortium |
| EPI_ISL_2659498, EPI_ISL_2659499 | Santa Clara County Public Health Laboratory | Chan-Zuckerberg Biohub | CZB Cliahub Consortium |
| EPI_ISL_2659500 | Humboldt County Public Health Laboratory | Chan-Zuckerberg Biohub | CZB Cliahub Consortium |
| EPI_ISL_2659501 | Santa Clara County Public Health Laboratory | Chan-Zuckerberg Biohub | CZB Cliahub Consortium |
| EPI_ISL_2659502, EPI_ISL_2659503 | CA DPH Viral and Rickettsial Disease Laboratory | Chan-Zuckerberg Biohub | CZB Cliahub Consortium |
| EPI_ISL_2659504 | County of San Luis Obispo Public Health Laboratory | Chan-Zuckerberg Biohub | CZB Cliahub Consortium |
| EPI_ISL_2659505 | Alameda County Public Health Lab | Chan-Zuckerberg Biohub | CZB Cliahub Consortium |
| EPI_ISL_2659506 | Santa Clara County Public Health Laboratory | Chan-Zuckerberg Biohub | CZB Cliahub Consortium |
| EPI_ISL_2659507 | CA DPH Viral and Rickettsial Disease Laboratory | Chan-Zuckerberg Biohub | CZB Cliahub Consortium |
| EPI_ISL_2659508 | Alameda County Public Health Lab | Chan-Zuckerberg Biohub | CZB Cliahub Consortium |
| EPI_ISL_2659509 | CA DPH Viral and Rickettsial Disease Laboratory | Chan-Zuckerberg Biohub | CZB Cliahub Consortium |
| EPI_ISL_2659510, EPI_ISL_2659511, EPI_ISL_2659512, EPI_ISL_2659513 | Santa Clara County Public Health Laboratory | Chan-Zuckerberg Biohub | CZB Cliahub Consortium |
| EPI_ISL_2659514 | CA DPH Viral and Rickettsial Disease Laboratory | Chan-Zuckerberg Biohub | CZB Cliahub Consortium |
| EPI_ISL_2659515, EPI_ISL_2659516 | Santa Clara County Public Health Laboratory | Chan-Zuckerberg Biohub | CZB Cliahub Consortium |
| EPI_ISL_2659517 | County of San Luis Obispo Public Health Laboratory | Chan-Zuckerberg Biohub | CZB Cliahub Consortium |
| EPI_ISL_2659518 | Santa Clara County Public Health Laboratory | Chan-Zuckerberg Biohub | CZB Cliahub Consortium |
| EPI_ISL_2659519 | Alameda County Public Health Lab | Chan-Zuckerberg Biohub | CZB Cliahub Consortium |
| EPI_ISL_2659520, EPI_ISL_2659521 | Santa Clara County Public Health Laboratory | Chan-Zuckerberg Biohub | CZB Cliahub Consortium |
| EPI_ISL_2659522 | CA DPH Viral and Rickettsial Disease Laboratory | Chan-Zuckerberg Biohub | CZB Cliahub Consortium |
| EPI_ISL_2659523 | Santa Clara County Public Health Laboratory | Chan-Zuckerberg Biohub | CZB Cliahub Consortium |
| EPI_ISL_2659524 | CA DPH Viral and Rickettsial Disease Laboratory | Chan-Zuckerberg Biohub | CZB Cliahub Consortium |
| EPI_ISL_2659525 | Santa Clara County Public Health Laboratory | Chan-Zuckerberg Biohub | CZB Cliahub Consortium |
| EPI_ISL_2659526 | Humboldt County Public Health Laboratory | Chan-Zuckerberg Biohub | CZB Cliahub Consortium |
| EPI_ISL_2659527, EPI_ISL_2659528 | Santa Clara County Public Health Laboratory | Chan-Zuckerberg Biohub | CZB Cliahub Consortium |
| EPI_ISL_2659529 | Contra Costa County Public Health Lab | Chan-Zuckerberg Biohub | CZB Cliahub Consortium |
| EPI_ISL_2659530, EPI_ISL_2659531, EPI_ISL_2659532 | Santa Clara County Public Health Laboratory | Chan-Zuckerberg Biohub | CZB Cliahub Consortium |
| EPI_ISL_2659533 | Contra Costa County Public Health Lab | Chan-Zuckerberg Biohub | CZB Cliahub Consortium |
| EPI_ISL_2659534 | Alameda County Public Health Lab | Chan-Zuckerberg Biohub | CZB Cliahub Consortium |
| EPI_ISL_2659535, EPI_ISL_2659536 | Santa Clara County Public Health Laboratory | Chan-Zuckerberg Biohub | CZB Cliahub Consortium |
| EPI_ISL_2659537 | UCSF Clinical Microbiology Laboratory | Chan-Zuckerberg Biohub | CZB Cliahub Consortium |
| EPI_ISL_2659538, EPI_ISL_2659539 | Contra Costa County Public Health Lab | Chan-Zuckerberg Biohub | CZB Cliahub Consortium |
| EPI_ISL_2659540 | Orange County Public Health Lab | Chan-Zuckerberg Biohub | CZB Cliahub Consortium |
| EPI_ISL_2659541, EPI_ISL_2659542, EPI_ISL_2659543 | Santa Clara County Public Health Laboratory | Chan-Zuckerberg Biohub | CZB Cliahub Consortium |
| EPI_ISL_2659544 | Alameda County Public Health Lab | Chan-Zuckerberg Biohub | CZB Cliahub Consortium |
| EPI_ISL_2659546, EPI_ISL_2659548 | Santa Clara County Public Health Laboratory | Chan-Zuckerberg Biohub | CZB Cliahub Consortium |
| EPI_ISL_2659549 | Humboldt County Public Health Laboratory | Chan-Zuckerberg Biohub | CZB Cliahub Consortium |
| EPI_ISL_2659550 | Orange County Public Health Lab | Chan-Zuckerberg Biohub | CZB Cliahub Consortium |
| EPI_ISL_2659551 | CA DPH Viral and Rickettsial Disease Laboratory | Chan-Zuckerberg Biohub | CZB Cliahub Consortium |
| EPI_ISL_2659552 | Contra Costa County Public Health Lab | Chan-Zuckerberg Biohub | CZB Cliahub Consortium |
| EPI_ISL_2659553 | Orange County Public Health Lab | Chan-Zuckerberg Biohub | CZB Cliahub Consortium |
| EPI_ISL_2659554 | Contra Costa County Public Health Lab | Chan-Zuckerberg Biohub | CZB Cliahub Consortium |

| | | | |
|---|---|---|---|
| EPI_ISL_2659555 | Santa Clara County Public Health Laboratory | Chan-Zuckerberg Biohub | CZB Cliahub Consortium |
| EPI_ISL_2659556 | CA DPH Viral and Rickettsial Disease Laboratory | Chan-Zuckerberg Biohub | CZB Cliahub Consortium |
| EPI_ISL_2659557 | Humboldt County Public Health Laboratory | Chan-Zuckerberg Biohub | CZB Cliahub Consortium |
| EPI_ISL_2659558, EPI_ISL_2659559 | Santa Clara County Public Health Laboratory | Chan-Zuckerberg Biohub | CZB Cliahub Consortium |
| EPI_ISL_2659560 | Contra Costa County Public Health Lab | Chan-Zuckerberg Biohub | CZB Cliahub Consortium |
| EPI_ISL_2659561 | Orange County Public Health Lab | Chan-Zuckerberg Biohub | CZB Cliahub Consortium |
| EPI_ISL_2659562 | Contra Costa County Public Health Lab | Chan-Zuckerberg Biohub | CZB Cliahub Consortium |
| EPI_ISL_2659563, EPI_ISL_2659564 | Santa Clara County Public Health Laboratory | Chan-Zuckerberg Biohub | CZB Cliahub Consortium |
| EPI_ISL_2659565 | Napa-Solano-Yolo- Marin County (NSYM) Public Health Laboratories | Chan-Zuckerberg Biohub | CZB Cliahub Consortium |
| EPI_ISL_2659566 | CA DPH Viral and Rickettsial Disease Laboratory | Chan-Zuckerberg Biohub | CZB Cliahub Consortium |
| EPI_ISL_2659567 | Santa Clara County Public Health Laboratory | Chan-Zuckerberg Biohub | CZB Cliahub Consortium |
| EPI_ISL_2659568 | Contra Costa County Public Health Lab | Chan-Zuckerberg Biohub | CZB Cliahub Consortium |
| EPI_ISL_2659569 | Santa Clara County Public Health Laboratory | Chan-Zuckerberg Biohub | CZB Cliahub Consortium |
| EPI_ISL_2659570 | Contra Costa County Public Health Lab | Chan-Zuckerberg Biohub | CZB Cliahub Consortium |
| EPI_ISL_2659571, EPI_ISL_2659572, EPI_ISL_2659573, EPI_ISL_2659574, EPI_ISL_2659575, EPI_ISL_2659576, EPI_ISL_2659577 | Santa Clara County Public Health Laboratory | Chan-Zuckerberg Biohub | CZB Cliahub Consortium |
| EPI_ISL_2659578 | Orange County Public Health Lab | Chan-Zuckerberg Biohub | CZB Cliahub Consortium |
| EPI_ISL_2659579 | CA DPH Viral and Rickettsial Disease Laboratory | Chan-Zuckerberg Biohub | CZB Cliahub Consortium |
| EPI_ISL_2659580, EPI_ISL_2659581 | Santa Clara County Public Health Laboratory | Chan-Zuckerberg Biohub | CZB Cliahub Consortium |
| EPI_ISL_2659582 | Alameda County Public Health Lab | Chan-Zuckerberg Biohub | CZB Cliahub Consortium |
| EPI_ISL_2659583, EPI_ISL_2659584, EPI_ISL_2659587 | Santa Clara County Public Health Laboratory | Chan-Zuckerberg Biohub | CZB Cliahub Consortium |
| EPI_ISL_2659588 | Alameda County Public Health Lab | Chan-Zuckerberg Biohub | CZB Cliahub Consortium |
| EPI_ISL_2659589, EPI_ISL_2659590 | Santa Clara County Public Health Laboratory | Chan-Zuckerberg Biohub | CZB Cliahub Consortium |
| EPI_ISL_2659591 | Humboldt County Public Health Laboratory | Chan-Zuckerberg Biohub | CZB Cliahub Consortium |
| EPI_ISL_2659592, EPI_ISL_2659593 | CA DPH Viral and Rickettsial Disease Laboratory | Chan-Zuckerberg Biohub | CZB Cliahub Consortium |
| EPI_ISL_2659594 | Santa Clara County Public Health Laboratory | Chan-Zuckerberg Biohub | CZB Cliahub Consortium |
| EPI_ISL_2659595 | Alameda County Public Health Lab | Chan-Zuckerberg Biohub | CZB Cliahub Consortium |
| EPI_ISL_2659596, EPI_ISL_2659597 | Contra Costa County Public Health Lab | Chan-Zuckerberg Biohub | CZB Cliahub Consortium |
| EPI_ISL_2659598, EPI_ISL_2659599, EPI_ISL_2659600 | Santa Clara County Public Health Laboratory | Chan-Zuckerberg Biohub | CZB Cliahub Consortium |
| EPI_ISL_2659601 | Contra Costa County Public Health Lab | Chan-Zuckerberg Biohub | CZB Cliahub Consortium |
| EPI_ISL_2659602 | Santa Clara County Public Health Laboratory | Chan-Zuckerberg Biohub | CZB Cliahub Consortium |
| EPI_ISL_2659603 | Orange County Public Health Lab | Chan-Zuckerberg Biohub | CZB Cliahub Consortium |
| EPI_ISL_2659604 | CA DPH Viral and Rickettsial Disease Laboratory | Chan-Zuckerberg Biohub | CZB Cliahub Consortium |
| EPI_ISL_2659605 | Santa Clara County Public Health Laboratory | Chan-Zuckerberg Biohub | CZB Cliahub Consortium |
| EPI_ISL_2659606 | County of San Luis Obispo Public Health Laboratory | Chan-Zuckerberg Biohub | CZB Cliahub Consortium |
| EPI_ISL_2659607 | Santa Clara County Public Health Laboratory | Chan-Zuckerberg Biohub | CZB Cliahub Consortium |
| EPI_ISL_2659608 | CA DPH Viral and Rickettsial Disease Laboratory | Chan-Zuckerberg Biohub | CZB Cliahub Consortium |
| EPI_ISL_2659609 | Humboldt County Public Health Laboratory | Chan-Zuckerberg Biohub | CZB Cliahub Consortium |
| EPI_ISL_2659610 | Santa Clara County Public Health Laboratory | Chan-Zuckerberg Biohub | CZB Cliahub Consortium |
| EPI_ISL_2659611 | Alameda County Public Health Lab | Chan-Zuckerberg Biohub | CZB Cliahub Consortium |
| EPI_ISL_2659612 | CA DPH Viral and Rickettsial Disease Laboratory | Chan-Zuckerberg Biohub | CZB Cliahub Consortium |
| EPI_ISL_2659613 | Santa Clara County Public Health Laboratory | Chan-Zuckerberg Biohub | CZB Cliahub Consortium |
| EPI_ISL_2659614 | Alameda County Public Health Lab | Chan-Zuckerberg Biohub | CZB Cliahub Consortium |
| EPI_ISL_2659615, EPI_ISL_2659616, EPI_ISL_2659617, EPI_ISL_2659618, EPI_ISL_2659619 | Santa Clara County Public Health Laboratory | Chan-Zuckerberg Biohub | CZB Cliahub Consortium |
| EPI_ISL_2659620 | CA DPH Viral and Rickettsial Disease Laboratory | Chan-Zuckerberg Biohub | CZB Cliahub Consortium |
| EPI_ISL_2659621 | Santa Clara County Public Health Laboratory | Chan-Zuckerberg Biohub | CZB Cliahub Consortium |
| EPI_ISL_2659622 | Orange County Public Health Lab | Chan-Zuckerberg Biohub | CZB Cliahub Consortium |
| EPI_ISL_2659623 | Santa Clara County Public Health Laboratory | Chan-Zuckerberg Biohub | CZB Cliahub Consortium |

| | | | |
|---|---|---|---|
| EPI_ISL_2659624 | Contra Costa County Public Health Lab | Chan-Zuckerberg Biohub | CZB Cliahub Consortium |
| EPI_ISL_2659625, EPI_ISL_2659626, EPI_ISL_2659627 | Santa Clara County Public Health Laboratory | Chan-Zuckerberg Biohub | CZB Cliahub Consortium |
| EPI_ISL_2659628 | Alameda County Public Health Lab | Chan-Zuckerberg Biohub | CZB Cliahub Consortium |
| EPI_ISL_2659629 | Santa Clara County Public Health Laboratory | Chan-Zuckerberg Biohub | CZB Cliahub Consortium |
| EPI_ISL_2659630 | Contra Costa County Public Health Lab | Chan-Zuckerberg Biohub | CZB Cliahub Consortium |
| EPI_ISL_2659631, EPI_ISL_2659632, EPI_ISL_2659633, EPI_ISL_2659634 | Santa Clara County Public Health Laboratory | Chan-Zuckerberg Biohub | CZB Cliahub Consortium |
| EPI_ISL_2659635 | Humboldt County Public Health Laboratory | Chan-Zuckerberg Biohub | CZB Cliahub Consortium |
| EPI_ISL_2659636 | Alameda County Public Health Lab | Chan-Zuckerberg Biohub | CZB Cliahub Consortium |
| EPI_ISL_2659637 | Santa Clara County Public Health Laboratory | Chan-Zuckerberg Biohub | CZB Cliahub Consortium |
| EPI_ISL_2659638 | Humboldt County Public Health Laboratory | Chan-Zuckerberg Biohub | CZB Cliahub Consortium |
| EPI_ISL_2659639 | Santa Clara County Public Health Laboratory | Chan-Zuckerberg Biohub | CZB Cliahub Consortium |
| EPI_ISL_2659640 | Orange County Public Health Lab | Chan-Zuckerberg Biohub | CZB Cliahub Consortium |
| EPI_ISL_2659641 | Alameda County Public Health Lab | Chan-Zuckerberg Biohub | CZB Cliahub Consortium |
| EPI_ISL_2659642 | CA DPH Viral and Rickettsial Disease Laboratory | Chan-Zuckerberg Biohub | CZB Cliahub Consortium |
| EPI_ISL_2659643 | Santa Clara County Public Health Laboratory | Chan-Zuckerberg Biohub | CZB Cliahub Consortium |
| EPI_ISL_2659644 | Orange County Public Health Lab | Chan-Zuckerberg Biohub | CZB Cliahub Consortium |
| EPI_ISL_2659645, EPI_ISL_2659646, EPI_ISL_2659647, EPI_ISL_2659648, EPI_ISL_2659649 | Santa Clara County Public Health Laboratory | Chan-Zuckerberg Biohub | CZB Cliahub Consortium |
| EPI_ISL_2659650 | Orange County Public Health Lab | Chan-Zuckerberg Biohub | CZB Cliahub Consortium |
| EPI_ISL_2659651 | Santa Clara County Public Health Laboratory | Chan-Zuckerberg Biohub | CZB Cliahub Consortium |
| EPI_ISL_2659652 | CA DPH Viral and Rickettsial Disease Laboratory | Chan-Zuckerberg Biohub | CZB Cliahub Consortium |
| EPI_ISL_2659653, EPI_ISL_2659654 | Alameda County Public Health Lab | Chan-Zuckerberg Biohub | CZB Cliahub Consortium |
| EPI_ISL_2659655, EPI_ISL_2659656 | Santa Clara County Public Health Laboratory | Chan-Zuckerberg Biohub | CZB Cliahub Consortium |
| EPI_ISL_2660327, EPI_ISL_2660328, EPI_ISL_2660329, EPI_ISL_2660330, EPI_ISL_2660331, EPI_ISL_2660332, EPI_ISL_2660333, EPI_ISL_2660334, EPI_ISL_2660335, EPI_ISL_2660336, EPI_ISL_2660337, EPI_ISL_2660338, EPI_ISL_2660339, EPI_ISL_2660340, EPI_ISL_2660341, EPI_ISL_2660342, EPI_ISL_2660343, EPI_ISL_2660344, EPI_ISL_2660345, EPI_ISL_2660346, EPI_ISL_2660347, EPI_ISL_2660348, EPI_ISL_2660349, EPI_ISL_2660351, EPI_ISL_2660352, EPI_ISL_2660353, EPI_ISL_2660354, EPI_ISL_2660355, EPI_ISL_2660356, EPI_ISL_2660357, EPI_ISL_2660358, EPI_ISL_2660359, EPI_ISL_2660361, EPI_ISL_2660362, EPI_ISL_2660363, EPI_ISL_2660365, EPI_ISL_2660366, EPI_ISL_2660369, EPI_ISL_2660371, EPI_ISL_2660373, EPI_ISL_2660374, EPI_ISL_2660375, EPI_ISL_2660376, EPI_ISL_2660377, EPI_ISL_2660378, EPI_ISL_2660379, EPI_ISL_2660380, EPI_ISL_2660381, EPI_ISL_2660382, EPI_ISL_2660385, EPI_ISL_2660386, EPI_ISL_2660389, EPI_ISL_2660391, EPI_ISL_2660392, EPI_ISL_2660393, EPI_ISL_2660395, EPI_ISL_2660396, EPI_ISL_2660398, EPI_ISL_2660399, EPI_ISL_2660401, EPI_ISL_2660403, EPI_ISL_2660404, EPI_ISL_2660405, EPI_ISL_2660406, EPI_ISL_2660408, EPI_ISL_2660409, EPI_ISL_2660411, EPI_ISL_2660412, EPI_ISL_2660414, EPI_ISL_2660415, EPI_ISL_2660416, EPI_ISL_2660417, EPI_ISL_2660419, EPI_ISL_2660420, EPI_ISL_2660422, EPI_ISL_2660424, EPI_ISL_2660425, EPI_ISL_2660426, EPI_ISL_2660427, EPI_ISL_2660428, EPI_ISL_2660429, EPI_ISL_2660431, EPI_ISL_2660432, EPI_ISL_2660433, EPI_ISL_2660434, EPI_ISL_2660435, EPI_ISL_2660436, EPI_ISL_2660439, EPI_ISL_2660440, EPI_ISL_2660441, EPI_ISL_2660442, EPI_ISL_2660445, EPI_ISL_2660446, EPI_ISL_2660447, EPI_ISL_2660448, EPI_ISL_2660449, EPI_ISL_2660450, EPI_ISL_2660451 | | | |
| see above | Infectious Disease Program, Broad Institute of Harvard and MIT | Infectious Disease Program, Broad Institute of Harvard and MIT | Lemieux,J.E., Siddle,K.J., Shaw,B., Adams,G., Pierce,V., Turbett,S., Anahtar,M., Branda,J., Slater,D., Harris,J., Lin,A.E., Gladden-Young,A., Lagerborg,K., Rudy,M., DeRuff,K., Carter,A., Normandin,E., Bauer,M., Reilly,S., Tomkins-Tinch,C., Loreth,C., Chaluvadi,S., Neumann,A., Cusick,C., Chapman,S.B., Gnirke,A., Flowers,K., Cerrato,F., Birren,B.W., Gallagher,G., Smole,S., Park,D.J., MacInnis,B.L., Ryan,E., LaRocque,R., Rosenberg,E., Sabeti,P.C. |
| EPI_ISL_2661465, EPI_ISL_2661467, EPI_ISL_2661468, EPI_ISL_2661469, EPI_ISL_2661470, EPI_ISL_2661471, EPI_ISL_2661472, EPI_ISL_2661473, EPI_ISL_2661474, EPI_ISL_2661475, EPI_ISL_2661476, EPI_ISL_2661477, EPI_ISL_2661478, EPI_ISL_2661479, EPI_ISL_2661480, EPI_ISL_2661481, EPI_ISL_2661482, EPI_ISL_2661483, EPI_ISL_2661484, EPI_ISL_2661485, EPI_ISL_2661486, EPI_ISL_2661488, EPI_ISL_2661489, EPI_ISL_2661490 | | | |
| see above | US Air Force School of Aerospace Medicine | US Air Force School of Aerospace Medicine | Anthony Fries, Jennifer Meyer, William Gruner, Amanda Javorina, Carol Garrett, Sarah Purves, Clarise Starr, Elizabeth Macias |
| EPI_ISL_2661526, EPI_ISL_2661527, EPI_ISL_2661528, EPI_ISL_2661529, EPI_ISL_2661530, EPI_ISL_2661531, EPI_ISL_2661532, EPI_ISL_2661533, EPI_ISL_2661534, EPI_ISL_2661535, EPI_ISL_2661536, EPI_ISL_2661537, EPI_ISL_2661538, EPI_ISL_2661539, EPI_ISL_2661540, EPI_ISL_2661541, EPI_ISL_2661542, EPI_ISL_2661543, EPI_ISL_2661544, EPI_ISL_2661545, EPI_ISL_2661546, EPI_ISL_2661547, EPI_ISL_2661548, EPI_ISL_2661549 | | | |
| see above | UHTL, University Hospitals | UHTL, University Hospitals | Sadri,N., Alouani,D., Song,X. |
| EPI_ISL_2661610, EPI_ISL_2661611, EPI_ISL_2661612, EPI_ISL_2661614, EPI_ISL_2661615, EPI_ISL_2661616, EPI_ISL_2661617, EPI_ISL_2661619, EPI_ISL_2661620, EPI_ISL_2661622, EPI_ISL_2661623, EPI_ISL_2661626, EPI_ISL_2661627, EPI_ISL_2661628, EPI_ISL_2661629, EPI_ISL_2661630, EPI_ISL_2661631, EPI_ISL_2661633, EPI_ISL_2661634, EPI_ISL_2661635, EPI_ISL_2661636, EPI_ISL_2661637, EPI_ISL_2661638, EPI_ISL_2661639, EPI_ISL_2661640, EPI_ISL_2661641, EPI_ISL_2661642, EPI_ISL_2661643, EPI_ISL_2661644, EPI_ISL_2661645, EPI_ISL_2661646, EPI_ISL_2661647, EPI_ISL_2661648, EPI_ISL_2661649, EPI_ISL_2661650, EPI_ISL_2661651, EPI_ISL_2661652, EPI_ISL_2661653, EPI_ISL_2661654, EPI_ISL_2661656, EPI_ISL_2661657, EPI_ISL_2661658, EPI_ISL_2661659, EPI_ISL_2661660, EPI_ISL_2661661, EPI_ISL_2661662, EPI_ISL_2661663, EPI_ISL_2661664, EPI_ISL_2661665, EPI_ISL_2661666, EPI_ISL_2661667, EPI_ISL_2661668, EPI_ISL_2661669, EPI_ISL_2661672, EPI_ISL_2661674, EPI_ISL_2661675, EPI_ISL_2661676, EPI_ISL_2661677, EPI_ISL_2661678, EPI_ISL_2661679, EPI_ISL_2661680, EPI_ISL_2661681, EPI_ISL_2661682, EPI_ISL_2661683, EPI_ISL_2661684, EPI_ISL_2661685, EPI_ISL_2661686, EPI_ISL_2661688, EPI_ISL_2661689, EPI_ISL_2661691, EPI_ISL_2661692, EPI_ISL_2661693, EPI_ISL_2661694, EPI_ISL_2661695, EPI_ISL_2661698, EPI_ISL_2661701, EPI_ISL_2661702, EPI_ISL_2661704, EPI_ISL_2661705, EPI_ISL_2661706, EPI_ISL_2661707, EPI_ISL_2661708, EPI_ISL_2661709, EPI_ISL_2661710, EPI_ISL_2661711, EPI_ISL_2661712, EPI_ISL_2661713, EPI_ISL_2661714, EPI_ISL_2661715, EPI_ISL_2661716, EPI_ISL_2661717, EPI_ISL_2661718, EPI_ISL_2661719, EPI_ISL_2661720, EPI_ISL_2661721, EPI_ISL_2661722, EPI_ISL_2661723, EPI_ISL_2661724, EPI_ISL_2661725, EPI_ISL_2661726, EPI_ISL_2661727, EPI_ISL_2661730, EPI_ISL_2661731, EPI_ISL_2661732, EPI_ISL_2661733, EPI_ISL_2661734, EPI_ISL_2661735, EPI_ISL_2661736, EPI_ISL_2661737, EPI_ISL_2661738, EPI_ISL_2661739, EPI_ISL_2661741, EPI_ISL_2661742, EPI_ISL_2661744 | | | |
| see above | North Dakota Department of Health, Public Health Laboratory | North Dakota Department of Health, Public Health Laboratory | Lisa Wingerter |
| EPI_ISL_2661933, EPI_ISL_2661934, EPI_ISL_2661935, EPI_ISL_2661936, EPI_ISL_2661937, EPI_ISL_2661939, EPI_ISL_2661940, EPI_ISL_2661941, EPI_ISL_2661942, EPI_ISL_2661943, EPI_ISL_2661944, EPI_ISL_2661945, EPI_ISL_2661946, EPI_ISL_2661947, EPI_ISL_2661948, EPI_ISL_2661949, EPI_ISL_2661950, EPI_ISL_2661951, EPI_ISL_2661952, EPI_ISL_2661953, EPI_ISL_2661954, EPI_ISL_2661955, EPI_ISL_2661956, EPI_ISL_2661957, EPI_ISL_2661958, EPI_ISL_2661959, EPI_ISL_2661960, EPI_ISL_2661961, EPI_ISL_2661962, EPI_ISL_2661963, EPI_ISL_2661964, EPI_ISL_2661965, EPI_ISL_2661966, EPI_ISL_2661967, EPI_ISL_2661968, EPI_ISL_2661969, EPI_ISL_2661970, EPI_ISL_2661971, EPI_ISL_2661972, EPI_ISL_2661973, EPI_ISL_2661974, EPI_ISL_2661975, EPI_ISL_2661976, EPI_ISL_2661977, EPI_ISL_2661978, EPI_ISL_2661979, EPI_ISL_2661980, EPI_ISL_2661981, EPI_ISL_2661982, EPI_ISL_2661983, EPI_ISL_2661984, EPI_ISL_2661985, EPI_ISL_2661986, EPI_ISL_2661987, EPI_ISL_2661988, EPI_ISL_2661989, EPI_ISL_2661990, EPI_ISL_2661991, EPI_ISL_2661992, EPI_ISL_2661993, EPI_ISL_2661994, EPI_ISL_2661995, EPI_ISL_2661996, EPI_ISL_2661997, EPI_ISL_2661998, EPI_ISL_2661999, EPI_ISL_2662000, EPI_ISL_2662001, EPI_ISL_2662002, EPI_ISL_2662003, EPI_ISL_2662004, EPI_ISL_2662005, EPI_ISL_2662006, EPI_ISL_2662007, EPI_ISL_2662008, EPI_ISL_2662009, EPI_ISL_2662010, EPI_ISL_2662011, EPI_ISL_2662012, EPI_ISL_2662014, EPI_ISL_2662015, EPI_ISL_2662016, EPI_ISL_2662017, EPI_ISL_2662018, EPI_ISL_2662019, EPI_ISL_2662020, EPI_ISL_2662021, EPI_ISL_2662022, EPI_ISL_2662023, EPI_ISL_2662024, EPI_ISL_2662025, EPI_ISL_2662026, EPI_ISL_2662027, EPI_ISL_2662028, EPI_ISL_2662029, EPI_ISL_2662030, EPI_ISL_2662031, EPI_ISL_2662032, EPI_ISL_2662033, EPI_ISL_2662034, EPI_ISL_2662035, EPI_ISL_2662036, EPI_ISL_2662037, EPI_ISL_2662038, EPI_ISL_2662039, EPI_ISL_2662040, EPI_ISL_2662041, EPI_ISL_2662042, EPI_ISL_2662043, EPI_ISL_2662044, EPI_ISL_2662045, EPI_ISL_2662046, EPI_ISL_2662047, EPI_ISL_2662048, EPI_ISL_2662049, EPI_ISL_2662050, EPI_ISL_2662051, EPI_ISL_2662052, EPI_ISL_2662053, EPI_ISL_2662054, EPI_ISL_2662055, EPI_ISL_2662056, EPI_ISL_2662057, EPI_ISL_2662059, EPI_ISL_2662061, EPI_ISL_2662062, EPI_ISL_2662063, EPI_ISL_2662064, EPI_ISL_2662065, EPI_ISL_2662066, EPI_ISL_2662067, EPI_ISL_2662068, EPI_ISL_2662069, EPI_ISL_2662070, EPI_ISL_2662072, EPI_ISL_2662073, EPI_ISL_2662074, EPI_ISL_2662075, EPI_ISL_2662076, EPI_ISL_2662077, EPI_ISL_2662078, EPI_ISL_2662079, EPI_ISL_2662080, EPI_ISL_2662081, EPI_ISL_2662082, EPI_ISL_2662083, EPI_ISL_2662084, EPI_ISL_2662085, EPI_ISL_2662086, EPI_ISL_2662087, EPI_ISL_2662088, EPI_ISL_2662089, EPI_ISL_2662090, EPI_ISL_2662091, EPI_ISL_2662092, EPI_ISL_2662093, EPI_ISL_2662094, EPI_ISL_2662095, EPI_ISL_2662096, EPI_ISL_2662097, EPI_ISL_2662098, EPI_ISL_2662099, EPI_ISL_2662100, EPI_ISL_2662102, EPI_ISL_2662103, EPI_ISL_2662104, EPI_ISL_2662105, EPI_ISL_2662106, EPI_ISL_2662107, EPI_ISL_2662108, EPI_ISL_2662109, EPI_ISL_2662110, EPI_ISL_2662111, EPI_ISL_2662112, EPI_ISL_2662113, EPI_ISL_2662114, EPI_ISL_2662115, EPI_ISL_2662116, EPI_ISL_2662117, EPI_ISL_2662118, EPI_ISL_2662119, EPI_ISL_2662120, EPI_ISL_2662121, EPI_ISL_2662122, EPI_ISL_2662123, EPI_ISL_2662124, EPI_ISL_2662125, EPI_ISL_2662126, EPI_ISL_2662127, EPI_ISL_2662128, EPI_ISL_2662129, EPI_ISL_2662130, EPI_ISL_2662131, EPI_ISL_2662132, EPI_ISL_2662133, EPI_ISL_2662134, EPI_ISL_2662135, EPI_ISL_2662136, EPI_ISL_2662137, EPI_ISL_2662138, EPI_ISL_2662139, EPI_ISL_2662140, EPI_ISL_2662142, EPI_ISL_2662143, EPI_ISL_2662144, EPI_ISL_2662145, EPI_ISL_2662146, EPI_ISL_2662147, EPI_ISL_2662148, EPI_ISL_2662150, EPI_ISL_2662151, EPI_ISL_2662152, EPI_ISL_2662153, EPI_ISL_2662154, EPI_ISL_2662155, EPI_ISL_2662156, EPI_ISL_2662157, EPI_ISL_2662158, EPI_ISL_2662159, EPI_ISL_2662160, EPI_ISL_2662161, EPI_ISL_2662162, EPI_ISL_2662163, EPI_ISL_2662164, EPI_ISL_2662165, EPI_ISL_2662166, EPI_ISL_2662167, EPI_ISL_2662168, EPI_ISL_2662169, EPI_ISL_2662170, EPI_ISL_2662171, EPI_ISL_2662172, EPI_ISL_2662173, EPI_ISL_2662174, EPI_ISL_2662175, EPI_ISL_2662176, EPI_ISL_2662177, EPI_ISL_2662178, EPI_ISL_2662179, EPI_ISL_2662181, EPI_ISL_2662182, EPI_ISL_2662183, EPI_ISL_2662185, EPI_ISL_2662186, EPI_ISL_2662188, EPI_ISL_2662189, EPI_ISL_2662190, EPI_ISL_2662191, EPI_ISL_2662192, EPI_ISL_2662193, EPI_ISL_2662194, EPI_ISL_2662195, EPI_ISL_2662196, EPI_ISL_2662197, EPI_ISL_2662199, EPI_ISL_2662200, EPI_ISL_2662201, EPI_ISL_2662202, EPI_ISL_2662203, EPI_ISL_2662204, EPI_ISL_2662205, EPI_ISL_2662206, EPI_ISL_2662207, EPI_ISL_2662208, EPI_ISL_2662209, EPI_ISL_2662210, EPI_ISL_2662211, EPI_ISL_2662212, EPI_ISL_2662213, EPI_ISL_2662214, EPI_ISL_2662215, EPI_ISL_2662216, EPI_ISL_2662217, EPI_ISL_2662218, EPI_ISL_2662219, EPI_ISL_2662220, EPI_ISL_2662221, EPI_ISL_2662222, EPI_ISL_2662223, EPI_ISL_2662224, EPI_ISL_2662225, EPI_ISL_2662226, EPI_ISL_2662227, EPI_ISL_2662228, EPI_ISL_2662229, EPI_ISL_2662230, EPI_ISL_2662231, EPI_ISL_2662232, EPI_ISL_2662233, EPI_ISL_2662234, EPI_ISL_2662235, EPI_ISL_2662236, EPI_ISL_2662237, EPI_ISL_2662238, EPI_ISL_2662239, EPI_ISL_2662240, EPI_ISL_2662241, EPI_ISL_2662242, EPI_ISL_2662243, EPI_ISL_2662244, EPI_ISL_2662245, EPI_ISL_2662246, EPI_ISL_2662247, EPI_ISL_2662248, EPI_ISL_2662249, EPI_ISL_2662250, EPI_ISL_2662251, EPI_ISL_2662252, EPI_ISL_2662253, EPI_ISL_2662254, EPI_ISL_2662255, EPI_ISL_2662256, EPI_ISL_2662257, EPI_ISL_2662258, EPI_ISL_2662259, EPI_ISL_2662260, EPI_ISL_2662261, EPI_ISL_2662262, EPI_ISL_2662263, EPI_ISL_2662264, EPI_ISL_2662265, EPI_ISL_2662266, EPI_ISL_2662267, EPI_ISL_2662268, EPI_ISL_2662269, EPI_ISL_2662270, EPI_ISL_2662271, EPI_ISL_2662273, EPI_ISL_2662274, EPI_ISL_2662275, EPI_ISL_2662276, EPI_ISL_2662277, EPI_ISL_2662278, EPI_ISL_2662279, EPI_ISL_2662280, EPI_ISL_2662281, EPI_ISL_2662282, EPI_ISL_2662283, EPI_ISL_2662284, EPI_ISL_2662285, EPI_ISL_2662286, EPI_ISL_2662287 | | | |
| see above | Reditus Laboratories | Reditus Laboratories | Joshua J. Geltz, Ph.D., Robert M. Sgambelluri, Ph.D., Rex Dyer, Ph.D., Cassy Philips, M.S., Alexa Eichelberger, M.S. |
| EPI_ISL_2662730, EPI_ISL_2662732, EPI_ISL_2662733, EPI_ISL_2662734, EPI_ISL_2662735, EPI_ISL_2662736, EPI_ISL_2662738, EPI_ISL_2662740, EPI_ISL_2662741, EPI_ISL_2662745, EPI_ISL_2662746, EPI_ISL_2662747, EPI_ISL_2662748, EPI_ISL_2662749, EPI_ISL_2662750, EPI_ISL_2662751, EPI_ISL_2662752, EPI_ISL_2662753, EPI_ISL_2662754, EPI_ISL_2662755, EPI_ISL_2662756, EPI_ISL_2662757, EPI_ISL_2662759, EPI_ISL_2662760, EPI_ISL_2662762, EPI_ISL_2662763, EPI_ISL_2662764, EPI_ISL_2662765, EPI_ISL_2662766, EPI_ISL_2662767, EPI_ISL_2662769, EPI_ISL_2662770, EPI_ISL_2662772, EPI_ISL_2662773, EPI_ISL_2662775, EPI_ISL_2662776, EPI_ISL_2662777, EPI_ISL_2662778, EPI_ISL_2662779, EPI_ISL_2662780, EPI_ISL_2662781, EPI_ISL_2662782, EPI_ISL_2662783, EPI_ISL_2662784, EPI_ISL_2662785, EPI_ISL_2662786, EPI_ISL_2662787, EPI_ISL_2662789, EPI_ISL_2662791, EPI_ISL_2662794, EPI_ISL_2662795, EPI_ISL_2662796, EPI_ISL_2662797, EPI_ISL_2662798, EPI_ISL_2662799, EPI_ISL_2662800, EPI_ISL_2662801, EPI_ISL_2662802, EPI_ISL_2662803, EPI_ISL_2662804, EPI_ISL_2662806, EPI_ISL_2662807, EPI_ISL_2662808, EPI_ISL_2662809, EPI_ISL_2662810, EPI_ISL_2662811, EPI_ISL_2662814, EPI_ISL_2662815 | | | |
| see above | DPHL | Delaware Public Health Lab | Rebecca Savage |
| EPI_ISL_2662828, EPI_ISL_2662829, EPI_ISL_2662830, EPI_ISL_2662831 | Erie County Medical Center (ECMC) | University at Buffalo Genomics and Bioinformatics Core | Jonathan Bard, Natalie Lamb, Alyssa Pohlman, Brandon Marzullo, Amanda Boccolucci, Norma Nowak, Donald Yergeau, Jennifer Surtees |
| EPI_ISL_2662832, EPI_ISL_2662833, EPI_ISL_2662834, EPI_ISL_2662835, EPI_ISL_2662836, EPI_ISL_2662837, EPI_ISL_2662838, EPI_ISL_2662839, EPI_ISL_2662840, EPI_ISL_2662841, EPI_ISL_2662842, EPI_ISL_2662843, EPI_ISL_2662844, EPI_ISL_2662845, EPI_ISL_2662846, EPI_ISL_2662847, EPI_ISL_2662848, EPI_ISL_2662849, EPI_ISL_2662850, EPI_ISL_2662851, EPI_ISL_2662852, EPI_ISL_2662853, EPI_ISL_2662854, EPI_ISL_2662855, EPI_ISL_2662856, EPI_ISL_2662857, EPI_ISL_2662858, EPI_ISL_2662859, EPI_ISL_2662860, EPI_ISL_2662861, EPI_ISL_2662862, EPI_ISL_2662863, EPI_ISL_2662864, EPI_ISL_2662865, EPI_ISL_2662866, EPI_ISL_2662867, EPI_ISL_2662868, EPI_ISL_2662869, EPI_ISL_2662870, EPI_ISL_2662871, EPI_ISL_2662872, EPI_ISL_2662873, EPI_ISL_2662874, EPI_ISL_2662875, EPI_ISL_2662876, EPI_ISL_2662877, EPI_ISL_2662878, EPI_ISL_2662879, EPI_ISL_2662880, EPI_ISL_2662881, EPI_ISL_2662882, EPI_ISL_2662883, EPI_ISL_2662884, EPI_ISL_2662885, EPI_ISL_2662886, EPI_ISL_2662887, EPI_ISL_2662888, EPI_ISL_2662889, EPI_ISL_2662890, EPI_ISL_2662891, EPI_ISL_2662892, EPI_ISL_2662893, EPI_ISL_2662894, EPI_ISL_2662895, EPI_ISL_2662896, EPI_ISL_2662897, EPI_ISL_2662898, EPI_ISL_2662900, EPI_ISL_2662901, EPI_ISL_2662902, EPI_ISL_2662903, EPI_ISL_2662904, EPI_ISL_2662905, EPI_ISL_2662906, EPI_ISL_2662907, EPI_ISL_2662908, EPI_ISL_2662909, EPI_ISL_2662910, EPI_ISL_2662911, EPI_ISL_2662912, EPI_ISL_2662913, EPI_ISL_2662914, EPI_ISL_2662915, EPI_ISL_2662916, EPI_ISL_2662919, EPI_ISL_2662920, EPI_ISL_2662921, EPI_ISL_2662922, | | | |

| | | | |
|---|---|---|---|
| EPI_ISL_2662923, EPI_ISL_2662924 | | | |
| see above | Erie County Public Health (ECPHL) | University at Buffalo Genomics and Bioinformatics Core | Jonathan Bard, Natalie Lamb, Alyssa Pohlman, Brandon Marzullo, Amanda Boccolucci, Norma Nowak, Donald Yergeau, Jennifer Surtees |
| EPI_ISL_2662925, EPI_ISL_2662926, EPI_ISL_2662927, EPI_ISL_2662928, EPI_ISL_2662929, EPI_ISL_2662930, EPI_ISL_2662931, EPI_ISL_2662932, EPI_ISL_2662933, EPI_ISL_2662934, EPI_ISL_2662935, EPI_ISL_2662936, EPI_ISL_2662937, EPI_ISL_2662938, EPI_ISL_2662939, EPI_ISL_2662940, EPI_ISL_2662941, EPI_ISL_2662942, EPI_ISL_2662943, EPI_ISL_2662944, EPI_ISL_2662945, EPI_ISL_2662946, EPI_ISL_2662947, EPI_ISL_2662948, EPI_ISL_2662949, EPI_ISL_2662950, EPI_ISL_2662951, EPI_ISL_2662952, EPI_ISL_2662953, EPI_ISL_2662954, EPI_ISL_2662955, EPI_ISL_2662956, EPI_ISL_2662957, EPI_ISL_2662958, EPI_ISL_2662959, EPI_ISL_2662960, EPI_ISL_2662961, EPI_ISL_2662962, EPI_ISL_2662963, EPI_ISL_2662964, EPI_ISL_2662965, EPI_ISL_2662966, EPI_ISL_2662967, EPI_ISL_2662968, EPI_ISL_2662969, EPI_ISL_2662970, EPI_ISL_2662971, EPI_ISL_2662972, EPI_ISL_2662973, EPI_ISL_2662974, EPI_ISL_2662975, EPI_ISL_2662976, EPI_ISL_2662977, EPI_ISL_2662978, EPI_ISL_2662980, EPI_ISL_2662981, EPI_ISL_2662982, EPI_ISL_2662984, EPI_ISL_2662985, EPI_ISL_2662986, EPI_ISL_2662987, EPI_ISL_2662988, EPI_ISL_2662989, EPI_ISL_2662990, EPI_ISL_2662993, EPI_ISL_2662994 | | | |
| see above | Kaleida Health Laboratories | University at Buffalo Genomics and Bioinformatics Core | Jonathan Bard, Natalie Lamb, Alyssa Pohlman, Brandon Marzullo, Amanda Boccolucci, Norma Nowak, Donald Yergeau, Jennifer Surtees |
| EPI_ISL_2663115, EPI_ISL_2663117, EPI_ISL_2663118, EPI_ISL_2663120, EPI_ISL_2663121, EPI_ISL_2663123, EPI_ISL_2663124, EPI_ISL_2663126, EPI_ISL_2663128, EPI_ISL_2663129, EPI_ISL_2663131, EPI_ISL_2663132, EPI_ISL_2663134, EPI_ISL_2663135, EPI_ISL_2663137, EPI_ISL_2663139, EPI_ISL_2663140, EPI_ISL_2663142, EPI_ISL_2663143, EPI_ISL_2663145, EPI_ISL_2663146, EPI_ISL_2663148, EPI_ISL_2663149, EPI_ISL_2663151, EPI_ISL_2663152, EPI_ISL_2663154, EPI_ISL_2663155, EPI_ISL_2663157, EPI_ISL_2663158, EPI_ISL_2663160, EPI_ISL_2663162, EPI_ISL_2663163, EPI_ISL_2663165, EPI_ISL_2663166, EPI_ISL_2663168, EPI_ISL_2663169, EPI_ISL_2663171, EPI_ISL_2663173, EPI_ISL_2663174, EPI_ISL_2663176, EPI_ISL_2663177, EPI_ISL_2663179, EPI_ISL_2663181, EPI_ISL_2663182, EPI_ISL_2663184, EPI_ISL_2663185, EPI_ISL_2663187, EPI_ISL_2663188, EPI_ISL_2663190 | | | |
| see above | Ochsner Health | Bioinfoexperts, LLC | Judy Crabtree, Amy Feehan, Rebecca Rose, David J. Nolan, Tessa LaFleur, Julia-Garcia-Diaz, Lucio Miele, Susanna L. Lamers |
| EPI_ISL_2663194, EPI_ISL_2663195, EPI_ISL_2663196, EPI_ISL_2663197, EPI_ISL_2663198 | Minnesota Department of Health, Public Health Laboratory | Minnesota Department of Health, Public Health Laboratory | Alexandra Lorentz, Jacob Garfin, Matt Plumb, and Xiong Wang |
| EPI_ISL_2663199 | Maple Grove Hospital | Minnesota Department of Health, Public Health Laboratory | Alexandra Lorentz, Jacob Garfin, Matt Plumb, and Xiong Wang |
| EPI_ISL_2663200, EPI_ISL_2663201, EPI_ISL_2663202, EPI_ISL_2663203, EPI_ISL_2663204 | M Health Fairview | Minnesota Department of Health, Public Health Laboratory | Alexandra Lorentz, Jacob Garfin, Matt Plumb, and Xiong Wang |
| EPI_ISL_2663206, EPI_ISL_2663207, EPI_ISL_2663208, EPI_ISL_2663209, EPI_ISL_2663210, EPI_ISL_2663211, EPI_ISL_2663212, EPI_ISL_2663213, EPI_ISL_2663214, EPI_ISL_2663215, EPI_ISL_2663216, EPI_ISL_2663217, EPI_ISL_2663218, EPI_ISL_2663219, EPI_ISL_2663220, EPI_ISL_2663221, EPI_ISL_2663222, EPI_ISL_2663223, EPI_ISL_2663224, EPI_ISL_2663225, EPI_ISL_2663226, EPI_ISL_2663227 | | | |
| see above | HealthPartners Central Lab | Minnesota Department of Health, Public Health Laboratory | Alexandra Lorentz, Jacob Garfin, Matt Plumb, and Xiong Wang |
| EPI_ISL_2663228 | M Health Fairview | Minnesota Department of Health, Public Health Laboratory | Alexandra Lorentz, Jacob Garfin, Matt Plumb, and Xiong Wang |
| EPI_ISL_2663229, EPI_ISL_2663230, EPI_ISL_2663231, EPI_ISL_2663232, EPI_ISL_2663233, EPI_ISL_2663234, EPI_ISL_2663235, EPI_ISL_2663236, EPI_ISL_2663237, EPI_ISL_2663238, EPI_ISL_2663239 | | | |
| see above | Mayo Clinic & Mayo Clinic Laboratories | Minnesota Department of Health, Public Health Laboratory | Alexandra Lorentz, Jacob Garfin, Matt Plumb, and Xiong Wang |
| EPI_ISL_2663326 | LESP Guanajuato | Instituto de Diagnostico y Referencia Epidemiologicos (INDRE) | Claudia Wong-Arambula, Abril Rodriguez-Maldonado, Vanessa Rivero-Arredondo, Ariadna Medina-Benitez, Joaquin Quiroz-Mercado, Sergio Rangel-Guerrero, Natividad Cruz-Ortiz, Tatiana Nunez-Garcia, Gisela Barrera-Badillo, Lucia Hernandez-Rivas, Irma Lopez-Martinez, Ernesto Ramirez-Gonzalez. |
| EPI_ISL_2663327 | LESP Nuevo Leon | Instituto de Diagnostico y Referencia Epidemiologicos (INDRE) | Claudia Wong-Arambula, Abril Rodriguez-Maldonado, Vanessa Rivero-Arredondo, Ariadna Medina-Benitez, Joaquin Quiroz-Mercado, Sergio Rangel-Guerrero, Natividad Cruz-Ortiz, Tatiana Nunez-Garcia, Gisela Barrera-Badillo, Lucia Hernandez-Rivas, Irma Lopez-Martinez, Ernesto Ramirez-Gonzalez. |
| EPI_ISL_2663328, EPI_ISL_2663329, EPI_ISL_2663330 | LESP Ciudad de Mexico | Instituto de Diagnostico y Referencia Epidemiologicos (INDRE) | Claudia Wong-Arambula, Abril Rodriguez-Maldonado, Vanessa Rivero-Arredondo, Ariadna Medina-Benitez, Joaquin Quiroz-Mercado, Sergio Rangel-Guerrero, Natividad Cruz-Ortiz, Tatiana Nunez-Garcia, Gisela Barrera-Badillo, Lucia Hernandez-Rivas, Irma Lopez-Martinez, Ernesto Ramirez-Gonzalez. |
| EPI_ISL_2663331, EPI_ISL_2663332 | LESP Queretaro | Instituto de Diagnostico y Referencia Epidemiologicos (INDRE) | Claudia Wong-Arambula, Abril Rodriguez-Maldonado, Vanessa Rivero-Arredondo, Ariadna Medina-Benitez, Joaquin Quiroz-Mercado, Sergio Rangel-Guerrero, Natividad Cruz-Ortiz, Tatiana Nunez-Garcia, Gisela Barrera-Badillo, Lucia Hernandez-Rivas, Irma Lopez-Martinez, Ernesto Ramirez-Gonzalez. |
| EPI_ISL_2663333, EPI_ISL_2663334 | LESP Sinaloa | Instituto de Diagnostico y Referencia Epidemiologicos (INDRE) | Claudia Wong-Arambula, Abril Rodriguez-Maldonado, Vanessa Rivero-Arredondo, Ariadna Medina-Benitez, Joaquin Quiroz-Mercado, Sergio Rangel-Guerrero, Natividad Cruz-Ortiz, Tatiana Nunez-Garcia, Gisela Barrera-Badillo, Lucia Hernandez-Rivas, Irma Lopez-Martinez, Ernesto Ramirez-Gonzalez. |
| EPI_ISL_2663336, EPI_ISL_2663337, EPI_ISL_2663338, EPI_ISL_2663339, EPI_ISL_2663340, EPI_ISL_2663341 | LESP Tabasco | Instituto de Diagnostico y Referencia Epidemiologicos (INDRE) | Claudia Wong-Arambula, Abril Rodriguez-Maldonado, Vanessa Rivero-Arredondo, Ariadna Medina-Benitez, Joaquin Quiroz-Mercado, Sergio Rangel-Guerrero, Natividad Cruz-Ortiz, Tatiana Nunez-Garcia, Gisela Barrera-Badillo, Lucia Hernandez-Rivas, Irma Lopez-Martinez, Ernesto Ramirez-Gonzalez. |
| EPI_ISL_2663342, EPI_ISL_2663343, EPI_ISL_2663344 | LESP Ciudad de Mexico | Instituto de Diagnostico y Referencia Epidemiologicos (INDRE) | Claudia Wong-Arambula, Abril Rodriguez-Maldonado, Vanessa Rivero-Arredondo, Ariadna Medina-Benitez, Joaquin Quiroz-Mercado, Sergio Rangel-Guerrero, Natividad Cruz-Ortiz, Tatiana Nunez-Garcia, Gisela Barrera-Badillo, Lucia Hernandez-Rivas, Irma Lopez-Martinez, Ernesto Ramirez-Gonzalez. |
| EPI_ISL_2663345 | LESP Colima | Instituto de Diagnostico y Referencia Epidemiologicos (INDRE) | Claudia Wong-Arambula, Abril Rodriguez-Maldonado, Vanessa Rivero-Arredondo, Ariadna Medina-Benitez, Joaquin Quiroz-Mercado, Sergio Rangel-Guerrero, Natividad Cruz-Ortiz, Tatiana Nunez-Garcia, Gisela Barrera-Badillo, Lucia Hernandez-Rivas, Irma Lopez-Martinez, Ernesto Ramirez-Gonzalez. |
| EPI_ISL_2663346, EPI_ISL_2663347, EPI_ISL_2663348, EPI_ISL_2663349 | LESP Tabasco | Instituto de Diagnostico y Referencia Epidemiologicos (INDRE) | Claudia Wong-Arambula, Abril Rodriguez-Maldonado, Vanessa Rivero-Arredondo, Ariadna Medina-Benitez, Joaquin Quiroz-Mercado, Sergio Rangel-Guerrero, Natividad Cruz-Ortiz, Tatiana Nunez-Garcia, Gisela Barrera-Badillo, Lucia Hernandez-Rivas, Irma Lopez-Martinez, Ernesto Ramirez-Gonzalez. |
| EPI_ISL_2663350 | LESP Tamaulipas | Instituto de Diagnostico y Referencia Epidemiologicos (INDRE) | Claudia Wong-Arambula, Abril Rodriguez-Maldonado, Vanessa Rivero-Arredondo, Ariadna Medina-Benitez, Joaquin Quiroz-Mercado, Sergio Rangel-Guerrero, Natividad Cruz-Ortiz, Tatiana Nunez-Garcia, Gisela Barrera-Badillo, Lucia Hernandez-Rivas, Irma Lopez-Martinez, Ernesto Ramirez-Gonzalez. |
| EPI_ISL_2663351 | LESP Tabasco | Instituto de Diagnostico y Referencia Epidemiologicos (INDRE) | Claudia Wong-Arambula, Abril Rodriguez-Maldonado, Vanessa Rivero-Arredondo, Ariadna Medina-Benitez, Joaquin Quiroz-Mercado, Sergio Rangel-Guerrero, Natividad Cruz-Ortiz, Tatiana Nunez-Garcia, Gisela Barrera-Badillo, Lucia Hernandez-Rivas, Irma Lopez-Martinez, Ernesto Ramirez-Gonzalez. |
| EPI_ISL_2663352 | LESP Oaxaca | Instituto de Diagnostico y Referencia Epidemiologicos (INDRE) | Claudia Wong-Arambula, Abril Rodriguez-Maldonado, Vanessa Rivero-Arredondo, Ariadna Medina-Benitez, Joaquin Quiroz-Mercado, Sergio Rangel-Guerrero, Natividad Cruz-Ortiz, Tatiana Nunez-Garcia, Gisela Barrera-Badillo, Lucia Hernandez-Rivas, Irma Lopez-Martinez, Ernesto Ramirez-Gonzalez. |
| EPI_ISL_2663353 | LESP Baja California Sur | Instituto de Diagnostico y Referencia Epidemiologicos (INDRE) | Claudia Wong-Arambula, Abril Rodriguez-Maldonado, Vanessa Rivero-Arredondo, Ariadna Medina-Benitez, Joaquin Quiroz-Mercado, Sergio Rangel-Guerrero, Natividad Cruz-Ortiz, Tatiana Nunez-Garcia, Gisela Barrera-Badillo, Lucia Hernandez-Rivas, Irma Lopez-Martinez, Ernesto Ramirez-Gonzalez. |
| EPI_ISL_2663354, EPI_ISL_2663355 | LESP Sonora | Instituto de Diagnostico y Referencia Epidemiologicos (INDRE) | Claudia Wong-Arambula, Abril Rodriguez-Maldonado, Vanessa Rivero-Arredondo, Ariadna Medina-Benitez, Joaquin Quiroz-Mercado, Sergio Rangel-Guerrero, Natividad Cruz-Ortiz, Tatiana Nunez-Garcia, Gisela Barrera-Badillo, Lucia Hernandez-Rivas, Irma Lopez-Martinez, Ernesto Ramirez-Gonzalez. |
| EPI_ISL_2663356, EPI_ISL_2663357 | LESP Quintana Roo | Instituto de Diagnostico y Referencia Epidemiologicos (INDRE) | Claudia Wong-Arambula, Abril Rodriguez-Maldonado, Vanessa Rivero-Arredondo, Ariadna Medina-Benitez, Joaquin Quiroz-Mercado, Sergio Rangel-Guerrero, Natividad Cruz-Ortiz, Tatiana Nunez-Garcia, Gisela Barrera-Badillo, Lucia Hernandez-Rivas, Irma Lopez-Martinez, Ernesto Ramirez-Gonzalez. |
| EPI_ISL_2663358 | LESP Ciudad de Mexico | Instituto de Diagnostico y Referencia Epidemiologicos (INDRE) | Claudia Wong-Arambula, Abril Rodriguez-Maldonado, Vanessa Rivero-Arredondo, Ariadna Medina-Benitez, Joaquin Quiroz-Mercado, Sergio Rangel-Guerrero, Natividad Cruz-Ortiz, Tatiana Nunez-Garcia, Gisela Barrera-Badillo, Lucia Hernandez-Rivas, Irma Lopez-Martinez, Ernesto Ramirez-Gonzalez. |
| EPI_ISL_2663359 | LESP Sonora | Instituto de Diagnostico y Referencia Epidemiologicos (INDRE) | Claudia Wong-Arambula, Abril Rodriguez-Maldonado, Vanessa Rivero-Arredondo, Ariadna Medina-Benitez, Joaquin Quiroz-Mercado, Sergio Rangel-Guerrero, Natividad Cruz-Ortiz, Tatiana Nunez-Garcia, Gisela Barrera-Badillo, Lucia Hernandez-Rivas, Irma Lopez-Martinez, Ernesto Ramirez-Gonzalez. |
| EPI_ISL_2663360, EPI_ISL_2663361, EPI_ISL_2663362, EPI_ISL_2663363, EPI_ISL_2663364, EPI_ISL_2663365, EPI_ISL_2663366, EPI_ISL_2663367, EPI_ISL_2663368, EPI_ISL_2663369, EPI_ISL_2663370, EPI_ISL_2663371, EPI_ISL_2663372, EPI_ISL_2663373, EPI_ISL_2663374, EPI_ISL_2663375 | | | |
| see above | LESP Hidalgo | Instituto de Diagnostico y Referencia Epidemiologicos (INDRE) | Claudia Wong-Arambula, Abril Rodriguez-Maldonado, Vanessa Rivero-Arredondo, Ariadna Medina-Benitez, Joaquin Quiroz-Mercado, Sergio Rangel-Guerrero, Natividad Cruz-Ortiz, Tatiana Nunez-Garcia, Gisela Barrera-Badillo, Lucia Hernandez-Rivas, Irma Lopez-Martinez, Ernesto Ramirez-Gonzalez. |
| EPI_ISL_2663376, EPI_ISL_2663377 | LESP Queretaro | Instituto de Diagnostico y Referencia Epidemiologicos (INDRE) | Claudia Wong-Arambula, Abril Rodriguez-Maldonado, Vanessa Rivero-Arredondo, Ariadna Medina-Benitez, Joaquin Quiroz-Mercado, Sergio Rangel-Guerrero, Natividad Cruz-Ortiz, Tatiana Nunez-Garcia, Gisela Barrera-Badillo, Lucia Hernandez-Rivas, Irma Lopez-Martinez, Ernesto Ramirez-Gonzalez. |
| EPI_ISL_2663378, EPI_ISL_2663379 | LESP Yucatan | Instituto de Diagnostico y Referencia Epidemiologicos (INDRE) | Claudia Wong-Arambula, Abril Rodriguez-Maldonado, Vanessa Rivero-Arredondo, Ariadna Medina-Benitez, Joaquin Quiroz-Mercado, Sergio Rangel-Guerrero, Natividad Cruz-Ortiz, Tatiana Nunez-Garcia, Gisela Barrera-Badillo, Lucia Hernandez-Rivas, Irma Lopez-Martinez, Ernesto Ramirez-Gonzalez. |
| EPI_ISL_2663380, EPI_ISL_2663381 | LESP Queretaro | Instituto de Diagnostico y Referencia Epidemiologicos (INDRE) | Claudia Wong-Arambula, Abril Rodriguez-Maldonado, Vanessa Rivero-Arredondo, Ariadna Medina-Benitez, Joaquin Quiroz-Mercado, Sergio Rangel-Guerrero, Natividad Cruz-Ortiz, Tatiana Nunez-Garcia, Gisela Barrera-Badillo, Lucia Hernandez-Rivas, Irma Lopez-Martinez, Ernesto Ramirez-Gonzalez. |
| EPI_ISL_2663382 | LESP Morelos | Instituto de Diagnostico y Referencia Epidemiologicos (INDRE) | Claudia Wong-Arambula, Abril Rodriguez-Maldonado, Vanessa Rivero-Arredondo, Ariadna Medina-Benitez, Joaquin Quiroz-Mercado, Sergio Rangel-Guerrero, Natividad Cruz-Ortiz, Tatiana Nunez-Garcia, Gisela Barrera-Badillo, Lucia Hernandez-Rivas, Irma Lopez-Martinez, Ernesto Ramirez-Gonzalez. |
| EPI_ISL_2663383, EPI_ISL_2663384, EPI_ISL_2663385, EPI_ISL_2663386, EPI_ISL_2663387, EPI_ISL_2663388 | LESP Guanajuato | Instituto de Diagnostico y Referencia Epidemiologicos (INDRE) | Claudia Wong-Arambula, Abril Rodriguez-Maldonado, Vanessa Rivero-Arredondo, Ariadna Medina-Benitez, Joaquin Quiroz-Mercado, Sergio Rangel-Guerrero, Natividad Cruz-Ortiz, Tatiana Nunez-Garcia, Gisela Barrera-Badillo, Lucia Hernandez-Rivas, Irma Lopez-Martinez, Ernesto Ramirez-Gonzalez. |
| EPI_ISL_2663389, EPI_ISL_2663390, | LESP Nuevo Leon | Instituto de Diagnostico y Referencia Epidemiologicos (INDRE) | Claudia Wong-Arambula, Abril Rodriguez-Maldonado, Vanessa Rivero-Arredondo, Ariadna Medina-Benitez, Joaquin Quiroz-Mercado, Sergio Rangel-Guerrero, Natividad Cruz-Ortiz, Tatiana Nunez-Garcia, Gisela Barrera-Badillo, Lucia Hernandez-Rivas, Irma Lopez-Martinez, Ernesto Ramirez-Gonzalez. |

| | | | |
|---|---|---|---|
| EPI_ISL_2663391, EPI_ISL_2663392 | | | |
| EPI_ISL_2663393 | LESP Ciudad de Mexico | Instituto de Diagnostico y Referencia Epidemiologicos (INDRE) | Claudia Wong-Arambula, Abril Rodriguez-Maldonado, Vanessa Rivero-Arredondo, Ariadna Medina-Benitez, Joaquin Quiroz-Mercado, Sergio Rangel-Guerrero, Natividad Cruz-Ortiz, Tatiana Nunez-Garcia, Gisela Barrera-Badillo, Lucia Hernandez-Rivas, Irma Lopez-Martinez, Ernesto Ramirez-Gonzalez. |
| EPI_ISL_2663394, EPI_ISL_2663395, EPI_ISL_2663396, EPI_ISL_2663397, EPI_ISL_2663398 | LESP Chihuahua | Instituto de Diagnostico y Referencia Epidemiologicos (INDRE) | Claudia Wong-Arambula, Abril Rodriguez-Maldonado, Vanessa Rivero-Arredondo, Ariadna Medina-Benitez, Joaquin Quiroz-Mercado, Sergio Rangel-Guerrero, Natividad Cruz-Ortiz, Tatiana Nunez-Garcia, Gisela Barrera-Badillo, Lucia Hernandez-Rivas, Irma Lopez-Martinez, Ernesto Ramirez-Gonzalez. |
| EPI_ISL_2663399 | LESP Puebla | Instituto de Diagnostico y Referencia Epidemiologicos (INDRE) | Claudia Wong-Arambula, Abril Rodriguez-Maldonado, Vanessa Rivero-Arredondo, Ariadna Medina-Benitez, Joaquin Quiroz-Mercado, Sergio Rangel-Guerrero, Natividad Cruz-Ortiz, Tatiana Nunez-Garcia, Gisela Barrera-Badillo, Lucia Hernandez-Rivas, Irma Lopez-Martinez, Ernesto Ramirez-Gonzalez. |
| EPI_ISL_2663400, EPI_ISL_2663401 | LESP Tamaulipas | Instituto de Diagnostico y Referencia Epidemiologicos (INDRE) | Claudia Wong-Arambula, Abril Rodriguez-Maldonado, Vanessa Rivero-Arredondo, Ariadna Medina-Benitez, Joaquin Quiroz-Mercado, Sergio Rangel-Guerrero, Natividad Cruz-Ortiz, Tatiana Nunez-Garcia, Gisela Barrera-Badillo, Lucia Hernandez-Rivas, Irma Lopez-Martinez, Ernesto Ramirez-Gonzalez. |
| EPI_ISL_2663402 | LESP Yucatan | Instituto de Diagnostico y Referencia Epidemiologicos (INDRE) | Claudia Wong-Arambula, Abril Rodriguez-Maldonado, Vanessa Rivero-Arredondo, Ariadna Medina-Benitez, Joaquin Quiroz-Mercado, Sergio Rangel-Guerrero, Natividad Cruz-Ortiz, Tatiana Nunez-Garcia, Gisela Barrera-Badillo, Lucia Hernandez-Rivas, Irma Lopez-Martinez, Ernesto Ramirez-Gonzalez. |
| EPI_ISL_2663403 | LESP Sonora | Instituto de Diagnostico y Referencia Epidemiologicos (INDRE) | Claudia Wong-Arambula, Abril Rodriguez-Maldonado, Vanessa Rivero-Arredondo, Ariadna Medina-Benitez, Joaquin Quiroz-Mercado, Sergio Rangel-Guerrero, Natividad Cruz-Ortiz, Tatiana Nunez-Garcia, Gisela Barrera-Badillo, Lucia Hernandez-Rivas, Irma Lopez-Martinez, Ernesto Ramirez-Gonzalez. |
| EPI_ISL_2663470, EPI_ISL_2663471, EPI_ISL_2663472, EPI_ISL_2663473, EPI_ISL_2663474, EPI_ISL_2663475, EPI_ISL_2663476, EPI_ISL_2663477, EPI_ISL_2663478, EPI_ISL_2663479, EPI_ISL_2663481, EPI_ISL_2663483, EPI_ISL_2663484, EPI_ISL_2663485, EPI_ISL_2663486, EPI_ISL_2663487, EPI_ISL_2663488, EPI_ISL_2663489, EPI_ISL_2663490, EPI_ISL_2663491, EPI_ISL_2663493, EPI_ISL_2663494, EPI_ISL_2663495, EPI_ISL_2663496, EPI_ISL_2663497, EPI_ISL_2663498, EPI_ISL_2663499, EPI_ISL_2663500, EPI_ISL_2663501, EPI_ISL_2663502, EPI_ISL_2663503, EPI_ISL_2663504, EPI_ISL_2663505, EPI_ISL_2663506, EPI_ISL_2663507, EPI_ISL_2663508, EPI_ISL_2663509, EPI_ISL_2663510 | | | |
| see above | Virginia Division of Consolidated Laboratory Services | Virginia Division of Consolidated Laboratory Services | Virginia Division of Consolidated Laboratory Services |
| EPI_ISL_2663512, EPI_ISL_2663514, EPI_ISL_2663515, EPI_ISL_2663516, EPI_ISL_2663517, EPI_ISL_2663521, EPI_ISL_2663522, EPI_ISL_2663523, EPI_ISL_2663525, EPI_ISL_2663526, EPI_ISL_2663527, EPI_ISL_2663529, EPI_ISL_2663531, EPI_ISL_2663533, EPI_ISL_2663534, EPI_ISL_2663535, EPI_ISL_2663536, EPI_ISL_2663537, EPI_ISL_2663538, EPI_ISL_2663539, EPI_ISL_2663540, EPI_ISL_2663541, EPI_ISL_2663542, EPI_ISL_2663543, EPI_ISL_2663544, EPI_ISL_2663545, EPI_ISL_2663546, EPI_ISL_2663547, EPI_ISL_2663548, EPI_ISL_2663549, EPI_ISL_2663550, EPI_ISL_2663551, EPI_ISL_2663552, EPI_ISL_2663553 | | | |
| see above | NYU Langone Health | Departments of Pathology and Medicine, New York University School of Medicine | Adriana Heguy, Dacia Dimartino, Emily Guzman, Christian Marier, Peter Meyn, Sitharam Ramaswami, Gael Westby, Paul Zappile, Yutong Zhang, Paolo Cotzia, Guiqing Wang |
| EPI_ISL_2663619, EPI_ISL_2663620, EPI_ISL_2663622, EPI_ISL_2663623, EPI_ISL_2663624, EPI_ISL_2663625, EPI_ISL_2663626, EPI_ISL_2663629, EPI_ISL_2663630, EPI_ISL_2663631, EPI_ISL_2663632, EPI_ISL_2663633, EPI_ISL_2663634, EPI_ISL_2663635, EPI_ISL_2663636, EPI_ISL_2663637, EPI_ISL_2663638, EPI_ISL_2663639, EPI_ISL_2663640, EPI_ISL_2663641, EPI_ISL_2663642, EPI_ISL_2663643, EPI_ISL_2663644, EPI_ISL_2663645, EPI_ISL_2663646, EPI_ISL_2663647, EPI_ISL_2663648, EPI_ISL_2663649, EPI_ISL_2663651, EPI_ISL_2663652, EPI_ISL_2663653, EPI_ISL_2663655, EPI_ISL_2663656, EPI_ISL_2663657, EPI_ISL_2663658, EPI_ISL_2663659, EPI_ISL_2663660, EPI_ISL_2663662, EPI_ISL_2663663, EPI_ISL_2663664, EPI_ISL_2663665, EPI_ISL_2663666, EPI_ISL_2663667, EPI_ISL_2663668, EPI_ISL_2663669, EPI_ISL_2663670, EPI_ISL_2663671, EPI_ISL_2663672, EPI_ISL_2663673, EPI_ISL_2663674, EPI_ISL_2663675, EPI_ISL_2663676, EPI_ISL_2663677, EPI_ISL_2663678, EPI_ISL_2663679, EPI_ISL_2663680, EPI_ISL_2663681, EPI_ISL_2663682, EPI_ISL_2663683, EPI_ISL_2663684, EPI_ISL_2663685, EPI_ISL_2663686, EPI_ISL_2663687, EPI_ISL_2663688, EPI_ISL_2663689, EPI_ISL_2663690, EPI_ISL_2663692, EPI_ISL_2663693, EPI_ISL_2663694, EPI_ISL_2663695, EPI_ISL_2663696, EPI_ISL_2663697, EPI_ISL_2663698, EPI_ISL_2663699, EPI_ISL_2663700, EPI_ISL_2663701, EPI_ISL_2663702, EPI_ISL_2663703, EPI_ISL_2663704, EPI_ISL_2663705, EPI_ISL_2663706, EPI_ISL_2663707, EPI_ISL_2663708, EPI_ISL_2663709, EPI_ISL_2663710, EPI_ISL_2663712, EPI_ISL_2663714, EPI_ISL_2663715, EPI_ISL_2663716, EPI_ISL_2663717, EPI_ISL_2663718, EPI_ISL_2663719, EPI_ISL_2663720, EPI_ISL_2663721, EPI_ISL_2663722, EPI_ISL_2663723, EPI_ISL_2663724, EPI_ISL_2663726, EPI_ISL_2663728, EPI_ISL_2663729, EPI_ISL_2663730, EPI_ISL_2663731, EPI_ISL_2663732, EPI_ISL_2663733, EPI_ISL_2663734, EPI_ISL_2663735, EPI_ISL_2663736, EPI_ISL_2663737, EPI_ISL_2663738, EPI_ISL_2663740, EPI_ISL_2663741, EPI_ISL_2663742, EPI_ISL_2663743, EPI_ISL_2663744, EPI_ISL_2663745, EPI_ISL_2663746, EPI_ISL_2663747, EPI_ISL_2663748, EPI_ISL_2663749, EPI_ISL_2663750, EPI_ISL_2663751, EPI_ISL_2663752, EPI_ISL_2663753, EPI_ISL_2663755, EPI_ISL_2663756, EPI_ISL_2663757, EPI_ISL_2663758, EPI_ISL_2663759, EPI_ISL_2663760, EPI_ISL_2663761, EPI_ISL_2663764, EPI_ISL_2663765, EPI_ISL_2663766, EPI_ISL_2663767, EPI_ISL_2663768, EPI_ISL_2663769, EPI_ISL_2663770, EPI_ISL_2663771, EPI_ISL_2663773, EPI_ISL_2663774, EPI_ISL_2663775, EPI_ISL_2663777, EPI_ISL_2663778, EPI_ISL_2663779, EPI_ISL_2663780, EPI_ISL_2663781 | | | |
| see above | OHSU Lab Services Molecular Microbiology Lab | Oregon SARS-CoV-2 Genome Sequencing Center | Brendan L. O'Connell, Sonia Acharya, Cierra LaBlanc, Ruth V. Nichols, Alec J. Hirsch, Donna Hansel, Guang Fan, Xuan Qin, Daniel N. Streblow, William B. Messer, Andrew C. Adey, Benjamin N. Bimber, Brian J. O'Roak |
| EPI_ISL_2665396 | Baylor University Medical Center | Baylor Scott & White Medical Center - Temple | Ari Rao, Linden Morales, Kimberly Walker, Marcus Volz, Shelby Johnson |
| EPI_ISL_2671483 | Unidad de Investigacion Biomedica de Zacatecas (UIBZ) | Unidad de Genomica Avanzada | Consorcio Mexicano de Vigilancia Genomica (CoViGen-Mex). Authors (in alphabetical order): Julio Elias Alvarado-Yaah, Carlos F. Arias, Santiago avila-Rios, Eduardo Becerril-Vargas, Victor Hugo Borja-Aburto, Celia Boukadida, Cristobal Chaidez-Quiroz, Juan Bautista Chale-Dzul, Ricardo Ciria Merce, Andreu Comas-Garcia, Celida Duque Molina, Julissa Enciso-Ibarra, Jose Antonio Enciso-Moreno, Gloria Elena Espinosa-Ayala, Fernando Fontove-Herrera, Daniel Fregoso-Rueda, Victor Eduardo Garcia-Arias, Alejandra Garcia-Gasca, Bruno Gomez-Gil, Jean Pierre Gonzalez, Irvin Gonzalez-Lopez, Concepcion Grajales-Muñiz, Ricardo Grande, Rosa Maria Gutierrez Rios, Jesus Hernandez, Alejandra Hernandez-Teran, Alfredo Herrera-Estrella, Carla Ivon Herrera-Najera, Pavel Isa, Veronica Mata-Haro, Daniel Lira Morales, Susana Lopez, Antonio Loza Roman, Brenda Irasema Maldonado-Meza, Bernardo Martinez-Miguel, Jose Arturo Martinez-Orozco, Celida Martinez- Rodriguez, Margarita Matias-Florentino, Fidencio Mejia-Nepomuceno, Maria Guadalupe de Jesus Mireles-Rivera, Gloria Maria Molina-Salinas, Hector Montoya-Fuentes, , Mario Mujica-Sanchez, Jose Esteban Muñoz-Medina, Jose de Jesus Nuñez-Contreras, Alicia Ocaña-Mondragon, Luis Alberto Ochoa-Carrera, Hector Esteban Paz-Juarez, Marissa Perez-Garcia, Francisco Pulido, Helen Haydee Fernanda Ramirez-Plascencia, Jorge Salas-Hernandez, Angel Gustavo Salas-Lais, Alejandro Sanchez-Flores, Clara Esperanza Santacruz-Tinoco, Maria Guadalupe Santiago-Mauricio, Selene Zarate, Nelly Selem-Mojica, Blanca Taboada, Gloria Vazquez, Joel Armando Vazquez-Perez. |
| EPI_ISL_2671484 | Centro de Investigacion Biomedica de Occidente (CIBO) | Unidad de Genomica Avanzada | Consorcio Mexicano de Vigilancia Genomica (CoViGen-Mex). Authors (in alphabetical order): Julio Elias Alvarado-Yaah, Carlos F. Arias, Santiago avila-Rios, Eduardo Becerril-Vargas, Victor Hugo Borja-Aburto, Celia Boukadida, Cristobal Chaidez-Quiroz, Juan Bautista Chale-Dzul, Ricardo Ciria Merce, Andreu Comas-Garcia, Celida Duque Molina, Julissa Enciso-Ibarra, Jose Antonio Enciso-Moreno, Gloria Elena Espinosa-Ayala, Fernando Fontove-Herrera, Daniel Fregoso-Rueda, Victor Eduardo Garcia-Arias, Alejandra Garcia-Gasca, Bruno Gomez-Gil, Jean Pierre Gonzalez, Irvin Gonzalez-Lopez, Concepcion Grajales-Muñiz, Ricardo Grande, Rosa Maria Gutierrez Rios, Jesus Hernandez, Alejandra Hernandez-Teran, Alfredo Herrera-Estrella, Carla Ivon Herrera-Najera, Pavel Isa, Veronica Mata-Haro, Daniel Lira Morales, Susana Lopez, Antonio Loza Roman, Brenda Irasema Maldonado-Meza, Bernardo Martinez-Miguel, Jose Arturo Martinez-Orozco, Celida Martinez- Rodriguez, Margarita Matias-Florentino, Fidencio Mejia-Nepomuceno, Maria Guadalupe de Jesus Mireles-Rivera, Gloria Maria Molina-Salinas, Hector Montoya-Fuentes, , Mario Mujica-Sanchez, Jose Esteban Muñoz-Medina, Jose de Jesus Nuñez-Contreras, Alicia Ocaña-Mondragon, Luis Alberto Ochoa-Carrera, Hector Esteban Paz-Juarez, Marissa Perez-Garcia, Francisco Pulido, Helen Haydee Fernanda Ramirez-Plascencia, Jorge Salas-Hernandez, Angel Gustavo Salas-Lais, Alejandro Sanchez-Flores, Clara Esperanza Santacruz-Tinoco, Maria Guadalupe Santiago-Mauricio, Selene Zarate, Nelly Selem-Mojica, Blanca Taboada, Gloria Vazquez, Joel Armando Vazquez-Perez. |
| EPI_ISL_2671485, EPI_ISL_2671486, EPI_ISL_2671487, EPI_ISL_2671488, EPI_ISL_2671489, EPI_ISL_2671490, EPI_ISL_2671491, EPI_ISL_2671492 | Laboratorio Central de Epidemiologia (LCE) | Unidad de Genomica Avanzada | Consorcio Mexicano de Vigilancia Genomica (CoViGen-Mex). Authors (in alphabetical order): Julio Elias Alvarado-Yaah, Carlos F. Arias, Santiago avila-Rios, Eduardo Becerril-Vargas, Victor Hugo Borja-Aburto, Celia Boukadida, Cristobal Chaidez-Quiroz, Juan Bautista Chale-Dzul, Ricardo Ciria Merce, Andreu Comas-Garcia, Celida Duque Molina, Julissa Enciso-Ibarra, Jose Antonio Enciso-Moreno, Gloria Elena Espinosa-Ayala, Fernando Fontove-Herrera, Daniel Fregoso-Rueda, Victor Eduardo Garcia-Arias, Alejandra Garcia-Gasca, Bruno Gomez-Gil, Jean Pierre Gonzalez, Irvin Gonzalez-Lopez, Concepcion Grajales-Muñiz, Ricardo Grande, Rosa Maria Gutierrez Rios, Jesus Hernandez, Alejandra Hernandez-Teran, Alfredo Herrera-Estrella, Carla Ivon Herrera-Najera, Pavel Isa, Veronica Mata-Haro, Daniel Lira Morales, Susana Lopez, Antonio Loza Roman, Brenda Irasema Maldonado-Meza, Bernardo Martinez-Miguel, Jose Arturo Martinez-Orozco, Celida Martinez- Rodriguez, Margarita Matias-Florentino, Fidencio Mejia-Nepomuceno, Maria Guadalupe de Jesus Mireles-Rivera, Gloria Maria Molina-Salinas, Hector Montoya-Fuentes, , Mario Mujica-Sanchez, Jose Esteban Muñoz-Medina, Jose de Jesus Nuñez-Contreras, Alicia Ocaña-Mondragon, Luis Alberto Ochoa-Carrera, Hector Esteban Paz-Juarez, Marissa Perez-Garcia, Francisco Pulido, Helen Haydee Fernanda Ramirez-Plascencia, Jorge Salas-Hernandez, Angel Gustavo Salas-Lais, Alejandro Sanchez-Flores, Clara Esperanza Santacruz-Tinoco, Maria Guadalupe Santiago-Mauricio, Selene Zarate, Nelly Selem-Mojica, Blanca Taboada, Gloria Vazquez, Joel Armando Vazquez-Perez. |
| EPI_ISL_2671493, EPI_ISL_2671494, EPI_ISL_2671495, EPI_ISL_2671496, EPI_ISL_2671497, EPI_ISL_2671498, EPI_ISL_2671499, EPI_ISL_2671500, EPI_ISL_2671501, EPI_ISL_2671502, EPI_ISL_2671503, EPI_ISL_2671504 | | | |
| see above | Unidad de Investigacion Medica de Yucatan (UIMY) | Unidad de Genomica Avanzada | Consorcio Mexicano de Vigilancia Genomica (CoViGen-Mex). Authors (in alphabetical order): Julio Elias Alvarado-Yaah, Carlos F. Arias, Santiago avila-Rios, Eduardo Becerril-Vargas, Victor Hugo Borja-Aburto, Celia Boukadida, Cristobal Chaidez-Quiroz, Juan Bautista Chale-Dzul, Ricardo Ciria Merce, Andreu Comas-Garcia, Celida Duque Molina, Julissa Enciso-Ibarra, Jose Antonio Enciso-Moreno, Gloria Elena Espinosa-Ayala, Fernando Fontove-Herrera, Daniel Fregoso-Rueda, Victor Eduardo Garcia-Arias, Alejandra Garcia-Gasca, Bruno Gomez-Gil, Jean Pierre Gonzalez, Irvin Gonzalez-Lopez, Concepcion Grajales-Muñiz, Ricardo Grande, Rosa Maria Gutierrez Rios, Jesus Hernandez, Alejandra Hernandez-Teran, Alfredo Herrera-Estrella, Carla Ivon Herrera-Najera, Pavel Isa, Veronica Mata-Haro, Daniel Lira Morales, Susana Lopez, Antonio Loza Roman, Brenda Irasema Maldonado-Meza, Bernardo Martinez-Miguel, Jose Arturo Martinez-Orozco, Celida Martinez- Rodriguez, Margarita Matias-Florentino, Fidencio Mejia-Nepomuceno, Maria Guadalupe de Jesus Mireles-Rivera, Gloria Maria Molina-Salinas, Hector Montoya-Fuentes, , Mario Mujica-Sanchez, Jose Esteban Muñoz-Medina, Jose de Jesus Nuñez-Contreras, Alicia Ocaña-Mondragon, Luis Alberto Ochoa-Carrera, Hector Esteban Paz-Juarez, Marissa Perez-Garcia, Francisco Pulido, Helen Haydee Fernanda Ramirez-Plascencia, Jorge Salas-Hernandez, Angel Gustavo Salas-Lais, Alejandro Sanchez-Flores, Clara Esperanza Santacruz-Tinoco, Maria Guadalupe Santiago-Mauricio, Selene Zarate, Nelly Selem-Mojica, Blanca Taboada, Gloria Vazquez, Joel Armando Vazquez-Perez. |
| EPI_ISL_2671505, EPI_ISL_2671506, EPI_ISL_2671507, EPI_ISL_2671508, EPI_ISL_2671509, EPI_ISL_2671510, EPI_ISL_2671511, EPI_ISL_2671512, EPI_ISL_2671513, EPI_ISL_2671514, EPI_ISL_2671515, EPI_ISL_2671516, EPI_ISL_2671517, EPI_ISL_2671518, EPI_ISL_2671519, EPI_ISL_2671520, EPI_ISL_2671521, EPI_ISL_2671522, EPI_ISL_2671523, EPI_ISL_2671524, EPI_ISL_2671525 | | | |
| see above | Centro de Investigacion Biomedica del Noreste (CIBIN) | Unidad de Genomica Avanzada | Consorcio Mexicano de Vigilancia Genomica (CoViGen-Mex). Authors (in alphabetical order): Julio Elias Alvarado-Yaah, Carlos F. Arias, Santiago avila-Rios, Eduardo Becerril-Vargas, Victor Hugo Borja-Aburto, Celia Boukadida, Cristobal Chaidez-Quiroz, Juan Bautista Chale-Dzul, Ricardo Ciria Merce, Andreu Comas-Garcia, Celida Duque Molina, Julissa Enciso-Ibarra, Jose Antonio Enciso-Moreno, Gloria Elena Espinosa-Ayala, Fernando Fontove-Herrera, Daniel Fregoso-Rueda, Victor Eduardo Garcia-Arias, Alejandra Garcia-Gasca, Bruno Gomez-Gil, Jean Pierre Gonzalez, Irvin Gonzalez-Lopez, Concepcion Grajales-Muñiz, Ricardo Grande, Rosa Maria Gutierrez Rios, Jesus Hernandez, Alejandra Hernandez-Teran, Alfredo Herrera-Estrella, Carla Ivon Herrera-Najera, Pavel Isa, Veronica Mata-Haro, Daniel Lira Morales, Susana Lopez, Antonio Loza Roman, Brenda Irasema Maldonado-Meza, Bernardo Martinez-Miguel, Jose Arturo Martinez-Orozco, Celida Martinez- Rodriguez, Margarita Matias-Florentino, Fidencio Mejia-Nepomuceno, Maria Guadalupe de Jesus Mireles-Rivera, Gloria Maria Molina-Salinas, Hector Montoya-Fuentes, , Mario Mujica-Sanchez, Jose Esteban Muñoz-Medina, Jose de Jesus Nuñez-Contreras, Alicia Ocaña-Mondragon, Luis Alberto Ochoa-Carrera, Hector Esteban Paz-Juarez, Marissa Perez-Garcia, Francisco Pulido, Helen Haydee Fernanda Ramirez-Plascencia, Jorge Salas-Hernandez, Angel Gustavo Salas-Lais, Alejandro Sanchez-Flores, Clara Esperanza Santacruz-Tinoco, Maria Guadalupe Santiago-Mauricio, Selene Zarate, Nelly Selem-Mojica, Blanca Taboada, Gloria Vazquez, Joel Armando Vazquez-Perez. |
| EPI_ISL_2671526, EPI_ISL_2671527, EPI_ISL_2671528, EPI_ISL_2671529, EPI_ISL_2671530, EPI_ISL_2671531, EPI_ISL_2671532, EPI_ISL_2671533, EPI_ISL_2671534, EPI_ISL_2671535, EPI_ISL_2671536, EPI_ISL_2671537, EPI_ISL_2671538, EPI_ISL_2671539, EPI_ISL_2671540, EPI_ISL_2671541, EPI_ISL_2671542 | | | |
| see above | Laboratorio Central de Epidemiologia (LCE) | Unidad de Genomica Avanzada | Consorcio Mexicano de Vigilancia Genomica (CoViGen-Mex). Authors (in alphabetical order): Julio Elias Alvarado-Yaah, Carlos F. Arias, Santiago avila-Rios, Eduardo Becerril-Vargas, Victor Hugo Borja-Aburto, Celia Boukadida, Cristobal Chaidez-Quiroz, Juan Bautista Chale-Dzul, Ricardo Ciria Merce, Andreu Comas-Garcia, Celida Duque Molina, Julissa Enciso-Ibarra, Jose Antonio Enciso-Moreno, Gloria Elena Espinosa-Ayala, Fernando Fontove-Herrera, Daniel Fregoso-Rueda, Victor Eduardo Garcia-Arias, Alejandra Garcia-Gasca, Bruno Gomez-Gil, Jean Pierre Gonzalez, Irvin Gonzalez-Lopez, Concepcion Grajales-Muñiz, Ricardo Grande, Rosa Maria Gutierrez Rios, Jesus Hernandez, Alejandra Hernandez-Teran, Alfredo Herrera-Estrella, Carla Ivon Herrera-Najera, Pavel Isa, Veronica Mata-Haro, Daniel Lira Morales, Susana Lopez, Antonio Loza Roman, Brenda Irasema Maldonado-Meza, Bernardo Martinez-Miguel, Jose Arturo Martinez-Orozco, Celida Martinez- Rodriguez, Margarita Matias-Florentino, Fidencio Mejia-Nepomuceno, Maria Guadalupe de Jesus Mireles-Rivera, Gloria Maria Molina-Salinas, Hector Montoya-Fuentes, , Mario Mujica-Sanchez, Jose Esteban Muñoz-Medina, Jose de Jesus Nuñez-Contreras, Alicia Ocaña-Mondragon, Luis Alberto Ochoa-Carrera, Hector Esteban Paz-Juarez, Marissa Perez-Garcia, Francisco Pulido, Helen Haydee Fernanda Ramirez-Plascencia, Jorge Salas-Hernandez, Angel Gustavo Salas-Lais, Alejandro Sanchez-Flores, Clara Esperanza Santacruz-Tinoco, Maria Guadalupe Santiago-Mauricio, Selene Zarate, Nelly Selem-Mojica, Blanca Taboada, Gloria Vazquez, Joel Armando Vazquez-Perez. |
| EPI_ISL_2671543, EPI_ISL_2671544, EPI_ISL_2671545, EPI_ISL_2671546, EPI_ISL_2671547 | Centro de Investigacion Biomedica del Noreste (CIBIN) | Unidad de Genomica Avanzada | Consorcio Mexicano de Vigilancia Genomica (CoViGen-Mex). Authors (in alphabetical order): Julio Elias Alvarado-Yaah, Carlos F. Arias, Santiago avila-Rios, Eduardo Becerril-Vargas, Victor Hugo Borja-Aburto, Celia Boukadida, Cristobal Chaidez-Quiroz, Juan Bautista Chale-Dzul, Ricardo Ciria Merce, Andreu Comas-Garcia, Celida Duque Molina, Julissa Enciso-Ibarra, Jose Antonio Enciso-Moreno, Gloria Elena Espinosa-Ayala, Fernando Fontove-Herrera, Daniel Fregoso-Rueda, Victor Eduardo Garcia-Arias, Alejandra Garcia-Gasca, Bruno Gomez-Gil, Jean Pierre Gonzalez, Irvin Gonzalez-Lopez, Concepcion Grajales-Muñiz, Ricardo Grande, Rosa Maria Gutierrez Rios, Jesus Hernandez, Alejandra Hernandez-Teran, Alfredo Herrera-Estrella, Carla Ivon Herrera-Najera, Pavel Isa, Veronica Mata-Haro, Daniel Lira Morales, Susana Lopez, Antonio Loza Roman, Brenda Irasema Maldonado-Meza, Bernardo Martinez-Miguel, Jose Arturo Martinez-Orozco, Celida Martinez- Rodriguez, Margarita Matias-Florentino, Fidencio Mejia-Nepomuceno, Maria Guadalupe de Jesus Mireles-Rivera, Gloria Maria Molina-Salinas, Hector Montoya-Fuentes, , Mario Mujica-Sanchez, Jose Esteban Muñoz-Medina, Jose de Jesus Nuñez-Contreras, Alicia Ocaña-Mondragon, Luis Alberto Ochoa-Carrera, Hector Esteban Paz-Juarez, Marissa Perez-Garcia, Francisco Pulido, Helen Haydee Fernanda Ramirez-Plascencia, Jorge Salas-Hernandez, Angel Gustavo Salas-Lais, Alejandro Sanchez-Flores, Clara Esperanza Santacruz-Tinoco, Maria Guadalupe Santiago-Mauricio, Selene Zarate, Nelly Selem-Mojica, Blanca Taboada, Gloria Vazquez, Joel Armando Vazquez-Perez. |
| EPI_ISL_2671548, EPI_ISL_2671549, EPI_ISL_2671550, EPI_ISL_2671551, EPI_ISL_2671552, EPI_ISL_2671553, EPI_ISL_2671554, EPI_ISL_2671555, EPI_ISL_2671556, EPI_ISL_2671557 | Centro de Investigacion Biomedica de Occidente (CIBO) | Unidad de Genomica Avanzada | Consorcio Mexicano de Vigilancia Genomica (CoViGen-Mex). Authors (in alphabetical order): Julio Elias Alvarado-Yaah, Carlos F. Arias, Santiago avila-Rios, Eduardo Becerril-Vargas, Victor Hugo Borja-Aburto, Celia Boukadida, Cristobal Chaidez-Quiroz, Juan Bautista Chale-Dzul, Ricardo Ciria Merce, Andreu Comas-Garcia, Celida Duque Molina, Julissa Enciso-Ibarra, Jose Antonio Enciso-Moreno, Gloria Elena Espinosa-Ayala, Fernando Fontove-Herrera, Daniel Fregoso-Rueda, Victor Eduardo Garcia-Arias, Alejandra Garcia-Gasca, Bruno Gomez-Gil, Jean Pierre Gonzalez, Irvin Gonzalez-Lopez, Concepcion Grajales-Muñiz, Ricardo Grande, Rosa Maria Gutierrez Rios, Jesus Hernandez, Alejandra Hernandez-Teran, Alfredo Herrera-Estrella, Carla Ivon Herrera-Najera, Pavel Isa, Veronica Mata-Haro, Daniel Lira Morales, Susana Lopez, Antonio Loza Roman, Brenda Irasema Maldonado-Meza, Bernardo Martinez-Miguel, Jose Arturo Martinez-Orozco, Celida Martinez- Rodriguez, Margarita Matias-Florentino, Fidencio Mejia-Nepomuceno, Maria Guadalupe de Jesus Mireles-Rivera, Gloria Maria Molina-Salinas, Hector Montoya-Fuentes, , Mario Mujica-Sanchez, Jose Esteban Muñoz-Medina, Jose de Jesus Nuñez-Contreras, Alicia Ocaña-Mondragon, Luis Alberto Ochoa-Carrera, Hector Esteban Paz-Juarez, Marissa Perez-Garcia, Francisco Pulido, Helen Haydee Fernanda Ramirez-Plascencia, Jorge Salas-Hernandez, Angel Gustavo Salas-Lais, Alejandro Sanchez-Flores, Clara Esperanza Santacruz-Tinoco, Maria Guadalupe Santiago-Mauricio, Selene Zarate, Nelly Selem-Mojica, Blanca Taboada, Gloria Vazquez, Joel Armando Vazquez-Perez. |
| EPI_ISL_2671558, EPI_ISL_2671559, EPI_ISL_2671560 | Centro de Investigacion Biomedica del Noreste (CIBIN) | Unidad de Genomica Avanzada | Consorcio Mexicano de Vigilancia Genomica (CoViGen-Mex). Authors (in alphabetical order): Julio Elias Alvarado-Yaah, Carlos F. Arias, Santiago avila-Rios, Eduardo Becerril-Vargas, Victor Hugo Borja-Aburto, Celia Boukadida, Cristobal Chaidez-Quiroz, Juan Bautista Chale-Dzul, Ricardo Ciria Merce, Andreu Comas-Garcia, Celida Duque Molina, Julissa Enciso-Ibarra, Jose Antonio Enciso-Moreno, Gloria Elena Espinosa-Ayala, Fernando Fontove-Herrera, Daniel Fregoso-Rueda, Victor Eduardo Garcia-Arias, Alejandra Garcia-Gasca, Bruno Gomez-Gil, Jean Pierre Gonzalez, Irvin Gonzalez-Lopez, Concepcion Grajales-Muñiz, Ricardo Grande, Rosa Maria Gutierrez Rios, Jesus Hernandez, Alejandra Hernandez-Teran, Alfredo Herrera-Estrella, Carla Ivon Herrera-Najera, Pavel Isa, Veronica Mata-Haro, Daniel Lira Morales, Susana Lopez, Antonio Loza Roman, Brenda Irasema Maldonado-Meza, Bernardo Martinez-Miguel, Jose Arturo Martinez-Orozco, Celida Martinez- Rodriguez, Margarita Matias-Florentino, Fidencio Mejia-Nepomuceno, Maria Guadalupe de Jesus Mireles-Rivera, Gloria Maria Molina-Salinas, Hector Montoya-Fuentes, , Mario Mujica-Sanchez, Jose Esteban Muñoz-Medina, Jose de Jesus Nuñez-Contreras, Alicia Ocaña-Mondragon, Luis Alberto Ochoa-Carrera, Hector Esteban Paz-Juarez, Marissa Perez-Garcia, Francisco Pulido, Helen Haydee Fernanda Ramirez-Plascencia, Jorge Salas-Hernandez, Angel Gustavo Salas- |

| | | | |
|---|---|---|---|
| | | | Lais, Alejandro Sanchez-Flores, Clara Esperanza Santacruz-Tinoco, Maria Guadalupe Santiago-Mauricio, Selene Zarate, Nelly Selem-Mojica, Blanca Taboada, Gloria Vazquez, Joel Armando Vazquez-Perez. |
| EPI_ISL_2671561, EPI_ISL_2671563 | Laboratorio Central de Epidemiologia (LCE) | Unidad de Genomica Avanzada | Consorcio Mexicano de Vigilancia Genomica (CoViGen-Mex). Authors (in alphabetical order): Julio Elias Alvarado-Yaah, Carlos F. Arias, Santiago avila-Rios, Eduardo Becerril-Vargas, Victor Hugo Borja-Aburto, Celia Boukadida, Cristobal Chaidez-Quiroz, Juan Bautista Chale-Dzul, Ricardo Ciria Merce, Andreu Comas-Garcia, Celida Duque Molina, Julissa Enciso-Ibarra, Jose Antonio Enciso-Moreno, Gloria Elena Espinosa-Ayala, Fernando Fontove-Herrera, Daniel Fregoso-Rueda, Victor Eduardo Garcia-Arias, Alejandra Garcia-Gasca, Bruno Gomez-Gil, Jean Pierre Gonzalez, Irvin Gonzalez-Lopez, Concepcion Grajales-Muñiz, Ricardo Grande, Rosa Maria Gutierrez Rios, Jesus Hernandez, Alejandra Hernandez-Teran, Alfredo Herrera-Estrella, Carla Ivon Herrera-Najera, Pavel Isa, Veronica Mata-Haro, Daniel Lira Morales, Susana Lopez, Antonio Loza Roman, Brenda Irasema Maldonado-Meza, Bernardo Martinez-Miguel, Jose Arturo Martinez-Orozco, Celida Martinez- Rodriguez, Margarita Matias-Florentino, Fidencio Mejia-Nepomuceno, Maria Guadalupe de Jesus Mireles-Rivera, Gloria Maria Molina-Salinas, Hector Montoya-Fuentes, , Mario Mujica-Sanchez, Jose Esteban Muñoz-Medina, Jose de Jesus Nuñez-Contreras, Alicia Ocaña-Mondragon, Luis Alberto Ochoa-Carrera, Hector Esteban Paz-Juarez, Marissa Perez-Garcia, Francisco Pulido, Helen Haydee Fernanda Ramirez-Plascencia, Jorge Salas-Hernandez, Angel Gustavo Salas-Lais, Alejandro Sanchez-Flores, Clara Esperanza Santacruz-Tinoco, Maria Guadalupe Santiago-Mauricio, Selene Zarate, Nelly Selem-Mojica, Blanca Taboada, Gloria Vazquez, Joel Armando Vazquez-Perez. |
| EPI_ISL_2671564, EPI_ISL_2671565, EPI_ISL_2671566, EPI_ISL_2671567, EPI_ISL_2671568 | Centro de Investigacion Biomedica de Occidente (CIBO) | Unidad de Genomica Avanzada | Consorcio Mexicano de Vigilancia Genomica (CoViGen-Mex). Authors (in alphabetical order): Julio Elias Alvarado-Yaah, Carlos F. Arias, Santiago avila-Rios, Eduardo Becerril-Vargas, Victor Hugo Borja-Aburto, Celia Boukadida, Cristobal Chaidez-Quiroz, Juan Bautista Chale-Dzul, Ricardo Ciria Merce, Andreu Comas-Garcia, Celida Duque Molina, Julissa Enciso-Ibarra, Jose Antonio Enciso-Moreno, Gloria Elena Espinosa-Ayala, Fernando Fontove-Herrera, Daniel Fregoso-Rueda, Victor Eduardo Garcia-Arias, Alejandra Garcia-Gasca, Bruno Gomez-Gil, Jean Pierre Gonzalez, Irvin Gonzalez-Lopez, Concepcion Grajales-Muñiz, Ricardo Grande, Rosa Maria Gutierrez Rios, Jesus Hernandez, Alejandra Hernandez-Teran, Alfredo Herrera-Estrella, Carla Ivon Herrera-Najera, Pavel Isa, Veronica Mata-Haro, Daniel Lira Morales, Susana Lopez, Antonio Loza Roman, Brenda Irasema Maldonado-Meza, Bernardo Martinez-Miguel, Jose Arturo Martinez-Orozco, Celida Martinez- Rodriguez, Margarita Matias-Florentino, Fidencio Mejia-Nepomuceno, Maria Guadalupe de Jesus Mireles-Rivera, Gloria Maria Molina-Salinas, Hector Montoya-Fuentes, , Mario Mujica-Sanchez, Jose Esteban Muñoz-Medina, Jose de Jesus Nuñez-Contreras, Alicia Ocaña-Mondragon, Luis Alberto Ochoa-Carrera, Hector Esteban Paz-Juarez, Marissa Perez-Garcia, Francisco Pulido, Helen Haydee Fernanda Ramirez-Plascencia, Jorge Salas-Hernandez, Angel Gustavo Salas-Lais, Alejandro Sanchez-Flores, Clara Esperanza Santacruz-Tinoco, Maria Guadalupe Santiago-Mauricio, Selene Zarate, Nelly Selem-Mojica, Blanca Taboada, Gloria Vazquez, Joel Armando Vazquez-Perez. |
| EPI_ISL_2671569 | Laboratorio Central de Epidemiologia (LCE) | Unidad de Genomica Avanzada | Consorcio Mexicano de Vigilancia Genomica (CoViGen-Mex). Authors (in alphabetical order): Julio Elias Alvarado-Yaah, Carlos F. Arias, Santiago avila-Rios, Eduardo Becerril-Vargas, Victor Hugo Borja-Aburto, Celia Boukadida, Cristobal Chaidez-Quiroz, Juan Bautista Chale-Dzul, Ricardo Ciria Merce, Andreu Comas-Garcia, Celida Duque Molina, Julissa Enciso-Ibarra, Jose Antonio Enciso-Moreno, Gloria Elena Espinosa-Ayala, Fernando Fontove-Herrera, Daniel Fregoso-Rueda, Victor Eduardo Garcia-Arias, Alejandra Garcia-Gasca, Bruno Gomez-Gil, Jean Pierre Gonzalez, Irvin Gonzalez-Lopez, Concepcion Grajales-Muñiz, Ricardo Grande, Rosa Maria Gutierrez Rios, Jesus Hernandez, Alejandra Hernandez-Teran, Alfredo Herrera-Estrella, Carla Ivon Herrera-Najera, Pavel Isa, Veronica Mata-Haro, Daniel Lira Morales, Susana Lopez, Antonio Loza Roman, Brenda Irasema Maldonado-Meza, Bernardo Martinez-Miguel, Jose Arturo Martinez-Orozco, Celida Martinez- Rodriguez, Margarita Matias-Florentino, Fidencio Mejia-Nepomuceno, Maria Guadalupe de Jesus Mireles-Rivera, Gloria Maria Molina-Salinas, Hector Montoya-Fuentes, , Mario Mujica-Sanchez, Jose Esteban Muñoz-Medina, Jose de Jesus Nuñez-Contreras, Alicia Ocaña-Mondragon, Luis Alberto Ochoa-Carrera, Hector Esteban Paz-Juarez, Marissa Perez-Garcia, Francisco Pulido, Helen Haydee Fernanda Ramirez-Plascencia, Jorge Salas-Hernandez, Angel Gustavo Salas-Lais, Alejandro Sanchez-Flores, Clara Esperanza Santacruz-Tinoco, Maria Guadalupe Santiago-Mauricio, Selene Zarate, Nelly Selem-Mojica, Blanca Taboada, Gloria Vazquez, Joel Armando Vazquez-Perez. |
| EPI_ISL_2671570, EPI_ISL_2671571, EPI_ISL_2671572, EPI_ISL_2671573, EPI_ISL_2671574, EPI_ISL_2671575, EPI_ISL_2671576, EPI_ISL_2671577, EPI_ISL_2671578, EPI_ISL_2671579, EPI_ISL_2671580, EPI_ISL_2671581, EPI_ISL_2671582, EPI_ISL_2671583, EPI_ISL_2671584, EPI_ISL_2671585, EPI_ISL_2671586 | | | |
| see above | Centro de Investigacion Biomedica de Occidente (CIBO) | Unidad de Genomica Avanzada | Consorcio Mexicano de Vigilancia Genomica (CoViGen-Mex). Authors (in alphabetical order): Julio Elias Alvarado-Yaah, Carlos F. Arias, Santiago avila-Rios, Eduardo Becerril-Vargas, Victor Hugo Borja-Aburto, Celia Boukadida, Cristobal Chaidez-Quiroz, Juan Bautista Chale-Dzul, Ricardo Ciria Merce, Andreu Comas-Garcia, Celida Duque Molina, Julissa Enciso-Ibarra, Jose Antonio Enciso-Moreno, Gloria Elena Espinosa-Ayala, Fernando Fontove-Herrera, Daniel Fregoso-Rueda, Victor Eduardo Garcia-Arias, Alejandra Garcia-Gasca, Bruno Gomez-Gil, Jean Pierre Gonzalez, Irvin Gonzalez-Lopez, Concepcion Grajales-Muñiz, Ricardo Grande, Rosa Maria Gutierrez Rios, Jesus Hernandez, Alejandra Hernandez-Teran, Alfredo Herrera-Estrella, Carla Ivon Herrera-Najera, Pavel Isa, Veronica Mata-Haro, Daniel Lira Morales, Susana Lopez, Antonio Loza Roman, Brenda Irasema Maldonado-Meza, Bernardo Martinez-Miguel, Jose Arturo Martinez-Orozco, Celida Martinez- Rodriguez, Margarita Matias-Florentino, Fidencio Mejia-Nepomuceno, Maria Guadalupe de Jesus Mireles-Rivera, Gloria Maria Molina-Salinas, Hector Montoya-Fuentes, , Mario Mujica-Sanchez, Jose Esteban Muñoz-Medina, Jose de Jesus Nuñez-Contreras, Alicia Ocaña-Mondragon, Luis Alberto Ochoa-Carrera, Hector Esteban Paz-Juarez, Marissa Perez-Garcia, Francisco Pulido, Helen Haydee Fernanda Ramirez-Plascencia, Jorge Salas-Hernandez, Angel Gustavo Salas-Lais, Alejandro Sanchez-Flores, Clara Esperanza Santacruz-Tinoco, Maria Guadalupe Santiago-Mauricio, Selene Zarate, Nelly Selem-Mojica, Blanca Taboada, Gloria Vazquez, Joel Armando Vazquez-Perez. |
| EPI_ISL_2671587, EPI_ISL_2671588, EPI_ISL_2671589, EPI_ISL_2671590, EPI_ISL_2671591, EPI_ISL_2671592, EPI_ISL_2671593, EPI_ISL_2671594, EPI_ISL_2671595, EPI_ISL_2671596, EPI_ISL_2671597, EPI_ISL_2671598, EPI_ISL_2671599, EPI_ISL_2671600, EPI_ISL_2671601, EPI_ISL_2671602, EPI_ISL_2671603, EPI_ISL_2671604, EPI_ISL_2671605, EPI_ISL_2671606, EPI_ISL_2671607 | | | |
| see above | Laboratorio Central de Epidemiologia (LCE) | Unidad de Genomica Avanzada | Consorcio Mexicano de Vigilancia Genomica (CoViGen-Mex). Authors (in alphabetical order): Julio Elias Alvarado-Yaah, Carlos F. Arias, Santiago avila-Rios, Eduardo Becerril-Vargas, Victor Hugo Borja-Aburto, Celia Boukadida, Cristobal Chaidez-Quiroz, Juan Bautista Chale-Dzul, Ricardo Ciria Merce, Andreu Comas-Garcia, Celida Duque Molina, Julissa Enciso-Ibarra, Jose Antonio Enciso-Moreno, Gloria Elena Espinosa-Ayala, Fernando Fontove-Herrera, Daniel Fregoso-Rueda, Victor Eduardo Garcia-Arias, Alejandra Garcia-Gasca, Bruno Gomez-Gil, Jean Pierre Gonzalez, Irvin Gonzalez-Lopez, Concepcion Grajales-Muñiz, Ricardo Grande, Rosa Maria Gutierrez Rios, Jesus Hernandez, Alejandra Hernandez-Teran, Alfredo Herrera-Estrella, Carla Ivon Herrera-Najera, Pavel Isa, Veronica Mata-Haro, Daniel Lira Morales, Susana Lopez, Antonio Loza Roman, Brenda Irasema Maldonado-Meza, Bernardo Martinez-Miguel, Jose Arturo Martinez-Orozco, Celida Martinez- Rodriguez, Margarita Matias-Florentino, Fidencio Mejia-Nepomuceno, Maria Guadalupe de Jesus Mireles-Rivera, Gloria Maria Molina-Salinas, Hector Montoya-Fuentes, , Mario Mujica-Sanchez, Jose Esteban Muñoz-Medina, Jose de Jesus Nuñez-Contreras, Alicia Ocaña-Mondragon, Luis Alberto Ochoa-Carrera, Hector Esteban Paz-Juarez, Marissa Perez-Garcia, Francisco Pulido, Helen Haydee Fernanda Ramirez-Plascencia, Jorge Salas-Hernandez, Angel Gustavo Salas-Lais, Alejandro Sanchez-Flores, Clara Esperanza Santacruz-Tinoco, Maria Guadalupe Santiago-Mauricio, Selene Zarate, Nelly Selem-Mojica, Blanca Taboada, Gloria Vazquez, Joel Armando Vazquez-Perez. |
| EPI_ISL_2671608, EPI_ISL_2671609, EPI_ISL_2671610, EPI_ISL_2671611, EPI_ISL_2671612 | Centro de Investigacion Biomedica de Occidente (CIBO) | Unidad de Genomica Avanzada | Consorcio Mexicano de Vigilancia Genomica (CoViGen-Mex). Authors (in alphabetical order): Julio Elias Alvarado-Yaah, Carlos F. Arias, Santiago avila-Rios, Eduardo Becerril-Vargas, Victor Hugo Borja-Aburto, Celia Boukadida, Cristobal Chaidez-Quiroz, Juan Bautista Chale-Dzul, Ricardo Ciria Merce, Andreu Comas-Garcia, Celida Duque Molina, Julissa Enciso-Ibarra, Jose Antonio Enciso-Moreno, Gloria Elena Espinosa-Ayala, Fernando Fontove-Herrera, Daniel Fregoso-Rueda, Victor Eduardo Garcia-Arias, Alejandra Garcia-Gasca, Bruno Gomez-Gil, Jean Pierre Gonzalez, Irvin Gonzalez-Lopez, Concepcion Grajales-Muñiz, Ricardo Grande, Rosa Maria Gutierrez Rios, Jesus Hernandez, Alejandra Hernandez-Teran, Alfredo Herrera-Estrella, Carla Ivon Herrera-Najera, Pavel Isa, Veronica Mata-Haro, Daniel Lira Morales, Susana Lopez, Antonio Loza Roman, Brenda Irasema Maldonado-Meza, Bernardo Martinez-Miguel, Jose Arturo Martinez-Orozco, Celida Martinez- Rodriguez, Margarita Matias-Florentino, Fidencio Mejia-Nepomuceno, Maria Guadalupe de Jesus Mireles-Rivera, Gloria Maria Molina-Salinas, Hector Montoya-Fuentes, , Mario Mujica-Sanchez, Jose Esteban Muñoz-Medina, Jose de Jesus Nuñez-Contreras, Alicia Ocaña-Mondragon, Luis Alberto Ochoa-Carrera, Hector Esteban Paz-Juarez, Marissa Perez-Garcia, Francisco Pulido, Helen Haydee Fernanda Ramirez-Plascencia, Jorge Salas-Hernandez, Angel Gustavo Salas-Lais, Alejandro Sanchez-Flores, Clara Esperanza Santacruz-Tinoco, Maria Guadalupe Santiago-Mauricio, Selene Zarate, Nelly Selem-Mojica, Blanca Taboada, Gloria Vazquez, Joel Armando Vazquez-Perez. |
| EPI_ISL_2671613, EPI_ISL_2671614, EPI_ISL_2671615 | Laboratorio Central de Epidemiologia (LCE) | Unidad de Genomica Avanzada | Consorcio Mexicano de Vigilancia Genomica (CoViGen-Mex). Authors (in alphabetical order): Julio Elias Alvarado-Yaah, Carlos F. Arias, Santiago avila-Rios, Eduardo Becerril-Vargas, Victor Hugo Borja-Aburto, Celia Boukadida, Cristobal Chaidez-Quiroz, Juan Bautista Chale-Dzul, Ricardo Ciria Merce, Andreu Comas-Garcia, Celida Duque Molina, Julissa Enciso-Ibarra, Jose Antonio Enciso-Moreno, Gloria Elena Espinosa-Ayala, Fernando Fontove-Herrera, Daniel Fregoso-Rueda, Victor Eduardo Garcia-Arias, Alejandra Garcia-Gasca, Bruno Gomez-Gil, Jean Pierre Gonzalez, Irvin Gonzalez-Lopez, Concepcion Grajales-Muñiz, Ricardo Grande, Rosa Maria Gutierrez Rios, Jesus Hernandez, Alejandra Hernandez-Teran, Alfredo Herrera-Estrella, Carla Ivon Herrera-Najera, Pavel Isa, Veronica Mata-Haro, Daniel Lira Morales, Susana Lopez, Antonio Loza Roman, Brenda Irasema Maldonado-Meza, Bernardo Martinez-Miguel, Jose Arturo Martinez-Orozco, Celida Martinez- Rodriguez, Margarita Matias-Florentino, Fidencio Mejia-Nepomuceno, Maria Guadalupe de Jesus Mireles-Rivera, Gloria Maria Molina-Salinas, Hector Montoya-Fuentes, , Mario Mujica-Sanchez, Jose Esteban Muñoz-Medina, Jose de Jesus Nuñez-Contreras, Alicia Ocaña-Mondragon, Luis Alberto Ochoa-Carrera, Hector Esteban Paz-Juarez, Marissa Perez-Garcia, Francisco Pulido, Helen Haydee Fernanda Ramirez-Plascencia, Jorge Salas-Hernandez, Angel Gustavo Salas-Lais, Alejandro Sanchez-Flores, Clara Esperanza Santacruz-Tinoco, Maria Guadalupe Santiago-Mauricio, Selene Zarate, Nelly Selem-Mojica, Blanca Taboada, Gloria Vazquez, Joel Armando Vazquez-Perez. |
| EPI_ISL_2671616, EPI_ISL_2671617, EPI_ISL_2671618 | Centro de Investigacion Biomedica de Occidente (CIBO) | Unidad de Genomica Avanzada | Consorcio Mexicano de Vigilancia Genomica (CoViGen-Mex). Authors (in alphabetical order): Julio Elias Alvarado-Yaah, Carlos F. Arias, Santiago avila-Rios, Eduardo Becerril-Vargas, Victor Hugo Borja-Aburto, Celia Boukadida, Cristobal Chaidez-Quiroz, Juan Bautista Chale-Dzul, Ricardo Ciria Merce, Andreu Comas-Garcia, Celida Duque Molina, Julissa Enciso-Ibarra, Jose Antonio Enciso-Moreno, Gloria Elena Espinosa-Ayala, Fernando Fontove-Herrera, Daniel Fregoso-Rueda, Victor Eduardo Garcia-Arias, Alejandra Garcia-Gasca, Bruno Gomez-Gil, Jean Pierre Gonzalez, Irvin Gonzalez-Lopez, Concepcion Grajales-Muñiz, Ricardo Grande, Rosa Maria Gutierrez Rios, Jesus Hernandez, Alejandra Hernandez-Teran, Alfredo Herrera-Estrella, Carla Ivon Herrera-Najera, Pavel Isa, Veronica Mata-Haro, Daniel Lira Morales, Susana Lopez, Antonio Loza Roman, Brenda Irasema Maldonado-Meza, Bernardo Martinez-Miguel, Jose Arturo Martinez-Orozco, Celida Martinez- Rodriguez, Margarita Matias-Florentino, Fidencio Mejia-Nepomuceno, Maria Guadalupe de Jesus Mireles-Rivera, Gloria Maria Molina-Salinas, Hector Montoya-Fuentes, , Mario Mujica-Sanchez, Jose Esteban Muñoz-Medina, Jose de Jesus Nuñez-Contreras, Alicia Ocaña-Mondragon, Luis Alberto Ochoa-Carrera, Hector Esteban Paz-Juarez, Marissa Perez-Garcia, Francisco Pulido, Helen Haydee Fernanda Ramirez-Plascencia, Jorge Salas-Hernandez, Angel Gustavo Salas-Lais, Alejandro Sanchez-Flores, Clara Esperanza Santacruz-Tinoco, Maria Guadalupe Santiago-Mauricio, Selene Zarate, Nelly Selem-Mojica, Blanca Taboada, Gloria Vazquez, Joel Armando Vazquez-Perez. |
| EPI_ISL_2671619, EPI_ISL_2671620, EPI_ISL_2671621, EPI_ISL_2671622, EPI_ISL_2671623, EPI_ISL_2671624, EPI_ISL_2671625, EPI_ISL_2671626, EPI_ISL_2671627, EPI_ISL_2671628, EPI_ISL_2671629, EPI_ISL_2671630, EPI_ISL_2671631, EPI_ISL_2671632, EPI_ISL_2671633, EPI_ISL_2671634, EPI_ISL_2671635, EPI_ISL_2671636 | | | |
| see above | Centro de Investigacion Biomedica del Noreste (CIBIN) | Unidad de Genomica Avanzada | Consorcio Mexicano de Vigilancia Genomica (CoViGen-Mex). Authors (in alphabetical order): Julio Elias Alvarado-Yaah, Carlos F. Arias, Santiago avila-Rios, Eduardo Becerril-Vargas, Victor Hugo Borja-Aburto, Celia Boukadida, Cristobal Chaidez-Quiroz, Juan Bautista Chale-Dzul, Ricardo Ciria Merce, Andreu Comas-Garcia, Celida Duque Molina, Julissa Enciso-Ibarra, Jose Antonio Enciso-Moreno, Gloria Elena Espinosa-Ayala, Fernando Fontove-Herrera, Daniel Fregoso-Rueda, Victor Eduardo Garcia-Arias, Alejandra Garcia-Gasca, Bruno Gomez-Gil, Jean Pierre Gonzalez, Irvin Gonzalez-Lopez, Concepcion Grajales-Muñiz, Ricardo Grande, Rosa Maria Gutierrez Rios, Jesus Hernandez, Alejandra Hernandez-Teran, Alfredo Herrera-Estrella, Carla Ivon Herrera-Najera, Pavel Isa, Veronica Mata-Haro, Daniel Lira Morales, Susana Lopez, Antonio Loza Roman, Brenda Irasema Maldonado-Meza, Bernardo Martinez-Miguel, Jose Arturo Martinez-Orozco, Celida Martinez- Rodriguez, Margarita Matias-Florentino, Fidencio Mejia-Nepomuceno, Maria Guadalupe de Jesus Mireles-Rivera, Gloria Maria Molina-Salinas, Hector Montoya-Fuentes, , Mario Mujica-Sanchez, Jose Esteban Muñoz-Medina, Jose de Jesus Nuñez-Contreras, Alicia Ocaña-Mondragon, Luis Alberto Ochoa-Carrera, Hector Esteban Paz-Juarez, Marissa Perez-Garcia, Francisco Pulido, Helen Haydee Fernanda Ramirez-Plascencia, Jorge Salas-Hernandez, Angel Gustavo Salas-Lais, Alejandro Sanchez-Flores, Clara Esperanza Santacruz-Tinoco, Maria Guadalupe Santiago-Mauricio, Selene Zarate, Nelly Selem-Mojica, Blanca Taboada, Gloria Vazquez, Joel Armando Vazquez-Perez. |
| EPI_ISL_2671637, EPI_ISL_2671638, EPI_ISL_2671639, EPI_ISL_2671640, EPI_ISL_2671641, EPI_ISL_2671642, EPI_ISL_2671643, EPI_ISL_2671644, EPI_ISL_2671645, EPI_ISL_2671646, EPI_ISL_2671647, EPI_ISL_2671648, EPI_ISL_2671649, EPI_ISL_2671650, EPI_ISL_2671651, EPI_ISL_2671652 | | | |
| see above | Laboratorio Central de Epidemiologia (LCE) | Unidad de Genomica Avanzada | Consorcio Mexicano de Vigilancia Genomica (CoViGen-Mex). Authors (in alphabetical order): Julio Elias Alvarado-Yaah, Carlos F. Arias, Santiago avila-Rios, Eduardo Becerril-Vargas, Victor Hugo Borja-Aburto, Celia Boukadida, Cristobal Chaidez-Quiroz, Juan Bautista Chale-Dzul, Ricardo Ciria Merce, Andreu Comas-Garcia, Celida Duque Molina, Julissa Enciso-Ibarra, Jose Antonio Enciso-Moreno, Gloria Elena Espinosa-Ayala, Fernando Fontove-Herrera, Daniel Fregoso-Rueda, Victor Eduardo Garcia-Arias, Alejandra Garcia-Gasca, Bruno Gomez-Gil, Jean Pierre Gonzalez, Irvin Gonzalez-Lopez, Concepcion Grajales-Muñiz, Ricardo Grande, Rosa Maria Gutierrez Rios, Jesus Hernandez, Alejandra Hernandez-Teran, Alfredo Herrera-Estrella, Carla Ivon Herrera-Najera, Pavel Isa, Veronica Mata-Haro, Daniel Lira Morales, Susana Lopez, Antonio Loza Roman, Brenda Irasema Maldonado-Meza, Bernardo Martinez-Miguel, Jose Arturo Martinez-Orozco, Celida Martinez- Rodriguez, Margarita Matias-Florentino, Fidencio Mejia-Nepomuceno, Maria Guadalupe de Jesus Mireles-Rivera, Gloria Maria Molina-Salinas, Hector Montoya-Fuentes, , Mario Mujica-Sanchez, Jose Esteban Muñoz-Medina, Jose de Jesus Nuñez-Contreras, Alicia Ocaña-Mondragon, Luis Alberto Ochoa-Carrera, Hector Esteban Paz-Juarez, Marissa Perez-Garcia, Francisco Pulido, Helen Haydee Fernanda Ramirez-Plascencia, Jorge Salas-Hernandez, Angel Gustavo Salas-Lais, Alejandro Sanchez-Flores, Clara Esperanza Santacruz-Tinoco, Maria Guadalupe Santiago-Mauricio, Selene Zarate, Nelly Selem-Mojica, Blanca Taboada, Gloria Vazquez, Joel Armando Vazquez-Perez. |
| EPI_ISL_2671653, EPI_ISL_2671654, EPI_ISL_2671655, EPI_ISL_2671656, EPI_ISL_2671657, EPI_ISL_2671658, EPI_ISL_2671659, EPI_ISL_2671660, EPI_ISL_2671661, EPI_ISL_2671662, EPI_ISL_2671663, EPI_ISL_2671664, EPI_ISL_2671665, EPI_ISL_2671666, EPI_ISL_2671667, EPI_ISL_2671668, EPI_ISL_2671669, EPI_ISL_2671670, EPI_ISL_2671671, EPI_ISL_2671672, EPI_ISL_2671673, EPI_ISL_2671674, EPI_ISL_2671675, EPI_ISL_2671676, EPI_ISL_2671677, EPI_ISL_2671678, EPI_ISL_2671679, EPI_ISL_2671680, EPI_ISL_2671681, EPI_ISL_2671682, EPI_ISL_2671683, EPI_ISL_2671684, EPI_ISL_2671685, EPI_ISL_2671686, EPI_ISL_2671687, EPI_ISL_2671688, EPI_ISL_2671689, EPI_ISL_2671690, EPI_ISL_2671691, EPI_ISL_2671692, EPI_ISL_2671693, EPI_ISL_2671694, EPI_ISL_2671695, EPI_ISL_2671696, EPI_ISL_2671697 | | | |
| see above | Unidad de Investigacion Medica de Yucatan (UIMY) | Unidad de Genomica Avanzada | Consorcio Mexicano de Vigilancia Genomica (CoViGen-Mex). Authors (in alphabetical order): Julio Elias Alvarado-Yaah, Carlos F. Arias, Santiago avila-Rios, Eduardo Becerril-Vargas, Victor Hugo Borja-Aburto, Celia Boukadida, Cristobal Chaidez-Quiroz, Juan Bautista Chale-Dzul, Ricardo Ciria Merce, Andreu Comas-Garcia, Celida Duque Molina, Julissa Enciso-Ibarra, Jose Antonio Enciso-Moreno, Gloria Elena Espinosa-Ayala, Fernando Fontove-Herrera, Daniel Fregoso-Rueda, Victor Eduardo Garcia-Arias, Alejandra Garcia-Gasca, Bruno Gomez-Gil, Jean Pierre Gonzalez, Irvin Gonzalez-Lopez, Concepcion Grajales-Muñiz, Ricardo Grande, Rosa Maria Gutierrez Rios, Jesus Hernandez, Alejandra Hernandez-Teran, Alfredo Herrera-Estrella, Carla Ivon Herrera-Najera, Pavel Isa, Veronica Mata-Haro, Daniel Lira Morales, Susana Lopez, Antonio Loza Roman, Brenda Irasema Maldonado-Meza, Bernardo Martinez-Miguel, Jose Arturo Martinez-Orozco, Celida Martinez- Rodriguez, Margarita Matias-Florentino, Fidencio Mejia-Nepomuceno, Maria Guadalupe de Jesus Mireles-Rivera, Gloria Maria Molina-Salinas, Hector Montoya-Fuentes, , Mario Mujica-Sanchez, Jose Esteban Muñoz-Medina, Jose de Jesus Nuñez-Contreras, Alicia Ocaña-Mondragon, Luis Alberto Ochoa-Carrera, Hector Esteban Paz-Juarez, Marissa Perez-Garcia, Francisco Pulido, Helen Haydee Fernanda Ramirez-Plascencia, Jorge Salas-Hernandez, Angel Gustavo Salas-Lais, Alejandro Sanchez-Flores, Clara Esperanza Santacruz-Tinoco, Maria Guadalupe Santiago-Mauricio, Selene Zarate, Nelly Selem-Mojica, Blanca Taboada, Gloria Vazquez, Joel Armando Vazquez-Perez. |
| EPI_ISL_2671698, EPI_ISL_2671699 | Centro de Investigacion Biomedica de Occidente (CIBO) | Unidad de Genomica Avanzada | Consorcio Mexicano de Vigilancia Genomica (CoViGen-Mex). Authors (in alphabetical order): Julio Elias Alvarado-Yaah, Carlos F. Arias, Santiago avila-Rios, Eduardo Becerril-Vargas, Victor Hugo Borja-Aburto, Celia Boukadida, Cristobal Chaidez-Quiroz, Juan Bautista Chale-Dzul, Ricardo Ciria Merce, Andreu Comas-Garcia, Celida Duque Molina, Julissa Enciso-Ibarra, Jose Antonio Enciso-Moreno, Gloria Elena Espinosa-Ayala, Fernando Fontove-Herrera, Daniel Fregoso-Rueda, Victor Eduardo Garcia-Arias, Alejandra Garcia-Gasca, Bruno Gomez-Gil, Jean Pierre Gonzalez, Irvin Gonzalez-Lopez, Concepcion Grajales-Muñiz, Ricardo Grande, Rosa Maria Gutierrez Rios, Jesus Hernandez, Alejandra Hernandez-Teran, Alfredo Herrera-Estrella, Carla Ivon Herrera-Najera, Pavel Isa, Veronica Mata-Haro, Daniel Lira Morales, Susana Lopez, Antonio Loza Roman, Brenda Irasema Maldonado-Meza, Bernardo Martinez-Miguel, Jose Arturo Martinez-Orozco, Celida Martinez- Rodriguez, Margarita Matias-Florentino, Fidencio Mejia-Nepomuceno, Maria Guadalupe de Jesus Mireles-Rivera, Gloria Maria Molina-Salinas, Hector Montoya-Fuentes, , Mario Mujica-Sanchez, Jose Esteban Muñoz-Medina, Jose de Jesus Nuñez-Contreras, Alicia Ocaña-Mondragon, Luis Alberto Ochoa-Carrera, Hector Esteban Paz-Juarez, Marissa Perez-Garcia, Francisco Pulido, Helen Haydee Fernanda Ramirez-Plascencia, Jorge Salas-Hernandez, Angel Gustavo Salas-Lais, Alejandro Sanchez-Flores, Clara Esperanza Santacruz-Tinoco, Maria Guadalupe Santiago-Mauricio, Selene Zarate, Nelly Selem-Mojica, Blanca Taboada, Gloria Vazquez, Joel Armando Vazquez-Perez. |
| EPI_ISL_2671700, EPI_ISL_2671701, EPI_ISL_2671702 | Unidad de Investigacion Biomedica de Zacatecas (UIBZ) | Unidad de Genomica Avanzada | Consorcio Mexicano de Vigilancia Genomica (CoViGen-Mex). Authors (in alphabetical order): Julio Elias Alvarado-Yaah, Carlos F. Arias, Santiago avila-Rios, Eduardo Becerril-Vargas, Victor Hugo Borja-Aburto, Celia Boukadida, Cristobal Chaidez-Quiroz, Juan Bautista Chale-Dzul, Ricardo Ciria Merce, Andreu Comas-Garcia, Celida Duque Molina, Julissa Enciso-Ibarra, Jose Antonio Enciso-Moreno, Gloria Elena Espinosa-Ayala, Fernando Fontove-Herrera, Daniel Fregoso-Rueda, Victor Eduardo Garcia-Arias, Alejandra Garcia-Gasca, Bruno Gomez-Gil, Jean Pierre Gonzalez, Irvin Gonzalez-Lopez, Concepcion Grajales-Muñiz, Ricardo Grande, Rosa Maria Gutierrez Rios, Jesus Hernandez, Alejandra Hernandez-Teran, Alfredo Herrera-Estrella, Carla Ivon Herrera-Najera, Pavel Isa, Veronica Mata-Haro, Daniel Lira Morales, Susana Lopez, Antonio Loza Roman, Brenda Irasema Maldonado-Meza, Bernardo Martinez-Miguel, Jose Arturo Martinez-Orozco, Celida Martinez- Rodriguez, Margarita Matias-Florentino, Fidencio Mejia-Nepomuceno, Maria Guadalupe de Jesus Mireles-Rivera, Gloria Maria Molina-Salinas, Hector Montoya-Fuentes, , Mario Mujica-Sanchez, Jose Esteban Muñoz-Medina, Jose de Jesus Nuñez-Contreras, Alicia Ocaña-Mondragon, Luis Alberto Ochoa-Carrera, Hector Esteban Paz-Juarez, Marissa Perez-Garcia, Francisco Pulido, Helen Haydee Fernanda Ramirez-Plascencia, Jorge Salas-Hernandez, Angel Gustavo Salas-Lais, Alejandro Sanchez-Flores, Clara Esperanza Santacruz-Tinoco, Maria Guadalupe Santiago-Mauricio, Selene Zarate, Nelly Selem-Mojica, Blanca Taboada, Gloria Vazquez, Joel Armando Vazquez-Perez. |
| EPI_ISL_2671703 | Centro de Investigacion Biomedica de Occidente (CIBO) | Unidad de Genomica Avanzada | Consorcio Mexicano de Vigilancia Genomica (CoViGen-Mex). Authors (in alphabetical order): Julio Elias Alvarado-Yaah, Carlos F. Arias, Santiago avila-Rios, Eduardo Becerril-Vargas, Victor Hugo Borja-Aburto, Celia Boukadida, Cristobal Chaidez-Quiroz, Juan Bautista Chale-Dzul, Ricardo Ciria Merce, Andreu Comas-Garcia, Celida Duque Molina, Julissa Enciso-Ibarra, Jose Antonio Enciso-Moreno, Gloria Elena Espinosa-Ayala, Fernando Fontove-Herrera, Daniel Fregoso-Rueda, Victor Eduardo Garcia-Arias, Alejandra Garcia-Gasca, Bruno Gomez-Gil, Jean Pierre Gonzalez, Irvin Gonzalez-Lopez, Concepcion Grajales-Muñiz, Ricardo Grande, Rosa Maria Gutierrez Rios, Jesus Hernandez, Alejandra Hernandez-Teran, Alfredo Herrera-Estrella, Carla Ivon Herrera-Najera, Pavel Isa, Veronica Mata-Haro, Daniel Lira Morales, Susana Lopez, Antonio Loza Roman, Brenda Irasema Maldonado-Meza, Bernardo Martinez-Miguel, Jose Arturo Martinez-Orozco, Celida Martinez- Rodriguez, Margarita Matias-Florentino, Fidencio Mejia-Nepomuceno, Maria Guadalupe de Jesus Mireles-Rivera, Gloria Maria Molina-Salinas, Hector Montoya-Fuentes, , Mario Mujica-Sanchez, Jose Esteban Muñoz-Medina, Jose de Jesus Nuñez-Contreras, Alicia Ocaña-Mondragon, Luis Alberto Ochoa-Carrera, Hector Esteban Paz-Juarez, Marissa Perez-Garcia, Francisco Pulido, Helen Haydee Fernanda Ramirez-Plascencia, Jorge Salas-Hernandez, Angel Gustavo Salas-Lais, Alejandro Sanchez-Flores, Clara Esperanza Santacruz-Tinoco, Maria Guadalupe Santiago-Mauricio, Selene Zarate, Nelly Selem-Mojica, Blanca Taboada, Gloria Vazquez, Joel Armando Vazquez-Perez. |
| EPI_ISL_2671704, EPI_ISL_2671705, EPI_ISL_2671706, EPI_ISL_2671707, EPI_ISL_2671708, EPI_ISL_2671709, EPI_ISL_2671710, EPI_ISL_2671711, EPI_ISL_2671712, EPI_ISL_2671713, EPI_ISL_2671714, EPI_ISL_2671715 | | | |
| see above | Unidad de Investigacion Medica de Yucatan (UIMY) | Unidad de Genomica Avanzada | Consorcio Mexicano de Vigilancia Genomica (CoViGen-Mex). Authors (in alphabetical order): Julio Elias Alvarado-Yaah, Carlos F. Arias, Santiago avila-Rios, Eduardo Becerril-Vargas, Victor Hugo Borja-Aburto, Celia Boukadida, Cristobal Chaidez-Quiroz, Juan Bautista Chale-Dzul, Ricardo Ciria Merce, Andreu Comas-Garcia, Celida Duque Molina, Julissa Enciso-Ibarra, Jose Antonio Enciso-Moreno, Gloria Elena Espinosa-Ayala, Fernando Fontove-Herrera, Daniel Fregoso-Rueda, Victor Eduardo Garcia-Arias, Alejandra Garcia-Gasca, Bruno Gomez-Gil, Jean Pierre Gonzalez, Irvin Gonzalez-Lopez, Concepcion Grajales-Muñiz, Ricardo Grande, Rosa Maria Gutierrez Rios, Jesus Hernandez, Alejandra Hernandez-Teran, Alfredo Herrera-Estrella, Carla Ivon Herrera-Najera, Pavel Isa, Veronica Mata-Haro, Daniel Lira Morales, Susana Lopez, Antonio Loza Roman, Brenda Irasema Maldonado-Meza, Bernardo Martinez-Miguel, Jose Arturo Martinez-Orozco, Celida Martinez- Rodriguez, Margarita Matias-Florentino, Fidencio Mejia-Nepomuceno, Maria Guadalupe de Jesus Mireles-Rivera, Gloria Maria Molina-Salinas, Hector Montoya-Fuentes, , Mario Mujica-Sanchez, Jose Esteban Muñoz-Medina, Jose de Jesus Nuñez-Contreras, Alicia Ocaña-Mondragon, Luis Alberto Ochoa-Carrera, Hector Esteban Paz-Juarez, Marissa Perez-Garcia, Francisco Pulido, Helen Haydee Fernanda Ramirez-Plascencia, Jorge Salas-Hernandez, Angel Gustavo Salas-Lais, Alejandro Sanchez-Flores, Clara Esperanza Santacruz-Tinoco, Maria Guadalupe Santiago-Mauricio, Selene Zarate, Nelly Selem-Mojica, Blanca Taboada, Gloria Vazquez, Joel Armando Vazquez-Perez. |
| EPI_ISL_2671716, | Centro de Investigacion Biomedica del | Unidad de Genomica Avanzada | Consorcio Mexicano de Vigilancia Genomica (CoViGen-Mex). Authors (in alphabetical order): Julio Elias Alvarado-Yaah, Carlos F. Arias, Santiago avila-Rios, Eduardo Becerril-Vargas, Victor Hugo Borja-Aburto, Celia Boukadida, Cristobal Chaidez-Quiroz, Juan Bautista Chale-Dzul, Ricardo |

| | | | |
|---|---|---|---|
| EPI_ISL_2671717, EPI_ISL_2671718, EPI_ISL_2671719, EPI_ISL_2671720, EPI_ISL_2671721, EPI_ISL_2671722 | Noreste (CIBIN) | | Ciria Merce, Andreu Comas-Garcia, Celida Duque Molina, Julissa Enciso-Ibarra, Jose Antonio Enciso-Moreno, Gloria Elena Espinosa-Ayala, Fernando Fontove-Herrera, Daniel Fregoso-Rueda, Victor Eduardo Garcia-Arias, Alejandra Garcia-Gasca, Bruno Gomez-Gil, Jean Pierre Gonzalez, Irvin Gonzalez-Lopez, Concepcion Grajales-Muñiz, Ricardo Grande, Rosa Maria Gutierrez Rios, Jesus Hernandez, Alejandra Hernandez-Teran, Alfredo Herrera-Estrella, Carla Ivon Herrera-Najera, Pavel Isa, Veronica Mata-Haro, Daniel Lira Morales, Susana Lopez, Antonio Loza Roman, Brenda Irasema Maldonado-Meza, Bernardo Martinez-Miguel, Jose Arturo Martinez-Orozco, Celida Martinez- Rodriguez, Margarita Matias-Florentino, Fidencio Mejia-Nepomuceno, Maria Guadalupe de Jesus Mireles-Rivera, Gloria Maria Molina-Salinas, Hector Montoya-Fuentes, , Mario Mujica-Sanchez, Jose Esteban Muñoz-Medina, Jose de Jesus Nuñez-Contreras, Alicia Ocaña-Mondragon, Luis Alberto Ochoa-Carrera, Hector Esteban Paz-Juarez, Marissa Perez-Garcia, Francisco Pulido, Helen Haydee Fernanda Ramirez-Plascencia, Jorge Salas-Hernandez, Angel Gustavo Salas-Lais, Alejandro Sanchez-Flores, Clara Esperanza Santacruz-Tinoco, Maria Guadalupe Santiago-Mauricio, Selene Zarate, Nelly Selem-Mojica, Blanca Taboada, Gloria Vazquez, Joel Armando Vazquez-Perez. |
| EPI_ISL_2671723, EPI_ISL_2671724, EPI_ISL_2671725, EPI_ISL_2671726, EPI_ISL_2671727, EPI_ISL_2671728, EPI_ISL_2671729, EPI_ISL_2671730, EPI_ISL_2671731, EPI_ISL_2671732, EPI_ISL_2671733, EPI_ISL_2671734, EPI_ISL_2671735, EPI_ISL_2671736, EPI_ISL_2671737, EPI_ISL_2671738, EPI_ISL_2671739, EPI_ISL_2671740, EPI_ISL_2671741, EPI_ISL_2671742, EPI_ISL_2671743, EPI_ISL_2671744, EPI_ISL_2671745 | | | |
| see above | Laboratorio Central de Epidemiologia (LCE) | Unidad de Genomica Avanzada | Consorcio Mexicano de Vigilancia Genomica (CoViGen-Mex). Authors (in alphabetical order): Julio Elias Alvarado-Yaah, Carlos F. Arias, Santiago avila-Rios, Eduardo Becerril-Vargas, Victor Hugo Borja-Aburto, Celia Boukadida, Cristobal Chaidez-Quiroz, Juan Bautista Chale-Dzul, Ricardo Ciria Merce, Andreu Comas-Garcia, Celida Duque Molina, Julissa Enciso-Ibarra, Jose Antonio Enciso-Moreno, Gloria Elena Espinosa-Ayala, Fernando Fontove-Herrera, Daniel Fregoso-Rueda, Victor Eduardo Garcia-Arias, Alejandra Garcia-Gasca, Bruno Gomez-Gil, Jean Pierre Gonzalez, Irvin Gonzalez-Lopez, Concepcion Grajales-Muñiz, Ricardo Grande, Rosa Maria Gutierrez Rios, Jesus Hernandez, Alejandra Hernandez-Teran, Alfredo Herrera-Estrella, Carla Ivon Herrera-Najera, Pavel Isa, Veronica Mata-Haro, Daniel Lira Morales, Susana Lopez, Antonio Loza Roman, Brenda Irasema Maldonado-Meza, Bernardo Martinez-Miguel, Jose Arturo Martinez-Orozco, Celida Martinez- Rodriguez, Margarita Matias-Florentino, Fidencio Mejia-Nepomuceno, Maria Guadalupe de Jesus Mireles-Rivera, Gloria Maria Molina-Salinas, Hector Montoya-Fuentes, , Mario Mujica-Sanchez, Jose Esteban Muñoz-Medina, Jose de Jesus Nuñez-Contreras, Alicia Ocaña-Mondragon, Luis Alberto Ochoa-Carrera, Hector Esteban Paz-Juarez, Marissa Perez-Garcia, Francisco Pulido, Helen Haydee Fernanda Ramirez-Plascencia, Jorge Salas-Hernandez, Angel Gustavo Salas-Lais, Alejandro Sanchez-Flores, Clara Esperanza Santacruz-Tinoco, Maria Guadalupe Santiago-Mauricio, Selene Zarate, Nelly Selem-Mojica, Blanca Taboada, Gloria Vazquez, Joel Armando Vazquez-Perez. |
| EPI_ISL_2671746, EPI_ISL_2671747, EPI_ISL_2671748, EPI_ISL_2671749, EPI_ISL_2671750, EPI_ISL_2671751, EPI_ISL_2671752, EPI_ISL_2671753, EPI_ISL_2671754, EPI_ISL_2671755, EPI_ISL_2671756, EPI_ISL_2671757 | | | |
| see above | Unidad de Investigacion Medica de Yucatan (UIMY) | Unidad de Genomica Avanzada | Consorcio Mexicano de Vigilancia Genomica (CoViGen-Mex). Authors (in alphabetical order): Julio Elias Alvarado-Yaah, Carlos F. Arias, Santiago avila-Rios, Eduardo Becerril-Vargas, Victor Hugo Borja-Aburto, Celia Boukadida, Cristobal Chaidez-Quiroz, Juan Bautista Chale-Dzul, Ricardo Ciria Merce, Andreu Comas-Garcia, Celida Duque Molina, Julissa Enciso-Ibarra, Jose Antonio Enciso-Moreno, Gloria Elena Espinosa-Ayala, Fernando Fontove-Herrera, Daniel Fregoso-Rueda, Victor Eduardo Garcia-Arias, Alejandra Garcia-Gasca, Bruno Gomez-Gil, Jean Pierre Gonzalez, Irvin Gonzalez-Lopez, Concepcion Grajales-Muñiz, Ricardo Grande, Rosa Maria Gutierrez Rios, Jesus Hernandez, Alejandra Hernandez-Teran, Alfredo Herrera-Estrella, Carla Ivon Herrera-Najera, Pavel Isa, Veronica Mata-Haro, Daniel Lira Morales, Susana Lopez, Antonio Loza Roman, Brenda Irasema Maldonado-Meza, Bernardo Martinez-Miguel, Jose Arturo Martinez-Orozco, Celida Martinez- Rodriguez, Margarita Matias-Florentino, Fidencio Mejia-Nepomuceno, Maria Guadalupe de Jesus Mireles-Rivera, Gloria Maria Molina-Salinas, Hector Montoya-Fuentes, , Mario Mujica-Sanchez, Jose Esteban Muñoz-Medina, Jose de Jesus Nuñez-Contreras, Alicia Ocaña-Mondragon, Luis Alberto Ochoa-Carrera, Hector Esteban Paz-Juarez, Marissa Perez-Garcia, Francisco Pulido, Helen Haydee Fernanda Ramirez-Plascencia, Jorge Salas-Hernandez, Angel Gustavo Salas-Lais, Alejandro Sanchez-Flores, Clara Esperanza Santacruz-Tinoco, Maria Guadalupe Santiago-Mauricio, Selene Zarate, Nelly Selem-Mojica, Blanca Taboada, Gloria Vazquez, Joel Armando Vazquez-Perez. |
| EPI_ISL_2671758, EPI_ISL_2671759, EPI_ISL_2671760, EPI_ISL_2671761, EPI_ISL_2671762, EPI_ISL_2671763, EPI_ISL_2671764, EPI_ISL_2671765, EPI_ISL_2671766, EPI_ISL_2671767, EPI_ISL_2671768, EPI_ISL_2671769, EPI_ISL_2671770, EPI_ISL_2671771, EPI_ISL_2671772, EPI_ISL_2671773, EPI_ISL_2671774, EPI_ISL_2671775, EPI_ISL_2671776, EPI_ISL_2671777, EPI_ISL_2671778, EPI_ISL_2671779, EPI_ISL_2671780 | | | |
| see above | Unidad de Investigacion Biomedica de Zacatecas (UIBZ) | Unidad de Genomica Avanzada | Consorcio Mexicano de Vigilancia Genomica (CoViGen-Mex). Authors (in alphabetical order): Julio Elias Alvarado-Yaah, Carlos F. Arias, Santiago avila-Rios, Eduardo Becerril-Vargas, Victor Hugo Borja-Aburto, Celia Boukadida, Cristobal Chaidez-Quiroz, Juan Bautista Chale-Dzul, Ricardo Ciria Merce, Andreu Comas-Garcia, Celida Duque Molina, Julissa Enciso-Ibarra, Jose Antonio Enciso-Moreno, Gloria Elena Espinosa-Ayala, Fernando Fontove-Herrera, Daniel Fregoso-Rueda, Victor Eduardo Garcia-Arias, Alejandra Garcia-Gasca, Bruno Gomez-Gil, Jean Pierre Gonzalez, Irvin Gonzalez-Lopez, Concepcion Grajales-Muñiz, Ricardo Grande, Rosa Maria Gutierrez Rios, Jesus Hernandez, Alejandra Hernandez-Teran, Alfredo Herrera-Estrella, Carla Ivon Herrera-Najera, Pavel Isa, Veronica Mata-Haro, Daniel Lira Morales, Susana Lopez, Antonio Loza Roman, Brenda Irasema Maldonado-Meza, Bernardo Martinez-Miguel, Jose Arturo Martinez-Orozco, Celida Martinez- Rodriguez, Margarita Matias-Florentino, Fidencio Mejia-Nepomuceno, Maria Guadalupe de Jesus Mireles-Rivera, Gloria Maria Molina-Salinas, Hector Montoya-Fuentes, , Mario Mujica-Sanchez, Jose Esteban Muñoz-Medina, Jose de Jesus Nuñez-Contreras, Alicia Ocaña-Mondragon, Luis Alberto Ochoa-Carrera, Hector Esteban Paz-Juarez, Marissa Perez-Garcia, Francisco Pulido, Helen Haydee Fernanda Ramirez-Plascencia, Jorge Salas-Hernandez, Angel Gustavo Salas-Lais, Alejandro Sanchez-Flores, Clara Esperanza Santacruz-Tinoco, Maria Guadalupe Santiago-Mauricio, Selene Zarate, Nelly Selem-Mojica, Blanca Taboada, Gloria Vazquez, Joel Armando Vazquez-Perez. |
| EPI_ISL_2671781, EPI_ISL_2671782, EPI_ISL_2671783, EPI_ISL_2671784, EPI_ISL_2671785 | Unidad de Investigacion Medica de Yucatan (UIMY) | Unidad de Genomica Avanzada | Consorcio Mexicano de Vigilancia Genomica (CoViGen-Mex). Authors (in alphabetical order): Julio Elias Alvarado-Yaah, Carlos F. Arias, Santiago avila-Rios, Eduardo Becerril-Vargas, Victor Hugo Borja-Aburto, Celia Boukadida, Cristobal Chaidez-Quiroz, Juan Bautista Chale-Dzul, Ricardo Ciria Merce, Andreu Comas-Garcia, Celida Duque Molina, Julissa Enciso-Ibarra, Jose Antonio Enciso-Moreno, Gloria Elena Espinosa-Ayala, Fernando Fontove-Herrera, Daniel Fregoso-Rueda, Victor Eduardo Garcia-Arias, Alejandra Garcia-Gasca, Bruno Gomez-Gil, Jean Pierre Gonzalez, Irvin Gonzalez-Lopez, Concepcion Grajales-Muñiz, Ricardo Grande, Rosa Maria Gutierrez Rios, Jesus Hernandez, Alejandra Hernandez-Teran, Alfredo Herrera-Estrella, Carla Ivon Herrera-Najera, Pavel Isa, Veronica Mata-Haro, Daniel Lira Morales, Susana Lopez, Antonio Loza Roman, Brenda Irasema Maldonado-Meza, Bernardo Martinez-Miguel, Jose Arturo Martinez-Orozco, Celida Martinez- Rodriguez, Margarita Matias-Florentino, Fidencio Mejia-Nepomuceno, Maria Guadalupe de Jesus Mireles-Rivera, Gloria Maria Molina-Salinas, Hector Montoya-Fuentes, , Mario Mujica-Sanchez, Jose Esteban Muñoz-Medina, Jose de Jesus Nuñez-Contreras, Alicia Ocaña-Mondragon, Luis Alberto Ochoa-Carrera, Hector Esteban Paz-Juarez, Marissa Perez-Garcia, Francisco Pulido, Helen Haydee Fernanda Ramirez-Plascencia, Jorge Salas-Hernandez, Angel Gustavo Salas-Lais, Alejandro Sanchez-Flores, Clara Esperanza Santacruz-Tinoco, Maria Guadalupe Santiago-Mauricio, Selene Zarate, Nelly Selem-Mojica, Blanca Taboada, Gloria Vazquez, Joel Armando Vazquez-Perez. |
| EPI_ISL_2671903, EPI_ISL_2671906, EPI_ISL_2671907, EPI_ISL_2671911, EPI_ISL_2671913, EPI_ISL_2671915, EPI_ISL_2671919, EPI_ISL_2671920, EPI_ISL_2671923, EPI_ISL_2671925, EPI_ISL_2671928, EPI_ISL_2671929, EPI_ISL_2671930, EPI_ISL_2671932, EPI_ISL_2671933, EPI_ISL_2671935, EPI_ISL_2671938, EPI_ISL_2671939, EPI_ISL_2671941, EPI_ISL_2671942, EPI_ISL_2671944, EPI_ISL_2671946, EPI_ISL_2671947, EPI_ISL_2671948, EPI_ISL_2671952, EPI_ISL_2671954, EPI_ISL_2671957, EPI_ISL_2671959, EPI_ISL_2671961, EPI_ISL_2671962, EPI_ISL_2671963, EPI_ISL_2671965, EPI_ISL_2671967, EPI_ISL_2671969, EPI_ISL_2671973, EPI_ISL_2671976, EPI_ISL_2671979, EPI_ISL_2671980, EPI_ISL_2671981, EPI_ISL_2671982, EPI_ISL_2671984, EPI_ISL_2671985, EPI_ISL_2671988, EPI_ISL_2671993, EPI_ISL_2671994, EPI_ISL_2671996, EPI_ISL_2671998, EPI_ISL_2671999, EPI_ISL_2672000, EPI_ISL_2672001, EPI_ISL_2672005, EPI_ISL_2672008, EPI_ISL_2672011, EPI_ISL_2672016, EPI_ISL_2672018, EPI_ISL_2672020, EPI_ISL_2672024, EPI_ISL_2672025, EPI_ISL_2672027, EPI_ISL_2672034, EPI_ISL_2672035, EPI_ISL_2672040, EPI_ISL_2672041, EPI_ISL_2672044, EPI_ISL_2672049, EPI_ISL_2672052, EPI_ISL_2672053, EPI_ISL_2672055, EPI_ISL_2672056, EPI_ISL_2672060, EPI_ISL_2672062, EPI_ISL_2672064, EPI_ISL_2672066, EPI_ISL_2672067, EPI_ISL_2672071, EPI_ISL_2672073, EPI_ISL_2672075, EPI_ISL_2672080, EPI_ISL_2672082, EPI_ISL_2672084, EPI_ISL_2672087, EPI_ISL_2672088, EPI_ISL_2672090 | | | |
| see above | TGen North | TGen North | Jolene Bowers, Heather Centner, Chris French, Hayley Yaglom, Ashlyn Pfeiffer, Darrin Lemmer, Dave Engelthaler, The Arizona COVID Genomics Union (ACGU) |
| EPI_ISL_2672091, EPI_ISL_2672092 | IN State Department of Health Laboratory Services | IN State Department of Health Laboratory Services | Cassandra Campion, Jamie Yeadon, Brian Pope, Lixia Liu, Kyle Brownlee, Melissa Hindenlang, Mark Glazier |
| EPI_ISL_2672093 | TGen North | TGen North | Jolene Bowers, Heather Centner, Chris French, Hayley Yaglom, Ashlyn Pfeiffer, Darrin Lemmer, Dave Engelthaler, The Arizona COVID Genomics Union (ACGU) |
| EPI_ISL_2672094, EPI_ISL_2672096, EPI_ISL_2672097, EPI_ISL_2672099 | IN State Department of Health Laboratory Services | IN State Department of Health Laboratory Services | Cassandra Campion, Jamie Yeadon, Brian Pope, Lixia Liu, Kyle Brownlee, Melissa Hindenlang, Mark Glazier |
| EPI_ISL_2672100 | TGen North | TGen North | Jolene Bowers, Heather Centner, Chris French, Hayley Yaglom, Ashlyn Pfeiffer, Darrin Lemmer, Dave Engelthaler, The Arizona COVID Genomics Union (ACGU) |
| EPI_ISL_2672101 | IN State Department of Health Laboratory Services | IN State Department of Health Laboratory Services | Cassandra Campion, Jamie Yeadon, Brian Pope, Lixia Liu, Kyle Brownlee, Melissa Hindenlang, Mark Glazier |
| EPI_ISL_2672102 | TGen North | TGen North | Jolene Bowers, Heather Centner, Chris French, Hayley Yaglom, Ashlyn Pfeiffer, Darrin Lemmer, Dave Engelthaler, The Arizona COVID Genomics Union (ACGU) |
| EPI_ISL_2672103, EPI_ISL_2672104, EPI_ISL_2672106, EPI_ISL_2672108 | IN State Department of Health Laboratory Services | IN State Department of Health Laboratory Services | Cassandra Campion, Jamie Yeadon, Brian Pope, Lixia Liu, Kyle Brownlee, Melissa Hindenlang, Mark Glazier |
| EPI_ISL_2672109 | TGen North | TGen North | Jolene Bowers, Heather Centner, Chris French, Hayley Yaglom, Ashlyn Pfeiffer, Darrin Lemmer, Dave Engelthaler, The Arizona COVID Genomics Union (ACGU) |
| EPI_ISL_2672110 | IN State Department of Health Laboratory Services | IN State Department of Health Laboratory Services | Cassandra Campion, Jamie Yeadon, Brian Pope, Lixia Liu, Kyle Brownlee, Melissa Hindenlang, Mark Glazier |
| EPI_ISL_2672112 | TGen North | TGen North | Jolene Bowers, Heather Centner, Chris French, Hayley Yaglom, Ashlyn Pfeiffer, Darrin Lemmer, Dave Engelthaler, The Arizona COVID Genomics Union (ACGU) |
| EPI_ISL_2672113, EPI_ISL_2672115, EPI_ISL_2672117, EPI_ISL_2672120, EPI_ISL_2672122 | IN State Department of Health Laboratory Services | IN State Department of Health Laboratory Services | Cassandra Campion, Jamie Yeadon, Brian Pope, Lixia Liu, Kyle Brownlee, Melissa Hindenlang, Mark Glazier |
| EPI_ISL_2672123 | TGen North | TGen North | Jolene Bowers, Heather Centner, Chris French, Hayley Yaglom, Ashlyn Pfeiffer, Darrin Lemmer, Dave Engelthaler, The Arizona COVID Genomics Union (ACGU) |
| EPI_ISL_2672124, EPI_ISL_2672125, EPI_ISL_2672127, EPI_ISL_2672129, EPI_ISL_2672130, EPI_ISL_2672132 | IN State Department of Health Laboratory Services | IN State Department of Health Laboratory Services | Cassandra Campion, Jamie Yeadon, Brian Pope, Lixia Liu, Kyle Brownlee, Melissa Hindenlang, Mark Glazier |
| EPI_ISL_2672133 | TGen North | TGen North | Jolene Bowers, Heather Centner, Chris French, Hayley Yaglom, Ashlyn Pfeiffer, Darrin Lemmer, Dave Engelthaler, The Arizona COVID Genomics Union (ACGU) |
| EPI_ISL_2672134, EPI_ISL_2672137, EPI_ISL_2672139, EPI_ISL_2672141, EPI_ISL_2672143, EPI_ISL_2672144 | IN State Department of Health Laboratory Services | IN State Department of Health Laboratory Services | Cassandra Campion, Jamie Yeadon, Brian Pope, Lixia Liu, Kyle Brownlee, Melissa Hindenlang, Mark Glazier |
| EPI_ISL_2672148, EPI_ISL_2672154, EPI_ISL_2672155, EPI_ISL_2672156, EPI_ISL_2672157, EPI_ISL_2672159, EPI_ISL_2672160, EPI_ISL_2672162, EPI_ISL_2672163, EPI_ISL_2672164, EPI_ISL_2672166, EPI_ISL_2672169, EPI_ISL_2672171, EPI_ISL_2672172, EPI_ISL_2672173, EPI_ISL_2672176, EPI_ISL_2672177, EPI_ISL_2672179, EPI_ISL_2672182, EPI_ISL_2672185, EPI_ISL_2672187, EPI_ISL_2672190, EPI_ISL_2672192, EPI_ISL_2672196, EPI_ISL_2672199, EPI_ISL_2672201, EPI_ISL_2672203, EPI_ISL_2672206, EPI_ISL_2672207, EPI_ISL_2672209, EPI_ISL_2672213 | | | |
| see above | TGen North | TGen North | Jolene Bowers, Heather Centner, Chris French, Hayley Yaglom, Ashlyn Pfeiffer, Darrin Lemmer, Dave Engelthaler, The Arizona COVID Genomics Union (ACGU) |
| EPI_ISL_2673683 | AR Dept. of Health-PHL, Molecular Diagnostics | Genomics and Discovery, Respiratory Viruses Branch, Division of Viral Diseases, Centers for Disease Control and Prevention | Jing Zhang, Anna Kelleher, Ying Tao, Yan Li, Brian Lynch, Krista Queen, Anna Uehara, Peter Cook, Han Jia Justin Ng, Rachel Marine, Clinton R. Paden, Dhwani Batra, Haibin Wang, Tara Coalter, Jasmine Padilla, Morgan Davis, Mili Sheth, Sarah Nobles, Mark Burroughs, Justin Lee, Adam Retchless, Suxiang Tong |
| EPI_ISL_2673684 | TN Division of Laboratory Services | Genomics and Discovery, Respiratory Viruses Branch, Division of Viral Diseases, Centers for Disease Control and Prevention | Jing Zhang, Anna Kelleher, Ying Tao, Yan Li, Brian Lynch, Krista Queen, Anna Uehara, Peter Cook, Han Jia Justin Ng, Rachel Marine, Clinton R. Paden, Dhwani Batra, Haibin Wang, Tara Coalter, Jasmine Padilla, Morgan Davis, Mili Sheth, Sarah Nobles, Mark Burroughs, Justin Lee, Adam Retchless, Suxiang Tong |
| EPI_ISL_2673690, EPI_ISL_2673691, EPI_ISL_2673692, EPI_ISL_2673693 | Washington State Department of Health Public Health Laboratories | Washington State Department of Health Public Health Laboratories | Drew MacKellar, Philip Dykema, Denny Russell, Joenice Gonzalez, Hannah Gray, Geoff Melly, Vanessa De Los Santos, Darren Lucas, JohnAric Peterson, Avi Singh, Rebecca Cao |
| EPI_ISL_2673696 | Instituto Nacional de Enfermedades Respiratorias (INER) | Centro de Investigación en Enfermedades Infecciosas (CIENI), Instituto Nacional de Enfermedades Respiratorias (INER) | Consorcio Mexicano de Vigilancia Genómica (CoViGen-Mex). Authors (in alphabetical order): Julio Elias Alvarado-Yaah, Carlos F. Arias, Santiago Ávila-Ríos, Eduardo Becerril-Vargas, Víctor Hugo Borja-Aburto, Celia Boukadida, Cristóbal Cháidez-Quiróz, Juan Bautista Chale-Dzul, Ricardo Ciria Merce, Andreu Comas-García, Célida Duque Molina, Julissa Enciso-Ibarra, José Antonio Enciso-Moreno, Gloria Elena Espinosa-Ayala, Fernando Fontove-Herrera, Daniel Fregoso-Rueda, Victor Eduardo Garcia-Arias, Alejandra Garcia-Gasca, Bruno Gomez-Gil, Jean Pierre González, Irvin González-López, Concepción Grajales-Muñiz, Ricardo Grande, Rosa María Gutierrez Rios, Alejandra Hernández-Terán, Alfredo Herrera-Estrella, Carla Ivón Herrera-Najera, Pavel Isa, Daniel Lira Morales, Susana Lopez, Antonio Loza Román, Brenda Irasema Maldonado-Meza, Bernardo Martínez-Miguel, José Arturo Martínez-Orozco, Célida Martínez- Rodríguez, Margarita Matías-Florentino, Fidencio Mejía-Nepomuceno, María Guadalupe de Jesús Mireles-Rivera, Gloria María Molina-Salinas, Hector Montoya-Fuentes, , Mario Mújica-Sánchez, José Esteban Muñoz-Medina, José de Jesús Nuñez-Contreras, Alicia Ocaña-Mondragón, Luis Alberto Ochoa-Carrera, Hector Esteban Paz-Juárez, Marissa Pérez-García, Francisco Pulido, Helen Haydee Fernanda Ramírez-Plascencia, Jorge Salas-Hernández, Angel Gustavo Salas-Lais, Alejandro Sanchez-Flores, Clara Esperanza |

| | | | |
|---|---|---|---|
| | | | Santacruz-Tinoco, María Guadalupe Santiago-Mauricio, Selene Zárate, Nelly Sélem-Mojica, Blanca Taboada, Gloria Vazquez, Joel Armando Vázquez-Pérez. |
| EPI_ISL_2673772, EPI_ISL_2673773, EPI_ISL_2673774, EPI_ISL_2673775, EPI_ISL_2673776, EPI_ISL_2673777, EPI_ISL_2673778, EPI_ISL_2673779, EPI_ISL_2673780, EPI_ISL_2673781, EPI_ISL_2673782 | | | |
| see above | Kaiser Permanente Washington Health Research Institute | Genomics and Discovery, Respiratory Viruses Branch, Division of Viral Diseases, Centers for Disease Control and Prevention | Jing Zhang, Anna Kelleher, Ying Tao, Yan Li, Brian Lynch, Krista Queen, Anna Uehara, Peter Cook, Han Jia Justin Ng, Rachel Marine, Clinton R. Paden, Dhwani Batra, Haibin Wang, Tara Coalter, Jasmine Padilla, Morgan Davis, Mili Sheth, Sarah Nobles, Mark Burroughs, Justin Lee, Adam Retchless, Suxiang Tong |
| EPI_ISL_2673861, EPI_ISL_2673862, EPI_ISL_2673863, EPI_ISL_2673864, EPI_ISL_2673865, EPI_ISL_2673866, EPI_ISL_2673867, EPI_ISL_2673868, EPI_ISL_2673869, EPI_ISL_2673870, EPI_ISL_2673871, EPI_ISL_2673872, EPI_ISL_2673873, EPI_ISL_2673874, EPI_ISL_2673875, EPI_ISL_2673876, EPI_ISL_2673877, EPI_ISL_2673878, EPI_ISL_2673879, EPI_ISL_2673880, EPI_ISL_2673881, EPI_ISL_2673882, EPI_ISL_2673883, EPI_ISL_2673884, EPI_ISL_2673885, EPI_ISL_2673886, EPI_ISL_2673887, EPI_ISL_2673888, EPI_ISL_2673889, EPI_ISL_2673890, EPI_ISL_2673891, EPI_ISL_2674369, EPI_ISL_2674375, EPI_ISL_2674376, EPI_ISL_2674378, EPI_ISL_2674379, EPI_ISL_2674380, EPI_ISL_2674381, EPI_ISL_2674382 | | | |
| see above | Wyoming Public Health Laboratory | Wyoming Public Health Laboratory | Jim Mildenberger, Taylor Fearing, Channing Weber, Ashley Norberg, Chayse Rowley, Marley Goetz, Brian Dominguez, Elliot Thomasson, Sam Britz, Cari Sloma, Robert Petit, and Rob Christensen |
| EPI_ISL_2674545, EPI_ISL_2674546, EPI_ISL_2674547, EPI_ISL_2674548, EPI_ISL_2674555, EPI_ISL_2674556, EPI_ISL_2674558, EPI_ISL_2674559, EPI_ISL_2674560, EPI_ISL_2674561, EPI_ISL_2674562, EPI_ISL_2674563, EPI_ISL_2674564, EPI_ISL_2674565, EPI_ISL_2674566, EPI_ISL_2674568, EPI_ISL_2674570, EPI_ISL_2674571, EPI_ISL_2674572, EPI_ISL_2674573, EPI_ISL_2674575, EPI_ISL_2674576, EPI_ISL_2674579, EPI_ISL_2674580, EPI_ISL_2674581, EPI_ISL_2674582, EPI_ISL_2674583, EPI_ISL_2674585, EPI_ISL_2674587, EPI_ISL_2674588, EPI_ISL_2674589, EPI_ISL_2674590, EPI_ISL_2674591, EPI_ISL_2674592, EPI_ISL_2674593, EPI_ISL_2674594, EPI_ISL_2674595, EPI_ISL_2674596, EPI_ISL_2674598, EPI_ISL_2674600, EPI_ISL_2674601, EPI_ISL_2674602, EPI_ISL_2674604, EPI_ISL_2674608, EPI_ISL_2674610, EPI_ISL_2674611, EPI_ISL_2674613, EPI_ISL_2674614, EPI_ISL_2674615, EPI_ISL_2674617, EPI_ISL_2674618, EPI_ISL_2674619, EPI_ISL_2674621, EPI_ISL_2674623, EPI_ISL_2674625, EPI_ISL_2674626, EPI_ISL_2674627 | | | |
| see above | Curative | New Mexico Department of Health Scientific Laboratory | Ellie Johnson, D'eldra Malone, Jennifer Benoit, Linda Salazar, Ratheesh Rajan, Keila Gutierrez, Anastacia Griego-Fisher |
| EPI_ISL_2674628 | LESP Guanajuato | Instituto de Diagnostico y Referencia Epidemiologicos (INDRE) | Claudia Wong-Arambula, Abril Rodriguez-Maldonado, Vanessa Rivero-Arredondo, Ariadna Medina-Benitez, Joaquin Quiroz-Mercado, Sergio Rangel-Guerrero, Natividad Cruz-Ortiz, Tatiana Nunez-Garcia, Gisela Barrera-Badillo, Lucia Hernandez-Rivas, Irma Lopez-Martinez, Ernesto Ramirez-Gonzalez. |
| EPI_ISL_2674629, EPI_ISL_2674630, EPI_ISL_2674631 | LESP Nuevo Leon | Instituto de Diagnostico y Referencia Epidemiologicos (INDRE) | Claudia Wong-Arambula, Abril Rodriguez-Maldonado, Vanessa Rivero-Arredondo, Ariadna Medina-Benitez, Joaquin Quiroz-Mercado, Sergio Rangel-Guerrero, Natividad Cruz-Ortiz, Tatiana Nunez-Garcia, Gisela Barrera-Badillo, Lucia Hernandez-Rivas, Irma Lopez-Martinez, Ernesto Ramirez-Gonzalez. |
| EPI_ISL_2674632 | LESP Ciudad de Mexico | Instituto de Diagnostico y Referencia Epidemiologicos (INDRE) | Claudia Wong-Arambula, Abril Rodriguez-Maldonado, Vanessa Rivero-Arredondo, Ariadna Medina-Benitez, Joaquin Quiroz-Mercado, Sergio Rangel-Guerrero, Natividad Cruz-Ortiz, Tatiana Nunez-Garcia, Gisela Barrera-Badillo, Lucia Hernandez-Rivas, Irma Lopez-Martinez, Ernesto Ramirez-Gonzalez. |
| EPI_ISL_2674633, EPI_ISL_2674634 | LESP Sinaloa | Instituto de Diagnostico y Referencia Epidemiologicos (INDRE) | Claudia Wong-Arambula, Abril Rodriguez-Maldonado, Vanessa Rivero-Arredondo, Ariadna Medina-Benitez, Joaquin Quiroz-Mercado, Sergio Rangel-Guerrero, Natividad Cruz-Ortiz, Tatiana Nunez-Garcia, Gisela Barrera-Badillo, Lucia Hernandez-Rivas, Irma Lopez-Martinez, Ernesto Ramirez-Gonzalez. |
| EPI_ISL_2674635, EPI_ISL_2674636, EPI_ISL_2674637, EPI_ISL_2674638, EPI_ISL_2674639, EPI_ISL_2674640 | LESP Tabasco | Instituto de Diagnostico y Referencia Epidemiologicos (INDRE) | Claudia Wong-Arambula, Abril Rodriguez-Maldonado, Vanessa Rivero-Arredondo, Ariadna Medina-Benitez, Joaquin Quiroz-Mercado, Sergio Rangel-Guerrero, Natividad Cruz-Ortiz, Tatiana Nunez-Garcia, Gisela Barrera-Badillo, Lucia Hernandez-Rivas, Irma Lopez-Martinez, Ernesto Ramirez-Gonzalez. |
| EPI_ISL_2674641 | LESP Michoacan | Instituto de Diagnostico y Referencia Epidemiologicos (INDRE) | Claudia Wong-Arambula, Abril Rodriguez-Maldonado, Vanessa Rivero-Arredondo, Ariadna Medina-Benitez, Joaquin Quiroz-Mercado, Sergio Rangel-Guerrero, Natividad Cruz-Ortiz, Tatiana Nunez-Garcia, Gisela Barrera-Badillo, Lucia Hernandez-Rivas, Irma Lopez-Martinez, Ernesto Ramirez-Gonzalez. |
| EPI_ISL_2674642 | LESP Durango | Instituto de Diagnostico y Referencia Epidemiologicos (INDRE) | Claudia Wong-Arambula, Abril Rodriguez-Maldonado, Vanessa Rivero-Arredondo, Ariadna Medina-Benitez, Joaquin Quiroz-Mercado, Sergio Rangel-Guerrero, Natividad Cruz-Ortiz, Tatiana Nunez-Garcia, Gisela Barrera-Badillo, Lucia Hernandez-Rivas, Irma Lopez-Martinez, Ernesto Ramirez-Gonzalez. |
| EPI_ISL_2674643, EPI_ISL_2674644, EPI_ISL_2674645, EPI_ISL_2674646, EPI_ISL_2674647, EPI_ISL_2674648, EPI_ISL_2674649 | LESP Sonora | Instituto de Diagnostico y Referencia Epidemiologicos (INDRE) | Claudia Wong-Arambula, Abril Rodriguez-Maldonado, Vanessa Rivero-Arredondo, Ariadna Medina-Benitez, Joaquin Quiroz-Mercado, Sergio Rangel-Guerrero, Natividad Cruz-Ortiz, Tatiana Nunez-Garcia, Gisela Barrera-Badillo, Lucia Hernandez-Rivas, Irma Lopez-Martinez, Ernesto Ramirez-Gonzalez. |
| EPI_ISL_2674650 | LESP Jalisco | Instituto de Diagnostico y Referencia Epidemiologicos (INDRE) | Claudia Wong-Arambula, Abril Rodriguez-Maldonado, Vanessa Rivero-Arredondo, Ariadna Medina-Benitez, Joaquin Quiroz-Mercado, Sergio Rangel-Guerrero, Natividad Cruz-Ortiz, Tatiana Nunez-Garcia, Gisela Barrera-Badillo, Lucia Hernandez-Rivas, Irma Lopez-Martinez, Ernesto Ramirez-Gonzalez. |
| EPI_ISL_2674651, EPI_ISL_2674652, EPI_ISL_2674653, EPI_ISL_2674654, EPI_ISL_2674655 | LESP Quintana Roo | Instituto de Diagnostico y Referencia Epidemiologicos (INDRE) | Claudia Wong-Arambula, Abril Rodriguez-Maldonado, Vanessa Rivero-Arredondo, Ariadna Medina-Benitez, Joaquin Quiroz-Mercado, Sergio Rangel-Guerrero, Natividad Cruz-Ortiz, Tatiana Nunez-Garcia, Gisela Barrera-Badillo, Lucia Hernandez-Rivas, Irma Lopez-Martinez, Ernesto Ramirez-Gonzalez. |
| EPI_ISL_2674656, EPI_ISL_2674657 | LESP Guanajuato | Instituto de Diagnostico y Referencia Epidemiologicos (INDRE) | Claudia Wong-Arambula, Abril Rodriguez-Maldonado, Vanessa Rivero-Arredondo, Ariadna Medina-Benitez, Joaquin Quiroz-Mercado, Sergio Rangel-Guerrero, Natividad Cruz-Ortiz, Tatiana Nunez-Garcia, Gisela Barrera-Badillo, Lucia Hernandez-Rivas, Irma Lopez-Martinez, Ernesto Ramirez-Gonzalez. |
| EPI_ISL_2674658, EPI_ISL_2674659, EPI_ISL_2674660, EPI_ISL_2674661, EPI_ISL_2674662, EPI_ISL_2674663 | LESP Hidalgo | Instituto de Diagnostico y Referencia Epidemiologicos (INDRE) | Claudia Wong-Arambula, Abril Rodriguez-Maldonado, Vanessa Rivero-Arredondo, Ariadna Medina-Benitez, Joaquin Quiroz-Mercado, Sergio Rangel-Guerrero, Natividad Cruz-Ortiz, Tatiana Nunez-Garcia, Gisela Barrera-Badillo, Lucia Hernandez-Rivas, Irma Lopez-Martinez, Ernesto Ramirez-Gonzalez. |
| EPI_ISL_2674664 | LESP Estado de Mexico | Instituto de Diagnostico y Referencia Epidemiologicos (INDRE) | Claudia Wong-Arambula, Abril Rodriguez-Maldonado, Vanessa Rivero-Arredondo, Ariadna Medina-Benitez, Joaquin Quiroz-Mercado, Sergio Rangel-Guerrero, Natividad Cruz-Ortiz, Tatiana Nunez-Garcia, Gisela Barrera-Badillo, Lucia Hernandez-Rivas, Irma Lopez-Martinez, Ernesto Ramirez-Gonzalez. |
| EPI_ISL_2674665, EPI_ISL_2674666, EPI_ISL_2674667 | LESP Hidalgo | Instituto de Diagnostico y Referencia Epidemiologicos (INDRE) | Claudia Wong-Arambula, Abril Rodriguez-Maldonado, Vanessa Rivero-Arredondo, Ariadna Medina-Benitez, Joaquin Quiroz-Mercado, Sergio Rangel-Guerrero, Natividad Cruz-Ortiz, Tatiana Nunez-Garcia, Gisela Barrera-Badillo, Lucia Hernandez-Rivas, Irma Lopez-Martinez, Ernesto Ramirez-Gonzalez. |
| EPI_ISL_2674668, EPI_ISL_2674669, EPI_ISL_2674670 | LESP Estado de Mexico | Instituto de Diagnostico y Referencia Epidemiologicos (INDRE) | Claudia Wong-Arambula, Abril Rodriguez-Maldonado, Vanessa Rivero-Arredondo, Ariadna Medina-Benitez, Joaquin Quiroz-Mercado, Sergio Rangel-Guerrero, Natividad Cruz-Ortiz, Tatiana Nunez-Garcia, Gisela Barrera-Badillo, Lucia Hernandez-Rivas, Irma Lopez-Martinez, Ernesto Ramirez-Gonzalez. |
| EPI_ISL_2674671, EPI_ISL_2674672 | LESP Hidalgo | Instituto de Diagnostico y Referencia Epidemiologicos (INDRE) | Claudia Wong-Arambula, Abril Rodriguez-Maldonado, Vanessa Rivero-Arredondo, Ariadna Medina-Benitez, Joaquin Quiroz-Mercado, Sergio Rangel-Guerrero, Natividad Cruz-Ortiz, Tatiana Nunez-Garcia, Gisela Barrera-Badillo, Lucia Hernandez-Rivas, Irma Lopez-Martinez, Ernesto Ramirez-Gonzalez. |
| EPI_ISL_2674673 | LESP Puebla | Instituto de Diagnostico y Referencia Epidemiologicos (INDRE) | Claudia Wong-Arambula, Abril Rodriguez-Maldonado, Vanessa Rivero-Arredondo, Ariadna Medina-Benitez, Joaquin Quiroz-Mercado, Sergio Rangel-Guerrero, Natividad Cruz-Ortiz, Tatiana Nunez-Garcia, Gisela Barrera-Badillo, Lucia Hernandez-Rivas, Irma Lopez-Martinez, Ernesto Ramirez-Gonzalez. |
| EPI_ISL_2674674, EPI_ISL_2674675, EPI_ISL_2674676, EPI_ISL_2674677, EPI_ISL_2674678, EPI_ISL_2674679, EPI_ISL_2674680, EPI_ISL_2674681, EPI_ISL_2674682 | LESP Hidalgo | Instituto de Diagnostico y Referencia Epidemiologicos (INDRE) | Claudia Wong-Arambula, Abril Rodriguez-Maldonado, Vanessa Rivero-Arredondo, Ariadna Medina-Benitez, Joaquin Quiroz-Mercado, Sergio Rangel-Guerrero, Natividad Cruz-Ortiz, Tatiana Nunez-Garcia, Gisela Barrera-Badillo, Lucia Hernandez-Rivas, Irma Lopez-Martinez, Ernesto Ramirez-Gonzalez. |
| EPI_ISL_2674683 | LESP Queretaro | Instituto de Diagnostico y Referencia Epidemiologicos (INDRE) | Claudia Wong-Arambula, Abril Rodriguez-Maldonado, Vanessa Rivero-Arredondo, Ariadna Medina-Benitez, Joaquin Quiroz-Mercado, Sergio Rangel-Guerrero, Natividad Cruz-Ortiz, Tatiana Nunez-Garcia, Gisela Barrera-Badillo, Lucia Hernandez-Rivas, Irma Lopez-Martinez, Ernesto Ramirez-Gonzalez. |
| EPI_ISL_2674684, EPI_ISL_2674685, EPI_ISL_2674686, EPI_ISL_2674687, EPI_ISL_2674688 | LESP Yucatan | Instituto de Diagnostico y Referencia Epidemiologicos (INDRE) | Claudia Wong-Arambula, Abril Rodriguez-Maldonado, Vanessa Rivero-Arredondo, Ariadna Medina-Benitez, Joaquin Quiroz-Mercado, Sergio Rangel-Guerrero, Natividad Cruz-Ortiz, Tatiana Nunez-Garcia, Gisela Barrera-Badillo, Lucia Hernandez-Rivas, Irma Lopez-Martinez, Ernesto Ramirez-Gonzalez. |
| EPI_ISL_2674689 | LESP Coahuila | Instituto de Diagnostico y Referencia Epidemiologicos (INDRE) | Claudia Wong-Arambula, Abril Rodriguez-Maldonado, Vanessa Rivero-Arredondo, Ariadna Medina-Benitez, Joaquin Quiroz-Mercado, Sergio Rangel-Guerrero, Natividad Cruz-Ortiz, Tatiana Nunez-Garcia, Gisela Barrera-Badillo, Lucia Hernandez-Rivas, Irma Lopez-Martinez, Ernesto Ramirez-Gonzalez. |
| EPI_ISL_2674690 | LESP Hidalgo | Instituto de Diagnostico y Referencia Epidemiologicos (INDRE) | Claudia Wong-Arambula, Abril Rodriguez-Maldonado, Vanessa Rivero-Arredondo, Ariadna Medina-Benitez, Joaquin Quiroz-Mercado, Sergio Rangel-Guerrero, Natividad Cruz-Ortiz, Tatiana Nunez-Garcia, Gisela Barrera-Badillo, Lucia Hernandez-Rivas, Irma Lopez-Martinez, Ernesto Ramirez-Gonzalez. |
| EPI_ISL_2674691, EPI_ISL_2674692, EPI_ISL_2674693, EPI_ISL_2674694, EPI_ISL_2674695, EPI_ISL_2674696 | LESP Guanajuato | Instituto de Diagnostico y Referencia Epidemiologicos (INDRE) | Claudia Wong-Arambula, Abril Rodriguez-Maldonado, Vanessa Rivero-Arredondo, Ariadna Medina-Benitez, Joaquin Quiroz-Mercado, Sergio Rangel-Guerrero, Natividad Cruz-Ortiz, Tatiana Nunez-Garcia, Gisela Barrera-Badillo, Lucia Hernandez-Rivas, Irma Lopez-Martinez, Ernesto Ramirez-Gonzalez. |
| EPI_ISL_2674697 | LESP Nuevo Leon | Instituto de Diagnostico y Referencia Epidemiologicos (INDRE) | Claudia Wong-Arambula, Abril Rodriguez-Maldonado, Vanessa Rivero-Arredondo, Ariadna Medina-Benitez, Joaquin Quiroz-Mercado, Sergio Rangel-Guerrero, Natividad Cruz-Ortiz, Tatiana Nunez-Garcia, Gisela Barrera-Badillo, Lucia Hernandez-Rivas, Irma Lopez-Martinez, Ernesto Ramirez-Gonzalez. |
| EPI_ISL_2674698 | LESP Ciudad de Mexico | Instituto de Diagnostico y Referencia Epidemiologicos (INDRE) | Claudia Wong-Arambula, Abril Rodriguez-Maldonado, Vanessa Rivero-Arredondo, Ariadna Medina-Benitez, Joaquin Quiroz-Mercado, Sergio Rangel-Guerrero, Natividad Cruz-Ortiz, Tatiana Nunez-Garcia, Gisela Barrera-Badillo, Lucia Hernandez-Rivas, Irma Lopez-Martinez, Ernesto Ramirez-Gonzalez. |

| | | | |
|---|---|---|---|
| EPI_ISL_2674699 | LESP Chihuahua | Instituto de Diagnostico y Referencia Epidemiologicos (INDRE) | Claudia Wong-Arambula, Abril Rodriguez-Maldonado, Vanessa Rivero-Arredondo, Ariadna Medina-Benitez, Joaquin Quiroz-Mercado, Sergio Rangel-Guerrero, Natividad Cruz-Ortiz, Tatiana Nunez-Garcia, Gisela Barrera-Badillo, Lucia Hernandez-Rivas, Irma Lopez-Martinez, Ernesto Ramirez-Gonzalez. |
| EPI_ISL_2674700 | LESP Puebla | Instituto de Diagnostico y Referencia Epidemiologicos (INDRE) | Claudia Wong-Arambula, Abril Rodriguez-Maldonado, Vanessa Rivero-Arredondo, Ariadna Medina-Benitez, Joaquin Quiroz-Mercado, Sergio Rangel-Guerrero, Natividad Cruz-Ortiz, Tatiana Nunez-Garcia, Gisela Barrera-Badillo, Lucia Hernandez-Rivas, Irma Lopez-Martinez, Ernesto Ramirez-Gonzalez. |
| EPI_ISL_2674701, EPI_ISL_2674702, EPI_ISL_2674703, EPI_ISL_2674704, EPI_ISL_2674705, EPI_ISL_2674706, EPI_ISL_2674707, EPI_ISL_2674708, EPI_ISL_2674709, EPI_ISL_2674710, EPI_ISL_2674711, EPI_ISL_2674712, EPI_ISL_2674713 | | | |
| see above | LESP Yucatan | Instituto de Diagnostico y Referencia Epidemiologicos (INDRE) | Claudia Wong-Arambula, Abril Rodriguez-Maldonado, Vanessa Rivero-Arredondo, Ariadna Medina-Benitez, Joaquin Quiroz-Mercado, Sergio Rangel-Guerrero, Natividad Cruz-Ortiz, Tatiana Nunez-Garcia, Gisela Barrera-Badillo, Lucia Hernandez-Rivas, Irma Lopez-Martinez, Ernesto Ramirez-Gonzalez. |
| EPI_ISL_2674714 | LESP Quintana Roo | Instituto de Diagnostico y Referencia Epidemiologicos (INDRE) | Claudia Wong-Arambula, Abril Rodriguez-Maldonado, Vanessa Rivero-Arredondo, Ariadna Medina-Benitez, Joaquin Quiroz-Mercado, Sergio Rangel-Guerrero, Natividad Cruz-Ortiz, Tatiana Nunez-Garcia, Gisela Barrera-Badillo, Lucia Hernandez-Rivas, Irma Lopez-Martinez, Ernesto Ramirez-Gonzalez. |
| EPI_ISL_2674715 | LESP Yucatan | Instituto de Diagnostico y Referencia Epidemiologicos (INDRE) | Claudia Wong-Arambula, Abril Rodriguez-Maldonado, Vanessa Rivero-Arredondo, Ariadna Medina-Benitez, Joaquin Quiroz-Mercado, Sergio Rangel-Guerrero, Natividad Cruz-Ortiz, Tatiana Nunez-Garcia, Gisela Barrera-Badillo, Lucia Hernandez-Rivas, Irma Lopez-Martinez, Ernesto Ramirez-Gonzalez. |
| EPI_ISL_2674716 | LESP San Luis Potosi | Instituto de Diagnostico y Referencia Epidemiologicos (INDRE) | Claudia Wong-Arambula, Abril Rodriguez-Maldonado, Vanessa Rivero-Arredondo, Ariadna Medina-Benitez, Joaquin Quiroz-Mercado, Sergio Rangel-Guerrero, Natividad Cruz-Ortiz, Tatiana Nunez-Garcia, Gisela Barrera-Badillo, Lucia Hernandez-Rivas, Irma Lopez-Martinez, Ernesto Ramirez-Gonzalez. |
| EPI_ISL_2674717, EPI_ISL_2674718, EPI_ISL_2674719 | LESP Sonora | Instituto de Diagnostico y Referencia Epidemiologicos (INDRE) | Claudia Wong-Arambula, Abril Rodriguez-Maldonado, Vanessa Rivero-Arredondo, Ariadna Medina-Benitez, Joaquin Quiroz-Mercado, Sergio Rangel-Guerrero, Natividad Cruz-Ortiz, Tatiana Nunez-Garcia, Gisela Barrera-Badillo, Lucia Hernandez-Rivas, Irma Lopez-Martinez, Ernesto Ramirez-Gonzalez. |
| EPI_ISL_2674900, EPI_ISL_2674901, EPI_ISL_2674902 | LESP Ciudad de Mexico | Instituto de Diagnostico y Referencia Epidemiologicos (INDRE) | Claudia Wong-Arambula, Abril Rodriguez-Maldonado, Vanessa Rivero-Arredondo, Ariadna Medina-Benitez, Joaquin Quiroz-Mercado, Sergio Rangel-Guerrero, Natividad Cruz-Ortiz, Tatiana Nunez-Garcia, Gisela Barrera-Badillo, Lucia Hernandez-Rivas, Irma Lopez-Martinez, Ernesto Ramirez-Gonzalez. |
| EPI_ISL_2674903 | LESP Jalisco | Instituto de Diagnostico y Referencia Epidemiologicos (INDRE) | Claudia Wong-Arambula, Abril Rodriguez-Maldonado, Vanessa Rivero-Arredondo, Ariadna Medina-Benitez, Joaquin Quiroz-Mercado, Sergio Rangel-Guerrero, Natividad Cruz-Ortiz, Tatiana Nunez-Garcia, Gisela Barrera-Badillo, Lucia Hernandez-Rivas, Irma Lopez-Martinez, Ernesto Ramirez-Gonzalez. |
| EPI_ISL_2674904 | LESP Ciudad de Mexico | Instituto de Diagnostico y Referencia Epidemiologicos (INDRE) | Claudia Wong-Arambula, Abril Rodriguez-Maldonado, Vanessa Rivero-Arredondo, Ariadna Medina-Benitez, Joaquin Quiroz-Mercado, Sergio Rangel-Guerrero, Natividad Cruz-Ortiz, Tatiana Nunez-Garcia, Gisela Barrera-Badillo, Lucia Hernandez-Rivas, Irma Lopez-Martinez, Ernesto Ramirez-Gonzalez. |
| EPI_ISL_2674905 | LESP Yucatan | Instituto de Diagnostico y Referencia Epidemiologicos (INDRE) | Claudia Wong-Arambula, Abril Rodriguez-Maldonado, Vanessa Rivero-Arredondo, Ariadna Medina-Benitez, Joaquin Quiroz-Mercado, Sergio Rangel-Guerrero, Natividad Cruz-Ortiz, Tatiana Nunez-Garcia, Gisela Barrera-Badillo, Lucia Hernandez-Rivas, Irma Lopez-Martinez, Ernesto Ramirez-Gonzalez. |
| EPI_ISL_2675189, EPI_ISL_2675190, EPI_ISL_2675191, EPI_ISL_2675192, EPI_ISL_2675193, EPI_ISL_2675194, EPI_ISL_2675195, EPI_ISL_2675196, EPI_ISL_2675197, EPI_ISL_2675198, EPI_ISL_2675199, EPI_ISL_2675200 | | | |
| see above | Wichita State University - Molecular Diagnostics Lab | Kansas Health and Environmental Lab | Mike Grose, Katherine Wiggins, Jonathan Barnell, Ben Olsen, and Phil Adam |
| EPI_ISL_2675203, EPI_ISL_2675204, EPI_ISL_2675205, EPI_ISL_2675206, EPI_ISL_2675207, EPI_ISL_2675208, EPI_ISL_2675209, EPI_ISL_2675210, EPI_ISL_2675211 | Houston Health Dept. | Houston Health Dept. | Ryker Penn, Pamela Brown, Adolpho Lara, Yanlai Lai |
| EPI_ISL_2675213, EPI_ISL_2675214, EPI_ISL_2675216, EPI_ISL_2675217, EPI_ISL_2675218, EPI_ISL_2675220, EPI_ISL_2675221, EPI_ISL_2675222, EPI_ISL_2675223, EPI_ISL_2675225, EPI_ISL_2675226, EPI_ISL_2675227, EPI_ISL_2675228, EPI_ISL_2675229, EPI_ISL_2675230, EPI_ISL_2675231, EPI_ISL_2675232, EPI_ISL_2675233, EPI_ISL_2675234, EPI_ISL_2675235, EPI_ISL_2675236, EPI_ISL_2675237, EPI_ISL_2675238, EPI_ISL_2675239, EPI_ISL_2675240 | | | |
| see above | South Dakota Public Health Laboratory | South Dakota Public Health Laboratory | Jacob Garfin and Chris Carlson |
| EPI_ISL_2675241, EPI_ISL_2675242, EPI_ISL_2675243, EPI_ISL_2675244, EPI_ISL_2675245, EPI_ISL_2675246, EPI_ISL_2675247, EPI_ISL_2675248, EPI_ISL_2675249, EPI_ISL_2675250 | Alabama Department of Public Health (ADPH) | Alabama Department of Public Health (ADPH) | Adrianna Walker |
| EPI_ISL_2676406, EPI_ISL_2676407, EPI_ISL_2676410, EPI_ISL_2676411, EPI_ISL_2676417, EPI_ISL_2676424, EPI_ISL_2676427 | DC Public Health Lab/ Dept. of Forensic Sciences | DC Public Health Lab/ Dept. of Forensic Sciences | Janis Doss, Scott Nguyen, Elizabeth Zelaya, Sarah Scott, Connie Maza, Monica Mann, Brittany Hamilton, David Payne, Jocelyn Hauser |
| EPI_ISL_2676450 | Wyoming Public Health Laboratory | Wyoming Public Health Laboratory | Jim Mildenberger, Taylor Fearing, Channing Weber, Ashley Norberg, Chayse Rowley, Marley Goetz, Brian Dominguez, Elliot Thomasson, Sam Britz, Cari Sloma, Robert Petit, and Rob Christensen |
| EPI_ISL_2676884, EPI_ISL_2676885, EPI_ISL_2676886, EPI_ISL_2676887, EPI_ISL_2676888, EPI_ISL_2676889, EPI_ISL_2676890, EPI_ISL_2676891, EPI_ISL_2676892, EPI_ISL_2676893, EPI_ISL_2676894, EPI_ISL_2676895, EPI_ISL_2676896, EPI_ISL_2676897, EPI_ISL_2676898, EPI_ISL_2676899, EPI_ISL_2676900, EPI_ISL_2676901, EPI_ISL_2676902, EPI_ISL_2676903, EPI_ISL_2676904, EPI_ISL_2676905, EPI_ISL_2676906, EPI_ISL_2676907, EPI_ISL_2676908, EPI_ISL_2676909, EPI_ISL_2676910, EPI_ISL_2676911, EPI_ISL_2676912, EPI_ISL_2676913, EPI_ISL_2676914, EPI_ISL_2676915, EPI_ISL_2676916, EPI_ISL_2676917, EPI_ISL_2676918, EPI_ISL_2676919, EPI_ISL_2676920, EPI_ISL_2676921, EPI_ISL_2676922, EPI_ISL_2676923, EPI_ISL_2676924, EPI_ISL_2676925, EPI_ISL_2676926, EPI_ISL_2676927, EPI_ISL_2676928, EPI_ISL_2676929, EPI_ISL_2676930, EPI_ISL_2676931, EPI_ISL_2676932, EPI_ISL_2676933, EPI_ISL_2676934, EPI_ISL_2676935, EPI_ISL_2676936, EPI_ISL_2676937, EPI_ISL_2676938, EPI_ISL_2676939, EPI_ISL_2676940, EPI_ISL_2676941, EPI_ISL_2676942, EPI_ISL_2676943, EPI_ISL_2676944, EPI_ISL_2676945, EPI_ISL_2676946, EPI_ISL_2676947, EPI_ISL_2676948, EPI_ISL_2676949, EPI_ISL_2676950, EPI_ISL_2676951, EPI_ISL_2676952, EPI_ISL_2676953, EPI_ISL_2676954, EPI_ISL_2676955, EPI_ISL_2676956, EPI_ISL_2676957, EPI_ISL_2676958, EPI_ISL_2676959, EPI_ISL_2676960, EPI_ISL_2676961, EPI_ISL_2676962, EPI_ISL_2676963, EPI_ISL_2676964, EPI_ISL_2676965, EPI_ISL_2676966, EPI_ISL_2676967, EPI_ISL_2676968, EPI_ISL_2676969, EPI_ISL_2676970, EPI_ISL_2676971, EPI_ISL_2676972, EPI_ISL_2676973, EPI_ISL_2676974, EPI_ISL_2676975, EPI_ISL_2676976, EPI_ISL_2676977, EPI_ISL_2676978, EPI_ISL_2676979, EPI_ISL_2676980, EPI_ISL_2676981, EPI_ISL_2676982, EPI_ISL_2676983, EPI_ISL_2676984, EPI_ISL_2676985, EPI_ISL_2676986, EPI_ISL_2676987, EPI_ISL_2676988, EPI_ISL_2676989, EPI_ISL_2676990, EPI_ISL_2676991, EPI_ISL_2676992, EPI_ISL_2676993, EPI_ISL_2676994, EPI_ISL_2676995, EPI_ISL_2676996, EPI_ISL_2676997, EPI_ISL_2676998, EPI_ISL_2676999, EPI_ISL_2677000, EPI_ISL_2677001, EPI_ISL_2677002, EPI_ISL_2677003 | | | |
| see above | Utah Public Health Laboratory | Utah Public Health Laboratory | Erin L. Young, Kelly F. Oakeson, Tara Gallagher |
| EPI_ISL_2677633, EPI_ISL_2677635, EPI_ISL_2677636, EPI_ISL_2677637, EPI_ISL_2677639, EPI_ISL_2677640, EPI_ISL_2677642, EPI_ISL_2677643, EPI_ISL_2677644 | Wichita State University - Molecular Diagnostics Lab | Kansas Health and Environmental Lab | Katherine Wiggins, Mike Grose, Jonathan Barnell, Ben Olsen, and Phil Adam |
| EPI_ISL_2677645, EPI_ISL_2677646 | Kansas Health and Environmental Lab | Kansas Health and Environmental Lab | Katherine Wiggins, Mike Grose, Jonathan Barnell, Ben Olsen, and Phil Adam |
| EPI_ISL_2677668, EPI_ISL_2677669, EPI_ISL_2677670, EPI_ISL_2677671, EPI_ISL_2677672, EPI_ISL_2677673, EPI_ISL_2677674, EPI_ISL_2677675, EPI_ISL_2677676, EPI_ISL_2677677, EPI_ISL_2677678, EPI_ISL_2677679, EPI_ISL_2677680, EPI_ISL_2677681, EPI_ISL_2677682, EPI_ISL_2677683, EPI_ISL_2677684, EPI_ISL_2677685, EPI_ISL_2677686, EPI_ISL_2677687, EPI_ISL_2677688, EPI_ISL_2677689, EPI_ISL_2677690, EPI_ISL_2677691, EPI_ISL_2677692, EPI_ISL_2677693, EPI_ISL_2677694, EPI_ISL_2677695, EPI_ISL_2677696, EPI_ISL_2677697, EPI_ISL_2677698, EPI_ISL_2677699, EPI_ISL_2677700, EPI_ISL_2677701, EPI_ISL_2677702, EPI_ISL_2677703, EPI_ISL_2677704, EPI_ISL_2677705, EPI_ISL_2677706, EPI_ISL_2677707, EPI_ISL_2677708, EPI_ISL_2677709, EPI_ISL_2677710, EPI_ISL_2677711, EPI_ISL_2677712, EPI_ISL_2677713, EPI_ISL_2677714, EPI_ISL_2677715, EPI_ISL_2677716, EPI_ISL_2677717, EPI_ISL_2677718, EPI_ISL_2677719, EPI_ISL_2677720, EPI_ISL_2677722, EPI_ISL_2677723, EPI_ISL_2677724, EPI_ISL_2677725, EPI_ISL_2677726, EPI_ISL_2677727, EPI_ISL_2677728, EPI_ISL_2677729, EPI_ISL_2677730, EPI_ISL_2677731, EPI_ISL_2677732, EPI_ISL_2677733, EPI_ISL_2677734, EPI_ISL_2677735, EPI_ISL_2677736, EPI_ISL_2677737, EPI_ISL_2677738, EPI_ISL_2677739, EPI_ISL_2677740, EPI_ISL_2677741, EPI_ISL_2677742, EPI_ISL_2677743, EPI_ISL_2677744, EPI_ISL_2677745, EPI_ISL_2677746, EPI_ISL_2677747, EPI_ISL_2677748, EPI_ISL_2677749, EPI_ISL_2677750, EPI_ISL_2677751, EPI_ISL_2677752, EPI_ISL_2677753, EPI_ISL_2677754, EPI_ISL_2677755, EPI_ISL_2677756, EPI_ISL_2677757, EPI_ISL_2677758, EPI_ISL_2677759, EPI_ISL_2677760, EPI_ISL_2677761, EPI_ISL_2677762, EPI_ISL_2677763, EPI_ISL_2677764, EPI_ISL_2677765, EPI_ISL_2677766, EPI_ISL_2677767, EPI_ISL_2677768, EPI_ISL_2677769, EPI_ISL_2677770, EPI_ISL_2677771, EPI_ISL_2677772, EPI_ISL_2677773, EPI_ISL_2677774, EPI_ISL_2677775, EPI_ISL_2677776, EPI_ISL_2677777, EPI_ISL_2677778, EPI_ISL_2677779, EPI_ISL_2677780, EPI_ISL_2677781, EPI_ISL_2677782, EPI_ISL_2677783, EPI_ISL_2677784, EPI_ISL_2677785, EPI_ISL_2677786, EPI_ISL_2677787, EPI_ISL_2677788, EPI_ISL_2677789, EPI_ISL_2677790, EPI_ISL_2677791, EPI_ISL_2677792, EPI_ISL_2677793, EPI_ISL_2677794, EPI_ISL_2677795, EPI_ISL_2677796, EPI_ISL_2677797, EPI_ISL_2677798, EPI_ISL_2677799, EPI_ISL_2677800, EPI_ISL_2677801, EPI_ISL_2677802, EPI_ISL_2677803, EPI_ISL_2677804, EPI_ISL_2677805, EPI_ISL_2677806, EPI_ISL_2677807, EPI_ISL_2677808, EPI_ISL_2677809, EPI_ISL_2677810, EPI_ISL_2677811, EPI_ISL_2677812, EPI_ISL_2677813, EPI_ISL_2677814, EPI_ISL_2677815, EPI_ISL_2677816, EPI_ISL_2677817, EPI_ISL_2677818, EPI_ISL_2677819, EPI_ISL_2677820, EPI_ISL_2677821, EPI_ISL_2677822, EPI_ISL_2677823, EPI_ISL_2677824, EPI_ISL_2677825, EPI_ISL_2677826, EPI_ISL_2677827, EPI_ISL_2677828, EPI_ISL_2677829, EPI_ISL_2677830, EPI_ISL_2677831, EPI_ISL_2677832, EPI_ISL_2677833, EPI_ISL_2677834, EPI_ISL_2677835, EPI_ISL_2677836, EPI_ISL_2677837, EPI_ISL_2677838, EPI_ISL_2677839, EPI_ISL_2677840, EPI_ISL_2677841, EPI_ISL_2677842, EPI_ISL_2677843, EPI_ISL_2677844, EPI_ISL_2677845, EPI_ISL_2677846, EPI_ISL_2677847, EPI_ISL_2677848, EPI_ISL_2677849, EPI_ISL_2677850, EPI_ISL_2677851, EPI_ISL_2677852, EPI_ISL_2677853, EPI_ISL_2677854, EPI_ISL_2677855, EPI_ISL_2677856, EPI_ISL_2677857, EPI_ISL_2677858, EPI_ISL_2677859, EPI_ISL_2677860, EPI_ISL_2677861, EPI_ISL_2677862, EPI_ISL_2677863, EPI_ISL_2677864, EPI_ISL_2677865, EPI_ISL_2677866, EPI_ISL_2677867, EPI_ISL_2677868, EPI_ISL_2677869, EPI_ISL_2677870, EPI_ISL_2677871, EPI_ISL_2677872, EPI_ISL_2677873, EPI_ISL_2677874, EPI_ISL_2677875, EPI_ISL_2677876, EPI_ISL_2677877, EPI_ISL_2677878, EPI_ISL_2677879, EPI_ISL_2677880, EPI_ISL_2677881, EPI_ISL_2677882, EPI_ISL_2677883, EPI_ISL_2677884, EPI_ISL_2677885, EPI_ISL_2677886, EPI_ISL_2677887, EPI_ISL_2677888, EPI_ISL_2677889, EPI_ISL_2677890, EPI_ISL_2677891, EPI_ISL_2677892, EPI_ISL_2677893, EPI_ISL_2677894, EPI_ISL_2677895, EPI_ISL_2677896, EPI_ISL_2677897, EPI_ISL_2677898, EPI_ISL_2677899, EPI_ISL_2677900, EPI_ISL_2677901, EPI_ISL_2677902, EPI_ISL_2677903, EPI_ISL_2677904, EPI_ISL_2677905, EPI_ISL_2677906, EPI_ISL_2677907, EPI_ISL_2677908, EPI_ISL_2677909, EPI_ISL_2677910, EPI_ISL_2677911, EPI_ISL_2677912, EPI_ISL_2677913, EPI_ISL_2677914, EPI_ISL_2677915, EPI_ISL_2677916, EPI_ISL_2677917, EPI_ISL_2677918, EPI_ISL_2677919, EPI_ISL_2677920, EPI_ISL_2677921, EPI_ISL_2677922, EPI_ISL_2677923, EPI_ISL_2677924, EPI_ISL_2677925, EPI_ISL_2677926, EPI_ISL_2677927, EPI_ISL_2677928, EPI_ISL_2677929, EPI_ISL_2677930, EPI_ISL_2677931, EPI_ISL_2677932, EPI_ISL_2677933, EPI_ISL_2677934, EPI_ISL_2677935, EPI_ISL_2677936, EPI_ISL_2677937, EPI_ISL_2677938, EPI_ISL_2677939, EPI_ISL_2677940, EPI_ISL_2677955, EPI_ISL_2677956, EPI_ISL_2677957, EPI_ISL_2677958, EPI_ISL_2677959, EPI_ISL_2677960, EPI_ISL_2677961, EPI_ISL_2677962, EPI_ISL_2677963, EPI_ISL_2677964, EPI_ISL_2677965, EPI_ISL_2677966, EPI_ISL_2677967, EPI_ISL_2677968, EPI_ISL_2677969, EPI_ISL_2677970, EPI_ISL_2677971, EPI_ISL_2677972, EPI_ISL_2677973, EPI_ISL_2677974, EPI_ISL_2677975, EPI_ISL_2677976, EPI_ISL_2677977, EPI_ISL_2677978, EPI_ISL_2677979, EPI_ISL_2677980, EPI_ISL_2677981, EPI_ISL_2677982, EPI_ISL_2677983, EPI_ISL_2677984, EPI_ISL_2677985, EPI_ISL_2677986, EPI_ISL_2677987, EPI_ISL_2677988, EPI_ISL_2677989, EPI_ISL_2677990, EPI_ISL_2677991, EPI_ISL_2677992, EPI_ISL_2677993, EPI_ISL_2677994, EPI_ISL_2677995, EPI_ISL_2677996, EPI_ISL_2677997, EPI_ISL_2677998, EPI_ISL_2677999, EPI_ISL_2678000, EPI_ISL_2678001, EPI_ISL_2678002, EPI_ISL_2678003, EPI_ISL_2678004, EPI_ISL_2678005, EPI_ISL_2678006, EPI_ISL_2678007, EPI_ISL_2678008, EPI_ISL_2678009, EPI_ISL_2678010, EPI_ISL_2678011, EPI_ISL_2678012, EPI_ISL_2678013, EPI_ISL_2678014, EPI_ISL_2678015, EPI_ISL_2678016, EPI_ISL_2678017, EPI_ISL_2678018, EPI_ISL_2678019, EPI_ISL_2678020, EPI_ISL_2678021, EPI_ISL_2678022, EPI_ISL_2678023, EPI_ISL_2678024, EPI_ISL_2678025, EPI_ISL_2678026, EPI_ISL_2678027, EPI_ISL_2678028, EPI_ISL_2678029, EPI_ISL_2678030, EPI_ISL_2678031, EPI_ISL_2678032, EPI_ISL_2678033, EPI_ISL_2678034, EPI_ISL_2678035, EPI_ISL_2678036, EPI_ISL_2678037, EPI_ISL_2678038, EPI_ISL_2678039, EPI_ISL_2678040, EPI_ISL_2678041, EPI_ISL_2678042, EPI_ISL_2678043, EPI_ISL_2678044, EPI_ISL_2678045, EPI_ISL_2678046, EPI_ISL_2678047, EPI_ISL_2678048, EPI_ISL_2678049, EPI_ISL_2678050, EPI_ISL_2678051, EPI_ISL_2678052, EPI_ISL_2678053, EPI_ISL_2678054, EPI_ISL_2678056, EPI_ISL_2678057, EPI_ISL_2678058, EPI_ISL_2678059, EPI_ISL_2678060, EPI_ISL_2678061, EPI_ISL_2678062, EPI_ISL_2678063, EPI_ISL_2678064, EPI_ISL_2678065, EPI_ISL_2678066, EPI_ISL_2678067, EPI_ISL_2678068, EPI_ISL_2678069, EPI_ISL_2678070, EPI_ISL_2678071, EPI_ISL_2678072, EPI_ISL_2678073, EPI_ISL_2678074, EPI_ISL_2678075, EPI_ISL_2678076, EPI_ISL_2678077, EPI_ISL_2678078, EPI_ISL_2678079, EPI_ISL_2678080, EPI_ISL_2678081, EPI_ISL_2678082, EPI_ISL_2678083, EPI_ISL_2678084, EPI_ISL_2678085, EPI_ISL_2678086, EPI_ISL_2678087, EPI_ISL_2678088, EPI_ISL_2678089, EPI_ISL_2678090, EPI_ISL_2678091, EPI_ISL_2678206, EPI_ISL_2678207, EPI_ISL_2678208, EPI_ISL_2678209, EPI_ISL_2678210, EPI_ISL_2678211, EPI_ISL_2678212, EPI_ISL_2678213, EPI_ISL_2678214, EPI_ISL_2678215, EPI_ISL_2678216, EPI_ISL_2678217, EPI_ISL_2678218, EPI_ISL_2678219, EPI_ISL_2678220, EPI_ISL_2678221, EPI_ISL_2678222, EPI_ISL_2678223, EPI_ISL_2678224, EPI_ISL_2678225, EPI_ISL_2678226, EPI_ISL_2678227, EPI_ISL_2678228, EPI_ISL_2678229, EPI_ISL_2678230, EPI_ISL_2678231, EPI_ISL_2678232, EPI_ISL_2678233, EPI_ISL_2678234, EPI_ISL_2678235, EPI_ISL_2678236, EPI_ISL_2678237, EPI_ISL_2678238, EPI_ISL_2678239, EPI_ISL_2678240, EPI_ISL_2678241, EPI_ISL_2678242, EPI_ISL_2678243, EPI_ISL_2678244, EPI_ISL_2678245, EPI_ISL_2678246, EPI_ISL_2678247, EPI_ISL_2678248, EPI_ISL_2678249, EPI_ISL_2678250, EPI_ISL_2678251, EPI_ISL_2678252, EPI_ISL_2678253, EPI_ISL_2678254, EPI_ISL_2678255, EPI_ISL_2678256, EPI_ISL_2678257, EPI_ISL_2678258, EPI_ISL_2678259, EPI_ISL_2678260, EPI_ISL_2678261, EPI_ISL_2678262, EPI_ISL_2678263, EPI_ISL_2678264, EPI_ISL_2678265, EPI_ISL_2678266, EPI_ISL_2678267, EPI_ISL_2678268, EPI_ISL_2678269, EPI_ISL_2678270, EPI_ISL_2678271, EPI_ISL_2678272, EPI_ISL_2678273, EPI_ISL_2678274, EPI_ISL_2678319, EPI_ISL_2678320, EPI_ISL_2678322, EPI_ISL_2678326, EPI_ISL_2678327, EPI_ISL_2678328, EPI_ISL_2678329, EPI_ISL_2678330, EPI_ISL_2678331, EPI_ISL_2678332, EPI_ISL_2678333, EPI_ISL_2678334, EPI_ISL_2678336, EPI_ISL_2678337, EPI_ISL_2678338, EPI_ISL_2678339, EPI_ISL_2678342, EPI_ISL_2678344, EPI_ISL_2678345, EPI_ISL_2678346, EPI_ISL_2678347, EPI_ISL_2678348, EPI_ISL_2678352, EPI_ISL_2678353, EPI_ISL_2678355, EPI_ISL_2678356, EPI_ISL_2678357, EPI_ISL_2678358, EPI_ISL_2678361, | | | |

| | | | |
|---|---|---|---|
| EPI_ISL_2678363, EPI_ISL_2678366, EPI_ISL_2678831, EPI_ISL_2678834, EPI_ISL_2678835, EPI_ISL_2678837, EPI_ISL_2678841, EPI_ISL_2678843, EPI_ISL_2678845, EPI_ISL_2678848, EPI_ISL_2678850, EPI_ISL_2678852, EPI_ISL_2678854, EPI_ISL_2678856, EPI_ISL_2678859, EPI_ISL_2678863, EPI_ISL_2678865, EPI_ISL_2678866, EPI_ISL_2678868, EPI_ISL_2678872, EPI_ISL_2678876, EPI_ISL_2678877, EPI_ISL_2678879, EPI_ISL_2678881, EPI_ISL_2678887, EPI_ISL_2678890, EPI_ISL_2678892, EPI_ISL_2678894, EPI_ISL_2678895, EPI_ISL_2678896, EPI_ISL_2678897, EPI_ISL_2678898, EPI_ISL_2678899, EPI_ISL_2678900, EPI_ISL_2678901, EPI_ISL_2678902, EPI_ISL_2678903, EPI_ISL_2678904, EPI_ISL_2678905, EPI_ISL_2678906, EPI_ISL_2678907, EPI_ISL_2678908, EPI_ISL_2678909, EPI_ISL_2678910, EPI_ISL_2678911, EPI_ISL_2678913, EPI_ISL_2678914, EPI_ISL_2678915, EPI_ISL_2678916, EPI_ISL_2678917, EPI_ISL_2678918, EPI_ISL_2678919, EPI_ISL_2678920, EPI_ISL_2678921, EPI_ISL_2678922, EPI_ISL_2678923, EPI_ISL_2678924, EPI_ISL_2678925, EPI_ISL_2678926, EPI_ISL_2678927, EPI_ISL_2678928, EPI_ISL_2678929, EPI_ISL_2678930, EPI_ISL_2678931, EPI_ISL_2678932, EPI_ISL_2678933, EPI_ISL_2678935, EPI_ISL_2678936, EPI_ISL_2678938, EPI_ISL_2678939, EPI_ISL_2678940, EPI_ISL_2678941, EPI_ISL_2678942, EPI_ISL_2678943, EPI_ISL_2678944, EPI_ISL_2678945, EPI_ISL_2678946, EPI_ISL_2678947, EPI_ISL_2678948, EPI_ISL_2678949, EPI_ISL_2678950, EPI_ISL_2678951, EPI_ISL_2678952, EPI_ISL_2678953, EPI_ISL_2678954, EPI_ISL_2678955, EPI_ISL_2678956, EPI_ISL_2678957, EPI_ISL_2678958, EPI_ISL_2678959, EPI_ISL_2678960, EPI_ISL_2678961, EPI_ISL_2678962, EPI_ISL_2678963, EPI_ISL_2678965, EPI_ISL_2678966, EPI_ISL_2678967, EPI_ISL_2678968, EPI_ISL_2678970, EPI_ISL_2678971, EPI_ISL_2678972, EPI_ISL_2678973, EPI_ISL_2678974, EPI_ISL_2678975, EPI_ISL_2678976, EPI_ISL_2678977, EPI_ISL_2678980, EPI_ISL_2678981, EPI_ISL_2678982, EPI_ISL_2678983, EPI_ISL_2678984, EPI_ISL_2678985, EPI_ISL_2678987, EPI_ISL_2678988, EPI_ISL_2678989, EPI_ISL_2678990, EPI_ISL_2678991, EPI_ISL_2678992, EPI_ISL_2678993, EPI_ISL_2678994, EPI_ISL_2678995, EPI_ISL_2678996, EPI_ISL_2678997, EPI_ISL_2678998, EPI_ISL_2678999, EPI_ISL_2679000, EPI_ISL_2679001, EPI_ISL_2679002, EPI_ISL_2679003, EPI_ISL_2679004, EPI_ISL_2679005, EPI_ISL_2679006, EPI_ISL_2679007, EPI_ISL_2679008, EPI_ISL_2679009, EPI_ISL_2679010, EPI_ISL_2679011, EPI_ISL_2679012, EPI_ISL_2679013, EPI_ISL_2679014, EPI_ISL_2679015, EPI_ISL_2679017, EPI_ISL_2679018, EPI_ISL_2679019, EPI_ISL_2679020, EPI_ISL_2679021, EPI_ISL_2679022, EPI_ISL_2679023, EPI_ISL_2679024, EPI_ISL_2679025, EPI_ISL_2679026, EPI_ISL_2679027, EPI_ISL_2679028, EPI_ISL_2679029, EPI_ISL_2679030, EPI_ISL_2679031, EPI_ISL_2679032, EPI_ISL_2679033, EPI_ISL_2679034, EPI_ISL_2679035, EPI_ISL_2679036, EPI_ISL_2679037, EPI_ISL_2679038, EPI_ISL_2679039, EPI_ISL_2679040, EPI_ISL_2679041, EPI_ISL_2679042, EPI_ISL_2679043, EPI_ISL_2679045, EPI_ISL_2679048, EPI_ISL_2679050, EPI_ISL_2679053, EPI_ISL_2679055, EPI_ISL_2679058, EPI_ISL_2679060, EPI_ISL_2679063, EPI_ISL_2679066, EPI_ISL_2679068, EPI_ISL_2679070, EPI_ISL_2679072, EPI_ISL_2679074, EPI_ISL_2679075, EPI_ISL_2679077, EPI_ISL_2679079, EPI_ISL_2679080, EPI_ISL_2679082, EPI_ISL_2679084, EPI_ISL_2679086, EPI_ISL_2679088, EPI_ISL_2679090, EPI_ISL_2679092, EPI_ISL_2679093, EPI_ISL_2679095, EPI_ISL_2679097, EPI_ISL_2679099, EPI_ISL_2679101, EPI_ISL_2679103, EPI_ISL_2679105, EPI_ISL_2679107, EPI_ISL_2679109, EPI_ISL_2679110, EPI_ISL_2679112, EPI_ISL_2679114, EPI_ISL_2679116, EPI_ISL_2679117, EPI_ISL_2679119, EPI_ISL_2679121, EPI_ISL_2679125, EPI_ISL_2679127, EPI_ISL_2679129, EPI_ISL_2679131, EPI_ISL_2679133, EPI_ISL_2679135, EPI_ISL_2679140, EPI_ISL_2679142, EPI_ISL_2679144, EPI_ISL_2679145, EPI_ISL_2679147, EPI_ISL_2679149, EPI_ISL_2679151, EPI_ISL_2679152, EPI_ISL_2679154, EPI_ISL_2679156, EPI_ISL_2679158, EPI_ISL_2679160, EPI_ISL_2679161, EPI_ISL_2679163, EPI_ISL_2679164, EPI_ISL_2679166, EPI_ISL_2679168, EPI_ISL_2679170, EPI_ISL_2679171, EPI_ISL_2679173, EPI_ISL_2679175, EPI_ISL_2679176, EPI_ISL_2679177, EPI_ISL_2679179, EPI_ISL_2679181, EPI_ISL_2679183, EPI_ISL_2679185, EPI_ISL_2679187, EPI_ISL_2679189, EPI_ISL_2679190, EPI_ISL_2679192, EPI_ISL_2679193, EPI_ISL_2679195, EPI_ISL_2679197, EPI_ISL_2679199, EPI_ISL_2679200, EPI_ISL_2679201, EPI_ISL_2679203, EPI_ISL_2679204, EPI_ISL_2679206, EPI_ISL_2679207, EPI_ISL_2679211, EPI_ISL_2679217, EPI_ISL_2679218, EPI_ISL_2679220, EPI_ISL_2679222, EPI_ISL_2679224, EPI_ISL_2679226, EPI_ISL_2679227, EPI_ISL_2679229, EPI_ISL_2679231, EPI_ISL_2679232, EPI_ISL_2679234, EPI_ISL_2679236, EPI_ISL_2679237, EPI_ISL_2679239, EPI_ISL_2679241, EPI_ISL_2679244, EPI_ISL_2679246, EPI_ISL_2679247, EPI_ISL_2679249, EPI_ISL_2679251, EPI_ISL_2679253, EPI_ISL_2679255, EPI_ISL_2679257, EPI_ISL_2679258, EPI_ISL_2679260, EPI_ISL_2679262, EPI_ISL_2679263, EPI_ISL_2679267, EPI_ISL_2679269, EPI_ISL_2679271, EPI_ISL_2679273, EPI_ISL_2679275, EPI_ISL_2679276, EPI_ISL_2679278, EPI_ISL_2679281, EPI_ISL_2679283, EPI_ISL_2679285, EPI_ISL_2679286, EPI_ISL_2679288, EPI_ISL_2679289, EPI_ISL_2679291, EPI_ISL_2679293, EPI_ISL_2679294, EPI_ISL_2679297, EPI_ISL_2679301, EPI_ISL_2679303, EPI_ISL_2679305, EPI_ISL_2679307, EPI_ISL_2679309, EPI_ISL_2679311, EPI_ISL_2679313, EPI_ISL_2679315, EPI_ISL_2679317, EPI_ISL_2679318, EPI_ISL_2679320, EPI_ISL_2679324, EPI_ISL_2679328, EPI_ISL_2679332, EPI_ISL_2679334, EPI_ISL_2679336, EPI_ISL_2679337, EPI_ISL_2679339, EPI_ISL_2679341, EPI_ISL_2679342, EPI_ISL_2679344, EPI_ISL_2679346, EPI_ISL_2679348, EPI_ISL_2679350, EPI_ISL_2679351, EPI_ISL_2679353, EPI_ISL_2679354, EPI_ISL_2679356, EPI_ISL_2679358, EPI_ISL_2679359, EPI_ISL_2679362, EPI_ISL_2679364, EPI_ISL_2679366, EPI_ISL_2679368, EPI_ISL_2679369, EPI_ISL_2679371, EPI_ISL_2679373, EPI_ISL_2679374, EPI_ISL_2679376, EPI_ISL_2679380, EPI_ISL_2679382, EPI_ISL_2679384, EPI_ISL_2679386, EPI_ISL_2679388, EPI_ISL_2679389, EPI_ISL_2679391, EPI_ISL_2679392, EPI_ISL_2679395, EPI_ISL_2679397, EPI_ISL_2679399, EPI_ISL_2679401, EPI_ISL_2679403, EPI_ISL_2679405, EPI_ISL_2679406, EPI_ISL_2679408, EPI_ISL_2679409, EPI_ISL_2679411, EPI_ISL_2679413, EPI_ISL_2679415, EPI_ISL_2679418, EPI_ISL_2679420, EPI_ISL_2679421, EPI_ISL_2679422, EPI_ISL_2679424, EPI_ISL_2679426, EPI_ISL_2679430, EPI_ISL_2679432, EPI_ISL_2679434, EPI_ISL_2679436, EPI_ISL_2679437, EPI_ISL_2679439, EPI_ISL_2679441, EPI_ISL_2679442, EPI_ISL_2679444, EPI_ISL_2679446, EPI_ISL_2679448, EPI_ISL_2679450, EPI_ISL_2679452, EPI_ISL_2679454, EPI_ISL_2679456, EPI_ISL_2679457, EPI_ISL_2679459, EPI_ISL_2679461, EPI_ISL_2679462, EPI_ISL_2679466, EPI_ISL_2679468, EPI_ISL_2679470, EPI_ISL_2679471, EPI_ISL_2679473, EPI_ISL_2679475, EPI_ISL_2679477, EPI_ISL_2679478, EPI_ISL_2679480, EPI_ISL_2679481, EPI_ISL_2679483, EPI_ISL_2679484, EPI_ISL_2679486, EPI_ISL_2679488, EPI_ISL_2679490, EPI_ISL_2679493, EPI_ISL_2679494, EPI_ISL_2679496, EPI_ISL_2679498, EPI_ISL_2679500, EPI_ISL_2679502, EPI_ISL_2679506, EPI_ISL_2679508, EPI_ISL_2679510, EPI_ISL_2679512, EPI_ISL_2679514, EPI_ISL_2679516, EPI_ISL_2679518, EPI_ISL_2679520, EPI_ISL_2679521, EPI_ISL_2679523, EPI_ISL_2679524, EPI_ISL_2679526, EPI_ISL_2679527, EPI_ISL_2679530, EPI_ISL_2679531, EPI_ISL_2679533, EPI_ISL_2679534, EPI_ISL_2679536, EPI_ISL_2679539, EPI_ISL_2679541, EPI_ISL_2679543, EPI_ISL_2679545, EPI_ISL_2679547, EPI_ISL_2679549, EPI_ISL_2679550, EPI_ISL_2679552, EPI_ISL_2679554, EPI_ISL_2679556, EPI_ISL_2679558, EPI_ISL_2679560, EPI_ISL_2679562, EPI_ISL_2679564, EPI_ISL_2679566, EPI_ISL_2679568, EPI_ISL_2679570, EPI_ISL_2679574, EPI_ISL_2679576, EPI_ISL_2679577, EPI_ISL_2679579, EPI_ISL_2679583, EPI_ISL_2679585, EPI_ISL_2679587, EPI_ISL_2679588, EPI_ISL_2679590, EPI_ISL_2679593, EPI_ISL_2679594, EPI_ISL_2679596, EPI_ISL_2679598, EPI_ISL_2679601, EPI_ISL_2679603, EPI_ISL_2679605, EPI_ISL_2679607, EPI_ISL_2679609, EPI_ISL_2679611, EPI_ISL_2679613, EPI_ISL_2679614, EPI_ISL_2679616, EPI_ISL_2679617, EPI_ISL_2679621, EPI_ISL_2679622, EPI_ISL_2679624, EPI_ISL_2679626, EPI_ISL_2679628, EPI_ISL_2679630, EPI_ISL_2679632, EPI_ISL_2679634, EPI_ISL_2679636, EPI_ISL_2679637, EPI_ISL_2679639, EPI_ISL_2679641, EPI_ISL_2679643, EPI_ISL_2679645, EPI_ISL_2679647, EPI_ISL_2679649, EPI_ISL_2679650, EPI_ISL_2679652, EPI_ISL_2679654, EPI_ISL_2679656, EPI_ISL_2679658, EPI_ISL_2679660, EPI_ISL_2679661, EPI_ISL_2679664, EPI_ISL_2679666, EPI_ISL_2679668, EPI_ISL_2679669, EPI_ISL_2679671, EPI_ISL_2679673, EPI_ISL_2679675, EPI_ISL_2679677, EPI_ISL_2679679, EPI_ISL_2679681, EPI_ISL_2679682, EPI_ISL_2679684, EPI_ISL_2679686, EPI_ISL_2679688, EPI_ISL_2679690, EPI_ISL_2679691, EPI_ISL_2679693, EPI_ISL_2679695, EPI_ISL_2679697, EPI_ISL_2679699, EPI_ISL_2679700, EPI_ISL_2679701, EPI_ISL_2679703, EPI_ISL_2679705, EPI_ISL_2679706, EPI_ISL_2679708, EPI_ISL_2679711, EPI_ISL_2679713, EPI_ISL_2679715, EPI_ISL_2679716, EPI_ISL_2679718, EPI_ISL_2679720, EPI_ISL_2679722, EPI_ISL_2679724, EPI_ISL_2679725, EPI_ISL_2679727, EPI_ISL_2679729, EPI_ISL_2679731, EPI_ISL_2679733, EPI_ISL_2679735, EPI_ISL_2679737, EPI_ISL_2679739, EPI_ISL_2679740, EPI_ISL_2679742, EPI_ISL_2679744, EPI_ISL_2679746, EPI_ISL_2679747, EPI_ISL_2679749, EPI_ISL_2679751, EPI_ISL_2679753, EPI_ISL_2679754, EPI_ISL_2679756, EPI_ISL_2679758, EPI_ISL_2679760, EPI_ISL_2679762, EPI_ISL_2679764, EPI_ISL_2679766, EPI_ISL_2679768, EPI_ISL_2679770, EPI_ISL_2679771, EPI_ISL_2679772, EPI_ISL_2679774, EPI_ISL_2679776 | | | |
| see above | Utah Public Health Laboratory | Utah Public Health Laboratory | Erin L. Young, Kelly F. Oakeson, Tara Gallagher |
| EPI_ISL_2680469, EPI_ISL_2680471, EPI_ISL_2680473, EPI_ISL_2680474, EPI_ISL_2680476, EPI_ISL_2680478, EPI_ISL_2680480, EPI_ISL_2680482, EPI_ISL_2680483, EPI_ISL_2680485, EPI_ISL_2680487, EPI_ISL_2680488, EPI_ISL_2680490, EPI_ISL_2680492 | | | |
| see above | Humboldt County Public Health Laboratory | Humboldt County Public Health Laboratory | Jeremy Corrigan |
| EPI_ISL_2680916, EPI_ISL_2680917, EPI_ISL_2680918, EPI_ISL_2680919, EPI_ISL_2680920, EPI_ISL_2680921, EPI_ISL_2680922, EPI_ISL_2680923, EPI_ISL_2680924, EPI_ISL_2680925, EPI_ISL_2680926, EPI_ISL_2680927 | | | |
| see above | Laboratorios Delia Barraza | Laboratorio nacional-LANIIA-CIAD | Consorcio Mexicano de Vigilancia Genómica (CoViGen-Mex). Authors (in alphabetical order): Julio Elias Alvarado-Yaah, Carlos F. Arias, Santiago Ávila-Ríos, Eduardo Becerril-Vargas, Víctor Hugo Borja-Aburto, Celia Boukadida, Cristóbal Cháidez-Quiróz, Juan Bautista Chale-Dzul, Ricardo Ciria Merce, Andreu Comas-García, Célida Duque Molina, Julissa Enciso-Ibarra, José Antonio Enciso-Moreno, Gloria Elena Espinosa-Ayala, Fernando Fontove-Herrera, Daniel Fregoso-Rueda, Victor Eduardo García-Arias, Alejandra García-Gasca, Bruno Gómez-Gil, Jean Pierre González, Irvin González-López, Concepción Grajales-Muñiz, Ricardo Grande, Rosa María Gutiérrez Rios, Jesús Hernández, Alejandra Hernández-Terán, Alfredo Herrera-Estrella, Carla Ivón Herrera-Najera, Pavel Isa, Verónica Mata-Haro, Daniel Lira Morales, Susana Lopez, Antonio Loza Román, Brenda Irasema Maldonado-Meza, Bernardo Martínez-Miguel, José Arturo Martínez-Orozco, Célida Martínez- Rodríguez, Margarita Matías-Florentino, Fidencio Mejía-Nepomuceno, María Guadalupe de Jesús Mireles-Rivera, Gloria María Molina-Salinas, Hector Montoya-Fuentes, , Mario Mújica-Sánchez, José Esteban Muñoz-Medina, José de Jesús Nuñez-Contreras, Alicia Ocaña-Mondragón, Luis Alberto Ochoa-Carrera, Hector Esteban Paz-Juárez, Marissa Perez-Garcia, Francisco Pulido, Helen Haydee Fernanda Ramírez-Plascencia, Jorge Salas-Hernández, Angel Gustavo Salas-Lais, Alejandro Sánchez-Flores, Clara Esperanza Santacruz-Tinoco, María Guadalupe Santiago-Mauricio, Selene Zárate, Nelly Sélem-Mojica, Blanca Taboada, Gloria Vazquez, Joel Armando Vázquez-Pérez. Laboratorios Delia Barraza: María Delia Barraza Sámano, Claudia Soto Félix, Melissa García Angulo. |
| EPI_ISL_2680928, EPI_ISL_2680929 | Hospital Fátima S.A. de C.V. | Laboratorio nacional-LANIIA-CIAD | Consorcio Mexicano de Vigilancia Genómica (CoViGen-Mex). Authors (in alphabetical order): Julio Elias Alvarado-Yaah, Carlos F. Arias, Santiago Ávila-Ríos, Eduardo Becerril-Vargas, Víctor Hugo Borja-Aburto, Celia Boukadida, Cristóbal Cháidez-Quiróz, Juan Bautista Chale-Dzul, Ricardo Ciria Merce, Andreu Comas-García, Célida Duque Molina, Julissa Enciso-Ibarra, José Antonio Enciso-Moreno, Gloria Elena Espinosa-Ayala, Fernando Fontove-Herrera, Daniel Fregoso-Rueda, Victor Eduardo García-Arias, Alejandra García-Gasca, Bruno Gómez-Gil, Jean Pierre González, Irvin González-López, Concepción Grajales-Muñiz, Ricardo Grande, Rosa María Gutiérrez Rios, Jesús Hernández, Alejandra Hernández-Terán, Alfredo Herrera-Estrella, Carla Ivón Herrera-Najera, Pavel Isa, Verónica Mata-Haro, Daniel Lira Morales, Susana Lopez, Antonio Loza Román, Brenda Irasema Maldonado-Meza, Bernardo Martínez-Miguel, José Arturo Martínez-Orozco, Célida Martínez- Rodríguez, Margarita Matías-Florentino, Fidencio Mejía-Nepomuceno, María Guadalupe de Jesús Mireles-Rivera, Gloria María Molina-Salinas, Hector Montoya-Fuentes, , Mario Mújica-Sánchez, José Esteban Muñoz-Medina, José de Jesús Nuñez-Contreras, Alicia Ocaña-Mondragón, Luis Alberto Ochoa-Carrera, Hector Esteban Paz-Juárez, Marissa Perez-Garcia, Francisco Pulido, Helen Haydee Fernanda Ramírez-Plascencia, Jorge Salas-Hernández, Angel Gustavo Salas-Lais, Alejandro Sánchez-Flores, Clara Esperanza Santacruz-Tinoco, María Guadalupe Santiago-Mauricio, Selene Zárate, Nelly Sélem-Mojica, Blanca Taboada, Gloria Vazquez, Joel Armando Vázquez-Pérez. |
| EPI_ISL_2680930 | Servicio Diagnóstico Clínico | Laboratorio nacional-LANIIA-CIAD | Consorcio Mexicano de Vigilancia Genómica (CoViGen-Mex). Authors (in alphabetical order): Julio Elias Alvarado-Yaah, Carlos F. Arias, Santiago Ávila-Ríos, Eduardo Becerril-Vargas, Víctor Hugo Borja-Aburto, Celia Boukadida, Cristóbal Cháidez-Quiróz, Juan Bautista Chale-Dzul, Ricardo Ciria Merce, Andreu Comas-García, Célida Duque Molina, Julissa Enciso-Ibarra, José Antonio Enciso-Moreno, Gloria Elena Espinosa-Ayala, Fernando Fontove-Herrera, Daniel Fregoso-Rueda, Victor Eduardo García-Arias, Alejandra García-Gasca, Bruno Gómez-Gil, Jean Pierre González, Irvin González-López, Concepción Grajales-Muñiz, Ricardo Grande, Rosa María Gutiérrez Rios, Jesús Hernández, Alejandra Hernández-Terán, Alfredo Herrera-Estrella, Carla Ivón Herrera-Najera, Pavel Isa, Verónica Mata-Haro, Daniel Lira Morales, Susana Lopez, Antonio Loza Román, Brenda Irasema Maldonado-Meza, Bernardo Martínez-Miguel, José Arturo Martínez-Orozco, Célida Martínez- Rodríguez, Margarita Matías-Florentino, Fidencio Mejía-Nepomuceno, María Guadalupe de Jesús Mireles-Rivera, Gloria María Molina-Salinas, Hector Montoya-Fuentes, , Mario Mújica-Sánchez, José Esteban Muñoz-Medina, José de Jesús Nuñez-Contreras, Alicia Ocaña-Mondragón, Luis Alberto Ochoa-Carrera, Hector Esteban Paz-Juárez, Marissa Perez-Garcia, Francisco Pulido, Helen Haydee Fernanda Ramírez-Plascencia, Jorge Salas-Hernández, Angel Gustavo Salas-Lais, Alejandro Sánchez-Flores, Clara Esperanza Santacruz-Tinoco, María Guadalupe Santiago-Mauricio, Selene Zárate, Nelly Sélem-Mojica, Blanca Taboada, Gloria Vazquez, Joel Armando Vázquez-Pérez. |
| EPI_ISL_2680931, EPI_ISL_2680932, EPI_ISL_2680933 | Hospital Margarita Maza de Juárez | Microbial Genomics Laboratory | Consorcio Mexicano de Vigilancia Genómica (CoViGen-Mex). Authors (in alphabetical order): Julio Elias Alvarado-Yaah, Carlos F. Arias, Santiago Ávila-Ríos, Eduardo Becerril-Vargas, Víctor Hugo Borja-Aburto, Celia Boukadida, Cristóbal Cháidez-Quiróz, Juan Bautista Chale-Dzul, Ricardo Ciria Merce, Andreu Comas-García, Célida Duque Molina, Julissa Enciso-Ibarra, José Antonio Enciso-Moreno, Gloria Elena Espinosa-Ayala, Fernando Fontove-Herrera, Daniel Fregoso-Rueda, Victor Eduardo García-Arias, Alejandra García-Gasca, Bruno Gómez-Gil, Jean Pierre González, Irvin González-López, Concepción Grajales-Muñiz, Ricardo Grande, Rosa María Gutiérrez Rios, Jesús Hernández, Alejandra Hernández-Terán, Alfredo Herrera-Estrella, Carla Ivón Herrera-Najera, Pavel Isa, Verónica Mata-Haro, Daniel Lira Morales, Susana Lopez, Antonio Loza Román, Brenda Irasema Maldonado-Meza, Bernardo Martínez-Miguel, José Arturo Martínez-Orozco, Célida Martínez- Rodríguez, Margarita Matías-Florentino, Fidencio Mejía-Nepomuceno, María Guadalupe de Jesús Mireles-Rivera, Gloria María Molina-Salinas, Hector Montoya-Fuentes, , Mario Mújica-Sánchez, José Esteban Muñoz-Medina, José de Jesús Nuñez-Contreras, Alicia Ocaña-Mondragón, Luis Alberto Ochoa-Carrera, Hector Esteban Paz-Juárez, Marissa Perez-Garcia, Francisco Pulido, Helen Haydee Fernanda Ramírez-Plascencia, Jorge Salas-Hernández, Angel Gustavo Salas-Lais, Alejandro Sánchez-Flores, Clara Esperanza Santacruz-Tinoco, María Guadalupe Santiago-Mauricio, Selene Zárate, Nelly Sélem-Mojica, Blanca Taboada, Gloria Vazquez, Joel Armando Vázquez-Pérez. |
| EPI_ISL_2680934 | Imágenes Diagnósticas de Mazatlán | Microbial Genomics Laboratory | Consorcio Mexicano de Vigilancia Genómica (CoViGen-Mex). Authors (in alphabetical order): Julio Elias Alvarado-Yaah, Carlos F. Arias, Santiago Ávila-Ríos, Eduardo Becerril-Vargas, Víctor Hugo Borja-Aburto, Celia Boukadida, Cristóbal Cháidez-Quiróz, Juan Bautista Chale-Dzul, Ricardo Ciria Merce, Andreu Comas-García, Célida Duque Molina, Julissa Enciso-Ibarra, José Antonio Enciso-Moreno, Gloria Elena Espinosa-Ayala, Fernando Fontove-Herrera, Daniel Fregoso-Rueda, Victor Eduardo García-Arias, Alejandra García-Gasca, Bruno Gómez-Gil, Jean Pierre González, Irvin González-López, Concepción Grajales-Muñiz, Ricardo Grande, Rosa María Gutiérrez Rios, Jesús Hernández, Alejandra Hernández-Terán, Alfredo Herrera-Estrella, Carla Ivón Herrera-Najera, Pavel Isa, Verónica Mata-Haro, Daniel Lira Morales, Susana Lopez, Antonio Loza Román, Brenda Irasema Maldonado-Meza, Bernardo Martínez-Miguel, José Arturo Martínez-Orozco, Célida Martínez- Rodríguez, Margarita Matías-Florentino, Fidencio Mejía-Nepomuceno, María Guadalupe de Jesús Mireles-Rivera, Gloria María Molina-Salinas, Hector Montoya-Fuentes, , Mario Mújica-Sánchez, José Esteban Muñoz-Medina, José de Jesús Nuñez-Contreras, Alicia Ocaña-Mondragón, Luis Alberto Ochoa-Carrera, Hector Esteban Paz-Juárez, Marissa Perez-Garcia, Francisco Pulido, Helen Haydee Fernanda Ramírez-Plascencia, Jorge Salas-Hernández, Angel Gustavo Salas-Lais, Alejandro Sánchez-Flores, Clara Esperanza Santacruz-Tinoco, María Guadalupe Santiago-Mauricio, Selene Zárate, Nelly Sélem-Mojica, Blanca Taboada, Gloria Vazquez, Joel Armando Vázquez-Pérez. |
| EPI_ISL_2680935 | Hospital Sharp | Microbial Genomics Laboratory | Consorcio Mexicano de Vigilancia Genómica (CoViGen-Mex). Authors (in alphabetical order): Julio Elias Alvarado-Yaah, Carlos F. Arias, Santiago Ávila-Ríos, Eduardo Becerril-Vargas, Víctor Hugo Borja-Aburto, Celia Boukadida, Cristóbal Cháidez-Quiróz, Juan Bautista Chale-Dzul, Ricardo Ciria Merce, Andreu Comas-García, Célida Duque Molina, Julissa Enciso-Ibarra, José Antonio Enciso-Moreno, Gloria Elena Espinosa-Ayala, Fernando Fontove-Herrera, Daniel Fregoso-Rueda, Victor Eduardo García-Arias, Alejandra García-Gasca, Bruno Gómez-Gil, Jean Pierre González, Irvin González-López, Concepción Grajales-Muñiz, Ricardo Grande, Rosa María Gutiérrez Rios, Jesús Hernández, Alejandra Hernández-Terán, Alfredo Herrera-Estrella, Carla Ivón Herrera-Najera, Pavel Isa, Verónica Mata-Haro, Daniel Lira Morales, Susana Lopez, Antonio Loza Román, Brenda Irasema Maldonado-Meza, Bernardo Martínez-Miguel, José Arturo Martínez-Orozco, Célida Martínez- Rodríguez, Margarita Matías-Florentino, Fidencio Mejía-Nepomuceno, María Guadalupe de Jesús Mireles-Rivera, Gloria María Molina-Salinas, Hector Montoya-Fuentes, , Mario Mújica-Sánchez, José Esteban Muñoz-Medina, José de Jesús Nuñez-Contreras, Alicia Ocaña-Mondragón, Luis Alberto Ochoa-Carrera, Hector Esteban Paz-Juárez, Marissa Perez-Garcia, Francisco Pulido, Helen Haydee Fernanda Ramírez-Plascencia, Jorge Salas-Hernández, Angel Gustavo Salas-Lais, Alejandro Sánchez-Flores, Clara Esperanza Santacruz-Tinoco, María Guadalupe Santiago-Mauricio, Selene Zárate, Nelly Sélem-Mojica, Blanca Taboada, Gloria Vazquez, Joel Armando Vázquez-Pérez. |
| EPI_ISL_2680936, EPI_ISL_2680937 | Hospital Margarita Maza de Juárez | Microbial Genomics Laboratory | Consorcio Mexicano de Vigilancia Genómica (CoViGen-Mex). Authors (in alphabetical order): Julio Elias Alvarado-Yaah, Carlos F. Arias, Santiago Ávila-Ríos, Eduardo Becerril-Vargas, Víctor Hugo Borja-Aburto, Celia Boukadida, Cristóbal Cháidez-Quiróz, Juan Bautista Chale-Dzul, Ricardo Ciria Merce, Andreu Comas-García, Célida Duque Molina, Julissa Enciso-Ibarra, José Antonio Enciso-Moreno, Gloria Elena Espinosa-Ayala, Fernando Fontove-Herrera, Daniel Fregoso-Rueda, Victor Eduardo García-Arias, Alejandra García-Gasca, Bruno Gómez-Gil, Jean Pierre González, Irvin González-López, Concepción Grajales-Muñiz, Ricardo Grande, Rosa María Gutiérrez Rios, Jesús Hernández, Alejandra Hernández-Terán, Alfredo Herrera-Estrella, Carla Ivón Herrera-Najera, Pavel Isa, Verónica Mata-Haro, Daniel Lira Morales, Susana Lopez, Antonio Loza Román, Brenda Irasema Maldonado-Meza, Bernardo Martínez-Miguel, José Arturo Martínez-Orozco, Célida Martínez- Rodríguez, Margarita Matías-Florentino, Fidencio Mejía-Nepomuceno, María Guadalupe de Jesús Mireles-Rivera, Gloria María Molina-Salinas, Hector Montoya-Fuentes, , Mario Mújica-Sánchez, José Esteban Muñoz-Medina, José de Jesús Nuñez-Contreras, Alicia Ocaña-Mondragón, Luis Alberto Ochoa-Carrera, Hector Esteban Paz-Juárez, Marissa Perez-Garcia, Francisco Pulido, Helen Haydee Fernanda Ramírez-Plascencia, Jorge Salas-Hernández, Angel Gustavo Salas-Lais, Alejandro Sánchez-Flores, Clara Esperanza Santacruz-Tinoco, María Guadalupe Santiago-Mauricio, Selene Zárate, Nelly Sélem-Mojica, Blanca Taboada, Gloria Vazquez, Joel Armando Vázquez-Pérez. |
| EPI_ISL_2680938, EPI_ISL_2680939, EPI_ISL_2680940, EPI_ISL_2680942 | Hospital Sharp | Microbial Genomics Laboratory | Consorcio Mexicano de Vigilancia Genómica (CoViGen-Mex). Authors (in alphabetical order): Julio Elias Alvarado-Yaah, Carlos F. Arias, Santiago Ávila-Ríos, Eduardo Becerril-Vargas, Víctor Hugo Borja-Aburto, Celia Boukadida, Cristóbal Cháidez-Quiróz, Juan Bautista Chale-Dzul, Ricardo Ciria Merce, Andreu Comas-García, Célida Duque Molina, Julissa Enciso-Ibarra, José Antonio Enciso-Moreno, Gloria Elena Espinosa-Ayala, Fernando Fontove-Herrera, Daniel Fregoso-Rueda, Victor Eduardo García-Arias, Alejandra García-Gasca, Bruno Gómez-Gil, Jean Pierre González, Irvin González-López, Concepción Grajales-Muñiz, Ricardo Grande, Rosa María Gutiérrez Rios, Jesús Hernández, Alejandra Hernández-Terán, Alfredo Herrera-Estrella, Carla Ivón Herrera-Najera, Pavel Isa, Verónica Mata-Haro, Daniel Lira Morales, Susana Lopez, Antonio Loza Román, Brenda Irasema Maldonado-Meza, Bernardo Martínez-Miguel, José Arturo Martínez-Orozco, Célida Martínez- Rodríguez, Margarita Matías-Florentino, Fidencio Mejía-Nepomuceno, María Guadalupe de Jesús Mireles-Rivera, Gloria María Molina-Salinas, Hector Montoya-Fuentes, , Mario Mújica-Sánchez, José Esteban Muñoz-Medina, José de Jesús Nuñez-Contreras, Alicia Ocaña-Mondragón, Luis Alberto Ochoa-Carrera, Hector Esteban Paz-Juárez, Marissa Perez-Garcia, Francisco Pulido, Helen Haydee Fernanda Ramírez-Plascencia, Jorge Salas-Hernández, Angel Gustavo Salas-Lais, Alejandro Sánchez-Flores, Clara Esperanza Santacruz-Tinoco, María Guadalupe Santiago-Mauricio, Selene Zárate, Nelly Sélem-Mojica, Blanca Taboada, Gloria Vazquez, Joel Armando Vázquez-Pérez. |
| EPI_ISL_2681046 | Laboratorio Central de Epidemiología (LCE) | Instituto de Biotecnología de la UNAM | Consorcio Mexicano de Vigilancia Genómica (CoViGen-Mex). Authors (in alphabetical order): Julio Elias Alvarado-Yaah, Carlos F. Arias, Santiago Ávila-Ríos, Eduardo Becerril-Vargas, Víctor Hugo Borja-Aburto, Celia Boukadida, Cristóbal Cháidez-Quiróz, Juan Bautista Chale-Dzul, Ricardo Ciria Merce, Andreu Comas-García, Célida Duque Molina, Julissa Enciso-Ibarra, José Antonio Enciso-Moreno, Gloria Elena Espinosa-Ayala, Fernando Fontove-Herrera, Daniel Fregoso-Rueda, Victor Eduardo García-Arias, Alejandra García-Gasca, Bruno Gómez-Gil, Jean Pierre González, Irvin González-López, Concepción Grajales-Muñiz, Ricardo Grande, Rosa María Gutiérrez Rios, Jesús Hernández, Alejandra Hernández-Terán, Alfredo Herrera-Estrella, Carla Ivón Herrera-Najera, Pavel Isa, Verónica Mata-Haro, Daniel Lira Morales, Susana Lopez, Antonio Loza Román, Brenda Irasema Maldonado-Meza, Bernardo Martínez-Miguel, José Arturo Martínez-Orozco, Célida Martínez- Rodríguez, Margarita Matías-Florentino, Fidencio Mejía-Nepomuceno, María Guadalupe de Jesús Mireles-Rivera, Gloria María Molina-Salinas, Hector Montoya-Fuentes, , Mario Mújica-Sánchez, José Esteban Muñoz-Medina, José de Jesús Nuñez-Contreras, Alicia Ocaña-Mondragón, Luis Alberto Ochoa-Carrera, Hector Esteban Paz-Juárez, Marissa Perez-Garcia, Francisco Pulido, Helen Haydee Fernanda Ramírez-Plascencia, Jorge Salas-Hernández, Angel Gustavo Salas-Lais, Alejandro Sánchez-Flores, Clara Esperanza Santacruz-Tinoco, María Guadalupe Santiago-Mauricio, Selene Zárate, Nelly Sélem-Mojica, Blanca Taboada, Gloria Vazquez, Joel Armando Vázquez-Pérez. |
| EPI_ISL_2681047, EPI_ISL_2681048, EPI_ISL_2681049 | Unidad de Investigación Biomédica de Zacatecas (UIBZ) | Instituto de Biotecnología de la UNAM | Consorcio Mexicano de Vigilancia Genómica (CoViGen-Mex). Authors (in alphabetical order): Julio Elias Alvarado-Yaah, Carlos F. Arias, Santiago Ávila-Ríos, Eduardo Becerril-Vargas, Víctor Hugo Borja-Aburto, Celia Boukadida, Cristóbal Cháidez-Quiróz, Juan Bautista Chale-Dzul, Ricardo Ciria Merce, Andreu Comas-García, Célida Duque Molina, Julissa Enciso-Ibarra, José Antonio Enciso-Moreno, Gloria Elena Espinosa-Ayala, Fernando Fontove-Herrera, Daniel Fregoso-Rueda, Victor Eduardo García-Arias, Alejandra García-Gasca, Bruno Gómez-Gil, Jean Pierre González, Irvin González-López, Concepción Grajales-Muñiz, Ricardo Grande, Rosa María Gutiérrez Rios, Jesús Hernández, Alejandra Hernández-Terán, Alfredo Herrera-Estrella, Carla Ivón Herrera-Najera, Pavel Isa, Verónica Mata-Haro, Daniel Lira Morales, Susana Lopez, Antonio Loza Román, Brenda Irasema Maldonado-Meza, Bernardo Martínez-Miguel, José Arturo Martínez-Orozco, Célida Martínez- Rodríguez, Margarita Matías-Florentino, Fidencio Mejía-Nepomuceno, María Guadalupe de Jesús Mireles-Rivera, Gloria María Molina-Salinas, Hector Montoya-Fuentes, , Mario Mújica-Sánchez, José Esteban Muñoz-Medina, José de Jesús Nuñez-Contreras, Alicia Ocaña-Mondragón, Luis Alberto Ochoa-Carrera, Hector Esteban Paz-Juárez, Marissa Perez-Garcia, Francisco Pulido, Helen Haydee Fernanda Ramírez-Plascencia, Jorge Salas-Hernández, Angel Gustavo Salas-Lais, Alejandro Sánchez-Flores, Clara Esperanza Santacruz-Tinoco, María Guadalupe Santiago-Mauricio, Selene Zárate, Nelly Sélem-Mojica, Blanca Taboada, Gloria Vazquez, Joel Armando Vázquez-Pérez. |
| EPI_ISL_2681050 | Unidad de Investigación Médica de Yucatán (UIMY) | Instituto de Biotecnología de la UNAM | Consorcio Mexicano de Vigilancia Genómica (CoViGen-Mex). Authors (in alphabetical order): Julio Elias Alvarado-Yaah, Carlos F. Arias, Santiago Ávila-Ríos, Eduardo Becerril-Vargas, Víctor Hugo Borja-Aburto, Celia Boukadida, Cristóbal Cháidez-Quiróz, Juan Bautista Chale-Dzul, Ricardo Ciria Merce, Andreu Comas-García, Célida Duque Molina, Julissa Enciso-Ibarra, José Antonio Enciso-Moreno, Gloria Elena Espinosa-Ayala, Fernando Fontove-Herrera, Daniel Fregoso-Rueda, Victor Eduardo García-Arias, Alejandra García-Gasca, Bruno Gómez-Gil, Jean Pierre González, Irvin González-López, Concepción Grajales-Muñiz, Ricardo Grande, Rosa María Gutiérrez Rios, Jesús Hernández, Alejandra Hernández-Terán, Alfredo Herrera-Estrella, Carla Ivón Herrera-Najera, Pavel Isa, Verónica Mata-Haro, Daniel Lira Morales, Susana Lopez, Antonio Loza Román, Brenda Irasema Maldonado-Meza, Bernardo Martínez-Miguel, José Arturo Martínez-Orozco, Célida Martínez- Rodríguez, Margarita Matías-Florentino, Fidencio Mejía-Nepomuceno, María Guadalupe de Jesús Mireles-Rivera, Gloria María Molina-Salinas, Hector Montoya-Fuentes, , Mario Mújica-Sánchez, José Esteban Muñoz-Medina, José de Jesús Nuñez-Contreras, Alicia Ocaña-Mondragón, Luis Alberto Ochoa-Carrera, Hector Esteban Paz-Juárez, Marissa Perez-Garcia, Francisco Pulido, Helen Haydee Fernanda Ramírez-Plascencia, Jorge Salas-Hernández, Angel Gustavo Salas-Lais, Alejandro Sánchez-Flores, Clara Esperanza Santacruz-Tinoco, María Guadalupe Santiago-Mauricio, Selene Zárate, Nelly Sélem-Mojica, Blanca Taboada, Gloria Vazquez, Joel Armando Vázquez-Pérez. |
| EPI_ISL_2681051 | Laboratorio Central de Epidemiología (LCE) | Instituto de Biotecnología de la UNAM | Consorcio Mexicano de Vigilancia Genómica (CoViGen-Mex). Authors (in alphabetical order): Julio Elias Alvarado-Yaah, Carlos F. Arias, Santiago Ávila-Ríos, Eduardo Becerril-Vargas, Víctor Hugo Borja-Aburto, Celia Boukadida, Cristóbal Cháidez-Quiróz, Juan Bautista Chale-Dzul, Ricardo Ciria Merce, Andreu Comas-García, Célida Duque Molina, Julissa Enciso-Ibarra, José Antonio Enciso-Moreno, Gloria Elena Espinosa-Ayala, Fernando Fontove-Herrera, Daniel Fregoso-Rueda, Victor Eduardo García-Arias, Alejandra García-Gasca, Bruno Gómez-Gil, Jean Pierre González, Irvin González-López, Concepción Grajales-Muñiz, Ricardo Grande, Rosa María Gutiérrez Rios, Jesús Hernández, Alejandra Hernández-Terán, Alfredo Herrera-Estrella, Carla Ivón Herrera-Najera, Pavel Isa, Verónica Mata-Haro, Daniel Lira Morales, Susana Lopez, Antonio Loza Román, Brenda Irasema Maldonado-Meza, Bernardo Martínez-Miguel, José Arturo Martínez-Orozco, Célida Martínez- Rodríguez, Margarita Matías-Florentino, Fidencio Mejía-Nepomuceno, María Guadalupe de Jesús Mireles-Rivera, Gloria María Molina-Salinas, Hector Montoya-Fuentes, , Mario Mújica-Sánchez, José Esteban Muñoz-Medina, José de Jesús Nuñez-Contreras, Alicia Ocaña-Mondragón, Luis Alberto Ochoa-Carrera, Hector Esteban Paz-Juárez, Marissa Perez-Garcia, Francisco Pulido, Helen Haydee Fernanda Ramírez-Plascencia, Jorge Salas-Hernández, Angel Gustavo Salas-Lais, Alejandro Sánchez-Flores, Clara Esperanza Santacruz-Tinoco, María Guadalupe Santiago-Mauricio, Selene Zárate, Nelly Sélem-Mojica, Blanca Taboada, Gloria Vazquez, Joel Armando Vázquez-Pérez. |

| | | | |
|---|---|---|---|
| EPI_ISL_2681052 | Unidad de Investigación Biomédica de Zacatecas (UIBZ) | Instituto de Biotecnología de la UNAM | Consorcio Mexicano de Vigilancia Genómica (CoViGen-Mex). Authors (in alphabetical order): Julio Elias Alvarado-Yaah, Carlos F. Arias, Santiago Ávila-Ríos, Eduardo Becerril-Vargas, Víctor Hugo Borja-Aburto, Celia Boukadida, Cristóbal Cháidez-Quiróz, Juan Bautista Chale-Dzul, Ricardo Ciria Merce, Andreu Comas-García, Célida Duque Molina, Julissa Enciso-Ibarra, José Antonio Enciso-Moreno, Gloria Elena Espinosa-Ayala, Fernando Fontove-Herrera, Daniel Fregoso-Rueda, Victor Eduardo García-Arias, Alejandra García-Gasca, Bruno Gómez-Gil, Jean Pierre González, Irvin González-López, Concepción Grajales-Muñiz, Ricardo Grande, Rosa María Gutiérrez Rios, Jesús Hernández, Alejandra Hernández-Terán, Alfredo Herrera-Estrella, Carla Ivón Herrera-Najera, Pavel Isa, Verónica Mata-Haro, Daniel Lira Morales, Susana Lopez, Antonio Loza Román, Brenda Irasema Maldonado-Meza, Bernardo Martínez-Miguel, José Arturo Martínez-Orozco, Célida Martínez- Rodríguez, Margarita Matías-Florentino, Fidencio Mejía-Nepomuceno, María Guadalupe de Jesús Mireles-Rivera, Gloria María Molina-Salinas, Hector Montoya-Fuentes, , Mario Mújica-Sánchez, José Esteban Muñoz-Medina, José de Jesús Nuñez-Contreras, Alicia Ocaña-Mondragón, Luis Alberto Ochoa-Carrera, Hector Esteban Paz-Juárez, Marissa Perez-Garcia, Francisco Pulido, Helen Haydee Fernanda Ramírez-Plascencia, Jorge Salas-Hernández, Angel Gustavo Salas-Lais, Alejandro Sánchez-Flores, Clara Esperanza Santacruz-Tinoco, María Guadalupe Santiago-Mauricio, Selene Zárate, Nelly Sélem-Mojica, Blanca Taboada, Gloria Vazquez, Joel Armando Vázquez-Pérez. |
| EPI_ISL_2681053 | Centro de Investigación Biomédica del Noreste (CIBIN) | Instituto de Biotecnología de la UNAM | Consorcio Mexicano de Vigilancia Genómica (CoViGen-Mex). Authors (in alphabetical order): Julio Elias Alvarado-Yaah, Carlos F. Arias, Santiago Ávila-Ríos, Eduardo Becerril-Vargas, Víctor Hugo Borja-Aburto, Celia Boukadida, Cristóbal Cháidez-Quiróz, Juan Bautista Chale-Dzul, Ricardo Ciria Merce, Andreu Comas-García, Célida Duque Molina, Julissa Enciso-Ibarra, José Antonio Enciso-Moreno, Gloria Elena Espinosa-Ayala, Fernando Fontove-Herrera, Daniel Fregoso-Rueda, Victor Eduardo García-Arias, Alejandra García-Gasca, Bruno Gómez-Gil, Jean Pierre González, Irvin González-López, Concepción Grajales-Muñiz, Ricardo Grande, Rosa María Gutiérrez Rios, Jesús Hernández, Alejandra Hernández-Terán, Alfredo Herrera-Estrella, Carla Ivón Herrera-Najera, Pavel Isa, Verónica Mata-Haro, Daniel Lira Morales, Susana Lopez, Antonio Loza Román, Brenda Irasema Maldonado-Meza, Bernardo Martínez-Miguel, José Arturo Martínez-Orozco, Célida Martínez- Rodríguez, Margarita Matías-Florentino, Fidencio Mejía-Nepomuceno, María Guadalupe de Jesús Mireles-Rivera, Gloria María Molina-Salinas, Hector Montoya-Fuentes, , Mario Mújica-Sánchez, José Esteban Muñoz-Medina, José de Jesús Nuñez-Contreras, Alicia Ocaña-Mondragón, Luis Alberto Ochoa-Carrera, Hector Esteban Paz-Juárez, Marissa Perez-Garcia, Francisco Pulido, Helen Haydee Fernanda Ramírez-Plascencia, Jorge Salas-Hernández, Angel Gustavo Salas-Lais, Alejandro Sánchez-Flores, Clara Esperanza Santacruz-Tinoco, María Guadalupe Santiago-Mauricio, Selene Zárate, Nelly Sélem-Mojica, Blanca Taboada, Gloria Vazquez, Joel Armando Vázquez-Pérez. |
| EPI_ISL_2681054 | Laboratorio Central de Epidemiología (LCE) | Instituto de Biotecnología de la UNAM | Consorcio Mexicano de Vigilancia Genómica (CoViGen-Mex). Authors (in alphabetical order): Julio Elias Alvarado-Yaah, Carlos F. Arias, Santiago Ávila-Ríos, Eduardo Becerril-Vargas, Víctor Hugo Borja-Aburto, Celia Boukadida, Cristóbal Cháidez-Quiróz, Juan Bautista Chale-Dzul, Ricardo Ciria Merce, Andreu Comas-García, Célida Duque Molina, Julissa Enciso-Ibarra, José Antonio Enciso-Moreno, Gloria Elena Espinosa-Ayala, Fernando Fontove-Herrera, Daniel Fregoso-Rueda, Victor Eduardo García-Arias, Alejandra García-Gasca, Bruno Gómez-Gil, Jean Pierre González, Irvin González-López, Concepción Grajales-Muñiz, Ricardo Grande, Rosa María Gutiérrez Rios, Jesús Hernández, Alejandra Hernández-Terán, Alfredo Herrera-Estrella, Carla Ivón Herrera-Najera, Pavel Isa, Verónica Mata-Haro, Daniel Lira Morales, Susana Lopez, Antonio Loza Román, Brenda Irasema Maldonado-Meza, Bernardo Martínez-Miguel, José Arturo Martínez-Orozco, Célida Martínez- Rodríguez, Margarita Matías-Florentino, Fidencio Mejía-Nepomuceno, María Guadalupe de Jesús Mireles-Rivera, Gloria María Molina-Salinas, Hector Montoya-Fuentes, , Mario Mújica-Sánchez, José Esteban Muñoz-Medina, José de Jesús Nuñez-Contreras, Alicia Ocaña-Mondragón, Luis Alberto Ochoa-Carrera, Hector Esteban Paz-Juárez, Marissa Perez-Garcia, Francisco Pulido, Helen Haydee Fernanda Ramírez-Plascencia, Jorge Salas-Hernández, Angel Gustavo Salas-Lais, Alejandro Sánchez-Flores, Clara Esperanza Santacruz-Tinoco, María Guadalupe Santiago-Mauricio, Selene Zárate, Nelly Sélem-Mojica, Blanca Taboada, Gloria Vazquez, Joel Armando Vázquez-Pérez. |
| EPI_ISL_2681055 | Unidad de Investigación Médica de Yucatán (UIMY) | Instituto de Biotecnología de la UNAM | Consorcio Mexicano de Vigilancia Genómica (CoViGen-Mex). Authors (in alphabetical order): Julio Elias Alvarado-Yaah, Carlos F. Arias, Santiago Ávila-Ríos, Eduardo Becerril-Vargas, Víctor Hugo Borja-Aburto, Celia Boukadida, Cristóbal Cháidez-Quiróz, Juan Bautista Chale-Dzul, Ricardo Ciria Merce, Andreu Comas-García, Célida Duque Molina, Julissa Enciso-Ibarra, José Antonio Enciso-Moreno, Gloria Elena Espinosa-Ayala, Fernando Fontove-Herrera, Daniel Fregoso-Rueda, Victor Eduardo García-Arias, Alejandra García-Gasca, Bruno Gómez-Gil, Jean Pierre González, Irvin González-López, Concepción Grajales-Muñiz, Ricardo Grande, Rosa María Gutiérrez Rios, Jesús Hernández, Alejandra Hernández-Terán, Alfredo Herrera-Estrella, Carla Ivón Herrera-Najera, Pavel Isa, Verónica Mata-Haro, Daniel Lira Morales, Susana Lopez, Antonio Loza Román, Brenda Irasema Maldonado-Meza, Bernardo Martínez-Miguel, José Arturo Martínez-Orozco, Célida Martínez- Rodríguez, Margarita Matías-Florentino, Fidencio Mejía-Nepomuceno, María Guadalupe de Jesús Mireles-Rivera, Gloria María Molina-Salinas, Hector Montoya-Fuentes, , Mario Mújica-Sánchez, José Esteban Muñoz-Medina, José de Jesús Nuñez-Contreras, Alicia Ocaña-Mondragón, Luis Alberto Ochoa-Carrera, Hector Esteban Paz-Juárez, Marissa Perez-Garcia, Francisco Pulido, Helen Haydee Fernanda Ramírez-Plascencia, Jorge Salas-Hernández, Angel Gustavo Salas-Lais, Alejandro Sánchez-Flores, Clara Esperanza Santacruz-Tinoco, María Guadalupe Santiago-Mauricio, Selene Zárate, Nelly Sélem-Mojica, Blanca Taboada, Gloria Vazquez, Joel Armando Vázquez-Pérez. |
| EPI_ISL_2681056 | Laboratorio Central de Epidemiología (LCE) | Instituto de Biotecnología de la UNAM | Consorcio Mexicano de Vigilancia Genómica (CoViGen-Mex). Authors (in alphabetical order): Julio Elias Alvarado-Yaah, Carlos F. Arias, Santiago Ávila-Ríos, Eduardo Becerril-Vargas, Víctor Hugo Borja-Aburto, Celia Boukadida, Cristóbal Cháidez-Quiróz, Juan Bautista Chale-Dzul, Ricardo Ciria Merce, Andreu Comas-García, Célida Duque Molina, Julissa Enciso-Ibarra, José Antonio Enciso-Moreno, Gloria Elena Espinosa-Ayala, Fernando Fontove-Herrera, Daniel Fregoso-Rueda, Victor Eduardo García-Arias, Alejandra García-Gasca, Bruno Gómez-Gil, Jean Pierre González, Irvin González-López, Concepción Grajales-Muñiz, Ricardo Grande, Rosa María Gutiérrez Rios, Jesús Hernández, Alejandra Hernández-Terán, Alfredo Herrera-Estrella, Carla Ivón Herrera-Najera, Pavel Isa, Verónica Mata-Haro, Daniel Lira Morales, Susana Lopez, Antonio Loza Román, Brenda Irasema Maldonado-Meza, Bernardo Martínez-Miguel, José Arturo Martínez-Orozco, Célida Martínez- Rodríguez, Margarita Matías-Florentino, Fidencio Mejía-Nepomuceno, María Guadalupe de Jesús Mireles-Rivera, Gloria María Molina-Salinas, Hector Montoya-Fuentes, , Mario Mújica-Sánchez, José Esteban Muñoz-Medina, José de Jesús Nuñez-Contreras, Alicia Ocaña-Mondragón, Luis Alberto Ochoa-Carrera, Hector Esteban Paz-Juárez, Marissa Perez-Garcia, Francisco Pulido, Helen Haydee Fernanda Ramírez-Plascencia, Jorge Salas-Hernández, Angel Gustavo Salas-Lais, Alejandro Sánchez-Flores, Clara Esperanza Santacruz-Tinoco, María Guadalupe Santiago-Mauricio, Selene Zárate, Nelly Sélem-Mojica, Blanca Taboada, Gloria Vazquez, Joel Armando Vázquez-Pérez. |
| EPI_ISL_2681057 | Unidad de Investigación Biomédica de Zacatecas (UIBZ) | Instituto de Biotecnología de la UNAM | Consorcio Mexicano de Vigilancia Genómica (CoViGen-Mex). Authors (in alphabetical order): Julio Elias Alvarado-Yaah, Carlos F. Arias, Santiago Ávila-Ríos, Eduardo Becerril-Vargas, Víctor Hugo Borja-Aburto, Celia Boukadida, Cristóbal Cháidez-Quiróz, Juan Bautista Chale-Dzul, Ricardo Ciria Merce, Andreu Comas-García, Célida Duque Molina, Julissa Enciso-Ibarra, José Antonio Enciso-Moreno, Gloria Elena Espinosa-Ayala, Fernando Fontove-Herrera, Daniel Fregoso-Rueda, Victor Eduardo García-Arias, Alejandra García-Gasca, Bruno Gómez-Gil, Jean Pierre González, Irvin González-López, Concepción Grajales-Muñiz, Ricardo Grande, Rosa María Gutiérrez Rios, Jesús Hernández, Alejandra Hernández-Terán, Alfredo Herrera-Estrella, Carla Ivón Herrera-Najera, Pavel Isa, Verónica Mata-Haro, Daniel Lira Morales, Susana Lopez, Antonio Loza Román, Brenda Irasema Maldonado-Meza, Bernardo Martínez-Miguel, José Arturo Martínez-Orozco, Célida Martínez- Rodríguez, Margarita Matías-Florentino, Fidencio Mejía-Nepomuceno, María Guadalupe de Jesús Mireles-Rivera, Gloria María Molina-Salinas, Hector Montoya-Fuentes, , Mario Mújica-Sánchez, José Esteban Muñoz-Medina, José de Jesús Nuñez-Contreras, Alicia Ocaña-Mondragón, Luis Alberto Ochoa-Carrera, Hector Esteban Paz-Juárez, Marissa Perez-Garcia, Francisco Pulido, Helen Haydee Fernanda Ramírez-Plascencia, Jorge Salas-Hernández, Angel Gustavo Salas-Lais, Alejandro Sánchez-Flores, Clara Esperanza Santacruz-Tinoco, María Guadalupe Santiago-Mauricio, Selene Zárate, Nelly Sélem-Mojica, Blanca Taboada, Gloria Vazquez, Joel Armando Vázquez-Pérez. |
| EPI_ISL_2681058, EPI_ISL_2681059, EPI_ISL_2681060, EPI_ISL_2681061, EPI_ISL_2681062, EPI_ISL_2681063, EPI_ISL_2681064, EPI_ISL_2681065, EPI_ISL_2681066 | Laboratorio Central de Epidemiología (LCE) | Instituto de Biotecnología de la UNAM | Consorcio Mexicano de Vigilancia Genómica (CoViGen-Mex). Authors (in alphabetical order): Julio Elias Alvarado-Yaah, Carlos F. Arias, Santiago Ávila-Ríos, Eduardo Becerril-Vargas, Víctor Hugo Borja-Aburto, Celia Boukadida, Cristóbal Cháidez-Quiróz, Juan Bautista Chale-Dzul, Ricardo Ciria Merce, Andreu Comas-García, Célida Duque Molina, Julissa Enciso-Ibarra, José Antonio Enciso-Moreno, Gloria Elena Espinosa-Ayala, Fernando Fontove-Herrera, Daniel Fregoso-Rueda, Victor Eduardo García-Arias, Alejandra García-Gasca, Bruno Gómez-Gil, Jean Pierre González, Irvin González-López, Concepción Grajales-Muñiz, Ricardo Grande, Rosa María Gutiérrez Rios, Jesús Hernández, Alejandra Hernández-Terán, Alfredo Herrera-Estrella, Carla Ivón Herrera-Najera, Pavel Isa, Verónica Mata-Haro, Daniel Lira Morales, Susana Lopez, Antonio Loza Román, Brenda Irasema Maldonado-Meza, Bernardo Martínez-Miguel, José Arturo Martínez-Orozco, Célida Martínez- Rodríguez, Margarita Matías-Florentino, Fidencio Mejía-Nepomuceno, María Guadalupe de Jesús Mireles-Rivera, Gloria María Molina-Salinas, Hector Montoya-Fuentes, , Mario Mújica-Sánchez, José Esteban Muñoz-Medina, José de Jesús Nuñez-Contreras, Alicia Ocaña-Mondragón, Luis Alberto Ochoa-Carrera, Hector Esteban Paz-Juárez, Marissa Perez-Garcia, Francisco Pulido, Helen Haydee Fernanda Ramírez-Plascencia, Jorge Salas-Hernández, Angel Gustavo Salas-Lais, Alejandro Sánchez-Flores, Clara Esperanza Santacruz-Tinoco, María Guadalupe Santiago-Mauricio, Selene Zárate, Nelly Sélem-Mojica, Blanca Taboada, Gloria Vazquez, Joel Armando Vázquez-Pérez. |
| EPI_ISL_2681067 | Unidad de Investigación Médica de Yucatán (UIMY) | Instituto de Biotecnología de la UNAM | Consorcio Mexicano de Vigilancia Genómica (CoViGen-Mex). Authors (in alphabetical order): Julio Elias Alvarado-Yaah, Carlos F. Arias, Santiago Ávila-Ríos, Eduardo Becerril-Vargas, Víctor Hugo Borja-Aburto, Celia Boukadida, Cristóbal Cháidez-Quiróz, Juan Bautista Chale-Dzul, Ricardo Ciria Merce, Andreu Comas-García, Célida Duque Molina, Julissa Enciso-Ibarra, José Antonio Enciso-Moreno, Gloria Elena Espinosa-Ayala, Fernando Fontove-Herrera, Daniel Fregoso-Rueda, Victor Eduardo García-Arias, Alejandra García-Gasca, Bruno Gómez-Gil, Jean Pierre González, Irvin González-López, Concepción Grajales-Muñiz, Ricardo Grande, Rosa María Gutiérrez Rios, Jesús Hernández, Alejandra Hernández-Terán, Alfredo Herrera-Estrella, Carla Ivón Herrera-Najera, Pavel Isa, Verónica Mata-Haro, Daniel Lira Morales, Susana Lopez, Antonio Loza Román, Brenda Irasema Maldonado-Meza, Bernardo Martínez-Miguel, José Arturo Martínez-Orozco, Célida Martínez- Rodríguez, Margarita Matías-Florentino, Fidencio Mejía-Nepomuceno, María Guadalupe de Jesús Mireles-Rivera, Gloria María Molina-Salinas, Hector Montoya-Fuentes, , Mario Mújica-Sánchez, José Esteban Muñoz-Medina, José de Jesús Nuñez-Contreras, Alicia Ocaña-Mondragón, Luis Alberto Ochoa-Carrera, Hector Esteban Paz-Juárez, Marissa Perez-Garcia, Francisco Pulido, Helen Haydee Fernanda Ramírez-Plascencia, Jorge Salas-Hernández, Angel Gustavo Salas-Lais, Alejandro Sánchez-Flores, Clara Esperanza Santacruz-Tinoco, María Guadalupe Santiago-Mauricio, Selene Zárate, Nelly Sélem-Mojica, Blanca Taboada, Gloria Vazquez, Joel Armando Vázquez-Pérez. |
| EPI_ISL_2681068 | Unidad de Investigación Biomédica de Zacatecas (UIBZ) | Instituto de Biotecnología de la UNAM | Consorcio Mexicano de Vigilancia Genómica (CoViGen-Mex). Authors (in alphabetical order): Julio Elias Alvarado-Yaah, Carlos F. Arias, Santiago Ávila-Ríos, Eduardo Becerril-Vargas, Víctor Hugo Borja-Aburto, Celia Boukadida, Cristóbal Cháidez-Quiróz, Juan Bautista Chale-Dzul, Ricardo Ciria Merce, Andreu Comas-García, Célida Duque Molina, Julissa Enciso-Ibarra, José Antonio Enciso-Moreno, Gloria Elena Espinosa-Ayala, Fernando Fontove-Herrera, Daniel Fregoso-Rueda, Victor Eduardo García-Arias, Alejandra García-Gasca, Bruno Gómez-Gil, Jean Pierre González, Irvin González-López, Concepción Grajales-Muñiz, Ricardo Grande, Rosa María Gutiérrez Rios, Jesús Hernández, Alejandra Hernández-Terán, Alfredo Herrera-Estrella, Carla Ivón Herrera-Najera, Pavel Isa, Verónica Mata-Haro, Daniel Lira Morales, Susana Lopez, Antonio Loza Román, Brenda Irasema Maldonado-Meza, Bernardo Martínez-Miguel, José Arturo Martínez-Orozco, Célida Martínez- Rodríguez, Margarita Matías-Florentino, Fidencio Mejía-Nepomuceno, María Guadalupe de Jesús Mireles-Rivera, Gloria María Molina-Salinas, Hector Montoya-Fuentes, , Mario Mújica-Sánchez, José Esteban Muñoz-Medina, José de Jesús Nuñez-Contreras, Alicia Ocaña-Mondragón, Luis Alberto Ochoa-Carrera, Hector Esteban Paz-Juárez, Marissa Perez-Garcia, Francisco Pulido, Helen Haydee Fernanda Ramírez-Plascencia, Jorge Salas-Hernández, Angel Gustavo Salas-Lais, Alejandro Sánchez-Flores, Clara Esperanza Santacruz-Tinoco, María Guadalupe Santiago-Mauricio, Selene Zárate, Nelly Sélem-Mojica, Blanca Taboada, Gloria Vazquez, Joel Armando Vázquez-Pérez. |
| EPI_ISL_2681069, EPI_ISL_2681070, EPI_ISL_2681071, EPI_ISL_2681072, EPI_ISL_2681073 | Laboratorio Central de Epidemiología (LCE) | Instituto de Biotecnología de la UNAM | Consorcio Mexicano de Vigilancia Genómica (CoViGen-Mex). Authors (in alphabetical order): Julio Elias Alvarado-Yaah, Carlos F. Arias, Santiago Ávila-Ríos, Eduardo Becerril-Vargas, Víctor Hugo Borja-Aburto, Celia Boukadida, Cristóbal Cháidez-Quiróz, Juan Bautista Chale-Dzul, Ricardo Ciria Merce, Andreu Comas-García, Célida Duque Molina, Julissa Enciso-Ibarra, José Antonio Enciso-Moreno, Gloria Elena Espinosa-Ayala, Fernando Fontove-Herrera, Daniel Fregoso-Rueda, Victor Eduardo García-Arias, Alejandra García-Gasca, Bruno Gómez-Gil, Jean Pierre González, Irvin González-López, Concepción Grajales-Muñiz, Ricardo Grande, Rosa María Gutiérrez Rios, Jesús Hernández, Alejandra Hernández-Terán, Alfredo Herrera-Estrella, Carla Ivón Herrera-Najera, Pavel Isa, Verónica Mata-Haro, Daniel Lira Morales, Susana Lopez, Antonio Loza Román, Brenda Irasema Maldonado-Meza, Bernardo Martínez-Miguel, José Arturo Martínez-Orozco, Célida Martínez- Rodríguez, Margarita Matías-Florentino, Fidencio Mejía-Nepomuceno, María Guadalupe de Jesús Mireles-Rivera, Gloria María Molina-Salinas, Hector Montoya-Fuentes, , Mario Mújica-Sánchez, José Esteban Muñoz-Medina, José de Jesús Nuñez-Contreras, Alicia Ocaña-Mondragón, Luis Alberto Ochoa-Carrera, Hector Esteban Paz-Juárez, Marissa Perez-Garcia, Francisco Pulido, Helen Haydee Fernanda Ramírez-Plascencia, Jorge Salas-Hernández, Angel Gustavo Salas-Lais, Alejandro Sánchez-Flores, Clara Esperanza Santacruz-Tinoco, María Guadalupe Santiago-Mauricio, Selene Zárate, Nelly Sélem-Mojica, Blanca Taboada, Gloria Vazquez, Joel Armando Vázquez-Pérez. |
| EPI_ISL_2681074, EPI_ISL_2681075 | Unidad de Investigación Médica de Yucatán (UIMY) | Instituto de Biotecnología de la UNAM | Consorcio Mexicano de Vigilancia Genómica (CoViGen-Mex). Authors (in alphabetical order): Julio Elias Alvarado-Yaah, Carlos F. Arias, Santiago Ávila-Ríos, Eduardo Becerril-Vargas, Víctor Hugo Borja-Aburto, Celia Boukadida, Cristóbal Cháidez-Quiróz, Juan Bautista Chale-Dzul, Ricardo Ciria Merce, Andreu Comas-García, Célida Duque Molina, Julissa Enciso-Ibarra, José Antonio Enciso-Moreno, Gloria Elena Espinosa-Ayala, Fernando Fontove-Herrera, Daniel Fregoso-Rueda, Victor Eduardo García-Arias, Alejandra García-Gasca, Bruno Gómez-Gil, Jean Pierre González, Irvin González-López, Concepción Grajales-Muñiz, Ricardo Grande, Rosa María Gutiérrez Rios, Jesús Hernández, Alejandra Hernández-Terán, Alfredo Herrera-Estrella, Carla Ivón Herrera-Najera, Pavel Isa, Verónica Mata-Haro, Daniel Lira Morales, Susana Lopez, Antonio Loza Román, Brenda Irasema Maldonado-Meza, Bernardo Martínez-Miguel, José Arturo Martínez-Orozco, Célida Martínez- Rodríguez, Margarita Matías-Florentino, Fidencio Mejía-Nepomuceno, María Guadalupe de Jesús Mireles-Rivera, Gloria María Molina-Salinas, Hector Montoya-Fuentes, , Mario Mújica-Sánchez, José Esteban Muñoz-Medina, José de Jesús Nuñez-Contreras, Alicia Ocaña-Mondragón, Luis Alberto Ochoa-Carrera, Hector Esteban Paz-Juárez, Marissa Perez-Garcia, Francisco Pulido, Helen Haydee Fernanda Ramírez-Plascencia, Jorge Salas-Hernández, Angel Gustavo Salas-Lais, Alejandro Sánchez-Flores, Clara Esperanza Santacruz-Tinoco, María Guadalupe Santiago-Mauricio, Selene Zárate, Nelly Sélem-Mojica, Blanca Taboada, Gloria Vazquez, Joel Armando Vázquez-Pérez. |
| EPI_ISL_2681076 | Laboratorio Central de Epidemiología (LCE) | Instituto de Biotecnología de la UNAM | Consorcio Mexicano de Vigilancia Genómica (CoViGen-Mex). Authors (in alphabetical order): Julio Elias Alvarado-Yaah, Carlos F. Arias, Santiago Ávila-Ríos, Eduardo Becerril-Vargas, Víctor Hugo Borja-Aburto, Celia Boukadida, Cristóbal Cháidez-Quiróz, Juan Bautista Chale-Dzul, Ricardo Ciria Merce, Andreu Comas-García, Célida Duque Molina, Julissa Enciso-Ibarra, José Antonio Enciso-Moreno, Gloria Elena Espinosa-Ayala, Fernando Fontove-Herrera, Daniel Fregoso-Rueda, Victor Eduardo García-Arias, Alejandra García-Gasca, Bruno Gómez-Gil, Jean Pierre González, Irvin González-López, Concepción Grajales-Muñiz, Ricardo Grande, Rosa María Gutiérrez Rios, Jesús Hernández, Alejandra Hernández-Terán, Alfredo Herrera-Estrella, Carla Ivón Herrera-Najera, Pavel Isa, Verónica Mata-Haro, Daniel Lira Morales, Susana Lopez, Antonio Loza Román, Brenda Irasema Maldonado-Meza, Bernardo Martínez-Miguel, José Arturo Martínez-Orozco, Célida Martínez- Rodríguez, Margarita Matías-Florentino, Fidencio Mejía-Nepomuceno, María Guadalupe de Jesús Mireles-Rivera, Gloria María Molina-Salinas, Hector Montoya-Fuentes, , Mario Mújica-Sánchez, José Esteban Muñoz-Medina, José de Jesús Nuñez-Contreras, Alicia Ocaña-Mondragón, Luis Alberto Ochoa-Carrera, Hector Esteban Paz-Juárez, Marissa Perez-Garcia, Francisco Pulido, Helen Haydee Fernanda Ramírez-Plascencia, Jorge Salas-Hernández, Angel Gustavo Salas-Lais, Alejandro Sánchez-Flores, Clara Esperanza Santacruz-Tinoco, María Guadalupe Santiago-Mauricio, Selene Zárate, Nelly Sélem-Mojica, Blanca Taboada, Gloria Vazquez, Joel Armando Vázquez-Pérez. |
| EPI_ISL_2681077, EPI_ISL_2681078, EPI_ISL_2681079, EPI_ISL_2681080 | Unidad de Investigación Médica de Yucatán (UIMY) | Instituto de Biotecnología de la UNAM | Consorcio Mexicano de Vigilancia Genómica (CoViGen-Mex). Authors (in alphabetical order): Julio Elias Alvarado-Yaah, Carlos F. Arias, Santiago Ávila-Ríos, Eduardo Becerril-Vargas, Víctor Hugo Borja-Aburto, Celia Boukadida, Cristóbal Cháidez-Quiróz, Juan Bautista Chale-Dzul, Ricardo Ciria Merce, Andreu Comas-García, Célida Duque Molina, Julissa Enciso-Ibarra, José Antonio Enciso-Moreno, Gloria Elena Espinosa-Ayala, Fernando Fontove-Herrera, Daniel Fregoso-Rueda, Victor Eduardo García-Arias, Alejandra García-Gasca, Bruno Gómez-Gil, Jean Pierre González, Irvin González-López, Concepción Grajales-Muñiz, Ricardo Grande, Rosa María Gutiérrez Rios, Jesús Hernández, Alejandra Hernández-Terán, Alfredo Herrera-Estrella, Carla Ivón Herrera-Najera, Pavel Isa, Verónica Mata-Haro, Daniel Lira Morales, Susana Lopez, Antonio Loza Román, Brenda Irasema Maldonado-Meza, Bernardo Martínez-Miguel, José Arturo Martínez-Orozco, Célida Martínez- Rodríguez, Margarita Matías-Florentino, Fidencio Mejía-Nepomuceno, María Guadalupe de Jesús Mireles-Rivera, Gloria María Molina-Salinas, Hector Montoya-Fuentes, , Mario Mújica-Sánchez, José Esteban Muñoz-Medina, José de Jesús Nuñez-Contreras, Alicia Ocaña-Mondragón, Luis Alberto Ochoa-Carrera, Hector Esteban Paz-Juárez, Marissa Perez-Garcia, Francisco Pulido, Helen Haydee Fernanda Ramírez-Plascencia, Jorge Salas-Hernández, Angel Gustavo Salas-Lais, Alejandro Sánchez-Flores, Clara Esperanza Santacruz-Tinoco, María Guadalupe Santiago-Mauricio, Selene Zárate, Nelly Sélem-Mojica, Blanca Taboada, Gloria Vazquez, Joel Armando Vázquez-Pérez. |
| EPI_ISL_2681081 | Laboratorio Central de Epidemiología (LCE) | Instituto de Biotecnología de la UNAM | Consorcio Mexicano de Vigilancia Genómica (CoViGen-Mex). Authors (in alphabetical order): Julio Elias Alvarado-Yaah, Carlos F. Arias, Santiago Ávila-Ríos, Eduardo Becerril-Vargas, Víctor Hugo Borja-Aburto, Celia Boukadida, Cristóbal Cháidez-Quiróz, Juan Bautista Chale-Dzul, Ricardo Ciria Merce, Andreu Comas-García, Célida Duque Molina, Julissa Enciso-Ibarra, José Antonio Enciso-Moreno, Gloria Elena Espinosa-Ayala, Fernando Fontove-Herrera, Daniel Fregoso-Rueda, Victor Eduardo García-Arias, Alejandra García-Gasca, Bruno Gómez-Gil, Jean Pierre González, Irvin González-López, Concepción Grajales-Muñiz, Ricardo Grande, Rosa María Gutiérrez Rios, Jesús Hernández, Alejandra Hernández-Terán, Alfredo Herrera-Estrella, Carla Ivón Herrera-Najera, Pavel Isa, Verónica Mata-Haro, Daniel Lira Morales, Susana Lopez, Antonio Loza Román, Brenda Irasema Maldonado-Meza, Bernardo Martínez-Miguel, José Arturo Martínez-Orozco, Célida Martínez- Rodríguez, Margarita Matías-Florentino, Fidencio Mejía-Nepomuceno, María Guadalupe de Jesús Mireles-Rivera, Gloria María Molina-Salinas, Hector Montoya-Fuentes, , Mario Mújica-Sánchez, José Esteban Muñoz-Medina, José de Jesús Nuñez-Contreras, Alicia Ocaña-Mondragón, Luis Alberto Ochoa-Carrera, Hector Esteban Paz-Juárez, Marissa Perez-Garcia, Francisco Pulido, Helen Haydee Fernanda Ramírez-Plascencia, Jorge Salas-Hernández, Angel Gustavo Salas-Lais, Alejandro Sánchez-Flores, Clara Esperanza Santacruz-Tinoco, María Guadalupe Santiago-Mauricio, Selene Zárate, Nelly Sélem-Mojica, Blanca Taboada, Gloria Vazquez, Joel Armando Vázquez-Pérez. |
| EPI_ISL_2681082, EPI_ISL_2681083, EPI_ISL_2681084, EPI_ISL_2681085 | Unidad de Investigación Médica de Yucatán (UIMY) | Instituto de Biotecnología de la UNAM | Consorcio Mexicano de Vigilancia Genómica (CoViGen-Mex). Authors (in alphabetical order): Julio Elias Alvarado-Yaah, Carlos F. Arias, Santiago Ávila-Ríos, Eduardo Becerril-Vargas, Víctor Hugo Borja-Aburto, Celia Boukadida, Cristóbal Cháidez-Quiróz, Juan Bautista Chale-Dzul, Ricardo Ciria Merce, Andreu Comas-García, Célida Duque Molina, Julissa Enciso-Ibarra, José Antonio Enciso-Moreno, Gloria Elena Espinosa-Ayala, Fernando Fontove-Herrera, Daniel Fregoso-Rueda, Victor Eduardo García-Arias, Alejandra García-Gasca, Bruno Gómez-Gil, Jean Pierre González, Irvin González-López, Concepción Grajales-Muñiz, Ricardo Grande, Rosa María Gutiérrez Rios, Jesús Hernández, Alejandra Hernández-Terán, Alfredo Herrera-Estrella, Carla Ivón Herrera-Najera, Pavel Isa, Verónica Mata-Haro, Daniel Lira Morales, Susana Lopez, Antonio Loza Román, Brenda Irasema Maldonado-Meza, Bernardo Martínez-Miguel, José Arturo Martínez-Orozco, Célida Martínez- Rodríguez, Margarita Matías-Florentino, Fidencio Mejía-Nepomuceno, María Guadalupe de Jesús Mireles-Rivera, Gloria María Molina-Salinas, Hector Montoya-Fuentes, , Mario Mújica-Sánchez, José Esteban Muñoz-Medina, José de Jesús Nuñez-Contreras, Alicia Ocaña-Mondragón, Luis Alberto Ochoa-Carrera, Hector Esteban Paz-Juárez, Marissa Perez-Garcia, Francisco Pulido, Helen Haydee Fernanda Ramírez-Plascencia, Jorge Salas-Hernández, Angel Gustavo Salas-Lais, Alejandro Sánchez-Flores, Clara Esperanza Santacruz-Tinoco, María Guadalupe Santiago-Mauricio, Selene Zárate, Nelly Sélem-Mojica, Blanca Taboada, Gloria Vazquez, Joel Armando Vázquez-Pérez. |
| EPI_ISL_2681086 | Centro de Investigación Biomédica del Noreste (CIBIN) | Instituto de Biotecnología de la UNAM | Consorcio Mexicano de Vigilancia Genómica (CoViGen-Mex). Authors (in alphabetical order): Julio Elias Alvarado-Yaah, Carlos F. Arias, Santiago Ávila-Ríos, Eduardo Becerril-Vargas, Víctor Hugo Borja-Aburto, Celia Boukadida, Cristóbal Cháidez-Quiróz, Juan Bautista Chale-Dzul, Ricardo Ciria Merce, Andreu Comas-García, Célida Duque Molina, Julissa Enciso-Ibarra, José Antonio Enciso-Moreno, Gloria Elena Espinosa-Ayala, Fernando Fontove-Herrera, Daniel Fregoso-Rueda, Victor Eduardo García-Arias, Alejandra García-Gasca, Bruno Gómez-Gil, Jean Pierre González, Irvin González-López, Concepción Grajales-Muñiz, Ricardo Grande, Rosa María Gutiérrez Rios, Jesús Hernández, Alejandra Hernández-Terán, Alfredo Herrera-Estrella, Carla Ivón Herrera-Najera, Pavel Isa, Verónica Mata-Haro, Daniel Lira Morales, Susana Lopez, Antonio Loza Román, Brenda Irasema Maldonado-Meza, Bernardo Martínez-Miguel, José Arturo Martínez-Orozco, Célida Martínez- Rodríguez, Margarita Matías-Florentino, Fidencio Mejía-Nepomuceno, María Guadalupe de Jesús Mireles-Rivera, Gloria María Molina-Salinas, Hector Montoya-Fuentes, , Mario Mújica-Sánchez, José Esteban Muñoz-Medina, José de Jesús Nuñez-Contreras, Alicia Ocaña-Mondragón, Luis Alberto Ochoa-Carrera, Hector Esteban Paz-Juárez, Marissa Perez-Garcia, Francisco Pulido, Helen Haydee Fernanda Ramírez-Plascencia, Jorge Salas-Hernández, Angel Gustavo Salas-Lais, Alejandro Sánchez-Flores, Clara Esperanza Santacruz-Tinoco, María Guadalupe Santiago-Mauricio, Selene Zárate, Nelly Sélem-Mojica, Blanca Taboada, Gloria Vazquez, Joel Armando Vázquez-Pérez. |
| EPI_ISL_2681087 | Laboratorio Central de Epidemiología (LCE) | Instituto de Biotecnología de la UNAM | Consorcio Mexicano de Vigilancia Genómica (CoViGen-Mex). Authors (in alphabetical order): Julio Elias Alvarado-Yaah, Carlos F. Arias, Santiago Ávila-Ríos, Eduardo Becerril-Vargas, Víctor Hugo Borja-Aburto, Celia Boukadida, Cristóbal Cháidez-Quiróz, Juan Bautista Chale-Dzul, Ricardo Ciria Merce, Andreu Comas-García, Célida Duque Molina, Julissa Enciso-Ibarra, José Antonio Enciso-Moreno, Gloria Elena Espinosa-Ayala, Fernando Fontove-Herrera, Daniel Fregoso-Rueda, Victor Eduardo García-Arias, Alejandra García-Gasca, Bruno Gómez-Gil, Jean Pierre González, Irvin González-López, Concepción Grajales-Muñiz, Ricardo Grande, Rosa María Gutiérrez Rios, Jesús Hernández, Alejandra Hernández-Terán, Alfredo Herrera-Estrella, Carla Ivón Herrera-Najera, Pavel Isa, Verónica Mata-Haro, Daniel Lira Morales, Susana Lopez, Antonio Loza Román, Brenda Irasema Maldonado-Meza, Bernardo Martínez-Miguel, José Arturo Martínez-Orozco, Célida Martínez- Rodríguez, Margarita Matías-Florentino, Fidencio Mejía-Nepomuceno, María Guadalupe de Jesús Mireles-Rivera, Gloria María Molina-Salinas, Hector Montoya-Fuentes, , Mario Mújica- |

| | | | |
|---|---|---|---|
| | | | Sánchez, José Esteban Muñoz-Medina, José de Jesús Nuñez-Contreras, Alicia Ocaña-Mondragón, Luis Alberto Ochoa-Carrera, Hector Esteban Paz-Juárez, Marissa Perez-Garcia, Francisco Pulido, Helen Haydee Fernanda Ramírez-Plascencia, Jorge Salas-Hernández, Angel Gustavo Salas-Lais, Alejandro Sánchez-Flores, Clara Esperanza Santacruz-Tinoco, María Guadalupe Santiago-Mauricio, Selene Zárate, Nelly Sélem-Mojica, Blanca Taboada, Gloria Vazquez, Joel Armando Vázquez-Pérez. |
| EPI_ISL_2681088, EPI_ISL_2681089, EPI_ISL_2681090, EPI_ISL_2681091 | Unidad de Investigación Médica de Yucatán (UIMY) | Instituto de Biotecnología de la UNAM | Consorcio Mexicano de Vigilancia Genómica (CoViGen-Mex). Authors (in alphabetical order): Julio Elias Alvarado-Yaah, Carlos F. Arias, Santiago Ávila-Ríos, Eduardo Becerril-Vargas, Víctor Hugo Borja-Aburto, Celia Boukadida, Cristóbal Cháidez-Quiróz, Juan Bautista Chale-Dzul, Ricardo Ciria Merce, Andreu Comas-García, Célida Duque Molina, Julissa Enciso-Ibarra, José Antonio Enciso-Moreno, Gloria Elena Espinosa-Ayala, Fernando Fontove-Herrera, Daniel Fregoso-Rueda, Victor Eduardo García-Arias, Alejandra García-Gasca, Bruno Gómez-Gil, Jean Pierre González, Irvin González-López, Concepción Grajales-Muñiz, Ricardo Grande, Rosa María Gutiérrez Rios, Jesús Hernández, Alejandra Hernández-Terán, Alfredo Herrera-Estrella, Carla Ivón Herrera-Najera, Pavel Isa, Verónica Mata-Haro, Daniel Lira Morales, Susana Lopez, Antonio Loza Román, Brenda Irasema Maldonado-Meza, Bernardo Martínez-Miguel, José Arturo Martínez-Orozco, Célida Martínez- Rodríguez, Margarita Matías-Florentino, Fidencio Mejía-Nepomuceno, María Guadalupe de Jesús Mireles-Rivera, Gloria María Molina-Salinas, Hector Montoya-Fuentes, , Mario Mújica-Sánchez, José Esteban Muñoz-Medina, José de Jesús Nuñez-Contreras, Alicia Ocaña-Mondragón, Luis Alberto Ochoa-Carrera, Hector Esteban Paz-Juárez, Marissa Perez-Garcia, Francisco Pulido, Helen Haydee Fernanda Ramírez-Plascencia, Jorge Salas-Hernández, Angel Gustavo Salas-Lais, Alejandro Sánchez-Flores, Clara Esperanza Santacruz-Tinoco, María Guadalupe Santiago-Mauricio, Selene Zárate, Nelly Sélem-Mojica, Blanca Taboada, Gloria Vazquez, Joel Armando Vázquez-Pérez. |
| EPI_ISL_2681092 | Unidad de Investigación Biomédica de Zacatecas (UIBZ) | Instituto de Biotecnología de la UNAM | Consorcio Mexicano de Vigilancia Genómica (CoViGen-Mex). Authors (in alphabetical order): Julio Elias Alvarado-Yaah, Carlos F. Arias, Santiago Ávila-Ríos, Eduardo Becerril-Vargas, Víctor Hugo Borja-Aburto, Celia Boukadida, Cristóbal Cháidez-Quiróz, Juan Bautista Chale-Dzul, Ricardo Ciria Merce, Andreu Comas-García, Célida Duque Molina, Julissa Enciso-Ibarra, José Antonio Enciso-Moreno, Gloria Elena Espinosa-Ayala, Fernando Fontove-Herrera, Daniel Fregoso-Rueda, Victor Eduardo García-Arias, Alejandra García-Gasca, Bruno Gómez-Gil, Jean Pierre González, Irvin González-López, Concepción Grajales-Muñiz, Ricardo Grande, Rosa María Gutiérrez Rios, Jesús Hernández, Alejandra Hernández-Terán, Alfredo Herrera-Estrella, Carla Ivón Herrera-Najera, Pavel Isa, Verónica Mata-Haro, Daniel Lira Morales, Susana Lopez, Antonio Loza Román, Brenda Irasema Maldonado-Meza, Bernardo Martínez-Miguel, José Arturo Martínez-Orozco, Célida Martínez- Rodríguez, Margarita Matías-Florentino, Fidencio Mejía-Nepomuceno, María Guadalupe de Jesús Mireles-Rivera, Gloria María Molina-Salinas, Hector Montoya-Fuentes, , Mario Mújica-Sánchez, José Esteban Muñoz-Medina, José de Jesús Nuñez-Contreras, Alicia Ocaña-Mondragón, Luis Alberto Ochoa-Carrera, Hector Esteban Paz-Juárez, Marissa Perez-Garcia, Francisco Pulido, Helen Haydee Fernanda Ramírez-Plascencia, Jorge Salas-Hernández, Angel Gustavo Salas-Lais, Alejandro Sánchez-Flores, Clara Esperanza Santacruz-Tinoco, María Guadalupe Santiago-Mauricio, Selene Zárate, Nelly Sélem-Mojica, Blanca Taboada, Gloria Vazquez, Joel Armando Vázquez-Pérez. |
| EPI_ISL_2681093 | Laboratorio Central de Epidemiología (LCE) | Instituto de Biotecnología de la UNAM | Consorcio Mexicano de Vigilancia Genómica (CoViGen-Mex). Authors (in alphabetical order): Julio Elias Alvarado-Yaah, Carlos F. Arias, Santiago Ávila-Ríos, Eduardo Becerril-Vargas, Víctor Hugo Borja-Aburto, Celia Boukadida, Cristóbal Cháidez-Quiróz, Juan Bautista Chale-Dzul, Ricardo Ciria Merce, Andreu Comas-García, Célida Duque Molina, Julissa Enciso-Ibarra, José Antonio Enciso-Moreno, Gloria Elena Espinosa-Ayala, Fernando Fontove-Herrera, Daniel Fregoso-Rueda, Victor Eduardo García-Arias, Alejandra García-Gasca, Bruno Gómez-Gil, Jean Pierre González, Irvin González-López, Concepción Grajales-Muñiz, Ricardo Grande, Rosa María Gutiérrez Rios, Jesús Hernández, Alejandra Hernández-Terán, Alfredo Herrera-Estrella, Carla Ivón Herrera-Najera, Pavel Isa, Verónica Mata-Haro, Daniel Lira Morales, Susana Lopez, Antonio Loza Román, Brenda Irasema Maldonado-Meza, Bernardo Martínez-Miguel, José Arturo Martínez-Orozco, Célida Martínez- Rodríguez, Margarita Matías-Florentino, Fidencio Mejía-Nepomuceno, María Guadalupe de Jesús Mireles-Rivera, Gloria María Molina-Salinas, Hector Montoya-Fuentes, , Mario Mújica-Sánchez, José Esteban Muñoz-Medina, José de Jesús Nuñez-Contreras, Alicia Ocaña-Mondragón, Luis Alberto Ochoa-Carrera, Hector Esteban Paz-Juárez, Marissa Perez-Garcia, Francisco Pulido, Helen Haydee Fernanda Ramírez-Plascencia, Jorge Salas-Hernández, Angel Gustavo Salas-Lais, Alejandro Sánchez-Flores, Clara Esperanza Santacruz-Tinoco, María Guadalupe Santiago-Mauricio, Selene Zárate, Nelly Sélem-Mojica, Blanca Taboada, Gloria Vazquez, Joel Armando Vázquez-Pérez. |
| EPI_ISL_2681094 | Unidad de Investigación Médica de Yucatán (UIMY) | Instituto de Biotecnología de la UNAM | Consorcio Mexicano de Vigilancia Genómica (CoViGen-Mex). Authors (in alphabetical order): Julio Elias Alvarado-Yaah, Carlos F. Arias, Santiago Ávila-Ríos, Eduardo Becerril-Vargas, Víctor Hugo Borja-Aburto, Celia Boukadida, Cristóbal Cháidez-Quiróz, Juan Bautista Chale-Dzul, Ricardo Ciria Merce, Andreu Comas-García, Célida Duque Molina, Julissa Enciso-Ibarra, José Antonio Enciso-Moreno, Gloria Elena Espinosa-Ayala, Fernando Fontove-Herrera, Daniel Fregoso-Rueda, Victor Eduardo García-Arias, Alejandra García-Gasca, Bruno Gómez-Gil, Jean Pierre González, Irvin González-López, Concepción Grajales-Muñiz, Ricardo Grande, Rosa María Gutiérrez Rios, Jesús Hernández, Alejandra Hernández-Terán, Alfredo Herrera-Estrella, Carla Ivón Herrera-Najera, Pavel Isa, Verónica Mata-Haro, Daniel Lira Morales, Susana Lopez, Antonio Loza Román, Brenda Irasema Maldonado-Meza, Bernardo Martínez-Miguel, José Arturo Martínez-Orozco, Célida Martínez- Rodríguez, Margarita Matías-Florentino, Fidencio Mejía-Nepomuceno, María Guadalupe de Jesús Mireles-Rivera, Gloria María Molina-Salinas, Hector Montoya-Fuentes, , Mario Mújica-Sánchez, José Esteban Muñoz-Medina, José de Jesús Nuñez-Contreras, Alicia Ocaña-Mondragón, Luis Alberto Ochoa-Carrera, Hector Esteban Paz-Juárez, Marissa Perez-Garcia, Francisco Pulido, Helen Haydee Fernanda Ramírez-Plascencia, Jorge Salas-Hernández, Angel Gustavo Salas-Lais, Alejandro Sánchez-Flores, Clara Esperanza Santacruz-Tinoco, María Guadalupe Santiago-Mauricio, Selene Zárate, Nelly Sélem-Mojica, Blanca Taboada, Gloria Vazquez, Joel Armando Vázquez-Pérez. |
| EPI_ISL_2681095, EPI_ISL_2681096 | Centro de Investigación Biomédica del Noreste (CIBIN) | Instituto de Biotecnología de la UNAM | Consorcio Mexicano de Vigilancia Genómica (CoViGen-Mex). Authors (in alphabetical order): Julio Elias Alvarado-Yaah, Carlos F. Arias, Santiago Ávila-Ríos, Eduardo Becerril-Vargas, Víctor Hugo Borja-Aburto, Celia Boukadida, Cristóbal Cháidez-Quiróz, Juan Bautista Chale-Dzul, Ricardo Ciria Merce, Andreu Comas-García, Célida Duque Molina, Julissa Enciso-Ibarra, José Antonio Enciso-Moreno, Gloria Elena Espinosa-Ayala, Fernando Fontove-Herrera, Daniel Fregoso-Rueda, Victor Eduardo García-Arias, Alejandra García-Gasca, Bruno Gómez-Gil, Jean Pierre González, Irvin González-López, Concepción Grajales-Muñiz, Ricardo Grande, Rosa María Gutiérrez Rios, Jesús Hernández, Alejandra Hernández-Terán, Alfredo Herrera-Estrella, Carla Ivón Herrera-Najera, Pavel Isa, Verónica Mata-Haro, Daniel Lira Morales, Susana Lopez, Antonio Loza Román, Brenda Irasema Maldonado-Meza, Bernardo Martínez-Miguel, José Arturo Martínez-Orozco, Célida Martínez- Rodríguez, Margarita Matías-Florentino, Fidencio Mejía-Nepomuceno, María Guadalupe de Jesús Mireles-Rivera, Gloria María Molina-Salinas, Hector Montoya-Fuentes, , Mario Mújica-Sánchez, José Esteban Muñoz-Medina, José de Jesús Nuñez-Contreras, Alicia Ocaña-Mondragón, Luis Alberto Ochoa-Carrera, Hector Esteban Paz-Juárez, Marissa Perez-Garcia, Francisco Pulido, Helen Haydee Fernanda Ramírez-Plascencia, Jorge Salas-Hernández, Angel Gustavo Salas-Lais, Alejandro Sánchez-Flores, Clara Esperanza Santacruz-Tinoco, María Guadalupe Santiago-Mauricio, Selene Zárate, Nelly Sélem-Mojica, Blanca Taboada, Gloria Vazquez, Joel Armando Vázquez-Pérez. |
| EPI_ISL_2681097 | Unidad de Investigación Biomédica de Zacatecas (UIBZ) | Instituto de Biotecnología de la UNAM | Consorcio Mexicano de Vigilancia Genómica (CoViGen-Mex). Authors (in alphabetical order): Julio Elias Alvarado-Yaah, Carlos F. Arias, Santiago Ávila-Ríos, Eduardo Becerril-Vargas, Víctor Hugo Borja-Aburto, Celia Boukadida, Cristóbal Cháidez-Quiróz, Juan Bautista Chale-Dzul, Ricardo Ciria Merce, Andreu Comas-García, Célida Duque Molina, Julissa Enciso-Ibarra, José Antonio Enciso-Moreno, Gloria Elena Espinosa-Ayala, Fernando Fontove-Herrera, Daniel Fregoso-Rueda, Victor Eduardo García-Arias, Alejandra García-Gasca, Bruno Gómez-Gil, Jean Pierre González, Irvin González-López, Concepción Grajales-Muñiz, Ricardo Grande, Rosa María Gutiérrez Rios, Jesús Hernández, Alejandra Hernández-Terán, Alfredo Herrera-Estrella, Carla Ivón Herrera-Najera, Pavel Isa, Verónica Mata-Haro, Daniel Lira Morales, Susana Lopez, Antonio Loza Román, Brenda Irasema Maldonado-Meza, Bernardo Martínez-Miguel, José Arturo Martínez-Orozco, Célida Martínez- Rodríguez, Margarita Matías-Florentino, Fidencio Mejía-Nepomuceno, María Guadalupe de Jesús Mireles-Rivera, Gloria María Molina-Salinas, Hector Montoya-Fuentes, , Mario Mújica-Sánchez, José Esteban Muñoz-Medina, José de Jesús Nuñez-Contreras, Alicia Ocaña-Mondragón, Luis Alberto Ochoa-Carrera, Hector Esteban Paz-Juárez, Marissa Perez-Garcia, Francisco Pulido, Helen Haydee Fernanda Ramírez-Plascencia, Jorge Salas-Hernández, Angel Gustavo Salas-Lais, Alejandro Sánchez-Flores, Clara Esperanza Santacruz-Tinoco, María Guadalupe Santiago-Mauricio, Selene Zárate, Nelly Sélem-Mojica, Blanca Taboada, Gloria Vazquez, Joel Armando Vázquez-Pérez. |
| EPI_ISL_2681098, EPI_ISL_2681099, EPI_ISL_2681100, EPI_ISL_2681101 | Unidad de Investigación Médica de Yucatán (UIMY) | Instituto de Biotecnología de la UNAM | Consorcio Mexicano de Vigilancia Genómica (CoViGen-Mex). Authors (in alphabetical order): Julio Elias Alvarado-Yaah, Carlos F. Arias, Santiago Ávila-Ríos, Eduardo Becerril-Vargas, Víctor Hugo Borja-Aburto, Celia Boukadida, Cristóbal Cháidez-Quiróz, Juan Bautista Chale-Dzul, Ricardo Ciria Merce, Andreu Comas-García, Célida Duque Molina, Julissa Enciso-Ibarra, José Antonio Enciso-Moreno, Gloria Elena Espinosa-Ayala, Fernando Fontove-Herrera, Daniel Fregoso-Rueda, Victor Eduardo García-Arias, Alejandra García-Gasca, Bruno Gómez-Gil, Jean Pierre González, Irvin González-López, Concepción Grajales-Muñiz, Ricardo Grande, Rosa María Gutiérrez Rios, Jesús Hernández, Alejandra Hernández-Terán, Alfredo Herrera-Estrella, Carla Ivón Herrera-Najera, Pavel Isa, Verónica Mata-Haro, Daniel Lira Morales, Susana Lopez, Antonio Loza Román, Brenda Irasema Maldonado-Meza, Bernardo Martínez-Miguel, José Arturo Martínez-Orozco, Célida Martínez- Rodríguez, Margarita Matías-Florentino, Fidencio Mejía-Nepomuceno, María Guadalupe de Jesús Mireles-Rivera, Gloria María Molina-Salinas, Hector Montoya-Fuentes, , Mario Mújica-Sánchez, José Esteban Muñoz-Medina, José de Jesús Nuñez-Contreras, Alicia Ocaña-Mondragón, Luis Alberto Ochoa-Carrera, Hector Esteban Paz-Juárez, Marissa Perez-Garcia, Francisco Pulido, Helen Haydee Fernanda Ramírez-Plascencia, Jorge Salas-Hernández, Angel Gustavo Salas-Lais, Alejandro Sánchez-Flores, Clara Esperanza Santacruz-Tinoco, María Guadalupe Santiago-Mauricio, Selene Zárate, Nelly Sélem-Mojica, Blanca Taboada, Gloria Vazquez, Joel Armando Vázquez-Pérez. |
| EPI_ISL_2681102, EPI_ISL_2681103, EPI_ISL_2681104 | Centro de Investigación Biomédica del Noreste (CIBIN) | Instituto de Biotecnología de la UNAM | Consorcio Mexicano de Vigilancia Genómica (CoViGen-Mex). Authors (in alphabetical order): Julio Elias Alvarado-Yaah, Carlos F. Arias, Santiago Ávila-Ríos, Eduardo Becerril-Vargas, Víctor Hugo Borja-Aburto, Celia Boukadida, Cristóbal Cháidez-Quiróz, Juan Bautista Chale-Dzul, Ricardo Ciria Merce, Andreu Comas-García, Célida Duque Molina, Julissa Enciso-Ibarra, José Antonio Enciso-Moreno, Gloria Elena Espinosa-Ayala, Fernando Fontove-Herrera, Daniel Fregoso-Rueda, Victor Eduardo García-Arias, Alejandra García-Gasca, Bruno Gómez-Gil, Jean Pierre González, Irvin González-López, Concepción Grajales-Muñiz, Ricardo Grande, Rosa María Gutiérrez Rios, Jesús Hernández, Alejandra Hernández-Terán, Alfredo Herrera-Estrella, Carla Ivón Herrera-Najera, Pavel Isa, Verónica Mata-Haro, Daniel Lira Morales, Susana Lopez, Antonio Loza Román, Brenda Irasema Maldonado-Meza, Bernardo Martínez-Miguel, José Arturo Martínez-Orozco, Célida Martínez- Rodríguez, Margarita Matías-Florentino, Fidencio Mejía-Nepomuceno, María Guadalupe de Jesús Mireles-Rivera, Gloria María Molina-Salinas, Hector Montoya-Fuentes, , Mario Mújica-Sánchez, José Esteban Muñoz-Medina, José de Jesús Nuñez-Contreras, Alicia Ocaña-Mondragón, Luis Alberto Ochoa-Carrera, Hector Esteban Paz-Juárez, Marissa Perez-Garcia, Francisco Pulido, Helen Haydee Fernanda Ramírez-Plascencia, Jorge Salas-Hernández, Angel Gustavo Salas-Lais, Alejandro Sánchez-Flores, Clara Esperanza Santacruz-Tinoco, María Guadalupe Santiago-Mauricio, Selene Zárate, Nelly Sélem-Mojica, Blanca Taboada, Gloria Vazquez, Joel Armando Vázquez-Pérez. |
| EPI_ISL_2681105 | Laboratorio Central de Epidemiología (LCE) | Instituto de Biotecnología de la UNAM | Consorcio Mexicano de Vigilancia Genómica (CoViGen-Mex). Authors (in alphabetical order): Julio Elias Alvarado-Yaah, Carlos F. Arias, Santiago Ávila-Ríos, Eduardo Becerril-Vargas, Víctor Hugo Borja-Aburto, Celia Boukadida, Cristóbal Cháidez-Quiróz, Juan Bautista Chale-Dzul, Ricardo Ciria Merce, Andreu Comas-García, Célida Duque Molina, Julissa Enciso-Ibarra, José Antonio Enciso-Moreno, Gloria Elena Espinosa-Ayala, Fernando Fontove-Herrera, Daniel Fregoso-Rueda, Victor Eduardo García-Arias, Alejandra García-Gasca, Bruno Gómez-Gil, Jean Pierre González, Irvin González-López, Concepción Grajales-Muñiz, Ricardo Grande, Rosa María Gutiérrez Rios, Jesús Hernández, Alejandra Hernández-Terán, Alfredo Herrera-Estrella, Carla Ivón Herrera-Najera, Pavel Isa, Verónica Mata-Haro, Daniel Lira Morales, Susana Lopez, Antonio Loza Román, Brenda Irasema Maldonado-Meza, Bernardo Martínez-Miguel, José Arturo Martínez-Orozco, Célida Martínez- Rodríguez, Margarita Matías-Florentino, Fidencio Mejía-Nepomuceno, María Guadalupe de Jesús Mireles-Rivera, Gloria María Molina-Salinas, Hector Montoya-Fuentes, , Mario Mújica-Sánchez, José Esteban Muñoz-Medina, José de Jesús Nuñez-Contreras, Alicia Ocaña-Mondragón, Luis Alberto Ochoa-Carrera, Hector Esteban Paz-Juárez, Marissa Perez-Garcia, Francisco Pulido, Helen Haydee Fernanda Ramírez-Plascencia, Jorge Salas-Hernández, Angel Gustavo Salas-Lais, Alejandro Sánchez-Flores, Clara Esperanza Santacruz-Tinoco, María Guadalupe Santiago-Mauricio, Selene Zárate, Nelly Sélem-Mojica, Blanca Taboada, Gloria Vazquez, Joel Armando Vázquez-Pérez. |
| EPI_ISL_2681106, EPI_ISL_2681107, EPI_ISL_2681108, EPI_ISL_2681109 | Unidad de Investigación Médica de Yucatán (UIMY) | Instituto de Biotecnología de la UNAM | Consorcio Mexicano de Vigilancia Genómica (CoViGen-Mex). Authors (in alphabetical order): Julio Elias Alvarado-Yaah, Carlos F. Arias, Santiago Ávila-Ríos, Eduardo Becerril-Vargas, Víctor Hugo Borja-Aburto, Celia Boukadida, Cristóbal Cháidez-Quiróz, Juan Bautista Chale-Dzul, Ricardo Ciria Merce, Andreu Comas-García, Célida Duque Molina, Julissa Enciso-Ibarra, José Antonio Enciso-Moreno, Gloria Elena Espinosa-Ayala, Fernando Fontove-Herrera, Daniel Fregoso-Rueda, Victor Eduardo García-Arias, Alejandra García-Gasca, Bruno Gómez-Gil, Jean Pierre González, Irvin González-López, Concepción Grajales-Muñiz, Ricardo Grande, Rosa María Gutiérrez Rios, Jesús Hernández, Alejandra Hernández-Terán, Alfredo Herrera-Estrella, Carla Ivón Herrera-Najera, Pavel Isa, Verónica Mata-Haro, Daniel Lira Morales, Susana Lopez, Antonio Loza Román, Brenda Irasema Maldonado-Meza, Bernardo Martínez-Miguel, José Arturo Martínez-Orozco, Célida Martínez- Rodríguez, Margarita Matías-Florentino, Fidencio Mejía-Nepomuceno, María Guadalupe de Jesús Mireles-Rivera, Gloria María Molina-Salinas, Hector Montoya-Fuentes, , Mario Mújica-Sánchez, José Esteban Muñoz-Medina, José de Jesús Nuñez-Contreras, Alicia Ocaña-Mondragón, Luis Alberto Ochoa-Carrera, Hector Esteban Paz-Juárez, Marissa Perez-Garcia, Francisco Pulido, Helen Haydee Fernanda Ramírez-Plascencia, Jorge Salas-Hernández, Angel Gustavo Salas-Lais, Alejandro Sánchez-Flores, Clara Esperanza Santacruz-Tinoco, María Guadalupe Santiago-Mauricio, Selene Zárate, Nelly Sélem-Mojica, Blanca Taboada, Gloria Vazquez, Joel Armando Vázquez-Pérez. |
| EPI_ISL_2681110 | Centro de Investigación Biomédica del Noreste (CIBIN) | Instituto de Biotecnología de la UNAM | Consorcio Mexicano de Vigilancia Genómica (CoViGen-Mex). Authors (in alphabetical order): Julio Elias Alvarado-Yaah, Carlos F. Arias, Santiago Ávila-Ríos, Eduardo Becerril-Vargas, Víctor Hugo Borja-Aburto, Celia Boukadida, Cristóbal Cháidez-Quiróz, Juan Bautista Chale-Dzul, Ricardo Ciria Merce, Andreu Comas-García, Célida Duque Molina, Julissa Enciso-Ibarra, José Antonio Enciso-Moreno, Gloria Elena Espinosa-Ayala, Fernando Fontove-Herrera, Daniel Fregoso-Rueda, Victor Eduardo García-Arias, Alejandra García-Gasca, Bruno Gómez-Gil, Jean Pierre González, Irvin González-López, Concepción Grajales-Muñiz, Ricardo Grande, Rosa María Gutiérrez Rios, Jesús Hernández, Alejandra Hernández-Terán, Alfredo Herrera-Estrella, Carla Ivón Herrera-Najera, Pavel Isa, Verónica Mata-Haro, Daniel Lira Morales, Susana Lopez, Antonio Loza Román, Brenda Irasema Maldonado-Meza, Bernardo Martínez-Miguel, José Arturo Martínez-Orozco, Célida Martínez- Rodríguez, Margarita Matías-Florentino, Fidencio Mejía-Nepomuceno, María Guadalupe de Jesús Mireles-Rivera, Gloria María Molina-Salinas, Hector Montoya-Fuentes, , Mario Mújica-Sánchez, José Esteban Muñoz-Medina, José de Jesús Nuñez-Contreras, Alicia Ocaña-Mondragón, Luis Alberto Ochoa-Carrera, Hector Esteban Paz-Juárez, Marissa Perez-Garcia, Francisco Pulido, Helen Haydee Fernanda Ramírez-Plascencia, Jorge Salas-Hernández, Angel Gustavo Salas-Lais, Alejandro Sánchez-Flores, Clara Esperanza Santacruz-Tinoco, María Guadalupe Santiago-Mauricio, Selene Zárate, Nelly Sélem-Mojica, Blanca Taboada, Gloria Vazquez, Joel Armando Vázquez-Pérez. |
| EPI_ISL_2681111, EPI_ISL_2681112, EPI_ISL_2681113 | Laboratorio Central de Epidemiología (LCE) | Instituto de Biotecnología de la UNAM | Consorcio Mexicano de Vigilancia Genómica (CoViGen-Mex). Authors (in alphabetical order): Julio Elias Alvarado-Yaah, Carlos F. Arias, Santiago Ávila-Ríos, Eduardo Becerril-Vargas, Víctor Hugo Borja-Aburto, Celia Boukadida, Cristóbal Cháidez-Quiróz, Juan Bautista Chale-Dzul, Ricardo Ciria Merce, Andreu Comas-García, Célida Duque Molina, Julissa Enciso-Ibarra, José Antonio Enciso-Moreno, Gloria Elena Espinosa-Ayala, Fernando Fontove-Herrera, Daniel Fregoso-Rueda, Victor Eduardo García-Arias, Alejandra García-Gasca, Bruno Gómez-Gil, Jean Pierre González, Irvin González-López, Concepción Grajales-Muñiz, Ricardo Grande, Rosa María Gutiérrez Rios, Jesús Hernández, Alejandra Hernández-Terán, Alfredo Herrera-Estrella, Carla Ivón Herrera-Najera, Pavel Isa, Verónica Mata-Haro, Daniel Lira Morales, Susana Lopez, Antonio Loza Román, Brenda Irasema Maldonado-Meza, Bernardo Martínez-Miguel, José Arturo Martínez-Orozco, Célida Martínez- Rodríguez, Margarita Matías-Florentino, Fidencio Mejía-Nepomuceno, María Guadalupe de Jesús Mireles-Rivera, Gloria María Molina-Salinas, Hector Montoya-Fuentes, , Mario Mújica-Sánchez, José Esteban Muñoz-Medina, José de Jesús Nuñez-Contreras, Alicia Ocaña-Mondragón, Luis Alberto Ochoa-Carrera, Hector Esteban Paz-Juárez, Marissa Perez-Garcia, Francisco Pulido, Helen Haydee Fernanda Ramírez-Plascencia, Jorge Salas-Hernández, Angel Gustavo Salas-Lais, Alejandro Sánchez-Flores, Clara Esperanza Santacruz-Tinoco, María Guadalupe Santiago-Mauricio, Selene Zárate, Nelly Sélem-Mojica, Blanca Taboada, Gloria Vazquez, Joel Armando Vázquez-Pérez. |
| EPI_ISL_2681114, EPI_ISL_2681115, EPI_ISL_2681116, EPI_ISL_2681117, EPI_ISL_2681118 | Unidad de Investigación Médica de Yucatán (UIMY) | Instituto de Biotecnología de la UNAM | Consorcio Mexicano de Vigilancia Genómica (CoViGen-Mex). Authors (in alphabetical order): Julio Elias Alvarado-Yaah, Carlos F. Arias, Santiago Ávila-Ríos, Eduardo Becerril-Vargas, Víctor Hugo Borja-Aburto, Celia Boukadida, Cristóbal Cháidez-Quiróz, Juan Bautista Chale-Dzul, Ricardo Ciria Merce, Andreu Comas-García, Célida Duque Molina, Julissa Enciso-Ibarra, José Antonio Enciso-Moreno, Gloria Elena Espinosa-Ayala, Fernando Fontove-Herrera, Daniel Fregoso-Rueda, Victor Eduardo García-Arias, Alejandra García-Gasca, Bruno Gómez-Gil, Jean Pierre González, Irvin González-López, Concepción Grajales-Muñiz, Ricardo Grande, Rosa María Gutiérrez Rios, Jesús Hernández, Alejandra Hernández-Terán, Alfredo Herrera-Estrella, Carla Ivón Herrera-Najera, Pavel Isa, Verónica Mata-Haro, Daniel Lira Morales, Susana Lopez, Antonio Loza Román, Brenda Irasema Maldonado-Meza, Bernardo Martínez-Miguel, José Arturo Martínez-Orozco, Célida Martínez- Rodríguez, Margarita Matías-Florentino, Fidencio Mejía-Nepomuceno, María Guadalupe de Jesús Mireles-Rivera, Gloria María Molina-Salinas, Hector Montoya-Fuentes, , Mario Mújica-Sánchez, José Esteban Muñoz-Medina, José de Jesús Nuñez-Contreras, Alicia Ocaña-Mondragón, Luis Alberto Ochoa-Carrera, Hector Esteban Paz-Juárez, Marissa Perez-Garcia, Francisco Pulido, Helen Haydee Fernanda Ramírez-Plascencia, Jorge Salas-Hernández, Angel Gustavo Salas-Lais, Alejandro Sánchez-Flores, Clara Esperanza Santacruz-Tinoco, María Guadalupe Santiago-Mauricio, Selene Zárate, Nelly Sélem-Mojica, Blanca Taboada, Gloria Vazquez, Joel Armando Vázquez-Pérez. |
| EPI_ISL_2681119, EPI_ISL_2681120, EPI_ISL_2681121, EPI_ISL_2681122, EPI_ISL_2681123 | Unidad de Investigación Biomédica de Zacatecas (UIBZ) | Instituto de Biotecnología de la UNAM | Consorcio Mexicano de Vigilancia Genómica (CoViGen-Mex). Authors (in alphabetical order): Julio Elias Alvarado-Yaah, Carlos F. Arias, Santiago Ávila-Ríos, Eduardo Becerril-Vargas, Víctor Hugo Borja-Aburto, Celia Boukadida, Cristóbal Cháidez-Quiróz, Juan Bautista Chale-Dzul, Ricardo Ciria Merce, Andreu Comas-García, Célida Duque Molina, Julissa Enciso-Ibarra, José Antonio Enciso-Moreno, Gloria Elena Espinosa-Ayala, Fernando Fontove-Herrera, Daniel Fregoso-Rueda, Victor Eduardo García-Arias, Alejandra García-Gasca, Bruno Gómez-Gil, Jean Pierre González, Irvin González-López, Concepción Grajales-Muñiz, Ricardo Grande, Rosa María Gutiérrez Rios, Jesús Hernández, Alejandra Hernández-Terán, Alfredo Herrera-Estrella, Carla Ivón Herrera-Najera, Pavel Isa, Verónica Mata-Haro, Daniel Lira Morales, Susana Lopez, Antonio Loza Román, Brenda Irasema Maldonado-Meza, Bernardo Martínez-Miguel, José Arturo Martínez-Orozco, Célida Martínez- Rodríguez, Margarita Matías-Florentino, Fidencio Mejía-Nepomuceno, María Guadalupe de Jesús Mireles-Rivera, Gloria María Molina-Salinas, Hector Montoya-Fuentes, , Mario Mújica-Sánchez, José Esteban Muñoz-Medina, José de Jesús Nuñez-Contreras, Alicia Ocaña-Mondragón, Luis Alberto Ochoa-Carrera, Hector Esteban Paz-Juárez, Marissa Perez-Garcia, Francisco Pulido, Helen Haydee Fernanda Ramírez-Plascencia, Jorge Salas-Hernández, Angel Gustavo Salas-Lais, Alejandro Sánchez-Flores, Clara Esperanza Santacruz-Tinoco, María Guadalupe Santiago-Mauricio, Selene Zárate, Nelly Sélem-Mojica, Blanca Taboada, Gloria Vazquez, Joel Armando Vázquez-Pérez. |
| EPI_ISL_2681124, EPI_ISL_2681125 | Laboratorio Central de Epidemiología (LCE) | Instituto de Biotecnología de la UNAM | Consorcio Mexicano de Vigilancia Genómica (CoViGen-Mex). Authors (in alphabetical order): Julio Elias Alvarado-Yaah, Carlos F. Arias, Santiago Ávila-Ríos, Eduardo Becerril-Vargas, Víctor Hugo Borja-Aburto, Celia Boukadida, Cristóbal Cháidez-Quiróz, Juan Bautista Chale-Dzul, Ricardo Ciria Merce, Andreu Comas-García, Célida Duque Molina, Julissa Enciso-Ibarra, José Antonio Enciso-Moreno, Gloria Elena Espinosa-Ayala, Fernando Fontove-Herrera, Daniel Fregoso-Rueda, Victor Eduardo García-Arias, Alejandra García-Gasca, Bruno Gómez-Gil, Jean Pierre González, Irvin González-López, Concepción Grajales-Muñiz, Ricardo Grande, Rosa María Gutiérrez Rios, Jesús Hernández, Alejandra Hernández-Terán, Alfredo Herrera-Estrella, Carla Ivón Herrera-Najera, Pavel Isa, Verónica Mata-Haro, Daniel Lira Morales, Susana Lopez, Antonio Loza Román, Brenda Irasema Maldonado-Meza, Bernardo Martínez-Miguel, José Arturo Martínez-Orozco, Célida Martínez- Rodríguez, Margarita Matías-Florentino, Fidencio Mejía-Nepomuceno, María Guadalupe de Jesús Mireles-Rivera, Gloria María Molina-Salinas, Hector Montoya-Fuentes, , Mario Mújica-Sánchez, José Esteban Muñoz-Medina, José de Jesús Nuñez-Contreras, Alicia Ocaña-Mondragón, Luis Alberto Ochoa-Carrera, Hector Esteban Paz-Juárez, Marissa Perez-Garcia, Francisco Pulido, Helen Haydee Fernanda Ramírez-Plascencia, Jorge Salas-Hernández, Angel Gustavo Salas-Lais, Alejandro Sánchez-Flores, Clara Esperanza Santacruz-Tinoco, María Guadalupe Santiago-Mauricio, Selene Zárate, Nelly Sélem-Mojica, Blanca Taboada, Gloria Vazquez, Joel Armando Vázquez-Pérez. |
| EPI_ISL_2681126 | Unidad de Investigación Médica de Yucatán (UIMY) | Instituto de Biotecnología de la UNAM | Consorcio Mexicano de Vigilancia Genómica (CoViGen-Mex). Authors (in alphabetical order): Julio Elias Alvarado-Yaah, Carlos F. Arias, Santiago Ávila-Ríos, Eduardo Becerril-Vargas, Víctor Hugo Borja-Aburto, Celia Boukadida, Cristóbal Cháidez-Quiróz, Juan Bautista Chale-Dzul, Ricardo Ciria Merce, Andreu Comas-García, Célida Duque Molina, Julissa Enciso-Ibarra, José Antonio Enciso-Moreno, Gloria Elena Espinosa-Ayala, Fernando Fontove-Herrera, Daniel Fregoso-Rueda, Victor Eduardo García-Arias, Alejandra García-Gasca, Bruno Gómez-Gil, Jean Pierre González, Irvin González-López, Concepción Grajales-Muñiz, Ricardo Grande, Rosa María Gutiérrez Rios, Jesús Hernández, Alejandra Hernández-Terán, Alfredo Herrera-Estrella, Carla Ivón Herrera-Najera, Pavel Isa, Verónica Mata-Haro, Daniel Lira Morales, Susana Lopez, Antonio Loza Román, Brenda Irasema Maldonado-Meza, Bernardo Martínez-Miguel, José Arturo Martínez-Orozco, Célida Martínez- Rodríguez, Margarita Matías-Florentino, Fidencio Mejía-Nepomuceno, María Guadalupe de Jesús Mireles-Rivera, Gloria María Molina-Salinas, Hector Montoya-Fuentes, , Mario Mújica-Sánchez, José Esteban Muñoz-Medina, José de Jesús Nuñez-Contreras, Alicia Ocaña-Mondragón, Luis Alberto Ochoa-Carrera, Hector Esteban Paz-Juárez, Marissa Perez-Garcia, Francisco Pulido, Helen Haydee Fernanda Ramírez-Plascencia, Jorge Salas-Hernández, Angel Gustavo Salas-Lais, Alejandro Sánchez-Flores, Clara Esperanza Santacruz-Tinoco, María Guadalupe Santiago-Mauricio, Selene Zárate, Nelly Sélem-Mojica, Blanca Taboada, Gloria Vazquez, Joel Armando Vázquez-Pérez. |
| EPI_ISL_2681127 | Centro de Investigación Biomédica del Noreste (CIBIN) | Instituto de Biotecnología de la UNAM | Consorcio Mexicano de Vigilancia Genómica (CoViGen-Mex). Authors (in alphabetical order): Julio Elias Alvarado-Yaah, Carlos F. Arias, Santiago Ávila-Ríos, Eduardo Becerril-Vargas, Víctor Hugo Borja-Aburto, Celia Boukadida, Cristóbal Cháidez-Quiróz, Juan Bautista Chale-Dzul, Ricardo Ciria Merce, Andreu Comas-García, Célida Duque Molina, Julissa Enciso-Ibarra, José Antonio Enciso-Moreno, Gloria Elena Espinosa-Ayala, Fernando Fontove-Herrera, Daniel Fregoso-Rueda, Victor Eduardo García-Arias, Alejandra García-Gasca, Bruno Gómez-Gil, Jean Pierre González, Irvin González-López, Concepción Grajales-Muñiz, Ricardo Grande, Rosa María Gutiérrez Rios, Jesús Hernández, Alejandra Hernández-Terán, Alfredo Herrera-Estrella, Carla Ivón Herrera-Najera, Pavel Isa, Verónica Mata-Haro, Daniel Lira Morales, Susana Lopez, Antonio Loza Román, Brenda Irasema Maldonado-Meza, Bernardo Martínez-Miguel, José Arturo Martínez-Orozco, Célida Martínez- Rodríguez, Margarita Matías-Florentino, Fidencio Mejía-Nepomuceno, María Guadalupe de Jesús Mireles-Rivera, Gloria María Molina-Salinas, Hector Montoya-Fuentes, , Mario Mújica-Sánchez, José Esteban Muñoz-Medina, José de Jesús Nuñez-Contreras, Alicia Ocaña-Mondragón, Luis Alberto Ochoa-Carrera, Hector Esteban Paz-Juárez, Marissa Perez-Garcia, Francisco Pulido, Helen Haydee Fernanda Ramírez-Plascencia, Jorge Salas-Hernández, Angel Gustavo Salas- |

| | | | |
|---|---|---|---|
| | | | Lais, Alejandro Sánchez-Flores, Clara Esperanza Santacruz-Tinoco, María Guadalupe Santiago-Mauricio, Selene Zárate, Nelly Sélem-Mojica, Blanca Taboada, Gloria Vazquez, Joel Armando Vázquez-Pérez. |
| EPI_ISL_2681128, EPI_ISL_2681129 | Laboratorio Central de Epidemiología (LCE) | Instituto de Biotecnología de la UNAM | Consorcio Mexicano de Vigilancia Genómica (CoViGen-Mex). Authors (in alphabetical order): Julio Elias Alvarado-Yaah, Carlos F. Arias, Santiago Ávila-Ríos, Eduardo Becerril-Vargas, Víctor Hugo Borja-Aburto, Celia Boukadida, Cristóbal Cháidez-Quiróz, Juan Bautista Chale-Dzul, Ricardo Ciria Merce, Andreu Comas-García, Célida Duque Molina, Julissa Enciso-Ibarra, José Antonio Enciso-Moreno, Gloria Elena Espinosa-Ayala, Fernando Fontove-Herrera, Daniel Fregoso-Rueda, Victor Eduardo García-Arias, Alejandra García-Gasca, Bruno Gómez-Gil, Jean Pierre González, Irvin González-López, Concepción Grajales-Muñiz, Ricardo Grande, Rosa María Gutiérrez Rios, Jesús Hernández, Alejandra Hernández-Terán, Alfredo Herrera-Estrella, Carla Ivón Herrera-Najera, Pavel Isa, Verónica Mata-Haro, Daniel Lira Morales, Susana Lopez, Antonio Loza Román, Brenda Irasema Maldonado-Meza, Bernardo Martínez-Miguel, José Arturo Martínez-Orozco, Célida Martínez- Rodríguez, Margarita Matías-Florentino, Fidencio Mejía-Nepomuceno, María Guadalupe de Jesús Mireles-Rivera, Gloria María Molina-Salinas, Hector Montoya-Fuentes, , Mario Mújica-Sánchez, José Esteban Muñoz-Medina, José de Jesús Nuñez-Contreras, Alicia Ocaña-Mondragón, Luis Alberto Ochoa-Carrera, Hector Esteban Paz-Juárez, Marissa Perez-Garcia, Francisco Pulido, Helen Haydee Fernanda Ramírez-Plascencia, Jorge Salas-Hernández, Angel Gustavo Salas-Lais, Alejandro Sánchez-Flores, Clara Esperanza Santacruz-Tinoco, María Guadalupe Santiago-Mauricio, Selene Zárate, Nelly Sélem-Mojica, Blanca Taboada, Gloria Vazquez, Joel Armando Vázquez-Pérez. |
| EPI_ISL_2681130 | Unidad de Investigación Médica de Yucatán (UIMY) | Instituto de Biotecnología de la UNAM | Consorcio Mexicano de Vigilancia Genómica (CoViGen-Mex). Authors (in alphabetical order): Julio Elias Alvarado-Yaah, Carlos F. Arias, Santiago Ávila-Ríos, Eduardo Becerril-Vargas, Víctor Hugo Borja-Aburto, Celia Boukadida, Cristóbal Cháidez-Quiróz, Juan Bautista Chale-Dzul, Ricardo Ciria Merce, Andreu Comas-García, Célida Duque Molina, Julissa Enciso-Ibarra, José Antonio Enciso-Moreno, Gloria Elena Espinosa-Ayala, Fernando Fontove-Herrera, Daniel Fregoso-Rueda, Victor Eduardo García-Arias, Alejandra García-Gasca, Bruno Gómez-Gil, Jean Pierre González, Irvin González-López, Concepción Grajales-Muñiz, Ricardo Grande, Rosa María Gutiérrez Rios, Jesús Hernández, Alejandra Hernández-Terán, Alfredo Herrera-Estrella, Carla Ivón Herrera-Najera, Pavel Isa, Verónica Mata-Haro, Daniel Lira Morales, Susana Lopez, Antonio Loza Román, Brenda Irasema Maldonado-Meza, Bernardo Martínez-Miguel, José Arturo Martínez-Orozco, Célida Martínez- Rodríguez, Margarita Matías-Florentino, Fidencio Mejía-Nepomuceno, María Guadalupe de Jesús Mireles-Rivera, Gloria María Molina-Salinas, Hector Montoya-Fuentes, , Mario Mújica-Sánchez, José Esteban Muñoz-Medina, José de Jesús Nuñez-Contreras, Alicia Ocaña-Mondragón, Luis Alberto Ochoa-Carrera, Hector Esteban Paz-Juárez, Marissa Perez-Garcia, Francisco Pulido, Helen Haydee Fernanda Ramírez-Plascencia, Jorge Salas-Hernández, Angel Gustavo Salas-Lais, Alejandro Sánchez-Flores, Clara Esperanza Santacruz-Tinoco, María Guadalupe Santiago-Mauricio, Selene Zárate, Nelly Sélem-Mojica, Blanca Taboada, Gloria Vazquez, Joel Armando Vázquez-Pérez. |
| EPI_ISL_2681131, EPI_ISL_2681132 | Laboratorio Central de Epidemiología (LCE) | Instituto de Biotecnología de la UNAM | Consorcio Mexicano de Vigilancia Genómica (CoViGen-Mex). Authors (in alphabetical order): Julio Elias Alvarado-Yaah, Carlos F. Arias, Santiago Ávila-Ríos, Eduardo Becerril-Vargas, Víctor Hugo Borja-Aburto, Celia Boukadida, Cristóbal Cháidez-Quiróz, Juan Bautista Chale-Dzul, Ricardo Ciria Merce, Andreu Comas-García, Célida Duque Molina, Julissa Enciso-Ibarra, José Antonio Enciso-Moreno, Gloria Elena Espinosa-Ayala, Fernando Fontove-Herrera, Daniel Fregoso-Rueda, Victor Eduardo García-Arias, Alejandra García-Gasca, Bruno Gómez-Gil, Jean Pierre González, Irvin González-López, Concepción Grajales-Muñiz, Ricardo Grande, Rosa María Gutiérrez Rios, Jesús Hernández, Alejandra Hernández-Terán, Alfredo Herrera-Estrella, Carla Ivón Herrera-Najera, Pavel Isa, Verónica Mata-Haro, Daniel Lira Morales, Susana Lopez, Antonio Loza Román, Brenda Irasema Maldonado-Meza, Bernardo Martínez-Miguel, José Arturo Martínez-Orozco, Célida Martínez- Rodríguez, Margarita Matías-Florentino, Fidencio Mejía-Nepomuceno, María Guadalupe de Jesús Mireles-Rivera, Gloria María Molina-Salinas, Hector Montoya-Fuentes, , Mario Mújica-Sánchez, José Esteban Muñoz-Medina, José de Jesús Nuñez-Contreras, Alicia Ocaña-Mondragón, Luis Alberto Ochoa-Carrera, Hector Esteban Paz-Juárez, Marissa Perez-Garcia, Francisco Pulido, Helen Haydee Fernanda Ramírez-Plascencia, Jorge Salas-Hernández, Angel Gustavo Salas-Lais, Alejandro Sánchez-Flores, Clara Esperanza Santacruz-Tinoco, María Guadalupe Santiago-Mauricio, Selene Zárate, Nelly Sélem-Mojica, Blanca Taboada, Gloria Vazquez, Joel Armando Vázquez-Pérez. |
| EPI_ISL_2681133 | Unidad de Investigación Médica de Yucatán (UIMY) | Instituto de Biotecnología de la UNAM | Consorcio Mexicano de Vigilancia Genómica (CoViGen-Mex). Authors (in alphabetical order): Julio Elias Alvarado-Yaah, Carlos F. Arias, Santiago Ávila-Ríos, Eduardo Becerril-Vargas, Víctor Hugo Borja-Aburto, Celia Boukadida, Cristóbal Cháidez-Quiróz, Juan Bautista Chale-Dzul, Ricardo Ciria Merce, Andreu Comas-García, Célida Duque Molina, Julissa Enciso-Ibarra, José Antonio Enciso-Moreno, Gloria Elena Espinosa-Ayala, Fernando Fontove-Herrera, Daniel Fregoso-Rueda, Victor Eduardo García-Arias, Alejandra García-Gasca, Bruno Gómez-Gil, Jean Pierre González, Irvin González-López, Concepción Grajales-Muñiz, Ricardo Grande, Rosa María Gutiérrez Rios, Jesús Hernández, Alejandra Hernández-Terán, Alfredo Herrera-Estrella, Carla Ivón Herrera-Najera, Pavel Isa, Verónica Mata-Haro, Daniel Lira Morales, Susana Lopez, Antonio Loza Román, Brenda Irasema Maldonado-Meza, Bernardo Martínez-Miguel, José Arturo Martínez-Orozco, Célida Martínez- Rodríguez, Margarita Matías-Florentino, Fidencio Mejía-Nepomuceno, María Guadalupe de Jesús Mireles-Rivera, Gloria María Molina-Salinas, Hector Montoya-Fuentes, , Mario Mújica-Sánchez, José Esteban Muñoz-Medina, José de Jesús Nuñez-Contreras, Alicia Ocaña-Mondragón, Luis Alberto Ochoa-Carrera, Hector Esteban Paz-Juárez, Marissa Perez-Garcia, Francisco Pulido, Helen Haydee Fernanda Ramírez-Plascencia, Jorge Salas-Hernández, Angel Gustavo Salas-Lais, Alejandro Sánchez-Flores, Clara Esperanza Santacruz-Tinoco, María Guadalupe Santiago-Mauricio, Selene Zárate, Nelly Sélem-Mojica, Blanca Taboada, Gloria Vazquez, Joel Armando Vázquez-Pérez. |
| EPI_ISL_2681134, EPI_ISL_2681135 | Laboratorio Central de Epidemiología (LCE) | Instituto de Biotecnología de la UNAM | Consorcio Mexicano de Vigilancia Genómica (CoViGen-Mex). Authors (in alphabetical order): Julio Elias Alvarado-Yaah, Carlos F. Arias, Santiago Ávila-Ríos, Eduardo Becerril-Vargas, Víctor Hugo Borja-Aburto, Celia Boukadida, Cristóbal Cháidez-Quiróz, Juan Bautista Chale-Dzul, Ricardo Ciria Merce, Andreu Comas-García, Célida Duque Molina, Julissa Enciso-Ibarra, José Antonio Enciso-Moreno, Gloria Elena Espinosa-Ayala, Fernando Fontove-Herrera, Daniel Fregoso-Rueda, Victor Eduardo García-Arias, Alejandra García-Gasca, Bruno Gómez-Gil, Jean Pierre González, Irvin González-López, Concepción Grajales-Muñiz, Ricardo Grande, Rosa María Gutiérrez Rios, Jesús Hernández, Alejandra Hernández-Terán, Alfredo Herrera-Estrella, Carla Ivón Herrera-Najera, Pavel Isa, Verónica Mata-Haro, Daniel Lira Morales, Susana Lopez, Antonio Loza Román, Brenda Irasema Maldonado-Meza, Bernardo Martínez-Miguel, José Arturo Martínez-Orozco, Célida Martínez- Rodríguez, Margarita Matías-Florentino, Fidencio Mejía-Nepomuceno, María Guadalupe de Jesús Mireles-Rivera, Gloria María Molina-Salinas, Hector Montoya-Fuentes, , Mario Mújica-Sánchez, José Esteban Muñoz-Medina, José de Jesús Nuñez-Contreras, Alicia Ocaña-Mondragón, Luis Alberto Ochoa-Carrera, Hector Esteban Paz-Juárez, Marissa Perez-Garcia, Francisco Pulido, Helen Haydee Fernanda Ramírez-Plascencia, Jorge Salas-Hernández, Angel Gustavo Salas-Lais, Alejandro Sánchez-Flores, Clara Esperanza Santacruz-Tinoco, María Guadalupe Santiago-Mauricio, Selene Zárate, Nelly Sélem-Mojica, Blanca Taboada, Gloria Vazquez, Joel Armando Vázquez-Pérez. |
| EPI_ISL_2681136 | Unidad de Investigación Médica de Yucatán (UIMY) | Instituto de Biotecnología de la UNAM | Consorcio Mexicano de Vigilancia Genómica (CoViGen-Mex). Authors (in alphabetical order): Julio Elias Alvarado-Yaah, Carlos F. Arias, Santiago Ávila-Ríos, Eduardo Becerril-Vargas, Víctor Hugo Borja-Aburto, Celia Boukadida, Cristóbal Cháidez-Quiróz, Juan Bautista Chale-Dzul, Ricardo Ciria Merce, Andreu Comas-García, Célida Duque Molina, Julissa Enciso-Ibarra, José Antonio Enciso-Moreno, Gloria Elena Espinosa-Ayala, Fernando Fontove-Herrera, Daniel Fregoso-Rueda, Victor Eduardo García-Arias, Alejandra García-Gasca, Bruno Gómez-Gil, Jean Pierre González, Irvin González-López, Concepción Grajales-Muñiz, Ricardo Grande, Rosa María Gutiérrez Rios, Jesús Hernández, Alejandra Hernández-Terán, Alfredo Herrera-Estrella, Carla Ivón Herrera-Najera, Pavel Isa, Verónica Mata-Haro, Daniel Lira Morales, Susana Lopez, Antonio Loza Román, Brenda Irasema Maldonado-Meza, Bernardo Martínez-Miguel, José Arturo Martínez-Orozco, Célida Martínez- Rodríguez, Margarita Matías-Florentino, Fidencio Mejía-Nepomuceno, María Guadalupe de Jesús Mireles-Rivera, Gloria María Molina-Salinas, Hector Montoya-Fuentes, , Mario Mújica-Sánchez, José Esteban Muñoz-Medina, José de Jesús Nuñez-Contreras, Alicia Ocaña-Mondragón, Luis Alberto Ochoa-Carrera, Hector Esteban Paz-Juárez, Marissa Perez-Garcia, Francisco Pulido, Helen Haydee Fernanda Ramírez-Plascencia, Jorge Salas-Hernández, Angel Gustavo Salas-Lais, Alejandro Sánchez-Flores, Clara Esperanza Santacruz-Tinoco, María Guadalupe Santiago-Mauricio, Selene Zárate, Nelly Sélem-Mojica, Blanca Taboada, Gloria Vazquez, Joel Armando Vázquez-Pérez. |
| EPI_ISL_2681137 | Centro de Investigación Biomédica del Noreste (CIBIN) | Instituto de Biotecnología de la UNAM | Consorcio Mexicano de Vigilancia Genómica (CoViGen-Mex). Authors (in alphabetical order): Julio Elias Alvarado-Yaah, Carlos F. Arias, Santiago Ávila-Ríos, Eduardo Becerril-Vargas, Víctor Hugo Borja-Aburto, Celia Boukadida, Cristóbal Cháidez-Quiróz, Juan Bautista Chale-Dzul, Ricardo Ciria Merce, Andreu Comas-García, Célida Duque Molina, Julissa Enciso-Ibarra, José Antonio Enciso-Moreno, Gloria Elena Espinosa-Ayala, Fernando Fontove-Herrera, Daniel Fregoso-Rueda, Victor Eduardo García-Arias, Alejandra García-Gasca, Bruno Gómez-Gil, Jean Pierre González, Irvin González-López, Concepción Grajales-Muñiz, Ricardo Grande, Rosa María Gutiérrez Rios, Jesús Hernández, Alejandra Hernández-Terán, Alfredo Herrera-Estrella, Carla Ivón Herrera-Najera, Pavel Isa, Verónica Mata-Haro, Daniel Lira Morales, Susana Lopez, Antonio Loza Román, Brenda Irasema Maldonado-Meza, Bernardo Martínez-Miguel, José Arturo Martínez-Orozco, Célida Martínez- Rodríguez, Margarita Matías-Florentino, Fidencio Mejía-Nepomuceno, María Guadalupe de Jesús Mireles-Rivera, Gloria María Molina-Salinas, Hector Montoya-Fuentes, , Mario Mújica-Sánchez, José Esteban Muñoz-Medina, José de Jesús Nuñez-Contreras, Alicia Ocaña-Mondragón, Luis Alberto Ochoa-Carrera, Hector Esteban Paz-Juárez, Marissa Perez-Garcia, Francisco Pulido, Helen Haydee Fernanda Ramírez-Plascencia, Jorge Salas-Hernández, Angel Gustavo Salas-Lais, Alejandro Sánchez-Flores, Clara Esperanza Santacruz-Tinoco, María Guadalupe Santiago-Mauricio, Selene Zárate, Nelly Sélem-Mojica, Blanca Taboada, Gloria Vazquez, Joel Armando Vázquez-Pérez. |
| EPI_ISL_2681138, EPI_ISL_2681139, EPI_ISL_2681140 | Laboratorio Central de Epidemiología (LCE) | Instituto de Biotecnología de la UNAM | Consorcio Mexicano de Vigilancia Genómica (CoViGen-Mex). Authors (in alphabetical order): Julio Elias Alvarado-Yaah, Carlos F. Arias, Santiago Ávila-Ríos, Eduardo Becerril-Vargas, Víctor Hugo Borja-Aburto, Celia Boukadida, Cristóbal Cháidez-Quiróz, Juan Bautista Chale-Dzul, Ricardo Ciria Merce, Andreu Comas-García, Célida Duque Molina, Julissa Enciso-Ibarra, José Antonio Enciso-Moreno, Gloria Elena Espinosa-Ayala, Fernando Fontove-Herrera, Daniel Fregoso-Rueda, Victor Eduardo García-Arias, Alejandra García-Gasca, Bruno Gómez-Gil, Jean Pierre González, Irvin González-López, Concepción Grajales-Muñiz, Ricardo Grande, Rosa María Gutiérrez Rios, Jesús Hernández, Alejandra Hernández-Terán, Alfredo Herrera-Estrella, Carla Ivón Herrera-Najera, Pavel Isa, Verónica Mata-Haro, Daniel Lira Morales, Susana Lopez, Antonio Loza Román, Brenda Irasema Maldonado-Meza, Bernardo Martínez-Miguel, José Arturo Martínez-Orozco, Célida Martínez- Rodríguez, Margarita Matías-Florentino, Fidencio Mejía-Nepomuceno, María Guadalupe de Jesús Mireles-Rivera, Gloria María Molina-Salinas, Hector Montoya-Fuentes, , Mario Mújica-Sánchez, José Esteban Muñoz-Medina, José de Jesús Nuñez-Contreras, Alicia Ocaña-Mondragón, Luis Alberto Ochoa-Carrera, Hector Esteban Paz-Juárez, Marissa Perez-Garcia, Francisco Pulido, Helen Haydee Fernanda Ramírez-Plascencia, Jorge Salas-Hernández, Angel Gustavo Salas-Lais, Alejandro Sánchez-Flores, Clara Esperanza Santacruz-Tinoco, María Guadalupe Santiago-Mauricio, Selene Zárate, Nelly Sélem-Mojica, Blanca Taboada, Gloria Vazquez, Joel Armando Vázquez-Pérez. |
| EPI_ISL_2681141 | Unidad de Investigación Médica de Yucatán (UIMY) | Instituto de Biotecnología de la UNAM | Consorcio Mexicano de Vigilancia Genómica (CoViGen-Mex). Authors (in alphabetical order): Julio Elias Alvarado-Yaah, Carlos F. Arias, Santiago Ávila-Ríos, Eduardo Becerril-Vargas, Víctor Hugo Borja-Aburto, Celia Boukadida, Cristóbal Cháidez-Quiróz, Juan Bautista Chale-Dzul, Ricardo Ciria Merce, Andreu Comas-García, Célida Duque Molina, Julissa Enciso-Ibarra, José Antonio Enciso-Moreno, Gloria Elena Espinosa-Ayala, Fernando Fontove-Herrera, Daniel Fregoso-Rueda, Victor Eduardo García-Arias, Alejandra García-Gasca, Bruno Gómez-Gil, Jean Pierre González, Irvin González-López, Concepción Grajales-Muñiz, Ricardo Grande, Rosa María Gutiérrez Rios, Jesús Hernández, Alejandra Hernández-Terán, Alfredo Herrera-Estrella, Carla Ivón Herrera-Najera, Pavel Isa, Verónica Mata-Haro, Daniel Lira Morales, Susana Lopez, Antonio Loza Román, Brenda Irasema Maldonado-Meza, Bernardo Martínez-Miguel, José Arturo Martínez-Orozco, Célida Martínez- Rodríguez, Margarita Matías-Florentino, Fidencio Mejía-Nepomuceno, María Guadalupe de Jesús Mireles-Rivera, Gloria María Molina-Salinas, Hector Montoya-Fuentes, , Mario Mújica-Sánchez, José Esteban Muñoz-Medina, José de Jesús Nuñez-Contreras, Alicia Ocaña-Mondragón, Luis Alberto Ochoa-Carrera, Hector Esteban Paz-Juárez, Marissa Perez-Garcia, Francisco Pulido, Helen Haydee Fernanda Ramírez-Plascencia, Jorge Salas-Hernández, Angel Gustavo Salas-Lais, Alejandro Sánchez-Flores, Clara Esperanza Santacruz-Tinoco, María Guadalupe Santiago-Mauricio, Selene Zárate, Nelly Sélem-Mojica, Blanca Taboada, Gloria Vazquez, Joel Armando Vázquez-Pérez. |
| EPI_ISL_2681142 | Unidad de Investigación Biomédica de Zacatecas (UIBZ) | Instituto de Biotecnología de la UNAM | Consorcio Mexicano de Vigilancia Genómica (CoViGen-Mex). Authors (in alphabetical order): Julio Elias Alvarado-Yaah, Carlos F. Arias, Santiago Ávila-Ríos, Eduardo Becerril-Vargas, Víctor Hugo Borja-Aburto, Celia Boukadida, Cristóbal Cháidez-Quiróz, Juan Bautista Chale-Dzul, Ricardo Ciria Merce, Andreu Comas-García, Célida Duque Molina, Julissa Enciso-Ibarra, José Antonio Enciso-Moreno, Gloria Elena Espinosa-Ayala, Fernando Fontove-Herrera, Daniel Fregoso-Rueda, Victor Eduardo García-Arias, Alejandra García-Gasca, Bruno Gómez-Gil, Jean Pierre González, Irvin González-López, Concepción Grajales-Muñiz, Ricardo Grande, Rosa María Gutiérrez Rios, Jesús Hernández, Alejandra Hernández-Terán, Alfredo Herrera-Estrella, Carla Ivón Herrera-Najera, Pavel Isa, Verónica Mata-Haro, Daniel Lira Morales, Susana Lopez, Antonio Loza Román, Brenda Irasema Maldonado-Meza, Bernardo Martínez-Miguel, José Arturo Martínez-Orozco, Célida Martínez- Rodríguez, Margarita Matías-Florentino, Fidencio Mejía-Nepomuceno, María Guadalupe de Jesús Mireles-Rivera, Gloria María Molina-Salinas, Hector Montoya-Fuentes, , Mario Mújica-Sánchez, José Esteban Muñoz-Medina, José de Jesús Nuñez-Contreras, Alicia Ocaña-Mondragón, Luis Alberto Ochoa-Carrera, Hector Esteban Paz-Juárez, Marissa Perez-Garcia, Francisco Pulido, Helen Haydee Fernanda Ramírez-Plascencia, Jorge Salas-Hernández, Angel Gustavo Salas-Lais, Alejandro Sánchez-Flores, Clara Esperanza Santacruz-Tinoco, María Guadalupe Santiago-Mauricio, Selene Zárate, Nelly Sélem-Mojica, Blanca Taboada, Gloria Vazquez, Joel Armando Vázquez-Pérez. |
| EPI_ISL_2681143, EPI_ISL_2681144, EPI_ISL_2681145, EPI_ISL_2681146, EPI_ISL_2681147, EPI_ISL_2681148, EPI_ISL_2681149, EPI_ISL_2681150, EPI_ISL_2681151, EPI_ISL_2681152, EPI_ISL_2681153, EPI_ISL_2681154, EPI_ISL_2681155, EPI_ISL_2681156, EPI_ISL_2681157, EPI_ISL_2681158, EPI_ISL_2681159 | | | |
| see above | Laboratorio Central de Epidemiología (LCE) | Instituto de Biotecnología de la UNAM | Consorcio Mexicano de Vigilancia Genómica (CoViGen-Mex). Authors (in alphabetical order): Julio Elias Alvarado-Yaah, Carlos F. Arias, Santiago Ávila-Ríos, Eduardo Becerril-Vargas, Víctor Hugo Borja-Aburto, Celia Boukadida, Cristóbal Cháidez-Quiróz, Juan Bautista Chale-Dzul, Ricardo Ciria Merce, Andreu Comas-García, Célida Duque Molina, Julissa Enciso-Ibarra, José Antonio Enciso-Moreno, Gloria Elena Espinosa-Ayala, Fernando Fontove-Herrera, Daniel Fregoso-Rueda, Victor Eduardo García-Arias, Alejandra García-Gasca, Bruno Gómez-Gil, Jean Pierre González, Irvin González-López, Concepción Grajales-Muñiz, Ricardo Grande, Rosa María Gutiérrez Rios, Jesús Hernández, Alejandra Hernández-Terán, Alfredo Herrera-Estrella, Carla Ivón Herrera-Najera, Pavel Isa, Verónica Mata-Haro, Daniel Lira Morales, Susana Lopez, Antonio Loza Román, Brenda Irasema Maldonado-Meza, Bernardo Martínez-Miguel, José Arturo Martínez-Orozco, Célida Martínez- Rodríguez, Margarita Matías-Florentino, Fidencio Mejía-Nepomuceno, María Guadalupe de Jesús Mireles-Rivera, Gloria María Molina-Salinas, Hector Montoya-Fuentes, , Mario Mújica-Sánchez, José Esteban Muñoz-Medina, José de Jesús Nuñez-Contreras, Alicia Ocaña-Mondragón, Luis Alberto Ochoa-Carrera, Hector Esteban Paz-Juárez, Marissa Perez-Garcia, Francisco Pulido, Helen Haydee Fernanda Ramírez-Plascencia, Jorge Salas-Hernández, Angel Gustavo Salas-Lais, Alejandro Sánchez-Flores, Clara Esperanza Santacruz-Tinoco, María Guadalupe Santiago-Mauricio, Selene Zárate, Nelly Sélem-Mojica, Blanca Taboada, Gloria Vazquez, Joel Armando Vázquez-Pérez. |
| EPI_ISL_2681160 | Unidad de Investigación Biomédica de Zacatecas (UIBZ) | Instituto de Biotecnología de la UNAM | Consorcio Mexicano de Vigilancia Genómica (CoViGen-Mex). Authors (in alphabetical order): Julio Elias Alvarado-Yaah, Carlos F. Arias, Santiago Ávila-Ríos, Eduardo Becerril-Vargas, Víctor Hugo Borja-Aburto, Celia Boukadida, Cristóbal Cháidez-Quiróz, Juan Bautista Chale-Dzul, Ricardo Ciria Merce, Andreu Comas-García, Célida Duque Molina, Julissa Enciso-Ibarra, José Antonio Enciso-Moreno, Gloria Elena Espinosa-Ayala, Fernando Fontove-Herrera, Daniel Fregoso-Rueda, Victor Eduardo García-Arias, Alejandra García-Gasca, Bruno Gómez-Gil, Jean Pierre González, Irvin González-López, Concepción Grajales-Muñiz, Ricardo Grande, Rosa María Gutiérrez Rios, Jesús Hernández, Alejandra Hernández-Terán, Alfredo Herrera-Estrella, Carla Ivón Herrera-Najera, Pavel Isa, Verónica Mata-Haro, Daniel Lira Morales, Susana Lopez, Antonio Loza Román, Brenda Irasema Maldonado-Meza, Bernardo Martínez-Miguel, José Arturo Martínez-Orozco, Célida Martínez- Rodríguez, Margarita Matías-Florentino, Fidencio Mejía-Nepomuceno, María Guadalupe de Jesús Mireles-Rivera, Gloria María Molina-Salinas, Hector Montoya-Fuentes, , Mario Mújica-Sánchez, José Esteban Muñoz-Medina, José de Jesús Nuñez-Contreras, Alicia Ocaña-Mondragón, Luis Alberto Ochoa-Carrera, Hector Esteban Paz-Juárez, Marissa Perez-Garcia, Francisco Pulido, Helen Haydee Fernanda Ramírez-Plascencia, Jorge Salas-Hernández, Angel Gustavo Salas-Lais, Alejandro Sánchez-Flores, Clara Esperanza Santacruz-Tinoco, María Guadalupe Santiago-Mauricio, Selene Zárate, Nelly Sélem-Mojica, Blanca Taboada, Gloria Vazquez, Joel Armando Vázquez-Pérez. |
| EPI_ISL_2681161, EPI_ISL_2681162, EPI_ISL_2681163, EPI_ISL_2681164, EPI_ISL_2681165, EPI_ISL_2681166, EPI_ISL_2681167, EPI_ISL_2681168, EPI_ISL_2681169, EPI_ISL_2681170, EPI_ISL_2681171, EPI_ISL_2681172, EPI_ISL_2681173, EPI_ISL_2681174, EPI_ISL_2681175 | | | |
| see above | Laboratorio Central de Epidemiología (LCE) | Instituto de Biotecnología de la UNAM | Consorcio Mexicano de Vigilancia Genómica (CoViGen-Mex). Authors (in alphabetical order): Julio Elias Alvarado-Yaah, Carlos F. Arias, Santiago Ávila-Ríos, Eduardo Becerril-Vargas, Víctor Hugo Borja-Aburto, Celia Boukadida, Cristóbal Cháidez-Quiróz, Juan Bautista Chale-Dzul, Ricardo Ciria Merce, Andreu Comas-García, Célida Duque Molina, Julissa Enciso-Ibarra, José Antonio Enciso-Moreno, Gloria Elena Espinosa-Ayala, Fernando Fontove-Herrera, Daniel Fregoso-Rueda, Victor Eduardo García-Arias, Alejandra García-Gasca, Bruno Gómez-Gil, Jean Pierre González, Irvin González-López, Concepción Grajales-Muñiz, Ricardo Grande, Rosa María Gutiérrez Rios, Jesús Hernández, Alejandra Hernández-Terán, Alfredo Herrera-Estrella, Carla Ivón Herrera-Najera, Pavel Isa, Verónica Mata-Haro, Daniel Lira Morales, Susana Lopez, Antonio Loza Román, Brenda Irasema Maldonado-Meza, Bernardo Martínez-Miguel, José Arturo Martínez-Orozco, Célida Martínez- Rodríguez, Margarita Matías-Florentino, Fidencio Mejía-Nepomuceno, María Guadalupe de Jesús Mireles-Rivera, Gloria María Molina-Salinas, Hector Montoya-Fuentes, , Mario Mújica-Sánchez, José Esteban Muñoz-Medina, José de Jesús Nuñez-Contreras, Alicia Ocaña-Mondragón, Luis Alberto Ochoa-Carrera, Hector Esteban Paz-Juárez, Marissa Perez-Garcia, Francisco Pulido, Helen Haydee Fernanda Ramírez-Plascencia, Jorge Salas-Hernández, Angel Gustavo Salas-Lais, Alejandro Sánchez-Flores, Clara Esperanza Santacruz-Tinoco, María Guadalupe Santiago-Mauricio, Selene Zárate, Nelly Sélem-Mojica, Blanca Taboada, Gloria Vazquez, Joel Armando Vázquez-Pérez. |
| EPI_ISL_2681176 | Unidad de Investigación Médica de Yucatán (UIMY) | Instituto de Biotecnología de la UNAM | Consorcio Mexicano de Vigilancia Genómica (CoViGen-Mex). Authors (in alphabetical order): Julio Elias Alvarado-Yaah, Carlos F. Arias, Santiago Ávila-Ríos, Eduardo Becerril-Vargas, Víctor Hugo Borja-Aburto, Celia Boukadida, Cristóbal Cháidez-Quiróz, Juan Bautista Chale-Dzul, Ricardo Ciria Merce, Andreu Comas-García, Célida Duque Molina, Julissa Enciso-Ibarra, José Antonio Enciso-Moreno, Gloria Elena Espinosa-Ayala, Fernando Fontove-Herrera, Daniel Fregoso-Rueda, Victor Eduardo García-Arias, Alejandra García-Gasca, Bruno Gómez-Gil, Jean Pierre González, Irvin González-López, Concepción Grajales-Muñiz, Ricardo Grande, Rosa María Gutiérrez Rios, Jesús Hernández, Alejandra Hernández-Terán, Alfredo Herrera-Estrella, Carla Ivón Herrera-Najera, Pavel Isa, Verónica Mata-Haro, Daniel Lira Morales, Susana Lopez, Antonio Loza Román, Brenda Irasema Maldonado-Meza, Bernardo Martínez-Miguel, José Arturo Martínez-Orozco, Célida Martínez- Rodríguez, Margarita Matías-Florentino, Fidencio Mejía-Nepomuceno, María Guadalupe de Jesús Mireles-Rivera, Gloria María Molina-Salinas, Hector Montoya-Fuentes, , Mario Mújica-Sánchez, José Esteban Muñoz-Medina, José de Jesús Nuñez-Contreras, Alicia Ocaña-Mondragón, Luis Alberto Ochoa-Carrera, Hector Esteban Paz-Juárez, Marissa Perez-Garcia, Francisco Pulido, Helen Haydee Fernanda Ramírez-Plascencia, Jorge Salas-Hernández, Angel Gustavo Salas-Lais, Alejandro Sánchez-Flores, Clara Esperanza Santacruz-Tinoco, María Guadalupe Santiago-Mauricio, Selene Zárate, Nelly Sélem-Mojica, Blanca Taboada, Gloria Vazquez, Joel Armando Vázquez-Pérez. |
| EPI_ISL_2681177, EPI_ISL_2681178, EPI_ISL_2681179, EPI_ISL_2681180, EPI_ISL_2681181, EPI_ISL_2681182, EPI_ISL_2681183, EPI_ISL_2681184, EPI_ISL_2681185, EPI_ISL_2681186, EPI_ISL_2681187, EPI_ISL_2681188, EPI_ISL_2681189 | | | |
| see above | Laboratorio Central de Epidemiología (LCE) | Instituto de Biotecnología de la UNAM | Consorcio Mexicano de Vigilancia Genómica (CoViGen-Mex). Authors (in alphabetical order): Julio Elias Alvarado-Yaah, Carlos F. Arias, Santiago Ávila-Ríos, Eduardo Becerril-Vargas, Víctor Hugo Borja-Aburto, Celia Boukadida, Cristóbal Cháidez-Quiróz, Juan Bautista Chale-Dzul, Ricardo Ciria Merce, Andreu Comas-García, Célida Duque Molina, Julissa Enciso-Ibarra, José Antonio Enciso-Moreno, Gloria Elena Espinosa-Ayala, Fernando Fontove-Herrera, Daniel Fregoso-Rueda, Victor Eduardo García-Arias, Alejandra García-Gasca, Bruno Gómez-Gil, Jean Pierre González, Irvin González-López, Concepción Grajales-Muñiz, Ricardo Grande, Rosa María Gutiérrez Rios, Jesús Hernández, Alejandra Hernández-Terán, Alfredo Herrera-Estrella, Carla Ivón Herrera-Najera, Pavel Isa, Verónica Mata-Haro, Daniel Lira Morales, Susana Lopez, Antonio Loza Román, Brenda Irasema Maldonado-Meza, Bernardo Martínez-Miguel, José Arturo Martínez-Orozco, Célida Martínez- Rodríguez, Margarita Matías-Florentino, Fidencio Mejía-Nepomuceno, María Guadalupe de Jesús Mireles-Rivera, Gloria María Molina-Salinas, Hector Montoya-Fuentes, , Mario Mújica-Sánchez, José Esteban Muñoz-Medina, José de Jesús Nuñez-Contreras, Alicia Ocaña-Mondragón, Luis Alberto Ochoa-Carrera, Hector Esteban Paz-Juárez, Marissa Perez-Garcia, Francisco Pulido, Helen Haydee Fernanda Ramírez-Plascencia, Jorge Salas-Hernández, Angel Gustavo Salas-Lais, Alejandro Sánchez-Flores, Clara Esperanza Santacruz-Tinoco, María Guadalupe Santiago-Mauricio, Selene Zárate, Nelly Sélem-Mojica, Blanca Taboada, Gloria Vazquez, Joel Armando Vázquez-Pérez. |
| EPI_ISL_2681190 | Unidad de Investigación Biomédica de Zacatecas (UIBZ) | Instituto de Biotecnología de la UNAM | Consorcio Mexicano de Vigilancia Genómica (CoViGen-Mex). Authors (in alphabetical order): Julio Elias Alvarado-Yaah, Carlos F. Arias, Santiago Ávila-Ríos, Eduardo Becerril-Vargas, Víctor Hugo Borja-Aburto, Celia Boukadida, Cristóbal Cháidez-Quiróz, Juan Bautista Chale-Dzul, Ricardo Ciria Merce, Andreu Comas-García, Célida Duque Molina, Julissa Enciso-Ibarra, José Antonio Enciso-Moreno, Gloria Elena Espinosa-Ayala, Fernando Fontove-Herrera, Daniel Fregoso-Rueda, Victor Eduardo García-Arias, Alejandra García-Gasca, Bruno Gómez-Gil, Jean Pierre González, Irvin González-López, Concepción Grajales-Muñiz, Ricardo Grande, Rosa María Gutiérrez Rios, Jesús Hernández, Alejandra Hernández-Terán, Alfredo Herrera-Estrella, Carla Ivón Herrera-Najera, Pavel Isa, Verónica Mata-Haro, Daniel Lira Morales, Susana Lopez, Antonio Loza Román, Brenda Irasema Maldonado-Meza, Bernardo Martínez-Miguel, José Arturo Martínez-Orozco, Célida Martínez- Rodríguez, Margarita Matías-Florentino, Fidencio Mejía-Nepomuceno, María Guadalupe de Jesús Mireles-Rivera, Gloria María Molina-Salinas, Hector Montoya-Fuentes, , Mario Mújica-Sánchez, José Esteban Muñoz-Medina, José de Jesús Nuñez-Contreras, Alicia Ocaña-Mondragón, Luis Alberto Ochoa-Carrera, Hector Esteban Paz-Juárez, Marissa Perez-Garcia, Francisco Pulido, Helen Haydee Fernanda Ramírez-Plascencia, Jorge Salas-Hernández, Angel Gustavo Salas-Lais, Alejandro Sánchez-Flores, Clara Esperanza Santacruz-Tinoco, María Guadalupe Santiago-Mauricio, Selene Zárate, Nelly Sélem-Mojica, Blanca Taboada, Gloria Vazquez, Joel Armando Vázquez-Pérez. |
| EPI_ISL_2681191, | Laboratorio Central de Epidemiología | Instituto de Biotecnología de la UNAM | Consorcio Mexicano de Vigilancia Genómica (CoViGen-Mex). Authors (in alphabetical order): Julio Elias Alvarado-Yaah, Carlos F. Arias, Santiago Ávila-Ríos, Eduardo Becerril-Vargas, Víctor Hugo Borja-Aburto, Celia Boukadida, Cristóbal Cháidez-Quiróz, Juan Bautista Chale-Dzul, Ricardo |

| | | | |
|---|---|---|---|
| EPI_ISL_2681192, EPI_ISL_2681193, EPI_ISL_2681194, EPI_ISL_2681195, EPI_ISL_2681196 | (LCE) | | Ciria Merce, Andreu Comas-García, Célida Duque Molina, Julissa Enciso-Ibarra, José Antonio Enciso-Moreno, Gloria Elena Espinosa-Ayala, Fernando Fontove-Herrera, Daniel Fregoso-Rueda, Victor Eduardo García-Arias, Alejandra García-Gasca, Bruno Gómez-Gil, Jean Pierre González, Irvin González-López, Concepción Grajales-Muñiz, Ricardo Grande, Rosa María Gutiérrez Rios, Jesús Hernández, Alejandra Hernández-Terán, Alfredo Herrera-Estrella, Carla Ivón Herrera-Najera, Pavel Isa, Verónica Mata-Haro, Daniel Lira Morales, Susana Lopez, Antonio Loza Román, Brenda Irasema Maldonado-Meza, Bernardo Martínez-Miguel, José Arturo Martínez-Orozco, Célida Martínez- Rodríguez, Margarita Matías-Florentino, Fidencio Mejía-Nepomuceno, María Guadalupe de Jesús Mireles-Rivera, Gloria María Molina-Salinas, Hector Montoya-Fuentes, , Mario Mújica-Sánchez, José Esteban Muñoz-Medina, José de Jesús Nuñez-Contreras, Alicia Ocaña-Mondragón, Luis Alberto Ochoa-Carrera, Hector Esteban Paz-Juárez, Marissa Perez-Garcia, Francisco Pulido, Helen Haydee Fernanda Ramírez-Plascencia, Jorge Salas-Hernández, Angel Gustavo Salas-Lais, Alejandro Sánchez-Flores, Clara Esperanza Santacruz-Tinoco, María Guadalupe Santiago-Mauricio, Selene Zárate, Nelly Sélem-Mojica, Blanca Taboada, Gloria Vazquez, Joel Armando Vázquez-Pérez. |
| EPI_ISL_2681197 | Centro de Investigación Biomédica del Noreste (CIBIN) | Instituto de Biotecnología de la UNAM | Consorcio Mexicano de Vigilancia Genómica (CoViGen-Mex). Authors (in alphabetical order): Julio Elias Alvarado-Yaah, Carlos F. Arias, Santiago Ávila-Ríos, Eduardo Becerril-Vargas, Víctor Hugo Borja-Aburto, Celia Boukadida, Cristóbal Cháidez-Quiróz, Juan Bautista Chale-Dzul, Ricardo Ciria Merce, Andreu Comas-García, Célida Duque Molina, Julissa Enciso-Ibarra, José Antonio Enciso-Moreno, Gloria Elena Espinosa-Ayala, Fernando Fontove-Herrera, Daniel Fregoso-Rueda, Victor Eduardo García-Arias, Alejandra García-Gasca, Bruno Gómez-Gil, Jean Pierre González, Irvin González-López, Concepción Grajales-Muñiz, Ricardo Grande, Rosa María Gutiérrez Rios, Jesús Hernández, Alejandra Hernández-Terán, Alfredo Herrera-Estrella, Carla Ivón Herrera-Najera, Pavel Isa, Verónica Mata-Haro, Daniel Lira Morales, Susana Lopez, Antonio Loza Román, Brenda Irasema Maldonado-Meza, Bernardo Martínez-Miguel, José Arturo Martínez-Orozco, Célida Martínez- Rodríguez, Margarita Matías-Florentino, Fidencio Mejía-Nepomuceno, María Guadalupe de Jesús Mireles-Rivera, Gloria María Molina-Salinas, Hector Montoya-Fuentes, , Mario Mújica-Sánchez, José Esteban Muñoz-Medina, José de Jesús Nuñez-Contreras, Alicia Ocaña-Mondragón, Luis Alberto Ochoa-Carrera, Hector Esteban Paz-Juárez, Marissa Perez-Garcia, Francisco Pulido, Helen Haydee Fernanda Ramírez-Plascencia, Jorge Salas-Hernández, Angel Gustavo Salas-Lais, Alejandro Sánchez-Flores, Clara Esperanza Santacruz-Tinoco, María Guadalupe Santiago-Mauricio, Selene Zárate, Nelly Sélem-Mojica, Blanca Taboada, Gloria Vazquez, Joel Armando Vázquez-Pérez. |
| EPI_ISL_2681198, EPI_ISL_2681199, EPI_ISL_2681200 | Laboratorio Central de Epidemiología (LCE) | Instituto de Biotecnología de la UNAM | Consorcio Mexicano de Vigilancia Genómica (CoViGen-Mex). Authors (in alphabetical order): Julio Elias Alvarado-Yaah, Carlos F. Arias, Santiago Ávila-Ríos, Eduardo Becerril-Vargas, Víctor Hugo Borja-Aburto, Celia Boukadida, Cristóbal Cháidez-Quiróz, Juan Bautista Chale-Dzul, Ricardo Ciria Merce, Andreu Comas-García, Célida Duque Molina, Julissa Enciso-Ibarra, José Antonio Enciso-Moreno, Gloria Elena Espinosa-Ayala, Fernando Fontove-Herrera, Daniel Fregoso-Rueda, Victor Eduardo García-Arias, Alejandra García-Gasca, Bruno Gómez-Gil, Jean Pierre González, Irvin González-López, Concepción Grajales-Muñiz, Ricardo Grande, Rosa María Gutiérrez Rios, Jesús Hernández, Alejandra Hernández-Terán, Alfredo Herrera-Estrella, Carla Ivón Herrera-Najera, Pavel Isa, Verónica Mata-Haro, Daniel Lira Morales, Susana Lopez, Antonio Loza Román, Brenda Irasema Maldonado-Meza, Bernardo Martínez-Miguel, José Arturo Martínez-Orozco, Célida Martínez- Rodríguez, Margarita Matías-Florentino, Fidencio Mejía-Nepomuceno, María Guadalupe de Jesús Mireles-Rivera, Gloria María Molina-Salinas, Hector Montoya-Fuentes, , Mario Mújica-Sánchez, José Esteban Muñoz-Medina, José de Jesús Nuñez-Contreras, Alicia Ocaña-Mondragón, Luis Alberto Ochoa-Carrera, Hector Esteban Paz-Juárez, Marissa Perez-Garcia, Francisco Pulido, Helen Haydee Fernanda Ramírez-Plascencia, Jorge Salas-Hernández, Angel Gustavo Salas-Lais, Alejandro Sánchez-Flores, Clara Esperanza Santacruz-Tinoco, María Guadalupe Santiago-Mauricio, Selene Zárate, Nelly Sélem-Mojica, Blanca Taboada, Gloria Vazquez, Joel Armando Vázquez-Pérez. |
| EPI_ISL_2681201 | Centro de Investigación Biomédica del Noreste (CIBIN) | Instituto de Biotecnología de la UNAM | Consorcio Mexicano de Vigilancia Genómica (CoViGen-Mex). Authors (in alphabetical order): Julio Elias Alvarado-Yaah, Carlos F. Arias, Santiago Ávila-Ríos, Eduardo Becerril-Vargas, Víctor Hugo Borja-Aburto, Celia Boukadida, Cristóbal Cháidez-Quiróz, Juan Bautista Chale-Dzul, Ricardo Ciria Merce, Andreu Comas-García, Célida Duque Molina, Julissa Enciso-Ibarra, José Antonio Enciso-Moreno, Gloria Elena Espinosa-Ayala, Fernando Fontove-Herrera, Daniel Fregoso-Rueda, Victor Eduardo García-Arias, Alejandra García-Gasca, Bruno Gómez-Gil, Jean Pierre González, Irvin González-López, Concepción Grajales-Muñiz, Ricardo Grande, Rosa María Gutiérrez Rios, Jesús Hernández, Alejandra Hernández-Terán, Alfredo Herrera-Estrella, Carla Ivón Herrera-Najera, Pavel Isa, Verónica Mata-Haro, Daniel Lira Morales, Susana Lopez, Antonio Loza Román, Brenda Irasema Maldonado-Meza, Bernardo Martínez-Miguel, José Arturo Martínez-Orozco, Célida Martínez- Rodríguez, Margarita Matías-Florentino, Fidencio Mejía-Nepomuceno, María Guadalupe de Jesús Mireles-Rivera, Gloria María Molina-Salinas, Hector Montoya-Fuentes, , Mario Mújica-Sánchez, José Esteban Muñoz-Medina, José de Jesús Nuñez-Contreras, Alicia Ocaña-Mondragón, Luis Alberto Ochoa-Carrera, Hector Esteban Paz-Juárez, Marissa Perez-Garcia, Francisco Pulido, Helen Haydee Fernanda Ramírez-Plascencia, Jorge Salas-Hernández, Angel Gustavo Salas-Lais, Alejandro Sánchez-Flores, Clara Esperanza Santacruz-Tinoco, María Guadalupe Santiago-Mauricio, Selene Zárate, Nelly Sélem-Mojica, Blanca Taboada, Gloria Vazquez, Joel Armando Vázquez-Pérez. |
| EPI_ISL_2681202, EPI_ISL_2681203, EPI_ISL_2681204, EPI_ISL_2681205, EPI_ISL_2681206 | Laboratorio Central de Epidemiología (LCE) | Instituto de Biotecnología de la UNAM | Consorcio Mexicano de Vigilancia Genómica (CoViGen-Mex). Authors (in alphabetical order): Julio Elias Alvarado-Yaah, Carlos F. Arias, Santiago Ávila-Ríos, Eduardo Becerril-Vargas, Víctor Hugo Borja-Aburto, Celia Boukadida, Cristóbal Cháidez-Quiróz, Juan Bautista Chale-Dzul, Ricardo Ciria Merce, Andreu Comas-García, Célida Duque Molina, Julissa Enciso-Ibarra, José Antonio Enciso-Moreno, Gloria Elena Espinosa-Ayala, Fernando Fontove-Herrera, Daniel Fregoso-Rueda, Victor Eduardo García-Arias, Alejandra García-Gasca, Bruno Gómez-Gil, Jean Pierre González, Irvin González-López, Concepción Grajales-Muñiz, Ricardo Grande, Rosa María Gutiérrez Rios, Jesús Hernández, Alejandra Hernández-Terán, Alfredo Herrera-Estrella, Carla Ivón Herrera-Najera, Pavel Isa, Verónica Mata-Haro, Daniel Lira Morales, Susana Lopez, Antonio Loza Román, Brenda Irasema Maldonado-Meza, Bernardo Martínez-Miguel, José Arturo Martínez-Orozco, Célida Martínez- Rodríguez, Margarita Matías-Florentino, Fidencio Mejía-Nepomuceno, María Guadalupe de Jesús Mireles-Rivera, Gloria María Molina-Salinas, Hector Montoya-Fuentes, , Mario Mújica-Sánchez, José Esteban Muñoz-Medina, José de Jesús Nuñez-Contreras, Alicia Ocaña-Mondragón, Luis Alberto Ochoa-Carrera, Hector Esteban Paz-Juárez, Marissa Perez-Garcia, Francisco Pulido, Helen Haydee Fernanda Ramírez-Plascencia, Jorge Salas-Hernández, Angel Gustavo Salas-Lais, Alejandro Sánchez-Flores, Clara Esperanza Santacruz-Tinoco, María Guadalupe Santiago-Mauricio, Selene Zárate, Nelly Sélem-Mojica, Blanca Taboada, Gloria Vazquez, Joel Armando Vázquez-Pérez. |
| EPI_ISL_2681207 | Unidad de Investigación Biomédica de Zacatecas (UIBZ) | Instituto de Biotecnología de la UNAM | Consorcio Mexicano de Vigilancia Genómica (CoViGen-Mex). Authors (in alphabetical order): Julio Elias Alvarado-Yaah, Carlos F. Arias, Santiago Ávila-Ríos, Eduardo Becerril-Vargas, Víctor Hugo Borja-Aburto, Celia Boukadida, Cristóbal Cháidez-Quiróz, Juan Bautista Chale-Dzul, Ricardo Ciria Merce, Andreu Comas-García, Célida Duque Molina, Julissa Enciso-Ibarra, José Antonio Enciso-Moreno, Gloria Elena Espinosa-Ayala, Fernando Fontove-Herrera, Daniel Fregoso-Rueda, Victor Eduardo García-Arias, Alejandra García-Gasca, Bruno Gómez-Gil, Jean Pierre González, Irvin González-López, Concepción Grajales-Muñiz, Ricardo Grande, Rosa María Gutiérrez Rios, Jesús Hernández, Alejandra Hernández-Terán, Alfredo Herrera-Estrella, Carla Ivón Herrera-Najera, Pavel Isa, Verónica Mata-Haro, Daniel Lira Morales, Susana Lopez, Antonio Loza Román, Brenda Irasema Maldonado-Meza, Bernardo Martínez-Miguel, José Arturo Martínez-Orozco, Célida Martínez- Rodríguez, Margarita Matías-Florentino, Fidencio Mejía-Nepomuceno, María Guadalupe de Jesús Mireles-Rivera, Gloria María Molina-Salinas, Hector Montoya-Fuentes, , Mario Mújica-Sánchez, José Esteban Muñoz-Medina, José de Jesús Nuñez-Contreras, Alicia Ocaña-Mondragón, Luis Alberto Ochoa-Carrera, Hector Esteban Paz-Juárez, Marissa Perez-Garcia, Francisco Pulido, Helen Haydee Fernanda Ramírez-Plascencia, Jorge Salas-Hernández, Angel Gustavo Salas-Lais, Alejandro Sánchez-Flores, Clara Esperanza Santacruz-Tinoco, María Guadalupe Santiago-Mauricio, Selene Zárate, Nelly Sélem-Mojica, Blanca Taboada, Gloria Vazquez, Joel Armando Vázquez-Pérez. |
| EPI_ISL_2681208 | Laboratorio Central de Epidemiología (LCE) | Instituto de Biotecnología de la UNAM | Consorcio Mexicano de Vigilancia Genómica (CoViGen-Mex). Authors (in alphabetical order): Julio Elias Alvarado-Yaah, Carlos F. Arias, Santiago Ávila-Ríos, Eduardo Becerril-Vargas, Víctor Hugo Borja-Aburto, Celia Boukadida, Cristóbal Cháidez-Quiróz, Juan Bautista Chale-Dzul, Ricardo Ciria Merce, Andreu Comas-García, Célida Duque Molina, Julissa Enciso-Ibarra, José Antonio Enciso-Moreno, Gloria Elena Espinosa-Ayala, Fernando Fontove-Herrera, Daniel Fregoso-Rueda, Victor Eduardo García-Arias, Alejandra García-Gasca, Bruno Gómez-Gil, Jean Pierre González, Irvin González-López, Concepción Grajales-Muñiz, Ricardo Grande, Rosa María Gutiérrez Rios, Jesús Hernández, Alejandra Hernández-Terán, Alfredo Herrera-Estrella, Carla Ivón Herrera-Najera, Pavel Isa, Verónica Mata-Haro, Daniel Lira Morales, Susana Lopez, Antonio Loza Román, Brenda Irasema Maldonado-Meza, Bernardo Martínez-Miguel, José Arturo Martínez-Orozco, Célida Martínez- Rodríguez, Margarita Matías-Florentino, Fidencio Mejía-Nepomuceno, María Guadalupe de Jesús Mireles-Rivera, Gloria María Molina-Salinas, Hector Montoya-Fuentes, , Mario Mújica-Sánchez, José Esteban Muñoz-Medina, José de Jesús Nuñez-Contreras, Alicia Ocaña-Mondragón, Luis Alberto Ochoa-Carrera, Hector Esteban Paz-Juárez, Marissa Perez-Garcia, Francisco Pulido, Helen Haydee Fernanda Ramírez-Plascencia, Jorge Salas-Hernández, Angel Gustavo Salas-Lais, Alejandro Sánchez-Flores, Clara Esperanza Santacruz-Tinoco, María Guadalupe Santiago-Mauricio, Selene Zárate, Nelly Sélem-Mojica, Blanca Taboada, Gloria Vazquez, Joel Armando Vázquez-Pérez. |
| EPI_ISL_2681209 | Unidad de Investigación Biomédica de Zacatecas (UIBZ) | Instituto de Biotecnología de la UNAM | Consorcio Mexicano de Vigilancia Genómica (CoViGen-Mex). Authors (in alphabetical order): Julio Elias Alvarado-Yaah, Carlos F. Arias, Santiago Ávila-Ríos, Eduardo Becerril-Vargas, Víctor Hugo Borja-Aburto, Celia Boukadida, Cristóbal Cháidez-Quiróz, Juan Bautista Chale-Dzul, Ricardo Ciria Merce, Andreu Comas-García, Célida Duque Molina, Julissa Enciso-Ibarra, José Antonio Enciso-Moreno, Gloria Elena Espinosa-Ayala, Fernando Fontove-Herrera, Daniel Fregoso-Rueda, Victor Eduardo García-Arias, Alejandra García-Gasca, Bruno Gómez-Gil, Jean Pierre González, Irvin González-López, Concepción Grajales-Muñiz, Ricardo Grande, Rosa María Gutiérrez Rios, Jesús Hernández, Alejandra Hernández-Terán, Alfredo Herrera-Estrella, Carla Ivón Herrera-Najera, Pavel Isa, Verónica Mata-Haro, Daniel Lira Morales, Susana Lopez, Antonio Loza Román, Brenda Irasema Maldonado-Meza, Bernardo Martínez-Miguel, José Arturo Martínez-Orozco, Célida Martínez- Rodríguez, Margarita Matías-Florentino, Fidencio Mejía-Nepomuceno, María Guadalupe de Jesús Mireles-Rivera, Gloria María Molina-Salinas, Hector Montoya-Fuentes, , Mario Mújica-Sánchez, José Esteban Muñoz-Medina, José de Jesús Nuñez-Contreras, Alicia Ocaña-Mondragón, Luis Alberto Ochoa-Carrera, Hector Esteban Paz-Juárez, Marissa Perez-Garcia, Francisco Pulido, Helen Haydee Fernanda Ramírez-Plascencia, Jorge Salas-Hernández, Angel Gustavo Salas-Lais, Alejandro Sánchez-Flores, Clara Esperanza Santacruz-Tinoco, María Guadalupe Santiago-Mauricio, Selene Zárate, Nelly Sélem-Mojica, Blanca Taboada, Gloria Vazquez, Joel Armando Vázquez-Pérez. |
| EPI_ISL_2681210 | Laboratorio Central de Epidemiología (LCE) | Instituto de Biotecnología de la UNAM | Consorcio Mexicano de Vigilancia Genómica (CoViGen-Mex). Authors (in alphabetical order): Julio Elias Alvarado-Yaah, Carlos F. Arias, Santiago Ávila-Ríos, Eduardo Becerril-Vargas, Víctor Hugo Borja-Aburto, Celia Boukadida, Cristóbal Cháidez-Quiróz, Juan Bautista Chale-Dzul, Ricardo Ciria Merce, Andreu Comas-García, Célida Duque Molina, Julissa Enciso-Ibarra, José Antonio Enciso-Moreno, Gloria Elena Espinosa-Ayala, Fernando Fontove-Herrera, Daniel Fregoso-Rueda, Victor Eduardo García-Arias, Alejandra García-Gasca, Bruno Gómez-Gil, Jean Pierre González, Irvin González-López, Concepción Grajales-Muñiz, Ricardo Grande, Rosa María Gutiérrez Rios, Jesús Hernández, Alejandra Hernández-Terán, Alfredo Herrera-Estrella, Carla Ivón Herrera-Najera, Pavel Isa, Verónica Mata-Haro, Daniel Lira Morales, Susana Lopez, Antonio Loza Román, Brenda Irasema Maldonado-Meza, Bernardo Martínez-Miguel, José Arturo Martínez-Orozco, Célida Martínez- Rodríguez, Margarita Matías-Florentino, Fidencio Mejía-Nepomuceno, María Guadalupe de Jesús Mireles-Rivera, Gloria María Molina-Salinas, Hector Montoya-Fuentes, , Mario Mújica-Sánchez, José Esteban Muñoz-Medina, José de Jesús Nuñez-Contreras, Alicia Ocaña-Mondragón, Luis Alberto Ochoa-Carrera, Hector Esteban Paz-Juárez, Marissa Perez-Garcia, Francisco Pulido, Helen Haydee Fernanda Ramírez-Plascencia, Jorge Salas-Hernández, Angel Gustavo Salas-Lais, Alejandro Sánchez-Flores, Clara Esperanza Santacruz-Tinoco, María Guadalupe Santiago-Mauricio, Selene Zárate, Nelly Sélem-Mojica, Blanca Taboada, Gloria Vazquez, Joel Armando Vázquez-Pérez. |
| EPI_ISL_2681211 | Unidad de Investigación Médica de Yucatán (UIMY) | Instituto de Biotecnología de la UNAM | Consorcio Mexicano de Vigilancia Genómica (CoViGen-Mex). Authors (in alphabetical order): Julio Elias Alvarado-Yaah, Carlos F. Arias, Santiago Ávila-Ríos, Eduardo Becerril-Vargas, Víctor Hugo Borja-Aburto, Celia Boukadida, Cristóbal Cháidez-Quiróz, Juan Bautista Chale-Dzul, Ricardo Ciria Merce, Andreu Comas-García, Célida Duque Molina, Julissa Enciso-Ibarra, José Antonio Enciso-Moreno, Gloria Elena Espinosa-Ayala, Fernando Fontove-Herrera, Daniel Fregoso-Rueda, Victor Eduardo García-Arias, Alejandra García-Gasca, Bruno Gómez-Gil, Jean Pierre González, Irvin González-López, Concepción Grajales-Muñiz, Ricardo Grande, Rosa María Gutiérrez Rios, Jesús Hernández, Alejandra Hernández-Terán, Alfredo Herrera-Estrella, Carla Ivón Herrera-Najera, Pavel Isa, Verónica Mata-Haro, Daniel Lira Morales, Susana Lopez, Antonio Loza Román, Brenda Irasema Maldonado-Meza, Bernardo Martínez-Miguel, José Arturo Martínez-Orozco, Célida Martínez- Rodríguez, Margarita Matías-Florentino, Fidencio Mejía-Nepomuceno, María Guadalupe de Jesús Mireles-Rivera, Gloria María Molina-Salinas, Hector Montoya-Fuentes, , Mario Mújica-Sánchez, José Esteban Muñoz-Medina, José de Jesús Nuñez-Contreras, Alicia Ocaña-Mondragón, Luis Alberto Ochoa-Carrera, Hector Esteban Paz-Juárez, Marissa Perez-Garcia, Francisco Pulido, Helen Haydee Fernanda Ramírez-Plascencia, Jorge Salas-Hernández, Angel Gustavo Salas-Lais, Alejandro Sánchez-Flores, Clara Esperanza Santacruz-Tinoco, María Guadalupe Santiago-Mauricio, Selene Zárate, Nelly Sélem-Mojica, Blanca Taboada, Gloria Vazquez, Joel Armando Vázquez-Pérez. |
| EPI_ISL_2681212 | Unidad de Investigación Biomédica de Zacatecas (UIBZ) | Instituto de Biotecnología de la UNAM | Consorcio Mexicano de Vigilancia Genómica (CoViGen-Mex). Authors (in alphabetical order): Julio Elias Alvarado-Yaah, Carlos F. Arias, Santiago Ávila-Ríos, Eduardo Becerril-Vargas, Víctor Hugo Borja-Aburto, Celia Boukadida, Cristóbal Cháidez-Quiróz, Juan Bautista Chale-Dzul, Ricardo Ciria Merce, Andreu Comas-García, Célida Duque Molina, Julissa Enciso-Ibarra, José Antonio Enciso-Moreno, Gloria Elena Espinosa-Ayala, Fernando Fontove-Herrera, Daniel Fregoso-Rueda, Victor Eduardo García-Arias, Alejandra García-Gasca, Bruno Gómez-Gil, Jean Pierre González, Irvin González-López, Concepción Grajales-Muñiz, Ricardo Grande, Rosa María Gutiérrez Rios, Jesús Hernández, Alejandra Hernández-Terán, Alfredo Herrera-Estrella, Carla Ivón Herrera-Najera, Pavel Isa, Verónica Mata-Haro, Daniel Lira Morales, Susana Lopez, Antonio Loza Román, Brenda Irasema Maldonado-Meza, Bernardo Martínez-Miguel, José Arturo Martínez-Orozco, Célida Martínez- Rodríguez, Margarita Matías-Florentino, Fidencio Mejía-Nepomuceno, María Guadalupe de Jesús Mireles-Rivera, Gloria María Molina-Salinas, Hector Montoya-Fuentes, , Mario Mújica-Sánchez, José Esteban Muñoz-Medina, José de Jesús Nuñez-Contreras, Alicia Ocaña-Mondragón, Luis Alberto Ochoa-Carrera, Hector Esteban Paz-Juárez, Marissa Perez-Garcia, Francisco Pulido, Helen Haydee Fernanda Ramírez-Plascencia, Jorge Salas-Hernández, Angel Gustavo Salas-Lais, Alejandro Sánchez-Flores, Clara Esperanza Santacruz-Tinoco, María Guadalupe Santiago-Mauricio, Selene Zárate, Nelly Sélem-Mojica, Blanca Taboada, Gloria Vazquez, Joel Armando Vázquez-Pérez. |
| EPI_ISL_2681213, EPI_ISL_2681214, EPI_ISL_2681215, EPI_ISL_2681216, EPI_ISL_2681217 | Laboratorio Central de Epidemiología (LCE) | Instituto de Biotecnología de la UNAM | Consorcio Mexicano de Vigilancia Genómica (CoViGen-Mex). Authors (in alphabetical order): Julio Elias Alvarado-Yaah, Carlos F. Arias, Santiago Ávila-Ríos, Eduardo Becerril-Vargas, Víctor Hugo Borja-Aburto, Celia Boukadida, Cristóbal Cháidez-Quiróz, Juan Bautista Chale-Dzul, Ricardo Ciria Merce, Andreu Comas-García, Célida Duque Molina, Julissa Enciso-Ibarra, José Antonio Enciso-Moreno, Gloria Elena Espinosa-Ayala, Fernando Fontove-Herrera, Daniel Fregoso-Rueda, Victor Eduardo García-Arias, Alejandra García-Gasca, Bruno Gómez-Gil, Jean Pierre González, Irvin González-López, Concepción Grajales-Muñiz, Ricardo Grande, Rosa María Gutiérrez Rios, Jesús Hernández, Alejandra Hernández-Terán, Alfredo Herrera-Estrella, Carla Ivón Herrera-Najera, Pavel Isa, Verónica Mata-Haro, Daniel Lira Morales, Susana Lopez, Antonio Loza Román, Brenda Irasema Maldonado-Meza, Bernardo Martínez-Miguel, José Arturo Martínez-Orozco, Célida Martínez- Rodríguez, Margarita Matías-Florentino, Fidencio Mejía-Nepomuceno, María Guadalupe de Jesús Mireles-Rivera, Gloria María Molina-Salinas, Hector Montoya-Fuentes, , Mario Mújica-Sánchez, José Esteban Muñoz-Medina, José de Jesús Nuñez-Contreras, Alicia Ocaña-Mondragón, Luis Alberto Ochoa-Carrera, Hector Esteban Paz-Juárez, Marissa Perez-Garcia, Francisco Pulido, Helen Haydee Fernanda Ramírez-Plascencia, Jorge Salas-Hernández, Angel Gustavo Salas-Lais, Alejandro Sánchez-Flores, Clara Esperanza Santacruz-Tinoco, María Guadalupe Santiago-Mauricio, Selene Zárate, Nelly Sélem-Mojica, Blanca Taboada, Gloria Vazquez, Joel Armando Vázquez-Pérez. |
| EPI_ISL_2681218 | Unidad de Investigación Médica de Yucatán (UIMY) | Instituto de Biotecnología de la UNAM | Consorcio Mexicano de Vigilancia Genómica (CoViGen-Mex). Authors (in alphabetical order): Julio Elias Alvarado-Yaah, Carlos F. Arias, Santiago Ávila-Ríos, Eduardo Becerril-Vargas, Víctor Hugo Borja-Aburto, Celia Boukadida, Cristóbal Cháidez-Quiróz, Juan Bautista Chale-Dzul, Ricardo Ciria Merce, Andreu Comas-García, Célida Duque Molina, Julissa Enciso-Ibarra, José Antonio Enciso-Moreno, Gloria Elena Espinosa-Ayala, Fernando Fontove-Herrera, Daniel Fregoso-Rueda, Victor Eduardo García-Arias, Alejandra García-Gasca, Bruno Gómez-Gil, Jean Pierre González, Irvin González-López, Concepción Grajales-Muñiz, Ricardo Grande, Rosa María Gutiérrez Rios, Jesús Hernández, Alejandra Hernández-Terán, Alfredo Herrera-Estrella, Carla Ivón Herrera-Najera, Pavel Isa, Verónica Mata-Haro, Daniel Lira Morales, Susana Lopez, Antonio Loza Román, Brenda Irasema Maldonado-Meza, Bernardo Martínez-Miguel, José Arturo Martínez-Orozco, Célida Martínez- Rodríguez, Margarita Matías-Florentino, Fidencio Mejía-Nepomuceno, María Guadalupe de Jesús Mireles-Rivera, Gloria María Molina-Salinas, Hector Montoya-Fuentes, , Mario Mújica-Sánchez, José Esteban Muñoz-Medina, José de Jesús Nuñez-Contreras, Alicia Ocaña-Mondragón, Luis Alberto Ochoa-Carrera, Hector Esteban Paz-Juárez, Marissa Perez-Garcia, Francisco Pulido, Helen Haydee Fernanda Ramírez-Plascencia, Jorge Salas-Hernández, Angel Gustavo Salas-Lais, Alejandro Sánchez-Flores, Clara Esperanza Santacruz-Tinoco, María Guadalupe Santiago-Mauricio, Selene Zárate, Nelly Sélem-Mojica, Blanca Taboada, Gloria Vazquez, Joel Armando Vázquez-Pérez. |
| EPI_ISL_2681219 | Centro de Investigación Biomédica del Noreste (CIBIN) | Instituto de Biotecnología de la UNAM | Consorcio Mexicano de Vigilancia Genómica (CoViGen-Mex). Authors (in alphabetical order): Julio Elias Alvarado-Yaah, Carlos F. Arias, Santiago Ávila-Ríos, Eduardo Becerril-Vargas, Víctor Hugo Borja-Aburto, Celia Boukadida, Cristóbal Cháidez-Quiróz, Juan Bautista Chale-Dzul, Ricardo Ciria Merce, Andreu Comas-García, Célida Duque Molina, Julissa Enciso-Ibarra, José Antonio Enciso-Moreno, Gloria Elena Espinosa-Ayala, Fernando Fontove-Herrera, Daniel Fregoso-Rueda, Victor Eduardo García-Arias, Alejandra García-Gasca, Bruno Gómez-Gil, Jean Pierre González, Irvin González-López, Concepción Grajales-Muñiz, Ricardo Grande, Rosa María Gutiérrez Rios, Jesús Hernández, Alejandra Hernández-Terán, Alfredo Herrera-Estrella, Carla Ivón Herrera-Najera, Pavel Isa, Verónica Mata-Haro, Daniel Lira Morales, Susana Lopez, Antonio Loza Román, Brenda Irasema Maldonado-Meza, Bernardo Martínez-Miguel, José Arturo Martínez-Orozco, Célida Martínez- Rodríguez, Margarita Matías-Florentino, Fidencio Mejía-Nepomuceno, María Guadalupe de Jesús Mireles-Rivera, Gloria María Molina-Salinas, Hector Montoya-Fuentes, , Mario Mújica-Sánchez, José Esteban Muñoz-Medina, José de Jesús Nuñez-Contreras, Alicia Ocaña-Mondragón, Luis Alberto Ochoa-Carrera, Hector Esteban Paz-Juárez, Marissa Perez-Garcia, Francisco Pulido, Helen Haydee Fernanda Ramírez-Plascencia, Jorge Salas-Hernández, Angel Gustavo Salas-Lais, Alejandro Sánchez-Flores, Clara Esperanza Santacruz-Tinoco, María Guadalupe Santiago-Mauricio, Selene Zárate, Nelly Sélem-Mojica, Blanca Taboada, Gloria Vazquez, Joel Armando Vázquez-Pérez. |
| EPI_ISL_2681220 | Laboratorio Central de Epidemiología (LCE) | Instituto de Biotecnología de la UNAM | Consorcio Mexicano de Vigilancia Genómica (CoViGen-Mex). Authors (in alphabetical order): Julio Elias Alvarado-Yaah, Carlos F. Arias, Santiago Ávila-Ríos, Eduardo Becerril-Vargas, Víctor Hugo Borja-Aburto, Celia Boukadida, Cristóbal Cháidez-Quiróz, Juan Bautista Chale-Dzul, Ricardo Ciria Merce, Andreu Comas-García, Célida Duque Molina, Julissa Enciso-Ibarra, José Antonio Enciso-Moreno, Gloria Elena Espinosa-Ayala, Fernando Fontove-Herrera, Daniel Fregoso-Rueda, Victor Eduardo García-Arias, Alejandra García-Gasca, Bruno Gómez-Gil, Jean Pierre González, Irvin González-López, Concepción Grajales-Muñiz, Ricardo Grande, Rosa María Gutiérrez Rios, Jesús Hernández, Alejandra Hernández-Terán, Alfredo Herrera-Estrella, Carla Ivón Herrera-Najera, Pavel Isa, Verónica Mata-Haro, Daniel Lira Morales, Susana Lopez, Antonio Loza Román, Brenda Irasema Maldonado-Meza, Bernardo Martínez-Miguel, José Arturo Martínez-Orozco, Célida Martínez- Rodríguez, Margarita Matías-Florentino, Fidencio Mejía-Nepomuceno, María Guadalupe de Jesús Mireles-Rivera, Gloria María Molina-Salinas, Hector Montoya-Fuentes, , Mario Mújica-Sánchez, José Esteban Muñoz-Medina, José de Jesús Nuñez-Contreras, Alicia Ocaña-Mondragón, Luis Alberto Ochoa-Carrera, Hector Esteban Paz-Juárez, Marissa Perez-Garcia, Francisco Pulido, Helen Haydee Fernanda Ramírez-Plascencia, Jorge Salas-Hernández, Angel Gustavo Salas-Lais, Alejandro Sánchez-Flores, Clara Esperanza Santacruz-Tinoco, María Guadalupe Santiago-Mauricio, Selene Zárate, Nelly Sélem-Mojica, Blanca Taboada, Gloria Vazquez, Joel Armando Vázquez-Pérez. |
| EPI_ISL_2681221 | Unidad de Investigación Biomédica de Zacatecas (UIBZ) | Instituto de Biotecnología de la UNAM | Consorcio Mexicano de Vigilancia Genómica (CoViGen-Mex). Authors (in alphabetical order): Julio Elias Alvarado-Yaah, Carlos F. Arias, Santiago Ávila-Ríos, Eduardo Becerril-Vargas, Víctor Hugo Borja-Aburto, Celia Boukadida, Cristóbal Cháidez-Quiróz, Juan Bautista Chale-Dzul, Ricardo Ciria Merce, Andreu Comas-García, Célida Duque Molina, Julissa Enciso-Ibarra, José Antonio Enciso-Moreno, Gloria Elena Espinosa-Ayala, Fernando Fontove-Herrera, Daniel Fregoso-Rueda, Victor Eduardo García-Arias, Alejandra García-Gasca, Bruno Gómez-Gil, Jean Pierre González, Irvin González-López, Concepción Grajales-Muñiz, Ricardo Grande, Rosa María Gutiérrez Rios, Jesús Hernández, Alejandra Hernández-Terán, Alfredo Herrera-Estrella, Carla Ivón Herrera-Najera, Pavel Isa, Verónica Mata-Haro, Daniel Lira Morales, Susana Lopez, Antonio Loza Román, Brenda Irasema Maldonado-Meza, Bernardo Martínez-Miguel, José Arturo Martínez-Orozco, Célida Martínez- Rodríguez, Margarita Matías-Florentino, Fidencio Mejía-Nepomuceno, María Guadalupe de Jesús Mireles-Rivera, Gloria María Molina-Salinas, Hector Montoya-Fuentes, , Mario Mújica-Sánchez, José Esteban Muñoz-Medina, José de Jesús Nuñez-Contreras, Alicia Ocaña-Mondragón, Luis Alberto Ochoa-Carrera, Hector Esteban Paz-Juárez, Marissa Perez-Garcia, Francisco Pulido, Helen Haydee Fernanda Ramírez-Plascencia, Jorge Salas-Hernández, Angel Gustavo Salas-Lais, Alejandro Sánchez-Flores, Clara Esperanza Santacruz-Tinoco, María Guadalupe Santiago-Mauricio, Selene Zárate, Nelly Sélem-Mojica, Blanca Taboada, Gloria Vazquez, Joel Armando Vázquez-Pérez. |
| EPI_ISL_2681222 | Laboratorio Central de Epidemiología (LCE) | Instituto de Biotecnología de la UNAM | Consorcio Mexicano de Vigilancia Genómica (CoViGen-Mex). Authors (in alphabetical order): Julio Elias Alvarado-Yaah, Carlos F. Arias, Santiago Ávila-Ríos, Eduardo Becerril-Vargas, Víctor Hugo Borja-Aburto, Celia Boukadida, Cristóbal Cháidez-Quiróz, Juan Bautista Chale-Dzul, Ricardo Ciria Merce, Andreu Comas-García, Célida Duque Molina, Julissa Enciso-Ibarra, José Antonio Enciso-Moreno, Gloria Elena Espinosa-Ayala, Fernando Fontove-Herrera, Daniel Fregoso-Rueda, Victor Eduardo García-Arias, Alejandra García-Gasca, Bruno Gómez-Gil, Jean Pierre González, Irvin González-López, Concepción Grajales-Muñiz, Ricardo Grande, Rosa María Gutiérrez Rios, Jesús Hernández, Alejandra Hernández-Terán, Alfredo Herrera-Estrella, Carla Ivón Herrera-Najera, Pavel Isa, Verónica Mata-Haro, Daniel Lira Morales, Susana Lopez, Antonio Loza Román, Brenda Irasema Maldonado-Meza, Bernardo Martínez-Miguel, José Arturo Martínez-Orozco, Célida Martínez- Rodríguez, Margarita Matías-Florentino, Fidencio Mejía-Nepomuceno, María Guadalupe de Jesús Mireles-Rivera, Gloria María Molina-Salinas, Hector Montoya-Fuentes, , Mario Mújica-Sánchez, José Esteban Muñoz-Medina, José de Jesús Nuñez-Contreras, Alicia Ocaña-Mondragón, Luis Alberto Ochoa-Carrera, Hector Esteban Paz-Juárez, Marissa Perez-Garcia, Francisco Pulido, Helen Haydee Fernanda Ramírez-Plascencia, Jorge Salas-Hernández, Angel Gustavo Salas-Lais, Alejandro Sánchez-Flores, Clara Esperanza Santacruz-Tinoco, María Guadalupe Santiago-Mauricio, Selene Zárate, Nelly Sélem-Mojica, Blanca Taboada, Gloria Vazquez, Joel Armando Vázquez-Pérez. |
| EPI_ISL_2681223 | Unidad de Investigación Biomédica de | Instituto de Biotecnología de la UNAM | Consorcio Mexicano de Vigilancia Genómica (CoViGen-Mex). Authors (in alphabetical order): Julio Elias Alvarado-Yaah, Carlos F. Arias, Santiago Ávila-Ríos, Eduardo Becerril-Vargas, Víctor Hugo Borja-Aburto, Celia Boukadida, Cristóbal Cháidez-Quiróz, Juan Bautista Chale-Dzul, Ricardo |

| | | | |
|---|---|---|---|
| | Zacatecas (UIBZ) | | Ciria Merce, Andreu Comas-García, Célida Duque Molina, Julissa Enciso-Ibarra, José Antonio Enciso-Moreno, Gloria Elena Espinosa-Ayala, Fernando Fontove-Herrera, Daniel Fregoso-Rueda, Victor Eduardo García-Arias, Alejandra García-Gasca, Bruno Gómez-Gil, Jean Pierre González, Irvin González-López, Concepción Grajales-Muñiz, Ricardo Grande, Rosa María Gutiérrez Rios, Jesús Hernández, Alejandra Hernández-Terán, Alfredo Herrera-Estrella, Carla Ivón Herrera-Najera, Pavel Isa, Verónica Mata-Haro, Daniel Lira Morales, Susana Lopez, Antonio Loza Román, Brenda Irasema Maldonado-Meza, Bernardo Martínez-Miguel, José Arturo Martínez-Orozco, Célida Martínez- Rodríguez, Margarita Matías-Florentino, Fidencio Mejía-Nepomuceno, María Guadalupe de Jesús Mireles-Rivera, Gloria María Molina-Salinas, Hector Montoya-Fuentes, , Mario Mújica-Sánchez, José Esteban Muñoz-Medina, José de Jesús Nuñez-Contreras, Alicia Ocaña-Mondragón, Luis Alberto Ochoa-Carrera, Hector Esteban Paz-Juárez, Marissa Perez-Garcia, Francisco Pulido, Helen Haydee Fernanda Ramírez-Plascencia, Jorge Salas-Hernández, Angel Gustavo Salas-Lais, Alejandro Sánchez-Flores, Clara Esperanza Santacruz-Tinoco, María Guadalupe Santiago-Mauricio, Selene Zárate, Nelly Sélem-Mojica, Blanca Taboada, Gloria Vazquez, Joel Armando Vázquez-Pérez. |
| EPI_ISL_2681224, EPI_ISL_2681225, EPI_ISL_2681226 | Laboratorio Central de Epidemiología (LCE) | Instituto de Biotecnología de la UNAM | Consorcio Mexicano de Vigilancia Genómica (CoViGen-Mex). Authors (in alphabetical order): Julio Elias Alvarado-Yaah, Carlos F. Arias, Santiago Ávila-Ríos, Eduardo Becerril-Vargas, Víctor Hugo Borja-Aburto, Celia Boukadida, Cristóbal Cháidez-Quiróz, Juan Bautista Chale-Dzul, Ricardo Ciria Merce, Andreu Comas-García, Célida Duque Molina, Julissa Enciso-Ibarra, José Antonio Enciso-Moreno, Gloria Elena Espinosa-Ayala, Fernando Fontove-Herrera, Daniel Fregoso-Rueda, Victor Eduardo García-Arias, Alejandra García-Gasca, Bruno Gómez-Gil, Jean Pierre González, Irvin González-López, Concepción Grajales-Muñiz, Ricardo Grande, Rosa María Gutiérrez Rios, Jesús Hernández, Alejandra Hernández-Terán, Alfredo Herrera-Estrella, Carla Ivón Herrera-Najera, Pavel Isa, Verónica Mata-Haro, Daniel Lira Morales, Susana Lopez, Antonio Loza Román, Brenda Irasema Maldonado-Meza, Bernardo Martínez-Miguel, José Arturo Martínez-Orozco, Célida Martínez- Rodríguez, Margarita Matías-Florentino, Fidencio Mejía-Nepomuceno, María Guadalupe de Jesús Mireles-Rivera, Gloria María Molina-Salinas, Hector Montoya-Fuentes, , Mario Mújica-Sánchez, José Esteban Muñoz-Medina, José de Jesús Nuñez-Contreras, Alicia Ocaña-Mondragón, Luis Alberto Ochoa-Carrera, Hector Esteban Paz-Juárez, Marissa Perez-Garcia, Francisco Pulido, Helen Haydee Fernanda Ramírez-Plascencia, Jorge Salas-Hernández, Angel Gustavo Salas-Lais, Alejandro Sánchez-Flores, Clara Esperanza Santacruz-Tinoco, María Guadalupe Santiago-Mauricio, Selene Zárate, Nelly Sélem-Mojica, Blanca Taboada, Gloria Vazquez, Joel Armando Vázquez-Pérez. |
| EPI_ISL_2681227, EPI_ISL_2681228 | Centro de Investigación Biomédica del Noreste (CIBIN) | Instituto de Biotecnología de la UNAM | Consorcio Mexicano de Vigilancia Genómica (CoViGen-Mex). Authors (in alphabetical order): Julio Elias Alvarado-Yaah, Carlos F. Arias, Santiago Ávila-Ríos, Eduardo Becerril-Vargas, Víctor Hugo Borja-Aburto, Celia Boukadida, Cristóbal Cháidez-Quiróz, Juan Bautista Chale-Dzul, Ricardo Ciria Merce, Andreu Comas-García, Célida Duque Molina, Julissa Enciso-Ibarra, José Antonio Enciso-Moreno, Gloria Elena Espinosa-Ayala, Fernando Fontove-Herrera, Daniel Fregoso-Rueda, Victor Eduardo García-Arias, Alejandra García-Gasca, Bruno Gómez-Gil, Jean Pierre González, Irvin González-López, Concepción Grajales-Muñiz, Ricardo Grande, Rosa María Gutiérrez Rios, Jesús Hernández, Alejandra Hernández-Terán, Alfredo Herrera-Estrella, Carla Ivón Herrera-Najera, Pavel Isa, Verónica Mata-Haro, Daniel Lira Morales, Susana Lopez, Antonio Loza Román, Brenda Irasema Maldonado-Meza, Bernardo Martínez-Miguel, José Arturo Martínez-Orozco, Célida Martínez- Rodríguez, Margarita Matías-Florentino, Fidencio Mejía-Nepomuceno, María Guadalupe de Jesús Mireles-Rivera, Gloria María Molina-Salinas, Hector Montoya-Fuentes, , Mario Mújica-Sánchez, José Esteban Muñoz-Medina, José de Jesús Nuñez-Contreras, Alicia Ocaña-Mondragón, Luis Alberto Ochoa-Carrera, Hector Esteban Paz-Juárez, Marissa Perez-Garcia, Francisco Pulido, Helen Haydee Fernanda Ramírez-Plascencia, Jorge Salas-Hernández, Angel Gustavo Salas-Lais, Alejandro Sánchez-Flores, Clara Esperanza Santacruz-Tinoco, María Guadalupe Santiago-Mauricio, Selene Zárate, Nelly Sélem-Mojica, Blanca Taboada, Gloria Vazquez, Joel Armando Vázquez-Pérez. |
| EPI_ISL_2681229, EPI_ISL_2681230, EPI_ISL_2681231 | Laboratorio Central de Epidemiología (LCE) | Instituto de Biotecnología de la UNAM | Consorcio Mexicano de Vigilancia Genómica (CoViGen-Mex). Authors (in alphabetical order): Julio Elias Alvarado-Yaah, Carlos F. Arias, Santiago Ávila-Ríos, Eduardo Becerril-Vargas, Víctor Hugo Borja-Aburto, Celia Boukadida, Cristóbal Cháidez-Quiróz, Juan Bautista Chale-Dzul, Ricardo Ciria Merce, Andreu Comas-García, Célida Duque Molina, Julissa Enciso-Ibarra, José Antonio Enciso-Moreno, Gloria Elena Espinosa-Ayala, Fernando Fontove-Herrera, Daniel Fregoso-Rueda, Victor Eduardo García-Arias, Alejandra García-Gasca, Bruno Gómez-Gil, Jean Pierre González, Irvin González-López, Concepción Grajales-Muñiz, Ricardo Grande, Rosa María Gutiérrez Rios, Jesús Hernández, Alejandra Hernández-Terán, Alfredo Herrera-Estrella, Carla Ivón Herrera-Najera, Pavel Isa, Verónica Mata-Haro, Daniel Lira Morales, Susana Lopez, Antonio Loza Román, Brenda Irasema Maldonado-Meza, Bernardo Martínez-Miguel, José Arturo Martínez-Orozco, Célida Martínez- Rodríguez, Margarita Matías-Florentino, Fidencio Mejía-Nepomuceno, María Guadalupe de Jesús Mireles-Rivera, Gloria María Molina-Salinas, Hector Montoya-Fuentes, , Mario Mújica-Sánchez, José Esteban Muñoz-Medina, José de Jesús Nuñez-Contreras, Alicia Ocaña-Mondragón, Luis Alberto Ochoa-Carrera, Hector Esteban Paz-Juárez, Marissa Perez-Garcia, Francisco Pulido, Helen Haydee Fernanda Ramírez-Plascencia, Jorge Salas-Hernández, Angel Gustavo Salas-Lais, Alejandro Sánchez-Flores, Clara Esperanza Santacruz-Tinoco, María Guadalupe Santiago-Mauricio, Selene Zárate, Nelly Sélem-Mojica, Blanca Taboada, Gloria Vazquez, Joel Armando Vázquez-Pérez. |
| EPI_ISL_2681232, EPI_ISL_2681233 | Unidad de Investigación Médica de Yucatán (UIMY) | Instituto de Biotecnología de la UNAM | Consorcio Mexicano de Vigilancia Genómica (CoViGen-Mex). Authors (in alphabetical order): Julio Elias Alvarado-Yaah, Carlos F. Arias, Santiago Ávila-Ríos, Eduardo Becerril-Vargas, Víctor Hugo Borja-Aburto, Celia Boukadida, Cristóbal Cháidez-Quiróz, Juan Bautista Chale-Dzul, Ricardo Ciria Merce, Andreu Comas-García, Célida Duque Molina, Julissa Enciso-Ibarra, José Antonio Enciso-Moreno, Gloria Elena Espinosa-Ayala, Fernando Fontove-Herrera, Daniel Fregoso-Rueda, Victor Eduardo García-Arias, Alejandra García-Gasca, Bruno Gómez-Gil, Jean Pierre González, Irvin González-López, Concepción Grajales-Muñiz, Ricardo Grande, Rosa María Gutiérrez Rios, Jesús Hernández, Alejandra Hernández-Terán, Alfredo Herrera-Estrella, Carla Ivón Herrera-Najera, Pavel Isa, Verónica Mata-Haro, Daniel Lira Morales, Susana Lopez, Antonio Loza Román, Brenda Irasema Maldonado-Meza, Bernardo Martínez-Miguel, José Arturo Martínez-Orozco, Célida Martínez- Rodríguez, Margarita Matías-Florentino, Fidencio Mejía-Nepomuceno, María Guadalupe de Jesús Mireles-Rivera, Gloria María Molina-Salinas, Hector Montoya-Fuentes, , Mario Mújica-Sánchez, José Esteban Muñoz-Medina, José de Jesús Nuñez-Contreras, Alicia Ocaña-Mondragón, Luis Alberto Ochoa-Carrera, Hector Esteban Paz-Juárez, Marissa Perez-Garcia, Francisco Pulido, Helen Haydee Fernanda Ramírez-Plascencia, Jorge Salas-Hernández, Angel Gustavo Salas-Lais, Alejandro Sánchez-Flores, Clara Esperanza Santacruz-Tinoco, María Guadalupe Santiago-Mauricio, Selene Zárate, Nelly Sélem-Mojica, Blanca Taboada, Gloria Vazquez, Joel Armando Vázquez-Pérez. |
| EPI_ISL_2681234, EPI_ISL_2681235 | Unidad de Investigación Biomédica de Zacatecas (UIBZ) | Instituto de Biotecnología de la UNAM | Consorcio Mexicano de Vigilancia Genómica (CoViGen-Mex). Authors (in alphabetical order): Julio Elias Alvarado-Yaah, Carlos F. Arias, Santiago Ávila-Ríos, Eduardo Becerril-Vargas, Víctor Hugo Borja-Aburto, Celia Boukadida, Cristóbal Cháidez-Quiróz, Juan Bautista Chale-Dzul, Ricardo Ciria Merce, Andreu Comas-García, Célida Duque Molina, Julissa Enciso-Ibarra, José Antonio Enciso-Moreno, Gloria Elena Espinosa-Ayala, Fernando Fontove-Herrera, Daniel Fregoso-Rueda, Victor Eduardo García-Arias, Alejandra García-Gasca, Bruno Gómez-Gil, Jean Pierre González, Irvin González-López, Concepción Grajales-Muñiz, Ricardo Grande, Rosa María Gutiérrez Rios, Jesús Hernández, Alejandra Hernández-Terán, Alfredo Herrera-Estrella, Carla Ivón Herrera-Najera, Pavel Isa, Verónica Mata-Haro, Daniel Lira Morales, Susana Lopez, Antonio Loza Román, Brenda Irasema Maldonado-Meza, Bernardo Martínez-Miguel, José Arturo Martínez-Orozco, Célida Martínez- Rodríguez, Margarita Matías-Florentino, Fidencio Mejía-Nepomuceno, María Guadalupe de Jesús Mireles-Rivera, Gloria María Molina-Salinas, Hector Montoya-Fuentes, , Mario Mújica-Sánchez, José Esteban Muñoz-Medina, José de Jesús Nuñez-Contreras, Alicia Ocaña-Mondragón, Luis Alberto Ochoa-Carrera, Hector Esteban Paz-Juárez, Marissa Perez-Garcia, Francisco Pulido, Helen Haydee Fernanda Ramírez-Plascencia, Jorge Salas-Hernández, Angel Gustavo Salas-Lais, Alejandro Sánchez-Flores, Clara Esperanza Santacruz-Tinoco, María Guadalupe Santiago-Mauricio, Selene Zárate, Nelly Sélem-Mojica, Blanca Taboada, Gloria Vazquez, Joel Armando Vázquez-Pérez. |
| EPI_ISL_2681236 | Unidad de Investigación Médica de Yucatán (UIMY) | Instituto de Biotecnología de la UNAM | Consorcio Mexicano de Vigilancia Genómica (CoViGen-Mex). Authors (in alphabetical order): Julio Elias Alvarado-Yaah, Carlos F. Arias, Santiago Ávila-Ríos, Eduardo Becerril-Vargas, Víctor Hugo Borja-Aburto, Celia Boukadida, Cristóbal Cháidez-Quiróz, Juan Bautista Chale-Dzul, Ricardo Ciria Merce, Andreu Comas-García, Célida Duque Molina, Julissa Enciso-Ibarra, José Antonio Enciso-Moreno, Gloria Elena Espinosa-Ayala, Fernando Fontove-Herrera, Daniel Fregoso-Rueda, Victor Eduardo García-Arias, Alejandra García-Gasca, Bruno Gómez-Gil, Jean Pierre González, Irvin González-López, Concepción Grajales-Muñiz, Ricardo Grande, Rosa María Gutiérrez Rios, Jesús Hernández, Alejandra Hernández-Terán, Alfredo Herrera-Estrella, Carla Ivón Herrera-Najera, Pavel Isa, Verónica Mata-Haro, Daniel Lira Morales, Susana Lopez, Antonio Loza Román, Brenda Irasema Maldonado-Meza, Bernardo Martínez-Miguel, José Arturo Martínez-Orozco, Célida Martínez- Rodríguez, Margarita Matías-Florentino, Fidencio Mejía-Nepomuceno, María Guadalupe de Jesús Mireles-Rivera, Gloria María Molina-Salinas, Hector Montoya-Fuentes, , Mario Mújica-Sánchez, José Esteban Muñoz-Medina, José de Jesús Nuñez-Contreras, Alicia Ocaña-Mondragón, Luis Alberto Ochoa-Carrera, Hector Esteban Paz-Juárez, Marissa Perez-Garcia, Francisco Pulido, Helen Haydee Fernanda Ramírez-Plascencia, Jorge Salas-Hernández, Angel Gustavo Salas-Lais, Alejandro Sánchez-Flores, Clara Esperanza Santacruz-Tinoco, María Guadalupe Santiago-Mauricio, Selene Zárate, Nelly Sélem-Mojica, Blanca Taboada, Gloria Vazquez, Joel Armando Vázquez-Pérez. |
| EPI_ISL_2681237, EPI_ISL_2681238 | Laboratorio Central de Epidemiología (LCE) | Instituto de Biotecnología de la UNAM | Consorcio Mexicano de Vigilancia Genómica (CoViGen-Mex). Authors (in alphabetical order): Julio Elias Alvarado-Yaah, Carlos F. Arias, Santiago Ávila-Ríos, Eduardo Becerril-Vargas, Víctor Hugo Borja-Aburto, Celia Boukadida, Cristóbal Cháidez-Quiróz, Juan Bautista Chale-Dzul, Ricardo Ciria Merce, Andreu Comas-García, Célida Duque Molina, Julissa Enciso-Ibarra, José Antonio Enciso-Moreno, Gloria Elena Espinosa-Ayala, Fernando Fontove-Herrera, Daniel Fregoso-Rueda, Victor Eduardo García-Arias, Alejandra García-Gasca, Bruno Gómez-Gil, Jean Pierre González, Irvin González-López, Concepción Grajales-Muñiz, Ricardo Grande, Rosa María Gutiérrez Rios, Jesús Hernández, Alejandra Hernández-Terán, Alfredo Herrera-Estrella, Carla Ivón Herrera-Najera, Pavel Isa, Verónica Mata-Haro, Daniel Lira Morales, Susana Lopez, Antonio Loza Román, Brenda Irasema Maldonado-Meza, Bernardo Martínez-Miguel, José Arturo Martínez-Orozco, Célida Martínez- Rodríguez, Margarita Matías-Florentino, Fidencio Mejía-Nepomuceno, María Guadalupe de Jesús Mireles-Rivera, Gloria María Molina-Salinas, Hector Montoya-Fuentes, , Mario Mújica-Sánchez, José Esteban Muñoz-Medina, José de Jesús Nuñez-Contreras, Alicia Ocaña-Mondragón, Luis Alberto Ochoa-Carrera, Hector Esteban Paz-Juárez, Marissa Perez-Garcia, Francisco Pulido, Helen Haydee Fernanda Ramírez-Plascencia, Jorge Salas-Hernández, Angel Gustavo Salas-Lais, Alejandro Sánchez-Flores, Clara Esperanza Santacruz-Tinoco, María Guadalupe Santiago-Mauricio, Selene Zárate, Nelly Sélem-Mojica, Blanca Taboada, Gloria Vazquez, Joel Armando Vázquez-Pérez. |
| EPI_ISL_2681239 | Unidad de Investigación Biomédica de Zacatecas (UIBZ) | Instituto de Biotecnología de la UNAM | Consorcio Mexicano de Vigilancia Genómica (CoViGen-Mex). Authors (in alphabetical order): Julio Elias Alvarado-Yaah, Carlos F. Arias, Santiago Ávila-Ríos, Eduardo Becerril-Vargas, Víctor Hugo Borja-Aburto, Celia Boukadida, Cristóbal Cháidez-Quiróz, Juan Bautista Chale-Dzul, Ricardo Ciria Merce, Andreu Comas-García, Célida Duque Molina, Julissa Enciso-Ibarra, José Antonio Enciso-Moreno, Gloria Elena Espinosa-Ayala, Fernando Fontove-Herrera, Daniel Fregoso-Rueda, Victor Eduardo García-Arias, Alejandra García-Gasca, Bruno Gómez-Gil, Jean Pierre González, Irvin González-López, Concepción Grajales-Muñiz, Ricardo Grande, Rosa María Gutiérrez Rios, Jesús Hernández, Alejandra Hernández-Terán, Alfredo Herrera-Estrella, Carla Ivón Herrera-Najera, Pavel Isa, Verónica Mata-Haro, Daniel Lira Morales, Susana Lopez, Antonio Loza Román, Brenda Irasema Maldonado-Meza, Bernardo Martínez-Miguel, José Arturo Martínez-Orozco, Célida Martínez- Rodríguez, Margarita Matías-Florentino, Fidencio Mejía-Nepomuceno, María Guadalupe de Jesús Mireles-Rivera, Gloria María Molina-Salinas, Hector Montoya-Fuentes, , Mario Mújica-Sánchez, José Esteban Muñoz-Medina, José de Jesús Nuñez-Contreras, Alicia Ocaña-Mondragón, Luis Alberto Ochoa-Carrera, Hector Esteban Paz-Juárez, Marissa Perez-Garcia, Francisco Pulido, Helen Haydee Fernanda Ramírez-Plascencia, Jorge Salas-Hernández, Angel Gustavo Salas-Lais, Alejandro Sánchez-Flores, Clara Esperanza Santacruz-Tinoco, María Guadalupe Santiago-Mauricio, Selene Zárate, Nelly Sélem-Mojica, Blanca Taboada, Gloria Vazquez, Joel Armando Vázquez-Pérez. |
| EPI_ISL_2681240 | Laboratorio Central de Epidemiología (LCE) | Instituto de Biotecnología de la UNAM | Consorcio Mexicano de Vigilancia Genómica (CoViGen-Mex). Authors (in alphabetical order): Julio Elias Alvarado-Yaah, Carlos F. Arias, Santiago Ávila-Ríos, Eduardo Becerril-Vargas, Víctor Hugo Borja-Aburto, Celia Boukadida, Cristóbal Cháidez-Quiróz, Juan Bautista Chale-Dzul, Ricardo Ciria Merce, Andreu Comas-García, Célida Duque Molina, Julissa Enciso-Ibarra, José Antonio Enciso-Moreno, Gloria Elena Espinosa-Ayala, Fernando Fontove-Herrera, Daniel Fregoso-Rueda, Victor Eduardo García-Arias, Alejandra García-Gasca, Bruno Gómez-Gil, Jean Pierre González, Irvin González-López, Concepción Grajales-Muñiz, Ricardo Grande, Rosa María Gutiérrez Rios, Jesús Hernández, Alejandra Hernández-Terán, Alfredo Herrera-Estrella, Carla Ivón Herrera-Najera, Pavel Isa, Verónica Mata-Haro, Daniel Lira Morales, Susana Lopez, Antonio Loza Román, Brenda Irasema Maldonado-Meza, Bernardo Martínez-Miguel, José Arturo Martínez-Orozco, Célida Martínez- Rodríguez, Margarita Matías-Florentino, Fidencio Mejía-Nepomuceno, María Guadalupe de Jesús Mireles-Rivera, Gloria María Molina-Salinas, Hector Montoya-Fuentes, , Mario Mújica-Sánchez, José Esteban Muñoz-Medina, José de Jesús Nuñez-Contreras, Alicia Ocaña-Mondragón, Luis Alberto Ochoa-Carrera, Hector Esteban Paz-Juárez, Marissa Perez-Garcia, Francisco Pulido, Helen Haydee Fernanda Ramírez-Plascencia, Jorge Salas-Hernández, Angel Gustavo Salas-Lais, Alejandro Sánchez-Flores, Clara Esperanza Santacruz-Tinoco, María Guadalupe Santiago-Mauricio, Selene Zárate, Nelly Sélem-Mojica, Blanca Taboada, Gloria Vazquez, Joel Armando Vázquez-Pérez. |
| EPI_ISL_2681241 | Unidad de Investigación Médica de Yucatán (UIMY) | Instituto de Biotecnología de la UNAM | Consorcio Mexicano de Vigilancia Genómica (CoViGen-Mex). Authors (in alphabetical order): Julio Elias Alvarado-Yaah, Carlos F. Arias, Santiago Ávila-Ríos, Eduardo Becerril-Vargas, Víctor Hugo Borja-Aburto, Celia Boukadida, Cristóbal Cháidez-Quiróz, Juan Bautista Chale-Dzul, Ricardo Ciria Merce, Andreu Comas-García, Célida Duque Molina, Julissa Enciso-Ibarra, José Antonio Enciso-Moreno, Gloria Elena Espinosa-Ayala, Fernando Fontove-Herrera, Daniel Fregoso-Rueda, Victor Eduardo García-Arias, Alejandra García-Gasca, Bruno Gómez-Gil, Jean Pierre González, Irvin González-López, Concepción Grajales-Muñiz, Ricardo Grande, Rosa María Gutiérrez Rios, Jesús Hernández, Alejandra Hernández-Terán, Alfredo Herrera-Estrella, Carla Ivón Herrera-Najera, Pavel Isa, Verónica Mata-Haro, Daniel Lira Morales, Susana Lopez, Antonio Loza Román, Brenda Irasema Maldonado-Meza, Bernardo Martínez-Miguel, José Arturo Martínez-Orozco, Célida Martínez- Rodríguez, Margarita Matías-Florentino, Fidencio Mejía-Nepomuceno, María Guadalupe de Jesús Mireles-Rivera, Gloria María Molina-Salinas, Hector Montoya-Fuentes, , Mario Mújica-Sánchez, José Esteban Muñoz-Medina, José de Jesús Nuñez-Contreras, Alicia Ocaña-Mondragón, Luis Alberto Ochoa-Carrera, Hector Esteban Paz-Juárez, Marissa Perez-Garcia, Francisco Pulido, Helen Haydee Fernanda Ramírez-Plascencia, Jorge Salas-Hernández, Angel Gustavo Salas-Lais, Alejandro Sánchez-Flores, Clara Esperanza Santacruz-Tinoco, María Guadalupe Santiago-Mauricio, Selene Zárate, Nelly Sélem-Mojica, Blanca Taboada, Gloria Vazquez, Joel Armando Vázquez-Pérez. |
| EPI_ISL_2681242 | Unidad de Investigación Biomédica de Zacatecas (UIBZ) | Instituto de Biotecnología de la UNAM | Consorcio Mexicano de Vigilancia Genómica (CoViGen-Mex). Authors (in alphabetical order): Julio Elias Alvarado-Yaah, Carlos F. Arias, Santiago Ávila-Ríos, Eduardo Becerril-Vargas, Víctor Hugo Borja-Aburto, Celia Boukadida, Cristóbal Cháidez-Quiróz, Juan Bautista Chale-Dzul, Ricardo Ciria Merce, Andreu Comas-García, Célida Duque Molina, Julissa Enciso-Ibarra, José Antonio Enciso-Moreno, Gloria Elena Espinosa-Ayala, Fernando Fontove-Herrera, Daniel Fregoso-Rueda, Victor Eduardo García-Arias, Alejandra García-Gasca, Bruno Gómez-Gil, Jean Pierre González, Irvin González-López, Concepción Grajales-Muñiz, Ricardo Grande, Rosa María Gutiérrez Rios, Jesús Hernández, Alejandra Hernández-Terán, Alfredo Herrera-Estrella, Carla Ivón Herrera-Najera, Pavel Isa, Verónica Mata-Haro, Daniel Lira Morales, Susana Lopez, Antonio Loza Román, Brenda Irasema Maldonado-Meza, Bernardo Martínez-Miguel, José Arturo Martínez-Orozco, Célida Martínez- Rodríguez, Margarita Matías-Florentino, Fidencio Mejía-Nepomuceno, María Guadalupe de Jesús Mireles-Rivera, Gloria María Molina-Salinas, Hector Montoya-Fuentes, , Mario Mújica-Sánchez, José Esteban Muñoz-Medina, José de Jesús Nuñez-Contreras, Alicia Ocaña-Mondragón, Luis Alberto Ochoa-Carrera, Hector Esteban Paz-Juárez, Marissa Perez-Garcia, Francisco Pulido, Helen Haydee Fernanda Ramírez-Plascencia, Jorge Salas-Hernández, Angel Gustavo Salas-Lais, Alejandro Sánchez-Flores, Clara Esperanza Santacruz-Tinoco, María Guadalupe Santiago-Mauricio, Selene Zárate, Nelly Sélem-Mojica, Blanca Taboada, Gloria Vazquez, Joel Armando Vázquez-Pérez. |
| EPI_ISL_2681243 | Laboratorio Central de Epidemiología (LCE) | Instituto de Biotecnología de la UNAM | Consorcio Mexicano de Vigilancia Genómica (CoViGen-Mex). Authors (in alphabetical order): Julio Elias Alvarado-Yaah, Carlos F. Arias, Santiago Ávila-Ríos, Eduardo Becerril-Vargas, Víctor Hugo Borja-Aburto, Celia Boukadida, Cristóbal Cháidez-Quiróz, Juan Bautista Chale-Dzul, Ricardo Ciria Merce, Andreu Comas-García, Célida Duque Molina, Julissa Enciso-Ibarra, José Antonio Enciso-Moreno, Gloria Elena Espinosa-Ayala, Fernando Fontove-Herrera, Daniel Fregoso-Rueda, Victor Eduardo García-Arias, Alejandra García-Gasca, Bruno Gómez-Gil, Jean Pierre González, Irvin González-López, Concepción Grajales-Muñiz, Ricardo Grande, Rosa María Gutiérrez Rios, Jesús Hernández, Alejandra Hernández-Terán, Alfredo Herrera-Estrella, Carla Ivón Herrera-Najera, Pavel Isa, Verónica Mata-Haro, Daniel Lira Morales, Susana Lopez, Antonio Loza Román, Brenda Irasema Maldonado-Meza, Bernardo Martínez-Miguel, José Arturo Martínez-Orozco, Célida Martínez- Rodríguez, Margarita Matías-Florentino, Fidencio Mejía-Nepomuceno, María Guadalupe de Jesús Mireles-Rivera, Gloria María Molina-Salinas, Hector Montoya-Fuentes, , Mario Mújica-Sánchez, José Esteban Muñoz-Medina, José de Jesús Nuñez-Contreras, Alicia Ocaña-Mondragón, Luis Alberto Ochoa-Carrera, Hector Esteban Paz-Juárez, Marissa Perez-Garcia, Francisco Pulido, Helen Haydee Fernanda Ramírez-Plascencia, Jorge Salas-Hernández, Angel Gustavo Salas-Lais, Alejandro Sánchez-Flores, Clara Esperanza Santacruz-Tinoco, María Guadalupe Santiago-Mauricio, Selene Zárate, Nelly Sélem-Mojica, Blanca Taboada, Gloria Vazquez, Joel Armando Vázquez-Pérez. |
| EPI_ISL_2681244 | Unidad de Investigación Médica de Yucatán (UIMY) | Instituto de Biotecnología de la UNAM | Consorcio Mexicano de Vigilancia Genómica (CoViGen-Mex). Authors (in alphabetical order): Julio Elias Alvarado-Yaah, Carlos F. Arias, Santiago Ávila-Ríos, Eduardo Becerril-Vargas, Víctor Hugo Borja-Aburto, Celia Boukadida, Cristóbal Cháidez-Quiróz, Juan Bautista Chale-Dzul, Ricardo Ciria Merce, Andreu Comas-García, Célida Duque Molina, Julissa Enciso-Ibarra, José Antonio Enciso-Moreno, Gloria Elena Espinosa-Ayala, Fernando Fontove-Herrera, Daniel Fregoso-Rueda, Victor Eduardo García-Arias, Alejandra García-Gasca, Bruno Gómez-Gil, Jean Pierre González, Irvin González-López, Concepción Grajales-Muñiz, Ricardo Grande, Rosa María Gutiérrez Rios, Jesús Hernández, Alejandra Hernández-Terán, Alfredo Herrera-Estrella, Carla Ivón Herrera-Najera, Pavel Isa, Verónica Mata-Haro, Daniel Lira Morales, Susana Lopez, Antonio Loza Román, Brenda Irasema Maldonado-Meza, Bernardo Martínez-Miguel, José Arturo Martínez-Orozco, Célida Martínez- Rodríguez, Margarita Matías-Florentino, Fidencio Mejía-Nepomuceno, María Guadalupe de Jesús Mireles-Rivera, Gloria María Molina-Salinas, Hector Montoya-Fuentes, , Mario Mújica-Sánchez, José Esteban Muñoz-Medina, José de Jesús Nuñez-Contreras, Alicia Ocaña-Mondragón, Luis Alberto Ochoa-Carrera, Hector Esteban Paz-Juárez, Marissa Perez-Garcia, Francisco Pulido, Helen Haydee Fernanda Ramírez-Plascencia, Jorge Salas-Hernández, Angel Gustavo Salas-Lais, Alejandro Sánchez-Flores, Clara Esperanza Santacruz-Tinoco, María Guadalupe Santiago-Mauricio, Selene Zárate, Nelly Sélem-Mojica, Blanca Taboada, Gloria Vazquez, Joel Armando Vázquez-Pérez. |
| EPI_ISL_2681245 | Laboratorio Central de Epidemiología (LCE) | Instituto de Biotecnología de la UNAM | Consorcio Mexicano de Vigilancia Genómica (CoViGen-Mex). Authors (in alphabetical order): Julio Elias Alvarado-Yaah, Carlos F. Arias, Santiago Ávila-Ríos, Eduardo Becerril-Vargas, Víctor Hugo Borja-Aburto, Celia Boukadida, Cristóbal Cháidez-Quiróz, Juan Bautista Chale-Dzul, Ricardo Ciria Merce, Andreu Comas-García, Célida Duque Molina, Julissa Enciso-Ibarra, José Antonio Enciso-Moreno, Gloria Elena Espinosa-Ayala, Fernando Fontove-Herrera, Daniel Fregoso-Rueda, Victor Eduardo García-Arias, Alejandra García-Gasca, Bruno Gómez-Gil, Jean Pierre González, Irvin González-López, Concepción Grajales-Muñiz, Ricardo Grande, Rosa María Gutiérrez Rios, Jesús Hernández, Alejandra Hernández-Terán, Alfredo Herrera-Estrella, Carla Ivón Herrera-Najera, Pavel Isa, Verónica Mata-Haro, Daniel Lira Morales, Susana Lopez, Antonio Loza Román, Brenda Irasema Maldonado-Meza, Bernardo Martínez-Miguel, José Arturo Martínez-Orozco, Célida Martínez- Rodríguez, Margarita Matías-Florentino, Fidencio Mejía-Nepomuceno, María Guadalupe de Jesús Mireles-Rivera, Gloria María Molina-Salinas, Hector Montoya-Fuentes, , Mario Mújica-Sánchez, José Esteban Muñoz-Medina, José de Jesús Nuñez-Contreras, Alicia Ocaña-Mondragón, Luis Alberto Ochoa-Carrera, Hector Esteban Paz-Juárez, Marissa Perez-Garcia, Francisco Pulido, Helen Haydee Fernanda Ramírez-Plascencia, Jorge Salas-Hernández, Angel Gustavo Salas-Lais, Alejandro Sánchez-Flores, Clara Esperanza Santacruz-Tinoco, María Guadalupe Santiago-Mauricio, Selene Zárate, Nelly Sélem-Mojica, Blanca Taboada, Gloria Vazquez, Joel Armando Vázquez-Pérez. |
| EPI_ISL_2681246 | Unidad de Investigación Biomédica de Zacatecas (UIBZ) | Instituto de Biotecnología de la UNAM | Consorcio Mexicano de Vigilancia Genómica (CoViGen-Mex). Authors (in alphabetical order): Julio Elias Alvarado-Yaah, Carlos F. Arias, Santiago Ávila-Ríos, Eduardo Becerril-Vargas, Víctor Hugo Borja-Aburto, Celia Boukadida, Cristóbal Cháidez-Quiróz, Juan Bautista Chale-Dzul, Ricardo Ciria Merce, Andreu Comas-García, Célida Duque Molina, Julissa Enciso-Ibarra, José Antonio Enciso-Moreno, Gloria Elena Espinosa-Ayala, Fernando Fontove-Herrera, Daniel Fregoso-Rueda, Victor Eduardo García-Arias, Alejandra García-Gasca, Bruno Gómez-Gil, Jean Pierre González, Irvin González-López, Concepción Grajales-Muñiz, Ricardo Grande, Rosa María Gutiérrez Rios, Jesús Hernández, Alejandra Hernández-Terán, Alfredo Herrera-Estrella, Carla Ivón Herrera-Najera, Pavel Isa, Verónica Mata-Haro, Daniel Lira Morales, Susana Lopez, Antonio Loza Román, Brenda Irasema Maldonado-Meza, Bernardo Martínez-Miguel, José Arturo Martínez-Orozco, Célida Martínez- Rodríguez, Margarita Matías-Florentino, Fidencio Mejía-Nepomuceno, María Guadalupe de Jesús Mireles-Rivera, Gloria María Molina-Salinas, Hector Montoya-Fuentes, , Mario Mújica-Sánchez, José Esteban Muñoz-Medina, José de Jesús Nuñez-Contreras, Alicia Ocaña-Mondragón, Luis Alberto Ochoa-Carrera, Hector Esteban Paz-Juárez, Marissa Perez-Garcia, Francisco Pulido, Helen Haydee Fernanda Ramírez-Plascencia, Jorge Salas-Hernández, Angel Gustavo Salas-Lais, Alejandro Sánchez-Flores, Clara Esperanza Santacruz-Tinoco, María Guadalupe Santiago-Mauricio, Selene Zárate, Nelly Sélem-Mojica, Blanca Taboada, Gloria Vazquez, Joel Armando Vázquez-Pérez. |
| EPI_ISL_2681247 | Laboratorio Central de Epidemiología (LCE) | Instituto de Biotecnología de la UNAM | Consorcio Mexicano de Vigilancia Genómica (CoViGen-Mex). Authors (in alphabetical order): Julio Elias Alvarado-Yaah, Carlos F. Arias, Santiago Ávila-Ríos, Eduardo Becerril-Vargas, Víctor Hugo Borja-Aburto, Celia Boukadida, Cristóbal Cháidez-Quiróz, Juan Bautista Chale-Dzul, Ricardo Ciria Merce, Andreu Comas-García, Célida Duque Molina, Julissa Enciso-Ibarra, José Antonio Enciso-Moreno, Gloria Elena Espinosa-Ayala, Fernando Fontove-Herrera, Daniel Fregoso-Rueda, Victor Eduardo García-Arias, Alejandra García-Gasca, Bruno Gómez-Gil, Jean Pierre González, Irvin González-López, Concepción Grajales-Muñiz, Ricardo Grande, Rosa María Gutiérrez Rios, Jesús Hernández, Alejandra Hernández-Terán, Alfredo Herrera-Estrella, Carla Ivón Herrera-Najera, Pavel Isa, Verónica Mata-Haro, Daniel Lira Morales, Susana Lopez, Antonio Loza Román, Brenda Irasema Maldonado-Meza, Bernardo Martínez-Miguel, José Arturo Martínez-Orozco, Célida Martínez- Rodríguez, Margarita Matías-Florentino, Fidencio Mejía-Nepomuceno, María Guadalupe de Jesús Mireles-Rivera, Gloria María Molina-Salinas, Hector Montoya-Fuentes, , Mario Mújica-Sánchez, José Esteban Muñoz-Medina, José de Jesús Nuñez-Contreras, Alicia Ocaña-Mondragón, Luis Alberto Ochoa-Carrera, Hector Esteban Paz-Juárez, Marissa Perez-Garcia, Francisco Pulido, Helen Haydee Fernanda Ramírez-Plascencia, Jorge Salas-Hernández, Angel Gustavo Salas-Lais, Alejandro Sánchez-Flores, Clara Esperanza Santacruz-Tinoco, María Guadalupe Santiago-Mauricio, Selene Zárate, Nelly Sélem-Mojica, Blanca Taboada, Gloria Vazquez, Joel Armando Vázquez-Pérez. |
| EPI_ISL_2681248 | Centro de Investigación Biomédica del Noreste (CIBIN) | Instituto de Biotecnología de la UNAM | Consorcio Mexicano de Vigilancia Genómica (CoViGen-Mex). Authors (in alphabetical order): Julio Elias Alvarado-Yaah, Carlos F. Arias, Santiago Ávila-Ríos, Eduardo Becerril-Vargas, Víctor Hugo Borja-Aburto, Celia Boukadida, Cristóbal Cháidez-Quiróz, Juan Bautista Chale-Dzul, Ricardo Ciria Merce, Andreu Comas-García, Célida Duque Molina, Julissa Enciso-Ibarra, José Antonio Enciso-Moreno, Gloria Elena Espinosa-Ayala, Fernando Fontove-Herrera, Daniel Fregoso-Rueda, Victor Eduardo García-Arias, Alejandra García-Gasca, Bruno Gómez-Gil, Jean Pierre González, Irvin |

| | | | |
|---|---|---|---|
| | | | González-López, Concepción Grajales-Muñiz, Ricardo Grande, Rosa María Gutiérrez Rios, Jesús Hernández, Alejandra Hernández-Terán, Alfredo Herrera-Estrella, Carla Ivón Herrera-Najera, Pavel Isa, Verónica Mata-Haro, Daniel Lira Morales, Susana Lopez, Antonio Loza Román, Brenda Irasema Maldonado-Meza, Bernardo Martínez-Miguel, José Arturo Martínez-Orozco, Célida Martínez- Rodríguez, Margarita Matías-Florentino, Fidencio Mejía-Nepomuceno, María Guadalupe de Jesús Mireles-Rivera, Gloria María Molina-Salinas, Hector Montoya-Fuentes, , Mario Mújica-Sánchez, José Esteban Muñoz-Medina, José de Jesús Nuñez-Contreras, Alicia Ocaña-Mondragón, Luis Alberto Ochoa-Carrera, Hector Esteban Paz-Juárez, Marissa Perez-Garcia, Francisco Pulido, Helen Haydee Fernanda Ramírez-Plascencia, Jorge Salas-Hernández, Angel Gustavo Salas-Lais, Alejandro Sánchez-Flores, Clara Esperanza Santacruz-Tinoco, María Guadalupe Santiago-Mauricio, Selene Zárate, Nelly Sélem-Mojica, Blanca Taboada, Gloria Vazquez, Joel Armando Vázquez-Pérez. |
| EPI_ISL_2681249, EPI_ISL_2681250, EPI_ISL_2681251, EPI_ISL_2681252, EPI_ISL_2681253, EPI_ISL_2681254, EPI_ISL_2681255, EPI_ISL_2681256, EPI_ISL_2681257, EPI_ISL_2681258, EPI_ISL_2681259 | | | |
| see above | Laboratorio Central de Epidemiología (LCE) | Instituto de Biotecnología de la UNAM | Consorcio Mexicano de Vigilancia Genómica (CoViGen-Mex). Authors (in alphabetical order): Julio Elias Alvarado-Yaah, Carlos F. Arias, Santiago Ávila-Ríos, Eduardo Becerril-Vargas, Víctor Hugo Borja-Aburto, Celia Boukadida, Cristóbal Cháidez-Quiróz, Juan Bautista Chale-Dzul, Ricardo Ciria Merce, Andreu Comas-García, Célida Duque Molina, Julissa Enciso-Ibarra, José Antonio Enciso-Moreno, Gloria Elena Espinosa-Ayala, Fernando Fontove-Herrera, Daniel Fregoso-Rueda, Victor Eduardo García-Arias, Alejandra García-Gasca, Bruno Gómez-Gil, Jean Pierre González, Irvin González-López, Concepción Grajales-Muñiz, Ricardo Grande, Rosa María Gutiérrez Rios, Jesús Hernández, Alejandra Hernández-Terán, Alfredo Herrera-Estrella, Carla Ivón Herrera-Najera, Pavel Isa, Verónica Mata-Haro, Daniel Lira Morales, Susana Lopez, Antonio Loza Román, Brenda Irasema Maldonado-Meza, Bernardo Martínez-Miguel, José Arturo Martínez-Orozco, Célida Martínez- Rodríguez, Margarita Matías-Florentino, Fidencio Mejía-Nepomuceno, María Guadalupe de Jesús Mireles-Rivera, Gloria María Molina-Salinas, Hector Montoya-Fuentes, , Mario Mújica-Sánchez, José Esteban Muñoz-Medina, José de Jesús Nuñez-Contreras, Alicia Ocaña-Mondragón, Luis Alberto Ochoa-Carrera, Hector Esteban Paz-Juárez, Marissa Perez-Garcia, Francisco Pulido, Helen Haydee Fernanda Ramírez-Plascencia, Jorge Salas-Hernández, Angel Gustavo Salas-Lais, Alejandro Sánchez-Flores, Clara Esperanza Santacruz-Tinoco, María Guadalupe Santiago-Mauricio, Selene Zárate, Nelly Sélem-Mojica, Blanca Taboada, Gloria Vazquez, Joel Armando Vázquez-Pérez. |
| EPI_ISL_2681260 | Centro de Investigación Biomédica del Noreste (CIBIN) | Instituto de Biotecnología de la UNAM | Consorcio Mexicano de Vigilancia Genómica (CoViGen-Mex). Authors (in alphabetical order): Julio Elias Alvarado-Yaah, Carlos F. Arias, Santiago Ávila-Ríos, Eduardo Becerril-Vargas, Víctor Hugo Borja-Aburto, Celia Boukadida, Cristóbal Cháidez-Quiróz, Juan Bautista Chale-Dzul, Ricardo Ciria Merce, Andreu Comas-García, Célida Duque Molina, Julissa Enciso-Ibarra, José Antonio Enciso-Moreno, Gloria Elena Espinosa-Ayala, Fernando Fontove-Herrera, Daniel Fregoso-Rueda, Victor Eduardo García-Arias, Alejandra García-Gasca, Bruno Gómez-Gil, Jean Pierre González, Irvin González-López, Concepción Grajales-Muñiz, Ricardo Grande, Rosa María Gutiérrez Rios, Jesús Hernández, Alejandra Hernández-Terán, Alfredo Herrera-Estrella, Carla Ivón Herrera-Najera, Pavel Isa, Verónica Mata-Haro, Daniel Lira Morales, Susana Lopez, Antonio Loza Román, Brenda Irasema Maldonado-Meza, Bernardo Martínez-Miguel, José Arturo Martínez-Orozco, Célida Martínez- Rodríguez, Margarita Matías-Florentino, Fidencio Mejía-Nepomuceno, María Guadalupe de Jesús Mireles-Rivera, Gloria María Molina-Salinas, Hector Montoya-Fuentes, , Mario Mújica-Sánchez, José Esteban Muñoz-Medina, José de Jesús Nuñez-Contreras, Alicia Ocaña-Mondragón, Luis Alberto Ochoa-Carrera, Hector Esteban Paz-Juárez, Marissa Perez-Garcia, Francisco Pulido, Helen Haydee Fernanda Ramírez-Plascencia, Jorge Salas-Hernández, Angel Gustavo Salas-Lais, Alejandro Sánchez-Flores, Clara Esperanza Santacruz-Tinoco, María Guadalupe Santiago-Mauricio, Selene Zárate, Nelly Sélem-Mojica, Blanca Taboada, Gloria Vazquez, Joel Armando Vázquez-Pérez. |
| EPI_ISL_2681261 | Unidad de Investigación Médica de Yucatán (UIMY) | Instituto de Biotecnología de la UNAM | Consorcio Mexicano de Vigilancia Genómica (CoViGen-Mex). Authors (in alphabetical order): Julio Elias Alvarado-Yaah, Carlos F. Arias, Santiago Ávila-Ríos, Eduardo Becerril-Vargas, Víctor Hugo Borja-Aburto, Celia Boukadida, Cristóbal Cháidez-Quiróz, Juan Bautista Chale-Dzul, Ricardo Ciria Merce, Andreu Comas-García, Célida Duque Molina, Julissa Enciso-Ibarra, José Antonio Enciso-Moreno, Gloria Elena Espinosa-Ayala, Fernando Fontove-Herrera, Daniel Fregoso-Rueda, Victor Eduardo García-Arias, Alejandra García-Gasca, Bruno Gómez-Gil, Jean Pierre González, Irvin González-López, Concepción Grajales-Muñiz, Ricardo Grande, Rosa María Gutiérrez Rios, Jesús Hernández, Alejandra Hernández-Terán, Alfredo Herrera-Estrella, Carla Ivón Herrera-Najera, Pavel Isa, Verónica Mata-Haro, Daniel Lira Morales, Susana Lopez, Antonio Loza Román, Brenda Irasema Maldonado-Meza, Bernardo Martínez-Miguel, José Arturo Martínez-Orozco, Célida Martínez- Rodríguez, Margarita Matías-Florentino, Fidencio Mejía-Nepomuceno, María Guadalupe de Jesús Mireles-Rivera, Gloria María Molina-Salinas, Hector Montoya-Fuentes, , Mario Mújica-Sánchez, José Esteban Muñoz-Medina, José de Jesús Nuñez-Contreras, Alicia Ocaña-Mondragón, Luis Alberto Ochoa-Carrera, Hector Esteban Paz-Juárez, Marissa Perez-Garcia, Francisco Pulido, Helen Haydee Fernanda Ramírez-Plascencia, Jorge Salas-Hernández, Angel Gustavo Salas-Lais, Alejandro Sánchez-Flores, Clara Esperanza Santacruz-Tinoco, María Guadalupe Santiago-Mauricio, Selene Zárate, Nelly Sélem-Mojica, Blanca Taboada, Gloria Vazquez, Joel Armando Vázquez-Pérez. |
| EPI_ISL_2681262, EPI_ISL_2681263, EPI_ISL_2681264 | Laboratorio Central de Epidemiología (LCE) | Instituto de Biotecnología de la UNAM | Consorcio Mexicano de Vigilancia Genómica (CoViGen-Mex). Authors (in alphabetical order): Julio Elias Alvarado-Yaah, Carlos F. Arias, Santiago Ávila-Ríos, Eduardo Becerril-Vargas, Víctor Hugo Borja-Aburto, Celia Boukadida, Cristóbal Cháidez-Quiróz, Juan Bautista Chale-Dzul, Ricardo Ciria Merce, Andreu Comas-García, Célida Duque Molina, Julissa Enciso-Ibarra, José Antonio Enciso-Moreno, Gloria Elena Espinosa-Ayala, Fernando Fontove-Herrera, Daniel Fregoso-Rueda, Victor Eduardo García-Arias, Alejandra García-Gasca, Bruno Gómez-Gil, Jean Pierre González, Irvin González-López, Concepción Grajales-Muñiz, Ricardo Grande, Rosa María Gutiérrez Rios, Jesús Hernández, Alejandra Hernández-Terán, Alfredo Herrera-Estrella, Carla Ivón Herrera-Najera, Pavel Isa, Verónica Mata-Haro, Daniel Lira Morales, Susana Lopez, Antonio Loza Román, Brenda Irasema Maldonado-Meza, Bernardo Martínez-Miguel, José Arturo Martínez-Orozco, Célida Martínez- Rodríguez, Margarita Matías-Florentino, Fidencio Mejía-Nepomuceno, María Guadalupe de Jesús Mireles-Rivera, Gloria María Molina-Salinas, Hector Montoya-Fuentes, , Mario Mújica-Sánchez, José Esteban Muñoz-Medina, José de Jesús Nuñez-Contreras, Alicia Ocaña-Mondragón, Luis Alberto Ochoa-Carrera, Hector Esteban Paz-Juárez, Marissa Perez-Garcia, Francisco Pulido, Helen Haydee Fernanda Ramírez-Plascencia, Jorge Salas-Hernández, Angel Gustavo Salas-Lais, Alejandro Sánchez-Flores, Clara Esperanza Santacruz-Tinoco, María Guadalupe Santiago-Mauricio, Selene Zárate, Nelly Sélem-Mojica, Blanca Taboada, Gloria Vazquez, Joel Armando Vázquez-Pérez. |
| EPI_ISL_2681265 | Centro de Investigación Biomédica del Noreste (CIBIN) | Instituto de Biotecnología de la UNAM | Consorcio Mexicano de Vigilancia Genómica (CoViGen-Mex). Authors (in alphabetical order): Julio Elias Alvarado-Yaah, Carlos F. Arias, Santiago Ávila-Ríos, Eduardo Becerril-Vargas, Víctor Hugo Borja-Aburto, Celia Boukadida, Cristóbal Cháidez-Quiróz, Juan Bautista Chale-Dzul, Ricardo Ciria Merce, Andreu Comas-García, Célida Duque Molina, Julissa Enciso-Ibarra, José Antonio Enciso-Moreno, Gloria Elena Espinosa-Ayala, Fernando Fontove-Herrera, Daniel Fregoso-Rueda, Victor Eduardo García-Arias, Alejandra García-Gasca, Bruno Gómez-Gil, Jean Pierre González, Irvin González-López, Concepción Grajales-Muñiz, Ricardo Grande, Rosa María Gutiérrez Rios, Jesús Hernández, Alejandra Hernández-Terán, Alfredo Herrera-Estrella, Carla Ivón Herrera-Najera, Pavel Isa, Verónica Mata-Haro, Daniel Lira Morales, Susana Lopez, Antonio Loza Román, Brenda Irasema Maldonado-Meza, Bernardo Martínez-Miguel, José Arturo Martínez-Orozco, Célida Martínez- Rodríguez, Margarita Matías-Florentino, Fidencio Mejía-Nepomuceno, María Guadalupe de Jesús Mireles-Rivera, Gloria María Molina-Salinas, Hector Montoya-Fuentes, , Mario Mújica-Sánchez, José Esteban Muñoz-Medina, José de Jesús Nuñez-Contreras, Alicia Ocaña-Mondragón, Luis Alberto Ochoa-Carrera, Hector Esteban Paz-Juárez, Marissa Perez-Garcia, Francisco Pulido, Helen Haydee Fernanda Ramírez-Plascencia, Jorge Salas-Hernández, Angel Gustavo Salas-Lais, Alejandro Sánchez-Flores, Clara Esperanza Santacruz-Tinoco, María Guadalupe Santiago-Mauricio, Selene Zárate, Nelly Sélem-Mojica, Blanca Taboada, Gloria Vazquez, Joel Armando Vázquez-Pérez. |
| EPI_ISL_2681266, EPI_ISL_2681267, EPI_ISL_2681268, EPI_ISL_2681269, EPI_ISL_2681270 | Laboratorio Central de Epidemiología (LCE) | Instituto de Biotecnología de la UNAM | Consorcio Mexicano de Vigilancia Genómica (CoViGen-Mex). Authors (in alphabetical order): Julio Elias Alvarado-Yaah, Carlos F. Arias, Santiago Ávila-Ríos, Eduardo Becerril-Vargas, Víctor Hugo Borja-Aburto, Celia Boukadida, Cristóbal Cháidez-Quiróz, Juan Bautista Chale-Dzul, Ricardo Ciria Merce, Andreu Comas-García, Célida Duque Molina, Julissa Enciso-Ibarra, José Antonio Enciso-Moreno, Gloria Elena Espinosa-Ayala, Fernando Fontove-Herrera, Daniel Fregoso-Rueda, Victor Eduardo García-Arias, Alejandra García-Gasca, Bruno Gómez-Gil, Jean Pierre González, Irvin González-López, Concepción Grajales-Muñiz, Ricardo Grande, Rosa María Gutiérrez Rios, Jesús Hernández, Alejandra Hernández-Terán, Alfredo Herrera-Estrella, Carla Ivón Herrera-Najera, Pavel Isa, Verónica Mata-Haro, Daniel Lira Morales, Susana Lopez, Antonio Loza Román, Brenda Irasema Maldonado-Meza, Bernardo Martínez-Miguel, José Arturo Martínez-Orozco, Célida Martínez- Rodríguez, Margarita Matías-Florentino, Fidencio Mejía-Nepomuceno, María Guadalupe de Jesús Mireles-Rivera, Gloria María Molina-Salinas, Hector Montoya-Fuentes, , Mario Mújica-Sánchez, José Esteban Muñoz-Medina, José de Jesús Nuñez-Contreras, Alicia Ocaña-Mondragón, Luis Alberto Ochoa-Carrera, Hector Esteban Paz-Juárez, Marissa Perez-Garcia, Francisco Pulido, Helen Haydee Fernanda Ramírez-Plascencia, Jorge Salas-Hernández, Angel Gustavo Salas-Lais, Alejandro Sánchez-Flores, Clara Esperanza Santacruz-Tinoco, María Guadalupe Santiago-Mauricio, Selene Zárate, Nelly Sélem-Mojica, Blanca Taboada, Gloria Vazquez, Joel Armando Vázquez-Pérez. |
| EPI_ISL_2681271 | Centro de Investigación Biomédica del Noreste (CIBIN) | Instituto de Biotecnología de la UNAM | Consorcio Mexicano de Vigilancia Genómica (CoViGen-Mex). Authors (in alphabetical order): Julio Elias Alvarado-Yaah, Carlos F. Arias, Santiago Ávila-Ríos, Eduardo Becerril-Vargas, Víctor Hugo Borja-Aburto, Celia Boukadida, Cristóbal Cháidez-Quiróz, Juan Bautista Chale-Dzul, Ricardo Ciria Merce, Andreu Comas-García, Célida Duque Molina, Julissa Enciso-Ibarra, José Antonio Enciso-Moreno, Gloria Elena Espinosa-Ayala, Fernando Fontove-Herrera, Daniel Fregoso-Rueda, Victor Eduardo García-Arias, Alejandra García-Gasca, Bruno Gómez-Gil, Jean Pierre González, Irvin González-López, Concepción Grajales-Muñiz, Ricardo Grande, Rosa María Gutiérrez Rios, Jesús Hernández, Alejandra Hernández-Terán, Alfredo Herrera-Estrella, Carla Ivón Herrera-Najera, Pavel Isa, Verónica Mata-Haro, Daniel Lira Morales, Susana Lopez, Antonio Loza Román, Brenda Irasema Maldonado-Meza, Bernardo Martínez-Miguel, José Arturo Martínez-Orozco, Célida Martínez- Rodríguez, Margarita Matías-Florentino, Fidencio Mejía-Nepomuceno, María Guadalupe de Jesús Mireles-Rivera, Gloria María Molina-Salinas, Hector Montoya-Fuentes, , Mario Mújica-Sánchez, José Esteban Muñoz-Medina, José de Jesús Nuñez-Contreras, Alicia Ocaña-Mondragón, Luis Alberto Ochoa-Carrera, Hector Esteban Paz-Juárez, Marissa Perez-Garcia, Francisco Pulido, Helen Haydee Fernanda Ramírez-Plascencia, Jorge Salas-Hernández, Angel Gustavo Salas-Lais, Alejandro Sánchez-Flores, Clara Esperanza Santacruz-Tinoco, María Guadalupe Santiago-Mauricio, Selene Zárate, Nelly Sélem-Mojica, Blanca Taboada, Gloria Vazquez, Joel Armando Vázquez-Pérez. |
| EPI_ISL_2681272 | Unidad de Investigación Biomédica de Zacatecas (UIBZ) | Instituto de Biotecnología de la UNAM | Consorcio Mexicano de Vigilancia Genómica (CoViGen-Mex). Authors (in alphabetical order): Julio Elias Alvarado-Yaah, Carlos F. Arias, Santiago Ávila-Ríos, Eduardo Becerril-Vargas, Víctor Hugo Borja-Aburto, Celia Boukadida, Cristóbal Cháidez-Quiróz, Juan Bautista Chale-Dzul, Ricardo Ciria Merce, Andreu Comas-García, Célida Duque Molina, Julissa Enciso-Ibarra, José Antonio Enciso-Moreno, Gloria Elena Espinosa-Ayala, Fernando Fontove-Herrera, Daniel Fregoso-Rueda, Victor Eduardo García-Arias, Alejandra García-Gasca, Bruno Gómez-Gil, Jean Pierre González, Irvin González-López, Concepción Grajales-Muñiz, Ricardo Grande, Rosa María Gutiérrez Rios, Jesús Hernández, Alejandra Hernández-Terán, Alfredo Herrera-Estrella, Carla Ivón Herrera-Najera, Pavel Isa, Verónica Mata-Haro, Daniel Lira Morales, Susana Lopez, Antonio Loza Román, Brenda Irasema Maldonado-Meza, Bernardo Martínez-Miguel, José Arturo Martínez-Orozco, Célida Martínez- Rodríguez, Margarita Matías-Florentino, Fidencio Mejía-Nepomuceno, María Guadalupe de Jesús Mireles-Rivera, Gloria María Molina-Salinas, Hector Montoya-Fuentes, , Mario Mújica-Sánchez, José Esteban Muñoz-Medina, José de Jesús Nuñez-Contreras, Alicia Ocaña-Mondragón, Luis Alberto Ochoa-Carrera, Hector Esteban Paz-Juárez, Marissa Perez-Garcia, Francisco Pulido, Helen Haydee Fernanda Ramírez-Plascencia, Jorge Salas-Hernández, Angel Gustavo Salas-Lais, Alejandro Sánchez-Flores, Clara Esperanza Santacruz-Tinoco, María Guadalupe Santiago-Mauricio, Selene Zárate, Nelly Sélem-Mojica, Blanca Taboada, Gloria Vazquez, Joel Armando Vázquez-Pérez. |
| EPI_ISL_2681273 | Laboratorio Central de Epidemiología (LCE) | Instituto de Biotecnología de la UNAM | Consorcio Mexicano de Vigilancia Genómica (CoViGen-Mex). Authors (in alphabetical order): Julio Elias Alvarado-Yaah, Carlos F. Arias, Santiago Ávila-Ríos, Eduardo Becerril-Vargas, Víctor Hugo Borja-Aburto, Celia Boukadida, Cristóbal Cháidez-Quiróz, Juan Bautista Chale-Dzul, Ricardo Ciria Merce, Andreu Comas-García, Célida Duque Molina, Julissa Enciso-Ibarra, José Antonio Enciso-Moreno, Gloria Elena Espinosa-Ayala, Fernando Fontove-Herrera, Daniel Fregoso-Rueda, Victor Eduardo García-Arias, Alejandra García-Gasca, Bruno Gómez-Gil, Jean Pierre González, Irvin González-López, Concepción Grajales-Muñiz, Ricardo Grande, Rosa María Gutiérrez Rios, Jesús Hernández, Alejandra Hernández-Terán, Alfredo Herrera-Estrella, Carla Ivón Herrera-Najera, Pavel Isa, Verónica Mata-Haro, Daniel Lira Morales, Susana Lopez, Antonio Loza Román, Brenda Irasema Maldonado-Meza, Bernardo Martínez-Miguel, José Arturo Martínez-Orozco, Célida Martínez- Rodríguez, Margarita Matías-Florentino, Fidencio Mejía-Nepomuceno, María Guadalupe de Jesús Mireles-Rivera, Gloria María Molina-Salinas, Hector Montoya-Fuentes, , Mario Mújica-Sánchez, José Esteban Muñoz-Medina, José de Jesús Nuñez-Contreras, Alicia Ocaña-Mondragón, Luis Alberto Ochoa-Carrera, Hector Esteban Paz-Juárez, Marissa Perez-Garcia, Francisco Pulido, Helen Haydee Fernanda Ramírez-Plascencia, Jorge Salas-Hernández, Angel Gustavo Salas-Lais, Alejandro Sánchez-Flores, Clara Esperanza Santacruz-Tinoco, María Guadalupe Santiago-Mauricio, Selene Zárate, Nelly Sélem-Mojica, Blanca Taboada, Gloria Vazquez, Joel Armando Vázquez-Pérez. |
| EPI_ISL_2681274, EPI_ISL_2681275 | Unidad de Investigación Médica de Yucatán (UIMY) | Instituto de Biotecnología de la UNAM | Consorcio Mexicano de Vigilancia Genómica (CoViGen-Mex). Authors (in alphabetical order): Julio Elias Alvarado-Yaah, Carlos F. Arias, Santiago Ávila-Ríos, Eduardo Becerril-Vargas, Víctor Hugo Borja-Aburto, Celia Boukadida, Cristóbal Cháidez-Quiróz, Juan Bautista Chale-Dzul, Ricardo Ciria Merce, Andreu Comas-García, Célida Duque Molina, Julissa Enciso-Ibarra, José Antonio Enciso-Moreno, Gloria Elena Espinosa-Ayala, Fernando Fontove-Herrera, Daniel Fregoso-Rueda, Victor Eduardo García-Arias, Alejandra García-Gasca, Bruno Gómez-Gil, Jean Pierre González, Irvin González-López, Concepción Grajales-Muñiz, Ricardo Grande, Rosa María Gutiérrez Rios, Jesús Hernández, Alejandra Hernández-Terán, Alfredo Herrera-Estrella, Carla Ivón Herrera-Najera, Pavel Isa, Verónica Mata-Haro, Daniel Lira Morales, Susana Lopez, Antonio Loza Román, Brenda Irasema Maldonado-Meza, Bernardo Martínez-Miguel, José Arturo Martínez-Orozco, Célida Martínez- Rodríguez, Margarita Matías-Florentino, Fidencio Mejía-Nepomuceno, María Guadalupe de Jesús Mireles-Rivera, Gloria María Molina-Salinas, Hector Montoya-Fuentes, , Mario Mújica-Sánchez, José Esteban Muñoz-Medina, José de Jesús Nuñez-Contreras, Alicia Ocaña-Mondragón, Luis Alberto Ochoa-Carrera, Hector Esteban Paz-Juárez, Marissa Perez-Garcia, Francisco Pulido, Helen Haydee Fernanda Ramírez-Plascencia, Jorge Salas-Hernández, Angel Gustavo Salas-Lais, Alejandro Sánchez-Flores, Clara Esperanza Santacruz-Tinoco, María Guadalupe Santiago-Mauricio, Selene Zárate, Nelly Sélem-Mojica, Blanca Taboada, Gloria Vazquez, Joel Armando Vázquez-Pérez. |
| EPI_ISL_2681276 | Laboratorio Central de Epidemiología (LCE) | Instituto de Biotecnología de la UNAM | Consorcio Mexicano de Vigilancia Genómica (CoViGen-Mex). Authors (in alphabetical order): Julio Elias Alvarado-Yaah, Carlos F. Arias, Santiago Ávila-Ríos, Eduardo Becerril-Vargas, Víctor Hugo Borja-Aburto, Celia Boukadida, Cristóbal Cháidez-Quiróz, Juan Bautista Chale-Dzul, Ricardo Ciria Merce, Andreu Comas-García, Célida Duque Molina, Julissa Enciso-Ibarra, José Antonio Enciso-Moreno, Gloria Elena Espinosa-Ayala, Fernando Fontove-Herrera, Daniel Fregoso-Rueda, Victor Eduardo García-Arias, Alejandra García-Gasca, Bruno Gómez-Gil, Jean Pierre González, Irvin González-López, Concepción Grajales-Muñiz, Ricardo Grande, Rosa María Gutiérrez Rios, Jesús Hernández, Alejandra Hernández-Terán, Alfredo Herrera-Estrella, Carla Ivón Herrera-Najera, Pavel Isa, Verónica Mata-Haro, Daniel Lira Morales, Susana Lopez, Antonio Loza Román, Brenda Irasema Maldonado-Meza, Bernardo Martínez-Miguel, José Arturo Martínez-Orozco, Célida Martínez- Rodríguez, Margarita Matías-Florentino, Fidencio Mejía-Nepomuceno, María Guadalupe de Jesús Mireles-Rivera, Gloria María Molina-Salinas, Hector Montoya-Fuentes, , Mario Mújica-Sánchez, José Esteban Muñoz-Medina, José de Jesús Nuñez-Contreras, Alicia Ocaña-Mondragón, Luis Alberto Ochoa-Carrera, Hector Esteban Paz-Juárez, Marissa Perez-Garcia, Francisco Pulido, Helen Haydee Fernanda Ramírez-Plascencia, Jorge Salas-Hernández, Angel Gustavo Salas-Lais, Alejandro Sánchez-Flores, Clara Esperanza Santacruz-Tinoco, María Guadalupe Santiago-Mauricio, Selene Zárate, Nelly Sélem-Mojica, Blanca Taboada, Gloria Vazquez, Joel Armando Vázquez-Pérez. |
| EPI_ISL_2681277 | Centro de Investigación Biomédica del Noreste (CIBIN) | Instituto de Biotecnología de la UNAM | Consorcio Mexicano de Vigilancia Genómica (CoViGen-Mex). Authors (in alphabetical order): Julio Elias Alvarado-Yaah, Carlos F. Arias, Santiago Ávila-Ríos, Eduardo Becerril-Vargas, Víctor Hugo Borja-Aburto, Celia Boukadida, Cristóbal Cháidez-Quiróz, Juan Bautista Chale-Dzul, Ricardo Ciria Merce, Andreu Comas-García, Célida Duque Molina, Julissa Enciso-Ibarra, José Antonio Enciso-Moreno, Gloria Elena Espinosa-Ayala, Fernando Fontove-Herrera, Daniel Fregoso-Rueda, Victor Eduardo García-Arias, Alejandra García-Gasca, Bruno Gómez-Gil, Jean Pierre González, Irvin González-López, Concepción Grajales-Muñiz, Ricardo Grande, Rosa María Gutiérrez Rios, Jesús Hernández, Alejandra Hernández-Terán, Alfredo Herrera-Estrella, Carla Ivón Herrera-Najera, Pavel Isa, Verónica Mata-Haro, Daniel Lira Morales, Susana Lopez, Antonio Loza Román, Brenda Irasema Maldonado-Meza, Bernardo Martínez-Miguel, José Arturo Martínez-Orozco, Célida Martínez- Rodríguez, Margarita Matías-Florentino, Fidencio Mejía-Nepomuceno, María Guadalupe de Jesús Mireles-Rivera, Gloria María Molina-Salinas, Hector Montoya-Fuentes, , Mario Mújica-Sánchez, José Esteban Muñoz-Medina, José de Jesús Nuñez-Contreras, Alicia Ocaña-Mondragón, Luis Alberto Ochoa-Carrera, Hector Esteban Paz-Juárez, Marissa Perez-Garcia, Francisco Pulido, Helen Haydee Fernanda Ramírez-Plascencia, Jorge Salas-Hernández, Angel Gustavo Salas-Lais, Alejandro Sánchez-Flores, Clara Esperanza Santacruz-Tinoco, María Guadalupe Santiago-Mauricio, Selene Zárate, Nelly Sélem-Mojica, Blanca Taboada, Gloria Vazquez, Joel Armando Vázquez-Pérez. |
| EPI_ISL_2681278, EPI_ISL_2681279, EPI_ISL_2681280, EPI_ISL_2681281, EPI_ISL_2681282, EPI_ISL_2681283, EPI_ISL_2681284 | Laboratorio Central de Epidemiología (LCE) | Instituto de Biotecnología de la UNAM | Consorcio Mexicano de Vigilancia Genómica (CoViGen-Mex). Authors (in alphabetical order): Julio Elias Alvarado-Yaah, Carlos F. Arias, Santiago Ávila-Ríos, Eduardo Becerril-Vargas, Víctor Hugo Borja-Aburto, Celia Boukadida, Cristóbal Cháidez-Quiróz, Juan Bautista Chale-Dzul, Ricardo Ciria Merce, Andreu Comas-García, Célida Duque Molina, Julissa Enciso-Ibarra, José Antonio Enciso-Moreno, Gloria Elena Espinosa-Ayala, Fernando Fontove-Herrera, Daniel Fregoso-Rueda, Victor Eduardo García-Arias, Alejandra García-Gasca, Bruno Gómez-Gil, Jean Pierre González, Irvin González-López, Concepción Grajales-Muñiz, Ricardo Grande, Rosa María Gutiérrez Rios, Jesús Hernández, Alejandra Hernández-Terán, Alfredo Herrera-Estrella, Carla Ivón Herrera-Najera, Pavel Isa, Verónica Mata-Haro, Daniel Lira Morales, Susana Lopez, Antonio Loza Román, Brenda Irasema Maldonado-Meza, Bernardo Martínez-Miguel, José Arturo Martínez-Orozco, Célida Martínez- Rodríguez, Margarita Matías-Florentino, Fidencio Mejía-Nepomuceno, María Guadalupe de Jesús Mireles-Rivera, Gloria María Molina-Salinas, Hector Montoya-Fuentes, , Mario Mújica-Sánchez, José Esteban Muñoz-Medina, José de Jesús Nuñez-Contreras, Alicia Ocaña-Mondragón, Luis Alberto Ochoa-Carrera, Hector Esteban Paz-Juárez, Marissa Perez-Garcia, Francisco Pulido, Helen Haydee Fernanda Ramírez-Plascencia, Jorge Salas-Hernández, Angel Gustavo Salas-Lais, Alejandro Sánchez-Flores, Clara Esperanza Santacruz-Tinoco, María Guadalupe Santiago-Mauricio, Selene Zárate, Nelly Sélem-Mojica, Blanca Taboada, Gloria Vazquez, Joel Armando Vázquez-Pérez. |
| EPI_ISL_2681285 | Centro de Investigación Biomédica del Noreste (CIBIN) | Instituto de Biotecnología de la UNAM | Consorcio Mexicano de Vigilancia Genómica (CoViGen-Mex). Authors (in alphabetical order): Julio Elias Alvarado-Yaah, Carlos F. Arias, Santiago Ávila-Ríos, Eduardo Becerril-Vargas, Víctor Hugo Borja-Aburto, Celia Boukadida, Cristóbal Cháidez-Quiróz, Juan Bautista Chale-Dzul, Ricardo Ciria Merce, Andreu Comas-García, Célida Duque Molina, Julissa Enciso-Ibarra, José Antonio Enciso-Moreno, Gloria Elena Espinosa-Ayala, Fernando Fontove-Herrera, Daniel Fregoso-Rueda, Victor Eduardo García-Arias, Alejandra García-Gasca, Bruno Gómez-Gil, Jean Pierre González, Irvin González-López, Concepción Grajales-Muñiz, Ricardo Grande, Rosa María Gutiérrez Rios, Jesús Hernández, Alejandra Hernández-Terán, Alfredo Herrera-Estrella, Carla Ivón Herrera-Najera, Pavel Isa, Verónica Mata-Haro, Daniel Lira Morales, Susana Lopez, Antonio Loza Román, Brenda Irasema Maldonado-Meza, Bernardo Martínez-Miguel, José Arturo Martínez-Orozco, Célida Martínez- Rodríguez, Margarita Matías-Florentino, Fidencio Mejía-Nepomuceno, María Guadalupe de Jesús Mireles-Rivera, Gloria María Molina-Salinas, Hector Montoya-Fuentes, , Mario Mújica-Sánchez, José Esteban Muñoz-Medina, José de Jesús Nuñez-Contreras, Alicia Ocaña-Mondragón, Luis Alberto Ochoa-Carrera, Hector Esteban Paz-Juárez, Marissa Perez-Garcia, Francisco Pulido, Helen Haydee Fernanda Ramírez-Plascencia, Jorge Salas-Hernández, Angel Gustavo Salas-Lais, Alejandro Sánchez-Flores, Clara Esperanza Santacruz-Tinoco, María Guadalupe Santiago-Mauricio, Selene Zárate, Nelly Sélem-Mojica, Blanca Taboada, Gloria Vazquez, Joel Armando Vázquez-Pérez. |
| EPI_ISL_2681286 | Unidad de Investigación Biomédica de Zacatecas (UIBZ) | Instituto de Biotecnología de la UNAM | Consorcio Mexicano de Vigilancia Genómica (CoViGen-Mex). Authors (in alphabetical order): Julio Elias Alvarado-Yaah, Carlos F. Arias, Santiago Ávila-Ríos, Eduardo Becerril-Vargas, Víctor Hugo Borja-Aburto, Celia Boukadida, Cristóbal Cháidez-Quiróz, Juan Bautista Chale-Dzul, Ricardo Ciria Merce, Andreu Comas-García, Célida Duque Molina, Julissa Enciso-Ibarra, José Antonio Enciso-Moreno, Gloria Elena Espinosa-Ayala, Fernando Fontove-Herrera, Daniel Fregoso-Rueda, Victor Eduardo García-Arias, Alejandra García-Gasca, Bruno Gómez-Gil, Jean Pierre González, Irvin González-López, Concepción Grajales-Muñiz, Ricardo Grande, Rosa María Gutiérrez Rios, Jesús Hernández, Alejandra Hernández-Terán, Alfredo Herrera-Estrella, Carla Ivón Herrera-Najera, Pavel Isa, Verónica Mata-Haro, Daniel Lira Morales, Susana Lopez, Antonio Loza Román, Brenda Irasema Maldonado-Meza, Bernardo Martínez-Miguel, José Arturo Martínez-Orozco, Célida Martínez- Rodríguez, Margarita Matías-Florentino, Fidencio Mejía-Nepomuceno, María Guadalupe de Jesús Mireles-Rivera, Gloria María Molina-Salinas, Hector Montoya-Fuentes, , Mario Mújica-Sánchez, José Esteban Muñoz-Medina, José de Jesús Nuñez-Contreras, Alicia Ocaña-Mondragón, Luis Alberto Ochoa-Carrera, Hector Esteban Paz-Juárez, Marissa Perez-Garcia, Francisco Pulido, Helen Haydee Fernanda Ramírez-Plascencia, Jorge Salas-Hernández, Angel Gustavo Salas-Lais, Alejandro Sánchez-Flores, Clara Esperanza Santacruz-Tinoco, María Guadalupe Santiago-Mauricio, Selene Zárate, Nelly Sélem-Mojica, Blanca Taboada, Gloria Vazquez, Joel Armando Vázquez-Pérez. |
| EPI_ISL_2681287, EPI_ISL_2681288 | Laboratorio Central de Epidemiología (LCE) | Instituto de Biotecnología de la UNAM | Consorcio Mexicano de Vigilancia Genómica (CoViGen-Mex). Authors (in alphabetical order): Julio Elias Alvarado-Yaah, Carlos F. Arias, Santiago Ávila-Ríos, Eduardo Becerril-Vargas, Víctor Hugo Borja-Aburto, Celia Boukadida, Cristóbal Cháidez-Quiróz, Juan Bautista Chale-Dzul, Ricardo Ciria Merce, Andreu Comas-García, Célida Duque Molina, Julissa Enciso-Ibarra, José Antonio Enciso-Moreno, Gloria Elena Espinosa-Ayala, Fernando Fontove-Herrera, Daniel Fregoso-Rueda, Victor Eduardo García-Arias, Alejandra García-Gasca, Bruno Gómez-Gil, Jean Pierre González, Irvin González-López, Concepción Grajales-Muñiz, Ricardo Grande, Rosa María Gutiérrez Rios, Jesús Hernández, Alejandra Hernández-Terán, Alfredo Herrera-Estrella, Carla Ivón Herrera-Najera, Pavel Isa, Verónica Mata-Haro, Daniel Lira Morales, Susana Lopez, Antonio Loza Román, Brenda Irasema Maldonado-Meza, Bernardo Martínez-Miguel, José Arturo Martínez-Orozco, Célida Martínez- Rodríguez, Margarita Matías-Florentino, Fidencio Mejía-Nepomuceno, María Guadalupe de Jesús Mireles-Rivera, Gloria María Molina-Salinas, Hector Montoya-Fuentes, , Mario Mújica-Sánchez, José Esteban Muñoz-Medina, José de Jesús Nuñez-Contreras, Alicia Ocaña-Mondragón, Luis Alberto Ochoa-Carrera, Hector Esteban Paz-Juárez, Marissa Perez-Garcia, Francisco Pulido, Helen Haydee Fernanda Ramírez-Plascencia, Jorge Salas-Hernández, Angel Gustavo Salas-Lais, Alejandro Sánchez-Flores, Clara Esperanza Santacruz-Tinoco, María Guadalupe Santiago-Mauricio, Selene Zárate, Nelly Sélem-Mojica, Blanca Taboada, Gloria Vazquez, Joel Armando Vázquez-Pérez. |

| | | | |
|---|---|---|---|
| EPI_ISL_2681289 | Centro de Investigación Biomédica del Noreste (CIBIN) | Instituto de Biotecnología de la UNAM | Consorcio Mexicano de Vigilancia Genómica (CoViGen-Mex). Authors (in alphabetical order): Julio Elias Alvarado-Yaah, Carlos F. Arias, Santiago Ávila-Ríos, Eduardo Becerril-Vargas, Víctor Hugo Borja-Aburto, Celia Boukadida, Cristóbal Cháidez-Quiróz, Juan Bautista Chale-Dzul, Ricardo Ciria Merce, Andreu Comas-García, Célida Duque Molina, Julissa Enciso-Ibarra, José Antonio Enciso-Moreno, Gloria Elena Espinosa-Ayala, Fernando Fontove-Herrera, Daniel Fregoso-Rueda, Victor Eduardo García-Arias, Alejandra García-Gasca, Bruno Gómez-Gil, Jean Pierre González, Irvin González-López, Concepción Grajales-Muñiz, Ricardo Grande, Rosa María Gutiérrez Rios, Jesús Hernández, Alejandra Hernández-Terán, Alfredo Herrera-Estrella, Carla Ivón Herrera-Najera, Pavel Isa, Verónica Mata-Haro, Daniel Lira Morales, Susana Lopez, Antonio Loza Román, Brenda Irasema Maldonado-Meza, Bernardo Martínez-Miguel, José Arturo Martínez-Orozco, Célida Martínez- Rodríguez, Margarita Matías-Florentino, Fidencio Mejía-Nepomuceno, María Guadalupe de Jesús Mireles-Rivera, Gloria María Molina-Salinas, Hector Montoya-Fuentes, , Mario Mújica-Sánchez, José Esteban Muñoz-Medina, José de Jesús Nuñez-Contreras, Alicia Ocaña-Mondragón, Luis Alberto Ochoa-Carrera, Hector Esteban Paz-Juárez, Marissa Perez-Garcia, Francisco Pulido, Helen Haydee Fernanda Ramírez-Plascencia, Jorge Salas-Hernández, Angel Gustavo Salas-Lais, Alejandro Sánchez-Flores, Clara Esperanza Santacruz-Tinoco, María Guadalupe Santiago-Mauricio, Selene Zárate, Nelly Sélem-Mojica, Blanca Taboada, Gloria Vazquez, Joel Armando Vázquez-Pérez. |
| EPI_ISL_2681290 | Laboratorio Central de Epidemiología (LCE) | Instituto de Biotecnología de la UNAM | Consorcio Mexicano de Vigilancia Genómica (CoViGen-Mex). Authors (in alphabetical order): Julio Elias Alvarado-Yaah, Carlos F. Arias, Santiago Ávila-Ríos, Eduardo Becerril-Vargas, Víctor Hugo Borja-Aburto, Celia Boukadida, Cristóbal Cháidez-Quiróz, Juan Bautista Chale-Dzul, Ricardo Ciria Merce, Andreu Comas-García, Célida Duque Molina, Julissa Enciso-Ibarra, José Antonio Enciso-Moreno, Gloria Elena Espinosa-Ayala, Fernando Fontove-Herrera, Daniel Fregoso-Rueda, Victor Eduardo García-Arias, Alejandra García-Gasca, Bruno Gómez-Gil, Jean Pierre González, Irvin González-López, Concepción Grajales-Muñiz, Ricardo Grande, Rosa María Gutiérrez Rios, Jesús Hernández, Alejandra Hernández-Terán, Alfredo Herrera-Estrella, Carla Ivón Herrera-Najera, Pavel Isa, Verónica Mata-Haro, Daniel Lira Morales, Susana Lopez, Antonio Loza Román, Brenda Irasema Maldonado-Meza, Bernardo Martínez-Miguel, José Arturo Martínez-Orozco, Célida Martínez- Rodríguez, Margarita Matías-Florentino, Fidencio Mejía-Nepomuceno, María Guadalupe de Jesús Mireles-Rivera, Gloria María Molina-Salinas, Hector Montoya-Fuentes, , Mario Mújica-Sánchez, José Esteban Muñoz-Medina, José de Jesús Nuñez-Contreras, Alicia Ocaña-Mondragón, Luis Alberto Ochoa-Carrera, Hector Esteban Paz-Juárez, Marissa Perez-Garcia, Francisco Pulido, Helen Haydee Fernanda Ramírez-Plascencia, Jorge Salas-Hernández, Angel Gustavo Salas-Lais, Alejandro Sánchez-Flores, Clara Esperanza Santacruz-Tinoco, María Guadalupe Santiago-Mauricio, Selene Zárate, Nelly Sélem-Mojica, Blanca Taboada, Gloria Vazquez, Joel Armando Vázquez-Pérez. |
| EPI_ISL_2681291 | Unidad de Investigación Biomédica de Zacatecas (UIBZ) | Instituto de Biotecnología de la UNAM | Consorcio Mexicano de Vigilancia Genómica (CoViGen-Mex). Authors (in alphabetical order): Julio Elias Alvarado-Yaah, Carlos F. Arias, Santiago Ávila-Ríos, Eduardo Becerril-Vargas, Víctor Hugo Borja-Aburto, Celia Boukadida, Cristóbal Cháidez-Quiróz, Juan Bautista Chale-Dzul, Ricardo Ciria Merce, Andreu Comas-García, Célida Duque Molina, Julissa Enciso-Ibarra, José Antonio Enciso-Moreno, Gloria Elena Espinosa-Ayala, Fernando Fontove-Herrera, Daniel Fregoso-Rueda, Victor Eduardo García-Arias, Alejandra García-Gasca, Bruno Gómez-Gil, Jean Pierre González, Irvin González-López, Concepción Grajales-Muñiz, Ricardo Grande, Rosa María Gutiérrez Rios, Jesús Hernández, Alejandra Hernández-Terán, Alfredo Herrera-Estrella, Carla Ivón Herrera-Najera, Pavel Isa, Verónica Mata-Haro, Daniel Lira Morales, Susana Lopez, Antonio Loza Román, Brenda Irasema Maldonado-Meza, Bernardo Martínez-Miguel, José Arturo Martínez-Orozco, Célida Martínez- Rodríguez, Margarita Matías-Florentino, Fidencio Mejía-Nepomuceno, María Guadalupe de Jesús Mireles-Rivera, Gloria María Molina-Salinas, Hector Montoya-Fuentes, , Mario Mújica-Sánchez, José Esteban Muñoz-Medina, José de Jesús Nuñez-Contreras, Alicia Ocaña-Mondragón, Luis Alberto Ochoa-Carrera, Hector Esteban Paz-Juárez, Marissa Perez-Garcia, Francisco Pulido, Helen Haydee Fernanda Ramírez-Plascencia, Jorge Salas-Hernández, Angel Gustavo Salas-Lais, Alejandro Sánchez-Flores, Clara Esperanza Santacruz-Tinoco, María Guadalupe Santiago-Mauricio, Selene Zárate, Nelly Sélem-Mojica, Blanca Taboada, Gloria Vazquez, Joel Armando Vázquez-Pérez. |
| EPI_ISL_2681292 | Unidad de Investigación Médica de Yucatán (UIMY) | Instituto de Biotecnología de la UNAM | Consorcio Mexicano de Vigilancia Genómica (CoViGen-Mex). Authors (in alphabetical order): Julio Elias Alvarado-Yaah, Carlos F. Arias, Santiago Ávila-Ríos, Eduardo Becerril-Vargas, Víctor Hugo Borja-Aburto, Celia Boukadida, Cristóbal Cháidez-Quiróz, Juan Bautista Chale-Dzul, Ricardo Ciria Merce, Andreu Comas-García, Célida Duque Molina, Julissa Enciso-Ibarra, José Antonio Enciso-Moreno, Gloria Elena Espinosa-Ayala, Fernando Fontove-Herrera, Daniel Fregoso-Rueda, Victor Eduardo García-Arias, Alejandra García-Gasca, Bruno Gómez-Gil, Jean Pierre González, Irvin González-López, Concepción Grajales-Muñiz, Ricardo Grande, Rosa María Gutiérrez Rios, Jesús Hernández, Alejandra Hernández-Terán, Alfredo Herrera-Estrella, Carla Ivón Herrera-Najera, Pavel Isa, Verónica Mata-Haro, Daniel Lira Morales, Susana Lopez, Antonio Loza Román, Brenda Irasema Maldonado-Meza, Bernardo Martínez-Miguel, José Arturo Martínez-Orozco, Célida Martínez- Rodríguez, Margarita Matías-Florentino, Fidencio Mejía-Nepomuceno, María Guadalupe de Jesús Mireles-Rivera, Gloria María Molina-Salinas, Hector Montoya-Fuentes, , Mario Mújica-Sánchez, José Esteban Muñoz-Medina, José de Jesús Nuñez-Contreras, Alicia Ocaña-Mondragón, Luis Alberto Ochoa-Carrera, Hector Esteban Paz-Juárez, Marissa Perez-Garcia, Francisco Pulido, Helen Haydee Fernanda Ramírez-Plascencia, Jorge Salas-Hernández, Angel Gustavo Salas-Lais, Alejandro Sánchez-Flores, Clara Esperanza Santacruz-Tinoco, María Guadalupe Santiago-Mauricio, Selene Zárate, Nelly Sélem-Mojica, Blanca Taboada, Gloria Vazquez, Joel Armando Vázquez-Pérez. |
| EPI_ISL_2681293 | Unidad de Investigación Biomédica de Zacatecas (UIBZ) | Instituto de Biotecnología de la UNAM | Consorcio Mexicano de Vigilancia Genómica (CoViGen-Mex). Authors (in alphabetical order): Julio Elias Alvarado-Yaah, Carlos F. Arias, Santiago Ávila-Ríos, Eduardo Becerril-Vargas, Víctor Hugo Borja-Aburto, Celia Boukadida, Cristóbal Cháidez-Quiróz, Juan Bautista Chale-Dzul, Ricardo Ciria Merce, Andreu Comas-García, Célida Duque Molina, Julissa Enciso-Ibarra, José Antonio Enciso-Moreno, Gloria Elena Espinosa-Ayala, Fernando Fontove-Herrera, Daniel Fregoso-Rueda, Victor Eduardo García-Arias, Alejandra García-Gasca, Bruno Gómez-Gil, Jean Pierre González, Irvin González-López, Concepción Grajales-Muñiz, Ricardo Grande, Rosa María Gutiérrez Rios, Jesús Hernández, Alejandra Hernández-Terán, Alfredo Herrera-Estrella, Carla Ivón Herrera-Najera, Pavel Isa, Verónica Mata-Haro, Daniel Lira Morales, Susana Lopez, Antonio Loza Román, Brenda Irasema Maldonado-Meza, Bernardo Martínez-Miguel, José Arturo Martínez-Orozco, Célida Martínez- Rodríguez, Margarita Matías-Florentino, Fidencio Mejía-Nepomuceno, María Guadalupe de Jesús Mireles-Rivera, Gloria María Molina-Salinas, Hector Montoya-Fuentes, , Mario Mújica-Sánchez, José Esteban Muñoz-Medina, José de Jesús Nuñez-Contreras, Alicia Ocaña-Mondragón, Luis Alberto Ochoa-Carrera, Hector Esteban Paz-Juárez, Marissa Perez-Garcia, Francisco Pulido, Helen Haydee Fernanda Ramírez-Plascencia, Jorge Salas-Hernández, Angel Gustavo Salas-Lais, Alejandro Sánchez-Flores, Clara Esperanza Santacruz-Tinoco, María Guadalupe Santiago-Mauricio, Selene Zárate, Nelly Sélem-Mojica, Blanca Taboada, Gloria Vazquez, Joel Armando Vázquez-Pérez. |
| EPI_ISL_2681294 | Laboratorio Central de Epidemiología (LCE) | Instituto de Biotecnología de la UNAM | Consorcio Mexicano de Vigilancia Genómica (CoViGen-Mex). Authors (in alphabetical order): Julio Elias Alvarado-Yaah, Carlos F. Arias, Santiago Ávila-Ríos, Eduardo Becerril-Vargas, Víctor Hugo Borja-Aburto, Celia Boukadida, Cristóbal Cháidez-Quiróz, Juan Bautista Chale-Dzul, Ricardo Ciria Merce, Andreu Comas-García, Célida Duque Molina, Julissa Enciso-Ibarra, José Antonio Enciso-Moreno, Gloria Elena Espinosa-Ayala, Fernando Fontove-Herrera, Daniel Fregoso-Rueda, Victor Eduardo García-Arias, Alejandra García-Gasca, Bruno Gómez-Gil, Jean Pierre González, Irvin González-López, Concepción Grajales-Muñiz, Ricardo Grande, Rosa María Gutiérrez Rios, Jesús Hernández, Alejandra Hernández-Terán, Alfredo Herrera-Estrella, Carla Ivón Herrera-Najera, Pavel Isa, Verónica Mata-Haro, Daniel Lira Morales, Susana Lopez, Antonio Loza Román, Brenda Irasema Maldonado-Meza, Bernardo Martínez-Miguel, José Arturo Martínez-Orozco, Célida Martínez- Rodríguez, Margarita Matías-Florentino, Fidencio Mejía-Nepomuceno, María Guadalupe de Jesús Mireles-Rivera, Gloria María Molina-Salinas, Hector Montoya-Fuentes, , Mario Mújica-Sánchez, José Esteban Muñoz-Medina, José de Jesús Nuñez-Contreras, Alicia Ocaña-Mondragón, Luis Alberto Ochoa-Carrera, Hector Esteban Paz-Juárez, Marissa Perez-Garcia, Francisco Pulido, Helen Haydee Fernanda Ramírez-Plascencia, Jorge Salas-Hernández, Angel Gustavo Salas-Lais, Alejandro Sánchez-Flores, Clara Esperanza Santacruz-Tinoco, María Guadalupe Santiago-Mauricio, Selene Zárate, Nelly Sélem-Mojica, Blanca Taboada, Gloria Vazquez, Joel Armando Vázquez-Pérez. |
| EPI_ISL_2681295 | Unidad de Investigación Médica de Yucatán (UIMY) | Instituto de Biotecnología de la UNAM | Consorcio Mexicano de Vigilancia Genómica (CoViGen-Mex). Authors (in alphabetical order): Julio Elias Alvarado-Yaah, Carlos F. Arias, Santiago Ávila-Ríos, Eduardo Becerril-Vargas, Víctor Hugo Borja-Aburto, Celia Boukadida, Cristóbal Cháidez-Quiróz, Juan Bautista Chale-Dzul, Ricardo Ciria Merce, Andreu Comas-García, Célida Duque Molina, Julissa Enciso-Ibarra, José Antonio Enciso-Moreno, Gloria Elena Espinosa-Ayala, Fernando Fontove-Herrera, Daniel Fregoso-Rueda, Victor Eduardo García-Arias, Alejandra García-Gasca, Bruno Gómez-Gil, Jean Pierre González, Irvin González-López, Concepción Grajales-Muñiz, Ricardo Grande, Rosa María Gutiérrez Rios, Jesús Hernández, Alejandra Hernández-Terán, Alfredo Herrera-Estrella, Carla Ivón Herrera-Najera, Pavel Isa, Verónica Mata-Haro, Daniel Lira Morales, Susana Lopez, Antonio Loza Román, Brenda Irasema Maldonado-Meza, Bernardo Martínez-Miguel, José Arturo Martínez-Orozco, Célida Martínez- Rodríguez, Margarita Matías-Florentino, Fidencio Mejía-Nepomuceno, María Guadalupe de Jesús Mireles-Rivera, Gloria María Molina-Salinas, Hector Montoya-Fuentes, , Mario Mújica-Sánchez, José Esteban Muñoz-Medina, José de Jesús Nuñez-Contreras, Alicia Ocaña-Mondragón, Luis Alberto Ochoa-Carrera, Hector Esteban Paz-Juárez, Marissa Perez-Garcia, Francisco Pulido, Helen Haydee Fernanda Ramírez-Plascencia, Jorge Salas-Hernández, Angel Gustavo Salas-Lais, Alejandro Sánchez-Flores, Clara Esperanza Santacruz-Tinoco, María Guadalupe Santiago-Mauricio, Selene Zárate, Nelly Sélem-Mojica, Blanca Taboada, Gloria Vazquez, Joel Armando Vázquez-Pérez. |
| EPI_ISL_2681296 | Centro de Investigación Biomédica del Noreste (CIBIN) | Instituto de Biotecnología de la UNAM | Consorcio Mexicano de Vigilancia Genómica (CoViGen-Mex). Authors (in alphabetical order): Julio Elias Alvarado-Yaah, Carlos F. Arias, Santiago Ávila-Ríos, Eduardo Becerril-Vargas, Víctor Hugo Borja-Aburto, Celia Boukadida, Cristóbal Cháidez-Quiróz, Juan Bautista Chale-Dzul, Ricardo Ciria Merce, Andreu Comas-García, Célida Duque Molina, Julissa Enciso-Ibarra, José Antonio Enciso-Moreno, Gloria Elena Espinosa-Ayala, Fernando Fontove-Herrera, Daniel Fregoso-Rueda, Victor Eduardo García-Arias, Alejandra García-Gasca, Bruno Gómez-Gil, Jean Pierre González, Irvin González-López, Concepción Grajales-Muñiz, Ricardo Grande, Rosa María Gutiérrez Rios, Jesús Hernández, Alejandra Hernández-Terán, Alfredo Herrera-Estrella, Carla Ivón Herrera-Najera, Pavel Isa, Verónica Mata-Haro, Daniel Lira Morales, Susana Lopez, Antonio Loza Román, Brenda Irasema Maldonado-Meza, Bernardo Martínez-Miguel, José Arturo Martínez-Orozco, Célida Martínez- Rodríguez, Margarita Matías-Florentino, Fidencio Mejía-Nepomuceno, María Guadalupe de Jesús Mireles-Rivera, Gloria María Molina-Salinas, Hector Montoya-Fuentes, , Mario Mújica-Sánchez, José Esteban Muñoz-Medina, José de Jesús Nuñez-Contreras, Alicia Ocaña-Mondragón, Luis Alberto Ochoa-Carrera, Hector Esteban Paz-Juárez, Marissa Perez-Garcia, Francisco Pulido, Helen Haydee Fernanda Ramírez-Plascencia, Jorge Salas-Hernández, Angel Gustavo Salas-Lais, Alejandro Sánchez-Flores, Clara Esperanza Santacruz-Tinoco, María Guadalupe Santiago-Mauricio, Selene Zárate, Nelly Sélem-Mojica, Blanca Taboada, Gloria Vazquez, Joel Armando Vázquez-Pérez. |
| EPI_ISL_2681297 | Laboratorio Central de Epidemiología (LCE) | Instituto de Biotecnología de la UNAM | Consorcio Mexicano de Vigilancia Genómica (CoViGen-Mex). Authors (in alphabetical order): Julio Elias Alvarado-Yaah, Carlos F. Arias, Santiago Ávila-Ríos, Eduardo Becerril-Vargas, Víctor Hugo Borja-Aburto, Celia Boukadida, Cristóbal Cháidez-Quiróz, Juan Bautista Chale-Dzul, Ricardo Ciria Merce, Andreu Comas-García, Célida Duque Molina, Julissa Enciso-Ibarra, José Antonio Enciso-Moreno, Gloria Elena Espinosa-Ayala, Fernando Fontove-Herrera, Daniel Fregoso-Rueda, Victor Eduardo García-Arias, Alejandra García-Gasca, Bruno Gómez-Gil, Jean Pierre González, Irvin González-López, Concepción Grajales-Muñiz, Ricardo Grande, Rosa María Gutiérrez Rios, Jesús Hernández, Alejandra Hernández-Terán, Alfredo Herrera-Estrella, Carla Ivón Herrera-Najera, Pavel Isa, Verónica Mata-Haro, Daniel Lira Morales, Susana Lopez, Antonio Loza Román, Brenda Irasema Maldonado-Meza, Bernardo Martínez-Miguel, José Arturo Martínez-Orozco, Célida Martínez- Rodríguez, Margarita Matías-Florentino, Fidencio Mejía-Nepomuceno, María Guadalupe de Jesús Mireles-Rivera, Gloria María Molina-Salinas, Hector Montoya-Fuentes, , Mario Mújica-Sánchez, José Esteban Muñoz-Medina, José de Jesús Nuñez-Contreras, Alicia Ocaña-Mondragón, Luis Alberto Ochoa-Carrera, Hector Esteban Paz-Juárez, Marissa Perez-Garcia, Francisco Pulido, Helen Haydee Fernanda Ramírez-Plascencia, Jorge Salas-Hernández, Angel Gustavo Salas-Lais, Alejandro Sánchez-Flores, Clara Esperanza Santacruz-Tinoco, María Guadalupe Santiago-Mauricio, Selene Zárate, Nelly Sélem-Mojica, Blanca Taboada, Gloria Vazquez, Joel Armando Vázquez-Pérez. |
| EPI_ISL_2681298 | Unidad de Investigación Médica de Yucatán (UIMY) | Instituto de Biotecnología de la UNAM | Consorcio Mexicano de Vigilancia Genómica (CoViGen-Mex). Authors (in alphabetical order): Julio Elias Alvarado-Yaah, Carlos F. Arias, Santiago Ávila-Ríos, Eduardo Becerril-Vargas, Víctor Hugo Borja-Aburto, Celia Boukadida, Cristóbal Cháidez-Quiróz, Juan Bautista Chale-Dzul, Ricardo Ciria Merce, Andreu Comas-García, Célida Duque Molina, Julissa Enciso-Ibarra, José Antonio Enciso-Moreno, Gloria Elena Espinosa-Ayala, Fernando Fontove-Herrera, Daniel Fregoso-Rueda, Victor Eduardo García-Arias, Alejandra García-Gasca, Bruno Gómez-Gil, Jean Pierre González, Irvin González-López, Concepción Grajales-Muñiz, Ricardo Grande, Rosa María Gutiérrez Rios, Jesús Hernández, Alejandra Hernández-Terán, Alfredo Herrera-Estrella, Carla Ivón Herrera-Najera, Pavel Isa, Verónica Mata-Haro, Daniel Lira Morales, Susana Lopez, Antonio Loza Román, Brenda Irasema Maldonado-Meza, Bernardo Martínez-Miguel, José Arturo Martínez-Orozco, Célida Martínez- Rodríguez, Margarita Matías-Florentino, Fidencio Mejía-Nepomuceno, María Guadalupe de Jesús Mireles-Rivera, Gloria María Molina-Salinas, Hector Montoya-Fuentes, , Mario Mújica-Sánchez, José Esteban Muñoz-Medina, José de Jesús Nuñez-Contreras, Alicia Ocaña-Mondragón, Luis Alberto Ochoa-Carrera, Hector Esteban Paz-Juárez, Marissa Perez-Garcia, Francisco Pulido, Helen Haydee Fernanda Ramírez-Plascencia, Jorge Salas-Hernández, Angel Gustavo Salas-Lais, Alejandro Sánchez-Flores, Clara Esperanza Santacruz-Tinoco, María Guadalupe Santiago-Mauricio, Selene Zárate, Nelly Sélem-Mojica, Blanca Taboada, Gloria Vazquez, Joel Armando Vázquez-Pérez. |
| EPI_ISL_2681299 | Laboratorio Central de Epidemiología (LCE) | Instituto de Biotecnología de la UNAM | Consorcio Mexicano de Vigilancia Genómica (CoViGen-Mex). Authors (in alphabetical order): Julio Elias Alvarado-Yaah, Carlos F. Arias, Santiago Ávila-Ríos, Eduardo Becerril-Vargas, Víctor Hugo Borja-Aburto, Celia Boukadida, Cristóbal Cháidez-Quiróz, Juan Bautista Chale-Dzul, Ricardo Ciria Merce, Andreu Comas-García, Célida Duque Molina, Julissa Enciso-Ibarra, José Antonio Enciso-Moreno, Gloria Elena Espinosa-Ayala, Fernando Fontove-Herrera, Daniel Fregoso-Rueda, Victor Eduardo García-Arias, Alejandra García-Gasca, Bruno Gómez-Gil, Jean Pierre González, Irvin González-López, Concepción Grajales-Muñiz, Ricardo Grande, Rosa María Gutiérrez Rios, Jesús Hernández, Alejandra Hernández-Terán, Alfredo Herrera-Estrella, Carla Ivón Herrera-Najera, Pavel Isa, Verónica Mata-Haro, Daniel Lira Morales, Susana Lopez, Antonio Loza Román, Brenda Irasema Maldonado-Meza, Bernardo Martínez-Miguel, José Arturo Martínez-Orozco, Célida Martínez- Rodríguez, Margarita Matías-Florentino, Fidencio Mejía-Nepomuceno, María Guadalupe de Jesús Mireles-Rivera, Gloria María Molina-Salinas, Hector Montoya-Fuentes, , Mario Mújica-Sánchez, José Esteban Muñoz-Medina, José de Jesús Nuñez-Contreras, Alicia Ocaña-Mondragón, Luis Alberto Ochoa-Carrera, Hector Esteban Paz-Juárez, Marissa Perez-Garcia, Francisco Pulido, Helen Haydee Fernanda Ramírez-Plascencia, Jorge Salas-Hernández, Angel Gustavo Salas-Lais, Alejandro Sánchez-Flores, Clara Esperanza Santacruz-Tinoco, María Guadalupe Santiago-Mauricio, Selene Zárate, Nelly Sélem-Mojica, Blanca Taboada, Gloria Vazquez, Joel Armando Vázquez-Pérez. |
| EPI_ISL_2681300 | Unidad de Investigación Médica de Yucatán (UIMY) | Instituto de Biotecnología de la UNAM | Consorcio Mexicano de Vigilancia Genómica (CoViGen-Mex). Authors (in alphabetical order): Julio Elias Alvarado-Yaah, Carlos F. Arias, Santiago Ávila-Ríos, Eduardo Becerril-Vargas, Víctor Hugo Borja-Aburto, Celia Boukadida, Cristóbal Cháidez-Quiróz, Juan Bautista Chale-Dzul, Ricardo Ciria Merce, Andreu Comas-García, Célida Duque Molina, Julissa Enciso-Ibarra, José Antonio Enciso-Moreno, Gloria Elena Espinosa-Ayala, Fernando Fontove-Herrera, Daniel Fregoso-Rueda, Victor Eduardo García-Arias, Alejandra García-Gasca, Bruno Gómez-Gil, Jean Pierre González, Irvin González-López, Concepción Grajales-Muñiz, Ricardo Grande, Rosa María Gutiérrez Rios, Jesús Hernández, Alejandra Hernández-Terán, Alfredo Herrera-Estrella, Carla Ivón Herrera-Najera, Pavel Isa, Verónica Mata-Haro, Daniel Lira Morales, Susana Lopez, Antonio Loza Román, Brenda Irasema Maldonado-Meza, Bernardo Martínez-Miguel, José Arturo Martínez-Orozco, Célida Martínez- Rodríguez, Margarita Matías-Florentino, Fidencio Mejía-Nepomuceno, María Guadalupe de Jesús Mireles-Rivera, Gloria María Molina-Salinas, Hector Montoya-Fuentes, , Mario Mújica-Sánchez, José Esteban Muñoz-Medina, José de Jesús Nuñez-Contreras, Alicia Ocaña-Mondragón, Luis Alberto Ochoa-Carrera, Hector Esteban Paz-Juárez, Marissa Perez-Garcia, Francisco Pulido, Helen Haydee Fernanda Ramírez-Plascencia, Jorge Salas-Hernández, Angel Gustavo Salas-Lais, Alejandro Sánchez-Flores, Clara Esperanza Santacruz-Tinoco, María Guadalupe Santiago-Mauricio, Selene Zárate, Nelly Sélem-Mojica, Blanca Taboada, Gloria Vazquez, Joel Armando Vázquez-Pérez. |
| EPI_ISL_2681301, EPI_ISL_2681302 | Laboratorio Central de Epidemiología (LCE) | Instituto de Biotecnología de la UNAM | Consorcio Mexicano de Vigilancia Genómica (CoViGen-Mex). Authors (in alphabetical order): Julio Elias Alvarado-Yaah, Carlos F. Arias, Santiago Ávila-Ríos, Eduardo Becerril-Vargas, Víctor Hugo Borja-Aburto, Celia Boukadida, Cristóbal Cháidez-Quiróz, Juan Bautista Chale-Dzul, Ricardo Ciria Merce, Andreu Comas-García, Célida Duque Molina, Julissa Enciso-Ibarra, José Antonio Enciso-Moreno, Gloria Elena Espinosa-Ayala, Fernando Fontove-Herrera, Daniel Fregoso-Rueda, Victor Eduardo García-Arias, Alejandra García-Gasca, Bruno Gómez-Gil, Jean Pierre González, Irvin González-López, Concepción Grajales-Muñiz, Ricardo Grande, Rosa María Gutiérrez Rios, Jesús Hernández, Alejandra Hernández-Terán, Alfredo Herrera-Estrella, Carla Ivón Herrera-Najera, Pavel Isa, Verónica Mata-Haro, Daniel Lira Morales, Susana Lopez, Antonio Loza Román, Brenda Irasema Maldonado-Meza, Bernardo Martínez-Miguel, José Arturo Martínez-Orozco, Célida Martínez- Rodríguez, Margarita Matías-Florentino, Fidencio Mejía-Nepomuceno, María Guadalupe de Jesús Mireles-Rivera, Gloria María Molina-Salinas, Hector Montoya-Fuentes, , Mario Mújica-Sánchez, José Esteban Muñoz-Medina, José de Jesús Nuñez-Contreras, Alicia Ocaña-Mondragón, Luis Alberto Ochoa-Carrera, Hector Esteban Paz-Juárez, Marissa Perez-Garcia, Francisco Pulido, Helen Haydee Fernanda Ramírez-Plascencia, Jorge Salas-Hernández, Angel Gustavo Salas-Lais, Alejandro Sánchez-Flores, Clara Esperanza Santacruz-Tinoco, María Guadalupe Santiago-Mauricio, Selene Zárate, Nelly Sélem-Mojica, Blanca Taboada, Gloria Vazquez, Joel Armando Vázquez-Pérez. |
| EPI_ISL_2681303 | Centro de Investigación Biomédica del Noreste (CIBIN) | Instituto de Biotecnología de la UNAM | Consorcio Mexicano de Vigilancia Genómica (CoViGen-Mex). Authors (in alphabetical order): Julio Elias Alvarado-Yaah, Carlos F. Arias, Santiago Ávila-Ríos, Eduardo Becerril-Vargas, Víctor Hugo Borja-Aburto, Celia Boukadida, Cristóbal Cháidez-Quiróz, Juan Bautista Chale-Dzul, Ricardo Ciria Merce, Andreu Comas-García, Célida Duque Molina, Julissa Enciso-Ibarra, José Antonio Enciso-Moreno, Gloria Elena Espinosa-Ayala, Fernando Fontove-Herrera, Daniel Fregoso-Rueda, Victor Eduardo García-Arias, Alejandra García-Gasca, Bruno Gómez-Gil, Jean Pierre González, Irvin González-López, Concepción Grajales-Muñiz, Ricardo Grande, Rosa María Gutiérrez Rios, Jesús Hernández, Alejandra Hernández-Terán, Alfredo Herrera-Estrella, Carla Ivón Herrera-Najera, Pavel Isa, Verónica Mata-Haro, Daniel Lira Morales, Susana Lopez, Antonio Loza Román, Brenda Irasema Maldonado-Meza, Bernardo Martínez-Miguel, José Arturo Martínez-Orozco, Célida Martínez- Rodríguez, Margarita Matías-Florentino, Fidencio Mejía-Nepomuceno, María Guadalupe de Jesús Mireles-Rivera, Gloria María Molina-Salinas, Hector Montoya-Fuentes, , Mario Mújica-Sánchez, José Esteban Muñoz-Medina, José de Jesús Nuñez-Contreras, Alicia Ocaña-Mondragón, Luis Alberto Ochoa-Carrera, Hector Esteban Paz-Juárez, Marissa Perez-Garcia, Francisco Pulido, Helen Haydee Fernanda Ramírez-Plascencia, Jorge Salas-Hernández, Angel Gustavo Salas-Lais, Alejandro Sánchez-Flores, Clara Esperanza Santacruz-Tinoco, María Guadalupe Santiago-Mauricio, Selene Zárate, Nelly Sélem-Mojica, Blanca Taboada, Gloria Vazquez, Joel Armando Vázquez-Pérez. |
| EPI_ISL_2681304 | Unidad de Investigación Médica de Yucatán (UIMY) | Instituto de Biotecnología de la UNAM | Consorcio Mexicano de Vigilancia Genómica (CoViGen-Mex). Authors (in alphabetical order): Julio Elias Alvarado-Yaah, Carlos F. Arias, Santiago Ávila-Ríos, Eduardo Becerril-Vargas, Víctor Hugo Borja-Aburto, Celia Boukadida, Cristóbal Cháidez-Quiróz, Juan Bautista Chale-Dzul, Ricardo Ciria Merce, Andreu Comas-García, Célida Duque Molina, Julissa Enciso-Ibarra, José Antonio Enciso-Moreno, Gloria Elena Espinosa-Ayala, Fernando Fontove-Herrera, Daniel Fregoso-Rueda, Victor Eduardo García-Arias, Alejandra García-Gasca, Bruno Gómez-Gil, Jean Pierre González, Irvin González-López, Concepción Grajales-Muñiz, Ricardo Grande, Rosa María Gutiérrez Rios, Jesús Hernández, Alejandra Hernández-Terán, Alfredo Herrera-Estrella, Carla Ivón Herrera-Najera, Pavel Isa, Verónica Mata-Haro, Daniel Lira Morales, Susana Lopez, Antonio Loza Román, Brenda Irasema Maldonado-Meza, Bernardo Martínez-Miguel, José Arturo Martínez-Orozco, Célida Martínez- Rodríguez, Margarita Matías-Florentino, Fidencio Mejía-Nepomuceno, María Guadalupe de Jesús Mireles-Rivera, Gloria María Molina-Salinas, Hector Montoya-Fuentes, , Mario Mújica-Sánchez, José Esteban Muñoz-Medina, José de Jesús Nuñez-Contreras, Alicia Ocaña-Mondragón, Luis Alberto Ochoa-Carrera, Hector Esteban Paz-Juárez, Marissa Perez-Garcia, Francisco Pulido, Helen Haydee Fernanda Ramírez-Plascencia, Jorge Salas-Hernández, Angel Gustavo Salas-Lais, Alejandro Sánchez-Flores, Clara Esperanza Santacruz-Tinoco, María Guadalupe Santiago-Mauricio, Selene Zárate, Nelly Sélem-Mojica, Blanca Taboada, Gloria Vazquez, Joel Armando Vázquez-Pérez. |
| EPI_ISL_2681305, EPI_ISL_2681306, EPI_ISL_2681307 | Laboratorio Central de Epidemiología (LCE) | Instituto de Biotecnología de la UNAM | Consorcio Mexicano de Vigilancia Genómica (CoViGen-Mex). Authors (in alphabetical order): Julio Elias Alvarado-Yaah, Carlos F. Arias, Santiago Ávila-Ríos, Eduardo Becerril-Vargas, Víctor Hugo Borja-Aburto, Celia Boukadida, Cristóbal Cháidez-Quiróz, Juan Bautista Chale-Dzul, Ricardo Ciria Merce, Andreu Comas-García, Célida Duque Molina, Julissa Enciso-Ibarra, José Antonio Enciso-Moreno, Gloria Elena Espinosa-Ayala, Fernando Fontove-Herrera, Daniel Fregoso-Rueda, Victor Eduardo García-Arias, Alejandra García-Gasca, Bruno Gómez-Gil, Jean Pierre González, Irvin González-López, Concepción Grajales-Muñiz, Ricardo Grande, Rosa María Gutiérrez Rios, Jesús Hernández, Alejandra Hernández-Terán, Alfredo Herrera-Estrella, Carla Ivón Herrera-Najera, Pavel Isa, Verónica Mata-Haro, Daniel Lira Morales, Susana Lopez, Antonio Loza Román, Brenda Irasema Maldonado-Meza, Bernardo Martínez-Miguel, José Arturo Martínez-Orozco, Célida Martínez- Rodríguez, Margarita Matías-Florentino, Fidencio Mejía-Nepomuceno, María Guadalupe de Jesús Mireles-Rivera, Gloria María Molina-Salinas, Hector Montoya-Fuentes, , Mario Mújica-Sánchez, José Esteban Muñoz-Medina, José de Jesús Nuñez-Contreras, Alicia Ocaña-Mondragón, Luis Alberto Ochoa-Carrera, Hector Esteban Paz-Juárez, Marissa Perez-Garcia, Francisco Pulido, Helen Haydee Fernanda Ramírez-Plascencia, Jorge Salas-Hernández, Angel Gustavo Salas-Lais, Alejandro Sánchez-Flores, Clara Esperanza Santacruz-Tinoco, María Guadalupe Santiago-Mauricio, Selene Zárate, Nelly Sélem-Mojica, Blanca Taboada, Gloria Vazquez, Joel Armando Vázquez-Pérez. |
| EPI_ISL_2681308 | Centro de Investigación Biomédica del Noreste (CIBIN) | Instituto de Biotecnología de la UNAM | Consorcio Mexicano de Vigilancia Genómica (CoViGen-Mex). Authors (in alphabetical order): Julio Elias Alvarado-Yaah, Carlos F. Arias, Santiago Ávila-Ríos, Eduardo Becerril-Vargas, Víctor Hugo Borja-Aburto, Celia Boukadida, Cristóbal Cháidez-Quiróz, Juan Bautista Chale-Dzul, Ricardo Ciria Merce, Andreu Comas-García, Célida Duque Molina, Julissa Enciso-Ibarra, José Antonio Enciso-Moreno, Gloria Elena Espinosa-Ayala, Fernando Fontove-Herrera, Daniel Fregoso-Rueda, Victor Eduardo García-Arias, Alejandra García-Gasca, Bruno Gómez-Gil, Jean Pierre González, Irvin González-López, Concepción Grajales-Muñiz, Ricardo Grande, Rosa María Gutiérrez Rios, Jesús Hernández, Alejandra Hernández-Terán, Alfredo Herrera-Estrella, Carla Ivón Herrera-Najera, Pavel Isa, Verónica Mata-Haro, Daniel Lira Morales, Susana Lopez, Antonio Loza Román, Brenda Irasema Maldonado-Meza, Bernardo Martínez-Miguel, José Arturo Martínez-Orozco, Célida Martínez- Rodríguez, Margarita Matías-Florentino, Fidencio Mejía-Nepomuceno, María Guadalupe de Jesús Mireles-Rivera, Gloria María Molina-Salinas, Hector Montoya-Fuentes, , Mario Mújica-Sánchez, José Esteban Muñoz-Medina, José de Jesús Nuñez-Contreras, Alicia Ocaña-Mondragón, Luis Alberto Ochoa-Carrera, Hector Esteban Paz-Juárez, Marissa Perez-Garcia, Francisco Pulido, Helen Haydee Fernanda Ramírez-Plascencia, Jorge Salas-Hernández, Angel Gustavo Salas-Lais, Alejandro Sánchez-Flores, Clara Esperanza Santacruz-Tinoco, María Guadalupe Santiago-Mauricio, Selene Zárate, Nelly Sélem-Mojica, Blanca Taboada, Gloria Vazquez, Joel Armando Vázquez-Pérez. |
| EPI_ISL_2681309 | Unidad de Investigación Biomédica de | Instituto de Biotecnología de la UNAM | Consorcio Mexicano de Vigilancia Genómica (CoViGen-Mex). Authors (in alphabetical order): Julio Elias Alvarado-Yaah, Carlos F. Arias, Santiago Ávila-Ríos, Eduardo Becerril-Vargas, Víctor Hugo Borja-Aburto, Celia Boukadida, Cristóbal Cháidez-Quiróz, Juan Bautista Chale-Dzul, Ricardo |

| | | | |
|---|---|---|---|
| | Zacatecas (UIBZ) | | Ciria Merce, Andreu Comas-García, Célida Duque Molina, Julissa Enciso-Ibarra, José Antonio Enciso-Moreno, Gloria Elena Espinosa-Ayala, Fernando Fontove-Herrera, Daniel Fregoso-Rueda, Victor Eduardo García-Arias, Alejandra García-Gasca, Bruno Gómez-Gil, Jean Pierre González, Irvin González-López, Concepción Grajales-Muñiz, Ricardo Grande, Rosa María Gutiérrez Rios, Jesús Hernández, Alejandra Hernández-Terán, Alfredo Herrera-Estrella, Carla Ivón Herrera-Najera, Pavel Isa, Verónica Mata-Haro, Daniel Lira Morales, Susana Lopez, Antonio Loza Román, Brenda Irasema Maldonado-Meza, Bernardo Martínez-Miguel, José Arturo Martínez-Orozco, Célida Martínez- Rodríguez, Margarita Matías-Florentino, Fidencio Mejía-Nepomuceno, María Guadalupe de Jesús Mireles-Rivera, Gloria María Molina-Salinas, Hector Montoya-Fuentes, , Mario Mújica-Sánchez, José Esteban Muñoz-Medina, José de Jesús Nuñez-Contreras, Alicia Ocaña-Mondragón, Luis Alberto Ochoa-Carrera, Hector Esteban Paz-Juárez, Marissa Perez-Garcia, Francisco Pulido, Helen Haydee Fernanda Ramírez-Plascencia, Jorge Salas-Hernández, Angel Gustavo Salas-Lais, Alejandro Sánchez-Flores, Clara Esperanza Santacruz-Tinoco, María Guadalupe Santiago-Mauricio, Selene Zárate, Nelly Sélem-Mojica, Blanca Taboada, Gloria Vazquez, Joel Armando Vázquez-Pérez. |
| EPI_ISL_2681310 | Laboratorio Central de Epidemiología (LCE) | Instituto de Biotecnología de la UNAM | Consorcio Mexicano de Vigilancia Genómica (CoViGen-Mex). Authors (in alphabetical order): Julio Elias Alvarado-Yaah, Carlos F. Arias, Santiago Ávila-Ríos, Eduardo Becerril-Vargas, Víctor Hugo Borja-Aburto, Celia Boukadida, Cristóbal Cháidez-Quiróz, Juan Bautista Chale-Dzul, Ricardo Ciria Merce, Andreu Comas-García, Célida Duque Molina, Julissa Enciso-Ibarra, José Antonio Enciso-Moreno, Gloria Elena Espinosa-Ayala, Fernando Fontove-Herrera, Daniel Fregoso-Rueda, Victor Eduardo García-Arias, Alejandra García-Gasca, Bruno Gómez-Gil, Jean Pierre González, Irvin González-López, Concepción Grajales-Muñiz, Ricardo Grande, Rosa María Gutiérrez Rios, Jesús Hernández, Alejandra Hernández-Terán, Alfredo Herrera-Estrella, Carla Ivón Herrera-Najera, Pavel Isa, Verónica Mata-Haro, Daniel Lira Morales, Susana Lopez, Antonio Loza Román, Brenda Irasema Maldonado-Meza, Bernardo Martínez-Miguel, José Arturo Martínez-Orozco, Célida Martínez- Rodríguez, Margarita Matías-Florentino, Fidencio Mejía-Nepomuceno, María Guadalupe de Jesús Mireles-Rivera, Gloria María Molina-Salinas, Hector Montoya-Fuentes, , Mario Mújica-Sánchez, José Esteban Muñoz-Medina, José de Jesús Nuñez-Contreras, Alicia Ocaña-Mondragón, Luis Alberto Ochoa-Carrera, Hector Esteban Paz-Juárez, Marissa Perez-Garcia, Francisco Pulido, Helen Haydee Fernanda Ramírez-Plascencia, Jorge Salas-Hernández, Angel Gustavo Salas-Lais, Alejandro Sánchez-Flores, Clara Esperanza Santacruz-Tinoco, María Guadalupe Santiago-Mauricio, Selene Zárate, Nelly Sélem-Mojica, Blanca Taboada, Gloria Vazquez, Joel Armando Vázquez-Pérez. |
| EPI_ISL_2681311 | Unidad de Investigación Médica de Yucatán (UIMY) | Instituto de Biotecnología de la UNAM | Consorcio Mexicano de Vigilancia Genómica (CoViGen-Mex). Authors (in alphabetical order): Julio Elias Alvarado-Yaah, Carlos F. Arias, Santiago Ávila-Ríos, Eduardo Becerril-Vargas, Víctor Hugo Borja-Aburto, Celia Boukadida, Cristóbal Cháidez-Quiróz, Juan Bautista Chale-Dzul, Ricardo Ciria Merce, Andreu Comas-García, Célida Duque Molina, Julissa Enciso-Ibarra, José Antonio Enciso-Moreno, Gloria Elena Espinosa-Ayala, Fernando Fontove-Herrera, Daniel Fregoso-Rueda, Victor Eduardo García-Arias, Alejandra García-Gasca, Bruno Gómez-Gil, Jean Pierre González, Irvin González-López, Concepción Grajales-Muñiz, Ricardo Grande, Rosa María Gutiérrez Rios, Jesús Hernández, Alejandra Hernández-Terán, Alfredo Herrera-Estrella, Carla Ivón Herrera-Najera, Pavel Isa, Verónica Mata-Haro, Daniel Lira Morales, Susana Lopez, Antonio Loza Román, Brenda Irasema Maldonado-Meza, Bernardo Martínez-Miguel, José Arturo Martínez-Orozco, Célida Martínez- Rodríguez, Margarita Matías-Florentino, Fidencio Mejía-Nepomuceno, María Guadalupe de Jesús Mireles-Rivera, Gloria María Molina-Salinas, Hector Montoya-Fuentes, , Mario Mújica-Sánchez, José Esteban Muñoz-Medina, José de Jesús Nuñez-Contreras, Alicia Ocaña-Mondragón, Luis Alberto Ochoa-Carrera, Hector Esteban Paz-Juárez, Marissa Perez-Garcia, Francisco Pulido, Helen Haydee Fernanda Ramírez-Plascencia, Jorge Salas-Hernández, Angel Gustavo Salas-Lais, Alejandro Sánchez-Flores, Clara Esperanza Santacruz-Tinoco, María Guadalupe Santiago-Mauricio, Selene Zárate, Nelly Sélem-Mojica, Blanca Taboada, Gloria Vazquez, Joel Armando Vázquez-Pérez. |
| EPI_ISL_2681312, EPI_ISL_2681313, EPI_ISL_2681314 | Laboratorio Central de Epidemiología (LCE) | Instituto de Biotecnología de la UNAM | Consorcio Mexicano de Vigilancia Genómica (CoViGen-Mex). Authors (in alphabetical order): Julio Elias Alvarado-Yaah, Carlos F. Arias, Santiago Ávila-Ríos, Eduardo Becerril-Vargas, Víctor Hugo Borja-Aburto, Celia Boukadida, Cristóbal Cháidez-Quiróz, Juan Bautista Chale-Dzul, Ricardo Ciria Merce, Andreu Comas-García, Célida Duque Molina, Julissa Enciso-Ibarra, José Antonio Enciso-Moreno, Gloria Elena Espinosa-Ayala, Fernando Fontove-Herrera, Daniel Fregoso-Rueda, Victor Eduardo García-Arias, Alejandra García-Gasca, Bruno Gómez-Gil, Jean Pierre González, Irvin González-López, Concepción Grajales-Muñiz, Ricardo Grande, Rosa María Gutiérrez Rios, Jesús Hernández, Alejandra Hernández-Terán, Alfredo Herrera-Estrella, Carla Ivón Herrera-Najera, Pavel Isa, Verónica Mata-Haro, Daniel Lira Morales, Susana Lopez, Antonio Loza Román, Brenda Irasema Maldonado-Meza, Bernardo Martínez-Miguel, José Arturo Martínez-Orozco, Célida Martínez- Rodríguez, Margarita Matías-Florentino, Fidencio Mejía-Nepomuceno, María Guadalupe de Jesús Mireles-Rivera, Gloria María Molina-Salinas, Hector Montoya-Fuentes, , Mario Mújica-Sánchez, José Esteban Muñoz-Medina, José de Jesús Nuñez-Contreras, Alicia Ocaña-Mondragón, Luis Alberto Ochoa-Carrera, Hector Esteban Paz-Juárez, Marissa Perez-Garcia, Francisco Pulido, Helen Haydee Fernanda Ramírez-Plascencia, Jorge Salas-Hernández, Angel Gustavo Salas-Lais, Alejandro Sánchez-Flores, Clara Esperanza Santacruz-Tinoco, María Guadalupe Santiago-Mauricio, Selene Zárate, Nelly Sélem-Mojica, Blanca Taboada, Gloria Vazquez, Joel Armando Vázquez-Pérez. |
| EPI_ISL_2681315 | Unidad de Investigación Biomédica de Zacatecas (UIBZ) | Instituto de Biotecnología de la UNAM | Consorcio Mexicano de Vigilancia Genómica (CoViGen-Mex). Authors (in alphabetical order): Julio Elias Alvarado-Yaah, Carlos F. Arias, Santiago Ávila-Ríos, Eduardo Becerril-Vargas, Víctor Hugo Borja-Aburto, Celia Boukadida, Cristóbal Cháidez-Quiróz, Juan Bautista Chale-Dzul, Ricardo Ciria Merce, Andreu Comas-García, Célida Duque Molina, Julissa Enciso-Ibarra, José Antonio Enciso-Moreno, Gloria Elena Espinosa-Ayala, Fernando Fontove-Herrera, Daniel Fregoso-Rueda, Victor Eduardo García-Arias, Alejandra García-Gasca, Bruno Gómez-Gil, Jean Pierre González, Irvin González-López, Concepción Grajales-Muñiz, Ricardo Grande, Rosa María Gutiérrez Rios, Jesús Hernández, Alejandra Hernández-Terán, Alfredo Herrera-Estrella, Carla Ivón Herrera-Najera, Pavel Isa, Verónica Mata-Haro, Daniel Lira Morales, Susana Lopez, Antonio Loza Román, Brenda Irasema Maldonado-Meza, Bernardo Martínez-Miguel, José Arturo Martínez-Orozco, Célida Martínez- Rodríguez, Margarita Matías-Florentino, Fidencio Mejía-Nepomuceno, María Guadalupe de Jesús Mireles-Rivera, Gloria María Molina-Salinas, Hector Montoya-Fuentes, , Mario Mújica-Sánchez, José Esteban Muñoz-Medina, José de Jesús Nuñez-Contreras, Alicia Ocaña-Mondragón, Luis Alberto Ochoa-Carrera, Hector Esteban Paz-Juárez, Marissa Perez-Garcia, Francisco Pulido, Helen Haydee Fernanda Ramírez-Plascencia, Jorge Salas-Hernández, Angel Gustavo Salas-Lais, Alejandro Sánchez-Flores, Clara Esperanza Santacruz-Tinoco, María Guadalupe Santiago-Mauricio, Selene Zárate, Nelly Sélem-Mojica, Blanca Taboada, Gloria Vazquez, Joel Armando Vázquez-Pérez. |
| EPI_ISL_2681316 | Centro de Investigación Biomédica del Noreste (CIBIN) | Instituto de Biotecnología de la UNAM | Consorcio Mexicano de Vigilancia Genómica (CoViGen-Mex). Authors (in alphabetical order): Julio Elias Alvarado-Yaah, Carlos F. Arias, Santiago Ávila-Ríos, Eduardo Becerril-Vargas, Víctor Hugo Borja-Aburto, Celia Boukadida, Cristóbal Cháidez-Quiróz, Juan Bautista Chale-Dzul, Ricardo Ciria Merce, Andreu Comas-García, Célida Duque Molina, Julissa Enciso-Ibarra, José Antonio Enciso-Moreno, Gloria Elena Espinosa-Ayala, Fernando Fontove-Herrera, Daniel Fregoso-Rueda, Victor Eduardo García-Arias, Alejandra García-Gasca, Bruno Gómez-Gil, Jean Pierre González, Irvin González-López, Concepción Grajales-Muñiz, Ricardo Grande, Rosa María Gutiérrez Rios, Jesús Hernández, Alejandra Hernández-Terán, Alfredo Herrera-Estrella, Carla Ivón Herrera-Najera, Pavel Isa, Verónica Mata-Haro, Daniel Lira Morales, Susana Lopez, Antonio Loza Román, Brenda Irasema Maldonado-Meza, Bernardo Martínez-Miguel, José Arturo Martínez-Orozco, Célida Martínez- Rodríguez, Margarita Matías-Florentino, Fidencio Mejía-Nepomuceno, María Guadalupe de Jesús Mireles-Rivera, Gloria María Molina-Salinas, Hector Montoya-Fuentes, , Mario Mújica-Sánchez, José Esteban Muñoz-Medina, José de Jesús Nuñez-Contreras, Alicia Ocaña-Mondragón, Luis Alberto Ochoa-Carrera, Hector Esteban Paz-Juárez, Marissa Perez-Garcia, Francisco Pulido, Helen Haydee Fernanda Ramírez-Plascencia, Jorge Salas-Hernández, Angel Gustavo Salas-Lais, Alejandro Sánchez-Flores, Clara Esperanza Santacruz-Tinoco, María Guadalupe Santiago-Mauricio, Selene Zárate, Nelly Sélem-Mojica, Blanca Taboada, Gloria Vazquez, Joel Armando Vázquez-Pérez. |
| EPI_ISL_2681317 | Unidad de Investigación Biomédica de Zacatecas (UIBZ) | Instituto de Biotecnología de la UNAM | Consorcio Mexicano de Vigilancia Genómica (CoViGen-Mex). Authors (in alphabetical order): Julio Elias Alvarado-Yaah, Carlos F. Arias, Santiago Ávila-Ríos, Eduardo Becerril-Vargas, Víctor Hugo Borja-Aburto, Celia Boukadida, Cristóbal Cháidez-Quiróz, Juan Bautista Chale-Dzul, Ricardo Ciria Merce, Andreu Comas-García, Célida Duque Molina, Julissa Enciso-Ibarra, José Antonio Enciso-Moreno, Gloria Elena Espinosa-Ayala, Fernando Fontove-Herrera, Daniel Fregoso-Rueda, Victor Eduardo García-Arias, Alejandra García-Gasca, Bruno Gómez-Gil, Jean Pierre González, Irvin González-López, Concepción Grajales-Muñiz, Ricardo Grande, Rosa María Gutiérrez Rios, Jesús Hernández, Alejandra Hernández-Terán, Alfredo Herrera-Estrella, Carla Ivón Herrera-Najera, Pavel Isa, Verónica Mata-Haro, Daniel Lira Morales, Susana Lopez, Antonio Loza Román, Brenda Irasema Maldonado-Meza, Bernardo Martínez-Miguel, José Arturo Martínez-Orozco, Célida Martínez- Rodríguez, Margarita Matías-Florentino, Fidencio Mejía-Nepomuceno, María Guadalupe de Jesús Mireles-Rivera, Gloria María Molina-Salinas, Hector Montoya-Fuentes, , Mario Mújica-Sánchez, José Esteban Muñoz-Medina, José de Jesús Nuñez-Contreras, Alicia Ocaña-Mondragón, Luis Alberto Ochoa-Carrera, Hector Esteban Paz-Juárez, Marissa Perez-Garcia, Francisco Pulido, Helen Haydee Fernanda Ramírez-Plascencia, Jorge Salas-Hernández, Angel Gustavo Salas-Lais, Alejandro Sánchez-Flores, Clara Esperanza Santacruz-Tinoco, María Guadalupe Santiago-Mauricio, Selene Zárate, Nelly Sélem-Mojica, Blanca Taboada, Gloria Vazquez, Joel Armando Vázquez-Pérez. |
| EPI_ISL_2681318 | Laboratorio Central de Epidemiología (LCE) | Instituto de Biotecnología de la UNAM | Consorcio Mexicano de Vigilancia Genómica (CoViGen-Mex). Authors (in alphabetical order): Julio Elias Alvarado-Yaah, Carlos F. Arias, Santiago Ávila-Ríos, Eduardo Becerril-Vargas, Víctor Hugo Borja-Aburto, Celia Boukadida, Cristóbal Cháidez-Quiróz, Juan Bautista Chale-Dzul, Ricardo Ciria Merce, Andreu Comas-García, Célida Duque Molina, Julissa Enciso-Ibarra, José Antonio Enciso-Moreno, Gloria Elena Espinosa-Ayala, Fernando Fontove-Herrera, Daniel Fregoso-Rueda, Victor Eduardo García-Arias, Alejandra García-Gasca, Bruno Gómez-Gil, Jean Pierre González, Irvin González-López, Concepción Grajales-Muñiz, Ricardo Grande, Rosa María Gutiérrez Rios, Jesús Hernández, Alejandra Hernández-Terán, Alfredo Herrera-Estrella, Carla Ivón Herrera-Najera, Pavel Isa, Verónica Mata-Haro, Daniel Lira Morales, Susana Lopez, Antonio Loza Román, Brenda Irasema Maldonado-Meza, Bernardo Martínez-Miguel, José Arturo Martínez-Orozco, Célida Martínez- Rodríguez, Margarita Matías-Florentino, Fidencio Mejía-Nepomuceno, María Guadalupe de Jesús Mireles-Rivera, Gloria María Molina-Salinas, Hector Montoya-Fuentes, , Mario Mújica-Sánchez, José Esteban Muñoz-Medina, José de Jesús Nuñez-Contreras, Alicia Ocaña-Mondragón, Luis Alberto Ochoa-Carrera, Hector Esteban Paz-Juárez, Marissa Perez-Garcia, Francisco Pulido, Helen Haydee Fernanda Ramírez-Plascencia, Jorge Salas-Hernández, Angel Gustavo Salas-Lais, Alejandro Sánchez-Flores, Clara Esperanza Santacruz-Tinoco, María Guadalupe Santiago-Mauricio, Selene Zárate, Nelly Sélem-Mojica, Blanca Taboada, Gloria Vazquez, Joel Armando Vázquez-Pérez. |
| EPI_ISL_2681319, EPI_ISL_2681320 | Unidad de Investigación Médica de Yucatán (UIMY) | Instituto de Biotecnología de la UNAM | Consorcio Mexicano de Vigilancia Genómica (CoViGen-Mex). Authors (in alphabetical order): Julio Elias Alvarado-Yaah, Carlos F. Arias, Santiago Ávila-Ríos, Eduardo Becerril-Vargas, Víctor Hugo Borja-Aburto, Celia Boukadida, Cristóbal Cháidez-Quiróz, Juan Bautista Chale-Dzul, Ricardo Ciria Merce, Andreu Comas-García, Célida Duque Molina, Julissa Enciso-Ibarra, José Antonio Enciso-Moreno, Gloria Elena Espinosa-Ayala, Fernando Fontove-Herrera, Daniel Fregoso-Rueda, Victor Eduardo García-Arias, Alejandra García-Gasca, Bruno Gómez-Gil, Jean Pierre González, Irvin González-López, Concepción Grajales-Muñiz, Ricardo Grande, Rosa María Gutiérrez Rios, Jesús Hernández, Alejandra Hernández-Terán, Alfredo Herrera-Estrella, Carla Ivón Herrera-Najera, Pavel Isa, Verónica Mata-Haro, Daniel Lira Morales, Susana Lopez, Antonio Loza Román, Brenda Irasema Maldonado-Meza, Bernardo Martínez-Miguel, José Arturo Martínez-Orozco, Célida Martínez- Rodríguez, Margarita Matías-Florentino, Fidencio Mejía-Nepomuceno, María Guadalupe de Jesús Mireles-Rivera, Gloria María Molina-Salinas, Hector Montoya-Fuentes, , Mario Mújica-Sánchez, José Esteban Muñoz-Medina, José de Jesús Nuñez-Contreras, Alicia Ocaña-Mondragón, Luis Alberto Ochoa-Carrera, Hector Esteban Paz-Juárez, Marissa Perez-Garcia, Francisco Pulido, Helen Haydee Fernanda Ramírez-Plascencia, Jorge Salas-Hernández, Angel Gustavo Salas-Lais, Alejandro Sánchez-Flores, Clara Esperanza Santacruz-Tinoco, María Guadalupe Santiago-Mauricio, Selene Zárate, Nelly Sélem-Mojica, Blanca Taboada, Gloria Vazquez, Joel Armando Vázquez-Pérez. |
| EPI_ISL_2681321 | Laboratorio Central de Epidemiología (LCE) | Instituto de Biotecnología de la UNAM | Consorcio Mexicano de Vigilancia Genómica (CoViGen-Mex). Authors (in alphabetical order): Julio Elias Alvarado-Yaah, Carlos F. Arias, Santiago Ávila-Ríos, Eduardo Becerril-Vargas, Víctor Hugo Borja-Aburto, Celia Boukadida, Cristóbal Cháidez-Quiróz, Juan Bautista Chale-Dzul, Ricardo Ciria Merce, Andreu Comas-García, Célida Duque Molina, Julissa Enciso-Ibarra, José Antonio Enciso-Moreno, Gloria Elena Espinosa-Ayala, Fernando Fontove-Herrera, Daniel Fregoso-Rueda, Victor Eduardo García-Arias, Alejandra García-Gasca, Bruno Gómez-Gil, Jean Pierre González, Irvin González-López, Concepción Grajales-Muñiz, Ricardo Grande, Rosa María Gutiérrez Rios, Jesús Hernández, Alejandra Hernández-Terán, Alfredo Herrera-Estrella, Carla Ivón Herrera-Najera, Pavel Isa, Verónica Mata-Haro, Daniel Lira Morales, Susana Lopez, Antonio Loza Román, Brenda Irasema Maldonado-Meza, Bernardo Martínez-Miguel, José Arturo Martínez-Orozco, Célida Martínez- Rodríguez, Margarita Matías-Florentino, Fidencio Mejía-Nepomuceno, María Guadalupe de Jesús Mireles-Rivera, Gloria María Molina-Salinas, Hector Montoya-Fuentes, , Mario Mújica-Sánchez, José Esteban Muñoz-Medina, José de Jesús Nuñez-Contreras, Alicia Ocaña-Mondragón, Luis Alberto Ochoa-Carrera, Hector Esteban Paz-Juárez, Marissa Perez-Garcia, Francisco Pulido, Helen Haydee Fernanda Ramírez-Plascencia, Jorge Salas-Hernández, Angel Gustavo Salas-Lais, Alejandro Sánchez-Flores, Clara Esperanza Santacruz-Tinoco, María Guadalupe Santiago-Mauricio, Selene Zárate, Nelly Sélem-Mojica, Blanca Taboada, Gloria Vazquez, Joel Armando Vázquez-Pérez. |
| EPI_ISL_2681322 | Unidad de Investigación Médica de Yucatán (UIMY) | Instituto de Biotecnología de la UNAM | Consorcio Mexicano de Vigilancia Genómica (CoViGen-Mex). Authors (in alphabetical order): Julio Elias Alvarado-Yaah, Carlos F. Arias, Santiago Ávila-Ríos, Eduardo Becerril-Vargas, Víctor Hugo Borja-Aburto, Celia Boukadida, Cristóbal Cháidez-Quiróz, Juan Bautista Chale-Dzul, Ricardo Ciria Merce, Andreu Comas-García, Célida Duque Molina, Julissa Enciso-Ibarra, José Antonio Enciso-Moreno, Gloria Elena Espinosa-Ayala, Fernando Fontove-Herrera, Daniel Fregoso-Rueda, Victor Eduardo García-Arias, Alejandra García-Gasca, Bruno Gómez-Gil, Jean Pierre González, Irvin González-López, Concepción Grajales-Muñiz, Ricardo Grande, Rosa María Gutiérrez Rios, Jesús Hernández, Alejandra Hernández-Terán, Alfredo Herrera-Estrella, Carla Ivón Herrera-Najera, Pavel Isa, Verónica Mata-Haro, Daniel Lira Morales, Susana Lopez, Antonio Loza Román, Brenda Irasema Maldonado-Meza, Bernardo Martínez-Miguel, José Arturo Martínez-Orozco, Célida Martínez- Rodríguez, Margarita Matías-Florentino, Fidencio Mejía-Nepomuceno, María Guadalupe de Jesús Mireles-Rivera, Gloria María Molina-Salinas, Hector Montoya-Fuentes, , Mario Mújica-Sánchez, José Esteban Muñoz-Medina, José de Jesús Nuñez-Contreras, Alicia Ocaña-Mondragón, Luis Alberto Ochoa-Carrera, Hector Esteban Paz-Juárez, Marissa Perez-Garcia, Francisco Pulido, Helen Haydee Fernanda Ramírez-Plascencia, Jorge Salas-Hernández, Angel Gustavo Salas-Lais, Alejandro Sánchez-Flores, Clara Esperanza Santacruz-Tinoco, María Guadalupe Santiago-Mauricio, Selene Zárate, Nelly Sélem-Mojica, Blanca Taboada, Gloria Vazquez, Joel Armando Vázquez-Pérez. |
| EPI_ISL_2681323 | Laboratorio Central de Epidemiología (LCE) | Instituto de Biotecnología de la UNAM | Consorcio Mexicano de Vigilancia Genómica (CoViGen-Mex). Authors (in alphabetical order): Julio Elias Alvarado-Yaah, Carlos F. Arias, Santiago Ávila-Ríos, Eduardo Becerril-Vargas, Víctor Hugo Borja-Aburto, Celia Boukadida, Cristóbal Cháidez-Quiróz, Juan Bautista Chale-Dzul, Ricardo Ciria Merce, Andreu Comas-García, Célida Duque Molina, Julissa Enciso-Ibarra, José Antonio Enciso-Moreno, Gloria Elena Espinosa-Ayala, Fernando Fontove-Herrera, Daniel Fregoso-Rueda, Victor Eduardo García-Arias, Alejandra García-Gasca, Bruno Gómez-Gil, Jean Pierre González, Irvin González-López, Concepción Grajales-Muñiz, Ricardo Grande, Rosa María Gutiérrez Rios, Jesús Hernández, Alejandra Hernández-Terán, Alfredo Herrera-Estrella, Carla Ivón Herrera-Najera, Pavel Isa, Verónica Mata-Haro, Daniel Lira Morales, Susana Lopez, Antonio Loza Román, Brenda Irasema Maldonado-Meza, Bernardo Martínez-Miguel, José Arturo Martínez-Orozco, Célida Martínez- Rodríguez, Margarita Matías-Florentino, Fidencio Mejía-Nepomuceno, María Guadalupe de Jesús Mireles-Rivera, Gloria María Molina-Salinas, Hector Montoya-Fuentes, , Mario Mújica-Sánchez, José Esteban Muñoz-Medina, José de Jesús Nuñez-Contreras, Alicia Ocaña-Mondragón, Luis Alberto Ochoa-Carrera, Hector Esteban Paz-Juárez, Marissa Perez-Garcia, Francisco Pulido, Helen Haydee Fernanda Ramírez-Plascencia, Jorge Salas-Hernández, Angel Gustavo Salas-Lais, Alejandro Sánchez-Flores, Clara Esperanza Santacruz-Tinoco, María Guadalupe Santiago-Mauricio, Selene Zárate, Nelly Sélem-Mojica, Blanca Taboada, Gloria Vazquez, Joel Armando Vázquez-Pérez. |
| EPI_ISL_2681324 | Centro de Investigación Biomédica del Noreste (CIBIN) | Instituto de Biotecnología de la UNAM | Consorcio Mexicano de Vigilancia Genómica (CoViGen-Mex). Authors (in alphabetical order): Julio Elias Alvarado-Yaah, Carlos F. Arias, Santiago Ávila-Ríos, Eduardo Becerril-Vargas, Víctor Hugo Borja-Aburto, Celia Boukadida, Cristóbal Cháidez-Quiróz, Juan Bautista Chale-Dzul, Ricardo Ciria Merce, Andreu Comas-García, Célida Duque Molina, Julissa Enciso-Ibarra, José Antonio Enciso-Moreno, Gloria Elena Espinosa-Ayala, Fernando Fontove-Herrera, Daniel Fregoso-Rueda, Victor Eduardo García-Arias, Alejandra García-Gasca, Bruno Gómez-Gil, Jean Pierre González, Irvin González-López, Concepción Grajales-Muñiz, Ricardo Grande, Rosa María Gutiérrez Rios, Jesús Hernández, Alejandra Hernández-Terán, Alfredo Herrera-Estrella, Carla Ivón Herrera-Najera, Pavel Isa, Verónica Mata-Haro, Daniel Lira Morales, Susana Lopez, Antonio Loza Román, Brenda Irasema Maldonado-Meza, Bernardo Martínez-Miguel, José Arturo Martínez-Orozco, Célida Martínez- Rodríguez, Margarita Matías-Florentino, Fidencio Mejía-Nepomuceno, María Guadalupe de Jesús Mireles-Rivera, Gloria María Molina-Salinas, Hector Montoya-Fuentes, , Mario Mújica-Sánchez, José Esteban Muñoz-Medina, José de Jesús Nuñez-Contreras, Alicia Ocaña-Mondragón, Luis Alberto Ochoa-Carrera, Hector Esteban Paz-Juárez, Marissa Perez-Garcia, Francisco Pulido, Helen Haydee Fernanda Ramírez-Plascencia, Jorge Salas-Hernández, Angel Gustavo Salas-Lais, Alejandro Sánchez-Flores, Clara Esperanza Santacruz-Tinoco, María Guadalupe Santiago-Mauricio, Selene Zárate, Nelly Sélem-Mojica, Blanca Taboada, Gloria Vazquez, Joel Armando Vázquez-Pérez. |
| EPI_ISL_2681325, EPI_ISL_2681326, EPI_ISL_2681327, EPI_ISL_2681328 | Unidad de Investigación Médica de Yucatán (UIMY) | Instituto de Biotecnología de la UNAM | Consorcio Mexicano de Vigilancia Genómica (CoViGen-Mex). Authors (in alphabetical order): Julio Elias Alvarado-Yaah, Carlos F. Arias, Santiago Ávila-Ríos, Eduardo Becerril-Vargas, Víctor Hugo Borja-Aburto, Celia Boukadida, Cristóbal Cháidez-Quiróz, Juan Bautista Chale-Dzul, Ricardo Ciria Merce, Andreu Comas-García, Célida Duque Molina, Julissa Enciso-Ibarra, José Antonio Enciso-Moreno, Gloria Elena Espinosa-Ayala, Fernando Fontove-Herrera, Daniel Fregoso-Rueda, Victor Eduardo García-Arias, Alejandra García-Gasca, Bruno Gómez-Gil, Jean Pierre González, Irvin González-López, Concepción Grajales-Muñiz, Ricardo Grande, Rosa María Gutiérrez Rios, Jesús Hernández, Alejandra Hernández-Terán, Alfredo Herrera-Estrella, Carla Ivón Herrera-Najera, Pavel Isa, Verónica Mata-Haro, Daniel Lira Morales, Susana Lopez, Antonio Loza Román, Brenda Irasema Maldonado-Meza, Bernardo Martínez-Miguel, José Arturo Martínez-Orozco, Célida Martínez- Rodríguez, Margarita Matías-Florentino, Fidencio Mejía-Nepomuceno, María Guadalupe de Jesús Mireles-Rivera, Gloria María Molina-Salinas, Hector Montoya-Fuentes, , Mario Mújica-Sánchez, José Esteban Muñoz-Medina, José de Jesús Nuñez-Contreras, Alicia Ocaña-Mondragón, Luis Alberto Ochoa-Carrera, Hector Esteban Paz-Juárez, Marissa Perez-Garcia, Francisco Pulido, Helen Haydee Fernanda Ramírez-Plascencia, Jorge Salas-Hernández, Angel Gustavo Salas-Lais, Alejandro Sánchez-Flores, Clara Esperanza Santacruz-Tinoco, María Guadalupe Santiago-Mauricio, Selene Zárate, Nelly Sélem-Mojica, Blanca Taboada, Gloria Vazquez, Joel Armando Vázquez-Pérez. |
| EPI_ISL_2681329, EPI_ISL_2681330 | Laboratorio Central de Epidemiología (LCE) | Instituto de Biotecnología de la UNAM | Consorcio Mexicano de Vigilancia Genómica (CoViGen-Mex). Authors (in alphabetical order): Julio Elias Alvarado-Yaah, Carlos F. Arias, Santiago Ávila-Ríos, Eduardo Becerril-Vargas, Víctor Hugo Borja-Aburto, Celia Boukadida, Cristóbal Cháidez-Quiróz, Juan Bautista Chale-Dzul, Ricardo Ciria Merce, Andreu Comas-García, Célida Duque Molina, Julissa Enciso-Ibarra, José Antonio Enciso-Moreno, Gloria Elena Espinosa-Ayala, Fernando Fontove-Herrera, Daniel Fregoso-Rueda, Victor Eduardo García-Arias, Alejandra García-Gasca, Bruno Gómez-Gil, Jean Pierre González, Irvin González-López, Concepción Grajales-Muñiz, Ricardo Grande, Rosa María Gutiérrez Rios, Jesús Hernández, Alejandra Hernández-Terán, Alfredo Herrera-Estrella, Carla Ivón Herrera-Najera, Pavel Isa, Verónica Mata-Haro, Daniel Lira Morales, Susana Lopez, Antonio Loza Román, Brenda Irasema Maldonado-Meza, Bernardo Martínez-Miguel, José Arturo Martínez-Orozco, Célida Martínez- Rodríguez, Margarita Matías-Florentino, Fidencio Mejía-Nepomuceno, María Guadalupe de Jesús Mireles-Rivera, Gloria María Molina-Salinas, Hector Montoya-Fuentes, , Mario Mújica-Sánchez, José Esteban Muñoz-Medina, José de Jesús Nuñez-Contreras, Alicia Ocaña-Mondragón, Luis Alberto Ochoa-Carrera, Hector Esteban Paz-Juárez, Marissa Perez-Garcia, Francisco Pulido, Helen Haydee Fernanda Ramírez-Plascencia, Jorge Salas-Hernández, Angel Gustavo Salas-Lais, Alejandro Sánchez-Flores, Clara Esperanza Santacruz-Tinoco, María Guadalupe Santiago-Mauricio, Selene Zárate, Nelly Sélem-Mojica, Blanca Taboada, Gloria Vazquez, Joel Armando Vázquez-Pérez. |
| EPI_ISL_2681331 | Centro de Investigación Biomédica del Noreste (CIBIN) | Instituto de Biotecnología de la UNAM | Consorcio Mexicano de Vigilancia Genómica (CoViGen-Mex). Authors (in alphabetical order): Julio Elias Alvarado-Yaah, Carlos F. Arias, Santiago Ávila-Ríos, Eduardo Becerril-Vargas, Víctor Hugo Borja-Aburto, Celia Boukadida, Cristóbal Cháidez-Quiróz, Juan Bautista Chale-Dzul, Ricardo Ciria Merce, Andreu Comas-García, Célida Duque Molina, Julissa Enciso-Ibarra, José Antonio Enciso-Moreno, Gloria Elena Espinosa-Ayala, Fernando Fontove-Herrera, Daniel Fregoso-Rueda, Victor Eduardo García-Arias, Alejandra García-Gasca, Bruno Gómez-Gil, Jean Pierre González, Irvin González-López, Concepción Grajales-Muñiz, Ricardo Grande, Rosa María Gutiérrez Rios, Jesús Hernández, Alejandra Hernández-Terán, Alfredo Herrera-Estrella, Carla Ivón Herrera-Najera, Pavel Isa, Verónica Mata-Haro, Daniel Lira Morales, Susana Lopez, Antonio Loza Román, Brenda Irasema Maldonado-Meza, Bernardo Martínez-Miguel, José Arturo Martínez-Orozco, Célida Martínez- Rodríguez, Margarita Matías-Florentino, Fidencio Mejía-Nepomuceno, María Guadalupe de Jesús Mireles-Rivera, Gloria María Molina-Salinas, Hector Montoya-Fuentes, , Mario Mújica-Sánchez, José Esteban Muñoz-Medina, José de Jesús Nuñez-Contreras, Alicia Ocaña-Mondragón, Luis Alberto Ochoa-Carrera, Hector Esteban Paz-Juárez, Marissa Perez-Garcia, Francisco Pulido, Helen Haydee Fernanda Ramírez-Plascencia, Jorge Salas-Hernández, Angel Gustavo Salas-Lais, Alejandro Sánchez-Flores, Clara Esperanza Santacruz-Tinoco, María Guadalupe Santiago-Mauricio, Selene Zárate, Nelly Sélem-Mojica, Blanca Taboada, Gloria Vazquez, Joel Armando Vázquez-Pérez. |
| EPI_ISL_2681332 | Laboratorio Central de Epidemiología (LCE) | Instituto de Biotecnología de la UNAM | Consorcio Mexicano de Vigilancia Genómica (CoViGen-Mex). Authors (in alphabetical order): Julio Elias Alvarado-Yaah, Carlos F. Arias, Santiago Ávila-Ríos, Eduardo Becerril-Vargas, Víctor Hugo Borja-Aburto, Celia Boukadida, Cristóbal Cháidez-Quiróz, Juan Bautista Chale-Dzul, Ricardo Ciria Merce, Andreu Comas-García, Célida Duque Molina, Julissa Enciso-Ibarra, José Antonio Enciso-Moreno, Gloria Elena Espinosa-Ayala, Fernando Fontove-Herrera, Daniel Fregoso-Rueda, Victor Eduardo García-Arias, Alejandra García-Gasca, Bruno Gómez-Gil, Jean Pierre González, Irvin González-López, Concepción Grajales-Muñiz, Ricardo Grande, Rosa María Gutiérrez Rios, Jesús Hernández, Alejandra Hernández-Terán, Alfredo Herrera-Estrella, Carla Ivón Herrera-Najera, Pavel Isa, Verónica Mata-Haro, Daniel Lira Morales, Susana Lopez, Antonio Loza Román, Brenda Irasema Maldonado-Meza, Bernardo Martínez-Miguel, José Arturo Martínez-Orozco, Célida Martínez- Rodríguez, Margarita Matías-Florentino, Fidencio Mejía-Nepomuceno, María Guadalupe de Jesús Mireles-Rivera, Gloria María Molina-Salinas, Hector Montoya-Fuentes, , Mario Mújica-Sánchez, José Esteban Muñoz-Medina, José de Jesús Nuñez-Contreras, Alicia Ocaña-Mondragón, Luis Alberto Ochoa-Carrera, Hector Esteban Paz-Juárez, Marissa Perez-Garcia, Francisco Pulido, Helen Haydee Fernanda Ramírez-Plascencia, Jorge Salas-Hernández, Angel Gustavo Salas-Lais, Alejandro Sánchez-Flores, Clara Esperanza Santacruz-Tinoco, María Guadalupe Santiago-Mauricio, Selene Zárate, Nelly Sélem-Mojica, Blanca Taboada, Gloria Vazquez, Joel Armando Vázquez-Pérez. |
| EPI_ISL_2681333 | Unidad de Investigación Médica de Yucatán (UIMY) | Instituto de Biotecnología de la UNAM | Consorcio Mexicano de Vigilancia Genómica (CoViGen-Mex). Authors (in alphabetical order): Julio Elias Alvarado-Yaah, Carlos F. Arias, Santiago Ávila-Ríos, Eduardo Becerril-Vargas, Víctor Hugo Borja-Aburto, Celia Boukadida, Cristóbal Cháidez-Quiróz, Juan Bautista Chale-Dzul, Ricardo Ciria Merce, Andreu Comas-García, Célida Duque Molina, Julissa Enciso-Ibarra, José Antonio Enciso-Moreno, Gloria Elena Espinosa-Ayala, Fernando Fontove-Herrera, Daniel Fregoso-Rueda, Victor Eduardo García-Arias, Alejandra García-Gasca, Bruno Gómez-Gil, Jean Pierre González, Irvin |

| | | | |
|---|---|---|---|
| | | | González-López, Concepción Grajales-Muñiz, Ricardo Grande, Rosa María Gutiérrez Rios, Jesús Hernández, Alejandra Hernández-Terán, Alfredo Herrera-Estrella, Carla Ivón Herrera-Najera, Pavel Isa, Verónica Mata-Haro, Daniel Lira Morales, Susana Lopez, Antonio Loza Román, Brenda Irasema Maldonado-Meza, Bernardo Martínez-Miguel, José Arturo Martínez-Orozco, Célida Martínez- Rodríguez, Margarita Matías-Florentino, Fidencio Mejía-Nepomuceno, María Guadalupe de Jesús Mireles-Rivera, Gloria María Molina-Salinas, Hector Montoya-Fuentes, , Mario Mújica-Sánchez, José Esteban Muñoz-Medina, José de Jesús Nuñez-Contreras, Alicia Ocaña-Mondragón, Luis Alberto Ochoa-Carrera, Hector Esteban Paz-Juárez, Marissa Perez-Garcia, Francisco Pulido, Helen Haydee Fernanda Ramírez-Plascencia, Jorge Salas-Hernández, Angel Gustavo Salas-Lais, Alejandro Sánchez-Flores, Clara Esperanza Santacruz-Tinoco, María Guadalupe Santiago-Mauricio, Selene Zárate, Nelly Sélem-Mojica, Blanca Taboada, Gloria Vazquez, Joel Armando Vázquez-Pérez. |
| EPI_ISL_2681334, EPI_ISL_2681335 | Laboratorio Central de Epidemiología (LCE) | Instituto de Biotecnología de la UNAM | Consorcio Mexicano de Vigilancia Genómica (CoViGen-Mex). Authors (in alphabetical order): Julio Elias Alvarado-Yaah, Carlos F. Arias, Santiago Ávila-Ríos, Eduardo Becerril-Vargas, Víctor Hugo Borja-Aburto, Celia Boukadida, Cristóbal Cháidez-Quiróz, Juan Bautista Chale-Dzul, Ricardo Ciria Merce, Andreu Comas-García, Célida Duque Molina, Julissa Enciso-Ibarra, José Antonio Enciso-Moreno, Gloria Elena Espinosa-Ayala, Fernando Fontove-Herrera, Daniel Fregoso-Rueda, Victor Eduardo García-Arias, Alejandra García-Gasca, Bruno Gómez-Gil, Jean Pierre González, Irvin González-López, Concepción Grajales-Muñiz, Ricardo Grande, Rosa María Gutiérrez Rios, Jesús Hernández, Alejandra Hernández-Terán, Alfredo Herrera-Estrella, Carla Ivón Herrera-Najera, Pavel Isa, Verónica Mata-Haro, Daniel Lira Morales, Susana Lopez, Antonio Loza Román, Brenda Irasema Maldonado-Meza, Bernardo Martínez-Miguel, José Arturo Martínez-Orozco, Célida Martínez- Rodríguez, Margarita Matías-Florentino, Fidencio Mejía-Nepomuceno, María Guadalupe de Jesús Mireles-Rivera, Gloria María Molina-Salinas, Hector Montoya-Fuentes, , Mario Mújica-Sánchez, José Esteban Muñoz-Medina, José de Jesús Nuñez-Contreras, Alicia Ocaña-Mondragón, Luis Alberto Ochoa-Carrera, Hector Esteban Paz-Juárez, Marissa Perez-Garcia, Francisco Pulido, Helen Haydee Fernanda Ramírez-Plascencia, Jorge Salas-Hernández, Angel Gustavo Salas-Lais, Alejandro Sánchez-Flores, Clara Esperanza Santacruz-Tinoco, María Guadalupe Santiago-Mauricio, Selene Zárate, Nelly Sélem-Mojica, Blanca Taboada, Gloria Vazquez, Joel Armando Vázquez-Pérez. |
| EPI_ISL_2681336 | Centro de Investigación Biomédica del Noreste (CIBIN) | Instituto de Biotecnología de la UNAM | Consorcio Mexicano de Vigilancia Genómica (CoViGen-Mex). Authors (in alphabetical order): Julio Elias Alvarado-Yaah, Carlos F. Arias, Santiago Ávila-Ríos, Eduardo Becerril-Vargas, Víctor Hugo Borja-Aburto, Celia Boukadida, Cristóbal Cháidez-Quiróz, Juan Bautista Chale-Dzul, Ricardo Ciria Merce, Andreu Comas-García, Célida Duque Molina, Julissa Enciso-Ibarra, José Antonio Enciso-Moreno, Gloria Elena Espinosa-Ayala, Fernando Fontove-Herrera, Daniel Fregoso-Rueda, Victor Eduardo García-Arias, Alejandra García-Gasca, Bruno Gómez-Gil, Jean Pierre González, Irvin González-López, Concepción Grajales-Muñiz, Ricardo Grande, Rosa María Gutiérrez Rios, Jesús Hernández, Alejandra Hernández-Terán, Alfredo Herrera-Estrella, Carla Ivón Herrera-Najera, Pavel Isa, Verónica Mata-Haro, Daniel Lira Morales, Susana Lopez, Antonio Loza Román, Brenda Irasema Maldonado-Meza, Bernardo Martínez-Miguel, José Arturo Martínez-Orozco, Célida Martínez- Rodríguez, Margarita Matías-Florentino, Fidencio Mejía-Nepomuceno, María Guadalupe de Jesús Mireles-Rivera, Gloria María Molina-Salinas, Hector Montoya-Fuentes, , Mario Mújica-Sánchez, José Esteban Muñoz-Medina, José de Jesús Nuñez-Contreras, Alicia Ocaña-Mondragón, Luis Alberto Ochoa-Carrera, Hector Esteban Paz-Juárez, Marissa Perez-Garcia, Francisco Pulido, Helen Haydee Fernanda Ramírez-Plascencia, Jorge Salas-Hernández, Angel Gustavo Salas-Lais, Alejandro Sánchez-Flores, Clara Esperanza Santacruz-Tinoco, María Guadalupe Santiago-Mauricio, Selene Zárate, Nelly Sélem-Mojica, Blanca Taboada, Gloria Vazquez, Joel Armando Vázquez-Pérez. |
| EPI_ISL_2681337 | Laboratorio Central de Epidemiología (LCE) | Instituto de Biotecnología de la UNAM | Consorcio Mexicano de Vigilancia Genómica (CoViGen-Mex). Authors (in alphabetical order): Julio Elias Alvarado-Yaah, Carlos F. Arias, Santiago Ávila-Ríos, Eduardo Becerril-Vargas, Víctor Hugo Borja-Aburto, Celia Boukadida, Cristóbal Cháidez-Quiróz, Juan Bautista Chale-Dzul, Ricardo Ciria Merce, Andreu Comas-García, Célida Duque Molina, Julissa Enciso-Ibarra, José Antonio Enciso-Moreno, Gloria Elena Espinosa-Ayala, Fernando Fontove-Herrera, Daniel Fregoso-Rueda, Victor Eduardo García-Arias, Alejandra García-Gasca, Bruno Gómez-Gil, Jean Pierre González, Irvin González-López, Concepción Grajales-Muñiz, Ricardo Grande, Rosa María Gutiérrez Rios, Jesús Hernández, Alejandra Hernández-Terán, Alfredo Herrera-Estrella, Carla Ivón Herrera-Najera, Pavel Isa, Verónica Mata-Haro, Daniel Lira Morales, Susana Lopez, Antonio Loza Román, Brenda Irasema Maldonado-Meza, Bernardo Martínez-Miguel, José Arturo Martínez-Orozco, Célida Martínez- Rodríguez, Margarita Matías-Florentino, Fidencio Mejía-Nepomuceno, María Guadalupe de Jesús Mireles-Rivera, Gloria María Molina-Salinas, Hector Montoya-Fuentes, , Mario Mújica-Sánchez, José Esteban Muñoz-Medina, José de Jesús Nuñez-Contreras, Alicia Ocaña-Mondragón, Luis Alberto Ochoa-Carrera, Hector Esteban Paz-Juárez, Marissa Perez-Garcia, Francisco Pulido, Helen Haydee Fernanda Ramírez-Plascencia, Jorge Salas-Hernández, Angel Gustavo Salas-Lais, Alejandro Sánchez-Flores, Clara Esperanza Santacruz-Tinoco, María Guadalupe Santiago-Mauricio, Selene Zárate, Nelly Sélem-Mojica, Blanca Taboada, Gloria Vazquez, Joel Armando Vázquez-Pérez. |
| EPI_ISL_2681338 | Unidad de Investigación Médica de Yucatán (UIMY) | Instituto de Biotecnología de la UNAM | Consorcio Mexicano de Vigilancia Genómica (CoViGen-Mex). Authors (in alphabetical order): Julio Elias Alvarado-Yaah, Carlos F. Arias, Santiago Ávila-Ríos, Eduardo Becerril-Vargas, Víctor Hugo Borja-Aburto, Celia Boukadida, Cristóbal Cháidez-Quiróz, Juan Bautista Chale-Dzul, Ricardo Ciria Merce, Andreu Comas-García, Célida Duque Molina, Julissa Enciso-Ibarra, José Antonio Enciso-Moreno, Gloria Elena Espinosa-Ayala, Fernando Fontove-Herrera, Daniel Fregoso-Rueda, Victor Eduardo García-Arias, Alejandra García-Gasca, Bruno Gómez-Gil, Jean Pierre González, Irvin González-López, Concepción Grajales-Muñiz, Ricardo Grande, Rosa María Gutiérrez Rios, Jesús Hernández, Alejandra Hernández-Terán, Alfredo Herrera-Estrella, Carla Ivón Herrera-Najera, Pavel Isa, Verónica Mata-Haro, Daniel Lira Morales, Susana Lopez, Antonio Loza Román, Brenda Irasema Maldonado-Meza, Bernardo Martínez-Miguel, José Arturo Martínez-Orozco, Célida Martínez- Rodríguez, Margarita Matías-Florentino, Fidencio Mejía-Nepomuceno, María Guadalupe de Jesús Mireles-Rivera, Gloria María Molina-Salinas, Hector Montoya-Fuentes, , Mario Mújica-Sánchez, José Esteban Muñoz-Medina, José de Jesús Nuñez-Contreras, Alicia Ocaña-Mondragón, Luis Alberto Ochoa-Carrera, Hector Esteban Paz-Juárez, Marissa Perez-Garcia, Francisco Pulido, Helen Haydee Fernanda Ramírez-Plascencia, Jorge Salas-Hernández, Angel Gustavo Salas-Lais, Alejandro Sánchez-Flores, Clara Esperanza Santacruz-Tinoco, María Guadalupe Santiago-Mauricio, Selene Zárate, Nelly Sélem-Mojica, Blanca Taboada, Gloria Vazquez, Joel Armando Vázquez-Pérez. |
| EPI_ISL_2681339, EPI_ISL_2681340 | Unidad de Investigación Biomédica de Zacatecas (UIBZ) | Instituto de Biotecnología de la UNAM | Consorcio Mexicano de Vigilancia Genómica (CoViGen-Mex). Authors (in alphabetical order): Julio Elias Alvarado-Yaah, Carlos F. Arias, Santiago Ávila-Ríos, Eduardo Becerril-Vargas, Víctor Hugo Borja-Aburto, Celia Boukadida, Cristóbal Cháidez-Quiróz, Juan Bautista Chale-Dzul, Ricardo Ciria Merce, Andreu Comas-García, Célida Duque Molina, Julissa Enciso-Ibarra, José Antonio Enciso-Moreno, Gloria Elena Espinosa-Ayala, Fernando Fontove-Herrera, Daniel Fregoso-Rueda, Victor Eduardo García-Arias, Alejandra García-Gasca, Bruno Gómez-Gil, Jean Pierre González, Irvin González-López, Concepción Grajales-Muñiz, Ricardo Grande, Rosa María Gutiérrez Rios, Jesús Hernández, Alejandra Hernández-Terán, Alfredo Herrera-Estrella, Carla Ivón Herrera-Najera, Pavel Isa, Verónica Mata-Haro, Daniel Lira Morales, Susana Lopez, Antonio Loza Román, Brenda Irasema Maldonado-Meza, Bernardo Martínez-Miguel, José Arturo Martínez-Orozco, Célida Martínez- Rodríguez, Margarita Matías-Florentino, Fidencio Mejía-Nepomuceno, María Guadalupe de Jesús Mireles-Rivera, Gloria María Molina-Salinas, Hector Montoya-Fuentes, , Mario Mújica-Sánchez, José Esteban Muñoz-Medina, José de Jesús Nuñez-Contreras, Alicia Ocaña-Mondragón, Luis Alberto Ochoa-Carrera, Hector Esteban Paz-Juárez, Marissa Perez-Garcia, Francisco Pulido, Helen Haydee Fernanda Ramírez-Plascencia, Jorge Salas-Hernández, Angel Gustavo Salas-Lais, Alejandro Sánchez-Flores, Clara Esperanza Santacruz-Tinoco, María Guadalupe Santiago-Mauricio, Selene Zárate, Nelly Sélem-Mojica, Blanca Taboada, Gloria Vazquez, Joel Armando Vázquez-Pérez. |
| EPI_ISL_2681341, EPI_ISL_2681342 | Unidad de Investigación Médica de Yucatán (UIMY) | Instituto de Biotecnología de la UNAM | Consorcio Mexicano de Vigilancia Genómica (CoViGen-Mex). Authors (in alphabetical order): Julio Elias Alvarado-Yaah, Carlos F. Arias, Santiago Ávila-Ríos, Eduardo Becerril-Vargas, Víctor Hugo Borja-Aburto, Celia Boukadida, Cristóbal Cháidez-Quiróz, Juan Bautista Chale-Dzul, Ricardo Ciria Merce, Andreu Comas-García, Célida Duque Molina, Julissa Enciso-Ibarra, José Antonio Enciso-Moreno, Gloria Elena Espinosa-Ayala, Fernando Fontove-Herrera, Daniel Fregoso-Rueda, Victor Eduardo García-Arias, Alejandra García-Gasca, Bruno Gómez-Gil, Jean Pierre González, Irvin González-López, Concepción Grajales-Muñiz, Ricardo Grande, Rosa María Gutiérrez Rios, Jesús Hernández, Alejandra Hernández-Terán, Alfredo Herrera-Estrella, Carla Ivón Herrera-Najera, Pavel Isa, Verónica Mata-Haro, Daniel Lira Morales, Susana Lopez, Antonio Loza Román, Brenda Irasema Maldonado-Meza, Bernardo Martínez-Miguel, José Arturo Martínez-Orozco, Célida Martínez- Rodríguez, Margarita Matías-Florentino, Fidencio Mejía-Nepomuceno, María Guadalupe de Jesús Mireles-Rivera, Gloria María Molina-Salinas, Hector Montoya-Fuentes, , Mario Mújica-Sánchez, José Esteban Muñoz-Medina, José de Jesús Nuñez-Contreras, Alicia Ocaña-Mondragón, Luis Alberto Ochoa-Carrera, Hector Esteban Paz-Juárez, Marissa Perez-Garcia, Francisco Pulido, Helen Haydee Fernanda Ramírez-Plascencia, Jorge Salas-Hernández, Angel Gustavo Salas-Lais, Alejandro Sánchez-Flores, Clara Esperanza Santacruz-Tinoco, María Guadalupe Santiago-Mauricio, Selene Zárate, Nelly Sélem-Mojica, Blanca Taboada, Gloria Vazquez, Joel Armando Vázquez-Pérez. |
| EPI_ISL_2681343, EPI_ISL_2681344 | Laboratorio Central de Epidemiología (LCE) | Instituto de Biotecnología de la UNAM | Consorcio Mexicano de Vigilancia Genómica (CoViGen-Mex). Authors (in alphabetical order): Julio Elias Alvarado-Yaah, Carlos F. Arias, Santiago Ávila-Ríos, Eduardo Becerril-Vargas, Víctor Hugo Borja-Aburto, Celia Boukadida, Cristóbal Cháidez-Quiróz, Juan Bautista Chale-Dzul, Ricardo Ciria Merce, Andreu Comas-García, Célida Duque Molina, Julissa Enciso-Ibarra, José Antonio Enciso-Moreno, Gloria Elena Espinosa-Ayala, Fernando Fontove-Herrera, Daniel Fregoso-Rueda, Victor Eduardo García-Arias, Alejandra García-Gasca, Bruno Gómez-Gil, Jean Pierre González, Irvin González-López, Concepción Grajales-Muñiz, Ricardo Grande, Rosa María Gutiérrez Rios, Jesús Hernández, Alejandra Hernández-Terán, Alfredo Herrera-Estrella, Carla Ivón Herrera-Najera, Pavel Isa, Verónica Mata-Haro, Daniel Lira Morales, Susana Lopez, Antonio Loza Román, Brenda Irasema Maldonado-Meza, Bernardo Martínez-Miguel, José Arturo Martínez-Orozco, Célida Martínez- Rodríguez, Margarita Matías-Florentino, Fidencio Mejía-Nepomuceno, María Guadalupe de Jesús Mireles-Rivera, Gloria María Molina-Salinas, Hector Montoya-Fuentes, , Mario Mújica-Sánchez, José Esteban Muñoz-Medina, José de Jesús Nuñez-Contreras, Alicia Ocaña-Mondragón, Luis Alberto Ochoa-Carrera, Hector Esteban Paz-Juárez, Marissa Perez-Garcia, Francisco Pulido, Helen Haydee Fernanda Ramírez-Plascencia, Jorge Salas-Hernández, Angel Gustavo Salas-Lais, Alejandro Sánchez-Flores, Clara Esperanza Santacruz-Tinoco, María Guadalupe Santiago-Mauricio, Selene Zárate, Nelly Sélem-Mojica, Blanca Taboada, Gloria Vazquez, Joel Armando Vázquez-Pérez. |
| EPI_ISL_2681345 | Unidad de Investigación Médica de Yucatán (UIMY) | Instituto de Biotecnología de la UNAM | Consorcio Mexicano de Vigilancia Genómica (CoViGen-Mex). Authors (in alphabetical order): Julio Elias Alvarado-Yaah, Carlos F. Arias, Santiago Ávila-Ríos, Eduardo Becerril-Vargas, Víctor Hugo Borja-Aburto, Celia Boukadida, Cristóbal Cháidez-Quiróz, Juan Bautista Chale-Dzul, Ricardo Ciria Merce, Andreu Comas-García, Célida Duque Molina, Julissa Enciso-Ibarra, José Antonio Enciso-Moreno, Gloria Elena Espinosa-Ayala, Fernando Fontove-Herrera, Daniel Fregoso-Rueda, Victor Eduardo García-Arias, Alejandra García-Gasca, Bruno Gómez-Gil, Jean Pierre González, Irvin González-López, Concepción Grajales-Muñiz, Ricardo Grande, Rosa María Gutiérrez Rios, Jesús Hernández, Alejandra Hernández-Terán, Alfredo Herrera-Estrella, Carla Ivón Herrera-Najera, Pavel Isa, Verónica Mata-Haro, Daniel Lira Morales, Susana Lopez, Antonio Loza Román, Brenda Irasema Maldonado-Meza, Bernardo Martínez-Miguel, José Arturo Martínez-Orozco, Célida Martínez- Rodríguez, Margarita Matías-Florentino, Fidencio Mejía-Nepomuceno, María Guadalupe de Jesús Mireles-Rivera, Gloria María Molina-Salinas, Hector Montoya-Fuentes, , Mario Mújica-Sánchez, José Esteban Muñoz-Medina, José de Jesús Nuñez-Contreras, Alicia Ocaña-Mondragón, Luis Alberto Ochoa-Carrera, Hector Esteban Paz-Juárez, Marissa Perez-Garcia, Francisco Pulido, Helen Haydee Fernanda Ramírez-Plascencia, Jorge Salas-Hernández, Angel Gustavo Salas-Lais, Alejandro Sánchez-Flores, Clara Esperanza Santacruz-Tinoco, María Guadalupe Santiago-Mauricio, Selene Zárate, Nelly Sélem-Mojica, Blanca Taboada, Gloria Vazquez, Joel Armando Vázquez-Pérez. |
| EPI_ISL_2681346 | Unidad de Investigación Biomédica de Zacatecas (UIBZ) | Instituto de Biotecnología de la UNAM | Consorcio Mexicano de Vigilancia Genómica (CoViGen-Mex). Authors (in alphabetical order): Julio Elias Alvarado-Yaah, Carlos F. Arias, Santiago Ávila-Ríos, Eduardo Becerril-Vargas, Víctor Hugo Borja-Aburto, Celia Boukadida, Cristóbal Cháidez-Quiróz, Juan Bautista Chale-Dzul, Ricardo Ciria Merce, Andreu Comas-García, Célida Duque Molina, Julissa Enciso-Ibarra, José Antonio Enciso-Moreno, Gloria Elena Espinosa-Ayala, Fernando Fontove-Herrera, Daniel Fregoso-Rueda, Victor Eduardo García-Arias, Alejandra García-Gasca, Bruno Gómez-Gil, Jean Pierre González, Irvin González-López, Concepción Grajales-Muñiz, Ricardo Grande, Rosa María Gutiérrez Rios, Jesús Hernández, Alejandra Hernández-Terán, Alfredo Herrera-Estrella, Carla Ivón Herrera-Najera, Pavel Isa, Verónica Mata-Haro, Daniel Lira Morales, Susana Lopez, Antonio Loza Román, Brenda Irasema Maldonado-Meza, Bernardo Martínez-Miguel, José Arturo Martínez-Orozco, Célida Martínez- Rodríguez, Margarita Matías-Florentino, Fidencio Mejía-Nepomuceno, María Guadalupe de Jesús Mireles-Rivera, Gloria María Molina-Salinas, Hector Montoya-Fuentes, , Mario Mújica-Sánchez, José Esteban Muñoz-Medina, José de Jesús Nuñez-Contreras, Alicia Ocaña-Mondragón, Luis Alberto Ochoa-Carrera, Hector Esteban Paz-Juárez, Marissa Perez-Garcia, Francisco Pulido, Helen Haydee Fernanda Ramírez-Plascencia, Jorge Salas-Hernández, Angel Gustavo Salas-Lais, Alejandro Sánchez-Flores, Clara Esperanza Santacruz-Tinoco, María Guadalupe Santiago-Mauricio, Selene Zárate, Nelly Sélem-Mojica, Blanca Taboada, Gloria Vazquez, Joel Armando Vázquez-Pérez. |
| EPI_ISL_2681347 | Laboratorio Central de Epidemiología (LCE) | Instituto de Biotecnología de la UNAM | Consorcio Mexicano de Vigilancia Genómica (CoViGen-Mex). Authors (in alphabetical order): Julio Elias Alvarado-Yaah, Carlos F. Arias, Santiago Ávila-Ríos, Eduardo Becerril-Vargas, Víctor Hugo Borja-Aburto, Celia Boukadida, Cristóbal Cháidez-Quiróz, Juan Bautista Chale-Dzul, Ricardo Ciria Merce, Andreu Comas-García, Célida Duque Molina, Julissa Enciso-Ibarra, José Antonio Enciso-Moreno, Gloria Elena Espinosa-Ayala, Fernando Fontove-Herrera, Daniel Fregoso-Rueda, Victor Eduardo García-Arias, Alejandra García-Gasca, Bruno Gómez-Gil, Jean Pierre González, Irvin González-López, Concepción Grajales-Muñiz, Ricardo Grande, Rosa María Gutiérrez Rios, Jesús Hernández, Alejandra Hernández-Terán, Alfredo Herrera-Estrella, Carla Ivón Herrera-Najera, Pavel Isa, Verónica Mata-Haro, Daniel Lira Morales, Susana Lopez, Antonio Loza Román, Brenda Irasema Maldonado-Meza, Bernardo Martínez-Miguel, José Arturo Martínez-Orozco, Célida Martínez- Rodríguez, Margarita Matías-Florentino, Fidencio Mejía-Nepomuceno, María Guadalupe de Jesús Mireles-Rivera, Gloria María Molina-Salinas, Hector Montoya-Fuentes, , Mario Mújica-Sánchez, José Esteban Muñoz-Medina, José de Jesús Nuñez-Contreras, Alicia Ocaña-Mondragón, Luis Alberto Ochoa-Carrera, Hector Esteban Paz-Juárez, Marissa Perez-Garcia, Francisco Pulido, Helen Haydee Fernanda Ramírez-Plascencia, Jorge Salas-Hernández, Angel Gustavo Salas-Lais, Alejandro Sánchez-Flores, Clara Esperanza Santacruz-Tinoco, María Guadalupe Santiago-Mauricio, Selene Zárate, Nelly Sélem-Mojica, Blanca Taboada, Gloria Vazquez, Joel Armando Vázquez-Pérez. |
| EPI_ISL_2681348 | Centro de Investigación Biomédica del Noreste (CIBIN) | Instituto de Biotecnología de la UNAM | Consorcio Mexicano de Vigilancia Genómica (CoViGen-Mex). Authors (in alphabetical order): Julio Elias Alvarado-Yaah, Carlos F. Arias, Santiago Ávila-Ríos, Eduardo Becerril-Vargas, Víctor Hugo Borja-Aburto, Celia Boukadida, Cristóbal Cháidez-Quiróz, Juan Bautista Chale-Dzul, Ricardo Ciria Merce, Andreu Comas-García, Célida Duque Molina, Julissa Enciso-Ibarra, José Antonio Enciso-Moreno, Gloria Elena Espinosa-Ayala, Fernando Fontove-Herrera, Daniel Fregoso-Rueda, Victor Eduardo García-Arias, Alejandra García-Gasca, Bruno Gómez-Gil, Jean Pierre González, Irvin González-López, Concepción Grajales-Muñiz, Ricardo Grande, Rosa María Gutiérrez Rios, Jesús Hernández, Alejandra Hernández-Terán, Alfredo Herrera-Estrella, Carla Ivón Herrera-Najera, Pavel Isa, Verónica Mata-Haro, Daniel Lira Morales, Susana Lopez, Antonio Loza Román, Brenda Irasema Maldonado-Meza, Bernardo Martínez-Miguel, José Arturo Martínez-Orozco, Célida Martínez- Rodríguez, Margarita Matías-Florentino, Fidencio Mejía-Nepomuceno, María Guadalupe de Jesús Mireles-Rivera, Gloria María Molina-Salinas, Hector Montoya-Fuentes, , Mario Mújica-Sánchez, José Esteban Muñoz-Medina, José de Jesús Nuñez-Contreras, Alicia Ocaña-Mondragón, Luis Alberto Ochoa-Carrera, Hector Esteban Paz-Juárez, Marissa Perez-Garcia, Francisco Pulido, Helen Haydee Fernanda Ramírez-Plascencia, Jorge Salas-Hernández, Angel Gustavo Salas-Lais, Alejandro Sánchez-Flores, Clara Esperanza Santacruz-Tinoco, María Guadalupe Santiago-Mauricio, Selene Zárate, Nelly Sélem-Mojica, Blanca Taboada, Gloria Vazquez, Joel Armando Vázquez-Pérez. |
| EPI_ISL_2681349 | Unidad de Investigación Biomédica de Zacatecas (UIBZ) | Instituto de Biotecnología de la UNAM | Consorcio Mexicano de Vigilancia Genómica (CoViGen-Mex). Authors (in alphabetical order): Julio Elias Alvarado-Yaah, Carlos F. Arias, Santiago Ávila-Ríos, Eduardo Becerril-Vargas, Víctor Hugo Borja-Aburto, Celia Boukadida, Cristóbal Cháidez-Quiróz, Juan Bautista Chale-Dzul, Ricardo Ciria Merce, Andreu Comas-García, Célida Duque Molina, Julissa Enciso-Ibarra, José Antonio Enciso-Moreno, Gloria Elena Espinosa-Ayala, Fernando Fontove-Herrera, Daniel Fregoso-Rueda, Victor Eduardo García-Arias, Alejandra García-Gasca, Bruno Gómez-Gil, Jean Pierre González, Irvin González-López, Concepción Grajales-Muñiz, Ricardo Grande, Rosa María Gutiérrez Rios, Jesús Hernández, Alejandra Hernández-Terán, Alfredo Herrera-Estrella, Carla Ivón Herrera-Najera, Pavel Isa, Verónica Mata-Haro, Daniel Lira Morales, Susana Lopez, Antonio Loza Román, Brenda Irasema Maldonado-Meza, Bernardo Martínez-Miguel, José Arturo Martínez-Orozco, Célida Martínez- Rodríguez, Margarita Matías-Florentino, Fidencio Mejía-Nepomuceno, María Guadalupe de Jesús Mireles-Rivera, Gloria María Molina-Salinas, Hector Montoya-Fuentes, , Mario Mújica-Sánchez, José Esteban Muñoz-Medina, José de Jesús Nuñez-Contreras, Alicia Ocaña-Mondragón, Luis Alberto Ochoa-Carrera, Hector Esteban Paz-Juárez, Marissa Perez-Garcia, Francisco Pulido, Helen Haydee Fernanda Ramírez-Plascencia, Jorge Salas-Hernández, Angel Gustavo Salas-Lais, Alejandro Sánchez-Flores, Clara Esperanza Santacruz-Tinoco, María Guadalupe Santiago-Mauricio, Selene Zárate, Nelly Sélem-Mojica, Blanca Taboada, Gloria Vazquez, Joel Armando Vázquez-Pérez. |
| EPI_ISL_2681350 | Laboratorio Central de Epidemiología (LCE) | Instituto de Biotecnología de la UNAM | Consorcio Mexicano de Vigilancia Genómica (CoViGen-Mex). Authors (in alphabetical order): Julio Elias Alvarado-Yaah, Carlos F. Arias, Santiago Ávila-Ríos, Eduardo Becerril-Vargas, Víctor Hugo Borja-Aburto, Celia Boukadida, Cristóbal Cháidez-Quiróz, Juan Bautista Chale-Dzul, Ricardo Ciria Merce, Andreu Comas-García, Célida Duque Molina, Julissa Enciso-Ibarra, José Antonio Enciso-Moreno, Gloria Elena Espinosa-Ayala, Fernando Fontove-Herrera, Daniel Fregoso-Rueda, Victor Eduardo García-Arias, Alejandra García-Gasca, Bruno Gómez-Gil, Jean Pierre González, Irvin González-López, Concepción Grajales-Muñiz, Ricardo Grande, Rosa María Gutiérrez Rios, Jesús Hernández, Alejandra Hernández-Terán, Alfredo Herrera-Estrella, Carla Ivón Herrera-Najera, Pavel Isa, Verónica Mata-Haro, Daniel Lira Morales, Susana Lopez, Antonio Loza Román, Brenda Irasema Maldonado-Meza, Bernardo Martínez-Miguel, José Arturo Martínez-Orozco, Célida Martínez- Rodríguez, Margarita Matías-Florentino, Fidencio Mejía-Nepomuceno, María Guadalupe de Jesús Mireles-Rivera, Gloria María Molina-Salinas, Hector Montoya-Fuentes, , Mario Mújica-Sánchez, José Esteban Muñoz-Medina, José de Jesús Nuñez-Contreras, Alicia Ocaña-Mondragón, Luis Alberto Ochoa-Carrera, Hector Esteban Paz-Juárez, Marissa Perez-Garcia, Francisco Pulido, Helen Haydee Fernanda Ramírez-Plascencia, Jorge Salas-Hernández, Angel Gustavo Salas-Lais, Alejandro Sánchez-Flores, Clara Esperanza Santacruz-Tinoco, María Guadalupe Santiago-Mauricio, Selene Zárate, Nelly Sélem-Mojica, Blanca Taboada, Gloria Vazquez, Joel Armando Vázquez-Pérez. |
| EPI_ISL_2681351 | Unidad de Investigación Médica de Yucatán (UIMY) | Instituto de Biotecnología de la UNAM | Consorcio Mexicano de Vigilancia Genómica (CoViGen-Mex). Authors (in alphabetical order): Julio Elias Alvarado-Yaah, Carlos F. Arias, Santiago Ávila-Ríos, Eduardo Becerril-Vargas, Víctor Hugo Borja-Aburto, Celia Boukadida, Cristóbal Cháidez-Quiróz, Juan Bautista Chale-Dzul, Ricardo Ciria Merce, Andreu Comas-García, Célida Duque Molina, Julissa Enciso-Ibarra, José Antonio Enciso-Moreno, Gloria Elena Espinosa-Ayala, Fernando Fontove-Herrera, Daniel Fregoso-Rueda, Victor Eduardo García-Arias, Alejandra García-Gasca, Bruno Gómez-Gil, Jean Pierre González, Irvin González-López, Concepción Grajales-Muñiz, Ricardo Grande, Rosa María Gutiérrez Rios, Jesús Hernández, Alejandra Hernández-Terán, Alfredo Herrera-Estrella, Carla Ivón Herrera-Najera, Pavel Isa, Verónica Mata-Haro, Daniel Lira Morales, Susana Lopez, Antonio Loza Román, Brenda Irasema Maldonado-Meza, Bernardo Martínez-Miguel, José Arturo Martínez-Orozco, Célida Martínez- Rodríguez, Margarita Matías-Florentino, Fidencio Mejía-Nepomuceno, María Guadalupe de Jesús Mireles-Rivera, Gloria María Molina-Salinas, Hector Montoya-Fuentes, , Mario Mújica-Sánchez, José Esteban Muñoz-Medina, José de Jesús Nuñez-Contreras, Alicia Ocaña-Mondragón, Luis Alberto Ochoa-Carrera, Hector Esteban Paz-Juárez, Marissa Perez-Garcia, Francisco Pulido, Helen Haydee Fernanda Ramírez-Plascencia, Jorge Salas-Hernández, Angel Gustavo Salas-Lais, Alejandro Sánchez-Flores, Clara Esperanza Santacruz-Tinoco, María Guadalupe Santiago-Mauricio, Selene Zárate, Nelly Sélem-Mojica, Blanca Taboada, Gloria Vazquez, Joel Armando Vázquez-Pérez. |
| EPI_ISL_2681352 | Laboratorio Central de Epidemiología (LCE) | Instituto de Biotecnología de la UNAM | Consorcio Mexicano de Vigilancia Genómica (CoViGen-Mex). Authors (in alphabetical order): Julio Elias Alvarado-Yaah, Carlos F. Arias, Santiago Ávila-Ríos, Eduardo Becerril-Vargas, Víctor Hugo Borja-Aburto, Celia Boukadida, Cristóbal Cháidez-Quiróz, Juan Bautista Chale-Dzul, Ricardo Ciria Merce, Andreu Comas-García, Célida Duque Molina, Julissa Enciso-Ibarra, José Antonio Enciso-Moreno, Gloria Elena Espinosa-Ayala, Fernando Fontove-Herrera, Daniel Fregoso-Rueda, Victor Eduardo García-Arias, Alejandra García-Gasca, Bruno Gómez-Gil, Jean Pierre González, Irvin González-López, Concepción Grajales-Muñiz, Ricardo Grande, Rosa María Gutiérrez Rios, Jesús Hernández, Alejandra Hernández-Terán, Alfredo Herrera-Estrella, Carla Ivón Herrera-Najera, Pavel Isa, Verónica Mata-Haro, Daniel Lira Morales, Susana Lopez, Antonio Loza Román, Brenda Irasema Maldonado-Meza, Bernardo Martínez-Miguel, José Arturo Martínez-Orozco, Célida Martínez- Rodríguez, Margarita Matías-Florentino, Fidencio Mejía-Nepomuceno, María Guadalupe de Jesús Mireles-Rivera, Gloria María Molina-Salinas, Hector Montoya-Fuentes, , Mario Mújica-Sánchez, José Esteban Muñoz-Medina, José de Jesús Nuñez-Contreras, Alicia Ocaña-Mondragón, Luis Alberto Ochoa-Carrera, Hector Esteban Paz-Juárez, Marissa Perez-Garcia, Francisco Pulido, Helen Haydee Fernanda Ramírez-Plascencia, Jorge Salas-Hernández, Angel Gustavo Salas-Lais, Alejandro Sánchez-Flores, Clara Esperanza Santacruz-Tinoco, María Guadalupe Santiago-Mauricio, Selene Zárate, Nelly Sélem-Mojica, Blanca Taboada, Gloria Vazquez, Joel Armando Vázquez-Pérez. |
| EPI_ISL_2681353 | Unidad de Investigación Médica de Yucatán (UIMY) | Instituto de Biotecnología de la UNAM | Consorcio Mexicano de Vigilancia Genómica (CoViGen-Mex). Authors (in alphabetical order): Julio Elias Alvarado-Yaah, Carlos F. Arias, Santiago Ávila-Ríos, Eduardo Becerril-Vargas, Víctor Hugo Borja-Aburto, Celia Boukadida, Cristóbal Cháidez-Quiróz, Juan Bautista Chale-Dzul, Ricardo Ciria Merce, Andreu Comas-García, Célida Duque Molina, Julissa Enciso-Ibarra, José Antonio Enciso-Moreno, Gloria Elena Espinosa-Ayala, Fernando Fontove-Herrera, Daniel Fregoso-Rueda, Victor Eduardo García-Arias, Alejandra García-Gasca, Bruno Gómez-Gil, Jean Pierre González, Irvin González-López, Concepción Grajales-Muñiz, Ricardo Grande, Rosa María Gutiérrez Rios, Jesús Hernández, Alejandra Hernández-Terán, Alfredo Herrera-Estrella, Carla Ivón Herrera-Najera, Pavel Isa, Verónica Mata-Haro, Daniel Lira Morales, Susana Lopez, Antonio Loza Román, Brenda Irasema Maldonado-Meza, Bernardo Martínez-Miguel, José Arturo Martínez-Orozco, Célida Martínez- Rodríguez, Margarita Matías-Florentino, Fidencio Mejía-Nepomuceno, María Guadalupe de Jesús Mireles-Rivera, Gloria María Molina-Salinas, Hector Montoya-Fuentes, , Mario Mújica-Sánchez, José Esteban Muñoz-Medina, José de Jesús Nuñez-Contreras, Alicia Ocaña-Mondragón, Luis Alberto Ochoa-Carrera, Hector Esteban Paz-Juárez, Marissa Perez-Garcia, Francisco Pulido, Helen Haydee Fernanda Ramírez-Plascencia, Jorge Salas-Hernández, Angel Gustavo Salas-Lais, Alejandro Sánchez-Flores, Clara Esperanza Santacruz-Tinoco, María Guadalupe Santiago-Mauricio, Selene Zárate, Nelly Sélem-Mojica, Blanca Taboada, Gloria Vazquez, Joel Armando Vázquez-Pérez. |
| EPI_ISL_2681354, EPI_ISL_2681355 | Unidad de Investigación Biomédica de Zacatecas (UIBZ) | Instituto de Biotecnología de la UNAM | Consorcio Mexicano de Vigilancia Genómica (CoViGen-Mex). Authors (in alphabetical order): Julio Elias Alvarado-Yaah, Carlos F. Arias, Santiago Ávila-Ríos, Eduardo Becerril-Vargas, Víctor Hugo Borja-Aburto, Celia Boukadida, Cristóbal Cháidez-Quiróz, Juan Bautista Chale-Dzul, Ricardo Ciria Merce, Andreu Comas-García, Célida Duque Molina, Julissa Enciso-Ibarra, José Antonio Enciso-Moreno, Gloria Elena Espinosa-Ayala, Fernando Fontove-Herrera, Daniel Fregoso-Rueda, Victor Eduardo García-Arias, Alejandra García-Gasca, Bruno Gómez-Gil, Jean Pierre González, Irvin |

| | | | |
|---|---|---|---|
| | | | González-López, Concepción Grajales-Muñiz, Ricardo Grande, Rosa María Gutiérrez Rios, Jesús Hernández, Alejandra Hernández-Terán, Alfredo Herrera-Estrella, Carla Ivón Herrera-Najera, Pavel Isa, Verónica Mata-Haro, Daniel Lira Morales, Susana Lopez, Antonio Loza Román, Brenda Irasema Maldonado-Meza, Bernardo Martínez-Miguel, José Arturo Martínez-Orozco, Célida Martínez- Rodríguez, Margarita Matías-Florentino, Fidencio Mejía-Nepomuceno, María Guadalupe de Jesús Mireles-Rivera, Gloria María Molina-Salinas, Hector Montoya-Fuentes, , Mario Mújica-Sánchez, José Esteban Muñoz-Medina, José de Jesús Nuñez-Contreras, Alicia Ocaña-Mondragón, Luis Alberto Ochoa-Carrera, Hector Esteban Paz-Juárez, Marissa Perez-Garcia, Francisco Pulido, Helen Haydee Fernanda Ramírez-Plascencia, Jorge Salas-Hernández, Angel Gustavo Salas-Lais, Alejandro Sánchez-Flores, Clara Esperanza Santacruz-Tinoco, María Guadalupe Santiago-Mauricio, Selene Zárate, Nelly Sélem-Mojica, Blanca Taboada, Gloria Vazquez, Joel Armando Vázquez-Pérez. |
| EPI_ISL_2681356 | Unidad de Investigación Médica de Yucatán (UIMY) | Instituto de Biotecnología de la UNAM | Consorcio Mexicano de Vigilancia Genómica (CoViGen-Mex). Authors (in alphabetical order): Julio Elias Alvarado-Yaah, Carlos F. Arias, Santiago Ávila-Ríos, Eduardo Becerril-Vargas, Víctor Hugo Borja-Aburto, Celia Boukadida, Cristóbal Cháidez-Quiróz, Juan Bautista Chale-Dzul, Ricardo Ciria Merce, Andreu Comas-García, Célida Duque Molina, Julissa Enciso-Ibarra, José Antonio Enciso-Moreno, Gloria Elena Espinosa-Ayala, Fernando Fontove-Herrera, Daniel Fregoso-Rueda, Victor Eduardo García-Arias, Alejandra García-Gasca, Bruno Gómez-Gil, Jean Pierre González, Irvin González-López, Concepción Grajales-Muñiz, Ricardo Grande, Rosa María Gutiérrez Rios, Jesús Hernández, Alejandra Hernández-Terán, Alfredo Herrera-Estrella, Carla Ivón Herrera-Najera, Pavel Isa, Verónica Mata-Haro, Daniel Lira Morales, Susana Lopez, Antonio Loza Román, Brenda Irasema Maldonado-Meza, Bernardo Martínez-Miguel, José Arturo Martínez-Orozco, Célida Martínez- Rodríguez, Margarita Matías-Florentino, Fidencio Mejía-Nepomuceno, María Guadalupe de Jesús Mireles-Rivera, Gloria María Molina-Salinas, Hector Montoya-Fuentes, , Mario Mújica-Sánchez, José Esteban Muñoz-Medina, José de Jesús Nuñez-Contreras, Alicia Ocaña-Mondragón, Luis Alberto Ochoa-Carrera, Hector Esteban Paz-Juárez, Marissa Perez-Garcia, Francisco Pulido, Helen Haydee Fernanda Ramírez-Plascencia, Jorge Salas-Hernández, Angel Gustavo Salas-Lais, Alejandro Sánchez-Flores, Clara Esperanza Santacruz-Tinoco, María Guadalupe Santiago-Mauricio, Selene Zárate, Nelly Sélem-Mojica, Blanca Taboada, Gloria Vazquez, Joel Armando Vázquez-Pérez. |
| EPI_ISL_2681357, EPI_ISL_2681358, EPI_ISL_2681359 | Centro de Investigación Biomédica del Noreste (CIBIN) | Instituto de Biotecnología de la UNAM | Consorcio Mexicano de Vigilancia Genómica (CoViGen-Mex). Authors (in alphabetical order): Julio Elias Alvarado-Yaah, Carlos F. Arias, Santiago Ávila-Ríos, Eduardo Becerril-Vargas, Víctor Hugo Borja-Aburto, Celia Boukadida, Cristóbal Cháidez-Quiróz, Juan Bautista Chale-Dzul, Ricardo Ciria Merce, Andreu Comas-García, Célida Duque Molina, Julissa Enciso-Ibarra, José Antonio Enciso-Moreno, Gloria Elena Espinosa-Ayala, Fernando Fontove-Herrera, Daniel Fregoso-Rueda, Victor Eduardo García-Arias, Alejandra García-Gasca, Bruno Gómez-Gil, Jean Pierre González, Irvin González-López, Concepción Grajales-Muñiz, Ricardo Grande, Rosa María Gutiérrez Rios, Jesús Hernández, Alejandra Hernández-Terán, Alfredo Herrera-Estrella, Carla Ivón Herrera-Najera, Pavel Isa, Verónica Mata-Haro, Daniel Lira Morales, Susana Lopez, Antonio Loza Román, Brenda Irasema Maldonado-Meza, Bernardo Martínez-Miguel, José Arturo Martínez-Orozco, Célida Martínez- Rodríguez, Margarita Matías-Florentino, Fidencio Mejía-Nepomuceno, María Guadalupe de Jesús Mireles-Rivera, Gloria María Molina-Salinas, Hector Montoya-Fuentes, , Mario Mújica-Sánchez, José Esteban Muñoz-Medina, José de Jesús Nuñez-Contreras, Alicia Ocaña-Mondragón, Luis Alberto Ochoa-Carrera, Hector Esteban Paz-Juárez, Marissa Perez-Garcia, Francisco Pulido, Helen Haydee Fernanda Ramírez-Plascencia, Jorge Salas-Hernández, Angel Gustavo Salas-Lais, Alejandro Sánchez-Flores, Clara Esperanza Santacruz-Tinoco, María Guadalupe Santiago-Mauricio, Selene Zárate, Nelly Sélem-Mojica, Blanca Taboada, Gloria Vazquez, Joel Armando Vázquez-Pérez. |
| EPI_ISL_2681360, EPI_ISL_2681361 | Unidad de Investigación Médica de Yucatán (UIMY) | Instituto de Biotecnología de la UNAM | Consorcio Mexicano de Vigilancia Genómica (CoViGen-Mex). Authors (in alphabetical order): Julio Elias Alvarado-Yaah, Carlos F. Arias, Santiago Ávila-Ríos, Eduardo Becerril-Vargas, Víctor Hugo Borja-Aburto, Celia Boukadida, Cristóbal Cháidez-Quiróz, Juan Bautista Chale-Dzul, Ricardo Ciria Merce, Andreu Comas-García, Célida Duque Molina, Julissa Enciso-Ibarra, José Antonio Enciso-Moreno, Gloria Elena Espinosa-Ayala, Fernando Fontove-Herrera, Daniel Fregoso-Rueda, Victor Eduardo García-Arias, Alejandra García-Gasca, Bruno Gómez-Gil, Jean Pierre González, Irvin González-López, Concepción Grajales-Muñiz, Ricardo Grande, Rosa María Gutiérrez Rios, Jesús Hernández, Alejandra Hernández-Terán, Alfredo Herrera-Estrella, Carla Ivón Herrera-Najera, Pavel Isa, Verónica Mata-Haro, Daniel Lira Morales, Susana Lopez, Antonio Loza Román, Brenda Irasema Maldonado-Meza, Bernardo Martínez-Miguel, José Arturo Martínez-Orozco, Célida Martínez- Rodríguez, Margarita Matías-Florentino, Fidencio Mejía-Nepomuceno, María Guadalupe de Jesús Mireles-Rivera, Gloria María Molina-Salinas, Hector Montoya-Fuentes, , Mario Mújica-Sánchez, José Esteban Muñoz-Medina, José de Jesús Nuñez-Contreras, Alicia Ocaña-Mondragón, Luis Alberto Ochoa-Carrera, Hector Esteban Paz-Juárez, Marissa Perez-Garcia, Francisco Pulido, Helen Haydee Fernanda Ramírez-Plascencia, Jorge Salas-Hernández, Angel Gustavo Salas-Lais, Alejandro Sánchez-Flores, Clara Esperanza Santacruz-Tinoco, María Guadalupe Santiago-Mauricio, Selene Zárate, Nelly Sélem-Mojica, Blanca Taboada, Gloria Vazquez, Joel Armando Vázquez-Pérez. |
| EPI_ISL_2681362 | Unidad de Investigación Biomédica de Zacatecas (UIBZ) | Instituto de Biotecnología de la UNAM | Consorcio Mexicano de Vigilancia Genómica (CoViGen-Mex). Authors (in alphabetical order): Julio Elias Alvarado-Yaah, Carlos F. Arias, Santiago Ávila-Ríos, Eduardo Becerril-Vargas, Víctor Hugo Borja-Aburto, Celia Boukadida, Cristóbal Cháidez-Quiróz, Juan Bautista Chale-Dzul, Ricardo Ciria Merce, Andreu Comas-García, Célida Duque Molina, Julissa Enciso-Ibarra, José Antonio Enciso-Moreno, Gloria Elena Espinosa-Ayala, Fernando Fontove-Herrera, Daniel Fregoso-Rueda, Victor Eduardo García-Arias, Alejandra García-Gasca, Bruno Gómez-Gil, Jean Pierre González, Irvin González-López, Concepción Grajales-Muñiz, Ricardo Grande, Rosa María Gutiérrez Rios, Jesús Hernández, Alejandra Hernández-Terán, Alfredo Herrera-Estrella, Carla Ivón Herrera-Najera, Pavel Isa, Verónica Mata-Haro, Daniel Lira Morales, Susana Lopez, Antonio Loza Román, Brenda Irasema Maldonado-Meza, Bernardo Martínez-Miguel, José Arturo Martínez-Orozco, Célida Martínez- Rodríguez, Margarita Matías-Florentino, Fidencio Mejía-Nepomuceno, María Guadalupe de Jesús Mireles-Rivera, Gloria María Molina-Salinas, Hector Montoya-Fuentes, , Mario Mújica-Sánchez, José Esteban Muñoz-Medina, José de Jesús Nuñez-Contreras, Alicia Ocaña-Mondragón, Luis Alberto Ochoa-Carrera, Hector Esteban Paz-Juárez, Marissa Perez-Garcia, Francisco Pulido, Helen Haydee Fernanda Ramírez-Plascencia, Jorge Salas-Hernández, Angel Gustavo Salas-Lais, Alejandro Sánchez-Flores, Clara Esperanza Santacruz-Tinoco, María Guadalupe Santiago-Mauricio, Selene Zárate, Nelly Sélem-Mojica, Blanca Taboada, Gloria Vazquez, Joel Armando Vázquez-Pérez. |
| EPI_ISL_2681363, EPI_ISL_2681364, EPI_ISL_2681365, EPI_ISL_2681366 | Unidad de Investigación Médica de Yucatán (UIMY) | Instituto de Biotecnología de la UNAM | Consorcio Mexicano de Vigilancia Genómica (CoViGen-Mex). Authors (in alphabetical order): Julio Elias Alvarado-Yaah, Carlos F. Arias, Santiago Ávila-Ríos, Eduardo Becerril-Vargas, Víctor Hugo Borja-Aburto, Celia Boukadida, Cristóbal Cháidez-Quiróz, Juan Bautista Chale-Dzul, Ricardo Ciria Merce, Andreu Comas-García, Célida Duque Molina, Julissa Enciso-Ibarra, José Antonio Enciso-Moreno, Gloria Elena Espinosa-Ayala, Fernando Fontove-Herrera, Daniel Fregoso-Rueda, Victor Eduardo García-Arias, Alejandra García-Gasca, Bruno Gómez-Gil, Jean Pierre González, Irvin González-López, Concepción Grajales-Muñiz, Ricardo Grande, Rosa María Gutiérrez Rios, Jesús Hernández, Alejandra Hernández-Terán, Alfredo Herrera-Estrella, Carla Ivón Herrera-Najera, Pavel Isa, Verónica Mata-Haro, Daniel Lira Morales, Susana Lopez, Antonio Loza Román, Brenda Irasema Maldonado-Meza, Bernardo Martínez-Miguel, José Arturo Martínez-Orozco, Célida Martínez- Rodríguez, Margarita Matías-Florentino, Fidencio Mejía-Nepomuceno, María Guadalupe de Jesús Mireles-Rivera, Gloria María Molina-Salinas, Hector Montoya-Fuentes, , Mario Mújica-Sánchez, José Esteban Muñoz-Medina, José de Jesús Nuñez-Contreras, Alicia Ocaña-Mondragón, Luis Alberto Ochoa-Carrera, Hector Esteban Paz-Juárez, Marissa Perez-Garcia, Francisco Pulido, Helen Haydee Fernanda Ramírez-Plascencia, Jorge Salas-Hernández, Angel Gustavo Salas-Lais, Alejandro Sánchez-Flores, Clara Esperanza Santacruz-Tinoco, María Guadalupe Santiago-Mauricio, Selene Zárate, Nelly Sélem-Mojica, Blanca Taboada, Gloria Vazquez, Joel Armando Vázquez-Pérez. |
| EPI_ISL_2681367 | Unidad de Investigación Biomédica de Zacatecas (UIBZ) | Instituto de Biotecnología de la UNAM | Consorcio Mexicano de Vigilancia Genómica (CoViGen-Mex). Authors (in alphabetical order): Julio Elias Alvarado-Yaah, Carlos F. Arias, Santiago Ávila-Ríos, Eduardo Becerril-Vargas, Víctor Hugo Borja-Aburto, Celia Boukadida, Cristóbal Cháidez-Quiróz, Juan Bautista Chale-Dzul, Ricardo Ciria Merce, Andreu Comas-García, Célida Duque Molina, Julissa Enciso-Ibarra, José Antonio Enciso-Moreno, Gloria Elena Espinosa-Ayala, Fernando Fontove-Herrera, Daniel Fregoso-Rueda, Victor Eduardo García-Arias, Alejandra García-Gasca, Bruno Gómez-Gil, Jean Pierre González, Irvin González-López, Concepción Grajales-Muñiz, Ricardo Grande, Rosa María Gutiérrez Rios, Jesús Hernández, Alejandra Hernández-Terán, Alfredo Herrera-Estrella, Carla Ivón Herrera-Najera, Pavel Isa, Verónica Mata-Haro, Daniel Lira Morales, Susana Lopez, Antonio Loza Román, Brenda Irasema Maldonado-Meza, Bernardo Martínez-Miguel, José Arturo Martínez-Orozco, Célida Martínez- Rodríguez, Margarita Matías-Florentino, Fidencio Mejía-Nepomuceno, María Guadalupe de Jesús Mireles-Rivera, Gloria María Molina-Salinas, Hector Montoya-Fuentes, , Mario Mújica-Sánchez, José Esteban Muñoz-Medina, José de Jesús Nuñez-Contreras, Alicia Ocaña-Mondragón, Luis Alberto Ochoa-Carrera, Hector Esteban Paz-Juárez, Marissa Perez-Garcia, Francisco Pulido, Helen Haydee Fernanda Ramírez-Plascencia, Jorge Salas-Hernández, Angel Gustavo Salas-Lais, Alejandro Sánchez-Flores, Clara Esperanza Santacruz-Tinoco, María Guadalupe Santiago-Mauricio, Selene Zárate, Nelly Sélem-Mojica, Blanca Taboada, Gloria Vazquez, Joel Armando Vázquez-Pérez. |
| EPI_ISL_2681368 | Unidad de Investigación Médica de Yucatán (UIMY) | Instituto de Biotecnología de la UNAM | Consorcio Mexicano de Vigilancia Genómica (CoViGen-Mex). Authors (in alphabetical order): Julio Elias Alvarado-Yaah, Carlos F. Arias, Santiago Ávila-Ríos, Eduardo Becerril-Vargas, Víctor Hugo Borja-Aburto, Celia Boukadida, Cristóbal Cháidez-Quiróz, Juan Bautista Chale-Dzul, Ricardo Ciria Merce, Andreu Comas-García, Célida Duque Molina, Julissa Enciso-Ibarra, José Antonio Enciso-Moreno, Gloria Elena Espinosa-Ayala, Fernando Fontove-Herrera, Daniel Fregoso-Rueda, Victor Eduardo García-Arias, Alejandra García-Gasca, Bruno Gómez-Gil, Jean Pierre González, Irvin González-López, Concepción Grajales-Muñiz, Ricardo Grande, Rosa María Gutiérrez Rios, Jesús Hernández, Alejandra Hernández-Terán, Alfredo Herrera-Estrella, Carla Ivón Herrera-Najera, Pavel Isa, Verónica Mata-Haro, Daniel Lira Morales, Susana Lopez, Antonio Loza Román, Brenda Irasema Maldonado-Meza, Bernardo Martínez-Miguel, José Arturo Martínez-Orozco, Célida Martínez- Rodríguez, Margarita Matías-Florentino, Fidencio Mejía-Nepomuceno, María Guadalupe de Jesús Mireles-Rivera, Gloria María Molina-Salinas, Hector Montoya-Fuentes, , Mario Mújica-Sánchez, José Esteban Muñoz-Medina, José de Jesús Nuñez-Contreras, Alicia Ocaña-Mondragón, Luis Alberto Ochoa-Carrera, Hector Esteban Paz-Juárez, Marissa Perez-Garcia, Francisco Pulido, Helen Haydee Fernanda Ramírez-Plascencia, Jorge Salas-Hernández, Angel Gustavo Salas-Lais, Alejandro Sánchez-Flores, Clara Esperanza Santacruz-Tinoco, María Guadalupe Santiago-Mauricio, Selene Zárate, Nelly Sélem-Mojica, Blanca Taboada, Gloria Vazquez, Joel Armando Vázquez-Pérez. |
| EPI_ISL_2681369 | Laboratorio Central de Epidemiología (LCE) | Instituto de Biotecnología de la UNAM | Consorcio Mexicano de Vigilancia Genómica (CoViGen-Mex). Authors (in alphabetical order): Julio Elias Alvarado-Yaah, Carlos F. Arias, Santiago Ávila-Ríos, Eduardo Becerril-Vargas, Víctor Hugo Borja-Aburto, Celia Boukadida, Cristóbal Cháidez-Quiróz, Juan Bautista Chale-Dzul, Ricardo Ciria Merce, Andreu Comas-García, Célida Duque Molina, Julissa Enciso-Ibarra, José Antonio Enciso-Moreno, Gloria Elena Espinosa-Ayala, Fernando Fontove-Herrera, Daniel Fregoso-Rueda, Victor Eduardo García-Arias, Alejandra García-Gasca, Bruno Gómez-Gil, Jean Pierre González, Irvin González-López, Concepción Grajales-Muñiz, Ricardo Grande, Rosa María Gutiérrez Rios, Jesús Hernández, Alejandra Hernández-Terán, Alfredo Herrera-Estrella, Carla Ivón Herrera-Najera, Pavel Isa, Verónica Mata-Haro, Daniel Lira Morales, Susana Lopez, Antonio Loza Román, Brenda Irasema Maldonado-Meza, Bernardo Martínez-Miguel, José Arturo Martínez-Orozco, Célida Martínez- Rodríguez, Margarita Matías-Florentino, Fidencio Mejía-Nepomuceno, María Guadalupe de Jesús Mireles-Rivera, Gloria María Molina-Salinas, Hector Montoya-Fuentes, , Mario Mújica-Sánchez, José Esteban Muñoz-Medina, José de Jesús Nuñez-Contreras, Alicia Ocaña-Mondragón, Luis Alberto Ochoa-Carrera, Hector Esteban Paz-Juárez, Marissa Perez-Garcia, Francisco Pulido, Helen Haydee Fernanda Ramírez-Plascencia, Jorge Salas-Hernández, Angel Gustavo Salas-Lais, Alejandro Sánchez-Flores, Clara Esperanza Santacruz-Tinoco, María Guadalupe Santiago-Mauricio, Selene Zárate, Nelly Sélem-Mojica, Blanca Taboada, Gloria Vazquez, Joel Armando Vázquez-Pérez. |
| EPI_ISL_2681370 | Unidad de Investigación Médica de Yucatán (UIMY) | Instituto de Biotecnología de la UNAM | Consorcio Mexicano de Vigilancia Genómica (CoViGen-Mex). Authors (in alphabetical order): Julio Elias Alvarado-Yaah, Carlos F. Arias, Santiago Ávila-Ríos, Eduardo Becerril-Vargas, Víctor Hugo Borja-Aburto, Celia Boukadida, Cristóbal Cháidez-Quiróz, Juan Bautista Chale-Dzul, Ricardo Ciria Merce, Andreu Comas-García, Célida Duque Molina, Julissa Enciso-Ibarra, José Antonio Enciso-Moreno, Gloria Elena Espinosa-Ayala, Fernando Fontove-Herrera, Daniel Fregoso-Rueda, Victor Eduardo García-Arias, Alejandra García-Gasca, Bruno Gómez-Gil, Jean Pierre González, Irvin González-López, Concepción Grajales-Muñiz, Ricardo Grande, Rosa María Gutiérrez Rios, Jesús Hernández, Alejandra Hernández-Terán, Alfredo Herrera-Estrella, Carla Ivón Herrera-Najera, Pavel Isa, Verónica Mata-Haro, Daniel Lira Morales, Susana Lopez, Antonio Loza Román, Brenda Irasema Maldonado-Meza, Bernardo Martínez-Miguel, José Arturo Martínez-Orozco, Célida Martínez- Rodríguez, Margarita Matías-Florentino, Fidencio Mejía-Nepomuceno, María Guadalupe de Jesús Mireles-Rivera, Gloria María Molina-Salinas, Hector Montoya-Fuentes, , Mario Mújica-Sánchez, José Esteban Muñoz-Medina, José de Jesús Nuñez-Contreras, Alicia Ocaña-Mondragón, Luis Alberto Ochoa-Carrera, Hector Esteban Paz-Juárez, Marissa Perez-Garcia, Francisco Pulido, Helen Haydee Fernanda Ramírez-Plascencia, Jorge Salas-Hernández, Angel Gustavo Salas-Lais, Alejandro Sánchez-Flores, Clara Esperanza Santacruz-Tinoco, María Guadalupe Santiago-Mauricio, Selene Zárate, Nelly Sélem-Mojica, Blanca Taboada, Gloria Vazquez, Joel Armando Vázquez-Pérez. |
| EPI_ISL_2681371 | Laboratorio Central de Epidemiología (LCE) | Instituto de Biotecnología de la UNAM | Consorcio Mexicano de Vigilancia Genómica (CoViGen-Mex). Authors (in alphabetical order): Julio Elias Alvarado-Yaah, Carlos F. Arias, Santiago Ávila-Ríos, Eduardo Becerril-Vargas, Víctor Hugo Borja-Aburto, Celia Boukadida, Cristóbal Cháidez-Quiróz, Juan Bautista Chale-Dzul, Ricardo Ciria Merce, Andreu Comas-García, Célida Duque Molina, Julissa Enciso-Ibarra, José Antonio Enciso-Moreno, Gloria Elena Espinosa-Ayala, Fernando Fontove-Herrera, Daniel Fregoso-Rueda, Victor Eduardo García-Arias, Alejandra García-Gasca, Bruno Gómez-Gil, Jean Pierre González, Irvin González-López, Concepción Grajales-Muñiz, Ricardo Grande, Rosa María Gutiérrez Rios, Jesús Hernández, Alejandra Hernández-Terán, Alfredo Herrera-Estrella, Carla Ivón Herrera-Najera, Pavel Isa, Verónica Mata-Haro, Daniel Lira Morales, Susana Lopez, Antonio Loza Román, Brenda Irasema Maldonado-Meza, Bernardo Martínez-Miguel, José Arturo Martínez-Orozco, Célida Martínez- Rodríguez, Margarita Matías-Florentino, Fidencio Mejía-Nepomuceno, María Guadalupe de Jesús Mireles-Rivera, Gloria María Molina-Salinas, Hector Montoya-Fuentes, , Mario Mújica-Sánchez, José Esteban Muñoz-Medina, José de Jesús Nuñez-Contreras, Alicia Ocaña-Mondragón, Luis Alberto Ochoa-Carrera, Hector Esteban Paz-Juárez, Marissa Perez-Garcia, Francisco Pulido, Helen Haydee Fernanda Ramírez-Plascencia, Jorge Salas-Hernández, Angel Gustavo Salas-Lais, Alejandro Sánchez-Flores, Clara Esperanza Santacruz-Tinoco, María Guadalupe Santiago-Mauricio, Selene Zárate, Nelly Sélem-Mojica, Blanca Taboada, Gloria Vazquez, Joel Armando Vázquez-Pérez. |
| EPI_ISL_2681372 | Unidad de Investigación Médica de Yucatán (UIMY) | Instituto de Biotecnología de la UNAM | Consorcio Mexicano de Vigilancia Genómica (CoViGen-Mex). Authors (in alphabetical order): Julio Elias Alvarado-Yaah, Carlos F. Arias, Santiago Ávila-Ríos, Eduardo Becerril-Vargas, Víctor Hugo Borja-Aburto, Celia Boukadida, Cristóbal Cháidez-Quiróz, Juan Bautista Chale-Dzul, Ricardo Ciria Merce, Andreu Comas-García, Célida Duque Molina, Julissa Enciso-Ibarra, José Antonio Enciso-Moreno, Gloria Elena Espinosa-Ayala, Fernando Fontove-Herrera, Daniel Fregoso-Rueda, Victor Eduardo García-Arias, Alejandra García-Gasca, Bruno Gómez-Gil, Jean Pierre González, Irvin González-López, Concepción Grajales-Muñiz, Ricardo Grande, Rosa María Gutiérrez Rios, Jesús Hernández, Alejandra Hernández-Terán, Alfredo Herrera-Estrella, Carla Ivón Herrera-Najera, Pavel Isa, Verónica Mata-Haro, Daniel Lira Morales, Susana Lopez, Antonio Loza Román, Brenda Irasema Maldonado-Meza, Bernardo Martínez-Miguel, José Arturo Martínez-Orozco, Célida Martínez- Rodríguez, Margarita Matías-Florentino, Fidencio Mejía-Nepomuceno, María Guadalupe de Jesús Mireles-Rivera, Gloria María Molina-Salinas, Hector Montoya-Fuentes, , Mario Mújica-Sánchez, José Esteban Muñoz-Medina, José de Jesús Nuñez-Contreras, Alicia Ocaña-Mondragón, Luis Alberto Ochoa-Carrera, Hector Esteban Paz-Juárez, Marissa Perez-Garcia, Francisco Pulido, Helen Haydee Fernanda Ramírez-Plascencia, Jorge Salas-Hernández, Angel Gustavo Salas-Lais, Alejandro Sánchez-Flores, Clara Esperanza Santacruz-Tinoco, María Guadalupe Santiago-Mauricio, Selene Zárate, Nelly Sélem-Mojica, Blanca Taboada, Gloria Vazquez, Joel Armando Vázquez-Pérez. |
| EPI_ISL_2681373 | Laboratorio Central de Epidemiología (LCE) | Instituto de Biotecnología de la UNAM | Consorcio Mexicano de Vigilancia Genómica (CoViGen-Mex). Authors (in alphabetical order): Julio Elias Alvarado-Yaah, Carlos F. Arias, Santiago Ávila-Ríos, Eduardo Becerril-Vargas, Víctor Hugo Borja-Aburto, Celia Boukadida, Cristóbal Cháidez-Quiróz, Juan Bautista Chale-Dzul, Ricardo Ciria Merce, Andreu Comas-García, Célida Duque Molina, Julissa Enciso-Ibarra, José Antonio Enciso-Moreno, Gloria Elena Espinosa-Ayala, Fernando Fontove-Herrera, Daniel Fregoso-Rueda, Victor Eduardo García-Arias, Alejandra García-Gasca, Bruno Gómez-Gil, Jean Pierre González, Irvin González-López, Concepción Grajales-Muñiz, Ricardo Grande, Rosa María Gutiérrez Rios, Jesús Hernández, Alejandra Hernández-Terán, Alfredo Herrera-Estrella, Carla Ivón Herrera-Najera, Pavel Isa, Verónica Mata-Haro, Daniel Lira Morales, Susana Lopez, Antonio Loza Román, Brenda Irasema Maldonado-Meza, Bernardo Martínez-Miguel, José Arturo Martínez-Orozco, Célida Martínez- Rodríguez, Margarita Matías-Florentino, Fidencio Mejía-Nepomuceno, María Guadalupe de Jesús Mireles-Rivera, Gloria María Molina-Salinas, Hector Montoya-Fuentes, , Mario Mújica-Sánchez, José Esteban Muñoz-Medina, José de Jesús Nuñez-Contreras, Alicia Ocaña-Mondragón, Luis Alberto Ochoa-Carrera, Hector Esteban Paz-Juárez, Marissa Perez-Garcia, Francisco Pulido, Helen Haydee Fernanda Ramírez-Plascencia, Jorge Salas-Hernández, Angel Gustavo Salas-Lais, Alejandro Sánchez-Flores, Clara Esperanza Santacruz-Tinoco, María Guadalupe Santiago-Mauricio, Selene Zárate, Nelly Sélem-Mojica, Blanca Taboada, Gloria Vazquez, Joel Armando Vázquez-Pérez. |
| EPI_ISL_2681374, EPI_ISL_2681375 | Unidad de Investigación Médica de Yucatán (UIMY) | Instituto de Biotecnología de la UNAM | Consorcio Mexicano de Vigilancia Genómica (CoViGen-Mex). Authors (in alphabetical order): Julio Elias Alvarado-Yaah, Carlos F. Arias, Santiago Ávila-Ríos, Eduardo Becerril-Vargas, Víctor Hugo Borja-Aburto, Celia Boukadida, Cristóbal Cháidez-Quiróz, Juan Bautista Chale-Dzul, Ricardo Ciria Merce, Andreu Comas-García, Célida Duque Molina, Julissa Enciso-Ibarra, José Antonio Enciso-Moreno, Gloria Elena Espinosa-Ayala, Fernando Fontove-Herrera, Daniel Fregoso-Rueda, Victor Eduardo García-Arias, Alejandra García-Gasca, Bruno Gómez-Gil, Jean Pierre González, Irvin González-López, Concepción Grajales-Muñiz, Ricardo Grande, Rosa María Gutiérrez Rios, Jesús Hernández, Alejandra Hernández-Terán, Alfredo Herrera-Estrella, Carla Ivón Herrera-Najera, Pavel Isa, Verónica Mata-Haro, Daniel Lira Morales, Susana Lopez, Antonio Loza Román, Brenda Irasema Maldonado-Meza, Bernardo Martínez-Miguel, José Arturo Martínez-Orozco, Célida Martínez- Rodríguez, Margarita Matías-Florentino, Fidencio Mejía-Nepomuceno, María Guadalupe de Jesús Mireles-Rivera, Gloria María Molina-Salinas, Hector Montoya-Fuentes, , Mario Mújica-Sánchez, José Esteban Muñoz-Medina, José de Jesús Nuñez-Contreras, Alicia Ocaña-Mondragón, Luis Alberto Ochoa-Carrera, Hector Esteban Paz-Juárez, Marissa Perez-Garcia, Francisco Pulido, Helen Haydee Fernanda Ramírez-Plascencia, Jorge Salas-Hernández, Angel Gustavo Salas-Lais, Alejandro Sánchez-Flores, Clara Esperanza Santacruz-Tinoco, María Guadalupe Santiago-Mauricio, Selene Zárate, Nelly Sélem-Mojica, Blanca Taboada, Gloria Vazquez, Joel Armando Vázquez-Pérez. |
| EPI_ISL_2681376 | Unidad de Investigación Biomédica de Zacatecas (UIBZ) | Instituto de Biotecnología de la UNAM | Consorcio Mexicano de Vigilancia Genómica (CoViGen-Mex). Authors (in alphabetical order): Julio Elias Alvarado-Yaah, Carlos F. Arias, Santiago Ávila-Ríos, Eduardo Becerril-Vargas, Víctor Hugo Borja-Aburto, Celia Boukadida, Cristóbal Cháidez-Quiróz, Juan Bautista Chale-Dzul, Ricardo Ciria Merce, Andreu Comas-García, Célida Duque Molina, Julissa Enciso-Ibarra, José Antonio Enciso-Moreno, Gloria Elena Espinosa-Ayala, Fernando Fontove-Herrera, Daniel Fregoso-Rueda, Victor Eduardo García-Arias, Alejandra García-Gasca, Bruno Gómez-Gil, Jean Pierre González, Irvin González-López, Concepción Grajales-Muñiz, Ricardo Grande, Rosa María Gutiérrez Rios, Jesús Hernández, Alejandra Hernández-Terán, Alfredo Herrera-Estrella, Carla Ivón Herrera-Najera, Pavel Isa, Verónica Mata-Haro, Daniel Lira Morales, Susana Lopez, Antonio Loza Román, Brenda Irasema Maldonado-Meza, Bernardo Martínez-Miguel, José Arturo Martínez-Orozco, Célida Martínez- Rodríguez, Margarita Matías-Florentino, Fidencio Mejía-Nepomuceno, María Guadalupe de Jesús Mireles-Rivera, Gloria María Molina-Salinas, Hector Montoya-Fuentes, , Mario Mújica-Sánchez, José Esteban Muñoz-Medina, José de Jesús Nuñez-Contreras, Alicia Ocaña-Mondragón, Luis Alberto Ochoa-Carrera, Hector Esteban Paz-Juárez, Marissa Perez-Garcia, Francisco Pulido, Helen Haydee Fernanda Ramírez-Plascencia, Jorge Salas-Hernández, Angel Gustavo Salas-Lais, Alejandro Sánchez-Flores, Clara Esperanza Santacruz-Tinoco, María Guadalupe Santiago-Mauricio, Selene Zárate, Nelly Sélem-Mojica, Blanca Taboada, Gloria Vazquez, Joel Armando Vázquez-Pérez. |
| EPI_ISL_2681377, EPI_ISL_2681378, EPI_ISL_2681379 | Unidad de Investigación Médica de Yucatán (UIMY) | Instituto de Biotecnología de la UNAM | Consorcio Mexicano de Vigilancia Genómica (CoViGen-Mex). Authors (in alphabetical order): Julio Elias Alvarado-Yaah, Carlos F. Arias, Santiago Ávila-Ríos, Eduardo Becerril-Vargas, Víctor Hugo Borja-Aburto, Celia Boukadida, Cristóbal Cháidez-Quiróz, Juan Bautista Chale-Dzul, Ricardo Ciria Merce, Andreu Comas-García, Célida Duque Molina, Julissa Enciso-Ibarra, José Antonio Enciso-Moreno, Gloria Elena Espinosa-Ayala, Fernando Fontove-Herrera, Daniel Fregoso-Rueda, Victor Eduardo García-Arias, Alejandra García-Gasca, Bruno Gómez-Gil, Jean Pierre González, Irvin González-López, Concepción Grajales-Muñiz, Ricardo Grande, Rosa María Gutiérrez Rios, Jesús Hernández, Alejandra Hernández-Terán, Alfredo Herrera-Estrella, Carla Ivón Herrera-Najera, Pavel Isa, Verónica Mata-Haro, Daniel Lira Morales, Susana Lopez, Antonio Loza Román, Brenda Irasema Maldonado-Meza, Bernardo Martínez-Miguel, José Arturo Martínez-Orozco, Célida Martínez- Rodríguez, Margarita Matías-Florentino, Fidencio Mejía-Nepomuceno, María Guadalupe de Jesús Mireles-Rivera, Gloria María Molina-Salinas, Hector Montoya-Fuentes, , Mario Mújica-Sánchez, José Esteban Muñoz-Medina, José de Jesús Nuñez-Contreras, Alicia Ocaña-Mondragón, Luis Alberto Ochoa-Carrera, Hector Esteban Paz-Juárez, Marissa Perez-Garcia, Francisco Pulido, Helen Haydee Fernanda Ramírez-Plascencia, Jorge Salas-Hernández, Angel Gustavo Salas-Lais, Alejandro Sánchez-Flores, Clara Esperanza Santacruz-Tinoco, María Guadalupe Santiago-Mauricio, Selene Zárate, Nelly Sélem-Mojica, Blanca Taboada, Gloria Vazquez, Joel Armando Vázquez-Pérez. |
| EPI_ISL_2681380 | Centro de Investigación Biomédica del Noreste (CIBIN) | Instituto de Biotecnología de la UNAM | Consorcio Mexicano de Vigilancia Genómica (CoViGen-Mex). Authors (in alphabetical order): Julio Elias Alvarado-Yaah, Carlos F. Arias, Santiago Ávila-Ríos, Eduardo Becerril-Vargas, Víctor Hugo Borja-Aburto, Celia Boukadida, Cristóbal Cháidez-Quiróz, Juan Bautista Chale-Dzul, Ricardo Ciria Merce, Andreu Comas-García, Célida Duque Molina, Julissa Enciso-Ibarra, José Antonio Enciso-Moreno, Gloria Elena Espinosa-Ayala, Fernando Fontove-Herrera, Daniel Fregoso-Rueda, Victor Eduardo García-Arias, Alejandra García-Gasca, Bruno Gómez-Gil, Jean Pierre González, Irvin González-López, Concepción Grajales-Muñiz, Ricardo Grande, Rosa María Gutiérrez Rios, Jesús Hernández, Alejandra Hernández-Terán, Alfredo Herrera-Estrella, Carla Ivón Herrera-Najera, Pavel Isa, Verónica Mata-Haro, Daniel Lira Morales, Susana Lopez, Antonio Loza Román, Brenda Irasema Maldonado-Meza, Bernardo Martínez-Miguel, José Arturo Martínez-Orozco, Célida Martínez- Rodríguez, Margarita Matías-Florentino, Fidencio Mejía-Nepomuceno, María Guadalupe de Jesús Mireles-Rivera, Gloria María Molina-Salinas, Hector Montoya-Fuentes, , Mario Mújica-Sánchez, José Esteban Muñoz-Medina, José de Jesús Nuñez-Contreras, Alicia Ocaña-Mondragón, Luis Alberto Ochoa-Carrera, Hector Esteban Paz-Juárez, Marissa Perez-Garcia, Francisco Pulido, Helen Haydee Fernanda Ramírez-Plascencia, Jorge Salas-Hernández, Angel Gustavo Salas-Lais, Alejandro Sánchez-Flores, Clara Esperanza Santacruz-Tinoco, María Guadalupe Santiago-Mauricio, Selene Zárate, Nelly Sélem-Mojica, Blanca Taboada, Gloria Vazquez, Joel Armando Vázquez-Pérez. |
| EPI_ISL_2681381 | Unidad de Investigación Biomédica de Zacatecas (UIBZ) | Instituto de Biotecnología de la UNAM | Consorcio Mexicano de Vigilancia Genómica (CoViGen-Mex). Authors (in alphabetical order): Julio Elias Alvarado-Yaah, Carlos F. Arias, Santiago Ávila-Ríos, Eduardo Becerril-Vargas, Víctor Hugo Borja-Aburto, Celia Boukadida, Cristóbal Cháidez-Quiróz, Juan Bautista Chale-Dzul, Ricardo Ciria Merce, Andreu Comas-García, Célida Duque Molina, Julissa Enciso-Ibarra, José Antonio Enciso-Moreno, Gloria Elena Espinosa-Ayala, Fernando Fontove-Herrera, Daniel Fregoso-Rueda, Victor Eduardo García-Arias, Alejandra García-Gasca, Bruno Gómez-Gil, Jean Pierre González, Irvin González-López, Concepción Grajales-Muñiz, Ricardo Grande, Rosa María Gutiérrez Rios, Jesús Hernández, Alejandra Hernández-Terán, Alfredo Herrera-Estrella, Carla Ivón Herrera-Najera, Pavel Isa, Verónica Mata-Haro, Daniel Lira Morales, Susana Lopez, Antonio Loza Román, Brenda |

| | | | |
|---|---|---|---|
| | | | Irasema Maldonado-Meza, Bernardo Martínez-Miguel, José Arturo Martínez-Orozco, Célida Martínez- Rodríguez, Margarita Matías-Florentino, Fidencio Mejía-Nepomuceno, María Guadalupe de Jesús Mireles-Rivera, Gloria María Molina-Salinas, Hector Montoya-Fuentes, , Mario Mújica-Sánchez, José Esteban Muñoz-Medina, José de Jesús Nuñez-Contreras, Alicia Ocaña-Mondragón, Luis Alberto Ochoa-Carrera, Hector Esteban Paz-Juárez, Marissa Perez-Garcia, Francisco Pulido, Helen Haydee Fernanda Ramírez-Plascencia, Jorge Salas-Hernández, Angel Gustavo Salas-Lais, Alejandro Sánchez-Flores, Clara Esperanza Santacruz-Tinoco, María Guadalupe Santiago-Mauricio, Selene Zárate, Nelly Sélem-Mojica, Blanca Taboada, Gloria Vazquez, Joel Armando Vázquez-Pérez. |
| EPI_ISL_2681382 | Unidad de Investigación Médica de Yucatán (UIMY) | Instituto de Biotecnología de la UNAM | Consorcio Mexicano de Vigilancia Genómica (CoViGen-Mex). Authors (in alphabetical order): Julio Elias Alvarado-Yaah, Carlos F. Arias, Santiago Ávila-Ríos, Eduardo Becerril-Vargas, Víctor Hugo Borja-Aburto, Celia Boukadida, Cristóbal Cháidez-Quiróz, Juan Bautista Chale-Dzul, Ricardo Ciria Merce, Andreu Comas-García, Célida Duque Molina, Julissa Enciso-Ibarra, José Antonio Enciso-Moreno, Gloria Elena Espinosa-Ayala, Fernando Fontove-Herrera, Daniel Fregoso-Rueda, Victor Eduardo García-Arias, Alejandra García-Gasca, Bruno Gómez-Gil, Jean Pierre González, Irvin González-López, Concepción Grajales-Muñiz, Ricardo Grande, Rosa María Gutiérrez Rios, Jesús Hernández, Alejandra Hernández-Terán, Alfredo Herrera-Estrella, Carla Ivón Herrera-Najera, Pavel Isa, Verónica Mata-Haro, Daniel Lira Morales, Susana Lopez, Antonio Loza Román, Brenda Irasema Maldonado-Meza, Bernardo Martínez-Miguel, José Arturo Martínez-Orozco, Célida Martínez- Rodríguez, Margarita Matías-Florentino, Fidencio Mejía-Nepomuceno, María Guadalupe de Jesús Mireles-Rivera, Gloria María Molina-Salinas, Hector Montoya-Fuentes, , Mario Mújica-Sánchez, José Esteban Muñoz-Medina, José de Jesús Nuñez-Contreras, Alicia Ocaña-Mondragón, Luis Alberto Ochoa-Carrera, Hector Esteban Paz-Juárez, Marissa Perez-Garcia, Francisco Pulido, Helen Haydee Fernanda Ramírez-Plascencia, Jorge Salas-Hernández, Angel Gustavo Salas-Lais, Alejandro Sánchez-Flores, Clara Esperanza Santacruz-Tinoco, María Guadalupe Santiago-Mauricio, Selene Zárate, Nelly Sélem-Mojica, Blanca Taboada, Gloria Vazquez, Joel Armando Vázquez-Pérez. |
| EPI_ISL_2681386, EPI_ISL_2681387, EPI_ISL_2681388 | Laboratorio Central de Epidemiología (LCE) | Instituto de Biotecnología de la UNAM | Consorcio Mexicano de Vigilancia Genómica (CoViGen-Mex). Authors (in alphabetical order): Julio Elias Alvarado-Yaah, Carlos F. Arias, Santiago Ávila-Ríos, Eduardo Becerril-Vargas, Víctor Hugo Borja-Aburto, Celia Boukadida, Cristóbal Cháidez-Quiróz, Juan Bautista Chale-Dzul, Ricardo Ciria Merce, Andreu Comas-García, Célida Duque Molina, Julissa Enciso-Ibarra, José Antonio Enciso-Moreno, Gloria Elena Espinosa-Ayala, Fernando Fontove-Herrera, Daniel Fregoso-Rueda, Victor Eduardo García-Arias, Alejandra García-Gasca, Bruno Gómez-Gil, Jean Pierre González, Irvin González-López, Concepción Grajales-Muñiz, Ricardo Grande, Rosa María Gutiérrez Rios, Jesús Hernández, Alejandra Hernández-Terán, Alfredo Herrera-Estrella, Carla Ivón Herrera-Najera, Pavel Isa, Verónica Mata-Haro, Daniel Lira Morales, Susana Lopez, Antonio Loza Román, Brenda Irasema Maldonado-Meza, Bernardo Martínez-Miguel, José Arturo Martínez-Orozco, Célida Martínez- Rodríguez, Margarita Matías-Florentino, Fidencio Mejía-Nepomuceno, María Guadalupe de Jesús Mireles-Rivera, Gloria María Molina-Salinas, Hector Montoya-Fuentes, , Mario Mújica-Sánchez, José Esteban Muñoz-Medina, José de Jesús Nuñez-Contreras, Alicia Ocaña-Mondragón, Luis Alberto Ochoa-Carrera, Hector Esteban Paz-Juárez, Marissa Perez-Garcia, Francisco Pulido, Helen Haydee Fernanda Ramírez-Plascencia, Jorge Salas-Hernández, Angel Gustavo Salas-Lais, Alejandro Sánchez-Flores, Clara Esperanza Santacruz-Tinoco, María Guadalupe Santiago-Mauricio, Selene Zárate, Nelly Sélem-Mojica, Blanca Taboada, Gloria Vazquez, Joel Armando Vázquez-Pérez. |
| EPI_ISL_2681420 | Respiratory Viruses Branch, Centers for Disease Control and Prevention | Respiratory Viruses Branch, Centers for Disease Control and Prevention | Howard,D., Batra,D., Cook,P.W., Moser,K., Paskey,A., Caravas,J.,Rambo-Martin,B., Morrison,S., Gulvick,C., Sammons,S., Unoarumhi,Y.,Schmerer,M., Roychoudhury,P., Xie,H., Shrestha,L., Nguyen,T.V.,Bakhash,S.M., Lin,M., Baker,N.R., Perez,R., Ellis,S., Breit,N.,Livingston,R.J., Huang,M.-L., Jerome,K.R., Mathias,P.,Greninger,A., Paden,C.R. and MacCannell,D. |
| EPI_ISL_2686358 | Mako Medical | Centers for Disease Control and Prevention Division of Viral Diseases, Pathogen Discovery | Dakota Howard, Dhwani Batra, Peter W. Cook, Kara Moser, Adrian Paskey, Jason Caravas, Benjamin Rambo-Martin, Shatavia Morrison, Christopher Gulvick, Scott Sammons, Yvette Unoarumhi, Darlene Wagner, Matthew Schmerer,Matthew Tugwell, Lauren Moon, Clinton R. Paden, Duncan MacCannell |
| EPI_ISL_2686359, EPI_ISL_2686360, EPI_ISL_2686361, EPI_ISL_2686362, EPI_ISL_2686363, EPI_ISL_2686364, EPI_ISL_2686365, EPI_ISL_2686366, EPI_ISL_2686367, EPI_ISL_2686368, EPI_ISL_2686369, EPI_ISL_2686370, EPI_ISL_2686371, EPI_ISL_2686372, EPI_ISL_2686373, EPI_ISL_2686374, EPI_ISL_2686375, EPI_ISL_2686376, EPI_ISL_2686377, EPI_ISL_2686378, EPI_ISL_2686379, EPI_ISL_2686380, EPI_ISL_2686381, EPI_ISL_2686382, EPI_ISL_2686383, EPI_ISL_2686384, EPI_ISL_2686385, EPI_ISL_2686386, EPI_ISL_2686387, EPI_ISL_2686388 | | | |
| see above | Quest Diagnostics Incorporated | Centers for Disease Control and Prevention Division of Viral Diseases, Pathogen Discovery | Dakota Howard, Dhwani Batra, Peter W. Cook, Kara Moser, Adrian Paskey, Jason Caravas, Benjamin Rambo-Martin, Shatavia Morrison, Christopher Gulvick, Scott Sammons, Yvette Unoarumhi, Darlene Wagner, Matthew Schmerer, S. H. Rosenthal, A. Gerasimova, R. M. Kagan, B. Anderson, M. Hua, Y. Liu, L.E. Bernstein, K.E. Livingston, A. Perez, I. A. Shlyakhter, R. V. Rolando, R. Owen, P. Tanpaiboon, F. Lacbawan, Clinton R. Paden, Duncan MacCannell |
| EPI_ISL_2686394, EPI_ISL_2686395, EPI_ISL_2686396, EPI_ISL_2686397, EPI_ISL_2686398, EPI_ISL_2686399, EPI_ISL_2686400, EPI_ISL_2686401, EPI_ISL_2686404, EPI_ISL_2686405, EPI_ISL_2686406, EPI_ISL_2686407, EPI_ISL_2686408, EPI_ISL_2686409, EPI_ISL_2686410, EPI_ISL_2686411, EPI_ISL_2686412, EPI_ISL_2686413, EPI_ISL_2686414, EPI_ISL_2686415, EPI_ISL_2686416, EPI_ISL_2686417, EPI_ISL_2686418, EPI_ISL_2686419, EPI_ISL_2686420, EPI_ISL_2686421, EPI_ISL_2686422, EPI_ISL_2686424, EPI_ISL_2686425, EPI_ISL_2686426, EPI_ISL_2686427, EPI_ISL_2686428, EPI_ISL_2686429, EPI_ISL_2686430, EPI_ISL_2686431, EPI_ISL_2686432, EPI_ISL_2686433, EPI_ISL_2686434, EPI_ISL_2686435, EPI_ISL_2686436, EPI_ISL_2686437, EPI_ISL_2686438, EPI_ISL_2686439, EPI_ISL_2686440, EPI_ISL_2686442, EPI_ISL_2686443, EPI_ISL_2686444, EPI_ISL_2686445, EPI_ISL_2686446, EPI_ISL_2686447, EPI_ISL_2686448, EPI_ISL_2686449, EPI_ISL_2686450, EPI_ISL_2686451, EPI_ISL_2686452, EPI_ISL_2686453, EPI_ISL_2686454, EPI_ISL_2686455, EPI_ISL_2686456, EPI_ISL_2686457, EPI_ISL_2686458, EPI_ISL_2686459, EPI_ISL_2686460, EPI_ISL_2686462, EPI_ISL_2686463, EPI_ISL_2686464, EPI_ISL_2686465, EPI_ISL_2686466, EPI_ISL_2686467, EPI_ISL_2686468, EPI_ISL_2686469, EPI_ISL_2686470, EPI_ISL_2686471, EPI_ISL_2686473, EPI_ISL_2686474, EPI_ISL_2686475, EPI_ISL_2686476, EPI_ISL_2686477, EPI_ISL_2686478, EPI_ISL_2686479, EPI_ISL_2686480, EPI_ISL_2686481, EPI_ISL_2686482, EPI_ISL_2686483, EPI_ISL_2686484, EPI_ISL_2686485, EPI_ISL_2686486, EPI_ISL_2686487, EPI_ISL_2686488, EPI_ISL_2686489, EPI_ISL_2686490, EPI_ISL_2686491, EPI_ISL_2686492, EPI_ISL_2686494, EPI_ISL_2686495, EPI_ISL_2686496, EPI_ISL_2686497, EPI_ISL_2686499, EPI_ISL_2686500, EPI_ISL_2686501, EPI_ISL_2686502, EPI_ISL_2686503, EPI_ISL_2686504, EPI_ISL_2686505, EPI_ISL_2686506, EPI_ISL_2686507, EPI_ISL_2686508, EPI_ISL_2686509, EPI_ISL_2686510, EPI_ISL_2686511, EPI_ISL_2686512, EPI_ISL_2686513, EPI_ISL_2686514, EPI_ISL_2686515, EPI_ISL_2686516, EPI_ISL_2686517, EPI_ISL_2686518, EPI_ISL_2686519, EPI_ISL_2686520, EPI_ISL_2686522, EPI_ISL_2686523, EPI_ISL_2686524, EPI_ISL_2686525, EPI_ISL_2686526, EPI_ISL_2686527, EPI_ISL_2686528, EPI_ISL_2686529, EPI_ISL_2686530, EPI_ISL_2686531, EPI_ISL_2686532, EPI_ISL_2686533, EPI_ISL_2686534, EPI_ISL_2686535, EPI_ISL_2686536, EPI_ISL_2686537, EPI_ISL_2686538, EPI_ISL_2686541, EPI_ISL_2686542, EPI_ISL_2686543, EPI_ISL_2686545, EPI_ISL_2686546, EPI_ISL_2686547, EPI_ISL_2686548, EPI_ISL_2686549, EPI_ISL_2686550, EPI_ISL_2686551, EPI_ISL_2686552, EPI_ISL_2686554, EPI_ISL_2686555, EPI_ISL_2686556, EPI_ISL_2686557, EPI_ISL_2686558, EPI_ISL_2686559, EPI_ISL_2686560, EPI_ISL_2686561, EPI_ISL_2686563, EPI_ISL_2686564, EPI_ISL_2686565, EPI_ISL_2686566, EPI_ISL_2686567, EPI_ISL_2686568, EPI_ISL_2686569, EPI_ISL_2686570, EPI_ISL_2686571, EPI_ISL_2686572, EPI_ISL_2686573, EPI_ISL_2686574, EPI_ISL_2686575, EPI_ISL_2686576, EPI_ISL_2686577, EPI_ISL_2686578, EPI_ISL_2686579, EPI_ISL_2686580, EPI_ISL_2686581, EPI_ISL_2686582, EPI_ISL_2686583, EPI_ISL_2686584, EPI_ISL_2686585, EPI_ISL_2686586, EPI_ISL_2686587, EPI_ISL_2686588, EPI_ISL_2686590, EPI_ISL_2686591, EPI_ISL_2686592, EPI_ISL_2686593, EPI_ISL_2686594, EPI_ISL_2686595, EPI_ISL_2686596, EPI_ISL_2686597, EPI_ISL_2686598, EPI_ISL_2686599, EPI_ISL_2686600, EPI_ISL_2686601, EPI_ISL_2686602, EPI_ISL_2686603, EPI_ISL_2686604, EPI_ISL_2686605, EPI_ISL_2686606, EPI_ISL_2686608, EPI_ISL_2686609, EPI_ISL_2686610, EPI_ISL_2686612, EPI_ISL_2686613, EPI_ISL_2686614, EPI_ISL_2686615, EPI_ISL_2686617, EPI_ISL_2686618, EPI_ISL_2686619, EPI_ISL_2686620, EPI_ISL_2686621, EPI_ISL_2686622, EPI_ISL_2686623, EPI_ISL_2686624, EPI_ISL_2686625, EPI_ISL_2686626, EPI_ISL_2686627, EPI_ISL_2686628, EPI_ISL_2686629, EPI_ISL_2686630, EPI_ISL_2686632, EPI_ISL_2686633, EPI_ISL_2686634, EPI_ISL_2686635, EPI_ISL_2686637, EPI_ISL_2686638, EPI_ISL_2686639, EPI_ISL_2686640, EPI_ISL_2686641, EPI_ISL_2686642, EPI_ISL_2686643, EPI_ISL_2686644, EPI_ISL_2686645, EPI_ISL_2686646, EPI_ISL_2686647, EPI_ISL_2686648, EPI_ISL_2686649, EPI_ISL_2686650, EPI_ISL_2686651, EPI_ISL_2686652, EPI_ISL_2686653, EPI_ISL_2686654, EPI_ISL_2686655, EPI_ISL_2686656, EPI_ISL_2686657, EPI_ISL_2686658, EPI_ISL_2686659, EPI_ISL_2686660, EPI_ISL_2686661, EPI_ISL_2686662, EPI_ISL_2686663, EPI_ISL_2686664, EPI_ISL_2686665, EPI_ISL_2686666, EPI_ISL_2686667, EPI_ISL_2686668, EPI_ISL_2686669, EPI_ISL_2686670, EPI_ISL_2686671, EPI_ISL_2686672, EPI_ISL_2686673, EPI_ISL_2686674, EPI_ISL_2686675, EPI_ISL_2686676, EPI_ISL_2686677, EPI_ISL_2686678, EPI_ISL_2686679, EPI_ISL_2686680, EPI_ISL_2686681, EPI_ISL_2686682, EPI_ISL_2686683, EPI_ISL_2686684, EPI_ISL_2686685, EPI_ISL_2686686, EPI_ISL_2686687, EPI_ISL_2686688, EPI_ISL_2686689, EPI_ISL_2686690, EPI_ISL_2686691, EPI_ISL_2686692, EPI_ISL_2686693, EPI_ISL_2686694, EPI_ISL_2686695, EPI_ISL_2686696, EPI_ISL_2686697, EPI_ISL_2686698, EPI_ISL_2686699, EPI_ISL_2686700, EPI_ISL_2686701, EPI_ISL_2686702, EPI_ISL_2686703, EPI_ISL_2686704, EPI_ISL_2686705, EPI_ISL_2686706, EPI_ISL_2686707, EPI_ISL_2686708, EPI_ISL_2686709, EPI_ISL_2686710, EPI_ISL_2686711, EPI_ISL_2686712, EPI_ISL_2686713, EPI_ISL_2686714, EPI_ISL_2686715, EPI_ISL_2686716, EPI_ISL_2686717, EPI_ISL_2686718, EPI_ISL_2686720, EPI_ISL_2686721, EPI_ISL_2686722, EPI_ISL_2686723, EPI_ISL_2686724, EPI_ISL_2686725, EPI_ISL_2686726, EPI_ISL_2686727, EPI_ISL_2686728, EPI_ISL_2686729, EPI_ISL_2686730, EPI_ISL_2686731, EPI_ISL_2686732, EPI_ISL_2686733, EPI_ISL_2686734, EPI_ISL_2686735, EPI_ISL_2686736, EPI_ISL_2686737, EPI_ISL_2686738, EPI_ISL_2686739, EPI_ISL_2686740, EPI_ISL_2686741, EPI_ISL_2686742, EPI_ISL_2686743, EPI_ISL_2686744, EPI_ISL_2686745, EPI_ISL_2686746, EPI_ISL_2686747, EPI_ISL_2686748, EPI_ISL_2686749, EPI_ISL_2686750, EPI_ISL_2686751, EPI_ISL_2686752, EPI_ISL_2686753, EPI_ISL_2686754, EPI_ISL_2686756, EPI_ISL_2686757, EPI_ISL_2686758, EPI_ISL_2686759, EPI_ISL_2686761, EPI_ISL_2686762, EPI_ISL_2686763, EPI_ISL_2686764, EPI_ISL_2686765, EPI_ISL_2686766, EPI_ISL_2686767, EPI_ISL_2686768, EPI_ISL_2686769, EPI_ISL_2686771, EPI_ISL_2686772, EPI_ISL_2686773, EPI_ISL_2686774, EPI_ISL_2686775, EPI_ISL_2686776, EPI_ISL_2686777, EPI_ISL_2686778, EPI_ISL_2686779, EPI_ISL_2686780, EPI_ISL_2686781, EPI_ISL_2686782, EPI_ISL_2686784, EPI_ISL_2686785, EPI_ISL_2686786, EPI_ISL_2686787, EPI_ISL_2686788, EPI_ISL_2686789, EPI_ISL_2686790, EPI_ISL_2686791, EPI_ISL_2686792, EPI_ISL_2686793, EPI_ISL_2686795, EPI_ISL_2686796, EPI_ISL_2686797, EPI_ISL_2686798, EPI_ISL_2686799, EPI_ISL_2686800, EPI_ISL_2686801, EPI_ISL_2686802, EPI_ISL_2686803, EPI_ISL_2686804, EPI_ISL_2686805, EPI_ISL_2686806, EPI_ISL_2686807, EPI_ISL_2686808, EPI_ISL_2686809, EPI_ISL_2686810, EPI_ISL_2686811 | | | |
| see above | Fulgent Genetics | Centers for Disease Control and Prevention Division of Viral Diseases, Pathogen Discovery | Dakota Howard, Dhwani Batra, Peter W. Cook, Kara Moser, Adrian Paskey, Jason Caravas, Benjamin Rambo-Martin, Shatavia Morrison, Christopher Gulvick, Scott Sammons, Yvette Unoarumhi, Darlene Wagner, Matthew Schmerer, Harry Gao, Mickey Li, John Gao, Joseph Fierro, Benafsh Sapra, Becky Tsai, Yan Meng, Doreen Ng, James Xie, Clinton R. Paden, Duncan MacCannell |
| EPI_ISL_2686840, EPI_ISL_2686841, EPI_ISL_2686842, EPI_ISL_2686843, EPI_ISL_2686844, EPI_ISL_2686845, EPI_ISL_2686846, EPI_ISL_2686847, EPI_ISL_2686848, EPI_ISL_2686849, EPI_ISL_2686850, EPI_ISL_2686851, EPI_ISL_2686852, EPI_ISL_2686853, EPI_ISL_2686854, EPI_ISL_2686855, EPI_ISL_2686856, EPI_ISL_2686857, EPI_ISL_2686858, EPI_ISL_2686859, EPI_ISL_2686860, EPI_ISL_2686861, EPI_ISL_2686862, EPI_ISL_2686863, EPI_ISL_2686864, EPI_ISL_2686865, EPI_ISL_2686866, EPI_ISL_2686867, EPI_ISL_2686868, EPI_ISL_2686869, EPI_ISL_2686870, EPI_ISL_2686871, EPI_ISL_2686872, EPI_ISL_2686873, EPI_ISL_2686874, EPI_ISL_2686876, EPI_ISL_2686877, EPI_ISL_2686878, EPI_ISL_2686879, EPI_ISL_2686880, EPI_ISL_2686881, EPI_ISL_2686882, EPI_ISL_2686883, EPI_ISL_2686884, EPI_ISL_2686885, EPI_ISL_2686887, EPI_ISL_2686888, EPI_ISL_2686889, EPI_ISL_2686890, EPI_ISL_2686891, EPI_ISL_2686892, EPI_ISL_2686893, EPI_ISL_2686894, EPI_ISL_2686895, EPI_ISL_2686896, EPI_ISL_2686897, EPI_ISL_2686898, EPI_ISL_2686899, EPI_ISL_2686900 | | | |
| see above | Infinity Biologix | Centers for Disease Control and Prevention Division of Viral Diseases, Pathogen Discovery | Dakota Howard, Dhwani Batra, Peter W. Cook, Kara Moser, Adrian Paskey, Jason Caravas, Benjamin Rambo-Martin, Shatavia Morrison, Christopher Gulvick, Scott Sammons, Yvette Unoarumhi, Darlene Wagner, Matthew Schmerer, Christian Bixby, Yihe Wang, Jonathan Schultz, Chirayu Goswami, Russ Hager, Robin Grimwood, Clinton R. Paden, Duncan MacCannell |
| EPI_ISL_2686901, EPI_ISL_2686902, EPI_ISL_2686903, EPI_ISL_2686904, EPI_ISL_2686905, EPI_ISL_2686906, EPI_ISL_2686907, EPI_ISL_2686908, EPI_ISL_2686909, EPI_ISL_2686910, EPI_ISL_2686911, EPI_ISL_2686912, EPI_ISL_2686913, EPI_ISL_2686914, EPI_ISL_2686915, EPI_ISL_2686916, EPI_ISL_2686917, EPI_ISL_2686918, EPI_ISL_2686919, EPI_ISL_2686920, EPI_ISL_2686921, EPI_ISL_2686922, EPI_ISL_2686923, EPI_ISL_2686924, EPI_ISL_2686925, EPI_ISL_2686926, EPI_ISL_2686927, EPI_ISL_2686928, EPI_ISL_2686929, EPI_ISL_2686930, EPI_ISL_2686931, EPI_ISL_2686932, EPI_ISL_2686933, EPI_ISL_2686934, EPI_ISL_2686935, EPI_ISL_2686936, EPI_ISL_2686937, EPI_ISL_2686938, EPI_ISL_2686939, EPI_ISL_2686940, EPI_ISL_2686941, EPI_ISL_2686942, EPI_ISL_2686943, EPI_ISL_2686944, EPI_ISL_2686945, EPI_ISL_2686946, EPI_ISL_2686947, EPI_ISL_2686948, EPI_ISL_2686949, EPI_ISL_2686950, EPI_ISL_2686951, EPI_ISL_2686952, EPI_ISL_2686953, EPI_ISL_2686954, EPI_ISL_2686955, EPI_ISL_2686956, EPI_ISL_2686957, EPI_ISL_2686958, EPI_ISL_2686959, EPI_ISL_2686960, EPI_ISL_2686961, EPI_ISL_2686962, EPI_ISL_2686963, EPI_ISL_2686964, EPI_ISL_2686965, EPI_ISL_2686966, EPI_ISL_2686967, EPI_ISL_2686968, EPI_ISL_2686969, EPI_ISL_2686970, EPI_ISL_2686971, EPI_ISL_2686972, EPI_ISL_2686973, EPI_ISL_2686974, EPI_ISL_2686975, EPI_ISL_2686977, EPI_ISL_2686978, EPI_ISL_2686979, EPI_ISL_2686980, EPI_ISL_2686981, EPI_ISL_2686982, EPI_ISL_2686983, EPI_ISL_2686984, EPI_ISL_2686985, EPI_ISL_2686986, EPI_ISL_2686987, EPI_ISL_2686988, EPI_ISL_2686989, EPI_ISL_2686990, EPI_ISL_2686991, EPI_ISL_2686992, EPI_ISL_2686993, EPI_ISL_2686994, EPI_ISL_2686995, EPI_ISL_2686996, EPI_ISL_2686997, EPI_ISL_2686998, EPI_ISL_2686999, EPI_ISL_2687000, EPI_ISL_2687001, EPI_ISL_2687002, EPI_ISL_2687003, EPI_ISL_2687004, EPI_ISL_2687005, EPI_ISL_2687006, EPI_ISL_2687007, EPI_ISL_2687008, EPI_ISL_2687009, EPI_ISL_2687010, EPI_ISL_2687011, EPI_ISL_2687012, EPI_ISL_2687013, EPI_ISL_2687014, EPI_ISL_2687015, EPI_ISL_2687016, EPI_ISL_2687017, EPI_ISL_2687018, EPI_ISL_2687019, EPI_ISL_2687020, EPI_ISL_2687021, EPI_ISL_2687022, EPI_ISL_2687023, EPI_ISL_2687024, EPI_ISL_2687025, EPI_ISL_2687026, EPI_ISL_2687027, EPI_ISL_2687028, EPI_ISL_2687030, EPI_ISL_2687031, EPI_ISL_2687032, EPI_ISL_2687033, EPI_ISL_2687034, EPI_ISL_2687035, EPI_ISL_2687036, EPI_ISL_2687037, EPI_ISL_2687038, EPI_ISL_2687039, EPI_ISL_2687040, EPI_ISL_2687041, EPI_ISL_2687042, EPI_ISL_2687043, EPI_ISL_2687044, EPI_ISL_2687045, EPI_ISL_2687046, EPI_ISL_2687047, EPI_ISL_2687048, EPI_ISL_2687049, EPI_ISL_2687050, EPI_ISL_2687051, EPI_ISL_2687052, EPI_ISL_2687053, EPI_ISL_2687054, EPI_ISL_2687055, EPI_ISL_2687056, EPI_ISL_2687057, EPI_ISL_2687058, EPI_ISL_2687060, EPI_ISL_2687061, EPI_ISL_2687062, EPI_ISL_2687063, EPI_ISL_2687064, EPI_ISL_2687065, EPI_ISL_2687066, EPI_ISL_2687067, EPI_ISL_2687068, EPI_ISL_2687070, EPI_ISL_2687071, EPI_ISL_2687072, EPI_ISL_2687073, EPI_ISL_2687074, EPI_ISL_2687075, EPI_ISL_2687076, EPI_ISL_2687077, EPI_ISL_2687079, EPI_ISL_2687080, EPI_ISL_2687081, EPI_ISL_2687082, EPI_ISL_2687083, EPI_ISL_2687084, EPI_ISL_2687085, EPI_ISL_2687086, EPI_ISL_2687087, EPI_ISL_2687088, EPI_ISL_2687089, EPI_ISL_2687090, EPI_ISL_2687091, EPI_ISL_2687092, EPI_ISL_2687093, EPI_ISL_2687094, EPI_ISL_2687095, EPI_ISL_2687096, EPI_ISL_2687098, EPI_ISL_2687099, EPI_ISL_2687100, EPI_ISL_2687101, EPI_ISL_2687102, EPI_ISL_2687103, EPI_ISL_2687104, EPI_ISL_2687105, EPI_ISL_2687106, EPI_ISL_2687107, EPI_ISL_2687108, EPI_ISL_2687109, EPI_ISL_2687110, EPI_ISL_2687111, EPI_ISL_2687112, EPI_ISL_2687113, EPI_ISL_2687114, EPI_ISL_2687115, EPI_ISL_2687116, EPI_ISL_2687117, EPI_ISL_2687118, EPI_ISL_2687119, EPI_ISL_2687120, EPI_ISL_2687121, EPI_ISL_2687122, EPI_ISL_2687123, EPI_ISL_2687124, EPI_ISL_2687125, EPI_ISL_2687126, EPI_ISL_2687127, EPI_ISL_2687128, EPI_ISL_2687129, EPI_ISL_2687130, EPI_ISL_2687131, EPI_ISL_2687132, EPI_ISL_2687133, EPI_ISL_2687134, EPI_ISL_2687135, EPI_ISL_2687136, EPI_ISL_2687137, EPI_ISL_2687139, EPI_ISL_2687140, EPI_ISL_2687141, EPI_ISL_2687142, EPI_ISL_2687143, EPI_ISL_2687144, EPI_ISL_2687145, EPI_ISL_2687146, EPI_ISL_2687147, EPI_ISL_2687148, EPI_ISL_2687149, EPI_ISL_2687150, EPI_ISL_2687151, EPI_ISL_2687152, EPI_ISL_2687153, EPI_ISL_2687154, EPI_ISL_2687155, EPI_ISL_2687156, EPI_ISL_2687157, EPI_ISL_2687159, EPI_ISL_2687160, EPI_ISL_2687161, EPI_ISL_2687162, EPI_ISL_2687163, EPI_ISL_2687164, EPI_ISL_2687165, EPI_ISL_2687166, EPI_ISL_2687167, EPI_ISL_2687168, EPI_ISL_2687169, EPI_ISL_2687170, EPI_ISL_2687171, EPI_ISL_2687172, EPI_ISL_2687173, EPI_ISL_2687174, EPI_ISL_2687175, EPI_ISL_2687176, EPI_ISL_2687177, EPI_ISL_2687178, EPI_ISL_2687179, EPI_ISL_2687180, EPI_ISL_2687181, EPI_ISL_2687182, EPI_ISL_2687183, EPI_ISL_2687184, EPI_ISL_2687185, EPI_ISL_2687186, EPI_ISL_2687187, EPI_ISL_2687188, EPI_ISL_2687189, EPI_ISL_2687190, EPI_ISL_2687191, EPI_ISL_2687192, EPI_ISL_2687193, EPI_ISL_2687194, EPI_ISL_2687195, EPI_ISL_2687196, EPI_ISL_2687197, EPI_ISL_2687198, EPI_ISL_2687199, EPI_ISL_2687200, EPI_ISL_2687201, EPI_ISL_2687202, EPI_ISL_2687203, EPI_ISL_2687204, EPI_ISL_2687205, EPI_ISL_2687206, EPI_ISL_2687207, EPI_ISL_2687208, EPI_ISL_2687209, EPI_ISL_2687210, EPI_ISL_2687211, EPI_ISL_2687212, EPI_ISL_2687213, EPI_ISL_2687214, EPI_ISL_2687215, EPI_ISL_2687216, EPI_ISL_2687217, EPI_ISL_2687218, EPI_ISL_2687219, EPI_ISL_2687220, EPI_ISL_2687221, EPI_ISL_2687222, EPI_ISL_2687224, EPI_ISL_2687225, EPI_ISL_2687226, EPI_ISL_2687227, EPI_ISL_2687228, EPI_ISL_2687229, EPI_ISL_2687230, EPI_ISL_2687231, EPI_ISL_2687232, EPI_ISL_2687233, EPI_ISL_2687234, EPI_ISL_2687235, EPI_ISL_2687236, EPI_ISL_2687237, EPI_ISL_2687238, EPI_ISL_2687239, EPI_ISL_2687240, EPI_ISL_2687241, EPI_ISL_2687242, EPI_ISL_2687243, EPI_ISL_2687244, EPI_ISL_2687245, EPI_ISL_2687246, EPI_ISL_2687247, EPI_ISL_2687248, EPI_ISL_2687249, EPI_ISL_2687250, EPI_ISL_2687251, EPI_ISL_2687252, EPI_ISL_2687253, EPI_ISL_2687254, EPI_ISL_2687255, EPI_ISL_2687256, EPI_ISL_2687257, EPI_ISL_2687258, EPI_ISL_2687260, EPI_ISL_2687261, EPI_ISL_2687262, EPI_ISL_2687263, EPI_ISL_2687264, EPI_ISL_2687265, EPI_ISL_2687266, EPI_ISL_2687267, EPI_ISL_2687268, EPI_ISL_2687269, EPI_ISL_2687270, EPI_ISL_2687271, EPI_ISL_2687272, EPI_ISL_2687273, EPI_ISL_2687274, EPI_ISL_2687275, EPI_ISL_2687276, EPI_ISL_2687277, EPI_ISL_2687278, EPI_ISL_2687279, EPI_ISL_2687280, EPI_ISL_2687281, EPI_ISL_2687282, EPI_ISL_2687283, EPI_ISL_2687284, EPI_ISL_2687286, EPI_ISL_2687287, EPI_ISL_2687288, EPI_ISL_2687289, EPI_ISL_2687290, EPI_ISL_2687291, EPI_ISL_2687292, EPI_ISL_2687293, EPI_ISL_2687294, EPI_ISL_2687296, EPI_ISL_2687297, EPI_ISL_2687298, EPI_ISL_2687299, EPI_ISL_2687300, EPI_ISL_2687301, EPI_ISL_2687302, EPI_ISL_2687303, EPI_ISL_2687304, EPI_ISL_2687306, EPI_ISL_2687307, EPI_ISL_2687308, EPI_ISL_2687309, EPI_ISL_2687310, EPI_ISL_2687311, EPI_ISL_2687312, EPI_ISL_2687313, EPI_ISL_2687314, EPI_ISL_2687316, EPI_ISL_2687317, EPI_ISL_2687318, EPI_ISL_2687319, EPI_ISL_2687320, EPI_ISL_2687321, EPI_ISL_2687322, EPI_ISL_2687323, EPI_ISL_2687324, EPI_ISL_2687325, EPI_ISL_2687326, EPI_ISL_2687327, EPI_ISL_2687328, EPI_ISL_2687329, EPI_ISL_2687330, EPI_ISL_2687331, EPI_ISL_2687332, EPI_ISL_2687333, EPI_ISL_2687334, EPI_ISL_2687335, EPI_ISL_2687336, EPI_ISL_2687337, EPI_ISL_2687338, EPI_ISL_2687339, EPI_ISL_2687340, EPI_ISL_2687341, EPI_ISL_2687342, EPI_ISL_2687343, EPI_ISL_2687344, EPI_ISL_2687345, EPI_ISL_2687346, EPI_ISL_2687347, EPI_ISL_2687348, EPI_ISL_2687349, EPI_ISL_2687350, EPI_ISL_2687351, EPI_ISL_2687352, EPI_ISL_2687353, EPI_ISL_2687354, EPI_ISL_2687355, EPI_ISL_2687356, EPI_ISL_2687358, EPI_ISL_2687359, EPI_ISL_2687360, EPI_ISL_2687361, EPI_ISL_2687362, EPI_ISL_2687363, EPI_ISL_2687364, EPI_ISL_2687365, EPI_ISL_2687366, EPI_ISL_2687367, EPI_ISL_2687368, EPI_ISL_2687369, EPI_ISL_2687370, EPI_ISL_2687371, EPI_ISL_2687372, EPI_ISL_2687373, EPI_ISL_2687374, EPI_ISL_2687375, EPI_ISL_2687376, EPI_ISL_2687377, EPI_ISL_2687378, EPI_ISL_2687379, EPI_ISL_2687380, EPI_ISL_2687381, EPI_ISL_2687382, EPI_ISL_2687383, EPI_ISL_2687384, EPI_ISL_2687385, EPI_ISL_2687386, EPI_ISL_2687387, EPI_ISL_2687388, EPI_ISL_2687389, EPI_ISL_2687390, EPI_ISL_2687391, EPI_ISL_2687393, EPI_ISL_2687394, EPI_ISL_2687395, EPI_ISL_2687396, EPI_ISL_2687397, EPI_ISL_2687398, EPI_ISL_2687399, EPI_ISL_2687400, EPI_ISL_2687401, EPI_ISL_2687402, EPI_ISL_2687403, EPI_ISL_2687404, EPI_ISL_2687405, EPI_ISL_2687406, EPI_ISL_2687407, EPI_ISL_2687408, EPI_ISL_2687409, EPI_ISL_2687410, EPI_ISL_2687411, EPI_ISL_2687412, EPI_ISL_2687413, EPI_ISL_2687414, EPI_ISL_2687415, EPI_ISL_2687416, EPI_ISL_2687417, EPI_ISL_2687418, EPI_ISL_2687419, EPI_ISL_2687420, EPI_ISL_2687421, EPI_ISL_2687422, EPI_ISL_2687423, EPI_ISL_2687424, EPI_ISL_2687425, EPI_ISL_2687426, EPI_ISL_2687427, EPI_ISL_2687428, EPI_ISL_2687429, EPI_ISL_2687430, EPI_ISL_2687432, EPI_ISL_2687433, EPI_ISL_2687434, EPI_ISL_2687436, EPI_ISL_2687438, EPI_ISL_2687439, EPI_ISL_2687440, EPI_ISL_2687441, EPI_ISL_2687442, EPI_ISL_2687443, EPI_ISL_2687444, EPI_ISL_2687445, EPI_ISL_2687446, EPI_ISL_2687447, EPI_ISL_2687448, EPI_ISL_2687449, EPI_ISL_2687450, EPI_ISL_2687451, EPI_ISL_2687452, EPI_ISL_2687453, EPI_ISL_2687454, EPI_ISL_2687455, EPI_ISL_2687456, EPI_ISL_2687457, EPI_ISL_2687458, EPI_ISL_2687459, EPI_ISL_2687460, EPI_ISL_2687461, EPI_ISL_2687462, EPI_ISL_2687463, EPI_ISL_2687464, EPI_ISL_2687465, EPI_ISL_2687466, EPI_ISL_2687467, EPI_ISL_2687468, EPI_ISL_2687469, EPI_ISL_2687470, EPI_ISL_2687471, EPI_ISL_2687472, EPI_ISL_2687473, EPI_ISL_2687474, EPI_ISL_2687475, EPI_ISL_2687476, EPI_ISL_2687477, EPI_ISL_2687478, EPI_ISL_2687479, EPI_ISL_2687480, EPI_ISL_2687481, EPI_ISL_2687482, EPI_ISL_2687483, EPI_ISL_2687484, EPI_ISL_2687485, EPI_ISL_2687486, EPI_ISL_2687487, EPI_ISL_2687488, EPI_ISL_2687489, EPI_ISL_2687490, EPI_ISL_2687491, EPI_ISL_2687492, EPI_ISL_2687494, EPI_ISL_2687496, EPI_ISL_2687497, EPI_ISL_2687498, EPI_ISL_2687499, EPI_ISL_2687500, EPI_ISL_2687501, EPI_ISL_2687502, EPI_ISL_2687503, EPI_ISL_2687504, EPI_ISL_2687505, EPI_ISL_2687507, EPI_ISL_2687509, EPI_ISL_2687510, EPI_ISL_2687511, EPI_ISL_2687512, EPI_ISL_2687513, EPI_ISL_2687514, EPI_ISL_2687515, EPI_ISL_2687516, EPI_ISL_2687517, EPI_ISL_2687518, EPI_ISL_2687519, EPI_ISL_2687520, EPI_ISL_2687521, EPI_ISL_2687522, EPI_ISL_2687523, EPI_ISL_2687524, EPI_ISL_2687525, EPI_ISL_2687526, EPI_ISL_2687527, EPI_ISL_2687528, EPI_ISL_2687529, EPI_ISL_2687530, EPI_ISL_2687531, EPI_ISL_2687532, EPI_ISL_2687533, EPI_ISL_2687534, EPI_ISL_2687535, EPI_ISL_2687536, EPI_ISL_2687537, EPI_ISL_2687538, EPI_ISL_2687539, EPI_ISL_2687540, EPI_ISL_2687541, EPI_ISL_2687542, EPI_ISL_2687543, EPI_ISL_2687544, EPI_ISL_2687545, EPI_ISL_2687546, EPI_ISL_2687547, EPI_ISL_2687548, EPI_ISL_2687549, EPI_ISL_2687550, EPI_ISL_2687551, EPI_ISL_2687552, EPI_ISL_2687553, EPI_ISL_2687554, EPI_ISL_2687555, EPI_ISL_2687556, EPI_ISL_2687557, EPI_ISL_2687558, EPI_ISL_2687559, EPI_ISL_2687560, EPI_ISL_2687561, EPI_ISL_2687562, EPI_ISL_2687563, EPI_ISL_2687564, EPI_ISL_2687565, EPI_ISL_2687566, EPI_ISL_2687567, EPI_ISL_2687568, EPI_ISL_2687569, EPI_ISL_2687570, EPI_ISL_2687571, EPI_ISL_2687572, EPI_ISL_2687573, EPI_ISL_2687575, EPI_ISL_2687576, EPI_ISL_2687578, EPI_ISL_2687579, EPI_ISL_2687580, EPI_ISL_2687581, EPI_ISL_2687582, EPI_ISL_2687583 | | | |
| see above | Aegis Sciences Corporation | Centers for Disease Control and Prevention Division of Viral Diseases, Pathogen Discovery | Dakota Howard, Dhwani Batra, Peter W. Cook, Kara Moser, Adrian Paskey, Jason Caravas, Benjamin Rambo-Martin, Shatavia Morrison, Christopher Gulvick, Scott Sammons, Yvette Unoarumhi, Darlene Wagner, Matthew Schmerer, Cyndi Clark, Patrick Campbell, Rob Case, Vikramsinha Ghorpade, Holly Houdeshell, Ola Kvalvaag, Dillon Nall, Ethan Sanders, Alec Vest, Shaun Westlund, Matthew Hardison, Clinton R. Paden, Duncan MacCannell |
| EPI_ISL_2688001, EPI_ISL_2688002, EPI_ISL_2688003, EPI_ISL_2688004, EPI_ISL_2688005, EPI_ISL_2688006 | Sonic Reference Laboratory | Sonic Reference Laboratory | "Robert Moller, Bijaya Dhakal, Krishna Maddula, John Woody, Cora Lahey, Ellen Pizarek" |
| EPI_ISL_2688187, EPI_ISL_2688188, EPI_ISL_2688189 | IN State Department of Health Laboratory Services | IN State Department of Health Laboratory Services | Cassandra Campion, Jamie Yeadon, Brian Pope, Lixia Liu, Kyle Brownlee, Melissa Hindenlang, Mark Glazier |
| EPI_ISL_2688190, EPI_ISL_2688192, EPI_ISL_2688193 | Kansas Health and Environmental Lab | Kansas Health and Environmental Lab | Mike Grose, Katherine Wiggins, Jonathan Barnell, Ben Olsen, and Phil Adam |
| EPI_ISL_2688194 | IN State Department of Health Laboratory Services | IN State Department of Health Laboratory Services | Cassandra Campion, Jamie Yeadon, Brian Pope, Lixia Liu, Kyle Brownlee, Melissa Hindenlang, Mark Glazier |
| EPI_ISL_2688195, EPI_ISL_2688196 | Kansas Health and Environmental Lab | Kansas Health and Environmental Lab | Mike Grose, Katherine Wiggins, Jonathan Barnell, Ben Olsen, and Phil Adam |

| | | | |
|---|---|---|---|
| EPI_ISL_2688197 | IN State Department of Health Laboratory Services | IN State Department of Health Laboratory Services | Cassandra Campion, Jamie Yeadon, Brian Pope, Lixia Liu, Kyle Brownlee, Melissa Hindenlang, Mark Glazier |
| EPI_ISL_2688198, EPI_ISL_2688199 | Kansas Health and Environmental Lab | Kansas Health and Environmental Lab | Mike Grose, Katherine Wiggins, Jonathan Barnell, Ben Olsen, and Phil Adam |
| EPI_ISL_2688200 | IN State Department of Health Laboratory Services | IN State Department of Health Laboratory Services | Cassandra Campion, Jamie Yeadon, Brian Pope, Lixia Liu, Kyle Brownlee, Melissa Hindenlang, Mark Glazier |
| EPI_ISL_2688201, EPI_ISL_2688202 | Kansas Health and Environmental Lab | Kansas Health and Environmental Lab | Mike Grose, Katherine Wiggins, Jonathan Barnell, Ben Olsen, and Phil Adam |
| EPI_ISL_2688203 | IN State Department of Health Laboratory Services | IN State Department of Health Laboratory Services | Cassandra Campion, Jamie Yeadon, Brian Pope, Lixia Liu, Kyle Brownlee, Melissa Hindenlang, Mark Glazier |
| EPI_ISL_2688204, EPI_ISL_2688205 | Kansas Health and Environmental Lab | Kansas Health and Environmental Lab | Mike Grose, Katherine Wiggins, Jonathan Barnell, Ben Olsen, and Phil Adam |
| EPI_ISL_2688206 | IN State Department of Health Laboratory Services | IN State Department of Health Laboratory Services | Cassandra Campion, Jamie Yeadon, Brian Pope, Lixia Liu, Kyle Brownlee, Melissa Hindenlang, Mark Glazier |
| EPI_ISL_2688207, EPI_ISL_2688208 | Kansas Health and Environmental Lab | Kansas Health and Environmental Lab | Mike Grose, Katherine Wiggins, Jonathan Barnell, Ben Olsen, and Phil Adam |
| EPI_ISL_2688209 | IN State Department of Health Laboratory Services | IN State Department of Health Laboratory Services | Cassandra Campion, Jamie Yeadon, Brian Pope, Lixia Liu, Kyle Brownlee, Melissa Hindenlang, Mark Glazier |
| EPI_ISL_2688211 | Kansas Health and Environmental Lab | Kansas Health and Environmental Lab | Mike Grose, Katherine Wiggins, Jonathan Barnell, Ben Olsen, and Phil Adam |
| EPI_ISL_2688212 | IN State Department of Health Laboratory Services | IN State Department of Health Laboratory Services | Cassandra Campion, Jamie Yeadon, Brian Pope, Lixia Liu, Kyle Brownlee, Melissa Hindenlang, Mark Glazier |
| EPI_ISL_2688214 | Kansas Health and Environmental Lab | Kansas Health and Environmental Lab | Mike Grose, Katherine Wiggins, Jonathan Barnell, Ben Olsen, and Phil Adam |
| EPI_ISL_2688215 | IN State Department of Health Laboratory Services | IN State Department of Health Laboratory Services | Cassandra Campion, Jamie Yeadon, Brian Pope, Lixia Liu, Kyle Brownlee, Melissa Hindenlang, Mark Glazier |
| EPI_ISL_2688216, EPI_ISL_2688217 | Kansas Health and Environmental Lab | Kansas Health and Environmental Lab | Mike Grose, Katherine Wiggins, Jonathan Barnell, Ben Olsen, and Phil Adam |
| EPI_ISL_2688218 | IN State Department of Health Laboratory Services | IN State Department of Health Laboratory Services | Cassandra Campion, Jamie Yeadon, Brian Pope, Lixia Liu, Kyle Brownlee, Melissa Hindenlang, Mark Glazier |
| EPI_ISL_2688219, EPI_ISL_2688221, EPI_ISL_2688222 | Kansas Health and Environmental Lab | Kansas Health and Environmental Lab | Mike Grose, Katherine Wiggins, Jonathan Barnell, Ben Olsen, and Phil Adam |
| EPI_ISL_2688223 | IN State Department of Health Laboratory Services | IN State Department of Health Laboratory Services | Cassandra Campion, Jamie Yeadon, Brian Pope, Lixia Liu, Kyle Brownlee, Melissa Hindenlang, Mark Glazier |
| EPI_ISL_2688224 | Kansas Health and Environmental Lab | Kansas Health and Environmental Lab | Mike Grose, Katherine Wiggins, Jonathan Barnell, Ben Olsen, and Phil Adam |
| EPI_ISL_2688225 | IN State Department of Health Laboratory Services | IN State Department of Health Laboratory Services | Cassandra Campion, Jamie Yeadon, Brian Pope, Lixia Liu, Kyle Brownlee, Melissa Hindenlang, Mark Glazier |
| EPI_ISL_2688226 | Kansas Health and Environmental Lab | Kansas Health and Environmental Lab | Mike Grose, Katherine Wiggins, Jonathan Barnell, Ben Olsen, and Phil Adam |
| EPI_ISL_2688227 | IN State Department of Health Laboratory Services | IN State Department of Health Laboratory Services | Cassandra Campion, Jamie Yeadon, Brian Pope, Lixia Liu, Kyle Brownlee, Melissa Hindenlang, Mark Glazier |
| EPI_ISL_2688228, EPI_ISL_2688229 | Kansas Health and Environmental Lab | Kansas Health and Environmental Lab | Mike Grose, Katherine Wiggins, Jonathan Barnell, Ben Olsen, and Phil Adam |
| EPI_ISL_2688230 | IN State Department of Health Laboratory Services | IN State Department of Health Laboratory Services | Cassandra Campion, Jamie Yeadon, Brian Pope, Lixia Liu, Kyle Brownlee, Melissa Hindenlang, Mark Glazier |
| EPI_ISL_2688231, EPI_ISL_2688232 | Kansas Health and Environmental Lab | Kansas Health and Environmental Lab | Mike Grose, Katherine Wiggins, Jonathan Barnell, Ben Olsen, and Phil Adam |
| EPI_ISL_2688233 | IN State Department of Health Laboratory Services | IN State Department of Health Laboratory Services | Cassandra Campion, Jamie Yeadon, Brian Pope, Lixia Liu, Kyle Brownlee, Melissa Hindenlang, Mark Glazier |
| EPI_ISL_2688234, EPI_ISL_2688235 | Kansas Health and Environmental Lab | Kansas Health and Environmental Lab | Mike Grose, Katherine Wiggins, Jonathan Barnell, Ben Olsen, and Phil Adam |
| EPI_ISL_2688236 | IN State Department of Health Laboratory Services | IN State Department of Health Laboratory Services | Cassandra Campion, Jamie Yeadon, Brian Pope, Lixia Liu, Kyle Brownlee, Melissa Hindenlang, Mark Glazier |
| EPI_ISL_2688237 | Kansas Health and Environmental Lab | Kansas Health and Environmental Lab | Mike Grose, Katherine Wiggins, Jonathan Barnell, Ben Olsen, and Phil Adam |
| EPI_ISL_2688238, EPI_ISL_2688239, EPI_ISL_2688240, EPI_ISL_2688244, EPI_ISL_2688245, EPI_ISL_2688246, EPI_ISL_2688247, EPI_ISL_2688248 | IN State Department of Health Laboratory Services | IN State Department of Health Laboratory Services | Cassandra Campion, Jamie Yeadon, Brian Pope, Lixia Liu, Kyle Brownlee, Melissa Hindenlang, Mark Glazier |
| EPI_ISL_2688259, EPI_ISL_2688260, EPI_ISL_2688261, EPI_ISL_2688262, EPI_ISL_2688263, EPI_ISL_2688264, EPI_ISL_2688265, EPI_ISL_2688266, EPI_ISL_2688267, EPI_ISL_2688268, EPI_ISL_2688269, EPI_ISL_2688270, EPI_ISL_2688271, EPI_ISL_2688272, EPI_ISL_2688273, EPI_ISL_2688274, EPI_ISL_2688275, EPI_ISL_2688276, EPI_ISL_2688277 | | | |
| see above | Centers for Disease Control and Prevention, Dengue Branch | Centers for Disease Control and Prevention, Dengue Branch | Gilberto A. Santiago, Glenda Gonzalez, Betzabel Flores, Keyla Charriez, Gabriela Paz-Bailey, Jorge L. Munoz-Jordan |
| EPI_ISL_2688278, EPI_ISL_2688279, EPI_ISL_2688280, EPI_ISL_2688281, EPI_ISL_2688282, EPI_ISL_2688283, EPI_ISL_2688284, EPI_ISL_2688285, EPI_ISL_2688286, EPI_ISL_2688287, EPI_ISL_2688288, EPI_ISL_2688289, EPI_ISL_2688290, EPI_ISL_2688291, EPI_ISL_2688292, EPI_ISL_2688293, EPI_ISL_2688294, EPI_ISL_2688295, EPI_ISL_2688296, EPI_ISL_2688297, EPI_ISL_2688298, EPI_ISL_2688299, EPI_ISL_2688300, EPI_ISL_2688301, EPI_ISL_2688302, EPI_ISL_2688303, EPI_ISL_2688304, EPI_ISL_2688305, EPI_ISL_2688306, EPI_ISL_2688307, EPI_ISL_2688308, EPI_ISL_2688309, EPI_ISL_2688310, EPI_ISL_2688311, EPI_ISL_2688312, EPI_ISL_2688313, EPI_ISL_2688314, EPI_ISL_2688315, EPI_ISL_2688316, EPI_ISL_2688317, EPI_ISL_2688318, EPI_ISL_2688319 | | | |
| see above | Puerto Rico Department of Health | Centers for Disease Control and Prevention, Dengue Branch | Gilberto A. Santiago, Glenda Gonzalez, Betzabel Flores, Keyla Charriez, Gabriela Paz-Bailey, Jorge L. Munoz-Jordan |
| EPI_ISL_2688804, EPI_ISL_2688805, EPI_ISL_2688806, EPI_ISL_2688807, EPI_ISL_2688808, EPI_ISL_2688809, EPI_ISL_2688810, EPI_ISL_2688811, EPI_ISL_2688812, EPI_ISL_2688813, EPI_ISL_2688814, EPI_ISL_2688816, EPI_ISL_2688817, EPI_ISL_2688818, EPI_ISL_2688819, EPI_ISL_2688820 | | | |
| see above | Massachusetts State Public Health Laboratory | Massachusetts State Public Health Laboratory | Andrew Lang, Timelia Fink, Glen Gallagher, Sandra Smole |
| EPI_ISL_2688821, EPI_ISL_2688822, EPI_ISL_2688823 | Kaiser Permanente Washington Health Research Institute | Genomics and Discovery, Respiratory Viruses Branch, Division of Viral Diseases, Centers for Disease Control and Prevention | Jing Zhang, Anna Kelleher, Ying Tao, Yan Li, Brian Lynch, Krista Queen, Anna Uehara, Peter Cook, Han Jia Justin Ng, Rachel Marine, Clinton R. Paden, Dhwani Batra, Haibin Wang, Tara Coalter, Jasmine Padilla, Morgan Davis, Mili Sheth, Sarah Nobles, Mark Burroughs, Justin Lee, Adam Retchless, Suxiang Tong |
| EPI_ISL_2688824, EPI_ISL_2688825, EPI_ISL_2688826, EPI_ISL_2688827, EPI_ISL_2688828 | LESP Tabasco | Instituto de Diagnostico y Referencia Epidemiologicos (INDRE) | Claudia Wong-Arambula, Abril Rodriguez-Maldonado, Vanessa Rivero-Arredondo, Ariadna Medina-Benitez, Joaquin Quiroz-Mercado, Sergio Rangel-Guerrero, Natividad Cruz-Ortiz, Tatiana Nunez-Garcia, Gisela Barrera-Badillo, Lucia Hernandez-Rivas, Irma Lopez-Martinez, Ernesto Ramirez-Gonzalez. |
| EPI_ISL_2688829, EPI_ISL_2688830 | LESP Queretaro | Instituto de Diagnostico y Referencia Epidemiologicos (INDRE) | Claudia Wong-Arambula, Abril Rodriguez-Maldonado, Vanessa Rivero-Arredondo, Ariadna Medina-Benitez, Joaquin Quiroz-Mercado, Sergio Rangel-Guerrero, Natividad Cruz-Ortiz, Tatiana Nunez-Garcia, Gisela Barrera-Badillo, Lucia Hernandez-Rivas, Irma Lopez-Martinez, Ernesto Ramirez-Gonzalez. |
| EPI_ISL_2688831 | LESP Veracruz | Instituto de Diagnostico y Referencia Epidemiologicos (INDRE) | Claudia Wong-Arambula, Abril Rodriguez-Maldonado, Vanessa Rivero-Arredondo, Ariadna Medina-Benitez, Joaquin Quiroz-Mercado, Sergio Rangel-Guerrero, Natividad Cruz-Ortiz, Tatiana Nunez-Garcia, Gisela Barrera-Badillo, Lucia Hernandez-Rivas, Irma Lopez-Martinez, Ernesto Ramirez-Gonzalez. |
| EPI_ISL_2688900 | Kaiser Permanente Washington Health Research Institute | Genomics and Discovery, Respiratory Viruses Branch, Division of Viral Diseases, Centers for Disease Control and Prevention | Jing Zhang, Anna Kelleher, Ying Tao, Yan Li, Brian Lynch, Krista Queen, Anna Uehara, Peter Cook, Han Jia Justin Ng, Rachel Marine, Clinton R. Paden, Dhwani Batra, Haibin Wang, Tara Coalter, Jasmine Padilla, Morgan Davis, Mili Sheth, Sarah Nobles, Mark Burroughs, Justin Lee, Adam Retchless, Suxiang Tong |
| EPI_ISL_2688901, EPI_ISL_2688902, EPI_ISL_2688903, EPI_ISL_2688904, EPI_ISL_2688905, EPI_ISL_2688906, EPI_ISL_2688907, EPI_ISL_2688908, | University of Pittsburgh | Genomics and Discovery, Respiratory Viruses Branch, Division of Viral Diseases, Centers for Disease Control and Prevention | Jing Zhang, Anna Kelleher, Ying Tao, Yan Li, Brian Lynch, Krista Queen, Anna Uehara, Peter Cook, Han Jia Justin Ng, Rachel Marine, Clinton R. Paden, Dhwani Batra, Haibin Wang, Tara Coalter, Jasmine Padilla, Morgan Davis, Mili Sheth, Sarah Nobles, Mark Burroughs, Justin Lee, Adam Retchless, Suxiang Tong |

| | | | |
|---|---|---|---|
| EPI_ISL_2688909, EPI_ISL_2688910 | | | |
| EPI_ISL_2688911 | LESP Ciudad de Mexico | Instituto de Diagnostico y Referencia Epidemiologicos (INDRE) | Claudia Wong-Arambula, Abril Rodriguez-Maldonado, Vanessa Rivero-Arredondo, Ariadna Medina-Benitez, Joaquin Quiroz-Mercado, Sergio Rangel-Guerrero, Natividad Cruz-Ortiz, Tatiana Nunez-Garcia, Gisela Barrera-Badillo, Lucia Hernandez-Rivas, Irma Lopez-Martinez, Ernesto Ramirez-Gonzalez. |
| EPI_ISL_2688912 | LESP Nuevo Leon | Instituto de Diagnostico y Referencia Epidemiologicos (INDRE) | Claudia Wong-Arambula, Abril Rodriguez-Maldonado, Vanessa Rivero-Arredondo, Ariadna Medina-Benitez, Joaquin Quiroz-Mercado, Sergio Rangel-Guerrero, Natividad Cruz-Ortiz, Tatiana Nunez-Garcia, Gisela Barrera-Badillo, Lucia Hernandez-Rivas, Irma Lopez-Martinez, Ernesto Ramirez-Gonzalez. |
| EPI_ISL_2688913, EPI_ISL_2688914 | LESP Jalisco | Instituto de Diagnostico y Referencia Epidemiologicos (INDRE) | Claudia Wong-Arambula, Abril Rodriguez-Maldonado, Vanessa Rivero-Arredondo, Ariadna Medina-Benitez, Joaquin Quiroz-Mercado, Sergio Rangel-Guerrero, Natividad Cruz-Ortiz, Tatiana Nunez-Garcia, Gisela Barrera-Badillo, Lucia Hernandez-Rivas, Irma Lopez-Martinez, Ernesto Ramirez-Gonzalez. |
| EPI_ISL_2688915, EPI_ISL_2688916, EPI_ISL_2688917, EPI_ISL_2688918 | LESP Morelos | Instituto de Diagnostico y Referencia Epidemiologicos (INDRE) | Claudia Wong-Arambula, Abril Rodriguez-Maldonado, Vanessa Rivero-Arredondo, Ariadna Medina-Benitez, Joaquin Quiroz-Mercado, Sergio Rangel-Guerrero, Natividad Cruz-Ortiz, Tatiana Nunez-Garcia, Gisela Barrera-Badillo, Lucia Hernandez-Rivas, Irma Lopez-Martinez, Ernesto Ramirez-Gonzalez. |
| EPI_ISL_2688919, EPI_ISL_2688920, EPI_ISL_2688921, EPI_ISL_2688922, EPI_ISL_2688923 | LESP Ciudad de Mexico | Instituto de Diagnostico y Referencia Epidemiologicos (INDRE) | Claudia Wong-Arambula, Abril Rodriguez-Maldonado, Vanessa Rivero-Arredondo, Ariadna Medina-Benitez, Joaquin Quiroz-Mercado, Sergio Rangel-Guerrero, Natividad Cruz-Ortiz, Tatiana Nunez-Garcia, Gisela Barrera-Badillo, Lucia Hernandez-Rivas, Irma Lopez-Martinez, Ernesto Ramirez-Gonzalez. |
| EPI_ISL_2688924, EPI_ISL_2688925 | LESP Queretaro | Instituto de Diagnostico y Referencia Epidemiologicos (INDRE) | Claudia Wong-Arambula, Abril Rodriguez-Maldonado, Vanessa Rivero-Arredondo, Ariadna Medina-Benitez, Joaquin Quiroz-Mercado, Sergio Rangel-Guerrero, Natividad Cruz-Ortiz, Tatiana Nunez-Garcia, Gisela Barrera-Badillo, Lucia Hernandez-Rivas, Irma Lopez-Martinez, Ernesto Ramirez-Gonzalez. |
| EPI_ISL_2688926 | LESP Ciudad de Mexico | Instituto de Diagnostico y Referencia Epidemiologicos (INDRE) | Claudia Wong-Arambula, Abril Rodriguez-Maldonado, Vanessa Rivero-Arredondo, Ariadna Medina-Benitez, Joaquin Quiroz-Mercado, Sergio Rangel-Guerrero, Natividad Cruz-Ortiz, Tatiana Nunez-Garcia, Gisela Barrera-Badillo, Lucia Hernandez-Rivas, Irma Lopez-Martinez, Ernesto Ramirez-Gonzalez. |
| EPI_ISL_2688927, EPI_ISL_2688928, EPI_ISL_2688929, EPI_ISL_2688930, EPI_ISL_2688931, EPI_ISL_2688932, EPI_ISL_2688933, EPI_ISL_2688934, EPI_ISL_2688935, EPI_ISL_2688936, EPI_ISL_2688937, EPI_ISL_2688938, EPI_ISL_2688939, EPI_ISL_2688940, EPI_ISL_2688941, EPI_ISL_2688942 | | | |
| see above | LESP Tabasco | Instituto de Diagnostico y Referencia Epidemiologicos (INDRE) | Claudia Wong-Arambula, Abril Rodriguez-Maldonado, Vanessa Rivero-Arredondo, Ariadna Medina-Benitez, Joaquin Quiroz-Mercado, Sergio Rangel-Guerrero, Natividad Cruz-Ortiz, Tatiana Nunez-Garcia, Gisela Barrera-Badillo, Lucia Hernandez-Rivas, Irma Lopez-Martinez, Ernesto Ramirez-Gonzalez. |
| EPI_ISL_2688943, EPI_ISL_2688944, EPI_ISL_2688945, EPI_ISL_2688946, EPI_ISL_2688947 | LESP Michoacan | Instituto de Diagnostico y Referencia Epidemiologicos (INDRE) | Claudia Wong-Arambula, Abril Rodriguez-Maldonado, Vanessa Rivero-Arredondo, Ariadna Medina-Benitez, Joaquin Quiroz-Mercado, Sergio Rangel-Guerrero, Natividad Cruz-Ortiz, Tatiana Nunez-Garcia, Gisela Barrera-Badillo, Lucia Hernandez-Rivas, Irma Lopez-Martinez, Ernesto Ramirez-Gonzalez. |
| EPI_ISL_2688948, EPI_ISL_2688949, EPI_ISL_2688950, EPI_ISL_2688951, EPI_ISL_2688952, EPI_ISL_2688953, EPI_ISL_2688954, EPI_ISL_2688955, EPI_ISL_2688956 | LESP Tabasco | Instituto de Diagnostico y Referencia Epidemiologicos (INDRE) | Claudia Wong-Arambula, Abril Rodriguez-Maldonado, Vanessa Rivero-Arredondo, Ariadna Medina-Benitez, Joaquin Quiroz-Mercado, Sergio Rangel-Guerrero, Natividad Cruz-Ortiz, Tatiana Nunez-Garcia, Gisela Barrera-Badillo, Lucia Hernandez-Rivas, Irma Lopez-Martinez, Ernesto Ramirez-Gonzalez. |
| EPI_ISL_2688957 | LESP Chihuahua | Instituto de Diagnostico y Referencia Epidemiologicos (INDRE) | Claudia Wong-Arambula, Abril Rodriguez-Maldonado, Vanessa Rivero-Arredondo, Ariadna Medina-Benitez, Joaquin Quiroz-Mercado, Sergio Rangel-Guerrero, Natividad Cruz-Ortiz, Tatiana Nunez-Garcia, Gisela Barrera-Badillo, Lucia Hernandez-Rivas, Irma Lopez-Martinez, Ernesto Ramirez-Gonzalez. |
| EPI_ISL_2688958 | LESP Tabasco | Instituto de Diagnostico y Referencia Epidemiologicos (INDRE) | Claudia Wong-Arambula, Abril Rodriguez-Maldonado, Vanessa Rivero-Arredondo, Ariadna Medina-Benitez, Joaquin Quiroz-Mercado, Sergio Rangel-Guerrero, Natividad Cruz-Ortiz, Tatiana Nunez-Garcia, Gisela Barrera-Badillo, Lucia Hernandez-Rivas, Irma Lopez-Martinez, Ernesto Ramirez-Gonzalez. |
| EPI_ISL_2688959 | LESP Estado de Mexico | Instituto de Diagnostico y Referencia Epidemiologicos (INDRE) | Claudia Wong-Arambula, Abril Rodriguez-Maldonado, Vanessa Rivero-Arredondo, Ariadna Medina-Benitez, Joaquin Quiroz-Mercado, Sergio Rangel-Guerrero, Natividad Cruz-Ortiz, Tatiana Nunez-Garcia, Gisela Barrera-Badillo, Lucia Hernandez-Rivas, Irma Lopez-Martinez, Ernesto Ramirez-Gonzalez. |
| EPI_ISL_2688960, EPI_ISL_2688961, EPI_ISL_2688962, EPI_ISL_2688963, EPI_ISL_2688964 | LESP Ciudad de Mexico | Instituto de Diagnostico y Referencia Epidemiologicos (INDRE) | Claudia Wong-Arambula, Abril Rodriguez-Maldonado, Vanessa Rivero-Arredondo, Ariadna Medina-Benitez, Joaquin Quiroz-Mercado, Sergio Rangel-Guerrero, Natividad Cruz-Ortiz, Tatiana Nunez-Garcia, Gisela Barrera-Badillo, Lucia Hernandez-Rivas, Irma Lopez-Martinez, Ernesto Ramirez-Gonzalez. |
| EPI_ISL_2688965 | LESP Estado de Mexico | Instituto de Diagnostico y Referencia Epidemiologicos (INDRE) | Claudia Wong-Arambula, Abril Rodriguez-Maldonado, Vanessa Rivero-Arredondo, Ariadna Medina-Benitez, Joaquin Quiroz-Mercado, Sergio Rangel-Guerrero, Natividad Cruz-Ortiz, Tatiana Nunez-Garcia, Gisela Barrera-Badillo, Lucia Hernandez-Rivas, Irma Lopez-Martinez, Ernesto Ramirez-Gonzalez. |
| EPI_ISL_2688966, EPI_ISL_2688967, EPI_ISL_2688968 | LESP Ciudad de Mexico | Instituto de Diagnostico y Referencia Epidemiologicos (INDRE) | Claudia Wong-Arambula, Abril Rodriguez-Maldonado, Vanessa Rivero-Arredondo, Ariadna Medina-Benitez, Joaquin Quiroz-Mercado, Sergio Rangel-Guerrero, Natividad Cruz-Ortiz, Tatiana Nunez-Garcia, Gisela Barrera-Badillo, Lucia Hernandez-Rivas, Irma Lopez-Martinez, Ernesto Ramirez-Gonzalez. |
| EPI_ISL_2688969 | LESP Queretaro | Instituto de Diagnostico y Referencia Epidemiologicos (INDRE) | Claudia Wong-Arambula, Abril Rodriguez-Maldonado, Vanessa Rivero-Arredondo, Ariadna Medina-Benitez, Joaquin Quiroz-Mercado, Sergio Rangel-Guerrero, Natividad Cruz-Ortiz, Tatiana Nunez-Garcia, Gisela Barrera-Badillo, Lucia Hernandez-Rivas, Irma Lopez-Martinez, Ernesto Ramirez-Gonzalez. |
| EPI_ISL_2688970 | LESP Michoacan | Instituto de Diagnostico y Referencia Epidemiologicos (INDRE) | Claudia Wong-Arambula, Abril Rodriguez-Maldonado, Vanessa Rivero-Arredondo, Ariadna Medina-Benitez, Joaquin Quiroz-Mercado, Sergio Rangel-Guerrero, Natividad Cruz-Ortiz, Tatiana Nunez-Garcia, Gisela Barrera-Badillo, Lucia Hernandez-Rivas, Irma Lopez-Martinez, Ernesto Ramirez-Gonzalez. |
| EPI_ISL_2688971 | LESP Zacatecas | Instituto de Diagnostico y Referencia Epidemiologicos (INDRE) | Claudia Wong-Arambula, Abril Rodriguez-Maldonado, Vanessa Rivero-Arredondo, Ariadna Medina-Benitez, Joaquin Quiroz-Mercado, Sergio Rangel-Guerrero, Natividad Cruz-Ortiz, Tatiana Nunez-Garcia, Gisela Barrera-Badillo, Lucia Hernandez-Rivas, Irma Lopez-Martinez, Ernesto Ramirez-Gonzalez. |
| EPI_ISL_2688972 | LESP Hidalgo | Instituto de Diagnostico y Referencia Epidemiologicos (INDRE) | Claudia Wong-Arambula, Abril Rodriguez-Maldonado, Vanessa Rivero-Arredondo, Ariadna Medina-Benitez, Joaquin Quiroz-Mercado, Sergio Rangel-Guerrero, Natividad Cruz-Ortiz, Tatiana Nunez-Garcia, Gisela Barrera-Badillo, Lucia Hernandez-Rivas, Irma Lopez-Martinez, Ernesto Ramirez-Gonzalez. |
| EPI_ISL_2688973 | LESP Morelos | Instituto de Diagnostico y Referencia Epidemiologicos (INDRE) | Claudia Wong-Arambula, Abril Rodriguez-Maldonado, Vanessa Rivero-Arredondo, Ariadna Medina-Benitez, Joaquin Quiroz-Mercado, Sergio Rangel-Guerrero, Natividad Cruz-Ortiz, Tatiana Nunez-Garcia, Gisela Barrera-Badillo, Lucia Hernandez-Rivas, Irma Lopez-Martinez, Ernesto Ramirez-Gonzalez. |
| EPI_ISL_2688974 | LESP Guanajuato | Instituto de Diagnostico y Referencia Epidemiologicos (INDRE) | Claudia Wong-Arambula, Abril Rodriguez-Maldonado, Vanessa Rivero-Arredondo, Ariadna Medina-Benitez, Joaquin Quiroz-Mercado, Sergio Rangel-Guerrero, Natividad Cruz-Ortiz, Tatiana Nunez-Garcia, Gisela Barrera-Badillo, Lucia Hernandez-Rivas, Irma Lopez-Martinez, Ernesto Ramirez-Gonzalez. |
| EPI_ISL_2688975 | LESP Jalisco | Instituto de Diagnostico y Referencia Epidemiologicos (INDRE) | Claudia Wong-Arambula, Abril Rodriguez-Maldonado, Vanessa Rivero-Arredondo, Ariadna Medina-Benitez, Joaquin Quiroz-Mercado, Sergio Rangel-Guerrero, Natividad Cruz-Ortiz, Tatiana Nunez-Garcia, Gisela Barrera-Badillo, Lucia Hernandez-Rivas, Irma Lopez-Martinez, Ernesto Ramirez-Gonzalez. |
| EPI_ISL_2688976, EPI_ISL_2688977, EPI_ISL_2688978, EPI_ISL_2688979 | LESP Guanajuato | Instituto de Diagnostico y Referencia Epidemiologicos (INDRE) | Claudia Wong-Arambula, Abril Rodriguez-Maldonado, Vanessa Rivero-Arredondo, Ariadna Medina-Benitez, Joaquin Quiroz-Mercado, Sergio Rangel-Guerrero, Natividad Cruz-Ortiz, Tatiana Nunez-Garcia, Gisela Barrera-Badillo, Lucia Hernandez-Rivas, Irma Lopez-Martinez, Ernesto Ramirez-Gonzalez. |
| EPI_ISL_2688980, EPI_ISL_2688981, EPI_ISL_2688982, EPI_ISL_2688983, EPI_ISL_2688984 | LESP Chihuahua | Instituto de Diagnostico y Referencia Epidemiologicos (INDRE) | Claudia Wong-Arambula, Abril Rodriguez-Maldonado, Vanessa Rivero-Arredondo, Ariadna Medina-Benitez, Joaquin Quiroz-Mercado, Sergio Rangel-Guerrero, Natividad Cruz-Ortiz, Tatiana Nunez-Garcia, Gisela Barrera-Badillo, Lucia Hernandez-Rivas, Irma Lopez-Martinez, Ernesto Ramirez-Gonzalez. |
| EPI_ISL_2688985 | LESP Coahuila | Instituto de Diagnostico y Referencia Epidemiologicos (INDRE) | Claudia Wong-Arambula, Abril Rodriguez-Maldonado, Vanessa Rivero-Arredondo, Ariadna Medina-Benitez, Joaquin Quiroz-Mercado, Sergio Rangel-Guerrero, Natividad Cruz-Ortiz, Tatiana Nunez-Garcia, Gisela Barrera-Badillo, Lucia Hernandez-Rivas, Irma Lopez-Martinez, Ernesto Ramirez-Gonzalez. |
| EPI_ISL_2688986, EPI_ISL_2688987 | LESP Puebla | Instituto de Diagnostico y Referencia Epidemiologicos (INDRE) | Claudia Wong-Arambula, Abril Rodriguez-Maldonado, Vanessa Rivero-Arredondo, Ariadna Medina-Benitez, Joaquin Quiroz-Mercado, Sergio Rangel-Guerrero, Natividad Cruz-Ortiz, Tatiana Nunez-Garcia, Gisela Barrera-Badillo, Lucia Hernandez-Rivas, Irma Lopez-Martinez, Ernesto Ramirez-Gonzalez. |
| EPI_ISL_2688988, EPI_ISL_2688989 | LESP Veracruz | Instituto de Diagnostico y Referencia Epidemiologicos (INDRE) | Claudia Wong-Arambula, Abril Rodriguez-Maldonado, Vanessa Rivero-Arredondo, Ariadna Medina-Benitez, Joaquin Quiroz-Mercado, Sergio Rangel-Guerrero, Natividad Cruz-Ortiz, Tatiana Nunez-Garcia, Gisela Barrera-Badillo, Lucia Hernandez-Rivas, Irma Lopez-Martinez, Ernesto Ramirez-Gonzalez. |
| EPI_ISL_2688990 | LESP Yucatan | Instituto de Diagnostico y Referencia Epidemiologicos (INDRE) | Claudia Wong-Arambula, Abril Rodriguez-Maldonado, Vanessa Rivero-Arredondo, Ariadna Medina-Benitez, Joaquin Quiroz-Mercado, Sergio Rangel-Guerrero, Natividad Cruz-Ortiz, Tatiana Nunez-Garcia, Gisela Barrera-Badillo, Lucia Hernandez-Rivas, Irma Lopez-Martinez, Ernesto Ramirez-Gonzalez. |
| EPI_ISL_2688997, EPI_ISL_2688998, EPI_ISL_2688999, EPI_ISL_2689000, EPI_ISL_2689001, EPI_ISL_2689002, EPI_ISL_2689003, EPI_ISL_2689004, EPI_ISL_2689005, EPI_ISL_2689006, EPI_ISL_2689007, EPI_ISL_2689008, EPI_ISL_2689009, EPI_ISL_2689010, EPI_ISL_2689011, EPI_ISL_2689012, EPI_ISL_2689013, EPI_ISL_2689014, EPI_ISL_2689015, EPI_ISL_2689016, EPI_ISL_2689017 | | | |
| see above | DC Public Health Lab/ Dept. of Forensic Sciences | DC Public Health Lab/ Dept. of Forensic Sciences | Janis Doss, Scott Nguyen, Elizabeth Zelaya, Sarah Scott, Connie Maza, Monica Mann, Brittany Hamilton, David Payne, Jocelyn Hauser |
| EPI_ISL_2689021 | Minnesota Department of Health, Public Health Laboratory | Minnesota Department of Health, Public Health Laboratory | Alexandra Lorentz, Jacob Garfin, Matt Plumb, and Xiong Wang |
| EPI_ISL_2689022, EPI_ISL_2689023 | Allina Health Laboratory | Minnesota Department of Health, Public Health Laboratory | Alexandra Lorentz, Jacob Garfin, Matt Plumb, and Xiong Wang |
| EPI_ISL_2689024, EPI_ISL_2689025 | Minnesota Department of Health, Public Health Laboratory | Minnesota Department of Health, Public Health Laboratory | Alexandra Lorentz, Jacob Garfin, Matt Plumb, and Xiong Wang |

| | | | |
|---|---|---|---|
| EPI_ISL_2689026, EPI_ISL_2689027, EPI_ISL_2689028 | M Health Fairview | Minnesota Department of Health, Public Health Laboratory | Alexandra Lorentz, Jacob Garfin, Matt Plumb, and Xiong Wang |
| EPI_ISL_2689029, EPI_ISL_2689030, EPI_ISL_2689031, EPI_ISL_2689032, EPI_ISL_2689033, EPI_ISL_2689034, EPI_ISL_2689035, EPI_ISL_2689036, EPI_ISL_2689037, EPI_ISL_2689038, EPI_ISL_2689039, EPI_ISL_2689040, EPI_ISL_2689041, EPI_ISL_2689042, EPI_ISL_2689043 | | | |
| see above | Mayo Clinic & Mayo Clinic Laboratories | Minnesota Department of Health, Public Health Laboratory | Alexandra Lorentz, Jacob Garfin, Matt Plumb, and Xiong Wang |
| EPI_ISL_2689045, EPI_ISL_2689046, EPI_ISL_2689047, EPI_ISL_2689048, EPI_ISL_2689049, EPI_ISL_2689050, EPI_ISL_2689051, EPI_ISL_2689052, EPI_ISL_2689053, EPI_ISL_2689054, EPI_ISL_2689055, EPI_ISL_2689056, EPI_ISL_2689057, EPI_ISL_2689058, EPI_ISL_2689059, EPI_ISL_2689060, EPI_ISL_2689061, EPI_ISL_2689062, EPI_ISL_2689063, EPI_ISL_2689064, EPI_ISL_2689065, EPI_ISL_2689066, EPI_ISL_2689067, EPI_ISL_2689068, EPI_ISL_2689069, EPI_ISL_2689070, EPI_ISL_2689071, EPI_ISL_2689072, EPI_ISL_2689073, EPI_ISL_2689074, EPI_ISL_2689075, EPI_ISL_2689076, EPI_ISL_2689077, EPI_ISL_2689078, EPI_ISL_2689079, EPI_ISL_2689080, EPI_ISL_2689081, EPI_ISL_2689082, EPI_ISL_2689083, EPI_ISL_2689084, EPI_ISL_2689085, EPI_ISL_2689086, EPI_ISL_2689087, EPI_ISL_2689088, EPI_ISL_2689089, EPI_ISL_2689090, EPI_ISL_2689091, EPI_ISL_2689092, EPI_ISL_2689093, EPI_ISL_2689094, EPI_ISL_2689095, EPI_ISL_2689096, EPI_ISL_2689097, EPI_ISL_2689098, EPI_ISL_2689099, EPI_ISL_2689100, EPI_ISL_2689101, EPI_ISL_2689102, EPI_ISL_2689103, EPI_ISL_2689104, EPI_ISL_2689105, EPI_ISL_2689106, EPI_ISL_2689107, EPI_ISL_2689108, EPI_ISL_2689109, EPI_ISL_2689110, EPI_ISL_2689111, EPI_ISL_2689112, EPI_ISL_2689113, EPI_ISL_2689114, EPI_ISL_2689115 | | | |
| see above | MD PHL | MD PHL | Maryland Department of Health Laboratories Administration |
| EPI_ISL_2689116, EPI_ISL_2689117, EPI_ISL_2689118, EPI_ISL_2689119, EPI_ISL_2689120, EPI_ISL_2689121, EPI_ISL_2689122, EPI_ISL_2689123, EPI_ISL_2689124, EPI_ISL_2689125, EPI_ISL_2689127, EPI_ISL_2689128, EPI_ISL_2689129, EPI_ISL_2689130, EPI_ISL_2689131, EPI_ISL_2689132, EPI_ISL_2689133, EPI_ISL_2689134, EPI_ISL_2689135, EPI_ISL_2689136, EPI_ISL_2689137, EPI_ISL_2689139, EPI_ISL_2689140, EPI_ISL_2689141, EPI_ISL_2689142, EPI_ISL_2689143 | | | |
| see above | Nebraska Public Health Laboratory | NPHL COVID-19 Response Team | NPHL COVID-19 Response Team |
| EPI_ISL_2689144, EPI_ISL_2689145, EPI_ISL_2689146, EPI_ISL_2689147, EPI_ISL_2689148, EPI_ISL_2689149 | MSK Microbiology Lab | MSK Microbiology Lab | Esther Babady, Tracy McMillen, Krupa Jani |
| EPI_ISL_2689594, EPI_ISL_2689595, EPI_ISL_2689596, EPI_ISL_2689597, EPI_ISL_2689598, EPI_ISL_2689599, EPI_ISL_2689600, EPI_ISL_2689601 | GA Department of Public Health Laboratory | Genomics and Discovery, Respiratory Viruses Branch, Division of Viral Diseases, Centers for Disease Control and Prevention | Jing Zhang, Anna Kelleher, Ying Tao, Yan Li, Brian Lynch, Krista Queen, Anna Uehara, Peter Cook, Han Jia Justin Ng, Rachel Marine, Clinton R. Paden, Dhwani Batra, Haibin Wang, Tara Coalter, Jasmine Padilla, Morgan Davis, Mili Sheth, Sarah Nobles, Mark Burroughs, Justin Lee, Adam Retchless, Suxiang Tong |
| EPI_ISL_2689602 | NM Dept. Health, Scientific Laboratory Division | Genomics and Discovery, Respiratory Viruses Branch, Division of Viral Diseases, Centers for Disease Control and Prevention | Jing Zhang, Anna Kelleher, Ying Tao, Yan Li, Brian Lynch, Krista Queen, Anna Uehara, Peter Cook, Han Jia Justin Ng, Rachel Marine, Clinton R. Paden, Dhwani Batra, Haibin Wang, Tara Coalter, Jasmine Padilla, Morgan Davis, Mili Sheth, Sarah Nobles, Mark Burroughs, Justin Lee, Adam Retchless, Suxiang Tong |
| EPI_ISL_2689603 | OK Public Health Laboratory, Oklahoma State DOH | Genomics and Discovery, Respiratory Viruses Branch, Division of Viral Diseases, Centers for Disease Control and Prevention | Jing Zhang, Anna Kelleher, Ying Tao, Yan Li, Brian Lynch, Krista Queen, Anna Uehara, Peter Cook, Han Jia Justin Ng, Rachel Marine, Clinton R. Paden, Dhwani Batra, Haibin Wang, Tara Coalter, Jasmine Padilla, Morgan Davis, Mili Sheth, Sarah Nobles, Mark Burroughs, Justin Lee, Adam Retchless, Suxiang Tong |
| EPI_ISL_2689604 | AR Dept. of Health-PHL, Molecular Diagnostics | Genomics and Discovery, Respiratory Viruses Branch, Division of Viral Diseases, Centers for Disease Control and Prevention | Jing Zhang, Anna Kelleher, Ying Tao, Yan Li, Brian Lynch, Krista Queen, Anna Uehara, Peter Cook, Han Jia Justin Ng, Rachel Marine, Clinton R. Paden, Dhwani Batra, Haibin Wang, Tara Coalter, Jasmine Padilla, Morgan Davis, Mili Sheth, Sarah Nobles, Mark Burroughs, Justin Lee, Adam Retchless, Suxiang Tong |
| EPI_ISL_2689605, EPI_ISL_2689606 | VA-Division of Consolidated Laboratory Services | Genomics and Discovery, Respiratory Viruses Branch, Division of Viral Diseases, Centers for Disease Control and Prevention | Jing Zhang, Anna Kelleher, Ying Tao, Yan Li, Brian Lynch, Krista Queen, Anna Uehara, Peter Cook, Han Jia Justin Ng, Rachel Marine, Clinton R. Paden, Dhwani Batra, Haibin Wang, Tara Coalter, Jasmine Padilla, Morgan Davis, Mili Sheth, Sarah Nobles, Mark Burroughs, Justin Lee, Adam Retchless, Suxiang Tong |
| EPI_ISL_2689607 | NC State Laboratory of Public Health | Genomics and Discovery, Respiratory Viruses Branch, Division of Viral Diseases, Centers for Disease Control and Prevention | Jing Zhang, Anna Kelleher, Ying Tao, Yan Li, Brian Lynch, Krista Queen, Anna Uehara, Peter Cook, Han Jia Justin Ng, Rachel Marine, Clinton R. Paden, Dhwani Batra, Haibin Wang, Tara Coalter, Jasmine Padilla, Morgan Davis, Mili Sheth, Sarah Nobles, Mark Burroughs, Justin Lee, Adam Retchless, Suxiang Tong |
| EPI_ISL_2689661, EPI_ISL_2689662, EPI_ISL_2689663 | Kansas Health and Environmental Lab | Kansas Health and Environmental Lab | Katherine Wiggins, Mike Grose, Jonathan Barnell, Ben Olsen, and Phil Adam |
| EPI_ISL_2689665, EPI_ISL_2689666, EPI_ISL_2689667, EPI_ISL_2689668, EPI_ISL_2689669, EPI_ISL_2689670, EPI_ISL_2689671, EPI_ISL_2689672, EPI_ISL_2689673, EPI_ISL_2689674, EPI_ISL_2689675, EPI_ISL_2689676, EPI_ISL_2689677, EPI_ISL_2689678, EPI_ISL_2689679, EPI_ISL_2689680, EPI_ISL_2689681, EPI_ISL_2689682, EPI_ISL_2689683, EPI_ISL_2689684, EPI_ISL_2689685 | | | |
| see above | Nebraska Public Health Laboratory | NPHL COVID-19 Response Team | NPHL COVID-19 Response Team |
| EPI_ISL_2689737, EPI_ISL_2689738, EPI_ISL_2689739, EPI_ISL_2689740, EPI_ISL_2689741, EPI_ISL_2689742, EPI_ISL_2689743, EPI_ISL_2689744, EPI_ISL_2689745, EPI_ISL_2689746, EPI_ISL_2689747, EPI_ISL_2689748, EPI_ISL_2689749, EPI_ISL_2689750, EPI_ISL_2689751, EPI_ISL_2689752, EPI_ISL_2689753, EPI_ISL_2689754 | | | |
| see above | Idaho Bureau of Laboratories | Center for Global Health, University of New Mexico Health Sciences Center | Daryl Domman, Kurt Schwalm, Valerie Morley, Matthew Burns, Robert Voermans, Christopher Ball, Darrell Dinwiddie |
| EPI_ISL_2689755, EPI_ISL_2689756, EPI_ISL_2689757 | Kansas Health and Environmental Lab | Kansas Health and Environmental Lab | Katherine Wiggins, Mike Grose, Jonathan Barnell, Ben Olsen, and Phil Adam |
| EPI_ISL_2689758, EPI_ISL_2689760, EPI_ISL_2689761, EPI_ISL_2689762, EPI_ISL_2689763, EPI_ISL_2689764, EPI_ISL_2689765, EPI_ISL_2689766, EPI_ISL_2689767, EPI_ISL_2689768, EPI_ISL_2689769, EPI_ISL_2689770, EPI_ISL_2689772, EPI_ISL_2689773, EPI_ISL_2689774, EPI_ISL_2689775, EPI_ISL_2689776, EPI_ISL_2689777, EPI_ISL_2689778, EPI_ISL_2689779, EPI_ISL_2689780, EPI_ISL_2689781, EPI_ISL_2689782 | | | |
| see above | Wichita State University - Molecular Diagnostics Lab | Kansas Health and Environmental Lab | Katherine Wiggins, Mike Grose, Jonathan Barnell, Ben Olsen, and Phil Adam |
| EPI_ISL_2689794 | LESP Sonora | Instituto de Diagnostico y Referencia Epidemiologicos (INDRE) | Claudia Wong-Arambula, Abril Rodriguez-Maldonado, Vanessa Rivero-Arredondo, Ariadna Medina-Benitez, Joaquin Quiroz-Mercado, Sergio Rangel-Guerrero, Natividad Cruz-Ortiz, Tatiana Nunez-Garcia, Gisela Barrera-Badillo, Lucia Hernandez-Rivas, Irma Lopez-Martinez, Ernesto Ramirez-Gonzalez. |
| EPI_ISL_2689795 | LESP Chiapas | Instituto de Diagnostico y Referencia Epidemiologicos (INDRE) | Claudia Wong-Arambula, Abril Rodriguez-Maldonado, Vanessa Rivero-Arredondo, Ariadna Medina-Benitez, Joaquin Quiroz-Mercado, Sergio Rangel-Guerrero, Natividad Cruz-Ortiz, Tatiana Nunez-Garcia, Gisela Barrera-Badillo, Lucia Hernandez-Rivas, Irma Lopez-Martinez, Ernesto Ramirez-Gonzalez. |
| EPI_ISL_2689796, EPI_ISL_2689797, EPI_ISL_2689798 | LESP Ciudad de Mexico | Instituto de Diagnostico y Referencia Epidemiologicos (INDRE) | Claudia Wong-Arambula, Abril Rodriguez-Maldonado, Vanessa Rivero-Arredondo, Ariadna Medina-Benitez, Joaquin Quiroz-Mercado, Sergio Rangel-Guerrero, Natividad Cruz-Ortiz, Tatiana Nunez-Garcia, Gisela Barrera-Badillo, Lucia Hernandez-Rivas, Irma Lopez-Martinez, Ernesto Ramirez-Gonzalez. |
| EPI_ISL_2689799 | LESP Durango | Instituto de Diagnostico y Referencia Epidemiologicos (INDRE) | Claudia Wong-Arambula, Abril Rodriguez-Maldonado, Vanessa Rivero-Arredondo, Ariadna Medina-Benitez, Joaquin Quiroz-Mercado, Sergio Rangel-Guerrero, Natividad Cruz-Ortiz, Tatiana Nunez-Garcia, Gisela Barrera-Badillo, Lucia Hernandez-Rivas, Irma Lopez-Martinez, Ernesto Ramirez-Gonzalez. |
| EPI_ISL_2689800, EPI_ISL_2689801, EPI_ISL_2689802, EPI_ISL_2689803, EPI_ISL_2689804 | LESP Ciudad de Mexico | Instituto de Diagnostico y Referencia Epidemiologicos (INDRE) | Claudia Wong-Arambula, Abril Rodriguez-Maldonado, Vanessa Rivero-Arredondo, Ariadna Medina-Benitez, Joaquin Quiroz-Mercado, Sergio Rangel-Guerrero, Natividad Cruz-Ortiz, Tatiana Nunez-Garcia, Gisela Barrera-Badillo, Lucia Hernandez-Rivas, Irma Lopez-Martinez, Ernesto Ramirez-Gonzalez. |
| EPI_ISL_2689805, EPI_ISL_2689806 | LESP Sinaloa | Instituto de Diagnostico y Referencia Epidemiologicos (INDRE) | Claudia Wong-Arambula, Abril Rodriguez-Maldonado, Vanessa Rivero-Arredondo, Ariadna Medina-Benitez, Joaquin Quiroz-Mercado, Sergio Rangel-Guerrero, Natividad Cruz-Ortiz, Tatiana Nunez-Garcia, Gisela Barrera-Badillo, Lucia Hernandez-Rivas, Irma Lopez-Martinez, Ernesto Ramirez-Gonzalez. |
| EPI_ISL_2689807 | LESP Zacatecas | Instituto de Diagnostico y Referencia Epidemiologicos (INDRE) | Claudia Wong-Arambula, Abril Rodriguez-Maldonado, Vanessa Rivero-Arredondo, Ariadna Medina-Benitez, Joaquin Quiroz-Mercado, Sergio Rangel-Guerrero, Natividad Cruz-Ortiz, Tatiana Nunez-Garcia, Gisela Barrera-Badillo, Lucia Hernandez-Rivas, Irma Lopez-Martinez, Ernesto Ramirez-Gonzalez. |
| EPI_ISL_2689808, EPI_ISL_2689809, EPI_ISL_2689810 | LESP Tabasco | Instituto de Diagnostico y Referencia Epidemiologicos (INDRE) | Claudia Wong-Arambula, Abril Rodriguez-Maldonado, Vanessa Rivero-Arredondo, Ariadna Medina-Benitez, Joaquin Quiroz-Mercado, Sergio Rangel-Guerrero, Natividad Cruz-Ortiz, Tatiana Nunez-Garcia, Gisela Barrera-Badillo, Lucia Hernandez-Rivas, Irma Lopez-Martinez, Ernesto Ramirez-Gonzalez. |
| EPI_ISL_2689811, EPI_ISL_2689812 | LESP Michoacan | Instituto de Diagnostico y Referencia Epidemiologicos (INDRE) | Claudia Wong-Arambula, Abril Rodriguez-Maldonado, Vanessa Rivero-Arredondo, Ariadna Medina-Benitez, Joaquin Quiroz-Mercado, Sergio Rangel-Guerrero, Natividad Cruz-Ortiz, Tatiana Nunez-Garcia, Gisela Barrera-Badillo, Lucia Hernandez-Rivas, Irma Lopez-Martinez, Ernesto Ramirez-Gonzalez. |
| EPI_ISL_2689813 | LESP Tabasco | Instituto de Diagnostico y Referencia Epidemiologicos (INDRE) | Claudia Wong-Arambula, Abril Rodriguez-Maldonado, Vanessa Rivero-Arredondo, Ariadna Medina-Benitez, Joaquin Quiroz-Mercado, Sergio Rangel-Guerrero, Natividad Cruz-Ortiz, Tatiana Nunez-Garcia, Gisela Barrera-Badillo, Lucia Hernandez-Rivas, Irma Lopez-Martinez, Ernesto Ramirez-Gonzalez. |
| EPI_ISL_2689814, EPI_ISL_2689815, EPI_ISL_2689816, EPI_ISL_2689817, EPI_ISL_2689818, EPI_ISL_2689819 | LESP Ciudad de Mexico | Instituto de Diagnostico y Referencia Epidemiologicos (INDRE) | Claudia Wong-Arambula, Abril Rodriguez-Maldonado, Vanessa Rivero-Arredondo, Ariadna Medina-Benitez, Joaquin Quiroz-Mercado, Sergio Rangel-Guerrero, Natividad Cruz-Ortiz, Tatiana Nunez-Garcia, Gisela Barrera-Badillo, Lucia Hernandez-Rivas, Irma Lopez-Martinez, Ernesto Ramirez-Gonzalez. |
| EPI_ISL_2689820 | LESP Durango | Instituto de Diagnostico y Referencia Epidemiologicos (INDRE) | Claudia Wong-Arambula, Abril Rodriguez-Maldonado, Vanessa Rivero-Arredondo, Ariadna Medina-Benitez, Joaquin Quiroz-Mercado, Sergio Rangel-Guerrero, Natividad Cruz-Ortiz, Tatiana Nunez-Garcia, Gisela Barrera-Badillo, Lucia Hernandez-Rivas, Irma Lopez-Martinez, Ernesto Ramirez-Gonzalez. |
| EPI_ISL_2689821, EPI_ISL_2689822, EPI_ISL_2689823 | LESP Ciudad de Mexico | Instituto de Diagnostico y Referencia Epidemiologicos (INDRE) | Claudia Wong-Arambula, Abril Rodriguez-Maldonado, Vanessa Rivero-Arredondo, Ariadna Medina-Benitez, Joaquin Quiroz-Mercado, Sergio Rangel-Guerrero, Natividad Cruz-Ortiz, Tatiana Nunez-Garcia, Gisela Barrera-Badillo, Lucia Hernandez-Rivas, Irma Lopez-Martinez, Ernesto Ramirez-Gonzalez. |

| | | | |
|---|---|---|---|
| EPI_ISL_2689824 | LESP Queretaro | Instituto de Diagnostico y Referencia Epidemiologicos (INDRE) | Claudia Wong-Arambula, Abril Rodriguez-Maldonado, Vanessa Rivero-Arredondo, Ariadna Medina-Benitez, Joaquin Quiroz-Mercado, Sergio Rangel-Guerrero, Natividad Cruz-Ortiz, Tatiana Nunez-Garcia, Gisela Barrera-Badillo, Lucia Hernandez-Rivas, Irma Lopez-Martinez, Ernesto Ramirez-Gonzalez. |
| EPI_ISL_2689825 | LESP Michoacan | Instituto de Diagnostico y Referencia Epidemiologicos (INDRE) | Claudia Wong-Arambula, Abril Rodriguez-Maldonado, Vanessa Rivero-Arredondo, Ariadna Medina-Benitez, Joaquin Quiroz-Mercado, Sergio Rangel-Guerrero, Natividad Cruz-Ortiz, Tatiana Nunez-Garcia, Gisela Barrera-Badillo, Lucia Hernandez-Rivas, Irma Lopez-Martinez, Ernesto Ramirez-Gonzalez. |
| EPI_ISL_2689826 | LESP Veracruz | Instituto de Diagnostico y Referencia Epidemiologicos (INDRE) | Claudia Wong-Arambula, Abril Rodriguez-Maldonado, Vanessa Rivero-Arredondo, Ariadna Medina-Benitez, Joaquin Quiroz-Mercado, Sergio Rangel-Guerrero, Natividad Cruz-Ortiz, Tatiana Nunez-Garcia, Gisela Barrera-Badillo, Lucia Hernandez-Rivas, Irma Lopez-Martinez, Ernesto Ramirez-Gonzalez. |
| EPI_ISL_2689827, EPI_ISL_2689828 | LESP Chihuahua | Instituto de Diagnostico y Referencia Epidemiologicos (INDRE) | Claudia Wong-Arambula, Abril Rodriguez-Maldonado, Vanessa Rivero-Arredondo, Ariadna Medina-Benitez, Joaquin Quiroz-Mercado, Sergio Rangel-Guerrero, Natividad Cruz-Ortiz, Tatiana Nunez-Garcia, Gisela Barrera-Badillo, Lucia Hernandez-Rivas, Irma Lopez-Martinez, Ernesto Ramirez-Gonzalez. |
| EPI_ISL_2689829 | LESP Sonora | Instituto de Diagnostico y Referencia Epidemiologicos (INDRE) | Claudia Wong-Arambula, Abril Rodriguez-Maldonado, Vanessa Rivero-Arredondo, Ariadna Medina-Benitez, Joaquin Quiroz-Mercado, Sergio Rangel-Guerrero, Natividad Cruz-Ortiz, Tatiana Nunez-Garcia, Gisela Barrera-Badillo, Lucia Hernandez-Rivas, Irma Lopez-Martinez, Ernesto Ramirez-Gonzalez. |
| EPI_ISL_2689847, EPI_ISL_2689848, EPI_ISL_2689849, EPI_ISL_2689850, EPI_ISL_2689851, EPI_ISL_2689852, EPI_ISL_2689853, EPI_ISL_2689854, EPI_ISL_2689855, EPI_ISL_2689856, EPI_ISL_2689857, EPI_ISL_2689858, EPI_ISL_2689859, EPI_ISL_2689860, EPI_ISL_2689861, EPI_ISL_2689862, EPI_ISL_2689863, EPI_ISL_2689864, EPI_ISL_2689865 | | | |
| see above | Virginia Division of Consolidated Laboratory Services | Virginia Division of Consolidated Laboratory Services | Virginia Division of Consolidated Laboratory Services |
| EPI_ISL_2689866 | OH Department of Health Laboratory | Centers for Disease Control and Prevention Division of Viral Diseases, Pathogen Discovery | Mili Sheth, Sarah Nobles, Jasmine Padilla, Mark Burroughs, Shoshona Le, Katie Dillon, Peter Cook, Clinton R. Paden, Dhwani Batra, Krista Queen, Kristen Knipe, Dakota Howard, Yvette Unoarumhi, Darlene Wagner, Matthew Schmerer, Ben L. Rambo-Martin, Kristine Lacek, Sam Shepard, Alison Laufer Halpin, Dave Wentworth, Vivien Dugan, Suxiang Tong, Justin Lee |
| EPI_ISL_2689867 | MO State Public Health Laboratory | Centers for Disease Control and Prevention Division of Viral Diseases, Pathogen Discovery | Mili Sheth, Sarah Nobles, Jasmine Padilla, Mark Burroughs, Shoshona Le, Katie Dillon, Peter Cook, Clinton R. Paden, Dhwani Batra, Krista Queen, Kristen Knipe, Dakota Howard, Yvette Unoarumhi, Darlene Wagner, Matthew Schmerer, Ben L. Rambo-Martin, Kristine Lacek, Sam Shepard, Alison Laufer Halpin, Dave Wentworth, Vivien Dugan, Suxiang Tong, Justin Lee |
| EPI_ISL_2689868, EPI_ISL_2689869, EPI_ISL_2689870, EPI_ISL_2689871, EPI_ISL_2689872, EPI_ISL_2689873, EPI_ISL_2689874 | "AK State Public Health Lab, State Health Department" | Centers for Disease Control and Prevention Division of Viral Diseases, Pathogen Discovery | Mili Sheth, Sarah Nobles, Jasmine Padilla, Mark Burroughs, Shoshona Le, Katie Dillon, Peter Cook, Clinton R. Paden, Dhwani Batra, Krista Queen, Kristen Knipe, Dakota Howard, Yvette Unoarumhi, Darlene Wagner, Matthew Schmerer, Ben L. Rambo-Martin, Kristine Lacek, Sam Shepard, Alison Laufer Halpin, Dave Wentworth, Vivien Dugan, Suxiang Tong, Justin Lee |
| EPI_ISL_2689875, EPI_ISL_2689876, EPI_ISL_2689877, EPI_ISL_2689878, EPI_ISL_2689879, EPI_ISL_2689880, EPI_ISL_2689881, EPI_ISL_2689882, EPI_ISL_2689883, EPI_ISL_2689884, EPI_ISL_2689885, EPI_ISL_2689886 | | | |
| see above | MS Public Health Laboratory | Centers for Disease Control and Prevention Division of Viral Diseases, Pathogen Discovery | Mili Sheth, Sarah Nobles, Jasmine Padilla, Mark Burroughs, Shoshona Le, Katie Dillon, Peter Cook, Clinton R. Paden, Dhwani Batra, Krista Queen, Kristen Knipe, Dakota Howard, Yvette Unoarumhi, Darlene Wagner, Matthew Schmerer, Ben L. Rambo-Martin, Kristine Lacek, Sam Shepard, Alison Laufer Halpin, Dave Wentworth, Vivien Dugan, Suxiang Tong, Justin Lee |
| EPI_ISL_2689887, EPI_ISL_2689888, EPI_ISL_2689889, EPI_ISL_2689890, EPI_ISL_2689891, EPI_ISL_2689892, EPI_ISL_2689893 | IA State Hygienic Laboratory | Centers for Disease Control and Prevention Division of Viral Diseases, Pathogen Discovery | Mili Sheth, Sarah Nobles, Jasmine Padilla, Mark Burroughs, Shoshona Le, Katie Dillon, Peter Cook, Clinton R. Paden, Dhwani Batra, Krista Queen, Kristen Knipe, Dakota Howard, Yvette Unoarumhi, Darlene Wagner, Matthew Schmerer, Ben L. Rambo-Martin, Kristine Lacek, Sam Shepard, Alison Laufer Halpin, Dave Wentworth, Vivien Dugan, Suxiang Tong, Justin Lee |
| EPI_ISL_2689894, EPI_ISL_2689895, EPI_ISL_2689896, EPI_ISL_2689897, EPI_ISL_2689898, EPI_ISL_2689899, EPI_ISL_2689900, EPI_ISL_2689901, EPI_ISL_2689902, EPI_ISL_2689903, EPI_ISL_2689904, EPI_ISL_2689905, EPI_ISL_2689906, EPI_ISL_2689907 | | | |
| see above | WY Public Health Laboratory | Centers for Disease Control and Prevention Division of Viral Diseases, Pathogen Discovery | Mili Sheth, Sarah Nobles, Jasmine Padilla, Mark Burroughs, Shoshona Le, Katie Dillon, Peter Cook, Clinton R. Paden, Dhwani Batra, Krista Queen, Kristen Knipe, Dakota Howard, Yvette Unoarumhi, Darlene Wagner, Matthew Schmerer, Ben L. Rambo-Martin, Kristine Lacek, Sam Shepard, Alison Laufer Halpin, Dave Wentworth, Vivien Dugan, Suxiang Tong, Justin Lee |
| EPI_ISL_2689908, EPI_ISL_2689909, EPI_ISL_2689910, EPI_ISL_2689911, EPI_ISL_2689912, EPI_ISL_2689913, EPI_ISL_2689914, EPI_ISL_2689915 | RI State Health Laboratories | Centers for Disease Control and Prevention Division of Viral Diseases, Pathogen Discovery | Mili Sheth, Sarah Nobles, Jasmine Padilla, Mark Burroughs, Shoshona Le, Katie Dillon, Peter Cook, Clinton R. Paden, Dhwani Batra, Krista Queen, Kristen Knipe, Dakota Howard, Yvette Unoarumhi, Darlene Wagner, Matthew Schmerer, Ben L. Rambo-Martin, Kristine Lacek, Sam Shepard, Alison Laufer Halpin, Dave Wentworth, Vivien Dugan, Suxiang Tong, Justin Lee |
| EPI_ISL_2689916, EPI_ISL_2689917, EPI_ISL_2689918, EPI_ISL_2689919, EPI_ISL_2689920, EPI_ISL_2689921, EPI_ISL_2689922, EPI_ISL_2689923, EPI_ISL_2689924, EPI_ISL_2689925, EPI_ISL_2689926, EPI_ISL_2689927, EPI_ISL_2689928, EPI_ISL_2689929, EPI_ISL_2689930, EPI_ISL_2689931 | | | |
| see above | NE Public Health Laboratory | Centers for Disease Control and Prevention Division of Viral Diseases, Pathogen Discovery | Mili Sheth, Sarah Nobles, Jasmine Padilla, Mark Burroughs, Shoshona Le, Katie Dillon, Peter Cook, Clinton R. Paden, Dhwani Batra, Krista Queen, Kristen Knipe, Dakota Howard, Yvette Unoarumhi, Darlene Wagner, Matthew Schmerer, Ben L. Rambo-Martin, Kristine Lacek, Sam Shepard, Alison Laufer Halpin, Dave Wentworth, Vivien Dugan, Suxiang Tong, Justin Lee |
| EPI_ISL_2689932, EPI_ISL_2689933 | CA-Los Angeles County Public Health Laboratory | Centers for Disease Control and Prevention Division of Viral Diseases, Pathogen Discovery | Mili Sheth, Sarah Nobles, Jasmine Padilla, Mark Burroughs, Shoshona Le, Katie Dillon, Peter Cook, Clinton R. Paden, Dhwani Batra, Krista Queen, Kristen Knipe, Dakota Howard, Yvette Unoarumhi, Darlene Wagner, Matthew Schmerer, Ben L. Rambo-Martin, Kristine Lacek, Sam Shepard, Alison Laufer Halpin, Dave Wentworth, Vivien Dugan, Suxiang Tong, Justin Lee |
| EPI_ISL_2689934, EPI_ISL_2689935, EPI_ISL_2689936, EPI_ISL_2689937, EPI_ISL_2689938, EPI_ISL_2689939, EPI_ISL_2689940 | NJ Public Health and Environmental Laboratories | Centers for Disease Control and Prevention Division of Viral Diseases, Pathogen Discovery | Mili Sheth, Sarah Nobles, Jasmine Padilla, Mark Burroughs, Shoshona Le, Katie Dillon, Peter Cook, Clinton R. Paden, Dhwani Batra, Krista Queen, Kristen Knipe, Dakota Howard, Yvette Unoarumhi, Darlene Wagner, Matthew Schmerer, Ben L. Rambo-Martin, Kristine Lacek, Sam Shepard, Alison Laufer Halpin, Dave Wentworth, Vivien Dugan, Suxiang Tong, Justin Lee |
| EPI_ISL_2689941, EPI_ISL_2689942, EPI_ISL_2689943, EPI_ISL_2689944, EPI_ISL_2689945, EPI_ISL_2689946, EPI_ISL_2689947, EPI_ISL_2689948, EPI_ISL_2689949, EPI_ISL_2689950, EPI_ISL_2689951, EPI_ISL_2689952, EPI_ISL_2689953 | | | |
| see above | MI - Michigan Department of Health and Human Services - Bureau of Laboratories | Centers for Disease Control and Prevention Division of Viral Diseases, Pathogen Discovery | Mili Sheth, Sarah Nobles, Jasmine Padilla, Mark Burroughs, Shoshona Le, Katie Dillon, Peter Cook, Clinton R. Paden, Dhwani Batra, Krista Queen, Kristen Knipe, Dakota Howard, Yvette Unoarumhi, Darlene Wagner, Matthew Schmerer, Ben L. Rambo-Martin, Kristine Lacek, Sam Shepard, Alison Laufer Halpin, Dave Wentworth, Vivien Dugan, Suxiang Tong, Justin Lee |
| EPI_ISL_2689954, EPI_ISL_2689955, EPI_ISL_2689956, EPI_ISL_2689957, EPI_ISL_2689958, EPI_ISL_2689959, EPI_ISL_2689960, EPI_ISL_2689961, EPI_ISL_2689962, EPI_ISL_2689963 | LA Office of Public Health Laboratories | Centers for Disease Control and Prevention Division of Viral Diseases, Pathogen Discovery | Mili Sheth, Sarah Nobles, Jasmine Padilla, Mark Burroughs, Shoshona Le, Katie Dillon, Peter Cook, Clinton R. Paden, Dhwani Batra, Krista Queen, Kristen Knipe, Dakota Howard, Yvette Unoarumhi, Darlene Wagner, Matthew Schmerer, Ben L. Rambo-Martin, Kristine Lacek, Sam Shepard, Alison Laufer Halpin, Dave Wentworth, Vivien Dugan, Suxiang Tong, Justin Lee |
| EPI_ISL_2689964 | "OK Public Health Laboratory, Oklahoma State DOH" | Centers for Disease Control and Prevention Division of Viral Diseases, Pathogen Discovery | Mili Sheth, Sarah Nobles, Jasmine Padilla, Mark Burroughs, Shoshona Le, Katie Dillon, Peter Cook, Clinton R. Paden, Dhwani Batra, Krista Queen, Kristen Knipe, Dakota Howard, Yvette Unoarumhi, Darlene Wagner, Matthew Schmerer, Ben L. Rambo-Martin, Kristine Lacek, Sam Shepard, Alison Laufer Halpin, Dave Wentworth, Vivien Dugan, Suxiang Tong, Justin Lee |
| EPI_ISL_2689965, EPI_ISL_2689966, EPI_ISL_2689967, EPI_ISL_2689968, EPI_ISL_2689969, EPI_ISL_2689970, EPI_ISL_2689971, EPI_ISL_2689972, EPI_ISL_2689973, EPI_ISL_2689974, EPI_ISL_2689975, EPI_ISL_2689976, EPI_ISL_2689977 | | | |
| see above | "AR Dept. of Health-PHL, Molecular Diagnostics" | Centers for Disease Control and Prevention Division of Viral Diseases, Pathogen Discovery | Mili Sheth, Sarah Nobles, Jasmine Padilla, Mark Burroughs, Shoshona Le, Katie Dillon, Peter Cook, Clinton R. Paden, Dhwani Batra, Krista Queen, Kristen Knipe, Dakota Howard, Yvette Unoarumhi, Darlene Wagner, Matthew Schmerer, Ben L. Rambo-Martin, Kristine Lacek, Sam Shepard, Alison Laufer Halpin, Dave Wentworth, Vivien Dugan, Suxiang Tong, Justin Lee |
| EPI_ISL_2689978, EPI_ISL_2689979, EPI_ISL_2689980, EPI_ISL_2689981, EPI_ISL_2689982, EPI_ISL_2689983, EPI_ISL_2689984, EPI_ISL_2689985, EPI_ISL_2689986, EPI_ISL_2689987, EPI_ISL_2689988 | | | |
| see above | OR State PHL-Virology/Immunology Section | Centers for Disease Control and Prevention Division of Viral Diseases, Pathogen Discovery | Mili Sheth, Sarah Nobles, Jasmine Padilla, Mark Burroughs, Shoshona Le, Katie Dillon, Peter Cook, Clinton R. Paden, Dhwani Batra, Krista Queen, Kristen Knipe, Dakota Howard, Yvette Unoarumhi, Darlene Wagner, Matthew Schmerer, Ben L. Rambo-Martin, Kristine Lacek, Sam Shepard, Alison Laufer Halpin, Dave Wentworth, Vivien Dugan, Suxiang Tong, Justin Lee |
| EPI_ISL_2689989, EPI_ISL_2689990, EPI_ISL_2689991, EPI_ISL_2689992, EPI_ISL_2689993, EPI_ISL_2689994, EPI_ISL_2689995, EPI_ISL_2689996, EPI_ISL_2689997, EPI_ISL_2689998, EPI_ISL_2689999, EPI_ISL_2690000, EPI_ISL_2690001, EPI_ISL_2690002, EPI_ISL_2690003, EPI_ISL_2690004, EPI_ISL_2690005, EPI_ISL_2690006, EPI_ISL_2690007, EPI_ISL_2690008, EPI_ISL_2690009, EPI_ISL_2690010, EPI_ISL_2690011, EPI_ISL_2690012, EPI_ISL_2690013 | | | |
| see above | IN State Department of Health Laboratory Services | Centers for Disease Control and Prevention Division of Viral Diseases, Pathogen Discovery | Mili Sheth, Sarah Nobles, Jasmine Padilla, Mark Burroughs, Shoshona Le, Katie Dillon, Peter Cook, Clinton R. Paden, Dhwani Batra, Krista Queen, Kristen Knipe, Dakota Howard, Yvette Unoarumhi, Darlene Wagner, Matthew Schmerer, Ben L. Rambo-Martin, Kristine Lacek, Sam Shepard, Alison Laufer Halpin, Dave Wentworth, Vivien Dugan, Suxiang Tong, Justin Lee |
| EPI_ISL_2690014, EPI_ISL_2690015, EPI_ISL_2690016 | NC State Laboratory of Public Health | Centers for Disease Control and Prevention Division of Viral Diseases, Pathogen Discovery | Mili Sheth, Sarah Nobles, Jasmine Padilla, Mark Burroughs, Shoshona Le, Katie Dillon, Peter Cook, Clinton R. Paden, Dhwani Batra, Krista Queen, Kristen Knipe, Dakota Howard, Yvette Unoarumhi, Darlene Wagner, Matthew Schmerer, Ben L. Rambo-Martin, Kristine Lacek, Sam Shepard, Alison Laufer Halpin, Dave Wentworth, Vivien Dugan, Suxiang Tong, Justin Lee |
| EPI_ISL_2690017, | VI-US Virgin Islands Department of | Centers for Disease Control and Prevention | Mili Sheth, Sarah Nobles, Jasmine Padilla, Mark Burroughs, Shoshona Le, Katie Dillon, Peter Cook, Clinton R. Paden, Dhwani Batra, Krista Queen, Kristen Knipe, Dakota Howard, Yvette Unoarumhi, Darlene Wagner, Matthew Schmerer, Ben L. Rambo-Martin, Kristine Lacek, Sam Shepard, |

| | | | |
|---|---|---|---|
| EPI_ISL_2690018, EPI_ISL_2690019, EPI_ISL_2690020, EPI_ISL_2690021 | Health | Division of Viral Diseases, Pathogen Discovery | Alison Laufer Halpin, Dave Wentworth, Vivien Dugan, Suxiang Tong, Justin Lee |
| EPI_ISL_2690022, EPI_ISL_2690023, EPI_ISL_2690024, EPI_ISL_2690025, EPI_ISL_2690026 | UT-Unified State Labs: Public Health Utah DOH | Centers for Disease Control and Prevention Division of Viral Diseases, Pathogen Discovery | Mili Sheth, Sarah Nobles, Jasmine Padilla, Mark Burroughs, Shoshona Le, Katie Dillon, Peter Cook, Clinton R. Paden, Dhwani Batra, Krista Queen, Kristen Knipe, Dakota Howard, Yvette Unoarumhi, Darlene Wagner, Matthew Schmerer, Ben L. Rambo-Martin, Kristine Lacek, Sam Shepard, Alison Laufer Halpin, Dave Wentworth, Vivien Dugan, Suxiang Tong, Justin Lee |
| EPI_ISL_2690027, EPI_ISL_2690028, EPI_ISL_2690029, EPI_ISL_2690030, EPI_ISL_2690031, EPI_ISL_2690032, EPI_ISL_2690033, EPI_ISL_2690034 | GA Department of Public Health Laboratory | Centers for Disease Control and Prevention Division of Viral Diseases, Pathogen Discovery | Mili Sheth, Sarah Nobles, Jasmine Padilla, Mark Burroughs, Shoshona Le, Katie Dillon, Peter Cook, Clinton R. Paden, Dhwani Batra, Krista Queen, Kristen Knipe, Dakota Howard, Yvette Unoarumhi, Darlene Wagner, Matthew Schmerer, Ben L. Rambo-Martin, Kristine Lacek, Sam Shepard, Alison Laufer Halpin, Dave Wentworth, Vivien Dugan, Suxiang Tong, Justin Lee |
| EPI_ISL_2690035, EPI_ISL_2690036, EPI_ISL_2690037, EPI_ISL_2690038, EPI_ISL_2690039, EPI_ISL_2690040, EPI_ISL_2690041, EPI_ISL_2690042 | NV-Southern Nevada Public Health Laboratory | Centers for Disease Control and Prevention Division of Viral Diseases, Pathogen Discovery | Mili Sheth, Sarah Nobles, Jasmine Padilla, Mark Burroughs, Shoshona Le, Katie Dillon, Peter Cook, Clinton R. Paden, Dhwani Batra, Krista Queen, Kristen Knipe, Dakota Howard, Yvette Unoarumhi, Darlene Wagner, Matthew Schmerer, Ben L. Rambo-Martin, Kristine Lacek, Sam Shepard, Alison Laufer Halpin, Dave Wentworth, Vivien Dugan, Suxiang Tong, Justin Lee |
| EPI_ISL_2690043, EPI_ISL_2690044, EPI_ISL_2690045, EPI_ISL_2690046 | "NM Dept. Health, Scientific Laboratory Division " | Centers for Disease Control and Prevention Division of Viral Diseases, Pathogen Discovery | Mili Sheth, Sarah Nobles, Jasmine Padilla, Mark Burroughs, Shoshona Le, Katie Dillon, Peter Cook, Clinton R. Paden, Dhwani Batra, Krista Queen, Kristen Knipe, Dakota Howard, Yvette Unoarumhi, Darlene Wagner, Matthew Schmerer, Ben L. Rambo-Martin, Kristine Lacek, Sam Shepard, Alison Laufer Halpin, Dave Wentworth, Vivien Dugan, Suxiang Tong, Justin Lee |
| EPI_ISL_2690047, EPI_ISL_2690048, EPI_ISL_2690049, EPI_ISL_2690050, EPI_ISL_2690051, EPI_ISL_2690052, EPI_ISL_2690053, EPI_ISL_2690054, EPI_ISL_2690055, EPI_ISL_2690056 | KS Health and Environmental Laboratories | Centers for Disease Control and Prevention Division of Viral Diseases, Pathogen Discovery | Mili Sheth, Sarah Nobles, Jasmine Padilla, Mark Burroughs, Shoshona Le, Katie Dillon, Peter Cook, Clinton R. Paden, Dhwani Batra, Krista Queen, Kristen Knipe, Dakota Howard, Yvette Unoarumhi, Darlene Wagner, Matthew Schmerer, Ben L. Rambo-Martin, Kristine Lacek, Sam Shepard, Alison Laufer Halpin, Dave Wentworth, Vivien Dugan, Suxiang Tong, Justin Lee |
| EPI_ISL_2690057, EPI_ISL_2690058 | NYC Department of Health and Mental Hygiene | Centers for Disease Control and Prevention Division of Viral Diseases, Pathogen Discovery | Mili Sheth, Sarah Nobles, Jasmine Padilla, Mark Burroughs, Shoshona Le, Katie Dillon, Peter Cook, Clinton R. Paden, Dhwani Batra, Krista Queen, Kristen Knipe, Dakota Howard, Yvette Unoarumhi, Darlene Wagner, Matthew Schmerer, Ben L. Rambo-Martin, Kristine Lacek, Sam Shepard, Alison Laufer Halpin, Dave Wentworth, Vivien Dugan, Suxiang Tong, Justin Lee |
| EPI_ISL_2690059, EPI_ISL_2690060, EPI_ISL_2690061, EPI_ISL_2690062, EPI_ISL_2690063, EPI_ISL_2690064, EPI_ISL_2690065, EPI_ISL_2690066, EPI_ISL_2690068, EPI_ISL_2690069, EPI_ISL_2690070, EPI_ISL_2690071, EPI_ISL_2690072, EPI_ISL_2690073, EPI_ISL_2690074, EPI_ISL_2690075, EPI_ISL_2690076, EPI_ISL_2690077, EPI_ISL_2690078, EPI_ISL_2690079, EPI_ISL_2690080, EPI_ISL_2690081, EPI_ISL_2690082, EPI_ISL_2690083, EPI_ISL_2690084, EPI_ISL_2690085, EPI_ISL_2690086 | | | |
| see above | ID Bureau of Laboratories | Centers for Disease Control and Prevention Division of Viral Diseases, Pathogen Discovery | Mili Sheth, Sarah Nobles, Jasmine Padilla, Mark Burroughs, Shoshona Le, Katie Dillon, Peter Cook, Clinton R. Paden, Dhwani Batra, Krista Queen, Kristen Knipe, Dakota Howard, Yvette Unoarumhi, Darlene Wagner, Matthew Schmerer, Ben L. Rambo-Martin, Kristine Lacek, Sam Shepard, Alison Laufer Halpin, Dave Wentworth, Vivien Dugan, Suxiang Tong, Justin Lee |
| EPI_ISL_2690088, EPI_ISL_2690089, EPI_ISL_2690090, EPI_ISL_2690091, EPI_ISL_2690092 | OH Department of Health Laboratory | Centers for Disease Control and Prevention Division of Viral Diseases, Pathogen Discovery | Mili Sheth, Sarah Nobles, Jasmine Padilla, Mark Burroughs, Shoshona Le, Katie Dillon, Peter Cook, Clinton R. Paden, Dhwani Batra, Krista Queen, Kristen Knipe, Dakota Howard, Yvette Unoarumhi, Darlene Wagner, Matthew Schmerer, Ben L. Rambo-Martin, Kristine Lacek, Sam Shepard, Alison Laufer Halpin, Dave Wentworth, Vivien Dugan, Suxiang Tong, Justin Lee |
| EPI_ISL_2690093, EPI_ISL_2690094 | "MN PHL Division, Minnesota Department of Health" | Centers for Disease Control and Prevention Division of Viral Diseases, Pathogen Discovery | Mili Sheth, Sarah Nobles, Jasmine Padilla, Mark Burroughs, Shoshona Le, Katie Dillon, Peter Cook, Clinton R. Paden, Dhwani Batra, Krista Queen, Kristen Knipe, Dakota Howard, Yvette Unoarumhi, Darlene Wagner, Matthew Schmerer, Ben L. Rambo-Martin, Kristine Lacek, Sam Shepard, Alison Laufer Halpin, Dave Wentworth, Vivien Dugan, Suxiang Tong, Justin Lee |
| EPI_ISL_2690095, EPI_ISL_2690096, EPI_ISL_2690097, EPI_ISL_2690098, EPI_ISL_2690099, EPI_ISL_2690100, EPI_ISL_2690101, EPI_ISL_2690102, EPI_ISL_2690103, EPI_ISL_2690104, EPI_ISL_2690105, EPI_ISL_2690106, EPI_ISL_2690107, EPI_ISL_2690108, EPI_ISL_2690109, EPI_ISL_2690110, EPI_ISL_2690111, EPI_ISL_2690112, EPI_ISL_2690113, EPI_ISL_2690114 | | | |
| see above | WA State Department of Health | Centers for Disease Control and Prevention Division of Viral Diseases, Pathogen Discovery | Mili Sheth, Sarah Nobles, Jasmine Padilla, Mark Burroughs, Shoshona Le, Katie Dillon, Peter Cook, Clinton R. Paden, Dhwani Batra, Krista Queen, Kristen Knipe, Dakota Howard, Yvette Unoarumhi, Darlene Wagner, Matthew Schmerer, Ben L. Rambo-Martin, Kristine Lacek, Sam Shepard, Alison Laufer Halpin, Dave Wentworth, Vivien Dugan, Suxiang Tong, Justin Lee |
| EPI_ISL_2690115, EPI_ISL_2690116, EPI_ISL_2690117, EPI_ISL_2690118, EPI_ISL_2690119 | PR Public Health Lab | Centers for Disease Control and Prevention Division of Viral Diseases, Pathogen Discovery | Mili Sheth, Sarah Nobles, Jasmine Padilla, Mark Burroughs, Shoshona Le, Katie Dillon, Peter Cook, Clinton R. Paden, Dhwani Batra, Krista Queen, Kristen Knipe, Dakota Howard, Yvette Unoarumhi, Darlene Wagner, Matthew Schmerer, Ben L. Rambo-Martin, Kristine Lacek, Sam Shepard, Alison Laufer Halpin, Dave Wentworth, Vivien Dugan, Suxiang Tong, Justin Lee |
| EPI_ISL_2690120 | Philadelphia Department of Public Health | Centers for Disease Control and Prevention Division of Viral Diseases, Pathogen Discovery | Mili Sheth, Sarah Nobles, Jasmine Padilla, Mark Burroughs, Shoshona Le, Katie Dillon, Peter Cook, Clinton R. Paden, Dhwani Batra, Krista Queen, Kristen Knipe, Dakota Howard, Yvette Unoarumhi, Darlene Wagner, Matthew Schmerer, Ben L. Rambo-Martin, Kristine Lacek, Sam Shepard, Alison Laufer Halpin, Dave Wentworth, Vivien Dugan, Suxiang Tong, Justin Lee |
| EPI_ISL_2690121, EPI_ISL_2690122, EPI_ISL_2690123, EPI_ISL_2690124, EPI_ISL_2690125, EPI_ISL_2690126, EPI_ISL_2690127, EPI_ISL_2690128 | MT Public Health Laboratory | Centers for Disease Control and Prevention Division of Viral Diseases, Pathogen Discovery | Mili Sheth, Sarah Nobles, Jasmine Padilla, Mark Burroughs, Shoshona Le, Katie Dillon, Peter Cook, Clinton R. Paden, Dhwani Batra, Krista Queen, Kristen Knipe, Dakota Howard, Yvette Unoarumhi, Darlene Wagner, Matthew Schmerer, Ben L. Rambo-Martin, Kristine Lacek, Sam Shepard, Alison Laufer Halpin, Dave Wentworth, Vivien Dugan, Suxiang Tong, Justin Lee |
| EPI_ISL_2690129, EPI_ISL_2690130, EPI_ISL_2690131, EPI_ISL_2690132, EPI_ISL_2690133, EPI_ISL_2690134, EPI_ISL_2690135, EPI_ISL_2690136, EPI_ISL_2690137, EPI_ISL_2690138, EPI_ISL_2690139, EPI_ISL_2690140, EPI_ISL_2690141, EPI_ISL_2690142, EPI_ISL_2690143, EPI_ISL_2690144, EPI_ISL_2690145, EPI_ISL_2690146, EPI_ISL_2690147, EPI_ISL_2690148, EPI_ISL_2690149, EPI_ISL_2690150, EPI_ISL_2690151, EPI_ISL_2690152, EPI_ISL_2690153, EPI_ISL_2690154, EPI_ISL_2690155, EPI_ISL_2690156, EPI_ISL_2690157, EPI_ISL_2690158, EPI_ISL_2690159, EPI_ISL_2690160, EPI_ISL_2690161, EPI_ISL_2690162, EPI_ISL_2690163, EPI_ISL_2690164, EPI_ISL_2690165, EPI_ISL_2690166, EPI_ISL_2690167, EPI_ISL_2690168, EPI_ISL_2690169, EPI_ISL_2690170, EPI_ISL_2690171, EPI_ISL_2690172, EPI_ISL_2690173, EPI_ISL_2690174, EPI_ISL_2690175, EPI_ISL_2690176, EPI_ISL_2690177, EPI_ISL_2690178, EPI_ISL_2690179, EPI_ISL_2690180, EPI_ISL_2690181, EPI_ISL_2690182, EPI_ISL_2690183, EPI_ISL_2690184, EPI_ISL_2690185, EPI_ISL_2690186, EPI_ISL_2690187, EPI_ISL_2690188, EPI_ISL_2690189, EPI_ISL_2690190, EPI_ISL_2690191, EPI_ISL_2690192, EPI_ISL_2690193 | | | |
| see above | Ochsner Health | Bioinfoexperts, LLC | Judy Crabtree, Amy Feehan, Rebecca Rose, David J. Nolan, Tessa LaFleur, Julia-Garcia-Diaz, Lucio Miele, Susanna L. Lamers |
| EPI_ISL_2690243, EPI_ISL_2690244 | Kansas Health and Environmental Lab | Kansas Health and Environmental Lab | Katherine Wiggins, Mike Grose, Jonathan Barnell, Ben Olsen, and Phil Adam |
| EPI_ISL_2690248, EPI_ISL_2690249, EPI_ISL_2690250, EPI_ISL_2690251, EPI_ISL_2690252, EPI_ISL_2690253, EPI_ISL_2690254, EPI_ISL_2690255 | University of Mississippi Medical Center, Department of Pathology | University of Mississippi Medical Center, Molecular and Genomics Core Facility | Ashley C. Johnson, Wenjie Wu, Ithiel J. Frame, Krishna K. Ayyalasomayajula, Michael R. Garrett, D. Ashley Robinson |
| EPI_ISL_2690256, EPI_ISL_2690257, EPI_ISL_2690258, EPI_ISL_2690259, EPI_ISL_2690260, EPI_ISL_2690261, EPI_ISL_2690262, EPI_ISL_2690263, EPI_ISL_2690264, EPI_ISL_2690265, EPI_ISL_2690266, EPI_ISL_2690267, EPI_ISL_2690268, EPI_ISL_2690269, EPI_ISL_2690270 | | | |
| see above | MS Public Health Laboratory | University of Mississippi Medical Center, Molecular and Genomics Core Facility | Ashley C. Johnson, Wenjie Wu, Ithiel J. Frame, Krishna K. Ayyalasomayajula, Michael R. Garrett, D. Ashley Robinson |
| EPI_ISL_2690271, EPI_ISL_2690272, EPI_ISL_2690273, EPI_ISL_2690274, EPI_ISL_2690275, EPI_ISL_2690276, EPI_ISL_2690277, EPI_ISL_2690278, EPI_ISL_2690279, EPI_ISL_2690280, EPI_ISL_2690281, EPI_ISL_2690282, EPI_ISL_2690283, EPI_ISL_2690284, EPI_ISL_2690285, EPI_ISL_2690286, EPI_ISL_2690287, EPI_ISL_2690288, EPI_ISL_2690290, EPI_ISL_2690291, EPI_ISL_2690292, EPI_ISL_2690293, EPI_ISL_2690294, EPI_ISL_2690295, EPI_ISL_2690296 | | | |
| see above | Nevada State Public Health Laboratory | Nevada State Public Health Laboratory | Andrew Gorzalski, Mark Pandori |
| EPI_ISL_2690297, EPI_ISL_2690300, EPI_ISL_2690303, EPI_ISL_2690304, EPI_ISL_2690305, EPI_ISL_2690308, EPI_ISL_2690309, EPI_ISL_2690310, EPI_ISL_2690311, EPI_ISL_2690312, EPI_ISL_2690317, EPI_ISL_2690318, EPI_ISL_2690320, EPI_ISL_2690321, EPI_ISL_2690326, EPI_ISL_2690328, EPI_ISL_2690329 | | | |
| see above | MSK Microbiology Lab | MSK Microbiology Lab | Esther Babady, Tracy McMillen, Krupa Jani |
| EPI_ISL_2690332, EPI_ISL_2690333, EPI_ISL_2690335, EPI_ISL_2690336, EPI_ISL_2690338, EPI_ISL_2690339, EPI_ISL_2690340, EPI_ISL_2690341, EPI_ISL_2690342, EPI_ISL_2690343, EPI_ISL_2690345, EPI_ISL_2690347, EPI_ISL_2690349, EPI_ISL_2690350, EPI_ISL_2690351, EPI_ISL_2690353, EPI_ISL_2690354, EPI_ISL_2690355, EPI_ISL_2690356, EPI_ISL_2690357, EPI_ISL_2690358, EPI_ISL_2690359, EPI_ISL_2690360, EPI_ISL_2690361, EPI_ISL_2690362, EPI_ISL_2690363, EPI_ISL_2690364, EPI_ISL_2690365, EPI_ISL_2690367, EPI_ISL_2690368, EPI_ISL_2690369, EPI_ISL_2690370, EPI_ISL_2690371, EPI_ISL_2690372, EPI_ISL_2690373, EPI_ISL_2690374, EPI_ISL_2690375, EPI_ISL_2690376, EPI_ISL_2690378, EPI_ISL_2690379, EPI_ISL_2690380, EPI_ISL_2690381, EPI_ISL_2690382, EPI_ISL_2690384, | | | |

| | | | |
|---|---|---|---|
| EPI_ISL_2690385, EPI_ISL_2690386, EPI_ISL_2690387, EPI_ISL_2690388, EPI_ISL_2690389, EPI_ISL_2690390, EPI_ISL_2690391, EPI_ISL_2690392, EPI_ISL_2690393, EPI_ISL_2690394, EPI_ISL_2690395, EPI_ISL_2690396, EPI_ISL_2690397, EPI_ISL_2690398, EPI_ISL_2690399, EPI_ISL_2690400 | | | |
| see above | Arizona State University | Arizona State University | Peter T. Skidmore, LaRinda A. Holland, Matthew F. Smith, Rabia Maqsood, Nicholas J. Mellor, Joy M. Blain, Valerie Harris, Joshua LaBaer, Vel Murugan, Efrem S. Lim |
| EPI_ISL_2690401, EPI_ISL_2690402, EPI_ISL_2690404, EPI_ISL_2690405, EPI_ISL_2690406, EPI_ISL_2690407, EPI_ISL_2690412, EPI_ISL_2690418, EPI_ISL_2690420, EPI_ISL_2690422, EPI_ISL_2690423, EPI_ISL_2690426, EPI_ISL_2690427, EPI_ISL_2690428, EPI_ISL_2690429, EPI_ISL_2690432, EPI_ISL_2690433, EPI_ISL_2690434, EPI_ISL_2690436, EPI_ISL_2690437, EPI_ISL_2690438, EPI_ISL_2690439, EPI_ISL_2690440, EPI_ISL_2690441, EPI_ISL_2690442, EPI_ISL_2690444, EPI_ISL_2690568, EPI_ISL_2690570, EPI_ISL_2690572, EPI_ISL_2690573, EPI_ISL_2690575, EPI_ISL_2690576, EPI_ISL_2690578, EPI_ISL_2690579, EPI_ISL_2690580, EPI_ISL_2690583, EPI_ISL_2690586, EPI_ISL_2690588, EPI_ISL_2690590, EPI_ISL_2690592, EPI_ISL_2690593, EPI_ISL_2690595, EPI_ISL_2690596, EPI_ISL_2690599, EPI_ISL_2690601, EPI_ISL_2690602, EPI_ISL_2690604, EPI_ISL_2690611, EPI_ISL_2690612, EPI_ISL_2690624, EPI_ISL_2690627, EPI_ISL_2690628, EPI_ISL_2690629, EPI_ISL_2690632, EPI_ISL_2690635, EPI_ISL_2690641, EPI_ISL_2690642, EPI_ISL_2690644, EPI_ISL_2690645, EPI_ISL_2690647, EPI_ISL_2690651, EPI_ISL_2690655, EPI_ISL_2690657, EPI_ISL_2690658, EPI_ISL_2690659, EPI_ISL_2690662, EPI_ISL_2690664, EPI_ISL_2690665, EPI_ISL_2690667, EPI_ISL_2690668, EPI_ISL_2690670, EPI_ISL_2690671, EPI_ISL_2690674, EPI_ISL_2690677, EPI_ISL_2690680, EPI_ISL_2690681, EPI_ISL_2690683, EPI_ISL_2690684, EPI_ISL_2690685, EPI_ISL_2690690, EPI_ISL_2690691, EPI_ISL_2690693, EPI_ISL_2690694, EPI_ISL_2690695, EPI_ISL_2690698, EPI_ISL_2690997, EPI_ISL_2690998, EPI_ISL_2691002, EPI_ISL_2691003, EPI_ISL_2691005, EPI_ISL_2691009, EPI_ISL_2691010, EPI_ISL_2691012, EPI_ISL_2691013, EPI_ISL_2691014, EPI_ISL_2691015, EPI_ISL_2691016, EPI_ISL_2691017, EPI_ISL_2691018, EPI_ISL_2691019, EPI_ISL_2691020, EPI_ISL_2691021, EPI_ISL_2691022, EPI_ISL_2691023, EPI_ISL_2691026, EPI_ISL_2691027, EPI_ISL_2691028, EPI_ISL_2691029, EPI_ISL_2691030, EPI_ISL_2691032, EPI_ISL_2691034, EPI_ISL_2691035, EPI_ISL_2691036, EPI_ISL_2691038, EPI_ISL_2691041, EPI_ISL_2691042, EPI_ISL_2691044, EPI_ISL_2691046, EPI_ISL_2691047, EPI_ISL_2691048, EPI_ISL_2691049, EPI_ISL_2691050, EPI_ISL_2691051, EPI_ISL_2691052, EPI_ISL_2691053, EPI_ISL_2691055, EPI_ISL_2691056, EPI_ISL_2691057, EPI_ISL_2691058, EPI_ISL_2691059, EPI_ISL_2691060, EPI_ISL_2691061, EPI_ISL_2691063, EPI_ISL_2691065, EPI_ISL_2691066, EPI_ISL_2691067, EPI_ISL_2691069, EPI_ISL_2691072, EPI_ISL_2691074, EPI_ISL_2691075, EPI_ISL_2691076, EPI_ISL_2691077, EPI_ISL_2691078, EPI_ISL_2691079, EPI_ISL_2691080, EPI_ISL_2691081, EPI_ISL_2691082, EPI_ISL_2691083, EPI_ISL_2691084, EPI_ISL_2691085, EPI_ISL_2691086, EPI_ISL_2691087, EPI_ISL_2691088 | | | |
| see above | MSK Microbiology Lab | MSK Microbiology Lab | Esther Babady, Tracy McMillen, Krupa Jani |
| EPI_ISL_2691220, EPI_ISL_2691221, EPI_ISL_2691222, EPI_ISL_2691223, EPI_ISL_2691224, EPI_ISL_2691225, EPI_ISL_2691226, EPI_ISL_2691227, EPI_ISL_2691228, EPI_ISL_2691230, EPI_ISL_2691231, EPI_ISL_2691232, EPI_ISL_2691233, EPI_ISL_2691234, EPI_ISL_2691235, EPI_ISL_2691237, EPI_ISL_2691238, EPI_ISL_2691240, EPI_ISL_2691241, EPI_ISL_2691242, EPI_ISL_2691243, EPI_ISL_2691244, EPI_ISL_2691245, EPI_ISL_2691246, EPI_ISL_2691247, EPI_ISL_2691250, EPI_ISL_2691251, EPI_ISL_2691252, EPI_ISL_2691254, EPI_ISL_2691255, EPI_ISL_2691256, EPI_ISL_2691257, EPI_ISL_2691258, EPI_ISL_2691259, EPI_ISL_2691260, EPI_ISL_2691261, EPI_ISL_2691262, EPI_ISL_2691263, EPI_ISL_2691264, EPI_ISL_2691265, EPI_ISL_2691266, EPI_ISL_2691268, EPI_ISL_2691269, EPI_ISL_2691270, EPI_ISL_2691272, EPI_ISL_2691273, EPI_ISL_2691274, EPI_ISL_2691275, EPI_ISL_2691276, EPI_ISL_2691277, EPI_ISL_2691278, EPI_ISL_2691279, EPI_ISL_2691281, EPI_ISL_2691282, EPI_ISL_2691283, EPI_ISL_2691284, EPI_ISL_2691285, EPI_ISL_2691286, EPI_ISL_2691287, EPI_ISL_2691288, EPI_ISL_2691289, EPI_ISL_2691290, EPI_ISL_2691291, EPI_ISL_2691292, EPI_ISL_2691293, EPI_ISL_2691294 | | | |
| see above | Santa Clara Valley Medical Center | Santa Clara County Public Health Laboratory | Santa Clara County Public Health Department |
| EPI_ISL_2691295 | Kaiser San Jose | Santa Clara County Public Health Laboratory | Santa Clara County Public Health Department |
| EPI_ISL_2691296 | PAMF Palo Alto | Santa Clara County Public Health Laboratory | Santa Clara County Public Health Department |
| EPI_ISL_2691299, EPI_ISL_2691303, EPI_ISL_2691305, EPI_ISL_2691307, EPI_ISL_2691308, EPI_ISL_2691310, EPI_ISL_2691311, EPI_ISL_2691312, EPI_ISL_2691313, EPI_ISL_2691314 | Santa Clara Valley Medical Center | Santa Clara County Public Health Laboratory | Santa Clara County Public Health Department |
| EPI_ISL_2691711, EPI_ISL_2691712, EPI_ISL_2691713, EPI_ISL_2691714, EPI_ISL_2691716, EPI_ISL_2691717, EPI_ISL_2691718, EPI_ISL_2691719, EPI_ISL_2691720, EPI_ISL_2691721, EPI_ISL_2691722, EPI_ISL_2691723, EPI_ISL_2691724, EPI_ISL_2691725, EPI_ISL_2691726, EPI_ISL_2691727, EPI_ISL_2691728, EPI_ISL_2691729, EPI_ISL_2691730, EPI_ISL_2691732, EPI_ISL_2691734, EPI_ISL_2691735, EPI_ISL_2691736, EPI_ISL_2691737, EPI_ISL_2691738, EPI_ISL_2691739, EPI_ISL_2691740, EPI_ISL_2691741, EPI_ISL_2691742, EPI_ISL_2691743, EPI_ISL_2691744, EPI_ISL_2691745, EPI_ISL_2691746, EPI_ISL_2691747, EPI_ISL_2691748, EPI_ISL_2691749, EPI_ISL_2691750, EPI_ISL_2691751, EPI_ISL_2691752, EPI_ISL_2691753, EPI_ISL_2691755, EPI_ISL_2691756, EPI_ISL_2691758, EPI_ISL_2691759, EPI_ISL_2691760, EPI_ISL_2691761, EPI_ISL_2691762, EPI_ISL_2691763, EPI_ISL_2691764, EPI_ISL_2691765, EPI_ISL_2691766, EPI_ISL_2691767, EPI_ISL_2691768, EPI_ISL_2691769, EPI_ISL_2691770, EPI_ISL_2691771, EPI_ISL_2691772, EPI_ISL_2691773, EPI_ISL_2691774, EPI_ISL_2691775, EPI_ISL_2691776, EPI_ISL_2691777, EPI_ISL_2691778, EPI_ISL_2691780, EPI_ISL_2691781, EPI_ISL_2691782, EPI_ISL_2691783, EPI_ISL_2691784, EPI_ISL_2691785, EPI_ISL_2691786, EPI_ISL_2691787, EPI_ISL_2691788, EPI_ISL_2691789, EPI_ISL_2691790, EPI_ISL_2691791, EPI_ISL_2691793, EPI_ISL_2691794, EPI_ISL_2691795, EPI_ISL_2691796, EPI_ISL_2691797, EPI_ISL_2691798, EPI_ISL_2691799, EPI_ISL_2691800, EPI_ISL_2691801, EPI_ISL_2691802, EPI_ISL_2691803, EPI_ISL_2691804, EPI_ISL_2691805, EPI_ISL_2691806, EPI_ISL_2691807, EPI_ISL_2691810, EPI_ISL_2691811, EPI_ISL_2691812, EPI_ISL_2691813, EPI_ISL_2691814, EPI_ISL_2691815, EPI_ISL_2691816, EPI_ISL_2691817, EPI_ISL_2691818, EPI_ISL_2691819, EPI_ISL_2691820, EPI_ISL_2691821, EPI_ISL_2691822, EPI_ISL_2691823, EPI_ISL_2691824, EPI_ISL_2691825, EPI_ISL_2691826, EPI_ISL_2691827, EPI_ISL_2691828, EPI_ISL_2691829, EPI_ISL_2691830, EPI_ISL_2691831, EPI_ISL_2691832, EPI_ISL_2691833, EPI_ISL_2691834, EPI_ISL_2691835, EPI_ISL_2691836, EPI_ISL_2691837, EPI_ISL_2691838, EPI_ISL_2691839, EPI_ISL_2691840, EPI_ISL_2691841, EPI_ISL_2691842, EPI_ISL_2691843, EPI_ISL_2691844, EPI_ISL_2691845, EPI_ISL_2691846, EPI_ISL_2691847, EPI_ISL_2691848, EPI_ISL_2691849, EPI_ISL_2691850, EPI_ISL_2691851, EPI_ISL_2691852, EPI_ISL_2691853, EPI_ISL_2691854, EPI_ISL_2691855, EPI_ISL_2691856, EPI_ISL_2691857, EPI_ISL_2691858, EPI_ISL_2691859, EPI_ISL_2691860, EPI_ISL_2691861, EPI_ISL_2691862, EPI_ISL_2691863, EPI_ISL_2691864, EPI_ISL_2691865, EPI_ISL_2691866, EPI_ISL_2691867, EPI_ISL_2691868, EPI_ISL_2691869, EPI_ISL_2691870, EPI_ISL_2691871, EPI_ISL_2691872, EPI_ISL_2691873, EPI_ISL_2691874, EPI_ISL_2691875, EPI_ISL_2691876, EPI_ISL_2691877, EPI_ISL_2691878, EPI_ISL_2691879, EPI_ISL_2691880, EPI_ISL_2691881, EPI_ISL_2691882, EPI_ISL_2691883, EPI_ISL_2691884, EPI_ISL_2691885, EPI_ISL_2691886, EPI_ISL_2691887, EPI_ISL_2691888, EPI_ISL_2691889, EPI_ISL_2691890, EPI_ISL_2691891, EPI_ISL_2691892, EPI_ISL_2691893, EPI_ISL_2691894, EPI_ISL_2691895, EPI_ISL_2691896, EPI_ISL_2691897, EPI_ISL_2691898, EPI_ISL_2691899, EPI_ISL_2691900, EPI_ISL_2691901, EPI_ISL_2691902, EPI_ISL_2691903, EPI_ISL_2691904, EPI_ISL_2691905, EPI_ISL_2691906, EPI_ISL_2691907, EPI_ISL_2691908, EPI_ISL_2691909, EPI_ISL_2691910, EPI_ISL_2691911 | | | |
| see above | Johns Hopkins Hospital Department of Pathology | Johns Hopkins Hospital Department of Pathology | C. Paul Morris, Chun Huai Luo, Adannaya Amadi, Matthew Schwartz, Nicholas Gallagher, Heba H. Mostafa |
| EPI_ISL_2691913, EPI_ISL_2691914, EPI_ISL_2691915, EPI_ISL_2691916, EPI_ISL_2691917, EPI_ISL_2691918, EPI_ISL_2691920, EPI_ISL_2691921, EPI_ISL_2691922, EPI_ISL_2691923, EPI_ISL_2691924, EPI_ISL_2691925, EPI_ISL_2691926, EPI_ISL_2691927, EPI_ISL_2691928, EPI_ISL_2691929, EPI_ISL_2691930, EPI_ISL_2691931, EPI_ISL_2691932, EPI_ISL_2691933, EPI_ISL_2691934, EPI_ISL_2691935, EPI_ISL_2691936, EPI_ISL_2691937, EPI_ISL_2691938, EPI_ISL_2691939, EPI_ISL_2691940 | | | |
| see above | Rush University Medical Center | RIPHL at Rush University Medical Center | Stefan Green, Kevin Kunstman, Max Kolton, Marieta Hyde, Laura Furtado, Felix Araujo Perez, Mary Hayden, Joyce Houlihan, Diane Springer |
| EPI_ISL_2691941 | Rush University Medical Center | RIPHL at Rush University Medical Center | Stefan Green, Kevin Kunstman, Max Kolton, Marieta Hyde, Laura Furtado, Felix Araujo Perez, Mary Hayden, Joyce Houlihan, Diane Springer, Do Young Kim |
| EPI_ISL_2691942, EPI_ISL_2691943, EPI_ISL_2691944, EPI_ISL_2691945, EPI_ISL_2691946, EPI_ISL_2691947, EPI_ISL_2691948, EPI_ISL_2691949, EPI_ISL_2691950, EPI_ISL_2691951, EPI_ISL_2691952, EPI_ISL_2691953, EPI_ISL_2691954, EPI_ISL_2691955, EPI_ISL_2691956, EPI_ISL_2691957 | | | |
| see above | Rush University Medical Center | RIPHL at Rush University Medical Center | Stefan Green, Kevin Kunstman, Max Kolton, Marieta Hyde, Laura Furtado, Felix Araujo Perez, Mary Hayden, Joyce Houlihan, Diane Springer |
| EPI_ISL_2691958, EPI_ISL_2691959, EPI_ISL_2691960, EPI_ISL_2691961, EPI_ISL_2691962, EPI_ISL_2691963, EPI_ISL_2691964, EPI_ISL_2691965, EPI_ISL_2691966, EPI_ISL_2691967, EPI_ISL_2691968, EPI_ISL_2691969, EPI_ISL_2691970, EPI_ISL_2691971, EPI_ISL_2691972, EPI_ISL_2691973, EPI_ISL_2691974, EPI_ISL_2691975, EPI_ISL_2691976, EPI_ISL_2691977, EPI_ISL_2691978, EPI_ISL_2691979, EPI_ISL_2691980, EPI_ISL_2691981, EPI_ISL_2691982, EPI_ISL_2691983, EPI_ISL_2691984, EPI_ISL_2691985, EPI_ISL_2691987, EPI_ISL_2691988, EPI_ISL_2691989, EPI_ISL_2691990, EPI_ISL_2691991, EPI_ISL_2691992, EPI_ISL_2691993, EPI_ISL_2691994, EPI_ISL_2691995, EPI_ISL_2691996, EPI_ISL_2691997, EPI_ISL_2691998, EPI_ISL_2691999, EPI_ISL_2692000, EPI_ISL_2692001, EPI_ISL_2692002, EPI_ISL_2692003, EPI_ISL_2692004, EPI_ISL_2692005, EPI_ISL_2692006, EPI_ISL_2692007, EPI_ISL_2692008, EPI_ISL_2692009, EPI_ISL_2692010, EPI_ISL_2692011, EPI_ISL_2692012, EPI_ISL_2692013, EPI_ISL_2692014, EPI_ISL_2692015, EPI_ISL_2692016, EPI_ISL_2692017, EPI_ISL_2692018, EPI_ISL_2692019, EPI_ISL_2692020, EPI_ISL_2692021, EPI_ISL_2692022, EPI_ISL_2692023, EPI_ISL_2692024, EPI_ISL_2692025, EPI_ISL_2692026 | | | |
| see above | University of Chicago Medicine | RIPHL at Rush University Medical Center | Stefan Green, Kevin Kunstman, Max Kolton, Marieta Hyde, Laura Furtado, Felix Araujo Perez, Lauren Megger |
| EPI_ISL_2692027, EPI_ISL_2692028, EPI_ISL_2692029, EPI_ISL_2692030, EPI_ISL_2692031, EPI_ISL_2692032, EPI_ISL_2692033, EPI_ISL_2692034, EPI_ISL_2692035, EPI_ISL_2692036, EPI_ISL_2692037, EPI_ISL_2692038, EPI_ISL_2692039, EPI_ISL_2692040, EPI_ISL_2692041, EPI_ISL_2692042, EPI_ISL_2692043, EPI_ISL_2692044, EPI_ISL_2692045, EPI_ISL_2692046, EPI_ISL_2692047, EPI_ISL_2692048, EPI_ISL_2692049, EPI_ISL_2692050, EPI_ISL_2692051 | | | |
| see above | Northwestern Memorial Hospital | RIPHL at Rush University Medical Center | Stefan Green, Kevin Kunstman, Max Kolton, Marieta Hyde, Laura Furtado, Felix Araujo Perez, Chao Qi, Amber Kimble |
| EPI_ISL_2692052, EPI_ISL_2692053, EPI_ISL_2692054, EPI_ISL_2692055, EPI_ISL_2692056, EPI_ISL_2692057, EPI_ISL_2692058, EPI_ISL_2692059, EPI_ISL_2692060, EPI_ISL_2692061, EPI_ISL_2692062, EPI_ISL_2692063, EPI_ISL_2692064, EPI_ISL_2692065, EPI_ISL_2692066, EPI_ISL_2692067, EPI_ISL_2692068, EPI_ISL_2692069, EPI_ISL_2692070, EPI_ISL_2692071, EPI_ISL_2692072, EPI_ISL_2692073, EPI_ISL_2692074, EPI_ISL_2692075, EPI_ISL_2692076, EPI_ISL_2692077, EPI_ISL_2692078, EPI_ISL_2692079, EPI_ISL_2692080, EPI_ISL_2692081, EPI_ISL_2692082, EPI_ISL_2692083, EPI_ISL_2692084 | | | |
| see above | John H Stroger Jr Hospital of Cook County | RIPHL at Rush University Medical Center | Stefan Green, Kevin Kunstman, Max Kolton, Marieta Hyde, Laura Furtado, Felix Araujo Perez, Ah-Young Sung, Sharon Welbel |
| EPI_ISL_2692085, EPI_ISL_2692086, EPI_ISL_2692087, EPI_ISL_2692088, EPI_ISL_2692089, EPI_ISL_2692090, EPI_ISL_2692091, EPI_ISL_2692092, EPI_ISL_2692093, EPI_ISL_2692094, EPI_ISL_2692095, EPI_ISL_2692096, EPI_ISL_2692097, EPI_ISL_2692098, EPI_ISL_2692099, EPI_ISL_2692100, EPI_ISL_2692101, EPI_ISL_2692102, EPI_ISL_2692103, EPI_ISL_2692104, EPI_ISL_2692105, EPI_ISL_2692106, EPI_ISL_2692107, EPI_ISL_2692108, EPI_ISL_2692109, EPI_ISL_2692110, EPI_ISL_2692111, EPI_ISL_2692112, EPI_ISL_2692113, EPI_ISL_2692114, EPI_ISL_2692115, EPI_ISL_2692116, EPI_ISL_2692117, EPI_ISL_2692118, EPI_ISL_2692119, EPI_ISL_2692120, EPI_ISL_2692121, EPI_ISL_2692122, EPI_ISL_2692123, EPI_ISL_2692124, EPI_ISL_2692125, EPI_ISL_2692126, EPI_ISL_2692127, EPI_ISL_2692128, EPI_ISL_2692129, EPI_ISL_2692130, EPI_ISL_2692131, EPI_ISL_2692132, EPI_ISL_2692133 | | | |
| see above | University of Illinois at Chicago | RIPHL at Rush University Medical Center | Stefan Green, Kevin Kunstman, Max Kolton, Marieta Hyde, Laura Furtado, Felix Araujo Perez, Julie Bentea, Omar Perez |
| EPI_ISL_2692134 | Quest Diagnostics | RIPHL at Rush University Medical Center | Stefan Green, Kevin Kunstman, Max Kolton, Marieta Hyde, Laura Furtado, Felix Araujo Perez, Chicago Department of Public Health |
| EPI_ISL_2692135 | Swedish Covenant Hospital | RIPHL at Rush University Medical Center | Stefan Green, Kevin Kunstman, Max Kolton, Marieta Hyde, Laura Furtado, Felix Araujo Perez, Chicago Department of Public Health |
| EPI_ISL_2692136, EPI_ISL_2692138, EPI_ISL_2692139, EPI_ISL_2692140, EPI_ISL_2692141 | Lifescan | RIPHL at Rush University Medical Center | Stefan Green, Kevin Kunstman, Max Kolton, Marieta Hyde, Laura Furtado, Felix Araujo Perez, Chicago Department of Public Health |
| EPI_ISL_2692145, EPI_ISL_2692146, EPI_ISL_2692147, EPI_ISL_2692148, EPI_ISL_2692149, EPI_ISL_2692150, EPI_ISL_2692151, EPI_ISL_2692152, EPI_ISL_2692153, EPI_ISL_2692154, EPI_ISL_2692155, EPI_ISL_2692156, EPI_ISL_2692157, EPI_ISL_2692158, EPI_ISL_2692159, EPI_ISL_2692160, EPI_ISL_2692161, EPI_ISL_2692162, EPI_ISL_2692163, EPI_ISL_2692164, EPI_ISL_2692165, EPI_ISL_2692166, EPI_ISL_2692167, EPI_ISL_2692168 | | | |
| see above | Montana Public Health Laboratory | Montana Public Health Laboratory | Joy Ritter, Michelle Mozer, Carrie Biskupiak, Deborah Gibson, Michael Dills |
| EPI_ISL_2692172, EPI_ISL_2692173, EPI_ISL_2692174, EPI_ISL_2692175, EPI_ISL_2692176, EPI_ISL_2692177, EPI_ISL_2692178, EPI_ISL_2692180, EPI_ISL_2692181, EPI_ISL_2692182, EPI_ISL_2692183, EPI_ISL_2692184, EPI_ISL_2692185, EPI_ISL_2692186, EPI_ISL_2692187, EPI_ISL_2692188, EPI_ISL_2692189, EPI_ISL_2692190, EPI_ISL_2692191, EPI_ISL_2692192, EPI_ISL_2692193, EPI_ISL_2692194, EPI_ISL_2692195, EPI_ISL_2692196, EPI_ISL_2692197, EPI_ISL_2692198, EPI_ISL_2692199, EPI_ISL_2692200, EPI_ISL_2692201, EPI_ISL_2692202, EPI_ISL_2692203, EPI_ISL_2692204 | | | |
| see above | Wyoming Public Health Laboratory | Wyoming Public Health Laboratory | Jim Mildenberger, Taylor Fearing, Channing Weber, Ashley Norberg, Chayse Rowley, Marley Goetz, Brian Dominguez, Elliot Thomasson, Sam Britz, Cari Sloma, Robert Petit, and Rob Christensen |
| EPI_ISL_2692208, EPI_ISL_2692209, EPI_ISL_2692210, EPI_ISL_2692211, EPI_ISL_2692212, EPI_ISL_2692213, EPI_ISL_2692214, EPI_ISL_2692215, EPI_ISL_2692216, EPI_ISL_2692217, EPI_ISL_2692218 | | | |
| see above | Seattle Flu Study | Seattle Flu Study | Deborah A. Nickerson, Chris D. Frazar, Jover Lee, Benjamin Pelle, Erica Ryke, Matthew Richardson, Amanda Adler, Elisabeth Brandstetter, Peter D. Han, Kairsten Fay, Misja Ilcisin, Kirsten Lacombe, Thomas R. Sibley, Melissa Truong, Caitlin R. Wolf, Karen Cowgill, Stephanie Schrag, Jeff Duchin, Michael Boeckh, Janet A. Englund, Michael Famulare, Barry R. Lutz, Mark J. Rieder, Lea M. Starita, Matthew Thompson, Helen Y. Chu, Trevor Bedford, Jay Shendure |
| EPI_ISL_2692219 | Seattle Flu Study | Seattle Flu Study | Deborah A. Nickerson, Chris D. Frazar, Jover Lee, Benjamin Pelle, Erica Ryke, Matthew Richardson, Amanda Adler, Elisabeth Brandstetter, Peter D. Han, Kairsten Fay, Misja Ilcisin, Kirsten Lacombe, Thomas R. Sibley, Melissa Truong, Caitlin R. Wolf, Michael Boeckh, Janet A. Englund, Michael Famulare, Barry R. Lutz, Mark J. Rieder, Lea M. Starita, Matthew Thompson, Jay Shendure, Trevor Bedford, Helen Y. Chu |
| EPI_ISL_2692220, EPI_ISL_2692221, EPI_ISL_2692222, EPI_ISL_2692223, EPI_ISL_2692224, EPI_ISL_2692225 | Seattle Flu Study | Seattle Flu Study | Deborah A. Nickerson, Chris D. Frazar, Jover Lee, Benjamin Pelle, Erica Ryke, Matthew Richardson, Amanda Adler, Elisabeth Brandstetter, Peter D. Han, Kairsten Fay, Misja Ilcisin, Kirsten Lacombe, Thomas R. Sibley, Melissa Truong, Caitlin R. Wolf, Karen Cowgill, Stephanie Schrag, Jeff Duchin, Michael Boeckh, Janet A. Englund, Michael Famulare, Barry R. Lutz, Mark J. Rieder, Lea M. Starita, Matthew Thompson, Helen Y. Chu, Trevor Bedford, Jay Shendure |
| EPI_ISL_2692226 | Seattle Flu Study | Seattle Flu Study | Deborah A. Nickerson, Chris D. Frazar, Jover Lee, Benjamin Pelle, Erica Ryke, Matthew Richardson, Amanda Adler, Elisabeth Brandstetter, Peter D. Han, Kairsten Fay, Misja Ilcisin, Kirsten Lacombe, Thomas R. Sibley, Melissa Truong, Caitlin R. Wolf, Michael Boeckh, Janet A. Englund, Michael Famulare, Barry R. Lutz, Mark J. Rieder, Lea M. Starita, Matthew Thompson, Jay Shendure, Trevor Bedford, Helen Y. Chu |
| EPI_ISL_2692227, EPI_ISL_2692228, EPI_ISL_2692229, EPI_ISL_2692230 | Seattle Flu Study | Seattle Flu Study | Deborah A. Nickerson, Chris D. Frazar, Jover Lee, Benjamin Pelle, Erica Ryke, Matthew Richardson, Amanda Adler, Elisabeth Brandstetter, Peter D. Han, Kairsten Fay, Misja Ilcisin, Kirsten Lacombe, Thomas R. Sibley, Melissa Truong, Caitlin R. Wolf, Karen Cowgill, Stephanie Schrag, Jeff Duchin, Michael Boeckh, Janet A. Englund, Michael Famulare, Barry R. Lutz, Mark J. Rieder, Lea M. Starita, Matthew Thompson, Helen Y. Chu, Trevor Bedford, Jay Shendure |
| EPI_ISL_2692231, EPI_ISL_2692232 | Seattle Flu Study | Seattle Flu Study | Deborah A. Nickerson, Chris D. Frazar, Jover Lee, Benjamin Pelle, Erica Ryke, Matthew Richardson, Amanda Adler, Elisabeth Brandstetter, Peter D. Han, Kairsten Fay, Misja Ilcisin, Kirsten Lacombe, Thomas R. Sibley, Melissa Truong, Caitlin R. Wolf, Michael Boeckh, Janet A. Englund, Michael Famulare, Barry R. Lutz, Mark J. Rieder, Lea M. Starita, Matthew Thompson, Jay Shendure, Trevor Bedford, Helen Y. Chu |
| EPI_ISL_2692233, EPI_ISL_2692234, EPI_ISL_2692235, EPI_ISL_2692236, EPI_ISL_2692237, EPI_ISL_2692238, EPI_ISL_2692239, EPI_ISL_2692241, EPI_ISL_2692242, EPI_ISL_2692243, EPI_ISL_2692244, EPI_ISL_2692245, EPI_ISL_2692246, EPI_ISL_2692247, EPI_ISL_2692248, EPI_ISL_2692249, EPI_ISL_2692250, EPI_ISL_2692251, EPI_ISL_2692252, EPI_ISL_2692253, EPI_ISL_2692254, EPI_ISL_2692255, EPI_ISL_2692256, EPI_ISL_2692257, EPI_ISL_2692258, EPI_ISL_2692259, EPI_ISL_2692260, EPI_ISL_2692261, EPI_ISL_2692262, EPI_ISL_2692263, EPI_ISL_2692264, EPI_ISL_2692265, EPI_ISL_2692266, EPI_ISL_2692267, EPI_ISL_2692268, EPI_ISL_2692269, EPI_ISL_2692270, EPI_ISL_2692271, EPI_ISL_2692272, EPI_ISL_2692273, EPI_ISL_2692274, EPI_ISL_2692275, EPI_ISL_2692277, EPI_ISL_2692278, | | | |

| | | | |
|---|---|---|---|
| EPI_ISL_2692279, EPI_ISL_2692280, EPI_ISL_2692282, EPI_ISL_2692283, EPI_ISL_2692284, EPI_ISL_2692285, EPI_ISL_2692286, EPI_ISL_2692287, EPI_ISL_2692288, EPI_ISL_2692289, EPI_ISL_2692290, EPI_ISL_2692291, EPI_ISL_2692292, EPI_ISL_2692293, EPI_ISL_2692294, EPI_ISL_2692295, EPI_ISL_2692296, EPI_ISL_2692297, EPI_ISL_2692299, EPI_ISL_2692300, EPI_ISL_2692301, EPI_ISL_2692304, EPI_ISL_2692305, EPI_ISL_2692308 | | | |
| see above | Atlas Genomics | Seattle Flu Study | Deborah A. Nickerson, Chris D. Frazar, Jover Lee, Benjamin Pelle, Erica Ryke, Matthew Richardson, Amanda Adler, Elisabeth Brandstetter, Peter D. Han, Kairsten Fay, Misja Ilcisin, Kirsten Lacombe, Thomas R. Sibley, Melissa Truong, Caitlin R. Wolf, Romesh Gautom, Geoff Melly, Brian Hiatt, Philip Dykema, Scott Lindquist, Michael Boeckh, Janet A. Englund, Michael Famulare, Barry R. Lutz, Mark J. Rieder, Lea M. Starita, Matthew Thompson, Helen Y. Chu, Jay Shendure, Trevor Bedford |
| EPI_ISL_2692309, EPI_ISL_2692310, EPI_ISL_2692311, EPI_ISL_2692313, EPI_ISL_2692314, EPI_ISL_2692315, EPI_ISL_2692317, EPI_ISL_2692318, EPI_ISL_2692319 | Seattle Flu Study | Seattle Flu Study | Deborah A. Nickerson, Chris D. Frazar, Jover Lee, Benjamin Pelle, Erica Ryke, Matthew Richardson, Amanda Adler, Elisabeth Brandstetter, Peter D. Han, Kairsten Fay, Misja Ilcisin, Kirsten Lacombe, Thomas R. Sibley, Melissa Truong, Caitlin R. Wolf, Karen Cowgill, Stephanie Schrag, Jeff Duchin, Michael Boeckh, Janet A. Englund, Michael Famulare, Barry R. Lutz, Mark J. Rieder, Lea M. Starita, Matthew Thompson, Helen Y. Chu, Trevor Bedford, Jay Shendure |
| EPI_ISL_2692320, EPI_ISL_2692321 | Seattle Flu Study | Seattle Flu Study | Deborah A. Nickerson, Chris D. Frazar, Jover Lee, Benjamin Pelle, Erica Ryke, Matthew Richardson, Amanda Adler, Elisabeth Brandstetter, Peter D. Han, Kairsten Fay, Misja Ilcisin, Kirsten Lacombe, Thomas R. Sibley, Melissa Truong, Caitlin R. Wolf, Michael Boeckh, Janet A. Englund, Michael Famulare, Barry R. Lutz, Mark J. Rieder, Lea M. Starita, Matthew Thompson, Jay Shendure, Trevor Bedford, Helen Y. Chu |
| EPI_ISL_2692322, EPI_ISL_2692323, EPI_ISL_2692324, EPI_ISL_2692325, EPI_ISL_2692326, EPI_ISL_2692327, EPI_ISL_2692328, EPI_ISL_2692330 | Seattle Flu Study | Seattle Flu Study | Deborah A. Nickerson, Chris D. Frazar, Jover Lee, Benjamin Pelle, Erica Ryke, Matthew Richardson, Amanda Adler, Elisabeth Brandstetter, Peter D. Han, Kairsten Fay, Misja Ilcisin, Kirsten Lacombe, Thomas R. Sibley, Melissa Truong, Caitlin R. Wolf, Karen Cowgill, Stephanie Schrag, Jeff Duchin, Michael Boeckh, Janet A. Englund, Michael Famulare, Barry R. Lutz, Mark J. Rieder, Lea M. Starita, Matthew Thompson, Helen Y. Chu, Trevor Bedford, Jay Shendure |
| EPI_ISL_2692332, EPI_ISL_2692333, EPI_ISL_2692334, EPI_ISL_2692336, EPI_ISL_2692337, EPI_ISL_2692338, EPI_ISL_2692339, EPI_ISL_2692340, EPI_ISL_2692341, EPI_ISL_2692342, EPI_ISL_2692343, EPI_ISL_2692344, EPI_ISL_2692345, EPI_ISL_2692346, EPI_ISL_2692347, EPI_ISL_2692348, EPI_ISL_2692349, EPI_ISL_2692350, EPI_ISL_2692352, EPI_ISL_2692353, EPI_ISL_2692354, EPI_ISL_2692355, EPI_ISL_2692356, EPI_ISL_2692357, EPI_ISL_2692358, EPI_ISL_2692359, EPI_ISL_2692360, EPI_ISL_2692361, EPI_ISL_2692362, EPI_ISL_2692364, EPI_ISL_2692365, EPI_ISL_2692366, EPI_ISL_2692367, EPI_ISL_2692368, EPI_ISL_2692369, EPI_ISL_2692370, EPI_ISL_2692371, EPI_ISL_2692372, EPI_ISL_2692374, EPI_ISL_2692375, EPI_ISL_2692377, EPI_ISL_2692378, EPI_ISL_2692379, EPI_ISL_2692380, EPI_ISL_2692381, EPI_ISL_2692382, EPI_ISL_2692383, EPI_ISL_2692384, EPI_ISL_2692385, EPI_ISL_2692386, EPI_ISL_2692388, EPI_ISL_2692390, EPI_ISL_2692391, EPI_ISL_2692392, EPI_ISL_2692393, EPI_ISL_2692394, EPI_ISL_2692395, EPI_ISL_2692396, EPI_ISL_2692397, EPI_ISL_2692398, EPI_ISL_2692399, EPI_ISL_2692400, EPI_ISL_2692403, EPI_ISL_2692404, EPI_ISL_2692405, EPI_ISL_2692406, EPI_ISL_2692407, EPI_ISL_2692408, EPI_ISL_2692409, EPI_ISL_2692410, EPI_ISL_2692411, EPI_ISL_2692412, EPI_ISL_2692413, EPI_ISL_2692414, EPI_ISL_2692415, EPI_ISL_2692416, EPI_ISL_2692417, EPI_ISL_2692418 | | | |
| see above | Northwest Laboratory | Seattle Flu Study | Deborah A. Nickerson, Chris D. Frazar, Jover Lee, Benjamin Pelle, Erica Ryke, Matthew Richardson, Amanda Adler, Elisabeth Brandstetter, Peter D. Han, Kairsten Fay, Misja Ilcisin, Kirsten Lacombe, Thomas R. Sibley, Melissa Truong, Caitlin R. Wolf, Romesh Gautom, Geoff Melly, Brian Hiatt, Philip Dykema, Scott Lindquist, Michael Boeckh, Janet A. Englund, Michael Famulare, Barry R. Lutz, Mark J. Rieder, Lea M. Starita, Matthew Thompson, Helen Y. Chu, Jay Shendure, Trevor Bedford |
| EPI_ISL_2692419, EPI_ISL_2692420, EPI_ISL_2692421, EPI_ISL_2692422, EPI_ISL_2692423, EPI_ISL_2692424, EPI_ISL_2692425, EPI_ISL_2692426, EPI_ISL_2692427, EPI_ISL_2692428, EPI_ISL_2692429, EPI_ISL_2692430, EPI_ISL_2692431, EPI_ISL_2692432, EPI_ISL_2692433, EPI_ISL_2692434, EPI_ISL_2692435, EPI_ISL_2692436, EPI_ISL_2692438 | | | |
| see above | Incyte Diagnostics Spokane | Seattle Flu Study | Deborah A. Nickerson, Chris D. Frazar, Jover Lee, Benjamin Pelle, Erica Ryke, Matthew Richardson, Amanda Adler, Elisabeth Brandstetter, Peter D. Han, Kairsten Fay, Misja Ilcisin, Kirsten Lacombe, Thomas R. Sibley, Melissa Truong, Caitlin R. Wolf, Romesh Gautom, Geoff Melly, Brian Hiatt, Philip Dykema, Scott Lindquist, Michael Boeckh, Janet A. Englund, Michael Famulare, Barry R. Lutz, Mark J. Rieder, Lea M. Starita, Matthew Thompson, Helen Y. Chu, Jay Shendure, Trevor Bedford |
| EPI_ISL_2692439, EPI_ISL_2692440, EPI_ISL_2692441, EPI_ISL_2692442, EPI_ISL_2692443, EPI_ISL_2692444, EPI_ISL_2692445, EPI_ISL_2692446, EPI_ISL_2692447, EPI_ISL_2692448, EPI_ISL_2692449, EPI_ISL_2692450, EPI_ISL_2692451, EPI_ISL_2692452, EPI_ISL_2692453, EPI_ISL_2692454, EPI_ISL_2692455, EPI_ISL_2692457, EPI_ISL_2692458, EPI_ISL_2692460, EPI_ISL_2692461, EPI_ISL_2692462, EPI_ISL_2692463, EPI_ISL_2692464, EPI_ISL_2692465, EPI_ISL_2692466, EPI_ISL_2692467, EPI_ISL_2692468, EPI_ISL_2692469, EPI_ISL_2692470, EPI_ISL_2692471, EPI_ISL_2692473, EPI_ISL_2692474, EPI_ISL_2692475, EPI_ISL_2692476, EPI_ISL_2692477, EPI_ISL_2692478, EPI_ISL_2692479, EPI_ISL_2692480, EPI_ISL_2692481, EPI_ISL_2692482, EPI_ISL_2692483, EPI_ISL_2692484, EPI_ISL_2692485, EPI_ISL_2692487, EPI_ISL_2692488, EPI_ISL_2692489, EPI_ISL_2692490, EPI_ISL_2692491, EPI_ISL_2692492, EPI_ISL_2692493, EPI_ISL_2692494, EPI_ISL_2692495, EPI_ISL_2692496, EPI_ISL_2692497 | | | |
| see above | Atlas Genomics | Seattle Flu Study | Deborah A. Nickerson, Chris D. Frazar, Jover Lee, Benjamin Pelle, Erica Ryke, Matthew Richardson, Amanda Adler, Elisabeth Brandstetter, Peter D. Han, Kairsten Fay, Misja Ilcisin, Kirsten Lacombe, Thomas R. Sibley, Melissa Truong, Caitlin R. Wolf, Romesh Gautom, Geoff Melly, Brian Hiatt, Philip Dykema, Scott Lindquist, Michael Boeckh, Janet A. Englund, Michael Famulare, Barry R. Lutz, Mark J. Rieder, Lea M. Starita, Matthew Thompson, Helen Y. Chu, Jay Shendure, Trevor Bedford |
| EPI_ISL_2692499 | Seattle Flu Study | Seattle Flu Study | Deborah A. Nickerson, Chris D. Frazar, Jover Lee, Benjamin Pelle, Erica Ryke, Matthew Richardson, Amanda Adler, Elisabeth Brandstetter, Peter D. Han, Kairsten Fay, Misja Ilcisin, Kirsten Lacombe, Thomas R. Sibley, Melissa Truong, Caitlin R. Wolf, Karen Cowgill, Stephanie Schrag, Jeff Duchin, Michael Boeckh, Janet A. Englund, Michael Famulare, Barry R. Lutz, Mark J. Rieder, Lea M. Starita, Matthew Thompson, Helen Y. Chu, Trevor Bedford, Jay Shendure |
| EPI_ISL_2692501, EPI_ISL_2692502, EPI_ISL_2692503, EPI_ISL_2692504, EPI_ISL_2692505, EPI_ISL_2692506, EPI_ISL_2692507, EPI_ISL_2692508 | Seattle Flu Study | Seattle Flu Study | Deborah A. Nickerson, Chris D. Frazar, Jover Lee, Benjamin Pelle, Erica Ryke, Matthew Richardson, Amanda Adler, Elisabeth Brandstetter, Peter D. Han, Kairsten Fay, Misja Ilcisin, Kirsten Lacombe, Thomas R. Sibley, Melissa Truong, Caitlin R. Wolf, Michael Boeckh, Janet A. Englund, Michael Famulare, Barry R. Lutz, Mark J. Rieder, Lea M. Starita, Matthew Thompson, Jay Shendure, Trevor Bedford, Helen Y. Chu |
| EPI_ISL_2692509, EPI_ISL_2692510, EPI_ISL_2692511, EPI_ISL_2692512, EPI_ISL_2692513, EPI_ISL_2692514, EPI_ISL_2692515, EPI_ISL_2692516, EPI_ISL_2692517, EPI_ISL_2692518, EPI_ISL_2692519, EPI_ISL_2692521, EPI_ISL_2692522, EPI_ISL_2692523, EPI_ISL_2692524, EPI_ISL_2692525, EPI_ISL_2692526, EPI_ISL_2692527, EPI_ISL_2692528, EPI_ISL_2692529 | | | |
| see above | Overlake Hospital | Seattle Flu Study | Deborah A. Nickerson, Chris D. Frazar, Jover Lee, Benjamin Pelle, Erica Ryke, Matthew Richardson, Amanda Adler, Elisabeth Brandstetter, Peter D. Han, Kairsten Fay, Misja Ilcisin, Kirsten Lacombe, Thomas R. Sibley, Melissa Truong, Caitlin R. Wolf, Romesh Gautom, Geoff Melly, Brian Hiatt, Philip Dykema, Scott Lindquist, Michael Boeckh, Janet A. Englund, Michael Famulare, Barry R. Lutz, Mark J. Rieder, Lea M. Starita, Matthew Thompson, Helen Y. Chu, Jay Shendure, Trevor Bedford |
| EPI_ISL_2692532 | GA Department of Public Health Laboratory | Genomics and Discovery, Respiratory Viruses Branch, Division of Viral Diseases, Centers for Disease Control and Prevention | Jing Zhang, Anna Kelleher, Ying Tao, Yan Li, Brian Lynch, Krista Queen, Anna Uehara, Peter Cook, Han Jia Justin Ng, Rachel Marine, Clinton R. Paden, Dhwani Batra, Haibin Wang, Tara Coalter, Jasmine Padilla, Morgan Davis, Mili Sheth, Sarah Nobles, Mark Burroughs, Justin Lee, Adam Retchless, Suxiang Tong |
| EPI_ISL_2692535 | GA Department of Public Health Laboratory | Genomics and Discovery, Respiratory Viruses Branch, Division of Viral Diseases, Centers for Disease Control and Prevention | Jing Zhang, Anna Kelleher, Ying Tao, Yan Li, Brian Lynch, Krista Queen, Anna Uehara, Peter Cook, Han Jia Justin Ng, Rachel Marine, Clinton R. Paden, Dhwani Batra, Haibin Wang, Tara Coalter, Jasmine Padilla, Morgan Davis, Mili Sheth, Sarah Nobles, Mark Burroughs, Justin Lee, Adam Retchless, Suxiang Tong |
| EPI_ISL_2692536, EPI_ISL_2692537, EPI_ISL_2692538 | OK Public Health Laboratory, Oklahoma State DOH | Genomics and Discovery, Respiratory Viruses Branch, Division of Viral Diseases, Centers for Disease Control and Prevention | Jing Zhang, Anna Kelleher, Ying Tao, Yan Li, Brian Lynch, Krista Queen, Anna Uehara, Peter Cook, Han Jia Justin Ng, Rachel Marine, Clinton R. Paden, Dhwani Batra, Haibin Wang, Tara Coalter, Jasmine Padilla, Morgan Davis, Mili Sheth, Sarah Nobles, Mark Burroughs, Justin Lee, Adam Retchless, Suxiang Tong |
| EPI_ISL_2692539 | Instituto Nacional de Medicina Genomica | Instituto Nacional de Medicina Genomica | Pendiente |
| EPI_ISL_2692540, EPI_ISL_2692541, EPI_ISL_2692542, EPI_ISL_2692544, EPI_ISL_2692545 | Salud Digna | Instituto Nacional de Medicina Genomica | Cedro-Tanda A, Hidalgo-Miranda A, Mendoza-Vargas A, Reyes-Grajeda JP, Abraham Campos-Romero, Moreno-Camacho José Luis, Rodriguez-Gallegos Jorge, Luna-Ruiz Marco, Gonzalez-Barrera D, Rangel-DeLeon D, Munguia-Garza P, Ramirez-Vega O, Escobar-Arrazola, M, Herrera-Montalvo LA. |
| EPI_ISL_2692546 | Instituto Nacional de Medicina Genomica | Instituto Nacional de Medicina Genomica | Cedro-Tanda A, Hidalgo-Miranda A, Mendoza-Vargas A, Reyes-Grajeda JP, Abraham Campos-Romero, Moreno-Camacho José Luis, Rodriguez-Gallegos Jorge, Luna-Ruiz Marco, Gonzalez-Barrera D, Rangel-DeLeon D, Munguia-Garza P, Ramirez-Vega O, Escobar-Arrazola, M, Herrera-Montalvo LA. |
| EPI_ISL_2692547, EPI_ISL_2692548, EPI_ISL_2692549, EPI_ISL_2692550, EPI_ISL_2692551, EPI_ISL_2692552, EPI_ISL_2692553, EPI_ISL_2692554, EPI_ISL_2692555, EPI_ISL_2692556, EPI_ISL_2692557, EPI_ISL_2692558, EPI_ISL_2692560, EPI_ISL_2692561, EPI_ISL_2692562, EPI_ISL_2692563, EPI_ISL_2692564, EPI_ISL_2692565, EPI_ISL_2692566, EPI_ISL_2692567, EPI_ISL_2692569, EPI_ISL_2692570, EPI_ISL_2692571, EPI_ISL_2692572, EPI_ISL_2692573, EPI_ISL_2692574, EPI_ISL_2692575, EPI_ISL_2692576, EPI_ISL_2692577, EPI_ISL_2692578, EPI_ISL_2692579, EPI_ISL_2692580, EPI_ISL_2692581, EPI_ISL_2692582, EPI_ISL_2692583, EPI_ISL_2692584, EPI_ISL_2692585, EPI_ISL_2692586, EPI_ISL_2692587, EPI_ISL_2692588, EPI_ISL_2692589, EPI_ISL_2692590, EPI_ISL_2692592, EPI_ISL_2692593, EPI_ISL_2692595, EPI_ISL_2692596, EPI_ISL_2692597, EPI_ISL_2692598, EPI_ISL_2692599, EPI_ISL_2692600, EPI_ISL_2692601, EPI_ISL_2692602, EPI_ISL_2692603, EPI_ISL_2692605, EPI_ISL_2692606, EPI_ISL_2692607, EPI_ISL_2692608, EPI_ISL_2692609, EPI_ISL_2692610, EPI_ISL_2692611, EPI_ISL_2692614, EPI_ISL_2692615, EPI_ISL_2692618, EPI_ISL_2692619, EPI_ISL_2692620, EPI_ISL_2692621, EPI_ISL_2692622, EPI_ISL_2692623, EPI_ISL_2692624, EPI_ISL_2692625, EPI_ISL_2692626, EPI_ISL_2692627, EPI_ISL_2692628, EPI_ISL_2692629, EPI_ISL_2692630, EPI_ISL_2692631, EPI_ISL_2692632, EPI_ISL_2692633, EPI_ISL_2692634, EPI_ISL_2692636, EPI_ISL_2692637, EPI_ISL_2692638, EPI_ISL_2692640, EPI_ISL_2692642, EPI_ISL_2692643, EPI_ISL_2692644, EPI_ISL_2692645, EPI_ISL_2692647, EPI_ISL_2692648, EPI_ISL_2692649, EPI_ISL_2692650, EPI_ISL_2692651, EPI_ISL_2692652, EPI_ISL_2692653, EPI_ISL_2692654, EPI_ISL_2692655, EPI_ISL_2692656, EPI_ISL_2692657, EPI_ISL_2692658, EPI_ISL_2692659, EPI_ISL_2692660, EPI_ISL_2692661, EPI_ISL_2692662, EPI_ISL_2692663, EPI_ISL_2692664, EPI_ISL_2692665, EPI_ISL_2692666, EPI_ISL_2692667, EPI_ISL_2692668, EPI_ISL_2692669, EPI_ISL_2692670, EPI_ISL_2692671, EPI_ISL_2692673, EPI_ISL_2692674, EPI_ISL_2692675, EPI_ISL_2692676, EPI_ISL_2692677, EPI_ISL_2692678, EPI_ISL_2692679, EPI_ISL_2692680, EPI_ISL_2692681, EPI_ISL_2692682, EPI_ISL_2692683, EPI_ISL_2692684, EPI_ISL_2692685, EPI_ISL_2692686, EPI_ISL_2692687, EPI_ISL_2692688, EPI_ISL_2692689, EPI_ISL_2692690, EPI_ISL_2692691, EPI_ISL_2692692, EPI_ISL_2692693, EPI_ISL_2692694, EPI_ISL_2692695, EPI_ISL_2692696, EPI_ISL_2692697, EPI_ISL_2692698, EPI_ISL_2692699, EPI_ISL_2692700, EPI_ISL_2692701, EPI_ISL_2692702, EPI_ISL_2692703, EPI_ISL_2692704, EPI_ISL_2692705, EPI_ISL_2692707, EPI_ISL_2692708, EPI_ISL_2692709, EPI_ISL_2692710, EPI_ISL_2692711, EPI_ISL_2692712, EPI_ISL_2692713, EPI_ISL_2692714, EPI_ISL_2692715, EPI_ISL_2692716, EPI_ISL_2692717, EPI_ISL_2692718, EPI_ISL_2692720, EPI_ISL_2692721, EPI_ISL_2692722, EPI_ISL_2692723, EPI_ISL_2692724, EPI_ISL_2692726, EPI_ISL_2692727, EPI_ISL_2692728, EPI_ISL_2692729, EPI_ISL_2692730, EPI_ISL_2692731, EPI_ISL_2692732, EPI_ISL_2692734, EPI_ISL_2692736, EPI_ISL_2692737, EPI_ISL_2692738, EPI_ISL_2692739, EPI_ISL_2692740, EPI_ISL_2692741, EPI_ISL_2692742, EPI_ISL_2692743, EPI_ISL_2692744, EPI_ISL_2692745, EPI_ISL_2692746, EPI_ISL_2692747, EPI_ISL_2692748, EPI_ISL_2692749, EPI_ISL_2692750, EPI_ISL_2692752, EPI_ISL_2692753, EPI_ISL_2692754, EPI_ISL_2692755, EPI_ISL_2692756, EPI_ISL_2692757, EPI_ISL_2692758, EPI_ISL_2692759, EPI_ISL_2692760, EPI_ISL_2692761, EPI_ISL_2692762, EPI_ISL_2692763, EPI_ISL_2692764, EPI_ISL_2692765, EPI_ISL_2692766, EPI_ISL_2692767, EPI_ISL_2692768, EPI_ISL_2692769, EPI_ISL_2692771, EPI_ISL_2692772, EPI_ISL_2692774, EPI_ISL_2692775, EPI_ISL_2692776, EPI_ISL_2692778, EPI_ISL_2692779, EPI_ISL_2692780, EPI_ISL_2692781, EPI_ISL_2692782, EPI_ISL_2692783, EPI_ISL_2692784, EPI_ISL_2692785, EPI_ISL_2692786, EPI_ISL_2692787, EPI_ISL_2692788, EPI_ISL_2692789, EPI_ISL_2692790, EPI_ISL_2692791, EPI_ISL_2692792, EPI_ISL_2692793, EPI_ISL_2692794, EPI_ISL_2692795, EPI_ISL_2692796, EPI_ISL_2692797, EPI_ISL_2692798, EPI_ISL_2692801, EPI_ISL_2692803, EPI_ISL_2692804, EPI_ISL_2692806, EPI_ISL_2692807, EPI_ISL_2692808, EPI_ISL_2692809, EPI_ISL_2692810, EPI_ISL_2692811, EPI_ISL_2692812, EPI_ISL_2692813, EPI_ISL_2692814, EPI_ISL_2692815, EPI_ISL_2692816, EPI_ISL_2692817, EPI_ISL_2692818, EPI_ISL_2692819, EPI_ISL_2692820, EPI_ISL_2692821, EPI_ISL_2692822, EPI_ISL_2692823 | | | |
| see above | Salud Digna | Instituto Nacional de Medicina Genomica | Cedro-Tanda A, Hidalgo-Miranda A, Mendoza-Vargas A, Reyes-Grajeda JP, Abraham Campos-Romero, Moreno-Camacho José Luis, Rodriguez-Gallegos Jorge, Luna-Ruiz Marco, Gonzalez-Barrera D, Rangel-DeLeon D, Munguia-Garza P, Ramirez-Vega O, Escobar-Arrazola, M, Herrera-Montalvo LA. |
| EPI_ISL_2692825 | Instituto Nacional de Medicina Genomica | Instituto Nacional de Medicina Genomica | Cedro-Tanda A, Hidalgo-Miranda A, Mendoza-Vargas A, Reyes-Grajeda JP, Abraham Campos-Romero, Moreno-Camacho José Luis, Rodriguez-Gallegos Jorge, Luna-Ruiz Marco, Gonzalez-Barrera D, Rangel-DeLeon D, Munguia-Garza P, Ramirez-Vega O, Escobar-Arrazola, M, Herrera-Montalvo LA. |
| EPI_ISL_2692826, EPI_ISL_2692827, EPI_ISL_2692828, EPI_ISL_2692829, EPI_ISL_2692831, EPI_ISL_2692832 | Salud Digna | Instituto Nacional de Medicina Genomica | Cedro-Tanda A, Hidalgo-Miranda A, Mendoza-Vargas A, Reyes-Grajeda JP, Abraham Campos-Romero, Moreno-Camacho José Luis, Rodriguez-Gallegos Jorge, Luna-Ruiz Marco, Gonzalez-Barrera D, Rangel-DeLeon D, Munguia-Garza P, Ramirez-Vega O, Escobar-Arrazola, M, Herrera-Montalvo LA. |
| EPI_ISL_2692833, EPI_ISL_2692834, EPI_ISL_2692835, EPI_ISL_2692836, EPI_ISL_2692837, EPI_ISL_2692838, EPI_ISL_2692839, EPI_ISL_2692840, EPI_ISL_2692841, EPI_ISL_2692842, EPI_ISL_2692843, EPI_ISL_2692844, EPI_ISL_2692845, EPI_ISL_2692847 | | | |
| see above | Instituto Nacional de Medicina Genomica | Instituto Nacional de Medicina Genomica | Cedro-Tanda A, Hidalgo-Miranda A, Mendoza-Vargas A, Reyes-Grajeda JP, Abraham Campos-Romero, Moreno-Camacho José Luis, Rodriguez-Gallegos Jorge, Luna-Ruiz Marco, Gonzalez-Barrera D, Rangel-DeLeon D, Munguia-Garza P, Ramirez-Vega O, Escobar-Arrazola, M, Herrera-Montalvo LA. |
| EPI_ISL_2692848, EPI_ISL_2692849, EPI_ISL_2692850, EPI_ISL_2692851, EPI_ISL_2692852 | Salud Digna | Instituto Nacional de Medicina Genomica | Cedro-Tanda A, Hidalgo-Miranda A, Mendoza-Vargas A, Reyes-Grajeda JP, Abraham Campos-Romero, Moreno-Camacho José Luis, Rodriguez-Gallegos Jorge, Luna-Ruiz Marco, Gonzalez-Barrera D, Rangel-DeLeon D, Munguia-Garza P, Ramirez-Vega O, Escobar-Arrazola, M, Herrera-Montalvo LA. |

EPI_ISL_2693007, EPI_ISL_2693008, EPI_ISL_2693009, EPI_ISL_2693010, EPI_ISL_2693011, EPI_ISL_2693012, EPI_ISL_2693013, EPI_ISL_2693014, EPI_ISL_2693015, EPI_ISL_2693016, EPI_ISL_2693017, EPI_ISL_2693018, EPI_ISL_2693019, EPI_ISL_2693020, EPI_ISL_2693021, EPI_ISL_2693022, EPI_ISL_2693023, EPI_ISL_2693024, EPI_ISL_2693025, EPI_ISL_2693026, EPI_ISL_2693027, EPI_ISL_2693028, EPI_ISL_2693029, EPI_ISL_2693030, EPI_ISL_2693031, EPI_ISL_2693032, EPI_ISL_2693033, EPI_ISL_2693034, EPI_ISL_2693035

| | | | |
|---|---|---|---|
| see above | Oregon State Public Health Laboratory | Oregon State Public Health Laboratory | Rafia Razzaque, Eugene Yeboah, Vanda Makris, Laura Tsaknaridis, John Fontana and Shane Sevey |

EPI_ISL_2693113, EPI_ISL_2693114, EPI_ISL_2693115, EPI_ISL_2693116, EPI_ISL_2693117, EPI_ISL_2693118, EPI_ISL_2693119, EPI_ISL_2693120, EPI_ISL_2693121, EPI_ISL_2693123, EPI_ISL_2693124, EPI_ISL_2693125, EPI_ISL_2693126, EPI_ISL_2693127, EPI_ISL_2693128, EPI_ISL_2693129, EPI_ISL_2693131, EPI_ISL_2693132, EPI_ISL_2693133, EPI_ISL_2693134, EPI_ISL_2693135, EPI_ISL_2693136, EPI_ISL_2693137, EPI_ISL_2693139, EPI_ISL_2693140, EPI_ISL_2693141, EPI_ISL_2693142, EPI_ISL_2693144, EPI_ISL_2693145, EPI_ISL_2693147, EPI_ISL_2693150, EPI_ISL_2693151, EPI_ISL_2693152, EPI_ISL_2693153, EPI_ISL_2693154, EPI_ISL_2693155, EPI_ISL_2693156, EPI_ISL_2693157, EPI_ISL_2693158, EPI_ISL_2693159, EPI_ISL_2693160, EPI_ISL_2693161, EPI_ISL_2693162, EPI_ISL_2693163, EPI_ISL_2693164, EPI_ISL_2693166, EPI_ISL_2693167, EPI_ISL_2693168, EPI_ISL_2693169, EPI_ISL_2693170, EPI_ISL_2693171, EPI_ISL_2693172, EPI_ISL_2693173, EPI_ISL_2693174, EPI_ISL_2693175, EPI_ISL_2693176, EPI_ISL_2693177, EPI_ISL_2693178, EPI_ISL_2693179, EPI_ISL_2693180, EPI_ISL_2693181, EPI_ISL_2693182, EPI_ISL_2693183, EPI_ISL_2693184, EPI_ISL_2693185, EPI_ISL_2693186, EPI_ISL_2693187, EPI_ISL_2693188, EPI_ISL_2693189, EPI_ISL_2693190, EPI_ISL_2693191, EPI_ISL_2693192, EPI_ISL_2693193, EPI_ISL_2693194, EPI_ISL_2693195, EPI_ISL_2693196, EPI_ISL_2693197, EPI_ISL_2693198, EPI_ISL_2693199, EPI_ISL_2693200, EPI_ISL_2693201, EPI_ISL_2693202, EPI_ISL_2693203, EPI_ISL_2693204, EPI_ISL_2693205, EPI_ISL_2693206, EPI_ISL_2693207, EPI_ISL_2693208, EPI_ISL_2693209, EPI_ISL_2693210, EPI_ISL_2693211, EPI_ISL_2693212, EPI_ISL_2693213, EPI_ISL_2693214, EPI_ISL_2693215, EPI_ISL_2693216, EPI_ISL_2693217, EPI_ISL_2693219, EPI_ISL_2693220, EPI_ISL_2693221, EPI_ISL_2693222, EPI_ISL_2693223, EPI_ISL_2693224, EPI_ISL_2693225, EPI_ISL_2693226, EPI_ISL_2693227, EPI_ISL_2693228, EPI_ISL_2693229, EPI_ISL_2693230, EPI_ISL_2693231, EPI_ISL_2693232, EPI_ISL_2693233, EPI_ISL_2693234, EPI_ISL_2693235, EPI_ISL_2693236, EPI_ISL_2693237, EPI_ISL_2693238, EPI_ISL_2693239, EPI_ISL_2693240, EPI_ISL_2693241, EPI_ISL_2693242, EPI_ISL_2693243, EPI_ISL_2693244, EPI_ISL_2693245, EPI_ISL_2693246, EPI_ISL_2693247, EPI_ISL_2693249, EPI_ISL_2693250, EPI_ISL_2693251, EPI_ISL_2693253, EPI_ISL_2693254, EPI_ISL_2693255, EPI_ISL_2693256, EPI_ISL_2693257, EPI_ISL_2693258, EPI_ISL_2693259, EPI_ISL_2693260, EPI_ISL_2693261, EPI_ISL_2693262, EPI_ISL_2693263, EPI_ISL_2693264, EPI_ISL_2693265, EPI_ISL_2693266, EPI_ISL_2693267, EPI_ISL_2693268, EPI_ISL_2693269, EPI_ISL_2693270, EPI_ISL_2693271, EPI_ISL_2693272, EPI_ISL_2693273, EPI_ISL_2693274, EPI_ISL_2693275, EPI_ISL_2693276, EPI_ISL_2693277, EPI_ISL_2693278, EPI_ISL_2693279, EPI_ISL_2693280, EPI_ISL_2693281, EPI_ISL_2693282, EPI_ISL_2693283, EPI_ISL_2693284, EPI_ISL_2693285, EPI_ISL_2693286, EPI_ISL_2693287, EPI_ISL_2693288, EPI_ISL_2693289, EPI_ISL_2693290, EPI_ISL_2693291, EPI_ISL_2693292, EPI_ISL_2693293, EPI_ISL_2693294, EPI_ISL_2693295, EPI_ISL_2693296, EPI_ISL_2693297, EPI_ISL_2693298, EPI_ISL_2693299, EPI_ISL_2693300, EPI_ISL_2693301, EPI_ISL_2693302, EPI_ISL_2693303, EPI_ISL_2693304, EPI_ISL_2693305, EPI_ISL_2693306, EPI_ISL_2693307, EPI_ISL_2693308, EPI_ISL_2693309, EPI_ISL_2693310, EPI_ISL_2693311, EPI_ISL_2693312, EPI_ISL_2693313, EPI_ISL_2693314, EPI_ISL_2693315

| | | | |
|---|---|---|---|
| see above | UW Virology Lab | UW Virology Lab | Pavitra Roychoudhury, Hong Xie, Lasata Shrestha, Tien V. Nguyen, Shah Mohamed Bakhash, Michelle Lin, Noah R. Baker, Ricardo Perez, Sean Ellis, Nathan Breit, Robert J. Livingston, Meei-Li Huang, Keith R Jerome, Patrick Mathias, Alexander Greninger |

EPI_ISL_2693333, EPI_ISL_2693334, EPI_ISL_2693335, EPI_ISL_2693336, EPI_ISL_2693337, EPI_ISL_2693338, EPI_ISL_2693339, EPI_ISL_2693340, EPI_ISL_2693341, EPI_ISL_2693342, EPI_ISL_2693343, EPI_ISL_2693344, EPI_ISL_2693345, EPI_ISL_2693346, EPI_ISL_2693347, EPI_ISL_2693348, EPI_ISL_2693349, EPI_ISL_2693350, EPI_ISL_2693351, EPI_ISL_2693352, EPI_ISL_2693353, EPI_ISL_2693354, EPI_ISL_2693355, EPI_ISL_2693356, EPI_ISL_2693357, EPI_ISL_2693358, EPI_ISL_2693359, EPI_ISL_2693360, EPI_ISL_2693361, EPI_ISL_2693362, EPI_ISL_2693363, EPI_ISL_2693364, EPI_ISL_2693365, EPI_ISL_2693366, EPI_ISL_2693367, EPI_ISL_2693368, EPI_ISL_2693369, EPI_ISL_2693370, EPI_ISL_2693371, EPI_ISL_2693372, EPI_ISL_2693373

| | | | |
|---|---|---|---|
| see above | ISDH | Quantigen Biosciences | Paul J. Childress |

EPI_ISL_2693677, EPI_ISL_2693678, EPI_ISL_2693679, EPI_ISL_2693680, EPI_ISL_2693681, EPI_ISL_2693682, EPI_ISL_2693683, EPI_ISL_2693684, EPI_ISL_2693685, EPI_ISL_2693686, EPI_ISL_2693687, EPI_ISL_2693688, EPI_ISL_2693689, EPI_ISL_2693690, EPI_ISL_2693691, EPI_ISL_2693692, EPI_ISL_2693693, EPI_ISL_2693694, EPI_ISL_2693695, EPI_ISL_2693696, EPI_ISL_2693697, EPI_ISL_2693698, EPI_ISL_2693699, EPI_ISL_2693700, EPI_ISL_2693701, EPI_ISL_2693702, EPI_ISL_2693703, EPI_ISL_2693704, EPI_ISL_2693705, EPI_ISL_2693706, EPI_ISL_2693707, EPI_ISL_2693708, EPI_ISL_2693709, EPI_ISL_2693710, EPI_ISL_2693711, EPI_ISL_2693712, EPI_ISL_2693713, EPI_ISL_2693714, EPI_ISL_2693715, EPI_ISL_2693716, EPI_ISL_2693717, EPI_ISL_2693718, EPI_ISL_2693719, EPI_ISL_2693720, EPI_ISL_2693721, EPI_ISL_2693722, EPI_ISL_2693723, EPI_ISL_2693724, EPI_ISL_2693725, EPI_ISL_2693726, EPI_ISL_2693727, EPI_ISL_2693728, EPI_ISL_2693729, EPI_ISL_2693730, EPI_ISL_2693731, EPI_ISL_2693732, EPI_ISL_2693733, EPI_ISL_2693734, EPI_ISL_2693735, EPI_ISL_2693736, EPI_ISL_2693737, EPI_ISL_2693738, EPI_ISL_2693739, EPI_ISL_2693740, EPI_ISL_2693741, EPI_ISL_2693742, EPI_ISL_2693743, EPI_ISL_2693744, EPI_ISL_2693745, EPI_ISL_2693746, EPI_ISL_2693747, EPI_ISL_2693748, EPI_ISL_2693749, EPI_ISL_2693750, EPI_ISL_2693751, EPI_ISL_2693753, EPI_ISL_2693754, EPI_ISL_2693755, EPI_ISL_2693756, EPI_ISL_2693757, EPI_ISL_2693758, EPI_ISL_2693759, EPI_ISL_2693760, EPI_ISL_2693761, EPI_ISL_2693762, EPI_ISL_2693763, EPI_ISL_2693764, EPI_ISL_2693765, EPI_ISL_2693766, EPI_ISL_2693767, EPI_ISL_2693768, EPI_ISL_2693769, EPI_ISL_2693770, EPI_ISL_2693771, EPI_ISL_2693772, EPI_ISL_2693773, EPI_ISL_2693774, EPI_ISL_2693775, EPI_ISL_2693776, EPI_ISL_2693777, EPI_ISL_2693778, EPI_ISL_2693779, EPI_ISL_2693780, EPI_ISL_2693781, EPI_ISL_2693782, EPI_ISL_2693783, EPI_ISL_2693784, EPI_ISL_2693785, EPI_ISL_2693786, EPI_ISL_2693787, EPI_ISL_2693788, EPI_ISL_2693789, EPI_ISL_2693790, EPI_ISL_2693791, EPI_ISL_2693792, EPI_ISL_2693793, EPI_ISL_2693794, EPI_ISL_2693795, EPI_ISL_2693796, EPI_ISL_2693797, EPI_ISL_2693798, EPI_ISL_2693799, EPI_ISL_2693800, EPI_ISL_2693801, EPI_ISL_2693802, EPI_ISL_2693803, EPI_ISL_2693804, EPI_ISL_2693805, EPI_ISL_2693806, EPI_ISL_2693807, EPI_ISL_2693808, EPI_ISL_2693809, EPI_ISL_2693810, EPI_ISL_2693811, EPI_ISL_2693812, EPI_ISL_2693813, EPI_ISL_2693815, EPI_ISL_2693816, EPI_ISL_2693817, EPI_ISL_2693818, EPI_ISL_2693819, EPI_ISL_2693820, EPI_ISL_2693821, EPI_ISL_2693822, EPI_ISL_2693823

| | | | |
|---|---|---|---|
| see above | UW Virology Lab | UW Virology Lab | Pavitra Roychoudhury, Hong Xie, Lasata Shrestha, Shah Mohamed Bakhash, Tien V. Nguyen, Noah R. Baker, Sean Ellis, Meei-Li Huang, Keith R Jerome, Alexander Greninger |

EPI_ISL_2696299, EPI_ISL_2696300, EPI_ISL_2696301, EPI_ISL_2696302, EPI_ISL_2696303, EPI_ISL_2696304, EPI_ISL_2696305, EPI_ISL_2696306, EPI_ISL_2696307, EPI_ISL_2696308, EPI_ISL_2696309, EPI_ISL_2696310, EPI_ISL_2696311, EPI_ISL_2696312, EPI_ISL_2696313, EPI_ISL_2696314, EPI_ISL_2696315, EPI_ISL_2696316, EPI_ISL_2696317, EPI_ISL_2696318, EPI_ISL_2696319, EPI_ISL_2696320, EPI_ISL_2696321, EPI_ISL_2696322, EPI_ISL_2696323, EPI_ISL_2696324, EPI_ISL_2696325, EPI_ISL_2696326, EPI_ISL_2696327, EPI_ISL_2696328, EPI_ISL_2696329, EPI_ISL_2696330, EPI_ISL_2696331, EPI_ISL_2696332, EPI_ISL_2696333, EPI_ISL_2696334, EPI_ISL_2696335, EPI_ISL_2696336, EPI_ISL_2696337, EPI_ISL_2696338, EPI_ISL_2696339, EPI_ISL_2696340, EPI_ISL_2696341, EPI_ISL_2696342, EPI_ISL_2696343, EPI_ISL_2696344, EPI_ISL_2696345, EPI_ISL_2696346, EPI_ISL_2696347, EPI_ISL_2696348, EPI_ISL_2696349, EPI_ISL_2696350, EPI_ISL_2696351, EPI_ISL_2696352, EPI_ISL_2696353, EPI_ISL_2696354, EPI_ISL_2696355, EPI_ISL_2696356, EPI_ISL_2696357, EPI_ISL_2696358, EPI_ISL_2696359, EPI_ISL_2696360, EPI_ISL_2696361, EPI_ISL_2696362, EPI_ISL_2696363, EPI_ISL_2696365, EPI_ISL_2696366, EPI_ISL_2696367, EPI_ISL_2696368, EPI_ISL_2696369, EPI_ISL_2696370, EPI_ISL_2696371, EPI_ISL_2696372, EPI_ISL_2696373, EPI_ISL_2696374, EPI_ISL_2696375, EPI_ISL_2696376, EPI_ISL_2696377, EPI_ISL_2696378, EPI_ISL_2696379, EPI_ISL_2696380, EPI_ISL_2696381, EPI_ISL_2696382, EPI_ISL_2696383, EPI_ISL_2696384, EPI_ISL_2696385, EPI_ISL_2696386, EPI_ISL_2696387, EPI_ISL_2696388, EPI_ISL_2696389, EPI_ISL_2696390, EPI_ISL_2696391, EPI_ISL_2696392, EPI_ISL_2696393, EPI_ISL_2696394, EPI_ISL_2696395, EPI_ISL_2696396, EPI_ISL_2696397, EPI_ISL_2696399, EPI_ISL_2696400, EPI_ISL_2696401, EPI_ISL_2696402, EPI_ISL_2696403, EPI_ISL_2696404, EPI_ISL_2696405, EPI_ISL_2696406, EPI_ISL_2696407, EPI_ISL_2696408, EPI_ISL_2696409, EPI_ISL_2696410, EPI_ISL_2696411, EPI_ISL_2696412, EPI_ISL_2696413, EPI_ISL_2696414, EPI_ISL_2696415, EPI_ISL_2696417, EPI_ISL_2696418, EPI_ISL_2696419, EPI_ISL_2696420, EPI_ISL_2696421, EPI_ISL_2696422, EPI_ISL_2696423, EPI_ISL_2696424, EPI_ISL_2696425, EPI_ISL_2696426, EPI_ISL_2696427, EPI_ISL_2696428, EPI_ISL_2696429, EPI_ISL_2696430, EPI_ISL_2696431, EPI_ISL_2696432, EPI_ISL_2696433, EPI_ISL_2696434, EPI_ISL_2696435, EPI_ISL_2696436, EPI_ISL_2696437, EPI_ISL_2696438, EPI_ISL_2696440, EPI_ISL_2696441, EPI_ISL_2696442, EPI_ISL_2696443, EPI_ISL_2696444, EPI_ISL_2696445, EPI_ISL_2696446, EPI_ISL_2696447, EPI_ISL_2696448, EPI_ISL_2696449, EPI_ISL_2696450, EPI_ISL_2696451, EPI_ISL_2696452, EPI_ISL_2696453, EPI_ISL_2696454, EPI_ISL_2696455, EPI_ISL_2696456, EPI_ISL_2696457, EPI_ISL_2696459, EPI_ISL_2696460, EPI_ISL_2696461, EPI_ISL_2696462, EPI_ISL_2696463, EPI_ISL_2696464, EPI_ISL_2696465, EPI_ISL_2696466, EPI_ISL_2696467, EPI_ISL_2696468, EPI_ISL_2696469, EPI_ISL_2696470, EPI_ISL_2696471, EPI_ISL_2696472, EPI_ISL_2696473, EPI_ISL_2696474, EPI_ISL_2696475, EPI_ISL_2696476, EPI_ISL_2696477, EPI_ISL_2696478, EPI_ISL_2696479, EPI_ISL_2696480, EPI_ISL_2696481, EPI_ISL_2696482, EPI_ISL_2696483, EPI_ISL_2696484, EPI_ISL_2696485, EPI_ISL_2696486, EPI_ISL_2696487, EPI_ISL_2696488, EPI_ISL_2696489, EPI_ISL_2696490, EPI_ISL_2696491, EPI_ISL_2696492, EPI_ISL_2696493, EPI_ISL_2696494, EPI_ISL_2696495, EPI_ISL_2696496, EPI_ISL_2696497, EPI_ISL_2696498, EPI_ISL_2696499, EPI_ISL_2696500, EPI_ISL_2696501, EPI_ISL_2696502, EPI_ISL_2696503, EPI_ISL_2696504, EPI_ISL_2696505, EPI_ISL_2696506, EPI_ISL_2696507, EPI_ISL_2696508, EPI_ISL_2696509, EPI_ISL_2696510, EPI_ISL_2696511, EPI_ISL_2696512, EPI_ISL_2696513, EPI_ISL_2696514, EPI_ISL_2696515, EPI_ISL_2696516, EPI_ISL_2696517, EPI_ISL_2696518, EPI_ISL_2696519, EPI_ISL_2696520, EPI_ISL_2696521, EPI_ISL_2696522, EPI_ISL_2696523, EPI_ISL_2696524, EPI_ISL_2696525, EPI_ISL_2696526, EPI_ISL_2696527, EPI_ISL_2696528, EPI_ISL_2696529, EPI_ISL_2696530, EPI_ISL_2696531, EPI_ISL_2696532, EPI_ISL_2696533, EPI_ISL_2696534, EPI_ISL_2696536, EPI_ISL_2696538, EPI_ISL_2696539, EPI_ISL_2696540, EPI_ISL_2696541, EPI_ISL_2696542, EPI_ISL_2696543, EPI_ISL_2696544, EPI_ISL_2696545, EPI_ISL_2696546, EPI_ISL_2696547, EPI_ISL_2696548, EPI_ISL_2696549, EPI_ISL_2696550, EPI_ISL_2696551, EPI_ISL_2696552, EPI_ISL_2696553, EPI_ISL_2696554, EPI_ISL_2696555, EPI_ISL_2696556, EPI_ISL_2696557, EPI_ISL_2696558, EPI_ISL_2696559, EPI_ISL_2696560, EPI_ISL_2696561, EPI_ISL_2696562, EPI_ISL_2696563, EPI_ISL_2696564, EPI_ISL_2696565, EPI_ISL_2696566, EPI_ISL_2696567, EPI_ISL_2696568, EPI_ISL_2696569, EPI_ISL_2696570, EPI_ISL_2696571, EPI_ISL_2696572, EPI_ISL_2696573, EPI_ISL_2696574, EPI_ISL_2696575, EPI_ISL_2696576, EPI_ISL_2696577, EPI_ISL_2696578, EPI_ISL_2696579, EPI_ISL_2696580, EPI_ISL_2696581, EPI_ISL_2696582, EPI_ISL_2696583, EPI_ISL_2696584, EPI_ISL_2696585, EPI_ISL_2696586, EPI_ISL_2696587, EPI_ISL_2696588, EPI_ISL_2696589, EPI_ISL_2696590, EPI_ISL_2696591, EPI_ISL_2696592, EPI_ISL_2696593, EPI_ISL_2696594, EPI_ISL_2696595, EPI_ISL_2696596, EPI_ISL_2696597, EPI_ISL_2696598, EPI_ISL_2696599, EPI_ISL_2696600, EPI_ISL_2696601, EPI_ISL_2696602, EPI_ISL_2696603, EPI_ISL_2696604, EPI_ISL_2696605, EPI_ISL_2696606, EPI_ISL_2696607, EPI_ISL_2696608, EPI_ISL_2696609, EPI_ISL_2696610, EPI_ISL_2696611, EPI_ISL_2696612, EPI_ISL_2696613, EPI_ISL_2696615, EPI_ISL_2696616, EPI_ISL_2696617, EPI_ISL_2696618, EPI_ISL_2696619, EPI_ISL_2696620, EPI_ISL_2696621, EPI_ISL_2696623, EPI_ISL_2696624, EPI_ISL_2696625, EPI_ISL_2696626, EPI_ISL_2696627, EPI_ISL_2696628, EPI_ISL_2696629, EPI_ISL_2696630, EPI_ISL_2696631, EPI_ISL_2696632, EPI_ISL_2696633, EPI_ISL_2696634, EPI_ISL_2696635, EPI_ISL_2696636, EPI_ISL_2696637, EPI_ISL_2696638, EPI_ISL_2696639, EPI_ISL_2696640, EPI_ISL_2696641, EPI_ISL_2696642, EPI_ISL_2696643, EPI_ISL_2696644, EPI_ISL_2696645, EPI_ISL_2696646, EPI_ISL_2696647, EPI_ISL_2696648, EPI_ISL_2696650, EPI_ISL_2696651, EPI_ISL_2696652, EPI_ISL_2696653, EPI_ISL_2696654, EPI_ISL_2696655, EPI_ISL_2696656, EPI_ISL_2696657, EPI_ISL_2696658, EPI_ISL_2696659, EPI_ISL_2696660, EPI_ISL_2696661, EPI_ISL_2696662, EPI_ISL_2696663, EPI_ISL_2696664, EPI_ISL_2696666, EPI_ISL_2696667, EPI_ISL_2696668, EPI_ISL_2696669, EPI_ISL_2696670, EPI_ISL_2696671, EPI_ISL_2696672, EPI_ISL_2696673, EPI_ISL_2696674, EPI_ISL_2696675, EPI_ISL_2696676, EPI_ISL_2696677, EPI_ISL_2696678, EPI_ISL_2696679, EPI_ISL_2696680, EPI_ISL_2696681, EPI_ISL_2696682, EPI_ISL_2696683, EPI_ISL_2696684, EPI_ISL_2696685, EPI_ISL_2696686, EPI_ISL_2696687, EPI_ISL_2696688, EPI_ISL_2696689, EPI_ISL_2696690, EPI_ISL_2696691, EPI_ISL_2696692, EPI_ISL_2696693, EPI_ISL_2696694, EPI_ISL_2696695, EPI_ISL_2696696, EPI_ISL_2696697, EPI_ISL_2696698, EPI_ISL_2696699, EPI_ISL_2696700, EPI_ISL_2696701, EPI_ISL_2696702, EPI_ISL_2696703, EPI_ISL_2696704, EPI_ISL_2696705, EPI_ISL_2696706, EPI_ISL_2696707, EPI_ISL_2696708, EPI_ISL_2696709, EPI_ISL_2696711, EPI_ISL_2696712, EPI_ISL_2696713, EPI_ISL_2696715, EPI_ISL_2696716, EPI_ISL_2696717, EPI_ISL_2696718, EPI_ISL_2696719, EPI_ISL_2696720, EPI_ISL_2696721, EPI_ISL_2696722, EPI_ISL_2696723, EPI_ISL_2696724, EPI_ISL_2696725, EPI_ISL_2696726, EPI_ISL_2696727, EPI_ISL_2696728, EPI_ISL_2696729, EPI_ISL_2696730, EPI_ISL_2696731, EPI_ISL_2696732, EPI_ISL_2696733, EPI_ISL_2696734, EPI_ISL_2696735, EPI_ISL_2696736, EPI_ISL_2696737, EPI_ISL_2696738, EPI_ISL_2696740, EPI_ISL_2696741, EPI_ISL_2696742, EPI_ISL_2696743, EPI_ISL_2696744, EPI_ISL_2696745, EPI_ISL_2696746, EPI_ISL_2696747, EPI_ISL_2696748, EPI_ISL_2696749, EPI_ISL_2696750, EPI_ISL_2696751, EPI_ISL_2696752, EPI_ISL_2696753, EPI_ISL_2696754, EPI_ISL_2696755, EPI_ISL_2696756, EPI_ISL_2696757, EPI_ISL_2696758, EPI_ISL_2696759, EPI_ISL_2696760, EPI_ISL_2696761, EPI_ISL_2696762, EPI_ISL_2696763, EPI_ISL_2696764, EPI_ISL_2696766, EPI_ISL_2696767, EPI_ISL_2696768, EPI_ISL_2696769, EPI_ISL_2696770, EPI_ISL_2696771, EPI_ISL_2696772, EPI_ISL_2696773, EPI_ISL_2696774, EPI_ISL_2696775, EPI_ISL_2696776, EPI_ISL_2696777, EPI_ISL_2696778, EPI_ISL_2696779, EPI_ISL_2696780, EPI_ISL_2696781, EPI_ISL_2696782, EPI_ISL_2696783, EPI_ISL_2696784, EPI_ISL_2696785, EPI_ISL_2696786, EPI_ISL_2696787, EPI_ISL_2696788, EPI_ISL_2696789, EPI_ISL_2696790, EPI_ISL_2696791, EPI_ISL_2696792, EPI_ISL_2696793, EPI_ISL_2696794, EPI_ISL_2696795, EPI_ISL_2696796, EPI_ISL_2696797, EPI_ISL_2696798, EPI_ISL_2696799, EPI_ISL_2696800, EPI_ISL_2696801, EPI_ISL_2696802, EPI_ISL_2696803, EPI_ISL_2696804, EPI_ISL_2696805, EPI_ISL_2696806, EPI_ISL_2696807, EPI_ISL_2696808, EPI_ISL_2696809, EPI_ISL_2696810, EPI_ISL_2696811, EPI_ISL_2696812, EPI_ISL_2696813, EPI_ISL_2696814, EPI_ISL_2696815, EPI_ISL_2696816, EPI_ISL_2696817, EPI_ISL_2696818, EPI_ISL_2696819, EPI_ISL_2696820, EPI_ISL_2696821, EPI_ISL_2696822, EPI_ISL_2696823, EPI_ISL_2696824, EPI_ISL_2696825, EPI_ISL_2696826, EPI_ISL_2696827, EPI_ISL_2696828, EPI_ISL_2696829, EPI_ISL_2696830, EPI_ISL_2696831, EPI_ISL_2696832, EPI_ISL_2696833, EPI_ISL_2696834, EPI_ISL_2696835, EPI_ISL_2696836, EPI_ISL_2696837, EPI_ISL_2696838, EPI_ISL_2696839, EPI_ISL_2696840, EPI_ISL_2696841, EPI_ISL_2696842, EPI_ISL_2696843, EPI_ISL_2696844, EPI_ISL_2696845, EPI_ISL_2696846, EPI_ISL_2696847, EPI_ISL_2696848, EPI_ISL_2696849, EPI_ISL_2696850, EPI_ISL_2696851, EPI_ISL_2696852, EPI_ISL_2696853, EPI_ISL_2696854, EPI_ISL_2696855, EPI_ISL_2696856, EPI_ISL_2696857, EPI_ISL_2696858, EPI_ISL_2696860, EPI_ISL_2696861, EPI_ISL_2696862, EPI_ISL_2696863, EPI_ISL_2696864, EPI_ISL_2696865, EPI_ISL_2696866, EPI_ISL_2696867, EPI_ISL_2696868, EPI_ISL_2696869, EPI_ISL_2696870, EPI_ISL_2696871, EPI_ISL_2696872, EPI_ISL_2696873, EPI_ISL_2696874, EPI_ISL_2696875, EPI_ISL_2696876, EPI_ISL_2696877, EPI_ISL_2696878, EPI_ISL_2696879, EPI_ISL_2696880, EPI_ISL_2696881, EPI_ISL_2696882, EPI_ISL_2696883, EPI_ISL_2696884, EPI_ISL_2696885, EPI_ISL_2696886, EPI_ISL_2696887, EPI_ISL_2696888, EPI_ISL_2696889, EPI_ISL_2696890, EPI_ISL_2696891, EPI_ISL_2696892, EPI_ISL_2696893, EPI_ISL_2696894, EPI_ISL_2696895

| | | | |
|---|---|---|---|
| see above | Laboratory, Gravity Diagnostics | Laboratory, Gravity Diagnostics | Diagnostics,G. |

EPI_ISL_2696947, EPI_ISL_2696949, EPI_ISL_2696950, EPI_ISL_2696952, EPI_ISL_2696953, EPI_ISL_2696954, EPI_ISL_2696955, EPI_ISL_2696957, EPI_ISL_2696958, EPI_ISL_2696959, EPI_ISL_2696960, EPI_ISL_2696962, EPI_ISL_2696964, EPI_ISL_2696965, EPI_ISL_2696966, EPI_ISL_2696967, EPI_ISL_2696968, EPI_ISL_2696969, EPI_ISL_2696970, EPI_ISL_2696971, EPI_ISL_2696972, EPI_ISL_2696973, EPI_ISL_2696975, EPI_ISL_2696976, EPI_ISL_2696977, EPI_ISL_2696978, EPI_ISL_2696980, EPI_ISL_2696981, EPI_ISL_2696984, EPI_ISL_2696985, EPI_ISL_2696986, EPI_ISL_2696987, EPI_ISL_2696988, EPI_ISL_2696989, EPI_ISL_2696990, EPI_ISL_2696991, EPI_ISL_2696992, EPI_ISL_2696993, EPI_ISL_2696994, EPI_ISL_2696995, EPI_ISL_2696998, EPI_ISL_2696999, EPI_ISL_2697001, EPI_ISL_2697003, EPI_ISL_2697004, EPI_ISL_2697005, EPI_ISL_2697006, EPI_ISL_2697007, EPI_ISL_2697008, EPI_ISL_2697010, EPI_ISL_2697011, EPI_ISL_2697012, EPI_ISL_2697013, EPI_ISL_2697014, EPI_ISL_2697015, EPI_ISL_2697016, EPI_ISL_2697017, EPI_ISL_2697019, EPI_ISL_2697020, EPI_ISL_2697021, EPI_ISL_2697023, EPI_ISL_2697024, EPI_ISL_2697025, EPI_ISL_2697026, EPI_ISL_2697027, EPI_ISL_2697028, EPI_ISL_2697029, EPI_ISL_2697030, EPI_ISL_2697031, EPI_ISL_2697032, EPI_ISL_2697034, EPI_ISL_2697035, EPI_ISL_2697037, EPI_ISL_2697038, EPI_ISL_2697039, EPI_ISL_2697040, EPI_ISL_2697041, EPI_ISL_2697042, EPI_ISL_2697043, EPI_ISL_2697044, EPI_ISL_2697045, EPI_ISL_2697046, EPI_ISL_2697047, EPI_ISL_2697048, EPI_ISL_2697050, EPI_ISL_2697052, EPI_ISL_2697053, EPI_ISL_2697054, EPI_ISL_2697055, EPI_ISL_2697056, EPI_ISL_2697057, EPI_ISL_2697058, EPI_ISL_2697059, EPI_ISL_2697062, EPI_ISL_2697063, EPI_ISL_2697065, EPI_ISL_2697066, EPI_ISL_2697068, EPI_ISL_2697070, EPI_ISL_2697071, EPI_ISL_2697072, EPI_ISL_2697073, EPI_ISL_2697074, EPI_ISL_2697075, EPI_ISL_2697076, EPI_ISL_2697078, EPI_ISL_2697079, EPI_ISL_2697080, EPI_ISL_2697081, EPI_ISL_2697083, EPI_ISL_2697085, EPI_ISL_2697086, EPI_ISL_2697087, EPI_ISL_2697088, EPI_ISL_2697089, EPI_ISL_2697090, EPI_ISL_2697092, EPI_ISL_2697095, EPI_ISL_2697097, EPI_ISL_2697098, EPI_ISL_2697099, EPI_ISL_2697100, EPI_ISL_2697101, EPI_ISL_2697102, EPI_ISL_2697103, EPI_ISL_2697104, EPI_ISL_2697106, EPI_ISL_2697107, EPI_ISL_2697109, EPI_ISL_2697110, EPI_ISL_2697111, EPI_ISL_2697112, EPI_ISL_2697114, EPI_ISL_2697115, EPI_ISL_2697116, EPI_ISL_2697117, EPI_ISL_2697118, EPI_ISL_2697119, EPI_ISL_2697120, EPI_ISL_2697121, EPI_ISL_2697122, EPI_ISL_2697124, EPI_ISL_2697125, EPI_ISL_2697126, EPI_ISL_2697127, EPI_ISL_2697128, EPI_ISL_2697129, EPI_ISL_2697130, EPI_ISL_2697131, EPI_ISL_2697132, EPI_ISL_2697133, EPI_ISL_2697134, EPI_ISL_2697136, EPI_ISL_2697137, EPI_ISL_2697138, EPI_ISL_2697139, EPI_ISL_2697140, EPI_ISL_2697141, EPI_ISL_2697142, EPI_ISL_2697143, EPI_ISL_2697145, EPI_ISL_2697147, EPI_ISL_2697148, EPI_ISL_2697149, EPI_ISL_2697150, EPI_ISL_2697152, EPI_ISL_2697153, EPI_ISL_2697154, EPI_ISL_2697156, EPI_ISL_2697157, EPI_ISL_2697158, EPI_ISL_2697160, EPI_ISL_2697161, EPI_ISL_2697167, EPI_ISL_2697171, EPI_ISL_2697174, EPI_ISL_2697180, EPI_ISL_2697183, EPI_ISL_2697184, EPI_ISL_2697187, EPI_ISL_2697190, EPI_ISL_2697191, EPI_ISL_2697194, EPI_ISL_2697195, EPI_ISL_2697196, EPI_ISL_2697200, EPI_ISL_2697202, EPI_ISL_2697206, EPI_ISL_2697210, EPI_ISL_2697211, EPI_ISL_2697224, EPI_ISL_2697228, EPI_ISL_2697235, EPI_ISL_2697240, EPI_ISL_2697245, EPI_ISL_2697250, EPI_ISL_2697252, EPI_ISL_2697258, EPI_ISL_2697265, EPI_ISL_2697266, EPI_ISL_2697277, EPI_ISL_2697278, EPI_ISL_2697281, EPI_ISL_2697290, EPI_ISL_2697293, EPI_ISL_2697295, EPI_ISL_2697296, EPI_ISL_2697298, EPI_ISL_2697299, EPI_ISL_2697302, EPI_ISL_2697303, EPI_ISL_2697305, EPI_ISL_2697306, EPI_ISL_2697310, EPI_ISL_2697311, EPI_ISL_2697314, EPI_ISL_2697318, EPI_ISL_2697320, EPI_ISL_2697321, EPI_ISL_2697323, EPI_ISL_2697324, EPI_ISL_2697325, EPI_ISL_2697326, EPI_ISL_2697328, EPI_ISL_2697329, EPI_ISL_2697331, EPI_ISL_2697332, EPI_ISL_2697333, EPI_ISL_2697335, EPI_ISL_2697337, EPI_ISL_2697338, EPI_ISL_2697341, EPI_ISL_2697343, EPI_ISL_2697344, EPI_ISL_2697346, EPI_ISL_2697348, EPI_ISL_2697350, EPI_ISL_2697351, EPI_ISL_2697352, EPI_ISL_2697353, EPI_ISL_2697355, EPI_ISL_2697360, EPI_ISL_2697361, EPI_ISL_2697367, EPI_ISL_2697368, EPI_ISL_2697370, EPI_ISL_2697373, EPI_ISL_2697374, EPI_ISL_2697378, EPI_ISL_2697380, EPI_ISL_2697382, EPI_ISL_2697383, EPI_ISL_2697384, EPI_ISL_2697385, EPI_ISL_2697387, EPI_ISL_2697388, EPI_ISL_2697389, EPI_ISL_2697390, EPI_ISL_2697393, EPI_ISL_2697394, EPI_ISL_2697396, EPI_ISL_2697398, EPI_ISL_2697399, EPI_ISL_2697400, EPI_ISL_2697403, EPI_ISL_2697406, EPI_ISL_2697407, EPI_ISL_2697408, EPI_ISL_2697409, EPI_ISL_2697410, EPI_ISL_2697415, EPI_ISL_2697416, EPI_ISL_2697419, EPI_ISL_2697420, EPI_ISL_2697423, EPI_ISL_2697424, EPI_ISL_2697425, EPI_ISL_2697428, EPI_ISL_2697429, EPI_ISL_2697432, EPI_ISL_2697433, EPI_ISL_2697434, EPI_ISL_2697435, EPI_ISL_2697437, EPI_ISL_2697442, EPI_ISL_2697444, EPI_ISL_2697445, EPI_ISL_2697446, EPI_ISL_2697447, EPI_ISL_2697448, EPI_ISL_2697450, EPI_ISL_2697453, EPI_ISL_2697458, EPI_ISL_2697460, EPI_ISL_2697462, EPI_ISL_2697469, EPI_ISL_2697470, EPI_ISL_2697472, EPI_ISL_2697475, EPI_ISL_2697476, EPI_ISL_2697477, EPI_ISL_2697478, EPI_ISL_2697479, EPI_ISL_2697480, EPI_ISL_2697481, EPI_ISL_2697482, EPI_ISL_2697483, EPI_ISL_2697484, EPI_ISL_2697485, EPI_ISL_2697486, EPI_ISL_2697487, EPI_ISL_2697488, EPI_ISL_2697489, EPI_ISL_2697490, EPI_ISL_2697491, EPI_ISL_2697492, EPI_ISL_2697493, EPI_ISL_2697494, EPI_ISL_2697495, EPI_ISL_2697496, EPI_ISL_2697497, EPI_ISL_2697498, EPI_ISL_2697499, EPI_ISL_2697500, EPI_ISL_2697501, EPI_ISL_2697502, EPI_ISL_2697503, EPI_ISL_2697504, EPI_ISL_2697505, EPI_ISL_2697506, EPI_ISL_2697507, EPI_ISL_2697508, EPI_ISL_2697509, EPI_ISL_2697510, EPI_ISL_2697511, EPI_ISL_2697512, EPI_ISL_2697513, EPI_ISL_2697514, EPI_ISL_2697515, EPI_ISL_2697516, EPI_ISL_2697517, EPI_ISL_2697518, EPI_ISL_2697519, EPI_ISL_2697520, EPI_ISL_2697521, EPI_ISL_2697522, EPI_ISL_2697523, EPI_ISL_2697524, EPI_ISL_2697525, EPI_ISL_2697526, EPI_ISL_2697527, EPI_ISL_2697528, EPI_ISL_2697529, EPI_ISL_2697530, EPI_ISL_2697531, EPI_ISL_2697532, EPI_ISL_2697533, EPI_ISL_2697534, EPI_ISL_2697535, EPI_ISL_2697536, EPI_ISL_2697537, EPI_ISL_2697538, EPI_ISL_2697539, EPI_ISL_2697540, EPI_ISL_2697541, EPI_ISL_2697542, EPI_ISL_2697543, EPI_ISL_2697544, EPI_ISL_2697545, EPI_ISL_2697546, EPI_ISL_2697547, EPI_ISL_2697548, EPI_ISL_2697549, EPI_ISL_2697550, EPI_ISL_2697551, EPI_ISL_2697552, EPI_ISL_2697553, EPI_ISL_2697554, EPI_ISL_2697555, EPI_ISL_2697556, EPI_ISL_2697557, EPI_ISL_2697558, EPI_ISL_2697559, EPI_ISL_2697560, EPI_ISL_2697561, EPI_ISL_2697562, EPI_ISL_2697563, EPI_ISL_2697564, EPI_ISL_2697565, EPI_ISL_2697566, EPI_ISL_2697567, EPI_ISL_2697568, EPI_ISL_2697569, EPI_ISL_2697570, EPI_ISL_2697571, EPI_ISL_2697572, EPI_ISL_2697573, EPI_ISL_2697574, EPI_ISL_2697575, EPI_ISL_2697576, EPI_ISL_2697577, EPI_ISL_2697578, EPI_ISL_2697579, EPI_ISL_2697580, EPI_ISL_2697581, EPI_ISL_2697582, EPI_ISL_2697583, EPI_ISL_2697584, EPI_ISL_2697585, EPI_ISL_2697586, EPI_ISL_2697587, EPI_ISL_2697588, EPI_ISL_2697589, EPI_ISL_2697590, EPI_ISL_2697591, EPI_ISL_2697592, EPI_ISL_2697593, EPI_ISL_2697594, EPI_ISL_2697595, EPI_ISL_2697596, EPI_ISL_2697597, EPI_ISL_2697598, EPI_ISL_2697599, EPI_ISL_2697600, EPI_ISL_2697601, EPI_ISL_2697602, EPI_ISL_2697603, EPI_ISL_2697604, EPI_ISL_2697605, EPI_ISL_2697606, EPI_ISL_2697607, EPI_ISL_2697608, EPI_ISL_2697609, EPI_ISL_2697610, EPI_ISL_2697611, EPI_ISL_2697612, EPI_ISL_2697613, EPI_ISL_2697614, EPI_ISL_2697615, EPI_ISL_2697616, EPI_ISL_2697617, EPI_ISL_2697618, EPI_ISL_2697619, EPI_ISL_2697620, EPI_ISL_2697621, EPI_ISL_2697622, EPI_ISL_2697623, EPI_ISL_2697624, EPI_ISL_2697625, EPI_ISL_2697626, EPI_ISL_2697627, EPI_ISL_2697628, EPI_ISL_2697629, EPI_ISL_2697630, EPI_ISL_2697631, EPI_ISL_2697632, EPI_ISL_2697633, EPI_ISL_2697634, EPI_ISL_2697635, EPI_ISL_2697636, EPI_ISL_2697637, EPI_ISL_2697638, EPI_ISL_2697639, EPI_ISL_2697640, EPI_ISL_2697641, EPI_ISL_2697642, EPI_ISL_2697643, EPI_ISL_2697644, EPI_ISL_2697645, EPI_ISL_2697646, EPI_ISL_2697647, EPI_ISL_2697648, EPI_ISL_2697649, EPI_ISL_2697650, EPI_ISL_2697651, EPI_ISL_2697652, EPI_ISL_2697653, EPI_ISL_2697654, EPI_ISL_2697655, EPI_ISL_2697656, EPI_ISL_2697657, EPI_ISL_2697658, EPI_ISL_2697659, EPI_ISL_2697660, EPI_ISL_2697661, EPI_ISL_2697662, EPI_ISL_2697663, EPI_ISL_2697664, EPI_ISL_2697665, EPI_ISL_2697666, EPI_ISL_2697667, EPI_ISL_2697668, EPI_ISL_2697669, EPI_ISL_2697670, EPI_ISL_2697671, EPI_ISL_2697672, EPI_ISL_2697678, EPI_ISL_2697679, EPI_ISL_2697680, EPI_ISL_2697681, EPI_ISL_2697682, EPI_ISL_2697683, EPI_ISL_2697684, EPI_ISL_2697685, EPI_ISL_2697686, EPI_ISL_2697687, EPI_ISL_2697688, EPI_ISL_2697689, EPI_ISL_2697690, EPI_ISL_2697691, EPI_ISL_2697692, EPI_ISL_2697693, EPI_ISL_2697694, EPI_ISL_2697695, EPI_ISL_2697696, EPI_ISL_2697697, EPI_ISL_2697698, EPI_ISL_2697699, EPI_ISL_2697700, EPI_ISL_2697701, EPI_ISL_2697702, EPI_ISL_2697703, EPI_ISL_2697704, EPI_ISL_2697705, EPI_ISL_2697706, EPI_ISL_2697707, EPI_ISL_2697708, EPI_ISL_2697709, EPI_ISL_2697710, EPI_ISL_2697711, EPI_ISL_2697712, EPI_ISL_2697713, EPI_ISL_2697714, EPI_ISL_2697715, EPI_ISL_2697716, EPI_ISL_2697717, EPI_ISL_2697718, EPI_ISL_2697719, EPI_ISL_2697720, EPI_ISL_2697721, EPI_ISL_2697722, EPI_ISL_2697723, EPI_ISL_2697724, EPI_ISL_2697725, EPI_ISL_2697726, EPI_ISL_2697727, EPI_ISL_2697728, EPI_ISL_2697729, EPI_ISL_2697730, EPI_ISL_2697731, EPI_ISL_2697732, EPI_ISL_2697733, EPI_ISL_2697734, EPI_ISL_2697735, EPI_ISL_2697736, EPI_ISL_2697737, EPI_ISL_2697738, EPI_ISL_2697739, EPI_ISL_2697740, EPI_ISL_2697741, EPI_ISL_2697742, EPI_ISL_2697743, EPI_ISL_2697744, EPI_ISL_2697745, EPI_ISL_2697746, EPI_ISL_2697747, EPI_ISL_2697748, EPI_ISL_2697749, EPI_ISL_2697750, EPI_ISL_2697751, EPI_ISL_2697752, EPI_ISL_2697753, EPI_ISL_2697754, EPI_ISL_2697755, EPI_ISL_2697756, EPI_ISL_2697757, EPI_ISL_2697758, EPI_ISL_2697759, EPI_ISL_2697760, EPI_ISL_2697761, EPI_ISL_2697762, EPI_ISL_2697763, EPI_ISL_2697764, EPI_ISL_2697765, EPI_ISL_2697766, EPI_ISL_2697767, EPI_ISL_2697769, EPI_ISL_2697770, EPI_ISL_2697771, EPI_ISL_2697772, EPI_ISL_2697773, EPI_ISL_2697774, EPI_ISL_2697775, EPI_ISL_2697776, EPI_ISL_2697777, EPI_ISL_2697778, EPI_ISL_2697779, EPI_ISL_2697780, EPI_ISL_2697781, EPI_ISL_2697782, EPI_ISL_2697783, EPI_ISL_2697784, EPI_ISL_2697785, EPI_ISL_2697786, EPI_ISL_2697787, EPI_ISL_2697788, EPI_ISL_2697789, EPI_ISL_2697790, EPI_ISL_2697791, EPI_ISL_2697792, EPI_ISL_2697793, EPI_ISL_2697794, EPI_ISL_2697795, EPI_ISL_2697796, EPI_ISL_2697798, EPI_ISL_2697799, EPI_ISL_2697800, EPI_ISL_2697801, EPI_ISL_2697802, EPI_ISL_2697803, EPI_ISL_2697804, EPI_ISL_2697805, EPI_ISL_2697806, EPI_ISL_2697807, EPI_ISL_2697808, EPI_ISL_2697809, EPI_ISL_2697810, EPI_ISL_2697811, EPI_ISL_2697812, EPI_ISL_2697813, EPI_ISL_2697814, EPI_ISL_2697815, EPI_ISL_2697816, EPI_ISL_2697817, EPI_ISL_2697818, EPI_ISL_2697819, EPI_ISL_2697820, EPI_ISL_2697821, EPI_ISL_2697822, EPI_ISL_2697823, EPI_ISL_2697824, EPI_ISL_2697825, EPI_ISL_2697826, EPI_ISL_2697827, EPI_ISL_2697828, EPI_ISL_2697829, EPI_ISL_2697830, EPI_ISL_2697831, EPI_ISL_2697832, EPI_ISL_2697833, EPI_ISL_2697834, EPI_ISL_2697835, EPI_ISL_2697836, EPI_ISL_2697837, EPI_ISL_2697838, EPI_ISL_2697839, EPI_ISL_2697840, EPI_ISL_2697841, EPI_ISL_2697842, EPI_ISL_2697843, EPI_ISL_2697844, EPI_ISL_2697845, EPI_ISL_2697846, EPI_ISL_2697847, EPI_ISL_2697848, EPI_ISL_2697849, EPI_ISL_2697850, EPI_ISL_2697851, EPI_ISL_2697852, EPI_ISL_2697853, EPI_ISL_2697854, EPI_ISL_2697855, EPI_ISL_2697856, EPI_ISL_2697857, EPI_ISL_2697858, EPI_ISL_2697859, EPI_ISL_2697860, EPI_ISL_2697861, EPI_ISL_2697862, EPI_ISL_2697863, EPI_ISL_2697864, EPI_ISL_2697865, EPI_ISL_2697866, EPI_ISL_2697867, EPI_ISL_2697868, EPI_ISL_2697869, EPI_ISL_2697870, EPI_ISL_2697871, EPI_ISL_2697872, EPI_ISL_2697873, EPI_ISL_2697874, EPI_ISL_2697875, EPI_ISL_2697876, EPI_ISL_2697877, EPI_ISL_2697878, EPI_ISL_2697879, EPI_ISL_2697880, EPI_ISL_2697881, EPI_ISL_2698108, EPI_ISL_2698109, EPI_ISL_2698110, EPI_ISL_2698111, EPI_ISL_2698112, EPI_ISL_2698113, EPI_ISL_2698114, EPI_ISL_2698115, EPI_ISL_2698116, EPI_ISL_2698117, EPI_ISL_2698118, EPI_ISL_2698119, EPI_ISL_2698120, EPI_ISL_2698121, EPI_ISL_2698122, EPI_ISL_2698123, EPI_ISL_2698124, EPI_ISL_2698125, EPI_ISL_2698127, EPI_ISL_2698128, EPI_ISL_2698129, EPI_ISL_2698130, EPI_ISL_2698131, EPI_ISL_2698132, EPI_ISL_2698133, EPI_ISL_2698134, EPI_ISL_2698135, EPI_ISL_2698136, EPI_ISL_2698137, EPI_ISL_2698138, EPI_ISL_2698139, EPI_ISL_2698140, EPI_ISL_2698141, EPI_ISL_2698142, EPI_ISL_2698143, EPI_ISL_2698144, EPI_ISL_2698145, EPI_ISL_2698146, EPI_ISL_2698147, EPI_ISL_2698148, EPI_ISL_2698149, EPI_ISL_2698150, EPI_ISL_2698151, EPI_ISL_2698152, EPI_ISL_2698153, EPI_ISL_2698154, EPI_ISL_2698155, EPI_ISL_2698156, EPI_ISL_2698157, EPI_ISL_2698158, EPI_ISL_2698159, EPI_ISL_2698160, EPI_ISL_2698161, EPI_ISL_2698162, EPI_ISL_2698163, EPI_ISL_2698164, EPI_ISL_2698165, EPI_ISL_2698166, EPI_ISL_2698167, EPI_ISL_2698168, EPI_ISL_2698169, EPI_ISL_2698170, EPI_ISL_2698171, EPI_ISL_2698172, EPI_ISL_2698173, EPI_ISL_2698174, EPI_ISL_2698175, EPI_ISL_2698176, EPI_ISL_2698177, EPI_ISL_2698178, EPI_ISL_2698179, EPI_ISL_2698180, EPI_ISL_2698181, EPI_ISL_2698182, EPI_ISL_2698183, EPI_ISL_2698184, EPI_ISL_2698185, EPI_ISL_2698186, EPI_ISL_2698187, EPI_ISL_2698188, EPI_ISL_2698189, EPI_ISL_2698190, EPI_ISL_2698191, EPI_ISL_2698192, EPI_ISL_2698193, EPI_ISL_2698194, EPI_ISL_2698195, EPI_ISL_2698196, EPI_ISL_2698197, EPI_ISL_2698198, EPI_ISL_2698199, EPI_ISL_2698200, EPI_ISL_2698201, EPI_ISL_2698202, EPI_ISL_2698203, EPI_ISL_2698204, EPI_ISL_2698205, EPI_ISL_2698206, EPI_ISL_2698207, EPI_ISL_2698208, EPI_ISL_2698209, EPI_ISL_2698210, EPI_ISL_2698211, EPI_ISL_2698212, EPI_ISL_2698213, EPI_ISL_2698214, EPI_ISL_2698215, EPI_ISL_2698216, EPI_ISL_2698217, EPI_ISL_2698218, EPI_ISL_2698219, EPI_ISL_2698220, EPI_ISL_2698221, EPI_ISL_2698222, EPI_ISL_2698223, EPI_ISL_2698224, EPI_ISL_2698225, EPI_ISL_2698226, EPI_ISL_2698227, EPI_ISL_2698228, EPI_ISL_2698229, EPI_ISL_2698230, EPI_ISL_2698231, EPI_ISL_2698232, EPI_ISL_2698233, EPI_ISL_2698234, EPI_ISL_2698235, EPI_ISL_2698236, EPI_ISL_2698237, EPI_ISL_2698238, EPI_ISL_2698239, EPI_ISL_2698240, EPI_ISL_2698241, EPI_ISL_2698242, EPI_ISL_2698243, EPI_ISL_2698244, EPI_ISL_2698245, EPI_ISL_2698246, EPI_ISL_2698247, EPI_ISL_2698248, EPI_ISL_2698249, EPI_ISL_2698250, EPI_ISL_2698251, EPI_ISL_2698252, EPI_ISL_2698253, EPI_ISL_2698254, EPI_ISL_2698255, EPI_ISL_2698256, EPI_ISL_2698257, EPI_ISL_2698258, EPI_ISL_2698259, EPI_ISL_2698260, EPI_ISL_2698261, EPI_ISL_2698262, EPI_ISL_2698263, EPI_ISL_2698264, EPI_ISL_2698265, EPI_ISL_2698266, EPI_ISL_2698267, EPI_ISL_2698268, EPI_ISL_2698269, EPI_ISL_2698270, EPI_ISL_2698271, EPI_ISL_2698272, EPI_ISL_2698273, EPI_ISL_2698274, EPI_ISL_2698275, EPI_ISL_2698276, EPI_ISL_2698277, EPI_ISL_2698278, EPI_ISL_2698279, EPI_ISL_2698280, EPI_ISL_2698281, EPI_ISL_2698282, EPI_ISL_2698283, EPI_ISL_2698284, EPI_ISL_2698285, EPI_ISL_2698286, EPI_ISL_2698287, EPI_ISL_2698288, EPI_ISL_2698289, EPI_ISL_2698290, EPI_ISL_2698291, EPI_ISL_2698292, EPI_ISL_2698293, EPI_ISL_2698294, EPI_ISL_2698295, EPI_ISL_2698296, EPI_ISL_2698297, EPI_ISL_2698298, EPI_ISL_2698299, EPI_ISL_2698300, EPI_ISL_2698301, EPI_ISL_2698302, EPI_ISL_2698303, EPI_ISL_2698304, EPI_ISL_2698305, EPI_ISL_2698306, EPI_ISL_2698307, EPI_ISL_2698308, EPI_ISL_2698309, EPI_ISL_2698310, EPI_ISL_2698311, EPI_ISL_2698312, EPI_ISL_2698313, EPI_ISL_2698314, EPI_ISL_2698315, EPI_ISL_2698316, EPI_ISL_2698317, EPI_ISL_2698326, EPI_ISL_2698327, EPI_ISL_2698328, EPI_ISL_2698329, EPI_ISL_2698330, EPI_ISL_2698331, EPI_ISL_2698332, EPI_ISL_2698333, EPI_ISL_2698334, EPI_ISL_2698335, EPI_ISL_2698336, EPI_ISL_2698337, EPI_ISL_2698338, EPI_ISL_2698339, EPI_ISL_2698340, EPI_ISL_2698341, EPI_ISL_2698342, EPI_ISL_2698343, EPI_ISL_2698344, EPI_ISL_2698345, EPI_ISL_2698346, EPI_ISL_2698347, EPI_ISL_2698348, EPI_ISL_2698349, EPI_ISL_2698350, EPI_ISL_2698351, EPI_ISL_2698352, EPI_ISL_2698353, EPI_ISL_2698354, EPI_ISL_2698355, EPI_ISL_2698356, EPI_ISL_2698357, EPI_ISL_2698358, EPI_ISL_2698359, EPI_ISL_2698360, EPI_ISL_2698361, EPI_ISL_2698362, EPI_ISL_2698363, EPI_ISL_2698364, EPI_ISL_2698365, EPI_ISL_2698366, EPI_ISL_2698367, EPI_ISL_2698368, EPI_ISL_2698369, EPI_ISL_2698370, EPI_ISL_2698371, EPI_ISL_2698372, EPI_ISL_2698373, EPI_ISL_2698374, EPI_ISL_2698375, EPI_ISL_2698376, EPI_ISL_2698377, EPI_ISL_2698378, EPI_ISL_2698379, EPI_ISL_2698380, EPI_ISL_2698381, EPI_ISL_2698382, EPI_ISL_2698383, EPI_ISL_2698384, EPI_ISL_2698385, EPI_ISL_2698386, EPI_ISL_2698387, EPI_ISL_2698388, EPI_ISL_2698389, EPI_ISL_2698390, EPI_ISL_2698391, EPI_ISL_2698392, EPI_ISL_2698393, EPI_ISL_2698394, EPI_ISL_2698395, EPI_ISL_2698396, EPI_ISL_2698397, EPI_ISL_2698398, EPI_ISL_2698399, EPI_ISL_2698400, EPI_ISL_2698401, EPI_ISL_2698402, EPI_ISL_2698403, EPI_ISL_2698404, EPI_ISL_2698405, EPI_ISL_2698406, EPI_ISL_2698407, EPI_ISL_2698408, EPI_ISL_2698409, EPI_ISL_2698410, EPI_ISL_2698411, EPI_ISL_2698412, EPI_ISL_2698413, EPI_ISL_2698414, EPI_ISL_2698415, EPI_ISL_2698416, EPI_ISL_2698417, EPI_ISL_2698418, EPI_ISL_2698419, EPI_ISL_2698420, EPI_ISL_2698421, EPI_ISL_2698422, EPI_ISL_2698423, EPI_ISL_2698426, EPI_ISL_2698427, EPI_ISL_2698428, EPI_ISL_2698429, EPI_ISL_2698430, EPI_ISL_2698431, EPI_ISL_2698432, EPI_ISL_2698434, EPI_ISL_2698435, EPI_ISL_2698436, EPI_ISL_2698437, EPI_ISL_2698438, EPI_ISL_2698440, EPI_ISL_2698441, EPI_ISL_2698443, EPI_ISL_2698445, EPI_ISL_2698446, EPI_ISL_2698448, EPI_ISL_2698449, EPI_ISL_2698452, EPI_ISL_2698453, EPI_ISL_2698454, EPI_ISL_2698456, EPI_ISL_2698457, EPI_ISL_2698459, EPI_ISL_2698460, EPI_ISL_2698462, EPI_ISL_2698463, EPI_ISL_2698464, EPI_ISL_2698465, EPI_ISL_2698466, EPI_ISL_2698468, EPI_ISL_2698469, EPI_ISL_2698472, EPI_ISL_2698474, EPI_ISL_2698475, EPI_ISL_2698477, EPI_ISL_2698479, EPI_ISL_2698480, EPI_ISL_2698482, EPI_ISL_2698483, EPI_ISL_2698485, EPI_ISL_2698486, EPI_ISL_2698488, EPI_ISL_2698489, EPI_ISL_2698491, EPI_ISL_2698492, EPI_ISL_2698494, EPI_ISL_2698495, EPI_ISL_2698497, EPI_ISL_2698500, EPI_ISL_2698503, EPI_ISL_2698505, EPI_ISL_2698506, EPI_ISL_2698508, EPI_ISL_2698510, EPI_ISL_2698511, EPI_ISL_2698513, EPI_ISL_2698514, EPI_ISL_2698516, EPI_ISL_2698518, EPI_ISL_2698519, EPI_ISL_2698521, EPI_ISL_2698523, EPI_ISL_2698525, EPI_ISL_2698526, EPI_ISL_2698528, EPI_ISL_2698529, EPI_ISL_2698531, EPI_ISL_2698533, EPI_ISL_2698534, EPI_ISL_2698536, EPI_ISL_2698538, EPI_ISL_2698540, EPI_ISL_2698541, EPI_ISL_2698543, EPI_ISL_2698544, EPI_ISL_2698546, EPI_ISL_2698548, EPI_ISL_2698551, EPI_ISL_2698553, EPI_ISL_2698555, EPI_ISL_2698556, EPI_ISL_2698558, EPI_ISL_2698560, EPI_ISL_2698561, EPI_ISL_2698563, EPI_ISL_2698565, EPI_ISL_2698566, EPI_ISL_2698568, EPI_ISL_2698569, EPI_ISL_2698571, EPI_ISL_2698573, EPI_ISL_2698574, EPI_ISL_2698576, EPI_ISL_2698578, EPI_ISL_2698580, EPI_ISL_2698581, EPI_ISL_2698583, EPI_ISL_2698584, EPI_ISL_2698587, EPI_ISL_2698589, EPI_ISL_2698592, EPI_ISL_2698594, EPI_ISL_2698596, EPI_ISL_2698597, EPI_ISL_2698599, EPI_ISL_2698600, EPI_ISL_2698602, EPI_ISL_2698603, EPI_ISL_2698605, EPI_ISL_2698606, EPI_ISL_2698608, EPI_ISL_2698609, EPI_ISL_2698611, EPI_ISL_2698613, EPI_ISL_2698614, EPI_ISL_2698616, EPI_ISL_2698618, EPI_ISL_2698619, EPI_ISL_2698621, EPI_ISL_2698623, EPI_ISL_2698624, EPI_ISL_2698626, EPI_ISL_2698627, EPI_ISL_2698629, EPI_ISL_2698630, EPI_ISL_2698632, EPI_ISL_2698633, EPI_ISL_2698635, EPI_ISL_2698637, EPI_ISL_2698638, EPI_ISL_2698640, EPI_ISL_2698642, EPI_ISL_2698643, EPI_ISL_2698645, EPI_ISL_2698647, EPI_ISL_2698649, EPI_ISL_2698650, EPI_ISL_2698652, EPI_ISL_2698654, EPI_ISL_2698656, EPI_ISL_2698657, EPI_ISL_2698659, EPI_ISL_2698660, EPI_ISL_2698662, EPI_ISL_2698663, EPI_ISL_2698665, EPI_ISL_2698667, EPI_ISL_2698668, EPI_ISL_2698670, EPI_ISL_2698671, EPI_ISL_2698673, EPI_ISL_2698674, EPI_ISL_2698676, EPI_ISL_2698677, EPI_ISL_2698679, EPI_ISL_2698681, EPI_ISL_2698682, EPI_ISL_2698683, EPI_ISL_2698684, EPI_ISL_2698685, EPI_ISL_2698686, EPI_ISL_2698687, EPI_ISL_2698688, EPI_ISL_2698689, EPI_ISL_2698690, EPI_ISL_2698691, EPI_ISL_2698692, EPI_ISL_2698693, EPI_ISL_2698694, EPI_ISL_2698695, EPI_ISL_2698696, EPI_ISL_2698697, EPI_ISL_2698698, EPI_ISL_2698699, EPI_ISL_2698702, EPI_ISL_2698704, EPI_ISL_2698705, EPI_ISL_2698706, EPI_ISL_2698707, EPI_ISL_2698708, EPI_ISL_2698709,

| | | | |
|---|---|---|---|
| EPI_ISL_2698710, EPI_ISL_2698711, EPI_ISL_2698712, EPI_ISL_2698713, EPI_ISL_2698714, EPI_ISL_2698715, EPI_ISL_2698716, EPI_ISL_2698717, EPI_ISL_2698718, EPI_ISL_2698719, EPI_ISL_2698720, EPI_ISL_2698721, EPI_ISL_2698722, EPI_ISL_2698723, EPI_ISL_2698724, EPI_ISL_2698725, EPI_ISL_2698726, EPI_ISL_2698727, EPI_ISL_2698728, EPI_ISL_2698729, EPI_ISL_2698730, EPI_ISL_2698731, EPI_ISL_2698732, EPI_ISL_2698733, EPI_ISL_2698734, EPI_ISL_2698735, EPI_ISL_2698736, EPI_ISL_2698737, EPI_ISL_2698738, EPI_ISL_2698739, EPI_ISL_2698740, EPI_ISL_2698741, EPI_ISL_2698742, EPI_ISL_2698743, EPI_ISL_2698744, EPI_ISL_2698745, EPI_ISL_2698746, EPI_ISL_2698747, EPI_ISL_2698748, EPI_ISL_2698749, EPI_ISL_2698750, EPI_ISL_2698751, EPI_ISL_2698752, EPI_ISL_2698753, EPI_ISL_2698754, EPI_ISL_2698755, EPI_ISL_2698756, EPI_ISL_2698757, EPI_ISL_2698758, EPI_ISL_2698759, EPI_ISL_2698760, EPI_ISL_2698761, EPI_ISL_2698762, EPI_ISL_2698763, EPI_ISL_2698764, EPI_ISL_2698765, EPI_ISL_2698766, EPI_ISL_2698767, EPI_ISL_2698768, EPI_ISL_2698769, EPI_ISL_2698770, EPI_ISL_2698771, EPI_ISL_2698772, EPI_ISL_2698773, EPI_ISL_2698774, EPI_ISL_2698775, EPI_ISL_2698776, EPI_ISL_2698777, EPI_ISL_2698778, EPI_ISL_2698779, EPI_ISL_2698780, EPI_ISL_2698781, EPI_ISL_2698782, EPI_ISL_2698783, EPI_ISL_2698784, EPI_ISL_2698785, EPI_ISL_2698786, EPI_ISL_2698787, EPI_ISL_2698788, EPI_ISL_2698789, EPI_ISL_2698790, EPI_ISL_2698791, EPI_ISL_2698792, EPI_ISL_2698793, EPI_ISL_2698794, EPI_ISL_2698795, EPI_ISL_2698796, EPI_ISL_2698797, EPI_ISL_2698798, EPI_ISL_2698799, EPI_ISL_2698800, EPI_ISL_2698801, EPI_ISL_2698803, EPI_ISL_2698804, EPI_ISL_2698805, EPI_ISL_2698806, EPI_ISL_2698807, EPI_ISL_2698808, EPI_ISL_2698809, EPI_ISL_2698810, EPI_ISL_2698811, EPI_ISL_2698812, EPI_ISL_2698813, EPI_ISL_2698815, EPI_ISL_2698816, EPI_ISL_2698817, EPI_ISL_2698818, EPI_ISL_2698819, EPI_ISL_2698820, EPI_ISL_2698821, EPI_ISL_2698822, EPI_ISL_2698823, EPI_ISL_2698824, EPI_ISL_2698825, EPI_ISL_2698826, EPI_ISL_2698827, EPI_ISL_2698828, EPI_ISL_2698829, EPI_ISL_2698830, EPI_ISL_2698831, EPI_ISL_2698832, EPI_ISL_2698833, EPI_ISL_2698834, EPI_ISL_2698835, EPI_ISL_2698836, EPI_ISL_2698837, EPI_ISL_2698838, EPI_ISL_2698840, EPI_ISL_2698841, EPI_ISL_2698842, EPI_ISL_2698843, EPI_ISL_2698844, EPI_ISL_2698846, EPI_ISL_2698847, EPI_ISL_2698848, EPI_ISL_2698849, EPI_ISL_2698851, EPI_ISL_2698852, EPI_ISL_2698853, EPI_ISL_2698854, EPI_ISL_2698857, EPI_ISL_2698858, EPI_ISL_2698859, EPI_ISL_2698860, EPI_ISL_2698861, EPI_ISL_2698862, EPI_ISL_2698864, EPI_ISL_2698865, EPI_ISL_2698866, EPI_ISL_2698867, EPI_ISL_2698868, EPI_ISL_2698869, EPI_ISL_2698870, EPI_ISL_2698871, EPI_ISL_2698873, EPI_ISL_2698874, EPI_ISL_2698875, EPI_ISL_2698876, EPI_ISL_2698877, EPI_ISL_2698878, EPI_ISL_2698879, EPI_ISL_2698880, EPI_ISL_2698881, EPI_ISL_2698882, EPI_ISL_2698884, EPI_ISL_2698885, EPI_ISL_2698886, EPI_ISL_2698887, EPI_ISL_2698888, EPI_ISL_2698889, EPI_ISL_2698890, EPI_ISL_2698891, EPI_ISL_2698892, EPI_ISL_2698893, EPI_ISL_2698895, EPI_ISL_2698896, EPI_ISL_2698897, EPI_ISL_2698898, EPI_ISL_2698899, EPI_ISL_2698900, EPI_ISL_2698901, EPI_ISL_2698902, EPI_ISL_2698904, EPI_ISL_2698905, EPI_ISL_2698906, EPI_ISL_2698907, EPI_ISL_2698908, EPI_ISL_2698909, EPI_ISL_2698910, EPI_ISL_2698911, EPI_ISL_2698912, EPI_ISL_2698914, EPI_ISL_2698915, EPI_ISL_2698916, EPI_ISL_2698917, EPI_ISL_2698918, EPI_ISL_2698919, EPI_ISL_2698920, EPI_ISL_2698921, EPI_ISL_2698922, EPI_ISL_2698923, EPI_ISL_2698925, EPI_ISL_2698926, EPI_ISL_2698927, EPI_ISL_2698928, EPI_ISL_2698930, EPI_ISL_2698931, EPI_ISL_2698932, EPI_ISL_2698933, EPI_ISL_2698934, EPI_ISL_2698935, EPI_ISL_2698936, EPI_ISL_2698937, EPI_ISL_2698938, EPI_ISL_2698939, EPI_ISL_2698940, EPI_ISL_2698941, EPI_ISL_2698944, EPI_ISL_2698945, EPI_ISL_2698946, EPI_ISL_2698947, EPI_ISL_2698948, EPI_ISL_2698949, EPI_ISL_2698950, EPI_ISL_2698951, EPI_ISL_2698952, EPI_ISL_2698953, EPI_ISL_2698954, EPI_ISL_2698955, EPI_ISL_2698956, EPI_ISL_2698957, EPI_ISL_2698958, EPI_ISL_2698959, EPI_ISL_2698960, EPI_ISL_2698961, EPI_ISL_2698962, EPI_ISL_2698963, EPI_ISL_2698964, EPI_ISL_2698965, EPI_ISL_2698966, EPI_ISL_2698967, EPI_ISL_2698968, EPI_ISL_2698969, EPI_ISL_2698970, EPI_ISL_2698971, EPI_ISL_2698972, EPI_ISL_2698974, EPI_ISL_2698975, EPI_ISL_2698976, EPI_ISL_2698977, EPI_ISL_2698978, EPI_ISL_2698979, EPI_ISL_2698980, EPI_ISL_2698981, EPI_ISL_2698982, EPI_ISL_2698983, EPI_ISL_2698984, EPI_ISL_2698985, EPI_ISL_2698986, EPI_ISL_2698988, EPI_ISL_2698989, EPI_ISL_2698991, EPI_ISL_2698992, EPI_ISL_2698993, EPI_ISL_2698994, EPI_ISL_2698995, EPI_ISL_2698996, EPI_ISL_2698997, EPI_ISL_2698998, EPI_ISL_2698999, EPI_ISL_2699000, EPI_ISL_2699001, EPI_ISL_2699002, EPI_ISL_2699003, EPI_ISL_2699004, EPI_ISL_2699005, EPI_ISL_2699006, EPI_ISL_2699007, EPI_ISL_2699008, EPI_ISL_2699009, EPI_ISL_2699010, EPI_ISL_2699011, EPI_ISL_2699012, EPI_ISL_2699013, EPI_ISL_2699014, EPI_ISL_2699015, EPI_ISL_2699016, EPI_ISL_2699017, EPI_ISL_2699018, EPI_ISL_2699019, EPI_ISL_2699020, EPI_ISL_2699021, EPI_ISL_2699022, EPI_ISL_2699023, EPI_ISL_2699025, EPI_ISL_2699026, EPI_ISL_2699027, EPI_ISL_2699028, EPI_ISL_2699029, EPI_ISL_2699030, EPI_ISL_2699031, EPI_ISL_2699032, EPI_ISL_2699033, EPI_ISL_2699034, EPI_ISL_2699035, EPI_ISL_2699036, EPI_ISL_2699037, EPI_ISL_2699038, EPI_ISL_2699041, EPI_ISL_2699042, EPI_ISL_2699043, EPI_ISL_2699045, EPI_ISL_2699049, EPI_ISL_2699050, EPI_ISL_2699051, EPI_ISL_2699052, EPI_ISL_2699053, EPI_ISL_2699054, EPI_ISL_2699056, EPI_ISL_2699057, EPI_ISL_2699058, EPI_ISL_2699059, EPI_ISL_2699060, EPI_ISL_2699061, EPI_ISL_2699062, EPI_ISL_2699063, EPI_ISL_2699064, EPI_ISL_2699065, EPI_ISL_2699067, EPI_ISL_2699068, EPI_ISL_2699069, EPI_ISL_2699070, EPI_ISL_2699071, EPI_ISL_2699072, EPI_ISL_2699073, EPI_ISL_2699074, EPI_ISL_2699075, EPI_ISL_2699076, EPI_ISL_2699077, EPI_ISL_2699078, EPI_ISL_2699079, EPI_ISL_2699080, EPI_ISL_2699081, EPI_ISL_2699082, EPI_ISL_2699083, EPI_ISL_2699084, EPI_ISL_2699085, EPI_ISL_2699086, EPI_ISL_2699087, EPI_ISL_2699088, EPI_ISL_2699089, EPI_ISL_2699090, EPI_ISL_2699091, EPI_ISL_2699092, EPI_ISL_2699093, EPI_ISL_2699094, EPI_ISL_2699095, EPI_ISL_2699096, EPI_ISL_2699097, EPI_ISL_2699098, EPI_ISL_2699099, EPI_ISL_2699100, EPI_ISL_2699101, EPI_ISL_2699102, EPI_ISL_2699103, EPI_ISL_2699104, EPI_ISL_2699105, EPI_ISL_2699106, EPI_ISL_2699107, EPI_ISL_2699108, EPI_ISL_2699109, EPI_ISL_2699110, EPI_ISL_2699111, EPI_ISL_2699112, EPI_ISL_2699113, EPI_ISL_2699114, EPI_ISL_2699115, EPI_ISL_2699116, EPI_ISL_2699117, EPI_ISL_2699118, EPI_ISL_2699119, EPI_ISL_2699120, EPI_ISL_2699121, EPI_ISL_2699122, EPI_ISL_2699124, EPI_ISL_2699125, EPI_ISL_2699126, EPI_ISL_2699127, EPI_ISL_2699128, EPI_ISL_2699129, EPI_ISL_2699130, EPI_ISL_2699131, EPI_ISL_2699132, EPI_ISL_2699134, EPI_ISL_2699135, EPI_ISL_2699136, EPI_ISL_2699137, EPI_ISL_2699139, EPI_ISL_2699140, EPI_ISL_2699141, EPI_ISL_2699142, EPI_ISL_2699144, EPI_ISL_2699145, EPI_ISL_2699146, EPI_ISL_2699147, EPI_ISL_2699148, EPI_ISL_2699149, EPI_ISL_2699150, EPI_ISL_2699151, EPI_ISL_2699152, EPI_ISL_2699153, EPI_ISL_2699154, EPI_ISL_2699155, EPI_ISL_2699158, EPI_ISL_2699159, EPI_ISL_2699160, EPI_ISL_2699161, EPI_ISL_2699162, EPI_ISL_2699163, EPI_ISL_2699164, EPI_ISL_2699165, EPI_ISL_2699166, EPI_ISL_2699167, EPI_ISL_2699168, EPI_ISL_2699169, EPI_ISL_2699170, EPI_ISL_2699171, EPI_ISL_2699173, EPI_ISL_2699174, EPI_ISL_2699175, EPI_ISL_2699176, EPI_ISL_2699177, EPI_ISL_2699178, EPI_ISL_2699179, EPI_ISL_2699180, EPI_ISL_2699181, EPI_ISL_2699182, EPI_ISL_2699183, EPI_ISL_2699184, EPI_ISL_2699185, EPI_ISL_2699187, EPI_ISL_2699188, EPI_ISL_2699189, EPI_ISL_2699190, EPI_ISL_2699191, EPI_ISL_2699192, EPI_ISL_2699194, EPI_ISL_2699195, EPI_ISL_2699196, EPI_ISL_2699197, EPI_ISL_2699198, EPI_ISL_2699199, EPI_ISL_2699200, EPI_ISL_2699201, EPI_ISL_2699202, EPI_ISL_2699203, EPI_ISL_2699204, EPI_ISL_2699205, EPI_ISL_2699206, EPI_ISL_2699207, EPI_ISL_2699208, EPI_ISL_2699209, EPI_ISL_2699210, EPI_ISL_2699211, EPI_ISL_2699213, EPI_ISL_2699215, EPI_ISL_2699216, EPI_ISL_2699217, EPI_ISL_2699218, EPI_ISL_2699220, EPI_ISL_2699222, EPI_ISL_2699223, EPI_ISL_2699224, EPI_ISL_2699225, EPI_ISL_2699226, EPI_ISL_2699227, EPI_ISL_2699228, EPI_ISL_2699229, EPI_ISL_2699230, EPI_ISL_2699231, EPI_ISL_2699232, EPI_ISL_2699233, EPI_ISL_2699234, EPI_ISL_2699235, EPI_ISL_2699236, EPI_ISL_2699237, EPI_ISL_2699238, EPI_ISL_2699239, EPI_ISL_2699240, EPI_ISL_2699241, EPI_ISL_2699242, EPI_ISL_2699243, EPI_ISL_2699244, EPI_ISL_2699245, EPI_ISL_2699246, EPI_ISL_2699247, EPI_ISL_2699248, EPI_ISL_2699249, EPI_ISL_2699250, EPI_ISL_2699251, EPI_ISL_2699252, EPI_ISL_2699253, EPI_ISL_2699254, EPI_ISL_2699255, EPI_ISL_2699256, EPI_ISL_2699257, EPI_ISL_2699258, EPI_ISL_2699259, EPI_ISL_2699260, EPI_ISL_2699261, EPI_ISL_2699262, EPI_ISL_2699263, EPI_ISL_2699264, EPI_ISL_2699265, EPI_ISL_2699266, EPI_ISL_2699267, EPI_ISL_2699268, EPI_ISL_2699269, EPI_ISL_2699270, EPI_ISL_2699271, EPI_ISL_2699272, EPI_ISL_2699273, EPI_ISL_2699274, EPI_ISL_2699275, EPI_ISL_2699276, EPI_ISL_2699277, EPI_ISL_2699278, EPI_ISL_2699279, EPI_ISL_2699280, EPI_ISL_2699281, EPI_ISL_2699282, EPI_ISL_2699283, EPI_ISL_2699284, EPI_ISL_2699285, EPI_ISL_2699286, EPI_ISL_2699287, EPI_ISL_2699288, EPI_ISL_2699289, EPI_ISL_2699290, EPI_ISL_2699291, EPI_ISL_2699292, EPI_ISL_2699293, EPI_ISL_2699294, EPI_ISL_2699295, EPI_ISL_2699296, EPI_ISL_2699297, EPI_ISL_2699298, EPI_ISL_2699299, EPI_ISL_2699300, EPI_ISL_2699301, EPI_ISL_2699302, EPI_ISL_2699303, EPI_ISL_2699304, EPI_ISL_2699305, EPI_ISL_2699306, EPI_ISL_2699307, EPI_ISL_2699308, EPI_ISL_2699309, EPI_ISL_2699310, EPI_ISL_2699311, EPI_ISL_2699312, EPI_ISL_2699313, EPI_ISL_2699314, EPI_ISL_2699315, EPI_ISL_2699316, EPI_ISL_2699317, EPI_ISL_2699318, EPI_ISL_2699319, EPI_ISL_2699320, EPI_ISL_2699321, EPI_ISL_2699322, EPI_ISL_2699323, EPI_ISL_2699324, EPI_ISL_2699325, EPI_ISL_2699326, EPI_ISL_2699327, EPI_ISL_2699328, EPI_ISL_2699329, EPI_ISL_2699330, EPI_ISL_2699331, EPI_ISL_2699332, EPI_ISL_2699333, EPI_ISL_2699334, EPI_ISL_2699335, EPI_ISL_2699336, EPI_ISL_2699337, EPI_ISL_2699338, EPI_ISL_2699339, EPI_ISL_2699340, EPI_ISL_2699341, EPI_ISL_2699342, EPI_ISL_2699343, EPI_ISL_2699344, EPI_ISL_2699345, EPI_ISL_2699346, EPI_ISL_2699347, EPI_ISL_2699348, EPI_ISL_2699349, EPI_ISL_2699350, EPI_ISL_2699351, EPI_ISL_2699352, EPI_ISL_2699353, EPI_ISL_2699354, EPI_ISL_2699355, EPI_ISL_2699356, EPI_ISL_2699357, EPI_ISL_2699358, EPI_ISL_2699359, EPI_ISL_2699360, EPI_ISL_2699361, EPI_ISL_2699362, EPI_ISL_2699363, EPI_ISL_2699364, EPI_ISL_2699365, EPI_ISL_2699366, EPI_ISL_2699367, EPI_ISL_2699368, EPI_ISL_2699369, EPI_ISL_2699370, EPI_ISL_2699371, EPI_ISL_2699372, EPI_ISL_2699373, EPI_ISL_2699374, EPI_ISL_2699375, EPI_ISL_2699376, EPI_ISL_2699377, EPI_ISL_2699378, EPI_ISL_2699379, EPI_ISL_2699380, EPI_ISL_2699381, EPI_ISL_2699382, EPI_ISL_2699383, EPI_ISL_2699384, EPI_ISL_2699385, EPI_ISL_2699386, EPI_ISL_2699387, EPI_ISL_2699388, EPI_ISL_2699389, EPI_ISL_2699390, EPI_ISL_2699391, EPI_ISL_2699392, EPI_ISL_2699393, EPI_ISL_2699394, EPI_ISL_2699395, EPI_ISL_2699396, EPI_ISL_2699397, EPI_ISL_2699398, EPI_ISL_2699399, EPI_ISL_2699400, EPI_ISL_2699401, EPI_ISL_2699402, EPI_ISL_2699403, EPI_ISL_2699404, EPI_ISL_2699405, EPI_ISL_2699406, EPI_ISL_2699407, EPI_ISL_2699408, EPI_ISL_2699409, EPI_ISL_2699410, EPI_ISL_2699411, EPI_ISL_2699412, EPI_ISL_2699413, EPI_ISL_2699414, EPI_ISL_2699415, EPI_ISL_2699416, EPI_ISL_2699417, EPI_ISL_2699418, EPI_ISL_2699419, EPI_ISL_2699420, EPI_ISL_2699421, EPI_ISL_2699422, EPI_ISL_2699423, EPI_ISL_2699424, EPI_ISL_2699425 | | | |
| see above | Public Health Laboratory, Minnesota Department of Health | University of Minnesota Genomics Center | Daryl M. Gohl, John Garbe, Jaquelyn Kuriger-Laber, Corbin Dirkx, and Sean Wang |
| EPI_ISL_2699427, EPI_ISL_2699428, EPI_ISL_2699429, EPI_ISL_2699430, EPI_ISL_2699431, EPI_ISL_2699432, EPI_ISL_2699433, EPI_ISL_2699434, EPI_ISL_2699435 | Laboratory, Gravity Diagnostics | Laboratory, Gravity Diagnostics | Diagnostics,G. |
| EPI_ISL_2699436, EPI_ISL_2699437, EPI_ISL_2699438, EPI_ISL_2699439, EPI_ISL_2699440, EPI_ISL_2699441, EPI_ISL_2699442, EPI_ISL_2699443, EPI_ISL_2699444, EPI_ISL_2699445, EPI_ISL_2699446, EPI_ISL_2699447, EPI_ISL_2699448, EPI_ISL_2699449, EPI_ISL_2699450, EPI_ISL_2699451, EPI_ISL_2699452, EPI_ISL_2699453, EPI_ISL_2699454, EPI_ISL_2699455, EPI_ISL_2699456, EPI_ISL_2699457, EPI_ISL_2699458, EPI_ISL_2699459, EPI_ISL_2699460, EPI_ISL_2699461, EPI_ISL_2699462, EPI_ISL_2699463, EPI_ISL_2699464, EPI_ISL_2699465, EPI_ISL_2699466, EPI_ISL_2699467, EPI_ISL_2699468, EPI_ISL_2699469, EPI_ISL_2699470, EPI_ISL_2699471, EPI_ISL_2699472, EPI_ISL_2699473, EPI_ISL_2699474, EPI_ISL_2699475, EPI_ISL_2699476, EPI_ISL_2699477, EPI_ISL_2699478, EPI_ISL_2699479, EPI_ISL_2699480, EPI_ISL_2699481, EPI_ISL_2699482, EPI_ISL_2699483, EPI_ISL_2699484, EPI_ISL_2699485, EPI_ISL_2699486, EPI_ISL_2699487, EPI_ISL_2699488, EPI_ISL_2699489, EPI_ISL_2699490, EPI_ISL_2699491, EPI_ISL_2699492, EPI_ISL_2699493, EPI_ISL_2699494, EPI_ISL_2699495, EPI_ISL_2699496, EPI_ISL_2699497, EPI_ISL_2699498, EPI_ISL_2699499, EPI_ISL_2699500, EPI_ISL_2699501, EPI_ISL_2699502, EPI_ISL_2699503, EPI_ISL_2699504, EPI_ISL_2699505, EPI_ISL_2699506, EPI_ISL_2699507, EPI_ISL_2699508, EPI_ISL_2699509, EPI_ISL_2699510, EPI_ISL_2699512, EPI_ISL_2699513, EPI_ISL_2699514, EPI_ISL_2699515, EPI_ISL_2699516, EPI_ISL_2699517, EPI_ISL_2699518, EPI_ISL_2699519, EPI_ISL_2709021, EPI_ISL_2709022, EPI_ISL_2709023, EPI_ISL_2709024, EPI_ISL_2709025, EPI_ISL_2709026, EPI_ISL_2709027, EPI_ISL_2709028, EPI_ISL_2709029, EPI_ISL_2709030, EPI_ISL_2709031, EPI_ISL_2709032, EPI_ISL_2709033, EPI_ISL_2709034, EPI_ISL_2709035, EPI_ISL_2709036, EPI_ISL_2709037, EPI_ISL_2709038, EPI_ISL_2709039, EPI_ISL_2709040, EPI_ISL_2709041, EPI_ISL_2709042, EPI_ISL_2709043, EPI_ISL_2709044, EPI_ISL_2709045, EPI_ISL_2709046, EPI_ISL_2709047, EPI_ISL_2709048, EPI_ISL_2709049, EPI_ISL_2709050, EPI_ISL_2709051, EPI_ISL_2709052, EPI_ISL_2709053, EPI_ISL_2709054, EPI_ISL_2709055, EPI_ISL_2709056, EPI_ISL_2709057, EPI_ISL_2709058, EPI_ISL_2709059, EPI_ISL_2709060, EPI_ISL_2709061, EPI_ISL_2709062, EPI_ISL_2709063, EPI_ISL_2709064, EPI_ISL_2709065, EPI_ISL_2709066, EPI_ISL_2709067, EPI_ISL_2709068, EPI_ISL_2709069, EPI_ISL_2709070, EPI_ISL_2709071, EPI_ISL_2709072, EPI_ISL_2709073, EPI_ISL_2709074, EPI_ISL_2709075, EPI_ISL_2709076, EPI_ISL_2709077, EPI_ISL_2709078, EPI_ISL_2709080, EPI_ISL_2709082, EPI_ISL_2709083, EPI_ISL_2709084, EPI_ISL_2709085, EPI_ISL_2709086, EPI_ISL_2709087, EPI_ISL_2709088, EPI_ISL_2709089, EPI_ISL_2709090, EPI_ISL_2709091, EPI_ISL_2709092, EPI_ISL_2709093, EPI_ISL_2709094, EPI_ISL_2709095, EPI_ISL_2709096, EPI_ISL_2709097, EPI_ISL_2709098, EPI_ISL_2709099, EPI_ISL_2709100, EPI_ISL_2709101, EPI_ISL_2709102, EPI_ISL_2709103, EPI_ISL_2709104, EPI_ISL_2709105, EPI_ISL_2709106, EPI_ISL_2709107, EPI_ISL_2709108, EPI_ISL_2709109, EPI_ISL_2709110, EPI_ISL_2709111, EPI_ISL_2709112, EPI_ISL_2709113, EPI_ISL_2709114, EPI_ISL_2709115, EPI_ISL_2709116, EPI_ISL_2709117, EPI_ISL_2709118, EPI_ISL_2709119, EPI_ISL_2709120, EPI_ISL_2709121, EPI_ISL_2709122, EPI_ISL_2709123, EPI_ISL_2709124, EPI_ISL_2709125, EPI_ISL_2709126, EPI_ISL_2709127, EPI_ISL_2709128, EPI_ISL_2709129, EPI_ISL_2709130, EPI_ISL_2709131, EPI_ISL_2709132, EPI_ISL_2709133, EPI_ISL_2709134, EPI_ISL_2709135, EPI_ISL_2709136, EPI_ISL_2709137, EPI_ISL_2709138, EPI_ISL_2709139, EPI_ISL_2709140, EPI_ISL_2709141, EPI_ISL_2709142, EPI_ISL_2709143, EPI_ISL_2709144, EPI_ISL_2709145, EPI_ISL_2709146, EPI_ISL_2709147, EPI_ISL_2709148, EPI_ISL_2709149, EPI_ISL_2709150, EPI_ISL_2709151, EPI_ISL_2709152, EPI_ISL_2709153, EPI_ISL_2709154, EPI_ISL_2709155, EPI_ISL_2709156, EPI_ISL_2709157, EPI_ISL_2709158, EPI_ISL_2709159, EPI_ISL_2709160, EPI_ISL_2709161, EPI_ISL_2709162, EPI_ISL_2709163, EPI_ISL_2709164, EPI_ISL_2709165, EPI_ISL_2709166, EPI_ISL_2709167, EPI_ISL_2709168, EPI_ISL_2709169, EPI_ISL_2709170, EPI_ISL_2709171, EPI_ISL_2709172, EPI_ISL_2709173, EPI_ISL_2709174, EPI_ISL_2709175, EPI_ISL_2709176, EPI_ISL_2709177, EPI_ISL_2709178, EPI_ISL_2709179, EPI_ISL_2709180, EPI_ISL_2709181, EPI_ISL_2709182, EPI_ISL_2709183, EPI_ISL_2709184, EPI_ISL_2709185, EPI_ISL_2709186, EPI_ISL_2709187, EPI_ISL_2709188, EPI_ISL_2709189, EPI_ISL_2709190, EPI_ISL_2709191, EPI_ISL_2709192, EPI_ISL_2709193, EPI_ISL_2709194, EPI_ISL_2709195, EPI_ISL_2709196, EPI_ISL_2709197, EPI_ISL_2709198, EPI_ISL_2709199, EPI_ISL_2709200, EPI_ISL_2709201, EPI_ISL_2709202, EPI_ISL_2709203, EPI_ISL_2709204, EPI_ISL_2709205, EPI_ISL_2709206, EPI_ISL_2709207, EPI_ISL_2709208, EPI_ISL_2709209, EPI_ISL_2709210, EPI_ISL_2709211, EPI_ISL_2709214, EPI_ISL_2709216, EPI_ISL_2709217, EPI_ISL_2709220, EPI_ISL_2709221, EPI_ISL_2709224, EPI_ISL_2709225, EPI_ISL_2709230, EPI_ISL_2709231, EPI_ISL_2709234, EPI_ISL_2709235, EPI_ISL_2709236, EPI_ISL_2709238, EPI_ISL_2709239, EPI_ISL_2709242, EPI_ISL_2709244, EPI_ISL_2709245 | | | |
| see above | Public Health Laboratory, Minnesota Department of Health | University of Minnesota Genomics Center | Daryl M. Gohl, John Garbe, Jaquelyn Kuriger-Laber, Corbin Dirkx, and Sean Wang |
| EPI_ISL_2709448, EPI_ISL_2709449, EPI_ISL_2709450, EPI_ISL_2709451, EPI_ISL_2709452, EPI_ISL_2709453, EPI_ISL_2709454, EPI_ISL_2709456, EPI_ISL_2709457, EPI_ISL_2709458, EPI_ISL_2709459, EPI_ISL_2709460, EPI_ISL_2709461, EPI_ISL_2709462, EPI_ISL_2709465, EPI_ISL_2709466, EPI_ISL_2709468, EPI_ISL_2709469, EPI_ISL_2709472, EPI_ISL_2709473, EPI_ISL_2709474, EPI_ISL_2709475 | | | |
| see above | IN State Department of Health Laboratory Services | IN State Department of Health Laboratory Services | Cassandra Campion, Jamie Yeadon, Brian Pope, Lixia Liu, Kyle Brownlee, Melissa Hindenlang, Mark Glazier |
| EPI_ISL_2709495, EPI_ISL_2709496 | Fulgent Genetics | Centers for Disease Control and Prevention Division of Viral Diseases, Pathogen Discovery | Dakota Howard, Dhwani Batra, Peter W. Cook, Kara Moser, Adrian Paskey, Jason Caravas, Benjamin Rambo-Martin, Shatavia Morrison, Christopher Gulvick, Scott Sammons, Yvette Unoarumhi, Darlene Wagner, Matthew Schmerer, Harry Gao, Mickey Li, John Gao, Joseph Fierro, Benafsh Sapra, Becky Tsai, Yan Meng, Doreen Ng, James Xie, Clinton R. Paden, Duncan MacCannell |
| EPI_ISL_2709497, EPI_ISL_2709498, EPI_ISL_2709499, EPI_ISL_2709500, EPI_ISL_2709501, EPI_ISL_2709502 | Aegis Sciences Corporation | Centers for Disease Control and Prevention Division of Viral Diseases, Pathogen Discovery | Dakota Howard, Dhwani Batra, Peter W. Cook, Kara Moser, Adrian Paskey, Jason Caravas, Benjamin Rambo-Martin, Shatavia Morrison, Christopher Gulvick, Scott Sammons, Yvette Unoarumhi, Darlene Wagner, Matthew Schmerer, Cyndi Clark, Patrick Campbell, Rob Case, Vikramsinha Ghorpade, Holly Houdeshell, Ola Kvalvaag, Dillon Nall, Ethan Sanders, Alec Vest, Shaun Westlund, Matthew Hardison, Clinton R. Paden, Duncan MacCannell |
| EPI_ISL_2709503, EPI_ISL_2709504, EPI_ISL_2709506, EPI_ISL_2709507, EPI_ISL_2709508, EPI_ISL_2709509, EPI_ISL_2709510, EPI_ISL_2709511 | Infinity Biologix | Centers for Disease Control and Prevention Division of Viral Diseases, Pathogen Discovery | Dakota Howard, Dhwani Batra, Peter W. Cook, Kara Moser, Adrian Paskey, Jason Caravas, Benjamin Rambo-Martin, Shatavia Morrison, Christopher Gulvick, Scott Sammons, Yvette Unoarumhi, Darlene Wagner, Matthew Schmerer, Christian Bixby, Yihe Wang, Jonathan Schultz, Chirayu Goswami, Russ Hager, Robin Grimwood, Clinton R. Paden, Duncan MacCannell |
| EPI_ISL_2709512 | Fulgent Genetics | Centers for Disease Control and Prevention Division of Viral Diseases, Pathogen Discovery | Dakota Howard, Dhwani Batra, Peter W. Cook, Kara Moser, Adrian Paskey, Jason Caravas, Benjamin Rambo-Martin, Shatavia Morrison, Christopher Gulvick, Scott Sammons, Yvette Unoarumhi, Darlene Wagner, Matthew Schmerer, Harry Gao, Mickey Li, John Gao, Joseph Fierro, Benafsh Sapra, Becky Tsai, Yan Meng, Doreen Ng, James Xie, Clinton R. Paden, Duncan MacCannell |
| EPI_ISL_2709513, EPI_ISL_2709514, EPI_ISL_2709515, EPI_ISL_2709516, EPI_ISL_2709517, EPI_ISL_2709518, EPI_ISL_2709519, EPI_ISL_2709520, EPI_ISL_2709521, EPI_ISL_2709522, EPI_ISL_2709523, EPI_ISL_2709524, EPI_ISL_2709525, EPI_ISL_2709526, EPI_ISL_2709527, EPI_ISL_2709528, EPI_ISL_2709529, EPI_ISL_2709530, EPI_ISL_2709531, EPI_ISL_2709532, EPI_ISL_2709533, EPI_ISL_2709534, EPI_ISL_2709535, EPI_ISL_2709536, EPI_ISL_2709537, EPI_ISL_2709538, EPI_ISL_2709539, EPI_ISL_2709540, EPI_ISL_2709541, EPI_ISL_2709542, EPI_ISL_2709543, EPI_ISL_2709544, EPI_ISL_2709545, EPI_ISL_2709546, EPI_ISL_2709547, EPI_ISL_2709548, EPI_ISL_2709549, EPI_ISL_2709550, EPI_ISL_2709551, EPI_ISL_2709552 | | | |
| see above | Aegis Sciences Corporation | Centers for Disease Control and Prevention Division of Viral Diseases, Pathogen Discovery | Dakota Howard, Dhwani Batra, Peter W. Cook, Kara Moser, Adrian Paskey, Jason Caravas, Benjamin Rambo-Martin, Shatavia Morrison, Christopher Gulvick, Scott Sammons, Yvette Unoarumhi, Darlene Wagner, Matthew Schmerer, Cyndi Clark, Patrick Campbell, Rob Case, Vikramsinha Ghorpade, Holly Houdeshell, Ola Kvalvaag, Dillon Nall, Ethan Sanders, Alec Vest, Shaun Westlund, Matthew Hardison, Clinton R. Paden, Duncan MacCannell |
| EPI_ISL_2709553 | MS Public Health Laboratory | Centers for Disease Control and Prevention Division of Viral Diseases, Pathogen Discovery | Mili Sheth, Sarah Nobles, Jasmine Padilla, Mark Burroughs, Shoshona Le, Katie Dillon, Peter Cook, Clinton R. Paden, Dhwani Batra, Krista Queen, Kristen Knipe, Dakota Howard, Yvette Unoarumhi, Darlene Wagner, Matthew Schmerer, Ben L. Rambo-Martin, Kristine Lacek, Sam Shepard, Alison Laufer Halpin, Dave Wentworth, Vivien Dugan, Suxiang Tong, Justin Lee |
| EPI_ISL_2709554 | IA State Hygienic Laboratory | Centers for Disease Control and Prevention Division of Viral Diseases, Pathogen Discovery | Mili Sheth, Sarah Nobles, Jasmine Padilla, Mark Burroughs, Shoshona Le, Katie Dillon, Peter Cook, Clinton R. Paden, Dhwani Batra, Krista Queen, Kristen Knipe, Dakota Howard, Yvette Unoarumhi, Darlene Wagner, Matthew Schmerer, Ben L. Rambo-Martin, Kristine Lacek, Sam Shepard, Alison Laufer Halpin, Dave Wentworth, Vivien Dugan, Suxiang Tong, Justin Lee |
| EPI_ISL_2709555 | WY Public Health Laboratory | Centers for Disease Control and Prevention Division of Viral Diseases, Pathogen Discovery | Mili Sheth, Sarah Nobles, Jasmine Padilla, Mark Burroughs, Shoshona Le, Katie Dillon, Peter Cook, Clinton R. Paden, Dhwani Batra, Krista Queen, Kristen Knipe, Dakota Howard, Yvette Unoarumhi, Darlene Wagner, Matthew Schmerer, Ben L. Rambo-Martin, Kristine Lacek, Sam Shepard, Alison Laufer Halpin, Dave Wentworth, Vivien Dugan, Suxiang Tong, Justin Lee |
| EPI_ISL_2709556 | NE Public Health Laboratory | Centers for Disease Control and Prevention Division of Viral Diseases, Pathogen Discovery | Mili Sheth, Sarah Nobles, Jasmine Padilla, Mark Burroughs, Shoshona Le, Katie Dillon, Peter Cook, Clinton R. Paden, Dhwani Batra, Krista Queen, Kristen Knipe, Dakota Howard, Yvette Unoarumhi, Darlene Wagner, Matthew Schmerer, Ben L. Rambo-Martin, Kristine Lacek, Sam Shepard, Alison Laufer Halpin, Dave Wentworth, Vivien Dugan, Suxiang Tong, Justin Lee |
| EPI_ISL_2709557 | CA-Los Angeles County Public Health Laboratory | Centers for Disease Control and Prevention Division of Viral Diseases, Pathogen Discovery | Mili Sheth, Sarah Nobles, Jasmine Padilla, Mark Burroughs, Shoshona Le, Katie Dillon, Peter Cook, Clinton R. Paden, Dhwani Batra, Krista Queen, Kristen Knipe, Dakota Howard, Yvette Unoarumhi, Darlene Wagner, Matthew Schmerer, Ben L. Rambo-Martin, Kristine Lacek, Sam Shepard, Alison Laufer Halpin, Dave Wentworth, Vivien Dugan, Suxiang Tong, Justin Lee |
| EPI_ISL_2709558 | NJ Public Health and Environmental | Centers for Disease Control and Prevention | Mili Sheth, Sarah Nobles, Jasmine Padilla, Mark Burroughs, Shoshona Le, Katie Dillon, Peter Cook, Clinton R. Paden, Dhwani Batra, Krista Queen, Kristen Knipe, Dakota Howard, Yvette Unoarumhi, Darlene Wagner, Matthew Schmerer, Ben L. Rambo-Martin, Kristine Lacek, Sam Shepard, |

| | | | |
|---|---|---|---|
| | Laboratories | Division of Viral Diseases, Pathogen Discovery | Alison Laufer Halpin, Dave Wentworth, Vivien Dugan, Suxiang Tong, Justin Lee |
| EPI_ISL_2709559 | MI - Michigan Department of Health and Human Services - Bureau of Laboratories | Centers for Disease Control and Prevention Division of Viral Diseases, Pathogen Discovery | Mili Sheth, Sarah Nobles, Jasmine Padilla, Mark Burroughs, Shoshona Le, Katie Dillon, Peter Cook, Clinton R. Paden, Dhwani Batra, Krista Queen, Kristen Knipe, Dakota Howard, Yvette Unoarumhi, Darlene Wagner, Matthew Schmerer, Ben L. Rambo-Martin, Kristine Lacek, Sam Shepard, Alison Laufer Halpin, Dave Wentworth, Vivien Dugan, Suxiang Tong, Justin Lee |
| EPI_ISL_2709560, EPI_ISL_2709561 | NV-Southern Nevada Public Health Laboratory | Centers for Disease Control and Prevention Division of Viral Diseases, Pathogen Discovery | Mili Sheth, Sarah Nobles, Jasmine Padilla, Mark Burroughs, Shoshona Le, Katie Dillon, Peter Cook, Clinton R. Paden, Dhwani Batra, Krista Queen, Kristen Knipe, Dakota Howard, Yvette Unoarumhi, Darlene Wagner, Matthew Schmerer, Ben L. Rambo-Martin, Kristine Lacek, Sam Shepard, Alison Laufer Halpin, Dave Wentworth, Vivien Dugan, Suxiang Tong, Justin Lee |
| EPI_ISL_2709562 | "NM Dept. Health, Scientific Laboratory Division " | Centers for Disease Control and Prevention Division of Viral Diseases, Pathogen Discovery | Mili Sheth, Sarah Nobles, Jasmine Padilla, Mark Burroughs, Shoshona Le, Katie Dillon, Peter Cook, Clinton R. Paden, Dhwani Batra, Krista Queen, Kristen Knipe, Dakota Howard, Yvette Unoarumhi, Darlene Wagner, Matthew Schmerer, Ben L. Rambo-Martin, Kristine Lacek, Sam Shepard, Alison Laufer Halpin, Dave Wentworth, Vivien Dugan, Suxiang Tong, Justin Lee |
| EPI_ISL_2709563, EPI_ISL_2709564 | ID Bureau of Laboratories | Centers for Disease Control and Prevention Division of Viral Diseases, Pathogen Discovery | Mili Sheth, Sarah Nobles, Jasmine Padilla, Mark Burroughs, Shoshona Le, Katie Dillon, Peter Cook, Clinton R. Paden, Dhwani Batra, Krista Queen, Kristen Knipe, Dakota Howard, Yvette Unoarumhi, Darlene Wagner, Matthew Schmerer, Ben L. Rambo-Martin, Kristine Lacek, Sam Shepard, Alison Laufer Halpin, Dave Wentworth, Vivien Dugan, Suxiang Tong, Justin Lee |
| EPI_ISL_2709565 | WA State Department of Health | Centers for Disease Control and Prevention Division of Viral Diseases, Pathogen Discovery | Mili Sheth, Sarah Nobles, Jasmine Padilla, Mark Burroughs, Shoshona Le, Katie Dillon, Peter Cook, Clinton R. Paden, Dhwani Batra, Krista Queen, Kristen Knipe, Dakota Howard, Yvette Unoarumhi, Darlene Wagner, Matthew Schmerer, Ben L. Rambo-Martin, Kristine Lacek, Sam Shepard, Alison Laufer Halpin, Dave Wentworth, Vivien Dugan, Suxiang Tong, Justin Lee |
| EPI_ISL_2709566 | PR Public Health Lab | Centers for Disease Control and Prevention Division of Viral Diseases, Pathogen Discovery | Mili Sheth, Sarah Nobles, Jasmine Padilla, Mark Burroughs, Shoshona Le, Katie Dillon, Peter Cook, Clinton R. Paden, Dhwani Batra, Krista Queen, Kristen Knipe, Dakota Howard, Yvette Unoarumhi, Darlene Wagner, Matthew Schmerer, Ben L. Rambo-Martin, Kristine Lacek, Sam Shepard, Alison Laufer Halpin, Dave Wentworth, Vivien Dugan, Suxiang Tong, Justin Lee |
| EPI_ISL_2709567 | Philadelphia Department of Public Health | Centers for Disease Control and Prevention Division of Viral Diseases, Pathogen Discovery | Mili Sheth, Sarah Nobles, Jasmine Padilla, Mark Burroughs, Shoshona Le, Katie Dillon, Peter Cook, Clinton R. Paden, Dhwani Batra, Krista Queen, Kristen Knipe, Dakota Howard, Yvette Unoarumhi, Darlene Wagner, Matthew Schmerer, Ben L. Rambo-Martin, Kristine Lacek, Sam Shepard, Alison Laufer Halpin, Dave Wentworth, Vivien Dugan, Suxiang Tong, Justin Lee |
| EPI_ISL_2709686 | LESP Queretaro | Instituto de Diagnostico y Referencia Epidemiologicos (INDRE) | Claudia Wong-Arambula, Abril Rodriguez-Maldonado, Vanessa Rivero-Arredondo, Ariadna Medina-Benitez, Joaquin Quiroz-Mercado, Sergio Rangel-Guerrero, Natividad Cruz-Ortiz, Tatiana Nunez-Garcia, Gisela Barrera-Badillo, Lucia Hernandez-Rivas, Irma Lopez-Martinez, Ernesto Ramirez-Gonzalez. |
| EPI_ISL_2709687 | LESP Baja California | Instituto de Diagnostico y Referencia Epidemiologicos (INDRE) | Claudia Wong-Arambula, Abril Rodriguez-Maldonado, Vanessa Rivero-Arredondo, Ariadna Medina-Benitez, Joaquin Quiroz-Mercado, Sergio Rangel-Guerrero, Natividad Cruz-Ortiz, Tatiana Nunez-Garcia, Gisela Barrera-Badillo, Lucia Hernandez-Rivas, Irma Lopez-Martinez, Ernesto Ramirez-Gonzalez. |
| EPI_ISL_2709688 | LESP Michoacan | Instituto de Diagnostico y Referencia Epidemiologicos (INDRE) | Claudia Wong-Arambula, Abril Rodriguez-Maldonado, Vanessa Rivero-Arredondo, Ariadna Medina-Benitez, Joaquin Quiroz-Mercado, Sergio Rangel-Guerrero, Natividad Cruz-Ortiz, Tatiana Nunez-Garcia, Gisela Barrera-Badillo, Lucia Hernandez-Rivas, Irma Lopez-Martinez, Ernesto Ramirez-Gonzalez. |
| EPI_ISL_2709689, EPI_ISL_2709690, EPI_ISL_2709691, EPI_ISL_2709692, EPI_ISL_2709693, EPI_ISL_2709694, EPI_ISL_2709695, EPI_ISL_2709696, EPI_ISL_2709697, EPI_ISL_2709698 | LESP Nuevo Leon | Instituto de Diagnostico y Referencia Epidemiologicos (INDRE) | Claudia Wong-Arambula, Abril Rodriguez-Maldonado, Vanessa Rivero-Arredondo, Ariadna Medina-Benitez, Joaquin Quiroz-Mercado, Sergio Rangel-Guerrero, Natividad Cruz-Ortiz, Tatiana Nunez-Garcia, Gisela Barrera-Badillo, Lucia Hernandez-Rivas, Irma Lopez-Martinez, Ernesto Ramirez-Gonzalez. |
| EPI_ISL_2709699 | LESP Jalisco | Instituto de Diagnostico y Referencia Epidemiologicos (INDRE) | Claudia Wong-Arambula, Abril Rodriguez-Maldonado, Vanessa Rivero-Arredondo, Ariadna Medina-Benitez, Joaquin Quiroz-Mercado, Sergio Rangel-Guerrero, Natividad Cruz-Ortiz, Tatiana Nunez-Garcia, Gisela Barrera-Badillo, Lucia Hernandez-Rivas, Irma Lopez-Martinez, Ernesto Ramirez-Gonzalez. |
| EPI_ISL_2709700, EPI_ISL_2709701, EPI_ISL_2709702 | LESP Morelos | Instituto de Diagnostico y Referencia Epidemiologicos (INDRE) | Claudia Wong-Arambula, Abril Rodriguez-Maldonado, Vanessa Rivero-Arredondo, Ariadna Medina-Benitez, Joaquin Quiroz-Mercado, Sergio Rangel-Guerrero, Natividad Cruz-Ortiz, Tatiana Nunez-Garcia, Gisela Barrera-Badillo, Lucia Hernandez-Rivas, Irma Lopez-Martinez, Ernesto Ramirez-Gonzalez. |
| EPI_ISL_2709703 | LESP Ciudad de Mexico | Instituto de Diagnostico y Referencia Epidemiologicos (INDRE) | Claudia Wong-Arambula, Abril Rodriguez-Maldonado, Vanessa Rivero-Arredondo, Ariadna Medina-Benitez, Joaquin Quiroz-Mercado, Sergio Rangel-Guerrero, Natividad Cruz-Ortiz, Tatiana Nunez-Garcia, Gisela Barrera-Badillo, Lucia Hernandez-Rivas, Irma Lopez-Martinez, Ernesto Ramirez-Gonzalez. |
| EPI_ISL_2709704, EPI_ISL_2709705 | LESP Estado de Mexico | Instituto de Diagnostico y Referencia Epidemiologicos (INDRE) | Claudia Wong-Arambula, Abril Rodriguez-Maldonado, Vanessa Rivero-Arredondo, Ariadna Medina-Benitez, Joaquin Quiroz-Mercado, Sergio Rangel-Guerrero, Natividad Cruz-Ortiz, Tatiana Nunez-Garcia, Gisela Barrera-Badillo, Lucia Hernandez-Rivas, Irma Lopez-Martinez, Ernesto Ramirez-Gonzalez. |
| EPI_ISL_2709706, EPI_ISL_2709707, EPI_ISL_2709708 | LESP Queretaro | Instituto de Diagnostico y Referencia Epidemiologicos (INDRE) | Claudia Wong-Arambula, Abril Rodriguez-Maldonado, Vanessa Rivero-Arredondo, Ariadna Medina-Benitez, Joaquin Quiroz-Mercado, Sergio Rangel-Guerrero, Natividad Cruz-Ortiz, Tatiana Nunez-Garcia, Gisela Barrera-Badillo, Lucia Hernandez-Rivas, Irma Lopez-Martinez, Ernesto Ramirez-Gonzalez. |
| EPI_ISL_2709709, EPI_ISL_2709710, EPI_ISL_2709711 | LESP Tabasco | Instituto de Diagnostico y Referencia Epidemiologicos (INDRE) | Claudia Wong-Arambula, Abril Rodriguez-Maldonado, Vanessa Rivero-Arredondo, Ariadna Medina-Benitez, Joaquin Quiroz-Mercado, Sergio Rangel-Guerrero, Natividad Cruz-Ortiz, Tatiana Nunez-Garcia, Gisela Barrera-Badillo, Lucia Hernandez-Rivas, Irma Lopez-Martinez, Ernesto Ramirez-Gonzalez. |
| EPI_ISL_2709712, EPI_ISL_2709713, EPI_ISL_2709714, EPI_ISL_2709715, EPI_ISL_2709716 | LESP Ciudad de Mexico | Instituto de Diagnostico y Referencia Epidemiologicos (INDRE) | Claudia Wong-Arambula, Abril Rodriguez-Maldonado, Vanessa Rivero-Arredondo, Ariadna Medina-Benitez, Joaquin Quiroz-Mercado, Sergio Rangel-Guerrero, Natividad Cruz-Ortiz, Tatiana Nunez-Garcia, Gisela Barrera-Badillo, Lucia Hernandez-Rivas, Irma Lopez-Martinez, Ernesto Ramirez-Gonzalez. |
| EPI_ISL_2709717 | LESP Jalisco | Instituto de Diagnostico y Referencia Epidemiologicos (INDRE) | Claudia Wong-Arambula, Abril Rodriguez-Maldonado, Vanessa Rivero-Arredondo, Ariadna Medina-Benitez, Joaquin Quiroz-Mercado, Sergio Rangel-Guerrero, Natividad Cruz-Ortiz, Tatiana Nunez-Garcia, Gisela Barrera-Badillo, Lucia Hernandez-Rivas, Irma Lopez-Martinez, Ernesto Ramirez-Gonzalez. |
| EPI_ISL_2709718 | LESP Sonora | Instituto de Diagnostico y Referencia Epidemiologicos (INDRE) | Claudia Wong-Arambula, Abril Rodriguez-Maldonado, Vanessa Rivero-Arredondo, Ariadna Medina-Benitez, Joaquin Quiroz-Mercado, Sergio Rangel-Guerrero, Natividad Cruz-Ortiz, Tatiana Nunez-Garcia, Gisela Barrera-Badillo, Lucia Hernandez-Rivas, Irma Lopez-Martinez, Ernesto Ramirez-Gonzalez. |
| EPI_ISL_2709719 | LESP Estado de Mexico | Instituto de Diagnostico y Referencia Epidemiologicos (INDRE) | Claudia Wong-Arambula, Abril Rodriguez-Maldonado, Vanessa Rivero-Arredondo, Ariadna Medina-Benitez, Joaquin Quiroz-Mercado, Sergio Rangel-Guerrero, Natividad Cruz-Ortiz, Tatiana Nunez-Garcia, Gisela Barrera-Badillo, Lucia Hernandez-Rivas, Irma Lopez-Martinez, Ernesto Ramirez-Gonzalez. |
| EPI_ISL_2709720 | LESP Ciudad de Mexico | Instituto de Diagnostico y Referencia Epidemiologicos (INDRE) | Claudia Wong-Arambula, Abril Rodriguez-Maldonado, Vanessa Rivero-Arredondo, Ariadna Medina-Benitez, Joaquin Quiroz-Mercado, Sergio Rangel-Guerrero, Natividad Cruz-Ortiz, Tatiana Nunez-Garcia, Gisela Barrera-Badillo, Lucia Hernandez-Rivas, Irma Lopez-Martinez, Ernesto Ramirez-Gonzalez. |
| EPI_ISL_2709721 | LESP Estado de Mexico | Instituto de Diagnostico y Referencia Epidemiologicos (INDRE) | Claudia Wong-Arambula, Abril Rodriguez-Maldonado, Vanessa Rivero-Arredondo, Ariadna Medina-Benitez, Joaquin Quiroz-Mercado, Sergio Rangel-Guerrero, Natividad Cruz-Ortiz, Tatiana Nunez-Garcia, Gisela Barrera-Badillo, Lucia Hernandez-Rivas, Irma Lopez-Martinez, Ernesto Ramirez-Gonzalez. |
| EPI_ISL_2709722 | LESP Hidalgo | Instituto de Diagnostico y Referencia Epidemiologicos (INDRE) | Claudia Wong-Arambula, Abril Rodriguez-Maldonado, Vanessa Rivero-Arredondo, Ariadna Medina-Benitez, Joaquin Quiroz-Mercado, Sergio Rangel-Guerrero, Natividad Cruz-Ortiz, Tatiana Nunez-Garcia, Gisela Barrera-Badillo, Lucia Hernandez-Rivas, Irma Lopez-Martinez, Ernesto Ramirez-Gonzalez. |
| EPI_ISL_2709723 | LESP Veracruz | Instituto de Diagnostico y Referencia Epidemiologicos (INDRE) | Claudia Wong-Arambula, Abril Rodriguez-Maldonado, Vanessa Rivero-Arredondo, Ariadna Medina-Benitez, Joaquin Quiroz-Mercado, Sergio Rangel-Guerrero, Natividad Cruz-Ortiz, Tatiana Nunez-Garcia, Gisela Barrera-Badillo, Lucia Hernandez-Rivas, Irma Lopez-Martinez, Ernesto Ramirez-Gonzalez. |
| EPI_ISL_2709724, EPI_ISL_2709725 | LESP Guanajuato | Instituto de Diagnostico y Referencia Epidemiologicos (INDRE) | Claudia Wong-Arambula, Abril Rodriguez-Maldonado, Vanessa Rivero-Arredondo, Ariadna Medina-Benitez, Joaquin Quiroz-Mercado, Sergio Rangel-Guerrero, Natividad Cruz-Ortiz, Tatiana Nunez-Garcia, Gisela Barrera-Badillo, Lucia Hernandez-Rivas, Irma Lopez-Martinez, Ernesto Ramirez-Gonzalez. |
| EPI_ISL_2709726, EPI_ISL_2709727, EPI_ISL_2709728, EPI_ISL_2709729 | LESP Nuevo Leon | Instituto de Diagnostico y Referencia Epidemiologicos (INDRE) | Claudia Wong-Arambula, Abril Rodriguez-Maldonado, Vanessa Rivero-Arredondo, Ariadna Medina-Benitez, Joaquin Quiroz-Mercado, Sergio Rangel-Guerrero, Natividad Cruz-Ortiz, Tatiana Nunez-Garcia, Gisela Barrera-Badillo, Lucia Hernandez-Rivas, Irma Lopez-Martinez, Ernesto Ramirez-Gonzalez. |
| EPI_ISL_2709730 | LESP Ciudad de Mexico | Instituto de Diagnostico y Referencia Epidemiologicos (INDRE) | Claudia Wong-Arambula, Abril Rodriguez-Maldonado, Vanessa Rivero-Arredondo, Ariadna Medina-Benitez, Joaquin Quiroz-Mercado, Sergio Rangel-Guerrero, Natividad Cruz-Ortiz, Tatiana Nunez-Garcia, Gisela Barrera-Badillo, Lucia Hernandez-Rivas, Irma Lopez-Martinez, Ernesto Ramirez-Gonzalez. |
| EPI_ISL_2709731 | LESP Coahuila | Instituto de Diagnostico y Referencia Epidemiologicos (INDRE) | Claudia Wong-Arambula, Abril Rodriguez-Maldonado, Vanessa Rivero-Arredondo, Ariadna Medina-Benitez, Joaquin Quiroz-Mercado, Sergio Rangel-Guerrero, Natividad Cruz-Ortiz, Tatiana Nunez-Garcia, Gisela Barrera-Badillo, Lucia Hernandez-Rivas, Irma Lopez-Martinez, Ernesto Ramirez-Gonzalez. |
| EPI_ISL_2709732 | LESP Chiapas | Instituto de Diagnostico y Referencia Epidemiologicos (INDRE) | Claudia Wong-Arambula, Abril Rodriguez-Maldonado, Vanessa Rivero-Arredondo, Ariadna Medina-Benitez, Joaquin Quiroz-Mercado, Sergio Rangel-Guerrero, Natividad Cruz-Ortiz, Tatiana Nunez-Garcia, Gisela Barrera-Badillo, Lucia Hernandez-Rivas, Irma Lopez-Martinez, Ernesto Ramirez-Gonzalez. |
| EPI_ISL_2709733 | LESP Veracruz | Instituto de Diagnostico y Referencia Epidemiologicos (INDRE) | Claudia Wong-Arambula, Abril Rodriguez-Maldonado, Vanessa Rivero-Arredondo, Ariadna Medina-Benitez, Joaquin Quiroz-Mercado, Sergio Rangel-Guerrero, Natividad Cruz-Ortiz, Tatiana Nunez-Garcia, Gisela Barrera-Badillo, Lucia Hernandez-Rivas, Irma Lopez-Martinez, Ernesto Ramirez-Gonzalez. |
| EPI_ISL_2709800, EPI_ISL_2709801, EPI_ISL_2709802, EPI_ISL_2709803, EPI_ISL_2709804, EPI_ISL_2709805, EPI_ISL_2709806, EPI_ISL_2709807, EPI_ISL_2709808, EPI_ISL_2709809, EPI_ISL_2709810, EPI_ISL_2709811, EPI_ISL_2709812, EPI_ISL_2709813, EPI_ISL_2709814, EPI_ISL_2709815, EPI_ISL_2709816, EPI_ISL_2709817, EPI_ISL_2709818, EPI_ISL_2709819, EPI_ISL_2709820, EPI_ISL_2709821, EPI_ISL_2709822, EPI_ISL_2709823, EPI_ISL_2709824, EPI_ISL_2709825, EPI_ISL_2709826, EPI_ISL_2709827, EPI_ISL_2709828, EPI_ISL_2709829, EPI_ISL_2709830, EPI_ISL_2709831, EPI_ISL_2709832, EPI_ISL_2709833, EPI_ISL_2709834, EPI_ISL_2709835, EPI_ISL_2709836, EPI_ISL_2709837, EPI_ISL_2709838, EPI_ISL_2709839, EPI_ISL_2709840, EPI_ISL_2709841, EPI_ISL_2709842, EPI_ISL_2709843, EPI_ISL_2709844, EPI_ISL_2709845, EPI_ISL_2709846, EPI_ISL_2709847 | | | |
| see above | MD PHL | MD PHL | Maryland Department of Health Laboratories Administration |
| EPI_ISL_2710012, EPI_ISL_2710013, EPI_ISL_2710014, EPI_ISL_2710015, EPI_ISL_2710016, EPI_ISL_2710017, EPI_ISL_2710018, EPI_ISL_2710019, EPI_ISL_2710020, EPI_ISL_2710021, EPI_ISL_2710024, EPI_ISL_2710025, EPI_ISL_2710026 | | | |
| see above | Ochsner Health | Bioinfoexperts, LLC | Judy Crabtree, Amy Feehan, Rebecca Rose, David J. Nolan, Tessa LaFleur, Julia-Garcia-Diaz, Lucio Miele, Susanna L. Lamers |
| EPI_ISL_2710027, EPI_ISL_2710028, EPI_ISL_2710029, EPI_ISL_2710030, EPI_ISL_2710031, EPI_ISL_2710032, EPI_ISL_2710033, EPI_ISL_2710034, EPI_ISL_2710035, EPI_ISL_2710036, EPI_ISL_2710037, EPI_ISL_2710039, EPI_ISL_2710041, EPI_ISL_2710042, EPI_ISL_2710043, EPI_ISL_2710044, EPI_ISL_2710045, EPI_ISL_2710046, EPI_ISL_2710047, EPI_ISL_2710048, EPI_ISL_2710049, EPI_ISL_2710050, | | | |

| | | | |
|---|---|---|---|
| EPI_ISL_2710051, EPI_ISL_2710052, EPI_ISL_2710053, EPI_ISL_2710054, EPI_ISL_2710055, EPI_ISL_2710056, EPI_ISL_2710057, EPI_ISL_2710058, EPI_ISL_2710059, EPI_ISL_2710060, EPI_ISL_2710061, EPI_ISL_2710062, EPI_ISL_2710063, EPI_ISL_2710064, EPI_ISL_2710065, EPI_ISL_2710066, EPI_ISL_2710067, EPI_ISL_2710068, EPI_ISL_2710069, EPI_ISL_2710070 | | | |
| see above | Vanderbilt University Medical Center | Genomics and Discovery, Respiratory Viruses Branch, Division of Viral Diseases, Centers for Disease Control and Prevention | Anna Kelleher, Ying Tao, Yan Li, Jing Zhang, Brian Lynch, Krista Queen, Anna Uehara, Peter Cook, Han Jia Justin Ng, Rachel Marine, Clinton R. Paden, Dhwani Batra, Haibin Wang, Tara Coalter, Jasmine Padilla, Morgan Davis, Mili Sheth, Sarah Nobles, Mark Burroughs, Justin Lee, Adam Retchless, Suxiang Tong |
| EPI_ISL_2710323 | OK Public Health Laboratory, Oklahoma State DOH | Genomics and Discovery, Respiratory Viruses Branch, Division of Viral Diseases, Centers for Disease Control and Prevention | Jing Zhang, Anna Kelleher, Ying Tao, Yan Li, Brian Lynch, Krista Queen, Anna Uehara, Peter Cook, Han Jia Justin Ng, Rachel Marine, Clinton R. Paden, Dhwani Batra, Haibin Wang, Tara Coalter, Jasmine Padilla, Morgan Davis, Mili Sheth, Sarah Nobles, Mark Burroughs, Justin Lee, Adam Retchless, Suxiang Tong |
| EPI_ISL_2710324, EPI_ISL_2710325, EPI_ISL_2710327, EPI_ISL_2710328, EPI_ISL_2710329, EPI_ISL_2710330, EPI_ISL_2710331, EPI_ISL_2710332, EPI_ISL_2710333, EPI_ISL_2710334, EPI_ISL_2710335, EPI_ISL_2710336 | | | |
| see above | Kansas Health and Environmental Lab | Kansas Health and Environmental Lab | Mike Grose, Katherine Wiggins, Jonathan Barnell, Ben Olsen, and Phil Adam |
| EPI_ISL_2710337, EPI_ISL_2710338, EPI_ISL_2710339, EPI_ISL_2710340, EPI_ISL_2710341, EPI_ISL_2710342, EPI_ISL_2710343, EPI_ISL_2710344, EPI_ISL_2710345, EPI_ISL_2710346, EPI_ISL_2710347, EPI_ISL_2710348, EPI_ISL_2710349, EPI_ISL_2710350, EPI_ISL_2710351, EPI_ISL_2710352, EPI_ISL_2710353, EPI_ISL_2710354, EPI_ISL_2710355, EPI_ISL_2710356, EPI_ISL_2710357, EPI_ISL_2710359, EPI_ISL_2710360, EPI_ISL_2710361 | | | |
| see above | Montana Public Health Laboratory | Montana Public Health Laboratory | Joy Ritter, Michelle Mozer, Carrie Biskupiak, Deborah Gibson, Michael Dills |
| EPI_ISL_2710362, EPI_ISL_2710363, EPI_ISL_2710364, EPI_ISL_2710365, EPI_ISL_2710366, EPI_ISL_2710367, EPI_ISL_2710368, EPI_ISL_2710369, EPI_ISL_2710370, EPI_ISL_2710371, EPI_ISL_2710372, EPI_ISL_2710373, EPI_ISL_2710374, EPI_ISL_2710375, EPI_ISL_2710376, EPI_ISL_2710377, EPI_ISL_2710378 | | | |
| see above | Kansas Health and Environmental Lab | Kansas Health and Environmental Lab | Mike Grose, Katherine Wiggins, Jonathan Barnell, Ben Olsen, and Phil Adam |
| EPI_ISL_2710379, EPI_ISL_2710380 | CT Department of Public Health | CT Department of Public Health | Claire Pearson, Tu N. Nguyen |
| EPI_ISL_2712451, EPI_ISL_2712452, EPI_ISL_2712453, EPI_ISL_2712454, EPI_ISL_2712455, EPI_ISL_2712456, EPI_ISL_2712457, EPI_ISL_2712458, EPI_ISL_2712459, EPI_ISL_2712460, EPI_ISL_2712461, EPI_ISL_2712462, EPI_ISL_2712463, EPI_ISL_2712464, EPI_ISL_2712465, EPI_ISL_2712466, EPI_ISL_2712467, EPI_ISL_2712468, EPI_ISL_2712469, EPI_ISL_2712470, EPI_ISL_2712471, EPI_ISL_2712472, EPI_ISL_2712473, EPI_ISL_2712474, EPI_ISL_2712475, EPI_ISL_2712476, EPI_ISL_2712477, EPI_ISL_2712478, EPI_ISL_2712479, EPI_ISL_2712480, EPI_ISL_2712481, EPI_ISL_2712482, EPI_ISL_2712483, EPI_ISL_2712484, EPI_ISL_2712485, EPI_ISL_2712486, EPI_ISL_2712487, EPI_ISL_2712488, EPI_ISL_2712489, EPI_ISL_2712490, EPI_ISL_2712491, EPI_ISL_2712492, EPI_ISL_2712493, EPI_ISL_2712494, EPI_ISL_2712495, EPI_ISL_2712496, EPI_ISL_2712497, EPI_ISL_2712498, EPI_ISL_2712499, EPI_ISL_2712500, EPI_ISL_2712501, EPI_ISL_2712502, EPI_ISL_2712503, EPI_ISL_2712504, EPI_ISL_2712505, EPI_ISL_2712506, EPI_ISL_2712507, EPI_ISL_2712508, EPI_ISL_2712509, EPI_ISL_2712510, EPI_ISL_2712511, EPI_ISL_2712512, EPI_ISL_2712513, EPI_ISL_2712514, EPI_ISL_2712515, EPI_ISL_2712516, EPI_ISL_2712517, EPI_ISL_2712518, EPI_ISL_2712519, EPI_ISL_2712520, EPI_ISL_2712521, EPI_ISL_2712522, EPI_ISL_2712523, EPI_ISL_2712524, EPI_ISL_2712525, EPI_ISL_2712526, EPI_ISL_2712527, EPI_ISL_2712528, EPI_ISL_2712529, EPI_ISL_2712530, EPI_ISL_2712531, EPI_ISL_2712532, EPI_ISL_2712533, EPI_ISL_2712534, EPI_ISL_2712535, EPI_ISL_2712536, EPI_ISL_2712537, EPI_ISL_2712538, EPI_ISL_2712539, EPI_ISL_2712540, EPI_ISL_2712541, EPI_ISL_2712542, EPI_ISL_2712543, EPI_ISL_2712544, EPI_ISL_2712545, EPI_ISL_2712546, EPI_ISL_2712547, EPI_ISL_2712548, EPI_ISL_2712549, EPI_ISL_2712550, EPI_ISL_2712551, EPI_ISL_2712552, EPI_ISL_2712553, EPI_ISL_2712554, EPI_ISL_2712555, EPI_ISL_2712556, EPI_ISL_2712557, EPI_ISL_2712558, EPI_ISL_2712559, EPI_ISL_2712560, EPI_ISL_2712561, EPI_ISL_2712562, EPI_ISL_2712563, EPI_ISL_2712564, EPI_ISL_2712565, EPI_ISL_2712566, EPI_ISL_2712567, EPI_ISL_2712568, EPI_ISL_2712569, EPI_ISL_2712570, EPI_ISL_2712571, EPI_ISL_2712572, EPI_ISL_2712573, EPI_ISL_2712574, EPI_ISL_2712575, EPI_ISL_2712576, EPI_ISL_2712577, EPI_ISL_2712578, EPI_ISL_2712579, EPI_ISL_2712580, EPI_ISL_2712581, EPI_ISL_2712582, EPI_ISL_2712583, EPI_ISL_2712584, EPI_ISL_2712585, EPI_ISL_2712586, EPI_ISL_2712587, EPI_ISL_2712588, EPI_ISL_2712589, EPI_ISL_2712590, EPI_ISL_2712591, EPI_ISL_2712592, EPI_ISL_2712593, EPI_ISL_2712594, EPI_ISL_2712595, EPI_ISL_2712596, EPI_ISL_2712597, EPI_ISL_2712598, EPI_ISL_2712599, EPI_ISL_2712600, EPI_ISL_2712601, EPI_ISL_2712602, EPI_ISL_2712603, EPI_ISL_2712604, EPI_ISL_2712605, EPI_ISL_2712606, EPI_ISL_2712607, EPI_ISL_2712608, EPI_ISL_2712609, EPI_ISL_2712610, EPI_ISL_2712611, EPI_ISL_2712612 | | | |
| see above | Sharp HealthCare Laboratory | Andersen lab at Scripps Research | SEARCH Alliance San Diego with Aaron Harding, Jacquelyn Berumen, Cathy Woerle, Liam McGinnis, Art Mendoza, Omid Bakhtar |
| EPI_ISL_2712613, EPI_ISL_2712614, EPI_ISL_2712615, EPI_ISL_2712616, EPI_ISL_2712617, EPI_ISL_2712618, EPI_ISL_2712619, EPI_ISL_2712620, EPI_ISL_2712621, EPI_ISL_2712622, EPI_ISL_2712623, EPI_ISL_2712624, EPI_ISL_2712625, EPI_ISL_2712626, EPI_ISL_2712627, EPI_ISL_2712628, EPI_ISL_2712629, EPI_ISL_2712630, EPI_ISL_2712631, EPI_ISL_2712632, EPI_ISL_2712633, EPI_ISL_2712634, EPI_ISL_2712635, EPI_ISL_2712636, EPI_ISL_2712637, EPI_ISL_2712638, EPI_ISL_2712639, EPI_ISL_2712640, EPI_ISL_2712641, EPI_ISL_2712642, EPI_ISL_2712643, EPI_ISL_2712644, EPI_ISL_2712645, EPI_ISL_2712646, EPI_ISL_2712647, EPI_ISL_2712648, EPI_ISL_2712649, EPI_ISL_2712650, EPI_ISL_2712651, EPI_ISL_2712652, EPI_ISL_2712653, EPI_ISL_2712654, EPI_ISL_2712655, EPI_ISL_2712656, EPI_ISL_2712657, EPI_ISL_2712658, EPI_ISL_2712659, EPI_ISL_2712660, EPI_ISL_2712661, EPI_ISL_2712662, EPI_ISL_2712663, EPI_ISL_2712664, EPI_ISL_2712665, EPI_ISL_2712666, EPI_ISL_2712667, EPI_ISL_2712668, EPI_ISL_2712669, EPI_ISL_2712670, EPI_ISL_2712671, EPI_ISL_2712672, EPI_ISL_2712673, EPI_ISL_2712674, EPI_ISL_2712675, EPI_ISL_2712676, EPI_ISL_2712677, EPI_ISL_2712678, EPI_ISL_2712679, EPI_ISL_2712680, EPI_ISL_2712681, EPI_ISL_2712682, EPI_ISL_2712683, EPI_ISL_2712684, EPI_ISL_2712685, EPI_ISL_2712686, EPI_ISL_2712687, EPI_ISL_2712688, EPI_ISL_2712689, EPI_ISL_2712690, EPI_ISL_2712691, EPI_ISL_2712692, EPI_ISL_2712693, EPI_ISL_2712694, EPI_ISL_2712695, EPI_ISL_2712696, EPI_ISL_2712697, EPI_ISL_2712698, EPI_ISL_2712699, EPI_ISL_2712700, EPI_ISL_2712701, EPI_ISL_2712702, EPI_ISL_2712703, EPI_ISL_2712704, EPI_ISL_2712705, EPI_ISL_2712706, EPI_ISL_2712707, EPI_ISL_2712708, EPI_ISL_2712709, EPI_ISL_2712710, EPI_ISL_2712711, EPI_ISL_2712712, EPI_ISL_2712713, EPI_ISL_2712714, EPI_ISL_2712715, EPI_ISL_2712716, EPI_ISL_2712717, EPI_ISL_2712718, EPI_ISL_2712719, EPI_ISL_2712720, EPI_ISL_2712721, EPI_ISL_2712722, EPI_ISL_2712723, EPI_ISL_2712724, EPI_ISL_2712725, EPI_ISL_2712726, EPI_ISL_2712727, EPI_ISL_2712728, EPI_ISL_2712729, EPI_ISL_2712730, EPI_ISL_2712731, EPI_ISL_2712732, EPI_ISL_2712733, EPI_ISL_2712734, EPI_ISL_2712735, EPI_ISL_2712736, EPI_ISL_2712737, EPI_ISL_2712738, EPI_ISL_2712739, EPI_ISL_2712740, EPI_ISL_2712741, EPI_ISL_2712742, EPI_ISL_2712743, EPI_ISL_2712744, EPI_ISL_2712745, EPI_ISL_2712746, EPI_ISL_2712747, EPI_ISL_2712748, EPI_ISL_2712749, EPI_ISL_2712750, EPI_ISL_2712751, EPI_ISL_2712752, EPI_ISL_2712753, EPI_ISL_2712754, EPI_ISL_2712755, EPI_ISL_2712756, EPI_ISL_2712757, EPI_ISL_2712758, EPI_ISL_2712759, EPI_ISL_2712760, EPI_ISL_2712761, EPI_ISL_2712762, EPI_ISL_2712763, EPI_ISL_2712764, EPI_ISL_2712765, EPI_ISL_2712766, EPI_ISL_2712767, EPI_ISL_2712768, EPI_ISL_2712769 | | | |
| see above | Instituto de Diagnostico y Referencia Epidemiologicos (InDRE) | Andersen lab at Scripps Research | SEARCH Alliance with Gisela Barrera Badillo, Jose Ernesto Ramirez Gonzalez, Maria Natividad Cruz Ortiz, Nervain Benjamin Contreras Gonzalez, Tatiana Ernestina Nunez Garcia, Dayanira Sarith Arellano Suarez, Irma Lopez Martinez, Rita Flores Leon, Beatriz Olivares Flores |
| EPI_ISL_2712770, EPI_ISL_2712771, EPI_ISL_2712772, EPI_ISL_2712773, EPI_ISL_2712774, EPI_ISL_2712775, EPI_ISL_2712776, EPI_ISL_2712777, EPI_ISL_2712778, EPI_ISL_2712779, EPI_ISL_2712780, EPI_ISL_2712781, EPI_ISL_2712782, EPI_ISL_2712783, EPI_ISL_2712784, EPI_ISL_2712785, EPI_ISL_2712786, EPI_ISL_2712787, EPI_ISL_2712788, EPI_ISL_2712789, EPI_ISL_2712790, EPI_ISL_2712791, EPI_ISL_2712792, EPI_ISL_2712793, EPI_ISL_2712794, EPI_ISL_2712795, EPI_ISL_2712796, EPI_ISL_2712797, EPI_ISL_2712798, EPI_ISL_2712799, EPI_ISL_2712800, EPI_ISL_2712801, EPI_ISL_2712802, EPI_ISL_2712803, EPI_ISL_2712804, EPI_ISL_2712805, EPI_ISL_2712806, EPI_ISL_2712807, EPI_ISL_2712808, EPI_ISL_2712809, EPI_ISL_2712810, EPI_ISL_2712811, EPI_ISL_2712812, EPI_ISL_2712813, EPI_ISL_2712814, EPI_ISL_2712816, EPI_ISL_2712817, EPI_ISL_2712818, EPI_ISL_2712819, EPI_ISL_2712820, EPI_ISL_2712821, EPI_ISL_2712822, EPI_ISL_2712823, EPI_ISL_2712824, EPI_ISL_2712825, EPI_ISL_2712826, EPI_ISL_2712827, EPI_ISL_2712828, EPI_ISL_2712829, EPI_ISL_2712830, EPI_ISL_2712831, EPI_ISL_2712832, EPI_ISL_2712833, EPI_ISL_2712834, EPI_ISL_2712835, EPI_ISL_2712836, EPI_ISL_2712837, EPI_ISL_2712838, EPI_ISL_2712839, EPI_ISL_2712840, EPI_ISL_2712841 | | | |
| see above | San Diego County Public Health Laboratory | Andersen lab at Scripps Research | SEARCH Alliance San Diego with Ashleigh Murphy, Jovan Shephard, Brett Austin |
| EPI_ISL_2712914, EPI_ISL_2712916, EPI_ISL_2712917, EPI_ISL_2712918, EPI_ISL_2712919, EPI_ISL_2712920, EPI_ISL_2712921, EPI_ISL_2712922, EPI_ISL_2712923, EPI_ISL_2712924, EPI_ISL_2712925, EPI_ISL_2712926, EPI_ISL_2712927, EPI_ISL_2712928, EPI_ISL_2712929, EPI_ISL_2712930, EPI_ISL_2712932, EPI_ISL_2712933, EPI_ISL_2712934, EPI_ISL_2712935, EPI_ISL_2712936, EPI_ISL_2712937, EPI_ISL_2712938, EPI_ISL_2712939, EPI_ISL_2712940, EPI_ISL_2712941, EPI_ISL_2712942, EPI_ISL_2712943, EPI_ISL_2712944, EPI_ISL_2712945, EPI_ISL_2712946, EPI_ISL_2712947, EPI_ISL_2712948, EPI_ISL_2712949, EPI_ISL_2712950, EPI_ISL_2712951, EPI_ISL_2712952, EPI_ISL_2712953, EPI_ISL_2712954, EPI_ISL_2712956, EPI_ISL_2712957, EPI_ISL_2712958, EPI_ISL_2712959, EPI_ISL_2712960, EPI_ISL_2712961, EPI_ISL_2712962, EPI_ISL_2712963, EPI_ISL_2712964, EPI_ISL_2712965, EPI_ISL_2712966, EPI_ISL_2712967, EPI_ISL_2712970, EPI_ISL_2712971, EPI_ISL_2712972, EPI_ISL_2712973, EPI_ISL_2712975 | | | |
| see above | Wyoming Public Health Laboratory | Wyoming Public Health Laboratory | Jim Mildenberger, Taylor Fearing, Channing Weber, Ashley Norberg, Chayse Rowley, Marley Goetz, Brian Dominguez, Elliot Thomasson, Sam Britz, Cari Sloma, Robert Petit, and Rob Christensen |
| EPI_ISL_2712976, EPI_ISL_2712977, EPI_ISL_2712978, EPI_ISL_2712979, EPI_ISL_2712980, EPI_ISL_2712981, EPI_ISL_2712982, EPI_ISL_2712983, EPI_ISL_2712984, EPI_ISL_2712985, EPI_ISL_2712986, EPI_ISL_2712987, EPI_ISL_2712988, EPI_ISL_2712989, EPI_ISL_2712990, EPI_ISL_2712991 | | | |
| see above | ISDH | Quantigen Biosciences | Paul J. Childress |
| EPI_ISL_2712992, EPI_ISL_2712993, EPI_ISL_2712994, EPI_ISL_2712995, EPI_ISL_2712996, EPI_ISL_2712997, EPI_ISL_2712998, EPI_ISL_2712999, EPI_ISL_2713000, EPI_ISL_2713001, EPI_ISL_2713002, EPI_ISL_2713003, EPI_ISL_2713004 | | | |
| see above | TXDSHS | TXDSHS | Rashmi Tuladhar, Bonnie Oh, Jenny Zhang, Maliha Rahman, Mayela Pedrueza, Anita Pokharel, Karen Bobier, Lorraine Rodriguez, Myong Koag, Chun Wang, Rachel Lee, Grace Kubin |
| EPI_ISL_2713005, EPI_ISL_2713006, EPI_ISL_2713007, EPI_ISL_2713008, EPI_ISL_2713009, EPI_ISL_2713010, EPI_ISL_2713011, EPI_ISL_2713012, EPI_ISL_2713013, EPI_ISL_2713014, EPI_ISL_2713015, EPI_ISL_2713016, EPI_ISL_2713017, EPI_ISL_2713018, EPI_ISL_2713019, EPI_ISL_2713020, EPI_ISL_2713021, EPI_ISL_2713022, EPI_ISL_2713023, EPI_ISL_2713024, EPI_ISL_2713025, EPI_ISL_2713026 | | | |
| see above | Wyoming Public Health Laboratory | Wyoming Public Health Laboratory | Jim Mildenberger, Taylor Fearing, Channing Weber, Ashley Norberg, Chayse Rowley, Marley Goetz, Brian Dominguez, Elliot Thomasson, Sam Britz, Cari Sloma, Robert Petit, and Rob Christensen |
| EPI_ISL_2713027, EPI_ISL_2713028 | University of Wisconsin-Madison AIDS Vaccine Research Laboratories | University of Wisconsin-Madison AIDS Vaccine Research Laboratories | Gage Moreno, Katarina Braun, et al. AIDS Vaccine Research Laboratories |
| EPI_ISL_2713029, EPI_ISL_2713030, EPI_ISL_2713031, EPI_ISL_2713032, EPI_ISL_2713033, EPI_ISL_2713034, EPI_ISL_2713035, EPI_ISL_2713036 | CO Dept. of Public Health and Environment, Lab Services Division | Genomics and Discovery, Respiratory Viruses Branch, Division of Viral Diseases, Centers for Disease Control and Prevention | Anna Kelleher, Ying Tao, Yan Li, Jing Zhang, Brian Lynch, Krista Queen, Anna Uehara, Peter Cook, Han Jia Justin Ng, Rachel Marine, Clinton R. Paden, Dhwani Batra, Haibin Wang, Tara Coalter, Jasmine Padilla, Morgan Davis, Mili Sheth, Sarah Nobles, Mark Burroughs, Justin Lee, Adam Retchless, Suxiang Tong |
| EPI_ISL_2713037, EPI_ISL_2713038, EPI_ISL_2713039 | Wyoming Public Health Laboratory | Wyoming Public Health Laboratory | Jim Mildenberger, Taylor Fearing, Channing Weber, Ashley Norberg, Chayse Rowley, Marley Goetz, Brian Dominguez, Elliot Thomasson, Sam Britz, Cari Sloma, Robert Petit, and Rob Christensen |
| EPI_ISL_2713041 | Vanderbilt University Medical Center | Genomics and Discovery, Respiratory Viruses Branch, Division of Viral Diseases, Centers for Disease Control and Prevention | Anna Kelleher, Ying Tao, Yan Li, Jing Zhang, Brian Lynch, Krista Queen, Anna Uehara, Peter Cook, Han Jia Justin Ng, Rachel Marine, Clinton R. Paden, Dhwani Batra, Haibin Wang, Tara Coalter, Jasmine Padilla, Morgan Davis, Mili Sheth, Sarah Nobles, Mark Burroughs, Justin Lee, Adam Retchless, Suxiang Tong |
| EPI_ISL_2713042, EPI_ISL_2713043, EPI_ISL_2713044, EPI_ISL_2713045, EPI_ISL_2713046, EPI_ISL_2713047, EPI_ISL_2713048, EPI_ISL_2713049, EPI_ISL_2713050, EPI_ISL_2713051, EPI_ISL_2713052, EPI_ISL_2713053, EPI_ISL_2713054, EPI_ISL_2713055, EPI_ISL_2713056, EPI_ISL_2713058, EPI_ISL_2713059 | | | |
| see above | University of Wisconsin-Madison AIDS Vaccine Research Laboratories | University of Wisconsin-Madison AIDS Vaccine Research Laboratories | Gage Moreno, Katarina Braun, et al. AIDS Vaccine Research Laboratories |
| EPI_ISL_2713060, EPI_ISL_2713061, EPI_ISL_2713062, EPI_ISL_2713063, EPI_ISL_2713064, EPI_ISL_2713065, EPI_ISL_2713066, EPI_ISL_2713067 | Oregon State Public Health Laboratory | Oregon State Public Health Laboratory | Rafia Razzaque, Eugene Yeboah, Vanda Makris, Laura Tsaknaridis, John Fontana and Shane Sevey |
| EPI_ISL_2713068, EPI_ISL_2713069, EPI_ISL_2713070, EPI_ISL_2713071, EPI_ISL_2713072, EPI_ISL_2713073 | ADPH | ADPH | Adrianna Walker |
| EPI_ISL_2713074 | Houston Health Department, Disease Prevention and Control | Houston Health Department, Disease Prevention and Control | Penn,R., Brown,P., Lara,A. and Lai,Y. |
| EPI_ISL_2713272, EPI_ISL_2713273, EPI_ISL_2713274, EPI_ISL_2713275, EPI_ISL_2713276, EPI_ISL_2713277, EPI_ISL_2713278, EPI_ISL_2713279, EPI_ISL_2713280, EPI_ISL_2713281, EPI_ISL_2713282, EPI_ISL_2713283, EPI_ISL_2713285, EPI_ISL_2713286, EPI_ISL_2713287, EPI_ISL_2713288, EPI_ISL_2713289, EPI_ISL_2713291, EPI_ISL_2713292, EPI_ISL_2713298, EPI_ISL_2713299, EPI_ISL_2713300, EPI_ISL_2713301 | | | |
| see above | Infectious Disease Program, Broad Institute of Harvard and MIT | Infectious Disease Program, Broad Institute of Harvard and MIT | Lemieux,J.E., Siddle,K.J., Shaw,B., Adams,G., Pierce,V., Turbett,S., Anahtar,M., Branda,J., Slater,D., Harris,J., Lin,A.E., Gladden-Young,A., Lagerborg,K., Rudy,M., DeRuff,K., Carter,A., Normandin,E., Bauer,M., Reilly,S., Tomkins-Tinch,C., Loreth,C., Chaluvadi,S., Neumann,A., Cusick,C., Chapman,S.B., Gnirke,A., Flowers,K., Cerrato,F., Birren,B.W., Gallagher,G., Smole,S., Park,D.J., MacInnis,B.L., Ryan,E., LaRocque,R., Rosenberg,E., Sabeti,P.C. |
| EPI_ISL_2714563, EPI_ISL_2714564, EPI_ISL_2714566, EPI_ISL_2714567, EPI_ISL_2714568, EPI_ISL_2714569, EPI_ISL_2714571, EPI_ISL_2714572, EPI_ISL_2714573, EPI_ISL_2714574, EPI_ISL_2714575, EPI_ISL_2714577, EPI_ISL_2714578, EPI_ISL_2714579, EPI_ISL_2714580, EPI_ISL_2714582, EPI_ISL_2714584, EPI_ISL_2714585, EPI_ISL_2714586, EPI_ISL_2714587, EPI_ISL_2714588, EPI_ISL_2714589, EPI_ISL_2714590, EPI_ISL_2714593, EPI_ISL_2714595, EPI_ISL_2714596, EPI_ISL_2714598, EPI_ISL_2714599, EPI_ISL_2714600, EPI_ISL_2714602, EPI_ISL_2714603, EPI_ISL_2714604, EPI_ISL_2714605, EPI_ISL_2714606, EPI_ISL_2714607, EPI_ISL_2714608, EPI_ISL_2714609, EPI_ISL_2714610, EPI_ISL_2714611, EPI_ISL_2714612, EPI_ISL_2714613, EPI_ISL_2714614, EPI_ISL_2714615, EPI_ISL_2714616, EPI_ISL_2714617, EPI_ISL_2714618, EPI_ISL_2714619, EPI_ISL_2714620, EPI_ISL_2714621, EPI_ISL_2714622, EPI_ISL_2714623, EPI_ISL_2714624, EPI_ISL_2714625, EPI_ISL_2714626, EPI_ISL_2714627, EPI_ISL_2714628 | | | |
| see above | North Dakota Department of Health, | North Dakota Department of Health, Public | Lisa Wingerter |

| | | | |
|---|---|---|---|
| | Public Health Laboratory | Health Laboratory | |
| EPI_ISL_2714918, EPI_ISL_2714919, EPI_ISL_2714920, EPI_ISL_2714921, EPI_ISL_2714922, EPI_ISL_2714923, EPI_ISL_2714924, EPI_ISL_2714925, EPI_ISL_2714926, EPI_ISL_2714927, EPI_ISL_2714928, EPI_ISL_2714929, EPI_ISL_2714930, EPI_ISL_2714931, EPI_ISL_2714932, EPI_ISL_2714933, EPI_ISL_2714934, EPI_ISL_2714935, EPI_ISL_2714936, EPI_ISL_2714937, EPI_ISL_2714938, EPI_ISL_2714939, EPI_ISL_2714940, EPI_ISL_2714941, EPI_ISL_2714942, EPI_ISL_2714943, EPI_ISL_2714944 | | | |
| see above | Incyte Diagnostics Spokane | Seattle Flu Study | Deborah A. Nickerson, Chris D. Frazar, Jover Lee, Benjamin Pelle, Erica Ryke, Matthew Richardson, Amanda Adler, Elisabeth Brandstetter, Peter D. Han, Kairsten Fay, Misja Ilcisin, Kirsten Lacombe, Thomas R. Sibley, Melissa Truong, Caitlin R. Wolf, Romesh Gautom, Geoff Melly, Brian Hiatt, Philip Dykema, Scott Lindquist, Michael Boeckh, Janet A. Englund, Michael Famulare, Barry R. Lutz, Mark J. Rieder, Lea M. Starita, Matthew Thompson, Helen Y. Chu, Jay Shendure, Trevor Bedford |
| EPI_ISL_2714945, EPI_ISL_2714946, EPI_ISL_2714947, EPI_ISL_2714948, EPI_ISL_2714949, EPI_ISL_2714950, EPI_ISL_2714951, EPI_ISL_2714953, EPI_ISL_2714954, EPI_ISL_2714956, EPI_ISL_2714957, EPI_ISL_2714958, EPI_ISL_2714959, EPI_ISL_2714960, EPI_ISL_2714961, EPI_ISL_2714962, EPI_ISL_2714963, EPI_ISL_2714964, EPI_ISL_2714965, EPI_ISL_2714966, EPI_ISL_2714967, EPI_ISL_2714968, EPI_ISL_2714969, EPI_ISL_2714970, EPI_ISL_2714971, EPI_ISL_2714972, EPI_ISL_2714973, EPI_ISL_2714974, EPI_ISL_2714975, EPI_ISL_2714976, EPI_ISL_2714977, EPI_ISL_2714978, EPI_ISL_2714979, EPI_ISL_2714980, EPI_ISL_2714981, EPI_ISL_2714982, EPI_ISL_2714983, EPI_ISL_2714984, EPI_ISL_2714985, EPI_ISL_2714986, EPI_ISL_2714987, EPI_ISL_2714988, EPI_ISL_2714989, EPI_ISL_2714990, EPI_ISL_2714992, EPI_ISL_2714993, EPI_ISL_2714994, EPI_ISL_2714995, EPI_ISL_2714996, EPI_ISL_2714997, EPI_ISL_2714999, EPI_ISL_2715000, EPI_ISL_2715001, EPI_ISL_2715002, EPI_ISL_2715003, EPI_ISL_2715005, EPI_ISL_2715006, EPI_ISL_2715007, EPI_ISL_2715008 | | | |
| see above | Interpath Laboratory | Seattle Flu Study | Deborah A. Nickerson, Chris D. Frazar, Jover Lee, Benjamin Pelle, Erica Ryke, Matthew Richardson, Amanda Adler, Elisabeth Brandstetter, Peter D. Han, Kairsten Fay, Misja Ilcisin, Kirsten Lacombe, Thomas R. Sibley, Melissa Truong, Caitlin R. Wolf, Romesh Gautom, Geoff Melly, Brian Hiatt, Philip Dykema, Scott Lindquist, Michael Boeckh, Janet A. Englund, Michael Famulare, Barry R. Lutz, Mark J. Rieder, Lea M. Starita, Matthew Thompson, Helen Y. Chu, Jay Shendure, Trevor Bedford |
| EPI_ISL_2715010, EPI_ISL_2715011, EPI_ISL_2715012, EPI_ISL_2715013, EPI_ISL_2715014, EPI_ISL_2715015, EPI_ISL_2715016, EPI_ISL_2715017, EPI_ISL_2715018, EPI_ISL_2715019, EPI_ISL_2715020, EPI_ISL_2715021, EPI_ISL_2715022, EPI_ISL_2715023, EPI_ISL_2715024, EPI_ISL_2715025, EPI_ISL_2715026, EPI_ISL_2715027, EPI_ISL_2715029, EPI_ISL_2715033, EPI_ISL_2715034, EPI_ISL_2715035, EPI_ISL_2715036, EPI_ISL_2715037, EPI_ISL_2715039, EPI_ISL_2715040, EPI_ISL_2715041, EPI_ISL_2715043, EPI_ISL_2715044, EPI_ISL_2715045, EPI_ISL_2715046, EPI_ISL_2715047, EPI_ISL_2715048, EPI_ISL_2715049, EPI_ISL_2715050, EPI_ISL_2715051, EPI_ISL_2715052, EPI_ISL_2715053, EPI_ISL_2715055, EPI_ISL_2715056, EPI_ISL_2715057, EPI_ISL_2715058, EPI_ISL_2715059, EPI_ISL_2715061, EPI_ISL_2715062, EPI_ISL_2715063, EPI_ISL_2715064, EPI_ISL_2715065, EPI_ISL_2715066, EPI_ISL_2715067, EPI_ISL_2715069, EPI_ISL_2715070, EPI_ISL_2715071, EPI_ISL_2715073, EPI_ISL_2715074, EPI_ISL_2715075, EPI_ISL_2715076, EPI_ISL_2715077, EPI_ISL_2715078, EPI_ISL_2715079, EPI_ISL_2715080, EPI_ISL_2715081, EPI_ISL_2715082 | | | |
| see above | Atlas Genomics | Seattle Flu Study | Deborah A. Nickerson, Chris D. Frazar, Jover Lee, Benjamin Pelle, Erica Ryke, Matthew Richardson, Amanda Adler, Elisabeth Brandstetter, Peter D. Han, Kairsten Fay, Misja Ilcisin, Kirsten Lacombe, Thomas R. Sibley, Melissa Truong, Caitlin R. Wolf, Romesh Gautom, Geoff Melly, Brian Hiatt, Philip Dykema, Scott Lindquist, Michael Boeckh, Janet A. Englund, Michael Famulare, Barry R. Lutz, Mark J. Rieder, Lea M. Starita, Matthew Thompson, Helen Y. Chu, Jay Shendure, Trevor Bedford |
| EPI_ISL_2715083, EPI_ISL_2715084, EPI_ISL_2715086, EPI_ISL_2715087, EPI_ISL_2715088, EPI_ISL_2715089, EPI_ISL_2715090, EPI_ISL_2715091, EPI_ISL_2715092, EPI_ISL_2715093, EPI_ISL_2715094, EPI_ISL_2715095, EPI_ISL_2715096, EPI_ISL_2715097, EPI_ISL_2715098 | | | |
| see above | Incyte Diagnostics Spokane | Seattle Flu Study | Deborah A. Nickerson, Chris D. Frazar, Jover Lee, Benjamin Pelle, Erica Ryke, Matthew Richardson, Amanda Adler, Elisabeth Brandstetter, Peter D. Han, Kairsten Fay, Misja Ilcisin, Kirsten Lacombe, Thomas R. Sibley, Melissa Truong, Caitlin R. Wolf, Romesh Gautom, Geoff Melly, Brian Hiatt, Philip Dykema, Scott Lindquist, Michael Boeckh, Janet A. Englund, Michael Famulare, Barry R. Lutz, Mark J. Rieder, Lea M. Starita, Matthew Thompson, Helen Y. Chu, Jay Shendure, Trevor Bedford |
| EPI_ISL_2715099, EPI_ISL_2715100, EPI_ISL_2715101, EPI_ISL_2715102, EPI_ISL_2715103, EPI_ISL_2715104, EPI_ISL_2715106, EPI_ISL_2715107, EPI_ISL_2715108, EPI_ISL_2715109, EPI_ISL_2715110, EPI_ISL_2715111, EPI_ISL_2715112, EPI_ISL_2715113, EPI_ISL_2715114, EPI_ISL_2715115, EPI_ISL_2715116, EPI_ISL_2715117, EPI_ISL_2715118, EPI_ISL_2715119, EPI_ISL_2715120, EPI_ISL_2715121, EPI_ISL_2715122, EPI_ISL_2715123, EPI_ISL_2715124, EPI_ISL_2715125, EPI_ISL_2715127, EPI_ISL_2715128, EPI_ISL_2715129, EPI_ISL_2715130, EPI_ISL_2715131, EPI_ISL_2715132, EPI_ISL_2715133, EPI_ISL_2715134, EPI_ISL_2715135, EPI_ISL_2715136, EPI_ISL_2715137, EPI_ISL_2715139, EPI_ISL_2715140, EPI_ISL_2715141, EPI_ISL_2715142, EPI_ISL_2715143, EPI_ISL_2715144, EPI_ISL_2715145, EPI_ISL_2715146, EPI_ISL_2715147, EPI_ISL_2715148, EPI_ISL_2715149, EPI_ISL_2715150, EPI_ISL_2715151, EPI_ISL_2715153, EPI_ISL_2715154, EPI_ISL_2715155, EPI_ISL_2715156, EPI_ISL_2715157, EPI_ISL_2715158, EPI_ISL_2715160, EPI_ISL_2715161, EPI_ISL_2715162, EPI_ISL_2715163, EPI_ISL_2715164, EPI_ISL_2715166, EPI_ISL_2715168, EPI_ISL_2715169, EPI_ISL_2715170, EPI_ISL_2715171, EPI_ISL_2715172, EPI_ISL_2715173, EPI_ISL_2715174, EPI_ISL_2715175, EPI_ISL_2715176 | | | |
| see above | Northwest Laboratory | Seattle Flu Study | Deborah A. Nickerson, Chris D. Frazar, Jover Lee, Benjamin Pelle, Erica Ryke, Matthew Richardson, Amanda Adler, Elisabeth Brandstetter, Peter D. Han, Kairsten Fay, Misja Ilcisin, Kirsten Lacombe, Thomas R. Sibley, Melissa Truong, Caitlin R. Wolf, Romesh Gautom, Geoff Melly, Brian Hiatt, Philip Dykema, Scott Lindquist, Michael Boeckh, Janet A. Englund, Michael Famulare, Barry R. Lutz, Mark J. Rieder, Lea M. Starita, Matthew Thompson, Helen Y. Chu, Jay Shendure, Trevor Bedford |
| EPI_ISL_2715178, EPI_ISL_2715179, EPI_ISL_2715180, EPI_ISL_2715181, EPI_ISL_2715182, EPI_ISL_2715184, EPI_ISL_2715185, EPI_ISL_2715186, EPI_ISL_2715187, EPI_ISL_2715188, EPI_ISL_2715189, EPI_ISL_2715190, EPI_ISL_2715192, EPI_ISL_2715193, EPI_ISL_2715194, EPI_ISL_2715195, EPI_ISL_2715196, EPI_ISL_2715197, EPI_ISL_2715198, EPI_ISL_2715199, EPI_ISL_2715200, EPI_ISL_2715201, EPI_ISL_2715202, EPI_ISL_2715203, EPI_ISL_2715204, EPI_ISL_2715205, EPI_ISL_2715206, EPI_ISL_2715207, EPI_ISL_2715208, EPI_ISL_2715209, EPI_ISL_2715212, EPI_ISL_2715213, EPI_ISL_2715214, EPI_ISL_2715215, EPI_ISL_2715217 | | | |
| see above | Atlas Genomics | Seattle Flu Study | Deborah A. Nickerson, Chris D. Frazar, Jover Lee, Benjamin Pelle, Erica Ryke, Matthew Richardson, Amanda Adler, Elisabeth Brandstetter, Peter D. Han, Kairsten Fay, Misja Ilcisin, Kirsten Lacombe, Thomas R. Sibley, Melissa Truong, Caitlin R. Wolf, Romesh Gautom, Geoff Melly, Brian Hiatt, Philip Dykema, Scott Lindquist, Michael Boeckh, Janet A. Englund, Michael Famulare, Barry R. Lutz, Mark J. Rieder, Lea M. Starita, Matthew Thompson, Helen Y. Chu, Jay Shendure, Trevor Bedford |
| EPI_ISL_2715218, EPI_ISL_2715220, EPI_ISL_2715221, EPI_ISL_2715222, EPI_ISL_2715223, EPI_ISL_2715224, EPI_ISL_2715225, EPI_ISL_2715226, EPI_ISL_2715227, EPI_ISL_2715228, EPI_ISL_2715229, EPI_ISL_2715230 | | | |
| see above | Incyte Diagnostics Spokane | Seattle Flu Study | Deborah A. Nickerson, Chris D. Frazar, Jover Lee, Benjamin Pelle, Erica Ryke, Matthew Richardson, Amanda Adler, Elisabeth Brandstetter, Peter D. Han, Kairsten Fay, Misja Ilcisin, Kirsten Lacombe, Thomas R. Sibley, Melissa Truong, Caitlin R. Wolf, Romesh Gautom, Geoff Melly, Brian Hiatt, Philip Dykema, Scott Lindquist, Michael Boeckh, Janet A. Englund, Michael Famulare, Barry R. Lutz, Mark J. Rieder, Lea M. Starita, Matthew Thompson, Helen Y. Chu, Jay Shendure, Trevor Bedford |
| EPI_ISL_2715231, EPI_ISL_2715232, EPI_ISL_2715233, EPI_ISL_2715234, EPI_ISL_2715235, EPI_ISL_2715236, EPI_ISL_2715237, EPI_ISL_2715238, EPI_ISL_2715239, EPI_ISL_2715240, EPI_ISL_2715241, EPI_ISL_2715242, EPI_ISL_2715243, EPI_ISL_2715244, EPI_ISL_2715245, EPI_ISL_2715246, EPI_ISL_2715247, EPI_ISL_2715249, EPI_ISL_2715250, EPI_ISL_2715251, EPI_ISL_2715252, EPI_ISL_2715253, EPI_ISL_2715254, EPI_ISL_2715255, EPI_ISL_2715256, EPI_ISL_2715258, EPI_ISL_2715259, EPI_ISL_2715260, EPI_ISL_2715261, EPI_ISL_2715263, EPI_ISL_2715264, EPI_ISL_2715268, EPI_ISL_2715270, EPI_ISL_2715271, EPI_ISL_2715272, EPI_ISL_2715273, EPI_ISL_2715274, EPI_ISL_2715275, EPI_ISL_2715277, EPI_ISL_2715278, EPI_ISL_2715279, EPI_ISL_2715281, EPI_ISL_2715283 | | | |
| see above | North Dakota Department of Health, Public Health Laboratory | North Dakota Department of Health, Public Health Laboratory | Lisa Wingerter |
| EPI_ISL_2715290, EPI_ISL_2715291, EPI_ISL_2715292, EPI_ISL_2715293, EPI_ISL_2715294, EPI_ISL_2715295, EPI_ISL_2715296, EPI_ISL_2715297, EPI_ISL_2715298, EPI_ISL_2715299, EPI_ISL_2715300, EPI_ISL_2715301, EPI_ISL_2715304, EPI_ISL_2715305, EPI_ISL_2715306, EPI_ISL_2715307, EPI_ISL_2715308, EPI_ISL_2715309, EPI_ISL_2715310, EPI_ISL_2715311, EPI_ISL_2715314, EPI_ISL_2715315, EPI_ISL_2715316, EPI_ISL_2715317, EPI_ISL_2715318, EPI_ISL_2715319, EPI_ISL_2715320, EPI_ISL_2715321, EPI_ISL_2715322, EPI_ISL_2715323, EPI_ISL_2715324, EPI_ISL_2715325, EPI_ISL_2715326, EPI_ISL_2715327, EPI_ISL_2715328, EPI_ISL_2715329, EPI_ISL_2715330, EPI_ISL_2715331, EPI_ISL_2715332, EPI_ISL_2715333, EPI_ISL_2715335, EPI_ISL_2715336, EPI_ISL_2715337, EPI_ISL_2715338, EPI_ISL_2715339, EPI_ISL_2715340, EPI_ISL_2715341, EPI_ISL_2715342, EPI_ISL_2715343, EPI_ISL_2715344, EPI_ISL_2715345, EPI_ISL_2715346, EPI_ISL_2715347, EPI_ISL_2715348, EPI_ISL_2715349, EPI_ISL_2715350, EPI_ISL_2715351, EPI_ISL_2715352 | | | |
| see above | US Air Force School of Aerospace Medicine | US Air Force School of Aerospace Medicine | Anthony Fries, Jennifer Meyer, William Gruner, Amanda Javorina, Carol Garrett, Sarah Purves, Clarise Starr, Elizabeth Macias |
| EPI_ISL_2715369, EPI_ISL_2715370, EPI_ISL_2715371 | Respiratory Viruses Branch, Centers for Disease Control and Prevention | Respiratory Viruses Branch, Centers for Disease Control and Prevention | Sheth,M., Nobles,S., Padilla,J., Burroughs,M., Le,S., Dillon,K., Cook,P., Paden,C.R., Batra,D., Queen,K., Knipe,K., Howard,D., Unoarumhi,Y., Wagner,D., Schmerer,M., Rambo-Martin,B.L., Lacek,K., Shepard,S., Halpin,A.L., Wentworth,D., Dugan,V., Tong,S., Lee,J. |
| EPI_ISL_2715373, EPI_ISL_2715374, EPI_ISL_2715375, EPI_ISL_2715376, EPI_ISL_2715377, EPI_ISL_2715378, EPI_ISL_2715379, EPI_ISL_2715380, EPI_ISL_2715381, EPI_ISL_2715382, EPI_ISL_2715384, EPI_ISL_2715385, EPI_ISL_2715386, EPI_ISL_2715387, EPI_ISL_2715388, EPI_ISL_2715389, EPI_ISL_2715390, EPI_ISL_2715391, EPI_ISL_2715392, EPI_ISL_2715393, EPI_ISL_2715396, EPI_ISL_2715397, EPI_ISL_2715398, EPI_ISL_2715399, EPI_ISL_2715401, EPI_ISL_2715402, EPI_ISL_2715403, EPI_ISL_2715404, EPI_ISL_2715405, EPI_ISL_2715406, EPI_ISL_2715408, EPI_ISL_2715409, EPI_ISL_2715410, EPI_ISL_2715411, EPI_ISL_2715412, EPI_ISL_2715413, EPI_ISL_2715414, EPI_ISL_2715415, EPI_ISL_2715416, EPI_ISL_2715417, EPI_ISL_2715418, EPI_ISL_2715419, EPI_ISL_2715420, EPI_ISL_2715422, EPI_ISL_2715423, EPI_ISL_2715424, EPI_ISL_2715425, EPI_ISL_2715426, EPI_ISL_2715427, EPI_ISL_2715428, EPI_ISL_2715429, EPI_ISL_2715430, EPI_ISL_2715432, EPI_ISL_2715433, EPI_ISL_2715434, EPI_ISL_2715435, EPI_ISL_2715436, EPI_ISL_2715437, EPI_ISL_2715438, EPI_ISL_2715440, EPI_ISL_2715442, EPI_ISL_2715443, EPI_ISL_2715444, EPI_ISL_2715445, EPI_ISL_2715446, EPI_ISL_2715447, EPI_ISL_2715448, EPI_ISL_2715449, EPI_ISL_2715450, EPI_ISL_2715452, EPI_ISL_2715453, EPI_ISL_2715454, EPI_ISL_2715455, EPI_ISL_2715456, EPI_ISL_2715457, EPI_ISL_2715458, EPI_ISL_2715459, EPI_ISL_2715460, EPI_ISL_2715461, EPI_ISL_2715462, EPI_ISL_2715463, EPI_ISL_2715464, EPI_ISL_2715465, EPI_ISL_2715466, EPI_ISL_2715467, EPI_ISL_2715468, EPI_ISL_2715469, EPI_ISL_2715470, EPI_ISL_2715471, EPI_ISL_2715472, EPI_ISL_2715473, EPI_ISL_2715474, EPI_ISL_2715475, EPI_ISL_2715476, EPI_ISL_2715477, EPI_ISL_2715478, EPI_ISL_2715479, EPI_ISL_2715480, EPI_ISL_2715481, EPI_ISL_2715482, EPI_ISL_2715483, EPI_ISL_2715484, EPI_ISL_2715485, EPI_ISL_2715486, EPI_ISL_2715487, EPI_ISL_2715488, EPI_ISL_2715489, EPI_ISL_2715490, EPI_ISL_2715491, EPI_ISL_2715492, EPI_ISL_2715493, EPI_ISL_2715494, EPI_ISL_2715495, EPI_ISL_2715496, EPI_ISL_2715497, EPI_ISL_2715498, EPI_ISL_2715499, EPI_ISL_2715500, EPI_ISL_2715501, EPI_ISL_2715502, EPI_ISL_2715503, EPI_ISL_2715505, EPI_ISL_2715506, EPI_ISL_2715507, EPI_ISL_2715508, EPI_ISL_2715510, EPI_ISL_2715511, EPI_ISL_2715512, EPI_ISL_2715513, EPI_ISL_2715514, EPI_ISL_2715515, EPI_ISL_2715517, EPI_ISL_2715518, EPI_ISL_2715519, EPI_ISL_2715520, EPI_ISL_2715521, EPI_ISL_2715523, EPI_ISL_2715524, EPI_ISL_2715525, EPI_ISL_2715527, EPI_ISL_2715528, EPI_ISL_2715529, EPI_ISL_2715530, EPI_ISL_2715532, EPI_ISL_2715533, EPI_ISL_2715534, EPI_ISL_2715535, EPI_ISL_2715536, EPI_ISL_2715537, EPI_ISL_2715538, EPI_ISL_2715539, EPI_ISL_2715540, EPI_ISL_2715541, EPI_ISL_2715542, EPI_ISL_2715543, EPI_ISL_2715544, EPI_ISL_2715545, EPI_ISL_2715546, EPI_ISL_2715547, EPI_ISL_2715548, EPI_ISL_2715549, EPI_ISL_2715550, EPI_ISL_2715551, EPI_ISL_2715552, EPI_ISL_2715553, EPI_ISL_2715554, EPI_ISL_2715555, EPI_ISL_2715556, EPI_ISL_2715557, EPI_ISL_2715558, EPI_ISL_2715559, EPI_ISL_2715560, EPI_ISL_2715561, EPI_ISL_2715562, EPI_ISL_2715563, EPI_ISL_2715564, EPI_ISL_2715565, EPI_ISL_2715566, EPI_ISL_2715567, EPI_ISL_2715568, EPI_ISL_2715569, EPI_ISL_2715570, EPI_ISL_2715571, EPI_ISL_2715572, EPI_ISL_2715574, EPI_ISL_2715575, EPI_ISL_2715576, EPI_ISL_2715578, EPI_ISL_2715579, EPI_ISL_2715580, EPI_ISL_2715581, EPI_ISL_2715582, EPI_ISL_2715583, EPI_ISL_2715584, EPI_ISL_2715585, EPI_ISL_2715586, EPI_ISL_2715587, EPI_ISL_2715588, EPI_ISL_2715589, EPI_ISL_2715590, EPI_ISL_2715591, EPI_ISL_2715592, EPI_ISL_2715593, EPI_ISL_2715594, EPI_ISL_2715595, EPI_ISL_2715596, EPI_ISL_2715597, EPI_ISL_2715598, EPI_ISL_2715599, EPI_ISL_2715600, EPI_ISL_2715601, EPI_ISL_2715602, EPI_ISL_2715603, EPI_ISL_2715604, EPI_ISL_2715605, EPI_ISL_2715606, EPI_ISL_2715608, EPI_ISL_2715609, EPI_ISL_2715610, EPI_ISL_2715611, EPI_ISL_2715612, EPI_ISL_2715613, EPI_ISL_2715614, EPI_ISL_2715615, EPI_ISL_2715616, EPI_ISL_2715617, EPI_ISL_2715618, EPI_ISL_2715619, EPI_ISL_2715620, EPI_ISL_2715621, EPI_ISL_2715622, EPI_ISL_2715623, EPI_ISL_2715624, EPI_ISL_2715625, EPI_ISL_2715626, EPI_ISL_2715627, EPI_ISL_2715628, EPI_ISL_2715629, EPI_ISL_2715630, EPI_ISL_2715631, EPI_ISL_2715632, EPI_ISL_2715633, EPI_ISL_2715634, EPI_ISL_2715635, EPI_ISL_2715636, EPI_ISL_2715637, EPI_ISL_2715638, EPI_ISL_2715640, EPI_ISL_2715641, EPI_ISL_2715642, EPI_ISL_2715643, EPI_ISL_2715644, EPI_ISL_2715645, EPI_ISL_2715646, EPI_ISL_2715647, EPI_ISL_2715648, EPI_ISL_2715649, EPI_ISL_2715650, EPI_ISL_2715651, EPI_ISL_2715652, EPI_ISL_2715654, EPI_ISL_2715655, EPI_ISL_2715656, EPI_ISL_2715657, EPI_ISL_2715658, EPI_ISL_2715659, EPI_ISL_2715660, EPI_ISL_2715661, EPI_ISL_2715662, EPI_ISL_2715663, EPI_ISL_2715664, EPI_ISL_2715665, EPI_ISL_2715666, EPI_ISL_2715667, EPI_ISL_2715668, EPI_ISL_2715669, EPI_ISL_2715670, EPI_ISL_2715671, EPI_ISL_2715672, EPI_ISL_2715673, EPI_ISL_2715674, EPI_ISL_2715676, EPI_ISL_2715677, EPI_ISL_2715679, EPI_ISL_2715680, EPI_ISL_2715681, EPI_ISL_2715682, EPI_ISL_2715685, EPI_ISL_2715686, EPI_ISL_2715687, EPI_ISL_2715688, EPI_ISL_2715690, EPI_ISL_2715692, EPI_ISL_2715694, EPI_ISL_2715695, EPI_ISL_2715696, EPI_ISL_2715697, EPI_ISL_2715698, EPI_ISL_2715699, EPI_ISL_2715700, EPI_ISL_2715701, EPI_ISL_2715702, EPI_ISL_2715703, EPI_ISL_2715704, EPI_ISL_2715705, EPI_ISL_2715706, EPI_ISL_2715707, EPI_ISL_2715708, EPI_ISL_2715709, EPI_ISL_2715710, EPI_ISL_2715712, EPI_ISL_2715713, EPI_ISL_2715714, EPI_ISL_2715715, EPI_ISL_2715716, EPI_ISL_2715717, EPI_ISL_2715718, EPI_ISL_2715720, EPI_ISL_2715721, EPI_ISL_2715722, EPI_ISL_2715723, EPI_ISL_2715724, EPI_ISL_2715725, EPI_ISL_2715726, EPI_ISL_2715728, EPI_ISL_2715729, EPI_ISL_2715730, EPI_ISL_2715731, EPI_ISL_2715732, EPI_ISL_2715733, EPI_ISL_2715734, EPI_ISL_2715735, EPI_ISL_2715736, EPI_ISL_2715737, EPI_ISL_2715738, EPI_ISL_2715739, EPI_ISL_2715741, EPI_ISL_2715743, EPI_ISL_2715744, EPI_ISL_2715745, EPI_ISL_2715746, EPI_ISL_2715747, EPI_ISL_2715748, EPI_ISL_2715751, EPI_ISL_2715753, EPI_ISL_2715754, EPI_ISL_2715755, EPI_ISL_2715756, EPI_ISL_2715757, EPI_ISL_2715758, EPI_ISL_2715759, EPI_ISL_2715760, EPI_ISL_2715763, EPI_ISL_2715764, EPI_ISL_2715765, EPI_ISL_2715766, EPI_ISL_2715767, EPI_ISL_2715769, EPI_ISL_2715770, EPI_ISL_2715772, EPI_ISL_2715773, EPI_ISL_2715774, EPI_ISL_2715775, EPI_ISL_2715777, EPI_ISL_2715778, EPI_ISL_2715779, EPI_ISL_2715781, EPI_ISL_2715782, EPI_ISL_2715784, EPI_ISL_2715785, EPI_ISL_2715786, EPI_ISL_2715787, EPI_ISL_2715788, EPI_ISL_2715789, EPI_ISL_2715790, EPI_ISL_2715791, EPI_ISL_2715792, EPI_ISL_2715793, EPI_ISL_2715794, EPI_ISL_2715795, EPI_ISL_2715796, EPI_ISL_2715797, EPI_ISL_2715798, EPI_ISL_2715799, EPI_ISL_2715800, EPI_ISL_2715801, EPI_ISL_2715802, EPI_ISL_2715803, EPI_ISL_2715805, EPI_ISL_2715806, EPI_ISL_2715807, EPI_ISL_2715808, EPI_ISL_2715810, EPI_ISL_2715811, EPI_ISL_2715813, EPI_ISL_2715814, EPI_ISL_2715815, EPI_ISL_2715816, EPI_ISL_2715817, EPI_ISL_2715818, EPI_ISL_2715819, EPI_ISL_2715820, EPI_ISL_2715821, EPI_ISL_2715822, EPI_ISL_2715823, EPI_ISL_2715824, EPI_ISL_2715825, EPI_ISL_2715827, EPI_ISL_2715828, EPI_ISL_2715829, EPI_ISL_2715830, EPI_ISL_2715831, EPI_ISL_2715832, EPI_ISL_2715833, EPI_ISL_2715834, EPI_ISL_2715835, EPI_ISL_2715836, EPI_ISL_2715837, EPI_ISL_2715838, EPI_ISL_2715839, EPI_ISL_2715840, EPI_ISL_2715841, EPI_ISL_2715842, EPI_ISL_2715843, EPI_ISL_2715844, EPI_ISL_2715845, EPI_ISL_2715846, EPI_ISL_2715847, EPI_ISL_2715848, EPI_ISL_2715849, EPI_ISL_2715850, EPI_ISL_2715851, EPI_ISL_2715852, EPI_ISL_2715853, EPI_ISL_2715855, EPI_ISL_2715856, EPI_ISL_2715857, EPI_ISL_2715858, EPI_ISL_2715859, EPI_ISL_2715860, EPI_ISL_2715861, EPI_ISL_2715862, EPI_ISL_2715863, EPI_ISL_2715864, EPI_ISL_2715865, EPI_ISL_2715866, EPI_ISL_2715867, EPI_ISL_2715868, EPI_ISL_2715869, EPI_ISL_2715870, EPI_ISL_2715872, EPI_ISL_2715873, EPI_ISL_2715874, EPI_ISL_2715875, EPI_ISL_2715876, EPI_ISL_2715877, EPI_ISL_2715878, EPI_ISL_2715880, EPI_ISL_2715881, EPI_ISL_2715882, EPI_ISL_2715883, EPI_ISL_2715884, EPI_ISL_2715885, EPI_ISL_2715886, EPI_ISL_2715888, EPI_ISL_2715889, EPI_ISL_2715890, EPI_ISL_2715892, EPI_ISL_2715893, EPI_ISL_2715895, EPI_ISL_2715896, EPI_ISL_2715897, EPI_ISL_2715898, EPI_ISL_2715899, EPI_ISL_2715900, EPI_ISL_2715901, EPI_ISL_2715902, EPI_ISL_2715903, EPI_ISL_2715904, EPI_ISL_2715905, EPI_ISL_2715906, EPI_ISL_2715907, EPI_ISL_2715909, EPI_ISL_2715911, EPI_ISL_2715912, EPI_ISL_2715914, EPI_ISL_2715915, EPI_ISL_2715916, EPI_ISL_2715917, EPI_ISL_2715918, EPI_ISL_2715920, EPI_ISL_2715921, EPI_ISL_2715922, EPI_ISL_2715923, EPI_ISL_2715924, EPI_ISL_2715925, EPI_ISL_2715927, EPI_ISL_2715928, EPI_ISL_2715929, EPI_ISL_2715930, EPI_ISL_2715931, EPI_ISL_2715933, EPI_ISL_2715934, EPI_ISL_2715935, EPI_ISL_2715937, EPI_ISL_2715939, EPI_ISL_2715940, EPI_ISL_2715941, EPI_ISL_2715942, EPI_ISL_2715943, EPI_ISL_2715944, EPI_ISL_2715947, EPI_ISL_2715948, EPI_ISL_2715949, EPI_ISL_2715950, EPI_ISL_2715951, EPI_ISL_2715952, EPI_ISL_2715953, EPI_ISL_2715954, EPI_ISL_2715955, EPI_ISL_2715956, EPI_ISL_2715957, EPI_ISL_2715958, EPI_ISL_2715960, EPI_ISL_2715961, EPI_ISL_2715962, EPI_ISL_2715963, EPI_ISL_2715965, EPI_ISL_2715967, EPI_ISL_2715968, EPI_ISL_2715969, EPI_ISL_2715970, EPI_ISL_2715971, EPI_ISL_2715972, EPI_ISL_2715973, EPI_ISL_2715974, EPI_ISL_2715975, EPI_ISL_2715976, EPI_ISL_2715977, EPI_ISL_2715978, EPI_ISL_2715979, EPI_ISL_2715980, EPI_ISL_2715981, EPI_ISL_2715983, EPI_ISL_2715984, EPI_ISL_2715985, EPI_ISL_2715986, EPI_ISL_2715987, EPI_ISL_2715988, EPI_ISL_2715989, EPI_ISL_2715990, EPI_ISL_2715991, EPI_ISL_2715992, EPI_ISL_2715993, EPI_ISL_2715994, EPI_ISL_2715995, EPI_ISL_2715996, EPI_ISL_2715997, EPI_ISL_2715998, EPI_ISL_2715999, EPI_ISL_2716000, EPI_ISL_2716001, EPI_ISL_2716002, EPI_ISL_2716003, EPI_ISL_2716004, EPI_ISL_2716007, EPI_ISL_2716008, EPI_ISL_2716009, EPI_ISL_2716010, EPI_ISL_2716011, EPI_ISL_2716012, EPI_ISL_2716013, EPI_ISL_2716014, EPI_ISL_2716015, EPI_ISL_2716016, EPI_ISL_2716017, EPI_ISL_2716018, EPI_ISL_2716019, EPI_ISL_2716020, EPI_ISL_2716021, EPI_ISL_2716022, EPI_ISL_2716023, EPI_ISL_2716024, EPI_ISL_2716025, EPI_ISL_2716026, EPI_ISL_2716028, EPI_ISL_2716029, EPI_ISL_2716030, EPI_ISL_2716031, EPI_ISL_2716032, EPI_ISL_2716033, EPI_ISL_2716035, EPI_ISL_2716036, EPI_ISL_2716038, EPI_ISL_2716039, EPI_ISL_2716040, EPI_ISL_2716041, EPI_ISL_2716042, EPI_ISL_2716043, EPI_ISL_2716045, EPI_ISL_2716046, EPI_ISL_2716047, EPI_ISL_2716049, EPI_ISL_2716050, EPI_ISL_2716052, EPI_ISL_2716053, EPI_ISL_2716054, EPI_ISL_2716055, EPI_ISL_2716056, EPI_ISL_2716057, EPI_ISL_2716058, EPI_ISL_2716059, EPI_ISL_2716060, EPI_ISL_2716061, EPI_ISL_2716062, EPI_ISL_2716063, EPI_ISL_2716064, EPI_ISL_2716065, EPI_ISL_2716066, EPI_ISL_2716067, EPI_ISL_2716068, EPI_ISL_2716069, EPI_ISL_2716070, EPI_ISL_2716071, EPI_ISL_2716072, EPI_ISL_2716073, EPI_ISL_2716074, EPI_ISL_2716075, EPI_ISL_2716076, EPI_ISL_2716078, EPI_ISL_2716079, EPI_ISL_2716081, EPI_ISL_2716082, EPI_ISL_2716083, EPI_ISL_2716084, EPI_ISL_2716085, EPI_ISL_2716086, EPI_ISL_2716087, EPI_ISL_2716088, EPI_ISL_2716089, EPI_ISL_2716090, EPI_ISL_2716091, EPI_ISL_2716092, EPI_ISL_2716093, EPI_ISL_2716094, EPI_ISL_2716095, EPI_ISL_2716096, EPI_ISL_2716097, EPI_ISL_2716098, EPI_ISL_2716099, EPI_ISL_2716100, EPI_ISL_2716101, EPI_ISL_2716102, EPI_ISL_2716104, EPI_ISL_2716105, EPI_ISL_2716106, EPI_ISL_2716107, EPI_ISL_2716108, EPI_ISL_2716110, EPI_ISL_2716111, EPI_ISL_2716112, EPI_ISL_2716113, EPI_ISL_2716114, EPI_ISL_2716115, EPI_ISL_2716116, EPI_ISL_2716117, EPI_ISL_2716118, EPI_ISL_2716119, EPI_ISL_2716120, EPI_ISL_2716121, EPI_ISL_2716122, EPI_ISL_2716123, EPI_ISL_2716124, EPI_ISL_2716125, EPI_ISL_2716126, EPI_ISL_2716128, EPI_ISL_2716130, EPI_ISL_2716131, EPI_ISL_2716132, EPI_ISL_2716133, EPI_ISL_2716134, EPI_ISL_2716135, EPI_ISL_2716137, EPI_ISL_2716139, EPI_ISL_2716140, EPI_ISL_2716141, EPI_ISL_2716142, EPI_ISL_2716143, EPI_ISL_2716144, EPI_ISL_2716145, EPI_ISL_2716146, EPI_ISL_2716147, EPI_ISL_2716148, EPI_ISL_2716149, EPI_ISL_2716150, EPI_ISL_2716151, EPI_ISL_2716152, EPI_ISL_2716153, EPI_ISL_2716155, EPI_ISL_2716156, EPI_ISL_2716157, EPI_ISL_2716158, EPI_ISL_2716159, EPI_ISL_2716160, EPI_ISL_2716161, EPI_ISL_2716162, EPI_ISL_2716163, EPI_ISL_2716164, EPI_ISL_2716165, EPI_ISL_2716166, EPI_ISL_2716167, EPI_ISL_2716168, EPI_ISL_2716169, EPI_ISL_2716170, EPI_ISL_2716171, EPI_ISL_2716172, EPI_ISL_2716173, EPI_ISL_2716174, EPI_ISL_2716175, EPI_ISL_2716176, EPI_ISL_2716177, EPI_ISL_2716178, EPI_ISL_2716179, EPI_ISL_2716180, EPI_ISL_2716181, EPI_ISL_2716182, EPI_ISL_2716183, EPI_ISL_2716184, EPI_ISL_2716185, EPI_ISL_2716186, EPI_ISL_2716187, EPI_ISL_2716188, EPI_ISL_2716189, EPI_ISL_2716190, EPI_ISL_2716191, EPI_ISL_2716192, EPI_ISL_2716193, EPI_ISL_2716194, EPI_ISL_2716195, EPI_ISL_2716196, EPI_ISL_2716197, EPI_ISL_2716198, EPI_ISL_2716199, EPI_ISL_2716200, EPI_ISL_2716201, EPI_ISL_2716202, EPI_ISL_2716203, EPI_ISL_2716204, EPI_ISL_2716205, EPI_ISL_2716206 | | | |
| see above | Pandemic Response Lab - NYC | Pandemic Response Lab, R&D | Henry Lee, Michael Hammerling, Melissa Hopkins, Cybill del Castillo, Shinyoung Clair Kang, William Ward, Pradeep Bugga, Sol Rey, Dylan Law, Katharine Nelson, Haiping Hao, Jon Laurent |
| EPI_ISL_2716208, EPI_ISL_2716209, EPI_ISL_2716210, EPI_ISL_2716211, EPI_ISL_2716212, EPI_ISL_2716213, EPI_ISL_2716214, EPI_ISL_2716215, EPI_ISL_2716216, EPI_ISL_2716217, EPI_ISL_2716218, EPI_ISL_2716219, EPI_ISL_2716220 | | | |
| see above | The Jackson Laboratory | The Jackson Laboratory | Long J, Renzette N, Adams M, Omerza G, Kelly K, Li L |
| EPI_ISL_2716221 | Respiratory Viruses Branch, Centers for Disease Control and Prevention | Respiratory Viruses Branch, Centers for Disease Control and Prevention | Howard,D., Batra,D., Cook,P.W., Moser,K., Paskey,A., Caravas,J.,Rambo-Martin,B., Morrison,S., Gulvick,C., Sammons,S., Unoarumhi,Y.,Wagner,D., Tugwell,M.S., Moon,L., Paden,C.R. and MacCannell,D. |
| EPI_ISL_2716222, EPI_ISL_2716223, EPI_ISL_2716224, EPI_ISL_2716225, EPI_ISL_2716226, EPI_ISL_2716227, EPI_ISL_2716228, EPI_ISL_2716229, EPI_ISL_2716230, EPI_ISL_2716231, EPI_ISL_2716232, EPI_ISL_2716233, EPI_ISL_2716234, EPI_ISL_2716235, EPI_ISL_2716236, EPI_ISL_2716237, EPI_ISL_2716238, EPI_ISL_2716239, EPI_ISL_2716240, EPI_ISL_2716241, EPI_ISL_2716242, EPI_ISL_2716243, EPI_ISL_2716244, EPI_ISL_2716245, EPI_ISL_2716246, EPI_ISL_2716247, EPI_ISL_2716248, EPI_ISL_2716249, EPI_ISL_2716250, EPI_ISL_2716251, EPI_ISL_2716252, EPI_ISL_2716253, EPI_ISL_2716254 | | | |
| see above | Yale Clinical Virology Lab | Grubaugh Lab - Yale School of Public Health | Joseph Fauver, Mallery Breban, Isabel Ott, Tara Alpert, Mary Petrone, Anderson Brito, Chantal Vogels, Annie Watkins, Chaney Kalinich, Jessica Rothman, Marie L. Landry, Nathan Grubaugh |
| EPI_ISL_2716255, EPI_ISL_2716256, EPI_ISL_2716257, EPI_ISL_2716258, EPI_ISL_2716259, EPI_ISL_2716260, EPI_ISL_2716261, EPI_ISL_2716262, EPI_ISL_2716263, EPI_ISL_2716264, EPI_ISL_2716265, EPI_ISL_2716266, EPI_ISL_2716267, EPI_ISL_2716268, EPI_ISL_2716269, EPI_ISL_2716270, EPI_ISL_2716271, EPI_ISL_2716272, EPI_ISL_2716273, EPI_ISL_2716274, EPI_ISL_2716275, EPI_ISL_2716276, EPI_ISL_2716277, EPI_ISL_2716278, EPI_ISL_2716279, EPI_ISL_2716280, EPI_ISL_2716282, EPI_ISL_2716283 | | | |
| see above | Wyoming Public Health Laboratory | Wyoming Public Health Laboratory | Jim Mildenberger, Taylor Fearing, Channing Weber, Ashley Norberg, Chayse Rowley, Marley Goetz, Brian Dominguez, Elliot Thomasson, Sam Britz, Cari Sloma, Robert Petit, and Rob Christensen |
| EPI_ISL_2716284, EPI_ISL_2716285, EPI_ISL_2716286, EPI_ISL_2716287, EPI_ISL_2716288, EPI_ISL_2716289, EPI_ISL_2716290, EPI_ISL_2716291, EPI_ISL_2716292, EPI_ISL_2716293, EPI_ISL_2716294, EPI_ISL_2716295, EPI_ISL_2716297, EPI_ISL_2716298, EPI_ISL_2716299, EPI_ISL_2716300, EPI_ISL_2716301, EPI_ISL_2716302, EPI_ISL_2716303, EPI_ISL_2716304, EPI_ISL_2716305, EPI_ISL_2716306, EPI_ISL_2716307, EPI_ISL_2716308, EPI_ISL_2716309, EPI_ISL_2716310 | | | |
| see above | Kansas Health and Environmental Lab | Kansas Health and Environmental Lab | Katherine Wiggins, Mike Grose, Jonathan Barnell, Ben Olsen, and Phil Adam |
| EPI_ISL_2716323, EPI_ISL_2716325, EPI_ISL_2716326, EPI_ISL_2716327, EPI_ISL_2716328, EPI_ISL_2716329, EPI_ISL_2716330, EPI_ISL_2716331, EPI_ISL_2716332, EPI_ISL_2716333, EPI_ISL_2716334, EPI_ISL_2716335, EPI_ISL_2716336, EPI_ISL_2716337, EPI_ISL_2716338, EPI_ISL_2716339, EPI_ISL_2716340, EPI_ISL_2716341, EPI_ISL_2716342, EPI_ISL_2716343, EPI_ISL_2716344, EPI_ISL_2716345, EPI_ISL_2716346, EPI_ISL_2716347, EPI_ISL_2716348, EPI_ISL_2716349, EPI_ISL_2716350, EPI_ISL_2716351, EPI_ISL_2716352, EPI_ISL_2716353, EPI_ISL_2716354, EPI_ISL_2716355, EPI_ISL_2716356, EPI_ISL_2716357, EPI_ISL_2716358, EPI_ISL_2716359, EPI_ISL_2716360, EPI_ISL_2716361, EPI_ISL_2716362, EPI_ISL_2716363, EPI_ISL_2716364, EPI_ISL_2716365, EPI_ISL_2716366, EPI_ISL_2716367, EPI_ISL_2716368, EPI_ISL_2716369, EPI_ISL_2716370, EPI_ISL_2716371, EPI_ISL_2716372, EPI_ISL_2716373, EPI_ISL_2716374, EPI_ISL_2716375, EPI_ISL_2716376, EPI_ISL_2716377, EPI_ISL_2716378, EPI_ISL_2716379, EPI_ISL_2716380, EPI_ISL_2716381, EPI_ISL_2716382, EPI_ISL_2716383, EPI_ISL_2716384, EPI_ISL_2716385, EPI_ISL_2716386, EPI_ISL_2716387, EPI_ISL_2716388, EPI_ISL_2716389, EPI_ISL_2716390, EPI_ISL_2716392, EPI_ISL_2716393, EPI_ISL_2716394, EPI_ISL_2716395, EPI_ISL_2716396, EPI_ISL_2716397, EPI_ISL_2716398, EPI_ISL_2716399, EPI_ISL_2716400, EPI_ISL_2716401, EPI_ISL_2716402, EPI_ISL_2716403, EPI_ISL_2716404, EPI_ISL_2716405, EPI_ISL_2716406, EPI_ISL_2716407, EPI_ISL_2716408, EPI_ISL_2716409, EPI_ISL_2716410, EPI_ISL_2716411, EPI_ISL_2716412 | | | |

| | | | |
|---|---|---|---|
| see above | Arizona State Public Health Laboratory | Arizona State Public Health Laboratory | Trung Huynh, Jessica Escobar, Katherine Fullerton, Nobuko Fukushima, Matthew Contursi, Stacy White, Linda Getsinger, Victor Waddell |
| EPI_ISL_2716428, EPI_ISL_2716429, EPI_ISL_2716430, EPI_ISL_2716431, EPI_ISL_2716432, EPI_ISL_2716433, EPI_ISL_2716434, EPI_ISL_2716435, EPI_ISL_2716436, EPI_ISL_2716437, EPI_ISL_2716438, EPI_ISL_2716439, EPI_ISL_2716440, EPI_ISL_2716441, EPI_ISL_2716442, EPI_ISL_2716443, EPI_ISL_2716444, EPI_ISL_2716445, EPI_ISL_2716446, EPI_ISL_2716447, EPI_ISL_2716448 | | | |
| see above | Virology, University of Washington | Virology, University of Washington | Greninger,A.L., Lin,M.J., Cox,R.M., Plemper,R.K. |
| EPI_ISL_2716450, EPI_ISL_2716451, EPI_ISL_2716452, EPI_ISL_2716453, EPI_ISL_2716454, EPI_ISL_2716455, EPI_ISL_2716456, EPI_ISL_2716457, EPI_ISL_2716458, EPI_ISL_2716459, EPI_ISL_2716460, EPI_ISL_2716461, EPI_ISL_2716462, EPI_ISL_2716463, EPI_ISL_2716464, EPI_ISL_2716465, EPI_ISL_2716466, EPI_ISL_2716467, EPI_ISL_2716468, EPI_ISL_2716469, EPI_ISL_2716470, EPI_ISL_2716471, EPI_ISL_2716472 | | | |
| see above | Washington State Department of Health Public Health Laboratories | Washington State Department of Health Public Health Laboratories | Drew MacKellar, Philip Dykema, Denny Russell, Joenice Gonzalez, Hannah Gray, Geoff Melly, Vanessa De Los Santos, Darren Lucas, JohnAric Peterson, Avi Singh, Rebecca Cao |
| EPI_ISL_2716477, EPI_ISL_2716478, EPI_ISL_2716479, EPI_ISL_2716480, EPI_ISL_2716481, EPI_ISL_2716482, EPI_ISL_2716483, EPI_ISL_2716484, EPI_ISL_2716485, EPI_ISL_2716486, EPI_ISL_2716487, EPI_ISL_2716488, EPI_ISL_2716489, EPI_ISL_2716490, EPI_ISL_2716491, EPI_ISL_2716492, EPI_ISL_2716493, EPI_ISL_2716494, EPI_ISL_2716495, EPI_ISL_2716496, EPI_ISL_2716497, EPI_ISL_2716498, EPI_ISL_2716499, EPI_ISL_2716500, EPI_ISL_2716501, EPI_ISL_2716502, EPI_ISL_2716503, EPI_ISL_2716504, EPI_ISL_2716505, EPI_ISL_2716506, EPI_ISL_2716507, EPI_ISL_2716508, EPI_ISL_2716509, EPI_ISL_2716510, EPI_ISL_2716511, EPI_ISL_2716512, EPI_ISL_2716513, EPI_ISL_2716514, EPI_ISL_2716515, EPI_ISL_2716516, EPI_ISL_2716517, EPI_ISL_2716518, EPI_ISL_2716519, EPI_ISL_2716520, EPI_ISL_2716521, EPI_ISL_2716522, EPI_ISL_2716523, EPI_ISL_2716524, EPI_ISL_2716525, EPI_ISL_2716526, EPI_ISL_2716527, EPI_ISL_2716528, EPI_ISL_2716529, EPI_ISL_2716530, EPI_ISL_2716531, EPI_ISL_2716532, EPI_ISL_2716533 | | | |
| see above | UW Virology Lab | UW Virology Lab | Pavitra Roychoudhury, Hong Xie, Lasata Shrestha, Shah Mohamed Bakhash, Tien V. Nguyen, Noah R. Baker, Sean Ellis, Meei-Li Huang, Keith R Jerome, Alexander Greninger |
| EPI_ISL_2716643, EPI_ISL_2716644, EPI_ISL_2716645, EPI_ISL_2716646, EPI_ISL_2716647, EPI_ISL_2716648, EPI_ISL_2716649, EPI_ISL_2716650, EPI_ISL_2716652, EPI_ISL_2716653, EPI_ISL_2716654, EPI_ISL_2716657, EPI_ISL_2716658, EPI_ISL_2716659, EPI_ISL_2716660, EPI_ISL_2716661, EPI_ISL_2716662, EPI_ISL_2716663, EPI_ISL_2716666, EPI_ISL_2716667, EPI_ISL_2716669, EPI_ISL_2716671, EPI_ISL_2716672, EPI_ISL_2716673, EPI_ISL_2716674, EPI_ISL_2716675, EPI_ISL_2716676, EPI_ISL_2716677, EPI_ISL_2716678, EPI_ISL_2716680, EPI_ISL_2716681, EPI_ISL_2716683, EPI_ISL_2716684, EPI_ISL_2716685, EPI_ISL_2716686, EPI_ISL_2716687, EPI_ISL_2716688, EPI_ISL_2716689, EPI_ISL_2716690, EPI_ISL_2716691, EPI_ISL_2716692, EPI_ISL_2716694, EPI_ISL_2716695, EPI_ISL_2716696, EPI_ISL_2716697, EPI_ISL_2716698, EPI_ISL_2716701, EPI_ISL_2716702, EPI_ISL_2716703, EPI_ISL_2716704, EPI_ISL_2716705, EPI_ISL_2716706, EPI_ISL_2716707, EPI_ISL_2716708, EPI_ISL_2716709, EPI_ISL_2716710, EPI_ISL_2716711, EPI_ISL_2716712, EPI_ISL_2716713, EPI_ISL_2716717, EPI_ISL_2716718, EPI_ISL_2716719, EPI_ISL_2716720, EPI_ISL_2716721, EPI_ISL_2716722, EPI_ISL_2716723 | | | |
| see above | Arizona State University | Arizona State University | Peter T. Skidmore, LaRinda A. Holland, Matthew F. Smith, Rabia Maqsood, Nicholas J. Mellor, Joy M. Blain, Valerie Harris, Joshua LaBaer, Vel Murugan, Efrem S. Lim |
| EPI_ISL_2716726, EPI_ISL_2716727, EPI_ISL_2716728, EPI_ISL_2716729, EPI_ISL_2716730, EPI_ISL_2716731, EPI_ISL_2716732, EPI_ISL_2716733, EPI_ISL_2716734, EPI_ISL_2716735, EPI_ISL_2716736, EPI_ISL_2716737, EPI_ISL_2716738, EPI_ISL_2716739 | | | |
| see above | UW Virology Lab | UW Virology Lab | Pavitra Roychoudhury, Hong Xie, Lasata Shrestha, Shah Mohamed Bakhash, Tien V. Nguyen, Noah R. Baker, Sean Ellis, Meei-Li Huang, Keith R Jerome, Alexander Greninger |
| EPI_ISL_2716839, EPI_ISL_2716840, EPI_ISL_2716841, EPI_ISL_2716842, EPI_ISL_2716843, EPI_ISL_2716844, EPI_ISL_2716845, EPI_ISL_2716846, EPI_ISL_2716847, EPI_ISL_2716848, EPI_ISL_2716849, EPI_ISL_2716850, EPI_ISL_2716851, EPI_ISL_2716852, EPI_ISL_2716853, EPI_ISL_2716854, EPI_ISL_2716855, EPI_ISL_2716856, EPI_ISL_2716857, EPI_ISL_2716858, EPI_ISL_2716859, EPI_ISL_2716860, EPI_ISL_2716861, EPI_ISL_2716862, EPI_ISL_2716863, EPI_ISL_2716864, EPI_ISL_2716865, EPI_ISL_2716866, EPI_ISL_2716867, EPI_ISL_2716868, EPI_ISL_2716869 | | | |
| see above | Alaska State Virology Laboratory | Alaska State Virology Laboratory | Stephanie DeRonde, Elva House, Jacob Zidek, Lisa Smith, Ph.D., Jack Chen, Ph.D. |
| EPI_ISL_2716871, EPI_ISL_2716872, EPI_ISL_2716873 | Advanced Molecular Diagnostics, Arizona Department of Health Services | Advanced Molecular Diagnostics, Arizona Department of Health Services | Huynh,T., Escobar,J., Fukushima,N., Fullerton,K., Contursi,M., Tesman,N. |
| EPI_ISL_2716876, EPI_ISL_2716877, EPI_ISL_2716878, EPI_ISL_2716879, EPI_ISL_2716880, EPI_ISL_2716881, EPI_ISL_2716882, EPI_ISL_2716883, EPI_ISL_2716884, EPI_ISL_2716885, EPI_ISL_2716886, EPI_ISL_2716887, EPI_ISL_2716888, EPI_ISL_2716889 | | | |
| see above | Monterey County Public Health Laboratory | Monterey County Public Health Laboratory | Monterey County Public Health Laboratory |
| EPI_ISL_2716890, EPI_ISL_2716891, EPI_ISL_2716892, EPI_ISL_2716893, EPI_ISL_2716894, EPI_ISL_2716895, EPI_ISL_2716896, EPI_ISL_2716897, EPI_ISL_2716898, EPI_ISL_2716899, EPI_ISL_2716900, EPI_ISL_2716901, EPI_ISL_2716902, EPI_ISL_2716903, EPI_ISL_2716904, EPI_ISL_2716905, EPI_ISL_2716906, EPI_ISL_2716907, EPI_ISL_2716908, EPI_ISL_2716909, EPI_ISL_2716910, EPI_ISL_2716912, EPI_ISL_2716913, EPI_ISL_2716914, EPI_ISL_2716915, EPI_ISL_2716916, EPI_ISL_2716917, EPI_ISL_2716918, EPI_ISL_2716919, EPI_ISL_2716920, EPI_ISL_2716921, EPI_ISL_2716922, EPI_ISL_2716923, EPI_ISL_2716924, EPI_ISL_2716925, EPI_ISL_2716926, EPI_ISL_2716927, EPI_ISL_2716928, EPI_ISL_2716929, EPI_ISL_2716930, EPI_ISL_2716931, EPI_ISL_2716932, EPI_ISL_2716933, EPI_ISL_2716934, EPI_ISL_2716935, EPI_ISL_2716936, EPI_ISL_2716937, EPI_ISL_2716938, EPI_ISL_2716939, EPI_ISL_2716940, EPI_ISL_2716941, EPI_ISL_2716942, EPI_ISL_2716943, EPI_ISL_2716944, EPI_ISL_2716945, EPI_ISL_2716946, EPI_ISL_2716947, EPI_ISL_2716948, EPI_ISL_2716949, EPI_ISL_2716950, EPI_ISL_2716952, EPI_ISL_2716953 | | | |
| see above | Southern Nevada Public Health Laboratory | Southern Nevada Public Health Laboratory | Michael Picker |
| EPI_ISL_2717676 | Public Health Laboratory - Infectious Disease Lab, Minnesota Department of Health Infectious Disease Laboratory Submission Group | Public Health Laboratory - Infectious Disease Lab, Minnesota Department of Health Infectious Disease Laboratory Submission Group | Lorentz,A., Garfin,J., Plumb,M., Wang,X. |
| EPI_ISL_2718532, EPI_ISL_2718533, EPI_ISL_2718534 | Laboratory Medicine, University of Washington | Laboratory Medicine, University of Washington | Roychoudhury,P., Xie,H., Shrestha,L., Bakhash,S.M., Nguyen,T.V., Baker,N.R., Ellis,S., Huang,M.-L., Jerome,K.R., Greninger,A. |
| EPI_ISL_2718535, EPI_ISL_2718536 | Laboratory Medicine, University of Washington | Laboratory Medicine, University of Washington | Castor,J.L. |
| EPI_ISL_2723649, EPI_ISL_2723650, EPI_ISL_2723651, EPI_ISL_2723652, EPI_ISL_2723653, EPI_ISL_2723654, EPI_ISL_2723656, EPI_ISL_2723657, EPI_ISL_2723658, EPI_ISL_2723659, EPI_ISL_2723660, EPI_ISL_2723661, EPI_ISL_2723662, EPI_ISL_2723663, EPI_ISL_2723664, EPI_ISL_2723665, EPI_ISL_2723666, EPI_ISL_2723667, EPI_ISL_2723668, EPI_ISL_2723669, EPI_ISL_2723671, EPI_ISL_2723672, EPI_ISL_2723673, EPI_ISL_2723675, EPI_ISL_2723676, EPI_ISL_2723677, EPI_ISL_2723678, EPI_ISL_2723679, EPI_ISL_2723680, EPI_ISL_2723681, EPI_ISL_2723682, EPI_ISL_2723683, EPI_ISL_2723685, EPI_ISL_2723686, EPI_ISL_2723687, EPI_ISL_2723688, EPI_ISL_2723689, EPI_ISL_2723690, EPI_ISL_2723691, EPI_ISL_2723692, EPI_ISL_2723694, EPI_ISL_2723696, EPI_ISL_2723697, EPI_ISL_2723698, EPI_ISL_2723699, EPI_ISL_2723700, EPI_ISL_2723701, EPI_ISL_2723702, EPI_ISL_2723703, EPI_ISL_2723704, EPI_ISL_2723705, EPI_ISL_2723709, EPI_ISL_2723710, EPI_ISL_2723711, EPI_ISL_2723713, EPI_ISL_2723714, EPI_ISL_2723715, EPI_ISL_2723716, EPI_ISL_2723717, EPI_ISL_2723718, EPI_ISL_2723720, EPI_ISL_2723721, EPI_ISL_2723725, EPI_ISL_2723726, EPI_ISL_2723727, EPI_ISL_2723728, EPI_ISL_2723731, EPI_ISL_2723733, EPI_ISL_2723734, EPI_ISL_2723736, EPI_ISL_2723737, EPI_ISL_2723738, EPI_ISL_2723739, EPI_ISL_2724007 | | | |
| see above | Curative | New Mexico Department of Health Scientific Laboratory | Ellie Johnson, D'eldra Malone, Jennifer Benoit, Ratheesh Rajan, Linda Salazar, Keila Gutierrez, Anastacia Griego-Fisher |
| EPI_ISL_2724009 | Molecular Biology Laboratory, Infectious Diseases Department, Instituto Nacional de Ciencias Medicas y Nutricion Salvador Zubiran | Instituto Nacional de Ciencias Medicas y Nutricion Infectious Diseases | Pilar Ramos Cervantes, Fernando Ledesma Barrientos, Violeta Ibarra Gonzalez, Patricia Leal, Eric Ochoa, Arturo Galindo, Guillermo Ruiz-Palacios y Santos |
| EPI_ISL_2724513, EPI_ISL_2724514, EPI_ISL_2724515, EPI_ISL_2724516, EPI_ISL_2724517, EPI_ISL_2724518, EPI_ISL_2724519, EPI_ISL_2724520, EPI_ISL_2724521, EPI_ISL_2724522, EPI_ISL_2724523, EPI_ISL_2724524, EPI_ISL_2724525, EPI_ISL_2724526, EPI_ISL_2724527, EPI_ISL_2724528, EPI_ISL_2724529, EPI_ISL_2724530, EPI_ISL_2724531, EPI_ISL_2724532, EPI_ISL_2724533, EPI_ISL_2724534, EPI_ISL_2724535, EPI_ISL_2724536, EPI_ISL_2724537, EPI_ISL_2724538, EPI_ISL_2724539, EPI_ISL_2724540, EPI_ISL_2724541, EPI_ISL_2724542, EPI_ISL_2724543, EPI_ISL_2724544, EPI_ISL_2724545, EPI_ISL_2724546, EPI_ISL_2724547, EPI_ISL_2724548, EPI_ISL_2724549, EPI_ISL_2724550, EPI_ISL_2724551, EPI_ISL_2724552, EPI_ISL_2724553, EPI_ISL_2724554, EPI_ISL_2724555, EPI_ISL_2724556, EPI_ISL_2724557, EPI_ISL_2724558, EPI_ISL_2724559, EPI_ISL_2724560, EPI_ISL_2724561, EPI_ISL_2724562, EPI_ISL_2724563, EPI_ISL_2724564, EPI_ISL_2724565, EPI_ISL_2724566, EPI_ISL_2724567, EPI_ISL_2724568, EPI_ISL_2724569, EPI_ISL_2724570, EPI_ISL_2724571, EPI_ISL_2724572, EPI_ISL_2724573, EPI_ISL_2724574, EPI_ISL_2724575, EPI_ISL_2724576, EPI_ISL_2724577, EPI_ISL_2724578, EPI_ISL_2724579, EPI_ISL_2724580, EPI_ISL_2724581, EPI_ISL_2724582, EPI_ISL_2724583, EPI_ISL_2724584, EPI_ISL_2724585, EPI_ISL_2724586, EPI_ISL_2724587, EPI_ISL_2724588, EPI_ISL_2724589, EPI_ISL_2724590, EPI_ISL_2724591, EPI_ISL_2724592, EPI_ISL_2724593, EPI_ISL_2724594, EPI_ISL_2724595, EPI_ISL_2724596, EPI_ISL_2724597, EPI_ISL_2724598, EPI_ISL_2724599, EPI_ISL_2724600, EPI_ISL_2724601, EPI_ISL_2724602, EPI_ISL_2724603, EPI_ISL_2724604, EPI_ISL_2724605, EPI_ISL_2724606, EPI_ISL_2724607, EPI_ISL_2724608, EPI_ISL_2724609, EPI_ISL_2724610, EPI_ISL_2724611, EPI_ISL_2724613, EPI_ISL_2724614, EPI_ISL_2724615, EPI_ISL_2724616, EPI_ISL_2724617, EPI_ISL_2724618, EPI_ISL_2724619, EPI_ISL_2724620, EPI_ISL_2724621, EPI_ISL_2724622, EPI_ISL_2724623, EPI_ISL_2724624, EPI_ISL_2724625, EPI_ISL_2724626, EPI_ISL_2724627, EPI_ISL_2724628, EPI_ISL_2724629, EPI_ISL_2724630, EPI_ISL_2724631, EPI_ISL_2724632, EPI_ISL_2724633, EPI_ISL_2724634, EPI_ISL_2724635, EPI_ISL_2724636, EPI_ISL_2724637, EPI_ISL_2724638, EPI_ISL_2724639, EPI_ISL_2724640, EPI_ISL_2724641, EPI_ISL_2724642, EPI_ISL_2724643, EPI_ISL_2724644, EPI_ISL_2724645, EPI_ISL_2724646, EPI_ISL_2724647, EPI_ISL_2724648, EPI_ISL_2724649, EPI_ISL_2724650, EPI_ISL_2724651, EPI_ISL_2724652, EPI_ISL_2724653, EPI_ISL_2724654, EPI_ISL_2724655, EPI_ISL_2724656, EPI_ISL_2724657, EPI_ISL_2724658, EPI_ISL_2724659, EPI_ISL_2724660, EPI_ISL_2724661, EPI_ISL_2724662, EPI_ISL_2724663, EPI_ISL_2724664, EPI_ISL_2724665, EPI_ISL_2724666, EPI_ISL_2724667, EPI_ISL_2724668, EPI_ISL_2724669, EPI_ISL_2724670, EPI_ISL_2724671, EPI_ISL_2724672, EPI_ISL_2724673, EPI_ISL_2724674, EPI_ISL_2724675, EPI_ISL_2724676, EPI_ISL_2724677, EPI_ISL_2724678, EPI_ISL_2724679, EPI_ISL_2724680, EPI_ISL_2724681, EPI_ISL_2724682, EPI_ISL_2724683, EPI_ISL_2724684, EPI_ISL_2724685, EPI_ISL_2724686, EPI_ISL_2724687, EPI_ISL_2724688, EPI_ISL_2724689, EPI_ISL_2724690, EPI_ISL_2724691, EPI_ISL_2724692, EPI_ISL_2724693, EPI_ISL_2724694, EPI_ISL_2724695, EPI_ISL_2724696, EPI_ISL_2724697, EPI_ISL_2724698, EPI_ISL_2724699, EPI_ISL_2724700, EPI_ISL_2724701, EPI_ISL_2724702, EPI_ISL_2724703, EPI_ISL_2724704, EPI_ISL_2724705, EPI_ISL_2724706, EPI_ISL_2724707, EPI_ISL_2724708, EPI_ISL_2724709, EPI_ISL_2724710, EPI_ISL_2724711, EPI_ISL_2724712, EPI_ISL_2724713, EPI_ISL_2724714, EPI_ISL_2724715, EPI_ISL_2724716, EPI_ISL_2724717, EPI_ISL_2724718, EPI_ISL_2724719, EPI_ISL_2724720, EPI_ISL_2724721, EPI_ISL_2724722, EPI_ISL_2724723, EPI_ISL_2724724, EPI_ISL_2724725, EPI_ISL_2724726, EPI_ISL_2724727, EPI_ISL_2724728, EPI_ISL_2724729, EPI_ISL_2724730, EPI_ISL_2724731, EPI_ISL_2724732, EPI_ISL_2724733, EPI_ISL_2724734, EPI_ISL_2724735, EPI_ISL_2724736, EPI_ISL_2724737, EPI_ISL_2724738, EPI_ISL_2724739, EPI_ISL_2724740, EPI_ISL_2724741, EPI_ISL_2724742, EPI_ISL_2724743, EPI_ISL_2724744, EPI_ISL_2724745, EPI_ISL_2724746, EPI_ISL_2724747, EPI_ISL_2724748, EPI_ISL_2724749, EPI_ISL_2724750, EPI_ISL_2724751, EPI_ISL_2724752, EPI_ISL_2724753, EPI_ISL_2724754, EPI_ISL_2724755, EPI_ISL_2724756, EPI_ISL_2724757, EPI_ISL_2724758, EPI_ISL_2724759, EPI_ISL_2724760, EPI_ISL_2724761, EPI_ISL_2724762, EPI_ISL_2724763, EPI_ISL_2724764, EPI_ISL_2724765, EPI_ISL_2724766, EPI_ISL_2724767, EPI_ISL_2724768, EPI_ISL_2724769, EPI_ISL_2724770, EPI_ISL_2724771, EPI_ISL_2724772, EPI_ISL_2724773, EPI_ISL_2724774, EPI_ISL_2724775, EPI_ISL_2724776, EPI_ISL_2724777, EPI_ISL_2724778, EPI_ISL_2724779, EPI_ISL_2724780, EPI_ISL_2724781, EPI_ISL_2724782, EPI_ISL_2724783, EPI_ISL_2724784, EPI_ISL_2724785, EPI_ISL_2724786, EPI_ISL_2724787, EPI_ISL_2724788, EPI_ISL_2724789, EPI_ISL_2724790, EPI_ISL_2724791, EPI_ISL_2724792, EPI_ISL_2724793, EPI_ISL_2724794, EPI_ISL_2724795, EPI_ISL_2724796, EPI_ISL_2724797, EPI_ISL_2724798, EPI_ISL_2724799, EPI_ISL_2724800, EPI_ISL_2724801, EPI_ISL_2724802, EPI_ISL_2724803, EPI_ISL_2724804, EPI_ISL_2724805, EPI_ISL_2724806, EPI_ISL_2724807, EPI_ISL_2724808, EPI_ISL_2724809, EPI_ISL_2724810, EPI_ISL_2724811, EPI_ISL_2724812, EPI_ISL_2724813, EPI_ISL_2724814, EPI_ISL_2724815, EPI_ISL_2724816, EPI_ISL_2724817, EPI_ISL_2724818, EPI_ISL_2724819, EPI_ISL_2724820, EPI_ISL_2724821, EPI_ISL_2724822, EPI_ISL_2724823, EPI_ISL_2724824, EPI_ISL_2724825, EPI_ISL_2724826, EPI_ISL_2724827, EPI_ISL_2724828, EPI_ISL_2724829, EPI_ISL_2724830, EPI_ISL_2724831, EPI_ISL_2724832, EPI_ISL_2724833, EPI_ISL_2724834, EPI_ISL_2724835, EPI_ISL_2724836, EPI_ISL_2724837, EPI_ISL_2724838, EPI_ISL_2724839, EPI_ISL_2724840, EPI_ISL_2724841, EPI_ISL_2724842, EPI_ISL_2724843, EPI_ISL_2724844, EPI_ISL_2724845, EPI_ISL_2724846, EPI_ISL_2724847, EPI_ISL_2724848, EPI_ISL_2724849, EPI_ISL_2724850, EPI_ISL_2724851, EPI_ISL_2724852, EPI_ISL_2724853, EPI_ISL_2724854, EPI_ISL_2724855, EPI_ISL_2724856, EPI_ISL_2724857, EPI_ISL_2724858, EPI_ISL_2724859, EPI_ISL_2724860, EPI_ISL_2724861, EPI_ISL_2724862, EPI_ISL_2724863, EPI_ISL_2724864, EPI_ISL_2724865, EPI_ISL_2724866, EPI_ISL_2724867, EPI_ISL_2724868, EPI_ISL_2724869, EPI_ISL_2724870, EPI_ISL_2724871, EPI_ISL_2724872, EPI_ISL_2724873, EPI_ISL_2724874, EPI_ISL_2724875, EPI_ISL_2724876, EPI_ISL_2724877, EPI_ISL_2724878, EPI_ISL_2724879, EPI_ISL_2724880, EPI_ISL_2724881, EPI_ISL_2724882, EPI_ISL_2724883, EPI_ISL_2724884, EPI_ISL_2724885, EPI_ISL_2724886, EPI_ISL_2724887, EPI_ISL_2724888, EPI_ISL_2724889, EPI_ISL_2724890, EPI_ISL_2724891, EPI_ISL_2724892, EPI_ISL_2724893, EPI_ISL_2724894, EPI_ISL_2724895, EPI_ISL_2724896, EPI_ISL_2724897, EPI_ISL_2724898, EPI_ISL_2724899, EPI_ISL_2724900, EPI_ISL_2724901, EPI_ISL_2724902, EPI_ISL_2724903, EPI_ISL_2724904, EPI_ISL_2724905, EPI_ISL_2724906, EPI_ISL_2724907, EPI_ISL_2724908, EPI_ISL_2724909, EPI_ISL_2724910, EPI_ISL_2724911, EPI_ISL_2724912 | | | |
| see above | State Laboratories Division, Hawaii State Department of Health | State Laboratories Division, Hawaii State Department of Health | Pamela O'Brien, Drew Kuwazaki, Ayana Garnet, Razvan Sultana, Edward Desmond |
| EPI_ISL_2725041, EPI_ISL_2725042, EPI_ISL_2725043, EPI_ISL_2725045, EPI_ISL_2725046, EPI_ISL_2725047, EPI_ISL_2725048, EPI_ISL_2725049, EPI_ISL_2725050, EPI_ISL_2725051, EPI_ISL_2725052, EPI_ISL_2725053, EPI_ISL_2725054, EPI_ISL_2725055, EPI_ISL_2725056, EPI_ISL_2725057, EPI_ISL_2725058, EPI_ISL_2725059, EPI_ISL_2725060, EPI_ISL_2725062, EPI_ISL_2725063, EPI_ISL_2725064, EPI_ISL_2725065, EPI_ISL_2725066, EPI_ISL_2725067, EPI_ISL_2725068, EPI_ISL_2725069, EPI_ISL_2725070, EPI_ISL_2725071 | | | |
| see above | IN State Department of Health Laboratory Services | IN State Department of Health Laboratory Services | Cassandra Campion, Jamie Yeadon, Brian Pope, Lixia Liu, Kyle Brownlee, Melissa Hindenlang, Mark Glazier |
| EPI_ISL_2725450, EPI_ISL_2725451 | Atlanta VA Medical Center | Genomics and Discovery, Respiratory Viruses Branch, Division of Viral Diseases, Centers for Disease Control and Prevention | Anna Kelleher, Ying Tao, Yan Li, Jing Zhang, Brian Lynch, Krista Queen, Anna Uehara, Peter Cook, Han Jia Justin Ng, Rachel Marine, Clinton R. Paden, Dhwani Batra, Haibin Wang, Tara Coalter, Jasmine Padilla, Morgan Davis, Mili Sheth, Sarah Nobles, Mark Burroughs, Justin Lee, Adam Retchless, Suxiang Tong |
| EPI_ISL_2725452, EPI_ISL_2725453, EPI_ISL_2725454, EPI_ISL_2725455, EPI_ISL_2725456, EPI_ISL_2725457, EPI_ISL_2725458, EPI_ISL_2725459, EPI_ISL_2725460, EPI_ISL_2725461, EPI_ISL_2725462, EPI_ISL_2725463, EPI_ISL_2725464, EPI_ISL_2725465, EPI_ISL_2725466, EPI_ISL_2725467, EPI_ISL_2725468, EPI_ISL_2725469, EPI_ISL_2725470, EPI_ISL_2725471, EPI_ISL_2725473, EPI_ISL_2725474, EPI_ISL_2725475, EPI_ISL_2725476, EPI_ISL_2725477, EPI_ISL_2725478, EPI_ISL_2725479, EPI_ISL_2725480, EPI_ISL_2725481, EPI_ISL_2725482, EPI_ISL_2725483 | | | |
| see above | DC Public Health Lab/ Dept. of Forensic Sciences | DC Public Health Lab/ Dept. of Forensic Sciences | Janis Doss, Scott Nguyen, Elizabeth Zelaya, Sarah Scott, Connie Maza, Monica Mann, Brittany Hamilton, David Payne, Jocelyn Hauser |
| EPI_ISL_2725517 | CA DPH Viral and Rickettsial Disease Laboratory | Chan-Zuckerberg Biohub | CZB Cliahub Consortium |
| EPI_ISL_2725518 | Santa Clara County Public Health Laboratory | Chan-Zuckerberg Biohub | CZB Cliahub Consortium |
| EPI_ISL_2725519 | Alameda County Public Health Lab | Chan-Zuckerberg Biohub | CZB Cliahub Consortium |
| EPI_ISL_2725520, EPI_ISL_2725521, EPI_ISL_2725522, EPI_ISL_2725523 | Contra Costa County Public Health Lab | Chan-Zuckerberg Biohub | CZB Cliahub Consortium |
| EPI_ISL_2725524 | Santa Clara County Public Health Laboratory | Chan-Zuckerberg Biohub | CZB Cliahub Consortium |
| EPI_ISL_2725558, EPI_ISL_2725559, EPI_ISL_2725560, EPI_ISL_2725561, EPI_ISL_2725562, EPI_ISL_2725563, EPI_ISL_2725564, EPI_ISL_2725565, EPI_ISL_2725566, EPI_ISL_2725567, EPI_ISL_2725568, EPI_ISL_2725569, EPI_ISL_2725570, EPI_ISL_2725571, EPI_ISL_2725572, EPI_ISL_2725573, EPI_ISL_2725574, EPI_ISL_2725575, EPI_ISL_2725576, EPI_ISL_2725577, EPI_ISL_2725578, EPI_ISL_2725579, | | | |

| | | | |
|---|---|---|---|
| EPI_ISL_2725580, EPI_ISL_2725581, EPI_ISL_2725582, EPI_ISL_2725583, EPI_ISL_2725584, EPI_ISL_2725585, EPI_ISL_2725586, EPI_ISL_2725587, EPI_ISL_2725588, EPI_ISL_2725589, EPI_ISL_2725590, EPI_ISL_2725591, EPI_ISL_2725592, EPI_ISL_2725593, EPI_ISL_2725594 | | | |
| see above | Rhode Island State Health Laboratory | Rhode Island State Health Laboratory | Ewa King, Richard C. Huard, Kristin Carpenter-Azevedo |
| EPI_ISL_2726533, EPI_ISL_2726534, EPI_ISL_2726535, EPI_ISL_2726536, EPI_ISL_2726537, EPI_ISL_2726538, EPI_ISL_2726539, EPI_ISL_2726540, EPI_ISL_2726541, EPI_ISL_2726544, EPI_ISL_2726545, EPI_ISL_2726547, EPI_ISL_2726548, EPI_ISL_2726549, EPI_ISL_2726550, EPI_ISL_2726551, EPI_ISL_2726552, EPI_ISL_2726553, EPI_ISL_2726554, EPI_ISL_2726555, EPI_ISL_2726556, EPI_ISL_2726557, EPI_ISL_2726558, EPI_ISL_2726559, EPI_ISL_2726560, EPI_ISL_2726561, EPI_ISL_2726562, EPI_ISL_2726563, EPI_ISL_2726564, EPI_ISL_2726565, EPI_ISL_2726566, EPI_ISL_2726567, EPI_ISL_2726568, EPI_ISL_2726569, EPI_ISL_2726571, EPI_ISL_2726574, EPI_ISL_2726575 | | | |
| see above | Wyoming Public Health Laboratory | Wyoming Public Health Laboratory | Jim Mildenberger, Taylor Fearing, Channing Weber, Ashley Norberg, Chayse Rowley, Marley Goetz, Brian Dominguez, Elliot Thomasson, Sam Britz, Cari Sloma, Robert Petit, and Rob Christensen |
| EPI_ISL_2726672, EPI_ISL_2726673, EPI_ISL_2726674, EPI_ISL_2726675, EPI_ISL_2726676, EPI_ISL_2726677, EPI_ISL_2726678, EPI_ISL_2726680, EPI_ISL_2726681, EPI_ISL_2726682, EPI_ISL_2726683, EPI_ISL_2726684, EPI_ISL_2726685, EPI_ISL_2726686, EPI_ISL_2726687, EPI_ISL_2726688, EPI_ISL_2726689, EPI_ISL_2726690, EPI_ISL_2726691, EPI_ISL_2726692, EPI_ISL_2726693, EPI_ISL_2726694, EPI_ISL_2726695, EPI_ISL_2726696 | | | |
| see above | Kansas Health and Environmental Lab | Kansas Health and Environmental Lab | Mike Grose, Katherine Wiggins, Jonathan Barnell, Ben Olsen, and Phil Adam |
| EPI_ISL_2726711, EPI_ISL_2726712, EPI_ISL_2726713, EPI_ISL_2726714, EPI_ISL_2726715, EPI_ISL_2726716, EPI_ISL_2726718, EPI_ISL_2726719, EPI_ISL_2726721, EPI_ISL_2726722, EPI_ISL_2726723, EPI_ISL_2726724, EPI_ISL_2726725, EPI_ISL_2726726, EPI_ISL_2726727, EPI_ISL_2726729, EPI_ISL_2726730, EPI_ISL_2726731, EPI_ISL_2726732, EPI_ISL_2726733, EPI_ISL_2726734, EPI_ISL_2726736, EPI_ISL_2726737, EPI_ISL_2726738, EPI_ISL_2726739, EPI_ISL_2726740, EPI_ISL_2726741, EPI_ISL_2726742, EPI_ISL_2726744, EPI_ISL_2726745, EPI_ISL_2726746, EPI_ISL_2726747, EPI_ISL_2726748, EPI_ISL_2726749, EPI_ISL_2726750, EPI_ISL_2726751, EPI_ISL_2726752, EPI_ISL_2726753, EPI_ISL_2726754, EPI_ISL_2726755, EPI_ISL_2726756, EPI_ISL_2726817, EPI_ISL_2726818, EPI_ISL_2726819, EPI_ISL_2726820, EPI_ISL_2726822, EPI_ISL_2726823, EPI_ISL_2726824, EPI_ISL_2726825, EPI_ISL_2726826, EPI_ISL_2726828, EPI_ISL_2726829, EPI_ISL_2726830, EPI_ISL_2726831, EPI_ISL_2726832, EPI_ISL_2726833, EPI_ISL_2726834, EPI_ISL_2726835, EPI_ISL_2726836, EPI_ISL_2726839, EPI_ISL_2726840, EPI_ISL_2726841, EPI_ISL_2726842, EPI_ISL_2726843, EPI_ISL_2726844, EPI_ISL_2726845, EPI_ISL_2726846, EPI_ISL_2726847, EPI_ISL_2726848, EPI_ISL_2726849, EPI_ISL_2726850, EPI_ISL_2726851, EPI_ISL_2726852, EPI_ISL_2726853 | | | |
| see above | Wyoming Public Health Laboratory | Wyoming Public Health Laboratory | Jim Mildenberger, Taylor Fearing, Channing Weber, Ashley Norberg, Chayse Rowley, Marley Goetz, Brian Dominguez, Elliot Thomasson, Sam Britz, Cari Sloma, Robert Petit, and Rob Christensen |
| EPI_ISL_2726930, EPI_ISL_2726931, EPI_ISL_2726932, EPI_ISL_2726933, EPI_ISL_2726934, EPI_ISL_2726935, EPI_ISL_2726936 | UW Virology Lab | UW Virology Lab | Pavitra Roychoudhury, Hong Xie, Lasata Shrestha, Shah Mohamed Bakhash, Tien V. Nguyen, Noah R. Baker, Sean Ellis, Meei-Li Huang, Keith R Jerome, Alexander Greninger |
| EPI_ISL_2726937, EPI_ISL_2726938, EPI_ISL_2726939, EPI_ISL_2726940, EPI_ISL_2726941, EPI_ISL_2726942, EPI_ISL_2726943, EPI_ISL_2726944 | ISDH | Quantigen Biosciences | Paul J. Childress |
| EPI_ISL_2726945, EPI_ISL_2726946, EPI_ISL_2726947, EPI_ISL_2726948, EPI_ISL_2726950, EPI_ISL_2726951, EPI_ISL_2726952, EPI_ISL_2726953, EPI_ISL_2726957, EPI_ISL_2726958, EPI_ISL_2726961, EPI_ISL_2726963, EPI_ISL_2726964, EPI_ISL_2726965, EPI_ISL_2726966 | | | |
| see above | Wyoming Public Health Laboratory | Wyoming Public Health Laboratory | Jim Mildenberger, Taylor Fearing, Channing Weber, Ashley Norberg, Chayse Rowley, Marley Goetz, Brian Dominguez, Elliot Thomasson, Sam Britz, Cari Sloma, Robert Petit, and Rob Christensen |
| EPI_ISL_2727055, EPI_ISL_2727056, EPI_ISL_2727057, EPI_ISL_2727058, EPI_ISL_2727059, EPI_ISL_2727060, EPI_ISL_2727061, EPI_ISL_2727062, EPI_ISL_2727063, EPI_ISL_2727064, EPI_ISL_2727065, EPI_ISL_2727066, EPI_ISL_2727067, EPI_ISL_2727068, EPI_ISL_2727069, EPI_ISL_2727070, EPI_ISL_2727071, EPI_ISL_2727072, EPI_ISL_2727073, EPI_ISL_2727074, EPI_ISL_2727075, EPI_ISL_2727076, EPI_ISL_2727077, EPI_ISL_2727078, EPI_ISL_2727079, EPI_ISL_2727080, EPI_ISL_2727081, EPI_ISL_2727082, EPI_ISL_2727083, EPI_ISL_2727084, EPI_ISL_2727085, EPI_ISL_2727086, EPI_ISL_2727087, EPI_ISL_2727088, EPI_ISL_2727089, EPI_ISL_2727090, EPI_ISL_2727091, EPI_ISL_2727092, EPI_ISL_2727093, EPI_ISL_2727094, EPI_ISL_2727095, EPI_ISL_2727096, EPI_ISL_2727097, EPI_ISL_2727098, EPI_ISL_2727099, EPI_ISL_2727100, EPI_ISL_2727101 | | | |
| see above | MD PHL | MD PHL | Maryland Department of Health Laboratories Administration |
| EPI_ISL_2727102 | URMC LABS | Wadsworth Center, New York State Department of Health | Kirsten St. George, Daryl M. Lamson, Alexis Russell, Matthew Shudt, Melissa A Leisner, Jonathan Plitnick, Catharine Prussing, Navjot Singh, John Kelly, Erasmus Schneider, Erica Lasek-Nesselquist |
| EPI_ISL_2727103 | WESTCHESTER MEDICAL CENTER | Wadsworth Center, New York State Department of Health | Kirsten St. George, Daryl M. Lamson, Alexis Russell, Matthew Shudt, Melissa A Leisner, Jonathan Plitnick, Catharine Prussing, Navjot Singh, John Kelly, Erasmus Schneider, Erica Lasek-Nesselquist |
| EPI_ISL_2727104 | ALBANY MEDICAL CENTER | Wadsworth Center, New York State Department of Health | Kirsten St. George, Daryl M. Lamson, Alexis Russell, Matthew Shudt, Melissa A Leisner, Jonathan Plitnick, Catharine Prussing, Navjot Singh, John Kelly, Erasmus Schneider, Erica Lasek-Nesselquist |
| EPI_ISL_2727105 | NORTHWELL HEALTH LABORATORIES | Wadsworth Center, New York State Department of Health | Kirsten St. George, Daryl M. Lamson, Alexis Russell, Matthew Shudt, Melissa A Leisner, Jonathan Plitnick, Catharine Prussing, Navjot Singh, John Kelly, Erasmus Schneider, Erica Lasek-Nesselquist |
| EPI_ISL_2727106, EPI_ISL_2727107, EPI_ISL_2727108, EPI_ISL_2727109 | URMC LABS | Wadsworth Center, New York State Department of Health | Kirsten St. George, Daryl M. Lamson, Alexis Russell, Matthew Shudt, Melissa A Leisner, Jonathan Plitnick, Catharine Prussing, Navjot Singh, John Kelly, Erasmus Schneider, Erica Lasek-Nesselquist |
| EPI_ISL_2727110 | MONTEFIORE MEDICAL CENTER LABORATORIES | Wadsworth Center, New York State Department of Health | Kirsten St. George, Daryl M. Lamson, Alexis Russell, Matthew Shudt, Melissa A Leisner, Jonathan Plitnick, Catharine Prussing, Navjot Singh, John Kelly, Erasmus Schneider, Erica Lasek-Nesselquist |
| EPI_ISL_2727111 | ALBANY MEDICAL CENTER | Wadsworth Center, New York State Department of Health | Kirsten St. George, Daryl M. Lamson, Alexis Russell, Matthew Shudt, Melissa A Leisner, Jonathan Plitnick, Catharine Prussing, Navjot Singh, John Kelly, Erasmus Schneider, Erica Lasek-Nesselquist |
| EPI_ISL_2727112 | TEMPUS LABS INC | Wadsworth Center, New York State Department of Health | Kirsten St. George, Daryl M. Lamson, Alexis Russell, Matthew Shudt, Melissa A Leisner, Jonathan Plitnick, Catharine Prussing, Navjot Singh, John Kelly, Erasmus Schneider, Erica Lasek-Nesselquist |
| EPI_ISL_2727113, EPI_ISL_2727114 | ALBANY MEDICAL CENTER | Wadsworth Center, New York State Department of Health | Kirsten St. George, Daryl M. Lamson, Alexis Russell, Matthew Shudt, Melissa A Leisner, Jonathan Plitnick, Catharine Prussing, Navjot Singh, John Kelly, Erasmus Schneider, Erica Lasek-Nesselquist |
| EPI_ISL_2727405, EPI_ISL_2727406, EPI_ISL_2727407, EPI_ISL_2727408, EPI_ISL_2727409, EPI_ISL_2727410, EPI_ISL_2727411, EPI_ISL_2727412, EPI_ISL_2727413, EPI_ISL_2727414, EPI_ISL_2727415, EPI_ISL_2727416, EPI_ISL_2727417, EPI_ISL_2727418, EPI_ISL_2727419, EPI_ISL_2727421, EPI_ISL_2727422, EPI_ISL_2727423, EPI_ISL_2727424, EPI_ISL_2727425, EPI_ISL_2727426, EPI_ISL_2727428, EPI_ISL_2727429, EPI_ISL_2727430, EPI_ISL_2727431, EPI_ISL_2727432, EPI_ISL_2727433, EPI_ISL_2727434, EPI_ISL_2727435, EPI_ISL_2727436, EPI_ISL_2727437, EPI_ISL_2727438, EPI_ISL_2727439, EPI_ISL_2727440, EPI_ISL_2727441, EPI_ISL_2727442 | | | |
| see above | Tampa General Hospital Esoteric Lab | Tampa General Hospital Esoteric Research & Development Lab | Amorce Lima, Jon Faughn, Deanna Becker, Suzane Silbert |
| EPI_ISL_2727443 | Atlas Genomics | Seattle Flu Study | Deborah A. Nickerson, Chris D. Frazar, Jover Lee, Benjamin Pelle, Erica Ryke, Matthew Richardson, Amanda Adler, Elisabeth Brandstetter, Peter D. Han, Kairsten Fay, Misja Ilcisin, Kirsten Lacombe, Thomas R. Sibley, Melissa Truong, Caitlin R. Wolf, Romesh Gautom, Geoff Melly, Brian Hiatt, Philip Dykema, Scott Lindquist, Michael Boeckh, Janet A. Englund, Michael Famulare, Barry R. Lutz, Mark J. Rieder, Lea M. Starita, Matthew Thompson, Helen Y. Chu, Jay Shendure, Trevor Bedford |
| EPI_ISL_2727444, EPI_ISL_2727445, EPI_ISL_2727446, EPI_ISL_2727447, EPI_ISL_2727448, EPI_ISL_2727450, EPI_ISL_2727452, EPI_ISL_2727453, EPI_ISL_2727454, EPI_ISL_2727455, EPI_ISL_2727456, EPI_ISL_2727458, EPI_ISL_2727461 | | | |
| see above | Wyoming Public Health Laboratory | Wyoming Public Health Laboratory | Jim Mildenberger, Taylor Fearing, Channing Weber, Ashley Norberg, Chayse Rowley, Marley Goetz, Brian Dominguez, Elliot Thomasson, Sam Britz, Cari Sloma, Robert Petit, and Rob Christensen |
| EPI_ISL_2727463, EPI_ISL_2727464, EPI_ISL_2727465, EPI_ISL_2727466, EPI_ISL_2727467, EPI_ISL_2727468, EPI_ISL_2727469, EPI_ISL_2727470, EPI_ISL_2727471, EPI_ISL_2727472, EPI_ISL_2727473, EPI_ISL_2727474, EPI_ISL_2727475, EPI_ISL_2727476, EPI_ISL_2727477, EPI_ISL_2727478, EPI_ISL_2727479, EPI_ISL_2727480, EPI_ISL_2727481, EPI_ISL_2727482, EPI_ISL_2727483, EPI_ISL_2727484, EPI_ISL_2727485, EPI_ISL_2727486, EPI_ISL_2727487, EPI_ISL_2727488, EPI_ISL_2727489, EPI_ISL_2727490, EPI_ISL_2727491, EPI_ISL_2727492, EPI_ISL_2727493, EPI_ISL_2727494, EPI_ISL_2727495, EPI_ISL_2727496, EPI_ISL_2727497, EPI_ISL_2727498 | | | |
| see above | Oregon State Public Health Laboratory | Oregon State Public Health Laboratory | Rafia Razzaque, Eugene Yeboah, Vanda Makris, Laura Tsaknaridis, John Fontana and Shane Sevey |
| EPI_ISL_2727522, EPI_ISL_2727523, EPI_ISL_2727524, EPI_ISL_2727525, EPI_ISL_2727526, EPI_ISL_2727527, EPI_ISL_2727528, EPI_ISL_2727529, EPI_ISL_2727530, EPI_ISL_2727531, EPI_ISL_2727532, EPI_ISL_2727533, EPI_ISL_2727534, EPI_ISL_2727535, EPI_ISL_2727536, EPI_ISL_2727537, EPI_ISL_2727538, EPI_ISL_2727539, EPI_ISL_2727540, EPI_ISL_2727541, EPI_ISL_2727542, EPI_ISL_2727543, EPI_ISL_2727544, EPI_ISL_2727545, EPI_ISL_2727546, EPI_ISL_2727547, EPI_ISL_2727548, EPI_ISL_2727549, EPI_ISL_2727550, EPI_ISL_2727551, EPI_ISL_2727552, EPI_ISL_2727553, EPI_ISL_2727554, EPI_ISL_2727555, EPI_ISL_2727556, EPI_ISL_2727557, EPI_ISL_2727558, EPI_ISL_2727559, EPI_ISL_2727560, EPI_ISL_2727561, EPI_ISL_2727562, EPI_ISL_2727563, EPI_ISL_2727564 | | | |
| see above | Tampa General Hospital Esoteric Lab | Tampa General Hospital Esoteric Research & Development Lab | Jon Faughn, Deanna Becker, Dominic Uy, Jorge Mecias-Frias, Elaine Vendrone, Vicki Healer, Amorce Lima, Suzane Silbert |
| EPI_ISL_2727570, EPI_ISL_2727571, EPI_ISL_2727572, EPI_ISL_2727573, EPI_ISL_2727574, EPI_ISL_2727575, EPI_ISL_2727577, EPI_ISL_2727579, EPI_ISL_2727580, EPI_ISL_2727581, EPI_ISL_2727582, EPI_ISL_2727585, EPI_ISL_2727586, EPI_ISL_2727587, EPI_ISL_2727588, EPI_ISL_2727589, EPI_ISL_2727590, EPI_ISL_2727591, EPI_ISL_2727592, EPI_ISL_2727593, EPI_ISL_2727594, EPI_ISL_2727595, EPI_ISL_2727596, EPI_ISL_2727598, EPI_ISL_2727599, EPI_ISL_2727600, EPI_ISL_2727601, EPI_ISL_2727602, EPI_ISL_2727603, EPI_ISL_2727604, EPI_ISL_2727605, EPI_ISL_2727606, EPI_ISL_2727607, EPI_ISL_2727608, EPI_ISL_2727609, EPI_ISL_2727610, EPI_ISL_2727611, EPI_ISL_2727612, EPI_ISL_2727613 | | | |
| see above | NJDOH, Public Health and Environmental Laboratories | NJ_PHEL | Lindsey Bodnar, Shiv K. Verma, Jacquelyn Deverell, Ryan Pachucki, Dana Woell, Allison Roder, Byeong Jeong |
| EPI_ISL_2727673, EPI_ISL_2727674, EPI_ISL_2727675, EPI_ISL_2727676, EPI_ISL_2727677, EPI_ISL_2727678, EPI_ISL_2727679, EPI_ISL_2727680, EPI_ISL_2727681, EPI_ISL_2727682, EPI_ISL_2727683, EPI_ISL_2727684, EPI_ISL_2727685, EPI_ISL_2727686, EPI_ISL_2727687, EPI_ISL_2727688, EPI_ISL_2727689, EPI_ISL_2727690, EPI_ISL_2727691, EPI_ISL_2727692, EPI_ISL_2727693, EPI_ISL_2727694, EPI_ISL_2727695, EPI_ISL_2727696, EPI_ISL_2727697, EPI_ISL_2727698, EPI_ISL_2727699, EPI_ISL_2727700, EPI_ISL_2727701, EPI_ISL_2727702, EPI_ISL_2727703, EPI_ISL_2727704, EPI_ISL_2727705, EPI_ISL_2727706, EPI_ISL_2727707, EPI_ISL_2727708, EPI_ISL_2727709, EPI_ISL_2727710, EPI_ISL_2727711, EPI_ISL_2727712, EPI_ISL_2727713, EPI_ISL_2727714, EPI_ISL_2727715, EPI_ISL_2727716, EPI_ISL_2727717, EPI_ISL_2727718, EPI_ISL_2727719, EPI_ISL_2727720, EPI_ISL_2727721, EPI_ISL_2727722, EPI_ISL_2727724, EPI_ISL_2727725, EPI_ISL_2727726, EPI_ISL_2727727, EPI_ISL_2727728, EPI_ISL_2727729, EPI_ISL_2727730, EPI_ISL_2727731, EPI_ISL_2727732, EPI_ISL_2727733, EPI_ISL_2727734, EPI_ISL_2727735, EPI_ISL_2727736, EPI_ISL_2727737, EPI_ISL_2727738, EPI_ISL_2727739, EPI_ISL_2727740, EPI_ISL_2727741, EPI_ISL_2727742, EPI_ISL_2727743, EPI_ISL_2727744, EPI_ISL_2727745, EPI_ISL_2727746, EPI_ISL_2727747, EPI_ISL_2727748, EPI_ISL_2727749, EPI_ISL_2727750, EPI_ISL_2727751, EPI_ISL_2727752, EPI_ISL_2727753, EPI_ISL_2727754, EPI_ISL_2727755, EPI_ISL_2727756, EPI_ISL_2727757, EPI_ISL_2727758, EPI_ISL_2727759, EPI_ISL_2727760, EPI_ISL_2727761, EPI_ISL_2727762, EPI_ISL_2727763, EPI_ISL_2727764, EPI_ISL_2727765, EPI_ISL_2727766, EPI_ISL_2727767, EPI_ISL_2727768, EPI_ISL_2727769, EPI_ISL_2727770, EPI_ISL_2727771, EPI_ISL_2727772, EPI_ISL_2727773, EPI_ISL_2727774, EPI_ISL_2727775, EPI_ISL_2727776, EPI_ISL_2727777, EPI_ISL_2727778, EPI_ISL_2727779, EPI_ISL_2727780, EPI_ISL_2727781, EPI_ISL_2727782, EPI_ISL_2727783, EPI_ISL_2727784, EPI_ISL_2727785, EPI_ISL_2727786, EPI_ISL_2727787, EPI_ISL_2727788, EPI_ISL_2727789, EPI_ISL_2727790, EPI_ISL_2727791, EPI_ISL_2727792, EPI_ISL_2727793, EPI_ISL_2727794, EPI_ISL_2727795, EPI_ISL_2727796, EPI_ISL_2727797, EPI_ISL_2727798, EPI_ISL_2727800, EPI_ISL_2727801, EPI_ISL_2727802, EPI_ISL_2727803, EPI_ISL_2727804, EPI_ISL_2727805, EPI_ISL_2727806, EPI_ISL_2727807, EPI_ISL_2727808, EPI_ISL_2727809, EPI_ISL_2727810, EPI_ISL_2727811, EPI_ISL_2727812, EPI_ISL_2727813, EPI_ISL_2727814, EPI_ISL_2727815, EPI_ISL_2727816, EPI_ISL_2727817, EPI_ISL_2727818, EPI_ISL_2727819, EPI_ISL_2727820, EPI_ISL_2727821, EPI_ISL_2727822, EPI_ISL_2727823, EPI_ISL_2727824, EPI_ISL_2727825, EPI_ISL_2727826, EPI_ISL_2727827, EPI_ISL_2727828, EPI_ISL_2727829, EPI_ISL_2727830, EPI_ISL_2727831, EPI_ISL_2727832, EPI_ISL_2727833, EPI_ISL_2727834 | | | |
| see above | Wisconsin State Laboratory of Hygiene Communicable Disease Division | Wisconsin State Laboratory of Hygiene Communicable Disease Division | Abigail C. Shockey, Alicia J. Mooney, Erika M. Hanson, Tonya Danz, Richard Griesser, Sara Wagner, Kelsey R. Florek |
| EPI_ISL_2727838, EPI_ISL_2727839, EPI_ISL_2727840, EPI_ISL_2727841, EPI_ISL_2727842, EPI_ISL_2727843, EPI_ISL_2727844, EPI_ISL_2727845, EPI_ISL_2727846, EPI_ISL_2727847, EPI_ISL_2727848, EPI_ISL_2727849, EPI_ISL_2727852, EPI_ISL_2727853, EPI_ISL_2727854, EPI_ISL_2727855, EPI_ISL_2727856, EPI_ISL_2727857 | | | |
| see above | Minnesota Department of Health, Public Health Laboratory | Minnesota Department of Health, Public Health Laboratory | Alexandra Lorentz, Jacob Garfin, Matt Plumb, and Xiong Wang |
| EPI_ISL_2727858, EPI_ISL_2727859, EPI_ISL_2727860, EPI_ISL_2727861, EPI_ISL_2727862, EPI_ISL_2727863, EPI_ISL_2727864, | M Health Fairview | Minnesota Department of Health, Public Health Laboratory | Alexandra Lorentz, Jacob Garfin, Matt Plumb, and Xiong Wang |

| | | | |
|---|---|---|---|
| EPI_ISL_2727865, EPI_ISL_2727866, EPI_ISL_2727867 | | | |
| EPI_ISL_2727868, EPI_ISL_2727869, EPI_ISL_2727870 | HealthPartners Central Lab | Minnesota Department of Health, Public Health Laboratory | Alexandra Lorentz, Jacob Garfin, Matt Plumb, and Xiong Wang |
| EPI_ISL_2727871, EPI_ISL_2727872, EPI_ISL_2727873 | Centracare Laboratory Services | Minnesota Department of Health, Public Health Laboratory | Alexandra Lorentz, Jacob Garfin, Matt Plumb, and Xiong Wang |
| EPI_ISL_2727929, EPI_ISL_2727930, EPI_ISL_2727932, EPI_ISL_2727934, EPI_ISL_2727935, EPI_ISL_2727936, EPI_ISL_2727937, EPI_ISL_2727938, EPI_ISL_2727939, EPI_ISL_2727941, EPI_ISL_2727942, EPI_ISL_2727943, EPI_ISL_2727944, EPI_ISL_2727945, EPI_ISL_2727946, EPI_ISL_2727947, EPI_ISL_2727949, EPI_ISL_2727950, EPI_ISL_2727952, EPI_ISL_2727953, EPI_ISL_2727954, EPI_ISL_2727955, EPI_ISL_2727956, EPI_ISL_2727957, EPI_ISL_2727958, EPI_ISL_2727959, EPI_ISL_2727960, EPI_ISL_2727961, EPI_ISL_2727962, EPI_ISL_2727963, EPI_ISL_2727964, EPI_ISL_2727965, EPI_ISL_2727966, EPI_ISL_2727967, EPI_ISL_2727968, EPI_ISL_2727969, EPI_ISL_2727970, EPI_ISL_2727971, EPI_ISL_2727972, EPI_ISL_2727973, EPI_ISL_2727975, EPI_ISL_2727977, EPI_ISL_2727978, EPI_ISL_2727981, EPI_ISL_2727982, EPI_ISL_2727983, EPI_ISL_2727987, EPI_ISL_2727988, EPI_ISL_2727989, EPI_ISL_2727991, EPI_ISL_2727992, EPI_ISL_2727993, EPI_ISL_2727994, EPI_ISL_2727995, EPI_ISL_2727997, EPI_ISL_2727999, EPI_ISL_2728000, EPI_ISL_2728001, EPI_ISL_2728002, EPI_ISL_2728003, EPI_ISL_2728004, EPI_ISL_2728005, EPI_ISL_2728006, EPI_ISL_2728007, EPI_ISL_2728008, EPI_ISL_2728009, EPI_ISL_2728010, EPI_ISL_2728011, EPI_ISL_2728012, EPI_ISL_2728013, EPI_ISL_2728014, EPI_ISL_2728015, EPI_ISL_2728016, EPI_ISL_2728017, EPI_ISL_2728018, EPI_ISL_2728019, EPI_ISL_2728020, EPI_ISL_2728021, EPI_ISL_2728022, EPI_ISL_2728023, EPI_ISL_2728024, EPI_ISL_2728025, EPI_ISL_2728026, EPI_ISL_2728027, EPI_ISL_2728028, EPI_ISL_2728030, EPI_ISL_2728031, EPI_ISL_2728032, EPI_ISL_2728033, EPI_ISL_2728034, EPI_ISL_2728035, EPI_ISL_2728036, EPI_ISL_2728037, EPI_ISL_2728038, EPI_ISL_2728039, EPI_ISL_2728040, EPI_ISL_2728041, EPI_ISL_2728042, EPI_ISL_2728043, EPI_ISL_2728044, EPI_ISL_2728045, EPI_ISL_2728046, EPI_ISL_2728047, EPI_ISL_2728048, EPI_ISL_2728049, EPI_ISL_2728050, EPI_ISL_2728051, EPI_ISL_2728052, EPI_ISL_2728053, EPI_ISL_2728054, EPI_ISL_2728055, EPI_ISL_2728056, EPI_ISL_2728057, EPI_ISL_2728058, EPI_ISL_2728059, EPI_ISL_2728060, EPI_ISL_2728061, EPI_ISL_2728062, EPI_ISL_2728063, EPI_ISL_2728064, EPI_ISL_2728065, EPI_ISL_2728066, EPI_ISL_2728067, EPI_ISL_2728068, EPI_ISL_2728069, EPI_ISL_2728070, EPI_ISL_2728071, EPI_ISL_2728072, EPI_ISL_2728073, EPI_ISL_2728074, EPI_ISL_2728075, EPI_ISL_2728076, EPI_ISL_2728077, EPI_ISL_2728078, EPI_ISL_2728079, EPI_ISL_2728080, EPI_ISL_2728081, EPI_ISL_2728082, EPI_ISL_2728083, EPI_ISL_2728084, EPI_ISL_2728085, EPI_ISL_2728086, EPI_ISL_2728087, EPI_ISL_2728088, EPI_ISL_2728089, EPI_ISL_2728090, EPI_ISL_2728091, EPI_ISL_2728092, EPI_ISL_2728093, EPI_ISL_2728094, EPI_ISL_2728095, EPI_ISL_2728096, EPI_ISL_2728097, EPI_ISL_2728098, EPI_ISL_2728099, EPI_ISL_2728100, EPI_ISL_2728101, EPI_ISL_2728102, EPI_ISL_2728103, EPI_ISL_2728104, EPI_ISL_2728105, EPI_ISL_2728106, EPI_ISL_2728109, EPI_ISL_2728110, EPI_ISL_2728112, EPI_ISL_2728114, EPI_ISL_2728115, EPI_ISL_2728116, EPI_ISL_2728117, EPI_ISL_2728118, EPI_ISL_2728119, EPI_ISL_2728120, EPI_ISL_2728121, EPI_ISL_2728122, EPI_ISL_2728123, EPI_ISL_2728124, EPI_ISL_2728125, EPI_ISL_2728127, EPI_ISL_2728128, EPI_ISL_2728130, EPI_ISL_2728131, EPI_ISL_2728133, EPI_ISL_2728135, EPI_ISL_2728136, EPI_ISL_2728137, EPI_ISL_2728138, EPI_ISL_2728139, EPI_ISL_2728140, EPI_ISL_2728141, EPI_ISL_2728143, EPI_ISL_2728144, EPI_ISL_2728146, EPI_ISL_2728149, EPI_ISL_2728154, EPI_ISL_2728155, EPI_ISL_2728156, EPI_ISL_2728157, EPI_ISL_2728160, EPI_ISL_2728162, EPI_ISL_2728163, EPI_ISL_2728166, EPI_ISL_2728168, EPI_ISL_2728169, EPI_ISL_2728171, EPI_ISL_2728174, EPI_ISL_2728175, EPI_ISL_2728176, EPI_ISL_2728177, EPI_ISL_2728178, EPI_ISL_2728179, EPI_ISL_2728180, EPI_ISL_2728182, EPI_ISL_2728183, EPI_ISL_2728185, EPI_ISL_2728186, EPI_ISL_2728187, EPI_ISL_2728189, EPI_ISL_2728190, EPI_ISL_2728193, EPI_ISL_2728194, EPI_ISL_2728195, EPI_ISL_2728196, EPI_ISL_2728197, EPI_ISL_2728198, EPI_ISL_2728201, EPI_ISL_2728202, EPI_ISL_2728203, EPI_ISL_2728204, EPI_ISL_2728205, EPI_ISL_2728206, EPI_ISL_2728207, EPI_ISL_2728210, EPI_ISL_2728211, EPI_ISL_2728213, EPI_ISL_2728214, EPI_ISL_2728215, EPI_ISL_2728216, EPI_ISL_2728218, EPI_ISL_2728219, EPI_ISL_2728220, EPI_ISL_2728221, EPI_ISL_2728222, EPI_ISL_2728223, EPI_ISL_2728224, EPI_ISL_2728225, EPI_ISL_2728226, EPI_ISL_2728227, EPI_ISL_2728228, EPI_ISL_2728229 | | | |
| see above | Wyoming Public Health Laboratory | Wyoming Public Health Laboratory | Jim Mildenberger, Taylor Fearing, Channing Weber, Ashley Norberg, Chayse Rowley, Marley Goetz, Brian Dominguez, Elliot Thomasson, Sam Britz, Cari Sloma, Robert Petit, and Rob Christensen |
| EPI_ISL_2731412, EPI_ISL_2731413, EPI_ISL_2731414, EPI_ISL_2731415, EPI_ISL_2731416, EPI_ISL_2731417, EPI_ISL_2731418, EPI_ISL_2731419, EPI_ISL_2731420, EPI_ISL_2731421, EPI_ISL_2731422, EPI_ISL_2731423, EPI_ISL_2731424, EPI_ISL_2731425, EPI_ISL_2731426, EPI_ISL_2731427, EPI_ISL_2731428, EPI_ISL_2731429, EPI_ISL_2731430, EPI_ISL_2731431, EPI_ISL_2731432, EPI_ISL_2731433, EPI_ISL_2731434, EPI_ISL_2731435, EPI_ISL_2731436, EPI_ISL_2731437 | | | |
| see above | Montana Public Health Laboratory | Montana Public Health Laboratory | Joy Ritter, Michelle Mozer, Carrie Biskupiak, Deborah Gibson, Michael Dills |
| EPI_ISL_2735848, EPI_ISL_2735850 | Seattle Flu Study | Seattle Flu Study | Deborah A. Nickerson, Chris D. Frazar, Jover Lee, Benjamin Pelle, Erica Ryke, Matthew Richardson, Amanda Adler, Elisabeth Brandstetter, Peter D. Han, Kairsten Fay, Misja Ilcisin, Kirsten Lacombe, Thomas R. Sibley, Melissa Truong, Caitlin R. Wolf, Michael Boeckh, Janet A. Englund, Michael Famulare, Barry R. Lutz, Mark J. Rieder, Lea M. Starita, Matthew Thompson, Jay Shendure, Trevor Bedford, Helen Y. Chu |
| EPI_ISL_2735852, EPI_ISL_2735854, EPI_ISL_2735856, EPI_ISL_2735858, EPI_ISL_2735860, EPI_ISL_2735861, EPI_ISL_2735863, EPI_ISL_2735865, EPI_ISL_2735867, EPI_ISL_2735869, EPI_ISL_2735871, EPI_ISL_2735873, EPI_ISL_2735874 | | | |
| see above | Seattle Flu Study | Seattle Flu Study | Deborah A. Nickerson, Chris D. Frazar, Jover Lee, Benjamin Pelle, Erica Ryke, Matthew Richardson, Amanda Adler, Elisabeth Brandstetter, Peter D. Han, Kairsten Fay, Misja Ilcisin, Kirsten Lacombe, Thomas R. Sibley, Melissa Truong, Caitlin R. Wolf, Karen Cowgill, Stephanie Schrag, Jeff Duchin, Michael Boeckh, Janet A. Englund, Michael Famulare, Barry R. Lutz, Mark J. Rieder, Lea M. Starita, Matthew Thompson, Helen Y. Chu, Trevor Bedford, Jay Shendure |
| EPI_ISL_2735878 | Seattle Flu Study | Seattle Flu Study | Deborah A. Nickerson, Chris D. Frazar, Jover Lee, Benjamin Pelle, Erica Ryke, Matthew Richardson, Amanda Adler, Elisabeth Brandstetter, Peter D. Han, Kairsten Fay, Misja Ilcisin, Kirsten Lacombe, Thomas R. Sibley, Melissa Truong, Caitlin R. Wolf, Michael Boeckh, Janet A. Englund, Michael Famulare, Barry R. Lutz, Mark J. Rieder, Lea M. Starita, Matthew Thompson, Jay Shendure, Trevor Bedford, Helen Y. Chu |
| EPI_ISL_2735880, EPI_ISL_2735882 | Seattle Flu Study | Seattle Flu Study | Deborah A. Nickerson, Chris D. Frazar, Jover Lee, Benjamin Pelle, Erica Ryke, Matthew Richardson, Amanda Adler, Elisabeth Brandstetter, Peter D. Han, Kairsten Fay, Misja Ilcisin, Kirsten Lacombe, Thomas R. Sibley, Melissa Truong, Caitlin R. Wolf, Karen Cowgill, Stephanie Schrag, Jeff Duchin, Michael Boeckh, Janet A. Englund, Michael Famulare, Barry R. Lutz, Mark J. Rieder, Lea M. Starita, Matthew Thompson, Helen Y. Chu, Trevor Bedford, Jay Shendure |
| EPI_ISL_2735884, EPI_ISL_2735886, EPI_ISL_2735887, EPI_ISL_2735891 | Seattle Flu Study | Seattle Flu Study | Deborah A. Nickerson, Chris D. Frazar, Jover Lee, Benjamin Pelle, Erica Ryke, Matthew Richardson, Amanda Adler, Elisabeth Brandstetter, Peter D. Han, Kairsten Fay, Misja Ilcisin, Kirsten Lacombe, Thomas R. Sibley, Melissa Truong, Caitlin R. Wolf, Michael Boeckh, Janet A. Englund, Michael Famulare, Barry R. Lutz, Mark J. Rieder, Lea M. Starita, Matthew Thompson, Jay Shendure, Trevor Bedford, Helen Y. Chu |
| EPI_ISL_2735895, EPI_ISL_2735897, EPI_ISL_2735899, EPI_ISL_2735900, EPI_ISL_2735902, EPI_ISL_2735904, EPI_ISL_2735906, EPI_ISL_2735908, EPI_ISL_2735910, EPI_ISL_2735911, EPI_ISL_2735913, EPI_ISL_2735915, EPI_ISL_2735916, EPI_ISL_2735918, EPI_ISL_2735924, EPI_ISL_2735926, EPI_ISL_2735928, EPI_ISL_2735929, EPI_ISL_2735931, EPI_ISL_2735933, EPI_ISL_2735935, EPI_ISL_2735936, EPI_ISL_2735938, EPI_ISL_2735940, EPI_ISL_2735941, EPI_ISL_2735943 | | | |
| see above | Seattle Flu Study | Seattle Flu Study | Deborah A. Nickerson, Chris D. Frazar, Jover Lee, Benjamin Pelle, Erica Ryke, Matthew Richardson, Amanda Adler, Elisabeth Brandstetter, Peter D. Han, Kairsten Fay, Misja Ilcisin, Kirsten Lacombe, Thomas R. Sibley, Melissa Truong, Caitlin R. Wolf, Karen Cowgill, Stephanie Schrag, Jeff Duchin, Michael Boeckh, Janet A. Englund, Michael Famulare, Barry R. Lutz, Mark J. Rieder, Lea M. Starita, Matthew Thompson, Helen Y. Chu, Trevor Bedford, Jay Shendure |
| EPI_ISL_2735945, EPI_ISL_2735947, EPI_ISL_2735949, EPI_ISL_2735950, EPI_ISL_2735952, EPI_ISL_2735954, EPI_ISL_2735955, EPI_ISL_2735958, EPI_ISL_2735959, EPI_ISL_2735962, EPI_ISL_2735963, EPI_ISL_2735965, EPI_ISL_2735967, EPI_ISL_2735969, EPI_ISL_2735971, EPI_ISL_2735972, EPI_ISL_2735975, EPI_ISL_2735976, EPI_ISL_2735978, EPI_ISL_2735980, EPI_ISL_2735982, EPI_ISL_2735984, EPI_ISL_2735985, EPI_ISL_2735987, EPI_ISL_2735991, EPI_ISL_2735992, EPI_ISL_2735994, EPI_ISL_2735996, EPI_ISL_2735998, EPI_ISL_2735999, EPI_ISL_2736001, EPI_ISL_2736003, EPI_ISL_2736005, EPI_ISL_2736006 | | | |
| see above | Incyte Diagnostics Spokane | Seattle Flu Study | Deborah A. Nickerson, Chris D. Frazar, Jover Lee, Benjamin Pelle, Erica Ryke, Matthew Richardson, Amanda Adler, Elisabeth Brandstetter, Peter D. Han, Kairsten Fay, Misja Ilcisin, Kirsten Lacombe, Thomas R. Sibley, Melissa Truong, Caitlin R. Wolf, Romesh Gautom, Geoff Melly, Brian Hiatt, Philip Dykema, Scott Lindquist, Michael Boeckh, Janet A. Englund, Michael Famulare, Barry R. Lutz, Mark J. Rieder, Lea M. Starita, Matthew Thompson, Helen Y. Chu, Jay Shendure, Trevor Bedford |
| EPI_ISL_2736010, EPI_ISL_2736012 | Seattle Flu Study | Seattle Flu Study | Deborah A. Nickerson, Chris D. Frazar, Jover Lee, Benjamin Pelle, Erica Ryke, Matthew Richardson, Amanda Adler, Elisabeth Brandstetter, Peter D. Han, Kairsten Fay, Misja Ilcisin, Kirsten Lacombe, Thomas R. Sibley, Melissa Truong, Caitlin R. Wolf, Karen Cowgill, Stephanie Schrag, Jeff Duchin, Michael Boeckh, Janet A. Englund, Michael Famulare, Barry R. Lutz, Mark J. Rieder, Lea M. Starita, Matthew Thompson, Helen Y. Chu, Trevor Bedford, Jay Shendure |
| EPI_ISL_2736013, EPI_ISL_2736015, EPI_ISL_2736017, EPI_ISL_2736019, EPI_ISL_2736021, EPI_ISL_2736022, EPI_ISL_2736024, EPI_ISL_2736026, EPI_ISL_2736028, EPI_ISL_2736029, EPI_ISL_2736031, EPI_ISL_2736033, EPI_ISL_2736040, EPI_ISL_2736044, EPI_ISL_2736051, EPI_ISL_2736055, EPI_ISL_2736058, EPI_ISL_2736060, EPI_ISL_2736064, EPI_ISL_2736065, EPI_ISL_2736070, EPI_ISL_2736073, EPI_ISL_2736077, EPI_ISL_2736079, EPI_ISL_2736080, EPI_ISL_2736081, EPI_ISL_2736083, EPI_ISL_2736086, EPI_ISL_2736088, EPI_ISL_2736090, EPI_ISL_2736091, EPI_ISL_2736095, EPI_ISL_2736098, EPI_ISL_2736101, EPI_ISL_2736106, EPI_ISL_2736108, EPI_ISL_2736109, EPI_ISL_2736111, EPI_ISL_2736115, EPI_ISL_2736118, EPI_ISL_2736125, EPI_ISL_2736128, EPI_ISL_2736131, EPI_ISL_2736134, EPI_ISL_2736137, EPI_ISL_2736141, EPI_ISL_2736144, EPI_ISL_2736147, EPI_ISL_2736150, EPI_ISL_2736154, EPI_ISL_2736157, EPI_ISL_2736160, EPI_ISL_2736162, EPI_ISL_2736163, EPI_ISL_2736165, EPI_ISL_2736169, EPI_ISL_2736172, EPI_ISL_2736174, EPI_ISL_2736176, EPI_ISL_2736179, EPI_ISL_2736182, EPI_ISL_2736186, EPI_ISL_2736189, EPI_ISL_2736192, EPI_ISL_2736196, EPI_ISL_2736198, EPI_ISL_2736199, EPI_ISL_2736201, EPI_ISL_2736205, EPI_ISL_2736207, EPI_ISL_2736208, EPI_ISL_2736210, EPI_ISL_2736212, EPI_ISL_2736214 | | | |
| see above | Northwest Laboratory | Seattle Flu Study | Deborah A. Nickerson, Chris D. Frazar, Jover Lee, Benjamin Pelle, Erica Ryke, Matthew Richardson, Amanda Adler, Elisabeth Brandstetter, Peter D. Han, Kairsten Fay, Misja Ilcisin, Kirsten Lacombe, Thomas R. Sibley, Melissa Truong, Caitlin R. Wolf, Romesh Gautom, Geoff Melly, Brian Hiatt, Philip Dykema, Scott Lindquist, Michael Boeckh, Janet A. Englund, Michael Famulare, Barry R. Lutz, Mark J. Rieder, Lea M. Starita, Matthew Thompson, Helen Y. Chu, Jay Shendure, Trevor Bedford |
| EPI_ISL_2736218, EPI_ISL_2736219, EPI_ISL_2736223, EPI_ISL_2736227, EPI_ISL_2736230, EPI_ISL_2736233, EPI_ISL_2736235, EPI_ISL_2736237, EPI_ISL_2736239, EPI_ISL_2736242, EPI_ISL_2736246, EPI_ISL_2736249, EPI_ISL_2736253, EPI_ISL_2736256, EPI_ISL_2736259, EPI_ISL_2736266, EPI_ISL_2736269, EPI_ISL_2736273, EPI_ISL_2736276, EPI_ISL_2736280, EPI_ISL_2736285, EPI_ISL_2736287, EPI_ISL_2736289, EPI_ISL_2736291, EPI_ISL_2736294, EPI_ISL_2736298, EPI_ISL_2736301, EPI_ISL_2736304, EPI_ISL_2736307, EPI_ISL_2736310, EPI_ISL_2736314, EPI_ISL_2736317, EPI_ISL_2736320, EPI_ISL_2736324, EPI_ISL_2736328, EPI_ISL_2736330, EPI_ISL_2736333, EPI_ISL_2736336 | | | |
| see above | Atlas Genomics | Seattle Flu Study | Deborah A. Nickerson, Chris D. Frazar, Jover Lee, Benjamin Pelle, Erica Ryke, Matthew Richardson, Amanda Adler, Elisabeth Brandstetter, Peter D. Han, Kairsten Fay, Misja Ilcisin, Kirsten Lacombe, Thomas R. Sibley, Melissa Truong, Caitlin R. Wolf, Romesh Gautom, Geoff Melly, Brian Hiatt, Philip Dykema, Scott Lindquist, Michael Boeckh, Janet A. Englund, Michael Famulare, Barry R. Lutz, Mark J. Rieder, Lea M. Starita, Matthew Thompson, Helen Y. Chu, Jay Shendure, Trevor Bedford |
| EPI_ISL_2736339, EPI_ISL_2736341, EPI_ISL_2736343, EPI_ISL_2736345, EPI_ISL_2736347, EPI_ISL_2736349, EPI_ISL_2736350, EPI_ISL_2736353, EPI_ISL_2736355, EPI_ISL_2736357, EPI_ISL_2736362, EPI_ISL_2736366, EPI_ISL_2736367, EPI_ISL_2736371, EPI_ISL_2736373, EPI_ISL_2736375, EPI_ISL_2736376, EPI_ISL_2736380, EPI_ISL_2736383, EPI_ISL_2736388, EPI_ISL_2736391, EPI_ISL_2736394, EPI_ISL_2736397, EPI_ISL_2736402, EPI_ISL_2736405, EPI_ISL_2736408, EPI_ISL_2736412, EPI_ISL_2736415, EPI_ISL_2736418, EPI_ISL_2736422, EPI_ISL_2736425, EPI_ISL_2736433, EPI_ISL_2736435, EPI_ISL_2736437, EPI_ISL_2736438, EPI_ISL_2736441, EPI_ISL_2736444, EPI_ISL_2736447, EPI_ISL_2736453, EPI_ISL_2736456, EPI_ISL_2736457, EPI_ISL_2736459, EPI_ISL_2736461, EPI_ISL_2736463, EPI_ISL_2736465, EPI_ISL_2736467, EPI_ISL_2736468, EPI_ISL_2736470, EPI_ISL_2736472, EPI_ISL_2736474, EPI_ISL_2736477, EPI_ISL_2736479, EPI_ISL_2736481, EPI_ISL_2736484, EPI_ISL_2736486, EPI_ISL_2736488, EPI_ISL_2736490, EPI_ISL_2736491, EPI_ISL_2736493, EPI_ISL_2736495, EPI_ISL_2736497, EPI_ISL_2736499, EPI_ISL_2736500, EPI_ISL_2736502, EPI_ISL_2736506, EPI_ISL_2736509, EPI_ISL_2736511, EPI_ISL_2736513, EPI_ISL_2736514, EPI_ISL_2736516, EPI_ISL_2736520, EPI_ISL_2736522, EPI_ISL_2736524, EPI_ISL_2736525, EPI_ISL_2736527, EPI_ISL_2736529, EPI_ISL_2736531, EPI_ISL_2736533, EPI_ISL_2736535, EPI_ISL_2736536, EPI_ISL_2736538, EPI_ISL_2736540, EPI_ISL_2736542, EPI_ISL_2736544, EPI_ISL_2736546, EPI_ISL_2736547, EPI_ISL_2736551, EPI_ISL_2736554, EPI_ISL_2736558, EPI_ISL_2736561, EPI_ISL_2736563, EPI_ISL_2736565, EPI_ISL_2736567, EPI_ISL_2736569, EPI_ISL_2736571, EPI_ISL_2736573, EPI_ISL_2736574, EPI_ISL_2736576 | | | |
| see above | Seattle Flu Study | Seattle Flu Study | Deborah A. Nickerson, Chris D. Frazar, Jover Lee, Benjamin Pelle, Erica Ryke, Matthew Richardson, Amanda Adler, Elisabeth Brandstetter, Peter D. Han, Kairsten Fay, Misja Ilcisin, Kirsten Lacombe, Thomas R. Sibley, Melissa Truong, Caitlin R. Wolf, Michael Boeckh, Janet A. Englund, Michael Famulare, Barry R. Lutz, Mark J. Rieder, Lea M. Starita, Matthew Thompson, Jay Shendure, Trevor Bedford, Helen Y. Chu |
| EPI_ISL_2736780, EPI_ISL_2736781, EPI_ISL_2736783 | LESP Sinaloa | Instituto de Diagnostico y Referencia Epidemiologicos (INDRE) | Claudia Wong-Arambula, Abril Rodriguez-Maldonado, Vanessa Rivero-Arredondo, Ariadna Medina-Benitez, Joaquin Quiroz-Mercado, Sergio Rangel-Guerrero, Natividad Cruz-Ortiz, Tatiana Nunez-Garcia, Gisela Barrera-Badillo, Lucia Hernandez-Rivas, Irma Lopez-Martinez, Ernesto Ramirez-Gonzalez. |
| EPI_ISL_2736784 | LESP Michoacan | Instituto de Diagnostico y Referencia Epidemiologicos (INDRE) | Claudia Wong-Arambula, Abril Rodriguez-Maldonado, Vanessa Rivero-Arredondo, Ariadna Medina-Benitez, Joaquin Quiroz-Mercado, Sergio Rangel-Guerrero, Natividad Cruz-Ortiz, Tatiana Nunez-Garcia, Gisela Barrera-Badillo, Lucia Hernandez-Rivas, Irma Lopez-Martinez, Ernesto Ramirez-Gonzalez. |
| EPI_ISL_2736785 | LESP Puebla | Instituto de Diagnostico y Referencia Epidemiologicos (INDRE) | Claudia Wong-Arambula, Abril Rodriguez-Maldonado, Vanessa Rivero-Arredondo, Ariadna Medina-Benitez, Joaquin Quiroz-Mercado, Sergio Rangel-Guerrero, Natividad Cruz-Ortiz, Tatiana Nunez-Garcia, Gisela Barrera-Badillo, Lucia Hernandez-Rivas, Irma Lopez-Martinez, Ernesto Ramirez-Gonzalez. |
| EPI_ISL_2736787 | LESP Guanajuato | Instituto de Diagnostico y Referencia Epidemiologicos (INDRE) | Claudia Wong-Arambula, Abril Rodriguez-Maldonado, Vanessa Rivero-Arredondo, Ariadna Medina-Benitez, Joaquin Quiroz-Mercado, Sergio Rangel-Guerrero, Natividad Cruz-Ortiz, Tatiana Nunez-Garcia, Gisela Barrera-Badillo, Lucia Hernandez-Rivas, Irma Lopez-Martinez, Ernesto Ramirez-Gonzalez. |
| EPI_ISL_2736790, EPI_ISL_2736792, EPI_ISL_2736795 | LESP Tabasco | Instituto de Diagnostico y Referencia Epidemiologicos (INDRE) | Claudia Wong-Arambula, Abril Rodriguez-Maldonado, Vanessa Rivero-Arredondo, Ariadna Medina-Benitez, Joaquin Quiroz-Mercado, Sergio Rangel-Guerrero, Natividad Cruz-Ortiz, Tatiana Nunez-Garcia, Gisela Barrera-Badillo, Lucia Hernandez-Rivas, Irma Lopez-Martinez, Ernesto Ramirez-Gonzalez. |
| EPI_ISL_2736797 | LESP Campeche | Instituto de Diagnostico y Referencia Epidemiologicos (INDRE) | Claudia Wong-Arambula, Abril Rodriguez-Maldonado, Vanessa Rivero-Arredondo, Ariadna Medina-Benitez, Joaquin Quiroz-Mercado, Sergio Rangel-Guerrero, Natividad Cruz-Ortiz, Tatiana Nunez-Garcia, Gisela Barrera-Badillo, Lucia Hernandez-Rivas, Irma Lopez-Martinez, Ernesto Ramirez-Gonzalez. |
| EPI_ISL_2736800, EPI_ISL_2736802, EPI_ISL_2736804, EPI_ISL_2736807, EPI_ISL_2736810, EPI_ISL_2736811, EPI_ISL_2736813, EPI_ISL_2736815, EPI_ISL_2736817, EPI_ISL_2736819, EPI_ISL_2736821, EPI_ISL_2736823 | | | |
| see above | LESP Tabasco | Instituto de Diagnostico y Referencia Epidemiologicos (INDRE) | Claudia Wong-Arambula, Abril Rodriguez-Maldonado, Vanessa Rivero-Arredondo, Ariadna Medina-Benitez, Joaquin Quiroz-Mercado, Sergio Rangel-Guerrero, Natividad Cruz-Ortiz, Tatiana Nunez-Garcia, Gisela Barrera-Badillo, Lucia Hernandez-Rivas, Irma Lopez-Martinez, Ernesto Ramirez-Gonzalez. |
| EPI_ISL_2736825 | LESP Michoacan | Instituto de Diagnostico y Referencia Epidemiologicos (INDRE) | Claudia Wong-Arambula, Abril Rodriguez-Maldonado, Vanessa Rivero-Arredondo, Ariadna Medina-Benitez, Joaquin Quiroz-Mercado, Sergio Rangel-Guerrero, Natividad Cruz-Ortiz, Tatiana Nunez-Garcia, Gisela Barrera-Badillo, Lucia Hernandez-Rivas, Irma Lopez-Martinez, Ernesto Ramirez-Gonzalez. |
| EPI_ISL_2736827 | LESP Guanajuato | Instituto de Diagnostico y Referencia Epidemiologicos (INDRE) | Claudia Wong-Arambula, Abril Rodriguez-Maldonado, Vanessa Rivero-Arredondo, Ariadna Medina-Benitez, Joaquin Quiroz-Mercado, Sergio Rangel-Guerrero, Natividad Cruz-Ortiz, Tatiana Nunez-Garcia, Gisela Barrera-Badillo, Lucia Hernandez-Rivas, Irma Lopez-Martinez, Ernesto Ramirez-Gonzalez. |
| EPI_ISL_2736829 | LESP Coahuila | Instituto de Diagnostico y Referencia Epidemiologicos (INDRE) | Claudia Wong-Arambula, Abril Rodriguez-Maldonado, Vanessa Rivero-Arredondo, Ariadna Medina-Benitez, Joaquin Quiroz-Mercado, Sergio Rangel-Guerrero, Natividad Cruz-Ortiz, Tatiana Nunez-Garcia, Gisela Barrera-Badillo, Lucia Hernandez-Rivas, Irma Lopez-Martinez, Ernesto Ramirez-Gonzalez. |
| EPI_ISL_2736832 | LESP Baja California | Instituto de Diagnostico y Referencia Epidemiologicos (INDRE) | Claudia Wong-Arambula, Abril Rodriguez-Maldonado, Vanessa Rivero-Arredondo, Ariadna Medina-Benitez, Joaquin Quiroz-Mercado, Sergio Rangel-Guerrero, Natividad Cruz-Ortiz, Tatiana Nunez-Garcia, Gisela Barrera-Badillo, Lucia Hernandez-Rivas, Irma Lopez-Martinez, Ernesto Ramirez-Gonzalez. |
| EPI_ISL_2736834 | LESP Michoacan | Instituto de Diagnostico y Referencia Epidemiologicos (INDRE) | Claudia Wong-Arambula, Abril Rodriguez-Maldonado, Vanessa Rivero-Arredondo, Ariadna Medina-Benitez, Joaquin Quiroz-Mercado, Sergio Rangel-Guerrero, Natividad Cruz-Ortiz, Tatiana Nunez-Garcia, Gisela Barrera-Badillo, Lucia Hernandez-Rivas, Irma Lopez-Martinez, Ernesto Ramirez-Gonzalez. |
| EPI_ISL_2736837, | LESP Tabasco | Instituto de Diagnostico y Referencia | Claudia Wong-Arambula, Abril Rodriguez-Maldonado, Vanessa Rivero-Arredondo, Ariadna Medina-Benitez, Joaquin Quiroz-Mercado, Sergio Rangel-Guerrero, Natividad Cruz-Ortiz, Tatiana Nunez-Garcia, Gisela Barrera-Badillo, Lucia Hernandez-Rivas, Irma Lopez-Martinez, Ernesto |

| | | | |
|---|---|---|---|
| EPI_ISL_2736839 | | Epidemiologicos (INDRE) | Ramirez-Gonzalez. |
| EPI_ISL_2736841 | LESP Guerrero | Instituto de Diagnostico y Referencia Epidemiologicos (INDRE) | Claudia Wong-Arambula, Abril Rodriguez-Maldonado, Vanessa Rivero-Arredondo, Ariadna Medina-Benitez, Joaquin Quiroz-Mercado, Sergio Rangel-Guerrero, Natividad Cruz-Ortiz, Tatiana Nunez-Garcia, Gisela Barrera-Badillo, Lucia Hernandez-Rivas, Irma Lopez-Martinez, Ernesto Ramirez-Gonzalez. |
| EPI_ISL_2736845 | LESP Yucatan | Instituto de Diagnostico y Referencia Epidemiologicos (INDRE) | Claudia Wong-Arambula, Abril Rodriguez-Maldonado, Vanessa Rivero-Arredondo, Ariadna Medina-Benitez, Joaquin Quiroz-Mercado, Sergio Rangel-Guerrero, Natividad Cruz-Ortiz, Tatiana Nunez-Garcia, Gisela Barrera-Badillo, Lucia Hernandez-Rivas, Irma Lopez-Martinez, Ernesto Ramirez-Gonzalez. |
| EPI_ISL_2736848, EPI_ISL_2736849, EPI_ISL_2736851 | LESP Michoacan | Instituto de Diagnostico y Referencia Epidemiologicos (INDRE) | Claudia Wong-Arambula, Abril Rodriguez-Maldonado, Vanessa Rivero-Arredondo, Ariadna Medina-Benitez, Joaquin Quiroz-Mercado, Sergio Rangel-Guerrero, Natividad Cruz-Ortiz, Tatiana Nunez-Garcia, Gisela Barrera-Badillo, Lucia Hernandez-Rivas, Irma Lopez-Martinez, Ernesto Ramirez-Gonzalez. |
| EPI_ISL_2736853 | LESP Tabasco | Instituto de Diagnostico y Referencia Epidemiologicos (INDRE) | Claudia Wong-Arambula, Abril Rodriguez-Maldonado, Vanessa Rivero-Arredondo, Ariadna Medina-Benitez, Joaquin Quiroz-Mercado, Sergio Rangel-Guerrero, Natividad Cruz-Ortiz, Tatiana Nunez-Garcia, Gisela Barrera-Badillo, Lucia Hernandez-Rivas, Irma Lopez-Martinez, Ernesto Ramirez-Gonzalez. |
| EPI_ISL_2736856 | LESP Sonora | Instituto de Diagnostico y Referencia Epidemiologicos (INDRE) | Claudia Wong-Arambula, Abril Rodriguez-Maldonado, Vanessa Rivero-Arredondo, Ariadna Medina-Benitez, Joaquin Quiroz-Mercado, Sergio Rangel-Guerrero, Natividad Cruz-Ortiz, Tatiana Nunez-Garcia, Gisela Barrera-Badillo, Lucia Hernandez-Rivas, Irma Lopez-Martinez, Ernesto Ramirez-Gonzalez. |
| EPI_ISL_2736857 | LESP Campeche | Instituto de Diagnostico y Referencia Epidemiologicos (INDRE) | Claudia Wong-Arambula, Abril Rodriguez-Maldonado, Vanessa Rivero-Arredondo, Ariadna Medina-Benitez, Joaquin Quiroz-Mercado, Sergio Rangel-Guerrero, Natividad Cruz-Ortiz, Tatiana Nunez-Garcia, Gisela Barrera-Badillo, Lucia Hernandez-Rivas, Irma Lopez-Martinez, Ernesto Ramirez-Gonzalez. |
| EPI_ISL_2736859, EPI_ISL_2736861, EPI_ISL_2736864 | LESP Guanajuato | Instituto de Diagnostico y Referencia Epidemiologicos (INDRE) | Claudia Wong-Arambula, Abril Rodriguez-Maldonado, Vanessa Rivero-Arredondo, Ariadna Medina-Benitez, Joaquin Quiroz-Mercado, Sergio Rangel-Guerrero, Natividad Cruz-Ortiz, Tatiana Nunez-Garcia, Gisela Barrera-Badillo, Lucia Hernandez-Rivas, Irma Lopez-Martinez, Ernesto Ramirez-Gonzalez. |
| EPI_ISL_2738387, EPI_ISL_2738389, EPI_ISL_2738391, EPI_ISL_2738393, EPI_ISL_2738395, EPI_ISL_2738397, EPI_ISL_2738398, EPI_ISL_2738400, EPI_ISL_2738401, EPI_ISL_2738402, EPI_ISL_2738403, EPI_ISL_2738404, EPI_ISL_2738406, EPI_ISL_2738407, EPI_ISL_2738409, EPI_ISL_2738411, EPI_ISL_2738413, EPI_ISL_2738415, EPI_ISL_2738417, EPI_ISL_2738419, EPI_ISL_2738420, EPI_ISL_2738422, EPI_ISL_2738424, EPI_ISL_2738426, EPI_ISL_2738428, EPI_ISL_2738430, EPI_ISL_2738431, EPI_ISL_2738433, EPI_ISL_2738435, EPI_ISL_2738437, EPI_ISL_2738441, EPI_ISL_2738444, EPI_ISL_2738447, EPI_ISL_2738450, EPI_ISL_2738454, EPI_ISL_2738458, EPI_ISL_2738462, EPI_ISL_2738464, EPI_ISL_2738466, EPI_ISL_2738469, EPI_ISL_2738471, EPI_ISL_2738473, EPI_ISL_2738475, EPI_ISL_2738478, EPI_ISL_2738480, EPI_ISL_2738482, EPI_ISL_2738484, EPI_ISL_2738485, EPI_ISL_2738486, EPI_ISL_2738488, EPI_ISL_2738490, EPI_ISL_2738492, EPI_ISL_2738493, EPI_ISL_2738495, EPI_ISL_2738497, EPI_ISL_2738499, EPI_ISL_2738504, EPI_ISL_2738508, EPI_ISL_2738510, EPI_ISL_2738512, EPI_ISL_2738513, EPI_ISL_2738515, EPI_ISL_2738517, EPI_ISL_2738519, EPI_ISL_2738520, EPI_ISL_2738525, EPI_ISL_2738528, EPI_ISL_2738532, EPI_ISL_2738536, EPI_ISL_2738540, EPI_ISL_2738541, EPI_ISL_2738543, EPI_ISL_2738545, EPI_ISL_2738548, EPI_ISL_2738552, EPI_ISL_2738553, EPI_ISL_2738555, EPI_ISL_2738557, EPI_ISL_2738559, EPI_ISL_2738561, EPI_ISL_2738563, EPI_ISL_2738565, EPI_ISL_2738567, EPI_ISL_2738571, EPI_ISL_2738574, EPI_ISL_2738578, EPI_ISL_2738581, EPI_ISL_2738583, EPI_ISL_2738585, EPI_ISL_2738587, EPI_ISL_2738589, EPI_ISL_2738591, EPI_ISL_2738593, EPI_ISL_2738595, EPI_ISL_2738597, EPI_ISL_2738599, EPI_ISL_2738603, EPI_ISL_2738608, EPI_ISL_2738611, EPI_ISL_2738614, EPI_ISL_2738618, EPI_ISL_2738622, EPI_ISL_2738626, EPI_ISL_2738629 | | | |
| see above | Hackensack Medical Center | New York Genome Center | Michael Zody, Andre Corvelo, Dayna M. Oschwald, Samantha Fennessey, Tom Maniatis, Liang Chen, Jose Mediavilla, Marcus Cunningham, Kaelea Composto, Kar Chow, David Perlin, Barry Kreiswirth |
| EPI_ISL_2747112, EPI_ISL_2747989, EPI_ISL_2748655, EPI_ISL_2749366, EPI_ISL_2751263, EPI_ISL_2751588, EPI_ISL_2752169, EPI_ISL_2753486 | Ohio Department of Health Laboratory | Ohio Department of Health Laboratory | Holmes, Jennifer; Eric Brandt, Keoni Omura, Glen McGillivary, Caitlin McDonnell, Jade Mowery, Stephanie Mccracken, Tyler Payne, Kirtana Ramadugu, Erica Leasure, Brent Lee, Kelsey Florek, Heather Blankenship, Allison Black, Quanta Brown, and Tammy Bannerman |
| EPI_ISL_2753615, EPI_ISL_2753616, EPI_ISL_2753617, EPI_ISL_2753618, EPI_ISL_2753619, EPI_ISL_2753620, EPI_ISL_2753621, EPI_ISL_2753623, EPI_ISL_2753624, EPI_ISL_2753625, EPI_ISL_2753627, EPI_ISL_2753628, EPI_ISL_2753629, EPI_ISL_2753630, EPI_ISL_2753631, EPI_ISL_2753632, EPI_ISL_2753633, EPI_ISL_2753634, EPI_ISL_2753635, EPI_ISL_2753636, EPI_ISL_2753637, EPI_ISL_2753638, EPI_ISL_2753639, EPI_ISL_2753640, EPI_ISL_2753641, EPI_ISL_2753642, EPI_ISL_2753643, EPI_ISL_2753644, EPI_ISL_2753645, EPI_ISL_2753646, EPI_ISL_2753647, EPI_ISL_2753648, EPI_ISL_2753649, EPI_ISL_2753650, EPI_ISL_2753651, EPI_ISL_2753652, EPI_ISL_2753653, EPI_ISL_2753654, EPI_ISL_2753655, EPI_ISL_2753656, EPI_ISL_2753657, EPI_ISL_2753658, EPI_ISL_2753659, EPI_ISL_2753660, EPI_ISL_2753661, EPI_ISL_2753662, EPI_ISL_2753663, EPI_ISL_2753664, EPI_ISL_2753665, EPI_ISL_2753666, EPI_ISL_2753667, EPI_ISL_2753668, EPI_ISL_2753669, EPI_ISL_2753671, EPI_ISL_2753673, EPI_ISL_2753674, EPI_ISL_2753676, EPI_ISL_2753677, EPI_ISL_2753678, EPI_ISL_2753679, EPI_ISL_2753680, EPI_ISL_2753681, EPI_ISL_2753682, EPI_ISL_2753683, EPI_ISL_2753684, EPI_ISL_2753685, EPI_ISL_2753686, EPI_ISL_2753688, EPI_ISL_2753689, EPI_ISL_2753690, EPI_ISL_2753691, EPI_ISL_2753692, EPI_ISL_2753693, EPI_ISL_2753694, EPI_ISL_2753695, EPI_ISL_2753696, EPI_ISL_2753697, EPI_ISL_2753698, EPI_ISL_2753699, EPI_ISL_2753700, EPI_ISL_2753701, EPI_ISL_2753702, EPI_ISL_2753703, EPI_ISL_2753704, EPI_ISL_2753705, EPI_ISL_2753706, EPI_ISL_2753707, EPI_ISL_2753708, EPI_ISL_2753709, EPI_ISL_2753710, EPI_ISL_2753711, EPI_ISL_2753712, EPI_ISL_2753713, EPI_ISL_2753715, EPI_ISL_2753716, EPI_ISL_2753717, EPI_ISL_2753718, EPI_ISL_2753719, EPI_ISL_2753720, EPI_ISL_2753721, EPI_ISL_2753723, EPI_ISL_2753724, EPI_ISL_2753726, EPI_ISL_2753727, EPI_ISL_2753728, EPI_ISL_2753729, EPI_ISL_2753730, EPI_ISL_2753731, EPI_ISL_2753732, EPI_ISL_2753733, EPI_ISL_2753734, EPI_ISL_2753735, EPI_ISL_2753736, EPI_ISL_2753737, EPI_ISL_2753738, EPI_ISL_2753739, EPI_ISL_2753740, EPI_ISL_2753742, EPI_ISL_2753743, EPI_ISL_2753745, EPI_ISL_2753746, EPI_ISL_2753747, EPI_ISL_2753748, EPI_ISL_2753749, EPI_ISL_2753751, EPI_ISL_2753752, EPI_ISL_2753753, EPI_ISL_2753755, EPI_ISL_2753756, EPI_ISL_2753757, EPI_ISL_2753758, EPI_ISL_2753759, EPI_ISL_2753761, EPI_ISL_2753762, EPI_ISL_2753764, EPI_ISL_2753765, EPI_ISL_2753766, EPI_ISL_2753767, EPI_ISL_2753768, EPI_ISL_2753769, EPI_ISL_2753770, EPI_ISL_2753771, EPI_ISL_2753772, EPI_ISL_2753774, EPI_ISL_2753775, EPI_ISL_2753776, EPI_ISL_2753777, EPI_ISL_2753778, EPI_ISL_2753779, EPI_ISL_2753780, EPI_ISL_2753781, EPI_ISL_2753782, EPI_ISL_2753783, EPI_ISL_2753784, EPI_ISL_2753786, EPI_ISL_2753788, EPI_ISL_2753789, EPI_ISL_2753791, EPI_ISL_2753792, EPI_ISL_2753793, EPI_ISL_2753794, EPI_ISL_2753795, EPI_ISL_2753796, EPI_ISL_2753798, EPI_ISL_2753799, EPI_ISL_2753800, EPI_ISL_2753801, EPI_ISL_2753802, EPI_ISL_2753803, EPI_ISL_2753804, EPI_ISL_2753805, EPI_ISL_2753806, EPI_ISL_2753807, EPI_ISL_2753808, EPI_ISL_2753810, EPI_ISL_2753811, EPI_ISL_2753812, EPI_ISL_2753813, EPI_ISL_2753814, EPI_ISL_2753815, EPI_ISL_2753816, EPI_ISL_2753817, EPI_ISL_2753818, EPI_ISL_2753819, EPI_ISL_2753820, EPI_ISL_2753821, EPI_ISL_2753822, EPI_ISL_2753823, EPI_ISL_2753824, EPI_ISL_2753825, EPI_ISL_2753826, EPI_ISL_2753827, EPI_ISL_2753828, EPI_ISL_2753829, EPI_ISL_2753830, EPI_ISL_2753831, EPI_ISL_2753832, EPI_ISL_2753833, EPI_ISL_2753834, EPI_ISL_2753835, EPI_ISL_2753836, EPI_ISL_2753837, EPI_ISL_2753838, EPI_ISL_2753839, EPI_ISL_2753840, EPI_ISL_2753841, EPI_ISL_2753842, EPI_ISL_2753843, EPI_ISL_2753844, EPI_ISL_2753845, EPI_ISL_2753846 | | | |
| see above | Missouri State Public Health Laboratory | Missouri State Public Health Laboratory | Matthew Sinn, Joshua Barry, Ashley New |
| EPI_ISL_2753847, EPI_ISL_2753851, EPI_ISL_2753852, EPI_ISL_2753853, EPI_ISL_2753856, EPI_ISL_2753857, EPI_ISL_2753858, EPI_ISL_2753859, EPI_ISL_2753860, EPI_ISL_2753861, EPI_ISL_2753862, EPI_ISL_2753863, EPI_ISL_2753865, EPI_ISL_2753866, EPI_ISL_2753867, EPI_ISL_2753876, EPI_ISL_2753877, EPI_ISL_2753879, EPI_ISL_2753880, EPI_ISL_2753881, EPI_ISL_2753882, EPI_ISL_2753883, EPI_ISL_2753884, EPI_ISL_2753886, EPI_ISL_2753887, EPI_ISL_2753888 | | | |
| see above | Wyoming Public Health Laboratory | Wyoming Public Health Laboratory | Jim Mildenberger, Taylor Fearing, Channing Weber, Ashley Norberg, Chayse Rowley, Marley Goetz, Brian Dominguez, Elliot Thomasson, Sam Britz, Cari Sloma, Robert Petit, and Rob Christensen |
| EPI_ISL_2753940, EPI_ISL_2753941, EPI_ISL_2753942, EPI_ISL_2753943, EPI_ISL_2753944, EPI_ISL_2753945, EPI_ISL_2753947, EPI_ISL_2753948, EPI_ISL_2753949, EPI_ISL_2753950, EPI_ISL_2753951 | | | |
| see above | NYU Langone Health | Departments of Pathology and Medicine, New York University School of Medicine | Adriana Heguy, Dacia Dimartino, Emily Guzman, Christian Marier, Peter Meyn, Sitharam Ramaswami, Gael Westby, Paul Zappile, Yutong Zhang, Paolo Cotzia, Guiqing Wang |
| EPI_ISL_2753958, EPI_ISL_2753961, EPI_ISL_2753965 | Ohio Department of Health Laboratory | Ohio Department of Health Laboratory | Holmes, Jennifer; Eric Brandt, Keoni Omura, Glen McGillivary, Caitlin McDonnell, Jade Mowery, Stephanie Mccracken, Tyler Payne, Kirtana Ramadugu, Erica Leasure, Brent Lee, Kelsey Florek, Heather Blankenship, Allison Black, Quanta Brown, and Tammy Bannerman |
| EPI_ISL_2753968, EPI_ISL_2753969, EPI_ISL_2753970, EPI_ISL_2753971, EPI_ISL_2753972 | Kaiser San Jose | Santa Clara County Public Health Laboratory | Santa Clara County Public Health Department |
| EPI_ISL_2753974 | PAMF Palo Alto | Santa Clara County Public Health Laboratory | Santa Clara County Public Health Department |
| EPI_ISL_2753975 | El Camino Hospital | Santa Clara County Public Health Laboratory | Santa Clara County Public Health Department |
| EPI_ISL_2753976 | Santa Clara Valley Medical Center | Santa Clara County Public Health Laboratory | Santa Clara County Public Health Department |
| EPI_ISL_2753978, EPI_ISL_2753979 | Kaiser Santa Clara | Santa Clara County Public Health Laboratory | Santa Clara County Public Health Department |
| EPI_ISL_2753981, EPI_ISL_2753982 | Kaiser San Jose | Santa Clara County Public Health Laboratory | Santa Clara County Public Health Department |
| EPI_ISL_2753983, EPI_ISL_2753985, EPI_ISL_2753986, EPI_ISL_2753987, EPI_ISL_2753988, EPI_ISL_2753989 | PAMF Palo Alto | Santa Clara County Public Health Laboratory | Santa Clara County Public Health Department |
| EPI_ISL_2753991, EPI_ISL_2753992, EPI_ISL_2753993, EPI_ISL_2753994, EPI_ISL_2753997, EPI_ISL_2753999, EPI_ISL_2754000, EPI_ISL_2754001, EPI_ISL_2754002, EPI_ISL_2754004, EPI_ISL_2754005, EPI_ISL_2754006, EPI_ISL_2754007, EPI_ISL_2754008, EPI_ISL_2754010, EPI_ISL_2754012, EPI_ISL_2754016, EPI_ISL_2754017, EPI_ISL_2754018, EPI_ISL_2754019, EPI_ISL_2754020 | | | |
| see above | Kaiser Regional | Santa Clara County Public Health Laboratory | Santa Clara County Public Health Department |
| EPI_ISL_2754022 | Good Samaritan Hospital | Santa Clara County Public Health Laboratory | Santa Clara County Public Health Department |
| EPI_ISL_2754036, EPI_ISL_2754037, EPI_ISL_2754038, EPI_ISL_2754039, EPI_ISL_2754040, EPI_ISL_2754042 | Monterey County Public Health Laboratory | Monterey County Public Health Laboratory | Monterey County Public Health Laboratory |
| EPI_ISL_2754045, EPI_ISL_2754046, EPI_ISL_2754047, EPI_ISL_2754048, EPI_ISL_2754049, EPI_ISL_2754050, EPI_ISL_2754051, EPI_ISL_2754052, EPI_ISL_2754055, EPI_ISL_2754056, EPI_ISL_2754057, EPI_ISL_2754058, EPI_ISL_2754059, EPI_ISL_2754060, EPI_ISL_2754061, EPI_ISL_2754062, EPI_ISL_2754064, EPI_ISL_2754065, EPI_ISL_2754066, EPI_ISL_2754067, EPI_ISL_2754068, EPI_ISL_2754069, EPI_ISL_2754070, EPI_ISL_2754071 | | | |
| see above | Wyoming Public Health Laboratory | Wyoming Public Health Laboratory | Jim Mildenberger, Taylor Fearing, Channing Weber, Ashley Norberg, Chayse Rowley, Marley Goetz, Brian Dominguez, Elliot Thomasson, Sam Britz, Cari Sloma, Robert Petit, and Rob Christensen |
| EPI_ISL_2754074, EPI_ISL_2754075, EPI_ISL_2754076, EPI_ISL_2754077, EPI_ISL_2754078, EPI_ISL_2754079, EPI_ISL_2754080, EPI_ISL_2754081, EPI_ISL_2754082, EPI_ISL_2754083, EPI_ISL_2754084, EPI_ISL_2754085, EPI_ISL_2754086, EPI_ISL_2754087, EPI_ISL_2754088, EPI_ISL_2754089, EPI_ISL_2754090, EPI_ISL_2754091, EPI_ISL_2754092, EPI_ISL_2754093, EPI_ISL_2754094 | | | |
| see above | Nebraska Public Health Laboratory | NPHL COVID-19 Response Team | NPHL COVID-19 Response Team |
| EPI_ISL_2754097, EPI_ISL_2754098 | Kansas Health and Environmental Lab | Wichita State University - Molecular Diagnostics Lab | Katherine Wiggins, Mike Grose, Jonathan Barnell, Ben Olsen, and Phil Adam |
| EPI_ISL_2754099, EPI_ISL_2754100, EPI_ISL_2754101, EPI_ISL_2754102, EPI_ISL_2754104, EPI_ISL_2754105, EPI_ISL_2754106, EPI_ISL_2754107, EPI_ISL_2754108, EPI_ISL_2754109, EPI_ISL_2754111, EPI_ISL_2754112, EPI_ISL_2754113, EPI_ISL_2754114 | | | |
| see above | Kansas Health and Environmental Lab | Clinical Reference Laboratory | Katherine Wiggins, Mike Grose, Jonathan Barnell, Ben Olsen, and Phil Adam |
| EPI_ISL_2754204 | Tampa General Hospital Esoteric Lab | Tampa General Hospital Esoteric Research & Development Lab | Jon Faughn, Deanna Becker, Dominic Uy, Jorge Mecias-Frias, Elaine Vendrone, Vicki Healer, Amorce Lima, Suzane Silbert |
| EPI_ISL_2754296, EPI_ISL_2754297, EPI_ISL_2754298, EPI_ISL_2754299, EPI_ISL_2754300, EPI_ISL_2754301, EPI_ISL_2754302, EPI_ISL_2754303, EPI_ISL_2754304, EPI_ISL_2754305, EPI_ISL_2754306, EPI_ISL_2754307, EPI_ISL_2754308, EPI_ISL_2754309 | | | |
| see above | Arizona State Public Health Laboratory | Arizona State Public Health Laboratory | Trung Huynh, Jessica Escobar, Katherine Fullerton, Nobuko Fukushima, Matthew Contursi, Stacy White, Linda Getsinger, Victor Waddell |
| EPI_ISL_2754803, | Ohio Department of Health Laboratory | Ohio Department of Health Laboratory | Holmes, Jennifer; Eric Brandt, Keoni Omura, Glen McGillivary, Caitlin McDonnell, Jade Mowery, Stephanie Mccracken, Tyler Payne, Kirtana Ramadugu, Erica Leasure, Brent Lee, Kelsey Florek, Heather Blankenship, Allison Black, Quanta Brown, and Tammy Bannerman |

| | | | |
|---|---|---|---|
| EPI_ISL_2754855 | | | |
| EPI_ISL_2754857, EPI_ISL_2754858, EPI_ISL_2754859, EPI_ISL_2754862, EPI_ISL_2754863, EPI_ISL_2754864, EPI_ISL_2754865, EPI_ISL_2754866, EPI_ISL_2754867, EPI_ISL_2754868, EPI_ISL_2754869, EPI_ISL_2754870, EPI_ISL_2754871, EPI_ISL_2754872, EPI_ISL_2754873, EPI_ISL_2754874, EPI_ISL_2754875, EPI_ISL_2754876, EPI_ISL_2754877, EPI_ISL_2754878, EPI_ISL_2754879, EPI_ISL_2754880, EPI_ISL_2754881, EPI_ISL_2754882, EPI_ISL_2754883, EPI_ISL_2754884, EPI_ISL_2754885, EPI_ISL_2754886, EPI_ISL_2754887, EPI_ISL_2754888, EPI_ISL_2754890, EPI_ISL_2754891, EPI_ISL_2754892, EPI_ISL_2754893, EPI_ISL_2754894, EPI_ISL_2754895, EPI_ISL_2754896, EPI_ISL_2754897, EPI_ISL_2754898, EPI_ISL_2754899, EPI_ISL_2754900, EPI_ISL_2754901, EPI_ISL_2754902 | | | |
| see above | Nebraska Public Health Laboratory | NPHL COVID-19 Response Team | NPHL COVID-19 Response Team |
| EPI_ISL_2754904, EPI_ISL_2754905 | LA Office of Public Health Laboratories | Genomics and Discovery, Respiratory Viruses Branch, Division of Viral Diseases, Centers for Disease Control and Prevention | Anna Kelleher, Ying Tao, Yan Li, Jing Zhang, Brian Lynch, Krista Queen, Anna Uehara, Peter Cook, Han Jia Justin Ng, Rachel Marine, Clinton R. Paden, Dhwani Batra, Haibin Wang, Tara Coalter, Jasmine Padilla, Morgan Davis, Mili Sheth, Sarah Nobles, Mark Burroughs, Justin Lee, Adam Retchless, Suxiang Tong |
| EPI_ISL_2754906 | Ohio Department of Health Laboratory | Ohio Department of Health Laboratory | Holmes, Jennifer; Eric Brandt, Keoni Omura, Glen McGillivary, Caitlin McDonnell, Jade Mowery, Stephanie Mccracken, Tyler Payne, Kirtana Ramadugu, Erica Leasure, Brent Lee, Kelsey Florek, Heather Blankenship, Allison Black, Quanta Brown, and Tammy Bannerman |
| EPI_ISL_2754907, EPI_ISL_2754908 | LA Office of Public Health Laboratories | Genomics and Discovery, Respiratory Viruses Branch, Division of Viral Diseases, Centers for Disease Control and Prevention | Anna Kelleher, Ying Tao, Yan Li, Jing Zhang, Brian Lynch, Krista Queen, Anna Uehara, Peter Cook, Han Jia Justin Ng, Rachel Marine, Clinton R. Paden, Dhwani Batra, Haibin Wang, Tara Coalter, Jasmine Padilla, Morgan Davis, Mili Sheth, Sarah Nobles, Mark Burroughs, Justin Lee, Adam Retchless, Suxiang Tong |
| EPI_ISL_2754909, EPI_ISL_2754910, EPI_ISL_2754911 | GA Department of Public Health Laboratory | Genomics and Discovery, Respiratory Viruses Branch, Division of Viral Diseases, Centers for Disease Control and Prevention | Anna Kelleher, Ying Tao, Yan Li, Jing Zhang, Brian Lynch, Krista Queen, Anna Uehara, Peter Cook, Han Jia Justin Ng, Rachel Marine, Clinton R. Paden, Dhwani Batra, Haibin Wang, Tara Coalter, Jasmine Padilla, Morgan Davis, Mili Sheth, Sarah Nobles, Mark Burroughs, Justin Lee, Adam Retchless, Suxiang Tong |
| EPI_ISL_2754912 | VA-Division of Consolidated Laboratory Services | Genomics and Discovery, Respiratory Viruses Branch, Division of Viral Diseases, Centers for Disease Control and Prevention | Anna Kelleher, Ying Tao, Yan Li, Jing Zhang, Brian Lynch, Krista Queen, Anna Uehara, Peter Cook, Han Jia Justin Ng, Rachel Marine, Clinton R. Paden, Dhwani Batra, Haibin Wang, Tara Coalter, Jasmine Padilla, Morgan Davis, Mili Sheth, Sarah Nobles, Mark Burroughs, Justin Lee, Adam Retchless, Suxiang Tong |
| EPI_ISL_2754913 | AR Dept. of Health-PHL, Molecular Diagnostics | Genomics and Discovery, Respiratory Viruses Branch, Division of Viral Diseases, Centers for Disease Control and Prevention | Anna Kelleher, Ying Tao, Yan Li, Jing Zhang, Brian Lynch, Krista Queen, Anna Uehara, Peter Cook, Han Jia Justin Ng, Rachel Marine, Clinton R. Paden, Dhwani Batra, Haibin Wang, Tara Coalter, Jasmine Padilla, Morgan Davis, Mili Sheth, Sarah Nobles, Mark Burroughs, Justin Lee, Adam Retchless, Suxiang Tong |
| EPI_ISL_2754914, EPI_ISL_2754915 | NJ Public Health and Environmental Laboratories | Genomics and Discovery, Respiratory Viruses Branch, Division of Viral Diseases, Centers for Disease Control and Prevention | Anna Kelleher, Ying Tao, Yan Li, Jing Zhang, Brian Lynch, Krista Queen, Anna Uehara, Peter Cook, Han Jia Justin Ng, Rachel Marine, Clinton R. Paden, Dhwani Batra, Haibin Wang, Tara Coalter, Jasmine Padilla, Morgan Davis, Mili Sheth, Sarah Nobles, Mark Burroughs, Justin Lee, Adam Retchless, Suxiang Tong |
| EPI_ISL_2754916 | NC State Laboratory of Public Health | Genomics and Discovery, Respiratory Viruses Branch, Division of Viral Diseases, Centers for Disease Control and Prevention | Anna Kelleher, Ying Tao, Yan Li, Jing Zhang, Brian Lynch, Krista Queen, Anna Uehara, Peter Cook, Han Jia Justin Ng, Rachel Marine, Clinton R. Paden, Dhwani Batra, Haibin Wang, Tara Coalter, Jasmine Padilla, Morgan Davis, Mili Sheth, Sarah Nobles, Mark Burroughs, Justin Lee, Adam Retchless, Suxiang Tong |
| EPI_ISL_2754917, EPI_ISL_2754918, EPI_ISL_2754919, EPI_ISL_2754920, EPI_ISL_2754921 | GA Department of Public Health Laboratory | Genomics and Discovery, Respiratory Viruses Branch, Division of Viral Diseases, Centers for Disease Control and Prevention | Anna Kelleher, Ying Tao, Yan Li, Jing Zhang, Brian Lynch, Krista Queen, Anna Uehara, Peter Cook, Han Jia Justin Ng, Rachel Marine, Clinton R. Paden, Dhwani Batra, Haibin Wang, Tara Coalter, Jasmine Padilla, Morgan Davis, Mili Sheth, Sarah Nobles, Mark Burroughs, Justin Lee, Adam Retchless, Suxiang Tong |
| EPI_ISL_2754922, EPI_ISL_2754923, EPI_ISL_2754924, EPI_ISL_2754925, EPI_ISL_2754926, EPI_ISL_2754927, EPI_ISL_2754928, EPI_ISL_2754929, EPI_ISL_2754930, EPI_ISL_2754931, EPI_ISL_2754932, EPI_ISL_2754933, EPI_ISL_2754934, EPI_ISL_2754935, EPI_ISL_2754936, EPI_ISL_2754937, EPI_ISL_2754938, EPI_ISL_2754939, EPI_ISL_2754940, EPI_ISL_2754941, EPI_ISL_2754942, EPI_ISL_2754943, EPI_ISL_2754944, EPI_ISL_2754945, EPI_ISL_2754946, EPI_ISL_2754947, EPI_ISL_2754948, EPI_ISL_2754949, EPI_ISL_2754950, EPI_ISL_2754951, EPI_ISL_2754953, EPI_ISL_2754954, EPI_ISL_2754955, EPI_ISL_2754956 | | | |
| see above | Baylor Scott and White Memorial Hospital | Genomics and Discovery, Respiratory Viruses Branch, Division of Viral Diseases, Centers for Disease Control and Prevention | Anna Kelleher, Ying Tao, Yan Li, Jing Zhang, Brian Lynch, Krista Queen, Anna Uehara, Peter Cook, Han Jia Justin Ng, Rachel Marine, Clinton R. Paden, Dhwani Batra, Haibin Wang, Tara Coalter, Jasmine Padilla, Morgan Davis, Mili Sheth, Sarah Nobles, Mark Burroughs, Justin Lee, Adam Retchless, Suxiang Tong |
| EPI_ISL_2754986, EPI_ISL_2754999, EPI_ISL_2755005, EPI_ISL_2755007, EPI_ISL_2755011, EPI_ISL_2755013, EPI_ISL_2755016, EPI_ISL_2755017, EPI_ISL_2755019, EPI_ISL_2755022, EPI_ISL_2755026, EPI_ISL_2755027, EPI_ISL_2755028, EPI_ISL_2755035, EPI_ISL_2755039, EPI_ISL_2755042, EPI_ISL_2755045, EPI_ISL_2755052, EPI_ISL_2755059, EPI_ISL_2755060, EPI_ISL_2755062, EPI_ISL_2755064, EPI_ISL_2755068, EPI_ISL_2755072, EPI_ISL_2755074, EPI_ISL_2755077, EPI_ISL_2755083, EPI_ISL_2755087, EPI_ISL_2755093, EPI_ISL_2755094, EPI_ISL_2755098, EPI_ISL_2755099 | | | |
| see above | Curative Labs | Curative Labs | Mikhail Hanewich-Hollatz, J.Cesar Ignacio-Espinoza, Elias L. Salfati, Nabjot Sandhu, George Way, Zhiyi Xie, Eugenia Khorosheva, Sophia Quasem, Janet Chen, Vladimir Slepnev |
| EPI_ISL_2755100 | Ohio Department of Health Laboratory | Ohio Department of Health Laboratory | Holmes, Jennifer; Eric Brandt, Keoni Omura, Glen McGillivary, Caitlin McDonnell, Jade Mowery, Stephanie Mccracken, Tyler Payne, Kirtana Ramadugu, Erica Leasure, Brent Lee, Kelsey Florek, Heather Blankenship, Allison Black, Quanta Brown, and Tammy Bannerman |
| EPI_ISL_2755103, EPI_ISL_2755104, EPI_ISL_2755108, EPI_ISL_2755110, EPI_ISL_2755117, EPI_ISL_2755121, EPI_ISL_2755122, EPI_ISL_2755154, EPI_ISL_2755159, EPI_ISL_2755163, EPI_ISL_2755165, EPI_ISL_2755177, EPI_ISL_2755183, EPI_ISL_2755185, EPI_ISL_2755191, EPI_ISL_2755203, EPI_ISL_2755231, EPI_ISL_2755239, EPI_ISL_2755251, EPI_ISL_2755255, EPI_ISL_2755262, EPI_ISL_2755269, EPI_ISL_2755273, EPI_ISL_2755289, EPI_ISL_2755294, EPI_ISL_2755305, EPI_ISL_2755306, EPI_ISL_2755310, EPI_ISL_2755317, EPI_ISL_2755318, EPI_ISL_2755321, EPI_ISL_2755325, EPI_ISL_2755329, EPI_ISL_2755331, EPI_ISL_2755339, EPI_ISL_2755341, EPI_ISL_2755342, EPI_ISL_2755344, EPI_ISL_2755349, EPI_ISL_2755354, EPI_ISL_2755355, EPI_ISL_2755360, EPI_ISL_2755362, EPI_ISL_2755366, EPI_ISL_2755367, EPI_ISL_2755372, EPI_ISL_2755373, EPI_ISL_2755376, EPI_ISL_2755381, EPI_ISL_2755390, EPI_ISL_2755401, EPI_ISL_2755408, EPI_ISL_2755410, EPI_ISL_2755417, EPI_ISL_2755438, EPI_ISL_2755443 | | | |
| see above | Curative Labs | Curative Labs | Mikhail Hanewich-Hollatz, J.Cesar Ignacio-Espinoza, Elias L. Salfati, Nabjot Sandhu, George Way, Zhiyi Xie, Eugenia Khorosheva, Sophia Quasem, Janet Chen, Vladimir Slepnev |
| EPI_ISL_2755444 | Ohio Department of Health Laboratory | Ohio Department of Health Laboratory | Holmes, Jennifer; Eric Brandt, Keoni Omura, Glen McGillivary, Caitlin McDonnell, Jade Mowery, Stephanie Mccracken, Tyler Payne, Kirtana Ramadugu, Erica Leasure, Brent Lee, Kelsey Florek, Heather Blankenship, Allison Black, Quanta Brown, and Tammy Bannerman |
| EPI_ISL_2755448, EPI_ISL_2755449, EPI_ISL_2755450, EPI_ISL_2755451, EPI_ISL_2755454, EPI_ISL_2755458, EPI_ISL_2755459, EPI_ISL_2755460, EPI_ISL_2755461, EPI_ISL_2755472, EPI_ISL_2755473, EPI_ISL_2755490, EPI_ISL_2755501, EPI_ISL_2755502, EPI_ISL_2755506, EPI_ISL_2755507, EPI_ISL_2755509, EPI_ISL_2755512, EPI_ISL_2755514, EPI_ISL_2755515, EPI_ISL_2755516, EPI_ISL_2755517, EPI_ISL_2755531, EPI_ISL_2755532, EPI_ISL_2755542, EPI_ISL_2755545, EPI_ISL_2755550, EPI_ISL_2755551, EPI_ISL_2755552, EPI_ISL_2755555, EPI_ISL_2755556, EPI_ISL_2755560, EPI_ISL_2755564, EPI_ISL_2755565, EPI_ISL_2755567, EPI_ISL_2755571, EPI_ISL_2755575, EPI_ISL_2755576, EPI_ISL_2755591, EPI_ISL_2755593, EPI_ISL_2755600, EPI_ISL_2755607, EPI_ISL_2755613, EPI_ISL_2755637, EPI_ISL_2755642, EPI_ISL_2755647, EPI_ISL_2755650, EPI_ISL_2755663, EPI_ISL_2755672, EPI_ISL_2755696, EPI_ISL_2755700, EPI_ISL_2755701, EPI_ISL_2755717, EPI_ISL_2755727, EPI_ISL_2755728, EPI_ISL_2755729, EPI_ISL_2755755, EPI_ISL_2755756, EPI_ISL_2755760, EPI_ISL_2755761, EPI_ISL_2755763, EPI_ISL_2755765, EPI_ISL_2755767, EPI_ISL_2755773, EPI_ISL_2755779, EPI_ISL_2755782, EPI_ISL_2755783, EPI_ISL_2755784, EPI_ISL_2755785, EPI_ISL_2755786, EPI_ISL_2755791, EPI_ISL_2755792, EPI_ISL_2755793, EPI_ISL_2755794, EPI_ISL_2755796, EPI_ISL_2755797, EPI_ISL_2755802, EPI_ISL_2755805, EPI_ISL_2755808, EPI_ISL_2755812, EPI_ISL_2755818, EPI_ISL_2755827, EPI_ISL_2755830, EPI_ISL_2755832, EPI_ISL_2755838, EPI_ISL_2755842, EPI_ISL_2755844, EPI_ISL_2755853, EPI_ISL_2755854, EPI_ISL_2755861, EPI_ISL_2755876, EPI_ISL_2755878, EPI_ISL_2755879, EPI_ISL_2755888, EPI_ISL_2755901, EPI_ISL_2755903, EPI_ISL_2755916, EPI_ISL_2755917, EPI_ISL_2755921, EPI_ISL_2755935, EPI_ISL_2755938, EPI_ISL_2755951, EPI_ISL_2755953, EPI_ISL_2755959, EPI_ISL_2755960, EPI_ISL_2755968, EPI_ISL_2755969, EPI_ISL_2755976, EPI_ISL_2755982, EPI_ISL_2755985, EPI_ISL_2755986, EPI_ISL_2755988 | | | |
| see above | Curative Labs | Curative Labs | Mikhail Hanewich-Hollatz, J.Cesar Ignacio-Espinoza, Elias L. Salfati, Nabjot Sandhu, George Way, Zhiyi Xie, Eugenia Khorosheva, Sophia Quasem, Janet Chen, Vladimir Slepnev |
| EPI_ISL_2755999 | Ohio Department of Health Laboratory | Ohio Department of Health Laboratory | Holmes, Jennifer; Eric Brandt, Keoni Omura, Glen McGillivary, Caitlin McDonnell, Jade Mowery, Stephanie Mccracken, Tyler Payne, Kirtana Ramadugu, Erica Leasure, Brent Lee, Kelsey Florek, Heather Blankenship, Allison Black, Quanta Brown, and Tammy Bannerman |
| EPI_ISL_2756002, EPI_ISL_2756004, EPI_ISL_2756006, EPI_ISL_2756010, EPI_ISL_2756013, EPI_ISL_2756014, EPI_ISL_2756016, EPI_ISL_2756019, EPI_ISL_2756021, EPI_ISL_2756022, EPI_ISL_2756023, EPI_ISL_2756026, EPI_ISL_2756030, EPI_ISL_2756031, EPI_ISL_2756034, EPI_ISL_2756035, EPI_ISL_2756037, EPI_ISL_2756038, EPI_ISL_2756039, EPI_ISL_2756041, EPI_ISL_2756045, EPI_ISL_2756046, EPI_ISL_2756048, EPI_ISL_2756049, EPI_ISL_2756052, EPI_ISL_2756056 | | | |
| see above | Curative Labs | Curative Labs | Mikhail Hanewich-Hollatz, J.Cesar Ignacio-Espinoza, Elias L. Salfati, Nabjot Sandhu, George Way, Zhiyi Xie, Eugenia Khorosheva, Sophia Quasem, Janet Chen, Vladimir Slepnev |
| EPI_ISL_2756127, EPI_ISL_2756219 | Ohio Department of Health Laboratory | Ohio Department of Health Laboratory | Holmes, Jennifer; Eric Brandt, Keoni Omura, Glen McGillivary, Caitlin McDonnell, Jade Mowery, Stephanie Mccracken, Tyler Payne, Kirtana Ramadugu, Erica Leasure, Brent Lee, Kelsey Florek, Heather Blankenship, Allison Black, Quanta Brown, and Tammy Bannerman |
| EPI_ISL_2756343, EPI_ISL_2756344, EPI_ISL_2756345, EPI_ISL_2756347, EPI_ISL_2756349, EPI_ISL_2756350, EPI_ISL_2756351, EPI_ISL_2756353, EPI_ISL_2756354, EPI_ISL_2756357, EPI_ISL_2756358, EPI_ISL_2756359, EPI_ISL_2756360, EPI_ISL_2756362, EPI_ISL_2756363 | | | |
| see above | Wyoming Public Health Laboratory | Wyoming Public Health Laboratory | Jim Mildenberger, Taylor Fearing, Channing Weber, Ashley Norberg, Chayse Rowley, Marley Goetz, Brian Dominguez, Elliot Thomasson, Sam Britz, Cari Sloma, Robert Petit, and Rob Christensen |
| EPI_ISL_2756422, EPI_ISL_2756423 | Baylor Scott and White Memorial Hospital | Genomics and Discovery, Respiratory Viruses Branch, Division of Viral Diseases, Centers for Disease Control and Prevention | Anna Kelleher, Ying Tao, Yan Li, Jing Zhang, Brian Lynch, Krista Queen, Anna Uehara, Peter Cook, Han Jia Justin Ng, Rachel Marine, Clinton R. Paden, Dhwani Batra, Haibin Wang, Tara Coalter, Jasmine Padilla, Morgan Davis, Mili Sheth, Sarah Nobles, Mark Burroughs, Justin Lee, Adam Retchless, Suxiang Tong |
| EPI_ISL_2756424 | VA-Division of Consolidated Laboratory Services | Genomics and Discovery, Respiratory Viruses Branch, Division of Viral Diseases, Centers for Disease Control and Prevention | Anna Kelleher, Ying Tao, Yan Li, Jing Zhang, Brian Lynch, Krista Queen, Anna Uehara, Peter Cook, Han Jia Justin Ng, Rachel Marine, Clinton R. Paden, Dhwani Batra, Haibin Wang, Tara Coalter, Jasmine Padilla, Morgan Davis, Mili Sheth, Sarah Nobles, Mark Burroughs, Justin Lee, Adam Retchless, Suxiang Tong |
| EPI_ISL_2756425, EPI_ISL_2756494, EPI_ISL_2756495, EPI_ISL_2756496, EPI_ISL_2756497, EPI_ISL_2756498, EPI_ISL_2756499, EPI_ISL_2756500, EPI_ISL_2756501, EPI_ISL_2756502, EPI_ISL_2756503, EPI_ISL_2756504, EPI_ISL_2756505, EPI_ISL_2756506, EPI_ISL_2756507, EPI_ISL_2756508, EPI_ISL_2756509, EPI_ISL_2756510, EPI_ISL_2756511, EPI_ISL_2756512, EPI_ISL_2756513, EPI_ISL_2756514, EPI_ISL_2756515, EPI_ISL_2756516, EPI_ISL_2756517, EPI_ISL_2756518, EPI_ISL_2756519, EPI_ISL_2756520 | | | |
| see above | Montana Public Health Laboratory | Montana Public Health Laboratory | Joy Ritter, Michelle Mozer, Carrie Biskupiak, Deborah Gibson, Michael Dills |
| EPI_ISL_2756522 | Tampa General Hospital Esoteric Lab | Tampa General Hospital Esoteric Research & Development Lab | Amorce Lima, Jon Faughn, Deanna Becker, Suzane Silbert |
| EPI_ISL_2756523, EPI_ISL_2756524, EPI_ISL_2756526, EPI_ISL_2756527, EPI_ISL_2756528, EPI_ISL_2756529, EPI_ISL_2756530, EPI_ISL_2756531, EPI_ISL_2756532, EPI_ISL_2756533, EPI_ISL_2756534, EPI_ISL_2756535, EPI_ISL_2756536, EPI_ISL_2756537, EPI_ISL_2756539, EPI_ISL_2756540 | | | |
| see above | Wichita State University - Molecular Diagnostics Lab | Kansas Health and Environmental Lab | Katherine Wiggins, Mike Grose, Jonathan Barnell, Ben Olsen, and Phil Adam |
| EPI_ISL_2756542, EPI_ISL_2756543, EPI_ISL_2756544, EPI_ISL_2756545, EPI_ISL_2756546, EPI_ISL_2756547, EPI_ISL_2756548, EPI_ISL_2756549, EPI_ISL_2756550, EPI_ISL_2756552, EPI_ISL_2756553, EPI_ISL_2756555 | | | |
| see above | Kansas Health and Environmental Lab | Kansas Health and Environmental Lab | Katherine Wiggins, Mike Grose, Jonathan Barnell, Ben Olsen, and Phil Adam |
| EPI_ISL_2756557 | Centers for Disease Control and Prevention, Dengue Branch | Centers for Disease Control and Prevention, Dengue Branch | Gilberto A. Santiago, Glenda Gonzalez, Betzabel Flores, Keyla Charriez, Gabriela Paz-Bailey, Jorge L. Munoz-Jordan |
| EPI_ISL_2757838, EPI_ISL_2757839, EPI_ISL_2757840, EPI_ISL_2757841, EPI_ISL_2757842, EPI_ISL_2757843, EPI_ISL_2757844, EPI_ISL_2757845, EPI_ISL_2757846, EPI_ISL_2757847, EPI_ISL_2757848, EPI_ISL_2757849 | | | |
| see above | CDPH VBL | California Department of Public Health | CDPH-COVIDNet |
| EPI_ISL_2757860, EPI_ISL_2757861 | Department of General Services Division of Consolidated Laboratory Services, Virginia Division of Consolidated Laboratory Services Sequencing Submission Group | Department of General Services Division of Consolidated Laboratory Services, Virginia Division of Consolidated Laboratory Services Sequencing Submission Group | Fink,L. |
| EPI_ISL_2757932, EPI_ISL_2757933, EPI_ISL_2757934, EPI_ISL_2757935, EPI_ISL_2757936, EPI_ISL_2757937, | ISDH | Quantigen Biosciences | Paul J. Childress |

| | | | |
|---|---|---|---|
| EPI_ISL_2757938, EPI_ISL_2757939, EPI_ISL_2757940 | | | |
| EPI_ISL_2758225, EPI_ISL_2758226, EPI_ISL_2758227, EPI_ISL_2758228, EPI_ISL_2758229, EPI_ISL_2758230, EPI_ISL_2758231, EPI_ISL_2758232, EPI_ISL_2758233, EPI_ISL_2758234, EPI_ISL_2758235 | | | |
| see above | Weill Cornell Medicine | New York Genome Center | Michael Zody, Andre Corvelo, Dayna M. Oschwald, Samantha Fennessey, Tom Maniatis, Melissa Cushing, Olivier Elemento, Margaret Elizabeth Ross, Chris Mason, Priya Velu, Hanna Rennert, Arryn Craney, Lars F Westblade |
| EPI_ISL_2758236, EPI_ISL_2758237 | Rush University Medical Center | RIPHL at Rush University Medical Center | Stefan Green, Kevin Kunstman, Max Kolton, Marieta Hyde, Laura Furtado, Felix Araujo Perez, Mary Hayden, Joyce Houlihan, Diane Springer |
| EPI_ISL_2758238 | University of Chicago Medicine | RIPHL at Rush University Medical Center | Stefan Green, Kevin Kunstman, Max Kolton, Marieta Hyde, Laura Furtado, Felix Araujo Perez, Lauren Megger |
| EPI_ISL_2758239 | Northwestern Memorial Hospital | RIPHL at Rush University Medical Center | Stefan Green, Kevin Kunstman, Max Kolton, Marieta Hyde, Laura Furtado, Felix Araujo Perez, Chao Qi, Amber Kimble |
| EPI_ISL_2758240, EPI_ISL_2758241, EPI_ISL_2758242, EPI_ISL_2758243, EPI_ISL_2758244 | John H Stroger Jr Hospital of Cook County | RIPHL at Rush University Medical Center | Stefan Green, Kevin Kunstman, Max Kolton, Marieta Hyde, Laura Furtado, Felix Araujo Perez, Ah-Young Sung, Sharon Welbel |
| EPI_ISL_2758245, EPI_ISL_2758246, EPI_ISL_2758247, EPI_ISL_2758248 | University of Illinois at Chicago | RIPHL at Rush University Medical Center | Stefan Green, Kevin Kunstman, Max Kolton, Marieta Hyde, Laura Furtado, Felix Araujo Perez, Julie Bentea, Omar Perez |
| EPI_ISL_2758249, EPI_ISL_2758250, EPI_ISL_2758251, EPI_ISL_2758252, EPI_ISL_2758253, EPI_ISL_2758254, EPI_ISL_2758255, EPI_ISL_2758256, EPI_ISL_2758257, EPI_ISL_2758258, EPI_ISL_2758259, EPI_ISL_2758260, EPI_ISL_2758261, EPI_ISL_2758262, EPI_ISL_2758263, EPI_ISL_2758264, EPI_ISL_2758265, EPI_ISL_2758267, EPI_ISL_2758268, EPI_ISL_2758269, EPI_ISL_2758270, EPI_ISL_2758271, EPI_ISL_2758272, EPI_ISL_2758273, EPI_ISL_2758274, EPI_ISL_2758275 | | | |
| see above | J.W. Ruby Memorial Hospital | WVU and Marshall University Combined Genomics Core Facilities | James Denvir, Peter Stoilov, Peter Perrotta, Wesley Kimble, Ryan Percifield |
| EPI_ISL_2758276, EPI_ISL_2758277, EPI_ISL_2758278 | Berkeley Medical Center | WVU and Marshall University Combined Genomics Core Facilities | James Denvir, Peter Stoilov, Peter Perrotta, Wesley Kimble, Ryan Percifield |
| EPI_ISL_2758279, EPI_ISL_2758280, EPI_ISL_2758281, EPI_ISL_2758282, EPI_ISL_2758283, EPI_ISL_2758284, EPI_ISL_2758285, EPI_ISL_2758286, EPI_ISL_2758287, EPI_ISL_2758288, EPI_ISL_2758289, EPI_ISL_2758290, EPI_ISL_2758291, EPI_ISL_2758292, EPI_ISL_2758293, EPI_ISL_2758294, EPI_ISL_2758295, EPI_ISL_2758296, EPI_ISL_2758297, EPI_ISL_2758298, EPI_ISL_2758299, EPI_ISL_2758300, EPI_ISL_2758301, EPI_ISL_2758302, EPI_ISL_2758303, EPI_ISL_2758304, EPI_ISL_2758305, EPI_ISL_2758307, EPI_ISL_2758308, EPI_ISL_2758309, EPI_ISL_2758310, EPI_ISL_2758311, EPI_ISL_2758312, EPI_ISL_2758313, EPI_ISL_2758314, EPI_ISL_2758315, EPI_ISL_2758316, EPI_ISL_2758317, EPI_ISL_2758318, EPI_ISL_2758319, EPI_ISL_2758320, EPI_ISL_2758321, EPI_ISL_2758322, EPI_ISL_2758323, EPI_ISL_2758324, EPI_ISL_2758325, EPI_ISL_2758326, EPI_ISL_2758327, EPI_ISL_2758328 | | | |
| see above | J.W. Ruby Memorial Hospital | WVU and Marshall University Combined Genomics Core Facilities | James Denvir, Peter Stoilov, Peter Perrotta, Wesley Kimble, Ryan Percifield |
| EPI_ISL_2758329, EPI_ISL_2758330, EPI_ISL_2758331 | Preston Taylor Community Health Center | WVU and Marshall University Combined Genomics Core Facilities | James Denvir, Peter Stoilov, Peter Perrotta, Wesley Kimble, Ryan Percifield |
| EPI_ISL_2758332, EPI_ISL_2758333, EPI_ISL_2758334, EPI_ISL_2758335, EPI_ISL_2758336, EPI_ISL_2758337, EPI_ISL_2758338, EPI_ISL_2758339, EPI_ISL_2758340, EPI_ISL_2758341, EPI_ISL_2758342, EPI_ISL_2758343, EPI_ISL_2758344, EPI_ISL_2758345, EPI_ISL_2758346, EPI_ISL_2758347, EPI_ISL_2758348, EPI_ISL_2758349, EPI_ISL_2758350, EPI_ISL_2758351, EPI_ISL_2758353, EPI_ISL_2758354, EPI_ISL_2758355, EPI_ISL_2758356, EPI_ISL_2758357, EPI_ISL_2758358, EPI_ISL_2758359, EPI_ISL_2758360, EPI_ISL_2758361, EPI_ISL_2758362, EPI_ISL_2758363, EPI_ISL_2758364, EPI_ISL_2758365, EPI_ISL_2758366, EPI_ISL_2758367 | | | |
| see above | QLabs | WVU and Marshall University Combined Genomics Core Facilities | James Denvir, Peter Stoilov, Peter Perrotta, Wesley Kimble, Ryan Percifield |
| EPI_ISL_2758368, EPI_ISL_2758369, EPI_ISL_2758371, EPI_ISL_2758372, EPI_ISL_2758373 | Berkeley Medical Center | WVU and Marshall University Combined Genomics Core Facilities | James Denvir, Peter Stoilov, Peter Perrotta, Wesley Kimble, Ryan Percifield |
| EPI_ISL_2758374, EPI_ISL_2758375 | Hampshire Memorial Hospital | WVU and Marshall University Combined Genomics Core Facilities | James Denvir, Peter Stoilov, Peter Perrotta, Wesley Kimble, Ryan Percifield |
| EPI_ISL_2758376, EPI_ISL_2758377, EPI_ISL_2758378 | Berkeley Medical Center | WVU and Marshall University Combined Genomics Core Facilities | James Denvir, Peter Stoilov, Peter Perrotta, Wesley Kimble, Ryan Percifield |
| EPI_ISL_2758379 | Camden Clark Medical Center Microbiology Department | WVU and Marshall University Combined Genomics Core Facilities | James Denvir, Peter Stoilov, Peter Perrotta, Wesley Kimble, Ryan Percifield |
| EPI_ISL_2758380, EPI_ISL_2758381, EPI_ISL_2758383, EPI_ISL_2758384, EPI_ISL_2758385, EPI_ISL_2758386, EPI_ISL_2758387, EPI_ISL_2758388, EPI_ISL_2758389, EPI_ISL_2758390, EPI_ISL_2758391, EPI_ISL_2758392, EPI_ISL_2758393, EPI_ISL_2758394, EPI_ISL_2758395, EPI_ISL_2758396, EPI_ISL_2758397, EPI_ISL_2758398, EPI_ISL_2758399, EPI_ISL_2758400 | | | |
| see above | QLabs | WVU and Marshall University Combined Genomics Core Facilities | James Denvir, Peter Stoilov, Peter Perrotta, Wesley Kimble, Ryan Percifield |
| EPI_ISL_2758407, EPI_ISL_2758408, EPI_ISL_2758409 | Humboldt County Public Health Laboratory | Chan-Zuckerberg Biohub | CZB Cliahub Consortium |
| EPI_ISL_2758410, EPI_ISL_2758411 | Orange County Public Health Lab | Chan-Zuckerberg Biohub | CZB Cliahub Consortium |
| EPI_ISL_2758413 | Santa Clara County Public Health Laboratory | Chan-Zuckerberg Biohub | CZB Cliahub Consortium |
| EPI_ISL_2758414, EPI_ISL_2758415 | Alameda County Public Health Lab | Chan-Zuckerberg Biohub | CZB Cliahub Consortium |
| EPI_ISL_2758416 | Orange County Public Health Lab | Chan-Zuckerberg Biohub | CZB Cliahub Consortium |
| EPI_ISL_2758417 | Alameda County Public Health Lab | Chan-Zuckerberg Biohub | CZB Cliahub Consortium |
| EPI_ISL_2758418, EPI_ISL_2758419, EPI_ISL_2758420 | Santa Clara County Public Health Laboratory | Chan-Zuckerberg Biohub | CZB Cliahub Consortium |
| EPI_ISL_2758421, EPI_ISL_2758422 | Orange County Public Health Lab | Chan-Zuckerberg Biohub | CZB Cliahub Consortium |
| EPI_ISL_2758423, EPI_ISL_2758424 | Santa Clara County Public Health Laboratory | Chan-Zuckerberg Biohub | CZB Cliahub Consortium |
| EPI_ISL_2758425 | Orange County Public Health Lab | Chan-Zuckerberg Biohub | CZB Cliahub Consortium |
| EPI_ISL_2758426, EPI_ISL_2758427, EPI_ISL_2758428 | Humboldt County Public Health Laboratory | Chan-Zuckerberg Biohub | CZB Cliahub Consortium |
| EPI_ISL_2758430 | Santa Clara County Public Health Laboratory | Chan-Zuckerberg Biohub | CZB Cliahub Consortium |
| EPI_ISL_2758431, EPI_ISL_2758432, EPI_ISL_2758433, EPI_ISL_2758434, EPI_ISL_2758435 | Orange County Public Health Lab | Chan-Zuckerberg Biohub | CZB Cliahub Consortium |
| EPI_ISL_2758436 | Contra Costa County Public Health Lab | Chan-Zuckerberg Biohub | CZB Cliahub Consortium |
| EPI_ISL_2758437, EPI_ISL_2758438 | Santa Clara County Public Health Laboratory | Chan-Zuckerberg Biohub | CZB Cliahub Consortium |
| EPI_ISL_2758439 | Orange County Public Health Lab | Chan-Zuckerberg Biohub | CZB Cliahub Consortium |
| EPI_ISL_2758440, EPI_ISL_2758441, EPI_ISL_2758442 | CA DPH Viral and Rickettsial Disease Laboratory | Chan-Zuckerberg Biohub | CZB Cliahub Consortium |
| EPI_ISL_2758443, EPI_ISL_2758444, EPI_ISL_2758445, EPI_ISL_2758446, EPI_ISL_2758447 | Alameda County Public Health Lab | Chan-Zuckerberg Biohub | CZB Cliahub Consortium |

| | | | |
|---|---|---|---|
| EPI_ISL_2758448, EPI_ISL_2758449 | CA DPH Viral and Rickettsial Disease Laboratory | Chan-Zuckerberg Biohub | CZB Cliahub Consortium |
| EPI_ISL_2758450, EPI_ISL_2758451 | Santa Clara County Public Health Laboratory | Chan-Zuckerberg Biohub | CZB Cliahub Consortium |
| EPI_ISL_2758452, EPI_ISL_2758453 | Humboldt County Public Health Laboratory | Chan-Zuckerberg Biohub | CZB Cliahub Consortium |
| EPI_ISL_2758454, EPI_ISL_2758455 | Alameda County Public Health Lab | Chan-Zuckerberg Biohub | CZB Cliahub Consortium |
| EPI_ISL_2758456, EPI_ISL_2758457, EPI_ISL_2758458, EPI_ISL_2758459 | Contra Costa County Public Health Lab | Chan-Zuckerberg Biohub | CZB Cliahub Consortium |
| EPI_ISL_2758460, EPI_ISL_2758461, EPI_ISL_2758462 | Humboldt County Public Health Laboratory | Chan-Zuckerberg Biohub | CZB Cliahub Consortium |
| EPI_ISL_2758463 | Alameda County Public Health Lab | Chan-Zuckerberg Biohub | CZB Cliahub Consortium |
| EPI_ISL_2758464, EPI_ISL_2758465, EPI_ISL_2758466, EPI_ISL_2758467 | CA DPH Viral and Rickettsial Disease Laboratory | Chan-Zuckerberg Biohub | CZB Cliahub Consortium |
| EPI_ISL_2758468, EPI_ISL_2758469 | Contra Costa County Public Health Lab | Chan-Zuckerberg Biohub | CZB Cliahub Consortium |
| EPI_ISL_2758470, EPI_ISL_2758471 | Alameda County Public Health Lab | Chan-Zuckerberg Biohub | CZB Cliahub Consortium |
| EPI_ISL_2758473 | Humboldt County Public Health Laboratory | Chan-Zuckerberg Biohub | CZB Cliahub Consortium |
| EPI_ISL_2758474 | County of San Luis Obispo Public Health Laboratory | Chan-Zuckerberg Biohub | CZB Cliahub Consortium |
| EPI_ISL_2758475 | Santa Clara County Public Health Laboratory | Chan-Zuckerberg Biohub | CZB Cliahub Consortium |
| EPI_ISL_2758476, EPI_ISL_2758477, EPI_ISL_2758478 | CA DPH Viral and Rickettsial Disease Laboratory | Chan-Zuckerberg Biohub | CZB Cliahub Consortium |
| EPI_ISL_2758479, EPI_ISL_2758480, EPI_ISL_2758481 | Contra Costa County Public Health Lab | Chan-Zuckerberg Biohub | CZB Cliahub Consortium |
| EPI_ISL_2758482 | County of San Luis Obispo Public Health Laboratory | Chan-Zuckerberg Biohub | CZB Cliahub Consortium |
| EPI_ISL_2758483, EPI_ISL_2758484, EPI_ISL_2758485, EPI_ISL_2758486 | CA DPH Viral and Rickettsial Disease Laboratory | Chan-Zuckerberg Biohub | CZB Cliahub Consortium |
| EPI_ISL_2758487 | Santa Clara County Public Health Laboratory | Chan-Zuckerberg Biohub | CZB Cliahub Consortium |
| EPI_ISL_2758488, EPI_ISL_2758489 | CA DPH Viral and Rickettsial Disease Laboratory | Chan-Zuckerberg Biohub | CZB Cliahub Consortium |
| EPI_ISL_2758490, EPI_ISL_2758491 | Orange County Public Health Lab | Chan-Zuckerberg Biohub | CZB Cliahub Consortium |
| EPI_ISL_2758492 | Santa Clara County Public Health Laboratory | Chan-Zuckerberg Biohub | CZB Cliahub Consortium |
| EPI_ISL_2758493, EPI_ISL_2758494 | CA DPH Viral and Rickettsial Disease Laboratory | Chan-Zuckerberg Biohub | CZB Cliahub Consortium |
| EPI_ISL_2758495, EPI_ISL_2758496, EPI_ISL_2758497, EPI_ISL_2758498, EPI_ISL_2758499, EPI_ISL_2758500 | County of San Luis Obispo Public Health Laboratory | Chan-Zuckerberg Biohub | CZB Cliahub Consortium |
| EPI_ISL_2758502 | Santa Clara County Public Health Laboratory | Chan-Zuckerberg Biohub | CZB Cliahub Consortium |
| EPI_ISL_2758503, EPI_ISL_2758504 | Contra Costa County Public Health Lab | Chan-Zuckerberg Biohub | CZB Cliahub Consortium |
| EPI_ISL_2758505, EPI_ISL_2758506, EPI_ISL_2758507, EPI_ISL_2758508, EPI_ISL_2758509, EPI_ISL_2758510, EPI_ISL_2758511, EPI_ISL_2758512, EPI_ISL_2758513 | CA DPH Viral and Rickettsial Disease Laboratory | Chan-Zuckerberg Biohub | CZB Cliahub Consortium |
| EPI_ISL_2758514 | County of San Luis Obispo Public Health Laboratory | Chan-Zuckerberg Biohub | CZB Cliahub Consortium |
| EPI_ISL_2758515 | Santa Clara County Public Health Laboratory | Chan-Zuckerberg Biohub | CZB Cliahub Consortium |
| EPI_ISL_2758516 | Orange County Public Health Lab | Chan-Zuckerberg Biohub | CZB Cliahub Consortium |
| EPI_ISL_2758517 | Humboldt County Public Health Laboratory | Chan-Zuckerberg Biohub | CZB Cliahub Consortium |
| EPI_ISL_2758518 | County of San Luis Obispo Public Health Laboratory | Chan-Zuckerberg Biohub | CZB Cliahub Consortium |
| EPI_ISL_2758519, EPI_ISL_2758520, EPI_ISL_2758521, EPI_ISL_2758522, EPI_ISL_2758523, EPI_ISL_2758524, EPI_ISL_2758525, EPI_ISL_2758526, EPI_ISL_2758527, EPI_ISL_2758528, EPI_ISL_2758529, EPI_ISL_2758530 | | | |
| see above | CA DPH Viral and Rickettsial Disease Laboratory | Chan-Zuckerberg Biohub | CZB Cliahub Consortium |
| EPI_ISL_2758531 | Humboldt County Public Health Laboratory | Chan-Zuckerberg Biohub | CZB Cliahub Consortium |
| EPI_ISL_2758532, EPI_ISL_2758533 | Contra Costa County Public Health Lab | Chan-Zuckerberg Biohub | CZB Cliahub Consortium |
| EPI_ISL_2758534, EPI_ISL_2758535, EPI_ISL_2758536, EPI_ISL_2758537, EPI_ISL_2758538, EPI_ISL_2758539, EPI_ISL_2758540 | CA DPH Viral and Rickettsial Disease Laboratory | Chan-Zuckerberg Biohub | CZB Cliahub Consortium |

| | | | |
|---|---|---|---|
| EPI_ISL_2758541 | Humboldt County Public Health Laboratory | Chan-Zuckerberg Biohub | CZB Cliahub Consortium |
| EPI_ISL_2758542 | Santa Clara County Public Health Laboratory | Chan-Zuckerberg Biohub | CZB Cliahub Consortium |
| EPI_ISL_2758543 | County of San Luis Obispo Public Health Laboratory | Chan-Zuckerberg Biohub | CZB Cliahub Consortium |
| EPI_ISL_2758544 | Humboldt County Public Health Laboratory | Chan-Zuckerberg Biohub | CZB Cliahub Consortium |
| EPI_ISL_2758545, EPI_ISL_2758546 | County of San Luis Obispo Public Health Laboratory | Chan-Zuckerberg Biohub | CZB Cliahub Consortium |
| EPI_ISL_2758547, EPI_ISL_2758548 | Santa Clara County Public Health Laboratory | Chan-Zuckerberg Biohub | CZB Cliahub Consortium |
| EPI_ISL_2758595, EPI_ISL_2758596, EPI_ISL_2758597, EPI_ISL_2758598, EPI_ISL_2758599, EPI_ISL_2758600, EPI_ISL_2758601, EPI_ISL_2758602, EPI_ISL_2758603, EPI_ISL_2758604, EPI_ISL_2758605, EPI_ISL_2758606 | | | |
| see above | County of San Luis Obispo Public Health Laboratory | County of San Luis Obispo Public Health Laboratory | Katrina Erwin, Frances Sidhu |
| EPI_ISL_2758607, EPI_ISL_2758608, EPI_ISL_2758609, EPI_ISL_2758612, EPI_ISL_2758613, EPI_ISL_2758614, EPI_ISL_2758615, EPI_ISL_2758616, EPI_ISL_2758617, EPI_ISL_2758618, EPI_ISL_2758619, EPI_ISL_2758620, EPI_ISL_2758621, EPI_ISL_2758622, EPI_ISL_2758623, EPI_ISL_2758624, EPI_ISL_2758625, EPI_ISL_2758626, EPI_ISL_2758627, EPI_ISL_2758628, EPI_ISL_2758629, EPI_ISL_2758630, EPI_ISL_2758631, EPI_ISL_2758632 | | | |
| see above | Microbiology Division, South Carolina Department of Health and Environmental Control | Microbiology Division, South Carolina Department of Health and Environmental Control | Freeman,J.S., Flores,H.V. |
| EPI_ISL_2758634 | Department of General Services Division of Consolidated Laboratory Services, Virginia Division of Consolidated Laboratory Services Sequencing Submission Group | Department of General Services Division of Consolidated Laboratory Services, Virginia Division of Consolidated Laboratory Services Sequencing Submission Group | Fink,L. |
| EPI_ISL_2758635 | DOHMH Morrisania | New York City Public Health Laboratory | Jade Wang, et al. |
| EPI_ISL_2758636 | DOHMH Fort Greene | New York City Public Health Laboratory | Jade Wang, et al. |
| EPI_ISL_2758637 | DOHMH Riverside | New York City Public Health Laboratory | Jade Wang, et al. |
| EPI_ISL_2758638 | DOHMH Chelsea | New York City Public Health Laboratory | Jade Wang, et al. |
| EPI_ISL_2758639 | DOHMH Riverside | New York City Public Health Laboratory | Jade Wang, et al. |
| EPI_ISL_2758640, EPI_ISL_2758641, EPI_ISL_2758642, EPI_ISL_2758643, EPI_ISL_2758644 | Quest Diagnostics | New York City Public Health Laboratory | Jade Wang, et al. |
| EPI_ISL_2758645 | OCME Office Of Chief Medical Examiner | New York City Public Health Laboratory | Jade Wang, et al. |
| EPI_ISL_2758834, EPI_ISL_2758835, EPI_ISL_2758836, EPI_ISL_2758837, EPI_ISL_2758838, EPI_ISL_2758839, EPI_ISL_2758840, EPI_ISL_2758841, EPI_ISL_2758842, EPI_ISL_2758843, EPI_ISL_2758844, EPI_ISL_2758845, EPI_ISL_2758846, EPI_ISL_2758847, EPI_ISL_2758848, EPI_ISL_2758849, EPI_ISL_2758850, EPI_ISL_2758852, EPI_ISL_2758853, EPI_ISL_2758855, EPI_ISL_2758856, EPI_ISL_2758857, EPI_ISL_2758858, EPI_ISL_2758860, EPI_ISL_2758861, EPI_ISL_2758864, EPI_ISL_2758865, EPI_ISL_2758867, EPI_ISL_2758868, EPI_ISL_2758869, EPI_ISL_2758870, EPI_ISL_2758871, EPI_ISL_2758873, EPI_ISL_2758874, EPI_ISL_2758875, EPI_ISL_2758876, EPI_ISL_2758878, EPI_ISL_2758879, EPI_ISL_2758880, EPI_ISL_2758882, EPI_ISL_2758883, EPI_ISL_2758884, EPI_ISL_2758887, EPI_ISL_2758889, EPI_ISL_2758890, EPI_ISL_2758891, EPI_ISL_2758892, EPI_ISL_2758893, EPI_ISL_2758894, EPI_ISL_2758895, EPI_ISL_2758896, EPI_ISL_2758897, EPI_ISL_2758899, EPI_ISL_2758900, EPI_ISL_2758901, EPI_ISL_2758902, EPI_ISL_2758903, EPI_ISL_2758904, EPI_ISL_2758905, EPI_ISL_2758906, EPI_ISL_2758907, EPI_ISL_2758908, EPI_ISL_2758909, EPI_ISL_2758910, EPI_ISL_2758911, EPI_ISL_2758912, EPI_ISL_2758913, EPI_ISL_2758914, EPI_ISL_2758915, EPI_ISL_2758917, EPI_ISL_2758918, EPI_ISL_2758919, EPI_ISL_2758920, EPI_ISL_2758921, EPI_ISL_2758922, EPI_ISL_2758923, EPI_ISL_2758925, EPI_ISL_2758926, EPI_ISL_2758927, EPI_ISL_2758928, EPI_ISL_2758929, EPI_ISL_2758930, EPI_ISL_2758931, EPI_ISL_2758933, EPI_ISL_2758934 | | | |
| see above | The Ohio State University Applied Microbiology Services Laboratory | The Ohio State University Applied Microbiology Services Laboratory | Seth A. Faith PhD |
| EPI_ISL_2759079, EPI_ISL_2759080, EPI_ISL_2759081, EPI_ISL_2759082, EPI_ISL_2759083, EPI_ISL_2759084, EPI_ISL_2759085, EPI_ISL_2759086, EPI_ISL_2759087, EPI_ISL_2759088 | Monterey County Public Health Laboratory | Monterey County Public Health Laboratory | Monterey County Public Health Laboratory |
| EPI_ISL_2759097, EPI_ISL_2759098, EPI_ISL_2759099 | Seattle Flu Study | Seattle Flu Study | Deborah A. Nickerson, Chris D. Frazar, Jover Lee, Benjamin Pelle, Erica Ryke, Matthew Richardson, Amanda Adler, Elisabeth Brandstetter, Peter D. Han, Kairsten Fay, Misja Ilcisin, Kirsten Lacombe, Thomas R. Sibley, Melissa Truong, Caitlin R. Wolf, Karen Cowgill, Stephanie Schrag, Jeff Duchin, Michael Boeckh, Janet A. Englund, Michael Famulare, Barry R. Lutz, Mark J. Rieder, Lea M. Starita, Matthew Thompson, Helen Y. Chu, Trevor Bedford, Jay Shendure |
| EPI_ISL_2759100 | Overlake Hospital | Seattle Flu Study | Deborah A. Nickerson, Chris D. Frazar, Jover Lee, Benjamin Pelle, Erica Ryke, Matthew Richardson, Amanda Adler, Elisabeth Brandstetter, Peter D. Han, Kairsten Fay, Misja Ilcisin, Kirsten Lacombe, Thomas R. Sibley, Melissa Truong, Caitlin R. Wolf, Romesh Gautom, Geoff Melly, Brian Hiatt, Philip Dykema, Scott Lindquist, Michael Boeckh, Janet A. Englund, Michael Famulare, Barry R. Lutz, Mark J. Rieder, Lea M. Starita, Matthew Thompson, Helen Y. Chu, Jay Shendure, Trevor Bedford |
| EPI_ISL_2759142, EPI_ISL_2759143, EPI_ISL_2759144, EPI_ISL_2759145, EPI_ISL_2759146, EPI_ISL_2759147, EPI_ISL_2759148, EPI_ISL_2759149, EPI_ISL_2759150, EPI_ISL_2759151, EPI_ISL_2759152, EPI_ISL_2759153, EPI_ISL_2759154, EPI_ISL_2759155, EPI_ISL_2759156, EPI_ISL_2759157, EPI_ISL_2759158, EPI_ISL_2759159, EPI_ISL_2759161, EPI_ISL_2759163, EPI_ISL_2759164, EPI_ISL_2759165, EPI_ISL_2759166, EPI_ISL_2759167, EPI_ISL_2759169, EPI_ISL_2759172, EPI_ISL_2759173, EPI_ISL_2759174, EPI_ISL_2759175, EPI_ISL_2759176, EPI_ISL_2759177, EPI_ISL_2759178, EPI_ISL_2759179, EPI_ISL_2759180, EPI_ISL_2759181, EPI_ISL_2759182, EPI_ISL_2759183, EPI_ISL_2759184, EPI_ISL_2759185, EPI_ISL_2759186, EPI_ISL_2759187, EPI_ISL_2759188, EPI_ISL_2759189, EPI_ISL_2759190, EPI_ISL_2759191, EPI_ISL_2759192, EPI_ISL_2759193, EPI_ISL_2759194, EPI_ISL_2759195, EPI_ISL_2759196, EPI_ISL_2759197, EPI_ISL_2759198, EPI_ISL_2759199, EPI_ISL_2759200, EPI_ISL_2759201, EPI_ISL_2759202, EPI_ISL_2759203, EPI_ISL_2759204, EPI_ISL_2759205, EPI_ISL_2759206, EPI_ISL_2759207, EPI_ISL_2759208, EPI_ISL_2759209, EPI_ISL_2759210, EPI_ISL_2759211, EPI_ISL_2759212, EPI_ISL_2759213, EPI_ISL_2759214, EPI_ISL_2759215, EPI_ISL_2759216, EPI_ISL_2759217, EPI_ISL_2759218, EPI_ISL_2759219, EPI_ISL_2759220, EPI_ISL_2759221, EPI_ISL_2759222, EPI_ISL_2759223, EPI_ISL_2759224, EPI_ISL_2759225, EPI_ISL_2759226, EPI_ISL_2759227, EPI_ISL_2759228, EPI_ISL_2759229, EPI_ISL_2759230, EPI_ISL_2759231, EPI_ISL_2759232, EPI_ISL_2759233, EPI_ISL_2759234, EPI_ISL_2759235, EPI_ISL_2759237, EPI_ISL_2759238, EPI_ISL_2759239, EPI_ISL_2759240, EPI_ISL_2759241, EPI_ISL_2759242, EPI_ISL_2759243, EPI_ISL_2759244, EPI_ISL_2759245, EPI_ISL_2759246, EPI_ISL_2759247, EPI_ISL_2759248, EPI_ISL_2759249, EPI_ISL_2759250, EPI_ISL_2759251, EPI_ISL_2759252, EPI_ISL_2759253, EPI_ISL_2759254, EPI_ISL_2759255, EPI_ISL_2759257, EPI_ISL_2759258, EPI_ISL_2759259, EPI_ISL_2759260, EPI_ISL_2759261, EPI_ISL_2759262, EPI_ISL_2759263, EPI_ISL_2759264, EPI_ISL_2759265, EPI_ISL_2759266, EPI_ISL_2759267, EPI_ISL_2759268, EPI_ISL_2759270, EPI_ISL_2759271, EPI_ISL_2759272, EPI_ISL_2759273, EPI_ISL_2759274, EPI_ISL_2759275, EPI_ISL_2759276, EPI_ISL_2759277, EPI_ISL_2759278, EPI_ISL_2759279, EPI_ISL_2759280, EPI_ISL_2759281, EPI_ISL_2759282, EPI_ISL_2759283, EPI_ISL_2759284, EPI_ISL_2759285, EPI_ISL_2759286, EPI_ISL_2759287, EPI_ISL_2759288, EPI_ISL_2759289, EPI_ISL_2759290, EPI_ISL_2759291, EPI_ISL_2759292, EPI_ISL_2759293, EPI_ISL_2759294, EPI_ISL_2759295, EPI_ISL_2759296, EPI_ISL_2759297, EPI_ISL_2759298, EPI_ISL_2759299, EPI_ISL_2759300, EPI_ISL_2759302, EPI_ISL_2759303, EPI_ISL_2759304, EPI_ISL_2759305, EPI_ISL_2759306, EPI_ISL_2759307, EPI_ISL_2759308, EPI_ISL_2759309, EPI_ISL_2759310, EPI_ISL_2759312, EPI_ISL_2759313, EPI_ISL_2759314, EPI_ISL_2759316, EPI_ISL_2759317, EPI_ISL_2759318, EPI_ISL_2759319, EPI_ISL_2759320, EPI_ISL_2759321, EPI_ISL_2759322, EPI_ISL_2759323, EPI_ISL_2759324, EPI_ISL_2759325, EPI_ISL_2759326, EPI_ISL_2759327, EPI_ISL_2759328, EPI_ISL_2759329, EPI_ISL_2759330, EPI_ISL_2759331, EPI_ISL_2759333, EPI_ISL_2759334, EPI_ISL_2759336, EPI_ISL_2759337, EPI_ISL_2759339, EPI_ISL_2759340, EPI_ISL_2759341, EPI_ISL_2759342, EPI_ISL_2759343, EPI_ISL_2759344, EPI_ISL_2759345, EPI_ISL_2759346, EPI_ISL_2759347, EPI_ISL_2759348, EPI_ISL_2759349, EPI_ISL_2759350, EPI_ISL_2759351, EPI_ISL_2759352, EPI_ISL_2759353, EPI_ISL_2759354, EPI_ISL_2759355, EPI_ISL_2759356, EPI_ISL_2759357, EPI_ISL_2759358, EPI_ISL_2759359, EPI_ISL_2759360, EPI_ISL_2759361, EPI_ISL_2759362, EPI_ISL_2759363, EPI_ISL_2759364, EPI_ISL_2759365, EPI_ISL_2759366, EPI_ISL_2759367, EPI_ISL_2759368, EPI_ISL_2759369, EPI_ISL_2759370, EPI_ISL_2759371, EPI_ISL_2759372, EPI_ISL_2759373, EPI_ISL_2759374, EPI_ISL_2759375, EPI_ISL_2759376, EPI_ISL_2759377, EPI_ISL_2759378, EPI_ISL_2759379, EPI_ISL_2759380, EPI_ISL_2759381, EPI_ISL_2759382, EPI_ISL_2759383, EPI_ISL_2759384, EPI_ISL_2759385, EPI_ISL_2759386, EPI_ISL_2759387, EPI_ISL_2759388, EPI_ISL_2759390, EPI_ISL_2759391, EPI_ISL_2759392, EPI_ISL_2759393, EPI_ISL_2759394, EPI_ISL_2759396, EPI_ISL_2759398, EPI_ISL_2759400, EPI_ISL_2759401, EPI_ISL_2759402, EPI_ISL_2759403, EPI_ISL_2759404, EPI_ISL_2759405, EPI_ISL_2759406, EPI_ISL_2759407, EPI_ISL_2759408, EPI_ISL_2759409, EPI_ISL_2759410, EPI_ISL_2759411, EPI_ISL_2759412, EPI_ISL_2759413, EPI_ISL_2759416, EPI_ISL_2759417, EPI_ISL_2759418, EPI_ISL_2759419, EPI_ISL_2759420, EPI_ISL_2759422, EPI_ISL_2759423, EPI_ISL_2759424, EPI_ISL_2759425, EPI_ISL_2759426, EPI_ISL_2759427, EPI_ISL_2759428, EPI_ISL_2759429, EPI_ISL_2759430, EPI_ISL_2759431, EPI_ISL_2759432, EPI_ISL_2759433, EPI_ISL_2759434, EPI_ISL_2759435, EPI_ISL_2759436, EPI_ISL_2759437, EPI_ISL_2759438, EPI_ISL_2759440, EPI_ISL_2759441, EPI_ISL_2759442, EPI_ISL_2759443, EPI_ISL_2759444, EPI_ISL_2759445, EPI_ISL_2759447, EPI_ISL_2759448, EPI_ISL_2759449, EPI_ISL_2759450, EPI_ISL_2759451, EPI_ISL_2759452, EPI_ISL_2759453, EPI_ISL_2759454, EPI_ISL_2759455, EPI_ISL_2759456, EPI_ISL_2759457, EPI_ISL_2759458, EPI_ISL_2759459, EPI_ISL_2759460, EPI_ISL_2759461, EPI_ISL_2759462, EPI_ISL_2759464, EPI_ISL_2759465, EPI_ISL_2759466, EPI_ISL_2759468, EPI_ISL_2759469, EPI_ISL_2759470, EPI_ISL_2759471, EPI_ISL_2759472, EPI_ISL_2759473, EPI_ISL_2759474, EPI_ISL_2759476, EPI_ISL_2759477, EPI_ISL_2759478, EPI_ISL_2759479, EPI_ISL_2759481, EPI_ISL_2759482, EPI_ISL_2759483, EPI_ISL_2759484, EPI_ISL_2759485, EPI_ISL_2759486, EPI_ISL_2759487, EPI_ISL_2759488, EPI_ISL_2759489, EPI_ISL_2759490, EPI_ISL_2759491, EPI_ISL_2759492, EPI_ISL_2759493, EPI_ISL_2759494, EPI_ISL_2759495, EPI_ISL_2759496, EPI_ISL_2759497, EPI_ISL_2759498, EPI_ISL_2759499, EPI_ISL_2759500, EPI_ISL_2759501, EPI_ISL_2759502, EPI_ISL_2759503, EPI_ISL_2759504, EPI_ISL_2759505, EPI_ISL_2759506, EPI_ISL_2759507, EPI_ISL_2759508, EPI_ISL_2759509, EPI_ISL_2759510, EPI_ISL_2759511, EPI_ISL_2759512, EPI_ISL_2759513, EPI_ISL_2759514, EPI_ISL_2759516, EPI_ISL_2759517, EPI_ISL_2759518, EPI_ISL_2759519, EPI_ISL_2759520, EPI_ISL_2759521, EPI_ISL_2759522, EPI_ISL_2759523, EPI_ISL_2759524, EPI_ISL_2759525, EPI_ISL_2759526, EPI_ISL_2759527, EPI_ISL_2759528, EPI_ISL_2759529, EPI_ISL_2759530, EPI_ISL_2759531, EPI_ISL_2759532, EPI_ISL_2759533, EPI_ISL_2759534, EPI_ISL_2759535, EPI_ISL_2759536, EPI_ISL_2759537, EPI_ISL_2759538, EPI_ISL_2759539, EPI_ISL_2759540, EPI_ISL_2759541, EPI_ISL_2759542, EPI_ISL_2759543, EPI_ISL_2759544, EPI_ISL_2759545, EPI_ISL_2759546, EPI_ISL_2759548, EPI_ISL_2759549, EPI_ISL_2759550, EPI_ISL_2759551, EPI_ISL_2759552, EPI_ISL_2759553, EPI_ISL_2759554, EPI_ISL_2759555, EPI_ISL_2759556, EPI_ISL_2759557, EPI_ISL_2759558, EPI_ISL_2759559, EPI_ISL_2759560, EPI_ISL_2759561, EPI_ISL_2759562, EPI_ISL_2759563, EPI_ISL_2759564, EPI_ISL_2759565, EPI_ISL_2759566, EPI_ISL_2759567, EPI_ISL_2759568, EPI_ISL_2759569, EPI_ISL_2759571, EPI_ISL_2759572, EPI_ISL_2759573, EPI_ISL_2759574, EPI_ISL_2759575, EPI_ISL_2759576, EPI_ISL_2759577, EPI_ISL_2759578, EPI_ISL_2759579, EPI_ISL_2759580, EPI_ISL_2759581, EPI_ISL_2759582, EPI_ISL_2759583, EPI_ISL_2759584, EPI_ISL_2759585, EPI_ISL_2759586, EPI_ISL_2759587, EPI_ISL_2759588, EPI_ISL_2759589, EPI_ISL_2759590, EPI_ISL_2759591, EPI_ISL_2759592, EPI_ISL_2759593, EPI_ISL_2759594, EPI_ISL_2759595, EPI_ISL_2759596, EPI_ISL_2759597, EPI_ISL_2759598, EPI_ISL_2759599, EPI_ISL_2759600, EPI_ISL_2759601, EPI_ISL_2759602, EPI_ISL_2759605, EPI_ISL_2759606, EPI_ISL_2759607, EPI_ISL_2759608, EPI_ISL_2759609, EPI_ISL_2759610, EPI_ISL_2759611, EPI_ISL_2759612, EPI_ISL_2759614, EPI_ISL_2759615, EPI_ISL_2759616, EPI_ISL_2759617, EPI_ISL_2759618, EPI_ISL_2759619, EPI_ISL_2759620, EPI_ISL_2759621, EPI_ISL_2759622, EPI_ISL_2759623, EPI_ISL_2759624, EPI_ISL_2759625, EPI_ISL_2759626, EPI_ISL_2759627, EPI_ISL_2759628, EPI_ISL_2759629, EPI_ISL_2759630, EPI_ISL_2759631, EPI_ISL_2759632, EPI_ISL_2759633, EPI_ISL_2759634, EPI_ISL_2759635, EPI_ISL_2759636, EPI_ISL_2759637, EPI_ISL_2759638, EPI_ISL_2759639, EPI_ISL_2759640, EPI_ISL_2759641, EPI_ISL_2759642, EPI_ISL_2759643, EPI_ISL_2759644 | | | |
| see above | Population Medicine and Diagnostic Sciences, Cornell University | Population Medicine and Diagnostic Sciences, Cornell University | Mitchell,P.K., Caserta,L.C., Cronk,B.D., Anderson,R.R., Laverack,M., Venugopalan,R., Plocharczyk,E., Diel,D.G. |
| EPI_ISL_2759876, EPI_ISL_2759877, EPI_ISL_2759878, EPI_ISL_2759879, EPI_ISL_2759880, EPI_ISL_2759881, EPI_ISL_2759882, EPI_ISL_2759883, EPI_ISL_2759884, EPI_ISL_2759885, EPI_ISL_2759886 | | | |
| see above | Washington State Department of Health Public Health Laboratories | Washington State Department of Health Public Health Laboratories | Drew MacKellar, Philip Dykema, Denny Russell, Joenice Gonzalez, Hannah Gray, Geoff Melly, Vanessa De Los Santos, Darren Lucas, JohnAric Peterson, Avi Singh, Rebecca Cao |
| EPI_ISL_2761672, EPI_ISL_2761674, EPI_ISL_2761675, EPI_ISL_2761676, EPI_ISL_2761678, EPI_ISL_2761680, EPI_ISL_2761681, EPI_ISL_2761683, EPI_ISL_2761685, EPI_ISL_2761686, EPI_ISL_2761688, EPI_ISL_2761690, EPI_ISL_2761691, EPI_ISL_2761693 | | | |
| see above | Pathology, University of Alabama at Birmingham | Pathology, University of Alabama at Birmingham | Moates,D., Hendrickson,R.C., Lefkowitz,E.J., Leal,S.M. |
| EPI_ISL_2768021, EPI_ISL_2768022, EPI_ISL_2768023, EPI_ISL_2768027, EPI_ISL_2768029 | Ohio Department of Health Laboratory | Ohio Department of Health Laboratory | Holmes, Jennifer; Eric Brandt, Keoni Omura, Glen McGillivary, Caitlin McDonnell, Jade Mowery, Stephanie Mccracken, Tyler Payne, Kirtana Ramadugu, Erica Leasure, Brent Lee, Kelsey Florek, Heather Blankenship, Allison Black, Quanta Brown, and Tammy Bannerman |
| EPI_ISL_2768051, EPI_ISL_2768052, EPI_ISL_2768054, EPI_ISL_2768055, EPI_ISL_2768056, EPI_ISL_2768057, EPI_ISL_2768058, EPI_ISL_2768060, EPI_ISL_2768061, EPI_ISL_2768062, EPI_ISL_2768063, EPI_ISL_2768064, EPI_ISL_2768065, EPI_ISL_2768066, EPI_ISL_2768067, EPI_ISL_2768068, EPI_ISL_2768069, EPI_ISL_2768070, EPI_ISL_2768071, EPI_ISL_2768072, EPI_ISL_2768073, EPI_ISL_2768074, EPI_ISL_2768076, EPI_ISL_2768077, EPI_ISL_2768078 | | | |
| see above | IN State Department of Health Laboratory Services | IN State Department of Health Laboratory Services | Cassandra Campion, Jamie Yeadon, Brian Pope, Lixia Liu, Kyle Brownlee, Melissa Hindenlang, Mark Glazier |

| | | | |
|---|---|---|---|
| EPI_ISL_2768080, EPI_ISL_2768088, EPI_ISL_2768100, EPI_ISL_2768101, EPI_ISL_2768102, EPI_ISL_2768103, EPI_ISL_2768104 | Ohio Department of Health Laboratory | Ohio Department of Health Laboratory | Holmes, Jennifer; Eric Brandt, Keoni Omura, Glen McGillivary, Caitlin McDonnell, Jade Mowery, Stephanie Mccracken, Tyler Payne, Kirtana Ramadugu, Erica Leasure, Brent Lee, Kelsey Florek, Heather Blankenship, Allison Black, Quanta Brown, and Tammy Bannerman |
| EPI_ISL_2769159, EPI_ISL_2769162, EPI_ISL_2769163, EPI_ISL_2769164, EPI_ISL_2769165, EPI_ISL_2769168, EPI_ISL_2769169, EPI_ISL_2769170, EPI_ISL_2769173, EPI_ISL_2769174, EPI_ISL_2769175, EPI_ISL_2769176, EPI_ISL_2769183, EPI_ISL_2769186, EPI_ISL_2769187, EPI_ISL_2769189, EPI_ISL_2769191, EPI_ISL_2769194, EPI_ISL_2769195, EPI_ISL_2769197, EPI_ISL_2769201, EPI_ISL_2769203, EPI_ISL_2769204, EPI_ISL_2769205, EPI_ISL_2769208, EPI_ISL_2769210, EPI_ISL_2769211, EPI_ISL_2769212, EPI_ISL_2769213, EPI_ISL_2769215, EPI_ISL_2769216, EPI_ISL_2769217, EPI_ISL_2769218, EPI_ISL_2769223, EPI_ISL_2769224, EPI_ISL_2769225, EPI_ISL_2769226, EPI_ISL_2769231, EPI_ISL_2769232, EPI_ISL_2769236, EPI_ISL_2769238, EPI_ISL_2769239, EPI_ISL_2769240, EPI_ISL_2769241, EPI_ISL_2769243, EPI_ISL_2769249, EPI_ISL_2769251, EPI_ISL_2769254, EPI_ISL_2769255, EPI_ISL_2769257, EPI_ISL_2769260, EPI_ISL_2769261, EPI_ISL_2769269, EPI_ISL_2769271, EPI_ISL_2769272, EPI_ISL_2769277, EPI_ISL_2769282, EPI_ISL_2769285, EPI_ISL_2769290, EPI_ISL_2769296, EPI_ISL_2769300, EPI_ISL_2769303, EPI_ISL_2769306, EPI_ISL_2769308, EPI_ISL_2769313, EPI_ISL_2769314, EPI_ISL_2769316, EPI_ISL_2769318, EPI_ISL_2769319, EPI_ISL_2769320, EPI_ISL_2769321, EPI_ISL_2769322, EPI_ISL_2769323, EPI_ISL_2769329, EPI_ISL_2769332, EPI_ISL_2769333, EPI_ISL_2769338, EPI_ISL_2769343, EPI_ISL_2769344, EPI_ISL_2769346, EPI_ISL_2769347, EPI_ISL_2769351, EPI_ISL_2769352 | | | |
| see above | TGen North | TGen North | "Jolene Bowers, Heather Centner, Brett Van Tassel, Chris French, Hayley Yaglom, Darrin Lemmer, Dave Engelthaler" |
| EPI_ISL_2769364, EPI_ISL_2769365, EPI_ISL_2769366, EPI_ISL_2769367, EPI_ISL_2769368 | Hackensack Medical Center | New York Genome Center | Michael Zody, Andre Corvelo, Dayna M. Oschwald, Samantha Fennessey, Tom Maniatis, Liang Chen, Jose Mediavilla, Marcus Cunningham, Kaelea Composto, Kar Chow, David Perlin, Barry Kreiswirth |
| EPI_ISL_2771492, EPI_ISL_2771493, EPI_ISL_2771494, EPI_ISL_2771495, EPI_ISL_2771496, EPI_ISL_2771499 | Ohio Department of Health Laboratory | Ohio Department of Health Laboratory | Holmes, Jennifer; Eric Brandt, Keoni Omura, Glen McGillivary, Caitlin McDonnell, Jade Mowery, Stephanie Mccracken, Tyler Payne, Kirtana Ramadugu, Erica Leasure, Brent Lee, Kelsey Florek, Heather Blankenship, Allison Black, Quanta Brown, and Tammy Bannerman |
| EPI_ISL_2772501 | Atlanta VA Medical Center | Genomics and Discovery, Respiratory Viruses Branch, Division of Viral Diseases, Centers for Disease Control and Prevention | Anna Kelleher, Ying Tao, Yan Li, Jing Zhang, Brian Lynch, Krista Queen, Anna Uehara, Peter Cook, Han Jia Justin Ng, Rachel Marine, Clinton R. Paden, Dhwani Batra, Haibin Wang, Tara Coalter, Jasmine Padilla, Morgan Davis, Mili Sheth, Sarah Nobles, Mark Burroughs, Justin Lee, Adam Retchless, Suxiang Tong |
| EPI_ISL_2772502, EPI_ISL_2772503, EPI_ISL_2772504 | California Hospital Medical Center | Los Angeles County Public Health Laboratories | P. Hemarajata et al. |
| EPI_ISL_2772505 | Pomona Valley Hospital Medical Center | Los Angeles County Public Health Laboratories | P. Hemarajata et al. |
| EPI_ISL_2772506, EPI_ISL_2772507, EPI_ISL_2772508, EPI_ISL_2772509, EPI_ISL_2772510, EPI_ISL_2772511, EPI_ISL_2772512, EPI_ISL_2772513, EPI_ISL_2772514, EPI_ISL_2772515, EPI_ISL_2772516, EPI_ISL_2772517, EPI_ISL_2772518, EPI_ISL_2772519, EPI_ISL_2772520, EPI_ISL_2772521 | | | |
| see above | UCLA Clinical Micro Lab | Los Angeles County Public Health Laboratories | P. Hemarajata et al. |
| EPI_ISL_2772522, EPI_ISL_2772523, EPI_ISL_2772524 | Flow Diagnostics Lab | Los Angeles County Public Health Laboratories | P. Hemarajata et al. |
| EPI_ISL_2772525 | Palmdale Regional Medical Center | Los Angeles County Public Health Laboratories | P. Hemarajata et al. |
| EPI_ISL_2772532, EPI_ISL_2772533, EPI_ISL_2772534, EPI_ISL_2772535, EPI_ISL_2772536, EPI_ISL_2772537, EPI_ISL_2772538, EPI_ISL_2772539, EPI_ISL_2772540, EPI_ISL_2772541, EPI_ISL_2772542, EPI_ISL_2772543, EPI_ISL_2772544, EPI_ISL_2772545, EPI_ISL_2772546, EPI_ISL_2772547, EPI_ISL_2772548, EPI_ISL_2772549, EPI_ISL_2772550, EPI_ISL_2772551, EPI_ISL_2772552, EPI_ISL_2772553, EPI_ISL_2772554, EPI_ISL_2772555, EPI_ISL_2772556, EPI_ISL_2772557, EPI_ISL_2772558, EPI_ISL_2772559, EPI_ISL_2772560, EPI_ISL_2772561, EPI_ISL_2772562, EPI_ISL_2772563, EPI_ISL_2772565, EPI_ISL_2772566, EPI_ISL_2772567, EPI_ISL_2772568, EPI_ISL_2772569, EPI_ISL_2772570, EPI_ISL_2772571, EPI_ISL_2772572, EPI_ISL_2772573, EPI_ISL_2772574, EPI_ISL_2772575, EPI_ISL_2772576, EPI_ISL_2772577, EPI_ISL_2772578, EPI_ISL_2772579, EPI_ISL_2772580, EPI_ISL_2772581, EPI_ISL_2772582 | | | |
| see above | Kansas Health and Environmental Lab | Kansas Health and Environmental Lab | Katherine Wiggins, Mike Grose, Jonathan Barnell, Ben Olsen, and Phil Adam |
| EPI_ISL_2772627 | UMGC | UMGC | Jeffrey Good, Nathanael Herrera, Gregg Thomas, Tim Thurman, David Xing |
| EPI_ISL_2772718, EPI_ISL_2772719, EPI_ISL_2772720, EPI_ISL_2772721, EPI_ISL_2772722, EPI_ISL_2772723 | Vanderbilt University Medical Center | Genomics and Discovery, Respiratory Viruses Branch, Division of Viral Diseases, Centers for Disease Control and Prevention | Anna Kelleher, Ying Tao, Yan Li, Jing Zhang, Brian Lynch, Krista Queen, Anna Uehara, Peter Cook, Han Jia Justin Ng, Rachel Marine, Clinton R. Paden, Dhwani Batra, Haibin Wang, Tara Coalter, Jasmine Padilla, Morgan Davis, Mili Sheth, Sarah Nobles, Mark Burroughs, Justin Lee, Adam Retchless, Suxiang Tong |
| EPI_ISL_2773872, EPI_ISL_2773873, EPI_ISL_2773874, EPI_ISL_2773875, EPI_ISL_2773876, EPI_ISL_2773877, EPI_ISL_2773878, EPI_ISL_2773879, EPI_ISL_2773880, EPI_ISL_2773881, EPI_ISL_2773882, EPI_ISL_2773883, EPI_ISL_2773884, EPI_ISL_2773885, EPI_ISL_2773886, EPI_ISL_2773887, EPI_ISL_2773888, EPI_ISL_2773889, EPI_ISL_2773890, EPI_ISL_2773891, EPI_ISL_2773892, EPI_ISL_2773893, EPI_ISL_2773894, EPI_ISL_2773895, EPI_ISL_2773896, EPI_ISL_2773897, EPI_ISL_2773898, EPI_ISL_2773899, EPI_ISL_2773900, EPI_ISL_2773901, EPI_ISL_2773902, EPI_ISL_2773903, EPI_ISL_2773904, EPI_ISL_2773905, EPI_ISL_2773906, EPI_ISL_2773907, EPI_ISL_2773908, EPI_ISL_2773909, EPI_ISL_2773910, EPI_ISL_2773911, EPI_ISL_2773912, EPI_ISL_2773913, EPI_ISL_2773914, EPI_ISL_2773915, EPI_ISL_2773916, EPI_ISL_2773917, EPI_ISL_2773918, EPI_ISL_2773919, EPI_ISL_2773920, EPI_ISL_2773921, EPI_ISL_2773922, EPI_ISL_2773923, EPI_ISL_2773924, EPI_ISL_2773925, EPI_ISL_2773926, EPI_ISL_2773927, EPI_ISL_2773928, EPI_ISL_2773929, EPI_ISL_2773930, EPI_ISL_2773931, EPI_ISL_2773932, EPI_ISL_2773933, EPI_ISL_2773934, EPI_ISL_2773935, EPI_ISL_2773936, EPI_ISL_2773937, EPI_ISL_2773938, EPI_ISL_2773939, EPI_ISL_2773940, EPI_ISL_2773941, EPI_ISL_2773942, EPI_ISL_2773943, EPI_ISL_2773944 | | | |
| see above | UW Virology Lab | UW Virology Lab | Pavitra Roychoudhury, Hong Xie, Lasata Shrestha, Shah Mohamed Bakhash, Margaret Mills, Noah Baker, Sean Ellis, Meei-Li Huang, Keith R Jerome, Alexander Greninger |
| EPI_ISL_2774241, EPI_ISL_2774242, EPI_ISL_2774243, EPI_ISL_2774244, EPI_ISL_2774245, EPI_ISL_2774246, EPI_ISL_2774247, EPI_ISL_2774248, EPI_ISL_2774249, EPI_ISL_2774252, EPI_ISL_2774253, EPI_ISL_2774254, EPI_ISL_2774255, EPI_ISL_2774256, EPI_ISL_2774257, EPI_ISL_2774258, EPI_ISL_2774259, EPI_ISL_2774260, EPI_ISL_2774261, EPI_ISL_2774263, EPI_ISL_2774264, EPI_ISL_2774265, EPI_ISL_2774266, EPI_ISL_2774267, EPI_ISL_2774268, EPI_ISL_2774269, EPI_ISL_2774270, EPI_ISL_2774271, EPI_ISL_2774272, EPI_ISL_2774273, EPI_ISL_2774274, EPI_ISL_2774275, EPI_ISL_2774276, EPI_ISL_2774277, EPI_ISL_2774278, EPI_ISL_2774279, EPI_ISL_2774281, EPI_ISL_2774282, EPI_ISL_2774283, EPI_ISL_2774284, EPI_ISL_2774285, EPI_ISL_2774286, EPI_ISL_2774287, EPI_ISL_2774288, EPI_ISL_2774289, EPI_ISL_2774290, EPI_ISL_2774291, EPI_ISL_2774292, EPI_ISL_2774293, EPI_ISL_2774294, EPI_ISL_2774295, EPI_ISL_2774296, EPI_ISL_2774297, EPI_ISL_2774298, EPI_ISL_2774299, EPI_ISL_2774300, EPI_ISL_2774301, EPI_ISL_2774302, EPI_ISL_2774303, EPI_ISL_2774304, EPI_ISL_2774305 | | | |
| see above | Baylor Scott & White-Temple | Baylor Scott & White-Temple | Ari Rao, Linden Morales, Kimberly Walker, Marcus Volz, Shelby Johnson |
| EPI_ISL_2774306, EPI_ISL_2774307, EPI_ISL_2774308, EPI_ISL_2774309, EPI_ISL_2774310, EPI_ISL_2774311 | Baylor Scott & White-Irving | Baylor Scott & White-Temple | Ari Rao, Linden Morales, Kimberly Walker, Marcus Volz, Shelby Johnson |
| EPI_ISL_2774312, EPI_ISL_2774313, EPI_ISL_2774314, EPI_ISL_2774315, EPI_ISL_2774316, EPI_ISL_2774317, EPI_ISL_2774318, EPI_ISL_2774319, EPI_ISL_2774320, EPI_ISL_2774323, EPI_ISL_2774324, EPI_ISL_2774325, EPI_ISL_2774326, EPI_ISL_2774327, EPI_ISL_2774328, EPI_ISL_2774329, EPI_ISL_2774330, EPI_ISL_2774331, EPI_ISL_2774332, EPI_ISL_2774333, EPI_ISL_2774334, EPI_ISL_2774335, EPI_ISL_2774336, EPI_ISL_2774337, EPI_ISL_2774338, EPI_ISL_2774339, EPI_ISL_2774340, EPI_ISL_2774341, EPI_ISL_2774343, EPI_ISL_2774344, EPI_ISL_2774345, EPI_ISL_2774346, EPI_ISL_2774347, EPI_ISL_2774348, EPI_ISL_2774349, EPI_ISL_2774350, EPI_ISL_2774351, EPI_ISL_2774352, EPI_ISL_2774353, EPI_ISL_2774354, EPI_ISL_2774355, EPI_ISL_2774356, EPI_ISL_2774357, EPI_ISL_2774358, EPI_ISL_2774360, EPI_ISL_2774361, EPI_ISL_2774362, EPI_ISL_2774363, EPI_ISL_2774364, EPI_ISL_2774365, EPI_ISL_2774367, EPI_ISL_2774368, EPI_ISL_2774369, EPI_ISL_2774370, EPI_ISL_2774371, EPI_ISL_2774372, EPI_ISL_2774373, EPI_ISL_2774374, EPI_ISL_2774375, EPI_ISL_2774376, EPI_ISL_2774377, EPI_ISL_2774378, EPI_ISL_2774379, EPI_ISL_2774380, EPI_ISL_2774381, EPI_ISL_2774382, EPI_ISL_2774383, EPI_ISL_2774384, EPI_ISL_2774385, EPI_ISL_2774386, EPI_ISL_2774387, EPI_ISL_2774388, EPI_ISL_2774389, EPI_ISL_2774390, EPI_ISL_2774391, EPI_ISL_2774392, EPI_ISL_2774393, EPI_ISL_2774394, EPI_ISL_2774395, EPI_ISL_2774396 | | | |
| see above | Illinois Department of Public Health - Springfield Lab | Illinois Department of Public Health - Springfield Lab | Bryan Sim, Gordon McCall |
| EPI_ISL_2774493 | Salud Digna | Instituto Nacional de Medicina Genomica | Hidalgo-Miranda A, Cedro-Tanda A, Cisneros-Villanueva M, Orjuela-Rodríguez M, Mendoza-Vargas A, Reyes-Grajeda JP, Abraham Campos-Romero, Moreno-Camacho José Luis, Rodriguez-Gallegos Jorge, Luna-Ruiz Marco, Gonzalez-Barrera D, Rangel-DeLeon D, Munguia-Garza P, Ramirez-Vega O, Escobar-Arrazola, M, Herrera-Montalvo LA. |
| EPI_ISL_2774495, EPI_ISL_2774496 | Weill Cornell Medicine | New York Genome Center | Michael Zody, Andre Corvelo, Dayna M. Oschwald, Samantha Fennessey, Tom Maniatis, Melissa Cushing, Olivier Elemento, Margaret Elizabeth Ross, Chris Mason, Priya Velu, Hanna Rennert, Arryn Craney, Lars F Westblade |
| EPI_ISL_2774497 | San Diego County Public Health Lab | San Diego County Public Health Lab | Ashleigh Murphy Schafer, Brett Austin |
| EPI_ISL_2774498, EPI_ISL_2774499, EPI_ISL_2774500, EPI_ISL_2774501, EPI_ISL_2774502, EPI_ISL_2774503, EPI_ISL_2774504 | Hackensack Medical Center | New York Genome Center | Michael Zody, Andre Corvelo, Dayna M. Oschwald, Samantha Fennessey, Tom Maniatis, Liang Chen, Jose Mediavilla, Marcus Cunningham, Kaelea Composto, Kar Chow, David Perlin, Barry Kreiswirth |
| EPI_ISL_2774512 | Monterey County Public Health Laboratory | Monterey County Public Health Laboratory | Monterey County Public Health Laboratory |
| EPI_ISL_2774516, EPI_ISL_2774517 | Interpath Laboratory | Seattle Flu Study | Deborah A. Nickerson, Chris D. Frazar, Jover Lee, Benjamin Pelle, Erica Ryke, Matthew Richardson, Amanda Adler, Elisabeth Brandstetter, Peter D. Han, Kairsten Fay, Misja Ilcisin, Kirsten Lacombe, Thomas R. Sibley, Melissa Truong, Caitlin R. Wolf, Romesh Gautom, Geoff Melly, Brian Hiatt, Philip Dykema, Scott Lindquist, Michael Boeckh, Janet A. Englund, Michael Famulare, Barry R. Lutz, Mark J. Rieder, Lea M. Starita, Matthew Thompson, Helen Y. Chu, Jay Shendure, Trevor Bedford |
| EPI_ISL_2774519, EPI_ISL_2774520, EPI_ISL_2774521, EPI_ISL_2774522, EPI_ISL_2774523 | Monterey County Public Health Laboratory | Monterey County Public Health Laboratory | Monterey County Public Health Laboratory |
| EPI_ISL_2774524, EPI_ISL_2774525, EPI_ISL_2774526 | Weill Cornell Medicine | New York Genome Center | Michael Zody, Andre Corvelo, Dayna M. Oschwald, Samantha Fennessey, Tom Maniatis, Melissa Cushing, Olivier Elemento, Margaret Elizabeth Ross, Chris Mason, Priya Velu, Hanna Rennert, Arryn Craney, Lars F Westblade |
| EPI_ISL_2774527, EPI_ISL_2774528, EPI_ISL_2774529, EPI_ISL_2774530, EPI_ISL_2774531, EPI_ISL_2774532, EPI_ISL_2774533, EPI_ISL_2774534, EPI_ISL_2774535, EPI_ISL_2774536, EPI_ISL_2774537, EPI_ISL_2774538, EPI_ISL_2774539, EPI_ISL_2774540, EPI_ISL_2774541, EPI_ISL_2774542, EPI_ISL_2774543, EPI_ISL_2774544, EPI_ISL_2774545, EPI_ISL_2774546, EPI_ISL_2774547, EPI_ISL_2774548, | | | |

| | | | |
|---|---|---|---|
| EPI_ISL_2774549, EPI_ISL_2774550, EPI_ISL_2774551, EPI_ISL_2774552, EPI_ISL_2774553, EPI_ISL_2774554 | | | |
| see above | Nebraska Public Health Laboratory | NPHL COVID-19 Response Team | NPHL COVID-19 Response Team |
| EPI_ISL_2774556 | Seattle Flu Study | Seattle Flu Study | Deborah A. Nickerson, Chris D. Frazar, Jover Lee, Benjamin Pelle, Erica Ryke, Matthew Richardson, Amanda Adler, Elisabeth Brandstetter, Peter D. Han, Kairsten Fay, Misja Ilcisin, Kirsten Lacombe, Thomas R. Sibley, Melissa Truong, Caitlin R. Wolf, Karen Cowgill, Stephanie Schrag, Jeff Duchin, Michael Boeckh, Janet A. Englund, Michael Famulare, Barry R. Lutz, Mark J. Rieder, Lea M. Starita, Matthew Thompson, Helen Y. Chu, Trevor Bedford, Jay Shendure |
| EPI_ISL_2774557, EPI_ISL_2774558 | Northwest Laboratory | Seattle Flu Study | Deborah A. Nickerson, Chris D. Frazar, Jover Lee, Benjamin Pelle, Erica Ryke, Matthew Richardson, Amanda Adler, Elisabeth Brandstetter, Peter D. Han, Kairsten Fay, Misja Ilcisin, Kirsten Lacombe, Thomas R. Sibley, Melissa Truong, Caitlin R. Wolf, Romesh Gautom, Geoff Melly, Brian Hiatt, Philip Dykema, Scott Lindquist, Michael Boeckh, Janet A. Englund, Michael Famulare, Barry R. Lutz, Mark J. Rieder, Lea M. Starita, Matthew Thompson, Helen Y. Chu, Jay Shendure, Trevor Bedford |
| EPI_ISL_2774559 | Incyte Diagnostics Spokane | Seattle Flu Study | Deborah A. Nickerson, Chris D. Frazar, Jover Lee, Benjamin Pelle, Erica Ryke, Matthew Richardson, Amanda Adler, Elisabeth Brandstetter, Peter D. Han, Kairsten Fay, Misja Ilcisin, Kirsten Lacombe, Thomas R. Sibley, Melissa Truong, Caitlin R. Wolf, Romesh Gautom, Geoff Melly, Brian Hiatt, Philip Dykema, Scott Lindquist, Michael Boeckh, Janet A. Englund, Michael Famulare, Barry R. Lutz, Mark J. Rieder, Lea M. Starita, Matthew Thompson, Helen Y. Chu, Jay Shendure, Trevor Bedford |
| EPI_ISL_2774560, EPI_ISL_2774561 | Atlas Genomics | Seattle Flu Study | Deborah A. Nickerson, Chris D. Frazar, Jover Lee, Benjamin Pelle, Erica Ryke, Matthew Richardson, Amanda Adler, Elisabeth Brandstetter, Peter D. Han, Kairsten Fay, Misja Ilcisin, Kirsten Lacombe, Thomas R. Sibley, Melissa Truong, Caitlin R. Wolf, Romesh Gautom, Geoff Melly, Brian Hiatt, Philip Dykema, Scott Lindquist, Michael Boeckh, Janet A. Englund, Michael Famulare, Barry R. Lutz, Mark J. Rieder, Lea M. Starita, Matthew Thompson, Helen Y. Chu, Jay Shendure, Trevor Bedford |
| EPI_ISL_2774562 | Martin Luther King Jr. Community Hospital | Los Angeles County Public Health Laboratories | P. Hemarajata et al. |
| EPI_ISL_2774563, EPI_ISL_2774564, EPI_ISL_2774565, EPI_ISL_2774566 | Los Angeles County Public Health Laboratories | Los Angeles County Public Health Laboratories | P. Hemarajata et al. |
| EPI_ISL_2774567 | USC Clinical Lab | Los Angeles County Public Health Laboratories | P. Hemarajata et al. |
| EPI_ISL_2774568 | Flow Diagnostics Lab | Los Angeles County Public Health Laboratories | P. Hemarajata et al. |
| EPI_ISL_2774570, EPI_ISL_2774571 | Los Angeles County Public Health Laboratories | Los Angeles County Public Health Laboratories | P. Hemarajata et al. |
| EPI_ISL_2774572 | Torrance Memorial Medical Center | Los Angeles County Public Health Laboratories | P. Hemarajata et al. |
| EPI_ISL_2774573, EPI_ISL_2774574 | Los Angeles County Public Health Laboratories | Los Angeles County Public Health Laboratories | P. Hemarajata et al. |
| EPI_ISL_2774575, EPI_ISL_2774576, EPI_ISL_2774577, EPI_ISL_2774579 | Santa Clara Valley Medical Center | Santa Clara County Public Health Laboratory | Santa Clara County Public Health Department |
| EPI_ISL_2774580, EPI_ISL_2774585, EPI_ISL_2774586, EPI_ISL_2774588 | PAMF Palo Alto | Santa Clara County Public Health Laboratory | Santa Clara County Public Health Department |
| EPI_ISL_2774591, EPI_ISL_2774592, EPI_ISL_2774593, EPI_ISL_2774594, EPI_ISL_2774595, EPI_ISL_2774596, EPI_ISL_2774598, EPI_ISL_2774599, EPI_ISL_2774600, EPI_ISL_2774601, EPI_ISL_2774602, EPI_ISL_2774604, EPI_ISL_2774605, EPI_ISL_2774607, EPI_ISL_2774608, EPI_ISL_2774609, EPI_ISL_2774610, EPI_ISL_2774613, EPI_ISL_2774614, EPI_ISL_2774615, EPI_ISL_2774618, EPI_ISL_2774620 | | | |
| see above | Santa Clara Valley Medical Center | Santa Clara County Public Health Laboratory | Santa Clara County Public Health Department |
| EPI_ISL_2774621 | El Camino Hospital | Santa Clara County Public Health Laboratory | Santa Clara County Public Health Department |
| EPI_ISL_2774622 | PAMF Palo Alto | Santa Clara County Public Health Laboratory | Santa Clara County Public Health Department |
| EPI_ISL_2774624, EPI_ISL_2774625, EPI_ISL_2774627, EPI_ISL_2774629, EPI_ISL_2774630, EPI_ISL_2774631, EPI_ISL_2774632, EPI_ISL_2774633, EPI_ISL_2774634 | Santa Clara Valley Medical Center | Santa Clara County Public Health Laboratory | Santa Clara County Public Health Department |
| EPI_ISL_2775501, EPI_ISL_2775502, EPI_ISL_2775503, EPI_ISL_2775506, EPI_ISL_2775508, EPI_ISL_2775510, EPI_ISL_2775511, EPI_ISL_2775512, EPI_ISL_2775517, EPI_ISL_2775518, EPI_ISL_2775519, EPI_ISL_2775520, EPI_ISL_2775521, EPI_ISL_2775522, EPI_ISL_2775523, EPI_ISL_2775524, EPI_ISL_2775526, EPI_ISL_2775527, EPI_ISL_2775528, EPI_ISL_2775530, EPI_ISL_2775532, EPI_ISL_2775538, EPI_ISL_2775539, EPI_ISL_2775541, EPI_ISL_2775542, EPI_ISL_2775543, EPI_ISL_2775544, EPI_ISL_2775545, EPI_ISL_2775548, EPI_ISL_2775549, EPI_ISL_2775550, EPI_ISL_2775551, EPI_ISL_2775552, EPI_ISL_2775553, EPI_ISL_2775554, EPI_ISL_2775555, EPI_ISL_2775558, EPI_ISL_2775559, EPI_ISL_2775560, EPI_ISL_2775561, EPI_ISL_2775564, EPI_ISL_2775565, EPI_ISL_2775566, EPI_ISL_2775567, EPI_ISL_2775568, EPI_ISL_2775569, EPI_ISL_2775571, EPI_ISL_2775572, EPI_ISL_2775573, EPI_ISL_2775575, EPI_ISL_2775576, EPI_ISL_2775579, EPI_ISL_2775580, EPI_ISL_2775581, EPI_ISL_2775582, EPI_ISL_2775583, EPI_ISL_2775584, EPI_ISL_2775585, EPI_ISL_2775586, EPI_ISL_2775587, EPI_ISL_2775588, EPI_ISL_2775589, EPI_ISL_2775590, EPI_ISL_2775591, EPI_ISL_2775592, EPI_ISL_2775593, EPI_ISL_2775596, EPI_ISL_2775597, EPI_ISL_2775599, EPI_ISL_2775600, EPI_ISL_2775601, EPI_ISL_2775602, EPI_ISL_2775603, EPI_ISL_2775606, EPI_ISL_2775607, EPI_ISL_2775608, EPI_ISL_2775611, EPI_ISL_2775612, EPI_ISL_2775613, EPI_ISL_2775614, EPI_ISL_2775615, EPI_ISL_2775616, EPI_ISL_2775617, EPI_ISL_2775618, EPI_ISL_2775619, EPI_ISL_2775620, EPI_ISL_2775622, EPI_ISL_2775623, EPI_ISL_2775624, EPI_ISL_2775626, EPI_ISL_2775628, EPI_ISL_2775629, EPI_ISL_2775630, EPI_ISL_2775631, EPI_ISL_2775632, EPI_ISL_2775635, EPI_ISL_2775636, EPI_ISL_2775637, EPI_ISL_2775638, EPI_ISL_2775639, EPI_ISL_2775640, EPI_ISL_2775641, EPI_ISL_2775642, EPI_ISL_2775643, EPI_ISL_2775644, EPI_ISL_2775645, EPI_ISL_2775646, EPI_ISL_2775648, EPI_ISL_2775649, EPI_ISL_2775651, EPI_ISL_2775652, EPI_ISL_2775653, EPI_ISL_2775654, EPI_ISL_2775655, EPI_ISL_2775656, EPI_ISL_2775657, EPI_ISL_2775658, EPI_ISL_2775659, EPI_ISL_2775660, EPI_ISL_2775661, EPI_ISL_2775662, EPI_ISL_2775663, EPI_ISL_2775664, EPI_ISL_2775665, EPI_ISL_2775666, EPI_ISL_2775667, EPI_ISL_2775668, EPI_ISL_2775669, EPI_ISL_2775670, EPI_ISL_2775671, EPI_ISL_2775673, EPI_ISL_2775675 | | | |
| see above | Wyoming Public Health Laboratory | Wyoming Public Health Laboratory | Jim Mildenberger, Taylor Fearing, Channing Weber, Ashley Norberg, Chayse Rowley, Marley Goetz, Brian Dominguez, Elliot Thomasson, Sam Britz, Cari Sloma, Robert Petit, and Rob Christensen |
| EPI_ISL_2775678, EPI_ISL_2775679, EPI_ISL_2775680, EPI_ISL_2775681, EPI_ISL_2775682, EPI_ISL_2775683, EPI_ISL_2775684, EPI_ISL_2775685, EPI_ISL_2775686, EPI_ISL_2775687, EPI_ISL_2775688, EPI_ISL_2775689, EPI_ISL_2775690, EPI_ISL_2775691, EPI_ISL_2775692, EPI_ISL_2775693, EPI_ISL_2775694, EPI_ISL_2775695, EPI_ISL_2775696, EPI_ISL_2775697, EPI_ISL_2775698, EPI_ISL_2775699, EPI_ISL_2775700, EPI_ISL_2775701, EPI_ISL_2775702, EPI_ISL_2775703, EPI_ISL_2775704, EPI_ISL_2775705, EPI_ISL_2775706, EPI_ISL_2775707, EPI_ISL_2775708, EPI_ISL_2775709, EPI_ISL_2775710, EPI_ISL_2775711, EPI_ISL_2775712, EPI_ISL_2775713, EPI_ISL_2775714, EPI_ISL_2775715, EPI_ISL_2775716, EPI_ISL_2775717, EPI_ISL_2775718, EPI_ISL_2775719, EPI_ISL_2775720, EPI_ISL_2775721, EPI_ISL_2775722, EPI_ISL_2775723, EPI_ISL_2775724, EPI_ISL_2775725, EPI_ISL_2775726, EPI_ISL_2775727, EPI_ISL_2775728, EPI_ISL_2775729, EPI_ISL_2775730, EPI_ISL_2775731, EPI_ISL_2775732, EPI_ISL_2775733, EPI_ISL_2775734, EPI_ISL_2775735, EPI_ISL_2775736, EPI_ISL_2775737, EPI_ISL_2775738, EPI_ISL_2775739, EPI_ISL_2775740, EPI_ISL_2775741, EPI_ISL_2775742, EPI_ISL_2775743, EPI_ISL_2775744, EPI_ISL_2775745, EPI_ISL_2775746, EPI_ISL_2775747, EPI_ISL_2775748, EPI_ISL_2775749, EPI_ISL_2775750, EPI_ISL_2775751, EPI_ISL_2775752, EPI_ISL_2775753, EPI_ISL_2775754, EPI_ISL_2775755, EPI_ISL_2775756, EPI_ISL_2775757, EPI_ISL_2775758, EPI_ISL_2775759, EPI_ISL_2775760, EPI_ISL_2775761, EPI_ISL_2775762, EPI_ISL_2775763, EPI_ISL_2775764, EPI_ISL_2775765, EPI_ISL_2775766, EPI_ISL_2775767, EPI_ISL_2775768, EPI_ISL_2775769, EPI_ISL_2775770, EPI_ISL_2775771, EPI_ISL_2775772, EPI_ISL_2775773, EPI_ISL_2775774, EPI_ISL_2775775, EPI_ISL_2775776, EPI_ISL_2775777, EPI_ISL_2775778, EPI_ISL_2775779, EPI_ISL_2775780, EPI_ISL_2775781, EPI_ISL_2775782, EPI_ISL_2775783, EPI_ISL_2775784, EPI_ISL_2775785, EPI_ISL_2775786, EPI_ISL_2775787, EPI_ISL_2775788, EPI_ISL_2775789, EPI_ISL_2775790, EPI_ISL_2775791, EPI_ISL_2775792, EPI_ISL_2775793, EPI_ISL_2775794, EPI_ISL_2775795, EPI_ISL_2775796, EPI_ISL_2775797, EPI_ISL_2775798, EPI_ISL_2775799, EPI_ISL_2775800, EPI_ISL_2775801, EPI_ISL_2775802, EPI_ISL_2775803, EPI_ISL_2775804, EPI_ISL_2775805, EPI_ISL_2775806, EPI_ISL_2775807, EPI_ISL_2775808, EPI_ISL_2775809, EPI_ISL_2775810, EPI_ISL_2775811, EPI_ISL_2775812, EPI_ISL_2775813, EPI_ISL_2775814, EPI_ISL_2775815, EPI_ISL_2775816, EPI_ISL_2775817, EPI_ISL_2775818, EPI_ISL_2775819, EPI_ISL_2775820, EPI_ISL_2775822, EPI_ISL_2775823, EPI_ISL_2775824, EPI_ISL_2775825, EPI_ISL_2775826, EPI_ISL_2775827, EPI_ISL_2775828, EPI_ISL_2775829, EPI_ISL_2775830, EPI_ISL_2775831, EPI_ISL_2775832, EPI_ISL_2775833, EPI_ISL_2775834, EPI_ISL_2775835, EPI_ISL_2775836, EPI_ISL_2775837, EPI_ISL_2775838, EPI_ISL_2775839, EPI_ISL_2775840, EPI_ISL_2775841, EPI_ISL_2775842, EPI_ISL_2775843, EPI_ISL_2775844, EPI_ISL_2775845, EPI_ISL_2775846, EPI_ISL_2775847, EPI_ISL_2775848, EPI_ISL_2775849, EPI_ISL_2775850, EPI_ISL_2775851, EPI_ISL_2775852, EPI_ISL_2775853, EPI_ISL_2775854, EPI_ISL_2775855, EPI_ISL_2775856, EPI_ISL_2775857, EPI_ISL_2775858, EPI_ISL_2775859, EPI_ISL_2775860, EPI_ISL_2775861, EPI_ISL_2775862, EPI_ISL_2775863, EPI_ISL_2775864, EPI_ISL_2775865, EPI_ISL_2775866, EPI_ISL_2775867, EPI_ISL_2775868, EPI_ISL_2775869, EPI_ISL_2775870, EPI_ISL_2775871, EPI_ISL_2775872, EPI_ISL_2775873, EPI_ISL_2775874, EPI_ISL_2775875, EPI_ISL_2775876 | | | |
| see above | UPMC Clinical Microbiology Laboratory | Microbial Genome Sequencing Center; Microbial Genomic Epidemiology Laboratory, University of Pittsburgh | Lee H. Harrison, Jane W. Marsh, Marissa P. Griffith, Vatsala R. Srinivasa, Kady D. Waggle, Daniel J. Snyder, Vaughn S. Cooper |
| EPI_ISL_2776014, EPI_ISL_2776015, EPI_ISL_2776016, EPI_ISL_2776017, EPI_ISL_2776018, EPI_ISL_2776019, EPI_ISL_2776020, EPI_ISL_2776021, EPI_ISL_2776022, EPI_ISL_2776023, EPI_ISL_2776024, EPI_ISL_2776025, EPI_ISL_2776026, EPI_ISL_2776027, EPI_ISL_2776028, EPI_ISL_2776029, EPI_ISL_2776030, EPI_ISL_2776031, EPI_ISL_2776032, EPI_ISL_2776033, EPI_ISL_2776034, EPI_ISL_2776035, EPI_ISL_2776036, EPI_ISL_2776037, EPI_ISL_2776038, EPI_ISL_2776039, EPI_ISL_2776040, EPI_ISL_2776041, EPI_ISL_2776042, EPI_ISL_2776043, EPI_ISL_2776044, EPI_ISL_2776045, EPI_ISL_2776046, EPI_ISL_2776047, EPI_ISL_2776048, EPI_ISL_2776049, EPI_ISL_2776050, EPI_ISL_2776051, EPI_ISL_2776052, EPI_ISL_2776053, EPI_ISL_2776054, EPI_ISL_2776055, EPI_ISL_2776056, EPI_ISL_2776057, EPI_ISL_2776058, EPI_ISL_2776059, EPI_ISL_2776060, EPI_ISL_2776061, EPI_ISL_2776062, EPI_ISL_2776063, EPI_ISL_2776064, EPI_ISL_2776065, EPI_ISL_2776066, EPI_ISL_2776067 | | | |
| see above | Virginia Division of Consolidated Laboratory Services | Virginia Division of Consolidated Laboratory Services | Virginia Division of Consolidated Laboratory Services |
| EPI_ISL_2776068, EPI_ISL_2776070, EPI_ISL_2776071, EPI_ISL_2776072, EPI_ISL_2776074, EPI_ISL_2776075, EPI_ISL_2776076, EPI_ISL_2776078, EPI_ISL_2776079, EPI_ISL_2776080, EPI_ISL_2776081, EPI_ISL_2776082, EPI_ISL_2776083, EPI_ISL_2776084, EPI_ISL_2776085, EPI_ISL_2776086, EPI_ISL_2776087, EPI_ISL_2776088, EPI_ISL_2776091, EPI_ISL_2776092, EPI_ISL_2776093, EPI_ISL_2776094, EPI_ISL_2776095, EPI_ISL_2776096, EPI_ISL_2776097, EPI_ISL_2776101, EPI_ISL_2776102, EPI_ISL_2776103, EPI_ISL_2776104, EPI_ISL_2776105, EPI_ISL_2776111, EPI_ISL_2776114, EPI_ISL_2776118 | | | |
| see above | Texas Children's Hospital | Texas Children's Microbiome Center | Ruth Ann Luna, Jennifer K. Spinler, James Dunn, James Versalovic, Ila Singh |
| EPI_ISL_2776126, EPI_ISL_2776137, EPI_ISL_2776138, EPI_ISL_2776139, EPI_ISL_2776143, EPI_ISL_2776144, EPI_ISL_2776147 | Wyoming Public Health Laboratory | Wyoming Public Health Laboratory | Jim Mildenberger, Taylor Fearing, Channing Weber, Ashley Norberg, Chayse Rowley, Marley Goetz, Brian Dominguez, Elliot Thomasson, Sam Britz, Cari Sloma, Robert Petit, and Rob Christensen |
| EPI_ISL_2776207, EPI_ISL_2776208, EPI_ISL_2776209, EPI_ISL_2776210, EPI_ISL_2776211, EPI_ISL_2776212, EPI_ISL_2776213 | Yale Clinical Virology Lab | Grubaugh Lab - Yale School of Public Health | Joseph Fauver, Mallery Breban, Isabel Ott, Tara Alpert, Mary Petrone, Anderson Brito, Chantal Vogels, Annie Watkins, Chaney Kalinich, Jessica Rothman, Kendall Billig, Tobias Koch, Marie L. Landry, Nathan Grubaugh |
| EPI_ISL_2776219 | Yale Pathology Lab | Grubaugh Lab - Yale School of Public Health | Joseph Fauver, Mallery Breban, Isabel Ott, Tara Alpert, Mary Petrone, Anderson Brito, Chantal Vogels, Annie Watkins, Chaney Kalinich, Jessica Rothman, Kendall Billig, Tobias Koch, Jianhui Wang, Chen Liu, Pei Hui, Nathan Grubaugh |
| EPI_ISL_2776221, EPI_ISL_2776222, EPI_ISL_2776223, EPI_ISL_2776224, EPI_ISL_2776225, EPI_ISL_2776226, EPI_ISL_2776227, EPI_ISL_2776228, EPI_ISL_2776229, EPI_ISL_2776230, EPI_ISL_2776232, EPI_ISL_2776233, EPI_ISL_2776234, EPI_ISL_2776235, EPI_ISL_2776236, EPI_ISL_2776237, EPI_ISL_2776238, EPI_ISL_2776239, EPI_ISL_2776240, EPI_ISL_2776241, EPI_ISL_2776242, EPI_ISL_2776243, EPI_ISL_2776244, EPI_ISL_2776245, EPI_ISL_2776246, EPI_ISL_2776247, EPI_ISL_2776248, EPI_ISL_2776249, EPI_ISL_2776250, EPI_ISL_2776251, EPI_ISL_2776252, EPI_ISL_2776253, EPI_ISL_2776254, EPI_ISL_2776255, EPI_ISL_2776256, EPI_ISL_2776257, EPI_ISL_2776258, EPI_ISL_2776259, EPI_ISL_2776260, EPI_ISL_2776261, EPI_ISL_2776262, EPI_ISL_2776263, EPI_ISL_2776264, EPI_ISL_2776265, EPI_ISL_2776266, EPI_ISL_2776267, EPI_ISL_2776268, EPI_ISL_2776270, EPI_ISL_2776271, EPI_ISL_2776272, EPI_ISL_2776273, EPI_ISL_2776274, EPI_ISL_2776275, EPI_ISL_2776276, EPI_ISL_2776277, EPI_ISL_2776278, EPI_ISL_2776279, EPI_ISL_2776280, EPI_ISL_2776281, EPI_ISL_2776283, EPI_ISL_2776284, EPI_ISL_2776285, EPI_ISL_2776286, EPI_ISL_2776287, EPI_ISL_2776288, EPI_ISL_2776289, EPI_ISL_2776290, EPI_ISL_2776292, EPI_ISL_2776293, EPI_ISL_2776294, EPI_ISL_2776295, EPI_ISL_2776296, EPI_ISL_2776297, EPI_ISL_2776298, EPI_ISL_2776299, EPI_ISL_2776300, EPI_ISL_2776301, EPI_ISL_2776302, EPI_ISL_2776303, EPI_ISL_2776304, EPI_ISL_2776305, EPI_ISL_2776306, EPI_ISL_2776307, EPI_ISL_2776308, EPI_ISL_2776309, EPI_ISL_2776310, EPI_ISL_2776311, EPI_ISL_2776312, EPI_ISL_2776313, EPI_ISL_2776314, EPI_ISL_2776315, EPI_ISL_2776316, EPI_ISL_2776317, EPI_ISL_2776318, EPI_ISL_2776319, EPI_ISL_2776320, EPI_ISL_2776321, EPI_ISL_2776322, EPI_ISL_2776323, EPI_ISL_2776324, EPI_ISL_2776325, EPI_ISL_2776327, EPI_ISL_2776329, EPI_ISL_2776330, EPI_ISL_2776331, EPI_ISL_2776332, EPI_ISL_2776333, EPI_ISL_2776334, EPI_ISL_2776335, EPI_ISL_2776336, EPI_ISL_2776337, EPI_ISL_2776338, EPI_ISL_2776339, EPI_ISL_2776341, EPI_ISL_2776342, EPI_ISL_2776343, EPI_ISL_2776344, EPI_ISL_2776345, EPI_ISL_2776346, EPI_ISL_2776347, EPI_ISL_2776348, EPI_ISL_2776349, EPI_ISL_2776350, EPI_ISL_2776351, EPI_ISL_2776352, EPI_ISL_2776353, EPI_ISL_2776354, EPI_ISL_2776355, EPI_ISL_2776356, EPI_ISL_2776357, EPI_ISL_2776358, EPI_ISL_2776359, EPI_ISL_2776360, EPI_ISL_2776361, EPI_ISL_2776362, EPI_ISL_2776363, EPI_ISL_2776364, EPI_ISL_2776365, EPI_ISL_2776366, EPI_ISL_2776367, EPI_ISL_2776368 | | | |

| | | | |
|---|---|---|---|
| see above | California Department of Public Health Valencia Branch Laboratory (CDPH VBL) | California Department of Public Health | CDPH-COVIDNet,Arias Lab at UCSB |
| EPI_ISL_2776371, EPI_ISL_2776372, EPI_ISL_2776373, EPI_ISL_2776374, EPI_ISL_2776375, EPI_ISL_2776377, EPI_ISL_2776378, EPI_ISL_2776380, EPI_ISL_2776383, EPI_ISL_2776384, EPI_ISL_2776385, EPI_ISL_2776386, EPI_ISL_2776387, EPI_ISL_2776389, EPI_ISL_2776390, EPI_ISL_2776391, EPI_ISL_2776392, EPI_ISL_2776393, EPI_ISL_2776394, EPI_ISL_2776395, EPI_ISL_2776396, EPI_ISL_2776397, EPI_ISL_2776399, EPI_ISL_2776400, EPI_ISL_2776401, EPI_ISL_2776402, EPI_ISL_2776404, EPI_ISL_2776405, EPI_ISL_2776406, EPI_ISL_2776407, EPI_ISL_2776408, EPI_ISL_2776409, EPI_ISL_2776410, EPI_ISL_2776411, EPI_ISL_2776412, EPI_ISL_2776413, EPI_ISL_2776414, EPI_ISL_2776415, EPI_ISL_2776416, EPI_ISL_2776417, EPI_ISL_2776419, EPI_ISL_2776421, EPI_ISL_2776422, EPI_ISL_2776423, EPI_ISL_2776425, EPI_ISL_2776426, EPI_ISL_2776427, EPI_ISL_2776428, EPI_ISL_2776429 | | | |
| see above | North Dakota Department of Health, Public Health Laboratory | North Dakota Department of Health, Public Health Laboratory | Lisa Wingerter |
| EPI_ISL_2777211, EPI_ISL_2777212, EPI_ISL_2777213, EPI_ISL_2777214 | Instituto Nacional de Enfermedades Respiratorias (INER) | Centro de Investigación en Enfermedades Infecciosas (CIENI), Instituto Nacional de Enfermedades Respiratorias (INER) | Consorcio Mexicano de Vigilancia Genómica (CoViGen-Mex). Authors (in alphabetical order): Julio Elias Alvarado-Yaah, Carlos F. Arias, Santiago Ávila-Ríos, Eduardo Becerril-Vargas, Víctor Hugo Borja-Aburto, Celia Boukadida, Cristóbal Cháidez-Quiróz, Juan Bautista Chale-Dzul, Ricardo Ciria Merce, Andreu Comas-García, Célida Duque Molina, Julissa Enciso-Ibarra, José Antonio Enciso-Moreno, Gloria Elena Espinosa-Ayala, Fernando Fontove-Herrera, Daniel Fregoso-Rueda, Victor Eduardo Garcia-Arias, Alejandra Garcia-Gasca, Bruno Gomez-Gil, Jean Pierre González, Irvin González-López, Concepción Grajales-Muñiz, Ricardo Grande, Rosa María Gutierrez Rios, Alejandra Hernández-Terán, Alfredo Herrera-Estrella, Carla Ivón Herrera-Najera, Pavel Isa, Daniel Lira Morales, Susana Lopez, Antonio Loza Román, Brenda Irasema Maldonado-Meza, Bernardo Martínez-Miguel, José Arturo Martínez-Orozco, Célida Martínez- Rodríguez, Margarita Matías-Florentino, Fidencio Mejía-Nepomuceno, María Guadalupe de Jesús Mireles-Rivera, Gloria María Molina-Salinas, Hector Montoya-Fuentes, Mario Mújica-Sánchez, José Esteban Muñoz-Medina, José de Jesús Nuñez-Contreras, Alicia Ocaña-Mondragón, Luis Alberto Ochoa-Carrera, Hector Esteban Paz-Juárez, Marissa Pérez-García, Francisco Pulido, Helen Haydee Fernanda Ramírez-Plascencia, Jorge Salas-Hernández, Angel Gustavo Salas-Lais, Alejandro Sanchez-Flores, Clara Esperanza Santacruz-Tinoco, María Guadalupe Santiago-Mauricio, Selene Zárate, Nelly Sélem-Mojica, Blanca Taboada, Kathia Elizabeth Tapia-Diaz, Gloria Vazquez, Joel Armando Vázquez-Pérez. |
| EPI_ISL_2777215 | Instituto Nacional de Cancerología | Centro de Investigación en Enfermedades Infecciosas (CIENI), Instituto Nacional de Enfermedades Respiratorias (INER) | Consorcio Mexicano de Vigilancia Genómica (CoViGen-Mex). Authors (in alphabetical order): Julio Elias Alvarado-Yaah, Carlos F. Arias, Santiago Ávila-Ríos, Eduardo Becerril-Vargas, Víctor Hugo Borja-Aburto, Celia Boukadida, Cristóbal Cháidez-Quiróz, Juan Bautista Chale-Dzul, Ricardo Ciria Merce, Andreu Comas-García, Célida Duque Molina, Julissa Enciso-Ibarra, José Antonio Enciso-Moreno, Gloria Elena Espinosa-Ayala, Fernando Fontove-Herrera, Daniel Fregoso-Rueda, Victor Eduardo Garcia-Arias, Alejandra Garcia-Gasca, Bruno Gomez-Gil, Jean Pierre González, Irvin González-López, Concepción Grajales-Muñiz, Ricardo Grande, Rosa María Gutierrez Rios, Alejandra Hernández-Terán, Alfredo Herrera-Estrella, Carla Ivón Herrera-Najera, Pavel Isa, Daniel Lira Morales, Susana Lopez, Antonio Loza Román, Brenda Irasema Maldonado-Meza, Bernardo Martínez-Miguel, José Arturo Martínez-Orozco, Célida Martínez- Rodríguez, Margarita Matías-Florentino, Fidencio Mejía-Nepomuceno, María Guadalupe de Jesús Mireles-Rivera, Gloria María Molina-Salinas, Hector Montoya-Fuentes, Mario Mújica-Sánchez, José Esteban Muñoz-Medina, José de Jesús Nuñez-Contreras, Alicia Ocaña-Mondragón, Luis Alberto Ochoa-Carrera, Hector Esteban Paz-Juárez, Marissa Pérez-García, Francisco Pulido, Helen Haydee Fernanda Ramírez-Plascencia, Jorge Salas-Hernández, Angel Gustavo Salas-Lais, Alejandro Sanchez-Flores, Clara Esperanza Santacruz-Tinoco, María Guadalupe Santiago-Mauricio, Selene Zárate, Nelly Sélem-Mojica, Blanca Taboada, Kathia Elizabeth Tapia-Diaz, Gloria Vazquez, Joel Armando Vázquez-Pérez. |
| EPI_ISL_2777216, EPI_ISL_2777217, EPI_ISL_2777218, EPI_ISL_2777219, EPI_ISL_2777220 | Instituto Nacional de Enfermedades Respiratorias (INER) | Centro de Investigación en Enfermedades Infecciosas (CIENI), Instituto Nacional de Enfermedades Respiratorias (INER) | Consorcio Mexicano de Vigilancia Genómica (CoViGen-Mex). Authors (in alphabetical order): Julio Elias Alvarado-Yaah, Carlos F. Arias, Santiago Ávila-Ríos, Eduardo Becerril-Vargas, Víctor Hugo Borja-Aburto, Celia Boukadida, Cristóbal Cháidez-Quiróz, Juan Bautista Chale-Dzul, Ricardo Ciria Merce, Andreu Comas-García, Célida Duque Molina, Julissa Enciso-Ibarra, José Antonio Enciso-Moreno, Gloria Elena Espinosa-Ayala, Fernando Fontove-Herrera, Daniel Fregoso-Rueda, Victor Eduardo Garcia-Arias, Alejandra Garcia-Gasca, Bruno Gomez-Gil, Jean Pierre González, Irvin González-López, Concepción Grajales-Muñiz, Ricardo Grande, Rosa María Gutierrez Rios, Alejandra Hernández-Terán, Alfredo Herrera-Estrella, Carla Ivón Herrera-Najera, Pavel Isa, Daniel Lira Morales, Susana Lopez, Antonio Loza Román, Brenda Irasema Maldonado-Meza, Bernardo Martínez-Miguel, José Arturo Martínez-Orozco, Célida Martínez- Rodríguez, Margarita Matías-Florentino, Fidencio Mejía-Nepomuceno, María Guadalupe de Jesús Mireles-Rivera, Gloria María Molina-Salinas, Hector Montoya-Fuentes, Mario Mújica-Sánchez, José Esteban Muñoz-Medina, José de Jesús Nuñez-Contreras, Alicia Ocaña-Mondragón, Luis Alberto Ochoa-Carrera, Hector Esteban Paz-Juárez, Marissa Pérez-García, Francisco Pulido, Helen Haydee Fernanda Ramírez-Plascencia, Jorge Salas-Hernández, Angel Gustavo Salas-Lais, Alejandro Sanchez-Flores, Clara Esperanza Santacruz-Tinoco, María Guadalupe Santiago-Mauricio, Selene Zárate, Nelly Sélem-Mojica, Blanca Taboada, Kathia Elizabeth Tapia-Diaz, Gloria Vazquez, Joel Armando Vázquez-Pérez. |
| EPI_ISL_2777221, EPI_ISL_2777222, EPI_ISL_2777223, EPI_ISL_2777224, EPI_ISL_2777225, EPI_ISL_2777226, EPI_ISL_2777227, EPI_ISL_2777228, EPI_ISL_2777229, EPI_ISL_2777230, EPI_ISL_2777231, EPI_ISL_2777232, EPI_ISL_2777233, EPI_ISL_2777234 | | | |
| see above | Instituto Nacional de Cancerología | Centro de Investigación en Enfermedades Infecciosas (CIENI), Instituto Nacional de Enfermedades Respiratorias (INER) | Consorcio Mexicano de Vigilancia Genómica (CoViGen-Mex). Authors (in alphabetical order): Julio Elias Alvarado-Yaah, Carlos F. Arias, Santiago Ávila-Ríos, Eduardo Becerril-Vargas, Víctor Hugo Borja-Aburto, Celia Boukadida, Cristóbal Cháidez-Quiróz, Juan Bautista Chale-Dzul, Ricardo Ciria Merce, Andreu Comas-García, Célida Duque Molina, Julissa Enciso-Ibarra, José Antonio Enciso-Moreno, Gloria Elena Espinosa-Ayala, Fernando Fontove-Herrera, Daniel Fregoso-Rueda, Victor Eduardo Garcia-Arias, Alejandra Garcia-Gasca, Bruno Gomez-Gil, Jean Pierre González, Irvin González-López, Concepción Grajales-Muñiz, Ricardo Grande, Rosa María Gutierrez Rios, Alejandra Hernández-Terán, Alfredo Herrera-Estrella, Carla Ivón Herrera-Najera, Pavel Isa, Daniel Lira Morales, Susana Lopez, Antonio Loza Román, Brenda Irasema Maldonado-Meza, Bernardo Martínez-Miguel, José Arturo Martínez-Orozco, Célida Martínez- Rodríguez, Margarita Matías-Florentino, Fidencio Mejía-Nepomuceno, María Guadalupe de Jesús Mireles-Rivera, Gloria María Molina-Salinas, Hector Montoya-Fuentes, Mario Mújica-Sánchez, José Esteban Muñoz-Medina, José de Jesús Nuñez-Contreras, Alicia Ocaña-Mondragón, Luis Alberto Ochoa-Carrera, Hector Esteban Paz-Juárez, Marissa Pérez-García, Francisco Pulido, Helen Haydee Fernanda Ramírez-Plascencia, Jorge Salas-Hernández, Angel Gustavo Salas-Lais, Alejandro Sanchez-Flores, Clara Esperanza Santacruz-Tinoco, María Guadalupe Santiago-Mauricio, Selene Zárate, Nelly Sélem-Mojica, Blanca Taboada, Kathia Elizabeth Tapia-Diaz, Gloria Vazquez, Joel Armando Vázquez-Pérez. |
| EPI_ISL_2778010, EPI_ISL_2778011, EPI_ISL_2778012, EPI_ISL_2778013, EPI_ISL_2778014, EPI_ISL_2778015, EPI_ISL_2778016, EPI_ISL_2778017 | Instituto Nacional de Medicina Genomica | Centro de Investigación en Enfermedades Infecciosas (CIENI), Instituto Nacional de Enfermedades Respiratorias (INER) | Matias-Florentino M, Perez-Garcia M, Hidalgo-Miranda A, Mendoza-Vargas A, Reyes-Grajeda JP, Cedro-Tanda A, Herrera-Montalvo LA, Reyes-Teran G, Avila-Rios S, Boukadida C |
| EPI_ISL_2778104, EPI_ISL_2778105, EPI_ISL_2778106, EPI_ISL_2778107, EPI_ISL_2778108, EPI_ISL_2778109, EPI_ISL_2778110, EPI_ISL_2778111, EPI_ISL_2778112, EPI_ISL_2778113, EPI_ISL_2778114, EPI_ISL_2778115, EPI_ISL_2778116, EPI_ISL_2778117, EPI_ISL_2778118, EPI_ISL_2778119, EPI_ISL_2778120, EPI_ISL_2778121, EPI_ISL_2778122, EPI_ISL_2778123, EPI_ISL_2778124, EPI_ISL_2778125, EPI_ISL_2778126, EPI_ISL_2778127, EPI_ISL_2778128, EPI_ISL_2778129, EPI_ISL_2778130, EPI_ISL_2778131, EPI_ISL_2778132, EPI_ISL_2778133, EPI_ISL_2778134, EPI_ISL_2778135, EPI_ISL_2778136, EPI_ISL_2778137, EPI_ISL_2778138, EPI_ISL_2778139, EPI_ISL_2778140, EPI_ISL_2778141, EPI_ISL_2778142, EPI_ISL_2778143, EPI_ISL_2778144, EPI_ISL_2778145, EPI_ISL_2778146, EPI_ISL_2778147, EPI_ISL_2778148, EPI_ISL_2778149, EPI_ISL_2778150, EPI_ISL_2778151, EPI_ISL_2778152, EPI_ISL_2778153, EPI_ISL_2778154, EPI_ISL_2778155, EPI_ISL_2778156, EPI_ISL_2778157, EPI_ISL_2778158, EPI_ISL_2778159, EPI_ISL_2778160, EPI_ISL_2778161, EPI_ISL_2778162, EPI_ISL_2778163, EPI_ISL_2778164, EPI_ISL_2778165, EPI_ISL_2778166, EPI_ISL_2778167, EPI_ISL_2778168, EPI_ISL_2778169, EPI_ISL_2778170, EPI_ISL_2778171, EPI_ISL_2778172, EPI_ISL_2778173, EPI_ISL_2778174, EPI_ISL_2778175, EPI_ISL_2778176, EPI_ISL_2778177, EPI_ISL_2778178, EPI_ISL_2778179, EPI_ISL_2778180, EPI_ISL_2778181, EPI_ISL_2778182, EPI_ISL_2778183, EPI_ISL_2778184, EPI_ISL_2778185, EPI_ISL_2778186, EPI_ISL_2778187, EPI_ISL_2778188, EPI_ISL_2778189, EPI_ISL_2778190, EPI_ISL_2778191, EPI_ISL_2778192, EPI_ISL_2778193, EPI_ISL_2778194, EPI_ISL_2778195, EPI_ISL_2778196, EPI_ISL_2778197, EPI_ISL_2778198, EPI_ISL_2778199, EPI_ISL_2778200, EPI_ISL_2778201, EPI_ISL_2778202, EPI_ISL_2778203, EPI_ISL_2778204, EPI_ISL_2778205, EPI_ISL_2778206, EPI_ISL_2778207, EPI_ISL_2778208, EPI_ISL_2778209, EPI_ISL_2778210, EPI_ISL_2778211, EPI_ISL_2778212, EPI_ISL_2778213, EPI_ISL_2778214, EPI_ISL_2778215, EPI_ISL_2778216, EPI_ISL_2778217, EPI_ISL_2778218, EPI_ISL_2778219, EPI_ISL_2778220, EPI_ISL_2778221, EPI_ISL_2778222, EPI_ISL_2778223, EPI_ISL_2778224, EPI_ISL_2778225, EPI_ISL_2778226, EPI_ISL_2778227, EPI_ISL_2778228, EPI_ISL_2778229, EPI_ISL_2778230, EPI_ISL_2778231, EPI_ISL_2778232, EPI_ISL_2778233, EPI_ISL_2778234, EPI_ISL_2778235, EPI_ISL_2778236, EPI_ISL_2778237, EPI_ISL_2778238, EPI_ISL_2778239, EPI_ISL_2778240, EPI_ISL_2778241, EPI_ISL_2778242, EPI_ISL_2778243, EPI_ISL_2778244, EPI_ISL_2778245, EPI_ISL_2778246, EPI_ISL_2778247, EPI_ISL_2778248, EPI_ISL_2778249, EPI_ISL_2778250, EPI_ISL_2778251, EPI_ISL_2778252, EPI_ISL_2778253, EPI_ISL_2778254, EPI_ISL_2778255, EPI_ISL_2778256, EPI_ISL_2778257, EPI_ISL_2778258, EPI_ISL_2778260, EPI_ISL_2778261, EPI_ISL_2778262, EPI_ISL_2778263, EPI_ISL_2778264, EPI_ISL_2778265, EPI_ISL_2778266, EPI_ISL_2778267, EPI_ISL_2778268, EPI_ISL_2778269, EPI_ISL_2778270, EPI_ISL_2778271, EPI_ISL_2778272, EPI_ISL_2778273, EPI_ISL_2778274, EPI_ISL_2778275, EPI_ISL_2778277, EPI_ISL_2778278, EPI_ISL_2778279, EPI_ISL_2778280, EPI_ISL_2778281, EPI_ISL_2778282, EPI_ISL_2778283, EPI_ISL_2778284, EPI_ISL_2778285, EPI_ISL_2778286, EPI_ISL_2778287, EPI_ISL_2778288, EPI_ISL_2778289, EPI_ISL_2778290, EPI_ISL_2778291, EPI_ISL_2778292, EPI_ISL_2778293, EPI_ISL_2778294, EPI_ISL_2778295, EPI_ISL_2778297, EPI_ISL_2778298, EPI_ISL_2778299, EPI_ISL_2778300, EPI_ISL_2778301, EPI_ISL_2778302, EPI_ISL_2778303, EPI_ISL_2778304, EPI_ISL_2778305, EPI_ISL_2778306, EPI_ISL_2778307, EPI_ISL_2778308, EPI_ISL_2778309, EPI_ISL_2778310, EPI_ISL_2778311, EPI_ISL_2778312, EPI_ISL_2778313, EPI_ISL_2778314, EPI_ISL_2778315, EPI_ISL_2778316, EPI_ISL_2778317, EPI_ISL_2778318, EPI_ISL_2778319, EPI_ISL_2778320, EPI_ISL_2778321, EPI_ISL_2778322, EPI_ISL_2778323, EPI_ISL_2778324, EPI_ISL_2778325, EPI_ISL_2778326, EPI_ISL_2778327, EPI_ISL_2778328, EPI_ISL_2778329, EPI_ISL_2778330, EPI_ISL_2778331, EPI_ISL_2778332, EPI_ISL_2778333, EPI_ISL_2778334, EPI_ISL_2778335, EPI_ISL_2778336, EPI_ISL_2778337, EPI_ISL_2778338, EPI_ISL_2778339, EPI_ISL_2778340, EPI_ISL_2778341, EPI_ISL_2778342, EPI_ISL_2778343, EPI_ISL_2778344, EPI_ISL_2778345, EPI_ISL_2778346, EPI_ISL_2778347, EPI_ISL_2778348, EPI_ISL_2778349, EPI_ISL_2778350, EPI_ISL_2778351, EPI_ISL_2778352, EPI_ISL_2778353, EPI_ISL_2778354, EPI_ISL_2778355, EPI_ISL_2778356, EPI_ISL_2778357, EPI_ISL_2778358, EPI_ISL_2778359, EPI_ISL_2778360, EPI_ISL_2778361, EPI_ISL_2778362, EPI_ISL_2778363, EPI_ISL_2778364, EPI_ISL_2778365, EPI_ISL_2778366, EPI_ISL_2778367, EPI_ISL_2778368, EPI_ISL_2778369, EPI_ISL_2778370, EPI_ISL_2778371, EPI_ISL_2778372, EPI_ISL_2778373, EPI_ISL_2778374, EPI_ISL_2778375, EPI_ISL_2778376, EPI_ISL_2778377, EPI_ISL_2778378, EPI_ISL_2778379, EPI_ISL_2778380, EPI_ISL_2778381, EPI_ISL_2778382, EPI_ISL_2778383, EPI_ISL_2778384, EPI_ISL_2778385, EPI_ISL_2778386, EPI_ISL_2778387, EPI_ISL_2778388, EPI_ISL_2778389, EPI_ISL_2778390, EPI_ISL_2778391, EPI_ISL_2778392, EPI_ISL_2778393, EPI_ISL_2778394, EPI_ISL_2778395, EPI_ISL_2778396, EPI_ISL_2778397, EPI_ISL_2778398, EPI_ISL_2778399, EPI_ISL_2778400, EPI_ISL_2778401, EPI_ISL_2778402, EPI_ISL_2778403, EPI_ISL_2778404, EPI_ISL_2778405, EPI_ISL_2778406, EPI_ISL_2778407, EPI_ISL_2778408, EPI_ISL_2778409, EPI_ISL_2778410, EPI_ISL_2778411, EPI_ISL_2778412, EPI_ISL_2778413, EPI_ISL_2778414, EPI_ISL_2778415, EPI_ISL_2778416, EPI_ISL_2778417, EPI_ISL_2778418, EPI_ISL_2778419, EPI_ISL_2778420, EPI_ISL_2778421, EPI_ISL_2778422, EPI_ISL_2778423, EPI_ISL_2778424, EPI_ISL_2778425, EPI_ISL_2778426, EPI_ISL_2778427, EPI_ISL_2778428, EPI_ISL_2778429, EPI_ISL_2778430, EPI_ISL_2778431, EPI_ISL_2778432, EPI_ISL_2778433, EPI_ISL_2778434, EPI_ISL_2778435, EPI_ISL_2778436, EPI_ISL_2778437, EPI_ISL_2778438, EPI_ISL_2778439, EPI_ISL_2778440, EPI_ISL_2778441, EPI_ISL_2778442, EPI_ISL_2778443, EPI_ISL_2778444, EPI_ISL_2778445, EPI_ISL_2778446, EPI_ISL_2778447, EPI_ISL_2778448, EPI_ISL_2778449, EPI_ISL_2778450, EPI_ISL_2778451, EPI_ISL_2778452, EPI_ISL_2778453, EPI_ISL_2778454, EPI_ISL_2778455, EPI_ISL_2778456, EPI_ISL_2778457, EPI_ISL_2778458, EPI_ISL_2778459, EPI_ISL_2778460, EPI_ISL_2778461, EPI_ISL_2778462, EPI_ISL_2778463, EPI_ISL_2778464, EPI_ISL_2778465, EPI_ISL_2778466, EPI_ISL_2778467, EPI_ISL_2778468, EPI_ISL_2778469, EPI_ISL_2778470, EPI_ISL_2778471, EPI_ISL_2778472, EPI_ISL_2778473, EPI_ISL_2778474, EPI_ISL_2778475, EPI_ISL_2778476, EPI_ISL_2778477, EPI_ISL_2778478, EPI_ISL_2778479, EPI_ISL_2778480, EPI_ISL_2778481, EPI_ISL_2778482, EPI_ISL_2778483, EPI_ISL_2778484, EPI_ISL_2778485, EPI_ISL_2778486, EPI_ISL_2778487, EPI_ISL_2778488, EPI_ISL_2778489, EPI_ISL_2778490, EPI_ISL_2778491, EPI_ISL_2778492, EPI_ISL_2778493, EPI_ISL_2778494, EPI_ISL_2778495, EPI_ISL_2778496, EPI_ISL_2778497, EPI_ISL_2778498, EPI_ISL_2778499, EPI_ISL_2778500, EPI_ISL_2778501, EPI_ISL_2778502, EPI_ISL_2778503, EPI_ISL_2778504, EPI_ISL_2778505, EPI_ISL_2778506, EPI_ISL_2778507, EPI_ISL_2778508, EPI_ISL_2778509, EPI_ISL_2778510, EPI_ISL_2778511, EPI_ISL_2778512, EPI_ISL_2778513, EPI_ISL_2778514, EPI_ISL_2778515, EPI_ISL_2778516, EPI_ISL_2778517, EPI_ISL_2778518, EPI_ISL_2778519, EPI_ISL_2778520, EPI_ISL_2778521, EPI_ISL_2778522, EPI_ISL_2778523, EPI_ISL_2778524, EPI_ISL_2778525, EPI_ISL_2778526, EPI_ISL_2778527, EPI_ISL_2778528, EPI_ISL_2778529, EPI_ISL_2778530, EPI_ISL_2778531, EPI_ISL_2778532, EPI_ISL_2778533, EPI_ISL_2778534, EPI_ISL_2778535, EPI_ISL_2778536, EPI_ISL_2778537, EPI_ISL_2778538, EPI_ISL_2778539, EPI_ISL_2778540, EPI_ISL_2778541, EPI_ISL_2778542, EPI_ISL_2778543, EPI_ISL_2778544, EPI_ISL_2778545, EPI_ISL_2778546, EPI_ISL_2778547, EPI_ISL_2778548, EPI_ISL_2778549, EPI_ISL_2778550, EPI_ISL_2778551, EPI_ISL_2778552, EPI_ISL_2778553, EPI_ISL_2778554, EPI_ISL_2778555, EPI_ISL_2778556, EPI_ISL_2778557, EPI_ISL_2778558, EPI_ISL_2778559, EPI_ISL_2778560, EPI_ISL_2778561, EPI_ISL_2778562, EPI_ISL_2778564, EPI_ISL_2778565, EPI_ISL_2778566, EPI_ISL_2778567, EPI_ISL_2778568, EPI_ISL_2778569, EPI_ISL_2778570, EPI_ISL_2778571, EPI_ISL_2778572, EPI_ISL_2778573, EPI_ISL_2778574, EPI_ISL_2778575, EPI_ISL_2778576, EPI_ISL_2778577, EPI_ISL_2778578, EPI_ISL_2778579, EPI_ISL_2778580, EPI_ISL_2778581, EPI_ISL_2778582, EPI_ISL_2778583, EPI_ISL_2778584, EPI_ISL_2778585, EPI_ISL_2778586, EPI_ISL_2778587, EPI_ISL_2778588, EPI_ISL_2778589, EPI_ISL_2778590, EPI_ISL_2778591, EPI_ISL_2778592, EPI_ISL_2778593, EPI_ISL_2778594, EPI_ISL_2778595, EPI_ISL_2778596, EPI_ISL_2778597, EPI_ISL_2778598, EPI_ISL_2778599, EPI_ISL_2778600, EPI_ISL_2778601, EPI_ISL_2778602, EPI_ISL_2778603, EPI_ISL_2778604, EPI_ISL_2778605, EPI_ISL_2778606, EPI_ISL_2778607, EPI_ISL_2778608, EPI_ISL_2778609, EPI_ISL_2778610, EPI_ISL_2778611, EPI_ISL_2778612, EPI_ISL_2778613, EPI_ISL_2778614, EPI_ISL_2778615, EPI_ISL_2778616, EPI_ISL_2778617, EPI_ISL_2778618, EPI_ISL_2778619, EPI_ISL_2778620, EPI_ISL_2778621, EPI_ISL_2778622, EPI_ISL_2778623, EPI_ISL_2778624, EPI_ISL_2778625, EPI_ISL_2778626, EPI_ISL_2778627, EPI_ISL_2778628, EPI_ISL_2778629, EPI_ISL_2778630, EPI_ISL_2778631, EPI_ISL_2778632, EPI_ISL_2778633, EPI_ISL_2778634, EPI_ISL_2778635, EPI_ISL_2778636, EPI_ISL_2778637, EPI_ISL_2778638, EPI_ISL_2778639, EPI_ISL_2778640, EPI_ISL_2778641, EPI_ISL_2778642, EPI_ISL_2778643, EPI_ISL_2778644, EPI_ISL_2778645, EPI_ISL_2778646, EPI_ISL_2778647, EPI_ISL_2778648, EPI_ISL_2778649, EPI_ISL_2778650, EPI_ISL_2778651, EPI_ISL_2778652, EPI_ISL_2778653, EPI_ISL_2778654, EPI_ISL_2778655, EPI_ISL_2778656, EPI_ISL_2778657, EPI_ISL_2778658, EPI_ISL_2778659, EPI_ISL_2778660, EPI_ISL_2778661, EPI_ISL_2778662, EPI_ISL_2778663, EPI_ISL_2778664, EPI_ISL_2778665, EPI_ISL_2778666, EPI_ISL_2778667, EPI_ISL_2778668, EPI_ISL_2778670, EPI_ISL_2778671, EPI_ISL_2778673, EPI_ISL_2778674, EPI_ISL_2778675, EPI_ISL_2778676, EPI_ISL_2778677, EPI_ISL_2778679, EPI_ISL_2778680, EPI_ISL_2778681, EPI_ISL_2778682, EPI_ISL_2778683, EPI_ISL_2778684, EPI_ISL_2778686, EPI_ISL_2778687, EPI_ISL_2778688, EPI_ISL_2778690, EPI_ISL_2778691, EPI_ISL_2778692, EPI_ISL_2778693, EPI_ISL_2778694, EPI_ISL_2778695, EPI_ISL_2778696, EPI_ISL_2778697, EPI_ISL_2778698, EPI_ISL_2778699, EPI_ISL_2778700, EPI_ISL_2778701, EPI_ISL_2778702, EPI_ISL_2778703, EPI_ISL_2778704, EPI_ISL_2778705, EPI_ISL_2778706, EPI_ISL_2778707, EPI_ISL_2778708, EPI_ISL_2778709, EPI_ISL_2778710, EPI_ISL_2778711, EPI_ISL_2778712, EPI_ISL_2778713, EPI_ISL_2778714, EPI_ISL_2778715, EPI_ISL_2778716, EPI_ISL_2778717, EPI_ISL_2778718, EPI_ISL_2778719, EPI_ISL_2778720, EPI_ISL_2778721, EPI_ISL_2778722, EPI_ISL_2778723, EPI_ISL_2778724, EPI_ISL_2778725, EPI_ISL_2778726, EPI_ISL_2778727, EPI_ISL_2778728, EPI_ISL_2778729, EPI_ISL_2778730, EPI_ISL_2778731, EPI_ISL_2778732, EPI_ISL_2778733, EPI_ISL_2778734, EPI_ISL_2778735, EPI_ISL_2778736, EPI_ISL_2778737, EPI_ISL_2778738, EPI_ISL_2778739, EPI_ISL_2778740, EPI_ISL_2778741, EPI_ISL_2778742, EPI_ISL_2778743, EPI_ISL_2778744, EPI_ISL_2778745, EPI_ISL_2778746, EPI_ISL_2778747, EPI_ISL_2778748, EPI_ISL_2778749, EPI_ISL_2778750, EPI_ISL_2778751, EPI_ISL_2778752, EPI_ISL_2778753, EPI_ISL_2778754, EPI_ISL_2778755, EPI_ISL_2778756, EPI_ISL_2778757, EPI_ISL_2778758, EPI_ISL_2778759, EPI_ISL_2778760, EPI_ISL_2778761, EPI_ISL_2778762, EPI_ISL_2778763, EPI_ISL_2778764, EPI_ISL_2778765, EPI_ISL_2778766, EPI_ISL_2778767, EPI_ISL_2778768, EPI_ISL_2778769, EPI_ISL_2778770, EPI_ISL_2778771, EPI_ISL_2778772, EPI_ISL_2778773, EPI_ISL_2778774, EPI_ISL_2778775, EPI_ISL_2778776, EPI_ISL_2778777, EPI_ISL_2778778, EPI_ISL_2778779, EPI_ISL_2778780, EPI_ISL_2778781, EPI_ISL_2778782, EPI_ISL_2778783, EPI_ISL_2778784, EPI_ISL_2778785, EPI_ISL_2778786, EPI_ISL_2778787, EPI_ISL_2778788, EPI_ISL_2778789, EPI_ISL_2778790, EPI_ISL_2778791, EPI_ISL_2778792, EPI_ISL_2778793, EPI_ISL_2778794, EPI_ISL_2778796, EPI_ISL_2778797, EPI_ISL_2778798, EPI_ISL_2778799, EPI_ISL_2778800, EPI_ISL_2778801, EPI_ISL_2778802, EPI_ISL_2778803, EPI_ISL_2778804, EPI_ISL_2778805, EPI_ISL_2778806, EPI_ISL_2778807, EPI_ISL_2778808, EPI_ISL_2778809, EPI_ISL_2778810, EPI_ISL_2778811, EPI_ISL_2778812, EPI_ISL_2778813, EPI_ISL_2778814, EPI_ISL_2778815, EPI_ISL_2778816, EPI_ISL_2778817, EPI_ISL_2778818, EPI_ISL_2778819, EPI_ISL_2778820, EPI_ISL_2778821, EPI_ISL_2778822, EPI_ISL_2778823, EPI_ISL_2778824, EPI_ISL_2778825, EPI_ISL_2778826, EPI_ISL_2778827, EPI_ISL_2778828, EPI_ISL_2778829, EPI_ISL_2778830, EPI_ISL_2778831, EPI_ISL_2778832, EPI_ISL_2778833, EPI_ISL_2778834, EPI_ISL_2778835, EPI_ISL_2778836, EPI_ISL_2778837, EPI_ISL_2778838, EPI_ISL_2778839, EPI_ISL_2778840, EPI_ISL_2778841, EPI_ISL_2778842, EPI_ISL_2778843, EPI_ISL_2778844, EPI_ISL_2778845, EPI_ISL_2778846, EPI_ISL_2778847, EPI_ISL_2778848, EPI_ISL_2778849, EPI_ISL_2778850, EPI_ISL_2778851, EPI_ISL_2778852, EPI_ISL_2778853, EPI_ISL_2778854, EPI_ISL_2778855, EPI_ISL_2778856, EPI_ISL_2778857, EPI_ISL_2778858, EPI_ISL_2778859, EPI_ISL_2778860, EPI_ISL_2778861, EPI_ISL_2778862, EPI_ISL_2778863, EPI_ISL_2778864, EPI_ISL_2778865, EPI_ISL_2778866, EPI_ISL_2778867, EPI_ISL_2778868, EPI_ISL_2778869, EPI_ISL_2778870, EPI_ISL_2778871, EPI_ISL_2778872, EPI_ISL_2778873, EPI_ISL_2778874, EPI_ISL_2778875, EPI_ISL_2778876, EPI_ISL_2778877, EPI_ISL_2778878, EPI_ISL_2778879, EPI_ISL_2778880, EPI_ISL_2778881, EPI_ISL_2778882, EPI_ISL_2778883, EPI_ISL_2778884, EPI_ISL_2778886, EPI_ISL_2778887, EPI_ISL_2778888, EPI_ISL_2778889, EPI_ISL_2778890, EPI_ISL_2778891, EPI_ISL_2778892, EPI_ISL_2778893, EPI_ISL_2778894, EPI_ISL_2778895, EPI_ISL_2778896, EPI_ISL_2778897, EPI_ISL_2778898, EPI_ISL_2778899, EPI_ISL_2778900, EPI_ISL_2778901, EPI_ISL_2778902, EPI_ISL_2778903, EPI_ISL_2778904, EPI_ISL_2778905, EPI_ISL_2778906, EPI_ISL_2778907, EPI_ISL_2778908, EPI_ISL_2778909, EPI_ISL_2778910, EPI_ISL_2778911, EPI_ISL_2778912, EPI_ISL_2778913, EPI_ISL_2778914, EPI_ISL_2778915, EPI_ISL_2778916, EPI_ISL_2778917, EPI_ISL_2778918, EPI_ISL_2778919, EPI_ISL_2778920, EPI_ISL_2778921, EPI_ISL_2778922, EPI_ISL_2778923, EPI_ISL_2778924, EPI_ISL_2778925, EPI_ISL_2778926, EPI_ISL_2778927, EPI_ISL_2778928, EPI_ISL_2778929, EPI_ISL_2778930, EPI_ISL_2778931, EPI_ISL_2778932, EPI_ISL_2778934, EPI_ISL_2778935, EPI_ISL_2778936, EPI_ISL_2778937, EPI_ISL_2778938, EPI_ISL_2778939, EPI_ISL_2778940, EPI_ISL_2778941, EPI_ISL_2778942, EPI_ISL_2778943, EPI_ISL_2778944, EPI_ISL_2778945, EPI_ISL_2778946, EPI_ISL_2778947, EPI_ISL_2778948, EPI_ISL_2778949, EPI_ISL_2778950, EPI_ISL_2778951, EPI_ISL_2778952, EPI_ISL_2778953, EPI_ISL_2778954 | | | |
| see above | California Department of Public Health Valencia Branch Laboratory (CDPH VBL) | California Department of Public Health | CDPH-COVIDNet,UCLA Technology Center for Genomics & Bioinformatics |
| EPI_ISL_2778955, EPI_ISL_2778957, EPI_ISL_2778958, EPI_ISL_2778959, EPI_ISL_2778960, EPI_ISL_2778961, EPI_ISL_2778962, EPI_ISL_2778963, EPI_ISL_2778964, EPI_ISL_2778965, EPI_ISL_2778966 | | | |
| see above | Salud Digna | Instituto Nacional de Medicina Genomica | Cedro-Tanda A, Hidalgo-Miranda A, Cisneros-Villanueva M, Orjuela-Rodríguez M, Mendoza-Vargas A, Reyes-Grajeda JP, Abraham Campos-Romero, Moreno-Camacho José Luis, Rodriguez-Gallegos Jorge, Luna-Ruiz Marco, Gonzalez-Barrera D, Rangel-DeLeon D, Munguia-Garza P, Ramirez-Vega O, Escobar-Arrazola, M, Herrera-Montalvo LA. |
| EPI_ISL_2778967, EPI_ISL_2778968 | Instituto Nacional de Medicina Genomica | Instituto Nacional de Medicina Genomica | Cedro-Tanda A, Hidalgo-Miranda A, Cisneros-Villanueva M, Orjuela-Rodríguez M, Mendoza-Vargas A, Reyes-Grajeda JP, Abraham Campos-Romero, Moreno-Camacho José Luis, Rodriguez-Gallegos Jorge, Luna-Ruiz Marco, Gonzalez-Barrera D, Rangel-DeLeon D, Munguia-Garza P, Ramirez-Vega O, Escobar-Arrazola, M, Herrera-Montalvo LA. |
| EPI_ISL_2778969, EPI_ISL_2778970, EPI_ISL_2778971, EPI_ISL_2778972, EPI_ISL_2778973, EPI_ISL_2778974, EPI_ISL_2778975, EPI_ISL_2778976, EPI_ISL_2778977, EPI_ISL_2778978, EPI_ISL_2778979, EPI_ISL_2778980, EPI_ISL_2778981, EPI_ISL_2778982, EPI_ISL_2778983 | | | |
| see above | Humboldt County Public Health Laboratory | Humboldt County Public Health Laboratory | Jeremy Corrigan |
| EPI_ISL_2778984, EPI_ISL_2778985, EPI_ISL_2778986, EPI_ISL_2778987, EPI_ISL_2778988 | LESP Tabasco | Instituto de Diagnostico y Referencia Epidemiologicos (INDRE) | Claudia Wong-Arambula, Abril Rodriguez-Maldonado, Vanessa Rivero-Arredondo, Ariadna Medina-Benitez, Joaquin Quiroz-Mercado, Sergio Rangel-Guerrero, Maribel Gonzalez-Villa, Natividad Cruz-Ortiz, Tatiana Nunez-Garcia, Gisela Barrera-Badillo, Lucia Hernandez-Rivas, Irma Lopez-Martinez, Ernesto Ramirez-Gonzalez. |
| EPI_ISL_2778989 | LESP Sonora | Instituto de Diagnostico y Referencia Epidemiologicos (INDRE) | Claudia Wong-Arambula, Abril Rodriguez-Maldonado, Vanessa Rivero-Arredondo, Ariadna Medina-Benitez, Joaquin Quiroz-Mercado, Sergio Rangel-Guerrero, Maribel Gonzalez-Villa, Natividad Cruz-Ortiz, Tatiana Nunez-Garcia, Gisela Barrera-Badillo, Lucia Hernandez-Rivas, Irma Lopez-Martinez, Ernesto Ramirez-Gonzalez. |
| EPI_ISL_2778990, EPI_ISL_2778991, EPI_ISL_2778992 | LESP Guanajuato | Instituto de Diagnostico y Referencia Epidemiologicos (INDRE) | Claudia Wong-Arambula, Abril Rodriguez-Maldonado, Vanessa Rivero-Arredondo, Ariadna Medina-Benitez, Joaquin Quiroz-Mercado, Sergio Rangel-Guerrero, Maribel Gonzalez-Villa, Natividad Cruz-Ortiz, Tatiana Nunez-Garcia, Gisela Barrera-Badillo, Lucia Hernandez-Rivas, Irma Lopez-Martinez, Ernesto Ramirez-Gonzalez. |
| EPI_ISL_2778993, EPI_ISL_2778994 | LESP Durango | Instituto de Diagnostico y Referencia Epidemiologicos (INDRE) | Claudia Wong-Arambula, Abril Rodriguez-Maldonado, Vanessa Rivero-Arredondo, Ariadna Medina-Benitez, Joaquin Quiroz-Mercado, Sergio Rangel-Guerrero, Maribel Gonzalez-Villa, Natividad Cruz-Ortiz, Tatiana Nunez-Garcia, Gisela Barrera-Badillo, Lucia Hernandez-Rivas, Irma Lopez-Martinez, Ernesto Ramirez-Gonzalez. |

| | | | |
|---|---|---|---|
| EPI_ISL_2778995 | LESP Nayarit | Instituto de Diagnostico y Referencia Epidemiologicos (INDRE) | Claudia Wong-Arambula, Abril Rodriguez-Maldonado, Vanessa Rivero-Arredondo, Ariadna Medina-Benitez, Joaquin Quiroz-Mercado, Sergio Rangel-Guerrero, Maribel Gonzalez-Villa, Natividad Cruz-Ortiz, Tatiana Nunez-Garcia, Gisela Barrera-Badillo, Lucia Hernandez-Rivas, Irma Lopez-Martinez, Ernesto Ramirez-Gonzalez. |
| EPI_ISL_2778996 | LESP Durango | Instituto de Diagnostico y Referencia Epidemiologicos (INDRE) | Claudia Wong-Arambula, Abril Rodriguez-Maldonado, Vanessa Rivero-Arredondo, Ariadna Medina-Benitez, Joaquin Quiroz-Mercado, Sergio Rangel-Guerrero, Maribel Gonzalez-Villa, Natividad Cruz-Ortiz, Tatiana Nunez-Garcia, Gisela Barrera-Badillo, Lucia Hernandez-Rivas, Irma Lopez-Martinez, Ernesto Ramirez-Gonzalez. |
| EPI_ISL_2778997 | LESP Chiapas | Instituto de Diagnostico y Referencia Epidemiologicos (INDRE) | Claudia Wong-Arambula, Abril Rodriguez-Maldonado, Vanessa Rivero-Arredondo, Ariadna Medina-Benitez, Joaquin Quiroz-Mercado, Sergio Rangel-Guerrero, Maribel Gonzalez-Villa, Natividad Cruz-Ortiz, Tatiana Nunez-Garcia, Gisela Barrera-Badillo, Lucia Hernandez-Rivas, Irma Lopez-Martinez, Ernesto Ramirez-Gonzalez. |
| EPI_ISL_2778998 | LESP Sinaloa | Instituto de Diagnostico y Referencia Epidemiologicos (INDRE) | Claudia Wong-Arambula, Abril Rodriguez-Maldonado, Vanessa Rivero-Arredondo, Ariadna Medina-Benitez, Joaquin Quiroz-Mercado, Sergio Rangel-Guerrero, Maribel Gonzalez-Villa, Natividad Cruz-Ortiz, Tatiana Nunez-Garcia, Gisela Barrera-Badillo, Lucia Hernandez-Rivas, Irma Lopez-Martinez, Ernesto Ramirez-Gonzalez. |
| EPI_ISL_2778999 | LESP Estado de Mexico | Instituto de Diagnostico y Referencia Epidemiologicos (INDRE) | Claudia Wong-Arambula, Abril Rodriguez-Maldonado, Vanessa Rivero-Arredondo, Ariadna Medina-Benitez, Joaquin Quiroz-Mercado, Sergio Rangel-Guerrero, Maribel Gonzalez-Villa, Natividad Cruz-Ortiz, Tatiana Nunez-Garcia, Gisela Barrera-Badillo, Lucia Hernandez-Rivas, Irma Lopez-Martinez, Ernesto Ramirez-Gonzalez. |
| EPI_ISL_2779000 | LESP Durango | Instituto de Diagnostico y Referencia Epidemiologicos (INDRE) | Claudia Wong-Arambula, Abril Rodriguez-Maldonado, Vanessa Rivero-Arredondo, Ariadna Medina-Benitez, Joaquin Quiroz-Mercado, Sergio Rangel-Guerrero, Maribel Gonzalez-Villa, Natividad Cruz-Ortiz, Tatiana Nunez-Garcia, Gisela Barrera-Badillo, Lucia Hernandez-Rivas, Irma Lopez-Martinez, Ernesto Ramirez-Gonzalez. |
| EPI_ISL_2779001 | LESP Campeche | Instituto de Diagnostico y Referencia Epidemiologicos (INDRE) | Claudia Wong-Arambula, Abril Rodriguez-Maldonado, Vanessa Rivero-Arredondo, Ariadna Medina-Benitez, Joaquin Quiroz-Mercado, Sergio Rangel-Guerrero, Maribel Gonzalez-Villa, Natividad Cruz-Ortiz, Tatiana Nunez-Garcia, Gisela Barrera-Badillo, Lucia Hernandez-Rivas, Irma Lopez-Martinez, Ernesto Ramirez-Gonzalez. |
| EPI_ISL_2779002, EPI_ISL_2779003, EPI_ISL_2779004 | LESP Tamaulipas | Instituto de Diagnostico y Referencia Epidemiologicos (INDRE) | Claudia Wong-Arambula, Abril Rodriguez-Maldonado, Vanessa Rivero-Arredondo, Ariadna Medina-Benitez, Joaquin Quiroz-Mercado, Sergio Rangel-Guerrero, Maribel Gonzalez-Villa, Natividad Cruz-Ortiz, Tatiana Nunez-Garcia, Gisela Barrera-Badillo, Lucia Hernandez-Rivas, Irma Lopez-Martinez, Ernesto Ramirez-Gonzalez. |
| EPI_ISL_2779005, EPI_ISL_2779006, EPI_ISL_2779007 | LESP Sinaloa | Instituto de Diagnostico y Referencia Epidemiologicos (INDRE) | Claudia Wong-Arambula, Abril Rodriguez-Maldonado, Vanessa Rivero-Arredondo, Ariadna Medina-Benitez, Joaquin Quiroz-Mercado, Sergio Rangel-Guerrero, Maribel Gonzalez-Villa, Natividad Cruz-Ortiz, Tatiana Nunez-Garcia, Gisela Barrera-Badillo, Lucia Hernandez-Rivas, Irma Lopez-Martinez, Ernesto Ramirez-Gonzalez. |
| EPI_ISL_2779008, EPI_ISL_2779009, EPI_ISL_2779010, EPI_ISL_2779011, EPI_ISL_2779012, EPI_ISL_2779013, EPI_ISL_2779014, EPI_ISL_2779015, EPI_ISL_2779016, EPI_ISL_2779017, EPI_ISL_2779018 | | | |
| see above | LESP Yucatan | Instituto de Diagnostico y Referencia Epidemiologicos (INDRE) | Claudia Wong-Arambula, Abril Rodriguez-Maldonado, Vanessa Rivero-Arredondo, Ariadna Medina-Benitez, Joaquin Quiroz-Mercado, Sergio Rangel-Guerrero, Maribel Gonzalez-Villa, Natividad Cruz-Ortiz, Tatiana Nunez-Garcia, Gisela Barrera-Badillo, Lucia Hernandez-Rivas, Irma Lopez-Martinez, Ernesto Ramirez-Gonzalez. |
| EPI_ISL_2779019 | LESP Guanajuato | Instituto de Diagnostico y Referencia Epidemiologicos (INDRE) | Claudia Wong-Arambula, Abril Rodriguez-Maldonado, Vanessa Rivero-Arredondo, Ariadna Medina-Benitez, Joaquin Quiroz-Mercado, Sergio Rangel-Guerrero, Maribel Gonzalez-Villa, Natividad Cruz-Ortiz, Tatiana Nunez-Garcia, Gisela Barrera-Badillo, Lucia Hernandez-Rivas, Irma Lopez-Martinez, Ernesto Ramirez-Gonzalez. |
| EPI_ISL_2779020, EPI_ISL_2779021, EPI_ISL_2779022, EPI_ISL_2779023 | LESP Yucatan | Instituto de Diagnostico y Referencia Epidemiologicos (INDRE) | Claudia Wong-Arambula, Abril Rodriguez-Maldonado, Vanessa Rivero-Arredondo, Ariadna Medina-Benitez, Joaquin Quiroz-Mercado, Sergio Rangel-Guerrero, Maribel Gonzalez-Villa, Natividad Cruz-Ortiz, Tatiana Nunez-Garcia, Gisela Barrera-Badillo, Lucia Hernandez-Rivas, Irma Lopez-Martinez, Ernesto Ramirez-Gonzalez. |
| EPI_ISL_2779024, EPI_ISL_2779025, EPI_ISL_2779026, EPI_ISL_2779027, EPI_ISL_2779028, EPI_ISL_2779029, EPI_ISL_2779030, EPI_ISL_2779031, EPI_ISL_2779032, EPI_ISL_2779033 | LESP Tabasco | Instituto de Diagnostico y Referencia Epidemiologicos (INDRE) | Claudia Wong-Arambula, Abril Rodriguez-Maldonado, Vanessa Rivero-Arredondo, Ariadna Medina-Benitez, Joaquin Quiroz-Mercado, Sergio Rangel-Guerrero, Maribel Gonzalez-Villa, Natividad Cruz-Ortiz, Tatiana Nunez-Garcia, Gisela Barrera-Badillo, Lucia Hernandez-Rivas, Irma Lopez-Martinez, Ernesto Ramirez-Gonzalez. |
| EPI_ISL_2779034 | LESP Coahuila | Instituto de Diagnostico y Referencia Epidemiologicos (INDRE) | Claudia Wong-Arambula, Abril Rodriguez-Maldonado, Vanessa Rivero-Arredondo, Ariadna Medina-Benitez, Joaquin Quiroz-Mercado, Sergio Rangel-Guerrero, Maribel Gonzalez-Villa, Natividad Cruz-Ortiz, Tatiana Nunez-Garcia, Gisela Barrera-Badillo, Lucia Hernandez-Rivas, Irma Lopez-Martinez, Ernesto Ramirez-Gonzalez. |
| EPI_ISL_2779035, EPI_ISL_2779036, EPI_ISL_2779037, EPI_ISL_2779038, EPI_ISL_2779039, EPI_ISL_2779040, EPI_ISL_2779041, EPI_ISL_2779042 | LESP Puebla | Instituto de Diagnostico y Referencia Epidemiologicos (INDRE) | Claudia Wong-Arambula, Abril Rodriguez-Maldonado, Vanessa Rivero-Arredondo, Ariadna Medina-Benitez, Joaquin Quiroz-Mercado, Sergio Rangel-Guerrero, Maribel Gonzalez-Villa, Natividad Cruz-Ortiz, Tatiana Nunez-Garcia, Gisela Barrera-Badillo, Lucia Hernandez-Rivas, Irma Lopez-Martinez, Ernesto Ramirez-Gonzalez. |
| EPI_ISL_2779043, EPI_ISL_2779044, EPI_ISL_2779045, EPI_ISL_2779046 | LESP Tabasco | Instituto de Diagnostico y Referencia Epidemiologicos (INDRE) | Claudia Wong-Arambula, Abril Rodriguez-Maldonado, Vanessa Rivero-Arredondo, Ariadna Medina-Benitez, Joaquin Quiroz-Mercado, Sergio Rangel-Guerrero, Maribel Gonzalez-Villa, Natividad Cruz-Ortiz, Tatiana Nunez-Garcia, Gisela Barrera-Badillo, Lucia Hernandez-Rivas, Irma Lopez-Martinez, Ernesto Ramirez-Gonzalez. |
| EPI_ISL_2779047, EPI_ISL_2779048 | LESP Guanajuato | Instituto de Diagnostico y Referencia Epidemiologicos (INDRE) | Claudia Wong-Arambula, Abril Rodriguez-Maldonado, Vanessa Rivero-Arredondo, Ariadna Medina-Benitez, Joaquin Quiroz-Mercado, Sergio Rangel-Guerrero, Maribel Gonzalez-Villa, Natividad Cruz-Ortiz, Tatiana Nunez-Garcia, Gisela Barrera-Badillo, Lucia Hernandez-Rivas, Irma Lopez-Martinez, Ernesto Ramirez-Gonzalez. |
| EPI_ISL_2779049 | LESP Queretaro | Instituto de Diagnostico y Referencia Epidemiologicos (INDRE) | Claudia Wong-Arambula, Abril Rodriguez-Maldonado, Vanessa Rivero-Arredondo, Ariadna Medina-Benitez, Joaquin Quiroz-Mercado, Sergio Rangel-Guerrero, Maribel Gonzalez-Villa, Natividad Cruz-Ortiz, Tatiana Nunez-Garcia, Gisela Barrera-Badillo, Lucia Hernandez-Rivas, Irma Lopez-Martinez, Ernesto Ramirez-Gonzalez. |
| EPI_ISL_2779050 | LESP Guerrero | Instituto de Diagnostico y Referencia Epidemiologicos (INDRE) | Claudia Wong-Arambula, Abril Rodriguez-Maldonado, Vanessa Rivero-Arredondo, Ariadna Medina-Benitez, Joaquin Quiroz-Mercado, Sergio Rangel-Guerrero, Maribel Gonzalez-Villa, Natividad Cruz-Ortiz, Tatiana Nunez-Garcia, Gisela Barrera-Badillo, Lucia Hernandez-Rivas, Irma Lopez-Martinez, Ernesto Ramirez-Gonzalez. |
| EPI_ISL_2779051, EPI_ISL_2779052 | LESP Quintana Roo | Instituto de Diagnostico y Referencia Epidemiologicos (INDRE) | Claudia Wong-Arambula, Abril Rodriguez-Maldonado, Vanessa Rivero-Arredondo, Ariadna Medina-Benitez, Joaquin Quiroz-Mercado, Sergio Rangel-Guerrero, Maribel Gonzalez-Villa, Natividad Cruz-Ortiz, Tatiana Nunez-Garcia, Gisela Barrera-Badillo, Lucia Hernandez-Rivas, Irma Lopez-Martinez, Ernesto Ramirez-Gonzalez. |
| EPI_ISL_2779053, EPI_ISL_2779054 | LESP Tabasco | Instituto de Diagnostico y Referencia Epidemiologicos (INDRE) | Claudia Wong-Arambula, Abril Rodriguez-Maldonado, Vanessa Rivero-Arredondo, Ariadna Medina-Benitez, Joaquin Quiroz-Mercado, Sergio Rangel-Guerrero, Maribel Gonzalez-Villa, Natividad Cruz-Ortiz, Tatiana Nunez-Garcia, Gisela Barrera-Badillo, Lucia Hernandez-Rivas, Irma Lopez-Martinez, Ernesto Ramirez-Gonzalez. |
| EPI_ISL_2779055 | LESP Sonora | Instituto de Diagnostico y Referencia Epidemiologicos (INDRE) | Claudia Wong-Arambula, Abril Rodriguez-Maldonado, Vanessa Rivero-Arredondo, Ariadna Medina-Benitez, Joaquin Quiroz-Mercado, Sergio Rangel-Guerrero, Maribel Gonzalez-Villa, Natividad Cruz-Ortiz, Tatiana Nunez-Garcia, Gisela Barrera-Badillo, Lucia Hernandez-Rivas, Irma Lopez-Martinez, Ernesto Ramirez-Gonzalez. |
| EPI_ISL_2779056, EPI_ISL_2779057, EPI_ISL_2779058 | LESP Guanajuato | Instituto de Diagnostico y Referencia Epidemiologicos (INDRE) | Claudia Wong-Arambula, Abril Rodriguez-Maldonado, Vanessa Rivero-Arredondo, Ariadna Medina-Benitez, Joaquin Quiroz-Mercado, Sergio Rangel-Guerrero, Maribel Gonzalez-Villa, Natividad Cruz-Ortiz, Tatiana Nunez-Garcia, Gisela Barrera-Badillo, Lucia Hernandez-Rivas, Irma Lopez-Martinez, Ernesto Ramirez-Gonzalez. |
| EPI_ISL_2779059 | LESP Campeche | Instituto de Diagnostico y Referencia Epidemiologicos (INDRE) | Claudia Wong-Arambula, Abril Rodriguez-Maldonado, Vanessa Rivero-Arredondo, Ariadna Medina-Benitez, Joaquin Quiroz-Mercado, Sergio Rangel-Guerrero, Maribel Gonzalez-Villa, Natividad Cruz-Ortiz, Tatiana Nunez-Garcia, Gisela Barrera-Badillo, Lucia Hernandez-Rivas, Irma Lopez-Martinez, Ernesto Ramirez-Gonzalez. |
| EPI_ISL_2779060, EPI_ISL_2779061, EPI_ISL_2779062, EPI_ISL_2779063, EPI_ISL_2779064, EPI_ISL_2779065 | LESP Tamaulipas | Instituto de Diagnostico y Referencia Epidemiologicos (INDRE) | Claudia Wong-Arambula, Abril Rodriguez-Maldonado, Vanessa Rivero-Arredondo, Ariadna Medina-Benitez, Joaquin Quiroz-Mercado, Sergio Rangel-Guerrero, Maribel Gonzalez-Villa, Natividad Cruz-Ortiz, Tatiana Nunez-Garcia, Gisela Barrera-Badillo, Lucia Hernandez-Rivas, Irma Lopez-Martinez, Ernesto Ramirez-Gonzalez. |
| EPI_ISL_2779066, EPI_ISL_2779067, EPI_ISL_2779068, EPI_ISL_2779069, EPI_ISL_2779070, EPI_ISL_2779071, EPI_ISL_2779072, EPI_ISL_2779073, EPI_ISL_2779074, EPI_ISL_2779075, EPI_ISL_2779076, EPI_ISL_2779077, EPI_ISL_2779078, EPI_ISL_2779079, EPI_ISL_2779080, EPI_ISL_2779081, EPI_ISL_2779082, EPI_ISL_2779083, EPI_ISL_2779084, EPI_ISL_2779085, EPI_ISL_2779086, EPI_ISL_2779087, EPI_ISL_2779088, EPI_ISL_2779089, EPI_ISL_2779090, EPI_ISL_2779091, EPI_ISL_2779092, EPI_ISL_2779093, EPI_ISL_2779094, EPI_ISL_2779095, EPI_ISL_2779096, EPI_ISL_2779097, EPI_ISL_2779098, EPI_ISL_2779099, EPI_ISL_2779100, EPI_ISL_2779101, EPI_ISL_2779102, EPI_ISL_2779103, EPI_ISL_2779104, EPI_ISL_2779105, EPI_ISL_2779106, EPI_ISL_2779107, EPI_ISL_2779108, EPI_ISL_2779109, EPI_ISL_2779110, EPI_ISL_2779111, EPI_ISL_2779112, EPI_ISL_2779113, EPI_ISL_2779114, EPI_ISL_2779115, EPI_ISL_2779116, EPI_ISL_2779117, EPI_ISL_2779118, EPI_ISL_2779119 | | | |
| see above | LESP Yucatan | Instituto de Diagnostico y Referencia Epidemiologicos (INDRE) | Claudia Wong-Arambula, Abril Rodriguez-Maldonado, Vanessa Rivero-Arredondo, Ariadna Medina-Benitez, Joaquin Quiroz-Mercado, Sergio Rangel-Guerrero, Maribel Gonzalez-Villa, Natividad Cruz-Ortiz, Tatiana Nunez-Garcia, Gisela Barrera-Badillo, Lucia Hernandez-Rivas, Irma Lopez-Martinez, Ernesto Ramirez-Gonzalez. |
| EPI_ISL_2779120 | LESP Guanajuato | Instituto de Diagnostico y Referencia Epidemiologicos (INDRE) | Claudia Wong-Arambula, Abril Rodriguez-Maldonado, Vanessa Rivero-Arredondo, Ariadna Medina-Benitez, Joaquin Quiroz-Mercado, Sergio Rangel-Guerrero, Maribel Gonzalez-Villa, Natividad Cruz-Ortiz, Tatiana Nunez-Garcia, Gisela Barrera-Badillo, Lucia Hernandez-Rivas, Irma Lopez-Martinez, Ernesto Ramirez-Gonzalez. |
| EPI_ISL_2779121, EPI_ISL_2779122, EPI_ISL_2779123, EPI_ISL_2779124, EPI_ISL_2779125, EPI_ISL_2779126, EPI_ISL_2779127, EPI_ISL_2779128, EPI_ISL_2779129, EPI_ISL_2779130, EPI_ISL_2779131, EPI_ISL_2779132, EPI_ISL_2779133, EPI_ISL_2779134, EPI_ISL_2779135, EPI_ISL_2779136, EPI_ISL_2779137, EPI_ISL_2779138, EPI_ISL_2779139, EPI_ISL_2779140, EPI_ISL_2779141, EPI_ISL_2779142, EPI_ISL_2779143, EPI_ISL_2779144, EPI_ISL_2779145, EPI_ISL_2779146, EPI_ISL_2779147, EPI_ISL_2779148, EPI_ISL_2779149, EPI_ISL_2779150, EPI_ISL_2779151, EPI_ISL_2779152, EPI_ISL_2779153, EPI_ISL_2779154, EPI_ISL_2779155, EPI_ISL_2779156, EPI_ISL_2779157, EPI_ISL_2779158 | | | |
| see above | LESP Yucatan | Instituto de Diagnostico y Referencia Epidemiologicos (INDRE) | Claudia Wong-Arambula, Abril Rodriguez-Maldonado, Vanessa Rivero-Arredondo, Ariadna Medina-Benitez, Joaquin Quiroz-Mercado, Sergio Rangel-Guerrero, Maribel Gonzalez-Villa, Natividad Cruz-Ortiz, Tatiana Nunez-Garcia, Gisela Barrera-Badillo, Lucia Hernandez-Rivas, Irma Lopez-Martinez, Ernesto Ramirez-Gonzalez. |
| EPI_ISL_2779159 | LESP San Luis Potosi | Instituto de Diagnostico y Referencia Epidemiologicos (INDRE) | Claudia Wong-Arambula, Abril Rodriguez-Maldonado, Vanessa Rivero-Arredondo, Ariadna Medina-Benitez, Joaquin Quiroz-Mercado, Sergio Rangel-Guerrero, Maribel Gonzalez-Villa, Natividad Cruz-Ortiz, Tatiana Nunez-Garcia, Gisela Barrera-Badillo, Lucia Hernandez-Rivas, Irma Lopez-Martinez, Ernesto Ramirez-Gonzalez. |
| EPI_ISL_2779160, EPI_ISL_2779161, EPI_ISL_2779162, EPI_ISL_2779163, EPI_ISL_2779164, EPI_ISL_2779165, EPI_ISL_2779166, EPI_ISL_2779167, EPI_ISL_2779168, EPI_ISL_2779169, EPI_ISL_2779170, EPI_ISL_2779171, EPI_ISL_2779172 | | | |
| see above | LESP Tabasco | Instituto de Diagnostico y Referencia Epidemiologicos (INDRE) | Claudia Wong-Arambula, Abril Rodriguez-Maldonado, Vanessa Rivero-Arredondo, Ariadna Medina-Benitez, Joaquin Quiroz-Mercado, Sergio Rangel-Guerrero, Maribel Gonzalez-Villa, Natividad Cruz-Ortiz, Tatiana Nunez-Garcia, Gisela Barrera-Badillo, Lucia Hernandez-Rivas, Irma Lopez-Martinez, Ernesto Ramirez-Gonzalez. |
| EPI_ISL_2779173 | LESP Ciudad de Mexico | Instituto de Diagnostico y Referencia Epidemiologicos (INDRE) | Claudia Wong-Arambula, Abril Rodriguez-Maldonado, Vanessa Rivero-Arredondo, Ariadna Medina-Benitez, Joaquin Quiroz-Mercado, Sergio Rangel-Guerrero, Maribel Gonzalez-Villa, Natividad Cruz-Ortiz, Tatiana Nunez-Garcia, Gisela Barrera-Badillo, Lucia Hernandez-Rivas, Irma Lopez-Martinez, Ernesto Ramirez-Gonzalez. |

| | | | |
|---|---|---|---|
| EPI_ISL_2779174 | LESP Puebla | Instituto de Diagnostico y Referencia Epidemiologicos (INDRE) | Claudia Wong-Arambula, Abril Rodriguez-Maldonado, Vanessa Rivero-Arredondo, Ariadna Medina-Benitez, Joaquin Quiroz-Mercado, Sergio Rangel-Guerrero, Maribel Gonzalez-Villa, Natividad Cruz-Ortiz, Tatiana Nunez-Garcia, Gisela Barrera-Badillo, Lucia Hernandez-Rivas, Irma Lopez-Martinez, Ernesto Ramirez-Gonzalez. |
| EPI_ISL_2779175 | LESP Jalisco | Instituto de Diagnostico y Referencia Epidemiologicos (INDRE) | Claudia Wong-Arambula, Abril Rodriguez-Maldonado, Vanessa Rivero-Arredondo, Ariadna Medina-Benitez, Joaquin Quiroz-Mercado, Sergio Rangel-Guerrero, Maribel Gonzalez-Villa, Natividad Cruz-Ortiz, Tatiana Nunez-Garcia, Gisela Barrera-Badillo, Lucia Hernandez-Rivas, Irma Lopez-Martinez, Ernesto Ramirez-Gonzalez. |
| EPI_ISL_2779176, EPI_ISL_2779177, EPI_ISL_2779178, EPI_ISL_2779179, EPI_ISL_2779180, EPI_ISL_2779181 | LESP Quintana Roo | Instituto de Diagnostico y Referencia Epidemiologicos (INDRE) | Claudia Wong-Arambula, Abril Rodriguez-Maldonado, Vanessa Rivero-Arredondo, Ariadna Medina-Benitez, Joaquin Quiroz-Mercado, Sergio Rangel-Guerrero, Maribel Gonzalez-Villa, Natividad Cruz-Ortiz, Tatiana Nunez-Garcia, Gisela Barrera-Badillo, Lucia Hernandez-Rivas, Irma Lopez-Martinez, Ernesto Ramirez-Gonzalez. |
| EPI_ISL_2779182, EPI_ISL_2779183 | LESP Tabasco | Instituto de Diagnostico y Referencia Epidemiologicos (INDRE) | Claudia Wong-Arambula, Abril Rodriguez-Maldonado, Vanessa Rivero-Arredondo, Ariadna Medina-Benitez, Joaquin Quiroz-Mercado, Sergio Rangel-Guerrero, Maribel Gonzalez-Villa, Natividad Cruz-Ortiz, Tatiana Nunez-Garcia, Gisela Barrera-Badillo, Lucia Hernandez-Rivas, Irma Lopez-Martinez, Ernesto Ramirez-Gonzalez. |
| EPI_ISL_2779184, EPI_ISL_2779185 | LESP Sonora | Instituto de Diagnostico y Referencia Epidemiologicos (INDRE) | Claudia Wong-Arambula, Abril Rodriguez-Maldonado, Vanessa Rivero-Arredondo, Ariadna Medina-Benitez, Joaquin Quiroz-Mercado, Sergio Rangel-Guerrero, Maribel Gonzalez-Villa, Natividad Cruz-Ortiz, Tatiana Nunez-Garcia, Gisela Barrera-Badillo, Lucia Hernandez-Rivas, Irma Lopez-Martinez, Ernesto Ramirez-Gonzalez. |
| EPI_ISL_2779186, EPI_ISL_2779187, EPI_ISL_2779188, EPI_ISL_2779189, EPI_ISL_2779190, EPI_ISL_2779191, EPI_ISL_2779192, EPI_ISL_2779193 | LESP Guanajuato | Instituto de Diagnostico y Referencia Epidemiologicos (INDRE) | Claudia Wong-Arambula, Abril Rodriguez-Maldonado, Vanessa Rivero-Arredondo, Ariadna Medina-Benitez, Joaquin Quiroz-Mercado, Sergio Rangel-Guerrero, Maribel Gonzalez-Villa, Natividad Cruz-Ortiz, Tatiana Nunez-Garcia, Gisela Barrera-Badillo, Lucia Hernandez-Rivas, Irma Lopez-Martinez, Ernesto Ramirez-Gonzalez. |
| EPI_ISL_2779194 | LESP Tabasco | Instituto de Diagnostico y Referencia Epidemiologicos (INDRE) | Claudia Wong-Arambula, Abril Rodriguez-Maldonado, Vanessa Rivero-Arredondo, Ariadna Medina-Benitez, Joaquin Quiroz-Mercado, Sergio Rangel-Guerrero, Maribel Gonzalez-Villa, Natividad Cruz-Ortiz, Tatiana Nunez-Garcia, Gisela Barrera-Badillo, Lucia Hernandez-Rivas, Irma Lopez-Martinez, Ernesto Ramirez-Gonzalez. |
| EPI_ISL_2779195 | LESP Campeche | Instituto de Diagnostico y Referencia Epidemiologicos (INDRE) | Claudia Wong-Arambula, Abril Rodriguez-Maldonado, Vanessa Rivero-Arredondo, Ariadna Medina-Benitez, Joaquin Quiroz-Mercado, Sergio Rangel-Guerrero, Maribel Gonzalez-Villa, Natividad Cruz-Ortiz, Tatiana Nunez-Garcia, Gisela Barrera-Badillo, Lucia Hernandez-Rivas, Irma Lopez-Martinez, Ernesto Ramirez-Gonzalez. |
| EPI_ISL_2779196 | LESP Guerrero | Instituto de Diagnostico y Referencia Epidemiologicos (INDRE) | Claudia Wong-Arambula, Abril Rodriguez-Maldonado, Vanessa Rivero-Arredondo, Ariadna Medina-Benitez, Joaquin Quiroz-Mercado, Sergio Rangel-Guerrero, Maribel Gonzalez-Villa, Natividad Cruz-Ortiz, Tatiana Nunez-Garcia, Gisela Barrera-Badillo, Lucia Hernandez-Rivas, Irma Lopez-Martinez, Ernesto Ramirez-Gonzalez. |
| EPI_ISL_2779197 | LESP Baja California Sur | Instituto de Diagnostico y Referencia Epidemiologicos (INDRE) | Claudia Wong-Arambula, Abril Rodriguez-Maldonado, Vanessa Rivero-Arredondo, Ariadna Medina-Benitez, Joaquin Quiroz-Mercado, Sergio Rangel-Guerrero, Maribel Gonzalez-Villa, Natividad Cruz-Ortiz, Tatiana Nunez-Garcia, Gisela Barrera-Badillo, Lucia Hernandez-Rivas, Irma Lopez-Martinez, Ernesto Ramirez-Gonzalez. |
| EPI_ISL_2779198, EPI_ISL_2779199, EPI_ISL_2779200, EPI_ISL_2779201, EPI_ISL_2779202, EPI_ISL_2779203, EPI_ISL_2779204, EPI_ISL_2779205, EPI_ISL_2779206, EPI_ISL_2779207, EPI_ISL_2779208, EPI_ISL_2779209, EPI_ISL_2779210 | | | |
| see above | LESP Tabasco | Instituto de Diagnostico y Referencia Epidemiologicos (INDRE) | Claudia Wong-Arambula, Abril Rodriguez-Maldonado, Vanessa Rivero-Arredondo, Ariadna Medina-Benitez, Joaquin Quiroz-Mercado, Sergio Rangel-Guerrero, Maribel Gonzalez-Villa, Natividad Cruz-Ortiz, Tatiana Nunez-Garcia, Gisela Barrera-Badillo, Lucia Hernandez-Rivas, Irma Lopez-Martinez, Ernesto Ramirez-Gonzalez. |
| EPI_ISL_2779211, EPI_ISL_2779212 | LESP Puebla | Instituto de Diagnostico y Referencia Epidemiologicos (INDRE) | Claudia Wong-Arambula, Abril Rodriguez-Maldonado, Vanessa Rivero-Arredondo, Ariadna Medina-Benitez, Joaquin Quiroz-Mercado, Sergio Rangel-Guerrero, Maribel Gonzalez-Villa, Natividad Cruz-Ortiz, Tatiana Nunez-Garcia, Gisela Barrera-Badillo, Lucia Hernandez-Rivas, Irma Lopez-Martinez, Ernesto Ramirez-Gonzalez. |
| EPI_ISL_2779213 | LESP Sonora | Instituto de Diagnostico y Referencia Epidemiologicos (INDRE) | Claudia Wong-Arambula, Abril Rodriguez-Maldonado, Vanessa Rivero-Arredondo, Ariadna Medina-Benitez, Joaquin Quiroz-Mercado, Sergio Rangel-Guerrero, Maribel Gonzalez-Villa, Natividad Cruz-Ortiz, Tatiana Nunez-Garcia, Gisela Barrera-Badillo, Lucia Hernandez-Rivas, Irma Lopez-Martinez, Ernesto Ramirez-Gonzalez. |
| EPI_ISL_2779214 | LESP Guerrero | Instituto de Diagnostico y Referencia Epidemiologicos (INDRE) | Claudia Wong-Arambula, Abril Rodriguez-Maldonado, Vanessa Rivero-Arredondo, Ariadna Medina-Benitez, Joaquin Quiroz-Mercado, Sergio Rangel-Guerrero, Maribel Gonzalez-Villa, Natividad Cruz-Ortiz, Tatiana Nunez-Garcia, Gisela Barrera-Badillo, Lucia Hernandez-Rivas, Irma Lopez-Martinez, Ernesto Ramirez-Gonzalez. |
| EPI_ISL_2779215, EPI_ISL_2779216, EPI_ISL_2779217, EPI_ISL_2779218 | LESP Guanajuato | Instituto de Diagnostico y Referencia Epidemiologicos (INDRE) | Claudia Wong-Arambula, Abril Rodriguez-Maldonado, Vanessa Rivero-Arredondo, Ariadna Medina-Benitez, Joaquin Quiroz-Mercado, Sergio Rangel-Guerrero, Maribel Gonzalez-Villa, Natividad Cruz-Ortiz, Tatiana Nunez-Garcia, Gisela Barrera-Badillo, Lucia Hernandez-Rivas, Irma Lopez-Martinez, Ernesto Ramirez-Gonzalez. |
| EPI_ISL_2779219 | LESP Tamaulipas | Instituto de Diagnostico y Referencia Epidemiologicos (INDRE) | Claudia Wong-Arambula, Abril Rodriguez-Maldonado, Vanessa Rivero-Arredondo, Ariadna Medina-Benitez, Joaquin Quiroz-Mercado, Sergio Rangel-Guerrero, Maribel Gonzalez-Villa, Natividad Cruz-Ortiz, Tatiana Nunez-Garcia, Gisela Barrera-Badillo, Lucia Hernandez-Rivas, Irma Lopez-Martinez, Ernesto Ramirez-Gonzalez. |
| EPI_ISL_2779220 | LESP Sinaloa | Instituto de Diagnostico y Referencia Epidemiologicos (INDRE) | Claudia Wong-Arambula, Abril Rodriguez-Maldonado, Vanessa Rivero-Arredondo, Ariadna Medina-Benitez, Joaquin Quiroz-Mercado, Sergio Rangel-Guerrero, Maribel Gonzalez-Villa, Natividad Cruz-Ortiz, Tatiana Nunez-Garcia, Gisela Barrera-Badillo, Lucia Hernandez-Rivas, Irma Lopez-Martinez, Ernesto Ramirez-Gonzalez. |
| EPI_ISL_2779221, EPI_ISL_2779222, EPI_ISL_2779223, EPI_ISL_2779224, EPI_ISL_2779225 | LESP Yucatan | Instituto de Diagnostico y Referencia Epidemiologicos (INDRE) | Claudia Wong-Arambula, Abril Rodriguez-Maldonado, Vanessa Rivero-Arredondo, Ariadna Medina-Benitez, Joaquin Quiroz-Mercado, Sergio Rangel-Guerrero, Maribel Gonzalez-Villa, Natividad Cruz-Ortiz, Tatiana Nunez-Garcia, Gisela Barrera-Badillo, Lucia Hernandez-Rivas, Irma Lopez-Martinez, Ernesto Ramirez-Gonzalez. |
| EPI_ISL_2779226 | LESP Michoacan | Instituto de Diagnostico y Referencia Epidemiologicos (INDRE) | Claudia Wong-Arambula, Abril Rodriguez-Maldonado, Vanessa Rivero-Arredondo, Ariadna Medina-Benitez, Joaquin Quiroz-Mercado, Sergio Rangel-Guerrero, Maribel Gonzalez-Villa, Natividad Cruz-Ortiz, Tatiana Nunez-Garcia, Gisela Barrera-Badillo, Lucia Hernandez-Rivas, Irma Lopez-Martinez, Ernesto Ramirez-Gonzalez. |
| EPI_ISL_2779227 | LESP Guerrero | Instituto de Diagnostico y Referencia Epidemiologicos (INDRE) | Claudia Wong-Arambula, Abril Rodriguez-Maldonado, Vanessa Rivero-Arredondo, Ariadna Medina-Benitez, Joaquin Quiroz-Mercado, Sergio Rangel-Guerrero, Maribel Gonzalez-Villa, Natividad Cruz-Ortiz, Tatiana Nunez-Garcia, Gisela Barrera-Badillo, Lucia Hernandez-Rivas, Irma Lopez-Martinez, Ernesto Ramirez-Gonzalez. |
| EPI_ISL_2779228, EPI_ISL_2779229, EPI_ISL_2779230, EPI_ISL_2779231 | LESP Yucatan | Instituto de Diagnostico y Referencia Epidemiologicos (INDRE) | Claudia Wong-Arambula, Abril Rodriguez-Maldonado, Vanessa Rivero-Arredondo, Ariadna Medina-Benitez, Joaquin Quiroz-Mercado, Sergio Rangel-Guerrero, Maribel Gonzalez-Villa, Natividad Cruz-Ortiz, Tatiana Nunez-Garcia, Gisela Barrera-Badillo, Lucia Hernandez-Rivas, Irma Lopez-Martinez, Ernesto Ramirez-Gonzalez. |
| EPI_ISL_2779232, EPI_ISL_2779233 | LESP Michoacan | Instituto de Diagnostico y Referencia Epidemiologicos (INDRE) | Claudia Wong-Arambula, Abril Rodriguez-Maldonado, Vanessa Rivero-Arredondo, Ariadna Medina-Benitez, Joaquin Quiroz-Mercado, Sergio Rangel-Guerrero, Maribel Gonzalez-Villa, Natividad Cruz-Ortiz, Tatiana Nunez-Garcia, Gisela Barrera-Badillo, Lucia Hernandez-Rivas, Irma Lopez-Martinez, Ernesto Ramirez-Gonzalez. |
| EPI_ISL_2779234, EPI_ISL_2779235, EPI_ISL_2779236, EPI_ISL_2779237, EPI_ISL_2779238, EPI_ISL_2779239, EPI_ISL_2779240, EPI_ISL_2779241, EPI_ISL_2779242, EPI_ISL_2779243, EPI_ISL_2779244, EPI_ISL_2779245, EPI_ISL_2779246, EPI_ISL_2779247, EPI_ISL_2779248, EPI_ISL_2779249, EPI_ISL_2779250, EPI_ISL_2779251, EPI_ISL_2779252, EPI_ISL_2779253, EPI_ISL_2779254, EPI_ISL_2779255, EPI_ISL_2779256 | | | |
| see above | LESP Tabasco | Instituto de Diagnostico y Referencia Epidemiologicos (INDRE) | Claudia Wong-Arambula, Abril Rodriguez-Maldonado, Vanessa Rivero-Arredondo, Ariadna Medina-Benitez, Joaquin Quiroz-Mercado, Sergio Rangel-Guerrero, Maribel Gonzalez-Villa, Natividad Cruz-Ortiz, Tatiana Nunez-Garcia, Gisela Barrera-Badillo, Lucia Hernandez-Rivas, Irma Lopez-Martinez, Ernesto Ramirez-Gonzalez. |
| EPI_ISL_2779257, EPI_ISL_2779258, EPI_ISL_2779259, EPI_ISL_2779260, EPI_ISL_2779261, EPI_ISL_2779262, EPI_ISL_2779263, EPI_ISL_2779264, EPI_ISL_2779265, EPI_ISL_2779266, EPI_ISL_2779267, EPI_ISL_2779268, EPI_ISL_2779269 | | | |
| see above | LESP Puebla | Instituto de Diagnostico y Referencia Epidemiologicos (INDRE) | Claudia Wong-Arambula, Abril Rodriguez-Maldonado, Vanessa Rivero-Arredondo, Ariadna Medina-Benitez, Joaquin Quiroz-Mercado, Sergio Rangel-Guerrero, Maribel Gonzalez-Villa, Natividad Cruz-Ortiz, Tatiana Nunez-Garcia, Gisela Barrera-Badillo, Lucia Hernandez-Rivas, Irma Lopez-Martinez, Ernesto Ramirez-Gonzalez. |
| EPI_ISL_2779271, EPI_ISL_2779272, EPI_ISL_2779273, EPI_ISL_2779274, EPI_ISL_2779275 | LESP Tabasco | Instituto de Diagnostico y Referencia Epidemiologicos (INDRE) | Claudia Wong-Arambula, Abril Rodriguez-Maldonado, Vanessa Rivero-Arredondo, Ariadna Medina-Benitez, Joaquin Quiroz-Mercado, Sergio Rangel-Guerrero, Maribel Gonzalez-Villa, Natividad Cruz-Ortiz, Tatiana Nunez-Garcia, Gisela Barrera-Badillo, Lucia Hernandez-Rivas, Irma Lopez-Martinez, Ernesto Ramirez-Gonzalez. |
| EPI_ISL_2779276 | LESP Guanajuato | Instituto de Diagnostico y Referencia Epidemiologicos (INDRE) | Claudia Wong-Arambula, Abril Rodriguez-Maldonado, Vanessa Rivero-Arredondo, Ariadna Medina-Benitez, Joaquin Quiroz-Mercado, Sergio Rangel-Guerrero, Maribel Gonzalez-Villa, Natividad Cruz-Ortiz, Tatiana Nunez-Garcia, Gisela Barrera-Badillo, Lucia Hernandez-Rivas, Irma Lopez-Martinez, Ernesto Ramirez-Gonzalez. |
| EPI_ISL_2779277, EPI_ISL_2779278, EPI_ISL_2779279 | LESP Tabasco | Instituto de Diagnostico y Referencia Epidemiologicos (INDRE) | Claudia Wong-Arambula, Abril Rodriguez-Maldonado, Vanessa Rivero-Arredondo, Ariadna Medina-Benitez, Joaquin Quiroz-Mercado, Sergio Rangel-Guerrero, Maribel Gonzalez-Villa, Natividad Cruz-Ortiz, Tatiana Nunez-Garcia, Gisela Barrera-Badillo, Lucia Hernandez-Rivas, Irma Lopez-Martinez, Ernesto Ramirez-Gonzalez. |
| EPI_ISL_2779280 | LESP Puebla | Instituto de Diagnostico y Referencia Epidemiologicos (INDRE) | Claudia Wong-Arambula, Abril Rodriguez-Maldonado, Vanessa Rivero-Arredondo, Ariadna Medina-Benitez, Joaquin Quiroz-Mercado, Sergio Rangel-Guerrero, Maribel Gonzalez-Villa, Natividad Cruz-Ortiz, Tatiana Nunez-Garcia, Gisela Barrera-Badillo, Lucia Hernandez-Rivas, Irma Lopez-Martinez, Ernesto Ramirez-Gonzalez. |
| EPI_ISL_2779281 | LESP Guanajuato | Instituto de Diagnostico y Referencia Epidemiologicos (INDRE) | Claudia Wong-Arambula, Abril Rodriguez-Maldonado, Vanessa Rivero-Arredondo, Ariadna Medina-Benitez, Joaquin Quiroz-Mercado, Sergio Rangel-Guerrero, Maribel Gonzalez-Villa, Natividad Cruz-Ortiz, Tatiana Nunez-Garcia, Gisela Barrera-Badillo, Lucia Hernandez-Rivas, Irma Lopez-Martinez, Ernesto Ramirez-Gonzalez. |
| EPI_ISL_2779551, EPI_ISL_2779556, EPI_ISL_2779557, EPI_ISL_2779558, EPI_ISL_2779559, EPI_ISL_2779561, EPI_ISL_2779562, EPI_ISL_2779566, EPI_ISL_2779568, EPI_ISL_2779570, EPI_ISL_2779574, EPI_ISL_2779575, EPI_ISL_2779578, EPI_ISL_2779579, EPI_ISL_2779580, EPI_ISL_2779582, EPI_ISL_2779584, EPI_ISL_2779587, EPI_ISL_2779588, EPI_ISL_2779589, EPI_ISL_2779590, EPI_ISL_2779592, EPI_ISL_2779593, EPI_ISL_2779594, EPI_ISL_2779597, EPI_ISL_2779602, EPI_ISL_2779603, EPI_ISL_2779604, EPI_ISL_2779608, EPI_ISL_2779609, EPI_ISL_2779612 | | | |
| see above | Santa Clara County Public Health Laboratory | Santa Clara County Public Health Laboratory | Santa Clara County Public Health Department |
| EPI_ISL_2779614, EPI_ISL_2779615, EPI_ISL_2779616, EPI_ISL_2779617, EPI_ISL_2779618, EPI_ISL_2779619, EPI_ISL_2779620, EPI_ISL_2779621, EPI_ISL_2779622, EPI_ISL_2779623, EPI_ISL_2779624, EPI_ISL_2779625, EPI_ISL_2779626, EPI_ISL_2779627, EPI_ISL_2779628, EPI_ISL_2779629, EPI_ISL_2779630, EPI_ISL_2779631, EPI_ISL_2779632, EPI_ISL_2779633, EPI_ISL_2779634, EPI_ISL_2779635, EPI_ISL_2779636, EPI_ISL_2779637 | | | |
| see above | Salud Digna | Instituto Nacional de Medicina Genomica | Cedro-Tanda A, Hidalgo-Miranda A, Cisneros-Villanueva M, Orjuela-Rodríguez M, Mendoza-Vargas A, Reyes-Grajeda JP, Abraham Campos-Romero, Moreno-Camacho José Luis, Rodriguez-Gallegos Jorge, Luna-Ruiz Marco, Gonzalez-Barrera D, Rangel-DeLeon D, Munguia-Garza P, Ramirez-Vega O, Escobar-Arrazola, M, Herrera-Montalvo LA. |
| EPI_ISL_2782383, EPI_ISL_2782384, EPI_ISL_2782385, EPI_ISL_2782386, EPI_ISL_2782387, EPI_ISL_2782388, EPI_ISL_2782389, EPI_ISL_2782390, EPI_ISL_2782391, EPI_ISL_2782392, EPI_ISL_2782393, EPI_ISL_2782394, EPI_ISL_2782395, EPI_ISL_2782396, EPI_ISL_2782397, EPI_ISL_2782398, EPI_ISL_2782399, EPI_ISL_2782400, EPI_ISL_2782401, EPI_ISL_2782402, EPI_ISL_2782403, EPI_ISL_2782404, EPI_ISL_2782405, EPI_ISL_2782406, EPI_ISL_2782407, EPI_ISL_2782408, EPI_ISL_2782409, EPI_ISL_2782410, EPI_ISL_2782411, EPI_ISL_2782412, EPI_ISL_2782413, EPI_ISL_2782414, EPI_ISL_2782415, EPI_ISL_2782416, EPI_ISL_2782417, EPI_ISL_2782418, EPI_ISL_2782419, EPI_ISL_2782420, EPI_ISL_2782421, EPI_ISL_2782422, EPI_ISL_2782423, EPI_ISL_2782424, EPI_ISL_2782425, EPI_ISL_2782426, EPI_ISL_2782427, EPI_ISL_2782428, EPI_ISL_2782429, EPI_ISL_2782430, EPI_ISL_2782431, EPI_ISL_2782432, EPI_ISL_2782433, EPI_ISL_2782434, EPI_ISL_2782435, EPI_ISL_2782436, EPI_ISL_2782437, EPI_ISL_2782438, EPI_ISL_2782439, EPI_ISL_2782440, EPI_ISL_2782441, EPI_ISL_2782442, EPI_ISL_2782443, EPI_ISL_2782444, EPI_ISL_2782445, EPI_ISL_2782446, EPI_ISL_2782447, EPI_ISL_2782448, | | | |

EPI_ISL_2782449, EPI_ISL_2782450, EPI_ISL_2782451, EPI_ISL_2782452, EPI_ISL_2782453, EPI_ISL_2782454, EPI_ISL_2782455, EPI_ISL_2782456, EPI_ISL_2782457, EPI_ISL_2782458, EPI_ISL_2782459, EPI_ISL_2782460, EPI_ISL_2782461, EPI_ISL_2782462, EPI_ISL_2782463, EPI_ISL_2782464, EPI_ISL_2782465, EPI_ISL_2782466, EPI_ISL_2782467, EPI_ISL_2782468, EPI_ISL_2782469, EPI_ISL_2782470, EPI_ISL_2782471, EPI_ISL_2782472, EPI_ISL_2782473, EPI_ISL_2782474, EPI_ISL_2782475, EPI_ISL_2782476, EPI_ISL_2782477, EPI_ISL_2782478, EPI_ISL_2782479, EPI_ISL_2782480, EPI_ISL_2782481, EPI_ISL_2782482, EPI_ISL_2782483, EPI_ISL_2782484, EPI_ISL_2782485, EPI_ISL_2782486, EPI_ISL_2782487, EPI_ISL_2782488, EPI_ISL_2782489, EPI_ISL_2782490, EPI_ISL_2782491, EPI_ISL_2782492, EPI_ISL_2782493, EPI_ISL_2782494, EPI_ISL_2782495, EPI_ISL_2782496, EPI_ISL_2782497, EPI_ISL_2782498, EPI_ISL_2782499, EPI_ISL_2782500, EPI_ISL_2782501, EPI_ISL_2782502, EPI_ISL_2782503, EPI_ISL_2782504, EPI_ISL_2782505, EPI_ISL_2782506, EPI_ISL_2782507, EPI_ISL_2782508, EPI_ISL_2782509, EPI_ISL_2782510, EPI_ISL_2782511, EPI_ISL_2782512, EPI_ISL_2782513, EPI_ISL_2782514, EPI_ISL_2782515, EPI_ISL_2782516, EPI_ISL_2782517, EPI_ISL_2782518, EPI_ISL_2782519, EPI_ISL_2782520, EPI_ISL_2782521, EPI_ISL_2782522, EPI_ISL_2782523, EPI_ISL_2782579, EPI_ISL_2782580, EPI_ISL_2782581, EPI_ISL_2782582, EPI_ISL_2782583, EPI_ISL_2782584, EPI_ISL_2782585, EPI_ISL_2782586, EPI_ISL_2782587, EPI_ISL_2782588, EPI_ISL_2782589, EPI_ISL_2782590, EPI_ISL_2782591, EPI_ISL_2782592, EPI_ISL_2782593, EPI_ISL_2782594, EPI_ISL_2782595, EPI_ISL_2782596, EPI_ISL_2782597, EPI_ISL_2782598, EPI_ISL_2782599, EPI_ISL_2782600, EPI_ISL_2782601, EPI_ISL_2782602, EPI_ISL_2782603, EPI_ISL_2782604, EPI_ISL_2782605, EPI_ISL_2782606, EPI_ISL_2782607, EPI_ISL_2782608, EPI_ISL_2782609, EPI_ISL_2782610, EPI_ISL_2782611, EPI_ISL_2782612, EPI_ISL_2782613, EPI_ISL_2782614, EPI_ISL_2782615, EPI_ISL_2782616, EPI_ISL_2782617, EPI_ISL_2782618, EPI_ISL_2782619, EPI_ISL_2782620, EPI_ISL_2782621, EPI_ISL_2782622, EPI_ISL_2782623, EPI_ISL_2782624, EPI_ISL_2782625, EPI_ISL_2782626, EPI_ISL_2782627, EPI_ISL_2782628, EPI_ISL_2782629, EPI_ISL_2782630, EPI_ISL_2782631, EPI_ISL_2782632, EPI_ISL_2782633, EPI_ISL_2782634, EPI_ISL_2782635, EPI_ISL_2782636, EPI_ISL_2782637, EPI_ISL_2782638, EPI_ISL_2782639, EPI_ISL_2782640, EPI_ISL_2782641, EPI_ISL_2782642, EPI_ISL_2782643, EPI_ISL_2782644, EPI_ISL_2782645, EPI_ISL_2782646, EPI_ISL_2782647, EPI_ISL_2782648, EPI_ISL_2782649, EPI_ISL_2782650, EPI_ISL_2782651, EPI_ISL_2782652, EPI_ISL_2782653, EPI_ISL_2782654, EPI_ISL_2782655, EPI_ISL_2782656, EPI_ISL_2782657, EPI_ISL_2782658, EPI_ISL_2782659, EPI_ISL_2782660, EPI_ISL_2782661, EPI_ISL_2782662, EPI_ISL_2782663, EPI_ISL_2782664, EPI_ISL_2782665, EPI_ISL_2782666, EPI_ISL_2782667, EPI_ISL_2782668, EPI_ISL_2782669, EPI_ISL_2782670, EPI_ISL_2782671, EPI_ISL_2782672, EPI_ISL_2782673, EPI_ISL_2782674, EPI_ISL_2782675, EPI_ISL_2782676, EPI_ISL_2782677, EPI_ISL_2782678, EPI_ISL_2782679, EPI_ISL_2782680, EPI_ISL_2782681, EPI_ISL_2782682, EPI_ISL_2782683, EPI_ISL_2782684, EPI_ISL_2782685, EPI_ISL_2782686, EPI_ISL_2782687, EPI_ISL_2782688, EPI_ISL_2782689, EPI_ISL_2782690, EPI_ISL_2782691, EPI_ISL_2782692, EPI_ISL_2782693, EPI_ISL_2782694, EPI_ISL_2782695, EPI_ISL_2782696, EPI_ISL_2782697, EPI_ISL_2782698, EPI_ISL_2782699, EPI_ISL_2782700, EPI_ISL_2782701, EPI_ISL_2782702, EPI_ISL_2782703, EPI_ISL_2782704, EPI_ISL_2782705, EPI_ISL_2782706, EPI_ISL_2782707, EPI_ISL_2782708, EPI_ISL_2782709, EPI_ISL_2782710, EPI_ISL_2782711, EPI_ISL_2782712, EPI_ISL_2782713, EPI_ISL_2782714, EPI_ISL_2782715, EPI_ISL_2782716, EPI_ISL_2782717, EPI_ISL_2782718, EPI_ISL_2782719, EPI_ISL_2782720, EPI_ISL_2782721, EPI_ISL_2782722, EPI_ISL_2782723, EPI_ISL_2782724, EPI_ISL_2782725, EPI_ISL_2782726, EPI_ISL_2782727, EPI_ISL_2782728, EPI_ISL_2782729, EPI_ISL_2782730, EPI_ISL_2782731, EPI_ISL_2782732, EPI_ISL_2782733, EPI_ISL_2782734, EPI_ISL_2782735, EPI_ISL_2782736, EPI_ISL_2782737, EPI_ISL_2782738, EPI_ISL_2782739, EPI_ISL_2782740, EPI_ISL_2782741, EPI_ISL_2782742, EPI_ISL_2782743, EPI_ISL_2782744, EPI_ISL_2782745, EPI_ISL_2782746, EPI_ISL_2782747, EPI_ISL_2782748, EPI_ISL_2782749, EPI_ISL_2782750, EPI_ISL_2782751, EPI_ISL_2782752, EPI_ISL_2782753, EPI_ISL_2782754, EPI_ISL_2782755, EPI_ISL_2782756, EPI_ISL_2782757, EPI_ISL_2782758, EPI_ISL_2782759, EPI_ISL_2782760, EPI_ISL_2782761, EPI_ISL_2782762, EPI_ISL_2782763, EPI_ISL_2782764, EPI_ISL_2782765, EPI_ISL_2782766, EPI_ISL_2782767, EPI_ISL_2782768, EPI_ISL_2782769, EPI_ISL_2782770, EPI_ISL_2782771, EPI_ISL_2782772, EPI_ISL_2782773, EPI_ISL_2782774, EPI_ISL_2782775, EPI_ISL_2782776, EPI_ISL_2782777, EPI_ISL_2782778, EPI_ISL_2782779, EPI_ISL_2782780, EPI_ISL_2782781, EPI_ISL_2782782, EPI_ISL_2782783, EPI_ISL_2782784, EPI_ISL_2782785, EPI_ISL_2782786, EPI_ISL_2782787, EPI_ISL_2782788, EPI_ISL_2782789, EPI_ISL_2782790, EPI_ISL_2782791, EPI_ISL_2782792, EPI_ISL_2782793, EPI_ISL_2782794, EPI_ISL_2782795, EPI_ISL_2782796, EPI_ISL_2782797, EPI_ISL_2782798, EPI_ISL_2782799, EPI_ISL_2782800, EPI_ISL_2782801, EPI_ISL_2782802, EPI_ISL_2782803, EPI_ISL_2782804, EPI_ISL_2782805, EPI_ISL_2782806, EPI_ISL_2782807, EPI_ISL_2782808, EPI_ISL_2782809, EPI_ISL_2782810, EPI_ISL_2782811, EPI_ISL_2782812, EPI_ISL_2782813, EPI_ISL_2782814, EPI_ISL_2782815, EPI_ISL_2782816, EPI_ISL_2782817, EPI_ISL_2782818, EPI_ISL_2782819, EPI_ISL_2782820, EPI_ISL_2782821, EPI_ISL_2782822, EPI_ISL_2782823, EPI_ISL_2782824, EPI_ISL_2782825, EPI_ISL_2782826, EPI_ISL_2782827, EPI_ISL_2782828, EPI_ISL_2782829, EPI_ISL_2782830, EPI_ISL_2782831, EPI_ISL_2782832, EPI_ISL_2782833, EPI_ISL_2782834, EPI_ISL_2782835, EPI_ISL_2782836, EPI_ISL_2782837, EPI_ISL_2782838, EPI_ISL_2782839, EPI_ISL_2782840, EPI_ISL_2782841, EPI_ISL_2782842, EPI_ISL_2782843, EPI_ISL_2782844, EPI_ISL_2782845, EPI_ISL_2782846, EPI_ISL_2782847, EPI_ISL_2782848, EPI_ISL_2782849, EPI_ISL_2782850, EPI_ISL_2782851, EPI_ISL_2782852, EPI_ISL_2782853, EPI_ISL_2782854, EPI_ISL_2782855, EPI_ISL_2782856, EPI_ISL_2782857, EPI_ISL_2782858, EPI_ISL_2782859, EPI_ISL_2782860, EPI_ISL_2782861, EPI_ISL_2782862, EPI_ISL_2782863, EPI_ISL_2782864, EPI_ISL_2782865, EPI_ISL_2782866, EPI_ISL_2782867, EPI_ISL_2782868, EPI_ISL_2782869, EPI_ISL_2782870, EPI_ISL_2782871, EPI_ISL_2782872, EPI_ISL_2782873, EPI_ISL_2782874, EPI_ISL_2782875, EPI_ISL_2782876, EPI_ISL_2782877, EPI_ISL_2782878, EPI_ISL_2782879, EPI_ISL_2782880, EPI_ISL_2782881, EPI_ISL_2782882, EPI_ISL_2782883, EPI_ISL_2782884, EPI_ISL_2782885, EPI_ISL_2782886, EPI_ISL_2782887, EPI_ISL_2782888, EPI_ISL_2782889, EPI_ISL_2782890, EPI_ISL_2782891, EPI_ISL_2782892, EPI_ISL_2782893, EPI_ISL_2782894, EPI_ISL_2782895, EPI_ISL_2782896, EPI_ISL_2782897, EPI_ISL_2782898, EPI_ISL_2782899, EPI_ISL_2782900, EPI_ISL_2782901, EPI_ISL_2782902, EPI_ISL_2782903, EPI_ISL_2782904, EPI_ISL_2782905, EPI_ISL_2782906, EPI_ISL_2782907, EPI_ISL_2782908, EPI_ISL_2782909, EPI_ISL_2782910, EPI_ISL_2782911, EPI_ISL_2782912, EPI_ISL_2782913, EPI_ISL_2782914, EPI_ISL_2782915, EPI_ISL_2782916, EPI_ISL_2782917, EPI_ISL_2782918, EPI_ISL_2782919, EPI_ISL_2782920, EPI_ISL_2782921, EPI_ISL_2782922, EPI_ISL_2782923, EPI_ISL_2782924, EPI_ISL_2782925, EPI_ISL_2782926, EPI_ISL_2782927, EPI_ISL_2782928, EPI_ISL_2782929, EPI_ISL_2782930, EPI_ISL_2782931, EPI_ISL_2782932, EPI_ISL_2782933, EPI_ISL_2782934, EPI_ISL_2782935, EPI_ISL_2782936, EPI_ISL_2782937, EPI_ISL_2782938, EPI_ISL_2782939, EPI_ISL_2782940, EPI_ISL_2782941, EPI_ISL_2782942, EPI_ISL_2782943, EPI_ISL_2782944, EPI_ISL_2782945, EPI_ISL_2782946, EPI_ISL_2782947, EPI_ISL_2782948, EPI_ISL_2782949, EPI_ISL_2782950, EPI_ISL_2782951, EPI_ISL_2782952, EPI_ISL_2782953, EPI_ISL_2782954, EPI_ISL_2782955, EPI_ISL_2782956, EPI_ISL_2782957, EPI_ISL_2782958, EPI_ISL_2782959, EPI_ISL_2782960, EPI_ISL_2782961, EPI_ISL_2782962, EPI_ISL_2782963, EPI_ISL_2782964, EPI_ISL_2782965, EPI_ISL_2782966, EPI_ISL_2782967, EPI_ISL_2782968, EPI_ISL_2782969, EPI_ISL_2782970, EPI_ISL_2782971, EPI_ISL_2782972, EPI_ISL_2782973, EPI_ISL_2782974, EPI_ISL_2782975, EPI_ISL_2782976, EPI_ISL_2782977, EPI_ISL_2782978, EPI_ISL_2782979, EPI_ISL_2782980, EPI_ISL_2782981, EPI_ISL_2782982, EPI_ISL_2782983, EPI_ISL_2782984, EPI_ISL_2782985, EPI_ISL_2782986, EPI_ISL_2782987, EPI_ISL_2782988, EPI_ISL_2782989, EPI_ISL_2782990, EPI_ISL_2782991, EPI_ISL_2782992, EPI_ISL_2782993, EPI_ISL_2782994, EPI_ISL_2782995, EPI_ISL_2782996, EPI_ISL_2782997, EPI_ISL_2782998, EPI_ISL_2782999, EPI_ISL_2783000, EPI_ISL_2783001, EPI_ISL_2783002, EPI_ISL_2783003, EPI_ISL_2783004, EPI_ISL_2783005, EPI_ISL_2783006, EPI_ISL_2783007, EPI_ISL_2783008, EPI_ISL_2783009, EPI_ISL_2783010, EPI_ISL_2783011, EPI_ISL_2783012, EPI_ISL_2783013, EPI_ISL_2783014, EPI_ISL_2783015, EPI_ISL_2783016, EPI_ISL_2783017, EPI_ISL_2783018, EPI_ISL_2783019, EPI_ISL_2783020, EPI_ISL_2783021, EPI_ISL_2783022, EPI_ISL_2783023, EPI_ISL_2783024, EPI_ISL_2783025, EPI_ISL_2783026, EPI_ISL_2783027, EPI_ISL_2783028, EPI_ISL_2783029, EPI_ISL_2783030, EPI_ISL_2783031, EPI_ISL_2783032, EPI_ISL_2783033, EPI_ISL_2783034, EPI_ISL_2783035, EPI_ISL_2783036, EPI_ISL_2783037, EPI_ISL_2783038, EPI_ISL_2783039, EPI_ISL_2783040, EPI_ISL_2783041, EPI_ISL_2783042, EPI_ISL_2783043, EPI_ISL_2783044, EPI_ISL_2783045, EPI_ISL_2783046, EPI_ISL_2783047, EPI_ISL_2783048, EPI_ISL_2783049, EPI_ISL_2783050, EPI_ISL_2783051, EPI_ISL_2783052, EPI_ISL_2783053, EPI_ISL_2783054, EPI_ISL_2783055, EPI_ISL_2783056, EPI_ISL_2783057, EPI_ISL_2783058, EPI_ISL_2783059, EPI_ISL_2783060, EPI_ISL_2783061, EPI_ISL_2783062, EPI_ISL_2783063, EPI_ISL_2783064, EPI_ISL_2783065, EPI_ISL_2783066, EPI_ISL_2783067, EPI_ISL_2783068, EPI_ISL_2783069, EPI_ISL_2783070, EPI_ISL_2783071, EPI_ISL_2783072, EPI_ISL_2783073, EPI_ISL_2783074, EPI_ISL_2783075, EPI_ISL_2783076, EPI_ISL_2783077, EPI_ISL_2783078, EPI_ISL_2783079, EPI_ISL_2783080, EPI_ISL_2783081, EPI_ISL_2783082, EPI_ISL_2783083, EPI_ISL_2783084, EPI_ISL_2783085, EPI_ISL_2783086, EPI_ISL_2783087, EPI_ISL_2783088, EPI_ISL_2783089, EPI_ISL_2783090, EPI_ISL_2783091, EPI_ISL_2783092, EPI_ISL_2783093, EPI_ISL_2783094, EPI_ISL_2783095, EPI_ISL_2783096, EPI_ISL_2783097, EPI_ISL_2783098, EPI_ISL_2783099, EPI_ISL_2783100, EPI_ISL_2783101, EPI_ISL_2783102, EPI_ISL_2783103, EPI_ISL_2783104, EPI_ISL_2783105, EPI_ISL_2783106, EPI_ISL_2783107, EPI_ISL_2783108, EPI_ISL_2783109, EPI_ISL_2783110, EPI_ISL_2783111, EPI_ISL_2783112, EPI_ISL_2783113, EPI_ISL_2783114, EPI_ISL_2783115, EPI_ISL_2783116, EPI_ISL_2783117, EPI_ISL_2783118, EPI_ISL_2783119, EPI_ISL_2783120, EPI_ISL_2783121, EPI_ISL_2783122, EPI_ISL_2783123, EPI_ISL_2783124, EPI_ISL_2783125, EPI_ISL_2783126, EPI_ISL_2783127, EPI_ISL_2783128, EPI_ISL_2783129, EPI_ISL_2783130, EPI_ISL_2783131, EPI_ISL_2783132, EPI_ISL_2783133, EPI_ISL_2783134, EPI_ISL_2783135, EPI_ISL_2783136, EPI_ISL_2783137, EPI_ISL_2783138, EPI_ISL_2783139, EPI_ISL_2783140, EPI_ISL_2783141, EPI_ISL_2783142, EPI_ISL_2783143, EPI_ISL_2783144, EPI_ISL_2783145, EPI_ISL_2783146, EPI_ISL_2783147, EPI_ISL_2783148, EPI_ISL_2783149, EPI_ISL_2783150, EPI_ISL_2783151, EPI_ISL_2783152, EPI_ISL_2783153, EPI_ISL_2783154, EPI_ISL_2783155, EPI_ISL_2783156, EPI_ISL_2783157, EPI_ISL_2783158, EPI_ISL_2783159, EPI_ISL_2783160, EPI_ISL_2783161, EPI_ISL_2783162, EPI_ISL_2783163, EPI_ISL_2783164, EPI_ISL_2783165, EPI_ISL_2783166, EPI_ISL_2783167, EPI_ISL_2783168, EPI_ISL_2783169, EPI_ISL_2783170, EPI_ISL_2783171, EPI_ISL_2783172, EPI_ISL_2783173, EPI_ISL_2783174, EPI_ISL_2783175, EPI_ISL_2783176, EPI_ISL_2783177, EPI_ISL_2783178, EPI_ISL_2783179, EPI_ISL_2783180, EPI_ISL_2783181, EPI_ISL_2783182, EPI_ISL_2783183, EPI_ISL_2783184, EPI_ISL_2783185, EPI_ISL_2783186, EPI_ISL_2783187, EPI_ISL_2783188, EPI_ISL_2783189, EPI_ISL_2783190, EPI_ISL_2783191, EPI_ISL_2783192, EPI_ISL_2783193, EPI_ISL_2783194, EPI_ISL_2783195, EPI_ISL_2783196, EPI_ISL_2783197, EPI_ISL_2783198, EPI_ISL_2783199, EPI_ISL_2783200, EPI_ISL_2783201, EPI_ISL_2783202, EPI_ISL_2783203, EPI_ISL_2783204, EPI_ISL_2783205, EPI_ISL_2783206, EPI_ISL_2783207, EPI_ISL_2783208, EPI_ISL_2783209, EPI_ISL_2783210, EPI_ISL_2783211, EPI_ISL_2783212, EPI_ISL_2783213, EPI_ISL_2783214, EPI_ISL_2783215, EPI_ISL_2783216, EPI_ISL_2783217, EPI_ISL_2783218, EPI_ISL_2783219, EPI_ISL_2783220, EPI_ISL_2783221, EPI_ISL_2783222, EPI_ISL_2783223, EPI_ISL_2783224, EPI_ISL_2783225, EPI_ISL_2783226, EPI_ISL_2783227, EPI_ISL_2783228, EPI_ISL_2783229, EPI_ISL_2783230, EPI_ISL_2783231, EPI_ISL_2783232, EPI_ISL_2783233, EPI_ISL_2783234, EPI_ISL_2783235, EPI_ISL_2783236, EPI_ISL_2783237, EPI_ISL_2783238, EPI_ISL_2783239, EPI_ISL_2783240, EPI_ISL_2783241, EPI_ISL_2783242, EPI_ISL_2783243

| | | | |
|---|---|---|---|
| see above | CDPH VBL | California Department of Public Health | CDPH-COVIDNet,UCSF Center for Advanced Technology |

EPI_ISL_2783244, EPI_ISL_2783245, EPI_ISL_2783246, EPI_ISL_2783247, EPI_ISL_2783248, EPI_ISL_2783249, EPI_ISL_2783250, EPI_ISL_2783251, EPI_ISL_2783252, EPI_ISL_2783253, EPI_ISL_2783254, EPI_ISL_2783255, EPI_ISL_2783256, EPI_ISL_2783257, EPI_ISL_2783258, EPI_ISL_2783259, EPI_ISL_2783260, EPI_ISL_2783261, EPI_ISL_2783262, EPI_ISL_2783263, EPI_ISL_2783264, EPI_ISL_2783265, EPI_ISL_2783266, EPI_ISL_2783267, EPI_ISL_2783268, EPI_ISL_2783269, EPI_ISL_2783270, EPI_ISL_2783271, EPI_ISL_2783272, EPI_ISL_2783273, EPI_ISL_2783274, EPI_ISL_2783275, EPI_ISL_2783276, EPI_ISL_2783277, EPI_ISL_2783278, EPI_ISL_2783279, EPI_ISL_2783280, EPI_ISL_2783281, EPI_ISL_2783282, EPI_ISL_2783283, EPI_ISL_2783284, EPI_ISL_2783285, EPI_ISL_2783286, EPI_ISL_2783287, EPI_ISL_2783288, EPI_ISL_2783289, EPI_ISL_2783290, EPI_ISL_2783291, EPI_ISL_2783292, EPI_ISL_2783293, EPI_ISL_2783294, EPI_ISL_2783295, EPI_ISL_2783296, EPI_ISL_2783297, EPI_ISL_2783298, EPI_ISL_2783299, EPI_ISL_2783300, EPI_ISL_2783301, EPI_ISL_2783302, EPI_ISL_2783303, EPI_ISL_2783304, EPI_ISL_2783305, EPI_ISL_2783306, EPI_ISL_2783307, EPI_ISL_2783308, EPI_ISL_2783309, EPI_ISL_2783310, EPI_ISL_2783311, EPI_ISL_2783312, EPI_ISL_2783313, EPI_ISL_2783314, EPI_ISL_2783315, EPI_ISL_2783316, EPI_ISL_2783317, EPI_ISL_2783318, EPI_ISL_2783319, EPI_ISL_2783320, EPI_ISL_2783321, EPI_ISL_2783322, EPI_ISL_2783323, EPI_ISL_2783324, EPI_ISL_2783325, EPI_ISL_2783326, EPI_ISL_2783327, EPI_ISL_2783328, EPI_ISL_2783329, EPI_ISL_2783330, EPI_ISL_2783331, EPI_ISL_2783332, EPI_ISL_2783333, EPI_ISL_2783334, EPI_ISL_2783335, EPI_ISL_2783336, EPI_ISL_2783337, EPI_ISL_2783338, EPI_ISL_2783339, EPI_ISL_2783340, EPI_ISL_2783341, EPI_ISL_2783342, EPI_ISL_2783343, EPI_ISL_2783344, EPI_ISL_2783345, EPI_ISL_2783346, EPI_ISL_2783347, EPI_ISL_2783348, EPI_ISL_2783349, EPI_ISL_2783350, EPI_ISL_2783351, EPI_ISL_2783352, EPI_ISL_2783353, EPI_ISL_2783354, EPI_ISL_2783355, EPI_ISL_2783356, EPI_ISL_2783357, EPI_ISL_2783358, EPI_ISL_2783359, EPI_ISL_2783360, EPI_ISL_2783361, EPI_ISL_2783362, EPI_ISL_2783363, EPI_ISL_2783364, EPI_ISL_2783365, EPI_ISL_2783366, EPI_ISL_2783367, EPI_ISL_2783368, EPI_ISL_2783369, EPI_ISL_2783370, EPI_ISL_2783371, EPI_ISL_2783372, EPI_ISL_2783373, EPI_ISL_2783374, EPI_ISL_2783375, EPI_ISL_2783376, EPI_ISL_2783377, EPI_ISL_2783378, EPI_ISL_2783379, EPI_ISL_2783380, EPI_ISL_2783381, EPI_ISL_2783382, EPI_ISL_2783383, EPI_ISL_2783384, EPI_ISL_2783385, EPI_ISL_2783386, EPI_ISL_2783387, EPI_ISL_2783388, EPI_ISL_2783389, EPI_ISL_2783390, EPI_ISL_2783391, EPI_ISL_2783392, EPI_ISL_2783393, EPI_ISL_2783394, EPI_ISL_2783395, EPI_ISL_2783396, EPI_ISL_2783397, EPI_ISL_2783398, EPI_ISL_2783399, EPI_ISL_2783400, EPI_ISL_2783401, EPI_ISL_2783402, EPI_ISL_2783403, EPI_ISL_2783404, EPI_ISL_2783405, EPI_ISL_2783406, EPI_ISL_2783407, EPI_ISL_2783408, EPI_ISL_2783409, EPI_ISL_2783410, EPI_ISL_2783411, EPI_ISL_2783412, EPI_ISL_2783413, EPI_ISL_2783414, EPI_ISL_2783415, EPI_ISL_2783416, EPI_ISL_2783417, EPI_ISL_2783418, EPI_ISL_2783419, EPI_ISL_2783420, EPI_ISL_2783421, EPI_ISL_2783422, EPI_ISL_2783423, EPI_ISL_2783424, EPI_ISL_2783425, EPI_ISL_2783426, EPI_ISL_2783427, EPI_ISL_2783428, EPI_ISL_2783429, EPI_ISL_2783430, EPI_ISL_2783431, EPI_ISL_2783432, EPI_ISL_2783433, EPI_ISL_2783434, EPI_ISL_2783435, EPI_ISL_2783436, EPI_ISL_2783437, EPI_ISL_2783438, EPI_ISL_2783439, EPI_ISL_2783440, EPI_ISL_2783441, EPI_ISL_2783442, EPI_ISL_2783443, EPI_ISL_2783444, EPI_ISL_2783445, EPI_ISL_2783446, EPI_ISL_2783447, EPI_ISL_2783448, EPI_ISL_2783449, EPI_ISL_2783450, EPI_ISL_2783451, EPI_ISL_2783452, EPI_ISL_2783453, EPI_ISL_2783454, EPI_ISL_2783455, EPI_ISL_2783456, EPI_ISL_2783457, EPI_ISL_2783458, EPI_ISL_2783459, EPI_ISL_2783460, EPI_ISL_2783461, EPI_ISL_2783462, EPI_ISL_2783463, EPI_ISL_2783464, EPI_ISL_2783465, EPI_ISL_2783466, EPI_ISL_2783467, EPI_ISL_2783468, EPI_ISL_2783469, EPI_ISL_2783470, EPI_ISL_2783471, EPI_ISL_2783472, EPI_ISL_2783473, EPI_ISL_2783474, EPI_ISL_2783475, EPI_ISL_2783476, EPI_ISL_2783477, EPI_ISL_2783478, EPI_ISL_2783479, EPI_ISL_2783480, EPI_ISL_2783481, EPI_ISL_2783482, EPI_ISL_2783483, EPI_ISL_2783484, EPI_ISL_2783485, EPI_ISL_2783486, EPI_ISL_2783487, EPI_ISL_2783488, EPI_ISL_2783489, EPI_ISL_2783490, EPI_ISL_2783491, EPI_ISL_2783492, EPI_ISL_2783493, EPI_ISL_2783494, EPI_ISL_2783495, EPI_ISL_2783496, EPI_ISL_2783497, EPI_ISL_2783498, EPI_ISL_2783499, EPI_ISL_2783500, EPI_ISL_2783501, EPI_ISL_2783502, EPI_ISL_2783503, EPI_ISL_2783504, EPI_ISL_2783505, EPI_ISL_2783506, EPI_ISL_2783507, EPI_ISL_2783508, EPI_ISL_2783509, EPI_ISL_2783510

| | | | |
|---|---|---|---|
| see above | CDPH VBL | California Department of Public Health | CDPH-COVIDNet,UCI Genome Sciences Center/GHTF |

EPI_ISL_2783515, EPI_ISL_2783518, EPI_ISL_2783527, EPI_ISL_2783530, EPI_ISL_2783532, EPI_ISL_2783533, EPI_ISL_2783536, EPI_ISL_2783539, EPI_ISL_2783542, EPI_ISL_2783543, EPI_ISL_2783550, EPI_ISL_2783560, EPI_ISL_2783580, EPI_ISL_2783581, EPI_ISL_2783585, EPI_ISL_2783594, EPI_ISL_2783599, EPI_ISL_2783601, EPI_ISL_2783604, EPI_ISL_2783608, EPI_ISL_2783615, EPI_ISL_2783616, EPI_ISL_2783618, EPI_ISL_2783624, EPI_ISL_2783631, EPI_ISL_2783632, EPI_ISL_2783634, EPI_ISL_2783642, EPI_ISL_2783643, EPI_ISL_2783644, EPI_ISL_2783652, EPI_ISL_2783662, EPI_ISL_2783665, EPI_ISL_2783681, EPI_ISL_2783693, EPI_ISL_2783703, EPI_ISL_2783718, EPI_ISL_2783747, EPI_ISL_2783759, EPI_ISL_2783769, EPI_ISL_2783771, EPI_ISL_2783774, EPI_ISL_2783782, EPI_ISL_2783785, EPI_ISL_2783788, EPI_ISL_2783789, EPI_ISL_2783791, EPI_ISL_2783809, EPI_ISL_2783818, EPI_ISL_2783827, EPI_ISL_2783834, EPI_ISL_2783860, EPI_ISL_2783881, EPI_ISL_2783888, EPI_ISL_2783897, EPI_ISL_2783901, EPI_ISL_2783905, EPI_ISL_2783906, EPI_ISL_2783907, EPI_ISL_2783908, EPI_ISL_2783909, EPI_ISL_2783915, EPI_ISL_2783924, EPI_ISL_2783927, EPI_ISL_2783928, EPI_ISL_2783933, EPI_ISL_2783937, EPI_ISL_2783939, EPI_ISL_2783943, EPI_ISL_2783945, EPI_ISL_2783946, EPI_ISL_2783949, EPI_ISL_2783951, EPI_ISL_2783952, EPI_ISL_2783953, EPI_ISL_2783954, EPI_ISL_2783964, EPI_ISL_2783967, EPI_ISL_2783972, EPI_ISL_2783973, EPI_ISL_2783975, EPI_ISL_2783977, EPI_ISL_2783979, EPI_ISL_2783980, EPI_ISL_2783981, EPI_ISL_2783982, EPI_ISL_2783983, EPI_ISL_2783985, EPI_ISL_2783991, EPI_ISL_2783992, EPI_ISL_2783996, EPI_ISL_2784004, EPI_ISL_2784008, EPI_ISL_2784013, EPI_ISL_2784018, EPI_ISL_2784023, EPI_ISL_2784026, EPI_ISL_2784032, EPI_ISL_2784033, EPI_ISL_2784036, EPI_ISL_2784037, EPI_ISL_2784039, EPI_ISL_2784042, EPI_ISL_2784050, EPI_ISL_2784051, EPI_ISL_2784052, EPI_ISL_2784059, EPI_ISL_2784061, EPI_ISL_2784070, EPI_ISL_2784071, EPI_ISL_2784072, EPI_ISL_2784075, EPI_ISL_2784078, EPI_ISL_2784083, EPI_ISL_2784091, EPI_ISL_2784092, EPI_ISL_2784102, EPI_ISL_2784104, EPI_ISL_2784106, EPI_ISL_2784107, EPI_ISL_2784123, EPI_ISL_2784127, EPI_ISL_2784132, EPI_ISL_2784135, EPI_ISL_2784147, EPI_ISL_2784155, EPI_ISL_2784167, EPI_ISL_2784169, EPI_ISL_2784172, EPI_ISL_2784175, EPI_ISL_2784177, EPI_ISL_2784178, EPI_ISL_2784181, EPI_ISL_2784193, EPI_ISL_2784196, EPI_ISL_2784198, EPI_ISL_2784216, EPI_ISL_2784217, EPI_ISL_2784221, EPI_ISL_2784225, EPI_ISL_2784226, EPI_ISL_2784252, EPI_ISL_2784255, EPI_ISL_2784256, EPI_ISL_2784259, EPI_ISL_2784260, EPI_ISL_2784274, EPI_ISL_2784277, EPI_ISL_2784278, EPI_ISL_2784286, EPI_ISL_2784291, EPI_ISL_2784303, EPI_ISL_2784305, EPI_ISL_2784307, EPI_ISL_2784311, EPI_ISL_2784316, EPI_ISL_2784320, EPI_ISL_2784321, EPI_ISL_2784322, EPI_ISL_2784324

| | | | |
|---|---|---|---|
| see above | Curative Labs | Curative Labs | Mikhail Hanewich-Hollatz, J.Cesar Ignacio-Espinoza, Elias L. Salfati, Nabjot Sandhu, George Way, Zhiyi Xie, Eugenia Khorosheva, Sophia Quasem, Janet Chen, Vladimir Slepnev |
| EPI_ISL_2784694, EPI_ISL_2784695 | Ohio Department of Health Laboratory | Ohio Department of Health Laboratory | Holmes, Jennifer; Eric Brandt, Keoni Omura, Glen McGillivary, Caitlin McDonnell, Jade Mowery, Stephanie Mccracken, Tyler Payne, Kirtana Ramadugu, Erica Leasure, Brent Lee, Kelsey Florek, Heather Blankenship, Allison Black, Quanta Brown, and Tammy Bannerman |

EPI_ISL_2784696, EPI_ISL_2784697, EPI_ISL_2784698, EPI_ISL_2784699, EPI_ISL_2784700, EPI_ISL_2784701, EPI_ISL_2784702, EPI_ISL_2784703, EPI_ISL_2784704, EPI_ISL_2784705, EPI_ISL_2784706, EPI_ISL_2784707, EPI_ISL_2784708, EPI_ISL_2784709, EPI_ISL_2784710, EPI_ISL_2784711, EPI_ISL_2784712, EPI_ISL_2784713, EPI_ISL_2784714, EPI_ISL_2784715

| | | | |
|---|---|---|---|
| see above | UW Virology Lab | UW Virology Lab | Pavitra Roychoudhury, Hong Xie, Lasata Shrestha, Shah Mohamed Bakhash, Margaret Mills, Noah Baker, Sean Ellis, Meei-Li Huang, Keith R Jerome, Alexander Greninger |
| EPI_ISL_2784716, EPI_ISL_2784717, EPI_ISL_2784718, EPI_ISL_2784723, EPI_ISL_2784725, EPI_ISL_2784726 | Ohio Department of Health Laboratory | Ohio Department of Health Laboratory | Holmes, Jennifer; Eric Brandt, Keoni Omura, Glen McGillivary, Caitlin McDonnell, Jade Mowery, Stephanie Mccracken, Tyler Payne, Kirtana Ramadugu, Erica Leasure, Brent Lee, Kelsey Florek, Heather Blankenship, Allison Black, Quanta Brown, and Tammy Bannerman |

EPI_ISL_2784728, EPI_ISL_2784729, EPI_ISL_2784730, EPI_ISL_2784731, EPI_ISL_2784732, EPI_ISL_2784733, EPI_ISL_2784734, EPI_ISL_2784735, EPI_ISL_2784736, EPI_ISL_2784737, EPI_ISL_2784738, EPI_ISL_2784739, EPI_ISL_2784740, EPI_ISL_2784741, EPI_ISL_2784742, EPI_ISL_2784743, EPI_ISL_2784744, EPI_ISL_2784745, EPI_ISL_2784746, EPI_ISL_2784747, EPI_ISL_2784748, EPI_ISL_2784749, EPI_ISL_2784750, EPI_ISL_2784751, EPI_ISL_2784752, EPI_ISL_2784753, EPI_ISL_2784754, EPI_ISL_2784755, EPI_ISL_2784756, EPI_ISL_2784757, EPI_ISL_2784758, EPI_ISL_2784759, EPI_ISL_2784760, EPI_ISL_2784761, EPI_ISL_2784762, EPI_ISL_2784763, EPI_ISL_2784764, EPI_ISL_2784765, EPI_ISL_2784766, EPI_ISL_2784767, EPI_ISL_2784768, EPI_ISL_2784769, EPI_ISL_2784770, EPI_ISL_2784771, EPI_ISL_2784772, EPI_ISL_2784773, EPI_ISL_2784774, EPI_ISL_2784776, EPI_ISL_2784777, EPI_ISL_2784778, EPI_ISL_2784779, EPI_ISL_2784780, EPI_ISL_2784781, EPI_ISL_2784782, EPI_ISL_2784783, EPI_ISL_2784785, EPI_ISL_2784786, EPI_ISL_2784787, EPI_ISL_2784788, EPI_ISL_2784789, EPI_ISL_2784790, EPI_ISL_2784791, EPI_ISL_2784793, EPI_ISL_2784794, EPI_ISL_2784795, EPI_ISL_2784796, EPI_ISL_2784797, EPI_ISL_2784798, EPI_ISL_2784799, EPI_ISL_2784800, EPI_ISL_2784801, EPI_ISL_2784802, EPI_ISL_2784803, EPI_ISL_2784804, EPI_ISL_2784806, EPI_ISL_2784807, EPI_ISL_2784808, EPI_ISL_2784809, EPI_ISL_2784810, EPI_ISL_2784811, EPI_ISL_2784812, EPI_ISL_2784813, EPI_ISL_2784814, EPI_ISL_2784815, EPI_ISL_2784816, EPI_ISL_2784817, EPI_ISL_2784818, EPI_ISL_2784819, EPI_ISL_2784820, EPI_ISL_2784821, EPI_ISL_2784822, EPI_ISL_2784823, EPI_ISL_2784824, EPI_ISL_2784825, EPI_ISL_2784826, EPI_ISL_2784827, EPI_ISL_2784828, EPI_ISL_2784829, EPI_ISL_2784830, EPI_ISL_2784831, EPI_ISL_2784832, EPI_ISL_2784834, EPI_ISL_2784835, EPI_ISL_2784836, EPI_ISL_2784837, EPI_ISL_2784839, EPI_ISL_2784840, EPI_ISL_2784841, EPI_ISL_2784842, EPI_ISL_2784843, EPI_ISL_2784844, EPI_ISL_2784847, EPI_ISL_2784848, EPI_ISL_2784849, EPI_ISL_2784850, EPI_ISL_2784851, EPI_ISL_2784852, EPI_ISL_2784854, EPI_ISL_2784855, EPI_ISL_2784856, EPI_ISL_2784857, EPI_ISL_2784858, EPI_ISL_2784859, EPI_ISL_2784860, EPI_ISL_2784861, EPI_ISL_2784862, EPI_ISL_2784863, EPI_ISL_2784864, EPI_ISL_2784865, EPI_ISL_2784866, EPI_ISL_2784867, EPI_ISL_2784868, EPI_ISL_2784870, EPI_ISL_2784871, EPI_ISL_2784872, EPI_ISL_2784873, EPI_ISL_2784874, EPI_ISL_2784875, EPI_ISL_2784876, EPI_ISL_2784877, EPI_ISL_2784878, EPI_ISL_2784879, EPI_ISL_2784880, EPI_ISL_2784881, EPI_ISL_2784882, EPI_ISL_2784883, EPI_ISL_2784884, EPI_ISL_2784885, EPI_ISL_2784886, EPI_ISL_2784887, EPI_ISL_2784889, EPI_ISL_2784890, EPI_ISL_2784891, EPI_ISL_2784892, EPI_ISL_2784893, EPI_ISL_2784894, EPI_ISL_2784895, EPI_ISL_2784896, EPI_ISL_2784897, EPI_ISL_2784898, EPI_ISL_2784899, EPI_ISL_2784900, EPI_ISL_2784901, EPI_ISL_2784902, EPI_ISL_2784903, EPI_ISL_2784904, EPI_ISL_2784905, EPI_ISL_2784906, EPI_ISL_2784907, EPI_ISL_2784908, EPI_ISL_2784909, EPI_ISL_2784910, EPI_ISL_2784911, EPI_ISL_2784912, EPI_ISL_2784913, EPI_ISL_2784914, EPI_ISL_2784915, EPI_ISL_2784916, EPI_ISL_2784917, EPI_ISL_2784918, EPI_ISL_2784919, EPI_ISL_2784920, EPI_ISL_2784921, EPI_ISL_2784922, EPI_ISL_2784923, EPI_ISL_2784924, EPI_ISL_2784925, EPI_ISL_2784926, EPI_ISL_2784927, EPI_ISL_2784929, EPI_ISL_2784930, EPI_ISL_2784931, EPI_ISL_2784933, EPI_ISL_2784934, EPI_ISL_2784935, EPI_ISL_2784936, EPI_ISL_2784937, EPI_ISL_2784938, EPI_ISL_2784939, EPI_ISL_2784940, EPI_ISL_2784941, EPI_ISL_2784942, EPI_ISL_2784943, EPI_ISL_2784944, EPI_ISL_2784945, EPI_ISL_2784946, EPI_ISL_2784947, EPI_ISL_2784948, EPI_ISL_2784949, EPI_ISL_2784950, EPI_ISL_2784951, EPI_ISL_2784952, EPI_ISL_2784953, EPI_ISL_2784954, EPI_ISL_2784955, EPI_ISL_2784956, EPI_ISL_2784957, EPI_ISL_2784958, EPI_ISL_2784959, EPI_ISL_2784960, EPI_ISL_2784961, EPI_ISL_2784962, EPI_ISL_2784963, EPI_ISL_2784964, EPI_ISL_2784965, EPI_ISL_2784966, EPI_ISL_2784967, EPI_ISL_2784968, EPI_ISL_2784969, EPI_ISL_2784970, EPI_ISL_2784971, EPI_ISL_2784972, EPI_ISL_2784973, EPI_ISL_2784974, EPI_ISL_2784975, EPI_ISL_2784976, EPI_ISL_2784978, EPI_ISL_2784979, EPI_ISL_2784980, EPI_ISL_2784981, EPI_ISL_2784982, EPI_ISL_2784983, EPI_ISL_2784985, EPI_ISL_2784986, EPI_ISL_2784987, EPI_ISL_2784988, EPI_ISL_2784989, EPI_ISL_2784990, EPI_ISL_2784991, EPI_ISL_2784992, EPI_ISL_2784993, EPI_ISL_2784994, EPI_ISL_2784995, EPI_ISL_2784996, EPI_ISL_2784997, EPI_ISL_2784998, EPI_ISL_2784999, EPI_ISL_2785000, EPI_ISL_2785001, EPI_ISL_2785002, EPI_ISL_2785003, EPI_ISL_2785004, EPI_ISL_2785005, EPI_ISL_2785006, EPI_ISL_2785007, EPI_ISL_2785008, EPI_ISL_2785009, EPI_ISL_2785010, EPI_ISL_2785011, EPI_ISL_2785012, EPI_ISL_2785013, EPI_ISL_2785014, EPI_ISL_2785015, EPI_ISL_2785016, EPI_ISL_2785017, EPI_ISL_2785018, EPI_ISL_2785019, EPI_ISL_2785020, EPI_ISL_2785021, EPI_ISL_2785022, EPI_ISL_2785023, EPI_ISL_2785024, EPI_ISL_2785025, EPI_ISL_2785026, EPI_ISL_2785027, EPI_ISL_2785028, EPI_ISL_2785029, EPI_ISL_2785030, EPI_ISL_2785031, EPI_ISL_2785032, EPI_ISL_2785033, EPI_ISL_2785034, EPI_ISL_2785035, EPI_ISL_2785036, EPI_ISL_2785037, EPI_ISL_2785038, EPI_ISL_2785039, EPI_ISL_2785040, EPI_ISL_2785041, EPI_ISL_2785042, EPI_ISL_2785043, EPI_ISL_2785044, EPI_ISL_2785045, EPI_ISL_2785046, EPI_ISL_2785047, EPI_ISL_2785048, EPI_ISL_2785049, EPI_ISL_2785050, EPI_ISL_2785051, EPI_ISL_2785052, EPI_ISL_2785053, EPI_ISL_2785054, EPI_ISL_2785055, EPI_ISL_2785056, EPI_ISL_2785057, EPI_ISL_2785058, EPI_ISL_2785059, EPI_ISL_2785060, EPI_ISL_2785061, EPI_ISL_2785062, EPI_ISL_2785063, EPI_ISL_2785064, EPI_ISL_2785065, EPI_ISL_2785066, EPI_ISL_2785067, EPI_ISL_2785068, EPI_ISL_2785069, EPI_ISL_2785070, EPI_ISL_2785071, EPI_ISL_2785072, EPI_ISL_2785073, EPI_ISL_2785074, EPI_ISL_2785075, EPI_ISL_2785077, EPI_ISL_2785078, EPI_ISL_2785079, EPI_ISL_2785080, EPI_ISL_2785081, EPI_ISL_2785082, EPI_ISL_2785083, EPI_ISL_2785084, EPI_ISL_2785085, EPI_ISL_2785086, EPI_ISL_2785087, EPI_ISL_2785088, EPI_ISL_2785089, EPI_ISL_2785090, EPI_ISL_2785091, EPI_ISL_2785092, EPI_ISL_2785093, EPI_ISL_2785094, EPI_ISL_2785095, EPI_ISL_2785096, EPI_ISL_2785097, EPI_ISL_2785098, EPI_ISL_2785099, EPI_ISL_2785100, EPI_ISL_2785101, EPI_ISL_2785102, EPI_ISL_2785103, EPI_ISL_2785104, EPI_ISL_2785105, EPI_ISL_2785106, EPI_ISL_2785107, EPI_ISL_2785108, EPI_ISL_2785109, EPI_ISL_2785110, EPI_ISL_2785111, EPI_ISL_2785112, EPI_ISL_2785113, EPI_ISL_2785114, EPI_ISL_2785115, EPI_ISL_2785118, EPI_ISL_2785119, EPI_ISL_2785120, EPI_ISL_2785121, EPI_ISL_2785122, EPI_ISL_2785123, EPI_ISL_2785124, EPI_ISL_2785125, EPI_ISL_2785126, EPI_ISL_2785127, EPI_ISL_2785128, EPI_ISL_2785129, EPI_ISL_2785130, EPI_ISL_2785132, EPI_ISL_2785133, EPI_ISL_2785134, EPI_ISL_2785135, EPI_ISL_2785136, EPI_ISL_2785137, EPI_ISL_2785138, EPI_ISL_2785139, EPI_ISL_2785140, EPI_ISL_2785141, EPI_ISL_2785142, EPI_ISL_2785143, EPI_ISL_2785144, EPI_ISL_2785145, EPI_ISL_2785146, EPI_ISL_2785147, EPI_ISL_2785148, EPI_ISL_2785149, EPI_ISL_2785150, EPI_ISL_2785151, EPI_ISL_2785152, EPI_ISL_2785153, EPI_ISL_2785154, EPI_ISL_2785155, EPI_ISL_2785156, EPI_ISL_2785157, EPI_ISL_2785158, EPI_ISL_2785159, EPI_ISL_2785161, EPI_ISL_2785162, EPI_ISL_2785163, EPI_ISL_2785164, EPI_ISL_2785165, EPI_ISL_2785166, EPI_ISL_2785167, EPI_ISL_2785168, EPI_ISL_2785169, EPI_ISL_2785170, EPI_ISL_2785171, EPI_ISL_2785172, EPI_ISL_2785173, EPI_ISL_2785174, EPI_ISL_2785175, EPI_ISL_2785177, EPI_ISL_2785178, EPI_ISL_2785179, EPI_ISL_2785180, EPI_ISL_2785181, EPI_ISL_2785182, EPI_ISL_2785183, EPI_ISL_2785184, EPI_ISL_2785185, EPI_ISL_2785186, EPI_ISL_2785187, EPI_ISL_2785188, EPI_ISL_2785189, EPI_ISL_2785190, EPI_ISL_2785191, EPI_ISL_2785192, EPI_ISL_2785193, EPI_ISL_2785194, EPI_ISL_2785195, EPI_ISL_2785196, EPI_ISL_2785197, EPI_ISL_2785198, EPI_ISL_2785199, EPI_ISL_2785200, EPI_ISL_2785201, EPI_ISL_2785202, EPI_ISL_2785203, EPI_ISL_2785204, EPI_ISL_2785205, EPI_ISL_2785206, EPI_ISL_2785207, EPI_ISL_2785208, EPI_ISL_2785209, EPI_ISL_2785210, EPI_ISL_2785211, EPI_ISL_2785212, EPI_ISL_2785213, EPI_ISL_2785214, EPI_ISL_2785215, EPI_ISL_2785216, EPI_ISL_2785217, EPI_ISL_2785218, EPI_ISL_2785219, EPI_ISL_2785220, EPI_ISL_2785221, EPI_ISL_2785222, EPI_ISL_2785223, EPI_ISL_2785224, EPI_ISL_2785225, EPI_ISL_2785226, EPI_ISL_2785227, EPI_ISL_2785228, EPI_ISL_2785230, EPI_ISL_2785231, EPI_ISL_2785232, EPI_ISL_2785233, EPI_ISL_2785234, EPI_ISL_2785235, EPI_ISL_2785236, EPI_ISL_2785237, EPI_ISL_2785238, EPI_ISL_2785239, EPI_ISL_2785240, EPI_ISL_2785241, EPI_ISL_2785242, EPI_ISL_2785243, EPI_ISL_2785244, EPI_ISL_2785245, EPI_ISL_2785246, EPI_ISL_2785247, EPI_ISL_2785248, EPI_ISL_2785249, EPI_ISL_2785250, EPI_ISL_2785251, EPI_ISL_2785252, EPI_ISL_2785253, EPI_ISL_2785254, EPI_ISL_2785255, EPI_ISL_2785256, EPI_ISL_2785257, EPI_ISL_2785258, EPI_ISL_2785259, EPI_ISL_2785261, EPI_ISL_2785262, EPI_ISL_2785263, EPI_ISL_2785264, EPI_ISL_2785265, EPI_ISL_2785266, EPI_ISL_2785267, EPI_ISL_2785268, EPI_ISL_2785269, EPI_ISL_2785270, EPI_ISL_2785271, EPI_ISL_2785272, EPI_ISL_2785273, EPI_ISL_2785274, EPI_ISL_2785275, EPI_ISL_2785276, EPI_ISL_2785277, EPI_ISL_2785278, EPI_ISL_2785279, EPI_ISL_2785280, EPI_ISL_2785281, EPI_ISL_2785282, EPI_ISL_2785283, EPI_ISL_2785284, EPI_ISL_2785285, EPI_ISL_2785286, EPI_ISL_2785287, EPI_ISL_2785288, EPI_ISL_2785289, EPI_ISL_2785290, EPI_ISL_2785291, EPI_ISL_2785292, EPI_ISL_2785293, EPI_ISL_2785294, EPI_ISL_2785295, EPI_ISL_2785296, EPI_ISL_2785297, EPI_ISL_2785298, EPI_ISL_2785299, EPI_ISL_2785300, EPI_ISL_2785301, EPI_ISL_2785302, EPI_ISL_2785303, EPI_ISL_2785304, EPI_ISL_2785305, EPI_ISL_2785306, EPI_ISL_2785307, EPI_ISL_2785308, EPI_ISL_2785309, EPI_ISL_2785310, EPI_ISL_2785311, EPI_ISL_2785312, EPI_ISL_2785313, EPI_ISL_2785314, EPI_ISL_2785315, EPI_ISL_2785316, EPI_ISL_2785317, EPI_ISL_2785318, EPI_ISL_2785319, EPI_ISL_2785320, EPI_ISL_2785321, EPI_ISL_2785322, EPI_ISL_2785323, EPI_ISL_2785324, EPI_ISL_2785325, EPI_ISL_2785326, EPI_ISL_2785327, EPI_ISL_2785328, EPI_ISL_2785329, EPI_ISL_2785330, EPI_ISL_2785331, EPI_ISL_2785332, EPI_ISL_2785333, EPI_ISL_2785334, EPI_ISL_2785335, EPI_ISL_2785336, EPI_ISL_2785337, EPI_ISL_2785338, EPI_ISL_2785339, EPI_ISL_2785340, EPI_ISL_2785341, EPI_ISL_2785342, EPI_ISL_2785343, EPI_ISL_2785344, EPI_ISL_2785345, EPI_ISL_2785346, EPI_ISL_2785347, EPI_ISL_2785348, EPI_ISL_2785349, EPI_ISL_2785350, EPI_ISL_2785351, EPI_ISL_2785352, EPI_ISL_2785353, EPI_ISL_2785354, EPI_ISL_2785355, EPI_ISL_2785356, EPI_ISL_2785357, EPI_ISL_2785358, EPI_ISL_2785359, EPI_ISL_2785360, EPI_ISL_2785361, EPI_ISL_2785362, EPI_ISL_2785363, EPI_ISL_2785364, EPI_ISL_2785365, EPI_ISL_2785366, EPI_ISL_2785367, EPI_ISL_2785368, EPI_ISL_2785369, EPI_ISL_2785370, EPI_ISL_2785371, EPI_ISL_2785372, EPI_ISL_2785373, EPI_ISL_2785374, EPI_ISL_2785375, EPI_ISL_2785376, EPI_ISL_2785377, EPI_ISL_2785378, EPI_ISL_2785379, EPI_ISL_2785380, EPI_ISL_2785381, EPI_ISL_2785382, EPI_ISL_2785383, EPI_ISL_2785384, EPI_ISL_2785385, EPI_ISL_2785386, EPI_ISL_2785387, EPI_ISL_2785388, EPI_ISL_2785389, EPI_ISL_2785390, EPI_ISL_2785391, EPI_ISL_2785392, EPI_ISL_2785393, EPI_ISL_2785394, EPI_ISL_2785395, EPI_ISL_2785396, EPI_ISL_2785397, EPI_ISL_2785398, EPI_ISL_2785399, EPI_ISL_2785400, EPI_ISL_2785401, EPI_ISL_2785402, EPI_ISL_2785403, EPI_ISL_2785404, EPI_ISL_2785405, EPI_ISL_2785406, EPI_ISL_2785407, EPI_ISL_2785409, EPI_ISL_2785410, EPI_ISL_2785411, EPI_ISL_2785412, EPI_ISL_2785413, EPI_ISL_2785414, EPI_ISL_2785415, EPI_ISL_2785416, EPI_ISL_2785417, EPI_ISL_2785418, EPI_ISL_2785419, EPI_ISL_2785420, EPI_ISL_2785421, EPI_ISL_2785422, EPI_ISL_2785423, EPI_ISL_2785424, EPI_ISL_2785425, EPI_ISL_2785426, EPI_ISL_2785427, EPI_ISL_2785428, EPI_ISL_2785429, EPI_ISL_2785430, EPI_ISL_2785431, EPI_ISL_2785432, EPI_ISL_2785433, EPI_ISL_2785434, EPI_ISL_2785435, EPI_ISL_2785436, EPI_ISL_2785437, EPI_ISL_2785438, EPI_ISL_2785439, EPI_ISL_2785440, EPI_ISL_2785441, EPI_ISL_2785442, EPI_ISL_2785443, EPI_ISL_2785444, EPI_ISL_2785445, EPI_ISL_2785446, EPI_ISL_2785447, EPI_ISL_2785448, EPI_ISL_2785449, EPI_ISL_2785450, EPI_ISL_2785451, EPI_ISL_2785452, EPI_ISL_2785453, EPI_ISL_2785454, EPI_ISL_2785455, EPI_ISL_2785456, EPI_ISL_2785457, EPI_ISL_2785458, EPI_ISL_2785459, EPI_ISL_2785460, EPI_ISL_2785461, EPI_ISL_2785462, EPI_ISL_2785463, EPI_ISL_2785464, EPI_ISL_2785465, EPI_ISL_2785466, EPI_ISL_2785467, EPI_ISL_2785468, EPI_ISL_2785469, EPI_ISL_2785471, EPI_ISL_2785472, EPI_ISL_2785473, EPI_ISL_2785474, EPI_ISL_2785475, EPI_ISL_2785476, EPI_ISL_2785477, EPI_ISL_2785478, EPI_ISL_2785479, EPI_ISL_2785480, EPI_ISL_2785481, EPI_ISL_2785482, EPI_ISL_2785483, EPI_ISL_2785484, EPI_ISL_2785485, EPI_ISL_2785486, EPI_ISL_2785487, EPI_ISL_2785488, EPI_ISL_2785489, EPI_ISL_2785490, EPI_ISL_2785491, EPI_ISL_2785492, EPI_ISL_2785493, EPI_ISL_2785494, EPI_ISL_2785495, EPI_ISL_2785496, EPI_ISL_2785497, EPI_ISL_2785499, EPI_ISL_2785500, EPI_ISL_2785501, EPI_ISL_2785502, EPI_ISL_2785503, EPI_ISL_2785504, EPI_ISL_2785505, EPI_ISL_2785506, EPI_ISL_2785507, EPI_ISL_2785508, EPI_ISL_2785509, EPI_ISL_2785510, EPI_ISL_2785511, EPI_ISL_2785512, EPI_ISL_2785513, EPI_ISL_2785514, EPI_ISL_2785515, EPI_ISL_2785516, EPI_ISL_2785517, EPI_ISL_2785518

| | | | |
|---|---|---|---|
| see above | Aegis Sciences Corporation | Centers for Disease Control and Prevention Division of Viral Diseases, Pathogen | Dakota Howard, Dhwani Batra, Peter W. Cook, Kara Moser, Adrian Paskey, Jason Caravas, Benjamin Rambo-Martin, Shatavia Morrison, Christopher Gulvick, Scott Sammons, Yvette Unoarumhi, Darlene Wagner, Matthew Schmerer, Cyndi Clark, Patrick Campbell, Rob Case, Vikramsinha Ghorpade, Holly Houdeshell, Ola Kvalvaag, Dillon Nall, Ethan Sanders, Alec Vest, Shaun Westlund, Matthew Hardison, Clinton R. Paden, Duncan MacCannell |

| | | | |
|---|---|---|---|
| | | Discovery | |
| EPI_ISL_2785519, EPI_ISL_2785520, EPI_ISL_2785521, EPI_ISL_2785522, EPI_ISL_2785523, EPI_ISL_2785524, EPI_ISL_2785526, EPI_ISL_2785528, EPI_ISL_2785529, EPI_ISL_2785530, EPI_ISL_2785531, EPI_ISL_2785532, EPI_ISL_2785533, EPI_ISL_2785534, EPI_ISL_2785535, EPI_ISL_2785536, EPI_ISL_2785538, EPI_ISL_2785539, EPI_ISL_2785540, EPI_ISL_2785541, EPI_ISL_2785542, EPI_ISL_2785543, EPI_ISL_2785544, EPI_ISL_2785545, EPI_ISL_2785546, EPI_ISL_2785547, EPI_ISL_2785548, EPI_ISL_2785549, EPI_ISL_2785550, EPI_ISL_2785551, EPI_ISL_2785552, EPI_ISL_2785553, EPI_ISL_2785554, EPI_ISL_2785555, EPI_ISL_2785556, EPI_ISL_2785557, EPI_ISL_2785558, EPI_ISL_2785559, EPI_ISL_2785560, EPI_ISL_2785561, EPI_ISL_2785562, EPI_ISL_2785563, EPI_ISL_2785564, EPI_ISL_2785565, EPI_ISL_2785566, EPI_ISL_2785567, EPI_ISL_2785568, EPI_ISL_2785569, EPI_ISL_2785570, EPI_ISL_2785571, EPI_ISL_2785572, EPI_ISL_2785573, EPI_ISL_2785574, EPI_ISL_2785575, EPI_ISL_2785576, EPI_ISL_2785577, EPI_ISL_2785578, EPI_ISL_2785579, EPI_ISL_2785580, EPI_ISL_2785581, EPI_ISL_2785583, EPI_ISL_2785584, EPI_ISL_2785585, EPI_ISL_2785586, EPI_ISL_2785587, EPI_ISL_2785588, EPI_ISL_2785589, EPI_ISL_2785590, EPI_ISL_2785591, EPI_ISL_2785592, EPI_ISL_2785593, EPI_ISL_2785594, EPI_ISL_2785595, EPI_ISL_2785596, EPI_ISL_2785597, EPI_ISL_2785598, EPI_ISL_2785599, EPI_ISL_2785600, EPI_ISL_2785601, EPI_ISL_2785602, EPI_ISL_2785603, EPI_ISL_2785604, EPI_ISL_2785605, EPI_ISL_2785606, EPI_ISL_2785607, EPI_ISL_2785608, EPI_ISL_2785609, EPI_ISL_2785610, EPI_ISL_2785611, EPI_ISL_2785612, EPI_ISL_2785613, EPI_ISL_2785614, EPI_ISL_2785615, EPI_ISL_2785616, EPI_ISL_2785617, EPI_ISL_2785619, EPI_ISL_2785621, EPI_ISL_2785622, EPI_ISL_2785623, EPI_ISL_2785624, EPI_ISL_2785625, EPI_ISL_2785626, EPI_ISL_2785627, EPI_ISL_2785628, EPI_ISL_2785629, EPI_ISL_2785630, EPI_ISL_2785631, EPI_ISL_2785632, EPI_ISL_2785633, EPI_ISL_2785634, EPI_ISL_2785635, EPI_ISL_2785636, EPI_ISL_2785637, EPI_ISL_2785638, EPI_ISL_2785639, EPI_ISL_2785640, EPI_ISL_2785642, EPI_ISL_2785643, EPI_ISL_2785644, EPI_ISL_2785645, EPI_ISL_2785646, EPI_ISL_2785647, EPI_ISL_2785648, EPI_ISL_2785649, EPI_ISL_2785650, EPI_ISL_2785651, EPI_ISL_2785652, EPI_ISL_2785653, EPI_ISL_2785654, EPI_ISL_2785655, EPI_ISL_2785656, EPI_ISL_2785658, EPI_ISL_2785659, EPI_ISL_2785660, EPI_ISL_2785661, EPI_ISL_2785662, EPI_ISL_2785664, EPI_ISL_2785665, EPI_ISL_2785666, EPI_ISL_2785668, EPI_ISL_2785669, EPI_ISL_2785670, EPI_ISL_2785672, EPI_ISL_2785673, EPI_ISL_2785674, EPI_ISL_2785675, EPI_ISL_2785676, EPI_ISL_2785677, EPI_ISL_2785678, EPI_ISL_2785679, EPI_ISL_2785680, EPI_ISL_2785681, EPI_ISL_2785683, EPI_ISL_2785685, EPI_ISL_2785687, EPI_ISL_2785690, EPI_ISL_2785691, EPI_ISL_2785692, EPI_ISL_2785694, EPI_ISL_2785695, EPI_ISL_2785698, EPI_ISL_2785699, EPI_ISL_2785700, EPI_ISL_2785701, EPI_ISL_2785702, EPI_ISL_2785703, EPI_ISL_2785704, EPI_ISL_2785705, EPI_ISL_2785706, EPI_ISL_2785707, EPI_ISL_2785708, EPI_ISL_2785709, EPI_ISL_2785710, EPI_ISL_2785711, EPI_ISL_2785712, EPI_ISL_2785713, EPI_ISL_2785714, EPI_ISL_2785715, EPI_ISL_2785717, EPI_ISL_2785718, EPI_ISL_2785719, EPI_ISL_2785720, EPI_ISL_2785721, EPI_ISL_2785722, EPI_ISL_2785723, EPI_ISL_2785724, EPI_ISL_2785725, EPI_ISL_2785726, EPI_ISL_2785727, EPI_ISL_2785728, EPI_ISL_2785729, EPI_ISL_2785730, EPI_ISL_2785731, EPI_ISL_2785732, EPI_ISL_2785733, EPI_ISL_2785734, EPI_ISL_2785735, EPI_ISL_2785736, EPI_ISL_2785737, EPI_ISL_2785739, EPI_ISL_2785740, EPI_ISL_2785741, EPI_ISL_2785742, EPI_ISL_2785744, EPI_ISL_2785745, EPI_ISL_2785746, EPI_ISL_2785747, EPI_ISL_2785748, EPI_ISL_2785749, EPI_ISL_2785750, EPI_ISL_2785751, EPI_ISL_2785752, EPI_ISL_2785753, EPI_ISL_2785754, EPI_ISL_2785755, EPI_ISL_2785756, EPI_ISL_2785757, EPI_ISL_2785758, EPI_ISL_2785759, EPI_ISL_2785760, EPI_ISL_2785761, EPI_ISL_2785762, EPI_ISL_2785763, EPI_ISL_2785764, EPI_ISL_2785765, EPI_ISL_2785766, EPI_ISL_2785767, EPI_ISL_2785768, EPI_ISL_2785769, EPI_ISL_2785770, EPI_ISL_2785771, EPI_ISL_2785772, EPI_ISL_2785773, EPI_ISL_2785774, EPI_ISL_2785775, EPI_ISL_2785776, EPI_ISL_2785777, EPI_ISL_2785778, EPI_ISL_2785779, EPI_ISL_2785780, EPI_ISL_2785781, EPI_ISL_2785782, EPI_ISL_2785783, EPI_ISL_2785784, EPI_ISL_2785785, EPI_ISL_2785786, EPI_ISL_2785787, EPI_ISL_2785788, EPI_ISL_2785789, EPI_ISL_2785790, EPI_ISL_2785791, EPI_ISL_2785792, EPI_ISL_2785794, EPI_ISL_2785795, EPI_ISL_2785796, EPI_ISL_2785797, EPI_ISL_2785798, EPI_ISL_2785799, EPI_ISL_2785800, EPI_ISL_2785802, EPI_ISL_2785803, EPI_ISL_2785805, EPI_ISL_2785809, EPI_ISL_2785811, EPI_ISL_2785812, EPI_ISL_2785813, EPI_ISL_2785815, EPI_ISL_2785816, EPI_ISL_2785817, EPI_ISL_2785818, EPI_ISL_2785819, EPI_ISL_2785821, EPI_ISL_2785822, EPI_ISL_2785823, EPI_ISL_2785824, EPI_ISL_2785826, EPI_ISL_2785827, EPI_ISL_2785828, EPI_ISL_2785829, EPI_ISL_2785830, EPI_ISL_2785831, EPI_ISL_2785832, EPI_ISL_2785833, EPI_ISL_2785835, EPI_ISL_2785836, EPI_ISL_2785837, EPI_ISL_2785839, EPI_ISL_2785840, EPI_ISL_2785841, EPI_ISL_2785842, EPI_ISL_2785843, EPI_ISL_2785844, EPI_ISL_2785845, EPI_ISL_2785846, EPI_ISL_2785847, EPI_ISL_2785848, EPI_ISL_2785849, EPI_ISL_2785850, EPI_ISL_2785851, EPI_ISL_2785852, EPI_ISL_2785853, EPI_ISL_2785854, EPI_ISL_2785855, EPI_ISL_2785856, EPI_ISL_2785857, EPI_ISL_2785858, EPI_ISL_2785859, EPI_ISL_2785860, EPI_ISL_2785861, EPI_ISL_2785862, EPI_ISL_2785863, EPI_ISL_2785864, EPI_ISL_2785865, EPI_ISL_2785866, EPI_ISL_2785867, EPI_ISL_2785868, EPI_ISL_2785869, EPI_ISL_2785870, EPI_ISL_2785871, EPI_ISL_2785872, EPI_ISL_2785873, EPI_ISL_2785874, EPI_ISL_2785875, EPI_ISL_2785876, EPI_ISL_2785877, EPI_ISL_2785879, EPI_ISL_2785880, EPI_ISL_2785881, EPI_ISL_2785882, EPI_ISL_2785883, EPI_ISL_2785884, EPI_ISL_2785885, EPI_ISL_2785887, EPI_ISL_2785888, EPI_ISL_2785889, EPI_ISL_2785890, EPI_ISL_2785891, EPI_ISL_2785893, EPI_ISL_2785894, EPI_ISL_2785895, EPI_ISL_2785896, EPI_ISL_2785897, EPI_ISL_2785898, EPI_ISL_2785899, EPI_ISL_2785900, EPI_ISL_2785902, EPI_ISL_2785903, EPI_ISL_2785904, EPI_ISL_2785905, EPI_ISL_2785906, EPI_ISL_2785907, EPI_ISL_2785908, EPI_ISL_2785909, EPI_ISL_2785910, EPI_ISL_2785912, EPI_ISL_2785913, EPI_ISL_2785914, EPI_ISL_2785915, EPI_ISL_2785916, EPI_ISL_2785917, EPI_ISL_2785919, EPI_ISL_2785921, EPI_ISL_2785922, EPI_ISL_2785923, EPI_ISL_2785924, EPI_ISL_2785925, EPI_ISL_2785926, EPI_ISL_2785927, EPI_ISL_2785928, EPI_ISL_2785929, EPI_ISL_2785930, EPI_ISL_2785932, EPI_ISL_2785933, EPI_ISL_2785934, EPI_ISL_2785935, EPI_ISL_2785936, EPI_ISL_2785937, EPI_ISL_2785938, EPI_ISL_2785939, EPI_ISL_2785940, EPI_ISL_2785941, EPI_ISL_2785942, EPI_ISL_2785943, EPI_ISL_2785944, EPI_ISL_2785945, EPI_ISL_2785946, EPI_ISL_2785947, EPI_ISL_2785948, EPI_ISL_2785949, EPI_ISL_2785950, EPI_ISL_2785951, EPI_ISL_2785952, EPI_ISL_2785953, EPI_ISL_2785954, EPI_ISL_2785955, EPI_ISL_2785956, EPI_ISL_2785957, EPI_ISL_2785958, EPI_ISL_2785959, EPI_ISL_2785960, EPI_ISL_2785961, EPI_ISL_2785962, EPI_ISL_2785963, EPI_ISL_2785964, EPI_ISL_2785965, EPI_ISL_2785966, EPI_ISL_2785967 | | | |
| see above | Fulgent Genetics | Centers for Disease Control and Prevention Division of Viral Diseases, Pathogen Discovery | Dakota Howard, Dhwani Batra, Peter W. Cook, Kara Moser, Adrian Paskey, Jason Caravas, Benjamin Rambo-Martin, Shatavia Morrison, Christopher Gulvick, Scott Sammons, Yvette Unoarumhi, Darlene Wagner, Matthew Schmerer, Harry Gao, Mickey Li, John Gao, Joseph Fierro, Benafsh Sapra, Becky Tsai, Yan Meng, Doreen Ng, James Xie, Clinton R. Paden, Duncan MacCannell |
| EPI_ISL_2785969, EPI_ISL_2785970, EPI_ISL_2785971, EPI_ISL_2785972, EPI_ISL_2785973, EPI_ISL_2785975, EPI_ISL_2785976, EPI_ISL_2785977, EPI_ISL_2785978, EPI_ISL_2785979, EPI_ISL_2785980, EPI_ISL_2785981, EPI_ISL_2785982, EPI_ISL_2785983, EPI_ISL_2785984, EPI_ISL_2785985, EPI_ISL_2785986, EPI_ISL_2785987, EPI_ISL_2785988, EPI_ISL_2785989, EPI_ISL_2785990, EPI_ISL_2785991, EPI_ISL_2785992, EPI_ISL_2785993, EPI_ISL_2785994, EPI_ISL_2785995, EPI_ISL_2785996, EPI_ISL_2785997, EPI_ISL_2785998, EPI_ISL_2785999, EPI_ISL_2786000, EPI_ISL_2786001, EPI_ISL_2786002, EPI_ISL_2786003, EPI_ISL_2786004, EPI_ISL_2786005, EPI_ISL_2786006, EPI_ISL_2786007, EPI_ISL_2786008, EPI_ISL_2786009, EPI_ISL_2786010, EPI_ISL_2786011, EPI_ISL_2786012, EPI_ISL_2786013, EPI_ISL_2786014, EPI_ISL_2786015, EPI_ISL_2786016, EPI_ISL_2786017, EPI_ISL_2786018, EPI_ISL_2786019, EPI_ISL_2786020, EPI_ISL_2786021, EPI_ISL_2786022, EPI_ISL_2786023, EPI_ISL_2786024, EPI_ISL_2786025, EPI_ISL_2786026, EPI_ISL_2786027, EPI_ISL_2786028, EPI_ISL_2786029, EPI_ISL_2786030, EPI_ISL_2786031, EPI_ISL_2786032, EPI_ISL_2786033, EPI_ISL_2786034, EPI_ISL_2786035, EPI_ISL_2786036, EPI_ISL_2786037, EPI_ISL_2786038, EPI_ISL_2786039, EPI_ISL_2786040, EPI_ISL_2786041, EPI_ISL_2786042, EPI_ISL_2786043, EPI_ISL_2786044, EPI_ISL_2786045, EPI_ISL_2786046, EPI_ISL_2786047, EPI_ISL_2786048, EPI_ISL_2786049, EPI_ISL_2786050, EPI_ISL_2786051, EPI_ISL_2786052, EPI_ISL_2786053, EPI_ISL_2786054, EPI_ISL_2786055, EPI_ISL_2786056, EPI_ISL_2786057, EPI_ISL_2786058, EPI_ISL_2786059, EPI_ISL_2786060, EPI_ISL_2786061, EPI_ISL_2786062, EPI_ISL_2786063, EPI_ISL_2786064, EPI_ISL_2786065, EPI_ISL_2786066, EPI_ISL_2786067, EPI_ISL_2786068, EPI_ISL_2786069, EPI_ISL_2786070, EPI_ISL_2786072, EPI_ISL_2786073, EPI_ISL_2786074, EPI_ISL_2786075, EPI_ISL_2786076, EPI_ISL_2786077, EPI_ISL_2786078, EPI_ISL_2786079, EPI_ISL_2786080, EPI_ISL_2786081, EPI_ISL_2786082, EPI_ISL_2786083, EPI_ISL_2786084, EPI_ISL_2786085, EPI_ISL_2786086, EPI_ISL_2786087, EPI_ISL_2786088, EPI_ISL_2786089, EPI_ISL_2786090, EPI_ISL_2786091, EPI_ISL_2786092, EPI_ISL_2786093, EPI_ISL_2786094, EPI_ISL_2786095, EPI_ISL_2786096, EPI_ISL_2786097, EPI_ISL_2786098, EPI_ISL_2786099, EPI_ISL_2786100, EPI_ISL_2786101, EPI_ISL_2786102, EPI_ISL_2786103, EPI_ISL_2786104, EPI_ISL_2786105, EPI_ISL_2786106, EPI_ISL_2786107, EPI_ISL_2786108, EPI_ISL_2786109, EPI_ISL_2786110, EPI_ISL_2786111, EPI_ISL_2786112, EPI_ISL_2786113, EPI_ISL_2786114, EPI_ISL_2786115, EPI_ISL_2786116, EPI_ISL_2786117, EPI_ISL_2786118, EPI_ISL_2786119, EPI_ISL_2786120, EPI_ISL_2786121, EPI_ISL_2786122, EPI_ISL_2786123, EPI_ISL_2786124, EPI_ISL_2786125, EPI_ISL_2786126, EPI_ISL_2786127, EPI_ISL_2786128, EPI_ISL_2786129, EPI_ISL_2786130, EPI_ISL_2786131, EPI_ISL_2786132, EPI_ISL_2786134, EPI_ISL_2786135, EPI_ISL_2786136, EPI_ISL_2786138, EPI_ISL_2786139, EPI_ISL_2786140, EPI_ISL_2786141, EPI_ISL_2786142, EPI_ISL_2786144, EPI_ISL_2786145, EPI_ISL_2786146, EPI_ISL_2786147, EPI_ISL_2786148, EPI_ISL_2786149, EPI_ISL_2786150, EPI_ISL_2786151, EPI_ISL_2786152, EPI_ISL_2786153, EPI_ISL_2786154, EPI_ISL_2786155, EPI_ISL_2786156, EPI_ISL_2786157, EPI_ISL_2786158, EPI_ISL_2786159, EPI_ISL_2786160, EPI_ISL_2786161, EPI_ISL_2786162, EPI_ISL_2786164, EPI_ISL_2786165, EPI_ISL_2786166, EPI_ISL_2786167, EPI_ISL_2786168, EPI_ISL_2786169, EPI_ISL_2786170, EPI_ISL_2786171, EPI_ISL_2786172, EPI_ISL_2786173, EPI_ISL_2786174, EPI_ISL_2786175, EPI_ISL_2786176, EPI_ISL_2786177, EPI_ISL_2786178, EPI_ISL_2786179, EPI_ISL_2786180, EPI_ISL_2786181, EPI_ISL_2786182, EPI_ISL_2786183, EPI_ISL_2786184, EPI_ISL_2786185, EPI_ISL_2786186, EPI_ISL_2786187, EPI_ISL_2786188, EPI_ISL_2786189, EPI_ISL_2786190, EPI_ISL_2786191, EPI_ISL_2786192, EPI_ISL_2786193, EPI_ISL_2786194, EPI_ISL_2786195, EPI_ISL_2786196, EPI_ISL_2786197, EPI_ISL_2786198, EPI_ISL_2786199, EPI_ISL_2786200, EPI_ISL_2786201, EPI_ISL_2786202, EPI_ISL_2786203, EPI_ISL_2786204, EPI_ISL_2786205, EPI_ISL_2786206, EPI_ISL_2786207, EPI_ISL_2786208, EPI_ISL_2786209, EPI_ISL_2786210, EPI_ISL_2786211, EPI_ISL_2786212, EPI_ISL_2786213, EPI_ISL_2786214, EPI_ISL_2786215, EPI_ISL_2786216, EPI_ISL_2786217, EPI_ISL_2786218, EPI_ISL_2786219, EPI_ISL_2786220, EPI_ISL_2786221, EPI_ISL_2786222, EPI_ISL_2786223, EPI_ISL_2786224, EPI_ISL_2786225, EPI_ISL_2786226, EPI_ISL_2786227, EPI_ISL_2786228, EPI_ISL_2786229, EPI_ISL_2786230, EPI_ISL_2786231, EPI_ISL_2786232, EPI_ISL_2786233, EPI_ISL_2786234, EPI_ISL_2786235, EPI_ISL_2786236, EPI_ISL_2786237, EPI_ISL_2786238, EPI_ISL_2786239, EPI_ISL_2786240, EPI_ISL_2786241, EPI_ISL_2786242, EPI_ISL_2786243, EPI_ISL_2786244, EPI_ISL_2786245, EPI_ISL_2786246, EPI_ISL_2786247, EPI_ISL_2786248, EPI_ISL_2786249, EPI_ISL_2786250, EPI_ISL_2786251, EPI_ISL_2786252, EPI_ISL_2786253, EPI_ISL_2786254, EPI_ISL_2786255, EPI_ISL_2786256, EPI_ISL_2786257, EPI_ISL_2786258, EPI_ISL_2786259, EPI_ISL_2786260, EPI_ISL_2786261, EPI_ISL_2786262, EPI_ISL_2786263, EPI_ISL_2786264, EPI_ISL_2786265, EPI_ISL_2786266, EPI_ISL_2786267, EPI_ISL_2786268, EPI_ISL_2786269, EPI_ISL_2786270, EPI_ISL_2786271, EPI_ISL_2786272, EPI_ISL_2786273, EPI_ISL_2786274, EPI_ISL_2786275, EPI_ISL_2786276, EPI_ISL_2786277, EPI_ISL_2786278, EPI_ISL_2786279, EPI_ISL_2786280, EPI_ISL_2786281, EPI_ISL_2786282, EPI_ISL_2786283, EPI_ISL_2786284, EPI_ISL_2786285, EPI_ISL_2786286, EPI_ISL_2786287, EPI_ISL_2786288, EPI_ISL_2786289, EPI_ISL_2786290, EPI_ISL_2786291, EPI_ISL_2786292, EPI_ISL_2786293, EPI_ISL_2786294, EPI_ISL_2786295, EPI_ISL_2786296, EPI_ISL_2786297, EPI_ISL_2786298, EPI_ISL_2786299, EPI_ISL_2786300, EPI_ISL_2786301, EPI_ISL_2786302, EPI_ISL_2786303, EPI_ISL_2786304, EPI_ISL_2786305, EPI_ISL_2786306, EPI_ISL_2786307, EPI_ISL_2786308, EPI_ISL_2786309, EPI_ISL_2786310, EPI_ISL_2786311, EPI_ISL_2786312, EPI_ISL_2786313, EPI_ISL_2786314, EPI_ISL_2786315, EPI_ISL_2786316, EPI_ISL_2786317, EPI_ISL_2786318, EPI_ISL_2786319, EPI_ISL_2786320, EPI_ISL_2786321, EPI_ISL_2786322, EPI_ISL_2786323, EPI_ISL_2786324, EPI_ISL_2786325, EPI_ISL_2786326, EPI_ISL_2786327, EPI_ISL_2786328, EPI_ISL_2786329, EPI_ISL_2786330, EPI_ISL_2786331, EPI_ISL_2786332, EPI_ISL_2786333, EPI_ISL_2786334, EPI_ISL_2786335, EPI_ISL_2786336, EPI_ISL_2786337, EPI_ISL_2786338, EPI_ISL_2786339, EPI_ISL_2786340, EPI_ISL_2786341, EPI_ISL_2786342, EPI_ISL_2786343, EPI_ISL_2786344, EPI_ISL_2786345, EPI_ISL_2786346, EPI_ISL_2786347, EPI_ISL_2786348, EPI_ISL_2786349, EPI_ISL_2786350, EPI_ISL_2786351, EPI_ISL_2786352, EPI_ISL_2786353, EPI_ISL_2786354, EPI_ISL_2786355, EPI_ISL_2786356, EPI_ISL_2786357, EPI_ISL_2786358, EPI_ISL_2786359, EPI_ISL_2786360, EPI_ISL_2786361, EPI_ISL_2786362, EPI_ISL_2786363, EPI_ISL_2786364, EPI_ISL_2786365, EPI_ISL_2786366, EPI_ISL_2786367, EPI_ISL_2786368, EPI_ISL_2786369, EPI_ISL_2786370, EPI_ISL_2786371, EPI_ISL_2786372, EPI_ISL_2786373, EPI_ISL_2786374, EPI_ISL_2786375, EPI_ISL_2786376, EPI_ISL_2786377, EPI_ISL_2786378, EPI_ISL_2786379, EPI_ISL_2786380, EPI_ISL_2786381, EPI_ISL_2786382, EPI_ISL_2786383, EPI_ISL_2786384, EPI_ISL_2786385, EPI_ISL_2786386, EPI_ISL_2786387, EPI_ISL_2786388, EPI_ISL_2786389, EPI_ISL_2786390, EPI_ISL_2786391, EPI_ISL_2786392, EPI_ISL_2786393, EPI_ISL_2786394, EPI_ISL_2786395, EPI_ISL_2786396, EPI_ISL_2786397, EPI_ISL_2786398, EPI_ISL_2786399, EPI_ISL_2786400, EPI_ISL_2786401, EPI_ISL_2786402, EPI_ISL_2786403, EPI_ISL_2786404, EPI_ISL_2786405, EPI_ISL_2786406, EPI_ISL_2786407, EPI_ISL_2786408, EPI_ISL_2786409, EPI_ISL_2786410, EPI_ISL_2786411, EPI_ISL_2786412, EPI_ISL_2786413, EPI_ISL_2786414, EPI_ISL_2786415, EPI_ISL_2786416, EPI_ISL_2786417, EPI_ISL_2786418, EPI_ISL_2786419, EPI_ISL_2786420, EPI_ISL_2786421, EPI_ISL_2786422, EPI_ISL_2786423, EPI_ISL_2786424, EPI_ISL_2786425, EPI_ISL_2786426, EPI_ISL_2786427, EPI_ISL_2786428, EPI_ISL_2786429, EPI_ISL_2786430, EPI_ISL_2786431, EPI_ISL_2786432, EPI_ISL_2786433, EPI_ISL_2786434, EPI_ISL_2786435, EPI_ISL_2786436, EPI_ISL_2786437, EPI_ISL_2786438, EPI_ISL_2786439, EPI_ISL_2786440, EPI_ISL_2786441, EPI_ISL_2786442, EPI_ISL_2786443, EPI_ISL_2786444, EPI_ISL_2786445, EPI_ISL_2786446, EPI_ISL_2786447, EPI_ISL_2786448, EPI_ISL_2786449, EPI_ISL_2786450, EPI_ISL_2786451, EPI_ISL_2786452, EPI_ISL_2786453, EPI_ISL_2786454, EPI_ISL_2786455, EPI_ISL_2786456, EPI_ISL_2786457, EPI_ISL_2786458, EPI_ISL_2786459, EPI_ISL_2786460, EPI_ISL_2786461, EPI_ISL_2786462, EPI_ISL_2786463, EPI_ISL_2786464, EPI_ISL_2786465, EPI_ISL_2786466, EPI_ISL_2786467, EPI_ISL_2786468, EPI_ISL_2786469, EPI_ISL_2786470, EPI_ISL_2786471, EPI_ISL_2786472, EPI_ISL_2786473, EPI_ISL_2786474, EPI_ISL_2786475, EPI_ISL_2786476, EPI_ISL_2786477, EPI_ISL_2786478, EPI_ISL_2786479, EPI_ISL_2786480, EPI_ISL_2786481, EPI_ISL_2786482, EPI_ISL_2786483, EPI_ISL_2786484, EPI_ISL_2786485, EPI_ISL_2786486, EPI_ISL_2786487, EPI_ISL_2786488, EPI_ISL_2786489, EPI_ISL_2786490, EPI_ISL_2786491, EPI_ISL_2786492, EPI_ISL_2786493, EPI_ISL_2786494, EPI_ISL_2786495, EPI_ISL_2786496, EPI_ISL_2786497, EPI_ISL_2786498, EPI_ISL_2786499, EPI_ISL_2786500, EPI_ISL_2786501, EPI_ISL_2786502, EPI_ISL_2786503, EPI_ISL_2786504, EPI_ISL_2786505, EPI_ISL_2786506, EPI_ISL_2786507, EPI_ISL_2786508, EPI_ISL_2786509, EPI_ISL_2786510, EPI_ISL_2786511, EPI_ISL_2786512, EPI_ISL_2786513, EPI_ISL_2786514, EPI_ISL_2786515, EPI_ISL_2786516, EPI_ISL_2786517, EPI_ISL_2786518, EPI_ISL_2786519, EPI_ISL_2786520, EPI_ISL_2786521, EPI_ISL_2786522, EPI_ISL_2786523, EPI_ISL_2786524, EPI_ISL_2786525, EPI_ISL_2786526, EPI_ISL_2786527, EPI_ISL_2786528, EPI_ISL_2786529, EPI_ISL_2786530, EPI_ISL_2786531, EPI_ISL_2786532, EPI_ISL_2786533, EPI_ISL_2786534, EPI_ISL_2786535, EPI_ISL_2786537, EPI_ISL_2786538, EPI_ISL_2786539, EPI_ISL_2786540, EPI_ISL_2786541, EPI_ISL_2786542, EPI_ISL_2786543, EPI_ISL_2786544, EPI_ISL_2786545, EPI_ISL_2786546, EPI_ISL_2786547, EPI_ISL_2786548, EPI_ISL_2786549, EPI_ISL_2786550, EPI_ISL_2786551, EPI_ISL_2786552, EPI_ISL_2786553, EPI_ISL_2786554, EPI_ISL_2786555, EPI_ISL_2786556, EPI_ISL_2786557, EPI_ISL_2786558, EPI_ISL_2786559, EPI_ISL_2786560, EPI_ISL_2786561, EPI_ISL_2786562, EPI_ISL_2786563, EPI_ISL_2786564, EPI_ISL_2786565, EPI_ISL_2786566, EPI_ISL_2786567, EPI_ISL_2786568, EPI_ISL_2786569, EPI_ISL_2786570, EPI_ISL_2786571, EPI_ISL_2786572, EPI_ISL_2786573, EPI_ISL_2786574, EPI_ISL_2786575, EPI_ISL_2786576, EPI_ISL_2786577, EPI_ISL_2786578, EPI_ISL_2786579, EPI_ISL_2786580, EPI_ISL_2786581, EPI_ISL_2786582, EPI_ISL_2786583, EPI_ISL_2786584, EPI_ISL_2786585, EPI_ISL_2786586, EPI_ISL_2786587, EPI_ISL_2786588, EPI_ISL_2786589, EPI_ISL_2786590, EPI_ISL_2786591, EPI_ISL_2786592, EPI_ISL_2786593, EPI_ISL_2786594, EPI_ISL_2786595, EPI_ISL_2786596, EPI_ISL_2786597, EPI_ISL_2786598, EPI_ISL_2786599, EPI_ISL_2786600, EPI_ISL_2786601, EPI_ISL_2786602, EPI_ISL_2786603, EPI_ISL_2786604, EPI_ISL_2786605, EPI_ISL_2786606, EPI_ISL_2786607, EPI_ISL_2786608, EPI_ISL_2786609, EPI_ISL_2786610, EPI_ISL_2786611, EPI_ISL_2786612, EPI_ISL_2786613, EPI_ISL_2786614, EPI_ISL_2786615, EPI_ISL_2786616, EPI_ISL_2786617, EPI_ISL_2786618, EPI_ISL_2786619, EPI_ISL_2786621, EPI_ISL_2786622, EPI_ISL_2786623, EPI_ISL_2786624, EPI_ISL_2786625, EPI_ISL_2786626, EPI_ISL_2786627, EPI_ISL_2786628, EPI_ISL_2786629, EPI_ISL_2786630 | | | |
| see above | Mako Medical | Centers for Disease Control and Prevention Division of Viral Diseases, Pathogen Discovery | Dakota Howard, Dhwani Batra, Peter W. Cook, Kara Moser, Adrian Paskey, Jason Caravas, Benjamin Rambo-Martin, Shatavia Morrison, Christopher Gulvick, Scott Sammons, Yvette Unoarumhi, Darlene Wagner, Matthew Schmerer,Matthew Tugwell, Lauren Moon, Clinton R. Paden, Duncan MacCannell |
| EPI_ISL_2786631, EPI_ISL_2786632, EPI_ISL_2786633, EPI_ISL_2786634, EPI_ISL_2786635, EPI_ISL_2786636, EPI_ISL_2786637, EPI_ISL_2786638, EPI_ISL_2786639, EPI_ISL_2786640, EPI_ISL_2786641, EPI_ISL_2786642, EPI_ISL_2786643, EPI_ISL_2786644, EPI_ISL_2786645, EPI_ISL_2786647, EPI_ISL_2786648, EPI_ISL_2786649, EPI_ISL_2786650, EPI_ISL_2786651, EPI_ISL_2786652, EPI_ISL_2786653, EPI_ISL_2786654, EPI_ISL_2786655, EPI_ISL_2786656, EPI_ISL_2786657, EPI_ISL_2786658, EPI_ISL_2786659, EPI_ISL_2786660, EPI_ISL_2786661, EPI_ISL_2786662, EPI_ISL_2786663, EPI_ISL_2786664, EPI_ISL_2786665, EPI_ISL_2786666, EPI_ISL_2786667, EPI_ISL_2786668, EPI_ISL_2786669, EPI_ISL_2786670, EPI_ISL_2786671, EPI_ISL_2786672, EPI_ISL_2786673, EPI_ISL_2786674, EPI_ISL_2786675, EPI_ISL_2786676, EPI_ISL_2786677, EPI_ISL_2786678, EPI_ISL_2786679, EPI_ISL_2786680, EPI_ISL_2786681, EPI_ISL_2786682, EPI_ISL_2786683, EPI_ISL_2786684, EPI_ISL_2786685, EPI_ISL_2786686, EPI_ISL_2786687, EPI_ISL_2786688, EPI_ISL_2786689, EPI_ISL_2786690, EPI_ISL_2786691, EPI_ISL_2786692, EPI_ISL_2786693, EPI_ISL_2786694, EPI_ISL_2786696, EPI_ISL_2786697, EPI_ISL_2786698, EPI_ISL_2786699, EPI_ISL_2786700 | | | |
| see above | Infinity Biologix | Centers for Disease Control and Prevention Division of Viral Diseases, Pathogen Discovery | Dakota Howard, Dhwani Batra, Peter W. Cook, Kara Moser, Adrian Paskey, Jason Caravas, Benjamin Rambo-Martin, Shatavia Morrison, Christopher Gulvick, Scott Sammons, Yvette Unoarumhi, Darlene Wagner, Matthew Schmerer, Christian Bixby, Yihe Wang, Jonathan Schultz, Chirayu Goswami, Russ Hager, Robin Grimwood, Clinton R. Paden, Duncan MacCannell |
| EPI_ISL_2786894 | Atlas Genomics | Seattle Flu Study | Deborah A. Nickerson, Chris D. Frazar, Jover Lee, Benjamin Pelle, Erica Ryke, Matthew Richardson, Amanda Adler, Elisabeth Brandstetter, Peter D. Han, Kairsten Fay, Misja Ilcisin, Kirsten Lacombe, Thomas R. Sibley, Melissa Truong, Caitlin R. Wolf, Romesh Gautom, Geoff Melly, Brian Hiatt, Philip Dykema, Scott Lindquist, Michael Boeckh, Janet A. Englund, Michael Famulare, Barry R. Lutz, Mark J. Rieder, Lea M. Starita, Matthew Thompson, Helen Y. Chu, Jay Shendure, Trevor Bedford |
| EPI_ISL_2786895 | Incyte Diagnostics Spokane | Seattle Flu Study | Deborah A. Nickerson, Chris D. Frazar, Jover Lee, Benjamin Pelle, Erica Ryke, Matthew Richardson, Amanda Adler, Elisabeth Brandstetter, Peter D. Han, Kairsten Fay, Misja Ilcisin, Kirsten Lacombe, Thomas R. Sibley, Melissa Truong, Caitlin R. Wolf, Romesh Gautom, Geoff Melly, Brian Hiatt, Philip Dykema, Scott Lindquist, Michael Boeckh, Janet A. Englund, Michael Famulare, Barry R. Lutz, Mark J. Rieder, Lea M. Starita, Matthew Thompson, Helen Y. Chu, Jay Shendure, Trevor Bedford |
| EPI_ISL_2786896 | Northwest Laboratory | Seattle Flu Study | Deborah A. Nickerson, Chris D. Frazar, Jover Lee, Benjamin Pelle, Erica Ryke, Matthew Richardson, Amanda Adler, Elisabeth Brandstetter, Peter D. Han, Kairsten Fay, Misja Ilcisin, Kirsten Lacombe, Thomas R. Sibley, Melissa Truong, Caitlin R. Wolf, Romesh Gautom, Geoff Melly, Brian Hiatt, Philip Dykema, Scott Lindquist, Michael Boeckh, Janet A. Englund, Michael Famulare, Barry R. Lutz, Mark J. Rieder, Lea M. Starita, Matthew Thompson, Helen Y. Chu, Jay Shendure, Trevor Bedford |
| EPI_ISL_2786899, EPI_ISL_2786900 | Seattle Flu Study | Seattle Flu Study | Deborah A. Nickerson, Chris D. Frazar, Jover Lee, Benjamin Pelle, Erica Ryke, Matthew Richardson, Amanda Adler, Elisabeth Brandstetter, Peter D. Han, Kairsten Fay, Misja Ilcisin, Kirsten Lacombe, Thomas R. Sibley, Melissa Truong, Caitlin R. Wolf, Michael Boeckh, Janet A. Englund, Michael Famulare, Barry R. Lutz, Mark J. Rieder, Lea M. Starita, Matthew Thompson, Jay Shendure, Trevor Bedford, Helen Y. Chu |
| EPI_ISL_2786901, EPI_ISL_2786902, EPI_ISL_2786903 | Northwest Laboratory | Seattle Flu Study | Deborah A. Nickerson, Chris D. Frazar, Jover Lee, Benjamin Pelle, Erica Ryke, Matthew Richardson, Amanda Adler, Elisabeth Brandstetter, Peter D. Han, Kairsten Fay, Misja Ilcisin, Kirsten Lacombe, Thomas R. Sibley, Melissa Truong, Caitlin R. Wolf, Romesh Gautom, Geoff Melly, Brian Hiatt, Philip Dykema, Scott Lindquist, Michael Boeckh, Janet A. Englund, Michael Famulare, Barry R. Lutz, Mark J. Rieder, Lea M. Starita, Matthew Thompson, Helen Y. Chu, Jay Shendure, Trevor Bedford |
| EPI_ISL_2786904, EPI_ISL_2786905, EPI_ISL_2786906 | Seattle Flu Study | Seattle Flu Study | Deborah A. Nickerson, Chris D. Frazar, Jover Lee, Benjamin Pelle, Erica Ryke, Matthew Richardson, Amanda Adler, Elisabeth Brandstetter, Peter D. Han, Kairsten Fay, Misja Ilcisin, Kirsten Lacombe, Thomas R. Sibley, Melissa Truong, Caitlin R. Wolf, Michael Boeckh, Janet A. Englund, Michael Famulare, Barry R. Lutz, Mark J. Rieder, Lea M. Starita, Matthew Thompson, Jay Shendure, Trevor Bedford, Helen Y. Chu |
| EPI_ISL_2786925, EPI_ISL_2786926, EPI_ISL_2786927, EPI_ISL_2787111 | Ohio Department of Health Laboratory | Ohio Department of Health Laboratory | Holmes, Jennifer; Eric Brandt, Keoni Omura, Glen McGillivary, Caitlin McDonnell, Jade Mowery, Stephanie Mccracken, Tyler Payne, Kirtana Ramadugu, Erica Leasure, Brent Lee, Kelsey Florek, Heather Blankenship, Allison Black, Quanta Brown, and Tammy Bannerman |
| EPI_ISL_2787209, EPI_ISL_2787212, EPI_ISL_2787215, EPI_ISL_2787217, EPI_ISL_2787220, EPI_ISL_2787223, EPI_ISL_2787226, EPI_ISL_2787228, EPI_ISL_2787231, EPI_ISL_2787234, EPI_ISL_2787236 | | | |
| see above | FL Bureau of Public Health Laboratories-Tampa | Centers for Disease Control and Prevention Division of Viral Diseases, Pathogen Discovery | Mili Sheth, Sarah Nobles, Jasmine Padilla, Mark Burroughs, Shoshona Le, Katie Dillon, Peter Cook, Clinton R. Paden, Dhwani Batra, Krista Queen, Kristen Knipe, Dakota Howard, Yvette Unoarumhi, Darlene Wagner, Matthew Schmerer, Ben L. Rambo-Martin, Kristine Lacek, Sam Shepard, Alison Laufer Halpin, Dave Wentworth, Vivien Dugan, Suxiang Tong, Justin Lee |
| EPI_ISL_2787240, EPI_ISL_2787243, EPI_ISL_2787245, EPI_ISL_2787248, EPI_ISL_2787251, EPI_ISL_2787254, EPI_ISL_2787257, EPI_ISL_2787260, EPI_ISL_2787263, EPI_ISL_2787266, EPI_ISL_2787270, EPI_ISL_2787272, EPI_ISL_2787275, EPI_ISL_2787277, EPI_ISL_2787280, EPI_ISL_2787283, EPI_ISL_2787285 | | | |
| see above | SC Dept of Health and Env. Control-Bureau of Laboratories | Centers for Disease Control and Prevention Division of Viral Diseases, Pathogen Discovery | Mili Sheth, Sarah Nobles, Jasmine Padilla, Mark Burroughs, Shoshona Le, Katie Dillon, Peter Cook, Clinton R. Paden, Dhwani Batra, Krista Queen, Kristen Knipe, Dakota Howard, Yvette Unoarumhi, Darlene Wagner, Matthew Schmerer, Ben L. Rambo-Martin, Kristine Lacek, Sam Shepard, Alison Laufer Halpin, Dave Wentworth, Vivien Dugan, Suxiang Tong, Justin Lee |
| EPI_ISL_2787288, EPI_ISL_2787290, EPI_ISL_2787293, EPI_ISL_2787296, EPI_ISL_2787299, EPI_ISL_2787302 | ND Dept. of Health Laboratory Services-Microbiology | Centers for Disease Control and Prevention Division of Viral Diseases, Pathogen Discovery | Mili Sheth, Sarah Nobles, Jasmine Padilla, Mark Burroughs, Shoshona Le, Katie Dillon, Peter Cook, Clinton R. Paden, Dhwani Batra, Krista Queen, Kristen Knipe, Dakota Howard, Yvette Unoarumhi, Darlene Wagner, Matthew Schmerer, Ben L. Rambo-Martin, Kristine Lacek, Sam Shepard, Alison Laufer Halpin, Dave Wentworth, Vivien Dugan, Suxiang Tong, Justin Lee |
| EPI_ISL_2787305 | "TX DSHS, Lab Services Section MC 1947" | Centers for Disease Control and Prevention Division of Viral Diseases, Pathogen Discovery | Mili Sheth, Sarah Nobles, Jasmine Padilla, Mark Burroughs, Shoshona Le, Katie Dillon, Peter Cook, Clinton R. Paden, Dhwani Batra, Krista Queen, Kristen Knipe, Dakota Howard, Yvette Unoarumhi, Darlene Wagner, Matthew Schmerer, Ben L. Rambo-Martin, Kristine Lacek, Sam Shepard, Alison Laufer Halpin, Dave Wentworth, Vivien Dugan, Suxiang Tong, Justin Lee |
| EPI_ISL_2787307, EPI_ISL_2787310, EPI_ISL_2787313, | FL Bureau of Public Health Laboratories-Miami | Centers for Disease Control and Prevention Division of Viral Diseases, Pathogen Discovery | Mili Sheth, Sarah Nobles, Jasmine Padilla, Mark Burroughs, Shoshona Le, Katie Dillon, Peter Cook, Clinton R. Paden, Dhwani Batra, Krista Queen, Kristen Knipe, Dakota Howard, Yvette Unoarumhi, Darlene Wagner, Matthew Schmerer, Ben L. Rambo-Martin, Kristine Lacek, Sam Shepard, Alison Laufer Halpin, Dave Wentworth, Vivien Dugan, Suxiang Tong, Justin Lee |

| | | | |
|---|---|---|---|
| EPI_ISL_2787315, EPI_ISL_2787318 | | | |
| EPI_ISL_2787321, EPI_ISL_2787323, EPI_ISL_2787325, EPI_ISL_2787328 | CA-Los Angeles County Public Health Laboratory | Centers for Disease Control and Prevention Division of Viral Diseases, Pathogen Discovery | Mili Sheth, Sarah Nobles, Jasmine Padilla, Mark Burroughs, Shoshona Le, Katie Dillon, Peter Cook, Clinton R. Paden, Dhwani Batra, Krista Queen, Kristen Knipe, Dakota Howard, Yvette Unoarumhi, Darlene Wagner, Matthew Schmerer, Ben L. Rambo-Martin, Kristine Lacek, Sam Shepard, Alison Laufer Halpin, Dave Wentworth, Vivien Dugan, Suxiang Tong, Justin Lee |
| EPI_ISL_2787331, EPI_ISL_2787334, EPI_ISL_2787336, EPI_ISL_2787339, EPI_ISL_2787342, EPI_ISL_2787345, EPI_ISL_2787348, EPI_ISL_2787351, EPI_ISL_2787354 | SD Public Health Laboratory | Centers for Disease Control and Prevention Division of Viral Diseases, Pathogen Discovery | Mili Sheth, Sarah Nobles, Jasmine Padilla, Mark Burroughs, Shoshona Le, Katie Dillon, Peter Cook, Clinton R. Paden, Dhwani Batra, Krista Queen, Kristen Knipe, Dakota Howard, Yvette Unoarumhi, Darlene Wagner, Matthew Schmerer, Ben L. Rambo-Martin, Kristine Lacek, Sam Shepard, Alison Laufer Halpin, Dave Wentworth, Vivien Dugan, Suxiang Tong, Justin Lee |
| EPI_ISL_2787356, EPI_ISL_2787359, EPI_ISL_2787361, EPI_ISL_2787364, EPI_ISL_2787367, EPI_ISL_2787369, EPI_ISL_2787372, EPI_ISL_2787375, EPI_ISL_2787377, EPI_ISL_2787380, EPI_ISL_2787383, EPI_ISL_2787386, EPI_ISL_2787389, EPI_ISL_2787392, EPI_ISL_2787395 | | | |
| see above | IA State Hygienic Laboratory | Centers for Disease Control and Prevention Division of Viral Diseases, Pathogen Discovery | Mili Sheth, Sarah Nobles, Jasmine Padilla, Mark Burroughs, Shoshona Le, Katie Dillon, Peter Cook, Clinton R. Paden, Dhwani Batra, Krista Queen, Kristen Knipe, Dakota Howard, Yvette Unoarumhi, Darlene Wagner, Matthew Schmerer, Ben L. Rambo-Martin, Kristine Lacek, Sam Shepard, Alison Laufer Halpin, Dave Wentworth, Vivien Dugan, Suxiang Tong, Justin Lee |
| EPI_ISL_2787397, EPI_ISL_2787400, EPI_ISL_2787403, EPI_ISL_2787406, EPI_ISL_2787408, EPI_ISL_2787411, EPI_ISL_2787414, EPI_ISL_2787416 | WY Public Health Laboratory | Centers for Disease Control and Prevention Division of Viral Diseases, Pathogen Discovery | Mili Sheth, Sarah Nobles, Jasmine Padilla, Mark Burroughs, Shoshona Le, Katie Dillon, Peter Cook, Clinton R. Paden, Dhwani Batra, Krista Queen, Kristen Knipe, Dakota Howard, Yvette Unoarumhi, Darlene Wagner, Matthew Schmerer, Ben L. Rambo-Martin, Kristine Lacek, Sam Shepard, Alison Laufer Halpin, Dave Wentworth, Vivien Dugan, Suxiang Tong, Justin Lee |
| EPI_ISL_2787419, EPI_ISL_2787421, EPI_ISL_2787424, EPI_ISL_2787427, EPI_ISL_2787430, EPI_ISL_2787433, EPI_ISL_2787436, EPI_ISL_2787439, EPI_ISL_2787442, EPI_ISL_2787444, EPI_ISL_2787447, EPI_ISL_2787450, EPI_ISL_2787453, EPI_ISL_2787456, EPI_ISL_2787459, EPI_ISL_2787462 | | | |
| see above | OR State PHL-Virology/Immunology Section | Centers for Disease Control and Prevention Division of Viral Diseases, Pathogen Discovery | Mili Sheth, Sarah Nobles, Jasmine Padilla, Mark Burroughs, Shoshona Le, Katie Dillon, Peter Cook, Clinton R. Paden, Dhwani Batra, Krista Queen, Kristen Knipe, Dakota Howard, Yvette Unoarumhi, Darlene Wagner, Matthew Schmerer, Ben L. Rambo-Martin, Kristine Lacek, Sam Shepard, Alison Laufer Halpin, Dave Wentworth, Vivien Dugan, Suxiang Tong, Justin Lee |
| EPI_ISL_2787464, EPI_ISL_2787467, EPI_ISL_2787471, EPI_ISL_2787474, EPI_ISL_2787477, EPI_ISL_2787480, EPI_ISL_2787483, EPI_ISL_2787485, EPI_ISL_2787488, EPI_ISL_2787491, EPI_ISL_2787494, EPI_ISL_2787497, EPI_ISL_2787500, EPI_ISL_2787503, EPI_ISL_2787506, EPI_ISL_2787508 | | | |
| see above | "NM Dept. Health, Scientific Laboratory Division " | Centers for Disease Control and Prevention Division of Viral Diseases, Pathogen Discovery | Mili Sheth, Sarah Nobles, Jasmine Padilla, Mark Burroughs, Shoshona Le, Katie Dillon, Peter Cook, Clinton R. Paden, Dhwani Batra, Krista Queen, Kristen Knipe, Dakota Howard, Yvette Unoarumhi, Darlene Wagner, Matthew Schmerer, Ben L. Rambo-Martin, Kristine Lacek, Sam Shepard, Alison Laufer Halpin, Dave Wentworth, Vivien Dugan, Suxiang Tong, Justin Lee |
| EPI_ISL_2787511 | "AK State Public Health Lab, State Health Department" | Centers for Disease Control and Prevention Division of Viral Diseases, Pathogen Discovery | Mili Sheth, Sarah Nobles, Jasmine Padilla, Mark Burroughs, Shoshona Le, Katie Dillon, Peter Cook, Clinton R. Paden, Dhwani Batra, Krista Queen, Kristen Knipe, Dakota Howard, Yvette Unoarumhi, Darlene Wagner, Matthew Schmerer, Ben L. Rambo-Martin, Kristine Lacek, Sam Shepard, Alison Laufer Halpin, Dave Wentworth, Vivien Dugan, Suxiang Tong, Justin Lee |
| EPI_ISL_2787514 | CT-Dr. Katherine A. Kelley State Public Health Lab | Centers for Disease Control and Prevention Division of Viral Diseases, Pathogen Discovery | Mili Sheth, Sarah Nobles, Jasmine Padilla, Mark Burroughs, Shoshona Le, Katie Dillon, Peter Cook, Clinton R. Paden, Dhwani Batra, Krista Queen, Kristen Knipe, Dakota Howard, Yvette Unoarumhi, Darlene Wagner, Matthew Schmerer, Ben L. Rambo-Martin, Kristine Lacek, Sam Shepard, Alison Laufer Halpin, Dave Wentworth, Vivien Dugan, Suxiang Tong, Justin Lee |
| EPI_ISL_2787517, EPI_ISL_2787520, EPI_ISL_2787522, EPI_ISL_2787525, EPI_ISL_2787528, EPI_ISL_2787531, EPI_ISL_2787534, EPI_ISL_2787536, EPI_ISL_2787539, EPI_ISL_2787542 | OH Department of Health Laboratory | Centers for Disease Control and Prevention Division of Viral Diseases, Pathogen Discovery | Mili Sheth, Sarah Nobles, Jasmine Padilla, Mark Burroughs, Shoshona Le, Katie Dillon, Peter Cook, Clinton R. Paden, Dhwani Batra, Krista Queen, Kristen Knipe, Dakota Howard, Yvette Unoarumhi, Darlene Wagner, Matthew Schmerer, Ben L. Rambo-Martin, Kristine Lacek, Sam Shepard, Alison Laufer Halpin, Dave Wentworth, Vivien Dugan, Suxiang Tong, Justin Lee |
| EPI_ISL_2787545, EPI_ISL_2787547, EPI_ISL_2787551, EPI_ISL_2787554, EPI_ISL_2787557, EPI_ISL_2787559, EPI_ISL_2787562, EPI_ISL_2787565, EPI_ISL_2787568, EPI_ISL_2787570, EPI_ISL_2787573, EPI_ISL_2787576 | | | |
| see above | MS Public Health Laboratory | Centers for Disease Control and Prevention Division of Viral Diseases, Pathogen Discovery | Mili Sheth, Sarah Nobles, Jasmine Padilla, Mark Burroughs, Shoshona Le, Katie Dillon, Peter Cook, Clinton R. Paden, Dhwani Batra, Krista Queen, Kristen Knipe, Dakota Howard, Yvette Unoarumhi, Darlene Wagner, Matthew Schmerer, Ben L. Rambo-Martin, Kristine Lacek, Sam Shepard, Alison Laufer Halpin, Dave Wentworth, Vivien Dugan, Suxiang Tong, Justin Lee |
| EPI_ISL_2787579, EPI_ISL_2787582, EPI_ISL_2787585 | WI State Laboratory of Hygiene | Centers for Disease Control and Prevention Division of Viral Diseases, Pathogen Discovery | Mili Sheth, Sarah Nobles, Jasmine Padilla, Mark Burroughs, Shoshona Le, Katie Dillon, Peter Cook, Clinton R. Paden, Dhwani Batra, Krista Queen, Kristen Knipe, Dakota Howard, Yvette Unoarumhi, Darlene Wagner, Matthew Schmerer, Ben L. Rambo-Martin, Kristine Lacek, Sam Shepard, Alison Laufer Halpin, Dave Wentworth, Vivien Dugan, Suxiang Tong, Justin Lee |
| EPI_ISL_2787588, EPI_ISL_2787591, EPI_ISL_2787593, EPI_ISL_2787596, EPI_ISL_2787599, EPI_ISL_2787602, EPI_ISL_2787605, EPI_ISL_2787607 | "PA Department of Health, Bureau of Laboratories" | Centers for Disease Control and Prevention Division of Viral Diseases, Pathogen Discovery | Mili Sheth, Sarah Nobles, Jasmine Padilla, Mark Burroughs, Shoshona Le, Katie Dillon, Peter Cook, Clinton R. Paden, Dhwani Batra, Krista Queen, Kristen Knipe, Dakota Howard, Yvette Unoarumhi, Darlene Wagner, Matthew Schmerer, Ben L. Rambo-Martin, Kristine Lacek, Sam Shepard, Alison Laufer Halpin, Dave Wentworth, Vivien Dugan, Suxiang Tong, Justin Lee |
| EPI_ISL_2787610, EPI_ISL_2787613, EPI_ISL_2787616, EPI_ISL_2787618, EPI_ISL_2787621, EPI_ISL_2787624, EPI_ISL_2787627, EPI_ISL_2787630, EPI_ISL_2787633, EPI_ISL_2787636, EPI_ISL_2787639, EPI_ISL_2787642, EPI_ISL_2787645, EPI_ISL_2787648, EPI_ISL_2787651, EPI_ISL_2787654, EPI_ISL_2787656, EPI_ISL_2787659, EPI_ISL_2787662 | | | |
| see above | Guam Public Health Laboratory | Centers for Disease Control and Prevention Division of Viral Diseases, Pathogen Discovery | Mili Sheth, Sarah Nobles, Jasmine Padilla, Mark Burroughs, Shoshona Le, Katie Dillon, Peter Cook, Clinton R. Paden, Dhwani Batra, Krista Queen, Kristen Knipe, Dakota Howard, Yvette Unoarumhi, Darlene Wagner, Matthew Schmerer, Ben L. Rambo-Martin, Kristine Lacek, Sam Shepard, Alison Laufer Halpin, Dave Wentworth, Vivien Dugan, Suxiang Tong, Justin Lee |
| EPI_ISL_2787665, EPI_ISL_2787668, EPI_ISL_2787670, EPI_ISL_2787674 | NH Dept. of Health and Human Services Public Health Labs | Centers for Disease Control and Prevention Division of Viral Diseases, Pathogen Discovery | Mili Sheth, Sarah Nobles, Jasmine Padilla, Mark Burroughs, Shoshona Le, Katie Dillon, Peter Cook, Clinton R. Paden, Dhwani Batra, Krista Queen, Kristen Knipe, Dakota Howard, Yvette Unoarumhi, Darlene Wagner, Matthew Schmerer, Ben L. Rambo-Martin, Kristine Lacek, Sam Shepard, Alison Laufer Halpin, Dave Wentworth, Vivien Dugan, Suxiang Tong, Justin Lee |
| EPI_ISL_2787676, EPI_ISL_2787679, EPI_ISL_2787682, EPI_ISL_2787685, EPI_ISL_2787688, EPI_ISL_2787691, EPI_ISL_2787694, EPI_ISL_2787696, EPI_ISL_2787699, EPI_ISL_2787702, EPI_ISL_2787705 | | | |
| see above | MI - Michigan Department of Health and Human Services - Bureau of Laboratories | Centers for Disease Control and Prevention Division of Viral Diseases, Pathogen Discovery | Mili Sheth, Sarah Nobles, Jasmine Padilla, Mark Burroughs, Shoshona Le, Katie Dillon, Peter Cook, Clinton R. Paden, Dhwani Batra, Krista Queen, Kristen Knipe, Dakota Howard, Yvette Unoarumhi, Darlene Wagner, Matthew Schmerer, Ben L. Rambo-Martin, Kristine Lacek, Sam Shepard, Alison Laufer Halpin, Dave Wentworth, Vivien Dugan, Suxiang Tong, Justin Lee |
| EPI_ISL_2787708, EPI_ISL_2787711, EPI_ISL_2787713, EPI_ISL_2787716, EPI_ISL_2787719, EPI_ISL_2787721, EPI_ISL_2787724, EPI_ISL_2787727, EPI_ISL_2787730, EPI_ISL_2787732, EPI_ISL_2787735 | | | |
| see above | "HI Dept. of Health, State Laboratories Division" | Centers for Disease Control and Prevention Division of Viral Diseases, Pathogen Discovery | Mili Sheth, Sarah Nobles, Jasmine Padilla, Mark Burroughs, Shoshona Le, Katie Dillon, Peter Cook, Clinton R. Paden, Dhwani Batra, Krista Queen, Kristen Knipe, Dakota Howard, Yvette Unoarumhi, Darlene Wagner, Matthew Schmerer, Ben L. Rambo-Martin, Kristine Lacek, Sam Shepard, Alison Laufer Halpin, Dave Wentworth, Vivien Dugan, Suxiang Tong, Justin Lee |
| EPI_ISL_2787738 | IN State Department of Health Laboratory Services | Centers for Disease Control and Prevention Division of Viral Diseases, Pathogen Discovery | Mili Sheth, Sarah Nobles, Jasmine Padilla, Mark Burroughs, Shoshona Le, Katie Dillon, Peter Cook, Clinton R. Paden, Dhwani Batra, Krista Queen, Kristen Knipe, Dakota Howard, Yvette Unoarumhi, Darlene Wagner, Matthew Schmerer, Ben L. Rambo-Martin, Kristine Lacek, Sam Shepard, Alison Laufer Halpin, Dave Wentworth, Vivien Dugan, Suxiang Tong, Justin Lee |
| EPI_ISL_2787741, EPI_ISL_2787744 | MO State Public Health Laboratory | Centers for Disease Control and Prevention Division of Viral Diseases, Pathogen Discovery | Mili Sheth, Sarah Nobles, Jasmine Padilla, Mark Burroughs, Shoshona Le, Katie Dillon, Peter Cook, Clinton R. Paden, Dhwani Batra, Krista Queen, Kristen Knipe, Dakota Howard, Yvette Unoarumhi, Darlene Wagner, Matthew Schmerer, Ben L. Rambo-Martin, Kristine Lacek, Sam Shepard, Alison Laufer Halpin, Dave Wentworth, Vivien Dugan, Suxiang Tong, Justin Lee |
| EPI_ISL_2787747, EPI_ISL_2787750, EPI_ISL_2787752, EPI_ISL_2787755, EPI_ISL_2787758, EPI_ISL_2787761, EPI_ISL_2787764 | AL Dept. of Public Health Bureau of Clinical Laboratories | Centers for Disease Control and Prevention Division of Viral Diseases, Pathogen Discovery | Mili Sheth, Sarah Nobles, Jasmine Padilla, Mark Burroughs, Shoshona Le, Katie Dillon, Peter Cook, Clinton R. Paden, Dhwani Batra, Krista Queen, Kristen Knipe, Dakota Howard, Yvette Unoarumhi, Darlene Wagner, Matthew Schmerer, Ben L. Rambo-Martin, Kristine Lacek, Sam Shepard, Alison Laufer Halpin, Dave Wentworth, Vivien Dugan, Suxiang Tong, Justin Lee |
| EPI_ISL_2787766 | Ohio Department of Health Laboratory | Ohio Department of Health Laboratory | Holmes, Jennifer; Eric Brandt, Keoni Omura, Glen McGillivary, Caitlin McDonnell, Jade Mowery, Stephanie Mccracken, Tyler Payne, Kirtana Ramadugu, Erica Leasure, Brent Lee, Kelsey Florek, Heather Blankenship, Allison Black, Quanta Brown, and Tammy Bannerman |
| EPI_ISL_2787767, EPI_ISL_2787769 | AL Dept. of Public Health Bureau of Clinical Laboratories | Centers for Disease Control and Prevention Division of Viral Diseases, Pathogen | Mili Sheth, Sarah Nobles, Jasmine Padilla, Mark Burroughs, Shoshona Le, Katie Dillon, Peter Cook, Clinton R. Paden, Dhwani Batra, Krista Queen, Kristen Knipe, Dakota Howard, Yvette Unoarumhi, Darlene Wagner, Matthew Schmerer, Ben L. Rambo-Martin, Kristine Lacek, Sam Shepard, Alison Laufer Halpin, Dave Wentworth, Vivien Dugan, Suxiang Tong, Justin Lee |

| | | | |
|---|---|---|---|
| | | Discovery | |
| EPI_ISL_2787770, EPI_ISL_2787771, EPI_ISL_2787772 | PR Public Health Lab | Centers for Disease Control and Prevention Division of Viral Diseases, Pathogen Discovery | Mili Sheth, Sarah Nobles, Jasmine Padilla, Mark Burroughs, Shoshona Le, Katie Dillon, Peter Cook, Clinton R. Paden, Dhwani Batra, Krista Queen, Kristen Knipe, Dakota Howard, Yvette Unoarumhi, Darlene Wagner, Matthew Schmerer, Ben L. Rambo-Martin, Kristine Lacek, Sam Shepard, Alison Laufer Halpin, Dave Wentworth, Vivien Dugan, Suxiang Tong, Justin Lee |
| EPI_ISL_2787774 | Philadelphia Department of Public Health | Centers for Disease Control and Prevention Division of Viral Diseases, Pathogen Discovery | Mili Sheth, Sarah Nobles, Jasmine Padilla, Mark Burroughs, Shoshona Le, Katie Dillon, Peter Cook, Clinton R. Paden, Dhwani Batra, Krista Queen, Kristen Knipe, Dakota Howard, Yvette Unoarumhi, Darlene Wagner, Matthew Schmerer, Ben L. Rambo-Martin, Kristine Lacek, Sam Shepard, Alison Laufer Halpin, Dave Wentworth, Vivien Dugan, Suxiang Tong, Justin Lee |
| EPI_ISL_2787777 | NYC Department of Health and Mental Hygiene | Centers for Disease Control and Prevention Division of Viral Diseases, Pathogen Discovery | Mili Sheth, Sarah Nobles, Jasmine Padilla, Mark Burroughs, Shoshona Le, Katie Dillon, Peter Cook, Clinton R. Paden, Dhwani Batra, Krista Queen, Kristen Knipe, Dakota Howard, Yvette Unoarumhi, Darlene Wagner, Matthew Schmerer, Ben L. Rambo-Martin, Kristine Lacek, Sam Shepard, Alison Laufer Halpin, Dave Wentworth, Vivien Dugan, Suxiang Tong, Justin Lee |
| EPI_ISL_2787780, EPI_ISL_2787782, EPI_ISL_2787785, EPI_ISL_2787788, EPI_ISL_2787791, EPI_ISL_2787794, EPI_ISL_2787797 | KS Health and Environmental Laboratories | Centers for Disease Control and Prevention Division of Viral Diseases, Pathogen Discovery | Mili Sheth, Sarah Nobles, Jasmine Padilla, Mark Burroughs, Shoshona Le, Katie Dillon, Peter Cook, Clinton R. Paden, Dhwani Batra, Krista Queen, Kristen Knipe, Dakota Howard, Yvette Unoarumhi, Darlene Wagner, Matthew Schmerer, Ben L. Rambo-Martin, Kristine Lacek, Sam Shepard, Alison Laufer Halpin, Dave Wentworth, Vivien Dugan, Suxiang Tong, Justin Lee |
| EPI_ISL_2787800, EPI_ISL_2787803 | NC State Laboratory of Public Health | Centers for Disease Control and Prevention Division of Viral Diseases, Pathogen Discovery | Mili Sheth, Sarah Nobles, Jasmine Padilla, Mark Burroughs, Shoshona Le, Katie Dillon, Peter Cook, Clinton R. Paden, Dhwani Batra, Krista Queen, Kristen Knipe, Dakota Howard, Yvette Unoarumhi, Darlene Wagner, Matthew Schmerer, Ben L. Rambo-Martin, Kristine Lacek, Sam Shepard, Alison Laufer Halpin, Dave Wentworth, Vivien Dugan, Suxiang Tong, Justin Lee |
| EPI_ISL_2787805, EPI_ISL_2787808, EPI_ISL_2787811, EPI_ISL_2787814, EPI_ISL_2787817, EPI_ISL_2787820, EPI_ISL_2787822 | MT Public Health Laboratory | Centers for Disease Control and Prevention Division of Viral Diseases, Pathogen Discovery | Mili Sheth, Sarah Nobles, Jasmine Padilla, Mark Burroughs, Shoshona Le, Katie Dillon, Peter Cook, Clinton R. Paden, Dhwani Batra, Krista Queen, Kristen Knipe, Dakota Howard, Yvette Unoarumhi, Darlene Wagner, Matthew Schmerer, Ben L. Rambo-Martin, Kristine Lacek, Sam Shepard, Alison Laufer Halpin, Dave Wentworth, Vivien Dugan, Suxiang Tong, Justin Lee |
| EPI_ISL_2787825, EPI_ISL_2787828, EPI_ISL_2787831, EPI_ISL_2787834, EPI_ISL_2787837, EPI_ISL_2787840, EPI_ISL_2787843, EPI_ISL_2787846, EPI_ISL_2787848 | NV-Southern Nevada Public Health Laboratory | Centers for Disease Control and Prevention Division of Viral Diseases, Pathogen Discovery | Mili Sheth, Sarah Nobles, Jasmine Padilla, Mark Burroughs, Shoshona Le, Katie Dillon, Peter Cook, Clinton R. Paden, Dhwani Batra, Krista Queen, Kristen Knipe, Dakota Howard, Yvette Unoarumhi, Darlene Wagner, Matthew Schmerer, Ben L. Rambo-Martin, Kristine Lacek, Sam Shepard, Alison Laufer Halpin, Dave Wentworth, Vivien Dugan, Suxiang Tong, Justin Lee |
| EPI_ISL_2787851, EPI_ISL_2787854, EPI_ISL_2787857, EPI_ISL_2787860, EPI_ISL_2787862, EPI_ISL_2787865, EPI_ISL_2787868, EPI_ISL_2787870, EPI_ISL_2787873, EPI_ISL_2787876, EPI_ISL_2787879, EPI_ISL_2787882, EPI_ISL_2787884, EPI_ISL_2787887, EPI_ISL_2787890, EPI_ISL_2787892, EPI_ISL_2787895, EPI_ISL_2787898, EPI_ISL_2787901, EPI_ISL_2787904, EPI_ISL_2787906, EPI_ISL_2787909, EPI_ISL_2787912, EPI_ISL_2787914, EPI_ISL_2787917, EPI_ISL_2787920, EPI_ISL_2787923, EPI_ISL_2787925, EPI_ISL_2787928 | | | |
| see above | NJ Public Health and Environmental Laboratories | Centers for Disease Control and Prevention Division of Viral Diseases, Pathogen Discovery | Mili Sheth, Sarah Nobles, Jasmine Padilla, Mark Burroughs, Shoshona Le, Katie Dillon, Peter Cook, Clinton R. Paden, Dhwani Batra, Krista Queen, Kristen Knipe, Dakota Howard, Yvette Unoarumhi, Darlene Wagner, Matthew Schmerer, Ben L. Rambo-Martin, Kristine Lacek, Sam Shepard, Alison Laufer Halpin, Dave Wentworth, Vivien Dugan, Suxiang Tong, Justin Lee |
| EPI_ISL_2787933, EPI_ISL_2787936, EPI_ISL_2787940, EPI_ISL_2787942, EPI_ISL_2787945, EPI_ISL_2787948, EPI_ISL_2787951, EPI_ISL_2787954, EPI_ISL_2787957, EPI_ISL_2787960 | KY State Public Health Lab | Centers for Disease Control and Prevention Division of Viral Diseases, Pathogen Discovery | Mili Sheth, Sarah Nobles, Jasmine Padilla, Mark Burroughs, Shoshona Le, Katie Dillon, Peter Cook, Clinton R. Paden, Dhwani Batra, Krista Queen, Kristen Knipe, Dakota Howard, Yvette Unoarumhi, Darlene Wagner, Matthew Schmerer, Ben L. Rambo-Martin, Kristine Lacek, Sam Shepard, Alison Laufer Halpin, Dave Wentworth, Vivien Dugan, Suxiang Tong, Justin Lee |
| EPI_ISL_2787962, EPI_ISL_2787965, EPI_ISL_2787967, EPI_ISL_2787970, EPI_ISL_2787973, EPI_ISL_2787976, EPI_ISL_2787979 | GA Department of Public Health Laboratory | Centers for Disease Control and Prevention Division of Viral Diseases, Pathogen Discovery | Mili Sheth, Sarah Nobles, Jasmine Padilla, Mark Burroughs, Shoshona Le, Katie Dillon, Peter Cook, Clinton R. Paden, Dhwani Batra, Krista Queen, Kristen Knipe, Dakota Howard, Yvette Unoarumhi, Darlene Wagner, Matthew Schmerer, Ben L. Rambo-Martin, Kristine Lacek, Sam Shepard, Alison Laufer Halpin, Dave Wentworth, Vivien Dugan, Suxiang Tong, Justin Lee |
| EPI_ISL_2787981, EPI_ISL_2787984, EPI_ISL_2787987, EPI_ISL_2787990, EPI_ISL_2787993, EPI_ISL_2787996, EPI_ISL_2787999, EPI_ISL_2788001, EPI_ISL_2788004 | RI State Health Laboratories | Centers for Disease Control and Prevention Division of Viral Diseases, Pathogen Discovery | Mili Sheth, Sarah Nobles, Jasmine Padilla, Mark Burroughs, Shoshona Le, Katie Dillon, Peter Cook, Clinton R. Paden, Dhwani Batra, Krista Queen, Kristen Knipe, Dakota Howard, Yvette Unoarumhi, Darlene Wagner, Matthew Schmerer, Ben L. Rambo-Martin, Kristine Lacek, Sam Shepard, Alison Laufer Halpin, Dave Wentworth, Vivien Dugan, Suxiang Tong, Justin Lee |
| EPI_ISL_2788007, EPI_ISL_2788010, EPI_ISL_2788013, EPI_ISL_2788015 | LA Office of Public Health Laboratories | Centers for Disease Control and Prevention Division of Viral Diseases, Pathogen Discovery | Mili Sheth, Sarah Nobles, Jasmine Padilla, Mark Burroughs, Shoshona Le, Katie Dillon, Peter Cook, Clinton R. Paden, Dhwani Batra, Krista Queen, Kristen Knipe, Dakota Howard, Yvette Unoarumhi, Darlene Wagner, Matthew Schmerer, Ben L. Rambo-Martin, Kristine Lacek, Sam Shepard, Alison Laufer Halpin, Dave Wentworth, Vivien Dugan, Suxiang Tong, Justin Lee |
| EPI_ISL_2788018, EPI_ISL_2788021, EPI_ISL_2788024, EPI_ISL_2788026, EPI_ISL_2788029, EPI_ISL_2788033, EPI_ISL_2788036, EPI_ISL_2788039, EPI_ISL_2788042 | WA State Department of Health | Centers for Disease Control and Prevention Division of Viral Diseases, Pathogen Discovery | Mili Sheth, Sarah Nobles, Jasmine Padilla, Mark Burroughs, Shoshona Le, Katie Dillon, Peter Cook, Clinton R. Paden, Dhwani Batra, Krista Queen, Kristen Knipe, Dakota Howard, Yvette Unoarumhi, Darlene Wagner, Matthew Schmerer, Ben L. Rambo-Martin, Kristine Lacek, Sam Shepard, Alison Laufer Halpin, Dave Wentworth, Vivien Dugan, Suxiang Tong, Justin Lee |
| EPI_ISL_2788045, EPI_ISL_2788048, EPI_ISL_2788051, EPI_ISL_2788053, EPI_ISL_2788056, EPI_ISL_2788059, EPI_ISL_2788061, EPI_ISL_2788064, EPI_ISL_2788070, EPI_ISL_2788073 | "AR Dept. of Health-PHL, Molecular Diagnostics" | Centers for Disease Control and Prevention Division of Viral Diseases, Pathogen Discovery | Mili Sheth, Sarah Nobles, Jasmine Padilla, Mark Burroughs, Shoshona Le, Katie Dillon, Peter Cook, Clinton R. Paden, Dhwani Batra, Krista Queen, Kristen Knipe, Dakota Howard, Yvette Unoarumhi, Darlene Wagner, Matthew Schmerer, Ben L. Rambo-Martin, Kristine Lacek, Sam Shepard, Alison Laufer Halpin, Dave Wentworth, Vivien Dugan, Suxiang Tong, Justin Lee |
| EPI_ISL_2788076, EPI_ISL_2788079, EPI_ISL_2788082, EPI_ISL_2788084, EPI_ISL_2788087, EPI_ISL_2788090, EPI_ISL_2788093 | VA-Division of Consolidated Laboratory Services | Centers for Disease Control and Prevention Division of Viral Diseases, Pathogen Discovery | Mili Sheth, Sarah Nobles, Jasmine Padilla, Mark Burroughs, Shoshona Le, Katie Dillon, Peter Cook, Clinton R. Paden, Dhwani Batra, Krista Queen, Kristen Knipe, Dakota Howard, Yvette Unoarumhi, Darlene Wagner, Matthew Schmerer, Ben L. Rambo-Martin, Kristine Lacek, Sam Shepard, Alison Laufer Halpin, Dave Wentworth, Vivien Dugan, Suxiang Tong, Justin Lee |
| EPI_ISL_2788097, EPI_ISL_2788100, EPI_ISL_2788103, EPI_ISL_2788106, EPI_ISL_2788109, EPI_ISL_2788112, | "CDPH, Viral and Rickettsial Disease Laboratory" | Centers for Disease Control and Prevention Division of Viral Diseases, Pathogen Discovery | Mili Sheth, Sarah Nobles, Jasmine Padilla, Mark Burroughs, Shoshona Le, Katie Dillon, Peter Cook, Clinton R. Paden, Dhwani Batra, Krista Queen, Kristen Knipe, Dakota Howard, Yvette Unoarumhi, Darlene Wagner, Matthew Schmerer, Ben L. Rambo-Martin, Kristine Lacek, Sam Shepard, Alison Laufer Halpin, Dave Wentworth, Vivien Dugan, Suxiang Tong, Justin Lee |

| | | | |
|---|---|---|---|
| EPI_ISL_2788115, EPI_ISL_2788118, EPI_ISL_2788121 | | | |
| EPI_ISL_2788124, EPI_ISL_2788127, EPI_ISL_2788130, EPI_ISL_2788133, EPI_ISL_2788135, EPI_ISL_2788138, EPI_ISL_2788141, EPI_ISL_2788144, EPI_ISL_2788147 | IL Dept. of Public Health Springfield Laboratory | Centers for Disease Control and Prevention Division of Viral Diseases, Pathogen Discovery | Mili Sheth, Sarah Nobles, Jasmine Padilla, Mark Burroughs, Shoshona Le, Katie Dillon, Peter Cook, Clinton R. Paden, Dhwani Batra, Krista Queen, Kristen Knipe, Dakota Howard, Yvette Unoarumhi, Darlene Wagner, Matthew Schmerer, Ben L. Rambo-Martin, Kristine Lacek, Sam Shepard, Alison Laufer Halpin, Dave Wentworth, Vivien Dugan, Suxiang Tong, Justin Lee |
| EPI_ISL_2788392, EPI_ISL_2788644, EPI_ISL_2788766 | Ohio Department of Health Laboratory | Ohio Department of Health Laboratory | Holmes, Jennifer; Eric Brandt, Keoni Omura, Glen McGillivary, Caitlin McDonnell, Jade Mowery, Stephanie Mccracken, Tyler Payne, Kirtana Ramadugu, Erica Leasure, Brent Lee, Kelsey Florek, Heather Blankenship, Allison Black, Quanta Brown, and Tammy Bannerman |
| EPI_ISL_2789879, EPI_ISL_2789880, EPI_ISL_2789881, EPI_ISL_2789882, EPI_ISL_2789884, EPI_ISL_2789885, EPI_ISL_2789886, EPI_ISL_2789887, EPI_ISL_2789888, EPI_ISL_2789889, EPI_ISL_2789899, EPI_ISL_2789900, EPI_ISL_2789901, EPI_ISL_2789902, EPI_ISL_2789903, EPI_ISL_2789904, EPI_ISL_2789906, EPI_ISL_2789907, EPI_ISL_2789908 | | | |
| see above | IN State Department of Health Laboratory Services | IN State Department of Health Laboratory Services | Cassandra Campion, Jamie Yeadon, Brian Pope, Lixia Liu, Kyle Brownlee, Melissa Hindenlang, Mark Glazier |
| EPI_ISL_2789909, EPI_ISL_2789910, EPI_ISL_2789911, EPI_ISL_2789912, EPI_ISL_2789913, EPI_ISL_2789914, EPI_ISL_2789915, EPI_ISL_2789916, EPI_ISL_2789917, EPI_ISL_2789919, EPI_ISL_2789920, EPI_ISL_2789921, EPI_ISL_2789922, EPI_ISL_2789923, EPI_ISL_2789924, EPI_ISL_2789925, EPI_ISL_2789926, EPI_ISL_2789927, EPI_ISL_2789929, EPI_ISL_2789930, EPI_ISL_2789931, EPI_ISL_2789932, EPI_ISL_2789933, EPI_ISL_2789934, EPI_ISL_2789935 | | | |
| see above | Kansas Health and Environmental Lab | Kansas Health and Environmental Lab | Katherine Wiggins, Mike Grose, Jonathan Barnell, Ben Olsen, and Phil Adam |
| EPI_ISL_2789937, EPI_ISL_2789939, EPI_ISL_2789940, EPI_ISL_2789941, EPI_ISL_2789942, EPI_ISL_2789943, EPI_ISL_2789945, EPI_ISL_2789946, EPI_ISL_2789947, EPI_ISL_2789948, EPI_ISL_2789949, EPI_ISL_2789950, EPI_ISL_2789951, EPI_ISL_2789953, EPI_ISL_2789954, EPI_ISL_2789955, EPI_ISL_2789957, EPI_ISL_2789958, EPI_ISL_2789959, EPI_ISL_2789960, EPI_ISL_2789961, EPI_ISL_2789962, EPI_ISL_2789963, EPI_ISL_2789964, EPI_ISL_2789965, EPI_ISL_2789966, EPI_ISL_2789968, EPI_ISL_2789969, EPI_ISL_2789970, EPI_ISL_2789971, EPI_ISL_2789972, EPI_ISL_2789973, EPI_ISL_2789974, EPI_ISL_2789975, EPI_ISL_2789976, EPI_ISL_2789977, EPI_ISL_2789978, EPI_ISL_2789979, EPI_ISL_2789980, EPI_ISL_2789981, EPI_ISL_2789982, EPI_ISL_2789983, EPI_ISL_2789984, EPI_ISL_2789986, EPI_ISL_2789987, EPI_ISL_2789988, EPI_ISL_2789989, EPI_ISL_2789990, EPI_ISL_2789991, EPI_ISL_2789992, EPI_ISL_2789993, EPI_ISL_2789994, EPI_ISL_2789995, EPI_ISL_2789996, EPI_ISL_2789997, EPI_ISL_2789998, EPI_ISL_2789999, EPI_ISL_2790000, EPI_ISL_2790001 | | | |
| see above | IVY3 Central Lab, Vanderbilt University Medical Center | Lauring Lab, University of Michigan, Department of Microbiology and Immunology | Valesano |
| EPI_ISL_2790034, EPI_ISL_2790035, EPI_ISL_2790036, EPI_ISL_2790037, EPI_ISL_2790038, EPI_ISL_2790039, EPI_ISL_2790040, EPI_ISL_2790041, EPI_ISL_2790042, EPI_ISL_2790043, EPI_ISL_2790044, EPI_ISL_2790045, EPI_ISL_2790046, EPI_ISL_2790047, EPI_ISL_2790049, EPI_ISL_2790050, EPI_ISL_2790051, EPI_ISL_2790052, EPI_ISL_2790053, EPI_ISL_2790054, EPI_ISL_2790055, EPI_ISL_2790056, EPI_ISL_2790057, EPI_ISL_2790058, EPI_ISL_2790059, EPI_ISL_2790060, EPI_ISL_2790061, EPI_ISL_2790062, EPI_ISL_2790063, EPI_ISL_2790064, EPI_ISL_2790065, EPI_ISL_2790066, EPI_ISL_2790067, EPI_ISL_2790068, EPI_ISL_2790069, EPI_ISL_2790070, EPI_ISL_2790071, EPI_ISL_2790072, EPI_ISL_2790073, EPI_ISL_2790074, EPI_ISL_2790075, EPI_ISL_2790076, EPI_ISL_2790077, EPI_ISL_2790078, EPI_ISL_2790079, EPI_ISL_2790080 | | | |
| see above | MD PHL | MD PHL | Maryland Department of Health Laboratories Administration |
| EPI_ISL_2790132, EPI_ISL_2790133, EPI_ISL_2790134, EPI_ISL_2790135, EPI_ISL_2790136 | Instituto Nacional de Cancerología | Centro de Investigación en Enfermedades Infecciosas (CIENI), Instituto Nacional de Enfermedades Respiratorias (INER) | Consorcio Mexicano de Vigilancia Genómica (CoViGen-Mex). Authors (in alphabetical order): Julio Elias Alvarado-Yaah, Carlos F. Arias, Santiago Ávila-Ríos, Eduardo Becerril-Vargas, Víctor Hugo Borja-Aburto, Celia Boukadida, Cristóbal Cháidez-Quiróz, Juan Bautista Chale-Dzul, Ricardo Ciria Merce, Andreu Comas-García, Célida Duque Molina, Julissa Enciso-Ibarra, José Antonio Enciso-Moreno, Gloria Elena Espinosa-Ayala, Fernando Fontove-Herrera, Daniel Fregoso-Rueda, Victor Eduardo Garcia-Arias, Alejandra Garcia-Gasca, Bruno Gomez-Gil, Jean Pierre González, Irvin González-López, Concepción Grajales-Muñiz, Ricardo Grande, Rosa María Gutierrez Rios, Alejandra Hernández-Terán, Alfredo Herrera-Estrella, Carla Ivón Herrera-Najera, Pavel Isa, Daniel Lira Morales, Susana Lopez, Antonio Loza Román, Brenda Irasema Maldonado-Meza, Bernardo Martínez-Miguel, José Arturo Martínez-Orozco, Célida Martínez- Rodríguez, Margarita Matías-Florentino, Fidencio Mejía-Nepomuceno, María Guadalupe de Jesús Mireles-Rivera, Gloria María Molina-Salinas, Hector Montoya-Fuentes, Mario Mújica-Sánchez, José Esteban Muñoz-Medina, José de Jesús Nuñez-Contreras, Alicia Ocaña-Mondragón, Luis Alberto Ochoa-Carrera, Hector Esteban Paz-Juárez, Marissa Pérez-García, Francisco Pulido, Helen Haydee Fernanda Ramírez-Plascencia, Jorge Salas-Hernández, Angel Gustavo Salas-Lais, Alejandro Sanchez-Flores, Clara Esperanza Santacruz-Tinoco, María Guadalupe Santiago-Mauricio, Selene Zárate, Nelly Sélem-Mojica, Blanca Taboada, Kathia Elizabeth Tapia-Diaz, Gloria Vazquez, Joel Armando Vázquez-Pérez. |
| EPI_ISL_2790175, EPI_ISL_2790176 | Instituto Nacional de Enfermedades Respiratorias (INER) | Centro de Investigación en Enfermedades Infecciosas (CIENI), Instituto Nacional de Enfermedades Respiratorias (INER) | Consorcio Mexicano de Vigilancia Genómica (CoViGen-Mex). Authors (in alphabetical order): Julio Elias Alvarado-Yaah, Carlos F. Arias, Santiago Ávila-Ríos, Eduardo Becerril-Vargas, Víctor Hugo Borja-Aburto, Celia Boukadida, Cristóbal Cháidez-Quiróz, Juan Bautista Chale-Dzul, Ricardo Ciria Merce, Andreu Comas-García, Célida Duque Molina, Julissa Enciso-Ibarra, José Antonio Enciso-Moreno, Gloria Elena Espinosa-Ayala, Fernando Fontove-Herrera, Daniel Fregoso-Rueda, Victor Eduardo Garcia-Arias, Alejandra Garcia-Gasca, Bruno Gomez-Gil, Jean Pierre González, Irvin González-López, Concepción Grajales-Muñiz, Ricardo Grande, Rosa María Gutierrez Rios, Alejandra Hernández-Terán, Alfredo Herrera-Estrella, Carla Ivón Herrera-Najera, Pavel Isa, Daniel Lira Morales, Susana Lopez, Antonio Loza Román, Brenda Irasema Maldonado-Meza, Bernardo Martínez-Miguel, José Arturo Martínez-Orozco, Célida Martínez- Rodríguez, Margarita Matías-Florentino, Fidencio Mejía-Nepomuceno, María Guadalupe de Jesús Mireles-Rivera, Gloria María Molina-Salinas, Hector Montoya-Fuentes, , Mario Mújica-Sánchez, José Esteban Muñoz-Medina, José de Jesús Nuñez-Contreras, Alicia Ocaña-Mondragón, Luis Alberto Ochoa-Carrera, Hector Esteban Paz-Juárez, Marissa Perez-Garcia, Francisco Pulido, Helen Haydee Fernanda Ramírez-Plascencia, Jorge Salas-Hernández, Angel Gustavo Salas-Lais, Alejandro Sanchez-Flores, Clara Esperanza Santacruz-Tinoco, María Guadalupe Santiago-Mauricio, Selene Zárate, Nelly Sélem-Mojica, Blanca Taboada, Gloria Vazquez, Joel Armando Vázquez-Pérez. |
| EPI_ISL_2790177, EPI_ISL_2790178, EPI_ISL_2790179 | Instituto Nacional de Medicina Genomica | Centro de Investigación en Enfermedades Infecciosas (CIENI), Instituto Nacional de Enfermedades Respiratorias (INER) | Matias-Florentino M, Perez-Garcia M, Hidalgo-Miranda A, Mendoza-Vargas A, Reyes-Grajeda JP, Cedro-Tanda A, Herrera-Montalvo LA, Reyes-Teran G, Avila-Rios S, Boukadida C |
| EPI_ISL_2790180 | Centracare Laboratory Services | Minnesota Department of Health, Public Health Laboratory | Alexandra Lorentz, Jacob Garfin, Matt Plumb, and Xiong Wang |
| EPI_ISL_2790181 | M Health Fairview | Minnesota Department of Health, Public Health Laboratory | Alexandra Lorentz, Jacob Garfin, Matt Plumb, and Xiong Wang |
| EPI_ISL_2790182 | Centracare Laboratory Services | Minnesota Department of Health, Public Health Laboratory | Alexandra Lorentz, Jacob Garfin, Matt Plumb, and Xiong Wang |
| EPI_ISL_2790183 | M Health Fairview | Minnesota Department of Health, Public Health Laboratory | Alexandra Lorentz, Jacob Garfin, Matt Plumb, and Xiong Wang |
| EPI_ISL_2790184 | Regions Hospital | Minnesota Department of Health, Public Health Laboratory | Alexandra Lorentz, Jacob Garfin, Matt Plumb, and Xiong Wang |
| EPI_ISL_2790185 | Minnesota Department of Health, Public Health Laboratory | Minnesota Department of Health, Public Health Laboratory | Alexandra Lorentz, Jacob Garfin, Matt Plumb, and Xiong Wang |
| EPI_ISL_2790186 | Centracare Laboratory Services | Minnesota Department of Health, Public Health Laboratory | Alexandra Lorentz, Jacob Garfin, Matt Plumb, and Xiong Wang |
| EPI_ISL_2790187 | Carris Health Rice Memorial Hospital | Minnesota Department of Health, Public Health Laboratory | Alexandra Lorentz, Jacob Garfin, Matt Plumb, and Xiong Wang |
| EPI_ISL_2790188, EPI_ISL_2790189 | M Health Fairview | Minnesota Department of Health, Public Health Laboratory | Alexandra Lorentz, Jacob Garfin, Matt Plumb, and Xiong Wang |
| EPI_ISL_2790190 | Regions Hospital | Minnesota Department of Health, Public Health Laboratory | Alexandra Lorentz, Jacob Garfin, Matt Plumb, and Xiong Wang |
| EPI_ISL_2790191, EPI_ISL_2790192, EPI_ISL_2790193 | M Health Fairview | Minnesota Department of Health, Public Health Laboratory | Alexandra Lorentz, Jacob Garfin, Matt Plumb, and Xiong Wang |
| EPI_ISL_2790194 | Maple Grove Hospital | Minnesota Department of Health, Public Health Laboratory | Alexandra Lorentz, Jacob Garfin, Matt Plumb, and Xiong Wang |
| EPI_ISL_2790195, EPI_ISL_2790196, EPI_ISL_2790197, EPI_ISL_2790198, EPI_ISL_2790199, EPI_ISL_2790200, EPI_ISL_2790201 | M Health Fairview | Minnesota Department of Health, Public Health Laboratory | Alexandra Lorentz, Jacob Garfin, Matt Plumb, and Xiong Wang |
| EPI_ISL_2790207 | Minnesota Department of Health, Public Health Laboratory | Minnesota Department of Health, Public Health Laboratory | Alexandra Lorentz, Jacob Garfin, Matt Plumb, and Xiong Wang |
| EPI_ISL_2790208 | M Health Fairview | Minnesota Department of Health, Public Health Laboratory | Alexandra Lorentz, Jacob Garfin, Matt Plumb, and Xiong Wang |
| EPI_ISL_2790209, EPI_ISL_2790210, EPI_ISL_2790211, EPI_ISL_2790212, EPI_ISL_2790213, EPI_ISL_2790214, EPI_ISL_2790215, EPI_ISL_2790216, EPI_ISL_2790217 | InDRE | Instituto Nacional de Medicina Genomica | Hidalgo-Miranda A, Mendoza-Vargas A, Reyes-Grajeda JP, Cedro-Tanda A, Cisneros-Villanueva M, Orjuela-Rodríguez M, Gisela Barrera-Badillo, Irma Lopez-Martinez, Jose Ernesto Ramirez González, Gonzalez-Barrera D, Rangel-DeLeon D, Munguia-Garza P, Garcia-Cardenas FJ, Gonzalez-Woge MA, Herrera-Montalvo LA. |
| EPI_ISL_2790218, EPI_ISL_2790219, EPI_ISL_2790220, EPI_ISL_2790221, EPI_ISL_2790222, EPI_ISL_2790223, EPI_ISL_2790224, EPI_ISL_2790225, EPI_ISL_2790226, EPI_ISL_2790227, EPI_ISL_2790228, EPI_ISL_2790229, EPI_ISL_2790230, EPI_ISL_2790231 | | | |
| see above | Salud Digna | Instituto Nacional de Medicina Genomica | Hidalgo-Miranda A, Cedro-Tanda A, Cisneros-Villanueva M, Orjuela-Rodríguez M, Mendoza-Vargas A, Reyes-Grajeda JP, Abraham Campos-Romero, Moreno-Camacho José Luis, Rodriguez-Gallegos Jorge, Luna-Ruiz Marco, Gonzalez-Barrera D, Rangel-DeLeon D, Munguia-Garza P, Ramirez-Vega O, Escobar-Arrazola, M, Herrera-Montalvo LA. |
| EPI_ISL_2790306, EPI_ISL_2790307, EPI_ISL_2790308 | Washington State Department of Health Public Health Laboratories | Washington State Department of Health Public Health Laboratories | Drew MacKellar, Philip Dykema, Denny Russell, Joenice Gonzalez, Hannah Gray, Geoff Melly, Vanessa De Los Santos, Darren Lucas, JohnAric Peterson, Avi Singh, Rebecca Cao |

| | | | |
|---|---|---|---|
| EPI_ISL_2790436, EPI_ISL_2790437, EPI_ISL_2790438, EPI_ISL_2790439, EPI_ISL_2790440, EPI_ISL_2790441, EPI_ISL_2790442, EPI_ISL_2790443, EPI_ISL_2790444, EPI_ISL_2790445, EPI_ISL_2790446, EPI_ISL_2790447, EPI_ISL_2790448, EPI_ISL_2790449, EPI_ISL_2790450, EPI_ISL_2790451, EPI_ISL_2790452, EPI_ISL_2790453, EPI_ISL_2790454, EPI_ISL_2790455, EPI_ISL_2790456, EPI_ISL_2790457, EPI_ISL_2790458, EPI_ISL_2790459, EPI_ISL_2790460, EPI_ISL_2790461, EPI_ISL_2790463, EPI_ISL_2790464, EPI_ISL_2790465 | | | |
| see above | UW Virology Lab | UW Virology Lab | Pavitra Roychoudhury, Hong Xie, Lasata Shrestha, Shah Mohamed Bakhash, Margaret Mills, Noah Baker, Sean Ellis, Meei-Li Huang, Keith R Jerome, Alexander Greninger |
| EPI_ISL_2790466, EPI_ISL_2790467, EPI_ISL_2790468, EPI_ISL_2790469, EPI_ISL_2790470, EPI_ISL_2790471, EPI_ISL_2790472, EPI_ISL_2790473, EPI_ISL_2790474, EPI_ISL_2790475, EPI_ISL_2790476, EPI_ISL_2790477, EPI_ISL_2790478, EPI_ISL_2790479, EPI_ISL_2790480, EPI_ISL_2790481, EPI_ISL_2790482, EPI_ISL_2790483, EPI_ISL_2790484, EPI_ISL_2790485, EPI_ISL_2790486, EPI_ISL_2790487, EPI_ISL_2790488, EPI_ISL_2790489, EPI_ISL_2790490, EPI_ISL_2790491, EPI_ISL_2790492, EPI_ISL_2790493, EPI_ISL_2790494, EPI_ISL_2790495, EPI_ISL_2790496, EPI_ISL_2790497, EPI_ISL_2790498, EPI_ISL_2790499, EPI_ISL_2790500, EPI_ISL_2790501, EPI_ISL_2790502, EPI_ISL_2790503, EPI_ISL_2790504, EPI_ISL_2790505, EPI_ISL_2790506, EPI_ISL_2790507, EPI_ISL_2790508, EPI_ISL_2790509, EPI_ISL_2790510, EPI_ISL_2790511, EPI_ISL_2790512, EPI_ISL_2790513, EPI_ISL_2790514, EPI_ISL_2790515, EPI_ISL_2790516, EPI_ISL_2790517, EPI_ISL_2790518, EPI_ISL_2790519, EPI_ISL_2790520, EPI_ISL_2790521, EPI_ISL_2790522, EPI_ISL_2790523, EPI_ISL_2790524, EPI_ISL_2790525, EPI_ISL_2790526, EPI_ISL_2790527, EPI_ISL_2790528, EPI_ISL_2790529, EPI_ISL_2790530, EPI_ISL_2790531, EPI_ISL_2790532, EPI_ISL_2790533, EPI_ISL_2790534, EPI_ISL_2790535, EPI_ISL_2790536, EPI_ISL_2790537, EPI_ISL_2790538, EPI_ISL_2790539, EPI_ISL_2790541, EPI_ISL_2790542, EPI_ISL_2790543, EPI_ISL_2790544 | | | |
| see above | Tampa General Hospital Esoteric Lab | Tampa General Hospital Esoteric Research & Development Lab | Jon Faughn, Deanna Becker, Dominic Uy, Jorge Mecias-Frias, Elaine Vendrone, Vicki Healer, Amorce Lima, Suzane Silbert |
| EPI_ISL_2790554, EPI_ISL_2790555, EPI_ISL_2790556, EPI_ISL_2790557, EPI_ISL_2790558, EPI_ISL_2790559, EPI_ISL_2790560, EPI_ISL_2790561, EPI_ISL_2790562, EPI_ISL_2790563, EPI_ISL_2790564, EPI_ISL_2790565, EPI_ISL_2790566, EPI_ISL_2790567, EPI_ISL_2790568, EPI_ISL_2790569, EPI_ISL_2790570, EPI_ISL_2790571, EPI_ISL_2790572, EPI_ISL_2790573, EPI_ISL_2790574, EPI_ISL_2790575, EPI_ISL_2790576, EPI_ISL_2790577, EPI_ISL_2790578, EPI_ISL_2790579, EPI_ISL_2790580, EPI_ISL_2790581, EPI_ISL_2790582, EPI_ISL_2790583, EPI_ISL_2790584, EPI_ISL_2790585, EPI_ISL_2790586, EPI_ISL_2790587, EPI_ISL_2790588, EPI_ISL_2790589, EPI_ISL_2790590, EPI_ISL_2790591, EPI_ISL_2790592, EPI_ISL_2790593, EPI_ISL_2790594, EPI_ISL_2790595, EPI_ISL_2790596, EPI_ISL_2790597, EPI_ISL_2790598, EPI_ISL_2790599, EPI_ISL_2790600, EPI_ISL_2790601, EPI_ISL_2790602, EPI_ISL_2790603, EPI_ISL_2790604, EPI_ISL_2790605, EPI_ISL_2790606, EPI_ISL_2790607, EPI_ISL_2790608, EPI_ISL_2790609, EPI_ISL_2790610, EPI_ISL_2790611, EPI_ISL_2790612, EPI_ISL_2790613, EPI_ISL_2790614, EPI_ISL_2790615, EPI_ISL_2790616, EPI_ISL_2790617 | | | |
| see above | UW Virology Lab | UW Virology Lab | Pavitra Roychoudhury, Hong Xie, Lasata Shrestha, Shah Mohamed Bakhash, Margaret Mills, Noah Baker, Sean Ellis, Meei-Li Huang, Keith R Jerome, Alexander Greninger |
| EPI_ISL_2790618, EPI_ISL_2790622, EPI_ISL_2790623, EPI_ISL_2790624, EPI_ISL_2790625, EPI_ISL_2790626, EPI_ISL_2790627, EPI_ISL_2790630, EPI_ISL_2790631, EPI_ISL_2790632, EPI_ISL_2790633, EPI_ISL_2790634, EPI_ISL_2790635, EPI_ISL_2790636, EPI_ISL_2790639, EPI_ISL_2790640, EPI_ISL_2790642, EPI_ISL_2790643, EPI_ISL_2790644, EPI_ISL_2790645, EPI_ISL_2790646, EPI_ISL_2790647, EPI_ISL_2790648, EPI_ISL_2790649, EPI_ISL_2790650, EPI_ISL_2790652, EPI_ISL_2790654, EPI_ISL_2790655, EPI_ISL_2790656, EPI_ISL_2790657, EPI_ISL_2790660, EPI_ISL_2790661, EPI_ISL_2790662, EPI_ISL_2790663, EPI_ISL_2790664, EPI_ISL_2790665, EPI_ISL_2790666, EPI_ISL_2790667, EPI_ISL_2790668, EPI_ISL_2790670, EPI_ISL_2790671, EPI_ISL_2790673, EPI_ISL_2790674, EPI_ISL_2790675, EPI_ISL_2790676, EPI_ISL_2790677, EPI_ISL_2790679, EPI_ISL_2790680, EPI_ISL_2790681, EPI_ISL_2790682, EPI_ISL_2790683, EPI_ISL_2790684, EPI_ISL_2790685, EPI_ISL_2790688, EPI_ISL_2790689, EPI_ISL_2790691, EPI_ISL_2790692, EPI_ISL_2790693 | | | |
| see above | Colorado Department of Public Health and Environment | Colorado Department of Public Health and Environment | Laura Bankers, Molly C. Hetherington-Rauth, Diana Ir, Alexandria Rossheim, Michael Martin, Mandy Waters, Shannon R. Matzinger, Sarah Elizabeth Totten, Emily A. Travanty |
| EPI_ISL_2790697, EPI_ISL_2790698, EPI_ISL_2790699, EPI_ISL_2790700, EPI_ISL_2790701, EPI_ISL_2790702, EPI_ISL_2790703, EPI_ISL_2790704, EPI_ISL_2790705, EPI_ISL_2790706, EPI_ISL_2790707, EPI_ISL_2790708, EPI_ISL_2790709, EPI_ISL_2790710, EPI_ISL_2790711 | | | |
| see above | Emory Molecular Diagnostics Laboratory, Emory Healthcare | Genomics and Discovery, Respiratory Viruses Branch, Division of Viral Diseases, Centers for Disease Control and Prevention | Anna Kelleher, Ying Tao, Yan Li, Jing Zhang, Brian Lynch, Krista Queen, Anna Uehara, Peter Cook, Han Jia Justin Ng, Rachel Marine, Clinton R. Paden, Dhwani Batra, Haibin Wang, Tara Coalter, Jasmine Padilla, Morgan Davis, Mili Sheth, Sarah Nobles, Mark Burroughs, Justin Lee, Adam Retchless, Suxiang Tong, Ahmed Babiker, Anne Piantadosi |
| EPI_ISL_2790732, EPI_ISL_2790733 | Monterey County Public Health Laboratory | Monterey County Public Health Laboratory | Monterey County Public Health Laboratory |
| EPI_ISL_2790734, EPI_ISL_2790735, EPI_ISL_2790736, EPI_ISL_2790737, EPI_ISL_2790738, EPI_ISL_2790739, EPI_ISL_2790740, EPI_ISL_2790741, EPI_ISL_2790742, EPI_ISL_2790743, EPI_ISL_2790744, EPI_ISL_2790745, EPI_ISL_2790746, EPI_ISL_2790747, EPI_ISL_2790748, EPI_ISL_2790749, EPI_ISL_2790750, EPI_ISL_2790751, EPI_ISL_2790752, EPI_ISL_2790753, EPI_ISL_2790754, EPI_ISL_2790755, EPI_ISL_2790756, EPI_ISL_2790757, EPI_ISL_2790758, EPI_ISL_2790759, EPI_ISL_2790760, EPI_ISL_2790761 | | | |
| see above | Oregon State Public Health Laboratory | Oregon State Public Health Laboratory | Rafia Razzaque, Eugene Yeboah, Vanda Makris, Laura Tsaknaridis, John Fontana and Shane Sevey |
| EPI_ISL_2790762, EPI_ISL_2790763, EPI_ISL_2790764, EPI_ISL_2790765, EPI_ISL_2790766, EPI_ISL_2790767, EPI_ISL_2790768, EPI_ISL_2790769, EPI_ISL_2790770, EPI_ISL_2790771, EPI_ISL_2790772, EPI_ISL_2790773, EPI_ISL_2790774, EPI_ISL_2790775, EPI_ISL_2790776, EPI_ISL_2790777, EPI_ISL_2790778, EPI_ISL_2790779, EPI_ISL_2790780, EPI_ISL_2790781, EPI_ISL_2790782, EPI_ISL_2790783, EPI_ISL_2790784, EPI_ISL_2790785, EPI_ISL_2790786, EPI_ISL_2790787, EPI_ISL_2790788, EPI_ISL_2790789, EPI_ISL_2790790, EPI_ISL_2790791, EPI_ISL_2790792, EPI_ISL_2790793, EPI_ISL_2790794, EPI_ISL_2790795, EPI_ISL_2790796, EPI_ISL_2790797, EPI_ISL_2790798, EPI_ISL_2790799 | | | |
| see above | Alaska State Virology Laboratory | Alaska State Virology Laboratory | Stephanie DeRonde, Elva House, Jacob Zidek, Lisa Smith, Ph.D., Jack Chen, Ph.D. |
| EPI_ISL_2791036, EPI_ISL_2791037, EPI_ISL_2791038, EPI_ISL_2791039, EPI_ISL_2791040, EPI_ISL_2791041, EPI_ISL_2791042, EPI_ISL_2791043, EPI_ISL_2791044, EPI_ISL_2791045, EPI_ISL_2791046, EPI_ISL_2791047, EPI_ISL_2791048, EPI_ISL_2791049, EPI_ISL_2791050, EPI_ISL_2791051, EPI_ISL_2791052, EPI_ISL_2791053, EPI_ISL_2791054, EPI_ISL_2791055, EPI_ISL_2791056, EPI_ISL_2791057, EPI_ISL_2791058, EPI_ISL_2791059, EPI_ISL_2791060, EPI_ISL_2791061, EPI_ISL_2791062, EPI_ISL_2791063, EPI_ISL_2791064, EPI_ISL_2791065, EPI_ISL_2791066, EPI_ISL_2791067, EPI_ISL_2791068, EPI_ISL_2791069, EPI_ISL_2791070, EPI_ISL_2791071, EPI_ISL_2791072, EPI_ISL_2791073, EPI_ISL_2791074, EPI_ISL_2791075, EPI_ISL_2791076, EPI_ISL_2791077, EPI_ISL_2791078, EPI_ISL_2791079, EPI_ISL_2791080, EPI_ISL_2791081, EPI_ISL_2791082, EPI_ISL_2791083, EPI_ISL_2791084, EPI_ISL_2791085, EPI_ISL_2791086, EPI_ISL_2791087, EPI_ISL_2791088, EPI_ISL_2791089, EPI_ISL_2791090, EPI_ISL_2791091, EPI_ISL_2791092, EPI_ISL_2791093, EPI_ISL_2791094, EPI_ISL_2791095, EPI_ISL_2791096, EPI_ISL_2791097, EPI_ISL_2791098, EPI_ISL_2791099, EPI_ISL_2791100, EPI_ISL_2791101, EPI_ISL_2791102, EPI_ISL_2791103, EPI_ISL_2791104, EPI_ISL_2791105, EPI_ISL_2791106, EPI_ISL_2791107, EPI_ISL_2791108, EPI_ISL_2791109, EPI_ISL_2791110, EPI_ISL_2791111, EPI_ISL_2791112, EPI_ISL_2791113, EPI_ISL_2791114, EPI_ISL_2791115, EPI_ISL_2791116, EPI_ISL_2791117, EPI_ISL_2791118, EPI_ISL_2791119, EPI_ISL_2791120, EPI_ISL_2791121, EPI_ISL_2791122, EPI_ISL_2791123, EPI_ISL_2791124, EPI_ISL_2791125, EPI_ISL_2791126, EPI_ISL_2791127, EPI_ISL_2791128, EPI_ISL_2791129, EPI_ISL_2791130, EPI_ISL_2791131, EPI_ISL_2791132, EPI_ISL_2791133, EPI_ISL_2791134, EPI_ISL_2791135, EPI_ISL_2791136, EPI_ISL_2791137, EPI_ISL_2791138, EPI_ISL_2791139, EPI_ISL_2791140, EPI_ISL_2791141, EPI_ISL_2791142, EPI_ISL_2791143, EPI_ISL_2791144, EPI_ISL_2791145, EPI_ISL_2791146, EPI_ISL_2791147, EPI_ISL_2791148, EPI_ISL_2791149, EPI_ISL_2791150, EPI_ISL_2791151, EPI_ISL_2791152, EPI_ISL_2791153, EPI_ISL_2791154, EPI_ISL_2791155, EPI_ISL_2791156, EPI_ISL_2791157, EPI_ISL_2791158, EPI_ISL_2791159, EPI_ISL_2791160, EPI_ISL_2791161, EPI_ISL_2791162, EPI_ISL_2791163, EPI_ISL_2791164, EPI_ISL_2791165, EPI_ISL_2791166, EPI_ISL_2791167, EPI_ISL_2791168, EPI_ISL_2791169, EPI_ISL_2791170, EPI_ISL_2791171, EPI_ISL_2791172, EPI_ISL_2791173, EPI_ISL_2791174, EPI_ISL_2791175, EPI_ISL_2791176, EPI_ISL_2791177, EPI_ISL_2791178, EPI_ISL_2791179, EPI_ISL_2791180, EPI_ISL_2791181, EPI_ISL_2791182, EPI_ISL_2791183, EPI_ISL_2791184, EPI_ISL_2791185, EPI_ISL_2791186, EPI_ISL_2791187, EPI_ISL_2791188, EPI_ISL_2791189, EPI_ISL_2791190, EPI_ISL_2791191, EPI_ISL_2791192, EPI_ISL_2791193, EPI_ISL_2791194, EPI_ISL_2791195, EPI_ISL_2791196, EPI_ISL_2791197, EPI_ISL_2791198, EPI_ISL_2791199, EPI_ISL_2791200, EPI_ISL_2791201, EPI_ISL_2791202, EPI_ISL_2791203, EPI_ISL_2791204, EPI_ISL_2791205, EPI_ISL_2791206, EPI_ISL_2791207, EPI_ISL_2791208, EPI_ISL_2791209, EPI_ISL_2791210, EPI_ISL_2791211, EPI_ISL_2791212, EPI_ISL_2791213, EPI_ISL_2791214, EPI_ISL_2791215, EPI_ISL_2791216, EPI_ISL_2791217, EPI_ISL_2791218, EPI_ISL_2791219, EPI_ISL_2791220, EPI_ISL_2791221, EPI_ISL_2791222, EPI_ISL_2791223 | | | |
| see above | UW Virology Lab | UW Virology Lab | Pavitra Roychoudhury, Hong Xie, Lasata Shrestha, Shah Mohamed Bakhash, Margaret Mills, Noah Baker, Sean Ellis, Meei-Li Huang, Keith R Jerome, Alexander Greninger |
| EPI_ISL_2791232, EPI_ISL_2791251 | Pathology and Laboratory Medicine Institute, Cleveland Clinic, Ohio, USA | Pathology and Laboratory Medicine Institute, Cleveland Clinic, Ohio, USA | Concetta Peck, Jennifer Starbuck, Joy Nakitandwe, Kristen McDonnell, David Plunkett, Zheng Jin Tu, Jay Brock, Yu-Wei Cheng, Gary Procop, Daniel Rhoads, Daniel H. Farkas, David Bosler |
| EPI_ISL_2791380, EPI_ISL_2791381, EPI_ISL_2791382, EPI_ISL_2791383, EPI_ISL_2791384, EPI_ISL_2791386, EPI_ISL_2791387, EPI_ISL_2791388, EPI_ISL_2791389, EPI_ISL_2791390, EPI_ISL_2791391, EPI_ISL_2791392, EPI_ISL_2791393, EPI_ISL_2791394, EPI_ISL_2791395, EPI_ISL_2791396, EPI_ISL_2791397, EPI_ISL_2791398, EPI_ISL_2791399, EPI_ISL_2791400, EPI_ISL_2791401, EPI_ISL_2791402, EPI_ISL_2791403, EPI_ISL_2791404, EPI_ISL_2791405, EPI_ISL_2791406, EPI_ISL_2791407, EPI_ISL_2791408, EPI_ISL_2791409, EPI_ISL_2791410, EPI_ISL_2791411, EPI_ISL_2791412, EPI_ISL_2791413, EPI_ISL_2791414, EPI_ISL_2791415, EPI_ISL_2791416, EPI_ISL_2791417, EPI_ISL_2791418, EPI_ISL_2791419, EPI_ISL_2791420, EPI_ISL_2791421, EPI_ISL_2791422, EPI_ISL_2791423, EPI_ISL_2791424, EPI_ISL_2791425, EPI_ISL_2791426, EPI_ISL_2791427, EPI_ISL_2791428, EPI_ISL_2791431, EPI_ISL_2791432, EPI_ISL_2791433, EPI_ISL_2791435, EPI_ISL_2791436, EPI_ISL_2791437, EPI_ISL_2791438, EPI_ISL_2791439, EPI_ISL_2791440, EPI_ISL_2791441, EPI_ISL_2791442, EPI_ISL_2791443, EPI_ISL_2791446, EPI_ISL_2791447, EPI_ISL_2791448, EPI_ISL_2791449, EPI_ISL_2791450, EPI_ISL_2791451, EPI_ISL_2791452, EPI_ISL_2791453, EPI_ISL_2791454 | | | |
| see above | UMGC | UMGC | Jeffrey Good, Nathanael Herrera, Gregg Thomas, Tim Thurman, David Xing |
| EPI_ISL_2791456, EPI_ISL_2791457, EPI_ISL_2791458, EPI_ISL_2791459, EPI_ISL_2791460, EPI_ISL_2791461, EPI_ISL_2791462, EPI_ISL_2791463, EPI_ISL_2791464, EPI_ISL_2791465, EPI_ISL_2791466, EPI_ISL_2791467, EPI_ISL_2791468, EPI_ISL_2791469, EPI_ISL_2791470, EPI_ISL_2791471, EPI_ISL_2791472, EPI_ISL_2791473, EPI_ISL_2791474, EPI_ISL_2791475, EPI_ISL_2791476, EPI_ISL_2791477, EPI_ISL_2791478, EPI_ISL_2791479, EPI_ISL_2791480, EPI_ISL_2791481, EPI_ISL_2791482, EPI_ISL_2791483, EPI_ISL_2791484, EPI_ISL_2791485, EPI_ISL_2791486, EPI_ISL_2791487 | | | |
| see above | Alaska State Virology Laboratory | Alaska State Virology Laboratory | Stephanie DeRonde, Elva House, Jacob Zidek, Lisa Smith, Ph.D., Jack Chen, Ph.D. |
| EPI_ISL_2795658 | LESP Hidalgo | LESP Hidalgo GENE2LIFE | HERRERA-GOMEZ CONNIE ALEJANDRA; CASTILLO-ORTEGA LAURA SOFIA; GARCIA-REYES MELITO; VIGUERAS-MORALES YAJAIRA SULIM; GARCIA-GONZALEZ OCTAVIO; VILLAFUENTES-TELLEZ HECTOR ALBERTO |
| EPI_ISL_2796532, EPI_ISL_2796533, EPI_ISL_2796534, EPI_ISL_2796535, EPI_ISL_2796536, EPI_ISL_2796537, EPI_ISL_2796538, EPI_ISL_2796539, EPI_ISL_2796540, EPI_ISL_2796541, EPI_ISL_2796542, EPI_ISL_2796543, EPI_ISL_2796544, EPI_ISL_2796545, EPI_ISL_2796546, EPI_ISL_2796547, EPI_ISL_2796548, EPI_ISL_2796549, EPI_ISL_2796550, EPI_ISL_2796551, EPI_ISL_2796552, EPI_ISL_2796553, EPI_ISL_2796555, EPI_ISL_2796556, EPI_ISL_2796557, EPI_ISL_2796559, EPI_ISL_2796560, EPI_ISL_2796561, EPI_ISL_2796564, EPI_ISL_2796565, EPI_ISL_2796566, EPI_ISL_2796567, EPI_ISL_2796568, EPI_ISL_2796569, EPI_ISL_2796570, EPI_ISL_2796572, EPI_ISL_2796573, EPI_ISL_2796574, EPI_ISL_2796575, EPI_ISL_2796576, EPI_ISL_2796577, EPI_ISL_2796578, EPI_ISL_2796579, EPI_ISL_2796580, EPI_ISL_2796581, EPI_ISL_2796582, EPI_ISL_2796583, EPI_ISL_2796584, EPI_ISL_2796585, EPI_ISL_2796586, EPI_ISL_2796587, EPI_ISL_2796588, EPI_ISL_2796589, EPI_ISL_2796590, EPI_ISL_2796591, EPI_ISL_2796592, EPI_ISL_2796593, EPI_ISL_2796594, EPI_ISL_2796595, EPI_ISL_2796596, EPI_ISL_2796597, EPI_ISL_2796598, EPI_ISL_2796599, EPI_ISL_2796600, EPI_ISL_2796601, EPI_ISL_2796602, EPI_ISL_2796603, EPI_ISL_2796606, EPI_ISL_2796608, EPI_ISL_2796609, EPI_ISL_2796610, EPI_ISL_2796611, EPI_ISL_2796613, EPI_ISL_2796614, EPI_ISL_2796615, EPI_ISL_2796616, EPI_ISL_2796617, EPI_ISL_2796618, EPI_ISL_2796619, EPI_ISL_2796620, EPI_ISL_2796621, EPI_ISL_2796623, EPI_ISL_2796624, EPI_ISL_2796625, EPI_ISL_2796626, EPI_ISL_2796627, EPI_ISL_2796628, EPI_ISL_2796629, EPI_ISL_2796630, EPI_ISL_2796631, EPI_ISL_2796632, EPI_ISL_2796633, EPI_ISL_2796634, EPI_ISL_2796636, EPI_ISL_2796637, EPI_ISL_2796638, EPI_ISL_2796639, EPI_ISL_2796640, EPI_ISL_2796643, EPI_ISL_2796644, EPI_ISL_2796645, EPI_ISL_2796646, EPI_ISL_2796647, EPI_ISL_2796648, EPI_ISL_2796649, EPI_ISL_2796650, EPI_ISL_2796651, EPI_ISL_2796652, EPI_ISL_2796653, EPI_ISL_2796655, EPI_ISL_2796656, EPI_ISL_2796657, EPI_ISL_2796658, EPI_ISL_2796659, EPI_ISL_2796660, EPI_ISL_2796661, EPI_ISL_2796662, EPI_ISL_2796664, EPI_ISL_2796665, EPI_ISL_2796666, EPI_ISL_2796667, EPI_ISL_2796668, EPI_ISL_2796669, EPI_ISL_2796670, EPI_ISL_2796671, EPI_ISL_2796672, EPI_ISL_2796673, EPI_ISL_2796674, EPI_ISL_2796675, EPI_ISL_2796677, EPI_ISL_2796678, EPI_ISL_2796679, EPI_ISL_2796680, EPI_ISL_2796681, EPI_ISL_2796682, EPI_ISL_2796683, EPI_ISL_2796684, EPI_ISL_2796685, EPI_ISL_2796686, EPI_ISL_2796687, EPI_ISL_2796688, EPI_ISL_2796689, EPI_ISL_2796690, EPI_ISL_2796691, EPI_ISL_2796692, EPI_ISL_2796693, EPI_ISL_2796695, EPI_ISL_2796696, EPI_ISL_2796697, EPI_ISL_2796698, EPI_ISL_2796699, EPI_ISL_2796700, EPI_ISL_2796701, EPI_ISL_2796702, EPI_ISL_2796703, EPI_ISL_2796704, EPI_ISL_2796705, EPI_ISL_2796706, EPI_ISL_2796707, EPI_ISL_2796708, EPI_ISL_2796712, EPI_ISL_2796713, EPI_ISL_2796714, EPI_ISL_2796715, EPI_ISL_2796716, EPI_ISL_2796717, EPI_ISL_2796718, EPI_ISL_2796719, EPI_ISL_2796720, EPI_ISL_2796721, EPI_ISL_2796722, EPI_ISL_2796723, EPI_ISL_2796724, EPI_ISL_2796725, EPI_ISL_2796726, EPI_ISL_2796727, EPI_ISL_2796728, EPI_ISL_2796729, EPI_ISL_2796730, EPI_ISL_2796731, EPI_ISL_2796732, EPI_ISL_2796733, EPI_ISL_2796734, EPI_ISL_2796735, EPI_ISL_2796736, EPI_ISL_2796737, EPI_ISL_2796738, EPI_ISL_2796739, EPI_ISL_2796740, EPI_ISL_2796741, EPI_ISL_2796742, EPI_ISL_2796743, EPI_ISL_2796744, EPI_ISL_2796745, EPI_ISL_2796746, EPI_ISL_2796747, EPI_ISL_2796748, EPI_ISL_2796749, EPI_ISL_2796750, EPI_ISL_2796751, EPI_ISL_2796752, EPI_ISL_2796754, EPI_ISL_2796755, EPI_ISL_2796756, EPI_ISL_2796757, EPI_ISL_2796758, EPI_ISL_2796759, EPI_ISL_2796760, EPI_ISL_2796761, EPI_ISL_2796762, EPI_ISL_2796763, EPI_ISL_2796764, EPI_ISL_2796765, EPI_ISL_2796766, EPI_ISL_2796767, EPI_ISL_2796768, EPI_ISL_2796769, EPI_ISL_2796770, EPI_ISL_2796771, EPI_ISL_2796772, EPI_ISL_2796773, EPI_ISL_2796774, EPI_ISL_2796775, EPI_ISL_2796776, EPI_ISL_2796777, EPI_ISL_2796779, EPI_ISL_2796780, EPI_ISL_2796781, EPI_ISL_2796782, EPI_ISL_2796783, EPI_ISL_2796784, EPI_ISL_2796785, EPI_ISL_2796786, EPI_ISL_2796787, EPI_ISL_2796788, EPI_ISL_2796789, EPI_ISL_2796790, EPI_ISL_2796791, EPI_ISL_2796792, EPI_ISL_2796793, EPI_ISL_2796794, EPI_ISL_2796795, EPI_ISL_2796796, EPI_ISL_2796797, EPI_ISL_2796798, EPI_ISL_2796799, EPI_ISL_2796800, EPI_ISL_2796801, EPI_ISL_2796802, EPI_ISL_2796803, EPI_ISL_2796804, EPI_ISL_2796805, EPI_ISL_2796806, EPI_ISL_2796807, EPI_ISL_2796808, EPI_ISL_2796809, EPI_ISL_2796810, EPI_ISL_2796811, EPI_ISL_2796812, EPI_ISL_2796813, EPI_ISL_2796814, EPI_ISL_2796816, EPI_ISL_2796817, EPI_ISL_2796818, EPI_ISL_2796819, EPI_ISL_2796820, EPI_ISL_2796821, EPI_ISL_2796822, EPI_ISL_2796823, EPI_ISL_2796824, EPI_ISL_2796825, EPI_ISL_2796827, EPI_ISL_2796828, EPI_ISL_2796829, EPI_ISL_2796830, EPI_ISL_2796831, EPI_ISL_2796832, EPI_ISL_2796833, EPI_ISL_2796834, EPI_ISL_2796836, EPI_ISL_2796837, EPI_ISL_2796838, EPI_ISL_2796839, EPI_ISL_2796840, EPI_ISL_2796841, EPI_ISL_2796842, EPI_ISL_2796843, EPI_ISL_2796845, EPI_ISL_2796846, EPI_ISL_2796847, EPI_ISL_2796848, EPI_ISL_2796849, EPI_ISL_2796850, EPI_ISL_2796851, EPI_ISL_2796852, EPI_ISL_2796853, EPI_ISL_2796854, EPI_ISL_2796855, EPI_ISL_2796856, EPI_ISL_2796857, EPI_ISL_2796858, EPI_ISL_2796859, EPI_ISL_2796860, EPI_ISL_2796861, EPI_ISL_2796863, EPI_ISL_2796864, EPI_ISL_2796865, EPI_ISL_2796866, EPI_ISL_2796867, EPI_ISL_2796868, EPI_ISL_2796869, EPI_ISL_2796870, EPI_ISL_2796871, EPI_ISL_2796872, EPI_ISL_2796873, EPI_ISL_2796874, EPI_ISL_2796876, EPI_ISL_2796877, EPI_ISL_2796878, EPI_ISL_2796879, EPI_ISL_2796880, EPI_ISL_2796881, EPI_ISL_2796882, EPI_ISL_2796883, EPI_ISL_2796884, EPI_ISL_2796885, EPI_ISL_2796886, EPI_ISL_2796887, EPI_ISL_2796890, EPI_ISL_2796891, EPI_ISL_2796892, EPI_ISL_2796893, EPI_ISL_2796894, EPI_ISL_2796895, EPI_ISL_2796896, EPI_ISL_2796897, EPI_ISL_2796898, EPI_ISL_2796899, EPI_ISL_2796900, EPI_ISL_2796901, EPI_ISL_2796903, EPI_ISL_2796904, EPI_ISL_2796905, EPI_ISL_2796907, EPI_ISL_2796908, EPI_ISL_2796909, EPI_ISL_2796910, EPI_ISL_2796911, EPI_ISL_2796912, EPI_ISL_2796913, EPI_ISL_2796915, EPI_ISL_2796916, EPI_ISL_2796917, EPI_ISL_2796918, EPI_ISL_2796919, EPI_ISL_2796920, EPI_ISL_2796921, EPI_ISL_2796922, EPI_ISL_2796923, EPI_ISL_2796924, EPI_ISL_2796925, EPI_ISL_2796926, EPI_ISL_2796927, EPI_ISL_2796928, EPI_ISL_2796929, EPI_ISL_2796930, EPI_ISL_2796931, EPI_ISL_2796932, EPI_ISL_2796933, EPI_ISL_2796934, EPI_ISL_2796935, EPI_ISL_2796936, EPI_ISL_2796937, EPI_ISL_2796938, EPI_ISL_2796939, EPI_ISL_2796940, EPI_ISL_2796941, EPI_ISL_2796942, EPI_ISL_2796943, EPI_ISL_2796944, EPI_ISL_2796945, EPI_ISL_2796946, EPI_ISL_2796947, EPI_ISL_2796948, EPI_ISL_2796949, EPI_ISL_2796950, EPI_ISL_2796951, EPI_ISL_2796952, EPI_ISL_2796953, EPI_ISL_2796954, EPI_ISL_2796955, EPI_ISL_2796956, EPI_ISL_2796957, EPI_ISL_2796958, EPI_ISL_2796959, EPI_ISL_2796960, EPI_ISL_2796961, EPI_ISL_2796962, EPI_ISL_2796963, EPI_ISL_2796964, EPI_ISL_2796965, EPI_ISL_2796966, EPI_ISL_2796967, EPI_ISL_2796968, EPI_ISL_2796969, EPI_ISL_2796970, EPI_ISL_2796971, EPI_ISL_2796972, EPI_ISL_2796973, EPI_ISL_2796974, EPI_ISL_2796975, EPI_ISL_2796976, EPI_ISL_2796977, EPI_ISL_2796978, EPI_ISL_2796979, EPI_ISL_2796980, EPI_ISL_2796981, EPI_ISL_2796982, EPI_ISL_2796983, EPI_ISL_2796984, EPI_ISL_2796985, EPI_ISL_2796986, EPI_ISL_2796987, EPI_ISL_2796988, EPI_ISL_2796989, EPI_ISL_2796990, EPI_ISL_2796991, EPI_ISL_2796992, EPI_ISL_2796993, EPI_ISL_2796994, EPI_ISL_2796995, EPI_ISL_2796996, EPI_ISL_2796997, EPI_ISL_2796998, EPI_ISL_2796999, EPI_ISL_2797000, EPI_ISL_2797001, EPI_ISL_2797002, EPI_ISL_2797003, EPI_ISL_2797004, EPI_ISL_2797005, EPI_ISL_2797006, EPI_ISL_2797007, EPI_ISL_2797008, EPI_ISL_2797009, EPI_ISL_2797010, EPI_ISL_2797011, EPI_ISL_2797012, EPI_ISL_2797013, EPI_ISL_2797014, EPI_ISL_2797015, EPI_ISL_2797016, EPI_ISL_2797017, EPI_ISL_2797019, EPI_ISL_2797020, EPI_ISL_2797021, EPI_ISL_2797022, EPI_ISL_2797023, EPI_ISL_2797024, EPI_ISL_2797025, EPI_ISL_2797026, EPI_ISL_2797027, EPI_ISL_2797028, EPI_ISL_2797029, EPI_ISL_2797030, EPI_ISL_2797031, EPI_ISL_2797032, EPI_ISL_2797033, EPI_ISL_2797034, EPI_ISL_2797035, EPI_ISL_2797036, EPI_ISL_2797037, EPI_ISL_2797038, EPI_ISL_2797039, EPI_ISL_2797041, EPI_ISL_2797042, EPI_ISL_2797043, EPI_ISL_2797044, EPI_ISL_2797045, EPI_ISL_2797046, EPI_ISL_2797047, EPI_ISL_2797048, EPI_ISL_2797049, EPI_ISL_2797050, EPI_ISL_2797051, EPI_ISL_2797052, EPI_ISL_2797053, EPI_ISL_2797054, EPI_ISL_2797056, EPI_ISL_2797057, EPI_ISL_2797058, EPI_ISL_2797059, EPI_ISL_2797060, EPI_ISL_2797061, EPI_ISL_2797062, EPI_ISL_2797063, EPI_ISL_2797064, EPI_ISL_2797065, EPI_ISL_2797066, EPI_ISL_2797067, EPI_ISL_2797068, EPI_ISL_2797069, EPI_ISL_2797070, EPI_ISL_2797071, EPI_ISL_2797072, EPI_ISL_2797073, EPI_ISL_2797074, EPI_ISL_2797075, EPI_ISL_2797076, EPI_ISL_2797077, EPI_ISL_2797078, EPI_ISL_2797079, EPI_ISL_2797080, EPI_ISL_2797081, EPI_ISL_2797082, EPI_ISL_2797084, EPI_ISL_2797085, EPI_ISL_2797086, EPI_ISL_2797087, EPI_ISL_2797088, EPI_ISL_2797089, EPI_ISL_2797090, EPI_ISL_2797091, EPI_ISL_2797092, EPI_ISL_2797093, EPI_ISL_2797094, EPI_ISL_2797095, EPI_ISL_2797096, EPI_ISL_2797097, EPI_ISL_2797098, EPI_ISL_2797099, EPI_ISL_2797100, EPI_ISL_2797101, EPI_ISL_2797102, EPI_ISL_2797104, EPI_ISL_2797105, EPI_ISL_2797106, EPI_ISL_2797107, EPI_ISL_2797108, EPI_ISL_2797109, EPI_ISL_2797111, EPI_ISL_2797112, EPI_ISL_2797113, EPI_ISL_2797114, EPI_ISL_2797115, EPI_ISL_2797116, EPI_ISL_2797117, EPI_ISL_2797118, EPI_ISL_2797119, EPI_ISL_2797120, EPI_ISL_2797122, EPI_ISL_2797124, EPI_ISL_2797125, EPI_ISL_2797126, EPI_ISL_2797127, EPI_ISL_2797128, EPI_ISL_2797129, EPI_ISL_2797130, EPI_ISL_2797131, EPI_ISL_2797132, EPI_ISL_2797134, EPI_ISL_2797135, EPI_ISL_2797136, EPI_ISL_2797139, EPI_ISL_2797140, EPI_ISL_2797141, EPI_ISL_2797142, EPI_ISL_2797143, EPI_ISL_2797144, EPI_ISL_2797145, EPI_ISL_2797146, EPI_ISL_2797147, EPI_ISL_2797149, EPI_ISL_2797150, EPI_ISL_2797151, EPI_ISL_2797152, EPI_ISL_2797153, EPI_ISL_2797154, EPI_ISL_2797155, EPI_ISL_2797156, EPI_ISL_2797157, EPI_ISL_2797158, EPI_ISL_2797159, EPI_ISL_2797160, EPI_ISL_2797161, EPI_ISL_2797162, EPI_ISL_2797163, EPI_ISL_2797164, EPI_ISL_2797165, EPI_ISL_2797166, EPI_ISL_2797167, EPI_ISL_2797168, EPI_ISL_2797169, EPI_ISL_2797170, EPI_ISL_2797171, EPI_ISL_2797172, EPI_ISL_2797173, EPI_ISL_2797174, EPI_ISL_2797175, EPI_ISL_2797178, EPI_ISL_2797179, EPI_ISL_2797181, EPI_ISL_2797182, EPI_ISL_2797183, EPI_ISL_2797184, EPI_ISL_2797185, EPI_ISL_2797186, EPI_ISL_2797187, EPI_ISL_2797188, EPI_ISL_2797189, EPI_ISL_2797190, EPI_ISL_2797191, EPI_ISL_2797192, EPI_ISL_2797193, EPI_ISL_2797194, EPI_ISL_2797195, EPI_ISL_2797197, EPI_ISL_2797198, EPI_ISL_2797199, EPI_ISL_2797200, EPI_ISL_2797201, EPI_ISL_2797203, EPI_ISL_2797204, EPI_ISL_2797205, EPI_ISL_2797206, EPI_ISL_2797207, EPI_ISL_2797208, EPI_ISL_2797209, EPI_ISL_2797210, EPI_ISL_2797211, EPI_ISL_2797212, EPI_ISL_2797213, EPI_ISL_2797214, EPI_ISL_2797215, EPI_ISL_2797216, EPI_ISL_2797218, EPI_ISL_2797219, EPI_ISL_2797220, EPI_ISL_2797221, EPI_ISL_2797222, EPI_ISL_2797223, EPI_ISL_2797224, EPI_ISL_2797225, EPI_ISL_2797226, EPI_ISL_2797227, EPI_ISL_2797229, EPI_ISL_2797231, EPI_ISL_2797232, EPI_ISL_2797233, EPI_ISL_2797234, EPI_ISL_2797235, EPI_ISL_2797236, EPI_ISL_2797237, EPI_ISL_2797238, EPI_ISL_2797239, EPI_ISL_2797240, EPI_ISL_2797241, EPI_ISL_2797242, EPI_ISL_2797243, EPI_ISL_2797244, EPI_ISL_2797245, EPI_ISL_2797246, EPI_ISL_2797247, EPI_ISL_2797248, EPI_ISL_2797249, EPI_ISL_2797250, EPI_ISL_2797251, EPI_ISL_2797252, EPI_ISL_2797253, EPI_ISL_2797254, EPI_ISL_2797255, EPI_ISL_2797256, EPI_ISL_2797257, EPI_ISL_2797258, EPI_ISL_2797259, EPI_ISL_2797260, EPI_ISL_2797261, EPI_ISL_2797262, EPI_ISL_2797263, EPI_ISL_2797264, EPI_ISL_2797265, EPI_ISL_2797266, EPI_ISL_2797267, EPI_ISL_2797268, EPI_ISL_2797269, EPI_ISL_2797270, EPI_ISL_2797271, EPI_ISL_2797272, EPI_ISL_2797273, EPI_ISL_2797274, EPI_ISL_2797275, EPI_ISL_2797276, EPI_ISL_2797277, EPI_ISL_2797278, EPI_ISL_2797279, EPI_ISL_2797280, EPI_ISL_2797281, EPI_ISL_2797282, EPI_ISL_2797283, EPI_ISL_2797284, EPI_ISL_2797285, EPI_ISL_2797286, EPI_ISL_2797287, EPI_ISL_2797288, EPI_ISL_2797289, EPI_ISL_2797290, EPI_ISL_2797291, EPI_ISL_2797292, EPI_ISL_2797293, EPI_ISL_2797294, EPI_ISL_2797295, EPI_ISL_2797296, EPI_ISL_2797297, EPI_ISL_2797299, EPI_ISL_2797300, EPI_ISL_2797301, EPI_ISL_2797302, EPI_ISL_2797303, EPI_ISL_2797304, EPI_ISL_2797305, EPI_ISL_2797306, EPI_ISL_2797307, EPI_ISL_2797308, EPI_ISL_2797309, EPI_ISL_2797310, EPI_ISL_2797311, EPI_ISL_2797312, EPI_ISL_2797313, EPI_ISL_2797314, EPI_ISL_2797315, EPI_ISL_2797316, EPI_ISL_2797317, EPI_ISL_2797318, EPI_ISL_2797319, EPI_ISL_2797321, EPI_ISL_2797322, EPI_ISL_2797323, EPI_ISL_2797324, EPI_ISL_2797325, EPI_ISL_2797326, EPI_ISL_2797327, EPI_ISL_2797328, EPI_ISL_2797329, EPI_ISL_2797330, EPI_ISL_2797331, EPI_ISL_2797332, EPI_ISL_2797333, EPI_ISL_2797334, EPI_ISL_2797335, EPI_ISL_2797336, EPI_ISL_2797337, EPI_ISL_2797338, EPI_ISL_2797339, EPI_ISL_2797340, EPI_ISL_2797341, EPI_ISL_2797342, EPI_ISL_2797343, EPI_ISL_2797344, EPI_ISL_2797345, EPI_ISL_2797346, EPI_ISL_2797347, EPI_ISL_2797348, EPI_ISL_2797349, EPI_ISL_2797350, EPI_ISL_2797351, EPI_ISL_2797353, EPI_ISL_2797354, EPI_ISL_2797355, EPI_ISL_2797356, EPI_ISL_2797357, EPI_ISL_2797358, EPI_ISL_2797359, EPI_ISL_2797360, EPI_ISL_2797361, EPI_ISL_2797362, EPI_ISL_2797363, EPI_ISL_2797364, EPI_ISL_2797365, EPI_ISL_2797366, EPI_ISL_2797367, EPI_ISL_2797368, EPI_ISL_2797369, EPI_ISL_2797371, EPI_ISL_2797373, EPI_ISL_2797374, EPI_ISL_2797375, EPI_ISL_2797376, EPI_ISL_2797377, EPI_ISL_2797379, EPI_ISL_2797380, EPI_ISL_2797381, EPI_ISL_2797383, EPI_ISL_2797384, EPI_ISL_2797385, EPI_ISL_2797386, EPI_ISL_2797387, EPI_ISL_2797388, EPI_ISL_2797389, EPI_ISL_2797390, EPI_ISL_2797391, EPI_ISL_2797392, EPI_ISL_2797393, EPI_ISL_2797394, EPI_ISL_2797396, EPI_ISL_2797398, EPI_ISL_2797399, EPI_ISL_2797400, EPI_ISL_2797401, EPI_ISL_2797402, EPI_ISL_2797403, EPI_ISL_2797404, EPI_ISL_2797405, EPI_ISL_2797406, EPI_ISL_2797407, EPI_ISL_2797408, EPI_ISL_2797409, EPI_ISL_2797410, EPI_ISL_2797411, EPI_ISL_2797412, EPI_ISL_2797413, EPI_ISL_2797414, EPI_ISL_2797415, EPI_ISL_2797416, EPI_ISL_2797417, EPI_ISL_2797418, EPI_ISL_2797419, EPI_ISL_2797420, EPI_ISL_2797421, EPI_ISL_2797423, EPI_ISL_2797424, EPI_ISL_2797425, EPI_ISL_2797426, EPI_ISL_2797427, EPI_ISL_2797428, EPI_ISL_2797429, EPI_ISL_2797430, EPI_ISL_2797431, EPI_ISL_2797432, EPI_ISL_2797434, EPI_ISL_2797435, EPI_ISL_2797436, EPI_ISL_2797437, EPI_ISL_2797438, EPI_ISL_2797439, EPI_ISL_2797440, EPI_ISL_2797441, EPI_ISL_2797442, EPI_ISL_2797444, EPI_ISL_2797445, EPI_ISL_2797446, EPI_ISL_2797447, EPI_ISL_2797448, EPI_ISL_2797449, EPI_ISL_2797451, EPI_ISL_2797452, EPI_ISL_2797453, EPI_ISL_2797454, EPI_ISL_2797456, EPI_ISL_2797457, EPI_ISL_2797458, EPI_ISL_2797462, EPI_ISL_2797464, EPI_ISL_2797465, EPI_ISL_2797466, EPI_ISL_2797467, EPI_ISL_2797468, EPI_ISL_2797469, EPI_ISL_2797470, EPI_ISL_2797471, EPI_ISL_2797472, EPI_ISL_2797473, EPI_ISL_2797474, EPI_ISL_2797475, EPI_ISL_2797478, EPI_ISL_2797479, EPI_ISL_2797480, EPI_ISL_2797481, EPI_ISL_2797482, EPI_ISL_2797483, EPI_ISL_2797484, EPI_ISL_2797485, EPI_ISL_2797486, EPI_ISL_2797488, EPI_ISL_2797489, EPI_ISL_2797490, EPI_ISL_2797491, EPI_ISL_2797492, EPI_ISL_2797493, EPI_ISL_2797494, EPI_ISL_2797495, EPI_ISL_2797496, EPI_ISL_2797497, EPI_ISL_2797498, EPI_ISL_2797500, EPI_ISL_2797501, EPI_ISL_2797503, EPI_ISL_2797504, EPI_ISL_2797505, EPI_ISL_2797506, EPI_ISL_2797507, EPI_ISL_2797508, EPI_ISL_2797509, EPI_ISL_2797510, EPI_ISL_2797511, EPI_ISL_2797512, EPI_ISL_2797515, EPI_ISL_2797516, EPI_ISL_2797517, EPI_ISL_2797518, EPI_ISL_2797519, EPI_ISL_2797520, EPI_ISL_2797521, EPI_ISL_2797522, EPI_ISL_2797523, EPI_ISL_2797524, EPI_ISL_2797525, EPI_ISL_2797526, EPI_ISL_2797527, EPI_ISL_2797528, EPI_ISL_2797529, EPI_ISL_2797530, EPI_ISL_2797531, EPI_ISL_2797533, EPI_ISL_2797534, EPI_ISL_2797535, EPI_ISL_2797536, EPI_ISL_2797537, EPI_ISL_2797538, EPI_ISL_2797539, EPI_ISL_2797540, EPI_ISL_2797541, EPI_ISL_2797542, EPI_ISL_2797544, EPI_ISL_2797546, EPI_ISL_2797547, EPI_ISL_2797548, EPI_ISL_2797549, EPI_ISL_2797550, EPI_ISL_2797551, EPI_ISL_2797552, EPI_ISL_2797553, EPI_ISL_2797554, EPI_ISL_2797555, EPI_ISL_2797556, EPI_ISL_2797557, EPI_ISL_2797558, EPI_ISL_2797559, EPI_ISL_2797560, EPI_ISL_2797561, EPI_ISL_2797562, EPI_ISL_2797563, EPI_ISL_2797564, EPI_ISL_2797565, EPI_ISL_2797566, EPI_ISL_2797567, EPI_ISL_2797568, EPI_ISL_2797569, EPI_ISL_2797570, EPI_ISL_2797571, EPI_ISL_2797572, EPI_ISL_2797573, EPI_ISL_2797574, EPI_ISL_2797575, EPI_ISL_2797576, EPI_ISL_2797578, EPI_ISL_2797579, EPI_ISL_2797580, EPI_ISL_2797581, EPI_ISL_2797582, EPI_ISL_2797583, EPI_ISL_2797584, EPI_ISL_2797585, EPI_ISL_2797586, EPI_ISL_2797587, EPI_ISL_2797588, EPI_ISL_2797589, EPI_ISL_2797590, EPI_ISL_2797591, EPI_ISL_2797592, EPI_ISL_2797593, EPI_ISL_2797594, EPI_ISL_2797595, EPI_ISL_2797596, EPI_ISL_2797597, EPI_ISL_2797598, EPI_ISL_2797599, EPI_ISL_2797600, EPI_ISL_2797601, EPI_ISL_2797602, EPI_ISL_2797603, EPI_ISL_2797604, EPI_ISL_2797605, EPI_ISL_2797606, EPI_ISL_2797607, EPI_ISL_2797608, EPI_ISL_2797609, EPI_ISL_2797610, EPI_ISL_2797611, EPI_ISL_2797612, EPI_ISL_2797613, EPI_ISL_2797614, EPI_ISL_2797616, EPI_ISL_2797618, EPI_ISL_2797619, EPI_ISL_2797620, EPI_ISL_2797621, EPI_ISL_2797622, EPI_ISL_2797623, EPI_ISL_2797624, EPI_ISL_2797625, EPI_ISL_2797626, EPI_ISL_2797627, EPI_ISL_2797628, EPI_ISL_2797629, EPI_ISL_2797630, EPI_ISL_2797631, EPI_ISL_2797632, EPI_ISL_2797633, EPI_ISL_2797634, EPI_ISL_2797635, EPI_ISL_2797636, EPI_ISL_2797637, EPI_ISL_2797638, EPI_ISL_2797639, EPI_ISL_2797642, EPI_ISL_2797643, EPI_ISL_2797644, EPI_ISL_2797645, EPI_ISL_2797646, EPI_ISL_2797647, EPI_ISL_2797648, EPI_ISL_2797649, EPI_ISL_2797650, EPI_ISL_2797651, EPI_ISL_2797652, EPI_ISL_2797653, EPI_ISL_2797654, EPI_ISL_2797655, EPI_ISL_2797656, EPI_ISL_2797657, EPI_ISL_2797658, EPI_ISL_2797659, EPI_ISL_2797660, EPI_ISL_2797661, EPI_ISL_2797662, EPI_ISL_2797663, EPI_ISL_2797664, EPI_ISL_2797665, EPI_ISL_2797666, EPI_ISL_2797667, EPI_ISL_2797669, EPI_ISL_2797670, EPI_ISL_2797671, EPI_ISL_2797672, EPI_ISL_2797673, EPI_ISL_2797674, EPI_ISL_2797675, EPI_ISL_2797676, EPI_ISL_2797677, EPI_ISL_2797678, EPI_ISL_2797679, EPI_ISL_2797680, EPI_ISL_2797681, EPI_ISL_2797682, EPI_ISL_2797683, EPI_ISL_2797684, EPI_ISL_2797685, EPI_ISL_2797686, EPI_ISL_2797687, EPI_ISL_2797688, EPI_ISL_2797689, EPI_ISL_2797690, EPI_ISL_2797691, EPI_ISL_2797692, EPI_ISL_2797693, EPI_ISL_2797694, EPI_ISL_2797695, EPI_ISL_2797696, | | | |

| | | | |
|---|---|---|---|
| EPI_ISL_2797697, EPI_ISL_2797698, EPI_ISL_2797699, EPI_ISL_2797700, EPI_ISL_2797701, EPI_ISL_2797702, EPI_ISL_2797703, EPI_ISL_2797704, EPI_ISL_2797705, EPI_ISL_2797706, EPI_ISL_2797707, EPI_ISL_2797708, EPI_ISL_2797709, EPI_ISL_2797710, EPI_ISL_2797711, EPI_ISL_2797712, EPI_ISL_2797713, EPI_ISL_2797714, EPI_ISL_2797715, EPI_ISL_2797716, EPI_ISL_2797717, EPI_ISL_2797718, EPI_ISL_2797719, EPI_ISL_2797720, EPI_ISL_2797721, EPI_ISL_2797722, EPI_ISL_2797723, EPI_ISL_2797724, EPI_ISL_2797725, EPI_ISL_2797726, EPI_ISL_2797727, EPI_ISL_2797728, EPI_ISL_2797729, EPI_ISL_2797730, EPI_ISL_2797731, EPI_ISL_2797732, EPI_ISL_2797735, EPI_ISL_2797736, EPI_ISL_2797737, EPI_ISL_2797738, EPI_ISL_2797739, EPI_ISL_2797740, EPI_ISL_2797741, EPI_ISL_2797743, EPI_ISL_2797744, EPI_ISL_2797745, EPI_ISL_2797746, EPI_ISL_2797747, EPI_ISL_2797748, EPI_ISL_2797749, EPI_ISL_2797750, EPI_ISL_2797751, EPI_ISL_2797752, EPI_ISL_2797753, EPI_ISL_2797755, EPI_ISL_2797756, EPI_ISL_2797757, EPI_ISL_2797758, EPI_ISL_2797759, EPI_ISL_2797760, EPI_ISL_2797761, EPI_ISL_2797762, EPI_ISL_2797764, EPI_ISL_2797765, EPI_ISL_2797766, EPI_ISL_2797767, EPI_ISL_2797768, EPI_ISL_2797769, EPI_ISL_2797770, EPI_ISL_2797771, EPI_ISL_2797772, EPI_ISL_2797773, EPI_ISL_2797774, EPI_ISL_2797775, EPI_ISL_2797776, EPI_ISL_2797777, EPI_ISL_2797778, EPI_ISL_2797779, EPI_ISL_2797780, EPI_ISL_2797781, EPI_ISL_2797782, EPI_ISL_2797783, EPI_ISL_2797784, EPI_ISL_2797785, EPI_ISL_2797786, EPI_ISL_2797787, EPI_ISL_2797788, EPI_ISL_2797789, EPI_ISL_2797790, EPI_ISL_2797791, EPI_ISL_2797792, EPI_ISL_2797793, EPI_ISL_2797795, EPI_ISL_2797796, EPI_ISL_2797797, EPI_ISL_2797798, EPI_ISL_2797799, EPI_ISL_2797800, EPI_ISL_2797801, EPI_ISL_2797802, EPI_ISL_2797803, EPI_ISL_2797804, EPI_ISL_2797805, EPI_ISL_2797806, EPI_ISL_2797807, EPI_ISL_2797808, EPI_ISL_2797809, EPI_ISL_2797811, EPI_ISL_2797812, EPI_ISL_2797813, EPI_ISL_2797814, EPI_ISL_2797816, EPI_ISL_2797817, EPI_ISL_2797818, EPI_ISL_2797819, EPI_ISL_2797821, EPI_ISL_2797822, EPI_ISL_2797823, EPI_ISL_2797824, EPI_ISL_2797825, EPI_ISL_2797826, EPI_ISL_2797827, EPI_ISL_2797828, EPI_ISL_2797829, EPI_ISL_2797830, EPI_ISL_2797831, EPI_ISL_2797832, EPI_ISL_2797833, EPI_ISL_2797834, EPI_ISL_2797835, EPI_ISL_2797836, EPI_ISL_2797837, EPI_ISL_2797838, EPI_ISL_2797839, EPI_ISL_2797840, EPI_ISL_2797841, EPI_ISL_2797842, EPI_ISL_2797843, EPI_ISL_2797844, EPI_ISL_2797845, EPI_ISL_2797846, EPI_ISL_2797847, EPI_ISL_2797848, EPI_ISL_2797849, EPI_ISL_2797850, EPI_ISL_2797851, EPI_ISL_2797852, EPI_ISL_2797853, EPI_ISL_2797854, EPI_ISL_2797855, EPI_ISL_2797856, EPI_ISL_2797857, EPI_ISL_2797858, EPI_ISL_2797859, EPI_ISL_2797860, EPI_ISL_2797861, EPI_ISL_2797864, EPI_ISL_2797865, EPI_ISL_2797866, EPI_ISL_2797867, EPI_ISL_2797869, EPI_ISL_2797870, EPI_ISL_2797871, EPI_ISL_2797872, EPI_ISL_2797874, EPI_ISL_2797875, EPI_ISL_2797876, EPI_ISL_2797877, EPI_ISL_2797878, EPI_ISL_2797879, EPI_ISL_2797880, EPI_ISL_2797881, EPI_ISL_2797882, EPI_ISL_2797883, EPI_ISL_2797884, EPI_ISL_2797885, EPI_ISL_2797886, EPI_ISL_2797887, EPI_ISL_2797888, EPI_ISL_2797890, EPI_ISL_2797891, EPI_ISL_2797892, EPI_ISL_2797893, EPI_ISL_2797894, EPI_ISL_2797895, EPI_ISL_2797896, EPI_ISL_2797897, EPI_ISL_2797898, EPI_ISL_2797899, EPI_ISL_2797900, EPI_ISL_2797901, EPI_ISL_2797902, EPI_ISL_2797903, EPI_ISL_2797904, EPI_ISL_2797905, EPI_ISL_2797906, EPI_ISL_2797907, EPI_ISL_2797909, EPI_ISL_2797910, EPI_ISL_2797911, EPI_ISL_2797912, EPI_ISL_2797913, EPI_ISL_2797914, EPI_ISL_2797915, EPI_ISL_2797916, EPI_ISL_2797917, EPI_ISL_2797918, EPI_ISL_2797919, EPI_ISL_2797920, EPI_ISL_2797921, EPI_ISL_2797922, EPI_ISL_2797923, EPI_ISL_2797924, EPI_ISL_2797925, EPI_ISL_2797926, EPI_ISL_2797927, EPI_ISL_2797929, EPI_ISL_2797930, EPI_ISL_2797931, EPI_ISL_2797932, EPI_ISL_2797933, EPI_ISL_2797934, EPI_ISL_2797935, EPI_ISL_2797936, EPI_ISL_2797937, EPI_ISL_2797938, EPI_ISL_2797939, EPI_ISL_2797940, EPI_ISL_2797941, EPI_ISL_2797943, EPI_ISL_2797947, EPI_ISL_2797948, EPI_ISL_2797949, EPI_ISL_2797950, EPI_ISL_2797951, EPI_ISL_2797952, EPI_ISL_2797953, EPI_ISL_2797954, EPI_ISL_2797955, EPI_ISL_2797957, EPI_ISL_2797959, EPI_ISL_2797960, EPI_ISL_2797961, EPI_ISL_2797962, EPI_ISL_2797963, EPI_ISL_2797964, EPI_ISL_2797966, EPI_ISL_2797967, EPI_ISL_2797968, EPI_ISL_2797969, EPI_ISL_2797970, EPI_ISL_2797971, EPI_ISL_2797973, EPI_ISL_2797974, EPI_ISL_2797975, EPI_ISL_2797976, EPI_ISL_2797977, EPI_ISL_2797979, EPI_ISL_2797980, EPI_ISL_2797981, EPI_ISL_2797982, EPI_ISL_2797983, EPI_ISL_2797984, EPI_ISL_2797985, EPI_ISL_2797986, EPI_ISL_2797987, EPI_ISL_2797988, EPI_ISL_2797989, EPI_ISL_2797990, EPI_ISL_2797991, EPI_ISL_2797992, EPI_ISL_2797993, EPI_ISL_2797994, EPI_ISL_2797995, EPI_ISL_2797996, EPI_ISL_2797997, EPI_ISL_2797998, EPI_ISL_2797999, EPI_ISL_2798000, EPI_ISL_2798001, EPI_ISL_2798002, EPI_ISL_2798003, EPI_ISL_2798004, EPI_ISL_2798005, EPI_ISL_2798006, EPI_ISL_2798007, EPI_ISL_2798008, EPI_ISL_2798009, EPI_ISL_2798010, EPI_ISL_2798011, EPI_ISL_2798012, EPI_ISL_2798013, EPI_ISL_2798014, EPI_ISL_2798015, EPI_ISL_2798016, EPI_ISL_2798017, EPI_ISL_2798018, EPI_ISL_2798019, EPI_ISL_2798020, EPI_ISL_2798021, EPI_ISL_2798023, EPI_ISL_2798024, EPI_ISL_2798025, EPI_ISL_2798026, EPI_ISL_2798027, EPI_ISL_2798028, EPI_ISL_2798029, EPI_ISL_2798030, EPI_ISL_2798031, EPI_ISL_2798032, EPI_ISL_2798033, EPI_ISL_2798034, EPI_ISL_2798035, EPI_ISL_2798036, EPI_ISL_2798037, EPI_ISL_2798038, EPI_ISL_2798039, EPI_ISL_2798040, EPI_ISL_2798041, EPI_ISL_2798042, EPI_ISL_2798043, EPI_ISL_2798044, EPI_ISL_2798045, EPI_ISL_2798046, EPI_ISL_2798047, EPI_ISL_2798048, EPI_ISL_2798049, EPI_ISL_2798050, EPI_ISL_2798051, EPI_ISL_2798052, EPI_ISL_2798053, EPI_ISL_2798054, EPI_ISL_2798055, EPI_ISL_2798056, EPI_ISL_2798057, EPI_ISL_2798058, EPI_ISL_2798059, EPI_ISL_2798060, EPI_ISL_2798061, EPI_ISL_2798062, EPI_ISL_2798063, EPI_ISL_2798064, EPI_ISL_2798065, EPI_ISL_2798066, EPI_ISL_2798067, EPI_ISL_2798068, EPI_ISL_2798069, EPI_ISL_2798070, EPI_ISL_2798071, EPI_ISL_2798072, EPI_ISL_2798073, EPI_ISL_2798074, EPI_ISL_2798075, EPI_ISL_2798076, EPI_ISL_2798077, EPI_ISL_2798078, EPI_ISL_2798079, EPI_ISL_2798080, EPI_ISL_2798081, EPI_ISL_2798082, EPI_ISL_2798083, EPI_ISL_2798085, EPI_ISL_2798086, EPI_ISL_2798087, EPI_ISL_2798088, EPI_ISL_2798089, EPI_ISL_2798090, EPI_ISL_2798091, EPI_ISL_2798092, EPI_ISL_2798093, EPI_ISL_2798095, EPI_ISL_2798096, EPI_ISL_2798097, EPI_ISL_2798098, EPI_ISL_2798099, EPI_ISL_2798100, EPI_ISL_2798101, EPI_ISL_2798102, EPI_ISL_2798103, EPI_ISL_2798104, EPI_ISL_2798105, EPI_ISL_2798106, EPI_ISL_2798107, EPI_ISL_2798108, EPI_ISL_2798109, EPI_ISL_2798110, EPI_ISL_2798111, EPI_ISL_2798112, EPI_ISL_2798113, EPI_ISL_2798114, EPI_ISL_2798115, EPI_ISL_2798116, EPI_ISL_2798117, EPI_ISL_2798118, EPI_ISL_2798120, EPI_ISL_2798121, EPI_ISL_2798122, EPI_ISL_2798123, EPI_ISL_2798125, EPI_ISL_2798126, EPI_ISL_2798127, EPI_ISL_2798128, EPI_ISL_2798129, EPI_ISL_2798130, EPI_ISL_2798131, EPI_ISL_2798132, EPI_ISL_2798133, EPI_ISL_2798134, EPI_ISL_2798135, EPI_ISL_2798137, EPI_ISL_2798138, EPI_ISL_2798139, EPI_ISL_2798140, EPI_ISL_2798141, EPI_ISL_2798142, EPI_ISL_2798143, EPI_ISL_2798144, EPI_ISL_2798146, EPI_ISL_2798147, EPI_ISL_2798148, EPI_ISL_2798149, EPI_ISL_2798150, EPI_ISL_2798151, EPI_ISL_2798152, EPI_ISL_2798153, EPI_ISL_2798154, EPI_ISL_2798155, EPI_ISL_2798156, EPI_ISL_2798157, EPI_ISL_2798158, EPI_ISL_2798159, EPI_ISL_2798160, EPI_ISL_2798162, EPI_ISL_2798163, EPI_ISL_2798165, EPI_ISL_2798166, EPI_ISL_2798167, EPI_ISL_2798168, EPI_ISL_2798169, EPI_ISL_2798170, EPI_ISL_2798171, EPI_ISL_2798172, EPI_ISL_2798173, EPI_ISL_2798174, EPI_ISL_2798175, EPI_ISL_2798176, EPI_ISL_2798177, EPI_ISL_2798178, EPI_ISL_2798179, EPI_ISL_2798180, EPI_ISL_2798181, EPI_ISL_2798182 | | | |
| see above | CDPH VBL | California Department of Public Health | CDPH-COVIDNet, UCSF Center for Advanced Technology |
| EPI_ISL_2800344 | LESP Hidalgo | LESP Hidalgo GENE2LIFE | HERRERA-GOMEZ CONNIE ALEJANDRA; CASTILLO-ORTEGA LAURA SOFIA; GARCIA-REYES MELITO; VIGUERAS-MORALES YAJAIRA SULIM; GARCIA-GONZALEZ OCTAVIO; VILLAFUENTES-TELLEZ HECTOR ALBERTO |
| EPI_ISL_2800347, EPI_ISL_2800348, EPI_ISL_2800349, EPI_ISL_2800351, EPI_ISL_2800352, EPI_ISL_2800353, EPI_ISL_2800355, EPI_ISL_2800356, EPI_ISL_2800357, EPI_ISL_2800358, EPI_ISL_2800359, EPI_ISL_2800360, EPI_ISL_2800361, EPI_ISL_2800362, EPI_ISL_2800363, EPI_ISL_2800364, EPI_ISL_2800365, EPI_ISL_2800366, EPI_ISL_2800367, EPI_ISL_2800368, EPI_ISL_2800369, EPI_ISL_2800370, EPI_ISL_2800371, EPI_ISL_2800372, EPI_ISL_2800373, EPI_ISL_2800374, EPI_ISL_2800375, EPI_ISL_2800376, EPI_ISL_2800377, EPI_ISL_2800378, EPI_ISL_2800379, EPI_ISL_2800380, EPI_ISL_2800381, EPI_ISL_2800382, EPI_ISL_2800383, EPI_ISL_2800384, EPI_ISL_2800385, EPI_ISL_2800386, EPI_ISL_2800387, EPI_ISL_2800388, EPI_ISL_2800389, EPI_ISL_2800390, EPI_ISL_2800391, EPI_ISL_2800392, EPI_ISL_2800393, EPI_ISL_2800394, EPI_ISL_2800395, EPI_ISL_2800396, EPI_ISL_2800397, EPI_ISL_2800398, EPI_ISL_2800399, EPI_ISL_2800400, EPI_ISL_2800401, EPI_ISL_2800402, EPI_ISL_2800403, EPI_ISL_2800404, EPI_ISL_2800405, EPI_ISL_2800406, EPI_ISL_2800407, EPI_ISL_2800408, EPI_ISL_2800409, EPI_ISL_2800410, EPI_ISL_2800411, EPI_ISL_2800412, EPI_ISL_2800413, EPI_ISL_2800414, EPI_ISL_2800415, EPI_ISL_2800416, EPI_ISL_2800417, EPI_ISL_2800418, EPI_ISL_2800419, EPI_ISL_2800420, EPI_ISL_2800421, EPI_ISL_2800422, EPI_ISL_2800423, EPI_ISL_2800424, EPI_ISL_2800425, EPI_ISL_2800426, EPI_ISL_2800427, EPI_ISL_2800428, EPI_ISL_2800429, EPI_ISL_2800430, EPI_ISL_2800431, EPI_ISL_2800432, EPI_ISL_2800433, EPI_ISL_2800434, EPI_ISL_2800435, EPI_ISL_2800436, EPI_ISL_2800437, EPI_ISL_2800438, EPI_ISL_2800439, EPI_ISL_2800440, EPI_ISL_2800441, EPI_ISL_2800442, EPI_ISL_2800443, EPI_ISL_2800444, EPI_ISL_2800445, EPI_ISL_2800446, EPI_ISL_2800447, EPI_ISL_2800448, EPI_ISL_2800449, EPI_ISL_2800450, EPI_ISL_2800451, EPI_ISL_2800452, EPI_ISL_2800453, EPI_ISL_2800454, EPI_ISL_2800455, EPI_ISL_2800456, EPI_ISL_2800457, EPI_ISL_2800458, EPI_ISL_2800459, EPI_ISL_2800460, EPI_ISL_2800461, EPI_ISL_2800462, EPI_ISL_2800463, EPI_ISL_2800464, EPI_ISL_2800465, EPI_ISL_2800466, EPI_ISL_2800467, EPI_ISL_2800468, EPI_ISL_2800469, EPI_ISL_2800470, EPI_ISL_2800471, EPI_ISL_2800472, EPI_ISL_2800473, EPI_ISL_2800474, EPI_ISL_2800475, EPI_ISL_2800476, EPI_ISL_2800477, EPI_ISL_2800478, EPI_ISL_2800479, EPI_ISL_2800480, EPI_ISL_2800481, EPI_ISL_2800482, EPI_ISL_2800483, EPI_ISL_2800484, EPI_ISL_2800485, EPI_ISL_2800486, EPI_ISL_2800487, EPI_ISL_2800488, EPI_ISL_2800489, EPI_ISL_2800490, EPI_ISL_2800491, EPI_ISL_2800492, EPI_ISL_2800493, EPI_ISL_2800494, EPI_ISL_2800495, EPI_ISL_2800496, EPI_ISL_2800497, EPI_ISL_2800498, EPI_ISL_2800499, EPI_ISL_2800500, EPI_ISL_2800501, EPI_ISL_2800502, EPI_ISL_2800503, EPI_ISL_2800504, EPI_ISL_2800505, EPI_ISL_2800506, EPI_ISL_2800507, EPI_ISL_2800508, EPI_ISL_2800509, EPI_ISL_2800510, EPI_ISL_2800511, EPI_ISL_2800512, EPI_ISL_2800513, EPI_ISL_2800514, EPI_ISL_2800515, EPI_ISL_2800516, EPI_ISL_2800517, EPI_ISL_2800518, EPI_ISL_2800519, EPI_ISL_2800520, EPI_ISL_2800521, EPI_ISL_2800522, EPI_ISL_2800523, EPI_ISL_2800524, EPI_ISL_2800525, EPI_ISL_2800526, EPI_ISL_2800527, EPI_ISL_2800528, EPI_ISL_2800529, EPI_ISL_2800530, EPI_ISL_2800531, EPI_ISL_2800532, EPI_ISL_2800533 | | | |
| see above | State Testing Facility | Altius Institute for Biomedical Research | Daniel Bates, Rebecca Bruders, Michael Buckley, Mark Frerker, Amanda Gale, Clem Green, Muhammad Halimun, Kneshay Harper, Matt Hartman, Alex Isner, Audra Johnson, Jessica Kunder, Lauren Mitchell, Jemma Nelson, Alex Nguyen, Sofia Olsson, Sadie Patraw, Tobias Ragoczy, Joshua Richards, Jean Robinston, Jacob Rodriguez, John Stamatoyannopoulos, Eric Thorland, Julia Wald |
| EPI_ISL_2800534 | Bertschi School | Altius Institute for Biomedical Research | Daniel Bates, Rebecca Bruders, Michael Buckley, Mark Frerker, Amanda Gale, Clem Green, Muhammad Halimun, Kneshay Harper, Matt Hartman, Alex Isner, Audra Johnson, Jessica Kunder, Lauren Mitchell, Jemma Nelson, Alex Nguyen, Sofia Olsson, Sadie Patraw, Tobias Ragoczy, Joshua Richards, Jean Robinston, Jacob Rodriguez, John Stamatoyannopoulos, Eric Thorland, Julia Wald |
| EPI_ISL_2800535, EPI_ISL_2800536, EPI_ISL_2800537, EPI_ISL_2800538, EPI_ISL_2800539, EPI_ISL_2800540, EPI_ISL_2800541, EPI_ISL_2800542, EPI_ISL_2800543, EPI_ISL_2800544, EPI_ISL_2800545, EPI_ISL_2800547, EPI_ISL_2800548, EPI_ISL_2800549, EPI_ISL_2800550, EPI_ISL_2800551, EPI_ISL_2800552, EPI_ISL_2800553, EPI_ISL_2800554, EPI_ISL_2800560, EPI_ISL_2800561, EPI_ISL_2800562, EPI_ISL_2800563, EPI_ISL_2800564, EPI_ISL_2800565, EPI_ISL_2800566, EPI_ISL_2800567, EPI_ISL_2800568, EPI_ISL_2800569, EPI_ISL_2800570, EPI_ISL_2800571, EPI_ISL_2800572, EPI_ISL_2800573, EPI_ISL_2800574, EPI_ISL_2800575, EPI_ISL_2800576, EPI_ISL_2800577, EPI_ISL_2800578, EPI_ISL_2800579, EPI_ISL_2800580, EPI_ISL_2800581, EPI_ISL_2800582, EPI_ISL_2800583, EPI_ISL_2800584, EPI_ISL_2800585, EPI_ISL_2800586, EPI_ISL_2800587, EPI_ISL_2800588, EPI_ISL_2800589, EPI_ISL_2800590, EPI_ISL_2800591, EPI_ISL_2800592, EPI_ISL_2800593, EPI_ISL_2800594, EPI_ISL_2800595, EPI_ISL_2800596, EPI_ISL_2800597, EPI_ISL_2800598, EPI_ISL_2800599, EPI_ISL_2800600 | | | |
| see above | State Testing Facility | Altius Institute for Biomedical Research | Daniel Bates, Rebecca Bruders, Michael Buckley, Mark Frerker, Amanda Gale, Clem Green, Muhammad Halimun, Kneshay Harper, Matt Hartman, Alex Isner, Audra Johnson, Jessica Kunder, Lauren Mitchell, Jemma Nelson, Alex Nguyen, Sofia Olsson, Sadie Patraw, Tobias Ragoczy, Joshua Richards, Jean Robinston, Jacob Rodriguez, John Stamatoyannopoulos, Eric Thorland, Julia Wald |
| EPI_ISL_2800601, EPI_ISL_2800602 | Seattle University Athletics | Altius Institute for Biomedical Research | Daniel Bates, Rebecca Bruders, Michael Buckley, Mark Frerker, Amanda Gale, Clem Green, Muhammad Halimun, Kneshay Harper, Matt Hartman, Alex Isner, Audra Johnson, Jessica Kunder, Lauren Mitchell, Jemma Nelson, Alex Nguyen, Sofia Olsson, Sadie Patraw, Tobias Ragoczy, Joshua Richards, Jean Robinston, Jacob Rodriguez, John Stamatoyannopoulos, Eric Thorland, Julia Wald |
| EPI_ISL_2800603, EPI_ISL_2800604, EPI_ISL_2800605, EPI_ISL_2800606, EPI_ISL_2800607, EPI_ISL_2800608, EPI_ISL_2800609, EPI_ISL_2800610, EPI_ISL_2800611, EPI_ISL_2800612, EPI_ISL_2800613, EPI_ISL_2800614, EPI_ISL_2800615, EPI_ISL_2800616, EPI_ISL_2800617, EPI_ISL_2800618, EPI_ISL_2800619, EPI_ISL_2800620, EPI_ISL_2800621, EPI_ISL_2800622, EPI_ISL_2800623, EPI_ISL_2800624, EPI_ISL_2800625, EPI_ISL_2800626, EPI_ISL_2800627, EPI_ISL_2800628, EPI_ISL_2800629, EPI_ISL_2800630, EPI_ISL_2800631, EPI_ISL_2800632, EPI_ISL_2800633, EPI_ISL_2800634, EPI_ISL_2800635, EPI_ISL_2800636, EPI_ISL_2800637, EPI_ISL_2800638, EPI_ISL_2800639, EPI_ISL_2800640, EPI_ISL_2800641, EPI_ISL_2800642, EPI_ISL_2800643, EPI_ISL_2800644, EPI_ISL_2800645, EPI_ISL_2800646, EPI_ISL_2800647, EPI_ISL_2800648, EPI_ISL_2800649, EPI_ISL_2800650, EPI_ISL_2800651, EPI_ISL_2800652, EPI_ISL_2800653, EPI_ISL_2800654, EPI_ISL_2800655, EPI_ISL_2800656, EPI_ISL_2800657, EPI_ISL_2800658, EPI_ISL_2800659, EPI_ISL_2800660, EPI_ISL_2800661, EPI_ISL_2800662, EPI_ISL_2800663, EPI_ISL_2800664, EPI_ISL_2800665, EPI_ISL_2800666, EPI_ISL_2800667, EPI_ISL_2800668, EPI_ISL_2800669, EPI_ISL_2800670, EPI_ISL_2800671, EPI_ISL_2800672, EPI_ISL_2800673, EPI_ISL_2800674, EPI_ISL_2800675, EPI_ISL_2800676, EPI_ISL_2800677, EPI_ISL_2800678, EPI_ISL_2800679, EPI_ISL_2800680, EPI_ISL_2800681, EPI_ISL_2800682, EPI_ISL_2800683, EPI_ISL_2800684, EPI_ISL_2800685, EPI_ISL_2800686, EPI_ISL_2800687, EPI_ISL_2800688, EPI_ISL_2800689, EPI_ISL_2800690, EPI_ISL_2800691, EPI_ISL_2800692, EPI_ISL_2800693, EPI_ISL_2800694, EPI_ISL_2800695, EPI_ISL_2800696, EPI_ISL_2800697, EPI_ISL_2800698, EPI_ISL_2800699, EPI_ISL_2800700, EPI_ISL_2800701, EPI_ISL_2800702, EPI_ISL_2800703, EPI_ISL_2800704, EPI_ISL_2800705, EPI_ISL_2800706, EPI_ISL_2800707, EPI_ISL_2800708 | | | |
| see above | State Testing Facility | Altius Institute for Biomedical Research | Daniel Bates, Rebecca Bruders, Michael Buckley, Mark Frerker, Amanda Gale, Clem Green, Muhammad Halimun, Kneshay Harper, Matt Hartman, Alex Isner, Audra Johnson, Jessica Kunder, Lauren Mitchell, Jemma Nelson, Alex Nguyen, Sofia Olsson, Sadie Patraw, Tobias Ragoczy, Joshua Richards, Jean Robinston, Jacob Rodriguez, John Stamatoyannopoulos, Eric Thorland, Julia Wald |
| EPI_ISL_2800709, EPI_ISL_2800710, EPI_ISL_2800711, EPI_ISL_2800712, EPI_ISL_2800713, EPI_ISL_2800714, EPI_ISL_2800715, EPI_ISL_2800716, EPI_ISL_2800717, EPI_ISL_2800718, EPI_ISL_2800719, EPI_ISL_2800720, EPI_ISL_2800721, EPI_ISL_2800722, EPI_ISL_2800723, EPI_ISL_2800724, EPI_ISL_2800725, EPI_ISL_2800726, EPI_ISL_2800727, EPI_ISL_2800728, EPI_ISL_2800729, EPI_ISL_2800730, EPI_ISL_2800731, EPI_ISL_2800732, EPI_ISL_2800733, EPI_ISL_2800734, EPI_ISL_2800735, EPI_ISL_2800736, EPI_ISL_2800737, EPI_ISL_2800738, EPI_ISL_2800739, EPI_ISL_2800740, EPI_ISL_2800741, EPI_ISL_2800742, EPI_ISL_2800743, EPI_ISL_2800744, EPI_ISL_2800745, EPI_ISL_2800746, EPI_ISL_2800747, EPI_ISL_2800748, EPI_ISL_2800749, EPI_ISL_2800750, EPI_ISL_2800751, EPI_ISL_2800752, EPI_ISL_2800753, EPI_ISL_2800754, EPI_ISL_2800755, EPI_ISL_2800756, EPI_ISL_2800757, EPI_ISL_2800758, EPI_ISL_2800759, EPI_ISL_2800760, EPI_ISL_2800761, EPI_ISL_2800762, EPI_ISL_2800763, EPI_ISL_2800764, EPI_ISL_2800765, EPI_ISL_2800766, EPI_ISL_2800767, EPI_ISL_2800768, EPI_ISL_2800769, EPI_ISL_2800770, EPI_ISL_2800771, EPI_ISL_2800772, EPI_ISL_2800773, EPI_ISL_2800774, EPI_ISL_2800775, EPI_ISL_2800776, EPI_ISL_2800777, EPI_ISL_2800778, EPI_ISL_2800779, EPI_ISL_2800780, EPI_ISL_2800781, EPI_ISL_2800782, EPI_ISL_2800783, EPI_ISL_2800784, EPI_ISL_2800785, EPI_ISL_2800786, EPI_ISL_2800787, EPI_ISL_2800788, EPI_ISL_2800789, EPI_ISL_2800790, EPI_ISL_2800791, EPI_ISL_2800792, EPI_ISL_2800793, EPI_ISL_2800794, EPI_ISL_2800795, EPI_ISL_2800796, EPI_ISL_2800797, EPI_ISL_2800798, EPI_ISL_2800799, EPI_ISL_2800800, EPI_ISL_2800801, EPI_ISL_2800802, EPI_ISL_2800803, EPI_ISL_2800804, EPI_ISL_2800805, EPI_ISL_2800806, EPI_ISL_2800807, EPI_ISL_2800808, EPI_ISL_2800809, EPI_ISL_2800810, EPI_ISL_2800811, EPI_ISL_2800812, EPI_ISL_2800813, EPI_ISL_2800814, EPI_ISL_2800815, EPI_ISL_2800816, EPI_ISL_2800817, EPI_ISL_2800818, EPI_ISL_2800819, EPI_ISL_2800820, EPI_ISL_2800821, EPI_ISL_2800846, EPI_ISL_2800847, EPI_ISL_2800848, EPI_ISL_2800849, EPI_ISL_2800850, EPI_ISL_2800851, EPI_ISL_2800852, EPI_ISL_2800853, EPI_ISL_2800854, EPI_ISL_2800855, EPI_ISL_2800856, EPI_ISL_2800857, EPI_ISL_2800858, EPI_ISL_2800859, EPI_ISL_2800860, EPI_ISL_2800861, EPI_ISL_2800862, EPI_ISL_2800863, EPI_ISL_2800864, EPI_ISL_2800865, EPI_ISL_2800866, EPI_ISL_2800867, EPI_ISL_2800868, EPI_ISL_2800869, EPI_ISL_2800870, EPI_ISL_2800871, EPI_ISL_2800872, EPI_ISL_2800873 | | | |
| see above | Illinois Department of Public Health | Illinois Department of Public Health - Chicago Lab | Vineet K. Dhiman, Ira Heimler, Joel Price |
| EPI_ISL_2800874, EPI_ISL_2800875, EPI_ISL_2800876, EPI_ISL_2800877, EPI_ISL_2800878, EPI_ISL_2800879, EPI_ISL_2800880, EPI_ISL_2800881, EPI_ISL_2800882, EPI_ISL_2800883, EPI_ISL_2800884, EPI_ISL_2800885, EPI_ISL_2800886, EPI_ISL_2800887, EPI_ISL_2800888, EPI_ISL_2800889, EPI_ISL_2800890, EPI_ISL_2800891, EPI_ISL_2800892, EPI_ISL_2800893, EPI_ISL_2800894, EPI_ISL_2800895, EPI_ISL_2800896, EPI_ISL_2800897, EPI_ISL_2800898, EPI_ISL_2800899, EPI_ISL_2800900, EPI_ISL_2800901, EPI_ISL_2800902, EPI_ISL_2800903, EPI_ISL_2800904, EPI_ISL_2800905, EPI_ISL_2800906, EPI_ISL_2800907, EPI_ISL_2800908, EPI_ISL_2800909, EPI_ISL_2800910, EPI_ISL_2800911, EPI_ISL_2800912 | | | |
| see above | Tampa General Hospital Esoteric Lab | Tampa General Hospital Esoteric Research & Development Lab | Jon Faughn, Deanna Becker, Dominic Uy, Jorge Mecias-Frias, Elaine Vendrone, Vicki Healer, Amorce Lima, Suzane Silbert |
| EPI_ISL_2800913, EPI_ISL_2800914, EPI_ISL_2800915 | LESP Guanajuato | Instituto de Diagnostico y Referencia Epidemiologicos (INDRE) | Claudia Wong-Arambula, Abril Rodriguez-Maldonado, Vanessa Rivero-Arredondo, Ariadna Medina-Benitez, Joaquin Quiroz-Mercado, Sergio Rangel-Guerrero, Maribel Gonzalez-Villa, Natividad Cruz-Ortiz, Tatiana Nunez-Garcia, Gisela Barrera-Badillo, Lucia Hernandez-Rivas, Irma Lopez-Martinez, Ernesto Ramirez-Gonzalez. |
| EPI_ISL_2800916 | LESP Yucatan | Instituto de Diagnostico y Referencia Epidemiologicos (INDRE) | Claudia Wong-Arambula, Abril Rodriguez-Maldonado, Vanessa Rivero-Arredondo, Ariadna Medina-Benitez, Joaquin Quiroz-Mercado, Sergio Rangel-Guerrero, Maribel Gonzalez-Villa, Natividad Cruz-Ortiz, Tatiana Nunez-Garcia, Gisela Barrera-Badillo, Lucia Hernandez-Rivas, Irma Lopez-Martinez, Ernesto Ramirez-Gonzalez. |
| EPI_ISL_2800917 | LESP Guanajuato | Instituto de Diagnostico y Referencia Epidemiologicos (INDRE) | Claudia Wong-Arambula, Abril Rodriguez-Maldonado, Vanessa Rivero-Arredondo, Ariadna Medina-Benitez, Joaquin Quiroz-Mercado, Sergio Rangel-Guerrero, Maribel Gonzalez-Villa, Natividad Cruz-Ortiz, Tatiana Nunez-Garcia, Gisela Barrera-Badillo, Lucia Hernandez-Rivas, Irma Lopez-Martinez, Ernesto Ramirez-Gonzalez. |
| EPI_ISL_2800918, EPI_ISL_2800919, EPI_ISL_2800920, EPI_ISL_2800921, EPI_ISL_2800922, EPI_ISL_2800923 | LESP Yucatan | Instituto de Diagnostico y Referencia Epidemiologicos (INDRE) | Claudia Wong-Arambula, Abril Rodriguez-Maldonado, Vanessa Rivero-Arredondo, Ariadna Medina-Benitez, Joaquin Quiroz-Mercado, Sergio Rangel-Guerrero, Maribel Gonzalez-Villa, Natividad Cruz-Ortiz, Tatiana Nunez-Garcia, Gisela Barrera-Badillo, Lucia Hernandez-Rivas, Irma Lopez-Martinez, Ernesto Ramirez-Gonzalez. |
| EPI_ISL_2800924 | LESP Guanajuato | Instituto de Diagnostico y Referencia Epidemiologicos (INDRE) | Claudia Wong-Arambula, Abril Rodriguez-Maldonado, Vanessa Rivero-Arredondo, Ariadna Medina-Benitez, Joaquin Quiroz-Mercado, Sergio Rangel-Guerrero, Maribel Gonzalez-Villa, Natividad Cruz-Ortiz, Tatiana Nunez-Garcia, Gisela Barrera-Badillo, Lucia Hernandez-Rivas, Irma Lopez-Martinez, Ernesto Ramirez-Gonzalez. |
| EPI_ISL_2800925 | LESP Tabasco | Instituto de Diagnostico y Referencia Epidemiologicos (INDRE) | Claudia Wong-Arambula, Abril Rodriguez-Maldonado, Vanessa Rivero-Arredondo, Ariadna Medina-Benitez, Joaquin Quiroz-Mercado, Sergio Rangel-Guerrero, Natividad Cruz-Ortiz, Tatiana Nunez-Garcia, Gisela Barrera-Badillo, Lucia Hernandez-Rivas, Irma Lopez-Martinez, Ernesto Ramirez-Gonzalez. |
| EPI_ISL_2800926 | LESP Guanajuato | Instituto de Diagnostico y Referencia Epidemiologicos (INDRE) | Claudia Wong-Arambula, Abril Rodriguez-Maldonado, Vanessa Rivero-Arredondo, Ariadna Medina-Benitez, Joaquin Quiroz-Mercado, Sergio Rangel-Guerrero, Natividad Cruz-Ortiz, Tatiana Nunez-Garcia, Gisela Barrera-Badillo, Lucia Hernandez-Rivas, Irma Lopez-Martinez, Ernesto Ramirez-Gonzalez. |
| EPI_ISL_2800927, EPI_ISL_2800928, EPI_ISL_2800929 | LESP Tabasco | Instituto de Diagnostico y Referencia Epidemiologicos (INDRE) | Claudia Wong-Arambula, Abril Rodriguez-Maldonado, Vanessa Rivero-Arredondo, Ariadna Medina-Benitez, Joaquin Quiroz-Mercado, Sergio Rangel-Guerrero, Natividad Cruz-Ortiz, Tatiana Nunez-Garcia, Gisela Barrera-Badillo, Lucia Hernandez-Rivas, Irma Lopez-Martinez, Ernesto Ramirez-Gonzalez. |
| EPI_ISL_2800930, EPI_ISL_2800931 | LESP Sonora | Instituto de Diagnostico y Referencia Epidemiologicos (INDRE) | Claudia Wong-Arambula, Abril Rodriguez-Maldonado, Vanessa Rivero-Arredondo, Ariadna Medina-Benitez, Joaquin Quiroz-Mercado, Sergio Rangel-Guerrero, Natividad Cruz-Ortiz, Tatiana Nunez-Garcia, Gisela Barrera-Badillo, Lucia Hernandez-Rivas, Irma Lopez-Martinez, Ernesto Ramirez-Gonzalez. |
| EPI_ISL_2800932 | LESP Guanajuato | Instituto de Diagnostico y Referencia Epidemiologicos (INDRE) | Claudia Wong-Arambula, Abril Rodriguez-Maldonado, Vanessa Rivero-Arredondo, Ariadna Medina-Benitez, Joaquin Quiroz-Mercado, Sergio Rangel-Guerrero, Natividad Cruz-Ortiz, Tatiana Nunez-Garcia, Gisela Barrera-Badillo, Lucia Hernandez-Rivas, Irma Lopez-Martinez, Ernesto Ramirez-Gonzalez. |
| EPI_ISL_2800933, EPI_ISL_2800934 | LESP Durango | Instituto de Diagnostico y Referencia Epidemiologicos (INDRE) | Claudia Wong-Arambula, Abril Rodriguez-Maldonado, Vanessa Rivero-Arredondo, Ariadna Medina-Benitez, Joaquin Quiroz-Mercado, Sergio Rangel-Guerrero, Natividad Cruz-Ortiz, Tatiana Nunez-Garcia, Gisela Barrera-Badillo, Lucia Hernandez-Rivas, Irma Lopez-Martinez, Ernesto Ramirez-Gonzalez. |
| EPI_ISL_2800935 | LESP Tamaulipas | Instituto de Diagnostico y Referencia Epidemiologicos (INDRE) | Claudia Wong-Arambula, Abril Rodriguez-Maldonado, Vanessa Rivero-Arredondo, Ariadna Medina-Benitez, Joaquin Quiroz-Mercado, Sergio Rangel-Guerrero, Natividad Cruz-Ortiz, Tatiana Nunez-Garcia, Gisela Barrera-Badillo, Lucia Hernandez-Rivas, Irma Lopez-Martinez, Ernesto Ramirez-Gonzalez. |
| EPI_ISL_2800936 | LESP Puebla | Instituto de Diagnostico y Referencia Epidemiologicos (INDRE) | Claudia Wong-Arambula, Abril Rodriguez-Maldonado, Vanessa Rivero-Arredondo, Ariadna Medina-Benitez, Joaquin Quiroz-Mercado, Sergio Rangel-Guerrero, Natividad Cruz-Ortiz, Tatiana Nunez-Garcia, Gisela Barrera-Badillo, Lucia Hernandez-Rivas, Irma Lopez-Martinez, Ernesto Ramirez-Gonzalez. |
| EPI_ISL_2800937 | LESP Sonora | Instituto de Diagnostico y Referencia Epidemiologicos (INDRE) | Claudia Wong-Arambula, Abril Rodriguez-Maldonado, Vanessa Rivero-Arredondo, Ariadna Medina-Benitez, Joaquin Quiroz-Mercado, Sergio Rangel-Guerrero, Natividad Cruz-Ortiz, Tatiana Nunez-Garcia, Gisela Barrera-Badillo, Lucia Hernandez-Rivas, Irma Lopez-Martinez, Ernesto Ramirez-Gonzalez. |

| | | | |
|---|---|---|---|
| EPI_ISL_2800938 | LESP Quintana Roo | Instituto de Diagnostico y Referencia Epidemiologicos (INDRE) | Claudia Wong-Arambula, Abril Rodriguez-Maldonado, Vanessa Rivero-Arredondo, Ariadna Medina-Benitez, Joaquin Quiroz-Mercado, Sergio Rangel-Guerrero, Natividad Cruz-Ortiz, Tatiana Nunez-Garcia, Gisela Barrera-Badillo, Lucia Hernandez-Rivas, Irma Lopez-Martinez, Ernesto Ramirez-Gonzalez. |
| EPI_ISL_2800939, EPI_ISL_2800940 | LESP Tabasco | Instituto de Diagnostico y Referencia Epidemiologicos (INDRE) | Claudia Wong-Arambula, Abril Rodriguez-Maldonado, Vanessa Rivero-Arredondo, Ariadna Medina-Benitez, Joaquin Quiroz-Mercado, Sergio Rangel-Guerrero, Natividad Cruz-Ortiz, Tatiana Nunez-Garcia, Gisela Barrera-Badillo, Lucia Hernandez-Rivas, Irma Lopez-Martinez, Ernesto Ramirez-Gonzalez. |
| EPI_ISL_2800941 | LESP Guanajuato | Instituto de Diagnostico y Referencia Epidemiologicos (INDRE) | Claudia Wong-Arambula, Abril Rodriguez-Maldonado, Vanessa Rivero-Arredondo, Ariadna Medina-Benitez, Joaquin Quiroz-Mercado, Sergio Rangel-Guerrero, Natividad Cruz-Ortiz, Tatiana Nunez-Garcia, Gisela Barrera-Badillo, Lucia Hernandez-Rivas, Irma Lopez-Martinez, Ernesto Ramirez-Gonzalez. |
| EPI_ISL_2800942 | LESP Tabasco | Instituto de Diagnostico y Referencia Epidemiologicos (INDRE) | Claudia Wong-Arambula, Abril Rodriguez-Maldonado, Vanessa Rivero-Arredondo, Ariadna Medina-Benitez, Joaquin Quiroz-Mercado, Sergio Rangel-Guerrero, Maribel Gonzalez-Villa, Natividad Cruz-Ortiz, Tatiana Nunez-Garcia, Gisela Barrera-Badillo, Lucia Hernandez-Rivas, Irma Lopez-Martinez, Ernesto Ramirez-Gonzalez. |
| EPI_ISL_2800943 | LESP Guanajuato | Instituto de Diagnostico y Referencia Epidemiologicos (INDRE) | Claudia Wong-Arambula, Abril Rodriguez-Maldonado, Vanessa Rivero-Arredondo, Ariadna Medina-Benitez, Joaquin Quiroz-Mercado, Sergio Rangel-Guerrero, Natividad Cruz-Ortiz, Tatiana Nunez-Garcia, Gisela Barrera-Badillo, Lucia Hernandez-Rivas, Irma Lopez-Martinez, Ernesto Ramirez-Gonzalez. |
| EPI_ISL_2800944 | LESP Yucatan | Instituto de Diagnostico y Referencia Epidemiologicos (INDRE) | Claudia Wong-Arambula, Abril Rodriguez-Maldonado, Vanessa Rivero-Arredondo, Ariadna Medina-Benitez, Joaquin Quiroz-Mercado, Sergio Rangel-Guerrero, Natividad Cruz-Ortiz, Tatiana Nunez-Garcia, Gisela Barrera-Badillo, Lucia Hernandez-Rivas, Irma Lopez-Martinez, Ernesto Ramirez-Gonzalez. |
| EPI_ISL_2800945 | LESP Michoacan | Instituto de Diagnostico y Referencia Epidemiologicos (INDRE) | Claudia Wong-Arambula, Abril Rodriguez-Maldonado, Vanessa Rivero-Arredondo, Ariadna Medina-Benitez, Joaquin Quiroz-Mercado, Sergio Rangel-Guerrero, Natividad Cruz-Ortiz, Tatiana Nunez-Garcia, Gisela Barrera-Badillo, Lucia Hernandez-Rivas, Irma Lopez-Martinez, Ernesto Ramirez-Gonzalez. |
| EPI_ISL_2800946, EPI_ISL_2800947, EPI_ISL_2800948 | Illinois Department of Public Health | Gagnon Lab, Southern Illinois University | Keith Gagnon |
| EPI_ISL_2800949 | Simple Laboratories | Gagnon Lab, Southern Illinois University | Keith Gagnon |
| EPI_ISL_2800950 | Illinois Department of Public Health | Gagnon Lab, Southern Illinois University | Keith Gagnon |
| EPI_ISL_2800951 | Simple Laboratories | Gagnon Lab, Southern Illinois University | Keith Gagnon |
| EPI_ISL_2800952, EPI_ISL_2800953, EPI_ISL_2800954, EPI_ISL_2800955, EPI_ISL_2800956, EPI_ISL_2800957 | Illinois Department of Public Health | Gagnon Lab, Southern Illinois University | Keith Gagnon |
| EPI_ISL_2800958 | Simple Laboratories | Gagnon Lab, Southern Illinois University | Keith Gagnon |
| EPI_ISL_2800959, EPI_ISL_2800960 | Illinois Department of Public Health | Gagnon Lab, Southern Illinois University | Keith Gagnon |
| EPI_ISL_2800961 | Simple Laboratories | Gagnon Lab, Southern Illinois University | Keith Gagnon |
| EPI_ISL_2800962, EPI_ISL_2800963 | Illinois Department of Public Health | Gagnon Lab, Southern Illinois University | Keith Gagnon |
| EPI_ISL_2800964, EPI_ISL_2800965 | Simple Laboratories | Gagnon Lab, Southern Illinois University | Keith Gagnon |
| EPI_ISL_2800966, EPI_ISL_2800967 | Illinois Department of Public Health | Gagnon Lab, Southern Illinois University | Keith Gagnon |
| EPI_ISL_2800968, EPI_ISL_2800969 | Simple Laboratories | Gagnon Lab, Southern Illinois University | Keith Gagnon |
| EPI_ISL_2800970, EPI_ISL_2800971, EPI_ISL_2800972 | Illinois Department of Public Health | Gagnon Lab, Southern Illinois University | Keith Gagnon |
| EPI_ISL_2800973 | Simple Laboratories | Gagnon Lab, Southern Illinois University | Keith Gagnon |
| EPI_ISL_2800974, EPI_ISL_2800975, EPI_ISL_2800976, EPI_ISL_2800977 | Illinois Department of Public Health | Gagnon Lab, Southern Illinois University | Keith Gagnon |
| EPI_ISL_2800978, EPI_ISL_2800979 | Simple Laboratories | Gagnon Lab, Southern Illinois University | Keith Gagnon |
| EPI_ISL_2800980 | Illinois Department of Public Health | Gagnon Lab, Southern Illinois University | Keith Gagnon |
| EPI_ISL_2800981 | Simple Laboratories | Gagnon Lab, Southern Illinois University | Keith Gagnon |
| EPI_ISL_2800982, EPI_ISL_2800983, EPI_ISL_2800984 | Illinois Department of Public Health | Gagnon Lab, Southern Illinois University | Keith Gagnon |
| EPI_ISL_2800985 | Simple Laboratories | Gagnon Lab, Southern Illinois University | Keith Gagnon |
| EPI_ISL_2800986, EPI_ISL_2800987 | Illinois Department of Public Health | Gagnon Lab, Southern Illinois University | Keith Gagnon |
| EPI_ISL_2800988 | Simple Laboratories | Gagnon Lab, Southern Illinois University | Keith Gagnon |
| EPI_ISL_2800989, EPI_ISL_2800990, EPI_ISL_2800991 | Illinois Department of Public Health | Gagnon Lab, Southern Illinois University | Keith Gagnon |
| EPI_ISL_2800992, EPI_ISL_2800993, EPI_ISL_2800994, EPI_ISL_2800995 | Simple Laboratories | Gagnon Lab, Southern Illinois University | Keith Gagnon |
| EPI_ISL_2800996 | Illinois Department of Public Health | Gagnon Lab, Southern Illinois University | Keith Gagnon |
| EPI_ISL_2800997 | Simple Laboratories | Gagnon Lab, Southern Illinois University | Keith Gagnon |
| EPI_ISL_2800998, EPI_ISL_2800999, EPI_ISL_2801000, EPI_ISL_2801001, EPI_ISL_2801002 | Illinois Department of Public Health | Gagnon Lab, Southern Illinois University | Keith Gagnon |
| EPI_ISL_2801003, EPI_ISL_2801004 | Simple Laboratories | Gagnon Lab, Southern Illinois University | Keith Gagnon |
| EPI_ISL_2801005, EPI_ISL_2801006, EPI_ISL_2801007, EPI_ISL_2801008 | Illinois Department of Public Health | Gagnon Lab, Southern Illinois University | Keith Gagnon |
| EPI_ISL_2801009, EPI_ISL_2801010, EPI_ISL_2801011 | Simple Laboratories | Gagnon Lab, Southern Illinois University | Keith Gagnon |
| EPI_ISL_2801012, EPI_ISL_2801013, EPI_ISL_2801014, EPI_ISL_2801015, EPI_ISL_2801016, EPI_ISL_2801017, EPI_ISL_2801018, EPI_ISL_2801019, EPI_ISL_2801020, EPI_ISL_2801021, EPI_ISL_2801022, EPI_ISL_2801023, EPI_ISL_2801024, EPI_ISL_2801025, EPI_ISL_2801026, EPI_ISL_2801027, EPI_ISL_2801028 | | | |
| see above | Illinois Department of Public Health | Gagnon Lab, Southern Illinois University | Keith Gagnon |
| EPI_ISL_2801029 | Simple Laboratories | Gagnon Lab, Southern Illinois University | Keith Gagnon |
| EPI_ISL_2801030 | Illinois Department of Public Health | Gagnon Lab, Southern Illinois University | Keith Gagnon |
| EPI_ISL_2801031 | Simple Laboratories | Gagnon Lab, Southern Illinois University | Keith Gagnon |
| EPI_ISL_2801032, EPI_ISL_2801033, EPI_ISL_2801034 | Illinois Department of Public Health | Gagnon Lab, Southern Illinois University | Keith Gagnon |
| EPI_ISL_2801035, EPI_ISL_2801036, EPI_ISL_2801037, | Simple Laboratories | Gagnon Lab, Southern Illinois University | Keith Gagnon |

| | | | |
|---|---|---|---|
| EPI_ISL_2801038 | | | |
| EPI_ISL_2801039 | Illinois Department of Public Health | Gagnon Lab, Southern Illinois University | Keith Gagnon |
| EPI_ISL_2801040 | Simple Laboratories | Gagnon Lab, Southern Illinois University | Keith Gagnon |
| EPI_ISL_2801041, EPI_ISL_2801042, EPI_ISL_2801043, EPI_ISL_2801044, EPI_ISL_2801045, EPI_ISL_2801046 | Illinois Department of Public Health | Gagnon Lab, Southern Illinois University | Keith Gagnon |
| EPI_ISL_2801047, EPI_ISL_2801048, EPI_ISL_2801049, EPI_ISL_2801050, EPI_ISL_2801051, EPI_ISL_2801052, EPI_ISL_2801053, EPI_ISL_2801054, EPI_ISL_2801055, EPI_ISL_2801056, EPI_ISL_2801057, EPI_ISL_2801058, EPI_ISL_2801059, EPI_ISL_2801060, EPI_ISL_2801061, EPI_ISL_2801062, EPI_ISL_2801063, EPI_ISL_2801064, EPI_ISL_2801065, EPI_ISL_2801066, EPI_ISL_2801067, EPI_ISL_2801068, EPI_ISL_2801069, EPI_ISL_2801070, EPI_ISL_2801071, EPI_ISL_2801072, EPI_ISL_2801073, EPI_ISL_2801074, EPI_ISL_2801075 | | | |
| see above | Simple Laboratories | Gagnon Lab, Southern Illinois University | Keith Gagnon |
| EPI_ISL_2801076, EPI_ISL_2801077, EPI_ISL_2801078, EPI_ISL_2801079, EPI_ISL_2801080, EPI_ISL_2801081, EPI_ISL_2801082, EPI_ISL_2801083, EPI_ISL_2801084, EPI_ISL_2801085, EPI_ISL_2801086, EPI_ISL_2801087, EPI_ISL_2801088 | | | |
| see above | Illinois Department of Public Health | Gagnon Lab, Southern Illinois University | Keith Gagnon |
| EPI_ISL_2801089, EPI_ISL_2801090, EPI_ISL_2801091, EPI_ISL_2801092, EPI_ISL_2801093, EPI_ISL_2801094, EPI_ISL_2801095, EPI_ISL_2801096, EPI_ISL_2801097, EPI_ISL_2801098 | Simple Laboratories | Gagnon Lab, Southern Illinois University | Keith Gagnon |
| EPI_ISL_2801099, EPI_ISL_2801100, EPI_ISL_2801135, EPI_ISL_2801136, EPI_ISL_2801137, EPI_ISL_2801138, EPI_ISL_2801139, EPI_ISL_2801140, EPI_ISL_2801141, EPI_ISL_2801142 | Illinois Department of Public Health | Gagnon Lab, Southern Illinois University | Keith Gagnon |
| EPI_ISL_2801143, EPI_ISL_2801144, EPI_ISL_2801145, EPI_ISL_2801146, EPI_ISL_2801147, EPI_ISL_2801148, EPI_ISL_2801149, EPI_ISL_2801150, EPI_ISL_2801151 | Simple Laboratories | Gagnon Lab, Southern Illinois University | Keith Gagnon |
| EPI_ISL_2801152, EPI_ISL_2801153, EPI_ISL_2801154, EPI_ISL_2801155, EPI_ISL_2801156, EPI_ISL_2801157, EPI_ISL_2801158, EPI_ISL_2801159, EPI_ISL_2801160 | Illinois Department of Public Health | Gagnon Lab, Southern Illinois University | Keith Gagnon |
| EPI_ISL_2801161, EPI_ISL_2801162 | Simple Laboratories | Gagnon Lab, Southern Illinois University | Keith Gagnon |
| EPI_ISL_2801163, EPI_ISL_2801164, EPI_ISL_2801165, EPI_ISL_2801166, EPI_ISL_2801167, EPI_ISL_2801168, EPI_ISL_2801169, EPI_ISL_2801170, EPI_ISL_2801171, EPI_ISL_2801172, EPI_ISL_2801173, EPI_ISL_2801174, EPI_ISL_2801175, EPI_ISL_2801176, EPI_ISL_2801177, EPI_ISL_2801178, EPI_ISL_2801179, EPI_ISL_2801180, EPI_ISL_2801181, EPI_ISL_2801182, EPI_ISL_2801183, EPI_ISL_2801184, EPI_ISL_2801185, EPI_ISL_2801186, EPI_ISL_2801187, EPI_ISL_2801188, EPI_ISL_2801189, EPI_ISL_2801190, EPI_ISL_2801191, EPI_ISL_2801192, EPI_ISL_2801193, EPI_ISL_2801194, EPI_ISL_2801195, EPI_ISL_2801196, EPI_ISL_2801197, EPI_ISL_2801198, EPI_ISL_2801199, EPI_ISL_2801200, EPI_ISL_2801201, EPI_ISL_2801202, EPI_ISL_2801203, EPI_ISL_2801204, EPI_ISL_2801205, EPI_ISL_2801206, EPI_ISL_2801207, EPI_ISL_2801208, EPI_ISL_2801209, EPI_ISL_2801210, EPI_ISL_2801211, EPI_ISL_2801212, EPI_ISL_2801213, EPI_ISL_2801214, EPI_ISL_2801215, EPI_ISL_2801216, EPI_ISL_2801217, EPI_ISL_2801218, EPI_ISL_2801219, EPI_ISL_2801220, EPI_ISL_2801221, EPI_ISL_2801222, EPI_ISL_2801223, EPI_ISL_2801224, EPI_ISL_2801225, EPI_ISL_2801226, EPI_ISL_2801227, EPI_ISL_2801228, EPI_ISL_2801229, EPI_ISL_2801230, EPI_ISL_2801231, EPI_ISL_2801232, EPI_ISL_2801233, EPI_ISL_2801234, EPI_ISL_2801235, EPI_ISL_2801236, EPI_ISL_2801237, EPI_ISL_2801238, EPI_ISL_2801239, EPI_ISL_2801240, EPI_ISL_2801241, EPI_ISL_2801242, EPI_ISL_2801243, EPI_ISL_2801244, EPI_ISL_2801245, EPI_ISL_2801246, EPI_ISL_2801247, EPI_ISL_2801248, EPI_ISL_2801249, EPI_ISL_2801250, EPI_ISL_2801251, EPI_ISL_2801252, EPI_ISL_2801253, EPI_ISL_2801254, EPI_ISL_2801255, EPI_ISL_2801256, EPI_ISL_2801257, EPI_ISL_2801258, EPI_ISL_2801259, EPI_ISL_2801260, EPI_ISL_2801261, EPI_ISL_2801262, EPI_ISL_2801263, EPI_ISL_2801264, EPI_ISL_2801265, EPI_ISL_2801266, EPI_ISL_2801267, EPI_ISL_2801268, EPI_ISL_2801269, EPI_ISL_2801270, EPI_ISL_2801271, EPI_ISL_2801272, EPI_ISL_2801273, EPI_ISL_2801274, EPI_ISL_2801275, EPI_ISL_2801276, EPI_ISL_2801277, EPI_ISL_2801278, EPI_ISL_2801279, EPI_ISL_2801280, EPI_ISL_2801281, EPI_ISL_2801282, EPI_ISL_2801283, EPI_ISL_2801284, EPI_ISL_2801285, EPI_ISL_2801286, EPI_ISL_2801287, EPI_ISL_2801288, EPI_ISL_2801289, EPI_ISL_2801290, EPI_ISL_2801291, EPI_ISL_2801292, EPI_ISL_2801293, EPI_ISL_2801294, EPI_ISL_2801295, EPI_ISL_2801296, EPI_ISL_2801297, EPI_ISL_2801298, EPI_ISL_2801299, EPI_ISL_2801300, EPI_ISL_2801301, EPI_ISL_2801302, EPI_ISL_2801303, EPI_ISL_2801304 | | | |
| see above | Illinois Department of Public Health | Gagnon Lab, Southern Illinois University | Keith Gagnon |
| EPI_ISL_2801433, EPI_ISL_2801435, EPI_ISL_2801436, EPI_ISL_2801437, EPI_ISL_2801438 | IVY3 Central Lab, Vanderbilt University Medical Center | Lauring Lab, University of Michigan, Department of Microbiology and Immunology | Valesano |
| EPI_ISL_2801439 | University of Michigan Clinical Microbiology Laboratory | Lauring Lab, University of Michigan, Department of Microbiology and Immunology | Valesano |
| EPI_ISL_2801440, EPI_ISL_2801441 | IVY3 Central Lab, Vanderbilt University Medical Center | Lauring Lab, University of Michigan, Department of Microbiology and Immunology | Valesano |
| EPI_ISL_2801442 | University of Michigan Clinical Microbiology Laboratory | Lauring Lab, University of Michigan, Department of Microbiology and Immunology | Valesano |
| EPI_ISL_2801443, EPI_ISL_2801444, EPI_ISL_2801445, EPI_ISL_2801446, EPI_ISL_2801447, EPI_ISL_2801448, EPI_ISL_2801449 | IVY3 Central Lab, Vanderbilt University Medical Center | Lauring Lab, University of Michigan, Department of Microbiology and Immunology | Valesano |
| EPI_ISL_2801450, EPI_ISL_2801451 | University of Michigan Clinical Microbiology Laboratory | Lauring Lab, University of Michigan, Department of Microbiology and Immunology | Valesano |
| EPI_ISL_2801452, EPI_ISL_2801453, EPI_ISL_2801454, EPI_ISL_2801455, EPI_ISL_2801456, EPI_ISL_2801457, EPI_ISL_2801458, EPI_ISL_2801459, EPI_ISL_2801465, EPI_ISL_2801466, EPI_ISL_2801467, EPI_ISL_2801468, EPI_ISL_2801469, EPI_ISL_2801470, EPI_ISL_2801471, EPI_ISL_2801472, EPI_ISL_2801473 | | | |
| see above | IVY3 Central Lab, Vanderbilt University Medical Center | Lauring Lab, University of Michigan, Department of Microbiology and Immunology | Valesano |
| EPI_ISL_2801474 | University of Michigan Clinical Microbiology Laboratory | Lauring Lab, University of Michigan, Department of Microbiology and Immunology | Valesano |
| EPI_ISL_2801475, EPI_ISL_2801476, EPI_ISL_2801477 | IVY3 Central Lab, Vanderbilt University Medical Center | Lauring Lab, University of Michigan, Department of Microbiology and Immunology | Valesano |
| EPI_ISL_2801478, EPI_ISL_2801479, EPI_ISL_2801480, EPI_ISL_2801481, EPI_ISL_2801482, EPI_ISL_2801483, EPI_ISL_2801484, EPI_ISL_2801485, EPI_ISL_2801486, EPI_ISL_2801487, EPI_ISL_2801488, EPI_ISL_2801489, EPI_ISL_2801490, EPI_ISL_2801491, EPI_ISL_2801492, EPI_ISL_2801493, EPI_ISL_2801494, EPI_ISL_2801495, EPI_ISL_2801496, EPI_ISL_2801508, EPI_ISL_2801509, EPI_ISL_2801510, EPI_ISL_2801511, EPI_ISL_2801512, EPI_ISL_2801513, EPI_ISL_2801514, EPI_ISL_2801515, EPI_ISL_2801516, EPI_ISL_2801517, EPI_ISL_2801518, EPI_ISL_2801519, EPI_ISL_2801520, EPI_ISL_2801521 | | | |
| see above | Nevada State Public Health Laboratory | Nevada State Public Health Laboratory | Andrew Gorzalski, Mark Pandori |
| EPI_ISL_2801522, EPI_ISL_2801523, EPI_ISL_2801524, EPI_ISL_2801525, EPI_ISL_2801526, | Oregon State Public Health Laboratory | Oregon State Public Health Laboratory | Rafia Razzaque, Eugene Yeboah, Vanda Makris, Laura Tsaknaridis, John Fontana and Shane Sevey |

| | | | |
|---|---|---|---|
| EPI_ISL_2801527 | | | |
| EPI_ISL_2801531, EPI_ISL_2801532, EPI_ISL_2801534, EPI_ISL_2801535, EPI_ISL_2801536, EPI_ISL_2801537, EPI_ISL_2801538, EPI_ISL_2801539, EPI_ISL_2801540, EPI_ISL_2801541, EPI_ISL_2801542, EPI_ISL_2801543, EPI_ISL_2801544, EPI_ISL_2801545, EPI_ISL_2801546, EPI_ISL_2801547, EPI_ISL_2801548, EPI_ISL_2801549, EPI_ISL_2801550, EPI_ISL_2801551, EPI_ISL_2801552, EPI_ISL_2801553, EPI_ISL_2801555 | | | |
| see above | Wyoming Public Health Laboratory | Wyoming Public Health Laboratory | Jim Mildenberger, Taylor Fearing, Channing Weber, Ashley Norberg, Chayse Rowley, Marley Goetz, Brian Dominguez, Elliot Thomasson, Sam Britz, Cari Sloma, Robert Petit, and Rob Christensen |
| EPI_ISL_2801556, EPI_ISL_2801558, EPI_ISL_2801562 | DeRisi Lab, University of California, San Francisco | DeRisi Lab, University of California, San Francisco | Sabrina Mann, Anthea Mitchell, Matthias Hapte-Selassie, Jamin Liu, Sara Sunshine, Matthew Laurie, Genay Pilarowski, Amy Kistler, Manu Vanaerschot, Patrick Ayscue, Lucy Li, Eric Chow, IDseq Team, James Peng, Carina Marquez, Luis Rubio, Gabriel Chamie, Diane Jones, Jon Jacobo, Susana Rojas, Susy Rojas, Valerie Tulier-Laiwa, Douglas Black, Jackie Martinez, Maya Petersen, Diane Havlir, and Joseph DeRisi |
| EPI_ISL_2801564, EPI_ISL_2801565, EPI_ISL_2801566, EPI_ISL_2801567, EPI_ISL_2801568, EPI_ISL_2801569, EPI_ISL_2801570, EPI_ISL_2801571, EPI_ISL_2801572, EPI_ISL_2801573, EPI_ISL_2801574, EPI_ISL_2801575, EPI_ISL_2801576, EPI_ISL_2801577, EPI_ISL_2801578, EPI_ISL_2801579, EPI_ISL_2801580, EPI_ISL_2801581, EPI_ISL_2801582, EPI_ISL_2801583, EPI_ISL_2801584, EPI_ISL_2801585, EPI_ISL_2801586, EPI_ISL_2801587, EPI_ISL_2801588, EPI_ISL_2801589, EPI_ISL_2801590, EPI_ISL_2801591, EPI_ISL_2801592, EPI_ISL_2801593, EPI_ISL_2801594, EPI_ISL_2801595, EPI_ISL_2801596, EPI_ISL_2801597, EPI_ISL_2801598, EPI_ISL_2801599, EPI_ISL_2801600, EPI_ISL_2801601, EPI_ISL_2801602, EPI_ISL_2801603, EPI_ISL_2801604, EPI_ISL_2801605, EPI_ISL_2801606, EPI_ISL_2801607, EPI_ISL_2801608, EPI_ISL_2801609, EPI_ISL_2801610, EPI_ISL_2801611, EPI_ISL_2801612, EPI_ISL_2801613, EPI_ISL_2801614, EPI_ISL_2801615, EPI_ISL_2801616, EPI_ISL_2801617, EPI_ISL_2801618, EPI_ISL_2801619, EPI_ISL_2801620, EPI_ISL_2801621, EPI_ISL_2801622, EPI_ISL_2801623, EPI_ISL_2801624, EPI_ISL_2801625, EPI_ISL_2801626, EPI_ISL_2801627, EPI_ISL_2801628, EPI_ISL_2801629, EPI_ISL_2801630, EPI_ISL_2801631, EPI_ISL_2801632, EPI_ISL_2801633, EPI_ISL_2801634, EPI_ISL_2801635, EPI_ISL_2801636, EPI_ISL_2801637, EPI_ISL_2801638, EPI_ISL_2801639, EPI_ISL_2801640, EPI_ISL_2801641, EPI_ISL_2801642, EPI_ISL_2801643, EPI_ISL_2801644, EPI_ISL_2801645, EPI_ISL_2801646, EPI_ISL_2801647, EPI_ISL_2801648, EPI_ISL_2801649, EPI_ISL_2801650, EPI_ISL_2801651, EPI_ISL_2801652, EPI_ISL_2801653, EPI_ISL_2801654, EPI_ISL_2801655, EPI_ISL_2801656, EPI_ISL_2801657 | | | |
| see above | Unidad de Investigación Médica de Yucatán (UIMY) | Instituto de Biotecnología de la UNAM | Consorcio Mexicano de Vigilancia Genómica (CoViGen-Mex). Authors (in alphabetical order): Julio Elias Alvarado-Yaah, Carlos F. Arias, Santiago Ávila-Ríos, Eduardo Becerril-Vargas, Víctor Hugo Borja-Aburto, Celia Boukadida, Cristóbal Cháidez-Quiróz, Juan Bautista Chale-Dzul, Ricardo Ciria Merce, Andreu Comas-García, Célida Duque Molina, Julissa Enciso-Ibarra, José Antonio Enciso-Moreno, Gloria Elena Espinosa-Ayala, Fernando Fontove-Herrera, Daniel Fregoso-Rueda, Victor Eduardo García-Arias, Alejandra García-Gasca, Bruno Gómez-Gil, Jean Pierre González, Irvin González-López, Concepción Grajales-Muñiz, Ricardo Grande, Rosa María Gutiérrez Rios, Jesús Hernández, Alejandra Hernández-Terán, Alfredo Herrera-Estrella, Carla Ivón Herrera-Najera, Pavel Isa, Verónica Mata-Haro, Daniel Lira Morales, Susana Lopez, Antonio Loza Román, Brenda Irasema Maldonado-Meza, Bernardo Martínez-Miguel, José Arturo Martínez-Orozco, Célida Martínez- Rodríguez, Margarita Matías-Florentino, Fidencio Mejía-Nepomuceno, María Guadalupe de Jesús Mireles-Rivera, Gloria María Molina-Salinas, Hector Montoya-Fuentes, , Mario Mújica-Sánchez, José Esteban Muñoz-Medina, José de Jesús Nuñez-Contreras, Alicia Ocaña-Mondragón, Luis Alberto Ochoa-Carrera, Hector Esteban Paz-Juárez, Marissa Perez-Garcia, Francisco Pulido, Helen Haydee Fernanda Ramírez-Plascencia, Jorge Salas-Hernández, Angel Gustavo Salas-Lais, Alejandro Sánchez-Flores, Clara Esperanza Santacruz-Tinoco, María Guadalupe Santiago-Mauricio, Selene Zárate, Nelly Sélem-Mojica, Blanca Taboada, Gloria Vazquez, Joel Armando Vázquez-Pérez. |
| EPI_ISL_2801658, EPI_ISL_2801659, EPI_ISL_2801660, EPI_ISL_2801661, EPI_ISL_2801662, EPI_ISL_2801663, EPI_ISL_2801664, EPI_ISL_2801665, EPI_ISL_2801666, EPI_ISL_2801667, EPI_ISL_2801668, EPI_ISL_2801669, EPI_ISL_2801670, EPI_ISL_2801671, EPI_ISL_2801672, EPI_ISL_2801673, EPI_ISL_2801674, EPI_ISL_2801675, EPI_ISL_2801676, EPI_ISL_2801677, EPI_ISL_2801678, EPI_ISL_2801679, EPI_ISL_2801680, EPI_ISL_2801681, EPI_ISL_2801682, EPI_ISL_2801683, EPI_ISL_2801684, EPI_ISL_2801685, EPI_ISL_2801686, EPI_ISL_2801687, EPI_ISL_2801688, EPI_ISL_2801689, EPI_ISL_2801691, EPI_ISL_2801692, EPI_ISL_2801693, EPI_ISL_2801694, EPI_ISL_2801695, EPI_ISL_2801696 | | | |
| see above | Centro de Investigación Biomédica del Noreste (CIBIN) | Instituto de Biotecnología de la UNAM | Consorcio Mexicano de Vigilancia Genómica (CoViGen-Mex). Authors (in alphabetical order): Julio Elias Alvarado-Yaah, Carlos F. Arias, Santiago Ávila-Ríos, Eduardo Becerril-Vargas, Víctor Hugo Borja-Aburto, Celia Boukadida, Cristóbal Cháidez-Quiróz, Juan Bautista Chale-Dzul, Ricardo Ciria Merce, Andreu Comas-García, Célida Duque Molina, Julissa Enciso-Ibarra, José Antonio Enciso-Moreno, Gloria Elena Espinosa-Ayala, Fernando Fontove-Herrera, Daniel Fregoso-Rueda, Victor Eduardo García-Arias, Alejandra García-Gasca, Bruno Gómez-Gil, Jean Pierre González, Irvin González-López, Concepción Grajales-Muñiz, Ricardo Grande, Rosa María Gutiérrez Rios, Jesús Hernández, Alejandra Hernández-Terán, Alfredo Herrera-Estrella, Carla Ivón Herrera-Najera, Pavel Isa, Verónica Mata-Haro, Daniel Lira Morales, Susana Lopez, Antonio Loza Román, Brenda Irasema Maldonado-Meza, Bernardo Martínez-Miguel, José Arturo Martínez-Orozco, Célida Martínez- Rodríguez, Margarita Matías-Florentino, Fidencio Mejía-Nepomuceno, María Guadalupe de Jesús Mireles-Rivera, Gloria María Molina-Salinas, Hector Montoya-Fuentes, , Mario Mújica-Sánchez, José Esteban Muñoz-Medina, José de Jesús Nuñez-Contreras, Alicia Ocaña-Mondragón, Luis Alberto Ochoa-Carrera, Hector Esteban Paz-Juárez, Marissa Perez-Garcia, Francisco Pulido, Helen Haydee Fernanda Ramírez-Plascencia, Jorge Salas-Hernández, Angel Gustavo Salas-Lais, Alejandro Sánchez-Flores, Clara Esperanza Santacruz-Tinoco, María Guadalupe Santiago-Mauricio, Selene Zárate, Nelly Sélem-Mojica, Blanca Taboada, Gloria Vazquez, Joel Armando Vázquez-Pérez. |
| EPI_ISL_2801697, EPI_ISL_2801698, EPI_ISL_2801699, EPI_ISL_2801700, EPI_ISL_2801701, EPI_ISL_2801702, EPI_ISL_2801703, EPI_ISL_2801704, EPI_ISL_2801705, EPI_ISL_2801706 | Unidad de Investigación Biomédica de Zacatecas (UIBZ) | Instituto de Biotecnología de la UNAM | Consorcio Mexicano de Vigilancia Genómica (CoViGen-Mex). Authors (in alphabetical order): Julio Elias Alvarado-Yaah, Carlos F. Arias, Santiago Ávila-Ríos, Eduardo Becerril-Vargas, Víctor Hugo Borja-Aburto, Celia Boukadida, Cristóbal Cháidez-Quiróz, Juan Bautista Chale-Dzul, Ricardo Ciria Merce, Andreu Comas-García, Célida Duque Molina, Julissa Enciso-Ibarra, José Antonio Enciso-Moreno, Gloria Elena Espinosa-Ayala, Fernando Fontove-Herrera, Daniel Fregoso-Rueda, Victor Eduardo García-Arias, Alejandra García-Gasca, Bruno Gómez-Gil, Jean Pierre González, Irvin González-López, Concepción Grajales-Muñiz, Ricardo Grande, Rosa María Gutiérrez Rios, Jesús Hernández, Alejandra Hernández-Terán, Alfredo Herrera-Estrella, Carla Ivón Herrera-Najera, Pavel Isa, Verónica Mata-Haro, Daniel Lira Morales, Susana Lopez, Antonio Loza Román, Brenda Irasema Maldonado-Meza, Bernardo Martínez-Miguel, José Arturo Martínez-Orozco, Célida Martínez- Rodríguez, Margarita Matías-Florentino, Fidencio Mejía-Nepomuceno, María Guadalupe de Jesús Mireles-Rivera, Gloria María Molina-Salinas, Hector Montoya-Fuentes, , Mario Mújica-Sánchez, José Esteban Muñoz-Medina, José de Jesús Nuñez-Contreras, Alicia Ocaña-Mondragón, Luis Alberto Ochoa-Carrera, Hector Esteban Paz-Juárez, Marissa Perez-Garcia, Francisco Pulido, Helen Haydee Fernanda Ramírez-Plascencia, Jorge Salas-Hernández, Angel Gustavo Salas-Lais, Alejandro Sánchez-Flores, Clara Esperanza Santacruz-Tinoco, María Guadalupe Santiago-Mauricio, Selene Zárate, Nelly Sélem-Mojica, Blanca Taboada, Gloria Vazquez, Joel Armando Vázquez-Pérez. |
| EPI_ISL_2801707, EPI_ISL_2801708, EPI_ISL_2801709, EPI_ISL_2801710, EPI_ISL_2801712, EPI_ISL_2801713, EPI_ISL_2801714, EPI_ISL_2801715, EPI_ISL_2801716, EPI_ISL_2801717, EPI_ISL_2801718, EPI_ISL_2801719, EPI_ISL_2801720, EPI_ISL_2801721, EPI_ISL_2801722, EPI_ISL_2801723, EPI_ISL_2801724, EPI_ISL_2801725, EPI_ISL_2801726, EPI_ISL_2801727, EPI_ISL_2801728, EPI_ISL_2801729, EPI_ISL_2801730, EPI_ISL_2801731, EPI_ISL_2801732, EPI_ISL_2801733, EPI_ISL_2801734, EPI_ISL_2801735, EPI_ISL_2801736, EPI_ISL_2801737, EPI_ISL_2801738, EPI_ISL_2801739, EPI_ISL_2801740, EPI_ISL_2801741, EPI_ISL_2801742, EPI_ISL_2801743, EPI_ISL_2801744, EPI_ISL_2801745, EPI_ISL_2801746, EPI_ISL_2801747, EPI_ISL_2801748, EPI_ISL_2801749, EPI_ISL_2801750, EPI_ISL_2801751, EPI_ISL_2801752 | | | |
| see above | Centro de Investigación Biomédica de Occidente (CIBO) | Instituto de Biotecnología de la UNAM | Consorcio Mexicano de Vigilancia Genómica (CoViGen-Mex). Authors (in alphabetical order): Julio Elias Alvarado-Yaah, Carlos F. Arias, Santiago Ávila-Ríos, Eduardo Becerril-Vargas, Víctor Hugo Borja-Aburto, Celia Boukadida, Cristóbal Cháidez-Quiróz, Juan Bautista Chale-Dzul, Ricardo Ciria Merce, Andreu Comas-García, Célida Duque Molina, Julissa Enciso-Ibarra, José Antonio Enciso-Moreno, Gloria Elena Espinosa-Ayala, Fernando Fontove-Herrera, Daniel Fregoso-Rueda, Victor Eduardo García-Arias, Alejandra García-Gasca, Bruno Gómez-Gil, Jean Pierre González, Irvin González-López, Concepción Grajales-Muñiz, Ricardo Grande, Rosa María Gutiérrez Rios, Jesús Hernández, Alejandra Hernández-Terán, Alfredo Herrera-Estrella, Carla Ivón Herrera-Najera, Pavel Isa, Verónica Mata-Haro, Daniel Lira Morales, Susana Lopez, Antonio Loza Román, Brenda Irasema Maldonado-Meza, Bernardo Martínez-Miguel, José Arturo Martínez-Orozco, Célida Martínez- Rodríguez, Margarita Matías-Florentino, Fidencio Mejía-Nepomuceno, María Guadalupe de Jesús Mireles-Rivera, Gloria María Molina-Salinas, Hector Montoya-Fuentes, , Mario Mújica-Sánchez, José Esteban Muñoz-Medina, José de Jesús Nuñez-Contreras, Alicia Ocaña-Mondragón, Luis Alberto Ochoa-Carrera, Hector Esteban Paz-Juárez, Marissa Perez-Garcia, Francisco Pulido, Helen Haydee Fernanda Ramírez-Plascencia, Jorge Salas-Hernández, Angel Gustavo Salas-Lais, Alejandro Sánchez-Flores, Clara Esperanza Santacruz-Tinoco, María Guadalupe Santiago-Mauricio, Selene Zárate, Nelly Sélem-Mojica, Blanca Taboada, Gloria Vazquez, Joel Armando Vázquez-Pérez. |
| EPI_ISL_2801753, EPI_ISL_2801754, EPI_ISL_2801755, EPI_ISL_2801756, EPI_ISL_2801757, EPI_ISL_2801758, EPI_ISL_2801759, EPI_ISL_2801760, EPI_ISL_2801761, EPI_ISL_2801762, EPI_ISL_2801763, EPI_ISL_2801764, EPI_ISL_2801765, EPI_ISL_2801766, EPI_ISL_2801767, EPI_ISL_2801768, EPI_ISL_2801769, EPI_ISL_2801770, EPI_ISL_2801771, EPI_ISL_2801772, EPI_ISL_2801773, EPI_ISL_2801774, EPI_ISL_2801775, EPI_ISL_2801776, EPI_ISL_2801777, EPI_ISL_2801778, EPI_ISL_2801779, EPI_ISL_2801780, EPI_ISL_2801781, EPI_ISL_2801782, EPI_ISL_2801783, EPI_ISL_2801784, EPI_ISL_2801785, EPI_ISL_2801786, EPI_ISL_2801787, EPI_ISL_2801788, EPI_ISL_2801789, EPI_ISL_2801790, EPI_ISL_2801791, EPI_ISL_2801792, EPI_ISL_2801793, EPI_ISL_2801794, EPI_ISL_2801795, EPI_ISL_2801796, EPI_ISL_2801797, EPI_ISL_2801798, EPI_ISL_2801799, EPI_ISL_2801800, EPI_ISL_2801801, EPI_ISL_2801802, EPI_ISL_2801803, EPI_ISL_2801804, EPI_ISL_2801805, EPI_ISL_2801806, EPI_ISL_2801807, EPI_ISL_2801808, EPI_ISL_2801809, EPI_ISL_2801810, EPI_ISL_2801811, EPI_ISL_2801812, EPI_ISL_2801813, EPI_ISL_2801814, EPI_ISL_2801815, EPI_ISL_2801816, EPI_ISL_2801817, EPI_ISL_2801818, EPI_ISL_2801819, EPI_ISL_2801820, EPI_ISL_2801821, EPI_ISL_2801822, EPI_ISL_2801823, EPI_ISL_2801824, EPI_ISL_2801825, EPI_ISL_2801826, EPI_ISL_2801827, EPI_ISL_2801828, EPI_ISL_2801829, EPI_ISL_2801830, EPI_ISL_2801831, EPI_ISL_2801832, EPI_ISL_2801833, EPI_ISL_2801834, EPI_ISL_2801835, EPI_ISL_2801836, EPI_ISL_2801837, EPI_ISL_2801838, EPI_ISL_2801839, EPI_ISL_2801840, EPI_ISL_2801841, EPI_ISL_2801842, EPI_ISL_2801843, EPI_ISL_2801844, EPI_ISL_2801845, EPI_ISL_2801846, EPI_ISL_2801847, EPI_ISL_2801848, EPI_ISL_2801849, EPI_ISL_2801850, EPI_ISL_2801851, EPI_ISL_2801852, EPI_ISL_2801853, EPI_ISL_2801854, EPI_ISL_2801855, EPI_ISL_2801856, EPI_ISL_2801857, EPI_ISL_2801858, EPI_ISL_2801859, EPI_ISL_2801860, EPI_ISL_2801861, EPI_ISL_2801862, EPI_ISL_2801863, EPI_ISL_2801864, EPI_ISL_2801865, EPI_ISL_2801866, EPI_ISL_2801867, EPI_ISL_2801868, EPI_ISL_2801869, EPI_ISL_2801870, EPI_ISL_2801871, EPI_ISL_2801872, EPI_ISL_2801873, EPI_ISL_2801874, EPI_ISL_2801875, EPI_ISL_2801876, EPI_ISL_2801877, EPI_ISL_2801878, EPI_ISL_2801879, EPI_ISL_2801880, EPI_ISL_2801881, EPI_ISL_2801882, EPI_ISL_2801883, EPI_ISL_2801884, EPI_ISL_2801885, EPI_ISL_2801886, EPI_ISL_2801887, EPI_ISL_2801888, EPI_ISL_2801889, EPI_ISL_2801890, EPI_ISL_2801891 | | | |
| see above | Laboratorio Central de Epidemiología (LCE) | Instituto de Biotecnología de la UNAM | Consorcio Mexicano de Vigilancia Genómica (CoViGen-Mex). Authors (in alphabetical order): Julio Elias Alvarado-Yaah, Carlos F. Arias, Santiago Ávila-Ríos, Eduardo Becerril-Vargas, Víctor Hugo Borja-Aburto, Celia Boukadida, Cristóbal Cháidez-Quiróz, Juan Bautista Chale-Dzul, Ricardo Ciria Merce, Andreu Comas-García, Célida Duque Molina, Julissa Enciso-Ibarra, José Antonio Enciso-Moreno, Gloria Elena Espinosa-Ayala, Fernando Fontove-Herrera, Daniel Fregoso-Rueda, Victor Eduardo García-Arias, Alejandra García-Gasca, Bruno Gómez-Gil, Jean Pierre González, Irvin González-López, Concepción Grajales-Muñiz, Ricardo Grande, Rosa María Gutiérrez Rios, Jesús Hernández, Alejandra Hernández-Terán, Alfredo Herrera-Estrella, Carla Ivón Herrera-Najera, Pavel Isa, Verónica Mata-Haro, Daniel Lira Morales, Susana Lopez, Antonio Loza Román, Brenda Irasema Maldonado-Meza, Bernardo Martínez-Miguel, José Arturo Martínez-Orozco, Célida Martínez- Rodríguez, Margarita Matías-Florentino, Fidencio Mejía-Nepomuceno, María Guadalupe de Jesús Mireles-Rivera, Gloria María Molina-Salinas, Hector Montoya-Fuentes, , Mario Mújica-Sánchez, José Esteban Muñoz-Medina, José de Jesús Nuñez-Contreras, Alicia Ocaña-Mondragón, Luis Alberto Ochoa-Carrera, Hector Esteban Paz-Juárez, Marissa Perez-Garcia, Francisco Pulido, Helen Haydee Fernanda Ramírez-Plascencia, Jorge Salas-Hernández, Angel Gustavo Salas-Lais, Alejandro Sánchez-Flores, Clara Esperanza Santacruz-Tinoco, María Guadalupe Santiago-Mauricio, Selene Zárate, Nelly Sélem-Mojica, Blanca Taboada, Gloria Vazquez, Joel Armando Vázquez-Pérez. |
